# Methadone Distribution Trends from 2017-2019 in the United States

**DOI:** 10.1101/2020.11.07.20227496

**Authors:** John A. Furst, Nicholas J. Mynarski, Kenneth L. McCall, Brian J. Piper

## Abstract

**Objective:** Methadone is an evidence based treatment for opioid use disorder and is also employed for acute pain. The primary objective of this study was to explore methadone distribution patterns between the years 2017 and 2019 across the United States (US). This study builds upon previous literature that has analyzed prior years of US distribution patterns, and further outlines regional and state specific methadone trends.

**Methods:** The Drug Enforcement Administration’s Automated Reports and Consolidated Ordering System (ARCOS) was used to acquire the number of narcotic treatment programs (NTPs) per state and methadone distribution weight in grams. Methadone distribution by weight, corrected for state populations, and number of NTPs were compared from 2017 to 2019 between states, within regions, and nationally.

**Results:** Between 2017 and 2019, the national distribution of methadone increased 12.30% for NTPs but decreased 34.57% for pain, for a total increase of 2.66%. While all states saw a decrease in distribution for pain, when compared regionally, the Northeast showed a significantly smaller decrease than all other regions. Additionally, the majority of states experienced an increase in distribution for NTPs and most states demonstrated a relatively stable or increasing number of NTPs, with an 11.49% increase in NTPs nationally. The number of NTPs per 100K in 2019 ranged from 2.08 in Rhode Island to 0.00 in Wyoming.

**Conclusion:** Although methadone distribution for OUD was increasing in the US, there were pronounced regional disparities.

## Introduction

Methadone (“Dolophine” and “Methadose”), a synthetic opioid, is considered a full agonist of the μ-opioid receptor, known for its long half-life relative to other opioids.^1^ After being identified as a potent pharmacotherapeutic option for those suffering from opioid use disorder (OUD), methadone decreased illicit opioid use, decreased HIV-associated risk behavior and reduced drug-related criminal behaviors.^2-4^ In addition to treatment for opioid substance use disorders, methadone has been used in the management of severe, chronic pain. No significant difference in pain relief between morphine and methadone in cancer patients was noted.^1,5^ Methadone is classified as a schedule II controlled substance in the United States (US) and is Food and Drug Administration (FDA) approved for detoxification treatment of opioid addiction and pain management in patients nonresponsive to non-narcotic analgesics.^1,3,5^

Methadone is the predominant prescription opioid in the US by morphine mg equivalents (MME).^6^ Despite methadone’s prominence, some research has shown there is a correlation between methadone distribution rate and overdose death rate.^7^ Both methadone distribution and methadone-associated deaths in the US increased between 2002-2006, an average of 25% and 22% per year, respectively.^8^ Following years of increasing distribution and associated deaths, a steady decline in methadone distribution occurred between 2007 and 2013, with a significant 4.8% decrease in methadone volume distribution occurring between 2015 and 2016. ^6^

Even with national measures being taken to decrease opioid misuse, deaths related to opioid overdoses, across all opioid formulations, continue to be a public health issue of serious concern. The most recent data shows the total number of opioid overdoses to be 47,600 and 46,802 in 2017 and 2018, respectively. These numbers demonstrate very large increases from the reported 21,088 in 2010, indicating the need for further efforts to combat the rise in opioid related mortality.^9^

Naloxone, an antagonist of opioid receptors, is a pharmacological therapy that has received considerable attention for its ability to reverse the adverse effects of opioids.^10^ Community accessibility of this therapy remains variable across many areas of the country, however, it has been shown to have significant impacts on the ability to curb opioid overdose deaths when pharmacists are enabled to distribute naloxone in the absence of prescription orders.^11^

In recent years, additional options for the treatment of OUD have emerged, such as buprenorphine, a semi-synthetic opioid that is, uniquely, a partial agonist of the μ-opioid receptor with a long half life.^12^ A recent publication analyzing trends in the use of methadone and buprenorphine for medication-assisted treatment from 2003-2015 demonstrated an increase in the number of Narcotic Treatment Programs (NTPs) and an increase in the number of clients receiving medication at those facilities within the US.^13^ However, regional disparities exist. Counties within the East North Central, South Atlantic, and Mountain division were more likely to have low availability of OUD medication providers and higher rates of opioid overdose mortality than the national average.^14^ Additionally, buprenorphine distribution has been on the rise from 2007-2017, however, distribution has been reported to be nonhomogeneous across the United States.^15^

Despite published data trends being clearly outlined for methadone for this earlier time frame and more recent data for buprenorphine trends, data of current methadone distribution as well as specifics pertaining to state and regional distribution trends is currently lacking. Investigation of recent methadone distribution trends will allow for a greater understanding of the most up-to-date pattern of use in treatment of OUD and recent geographic fluctuations of methadone use within the US.

## Methods

### Data Sources

Methadone and buprenorphine distribution amount (in grams) was extracted from the DEA’s Automated Reports and Consolidated Ordering System (ARCOS) yearly drug summary data reports for the years 2017-2019, which was the most recent available when data-analysis was conducted (10/2020). Mandated by the 1970 Controlled Substances Act, this federal program provides a comprehensive drug reporting system that tracks the distribution of controlled substances, in grams, to pharmacies, hospitals, clinical providers, and NTPs, and has been utilized in previous pharmacoepidemiological studies.^6,7,15-17^ Methadone distributed to NTPs was classified as treatment of OUD. Methadone distribution to non-NTP locations, i.e. hospitals, pharmacies, and providers, was considered to be for pain management. State population estimates were gathered from the decennial Census published by the US Census Bureau, Population Division, to normalize data between states. Procedures were approved by the Institutional Review Board from the University of New England and Geisinger.

### Data Analysis

The following analyses were completed: (1) US total methadone distribution to NTPs for management of OUD and distribution to other clinical locations for pain management, e.g. hospitals, pharmacies, practitioners, was calculated and converted to kg for each year; (2) US Census Bureau, Regions and Divisions, was used to group states into regions (Northeast, Midwest, South, West).^18^ Average percent change of total, NTP, and pain distribution of methadone as well as total number of NTPs were compared between each region and compared amongst states within the Northeast. SEM was calculated and regional comparisons to the Northeast region were analyzed using t-tests; (3) percent change of methadone distribution from 2017 to 2019 for OUD and pain management for each state per 100K population quotient; (4) percent change in number of NTPs from 2017 to 2019 for each state per 100K population quotient; (5) number of NTPs in 2019 for each state per 100K population quotient; (6) percent change in distribution of methadone compared to buprenorphine from 2017 to 2019. For analyses (3) through (5), statistical significance was defined as any state with a percent change larger than ±1.96 SD from the national average. A Z-score between 1.96 and 2.58 was indicated as p < 0.05 and greater than 2.58 as p < 0.01. Data analysis and figures were completed with GraphPad Prism, version 8.4.3. Heat maps were prepared with JMP, version 15.

## Results

Following a 6.34% decrease between 2017 and 2018, the US total methadone distribution increased 9.61% between 2018 and 2019, for an overall increase of 2.66% between the years 2017 and 2019. More specifically, distribution of methadone for NTPs increased by 12.30% between 2017 and 2019, in contrast to distribution for pain which decreased steadily across these years for an overall reduction of 34.57% (Figure 1).

**Figure 1:**
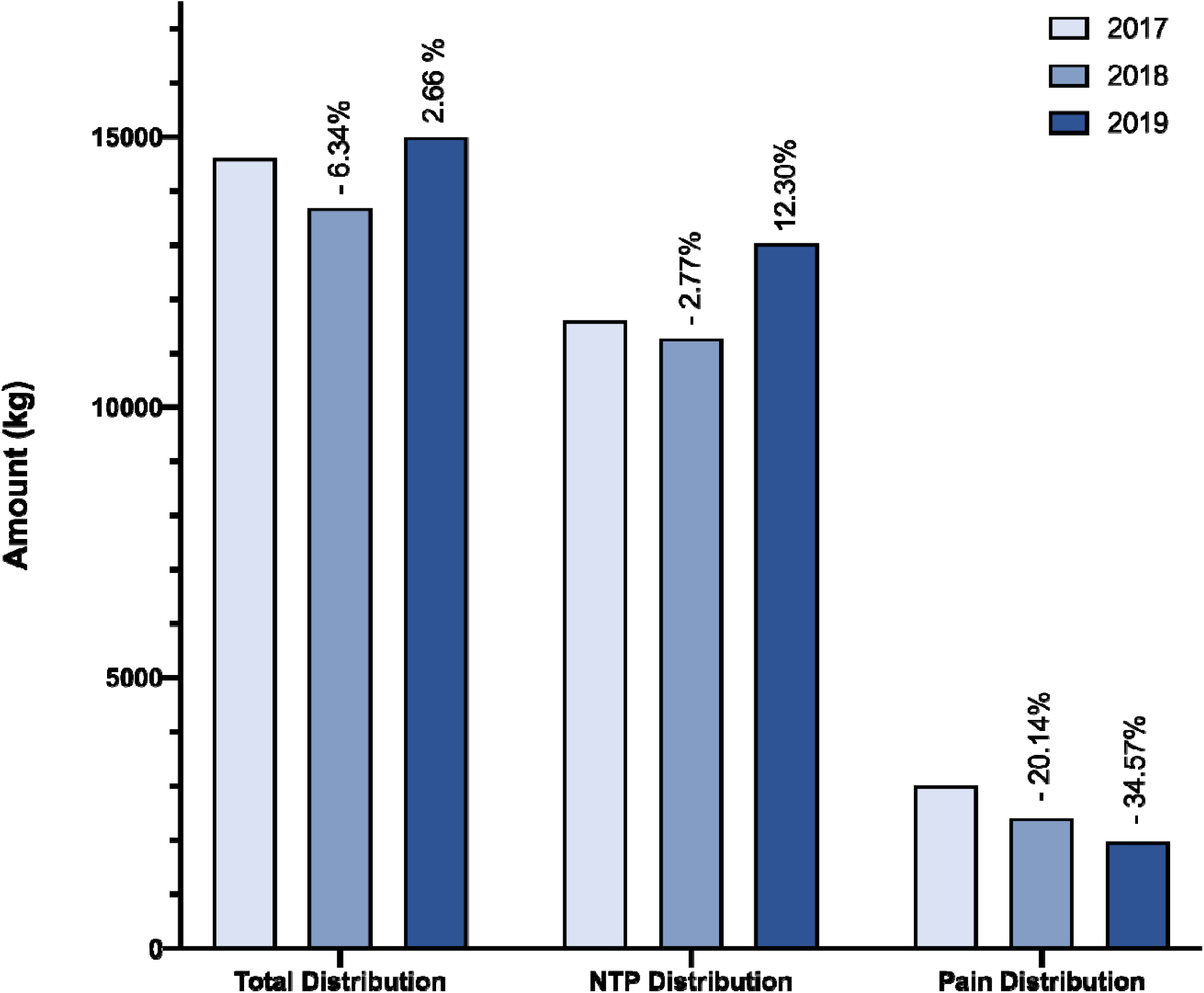
United States methadone distribution amounts as reported to the Drug Enforcement Administration’s Automated Reports and Consolidated Ordering System in kilograms for 2017, 2018, and 2019. Annotations denote percent change relative to 2017. NTP: Narcotic Treatment Program.

Regional analysis identified appreciable variation in pain distribution. When compared to the Northeast region, the South (p < 0.01) and West regions (p < 0.01) as well as the Midwest region (p < 0.05), experienced significantly greater declines. No other regional comparisons to the NE region were found to be statistically significant (Figure 2). A more focused view of the Northeast (NE) region shows wide variation amongst states in percent change for distribution types and number of NTPs (Figure 2). Correlation analysis demonstrated a strong positive association (r = 0.89) between total distribution and NTP distribution amongst NE states. This is in agreement with the finding that all NE states saw a decline in distribution for pain management despite all but one state that saw an increase in total distribution (Figure 2).

**Figure 2:**
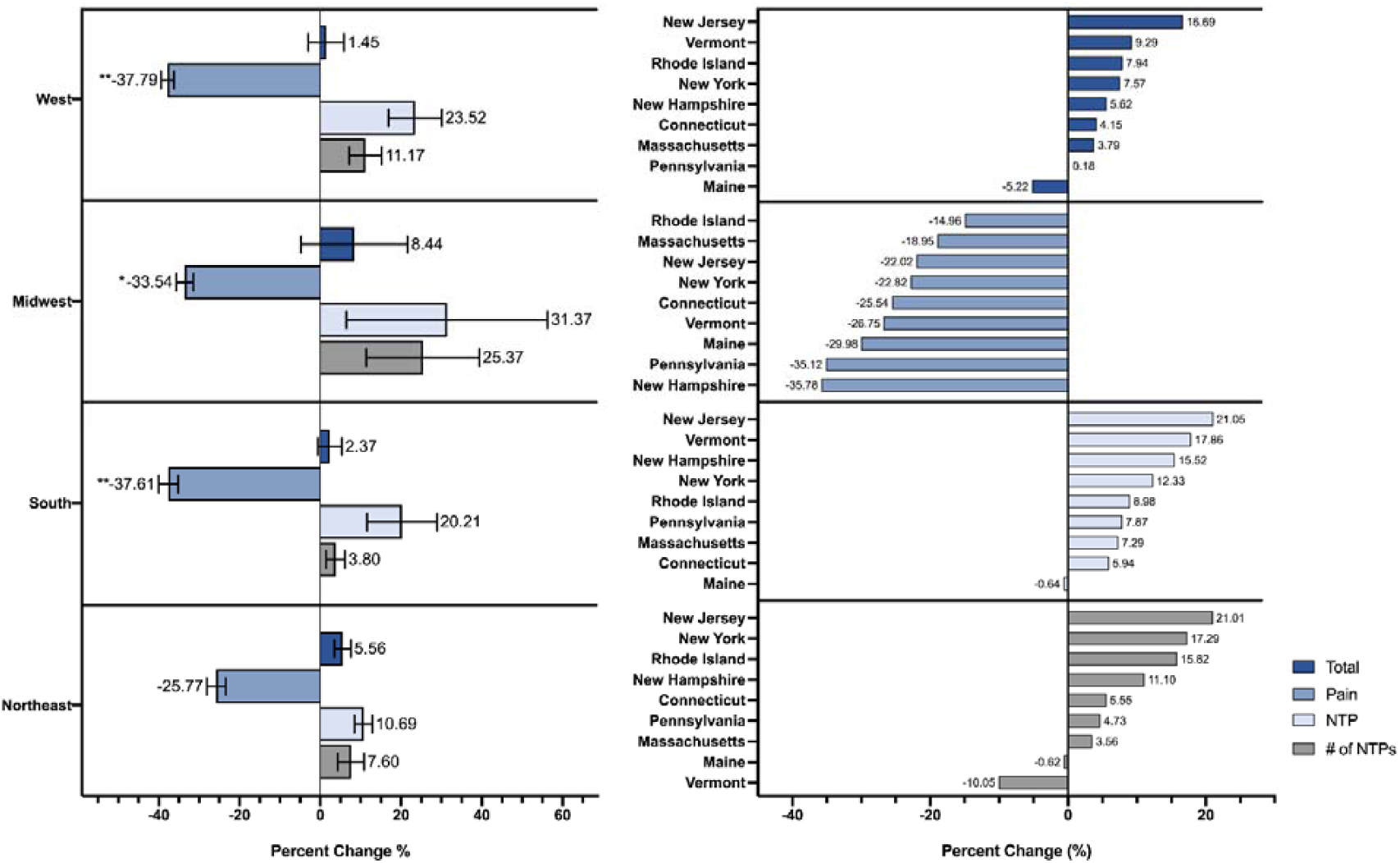
Regional comparison of percent change in number of methadone NTPs and methadone distribution as reported to the Drug Enforcement Administration’s Automated Reports and Consolidated Ordering System between 2017 and 2019. All regions were compared in reference to the NE region (left). Values and lines indicate SEM. Statistical analysis performed by unpaired t-tests (* p < 0.05, ** p < 0.01). Percent change in methadone distribution and number of Narcotic Treatment Programs (NTP) per 100K population between 2017 and 2019 in Northeast States (right).

To further analyze distribution of methadone during this time period, the total distribution for all US states was compared. Overall, approximately half of US states increased their total distribution of methadone between 2017 and 2019. Most notably, North Dakota experienced a significant increase (p < 0.00001) in comparison to the national average (Sup Figure 1). When total distribution was separated into distribution for pain and NTPs, per state analyses revealed additional trends. All states, including the District of Columbia (DC), showed a decrease in distribution for pain over this time period. Both Mississippi (p < 0.05) and Oklahoma (p < 0.05) had significantly larger decreases in pain distribution when compared to the national average. In contrast, Rhode Island (p < 0.05) had a significantly smaller decrease in pain distribution (Figure 3).

**Figure 3:**
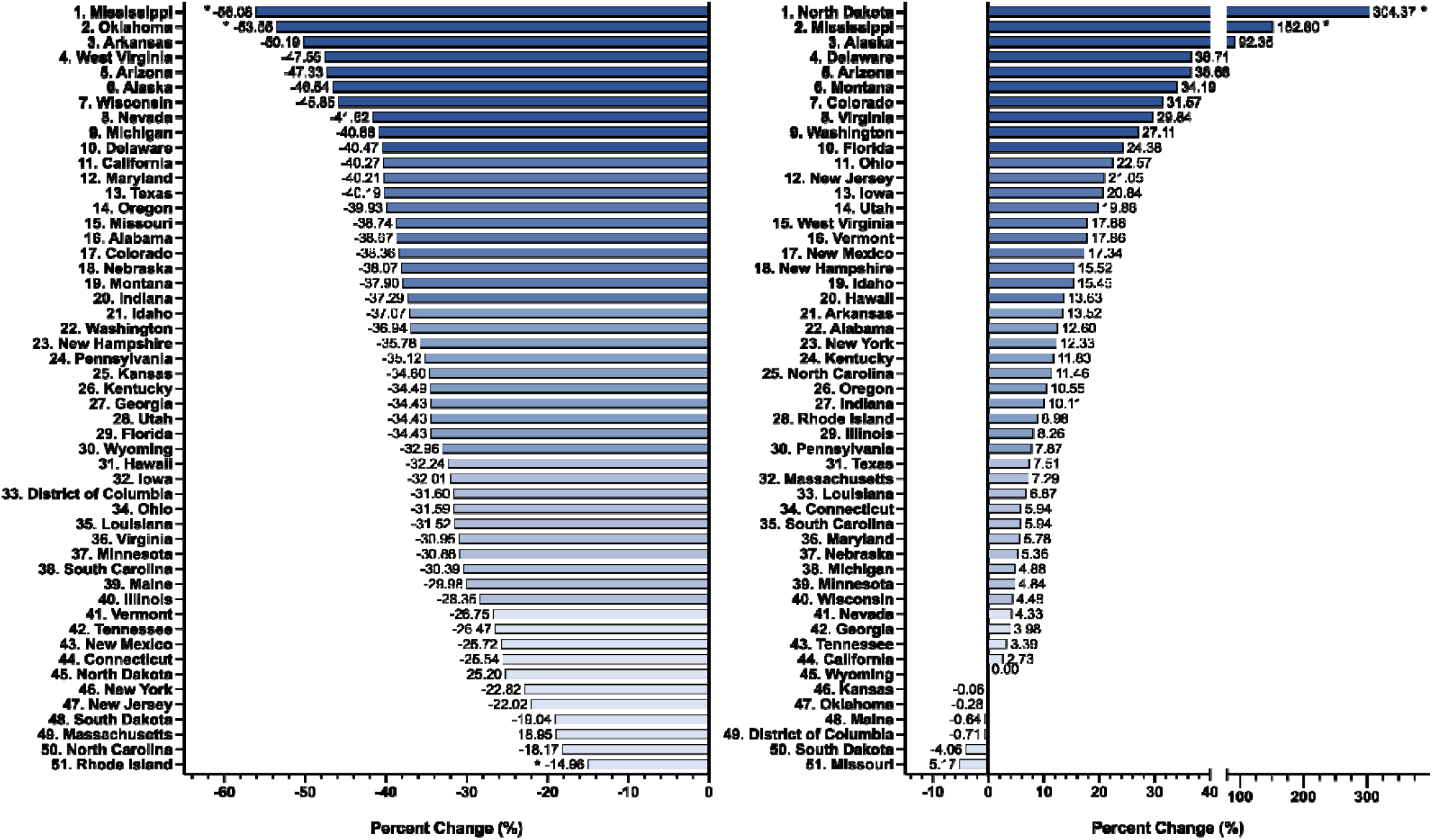
Percent change in methadone distribution as reported to the Drug Enforcement Administration’s Automated Reports and Consolidated Ordering System for pain (left) and OUD (right) per 100K population between 2017 and 2019. Percent change beyond 1.96 standard deviations from the US mean was considered significant (* p < .05).

Examination of methadone to NTPs showed increases in distribution across the majority of states, with North Dakota (p < 0.00001) and Mississippi (p < 0.01) experiencing a significantly greater increase in relation to the US average (Figure 3). Furthermore, percent changes in total methadone distribution were much smaller than changes in total buprenorphine distribution for the majority of states. As a whole, the US saw a 2.66% increase in total methadone distribution relative to a 31.60% increase in total buprenorphine distribution (Sup Figure 2). The average change in total buprenorphine distribution per state, 34.71%, was significantly larger than the average change in total methadone distribution per state, 4.13% (p < 0.0001).

Similar to the increase in methadone distribution for NTPs seen in most states, many states also experienced an increase in the number of NTPs during this period. Of note, Iowa (p < 0.00001) and Ohio (p < 0.01) demonstrated significantly greater increases in the number of NTPs in comparison to the nation’s average change (Sup Figure 3). Focusing specifically on 2019, Rhode Island (p < 0.0001), Delaware (p < 0.05), Vermont (p < 0.05), and Maryland (p < 0.05) all had a significantly greater number of NTPs in 2019 when compared to the US average. Notably, Rhode Island had over two NTPs per 100K population (2.08), representing the maximum number across all states. In contrast, Wyoming (population = 547,637) was without a single methadone NTP in 2019 (Figure 4).

**Figure 4:**
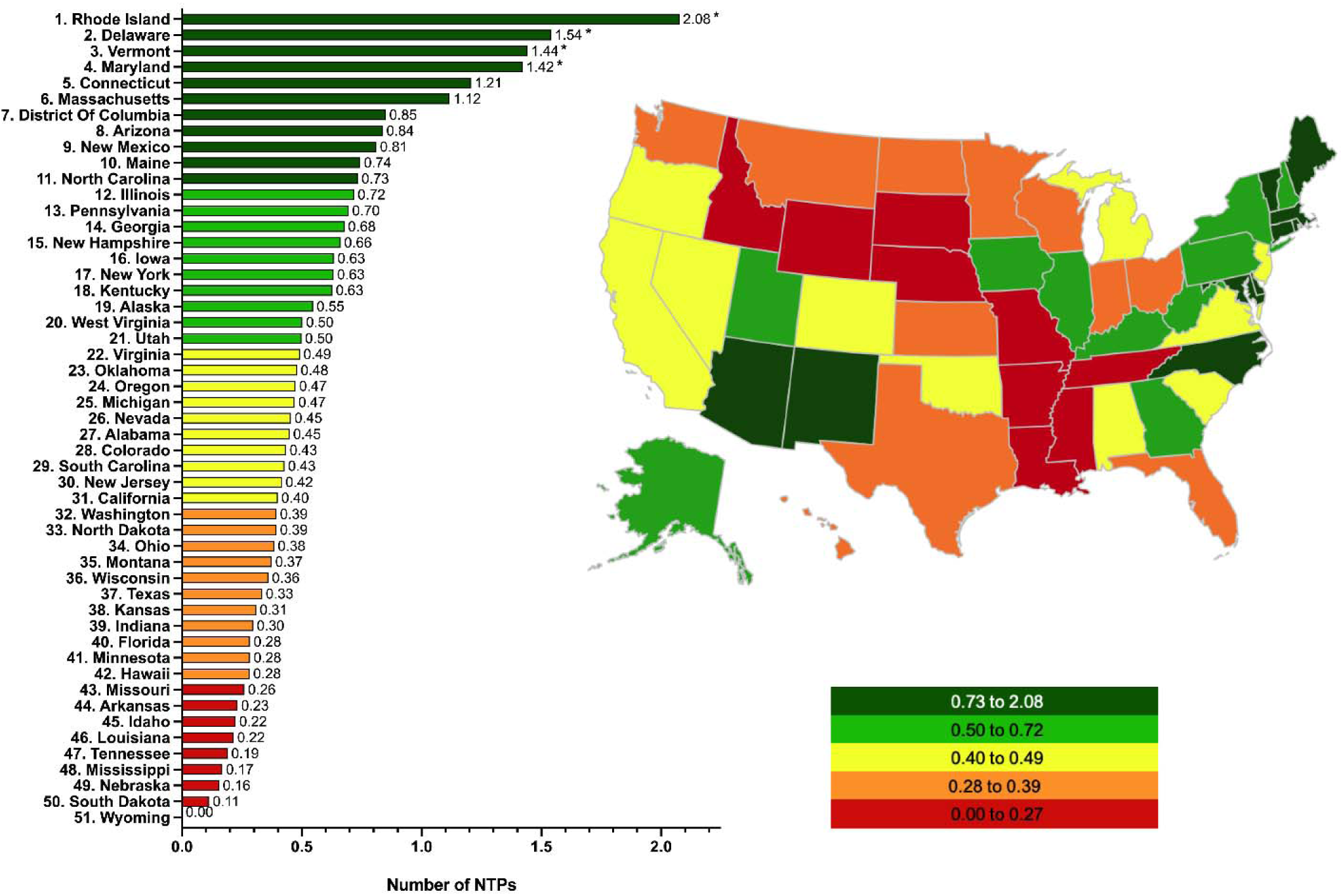
Number of Narcotic Treatment Programs as reported to the Drug Enforcement Administration’s Automated Reports and Consolidated Ordering System per 100K population in 2019. Values beyond 1.96 standard deviations from the US mean were considered significant (* p < .05).

## Discussion

This report demonstrated an increase in the distribution of methadone across the US between 2017-2019 and extends upon earlier research.^6,8^ This recent increase in methadone distribution is correlated with an increasing number of NTPs across the US. Between 2017 and 2019, 44 states increased their distribution of methadone to NTPs and over half of all states increased their total number of NTPs when adjusted for population. This data corroborates the 2019 Substance Abuse and Mental Health Services (SAMHSA) report; the number of individual clients receiving methadone from an NTP facility increased from 382,867 to 408,550 between 2017 and 2019.^19^ This may suggest that the national increase in methadone distribution is related to an increase in the number of individuals prescribed methadone from NTPs, rather than an increase in the amount prescribed per individual already receiving methadone treatment.^19^ Many of these patterns may be explained by legislation at the state and federal level, or other factors such as NTP distribution independent of barriers created by government policy.

As opposed to the trends in total distribution of methadone, analysis of distribution of methadone specifically for pain management demonstrated that all fifty states, and also the District of Columbia, have decreased in this regard. This is consistent with and extends upon previous reports demonstrating that opioid distribution amount and prescription rates have declined in the years leading up to 2017.^6,21^ It is likely that efforts from both the FDA and CDC have played significant roles in the widespread decreases seen across all states. In the wake of the opioid crisis, both the FDA and CDC have taken steps to eliminate unsafe opioid practices across the US which include improved provider education, appropriate drug labeling, increased naloxone access, and new CDC guidelines on prescribing opioids for chronic pain.^22,23^ Specifically, actions of the FDA between 2002-2014 resulted in a decrease use of methadone for pain and an associated decrease in methadone overdose deaths despite additional persons receiving methadone for OUD.^7^ Mississippi and Oklahoma had the largest decrease in methadone distribution for pain, demonstrating decreases significantly larger than the national average. Despite these large decreases, in 2018 Mississippi and Oklahoma providers wrote 76.8 and 79.1 opioid prescriptions for every 100 persons, respectively, in comparison to the US national average of 51.4 prescriptions per 100 persons.^24,25^ However, both of these prescription rates are the lowest either of these states have seen since this data became available in 2006 and could suggest that these states are making strides to improve their management of opioid misuse.^24,25^ Analyzing the same dataset revealed that Rhode Island had a significantly smaller decrease in the amount of methadone prescribed for pain in comparison to the national average, however, Rhode Island providers wrote 43.0 opioid prescriptions for every 100 persons in 2018.^26^ In alignment with this finding, the Northeast as a region saw a significantly smaller decrease in distribution for pain in comparison to the other three US regions. The methadone distribution increases to the Northeast are instead reflected by use in NTPs for patients with OUD.

As a region, the Northeast demonstrated an increase in the number of NTPs across nearly all states between the years 2017 and 2019. Many states experienced an increase in the number of NTPs and a state-by-state analysis showed that five out of the nine Northeast states are amongst the top ten states in the country based on the number of NTPs per 100k in 2019. Additionally, the most recently reported CDC statistics on opioid related overdose deaths demonstrate that some of the highest state opioid related overdose death rates across the country occurred in NE states in 2018.^9^ Considering this alongside the high number of NTPs consolidated within this region, this may help justify the need for the large number of NTPs in the NE and may even suggest that legislation within this region has been progressive in attempts to improve opioid treatment access. One member of this region, Rhode Island, was the only state in the US to have over 2 NTPs per 100K population in 2019, a number that increased 15.8% from 2017. While it is unclear if this expanded methadone access relative to other states was driven by state specific legislation, it should be noted that the 21st Century Cures Act passed by the federal government allocated roughly one billion dollars to be distributed across states and territories during 2017 and 2018 to expand access to evidence-based treatment for OUD and reduce unmet treatment needs.^27^ The 21st Century Cures Act may have enabled RI to reach some of their NTP goals, and has possibly driven the rise in NTP facilities observed in states, such as Iowa and Ohio, that saw significantly larger percent increases in the number of NTPs per 100K over this time period (149.26 and 95.14, respectively). Both Iowa and Ohio were reported to have used a majority of their grant money on treatment, as opposed to other measures such as prevention and administration, which could have contributed to their significant success in expanding methadone access in the form of NTPs.^28^ On the contrary, Wyoming received four million dollars in grant money through this act but remained without a single methadone NTP in 2019.^19,28^ This is likely due in part to the continued absence of Medicaid coverage for methadone within Wyoming.^29^

Almost all states increased methadone distribution to NTPs between 2017 and 2019. Specifically, North Dakota and Mississippi significantly increased in this distribution despite no increase in the number of NTPs. In Mississippi, which had a higher than average distribution of methadone for pain despite its significant decline in use as mentioned above, the increase in methadone distribution to NTPs may suggest that there is an increased need for NTPs in this region as a result of methadone continuing to be used for pain management. These significant increases in methadone distribution to rural states such as North Dakota and Mississippi may be because the majority of facilities that use medication assisted treatment for OUD reported that they did not offer the following services: (1) detoxification from opioids with lofexidine or clonidine (83 percent); (2) detoxification from opioids with methadone or buprenorphine (69 percent).^19^ The rural nature of these states decreases the likelihood that patients requiring methadone would have close access to an NTP supplying methadone. This is reflected in one-way driving times upwards of 120 minutes for patients to reach an NTP that can fill their prescription.^30^ The significant increase in methadone distribution to North Dakota and Mississippi could be due to a number of reasons. Namely, more people could be traveling to fill their prescriptions, physicians are providing more methadone per visit for these patients, and patients in neighboring states are traveling to these states to fill their methadone prescriptions because it is closer than the nearest NTP in their home state.

In addition to the recent methadone NTP distribution and use, buprenorphine distribution has been and continues to increase. From 2017 to 2019, national distribution of buprenorphine increased 31.60%, which is much larger than the 2.66% increase in methadone distribution seen during this time period. However, it is unlikely that this difference is driven by cost, as these drugs show little difference in treatment cost according to the US Department of Defense, in the setting of a treatment program. The Defense Health Agency TRICARE program estimates the weekly cost of buprenorphine and methadone treatment program cost to be $115 and $126, respectively, taking into consideration medication cost, as well as the number of visits and associated therapy required.^31^ The Comprehensive Addiction and Recovery Act that was passed in 2016 may help to explain this difference in distribution between the drugs, as it enabled qualified mid-level providers to prescribe buprenorphine, while also increasing the maximum number of patients that qualified providers can treat with buprenorphine, or other schedule III, IV, or V narcotics for the treatment of OUD, from 100 to 275 patients.^32^ These changes have expanded buprenorphine access and could be playing an appreciable role in the large increase in buprenorphine distribution as opposed to methadone. The 2019 National Survey of Substance Abuse Treatment Services (N-SSATS) supports this conclusion as the proportion of opioid treatment programs that provided methadone-only treatment decreased from 51% of all facilities in 2009 to 21% of all facilities in 2019.^19^ Additionally, it is important to recognize that both drugs are equally efficacious at controlling opioid misuse and show no significant difference in treatment related mortality, however, methadone is superior at retaining patients on treatment regimens.^33,34^

The use of ARCOS in the current study has provided a unique opportunity to make observations about the distribution patterns of opioid treatment medications nationally, which has emphasized the need for continued strides to improve methadone access in the care of those suffering from OUD. The federal regulation (42 CFR Part 2) prevented methadone from being entered into Prescription Drug Monitoring Programs until June, 2020 has been an impediment to pharmacoepidemiology research using this data source^35^. However, while ARCOS does overcome this limitation, the use of ARCOS as a data source does have some inherent limitations. Although changes in total distribution of methadone and buprenorphine could be compared, it was not possible to directly compare distribution of buprenorphine and methadone specifically for the treatment of OUD since both medications are also approved for pain, and in the case of buprenorphine, can be distributed in settings outside of certified NTPs. Furthermore, ARCOS does not report individual distribution amounts to specific NTPs, limiting the ability to compare distribution activity between individual NTP facilities. Although legislation at a federal level may help to explain some of the trends uncovered, variations in policy at the state level likely plays a significant role as well but lies outside the scope of the broader focus of this study. Future research on this topic is warranted to follow the response of the US to the opioid epidemic as it relates to implementation of opioid treatment programs and expanded access to pharmacological treatment therapies for OUD. Recent research demonstrating the importance of methadone treatment in vulnerable populations like prisoners, in addition to increased government funding for the treatment of OUD, may likely help shape new trends in the distribution of methadone and other OUD pharmacotherapies in upcoming years.^36^

## Conclusions

Opioid use disorder is a complex, chronic disease which requires an individualized and multifaceted approach to patient care. While both methadone and buprenorphine therapy are more effective than abstinence-based treatment, each modality has advantages.^8,34^ Methadone has higher treatment retention rates than buprenorphine and may be preferred for patients at higher risk of dropout such as persons who inject opioids. Methadone should also be considered if opioid use persists despite an optimal buprenorphine dose. While buprenorphine has a lower risk of overdose than methadone and may be preferred for patients who have work or family commitments that make it difficult to attend clinic daily or if their jobs require higher levels of cognitive function or psychomotor performance. Buprenorphine is also recommended for patients at higher risk of methadone toxicity such as the risk of serious arrhythmias in patients with a history of cardiac conduction abnormalities or taking medications affecting cardiac conduction. For these reasons, increased access to both methadone and buprenorphine are needed to individualize care and address the opioid crisis in regions of the country that continue to be disproportionately affected.

## Supporting information

Raw data from 2017

Raw data from 2018

Raw data from 2019

## Data Availability

Raw data in the form of pdfs are available at: https://www.deadiversion.usdoj.gov/arcos/retail_drug_summary/
Extracted data from these 5-1,100 page reports is included in the supplemental materials.

https://www.deadiversion.usdoj.gov/arcos/retail_drug_summary/report_yr_2019.pdf

## Acknowledgements

Iris Johnston provided technical support. The Drug Enforcement Administration staff is appreciated for making data publically available.

## Figure Legends

**Figure 1:** United States methadone distribution amounts as reported to the Drug Enforcement Administration’s Automated Reports and Consolidated Ordering System in kilograms for 2017, 2018, and 2019. Annotations denote percent change relative to 2017.

**Figure 2:** Regional comparison of percent change in number of methadone NTPs and methadone distribution as reported to the Drug Enforcement Administration’s Automated Reports and Consolidated Ordering System between 2017 and 2019. All regions were compared in reference to the NE region (left). Values and lines indicate SEM. Statistical analysis performed by unpaired t-tests (* p < 0.05, ** p < 0.01). Percent change in methadone distribution and number of Narcotic Treatment Programs (NTP) per 100K population between 2017 and 2019 in Northeast States (right).

**Figure 3:** Percent change in methadone distribution as reported to the Drug Enforcement Administration’s Automated Reports and Consolidated Ordering System for pain (left) and OUD (right) per 100K population between 2017 and 2019. Percent change beyond 1.96 standard deviations from the US mean was considered significant (* p < .05).

**Figure 4:** Number of Narcotic Treatment Programs as reported to the Drug Enforcement Administration’s Automated Reports and Consolidated Ordering System per 100K population in 2019. Values beyond 1.96 standard deviations from the US mean were considered significant (* p < .05).

**Sup Figure 1:**
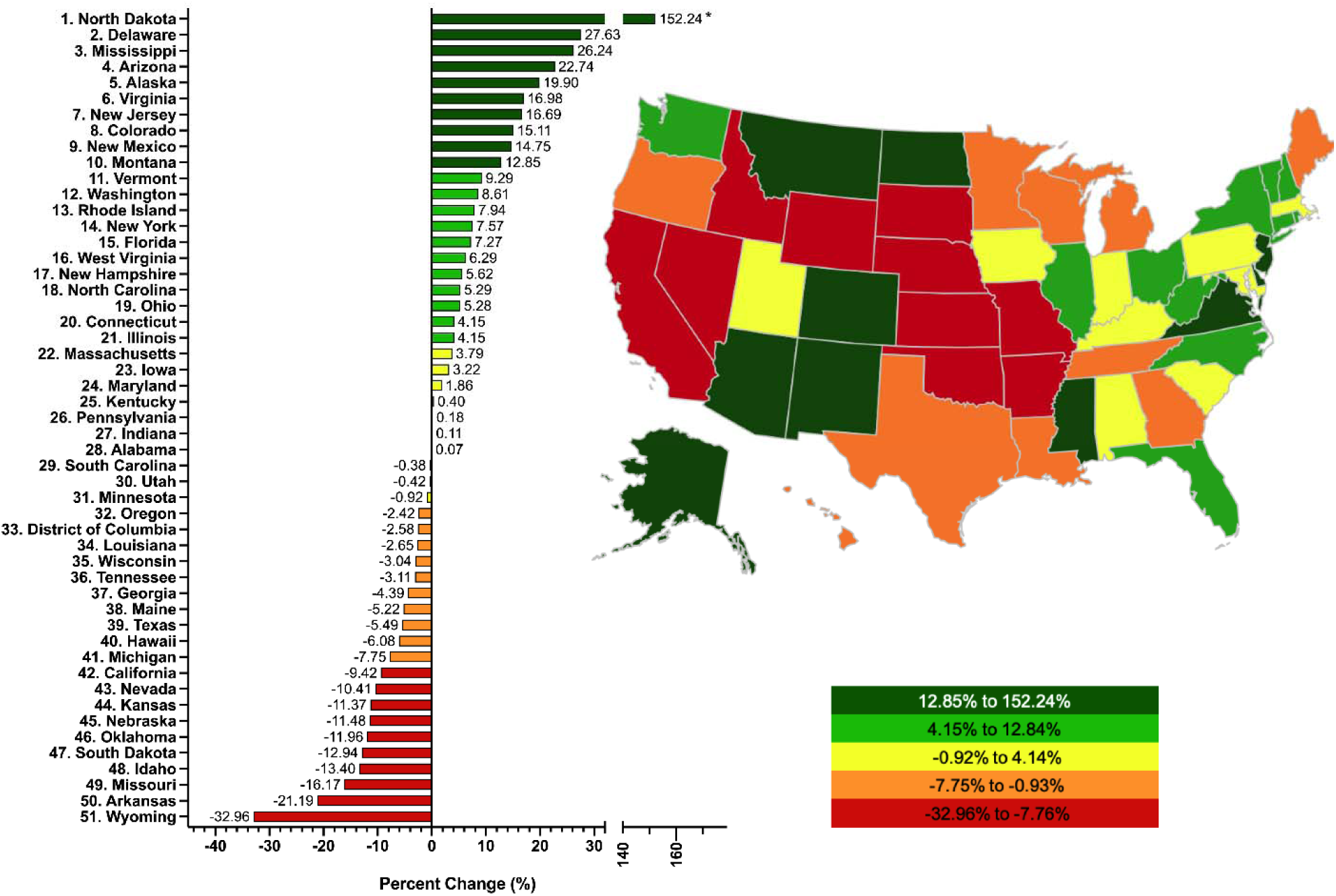
Percent change in total methadone distribution as reported to the Drug Enforcement Administration’s Automated Reports and Consolidated Ordering System per 100K population between 2017 and 2019. Percent change beyond 1.96 standard deviations from the US mean was considered significant (* p < .05).

**Sup Figure 2:**
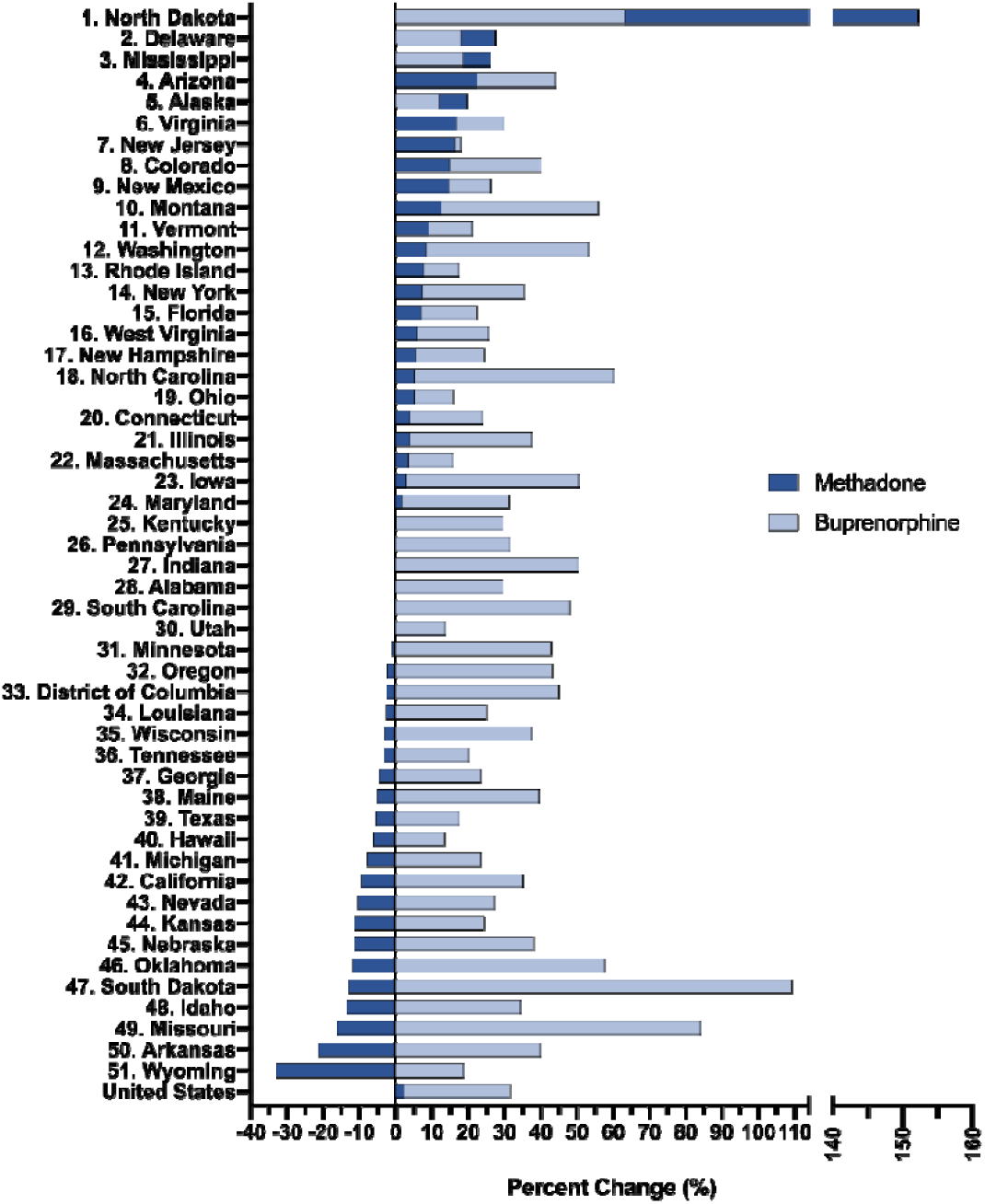
Percent change in total buprenorphine distribution (light blue) and total methadone distribution (dark blue) as reported to the Drug Enforcement Administration’s Automated Reports and Consolidated Ordering System per 100K population between 2017 and 2019. Also depicted, percent change of total methadone and buprenorphine distribution for the United States as a whole from 2017 to 2019.

**Sup Figure 3:**
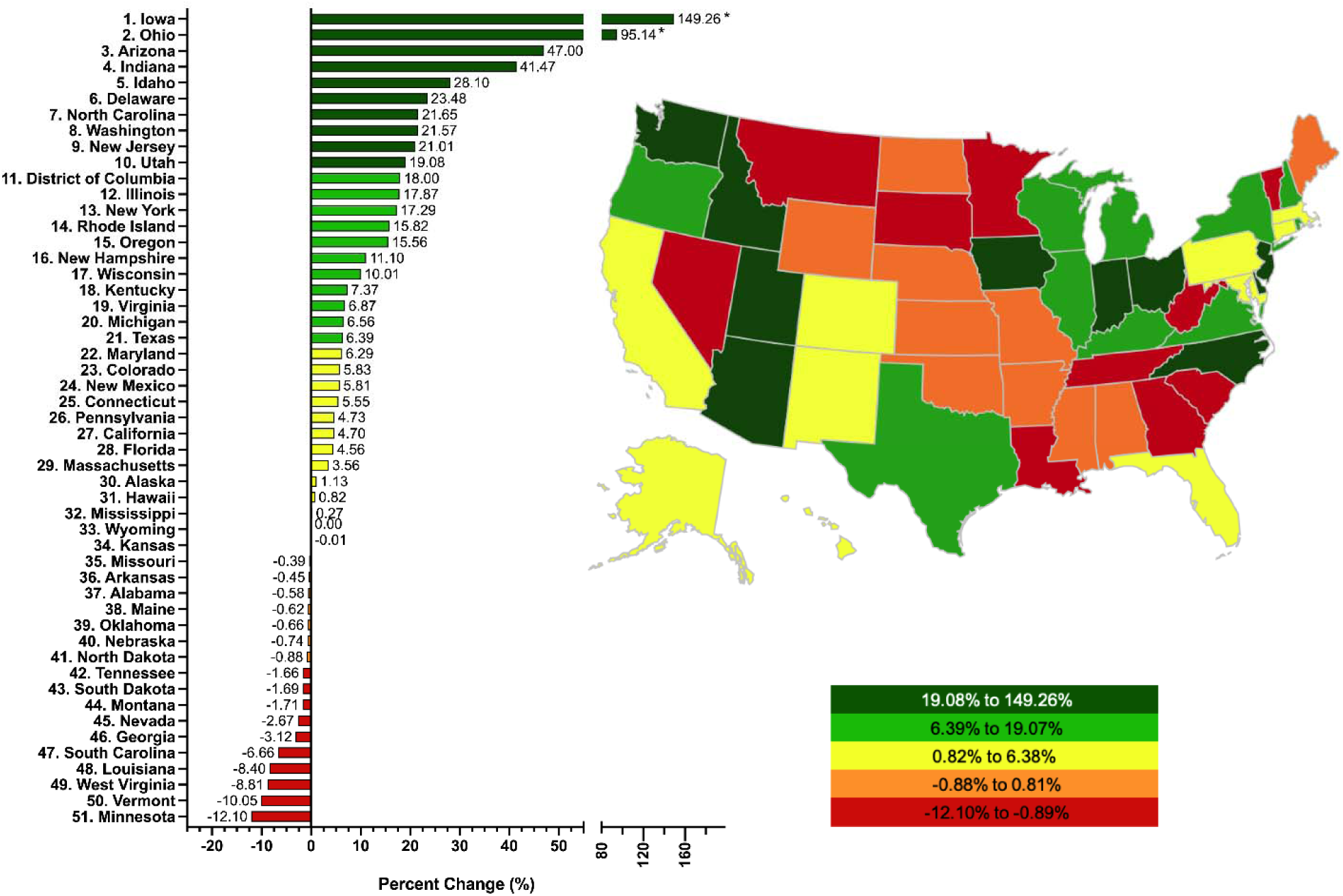
Percent change in number of Narcotic Treatment Programs (NTP) per 100K population as reported to the Drug Enforcement Administration’s Automated Reports and Consolidated Ordering System from 2017 to 2019. Percent change beyond 1.96 standard deviations from the US mean was considered significant (* p < .05).

