## Supplementary material for "Methadone Distribution Trends from 2017-2019 in the United States": Raw data from 2017

DATE RANGE: 01/01/2017 TO 12/31/2017

### ARCOS 3 - REPORT 1

Run Date: 07/03/2018

#### RETAIL DRUG DISTRIBUTION BY ZIP CODE WITHIN STATE BY GRAMS WT

DRUG CODE: 1100 DRUG NAME: AMPHETAMINE

STATE: ALABAMA

| ZIP CODE | QUARTER 1 | QUARTER 2 | QUARTER 3 | QUARTER 4 | TOTAL GRAMS |
|---|---|---|---|---|---|
| 350 | 17,317.77 | 17,592.62 | 17,601.30 | 18,026.63 | 70,538.32 |
| 351 | 8,853.04 | 8,730.36 | 8,871.69 | 8,885.75 | 35,340.84 |
| 352 | 19,011.67 | 18,775.54 | 19,012 | 19,869.23 | 76,668.44 |
| 354 | 8,415.59 | 8,008.47 | 7,903.15 | 8,576.03 | 32,903.24 |
| 355 | 3,437.61 | 3,592.54 | 3,505.06 | 3,423.91 | 13,959.12 |
| 356 | 9,419.78 | 9,504.50 | 9,896.92 | 10,055.83 | 38,877.03 |
| 357 | 4,092.23 | 4,275.89 | 4,362.04 | 4,447.32 | 17,177.48 |
| 358 | 5,351.36 | 5,555.98 | 5,650.23 | 5,681.07 | 22,238.64 |
| 359 | 4,819.79 | 4,836.55 | 4,619.88 | 5,088.01 | 19,364.23 |
| 360 | 6,516.37 | 6,441.69 | 6,372.90 | 6,768.46 | 26,099.42 |
| 361 | 5,920.66 | 5,825.58 | 5,225.22 | 5,243.17 | 22,214.63 |
| 362 | 2,442.19 | 2,350.13 | 2,347.92 | 2,345.96 | 9,486.20 |
| 363 | 5,219.18 | 5,291.86 | 5,210.23 | 5,424.80 | 21,146.07 |
| 364 | 1,273.24 | 1,315.80 | 1,144.04 | 1,299.77 | 5,032.85 |
| 365 | 11,488.92 | 11,634.86 | 12,049.97 | 12,420.28 | 47,594.03 |
| 366 | 7,815.82 | 7,687.68 | 7,886.41 | 7,953.48 | 31,343.39 |
| 367 | 1,557.50 | 1,463.38 | 1,478.39 | 1,572.70 | 6,071.97 |
| 368 | 6,415.23 | 6,190.82 | 6,134.74 | 6,677.12 | 25,417.91 |
| 369 | 154.58 | 123.62 | 127.6 | 143.08 | 548.88 |
| TOTAL | 129,522.53 | 129,197.87 | 129,399.69 | 133,902.60 | 522,022.69 |

STATE: ALASKA

| ZIP CODE | QUARTER 1 | QUARTER 2 | QUARTER 3 | QUARTER 4 | TOTAL GRAMS |
|---|---|---|---|---|---|
| 995 | 4,873.14 | 4,889.14 | 4,867.94 | 5,002.53 | 19,632.75 |
| 996 | 1,456.63 | 1,348.71 | 1,465.13 | 1,383.28 | 5,653.75 |
| 997 | 1,142.20 | 1,151.50 | 1,215.34 | 1,125.86 | 4,634.90 |
| 998 | 380.28 | 454.81 | 459.8 | 425.87 | 1,720.76 |
| 999 | 165.38 | 191.37 | 181.93 | 154.42 | 693.1 |
| TOTAL | 8,017.63 | 8,035.53 | 8,190.14 | 8,091.96 | 32,335.26 |

STATE: AMERICAN SAMOA

| ZIP CODE | QUARTER 1 | QUARTER 2 | QUARTER 3 | QUARTER 4 | TOTAL GRAMS |
|---|---|---|---|---|---|

| | | | | | | |
|---|---|---|---|---|---|---|
| | 967 | 1.25 | 1.25 | 0 | 0 | 2.5 |
| TOTAL | | 1.25 | 1.25 | 0 | 0 | 2.5 |

STATE: ARIZONA

| ZIP CODE | | QUARTER 1 | QUARTER 2 | QUARTER 3 | QUARTER 4 | TOTAL GRAMS |
|---|---|---|---|---|---|---|
| | 850 | 17,085.55 | 17,122.42 | 16,792.26 | 17,695.67 | 68,695.90 |
| | 851 | 2,949.78 | 3,116.14 | 3,116.29 | 3,382.39 | 12,564.60 |
| | 852 | 32,696.05 | 32,496.97 | 33,207.28 | 36,130.70 | 134,531 |
| | 853 | 11,110.41 | 10,528 | 10,559.23 | 10,991.41 | 43,189.05 |
| | 855 | 495.76 | 409.32 | 472.1 | 477.97 | 1,855.15 |
| | 856 | 1,667.39 | 1,636.98 | 1,527.61 | 1,696 | 6,527.98 |
| | 857 | 10,892.17 | 10,595.68 | 10,465.69 | 10,987.87 | 42,941.41 |
| | 859 | 529.34 | 520.01 | 497.23 | 472.8 | 2,019.38 |
| | 860 | 1,319.04 | 1,228.49 | 1,210.32 | 1,333.18 | 5,091.03 |
| | 863 | 1,569.48 | 1,587.04 | 1,464.17 | 1,718.05 | 6,338.74 |
| | 864 | 942.56 | 892.58 | 859.54 | 923.26 | 3,617.94 |
| | 865 | 54.23 | 51.09 | 27.5 | 64.94 | 197.76 |
| TOTAL | | 81,311.76 | 80,184.72 | 80,199.22 | 85,874.24 | 327,569.94 |

STATE: ARKANSAS

| ZIP CODE | | QUARTER 1 | QUARTER 2 | QUARTER 3 | QUARTER 4 | TOTAL GRAMS |
|---|---|---|---|---|---|---|
| | 716 | 2,788.22 | 2,809.06 | 2,886.64 | 2,919.10 | 11,403.02 |
| | 717 | 965.54 | 961.04 | 883.31 | 971.54 | 3,781.43 |
| | 718 | 959.19 | 832.46 | 896.19 | 951.83 | 3,639.67 |
| | 719 | 3,491.96 | 3,332.89 | 3,360.64 | 3,380.55 | 13,566.04 |
| | 720 | 7,605.38 | 7,648.33 | 7,746.66 | 8,031.25 | 31,031.62 |
| | 721 | 6,064.07 | 6,106.61 | 6,095.45 | 6,280.22 | 24,546.35 |
| | 722 | 7,089.99 | 7,013.36 | 7,301.47 | 7,287.35 | 28,692.17 |
| | 723 | 1,517.55 | 1,539.92 | 1,482.32 | 1,548.20 | 6,087.99 |
| | 724 | 2,812.94 | 2,734.62 | 2,673.57 | 2,854.60 | 11,075.73 |
| | 725 | 770.69 | 726.82 | 750.23 | 754.58 | 3,002.32 |
| | 726 | 1,540.53 | 1,469.13 | 1,554.03 | 1,537 | 6,100.69 |
| | 727 | 12,037.52 | 12,369.29 | 12,280.49 | 12,836.22 | 49,523.52 |
| | 728 | 986.65 | 986.5 | 934.46 | 944.66 | 3,852.27 |
| | 729 | 1,974.59 | 2,021.71 | 2,058.32 | 2,014.25 | 8,068.87 |

| | | | | | |
|---|---|---|---|---|---|
| TOTAL | 50,604.82 | 50,551.74 | 50,903.78 | 52,311.35 | 204,371.69 |

STATE: CALIFORNIA

| ZIP CODE | QUARTER 1 | QUARTER 2 | QUARTER 3 | QUARTER 4 | TOTAL GRAMS |
|---|---|---|---|---|---|
| 900 | 21,190.33 | 22,283.22 | 21,463.94 | 22,760.37 | 87,697.86 |
| 902 | 12,294.30 | 12,770.35 | 12,367.35 | 12,893.80 | 50,325.80 |
| 903 | 284.12 | 308.32 | 300.17 | 268.1 | 1,160.71 |
| 904 | 3,922.68 | 4,176.69 | 4,174.37 | 4,013.15 | 16,286.89 |
| 905 | 1,735.29 | 1,845.88 | 1,699.89 | 1,773.27 | 7,054.33 |
| 906 | 2,281.68 | 2,339.11 | 2,313.36 | 2,520.19 | 9,454.34 |
| 907 | 3,240.07 | 3,310.64 | 3,332.77 | 3,319.06 | 13,202.54 |
| 908 | 2,910.12 | 3,110.81 | 2,869.36 | 3,035.64 | 11,925.93 |
| 910 | 1,581.76 | 1,684.27 | 1,612.79 | 1,533.60 | 6,412.42 |
| 911 | 2,567.45 | 2,549.78 | 2,645.98 | 2,681.86 | 10,445.07 |
| 912 | 1,515.12 | 1,620.92 | 1,521.64 | 1,551.68 | 6,209.36 |
| 913 | 14,340.39 | 14,686.23 | 14,323.01 | 14,801.06 | 58,150.69 |
| 914 | 4,225.08 | 4,490.77 | 4,553.27 | 4,423.62 | 17,692.74 |
| 915 | 1,745.92 | 1,786.70 | 1,849.83 | 1,784.17 | 7,166.62 |
| 916 | 1,635.52 | 1,720.58 | 1,679.19 | 1,750.31 | 6,785.60 |
| 917 | 6,716.13 | 6,933.52 | 6,657.49 | 6,996.58 | 27,303.72 |
| 918 | 370.21 | 402.93 | 394.83 | 377.61 | 1,545.58 |
| 919 | 2,659.25 | 2,863.81 | 2,620.39 | 2,672.26 | 10,815.71 |
| 920 | 26,382.62 | 26,689.43 | 26,373.24 | 27,140.41 | 106,585.70 |
| 921 | 17,531.85 | 17,972.54 | 17,362.58 | 18,097.34 | 70,964.31 |
| 922 | 3,662.11 | 3,531.42 | 3,351.59 | 3,520.91 | 14,066.03 |
| 923 | 4,086.95 | 4,025.14 | 4,055.80 | 4,255.37 | 16,423.26 |
| 924 | 314.71 | 363.32 | 311.13 | 345.45 | 1,334.61 |
| 925 | 6,303.63 | 6,478.64 | 6,260.11 | 6,536.16 | 25,578.54 |
| 926 | 21,129.93 | 21,430.31 | 21,464.66 | 21,524.94 | 85,549.84 |
| 927 | 4,741.72 | 4,589.58 | 4,533.97 | 4,533.72 | 18,398.99 |
| 928 | 7,901.55 | 8,001.24 | 7,967.18 | 7,972.67 | 31,842.64 |
| 930 | 5,658 | 5,870.35 | 5,803.10 | 5,636.99 | 22,968.44 |
| 931 | 3,418.50 | 3,490.90 | 3,200.40 | 3,150.94 | 13,260.74 |
| 932 | 1,826.73 | 1,878.82 | 1,766.65 | 1,860.53 | 7,332.73 |
| 933 | 1,830.32 | 1,980.89 | 1,917.32 | 1,991.08 | 7,719.61 |
| 934 | 3,243.50 | 3,464.45 | 3,384.48 | 3,554.32 | 13,646.75 |

| | | | | | | |
|---|---|---|---|---|---|---|
| | 935 | 2,266.78 | 2,331.36 | 2,216.47 | 2,324.32 | 9,138.93 |
| | 936 | 2,191.61 | 2,238.72 | 2,220.73 | 2,267.07 | 8,918.13 |
| | 937 | 3,220.95 | 3,489.91 | 3,420.07 | 3,443.30 | 13,574.23 |
| | 939 | 1,782.44 | 1,517.15 | 1,647.58 | 1,661.16 | 6,608.33 |
| | 940 | 6,015.56 | 5,875.57 | 5,830.74 | 6,075.81 | 23,797.68 |
| | 941 | 11,324.71 | 11,891.46 | 11,453.66 | 12,133.52 | 46,803.35 |
| | 943 | 1,042.64 | 1,049.59 | 1,020.94 | 1,029.72 | 4,142.89 |
| | 944 | 1,300.21 | 1,214.39 | 1,265.94 | 1,289.23 | 5,069.77 |
| | 945 | 15,812.97 | 16,272.27 | 16,129.27 | 16,479.85 | 64,694.36 |
| | 946 | 3,512.79 | 3,749.70 | 3,607.58 | 3,806.18 | 14,676.25 |
| | 947 | 1,390.10 | 1,431.27 | 1,322.38 | 1,537.79 | 5,681.54 |
| | 948 | 728.19 | 668.91 | 659.4 | 804.53 | 2,861.03 |
| | 949 | 4,749.65 | 4,929.17 | 4,658.09 | 5,030.87 | 19,367.78 |
| | 950 | 7,343.05 | 7,224.69 | 7,112.19 | 7,503 | 29,182.93 |
| | 951 | 3,836.41 | 4,018.19 | 3,870.05 | 3,868.50 | 15,593.15 |
| | 952 | 1,954.64 | 1,911.29 | 1,927.89 | 2,044.15 | 7,837.97 |
| | 953 | 4,988.01 | 4,896.66 | 4,795.65 | 4,973.85 | 19,654.17 |
| | 954 | 3,788.85 | 3,787.57 | 3,910.37 | 3,706.14 | 15,192.93 |
| | 955 | 812.13 | 917.55 | 872.84 | 937.29 | 3,539.81 |
| | 956 | 10,543.11 | 9,969.77 | 10,392.98 | 10,298.08 | 41,203.94 |
| | 957 | 2,330.69 | 2,259.98 | 2,288.37 | 2,483.16 | 9,362.20 |
| | 958 | 4,801.01 | 4,939.51 | 4,870.09 | 5,178.32 | 19,788.93 |
| | 959 | 4,385.25 | 4,154.12 | 4,210.39 | 4,367.89 | 17,117.65 |
| | 960 | 1,650.51 | 1,617.02 | 1,718.10 | 1,743.77 | 6,729.40 |
| | 961 | 528.79 | 534 | 603.12 | 622.15 | 2,288.06 |
| TOTAL | | 295,548.59 | 301,541.38 | 296,156.73 | 304,886.81 | 1,198,133.51 |

STATE: COLORADO

| ZIP CODE | | QUARTER 1 | QUARTER 2 | QUARTER 3 | QUARTER 4 | TOTAL GRAMS |
|---|---|---|---|---|---|---|
| | 800 | 11,696.71 | 11,873.03 | 12,101.73 | 12,318.67 | 47,990.14 |
| | 801 | 14,129.25 | 14,264.28 | 14,925.21 | 15,316.41 | 58,635.15 |
| | 802 | 19,488.96 | 19,977.75 | 20,030.15 | 20,805.07 | 80,301.93 |
| | 803 | 3,425.91 | 3,432.56 | 3,287.27 | 3,594.31 | 13,740.05 |
| | 804 | 2,992.20 | 2,898.28 | 3,075.77 | 3,195.80 | 12,162.05 |
| | 805 | 7,533.98 | 7,640.99 | 7,691.15 | 7,985.39 | 30,851.51 |
| | 806 | 2,383.33 | 2,337.20 | 2,339.16 | 2,445.47 | 9,505.16 |

| | | | | | | |
|---|---|---|---|---|---|---|
| | 807 | 311.91 | 304.22 | 351.84 | 357.16 | 1,325.13 |
| | 808 | 1,157.90 | 1,131.96 | 1,196.88 | 1,111.36 | 4,598.10 |
| | 809 | 7,517.63 | 7,595.61 | 7,720.62 | 7,692.14 | 30,526 |
| | 810 | 1,344.58 | 1,397.99 | 1,443.49 | 1,503.88 | 5,689.94 |
| | 811 | 221.92 | 209.73 | 271.26 | 245.93 | 948.84 |
| | 812 | 541.97 | 549.05 | 584.12 | 599.06 | 2,274.20 |
| | 813 | 659.07 | 580.92 | 707.89 | 670.75 | 2,618.63 |
| | 814 | 541.33 | 554.16 | 592 | 590.08 | 2,277.57 |
| | 815 | 1,676.64 | 1,700.33 | 1,645.41 | 1,712.63 | 6,735.01 |
| | 816 | 2,338.52 | 2,314.90 | 2,366.51 | 2,254.82 | 9,274.75 |
| TOTAL | | 77,961.81 | 78,762.96 | 80,330.46 | 82,398.93 | 319,454.16 |

STATE: CONNECTICUT

| ZIP CODE | | QUARTER 1 | QUARTER 2 | QUARTER 3 | QUARTER 4 | TOTAL GRAMS |
|---|---|---|---|---|---|---|
| | 60 | 8,933.06 | 9,171.42 | 9,042.11 | 9,352.24 | 36,498.83 |
| | 61 | 3,307 | 3,319.39 | 3,222.41 | 3,337.42 | 13,186.22 |
| | 62 | 2,287.48 | 2,162.52 | 2,143.04 | 2,278.32 | 8,871.36 |
| | 63 | 4,877.18 | 5,125.23 | 5,472.97 | 5,471.01 | 20,946.39 |
| | 64 | 12,158.63 | 12,459.19 | 12,371.59 | 12,745.33 | 49,734.74 |
| | 65 | 3,984.48 | 4,079.48 | 4,003.77 | 4,186.85 | 16,254.58 |
| | 66 | 1,363.78 | 1,439.68 | 1,463.26 | 1,523.12 | 5,789.84 |
| | 67 | 3,421.25 | 3,383.24 | 3,466.52 | 3,534.35 | 13,805.36 |
| | 68 | 9,316.71 | 9,788.80 | 9,843.18 | 10,230.35 | 39,179.04 |
| | 69 | 1,642.23 | 1,833.29 | 1,777.89 | 1,752.73 | 7,006.14 |
| TOTAL | | 51,291.80 | 52,762.24 | 52,806.74 | 54,411.72 | 211,272.50 |

STATE: DELAWARE

| ZIP CODE | | QUARTER 1 | QUARTER 2 | QUARTER 3 | QUARTER 4 | TOTAL GRAMS |
|---|---|---|---|---|---|---|
| | 197 | 9,287.84 | 9,074.47 | 8,805.55 | 9,323.69 | 36,491.55 |
| | 198 | 6,799.40 | 6,770.01 | 6,609.31 | 7,015.87 | 27,194.59 |
| | 199 | 5,547.29 | 5,657.26 | 5,785.12 | 5,526.09 | 22,515.76 |
| TOTAL | | 21,634.53 | 21,501.74 | 21,199.98 | 21,865.65 | 86,201.90 |

DATE RANGE: 01/01/2017 TO 12/31/2017 — ARCOS 3 - REPORT 1 — Run Date: 07/03/2018

RETAIL DRUG DISTRIBUTION BY ZIP CODE WITHIN STATE BY GRAMS WT

STATE: DISTRICT OF COLUMBIA

| ZIP CODE | QUARTER 1 | QUARTER 2 | QUARTER 3 | QUARTER 4 | TOTAL GRAMS |
|---|---|---|---|---|---|
| 200 | 10,996.52 | 11,145.90 | 10,861.12 | 11,405.14 | 44,408.68 |
| 203 | 125.13 | 106.36 | 114.8 | 184.37 | 530.66 |
| 204 | 444.22 | 388.26 | 527.48 | 366.3 | 1,726.26 |
| 205 | 0 | 0 | 1.25 | 0 | 1.25 |
| TOTAL | 11,565.87 | 11,640.52 | 11,504.65 | 11,955.81 | 46,666.85 |

STATE: FLORIDA

| ZIP CODE | QUARTER 1 | QUARTER 2 | QUARTER 3 | QUARTER 4 | TOTAL GRAMS |
|---|---|---|---|---|---|
| 320 | 9,146.12 | 9,207.99 | 9,041.66 | 9,609.20 | 37,004.97 |
| 321 | 7,373.40 | 7,526.75 | 7,433.44 | 7,992.75 | 30,326.34 |
| 322 | 21,971.65 | 22,084.34 | 21,925.61 | 22,612.64 | 88,594.24 |
| 323 | 9,889.21 | 9,661.28 | 9,289.17 | 9,882.67 | 38,722.33 |
| 324 | 8,460.43 | 8,427.05 | 8,049.03 | 8,703.87 | 33,640.38 |
| 325 | 18,579.24 | 18,584.37 | 19,630.60 | 18,646.41 | 75,440.62 |
| 326 | 5,964 | 5,814.84 | 5,722.68 | 6,086.29 | 23,587.81 |
| 327 | 16,761.71 | 17,214.41 | 16,724.06 | 18,667.88 | 69,368.06 |
| 328 | 13,391.49 | 13,285.12 | 13,026 | 14,154.71 | 53,857.32 |
| 329 | 12,842.22 | 12,697.29 | 12,777.45 | 13,707.01 | 52,023.97 |
| 330 | 14,727.70 | 14,941.64 | 13,669.64 | 15,286.96 | 58,625.94 |
| 331 | 16,637.04 | 16,744.48 | 15,447.49 | 17,890.81 | 66,719.82 |
| 333 | 11,919.82 | 12,383.42 | 11,858.74 | 13,286.79 | 49,448.77 |
| 334 | 24,406.03 | 23,924.55 | 23,260.12 | 25,221.13 | 96,811.83 |
| 335 | 10,112.16 | 10,176.29 | 10,422.99 | 10,955.02 | 41,666.46 |
| 336 | 13,939.13 | 14,414.81 | 14,191.35 | 15,338.86 | 57,884.15 |
| 337 | 13,469.21 | 13,587.98 | 13,954.16 | 14,728.35 | 55,739.70 |
| 338 | 7,175.21 | 6,998.49 | 6,800.10 | 7,472.60 | 28,446.40 |
| 339 | 12,465.27 | 12,580.42 | 11,567.97 | 13,482.35 | 50,096.01 |
| 341 | 6,902.51 | 6,763.77 | 6,346.86 | 7,617.42 | 27,630.56 |
| 342 | 13,439.02 | 13,363.75 | 13,146.62 | 14,212.58 | 54,161.97 |
| 344 | 3,968.04 | 3,990.32 | 3,632.20 | 4,283.13 | 15,873.69 |
| 346 | 8,663.74 | 8,873.38 | 8,915.93 | 9,606.74 | 36,059.79 |
| 347 | 7,964.67 | 7,923.20 | 7,507.01 | 8,313.70 | 31,708.58 |
| 349 | 6,725.69 | 6,835.63 | 6,931.43 | 7,272.84 | 27,765.59 |

| | | | | | |
|---|---|---|---|---|---|
| TOTAL | 296,894.71 | 298,005.57 | 291,272.31 | 315,032.71 | 1,201,205.30 |

STATE: GEORGIA

| ZIP CODE | QUARTER 1 | QUARTER 2 | QUARTER 3 | QUARTER 4 | TOTAL GRAMS |
|---|---|---|---|---|---|
| 300 | 37,676.98 | 37,377.16 | 37,809.55 | 38,916.56 | 151,780.25 |
| 301 | 21,547.12 | 21,826.22 | 21,991.20 | 22,618.70 | 87,983.24 |
| 302 | 13,337.13 | 13,286.42 | 13,554.99 | 14,429.06 | 54,607.60 |
| 303 | 22,373.26 | 22,698.53 | 22,705.62 | 23,290.28 | 91,067.69 |
| 304 | 6,930.35 | 6,837.09 | 6,915.72 | 7,049.87 | 27,733.03 |
| 305 | 11,686.40 | 11,679.45 | 11,764.08 | 12,444.59 | 47,574.52 |
| 306 | 8,229.83 | 7,813.74 | 7,927.69 | 8,397.37 | 32,368.63 |
| 307 | 6,685.50 | 6,600.43 | 6,483.94 | 6,687.35 | 26,457.22 |
| 308 | 3,526.40 | 3,380.23 | 3,524.14 | 3,604.96 | 14,035.73 |
| 309 | 5,474.83 | 5,373.12 | 5,549.52 | 5,579.15 | 21,976.62 |
| 310 | 7,300.20 | 6,997.06 | 6,842.57 | 7,564.48 | 28,704.31 |
| 312 | 2,805.14 | 2,791.83 | 2,692.69 | 2,849.14 | 11,138.80 |
| 313 | 6,238.15 | 5,973.83 | 6,225.68 | 6,040.99 | 24,478.65 |
| 314 | 6,046.71 | 6,233.43 | 5,904.11 | 6,292.20 | 24,476.45 |
| 315 | 7,885.51 | 7,701.29 | 7,967.72 | 8,233.54 | 31,788.06 |
| 316 | 3,508.77 | 3,226.40 | 3,135.50 | 3,352.62 | 13,223.29 |
| 317 | 6,614.76 | 6,332.81 | 6,380.13 | 6,657.74 | 25,985.44 |
| 318 | 474.43 | 427.07 | 455.52 | 496.2 | 1,853.22 |
| 319 | 6,616.44 | 6,793.61 | 6,993.49 | 6,727.59 | 27,131.13 |
| 398 | 1,526.83 | 1,363 | 1,503.87 | 1,506.14 | 5,899.84 |
| TOTAL | 186,484.74 | 184,712.72 | 186,327.73 | 192,738.53 | 750,263.72 |

STATE: GUAM

| ZIP CODE | QUARTER 1 | QUARTER 2 | QUARTER 3 | QUARTER 4 | TOTAL GRAMS |
|---|---|---|---|---|---|
| 969 | 411.77 | 559.5 | 574.21 | 358.63 | 1,904.11 |
| TOTAL | 411.77 | 559.5 | 574.21 | 358.63 | 1,904.11 |

STATE: HAWAII

| ZIP CODE | QUARTER 1 | QUARTER 2 | QUARTER 3 | QUARTER 4 | TOTAL GRAMS |
|---|---|---|---|---|---|
| 967 | 4,677.70 | 4,633.36 | 4,693.58 | 4,785.69 | 18,790.33 |

| | | | | | | |
|---|---|---|---|---|---|---|
| | 968 | 4,891.50 | 5,201.06 | 5,246.38 | 4,790.81 | 20,129.75 |
| TOTAL | | 9,569.20 | 9,834.42 | 9,939.96 | 9,576.50 | 38,920.08 |

STATE: IDAHO

| ZIP CODE | | QUARTER 1 | QUARTER 2 | QUARTER 3 | QUARTER 4 | TOTAL GRAMS |
|---|---|---|---|---|---|---|
| | 832 | 1,936.65 | 2,020.93 | 2,030.11 | 2,205.96 | 8,193.65 |
| | 833 | 1,770.90 | 1,849.49 | 1,790.76 | 1,917.01 | 7,328.16 |
| | 834 | 4,511.05 | 4,455.38 | 4,542.72 | 4,865.26 | 18,374.41 |
| | 835 | 363.39 | 334.26 | 410.27 | 414.16 | 1,522.08 |
| | 836 | 7,113.85 | 7,341.79 | 7,319.31 | 7,549.85 | 29,324.80 |
| | 837 | 6,912.39 | 7,182.50 | 7,174.81 | 7,482.23 | 28,751.93 |
| | 838 | 2,000.15 | 2,056.63 | 2,067.54 | 2,145.21 | 8,269.53 |
| TOTAL | | 24,608.38 | 25,240.98 | 25,335.52 | 26,579.68 | 101,764.56 |

STATE: ILLINOIS

| ZIP CODE | | QUARTER 1 | QUARTER 2 | QUARTER 3 | QUARTER 4 | TOTAL GRAMS |
|---|---|---|---|---|---|---|
| | 600 | 32,258.21 | 32,583.18 | 32,973.54 | 33,987.59 | 131,802.52 |
| | 601 | 22,148.68 | 22,334.46 | 22,703.45 | 23,420.12 | 90,606.71 |
| | 602 | 1,470.60 | 1,483.08 | 1,394.87 | 1,495.24 | 5,843.79 |
| | 603 | 1,262.07 | 1,200.83 | 1,194.54 | 1,371.88 | 5,029.32 |
| | 604 | 18,245.11 | 17,922.09 | 18,210.04 | 18,869.25 | 73,246.49 |
| | 605 | 17,328.76 | 17,421.46 | 17,470.52 | 18,237.56 | 70,458.30 |
| | 606 | 36,207.78 | 36,728.69 | 36,788.13 | 38,577.71 | 148,302.31 |
| | 607 | 2,114.51 | 2,201.15 | 2,182.65 | 2,338.65 | 8,836.96 |
| | 608 | 618.01 | 648.01 | 707.04 | 687.3 | 2,660.36 |
| | 609 | 1,987.42 | 1,969.36 | 2,027.29 | 2,158.88 | 8,142.95 |
| | 610 | 3,460.11 | 3,426.85 | 3,553.35 | 3,525.47 | 13,965.78 |
| | 611 | 3,749.15 | 3,811.54 | 3,928.17 | 3,952.67 | 15,441.53 |
| | 612 | 3,160.71 | 3,104.18 | 3,048.40 | 3,088.29 | 12,401.58 |
| | 613 | 2,377.45 | 2,454.66 | 2,422.23 | 2,556.25 | 9,810.59 |
| | 614 | 1,968.80 | 1,927.23 | 1,925.05 | 1,954.03 | 7,775.11 |
| | 615 | 3,190.98 | 3,162.57 | 3,184.67 | 3,310.95 | 12,849.17 |
| | 616 | 4,008.57 | 3,971.24 | 4,095.63 | 4,225.31 | 16,300.75 |
| | 617 | 3,810.21 | 3,469.35 | 3,572.56 | 3,748.50 | 14,600.62 |
| | 618 | 3,926.67 | 3,904.98 | 3,668.10 | 3,922.61 | 15,422.36 |

| | | | | | |
|---|---|---|---|---|---|
| 619 | 1,314.31 | 1,222.32 | 1,127.18 | 1,304.52 | 4,968.33 |
| 620 | 3,596.67 | 3,534.07 | 3,495.65 | 3,728.19 | 14,354.58 |
| 622 | 5,212.19 | 5,019.72 | 5,295.60 | 5,495.74 | 21,023.25 |
| 623 | 1,931.39 | 1,934.56 | 1,867.40 | 1,993.46 | 7,726.81 |
| 624 | 1,256.72 | 1,183.47 | 1,176.22 | 1,180.58 | 4,796.99 |
| 625 | 1,802.72 | 1,684.66 | 1,605.38 | 1,721.83 | 6,814.59 |
| 626 | 1,533.48 | 1,512.41 | 1,490.88 | 1,636.76 | 6,173.53 |
| 627 | 2,433.51 | 2,394.49 | 2,326.37 | 2,470.36 | 9,624.73 |
| 628 | 2,382.42 | 2,337.55 | 2,368.47 | 2,402.20 | 9,490.64 |
| 629 | 3,971.66 | 3,830.07 | 3,855.80 | 3,838.99 | 15,496.52 |
| TOTAL | 188,728.87 | 188,378.23 | 189,659.18 | 197,200.89 | 763,967.17 |

STATE: INDIANA

| ZIP CODE | QUARTER 1 | QUARTER 2 | QUARTER 3 | QUARTER 4 | TOTAL GRAMS |
|---|---|---|---|---|---|
| 460 | 14,082.04 | 14,427.79 | 14,621.18 | 15,147.16 | 58,278.17 |
| 461 | 8,537.15 | 8,637.74 | 8,835.51 | 9,152.99 | 35,163.39 |
| 462 | 15,913.92 | 16,007.20 | 16,383.52 | 16,792.26 | 65,096.90 |
| 463 | 11,699.54 | 11,559.31 | 12,066.72 | 12,367.67 | 47,693.24 |
| 464 | 1,287.23 | 1,217.21 | 1,179.85 | 1,182.23 | 4,866.52 |
| 465 | 16,953.33 | 17,451.93 | 17,211.78 | 17,726.66 | 69,343.70 |
| 466 | 4,512.19 | 4,660 | 4,598.08 | 4,851.14 | 18,621.41 |
| 467 | 3,454.65 | 3,314.09 | 3,289.37 | 3,471.43 | 13,529.54 |
| 468 | 5,698.03 | 5,598.25 | 5,614.15 | 5,962.56 | 22,872.99 |
| 469 | 5,346.98 | 5,294.25 | 5,416.86 | 5,570.08 | 21,628.17 |
| 470 | 1,565.39 | 1,532.66 | 1,500.86 | 1,660.66 | 6,259.57 |
| 471 | 7,675.23 | 8,608.07 | 8,248.72 | 7,710.49 | 32,242.51 |
| 472 | 2,185.13 | 2,225.92 | 2,365.60 | 2,441.40 | 9,218.05 |
| 473 | 5,005.73 | 4,862.14 | 5,062.47 | 5,002.41 | 19,932.75 |
| 474 | 4,052.96 | 3,972.20 | 3,838.75 | 4,136.40 | 16,000.31 |
| 475 | 2,231.19 | 2,174.47 | 2,262.22 | 2,371.55 | 9,039.43 |
| 476 | 2,550.80 | 2,548 | 2,520.70 | 2,739.84 | 10,359.34 |
| 477 | 5,789.77 | 5,555.45 | 5,876.81 | 5,816.12 | 23,038.15 |
| 478 | 2,258.54 | 2,249.28 | 2,290.69 | 2,432.91 | 9,231.42 |
| 479 | 6,918.42 | 6,840.65 | 6,770.16 | 7,045.86 | 27,575.09 |
| TOTAL | 127,718.22 | 128,736.61 | 129,954 | 133,581.82 | 519,990.65 |

| STATE: | IOWA | | | | |
|---|---|---|---|---|---|
| ZIP CODE | QUARTER 1 | QUARTER 2 | QUARTER 3 | QUARTER 4 | TOTAL GRAMS |
| 500 | 4,007.25 | 3,989.62 | 4,087.29 | 4,321.12 | 16,405.28 |
| 501 | 1,758.57 | 1,643.92 | 1,837.44 | 1,917.16 | 7,157.09 |
| 502 | 2,882.65 | 2,972.32 | 2,942.58 | 3,054.74 | 11,852.29 |
| 503 | 5,567.10 | 5,762.03 | 5,860.45 | 6,020.78 | 23,210.36 |
| 504 | 816.87 | 850.76 | 811.09 | 837.64 | 3,316.36 |
| 505 | 1,125.31 | 1,149.99 | 1,138.33 | 1,218.90 | 4,632.53 |
| 506 | 1,471.68 | 1,466.43 | 1,379.27 | 1,636.21 | 5,953.59 |
| 507 | 1,124.64 | 1,208.32 | 1,183.53 | 1,233.66 | 4,750.15 |
| 508 | 168.56 | 173.34 | 171.79 | 148.28 | 661.97 |
| 510 | 923.41 | 950.13 | 947.23 | 961.13 | 3,781.90 |
| 511 | 2,530.50 | 2,644.26 | 2,688.89 | 2,698.10 | 10,561.75 |
| 512 | 433.75 | 385.45 | 406.02 | 472.05 | 1,697.27 |
| 513 | 681.41 | 713.7 | 726.62 | 720.64 | 2,842.37 |
| 514 | 410.67 | 381.67 | 393.64 | 412.03 | 1,598.01 |
| 515 | 1,992.99 | 2,066.72 | 2,092.30 | 2,098.33 | 8,250.34 |
| 516 | 190.95 | 184.42 | 187.6 | 192.62 | 755.59 |
| 520 | 3,062.20 | 3,144.39 | 3,217.16 | 3,377.22 | 12,800.97 |
| 521 | 482.58 | 426.14 | 471.72 | 522.79 | 1,903.23 |
| 522 | 4,806.13 | 4,885.96 | 4,786.67 | 4,995.88 | 19,474.64 |
| 523 | 2,248.97 | 2,263.87 | 2,342.46 | 2,567.90 | 9,423.20 |
| 524 | 3,762.67 | 3,876.31 | 3,941.52 | 4,263.08 | 15,843.58 |
| 525 | 928.9 | 904.04 | 925.1 | 976.7 | 3,734.74 |
| 526 | 1,574.98 | 1,660.19 | 1,609.89 | 1,595.20 | 6,440.26 |
| 527 | 3,153.56 | 3,122.25 | 3,195.28 | 3,273.03 | 12,744.12 |
| 528 | 2,329.13 | 2,568.24 | 2,473.54 | 2,607.58 | 9,978.49 |
| TOTAL | 48,435.43 | 49,394.47 | 49,817.41 | 52,122.77 | 199,770.08 |

| STATE: | KANSAS | | | | |
|---|---|---|---|---|---|
| ZIP CODE | QUARTER 1 | QUARTER 2 | QUARTER 3 | QUARTER 4 | TOTAL GRAMS |
| 660 | 13,760.24 | 14,306.76 | 14,433.43 | 14,969.69 | 57,470.12 |
| 661 | 1,682.82 | 1,874.74 | 1,751.35 | 1,872.93 | 7,181.84 |
| 662 | 17,745.17 | 18,500.59 | 18,647.34 | 19,424.40 | 74,317.50 |
| 664 | 966.74 | 924.22 | 1,103.54 | 785.16 | 3,779.66 |

| | | | | | | |
|---|---|---|---|---|---|---|
| | 665 | 1,535.75 | 1,433.16 | 1,461.22 | 1,609.66 | 6,039.79 |
| | 666 | 4,714.97 | 4,994.61 | 5,006.25 | 5,122.81 | 19,838.64 |
| | 667 | 1,709.61 | 1,672.62 | 1,703.08 | 1,704.10 | 6,789.41 |
| | 668 | 645.09 | 618.96 | 619.26 | 607.98 | 2,491.29 |
| | 669 | 104.15 | 108.91 | 89.56 | 116.87 | 419.49 |
| | 670 | 2,389.58 | 2,400.78 | 2,470.36 | 2,478.99 | 9,739.71 |
| | 671 | 910.24 | 929.37 | 976.74 | 1,086.33 | 3,902.68 |
| | 672 | 7,395.99 | 7,313.87 | 7,454.12 | 7,592.29 | 29,756.27 |
| | 673 | 813.9 | 761.67 | 794.99 | 782.72 | 3,153.28 |
| | 674 | 1,313.93 | 1,235.99 | 1,250.87 | 1,257.59 | 5,058.38 |
| | 675 | 1,094.34 | 1,010 | 1,035.38 | 1,182.57 | 4,322.29 |
| | 676 | 608.71 | 602.23 | 665.44 | 730.47 | 2,606.85 |
| | 677 | 351.49 | 337.56 | 333.83 | 356.52 | 1,379.40 |
| | 678 | 684.29 | 644.57 | 662.79 | 686.34 | 2,677.99 |
| | 679 | 213.79 | 216.18 | 200.61 | 203.54 | 834.12 |
| TOTAL | | 58,640.80 | 59,886.79 | 60,660.16 | 62,570.96 | 241,758.71 |
| STATE: | | KENTUCKY | | | | |
| ZIP CODE | | QUARTER 1 | QUARTER 2 | QUARTER 3 | QUARTER 4 | TOTAL GRAMS |
| | 400 | 3,799.62 | 3,561.18 | 3,803.46 | 3,845.21 | 15,009.47 |
| | 401 | 1,626.35 | 1,684.28 | 1,861.63 | 1,381.23 | 6,553.49 |
| | 402 | 16,526.98 | 16,030.78 | 16,159.74 | 16,825.22 | 65,542.72 |
| | 403 | 4,014.41 | 3,751.53 | 3,913.64 | 4,033.78 | 15,713.36 |
| | 404 | 1,626.28 | 1,650.46 | 1,606.80 | 1,828.94 | 6,712.48 |
| | 405 | 6,901.37 | 6,739.77 | 6,768.40 | 7,247.03 | 27,656.57 |
| | 406 | 733.18 | 750.61 | 712.82 | 765.58 | 2,962.19 |
| | 407 | 548.77 | 519.13 | 515.76 | 582.06 | 2,165.72 |
| | 408 | 150.05 | 185.11 | 160.31 | 173.37 | 668.84 |
| | 409 | 174.23 | 175.91 | 202.89 | 226.33 | 779.36 |
| | 410 | 9,460.87 | 9,428.55 | 9,257.37 | 10,049.20 | 38,195.99 |
| | 411 | 1,351.43 | 1,344.22 | 1,358.49 | 1,396.30 | 5,450.44 |
| | 412 | 383.97 | 327.08 | 303 | 350.08 | 1,364.13 |
| | 413 | 143.76 | 114.73 | 106.38 | 128.02 | 492.89 |
| | 414 | 146.43 | 138.74 | 119.86 | 140.52 | 545.55 |
| | 415 | 458.58 | 511.43 | 520.73 | 504.19 | 1,994.93 |
| | 416 | 333.06 | 309.02 | 333.96 | 337.28 | 1,313.32 |

| | | | | | | |
|---|---|---|---|---|---|---|
| | 417 | 368.09 | 319.9 | 349.19 | 369.16 | 1,406.34 |
| | 418 | 120.15 | 88.01 | 109.48 | 89.96 | 407.6 |
| | 420 | 4,204.70 | 4,332.25 | 4,347.94 | 4,491.43 | 17,376.32 |
| | 421 | 2,877.44 | 2,795.05 | 2,861.53 | 2,993.76 | 11,527.78 |
| | 422 | 2,169.96 | 2,073.88 | 2,316.23 | 2,102.13 | 8,662.20 |
| | 423 | 3,558.06 | 3,451.01 | 3,361.22 | 3,513.17 | 13,883.46 |
| | 424 | 3,457.80 | 3,433.88 | 3,319.88 | 3,471.92 | 13,683.48 |
| | 425 | 399.28 | 398.05 | 349.51 | 432.53 | 1,579.37 |
| | 426 | 222.87 | 191.4 | 209.92 | 211.68 | 835.87 |
| | 427 | 2,357.79 | 2,249.94 | 2,352.46 | 2,503.57 | 9,463.76 |
| TOTAL | | 68,115.48 | 66,555.90 | 67,282.60 | 69,993.65 | 271,947.63 |

STATE: LOUISIANA

| ZIP CODE | | QUARTER 1 | QUARTER 2 | QUARTER 3 | QUARTER 4 | TOTAL GRAMS |
|---|---|---|---|---|---|---|
| | 700 | 19,234.03 | 18,825.21 | 19,172.10 | 20,164.14 | 77,395.48 |
| | 701 | 9,828.23 | 9,744.46 | 9,810.42 | 10,601.66 | 39,984.77 |
| | 703 | 6,074.21 | 5,728.41 | 5,752.10 | 6,197.80 | 23,752.52 |
| | 704 | 18,262.36 | 18,193.06 | 18,838.30 | 19,606.50 | 74,900.22 |
| | 705 | 31,529.78 | 30,602.94 | 31,156.61 | 33,096.45 | 126,385.78 |
| | 706 | 10,912.52 | 10,783.84 | 10,932.92 | 11,738.63 | 44,367.91 |
| | 707 | 18,031.06 | 18,298.23 | 18,444.97 | 19,670.39 | 74,444.65 |
| | 708 | 15,974.03 | 15,780.14 | 15,668.68 | 16,706.46 | 64,129.31 |
| | 710 | 1,865.68 | 1,822.96 | 1,949.23 | 1,989.83 | 7,627.70 |
| | 711 | 7,227.87 | 7,146.95 | 7,343.48 | 7,418.68 | 29,136.98 |
| | 712 | 6,671.04 | 6,456.30 | 6,577.05 | 6,743.51 | 26,447.90 |
| | 713 | 5,312.33 | 5,305.65 | 5,246.79 | 5,668.09 | 21,532.86 |
| | 714 | 1,916.92 | 1,714.14 | 1,927.70 | 1,943.64 | 7,502.40 |
| TOTAL | | 152,840.06 | 150,402.29 | 152,820.35 | 161,545.78 | 617,608.48 |

STATE: MAINE

| ZIP CODE | | QUARTER 1 | QUARTER 2 | QUARTER 3 | QUARTER 4 | TOTAL GRAMS |
|---|---|---|---|---|---|---|
| | 39 | 811.63 | 834.75 | 912.81 | 901.4 | 3,460.59 |
| | 40 | 7,349.93 | 7,760.05 | 8,147.28 | 7,889.66 | 31,146.92 |
| | 41 | 3,797.64 | 4,161.47 | 4,221.02 | 4,061.28 | 16,241.41 |
| | 42 | 1,817.12 | 1,950.28 | 1,916.86 | 2,007.25 | 7,691.51 |

| | | | | | | |
|---|---|---|---|---|---|---|
| | 43 | 1,365.26 | 1,391.25 | 1,505.57 | 1,437.36 | 5,699.44 |
| | 44 | 2,155.53 | 2,146.54 | 2,163.66 | 2,264 | 8,729.73 |
| | 45 | 656.27 | 771.75 | 790.47 | 758.08 | 2,976.57 |
| | 46 | 679.18 | 744.43 | 820.01 | 746.88 | 2,990.50 |
| | 47 | 504.94 | 553.17 | 501.82 | 459.47 | 2,019.40 |
| | 48 | 508.01 | 601.72 | 581.36 | 568.4 | 2,259.49 |
| | 49 | 1,774.41 | 1,861.93 | 1,893.68 | 1,926.70 | 7,456.72 |
| TOTAL | | 21,419.92 | 22,777.34 | 23,454.54 | 23,020.48 | 90,672.28 |

STATE: MARYLAND

| ZIP CODE | | QUARTER 1 | QUARTER 2 | QUARTER 3 | QUARTER 4 | TOTAL GRAMS |
|---|---|---|---|---|---|---|
| | 206 | 4,305.63 | 4,261.36 | 4,411.87 | 4,322.56 | 17,301.42 |
| | 207 | 6,198.69 | 6,038.62 | 5,998.51 | 6,036.84 | 24,272.66 |
| | 208 | 14,360.66 | 14,503.05 | 14,634.05 | 14,455.43 | 57,953.19 |
| | 209 | 2,611.66 | 2,646.45 | 2,568.07 | 2,753.44 | 10,579.62 |
| | 210 | 16,700.18 | 17,201.82 | 17,225.21 | 17,953.79 | 69,081 |
| | 211 | 7,843.55 | 8,070.78 | 7,672.37 | 8,160.60 | 31,747.30 |
| | 212 | 16,051.43 | 16,095.77 | 16,134.42 | 16,944.78 | 65,226.40 |
| | 214 | 2,555.94 | 2,641.90 | 2,619.49 | 2,708.32 | 10,525.65 |
| | 215 | 1,748.91 | 1,843.08 | 1,862.29 | 1,933.06 | 7,387.34 |
| | 216 | 2,973.73 | 3,027.90 | 3,066.30 | 3,137.70 | 12,205.63 |
| | 217 | 10,277.06 | 10,213.21 | 10,200.74 | 10,770.95 | 41,461.96 |
| | 218 | 2,013.85 | 2,042.68 | 2,023.88 | 2,076.93 | 8,157.34 |
| | 219 | 2,539.02 | 2,521.03 | 2,623.55 | 2,680.27 | 10,363.87 |
| TOTAL | | 90,180.31 | 91,107.65 | 91,040.75 | 93,934.67 | 366,263.38 |

STATE: MASSACHUSETTS

| ZIP CODE | | QUARTER 1 | QUARTER 2 | QUARTER 3 | QUARTER 4 | TOTAL GRAMS |
|---|---|---|---|---|---|---|
| | 10 | 8,821.20 | 9,201.54 | 8,646.89 | 9,207 | 35,876.63 |
| | 11 | 2,263.63 | 2,164.77 | 2,071.93 | 2,264.63 | 8,764.96 |
| | 12 | 2,319.16 | 2,295.96 | 2,212.25 | 2,417.26 | 9,244.63 |
| | 13 | 2,465.56 | 2,520.33 | 2,559.08 | 2,668.17 | 10,213.14 |
| | 14 | 5,077.38 | 5,268.81 | 5,145.08 | 5,385.99 | 20,877.26 |
| | 15 | 7,214.75 | 7,316.36 | 6,879.90 | 7,332.50 | 28,743.51 |
| | 16 | 5,800.43 | 5,978.76 | 5,856.56 | 5,675.37 | 23,311.12 |

| | | | | | | |
|---|---|---|---|---|---|---|
| | 17 | 11,031.49 | 11,536.75 | 11,243.66 | 11,617 | 45,428.90 |
| | 18 | 19,618.17 | 20,333.38 | 20,003.70 | 20,565.59 | 80,520.84 |
| | 19 | 17,907.80 | 17,710.95 | 17,926.62 | 18,192.87 | 71,738.24 |
| | 20 | 13,411.10 | 13,807.91 | 13,524.27 | 14,111.91 | 54,855.19 |
| | 21 | 33,206.60 | 34,157.36 | 32,873.18 | 34,291.66 | 134,528.80 |
| | 22 | 1,714.26 | 1,685.24 | 1,492.41 | 1,582.23 | 6,474.14 |
| | 23 | 13,004.34 | 13,302.17 | 13,264.71 | 13,563.67 | 53,134.89 |
| | 24 | 11,168.54 | 11,327.45 | 10,511.87 | 11,251.79 | 44,259.65 |
| | 25 | 3,843.66 | 4,161.42 | 4,162.77 | 4,027.41 | 16,195.26 |
| | 26 | 4,546.35 | 4,979.82 | 5,299.21 | 4,916.52 | 19,741.90 |
| | 27 | 11,580.96 | 11,969.58 | 12,017.31 | 12,382.07 | 47,949.92 |
| TOTAL | | 174,995.38 | 179,718.56 | 175,691.40 | 181,453.64 | 711,858.98 |

STATE: MICHIGAN

| ZIP CODE | | QUARTER 1 | QUARTER 2 | QUARTER 3 | QUARTER 4 | TOTAL GRAMS |
|---|---|---|---|---|---|---|
| | 480 | 38,656.60 | 38,648.05 | 39,024.60 | 39,455.22 | 155,784.47 |
| | 481 | 39,332.73 | 39,905.80 | 39,748.89 | 41,263.84 | 160,251.26 |
| | 482 | 6,850.44 | 7,019.11 | 6,703.56 | 7,003.62 | 27,576.73 |
| | 483 | 32,245.21 | 32,071.50 | 32,229.85 | 32,989.82 | 129,536.38 |
| | 484 | 8,367.48 | 8,316.75 | 8,423.08 | 8,726.81 | 33,834.12 |
| | 485 | 2,865.71 | 2,810.55 | 2,800.97 | 2,753.72 | 11,230.95 |
| | 486 | 5,092.86 | 5,052.84 | 4,979.66 | 5,071.62 | 20,196.98 |
| | 487 | 2,974.80 | 3,087.54 | 2,912.99 | 2,944.34 | 11,919.67 |
| | 488 | 14,864.89 | 14,396.30 | 14,521.59 | 15,414.39 | 59,197.17 |
| | 489 | 4,268.94 | 4,142.66 | 4,247.53 | 4,255.99 | 16,915.12 |
| | 490 | 18,047.81 | 17,831.34 | 17,620.62 | 17,845.43 | 71,345.20 |
| | 491 | 2,198.71 | 2,237.25 | 2,193.20 | 2,214.41 | 8,843.57 |
| | 492 | 4,228.85 | 4,093.76 | 4,238.92 | 4,228.24 | 16,789.77 |
| | 493 | 7,137.22 | 6,984.17 | 6,939.20 | 7,169.91 | 28,230.50 |
| | 494 | 19,061.59 | 19,115.64 | 19,176.82 | 19,731.66 | 77,085.71 |
| | 495 | 19,011.11 | 19,421.45 | 19,183.13 | 19,649.02 | 77,264.71 |
| | 496 | 5,556.97 | 5,516.70 | 5,745.99 | 5,672.27 | 22,491.93 |
| | 497 | 3,247.40 | 3,415.96 | 3,606.06 | 3,490.72 | 13,760.14 |
| | 498 | 3,145.86 | 3,282.97 | 3,339.05 | 3,341.59 | 13,109.47 |
| | 499 | 1,732.95 | 1,668.29 | 1,584.75 | 1,591.47 | 6,577.46 |
| TOTAL | | 238,888.13 | 239,018.63 | 239,220.46 | 244,814.09 | 961,941.31 |

STATE: MINNESOTA

| ZIP CODE | QUARTER 1 | QUARTER 2 | QUARTER 3 | QUARTER 4 | TOTAL GRAMS |
|---|---|---|---|---|---|
| 550 | 12,269.97 | 12,406.39 | 12,449.60 | 12,896 | 50,021.96 |
| 551 | 17,582.18 | 17,522.84 | 17,539.11 | 18,256.55 | 70,900.68 |
| 553 | 22,333.84 | 22,337.21 | 22,514.02 | 23,228.86 | 90,413.93 |
| 554 | 25,523.27 | 26,036.67 | 25,618.70 | 26,664.52 | 103,843.16 |
| 556 | 115.42 | 102.78 | 121.52 | 107.42 | 447.14 |
| 557 | 3,059.16 | 3,086.96 | 3,034.53 | 3,101.94 | 12,282.59 |
| 558 | 2,063.74 | 2,014.90 | 1,931.05 | 2,140.05 | 8,149.74 |
| 559 | 5,234.76 | 5,273.28 | 5,150.75 | 5,327.15 | 20,985.94 |
| 560 | 5,190.94 | 5,166.22 | 5,069.61 | 5,297.46 | 20,724.23 |
| 561 | 776.7 | 835.56 | 735.28 | 832.42 | 3,179.96 |
| 562 | 1,672.96 | 1,590.17 | 1,553.49 | 1,715.47 | 6,532.09 |
| 563 | 4,197.23 | 4,174.58 | 4,154.96 | 4,346.07 | 16,872.84 |
| 564 | 2,198.72 | 2,168.91 | 2,287.62 | 2,255.63 | 8,910.88 |
| 565 | 2,553.22 | 2,587.72 | 2,638.54 | 2,808.72 | 10,588.20 |
| 566 | 1,209.33 | 1,237.78 | 1,242.86 | 1,299.88 | 4,989.85 |
| 567 | 658.95 | 677.45 | 701.69 | 770.25 | 2,808.34 |
| TOTAL | 106,640.39 | 107,219.42 | 106,743.33 | 111,048.39 | 431,651.53 |

STATE: MISSISSIPPI

| ZIP CODE | QUARTER 1 | QUARTER 2 | QUARTER 3 | QUARTER 4 | TOTAL GRAMS |
|---|---|---|---|---|---|
| 386 | 9,325.98 | 8,954.60 | 9,062.62 | 9,487.48 | 36,830.68 |
| 387 | 1,566.22 | 1,447.84 | 1,525.47 | 1,520.57 | 6,060.10 |
| 388 | 5,068.11 | 4,916.24 | 4,730.20 | 4,970.83 | 19,685.38 |
| 389 | 1,515.18 | 1,500.01 | 1,466.44 | 1,491.24 | 5,972.87 |
| 390 | 5,218.56 | 4,994.31 | 5,019.33 | 5,063.92 | 20,296.12 |
| 391 | 6,290.53 | 6,140.98 | 6,128.04 | 6,366.66 | 24,926.21 |
| 392 | 5,283.71 | 5,074.05 | 5,079.07 | 5,266.11 | 20,702.94 |
| 393 | 4,176.49 | 4,104.13 | 3,859.95 | 4,045.21 | 16,185.78 |
| 394 | 8,916.65 | 8,647.63 | 8,665.39 | 8,957.79 | 35,187.46 |
| 395 | 8,850.27 | 8,723.88 | 8,930.98 | 9,452.34 | 35,957.47 |
| 396 | 2,505.74 | 2,494.17 | 2,487.21 | 2,424.41 | 9,911.53 |
| 397 | 3,886.42 | 3,448.36 | 3,482.63 | 3,503.70 | 14,321.11 |

| | | | | | | |
|---|---|---|---|---|---|---|
| TOTAL | | 62,603.86 | 60,446.20 | 60,437.33 | 62,550.26 | 246,037.65 |

STATE: MISSOURI

| ZIP CODE | | QUARTER 1 | QUARTER 2 | QUARTER 3 | QUARTER 4 | TOTAL GRAMS |
|---|---|---|---|---|---|---|
| | 630 | 19,181.59 | 19,284.93 | 19,296.99 | 20,172.99 | 77,936.50 |
| | 631 | 38,390.63 | 37,857.57 | 39,594.90 | 40,418.54 | 156,261.64 |
| | 633 | 13,801.98 | 13,789.18 | 13,775.98 | 14,406.98 | 55,774.12 |
| | 634 | 1,140.37 | 1,115.66 | 1,143.29 | 1,248.91 | 4,648.23 |
| | 635 | 1,183.08 | 1,256.13 | 1,169.78 | 1,301.98 | 4,910.97 |
| | 636 | 1,997.66 | 2,048.85 | 1,956.21 | 2,068.95 | 8,071.67 |
| | 637 | 2,217.43 | 2,242.36 | 2,250.24 | 2,278.04 | 8,988.07 |
| | 638 | 1,612.84 | 1,507.82 | 1,556.13 | 1,645.27 | 6,322.06 |
| | 639 | 565.93 | 522.82 | 492.89 | 506.96 | 2,088.60 |
| | 640 | 13,719.85 | 14,107.80 | 14,160.35 | 14,531.50 | 56,519.50 |
| | 641 | 16,730.58 | 17,453.02 | 17,306.09 | 18,154.32 | 69,644.01 |
| | 644 | 689.6 | 646.84 | 626.9 | 679.84 | 2,643.18 |
| | 645 | 1,134 | 1,090.84 | 1,159.53 | 1,174.74 | 4,559.11 |
| | 646 | 426.64 | 443.96 | 422.91 | 456.25 | 1,749.76 |
| | 647 | 944.67 | 984.64 | 1,007.46 | 1,035.43 | 3,972.20 |
| | 648 | 1,555.05 | 1,453.38 | 1,499.07 | 1,466.53 | 5,974.03 |
| | 650 | 1,018.86 | 1,075.39 | 1,121.85 | 1,139.17 | 4,355.27 |
| | 651 | 1,176.32 | 1,158.61 | 1,151.90 | 1,144.43 | 4,631.26 |
| | 652 | 5,358.44 | 5,191.98 | 5,038.46 | 5,447.29 | 21,036.17 |
| | 653 | 998.82 | 974.58 | 961.19 | 984.39 | 3,918.98 |
| | 654 | 832.27 | 855.1 | 934.73 | 913.25 | 3,535.35 |
| | 655 | 684.21 | 637.07 | 691.81 | 634 | 2,647.09 |
| | 656 | 1,357.64 | 1,263.55 | 1,350.67 | 1,384.11 | 5,355.97 |
| | 657 | 1,725.62 | 1,775.44 | 1,776.91 | 1,893.37 | 7,171.34 |
| | 658 | 3,429.02 | 3,424.56 | 3,459.75 | 3,568.61 | 13,881.94 |
| TOTAL | | 131,873.10 | 132,162.08 | 133,905.99 | 138,655.85 | 536,597.02 |

STATE: MONTANA

| ZIP CODE | | QUARTER 1 | QUARTER 2 | QUARTER 3 | QUARTER 4 | TOTAL GRAMS |
|---|---|---|---|---|---|---|
| | 590 | 567.46 | 594.35 | 640.11 | 597.27 | 2,399.19 |
| | 591 | 2,343.69 | 2,356.78 | 2,317.45 | 2,419.18 | 9,437.10 |

| | | | | | | |
|---|---|---|---|---|---|---|
| | 592 | 366.39 | 350.56 | 347.69 | 356.48 | 1,421.12 |
| | 593 | 309.3 | 328.65 | 316.07 | 312.53 | 1,266.55 |
| | 594 | 1,105.99 | 1,142.66 | 1,163.40 | 1,117.01 | 4,529.06 |
| | 595 | 199.58 | 193.12 | 220.31 | 219.75 | 832.76 |
| | 596 | 1,078.58 | 1,069.82 | 1,080.63 | 1,137.37 | 4,366.40 |
| | 597 | 2,979.15 | 3,154.06 | 3,258.84 | 3,376.49 | 12,768.54 |
| | 598 | 3,476.33 | 3,486.02 | 3,562.76 | 3,602.68 | 14,127.79 |
| | 599 | 1,179.06 | 1,202.38 | 1,252.61 | 1,214.09 | 4,848.14 |
| TOTAL | | 13,605.53 | 13,878.40 | 14,159.87 | 14,352.85 | 55,996.65 |

| STATE: | NEBRASKA | | | | |
|---|---|---|---|---|---|
| ZIP CODE | QUARTER 1 | QUARTER 2 | QUARTER 3 | QUARTER 4 | TOTAL GRAMS |
| 680 | 2,240.59 | 2,316.89 | 2,295.69 | 2,347.71 | 9,200.88 |
| 681 | 11,977.48 | 12,024.90 | 12,377.83 | 12,739.13 | 49,119.34 |
| 683 | 520.51 | 503.38 | 531.87 | 506.4 | 2,062.16 |
| 684 | 450.57 | 424.11 | 470.27 | 460.84 | 1,805.79 |
| 685 | 5,322.50 | 5,188.87 | 5,377.41 | 5,620.76 | 21,509.54 |
| 686 | 445.86 | 490.58 | 466.69 | 474.83 | 1,877.96 |
| 687 | 1,024.39 | 1,023.16 | 1,046.33 | 1,131.23 | 4,225.11 |
| 688 | 1,448.08 | 1,404.51 | 1,447.51 | 1,620.31 | 5,920.41 |
| 689 | 476.83 | 467.89 | 510.8 | 484.47 | 1,939.99 |
| 690 | 181.53 | 176.25 | 197.56 | 168.88 | 724.22 |
| 691 | 678.64 | 637.9 | 696.39 | 672.44 | 2,685.37 |
| 692 | 28.83 | 26.73 | 41.47 | 39.16 | 136.19 |
| 693 | 527.25 | 540.12 | 557.75 | 484.2 | 2,109.32 |
| TOTAL | 25,323.06 | 25,225.29 | 26,017.57 | 26,750.36 | 103,316.28 |

| STATE: | NEVADA | | | | |
|---|---|---|---|---|---|
| ZIP CODE | QUARTER 1 | QUARTER 2 | QUARTER 3 | QUARTER 4 | TOTAL GRAMS |
| 890 | 5,755.79 | 5,737.56 | 5,701.19 | 5,840.85 | 23,035.39 |
| 891 | 14,646.21 | 15,020.56 | 14,888.37 | 15,247.04 | 59,802.18 |
| 893 | 60.72 | 55.73 | 52.7 | 45.63 | 214.78 |
| 894 | 1,537.89 | 1,547.57 | 1,569.84 | 1,726.76 | 6,382.06 |
| 895 | 2,604.83 | 2,713.18 | 2,823.42 | 2,851.37 | 10,992.80 |
| 897 | 495.33 | 484.57 | 504.33 | 533.57 | 2,017.80 |

| | | | | | |
|---|---|---|---|---|---|
| | 898 | 394.42 | 425.21 | 430.54 | 452.37 | 1,702.54 |
| TOTAL | | 25,495.19 | 25,984.38 | 25,970.39 | 26,697.59 | 104,147.55 |

STATE: NEW HAMPSHIRE

| ZIP CODE | | QUARTER 1 | QUARTER 2 | QUARTER 3 | QUARTER 4 | TOTAL GRAMS |
|---|---|---|---|---|---|---|
| | 30 | 11,366.07 | 11,964.81 | 11,848.87 | 12,047.21 | 47,226.96 |
| | 31 | 5,938.12 | 6,306.54 | 6,145.86 | 6,467.55 | 24,858.07 |
| | 32 | 2,642.38 | 2,809.01 | 2,831.69 | 2,834.78 | 11,117.86 |
| | 33 | 1,963.79 | 1,977.96 | 2,061.07 | 2,058.70 | 8,061.52 |
| | 34 | 1,473.37 | 1,537.42 | 1,496.38 | 1,598.25 | 6,105.42 |
| | 35 | 524.8 | 536.52 | 508.71 | 543.84 | 2,113.87 |
| | 36 | 95.36 | 91.15 | 94.54 | 96.88 | 377.93 |
| | 37 | 1,754.28 | 1,855.55 | 1,734.25 | 1,772.81 | 7,116.89 |
| | 38 | 11,510.33 | 11,662.69 | 11,754.65 | 11,962.92 | 46,890.59 |
| TOTAL | | 37,268.50 | 38,741.65 | 38,476.02 | 39,382.94 | 153,869.11 |

STATE: NEW JERSEY

| ZIP CODE | | QUARTER 1 | QUARTER 2 | QUARTER 3 | QUARTER 4 | TOTAL GRAMS |
|---|---|---|---|---|---|---|
| | 70 | 15,822.01 | 16,225.65 | 16,176.53 | 17,069.03 | 65,293.22 |
| | 71 | 1,362.78 | 1,351.99 | 1,209.23 | 1,389.54 | 5,313.54 |
| | 72 | 577.89 | 593.78 | 551.94 | 615.44 | 2,339.05 |
| | 73 | 1,418.64 | 1,502.29 | 1,486.48 | 1,586.56 | 5,993.97 |
| | 74 | 5,893.38 | 5,951.77 | 5,977.31 | 6,428.95 | 24,251.41 |
| | 75 | 601.72 | 641.84 | 614.53 | 568.53 | 2,426.62 |
| | 76 | 5,189.34 | 5,197.87 | 5,345.28 | 5,628.45 | 21,360.94 |
| | 77 | 7,943.89 | 8,216.46 | 8,314.21 | 8,677.09 | 33,151.65 |
| | 78 | 5,307.88 | 5,315.30 | 5,343.38 | 5,521.29 | 21,487.85 |
| | 79 | 4,524.63 | 4,595.77 | 4,762.45 | 4,712.85 | 18,595.70 |
| | 80 | 23,402.18 | 23,353.42 | 23,142.33 | 23,732.08 | 93,630.01 |
| | 81 | 1,613.60 | 1,684.18 | 1,700.54 | 1,741.29 | 6,739.61 |
| | 82 | 3,754.25 | 3,953.29 | 4,056.56 | 3,952.98 | 15,717.08 |
| | 83 | 2,608.24 | 2,490.07 | 2,481.96 | 2,590.39 | 10,170.66 |
| | 84 | 380.06 | 390.25 | 390.37 | 334.11 | 1,494.79 |
| | 85 | 3,460.34 | 3,549.86 | 3,487.46 | 3,588.23 | 14,085.89 |
| | 86 | 2,698.23 | 2,721.11 | 2,689.84 | 2,709.88 | 10,819.06 |

| | | | | | | |
|---|---|---|---|---|---|---|
| | 87 | 5,016.36 | 5,413.95 | 5,738.26 | 6,068.13 | 22,236.70 |
| | 88 | 9,402.33 | 9,540.22 | 9,105.88 | 9,549.85 | 37,598.28 |
| | 89 | 991.79 | 975.82 | 916.5 | 1,088.59 | 3,972.70 |
| TOTAL | | 101,969.54 | 103,664.89 | 103,491.04 | 107,553.26 | 416,678.73 |

STATE: NEW MEXICO

| ZIP CODE | | QUARTER 1 | QUARTER 2 | QUARTER 3 | QUARTER 4 | TOTAL GRAMS |
|---|---|---|---|---|---|---|
| | 870 | 752.21 | 657.43 | 659.95 | 616.09 | 2,685.68 |
| | 871 | 8,068.49 | 7,692.69 | 7,111.24 | 6,479.99 | 29,352.41 |
| | 873 | 129.82 | 123.92 | 126.73 | 126.8 | 507.27 |
| | 874 | 631.23 | 632.8 | 633.83 | 581.07 | 2,478.93 |
| | 875 | 1,944.24 | 1,958.64 | 1,952.37 | 1,919.33 | 7,774.58 |
| | 877 | 186.94 | 176.19 | 183.3 | 168.11 | 714.54 |
| | 878 | 62.71 | 48.82 | 40.93 | 45.49 | 197.95 |
| | 879 | 83.55 | 79.04 | 76.79 | 68.51 | 307.89 |
| | 880 | 1,963.32 | 1,922.91 | 1,753.11 | 2,000.27 | 7,639.61 |
| | 881 | 327.22 | 235.34 | 394.08 | 340.12 | 1,296.76 |
| | 882 | 1,741.97 | 1,797.58 | 1,785.25 | 1,772.94 | 7,097.74 |
| | 883 | 847.47 | 837.96 | 847.84 | 839.84 | 3,373.11 |
| | 884 | 62.88 | 63.04 | 57.24 | 50.7 | 233.86 |
| TOTAL | | 16,802.05 | 16,226.36 | 15,622.66 | 15,009.26 | 63,660.33 |

STATE: NEW YORK

| ZIP CODE | | QUARTER 1 | QUARTER 2 | QUARTER 3 | QUARTER 4 | TOTAL GRAMS |
|---|---|---|---|---|---|---|
| | 100 | 47,685.50 | 50,501.81 | 49,015.61 | 51,570.23 | 198,773.15 |
| | 101 | 2,854.39 | 2,719.06 | 2,563.27 | 2,918.06 | 11,054.78 |
| | 102 | 478.26 | 477.81 | 471.29 | 486.35 | 1,913.71 |
| | 103 | 3,111.01 | 3,307.45 | 3,231.45 | 3,407.75 | 13,057.66 |
| | 104 | 3,929 | 3,887.25 | 3,650.77 | 3,862.95 | 15,329.97 |
| | 105 | 8,901.30 | 9,158.57 | 9,351.09 | 9,614.99 | 37,025.95 |
| | 106 | 1,313.83 | 1,296.55 | 1,191.40 | 1,247.61 | 5,049.39 |
| | 107 | 1,627.88 | 1,781.05 | 1,772.28 | 1,852.64 | 7,033.85 |
| | 108 | 768.11 | 765.46 | 706.68 | 762.88 | 3,003.13 |
| | 109 | 6,258.25 | 6,351.17 | 6,242.36 | 6,741.89 | 25,593.67 |
| | 110 | 2,363.33 | 2,464.57 | 2,550.93 | 2,527.27 | 9,906.10 |

| | | | | | |
|---|---|---|---|---|---|
| 111 | 2,606.59 | 2,690.64 | 2,703.04 | 2,888.19 | 10,888.46 |
| 112 | 15,912.46 | 16,678.13 | 16,141.09 | 17,061.77 | 65,793.45 |
| 113 | 5,195.38 | 5,472.84 | 5,269.04 | 5,644.96 | 21,582.22 |
| 114 | 1,259.58 | 1,316.61 | 1,230.88 | 1,330.71 | 5,137.78 |
| 115 | 7,035.65 | 7,334.60 | 7,506.12 | 7,684.90 | 29,561.27 |
| 116 | 523.49 | 520.42 | 503.4 | 543.59 | 2,090.90 |
| 117 | 19,754.28 | 20,088.74 | 20,570.61 | 21,076.11 | 81,489.74 |
| 118 | 974.16 | 1,100.85 | 1,059.99 | 1,099.63 | 4,234.63 |
| 119 | 2,702.73 | 2,976.62 | 3,237.74 | 2,968.02 | 11,885.11 |
| 120 | 5,089.25 | 5,108.10 | 5,303.27 | 5,347.21 | 20,847.83 |
| 121 | 3,571.85 | 3,591.02 | 3,631.99 | 3,798.22 | 14,593.08 |
| 122 | 2,757.39 | 2,800.59 | 2,800.66 | 2,947.66 | 11,306.30 |
| 123 | 2,195.85 | 2,196.15 | 2,126.06 | 2,329.26 | 8,847.32 |
| 124 | 1,742.70 | 1,828.83 | 1,919.16 | 1,928.61 | 7,419.30 |
| 125 | 5,145.02 | 5,313.37 | 5,236.16 | 5,667.06 | 21,361.61 |
| 126 | 1,262.35 | 1,300.02 | 1,275.32 | 1,359.24 | 5,196.93 |
| 127 | 1,113.24 | 1,127.64 | 1,178.42 | 1,172.83 | 4,592.13 |
| 128 | 3,205.85 | 3,213.98 | 3,409.34 | 3,343.34 | 13,172.51 |
| 129 | 2,159.74 | 2,244.31 | 2,290.65 | 2,351.49 | 9,046.19 |
| 130 | 5,602.05 | 5,570.61 | 5,672.83 | 6,091.26 | 22,936.75 |
| 131 | 1,745.53 | 1,713.55 | 1,701.69 | 1,881.86 | 7,042.63 |
| 132 | 3,058.22 | 3,122.78 | 3,106.88 | 3,272.72 | 12,560.60 |
| 133 | 1,000.73 | 986.42 | 904.02 | 1,053.83 | 3,945 |
| 134 | 1,868.75 | 1,994.33 | 1,797.23 | 1,932.55 | 7,592.86 |
| 135 | 937.37 | 941.53 | 970 | 954.62 | 3,803.52 |
| 136 | 2,531.80 | 2,638.83 | 2,726.73 | 2,674.07 | 10,571.43 |
| 137 | 1,248.33 | 1,252.52 | 1,185.77 | 1,253.74 | 4,940.36 |
| 138 | 1,518.94 | 1,630.03 | 1,504.45 | 1,656.66 | 6,310.08 |
| 139 | 1,081.32 | 1,021.85 | 959.78 | 1,037.87 | 4,100.82 |
| 140 | 6,193.95 | 6,213.22 | 6,284.54 | 6,494.76 | 25,186.47 |
| 141 | 2,806.79 | 2,892.28 | 2,887.81 | 2,862.07 | 11,448.95 |
| 142 | 8,823.66 | 8,918.68 | 8,857.45 | 9,367.87 | 35,967.66 |
| 143 | 1,005.50 | 1,048.34 | 989.63 | 1,156.62 | 4,200.09 |
| 144 | 5,382.73 | 5,393.73 | 5,397.44 | 5,816.16 | 21,990.06 |
| 145 | 4,863.37 | 5,150.55 | 5,006.84 | 5,250.25 | 20,271.01 |
| 146 | 9,129.97 | 9,516.47 | 9,316.67 | 9,859.76 | 37,822.87 |
| 147 | 1,598.31 | 1,598.96 | 1,508.75 | 1,511.42 | 6,217.44 |

| | | | | | | |
|---|---|---|---|---|---|---|
| | 148 | 3,576.33 | 3,605.26 | 3,627.32 | 3,676.92 | 14,485.83 |
| | 149 | 906.85 | 874.03 | 880.18 | 890.33 | 3,551.39 |
| TOTAL | | 228,378.92 | 235,698.18 | 233,426.08 | 244,230.76 | 941,733.94 |

STATE: NORTH CAROLINA

| ZIP CODE | | QUARTER 1 | QUARTER 2 | QUARTER 3 | QUARTER 4 | TOTAL GRAMS |
|---|---|---|---|---|---|---|
| | 270 | 4,002.76 | 3,918.63 | 3,793.51 | 4,133.40 | 15,848.30 |
| | 271 | 5,610.48 | 5,417.73 | 5,595.30 | 5,774.14 | 22,397.65 |
| | 272 | 10,948.88 | 10,964.31 | 10,830.60 | 11,508.69 | 44,252.48 |
| | 273 | 5,801.83 | 5,874.17 | 5,856.59 | 6,040.14 | 23,572.73 |
| | 274 | 7,700.25 | 7,835.55 | 7,711.70 | 8,003.83 | 31,251.33 |
| | 275 | 23,093.80 | 23,304.62 | 23,170.67 | 24,480.88 | 94,049.97 |
| | 276 | 13,962.66 | 14,177.32 | 14,111.56 | 15,149.33 | 57,400.87 |
| | 277 | 5,653.65 | 5,879.62 | 5,765.86 | 5,889.22 | 23,188.35 |
| | 278 | 9,201.28 | 8,794.35 | 8,798.11 | 9,371.84 | 36,165.58 |
| | 279 | 2,981.77 | 3,179.85 | 3,149.41 | 3,313.22 | 12,624.25 |
| | 280 | 17,724.67 | 17,513.17 | 17,411.99 | 18,153.37 | 70,803.20 |
| | 281 | 11,947.85 | 11,931.06 | 12,097.88 | 12,292.27 | 48,269.06 |
| | 282 | 19,401.42 | 19,824.62 | 19,539.60 | 20,385.49 | 79,151.13 |
| | 283 | 14,806.90 | 14,563.26 | 14,686.99 | 14,952.25 | 59,009.40 |
| | 284 | 16,914.69 | 17,390.68 | 17,302.66 | 17,740.92 | 69,348.95 |
| | 285 | 11,357.36 | 11,417.50 | 11,732.43 | 10,965.96 | 45,473.25 |
| | 286 | 7,914.07 | 7,908 | 7,857.71 | 8,171.47 | 31,851.25 |
| | 287 | 7,119.54 | 7,173.23 | 7,063.36 | 7,359.54 | 28,715.67 |
| | 288 | 4,305.50 | 4,330.87 | 4,381.53 | 4,490.35 | 17,508.25 |
| | 289 | 385.46 | 406.43 | 436.57 | 421.36 | 1,649.82 |
| TOTAL | | 200,834.82 | 201,804.97 | 201,294.03 | 208,597.67 | 812,531.49 |

STATE: NORTH DAKOTA

| ZIP CODE | | QUARTER 1 | QUARTER 2 | QUARTER 3 | QUARTER 4 | TOTAL GRAMS |
|---|---|---|---|---|---|---|
| | 580 | 712.58 | 669.74 | 695.3 | 691.53 | 2,769.15 |
| | 581 | 2,147.83 | 2,115.42 | 2,219.63 | 2,236.90 | 8,719.78 |
| | 582 | 1,486.56 | 1,487.49 | 1,548.45 | 1,524.90 | 6,047.40 |
| | 583 | 582.59 | 512.39 | 529.29 | 494.65 | 2,118.92 |
| | 584 | 275.03 | 302.77 | 276.72 | 267.32 | 1,121.84 |

| | | | | | | |
|---|---|---|---|---|---|---|
| | 585 | 1,435.23 | 1,509.72 | 1,452.50 | 1,528.08 | 5,925.53 |
| | 586 | 350.79 | 374.91 | 383.07 | 377.83 | 1,486.60 |
| | 587 | 1,305.35 | 1,383.08 | 1,380.44 | 1,426.98 | 5,495.85 |
| | 588 | 512.95 | 531.38 | 585.45 | 649.46 | 2,279.24 |
| TOTAL | | 8,808.91 | 8,886.90 | 9,070.85 | 9,197.65 | 35,964.31 |

| STATE: | OHIO | | | | | |
|---|---|---|---|---|---|---|
| ZIP CODE | | QUARTER 1 | QUARTER 2 | QUARTER 3 | QUARTER 4 | TOTAL GRAMS |
| | 430 | 15,577.69 | 15,691.25 | 15,377.30 | 16,021.50 | 62,667.74 |
| | 431 | 5,802.53 | 5,925.26 | 5,910.77 | 6,090.54 | 23,729.10 |
| | 432 | 31,407.16 | 31,151.36 | 32,995.32 | 36,156.42 | 131,710.26 |
| | 433 | 2,368.10 | 2,299.59 | 2,279.74 | 2,399.05 | 9,346.48 |
| | 434 | 2,061.63 | 2,044.61 | 2,024.24 | 2,168.74 | 8,299.22 |
| | 435 | 5,118.27 | 5,192.30 | 5,074.72 | 5,421.81 | 20,807.10 |
| | 436 | 7,949.73 | 8,157.50 | 8,246.90 | 8,406.35 | 32,760.48 |
| | 437 | 1,537.62 | 1,601.49 | 1,587.40 | 1,705.34 | 6,431.85 |
| | 438 | 215.21 | 223.07 | 234.29 | 258.23 | 930.8 |
| | 439 | 1,926.51 | 1,999.75 | 1,987.25 | 2,039.04 | 7,952.55 |
| | 440 | 14,161.18 | 14,420.70 | 14,591.11 | 15,394.64 | 58,567.63 |
| | 441 | 19,830.89 | 20,202.49 | 20,483.60 | 21,321.46 | 81,838.44 |
| | 442 | 8,214 | 8,192.14 | 8,318.92 | 8,443.48 | 33,168.54 |
| | 443 | 3,957.60 | 4,033.68 | 3,833.38 | 3,935.89 | 15,760.55 |
| | 444 | 4,410.40 | 4,349.92 | 4,489.45 | 4,658.23 | 17,908 |
| | 445 | 4,264.52 | 4,327.17 | 4,567.68 | 4,766.48 | 17,925.85 |
| | 446 | 5,779.88 | 5,891.81 | 5,859.68 | 5,934.44 | 23,465.81 |
| | 447 | 5,141.28 | 5,207.49 | 5,223.87 | 5,293.16 | 20,865.80 |
| | 448 | 3,352.58 | 3,241.79 | 3,298.12 | 3,441.26 | 13,333.75 |
| | 449 | 881.48 | 891.29 | 825.07 | 879.31 | 3,477.15 |
| | 450 | 14,744.50 | 14,931.86 | 14,868.74 | 15,574.40 | 60,119.50 |
| | 451 | 5,284.21 | 5,197.16 | 5,168.43 | 5,630.70 | 21,280.50 |
| | 452 | 21,077.55 | 20,853.22 | 20,818.05 | 21,932.22 | 84,681.04 |
| | 453 | 5,013.22 | 5,000.12 | 4,918.38 | 5,165.61 | 20,097.33 |
| | 454 | 6,024.62 | 5,994.59 | 5,846 | 6,186.27 | 24,051.48 |
| | 455 | 1,610.94 | 1,440.29 | 1,391.31 | 1,445.84 | 5,888.38 |
| | 456 | 3,719.97 | 3,529.66 | 3,463.64 | 3,702.07 | 14,415.34 |
| | 457 | 3,170.28 | 2,949.72 | 2,885.08 | 3,070.89 | 12,075.97 |

| | | | | | | |
|---|---|---|---|---|---|---|
| | 458 | 3,338.24 | 3,259.81 | 3,344.34 | 3,520.82 | 13,463.21 |
| TOTAL | | 207,941.79 | 208,201.09 | 209,912.78 | 220,964.19 | 847,019.85 |

STATE: OKLAHOMA

| ZIP CODE | | QUARTER 1 | QUARTER 2 | QUARTER 3 | QUARTER 4 | TOTAL GRAMS |
|---|---|---|---|---|---|---|
| | 730 | 10,397.12 | 10,269.08 | 10,391.43 | 10,839.02 | 41,896.65 |
| | 731 | 9,047.19 | 9,215.82 | 8,922.56 | 9,401.65 | 36,587.22 |
| | 734 | 910.77 | 878.53 | 929.02 | 969.52 | 3,687.84 |
| | 735 | 2,452.25 | 2,497.80 | 2,713.73 | 2,285.70 | 9,949.48 |
| | 736 | 240.67 | 265.19 | 258.97 | 277.51 | 1,042.34 |
| | 737 | 909.53 | 995.94 | 986.48 | 1,052.38 | 3,944.33 |
| | 738 | 163.88 | 159.96 | 173.77 | 172.13 | 669.74 |
| | 739 | 73.34 | 75.8 | 70.78 | 58.19 | 278.11 |
| | 740 | 11,938.26 | 11,867.98 | 11,871.44 | 12,446.58 | 48,124.26 |
| | 741 | 9,406.92 | 9,695.19 | 9,520.66 | 9,940.79 | 38,563.56 |
| | 743 | 1,008.76 | 998.96 | 982.4 | 992.04 | 3,982.16 |
| | 744 | 2,599.40 | 2,651.18 | 2,682.03 | 2,620.17 | 10,552.78 |
| | 745 | 486.54 | 478.65 | 450.64 | 474.46 | 1,890.29 |
| | 746 | 265.01 | 259.01 | 274.66 | 263.01 | 1,061.69 |
| | 747 | 581.73 | 552.82 | 629.19 | 657.41 | 2,421.15 |
| | 748 | 1,364.83 | 1,301.56 | 1,256.71 | 1,366.84 | 5,289.94 |
| | 749 | 438.53 | 407.47 | 401.65 | 415.54 | 1,663.19 |
| TOTAL | | 52,284.73 | 52,570.94 | 52,516.12 | 54,232.94 | 211,604.73 |

STATE: OREGON

| ZIP CODE | | QUARTER 1 | QUARTER 2 | QUARTER 3 | QUARTER 4 | TOTAL GRAMS |
|---|---|---|---|---|---|---|
| | 970 | 12,457.94 | 12,606.28 | 12,605.77 | 12,828.90 | 50,498.89 |
| | 971 | 4,749.31 | 4,985.34 | 4,884.17 | 4,914.79 | 19,533.61 |
| | 972 | 17,358.93 | 18,610.34 | 18,357.45 | 18,514.58 | 72,841.30 |
| | 973 | 8,537.23 | 8,739.51 | 8,465.55 | 8,721.94 | 34,464.23 |
| | 974 | 6,683.65 | 6,741.94 | 6,648.55 | 6,618.30 | 26,692.44 |
| | 975 | 2,114.95 | 2,181.56 | 2,224.05 | 2,427.32 | 8,947.88 |
| | 976 | 486.81 | 420.59 | 437.81 | 457.61 | 1,802.82 |
| | 977 | 3,610.31 | 3,737.36 | 3,744.50 | 3,718.32 | 14,810.49 |
| | 978 | 826.36 | 939.42 | 854.07 | 933.33 | 3,553.18 |

| | | | | | | |
|---|---|---|---|---|---|---|
| | 979 | 360.11 | 310.05 | 347.04 | 402.15 | 1,419.35 |
| TOTAL | | 57,185.60 | 59,272.39 | 58,568.96 | 59,537.24 | 234,564.19 |

| STATE: | PENNSYLVANIA | | | | | |
|---|---|---|---|---|---|---|
| ZIP CODE | | QUARTER 1 | QUARTER 2 | QUARTER 3 | QUARTER 4 | TOTAL GRAMS |
| | 150 | 6,286.33 | 6,292.89 | 6,342.71 | 6,392.33 | 25,314.26 |
| | 151 | 5,093.39 | 4,999.04 | 5,115.10 | 5,217.30 | 20,424.83 |
| | 152 | 10,722.99 | 10,592.92 | 10,789.04 | 11,054.52 | 43,159.47 |
| | 153 | 2,383.27 | 2,429.01 | 2,455.01 | 2,470.31 | 9,737.60 |
| | 154 | 1,981.13 | 1,879.02 | 1,785.30 | 1,925.75 | 7,571.20 |
| | 155 | 894.99 | 862.51 | 892.49 | 972.73 | 3,622.72 |
| | 156 | 4,535.84 | 4,380.69 | 4,407.35 | 4,442.83 | 17,766.71 |
| | 157 | 1,479.39 | 1,269.26 | 1,326.14 | 1,442.14 | 5,516.93 |
| | 158 | 1,272.77 | 1,273.05 | 1,277.49 | 1,247.72 | 5,071.03 |
| | 159 | 1,395.40 | 1,373.64 | 1,399.31 | 1,445.92 | 5,614.27 |
| | 160 | 3,090.29 | 3,108.98 | 3,099.60 | 3,112.39 | 12,411.26 |
| | 161 | 2,468.82 | 2,376.40 | 2,529.17 | 2,728.85 | 10,103.24 |
| | 162 | 964.99 | 916.22 | 880.24 | 955.62 | 3,717.07 |
| | 163 | 2,055.05 | 2,017.23 | 1,881.06 | 1,978.75 | 7,932.09 |
| | 164 | 964.97 | 943.9 | 980.54 | 1,036.47 | 3,925.88 |
| | 165 | 4,014.10 | 4,090.38 | 4,092.97 | 4,047.28 | 16,244.73 |
| | 166 | 2,734.21 | 2,702.93 | 2,715.19 | 2,786.90 | 10,939.23 |
| | 167 | 609.27 | 574.59 | 589.98 | 589.46 | 2,363.30 |
| | 168 | 2,883.91 | 2,773.32 | 2,750.02 | 2,815.66 | 11,222.91 |
| | 169 | 467.32 | 467.82 | 469.61 | 480.79 | 1,885.54 |
| | 170 | 11,695.29 | 11,829.93 | 11,864.55 | 11,808.30 | 47,198.07 |
| | 171 | 2,868.87 | 2,955.66 | 3,030.41 | 3,090.65 | 11,945.59 |
| | 172 | 3,106.65 | 3,102.76 | 3,227.46 | 3,228.57 | 12,665.44 |
| | 173 | 5,274.53 | 5,339.07 | 5,189.58 | 5,404.58 | 21,207.76 |
| | 174 | 4,170.01 | 4,205.22 | 4,132.61 | 4,291.17 | 16,799.01 |
| | 175 | 3,720.63 | 3,610.40 | 3,782.78 | 3,698.35 | 14,812.16 |
| | 176 | 3,964.30 | 3,960.05 | 4,040.38 | 4,139.95 | 16,104.68 |
| | 177 | 2,099.91 | 2,029.04 | 2,006.73 | 2,162.58 | 8,298.26 |
| | 178 | 2,596.46 | 2,668.49 | 2,617.31 | 2,743.48 | 10,625.74 |
| | 179 | 962.74 | 945.43 | 905.74 | 955.84 | 3,769.75 |
| | 180 | 5,581.76 | 5,639.63 | 5,613.68 | 5,811.15 | 22,646.22 |

| | | | | | | |
|---|---|---|---|---|---|---|
| | 181 | 2,981.36 | 3,027.62 | 2,866.73 | 3,015.86 | 11,891.57 |
| | 182 | 932.5 | 1,008.12 | 938.18 | 1,024.51 | 3,903.31 |
| | 183 | 1,324.87 | 1,298.86 | 1,422.59 | 1,501.55 | 5,547.87 |
| | 184 | 2,091.83 | 2,352.68 | 2,274.44 | 2,423.51 | 9,142.46 |
| | 185 | 3,665.84 | 3,633.99 | 3,800 | 4,013.76 | 15,113.59 |
| | 186 | 1,757.25 | 1,777.49 | 1,761.16 | 1,893.87 | 7,189.77 |
| | 187 | 16,263.25 | 18,137.16 | 15,896.68 | 17,945.60 | 68,242.69 |
| | 188 | 657.78 | 630.93 | 574.36 | 599.41 | 2,462.48 |
| | 189 | 7,281.32 | 7,384.59 | 7,629.80 | 7,678.36 | 29,974.07 |
| | 190 | 22,526.70 | 23,093.92 | 23,116.35 | 23,352.81 | 92,089.78 |
| | 191 | 20,498.52 | 20,729.84 | 19,846.22 | 20,842.10 | 81,916.68 |
| | 193 | 10,193.07 | 10,256.68 | 10,220.75 | 10,266.77 | 40,937.27 |
| | 194 | 11,137.20 | 11,400.16 | 11,433.18 | 11,631.63 | 45,602.17 |
| | 195 | 1,907.18 | 1,802.08 | 1,866.09 | 1,887.42 | 7,462.77 |
| | 196 | 2,945.43 | 3,103.97 | 3,003.17 | 3,189 | 12,241.57 |
| TOTAL | | 208,503.68 | 211,247.57 | 208,839.25 | 215,744.50 | 844,335 |

STATE: PUERTO RICO

| ZIP CODE | QUARTER 1 | QUARTER 2 | QUARTER 3 | QUARTER 4 | TOTAL GRAMS |
|---|---|---|---|---|---|
| 6 | 2,036.97 | 1,883.50 | 1,884.21 | 1,821.97 | 7,626.65 |
| 7 | 2,215.78 | 2,035.05 | 1,758.58 | 1,764.10 | 7,773.51 |
| 9 | 4,823.87 | 4,523.81 | 4,202.79 | 4,164.60 | 17,715.07 |
| TOTAL | 9,076.62 | 8,442.36 | 7,845.58 | 7,750.67 | 33,115.23 |

STATE: RHODE ISLAND

| ZIP CODE | QUARTER 1 | QUARTER 2 | QUARTER 3 | QUARTER 4 | TOTAL GRAMS |
|---|---|---|---|---|---|
| 28 | 22,511.48 | 23,256.34 | 23,363.81 | 23,812.45 | 92,944.08 |
| 29 | 12,873.29 | 13,224.85 | 13,490.61 | 14,006.29 | 53,595.04 |
| TOTAL | 35,384.77 | 36,481.19 | 36,854.42 | 37,818.74 | 146,539.12 |

STATE: SOUTH CAROLINA

| ZIP CODE | QUARTER 1 | QUARTER 2 | QUARTER 3 | QUARTER 4 | TOTAL GRAMS |
|---|---|---|---|---|---|
| 290 | 12,157.26 | 11,987.72 | 11,757.93 | 12,317.68 | 48,220.59 |
| 291 | 9,839.22 | 9,812.74 | 9,574.48 | 9,939.63 | 39,166.07 |

| | | | | | | |
|---|---|---|---|---|---|---|
| | 292 | 10,443.79 | 10,172.93 | 10,163.75 | 10,346.59 | 41,127.06 |
| | 293 | 8,530.70 | 8,451.77 | 8,245.80 | 8,683.32 | 33,911.59 |
| | 294 | 33,171.24 | 32,863.04 | 32,244.56 | 34,065.76 | 132,344.60 |
| | 295 | 18,135.98 | 17,838.95 | 18,024.01 | 18,110.94 | 72,109.88 |
| | 296 | 28,183.57 | 28,062.82 | 27,985.60 | 29,606.06 | 113,838.05 |
| | 297 | 7,620.71 | 7,439.27 | 7,809.08 | 8,085.47 | 30,954.53 |
| | 298 | 4,826.42 | 4,508.63 | 4,191.87 | 4,577.69 | 18,104.61 |
| | 299 | 3,752.47 | 3,816.27 | 3,939.56 | 3,907.65 | 15,415.95 |
| TOTAL | | 136,661.36 | 134,954.14 | 133,936.64 | 139,640.79 | 545,192.93 |

STATE: SOUTH DAKOTA

| ZIP CODE | | QUARTER 1 | QUARTER 2 | QUARTER 3 | QUARTER 4 | TOTAL GRAMS |
|---|---|---|---|---|---|---|
| | 570 | 2,154.92 | 2,065.66 | 2,033.34 | 2,121.19 | 8,375.11 |
| | 571 | 9,124.64 | 8,230.04 | 7,772.28 | 8,240.94 | 33,367.90 |
| | 572 | 641.09 | 605.57 | 643.44 | 648.48 | 2,538.58 |
| | 573 | 604.76 | 637.5 | 656.83 | 609.2 | 2,508.29 |
| | 574 | 659.33 | 657.28 | 727.14 | 707.86 | 2,751.61 |
| | 575 | 244.23 | 255.81 | 252.02 | 282.3 | 1,034.36 |
| | 576 | 53.14 | 53.1 | 42.28 | 55.12 | 203.64 |
| | 577 | 2,501.35 | 2,410.86 | 2,528.44 | 2,592.09 | 10,032.74 |
| TOTAL | | 15,983.46 | 14,915.82 | 14,655.77 | 15,257.18 | 60,812.23 |

STATE: TENNESSEE

| ZIP CODE | | QUARTER 1 | QUARTER 2 | QUARTER 3 | QUARTER 4 | TOTAL GRAMS |
|---|---|---|---|---|---|---|
| | 370 | 19,914.15 | 20,446.43 | 20,113.64 | 21,212.23 | 81,686.45 |
| | 371 | 12,690.80 | 12,666.69 | 12,678.29 | 12,972.62 | 51,008.40 |
| | 372 | 13,795.91 | 14,113.90 | 14,207.19 | 14,692.42 | 56,809.42 |
| | 373 | 11,994.58 | 12,025.25 | 11,804.24 | 12,194.45 | 48,018.52 |
| | 374 | 8,955.75 | 8,882.51 | 8,749.13 | 9,356.93 | 35,944.32 |
| | 376 | 4,034.02 | 3,948.45 | 4,020.91 | 4,234.17 | 16,237.55 |
| | 377 | 4,367.01 | 4,456.34 | 4,381.72 | 4,506.26 | 17,711.33 |
| | 378 | 6,661.64 | 6,643.52 | 6,808.64 | 7,077.53 | 27,191.33 |
| | 379 | 11,142.59 | 10,967.09 | 11,197.71 | 11,645.90 | 44,953.29 |
| | 380 | 9,864.75 | 9,387.08 | 9,541.94 | 10,031.19 | 38,824.96 |
| | 381 | 11,785.36 | 11,673.54 | 11,744.76 | 12,407.81 | 47,611.47 |

| | | | | | | |
|---|---|---|---|---|---|---|
| | 382 | 3,312.02 | 3,217.44 | 3,223.76 | 3,205.07 | 12,958.29 |
| | 383 | 4,753.89 | 4,460.62 | 4,615.68 | 4,934.45 | 18,764.64 |
| | 384 | 3,479.16 | 3,450.80 | 3,370.36 | 3,484.59 | 13,784.91 |
| | 385 | 2,269.08 | 2,145.98 | 2,261.77 | 2,257.04 | 8,933.87 |
| TOTAL | | 129,020.71 | 128,485.64 | 128,719.74 | 134,212.66 | 520,438.75 |

STATE: TEXAS

| ZIP CODE | | QUARTER 1 | QUARTER 2 | QUARTER 3 | QUARTER 4 | TOTAL GRAMS |
|---|---|---|---|---|---|---|
| | 750 | 52,977.30 | 53,821.17 | 52,663.51 | 52,865.68 | 212,327.66 |
| | 751 | 8,796.92 | 8,947.83 | 8,878.34 | 9,416.20 | 36,039.29 |
| | 752 | 25,195.07 | 25,973.74 | 25,521.70 | 26,973.85 | 103,664.36 |
| | 753 | 16.46 | 23.47 | 31.01 | 34.92 | 105.86 |
| | 754 | 3,944.85 | 4,002.35 | 4,046.93 | 4,393.16 | 16,387.29 |
| | 755 | 1,189.32 | 1,219.66 | 1,167.56 | 1,261.41 | 4,837.95 |
| | 756 | 3,681.08 | 3,728.43 | 3,579.84 | 3,796.61 | 14,785.96 |
| | 757 | 4,673.25 | 4,869.17 | 4,698.22 | 4,980.23 | 19,220.87 |
| | 758 | 848.54 | 775.87 | 766.17 | 894 | 3,284.58 |
| | 759 | 2,483.80 | 2,423.55 | 2,507.10 | 2,477.30 | 9,891.75 |
| | 760 | 22,617.33 | 22,706.25 | 23,081.17 | 23,967.62 | 92,372.37 |
| | 761 | 13,578.94 | 13,772.70 | 13,737.87 | 14,827.14 | 55,916.65 |
| | 762 | 11,303.11 | 11,333.85 | 11,402.01 | 11,950.33 | 45,989.30 |
| | 763 | 3,172.93 | 3,311.83 | 3,379.12 | 3,449.63 | 13,313.51 |
| | 764 | 1,388.39 | 1,310.96 | 1,365.69 | 1,415.35 | 5,480.39 |
| | 765 | 5,730.29 | 5,722.54 | 6,196.89 | 5,541.52 | 23,191.24 |
| | 766 | 1,121.36 | 1,109.95 | 1,080.50 | 1,192.41 | 4,504.22 |
| | 767 | 3,475.59 | 3,373.16 | 3,275.31 | 3,610.29 | 13,734.35 |
| | 768 | 778.49 | 800.96 | 804.68 | 820 | 3,204.13 |
| | 769 | 1,629.92 | 1,654.43 | 1,622.73 | 1,832.75 | 6,739.83 |
| | 770 | 38,848.59 | 39,521.68 | 38,044.50 | 41,628.73 | 158,043.50 |
| | 772 | 38.16 | 49.11 | 32.22 | 49.89 | 169.38 |
| | 773 | 28,335.85 | 28,543.69 | 28,415.73 | 30,315.97 | 115,611.24 |
| | 774 | 20,162.21 | 20,297.03 | 19,828.38 | 21,359.97 | 81,647.59 |
| | 775 | 22,757.20 | 22,986.66 | 22,438.01 | 23,847.21 | 92,029.08 |
| | 776 | 4,828.73 | 4,819.71 | 4,778.33 | 5,054.03 | 19,480.80 |
| | 777 | 2,512.50 | 2,517.69 | 2,426.06 | 2,621.17 | 10,077.42 |
| | 778 | 6,977.41 | 6,735 | 6,715.97 | 7,131.18 | 27,559.56 |

| | | | | | | |
|---|---|---|---|---|---|---|
| | 779 | 2,177.86 | 2,153.85 | 2,004.27 | 2,213.75 | 8,549.73 |
| | 780 | 4,581.36 | 4,756.95 | 4,649.12 | 5,136.69 | 19,124.12 |
| | 781 | 5,936.02 | 5,925.33 | 6,221.20 | 6,441.24 | 24,523.79 |
| | 782 | 26,025.30 | 25,617.27 | 25,924.24 | 27,682.14 | 105,248.95 |
| | 783 | 2,060.57 | 2,015.38 | 1,982.18 | 1,890.07 | 7,948.20 |
| | 784 | 4,859.77 | 4,635.57 | 4,660.40 | 4,900.94 | 19,056.68 |
| | 785 | 3,078.25 | 2,792.13 | 2,838.76 | 3,174.33 | 11,883.47 |
| | 786 | 16,825.95 | 16,889.88 | 17,093.83 | 18,197.40 | 69,007.06 |
| | 787 | 28,082.76 | 28,823.78 | 28,213.16 | 30,580.73 | 115,700.43 |
| | 788 | 629.59 | 627.81 | 706.57 | 615.04 | 2,579.01 |
| | 789 | 690.71 | 656.82 | 694.38 | 730.93 | 2,772.84 |
| | 790 | 903.12 | 888.58 | 858.18 | 899.4 | 3,549.28 |
| | 791 | 1,688.17 | 1,812.12 | 1,755.97 | 1,921.51 | 7,177.77 |
| | 792 | 70.29 | 71.69 | 83.07 | 81.15 | 306.2 |
| | 793 | 466.52 | 455.64 | 466.39 | 481.19 | 1,869.74 |
| | 794 | 4,592.37 | 4,558.65 | 4,591.74 | 4,817.78 | 18,560.54 |
| | 795 | 464.01 | 478.06 | 516.82 | 522.06 | 1,980.95 |
| | 796 | 2,159.51 | 2,162.05 | 2,092.64 | 2,220.52 | 8,634.72 |
| | 797 | 5,707.66 | 6,055.47 | 6,110.70 | 6,384.55 | 24,258.38 |
| | 798 | 248 | 217.44 | 201.4 | 215.05 | 881.89 |
| | 799 | 3,159.62 | 3,078.47 | 3,566.71 | 2,831.02 | 12,635.82 |
| TOTAL | | 407,471 | 411,025.38 | 407,717.28 | 429,646.04 | 1,655,859.70 |

STATE: UTAH

| ZIP CODE | | QUARTER 1 | QUARTER 2 | QUARTER 3 | QUARTER 4 | TOTAL GRAMS |
|---|---|---|---|---|---|---|
| | 840 | 40,080.76 | 41,363.65 | 41,678.84 | 43,562.33 | 166,685.58 |
| | 841 | 15,987.90 | 16,400.75 | 16,333.30 | 16,999.48 | 65,721.43 |
| | 843 | 4,110.92 | 4,286.18 | 4,329.52 | 4,590.22 | 17,316.84 |
| | 844 | 3,671.30 | 3,827.05 | 3,777.10 | 4,159.28 | 15,434.73 |
| | 845 | 726.09 | 593.64 | 637.05 | 672.19 | 2,628.97 |
| | 846 | 8,307.69 | 8,328.13 | 8,538.70 | 8,860.93 | 34,035.45 |
| | 847 | 6,328.21 | 6,229.66 | 6,454.22 | 6,851.31 | 25,863.40 |
| TOTAL | | 79,212.87 | 81,029.06 | 81,748.73 | 85,695.74 | 327,686.40 |

STATE: VERMONT

| ZIP CODE | QUARTER 1 | QUARTER 2 | QUARTER 3 | QUARTER 4 | TOTAL GRAMS |
|---|---|---|---|---|---|
| 50 | 635.55 | 578.1 | 497.19 | 590.97 | 2,301.81 |
| 51 | 542.27 | 490.95 | 570.95 | 524.63 | 2,128.80 |
| 52 | 712.57 | 709.25 | 692.82 | 723.12 | 2,837.76 |
| 53 | 1,614.01 | 1,609.38 | 1,644.18 | 1,731.01 | 6,598.58 |
| 54 | 3,253.72 | 3,338.83 | 3,274.56 | 3,401.11 | 13,268.22 |
| 56 | 1,636.76 | 1,638.93 | 1,650.35 | 1,659.53 | 6,585.57 |
| 57 | 840.5 | 919.94 | 880 | 916.78 | 3,557.22 |
| 58 | 564.64 | 623.97 | 570.99 | 651.86 | 2,411.46 |
| TOTAL | 9,800.02 | 9,909.35 | 9,781.04 | 10,199.01 | 39,689.42 |

STATE: VIRGIN ISLANDS

| ZIP CODE | QUARTER 1 | QUARTER 2 | QUARTER 3 | QUARTER 4 | TOTAL GRAMS |
|---|---|---|---|---|---|
| 8 | 368.82 | 364.19 | 251.15 | 233.08 | 1,217.24 |
| TOTAL | 368.82 | 364.19 | 251.15 | 233.08 | 1,217.24 |

STATE: VIRGINIA

| ZIP CODE | QUARTER 1 | QUARTER 2 | QUARTER 3 | QUARTER 4 | TOTAL GRAMS |
|---|---|---|---|---|---|
| 201 | 18,061.29 | 17,971.69 | 17,923.90 | 18,716.86 | 72,673.74 |
| 220 | 10,528.57 | 10,214.45 | 10,448.90 | 10,454.96 | 41,646.88 |
| 221 | 6,912.54 | 7,062.72 | 6,868.53 | 7,331.99 | 28,175.78 |
| 222 | 4,791.73 | 5,029.40 | 4,714.86 | 4,629.21 | 19,165.20 |
| 223 | 3,933.01 | 4,032.93 | 4,011.68 | 4,275.69 | 16,253.31 |
| 224 | 4,542.77 | 4,681.80 | 4,481.74 | 4,700.12 | 18,406.43 |
| 225 | 2,059.65 | 2,106.04 | 1,978.13 | 2,128.30 | 8,272.12 |
| 226 | 3,036.79 | 3,128.81 | 3,094.84 | 3,327.86 | 12,588.30 |
| 227 | 1,083.35 | 1,052.26 | 1,080.33 | 1,065.64 | 4,281.58 |
| 228 | 3,183.47 | 3,097.28 | 2,897.85 | 3,154.29 | 12,332.89 |
| 229 | 6,345.28 | 6,249.85 | 6,318.83 | 6,503.63 | 25,417.59 |
| 230 | 4,907.31 | 4,799.23 | 4,802.70 | 5,076 | 19,585.24 |
| 231 | 11,468.86 | 11,681.76 | 11,567.48 | 12,200.32 | 46,918.42 |
| 232 | 15,970.95 | 16,035.99 | 16,040.45 | 16,221.24 | 64,268.63 |
| 233 | 9,643.61 | 9,850.78 | 9,844.15 | 9,987.53 | 39,326.07 |
| 234 | 24,154.23 | 24,390.20 | 24,181.45 | 24,883.07 | 97,608.95 |

| | | | | | | |
|---|---|---|---|---|---|---|
| | 235 | 6,124.39 | 6,373.08 | 6,014.14 | 6,313.24 | 24,824.85 |
| | 236 | 9,406.51 | 9,584.18 | 10,035.86 | 9,236.87 | 38,263.42 |
| | 237 | 4,110.91 | 4,169.09 | 3,971.07 | 4,184.49 | 16,435.56 |
| | 238 | 6,597.99 | 6,583.20 | 6,295.86 | 6,774.44 | 26,251.49 |
| | 239 | 1,029.59 | 1,027.78 | 1,050.80 | 1,032.35 | 4,140.52 |
| | 240 | 5,321.29 | 5,428.17 | 5,202.75 | 5,498.57 | 21,450.78 |
| | 241 | 2,753.62 | 2,723.67 | 2,620.40 | 2,783.92 | 10,881.61 |
| | 242 | 1,437.57 | 1,304.17 | 1,385.50 | 1,389.63 | 5,516.87 |
| | 243 | 906.76 | 886.96 | 830.7 | 849.8 | 3,474.22 |
| | 244 | 3,251.13 | 3,309.54 | 3,211.31 | 3,448.53 | 13,220.51 |
| | 245 | 5,239.40 | 5,229.88 | 5,329.16 | 5,409.06 | 21,207.50 |
| | 246 | 356.39 | 357.62 | 392.01 | 388.71 | 1,494.73 |
| TOTAL | | 177,158.96 | 178,362.53 | 176,595.38 | 181,966.32 | 714,083.19 |

STATE: WASHINGTON

| ZIP CODE | | QUARTER 1 | QUARTER 2 | QUARTER 3 | QUARTER 4 | TOTAL GRAMS |
|---|---|---|---|---|---|---|
| | 980 | 26,199.11 | 26,917.52 | 26,847.86 | 26,763.73 | 106,728.22 |
| | 981 | 17,081.65 | 17,494.90 | 17,264.76 | 17,933.80 | 69,775.11 |
| | 982 | 15,963.42 | 16,382.22 | 16,092.72 | 16,609.58 | 65,047.94 |
| | 983 | 10,153.15 | 10,528.89 | 10,394.54 | 10,587.39 | 41,663.97 |
| | 984 | 7,189.03 | 7,340.15 | 7,140.58 | 6,990.34 | 28,660.10 |
| | 985 | 5,105.72 | 5,292.46 | 5,248.30 | 5,368.19 | 21,014.67 |
| | 986 | 8,020.81 | 8,270.09 | 8,090.06 | 8,099.95 | 32,480.91 |
| | 988 | 1,818.96 | 1,749.07 | 1,800.57 | 1,894.11 | 7,262.71 |
| | 989 | 2,520.28 | 2,516.04 | 2,357.39 | 2,426.01 | 9,819.72 |
| | 990 | 897.67 | 929.36 | 979.18 | 1,000.32 | 3,806.53 |
| | 991 | 1,174.44 | 1,021.36 | 1,079.61 | 1,092.36 | 4,367.77 |
| | 992 | 5,401.87 | 5,452.08 | 5,430.31 | 5,717.49 | 22,001.75 |
| | 993 | 5,483.30 | 5,676.46 | 5,339.66 | 5,530.12 | 22,029.54 |
| | 994 | 186.99 | 176.27 | 182.49 | 196.41 | 742.16 |
| TOTAL | | 107,196.40 | 109,746.87 | 108,248.03 | 110,209.80 | 435,401.10 |

STATE: WEST VIRGINIA

| ZIP CODE | | QUARTER 1 | QUARTER 2 | QUARTER 3 | QUARTER 4 | TOTAL GRAMS |
|---|---|---|---|---|---|---|
| | 247 | 283.55 | 272.89 | 309.84 | 292 | 1,158.28 |

| | | | | | | |
|---|---|---|---|---|---|---|
| | 248 | 221.17 | 187.41 | 219.48 | 201.31 | 829.37 |
| | 249 | 411.88 | 436.06 | 426.43 | 394.08 | 1,668.45 |
| | 250 | 907.09 | 845.37 | 805.15 | 832.62 | 3,390.23 |
| | 251 | 676.9 | 670.38 | 702.5 | 713.54 | 2,763.32 |
| | 252 | 540.04 | 557.15 | 503.51 | 470.89 | 2,071.59 |
| | 253 | 3,663.21 | 3,640.36 | 3,707.73 | 3,485.15 | 14,496.45 |
| | 254 | 2,425.50 | 2,449.94 | 2,474.69 | 2,579.58 | 9,929.71 |
| | 255 | 2,397.40 | 2,230.80 | 2,227.81 | 2,453.65 | 9,309.66 |
| | 256 | 266.95 | 273.59 | 246.71 | 261.15 | 1,048.40 |
| | 257 | 1,749.10 | 1,661.37 | 1,769.47 | 1,784.52 | 6,964.46 |
| | 258 | 1,225.69 | 1,243.03 | 1,129.56 | 1,264.32 | 4,862.60 |
| | 259 | 329.28 | 265.6 | 276.74 | 260.2 | 1,131.82 |
| | 260 | 1,452.94 | 1,427.92 | 1,395.42 | 1,397.80 | 5,674.08 |
| | 261 | 2,315.91 | 2,288.21 | 2,206.75 | 2,217.07 | 9,027.94 |
| | 262 | 295.14 | 278.38 | 313.09 | 329.37 | 1,215.98 |
| | 263 | 954.76 | 960.65 | 996.3 | 1,004.76 | 3,916.47 |
| | 264 | 268.85 | 239.79 | 215.18 | 271.04 | 994.86 |
| | 265 | 2,842.17 | 2,705.95 | 2,660.63 | 2,752.79 | 10,961.54 |
| | 266 | 137.27 | 130.71 | 131.52 | 123.78 | 523.28 |
| | 267 | 256.58 | 276.47 | 281.04 | 290.63 | 1,104.72 |
| | 268 | 233.96 | 225.05 | 224.04 | 246.54 | 929.59 |
| TOTAL | | 23,855.34 | 23,267.08 | 23,223.59 | 23,626.79 | 93,972.80 |

STATE: WISCONSIN

| ZIP CODE | | QUARTER 1 | QUARTER 2 | QUARTER 3 | QUARTER 4 | TOTAL GRAMS |
|---|---|---|---|---|---|---|
| | 530 | 14,494.54 | 14,076.99 | 14,146.19 | 14,728.04 | 57,445.76 |
| | 531 | 15,111.66 | 14,688.09 | 14,682.48 | 15,265.22 | 59,747.45 |
| | 532 | 13,660.07 | 13,381.61 | 13,086.07 | 13,621.37 | 53,749.12 |
| | 534 | 4,307.56 | 4,086.75 | 3,988.51 | 4,108.37 | 16,491.19 |
| | 535 | 10,047.03 | 9,848.42 | 9,735.87 | 10,208.38 | 39,839.70 |
| | 537 | 8,599.88 | 8,268.59 | 8,065.53 | 8,300.65 | 33,234.65 |
| | 538 | 1,210.99 | 1,127.64 | 1,070.99 | 1,145.22 | 4,554.84 |
| | 539 | 3,680.43 | 3,490.13 | 3,510.22 | 3,600.40 | 14,281.18 |
| | 540 | 2,619.15 | 2,568.49 | 2,571.74 | 2,498.43 | 10,257.81 |
| | 541 | 6,814.01 | 6,845.83 | 6,814.58 | 6,861.29 | 27,335.71 |
| | 542 | 2,748.42 | 2,688.36 | 2,745.60 | 2,718.71 | 10,901.09 |

| | | | | | | |
|---|---|---|---|---|---|---|
| | 543 | 8,151.85 | 7,953.90 | 8,163.90 | 8,352.14 | 32,621.79 |
| | 544 | 8,807.30 | 8,498.18 | 8,326.06 | 8,338.27 | 33,969.81 |
| | 545 | 2,367.05 | 2,264.78 | 2,280.49 | 2,185.23 | 9,097.55 |
| | 546 | 4,561.95 | 4,500.34 | 4,274.53 | 4,469.03 | 17,805.85 |
| | 547 | 4,486.56 | 4,533.37 | 4,197.07 | 4,503.49 | 17,720.49 |
| | 548 | 2,525.77 | 2,396.82 | 2,396.23 | 2,355.12 | 9,673.94 |
| | 549 | 15,792.30 | 15,395.98 | 15,411.44 | 15,505.34 | 62,105.06 |
| TOTAL | | 129,986.52 | 126,614.27 | 125,467.50 | 128,764.70 | 510,832.99 |

STATE: WYOMING

| ZIP CODE | | QUARTER 1 | QUARTER 2 | QUARTER 3 | QUARTER 4 | TOTAL GRAMS |
|---|---|---|---|---|---|---|
| | 820 | 1,404.46 | 1,354.91 | 1,313.54 | 1,417.19 | 5,490.10 |
| | 822 | 87.49 | 80.74 | 79.08 | 79.35 | 326.66 |
| | 823 | 122.86 | 116.28 | 131.73 | 83.68 | 454.55 |
| | 824 | 899.32 | 845.86 | 803.52 | 809.94 | 3,358.64 |
| | 825 | 306.28 | 310.44 | 306.1 | 294.89 | 1,217.71 |
| | 826 | 1,063.62 | 970.14 | 937.79 | 999.83 | 3,971.38 |
| | 827 | 490.5 | 456.51 | 529.46 | 489.21 | 1,965.68 |
| | 828 | 609.67 | 564.28 | 670.01 | 598.46 | 2,442.42 |
| | 829 | 1,024.68 | 987.22 | 992.08 | 1,071.36 | 4,075.34 |
| | 830 | 469.62 | 436.05 | 533.86 | 511.34 | 1,950.87 |
| | 831 | 131.36 | 109.68 | 113.77 | 127.79 | 482.6 |
| TOTAL | | 6,609.86 | 6,232.11 | 6,410.94 | 6,483.04 | 25,735.95 |

DRUG CODE: 1105B DRUG NAME: DL-METHAMPHETAMINE RACEMIC BASE
STATE: ALABAMA

| ZIP CODE | | QUARTER 1 | QUARTER 2 | QUARTER 3 | QUARTER 4 | TOTAL GRAMS |
|---|---|---|---|---|---|---|
| | 350 | 1.61 | 2.41 | 0.8 | 1.21 | 6.03 |
| | 351 | 4.42 | 5.22 | 3.62 | 4.82 | 18.08 |
| | 352 | 5.22 | 4.42 | 4.42 | 3.62 | 17.68 |
| | 354 | 0 | 1.61 | 1.21 | 1.21 | 4.03 |
| | 355 | 3.62 | 4.02 | 8.04 | 4.42 | 20.1 |
| | 356 | 0 | 0.8 | 2.81 | 0 | 3.61 |
| | 358 | 0.4 | 0 | 0 | 0 | 0.4 |
| | 359 | 0 | 0 | 0.4 | 0.4 | 0.8 |

| | | | | | | |
|---|---|---|---|---|---|---|
| | 361 | 1.21 | 0.8 | 0.8 | 0 | 2.81 |
| | 362 | 4.42 | 4.02 | 4.42 | 4.42 | 17.28 |
| | 365 | 0.4 | 0 | 0 | 0 | 0.4 |
| | 366 | 1.21 | 1.21 | 1.21 | 1.21 | 4.84 |
| TOTAL | | 22.51 | 24.51 | 27.73 | 21.31 | 96.06 |

STATE: ALASKA

| ZIP CODE | | QUARTER 1 | QUARTER 2 | QUARTER 3 | QUARTER 4 | TOTAL GRAMS |
|---|---|---|---|---|---|---|
| | 995 | 0.8 | 3.21 | 2.41 | 0 | 6.42 |
| | 997 | 0 | 1.61 | 1.61 | 1.21 | 4.43 |
| TOTAL | | 0.8 | 4.82 | 4.02 | 1.21 | 10.85 |

STATE: ARIZONA

| ZIP CODE | | QUARTER 1 | QUARTER 2 | QUARTER 3 | QUARTER 4 | TOTAL GRAMS |
|---|---|---|---|---|---|---|
| | 850 | 2.81 | 2.01 | 2.01 | 2.01 | 8.84 |
| | 852 | 21.7 | 25.32 | 22.1 | 30.54 | 99.66 |
| | 853 | 5.22 | 2.81 | 2.81 | 2.41 | 13.25 |
| | 855 | 2.01 | 1.21 | 1.21 | 1.21 | 5.64 |
| | 856 | 2.41 | 0.8 | 0.4 | 2.41 | 6.02 |
| | 857 | 0 | 0 | 0 | 0.8 | 0.8 |
| TOTAL | | 34.15 | 32.15 | 28.53 | 39.38 | 134.21 |

STATE: ARKANSAS

| ZIP CODE | | QUARTER 1 | QUARTER 2 | QUARTER 3 | QUARTER 4 | TOTAL GRAMS |
|---|---|---|---|---|---|---|
| | 720 | 0.8 | 4.42 | 3.62 | 2.81 | 11.65 |
| | 721 | 2.01 | 1.61 | 0.4 | 0.4 | 4.42 |
| | 722 | 0.8 | 0.8 | 0.8 | 1.21 | 3.61 |
| | 727 | 1.61 | 0.4 | 2.01 | 1.61 | 5.63 |
| TOTAL | | 5.22 | 7.23 | 6.83 | 6.03 | 25.31 |

STATE: CALIFORNIA

| ZIP CODE | | QUARTER 1 | QUARTER 2 | QUARTER 3 | QUARTER 4 | TOTAL GRAMS |
|---|---|---|---|---|---|---|
| | 900 | 34.16 | 22.5 | 10.85 | 8.04 | 75.55 |

| | | | | | |
|---|---|---|---|---|---|
| 902 | 8.84 | 2.81 | 4.82 | 4.02 | 20.49 |
| 904 | 0 | 0.8 | 0 | 0 | 0.8 |
| 905 | 0.8 | 0 | 0 | 0 | 0.8 |
| 906 | 0 | 0.8 | 0 | 0 | 0.8 |
| 907 | 0 | 0 | 0 | 0.8 | 0.8 |
| 908 | 1.21 | 0.4 | 0.8 | 0.4 | 2.81 |
| 910 | 10.05 | 0.4 | 4.82 | 4.82 | 20.09 |
| 911 | 0.8 | 0 | 2.81 | 4.02 | 7.63 |
| 912 | 1.61 | 8.44 | 5.22 | 4.42 | 19.69 |
| 913 | 30.94 | 39.78 | 34.56 | 30.54 | 135.82 |
| 914 | 6.83 | 4.42 | 3.62 | 2.81 | 17.68 |
| 917 | 20.09 | 11.25 | 1.61 | 2.41 | 35.36 |
| 919 | 2.01 | 2.01 | 2.81 | 2.01 | 8.84 |
| 920 | 32.15 | 38.58 | 36.97 | 26.92 | 134.62 |
| 921 | 9.64 | 6.03 | 9.64 | 18.49 | 43.8 |
| 922 | 6.43 | 5.22 | 4.82 | 3.21 | 19.68 |
| 923 | 0 | 0 | 0 | 5.63 | 5.63 |
| 925 | 4.42 | 5.22 | 6.43 | 5.63 | 21.7 |
| 926 | 20.9 | 23.71 | 25.72 | 12.86 | 83.19 |
| 927 | 7.23 | 10.85 | 6.03 | 2.41 | 26.52 |
| 928 | 5.63 | 11.65 | 8.84 | 8.04 | 34.16 |
| 930 | 6.43 | 2.81 | 3.62 | 3.21 | 16.07 |
| 931 | 7.23 | 3.62 | 3.21 | 0.4 | 14.46 |
| 933 | 0 | 0 | 0 | 0.4 | 0.4 |
| 934 | 0 | 0.8 | 1.21 | 0.8 | 2.81 |
| 935 | 2.81 | 0.8 | 0.4 | 1.61 | 5.62 |
| 936 | 0.4 | 0 | 0 | 0 | 0.4 |
| 937 | 6.43 | 4.42 | 6.03 | 5.63 | 22.51 |
| 939 | 1.61 | 1.21 | 1.21 | 0.8 | 4.83 |
| 940 | 9.24 | 9.24 | 10.05 | 14.87 | 43.4 |
| 941 | 13.26 | 16.88 | 16.88 | 12.06 | 59.08 |
| 943 | 0 | 0.4 | 0 | 0 | 0.4 |
| 945 | 10.05 | 10.45 | 7.64 | 8.84 | 36.98 |
| 946 | 3.62 | 2.41 | 2.41 | 2.01 | 10.45 |
| 947 | 0.8 | 2.01 | 1.21 | 0 | 4.02 |
| 949 | 10.05 | 8.44 | 2.41 | 1.61 | 22.51 |
| 950 | 10.45 | 11.65 | 6.43 | 10.85 | 39.38 |

| | | | | | | |
|---|---|---|---|---|---|---|
| | 951 | 12.46 | 12.46 | 14.47 | 15.27 | 54.66 |
| | 952 | 0 | 0.4 | 0 | 0 | 0.4 |
| | 953 | 3.21 | 3.21 | 2.81 | 2.81 | 12.04 |
| | 954 | 0 | 0.8 | 0.8 | 0.4 | 2 |
| | 955 | 1.21 | 0.8 | 1.21 | 1.21 | 4.43 |
| | 956 | 2.01 | 4.82 | 4.42 | 5.63 | 16.88 |
| | 957 | 2.01 | 0.8 | 2.41 | 2.41 | 7.63 |
| | 958 | 7.23 | 6.03 | 4.82 | 4.02 | 22.1 |
| | 959 | 6.03 | 10.85 | 6.43 | 6.83 | 30.14 |
| | 960 | 0.4 | 0.8 | 0.8 | 0.8 | 2.8 |
| TOTAL | | 320.68 | 310.98 | 271.25 | 249.95 | 1,152.86 |

STATE: COLORADO

| ZIP CODE | | QUARTER 1 | QUARTER 2 | QUARTER 3 | QUARTER 4 | TOTAL GRAMS |
|---|---|---|---|---|---|---|
| | 800 | 9.64 | 9.64 | 11.65 | 8.04 | 38.97 |
| | 801 | 15.67 | 20.49 | 13.26 | 21.3 | 70.72 |
| | 802 | 8.44 | 5.22 | 10.45 | 12.46 | 36.57 |
| | 803 | 0.4 | 0 | 0 | 0 | 0.4 |
| | 804 | 1.21 | 1.21 | 0.4 | 3.62 | 6.44 |
| | 805 | 4.42 | 6.03 | 6.03 | 2.41 | 18.89 |
| | 806 | 0 | 0.4 | 0 | 0.4 | 0.8 |
| | 809 | 3.21 | 6.83 | 2.81 | 2.81 | 15.66 |
| | 810 | 1.61 | 0.8 | 2.81 | 0.8 | 6.02 |
| | 811 | 0 | 0 | 0.4 | 0.4 | 0.8 |
| | 812 | 2.01 | 1.61 | 0.4 | 0.8 | 4.82 |
| | 813 | 0 | 0 | 0 | 1.21 | 1.21 |
| | 815 | 1.21 | 1.21 | 0.4 | 0 | 2.82 |
| | 816 | 1.21 | 0.8 | 1.21 | 3.21 | 6.43 |
| TOTAL | | 49.03 | 54.24 | 49.82 | 57.46 | 210.55 |

STATE: CONNECTICUT

| ZIP CODE | | QUARTER 1 | QUARTER 2 | QUARTER 3 | QUARTER 4 | TOTAL GRAMS |
|---|---|---|---|---|---|---|
| | 60 | 4.42 | 6.03 | 5.22 | 6.03 | 21.7 |
| | 61 | 2.81 | 2.81 | 4.02 | 2.41 | 12.05 |
| | 64 | 0.8 | 1.61 | 0 | 0 | 2.41 |

| | | | | | |
|---|---|---|---|---|---|
| 65 | 4.42 | 6.03 | 1.21 | 3.21 | 14.87 |
| 66 | 0 | 0 | 0.4 | 0 | 0.4 |
| 67 | 1.21 | 0.4 | 2.41 | 4.82 | 8.84 |
| 68 | 6.43 | 5.63 | 5.63 | 6.03 | 23.72 |
| 69 | 0.4 | 2.01 | 1.21 | 2.01 | 5.63 |
| TOTAL | 20.49 | 24.52 | 20.1 | 24.51 | 89.62 |

STATE: DELAWARE

| ZIP CODE | QUARTER 1 | QUARTER 2 | QUARTER 3 | QUARTER 4 | TOTAL GRAMS |
|---|---|---|---|---|---|
| 197 | 0 | 0 | 0.4 | 0.8 | 1.2 |
| 198 | 1.21 | 2.01 | 3.21 | 2.01 | 8.44 |
| 199 | 1.21 | 1.61 | 1.21 | 1.61 | 5.64 |
| TOTAL | 2.42 | 3.62 | 4.82 | 4.42 | 15.28 |

STATE: DISTRICT OF COLUMBIA

| ZIP CODE | QUARTER 1 | QUARTER 2 | QUARTER 3 | QUARTER 4 | TOTAL GRAMS |
|---|---|---|---|---|---|
| 200 | 4.02 | 2.01 | 0 | 0.8 | 6.83 |
| TOTAL | 4.02 | 2.01 | 0 | 0.8 | 6.83 |

STATE: FLORIDA

| ZIP CODE | QUARTER 1 | QUARTER 2 | QUARTER 3 | QUARTER 4 | TOTAL GRAMS |
|---|---|---|---|---|---|
| 321 | 0 | 0 | 0.4 | 1.61 | 2.01 |
| 322 | 3.62 | 6.03 | 4.42 | 2.01 | 16.08 |
| 323 | 0.8 | 0.4 | 0 | 0 | 1.2 |
| 324 | 0 | 0.4 | 1.61 | 0 | 2.01 |
| 325 | 2.81 | 2.41 | 3.62 | 1.61 | 10.45 |
| 326 | 2.81 | 2.81 | 1.61 | 1.21 | 8.44 |
| 327 | 1.61 | 2.01 | 2.81 | 2.41 | 8.84 |
| 328 | 0 | 0 | 3.21 | 2.01 | 5.22 |
| 329 | 1.61 | 2.41 | 1.61 | 0.8 | 6.43 |
| 330 | 0.8 | 0 | 2.41 | 3.21 | 6.42 |
| 331 | 1.61 | 3.21 | 0 | 2.01 | 6.83 |
| 333 | 1.21 | 3.21 | 4.42 | 2.41 | 11.25 |
| 334 | 1.61 | 2.81 | 0.4 | 0.8 | 5.62 |

| | | | | | | |
|---|---|---|---|---|---|---|
| | 335 | 1.21 | 0.4 | 0.4 | 0.4 | 2.41 |
| | 336 | 0.8 | 0 | 0 | 0 | 0.8 |
| | 337 | 0.8 | 0.8 | 0 | 0.4 | 2 |
| | 338 | 0 | 4.42 | 1.61 | 3.21 | 9.24 |
| | 339 | 2.01 | 4.02 | 3.62 | 2.41 | 12.06 |
| | 341 | 0.4 | 0 | 0.4 | 1.21 | 2.01 |
| | 342 | 2.81 | 4.02 | 2.41 | 2.41 | 11.65 |
| | 346 | 0.4 | 0 | 1.61 | 1.61 | 3.62 |
| | 347 | 1.21 | 0 | 0 | 0 | 1.21 |
| TOTAL | | 28.13 | 39.36 | 36.57 | 31.74 | 135.8 |

STATE: GEORGIA

| ZIP CODE | | QUARTER 1 | QUARTER 2 | QUARTER 3 | QUARTER 4 | TOTAL GRAMS |
|---|---|---|---|---|---|---|
| | 300 | 5.22 | 4.42 | 5.22 | 6.83 | 21.69 |
| | 301 | 6.03 | 4.42 | 4.82 | 6.83 | 22.1 |
| | 303 | 6.43 | 5.22 | 2.81 | 2.41 | 16.87 |
| | 304 | 2.01 | 2.01 | 1.61 | 1.61 | 7.24 |
| | 305 | 1.61 | 0 | 0 | 0 | 1.61 |
| | 306 | 0.8 | 0.4 | 0.8 | 0 | 2 |
| | 309 | 3.21 | 2.81 | 0 | 0 | 6.02 |
| | 310 | 2.81 | 1.21 | 0 | 0 | 4.02 |
| | 313 | 1.21 | 1.21 | 0.8 | 2.41 | 5.63 |
| | 315 | 2.01 | 2.81 | 0 | 0 | 4.82 |
| | 316 | 0.4 | 0 | 0.4 | 0 | 0.8 |
| | 317 | 0.4 | 0 | 0.4 | 0.4 | 1.2 |
| | 398 | 0 | 0 | 0.8 | 0 | 0.8 |
| TOTAL | | 32.14 | 24.51 | 17.66 | 20.49 | 94.8 |

STATE: HAWAII

| ZIP CODE | | QUARTER 1 | QUARTER 2 | QUARTER 3 | QUARTER 4 | TOTAL GRAMS |
|---|---|---|---|---|---|---|
| | 967 | 3.21 | 5.63 | 0.8 | 0.4 | 10.04 |
| | 968 | 2.01 | 1.21 | 3.21 | 4.02 | 10.45 |
| TOTAL | | 5.22 | 6.84 | 4.01 | 4.42 | 20.49 |

STATE: IDAHO

| ZIP CODE | QUARTER 1 | QUARTER 2 | QUARTER 3 | QUARTER 4 | TOTAL GRAMS |
|---|---|---|---|---|---|
| 832 | 0 | 0 | 0.8 | 0 | 0.8 |
| 833 | 0.8 | 0 | 0 | 0 | 0.8 |
| 834 | 0.4 | 0 | 0.8 | 0.4 | 1.6 |
| 836 | 0.4 | 2.41 | 5.63 | 7.64 | 16.08 |
| 837 | 1.61 | 0.8 | 2.81 | 1.61 | 6.83 |
| 838 | 4.42 | 3.62 | 0.4 | 0.4 | 8.84 |
| TOTAL | 7.63 | 6.83 | 10.44 | 10.05 | 34.95 |

STATE: ILLINOIS

| ZIP CODE | QUARTER 1 | QUARTER 2 | QUARTER 3 | QUARTER 4 | TOTAL GRAMS |
|---|---|---|---|---|---|
| 600 | 20.49 | 21.3 | 16.48 | 20.9 | 79.17 |
| 601 | 4.42 | 4.42 | 4.82 | 4.82 | 18.48 |
| 602 | 0 | 0 | 0.4 | 0 | 0.4 |
| 603 | 0 | 0 | 1.21 | 0 | 1.21 |
| 604 | 1.61 | 2.41 | 2.41 | 4.42 | 10.85 |
| 605 | 19.29 | 16.07 | 17.68 | 18.49 | 71.53 |
| 606 | 11.25 | 6.83 | 10.05 | 6.43 | 34.56 |
| 607 | 0.4 | 0.4 | 0 | 0 | 0.8 |
| 611 | 1.21 | 0 | 1.21 | 0.8 | 3.22 |
| 612 | 0 | 0.8 | 2.41 | 0 | 3.21 |
| 614 | 1.21 | 2.01 | 1.21 | 1.21 | 5.64 |
| 615 | 0 | 0 | 0 | 0.8 | 0.8 |
| 617 | 0 | 0 | 0 | 0.4 | 0.4 |
| 618 | 0 | 0.4 | 0 | 0 | 0.4 |
| 620 | 0.4 | 0.4 | 1.61 | 0 | 2.41 |
| 623 | 0 | 0.4 | 0 | 0 | 0.4 |
| 624 | 0 | 0 | 0.8 | 0.4 | 1.2 |
| 627 | 3.62 | 2.81 | 3.21 | 3.62 | 13.26 |
| 629 | 0 | 0 | 0 | 0.4 | 0.4 |
| TOTAL | 63.9 | 58.25 | 63.5 | 62.69 | 248.34 |

STATE: INDIANA

| ZIP CODE | QUARTER 1 | QUARTER 2 | QUARTER 3 | QUARTER 4 | TOTAL GRAMS |
|---|---|---|---|---|---|

| | | | | | | |
|---|---|---|---|---|---|---|
| | 460 | 9.24 | 6.43 | 5.22 | 7.23 | 28.12 |
| | 462 | 2.41 | 1.21 | 0.4 | 0 | 4.02 |
| | 463 | 0 | 0 | 0.8 | 1.21 | 2.01 |
| | 469 | 7.23 | 9.64 | 14.47 | 6.43 | 37.77 |
| | 471 | 2.81 | 2.41 | 2.81 | 1.21 | 9.24 |
| | 474 | 6.43 | 6.03 | 4.82 | 1.61 | 18.89 |
| | 477 | 0.8 | 0.4 | 1.21 | 0.4 | 2.81 |
| | 478 | 0.8 | 0 | 0 | 0 | 0.8 |
| TOTAL | | 29.72 | 26.12 | 29.73 | 18.09 | 103.66 |

STATE: IOWA

| ZIP CODE | | QUARTER 1 | QUARTER 2 | QUARTER 3 | QUARTER 4 | TOTAL GRAMS |
|---|---|---|---|---|---|---|
| | 500 | 0 | 0 | 1.21 | 1.61 | 2.82 |
| | 501 | 0 | 0.4 | 0 | 0 | 0.4 |
| | 502 | 4.02 | 6.43 | 7.64 | 8.04 | 26.13 |
| | 503 | 2.01 | 2.81 | 1.61 | 2.41 | 8.84 |
| | 504 | 3.21 | 4.02 | 3.62 | 5.22 | 16.07 |
| | 505 | 0 | 0 | 0.4 | 0.4 | 0.8 |
| | 506 | 0.4 | 0.4 | 0 | 0 | 0.8 |
| | 515 | 1.61 | 1.61 | 2.41 | 0.8 | 6.43 |
| | 516 | 1.61 | 1.21 | 0 | 0.8 | 3.62 |
| | 520 | 0.4 | 0 | 0 | 0 | 0.4 |
| | 528 | 1.61 | 2.01 | 2.01 | 2.01 | 7.64 |
| TOTAL | | 14.87 | 18.89 | 18.9 | 21.29 | 73.95 |

STATE: KANSAS

| ZIP CODE | | QUARTER 1 | QUARTER 2 | QUARTER 3 | QUARTER 4 | TOTAL GRAMS |
|---|---|---|---|---|---|---|
| | 660 | 3.21 | 4.42 | 3.62 | 3.62 | 14.87 |
| | 661 | 0 | 0.8 | 0 | 0 | 0.8 |
| | 665 | 1.61 | 1.61 | 1.21 | 1.61 | 6.04 |
| | 666 | 0 | 0 | 0 | 1.21 | 1.21 |
| | 670 | 0 | 0 | 0 | 0.4 | 0.4 |
| | 672 | 6.03 | 5.63 | 3.21 | 3.62 | 18.49 |
| TOTAL | | 10.85 | 12.46 | 8.04 | 10.46 | 41.81 |

STATE: KENTUCKY

| ZIP CODE | QUARTER 1 | QUARTER 2 | QUARTER 3 | QUARTER 4 | TOTAL GRAMS |
|---|---|---|---|---|---|
| 400 | 0.4 | 0 | 0 | 0 | 0.4 |
| 402 | 2.01 | 0.4 | 0.4 | 0 | 2.81 |
| 403 | 2.81 | 2.01 | 3.21 | 1.61 | 9.64 |
| 405 | 0 | 2.41 | 2.81 | 5.22 | 10.44 |
| 406 | 0 | 0 | 0.8 | 0.4 | 1.2 |
| 408 | 2.41 | 1.21 | 0 | 0 | 3.62 |
| 410 | 0.4 | 0.4 | 0 | 0.4 | 1.2 |
| 417 | 0.4 | 0.4 | 0 | 0 | 0.8 |
| 420 | 0.8 | 0.8 | 0 | 1.21 | 2.81 |
| 424 | 1.61 | 1.61 | 2.01 | 0 | 5.23 |
| 425 | 0 | 0.8 | 0.4 | 0.4 | 1.6 |
| TOTAL | 10.84 | 10.04 | 9.63 | 9.24 | 39.75 |

STATE: LOUISIANA

| ZIP CODE | QUARTER 1 | QUARTER 2 | QUARTER 3 | QUARTER 4 | TOTAL GRAMS |
|---|---|---|---|---|---|
| 700 | 0 | 0.4 | 0 | 0 | 0.4 |
| 701 | 0.4 | 0 | 0 | 0 | 0.4 |
| 703 | 1.61 | 2.41 | 2.81 | 3.21 | 10.04 |
| 704 | 2.01 | 1.61 | 2.41 | 1.21 | 7.24 |
| 705 | 0.4 | 0 | 0 | 0 | 0.4 |
| 706 | 1.21 | 0.4 | 1.61 | 0.8 | 4.02 |
| 707 | 1.61 | 2.41 | 0.8 | 2.01 | 6.83 |
| 708 | 2.41 | 2.81 | 0.8 | 0 | 6.02 |
| 710 | 0.8 | 0 | 0 | 0 | 0.8 |
| 711 | 0.8 | 2.41 | 2.41 | 2.81 | 8.43 |
| 712 | 0 | 0.4 | 0.4 | 0 | 0.8 |
| 713 | 2.81 | 2.01 | 2.81 | 2.01 | 9.64 |
| TOTAL | 14.06 | 14.86 | 14.05 | 12.05 | 55.02 |

STATE: MAINE

| ZIP CODE | QUARTER 1 | QUARTER 2 | QUARTER 3 | QUARTER 4 | TOTAL GRAMS |
|---|---|---|---|---|---|
| 39 | 0 | 0 | 0.4 | 0 | 0.4 |

| | | | | | | |
|---|---|---|---|---|---|---|
| | 40 | 3.21 | 3.62 | 1.61 | 6.03 | 14.47 |
| | 41 | 0 | 0 | 0 | 2.01 | 2.01 |
| | 42 | 0 | 2.01 | 8.04 | 3.62 | 13.67 |
| | 48 | 0 | 1.21 | 0.4 | 1.21 | 2.82 |
| TOTAL | | 3.21 | 6.84 | 10.45 | 12.87 | 33.37 |

| STATE: | MARYLAND | | | | | |
|---|---|---|---|---|---|---|
| ZIP CODE | | QUARTER 1 | QUARTER 2 | QUARTER 3 | QUARTER 4 | TOTAL GRAMS |
| | 206 | 0 | 0.4 | 0 | 0 | 0.4 |
| | 207 | 0 | 0 | 0.4 | 0.8 | 1.2 |
| | 208 | 11.25 | 7.23 | 8.44 | 8.84 | 35.76 |
| | 210 | 10.45 | 7.23 | 7.23 | 7.64 | 32.55 |
| | 211 | 8.84 | 8.84 | 11.65 | 6.03 | 35.36 |
| | 212 | 9.64 | 18.89 | 8.04 | 15.67 | 52.24 |
| | 214 | 0.4 | 0 | 0.8 | 0 | 1.2 |
| | 217 | 0.8 | 2.01 | 1.61 | 1.21 | 5.63 |
| TOTAL | | 41.38 | 44.6 | 38.17 | 40.19 | 164.34 |

| STATE: | MASSACHUSETTS | | | | | |
|---|---|---|---|---|---|---|
| ZIP CODE | | QUARTER 1 | QUARTER 2 | QUARTER 3 | QUARTER 4 | TOTAL GRAMS |
| | 10 | 19.29 | 18.08 | 17.68 | 16.48 | 71.53 |
| | 12 | 2.41 | 2.01 | 2.41 | 1.61 | 8.44 |
| | 14 | 3.21 | 2.81 | 2.01 | 0.8 | 8.83 |
| | 16 | 0 | 0 | 0 | 1.21 | 1.21 |
| | 17 | 3.62 | 3.21 | 3.21 | 4.02 | 14.06 |
| | 18 | 2.81 | 2.81 | 4.42 | 6.43 | 16.47 |
| | 19 | 2.81 | 4.42 | 4.02 | 2.81 | 14.06 |
| | 20 | 0 | 0.8 | 0 | 0 | 0.8 |
| | 21 | 5.63 | 4.02 | 4.42 | 2.81 | 16.88 |
| | 23 | 0.8 | 0.4 | 0.8 | 0.4 | 2.4 |
| | 24 | 2.81 | 2.01 | 1.61 | 1.21 | 7.64 |
| | 27 | 1.21 | 0.8 | 2.41 | 1.61 | 6.03 |
| TOTAL | | 44.6 | 41.37 | 42.99 | 39.39 | 168.35 |

RETAIL DRUG DISTRIBUTION BY ZIP CODE WITHIN STATE BY GRAMS WT

STATE: MICHIGAN

| ZIP CODE | QUARTER 1 | QUARTER 2 | QUARTER 3 | QUARTER 4 | TOTAL GRAMS |
|---|---|---|---|---|---|
| 480 | 1.61 | 0.8 | 0.4 | 1.21 | 4.02 |
| 481 | 4.82 | 4.42 | 4.42 | 2.41 | 16.07 |
| 483 | 0.8 | 2.01 | 3.21 | 2.81 | 8.83 |
| 484 | 0 | 0.4 | 0 | 0 | 0.4 |
| 486 | 2.41 | 0 | 0 | 0 | 2.41 |
| 488 | 2.01 | 1.61 | 1.61 | 0.8 | 6.03 |
| 489 | 0 | 0 | 0 | 0.4 | 0.4 |
| 490 | 9.64 | 8.84 | 10.85 | 12.46 | 41.79 |
| 492 | 0 | 0 | 0 | 0.8 | 0.8 |
| 493 | 1.61 | 3.62 | 0 | 2.01 | 7.24 |
| 495 | 0 | 0.4 | 0.4 | 0.8 | 1.6 |
| 498 | 0 | 0.4 | 0.8 | 0.8 | 2 |
| TOTAL | 22.9 | 22.5 | 21.69 | 24.5 | 91.59 |

STATE: MINNESOTA

| ZIP CODE | QUARTER 1 | QUARTER 2 | QUARTER 3 | QUARTER 4 | TOTAL GRAMS |
|---|---|---|---|---|---|
| 550 | 0.8 | 0.4 | 5.22 | 3.62 | 10.04 |
| 551 | 13.26 | 12.06 | 5.63 | 8.04 | 38.99 |
| 553 | 3.62 | 5.22 | 7.64 | 5.63 | 22.11 |
| 554 | 27.73 | 34.96 | 18.08 | 14.47 | 95.24 |
| 557 | 1.21 | 4.02 | 2.41 | 4.42 | 12.06 |
| 559 | 21.7 | 24.91 | 19.69 | 19.29 | 85.59 |
| 560 | 2.41 | 2.41 | 1.21 | 1.61 | 7.64 |
| 562 | 2.81 | 0.4 | 1.61 | 0.8 | 5.62 |
| 563 | 1.21 | 0.8 | 0 | 0 | 2.01 |
| 565 | 2.41 | 2.01 | 2.41 | 1.21 | 8.04 |
| 566 | 1.21 | 0 | 0 | 0.8 | 2.01 |
| TOTAL | 78.37 | 87.19 | 63.9 | 59.89 | 289.35 |

STATE: MISSISSIPPI

| ZIP CODE | QUARTER 1 | QUARTER 2 | QUARTER 3 | QUARTER 4 | TOTAL GRAMS |
|---|---|---|---|---|---|
| 386 | 0 | 0 | 0 | 0.4 | 0.4 |

| | | | | | | |
|---|---|---|---|---|---|---|
| | 391 | 0.4 | 0 | 0 | 0 | 0.4 |
| | 392 | 6.43 | 6.43 | 7.64 | 5.63 | 26.13 |
| | 394 | 0 | 0 | 0 | 0.4 | 0.4 |
| | 396 | 1.21 | 0.8 | 1.21 | 1.21 | 4.43 |
| | 397 | 0.4 | 0.8 | 0 | 0 | 1.2 |
| TOTAL | | 8.44 | 8.03 | 8.85 | 7.64 | 32.96 |

STATE: MISSOURI

| ZIP CODE | | QUARTER 1 | QUARTER 2 | QUARTER 3 | QUARTER 4 | TOTAL GRAMS |
|---|---|---|---|---|---|---|
| | 630 | 3.62 | 2.41 | 2.41 | 3.21 | 11.65 |
| | 631 | 34.16 | 49.43 | 6.43 | 48.62 | 138.64 |
| | 634 | 0.8 | 1.21 | 0 | 2.01 | 4.02 |
| | 636 | 2.41 | 0 | 0 | 1.21 | 3.62 |
| | 639 | 1.61 | 1.21 | 1.61 | 1.61 | 6.04 |
| | 640 | 0.8 | 0.4 | 0.8 | 2.81 | 4.81 |
| | 641 | 1.61 | 1.61 | 2.01 | 1.21 | 6.44 |
| | 645 | 1.21 | 2.01 | 2.81 | 2.01 | 8.04 |
| | 652 | 1.21 | 2.41 | 0 | 0 | 3.62 |
| | 653 | 0.4 | 0.8 | 0 | 0 | 1.2 |
| | 655 | 0.4 | 0 | 0 | 0 | 0.4 |
| | 657 | 0.8 | 0 | 0 | 0 | 0.8 |
| | 658 | 0 | 0.8 | 0 | 0 | 0.8 |
| TOTAL | | 49.03 | 62.29 | 16.07 | 62.69 | 190.08 |

STATE: MONTANA

| ZIP CODE | | QUARTER 1 | QUARTER 2 | QUARTER 3 | QUARTER 4 | TOTAL GRAMS |
|---|---|---|---|---|---|---|
| | 593 | 0.4 | 0.4 | 0.4 | 0.4 | 1.6 |
| | 594 | 0.4 | 0 | 0 | 0 | 0.4 |
| | 597 | 0 | 0.4 | 0 | 0 | 0.4 |
| | 598 | 0.4 | 0 | 0 | 0 | 0.4 |
| TOTAL | | 1.2 | 0.8 | 0.4 | 0.4 | 2.8 |

STATE: NEBRASKA

| ZIP CODE | | QUARTER 1 | QUARTER 2 | QUARTER 3 | QUARTER 4 | TOTAL GRAMS |
|---|---|---|---|---|---|---|

| | | | | | |
|---|---|---|---|---|---|
| 680 | 0.4 | 1.61 | 0 | 0 | 2.01 |
| 681 | 4.82 | 6.03 | 5.63 | 5.63 | 22.11 |
| 684 | 0 | 0 | 1.21 | 2.01 | 3.22 |
| 685 | 0 | 0.8 | 1.21 | 0.8 | 2.81 |
| 687 | 0.8 | 0.4 | 0.8 | 0.8 | 2.8 |
| TOTAL | 6.02 | 8.84 | 8.85 | 9.24 | 32.95 |

STATE: NEVADA

| ZIP CODE | QUARTER 1 | QUARTER 2 | QUARTER 3 | QUARTER 4 | TOTAL GRAMS |
|---|---|---|---|---|---|
| 890 | 1.21 | 0.8 | 0 | 0 | 2.01 |
| 891 | 20.09 | 25.72 | 17.68 | 18.08 | 81.57 |
| 893 | 0.4 | 0.8 | 0.4 | 1.61 | 3.21 |
| 894 | 0 | 0.4 | 1.61 | 0 | 2.01 |
| 895 | 2.01 | 1.21 | 2.41 | 2.41 | 8.04 |
| 897 | 0 | 0 | 0 | 0.4 | 0.4 |
| 898 | 0 | 0.4 | 0 | 0 | 0.4 |
| TOTAL | 23.71 | 29.33 | 22.1 | 22.5 | 97.64 |

STATE: NEW HAMPSHIRE

| ZIP CODE | QUARTER 1 | QUARTER 2 | QUARTER 3 | QUARTER 4 | TOTAL GRAMS |
|---|---|---|---|---|---|
| 31 | 2.41 | 2.41 | 3.62 | 1.61 | 10.05 |
| 37 | 3.21 | 2.41 | 3.21 | 1.21 | 10.04 |
| 38 | 2.41 | 2.81 | 0 | 0.8 | 6.02 |
| TOTAL | 8.03 | 7.63 | 6.83 | 3.62 | 26.11 |

STATE: NEW JERSEY

| ZIP CODE | QUARTER 1 | QUARTER 2 | QUARTER 3 | QUARTER 4 | TOTAL GRAMS |
|---|---|---|---|---|---|
| 70 | 4.42 | 1.21 | 2.81 | 2.01 | 10.45 |
| 71 | 0 | 0 | 0 | 0.8 | 0.8 |
| 73 | 0.4 | 0 | 0 | 0 | 0.4 |
| 74 | 2.41 | 3.21 | 2.41 | 3.21 | 11.24 |
| 76 | 9.24 | 7.23 | 6.83 | 7.64 | 30.94 |
| 77 | 0 | 0 | 0 | 1.21 | 1.21 |

| | | | | | | |
|---|---|---|---|---|---|---|
| | 78 | 4.82 | 1.21 | 3.62 | 4.82 | 14.47 |
| | 79 | 1.21 | 1.21 | 1.61 | 0.8 | 4.83 |
| | 80 | 7.64 | 8.44 | 8.04 | 11.25 | 35.37 |
| | 81 | 1.21 | 1.21 | 2.01 | 0.8 | 5.23 |
| | 85 | 1.21 | 1.61 | 0.4 | 0.4 | 3.62 |
| | 86 | 0 | 0 | 0.4 | 1.21 | 1.61 |
| | 87 | 0.8 | 0 | 0 | 0 | 0.8 |
| | 88 | 4.82 | 5.22 | 3.62 | 3.62 | 17.28 |
| TOTAL | | 38.18 | 30.55 | 31.75 | 37.77 | 138.25 |

STATE: NEW MEXICO

| ZIP CODE | QUARTER 1 | QUARTER 2 | QUARTER 3 | QUARTER 4 | TOTAL GRAMS |
|---|---|---|---|---|---|
| 871 | 20.9 | 15.67 | 12.06 | 4.02 | 52.65 |
| 880 | 2.01 | 2.41 | 0 | 0.8 | 5.22 |
| 882 | 3.21 | 0.4 | 0.4 | 0 | 4.01 |
| 883 | 1.21 | 2.41 | 0.4 | 0 | 4.02 |
| TOTAL | 27.33 | 20.89 | 12.86 | 4.82 | 65.9 |

STATE: NEW YORK

| ZIP CODE | QUARTER 1 | QUARTER 2 | QUARTER 3 | QUARTER 4 | TOTAL GRAMS |
|---|---|---|---|---|---|
| 100 | 20.49 | 19.29 | 16.48 | 16.88 | 73.14 |
| 101 | 0.4 | 0.8 | 0 | 0.8 | 2 |
| 104 | 0.8 | 0.8 | 0.4 | 0.8 | 2.8 |
| 105 | 2.01 | 1.21 | 1.61 | 5.63 | 10.46 |
| 106 | 8.84 | 8.84 | 10.05 | 10.85 | 38.58 |
| 108 | 0 | 0 | 1.21 | 0 | 1.21 |
| 109 | 6.03 | 6.83 | 4.82 | 2.81 | 20.49 |
| 111 | 0 | 0.8 | 0 | 0 | 0.8 |
| 112 | 0.4 | 0.4 | 1.21 | 2.41 | 4.42 |
| 113 | 2.81 | 2.81 | 1.61 | 2.41 | 9.64 |
| 114 | 1.61 | 2.01 | 1.61 | 2.41 | 7.64 |
| 115 | 2.41 | 5.22 | 4.82 | 4.42 | 16.87 |
| 117 | 6.03 | 8.04 | 8.04 | 16.07 | 38.18 |
| 118 | 4.02 | 3.21 | 0 | 0 | 7.23 |
| 119 | 0.4 | 0 | 0 | 0 | 0.4 |

| | | | | | | |
|---|---|---|---|---|---|---|
| | 120 | 2.01 | 1.21 | 1.61 | 2.01 | 6.84 |
| | 122 | 0 | 0.8 | 1.61 | 0 | 2.41 |
| | 123 | 3.62 | 3.62 | 3.21 | 2.01 | 12.46 |
| | 125 | 0.8 | 0.4 | 0 | 1.21 | 2.41 |
| | 127 | 0 | 0 | 0.4 | 1.61 | 2.01 |
| | 128 | 0.8 | 1.61 | 1.21 | 0.8 | 4.42 |
| | 130 | 0.8 | 1.21 | 2.41 | 2.81 | 7.23 |
| | 131 | 0 | 0.4 | 0 | 0 | 0.4 |
| | 134 | 0.8 | 0 | 0.4 | 0.4 | 1.6 |
| | 136 | 0 | 0 | 0 | 2.01 | 2.01 |
| | 139 | 0 | 0.8 | 0.8 | 0.8 | 2.4 |
| | 140 | 0.8 | 1.21 | 3.62 | 6.03 | 11.66 |
| | 141 | 1.61 | 3.21 | 2.81 | 3.21 | 10.84 |
| | 142 | 2.81 | 3.62 | 2.41 | 1.61 | 10.45 |
| | 144 | 2.41 | 0 | 0 | 0 | 2.41 |
| | 148 | 1.61 | 2.41 | 4.42 | 3.62 | 12.06 |
| TOTAL | | 74.32 | 80.76 | 76.77 | 93.62 | 325.47 |

| STATE: | NORTH CAROLINA | | | | | |
|---|---|---|---|---|---|---|
| ZIP CODE | | QUARTER 1 | QUARTER 2 | QUARTER 3 | QUARTER 4 | TOTAL GRAMS |
| | 270 | 0 | 0 | 0 | 0.4 | 0.4 |
| | 271 | 0 | 0 | 0.8 | 0.8 | 1.6 |
| | 272 | 7.64 | 6.43 | 1.61 | 2.41 | 18.09 |
| | 273 | 0 | 0.8 | 0.8 | 0 | 1.6 |
| | 274 | 0 | 0.4 | 0 | 0.8 | 1.2 |
| | 275 | 11.25 | 22.5 | 10.85 | 10.85 | 55.45 |
| | 276 | 6.83 | 6.43 | 10.05 | 8.44 | 31.75 |
| | 277 | 2.81 | 4.02 | 4.02 | 3.21 | 14.06 |
| | 278 | 0.8 | 1.21 | 0 | 0 | 2.01 |
| | 280 | 2.01 | 0.8 | 0.4 | 2.41 | 5.62 |
| | 281 | 7.23 | 4.42 | 6.03 | 6.43 | 24.11 |
| | 282 | 2.01 | 2.01 | 2.41 | 2.01 | 8.44 |
| | 283 | 3.21 | 4.02 | 2.81 | 2.41 | 12.45 |
| | 284 | 0 | 0 | 0 | 1.61 | 1.61 |
| | 285 | 0.4 | 0 | 0 | 0 | 0.4 |
| | 286 | 2.01 | 1.21 | 0 | 0 | 3.22 |

| | | | | | | |
|---|---|---|---|---|---|---|
| | 287 | 0.4 | 1.21 | 1.21 | 0.8 | 3.62 |
| | 288 | 1.21 | 1.21 | 1.21 | 0 | 3.63 |
| TOTAL | | 47.81 | 56.67 | 42.2 | 42.58 | 189.26 |

STATE: NORTH DAKOTA

| ZIP CODE | | QUARTER 1 | QUARTER 2 | QUARTER 3 | QUARTER 4 | TOTAL GRAMS |
|---|---|---|---|---|---|---|
| | 581 | 1.21 | 0 | 0 | 0 | 1.21 |
| | 584 | 1.61 | 4.02 | 4.02 | 5.63 | 15.28 |
| | 587 | 0 | 3.62 | 2.01 | 1.61 | 7.24 |
| TOTAL | | 2.82 | 7.64 | 6.03 | 7.24 | 23.73 |

STATE: OHIO

| ZIP CODE | | QUARTER 1 | QUARTER 2 | QUARTER 3 | QUARTER 4 | TOTAL GRAMS |
|---|---|---|---|---|---|---|
| | 430 | 0 | 0 | 0.4 | 0.4 | 0.8 |
| | 431 | 0.4 | 1.21 | 1.61 | 1.21 | 4.43 |
| | 432 | 47.02 | 49.83 | 21.3 | 68.72 | 186.87 |
| | 434 | 0 | 0.8 | 0.8 | 0 | 1.6 |
| | 439 | 0 | 0.4 | 0 | 0 | 0.4 |
| | 440 | 0 | 0 | 0 | 0.4 | 0.4 |
| | 441 | 4.02 | 4.42 | 4.02 | 2.01 | 14.47 |
| | 442 | 0 | 0.4 | 0 | 0 | 0.4 |
| | 444 | 0 | 0.8 | 0 | 0.4 | 1.2 |
| | 445 | 2.01 | 3.21 | 2.41 | 2.01 | 9.64 |
| | 446 | 2.01 | 0.4 | 1.21 | 2.41 | 6.03 |
| | 447 | 1.21 | 1.61 | 1.61 | 1.21 | 5.64 |
| | 448 | 0 | 1.21 | 1.61 | 2.01 | 4.83 |
| | 450 | 9.64 | 9.24 | 8.84 | 6.83 | 34.55 |
| | 451 | 0.8 | 0 | 0 | 0 | 0.8 |
| | 452 | 2.01 | 0.8 | 2.01 | 1.61 | 6.43 |
| | 453 | 1.21 | 0.8 | 0.8 | 1.21 | 4.02 |
| | 454 | 5.22 | 4.42 | 2.01 | 2.81 | 14.46 |
| | 457 | 2.01 | 1.21 | 0 | 0 | 3.22 |
| TOTAL | | 77.56 | 80.76 | 48.63 | 93.24 | 300.19 |

DATE RANGE: 01/01/2017 TO 12/31/2017 ARCOS 3 - REPORT 1 Run Date: 07/03/2018

RETAIL DRUG DISTRIBUTION BY ZIP CODE WITHIN STATE BY GRAMS WT

STATE: OKLAHOMA

| ZIP CODE | QUARTER 1 | QUARTER 2 | QUARTER 3 | QUARTER 4 | TOTAL GRAMS |
|---|---|---|---|---|---|
| 730 | 4.02 | 3.62 | 3.21 | 1.61 | 12.46 |
| 731 | 3.21 | 4.02 | 5.22 | 1.61 | 14.06 |
| 735 | 2.01 | 1.61 | 2.81 | 2.81 | 9.24 |
| 737 | 0 | 0 | 0 | 1.21 | 1.21 |
| 740 | 7.23 | 3.21 | 6.83 | 4.02 | 21.29 |
| 741 | 5.63 | 3.62 | 0.8 | 2.41 | 12.46 |
| 745 | 0 | 0.4 | 0.8 | 1.61 | 2.81 |
| 747 | 1.21 | 0 | 0.8 | 0.4 | 2.41 |
| 748 | 0.4 | 0 | 0 | 0.8 | 1.2 |
| TOTAL | 23.71 | 16.48 | 20.47 | 16.48 | 77.14 |

STATE: OREGON

| ZIP CODE | QUARTER 1 | QUARTER 2 | QUARTER 3 | QUARTER 4 | TOTAL GRAMS |
|---|---|---|---|---|---|
| 970 | 5.22 | 4.82 | 2.81 | 6.83 | 19.68 |
| 971 | 8.04 | 0.4 | 0 | 0.4 | 8.84 |
| 972 | 10.05 | 7.64 | 6.43 | 9.64 | 33.76 |
| 973 | 6.03 | 2.81 | 6.03 | 9.64 | 24.51 |
| 974 | 0.8 | 0.4 | 1.21 | 2.81 | 5.22 |
| 975 | 5.63 | 7.23 | 1.61 | 0 | 14.47 |
| 977 | 0.4 | 1.61 | 1.21 | 2.01 | 5.23 |
| 978 | 8.04 | 8.04 | 0 | 0 | 16.08 |
| TOTAL | 44.21 | 32.95 | 19.3 | 31.33 | 127.79 |

STATE: PENNSYLVANIA

| ZIP CODE | QUARTER 1 | QUARTER 2 | QUARTER 3 | QUARTER 4 | TOTAL GRAMS |
|---|---|---|---|---|---|
| 150 | 6.83 | 2.81 | 6.43 | 5.63 | 21.7 |
| 151 | 3.62 | 2.81 | 3.21 | 2.41 | 12.05 |
| 152 | 16.07 | 20.9 | 22.1 | 18.49 | 77.56 |
| 156 | 0.8 | 1.21 | 1.21 | 0.4 | 3.62 |
| 160 | 2.41 | 3.21 | 1.21 | 3.21 | 10.04 |
| 168 | 2.81 | 5.22 | 3.62 | 4.82 | 16.47 |
| 170 | 4.02 | 2.81 | 3.21 | 2.41 | 12.45 |

| | | | | | | |
|---|---|---|---|---|---|---|
| | 171 | 0 | 1.21 | 1.21 | 4.02 | 6.44 |
| | 173 | 1.61 | 2.01 | 0.8 | 4.82 | 9.24 |
| | 174 | 0 | 0.4 | 1.61 | 0.8 | 2.81 |
| | 175 | 1.21 | 0.4 | 0 | 0 | 1.61 |
| | 176 | 0.8 | 0.8 | 0.4 | 0 | 2 |
| | 178 | 2.81 | 4.02 | 6.03 | 6.03 | 18.89 |
| | 180 | 1.61 | 0.4 | 0.4 | 3.21 | 5.62 |
| | 181 | 2.81 | 2.81 | 1.21 | 3.21 | 10.04 |
| | 183 | 1.61 | 0 | 0 | 0 | 1.61 |
| | 184 | 3.62 | 5.22 | 3.62 | 2.01 | 14.47 |
| | 185 | 4.42 | 4.02 | 2.81 | 0 | 11.25 |
| | 186 | 1.61 | 2.01 | 1.61 | 2.01 | 7.24 |
| | 187 | 32.95 | 45.81 | 32.55 | 30.54 | 141.85 |
| | 189 | 4.42 | 4.42 | 5.22 | 5.63 | 19.69 |
| | 190 | 8.84 | 7.64 | 5.22 | 5.22 | 26.92 |
| | 191 | 9.64 | 7.23 | 4.02 | 8.04 | 28.93 |
| | 193 | 1.21 | 2.81 | 4.02 | 3.62 | 11.66 |
| | 194 | 1.61 | 3.21 | 2.41 | 3.62 | 10.85 |
| | 195 | 0.4 | 0.8 | 1.21 | 0 | 2.41 |
| | 196 | 0.8 | 0 | 0.8 | 0 | 1.6 |
| TOTAL | | 118.54 | 134.19 | 116.14 | 120.15 | 489.02 |

STATE: RHODE ISLAND

| ZIP CODE | QUARTER 1 | QUARTER 2 | QUARTER 3 | QUARTER 4 | TOTAL GRAMS |
|---|---|---|---|---|---|
| 28 | 4.42 | 2.41 | 1.61 | 2.01 | 10.45 |
| 29 | 0.4 | 3.21 | 1.21 | 1.61 | 6.43 |
| TOTAL | 4.82 | 5.62 | 2.82 | 3.62 | 16.88 |

STATE: SOUTH CAROLINA

| ZIP CODE | QUARTER 1 | QUARTER 2 | QUARTER 3 | QUARTER 4 | TOTAL GRAMS |
|---|---|---|---|---|---|
| 290 | 0 | 0.4 | 1.61 | 0.8 | 2.81 |
| 292 | 4.02 | 2.81 | 2.01 | 0.8 | 9.64 |
| 293 | 5.22 | 0.8 | 0 | 0 | 6.02 |
| 294 | 0.8 | 0.8 | 3.21 | 2.41 | 7.22 |

| | | | | | |
|---|---|---|---|---|---|
| | 295 | 2.41 | 2.81 | 5.63 | 2.81 | 13.66 |
| | 296 | 6.03 | 4.02 | 4.02 | 5.63 | 19.7 |
| | 297 | 0 | 0 | 0 | 0.8 | 0.8 |
| | 298 | 0 | 0.4 | 0 | 0 | 0.4 |
| TOTAL | | 18.48 | 12.04 | 16.48 | 13.25 | 60.25 |

STATE: SOUTH DAKOTA

| ZIP CODE | QUARTER 1 | QUARTER 2 | QUARTER 3 | QUARTER 4 | TOTAL GRAMS |
|---|---|---|---|---|---|
| 571 | 11.25 | 18.89 | 16.07 | 13.66 | 59.87 |
| 573 | 0 | 0.8 | 0 | 0 | 0.8 |
| 574 | 0 | 0.4 | 0 | 0 | 0.4 |
| 577 | 4.42 | 2.01 | 1.21 | 0.8 | 8.44 |
| TOTAL | 15.67 | 22.1 | 17.28 | 14.46 | 69.51 |

STATE: TENNESSEE

| ZIP CODE | QUARTER 1 | QUARTER 2 | QUARTER 3 | QUARTER 4 | TOTAL GRAMS |
|---|---|---|---|---|---|
| 372 | 0 | 0.4 | 0 | 3.21 | 3.61 |
| 373 | 0 | 0 | 1.21 | 1.61 | 2.82 |
| 374 | 0.4 | 1.21 | 0.4 | 0.8 | 2.81 |
| 378 | 1.21 | 1.61 | 1.21 | 2.01 | 6.04 |
| 379 | 0.4 | 1.21 | 1.61 | 1.21 | 4.43 |
| 380 | 2.01 | 0.8 | 1.21 | 1.61 | 5.63 |
| 381 | 6.43 | 7.23 | 6.83 | 4.82 | 25.31 |
| TOTAL | 10.45 | 12.46 | 12.47 | 15.27 | 50.65 |

STATE: TEXAS

| ZIP CODE | QUARTER 1 | QUARTER 2 | QUARTER 3 | QUARTER 4 | TOTAL GRAMS |
|---|---|---|---|---|---|
| 750 | 24.91 | 27.73 | 24.11 | 24.51 | 101.26 |
| 751 | 0 | 0 | 0 | 0.8 | 0.8 |
| 752 | 17.68 | 18.49 | 13.66 | 15.27 | 65.1 |
| 754 | 2.01 | 2.41 | 3.21 | 2.81 | 10.44 |
| 755 | 0 | 0.8 | 0 | 0 | 0.8 |
| 756 | 1.21 | 2.81 | 0 | 0 | 4.02 |
| 757 | 2.01 | 0.8 | 0.8 | 0 | 3.61 |

| | | | | | | |
|---|---|---|---|---|---|---|
| | 759 | 0 | 0.4 | 0 | 0 | 0.4 |
| | 760 | 11.25 | 6.43 | 5.63 | 11.25 | 34.56 |
| | 761 | 0 | 0.4 | 1.21 | 0.8 | 2.41 |
| | 762 | 4.42 | 1.21 | 0.4 | 3.21 | 9.24 |
| | 763 | 0 | 0.8 | 0 | 0 | 0.8 |
| | 764 | 0 | 1.61 | 0.8 | 0.8 | 3.21 |
| | 765 | 0 | 0 | 0 | 0.8 | 0.8 |
| | 766 | 0 | 0 | 0 | 1.21 | 1.21 |
| | 769 | 0 | 0.8 | 0.8 | 0.8 | 2.4 |
| | 770 | 9.24 | 6.43 | 2.41 | 5.63 | 23.71 |
| | 773 | 3.21 | 1.21 | 0.8 | 1.61 | 6.83 |
| | 774 | 3.62 | 6.03 | 3.62 | 4.42 | 17.69 |
| | 775 | 3.62 | 3.62 | 3.21 | 0.8 | 11.25 |
| | 778 | 0 | 0 | 0 | 1.21 | 1.21 |
| | 780 | 0.4 | 0 | 0.4 | 0 | 0.8 |
| | 781 | 1.61 | 4.02 | 2.41 | 0 | 8.04 |
| | 782 | 4.02 | 3.21 | 2.01 | 2.41 | 11.65 |
| | 783 | 0 | 0.4 | 0 | 0 | 0.4 |
| | 784 | 2.41 | 2.41 | 1.21 | 1.21 | 7.24 |
| | 785 | 4.42 | 2.81 | 6.83 | 3.62 | 17.68 |
| | 786 | 3.21 | 2.41 | 4.02 | 4.82 | 14.46 |
| | 787 | 8.04 | 10.45 | 10.05 | 9.64 | 38.18 |
| | 788 | 2.01 | 3.62 | 0 | 0 | 5.63 |
| | 789 | 2.01 | 0.8 | 0.8 | 1.61 | 5.22 |
| | 790 | 1.61 | 1.61 | 1.61 | 1.21 | 6.04 |
| | 797 | 3.62 | 2.41 | 1.21 | 2.01 | 9.25 |
| | 799 | 0 | 0 | 0 | 0.4 | 0.4 |
| TOTAL | | 116.54 | 116.13 | 91.21 | 102.86 | 426.74 |

| STATE: | UTAH | | | | |
|---|---|---|---|---|---|
| ZIP CODE | QUARTER 1 | QUARTER 2 | QUARTER 3 | QUARTER 4 | TOTAL GRAMS |
| 840 | 6.83 | 6.83 | 7.23 | 4.82 | 25.71 |
| 841 | 4.42 | 2.41 | 9.24 | 9.24 | 25.31 |
| 846 | 6.43 | 5.63 | 4.42 | 5.63 | 22.11 |
| 847 | 7.64 | 8.44 | 5.63 | 9.24 | 30.95 |
| TOTAL | 25.32 | 23.31 | 26.52 | 28.93 | 104.08 |

STATE: VERMONT

| ZIP CODE | QUARTER 1 | QUARTER 2 | QUARTER 3 | QUARTER 4 | TOTAL GRAMS |
|---|---|---|---|---|---|
| 56 | 0 | 1.21 | 1.21 | 2.01 | 4.43 |
| 57 | 0.8 | 0.8 | 0.8 | 0.4 | 2.8 |
| TOTAL | 0.8 | 2.01 | 2.01 | 2.41 | 7.23 |

STATE: VIRGINIA

| ZIP CODE | QUARTER 1 | QUARTER 2 | QUARTER 3 | QUARTER 4 | TOTAL GRAMS |
|---|---|---|---|---|---|
| 201 | 8.04 | 7.23 | 4.02 | 7.64 | 26.93 |
| 220 | 4.42 | 3.21 | 3.62 | 4.02 | 15.27 |
| 221 | 0 | 0.4 | 0 | 1.21 | 1.61 |
| 222 | 1.61 | 2.01 | 1.21 | 3.21 | 8.04 |
| 223 | 1.61 | 1.21 | 0.8 | 2.01 | 5.63 |
| 224 | 1.61 | 0 | 0.8 | 1.61 | 4.02 |
| 228 | 0 | 0.8 | 0 | 0 | 0.8 |
| 229 | 0.4 | 0 | 0 | 1.61 | 2.01 |
| 231 | 4.02 | 2.81 | 3.21 | 0.8 | 10.84 |
| 232 | 7.23 | 5.22 | 5.22 | 4.82 | 22.49 |
| 233 | 1.61 | 0 | 0 | 0 | 1.61 |
| 234 | 7.64 | 5.22 | 4.82 | 4.42 | 22.1 |
| 235 | 3.21 | 2.81 | 2.41 | 1.61 | 10.04 |
| 237 | 0 | 0.4 | 1.21 | 1.21 | 2.82 |
| 238 | 4.02 | 3.21 | 4.02 | 4.42 | 15.67 |
| 240 | 3.21 | 3.21 | 5.63 | 3.21 | 15.26 |
| 241 | 0 | 1.61 | 0.8 | 2.01 | 4.42 |
| 245 | 2.41 | 1.21 | 2.01 | 0 | 5.63 |
| TOTAL | 51.04 | 40.56 | 39.78 | 43.81 | 175.19 |

STATE: WASHINGTON

| ZIP CODE | QUARTER 1 | QUARTER 2 | QUARTER 3 | QUARTER 4 | TOTAL GRAMS |
|---|---|---|---|---|---|
| 980 | 28.13 | 33.76 | 36.57 | 37.37 | 135.83 |
| 981 | 10.05 | 15.67 | 8.84 | 12.86 | 47.42 |
| 982 | 15.27 | 13.26 | 18.49 | 15.27 | 62.29 |

| | | | | | |
|---|---|---|---|---|---|
| 983 | 6.03 | 4.42 | 1.21 | 3.21 | 14.87 |
| 984 | 2.81 | 2.01 | 2.01 | 2.41 | 9.24 |
| 985 | 4.42 | 3.21 | 2.01 | 1.61 | 11.25 |
| 986 | 16.07 | 14.06 | 14.06 | 20.49 | 64.68 |
| 988 | 1.21 | 0.4 | 0 | 0 | 1.61 |
| 989 | 1.61 | 1.61 | 0.8 | 0.8 | 4.82 |
| 992 | 5.22 | 6.83 | 5.22 | 8.04 | 25.31 |
| 993 | 0 | 0.4 | 2.01 | 1.61 | 4.02 |
| TOTAL | 90.82 | 95.63 | 91.22 | 103.67 | 381.34 |

STATE: WEST VIRGINIA

| ZIP CODE | QUARTER 1 | QUARTER 2 | QUARTER 3 | QUARTER 4 | TOTAL GRAMS |
|---|---|---|---|---|---|
| 254 | 0 | 0 | 1.21 | 0 | 1.21 |
| 258 | 0.8 | 3.21 | 0.4 | 2.01 | 6.42 |
| 260 | 2.41 | 1.21 | 1.61 | 0 | 5.23 |
| 265 | 0 | 0 | 2.41 | 1.21 | 3.62 |
| TOTAL | 3.21 | 4.42 | 5.63 | 3.22 | 16.48 |

STATE: WISCONSIN

| ZIP CODE | QUARTER 1 | QUARTER 2 | QUARTER 3 | QUARTER 4 | TOTAL GRAMS |
|---|---|---|---|---|---|
| 530 | 3.21 | 1.61 | 0 | 0 | 4.82 |
| 531 | 2.01 | 4.02 | 2.81 | 4.42 | 13.26 |
| 532 | 9.64 | 15.67 | 9.24 | 3.62 | 38.17 |
| 535 | 2.41 | 3.62 | 0 | 0.4 | 6.43 |
| 537 | 16.88 | 14.06 | 12.86 | 7.64 | 51.44 |
| 539 | 2.41 | 0.4 | 1.21 | 1.61 | 5.63 |
| 540 | 1.61 | 2.01 | 2.81 | 1.21 | 7.64 |
| 541 | 0 | 0.4 | 2.01 | 1.61 | 4.02 |
| 542 | 7.23 | 6.03 | 8.44 | 8.04 | 29.74 |
| 543 | 4.82 | 1.21 | 1.21 | 2.81 | 10.05 |
| 544 | 0 | 0 | 1.61 | 0 | 1.61 |
| 549 | 11.25 | 13.26 | 0 | 0 | 24.51 |
| TOTAL | 61.47 | 62.29 | 42.2 | 31.36 | 197.32 |

STATE: WYOMING

| ZIP CODE | QUARTER 1 | QUARTER 2 | QUARTER 3 | QUARTER 4 | TOTAL GRAMS |
|---|---|---|---|---|---|
| 820 | 2.81 | 4.82 | 3.21 | 2.41 | 13.25 |
| 826 | 1.61 | 1.61 | 1.61 | 0.8 | 5.63 |
| 829 | 1.61 | 0.8 | 0 | 0 | 2.41 |
| TOTAL | 6.03 | 7.23 | 4.82 | 3.21 | 21.29 |

DRUG CODE: 1105D DRUG NAME: D-METHAMPHETAMINE

STATE: ALABAMA

| ZIP CODE | QUARTER 1 | QUARTER 2 | QUARTER 3 | QUARTER 4 | TOTAL GRAMS |
|---|---|---|---|---|---|
| 350 | 0 | 0 | 2.01 | 0.8 | 2.81 |
| 351 | 0 | 0.8 | 2.01 | 0.8 | 3.61 |
| 356 | 0 | 0 | 2.81 | 5.62 | 8.43 |
| TOTAL | 0 | 0.8 | 6.83 | 7.22 | 14.85 |

STATE: ALASKA

| ZIP CODE | QUARTER 1 | QUARTER 2 | QUARTER 3 | QUARTER 4 | TOTAL GRAMS |
|---|---|---|---|---|---|
| 996 | 0 | 0 | 0.8 | 0 | 0.8 |
| TOTAL | 0 | 0 | 0.8 | 0 | 0.8 |

STATE: ARIZONA

| ZIP CODE | QUARTER 1 | QUARTER 2 | QUARTER 3 | QUARTER 4 | TOTAL GRAMS |
|---|---|---|---|---|---|
| 852 | 0 | 4.42 | 0.8 | 9.64 | 14.86 |
| 853 | 0 | 0 | 3.21 | 1.61 | 4.82 |
| TOTAL | 0 | 4.42 | 4.01 | 11.25 | 19.68 |

STATE: CALIFORNIA

| ZIP CODE | QUARTER 1 | QUARTER 2 | QUARTER 3 | QUARTER 4 | TOTAL GRAMS |
|---|---|---|---|---|---|
| 900 | 0 | 8.84 | 16.87 | 17.27 | 42.98 |
| 902 | 0 | 0 | 0.8 | 0.4 | 1.2 |
| 914 | 0 | 1.21 | 0.8 | 0.8 | 2.81 |
| 917 | 0 | 8.03 | 22.09 | 21.69 | 51.81 |
| 920 | 0 | 1.21 | 6.83 | 4.82 | 12.86 |

| | | | | | |
|---|---|---|---|---|---|
| 921 | 0 | 0.8 | 4.02 | 1.21 | 6.03 |
| 923 | 0 | 0 | 1.21 | 2.81 | 4.02 |
| 926 | 0 | 0 | 1.61 | 2.01 | 3.62 |
| 927 | 0 | 0.8 | 1.61 | 0.8 | 3.21 |
| 928 | 0 | 0.8 | 2.41 | 2.41 | 5.62 |
| 931 | 0 | 0.8 | 0.8 | 2.01 | 3.61 |
| 940 | 0 | 0.4 | 2.01 | 2.41 | 4.82 |
| 943 | 0 | 0.16 | 0.08 | 0 | 0.24 |
| TOTAL | 0 | 23.05 | 61.14 | 58.64 | 142.83 |

STATE: CONNECTICUT

| ZIP CODE | QUARTER 1 | QUARTER 2 | QUARTER 3 | QUARTER 4 | TOTAL GRAMS |
|---|---|---|---|---|---|
| 65 | 0 | 0 | 3.62 | 4.02 | 7.64 |
| 66 | 0 | 0 | 0 | 0.4 | 0.4 |
| TOTAL | 0 | 0 | 3.62 | 4.42 | 8.04 |

STATE: DISTRICT OF COLUMBIA

| ZIP CODE | QUARTER 1 | QUARTER 2 | QUARTER 3 | QUARTER 4 | TOTAL GRAMS |
|---|---|---|---|---|---|
| 200 | 0 | 0.8 | 2.81 | 1.61 | 5.22 |
| TOTAL | 0 | 0.8 | 2.81 | 1.61 | 5.22 |

STATE: FLORIDA

| ZIP CODE | QUARTER 1 | QUARTER 2 | QUARTER 3 | QUARTER 4 | TOTAL GRAMS |
|---|---|---|---|---|---|
| 326 | 0 | 0.8 | 2.01 | 1.21 | 4.02 |
| 329 | 0 | 0 | 0 | 0.8 | 0.8 |
| 334 | 0 | 4.02 | 0 | 0 | 4.02 |
| 337 | 0 | 0 | 2.41 | 1.21 | 3.62 |
| TOTAL | 0 | 4.82 | 4.42 | 3.22 | 12.46 |

STATE: GEORGIA

| ZIP CODE | QUARTER 1 | QUARTER 2 | QUARTER 3 | QUARTER 4 | TOTAL GRAMS |
|---|---|---|---|---|---|
| 310 | 0 | 1.61 | 2.41 | 2.41 | 6.43 |
| 315 | 0 | 0 | 2.41 | 2.41 | 4.82 |

| | | | | | |
|---|---|---|---|---|---|
| TOTAL | 0 | 1.61 | 4.82 | 4.82 | 11.25 |

STATE: HAWAII

| ZIP CODE | QUARTER 1 | QUARTER 2 | QUARTER 3 | QUARTER 4 | TOTAL GRAMS |
|---|---|---|---|---|---|
| 967 | 0 | 0 | 1.61 | 1.61 | 3.22 |
| TOTAL | 0 | 0 | 1.61 | 1.61 | 3.22 |

STATE: IDAHO

| ZIP CODE | QUARTER 1 | QUARTER 2 | QUARTER 3 | QUARTER 4 | TOTAL GRAMS |
|---|---|---|---|---|---|
| 838 | 0 | 1.21 | 5.22 | 4.02 | 10.45 |
| TOTAL | 0 | 1.21 | 5.22 | 4.02 | 10.45 |

STATE: ILLINOIS

| ZIP CODE | QUARTER 1 | QUARTER 2 | QUARTER 3 | QUARTER 4 | TOTAL GRAMS |
|---|---|---|---|---|---|
| 606 | 0 | 0 | 2.41 | 1.61 | 4.02 |
| 607 | 0 | 0 | 1.21 | 0.8 | 2.01 |
| TOTAL | 0 | 0 | 3.62 | 2.41 | 6.03 |

STATE: KANSAS

| ZIP CODE | QUARTER 1 | QUARTER 2 | QUARTER 3 | QUARTER 4 | TOTAL GRAMS |
|---|---|---|---|---|---|
| 660 | 0 | 0 | 0 | 0.8 | 0.8 |
| 666 | 0 | 0 | 0 | 0.8 | 0.8 |
| 672 | 0 | 0.8 | 0.8 | 1.21 | 2.81 |
| TOTAL | 0 | 0.8 | 0.8 | 2.81 | 4.41 |

STATE: KENTUCKY

| ZIP CODE | QUARTER 1 | QUARTER 2 | QUARTER 3 | QUARTER 4 | TOTAL GRAMS |
|---|---|---|---|---|---|
| 408 | 0 | 0.4 | 1.61 | 1.61 | 3.62 |
| TOTAL | 0 | 0.4 | 1.61 | 1.61 | 3.62 |

STATE: LOUISIANA

| ZIP CODE | QUARTER 1 | QUARTER 2 | QUARTER 3 | QUARTER 4 | TOTAL GRAMS |
|---|---|---|---|---|---|
| 708 | 0 | 0 | 2.01 | 2.41 | 4.42 |
| TOTAL | 0 | 0 | 2.01 | 2.41 | 4.42 |

STATE: MARYLAND

| ZIP CODE | QUARTER 1 | QUARTER 2 | QUARTER 3 | QUARTER 4 | TOTAL GRAMS |
|---|---|---|---|---|---|
| 208 | 0 | 0 | 0.8 | 1.21 | 2.01 |
| 210 | 0 | 0 | 0.4 | 2.41 | 2.81 |
| TOTAL | 0 | 0 | 1.2 | 3.62 | 4.82 |

STATE: MASSACHUSETTS

| ZIP CODE | QUARTER 1 | QUARTER 2 | QUARTER 3 | QUARTER 4 | TOTAL GRAMS |
|---|---|---|---|---|---|
| 10 | 0 | 0.4 | 2.01 | 1.61 | 4.02 |
| 18 | 0 | 0 | 0.4 | 0 | 0.4 |
| 21 | 0 | 0.4 | 1.21 | 0.4 | 2.01 |
| 24 | 0 | 0 | 0 | 0.4 | 0.4 |
| TOTAL | 0 | 0.8 | 3.62 | 2.41 | 6.83 |

STATE: MINNESOTA

| ZIP CODE | QUARTER 1 | QUARTER 2 | QUARTER 3 | QUARTER 4 | TOTAL GRAMS |
|---|---|---|---|---|---|
| 551 | 0 | 0.8 | 4.42 | 3.62 | 8.84 |
| 554 | 0 | 2.81 | 20.89 | 15.26 | 38.96 |
| 559 | 0 | 0 | 0 | 4.02 | 4.02 |
| 563 | 0 | 0.8 | 0.4 | 0 | 1.2 |
| TOTAL | 0 | 4.41 | 25.71 | 22.9 | 53.02 |

STATE: MISSISSIPPI

| ZIP CODE | QUARTER 1 | QUARTER 2 | QUARTER 3 | QUARTER 4 | TOTAL GRAMS |
|---|---|---|---|---|---|
| 397 | 0 | 0 | 1.61 | 0.8 | 2.41 |
| TOTAL | 0 | 0 | 1.61 | 0.8 | 2.41 |

STATE: MISSOURI

| ZIP CODE | QUARTER 1 | QUARTER 2 | QUARTER 3 | QUARTER 4 | TOTAL GRAMS |
|---|---|---|---|---|---|
| 631 | 0 | 2.81 | 10.85 | 15.67 | 29.33 |
| 641 | 0 | 0 | 0 | 0.8 | 0.8 |
| 655 | 0 | 0.4 | 1.21 | 0.4 | 2.01 |
| TOTAL | 0 | 3.21 | 12.06 | 16.87 | 32.14 |

STATE: NEBRASKA

| ZIP CODE | QUARTER 1 | QUARTER 2 | QUARTER 3 | QUARTER 4 | TOTAL GRAMS |
|---|---|---|---|---|---|
| 684 | 0 | 0 | 0 | 0.8 | 0.8 |
| TOTAL | 0 | 0 | 0 | 0.8 | 0.8 |

STATE: NEVADA

| ZIP CODE | QUARTER 1 | QUARTER 2 | QUARTER 3 | QUARTER 4 | TOTAL GRAMS |
|---|---|---|---|---|---|
| 891 | 0 | 0 | 0.4 | 0 | 0.4 |
| TOTAL | 0 | 0 | 0.4 | 0 | 0.4 |

STATE: NEW JERSEY

| ZIP CODE | QUARTER 1 | QUARTER 2 | QUARTER 3 | QUARTER 4 | TOTAL GRAMS |
|---|---|---|---|---|---|
| 70 | 0 | 0.8 | 0.8 | 0 | 1.6 |
| 79 | 0 | 0 | 0.4 | 0.8 | 1.2 |
| 85 | 0 | 0 | 1.61 | 1.21 | 2.82 |
| TOTAL | 0 | 0.8 | 2.81 | 2.01 | 5.62 |

STATE: NEW YORK

| ZIP CODE | QUARTER 1 | QUARTER 2 | QUARTER 3 | QUARTER 4 | TOTAL GRAMS |
|---|---|---|---|---|---|
| 101 | 0 | 0 | 0 | 0.8 | 0.8 |
| 104 | 0 | 0 | 4.02 | 4.1 | 8.12 |
| 106 | 0 | 0 | 1.21 | 1.21 | 2.42 |
| 113 | 0 | 0 | 1.61 | 0.8 | 2.41 |
| 128 | 4.02 | 0 | 0 | 0 | 4.02 |
| TOTAL | 4.02 | 0 | 6.84 | 6.91 | 17.77 |

STATE: NORTH CAROLINA

| ZIP CODE | QUARTER 1 | QUARTER 2 | QUARTER 3 | QUARTER 4 | TOTAL GRAMS |
|---|---|---|---|---|---|
| 272 | 0 | 0 | 0.8 | 0 | 0.8 |
| 275 | 0 | 0 | 4.02 | 4.02 | 8.04 |
| 278 | 0 | 0 | 1.21 | 1.61 | 2.82 |
| 280 | 0 | 0 | 0.8 | 0.8 | 1.6 |
| 281 | 0 | 0.8 | 0.4 | 0 | 1.2 |
| 286 | 0 | 0.8 | 1.61 | 2.01 | 4.42 |
| 288 | 0 | 0.4 | 0.8 | 1.61 | 2.81 |
| TOTAL | 0 | 2 | 9.64 | 10.05 | 21.69 |

STATE: NORTH DAKOTA

| ZIP CODE | QUARTER 1 | QUARTER 2 | QUARTER 3 | QUARTER 4 | TOTAL GRAMS |
|---|---|---|---|---|---|
| 581 | 0 | 0 | 1.61 | 0.4 | 2.01 |
| 582 | 0 | 0 | 4.02 | 0 | 4.02 |
| TOTAL | 0 | 0 | 5.63 | 0.4 | 6.03 |

STATE: OHIO

| ZIP CODE | QUARTER 1 | QUARTER 2 | QUARTER 3 | QUARTER 4 | TOTAL GRAMS |
|---|---|---|---|---|---|
| 432 | 0 | 0 | 3.62 | 7.63 | 11.25 |
| 446 | 0 | 0 | 0.4 | 0 | 0.4 |
| 450 | 0 | 0 | 2.01 | 2.41 | 4.42 |
| 452 | 0 | 0 | 0.4 | 0 | 0.4 |
| 454 | 0 | 0 | 0 | 0.8 | 0.8 |
| 457 | 0 | 0.4 | 2.01 | 1.61 | 4.02 |
| TOTAL | 0 | 0.4 | 8.44 | 12.45 | 21.29 |

STATE: OKLAHOMA

| ZIP CODE | QUARTER 1 | QUARTER 2 | QUARTER 3 | QUARTER 4 | TOTAL GRAMS |
|---|---|---|---|---|---|
| 741 | 0 | 0.4 | 4.02 | 2.81 | 7.23 |
| TOTAL | 0 | 0.4 | 4.02 | 2.81 | 7.23 |

STATE: OREGON

| ZIP CODE | QUARTER 1 | QUARTER 2 | QUARTER 3 | QUARTER 4 | TOTAL GRAMS |
|---|---|---|---|---|---|
| 972 | 0 | 0.4 | 2.01 | 2.01 | 4.42 |
| 973 | 0 | 4.82 | 0 | 0 | 4.82 |
| 974 | 0 | 0 | 0 | 0.4 | 0.4 |
| 978 | 0 | 4.02 | 12.05 | 12.05 | 28.12 |
| TOTAL | 0 | 9.24 | 14.06 | 14.46 | 37.76 |

STATE: PENNSYLVANIA

| ZIP CODE | QUARTER 1 | QUARTER 2 | QUARTER 3 | QUARTER 4 | TOTAL GRAMS |
|---|---|---|---|---|---|
| 151 | 0 | 0 | 2.01 | 2.01 | 4.02 |
| 187 | 0 | 3.21 | 6.43 | 6.03 | 15.67 |
| 190 | 0 | 0 | 3.21 | 3.21 | 6.42 |
| TOTAL | 0 | 3.21 | 11.65 | 11.25 | 26.11 |

STATE: RHODE ISLAND

| ZIP CODE | QUARTER 1 | QUARTER 2 | QUARTER 3 | QUARTER 4 | TOTAL GRAMS |
|---|---|---|---|---|---|
| 28 | 0 | 0 | 1.61 | 2.01 | 3.62 |
| TOTAL | 0 | 0 | 1.61 | 2.01 | 3.62 |

STATE: SOUTH CAROLINA

| ZIP CODE | QUARTER 1 | QUARTER 2 | QUARTER 3 | QUARTER 4 | TOTAL GRAMS |
|---|---|---|---|---|---|
| 292 | 0 | 2.01 | 4.02 | 2.81 | 8.84 |
| TOTAL | 0 | 2.01 | 4.02 | 2.81 | 8.84 |

STATE: TENNESSEE

| ZIP CODE | QUARTER 1 | QUARTER 2 | QUARTER 3 | QUARTER 4 | TOTAL GRAMS |
|---|---|---|---|---|---|
| 370 | 0 | 0 | 0 | 0.4 | 0.4 |
| TOTAL | 0 | 0 | 0 | 0.4 | 0.4 |

STATE: TEXAS

| ZIP CODE | QUARTER 1 | QUARTER 2 | QUARTER 3 | QUARTER 4 | TOTAL GRAMS |
|---|---|---|---|---|---|
| 750 | 0 | 0 | 1.21 | 0 | 1.21 |

| | | | | | | |
|---|---|---|---|---|---|---|
| | 751 | 0 | 0 | 0 | 0.4 | 0.4 |
| | 752 | 0 | 0.8 | 4.02 | 1.61 | 6.43 |
| | 754 | 0 | 0 | 0.4 | 0 | 0.4 |
| | 756 | 0 | 0.4 | 2.01 | 2.01 | 4.42 |
| | 757 | 0 | 1.61 | 1.21 | 2.81 | 5.63 |
| | 759 | 0 | 0.8 | 0.8 | 1.61 | 3.21 |
| | 760 | 0 | 0 | 1.61 | 1.21 | 2.82 |
| | 761 | 0 | 0 | 0 | 0.4 | 0.4 |
| | 770 | 0 | 0.8 | 2.41 | 2.41 | 5.62 |
| | 782 | 0 | 0 | 0 | 1.21 | 1.21 |
| | 787 | 0 | 0 | 0.8 | 0 | 0.8 |
| | 788 | 0 | 0 | 3.62 | 3.21 | 6.83 |
| | 799 | 0 | 0 | 0 | 0.4 | 0.4 |
| TOTAL | | 0 | 4.41 | 18.09 | 17.28 | 39.78 |

STATE: WASHINGTON

| ZIP CODE | | QUARTER 1 | QUARTER 2 | QUARTER 3 | QUARTER 4 | TOTAL GRAMS |
|---|---|---|---|---|---|---|
| | 981 | 0 | 0 | 0.8 | 0.8 | 1.6 |
| | 986 | 0 | 0 | 0 | 0.8 | 0.8 |
| TOTAL | | 0 | 0 | 0.8 | 1.6 | 2.4 |

STATE: WEST VIRGINIA

| ZIP CODE | | QUARTER 1 | QUARTER 2 | QUARTER 3 | QUARTER 4 | TOTAL GRAMS |
|---|---|---|---|---|---|---|
| | 260 | 0 | 0 | 2.41 | 1.21 | 3.62 |
| TOTAL | | 0 | 0 | 2.41 | 1.21 | 3.62 |

STATE: WISCONSIN

| ZIP CODE | | QUARTER 1 | QUARTER 2 | QUARTER 3 | QUARTER 4 | TOTAL GRAMS |
|---|---|---|---|---|---|---|
| | 530 | 0 | 0.8 | 2.41 | 1.61 | 4.82 |
| | 531 | 0 | 0 | 0 | 1.21 | 1.21 |
| | 532 | 0 | 0 | 2.41 | 1.61 | 4.02 |
| | 535 | 0 | 0 | 2.81 | 2.01 | 4.82 |
| | 537 | 0 | 0.8 | 2.41 | 2.81 | 6.02 |
| | 549 | 0 | 0 | 3.62 | 3.62 | 7.24 |

| | | | | | | |
|---|---|---|---|---|---|---|
| TOTAL | | 0 | 1.6 | 13.66 | 12.87 | 28.13 |

DRUG CODE: 1205 DRUG NAME: LISDEXAMFETAMINE

STATE: ALABAMA

| ZIP CODE | QUARTER 1 | QUARTER 2 | QUARTER 3 | QUARTER 4 | TOTAL GRAMS |
|---|---|---|---|---|---|
| 350 | 6,887.48 | 6,568.95 | 6,546.98 | 6,970.15 | 26,973.56 |
| 351 | 4,517.85 | 4,297.02 | 4,361.19 | 4,520.74 | 17,696.80 |
| 352 | 9,847.93 | 9,235.73 | 9,262.90 | 9,830.01 | 38,176.57 |
| 354 | 4,604.57 | 4,298.75 | 4,232.85 | 4,773.95 | 17,910.12 |
| 355 | 1,366.05 | 1,297.26 | 1,296.68 | 1,356.22 | 5,316.21 |
| 356 | 3,860.55 | 3,854.19 | 3,853.04 | 4,056.53 | 15,624.31 |
| 357 | 2,342.46 | 2,232.04 | 2,347.09 | 2,297.95 | 9,219.54 |
| 358 | 2,611.86 | 2,548.26 | 2,550.58 | 2,700.31 | 10,411.01 |
| 359 | 2,580.64 | 2,403.74 | 2,330.32 | 2,428.60 | 9,743.30 |
| 360 | 4,000.45 | 3,571.50 | 3,784.24 | 3,859.40 | 15,215.59 |
| 361 | 2,972.59 | 2,653.48 | 2,614.17 | 2,881.83 | 11,122.07 |
| 362 | 1,970.74 | 1,784.59 | 1,801.36 | 1,932.59 | 7,489.28 |
| 363 | 4,388.94 | 4,171.57 | 4,181.51 | 4,271.19 | 17,013.21 |
| 364 | 987.39 | 1,018.03 | 1,027.86 | 1,064.28 | 4,097.56 |
| 365 | 7,968.53 | 7,737.87 | 7,954.08 | 8,354.70 | 32,015.18 |
| 366 | 6,723.88 | 6,346.38 | 6,400.72 | 6,445.24 | 25,916.22 |
| 367 | 816.86 | 637.07 | 719.73 | 790.84 | 2,964.50 |
| 368 | 4,518.43 | 4,223.02 | 4,349.05 | 4,723.08 | 17,813.58 |
| 369 | 157.24 | 128.92 | 156.09 | 164.18 | 606.43 |
| TOTAL | 73,124.44 | 69,008.37 | 69,770.44 | 73,421.79 | 285,325.04 |

STATE: ALASKA

| ZIP CODE | QUARTER 1 | QUARTER 2 | QUARTER 3 | QUARTER 4 | TOTAL GRAMS |
|---|---|---|---|---|---|
| 995 | 1,127.87 | 1,201.87 | 1,235.40 | 1,258.52 | 4,823.66 |
| 996 | 535.32 | 474.04 | 559.02 | 585.04 | 2,153.42 |
| 997 | 352.06 | 373.45 | 367.67 | 343.97 | 1,437.15 |
| 998 | 216.79 | 216.21 | 216.79 | 211.58 | 861.37 |
| 999 | 134.7 | 147.99 | 155.51 | 152.04 | 590.24 |
| TOTAL | 2,366.74 | 2,413.56 | 2,534.39 | 2,551.15 | 9,865.84 |

DATE RANGE: 01/01/2017 TO 12/31/2017 ARCOS 3 - REPORT 1 Run Date: 07/03/2018

RETAIL DRUG DISTRIBUTION BY ZIP CODE WITHIN STATE BY GRAMS WT

STATE: ARIZONA

| ZIP CODE | QUARTER 1 | QUARTER 2 | QUARTER 3 | QUARTER 4 | TOTAL GRAMS |
|---|---|---|---|---|---|
| 850 | 6,502.47 | 6,472.99 | 6,322.10 | 6,675.90 | 25,973.46 |
| 851 | 1,233.67 | 1,223.84 | 1,298.41 | 1,367.78 | 5,123.70 |
| 852 | 16,863.18 | 16,603.61 | 17,038.92 | 18,626.96 | 69,132.67 |
| 853 | 4,206.26 | 3,956.52 | 3,975.59 | 4,406.86 | 16,545.23 |
| 855 | 132.38 | 118.51 | 95.96 | 145.1 | 491.95 |
| 856 | 682.74 | 483.87 | 543.99 | 593.13 | 2,303.73 |
| 857 | 4,683.19 | 4,330.55 | 4,413.22 | 4,771.06 | 18,198.02 |
| 859 | 131.23 | 145.1 | 184.99 | 196.55 | 657.87 |
| 860 | 347.44 | 335.88 | 299.46 | 350.91 | 1,333.69 |
| 863 | 633.02 | 586.19 | 644.58 | 720.31 | 2,584.10 |
| 864 | 237.02 | 258.41 | 264.19 | 276.33 | 1,035.95 |
| TOTAL | 35,652.60 | 34,515.47 | 35,081.41 | 38,130.89 | 143,380.37 |

STATE: ARKANSAS

| ZIP CODE | QUARTER 1 | QUARTER 2 | QUARTER 3 | QUARTER 4 | TOTAL GRAMS |
|---|---|---|---|---|---|
| 716 | 1,601.34 | 1,442.94 | 1,438.31 | 1,527.92 | 6,010.51 |
| 717 | 805.87 | 624.93 | 607.01 | 783.9 | 2,821.71 |
| 718 | 764.25 | 709.33 | 662.5 | 789.68 | 2,925.76 |
| 719 | 2,129.14 | 1,882.29 | 1,967.85 | 2,043.01 | 8,022.29 |
| 720 | 4,554.27 | 4,096.99 | 4,297.60 | 4,655.44 | 17,604.30 |
| 721 | 3,426.98 | 3,054.10 | 3,112.49 | 3,387.67 | 12,981.24 |
| 722 | 3,597.52 | 3,545.49 | 3,517.16 | 3,646.39 | 14,306.56 |
| 723 | 1,250.43 | 1,045.78 | 1,030.17 | 1,175.28 | 4,501.66 |
| 724 | 3,029.82 | 2,538.44 | 2,658.68 | 2,813.61 | 11,040.55 |
| 725 | 711.64 | 689.1 | 608.16 | 659.61 | 2,668.51 |
| 726 | 812.81 | 752.11 | 771.76 | 797.78 | 3,134.46 |
| 727 | 5,346.85 | 4,933.51 | 5,017.33 | 5,497.73 | 20,795.42 |
| 728 | 1,204.18 | 1,005.32 | 1,035.96 | 1,186.26 | 4,431.72 |
| 729 | 2,699.73 | 2,412.41 | 2,282.34 | 2,564.45 | 9,958.93 |
| TOTAL | 31,934.83 | 28,732.74 | 29,007.32 | 31,528.73 | 121,203.62 |

STATE: CALIFORNIA

RETAIL DRUG DISTRIBUTION BY ZIP CODE WITHIN STATE BY GRAMS WT

| ZIP CODE | QUARTER 1 | QUARTER 2 | QUARTER 3 | QUARTER 4 | TOTAL GRAMS |
|---|---|---|---|---|---|
| 900 | 6,115.14 | 6,404.77 | 6,296.67 | 6,443.50 | 25,260.08 |
| 902 | 4,816.73 | 4,902.87 | 4,795.34 | 4,883.79 | 19,398.73 |
| 903 | 146.84 | 142.21 | 134.12 | 168.81 | 591.98 |
| 904 | 964.85 | 1,097.81 | 1,092.61 | 1,098.39 | 4,253.66 |
| 905 | 838.25 | 785.64 | 768.29 | 755 | 3,147.18 |
| 906 | 1,094.92 | 1,040 | 1,141.17 | 1,146.95 | 4,423.04 |
| 907 | 1,501.90 | 1,468.95 | 1,397.27 | 1,600.18 | 5,968.30 |
| 908 | 918.6 | 844.6 | 765.98 | 756.91 | 3,286.09 |
| 910 | 775.81 | 697.19 | 718 | 741.12 | 2,932.12 |
| 911 | 1,399 | 1,432.53 | 1,482.83 | 1,507.68 | 5,822.04 |
| 912 | 476.93 | 455.54 | 508.73 | 508.15 | 1,949.35 |
| 913 | 5,970.62 | 5,902.40 | 5,859.04 | 5,842.28 | 23,574.34 |
| 914 | 1,105.91 | 1,297.83 | 1,131.92 | 1,296.68 | 4,832.34 |
| 915 | 522.6 | 507.57 | 570.01 | 585.62 | 2,185.80 |
| 916 | 456.7 | 481.56 | 503.53 | 536.48 | 1,978.27 |
| 917 | 2,899.75 | 2,907.26 | 2,755.22 | 2,977.22 | 11,539.45 |
| 918 | 119.67 | 134.12 | 97.12 | 160.13 | 511.04 |
| 919 | 1,039.42 | 1,046.36 | 901.84 | 1,007.05 | 3,994.67 |
| 920 | 13,825.26 | 13,622.35 | 13,816.59 | 14,444.41 | 55,708.61 |
| 921 | 4,943.91 | 4,876.85 | 4,618.33 | 4,978.02 | 19,417.11 |
| 922 | 1,291.48 | 1,316.33 | 1,252.74 | 1,343.97 | 5,204.52 |
| 923 | 1,927.96 | 1,935.48 | 1,974.21 | 2,014.56 | 7,852.21 |
| 924 | 144.53 | 153.2 | 148.57 | 141.06 | 587.36 |
| 925 | 2,287.54 | 2,214.12 | 1,997.80 | 2,225.73 | 8,725.19 |
| 926 | 8,367.42 | 8,373.20 | 8,136.18 | 8,311.34 | 33,188.14 |
| 927 | 1,363.74 | 1,357.38 | 1,261.41 | 1,376.46 | 5,358.99 |
| 928 | 2,910.16 | 2,999.76 | 2,908.42 | 3,005.54 | 11,823.88 |
| 930 | 2,872 | 2,922.30 | 2,648.85 | 2,913.62 | 11,356.77 |
| 931 | 1,007.05 | 1,051.56 | 922.07 | 930.16 | 3,910.84 |
| 932 | 1,155.04 | 1,118.62 | 1,097.23 | 1,116.31 | 4,487.20 |
| 933 | 850.96 | 826.1 | 763.09 | 852.7 | 3,292.85 |
| 934 | 2,038.38 | 2,023.35 | 1,912.35 | 1,997.91 | 7,971.99 |
| 935 | 1,023.24 | 1,117.47 | 1,003.58 | 1,114 | 4,258.29 |
| 936 | 1,464.33 | 1,475.31 | 1,371.25 | 1,444.09 | 5,754.98 |
| 937 | 2,038.38 | 2,130.88 | 1,819.28 | 2,035.49 | 8,024.03 |
| 939 | 752.69 | 794.89 | 749.8 | 838.25 | 3,135.63 |

| | | | | | | |
|---|---|---|---|---|---|---|
| | 940 | 2,119.31 | 2,146.49 | 2,129.14 | 2,273.67 | 8,668.61 |
| | 941 | 3,562.25 | 3,602.14 | 3,848.99 | 4,002.19 | 15,015.57 |
| | 943 | 424.9 | 431.26 | 404.67 | 457.28 | 1,718.11 |
| | 944 | 476.93 | 471.15 | 501.21 | 466.53 | 1,915.82 |
| | 945 | 4,812.68 | 4,860.09 | 4,757.18 | 4,936.97 | 19,366.92 |
| | 946 | 855.01 | 1,040.58 | 965.43 | 1,035.38 | 3,896.40 |
| | 947 | 501.21 | 459.01 | 497.17 | 551.51 | 2,008.90 |
| | 948 | 68.79 | 75.15 | 97.7 | 104.06 | 345.7 |
| | 949 | 1,312.87 | 1,444.67 | 1,325.58 | 1,494.39 | 5,577.51 |
| | 950 | 2,199.67 | 2,379.46 | 2,085.78 | 2,447.68 | 9,112.59 |
| | 951 | 1,235.40 | 1,241.18 | 1,212.85 | 1,284.54 | 4,973.97 |
| | 952 | 938.83 | 916.87 | 926.69 | 993.18 | 3,775.57 |
| | 953 | 2,352.87 | 2,304.88 | 2,325.12 | 2,348.24 | 9,331.11 |
| | 954 | 661.35 | 634.75 | 628.97 | 685.05 | 2,610.12 |
| | 955 | 129.49 | 96.54 | 115.62 | 144.53 | 486.18 |
| | 956 | 3,328.70 | 3,114.22 | 2,881.25 | 3,078.96 | 12,403.13 |
| | 957 | 993.75 | 1,003 | 1,057.34 | 1,069.49 | 4,123.58 |
| | 958 | 1,405.36 | 1,470.69 | 1,352.75 | 1,615.79 | 5,844.59 |
| | 959 | 1,501.90 | 1,347.55 | 1,434.27 | 1,438.31 | 5,722.03 |
| | 960 | 831.31 | 744.59 | 787.37 | 811.65 | 3,174.92 |
| | 961 | 121.4 | 132.38 | 137.59 | 120.24 | 511.61 |
| TOTAL | | 111,261.69 | 111,775.01 | 108,794.11 | 114,459.20 | 446,290.01 |

| STATE: | COLORADO | | | | | |
|---|---|---|---|---|---|---|
| ZIP CODE | | QUARTER 1 | QUARTER 2 | QUARTER 3 | QUARTER 4 | TOTAL GRAMS |
| | 800 | 4,064.62 | 4,066.36 | 3,838.58 | 4,238.63 | 16,208.19 |
| | 801 | 5,368.24 | 5,329.50 | 5,487.90 | 5,560.17 | 21,745.81 |
| | 802 | 5,706.43 | 5,782.16 | 5,815.11 | 6,117.45 | 23,421.15 |
| | 803 | 1,530.23 | 1,681.11 | 1,581.68 | 1,703.08 | 6,496.10 |
| | 804 | 924.96 | 861.37 | 829 | 931.9 | 3,547.23 |
| | 805 | 2,823.44 | 2,891.66 | 2,902.06 | 3,180.71 | 11,797.87 |
| | 806 | 1,261.41 | 1,145.22 | 1,155.62 | 1,190.31 | 4,752.56 |
| | 807 | 116.2 | 142.79 | 114.46 | 149.73 | 523.18 |
| | 808 | 431.84 | 446.29 | 456.12 | 436.47 | 1,770.72 |
| | 809 | 2,758.12 | 2,822.86 | 2,923.45 | 2,989.36 | 11,493.79 |
| | 810 | 580.99 | 546.88 | 520.87 | 609.32 | 2,258.06 |

| | | | | | | |
|---|---|---|---|---|---|---|
| | 811 | 86.72 | 83.25 | 102.9 | 95.39 | 368.26 |
| | 812 | 116.78 | 108.1 | 127.76 | 135.28 | 487.92 |
| | 813 | 169.38 | 195.98 | 216.79 | 180.95 | 763.1 |
| | 814 | 127.76 | 115.62 | 129.49 | 145.1 | 517.97 |
| | 815 | 491.96 | 419.7 | 437.04 | 417.39 | 1,766.09 |
| | 816 | 760.2 | 756.73 | 856.74 | 741.12 | 3,114.79 |
| TOTAL | | 27,319.28 | 27,395.58 | 27,495.57 | 28,822.36 | 111,032.79 |

STATE: CONNECTICUT

| ZIP CODE | | QUARTER 1 | QUARTER 2 | QUARTER 3 | QUARTER 4 | TOTAL GRAMS |
|---|---|---|---|---|---|---|
| | 60 | 5,839.97 | 6,112.25 | 5,872.92 | 6,278.17 | 24,103.31 |
| | 61 | 2,263.84 | 2,181.75 | 1,933.17 | 2,218.75 | 8,597.51 |
| | 62 | 1,691.52 | 1,708.86 | 1,589.78 | 1,799.05 | 6,789.21 |
| | 63 | 3,620.64 | 3,561.10 | 3,574.39 | 3,711.40 | 14,467.53 |
| | 64 | 7,263.25 | 7,562.13 | 7,042.41 | 7,616.47 | 29,484.26 |
| | 65 | 1,826.22 | 1,837.78 | 1,666.08 | 1,973.06 | 7,303.14 |
| | 66 | 1,385.71 | 1,465.48 | 1,382.82 | 1,592.67 | 5,826.68 |
| | 67 | 3,033.87 | 3,005.54 | 2,804.94 | 3,046.59 | 11,890.94 |
| | 68 | 7,982.98 | 8,079.53 | 8,050.04 | 8,086.46 | 32,199.01 |
| | 69 | 1,038.27 | 1,013.41 | 986.82 | 1,051.56 | 4,090.06 |
| TOTAL | | 35,946.27 | 36,527.83 | 34,903.37 | 37,374.18 | 144,751.65 |

STATE: DELAWARE

| ZIP CODE | | QUARTER 1 | QUARTER 2 | QUARTER 3 | QUARTER 4 | TOTAL GRAMS |
|---|---|---|---|---|---|---|
| | 197 | 5,331.24 | 5,245.10 | 5,080.34 | 5,646.88 | 21,303.56 |
| | 198 | 3,632.20 | 3,540.28 | 3,472.07 | 3,728.75 | 14,373.30 |
| | 199 | 5,426.62 | 5,445.70 | 5,190.76 | 5,476.92 | 21,540 |
| TOTAL | | 14,390.06 | 14,231.08 | 13,743.17 | 14,852.55 | 57,216.86 |

STATE: DISTRICT OF COLUMBIA

| ZIP CODE | | QUARTER 1 | QUARTER 2 | QUARTER 3 | QUARTER 4 | TOTAL GRAMS |
|---|---|---|---|---|---|---|
| | 200 | 5,067.62 | 5,111.56 | 5,134.11 | 5,428.94 | 20,742.23 |
| | 203 | 5.2 | 17.34 | 71.11 | 0 | 93.65 |
| | 204 | 54.34 | 37.58 | 56.08 | 26.01 | 174.01 |

| | | | | | | |
|---|---|---|---|---|---|---|
| TOTAL | | 5,127.16 | 5,166.48 | 5,261.30 | 5,454.95 | 21,009.89 |

| STATE: | FLORIDA | | | | | |
|---|---|---|---|---|---|---|
| ZIP CODE | | QUARTER 1 | QUARTER 2 | QUARTER 3 | QUARTER 4 | TOTAL GRAMS |
| | 320 | 4,714.41 | 4,438.65 | 4,289.50 | 4,597.63 | 18,040.19 |
| | 321 | 3,183.02 | 2,947.73 | 2,809.57 | 3,101.51 | 12,041.83 |
| | 322 | 8,924.13 | 8,328.69 | 8,162.66 | 8,677.43 | 34,092.91 |
| | 323 | 5,196.54 | 4,936.40 | 4,728.86 | 5,321.99 | 20,183.79 |
| | 324 | 3,543.17 | 3,189.38 | 3,231 | 3,143.13 | 13,106.68 |
| | 325 | 7,840.19 | 7,552.30 | 7,503.16 | 7,640.17 | 30,535.82 |
| | 326 | 3,334.48 | 3,280.72 | 3,038.67 | 3,324.08 | 12,977.95 |
| | 327 | 9,707.46 | 9,268.10 | 8,708.27 | 9,508.47 | 37,192.30 |
| | 328 | 8,386.50 | 7,861.58 | 7,389.62 | 8,239.34 | 31,877.04 |
| | 329 | 7,046.46 | 6,532.53 | 6,304.18 | 6,934.31 | 26,817.48 |
| | 330 | 7,102.54 | 6,563.75 | 5,700.64 | 6,858.58 | 26,225.51 |
| | 331 | 8,654.74 | 7,758.10 | 7,364.42 | 8,572.64 | 32,349.90 |
| | 333 | 3,713.71 | 3,679.61 | 3,257.02 | 3,695.79 | 14,346.13 |
| | 334 | 9,582.01 | 9,068.08 | 8,299.20 | 9,274.46 | 36,223.75 |
| | 335 | 7,008.31 | 6,646.99 | 6,686.88 | 7,015.24 | 27,357.42 |
| | 336 | 6,242.32 | 5,952.12 | 5,708.74 | 6,374.13 | 24,277.31 |
| | 337 | 5,224.29 | 4,665.27 | 4,723.08 | 4,933.30 | 19,545.94 |
| | 338 | 6,238.86 | 5,828.98 | 5,356.67 | 6,055.60 | 23,480.11 |
| | 339 | 5,076.30 | 4,714.98 | 4,472.18 | 4,919.63 | 19,183.09 |
| | 341 | 2,322.23 | 2,219.90 | 1,941.84 | 2,085.78 | 8,569.75 |
| | 342 | 5,899.51 | 5,728.97 | 5,482.12 | 6,053.86 | 23,164.46 |
| | 344 | 2,662.15 | 2,458.08 | 2,525.14 | 2,763.90 | 10,409.27 |
| | 346 | 4,411.48 | 4,025.31 | 3,871.54 | 4,031.67 | 16,340 |
| | 347 | 5,349.16 | 4,982.07 | 4,778 | 5,261.29 | 20,370.52 |
| | 349 | 2,610.12 | 2,455.77 | 2,334.37 | 2,691.06 | 10,091.32 |
| TOTAL | | 143,974.09 | 135,084.06 | 128,667.33 | 141,074.99 | 548,800.47 |

| STATE: | GEORGIA | | | | | |
|---|---|---|---|---|---|---|
| ZIP CODE | | QUARTER 1 | QUARTER 2 | QUARTER 3 | QUARTER 4 | TOTAL GRAMS |
| | 300 | 25,595.38 | 24,502.19 | 24,767.54 | 25,312.08 | 100,177.19 |
| | 301 | 12,216.99 | 11,183.34 | 11,857.99 | 12,042.40 | 47,300.72 |

| | | | | | | |
|---|---|---|---|---|---|---|
| | 302 | 8,004.95 | 7,567.33 | 7,695.09 | 7,929.80 | 31,197.17 |
| | 303 | 12,388.68 | 12,419.32 | 12,179.41 | 12,658.08 | 49,645.49 |
| | 304 | 3,139.08 | 2,983.57 | 2,968.54 | 3,152.38 | 12,243.57 |
| | 305 | 6,744.11 | 5,964.84 | 6,381.65 | 6,642.95 | 25,733.55 |
| | 306 | 5,142.78 | 4,717.87 | 4,962.99 | 5,014.44 | 19,838.08 |
| | 307 | 2,650.59 | 2,475.42 | 2,498.55 | 2,593.36 | 10,217.92 |
| | 308 | 2,288.70 | 2,038.38 | 2,052.83 | 2,067.86 | 8,447.77 |
| | 309 | 3,040.81 | 2,618.21 | 2,830.96 | 3,113.65 | 11,603.63 |
| | 310 | 5,160.70 | 4,705.73 | 4,770.48 | 4,970.50 | 19,607.41 |
| | 312 | 2,192.16 | 2,054.57 | 2,043.01 | 2,213.54 | 8,503.28 |
| | 313 | 2,608.39 | 2,388.71 | 2,375.41 | 2,549.42 | 9,921.93 |
| | 314 | 3,428.13 | 3,151.80 | 3,061.62 | 3,226.38 | 12,867.93 |
| | 315 | 3,354.14 | 2,949.47 | 3,142.55 | 3,056.99 | 12,503.15 |
| | 316 | 1,680.54 | 1,393.22 | 1,625.62 | 1,566.07 | 6,265.45 |
| | 317 | 3,070.87 | 2,896.86 | 2,922.30 | 3,024.04 | 11,914.07 |
| | 318 | 232.97 | 235.86 | 234.71 | 235.86 | 939.4 |
| | 319 | 2,228.58 | 2,198.51 | 2,287.54 | 2,158.63 | 8,873.26 |
| | 398 | 442.82 | 461.9 | 450.34 | 417.97 | 1,773.03 |
| TOTAL | | 105,611.37 | 98,907.10 | 101,109.13 | 103,946.40 | 409,574 |

| STATE: | GUAM | | | | | |
|---|---|---|---|---|---|---|
| ZIP CODE | | QUARTER 1 | QUARTER 2 | QUARTER 3 | QUARTER 4 | TOTAL GRAMS |
| | 969 | 37.58 | 27.17 | 12.14 | 13.87 | 90.76 |
| TOTAL | | 37.58 | 27.17 | 12.14 | 13.87 | 90.76 |

| STATE: | HAWAII | | | | | |
|---|---|---|---|---|---|---|
| ZIP CODE | | QUARTER 1 | QUARTER 2 | QUARTER 3 | QUARTER 4 | TOTAL GRAMS |
| | 967 | 760.78 | 686.78 | 705.28 | 663.08 | 2,815.92 |
| | 968 | 587.93 | 617.99 | 626.66 | 709.91 | 2,542.49 |
| TOTAL | | 1,348.71 | 1,304.77 | 1,331.94 | 1,372.99 | 5,358.41 |

| STATE: | IDAHO | | | | | |
|---|---|---|---|---|---|---|
| ZIP CODE | | QUARTER 1 | QUARTER 2 | QUARTER 3 | QUARTER 4 | TOTAL GRAMS |
| | 832 | 450.34 | 497.17 | 502.37 | 562.49 | 2,012.37 |

| | | | | | | |
|---|---|---|---|---|---|---|
| | 833 | 721.47 | 774.65 | 810.5 | 815.12 | 3,121.74 |
| | 834 | 1,438.31 | 1,387.44 | 1,389.75 | 1,481.67 | 5,697.17 |
| | 835 | 157.82 | 156.67 | 184.41 | 174.01 | 672.91 |
| | 836 | 2,739.62 | 2,900.91 | 2,872.58 | 3,217.13 | 11,730.24 |
| | 837 | 2,004.27 | 2,086.36 | 1,977.10 | 2,147.64 | 8,215.37 |
| | 838 | 980.46 | 945.77 | 1,045.78 | 975.25 | 3,947.26 |
| TOTAL | | 8,492.29 | 8,748.97 | 8,782.49 | 9,373.31 | 35,397.06 |

STATE: ILLINOIS

| ZIP CODE | | QUARTER 1 | QUARTER 2 | QUARTER 3 | QUARTER 4 | TOTAL GRAMS |
|---|---|---|---|---|---|---|
| | 600 | 16,113.96 | 15,945.15 | 15,856.70 | 16,303.58 | 64,219.39 |
| | 601 | 9,868.17 | 9,607.44 | 9,731.74 | 9,908.06 | 39,115.41 |
| | 602 | 741.7 | 770.61 | 669.44 | 733.03 | 2,914.78 |
| | 603 | 633.02 | 647.47 | 614.52 | 617.41 | 2,512.42 |
| | 604 | 7,208.33 | 7,077.10 | 7,076.52 | 7,409.51 | 28,771.46 |
| | 605 | 8,826.43 | 8,983.10 | 8,632.77 | 9,198.15 | 35,640.45 |
| | 606 | 10,593.10 | 10,353.19 | 10,498.87 | 11,028.99 | 42,474.15 |
| | 607 | 702.97 | 716.27 | 633.02 | 685.63 | 2,737.89 |
| | 608 | 165.34 | 184.99 | 141.63 | 184.41 | 676.37 |
| | 609 | 1,088.56 | 1,016.88 | 1,029.02 | 1,101.86 | 4,236.32 |
| | 610 | 931.32 | 849.81 | 874.67 | 912.24 | 3,568.04 |
| | 611 | 1,061.97 | 952.13 | 924.38 | 1,052.14 | 3,990.62 |
| | 612 | 679.27 | 596.02 | 624.35 | 649.21 | 2,548.85 |
| | 613 | 557.29 | 570.58 | 560.76 | 615.1 | 2,303.73 |
| | 614 | 675.22 | 649.78 | 638.8 | 727.83 | 2,691.63 |
| | 615 | 682.16 | 716.27 | 646.89 | 728.98 | 2,774.30 |
| | 616 | 1,122.67 | 1,072.95 | 1,090.30 | 1,207.07 | 4,492.99 |
| | 617 | 1,412.30 | 1,352.18 | 1,397.85 | 1,517.51 | 5,679.84 |
| | 618 | 1,352.75 | 1,253.32 | 1,207.07 | 1,517.51 | 5,330.65 |
| | 619 | 244.54 | 201.76 | 240.49 | 249.16 | 935.95 |
| | 620 | 937.68 | 848.65 | 946.35 | 1,004.16 | 3,736.84 |
| | 622 | 1,611.74 | 1,462.59 | 1,596.71 | 1,799.05 | 6,470.09 |
| | 623 | 768.29 | 778.7 | 809.92 | 898.95 | 3,255.86 |
| | 624 | 260.72 | 253.79 | 277.49 | 250.9 | 1,042.90 |
| | 625 | 486.18 | 424.9 | 456.7 | 530.12 | 1,897.90 |
| | 626 | 378.66 | 345.7 | 342.24 | 391.37 | 1,457.97 |

| | | | | | | |
|---|---|---|---|---|---|---|
| | 627 | 645.74 | 564.23 | 593.13 | 624.93 | 2,428.03 |
| | 628 | 413.92 | 360.16 | 383.28 | 422.01 | 1,579.37 |
| | 629 | 1,020.35 | 927.27 | 973.52 | 1,034.80 | 3,955.94 |
| TOTAL | | 71,184.35 | 69,482.99 | 69,469.13 | 73,303.67 | 283,440.14 |

STATE: INDIANA

| ZIP CODE | | QUARTER 1 | QUARTER 2 | QUARTER 3 | QUARTER 4 | TOTAL GRAMS |
|---|---|---|---|---|---|---|
| | 460 | 10,186.70 | 10,034.66 | 10,127.16 | 10,645.71 | 40,994.23 |
| | 461 | 6,612.89 | 6,253.89 | 6,496.11 | 6,775.33 | 26,138.22 |
| | 462 | 9,568.71 | 9,423.03 | 9,494.14 | 9,862.96 | 38,348.84 |
| | 463 | 8,215.96 | 7,985.87 | 7,985.30 | 8,460.49 | 32,647.62 |
| | 464 | 805.87 | 704.7 | 579.26 | 712.22 | 2,802.05 |
| | 465 | 8,154.10 | 8,155.26 | 7,595.08 | 7,955.81 | 31,860.25 |
| | 466 | 2,615.90 | 2,568.50 | 2,434.96 | 2,670.82 | 10,290.18 |
| | 467 | 2,678.34 | 2,443.05 | 2,637.29 | 2,574.86 | 10,333.54 |
| | 468 | 3,424.66 | 3,425.82 | 3,437.96 | 3,685.39 | 13,973.83 |
| | 469 | 6,778.80 | 6,544.67 | 6,485.70 | 6,817.53 | 26,626.70 |
| | 470 | 653.83 | 670.6 | 620.3 | 692.56 | 2,637.29 |
| | 471 | 4,964.72 | 5,537.62 | 5,510.45 | 5,182.09 | 21,194.88 |
| | 472 | 1,390.91 | 1,314.02 | 1,330.21 | 1,530.23 | 5,565.37 |
| | 473 | 3,468.02 | 3,026.93 | 3,181.28 | 3,378.42 | 13,054.65 |
| | 474 | 3,046.01 | 2,684.70 | 2,809.57 | 3,011.32 | 11,551.60 |
| | 475 | 1,741.82 | 1,607.12 | 1,657.41 | 1,690.94 | 6,697.29 |
| | 476 | 2,147.06 | 2,054.57 | 2,114.69 | 2,288.12 | 8,604.44 |
| | 477 | 5,104.62 | 4,836.96 | 5,092.48 | 5,043.34 | 20,077.40 |
| | 478 | 1,327.90 | 1,167.18 | 1,164.29 | 1,218.63 | 4,878 |
| | 479 | 3,472.65 | 3,040.23 | 3,212.50 | 3,367.43 | 13,092.81 |
| TOTAL | | 86,359.47 | 83,479.38 | 83,966.14 | 87,564.20 | 341,369.19 |

STATE: IOWA

| ZIP CODE | | QUARTER 1 | QUARTER 2 | QUARTER 3 | QUARTER 4 | TOTAL GRAMS |
|---|---|---|---|---|---|---|
| | 500 | 2,621.68 | 2,391.60 | 2,456.35 | 2,544.22 | 10,013.85 |
| | 501 | 1,408.83 | 1,379.92 | 1,320.96 | 1,403.63 | 5,513.34 |
| | 502 | 1,560.29 | 1,499.01 | 1,468.95 | 1,613.48 | 6,141.73 |
| | 503 | 2,839.05 | 2,858.70 | 2,759.85 | 2,889.34 | 11,346.94 |

| | | | | | | |
|---|---|---|---|---|---|---|
| | 504 | 912.82 | 883.34 | 857.9 | 832.46 | 3,486.52 |
| | 505 | 1,510.58 | 1,502.48 | 1,497.28 | 1,515.78 | 6,026.12 |
| | 506 | 1,550.46 | 1,411.14 | 1,351.60 | 1,431.38 | 5,744.58 |
| | 507 | 1,182.79 | 1,156.78 | 1,094.34 | 1,142.90 | 4,576.81 |
| | 508 | 167.65 | 192.51 | 160.71 | 187.3 | 708.17 |
| | 510 | 1,194.35 | 1,152.15 | 1,057.34 | 1,145.22 | 4,549.06 |
| | 511 | 2,483.52 | 2,565.03 | 2,545.37 | 2,726.90 | 10,320.82 |
| | 512 | 445.14 | 386.75 | 437.04 | 476.93 | 1,745.86 |
| | 513 | 749.22 | 616.83 | 611.63 | 627.24 | 2,604.92 |
| | 514 | 365.36 | 323.74 | 308.13 | 327.2 | 1,324.43 |
| | 515 | 1,699.04 | 1,656.83 | 1,636.60 | 1,726.21 | 6,718.68 |
| | 516 | 191.93 | 201.76 | 174.01 | 186.15 | 753.85 |
| | 520 | 1,733.72 | 1,713.49 | 1,640.65 | 1,856.86 | 6,944.72 |
| | 521 | 208.69 | 189.62 | 238.76 | 242.8 | 879.87 |
| | 522 | 2,460.39 | 2,582.95 | 2,494.50 | 2,465.02 | 10,002.86 |
| | 523 | 1,798.47 | 1,678.22 | 1,649.90 | 1,782.28 | 6,908.87 |
| | 524 | 2,510.69 | 2,501.44 | 2,383.51 | 2,460.97 | 9,856.61 |
| | 525 | 1,126.72 | 1,139.44 | 1,067.17 | 1,195.51 | 4,528.84 |
| | 526 | 1,253.32 | 1,077 | 1,105.33 | 1,147.53 | 4,583.18 |
| | 527 | 1,688.63 | 1,546.42 | 1,581.68 | 1,731.41 | 6,548.14 |
| | 528 | 1,523.87 | 1,418.66 | 1,375.30 | 1,493.23 | 5,811.06 |
| TOTAL | | 35,187.21 | 34,025.81 | 33,274.86 | 35,151.95 | 137,639.83 |

STATE: KANSAS

| ZIP CODE | | QUARTER 1 | QUARTER 2 | QUARTER 3 | QUARTER 4 | TOTAL GRAMS |
|---|---|---|---|---|---|---|
| | 660 | 7,128.55 | 7,028.54 | 7,060.91 | 7,502.58 | 28,720.58 |
| | 661 | 885.65 | 864.84 | 805.29 | 961.96 | 3,517.74 |
| | 662 | 7,574.84 | 7,430.32 | 7,595.66 | 8,048.31 | 30,649.13 |
| | 664 | 509.31 | 433.58 | 472.31 | 490.81 | 1,906.01 |
| | 665 | 849.23 | 812.81 | 808.18 | 955.02 | 3,425.24 |
| | 666 | 3,448.37 | 3,455.30 | 3,489.99 | 3,646.08 | 14,039.74 |
| | 667 | 1,246.96 | 1,155.62 | 1,157.93 | 1,180.48 | 4,740.99 |
| | 668 | 487.34 | 495.43 | 470.57 | 539.95 | 1,993.29 |
| | 669 | 110.42 | 107.53 | 98.28 | 104.64 | 420.87 |
| | 670 | 1,505.95 | 1,447.56 | 1,502.48 | 1,629.09 | 6,085.08 |
| | 671 | 617.99 | 607.58 | 598.33 | 627.24 | 2,451.14 |

| | | | | | |
|---|---|---|---|---|---|
| 672 | 3,770.95 | 4,003.34 | 3,757.07 | 4,072.14 | 15,603.50 |
| 673 | 545.15 | 550.35 | 560.76 | 631.29 | 2,287.55 |
| 674 | 920.91 | 918.02 | 819.75 | 940.57 | 3,599.25 |
| 675 | 853.85 | 778.12 | 744.59 | 879.87 | 3,256.43 |
| 676 | 589.66 | 564.8 | 550.93 | 585.62 | 2,291.01 |
| 677 | 119.67 | 109.26 | 137.01 | 133.54 | 499.48 |
| 678 | 346.86 | 304.66 | 288.47 | 332.41 | 1,272.40 |
| 679 | 92.5 | 82.09 | 95.96 | 122.56 | 393.11 |
| TOTAL | 31,604.16 | 31,149.75 | 31,014.47 | 33,384.16 | 127,152.54 |

STATE: KENTUCKY

| ZIP CODE | QUARTER 1 | QUARTER 2 | QUARTER 3 | QUARTER 4 | TOTAL GRAMS |
|---|---|---|---|---|---|
| 400 | 3,260.48 | 3,109.60 | 3,059.31 | 3,136.19 | 12,565.58 |
| 401 | 990.86 | 920.34 | 884.49 | 886.23 | 3,681.92 |
| 402 | 10,683.87 | 10,215.61 | 9,907.48 | 10,216.76 | 41,023.72 |
| 403 | 2,515.89 | 2,276.56 | 2,389.29 | 2,523.41 | 9,705.15 |
| 404 | 1,027.86 | 962.54 | 1,020.92 | 1,071.22 | 4,082.54 |
| 405 | 3,834.53 | 3,618.90 | 3,757.62 | 4,072.14 | 15,283.19 |
| 406 | 464.79 | 492.54 | 451.5 | 408.14 | 1,816.97 |
| 407 | 299.46 | 275.75 | 332.99 | 353.22 | 1,261.42 |
| 408 | 117.35 | 94.23 | 101.17 | 91.92 | 404.67 |
| 409 | 143.95 | 147.42 | 130.65 | 150.88 | 572.9 |
| 410 | 4,657.75 | 4,442.12 | 4,352.51 | 4,464.09 | 17,916.47 |
| 411 | 698.92 | 620.88 | 659.03 | 696.61 | 2,675.44 |
| 412 | 280.96 | 237.6 | 196.55 | 231.82 | 946.93 |
| 413 | 51.45 | 42.78 | 42.78 | 52.61 | 189.62 |
| 414 | 101.75 | 91.92 | 88.45 | 91.92 | 374.04 |
| 415 | 391.37 | 368.25 | 455.54 | 448.61 | 1,663.77 |
| 416 | 279.22 | 258.99 | 265.93 | 259.57 | 1,063.71 |
| 417 | 205.8 | 165.34 | 163.6 | 187.88 | 722.62 |
| 418 | 47.4 | 33.53 | 34.69 | 40.47 | 156.09 |
| 420 | 4,264.64 | 4,066.36 | 4,087.75 | 4,229.96 | 16,648.71 |
| 421 | 1,708.29 | 1,630.24 | 1,710.02 | 1,665.51 | 6,714.06 |
| 422 | 988.55 | 865.42 | 1,077 | 824.37 | 3,755.34 |
| 423 | 2,134.92 | 1,943.57 | 1,879.40 | 1,890.39 | 7,848.28 |

| | | | | | | |
|---|---|---|---|---|---|---|
| | 424 | 2,896.86 | 2,652.90 | 2,534.39 | 2,827.49 | 10,911.64 |
| | 425 | 348.02 | 295.41 | 338.77 | 358.42 | 1,340.62 |
| | 426 | 105.21 | 90.76 | 106.95 | 84.4 | 387.32 |
| | 427 | 1,477.62 | 1,413.45 | 1,457.97 | 1,569.54 | 5,918.58 |
| TOTAL | | 43,977.77 | 41,333.01 | 41,486.75 | 42,833.77 | 169,631.30 |

| STATE: | LOUISIANA | | | | | |
|---|---|---|---|---|---|---|
| ZIP CODE | | QUARTER 1 | QUARTER 2 | QUARTER 3 | QUARTER 4 | TOTAL GRAMS |
| | 700 | 9,042.06 | 8,172.60 | 8,340.25 | 8,679.02 | 34,233.93 |
| | 701 | 4,853.73 | 4,475.65 | 4,389.51 | 4,582.60 | 18,301.49 |
| | 703 | 5,498.89 | 4,848.52 | 5,126.59 | 5,414.48 | 20,888.48 |
| | 704 | 9,648.49 | 8,777.29 | 9,028.19 | 9,412.62 | 36,866.59 |
| | 705 | 19,229.92 | 17,358.61 | 17,413.53 | 18,243.10 | 72,245.16 |
| | 706 | 9,675.08 | 8,771.51 | 8,823.54 | 9,339.09 | 36,609.22 |
| | 707 | 12,591.02 | 11,593.22 | 11,686.87 | 12,362.09 | 48,233.20 |
| | 708 | 10,027.72 | 8,866.90 | 8,954.19 | 9,740.99 | 37,589.80 |
| | 710 | 1,926.81 | 1,803.09 | 1,835.47 | 2,037.80 | 7,603.17 |
| | 711 | 6,286.26 | 5,799.50 | 5,967.73 | 6,007.62 | 24,061.11 |
| | 712 | 3,975.02 | 3,557.63 | 3,509.07 | 3,940.33 | 14,982.05 |
| | 713 | 3,566.30 | 3,228.69 | 3,203.25 | 3,507.91 | 13,506.15 |
| | 714 | 1,689.79 | 1,442.36 | 1,472.42 | 1,634.87 | 6,239.44 |
| TOTAL | | 98,011.09 | 88,695.57 | 89,750.61 | 94,902.52 | 371,359.79 |

| STATE: | MAINE | | | | | |
|---|---|---|---|---|---|---|
| ZIP CODE | | QUARTER 1 | QUARTER 2 | QUARTER 3 | QUARTER 4 | TOTAL GRAMS |
| | 39 | 600.07 | 623.77 | 645.16 | 643.43 | 2,512.43 |
| | 40 | 5,366.50 | 5,681.57 | 5,759.61 | 5,724.35 | 22,532.03 |
| | 41 | 2,079.43 | 2,155.73 | 2,203.14 | 2,075.38 | 8,513.68 |
| | 42 | 1,589.78 | 1,748.75 | 1,644.12 | 1,839.51 | 6,822.16 |
| | 43 | 708.17 | 776.39 | 808.76 | 824.95 | 3,118.27 |
| | 44 | 2,043.01 | 1,899.06 | 1,841.83 | 1,989.82 | 7,773.72 |
| | 45 | 402.36 | 414.5 | 400.62 | 402.94 | 1,620.42 |
| | 46 | 842.87 | 822.64 | 812.81 | 803.56 | 3,281.88 |
| | 47 | 922.07 | 848.07 | 919.76 | 911.09 | 3,600.99 |
| | 48 | 308.13 | 320.85 | 340.5 | 315.64 | 1,285.12 |

| | | | | | |
|---|---|---|---|---|---|
| 49 | 1,423.86 | 1,528.50 | 1,604.23 | 1,693.25 | 6,249.84 |
| TOTAL | 16,286.25 | 16,819.83 | 16,980.54 | 17,223.92 | 67,310.54 |

STATE: MARYLAND

| ZIP CODE | QUARTER 1 | QUARTER 2 | QUARTER 3 | QUARTER 4 | TOTAL GRAMS |
|---|---|---|---|---|---|
| 206 | 3,302.69 | 3,062.77 | 2,959.29 | 3,128.68 | 12,453.43 |
| 207 | 4,086.01 | 3,868.07 | 3,770.37 | 4,111.45 | 15,835.90 |
| 208 | 7,765.04 | 7,907.25 | 7,516.46 | 7,969.69 | 31,158.44 |
| 209 | 1,446.41 | 1,459.12 | 1,355.64 | 1,459.70 | 5,720.87 |
| 210 | 10,254.92 | 10,191.32 | 9,768.73 | 10,474.59 | 40,689.56 |
| 211 | 5,183.82 | 5,044.50 | 4,806.90 | 5,087.86 | 20,123.08 |
| 212 | 9,538.65 | 9,251.91 | 8,841.46 | 9,650.86 | 37,282.88 |
| 214 | 1,460.28 | 1,496.70 | 1,444.09 | 1,444.09 | 5,845.16 |
| 215 | 2,062.08 | 1,956.87 | 1,923.34 | 1,934.32 | 7,876.61 |
| 216 | 2,400.85 | 2,418.19 | 2,220.48 | 2,515.89 | 9,555.41 |
| 217 | 5,680.41 | 5,709.89 | 5,440.50 | 6,036.52 | 22,867.32 |
| 218 | 2,317.60 | 2,204.87 | 2,140.70 | 2,199.67 | 8,862.84 |
| 219 | 1,271.82 | 1,222.68 | 1,257.95 | 1,432.53 | 5,184.98 |
| TOTAL | 56,770.58 | 55,794.14 | 53,445.91 | 57,445.85 | 223,456.48 |

STATE: MASSACHUSETTS

| ZIP CODE | QUARTER 1 | QUARTER 2 | QUARTER 3 | QUARTER 4 | TOTAL GRAMS |
|---|---|---|---|---|---|
| 10 | 1,834.89 | 1,759.16 | 1,669.55 | 1,847.03 | 7,110.63 |
| 11 | 797.2 | 832.46 | 626.66 | 692.56 | 2,948.88 |
| 12 | 497.17 | 456.12 | 483.87 | 591.97 | 2,029.13 |
| 13 | 287.32 | 339.34 | 301.19 | 395.42 | 1,323.27 |
| 14 | 1,341.77 | 1,367.21 | 1,216.32 | 1,344.66 | 5,269.96 |
| 15 | 1,515.20 | 1,512.31 | 1,364.89 | 1,556.25 | 5,948.65 |
| 16 | 962.54 | 942.88 | 871.2 | 935.37 | 3,711.99 |
| 17 | 2,519.94 | 2,631.51 | 2,379.46 | 2,665.62 | 10,196.53 |
| 18 | 4,408.01 | 4,466.40 | 4,302.80 | 4,308.58 | 17,485.79 |
| 19 | 3,907.96 | 3,848.99 | 3,782.51 | 4,075.03 | 15,614.49 |
| 20 | 3,022.88 | 3,019.99 | 2,895.70 | 3,093.41 | 12,031.98 |
| 21 | 5,389.63 | 5,624.91 | 5,167.64 | 5,965.41 | 22,147.59 |
| 22 | 292.52 | 323.16 | 250.32 | 387.33 | 1,253.33 |

| | | | | | |
|---|---|---|---|---|---|
| 23 | 2,516.47 | 2,535.55 | 2,378.88 | 2,433.22 | 9,864.12 |
| 24 | 2,601.45 | 2,767.94 | 2,619.37 | 2,726.32 | 10,715.08 |
| 25 | 623.77 | 697.77 | 744.59 | 664.24 | 2,730.37 |
| 26 | 757.89 | 855.01 | 1,000.69 | 901.26 | 3,514.85 |
| 27 | 2,291.01 | 2,240.14 | 2,167.30 | 2,302.57 | 9,001.02 |
| TOTAL | 35,567.62 | 36,220.85 | 34,222.94 | 36,886.25 | 142,897.66 |

STATE: MICHIGAN

| ZIP CODE | QUARTER 1 | QUARTER 2 | QUARTER 3 | QUARTER 4 | TOTAL GRAMS |
|---|---|---|---|---|---|
| 480 | 7,469.05 | 7,510.10 | 7,097.33 | 7,806.66 | 29,883.14 |
| 481 | 7,124.45 | 6,782.85 | 6,649.31 | 7,405.46 | 27,962.07 |
| 482 | 1,644.69 | 1,608.27 | 1,437.73 | 1,652.21 | 6,342.90 |
| 483 | 6,638.32 | 6,420.38 | 6,383.84 | 6,852.10 | 26,294.64 |
| 484 | 2,098.50 | 1,927.39 | 1,855.70 | 2,034.91 | 7,916.50 |
| 485 | 814.54 | 822.06 | 744.59 | 811.07 | 3,192.26 |
| 486 | 2,552.31 | 2,387.55 | 2,360.38 | 2,528.61 | 9,828.85 |
| 487 | 1,012.25 | 1,097.81 | 956.76 | 1,029.02 | 4,095.84 |
| 488 | 4,685.50 | 4,432.87 | 4,524.21 | 4,666.42 | 18,309 |
| 489 | 1,121.51 | 1,113.42 | 1,188.57 | 1,104.75 | 4,528.25 |
| 490 | 6,018.60 | 5,516.23 | 5,459.58 | 5,863.09 | 22,857.50 |
| 491 | 869.46 | 893.74 | 848.65 | 912.82 | 3,524.67 |
| 492 | 1,610.59 | 1,450.45 | 1,448.72 | 1,627.93 | 6,137.69 |
| 493 | 1,921.60 | 1,900.21 | 1,766.67 | 2,021.04 | 7,609.52 |
| 494 | 5,005.77 | 5,046.81 | 5,040.45 | 5,376.33 | 20,469.36 |
| 495 | 4,228.80 | 4,140.35 | 4,158.27 | 4,247.30 | 16,774.72 |
| 496 | 2,494.50 | 2,641.92 | 2,490.45 | 2,620.53 | 10,247.40 |
| 497 | 1,345.82 | 1,431.95 | 1,312.87 | 1,425.59 | 5,516.23 |
| 498 | 1,230.77 | 1,182.21 | 1,151.58 | 1,263.73 | 4,828.29 |
| 499 | 386.75 | 371.14 | 362.47 | 413.34 | 1,533.70 |
| TOTAL | 60,273.78 | 58,677.71 | 57,238.13 | 61,662.91 | 237,852.53 |

STATE: MINNESOTA

| ZIP CODE | QUARTER 1 | QUARTER 2 | QUARTER 3 | QUARTER 4 | TOTAL GRAMS |
|---|---|---|---|---|---|
| 550 | 2,770.83 | 2,751.76 | 2,805.52 | 2,768.52 | 11,096.63 |
| 551 | 4,009.12 | 4,058.26 | 4,020.11 | 4,125.90 | 16,213.39 |
| 553 | 5,861.93 | 5,845.17 | 5,789.67 | 6,052.71 | 23,549.48 |

| | | | | | | |
|---|---|---|---|---|---|---|
| | 554 | 5,685.61 | 6,018.60 | 5,937.67 | 6,053.29 | 23,695.17 |
| | 556 | 17.92 | 10.98 | 19.08 | 13.3 | 61.28 |
| | 557 | 1,099.55 | 1,134.81 | 1,115.15 | 1,036.53 | 4,386.04 |
| | 558 | 372.3 | 433 | 383.28 | 408.72 | 1,597.30 |
| | 559 | 1,218.06 | 1,344.08 | 1,233.67 | 1,211.70 | 5,007.51 |
| | 560 | 3,013.06 | 2,981.26 | 3,141.97 | 2,877.78 | 12,014.07 |
| | 561 | 684.47 | 721.47 | 671.75 | 615.68 | 2,693.37 |
| | 562 | 853.28 | 909.93 | 870.62 | 620.88 | 3,254.71 |
| | 563 | 1,520.98 | 1,420.97 | 1,459.70 | 1,330.79 | 5,732.44 |
| | 564 | 1,106.48 | 1,087.98 | 1,027.28 | 890.85 | 4,112.59 |
| | 565 | 1,919.29 | 1,837.20 | 1,708.86 | 1,392.64 | 6,857.99 |
| | 566 | 896.63 | 878.13 | 905.88 | 894.9 | 3,575.54 |
| | 567 | 774.65 | 815.12 | 717.42 | 524.91 | 2,832.10 |
| TOTAL | | 31,804.16 | 32,248.72 | 31,807.63 | 30,819.10 | 126,679.61 |
| STATE: | MISSISSIPPI | | | | | |
| ZIP CODE | | QUARTER 1 | QUARTER 2 | QUARTER 3 | QUARTER 4 | TOTAL GRAMS |
| | 386 | 7,082.88 | 6,477.61 | 6,705.96 | 7,035.48 | 27,301.93 |
| | 387 | 630.71 | 508.15 | 550.93 | 628.39 | 2,318.18 |
| | 388 | 2,974.32 | 2,758.69 | 2,707.24 | 2,887.61 | 11,327.86 |
| | 389 | 905.88 | 867.73 | 806.45 | 848.65 | 3,428.71 |
| | 390 | 2,983.57 | 2,727.48 | 2,762.16 | 2,911.31 | 11,384.52 |
| | 391 | 3,481.32 | 3,259.33 | 3,354.71 | 3,431.02 | 13,526.38 |
| | 392 | 3,144.86 | 2,804.36 | 2,751.18 | 3,098.04 | 11,798.44 |
| | 393 | 2,283.50 | 2,150.53 | 2,296.79 | 2,352.87 | 9,083.69 |
| | 394 | 6,845.86 | 6,275.85 | 6,289.15 | 6,736.60 | 26,147.46 |
| | 395 | 6,278.74 | 5,785.05 | 5,926.68 | 6,296.09 | 24,286.56 |
| | 396 | 2,027.40 | 1,705.97 | 1,826.22 | 1,964.38 | 7,523.97 |
| | 397 | 2,101.97 | 1,794.42 | 1,884.61 | 2,012.37 | 7,793.37 |
| TOTAL | | 40,741.01 | 37,115.17 | 37,862.08 | 40,202.81 | 155,921.07 |
| STATE: | MISSOURI | | | | | |
| ZIP CODE | | QUARTER 1 | QUARTER 2 | QUARTER 3 | QUARTER 4 | TOTAL GRAMS |
| | 630 | 7,741.34 | 7,387.54 | 7,437.26 | 7,758.10 | 30,324.24 |
| | 631 | 21,788.01 | 21,029.54 | 20,719.10 | 21,471.79 | 85,008.44 |

| | | | | | | |
|---|---|---|---|---|---|---|
| | 633 | 7,036.63 | 6,884.01 | 7,040.10 | 7,345.34 | 28,306.08 |
| | 634 | 532.43 | 520.87 | 545.15 | 587.93 | 2,186.38 |
| | 635 | 613.36 | 530.7 | 497.17 | 557.29 | 2,198.52 |
| | 636 | 860.79 | 786.79 | 778.7 | 858.48 | 3,284.76 |
| | 637 | 572.9 | 524.34 | 531.27 | 528.96 | 2,157.47 |
| | 638 | 1,031.91 | 920.91 | 942.3 | 1,227.31 | 4,122.43 |
| | 639 | 426.06 | 354.95 | 346.86 | 367.67 | 1,495.54 |
| | 640 | 5,790.25 | 5,648.62 | 5,611.04 | 5,805.86 | 22,855.77 |
| | 641 | 9,015.47 | 8,793.48 | 8,739.14 | 9,095.25 | 35,643.34 |
| | 644 | 404.09 | 360.73 | 368.25 | 419.7 | 1,552.77 |
| | 645 | 1,209.39 | 1,143.48 | 1,216.32 | 1,241.18 | 4,810.37 |
| | 646 | 191.35 | 174.01 | 191.35 | 220.26 | 776.97 |
| | 647 | 794.89 | 640.53 | 692.56 | 697.19 | 2,825.17 |
| | 648 | 1,147.53 | 1,042.31 | 1,074.11 | 1,145.22 | 4,409.17 |
| | 650 | 493.12 | 467.1 | 538.21 | 518.56 | 2,016.99 |
| | 651 | 694.3 | 622.61 | 645.74 | 746.91 | 2,709.56 |
| | 652 | 2,259.21 | 2,074.22 | 2,020.46 | 2,381.77 | 8,735.66 |
| | 653 | 517.98 | 420.86 | 418.54 | 454.39 | 1,811.77 |
| | 654 | 382.7 | 335.3 | 428.95 | 471.15 | 1,618.10 |
| | 655 | 554.4 | 583.88 | 561.91 | 600.07 | 2,300.26 |
| | 656 | 794.89 | 634.75 | 656.14 | 746.91 | 2,832.69 |
| | 657 | 1,168.34 | 1,087.41 | 1,145.79 | 1,168.92 | 4,570.46 |
| | 658 | 1,888.07 | 1,851.08 | 1,918.71 | 2,001.96 | 7,659.82 |
| TOTAL | | 67,909.41 | 64,820.02 | 65,065.13 | 68,418.17 | 266,212.73 |

STATE: MONTANA

| ZIP CODE | | QUARTER 1 | QUARTER 2 | QUARTER 3 | QUARTER 4 | TOTAL GRAMS |
|---|---|---|---|---|---|---|
| | 590 | 282.11 | 192.51 | 216.21 | 200.02 | 890.85 |
| | 591 | 1,147.53 | 1,066.59 | 1,050.99 | 1,159.09 | 4,424.20 |
| | 592 | 116.2 | 102.32 | 105.79 | 115.04 | 439.35 |
| | 593 | 200.02 | 201.76 | 195.98 | 236.44 | 834.2 |
| | 594 | 746.91 | 657.88 | 637.64 | 674.06 | 2,716.49 |
| | 595 | 129.49 | 127.76 | 127.76 | 125.45 | 510.46 |
| | 596 | 304.66 | 323.74 | 300.03 | 396 | 1,324.43 |
| | 597 | 1,119.20 | 1,022.66 | 1,080.47 | 1,142.33 | 4,364.66 |
| | 598 | 1,389.75 | 1,360.27 | 1,363.16 | 1,428.49 | 5,541.67 |

| | | | | | |
|---|---|---|---|---|---|
| 599 | 408.72 | 479.82 | 490.81 | 484.45 | 1,863.80 |
| TOTAL | 5,844.59 | 5,535.31 | 5,568.84 | 5,961.37 | 22,910.11 |

STATE: NEBRASKA

| ZIP CODE | QUARTER 1 | QUARTER 2 | QUARTER 3 | QUARTER 4 | TOTAL GRAMS |
|---|---|---|---|---|---|
| 680 | 1,511.15 | 1,419.24 | 1,551.04 | 1,642.96 | 6,124.39 |
| 681 | 6,354.48 | 6,159.08 | 6,303.02 | 6,722.72 | 25,539.30 |
| 683 | 519.13 | 413.34 | 437.04 | 452.07 | 1,821.58 |
| 684 | 413.92 | 357.27 | 345.7 | 413.34 | 1,530.23 |
| 685 | 3,816.62 | 3,854.77 | 3,851.42 | 4,265.71 | 15,788.52 |
| 686 | 431.26 | 455.54 | 395.42 | 493.7 | 1,775.92 |
| 687 | 583.88 | 564.23 | 526.07 | 590.24 | 2,264.42 |
| 688 | 1,326.74 | 1,243.49 | 1,211.70 | 1,296.10 | 5,078.03 |
| 689 | 486.18 | 428.37 | 423.17 | 446.29 | 1,784.01 |
| 690 | 139.9 | 155.51 | 192.51 | 174.01 | 661.93 |
| 691 | 382.12 | 383.28 | 367.67 | 387.33 | 1,520.40 |
| 692 | 20.81 | 13.3 | 13.3 | 16.19 | 63.6 |
| 693 | 430.68 | 471.15 | 457.86 | 453.81 | 1,813.50 |
| TOTAL | 16,416.87 | 15,918.57 | 16,075.92 | 17,354.47 | 65,765.83 |

STATE: NEVADA

| ZIP CODE | QUARTER 1 | QUARTER 2 | QUARTER 3 | QUARTER 4 | TOTAL GRAMS |
|---|---|---|---|---|---|
| 890 | 1,891.54 | 1,881.14 | 1,723.89 | 1,878.25 | 7,374.82 |
| 891 | 3,993.51 | 3,779.62 | 3,708.97 | 3,832.22 | 15,314.32 |
| 893 | 22.55 | 6.36 | 15.03 | 7.52 | 51.46 |
| 894 | 666.55 | 644.58 | 626.66 | 690.25 | 2,628.04 |
| 895 | 952.13 | 939.41 | 900.68 | 1,001.27 | 3,793.49 |
| 897 | 221.99 | 245.69 | 171.7 | 242.22 | 881.6 |
| 898 | 51.45 | 54.92 | 60.12 | 68.22 | 234.71 |
| TOTAL | 7,799.72 | 7,551.72 | 7,207.05 | 7,719.95 | 30,278.44 |

STATE: NEW HAMPSHIRE

| ZIP CODE | QUARTER 1 | QUARTER 2 | QUARTER 3 | QUARTER 4 | TOTAL GRAMS |
|---|---|---|---|---|---|
| 30 | 4,035.14 | 4,136.31 | 3,943.80 | 3,962.88 | 16,078.13 |

| | | | | | | |
|---|---|---|---|---|---|---|
| | 31 | 2,226.84 | 2,322.23 | 2,161.52 | 2,202.56 | 8,913.15 |
| | 32 | 1,127.30 | 1,119.78 | 1,042.31 | 1,153.31 | 4,442.70 |
| | 33 | 889.12 | 809.34 | 851.54 | 883.91 | 3,433.91 |
| | 34 | 425.48 | 431.26 | 368.25 | 411.61 | 1,636.60 |
| | 35 | 310.44 | 289.63 | 302.35 | 304.66 | 1,207.08 |
| | 36 | 29.48 | 28.33 | 25.44 | 43.94 | 127.19 |
| | 37 | 756.15 | 794.89 | 722.05 | 752.11 | 3,025.20 |
| | 38 | 4,151.34 | 4,206.83 | 4,147.29 | 4,268.69 | 16,774.15 |
| TOTAL | | 13,951.29 | 14,138.60 | 13,564.55 | 13,983.67 | 55,638.11 |

STATE: NEW JERSEY

| ZIP CODE | | QUARTER 1 | QUARTER 2 | QUARTER 3 | QUARTER 4 | TOTAL GRAMS |
|---|---|---|---|---|---|---|
| | 70 | 8,994.08 | 8,970.96 | 8,282.44 | 8,923.55 | 35,171.03 |
| | 71 | 501.79 | 501.79 | 526.07 | 603.54 | 2,133.19 |
| | 72 | 386.17 | 354.38 | 303.5 | 377.5 | 1,421.55 |
| | 73 | 504.1 | 578.68 | 530.7 | 579.26 | 2,192.74 |
| | 74 | 3,320.03 | 3,465.71 | 3,297.48 | 3,399.23 | 13,482.45 |
| | 75 | 396 | 387.91 | 363.62 | 356.11 | 1,503.64 |
| | 76 | 3,296.90 | 3,292.28 | 3,130.41 | 3,431.60 | 13,151.19 |
| | 77 | 3,092.26 | 3,281.30 | 3,228.69 | 3,297.48 | 12,899.73 |
| | 78 | 3,009.59 | 2,909.58 | 2,829.22 | 2,932.70 | 11,681.09 |
| | 79 | 3,140.24 | 3,187.07 | 3,076.65 | 3,263.37 | 12,667.33 |
| | 80 | 9,025.30 | 9,121.84 | 8,727.58 | 8,733.36 | 35,608.08 |
| | 81 | 623.19 | 624.93 | 590.82 | 659.61 | 2,498.55 |
| | 82 | 1,115.73 | 1,309.40 | 1,296.10 | 1,190.31 | 4,911.54 |
| | 83 | 994.33 | 1,029.02 | 846.34 | 897.21 | 3,766.90 |
| | 84 | 90.18 | 126.6 | 149.73 | 115.04 | 481.55 |
| | 85 | 2,161.52 | 2,307.78 | 2,088.10 | 2,306.04 | 8,863.44 |
| | 86 | 1,415.19 | 1,521.56 | 1,414.03 | 1,492.08 | 5,842.86 |
| | 87 | 2,302.57 | 2,233.78 | 2,318.76 | 2,252.86 | 9,107.97 |
| | 88 | 4,222.44 | 4,163.48 | 4,001.03 | 4,291.24 | 16,678.19 |
| | 89 | 471.73 | 456.7 | 454.96 | 514.51 | 1,897.90 |
| TOTAL | | 49,063.34 | 49,824.75 | 47,456.23 | 49,616.60 | 195,960.92 |

STATE: NEW MEXICO

| ZIP CODE | | QUARTER 1 | QUARTER 2 | QUARTER 3 | QUARTER 4 | TOTAL GRAMS |
|---|---|---|---|---|---|---|
| | 870 | 273.44 | 239.33 | 254.36 | 263.61 | 1,030.74 |
| | 871 | 2,382.93 | 2,151.69 | 2,018.15 | 1,737.19 | 8,289.96 |
| | 873 | 30.06 | 24.28 | 17.92 | 32.37 | 104.63 |
| | 874 | 128.34 | 125.45 | 132.38 | 143.37 | 529.54 |
| | 875 | 402.94 | 412.76 | 396.58 | 386.17 | 1,598.45 |
| | 877 | 79.2 | 94.23 | 70.53 | 91.34 | 335.3 |
| | 878 | 40.47 | 45.67 | 39.89 | 42.2 | 168.23 |
| | 879 | 33.53 | 17.34 | 17.92 | 13.87 | 82.66 |
| | 880 | 437.04 | 401.78 | 366.52 | 433 | 1,638.34 |
| | 881 | 133.54 | 94.81 | 90.76 | 93.65 | 412.76 |
| | 882 | 556.71 | 536.48 | 497.17 | 580.41 | 2,170.77 |
| | 883 | 198.87 | 193.66 | 169.38 | 235.86 | 797.77 |
| | 884 | 14.45 | 31.22 | 30.06 | 24.86 | 100.59 |
| TOTAL | | 4,711.52 | 4,368.70 | 4,101.62 | 4,077.90 | 17,259.74 |

STATE: NEW YORK

| ZIP CODE | | QUARTER 1 | QUARTER 2 | QUARTER 3 | QUARTER 4 | TOTAL GRAMS |
|---|---|---|---|---|---|---|
| | 100 | 15,013.26 | 16,397.81 | 15,924.34 | 17,150.45 | 64,485.86 |
| | 101 | 1,001.85 | 1,003 | 902.99 | 933.05 | 3,840.89 |
| | 102 | 174.59 | 156.67 | 139.9 | 157.82 | 628.98 |
| | 103 | 1,139.44 | 1,146.37 | 1,018.61 | 1,164.87 | 4,469.29 |
| | 104 | 1,279.34 | 1,285.12 | 994.33 | 1,116.89 | 4,675.68 |
| | 105 | 5,071.67 | 5,125.43 | 5,021.95 | 5,334.71 | 20,553.76 |
| | 106 | 556.13 | 511.04 | 522.6 | 529.54 | 2,119.31 |
| | 107 | 679.27 | 692.56 | 672.33 | 681 | 2,725.16 |
| | 108 | 474.04 | 403.51 | 412.76 | 410.45 | 1,700.76 |
| | 109 | 2,903.80 | 2,614.17 | 2,565.03 | 2,607.23 | 10,690.23 |
| | 110 | 1,291.48 | 1,338.30 | 1,248.70 | 1,378.77 | 5,257.25 |
| | 111 | 737.66 | 815.12 | 764.83 | 814.54 | 3,132.15 |
| | 112 | 5,737.64 | 5,683.88 | 5,239.90 | 5,890.84 | 22,552.26 |
| | 113 | 1,627.35 | 1,641.80 | 1,514.62 | 1,714.07 | 6,497.84 |
| | 114 | 560.18 | 530.7 | 575.21 | 534.16 | 2,200.25 |
| | 115 | 4,342.69 | 4,370.44 | 4,141.51 | 4,382.58 | 17,237.22 |
| | 116 | 348.02 | 332.41 | 312.75 | 340.5 | 1,333.68 |
| | 117 | 8,655.31 | 8,550.10 | 8,174.33 | 8,763.42 | 34,143.16 |

| | | | | | | |
|---|---|---|---|---|---|---|
| | 118 | 503.53 | 552.66 | 522.02 | 626.66 | 2,204.87 |
| | 119 | 1,002.43 | 1,092.03 | 1,179.32 | 1,050.99 | 4,324.77 |
| | 120 | 1,895.59 | 1,804.25 | 1,899.64 | 1,934.90 | 7,534.38 |
| | 121 | 1,052.72 | 945.19 | 1,034.80 | 1,110.53 | 4,143.24 |
| | 122 | 718.58 | 746.91 | 675.8 | 800.09 | 2,941.38 |
| | 123 | 767.14 | 723.2 | 720.89 | 714.53 | 2,925.76 |
| | 124 | 456.12 | 437.62 | 485.03 | 474.62 | 1,853.39 |
| | 125 | 2,218.75 | 2,370.79 | 2,316.45 | 2,417.04 | 9,323.03 |
| | 126 | 502.95 | 490.81 | 437.62 | 487.92 | 1,919.30 |
| | 127 | 449.18 | 542.26 | 470 | 458.43 | 1,919.87 |
| | 128 | 1,385.68 | 1,403.60 | 1,346.97 | 1,446.98 | 5,583.23 |
| | 129 | 453.81 | 499.48 | 429.53 | 457.86 | 1,840.68 |
| | 130 | 1,834.89 | 1,839.51 | 1,821.02 | 1,915.25 | 7,410.67 |
| | 131 | 727.83 | 725.52 | 627.24 | 737.66 | 2,818.25 |
| | 132 | 913.98 | 867.15 | 931.32 | 980.46 | 3,692.91 |
| | 133 | 579.26 | 553.82 | 500.63 | 491.39 | 2,125.10 |
| | 134 | 659.61 | 698.34 | 675.22 | 701.24 | 2,734.41 |
| | 135 | 232.97 | 242.22 | 246.85 | 238.76 | 960.8 |
| | 136 | 1,170.65 | 1,095.50 | 1,167.18 | 1,282.80 | 4,716.13 |
| | 137 | 559.02 | 562.49 | 573.48 | 568.85 | 2,263.84 |
| | 138 | 799.51 | 768.29 | 753.26 | 848.07 | 3,169.13 |
| | 139 | 573.48 | 599.49 | 509.31 | 605.85 | 2,288.13 |
| | 140 | 1,589.20 | 1,468.37 | 1,454.50 | 1,533.70 | 6,045.77 |
| | 141 | 593.13 | 591.4 | 576.37 | 563.07 | 2,323.97 |
| | 142 | 1,936.64 | 2,030.87 | 1,865.53 | 2,122.21 | 7,955.25 |
| | 143 | 137.59 | 151.46 | 164.18 | 137.01 | 590.24 |
| | 144 | 2,037.22 | 1,978.26 | 1,955.71 | 2,278.29 | 8,249.48 |
| | 145 | 1,699.61 | 1,833.16 | 1,714.07 | 1,847.61 | 7,094.45 |
| | 146 | 2,645.39 | 2,617.64 | 2,684.12 | 2,837.89 | 10,785.04 |
| | 147 | 707.02 | 709.91 | 732.45 | 690.25 | 2,839.63 |
| | 148 | 1,139.44 | 1,066.02 | 1,071.22 | 1,100.12 | 4,376.80 |
| | 149 | 324.31 | 282.11 | 255.52 | 359 | 1,220.94 |
| TOTAL | | 83,860.95 | 84,888.76 | 81,943.94 | 87,724.92 | 338,418.57 |

| | | | | | |
|---|---|---|---|---|---|
| STATE: | NORTH CAROLINA | | | | |
| ZIP CODE | QUARTER 1 | QUARTER 2 | QUARTER 3 | QUARTER 4 | TOTAL GRAMS |

| | | | | | | |
|---|---|---|---|---|---|---|
| | 270 | 2,374.26 | 2,327.43 | 2,168.45 | 2,529.77 | 9,399.91 |
| | 271 | 3,333.90 | 3,076.65 | 3,211.35 | 3,335.06 | 12,956.96 |
| | 272 | 5,997.21 | 5,915.70 | 5,637.63 | 6,329.04 | 23,879.58 |
| | 273 | 3,554.16 | 3,438.54 | 3,220.02 | 3,530.46 | 13,743.18 |
| | 274 | 3,828.18 | 3,720.65 | 3,447.79 | 3,872.11 | 14,868.73 |
| | 275 | 11,765.49 | 11,805.38 | 11,125.53 | 12,384.06 | 47,080.46 |
| | 276 | 5,605.84 | 5,476.34 | 5,374.02 | 5,849.79 | 22,305.99 |
| | 277 | 2,215.89 | 2,186.37 | 2,190.42 | 2,209.04 | 8,801.72 |
| | 278 | 5,287.88 | 4,710.36 | 4,757.76 | 5,164.75 | 19,920.75 |
| | 279 | 1,271.82 | 1,286.27 | 1,182.21 | 1,261.99 | 5,002.29 |
| | 280 | 9,887.24 | 9,336.89 | 9,132.82 | 9,801.69 | 38,158.64 |
| | 281 | 7,577.16 | 7,320.48 | 6,774.18 | 7,589.30 | 29,261.12 |
| | 282 | 8,201.50 | 8,211.33 | 7,762.15 | 8,526.98 | 32,701.96 |
| | 283 | 8,161.62 | 7,671.97 | 7,293.31 | 8,018.25 | 31,145.15 |
| | 284 | 6,533.69 | 6,281.63 | 6,176.42 | 6,520.39 | 25,512.13 |
| | 285 | 5,372.86 | 5,316.79 | 5,422 | 5,496.57 | 21,608.22 |
| | 286 | 7,763.88 | 7,685.84 | 7,508.36 | 8,584.21 | 31,542.29 |
| | 287 | 3,830.49 | 3,703.31 | 3,783.66 | 3,922.41 | 15,239.87 |
| | 288 | 1,575.32 | 1,619.84 | 1,491.50 | 1,719.27 | 6,405.93 |
| | 289 | 262.46 | 297.72 | 237.02 | 239.91 | 1,037.11 |
| TOTAL | | 104,400.85 | 101,389.49 | 97,896.60 | 106,885.05 | 410,571.99 |

| STATE: | | NORTH DAKOTA | | | | |
|---|---|---|---|---|---|---|
| ZIP CODE | | QUARTER 1 | QUARTER 2 | QUARTER 3 | QUARTER 4 | TOTAL GRAMS |
| | 580 | 647.47 | 557.87 | 612.79 | 572.9 | 2,391.03 |
| | 581 | 1,770.72 | 1,614.63 | 1,624.46 | 1,668.97 | 6,678.78 |
| | 582 | 1,231.35 | 1,116.89 | 1,162.56 | 1,241.76 | 4,752.56 |
| | 583 | 313.91 | 314.49 | 266.5 | 294.25 | 1,189.15 |
| | 584 | 208.12 | 161.87 | 165.34 | 185.57 | 720.9 |
| | 585 | 550.93 | 455.54 | 496.59 | 527.81 | 2,030.87 |
| | 586 | 115.04 | 131.81 | 120.24 | 121.4 | 488.49 |
| | 587 | 873.51 | 959.07 | 941.72 | 987.97 | 3,762.27 |
| | 588 | 220.83 | 204.07 | 202.34 | 250.9 | 878.14 |
| TOTAL | | 5,931.88 | 5,516.24 | 5,592.54 | 5,851.53 | 22,892.19 |

STATE: OHIO

| ZIP CODE | QUARTER 1 | QUARTER 2 | QUARTER 3 | QUARTER 4 | TOTAL GRAMS |
|---|---|---|---|---|---|
| 430 | 9,625.94 | 9,287.75 | 9,142.65 | 9,550.21 | 37,606.55 |
| 431 | 3,734.53 | 3,619.48 | 3,610.81 | 3,836.27 | 14,801.09 |
| 432 | 22,553.99 | 19,659.45 | 20,684.42 | 20,626.03 | 83,523.89 |
| 433 | 900.68 | 926.69 | 861.95 | 904.73 | 3,594.05 |
| 434 | 1,119.20 | 1,053.88 | 972.36 | 1,118.62 | 4,264.06 |
| 435 | 2,432.64 | 2,286.39 | 2,199.09 | 2,352.29 | 9,270.41 |
| 436 | 3,063.35 | 3,003.23 | 2,807.25 | 2,797.43 | 11,671.26 |
| 437 | 1,084.52 | 1,051.56 | 909.93 | 991.44 | 4,037.45 |
| 438 | 182.1 | 212.16 | 181.52 | 189.62 | 765.4 |
| 439 | 1,106.48 | 1,140.59 | 1,083.94 | 1,163.14 | 4,494.15 |
| 440 | 9,544.43 | 9,318.39 | 9,278.51 | 9,517.26 | 37,658.59 |
| 441 | 10,734.97 | 10,393.66 | 10,153.29 | 10,925.86 | 42,207.78 |
| 442 | 4,800.54 | 4,778 | 4,752.56 | 4,984.38 | 19,315.48 |
| 443 | 1,799.63 | 1,716.38 | 1,725.63 | 1,795.58 | 7,037.22 |
| 444 | 2,199.09 | 2,184.06 | 2,189.84 | 2,263.84 | 8,836.83 |
| 445 | 2,023.93 | 1,927.96 | 1,899.64 | 2,000.23 | 7,851.76 |
| 446 | 3,395.76 | 3,320.03 | 3,235.63 | 3,341.42 | 13,292.84 |
| 447 | 2,806.10 | 2,661.57 | 2,557.51 | 2,719.96 | 10,745.14 |
| 448 | 1,842.40 | 1,655.10 | 1,704.24 | 1,712.33 | 6,914.07 |
| 449 | 766.56 | 737.08 | 656.14 | 745.17 | 2,904.95 |
| 450 | 6,795.57 | 6,327.88 | 6,314.01 | 7,048.02 | 26,485.48 |
| 451 | 2,386.97 | 2,306.04 | 2,200.25 | 2,432.64 | 9,325.90 |
| 452 | 7,726.88 | 7,364.42 | 7,287.53 | 7,759.26 | 30,138.09 |
| 453 | 3,432.76 | 3,297.48 | 3,348.93 | 3,355.87 | 13,435.04 |
| 454 | 3,682.50 | 3,718.92 | 3,533.93 | 3,843.79 | 14,779.14 |
| 455 | 837.67 | 744.01 | 776.39 | 824.37 | 3,182.44 |
| 456 | 1,827.95 | 1,702.50 | 1,673.60 | 1,760.89 | 6,964.94 |
| 457 | 1,694.99 | 1,559.71 | 1,496.70 | 1,669.55 | 6,420.95 |
| 458 | 1,238.87 | 1,256.79 | 1,138.86 | 1,354.49 | 4,989.01 |
| TOTAL | 115,341 | 109,211.16 | 108,377.11 | 113,584.69 | 446,513.96 |

STATE: OKLAHOMA

| ZIP CODE | QUARTER 1 | QUARTER 2 | QUARTER 3 | QUARTER 4 | TOTAL GRAMS |
|---|---|---|---|---|---|
| 730 | 8,606.17 | 8,441.42 | 8,430.43 | 8,830.48 | 34,308.50 |

| | | | | | | |
|---|---|---|---|---|---|---|
| | 731 | 7,066.05 | 6,684.57 | 6,756.13 | 7,066.15 | 27,572.90 |
| | 734 | 781.59 | 670.6 | 702.39 | 716.27 | 2,870.85 |
| | 735 | 1,215.74 | 1,221.53 | 1,312.29 | 1,309.40 | 5,058.96 |
| | 736 | 327.78 | 307.55 | 321.42 | 335.88 | 1,292.63 |
| | 737 | 831.31 | 846.34 | 805.87 | 882.76 | 3,366.28 |
| | 738 | 122.56 | 99.43 | 64.75 | 98.86 | 385.6 |
| | 739 | 70.53 | 60.7 | 37 | 36.42 | 204.65 |
| | 740 | 6,848.17 | 6,536.58 | 6,756.83 | 7,523.39 | 27,664.97 |
| | 741 | 4,449.06 | 4,211.46 | 4,343.27 | 4,523.05 | 17,526.84 |
| | 743 | 1,134.23 | 1,097.81 | 1,006.47 | 1,211.12 | 4,449.63 |
| | 744 | 1,816.97 | 1,639.49 | 1,732.57 | 1,864.37 | 7,053.40 |
| | 745 | 622.04 | 621.46 | 729.56 | 671.17 | 2,644.23 |
| | 746 | 428.95 | 359.58 | 394.26 | 426.64 | 1,609.43 |
| | 747 | 627.82 | 545.15 | 648.05 | 612.79 | 2,433.81 |
| | 748 | 1,553.93 | 1,471.84 | 1,318.65 | 1,644.69 | 5,989.11 |
| | 749 | 612.21 | 489.65 | 506.42 | 637.07 | 2,245.35 |
| TOTAL | | 37,115.11 | 35,305.16 | 35,866.36 | 38,390.51 | 146,677.14 |

| STATE: | OREGON | | | | | |
|---|---|---|---|---|---|---|
| ZIP CODE | | QUARTER 1 | QUARTER 2 | QUARTER 3 | QUARTER 4 | TOTAL GRAMS |
| | 970 | 3,696.95 | 3,765.17 | 3,661.11 | 3,802.13 | 14,925.36 |
| | 971 | 1,018.03 | 1,085.09 | 1,094.92 | 1,091.45 | 4,289.49 |
| | 972 | 4,454.26 | 4,639.83 | 4,473.34 | 4,714.98 | 18,282.41 |
| | 973 | 1,715.22 | 1,647.01 | 1,579.95 | 1,738.92 | 6,681.10 |
| | 974 | 1,479.36 | 1,433.69 | 1,396.69 | 1,565.49 | 5,875.23 |
| | 975 | 608.74 | 631.29 | 603.54 | 601.22 | 2,444.79 |
| | 976 | 102.32 | 91.92 | 67.64 | 119.67 | 381.55 |
| | 977 | 1,315.18 | 1,286.85 | 1,328.47 | 1,325.01 | 5,255.51 |
| | 978 | 397.15 | 422.59 | 406.98 | 432.42 | 1,659.14 |
| | 979 | 73.42 | 82.67 | 79.2 | 87.87 | 323.16 |
| TOTAL | | 14,860.63 | 15,086.11 | 14,691.84 | 15,479.16 | 60,117.74 |

| STATE: | PENNSYLVANIA | | | | | |
|---|---|---|---|---|---|---|
| ZIP CODE | | QUARTER 1 | QUARTER 2 | QUARTER 3 | QUARTER 4 | TOTAL GRAMS |
| | 150 | 2,642.50 | 2,462.13 | 2,489.30 | 2,728.05 | 10,321.98 |

| | | | | | |
|---|---|---|---|---|---|
| 151 | 1,852.23 | 1,866.11 | 1,822.75 | 1,967.27 | 7,508.36 |
| 152 | 4,263.49 | 3,936.28 | 3,947.84 | 4,344.42 | 16,492.03 |
| 153 | 939.99 | 920.91 | 982.77 | 970.63 | 3,814.30 |
| 154 | 730.72 | 757.89 | 744.01 | 821.48 | 3,054.10 |
| 155 | 286.16 | 278.64 | 270.55 | 308.13 | 1,143.48 |
| 156 | 1,659.73 | 1,690.94 | 1,673.60 | 1,827.95 | 6,852.22 |
| 157 | 1,289.16 | 1,244.65 | 1,189.15 | 1,313.44 | 5,036.40 |
| 158 | 1,096.08 | 1,079.31 | 1,129.03 | 1,152.73 | 4,457.15 |
| 159 | 757.89 | 789.68 | 760.2 | 781.59 | 3,089.36 |
| 160 | 1,330.21 | 1,349.86 | 1,384.55 | 1,463.17 | 5,527.79 |
| 161 | 1,608.27 | 1,531.97 | 1,515.20 | 1,677.65 | 6,333.09 |
| 162 | 571.16 | 637.07 | 596.6 | 641.11 | 2,445.94 |
| 163 | 1,205.34 | 1,080.47 | 1,013.41 | 1,128.45 | 4,427.67 |
| 164 | 639.38 | 609.32 | 659.61 | 686.78 | 2,595.09 |
| 165 | 2,848.88 | 2,840.78 | 2,806.10 | 2,826.33 | 11,322.09 |
| 166 | 1,324.43 | 1,363.16 | 1,314.02 | 1,438.89 | 5,440.50 |
| 167 | 479.82 | 449.76 | 501.79 | 470 | 1,901.37 |
| 168 | 1,513.47 | 1,334.83 | 1,483.40 | 1,493.81 | 5,825.51 |
| 169 | 417.97 | 381.55 | 352.06 | 417.97 | 1,569.55 |
| 170 | 4,490.68 | 4,461.78 | 4,439.23 | 4,573.93 | 17,965.62 |
| 171 | 1,296.10 | 1,235.40 | 1,274.13 | 1,356.80 | 5,162.43 |
| 172 | 1,150.42 | 1,115.73 | 1,199.56 | 1,301.30 | 4,767.01 |
| 173 | 2,170.19 | 2,100.82 | 1,934.90 | 2,052.26 | 8,258.17 |
| 174 | 1,186.84 | 1,274.13 | 1,127.87 | 1,326.16 | 4,915 |
| 175 | 1,367.21 | 1,365.47 | 1,349.29 | 1,431.95 | 5,513.92 |
| 176 | 1,500.17 | 1,631.98 | 1,486.30 | 1,629.66 | 6,248.11 |
| 177 | 739.39 | 702.39 | 707.59 | 770.61 | 2,919.98 |
| 178 | 636.49 | 664.24 | 671.17 | 855.59 | 2,827.49 |
| 179 | 256.1 | 300.61 | 289.05 | 280.38 | 1,126.14 |
| 180 | 2,293.32 | 2,193.31 | 2,113.53 | 2,151.11 | 8,751.27 |
| 181 | 902.99 | 825.53 | 844.6 | 975.25 | 3,548.37 |
| 182 | 339.92 | 309.86 | 272.29 | 301.19 | 1,223.26 |
| 183 | 517.4 | 511.62 | 513.35 | 551.51 | 2,093.88 |
| 184 | 592.55 | 586.19 | 548.62 | 637.64 | 2,365 |
| 185 | 398.89 | 418.54 | 463.06 | 427.22 | 1,707.71 |
| 186 | 454.96 | 426.06 | 359.58 | 386.75 | 1,627.35 |
| 187 | 10,886.78 | 10,912.79 | 10,891.98 | 11,118.02 | 43,809.57 |

| | | | | | | |
|---|---|---|---|---|---|---|
| | 188 | 200.02 | 231.82 | 176.32 | 180.37 | 788.53 |
| | 189 | 2,348.82 | 2,304.88 | 2,410.10 | 2,374.83 | 9,438.63 |
| | 190 | 8,091.67 | 8,115.95 | 7,782.38 | 8,362.79 | 32,352.79 |
| | 191 | 5,821.47 | 5,726.08 | 5,551.49 | 6,157.34 | 23,256.38 |
| | 193 | 4,832.92 | 4,594.16 | 4,469.29 | 4,748.51 | 18,644.88 |
| | 194 | 3,574.39 | 3,737.42 | 3,560.52 | 3,660.53 | 14,532.86 |
| | 195 | 965.43 | 935.94 | 832.46 | 937.68 | 3,671.51 |
| | 196 | 1,721.58 | 1,749.91 | 1,667.82 | 1,788.06 | 6,927.37 |
| TOTAL | | 86,193.58 | 85,037.92 | 83,572.42 | 88,797.29 | 343,601.21 |

STATE: PUERTO RICO

| ZIP CODE | | QUARTER 1 | QUARTER 2 | QUARTER 3 | QUARTER 4 | TOTAL GRAMS |
|---|---|---|---|---|---|---|
| | 6 | 304.08 | 285.58 | 286.74 | 317.96 | 1,194.36 |
| | 7 | 313.33 | 219.68 | 221.41 | 223.15 | 977.57 |
| | 9 | 1,503.06 | 1,360.27 | 1,222.10 | 1,348.13 | 5,433.56 |
| TOTAL | | 2,120.47 | 1,865.53 | 1,730.25 | 1,889.24 | 7,605.49 |

STATE: RHODE ISLAND

| ZIP CODE | | QUARTER 1 | QUARTER 2 | QUARTER 3 | QUARTER 4 | TOTAL GRAMS |
|---|---|---|---|---|---|---|
| | 28 | 5,021.95 | 5,241.63 | 4,904.02 | 5,330.08 | 20,497.68 |
| | 29 | 2,417.04 | 2,433.80 | 2,267.89 | 2,495.08 | 9,613.81 |
| TOTAL | | 7,438.99 | 7,675.43 | 7,171.91 | 7,825.16 | 30,111.49 |

STATE: SOUTH CAROLINA

| ZIP CODE | | QUARTER 1 | QUARTER 2 | QUARTER 3 | QUARTER 4 | TOTAL GRAMS |
|---|---|---|---|---|---|---|
| | 290 | 5,370.55 | 4,957.79 | 5,063.58 | 5,654.97 | 21,046.89 |
| | 291 | 3,658.79 | 3,538.55 | 3,169.72 | 3,646.08 | 14,013.14 |
| | 292 | 4,750.25 | 4,165.21 | 4,334.59 | 4,929.46 | 18,179.51 |
| | 293 | 4,939.29 | 4,854.31 | 4,455.99 | 5,212.15 | 19,461.74 |
| | 294 | 14,073.84 | 13,381.86 | 13,131.43 | 14,348.76 | 54,935.89 |
| | 295 | 7,828.63 | 7,261.51 | 7,385.81 | 7,682.95 | 30,158.90 |
| | 296 | 14,389.49 | 13,774.97 | 13,626.40 | 15,154.89 | 56,945.75 |
| | 297 | 3,287.65 | 3,104.40 | 3,008.43 | 3,474.96 | 12,875.44 |

| | | | | | |
|---|---|---|---|---|---|
| 298 | 2,540.75 | 2,211.23 | 2,127.41 | 2,411.83 | 9,291.22 |
| 299 | 1,975.37 | 1,857.44 | 1,758 | 1,932.01 | 7,522.82 |
| TOTAL | 62,814.61 | 59,107.27 | 58,061.36 | 64,448.06 | 244,431.30 |

STATE: SOUTH DAKOTA

| ZIP CODE | QUARTER 1 | QUARTER 2 | QUARTER 3 | QUARTER 4 | TOTAL GRAMS |
|---|---|---|---|---|---|
| 570 | 1,643.54 | 1,536.59 | 1,564.34 | 1,666.08 | 6,410.55 |
| 571 | 6,508.83 | 6,343.49 | 5,917.43 | 6,098.96 | 24,868.71 |
| 572 | 467.1 | 379.81 | 389.06 | 411.03 | 1,647 |
| 573 | 950.97 | 851.54 | 861.37 | 905.3 | 3,569.18 |
| 574 | 601.8 | 494.28 | 481.56 | 522.02 | 2,099.66 |
| 575 | 330.67 | 261.3 | 299.46 | 283.85 | 1,175.28 |
| 576 | 7.52 | 12.72 | 23.12 | 14.45 | 57.81 |
| 577 | 944.62 | 1,019.77 | 978.72 | 1,064.28 | 4,007.39 |
| TOTAL | 11,455.05 | 10,899.50 | 10,515.06 | 10,965.97 | 43,835.58 |

STATE: TENNESSEE

| ZIP CODE | QUARTER 1 | QUARTER 2 | QUARTER 3 | QUARTER 4 | TOTAL GRAMS |
|---|---|---|---|---|---|
| 370 | 12,078.82 | 11,279.31 | 11,620.39 | 11,893.83 | 46,872.35 |
| 371 | 7,747.70 | 7,414.71 | 7,182.31 | 7,544.21 | 29,888.93 |
| 372 | 7,357.48 | 7,256.31 | 7,077.68 | 7,317.01 | 29,008.48 |
| 373 | 5,731.28 | 5,057.22 | 5,291.35 | 5,364.19 | 21,444.04 |
| 374 | 3,478.43 | 3,401.54 | 3,198.63 | 3,322.92 | 13,401.52 |
| 376 | 2,428.02 | 2,291.59 | 2,185.22 | 2,419.93 | 9,324.76 |
| 377 | 2,804.94 | 2,664.46 | 2,752.91 | 2,870.27 | 11,092.58 |
| 378 | 4,287.77 | 4,158.27 | 4,302.80 | 4,413.79 | 17,162.63 |
| 379 | 5,912.23 | 5,758.45 | 5,821.47 | 6,108.20 | 23,600.35 |
| 380 | 5,972.93 | 5,508.14 | 5,511.61 | 5,722.03 | 22,714.71 |
| 381 | 7,825.16 | 7,374.82 | 7,397.48 | 7,741.05 | 30,338.51 |
| 382 | 913.4 | 673.49 | 796.04 | 864.26 | 3,247.19 |
| 383 | 2,474.85 | 2,143.02 | 2,264.42 | 2,441.32 | 9,323.61 |
| 384 | 1,734.30 | 1,684.58 | 1,731.99 | 1,807.72 | 6,958.59 |
| 385 | 1,667.82 | 1,426.17 | 1,507.11 | 1,619.84 | 6,220.94 |
| TOTAL | 72,415.13 | 68,092.08 | 68,641.41 | 71,450.57 | 280,599.19 |

RETAIL DRUG DISTRIBUTION BY ZIP CODE WITHIN STATE BY GRAMS WT

STATE: TEXAS

| ZIP CODE | QUARTER 1 | QUARTER 2 | QUARTER 3 | QUARTER 4 | TOTAL GRAMS |
|---|---|---|---|---|---|
| 750 | 34,490.60 | 32,915.86 | 31,851 | 31,782.78 | 131,040.24 |
| 751 | 8,307.88 | 7,787.59 | 7,426.85 | 7,870.83 | 31,393.15 |
| 752 | 15,162.41 | 15,187.27 | 14,685.47 | 15,337.57 | 60,372.72 |
| 753 | 16.19 | 10.98 | 10.41 | 20.81 | 58.39 |
| 754 | 2,737.30 | 2,375.41 | 2,393.91 | 2,703.77 | 10,210.39 |
| 755 | 939.99 | 771.76 | 856.74 | 902.41 | 3,470.90 |
| 756 | 3,690.59 | 3,440.85 | 3,411.95 | 3,669.78 | 14,213.17 |
| 757 | 3,737.42 | 3,592.31 | 3,582.49 | 3,874.43 | 14,786.65 |
| 758 | 750.37 | 686.2 | 694.3 | 801.82 | 2,932.69 |
| 759 | 2,306.04 | 2,189.84 | 1,974.79 | 2,356.91 | 8,827.58 |
| 760 | 14,021.24 | 13,860.53 | 13,611.36 | 14,301.62 | 55,794.75 |
| 761 | 7,353.43 | 7,073.63 | 6,756.83 | 7,175.96 | 28,359.85 |
| 762 | 6,072.94 | 6,026.11 | 5,881.59 | 6,342.34 | 24,322.98 |
| 763 | 1,888.07 | 1,853.39 | 1,870.73 | 1,889.23 | 7,501.42 |
| 764 | 605.85 | 548.62 | 579.83 | 593.71 | 2,328.01 |
| 765 | 3,162.79 | 2,629.78 | 2,674.29 | 2,641.34 | 11,108.20 |
| 766 | 825.53 | 700.66 | 693.72 | 757.89 | 2,977.80 |
| 767 | 2,995.71 | 2,720.54 | 2,650.59 | 2,756.38 | 11,123.22 |
| 768 | 401.2 | 385.01 | 391.37 | 399.47 | 1,577.05 |
| 769 | 1,696.15 | 1,532.54 | 1,451.61 | 1,549.89 | 6,230.19 |
| 770 | 28,521.72 | 28,156.94 | 25,092.20 | 27,585.20 | 109,356.06 |
| 772 | 11.56 | 16.19 | 24.28 | 10.98 | 63.01 |
| 773 | 20,837.61 | 20,003.42 | 19,122.39 | 20,841.08 | 80,804.50 |
| 774 | 17,066.67 | 16,825.60 | 14,930.01 | 16,550.42 | 65,372.70 |
| 775 | 13,783.64 | 12,850.01 | 11,964.94 | 12,523.38 | 51,121.97 |
| 776 | 3,392.29 | 3,343.15 | 3,187.64 | 3,666.89 | 13,589.97 |
| 777 | 1,775.35 | 1,796.73 | 1,615.79 | 1,815.81 | 7,003.68 |
| 778 | 6,240.01 | 5,759.03 | 5,626.65 | 6,394.36 | 24,020.05 |
| 779 | 2,244.18 | 1,985.20 | 1,796.16 | 1,978.84 | 8,004.38 |
| 780 | 5,341.64 | 4,904.60 | 4,729.44 | 5,397.14 | 20,372.82 |
| 781 | 3,970.39 | 3,543.17 | 3,543.17 | 4,041.50 | 15,098.23 |
| 782 | 16,416.31 | 16,111.65 | 15,574.94 | 16,656.85 | 64,759.75 |
| 783 | 2,316.45 | 2,047.05 | 2,032.60 | 2,128.56 | 8,524.66 |
| 784 | 4,151.34 | 3,670.36 | 3,458.19 | 3,864.60 | 15,144.49 |

| | | | | | | |
|---|---|---|---|---|---|---|
| | 785 | 7,868.52 | 6,755.10 | 6,770.71 | 7,408.35 | 28,802.68 |
| | 786 | 9,138.03 | 8,802.73 | 8,695.20 | 9,405.69 | 36,041.65 |
| | 787 | 13,569.74 | 13,800.40 | 13,377.23 | 14,704.38 | 55,451.75 |
| | 788 | 722.05 | 617.99 | 649.21 | 687.94 | 2,677.19 |
| | 789 | 411.61 | 418.54 | 376.92 | 464.21 | 1,671.28 |
| | 790 | 700.66 | 552.66 | 554.98 | 623.19 | 2,431.49 |
| | 791 | 1,178.17 | 1,198.98 | 1,194.93 | 1,368.94 | 4,941.02 |
| | 792 | 78.62 | 57.81 | 84.4 | 81.51 | 302.34 |
| | 793 | 411.61 | 383.86 | 317.96 | 365.36 | 1,478.79 |
| | 794 | 3,316.56 | 3,037.34 | 2,961.61 | 3,484.21 | 12,799.72 |
| | 795 | 414.5 | 380.39 | 353.8 | 459.59 | 1,608.28 |
| | 796 | 1,485.14 | 1,333.10 | 1,267.20 | 1,585.15 | 5,670.59 |
| | 797 | 2,864.49 | 2,805.52 | 2,792.22 | 2,939.06 | 11,401.29 |
| | 798 | 83.82 | 87.87 | 60.7 | 67.06 | 299.45 |
| | 799 | 3,095.73 | 2,782.97 | 2,719.38 | 2,933.86 | 11,531.94 |
| TOTAL | | 282,570.11 | 270,317.24 | 258,324.68 | 277,763.05 | 1,088,975.08 |

STATE: UTAH

| ZIP CODE | | QUARTER 1 | QUARTER 2 | QUARTER 3 | QUARTER 4 | TOTAL GRAMS |
|---|---|---|---|---|---|---|
| | 840 | 12,689.87 | 12,550.55 | 12,461.52 | 13,668.60 | 51,370.54 |
| | 841 | 3,898.71 | 3,920.67 | 3,903.33 | 4,133.99 | 15,856.70 |
| | 843 | 795.47 | 760.78 | 834.78 | 970.05 | 3,361.08 |
| | 844 | 1,083.94 | 1,005.89 | 988.55 | 1,005.89 | 4,084.27 |
| | 845 | 126.03 | 112.73 | 97.12 | 143.37 | 479.25 |
| | 846 | 1,952.82 | 1,955.71 | 2,016.41 | 2,115.27 | 8,040.21 |
| | 847 | 851.54 | 805.29 | 811.65 | 945.19 | 3,413.67 |
| TOTAL | | 21,398.38 | 21,111.62 | 21,113.36 | 22,982.36 | 86,605.72 |

STATE: VERMONT

| ZIP CODE | | QUARTER 1 | QUARTER 2 | QUARTER 3 | QUARTER 4 | TOTAL GRAMS |
|---|---|---|---|---|---|---|
| | 50 | 275.75 | 308.71 | 258.99 | 327.2 | 1,170.65 |

DRUG CODE:1205DRUG NAME:LISDEXAMFETAMINE
STATE:VERMONT

| ZIP CODE | QUARTER 1 | QUARTER 2 | QUARTER 3 | QUARTER 4 | TOTAL GRAMS |
|---|---|---|---|---|---|
| 51 | 298.3 | 276.33 | 253.79 | 261.88 | 1,090.30 |
| 52 | 563.07 | 631.86 | 619.15 | 694.88 | 2,508.96 |
| 53 | 696.61 | 686.78 | 678.69 | 696.03 | 2,758.11 |
| 54 | 2,720.54 | 2,583.53 | 2,584.11 | 2,809.57 | 10,697.75 |
| 56 | 649.21 | 648.63 | 668.28 | 698.34 | 2,664.46 |
| 57 | 890.85 | 884.49 | 914.55 | 926.12 | 3,616.01 |
| 58 | 781.01 | 772.34 | 809.34 | 835.35 | 3,198.04 |
| TOTAL | 6,875.34 | 6,792.67 | 6,786.90 | 7,249.37 | 27,704.28 |

STATE: VIRGIN ISLANDS

| ZIP CODE | QUARTER 1 | QUARTER 2 | QUARTER 3 | QUARTER 4 | TOTAL GRAMS |
|---|---|---|---|---|---|
| 8 | 78.62 | 68.22 | 47.98 | 72.84 | 267.66 |
| TOTAL | 78.62 | 68.22 | 47.98 | 72.84 | 267.66 |

STATE: VIRGINIA

| ZIP CODE | QUARTER 1 | QUARTER 2 | QUARTER 3 | QUARTER 4 | TOTAL GRAMS |
|---|---|---|---|---|---|
| 201 | 6,943.56 | 7,169.02 | 6,920.44 | 7,476.57 | 28,509.59 |
| 220 | 3,616.59 | 3,726.43 | 3,355.29 | 3,389.98 | 14,088.29 |
| 221 | 2,911.31 | 2,813.61 | 2,869.11 | 2,899.17 | 11,493.20 |
| 222 | 1,976.52 | 2,223.37 | 2,030.29 | 2,063.24 | 8,293.42 |
| 223 | 1,744.13 | 1,879.40 | 1,890.97 | 1,892.12 | 7,406.62 |
| 224 | 2,108.91 | 2,033.18 | 2,012.94 | 2,162.67 | 8,317.70 |
| 225 | 1,005.32 | 1,020.35 | 976.99 | 1,063.70 | 4,066.36 |
| 226 | 1,497.86 | 1,429.06 | 1,325.01 | 1,483.40 | 5,735.33 |
| 227 | 496.01 | 506.99 | 519.71 | 541.68 | 2,064.39 |
| 228 | 1,219.79 | 1,002.43 | 1,023.24 | 1,123.25 | 4,368.71 |
| 229 | 2,284.65 | 2,220.48 | 2,147.06 | 2,353.45 | 9,005.64 |
| 230 | 1,668.97 | 1,444.09 | 1,495.54 | 1,596.13 | 6,204.73 |
| 231 | 4,690.70 | 4,757.18 | 4,227.65 | 4,605.72 | 18,281.25 |
| 232 | 5,434.14 | 5,330.66 | 5,102.89 | 5,473.45 | 21,341.14 |
| 233 | 3,262.22 | 3,210.77 | 2,947.15 | 3,027.51 | 12,447.65 |
| 234 | 7,249.37 | 7,145.89 | 6,751.05 | 6,800.77 | 27,947.08 |
| 235 | 1,628.51 | 1,733.72 | 1,696.15 | 1,656.26 | 6,714.64 |
| 236 | 3,102.08 | 2,972.01 | 3,029.24 | 3,043.12 | 12,146.45 |

| | | | | | | |
|---|---|---|---|---|---|---|
| | 237 | 1,241.18 | 1,307.66 | 1,138.28 | 1,301.88 | 4,989 |
| | 238 | 2,201.40 | 2,180.02 | 1,982.88 | 2,068.21 | 8,432.51 |
| | 239 | 645.16 | 501.21 | 566.54 | 537.05 | 2,249.96 |
| | 240 | 3,248.92 | 3,130.99 | 2,872 | 3,412.52 | 12,664.43 |
| | 241 | 1,949.35 | 1,875.93 | 1,728.52 | 1,915.25 | 7,469.05 |
| | 242 | 970.05 | 865.42 | 896.63 | 918.6 | 3,650.70 |
| | 243 | 944.62 | 922.65 | 885.07 | 946.93 | 3,699.27 |
| | 244 | 1,425.02 | 1,355.07 | 1,345.82 | 1,335.99 | 5,461.90 |
| | 245 | 2,892.23 | 2,863.33 | 2,805.52 | 3,050.63 | 11,611.71 |
| | 246 | 679.27 | 542.84 | 535.32 | 563.65 | 2,321.08 |
| TOTAL | | 69,037.84 | 68,163.76 | 65,077.30 | 68,702.90 | 270,981.80 |

STATE: WASHINGTON

| ZIP CODE | | QUARTER 1 | QUARTER 2 | QUARTER 3 | QUARTER 4 | TOTAL GRAMS |
|---|---|---|---|---|---|---|
| | 980 | 9,739.25 | 9,952.57 | 9,669.18 | 9,888.31 | 39,249.31 |
| | 981 | 5,493.11 | 5,927.84 | 5,846.33 | 5,873.50 | 23,140.78 |
| | 982 | 3,336.79 | 3,399.23 | 3,603.88 | 3,526.99 | 13,866.89 |
| | 983 | 3,399.81 | 3,478.43 | 3,385.35 | 3,308.47 | 13,572.06 |
| | 984 | 2,076.54 | 2,073.07 | 1,987.51 | 1,962.07 | 8,099.19 |
| | 985 | 1,777.08 | 1,815.81 | 1,741.82 | 1,892.70 | 7,227.41 |
| | 986 | 2,270.20 | 2,322.23 | 2,201.40 | 2,336.68 | 9,130.51 |
| | 988 | 254.94 | 216.21 | 238.18 | 289.05 | 998.38 |
| | 989 | 488.49 | 569.43 | 539.37 | 647.47 | 2,244.76 |
| | 990 | 423.17 | 420.28 | 376.92 | 419.7 | 1,640.07 |
| | 991 | 265.93 | 234.71 | 301.19 | 337.03 | 1,138.86 |
| | 992 | 2,212.97 | 2,246.50 | 2,073.07 | 2,415.88 | 8,948.42 |
| | 993 | 1,659.15 | 1,681.69 | 1,666.66 | 1,745.28 | 6,752.78 |
| | 994 | 81.51 | 63.01 | 76.89 | 81.51 | 302.92 |
| TOTAL | | 33,478.94 | 34,401.01 | 33,707.75 | 34,724.64 | 136,312.34 |

STATE: WEST VIRGINIA

| ZIP CODE | | QUARTER 1 | QUARTER 2 | QUARTER 3 | QUARTER 4 | TOTAL GRAMS |
|---|---|---|---|---|---|---|
| | 247 | 378.66 | 346.86 | 367.09 | 375.19 | 1,467.80 |
| | 248 | 268.82 | 262.46 | 258.41 | 283.85 | 1,073.54 |
| | 249 | 379.81 | 373.45 | 351.48 | 389.06 | 1,493.80 |

| | | | | | | |
|---|---|---|---|---|---|---|
| | 250 | 821.48 | 697.19 | 634.75 | 693.14 | 2,846.56 |
| | 251 | 577.52 | 506.99 | 540.52 | 535.9 | 2,160.93 |
| | 252 | 406.4 | 380.39 | 415.08 | 389.06 | 1,590.93 |
| | 253 | 2,006.59 | 1,788.64 | 1,779.39 | 1,784.59 | 7,359.21 |
| | 254 | 1,333.68 | 1,375.30 | 1,291.48 | 1,376.46 | 5,376.92 |
| | 255 | 1,784.59 | 1,749.91 | 1,625.62 | 1,885.18 | 7,045.30 |
| | 256 | 235.29 | 239.33 | 218.52 | 232.97 | 926.11 |
| | 257 | 1,638.34 | 1,516.36 | 1,488.03 | 1,553.93 | 6,196.66 |
| | 258 | 1,023.82 | 1,026.71 | 997.22 | 1,058.50 | 4,106.25 |
| | 259 | 360.73 | 299.46 | 294.25 | 311.02 | 1,265.46 |
| | 260 | 1,125.56 | 1,080.47 | 1,055.61 | 1,075.84 | 4,337.48 |
| | 261 | 2,132.61 | 2,018.73 | 1,942.42 | 2,002.54 | 8,096.30 |
| | 262 | 607.01 | 604.11 | 559.02 | 653.25 | 2,423.39 |
| | 263 | 1,371.25 | 1,383.39 | 1,319.80 | 1,360.27 | 5,434.71 |
| | 264 | 376.92 | 341.66 | 330.67 | 359.58 | 1,408.83 |
| | 265 | 2,277.71 | 1,966.12 | 2,012.94 | 2,138.97 | 8,395.74 |
| | 266 | 285 | 224.88 | 231.82 | 250.32 | 992.02 |
| | 267 | 683.31 | 741.7 | 715.69 | 729.56 | 2,870.26 |
| | 268 | 445.72 | 440.51 | 433.58 | 399.47 | 1,719.28 |
| TOTAL | | 20,520.82 | 19,364.62 | 18,863.39 | 19,838.65 | 78,587.48 |

STATE: WISCONSIN

| ZIP CODE | | QUARTER 1 | QUARTER 2 | QUARTER 3 | QUARTER 4 | TOTAL GRAMS |
|---|---|---|---|---|---|---|
| | 530 | 6,955.12 | 7,067.27 | 7,077.10 | 7,714.74 | 28,814.23 |
| | 531 | 7,632.08 | 7,828.05 | 7,852.33 | 8,776.14 | 32,088.60 |
| | 532 | 8,535.65 | 8,441.99 | 8,433.90 | 9,302.79 | 34,714.33 |
| | 534 | 2,296.79 | 2,263.26 | 2,162.67 | 2,494.50 | 9,217.22 |
| | 535 | 4,726.55 | 5,050.28 | 4,825.40 | 5,315.63 | 19,917.86 |
| | 537 | 2,826.91 | 2,836.74 | 2,958.14 | 3,159.32 | 11,781.11 |
| | 538 | 808.76 | 804.14 | 834.78 | 954.44 | 3,402.12 |
| | 539 | 1,858.59 | 1,914.67 | 1,888.65 | 1,996.18 | 7,658.09 |
| | 540 | 721.47 | 670.02 | 649.78 | 767.14 | 2,808.41 |
| | 541 | 3,609.66 | 3,584.22 | 3,644.92 | 3,940.33 | 14,779.13 |
| | 542 | 1,641.23 | 1,683.43 | 1,653.94 | 1,645.85 | 6,624.45 |
| | 543 | 5,074.56 | 5,378.64 | 5,283.83 | 5,551.49 | 21,288.52 |
| | 544 | 5,360.72 | 5,341.07 | 5,144.51 | 5,479.81 | 21,326.11 |

| ZIP CODE | QUARTER 1 | QUARTER 2 | QUARTER 3 | QUARTER 4 | TOTAL GRAMS |
|---|---|---|---|---|---|
| 545 | 1,238.29 | 1,173.54 | 1,303.04 | 1,215.17 | 4,930.04 |
| 546 | 1,743.55 | 1,778.81 | 1,703.08 | 1,919.87 | 7,145.31 |
| 547 | 1,367.21 | 1,321.54 | 1,286.27 | 1,471.84 | 5,446.86 |
| 548 | 1,388.60 | 1,457.97 | 1,404.20 | 1,580.53 | 5,831.30 |
| 549 | 7,261.51 | 7,218.73 | 7,318.17 | 7,979.51 | 29,777.92 |
| TOTAL | 65,047.25 | 65,814.37 | 65,424.71 | 71,265.28 | 267,551.61 |

STATE: WYOMING

| ZIP CODE | QUARTER 1 | QUARTER 2 | QUARTER 3 | QUARTER 4 | TOTAL GRAMS |
|---|---|---|---|---|---|
| 820 | 580.99 | 603.54 | 633.6 | 628.97 | 2,447.10 |
| 822 | 104.06 | 83.82 | 77.47 | 72.84 | 338.19 |
| 823 | 35.26 | 31.8 | 24.86 | 27.75 | 119.67 |
| 824 | 459.01 | 443.98 | 477.51 | 536.48 | 1,916.98 |
| 825 | 111 | 60.12 | 84.4 | 61.86 | 317.38 |
| 826 | 357.84 | 335.88 | 295.41 | 345.7 | 1,334.83 |
| 827 | 291.94 | 276.91 | 279.22 | 256.1 | 1,104.17 |
| 828 | 220.26 | 195.98 | 190.77 | 171.12 | 778.13 |
| 829 | 391.37 | 382.12 | 335.3 | 412.19 | 1,520.98 |
| 830 | 137.59 | 107.53 | 142.21 | 94.81 | 482.14 |
| 831 | 12.14 | 37 | 27.75 | 13.87 | 90.76 |
| TOTAL | 2,701.46 | 2,558.68 | 2,568.50 | 2,621.69 | 10,450.33 |

DRUG CODE: 1724 DRUG NAME: METHYLPHENIDATE (DL;D;L;ISOMERS)

STATE: ALABAMA

| ZIP CODE | QUARTER 1 | QUARTER 2 | QUARTER 3 | QUARTER 4 | TOTAL GRAMS |
|---|---|---|---|---|---|
| 350 | 9,161.26 | 8,435.17 | 8,416.40 | 9,317.56 | 35,330.39 |
| 351 | 5,060.77 | 4,583.27 | 4,594.38 | 5,076.71 | 19,315.13 |
| 352 | 9,560.83 | 8,226.96 | 8,098.49 | 8,932.20 | 34,818.48 |
| 354 | 2,827.34 | 2,494.94 | 2,468.52 | 2,801.73 | 10,592.53 |
| 355 | 2,057.54 | 2,119.26 | 1,967.89 | 2,251.21 | 8,395.90 |
| 356 | 4,403.64 | 4,087.63 | 4,078.18 | 4,319.75 | 16,889.20 |
| 357 | 2,672.42 | 2,482.29 | 2,540.19 | 2,608.52 | 10,303.42 |
| 358 | 3,027.85 | 2,981.22 | 2,935.96 | 2,894.13 | 11,839.16 |
| 359 | 4,164.49 | 3,679.80 | 3,707.55 | 4,106.04 | 15,657.88 |
| 360 | 3,914.23 | 3,436.26 | 3,635.07 | 3,810.50 | 14,796.06 |

| | | | | | | |
|---|---|---|---|---|---|---|
| | 361 | 3,058.49 | 2,971.62 | 2,890.99 | 2,882.54 | 11,803.64 |
| | 362 | 2,082.46 | 2,015.89 | 1,885.83 | 2,084.16 | 8,068.34 |
| | 363 | 6,071.56 | 5,600.51 | 5,391.28 | 5,856.94 | 22,920.29 |
| | 364 | 1,490.17 | 1,262.12 | 1,282.66 | 1,472.50 | 5,507.45 |
| | 365 | 6,891.06 | 6,275.41 | 6,260.52 | 6,850.85 | 26,277.84 |
| | 366 | 5,343 | 5,146.06 | 4,940.50 | 5,170.31 | 20,599.87 |
| | 367 | 671.41 | 540.53 | 588.27 | 665.01 | 2,465.22 |
| | 368 | 3,785.78 | 3,425.56 | 3,413.17 | 3,927.82 | 14,552.33 |
| | 369 | 197.55 | 174.73 | 177.29 | 186.58 | 736.15 |
| TOTAL | | 76,441.85 | 69,939.23 | 69,273.14 | 75,215.06 | 290,869.28 |

| STATE: | ALASKA | | | | | |
|---|---|---|---|---|---|---|
| ZIP CODE | | QUARTER 1 | QUARTER 2 | QUARTER 3 | QUARTER 4 | TOTAL GRAMS |
| | 995 | 4,385.48 | 4,337.54 | 4,266.74 | 4,395.24 | 17,385 |
| | 996 | 1,533.45 | 1,411.27 | 1,400.01 | 1,431.88 | 5,776.61 |
| | 997 | 1,415.27 | 1,203.83 | 1,269.01 | 1,305.07 | 5,193.18 |
| | 998 | 524.28 | 528.32 | 480.22 | 524.61 | 2,057.43 |
| | 999 | 449.77 | 457.7 | 396.97 | 422.31 | 1,726.75 |
| TOTAL | | 8,308.25 | 7,938.66 | 7,812.95 | 8,079.11 | 32,138.97 |

| STATE: | AMERICAN SAMOA | | | | | |
|---|---|---|---|---|---|---|
| ZIP CODE | | QUARTER 1 | QUARTER 2 | QUARTER 3 | QUARTER 4 | TOTAL GRAMS |
| | 967 | 2.59 | 6.05 | 4.32 | 4.76 | 17.72 |
| TOTAL | | 2.59 | 6.05 | 4.32 | 4.76 | 17.72 |

| STATE: | ARIZONA | | | | | |
|---|---|---|---|---|---|---|
| ZIP CODE | | QUARTER 1 | QUARTER 2 | QUARTER 3 | QUARTER 4 | TOTAL GRAMS |
| | 850 | 12,674.64 | 11,752.78 | 11,633.75 | 12,168.87 | 48,230.04 |
| | 851 | 3,733.77 | 3,606.55 | 4,046.76 | 4,052.18 | 15,439.26 |
| | 852 | 25,509.39 | 24,715.80 | 24,777.61 | 28,175.65 | 103,178.45 |
| | 853 | 12,650.88 | 11,348.09 | 11,221.77 | 11,877.47 | 47,098.21 |
| | 855 | 819.52 | 731.32 | 713.93 | 789.63 | 3,054.40 |
| | 856 | 3,098.53 | 2,694.97 | 2,557.20 | 2,746.76 | 11,097.46 |
| | 857 | 12,366.76 | 11,394.05 | 11,433.69 | 12,123.75 | 47,318.25 |

| | | | | | | |
|---|---|---|---|---|---|---|
| | 859 | 828.97 | 761.45 | 798.21 | 739.94 | 3,128.57 |
| | 860 | 1,574.67 | 1,414.74 | 1,486.06 | 1,388.10 | 5,863.57 |
| | 863 | 2,115.81 | 2,044.57 | 1,879.15 | 2,205.66 | 8,245.19 |
| | 864 | 1,158.48 | 976.03 | 955.28 | 1,027.41 | 4,117.20 |
| | 865 | 311.98 | 300.74 | 349.51 | 387.47 | 1,349.70 |
| TOTAL | | 76,843.40 | 71,741.09 | 71,852.92 | 77,682.89 | 298,120.30 |

STATE: ARKANSAS

| ZIP CODE | | QUARTER 1 | QUARTER 2 | QUARTER 3 | QUARTER 4 | TOTAL GRAMS |
|---|---|---|---|---|---|---|
| | 716 | 1,691.17 | 1,425.22 | 1,517.44 | 1,641.14 | 6,274.97 |
| | 717 | 1,075.10 | 968.32 | 912.15 | 1,046.47 | 4,002.04 |
| | 718 | 1,091.60 | 985.17 | 950.16 | 955.58 | 3,982.51 |
| | 719 | 2,267.64 | 2,061.43 | 2,081.54 | 2,151.78 | 8,562.39 |
| | 720 | 6,507.87 | 6,102.11 | 5,955.99 | 6,418.92 | 24,984.89 |
| | 721 | 5,053.99 | 4,572.57 | 4,489.51 | 4,811.28 | 18,927.35 |
| | 722 | 4,476.63 | 4,052.98 | 4,095.46 | 4,238.70 | 16,863.77 |
| | 723 | 1,570.28 | 1,365.16 | 1,312.38 | 1,445.89 | 5,693.71 |
| | 724 | 4,157.76 | 3,379.63 | 3,351.49 | 3,668.83 | 14,557.71 |
| | 725 | 1,251.81 | 1,136.46 | 1,088.54 | 1,226.98 | 4,703.79 |
| | 726 | 1,535.92 | 1,317.23 | 1,375.66 | 1,353.38 | 5,582.19 |
| | 727 | 7,303.05 | 7,181.34 | 6,716.43 | 7,068.60 | 28,269.42 |
| | 728 | 1,540.58 | 1,394.98 | 1,290.67 | 1,436.42 | 5,662.65 |
| | 729 | 3,214.88 | 2,942.88 | 2,848.39 | 3,027.36 | 12,033.51 |
| TOTAL | | 42,738.28 | 38,885.48 | 37,985.81 | 40,491.33 | 160,100.90 |

STATE: CALIFORNIA

| ZIP CODE | | QUARTER 1 | QUARTER 2 | QUARTER 3 | QUARTER 4 | TOTAL GRAMS |
|---|---|---|---|---|---|---|
| | 900 | 10,209.38 | 10,222.70 | 10,051.88 | 10,540.13 | 41,024.09 |
| | 902 | 7,452.04 | 7,389.82 | 7,078.97 | 7,544.09 | 29,464.92 |
| | 903 | 396.6 | 482.04 | 345.21 | 415.31 | 1,639.16 |
| | 904 | 1,383.88 | 1,432.59 | 1,130.18 | 1,536.26 | 5,482.91 |
| | 905 | 1,374.39 | 1,361.06 | 1,127.23 | 1,157.36 | 5,020.04 |
| | 906 | 3,965.02 | 3,172.14 | 3,224.59 | 3,367 | 13,728.75 |
| | 907 | 3,489.71 | 3,526.11 | 3,162.86 | 3,498.31 | 13,676.99 |
| | 908 | 2,232.36 | 2,335.84 | 2,148.82 | 2,197.29 | 8,914.31 |
| | 910 | 1,463.52 | 1,561.19 | 1,397.90 | 1,500.26 | 5,922.87 |

| | | | | | |
|---|---|---|---|---|---|
| 911 | 2,011.74 | 2,014.24 | 2,154.49 | 2,325.17 | 8,505.64 |
| 912 | 956.45 | 1,029.31 | 963.56 | 983.76 | 3,933.08 |
| 913 | 12,460.26 | 12,134.13 | 11,856.17 | 12,167.19 | 48,617.75 |
| 914 | 2,798.67 | 2,819.43 | 2,672.01 | 2,796.15 | 11,086.26 |
| 915 | 840.56 | 834.67 | 774.99 | 849.01 | 3,299.23 |
| 916 | 825.31 | 773.85 | 693.73 | 814.7 | 3,107.59 |
| 917 | 8,532.29 | 8,437.93 | 7,830.01 | 8,466.61 | 33,266.84 |
| 918 | 239.68 | 223.74 | 235.31 | 266.62 | 965.35 |
| 919 | 2,657.02 | 2,779.33 | 2,475.68 | 2,791.50 | 10,703.53 |
| 920 | 28,871.62 | 27,565.48 | 26,836.29 | 29,386.35 | 112,659.74 |
| 921 | 11,815.40 | 11,520.47 | 11,053.14 | 11,823.78 | 46,212.79 |
| 922 | 4,642.07 | 4,257.95 | 3,826.10 | 4,432.21 | 17,158.33 |
| 923 | 7,783.63 | 7,616.34 | 7,174.54 | 7,810.92 | 30,385.43 |
| 924 | 844.85 | 830.64 | 798.58 | 841.04 | 3,315.11 |
| 925 | 9,237.73 | 9,276.17 | 7,867.55 | 9,149.07 | 35,530.52 |
| 926 | 12,364.70 | 11,923.46 | 11,485.25 | 12,024.84 | 47,798.25 |
| 927 | 3,105.90 | 2,866.14 | 2,938.86 | 3,125.76 | 12,036.66 |
| 928 | 7,931.58 | 7,886.57 | 7,648.69 | 8,078.30 | 31,545.14 |
| 930 | 6,125.24 | 6,006.09 | 5,996.03 | 5,788.31 | 23,915.67 |
| 931 | 1,592.40 | 1,648.08 | 1,513.51 | 1,552.16 | 6,306.15 |
| 932 | 3,441.34 | 3,118.61 | 3,078.55 | 3,375.15 | 13,013.65 |
| 933 | 3,083.14 | 2,891.72 | 2,798.86 | 2,895.44 | 11,669.16 |
| 934 | 3,619.09 | 3,388.01 | 3,172.83 | 3,281.67 | 13,461.60 |
| 935 | 3,424.96 | 3,361.87 | 3,387.89 | 3,385.04 | 13,559.76 |
| 936 | 3,864.47 | 3,727.37 | 3,487.68 | 3,931.27 | 15,010.79 |
| 937 | 5,438.25 | 5,452.37 | 4,707.94 | 5,097.53 | 20,696.09 |
| 939 | 2,289.70 | 2,107.07 | 2,014.63 | 2,124.17 | 8,535.57 |
| 940 | 5,424.77 | 5,465.80 | 5,229.20 | 5,577.43 | 21,697.20 |
| 941 | 6,099.54 | 6,327.67 | 5,818.78 | 6,463.91 | 24,709.90 |
| 943 | 1,187.03 | 1,230.51 | 1,038.51 | 1,154.88 | 4,610.93 |
| 944 | 1,191.84 | 1,126.99 | 1,135.94 | 1,068.82 | 4,523.59 |
| 945 | 19,319.78 | 18,557.26 | 17,727.88 | 18,720.63 | 74,325.55 |
| 946 | 3,101.03 | 3,087.88 | 2,873.16 | 2,994.28 | 12,056.35 |
| 947 | 770.92 | 854.04 | 797.82 | 987.96 | 3,410.74 |
| 948 | 693.28 | 586.8 | 640.64 | 676.17 | 2,596.89 |
| 949 | 3,887.74 | 3,943.73 | 3,607.36 | 4,075.49 | 15,514.32 |
| 950 | 7,080.76 | 6,577.84 | 6,374.56 | 6,947.83 | 26,980.99 |

| | | | | | | |
|---|---|---|---|---|---|---|
| | 951 | 4,098.17 | 4,144.28 | 3,744.79 | 4,236.89 | 16,224.13 |
| | 952 | 3,296.68 | 3,015.26 | 2,783.26 | 2,833.30 | 11,928.50 |
| | 953 | 6,040.33 | 5,697.10 | 5,638.32 | 5,988.12 | 23,363.87 |
| | 954 | 3,757.84 | 3,675.01 | 3,653.13 | 3,268.56 | 14,354.54 |
| | 955 | 978.96 | 1,007.67 | 946.17 | 1,126.42 | 4,059.22 |
| | 956 | 13,866.13 | 12,664.13 | 13,070.65 | 12,962.23 | 52,563.14 |
| | 957 | 3,361.14 | 3,016.93 | 3,038.52 | 3,311.47 | 12,728.06 |
| | 958 | 5,941.85 | 5,903.21 | 5,524.66 | 5,700.79 | 23,070.51 |
| | 959 | 3,772.06 | 3,709.17 | 3,610.31 | 3,978.47 | 15,070.01 |
| | 960 | 2,956.02 | 2,743.86 | 2,639.55 | 2,993.49 | 11,332.92 |
| | 961 | 426.79 | 481.12 | 365.02 | 419.29 | 1,692.22 |
| TOTAL | | 280,017.61 | 271,790.79 | 259,368.74 | 276,806.16 | 1,087,983.30 |

STATE: COLORADO

| ZIP CODE | | QUARTER 1 | QUARTER 2 | QUARTER 3 | QUARTER 4 | TOTAL GRAMS |
|---|---|---|---|---|---|---|
| | 800 | 10,250.66 | 9,652.78 | 9,466.71 | 10,162.74 | 39,532.89 |
| | 801 | 11,279.12 | 10,999.97 | 10,710.12 | 11,293.66 | 44,282.87 |
| | 802 | 10,796.10 | 10,462.18 | 10,340.02 | 11,267.79 | 42,866.09 |
| | 803 | 1,973.84 | 1,951.01 | 1,784.69 | 2,008.84 | 7,718.38 |
| | 804 | 1,781.16 | 1,574.34 | 1,695.62 | 1,808.15 | 6,859.27 |
| | 805 | 6,674.71 | 6,544.62 | 6,488.51 | 7,059.72 | 26,767.56 |
| | 806 | 2,376.41 | 2,223.50 | 2,180.97 | 2,497.51 | 9,278.39 |
| | 807 | 408.04 | 400.1 | 445.86 | 490.76 | 1,744.76 |
| | 808 | 1,447.62 | 1,418.35 | 1,587.05 | 1,314.79 | 5,767.81 |
| | 809 | 8,199.44 | 7,907.60 | 7,743.07 | 7,713.09 | 31,563.20 |
| | 810 | 2,275.29 | 2,079.49 | 2,031.18 | 2,275.41 | 8,661.37 |
| | 811 | 333.24 | 280.4 | 272.12 | 283.9 | 1,169.66 |
| | 812 | 660.61 | 650.1 | 569.74 | 649.83 | 2,530.28 |
| | 813 | 514.94 | 527.22 | 545.9 | 520.8 | 2,108.86 |
| | 814 | 751.17 | 718.59 | 612.08 | 793.48 | 2,875.32 |
| | 815 | 2,066.14 | 2,002.86 | 1,961.49 | 2,148.07 | 8,178.56 |
| | 816 | 1,173.24 | 1,017.97 | 1,125.70 | 1,173.29 | 4,490.20 |
| TOTAL | | 62,961.73 | 60,411.08 | 59,560.83 | 63,461.83 | 246,395.47 |

STATE: CONNECTICUT

| ZIP CODE | QUARTER 1 | QUARTER 2 | QUARTER 3 | QUARTER 4 | TOTAL GRAMS |
|---|---|---|---|---|---|
| 60 | 11,671.18 | 11,452.17 | 10,765.50 | 11,861.07 | 45,749.92 |
| 61 | 4,257.82 | 4,304.71 | 3,825.09 | 4,157.78 | 16,545.40 |
| 62 | 2,658.90 | 2,402.13 | 2,290.16 | 2,440.39 | 9,791.58 |
| 63 | 4,658.81 | 4,827.29 | 4,662.90 | 4,721.58 | 18,870.58 |
| 64 | 12,777.82 | 12,202.26 | 11,055.18 | 12,161.56 | 48,196.82 |
| 65 | 3,601.31 | 3,570.67 | 3,099.11 | 3,769.30 | 14,040.39 |
| 66 | 1,579.27 | 1,519.61 | 1,463.34 | 1,566.66 | 6,128.88 |
| 67 | 4,665.87 | 4,559.68 | 4,286.49 | 4,351.32 | 17,863.36 |
| 68 | 8,721.27 | 8,967.98 | 8,019.48 | 8,897.50 | 34,606.23 |
| 69 | 1,062.03 | 1,201.49 | 1,064.99 | 1,194.25 | 4,522.76 |
| TOTAL | 55,654.28 | 55,007.99 | 50,532.24 | 55,121.41 | 216,315.92 |

STATE: DELAWARE

| ZIP CODE | QUARTER 1 | QUARTER 2 | QUARTER 3 | QUARTER 4 | TOTAL GRAMS |
|---|---|---|---|---|---|
| 197 | 7,188.95 | 7,109.30 | 6,546.90 | 7,190.23 | 28,035.38 |
| 198 | 5,660.29 | 5,431.11 | 4,903.65 | 5,401.81 | 21,396.86 |
| 199 | 6,573.53 | 6,368.55 | 6,202.81 | 6,518.63 | 25,663.52 |
| TOTAL | 19,422.77 | 18,908.96 | 17,653.36 | 19,110.67 | 75,095.76 |

STATE: DISTRICT OF COLUMBIA

| ZIP CODE | QUARTER 1 | QUARTER 2 | QUARTER 3 | QUARTER 4 | TOTAL GRAMS |
|---|---|---|---|---|---|
| 200 | 7,364.24 | 7,699.14 | 6,667.48 | 7,396.72 | 29,127.58 |
| 203 | 308.75 | 126.94 | 331.19 | 378.56 | 1,145.44 |
| 204 | 476.45 | 450.08 | 548.31 | 452.15 | 1,926.99 |
| TOTAL | 8,149.44 | 8,276.16 | 7,546.98 | 8,227.43 | 32,200.01 |

STATE: FLORIDA

| ZIP CODE | QUARTER 1 | QUARTER 2 | QUARTER 3 | QUARTER 4 | TOTAL GRAMS |
|---|---|---|---|---|---|
| 320 | 8,790.46 | 8,289.69 | 7,480.66 | 8,455.25 | 33,016.06 |
| 321 | 5,479.50 | 5,095.65 | 4,620.43 | 5,019.06 | 20,214.64 |
| 322 | 14,034.63 | 12,371.23 | 11,660.22 | 12,823.21 | 50,889.29 |
| 323 | 5,454.22 | 4,862.46 | 4,675.48 | 5,155.33 | 20,147.49 |
| 324 | 6,714 | 6,192.87 | 5,833.93 | 6,343.86 | 25,084.66 |

| | | | | | | |
|---|---|---|---|---|---|---|
| | 325 | 14,120.03 | 12,968.02 | 12,990.17 | 12,603.16 | 52,681.38 |
| | 326 | 6,215.67 | 5,602.05 | 5,225.13 | 5,944.78 | 22,987.63 |
| | 327 | 12,829.80 | 12,286.62 | 10,870.66 | 12,346.74 | 48,333.82 |
| | 328 | 8,690.02 | 8,233.54 | 7,354 | 8,139.28 | 32,416.84 |
| | 329 | 10,532.68 | 9,297.86 | 8,789.62 | 10,026.71 | 38,646.87 |
| | 330 | 10,848.15 | 10,203.93 | 7,987.05 | 9,962.47 | 39,001.60 |
| | 331 | 12,026.96 | 10,966.96 | 8,922.27 | 11,140.70 | 43,056.89 |
| | 333 | 5,064.82 | 4,725.16 | 4,026.88 | 4,922.07 | 18,738.93 |
| | 334 | 14,209.52 | 13,411.36 | 11,403.27 | 13,464.87 | 52,489.02 |
| | 335 | 10,195.09 | 9,758.19 | 8,894.22 | 9,701.85 | 38,549.35 |
| | 336 | 10,038.92 | 9,307.28 | 8,364.88 | 9,830.23 | 37,541.31 |
| | 337 | 7,735.35 | 7,107.83 | 6,517.37 | 7,531.50 | 28,892.05 |
| | 338 | 9,032.23 | 8,332.74 | 7,545.04 | 8,531.81 | 33,441.82 |
| | 339 | 9,495.59 | 8,811.85 | 7,761.85 | 9,300.20 | 35,369.49 |
| | 341 | 3,737.43 | 3,300.67 | 2,519.19 | 3,219.06 | 12,776.35 |
| | 342 | 10,019.59 | 9,512.45 | 8,441.46 | 9,385.20 | 37,358.70 |
| | 344 | 6,000.37 | 5,591.14 | 5,111.28 | 5,473.52 | 22,176.31 |
| | 346 | 8,799.20 | 7,928.68 | 7,544.39 | 7,888.98 | 32,161.25 |
| | 347 | 7,397.04 | 6,800.24 | 6,116.50 | 6,899.48 | 27,213.26 |
| | 349 | 5,284.84 | 4,822.06 | 4,240.80 | 5,240.95 | 19,588.65 |
| TOTAL | | 222,746.11 | 205,780.53 | 184,896.75 | 209,350.27 | 822,773.66 |

| STATE: | GEORGIA | | | | | |
|---|---|---|---|---|---|---|
| ZIP CODE | | QUARTER 1 | QUARTER 2 | QUARTER 3 | QUARTER 4 | TOTAL GRAMS |
| | 300 | 32,780.44 | 29,362.49 | 29,800 | 31,634.78 | 123,577.71 |
| | 301 | 20,112.09 | 17,914.74 | 18,238.81 | 19,762.47 | 76,028.11 |
| | 302 | 15,750.58 | 13,406.16 | 13,547.95 | 14,751.67 | 57,456.36 |
| | 303 | 13,045.71 | 12,007.49 | 11,951.14 | 12,900.86 | 49,905.20 |
| | 304 | 4,586.83 | 4,377.83 | 4,197.83 | 4,203.94 | 17,366.43 |
| | 305 | 10,681.46 | 9,397.92 | 9,174.30 | 9,844.60 | 39,098.28 |
| | 306 | 6,314.23 | 5,548.90 | 5,731.62 | 6,106.45 | 23,701.20 |
| | 307 | 6,878.02 | 6,123.02 | 6,042.93 | 6,686.07 | 25,730.04 |
| | 308 | 3,584.09 | 2,765.33 | 3,213.02 | 3,308 | 12,870.44 |
| | 309 | 5,318.44 | 4,714.61 | 4,876.84 | 5,062.80 | 19,972.69 |
| | 310 | 12,287.26 | 10,688.73 | 10,625.51 | 11,570.59 | 45,172.09 |
| | 312 | 4,099.79 | 3,678.90 | 3,913.03 | 4,147.74 | 15,839.46 |

| | | | | | |
|---|---|---|---|---|---|
| 313 | 5,584.61 | 4,743.66 | 4,928.94 | 5,240.90 | 20,498.11 |
| 314 | 4,180.07 | 3,763.71 | 3,697.64 | 4,065.73 | 15,707.15 |
| 315 | 9,268.35 | 7,916.26 | 8,028.97 | 8,518.32 | 33,731.90 |
| 316 | 4,307.34 | 3,510.49 | 3,541.69 | 4,097.07 | 15,456.59 |
| 317 | 9,232.57 | 7,751.06 | 7,935.79 | 8,520.12 | 33,439.54 |
| 318 | 622.89 | 501.36 | 492.07 | 649.63 | 2,265.95 |
| 319 | 5,260.62 | 4,729.13 | 5,036.25 | 4,886.96 | 19,912.96 |
| 398 | 1,613.69 | 1,406.23 | 1,433.09 | 1,522.09 | 5,975.10 |
| TOTAL | 175,509.08 | 154,308.02 | 156,407.42 | 167,480.79 | 653,705.31 |

STATE: GUAM

| ZIP CODE | QUARTER 1 | QUARTER 2 | QUARTER 3 | QUARTER 4 | TOTAL GRAMS |
|---|---|---|---|---|---|
| 969 | 562.07 | 394.32 | 633.74 | 324.62 | 1,914.75 |
| TOTAL | 562.07 | 394.32 | 633.74 | 324.62 | 1,914.75 |

STATE: HAWAII

| ZIP CODE | QUARTER 1 | QUARTER 2 | QUARTER 3 | QUARTER 4 | TOTAL GRAMS |
|---|---|---|---|---|---|
| 967 | 5,932.28 | 5,642.34 | 5,182.28 | 5,809.31 | 22,566.21 |
| 968 | 4,961.27 | 5,290.64 | 5,600.54 | 5,217.64 | 21,070.09 |
| TOTAL | 10,893.55 | 10,932.98 | 10,782.82 | 11,026.95 | 43,636.30 |

STATE: IDAHO

| ZIP CODE | QUARTER 1 | QUARTER 2 | QUARTER 3 | QUARTER 4 | TOTAL GRAMS |
|---|---|---|---|---|---|
| 832 | 2,814.52 | 2,670.72 | 2,514.78 | 2,775.20 | 10,775.22 |
| 833 | 2,721.89 | 2,691.20 | 2,471.35 | 2,697.27 | 10,581.71 |
| 834 | 3,475.36 | 3,452.25 | 3,208.10 | 3,405.98 | 13,541.69 |
| 835 | 821.99 | 794.34 | 731 | 816.83 | 3,164.16 |
| 836 | 9,011.63 | 8,771.92 | 8,270.97 | 8,820.35 | 34,874.87 |
| 837 | 6,470.51 | 6,627.44 | 6,096.62 | 6,876.04 | 26,070.61 |
| 838 | 2,666.14 | 2,764.18 | 2,504.17 | 2,644.36 | 10,578.85 |
| TOTAL | 27,982.04 | 27,772.05 | 25,796.99 | 28,036.03 | 109,587.11 |

STATE: ILLINOIS

| ZIP CODE | QUARTER 1 | QUARTER 2 | QUARTER 3 | QUARTER 4 | TOTAL GRAMS |
|---|---|---|---|---|---|
| 600 | 27,702.37 | 26,558.32 | 25,087.75 | 27,264.35 | 106,612.79 |
| 601 | 19,455.07 | 18,852.84 | 17,840.15 | 19,338.96 | 75,487.02 |
| 602 | 1,348.23 | 1,399.17 | 1,215.32 | 1,347.51 | 5,310.23 |
| 603 | 1,364.29 | 1,282.89 | 1,175.78 | 1,277.31 | 5,100.27 |
| 604 | 14,190.91 | 12,930.42 | 12,629.21 | 13,519.58 | 53,270.12 |
| 605 | 16,594.64 | 15,887.38 | 14,773.61 | 15,842.07 | 63,097.70 |
| 606 | 16,991.14 | 16,843.74 | 14,869.40 | 17,106.37 | 65,810.65 |
| 607 | 1,356.06 | 1,317.43 | 1,110.60 | 1,244.14 | 5,028.23 |
| 608 | 680.19 | 646.44 | 550.53 | 690.07 | 2,567.23 |
| 609 | 2,354.96 | 2,149.79 | 2,179.07 | 2,388.45 | 9,072.27 |
| 610 | 3,950.59 | 3,681.85 | 3,651.37 | 3,860.92 | 15,144.73 |
| 611 | 4,272.64 | 4,061.16 | 3,789.22 | 4,138.66 | 16,261.68 |
| 612 | 3,321.01 | 3,101.73 | 2,981.19 | 3,130.88 | 12,534.81 |
| 613 | 2,586.04 | 2,335.25 | 2,284.12 | 2,379.92 | 9,585.33 |
| 614 | 2,325.54 | 2,217.50 | 1,994.54 | 2,175.63 | 8,713.21 |
| 615 | 4,314.42 | 3,932.42 | 3,997.96 | 4,170.24 | 16,415.04 |
| 616 | 5,367.43 | 5,025.45 | 4,960.73 | 5,219.05 | 20,572.66 |
| 617 | 6,387.51 | 5,860.14 | 5,998.44 | 6,338.34 | 24,584.43 |
| 618 | 6,788.93 | 6,384.30 | 5,888.13 | 6,595.76 | 25,657.12 |
| 619 | 2,387.90 | 2,148.11 | 2,042.04 | 2,478.07 | 9,056.12 |
| 620 | 5,831.98 | 5,091.08 | 5,207.25 | 5,761.98 | 21,892.29 |
| 622 | 7,838.04 | 6,945.33 | 7,077.19 | 7,490.16 | 29,350.72 |
| 623 | 2,283.02 | 2,094.15 | 2,127.43 | 2,176.36 | 8,680.96 |
| 624 | 2,271.81 | 2,038.44 | 2,171.39 | 2,270.75 | 8,752.39 |
| 625 | 3,158.43 | 2,604.47 | 2,609.39 | 2,704.02 | 11,076.31 |
| 626 | 4,218.11 | 3,773.58 | 3,881.66 | 3,885.27 | 15,758.62 |
| 627 | 5,064.49 | 4,539.57 | 4,494.50 | 5,007.90 | 19,106.46 |
| 628 | 3,910.89 | 3,423.38 | 3,550.21 | 3,676.86 | 14,561.34 |
| 629 | 4,745.40 | 4,010.40 | 4,196.46 | 4,292.46 | 17,244.72 |
| TOTAL | 183,062.04 | 171,136.73 | 164,334.64 | 177,772.04 | 696,305.45 |

STATE: INDIANA

| ZIP CODE | QUARTER 1 | QUARTER 2 | QUARTER 3 | QUARTER 4 | TOTAL GRAMS |
|---|---|---|---|---|---|
| 460 | 14,089.61 | 12,897.94 | 12,931.52 | 13,888.78 | 53,807.85 |
| 461 | 11,559.76 | 10,774.40 | 10,448.41 | 11,226.63 | 44,009.20 |

| | | | | | | |
|---|---|---|---|---|---|---|
| | 462 | 17,733.17 | 16,260.15 | 16,303.11 | 16,538.83 | 66,835.26 |
| | 463 | 9,991.41 | 9,536.88 | 9,182.13 | 9,795.27 | 38,505.69 |
| | 464 | 923.86 | 799.57 | 708.55 | 858.05 | 3,290.03 |
| | 465 | 9,134.39 | 8,397.91 | 7,878.03 | 8,279.19 | 33,689.52 |
| | 466 | 2,565.06 | 2,548.31 | 2,395.18 | 2,643.79 | 10,152.34 |
| | 467 | 5,243.64 | 4,726.91 | 4,573.06 | 4,851.75 | 19,395.36 |
| | 468 | 7,486.39 | 6,925.02 | 6,676.43 | 7,043.26 | 28,131.10 |
| | 469 | 5,286.85 | 4,969.14 | 4,735.14 | 5,166.53 | 20,157.66 |
| | 470 | 1,647.10 | 1,460.80 | 1,520.15 | 1,664.70 | 6,292.75 |
| | 471 | 7,881.05 | 8,823.19 | 7,789.15 | 6,740.26 | 31,233.65 |
| | 472 | 3,944.17 | 3,551.86 | 3,608.62 | 3,594.78 | 14,699.43 |
| | 473 | 5,826.24 | 5,110.13 | 5,196.34 | 5,140.20 | 21,272.91 |
| | 474 | 3,771.76 | 3,439.47 | 3,643.10 | 3,659.79 | 14,514.12 |
| | 475 | 3,416.83 | 2,730.67 | 2,823 | 2,892.48 | 11,862.98 |
| | 476 | 2,705.06 | 2,432.15 | 2,482.61 | 2,858.41 | 10,478.23 |
| | 477 | 5,853.05 | 5,153.65 | 5,296.97 | 5,504.49 | 21,808.16 |
| | 478 | 3,384.90 | 3,056.57 | 2,993.15 | 3,185.86 | 12,620.48 |
| | 479 | 6,060.76 | 5,425.83 | 5,276.90 | 5,778.93 | 22,542.42 |
| TOTAL | | 128,505.06 | 119,020.55 | 116,461.55 | 121,311.98 | 485,299.14 |

STATE: IOWA

| ZIP CODE | QUARTER 1 | QUARTER 2 | QUARTER 3 | QUARTER 4 | TOTAL GRAMS |
|---|---|---|---|---|---|
| 500 | 6,914.61 | 6,373.60 | 6,131.84 | 6,799.48 | 26,219.53 |
| 501 | 3,862.64 | 3,801.39 | 3,704.67 | 3,841.20 | 15,209.90 |
| 502 | 4,840.53 | 4,778.18 | 4,508.95 | 4,699.13 | 18,826.79 |
| 503 | 8,294.83 | 7,942.46 | 7,316.76 | 7,634.38 | 31,188.43 |
| 504 | 2,111.32 | 2,011.19 | 1,914.38 | 2,017.85 | 8,054.74 |
| 505 | 2,333.37 | 2,248.23 | 2,146 | 2,276.56 | 9,004.16 |
| 506 | 3,301.48 | 3,194.30 | 3,018.75 | 3,189.48 | 12,704.01 |
| 507 | 2,056.92 | 2,042.68 | 1,823.75 | 1,895.52 | 7,818.87 |
| 508 | 638.53 | 524.71 | 551.3 | 628.53 | 2,343.07 |
| 510 | 2,159.49 | 2,089.79 | 1,981.82 | 2,216.22 | 8,447.32 |
| 511 | 4,031.32 | 4,005.95 | 3,523.10 | 3,852.04 | 15,412.41 |
| 512 | 1,174.37 | 938.68 | 963.89 | 1,077.43 | 4,154.37 |
| 513 | 1,070.86 | 1,066.58 | 1,012.98 | 1,028.74 | 4,179.16 |
| 514 | 1,006.79 | 945.73 | 964.13 | 993.61 | 3,910.26 |

| | | | | | | |
|---|---|---|---|---|---|---|
| | 515 | 3,591.30 | 3,420.85 | 3,225.68 | 3,435.69 | 13,673.52 |
| | 516 | 528.61 | 494.58 | 466.33 | 510.43 | 1,999.95 |
| | 520 | 4,129.64 | 3,808.24 | 3,768.31 | 3,951.66 | 15,657.85 |
| | 521 | 1,164.14 | 1,037.37 | 1,138.13 | 1,107.32 | 4,446.96 |
| | 522 | 7,465.49 | 7,208.33 | 6,640.64 | 7,026.89 | 28,341.35 |
| | 523 | 4,053.20 | 3,929.50 | 3,705.80 | 4,187.41 | 15,875.91 |
| | 524 | 6,665 | 6,518.80 | 6,216.02 | 6,642.64 | 26,042.46 |
| | 525 | 2,431.97 | 2,222.97 | 2,079.12 | 2,282.52 | 9,016.58 |
| | 526 | 3,181.20 | 2,943.75 | 2,914.04 | 2,957.72 | 11,996.71 |
| | 527 | 4,767.76 | 4,441.09 | 4,229.68 | 4,572.23 | 18,010.76 |
| | 528 | 3,526.06 | 3,325.13 | 3,169.79 | 3,520.34 | 13,541.32 |
| TOTAL | | 85,301.43 | 81,314.08 | 77,115.86 | 82,345.02 | 326,076.39 |

| STATE: | KANSAS | | | | | |
|---|---|---|---|---|---|---|
| ZIP CODE | | QUARTER 1 | QUARTER 2 | QUARTER 3 | QUARTER 4 | TOTAL GRAMS |
| | 660 | 10,341.32 | 9,715.61 | 9,537.33 | 10,190.73 | 39,784.99 |
| | 661 | 1,344.29 | 1,240.22 | 1,077.36 | 1,254.88 | 4,916.75 |
| | 662 | 9,060.52 | 8,677.60 | 8,157.39 | 8,845.41 | 34,740.92 |
| | 664 | 1,703.39 | 1,510.85 | 1,791.67 | 1,498.25 | 6,504.16 |
| | 665 | 1,851.79 | 1,755.45 | 1,568.09 | 1,820.34 | 6,995.67 |
| | 666 | 5,750.58 | 5,641.77 | 5,335.29 | 5,404.63 | 22,132.27 |
| | 667 | 1,926.90 | 1,819.40 | 1,747.49 | 1,846.60 | 7,340.39 |
| | 668 | 807.55 | 750.56 | 694.56 | 720.71 | 2,973.38 |
| | 669 | 276.85 | 267.34 | 280.6 | 274.42 | 1,099.21 |
| | 670 | 2,868.64 | 2,968.55 | 2,843.22 | 2,780.68 | 11,461.09 |
| | 671 | 1,540.86 | 1,433.88 | 1,513.80 | 1,584.87 | 6,073.41 |
| | 672 | 6,939.78 | 6,406.41 | 6,153.12 | 6,525.37 | 26,024.68 |
| | 673 | 1,548.06 | 1,512.42 | 1,476.63 | 1,551.44 | 6,088.55 |
| | 674 | 2,114.07 | 1,866.14 | 1,819.18 | 1,976.15 | 7,775.54 |
| | 675 | 1,858.26 | 1,742.18 | 1,582.57 | 1,741.96 | 6,924.97 |
| | 676 | 1,069.50 | 968.36 | 943.33 | 1,053.38 | 4,034.57 |
| | 677 | 323.29 | 347.86 | 322.39 | 347.29 | 1,340.83 |
| | 678 | 884.5 | 883.68 | 833.99 | 902.74 | 3,504.91 |
| | 679 | 119.33 | 114.87 | 136.4 | 166.39 | 536.99 |
| TOTAL | | 52,329.48 | 49,623.15 | 47,814.41 | 50,486.24 | 200,253.28 |

STATE: KENTUCKY

| ZIP CODE | QUARTER 1 | QUARTER 2 | QUARTER 3 | QUARTER 4 | TOTAL GRAMS |
|---|---|---|---|---|---|
| 400 | 4,553.38 | 4,193.77 | 4,123.80 | 4,136.79 | 17,007.74 |
| 401 | 1,943.50 | 1,893.48 | 2,041.85 | 1,818.31 | 7,697.14 |
| 402 | 15,014.92 | 13,169.87 | 13,160.57 | 13,849.07 | 55,194.43 |
| 403 | 6,302.88 | 5,910.80 | 5,904.27 | 6,086.81 | 24,204.76 |
| 404 | 2,380.81 | 2,190.26 | 2,014.71 | 2,228.09 | 8,813.87 |
| 405 | 5,751.75 | 4,956.51 | 5,281.95 | 5,651.27 | 21,641.48 |
| 406 | 1,084.53 | 1,009.76 | 944.98 | 1,020.29 | 4,059.56 |
| 407 | 1,808.40 | 1,607.42 | 1,534.94 | 1,691.14 | 6,641.90 |
| 408 | 406.69 | 365.15 | 345.82 | 329.69 | 1,447.35 |
| 409 | 633.22 | 566.94 | 576.07 | 604.58 | 2,380.81 |
| 410 | 10,725.63 | 9,823.63 | 9,412.40 | 10,556.60 | 40,518.26 |
| 411 | 2,262.41 | 2,032.11 | 1,965.03 | 2,177.50 | 8,437.05 |
| 412 | 808.02 | 703.77 | 682.31 | 697.69 | 2,891.79 |
| 413 | 398.75 | 352.3 | 387.77 | 393.06 | 1,531.88 |
| 414 | 320.03 | 305.89 | 282.52 | 309.65 | 1,218.09 |
| 415 | 1,166.56 | 1,085.71 | 1,159.66 | 1,225.98 | 4,637.91 |
| 416 | 651.77 | 538.48 | 602.36 | 619.14 | 2,411.75 |
| 417 | 1,025.37 | 891.63 | 848.6 | 899.04 | 3,664.64 |
| 418 | 235.35 | 273.68 | 283.77 | 288.51 | 1,081.31 |
| 420 | 4,888.27 | 4,255.24 | 4,424.36 | 4,636.70 | 18,204.57 |
| 421 | 3,214.28 | 2,818.25 | 3,017.34 | 3,167.17 | 12,217.04 |
| 422 | 3,086.43 | 2,329.28 | 3,225.93 | 2,500.47 | 11,142.11 |
| 423 | 3,029.72 | 2,762.02 | 2,590.65 | 2,796.22 | 11,178.61 |
| 424 | 3,736.19 | 3,448.18 | 3,375.02 | 3,800.68 | 14,360.07 |
| 425 | 1,005.02 | 887.84 | 836.67 | 1,145.43 | 3,874.96 |
| 426 | 637.76 | 558.94 | 625.29 | 729.3 | 2,551.29 |
| 427 | 2,885.74 | 2,633.04 | 2,776.41 | 2,862.31 | 11,157.50 |
| TOTAL | 79,957.38 | 71,563.95 | 72,425.05 | 76,221.49 | 300,167.87 |

STATE: LOUISIANA

| ZIP CODE | QUARTER 1 | QUARTER 2 | QUARTER 3 | QUARTER 4 | TOTAL GRAMS |
|---|---|---|---|---|---|
| 700 | 10,721.37 | 9,198.57 | 8,870.38 | 9,867.30 | 38,657.62 |
| 701 | 4,809.75 | 4,300.68 | 4,108.35 | 4,791.81 | 18,010.59 |

| | | | | | | |
|---|---|---|---|---|---|---|
| | 703 | 5,247.83 | 4,485.31 | 4,358.12 | 4,623.12 | 18,714.38 |
| | 704 | 10,669.09 | 9,413.51 | 9,145.08 | 10,037.74 | 39,265.42 |
| | 705 | 14,506.78 | 11,539.41 | 12,188.45 | 13,519.16 | 51,753.80 |
| | 706 | 9,139.90 | 7,582.95 | 7,474.18 | 8,155.89 | 32,352.92 |
| | 707 | 10,712.30 | 9,164.39 | 9,438.20 | 10,438.40 | 39,753.29 |
| | 708 | 7,989.68 | 6,667.66 | 6,749.88 | 7,466.19 | 28,873.41 |
| | 710 | 1,976.36 | 1,666.23 | 1,763.02 | 2,064.32 | 7,469.93 |
| | 711 | 5,357.42 | 4,733 | 4,466.94 | 4,891.96 | 19,449.32 |
| | 712 | 5,323.33 | 4,519.17 | 4,606.46 | 5,077.61 | 19,526.57 |
| | 713 | 3,941.09 | 3,310.57 | 3,370.04 | 3,830.31 | 14,452.01 |
| | 714 | 2,817.04 | 2,290.34 | 2,695.61 | 2,638.17 | 10,441.16 |
| TOTAL | | 93,211.94 | 78,871.79 | 79,234.71 | 87,401.98 | 338,720.42 |
| STATE: | | MAINE | | | | |
| ZIP CODE | | QUARTER 1 | QUARTER 2 | QUARTER 3 | QUARTER 4 | TOTAL GRAMS |
| | 39 | 743.55 | 742.18 | 790.65 | 699.5 | 2,975.88 |
| | 40 | 7,783.99 | 7,807.19 | 7,733.55 | 7,627.75 | 30,952.48 |
| | 41 | 2,887.36 | 2,961.21 | 2,914.33 | 2,977.35 | 11,740.25 |
| | 42 | 3,236.98 | 3,120.45 | 2,907.68 | 3,207.46 | 12,472.57 |
| | 43 | 2,652.56 | 2,849.68 | 2,787.66 | 2,715.59 | 11,005.49 |
| | 44 | 6,744.14 | 6,455.98 | 6,164.10 | 6,465.26 | 25,829.48 |
| | 45 | 1,328.13 | 1,314.51 | 1,273.07 | 1,307.49 | 5,223.20 |
| | 46 | 2,020.29 | 1,956.39 | 1,982.23 | 2,048.10 | 8,007.01 |
| | 47 | 2,186.48 | 2,095.50 | 1,810.11 | 2,030.92 | 8,123.01 |
| | 48 | 1,147.72 | 1,252.25 | 1,242.19 | 1,150.67 | 4,792.83 |
| | 49 | 4,977.73 | 5,169.78 | 5,005.06 | 4,950.03 | 20,102.60 |
| TOTAL | | 35,708.93 | 35,725.12 | 34,610.63 | 35,180.12 | 141,224.80 |
| STATE: | | MARYLAND | | | | |
| ZIP CODE | | QUARTER 1 | QUARTER 2 | QUARTER 3 | QUARTER 4 | TOTAL GRAMS |
| | 206 | 4,394.87 | 3,845.12 | 3,706.63 | 3,890.24 | 15,836.86 |
| | 207 | 7,077.91 | 7,090.35 | 6,293.47 | 6,451.98 | 26,913.71 |
| | 208 | 15,914.19 | 15,509.18 | 14,473 | 15,800.60 | 61,696.97 |
| | 209 | 2,779.33 | 2,725.99 | 2,668.17 | 2,653.85 | 10,827.34 |
| | 210 | 17,456.93 | 17,128.60 | 15,573.82 | 17,318.05 | 67,477.40 |

| | | | | | | |
|---|---|---|---|---|---|---|
| | 211 | 8,814.42 | 8,398.21 | 7,681.10 | 8,627.15 | 33,520.88 |
| | 212 | 20,715.10 | 20,047.12 | 18,214.30 | 20,263.68 | 79,240.20 |
| | 214 | 2,053.58 | 1,930.34 | 1,728.49 | 1,898.44 | 7,610.85 |
| | 215 | 3,614.88 | 3,748.20 | 3,444.20 | 3,582.97 | 14,390.25 |
| | 216 | 4,128.04 | 4,025.62 | 3,664.30 | 3,840.95 | 15,658.91 |
| | 217 | 12,157.43 | 11,520 | 10,864.46 | 11,622.10 | 46,163.99 |
| | 218 | 3,700.59 | 3,287.41 | 3,052.48 | 3,358.68 | 13,399.16 |
| | 219 | 1,889.01 | 1,795.86 | 1,750 | 2,026.94 | 7,461.81 |
| TOTAL | | 104,696.28 | 101,052 | 93,114.42 | 101,335.63 | 400,198.33 |

STATE: MASSACHUSETTS

| ZIP CODE | QUARTER 1 | QUARTER 2 | QUARTER 3 | QUARTER 4 | TOTAL GRAMS |
|---|---|---|---|---|---|
| 10 | 10,696.65 | 10,812.48 | 10,048.63 | 10,574.22 | 42,131.98 |
| 11 | 4,630.97 | 4,540.66 | 3,784.24 | 4,490.52 | 17,446.39 |
| 12 | 4,428.59 | 4,286.89 | 3,992.21 | 4,396.11 | 17,103.80 |
| 13 | 2,166.20 | 2,081.19 | 2,113.34 | 2,089.39 | 8,450.12 |
| 14 | 6,176.46 | 6,193.40 | 5,767.51 | 5,890.35 | 24,027.72 |
| 15 | 8,351.34 | 7,967.56 | 7,777.96 | 8,192.24 | 32,289.10 |
| 16 | 4,678.74 | 4,442.30 | 4,041.47 | 4,469.38 | 17,631.89 |
| 17 | 10,737.19 | 11,049.44 | 9,752.55 | 10,783.77 | 42,322.95 |
| 18 | 16,054.06 | 16,104.72 | 14,322.75 | 15,340.88 | 61,822.41 |
| 19 | 11,945.15 | 11,738.55 | 10,622.94 | 11,755.43 | 46,062.07 |
| 20 | 10,240.17 | 10,113.76 | 9,083.04 | 10,132.71 | 39,569.68 |
| 21 | 20,996.92 | 21,486.97 | 19,808.94 | 21,908.75 | 84,201.58 |
| 22 | 1,216.59 | 1,158.78 | 782.09 | 1,017.86 | 4,175.32 |
| 23 | 9,473.26 | 8,970.71 | 8,450.54 | 9,138.49 | 36,033 |
| 24 | 11,568.60 | 11,820.92 | 10,667.24 | 11,942.44 | 45,999.20 |
| 25 | 3,043.92 | 3,061.48 | 2,968.55 | 2,975.96 | 12,049.91 |
| 26 | 3,898.22 | 3,959.74 | 4,158.11 | 4,062.84 | 16,078.91 |
| 27 | 9,420.18 | 9,691.30 | 8,689.68 | 9,453.08 | 37,254.24 |
| TOTAL | 149,723.21 | 149,480.85 | 136,831.79 | 148,614.42 | 584,650.27 |

STATE: MICHIGAN

| ZIP CODE | QUARTER 1 | QUARTER 2 | QUARTER 3 | QUARTER 4 | TOTAL GRAMS |
|---|---|---|---|---|---|
| 480 | 25,111.36 | 24,199.28 | 22,158.43 | 24,259.70 | 95,728.77 |

| | | | | | | |
|---|---|---|---|---|---|---|
| | 481 | 28,053.25 | 26,412.76 | 24,636.99 | 27,948.24 | 107,051.24 |
| | 482 | 5,624.20 | 5,426.41 | 4,579.84 | 5,387.08 | 21,017.53 |
| | 483 | 19,612.91 | 19,325.40 | 18,162.22 | 19,785.52 | 76,886.05 |
| | 484 | 6,722.05 | 6,610.79 | 6,020.22 | 6,906.12 | 26,259.18 |
| | 485 | 2,665.24 | 2,577.78 | 2,269.66 | 2,386 | 9,898.68 |
| | 486 | 7,634.85 | 7,454.12 | 7,009.19 | 7,315.87 | 29,414.03 |
| | 487 | 4,524.13 | 4,318.12 | 3,868.12 | 4,349.18 | 17,059.55 |
| | 488 | 12,981.63 | 12,354.69 | 11,760.95 | 12,970.57 | 50,067.84 |
| | 489 | 3,104.34 | 3,031.06 | 2,879.86 | 3,029.44 | 12,044.70 |
| | 490 | 14,042.78 | 13,395.76 | 12,976.79 | 13,899.57 | 54,314.90 |
| | 491 | 1,230.47 | 1,150.32 | 1,056.07 | 1,025.01 | 4,461.87 |
| | 492 | 6,004.54 | 5,507.74 | 5,506.89 | 6,022.77 | 23,041.94 |
| | 493 | 6,366.75 | 5,977.68 | 5,945.37 | 6,091.85 | 24,381.65 |
| | 494 | 18,020.11 | 17,049.84 | 15,998.55 | 17,553.27 | 68,621.77 |
| | 495 | 13,843.45 | 13,191.30 | 12,242.55 | 13,203.97 | 52,481.27 |
| | 496 | 7,405.14 | 7,113.64 | 7,021.67 | 7,231.88 | 28,772.33 |
| | 497 | 5,594.58 | 5,456.06 | 5,383.87 | 5,305.16 | 21,739.67 |
| | 498 | 6,576.51 | 6,385.74 | 6,353.84 | 6,559.10 | 25,875.19 |
| | 499 | 1,511.69 | 1,390.17 | 1,401.84 | 1,453.83 | 5,757.53 |
| TOTAL | | 196,629.98 | 188,328.66 | 177,232.92 | 192,684.13 | 754,875.69 |

STATE: MINNESOTA

| ZIP CODE | | QUARTER 1 | QUARTER 2 | QUARTER 3 | QUARTER 4 | TOTAL GRAMS |
|---|---|---|---|---|---|---|
| | 550 | 11,192.38 | 10,961.66 | 10,422.48 | 11,432.66 | 44,009.18 |
| | 551 | 15,086.57 | 14,592.36 | 13,771.80 | 15,475.54 | 58,926.27 |
| | 553 | 21,128.21 | 20,204.76 | 18,859.36 | 21,092.47 | 81,284.80 |
| | 554 | 20,216 | 20,102.71 | 18,539.43 | 20,787.54 | 79,645.68 |
| | 556 | 216.85 | 167.5 | 184.53 | 175.83 | 744.71 |
| | 557 | 2,306.59 | 2,145.72 | 2,125.12 | 2,434.79 | 9,012.22 |
| | 558 | 2,606.19 | 2,434.30 | 2,331.63 | 2,606.64 | 9,978.76 |
| | 559 | 8,028.16 | 8,053.74 | 7,633.88 | 8,390.54 | 32,106.32 |
| | 560 | 4,778.17 | 4,659.90 | 4,458.46 | 4,618.03 | 18,514.56 |
| | 561 | 1,448.76 | 1,385.71 | 1,337.86 | 1,447.65 | 5,619.98 |
| | 562 | 2,305.19 | 2,233.60 | 2,089.41 | 2,276.69 | 8,904.89 |
| | 563 | 6,182.01 | 5,963.94 | 5,479.34 | 6,064.84 | 23,690.13 |
| | 564 | 2,152.94 | 2,078.75 | 2,004.56 | 2,200.06 | 8,436.31 |

| | | | | | | |
|---|---|---|---|---|---|---|
| | 565 | 3,721.45 | 3,555.36 | 3,456.10 | 3,878.39 | 14,611.30 |
| | 566 | 1,022.61 | 1,128.05 | 943.24 | 1,000.60 | 4,094.50 |
| | 567 | 989.41 | 895.8 | 864.35 | 1,011.61 | 3,761.17 |
| TOTAL | | 103,381.49 | 100,563.86 | 94,501.55 | 104,893.88 | 403,340.78 |

STATE: MISSISSIPPI

| ZIP CODE | | QUARTER 1 | QUARTER 2 | QUARTER 3 | QUARTER 4 | TOTAL GRAMS |
|---|---|---|---|---|---|---|
| | 386 | 6,538.18 | 5,558.31 | 5,920.57 | 6,272.18 | 24,289.24 |
| | 387 | 631.97 | 506.44 | 605.83 | 571.65 | 2,315.89 |
| | 388 | 3,225.61 | 2,849.45 | 2,823.37 | 3,135.53 | 12,033.96 |
| | 389 | 1,155.63 | 1,042.58 | 1,001.66 | 1,132.75 | 4,332.62 |
| | 390 | 3,954.67 | 3,545.09 | 3,537.98 | 3,705.87 | 14,743.61 |
| | 391 | 4,101.81 | 3,486.17 | 3,535.60 | 3,555.50 | 14,679.08 |
| | 392 | 3,519.37 | 3,125.52 | 3,209.05 | 3,464.69 | 13,318.63 |
| | 393 | 2,822.32 | 2,565.60 | 2,501.34 | 2,749.05 | 10,638.31 |
| | 394 | 7,190.23 | 6,125.16 | 6,290.16 | 6,730.88 | 26,336.43 |
| | 395 | 9,636.51 | 8,213.61 | 8,477.45 | 9,077.15 | 35,404.72 |
| | 396 | 2,035.58 | 1,720.01 | 1,720.74 | 1,819.34 | 7,295.67 |
| | 397 | 1,652.68 | 1,355.88 | 1,492.44 | 1,430.56 | 5,931.56 |
| TOTAL | | 46,464.56 | 40,093.82 | 41,116.19 | 43,645.15 | 171,319.72 |

STATE: MISSOURI

| ZIP CODE | | QUARTER 1 | QUARTER 2 | QUARTER 3 | QUARTER 4 | TOTAL GRAMS |
|---|---|---|---|---|---|---|
| | 630 | 17,140.75 | 15,786.42 | 15,589.33 | 17,353.11 | 65,869.61 |
| | 631 | 45,187.48 | 42,161.05 | 45,910.07 | 48,018.76 | 181,277.36 |
| | 633 | 11,682.21 | 10,474.82 | 10,493.13 | 11,617.86 | 44,268.02 |
| | 634 | 1,395.50 | 1,383.65 | 1,281.37 | 1,343.49 | 5,404.01 |
| | 635 | 749.88 | 692.07 | 659.52 | 716.37 | 2,817.84 |
| | 636 | 2,252.05 | 2,209.74 | 2,064.06 | 2,240.11 | 8,765.96 |
| | 637 | 2,153.21 | 1,936.61 | 1,940.84 | 2,112.32 | 8,142.98 |
| | 638 | 2,409.83 | 2,054.49 | 2,085.47 | 2,430.76 | 8,980.55 |
| | 639 | 965.82 | 850.76 | 784.12 | 773.69 | 3,374.39 |
| | 640 | 10,054.42 | 9,618.55 | 9,284.59 | 10,068.10 | 39,025.66 |
| | 641 | 12,135.52 | 11,756.34 | 11,358.29 | 12,089.84 | 47,339.99 |
| | 644 | 1,077.13 | 968.88 | 919.15 | 1,048.22 | 4,013.38 |

| | | | | | | |
|---|---|---|---|---|---|---|
| | 645 | 2,036.98 | 1,902.20 | 1,924.44 | 1,933.26 | 7,796.88 |
| | 646 | 596.73 | 543.95 | 529.93 | 572.87 | 2,243.48 |
| | 647 | 1,006.12 | 927.83 | 981.84 | 1,053.03 | 3,968.82 |
| | 648 | 3,153.84 | 2,953.08 | 2,897.11 | 3,091.51 | 12,095.54 |
| | 650 | 1,410.91 | 1,288.43 | 1,304.83 | 1,425.80 | 5,429.97 |
| | 651 | 1,763.56 | 1,590.51 | 1,566.11 | 1,618.79 | 6,538.97 |
| | 652 | 4,013.86 | 3,605.60 | 3,587.63 | 3,894.97 | 15,102.06 |
| | 653 | 1,280.99 | 1,208.53 | 1,219.52 | 1,123.51 | 4,832.55 |
| | 654 | 1,576.43 | 1,428.33 | 1,574.27 | 1,466.21 | 6,045.24 |
| | 655 | 1,549.08 | 1,463.20 | 1,391.70 | 1,384.88 | 5,788.86 |
| | 656 | 1,662.12 | 1,580.08 | 1,656.45 | 1,680.38 | 6,579.03 |
| | 657 | 3,609.27 | 3,445.95 | 3,419.88 | 3,447.27 | 13,922.37 |
| | 658 | 4,320.40 | 4,312.97 | 4,066.67 | 4,051.40 | 16,751.44 |
| TOTAL | | 135,184.09 | 126,144.04 | 128,490.32 | 136,556.51 | 526,374.96 |

| STATE: | MONTANA | | | | | |
|---|---|---|---|---|---|---|
| ZIP CODE | | QUARTER 1 | QUARTER 2 | QUARTER 3 | QUARTER 4 | TOTAL GRAMS |
| | 590 | 1,056.93 | 807.39 | 946.4 | 946.61 | 3,757.33 |
| | 591 | 4,457.82 | 4,194.62 | 3,892.82 | 4,128.81 | 16,674.07 |
| | 592 | 658.65 | 545.99 | 585.97 | 626.06 | 2,416.67 |
| | 593 | 806.22 | 731.88 | 706.4 | 741.61 | 2,986.11 |
| | 594 | 2,599.61 | 2,453.46 | 2,324.19 | 2,439.07 | 9,816.33 |
| | 595 | 373.59 | 351.83 | 347.9 | 335.51 | 1,408.83 |
| | 596 | 1,330.35 | 1,219.30 | 1,196.20 | 1,286.68 | 5,032.53 |
| | 597 | 2,551.15 | 2,555 | 2,508.39 | 2,580.21 | 10,194.75 |
| | 598 | 3,185.29 | 3,071.54 | 3,092.23 | 3,002.27 | 12,351.33 |
| | 599 | 1,328.25 | 1,244.61 | 1,358.27 | 1,406.78 | 5,337.91 |
| TOTAL | | 18,347.86 | 17,175.62 | 16,958.77 | 17,493.61 | 69,975.86 |

| STATE: | NEBRASKA | | | | | |
|---|---|---|---|---|---|---|
| ZIP CODE | | QUARTER 1 | QUARTER 2 | QUARTER 3 | QUARTER 4 | TOTAL GRAMS |
| | 680 | 3,418.53 | 3,273.88 | 3,142.56 | 3,242.18 | 13,077.15 |
| | 681 | 12,992.39 | 12,181.43 | 11,631.17 | 12,841.65 | 49,646.64 |
| | 683 | 936.58 | 934.47 | 885.16 | 888.03 | 3,644.24 |
| | 684 | 760.14 | 754.32 | 735.4 | 668.47 | 2,918.33 |

| | | | | | | |
|---|---|---|---|---|---|---|
| | 685 | 6,079.33 | 5,641.24 | 5,623.48 | 5,836.69 | 23,180.74 |
| | 686 | 784.52 | 686.61 | 677.12 | 805.67 | 2,953.92 |
| | 687 | 1,638.50 | 1,411.70 | 1,464.06 | 1,560.89 | 6,075.15 |
| | 688 | 2,541.93 | 2,364.49 | 2,299.66 | 2,452.08 | 9,658.16 |
| | 689 | 1,214.93 | 1,082.34 | 1,047.70 | 1,156.18 | 4,501.15 |
| | 690 | 314.4 | 282.68 | 252.16 | 278.42 | 1,127.66 |
| | 691 | 1,320.32 | 1,112.53 | 1,145.91 | 1,130.32 | 4,709.08 |
| | 692 | 49.25 | 72.06 | 22.86 | 40.68 | 184.85 |
| | 693 | 975.72 | 817.59 | 815.47 | 854.5 | 3,463.28 |
| TOTAL | | 33,026.54 | 30,615.34 | 29,742.71 | 31,755.76 | 125,140.35 |

STATE: NEVADA

| ZIP CODE | | QUARTER 1 | QUARTER 2 | QUARTER 3 | QUARTER 4 | TOTAL GRAMS |
|---|---|---|---|---|---|---|
| | 890 | 3,271.20 | 3,154.43 | 3,135.03 | 3,373.52 | 12,934.18 |
| | 891 | 7,245.65 | 6,873.89 | 6,287.29 | 6,830.92 | 27,237.75 |
| | 893 | 28.79 | 27.41 | 28.45 | 25.34 | 109.99 |
| | 894 | 1,600.46 | 1,677.76 | 1,510.88 | 1,667.71 | 6,456.81 |
| | 895 | 2,355.30 | 2,182.47 | 2,176.33 | 2,243.50 | 8,957.60 |
| | 897 | 474.68 | 460.72 | 485.52 | 495.84 | 1,916.76 |
| | 898 | 430.89 | 422.14 | 353.97 | 421.19 | 1,628.19 |
| TOTAL | | 15,406.97 | 14,798.82 | 13,977.47 | 15,058.02 | 59,241.28 |

STATE: NEW HAMPSHIRE

| ZIP CODE | | QUARTER 1 | QUARTER 2 | QUARTER 3 | QUARTER 4 | TOTAL GRAMS |
|---|---|---|---|---|---|---|
| | 30 | 8,586.96 | 8,615.94 | 8,059.43 | 8,698.29 | 33,960.62 |
| | 31 | 5,283.71 | 5,391.13 | 4,938.43 | 5,011.45 | 20,624.72 |
| | 32 | 3,246.35 | 3,220.45 | 3,183.71 | 3,199.49 | 12,850 |
| | 33 | 2,306.91 | 2,169.36 | 2,193.93 | 2,221.47 | 8,891.67 |
| | 34 | 1,992.72 | 2,232.18 | 1,974.29 | 2,056.86 | 8,256.05 |
| | 35 | 1,750.93 | 1,961.06 | 1,706.04 | 1,778.44 | 7,196.47 |
| | 36 | 108.35 | 126.29 | 130.22 | 111.37 | 476.23 |
| | 37 | 3,278.82 | 3,322.71 | 3,071.33 | 3,403.98 | 13,076.84 |
| | 38 | 7,855.36 | 7,697.51 | 7,324.38 | 7,430.10 | 30,307.35 |
| TOTAL | | 34,410.11 | 34,736.63 | 32,581.76 | 33,911.45 | 135,639.95 |

DATE RANGE: 01/01/2017 TO 12/31/2017 ARCOS 3 - REPORT 1 Run Date: 07/03/2018

RETAIL DRUG DISTRIBUTION BY ZIP CODE WITHIN STATE BY GRAMS WT

STATE: NEW JERSEY

| ZIP CODE | QUARTER 1 | QUARTER 2 | QUARTER 3 | QUARTER 4 | TOTAL GRAMS |
|---|---|---|---|---|---|
| 70 | 16,536.10 | 16,440.24 | 14,547.20 | 16,476.76 | 64,000.30 |
| 71 | 1,987.73 | 1,780.36 | 1,590.30 | 1,861.64 | 7,220.03 |
| 72 | 1,091.77 | 1,007.62 | 899.08 | 958.32 | 3,956.79 |
| 73 | 1,120.78 | 1,102.32 | 953.88 | 977.99 | 4,154.97 |
| 74 | 5,508.84 | 5,111.87 | 4,500.91 | 5,246.84 | 20,368.46 |
| 75 | 1,052.81 | 955.1 | 842.07 | 889.87 | 3,739.85 |
| 76 | 4,927.11 | 4,877.05 | 3,968.49 | 4,798.82 | 18,571.47 |
| 77 | 6,581.40 | 6,213.08 | 5,795.21 | 6,417.06 | 25,006.75 |
| 78 | 4,976 | 4,960.23 | 4,451.94 | 4,667.55 | 19,055.72 |
| 79 | 4,938.93 | 4,718.32 | 4,501.52 | 4,911.93 | 19,070.70 |
| 80 | 18,279.24 | 17,353.24 | 16,116.39 | 17,379.24 | 69,128.11 |
| 81 | 1,672 | 1,637.20 | 1,316.27 | 1,556.50 | 6,181.97 |
| 82 | 2,789.98 | 2,681.01 | 2,790.22 | 2,771.31 | 11,032.52 |
| 83 | 2,499.24 | 2,436.85 | 2,045.85 | 2,435.23 | 9,417.17 |
| 84 | 217.88 | 198.26 | 214.83 | 212.9 | 843.87 |
| 85 | 4,166.33 | 4,156.27 | 3,833.62 | 4,094.62 | 16,250.84 |
| 86 | 3,088.52 | 2,923.02 | 2,788.08 | 3,012.34 | 11,811.96 |
| 87 | 4,908.75 | 4,913.10 | 4,529.38 | 4,899.13 | 19,250.36 |
| 88 | 9,040.92 | 8,902.67 | 7,684.57 | 8,704.96 | 34,333.12 |
| 89 | 878.11 | 849.04 | 770.51 | 887.72 | 3,385.38 |
| TOTAL | 96,262.44 | 93,216.85 | 84,140.32 | 93,160.73 | 366,780.34 |

STATE: NEW MEXICO

| ZIP CODE | QUARTER 1 | QUARTER 2 | QUARTER 3 | QUARTER 4 | TOTAL GRAMS |
|---|---|---|---|---|---|
| 870 | 1,540.19 | 1,495.40 | 1,411.24 | 1,450.44 | 5,897.27 |
| 871 | 9,261.90 | 8,581.27 | 8,683.35 | 8,352.82 | 34,879.34 |
| 873 | 580.83 | 486.9 | 540.96 | 579.23 | 2,187.92 |
| 874 | 1,616.84 | 1,526.76 | 1,383.11 | 1,406.68 | 5,933.39 |
| 875 | 1,919.46 | 1,899.94 | 1,877.54 | 1,899.17 | 7,596.11 |
| 877 | 269.53 | 305.69 | 290.95 | 299.88 | 1,166.05 |
| 878 | 122.1 | 126.09 | 118.12 | 135.2 | 501.51 |
| 879 | 84.05 | 78.79 | 119.92 | 86.76 | 369.52 |
| 880 | 1,804.04 | 1,703.67 | 1,704.25 | 1,691.27 | 6,903.23 |

| | | | | | | |
|---|---|---|---|---|---|---|
| | 881 | 532.68 | 408.65 | 354.56 | 418.27 | 1,714.16 |
| | 882 | 1,667.39 | 1,586.96 | 1,627.62 | 1,740.35 | 6,622.32 |
| | 883 | 527.2 | 552.49 | 463.92 | 566.85 | 2,110.46 |
| | 884 | 94.12 | 108.48 | 97.75 | 88.76 | 389.11 |
| TOTAL | | 20,020.33 | 18,861.09 | 18,673.29 | 18,715.68 | 76,270.39 |

| STATE: | NEW YORK | | | | | |
|---|---|---|---|---|---|---|
| ZIP CODE | QUARTER 1 | | QUARTER 2 | QUARTER 3 | QUARTER 4 | TOTAL GRAMS |
| | 100 | 21,492.48 | 22,215.87 | 19,529.45 | 21,921.89 | 85,159.69 |
| | 101 | 1,413.86 | 1,258.19 | 1,073.32 | 1,318.34 | 5,063.71 |
| | 102 | 117.7 | 162.69 | 141.56 | 128.19 | 550.14 |
| | 103 | 2,704.18 | 2,610.14 | 2,266.59 | 2,721.93 | 10,302.84 |
| | 104 | 8,180.02 | 8,250.37 | 6,942.63 | 7,847.02 | 31,220.04 |
| | 105 | 10,713.65 | 10,326.44 | 9,274.78 | 10,182.34 | 40,497.21 |
| | 106 | 1,192.57 | 1,227.06 | 1,051.85 | 1,199.93 | 4,671.41 |
| | 107 | 1,959.55 | 1,940.26 | 1,646.08 | 1,984.71 | 7,530.60 |
| | 108 | 999.51 | 977.61 | 912.11 | 960.91 | 3,850.14 |
| | 109 | 7,157.53 | 6,897.03 | 6,228.60 | 6,724.51 | 27,007.67 |
| | 110 | 2,929.86 | 2,918.88 | 2,554.90 | 2,806.08 | 11,209.72 |
| | 111 | 1,115.95 | 1,189.72 | 996.58 | 1,205.51 | 4,507.76 |
| | 112 | 12,541.73 | 12,561.70 | 10,429.42 | 11,866.55 | 47,399.40 |
| | 113 | 4,889.04 | 5,011.21 | 4,045.01 | 5,033.13 | 18,978.39 |
| | 114 | 2,627.97 | 2,481.26 | 1,919.45 | 2,382 | 9,410.68 |
| | 115 | 8,662.71 | 8,549.54 | 7,656.59 | 8,495.36 | 33,364.20 |
| | 116 | 865.48 | 833.45 | 686.85 | 831.66 | 3,217.44 |
| | 117 | 21,082.74 | 20,253.17 | 18,160.80 | 20,379.77 | 79,876.48 |
| | 118 | 1,386.54 | 1,351.45 | 1,206.56 | 1,352.87 | 5,297.42 |
| | 119 | 2,749.16 | 2,889.63 | 2,818.64 | 2,741.54 | 11,198.97 |
| | 120 | 7,789.82 | 7,511.86 | 6,964.89 | 7,728.51 | 29,995.08 |
| | 121 | 4,778.41 | 4,849.29 | 4,294.06 | 4,834.74 | 18,756.50 |
| | 122 | 2,879.35 | 2,665.11 | 2,591.22 | 2,907.72 | 11,043.40 |
| | 123 | 3,250.53 | 3,306.83 | 2,950.86 | 3,174.89 | 12,683.11 |
| | 124 | 1,781.58 | 1,857.07 | 1,807.56 | 1,918.34 | 7,364.55 |
| | 125 | 5,395.25 | 5,397.91 | 4,629.63 | 5,256.87 | 20,679.66 |
| | 126 | 1,100.39 | 1,105.85 | 926.08 | 1,231.28 | 4,363.60 |
| | 127 | 1,059.53 | 1,246.39 | 1,031.59 | 1,026.75 | 4,364.26 |

| | | | | | | |
|---|---|---|---|---|---|---|
| | 128 | 5,016.45 | 5,021.28 | 4,609.41 | 5,034.40 | 19,681.54 |
| | 129 | 4,024.96 | 3,945.93 | 3,758.58 | 4,180.72 | 15,910.19 |
| | 130 | 6,330.80 | 6,415.33 | 5,927.36 | 6,506.02 | 25,179.51 |
| | 131 | 2,333.46 | 2,339.73 | 1,986.41 | 2,314.08 | 8,973.68 |
| | 132 | 3,552.69 | 3,540.55 | 3,239.84 | 3,636.59 | 13,969.67 |
| | 133 | 2,001.54 | 1,986.60 | 1,804.64 | 2,066.61 | 7,859.39 |
| | 134 | 2,601.64 | 2,730.55 | 2,511.09 | 2,579.02 | 10,422.30 |
| | 135 | 1,043.62 | 1,082.11 | 958.12 | 1,020.15 | 4,104 |
| | 136 | 5,740.21 | 5,721.72 | 5,699.25 | 5,572.65 | 22,733.83 |
| | 137 | 1,400.38 | 1,372.61 | 1,138.09 | 1,366.13 | 5,277.21 |
| | 138 | 2,545.18 | 2,464.76 | 2,312.85 | 2,575.21 | 9,898 |
| | 139 | 1,271.89 | 1,130.74 | 973.63 | 1,172.55 | 4,548.81 |
| | 140 | 5,840.44 | 5,735.78 | 5,437.63 | 5,968.72 | 22,982.57 |
| | 141 | 2,833.62 | 2,887.75 | 2,636.20 | 2,739.21 | 11,096.78 |
| | 142 | 7,477.91 | 7,404.61 | 7,085.50 | 7,756.33 | 29,724.35 |
| | 143 | 628.39 | 604.59 | 607.52 | 669.09 | 2,509.59 |
| | 144 | 7,762.32 | 7,600.52 | 7,129.23 | 7,704.70 | 30,196.77 |
| | 145 | 8,090.46 | 8,096.18 | 7,595.74 | 7,771.65 | 31,554.03 |
| | 146 | 12,713.74 | 12,280.29 | 11,537.99 | 12,554.63 | 49,086.65 |
| | 147 | 2,214.19 | 2,218.18 | 1,995.66 | 2,213.90 | 8,641.93 |
| | 148 | 5,396.06 | 5,275.46 | 4,922.28 | 5,496.74 | 21,090.54 |
| | 149 | 1,320.82 | 1,311.19 | 1,195.38 | 1,247.70 | 5,075.09 |
| TOTAL | | 234,957.86 | 233,012.44 | 209,800.06 | 232,310.14 | 910,080.50 |

STATE: NORTH CAROLINA

| ZIP CODE | | QUARTER 1 | QUARTER 2 | QUARTER 3 | QUARTER 4 | TOTAL GRAMS |
|---|---|---|---|---|---|---|
| | 270 | 5,347.53 | 4,729.80 | 4,373.23 | 4,930.23 | 19,380.79 |
| | 271 | 5,801.83 | 5,109.41 | 4,868.43 | 5,521.21 | 21,300.88 |
| | 272 | 11,940.38 | 11,242.68 | 10,305.83 | 11,315.21 | 44,804.10 |
| | 273 | 7,595.65 | 6,882.43 | 6,128.73 | 6,867.53 | 27,474.34 |
| | 274 | 6,156.92 | 5,611.18 | 5,449.20 | 5,486.10 | 22,703.40 |
| | 275 | 25,913.54 | 24,308.45 | 22,580.13 | 24,637.95 | 97,440.07 |
| | 276 | 9,765.52 | 9,014.17 | 8,489.32 | 9,064.09 | 36,333.10 |
| | 277 | 6,095.73 | 6,022.85 | 5,659.28 | 6,038.51 | 23,816.37 |
| | 278 | 10,033.17 | 8,748.20 | 8,421.84 | 9,318.31 | 36,521.52 |
| | 279 | 2,573.30 | 2,550.91 | 2,164.54 | 2,630.78 | 9,919.53 |

| | | | | | | |
|---|---|---|---|---|---|---|
| | 280 | 19,201.70 | 17,638.76 | 16,003.66 | 17,903.19 | 70,747.31 |
| | 281 | 14,830.84 | 13,388.42 | 12,348.56 | 13,806.58 | 54,374.40 |
| | 282 | 14,011.10 | 13,173.14 | 11,974.27 | 14,002.90 | 53,161.41 |
| | 283 | 17,635.59 | 15,936.71 | 14,817.50 | 15,904 | 64,293.80 |
| | 284 | 9,552.92 | 8,954.21 | 7,734.92 | 8,687.68 | 34,929.73 |
| | 285 | 12,630.15 | 11,988.27 | 11,266.90 | 10,813.42 | 46,698.74 |
| | 286 | 12,064.47 | 11,152.39 | 10,348.70 | 11,321.38 | 44,886.94 |
| | 287 | 9,245.32 | 8,533.79 | 8,162.40 | 8,831.31 | 34,772.82 |
| | 288 | 4,170.53 | 3,951.81 | 3,687.15 | 3,916.05 | 15,725.54 |
| | 289 | 750.41 | 806.64 | 698.2 | 639.64 | 2,894.89 |
| TOTAL | | 205,316.60 | 189,744.22 | 175,482.79 | 191,636.07 | 762,179.68 |

STATE: NORTH DAKOTA

| ZIP CODE | | QUARTER 1 | QUARTER 2 | QUARTER 3 | QUARTER 4 | TOTAL GRAMS |
|---|---|---|---|---|---|---|
| | 580 | 1,450.21 | 1,458.94 | 1,365.79 | 1,533.25 | 5,808.19 |
| | 581 | 3,674.74 | 3,556.80 | 3,298.05 | 3,520.11 | 14,049.70 |
| | 582 | 2,437.98 | 2,325.69 | 2,238.03 | 2,317.92 | 9,319.62 |
| | 583 | 1,143.84 | 972.02 | 970.93 | 972.19 | 4,058.98 |
| | 584 | 719.88 | 663.34 | 615.92 | 663.49 | 2,662.63 |
| | 585 | 3,023.03 | 2,714.86 | 2,716.90 | 3,141.48 | 11,596.27 |
| | 586 | 734.6 | 729.22 | 678.16 | 713.27 | 2,855.25 |
| | 587 | 1,802.17 | 1,751.75 | 1,653.63 | 1,792.39 | 6,999.94 |
| | 588 | 707.01 | 698.35 | 663.02 | 709.63 | 2,778.01 |
| TOTAL | | 15,693.46 | 14,870.97 | 14,200.43 | 15,363.73 | 60,128.59 |

STATE: OHIO

| ZIP CODE | | QUARTER 1 | QUARTER 2 | QUARTER 3 | QUARTER 4 | TOTAL GRAMS |
|---|---|---|---|---|---|---|
| | 430 | 17,332.65 | 16,053.31 | 15,442.04 | 16,775.34 | 65,603.34 |
| | 431 | 8,352.38 | 8,065.09 | 7,596.62 | 8,276.50 | 32,290.59 |
| | 432 | 40,131.49 | 37,467.16 | 42,200.52 | 46,052.29 | 165,851.46 |
| | 433 | 2,848.10 | 2,849.32 | 2,697.17 | 2,836.56 | 11,231.15 |
| | 434 | 2,862.05 | 2,700.28 | 2,619.66 | 2,841.30 | 11,023.29 |
| | 435 | 7,048.03 | 6,202.97 | 6,115.83 | 6,443.44 | 25,810.27 |
| | 436 | 8,101.27 | 7,577.81 | 7,226.65 | 7,358.55 | 30,264.28 |
| | 437 | 2,718 | 2,494.03 | 2,406.14 | 2,543.28 | 10,161.45 |

| | | | | | | |
|---|---|---|---|---|---|---|
| | 438 | 428.48 | 563.17 | 558.54 | 475.68 | 2,025.87 |
| | 439 | 2,312.62 | 2,207.20 | 2,026.23 | 2,106.12 | 8,652.17 |
| | 440 | 14,280.30 | 13,460.02 | 12,523.41 | 14,058.56 | 54,322.29 |
| | 441 | 16,749.30 | 15,826.44 | 15,043.63 | 16,195.84 | 63,815.21 |
| | 442 | 8,537.39 | 7,901.79 | 7,928.47 | 8,163.59 | 32,531.24 |
| | 443 | 4,256.22 | 3,864.25 | 3,725.59 | 3,866.40 | 15,712.46 |
| | 444 | 3,818.23 | 3,687.54 | 3,425.86 | 3,727.82 | 14,659.45 |
| | 445 | 3,051.83 | 2,889.61 | 2,704.47 | 2,940.84 | 11,586.75 |
| | 446 | 6,410.24 | 5,817.78 | 5,813 | 6,241.29 | 24,282.31 |
| | 447 | 5,003.73 | 4,475.86 | 4,434.39 | 4,709.05 | 18,623.03 |
| | 448 | 4,793.69 | 4,197.03 | 4,252.97 | 4,484.37 | 17,728.06 |
| | 449 | 1,676.70 | 1,533.46 | 1,347.58 | 1,488.75 | 6,046.49 |
| | 450 | 16,851.57 | 15,417.48 | 15,103.85 | 16,417.80 | 63,790.70 |
| | 451 | 6,878.56 | 6,148.64 | 6,184.71 | 6,610.11 | 25,822.02 |
| | 452 | 20,998.37 | 19,307.71 | 18,377.16 | 20,758.17 | 79,441.41 |
| | 453 | 8,014.14 | 7,586.48 | 7,235.89 | 8,176.20 | 31,012.71 |
| | 454 | 9,972.49 | 9,466.86 | 8,898.73 | 9,441.29 | 37,779.37 |
| | 455 | 2,777 | 2,415.70 | 2,436.50 | 2,533.96 | 10,163.16 |
| | 456 | 6,877.73 | 6,326.05 | 6,361.71 | 6,779.51 | 26,345 |
| | 457 | 2,628.01 | 2,236.94 | 2,318.12 | 2,566.04 | 9,749.11 |
| | 458 | 5,358.81 | 5,091.06 | 4,722.91 | 5,397.68 | 20,570.46 |
| TOTAL | | 241,069.38 | 223,831.04 | 221,728.35 | 240,266.33 | 926,895.10 |

| STATE: | OKLAHOMA | | | | | |
|---|---|---|---|---|---|---|
| ZIP CODE | | QUARTER 1 | QUARTER 2 | QUARTER 3 | QUARTER 4 | TOTAL GRAMS |
| | 730 | 9,707.97 | 9,098.98 | 8,987.46 | 9,618.02 | 37,412.43 |
| | 731 | 7,160.20 | 6,628.87 | 6,654.63 | 7,058.33 | 27,502.03 |
| | 734 | 650.08 | 611.8 | 682.13 | 635.67 | 2,579.68 |
| | 735 | 3,626.02 | 3,401.31 | 3,475.03 | 2,999.04 | 13,501.40 |
| | 736 | 759.87 | 690.89 | 693.25 | 817.29 | 2,961.30 |
| | 737 | 1,326.94 | 1,110.31 | 1,126.22 | 1,242.17 | 4,805.64 |
| | 738 | 190.45 | 162.12 | 207.85 | 188.06 | 748.48 |
| | 739 | 103.67 | 48.39 | 55.61 | 55.21 | 262.88 |
| | 740 | 11,279.12 | 9,967.10 | 9,916.91 | 10,889.57 | 42,052.70 |
| | 741 | 6,502.51 | 5,885.16 | 5,810.75 | 5,926.96 | 24,125.38 |
| | 743 | 1,878.95 | 1,638.12 | 1,735.97 | 1,879.81 | 7,132.85 |

| | | | | | | |
|---|---|---|---|---|---|---|
| | 744 | 3,093.25 | 2,682.67 | 3,039.49 | 3,042.11 | 11,857.52 |
| | 745 | 945.61 | 827.47 | 726.11 | 767.01 | 3,266.20 |
| | 746 | 729.35 | 633.19 | 660.62 | 763.7 | 2,786.86 |
| | 747 | 791.46 | 689.67 | 671.33 | 795.76 | 2,948.22 |
| | 748 | 1,706.14 | 1,552.14 | 1,531.90 | 1,621.11 | 6,411.29 |
| | 749 | 936.2 | 763.62 | 712.6 | 745.3 | 3,157.72 |
| TOTAL | | 51,387.79 | 46,391.81 | 46,687.86 | 49,045.12 | 193,512.58 |

STATE: OREGON

| ZIP CODE | | QUARTER 1 | QUARTER 2 | QUARTER 3 | QUARTER 4 | TOTAL GRAMS |
|---|---|---|---|---|---|---|
| | 970 | 12,889.34 | 12,893.73 | 11,599.11 | 12,565.74 | 49,947.92 |
| | 971 | 5,241.58 | 5,507.11 | 5,103.33 | 5,569.05 | 21,421.07 |
| | 972 | 15,798.87 | 16,616.18 | 15,549.64 | 16,192.79 | 64,157.48 |
| | 973 | 11,533.67 | 11,573.52 | 10,364.94 | 10,980.14 | 44,452.27 |
| | 974 | 7,381.19 | 7,326.44 | 6,808.83 | 7,130.31 | 28,646.77 |
| | 975 | 3,201.75 | 3,141.60 | 2,958.02 | 3,228.84 | 12,530.21 |
| | 976 | 665.48 | 641.08 | 606.27 | 653.17 | 2,566 |
| | 977 | 3,255.04 | 3,388.50 | 3,093.79 | 3,274.52 | 13,011.85 |
| | 978 | 1,798.69 | 1,751.80 | 1,569.54 | 1,905.90 | 7,025.93 |
| | 979 | 562.37 | 510.22 | 523.74 | 548.4 | 2,144.73 |
| TOTAL | | 62,327.98 | 63,350.18 | 58,177.21 | 62,048.86 | 245,904.23 |

STATE: PENNSYLVANIA

| ZIP CODE | | QUARTER 1 | QUARTER 2 | QUARTER 3 | QUARTER 4 | TOTAL GRAMS |
|---|---|---|---|---|---|---|
| | 150 | 7,925.16 | 7,547.51 | 6,972.05 | 7,586.98 | 30,031.70 |
| | 151 | 6,674.83 | 6,407.39 | 6,037.05 | 6,670.25 | 25,789.52 |
| | 152 | 12,436.52 | 11,748.21 | 10,863.74 | 11,916.89 | 46,965.36 |
| | 153 | 3,655.46 | 3,446.85 | 3,276.07 | 3,315.26 | 13,693.64 |
| | 154 | 1,610.95 | 1,505.60 | 1,418.48 | 1,530.41 | 6,065.44 |
| | 155 | 1,087.15 | 1,048.28 | 1,119.43 | 1,133.56 | 4,388.42 |
| | 156 | 5,392.92 | 5,161.79 | 4,948.83 | 5,259.73 | 20,763.27 |
| | 157 | 2,404.06 | 2,367.21 | 2,253.68 | 2,287.89 | 9,312.84 |
| | 158 | 2,219.69 | 2,127.84 | 2,145.79 | 2,156.05 | 8,649.37 |
| | 159 | 2,225.53 | 2,247.33 | 2,016.08 | 2,221.61 | 8,710.55 |
| | 160 | 3,991.18 | 3,656.04 | 3,729.83 | 3,787.36 | 15,164.41 |

| | | | | | | |
|---|---|---|---|---|---|---|
| | 161 | 4,074.78 | 3,905.03 | 3,716.42 | 4,030.86 | 15,727.09 |
| | 162 | 1,752.41 | 1,547.09 | 1,469.27 | 1,577.21 | 6,345.98 |
| | 163 | 2,518.81 | 2,353.54 | 2,100.79 | 2,280.44 | 9,253.58 |
| | 164 | 1,198.15 | 1,078.94 | 1,161.43 | 1,183.85 | 4,622.37 |
| | 165 | 4,797.41 | 4,621.58 | 4,339.09 | 4,477.37 | 18,235.45 |
| | 166 | 3,873.90 | 3,646.45 | 3,358.61 | 3,731.46 | 14,610.42 |
| | 167 | 771.4 | 754.1 | 749.56 | 829.08 | 3,104.14 |
| | 168 | 3,337.31 | 3,126.39 | 3,288.88 | 3,150.79 | 12,903.37 |
| | 169 | 790.34 | 749.84 | 700.04 | 718.22 | 2,958.44 |
| | 170 | 15,911.07 | 14,764.73 | 14,565.48 | 15,403.46 | 60,644.74 |
| | 171 | 3,127.57 | 3,059.25 | 2,794.75 | 3,130.26 | 12,111.83 |
| | 172 | 4,035.27 | 3,732.71 | 3,623.45 | 3,767.48 | 15,158.91 |
| | 173 | 6,825.74 | 6,499.15 | 5,877.39 | 6,383.93 | 25,586.21 |
| | 174 | 4,804.17 | 4,913.56 | 4,396.63 | 4,847.50 | 18,961.86 |
| | 175 | 5,199.17 | 5,000.91 | 4,779.95 | 4,870.68 | 19,850.71 |
| | 176 | 4,639.80 | 4,560.60 | 4,259.44 | 4,384.13 | 17,843.97 |
| | 177 | 3,006.43 | 2,947.96 | 2,738.72 | 3,026.62 | 11,719.73 |
| | 178 | 3,512.76 | 3,429.54 | 3,173.57 | 3,581.26 | 13,697.13 |
| | 179 | 1,439.11 | 1,392.87 | 1,265.37 | 1,265.05 | 5,362.40 |
| | 180 | 6,047.87 | 5,975.95 | 5,671.98 | 5,822.20 | 23,518 |
| | 181 | 3,094.71 | 2,926.70 | 2,830.71 | 2,912.53 | 11,764.65 |
| | 182 | 1,400.64 | 1,313.18 | 1,156.81 | 1,397.34 | 5,267.97 |
| | 183 | 1,504.55 | 1,477.29 | 1,425.50 | 1,615.06 | 6,022.40 |
| | 184 | 2,223.63 | 2,122.78 | 2,061.21 | 2,238.57 | 8,646.19 |
| | 185 | 2,598.78 | 2,521.21 | 2,453.07 | 2,630.23 | 10,203.29 |
| | 186 | 2,388.58 | 2,252.64 | 1,965.19 | 2,302.33 | 8,908.74 |
| | 187 | 21,679.25 | 25,213.04 | 25,890.09 | 28,674.36 | 101,456.74 |
| | 188 | 1,034.65 | 998.19 | 1,023.31 | 1,053.55 | 4,109.70 |
| | 189 | 6,221.09 | 5,803.88 | 5,447.47 | 6,039.74 | 23,512.18 |
| | 190 | 18,505.54 | 17,926.45 | 17,052.87 | 17,891.22 | 71,376.08 |
| | 191 | 13,693.46 | 13,342.14 | 11,929.72 | 13,785.29 | 52,750.61 |
| | 193 | 9,191.35 | 8,783.61 | 8,399.69 | 9,111.26 | 35,485.91 |
| | 194 | 8,657.94 | 8,431.33 | 8,118.89 | 8,643.23 | 33,851.39 |
| | 195 | 2,527.26 | 2,430.46 | 2,356.19 | 2,567.79 | 9,881.70 |
| | 196 | 3,790.63 | 3,758.84 | 3,473.21 | 3,803.67 | 14,826.35 |
| TOTAL | | 229,798.98 | 224,625.98 | 214,395.78 | 230,994.01 | 899,814.75 |

STATE: PUERTO RICO

| ZIP CODE | QUARTER 1 | QUARTER 2 | QUARTER 3 | QUARTER 4 | TOTAL GRAMS |
|---|---|---|---|---|---|
| 6 | 5,504.94 | 4,176.44 | 3,558.11 | 3,489.28 | 16,728.77 |
| 7 | 7,153.45 | 5,488.30 | 4,081.09 | 3,636.42 | 20,359.26 |
| 9 | 9,873.94 | 8,198.92 | 6,410.97 | 6,292.94 | 30,776.77 |
| TOTAL | 22,532.33 | 17,863.66 | 14,050.17 | 13,418.64 | 67,864.80 |

STATE: RHODE ISLAND

| ZIP CODE | QUARTER 1 | QUARTER 2 | QUARTER 3 | QUARTER 4 | TOTAL GRAMS |
|---|---|---|---|---|---|
| 28 | 12,735.71 | 13,165.48 | 11,892.83 | 12,809.09 | 50,603.11 |
| 29 | 6,304.49 | 6,384.24 | 5,770.50 | 6,236.93 | 24,696.16 |
| TOTAL | 19,040.20 | 19,549.72 | 17,663.33 | 19,046.02 | 75,299.27 |

STATE: SOUTH CAROLINA

| ZIP CODE | QUARTER 1 | QUARTER 2 | QUARTER 3 | QUARTER 4 | TOTAL GRAMS |
|---|---|---|---|---|---|
| 290 | 7,502.84 | 6,642.26 | 6,351.30 | 7,067.53 | 27,563.93 |
| 291 | 6,225.30 | 5,607.08 | 5,218.87 | 5,749.86 | 22,801.11 |
| 292 | 6,906.67 | 6,042.33 | 6,277.32 | 6,162.62 | 25,388.94 |
| 293 | 7,948.24 | 7,131.80 | 7,019.13 | 7,852.10 | 29,951.27 |
| 294 | 19,877.30 | 17,810.55 | 17,669.92 | 19,312.20 | 74,669.97 |
| 295 | 11,965.68 | 10,673.47 | 10,312.30 | 11,198.43 | 44,149.88 |
| 296 | 20,608.20 | 18,544.67 | 17,910.25 | 19,764.99 | 76,828.11 |
| 297 | 7,698.83 | 6,800.71 | 6,602.60 | 7,366.84 | 28,468.98 |
| 298 | 3,348.03 | 2,937.34 | 2,697.97 | 3,104.92 | 12,088.26 |
| 299 | 3,160.56 | 2,754.48 | 2,986.74 | 2,959.95 | 11,861.73 |
| TOTAL | 95,241.65 | 84,944.69 | 83,046.40 | 90,539.44 | 353,772.18 |

STATE: SOUTH DAKOTA

| ZIP CODE | QUARTER 1 | QUARTER 2 | QUARTER 3 | QUARTER 4 | TOTAL GRAMS |
|---|---|---|---|---|---|
| 570 | 2,748.79 | 2,503.21 | 2,462.63 | 2,618.44 | 10,333.07 |
| 571 | 13,967.95 | 10,191.14 | 12,911.46 | 14,211.90 | 51,282.45 |
| 572 | 1,091.78 | 1,006.29 | 1,003.09 | 1,118.04 | 4,219.20 |
| 573 | 1,821.48 | 1,513.61 | 1,653.98 | 1,723.56 | 6,712.63 |

| | | | | | | |
|---|---|---|---|---|---|---|
| | 574 | 1,178.47 | 1,158.80 | 1,125.22 | 1,142.37 | 4,604.86 |
| | 575 | 948.5 | 841.9 | 891.01 | 942.9 | 3,624.31 |
| | 576 | 209.08 | 167.2 | 173.8 | 177.26 | 727.34 |
| | 577 | 3,019.16 | 2,797.93 | 2,747.61 | 2,916.60 | 11,481.30 |
| TOTAL | | 24,985.21 | 20,180.08 | 22,968.80 | 24,851.07 | 92,985.16 |
| STATE: | TENNESSEE | | | | | |
| ZIP CODE | | QUARTER 1 | QUARTER 2 | QUARTER 3 | QUARTER 4 | TOTAL GRAMS |
| | 370 | 13,202.28 | 12,023.41 | 12,222.70 | 12,591.06 | 50,039.45 |
| | 371 | 8,851.90 | 8,458.74 | 8,038.71 | 8,773.20 | 34,122.55 |
| | 372 | 6,234.62 | 5,664.03 | 5,511.56 | 5,873.17 | 23,283.38 |
| | 373 | 8,202.75 | 7,604.27 | 7,553 | 8,239.40 | 31,599.42 |
| | 374 | 4,014.57 | 3,374.73 | 3,439.03 | 3,423.71 | 14,252.04 |
| | 376 | 5,151.61 | 4,720.58 | 4,731.06 | 4,993.29 | 19,596.54 |
| | 377 | 4,905.12 | 4,522.40 | 4,552.80 | 4,721.48 | 18,701.80 |
| | 378 | 7,049.75 | 6,562.54 | 6,707.80 | 7,109.45 | 27,429.54 |
| | 379 | 6,631.83 | 6,149.37 | 5,935.75 | 6,523.70 | 25,240.65 |
| | 380 | 6,694.60 | 5,730.54 | 5,935.58 | 6,061.46 | 24,422.18 |
| | 381 | 7,931.67 | 7,261.14 | 7,307.20 | 7,684.88 | 30,184.89 |
| | 382 | 1,667.54 | 1,444.87 | 1,542.15 | 1,492.76 | 6,147.32 |
| | 383 | 3,782.91 | 3,266.70 | 3,515.50 | 3,628.19 | 14,193.30 |
| | 384 | 2,149.66 | 2,018.96 | 1,878.81 | 1,945.78 | 7,993.21 |
| | 385 | 2,224.31 | 2,103.86 | 2,023.21 | 2,085.18 | 8,436.56 |
| TOTAL | | 88,695.12 | 80,906.14 | 80,894.86 | 85,146.71 | 335,642.83 |
| STATE: | TEXAS | | | | | |
| ZIP CODE | | QUARTER 1 | QUARTER 2 | QUARTER 3 | QUARTER 4 | TOTAL GRAMS |
| | 750 | 37,816.45 | 34,580.49 | 30,803.86 | 32,122 | 135,322.80 |
| | 751 | 9,702.23 | 8,922.55 | 8,308.14 | 8,893.24 | 35,826.16 |
| | 752 | 12,081.40 | 11,712.06 | 10,308.33 | 11,846.52 | 45,948.31 |
| | 753 | 15.91 | 12.88 | 20.58 | 33.72 | 83.09 |
| | 754 | 5,641.70 | 5,080.66 | 4,882.98 | 5,515.57 | 21,120.91 |
| | 755 | 1,593.80 | 1,375.52 | 1,273.69 | 1,426.04 | 5,669.05 |
| | 756 | 4,316.40 | 4,102.66 | 3,894.61 | 4,349.17 | 16,662.84 |
| | 757 | 5,373.97 | 5,117.54 | 4,858.70 | 5,207 | 20,557.21 |

| | | | | | |
|---|---|---|---|---|---|
| 758 | 1,291.96 | 1,140.41 | 1,111.14 | 1,367.29 | 4,910.80 |
| 759 | 4,585.90 | 4,187.07 | 3,812.75 | 4,405.62 | 16,991.34 |
| 760 | 19,234.20 | 17,608.14 | 16,780.17 | 17,924.23 | 71,546.74 |
| 761 | 11,029.85 | 10,602.12 | 9,320.83 | 10,754.86 | 41,707.66 |
| 762 | 8,149.83 | 7,431.80 | 7,393.14 | 8,151.84 | 31,126.61 |
| 763 | 3,977.42 | 3,684.11 | 3,844.27 | 3,583.97 | 15,089.77 |
| 764 | 1,418.37 | 1,293.93 | 1,243.60 | 1,421.31 | 5,377.21 |
| 765 | 9,760.11 | 9,573.44 | 9,532.33 | 9,274.30 | 38,140.18 |
| 766 | 1,446.56 | 1,283.63 | 1,278.30 | 1,428.66 | 5,437.15 |
| 767 | 4,267.94 | 4,014.31 | 3,680.95 | 4,291.75 | 16,254.95 |
| 768 | 1,494.99 | 1,341.51 | 1,231.52 | 1,425.78 | 5,493.80 |
| 769 | 2,739.17 | 2,638.69 | 2,305.61 | 2,808.51 | 10,491.98 |
| 770 | 25,685.28 | 25,106.46 | 20,876.30 | 24,793.55 | 96,461.59 |
| 772 | 3.46 | 1.73 | 4.32 | 0.86 | 10.37 |
| 773 | 18,968.01 | 17,356.57 | 15,315.36 | 17,959.91 | 69,599.85 |
| 774 | 18,345.33 | 17,135.67 | 14,319.58 | 17,252.26 | 67,052.84 |
| 775 | 18,177.38 | 15,885.32 | 14,103.94 | 16,430.64 | 64,597.28 |
| 776 | 4,153.96 | 3,823.84 | 3,012.65 | 3,684.57 | 14,675.02 |
| 777 | 1,874.32 | 1,555.08 | 1,352.94 | 1,603.87 | 6,386.21 |
| 778 | 4,963.45 | 4,694.31 | 4,396.45 | 4,706.60 | 18,760.81 |
| 779 | 2,367.19 | 2,227.64 | 1,971.72 | 2,337.55 | 8,904.10 |
| 780 | 6,092.74 | 5,709.12 | 5,206.09 | 5,912.44 | 22,920.39 |
| 781 | 6,484.17 | 5,803.91 | 5,632.65 | 6,000.11 | 23,920.84 |
| 782 | 26,487.69 | 24,668.86 | 23,781.95 | 25,294.86 | 100,233.36 |
| 783 | 3,412.07 | 3,033.60 | 2,698.83 | 3,080.82 | 12,225.32 |
| 784 | 6,041.18 | 5,547.24 | 5,015.67 | 5,390.85 | 21,994.94 |
| 785 | 12,439.35 | 11,093.07 | 10,448.98 | 11,815.33 | 45,796.73 |
| 786 | 13,252.23 | 12,212.78 | 11,540.92 | 13,531.98 | 50,537.91 |
| 787 | 13,339.23 | 12,625.03 | 11,788.17 | 13,097.86 | 50,850.29 |
| 788 | 1,573.15 | 1,405.96 | 1,313.43 | 1,424.73 | 5,717.27 |
| 789 | 648.12 | 551.97 | 668.24 | 645.44 | 2,513.77 |
| 790 | 1,494.67 | 1,307.70 | 1,199.92 | 1,456.97 | 5,459.26 |
| 791 | 2,425.38 | 2,405.36 | 2,226.95 | 2,447.54 | 9,505.23 |
| 792 | 313.95 | 284.66 | 297.48 | 340.04 | 1,236.13 |
| 793 | 1,161.79 | 1,070 | 1,024.52 | 1,156.48 | 4,412.79 |
| 794 | 5,310.22 | 5,237.55 | 4,842.01 | 5,231.99 | 20,621.77 |
| 795 | 1,104.54 | 1,049.97 | 1,084.80 | 1,146.53 | 4,385.84 |

| | | | | | | |
|---|---|---|---|---|---|---|
| | 796 | 3,641.10 | 3,449.05 | 2,966.62 | 3,273.03 | 13,329.80 |
| | 797 | 4,578.67 | 4,266.79 | 4,012.20 | 4,442.87 | 17,300.53 |
| | 798 | 276.58 | 245.47 | 201.56 | 196.61 | 920.22 |
| | 799 | 8,857.38 | 8,203.81 | 8,007.53 | 7,617.61 | 32,686.33 |
| TOTAL | | 359,410.75 | 333,662.04 | 305,197.28 | 338,505.28 | 1,336,775.35 |

STATE: UTAH

| ZIP CODE | QUARTER 1 | QUARTER 2 | QUARTER 3 | QUARTER 4 | TOTAL GRAMS |
|---|---|---|---|---|---|
| 840 | 24,645.26 | 23,494.80 | 22,752.54 | 24,528.92 | 95,421.52 |
| 841 | 9,216.31 | 8,839.17 | 8,595.55 | 9,049.94 | 35,700.97 |
| 843 | 2,220.30 | 2,104.38 | 2,005.49 | 2,314.82 | 8,644.99 |
| 844 | 2,811.59 | 2,713.48 | 2,582.80 | 2,853.78 | 10,961.65 |
| 845 | 563.06 | 509.15 | 512.41 | 514.15 | 2,098.77 |
| 846 | 4,938.24 | 4,597.42 | 4,360.63 | 4,696.20 | 18,592.49 |
| 847 | 3,356.15 | 3,009.04 | 3,007.10 | 3,368.28 | 12,740.57 |
| TOTAL | 47,750.91 | 45,267.44 | 43,816.52 | 47,326.09 | 184,160.96 |

STATE: VERMONT

| ZIP CODE | QUARTER 1 | QUARTER 2 | QUARTER 3 | QUARTER 4 | TOTAL GRAMS |
|---|---|---|---|---|---|
| 50 | 1,555.46 | 1,636.53 | 1,315.32 | 1,445.10 | 5,952.41 |
| 51 | 790 | 880.22 | 798.03 | 852.02 | 3,320.27 |
| 52 | 1,257.68 | 1,238.29 | 1,301.11 | 1,315.21 | 5,112.29 |
| 53 | 1,164.39 | 1,200.30 | 1,194.69 | 1,281.78 | 4,841.16 |
| 54 | 6,413.43 | 6,308.38 | 6,172.16 | 6,679.94 | 25,573.91 |
| 56 | 3,521.61 | 3,513.80 | 3,400.69 | 3,443.07 | 13,879.17 |
| 57 | 2,525.43 | 2,488.16 | 2,540.25 | 2,401.13 | 9,954.97 |
| 58 | 2,023.54 | 2,109.98 | 1,939.70 | 2,219.95 | 8,293.17 |
| TOTAL | 19,251.54 | 19,375.66 | 18,661.95 | 19,638.20 | 76,927.35 |

STATE: VIRGIN ISLANDS

| ZIP CODE | QUARTER 1 | QUARTER 2 | QUARTER 3 | QUARTER 4 | TOTAL GRAMS |
|---|---|---|---|---|---|
| 8 | 210 | 180.55 | 99.13 | 107.51 | 597.19 |
| TOTAL | 210 | 180.55 | 99.13 | 107.51 | 597.19 |

STATE: VIRGINIA

| ZIP CODE | QUARTER 1 | QUARTER 2 | QUARTER 3 | QUARTER 4 | TOTAL GRAMS |
|---|---|---|---|---|---|
| 201 | 12,950.80 | 11,934.74 | 11,613.73 | 12,562.60 | 49,061.87 |
| 220 | 9,669.13 | 9,161.77 | 8,906.60 | 8,538.82 | 36,276.32 |
| 221 | 5,909.88 | 5,692.87 | 5,328.43 | 6,084.89 | 23,016.07 |
| 222 | 2,808.94 | 2,943.80 | 2,638.54 | 2,711.88 | 11,103.16 |
| 223 | 3,410.84 | 3,216.69 | 3,097.55 | 3,321.97 | 13,047.05 |
| 224 | 4,726.86 | 4,616.67 | 4,163.32 | 4,624.52 | 18,131.37 |
| 225 | 2,311.14 | 2,399.63 | 2,189.37 | 2,275.76 | 9,175.90 |
| 226 | 3,066.63 | 2,861.91 | 2,803.78 | 3,002.06 | 11,734.38 |
| 227 | 927.03 | 888.33 | 837.39 | 993.06 | 3,645.81 |
| 228 | 2,157.23 | 1,906.88 | 1,835.64 | 2,127.43 | 8,027.18 |
| 229 | 6,264.18 | 5,923.02 | 5,495.29 | 6,061.54 | 23,744.03 |
| 230 | 3,864.80 | 3,709.85 | 3,458.53 | 3,840.51 | 14,873.69 |
| 231 | 8,456.95 | 8,625.66 | 7,301.92 | 8,492.20 | 32,876.73 |
| 232 | 10,030.21 | 9,723.57 | 8,793.09 | 9,793.95 | 38,340.82 |
| 233 | 6,292.79 | 6,448.77 | 5,377.03 | 5,750.43 | 23,869.02 |
| 234 | 12,423.86 | 12,267.52 | 10,738.09 | 11,982.98 | 47,412.45 |
| 235 | 3,265.54 | 3,495.31 | 2,916.35 | 3,326.55 | 13,003.75 |
| 236 | 8,265.88 | 7,686 | 7,252.76 | 7,214.03 | 30,418.67 |
| 237 | 3,306.26 | 3,110.94 | 3,116.68 | 2,941.05 | 12,474.93 |
| 238 | 5,284.91 | 5,001.99 | 4,298.07 | 5,016.48 | 19,601.45 |
| 239 | 1,231.24 | 1,117.31 | 1,246.57 | 1,360.40 | 4,955.52 |
| 240 | 5,963.96 | 5,457.88 | 5,360.06 | 5,801.32 | 22,583.22 |
| 241 | 3,504.27 | 3,341.05 | 3,161.99 | 3,686.84 | 13,694.15 |
| 242 | 2,625.17 | 2,356.96 | 2,277.76 | 2,426.81 | 9,686.70 |
| 243 | 1,910.49 | 1,827.06 | 1,689.94 | 1,900.32 | 7,327.81 |
| 244 | 2,592.99 | 2,538.56 | 2,444.89 | 2,692.50 | 10,268.94 |
| 245 | 6,168.59 | 5,748.66 | 5,775.86 | 5,922.65 | 23,615.76 |
| 246 | 992.71 | 902.53 | 886 | 1,035.98 | 3,817.22 |
| TOTAL | 140,383.28 | 134,905.93 | 125,005.23 | 135,489.53 | 535,783.97 |

STATE: WASHINGTON

| ZIP CODE | QUARTER 1 | QUARTER 2 | QUARTER 3 | QUARTER 4 | TOTAL GRAMS |
|---|---|---|---|---|---|
| 980 | 24,450.66 | 24,686.77 | 22,586.62 | 24,583.06 | 96,307.11 |

| | | | | | | |
|---|---|---|---|---|---|---|
| | 981 | 12,850.13 | 13,229.23 | 12,331.70 | 13,434.65 | 51,845.71 |
| | 982 | 15,646.24 | 15,821.80 | 14,441.39 | 15,633.49 | 61,542.92 |
| | 983 | 12,544.26 | 12,514.69 | 11,552.82 | 12,226.06 | 48,837.83 |
| | 984 | 8,218.95 | 8,133.18 | 7,661.01 | 7,378.51 | 31,391.65 |
| | 985 | 7,361.37 | 7,245.98 | 6,703.06 | 7,404.30 | 28,714.71 |
| | 986 | 9,271.48 | 9,457.73 | 8,797.92 | 9,076.93 | 36,604.06 |
| | 988 | 2,546.93 | 2,626.66 | 2,288.91 | 2,570.10 | 10,032.60 |
| | 989 | 3,531.78 | 3,489.60 | 2,991.30 | 3,499.63 | 13,512.31 |
| | 990 | 1,811.15 | 1,735.95 | 1,732.45 | 1,942.43 | 7,221.98 |
| | 991 | 1,286.42 | 1,225.84 | 1,203.96 | 1,272.52 | 4,988.74 |
| | 992 | 8,251.36 | 8,213.21 | 7,525.95 | 8,188.63 | 32,179.15 |
| | 993 | 5,500.34 | 5,695.43 | 4,816.07 | 5,495.04 | 21,506.88 |
| | 994 | 515.18 | 562.41 | 512.01 | 581.27 | 2,170.87 |
| TOTAL | | 113,786.25 | 114,638.48 | 105,145.17 | 113,286.62 | 446,856.52 |

| STATE: | WEST VIRGINIA | | | | | |
|---|---|---|---|---|---|---|
| ZIP CODE | | QUARTER 1 | QUARTER 2 | QUARTER 3 | QUARTER 4 | TOTAL GRAMS |
| | 247 | 1,066.97 | 1,001.32 | 929.46 | 1,055.32 | 4,053.07 |
| | 248 | 452.45 | 407.79 | 431.35 | 408.42 | 1,700.01 |
| | 249 | 793.58 | 725.05 | 679.93 | 674.92 | 2,873.48 |
| | 250 | 1,191.92 | 1,059.08 | 1,019.90 | 1,114.21 | 4,385.11 |
| | 251 | 974.91 | 895.4 | 892.08 | 988.95 | 3,751.34 |
| | 252 | 823.85 | 763.56 | 667.6 | 754.5 | 3,009.51 |
| | 253 | 3,124.01 | 2,933.73 | 2,703.09 | 2,769.05 | 11,529.88 |
| | 254 | 3,232.72 | 2,965.08 | 2,962.49 | 3,008.45 | 12,168.74 |
| | 255 | 3,354.40 | 2,948.77 | 2,934.76 | 3,062.98 | 12,300.91 |
| | 256 | 762.63 | 670.96 | 699.95 | 735.36 | 2,868.90 |
| | 257 | 2,529.03 | 2,355.92 | 2,448.76 | 2,435.17 | 9,768.88 |
| | 258 | 2,294.74 | 2,218.79 | 2,055.66 | 2,093.42 | 8,662.61 |
| | 259 | 828.18 | 760.4 | 782.28 | 792.75 | 3,163.61 |
| | 260 | 2,213.89 | 1,903.48 | 1,973.91 | 1,990.34 | 8,081.62 |
| | 261 | 2,027.45 | 2,062.72 | 1,877.11 | 1,953.42 | 7,920.70 |
| | 262 | 894.27 | 873.52 | 829.45 | 979.76 | 3,577 |
| | 263 | 1,474.33 | 1,353.63 | 1,393 | 1,381.21 | 5,602.17 |
| | 264 | 479.51 | 482.05 | 450.37 | 498.15 | 1,910.08 |
| | 265 | 3,110.85 | 2,767.61 | 2,892.80 | 3,148.25 | 11,919.51 |

| | | | | | | |
|---|---|---|---|---|---|---|
| | 266 | 415.01 | 391.71 | 426.44 | 405.26 | 1,638.42 |
| | 267 | 890.18 | 814.56 | 748.96 | 800.07 | 3,253.77 |
| | 268 | 621.62 | 544.13 | 541.17 | 530.51 | 2,237.43 |
| TOTAL | | 33,556.50 | 30,899.26 | 30,340.52 | 31,580.47 | 126,376.75 |

STATE: WISCONSIN

| ZIP CODE | | QUARTER 1 | QUARTER 2 | QUARTER 3 | QUARTER 4 | TOTAL GRAMS |
|---|---|---|---|---|---|---|
| | 530 | 12,503.68 | 12,184.28 | 11,911.43 | 13,005.27 | 49,604.66 |
| | 531 | 11,274.08 | 10,940.33 | 10,161.70 | 11,282.45 | 43,658.56 |
| | 532 | 12,172.92 | 11,434.21 | 10,772.87 | 11,559.31 | 45,939.31 |
| | 534 | 2,381.45 | 2,223.07 | 2,099.56 | 2,219.18 | 8,923.26 |
| | 535 | 10,728.68 | 10,553.77 | 9,507.98 | 10,595.26 | 41,385.69 |
| | 537 | 6,767.53 | 6,397.71 | 6,119.76 | 6,466.38 | 25,751.38 |
| | 538 | 930.72 | 903.7 | 813.06 | 1,050.59 | 3,698.07 |
| | 539 | 3,714.14 | 3,484.39 | 3,208.81 | 3,664.07 | 14,071.41 |
| | 540 | 2,467.63 | 2,480.45 | 2,320.40 | 2,381.29 | 9,649.77 |
| | 541 | 3,947.60 | 3,920.34 | 3,848.70 | 3,988.37 | 15,705.01 |
| | 542 | 2,136.21 | 2,134.02 | 2,067.68 | 2,137.20 | 8,475.11 |
| | 543 | 3,801.18 | 3,601.28 | 3,516.05 | 3,717.73 | 14,636.24 |
| | 544 | 7,788.01 | 7,422.14 | 7,037.13 | 7,605.08 | 29,852.36 |
| | 545 | 1,587.44 | 1,575.06 | 1,567.32 | 1,580.85 | 6,310.67 |
| | 546 | 5,524.63 | 5,108.76 | 4,993.63 | 5,409.04 | 21,036.06 |
| | 547 | 5,755.74 | 5,442.96 | 5,061.03 | 5,602.30 | 21,862.03 |
| | 548 | 2,785.99 | 2,837.29 | 2,557.94 | 2,820.77 | 11,001.99 |
| | 549 | 10,842.89 | 10,398.16 | 9,871.44 | 10,990.63 | 42,103.12 |
| TOTAL | | 107,110.52 | 103,041.92 | 97,436.49 | 106,075.77 | 413,664.70 |

STATE: WYOMING

| ZIP CODE | | QUARTER 1 | QUARTER 2 | QUARTER 3 | QUARTER 4 | TOTAL GRAMS |
|---|---|---|---|---|---|---|
| | 820 | 2,345.31 | 2,184.91 | 2,208.88 | 2,270.66 | 9,009.76 |
| | 822 | 308.45 | 260.47 | 244.43 | 255.45 | 1,068.80 |
| | 823 | 131.41 | 111.72 | 95.15 | 101.21 | 439.49 |
| | 824 | 878.35 | 747.08 | 877.12 | 794.82 | 3,297.37 |
| | 825 | 548.5 | 506.04 | 399.74 | 476.29 | 1,930.57 |
| | 826 | 1,710.32 | 1,586.53 | 1,497 | 1,539.41 | 6,333.26 |

| | | | | | | |
|---|---|---|---|---|---|---|
| | 827 | 712.95 | 664.17 | 609.94 | 716.72 | 2,703.78 |
| | 828 | 653.68 | 615.53 | 588.83 | 613.52 | 2,471.56 |
| | 829 | 833.83 | 783.54 | 777.52 | 834.78 | 3,229.67 |
| | 830 | 362.48 | 325.16 | 294.86 | 311.61 | 1,294.11 |
| | 831 | 257.42 | 162.13 | 201.37 | 176.68 | 797.6 |
| TOTAL | | 8,742.70 | 7,947.28 | 7,794.84 | 8,091.15 | 32,575.97 |

DRUG CODE: 2012 DRUG NAME: GAMMA HYDROXYBUTYRIC ACID(FDA APPROV

STATE: CALIFORNIA

| ZIP CODE | | QUARTER 1 | QUARTER 2 | QUARTER 3 | QUARTER 4 | TOTAL GRAMS |
|---|---|---|---|---|---|---|
| | 908 | 594.43 | 520.13 | 520.13 | 445.82 | 2,080.51 |
| | 928 | 817.34 | 817.34 | 817.34 | 817.34 | 3,269.36 |
| | 940 | 148.61 | 0 | 0 | 2,526.34 | 2,674.95 |
| TOTAL | | 1,560.38 | 1,337.47 | 1,337.47 | 3,789.50 | 8,024.82 |

STATE: ILLINOIS

| ZIP CODE | | QUARTER 1 | QUARTER 2 | QUARTER 3 | QUARTER 4 | TOTAL GRAMS |
|---|---|---|---|---|---|---|
| | 606 | 520.13 | 743.04 | 668.74 | 743.04 | 2,674.95 |
| TOTAL | | 520.13 | 743.04 | 668.74 | 743.04 | 2,674.95 |

STATE: LOUISIANA

| ZIP CODE | | QUARTER 1 | QUARTER 2 | QUARTER 3 | QUARTER 4 | TOTAL GRAMS |
|---|---|---|---|---|---|---|
| | 700 | 297.22 | 148.61 | 0 | 0 | 445.83 |
| TOTAL | | 297.22 | 148.61 | 0 | 0 | 445.83 |

STATE: MASSACHUSETTS

| ZIP CODE | | QUARTER 1 | QUARTER 2 | QUARTER 3 | QUARTER 4 | TOTAL GRAMS |
|---|---|---|---|---|---|---|
| | 21 | 0 | 1,560.38 | 297.22 | 817.34 | 2,674.94 |
| TOTAL | | 0 | 1,560.38 | 297.22 | 817.34 | 2,674.94 |

STATE: MICHIGAN

| ZIP CODE | | QUARTER 1 | QUARTER 2 | QUARTER 3 | QUARTER 4 | TOTAL GRAMS |
|---|---|---|---|---|---|---|

| | | | | | | |
|---|---|---|---|---|---|---|
| | 481 | 594.43 | 0 | 297.22 | 445.82 | 1,337.47 |
| TOTAL | | 594.43 | 0 | 297.22 | 445.82 | 1,337.47 |

STATE: MISSOURI

| ZIP CODE | | QUARTER 1 | QUARTER 2 | QUARTER 3 | QUARTER 4 | TOTAL GRAMS |
|---|---|---|---|---|---|---|
| | 631 | 9,221,200.70 | 5,490,842.69 | 8,990,338.18 | 6,709,576.90 | 30,411,958.47 |
| TOTAL | | 9,221,200.70 | 5,490,842.69 | 8,990,338.18 | 6,709,576.90 | 30,411,958.47 |

STATE: NEW YORK

| ZIP CODE | | QUARTER 1 | QUARTER 2 | QUARTER 3 | QUARTER 4 | TOTAL GRAMS |
|---|---|---|---|---|---|---|
| | 104 | 0 | 0 | 2,972.16 | 4,978.37 | 7,950.53 |
| TOTAL | | 0 | 0 | 2,972.16 | 4,978.37 | 7,950.53 |

STATE: NORTH CAROLINA

| ZIP CODE | | QUARTER 1 | QUARTER 2 | QUARTER 3 | QUARTER 4 | TOTAL GRAMS |
|---|---|---|---|---|---|---|
| | 277 | 222.91 | 817.34 | 594.43 | 668.74 | 2,303.42 |
| | 280 | 0 | 594.43 | 1,411.78 | 668.74 | 2,674.95 |
| TOTAL | | 222.91 | 1,411.77 | 2,006.21 | 1,337.48 | 4,978.37 |

STATE: OHIO

| ZIP CODE | | QUARTER 1 | QUARTER 2 | QUARTER 3 | QUARTER 4 | TOTAL GRAMS |
|---|---|---|---|---|---|---|
| | 441 | 0 | 817.34 | 817.34 | 817.34 | 2,452.02 |
| | 452 | 1,188.86 | 1,263.17 | 1,114.56 | 1,411.78 | 4,978.37 |
| TOTAL | | 1,188.86 | 2,080.51 | 1,931.90 | 2,229.12 | 7,430.39 |

STATE: SOUTH CAROLINA

| ZIP CODE | | QUARTER 1 | QUARTER 2 | QUARTER 3 | QUARTER 4 | TOTAL GRAMS |
|---|---|---|---|---|---|---|
| | 292 | 0 | 1,337.47 | 371.52 | 743.04 | 2,452.03 |
| | 296 | 965.95 | 1,411.78 | 0 | 2,749.25 | 5,126.98 |
| TOTAL | | 965.95 | 2,749.25 | 371.52 | 3,492.29 | 7,579.01 |

STATE: TENNESSEE

| ZIP CODE | QUARTER 1 | QUARTER 2 | QUARTER 3 | QUARTER 4 | TOTAL GRAMS |
|---|---|---|---|---|---|
| 381 | 594.43 | 594.43 | 594.43 | 594.43 | 2,377.72 |
| TOTAL | 594.43 | 594.43 | 594.43 | 594.43 | 2,377.72 |

STATE: TEXAS

| ZIP CODE | QUARTER 1 | QUARTER 2 | QUARTER 3 | QUARTER 4 | TOTAL GRAMS |
|---|---|---|---|---|---|
| 752 | 1,486.08 | 1,263.17 | 520.13 | 0 | 3,269.38 |
| 770 | 817.34 | 222.91 | 520.13 | 668.74 | 2,229.12 |
| 782 | 1,040.26 | 594.43 | 1,188.86 | 743.04 | 3,566.59 |
| 787 | 817.34 | 891.65 | 0 | 371.52 | 2,080.51 |
| TOTAL | 4,161.02 | 2,972.16 | 2,229.12 | 1,783.30 | 11,145.60 |

STATE: VIRGINIA

| ZIP CODE | QUARTER 1 | QUARTER 2 | QUARTER 3 | QUARTER 4 | TOTAL GRAMS |
|---|---|---|---|---|---|
| 235 | 0 | 2,229.12 | 1,634.69 | 2,229.12 | 6,092.93 |
| TOTAL | 0 | 2,229.12 | 1,634.69 | 2,229.12 | 6,092.93 |

STATE: WASHINGTON

| ZIP CODE | QUARTER 1 | QUARTER 2 | QUARTER 3 | QUARTER 4 | TOTAL GRAMS |
|---|---|---|---|---|---|
| 981 | 371.52 | 817.34 | 594.43 | 445.82 | 2,229.11 |
| TOTAL | 371.52 | 817.34 | 594.43 | 445.82 | 2,229.11 |

DRUG CODE: 2100 DRUG NAME: BARBITURIC ACID DERIVIATIVE OR SALT [PER

STATE: ALABAMA

| ZIP CODE | QUARTER 1 | QUARTER 2 | QUARTER 3 | QUARTER 4 | TOTAL GRAMS |
|---|---|---|---|---|---|
| 350 | 250 | 180 | 205 | 150 | 785 |
| 351 | 75 | 80 | 60 | 50 | 265 |
| 352 | 195 | 135 | 100 | 170 | 600 |
| 354 | 60 | 30 | 55 | 70 | 215 |
| 355 | 15 | 35 | 50 | 45 | 145 |
| 356 | 170 | 85 | 95 | 80 | 430 |

| | | | | | |
|---|---|---|---|---|---|
| 357 | 80 | 75 | 5 | 35 | 195 |
| 358 | 100 | 75 | 80 | 55 | 310 |
| 359 | 175 | 135 | 130 | 110 | 550 |
| 360 | 80 | 35 | 10 | 65 | 190 |
| 361 | 30 | 20 | 20 | 15 | 85 |
| 362 | 95 | 65 | 60 | 80 | 300 |
| 363 | 100 | 60 | 75 | 45 | 280 |
| 364 | 75 | 70 | 80 | 50 | 275 |
| 365 | 130 | 125 | 160 | 230 | 645 |
| 366 | 70 | 65 | 70 | 55 | 260 |
| 367 | 15 | 20 | 20 | 20 | 75 |
| 368 | 150 | 125 | 145 | 120 | 540 |
| TOTAL | 1,865 | 1,415 | 1,420 | 1,445 | 6,145 |

STATE: ALASKA

| ZIP CODE | QUARTER 1 | QUARTER 2 | QUARTER 3 | QUARTER 4 | TOTAL GRAMS |
|---|---|---|---|---|---|
| 995 | 60 | 65 | 60 | 55 | 240 |
| 996 | 95 | 60 | 55 | 60 | 270 |
| 998 | 50 | 40 | 40 | 30 | 160 |
| 999 | 20 | 10 | 10 | 5 | 45 |
| TOTAL | 225 | 175 | 165 | 150 | 715 |

STATE: ARIZONA

| ZIP CODE | QUARTER 1 | QUARTER 2 | QUARTER 3 | QUARTER 4 | TOTAL GRAMS |
|---|---|---|---|---|---|
| 850 | 790 | 1,085 | 865 | 750 | 3,490 |
| 851 | 405 | 270 | 300 | 405 | 1,380 |
| 852 | 1,665 | 1,495 | 1,295 | 1,380 | 5,835 |
| 853 | 1,150 | 900 | 780 | 695 | 3,525 |
| 855 | 95 | 90 | 35 | 10 | 230 |
| 856 | 165 | 130 | 75 | 125 | 495 |
| 857 | 910 | 740 | 700 | 645 | 2,995 |
| 859 | 85 | 100 | 105 | 25 | 315 |
| 860 | 25 | 5 | 5 | 20 | 55 |
| 863 | 195 | 205 | 155 | 175 | 730 |
| 864 | 335 | 360 | 215 | 290 | 1,200 |

| | | | | | |
|---|---|---|---|---|---|
| TOTAL | 5,820 | 5,380 | 4,530 | 4,520 | 20,250 |

STATE: ARKANSAS

| ZIP CODE | QUARTER 1 | QUARTER 2 | QUARTER 3 | QUARTER 4 | TOTAL GRAMS |
|---|---|---|---|---|---|
| 716 | 120 | 100 | 160 | 100 | 480 |
| 717 | 70 | 40 | 55 | 55 | 220 |
| 718 | 40 | 45 | 45 | 30 | 160 |
| 719 | 275 | 190 | 95 | 80 | 640 |
| 720 | 255 | 120 | 170 | 180 | 725 |
| 721 | 140 | 180 | 155 | 190 | 665 |
| 722 | 165 | 130 | 90 | 55 | 440 |
| 723 | 55 | 80 | 50 | 95 | 280 |
| 724 | 130 | 110 | 90 | 80 | 410 |
| 725 | 80 | 75 | 75 | 70 | 300 |
| 726 | 40 | 15 | 10 | 5 | 70 |
| 727 | 110 | 95 | 130 | 75 | 410 |
| 728 | 25 | 0 | 10 | 10 | 45 |
| 729 | 135 | 155 | 130 | 60 | 480 |
| TOTAL | 1,640 | 1,335 | 1,265 | 1,085 | 5,325 |

STATE: CALIFORNIA

| ZIP CODE | QUARTER 1 | QUARTER 2 | QUARTER 3 | QUARTER 4 | TOTAL GRAMS |
|---|---|---|---|---|---|
| 900 | 365 | 305 | 360 | 350 | 1,380 |
| 902 | 585 | 585 | 630 | 620 | 2,420 |
| 903 | 40 | 20 | 35 | 35 | 130 |
| 904 | 25 | 5 | 10 | 45 | 85 |
| 905 | 145 | 125 | 60 | 110 | 440 |
| 906 | 305 | 360 | 385 | 330 | 1,380 |
| 907 | 355 | 395 | 370 | 330 | 1,450 |
| 908 | 140 | 235 | 205 | 145 | 725 |
| 910 | 70 | 100 | 155 | 50 | 375 |
| 911 | 65 | 30 | 90 | 25 | 210 |
| 912 | 30 | 20 | 0 | 45 | 95 |
| 913 | 695 | 735 | 835 | 755 | 3,020 |
| 914 | 105 | 175 | 130 | 105 | 515 |

| | | | | | |
|---|---|---|---|---|---|
| 915 | 130 | 65 | 140 | 85 | 420 |
| 916 | 15 | 80 | 55 | 40 | 190 |
| 917 | 505 | 485 | 740 | 435 | 2,165 |
| 918 | 15 | 0 | 10 | 5 | 30 |
| 919 | 40 | 45 | 105 | 55 | 245 |
| 920 | 720 | 545 | 525 | 915 | 2,705 |
| 921 | 315 | 325 | 450 | 290 | 1,380 |
| 922 | 325 | 330 | 405 | 370 | 1,430 |
| 923 | 585 | 430 | 690 | 405 | 2,110 |
| 924 | 45 | 20 | 45 | 25 | 135 |
| 925 | 645 | 500 | 630 | 454 | 2,229 |
| 926 | 905 | 995 | 980 | 944.5 | 3,824.50 |
| 927 | 250 | 220 | 165 | 215 | 850 |
| 928 | 450 | 420 | 660 | 465 | 1,995 |
| 930 | 725 | 740 | 600 | 520 | 2,585 |
| 931 | 80 | 75 | 65 | 50 | 270 |
| 932 | 280 | 275 | 360 | 270 | 1,185 |
| 933 | 275 | 260 | 170 | 150 | 855 |
| 934 | 190 | 195 | 225 | 240 | 850 |
| 935 | 295 | 160 | 250 | 220 | 925 |
| 936 | 260 | 145 | 220 | 235 | 860 |
| 937 | 215 | 170 | 170 | 170 | 725 |
| 939 | 160 | 220 | 290 | 165 | 835 |
| 940 | 170 | 240 | 295 | 210 | 915 |
| 941 | 215 | 115 | 215 | 165 | 710 |
| 943 | 55 | 70 | 95 | 35 | 255 |
| 944 | 25 | 30 | 20 | 15 | 90 |
| 945 | 760 | 650 | 815 | 735 | 2,960 |
| 946 | 105 | 65 | 85 | 45 | 300 |
| 947 | 30 | 40 | 30 | 20 | 120 |
| 948 | 25 | 10 | 20 | 10 | 65 |
| 949 | 160 | 175 | 175 | 135 | 645 |
| 950 | 415 | 350 | 420 | 380 | 1,565 |
| 951 | 270 | 270 | 355 | 300 | 1,195 |
| 952 | 170 | 175 | 140 | 240 | 725 |
| 953 | 570 | 525 | 545 | 500 | 2,140 |
| 954 | 80 | 65 | 165 | 75 | 385 |

| | | | | | | |
|---|---|---|---|---|---|---|
| | 955 | 75 | 60 | 40 | 50 | 225 |
| | 956 | 625 | 530 | 615 | 530 | 2,300 |
| | 957 | 160 | 170 | 175 | 165 | 670 |
| | 958 | 230 | 230 | 170 | 145 | 775 |
| | 959 | 435 | 370 | 350 | 385 | 1,540 |
| | 960 | 100 | 155 | 160 | 105 | 520 |
| | 961 | 20 | 15 | 10 | 15 | 60 |
| TOTAL | | 15,045 | 14,100 | 16,110 | 13,928.50 | 59,183.50 |

STATE: COLORADO

| ZIP CODE | | QUARTER 1 | QUARTER 2 | QUARTER 3 | QUARTER 4 | TOTAL GRAMS |
|---|---|---|---|---|---|---|
| | 800 | 250 | 185 | 95 | 125 | 655 |
| | 801 | 295 | 305 | 220 | 225 | 1,045 |
| | 802 | 230 | 250 | 220 | 235 | 935 |
| | 803 | 35 | 40 | 10 | 25 | 110 |
| | 804 | 75 | 105 | 60 | 60 | 300 |
| | 805 | 220 | 170 | 175 | 170 | 735 |
| | 806 | 100 | 105 | 60 | 40 | 305 |
| | 807 | 70 | 70 | 30 | 30 | 200 |
| | 808 | 85 | 60 | 60 | 80 | 285 |
| | 809 | 320 | 230 | 230 | 135 | 915 |
| | 810 | 110 | 155 | 145 | 105 | 515 |
| | 811 | 5 | 20 | 30 | 20 | 75 |
| | 812 | 50 | 40 | 20 | 15 | 125 |
| | 813 | 30 | 10 | 5 | 10 | 55 |
| | 814 | 20 | 45 | 15 | 30 | 110 |
| | 815 | 20 | 20 | 35 | 5 | 80 |
| | 816 | 60 | 30 | 30 | 20 | 140 |
| TOTAL | | 1,975 | 1,840 | 1,440 | 1,330 | 6,585 |

STATE: CONNECTICUT

| ZIP CODE | | QUARTER 1 | QUARTER 2 | QUARTER 3 | QUARTER 4 | TOTAL GRAMS |
|---|---|---|---|---|---|---|
| | 60 | 495 | 475 | 580 | 395 | 1,945 |
| | 61 | 80 | 75 | 175 | 80 | 410 |
| | 62 | 90 | 90 | 85 | 120 | 385 |

| | | | | | | |
|---|---|---|---|---|---|---|
| | 63 | 145 | 130 | 105 | 155 | 535 |
| | 64 | 580 | 515 | 610 | 560 | 2,265 |
| | 65 | 50 | 95 | 65 | 65 | 275 |
| | 66 | 85 | 90 | 100 | 95 | 370 |
| | 67 | 220 | 195 | 205 | 165 | 785 |
| | 68 | 395 | 390 | 480 | 320 | 1,585 |
| | 69 | 20 | 30 | 45 | 60 | 155 |
| TOTAL | | 2,160 | 2,085 | 2,450 | 2,015 | 8,710 |

STATE: DELAWARE

| ZIP CODE | QUARTER 1 | QUARTER 2 | QUARTER 3 | QUARTER 4 | TOTAL GRAMS |
|---|---|---|---|---|---|
| 197 | 130 | 90 | 100 | 110 | 430 |
| 198 | 170 | 175 | 190 | 200 | 735 |
| 199 | 180 | 115 | 200 | 180 | 675 |
| TOTAL | 480 | 380 | 490 | 490 | 1,840 |

STATE: DISTRICT OF COLUMBIA

| ZIP CODE | QUARTER 1 | QUARTER 2 | QUARTER 3 | QUARTER 4 | TOTAL GRAMS |
|---|---|---|---|---|---|
| 200 | 280 | 180 | 285 | 225 | 970 |
| TOTAL | 280 | 180 | 285 | 225 | 970 |

STATE: FLORIDA

| ZIP CODE | QUARTER 1 | QUARTER 2 | QUARTER 3 | QUARTER 4 | TOTAL GRAMS |
|---|---|---|---|---|---|
| 320 | 710 | 575 | 580 | 595 | 2,460 |
| 321 | 545 | 505 | 435 | 400 | 1,885 |
| 322 | 855 | 725 | 810 | 720 | 3,110 |
| 323 | 515 | 440 | 450 | 470 | 1,875 |
| 324 | 320 | 285 | 205 | 180 | 990 |
| 325 | 550 | 465 | 375 | 395 | 1,785 |
| 326 | 310 | 330 | 255 | 250 | 1,145 |
| 327 | 965 | 715 | 970 | 825 | 3,475 |
| 328 | 635 | 485 | 620 | 515 | 2,255 |
| 329 | 860 | 740 | 850 | 710 | 3,160 |
| 330 | 1,650 | 1,175 | 1,090 | 1,050 | 4,965 |

| | | | | | |
|---|---|---|---|---|---|
| 331 | 785 | 805 | 855 | 775 | 3,220 |
| 333 | 850 | 680 | 710 | 560 | 2,800 |
| 334 | 1,980 | 1,620 | 1,755 | 1,680 | 7,035 |
| 335 | 965 | 690 | 820 | 830 | 3,305 |
| 336 | 845 | 735 | 835 | 715 | 3,130 |
| 337 | 950 | 860 | 1,025 | 955 | 3,790 |
| 338 | 770 | 495 | 445 | 500 | 2,210 |
| 339 | 1,255 | 645 | 715 | 785 | 3,400 |
| 341 | 390 | 305 | 395 | 295 | 1,385 |
| 342 | 785 | 660 | 725 | 670 | 2,840 |
| 344 | 720 | 435 | 695 | 495 | 2,345 |
| 346 | 875 | 945 | 850 | 875 | 3,545 |
| 347 | 450 | 410 | 445 | 435 | 1,740 |
| 349 | 600 | 415 | 535 | 445 | 1,995 |
| TOTAL | 20,135 | 16,140 | 17,445 | 16,125 | 69,845 |

STATE: GEORGIA

| ZIP CODE | QUARTER 1 | QUARTER 2 | QUARTER 3 | QUARTER 4 | TOTAL GRAMS |
|---|---|---|---|---|---|
| 300 | 1,495 | 1,400 | 1,065 | 945 | 4,905 |
| 301 | 785 | 760 | 575 | 625 | 2,745 |
| 302 | 580 | 550 | 550 | 410 | 2,090 |
| 303 | 435 | 410 | 255 | 255 | 1,355 |
| 304 | 240 | 280 | 185 | 85 | 790 |
| 305 | 260 | 330 | 140 | 160 | 890 |
| 306 | 215 | 255 | 200 | 155 | 825 |
| 307 | 180 | 145 | 130 | 50 | 505 |
| 308 | 140 | 135 | 150 | 95 | 520 |
| 309 | 195 | 210 | 205 | 115 | 725 |
| 310 | 310 | 235 | 240 | 160 | 945 |
| 312 | 130 | 65 | 60 | 65 | 320 |
| 313 | 105 | 100 | 100 | 60 | 365 |
| 314 | 165 | 145 | 65 | 85 | 460 |
| 315 | 590 | 605 | 405 | 325 | 1,925 |
| 316 | 110 | 100 | 80 | 70 | 360 |
| 317 | 230 | 165 | 255 | 230 | 880 |
| 318 | 0 | 5 | 0 | 5 | 10 |

| | | | | | |
|---|---|---|---|---|---|
| | 319 | 85 | 45 | 65 | 55 | 250 |
| | 398 | 35 | 35 | 50 | 40 | 160 |
| TOTAL | | 6,285 | 5,975 | 4,775 | 3,990 | 21,025 |

STATE: GUAM

| ZIP CODE | QUARTER 1 | QUARTER 2 | QUARTER 3 | QUARTER 4 | TOTAL GRAMS |
|---|---|---|---|---|---|
| 969 | 0 | 10 | 0 | 0 | 10 |
| TOTAL | 0 | 10 | 0 | 0 | 10 |

STATE: HAWAII

| ZIP CODE | QUARTER 1 | QUARTER 2 | QUARTER 3 | QUARTER 4 | TOTAL GRAMS |
|---|---|---|---|---|---|
| 967 | 450 | 550 | 610 | 425 | 2,035 |
| 968 | 320 | 380 | 330 | 315 | 1,345 |
| TOTAL | 770 | 930 | 940 | 740 | 3,380 |

STATE: IDAHO

| ZIP CODE | QUARTER 1 | QUARTER 2 | QUARTER 3 | QUARTER 4 | TOTAL GRAMS |
|---|---|---|---|---|---|
| 832 | 60 | 40 | 25 | 30 | 155 |
| 833 | 115 | 140 | 40 | 25 | 320 |
| 834 | 135 | 130 | 120 | 105 | 490 |
| 835 | 65 | 90 | 40 | 55 | 250 |
| 836 | 125 | 125 | 90 | 65 | 405 |
| 837 | 85 | 85 | 110 | 130 | 410 |
| 838 | 55 | 35 | 20 | 25 | 135 |
| TOTAL | 640 | 645 | 445 | 435 | 2,165 |

STATE: ILLINOIS

| ZIP CODE | QUARTER 1 | QUARTER 2 | QUARTER 3 | QUARTER 4 | TOTAL GRAMS |
|---|---|---|---|---|---|
| 600 | 1,675 | 1,655 | 1,815 | 1,675 | 6,820 |
| 601 | 950 | 1,140 | 1,180 | 1,180 | 4,450 |
| 602 | 40 | 20 | 55 | 40 | 155 |
| 603 | 20 | 20 | 10 | 30 | 80 |
| 604 | 1,135 | 1,040 | 1,095 | 1,100 | 4,370 |

| | | | | | | |
|---|---|---|---|---|---|---|
| | 605 | 755 | 650 | 910 | 825 | 3,140 |
| | 606 | 730 | 665 | 760 | 735 | 2,890 |
| | 607 | 55 | 65 | 85 | 55 | 260 |
| | 608 | 15 | 0 | 5 | 35 | 55 |
| | 609 | 100 | 65 | 70 | 115 | 350 |
| | 610 | 100 | 115 | 95 | 70 | 380 |
| | 611 | 125 | 95 | 135 | 160 | 515 |
| | 612 | 15 | 30 | 40 | 40 | 125 |
| | 613 | 45 | 40 | 40 | 30 | 155 |
| | 614 | 65 | 60 | 75 | 80 | 280 |
| | 615 | 105 | 90 | 110 | 120 | 425 |
| | 616 | 100 | 105 | 155 | 115 | 475 |
| | 617 | 140 | 125 | 135 | 125 | 525 |
| | 618 | 135 | 215 | 295 | 240 | 885 |
| | 619 | 10 | 10 | 5 | 5 | 30 |
| | 620 | 190 | 225 | 220 | 170 | 805 |
| | 622 | 190 | 175 | 170 | 145 | 680 |
| | 623 | 25 | 30 | 35 | 65 | 155 |
| | 624 | 15 | 20 | 30 | 10 | 75 |
| | 625 | 55 | 110 | 70 | 55 | 290 |
| | 626 | 65 | 45 | 80 | 75 | 265 |
| | 627 | 170 | 185 | 155 | 165 | 675 |
| | 628 | 80 | 110 | 55 | 55 | 300 |
| | 629 | 250 | 185 | 90 | 100 | 625 |
| TOTAL | | 7,355 | 7,290 | 7,975 | 7,615 | 30,235 |

STATE: INDIANA

| ZIP CODE | | QUARTER 1 | QUARTER 2 | QUARTER 3 | QUARTER 4 | TOTAL GRAMS |
|---|---|---|---|---|---|---|
| | 460 | 270 | 195 | 185 | 160 | 810 |
| | 461 | 385 | 315 | 300 | 285 | 1,285 |
| | 462 | 440 | 435 | 395 | 295 | 1,565 |
| | 463 | 600 | 595 | 635 | 580 | 2,410 |
| | 464 | 100 | 75 | 95 | 105 | 375 |
| | 465 | 120 | 165 | 155 | 155 | 595 |
| | 466 | 15 | 30 | 30 | 60 | 135 |
| | 467 | 65 | 100 | 75 | 75 | 315 |

| | | | | | | |
|---|---|---|---|---|---|---|
| | 468 | 55 | 55 | 100 | 75 | 285 |
| | 469 | 120 | 110 | 80 | 70 | 380 |
| | 470 | 80 | 65 | 55 | 80 | 280 |
| | 471 | 180 | 225 | 225 | 205 | 835 |
| | 472 | 95 | 50 | 80 | 80 | 305 |
| | 473 | 90 | 150 | 85 | 105 | 430 |
| | 474 | 90 | 85 | 115 | 75 | 365 |
| | 475 | 160 | 165 | 205 | 175 | 705 |
| | 476 | 95 | 125 | 85 | 115 | 420 |
| | 477 | 310 | 190 | 230 | 260 | 990 |
| | 478 | 50 | 75 | 50 | 60 | 235 |
| | 479 | 105 | 95 | 135 | 105 | 440 |
| TOTAL | | 3,425 | 3,300 | 3,315 | 3,120 | 13,160 |

STATE: IOWA

| ZIP CODE | QUARTER 1 | QUARTER 2 | QUARTER 3 | QUARTER 4 | TOTAL GRAMS |
|---|---|---|---|---|---|
| 500 | 50 | 50 | 50 | 60 | 210 |
| 501 | 30 | 25 | 25 | 5 | 85 |
| 502 | 45 | 35 | 25 | 20 | 125 |
| 503 | 50 | 70 | 90 | 100 | 310 |
| 504 | 5 | 15 | 5 | 0 | 25 |
| 505 | 5 | 5 | 15 | 15 | 40 |
| 506 | 10 | 20 | 10 | 15 | 55 |
| 507 | 0 | 10 | 15 | 5 | 30 |
| 508 | 15 | 30 | 25 | 20 | 90 |
| 510 | 5 | 0 | 5 | 0 | 10 |
| 511 | 60 | 80 | 35 | 60 | 235 |
| 512 | 5 | 0 | 0 | 0 | 5 |
| 513 | 5 | 0 | 0 | 5 | 10 |
| 514 | 20 | 5 | 5 | 0 | 30 |
| 515 | 80 | 65 | 80 | 80 | 305 |
| 520 | 105 | 80 | 80 | 130 | 395 |
| 522 | 25 | 10 | 0 | 10 | 45 |
| 523 | 10 | 10 | 25 | 15 | 60 |
| 524 | 20 | 15 | 15 | 20 | 70 |
| 525 | 0 | 5 | 5 | 5 | 15 |

| | | | | | | |
|---|---|---|---|---|---|---|
| | 526 | 45 | 25 | 30 | 15 | 115 |
| | 527 | 40 | 10 | 10 | 15 | 75 |
| | 528 | 90 | 110 | 100 | 140 | 440 |
| TOTAL | | 720 | 675 | 650 | 735 | 2,780 |

| STATE: | KANSAS | | | | | |
|---|---|---|---|---|---|---|
| ZIP CODE | | QUARTER 1 | QUARTER 2 | QUARTER 3 | QUARTER 4 | TOTAL GRAMS |
| | 660 | 140 | 160 | 110 | 70 | 480 |
| | 661 | 140 | 80 | 95 | 95 | 410 |
| | 662 | 355 | 515 | 320 | 315 | 1,505 |
| | 664 | 0 | 0 | 0 | 5 | 5 |
| | 665 | 25 | 20 | 20 | 10 | 75 |
| | 666 | 50 | 50 | 95 | 90 | 285 |
| | 667 | 35 | 55 | 60 | 60 | 210 |
| | 668 | 45 | 30 | 15 | 10 | 100 |
| | 670 | 105 | 115 | 105 | 95 | 420 |
| | 671 | 25 | 25 | 15 | 5 | 70 |
| | 672 | 250 | 285 | 140 | 155 | 830 |
| | 673 | 15 | 20 | 0 | 0 | 35 |
| | 674 | 70 | 55 | 25 | 35 | 185 |
| | 675 | 10 | 25 | 10 | 5 | 50 |
| | 676 | 0 | 0 | 0 | 5 | 5 |
| | 677 | 5 | 5 | 0 | 0 | 10 |
| | 678 | 35 | 40 | 20 | 20 | 115 |
| | 679 | 0 | 10 | 0 | 20 | 30 |
| TOTAL | | 1,305 | 1,490 | 1,030 | 995 | 4,820 |

| STATE: | KENTUCKY | | | | | |
|---|---|---|---|---|---|---|
| ZIP CODE | | QUARTER 1 | QUARTER 2 | QUARTER 3 | QUARTER 4 | TOTAL GRAMS |
| | 400 | 135 | 170 | 85 | 75 | 465 |
| | 401 | 75 | 35 | 40 | 60 | 210 |
| | 402 | 640 | 535 | 505 | 410 | 2,090 |
| | 403 | 100 | 105 | 60 | 55 | 320 |
| | 404 | 40 | 30 | 25 | 30 | 125 |
| | 405 | 85 | 20 | 35 | 40 | 180 |

| | | | | | | |
|---|---|---|---|---|---|---|
| | 406 | 10 | 0 | 10 | 0 | 20 |
| | 407 | 65 | 60 | 55 | 65 | 245 |
| | 409 | 35 | 50 | 40 | 30 | 155 |
| | 410 | 325 | 325 | 255 | 225 | 1,130 |
| | 411 | 20 | 15 | 0 | 5 | 40 |
| | 412 | 10 | 5 | 0 | 5 | 20 |
| | 415 | 10 | 5 | 0 | 0 | 15 |
| | 417 | 5 | 0 | 0 | 0 | 5 |
| | 420 | 130 | 100 | 135 | 125 | 490 |
| | 421 | 105 | 120 | 120 | 70 | 415 |
| | 422 | 55 | 90 | 70 | 85 | 300 |
| | 423 | 40 | 50 | 60 | 40 | 190 |
| | 424 | 25 | 65 | 85 | 15 | 190 |
| | 425 | 10 | 35 | 25 | 30 | 100 |
| | 426 | 30 | 15 | 30 | 30 | 105 |
| | 427 | 110 | 165 | 104 | 70 | 449 |
| TOTAL | | 2,060 | 1,995 | 1,739 | 1,465 | 7,259 |

STATE: LOUISIANA

| ZIP CODE | | QUARTER 1 | QUARTER 2 | QUARTER 3 | QUARTER 4 | TOTAL GRAMS |
|---|---|---|---|---|---|---|
| | 700 | 745 | 845 | 780 | 610 | 2,980 |
| | 701 | 270 | 185 | 255 | 180 | 890 |
| | 703 | 95 | 70 | 75 | 70 | 310 |
| | 704 | 590 | 540 | 620 | 435 | 2,185 |
| | 705 | 410 | 360 | 420 | 430 | 1,620 |
| | 706 | 330 | 270 | 310 | 335 | 1,245 |
| | 707 | 150 | 185 | 210 | 145 | 690 |
| | 708 | 140 | 160 | 180 | 160 | 640 |
| | 710 | 30 | 60 | 95 | 15 | 200 |
| | 711 | 65 | 55 | 165 | 105 | 390 |
| | 712 | 205 | 175 | 195 | 110 | 685 |
| | 713 | 245 | 310 | 340 | 265 | 1,160 |
| | 714 | 95 | 105 | 140 | 90 | 430 |
| TOTAL | | 3,370 | 3,320 | 3,785 | 2,950 | 13,425 |

STATE: MAINE

| ZIP CODE | QUARTER 1 | QUARTER 2 | QUARTER 3 | QUARTER 4 | TOTAL GRAMS |
|---|---|---|---|---|---|
| 39 | 5 | 0 | 0 | 0 | 5 |
| 40 | 175 | 150 | 200 | 210 | 735 |
| 41 | 65 | 75 | 60 | 80 | 280 |
| 42 | 35 | 15 | 20 | 10 | 80 |
| 43 | 25 | 20 | 25 | 15 | 85 |
| 44 | 60 | 15 | 5 | 35 | 115 |
| 45 | 10 | 20 | 10 | 0 | 40 |
| 46 | 10 | 0 | 0 | 5 | 15 |
| 47 | 30 | 15 | 10 | 25 | 80 |
| 49 | 65 | 65 | 75 | 50 | 255 |
| TOTAL | 480 | 375 | 405 | 430 | 1,690 |

STATE: MARYLAND

| ZIP CODE | QUARTER 1 | QUARTER 2 | QUARTER 3 | QUARTER 4 | TOTAL GRAMS |
|---|---|---|---|---|---|
| 206 | 340 | 275 | 320 | 300 | 1,235 |
| 207 | 280 | 265 | 220 | 175 | 940 |
| 208 | 260 | 220 | 350 | 245 | 1,075 |
| 209 | 165 | 45 | 95 | 105 | 410 |
| 210 | 325 | 375 | 340 | 265 | 1,305 |
| 211 | 110 | 115 | 85 | 155 | 465 |
| 212 | 420 | 315 | 370 | 285 | 1,390 |
| 214 | 80 | 110 | 100 | 70 | 360 |
| 215 | 90 | 90 | 60 | 80 | 320 |
| 216 | 90 | 50 | 75 | 100 | 315 |
| 217 | 315 | 345 | 400 | 275 | 1,335 |
| 218 | 95 | 35 | 65 | 30 | 225 |
| 219 | 40 | 60 | 40 | 25 | 165 |
| TOTAL | 2,610 | 2,300 | 2,520 | 2,110 | 9,540 |

STATE: MASSACHUSETTS

| ZIP CODE | QUARTER 1 | QUARTER 2 | QUARTER 3 | QUARTER 4 | TOTAL GRAMS |
|---|---|---|---|---|---|
| 10 | 240 | 195 | 255 | 145 | 835 |
| 11 | 35 | 15 | 30 | 20 | 100 |

| | | | | | | |
|---|---|---|---|---|---|---|
| | 12 | 10 | 30 | 45 | 15 | 100 |
| | 13 | 10 | 0 | 35 | 15 | 60 |
| | 14 | 180 | 140 | 225 | 75 | 620 |
| | 15 | 200 | 155 | 180 | 155 | 690 |
| | 16 | 190 | 185 | 295 | 120 | 790 |
| | 17 | 190 | 185 | 215 | 225 | 815 |
| | 18 | 250 | 260 | 255 | 255 | 1,020 |
| | 19 | 225 | 275 | 275 | 300 | 1,075 |
| | 20 | 135 | 190 | 155 | 165 | 645 |
| | 21 | 365 | 355 | 465 | 340 | 1,525 |
| | 22 | 10 | 5 | 10 | 5 | 30 |
| | 23 | 130 | 145 | 200 | 180 | 655 |
| | 24 | 155 | 165 | 195 | 225 | 740 |
| | 25 | 35 | 25 | 10 | 25 | 95 |
| | 26 | 85 | 65 | 70 | 65 | 285 |
| | 27 | 180 | 165 | 270 | 125 | 740 |
| TOTAL | | 2,625 | 2,555 | 3,185 | 2,455 | 10,820 |

STATE: MICHIGAN

| ZIP CODE | | QUARTER 1 | QUARTER 2 | QUARTER 3 | QUARTER 4 | TOTAL GRAMS |
|---|---|---|---|---|---|---|
| | 480 | 1,590 | 1,685 | 1,615 | 1,400 | 6,290 |
| | 481 | 1,270 | 1,475 | 1,470 | 1,490 | 5,705 |
| | 482 | 225 | 325 | 350 | 285 | 1,185 |
| | 483 | 980 | 1,020 | 1,080 | 920 | 4,000 |
| | 484 | 300 | 435 | 200 | 210 | 1,145 |
| | 485 | 45 | 60 | 135 | 85 | 325 |
| | 486 | 130 | 145 | 150 | 145 | 570 |
| | 487 | 100 | 90 | 70 | 70 | 330 |
| | 488 | 205 | 400 | 385 | 405 | 1,395 |
| | 489 | 165 | 140 | 125 | 130 | 560 |
| | 490 | 260 | 310 | 385 | 300 | 1,255 |
| | 491 | 15 | 15 | 10 | 15 | 55 |
| | 492 | 220 | 180 | 205 | 210 | 815 |
| | 493 | 55 | 50 | 35 | 70 | 210 |
| | 494 | 275 | 385 | 400 | 455 | 1,515 |
| | 495 | 130 | 205 | 220 | 215 | 770 |

| | | | | | |
|---|---|---|---|---|---|
| 496 | 30 | 60 | 50 | 60 | 200 |
| 497 | 45 | 65 | 145 | 25 | 280 |
| 498 | 95 | 65 | 50 | 35 | 245 |
| 499 | 5 | 15 | 10 | 30 | 60 |
| TOTAL | 6,140 | 7,125 | 7,090 | 6,555 | 26,910 |

STATE: MINNESOTA

| ZIP CODE | QUARTER 1 | QUARTER 2 | QUARTER 3 | QUARTER 4 | TOTAL GRAMS |
|---|---|---|---|---|---|
| 550 | 170 | 130 | 165 | 125 | 590 |
| 551 | 260 | 280 | 280 | 235 | 1,055 |
| 553 | 310 | 265 | 280 | 265 | 1,120 |
| 554 | 195 | 220 | 210 | 225 | 850 |
| 556 | 0 | 0 | 5 | 0 | 5 |
| 557 | 95 | 100 | 85 | 100 | 380 |
| 558 | 25 | 30 | 25 | 50 | 130 |
| 559 | 0 | 5 | 0 | 5 | 10 |
| 560 | 70 | 70 | 40 | 45 | 225 |
| 561 | 15 | 15 | 15 | 25 | 70 |
| 562 | 60 | 20 | 70 | 55 | 205 |
| 563 | 85 | 55 | 45 | 55 | 240 |
| 564 | 50 | 80 | 60 | 15 | 205 |
| 565 | 30 | 25 | 25 | 35 | 115 |
| 566 | 25 | 20 | 35 | 25 | 105 |
| 567 | 0 | 5 | 5 | 0 | 10 |
| TOTAL | 1,390 | 1,320 | 1,345 | 1,260 | 5,315 |

STATE: MISSISSIPPI

| ZIP CODE | QUARTER 1 | QUARTER 2 | QUARTER 3 | QUARTER 4 | TOTAL GRAMS |
|---|---|---|---|---|---|
| 386 | 155 | 130 | 135 | 65 | 485 |
| 387 | 10 | 5 | 5 | 15 | 35 |
| 388 | 125 | 95 | 60 | 75 | 355 |
| 389 | 50 | 30 | 55 | 40 | 175 |
| 390 | 55 | 80 | 35 | 10 | 180 |
| 391 | 130 | 25 | 90 | 50 | 295 |
| 392 | 45 | 45 | 35 | 45 | 170 |

| | | | | | | |
|---|---|---|---|---|---|---|
| | 393 | 35 | 25 | 15 | 40 | 115 |
| | 394 | 215 | 195 | 250 | 105 | 765 |
| | 395 | 275 | 320 | 310 | 200 | 1,105 |
| | 396 | 65 | 50 | 45 | 30 | 190 |
| | 397 | 40 | 50 | 45 | 15 | 150 |
| TOTAL | | 1,200 | 1,050 | 1,080 | 690 | 4,020 |

STATE: MISSOURI

| ZIP CODE | | QUARTER 1 | QUARTER 2 | QUARTER 3 | QUARTER 4 | TOTAL GRAMS |
|---|---|---|---|---|---|---|
| | 630 | 660 | 590 | 725 | 570 | 2,545 |
| | 631 | 1,480 | 1,850 | 755 | 570 | 4,655 |
| | 633 | 625 | 520 | 615 | 400 | 2,160 |
| | 634 | 5 | 0 | 0 | 5 | 10 |
| | 635 | 20 | 10 | 15 | 15 | 60 |
| | 636 | 60 | 20 | 45 | 35 | 160 |
| | 637 | 75 | 65 | 75 | 75 | 290 |
| | 638 | 40 | 65 | 65 | 40 | 210 |
| | 639 | 85 | 95 | 65 | 80 | 325 |
| | 640 | 240 | 245 | 315 | 320 | 1,120 |
| | 641 | 385 | 465 | 460 | 580 | 1,890 |
| | 644 | 10 | 15 | 15 | 5 | 45 |
| | 645 | 10 | 30 | 60 | 45 | 145 |
| | 647 | 5 | 5 | 15 | 10 | 35 |
| | 648 | 65 | 100 | 95 | 75 | 335 |
| | 650 | 70 | 45 | 65 | 35 | 215 |
| | 651 | 55 | 55 | 75 | 60 | 245 |
| | 652 | 10 | 15 | 5 | 10 | 40 |
| | 653 | 0 | 0 | 15 | 10 | 25 |
| | 654 | 10 | 5 | 0 | 5 | 20 |
| | 655 | 0 | 15 | 5 | 10 | 30 |
| | 656 | 55 | 50 | 75 | 35 | 215 |
| | 657 | 70 | 115 | 155 | 110 | 450 |
| | 658 | 145 | 140 | 195 | 115 | 595 |
| TOTAL | | 4,180 | 4,515 | 3,910 | 3,215 | 15,820 |

STATE: MONTANA

| ZIP CODE | QUARTER 1 | QUARTER 2 | QUARTER 3 | QUARTER 4 | TOTAL GRAMS |
|---|---|---|---|---|---|
| 590 | 25 | 25 | 20 | 10 | 80 |
| 591 | 50 | 75 | 75 | 60 | 260 |
| 592 | 5 | 10 | 0 | 10 | 25 |
| 593 | 40 | 45 | 30 | 5 | 120 |
| 594 | 30 | 30 | 95 | 50 | 205 |
| 595 | 5 | 10 | 0 | 0 | 15 |
| 596 | 15 | 40 | 25 | 20 | 100 |
| 597 | 105 | 100 | 95 | 60 | 360 |
| 598 | 115 | 125 | 185 | 140 | 565 |
| 599 | 100 | 80 | 65 | 20 | 265 |
| TOTAL | 490 | 540 | 590 | 375 | 1,995 |

STATE: NEBRASKA

| ZIP CODE | QUARTER 1 | QUARTER 2 | QUARTER 3 | QUARTER 4 | TOTAL GRAMS |
|---|---|---|---|---|---|
| 680 | 100 | 75 | 75 | 110 | 360 |
| 681 | 305 | 285 | 340 | 310 | 1,240 |
| 684 | 15 | 15 | 25 | 20 | 75 |
| 685 | 120 | 160 | 155 | 140 | 575 |
| 686 | 20 | 40 | 40 | 20 | 120 |
| 687 | 25 | 20 | 50 | 45 | 140 |
| 688 | 35 | 25 | 20 | 20 | 100 |
| 689 | 10 | 5 | 15 | 10 | 40 |
| 691 | 10 | 0 | 5 | 15 | 30 |
| 692 | 0 | 0 | 15 | 0 | 15 |
| 693 | 75 | 80 | 60 | 60 | 275 |
| TOTAL | 715 | 705 | 800 | 750 | 2,970 |

STATE: NEVADA

| ZIP CODE | QUARTER 1 | QUARTER 2 | QUARTER 3 | QUARTER 4 | TOTAL GRAMS |
|---|---|---|---|---|---|
| 890 | 1,410 | 1,490 | 1,175 | 1,415 | 5,490 |
| 891 | 2,260 | 2,115 | 1,795 | 1,925 | 8,095 |
| 893 | 15 | 70 | 35 | 35 | 155 |

| | | | | | | |
|---|---|---|---|---|---|---|
| | 894 | 475 | 405 | 375 | 315 | 1,570 |
| | 895 | 535 | 470 | 510 | 315 | 1,830 |
| | 897 | 85 | 90 | 110 | 75 | 360 |
| | 898 | 240 | 195 | 195 | 145 | 775 |
| TOTAL | | 5,020 | 4,835 | 4,195 | 4,225 | 18,275 |

STATE: NEW HAMPSHIRE

| ZIP CODE | QUARTER 1 | QUARTER 2 | QUARTER 3 | QUARTER 4 | TOTAL GRAMS |
|---|---|---|---|---|---|
| 30 | 120 | 180 | 175 | 210 | 685 |
| 31 | 145 | 150 | 110 | 125 | 530 |
| 32 | 40 | 30 | 25 | 30 | 125 |
| 33 | 0 | 5 | 5 | 5 | 15 |
| 34 | 10 | 5 | 20 | 10 | 45 |
| 36 | 10 | 10 | 10 | 10 | 40 |
| 37 | 0 | 0 | 5 | 0 | 5 |
| 38 | 160 | 130 | 125 | 135 | 550 |
| TOTAL | 485 | 510 | 475 | 525 | 1,995 |

STATE: NEW JERSEY

| ZIP CODE | QUARTER 1 | QUARTER 2 | QUARTER 3 | QUARTER 4 | TOTAL GRAMS |
|---|---|---|---|---|---|
| 70 | 640 | 690 | 855 | 730 | 2,915 |
| 71 | 30 | 60 | 35 | 35 | 160 |
| 72 | 105 | 30 | 60 | 45 | 240 |
| 73 | 65 | 50 | 55 | 35 | 205 |
| 74 | 645 | 590 | 745 | 560 | 2,540 |
| 75 | 40 | 35 | 40 | 50 | 165 |
| 76 | 325 | 260 | 345 | 365 | 1,295 |
| 77 | 515 | 385 | 535 | 410 | 1,845 |
| 78 | 215 | 205 | 230 | 185 | 835 |
| 79 | 215 | 105 | 120 | 160 | 600 |
| 80 | 645 | 650 | 620 | 620 | 2,535 |
| 81 | 45 | 40 | 10 | 35 | 130 |
| 82 | 140 | 180 | 320 | 170 | 810 |
| 83 | 130 | 100 | 110 | 90 | 430 |
| 84 | 10 | 10 | 10 | 5 | 35 |

| | | | | | | |
|---|---|---|---|---|---|---|
| | 85 | 70 | 45 | 75 | 40 | 230 |
| | 86 | 60 | 50 | 55 | 70 | 235 |
| | 87 | 210 | 300 | 385 | 215 | 1,110 |
| | 88 | 425 | 400 | 435 | 400 | 1,660 |
| | 89 | 50 | 20 | 25 | 40 | 135 |
| TOTAL | | 4,580 | 4,205 | 5,065 | 4,260 | 18,110 |

STATE: NEW MEXICO

| ZIP CODE | | QUARTER 1 | QUARTER 2 | QUARTER 3 | QUARTER 4 | TOTAL GRAMS |
|---|---|---|---|---|---|---|
| | 870 | 100 | 105 | 80 | 90 | 375 |
| | 871 | 320 | 225 | 325 | 265 | 1,135 |
| | 873 | 0 | 0 | 0 | 5 | 5 |
| | 874 | 35 | 0 | 10 | 0 | 45 |
| | 875 | 90 | 115 | 115 | 95 | 415 |
| | 877 | 15 | 5 | 20 | 25 | 65 |
| | 879 | 5 | 5 | 15 | 35 | 60 |
| | 880 | 130 | 185 | 130 | 130 | 575 |
| | 881 | 10 | 10 | 15 | 0 | 35 |
| | 882 | 95 | 145 | 90 | 95 | 425 |
| | 883 | 35 | 40 | 55 | 45 | 175 |
| | 884 | 5 | 0 | 5 | 5 | 15 |
| TOTAL | | 840 | 835 | 860 | 790 | 3,325 |

STATE: NEW YORK

| ZIP CODE | | QUARTER 1 | QUARTER 2 | QUARTER 3 | QUARTER 4 | TOTAL GRAMS |
|---|---|---|---|---|---|---|
| | 100 | 450 | 300 | 385 | 315 | 1,450 |
| | 101 | 60 | 45 | 30 | 20 | 155 |
| | 103 | 200 | 130 | 185 | 130 | 645 |
| | 104 | 210 | 180 | 180 | 205 | 775 |
| | 105 | 210 | 190 | 130 | 175 | 705 |
| | 106 | 40 | 50 | 30 | 65 | 185 |
| | 107 | 110 | 110 | 60 | 40 | 320 |
| | 108 | 75 | 60 | 50 | 20 | 205 |
| | 109 | 315 | 175 | 255 | 265 | 1,010 |
| | 110 | 140 | 90 | 70 | 55 | 355 |

| | | | | | |
|---|---|---|---|---|---|
| 111 | 35 | 60 | 40 | 0 | 135 |
| 112 | 365 | 450 | 360 | 340 | 1,515 |
| 113 | 255 | 295 | 170 | 180 | 900 |
| 114 | 110 | 100 | 155 | 105 | 470 |
| 115 | 300 | 355 | 315 | 285 | 1,255 |
| 116 | 25 | 25 | 30 | 10 | 90 |
| 117 | 615 | 635 | 545 | 430 | 2,225 |
| 118 | 35 | 50 | 60 | 40 | 185 |
| 119 | 70 | 60 | 65 | 125 | 320 |
| 120 | 35 | 80 | 100 | 40 | 255 |
| 121 | 25 | 10 | 40 | 30 | 105 |
| 122 | 20 | 15 | 15 | 60 | 110 |
| 123 | 115 | 95 | 130 | 95 | 435 |
| 124 | 85 | 75 | 85 | 80 | 325 |
| 125 | 65 | 50 | 75 | 125 | 315 |
| 126 | 40 | 35 | 65 | 30 | 170 |
| 127 | 15 | 0 | 35 | 30 | 80 |
| 128 | 85 | 65 | 40 | 65 | 255 |
| 130 | 5 | 5 | 5 | 15 | 30 |
| 131 | 35 | 35 | 30 | 30 | 130 |
| 132 | 5 | 0 | 5 | 0 | 10 |
| 133 | 45 | 35 | 25 | 40 | 145 |
| 134 | 15 | 5 | 15 | 0 | 35 |
| 135 | 0 | 5 | 0 | 0 | 5 |
| 136 | 0 | 10 | 20 | 15 | 45 |
| 137 | 35 | 35 | 20 | 30 | 120 |
| 138 | 70 | 25 | 60 | 70 | 225 |
| 139 | 45 | 35 | 20 | 30 | 130 |
| 140 | 5 | 15 | 15 | 80 | 115 |
| 141 | 30 | 30 | 45 | 30 | 135 |
| 142 | 120 | 90 | 120 | 130 | 460 |
| 143 | 0 | 10 | 5 | 10 | 25 |
| 144 | 75 | 85 | 70 | 30 | 260 |
| 145 | 10 | 60 | 105 | 50 | 225 |
| 146 | 80 | 85 | 80 | 70 | 315 |
| 147 | 20 | 25 | 50 | 50 | 145 |
| 148 | 10 | 10 | 10 | 15 | 45 |

| | | | | | | |
|---|---|---|---|---|---|---|
| | 149 | 10 | 5 | 10 | 10 | 35 |
| TOTAL | | 4,720 | 4,390 | 4,410 | 4,065 | 17,585 |

| STATE: | NORTH CAROLINA | | | | | |
|---|---|---|---|---|---|---|
| ZIP CODE | | QUARTER 1 | QUARTER 2 | QUARTER 3 | QUARTER 4 | TOTAL GRAMS |
| | 270 | 75 | 110 | 90 | 85 | 360 |
| | 271 | 90 | 100 | 115 | 105 | 410 |
| | 272 | 300 | 220 | 325 | 225 | 1,070 |
| | 273 | 90 | 90 | 110 | 120 | 410 |
| | 274 | 45 | 20 | 45 | 95 | 205 |
| | 275 | 305 | 315 | 450 | 325 | 1,395 |
| | 276 | 185 | 205 | 200 | 205 | 795 |
| | 277 | 130 | 80 | 60 | 40 | 310 |
| | 278 | 185 | 165 | 200 | 130 | 680 |
| | 279 | 90 | 75 | 130 | 90 | 385 |
| | 280 | 240 | 340 | 285 | 195 | 1,060 |
| | 281 | 310 | 190 | 325 | 210 | 1,035 |
| | 282 | 265 | 285 | 260 | 265 | 1,075 |
| | 283 | 405 | 355 | 310 | 340 | 1,410 |
| | 284 | 320 | 330 | 350 | 305 | 1,305 |
| | 285 | 230 | 240 | 240 | 245 | 955 |
| | 286 | 245 | 250 | 205 | 310 | 1,010 |
| | 287 | 200 | 205 | 240 | 160 | 805 |
| | 288 | 80 | 100 | 105 | 105 | 390 |
| | 289 | 85 | 40 | 40 | 35 | 200 |
| TOTAL | | 3,875 | 3,715 | 4,085 | 3,590 | 15,265 |

| STATE: | NORTH DAKOTA | | | | | |
|---|---|---|---|---|---|---|
| ZIP CODE | | QUARTER 1 | QUARTER 2 | QUARTER 3 | QUARTER 4 | TOTAL GRAMS |
| | 580 | 0 | 5 | 0 | 0 | 5 |
| | 581 | 20 | 0 | 5 | 5 | 30 |
| | 582 | 25 | 25 | 5 | 15 | 70 |
| | 584 | 0 | 5 | 0 | 0 | 5 |
| | 585 | 35 | 50 | 55 | 40 | 180 |
| | 586 | 15 | 10 | 10 | 5 | 40 |

| | | | | | | |
|---|---|---|---|---|---|---|
| | 587 | 0 | 10 | 5 | 10 | 25 |
| | 588 | 75 | 65 | 45 | 25 | 210 |
| TOTAL | | 170 | 170 | 125 | 100 | 565 |

| STATE: | OHIO | | | | |
|---|---|---|---|---|---|
| ZIP CODE | QUARTER 1 | QUARTER 2 | QUARTER 3 | QUARTER 4 | TOTAL GRAMS |
| 430 | 375 | 330 | 220 | 200 | 1,125 |
| 431 | 295 | 260 | 175 | 150 | 880 |
| 432 | 1,285 | 1,455 | 285 | 270 | 3,295 |
| 433 | 30 | 35 | 25 | 5 | 95 |
| 434 | 75 | 65 | 50 | 55 | 245 |
| 435 | 195 | 210 | 45 | 30 | 480 |
| 436 | 270 | 255 | 140 | 170 | 835 |
| 437 | 30 | 20 | 10 | 5 | 65 |
| 438 | 15 | 0 | 5 | 0 | 20 |
| 439 | 25 | 20 | 45 | 30 | 120 |
| 440 | 245 | 275 | 320 | 200 | 1,040 |
| 441 | 460 | 400 | 410 | 235 | 1,505 |
| 442 | 175 | 110 | 180 | 120 | 585 |
| 443 | 65 | 70 | 45 | 45 | 225 |
| 444 | 40 | 55 | 40 | 30 | 165 |
| 445 | 25 | 35 | 85 | 35 | 180 |
| 446 | 120 | 190 | 145 | 110 | 565 |
| 447 | 145 | 5 | 85 | 95 | 330 |
| 448 | 50 | 15 | 15 | 30 | 110 |
| 449 | 40 | 35 | 55 | 20 | 150 |
| 450 | 275 | 270 | 265 | 175 | 985 |
| 451 | 140 | 75 | 80 | 60 | 355 |
| 452 | 545 | 510 | 330 | 295 | 1,680 |
| 453 | 185 | 155 | 100 | 95 | 535 |
| 454 | 90 | 145 | 85 | 90 | 410 |
| 455 | 40 | 45 | 35 | 70 | 190 |
| 456 | 125 | 110 | 30 | 50 | 315 |
| 457 | 45 | 40 | 60 | 40 | 185 |
| 458 | 15 | 30 | 60 | 20 | 125 |
| TOTAL | 5,420 | 5,220 | 3,425 | 2,730 | 16,795 |

STATE: OKLAHOMA

| ZIP CODE | QUARTER 1 | QUARTER 2 | QUARTER 3 | QUARTER 4 | TOTAL GRAMS |
|---|---|---|---|---|---|
| 730 | 520 | 530 | 555 | 550 | 2,155 |
| 731 | 670 | 610 | 760 | 615 | 2,655 |
| 734 | 80 | 70 | 105 | 65 | 320 |
| 735 | 80 | 90 | 135 | 145 | 450 |
| 736 | 25 | 10 | 5 | 15 | 55 |
| 737 | 35 | 40 | 20 | 55 | 150 |
| 738 | 20 | 60 | 35 | 25 | 140 |
| 740 | 560 | 590 | 575 | 610 | 2,335 |
| 741 | 215 | 320 | 280 | 315 | 1,130 |
| 743 | 75 | 20 | 25 | 25 | 145 |
| 744 | 185 | 135 | 265 | 250 | 835 |
| 745 | 50 | 40 | 45 | 40 | 175 |
| 746 | 30 | 30 | 45 | 30 | 135 |
| 747 | 50 | 50 | 35 | 30 | 165 |
| 748 | 50 | 85 | 155 | 65 | 355 |
| 749 | 20 | 20 | 15 | 0 | 55 |
| TOTAL | 2,665 | 2,700 | 3,055 | 2,835 | 11,255 |

STATE: OREGON

| ZIP CODE | QUARTER 1 | QUARTER 2 | QUARTER 3 | QUARTER 4 | TOTAL GRAMS |
|---|---|---|---|---|---|
| 970 | 240 | 235 | 100 | 100 | 675 |
| 971 | 115 | 105 | 55 | 30 | 305 |
| 972 | 180 | 230 | 175 | 130 | 715 |
| 973 | 255 | 210 | 190 | 220 | 875 |
| 974 | 240 | 190 | 210 | 135 | 775 |
| 975 | 100 | 95 | 45 | 55 | 295 |
| 977 | 65 | 40 | 25 | 40 | 170 |
| 978 | 10 | 10 | 10 | 5 | 35 |
| 979 | 0 | 0 | 20 | 0 | 20 |
| TOTAL | 1,205 | 1,115 | 830 | 715 | 3,865 |

DATE RANGE: 01/01/2017 TO 12/31/2017 ARCOS 3 - REPORT 1 Run Date: 07/03/2018

RETAIL DRUG DISTRIBUTION BY ZIP CODE WITHIN STATE BY GRAMS WT

STATE: PENNSYLVANIA

| ZIP CODE | QUARTER 1 | QUARTER 2 | QUARTER 3 | QUARTER 4 | TOTAL GRAMS |
|---|---|---|---|---|---|
| 150 | 245 | 180 | 200 | 160 | 785 |
| 151 | 105 | 95 | 150 | 30 | 380 |
| 152 | 245 | 165 | 185 | 100 | 695 |
| 153 | 60 | 35 | 55 | 35 | 185 |
| 154 | 85 | 75 | 45 | 25 | 230 |
| 155 | 35 | 15 | 25 | 15 | 90 |
| 156 | 75 | 105 | 110 | 20 | 310 |
| 157 | 75 | 60 | 55 | 30 | 220 |
| 158 | 15 | 20 | 10 | 0 | 45 |
| 159 | 95 | 100 | 60 | 65 | 320 |
| 160 | 50 | 70 | 25 | 35 | 180 |
| 161 | 60 | 35 | 15 | 20 | 130 |
| 162 | 30 | 40 | 30 | 20 | 120 |
| 163 | 15 | 15 | 20 | 15 | 65 |
| 164 | 25 | 20 | 5 | 5 | 55 |
| 165 | 25 | 45 | 40 | 25 | 135 |
| 166 | 35 | 25 | 35 | 5 | 100 |
| 167 | 15 | 10 | 15 | 10 | 50 |
| 168 | 15 | 15 | 10 | 10 | 50 |
| 169 | 5 | 5 | 5 | 5 | 20 |
| 170 | 265 | 360 | 310 | 285 | 1,220 |
| 171 | 55 | 50 | 50 | 60 | 215 |
| 172 | 75 | 100 | 60 | 55 | 290 |
| 173 | 130 | 105 | 90 | 130 | 455 |
| 174 | 75 | 95 | 130 | 145 | 445 |
| 175 | 180 | 155 | 180 | 125 | 640 |
| 176 | 95 | 80 | 95 | 65 | 335 |
| 177 | 115 | 105 | 90 | 55 | 365 |
| 178 | 50 | 75 | 90 | 60 | 275 |
| 179 | 20 | 60 | 25 | 35 | 140 |
| 180 | 110 | 140 | 140 | 145 | 535 |
| 181 | 105 | 85 | 165 | 90 | 445 |
| 182 | 25 | 35 | 30 | 40 | 130 |
| 183 | 80 | 70 | 145 | 35 | 330 |
| 184 | 55 | 50 | 45 | 15 | 165 |

| | | | | | | |
|---|---|---|---|---|---|---|
| | 185 | 155 | 115 | 160 | 140 | 570 |
| | 186 | 90 | 100 | 65 | 80 | 335 |
| | 187 | 575 | 780 | 610 | 430 | 2,395 |
| | 188 | 0 | 0 | 0 | 5 | 5 |
| | 189 | 195 | 145 | 190 | 170 | 700 |
| | 190 | 715 | 715 | 580 | 540 | 2,550 |
| | 191 | 695 | 590 | 630 | 550 | 2,465 |
| | 193 | 115 | 110 | 145 | 155 | 525 |
| | 194 | 270 | 360 | 270 | 330 | 1,230 |
| | 195 | 55 | 70 | 50 | 50 | 225 |
| | 196 | 130 | 165 | 120 | 110 | 525 |
| TOTAL | | 5,740 | 5,845 | 5,560 | 4,530 | 21,675 |

STATE: PUERTO RICO

| ZIP CODE | | QUARTER 1 | QUARTER 2 | QUARTER 3 | QUARTER 4 | TOTAL GRAMS |
|---|---|---|---|---|---|---|
| | 6 | 60 | 60 | 60 | 45 | 225 |
| | 7 | 15 | 10 | 30 | 25 | 80 |
| | 9 | 60 | 80 | 75 | 85 | 300 |
| TOTAL | | 135 | 150 | 165 | 155 | 605 |

STATE: RHODE ISLAND

| ZIP CODE | | QUARTER 1 | QUARTER 2 | QUARTER 3 | QUARTER 4 | TOTAL GRAMS |
|---|---|---|---|---|---|---|
| | 28 | 345 | 345 | 400 | 335 | 1,425 |
| | 29 | 290 | 255 | 290 | 260 | 1,095 |
| TOTAL | | 635 | 600 | 690 | 595 | 2,520 |

STATE: SOUTH CAROLINA

| ZIP CODE | | QUARTER 1 | QUARTER 2 | QUARTER 3 | QUARTER 4 | TOTAL GRAMS |
|---|---|---|---|---|---|---|
| | 290 | 200 | 200 | 115 | 130 | 645 |
| | 291 | 130 | 75 | 95 | 110 | 410 |
| | 292 | 105 | 140 | 165 | 85 | 495 |
| | 293 | 240 | 140 | 160 | 135 | 675 |
| | 294 | 545 | 500 | 660 | 515 | 2,220 |
| | 295 | 470 | 390 | 410 | 500 | 1,770 |

| | | | | | | |
|---|---|---|---|---|---|---|
| | 296 | 640 | 565 | 570 | 515 | 2,290 |
| | 297 | 110 | 70 | 55 | 70 | 305 |
| | 298 | 50 | 45 | 60 | 35 | 190 |
| | 299 | 130 | 80 | 85 | 100 | 395 |
| TOTAL | | 2,620 | 2,205 | 2,375 | 2,195 | 9,395 |

STATE: SOUTH DAKOTA

| ZIP CODE | | QUARTER 1 | QUARTER 2 | QUARTER 3 | QUARTER 4 | TOTAL GRAMS |
|---|---|---|---|---|---|---|
| | 570 | 35 | 15 | 5 | 10 | 65 |
| | 571 | 135 | 110 | 85 | 70 | 400 |
| | 572 | 5 | 0 | 0 | 15 | 20 |
| | 573 | 25 | 15 | 25 | 25 | 90 |
| | 575 | 30 | 25 | 25 | 25 | 105 |
| | 577 | 40 | 55 | 85 | 45 | 225 |
| TOTAL | | 270 | 220 | 225 | 190 | 905 |

STATE: TENNESSEE

| ZIP CODE | | QUARTER 1 | QUARTER 2 | QUARTER 3 | QUARTER 4 | TOTAL GRAMS |
|---|---|---|---|---|---|---|
| | 370 | 385 | 325 | 285 | 265 | 1,260 |
| | 371 | 260 | 225 | 315 | 195 | 995 |
| | 372 | 235 | 245 | 200 | 150 | 830 |
| | 373 | 195 | 280 | 305 | 285 | 1,065 |
| | 374 | 120 | 185 | 130 | 115 | 550 |
| | 376 | 275 | 345 | 290 | 280 | 1,190 |
| | 377 | 130 | 165 | 170 | 205 | 670 |
| | 378 | 285 | 275 | 250 | 240 | 1,050 |
| | 379 | 265 | 215 | 220 | 305 | 1,005 |
| | 380 | 110 | 145 | 180 | 110 | 545 |
| | 381 | 175 | 150 | 185 | 180 | 690 |
| | 382 | 35 | 20 | 35 | 30 | 120 |
| | 383 | 225 | 165 | 165 | 120 | 675 |
| | 384 | 50 | 30 | 50 | 30 | 160 |
| | 385 | 115 | 145 | 100 | 140 | 500 |
| TOTAL | | 2,860 | 2,915 | 2,880 | 2,650 | 11,305 |

STATE: TEXAS

| ZIP CODE | QUARTER 1 | QUARTER 2 | QUARTER 3 | QUARTER 4 | TOTAL GRAMS |
|---|---|---|---|---|---|
| 750 | 1,315 | 1,225 | 1,105 | 1,165 | 4,810 |
| 751 | 415 | 430 | 430 | 330 | 1,605 |
| 752 | 400 | 510 | 470 | 395 | 1,775 |
| 753 | 0 | 0 | 5 | 0 | 5 |
| 754 | 165 | 165 | 215 | 180 | 725 |
| 755 | 30 | 10 | 10 | 10 | 60 |
| 756 | 110 | 135 | 105 | 105 | 455 |
| 757 | 115 | 135 | 205 | 135 | 590 |
| 758 | 65 | 55 | 35 | 25 | 180 |
| 759 | 265 | 135 | 120 | 190 | 710 |
| 760 | 755 | 790 | 800 | 775 | 3,120 |
| 761 | 500 | 525 | 570 | 445 | 2,040 |
| 762 | 425 | 450 | 390 | 230 | 1,495 |
| 763 | 75 | 65 | 65 | 60 | 265 |
| 764 | 0 | 10 | 5 | 5 | 20 |
| 765 | 70 | 70 | 70 | 30 | 240 |
| 766 | 5 | 10 | 0 | 20 | 35 |
| 767 | 25 | 35 | 45 | 30 | 135 |
| 768 | 10 | 15 | 20 | 25 | 70 |
| 769 | 10 | 10 | 25 | 20 | 65 |
| 770 | 800 | 940 | 1,134.90 | 860 | 3,734.90 |
| 773 | 1,060 | 960 | 845 | 680 | 3,545 |
| 774 | 600 | 620 | 610 | 575 | 2,405 |
| 775 | 915 | 850 | 910 | 745 | 3,420 |
| 776 | 230 | 200 | 315 | 225 | 970 |
| 777 | 85 | 105 | 135 | 140 | 465 |
| 778 | 25 | 55 | 45 | 35 | 160 |
| 779 | 25 | 15 | 40 | 20 | 100 |
| 780 | 350 | 410 | 455 | 255 | 1,470 |
| 781 | 260 | 215 | 340 | 205 | 1,020 |
| 782 | 750 | 900 | 1,000 | 645 | 3,295 |
| 783 | 120 | 210 | 175 | 115 | 620 |
| 784 | 560 | 455 | 535 | 450 | 2,000 |
| 785 | 280 | 350 | 335 | 240 | 1,205 |

| | | | | | | |
|---|---|---|---|---|---|---|
| | 786 | 510 | 555 | 465 | 400 | 1,930 |
| | 787 | 480 | 385 | 570 | 385 | 1,820 |
| | 788 | 10 | 25 | 15 | 0 | 50 |
| | 789 | 5 | 0 | 10 | 0 | 15 |
| | 790 | 15 | 10 | 20 | 35 | 80 |
| | 791 | 130 | 120 | 80 | 70 | 400 |
| | 792 | 0 | 0 | 5 | 5 | 10 |
| | 793 | 10 | 20 | 10 | 5 | 45 |
| | 794 | 150 | 75 | 150 | 160 | 535 |
| | 795 | 0 | 0 | 0 | 5 | 5 |
| | 796 | 15 | 20 | 40 | 15 | 90 |
| | 797 | 180 | 180 | 170 | 155 | 685 |
| | 798 | 0 | 0 | 0 | 5 | 5 |
| | 799 | 235 | 185 | 195 | 210 | 825 |
| TOTAL | | 12,555 | 12,640 | 13,294.90 | 10,815 | 49,304.90 |

| STATE: | UTAH | | | | |
|---|---|---|---|---|---|
| ZIP CODE | QUARTER 1 | QUARTER 2 | QUARTER 3 | QUARTER 4 | TOTAL GRAMS |
| 840 | 745 | 915 | 575 | 635 | 2,870 |
| 841 | 160 | 175 | 140 | 160 | 635 |
| 843 | 70 | 50 | 15 | 5 | 140 |
| 844 | 155 | 150 | 65 | 65 | 435 |
| 845 | 15 | 10 | 5 | 10 | 40 |
| 846 | 120 | 100 | 95 | 115 | 430 |
| 847 | 365 | 310 | 230 | 230 | 1,135 |
| TOTAL | 1,630 | 1,710 | 1,125 | 1,220 | 5,685 |

| STATE: | VERMONT | | | | |
|---|---|---|---|---|---|
| ZIP CODE | QUARTER 1 | QUARTER 2 | QUARTER 3 | QUARTER 4 | TOTAL GRAMS |
| 51 | 5 | 0 | 0 | 0 | 5 |
| 52 | 10 | 0 | 5 | 0 | 15 |
| 53 | 5 | 0 | 0 | 0 | 5 |
| 54 | 15 | 0 | 0 | 5 | 20 |
| 56 | 5 | 0 | 10 | 0 | 15 |
| 57 | 55 | 60 | 70 | 35 | 220 |

| | | | | | |
|---|---|---|---|---|---|
| 58 | 5 | 0 | 0 | 0 | 5 |
| TOTAL | 100 | 60 | 85 | 40 | 285 |

STATE: VIRGIN ISLANDS

| ZIP CODE | QUARTER 1 | QUARTER 2 | QUARTER 3 | QUARTER 4 | TOTAL GRAMS |
|---|---|---|---|---|---|
| 8 | 20 | 45 | 25 | 15 | 105 |
| TOTAL | 20 | 45 | 25 | 15 | 105 |

STATE: VIRGINIA

| ZIP CODE | QUARTER 1 | QUARTER 2 | QUARTER 3 | QUARTER 4 | TOTAL GRAMS |
|---|---|---|---|---|---|
| 201 | 495 | 490 | 365 | 415 | 1,765 |
| 220 | 85 | 145 | 100 | 95 | 425 |
| 221 | 70 | 90 | 80 | 50 | 290 |
| 222 | 85 | 75 | 90 | 55 | 305 |
| 223 | 80 | 95 | 60 | 95 | 330 |
| 224 | 90 | 105 | 115 | 145 | 455 |
| 225 | 35 | 40 | 60 | 45 | 180 |
| 226 | 40 | 20 | 25 | 20 | 105 |
| 227 | 25 | 20 | 35 | 15 | 95 |
| 228 | 35 | 20 | 10 | 25 | 90 |
| 229 | 50 | 85 | 85 | 100 | 320 |
| 230 | 150 | 130 | 150 | 200 | 630 |
| 231 | 255 | 340 | 200 | 255 | 1,050 |
| 232 | 295 | 220 | 175 | 235 | 925 |
| 233 | 90 | 150 | 65 | 75 | 380 |
| 234 | 330 | 390 | 245 | 255 | 1,220 |
| 235 | 170 | 185 | 105 | 145 | 605 |
| 236 | 135 | 170 | 205 | 195 | 705 |
| 237 | 10 | 15 | 60 | 0 | 85 |
| 238 | 185 | 135 | 130 | 140 | 590 |
| 239 | 25 | 35 | 20 | 15 | 95 |
| 240 | 220 | 200 | 145 | 130 | 695 |
| 241 | 105 | 85 | 70 | 80 | 340 |
| 242 | 80 | 125 | 100 | 95 | 400 |
| 243 | 110 | 120 | 60 | 50 | 340 |

| | | | | | |
|---|---|---|---|---|---|
| 244 | 135 | 150 | 65 | 20 | 370 |
| 245 | 170 | 170 | 220 | 105 | 665 |
| 246 | 115 | 160 | 105 | 65 | 445 |
| TOTAL | 3,670 | 3,965 | 3,145 | 3,120 | 13,900 |

STATE: WASHINGTON

| ZIP CODE | QUARTER 1 | QUARTER 2 | QUARTER 3 | QUARTER 4 | TOTAL GRAMS |
|---|---|---|---|---|---|
| 980 | 455 | 460 | 405 | 260 | 1,580 |
| 981 | 350 | 330 | 215 | 150 | 1,045 |
| 982 | 350 | 315 | 330 | 185 | 1,180 |
| 983 | 320 | 370 | 220 | 185 | 1,095 |
| 984 | 280 | 300 | 225 | 240 | 1,045 |
| 985 | 135 | 90 | 60 | 115 | 400 |
| 986 | 410 | 320 | 205 | 255 | 1,190 |
| 988 | 5 | 15 | 25 | 10 | 55 |
| 989 | 90 | 95 | 110 | 60 | 355 |
| 990 | 10 | 25 | 20 | 25 | 80 |
| 991 | 25 | 40 | 30 | 15 | 110 |
| 992 | 95 | 155 | 125 | 65 | 440 |
| 993 | 145 | 160 | 140 | 130 | 575 |
| 994 | 40 | 25 | 20 | 5 | 90 |
| TOTAL | 2,710 | 2,700 | 2,130 | 1,700 | 9,240 |

STATE: WEST VIRGINIA

| ZIP CODE | QUARTER 1 | QUARTER 2 | QUARTER 3 | QUARTER 4 | TOTAL GRAMS |
|---|---|---|---|---|---|
| 247 | 5 | 10 | 10 | 5 | 30 |
| 248 | 5 | 0 | 5 | 0 | 10 |
| 249 | 25 | 20 | 25 | 0 | 70 |
| 250 | 35 | 20 | 10 | 15 | 80 |
| 251 | 25 | 5 | 20 | 25 | 75 |
| 252 | 0 | 0 | 0 | 5 | 5 |
| 253 | 55 | 55 | 55 | 40 | 205 |
| 254 | 90 | 75 | 15 | 65 | 245 |
| 255 | 70 | 30 | 55 | 55 | 210 |
| 257 | 35 | 40 | 30 | 10 | 115 |

| | | | | | | |
|---|---|---|---|---|---|---|
| | 258 | 45 | 35 | 60 | 25 | 165 |
| | 259 | 5 | 0 | 0 | 0 | 5 |
| | 260 | 90 | 85 | 30 | 40 | 245 |
| | 261 | 5 | 0 | 10 | 0 | 15 |
| | 262 | 20 | 20 | 20 | 20 | 80 |
| | 263 | 60 | 40 | 75 | 35 | 210 |
| | 265 | 45 | 65 | 50 | 55 | 215 |
| | 266 | 20 | 15 | 10 | 5 | 50 |
| | 267 | 30 | 35 | 20 | 5 | 90 |
| TOTAL | | 665 | 550 | 500 | 405 | 2,120 |

STATE: WISCONSIN

| ZIP CODE | | QUARTER 1 | QUARTER 2 | QUARTER 3 | QUARTER 4 | TOTAL GRAMS |
|---|---|---|---|---|---|---|
| | 530 | 210 | 205 | 265 | 245 | 925 |
| | 531 | 405 | 455 | 435 | 360 | 1,655 |
| | 532 | 280 | 210 | 250 | 285 | 1,025 |
| | 534 | 55 | 20 | 45 | 10 | 130 |
| | 535 | 205 | 150 | 265 | 200 | 820 |
| | 537 | 25 | 25 | 20 | 25 | 95 |
| | 538 | 5 | 0 | 0 | 0 | 5 |
| | 539 | 20 | 5 | 30 | 25 | 80 |
| | 540 | 25 | 35 | 55 | 35 | 150 |
| | 541 | 25 | 15 | 15 | 40 | 95 |
| | 542 | 10 | 10 | 30 | 0 | 50 |
| | 543 | 20 | 35 | 30 | 20 | 105 |
| | 544 | 55 | 20 | 30 | 25 | 130 |
| | 545 | 40 | 20 | 25 | 10 | 95 |
| | 546 | 30 | 20 | 20 | 15 | 85 |
| | 547 | 45 | 45 | 50 | 70 | 210 |
| | 548 | 50 | 70 | 25 | 40 | 185 |
| | 549 | 115 | 110 | 160 | 135 | 520 |
| TOTAL | | 1,620 | 1,450 | 1,750 | 1,540 | 6,360 |

STATE: WYOMING

| ZIP CODE | QUARTER 1 | QUARTER 2 | QUARTER 3 | QUARTER 4 | TOTAL GRAMS |
|---|---|---|---|---|---|

| | | | | | | |
|---|---|---|---|---|---|---|
| | 820 | 15 | 25 | 25 | 15 | 80 |
| | 822 | 0 | 0 | 0 | 10 | 10 |
| | 823 | 5 | 0 | 0 | 0 | 5 |
| | 824 | 45 | 55 | 95 | 70 | 265 |
| | 825 | 25 | 35 | 20 | 20 | 100 |
| | 826 | 65 | 50 | 30 | 30 | 175 |
| | 827 | 0 | 5 | 0 | 5 | 10 |
| | 828 | 180 | 150 | 170 | 180 | 680 |
| | 829 | 95 | 75 | 5 | 20 | 195 |
| | 830 | 0 | 0 | 5 | 0 | 5 |
| TOTAL | | 430 | 395 | 350 | 350 | 1,525 |

DRUG CODE: 2125 DRUG NAME: AMOBARBITAL (SCHEDULE 2)

STATE: ARIZONA

| ZIP CODE | QUARTER 1 | QUARTER 2 | QUARTER 3 | QUARTER 4 | TOTAL GRAMS |
|---|---|---|---|---|---|
| 850 | 11.85 | 11.39 | 12.76 | 8.2 | 44.2 |
| 857 | 0.91 | 0 | 0 | 0 | 0.91 |
| TOTAL | 12.76 | 11.39 | 12.76 | 8.2 | 45.11 |

STATE: ARKANSAS

| ZIP CODE | QUARTER 1 | QUARTER 2 | QUARTER 3 | QUARTER 4 | TOTAL GRAMS |
|---|---|---|---|---|---|
| 722 | 0 | 0.46 | 0.46 | 0 | 0.92 |
| TOTAL | 0 | 0.46 | 0.46 | 0 | 0.92 |

STATE: CALIFORNIA

| ZIP CODE | QUARTER 1 | QUARTER 2 | QUARTER 3 | QUARTER 4 | TOTAL GRAMS |
|---|---|---|---|---|---|
| 900 | 13.67 | 13.67 | 9.57 | 7.75 | 44.66 |
| 910 | 0.46 | 0 | 0 | 0.46 | 0.92 |
| 911 | 0 | 0 | 0.91 | 0 | 0.91 |
| 915 | 1.82 | 0 | 0.91 | 2.28 | 5.01 |
| 920 | 1.82 | 1.82 | 2.73 | 0.91 | 7.28 |
| 921 | 0 | 2.73 | 1.82 | 0 | 4.55 |
| 923 | 1.82 | 0.91 | 0.46 | 0.91 | 4.1 |
| 926 | 0 | 0 | 0.46 | 0.91 | 1.37 |

| | | | | | | |
|---|---|---|---|---|---|---|
| | 928 | 3.19 | 1.82 | 2.73 | 0 | 7.74 |
| | 941 | 0 | 0 | 4.56 | 7.29 | 11.85 |
| | 958 | 4.56 | 0 | 5.92 | 1.82 | 12.3 |
| TOTAL | | 27.34 | 20.95 | 30.07 | 22.33 | 100.69 |

STATE: COLORADO

| ZIP CODE | | QUARTER 1 | QUARTER 2 | QUARTER 3 | QUARTER 4 | TOTAL GRAMS |
|---|---|---|---|---|---|---|
| | 800 | 15.04 | 7.75 | 10.48 | 10.03 | 43.3 |
| | 801 | 6.84 | 4.56 | 0 | 0 | 11.4 |
| | 802 | 5.92 | 3.65 | 4.1 | 0 | 13.67 |
| TOTAL | | 27.8 | 15.96 | 14.58 | 10.03 | 68.37 |

STATE: CONNECTICUT

| ZIP CODE | | QUARTER 1 | QUARTER 2 | QUARTER 3 | QUARTER 4 | TOTAL GRAMS |
|---|---|---|---|---|---|---|
| | 61 | 0 | 0.46 | 0 | 1.37 | 1.83 |
| | 65 | 0 | 1.82 | 0 | 0 | 1.82 |
| TOTAL | | 0 | 2.28 | 0 | 1.37 | 3.65 |

STATE: DISTRICT OF COLUMBIA

| ZIP CODE | | QUARTER 1 | QUARTER 2 | QUARTER 3 | QUARTER 4 | TOTAL GRAMS |
|---|---|---|---|---|---|---|
| | 200 | 0.46 | 0 | 1.82 | 4.56 | 6.84 |
| TOTAL | | 0.46 | 0 | 1.82 | 4.56 | 6.84 |

STATE: FLORIDA

| ZIP CODE | | QUARTER 1 | QUARTER 2 | QUARTER 3 | QUARTER 4 | TOTAL GRAMS |
|---|---|---|---|---|---|---|
| | 322 | 0 | 0.91 | 5.92 | 0.46 | 7.29 |
| | 325 | 0.91 | 0 | 0 | 0 | 0.91 |
| | 328 | 3.19 | 1.37 | 1.37 | 5.47 | 11.4 |
| | 330 | 0 | 0 | 1.37 | 2.28 | 3.65 |
| TOTAL | | 4.1 | 2.28 | 8.66 | 8.21 | 23.25 |

STATE: GEORGIA

| ZIP CODE | QUARTER 1 | QUARTER 2 | QUARTER 3 | QUARTER 4 | TOTAL GRAMS |
|---|---|---|---|---|---|
| 301 | 0 | 0 | 0.46 | 0 | 0.46 |
| 303 | 1.82 | 0.91 | 7.75 | 0 | 10.48 |
| 309 | 0 | 0 | 2.28 | 13.22 | 15.5 |
| TOTAL | 1.82 | 0.91 | 10.49 | 13.22 | 26.44 |

STATE: ILLINOIS

| ZIP CODE | QUARTER 1 | QUARTER 2 | QUARTER 3 | QUARTER 4 | TOTAL GRAMS |
|---|---|---|---|---|---|
| 601 | 0.46 | 0.91 | 0.46 | 1.37 | 3.2 |
| 602 | 0.91 | 0 | 0.46 | 0 | 1.37 |
| 604 | 2.73 | 0 | 0 | 0 | 2.73 |
| 606 | 0 | 0 | 3.19 | 1.82 | 5.01 |
| 617 | 0 | 0 | 0.46 | 0 | 0.46 |
| 618 | 0.46 | 0 | 0 | 0 | 0.46 |
| 627 | 0 | 0 | 2.28 | 0.46 | 2.74 |
| TOTAL | 4.56 | 0.91 | 6.85 | 3.65 | 15.97 |

STATE: INDIANA

| ZIP CODE | QUARTER 1 | QUARTER 2 | QUARTER 3 | QUARTER 4 | TOTAL GRAMS |
|---|---|---|---|---|---|
| 462 | 0 | 0 | 2.28 | 1.82 | 4.1 |
| TOTAL | 0 | 0 | 2.28 | 1.82 | 4.1 |

STATE: IOWA

| ZIP CODE | QUARTER 1 | QUARTER 2 | QUARTER 3 | QUARTER 4 | TOTAL GRAMS |
|---|---|---|---|---|---|
| 522 | 0 | 0 | 0.46 | 0 | 0.46 |
| TOTAL | 0 | 0 | 0.46 | 0 | 0.46 |

STATE: KANSAS

| ZIP CODE | QUARTER 1 | QUARTER 2 | QUARTER 3 | QUARTER 4 | TOTAL GRAMS |
|---|---|---|---|---|---|
| 661 | 1.82 | 5.01 | 0 | 5.47 | 12.3 |
| 672 | 0 | 0 | 0.46 | 0.91 | 1.37 |
| TOTAL | 1.82 | 5.01 | 0.46 | 6.38 | 13.67 |

| STATE: | KENTUCKY | | | | |
|---|---|---|---|---|---|
| ZIP CODE | QUARTER 1 | QUARTER 2 | QUARTER 3 | QUARTER 4 | TOTAL GRAMS |
| 402 | 4.56 | 2.73 | 2.73 | 5.47 | 15.49 |
| 405 | 0 | 0.91 | 0.91 | 1.82 | 3.64 |
| TOTAL | 4.56 | 3.64 | 3.64 | 7.29 | 19.13 |

| STATE: | MAINE | | | | |
|---|---|---|---|---|---|
| ZIP CODE | QUARTER 1 | QUARTER 2 | QUARTER 3 | QUARTER 4 | TOTAL GRAMS |
| 41 | 0 | 0 | 0.46 | 2.28 | 2.74 |
| TOTAL | 0 | 0 | 0.46 | 2.28 | 2.74 |

| STATE: | MARYLAND | | | | |
|---|---|---|---|---|---|
| ZIP CODE | QUARTER 1 | QUARTER 2 | QUARTER 3 | QUARTER 4 | TOTAL GRAMS |
| 208 | 0.91 | 1.82 | 3.19 | 0 | 5.92 |
| 209 | 0 | 0 | 0 | 1.82 | 1.82 |
| 212 | 6.38 | 1.37 | 3.19 | 8.66 | 19.6 |
| TOTAL | 7.29 | 3.19 | 6.38 | 10.48 | 27.34 |

| STATE: | MASSACHUSETTS | | | | |
|---|---|---|---|---|---|
| ZIP CODE | QUARTER 1 | QUARTER 2 | QUARTER 3 | QUARTER 4 | TOTAL GRAMS |
| 16 | 0 | 0 | 0 | 1.82 | 1.82 |
| 21 | 7.29 | 1.82 | 0.46 | 2.73 | 12.3 |
| 22 | 2.28 | 0 | 0.46 | 3.19 | 5.93 |
| TOTAL | 9.57 | 1.82 | 0.92 | 7.74 | 20.05 |

| STATE: | MICHIGAN | | | | |
|---|---|---|---|---|---|
| ZIP CODE | QUARTER 1 | QUARTER 2 | QUARTER 3 | QUARTER 4 | TOTAL GRAMS |
| 480 | 0 | 0 | 0.91 | 0 | 0.91 |
| 481 | 0 | 0 | 0.46 | 0 | 0.46 |
| 482 | 1.37 | 0.91 | 3.19 | 0.46 | 5.93 |
| 483 | 0 | 0 | 0 | 1.37 | 1.37 |

| | | | | | | |
|---|---|---|---|---|---|---|
| | 489 | 0 | 0.91 | 0 | 0 | 0.91 |
| | 490 | 1.82 | 0 | 0 | 0 | 1.82 |
| | 495 | 2.73 | 2.28 | 1.37 | 0.91 | 7.29 |
| TOTAL | | 5.92 | 4.1 | 5.93 | 2.74 | 18.69 |

| STATE: | MINNESOTA | | | | |
|---|---|---|---|---|---|
| ZIP CODE | QUARTER 1 | QUARTER 2 | QUARTER 3 | QUARTER 4 | TOTAL GRAMS |
| 551 | 2.28 | 0 | 0.91 | 1.37 | 4.56 |
| 554 | 2.28 | 2.73 | 5.92 | 3.65 | 14.58 |
| 559 | 0.46 | 0.46 | 0.46 | 2.73 | 4.11 |
| TOTAL | 5.02 | 3.19 | 7.29 | 7.75 | 23.25 |

| STATE: | MISSISSIPPI | | | | |
|---|---|---|---|---|---|
| ZIP CODE | QUARTER 1 | QUARTER 2 | QUARTER 3 | QUARTER 4 | TOTAL GRAMS |
| 392 | 1.82 | 0 | 1.37 | 1.37 | 4.56 |
| TOTAL | 1.82 | 0 | 1.37 | 1.37 | 4.56 |

| STATE: | MISSOURI | | | | |
|---|---|---|---|---|---|
| ZIP CODE | QUARTER 1 | QUARTER 2 | QUARTER 3 | QUARTER 4 | TOTAL GRAMS |
| 631 | 0 | 0 | 2.73 | 0 | 2.73 |
| 641 | 0.46 | 0.91 | 0 | 0.91 | 2.28 |
| 652 | 0 | 0 | 0 | 1.82 | 1.82 |
| TOTAL | 0.46 | 0.91 | 2.73 | 2.73 | 6.83 |

| STATE: | NEBRASKA | | | | |
|---|---|---|---|---|---|
| ZIP CODE | QUARTER 1 | QUARTER 2 | QUARTER 3 | QUARTER 4 | TOTAL GRAMS |
| 681 | 2.73 | 0 | 0 | 0.91 | 3.64 |
| TOTAL | 2.73 | 0 | 0 | 0.91 | 3.64 |

| STATE: | NEW HAMPSHIRE | | | | |
|---|---|---|---|---|---|
| ZIP CODE | QUARTER 1 | QUARTER 2 | QUARTER 3 | QUARTER 4 | TOTAL GRAMS |
| 37 | 4.1 | 1.82 | 2.73 | 3.65 | 12.3 |

| | | | | | |
|---|---|---|---|---|---|
| TOTAL | 4.1 | 1.82 | 2.73 | 3.65 | 12.3 |

STATE: NEW JERSEY

| ZIP CODE | QUARTER 1 | QUARTER 2 | QUARTER 3 | QUARTER 4 | TOTAL GRAMS |
|---|---|---|---|---|---|
| 70 | 0 | 0.91 | 0 | 0 | 0.91 |
| 71 | 0 | 0 | 3.65 | 6.38 | 10.03 |
| 74 | 0 | 0 | 0.91 | 0 | 0.91 |
| 76 | 1.37 | 0 | 0 | 7.29 | 8.66 |
| 77 | 0 | 0 | 0 | 0.46 | 0.46 |
| 79 | 1.82 | 2.28 | 2.28 | 0.46 | 6.84 |
| 80 | 0 | 0 | 0.46 | 0 | 0.46 |
| 81 | 0.46 | 0 | 0 | 1.82 | 2.28 |
| TOTAL | 3.65 | 3.19 | 7.3 | 16.41 | 30.55 |

STATE: NEW YORK

| ZIP CODE | QUARTER 1 | QUARTER 2 | QUARTER 3 | QUARTER 4 | TOTAL GRAMS |
|---|---|---|---|---|---|
| 100 | 27.34 | 12.3 | 9.57 | 15.49 | 64.7 |
| 103 | 0 | 0 | 0.91 | 0 | 0.91 |
| 104 | 3.65 | 0 | 0 | 1.37 | 5.02 |
| 105 | 0 | 0 | 0 | 2.28 | 2.28 |
| 110 | 11.85 | 0.91 | 2.73 | 7.29 | 22.78 |
| 112 | 3.65 | 1.82 | 0.46 | 2.73 | 8.66 |
| 113 | 0 | 0 | 0 | 1.37 | 1.37 |
| 117 | 3.65 | 1.37 | 0 | 5.01 | 10.03 |
| 122 | 0 | 0.46 | 1.82 | 4.56 | 6.84 |
| 126 | 0 | 0 | 0.46 | 1.82 | 2.28 |
| 142 | 4.56 | 2.28 | 9.11 | 7.29 | 23.24 |
| 146 | 0 | 2.73 | 1.82 | 0 | 4.55 |
| TOTAL | 54.7 | 21.87 | 26.88 | 49.21 | 152.66 |

STATE: NORTH CAROLINA

| ZIP CODE | QUARTER 1 | QUARTER 2 | QUARTER 3 | QUARTER 4 | TOTAL GRAMS |
|---|---|---|---|---|---|
| 271 | 1.37 | 2.73 | 2.28 | 1.82 | 8.2 |
| 275 | 3.65 | 1.37 | 2.28 | 0.91 | 8.21 |

| | | | | | | |
|---|---|---|---|---|---|---|
| | 277 | 4.1 | 0 | 0.91 | 0 | 5.01 |
| | 280 | 3.65 | 0.91 | 0 | 4.1 | 8.66 |
| | 282 | 3.65 | 0 | 0 | 4.56 | 8.21 |
| TOTAL | | 16.42 | 5.01 | 5.47 | 11.39 | 38.29 |

STATE: OHIO

| ZIP CODE | | QUARTER 1 | QUARTER 2 | QUARTER 3 | QUARTER 4 | TOTAL GRAMS |
|---|---|---|---|---|---|---|
| | 432 | 0 | 0.46 | 0.46 | 0.46 | 1.38 |
| | 443 | 0 | 0 | 1.82 | 0 | 1.82 |
| | 452 | 2.28 | 1.37 | 3.19 | 4.1 | 10.94 |
| | 454 | 0.91 | 1.82 | 0 | 0.91 | 3.64 |
| TOTAL | | 3.19 | 3.65 | 5.47 | 5.47 | 17.78 |

STATE: OREGON

| ZIP CODE | | QUARTER 1 | QUARTER 2 | QUARTER 3 | QUARTER 4 | TOTAL GRAMS |
|---|---|---|---|---|---|---|
| | 972 | 1.37 | 1.37 | 1.37 | 2.73 | 6.84 |
| TOTAL | | 1.37 | 1.37 | 1.37 | 2.73 | 6.84 |

STATE: PENNSYLVANIA

| ZIP CODE | | QUARTER 1 | QUARTER 2 | QUARTER 3 | QUARTER 4 | TOTAL GRAMS |
|---|---|---|---|---|---|---|
| | 152 | 0 | 0 | 0 | 4.56 | 4.56 |
| | 170 | 0.46 | 0 | 0 | 0 | 0.46 |
| | 174 | 0.46 | 0.91 | 0.91 | 0 | 2.28 |
| | 178 | 5.47 | 0 | 0 | 0 | 5.47 |
| | 180 | 0 | 0 | 0 | 0.46 | 0.46 |
| | 187 | 0 | 0 | 0.91 | 0 | 0.91 |
| | 191 | 7.29 | 1.82 | 2.28 | 3.65 | 15.04 |
| TOTAL | | 13.68 | 2.73 | 4.1 | 8.67 | 29.18 |

STATE: RHODE ISLAND

| ZIP CODE | | QUARTER 1 | QUARTER 2 | QUARTER 3 | QUARTER 4 | TOTAL GRAMS |
|---|---|---|---|---|---|---|
| | 29 | 3.65 | 0 | 0 | 0 | 3.65 |

| | | | | | | |
|---|---|---|---|---|---|---|
| TOTAL | | 3.65 | 0 | 0 | 0 | 3.65 |

| STATE: | SOUTH CAROLINA | | | | | |
|---|---|---|---|---|---|---|
| ZIP CODE | | QUARTER 1 | QUARTER 2 | QUARTER 3 | QUARTER 4 | TOTAL GRAMS |
| | 292 | 0 | 0 | 2.73 | 0 | 2.73 |
| | 294 | 1.37 | 4.1 | 0.91 | 4.1 | 10.48 |
| | 296 | 0 | 0 | 1.82 | 0 | 1.82 |
| TOTAL | | 1.37 | 4.1 | 5.46 | 4.1 | 15.03 |

| STATE: | TENNESSEE | | | | | |
|---|---|---|---|---|---|---|
| ZIP CODE | | QUARTER 1 | QUARTER 2 | QUARTER 3 | QUARTER 4 | TOTAL GRAMS |
| | 372 | 4.1 | 3.19 | 4.56 | 1.37 | 13.22 |
| | 378 | 0 | 0.46 | 0 | 0 | 0.46 |
| | 381 | 0 | 1.37 | 0.46 | 0.46 | 2.29 |
| TOTAL | | 4.1 | 5.02 | 5.02 | 1.83 | 15.97 |

| STATE: | TEXAS | | | | | |
|---|---|---|---|---|---|---|
| ZIP CODE | | QUARTER 1 | QUARTER 2 | QUARTER 3 | QUARTER 4 | TOTAL GRAMS |
| | 750 | 0 | 0 | 0 | 0.46 | 0.46 |
| | 752 | 4.56 | 0 | 0 | 0 | 4.56 |
| | 753 | 0.46 | 0 | 0 | 0.46 | 0.92 |
| | 761 | 0 | 0 | 0 | 0.46 | 0.46 |
| | 765 | 0 | 0.46 | 4.1 | 1.82 | 6.38 |
| | 770 | 8.66 | 4.56 | 6.38 | 4.1 | 23.7 |
| | 782 | 4.56 | 4.56 | 8.66 | 4.56 | 22.34 |
| | 787 | 7.29 | 4.1 | 8.2 | 3.65 | 23.24 |
| | 794 | 0 | 0 | 0.46 | 0.91 | 1.37 |
| TOTAL | | 25.53 | 13.68 | 27.8 | 16.42 | 83.43 |

| STATE: | UTAH | | | | | |
|---|---|---|---|---|---|---|
| ZIP CODE | | QUARTER 1 | QUARTER 2 | QUARTER 3 | QUARTER 4 | TOTAL GRAMS |
| | 841 | 1.82 | 13.67 | 14.58 | 4.1 | 34.17 |
| TOTAL | | 1.82 | 13.67 | 14.58 | 4.1 | 34.17 |

STATE: VERMONT

| ZIP CODE | QUARTER 1 | QUARTER 2 | QUARTER 3 | QUARTER 4 | TOTAL GRAMS |
|---|---|---|---|---|---|
| 54 | 0 | 0 | 0.91 | 0 | 0.91 |
| TOTAL | 0 | 0 | 0.91 | 0 | 0.91 |

STATE: VIRGINIA

| ZIP CODE | QUARTER 1 | QUARTER 2 | QUARTER 3 | QUARTER 4 | TOTAL GRAMS |
|---|---|---|---|---|---|
| 229 | 0.46 | 0.46 | 1.37 | 1.37 | 3.66 |
| 232 | 3.19 | 1.82 | 0.91 | 0.91 | 6.83 |
| 240 | 0 | 0.46 | 0.46 | 0 | 0.92 |
| TOTAL | 3.65 | 2.74 | 2.74 | 2.28 | 11.41 |

STATE: WASHINGTON

| ZIP CODE | QUARTER 1 | QUARTER 2 | QUARTER 3 | QUARTER 4 | TOTAL GRAMS |
|---|---|---|---|---|---|
| 980 | 0.46 | 0 | 0.46 | 1.37 | 2.29 |
| 981 | 2.28 | 0 | 10.94 | 15.95 | 29.17 |
| 992 | 1.82 | 0 | 0 | 3.65 | 5.47 |
| TOTAL | 4.56 | 0 | 11.4 | 20.97 | 36.93 |

STATE: WEST VIRGINIA

| ZIP CODE | QUARTER 1 | QUARTER 2 | QUARTER 3 | QUARTER 4 | TOTAL GRAMS |
|---|---|---|---|---|---|
| 265 | 0 | 0 | 0 | 1.82 | 1.82 |
| TOTAL | 0 | 0 | 0 | 1.82 | 1.82 |

STATE: WISCONSIN

| ZIP CODE | QUARTER 1 | QUARTER 2 | QUARTER 3 | QUARTER 4 | TOTAL GRAMS |
|---|---|---|---|---|---|
| 532 | 5.01 | 2.28 | 2.73 | 6.38 | 16.4 |
| 537 | 1.37 | 0.91 | 4.1 | 3.65 | 10.03 |
| 543 | 0 | 0 | 0.46 | 0 | 0.46 |
| TOTAL | 6.38 | 3.19 | 7.29 | 10.03 | 26.89 |

DRUG CODE: 2165 DRUG NAME: BUTALBITAL

STATE: ALABAMA

| ZIP CODE | QUARTER 1 | QUARTER 2 | QUARTER 3 | QUARTER 4 | TOTAL GRAMS |
|---|---|---|---|---|---|
| 350 | 625 | 625 | 605 | 525 | 2,380 |
| 351 | 330 | 445 | 280 | 270 | 1,325 |
| 352 | 270 | 325 | 300 | 300 | 1,195 |
| 354 | 255 | 305 | 420 | 280 | 1,260 |
| 355 | 175 | 165 | 150 | 120 | 610 |
| 356 | 315 | 370 | 385 | 280 | 1,350 |
| 357 | 140 | 165 | 140 | 160 | 605 |
| 358 | 160 | 200 | 155 | 125 | 640 |
| 359 | 245 | 285 | 305 | 270 | 1,105 |
| 360 | 170 | 175 | 140 | 195 | 680 |
| 361 | 90 | 100 | 100 | 75 | 365 |
| 362 | 125 | 145 | 150 | 165 | 585 |
| 363 | 325 | 255 | 285 | 320 | 1,185 |
| 364 | 100 | 85 | 105 | 95 | 385 |
| 365 | 420 | 445 | 410 | 255 | 1,530 |
| 366 | 275 | 275 | 290 | 255 | 1,095 |
| 367 | 190 | 150 | 125 | 105 | 570 |
| 368 | 195 | 210 | 255 | 225 | 885 |
| 369 | 10 | 20 | 15 | 5 | 50 |
| TOTAL | 4,415 | 4,745 | 4,615 | 4,025 | 17,800 |

STATE: ALASKA

| ZIP CODE | QUARTER 1 | QUARTER 2 | QUARTER 3 | QUARTER 4 | TOTAL GRAMS |
|---|---|---|---|---|---|
| 995 | 85 | 130 | 105 | 60 | 380 |
| 996 | 80 | 75 | 110 | 100 | 365 |
| 997 | 0 | 15 | 30 | 25 | 70 |
| 998 | 15 | 45 | 30 | 10 | 100 |
| 999 | 0 | 0 | 0 | 10 | 10 |
| TOTAL | 180 | 265 | 275 | 205 | 925 |

STATE: AMERICAN SAMOA

| ZIP CODE | QUARTER 1 | QUARTER 2 | QUARTER 3 | QUARTER 4 | TOTAL GRAMS |
|---|---|---|---|---|---|
| 967 | 10 | 0 | 0 | 0 | 10 |
| TOTAL | 10 | 0 | 0 | 0 | 10 |

STATE: ARIZONA

| ZIP CODE | QUARTER 1 | QUARTER 2 | QUARTER 3 | QUARTER 4 | TOTAL GRAMS |
|---|---|---|---|---|---|
| 850 | 720 | 715 | 785 | 875 | 3,095 |
| 851 | 130 | 235 | 275 | 270 | 910 |
| 852 | 1,675 | 1,665 | 1,895 | 1,855 | 7,090 |
| 853 | 785 | 665 | 820 | 950 | 3,220 |
| 855 | 195 | 220 | 165 | 140 | 720 |
| 856 | 250 | 225 | 245 | 275 | 995 |
| 857 | 500 | 410 | 505 | 570 | 1,985 |
| 859 | 55 | 60 | 90 | 85 | 290 |
| 860 | 30 | 45 | 35 | 70 | 180 |
| 863 | 145 | 200 | 255 | 200 | 800 |
| 864 | 225 | 265 | 425 | 335 | 1,250 |
| TOTAL | 4,710 | 4,705 | 5,495 | 5,625 | 20,535 |

STATE: ARKANSAS

| ZIP CODE | QUARTER 1 | QUARTER 2 | QUARTER 3 | QUARTER 4 | TOTAL GRAMS |
|---|---|---|---|---|---|
| 716 | 100 | 105 | 120 | 120 | 445 |
| 717 | 115 | 70 | 95 | 85 | 365 |
| 718 | 120 | 170 | 140 | 105 | 535 |
| 719 | 290 | 235 | 360 | 325 | 1,210 |
| 720 | 365 | 470 | 465 | 425 | 1,725 |
| 721 | 365 | 310 | 275 | 270 | 1,220 |
| 722 | 295 | 265 | 305 | 325 | 1,190 |
| 723 | 145 | 165 | 195 | 205 | 710 |
| 724 | 305 | 300 | 355 | 340 | 1,300 |
| 725 | 120 | 155 | 180 | 195 | 650 |
| 726 | 160 | 235 | 185 | 185 | 765 |
| 727 | 605 | 705 | 565 | 635 | 2,510 |
| 728 | 90 | 85 | 130 | 75 | 380 |
| 729 | 220 | 165 | 150 | 225 | 760 |

| | | | | | |
|---|---|---|---|---|---|
| TOTAL | 3,295 | 3,435 | 3,520 | 3,515 | 13,765 |

STATE: CALIFORNIA

| ZIP CODE | QUARTER 1 | QUARTER 2 | QUARTER 3 | QUARTER 4 | TOTAL GRAMS |
|---|---|---|---|---|---|
| 900 | 910 | 790 | 1,005 | 820 | 3,525 |
| 902 | 935 | 1,080 | 1,030 | 885 | 3,930 |
| 903 | 45 | 30 | 20 | 20 | 115 |
| 904 | 140 | 120 | 110 | 105 | 475 |
| 905 | 220 | 175 | 215 | 215 | 825 |
| 906 | 410 | 495 | 485 | 415 | 1,805 |
| 907 | 265 | 260 | 290 | 310 | 1,125 |
| 908 | 180 | 180 | 180 | 150 | 690 |
| 910 | 275 | 230 | 285 | 250 | 1,040 |
| 911 | 95 | 140 | 105 | 140 | 480 |
| 912 | 155 | 110 | 85 | 80 | 430 |
| 913 | 1,275 | 1,195 | 1,310 | 1,145 | 4,925 |
| 914 | 275 | 275 | 295 | 365 | 1,210 |
| 915 | 30 | 40 | 70 | 110 | 250 |
| 916 | 90 | 125 | 135 | 135 | 485 |
| 917 | 1,165 | 1,085 | 1,060 | 865 | 4,175 |
| 918 | 45 | 60 | 60 | 55 | 220 |
| 919 | 115 | 155 | 110 | 95 | 475 |
| 920 | 1,195 | 1,245 | 1,005 | 1,205 | 4,650 |
| 921 | 365 | 325 | 375 | 420 | 1,485 |
| 922 | 470 | 295 | 330 | 390 | 1,485 |
| 923 | 785 | 895 | 800 | 780 | 3,260 |
| 924 | 50 | 60 | 20 | 15 | 145 |
| 925 | 1,025 | 1,160 | 1,095 | 870 | 4,150 |
| 926 | 1,350 | 1,270 | 1,320 | 1,145 | 5,085 |
| 927 | 195 | 230 | 240 | 300 | 965 |
| 928 | 890 | 1,020 | 995 | 910 | 3,815 |
| 930 | 795 | 690 | 710 | 690 | 2,885 |
| 931 | 115 | 170 | 145 | 190 | 620 |
| 932 | 515 | 480 | 420 | 465 | 1,880 |
| 933 | 415 | 495 | 395 | 445 | 1,750 |
| 934 | 510 | 710 | 590 | 520 | 2,330 |

| | | | | | | |
|---|---|---|---|---|---|---|
| | 935 | 495 | 485 | 470 | 370 | 1,820 |
| | 936 | 505 | 420 | 405 | 460 | 1,790 |
| | 937 | 245 | 255 | 295 | 265 | 1,060 |
| | 939 | 370 | 415 | 465 | 420 | 1,670 |
| | 940 | 220 | 280 | 335 | 300 | 1,135 |
| | 941 | 110 | 115 | 130 | 120 | 475 |
| | 943 | 20 | 15 | 10 | 5 | 50 |
| | 944 | 30 | 15 | 25 | 20 | 90 |
| | 945 | 1,725 | 1,720 | 1,495 | 1,425 | 6,365 |
| | 946 | 120 | 75 | 95 | 60 | 350 |
| | 947 | 50 | 35 | 60 | 55 | 200 |
| | 948 | 35 | 35 | 5 | 20 | 95 |
| | 949 | 345 | 340 | 455 | 425 | 1,565 |
| | 950 | 400 | 435 | 650 | 460 | 1,945 |
| | 951 | 250 | 305 | 310 | 280 | 1,145 |
| | 952 | 485 | 385 | 495 | 440 | 1,805 |
| | 953 | 895 | 970 | 1,155 | 1,120 | 4,140 |
| | 954 | 260 | 310 | 235 | 280 | 1,085 |
| | 955 | 95 | 85 | 110 | 125 | 415 |
| | 956 | 1,110 | 1,185 | 1,165 | 1,165 | 4,625 |
| | 957 | 255 | 200 | 225 | 125 | 805 |
| | 958 | 370 | 325 | 325 | 330 | 1,350 |
| | 959 | 665 | 665 | 585 | 670 | 2,585 |
| | 960 | 215 | 220 | 185 | 185 | 805 |
| | 961 | 60 | 60 | 85 | 50 | 255 |
| TOTAL | | 24,630 | 24,940 | 25,060 | 23,680 | 98,310 |

| STATE: | COLORADO | | | | | |
|---|---|---|---|---|---|---|
| ZIP CODE | | QUARTER 1 | QUARTER 2 | QUARTER 3 | QUARTER 4 | TOTAL GRAMS |
| | 800 | 270 | 245 | 255 | 340 | 1,110 |
| | 801 | 270 | 325 | 280 | 410 | 1,285 |
| | 802 | 200 | 180 | 230 | 265 | 875 |
| | 803 | 30 | 5 | 25 | 40 | 100 |
| | 804 | 25 | 40 | 65 | 55 | 185 |
| | 805 | 210 | 240 | 235 | 260 | 945 |
| | 806 | 40 | 110 | 55 | 80 | 285 |

| | | | | | | |
|---|---|---|---|---|---|---|
| | 807 | 30 | 5 | 0 | 0 | 35 |
| | 808 | 25 | 45 | 25 | 35 | 130 |
| | 809 | 215 | 195 | 270 | 245 | 925 |
| | 810 | 125 | 105 | 195 | 150 | 575 |
| | 811 | 30 | 45 | 40 | 20 | 135 |
| | 812 | 80 | 40 | 80 | 100 | 300 |
| | 813 | 5 | 5 | 0 | 15 | 25 |
| | 814 | 45 | 65 | 75 | 105 | 290 |
| | 815 | 80 | 115 | 100 | 100 | 395 |
| | 816 | 35 | 70 | 55 | 55 | 215 |
| TOTAL | | 1,715 | 1,835 | 1,985 | 2,275 | 7,810 |

STATE: CONNECTICUT

| ZIP CODE | QUARTER 1 | QUARTER 2 | QUARTER 3 | QUARTER 4 | TOTAL GRAMS |
|---|---|---|---|---|---|
| 60 | 920 | 810 | 880 | 625 | 3,235 |
| 61 | 80 | 145 | 205 | 75 | 505 |
| 62 | 135 | 65 | 105 | 65 | 370 |
| 63 | 310 | 280 | 240 | 245 | 1,075 |
| 64 | 695 | 685 | 865 | 695 | 2,940 |
| 65 | 70 | 90 | 85 | 85 | 330 |
| 66 | 120 | 65 | 90 | 80 | 355 |
| 67 | 375 | 375 | 365 | 350 | 1,465 |
| 68 | 460 | 410 | 360 | 370 | 1,600 |
| 69 | 90 | 115 | 155 | 65 | 425 |
| TOTAL | 3,255 | 3,040 | 3,350 | 2,655 | 12,300 |

STATE: DELAWARE

| ZIP CODE | QUARTER 1 | QUARTER 2 | QUARTER 3 | QUARTER 4 | TOTAL GRAMS |
|---|---|---|---|---|---|
| 197 | 255 | 215 | 220 | 230 | 920 |
| 198 | 170 | 175 | 185 | 185 | 715 |
| 199 | 135 | 160 | 90 | 90 | 475 |
| TOTAL | 560 | 550 | 495 | 505 | 2,110 |

STATE: DISTRICT OF COLUMBIA

RETAIL DRUG DISTRIBUTION BY ZIP CODE WITHIN STATE BY GRAMS WT

| ZIP CODE | QUARTER 1 | QUARTER 2 | QUARTER 3 | QUARTER 4 | TOTAL GRAMS |
|---|---|---|---|---|---|
| 200 | 315 | 350 | 395 | 340 | 1,400 |
| TOTAL | 315 | 350 | 395 | 340 | 1,400 |

STATE: FLORIDA

| ZIP CODE | QUARTER 1 | QUARTER 2 | QUARTER 3 | QUARTER 4 | TOTAL GRAMS |
|---|---|---|---|---|---|
| 320 | 480 | 635 | 855 | 730 | 2,700 |
| 321 | 350 | 530 | 605 | 630 | 2,115 |
| 322 | 850 | 895 | 920 | 750 | 3,415 |
| 323 | 330 | 345 | 355 | 475 | 1,505 |
| 324 | 640 | 595 | 575 | 645 | 2,455 |
| 325 | 525 | 370 | 440 | 405 | 1,740 |
| 326 | 290 | 345 | 400 | 385 | 1,420 |
| 327 | 785 | 770 | 680 | 735 | 2,970 |
| 328 | 475 | 525 | 580 | 640 | 2,220 |
| 329 | 725 | 815 | 945 | 810 | 3,295 |
| 330 | 1,000 | 1,240 | 1,255 | 1,065 | 4,560 |
| 331 | 350 | 350 | 375 | 340 | 1,415 |
| 333 | 530 | 605 | 735 | 780 | 2,650 |
| 334 | 1,480 | 1,615 | 1,530 | 1,790 | 6,415 |
| 335 | 900 | 990 | 1,120 | 1,185 | 4,195 |
| 336 | 525 | 575 | 660 | 565 | 2,325 |
| 337 | 955 | 890 | 710 | 730 | 3,285 |
| 338 | 295 | 410 | 465 | 435 | 1,605 |
| 339 | 715 | 955 | 1,080 | 990 | 3,740 |
| 341 | 320 | 310 | 265 | 265 | 1,160 |
| 342 | 590 | 780 | 660 | 670 | 2,700 |
| 344 | 350 | 560 | 575 | 485 | 1,970 |
| 346 | 750 | 775 | 600 | 720 | 2,845 |
| 347 | 310 | 425 | 415 | 465 | 1,615 |
| 349 | 315 | 320 | 415 | 355 | 1,405 |
| TOTAL | 14,835 | 16,625 | 17,215 | 17,045 | 65,720 |

STATE: GEORGIA

| ZIP CODE | QUARTER 1 | QUARTER 2 | QUARTER 3 | QUARTER 4 | TOTAL GRAMS |
|---|---|---|---|---|---|

| | | | | | | |
|---|---|---|---|---|---|---|
| | 300 | 1,285 | 1,325 | 1,620 | 1,570 | 5,800 |
| | 301 | 1,325 | 1,500 | 1,700 | 1,850 | 6,375 |
| | 302 | 1,065 | 1,150 | 1,150 | 1,205 | 4,570 |
| | 303 | 480 | 530 | 690 | 675 | 2,375 |
| | 304 | 285 | 315 | 335 | 305 | 1,240 |
| | 305 | 825 | 860 | 885 | 835 | 3,405 |
| | 306 | 330 | 390 | 435 | 475 | 1,630 |
| | 307 | 430 | 375 | 470 | 455 | 1,730 |
| | 308 | 155 | 200 | 205 | 165 | 725 |
| | 309 | 235 | 180 | 255 | 210 | 880 |
| | 310 | 360 | 325 | 375 | 365 | 1,425 |
| | 312 | 185 | 135 | 225 | 175 | 720 |
| | 313 | 205 | 275 | 270 | 160 | 910 |
| | 314 | 75 | 85 | 115 | 105 | 380 |
| | 315 | 760 | 770 | 685 | 685 | 2,900 |
| | 316 | 335 | 320 | 315 | 355 | 1,325 |
| | 317 | 335 | 445 | 415 | 355 | 1,550 |
| | 318 | 30 | 35 | 15 | 30 | 110 |
| | 319 | 365 | 380 | 355 | 315 | 1,415 |
| | 398 | 105 | 65 | 90 | 75 | 335 |
| TOTAL | | 9,170 | 9,660 | 10,605 | 10,365 | 39,800 |

STATE: GUAM

| ZIP CODE | | QUARTER 1 | QUARTER 2 | QUARTER 3 | QUARTER 4 | TOTAL GRAMS |
|---|---|---|---|---|---|---|
| | 969 | 25 | 5 | 5 | 5 | 40 |
| TOTAL | | 25 | 5 | 5 | 5 | 40 |

STATE: HAWAII

| ZIP CODE | | QUARTER 1 | QUARTER 2 | QUARTER 3 | QUARTER 4 | TOTAL GRAMS |
|---|---|---|---|---|---|---|
| | 967 | 810 | 820 | 740 | 670 | 3,040 |
| | 968 | 300 | 255 | 305 | 260 | 1,120 |
| TOTAL | | 1,110 | 1,075 | 1,045 | 930 | 4,160 |

STATE: IDAHO

| ZIP CODE | QUARTER 1 | QUARTER 2 | QUARTER 3 | QUARTER 4 | TOTAL GRAMS |
|---|---|---|---|---|---|
| 832 | 175 | 215 | 190 | 290 | 870 |
| 833 | 120 | 65 | 200 | 165 | 550 |
| 834 | 115 | 80 | 145 | 150 | 490 |
| 835 | 45 | 60 | 70 | 85 | 260 |
| 836 | 345 | 335 | 365 | 295 | 1,340 |
| 837 | 185 | 170 | 190 | 190 | 735 |
| 838 | 195 | 315 | 270 | 220 | 1,000 |
| TOTAL | 1,180 | 1,240 | 1,430 | 1,395 | 5,245 |

STATE: ILLINOIS

| ZIP CODE | QUARTER 1 | QUARTER 2 | QUARTER 3 | QUARTER 4 | TOTAL GRAMS |
|---|---|---|---|---|---|
| 600 | 1,925 | 1,755 | 1,620 | 1,670 | 6,970 |
| 601 | 1,010 | 1,190 | 1,045 | 985 | 4,230 |
| 602 | 35 | 30 | 30 | 45 | 140 |
| 603 | 0 | 35 | 40 | 30 | 105 |
| 604 | 930 | 1,025 | 955 | 920 | 3,830 |
| 605 | 580 | 640 | 595 | 480 | 2,295 |
| 606 | 475 | 485 | 420 | 490 | 1,870 |
| 607 | 30 | 20 | 15 | 40 | 105 |
| 608 | 20 | 0 | 25 | 15 | 60 |
| 609 | 50 | 115 | 160 | 130 | 455 |
| 610 | 105 | 50 | 90 | 95 | 340 |
| 611 | 175 | 220 | 180 | 195 | 770 |
| 612 | 50 | 30 | 20 | 20 | 120 |
| 613 | 95 | 65 | 85 | 70 | 315 |
| 614 | 70 | 95 | 80 | 105 | 350 |
| 615 | 125 | 135 | 245 | 150 | 655 |
| 616 | 90 | 105 | 85 | 80 | 360 |
| 617 | 150 | 150 | 150 | 95 | 545 |
| 618 | 220 | 175 | 175 | 135 | 705 |
| 619 | 25 | 40 | 30 | 30 | 125 |
| 620 | 220 | 265 | 250 | 240 | 975 |
| 622 | 185 | 220 | 255 | 195 | 855 |
| 623 | 60 | 80 | 80 | 70 | 290 |
| 624 | 110 | 150 | 145 | 85 | 490 |

| | | | | | | |
|---|---|---|---|---|---|---|
| | 625 | 135 | 175 | 100 | 125 | 535 |
| | 626 | 25 | 30 | 35 | 40 | 130 |
| | 627 | 135 | 120 | 125 | 150 | 530 |
| | 628 | 105 | 155 | 100 | 205 | 565 |
| | 629 | 165 | 185 | 315 | 280 | 945 |
| TOTAL | | 7,300 | 7,740 | 7,450 | 7,170 | 29,660 |

| STATE: | INDIANA | | | | |
|---|---|---|---|---|---|
| ZIP CODE | QUARTER 1 | QUARTER 2 | QUARTER 3 | QUARTER 4 | TOTAL GRAMS |
| 460 | 310 | 340 | 290 | 275 | 1,215 |
| 461 | 365 | 285 | 420 | 325 | 1,395 |
| 462 | 435 | 470 | 575 | 570 | 2,050 |
| 463 | 455 | 395 | 350 | 370 | 1,570 |
| 464 | 85 | 90 | 150 | 130 | 455 |
| 465 | 250 | 200 | 160 | 185 | 795 |
| 466 | 65 | 65 | 30 | 15 | 175 |
| 467 | 180 | 165 | 255 | 220 | 820 |
| 468 | 95 | 65 | 105 | 130 | 395 |
| 469 | 60 | 115 | 50 | 105 | 330 |
| 470 | 85 | 110 | 105 | 110 | 410 |
| 471 | 370 | 300 | 435 | 335 | 1,440 |
| 472 | 35 | 70 | 100 | 90 | 295 |
| 473 | 155 | 150 | 175 | 175 | 655 |
| 474 | 155 | 195 | 225 | 220 | 795 |
| 475 | 185 | 165 | 110 | 100 | 560 |
| 476 | 130 | 160 | 150 | 165 | 605 |
| 477 | 205 | 175 | 235 | 240 | 855 |
| 478 | 80 | 60 | 85 | 45 | 270 |
| 479 | 110 | 105 | 185 | 110 | 510 |
| TOTAL | 3,810 | 3,680 | 4,190 | 3,915 | 15,595 |

| STATE: | IOWA | | | | |
|---|---|---|---|---|---|
| ZIP CODE | QUARTER 1 | QUARTER 2 | QUARTER 3 | QUARTER 4 | TOTAL GRAMS |
| 500 | 80 | 80 | 75 | 55 | 290 |
| 501 | 65 | 65 | 50 | 30 | 210 |

| | | | | | | |
|---|---|---|---|---|---|---|
| | 502 | 95 | 85 | 80 | 105 | 365 |
| | 503 | 85 | 140 | 95 | 105 | 425 |
| | 504 | 45 | 45 | 40 | 30 | 160 |
| | 505 | 70 | 35 | 80 | 85 | 270 |
| | 506 | 55 | 30 | 55 | 70 | 210 |
| | 507 | 25 | 60 | 35 | 35 | 155 |
| | 508 | 5 | 0 | 0 | 20 | 25 |
| | 510 | 80 | 100 | 85 | 60 | 325 |
| | 511 | 25 | 60 | 55 | 35 | 175 |
| | 512 | 0 | 0 | 5 | 0 | 5 |
| | 513 | 5 | 15 | 20 | 0 | 40 |
| | 514 | 100 | 95 | 105 | 70 | 370 |
| | 515 | 35 | 15 | 15 | 25 | 90 |
| | 516 | 45 | 20 | 50 | 40 | 155 |
| | 520 | 65 | 105 | 105 | 90 | 365 |
| | 521 | 40 | 40 | 45 | 45 | 170 |
| | 522 | 20 | 60 | 10 | 30 | 120 |
| | 523 | 20 | 15 | 30 | 30 | 95 |
| | 524 | 20 | 15 | 30 | 20 | 85 |
| | 525 | 30 | 20 | 20 | 30 | 100 |
| | 526 | 65 | 45 | 100 | 60 | 270 |
| | 527 | 105 | 95 | 90 | 105 | 395 |
| | 528 | 50 | 50 | 60 | 70 | 230 |
| TOTAL | | 1,230 | 1,290 | 1,335 | 1,245 | 5,100 |

STATE: KANSAS

| ZIP CODE | | QUARTER 1 | QUARTER 2 | QUARTER 3 | QUARTER 4 | TOTAL GRAMS |
|---|---|---|---|---|---|---|
| | 660 | 390 | 250 | 230 | 320 | 1,190 |
| | 661 | 85 | 70 | 145 | 35 | 335 |
| | 662 | 340 | 345 | 595 | 325 | 1,605 |
| | 664 | 70 | 55 | 75 | 75 | 275 |
| | 665 | 65 | 15 | 50 | 15 | 145 |
| | 666 | 110 | 55 | 50 | 30 | 245 |
| | 667 | 130 | 145 | 135 | 115 | 525 |
| | 668 | 55 | 80 | 105 | 105 | 345 |
| | 669 | 0 | 0 | 5 | 0 | 5 |

| | | | | | | |
|---|---|---|---|---|---|---|
| | 670 | 70 | 85 | 110 | 140 | 405 |
| | 671 | 70 | 80 | 75 | 80 | 305 |
| | 672 | 170 | 140 | 280 | 295 | 885 |
| | 673 | 160 | 110 | 140 | 140 | 550 |
| | 674 | 285 | 205 | 245 | 220 | 955 |
| | 675 | 65 | 45 | 110 | 65 | 285 |
| | 676 | 10 | 15 | 30 | 10 | 65 |
| | 677 | 10 | 35 | 35 | 25 | 105 |
| | 678 | 50 | 35 | 60 | 25 | 170 |
| | 679 | 85 | 110 | 105 | 110 | 410 |
| TOTAL | | 2,220 | 1,875 | 2,580 | 2,130 | 8,805 |

| STATE: | KENTUCKY | | | | |
|---|---|---|---|---|---|
| ZIP CODE | QUARTER 1 | QUARTER 2 | QUARTER 3 | QUARTER 4 | TOTAL GRAMS |
| 400 | 80 | 155 | 190 | 195 | 620 |
| 401 | 20 | 15 | 30 | 20 | 85 |
| 402 | 355 | 300 | 440 | 375 | 1,470 |
| 403 | 175 | 165 | 215 | 240 | 795 |
| 404 | 20 | 20 | 50 | 45 | 135 |
| 405 | 75 | 90 | 120 | 105 | 390 |
| 406 | 5 | 10 | 30 | 5 | 50 |
| 407 | 100 | 75 | 95 | 85 | 355 |
| 408 | 75 | 45 | 25 | 50 | 195 |
| 409 | 25 | 40 | 35 | 65 | 165 |
| 410 | 330 | 250 | 385 | 410 | 1,375 |
| 411 | 50 | 45 | 80 | 50 | 225 |
| 412 | 15 | 10 | 10 | 10 | 45 |
| 413 | 0 | 35 | 0 | 0 | 35 |
| 414 | 0 | 10 | 5 | 5 | 20 |
| 415 | 55 | 65 | 30 | 20 | 170 |
| 416 | 30 | 35 | 30 | 40 | 135 |
| 417 | 35 | 30 | 45 | 0 | 110 |
| 418 | 45 | 20 | 40 | 55 | 160 |
| 420 | 245 | 245 | 275 | 255 | 1,020 |
| 421 | 110 | 95 | 130 | 80 | 415 |
| 422 | 30 | 90 | 70 | 70 | 260 |

| | | | | | | |
|---|---|---|---|---|---|---|
| | 423 | 205 | 190 | 195 | 195 | 785 |
| | 424 | 75 | 125 | 115 | 110 | 425 |
| | 425 | 30 | 35 | 30 | 35 | 130 |
| | 426 | 25 | 35 | 25 | 30 | 115 |
| | 427 | 175 | 180 | 135 | 190 | 680 |
| TOTAL | | 2,385 | 2,410 | 2,830 | 2,740 | 10,365 |

STATE: LOUISIANA

| ZIP CODE | QUARTER 1 | QUARTER 2 | QUARTER 3 | QUARTER 4 | TOTAL GRAMS |
|---|---|---|---|---|---|
| 700 | 1,225 | 1,160 | 990 | 980 | 4,355 |
| 701 | 200 | 290 | 325 | 265 | 1,080 |
| 703 | 60 | 105 | 70 | 95 | 330 |
| 704 | 610 | 630 | 635 | 525 | 2,400 |
| 705 | 345 | 295 | 280 | 265 | 1,185 |
| 706 | 220 | 205 | 220 | 215 | 860 |
| 707 | 245 | 280 | 280 | 250 | 1,055 |
| 708 | 120 | 175 | 115 | 135 | 545 |
| 710 | 55 | 20 | 40 | 105 | 220 |
| 711 | 155 | 165 | 170 | 155 | 645 |
| 712 | 315 | 275 | 310 | 370 | 1,270 |
| 713 | 200 | 170 | 115 | 130 | 615 |
| 714 | 400 | 280 | 285 | 225 | 1,190 |
| TOTAL | 4,150 | 4,050 | 3,835 | 3,715 | 15,750 |

STATE: MAINE

| ZIP CODE | QUARTER 1 | QUARTER 2 | QUARTER 3 | QUARTER 4 | TOTAL GRAMS |
|---|---|---|---|---|---|
| 39 | 15 | 15 | 15 | 5 | 50 |
| 40 | 90 | 130 | 130 | 120 | 470 |
| 41 | 45 | 35 | 30 | 30 | 140 |
| 42 | 75 | 50 | 60 | 25 | 210 |
| 43 | 100 | 60 | 55 | 65 | 280 |
| 44 | 25 | 30 | 35 | 10 | 100 |
| 45 | 5 | 10 | 20 | 0 | 35 |
| 46 | 40 | 10 | 25 | 55 | 130 |
| 47 | 170 | 185 | 115 | 125 | 595 |

| | | | | | | |
|---|---|---|---|---|---|---|
| | 48 | 0 | 10 | 0 | 10 | 20 |
| | 49 | 185 | 100 | 125 | 80 | 490 |
| TOTAL | | 750 | 635 | 610 | 525 | 2,520 |

STATE: MARYLAND

| ZIP CODE | | QUARTER 1 | QUARTER 2 | QUARTER 3 | QUARTER 4 | TOTAL GRAMS |
|---|---|---|---|---|---|---|
| | 206 | 420 | 445 | 445 | 560 | 1,870 |
| | 207 | 360 | 345 | 565 | 375 | 1,645 |
| | 208 | 585 | 555 | 715 | 575 | 2,430 |
| | 209 | 130 | 100 | 115 | 85 | 430 |
| | 210 | 825 | 785 | 790 | 730 | 3,130 |
| | 211 | 235 | 290 | 345 | 250 | 1,120 |
| | 212 | 645 | 605 | 610 | 765 | 2,625 |
| | 214 | 115 | 85 | 105 | 90 | 395 |
| | 215 | 195 | 210 | 200 | 145 | 750 |
| | 216 | 140 | 110 | 100 | 155 | 505 |
| | 217 | 945 | 765 | 680 | 790 | 3,180 |
| | 218 | 180 | 110 | 130 | 115 | 535 |
| | 219 | 75 | 85 | 95 | 120 | 375 |
| TOTAL | | 4,850 | 4,490 | 4,895 | 4,755 | 18,990 |

STATE: MASSACHUSETTS

| ZIP CODE | | QUARTER 1 | QUARTER 2 | QUARTER 3 | QUARTER 4 | TOTAL GRAMS |
|---|---|---|---|---|---|---|
| | 10 | 325 | 310 | 275 | 345 | 1,255 |
| | 11 | 60 | 40 | 35 | 20 | 155 |
| | 12 | 90 | 50 | 65 | 55 | 260 |
| | 13 | 50 | 25 | 70 | 10 | 155 |
| | 14 | 85 | 85 | 50 | 50 | 270 |
| | 15 | 180 | 165 | 180 | 150 | 675 |
| | 16 | 50 | 20 | 30 | 10 | 110 |
| | 17 | 95 | 85 | 115 | 115 | 410 |
| | 18 | 280 | 280 | 235 | 260 | 1,055 |
| | 19 | 135 | 150 | 150 | 65 | 500 |
| | 20 | 110 | 105 | 105 | 110 | 430 |
| | 21 | 465 | 380 | 455 | 295 | 1,595 |

| | | | | | | |
|---|---|---|---|---|---|---|
| | 22 | 0 | 10 | 10 | 0 | 20 |
| | 23 | 50 | 120 | 70 | 90 | 330 |
| | 24 | 75 | 70 | 80 | 60 | 285 |
| | 25 | 120 | 120 | 115 | 155 | 510 |
| | 26 | 150 | 215 | 125 | 115 | 605 |
| | 27 | 210 | 175 | 185 | 110 | 680 |
| TOTAL | | 2,530 | 2,405 | 2,350 | 2,015 | 9,300 |

STATE: MICHIGAN

| ZIP CODE | | QUARTER 1 | QUARTER 2 | QUARTER 3 | QUARTER 4 | TOTAL GRAMS |
|---|---|---|---|---|---|---|
| | 480 | 1,585 | 1,420 | 1,505 | 1,480 | 5,990 |
| | 481 | 1,595 | 1,430 | 1,630 | 1,650 | 6,305 |
| | 482 | 475 | 380 | 390 | 310 | 1,555 |
| | 483 | 1,060 | 1,115 | 1,170 | 1,205 | 4,550 |
| | 484 | 625 | 660 | 635 | 790 | 2,710 |
| | 485 | 280 | 225 | 340 | 175 | 1,020 |
| | 486 | 215 | 240 | 290 | 355 | 1,100 |
| | 487 | 140 | 155 | 180 | 100 | 575 |
| | 488 | 810 | 640 | 610 | 740 | 2,800 |
| | 489 | 275 | 280 | 280 | 235 | 1,070 |
| | 490 | 585 | 565 | 470 | 520 | 2,140 |
| | 491 | 20 | 0 | 10 | 40 | 70 |
| | 492 | 295 | 250 | 285 | 220 | 1,050 |
| | 493 | 130 | 120 | 160 | 90 | 500 |
| | 494 | 385 | 330 | 315 | 240 | 1,270 |
| | 495 | 205 | 185 | 120 | 130 | 640 |
| | 496 | 245 | 185 | 190 | 155 | 775 |
| | 497 | 185 | 185 | 200 | 140 | 710 |
| | 498 | 135 | 95 | 140 | 145 | 515 |
| | 499 | 20 | 35 | 20 | 100 | 175 |
| TOTAL | | 9,265 | 8,495 | 8,940 | 8,820 | 35,520 |

STATE: MINNESOTA

| ZIP CODE | | QUARTER 1 | QUARTER 2 | QUARTER 3 | QUARTER 4 | TOTAL GRAMS |
|---|---|---|---|---|---|---|
| | 550 | 145 | 140 | 160 | 200 | 645 |

| | | | | | | |
|---|---|---|---|---|---|---|
| | 551 | 235 | 270 | 280 | 355 | 1,140 |
| | 553 | 310 | 405 | 320 | 395 | 1,430 |
| | 554 | 240 | 135 | 205 | 195 | 775 |
| | 556 | 5 | 0 | 0 | 5 | 10 |
| | 557 | 95 | 110 | 115 | 190 | 510 |
| | 558 | 15 | 15 | 40 | 20 | 90 |
| | 559 | 35 | 40 | 20 | 20 | 115 |
| | 560 | 75 | 110 | 50 | 85 | 320 |
| | 561 | 0 | 10 | 0 | 5 | 15 |
| | 562 | 45 | 70 | 80 | 35 | 230 |
| | 563 | 105 | 110 | 60 | 125 | 400 |
| | 564 | 45 | 55 | 70 | 80 | 250 |
| | 565 | 110 | 90 | 95 | 125 | 420 |
| | 566 | 30 | 25 | 40 | 40 | 135 |
| | 567 | 45 | 55 | 60 | 60 | 220 |
| TOTAL | | 1,535 | 1,640 | 1,595 | 1,935 | 6,705 |

STATE: MISSISSIPPI

| ZIP CODE | | QUARTER 1 | QUARTER 2 | QUARTER 3 | QUARTER 4 | TOTAL GRAMS |
|---|---|---|---|---|---|---|
| | 386 | 240 | 215 | 230 | 275 | 960 |
| | 387 | 40 | 30 | 70 | 20 | 160 |
| | 388 | 185 | 265 | 215 | 170 | 835 |
| | 389 | 15 | 0 | 10 | 10 | 35 |
| | 390 | 150 | 155 | 160 | 175 | 640 |
| | 391 | 335 | 340 | 305 | 295 | 1,275 |
| | 392 | 70 | 55 | 150 | 65 | 340 |
| | 393 | 195 | 195 | 180 | 145 | 715 |
| | 394 | 275 | 300 | 270 | 200 | 1,045 |
| | 395 | 425 | 385 | 400 | 560 | 1,770 |
| | 396 | 85 | 45 | 35 | 5 | 170 |
| | 397 | 115 | 145 | 160 | 180 | 600 |
| TOTAL | | 2,130 | 2,130 | 2,185 | 2,100 | 8,545 |

STATE: MISSOURI

| ZIP CODE | | QUARTER 1 | QUARTER 2 | QUARTER 3 | QUARTER 4 | TOTAL GRAMS |
|---|---|---|---|---|---|---|

| | | | | | |
|---|---|---|---|---|---|
| | 630 | 630 | 645 | 665 | 835 | 2,775 |
| | 631 | 635 | 640 | 1,510 | 1,605 | 4,390 |
| | 633 | 420 | 450 | 480 | 630 | 1,980 |
| | 634 | 80 | 110 | 80 | 75 | 345 |
| | 635 | 10 | 40 | 35 | 10 | 95 |
| | 636 | 220 | 285 | 295 | 220 | 1,020 |
| | 637 | 340 | 295 | 295 | 250 | 1,180 |
| | 638 | 175 | 125 | 100 | 90 | 490 |
| | 639 | 60 | 65 | 60 | 50 | 235 |
| | 640 | 470 | 550 | 500 | 515 | 2,035 |
| | 641 | 775 | 775 | 645 | 790 | 2,985 |
| | 644 | 60 | 105 | 50 | 90 | 305 |
| | 645 | 75 | 100 | 95 | 70 | 340 |
| | 646 | 115 | 140 | 90 | 80 | 425 |
| | 647 | 135 | 140 | 150 | 95 | 520 |
| | 648 | 205 | 165 | 80 | 150 | 600 |
| | 650 | 95 | 75 | 90 | 55 | 315 |
| | 651 | 15 | 20 | 35 | 40 | 110 |
| | 652 | 75 | 150 | 110 | 60 | 395 |
| | 653 | 100 | 80 | 100 | 75 | 355 |
| | 654 | 100 | 110 | 110 | 90 | 410 |
| | 655 | 150 | 180 | 155 | 150 | 635 |
| | 656 | 110 | 90 | 120 | 120 | 440 |
| | 657 | 145 | 210 | 145 | 150 | 650 |
| | 658 | 210 | 180 | 110 | 220 | 720 |
| TOTAL | | 5,405 | 5,725 | 6,105 | 6,515 | 23,750 |

STATE: MONTANA

| ZIP CODE | QUARTER 1 | QUARTER 2 | QUARTER 3 | QUARTER 4 | TOTAL GRAMS |
|---|---|---|---|---|---|
| 590 | 80 | 115 | 65 | 35 | 295 |
| 591 | 185 | 140 | 245 | 115 | 685 |
| 592 | 15 | 15 | 10 | 10 | 50 |
| 593 | 55 | 65 | 35 | 105 | 260 |
| 594 | 30 | 15 | 30 | 25 | 100 |
| 595 | 10 | 20 | 5 | 10 | 45 |
| 596 | 40 | 55 | 30 | 40 | 165 |

| | | | | | | |
|---|---|---|---|---|---|---|
| | 597 | 150 | 145 | 220 | 135 | 650 |
| | 598 | 115 | 125 | 155 | 100 | 495 |
| | 599 | 55 | 75 | 85 | 120 | 335 |
| TOTAL | | 735 | 770 | 880 | 695 | 3,080 |

| STATE: | NEBRASKA | | | | | |
|---|---|---|---|---|---|---|
| ZIP CODE | | QUARTER 1 | QUARTER 2 | QUARTER 3 | QUARTER 4 | TOTAL GRAMS |
| | 680 | 45 | 30 | 25 | 35 | 135 |
| | 681 | 180 | 205 | 250 | 260 | 895 |
| | 683 | 45 | 80 | 75 | 85 | 285 |
| | 684 | 35 | 25 | 25 | 25 | 110 |
| | 685 | 240 | 250 | 230 | 225 | 945 |
| | 686 | 80 | 85 | 80 | 95 | 340 |
| | 687 | 45 | 45 | 25 | 50 | 165 |
| | 688 | 150 | 130 | 130 | 155 | 565 |
| | 689 | 10 | 5 | 10 | 25 | 50 |
| | 691 | 75 | 105 | 135 | 140 | 455 |
| | 692 | 0 | 0 | 10 | 5 | 15 |
| | 693 | 145 | 120 | 110 | 100 | 475 |
| TOTAL | | 1,050 | 1,080 | 1,105 | 1,200 | 4,435 |

| STATE: | NEVADA | | | | | |
|---|---|---|---|---|---|---|
| ZIP CODE | | QUARTER 1 | QUARTER 2 | QUARTER 3 | QUARTER 4 | TOTAL GRAMS |
| | 890 | 670 | 620 | 855 | 825 | 2,970 |
| | 891 | 1,425 | 1,465 | 1,620 | 1,690 | 6,200 |
| | 893 | 15 | 40 | 90 | 75 | 220 |
| | 894 | 530 | 460 | 565 | 650 | 2,205 |
| | 895 | 620 | 675 | 680 | 550 | 2,525 |
| | 897 | 140 | 155 | 175 | 120 | 590 |
| | 898 | 35 | 5 | 40 | 25 | 105 |
| TOTAL | | 3,435 | 3,420 | 4,025 | 3,935 | 14,815 |

| STATE: | NEW HAMPSHIRE | | | | | |
|---|---|---|---|---|---|---|
| ZIP CODE | | QUARTER 1 | QUARTER 2 | QUARTER 3 | QUARTER 4 | TOTAL GRAMS |

| | | | | | |
|---|---|---|---|---|---|
| | 30 | 165 | 145 | 185 | 170 | 665 |
| | 31 | 130 | 160 | 110 | 100 | 500 |
| | 32 | 120 | 125 | 115 | 135 | 495 |
| | 33 | 35 | 10 | 40 | 50 | 135 |
| | 34 | 45 | 15 | 60 | 25 | 145 |
| | 35 | 30 | 25 | 40 | 65 | 160 |
| | 37 | 20 | 10 | 15 | 45 | 90 |
| | 38 | 80 | 125 | 145 | 125 | 475 |
| TOTAL | | 625 | 615 | 710 | 715 | 2,665 |

STATE: NEW JERSEY

| ZIP CODE | | QUARTER 1 | QUARTER 2 | QUARTER 3 | QUARTER 4 | TOTAL GRAMS |
|---|---|---|---|---|---|---|
| | 70 | 940 | 845 | 950 | 910 | 3,645 |
| | 71 | 50 | 25 | 55 | 70 | 200 |
| | 72 | 5 | 5 | 0 | 5 | 15 |
| | 73 | 5 | 25 | 20 | 0 | 50 |
| | 74 | 550 | 620 | 565 | 515 | 2,250 |
| | 75 | 95 | 110 | 85 | 80 | 370 |
| | 76 | 290 | 190 | 220 | 260 | 960 |
| | 77 | 895 | 750 | 560 | 490 | 2,695 |
| | 78 | 330 | 355 | 295 | 220 | 1,200 |
| | 79 | 155 | 155 | 165 | 85 | 560 |
| | 80 | 1,200 | 1,350 | 1,410 | 1,320 | 5,280 |
| | 81 | 35 | 40 | 45 | 15 | 135 |
| | 82 | 300 | 265 | 380 | 250 | 1,195 |
| | 83 | 110 | 135 | 95 | 85 | 425 |
| | 84 | 5 | 15 | 5 | 5 | 30 |
| | 85 | 235 | 195 | 170 | 165 | 765 |
| | 86 | 260 | 240 | 210 | 215 | 925 |
| | 87 | 310 | 240 | 305 | 230 | 1,085 |
| | 88 | 745 | 595 | 700 | 625 | 2,665 |
| | 89 | 55 | 35 | 70 | 60 | 220 |
| TOTAL | | 6,570 | 6,190 | 6,305 | 5,605 | 24,670 |

STATE: NEW MEXICO

| ZIP CODE | QUARTER 1 | QUARTER 2 | QUARTER 3 | QUARTER 4 | TOTAL GRAMS |
|---|---|---|---|---|---|
| 870 | 95 | 90 | 105 | 125 | 415 |
| 871 | 280 | 320 | 365 | 295 | 1,260 |
| 873 | 0 | 0 | 5 | 0 | 5 |
| 874 | 15 | 60 | 40 | 40 | 155 |
| 875 | 50 | 85 | 105 | 90 | 330 |
| 877 | 25 | 20 | 15 | 20 | 80 |
| 878 | 0 | 5 | 0 | 0 | 5 |
| 879 | 30 | 55 | 65 | 50 | 200 |
| 880 | 150 | 155 | 110 | 60 | 475 |
| 881 | 15 | 5 | 30 | 5 | 55 |
| 882 | 150 | 185 | 130 | 155 | 620 |
| 883 | 5 | 10 | 20 | 20 | 55 |
| 884 | 5 | 20 | 5 | 15 | 45 |
| TOTAL | 820 | 1,010 | 995 | 875 | 3,700 |

STATE: NEW YORK

| ZIP CODE | QUARTER 1 | QUARTER 2 | QUARTER 3 | QUARTER 4 | TOTAL GRAMS |
|---|---|---|---|---|---|
| 100 | 295 | 310 | 330 | 355 | 1,290 |
| 101 | 45 | 45 | 55 | 35 | 180 |
| 103 | 260 | 160 | 245 | 145 | 810 |
| 104 | 125 | 125 | 85 | 70 | 405 |
| 105 | 210 | 260 | 190 | 125 | 785 |
| 106 | 25 | 30 | 55 | 75 | 185 |
| 107 | 65 | 35 | 90 | 75 | 265 |
| 108 | 10 | 10 | 10 | 0 | 30 |
| 109 | 270 | 315 | 320 | 280 | 1,185 |
| 110 | 40 | 15 | 45 | 25 | 125 |
| 111 | 15 | 5 | 15 | 0 | 35 |
| 112 | 240 | 265 | 220 | 135 | 860 |
| 113 | 90 | 65 | 65 | 45 | 265 |
| 114 | 110 | 105 | 105 | 130 | 450 |
| 115 | 265 | 255 | 330 | 260 | 1,110 |
| 116 | 15 | 0 | 0 | 10 | 25 |
| 117 | 855 | 900 | 710 | 775 | 3,240 |
| 118 | 5 | 10 | 5 | 0 | 20 |

| | | | | | | |
|---|---|---|---|---|---|---|
| | 119 | 120 | 110 | 130 | 70 | 430 |
| | 120 | 275 | 215 | 230 | 195 | 915 |
| | 121 | 25 | 65 | 45 | 45 | 180 |
| | 122 | 130 | 105 | 155 | 55 | 445 |
| | 123 | 55 | 100 | 60 | 50 | 265 |
| | 124 | 80 | 60 | 45 | 30 | 215 |
| | 125 | 95 | 165 | 190 | 90 | 540 |
| | 126 | 60 | 75 | 45 | 10 | 190 |
| | 127 | 90 | 165 | 70 | 80 | 405 |
| | 128 | 165 | 140 | 120 | 145 | 570 |
| | 129 | 60 | 70 | 40 | 80 | 250 |
| | 130 | 190 | 180 | 140 | 185 | 695 |
| | 131 | 70 | 90 | 65 | 45 | 270 |
| | 132 | 70 | 95 | 75 | 40 | 280 |
| | 133 | 80 | 75 | 40 | 50 | 245 |
| | 134 | 25 | 40 | 20 | 35 | 120 |
| | 135 | 25 | 25 | 30 | 35 | 115 |
| | 136 | 145 | 120 | 150 | 95 | 510 |
| | 137 | 0 | 20 | 20 | 15 | 55 |
| | 138 | 40 | 50 | 70 | 45 | 205 |
| | 139 | 45 | 50 | 45 | 35 | 175 |
| | 140 | 195 | 235 | 165 | 180 | 775 |
| | 141 | 125 | 105 | 95 | 65 | 390 |
| | 142 | 225 | 280 | 220 | 195 | 920 |
| | 143 | 0 | 5 | 15 | 5 | 25 |
| | 144 | 175 | 120 | 160 | 155 | 610 |
| | 145 | 100 | 125 | 120 | 110 | 455 |
| | 146 | 205 | 115 | 155 | 140 | 615 |
| | 147 | 60 | 65 | 75 | 55 | 255 |
| | 148 | 75 | 90 | 80 | 75 | 320 |
| | 149 | 20 | 10 | 20 | 20 | 70 |
| TOTAL | | 5,965 | 6,075 | 5,765 | 4,970 | 22,775 |

| STATE: | NORTH CAROLINA | | | | |
|---|---|---|---|---|---|
| ZIP CODE | QUARTER 1 | QUARTER 2 | QUARTER 3 | QUARTER 4 | TOTAL GRAMS |
| 270 | 220 | 185 | 185 | 185 | 775 |

| | | | | | |
|---|---|---|---|---|---|
| 271 | 145 | 95 | 140 | 105 | 485 |
| 272 | 200 | 130 | 180 | 215 | 725 |
| 273 | 90 | 110 | 95 | 75 | 370 |
| 274 | 120 | 140 | 100 | 60 | 420 |
| 275 | 430 | 350 | 325 | 455 | 1,560 |
| 276 | 185 | 170 | 300 | 215 | 870 |
| 277 | 10 | 30 | 50 | 35 | 125 |
| 278 | 155 | 155 | 165 | 260 | 735 |
| 279 | 230 | 225 | 255 | 210 | 920 |
| 280 | 480 | 500 | 410 | 495 | 1,885 |
| 281 | 225 | 295 | 285 | 305 | 1,110 |
| 282 | 205 | 225 | 300 | 185 | 915 |
| 283 | 535 | 375 | 430 | 490 | 1,830 |
| 284 | 340 | 345 | 480 | 425 | 1,590 |
| 285 | 390 | 380 | 290 | 265 | 1,325 |
| 286 | 420 | 525 | 425 | 390 | 1,760 |
| 287 | 560 | 610 | 570 | 600 | 2,340 |
| 288 | 160 | 120 | 75 | 100 | 455 |
| 289 | 40 | 65 | 45 | 45 | 195 |
| TOTAL | 5,140 | 5,030 | 5,105 | 5,115 | 20,390 |

STATE: NORTH DAKOTA

| ZIP CODE | QUARTER 1 | QUARTER 2 | QUARTER 3 | QUARTER 4 | TOTAL GRAMS |
|---|---|---|---|---|---|
| 580 | 50 | 55 | 50 | 50 | 205 |
| 581 | 40 | 35 | 20 | 25 | 120 |
| 582 | 35 | 30 | 30 | 10 | 105 |
| 583 | 0 | 25 | 15 | 0 | 40 |
| 584 | 15 | 25 | 0 | 5 | 45 |
| 585 | 35 | 35 | 25 | 45 | 140 |
| 586 | 15 | 0 | 0 | 5 | 20 |
| 587 | 25 | 25 | 35 | 35 | 120 |
| 588 | 95 | 105 | 130 | 160 | 490 |
| TOTAL | 310 | 335 | 305 | 335 | 1,285 |

STATE: OHIO

| ZIP CODE | QUARTER 1 | QUARTER 2 | QUARTER 3 | QUARTER 4 | TOTAL GRAMS |
|---|---|---|---|---|---|
| 430 | 310 | 315 | 445 | 370 | 1,440 |
| 431 | 225 | 135 | 295 | 290 | 945 |
| 432 | 445 | 300 | 1,260 | 1,570 | 3,575 |
| 433 | 85 | 60 | 95 | 80 | 320 |
| 434 | 85 | 85 | 105 | 60 | 335 |
| 435 | 345 | 365 | 520 | 400 | 1,630 |
| 436 | 430 | 290 | 375 | 455 | 1,550 |
| 437 | 175 | 145 | 150 | 105 | 575 |
| 438 | 5 | 10 | 15 | 15 | 45 |
| 439 | 40 | 60 | 100 | 65 | 265 |
| 440 | 500 | 450 | 535 | 570 | 2,055 |
| 441 | 450 | 345 | 375 | 465 | 1,635 |
| 442 | 270 | 245 | 190 | 285 | 990 |
| 443 | 90 | 95 | 65 | 65 | 315 |
| 444 | 230 | 250 | 200 | 135 | 815 |
| 445 | 35 | 70 | 40 | 70 | 215 |
| 446 | 200 | 255 | 235 | 245 | 935 |
| 447 | 230 | 210 | 190 | 255 | 885 |
| 448 | 80 | 125 | 120 | 140 | 465 |
| 449 | 40 | 75 | 100 | 40 | 255 |
| 450 | 160 | 205 | 350 | 390 | 1,105 |
| 451 | 105 | 85 | 95 | 135 | 420 |
| 452 | 310 | 225 | 440 | 400 | 1,375 |
| 453 | 290 | 255 | 380 | 300 | 1,225 |
| 454 | 305 | 285 | 335 | 325 | 1,250 |
| 455 | 35 | 55 | 50 | 55 | 195 |
| 456 | 190 | 150 | 175 | 165 | 680 |
| 457 | 70 | 55 | 80 | 70 | 275 |
| 458 | 180 | 255 | 180 | 245 | 860 |
| TOTAL | 5,915 | 5,455 | 7,495 | 7,765 | 26,630 |

STATE: OKLAHOMA

| ZIP CODE | QUARTER 1 | QUARTER 2 | QUARTER 3 | QUARTER 4 | TOTAL GRAMS |
|---|---|---|---|---|---|
| 730 | 1,050 | 1,070 | 1,020 | 870 | 4,010 |
| 731 | 950 | 955 | 910 | 765 | 3,580 |

| | | | | | | |
|---|---|---|---|---|---|---|
| | 734 | 125 | 105 | 115 | 110 | 455 |
| | 735 | 345 | 325 | 310 | 240 | 1,220 |
| | 736 | 100 | 60 | 60 | 55 | 275 |
| | 737 | 245 | 275 | 275 | 255 | 1,050 |
| | 738 | 95 | 45 | 25 | 25 | 190 |
| | 739 | 20 | 10 | 15 | 5 | 50 |
| | 740 | 995 | 915 | 700 | 705 | 3,315 |
| | 741 | 385 | 365 | 325 | 325 | 1,400 |
| | 743 | 195 | 205 | 245 | 235 | 880 |
| | 744 | 235 | 295 | 250 | 250 | 1,030 |
| | 745 | 35 | 40 | 25 | 35 | 135 |
| | 746 | 15 | 20 | 20 | 20 | 75 |
| | 747 | 265 | 240 | 180 | 180 | 865 |
| | 748 | 240 | 175 | 200 | 205 | 820 |
| | 749 | 150 | 175 | 150 | 165 | 640 |
| TOTAL | | 5,445 | 5,275 | 4,825 | 4,445 | 19,990 |

STATE: OREGON

| ZIP CODE | | QUARTER 1 | QUARTER 2 | QUARTER 3 | QUARTER 4 | TOTAL GRAMS |
|---|---|---|---|---|---|---|
| | 970 | 365 | 340 | 450 | 480 | 1,635 |
| | 971 | 235 | 160 | 220 | 270 | 885 |
| | 972 | 250 | 235 | 255 | 250 | 990 |
| | 973 | 385 | 265 | 320 | 380 | 1,350 |
| | 974 | 460 | 470 | 490 | 485 | 1,905 |
| | 975 | 275 | 245 | 275 | 330 | 1,125 |
| | 976 | 45 | 35 | 60 | 50 | 190 |
| | 977 | 170 | 125 | 160 | 135 | 590 |
| | 978 | 160 | 170 | 145 | 165 | 640 |
| | 979 | 25 | 30 | 15 | 5 | 75 |
| TOTAL | | 2,370 | 2,075 | 2,390 | 2,550 | 9,385 |

STATE: PENNSYLVANIA

| ZIP CODE | | QUARTER 1 | QUARTER 2 | QUARTER 3 | QUARTER 4 | TOTAL GRAMS |
|---|---|---|---|---|---|---|
| | 150 | 450 | 285 | 335 | 355 | 1,425 |
| | 151 | 205 | 120 | 190 | 250 | 765 |

| | | | | | |
|---|---|---|---|---|---|
| 152 | 435 | 430 | 445 | 600 | 1,910 |
| 153 | 145 | 95 | 130 | 200 | 570 |
| 154 | 150 | 145 | 215 | 155 | 665 |
| 155 | 45 | 25 | 35 | 30 | 135 |
| 156 | 330 | 205 | 325 | 390 | 1,250 |
| 157 | 105 | 80 | 160 | 135 | 480 |
| 158 | 95 | 100 | 125 | 110 | 430 |
| 159 | 145 | 220 | 265 | 190 | 820 |
| 160 | 90 | 95 | 125 | 80 | 390 |
| 161 | 55 | 70 | 70 | 80 | 275 |
| 162 | 35 | 45 | 50 | 85 | 215 |
| 163 | 15 | 55 | 10 | 20 | 100 |
| 164 | 125 | 125 | 160 | 130 | 540 |
| 165 | 230 | 185 | 165 | 125 | 705 |
| 166 | 75 | 50 | 70 | 95 | 290 |
| 167 | 30 | 50 | 40 | 25 | 145 |
| 168 | 45 | 25 | 45 | 35 | 150 |
| 169 | 45 | 50 | 55 | 40 | 190 |
| 170 | 420 | 460 | 420 | 425 | 1,725 |
| 171 | 120 | 80 | 170 | 170 | 540 |
| 172 | 180 | 185 | 190 | 145 | 700 |
| 173 | 350 | 415 | 395 | 285 | 1,445 |
| 174 | 150 | 140 | 185 | 105 | 580 |
| 175 | 115 | 50 | 60 | 80 | 305 |
| 176 | 210 | 260 | 220 | 185 | 875 |
| 177 | 165 | 120 | 130 | 135 | 550 |
| 178 | 155 | 180 | 110 | 120 | 565 |
| 179 | 130 | 125 | 190 | 125 | 570 |
| 180 | 295 | 280 | 330 | 250 | 1,155 |
| 181 | 120 | 45 | 90 | 30 | 285 |
| 182 | 175 | 235 | 180 | 140 | 730 |
| 183 | 170 | 155 | 215 | 160 | 700 |
| 184 | 380 | 345 | 410 | 330 | 1,465 |
| 185 | 150 | 140 | 105 | 140 | 535 |
| 186 | 215 | 220 | 250 | 220 | 905 |
| 187 | 690 | 590 | 485 | 505 | 2,270 |
| 188 | 15 | 65 | 55 | 60 | 195 |

| | | | | | |
|---|---|---|---|---|---|
| 189 | 475 | 445 | 400 | 445 | 1,765 |
| 190 | 1,705 | 1,745 | 1,895 | 1,460 | 6,805 |
| 191 | 1,130 | 1,185 | 1,075 | 1,105 | 4,495 |
| 193 | 370 | 325 | 360 | 430 | 1,485 |
| 194 | 515 | 540 | 525 | 360 | 1,940 |
| 195 | 340 | 265 | 235 | 305 | 1,145 |
| 196 | 430 | 395 | 410 | 385 | 1,620 |
| TOTAL | 12,020 | 11,445 | 12,105 | 11,230 | 46,800 |

STATE: RHODE ISLAND

| ZIP CODE | QUARTER 1 | QUARTER 2 | QUARTER 3 | QUARTER 4 | TOTAL GRAMS |
|---|---|---|---|---|---|
| 28 | 290 | 310 | 260 | 250 | 1,110 |
| 29 | 85 | 105 | 100 | 70 | 360 |
| TOTAL | 375 | 415 | 360 | 320 | 1,470 |

STATE: SOUTH CAROLINA

| ZIP CODE | QUARTER 1 | QUARTER 2 | QUARTER 3 | QUARTER 4 | TOTAL GRAMS |
|---|---|---|---|---|---|
| 290 | 285 | 285 | 245 | 315 | 1,130 |
| 291 | 265 | 305 | 230 | 230 | 1,030 |
| 292 | 270 | 280 | 245 | 235 | 1,030 |
| 293 | 200 | 115 | 285 | 245 | 845 |
| 294 | 500 | 575 | 425 | 515 | 2,015 |
| 295 | 545 | 615 | 625 | 680 | 2,465 |
| 296 | 910 | 905 | 1,040 | 790 | 3,645 |
| 297 | 175 | 145 | 125 | 170 | 615 |
| 298 | 60 | 75 | 80 | 50 | 265 |
| 299 | 85 | 105 | 130 | 120 | 440 |
| TOTAL | 3,295 | 3,405 | 3,430 | 3,350 | 13,480 |

STATE: SOUTH DAKOTA

| ZIP CODE | QUARTER 1 | QUARTER 2 | QUARTER 3 | QUARTER 4 | TOTAL GRAMS |
|---|---|---|---|---|---|
| 570 | 35 | 85 | 60 | 85 | 265 |
| 571 | 40 | 60 | 90 | 100 | 290 |
| 572 | 75 | 80 | 55 | 30 | 240 |

| | | | | | |
|---|---|---|---|---|---|
| 573 | 40 | 60 | 50 | 75 | 225 |
| 574 | 140 | 90 | 130 | 140 | 500 |
| 575 | 50 | 45 | 35 | 30 | 160 |
| 576 | 105 | 75 | 65 | 90 | 335 |
| 577 | 175 | 140 | 165 | 100 | 580 |
| TOTAL | 660 | 635 | 650 | 650 | 2,595 |

STATE: TENNESSEE

| ZIP CODE | QUARTER 1 | QUARTER 2 | QUARTER 3 | QUARTER 4 | TOTAL GRAMS |
|---|---|---|---|---|---|
| 370 | 370 | 475 | 510 | 510 | 1,865 |
| 371 | 590 | 650 | 565 | 630 | 2,435 |
| 372 | 215 | 235 | 300 | 165 | 915 |
| 373 | 395 | 445 | 465 | 410 | 1,715 |
| 374 | 225 | 160 | 160 | 150 | 695 |
| 376 | 135 | 180 | 225 | 250 | 790 |
| 377 | 140 | 130 | 170 | 215 | 655 |
| 378 | 330 | 275 | 295 | 345 | 1,245 |
| 379 | 150 | 190 | 245 | 250 | 835 |
| 380 | 185 | 160 | 185 | 270 | 800 |
| 381 | 95 | 115 | 180 | 140 | 530 |
| 382 | 130 | 145 | 170 | 160 | 605 |
| 383 | 315 | 240 | 310 | 265 | 1,130 |
| 384 | 90 | 60 | 80 | 155 | 385 |
| 385 | 165 | 165 | 165 | 170 | 665 |
| TOTAL | 3,530 | 3,625 | 4,025 | 4,085 | 15,265 |

STATE: TEXAS

| ZIP CODE | QUARTER 1 | QUARTER 2 | QUARTER 3 | QUARTER 4 | TOTAL GRAMS |
|---|---|---|---|---|---|
| 750 | 1,785 | 1,580 | 1,690 | 1,825 | 6,880 |
| 751 | 560 | 520 | 545 | 570 | 2,195 |
| 752 | 460 | 420 | 390 | 390 | 1,660 |
| 753 | 0 | 5 | 0 | 0 | 5 |
| 754 | 595 | 720 | 425 | 495 | 2,235 |
| 755 | 145 | 135 | 110 | 130 | 520 |
| 756 | 455 | 425 | 360 | 460 | 1,700 |

| | | | | | |
|---|---|---|---|---|---|
| 757 | 585 | 555 | 435 | 505 | 2,080 |
| 758 | 225 | 245 | 265 | 200 | 935 |
| 759 | 345 | 450 | 345 | 525 | 1,665 |
| 760 | 995 | 1,015 | 1,080 | 955 | 4,045 |
| 761 | 710 | 760 | 780 | 765 | 3,015 |
| 762 | 450 | 400 | 530 | 435 | 1,815 |
| 763 | 250 | 275 | 255 | 305 | 1,085 |
| 764 | 275 | 250 | 200 | 160 | 885 |
| 765 | 135 | 200 | 140 | 95 | 570 |
| 766 | 270 | 260 | 245 | 225 | 1,000 |
| 767 | 200 | 170 | 145 | 170 | 685 |
| 768 | 160 | 190 | 160 | 120 | 630 |
| 769 | 110 | 80 | 100 | 130 | 420 |
| 770 | 1,045 | 975 | 1,075 | 1,140 | 4,235 |
| 773 | 1,240 | 1,015 | 1,345 | 1,230 | 4,830 |
| 774 | 630 | 575 | 610 | 680 | 2,495 |
| 775 | 1,035 | 1,010 | 1,115 | 1,205 | 4,365 |
| 776 | 295 | 280 | 335 | 230 | 1,140 |
| 777 | 155 | 85 | 150 | 180 | 570 |
| 778 | 140 | 175 | 205 | 200 | 720 |
| 779 | 145 | 115 | 95 | 140 | 495 |
| 780 | 245 | 255 | 295 | 345 | 1,140 |
| 781 | 380 | 305 | 350 | 380 | 1,415 |
| 782 | 880 | 930 | 935 | 1,120 | 3,865 |
| 783 | 215 | 220 | 185 | 180 | 800 |
| 784 | 330 | 285 | 275 | 320 | 1,210 |
| 785 | 345 | 230 | 205 | 360 | 1,140 |
| 786 | 615 | 740 | 580 | 730 | 2,665 |
| 787 | 500 | 430 | 440 | 585 | 1,955 |
| 788 | 30 | 40 | 70 | 45 | 185 |
| 789 | 135 | 155 | 135 | 125 | 550 |
| 790 | 250 | 185 | 225 | 210 | 870 |
| 791 | 425 | 355 | 455 | 380 | 1,615 |
| 792 | 40 | 20 | 15 | 0 | 75 |
| 793 | 155 | 200 | 175 | 170 | 700 |
| 794 | 180 | 205 | 195 | 170 | 750 |
| 795 | 50 | 80 | 55 | 75 | 260 |

| ZIP CODE | QUARTER 1 | QUARTER 2 | QUARTER 3 | QUARTER 4 | TOTAL GRAMS |
|---|---|---|---|---|---|
| 796 | 140 | 190 | 180 | 165 | 675 |
| 797 | 315 | 255 | 390 | 410 | 1,370 |
| 798 | 10 | 15 | 30 | 30 | 85 |
| 799 | 150 | 170 | 125 | 150 | 595 |
| TOTAL | 18,785 | 18,150 | 18,445 | 19,415 | 74,795 |

STATE: UTAH

| ZIP CODE | QUARTER 1 | QUARTER 2 | QUARTER 3 | QUARTER 4 | TOTAL GRAMS |
|---|---|---|---|---|---|
| 840 | 1,265 | 1,080 | 1,495 | 1,645 | 5,485 |
| 841 | 385 | 420 | 375 | 410 | 1,590 |
| 843 | 210 | 175 | 200 | 240 | 825 |
| 844 | 270 | 295 | 440 | 335 | 1,340 |
| 845 | 155 | 60 | 125 | 150 | 490 |
| 846 | 390 | 415 | 485 | 390 | 1,680 |
| 847 | 380 | 290 | 310 | 330 | 1,310 |
| TOTAL | 3,055 | 2,735 | 3,430 | 3,500 | 12,720 |

STATE: VERMONT

| ZIP CODE | QUARTER 1 | QUARTER 2 | QUARTER 3 | QUARTER 4 | TOTAL GRAMS |
|---|---|---|---|---|---|
| 50 | 0 | 0 | 5 | 0 | 5 |
| 51 | 5 | 5 | 5 | 5 | 20 |
| 52 | 25 | 5 | 5 | 5 | 40 |
| 53 | 5 | 0 | 0 | 5 | 10 |
| 54 | 40 | 20 | 40 | 50 | 150 |
| 56 | 0 | 15 | 10 | 20 | 45 |
| 57 | 45 | 35 | 15 | 25 | 120 |
| 58 | 30 | 5 | 15 | 30 | 80 |
| TOTAL | 150 | 85 | 95 | 140 | 470 |

STATE: VIRGIN ISLANDS

| ZIP CODE | QUARTER 1 | QUARTER 2 | QUARTER 3 | QUARTER 4 | TOTAL GRAMS |
|---|---|---|---|---|---|
| 8 | 0 | 0 | 20 | 25 | 45 |
| TOTAL | 0 | 0 | 20 | 25 | 45 |

DATE RANGE: 01/01/2017 TO 12/31/2017 ARCOS 3 - REPORT 1 Run Date: 07/03/2018

RETAIL DRUG DISTRIBUTION BY ZIP CODE WITHIN STATE BY GRAMS WT

| STATE: | VIRGINIA | | | | |
|---|---|---|---|---|---|
| ZIP CODE | QUARTER 1 | QUARTER 2 | QUARTER 3 | QUARTER 4 | TOTAL GRAMS |
| 201 | 615 | 565 | 515 | 430 | 2,125 |
| 220 | 180 | 185 | 160 | 175 | 700 |
| 221 | 370 | 300 | 315 | 325 | 1,310 |
| 222 | 45 | 40 | 70 | 35 | 190 |
| 223 | 195 | 115 | 160 | 220 | 690 |
| 224 | 230 | 225 | 135 | 145 | 735 |
| 225 | 110 | 125 | 95 | 75 | 405 |
| 226 | 180 | 245 | 185 | 110 | 720 |
| 227 | 30 | 35 | 20 | 25 | 110 |
| 228 | 90 | 70 | 130 | 105 | 395 |
| 229 | 255 | 200 | 240 | 220 | 915 |
| 230 | 185 | 195 | 145 | 185 | 710 |
| 231 | 240 | 220 | 260 | 265 | 985 |
| 232 | 260 | 305 | 310 | 240 | 1,115 |
| 233 | 240 | 170 | 140 | 160 | 710 |
| 234 | 295 | 285 | 295 | 205 | 1,080 |
| 235 | 80 | 55 | 75 | 90 | 300 |
| 236 | 285 | 220 | 235 | 225 | 965 |
| 237 | 35 | 55 | 5 | 10 | 105 |
| 238 | 250 | 180 | 285 | 125 | 840 |
| 239 | 35 | 15 | 20 | 5 | 75 |
| 240 | 190 | 195 | 250 | 290 | 925 |
| 241 | 105 | 145 | 130 | 140 | 520 |
| 242 | 310 | 280 | 295 | 355 | 1,240 |
| 243 | 150 | 105 | 120 | 115 | 490 |
| 244 | 45 | 80 | 70 | 125 | 320 |
| 245 | 220 | 185 | 200 | 195 | 800 |
| 246 | 320 | 360 | 305 | 410 | 1,395 |
| TOTAL | 5,545 | 5,155 | 5,165 | 5,005 | 20,870 |

| STATE: | WASHINGTON | | | | |
|---|---|---|---|---|---|
| ZIP CODE | QUARTER 1 | QUARTER 2 | QUARTER 3 | QUARTER 4 | TOTAL GRAMS |
| 980 | 700 | 730 | 785 | 955 | 3,170 |

| | | | | | | |
|---|---|---|---|---|---|---|
| | 981 | 200 | 320 | 285 | 395 | 1,200 |
| | 982 | 540 | 630 | 665 | 745 | 2,580 |
| | 983 | 390 | 420 | 530 | 520 | 1,860 |
| | 984 | 175 | 155 | 235 | 250 | 815 |
| | 985 | 410 | 500 | 375 | 405 | 1,690 |
| | 986 | 155 | 250 | 300 | 330 | 1,035 |
| | 988 | 215 | 195 | 215 | 165 | 790 |
| | 989 | 175 | 165 | 185 | 185 | 710 |
| | 990 | 100 | 70 | 95 | 140 | 405 |
| | 991 | 75 | 120 | 125 | 135 | 455 |
| | 992 | 235 | 285 | 295 | 380 | 1,195 |
| | 993 | 370 | 360 | 300 | 315 | 1,345 |
| | 994 | 25 | 30 | 55 | 45 | 155 |
| TOTAL | | 3,765 | 4,230 | 4,445 | 4,965 | 17,405 |

STATE: WEST VIRGINIA

| ZIP CODE | | QUARTER 1 | QUARTER 2 | QUARTER 3 | QUARTER 4 | TOTAL GRAMS |
|---|---|---|---|---|---|---|
| | 247 | 60 | 70 | 45 | 55 | 230 |
| | 248 | 10 | 0 | 0 | 5 | 15 |
| | 249 | 35 | 40 | 25 | 70 | 170 |
| | 250 | 25 | 30 | 30 | 15 | 100 |
| | 251 | 0 | 20 | 15 | 30 | 65 |
| | 252 | 120 | 160 | 165 | 160 | 605 |
| | 253 | 85 | 80 | 75 | 65 | 305 |
| | 254 | 145 | 105 | 125 | 100 | 475 |
| | 255 | 50 | 75 | 95 | 50 | 270 |
| | 256 | 50 | 55 | 55 | 65 | 225 |
| | 257 | 80 | 80 | 75 | 55 | 290 |
| | 258 | 140 | 125 | 130 | 100 | 495 |
| | 259 | 0 | 0 | 0 | 5 | 5 |
| | 260 | 60 | 80 | 65 | 130 | 335 |
| | 261 | 45 | 45 | 80 | 60 | 230 |
| | 262 | 55 | 50 | 60 | 75 | 240 |
| | 263 | 55 | 60 | 75 | 60 | 250 |
| | 264 | 15 | 15 | 35 | 15 | 80 |
| | 265 | 95 | 90 | 105 | 125 | 415 |

| | | | | | | |
|---|---|---|---|---|---|---|
| | 266 | 5 | 0 | 0 | 0 | 5 |
| | 267 | 75 | 40 | 75 | 0 | 190 |
| | 268 | 10 | 20 | 25 | 20 | 75 |
| TOTAL | | 1,215 | 1,240 | 1,355 | 1,260 | 5,070 |

STATE: WISCONSIN

| ZIP CODE | | QUARTER 1 | QUARTER 2 | QUARTER 3 | QUARTER 4 | TOTAL GRAMS |
|---|---|---|---|---|---|---|
| | 530 | 345 | 275 | 340 | 180 | 1,140 |
| | 531 | 325 | 355 | 405 | 365 | 1,450 |
| | 532 | 130 | 145 | 230 | 190 | 695 |
| | 534 | 15 | 30 | 50 | 20 | 115 |
| | 535 | 125 | 145 | 130 | 140 | 540 |
| | 537 | 20 | 10 | 20 | 20 | 70 |
| | 538 | 10 | 10 | 10 | 5 | 35 |
| | 539 | 25 | 60 | 40 | 35 | 160 |
| | 540 | 40 | 20 | 0 | 20 | 80 |
| | 541 | 35 | 75 | 65 | 65 | 240 |
| | 542 | 55 | 30 | 55 | 25 | 165 |
| | 543 | 40 | 75 | 70 | 55 | 240 |
| | 544 | 50 | 80 | 125 | 85 | 340 |
| | 545 | 70 | 90 | 80 | 75 | 315 |
| | 546 | 35 | 40 | 25 | 35 | 135 |
| | 547 | 115 | 130 | 100 | 140 | 485 |
| | 548 | 60 | 40 | 50 | 30 | 180 |
| | 549 | 175 | 135 | 145 | 155 | 610 |
| TOTAL | | 1,670 | 1,745 | 1,940 | 1,640 | 6,995 |

STATE: WYOMING

| ZIP CODE | | QUARTER 1 | QUARTER 2 | QUARTER 3 | QUARTER 4 | TOTAL GRAMS |
|---|---|---|---|---|---|---|
| | 820 | 25 | 15 | 35 | 40 | 115 |
| | 822 | 55 | 70 | 60 | 50 | 235 |
| | 823 | 0 | 5 | 0 | 5 | 10 |
| | 824 | 140 | 155 | 130 | 130 | 555 |
| | 825 | 40 | 15 | 35 | 35 | 125 |
| | 826 | 30 | 45 | 100 | 80 | 255 |

| | | | | | | |
|---|---|---|---|---|---|---|
| | 827 | 5 | 0 | 10 | 5 | 20 |
| | 828 | 5 | 15 | 5 | 10 | 35 |
| | 829 | 35 | 55 | 95 | 90 | 275 |
| | 831 | 5 | 0 | 0 | 0 | 5 |
| TOTAL | | 340 | 375 | 470 | 445 | 1,630 |

DRUG CODE: 2270 DRUG NAME: PENTOBARBITAL (SCHEDULE 2)

STATE: ALABAMA

| ZIP CODE | | QUARTER 1 | QUARTER 2 | QUARTER 3 | QUARTER 4 | TOTAL GRAMS |
|---|---|---|---|---|---|---|
| | 350 | 1,510.65 | 1,599.51 | 3,287.88 | 2,399.26 | 8,797.30 |
| | 351 | 710.89 | 1,688.37 | 888.62 | 887.24 | 4,175.12 |
| | 352 | 394.54 | 2,772.10 | 2,464.87 | 926.8 | 6,558.31 |
| | 354 | 1,066.34 | 0 | 1,155.20 | 189.12 | 2,410.66 |
| | 355 | 0 | 533.17 | 533.17 | 533.17 | 1,599.51 |
| | 356 | 355.45 | 0 | 710.89 | 88.86 | 1,155.20 |
| | 357 | 177.72 | 710.89 | 2,310.40 | 888.62 | 4,087.63 |
| | 358 | 266.58 | 0 | 177.72 | 205.07 | 649.37 |
| | 359 | 888.62 | 977.48 | 1,777.23 | 1,154.37 | 4,797.70 |
| | 360 | 1,598.41 | 1,331.82 | 976.38 | 3,821.04 | 7,727.65 |
| | 361 | 88.86 | 0 | 2,754.71 | 88.86 | 2,932.43 |
| | 362 | 533.17 | 0 | 710.89 | 1,066.34 | 2,310.40 |
| | 363 | 1,066.34 | 88.86 | 1,244.06 | 355.45 | 2,754.71 |
| | 364 | 177.17 | 622.03 | 621.48 | 532.62 | 1,953.30 |
| | 365 | 1,244.06 | 2,574.78 | 266.58 | 3,554.46 | 7,639.88 |
| | 366 | 0 | 890.42 | 1,421.78 | 0 | 2,312.20 |
| | 367 | 533.17 | 0 | 177.72 | 355.45 | 1,066.34 |
| | 368 | 533.17 | 710.89 | 888.62 | 533.17 | 2,665.85 |
| TOTAL | | 11,145.14 | 14,500.32 | 22,368.20 | 17,579.90 | 65,593.56 |

STATE: ALASKA

| ZIP CODE | | QUARTER 1 | QUARTER 2 | QUARTER 3 | QUARTER 4 | TOTAL GRAMS |
|---|---|---|---|---|---|---|
| | 995 | 2,843.57 | 1,886.14 | 3,376.74 | 1,711.15 | 9,817.60 |
| | 996 | 622.03 | 1,421.78 | 977.48 | 2,488.12 | 5,509.41 |
| | 997 | 1,599.51 | 1,332.92 | 1,421.78 | 1,332.92 | 5,687.13 |
| | 998 | 622.03 | 355.45 | 444.31 | 355.45 | 1,777.24 |

| | | | | | | |
|---|---|---|---|---|---|---|
| TOTAL | | 5,687.14 | 4,996.29 | 6,220.31 | 5,887.64 | 22,791.38 |

STATE: ARIZONA

| ZIP CODE | | QUARTER 1 | QUARTER 2 | QUARTER 3 | QUARTER 4 | TOTAL GRAMS |
|---|---|---|---|---|---|---|
| | 850 | 6,799.04 | 3,520.74 | 9,301.38 | 10,704.39 | 30,325.55 |
| | 851 | 533.17 | 3,462.30 | 2,043.81 | 3,998.77 | 10,038.05 |
| | 852 | 12,253.22 | 10,837.80 | 11,285.77 | 16,172.24 | 50,549.03 |
| | 853 | 9,063.87 | 15,106.46 | 13,684.67 | 15,373.04 | 53,228.04 |
| | 855 | 1,155.20 | 0 | 0 | 1,421.78 | 2,576.98 |
| | 856 | 444.31 | 2,043.81 | 1,510.65 | 3,021.29 | 7,020.06 |
| | 857 | 8,944.94 | 5,493.46 | 9,201.49 | 10,510.26 | 34,150.15 |
| | 859 | 0 | 444.31 | 1,066.34 | 0 | 1,510.65 |
| | 860 | 537.73 | 401.02 | 1,952.11 | 2,071.16 | 4,962.02 |
| | 863 | 4,176.49 | 2,399.26 | 5,776 | 5,332.60 | 17,684.35 |
| | 864 | 977.48 | 888.62 | 622.03 | 533.17 | 3,021.30 |
| TOTAL | | 44,885.45 | 44,597.78 | 56,444.25 | 69,138.70 | 215,066.18 |

STATE: ARKANSAS

| ZIP CODE | | QUARTER 1 | QUARTER 2 | QUARTER 3 | QUARTER 4 | TOTAL GRAMS |
|---|---|---|---|---|---|---|
| | 717 | 0 | 533.17 | 0 | 444.31 | 977.48 |
| | 718 | 0 | 622.03 | 0 | 177.72 | 799.75 |
| | 719 | 799.48 | 1,066.34 | 799.75 | 710.89 | 3,376.46 |
| | 720 | 710.89 | 266.58 | 1,777.23 | 888.62 | 3,643.32 |
| | 721 | 888.62 | 2,754.71 | 2,756.53 | 1.82 | 6,401.68 |
| | 722 | 533.17 | 4,470.42 | 1,094.76 | 732.22 | 6,830.57 |
| | 723 | 0 | 177.72 | 888.62 | 533.17 | 1,599.51 |
| | 724 | 2,221.54 | 799.75 | 799.75 | 622.03 | 4,443.07 |
| | 726 | 533.17 | 1,155.20 | 266.58 | 890.44 | 2,845.39 |
| | 727 | 977.2 | 978.57 | 533.17 | 355.17 | 2,844.11 |
| | 728 | 0 | 1,066.34 | 0 | 0 | 1,066.34 |
| | 729 | 888.62 | 0 | 0 | 710.89 | 1,599.51 |
| TOTAL | | 7,552.69 | 13,890.83 | 8,916.39 | 6,067.28 | 36,427.19 |

STATE: CALIFORNIA

RETAIL DRUG DISTRIBUTION BY ZIP CODE WITHIN STATE BY GRAMS WT

| ZIP CODE | QUARTER 1 | QUARTER 2 | QUARTER 3 | QUARTER 4 | TOTAL GRAMS |
|---|---|---|---|---|---|
| 900 | 4,580.24 | 1,399.45 | 1,985.03 | 2,230.20 | 10,194.92 |
| 902 | 355.45 | 8,897.54 | 9,597.04 | 11.39 | 18,861.42 |
| 905 | 0 | 68.36 | 0 | 39.19 | 107.55 |
| 906 | 0 | 2.28 | 1,066.34 | 1,244.06 | 2,312.68 |
| 907 | 0 | 533.17 | 533.17 | 355.45 | 1,421.79 |
| 908 | 29.62 | 1,066.34 | 0 | 13.67 | 1,109.63 |
| 910 | 3,050.91 | 1,296.47 | 1,254.09 | 1,584.92 | 7,186.39 |
| 911 | 2,132.68 | 0 | 0 | 2,223.82 | 4,356.50 |
| 912 | 1,332.92 | 0 | 177.72 | 1,510.65 | 3,021.29 |
| 913 | 3,998.77 | 4,989 | 6,402.59 | 4,889.66 | 20,280.02 |
| 914 | 266.58 | 710.89 | 447.04 | 0 | 1,424.51 |
| 915 | 20.51 | 0 | 0 | 16.86 | 37.37 |
| 916 | 266.58 | 88.86 | 444.31 | 622.03 | 1,421.78 |
| 917 | 1,338.39 | 22,308.79 | 2,262.64 | 3,199.01 | 29,108.83 |
| 918 | 0 | 710.89 | 0 | 0 | 710.89 |
| 919 | 1,822.80 | 1,244.06 | 2,665.85 | 3,203.57 | 8,936.28 |
| 920 | 8,175.26 | 4,976.24 | 10,245.05 | 9,311.32 | 32,707.87 |
| 921 | 2,615.72 | 1,471 | 2,115.82 | 2,016.02 | 8,218.56 |
| 922 | 4,354.21 | 9,152.73 | 7,376.42 | 7,819.81 | 28,703.17 |
| 923 | 3,533.67 | 8,245.80 | 5,670.64 | 9,227.36 | 26,677.47 |
| 924 | 0 | 2,156.37 | 2,132.68 | 9.11 | 4,298.16 |
| 925 | 6,137.37 | 15,204.89 | 12,575.95 | 12,225.06 | 46,143.27 |
| 926 | 991.15 | 2,226.09 | 2,231.11 | 1,697.11 | 7,145.46 |
| 927 | 303.04 | 339.5 | 9.11 | 4,701.46 | 5,353.11 |
| 928 | 3,921.30 | 2,021.03 | 8,568.98 | 2,422.05 | 16,933.36 |
| 930 | 2,134.50 | 1,421.78 | 1,956.78 | 1,690.19 | 7,203.25 |
| 931 | 355.45 | 355.45 | 444.31 | 355.45 | 1,510.66 |
| 932 | 9,599.78 | 5,164.90 | 5,159.44 | 5,600.10 | 25,524.22 |
| 933 | 8,712.98 | 5,420.55 | 6,603.09 | 8,175.26 | 28,911.88 |
| 934 | 2,852.68 | 3,998.77 | 3,465.60 | 710.89 | 11,027.94 |
| 935 | 6,589.42 | 2,665.85 | 8,708.43 | 6,220.31 | 24,184.01 |
| 936 | 3,821.04 | 1,435.46 | 3,113.34 | 2,493.59 | 10,863.43 |
| 937 | 9,064.78 | 96.15 | 9,809.40 | 196.86 | 19,167.19 |
| 939 | 10,125.65 | 3,111.06 | 2,223.36 | 2,182.61 | 17,642.68 |
| 940 | 1,332.92 | 622.03 | 1,688.37 | 977.48 | 4,620.80 |
| 941 | 1,025.78 | 219.19 | 31.44 | 1,190.74 | 2,467.15 |

| | | | | | | |
|---|---|---|---|---|---|---|
| | 943 | 533.17 | 0.91 | 897.73 | 100.25 | 1,532.06 |
| | 944 | 977.48 | 177.72 | 888.62 | 355.45 | 2,399.27 |
| | 945 | 10,406.27 | 5,343.08 | 8,274.95 | 6,333.68 | 30,357.98 |
| | 946 | 1,102.79 | 1,510.65 | 534.99 | 1,252.63 | 4,401.06 |
| | 947 | 622.03 | 533.17 | 1,066.34 | 1,421.78 | 3,643.32 |
| | 948 | 0 | 0 | 0 | 266.58 | 266.58 |
| | 949 | 5,864.58 | 1,244.06 | 2,332 | 8,001.18 | 17,441.82 |
| | 950 | 1,874.93 | 1,244.06 | 5,603.74 | 3,375.91 | 12,098.64 |
| | 951 | 532.89 | 177.45 | 314.34 | 538.09 | 1,562.77 |
| | 952 | 3,021.29 | 2,754.71 | 1,777.23 | 3,554.46 | 11,107.69 |
| | 953 | 18,926.85 | 8,976.83 | 10,143.06 | 16,349.21 | 54,395.95 |
| | 954 | 1,954.95 | 4,268.53 | 3,554.46 | 988.59 | 10,766.53 |
| | 955 | 3,021.29 | 710.89 | 1,332.92 | 2,488.12 | 7,553.22 |
| | 956 | 5,254.22 | 10,762.08 | 9,921.96 | 13,523.62 | 39,461.88 |
| | 957 | 177.72 | 0 | 534.99 | 88.86 | 801.57 |
| | 958 | 2,274.40 | 8,781.34 | 988.87 | 2,335.46 | 14,380.07 |
| | 959 | 6,574.93 | 4,265.35 | 9,681.78 | 3,375.91 | 23,897.97 |
| | 960 | 5,101.56 | 7,642.09 | 9,152.73 | 6,590.33 | 28,486.71 |
| | 961 | 355.45 | 1,244.06 | 1,510.65 | 1,332.92 | 4,443.08 |
| TOTAL | | 173,418.95 | 173,257.92 | 189,476.50 | 172,640.29 | 708,793.66 |

STATE: COLORADO

| ZIP CODE | | QUARTER 1 | QUARTER 2 | QUARTER 3 | QUARTER 4 | TOTAL GRAMS |
|---|---|---|---|---|---|---|
| | 800 | 1,772.67 | 2,470.07 | 3,380.84 | 3,886.21 | 11,509.79 |
| | 801 | 4,891.39 | 3,287.88 | 5,470.68 | 9,931.98 | 23,581.93 |
| | 802 | 8,086.40 | 1,877.48 | 3,287.88 | 7,022.79 | 20,274.55 |
| | 803 | 1,688.37 | 355.45 | 177.72 | 1,521.58 | 3,743.12 |
| | 804 | 6,486.89 | 2,754.71 | 2,576.98 | 7,552.95 | 19,371.53 |
| | 805 | 5,776 | 7,309.43 | 7,123.05 | 7,197.78 | 27,406.26 |
| | 806 | 2,754.71 | 1,332.92 | 1,688.37 | 1,692.93 | 7,468.93 |
| | 807 | 266.58 | 533.17 | 266.58 | 0 | 1,066.33 |
| | 808 | 888.62 | 2,132.68 | 710.89 | 355.45 | 4,087.64 |
| | 809 | 2,932.43 | 3,021.29 | 383.7 | 3,554.46 | 9,891.88 |
| | 810 | 2,665.85 | 3,910.82 | 3,376.74 | 2,843.57 | 12,796.98 |
| | 811 | 177.72 | 710.89 | 0 | 710.07 | 1,598.68 |
| | 812 | 888.62 | 531.52 | 88.86 | 1,423.61 | 2,932.61 |

| | | | | | |
|---|---|---|---|---|---|
| 813 | 177.72 | 1,510.65 | 1,066.34 | 2,932.43 | 5,687.14 |
| 814 | 1,244.06 | 4,533.21 | 266.58 | 4,354.21 | 10,398.06 |
| 815 | 4,265.35 | 1,332.92 | 2,000.52 | 3,110.15 | 10,708.94 |
| 816 | 2,310.40 | 977.48 | 1,155.20 | 2,488.12 | 6,931.20 |
| TOTAL | 47,273.78 | 38,582.57 | 33,020.93 | 60,578.29 | 179,455.57 |

STATE: CONNECTICUT

| ZIP CODE | QUARTER 1 | QUARTER 2 | QUARTER 3 | QUARTER 4 | TOTAL GRAMS |
|---|---|---|---|---|---|
| 60 | 2,570.39 | 4,172.09 | 3,814.45 | 5,067.27 | 15,624.20 |
| 61 | 287.09 | 489.88 | 401.02 | 1,489.68 | 2,667.67 |
| 62 | 710.89 | 1,155.20 | 2,043.81 | 3,465.60 | 7,375.50 |
| 63 | 1,510.65 | 266.58 | 622.03 | 1,599.51 | 3,998.77 |
| 64 | 7,107.27 | 5,242.83 | 7,286.64 | 6,930.92 | 26,567.66 |
| 65 | 739.15 | 0 | 888.62 | 797.02 | 2,424.79 |
| 66 | 186.84 | 271.14 | 177.72 | 266.58 | 902.28 |
| 67 | 1,243.51 | 2,843.02 | 4,531.66 | 1,954.40 | 10,572.59 |
| 68 | 533.17 | 0 | 630.23 | 0 | 1,163.40 |
| 69 | 0 | 88.86 | 1,066.34 | 0 | 1,155.20 |
| TOTAL | 14,888.96 | 14,529.60 | 21,462.52 | 21,570.98 | 72,452.06 |

STATE: DELAWARE

| ZIP CODE | QUARTER 1 | QUARTER 2 | QUARTER 3 | QUARTER 4 | TOTAL GRAMS |
|---|---|---|---|---|---|
| 197 | 0 | 91.14 | 0 | 307.68 | 398.82 |
| 198 | 76.56 | 109.37 | 111.65 | 61.06 | 358.64 |
| 199 | 710.89 | 1,775.58 | 1,597.86 | 2,307.10 | 6,391.43 |
| TOTAL | 787.45 | 1,976.09 | 1,709.51 | 2,675.84 | 7,148.89 |

STATE: DISTRICT OF COLUMBIA

| ZIP CODE | QUARTER 1 | QUARTER 2 | QUARTER 3 | QUARTER 4 | TOTAL GRAMS |
|---|---|---|---|---|---|
| 200 | 195.95 | 9.11 | 871.3 | 1,476.47 | 2,552.83 |
| TOTAL | 195.95 | 9.11 | 871.3 | 1,476.47 | 2,552.83 |

STATE: FLORIDA

| ZIP CODE | QUARTER 1 | QUARTER 2 | QUARTER 3 | QUARTER 4 | TOTAL GRAMS |
|---|---|---|---|---|---|
| 320 | 2,842.37 | 1,422.24 | 2,935.62 | 2,399.26 | 9,599.49 |
| 321 | 716.36 | 4,909.71 | 1,156.11 | 3,909.91 | 10,692.09 |
| 322 | 1,244.69 | 426.08 | 4,096.92 | 2,441.81 | 8,209.50 |
| 323 | 1,244.06 | 1,421.78 | 6,131.44 | 2,406.55 | 11,203.83 |
| 324 | 1,421.78 | 3,999.22 | 2,135.41 | 3,110.61 | 10,667.02 |
| 325 | 5,152.59 | 9,963.42 | 621.21 | 1,244.06 | 16,981.28 |
| 326 | 1,453.68 | 1,066.34 | 1,424.06 | 2,091.66 | 6,035.74 |
| 327 | 3,648.79 | 4,195.44 | 5,874.23 | 4,479.15 | 18,197.61 |
| 328 | 430.64 | 710.34 | 5,142.76 | 2,328.71 | 8,612.45 |
| 329 | 5,804.79 | 3,465.60 | 4,175.67 | 3,287.33 | 16,733.39 |
| 330 | 629.78 | 991.15 | 1,531.78 | 1,642.34 | 4,795.05 |
| 331 | 116.2 | 474.56 | 401.02 | 861.27 | 1,853.05 |
| 333 | 3,998.77 | 5,243.28 | 4,443.08 | 3,547.62 | 17,232.75 |
| 334 | 318.72 | 7,247.63 | 4,022.83 | 713.44 | 12,302.62 |
| 335 | 2,579.34 | 2,221.54 | 2,314.04 | 1,687.82 | 8,802.74 |
| 336 | 5,566.38 | 588.58 | 1,211.71 | 5,826.58 | 13,193.25 |
| 337 | 1,371.66 | 4,958.02 | 3,519.73 | 3,065.95 | 12,915.36 |
| 338 | 3,529.85 | 8,120.30 | 1,690.83 | 9,125.39 | 22,466.37 |
| 339 | 2,487.30 | 2,276.22 | 3,749.59 | 1,692.01 | 10,205.12 |
| 341 | 653.93 | 266.58 | 444.31 | 3,380.38 | 4,745.20 |
| 342 | 622.03 | 822.54 | 1,512.47 | 2,595.21 | 5,552.25 |
| 344 | 12,350.10 | 3,819.94 | 5,330.32 | 3,496.67 | 24,997.03 |
| 346 | 2,399.26 | 2,488.12 | 2,487.30 | 2,842.74 | 10,217.42 |
| 347 | 2,310.40 | 2,398.71 | 3,554.19 | 266.31 | 8,529.61 |
| 349 | 2,221.54 | 2,665.85 | 535.45 | 4,443.08 | 9,865.92 |
| TOTAL | 65,115.01 | 76,163.19 | 70,442.08 | 72,885.86 | 284,606.14 |

STATE: GEORGIA

| ZIP CODE | QUARTER 1 | QUARTER 2 | QUARTER 3 | QUARTER 4 | TOTAL GRAMS |
|---|---|---|---|---|---|
| 300 | 4,192.95 | 7,280.59 | 5,595.13 | 5,819.18 | 22,887.85 |
| 301 | 4,892.58 | 3,466.51 | 5,336.06 | 4,533.48 | 18,228.63 |
| 302 | 1,596.76 | 1,332.92 | 1,154.92 | 3,195.44 | 7,280.04 |
| 303 | 857.89 | 2,591.57 | 457.98 | 1,476.47 | 5,383.91 |
| 304 | 0 | 0 | 444.31 | 2,843.57 | 3,287.88 |
| 305 | 1,656.47 | 1,332.92 | 1,786.07 | 3,296.71 | 8,072.17 |

| | | | | | | |
|---|---|---|---|---|---|---|
| | 306 | 2,487.57 | 3,376.74 | 2,664.75 | 5,153.97 | 13,683.03 |
| | 307 | 1,510.65 | 2,397.06 | 4,354.21 | 888.62 | 9,150.54 |
| | 308 | 4,265.35 | 177.72 | 0 | 0 | 4,443.07 |
| | 309 | 2,221.54 | 4.56 | 4,525.10 | 539.55 | 7,290.75 |
| | 310 | 177.72 | 710.89 | 443.21 | 977.48 | 2,309.30 |
| | 312 | 365.47 | 2,150.90 | 1,958.60 | 280.26 | 4,755.23 |
| | 313 | 1,155.20 | 1,510.65 | 355.45 | 5,864.86 | 8,886.16 |
| | 314 | 177.45 | 92.23 | 632.51 | 266.31 | 1,168.50 |
| | 315 | 3,375.09 | 1,509 | 3,021.29 | 2,238.12 | 10,143.50 |
| | 316 | 1,071.81 | 0 | 1,071.81 | 0 | 2,143.62 |
| | 317 | 6,309.17 | 4,176.49 | 3,998.77 | 5,776 | 20,260.43 |
| | 318 | 0 | 531.52 | 0 | 531.52 | 1,063.04 |
| | 319 | 622.03 | 1,155.20 | 799.75 | 533.17 | 3,110.15 |
| | 398 | 88.86 | 88.86 | 88.86 | 0 | 266.58 |
| TOTAL | | 37,024.56 | 33,886.33 | 38,688.78 | 44,214.71 | 153,814.38 |

STATE: GUAM

| ZIP CODE | | QUARTER 1 | QUARTER 2 | QUARTER 3 | QUARTER 4 | TOTAL GRAMS |
|---|---|---|---|---|---|---|
| | 969 | 0 | 3,215.42 | 0 | 3,215.42 | 6,430.84 |
| TOTAL | | 0 | 3,215.42 | 0 | 3,215.42 | 6,430.84 |

DRUG CODE:2270DRUG NAME:PENTOBARBITAL (SCHEDULE 2)

STATE: HAWAII

| ZIP CODE | | QUARTER 1 | QUARTER 2 | QUARTER 3 | QUARTER 4 | TOTAL GRAMS |
|---|---|---|---|---|---|---|
| | 967 | 4,287.23 | 888.62 | 8,204.42 | 4,545.61 | 17,925.88 |
| | 968 | 344.05 | 341.78 | 9,020.13 | 312.15 | 10,018.11 |
| TOTAL | | 4,631.28 | 1,230.40 | 17,224.55 | 4,857.76 | 27,943.99 |

STATE: IDAHO

| ZIP CODE | | QUARTER 1 | QUARTER 2 | QUARTER 3 | QUARTER 4 | TOTAL GRAMS |
|---|---|---|---|---|---|---|
| | 832 | 622.03 | 91.6 | 2,491.77 | 1,689.28 | 4,894.68 |
| | 833 | 355.45 | 5,687.14 | 3,199.01 | 1,510.65 | 10,752.25 |

| | | | | | | |
|---|---|---|---|---|---|---|
| | 834 | 1,510.65 | 3,397.24 | 4,354.21 | 4,087.63 | 13,349.73 |
| | 835 | 533.17 | 1,244.06 | 1,510.65 | 1,244.06 | 4,531.94 |
| | 836 | 9,073.90 | 4,535.13 | 8,624.01 | 12,447.90 | 34,680.94 |
| | 837 | 544.11 | 3,188.99 | 4,464.95 | 943.3 | 9,141.35 |
| | 838 | 3,502.05 | 1,954.95 | 3,820.49 | 3,821.04 | 13,098.53 |
| TOTAL | | 16,141.36 | 20,099.11 | 28,465.09 | 25,743.86 | 90,449.42 |

| STATE: | ILLINOIS | | | | | |
|---|---|---|---|---|---|---|
| ZIP CODE | | QUARTER 1 | QUARTER 2 | QUARTER 3 | QUARTER 4 | TOTAL GRAMS |
| | 600 | 10,347.57 | 12,501.90 | 10,929.96 | 14,190.77 | 47,970.20 |
| | 601 | 8,896.54 | 3,960.39 | 7,907.30 | 7,853.35 | 28,617.58 |
| | 602 | 265.76 | 88.86 | 532.34 | 93.42 | 980.38 |
| | 603 | 0 | 977.48 | 0 | 622.03 | 1,599.51 |
| | 604 | 5,174.02 | 5,927.37 | 1,600.78 | 10,685.89 | 23,388.06 |
| | 605 | 4,176.49 | 2,771.93 | 5,598.27 | 3,578.97 | 16,125.66 |
| | 606 | 3,534.86 | 3,015.37 | 5,026.83 | 23,400.64 | 34,977.70 |
| | 607 | 533.17 | 444.31 | 0 | 622.03 | 1,599.51 |
| | 608 | 0 | 88.86 | 0 | 0 | 88.86 |
| | 609 | 0 | 2,219.34 | 565.07 | 1,244.06 | 4,028.47 |
| | 610 | 2,665.85 | 4,709.66 | 2,043.81 | 5,420.55 | 14,839.87 |
| | 611 | 621.76 | 2,074.35 | 1,068.80 | 1,510.65 | 5,275.56 |
| | 612 | 0 | 0 | 977.48 | 533.17 | 1,510.65 |
| | 613 | 2,129.38 | 1,594.56 | 1,595.11 | 3,016.89 | 8,335.94 |
| | 614 | 1,954.95 | 2,576.98 | 2,851.77 | 1,244.06 | 8,627.76 |
| | 615 | 710.89 | 710.89 | 355.45 | 2,043.81 | 3,821.04 |
| | 616 | 2,497.24 | 2,791.16 | 0 | 3,558.11 | 8,846.51 |
| | 617 | 977.48 | 533.17 | 533.17 | 710.89 | 2,754.71 |
| | 618 | 2,052.93 | 4,623.53 | 3,471.07 | 3,914.19 | 14,061.72 |
| | 619 | 444.31 | 977.48 | 710.89 | 1,775.58 | 3,908.26 |
| | 620 | 888.62 | 177.72 | 2,754.71 | 1,155.20 | 4,976.25 |
| | 622 | 1,155.20 | 2,043.81 | 1,421.78 | 1,331.55 | 5,952.34 |
| | 623 | 88.86 | 1,155.20 | 1,244.06 | 355.45 | 2,843.57 |
| | 624 | 3,199.01 | 0 | 888.07 | 3,643.32 | 7,730.40 |
| | 625 | 710.89 | 533.17 | 622.03 | 888.62 | 2,754.71 |
| | 626 | 1,244.06 | 1,510.10 | 1,332.92 | 3,109.60 | 7,196.68 |
| | 627 | 199.96 | 461.99 | 2,132.68 | 371.3 | 3,165.93 |

| | | | | | | |
|---|---|---|---|---|---|---|
| | 628 | 533.17 | 710.89 | 355.45 | 1,421.23 | 3,020.74 |
| | 629 | 1,155.20 | 888.62 | 3,021.29 | 3,198.19 | 8,263.30 |
| TOTAL | | 56,158.17 | 60,069.09 | 59,541.09 | 101,493.52 | 277,261.87 |

STATE: INDIANA

| ZIP CODE | | QUARTER 1 | QUARTER 2 | QUARTER 3 | QUARTER 4 | TOTAL GRAMS |
|---|---|---|---|---|---|---|
| | 460 | 3,199.01 | 3,552.81 | 4,174.84 | 4,363.33 | 15,289.99 |
| | 461 | 1,155.20 | 2,930.51 | 4,178.22 | 3,287.88 | 11,551.81 |
| | 462 | 5,184.40 | 717.73 | 1,615.72 | 2,671.95 | 10,189.80 |
| | 463 | 3,109.60 | 4,798.52 | 2,220.71 | 8,527.95 | 18,656.78 |
| | 464 | 0 | 355.45 | 533.17 | 4.56 | 893.18 |
| | 465 | 4,176.49 | 3,466.51 | 4,089.45 | 4,444.90 | 16,177.35 |
| | 466 | 355.45 | 1,510.65 | 266.58 | 534.99 | 2,667.67 |
| | 467 | 5,598.27 | 3,021.29 | 3,110.15 | 5,425.11 | 17,154.82 |
| | 468 | 3,021.29 | 4,361.50 | 3,943.17 | 4,450.82 | 15,776.78 |
| | 469 | 2,399.26 | 1,332.92 | 3,373.99 | 4,210.67 | 11,316.84 |
| | 470 | 0 | 0 | 0 | 177.72 | 177.72 |
| | 471 | 1,510.65 | 1,957.23 | 2,315.87 | 2,045.82 | 7,829.57 |
| | 472 | 799.75 | 1,599.51 | 2,398.44 | 1,865.27 | 6,662.97 |
| | 473 | 3,293.34 | 977.48 | 1,331.27 | 2,400.17 | 8,002.26 |
| | 474 | 2,319.59 | 1,598.41 | 3,310.66 | 3,107.68 | 10,336.34 |
| | 475 | 710.89 | 0 | 1,510.65 | 1,243.79 | 3,465.33 |
| | 476 | 2,394.04 | 533.17 | 976.1 | 2,216.59 | 6,119.90 |
| | 477 | 9.11 | 2,942.17 | 1,070.44 | 1,601.79 | 5,623.51 |
| | 478 | 1,420.13 | 1,330.72 | 1,066.34 | 2,751.41 | 6,568.60 |
| | 479 | 621.21 | 1,063.86 | 1,953.30 | 1,685.07 | 5,323.44 |
| TOTAL | | 41,277.68 | 38,050.44 | 43,439.07 | 57,017.47 | 179,784.66 |

STATE: IOWA

| ZIP CODE | | QUARTER 1 | QUARTER 2 | QUARTER 3 | QUARTER 4 | TOTAL GRAMS |
|---|---|---|---|---|---|---|
| | 500 | 888.62 | 5,242.83 | 1,244.06 | 7,286.64 | 14,662.15 |
| | 501 | 88.86 | 624.76 | 977.48 | 891.35 | 2,582.45 |
| | 502 | 355.45 | 444.31 | 977.48 | 622.94 | 2,400.18 |
| | 503 | 1,519.76 | 1,337.48 | 2,132.68 | 892.26 | 5,882.18 |
| | 504 | 2.73 | 1.82 | 177.17 | 270.59 | 452.31 |
| | 505 | 1,066.34 | 0 | 355.45 | 888.62 | 2,310.41 |

| | | | | | | |
|---|---|---|---|---|---|---|
| | 506 | 266.58 | 0 | 266.58 | 1,782.70 | 2,315.86 |
| | 507 | 0 | 0 | 888.62 | 0 | 888.62 |
| | 508 | 0 | 0 | 355.45 | 355.45 | 710.9 |
| | 510 | 0 | 710.89 | 355.45 | 0 | 1,066.34 |
| | 511 | 533.17 | 710.89 | 266.58 | 888.62 | 2,399.26 |
| | 513 | 533.17 | 266.58 | 0 | 1,866.09 | 2,665.84 |
| | 515 | 533.17 | 0 | 177.72 | 2,049.28 | 2,760.17 |
| | 516 | 535.9 | 535.9 | 0 | 0 | 1,071.80 |
| | 520 | 0 | 177.72 | 1,155.20 | 444.31 | 1,777.23 |
| | 521 | 0 | 177.72 | 0 | 0 | 177.72 |
| | 522 | 872.67 | 36 | 66.08 | 1,220.36 | 2,195.11 |
| | 523 | 1,155.20 | 444.31 | 533.17 | 622.03 | 2,754.71 |
| | 524 | 444.31 | 1,599.23 | 2,310.40 | 1,903.18 | 6,257.12 |
| | 525 | 0 | 1,421.78 | 266.58 | 710.89 | 2,399.25 |
| | 526 | 1,599.51 | 1,777.23 | 799.75 | 4,620.80 | 8,797.29 |
| | 527 | 177.72 | 0 | 355.45 | 1,421.78 | 1,954.95 |
| | 528 | 710.89 | 1,066.34 | 533.17 | 710.89 | 3,021.29 |
| TOTAL | | 11,284.05 | 16,575.79 | 14,194.52 | 29,448.78 | 71,503.14 |

STATE: KANSAS

| ZIP CODE | | QUARTER 1 | QUARTER 2 | QUARTER 3 | QUARTER 4 | TOTAL GRAMS |
|---|---|---|---|---|---|---|
| | 660 | 1,777.69 | 2,132.68 | 3,643.32 | 5,153.97 | 12,707.66 |
| | 661 | 341.78 | 0 | 266.58 | 355.45 | 963.81 |
| | 662 | 453.42 | 444.31 | 355.45 | 6,494.18 | 7,747.36 |
| | 664 | 88.86 | 266.58 | 710.89 | 2,043.81 | 3,110.14 |
| | 665 | 7,108.92 | 177.72 | 4,620.80 | 888.62 | 12,796.06 |
| | 666 | 1,672.42 | 622.03 | 1,599.51 | 710.89 | 4,604.85 |
| | 667 | 0 | 0 | 710.89 | 2,488.12 | 3,199.01 |
| | 668 | 0 | 0 | 0 | 177.72 | 177.72 |
| | 669 | 0 | 177.72 | 0 | 0 | 177.72 |
| | 670 | 1,155.20 | 1,688.37 | 2,399.26 | 1,510.65 | 6,753.48 |
| | 671 | 1,066.34 | 1,244.06 | 1,866.09 | 1,954.95 | 6,131.44 |
| | 672 | 3,080.53 | 2,540.07 | 2,313.59 | 4,809.91 | 12,744.10 |
| | 673 | 533.17 | 622.03 | 0 | 888.62 | 2,043.82 |
| | 674 | 799.75 | 1,421.78 | 356.81 | 88.86 | 2,667.20 |
| | 675 | 1,954.95 | 533.17 | 533.17 | 3,998.77 | 7,020.06 |

| | | | | | | |
|---|---|---|---|---|---|---|
| | 676 | 177.72 | 533.17 | 267.5 | 888.62 | 1,867.01 |
| | 677 | 0 | 355.45 | 357.27 | 0 | 712.72 |
| | 678 | 1,510.65 | 1,688.37 | 1,332.92 | 533.17 | 5,065.11 |
| | 679 | 0 | 0 | 0.91 | 0 | 0.91 |
| TOTAL | | 21,721.40 | 14,447.51 | 21,334.96 | 32,986.31 | 90,490.18 |

STATE: KENTUCKY

| ZIP CODE | | QUARTER 1 | QUARTER 2 | QUARTER 3 | QUARTER 4 | TOTAL GRAMS |
|---|---|---|---|---|---|---|
| | 400 | 1,421.78 | 977.48 | 3,020.74 | 532.62 | 5,952.62 |
| | 401 | 0 | 886.97 | 0 | 177.72 | 1,064.69 |
| | 402 | 2,339 | 2,894.95 | 1,955.02 | 7,310.42 | 14,499.39 |
| | 403 | 1,332.92 | 4,172.35 | 5,597.71 | 1,155.20 | 12,258.18 |
| | 404 | 2,220.71 | 2,488.12 | 1,598.41 | 2,398.99 | 8,706.23 |
| | 405 | 1,083.19 | 0 | 4,228.74 | 2,962.19 | 8,274.12 |
| | 406 | 888.62 | 267.04 | 177.72 | 88.59 | 1,421.97 |
| | 407 | 177.72 | 1,066.34 | 1,066.34 | 1,244.06 | 3,554.46 |
| | 408 | 0 | 0 | 0 | 444.31 | 444.31 |
| | 409 | 88.86 | 444.31 | 444.31 | 177.72 | 1,155.20 |
| | 410 | 886.69 | 1,604.43 | 2,129.93 | 1,862.79 | 6,483.84 |
| | 411 | 0 | 0 | 0 | 0.91 | 0.91 |
| | 412 | 0 | 0 | 0 | 265.76 | 265.76 |
| | 417 | 0 | 0 | 0 | 355.45 | 355.45 |
| | 420 | 2,843.57 | 2,668.58 | 3,110.15 | 3,643.32 | 12,265.62 |
| | 421 | 2,576.98 | 1,510.65 | 4,354.21 | 1,065.24 | 9,507.08 |
| | 422 | 1,510.65 | 888.62 | 1,066.34 | 2,488.12 | 5,953.73 |
| | 423 | 1,331.82 | 443.76 | 1,154.92 | 2,660.62 | 5,591.12 |
| | 424 | 533.17 | 1,507.90 | 710.89 | 1,155.20 | 3,907.16 |
| | 425 | 354.35 | 0 | 531.52 | 88.86 | 974.73 |
| | 426 | 0 | 177.17 | 88.86 | 533.17 | 799.2 |
| | 427 | 886.97 | 888.62 | 3,999.40 | 2,132.68 | 7,907.67 |
| TOTAL | | 20,477 | 22,887.29 | 35,235.21 | 32,743.94 | 111,343.44 |

STATE: LOUISIANA

| ZIP CODE | QUARTER 1 | QUARTER 2 | QUARTER 3 | QUARTER 4 | TOTAL GRAMS |
|---|---|---|---|---|---|

| | | | | | |
|---|---|---|---|---|---|
| 700 | 2,491.03 | 3,109.88 | 2,501.79 | 3,921.75 | 12,024.45 |
| 701 | 476.21 | 1,421.78 | 2,079.81 | 808.87 | 4,786.67 |
| 703 | 888.62 | 3,200.65 | 1,332.92 | 2,131.85 | 7,554.04 |
| 704 | 4,706.91 | 4,354.21 | 5,965.11 | 5,864.86 | 20,891.09 |
| 705 | 3,590.68 | 4,411.57 | 1,785.14 | 3,707.99 | 13,495.38 |
| 706 | 531.52 | 0 | 5,154.14 | 1,336.57 | 7,022.23 |
| 707 | 177.17 | 710.89 | 266.58 | 444.31 | 1,598.95 |
| 708 | 2,178.25 | 0 | 1,421.78 | 1,648.81 | 5,248.84 |
| 710 | 1,063.04 | 0 | 265.76 | 1,420.96 | 2,749.76 |
| 711 | 5,952.07 | 354.35 | 2,754.71 | 1,474.54 | 10,535.67 |
| 712 | 1,433.99 | 989.69 | 1,426.98 | 1,163.03 | 5,013.69 |
| 713 | 0 | 2,131.85 | 1,510.10 | 1,332.92 | 4,974.87 |
| 714 | 1,331.27 | 799.75 | 799.75 | 1,509 | 4,439.77 |
| TOTAL | 24,820.76 | 21,484.62 | 27,264.57 | 26,765.46 | 100,335.41 |

STATE: MAINE

| ZIP CODE | QUARTER 1 | QUARTER 2 | QUARTER 3 | QUARTER 4 | TOTAL GRAMS |
|---|---|---|---|---|---|
| 39 | 1,154.92 | 0 | 1,332.92 | 0 | 2,487.84 |
| 40 | 4,087.63 | 4,621.25 | 4,354.21 | 5,242.83 | 18,305.92 |
| 41 | 1,421.78 | 856.26 | 1,545.28 | 2,043.81 | 5,867.13 |
| 42 | 5,065.11 | 1,332.92 | 3,732.18 | 3,109.60 | 13,239.81 |
| 43 | 1,599.51 | 533.17 | 888.62 | 533.17 | 3,554.47 |
| 44 | 4,354.21 | 4,354.21 | 3,287.88 | 8,015.76 | 20,012.06 |
| 45 | 266.58 | 88.59 | 88.59 | 976.65 | 1,420.41 |
| 46 | 1,155.20 | 2,576.98 | 177.72 | 1,244.06 | 5,153.96 |
| 47 | 355.45 | 355.45 | 533.17 | 266.58 | 1,510.65 |
| 48 | 266.31 | 1,154.10 | 1,332.92 | 621.21 | 3,374.54 |
| 49 | 1,599.51 | 1,244.06 | 2,132.68 | 977.48 | 5,953.73 |
| TOTAL | 21,326.21 | 17,116.99 | 19,406.17 | 23,031.15 | 80,880.52 |

STATE: MARYLAND

| ZIP CODE | QUARTER 1 | QUARTER 2 | QUARTER 3 | QUARTER 4 | TOTAL GRAMS |
|---|---|---|---|---|---|
| 206 | 2,232.93 | 443.48 | 2,410.65 | 1,687.54 | 6,774.60 |
| 207 | 2,499.06 | 1,155.20 | 4,128.64 | 799.75 | 8,582.65 |
| 208 | 904.56 | 445.86 | 1,515.66 | 1,155.20 | 4,021.28 |

| | | | | | | |
|---|---|---|---|---|---|---|
| | 209 | 0 | 0 | 0 | 355.45 | 355.45 |
| | 210 | 2,754.16 | 3,287.88 | 3,641.02 | 4,620.52 | 14,303.58 |
| | 211 | 1,510.65 | 888.62 | 2,132.68 | 7,211.45 | 11,743.40 |
| | 212 | 5,575.49 | 180 | 858.08 | 2,335.92 | 8,949.49 |
| | 214 | 533.17 | 0 | 1,066.34 | 0 | 1,599.51 |
| | 215 | 533.17 | 0 | 977.48 | 533.17 | 2,043.82 |
| | 216 | 1,953.30 | 531.52 | 88.86 | 531.52 | 3,105.20 |
| | 217 | 2,487.02 | 6,753.47 | 6,130.62 | 7,194.48 | 22,565.59 |
| | 218 | 620.66 | 533.17 | 444.31 | 444.31 | 2,042.45 |
| | 219 | 1,421.78 | 444.31 | 444.31 | 533.17 | 2,843.57 |
| TOTAL | | 23,025.95 | 14,663.51 | 23,838.65 | 27,402.48 | 88,930.59 |

STATE: MASSACHUSETTS

| ZIP CODE | | QUARTER 1 | QUARTER 2 | QUARTER 3 | QUARTER 4 | TOTAL GRAMS |
|---|---|---|---|---|---|---|
| | 010 | 1,776.96 | 4,353.66 | 2,043.26 | 1,598.13 | 9,772.01 |
| | 011 | 888.62 | 0 | 0 | 1,165.22 | 2,053.84 |
| | 012 | 710.89 | 1,332.92 | 444.31 | 1,155.20 | 3,643.32 |
| | 013 | 177.72 | 1,777.23 | 444.31 | 1,688.37 | 4,087.63 |
| | 014 | 977.48 | 444.31 | 3,643.32 | 1,599.51 | 6,664.62 |
| | 015 | 1,954.40 | 1,421.78 | 2,932.43 | 5,680.54 | 11,989.15 |
| | 016 | 22.79 | 13.67 | 931.07 | 633.78 | 1,601.31 |
| | 017 | 1,155.20 | 1,954.68 | 3,287.88 | 799.75 | 7,197.51 |
| | 018 | 444.31 | 4,265.35 | 1,155.20 | 2,576.98 | 8,441.84 |
| | 019 | 1,510.65 | 1,332.92 | 1,066.34 | 2,575.61 | 6,485.52 |
| | 020 | 5,172.20 | 1,297.38 | 2,521.84 | 1,534.80 | 10,526.22 |
| | 021 | 0 | 0 | 0 | 15.95 | 15.95 |
| | 022 | 1,599.51 | 444.31 | 2,310.40 | 2,221.54 | 6,575.76 |
| | 023 | 622.03 | 533.17 | 799.75 | 1,421.78 | 3,376.73 |
| | 024 | 533.17 | 1,332.92 | 1,066.34 | 1,332.92 | 4,265.35 |
| | 025 | 1,066.34 | 1,244.06 | 2,665.85 | 2,399.26 | 7,375.51 |
| | 026 | 710.89 | 2,132.68 | 2,043.81 | 1,599.51 | 6,486.89 |
| TOTAL | | 21,722.42 | 26,302.17 | 31,710.32 | 33,023.51 | 112,758.42 |

STATE: MICHIGAN

| ZIP CODE | QUARTER 1 | QUARTER 2 | QUARTER 3 | QUARTER 4 | TOTAL GRAMS |
|---|---|---|---|---|---|
| 480 | 9,441.83 | 8,642.35 | 13,620.14 | 9,670.41 | 41,374.73 |
| 481 | 14,898.45 | 11,543.23 | 14,289.01 | 17,603.68 | 58,334.37 |
| 482 | 3,618.26 | 4,536.04 | 3,271.93 | 8,144.73 | 19,570.96 |
| 483 | 5,889.92 | 4,271.91 | 7,914.41 | 7,642.54 | 25,718.78 |
| 484 | 5,418.63 | 5,507.49 | 9,237.28 | 7,466.64 | 27,630.04 |
| 485 | 2,132.68 | 1,244.06 | 3,465.60 | 183.19 | 7,025.53 |
| 486 | 2,142.49 | 2,579.44 | 1,872.02 | 1,272.76 | 7,866.71 |
| 487 | 533.17 | 533.17 | 888.62 | 3,998.77 | 5,953.73 |
| 488 | 11,465.32 | 6,219.76 | 16,083.11 | 10,573.42 | 44,341.61 |
| 489 | 4,553.81 | 799.75 | 8.2 | 1,435.46 | 6,797.22 |
| 490 | 4,870.88 | 14,304.42 | 13,642.56 | 17,229.47 | 50,047.33 |
| 491 | 1,244.06 | 622.03 | 2,221.54 | 533.17 | 4,620.80 |
| 492 | 2,310.40 | 810.32 | 3,020.19 | 1,333.83 | 7,474.74 |
| 493 | 1,954.95 | 3,199.01 | 5,599.19 | 3,554.46 | 14,307.61 |
| 494 | 4,621.71 | 3,825.60 | 3,379.47 | 5,871.24 | 17,698.02 |
| 495 | 6,266.79 | 3,340.74 | 4,198.36 | 3,919.02 | 17,724.91 |
| 496 | 2,132.68 | 2,403.82 | 3,020.74 | 1,887.23 | 9,444.47 |
| 497 | 3,199.01 | 622.03 | 1,866.09 | 1,421.78 | 7,108.91 |
| 498 | 2,840.27 | 1,421.78 | 2,132.68 | 1,954.95 | 8,349.68 |
| 499 | 0 | 1,686.72 | 354.9 | 1,509.55 | 3,551.17 |
| TOTAL | 89,535.31 | 78,113.67 | 110,086.04 | 107,206.30 | 384,941.32 |

STATE: MINNESOTA

| ZIP CODE | QUARTER 1 | QUARTER 2 | QUARTER 3 | QUARTER 4 | TOTAL GRAMS |
|---|---|---|---|---|---|
| 550 | 2,759.26 | 5,153.97 | 3,909.91 | 4,887.38 | 16,710.52 |
| 551 | 3,023.57 | 4,903.33 | 3,988.29 | 10,745.41 | 22,660.60 |
| 553 | 4,798.52 | 7,642.09 | 7,730.95 | 8,975.01 | 29,146.57 |
| 554 | 4,010.88 | 4,074.04 | 3,122.54 | 15,108.08 | 26,315.54 |
| 556 | 0 | 0 | 355.45 | 0 | 355.45 |
| 557 | 1,151.90 | 1,421.78 | 1,599.51 | 1,954.95 | 6,128.14 |
| 558 | 177.72 | 1,166.59 | 444.31 | 538.64 | 2,327.26 |
| 559 | 2,665.48 | 2,494.50 | 1,333.01 | 2,490.40 | 8,983.39 |
| 560 | 1,420.68 | 3,021.29 | 2,043.81 | 533.17 | 7,018.95 |
| 561 | 266.58 | 177.72 | 88.86 | 0 | 533.16 |
| 562 | 977.48 | 799.75 | 533.17 | 1,155.20 | 3,465.60 |

| | | | | | | |
|---|---|---|---|---|---|---|
| | 563 | 1,510.10 | 2,221.54 | 3,731.63 | 2,754.71 | 10,217.98 |
| | 564 | 355.45 | 266.58 | 1,777.23 | 0 | 2,399.26 |
| | 565 | 2,665.85 | 1,688.37 | 0 | 1,421.78 | 5,776 |
| | 566 | 444.31 | 0 | 0 | 444.31 | 888.62 |
| | 567 | 0 | 1,066.34 | 531.52 | 354.9 | 1,952.76 |
| TOTAL | | 26,227.78 | 36,097.89 | 31,190.19 | 51,363.94 | 144,879.80 |

STATE: MISSISSIPPI

| ZIP CODE | | QUARTER 1 | QUARTER 2 | QUARTER 3 | QUARTER 4 | TOTAL GRAMS |
|---|---|---|---|---|---|---|
| | 386 | 3,376.74 | 1,866.09 | 0 | 1,777.23 | 7,020.06 |
| | 387 | 0 | 0 | 355.45 | 0 | 355.45 |
| | 388 | 0 | 355.45 | 728.21 | 0 | 1,083.66 |
| | 389 | 0 | 533.17 | 0 | 0 | 533.17 |
| | 390 | 0 | 177.72 | 0 | 177.72 | 355.44 |
| | 391 | 355.45 | 265.76 | 0 | 355.45 | 976.66 |
| | 392 | 3,557.65 | 0 | 2,535.51 | 2,843.57 | 8,936.73 |
| | 393 | 974.73 | 0 | 177.72 | 1,063.04 | 2,215.49 |
| | 394 | 462.54 | 906.84 | 533.17 | 1,066.34 | 2,968.89 |
| | 395 | 2,310.40 | 888.62 | 1,420.13 | 799.75 | 5,418.90 |
| | 396 | 88.59 | 531.52 | 0 | 0 | 620.11 |
| | 397 | 0 | 0 | 531.52 | 1,507.35 | 2,038.87 |
| TOTAL | | 11,126.10 | 5,525.17 | 6,281.71 | 9,590.45 | 32,523.43 |

STATE: MISSOURI

| ZIP CODE | | QUARTER 1 | QUARTER 2 | QUARTER 3 | QUARTER 4 | TOTAL GRAMS |
|---|---|---|---|---|---|---|
| | 630 | 5,506.11 | 2,219.89 | 5,774.35 | 7,021.88 | 20,522.23 |
| | 631 | 3,101.49 | 2,084.83 | 5,736.81 | 3,764.44 | 14,687.57 |
| | 633 | 3,020.74 | 3,558.55 | 5,962.83 | 3,907.98 | 16,450.10 |
| | 634 | 0 | 0 | 1,421.78 | 1,066.34 | 2,488.12 |
| | 635 | 0 | 178.63 | 979.3 | 356.36 | 1,514.29 |
| | 636 | 1,155.20 | 2,488.12 | 1,510.65 | 2,399.26 | 7,553.23 |
| | 637 | 1,155.20 | 1,782.70 | 801.58 | 1,345.68 | 5,085.16 |
| | 638 | 88.86 | 177.72 | 177.72 | 355.45 | 799.75 |
| | 639 | 0 | 355.45 | 177.72 | 0 | 533.17 |
| | 640 | 6,664.61 | 4,620.80 | 7,127.15 | 11,374.27 | 29,786.83 |
| | 641 | 901.37 | 747.07 | 1,440.92 | 2,819.14 | 5,908.50 |

| | | | | | | |
|---|---|---|---|---|---|---|
| | 644 | 2,132.68 | 977.48 | 1,510.65 | 3,021.29 | 7,642.10 |
| | 646 | 355.45 | 0 | 0 | 0 | 355.45 |
| | 647 | 533.17 | 977.48 | 444.31 | 1,244.06 | 3,199.02 |
| | 648 | 977.48 | 1,599.51 | 622.03 | 2,310.40 | 5,509.42 |
| | 650 | 1,424.52 | 0 | 0 | 1,066.34 | 2,490.86 |
| | 651 | 1,599.51 | 88.86 | 1,332.92 | 88.86 | 3,110.15 |
| | 652 | 5,493.46 | 1,214.44 | 5,202.27 | 232.86 | 12,143.03 |
| | 653 | 0 | 0 | 888.62 | 0 | 888.62 |
| | 654 | 355.45 | 0 | 533.17 | 0 | 888.62 |
| | 655 | 620.38 | 886.97 | 1,333.38 | 886.97 | 3,727.70 |
| | 656 | 1,242.41 | 1,688.37 | 2,576.98 | 5,153.97 | 10,661.73 |
| | 657 | 533.17 | 266.58 | 1,332.92 | 1,155.20 | 3,287.87 |
| | 658 | 91.6 | 192.31 | 266.58 | 1,243.79 | 1,794.28 |
| TOTAL | | 36,952.86 | 26,105.76 | 47,154.64 | 50,814.54 | 161,027.80 |

| STATE: | MONTANA | | | | |
|---|---|---|---|---|---|
| ZIP CODE | QUARTER 1 | QUARTER 2 | QUARTER 3 | QUARTER 4 | TOTAL GRAMS |
| 590 | 1,510.10 | 1,155.20 | 889.53 | 5,598.27 | 9,153.10 |
| 591 | 1,246.80 | 800.66 | 1,868.83 | 1,156.11 | 5,072.40 |
| 592 | 0 | 533.17 | 177.72 | 710.89 | 1,421.78 |
| 593 | 0 | 0 | 622.03 | 3,465.60 | 4,087.63 |
| 594 | 1,244.06 | 799.75 | 2,310.40 | 1,510.65 | 5,864.86 |
| 595 | 533.17 | 0 | 533.17 | 0 | 1,066.34 |
| 596 | 1,688.37 | 177.72 | 355.45 | 2,221.54 | 4,443.08 |
| 597 | 1,421.78 | 3,643.32 | 3,554.46 | 8,441.84 | 17,061.40 |
| 598 | 3,821.04 | 2,315.87 | 5,509.41 | 5,598.27 | 17,244.59 |
| 599 | 977.48 | 1,066.34 | 5,688.96 | 977.48 | 8,710.26 |
| TOTAL | 12,442.80 | 10,492.03 | 21,509.96 | 29,680.65 | 74,125.44 |

| STATE: | NEBRASKA | | | | |
|---|---|---|---|---|---|
| ZIP CODE | QUARTER 1 | QUARTER 2 | QUARTER 3 | QUARTER 4 | TOTAL GRAMS |
| 680 | 1,066.34 | 1,777.23 | 2,310.40 | 4,265.35 | 9,419.32 |
| 681 | 2,306.30 | 2,807.11 | 4,499.58 | 6,284.01 | 15,897 |
| 683 | 1,422.70 | 799.75 | 1,692.01 | 1,421.78 | 5,336.24 |
| 684 | 0 | 799.75 | 1,155.20 | 1,066.34 | 3,021.29 |

| | | | | | | |
|---|---|---|---|---|---|---|
| | 685 | 3,643.32 | 3,110.15 | 4,000.59 | 4,620.80 | 15,374.86 |
| | 686 | 266.58 | 266.58 | 444.31 | 888.62 | 1,866.09 |
| | 687 | 0 | 1,332.92 | 2,488.12 | 1,421.78 | 5,242.82 |
| | 688 | 1,599.51 | 2,576.98 | 1,599.51 | 2,046.55 | 7,822.55 |
| | 689 | 533.17 | 710.89 | 799.75 | 2,843.57 | 4,887.38 |
| | 690 | 0 | 0 | 0 | 533.17 | 533.17 |
| | 691 | 801.12 | 2,310.40 | 1,245.43 | 2,311.77 | 6,668.72 |
| | 693 | 888.62 | 2,576.98 | 267.04 | 2,665.85 | 6,398.49 |
| TOTAL | | 12,527.66 | 19,068.74 | 20,501.94 | 30,369.59 | 82,467.93 |

STATE: NEVADA

| ZIP CODE | | QUARTER 1 | QUARTER 2 | QUARTER 3 | QUARTER 4 | TOTAL GRAMS |
|---|---|---|---|---|---|---|
| | 890 | 1,778.14 | 977.93 | 1,422.70 | 1,599.51 | 5,778.28 |
| | 891 | 3,355.41 | 6,113.67 | 9,279.87 | 9,916.12 | 28,665.07 |
| | 893 | 266.58 | 0 | 0 | 0 | 266.58 |
| | 894 | 4,354.67 | 2,932.43 | 4,443.08 | 4,443.08 | 16,173.26 |
| | 895 | 959.25 | 911.86 | 2,688.63 | 2,400.63 | 6,960.37 |
| | 897 | 710.89 | 622.03 | 180 | 799.75 | 2,312.67 |
| | 898 | 799.75 | 0 | 266.58 | 533.17 | 1,599.50 |
| TOTAL | | 12,224.69 | 11,557.92 | 18,280.86 | 19,692.26 | 61,755.73 |

DRUG CODE:2270DRUG NAME:PENTOBARBITAL (SCHEDULE 2)

STATE: NEW HAMPSHIRE

| ZIP CODE | | QUARTER 1 | QUARTER 2 | QUARTER 3 | QUARTER 4 | TOTAL GRAMS |
|---|---|---|---|---|---|---|
| | 030 | 2,310.40 | 5,239.53 | 4,264.25 | 4,620.42 | 16,434.60 |
| | 031 | 18.23 | 177.72 | 88.86 | 355.45 | 640.26 |
| | 032 | 2,665.85 | 4,976.24 | 5,418.90 | 5,242.83 | 18,303.82 |
| | 033 | 975.83 | 177.72 | 266.58 | 1,064.69 | 2,484.82 |
| | 034 | 1,421.78 | 799.75 | 1,599.51 | 2,665.85 | 6,486.89 |
| | 035 | 0 | 88.86 | 177.72 | 0 | 266.58 |
| | 036 | 88.86 | 266.58 | 0 | 355.45 | 710.89 |
| | 037 | 1,546.65 | 2,343.56 | 2,488.12 | 710.89 | 7,089.22 |
| | 038 | 2,843.57 | 2,576.98 | 2,488.12 | 1,599.51 | 9,508.18 |
| TOTAL | | 11,871.17 | 16,646.94 | 16,792.06 | 16,615.09 | 61,925.26 |

STATE: NEW JERSEY

| ZIP CODE | QUARTER 1 | QUARTER 2 | QUARTER 3 | QUARTER 4 | TOTAL GRAMS |
|---|---|---|---|---|---|
| 070 | 1,332.92 | 1,599.51 | 1,332.92 | 1,906.55 | 6,171.90 |
| 071 | 86.58 | 9.11 | 2,349.59 | 43.29 | 2,488.57 |
| 072 | 0 | 0 | 0.91 | 0 | 0.91 |
| 074 | 532.62 | 1,065.79 | 1,067.06 | 1,157.02 | 3,822.49 |
| 075 | 0 | 0 | 122.13 | 0 | 122.13 |
| 076 | 1,954.95 | 1,911.66 | 1,877.48 | 2,087.56 | 7,831.65 |
| 077 | 1,421.78 | 799.75 | 2,756.53 | 3,465.60 | 8,443.66 |
| 078 | 4,176.49 | 1,599.51 | 6,220.31 | 1,599.51 | 13,595.82 |
| 079 | 1,102.79 | 532.89 | 977.48 | 199.6 | 2,812.76 |
| 080 | 2,323.61 | 1,784.52 | 3,472.89 | 4,370.62 | 11,951.64 |
| 081 | 0 | 0 | 0 | 9.11 | 9.11 |
| 082 | 0 | 977.48 | 266.58 | 799.48 | 2,043.54 |
| 083 | 1,332.92 | 1,866.09 | 1,954.95 | 1,244.06 | 6,398.02 |
| 085 | 2,749.76 | 1,064.69 | 3,016.07 | 2,752.51 | 9,583.03 |
| 086 | 27.34 | 0 | 177.72 | 88.86 | 293.92 |
| 087 | 177.72 | 533.17 | 444.31 | 266.58 | 1,421.78 |
| 088 | 1,166.20 | 1,512.01 | 266.58 | 1,010.83 | 3,955.62 |
| 089 | 444.31 | 458.89 | 23.7 | 44.66 | 971.56 |
| TOTAL | 18,829.99 | 15,715.07 | 26,327.21 | 21,045.84 | 81,918.11 |

STATE: NEW MEXICO

| ZIP CODE | QUARTER 1 | QUARTER 2 | QUARTER 3 | QUARTER 4 | TOTAL GRAMS |
|---|---|---|---|---|---|
| 870 | 3,910.82 | 4,266.26 | 2,844.48 | 4,799.43 | 15,820.99 |
| 871 | 2,064.32 | 2,400.17 | 2,665.85 | 5,513.42 | 12,643.76 |
| 874 | 1,782.70 | 2,221.54 | 3,026.76 | 3,826.51 | 10,857.51 |
| 875 | 1,332.92 | 3,110.15 | 6,677.37 | 977.48 | 12,097.92 |
| 877 | 799.75 | 0 | 622.03 | 88.86 | 1,510.64 |
| 878 | 0 | 0 | 177.72 | 355.45 | 533.17 |
| 879 | 354.35 | 0 | 177.17 | 177.17 | 708.69 |
| 880 | 2,310.40 | 5,143.88 | 5,335.06 | 3,294.26 | 16,083.60 |
| 881 | 355.45 | 533.17 | 888.62 | 0 | 1,777.24 |
| 882 | 710.89 | 267.95 | 3,469.70 | 2,399.26 | 6,847.80 |

| | | | | | | |
|---|---|---|---|---|---|---|
| | 883 | 266.58 | 177.72 | 622.03 | 355.45 | 1,421.78 |
| TOTAL | | 13,888.18 | 18,120.84 | 26,506.79 | 21,787.29 | 80,303.10 |

STATE: NEW YORK

| ZIP CODE | | QUARTER 1 | QUARTER 2 | QUARTER 3 | QUARTER 4 | TOTAL GRAMS |
|---|---|---|---|---|---|---|
| | 100 | 91.14 | 2,148.17 | 682.64 | 477.12 | 3,399.07 |
| | 101 | 0 | 88.86 | 177.72 | 0 | 266.58 |
| | 103 | 0 | 0 | 355.45 | 355.45 | 710.9 |
| | 104 | 0 | 93.14 | 191.39 | 19.14 | 303.67 |
| | 105 | 975.2 | 1,778.14 | 982.03 | 1,599.51 | 5,334.88 |
| | 109 | 976.1 | 977.48 | 1,065.24 | 444.31 | 3,463.13 |
| | 110 | 0 | 0 | 88.86 | 407.12 | 495.98 |
| | 112 | 23.7 | 2.73 | 1,070.90 | 10.94 | 1,108.27 |
| | 113 | 2.28 | 8.2 | 0 | 5.47 | 15.95 |
| | 114 | 0 | 0 | 533.17 | 0 | 533.17 |
| | 115 | 799.75 | 442.93 | 444.31 | 799.75 | 2,486.74 |
| | 117 | 710.89 | 1,242.96 | 3,483.83 | 1,619.37 | 7,057.05 |
| | 118 | 0 | 0 | 88.59 | 0 | 88.59 |
| | 119 | 444.31 | 533.17 | 1,066.34 | 711.8 | 2,755.62 |
| | 120 | 444.31 | 1,420.13 | 1,065.79 | 1,331.55 | 4,261.78 |
| | 121 | 5,152.32 | 5,065.11 | 5,420.55 | 7,107.27 | 22,745.25 |
| | 122 | 1,197.47 | 88.86 | 88.86 | 444.31 | 1,819.50 |
| | 123 | 88.86 | 200.51 | 88.86 | 177.72 | 555.95 |
| | 124 | 0 | 1,688.37 | 0 | 533.17 | 2,221.54 |

DRUG CODE:2270DRUG NAME:PENTOBARBITAL (SCHEDULE 2)
STATE:NEW YORK

| ZIP CODE | | QUARTER 1 | QUARTER 2 | QUARTER 3 | QUARTER 4 | TOTAL GRAMS |
|---|---|---|---|---|---|---|
| | 125 | 7,552.49 | 5,772.70 | 9,062.22 | 7,015.11 | 29,402.52 |
| | 126 | 622.03 | 266.58 | 533.17 | 379.14 | 1,800.92 |
| | 127 | 0 | 0 | 0 | 88.86 | 88.86 |
| | 128 | 1,777.23 | 2,664.20 | 4,443.08 | 3,730.53 | 12,615.04 |
| | 129 | 1,599.51 | 1,688.37 | 1,599.51 | 3,732.18 | 8,619.57 |
| | 130 | 3,549.79 | 4,615.30 | 5,149.02 | 5,503.64 | 18,817.75 |

| | | | | | | |
|---|---|---|---|---|---|---|
| | 131 | 266.58 | 1,332.92 | 977.48 | 4,443.08 | 7,020.06 |
| | 132 | 747.35 | 736.58 | 1,082.46 | 1,273.68 | 3,840.07 |
| | 133 | 1,777.23 | 3,287.88 | 888.62 | 1,777.23 | 7,730.96 |
| | 134 | 2,310.40 | 3,373.44 | 1,510.65 | 2,043.81 | 9,238.30 |
| | 136 | 0 | 265.76 | 1,778.31 | 977.48 | 3,021.55 |
| | 137 | 442.93 | 708.69 | 531.52 | 531.52 | 2,214.66 |
| | 138 | 1,509.55 | 1,951.65 | 2,396.24 | 3,195.17 | 9,052.61 |
| | 139 | 0 | 177.72 | 177.72 | 88.86 | 444.3 |
| | 140 | 4,265.35 | 4,972.67 | 5,152.59 | 8,086.40 | 22,477.01 |
| | 141 | 4,627.63 | 3,287.88 | 3,110.15 | 3,465.32 | 14,490.98 |
| | 142 | 2,485.92 | 3,838.71 | 5,901.21 | 6,710.36 | 18,936.20 |
| | 143 | 177.17 | 266.31 | 266.31 | 1,154.92 | 1,864.71 |
| | 144 | 2,575.88 | 1,686.44 | 5,507.76 | 5,516.23 | 15,286.31 |
| | 145 | 4,876.94 | 11,444.44 | 10,829.83 | 5,232.38 | 32,383.59 |
| | 146 | 263.58 | 400.74 | 845.96 | 337.67 | 1,847.95 |
| | 147 | 1,155.20 | 975.83 | 1,597.86 | 977.48 | 4,706.37 |
| | 148 | 2,664.75 | 7,194.76 | 3,639.75 | 7,372.21 | 20,871.47 |
| TOTAL | | 56,153.84 | 76,688.33 | 83,875.95 | 89,677.26 | 306,395.38 |

STATE: NORTH CAROLINA

| ZIP CODE | | QUARTER 1 | QUARTER 2 | QUARTER 3 | QUARTER 4 | TOTAL GRAMS |
|---|---|---|---|---|---|---|
| | 270 | 1,954.95 | 2,488.12 | 2,219.89 | 1,774.48 | 8,437.44 |
| | 271 | 5,523.08 | 1,646.44 | 1,442.29 | 1,433.18 | 10,044.99 |
| | 272 | 1,332.92 | 2,843.57 | 3,913.10 | 3,998.77 | 12,088.36 |
| | 273 | 1,421.78 | 2,043.54 | 4,975.97 | 3,288.51 | 11,729.80 |
| | 274 | 1,884.32 | 1,442.29 | 2,148.17 | 2,155.01 | 7,629.79 |
| | 275 | 4,351 | 3,284.03 | 7,328.17 | 4,047.87 | 19,011.07 |
| | 276 | 97.98 | 4,798.52 | 533.17 | 5,509.41 | 10,939.08 |
| | 277 | 480.76 | 2,645.34 | 534.08 | 3,346.21 | 7,006.39 |
| | 278 | 2,266.28 | 2,780.95 | 2,689.54 | 3,448.82 | 11,185.59 |
| | 279 | 177.72 | 799.75 | 1,777.23 | 710.89 | 3,465.59 |
| | 280 | 2,220.99 | 1,687.82 | 2,843.02 | 5,686.95 | 12,438.78 |
| | 281 | 7,010.16 | 9,232.25 | 4,882.98 | 9,500.48 | 30,625.87 |
| | 282 | 3,643.32 | 1,089.12 | 3,839.27 | 3,009.90 | 11,581.61 |
| | 283 | 8,264.12 | 6,309.17 | 5,862.11 | 9,686.81 | 30,122.21 |
| | 284 | 1,102.79 | 635.43 | 1,626.85 | 1,525.59 | 4,890.66 |

| | | | | | | |
|---|---|---|---|---|---|---|
| | 285 | 2,129.38 | 1,866.09 | 1,155.20 | 3,462.30 | 8,612.97 |
| | 286 | 2,221.54 | 5,776 | 6,398.03 | 2,843.57 | 17,239.14 |
| | 287 | 3,110.15 | 1,954.95 | 3,821.04 | 6,931.20 | 15,817.34 |
| | 288 | 1,929.89 | 2,132.68 | 2,576.98 | 2,317.23 | 8,956.78 |
| | 289 | 88.86 | 177.72 | 177.72 | 266.58 | 710.88 |
| TOTAL | | 51,211.99 | 55,633.78 | 60,744.81 | 74,943.76 | 242,534.34 |

| STATE: | NORTH DAKOTA | | | | | |
|---|---|---|---|---|---|---|
| ZIP CODE | | QUARTER 1 | QUARTER 2 | QUARTER 3 | QUARTER 4 | TOTAL GRAMS |
| | 580 | 1,244.06 | 2,310.40 | 799.75 | 2,576.98 | 6,931.19 |
| | 581 | 906.84 | 2,232.47 | 862.64 | 1,954.95 | 5,956.90 |
| | 582 | 2,665.85 | 888.62 | 177.72 | 355.45 | 4,087.64 |
| | 584 | 2,132.68 | 533.17 | 2,132.68 | 1,066.34 | 5,864.87 |
| | 585 | 533.17 | 0 | 284.81 | 533.17 | 1,351.15 |
| | 586 | 0 | 0 | 0 | 888.62 | 888.62 |
| | 587 | 1,244.06 | 533.17 | 533.17 | 977.48 | 3,287.88 |
| TOTAL | | 8,726.66 | 6,497.83 | 4,790.77 | 8,352.99 | 28,368.25 |

| STATE: | OHIO | | | | | |
|---|---|---|---|---|---|---|
| | 430 | 3,462.84 | 3,467.51 | 3,643.86 | 6,398.37 | 16,972.58 |
| | 431 | 797.55 | 533.17 | 1,331.27 | 1,952.75 | 4,614.74 |
| | 432 | 791.27 | 4,148.50 | 2,836.45 | 908.84 | 8,685.06 |
| | 433 | 355.45 | 4,999.03 | 4,465.86 | 4,821.31 | 14,641.65 |
| | 434 | 177.17 | 977.48 | 443.76 | 354.9 | 1,953.31 |
| | 435 | 2,405.64 | 3,733.55 | 2,851.77 | 2,403.82 | 11,394.78 |
| | 436 | 1,691.10 | 669.88 | 1,717.99 | 1,066.34 | 5,145.31 |
| | 437 | 3,643.32 | 1,688.37 | 3,643.32 | 3,554.46 | 12,529.47 |
| | 438 | 0 | 1,599.51 | 888.62 | 1,066.34 | 3,554.47 |
| | 439 | 444.31 | 710.89 | 1,155.20 | 177.72 | 2,488.12 |
| | 440 | 3,999.86 | 5,508.86 | 3,998.22 | 10,928.59 | 24,435.53 |
| | 441 | 6,905.86 | 8,366.10 | 3,098.30 | 6,475.95 | 24,846.21 |
| | 442 | 6,134.44 | 1,954.95 | 5,153.97 | 6,139.65 | 19,383.01 |
| | 443 | 710.89 | 2,132.68 | 5,360.40 | 1,510.65 | 9,714.62 |
| | 444 | 444.31 | 799.75 | 4,887.38 | 4,709.66 | 10,841.10 |
| | 445 | 800.39 | 1,333.56 | 2,122.38 | 888.07 | 5,144.40 |

| | | | | | | |
|---|---|---|---|---|---|---|
| | 446 | 2,754.71 | 1,516.11 | 5,690.68 | 4,626.27 | 14,587.77 |
| | 447 | 3,733.27 | 975.83 | 1,775.58 | 1,420.68 | 7,905.36 |
| | 448 | 3,465.60 | 2,399.26 | 5,419.45 | 4,265.35 | 15,549.66 |
| | 449 | 0 | 532.07 | 9.11 | 444.31 | 985.49 |
| | 450 | 5,868.50 | 1,958.60 | 3,552.81 | 9,333.37 | 20,713.28 |
| | 451 | 622.03 | 2,132.68 | 1,688.09 | 3,109.88 | 7,552.68 |
| | 452 | 1,934.45 | 1,054.95 | 924.16 | 2,103.51 | 6,017.07 |
| | 453 | 3,819.40 | 4,263.98 | 6,307.24 | 2,665.85 | 17,056.47 |
| | 454 | 970.64 | 2,866.35 | 1,594.95 | 5,621.06 | 11,053 |
| | 455 | 710.89 | 799.75 | 1,510.65 | 355.45 | 3,376.74 |
| | 456 | 1,066.34 | 977.48 | 444.31 | 533.17 | 3,021.30 |
| | 457 | 1,155.20 | 1,777.23 | 1,155.20 | 444.31 | 4,531.94 |
| | 458 | 979.3 | 1,866.09 | 555.95 | 3,025.39 | 6,426.73 |
| TOTAL | | 59,844.73 | 65,744.17 | 78,226.93 | 91,306.02 | 295,121.85 |

STATE: OKLAHOMA

| ZIP CODE | | QUARTER 1 | QUARTER 2 | QUARTER 3 | QUARTER 4 | TOTAL GRAMS |
|---|---|---|---|---|---|---|
| | 730 | 4,176.49 | 2,844.48 | 4,093.10 | 7,551.58 | 18,665.65 |
| | 731 | 2,426.60 | 5,780.55 | 2,762 | 9,824.89 | 20,794.04 |
| | 734 | 1,332.92 | 1,777.23 | 0 | 1,421.78 | 4,531.93 |
| | 735 | 533.17 | 177.72 | 0 | 0 | 710.89 |
| | 736 | 0 | 355.45 | 0 | 0 | 355.45 |
| | 740 | 1,599.51 | 1,599.51 | 1,688.37 | 799.75 | 5,687.14 |
| | 741 | 2,371.92 | 7,559.78 | 3,514.81 | 5,795.14 | 19,241.65 |
| | 743 | 177.17 | 0 | 177.17 | 88.86 | 443.2 |
| | 744 | 533.17 | 0 | 977.48 | 533.17 | 2,043.82 |
| | 745 | 88.59 | 1,063.04 | 265.76 | 0 | 1,417.39 |
| | 748 | 444.31 | 355.45 | 622.03 | 799.75 | 2,221.54 |
| TOTAL | | 13,683.85 | 21,513.21 | 14,100.72 | 26,814.92 | 76,112.70 |

STATE: OREGON

| ZIP CODE | | QUARTER 1 | QUARTER 2 | QUARTER 3 | QUARTER 4 | TOTAL GRAMS |
|---|---|---|---|---|---|---|
| | 970 | 5,864.86 | 7,907.02 | 5,953.72 | 10,041.35 | 29,766.95 |
| | 971 | 4,709.66 | 533.17 | 4,087.63 | 1,688.37 | 11,018.83 |
| | 972 | 2,310.40 | 2,533.69 | 4,799.43 | 4,187.43 | 13,830.95 |

| | | | | | | |
|---|---|---|---|---|---|---|
| | 973 | 4,976.24 | 5,153.97 | 7,464.37 | 5,864.86 | 23,459.44 |
| | 974 | 4,531.94 | 7,464.37 | 5,864.86 | 6,753.47 | 24,614.64 |
| | 975 | 2,132.68 | 4,797.97 | 4,531.94 | 5,331.42 | 16,794.01 |
| | 976 | 888.62 | 710.89 | 2,843.57 | 1,421.78 | 5,864.86 |
| | 977 | 2,137.23 | 3,021.29 | 6,758.94 | 3,201.75 | 15,119.21 |
| | 978 | 1,066.34 | 2,576.98 | 1,332.92 | 6,931.20 | 11,907.44 |
| | 979 | 355.45 | 622.03 | 355.45 | 977.48 | 2,310.41 |
| TOTAL | | 28,973.42 | 35,321.38 | 43,992.83 | 46,399.11 | 154,686.74 |

STATE: PENNSYLVANIA

| ZIP CODE | QUARTER 1 | QUARTER 2 | QUARTER 3 | QUARTER 4 | TOTAL GRAMS |
|---|---|---|---|---|---|
| 150 | 1,421.78 | 2,488.12 | 266.58 | 2,754.71 | 6,931.19 |
| 151 | 2,396.24 | 1,537.99 | 2,753.33 | 1,599.51 | 8,287.07 |
| 152 | 623.85 | 4,137.30 | 4,625.17 | 3,462.85 | 12,849.17 |
| 153 | 1,332.92 | 0 | 5,590.03 | 0 | 6,922.95 |
| 154 | 0 | 88.86 | 1,687.27 | 177.72 | 1,953.85 |
| 155 | 355.45 | 710.89 | 710.89 | 444.31 | 2,221.54 |
| 156 | 3,554.19 | 1,066.34 | 1,599.51 | 4,265.08 | 10,485.12 |
| 159 | 288.54 | 446.59 | 43.29 | 378.23 | 1,156.65 |
| 160 | 266.58 | 0 | 1,597.86 | 533.17 | 2,397.61 |
| 161 | 1,066.34 | 2,132.68 | 0 | 1,688.37 | 4,887.39 |
| 162 | 0 | 0 | 266.58 | 177.72 | 444.3 |
| 163 | 1,244.06 | 1,066.34 | 2,132.68 | 533.62 | 4,976.70 |
| 164 | 2,043.81 | 1,954.95 | 2,399.26 | 1,244.06 | 7,642.08 |
| 165 | 2,111.60 | 2,932.43 | 290.28 | 1,622.10 | 6,956.41 |
| 166 | 533.17 | 1,155.20 | 710.89 | 0 | 2,399.26 |
| 168 | 2,932.43 | 3,110.15 | 710.89 | 1,244.06 | 7,997.53 |
| 169 | 355.45 | 1,421.78 | 2,043.81 | 533.17 | 4,354.21 |
| 170 | 3,109.60 | 2,842.74 | 4,620.25 | 4,043.51 | 14,616.10 |
| 171 | 798.65 | 977.48 | 533.17 | 821.44 | 3,130.74 |
| 172 | 2,398.16 | 2,132.68 | 3,020.74 | 2,043.54 | 9,595.12 |
| 173 | 888.62 | 177.72 | 710.89 | 888.62 | 2,665.85 |
| 174 | 3,155.72 | 2,064.32 | 988.87 | 2,399.26 | 8,608.17 |
| 175 | 1,066.34 | 1,244.06 | 3,554.46 | 799.75 | 6,664.61 |
| 176 | 1,827.36 | 1,245.88 | 0 | 894.08 | 3,967.32 |
| 177 | 533.17 | 622.03 | 1,599.51 | 1,599.51 | 4,354.22 |

| | | | | | | |
|---|---|---|---|---|---|---|
| | 178 | 2,157.74 | 2,932.43 | 3,998.77 | 1,954.95 | 11,043.89 |
| | 179 | 266.58 | 177.72 | 88.86 | 266.58 | 799.74 |
| | 180 | 888.62 | 2,498.15 | 2,310.40 | 3,749.67 | 9,446.84 |
| | 181 | 1,182.54 | 690.39 | 36.46 | 710.89 | 2,620.28 |
| | 182 | 710.89 | 0 | 888.62 | 355.45 | 1,954.96 |
| | 183 | 710.89 | 977.48 | 1,777.23 | 444.31 | 3,909.91 |
| | 184 | 622.03 | 1,599.51 | 1,954.95 | 1,599.51 | 5,776 |
| | 185 | 0 | 0 | 888.62 | 355.45 | 1,244.07 |
| | 186 | 0 | 177.72 | 888.62 | 1,244.06 | 2,310.40 |
| | 187 | 266.58 | 0 | 5,598.27 | 634.33 | 6,499.18 |
| | 188 | 88.86 | 88.86 | 266.58 | 0.91 | 445.21 |
| | 189 | 2,842.47 | 5,153.97 | 5,151.22 | 6,753.47 | 19,901.13 |
| | 190 | 977.48 | 1,797.74 | 1,970.81 | 1,538.90 | 6,284.93 |
| | 191 | 487.6 | 1,328.35 | 746.44 | 761.93 | 3,324.32 |
| | 193 | 3,909.63 | 2,222.45 | 3,998.77 | 7,553.23 | 17,684.08 |
| | 194 | 822.99 | 5,244.65 | 8,372.58 | 1,260.92 | 15,701.14 |
| | 195 | 1,332.92 | 355.45 | 2,754.71 | 1,155.20 | 5,598.28 |
| | 196 | 1,952.75 | 2,399.89 | 3,559.65 | 269.04 | 8,181.33 |
| TOTAL | | 53,524.60 | 63,201.29 | 87,707.77 | 64,757.19 | 269,190.85 |

| STATE: | PUERTO RICO | | | | |
|---|---|---|---|---|---|
| ZIP CODE | QUARTER 1 | QUARTER 2 | QUARTER 3 | QUARTER 4 | TOTAL GRAMS |
| 009 | 0 | 0 | 0.2 | 0 | 0.2 |
| TOTAL | 0 | 0 | 0.2 | 0 | 0.2 |

| STATE: | RHODE ISLAND | | | | |
|---|---|---|---|---|---|
| ZIP CODE | QUARTER 1 | QUARTER 2 | QUARTER 3 | QUARTER 4 | TOTAL GRAMS |
| 028 | 3,021.29 | 5,953.72 | 7,108.65 | 6,575.75 | 22,659.41 |
| 029 | 186.38 | 992.15 | 181.82 | 542.64 | 1,902.99 |
| TOTAL | 3,207.67 | 6,945.87 | 7,290.47 | 7,118.39 | 24,562.40 |

| STATE: | SOUTH CAROLINA | | | | |
|---|---|---|---|---|---|
| ZIP CODE | QUARTER 1 | QUARTER 2 | QUARTER 3 | QUARTER 4 | TOTAL GRAMS |
| 290 | 3,014.69 | 799.75 | 2,132.40 | 1,688.37 | 7,635.21 |

| | | | | | | |
|---|---|---|---|---|---|---|
| | 291 | 88.86 | 1,066.34 | 1,244.06 | 2,043.81 | 4,443.07 |
| | 292 | 1,191.66 | 3,235.47 | 1,266.39 | 3,502.05 | 9,195.57 |
| | 293 | 2,665.85 | 2,136.32 | 1,777.23 | 1,510.65 | 8,090.05 |
| | 294 | 1,510.65 | 1,332.92 | 2,221.54 | 2,537.34 | 7,602.45 |
| | 295 | 2,155.46 | 1,066.34 | 1,547.10 | 1,954.95 | 6,723.85 |
| | 296 | 885.87 | 995.7 | 355.45 | 1,256.82 | 3,493.84 |
| | 297 | 0 | 355.45 | 355.45 | 177.72 | 888.62 |
| | 298 | 533.17 | 1,599.51 | 3,021.29 | 1,155.20 | 6,309.17 |
| | 299 | 533.17 | 266.58 | 0 | 533.17 | 1,332.92 |
| TOTAL | | 12,579.38 | 12,854.38 | 13,920.91 | 16,360.08 | 55,714.75 |

| STATE: | SOUTH DAKOTA | | | | | |
|---|---|---|---|---|---|---|
| ZIP CODE | | QUARTER 1 | QUARTER 2 | QUARTER 3 | QUARTER 4 | TOTAL GRAMS |
| | 570 | 533.17 | 1,599.51 | 1,777.23 | 1,777.23 | 5,687.14 |
| | 571 | 711.35 | 1,244.97 | 533.17 | 1,600.42 | 4,089.91 |
| | 573 | 0 | 0 | 0 | 177.72 | 177.72 |
| | 575 | 533.17 | 0 | 0 | 1,066.34 | 1,599.51 |
| | 577 | 4,003.32 | 2,581.54 | 4,365.61 | 2,488.12 | 13,438.59 |
| TOTAL | | 5,781.01 | 5,426.02 | 6,676.01 | 7,109.83 | 24,992.87 |

| STATE: | TENNESSEE | | | | | |
|---|---|---|---|---|---|---|
| ZIP CODE | | QUARTER 1 | QUARTER 2 | QUARTER 3 | QUARTER 4 | TOTAL GRAMS |
| | 370 | 3,550.06 | 3,550.34 | 4,262.05 | 5,145.17 | 16,507.62 |
| | 371 | 2,485.92 | 3,992.72 | 3,107.40 | 2,487.85 | 12,073.89 |
| | 372 | 269.32 | 367.29 | 444.31 | 2,952.94 | 4,033.86 |
| | 373 | 2,665.02 | 2,575.61 | 4,441.43 | 6,662.96 | 16,345.02 |
| | 374 | 266.58 | 2,433.44 | 897.73 | 631.14 | 4,228.89 |
| | 376 | 3,474.24 | 5,193.70 | 4,797.97 | 2,494.68 | 15,960.59 |
| | 377 | 1,955.59 | 3,642.58 | 2,580.80 | 1,687.54 | 9,866.51 |
| | 378 | 3,198.54 | 2,668.56 | 3,110.23 | 4,714.82 | 13,692.15 |
| | 379 | 3,283.48 | 6,061.24 | 4,631.34 | 1,950.28 | 15,926.34 |
| | 380 | 1,953.58 | 1,066.34 | 799.75 | 1,688.37 | 5,508.04 |
| | 381 | 779.7 | 5,611.67 | 6.84 | 2,065.04 | 8,463.25 |
| | 382 | 355.45 | 266.58 | 1,599.51 | 1,155.20 | 3,376.74 |
| | 383 | 1,598.96 | 531.52 | 1,510.37 | 1,510.10 | 5,150.95 |

| | | | | | | |
|---|---|---|---|---|---|---|
| | 384 | 801.58 | 2,310.12 | 88.86 | 2,756.25 | 5,956.81 |
| | 385 | 0 | 1,066.34 | 2,925.83 | 1,510.65 | 5,502.82 |
| TOTAL | | 26,638.02 | 41,338.05 | 35,204.42 | 39,412.99 | 142,593.48 |

STATE: TEXAS

| ZIP CODE | QUARTER 1 | QUARTER 2 | QUARTER 3 | QUARTER 4 | TOTAL GRAMS |
|---|---|---|---|---|---|
| 750 | 5,466.12 | 6,058.08 | 9,508.35 | 14,851.07 | 35,883.62 |
| 751 | 2,843.57 | 4,620.80 | 2,929.13 | 4,620.80 | 15,014.30 |
| 752 | 3,866.61 | 1,637.33 | 2,135.41 | 4,070.04 | 11,709.39 |
| 753 | 0 | 0 | 32.81 | 106.63 | 139.44 |
| 754 | 2,399.26 | 1,689.28 | 2,042.17 | 3,021.65 | 9,152.36 |
| 755 | 444.31 | 355.45 | 355.45 | 888.62 | 2,043.83 |
| 756 | 2,049.74 | 1,420.68 | 2,486.47 | 4,537.40 | 10,494.29 |
| 757 | 3,019.72 | 2,798.63 | 1,662.12 | 2,501.52 | 9,981.99 |
| 758 | 2,857.24 | 533.17 | 0 | 1,421.78 | 4,812.19 |
| 759 | 2,308.20 | 5,059.61 | 3,017.99 | 1,064.96 | 11,450.76 |
| 760 | 3,021.29 | 3,998.77 | 9,508.18 | 7,730.95 | 24,259.19 |
| 761 | 14,221.49 | 9,158.20 | 1,055.86 | 11,713.77 | 36,149.32 |
| 762 | 2,401.54 | 1,777.23 | 4,443.99 | 3,379.47 | 12,002.23 |
| 763 | 888.62 | 4,443.08 | 977.48 | 2,043.81 | 8,352.99 |
| 764 | 177.72 | 177.72 | 1,599.51 | 1,244.06 | 3,199.01 |
| 765 | 1,041.27 | 2,390.60 | 3,126.56 | 1,335.47 | 7,893.90 |
| 766 | 88.86 | 177.72 | 177.72 | 177.72 | 622.02 |
| 767 | 2,754.71 | 266.58 | 1,331.82 | 799.75 | 5,152.86 |
| 768 | 888.62 | 177.72 | 0 | 710.89 | 1,777.23 |
| 769 | 0 | 0 | 2,321.34 | 0 | 2,321.34 |
| 770 | 1,229.02 | 20,176.12 | 19,353.58 | 4,133.65 | 44,892.37 |
| 773 | 3,021.75 | 1,264.47 | 4,483.99 | 4,458.11 | 13,228.32 |
| 774 | 2,310.40 | 710.89 | 2,932.43 | 977.48 | 6,931.20 |
| 775 | 1,066.34 | 3,475.06 | 7,287.10 | 6,875.49 | 18,703.99 |
| 776 | 354.35 | 444.31 | 2,040.79 | 620.66 | 3,460.11 |
| 777 | 2,043.81 | 0 | 0 | 1,598.68 | 3,642.49 |
| 778 | 2,222.07 | 3,727.78 | 2,841.90 | 3,999.30 | 12,791.05 |
| 779 | 177.72 | 444.31 | 622.03 | 622.03 | 1,866.09 |
| 780 | 4,887.38 | 88.86 | 1,155.20 | 4,442.53 | 10,573.97 |
| 781 | 888.62 | 622.03 | 2,043.81 | 3,376.74 | 6,931.20 |

| | | | | | | |
|---|---|---|---|---|---|---|
| | 782 | 1,723 | 398.74 | 3,021.02 | 2,780.23 | 7,922.99 |
| | 783 | 1,332.92 | 533.17 | 2,576.98 | 622.03 | 5,065.10 |
| | 784 | 544.11 | 0 | 0 | 11.85 | 555.96 |
| | 785 | 9,685.90 | 9,512.74 | 11,023.38 | 10,060.21 | 40,282.23 |
| | 786 | 8,708.43 | 3,021.29 | 3,821.04 | 7,997.54 | 23,548.30 |
| | 787 | 266.58 | 1,818.24 | 1,421.78 | 2,518.20 | 6,024.80 |
| | 788 | 1,866.09 | 0 | 710.89 | 1,599.51 | 4,176.49 |
| | 789 | 0 | 1,244.06 | 0 | 0 | 1,244.06 |
| | 790 | 1,066.34 | 3,554.46 | 1,777.23 | 2,932.43 | 9,330.46 |
| | 791 | 988.87 | 626.59 | 9,961.60 | 1,866.09 | 13,443.15 |
| | 793 | 710.89 | 1,157.02 | 1,954.95 | 266.58 | 4,089.44 |
| | 794 | 2,365.08 | 3,224.08 | 2,319.97 | 3,583.62 | 11,492.75 |
| | 795 | 355.45 | 355.45 | 533.17 | 531.52 | 1,775.59 |
| | 796 | 2,488.12 | 1,954.40 | 1,954.95 | 1,066.34 | 7,463.81 |
| | 797 | 3,820.22 | 2,221.54 | 6,432.83 | 5,248.01 | 17,722.60 |
| | 798 | 0 | 0 | 533.17 | 0 | 533.17 |
| | 799 | 551.4 | 640.26 | 2,861.80 | 2,435.72 | 6,489.18 |
| TOTAL | | 105,413.75 | 107,956.52 | 142,377.95 | 140,844.91 | 496,593.13 |

| STATE: | UTAH | | | | |
|---|---|---|---|---|---|
| ZIP CODE | QUARTER 1 | QUARTER 2 | QUARTER 3 | QUARTER 4 | TOTAL GRAMS |
| 840 | 622.03 | 3,381.29 | 4,880.78 | 3,199.01 | 12,083.11 |
| 841 | 1,736.22 | 1,491.96 | 4,158.26 | 5,987.90 | 13,374.34 |
| 843 | 0 | 0 | 355.45 | 88.86 | 444.31 |
| 844 | 1,777.23 | 444.31 | 444.31 | 1,066.34 | 3,732.19 |
| 845 | 1,244.06 | 533.17 | 177.72 | 3,287.88 | 5,242.83 |
| 846 | 2,399.26 | 3,465.60 | 1,066.34 | 3,732.18 | 10,663.38 |
| 847 | 1,866.09 | 1,955.41 | 2,221.54 | 5,331.69 | 11,374.73 |
| TOTAL | 9,644.89 | 11,271.74 | 13,304.40 | 22,693.86 | 56,914.89 |

| STATE: | VERMONT | | | | |
|---|---|---|---|---|---|
| ZIP CODE | QUARTER 1 | QUARTER 2 | QUARTER 3 | QUARTER 4 | TOTAL GRAMS |
| 050 | 1,421.78 | 977.2 | 2,665.85 | 710.62 | 5,775.45 |
| 051 | 355.45 | 532.07 | 0 | 444.31 | 1,331.83 |
| 052 | 533.17 | 622.03 | 1,688.37 | 799.75 | 3,643.32 |

| | | | | | | |
|---|---|---|---|---|---|---|
| | 053 | 88.86 | 355.45 | 88.86 | 355.45 | 888.62 |
| | 054 | 2,064.68 | 4,389.65 | 7,749.11 | 6,929.55 | 21,132.99 |
| | 056 | 3,199.01 | 2,043.81 | 1,066.34 | 5,509.41 | 11,818.57 |
| | 057 | 2,043.81 | 444.31 | 2,399.26 | 3,021.29 | 7,908.67 |
| | 058 | 88.86 | 0 | 1,773.93 | 0 | 1,862.79 |
| TOTAL | | 9,795.62 | 9,364.52 | 17,431.72 | 17,770.38 | 54,362.24 |

STATE: VIRGINIA

| ZIP CODE | QUARTER 1 | QUARTER 2 | QUARTER 3 | QUARTER 4 | TOTAL GRAMS |
|---|---|---|---|---|---|
| 201 | 3,198.74 | 1,243.79 | 2,044.73 | 5,776 | 12,263.26 |
| 220 | 3,732.18 | 2,665.85 | 2,399.26 | 3,305.19 | 12,102.48 |
| 221 | 891.35 | 799.75 | 1,599.51 | 622.03 | 3,912.64 |
| 222 | 0 | 0 | 88.86 | 0 | 88.86 |
| 223 | 446.59 | 893.17 | 444.31 | 977.48 | 2,761.55 |
| 224 | 88.86 | 976.1 | 1,155.20 | 976.1 | 3,196.26 |
| 225 | 1,242.41 | 977.48 | 1,686.72 | 1,777.23 | 5,683.84 |
| 226 | 1,155.20 | 2,223.36 | 1,874.29 | 3,379.47 | 8,632.32 |
| 227 | 620.38 | 0 | 2,042.17 | 0 | 2,662.55 |
| 228 | 1,510.65 | 710.89 | 2,665.85 | 1,066.34 | 5,953.73 |
| 229 | 5,513.06 | 6,053.97 | 6,073.57 | 4,537.40 | 22,178 |
| 230 | 2,132.68 | 3,287.88 | 3,465.32 | 2,843.57 | 11,729.45 |
| 231 | 622.03 | 888.62 | 533.17 | 1,243.24 | 3,287.06 |
| 232 | 196.41 | 332.21 | 90.68 | 983.86 | 1,603.16 |
| 233 | 710.89 | 710.89 | 1,244.06 | 1,155.20 | 3,821.04 |
| 234 | 8,620.43 | 5,363.37 | 5,065.11 | 4,272.19 | 23,321.10 |
| 235 | 2,132.68 | 1,157.93 | 2,138.14 | 21.87 | 5,450.62 |
| 236 | 1,066.34 | 1,954.95 | 2,754.71 | 2,843.57 | 8,619.57 |
| 237 | 266.58 | 0 | 271.14 | 4.56 | 542.28 |
| 238 | 1,688.37 | 3,732.18 | 1,777.23 | 1,066.34 | 8,264.12 |
| 239 | 0 | 444.31 | 977.48 | 1,155.20 | 2,576.99 |
| 240 | 0 | 3,652.16 | 2,852.68 | 3,310.39 | 9,815.23 |
| 241 | 710.89 | 799.75 | 533.17 | 533.17 | 2,576.98 |
| 242 | 0 | 0 | 0 | 2,665.85 | 2,665.85 |
| 243 | 2,132.68 | 888.62 | 355.45 | 2,308.20 | 5,684.95 |
| 244 | 1,155.20 | 0 | 888.62 | 2,310.40 | 4,354.22 |
| 245 | 3,021.29 | 6,844.16 | 1,067.25 | 3,998.77 | 14,931.47 |

| | | | | | | |
|---|---|---|---|---|---|---|
| | 246 | 0 | 622.03 | 1,066.34 | 622.03 | 2,310.40 |
| TOTAL | | 42,855.89 | 47,223.42 | 47,155.02 | 53,755.65 | 190,989.98 |

| STATE: | WASHINGTON | | | | | |
|---|---|---|---|---|---|---|
| ZIP CODE | | QUARTER 1 | QUARTER 2 | QUARTER 3 | QUARTER 4 | TOTAL GRAMS |
| | 980 | 4,265.35 | 5,949.60 | 6,309.17 | 4,620.80 | 21,144.92 |
| | 981 | 1,344.32 | 713.17 | 2,271.66 | 1,472.82 | 5,801.97 |
| | 982 | 14,132.17 | 11,467.69 | 11,114.07 | 19,343.55 | 56,057.48 |
| | 983 | 4,265.35 | 3,909.91 | 2,309.57 | 12,351.75 | 22,836.58 |
| | 984 | 1,927.61 | 2,932.43 | 2,316.78 | 1,688.37 | 8,865.19 |
| | 985 | 4,887.38 | 8,797.29 | 4,709.66 | 8,264.12 | 26,658.45 |
| | 986 | 7,380.85 | 4,709.66 | 8,877.90 | 6,839.04 | 27,807.45 |
| | 988 | 1,332.92 | 2,579.72 | 1,421.78 | 2,044.27 | 7,378.69 |
| | 989 | 3,554.46 | 2,132.68 | 2,402.91 | 3,021.29 | 11,111.34 |
| | 990 | 7,819.81 | 2,310.40 | 6,309.17 | 8,441.84 | 24,881.22 |
| | 991 | 0 | 2,046.55 | 1,066.34 | 1,866.09 | 4,978.98 |
| | 992 | 915.96 | 4,907.89 | 3,110.15 | 2,892.33 | 11,826.33 |
| | 993 | 3,554.46 | 888.62 | 4,443.08 | 1,074.54 | 9,960.70 |
| | 994 | 177.72 | 266.58 | 355.45 | 710.89 | 1,510.64 |
| TOTAL | | 55,558.36 | 53,612.19 | 57,017.69 | 74,631.70 | 240,819.94 |

| STATE: | WEST VIRGINIA | | | | | |
|---|---|---|---|---|---|---|
| ZIP CODE | | QUARTER 1 | QUARTER 2 | QUARTER 3 | QUARTER 4 | TOTAL GRAMS |
| | 247 | 0 | 1,066.34 | 0 | 0 | 1,066.34 |
| | 250 | 533.17 | 88.86 | 266.58 | 266.58 | 1,155.19 |
| | 251 | 0 | 266.58 | 0 | 0 | 266.58 |
| | 252 | 88.86 | 355.45 | 444.31 | 177.72 | 1,066.34 |
| | 253 | 266.58 | 2,665.30 | 922.79 | 1,599.51 | 5,454.18 |
| | 254 | 1,332.92 | 2,576.98 | 1,332.92 | 533.17 | 5,775.99 |
| | 255 | 355.45 | 710.89 | 1,066.34 | 710.89 | 2,843.57 |
| | 256 | 0 | 355.45 | 0 | 0 | 355.45 |
| | 257 | 929.35 | 0 | 1,787.26 | 24.15 | 2,740.76 |
| | 258 | 0 | 710.89 | 0 | 0 | 710.89 |
| | 259 | 177.72 | 355.45 | 888.62 | 266.58 | 1,688.37 |
| | 260 | 1,066.34 | 622.03 | 3,199.01 | 1,954.95 | 6,842.33 |

| | | | | | | |
|---|---|---|---|---|---|---|
| | 261 | 266.58 | 977.48 | 1,332.92 | 533.17 | 3,110.15 |
| | 262 | 177.72 | 266.58 | 0 | 177.72 | 622.02 |
| | 263 | 0 | 88.86 | 266.58 | 0 | 355.44 |
| | 264 | 0 | 444.31 | 0 | 88.59 | 532.9 |
| | 265 | 532.07 | 666.23 | 355.45 | 1,609.80 | 3,163.55 |
| | 266 | 1,066.34 | 0 | 885.87 | 0 | 1,952.21 |
| | 267 | 0 | 355.45 | 1,244.06 | 355.45 | 1,954.96 |
| | 268 | 177.72 | 0 | 266.58 | 0 | 444.3 |
| TOTAL | | 6,970.82 | 12,573.13 | 14,259.29 | 8,298.28 | 42,101.52 |

| STATE: | WISCONSIN | | | | | |
|---|---|---|---|---|---|---|
| ZIP CODE | | QUARTER 1 | QUARTER 2 | QUARTER 3 | QUARTER 4 | TOTAL GRAMS |
| | 530 | 2,129.93 | 3,463.38 | 3,997.12 | 5,943.82 | 15,534.25 |
| | 531 | 6,581.22 | 6,756.21 | 5,157.51 | 11,200.56 | 29,695.50 |
| | 532 | 2,141.79 | 6,777.05 | 4,846.37 | 5,718.67 | 19,483.88 |
| | 534 | 0 | 1,155.20 | 266.58 | 888.62 | 2,310.40 |
| | 535 | 7,374.40 | 8,620.93 | 6,309.17 | 14,838.22 | 37,142.72 |
| | 537 | 4,268.09 | 891.8 | 5,116.14 | 6,142.84 | 16,418.87 |
| | 538 | 1,063.04 | 88.86 | 1,329.62 | 444.31 | 2,925.83 |
| | 539 | 710.89 | 888.62 | 1,335.66 | 3,731.63 | 6,666.80 |
| | 540 | 177.72 | 1,421.78 | 1,866.09 | 1,155.20 | 4,620.79 |
| | 541 | 1,599.23 | 533.17 | 889.07 | 266.31 | 3,287.78 |
| | 542 | 531.52 | 1,332.10 | 1,420.13 | 443.21 | 3,726.96 |
| | 543 | 1,159.57 | 710.89 | 1,257.54 | 355.45 | 3,483.45 |
| | 544 | 1,775.58 | 1,330.72 | 3,199.72 | 4,572.92 | 10,878.94 |
| | 545 | 355.45 | 710.89 | 1,422.24 | 1,866.09 | 4,354.67 |
| | 546 | 710.89 | 799.75 | 1,164.31 | 733.68 | 3,408.63 |
| | 547 | 2,308.20 | 3,104.10 | 2,930.23 | 3,198.46 | 11,540.99 |
| | 548 | 2,488.12 | 1,066.34 | 3,909.91 | 2,843.57 | 10,307.94 |
| | 549 | 5,595.53 | 3,909.34 | 5,702.61 | 8,424.61 | 23,632.09 |
| TOTAL | | 40,971.17 | 43,561.13 | 52,120.02 | 72,768.17 | 209,420.49 |

| STATE: | WYOMING | | | | | |
|---|---|---|---|---|---|---|
| ZIP CODE | | QUARTER 1 | QUARTER 2 | QUARTER 3 | QUARTER 4 | TOTAL GRAMS |
| | 820 | 1,066.34 | 2,043.81 | 2,156.37 | 6,131.44 | 11,397.96 |

| | | | | | | |
|---|---|---|---|---|---|---|
| | 822 | 0 | 0 | 444.31 | 0 | 444.31 |
| | 823 | 0 | 0 | 1,066.34 | 0 | 1,066.34 |
| | 824 | 177.72 | 5,065.11 | 266.58 | 1,599.51 | 7,108.92 |
| | 826 | 1,066.34 | 2,404.73 | 9.11 | 2,843.57 | 6,323.75 |
| | 827 | 710.89 | 799.75 | 1,155.20 | 355.45 | 3,021.29 |
| | 828 | 1,510.65 | 1,066.34 | 2,754.71 | 1,688.37 | 7,020.07 |
| | 829 | 0 | 710.89 | 1,244.06 | 1,421.78 | 3,376.73 |
| | 830 | 0 | 0 | 0 | 177.72 | 177.72 |
| | 831 | 0 | 533.17 | 0 | 0 | 533.17 |
| TOTAL | | 4,531.94 | 12,623.80 | 9,096.68 | 14,217.84 | 40,470.26 |

DRUG CODE: 2315 DRUG NAME: SECOBARBITAL (SCHEDULE 2)
STATE: ARIZONA

| ZIP CODE | | QUARTER 1 | QUARTER 2 | QUARTER 3 | QUARTER 4 | TOTAL GRAMS |
|---|---|---|---|---|---|---|
| | 852 | 0 | 0 | 0 | 9.16 | 9.16 |
| | 853 | 9.16 | 0 | 0 | 0 | 9.16 |
| TOTAL | | 9.16 | 0 | 0 | 9.16 | 18.32 |

STATE: CALIFORNIA

| ZIP CODE | | QUARTER 1 | QUARTER 2 | QUARTER 3 | QUARTER 4 | TOTAL GRAMS |
|---|---|---|---|---|---|---|
| | 900 | 18.31 | 9.16 | 9.16 | 18.31 | 54.94 |
| | 902 | 0 | 9.16 | 9.16 | 0 | 18.32 |
| | 906 | 27.47 | 18.31 | 27.47 | 18.31 | 91.56 |
| | 910 | 27.47 | 9.16 | 9.16 | 27.47 | 73.26 |
| | 912 | 0 | 0 | 9.16 | 9.16 | 18.32 |
| | 913 | 9.16 | 0 | 9.16 | 0 | 18.32 |
| | 914 | 9.16 | 9.16 | 9.16 | 0 | 27.48 |
| | 920 | 119.02 | 73.24 | 201.41 | 265.5 | 659.17 |
| | 921 | 9.16 | 0 | 9.16 | 18.31 | 36.63 |
| | 926 | 0 | 36.62 | 18.31 | 18.31 | 73.24 |
| | 928 | 137.33 | 164.79 | 146.48 | 210.57 | 659.17 |
| | 930 | 9.16 | 0 | 18.31 | 18.31 | 45.78 |
| | 931 | 0 | 0 | 18.31 | 18.31 | 36.62 |
| | 934 | 0 | 9.16 | 0 | 0 | 9.16 |

| | | | | | | |
|---|---|---|---|---|---|---|
| | 935 | 0 | 9.16 | 0 | 0 | 9.16 |
| | 939 | 0 | 9.16 | 9.16 | 9.16 | 27.48 |
| | 941 | 421.13 | 311.27 | 347.89 | 320.43 | 1,400.72 |
| | 943 | 36.62 | 9.16 | 9.16 | 27.47 | 82.41 |
| | 944 | 9.16 | 45.78 | 45.78 | 54.93 | 155.65 |
| | 945 | 9.16 | 9.16 | 9.16 | 0 | 27.48 |
| | 949 | 18.31 | 0 | 9.16 | 0 | 27.47 |
| | 950 | 0 | 18.31 | 9.16 | 9.16 | 36.63 |
| | 956 | 9.16 | 183.1 | 173.95 | 283.81 | 650.02 |
| | 958 | 27.47 | 45.78 | 45.78 | 0 | 119.03 |
| | 961 | 0 | 9.16 | 9.16 | 0 | 18.32 |
| TOTAL | | 897.25 | 988.8 | 1,162.77 | 1,327.52 | 4,376.34 |

STATE: COLORADO

| ZIP CODE | | QUARTER 1 | QUARTER 2 | QUARTER 3 | QUARTER 4 | TOTAL GRAMS |
|---|---|---|---|---|---|---|
| | 800 | 45.78 | 18.31 | 64.09 | 27.47 | 155.65 |
| | 801 | 9.16 | 18.31 | 0 | 9.16 | 36.63 |
| | 802 | 0 | 0 | 18.31 | 0 | 18.31 |
| | 803 | 0 | 9.16 | 0 | 9.16 | 18.32 |
| | 804 | 0 | 0 | 0 | 9.16 | 9.16 |
| | 805 | 9.16 | 0 | 0 | 0 | 9.16 |
| | 809 | 36.62 | 36.62 | 9.16 | 0 | 82.4 |
| | 810 | 9.16 | 0 | 0 | 0 | 9.16 |
| | 812 | 0 | 9.16 | 0 | 0 | 9.16 |
| | 814 | 0 | 0 | 0 | 9.16 | 9.16 |
| | 815 | 0 | 0 | 9.16 | 0 | 9.16 |
| | 816 | 0 | 0 | 9.16 | 0 | 9.16 |
| TOTAL | | 109.88 | 91.56 | 109.88 | 64.11 | 375.43 |

STATE: CONNECTICUT

| ZIP CODE | | QUARTER 1 | QUARTER 2 | QUARTER 3 | QUARTER 4 | TOTAL GRAMS |
|---|---|---|---|---|---|---|
| | 63 | 9.16 | 0 | 0 | 0 | 9.16 |
| | 64 | 9.16 | 9.16 | 9.16 | 9.16 | 36.64 |
| | 65 | 0 | 0 | 0 | 9.16 | 9.16 |
| | 67 | 9.16 | 18.31 | 0 | 0 | 27.47 |

| | | | | | | |
|---|---|---|---|---|---|---|
| | 68 | 9.16 | 0 | 0 | 0 | 9.16 |
| TOTAL | | 36.64 | 27.47 | 9.16 | 18.32 | 91.59 |

STATE: DELAWARE

| ZIP CODE | | QUARTER 1 | QUARTER 2 | QUARTER 3 | QUARTER 4 | TOTAL GRAMS |
|---|---|---|---|---|---|---|
| | 197 | 0 | 9.16 | 9.16 | 9.16 | 27.48 |
| TOTAL | | 0 | 9.16 | 9.16 | 9.16 | 27.48 |

STATE: FLORIDA

| ZIP CODE | | QUARTER 1 | QUARTER 2 | QUARTER 3 | QUARTER 4 | TOTAL GRAMS |
|---|---|---|---|---|---|---|
| | 330 | 45.78 | 9.16 | 18.31 | 9.16 | 82.41 |
| | 331 | 18.31 | 18.31 | 9.16 | 18.31 | 64.09 |
| | 333 | 0 | 0 | 0 | 9.16 | 9.16 |
| | 334 | 27.47 | 36.62 | 54.93 | 36.62 | 155.64 |
| | 337 | 0 | 9.16 | 9.16 | 9.16 | 27.48 |
| | 341 | 9.16 | 0 | 0 | 0 | 9.16 |
| | 342 | 0 | 0 | 0 | 9.16 | 9.16 |
| | 349 | 0 | 0 | 9.16 | 0 | 9.16 |
| TOTAL | | 100.72 | 73.25 | 100.72 | 91.57 | 366.26 |

STATE: GEORGIA

| ZIP CODE | | QUARTER 1 | QUARTER 2 | QUARTER 3 | QUARTER 4 | TOTAL GRAMS |
|---|---|---|---|---|---|---|
| | 300 | 9.16 | 0 | 0 | 0 | 9.16 |
| | 302 | 27.47 | 9.16 | 0 | 0 | 36.63 |
| TOTAL | | 36.63 | 9.16 | 0 | 0 | 45.79 |

STATE: HAWAII

| ZIP CODE | | QUARTER 1 | QUARTER 2 | QUARTER 3 | QUARTER 4 | TOTAL GRAMS |
|---|---|---|---|---|---|---|
| | 968 | 9.16 | 0 | 0 | 0 | 9.16 |
| TOTAL | | 9.16 | 0 | 0 | 0 | 9.16 |

STATE: ILLINOIS

RETAIL DRUG DISTRIBUTION BY ZIP CODE WITHIN STATE BY GRAMS WT

| ZIP CODE | QUARTER 1 | QUARTER 2 | QUARTER 3 | QUARTER 4 | TOTAL GRAMS |
|---|---|---|---|---|---|
| 600 | 27.47 | 36.62 | 0 | 9.16 | 73.25 |
| 606 | 18.31 | 18.31 | 27.47 | 18.31 | 82.4 |
| 610 | 0 | 0 | 9.16 | 0 | 9.16 |
| TOTAL | 45.78 | 54.93 | 36.63 | 27.47 | 164.81 |

STATE: KANSAS

| ZIP CODE | QUARTER 1 | QUARTER 2 | QUARTER 3 | QUARTER 4 | TOTAL GRAMS |
|---|---|---|---|---|---|
| 662 | 0 | 0 | 0 | 9.16 | 9.16 |
| 667 | 27.47 | 18.31 | 18.31 | 18.31 | 82.4 |
| 672 | 9.16 | 0 | 0 | 0 | 9.16 |
| TOTAL | 36.63 | 18.31 | 18.31 | 27.47 | 100.72 |

STATE: MARYLAND

| ZIP CODE | QUARTER 1 | QUARTER 2 | QUARTER 3 | QUARTER 4 | TOTAL GRAMS |
|---|---|---|---|---|---|
| 208 | 0 | 9.16 | 0 | 0 | 9.16 |
| 209 | 18.31 | 18.31 | 18.31 | 18.31 | 73.24 |
| 210 | 0 | 9.16 | 0 | 0 | 9.16 |
| 217 | 45.78 | 27.47 | 27.47 | 27.47 | 128.19 |
| TOTAL | 64.09 | 64.1 | 45.78 | 45.78 | 219.75 |

STATE: MASSACHUSETTS

| ZIP CODE | QUARTER 1 | QUARTER 2 | QUARTER 3 | QUARTER 4 | TOTAL GRAMS |
|---|---|---|---|---|---|
| 10 | 9.16 | 9.16 | 0 | 0 | 18.32 |
| 16 | 18.31 | 9.16 | 18.31 | 18.31 | 64.09 |
| 17 | 0 | 18.31 | 9.16 | 9.16 | 36.63 |
| 18 | 9.16 | 0 | 0 | 0 | 9.16 |
| 24 | 18.31 | 9.16 | 9.16 | 9.16 | 45.79 |
| 26 | 0 | 0 | 0 | 18.31 | 18.31 |
| TOTAL | 54.94 | 45.79 | 36.63 | 54.94 | 192.3 |

STATE: MICHIGAN

| ZIP CODE | QUARTER 1 | QUARTER 2 | QUARTER 3 | QUARTER 4 | TOTAL GRAMS |
|---|---|---|---|---|---|

| | | | | | | |
|---|---|---|---|---|---|---|
| | 480 | 0 | 0 | 0 | 18.31 | 18.31 |
| TOTAL | | 0 | 0 | 0 | 18.31 | 18.31 |

| STATE: | MINNESOTA | | | | |
|---|---|---|---|---|---|
| ZIP CODE | QUARTER 1 | QUARTER 2 | QUARTER 3 | QUARTER 4 | TOTAL GRAMS |
| 554 | 9.16 | 0 | 0 | 9.16 | 18.32 |
| TOTAL | 9.16 | 0 | 0 | 9.16 | 18.32 |

| STATE: | MISSOURI | | | | |
|---|---|---|---|---|---|
| ZIP CODE | QUARTER 1 | QUARTER 2 | QUARTER 3 | QUARTER 4 | TOTAL GRAMS |
| 637 | 18.31 | 9.16 | 27.47 | 18.31 | 73.25 |
| 654 | 9.16 | 9.16 | 9.16 | 0 | 27.48 |
| TOTAL | 27.47 | 18.32 | 36.63 | 18.31 | 100.73 |

| STATE: | NEVADA | | | | |
|---|---|---|---|---|---|
| ZIP CODE | QUARTER 1 | QUARTER 2 | QUARTER 3 | QUARTER 4 | TOTAL GRAMS |
| 891 | 9.16 | 0 | 0 | 0 | 9.16 |
| TOTAL | 9.16 | 0 | 0 | 0 | 9.16 |

| STATE: | NEW HAMPSHIRE | | | | |
|---|---|---|---|---|---|
| ZIP CODE | QUARTER 1 | QUARTER 2 | QUARTER 3 | QUARTER 4 | TOTAL GRAMS |
| 30 | 9.16 | 0 | 0 | 0 | 9.16 |
| 33 | 9.16 | 0 | 0 | 0 | 9.16 |
| 37 | 0 | 9.16 | 0 | 0 | 9.16 |
| TOTAL | 18.32 | 9.16 | 0 | 0 | 27.48 |

| STATE: | NEW JERSEY | | | | |
|---|---|---|---|---|---|
| ZIP CODE | QUARTER 1 | QUARTER 2 | QUARTER 3 | QUARTER 4 | TOTAL GRAMS |
| 70 | 27.47 | 18.31 | 27.47 | 18.31 | 91.56 |
| 74 | 9.16 | 0 | 0 | 0 | 9.16 |
| 77 | 9.16 | 0 | 0 | 9.16 | 18.32 |
| 78 | 0 | 9.16 | 0 | 0 | 9.16 |

| | | | | | | |
|---|---|---|---|---|---|---|
| | 80 | 9.16 | 9.16 | 9.16 | 9.16 | 36.64 |
| | 85 | 9.16 | 0 | 0 | 0 | 9.16 |
| | 88 | 9.16 | 18.31 | 18.31 | 9.16 | 54.94 |
| TOTAL | | 73.27 | 54.94 | 54.94 | 45.79 | 228.94 |

DRUG CODE:2315DRUG NAME:SECOBARBITAL (SCHEDULE 2)

STATE: NEW YORK

| ZIP CODE | QUARTER 1 | QUARTER 2 | QUARTER 3 | QUARTER 4 | TOTAL GRAMS |
|---|---|---|---|---|---|
| 100 | 100.71 | 119.02 | 64.09 | 54.93 | 338.75 |
| 101 | 9.16 | 0 | 0 | 0 | 9.16 |
| 103 | 36.62 | 27.47 | 27.47 | 27.47 | 119.03 |
| 105 | 27.47 | 9.16 | 9.16 | 9.16 | 54.95 |
| 106 | 9.16 | 9.16 | 0 | 0 | 18.32 |
| 108 | 9.16 | 9.16 | 0 | 9.16 | 27.48 |
| 110 | 9.16 | 0 | 9.16 | 9.16 | 27.48 |
| 112 | 173.95 | 119.02 | 109.86 | 109.86 | 512.69 |
| 113 | 0 | 9.16 | 0 | 0 | 9.16 |
| 114 | 18.31 | 27.47 | 18.31 | 18.31 | 82.4 |
| 115 | 0 | 9.16 | 0 | 0 | 9.16 |
| 116 | 45.78 | 36.62 | 45.78 | 45.78 | 173.96 |
| 117 | 18.31 | 0 | 0 | 9.16 | 27.47 |
| 119 | 9.16 | 18.31 | 0 | 27.47 | 54.94 |
| 142 | 0 | 9.16 | 9.16 | 9.16 | 27.48 |
| 144 | 0 | 0 | 0 | 9.16 | 9.16 |
| TOTAL | 466.95 | 402.87 | 292.99 | 338.78 | 1,501.59 |

STATE: NORTH CAROLINA

| ZIP CODE | QUARTER 1 | QUARTER 2 | QUARTER 3 | QUARTER 4 | TOTAL GRAMS |
|---|---|---|---|---|---|
| 275 | 0 | 9.16 | 0 | 0 | 9.16 |
| TOTAL | 0 | 9.16 | 0 | 0 | 9.16 |

STATE: OHIO

| ZIP CODE | QUARTER 1 | QUARTER 2 | QUARTER 3 | QUARTER 4 | TOTAL GRAMS |
|---|---|---|---|---|---|
| 430 | 9.16 | 0 | 9.16 | 9.16 | 27.48 |

| | | | | | |
|---|---|---|---|---|---|
| 448 | 36.62 | 18.31 | 18.31 | 27.47 | 100.71 |
| 452 | 9.16 | 0 | 0 | 0 | 9.16 |
| TOTAL | 54.94 | 18.31 | 27.47 | 36.63 | 137.35 |

STATE: OKLAHOMA

| ZIP CODE | QUARTER 1 | QUARTER 2 | QUARTER 3 | QUARTER 4 | TOTAL GRAMS |
|---|---|---|---|---|---|
| 740 | 0 | 0 | 27.47 | 9.16 | 36.63 |
| TOTAL | 0 | 0 | 27.47 | 9.16 | 36.63 |

STATE: OREGON

| ZIP CODE | QUARTER 1 | QUARTER 2 | QUARTER 3 | QUARTER 4 | TOTAL GRAMS |
|---|---|---|---|---|---|
| 970 | 9.16 | 0 | 9.16 | 9.16 | 27.48 |
| 971 | 0 | 9.16 | 9.16 | 9.16 | 27.48 |
| 972 | 119.02 | 91.55 | 128.17 | 128.17 | 466.91 |
| 973 | 54.93 | 36.62 | 36.62 | 91.55 | 219.72 |
| 974 | 36.62 | 0 | 18.31 | 27.47 | 82.4 |
| 975 | 27.47 | 36.62 | 9.16 | 36.62 | 109.87 |
| 977 | 0 | 0 | 9.16 | 0 | 9.16 |
| 978 | 9.16 | 0 | 0 | 0 | 9.16 |
| TOTAL | 256.36 | 173.95 | 219.74 | 302.13 | 952.18 |

STATE: PENNSYLVANIA

| ZIP CODE | QUARTER 1 | QUARTER 2 | QUARTER 3 | QUARTER 4 | TOTAL GRAMS |
|---|---|---|---|---|---|
| 170 | 0 | 0 | 9.16 | 0 | 9.16 |
| 173 | 9.16 | 27.47 | 18.31 | 36.62 | 91.56 |
| 187 | 9.16 | 27.47 | 18.31 | 18.31 | 73.25 |
| 190 | 9.16 | 18.31 | 27.47 | 18.31 | 73.25 |
| 191 | 9.16 | 0 | 0 | 0 | 9.16 |
| 193 | 0 | 0 | 9.16 | 0 | 9.16 |
| TOTAL | 36.64 | 73.25 | 82.41 | 73.24 | 265.54 |

STATE: RHODE ISLAND

| ZIP CODE | QUARTER 1 | QUARTER 2 | QUARTER 3 | QUARTER 4 | TOTAL GRAMS |
|---|---|---|---|---|---|

| | | | | | | |
|---|---|---|---|---|---|---|
| | 28 | 9.16 | 9.16 | 0 | 18.31 | 36.63 |
| TOTAL | | 9.16 | 9.16 | 0 | 18.31 | 36.63 |

| STATE: | SOUTH CAROLINA | | | | | |
|---|---|---|---|---|---|---|
| ZIP CODE | QUARTER 1 | | QUARTER 2 | QUARTER 3 | QUARTER 4 | TOTAL GRAMS |
| | 296 | 0 | 0 | 18.31 | 18.31 | 36.62 |
| TOTAL | | 0 | 0 | 18.31 | 18.31 | 36.62 |

| STATE: | SOUTH DAKOTA | | | | | |
|---|---|---|---|---|---|---|
| ZIP CODE | QUARTER 1 | | QUARTER 2 | QUARTER 3 | QUARTER 4 | TOTAL GRAMS |
| | 571 | 0 | 0 | 0 | 9.16 | 9.16 |
| TOTAL | | 0 | 0 | 0 | 9.16 | 9.16 |

| STATE: | TENNESSEE | | | | | |
|---|---|---|---|---|---|---|
| ZIP CODE | QUARTER 1 | | QUARTER 2 | QUARTER 3 | QUARTER 4 | TOTAL GRAMS |
| | 381 | 0 | 9.16 | 0 | 0 | 9.16 |
| ZIP CODE | QUARTER 1 | | QUARTER 2 | QUARTER 3 | QUARTER 4 | TOTAL GRAMS |
| TOTAL | | 0 | 9.16 | 0 | 0 | 9.16 |

| STATE: | TEXAS | | | | | |
|---|---|---|---|---|---|---|
| ZIP CODE | QUARTER 1 | | QUARTER 2 | QUARTER 3 | QUARTER 4 | TOTAL GRAMS |
| | 750 | 9.16 | 0 | 9.16 | 18.31 | 36.63 |
| | 786 | 0 | 0 | 0 | 9.16 | 9.16 |
| TOTAL | | 9.16 | 0 | 9.16 | 27.47 | 45.79 |

| STATE: | VERMONT | | | | | |
|---|---|---|---|---|---|---|
| ZIP CODE | QUARTER 1 | | QUARTER 2 | QUARTER 3 | QUARTER 4 | TOTAL GRAMS |
| | 57 | 54.93 | 27.47 | 0 | 54.93 | 137.33 |
| TOTAL | | 54.93 | 27.47 | 0 | 54.93 | 137.33 |

STATE: VIRGINIA

| ZIP CODE | QUARTER 1 | QUARTER 2 | QUARTER 3 | QUARTER 4 | TOTAL GRAMS |
|---|---|---|---|---|---|
| 220 | 9.16 | 0 | 0 | 0 | 9.16 |
| 222 | 0 | 9.16 | 0 | 0 | 9.16 |
| 235 | 18.31 | 18.31 | 27.47 | 27.47 | 91.56 |
| TOTAL | 27.47 | 27.47 | 27.47 | 27.47 | 109.88 |

STATE: WASHINGTON

| ZIP CODE | QUARTER 1 | QUARTER 2 | QUARTER 3 | QUARTER 4 | TOTAL GRAMS |
|---|---|---|---|---|---|
| 980 | 0 | 0 | 18.31 | 9.16 | 27.47 |
| 981 | 128.17 | 128.17 | 109.86 | 128.17 | 494.37 |
| 982 | 9.16 | 9.16 | 0 | 0 | 18.32 |
| 983 | 45.78 | 0 | 9.16 | 0 | 54.94 |
| 984 | 0 | 0 | 0 | 9.16 | 9.16 |
| 985 | 9.16 | 9.16 | 0 | 0 | 18.32 |
| 986 | 18.31 | 18.31 | 0 | 9.16 | 45.78 |
| 988 | 9.16 | 9.16 | 9.16 | 9.16 | 36.64 |
| 992 | 9.16 | 0 | 0 | 0 | 9.16 |
| 993 | 0 | 0 | 9.16 | 0 | 9.16 |
| TOTAL | 228.9 | 173.96 | 155.65 | 164.81 | 723.32 |

STATE: WISCONSIN

| ZIP CODE | QUARTER 1 | QUARTER 2 | QUARTER 3 | QUARTER 4 | TOTAL GRAMS |
|---|---|---|---|---|---|
| 535 | 9.16 | 0 | 0 | 0 | 9.16 |
| TOTAL | 9.16 | 0 | 0 | 0 | 9.16 |

DRUG CODE: 7365 DRUG NAME: DRONABINOL IN AN ORAL SOLUTION IN FDA AP

STATE: ALABAMA

| ZIP CODE | QUARTER 1 | QUARTER 2 | QUARTER 3 | QUARTER 4 | TOTAL GRAMS |
|---|---|---|---|---|---|
| 352 | 0 | 0 | 0.75 | 0.15 | 0.9 |
| 354 | 0 | 0 | 0 | 0.9 | 0.9 |
| 356 | 0 | 0 | 0 | 0.3 | 0.3 |
| 358 | 0 | 0 | 0 | 0.3 | 0.3 |
| 363 | 0 | 0 | 0.75 | 0 | 0.75 |

| | | | | | | |
|---|---|---|---|---|---|---|
| | 366 | 0 | 0 | 0 | 1.05 | 1.05 |
| TOTAL | | 0 | 0 | 1.5 | 2.7 | 4.2 |

| STATE: | ARIZONA | | | | |
|---|---|---|---|---|---|
| ZIP CODE | QUARTER 1 | QUARTER 2 | QUARTER 3 | QUARTER 4 | TOTAL GRAMS |
| 850 | 0 | 0 | 12 | 7.2 | 19.2 |
| 852 | 0 | 0 | 2.4 | 0 | 2.4 |
| TOTAL | 0 | 0 | 14.4 | 7.2 | 21.6 |

| STATE: | ARKANSAS | | | | |
|---|---|---|---|---|---|
| ZIP CODE | QUARTER 1 | QUARTER 2 | QUARTER 3 | QUARTER 4 | TOTAL GRAMS |
| 720 | 0 | 0 | 0 | 0.6 | 0.6 |
| 727 | 0 | 0 | 0.6 | 0 | 0.6 |
| TOTAL | 0 | 0 | 0.6 | 0.6 | 1.2 |

| STATE: | CALIFORNIA | | | | |
|---|---|---|---|---|---|
| ZIP CODE | QUARTER 1 | QUARTER 2 | QUARTER 3 | QUARTER 4 | TOTAL GRAMS |
| 900 | 0 | 0 | 5.55 | 1.95 | 7.5 |
| 904 | 0 | 0 | 0.9 | 1.35 | 2.25 |
| 906 | 0 | 0 | 0 | 0.15 | 0.15 |
| 911 | 0 | 0 | 0 | 0.15 | 0.15 |
| 913 | 0 | 0 | 1.5 | 3 | 4.5 |
| 920 | 0 | 0 | 0 | 6.15 | 6.15 |
| 921 | 0 | 0 | 0.9 | 6.45 | 7.35 |
| 922 | 0 | 0 | 0 | 0.6 | 0.6 |
| 924 | 0 | 0 | 0.3 | 2.7 | 3 |
| 925 | 0 | 0 | 0 | 0.9 | 0.9 |
| 928 | 0 | 0 | 0 | 0.15 | 0.15 |
| 933 | 0 | 0 | 0.15 | 1.2 | 1.35 |
| 937 | 0 | 0 | 0 | 0.6 | 0.6 |
| 941 | 0 | 0 | 0.6 | 0 | 0.6 |
| 945 | 0 | 0 | 1.8 | 0.75 | 2.55 |
| 946 | 0 | 0 | 0.9 | 0 | 0.9 |

| | | | | | | |
|---|---|---|---|---|---|---|
| | 952 | 0 | 0 | 0 | 0.15 | 0.15 |
| | 956 | 0 | 0 | 0.15 | 0 | 0.15 |
| | 958 | 0 | 0 | 2.4 | 2.4 | 4.8 |
| TOTAL | | 0 | 0 | 15.15 | 28.65 | 43.8 |

STATE: COLORADO

| ZIP CODE | | QUARTER 1 | QUARTER 2 | QUARTER 3 | QUARTER 4 | TOTAL GRAMS |
|---|---|---|---|---|---|---|
| | 801 | 0 | 0 | 1.2 | 0 | 1.2 |
| | 802 | 0 | 0 | 2.4 | 0 | 2.4 |
| | 815 | 0 | 0 | 0 | 0.15 | 0.15 |
| TOTAL | | 0 | 0 | 3.6 | 0.15 | 3.75 |

STATE: CONNECTICUT

| ZIP CODE | | QUARTER 1 | QUARTER 2 | QUARTER 3 | QUARTER 4 | TOTAL GRAMS |
|---|---|---|---|---|---|---|
| | 60 | 0 | 0 | 0 | 0.15 | 0.15 |
| | 64 | 0 | 0 | 0 | 0.15 | 0.15 |
| | 65 | 0 | 0 | 0 | 0.15 | 0.15 |
| | 68 | 0 | 0 | 1.2 | 0 | 1.2 |
| TOTAL | | 0 | 0 | 1.2 | 0.45 | 1.65 |

STATE: DELAWARE

| ZIP CODE | | QUARTER 1 | QUARTER 2 | QUARTER 3 | QUARTER 4 | TOTAL GRAMS |
|---|---|---|---|---|---|---|
| | 198 | 0 | 0 | 0.15 | 0 | 0.15 |
| TOTAL | | 0 | 0 | 0.15 | 0 | 0.15 |

STATE: DISTRICT OF COLUMBIA

| ZIP CODE | | QUARTER 1 | QUARTER 2 | QUARTER 3 | QUARTER 4 | TOTAL GRAMS |
|---|---|---|---|---|---|---|
| | 200 | 0 | 0 | 0 | 0.75 | 0.75 |
| TOTAL | | 0 | 0 | 0 | 0.75 | 0.75 |

STATE: FLORIDA

| ZIP CODE | | QUARTER 1 | QUARTER 2 | QUARTER 3 | QUARTER 4 | TOTAL GRAMS |
|---|---|---|---|---|---|---|

| | | | | | |
|---|---|---|---|---|---|
| 323 | 0 | 0 | 0.3 | 0 | 0.3 |
| 325 | 0 | 0 | 0.15 | 0 | 0.15 |
| 327 | 0 | 0 | 2.4 | 2.4 | 4.8 |
| 328 | 0 | 0 | 0.9 | 1.5 | 2.4 |
| 330 | 0 | 0 | 0.15 | 0 | 0.15 |
| 331 | 0 | 0 | 0.3 | 0.15 | 0.45 |
| 333 | 0 | 0 | 0.3 | 0.3 | 0.6 |
| 334 | 0 | 0 | 0 | 0.3 | 0.3 |
| 335 | 0 | 0 | 4.8 | 4.8 | 9.6 |
| 336 | 0 | 0 | 0 | 1.2 | 1.2 |
| 337 | 0 | 0 | 0 | 1.8 | 1.8 |
| 338 | 0 | 0 | 0 | 0.45 | 0.45 |
| 342 | 0 | 0 | 4.65 | 9.75 | 14.4 |
| TOTAL | 0 | 0 | 13.95 | 22.65 | 36.6 |

STATE: GEORGIA

| ZIP CODE | QUARTER 1 | QUARTER 2 | QUARTER 3 | QUARTER 4 | TOTAL GRAMS |
|---|---|---|---|---|---|
| 300 | 0 | 0 | 0.3 | 1.05 | 1.35 |
| 301 | 0 | 0 | 0.75 | 0.15 | 0.9 |
| 303 | 0 | 0 | 0 | 1.95 | 1.95 |
| 314 | 0 | 0 | 0.3 | 0 | 0.3 |
| 319 | 0 | 0 | 0 | 0.15 | 0.15 |
| TOTAL | 0 | 0 | 1.35 | 3.3 | 4.65 |

STATE: ILLINOIS

| ZIP CODE | QUARTER 1 | QUARTER 2 | QUARTER 3 | QUARTER 4 | TOTAL GRAMS |
|---|---|---|---|---|---|
| 600 | 0 | 0 | 0.3 | 0.75 | 1.05 |
| TOTAL | 0 | 0 | 0.3 | 0.75 | 1.05 |

STATE: INDIANA

| ZIP CODE | QUARTER 1 | QUARTER 2 | QUARTER 3 | QUARTER 4 | TOTAL GRAMS |
|---|---|---|---|---|---|
| 460 | 0 | 0 | 0.9 | 0 | 0.9 |
| 467 | 0 | 0 | 0.45 | 0 | 0.45 |
| 473 | 0 | 0 | 0.3 | 0 | 0.3 |

| | | | | | |
|---|---|---|---|---|---|
| TOTAL | 0 | 0 | 1.65 | 0 | 1.65 |

STATE: KANSAS

| ZIP CODE | QUARTER 1 | QUARTER 2 | QUARTER 3 | QUARTER 4 | TOTAL GRAMS |
|---|---|---|---|---|---|
| 662 | 0 | 0 | 0 | 0.6 | 0.6 |
| TOTAL | 0 | 0 | 0 | 0.6 | 0.6 |

STATE: KENTUCKY

| ZIP CODE | QUARTER 1 | QUARTER 2 | QUARTER 3 | QUARTER 4 | TOTAL GRAMS |
|---|---|---|---|---|---|
| 402 | 0 | 0 | 0 | 0.15 | 0.15 |
| 403 | 0 | 0 | 0.15 | 0 | 0.15 |
| 405 | 0 | 0 | 0.6 | 0.6 | 1.2 |
| TOTAL | 0 | 0 | 0.75 | 0.75 | 1.5 |

STATE: LOUISIANA

| ZIP CODE | QUARTER 1 | QUARTER 2 | QUARTER 3 | QUARTER 4 | TOTAL GRAMS |
|---|---|---|---|---|---|
| 700 | 0 | 0 | 0.15 | 0.15 | 0.3 |
| TOTAL | 0 | 0 | 0.15 | 0.15 | 0.3 |

STATE: MAINE

| ZIP CODE | QUARTER 1 | QUARTER 2 | QUARTER 3 | QUARTER 4 | TOTAL GRAMS |
|---|---|---|---|---|---|
| 42 | 0 | 0 | 0.3 | 0 | 0.3 |
| TOTAL | 0 | 0 | 0.3 | 0 | 0.3 |

STATE: MARYLAND

| ZIP CODE | QUARTER 1 | QUARTER 2 | QUARTER 3 | QUARTER 4 | TOTAL GRAMS |
|---|---|---|---|---|---|
| 208 | 0 | 0 | 0 | 0.9 | 0.9 |
| 210 | 0 | 0 | 0 | 0.3 | 0.3 |
| 212 | 0 | 0 | 0.15 | 0.6 | 0.75 |
| TOTAL | 0 | 0 | 0.15 | 1.8 | 1.95 |

STATE: MASSACHUSETTS

| ZIP CODE | QUARTER 1 | QUARTER 2 | QUARTER 3 | QUARTER 4 | TOTAL GRAMS |
|---|---|---|---|---|---|
| 17 | 0 | 0 | 0 | 0.9 | 0.9 |
| 21 | 0 | 0 | 0.15 | 0 | 0.15 |
| 22 | 0 | 0 | 0.6 | 1.95 | 2.55 |
| 23 | 0 | 0 | 0 | 0.15 | 0.15 |
| TOTAL | 0 | 0 | 0.75 | 3 | 3.75 |

STATE: MICHIGAN

| ZIP CODE | QUARTER 1 | QUARTER 2 | QUARTER 3 | QUARTER 4 | TOTAL GRAMS |
|---|---|---|---|---|---|
| 480 | 0 | 0 | 0 | 0.15 | 0.15 |
| 481 | 0 | 0 | 0.3 | 0.45 | 0.75 |
| 482 | 0 | 0 | 0.3 | 0.15 | 0.45 |
| 483 | 0 | 0 | 0 | 0.6 | 0.6 |
| 484 | 0 | 0 | 0.15 | 0 | 0.15 |
| 488 | 0 | 0 | 0.15 | 0 | 0.15 |
| 494 | 0 | 0 | 0.15 | 0 | 0.15 |
| TOTAL | 0 | 0 | 1.05 | 1.35 | 2.4 |

STATE: MINNESOTA

| ZIP CODE | QUARTER 1 | QUARTER 2 | QUARTER 3 | QUARTER 4 | TOTAL GRAMS |
|---|---|---|---|---|---|
| 551 | 0 | 0 | 0 | 0.45 | 0.45 |
| 553 | 0 | 0 | 0 | 2.4 | 2.4 |
| TOTAL | 0 | 0 | 0 | 2.85 | 2.85 |

STATE: MISSISSIPPI

| ZIP CODE | QUARTER 1 | QUARTER 2 | QUARTER 3 | QUARTER 4 | TOTAL GRAMS |
|---|---|---|---|---|---|
| 392 | 0 | 0 | 0.6 | 0.15 | 0.75 |
| TOTAL | 0 | 0 | 0.6 | 0.15 | 0.75 |

STATE: MISSOURI

| ZIP CODE | QUARTER 1 | QUARTER 2 | QUARTER 3 | QUARTER 4 | TOTAL GRAMS |
|---|---|---|---|---|---|
| 630 | 0 | 0 | 0 | 0.15 | 0.15 |

| | | | | | | |
|---|---|---|---|---|---|---|
| | 631 | 0 | 0 | 4.2 | 2.55 | 6.75 |
| | 640 | 0 | 0 | 0 | 0.3 | 0.3 |
| | 641 | 0 | 0 | 0 | 0.6 | 0.6 |
| | 648 | 0 | 0 | 0.15 | 0 | 0.15 |
| | 657 | 0 | 0 | 0 | 0.6 | 0.6 |
| TOTAL | | 0 | 0 | 4.35 | 4.2 | 8.55 |

STATE: MONTANA

| ZIP CODE | | QUARTER 1 | QUARTER 2 | QUARTER 3 | QUARTER 4 | TOTAL GRAMS |
|---|---|---|---|---|---|---|
| | 591 | 0 | 0 | 0 | 0.3 | 0.3 |
| TOTAL | | 0 | 0 | 0 | 0.3 | 0.3 |

STATE: NEVADA

| ZIP CODE | | QUARTER 1 | QUARTER 2 | QUARTER 3 | QUARTER 4 | TOTAL GRAMS |
|---|---|---|---|---|---|---|
| | 891 | 0 | 0 | 4.8 | 6.6 | 11.4 |
| | 895 | 0 | 0 | 0.15 | 0 | 0.15 |
| TOTAL | | 0 | 0 | 4.95 | 6.6 | 11.55 |

DRUG CODE:7365DRUG NAME:DRONABINOL IN AN ORAL SOLUTION IN FDA AP

STATE: NEW HAMPSHIRE

| ZIP CODE | | QUARTER 1 | QUARTER 2 | QUARTER 3 | QUARTER 4 | TOTAL GRAMS |
|---|---|---|---|---|---|---|
| | 030 | 0 | 0 | 0.3 | 0.6 | 0.9 |
| | 031 | 0 | 0 | 0.45 | 0.45 | 0.9 |
| TOTAL | | 0 | 0 | 0.75 | 1.05 | 1.8 |

STATE: NEW JERSEY

| ZIP CODE | | QUARTER 1 | QUARTER 2 | QUARTER 3 | QUARTER 4 | TOTAL GRAMS |
|---|---|---|---|---|---|---|
| | 070 | 0 | 0 | 70.8 | 69.9 | 140.7 |
| | 073 | 0 | 0 | 0 | 0.15 | 0.15 |
| | 076 | 0 | 0 | 0 | 0.45 | 0.45 |
| | 080 | 0 | 0 | 0 | 0.9 | 0.9 |
| | 082 | 0 | 0 | 0 | 0.9 | 0.9 |
| | 088 | 0 | 0 | 0 | 0.45 | 0.45 |

| | | | | | | |
|---|---|---|---|---|---|---|
| TOTAL | | 0 | 0 | 70.8 | 72.75 | 143.55 |

| STATE: | NEW MEXICO | | | | | |
|---|---|---|---|---|---|---|
| ZIP CODE | | QUARTER 1 | QUARTER 2 | QUARTER 3 | QUARTER 4 | TOTAL GRAMS |
| | 871 | 0 | 0 | 0.15 | 0 | 0.15 |
| TOTAL | | 0 | 0 | 0.15 | 0 | 0.15 |

| STATE: | NEW YORK | | | | | |
|---|---|---|---|---|---|---|
| ZIP CODE | | QUARTER 1 | QUARTER 2 | QUARTER 3 | QUARTER 4 | TOTAL GRAMS |
| | 100 | 0 | 0 | 0 | 0.15 | 0.15 |
| | 103 | 0 | 0 | 0.15 | 0.45 | 0.6 |
| | 112 | 0 | 0 | 0.45 | 0 | 0.45 |
| | 115 | 0 | 0 | 0 | 0.3 | 0.3 |
| | 120 | 0 | 0 | 0.15 | 0 | 0.15 |
| | 140 | 0 | 0 | 0 | 0.15 | 0.15 |
| | 142 | 0 | 0 | 0 | 0.15 | 0.15 |
| TOTAL | | 0 | 0 | 0.75 | 1.2 | 1.95 |

| STATE: | NORTH CAROLINA | | | | | |
|---|---|---|---|---|---|---|
| ZIP CODE | | QUARTER 1 | QUARTER 2 | QUARTER 3 | QUARTER 4 | TOTAL GRAMS |
| | 271 | 0 | 0 | 0.15 | 0 | 0.15 |
| | 273 | 0 | 0 | 0.3 | 0 | 0.3 |
| | 274 | 0 | 0 | 0 | 0.3 | 0.3 |
| | 276 | 0 | 0 | 0 | 0.3 | 0.3 |
| TOTAL | | 0 | 0 | 0.45 | 0.6 | 1.05 |

| STATE: | OHIO | | | | | |
|---|---|---|---|---|---|---|
| ZIP CODE | | QUARTER 1 | QUARTER 2 | QUARTER 3 | QUARTER 4 | TOTAL GRAMS |
| | 432 | 0 | 0 | 0 | 5.1 | 5.1 |
| | 436 | 0 | 0 | 0.3 | 1.2 | 1.5 |
| | 441 | 0 | 0 | 0 | 1.8 | 1.8 |
| | 444 | 0 | 0 | 0 | 0.15 | 0.15 |
| | 448 | 0 | 0 | 0.15 | 0 | 0.15 |

| | | | | | | |
|---|---|---|---|---|---|---|
| TOTAL | | 0 | 0 | 0.45 | 8.25 | 8.7 |
| STATE: | PENNSYLVANIA | | | | | |
| ZIP CODE | | QUARTER 1 | QUARTER 2 | QUARTER 3 | QUARTER 4 | TOTAL GRAMS |
| | 151 | 0 | 0 | 0 | 0.3 | 0.3 |
| | 152 | 0 | 0 | 0 | 0.3 | 0.3 |
| | 157 | 0 | 0 | 0 | 0.15 | 0.15 |
| | 176 | 0 | 0 | 0 | 0.15 | 0.15 |
| | 179 | 0 | 0 | 0 | 0.15 | 0.15 |
| | 186 | 0 | 0 | 0 | 0.45 | 0.45 |
| | 190 | 0 | 0 | 14.1 | 0.3 | 14.4 |
| | 191 | 0 | 0 | 0.3 | 0 | 0.3 |
| | 193 | 0 | 0 | 0 | 0.15 | 0.15 |
| TOTAL | | 0 | 0 | 14.4 | 1.95 | 16.35 |
| STATE: | SOUTH CAROLINA | | | | | |
| ZIP CODE | | QUARTER 1 | QUARTER 2 | QUARTER 3 | QUARTER 4 | TOTAL GRAMS |
| | 294 | 0 | 0 | 0.3 | 0 | 0.3 |
| TOTAL | | 0 | 0 | 0.3 | 0 | 0.3 |
| STATE: | TENNESSEE | | | | | |
| ZIP CODE | | QUARTER 1 | QUARTER 2 | QUARTER 3 | QUARTER 4 | TOTAL GRAMS |
| | 372 | 0 | 0 | 0 | 0.6 | 0.6 |
| | 376 | 0 | 0 | 0 | 0.75 | 0.75 |
| | 378 | 0 | 0 | 0 | 0.15 | 0.15 |
| | 381 | 0 | 0 | 1.5 | 0.6 | 2.1 |
| TOTAL | | 0 | 0 | 1.5 | 2.1 | 3.6 |
| STATE: | TEXAS | | | | | |
| ZIP CODE | | QUARTER 1 | QUARTER 2 | QUARTER 3 | QUARTER 4 | TOTAL GRAMS |
| | 750 | 0 | 0 | 0 | 0.3 | 0.3 |
| | 753 | 0 | 0 | 0.6 | 1.5 | 2.1 |
| | 761 | 0 | 0 | 2.7 | 0 | 2.7 |

| | | | | | | |
|---|---|---|---|---|---|---|
| | 765 | 0 | 0 | 0 | 0.6 | 0.6 |
| | 770 | 0 | 0 | 0.9 | 0.9 | 1.8 |
| | 773 | 0 | 0 | 0 | 0.15 | 0.15 |
| | 775 | 0 | 0 | 0 | 1.35 | 1.35 |
| | 782 | 0 | 0 | 0 | 0.3 | 0.3 |
| | 787 | 0 | 0 | 2.4 | 0 | 2.4 |
| TOTAL | | 0 | 0 | 6.6 | 5.1 | 11.7 |

STATE: UTAH

| ZIP CODE | | QUARTER 1 | QUARTER 2 | QUARTER 3 | QUARTER 4 | TOTAL GRAMS |
|---|---|---|---|---|---|---|
| | 841 | 0 | 0 | 0.45 | 0.15 | 0.6 |
| TOTAL | | 0 | 0 | 0.45 | 0.15 | 0.6 |

STATE: VIRGINIA

| ZIP CODE | | QUARTER 1 | QUARTER 2 | QUARTER 3 | QUARTER 4 | TOTAL GRAMS |
|---|---|---|---|---|---|---|
| | 201 | 0 | 0 | 0 | 0.45 | 0.45 |
| | 220 | 0 | 0 | 0 | 0.15 | 0.15 |
| | 235 | 0 | 0 | 0 | 0.15 | 0.15 |
| TOTAL | | 0 | 0 | 0 | 0.75 | 0.75 |

STATE: WISCONSIN

| ZIP CODE | | QUARTER 1 | QUARTER 2 | QUARTER 3 | QUARTER 4 | TOTAL GRAMS |
|---|---|---|---|---|---|---|
| | 531 | 0 | 0 | 0.15 | 0 | 0.15 |
| TOTAL | | 0 | 0 | 0.15 | 0 | 0.15 |

DRUG CODE: 7379 DRUG NAME: NABILONE

STATE: ARIZONA

| ZIP CODE | | QUARTER 1 | QUARTER 2 | QUARTER 3 | QUARTER 4 | TOTAL GRAMS |
|---|---|---|---|---|---|---|
| | 852 | 0 | 0 | 0.15 | 0.05 | 0.2 |
| TOTAL | | 0 | 0 | 0.15 | 0.05 | 0.2 |

STATE: CALIFORNIA

| ZIP CODE | QUARTER 1 | QUARTER 2 | QUARTER 3 | QUARTER 4 | TOTAL GRAMS |
|---|---|---|---|---|---|
| 945 | 0 | 0.25 | 0.05 | 0.35 | 0.65 |
| TOTAL | 0 | 0.25 | 0.05 | 0.35 | 0.65 |

STATE: COLORADO

| ZIP CODE | QUARTER 1 | QUARTER 2 | QUARTER 3 | QUARTER 4 | TOTAL GRAMS |
|---|---|---|---|---|---|
| 801 | 0.1 | 0.05 | 0 | 0.15 | 0.3 |
| 815 | 0.05 | 0 | 0 | 0 | 0.05 |
| TOTAL | 0.15 | 0.05 | 0 | 0.15 | 0.35 |

STATE: CONNECTICUT

| ZIP CODE | QUARTER 1 | QUARTER 2 | QUARTER 3 | QUARTER 4 | TOTAL GRAMS |
|---|---|---|---|---|---|
| 60 | 0.1 | 0.1 | 0.1 | 0 | 0.3 |
| 64 | 0 | 0.05 | 0 | 0 | 0.05 |
| TOTAL | 0.1 | 0.15 | 0.1 | 0 | 0.35 |

STATE: FLORIDA

| ZIP CODE | QUARTER 1 | QUARTER 2 | QUARTER 3 | QUARTER 4 | TOTAL GRAMS |
|---|---|---|---|---|---|
| 322 | 0.05 | 0 | 0 | 0 | 0.05 |
| 330 | 0.25 | 0.2 | 0.05 | 0 | 0.5 |
| 331 | 0 | 0 | 0 | 0.15 | 0.15 |
| 335 | 0.4 | 0.15 | 0.2 | 0 | 0.75 |
| 336 | 0.15 | 0 | 0 | 0 | 0.15 |
| 337 | 0.05 | 0.1 | 0 | 0 | 0.15 |
| 338 | 0.15 | 0 | 0.1 | 0.05 | 0.3 |
| 346 | 0 | 0 | 0.1 | 0 | 0.1 |
| TOTAL | 1.05 | 0.45 | 0.45 | 0.2 | 2.15 |

STATE: HAWAII

| ZIP CODE | QUARTER 1 | QUARTER 2 | QUARTER 3 | QUARTER 4 | TOTAL GRAMS |
|---|---|---|---|---|---|
| 967 | 0 | 0 | 0.15 | 0 | 0.15 |
| 968 | 0.45 | 0 | 0 | 0 | 0.45 |

| | | | | | |
|---|---|---|---|---|---|
| TOTAL | 0.45 | 0 | 0.15 | 0 | 0.6 |

STATE: ILLINOIS

| ZIP CODE | QUARTER 1 | QUARTER 2 | QUARTER 3 | QUARTER 4 | TOTAL GRAMS |
|---|---|---|---|---|---|
| 605 | 0.1 | 0 | 0 | 0 | 0.1 |
| 606 | 0.1 | 0.05 | 0.1 | 0.1 | 0.35 |
| 619 | 0.05 | 0 | 0 | 0 | 0.05 |
| TOTAL | 0.25 | 0.05 | 0.1 | 0.1 | 0.5 |

STATE: IOWA

| ZIP CODE | QUARTER 1 | QUARTER 2 | QUARTER 3 | QUARTER 4 | TOTAL GRAMS |
|---|---|---|---|---|---|
| 500 | 0.05 | 0.05 | 0 | 0 | 0.1 |
| TOTAL | 0.05 | 0.05 | 0 | 0 | 0.1 |

STATE: KENTUCKY

| ZIP CODE | QUARTER 1 | QUARTER 2 | QUARTER 3 | QUARTER 4 | TOTAL GRAMS |
|---|---|---|---|---|---|
| 410 | 0.15 | 0.2 | 0 | 0 | 0.35 |
| TOTAL | 0.15 | 0.2 | 0 | 0 | 0.35 |

STATE: MAINE

| ZIP CODE | QUARTER 1 | QUARTER 2 | QUARTER 3 | QUARTER 4 | TOTAL GRAMS |
|---|---|---|---|---|---|
| 44 | 0.1 | 0.1 | 0.1 | 0 | 0.3 |
| TOTAL | 0.1 | 0.1 | 0.1 | 0 | 0.3 |

STATE: MARYLAND

| ZIP CODE | QUARTER 1 | QUARTER 2 | QUARTER 3 | QUARTER 4 | TOTAL GRAMS |
|---|---|---|---|---|---|
| 208 | 0.7 | 0.9 | 1 | 0.8 | 3.4 |
| 212 | 0.1 | 0 | 0 | 0 | 0.1 |
| 216 | 0.25 | 0.2 | 0.05 | 0.1 | 0.6 |
| TOTAL | 1.05 | 1.1 | 1.05 | 0.9 | 4.1 |

DATE RANGE: 01/01/2017 TO 12/31/2017 ARCOS 3 - REPORT 1 Run Date: 07/03/2018

RETAIL DRUG DISTRIBUTION BY ZIP CODE WITHIN STATE BY GRAMS WT

| STATE: | MINNESOTA | | | | |
|---|---|---|---|---|---|
| ZIP CODE | QUARTER 1 | QUARTER 2 | QUARTER 3 | QUARTER 4 | TOTAL GRAMS |
| 551 | 0.05 | 0 | 0 | 0 | 0.05 |
| 558 | 0.05 | 0 | 0 | 0 | 0.05 |
| TOTAL | 0.1 | 0 | 0 | 0 | 0.1 |

| STATE: | MISSOURI | | | | |
|---|---|---|---|---|---|
| ZIP CODE | QUARTER 1 | QUARTER 2 | QUARTER 3 | QUARTER 4 | TOTAL GRAMS |
| 631 | 0.2 | 0 | 0 | 0.05 | 0.25 |
| 641 | 0 | 0.1 | 0.1 | 0.1 | 0.3 |
| TOTAL | 0.2 | 0.1 | 0.1 | 0.15 | 0.55 |

| STATE: | NEBRASKA | | | | |
|---|---|---|---|---|---|
| ZIP CODE | QUARTER 1 | QUARTER 2 | QUARTER 3 | QUARTER 4 | TOTAL GRAMS |
| 681 | 0.15 | 0 | 0 | 0 | 0.15 |
| 684 | 0 | 0 | 0 | 0.15 | 0.15 |
| TOTAL | 0.15 | 0 | 0 | 0.15 | 0.3 |

| STATE: | NEW HAMPSHIRE | | | | |
|---|---|---|---|---|---|
| ZIP CODE | QUARTER 1 | QUARTER 2 | QUARTER 3 | QUARTER 4 | TOTAL GRAMS |
| 37 | 0 | 0 | 0 | 0.05 | 0.05 |
| TOTAL | 0 | 0 | 0 | 0.05 | 0.05 |

| STATE: | NEW JERSEY | | | | |
|---|---|---|---|---|---|
| ZIP CODE | QUARTER 1 | QUARTER 2 | QUARTER 3 | QUARTER 4 | TOTAL GRAMS |
| 80 | 0 | 0 | 0.1 | 0 | 0.1 |
| 83 | 0.1 | 0 | 0 | 0 | 0.1 |
| 87 | 0 | 0.1 | 0 | 0 | 0.1 |
| 88 | 0 | 0.05 | 0 | 0 | 0.05 |
| TOTAL | 0.1 | 0.15 | 0.1 | 0 | 0.35 |

DATE RANGE: 01/01/2017 TO 12/31/2017 ARCOS 3 - REPORT 1 Run Date: 07/03/2018

RETAIL DRUG DISTRIBUTION BY ZIP CODE WITHIN STATE BY GRAMS WT

STATE: NEW MEXICO

| ZIP CODE | QUARTER 1 | QUARTER 2 | QUARTER 3 | QUARTER 4 | TOTAL GRAMS |
|---|---|---|---|---|---|
| 877 | 0 | 0 | 0.1 | 0 | 0.1 |
| TOTAL | 0 | 0 | 0.1 | 0 | 0.1 |

STATE: NEW YORK

| ZIP CODE | QUARTER 1 | QUARTER 2 | QUARTER 3 | QUARTER 4 | TOTAL GRAMS |
|---|---|---|---|---|---|
| 100 | 0.15 | 0 | 0 | 0 | 0.15 |
| 110 | 0 | 0.05 | 0.05 | 0 | 0.1 |
| 117 | 0 | 0.05 | 0 | 0 | 0.05 |
| 132 | 0.05 | 0.05 | 0 | 0 | 0.1 |
| TOTAL | 0.2 | 0.15 | 0.05 | 0 | 0.4 |

STATE: NORTH CAROLINA

| ZIP CODE | QUARTER 1 | QUARTER 2 | QUARTER 3 | QUARTER 4 | TOTAL GRAMS |
|---|---|---|---|---|---|
| 277 | 0.2 | 0.15 | 0.15 | 0.25 | 0.75 |
| TOTAL | 0.2 | 0.15 | 0.15 | 0.25 | 0.75 |

STATE: OHIO

| ZIP CODE | QUARTER 1 | QUARTER 2 | QUARTER 3 | QUARTER 4 | TOTAL GRAMS |
|---|---|---|---|---|---|
| 430 | 0.1 | 0.1 | 0 | 0 | 0.2 |
| 432 | 0 | 0 | 0.1 | 0.7 | 0.8 |
| TOTAL | 0.1 | 0.1 | 0.1 | 0.7 | 1 |

STATE: OREGON

| ZIP CODE | QUARTER 1 | QUARTER 2 | QUARTER 3 | QUARTER 4 | TOTAL GRAMS |
|---|---|---|---|---|---|
| 972 | 0 | 0 | 0 | 0.05 | 0.05 |
| TOTAL | 0 | 0 | 0 | 0.05 | 0.05 |

DRUG CODE:7379DRUG NAME:NABILONE

STATE: PENNSYLVANIA

| ZIP CODE | QUARTER 1 | QUARTER 2 | QUARTER 3 | QUARTER 4 | TOTAL GRAMS |
|---|---|---|---|---|---|
| 184 | 0 | 0 | 0.15 | 0 | 0.15 |
| 187 | 0.05 | 0.05 | 0 | 0.1 | 0.2 |
| TOTAL | 0.05 | 0.05 | 0.15 | 0.1 | 0.35 |

STATE: VIRGINIA

| ZIP CODE | QUARTER 1 | QUARTER 2 | QUARTER 3 | QUARTER 4 | TOTAL GRAMS |
|---|---|---|---|---|---|
| 220 | 0.15 | 0.1 | 0.05 | 0 | 0.3 |
| 222 | 0 | 0.2 | 0.1 | 0 | 0.3 |
| 231 | 0 | 0.05 | 0.05 | 0 | 0.1 |
| 232 | 0.05 | 0 | 0 | 0 | 0.05 |
| TOTAL | 0.2 | 0.35 | 0.2 | 0 | 0.75 |

STATE: WASHINGTON

| ZIP CODE | QUARTER 1 | QUARTER 2 | QUARTER 3 | QUARTER 4 | TOTAL GRAMS |
|---|---|---|---|---|---|
| 980 | 0 | 0 | 0 | 0.05 | 0.05 |
| TOTAL | 0 | 0 | 0 | 0.05 | 0.05 |

DRUG CODE: 9041L DRUG NAME: COCAINE

STATE: ALABAMA

| ZIP CODE | QUARTER 1 | QUARTER 2 | QUARTER 3 | QUARTER 4 | TOTAL GRAMS |
|---|---|---|---|---|---|
| 350 | 0.71 | 0 | 0 | 0.29 | 1 |
| 351 | 1.29 | 2.29 | 1.29 | 1.29 | 6.16 |
| 352 | 14.28 | 13.28 | 11.28 | 7.86 | 46.7 |
| 354 | 0.43 | 1.14 | 1 | 0.71 | 3.28 |
| 356 | 2.57 | 3.71 | 2.29 | 4.14 | 12.71 |
| 357 | 1.14 | 0.57 | 0.86 | 0.86 | 3.43 |
| 358 | 4.86 | 3.57 | 3.71 | 1 | 13.14 |
| 359 | 0.43 | 2.29 | 1.86 | 1.43 | 6.01 |
| 360 | 0.29 | 1.71 | 0 | 1.57 | 3.57 |
| 361 | 20.28 | 22.71 | 15.71 | 16.43 | 75.13 |

| | | | | | | |
|---|---|---|---|---|---|---|
| | 362 | 2.29 | 0 | 0.71 | 1.14 | 4.14 |
| | 364 | 1.57 | 1.57 | 1.57 | 0.29 | 5 |
| | 365 | 6.86 | 8 | 6 | 8.29 | 29.15 |
| | 366 | 3.86 | 3 | 3 | 2.86 | 12.72 |
| | 367 | 0 | 0 | 0.29 | 0 | 0.29 |
| | 368 | 1.57 | 2 | 0.29 | 2.43 | 6.29 |
| TOTAL | | 62.43 | 65.84 | 49.86 | 50.59 | 228.72 |

STATE: ALASKA

| ZIP CODE | | QUARTER 1 | QUARTER 2 | QUARTER 3 | QUARTER 4 | TOTAL GRAMS |
|---|---|---|---|---|---|---|
| | 995 | 4.43 | 4.57 | 3.43 | 6 | 18.43 |
| | 996 | 0.29 | 1 | 1.29 | 0.29 | 2.87 |
| | 997 | 3.57 | 14.07 | 4.29 | 3.43 | 25.36 |
| | 998 | 0.29 | 0 | 0.14 | 0.29 | 0.72 |
| | 999 | 0 | 0.14 | 0.43 | 0 | 0.57 |
| TOTAL | | 8.58 | 19.78 | 9.58 | 10.01 | 47.95 |

STATE: ARIZONA

| ZIP CODE | | QUARTER 1 | QUARTER 2 | QUARTER 3 | QUARTER 4 | TOTAL GRAMS |
|---|---|---|---|---|---|---|
| | 850 | 42.71 | 42.46 | 39.71 | 39.14 | 164.02 |
| | 851 | 2.43 | 4.57 | 0.71 | 3.14 | 10.85 |
| | 852 | 36.35 | 27.78 | 29.57 | 25.43 | 119.13 |
| | 853 | 5.14 | 8.71 | 9.14 | 8.86 | 31.85 |
| | 855 | 0 | 0 | 0.29 | 0 | 0.29 |
| | 856 | 2.29 | 1.29 | 1.14 | 1.29 | 6.01 |
| | 857 | 9.71 | 13.71 | 11.43 | 16.14 | 50.99 |
| | 859 | 0.57 | 0 | 0.29 | 0.14 | 1 |
| | 860 | 7.43 | 5.71 | 3 | 3.29 | 19.43 |
| | 863 | 0.29 | 0 | 1 | 0.71 | 2 |
| | 864 | 0.71 | 2.57 | 3.14 | 1.71 | 8.13 |
| | 865 | 2.86 | 2.86 | 0.14 | 0 | 5.86 |
| TOTAL | | 110.49 | 109.66 | 99.56 | 99.85 | 419.56 |

STATE: ARKANSAS

| ZIP CODE | QUARTER 1 | QUARTER 2 | QUARTER 3 | QUARTER 4 | TOTAL GRAMS |
|---|---|---|---|---|---|
| 716 | 0 | 0 | 0.29 | 0 | 0.29 |
| 719 | 0.86 | 1.71 | 0.43 | 1.14 | 4.14 |
| 720 | 10 | 6 | 9 | 9.43 | 34.43 |
| 721 | 1.14 | 0.57 | 2.29 | 2.14 | 6.14 |
| 722 | 1.86 | 1.71 | 4.57 | 0.57 | 8.71 |
| 723 | 0 | 4.61 | 0.14 | 0 | 4.75 |
| 724 | 16.18 | 15.75 | 12.03 | 16.18 | 60.14 |
| 725 | 1.29 | 1.57 | 1 | 0.14 | 4 |
| 726 | 2.29 | 1.86 | 2.57 | 3.57 | 10.29 |
| 727 | 19.86 | 2.86 | 2.71 | 2.71 | 28.14 |
| 728 | 0 | 0.29 | 0.29 | 0.29 | 0.87 |
| 729 | 2.86 | 4 | 2.86 | 4 | 13.72 |
| TOTAL | 56.34 | 40.93 | 38.18 | 40.17 | 175.62 |

STATE: CALIFORNIA

| ZIP CODE | QUARTER 1 | QUARTER 2 | QUARTER 3 | QUARTER 4 | TOTAL GRAMS |
|---|---|---|---|---|---|
| 900 | 53.78 | 53.32 | 56.92 | 52.6 | 216.62 |
| 902 | 20.71 | 21 | 18 | 22.93 | 82.64 |
| 903 | 0.57 | 0 | 0.57 | 0.29 | 1.43 |
| 904 | 20.57 | 18.28 | 17.32 | 17.86 | 74.03 |
| 905 | 13.14 | 10.43 | 18.71 | 18.28 | 60.56 |
| 906 | 3.86 | 1.71 | 2.43 | 4.29 | 12.29 |
| 907 | 12.28 | 12.71 | 11.43 | 7.86 | 44.28 |
| 908 | 16.86 | 17.57 | 17.71 | 14 | 66.14 |
| 910 | 7.71 | 7.29 | 6.71 | 6.29 | 28 |
| 911 | 5 | 4 | 3.71 | 4.29 | 17 |
| 912 | 10.14 | 11.14 | 10.29 | 9 | 40.57 |
| 913 | 28.57 | 33.57 | 27.86 | 24 | 114 |
| 914 | 9.71 | 14.93 | 6.43 | 4.71 | 35.78 |
| 915 | 0 | 0.29 | 0 | 0 | 0.29 |
| 917 | 22.14 | 18.43 | 16.14 | 14 | 70.71 |
| 918 | 0.71 | 0.86 | 0 | 1.57 | 3.14 |
| 919 | 9.29 | 8.29 | 10.29 | 7.14 | 35.01 |
| 920 | 35.5 | 34.32 | 43.03 | 28.39 | 141.24 |
| 921 | 59 | 59.42 | 60.14 | 56.57 | 235.13 |

| | | | | | | |
|---|---|---|---|---|---|---|
| | 922 | 16.57 | 15.14 | 12.86 | 14.14 | 58.71 |
| | 923 | 13.14 | 12.14 | 13 | 9.71 | 47.99 |
| | 925 | 11.71 | 7.14 | 10 | 9.29 | 38.14 |
| | 926 | 19.14 | 20.14 | 24.14 | 24.86 | 88.28 |
| | 927 | 9.86 | 6.86 | 8.14 | 12.71 | 37.57 |
| | 928 | 13.14 | 19.86 | 13.86 | 13 | 59.86 |
| | 930 | 3.71 | 4.71 | 7.14 | 7 | 22.56 |
| | 931 | 6.43 | 13.86 | 8.71 | 10.14 | 39.14 |
| | 932 | 23.14 | 18.21 | 24.43 | 26.14 | 91.92 |
| | 933 | 17.71 | 11.28 | 14.14 | 12.14 | 55.27 |
| | 934 | 7.14 | 5.57 | 7.86 | 7.43 | 28 |
| | 935 | 4.57 | 1.86 | 5.71 | 5.14 | 17.28 |
| | 936 | 3.57 | 1.86 | 2.86 | 2.57 | 10.86 |
| | 937 | 11.43 | 7.14 | 8.86 | 12.28 | 39.71 |
| | 939 | 17.57 | 12.28 | 14.71 | 16.78 | 61.34 |
| | 940 | 25.82 | 27.61 | 19 | 21.14 | 93.57 |
| | 941 | 83.28 | 69.78 | 48.96 | 45.6 | 247.62 |
| | 943 | 24.18 | 14.71 | 14.28 | 24 | 77.17 |
| | 944 | 0 | 2.14 | 2.29 | 5.32 | 9.75 |
| | 945 | 37.14 | 44.93 | 36.35 | 35 | 153.42 |
| | 946 | 21.43 | 21.57 | 23 | 11.86 | 77.86 |
| | 947 | 3.43 | 1.29 | 1.14 | 2.29 | 8.15 |
| | 948 | 9.57 | 10.29 | 9 | 6.86 | 35.72 |
| | 949 | 10.29 | 12.43 | 11.57 | 14.86 | 49.15 |
| | 950 | 16.86 | 10.57 | 16.14 | 17.14 | 60.71 |
| | 951 | 24.86 | 16.28 | 25.28 | 15.14 | 81.56 |
| | 952 | 14.39 | 17.39 | 13.07 | 19.71 | 64.56 |
| | 953 | 35.35 | 26.57 | 24.71 | 25.21 | 111.84 |
| | 954 | 5.36 | 8.07 | 8.14 | 8 | 29.57 |
| | 955 | 2.43 | 0.71 | 1.86 | 5.29 | 10.29 |
| | 956 | 38.57 | 39.57 | 30 | 38.93 | 147.07 |
| | 957 | 0.29 | 1.29 | 0.86 | 0.57 | 3.01 |
| | 958 | 35.57 | 35.43 | 27 | 31.57 | 129.57 |
| | 959 | 14.28 | 16 | 10.86 | 14 | 55.14 |
| | 960 | 13.43 | 12.43 | 5.71 | 5.79 | 37.36 |
| | 961 | 2.71 | 4.43 | 2.43 | 2.43 | 12 |
| TOTAL | | 927.61 | 879.1 | 835.76 | 828.11 | 3,470.58 |

STATE: COLORADO

| ZIP CODE | QUARTER 1 | QUARTER 2 | QUARTER 3 | QUARTER 4 | TOTAL GRAMS |
|---|---|---|---|---|---|
| 800 | 8.29 | 11.71 | 6.14 | 8.29 | 34.43 |
| 801 | 16.43 | 28.18 | 17.78 | 22 | 84.39 |
| 802 | 19.25 | 13.86 | 9.86 | 12.14 | 55.11 |
| 803 | 0.86 | 1.86 | 1 | 1.86 | 5.58 |
| 804 | 0.57 | 0.71 | 0.29 | 2.29 | 3.86 |
| 805 | 24.57 | 40.28 | 20.57 | 24.86 | 110.28 |
| 806 | 0.71 | 1.14 | 3.29 | 1.57 | 6.71 |
| 807 | 0.71 | 1 | 1.43 | 1.29 | 4.43 |
| 809 | 1.57 | 0.86 | 1.71 | 1.86 | 6 |
| 810 | 6 | 7.14 | 5.43 | 1 | 19.57 |
| 813 | 0.29 | 0.29 | 0.57 | 0.43 | 1.58 |
| 814 | 1.71 | 1.14 | 1.71 | 0.86 | 5.42 |
| 816 | 1.14 | 1 | 1.43 | 2.14 | 5.71 |
| TOTAL | 82.1 | 109.17 | 71.21 | 80.59 | 343.07 |

STATE: CONNECTICUT

| ZIP CODE | QUARTER 1 | QUARTER 2 | QUARTER 3 | QUARTER 4 | TOTAL GRAMS |
|---|---|---|---|---|---|
| 060 | 0.14 | 0.43 | 0.57 | 0.57 | 1.71 |
| 061 | 1.07 | 0.71 | 1.71 | 0 | 3.49 |
| 062 | 2.57 | 1.43 | 2.57 | 0.86 | 7.43 |
| 063 | 6.57 | 4.86 | 1.43 | 3.14 | 16 |
| 064 | 4.86 | 1.43 | 3 | 1.43 | 10.72 |
| 065 | 8.14 | 9.29 | 10.14 | 8.29 | 35.86 |
| 066 | 1.43 | 1.43 | 1.43 | 1.43 | 5.72 |
| 067 | 0.86 | 2.29 | 2.57 | 1.29 | 7.01 |
| 068 | 13.43 | 10.43 | 26.71 | 12.43 | 63 |
| TOTAL | 39.07 | 32.3 | 50.13 | 29.44 | 150.94 |

STATE: DELAWARE

| ZIP CODE | QUARTER 1 | QUARTER 2 | QUARTER 3 | QUARTER 4 | TOTAL GRAMS |
|---|---|---|---|---|---|
| 197 | 0.57 | 0.57 | 1.71 | 0.57 | 3.42 |

| | | | | | | |
|---|---|---|---|---|---|---|
| | 198 | 0.43 | 0.86 | 1.14 | 0.71 | 3.14 |
| | 199 | 5.57 | 17.43 | 12 | 10.29 | 45.29 |
| TOTAL | | 6.57 | 18.86 | 14.85 | 11.57 | 51.85 |

STATE: DISTRICT OF COLUMBIA

| ZIP CODE | | QUARTER 1 | QUARTER 2 | QUARTER 3 | QUARTER 4 | TOTAL GRAMS |
|---|---|---|---|---|---|---|
| | 200 | 22.25 | 21.46 | 28.14 | 20.75 | 92.6 |
| | 204 | 0.29 | 0 | 0 | 0 | 0.29 |
| TOTAL | | 22.54 | 21.46 | 28.14 | 20.75 | 92.89 |

STATE: FLORIDA

| ZIP CODE | | QUARTER 1 | QUARTER 2 | QUARTER 3 | QUARTER 4 | TOTAL GRAMS |
|---|---|---|---|---|---|---|
| | 320 | 4.14 | 5.43 | 3.29 | 4.29 | 17.15 |
| | 321 | 1.57 | 0.14 | 1 | 1.57 | 4.28 |
| | 322 | 28.43 | 32.86 | 36.28 | 35.28 | 132.85 |
| | 323 | 3.86 | 4 | 2.86 | 3 | 13.72 |
| | 324 | 0.29 | 0.43 | 0.29 | 0.14 | 1.15 |
| | 325 | 3.14 | 5 | 4.43 | 0.71 | 13.28 |
| | 326 | 4.57 | 5.86 | 5.14 | 5.57 | 21.14 |
| | 327 | 3.79 | 8.5 | 6.86 | 2.43 | 21.58 |
| | 328 | 2.71 | 4.14 | 7 | 4 | 17.85 |
| | 329 | 5.86 | 6.43 | 10 | 10.57 | 32.86 |
| | 330 | 16.28 | 19 | 18.86 | 16.93 | 71.07 |
| | 331 | 4.57 | 7.71 | 9.43 | 18.89 | 40.6 |
| | 333 | 15.14 | 13.28 | 12 | 18.43 | 58.85 |
| | 334 | 28.39 | 31.61 | 29.43 | 26.14 | 115.57 |
| | 335 | 20.71 | 9.61 | 9.04 | 3.29 | 42.65 |
| | 336 | 74.57 | 85.78 | 67.5 | 68.78 | 296.63 |
| | 337 | 8.43 | 7.57 | 11.43 | 6.86 | 34.29 |
| | 338 | 6.86 | 2.14 | 4.14 | 3.43 | 16.57 |
| | 339 | 10.14 | 15.71 | 7 | 4.86 | 37.71 |
| | 341 | 13.71 | 6.57 | 18 | 6.29 | 44.57 |
| | 342 | 18.28 | 19.28 | 17.14 | 11 | 65.7 |
| | 344 | 1.29 | 1.57 | 1.14 | 0.57 | 4.57 |

| | | | | | | |
|---|---|---|---|---|---|---|
| | 346 | 3 | 5.14 | 4 | 7.43 | 19.57 |
| | 347 | 1 | 1.29 | 0.86 | 1.07 | 4.22 |
| | 349 | 8.43 | 7.71 | 7.57 | 10 | 33.71 |
| TOTAL | | 289.16 | 306.76 | 294.69 | 271.53 | 1,162.14 |

STATE: GEORGIA

DRUG CODE:9041LDRUG NAME:COCAINE
STATE:GEORGIA

| ZIP CODE | | QUARTER 1 | QUARTER 2 | QUARTER 3 | QUARTER 4 | TOTAL GRAMS |
|---|---|---|---|---|---|---|
| | 300 | 55.75 | 66 | 55.28 | 71.71 | 248.74 |
| | 301 | 24.43 | 22 | 18.86 | 23.86 | 89.15 |
| | 302 | 12.43 | 7.57 | 10.71 | 10 | 40.71 |
| | 303 | 37.28 | 40.85 | 36.57 | 48.28 | 162.98 |
| | 304 | 3.43 | 2.29 | 2.14 | 2 | 9.86 |
| | 305 | 1.29 | 3.43 | 2.86 | 5.29 | 12.87 |
| | 306 | 3.14 | 3.71 | 3.14 | 2.43 | 12.42 |
| | 307 | 1.29 | 1.29 | 0.43 | 0 | 3.01 |
| | 309 | 2.57 | 2.04 | 0.14 | 1.57 | 6.32 |
| | 310 | 0.43 | 0.43 | 1.29 | 1 | 3.15 |
| | 312 | 0.57 | 1.14 | 1.14 | 0.71 | 3.56 |
| | 314 | 1.14 | 4.14 | 2.14 | 2 | 9.42 |
| | 315 | 0.57 | 0.71 | 1.29 | 2.14 | 4.71 |
| | 316 | 2.57 | 3.43 | 1.14 | 0.29 | 7.43 |
| | 317 | 6.29 | 9.57 | 8.43 | 9.86 | 34.15 |
| | 319 | 1.29 | 2.57 | 1.57 | 3 | 8.43 |
| TOTAL | | 154.47 | 171.17 | 147.13 | 184.14 | 656.91 |

STATE: GUAM

| ZIP CODE | | QUARTER 1 | QUARTER 2 | QUARTER 3 | QUARTER 4 | TOTAL GRAMS |
|---|---|---|---|---|---|---|
| | 969 | 0 | 0 | 0.36 | 0 | 0.36 |
| TOTAL | | 0 | 0 | 0.36 | 0 | 0.36 |

STATE: HAWAII

| ZIP CODE | QUARTER 1 | QUARTER 2 | QUARTER 3 | QUARTER 4 | TOTAL GRAMS |
|---|---|---|---|---|---|
| 967 | 13.71 | 9.14 | 10.57 | 10.71 | 44.13 |
| 968 | 14.57 | 14.71 | 14.28 | 9.71 | 53.27 |
| TOTAL | 28.28 | 23.85 | 24.85 | 20.42 | 97.4 |

STATE: IDAHO

| ZIP CODE | QUARTER 1 | QUARTER 2 | QUARTER 3 | QUARTER 4 | TOTAL GRAMS |
|---|---|---|---|---|---|
| 832 | 10 | 1.43 | 6.86 | 6.57 | 24.86 |
| 833 | 10.64 | 10.21 | 10.5 | 15.11 | 46.46 |
| 834 | 30.03 | 6.71 | 8.43 | 29.57 | 74.74 |
| 835 | 0 | 2 | 0.57 | 0 | 2.57 |
| 836 | 9.43 | 11.28 | 8.71 | 11.28 | 40.7 |
| 837 | 4.07 | 5.79 | 10.36 | 5 | 25.22 |
| 838 | 0.43 | 0 | 1.43 | 0.14 | 2 |
| TOTAL | 64.6 | 37.42 | 46.86 | 67.67 | 216.55 |

STATE: ILLINOIS

| ZIP CODE | QUARTER 1 | QUARTER 2 | QUARTER 3 | QUARTER 4 | TOTAL GRAMS |
|---|---|---|---|---|---|
| 600 | 38.64 | 24.28 | 20.86 | 31 | 114.78 |
| 601 | 58.85 | 56.6 | 53.43 | 54 | 222.88 |
| 602 | 12.43 | 13.43 | 2.71 | 14.21 | 42.78 |
| 603 | 1.14 | 2.14 | 1.14 | 1.57 | 5.99 |
| 604 | 14.57 | 22.28 | 17.28 | 18.14 | 72.27 |
| 605 | 40.57 | 46.28 | 29.14 | 39.85 | 155.84 |
| 606 | 61.28 | 77.6 | 44.85 | 75.42 | 259.15 |
| 607 | 0.14 | 0 | 0 | 0 | 0.14 |
| 608 | 0 | 0 | 0.29 | 0 | 0.29 |
| 609 | 19.14 | 30.5 | 29.93 | 32.5 | 112.07 |
| 610 | 16 | 14.71 | 6.29 | 3.57 | 40.57 |
| 611 | 6.71 | 5.29 | 6.71 | 4.29 | 23 |
| 612 | 5 | 3.86 | 2.71 | 3 | 14.57 |
| 613 | 0.43 | 1.14 | 0.71 | 1 | 3.28 |
| 614 | 1 | 12.5 | 9.93 | 10.21 | 33.64 |

| | | | | | | |
|---|---|---|---|---|---|---|
| | 615 | 2.29 | 2.29 | 3.86 | 2.86 | 11.3 |
| | 616 | 5.43 | 5.29 | 4.71 | 4.14 | 19.57 |
| | 617 | 7.14 | 5.14 | 2.57 | 6.86 | 21.71 |
| | 618 | 1.57 | 2.71 | 1.57 | 2.86 | 8.71 |
| | 619 | 0.43 | 0 | 0.14 | 0 | 0.57 |
| | 620 | 11 | 5.57 | 9.14 | 1.14 | 26.85 |
| | 622 | 7.14 | 5.14 | 5.57 | 5.14 | 22.99 |
| | 623 | 1.43 | 1.14 | 1.14 | 5 | 8.71 |
| | 624 | 0 | 0.43 | 0.43 | 0.14 | 1 |
| | 625 | 5.43 | 7.86 | 6.43 | 6.86 | 26.58 |
| | 626 | 0.57 | 0.43 | 0.14 | 0.57 | 1.71 |
| | 627 | 1.71 | 3.71 | 2.14 | 1.71 | 9.27 |
| | 628 | 6.14 | 5.93 | 7.86 | 7.93 | 27.86 |
| | 629 | 0.43 | 0.57 | 1 | 0 | 2 |
| TOTAL | | 326.61 | 356.82 | 272.68 | 333.97 | 1,290.08 |

STATE: INDIANA

| ZIP CODE | | QUARTER 1 | QUARTER 2 | QUARTER 3 | QUARTER 4 | TOTAL GRAMS |
|---|---|---|---|---|---|---|
| | 460 | 24.64 | 25.5 | 24.78 | 21.07 | 95.99 |
| | 461 | 11.43 | 5.14 | 9.57 | 11 | 37.14 |
| | 462 | 93.99 | 53.85 | 42.14 | 56.57 | 246.55 |
| | 463 | 29.43 | 22.86 | 21.14 | 16.86 | 90.29 |
| | 464 | 4.43 | 4.14 | 3.57 | 4.71 | 16.85 |
| | 465 | 7.18 | 7.61 | 0.29 | 2.29 | 17.37 |
| | 466 | 14.89 | 17.5 | 8.57 | 20.36 | 61.32 |
| | 467 | 3 | 5.29 | 3.93 | 4.29 | 16.51 |
| | 468 | 1 | 1 | 3 | 1.14 | 6.14 |
| | 469 | 18.28 | 9.29 | 14.28 | 16.86 | 58.71 |
| | 471 | 9.29 | 5.86 | 6.57 | 16 | 37.72 |
| | 472 | 10.29 | 8.14 | 5.14 | 6.86 | 30.43 |
| | 473 | 8.71 | 14.14 | 2 | 4.43 | 29.28 |
| | 474 | 0 | 0 | 0.57 | 0 | 0.57 |
| | 475 | 1.29 | 1.14 | 0.86 | 1.14 | 4.43 |
| | 476 | 1.14 | 1 | 0.86 | 1 | 4 |
| | 477 | 2 | 3.29 | 0.29 | 1.71 | 7.29 |
| | 478 | 1 | 0.71 | 0.29 | 0.71 | 2.71 |

| | | | | | |
|---|---|---|---|---|---|
| 479 | 9.14 | 9.29 | 5.29 | 13.93 | 37.65 |
| TOTAL | 251.13 | 195.75 | 153.14 | 200.93 | 800.95 |

STATE: IOWA

| ZIP CODE | QUARTER 1 | QUARTER 2 | QUARTER 3 | QUARTER 4 | TOTAL GRAMS |
|---|---|---|---|---|---|
| 500 | 0.29 | 0.29 | 0.29 | 0.57 | 1.44 |
| 501 | 0.14 | 0.29 | 0.71 | 0 | 1.14 |
| 502 | 1.71 | 2.57 | 1.14 | 2.14 | 7.56 |
| 503 | 14.28 | 16.14 | 11.43 | 11.14 | 52.99 |
| 504 | 0.29 | 0 | 0.14 | 0.14 | 0.57 |
| 505 | 2.43 | 2 | 2 | 0 | 6.43 |
| 506 | 1.57 | 0.86 | 2.43 | 1.43 | 6.29 |
| 507 | 23.43 | 26 | 19.86 | 30.28 | 99.57 |
| 508 | 0 | 0 | 0.29 | 0.29 | 0.58 |
| 510 | 0.86 | 0 | 0.14 | 1.43 | 2.43 |
| 511 | 4.71 | 5.43 | 6.14 | 6.57 | 22.85 |
| 512 | 0.14 | 0 | 0.29 | 0 | 0.43 |
| 513 | 0.14 | 0 | 0.29 | 0.14 | 0.57 |
| 514 | 0.57 | 0 | 0.29 | 0 | 0.86 |
| 515 | 1.57 | 1.14 | 1.29 | 1 | 5 |
| 516 | 0 | 0 | 0.43 | 0.43 | 0.86 |
| 520 | 3.14 | 2.29 | 0.71 | 2.86 | 9 |
| 521 | 0.71 | 0.71 | 0.14 | 0.43 | 1.99 |
| 522 | 15.28 | 5.43 | 7.32 | 6.57 | 34.6 |
| 523 | 0 | 0 | 0.29 | 0.29 | 0.58 |
| 524 | 9.57 | 12.28 | 9.29 | 11.71 | 42.85 |
| 525 | 5.29 | 6.29 | 5.71 | 6.14 | 23.43 |
| 526 | 1.57 | 1.14 | 1.57 | 1.71 | 5.99 |
| 527 | 5.29 | 3.29 | 6.57 | 1.86 | 17.01 |
| 528 | 12.43 | 16.71 | 7.36 | 11.5 | 48 |
| TOTAL | 105.41 | 102.86 | 86.12 | 98.63 | 393.02 |

STATE: KANSAS

| ZIP CODE | QUARTER 1 | QUARTER 2 | QUARTER 3 | QUARTER 4 | TOTAL GRAMS |
|---|---|---|---|---|---|
| 660 | 7.57 | 7.14 | 9.29 | 7.43 | 31.43 |

| | | | | | | |
|---|---|---|---|---|---|---|
| | 661 | 2.43 | 4 | 2.57 | 3.57 | 12.57 |
| | 662 | 35.68 | 45.75 | 47.39 | 56.53 | 185.35 |
| | 664 | 0 | 0.14 | 0.29 | 0 | 0.43 |
| | 665 | 4.71 | 4.71 | 5.14 | 6.86 | 21.42 |
| | 666 | 3.71 | 9.71 | 9.79 | 9.21 | 32.42 |
| | 667 | 1.86 | 3 | 1.86 | 1.86 | 8.58 |
| | 670 | 1 | 1.14 | 0.71 | 0.86 | 3.71 |
| | 671 | 0 | 0.71 | 0.29 | 0 | 1 |
| | 672 | 4.57 | 5.43 | 3.14 | 8.86 | 22 |
| | 673 | 0.57 | 1.14 | 0 | 0.29 | 2 |
| | 674 | 0.86 | 0 | 0.71 | 1.57 | 3.14 |
| | 676 | 0 | 0.29 | 0.57 | 0.29 | 1.15 |
| | 677 | 0.29 | 0.57 | 0.29 | 0 | 1.15 |
| | 678 | 0 | 0.71 | 0.29 | 0 | 1 |
| | 679 | 0.29 | 0.43 | 0 | 0.57 | 1.29 |
| TOTAL | | 63.54 | 84.87 | 82.33 | 97.9 | 328.64 |

| STATE: | KENTUCKY | | | | | |
|---|---|---|---|---|---|---|
| ZIP CODE | | QUARTER 1 | QUARTER 2 | QUARTER 3 | QUARTER 4 | TOTAL GRAMS |
| | 400 | 1.71 | 1 | 1.57 | 1.29 | 5.57 |
| | 402 | 26.43 | 25.14 | 24 | 23.14 | 98.71 |
| | 403 | 4.86 | 4.14 | 2 | 2.57 | 13.57 |
| | 404 | 0.57 | 0.57 | 0 | 1.71 | 2.85 |
| | 405 | 18.53 | 20.96 | 11.89 | 12.75 | 64.13 |
| | 406 | 0 | 0 | 0.86 | 0 | 0.86 |
| | 407 | 2.71 | 1 | 1.57 | 1.43 | 6.71 |
| | 409 | 0 | 0.57 | 0.86 | 0.29 | 1.72 |
| | 410 | 11.86 | 14.86 | 6.29 | 10.46 | 43.47 |
| | 411 | 9.29 | 11.14 | 7.29 | 9.86 | 37.58 |
| | 412 | 0.86 | 0.14 | 0.14 | 0.29 | 1.43 |
| | 415 | 0.43 | 0 | 0.14 | 0.43 | 1 |
| | 416 | 2.43 | 2.86 | 2.29 | 4.14 | 11.72 |
| | 420 | 14.36 | 14.93 | 10.75 | 21.21 | 61.25 |
| | 421 | 4.57 | 8.71 | 7.57 | 11.43 | 32.28 |
| | 422 | 0 | 0.29 | 1 | 1.14 | 2.43 |
| | 423 | 4.29 | 5.14 | 2.29 | 6.29 | 18.01 |

| | | | | | | |
|---|---|---|---|---|---|---|
| | 424 | 0.86 | 0 | 0.29 | 1.71 | 2.86 |
| | 426 | 0 | 0 | 0.57 | 0.43 | 1 |
| | 427 | 6.14 | 6.43 | 7.14 | 7.57 | 27.28 |
| TOTAL | | 109.9 | 117.88 | 88.51 | 118.14 | 434.43 |

STATE: LOUISIANA

| ZIP CODE | | QUARTER 1 | QUARTER 2 | QUARTER 3 | QUARTER 4 | TOTAL GRAMS |
|---|---|---|---|---|---|---|
| | 700 | 5.61 | 1.71 | 2 | 1.29 | 10.61 |
| | 701 | 15.71 | 15.14 | 11.28 | 24 | 66.13 |
| | 703 | 1.43 | 3.86 | 3 | 5.29 | 13.58 |
| | 704 | 2 | 1.71 | 1.86 | 1.86 | 7.43 |
| | 705 | 15.86 | 18.71 | 17.57 | 20.71 | 72.85 |
| | 706 | 7.86 | 9 | 6.57 | 7.71 | 31.14 |
| | 707 | 0 | 0 | 0.43 | 0 | 0.43 |
| | 708 | 3 | 2.57 | 3.71 | 2.43 | 11.71 |
| | 710 | 0.43 | 0.14 | 0.29 | 0.29 | 1.15 |
| | 711 | 31 | 32.86 | 27.57 | 36.85 | 128.28 |
| | 712 | 0.86 | 1.71 | 0.57 | 0.57 | 3.71 |
| | 713 | 5.14 | 6.29 | 8 | 10.71 | 30.14 |
| | 714 | 3.43 | 3 | 3.86 | 2 | 12.29 |
| TOTAL | | 92.33 | 96.7 | 86.71 | 113.71 | 389.45 |

STATE: MAINE

| ZIP CODE | | QUARTER 1 | QUARTER 2 | QUARTER 3 | QUARTER 4 | TOTAL GRAMS |
|---|---|---|---|---|---|---|
| | 039 | 0.14 | 0.43 | 1.29 | 0.57 | 2.43 |
| | 040 | 0.71 | 0.43 | 0.57 | 0.29 | 2 |
| | 041 | 4.71 | 3.14 | 4 | 4.43 | 16.28 |
| | 042 | 0.29 | 0.71 | 0.29 | 0.14 | 1.43 |
| | 043 | 1.86 | 1.57 | 2 | 2.57 | 8 |
| | 044 | 8.71 | 13.36 | 6.29 | 3.07 | 31.43 |
| | 045 | 0 | 0 | 0.14 | 0.57 | 0.71 |
| | 046 | 1.14 | 0.86 | 0.71 | 1 | 3.71 |
| | 047 | 2.14 | 1.71 | 1.86 | 1 | 6.71 |
| | 048 | 2 | 2.14 | 3.14 | 2.43 | 9.71 |
| | 049 | 0.71 | 0.57 | 2.57 | 1.14 | 4.99 |

| | | | | | |
|---|---|---|---|---|---|
| TOTAL | 22.41 | 24.92 | 22.86 | 17.21 | 87.4 |

STATE: MARYLAND

| ZIP CODE | QUARTER 1 | QUARTER 2 | QUARTER 3 | QUARTER 4 | TOTAL GRAMS |
|---|---|---|---|---|---|
| 206 | 3.29 | 0.43 | 2.86 | 0.71 | 7.29 |
| 207 | 3.71 | 5.43 | 6.43 | 4.29 | 19.86 |
| 208 | 19.07 | 12 | 23.14 | 14.71 | 68.92 |
| 209 | 6.29 | 7.86 | 0 | 2 | 16.15 |
| 210 | 11.04 | 10.71 | 5.46 | 9.18 | 36.39 |
| 211 | 7.43 | 8.43 | 6.43 | 7 | 29.29 |
| 212 | 33.57 | 49.18 | 35.14 | 28.86 | 146.75 |
| 214 | 37.25 | 40.57 | 29.07 | 31.36 | 138.25 |
| 215 | 1.14 | 1.14 | 0.86 | 0.71 | 3.85 |
| 216 | 1.29 | 0.71 | 0.57 | 2.29 | 4.86 |
| 217 | 10.57 | 7.14 | 6.14 | 5.14 | 28.99 |
| 218 | 3.43 | 0.57 | 1 | 0.86 | 5.86 |
| 219 | 2.71 | 2.43 | 2.43 | 3.57 | 11.14 |
| TOTAL | 140.79 | 146.6 | 119.53 | 110.68 | 517.6 |

STATE: MASSACHUSETTS

| ZIP CODE | QUARTER 1 | QUARTER 2 | QUARTER 3 | QUARTER 4 | TOTAL GRAMS |
|---|---|---|---|---|---|
| 10 | 8.43 | 5.71 | 5.57 | 7.57 | 27.28 |
| 11 | 16.57 | 17 | 16.57 | 16.28 | 66.42 |
| 12 | 7.86 | 6.71 | 4 | 5.86 | 24.43 |
| 13 | 0.57 | 0.71 | 1 | 0.86 | 3.14 |
| 14 | 13 | 8.29 | 10.43 | 8.57 | 40.29 |
| 15 | 3.86 | 4.29 | 3.57 | 2.43 | 14.15 |
| 16 | 27.14 | 23.86 | 16.57 | 20.28 | 87.85 |
| 17 | 10.43 | 8.71 | 5 | 10.86 | 35 |
| 18 | 20.28 | 12.71 | 12.86 | 18.43 | 64.28 |
| 19 | 9.71 | 10.86 | 8.29 | 9.14 | 38 |
| 20 | 0 | 0.29 | 0 | 0.14 | 0.43 |
| 21 | 84.42 | 78.28 | 73.71 | 88.64 | 325.05 |
| 22 | 7.29 | 9.57 | 7.43 | 3.86 | 28.15 |
| 23 | 2.14 | 2.43 | 2.29 | 2.86 | 9.72 |

| | | | | | | |
|---|---|---|---|---|---|---|
| | 24 | 9.14 | 5.71 | 6.29 | 2.86 | 24 |
| | 25 | 3.86 | 4 | 3.43 | 2.71 | 14 |
| | 26 | 2.14 | 1.14 | 1.14 | 1 | 5.42 |
| | 27 | 1.57 | 2.43 | 2.43 | 1.43 | 7.86 |
| TOTAL | | 228.41 | 202.7 | 180.58 | 203.78 | 815.47 |

STATE: MICHIGAN

| ZIP CODE | | QUARTER 1 | QUARTER 2 | QUARTER 3 | QUARTER 4 | TOTAL GRAMS |
|---|---|---|---|---|---|---|
| | 480 | 43.35 | 56.71 | 56.35 | 59.5 | 215.91 |
| | 481 | 46.85 | 37.6 | 39.78 | 30 | 154.23 |
| | 482 | 17.57 | 18.57 | 19.43 | 16.86 | 72.43 |
| | 483 | 24.43 | 30.86 | 20.86 | 23.28 | 99.43 |
| | 484 | 6.29 | 7.14 | 3.29 | 4.57 | 21.29 |
| | 485 | 12 | 13.86 | 8.86 | 11.14 | 45.86 |
| | 486 | 4.14 | 3.86 | 4.57 | 8.29 | 20.86 |
| | 487 | 5.71 | 7.57 | 4.71 | 3.71 | 21.7 |
| | 488 | 8.86 | 7.71 | 12 | 10.14 | 38.71 |
| | 489 | 3.57 | 9.29 | 4.57 | 4.14 | 21.57 |
| | 490 | 38.57 | 43.6 | 50.39 | 53.64 | 186.2 |
| | 491 | 11.78 | 38.57 | 4.57 | 5.61 | 60.53 |
| | 492 | 5.43 | 2.71 | 2.43 | 3.71 | 14.28 |
| | 493 | 0.29 | 0.86 | 1.43 | 0.86 | 3.44 |
| | 494 | 63.14 | 74.35 | 52.35 | 63.71 | 253.55 |
| | 495 | 33.43 | 23.71 | 24.57 | 30 | 111.71 |
| | 496 | 16.86 | 16.14 | 13.71 | 14.43 | 61.14 |
| | 497 | 20 | 21.18 | 10.43 | 30 | 81.61 |
| | 498 | 4.71 | 7.14 | 1.71 | 7.14 | 20.7 |
| | 499 | 0.57 | 0.71 | 0.57 | 0.86 | 2.71 |
| TOTAL | | 367.55 | 422.14 | 336.58 | 381.59 | 1,507.86 |

STATE: MINNESOTA

| ZIP CODE | | QUARTER 1 | QUARTER 2 | QUARTER 3 | QUARTER 4 | TOTAL GRAMS |
|---|---|---|---|---|---|---|
| | 550 | 8 | 12.57 | 10.71 | 11.43 | 42.71 |
| | 551 | 15.43 | 11.57 | 16.14 | 18.43 | 61.57 |
| | 553 | 26.43 | 25.28 | 25.14 | 33.93 | 110.78 |

| | | | | | | |
|---|---|---|---|---|---|---|
| | 554 | 81.14 | 90.24 | 65.14 | 73.71 | 310.23 |
| | 557 | 5.71 | 6.57 | 7.29 | 6.43 | 26 |
| | 558 | 19.71 | 22.14 | 14.14 | 12.43 | 68.42 |
| | 559 | 38.96 | 46.78 | 41.39 | 34.96 | 162.09 |
| | 560 | 7.14 | 7.43 | 9.71 | 5.71 | 29.99 |
| | 561 | 0.14 | 0.43 | 0 | 0.86 | 1.43 |
| | 562 | 4.71 | 3.29 | 4.86 | 4.71 | 17.57 |
| | 563 | 0.57 | 1.71 | 0.29 | 0.57 | 3.14 |
| | 564 | 1 | 2.14 | 1.57 | 1.43 | 6.14 |
| | 565 | 0.86 | 0.43 | 0.57 | 0 | 1.86 |
| | 566 | 1.29 | 2.86 | 1 | 5 | 10.15 |
| | 567 | 0.43 | 0.14 | 1.29 | 1.57 | 3.43 |
| TOTAL | | 211.52 | 233.58 | 199.24 | 211.17 | 855.51 |

STATE: MISSISSIPPI

| ZIP CODE | | QUARTER 1 | QUARTER 2 | QUARTER 3 | QUARTER 4 | TOTAL GRAMS |
|---|---|---|---|---|---|---|
| | 386 | 0.29 | 0 | 4 | 0.29 | 4.58 |
| | 388 | 0.86 | 1 | 0.71 | 0.43 | 3 |
| | 390 | 1.57 | 0 | 0.14 | 0.43 | 2.14 |
| | 391 | 0 | 0.14 | 0 | 0.14 | 0.28 |
| | 392 | 10.57 | 14.86 | 15.71 | 9.29 | 50.43 |
| | 393 | 0 | 0 | 0.14 | 0 | 0.14 |
| | 394 | 19.71 | 20.46 | 23.89 | 21.14 | 85.2 |
| | 395 | 12.43 | 44 | 7.43 | 30.75 | 94.61 |
| | 396 | 13.39 | 8.93 | 8.93 | 8.93 | 40.18 |
| | 397 | 20.86 | 12.14 | 14.28 | 11.14 | 58.42 |
| TOTAL | | 79.68 | 101.53 | 75.23 | 82.54 | 338.98 |

STATE: MISSOURI

| ZIP CODE | | QUARTER 1 | QUARTER 2 | QUARTER 3 | QUARTER 4 | TOTAL GRAMS |
|---|---|---|---|---|---|---|
| | 630 | 5.43 | 6.71 | 4.29 | 2.29 | 18.72 |

| | | | | | | |
|---|---|---|---|---|---|---|
| | 631 | 17 | 26.14 | 20.71 | 25.57 | 89.42 |
| | 633 | 6.29 | 5.29 | 6.57 | 6.14 | 24.29 |
| | 634 | 0.29 | 2.14 | 1.86 | 0.57 | 4.86 |
| | 635 | 0.57 | 0.29 | 0.86 | 0.29 | 2.01 |
| | 636 | 6.43 | 4.14 | 7.14 | 7.86 | 25.57 |
| | 637 | 12.5 | 21 | 12.21 | 22 | 67.71 |
| | 638 | 0.14 | 0 | 0.14 | 0 | 0.28 |
| | 639 | 0.14 | 0 | 0.57 | 0 | 0.71 |
| | 640 | 27.75 | 33.07 | 38.53 | 19.46 | 118.81 |
| | 641 | 16.64 | 17.28 | 7.57 | 6.86 | 48.35 |
| | 644 | 0.57 | 1.86 | 0.29 | 1.07 | 3.79 |
| | 645 | 5.71 | 4.29 | 5.71 | 7.57 | 23.28 |
| | 647 | 2 | 2.14 | 2.43 | 0.71 | 7.28 |
| | 648 | 16.71 | 16.57 | 11.43 | 12.71 | 57.42 |
| | 650 | 0 | 0 | 0.14 | 0 | 0.14 |
| | 651 | 2.71 | 0.57 | 0.71 | 2.43 | 6.42 |
| | 652 | 14.18 | 12.18 | 10.32 | 11.82 | 48.5 |
| | 653 | 9 | 7.57 | 7 | 9.43 | 33 |
| | 654 | 0.71 | 0.71 | 1.29 | 1.93 | 4.64 |
| | 655 | 0 | 0.36 | 0 | 0 | 0.36 |
| | 656 | 8.43 | 4.57 | 4.86 | 4.57 | 22.43 |
| | 657 | 0 | 0.14 | 0.43 | 0 | 0.57 |
| | 658 | 11 | 13.86 | 14.86 | 11.28 | 51 |
| TOTAL | | 164.2 | 180.88 | 159.92 | 154.56 | 659.56 |

| STATE: | MONTANA | | | | |
|---|---|---|---|---|---|
| ZIP CODE | QUARTER 1 | QUARTER 2 | QUARTER 3 | QUARTER 4 | TOTAL GRAMS |
| 590 | 0 | 0 | 0.57 | 0.57 | 1.14 |
| 591 | 8.29 | 9.43 | 10 | 15.86 | 43.58 |
| 592 | 3.43 | 6.29 | 2.86 | 4.43 | 17.01 |
| 593 | 0.57 | 0 | 0.86 | 0 | 1.43 |
| 594 | 4.14 | 5.71 | 4.43 | 5.29 | 19.57 |
| 596 | 7.57 | 9.14 | 7.14 | 12.43 | 36.28 |
| 597 | 3.86 | 3.57 | 5.14 | 4.14 | 16.71 |
| 598 | 5.32 | 5.32 | 0.79 | 0.71 | 12.14 |
| 599 | 15.68 | 15.53 | 9.93 | 22.61 | 63.75 |

| | | | | | | |
|---|---|---|---|---|---|---|
| TOTAL | | 48.86 | 54.99 | 41.72 | 66.04 | 211.61 |

| STATE: | NEBRASKA | | | | | |
|---|---|---|---|---|---|---|
| ZIP CODE | | QUARTER 1 | QUARTER 2 | QUARTER 3 | QUARTER 4 | TOTAL GRAMS |
| | 680 | 0.29 | 1.71 | 2 | 1.29 | 5.29 |
| | 681 | 9.86 | 12.43 | 10.71 | 10.57 | 43.57 |
| | 683 | 0.14 | 0 | 0.14 | 0 | 0.28 |
| | 684 | 0.14 | 0 | 0.43 | 0 | 0.57 |
| | 685 | 0.29 | 2.86 | 1.86 | 1 | 6.01 |
| | 686 | 0.29 | 0.14 | 4.61 | 0 | 5.04 |
| | 687 | 0 | 0.71 | 2 | 0 | 2.71 |
| | 688 | 3.71 | 3.57 | 4.71 | 4.86 | 16.85 |
| | 689 | 1.29 | 0.86 | 2 | 3.14 | 7.29 |
| | 691 | 4 | 4.57 | 6.29 | 5.43 | 20.29 |
| | 693 | 1.57 | 4.14 | 2.29 | 3.29 | 11.29 |
| TOTAL | | 21.58 | 30.99 | 37.04 | 29.58 | 119.19 |

| STATE: | NEVADA | | | | | |
|---|---|---|---|---|---|---|
| ZIP CODE | | QUARTER 1 | QUARTER 2 | QUARTER 3 | QUARTER 4 | TOTAL GRAMS |
| | 890 | 0.86 | 1.14 | 0.29 | 2.14 | 4.43 |
| | 891 | 14.57 | 12 | 8.57 | 9.75 | 44.89 |
| | 894 | 1.29 | 1.71 | 1.57 | 0.71 | 5.28 |
| | 895 | 21.18 | 30.39 | 29.21 | 19.28 | 100.06 |
| | 897 | 0.57 | 2 | 0.86 | 0 | 3.43 |
| | 898 | 6.14 | 4.86 | 3.43 | 7.43 | 21.86 |
| TOTAL | | 44.61 | 52.1 | 43.93 | 39.31 | 179.95 |

| STATE: | NEW HAMPSHIRE | | | | | |
|---|---|---|---|---|---|---|
| ZIP CODE | | QUARTER 1 | QUARTER 2 | QUARTER 3 | QUARTER 4 | TOTAL GRAMS |
| | 30 | 7 | 11.04 | 7.29 | 13.43 | 38.76 |
| | 31 | 10.86 | 15.43 | 8.29 | 18 | 52.58 |
| | 32 | 1 | 2.43 | 1.43 | 1.71 | 6.57 |

| | | | | | |
|---|---|---|---|---|---|
| | 33 | 0.29 | 0 | 0.57 | 0.29 | 1.15 |
| | 34 | 0.29 | 1.86 | 0 | 1 | 3.15 |
| | 35 | 1.43 | 0.43 | 0.71 | 1 | 3.57 |
| | 37 | 5 | 9.14 | 5.71 | 5.57 | 25.42 |
| | 38 | 12.86 | 14.57 | 16.57 | 15.14 | 59.14 |
| TOTAL | | 38.73 | 54.9 | 40.57 | 56.14 | 190.34 |

STATE: NEW JERSEY

| ZIP CODE | QUARTER 1 | QUARTER 2 | QUARTER 3 | QUARTER 4 | TOTAL GRAMS |
|---|---|---|---|---|---|
| 070 | 19 | 24.43 | 22.43 | 14.43 | 80.29 |
| 071 | 10.43 | 12.71 | 8.71 | 9.43 | 41.28 |
| 072 | 2.57 | 0.43 | 0.86 | 0.57 | 4.43 |
| 073 | 1.71 | 1.29 | 1 | 0.43 | 4.43 |
| 074 | 36.57 | 42.57 | 31.28 | 35.71 | 146.13 |
| 075 | 4.57 | 1.71 | 1 | 1 | 8.28 |
| 076 | 11.86 | 11.57 | 14.57 | 12 | 50 |
| 077 | 6.43 | 8.86 | 11.43 | 7 | 33.72 |
| 078 | 8.14 | 6.86 | 5.14 | 9.43 | 29.57 |
| 079 | 3.43 | 3.71 | 4.14 | 5.29 | 16.57 |
| 080 | 17.86 | 14 | 14.14 | 12 | 58 |
| 081 | 2.86 | 1.43 | 2.86 | 1.43 | 8.58 |
| 082 | 9.18 | 2.71 | 4.14 | 3.14 | 19.17 |
| 083 | 2.43 | 1.14 | 1.29 | 1.43 | 6.29 |
| 084 | 0.71 | 2.86 | 1.71 | 2.29 | 7.57 |
| 085 | 2.86 | 2.29 | 2.29 | 2.57 | 10.01 |
| 086 | 4.14 | 2.14 | 1.18 | 4.14 | 11.6 |
| 087 | 6 | 8 | 2.57 | 8.43 | 25 |
| 088 | 14.57 | 12.57 | 8.14 | 12.86 | 48.14 |
| 089 | 10.71 | 7.43 | 10 | 8.57 | 36.71 |
| TOTAL | 176.03 | 168.71 | 148.88 | 152.15 | 645.77 |

STATE: NEW MEXICO

| ZIP CODE | QUARTER 1 | QUARTER 2 | QUARTER 3 | QUARTER 4 | TOTAL GRAMS |
|---|---|---|---|---|---|

| | | | | | | |
|---|---|---|---|---|---|---|
| | 871 | 65.92 | 61.57 | 65.92 | 59.71 | 253.12 |
| | 873 | 0 | 0.43 | 0.86 | 0 | 1.29 |
| | 874 | 0 | 0.43 | 2.71 | 0.71 | 3.85 |
| | 875 | 4.57 | 3.57 | 2.43 | 4.14 | 14.71 |
| | 880 | 1.43 | 6.29 | 0 | 0.57 | 8.29 |
| | 882 | 4.43 | 1.29 | 1.71 | 1.43 | 8.86 |
| | 883 | 0 | 0.14 | 0.14 | 0 | 0.28 |
| TOTAL | | 76.35 | 73.72 | 73.77 | 66.56 | 290.4 |

STATE: NEW YORK

| ZIP CODE | | QUARTER 1 | QUARTER 2 | QUARTER 3 | QUARTER 4 | TOTAL GRAMS |
|---|---|---|---|---|---|---|
| | 100 | 138.1 | 124.92 | 76.14 | 125.99 | 465.15 |
| | 101 | 0.29 | 1.86 | 0.43 | 0.43 | 3.01 |
| | 103 | 1 | 0.86 | 0.57 | 1 | 3.43 |
| | 104 | 6.29 | 9.14 | 4.43 | 4.57 | 24.43 |
| | 105 | 2.86 | 3.14 | 7.29 | 4.57 | 17.86 |
| | 106 | 0.71 | 2.14 | 0 | 0 | 2.85 |
| | 107 | 1.43 | 3.14 | 1.43 | 1.14 | 7.14 |
| | 108 | 0.57 | 1.14 | 0 | 0 | 1.71 |
| | 109 | 5.71 | 6.86 | 6.43 | 7 | 26 |
| | 110 | 15.43 | 8.29 | 16.5 | 11.71 | 51.93 |
| | 111 | 0.43 | 1 | 0 | 0.29 | 1.72 |
| | 112 | 14.57 | 21.78 | 29.71 | 8.61 | 74.67 |
| | 113 | 2.86 | 1.86 | 5.14 | 2 | 11.86 |
| | 115 | 6.43 | 6.14 | 17.57 | 8.43 | 38.57 |
| | 116 | 1.71 | 2.14 | 1.14 | 1.71 | 6.7 |
| | 117 | 5.29 | 4.43 | 7.43 | 5.71 | 22.86 |
| | 118 | 2.29 | 3.14 | 3.14 | 1.43 | 10 |
| | 119 | 1.57 | 3.5 | 1.71 | 1.14 | 7.92 |
| | 121 | 0.14 | 0 | 0.86 | 0.14 | 1.14 |
| | 122 | 2.57 | 3.71 | 2.57 | 1 | 9.85 |
| | 123 | 0.86 | 1.43 | 0.14 | 1 | 3.43 |
| | 125 | 0.64 | 0.36 | 1.14 | 3 | 5.14 |
| | 126 | 9.86 | 7.71 | 6.14 | 6.14 | 29.85 |
| | 127 | 2.57 | 0 | 2.14 | 0.57 | 5.28 |
| | 128 | 10.14 | 11 | 8.43 | 9 | 38.57 |

| | | | | | | |
|---|---|---|---|---|---|---|
| | 129 | 0.57 | 0.86 | 0.57 | 0 | 2 |
| | 130 | 3.29 | 2.07 | 1.57 | 2 | 8.93 |
| | 131 | 4.29 | 3.14 | 1.71 | 2.29 | 11.43 |
| | 132 | 6.79 | 9.64 | 11.21 | 7.79 | 35.43 |
| | 133 | 0 | 1 | 0 | 0.29 | 1.29 |
| | 134 | 3.71 | 1.29 | 2.29 | 2.14 | 9.43 |
| | 135 | 2 | 0.29 | 1.43 | 1.71 | 5.43 |
| | 136 | 2.29 | 1.57 | 3.57 | 4.57 | 12 |
| | 137 | 0.71 | 0.57 | 0.71 | 1 | 2.99 |
| | 138 | 0.57 | 0.57 | 1.14 | 1 | 3.28 |
| | 139 | 2.86 | 2.29 | 3.29 | 1.57 | 10.01 |
| | 140 | 2 | 2.43 | 1.86 | 2.14 | 8.43 |
| | 141 | 1.86 | 4.14 | 2.43 | 0 | 8.43 |
| | 142 | 48.75 | 60.75 | 44.71 | 46.85 | 201.06 |
| | 144 | 4.71 | 6.29 | 3.86 | 4.71 | 19.57 |
| | 145 | 0 | 1.14 | 1.43 | 0.43 | 3 |
| | 146 | 36.68 | 32.14 | 23 | 30.57 | 122.39 |
| | 148 | 4.43 | 7.29 | 11 | 9.43 | 32.15 |
| | 149 | 2 | 3.86 | 3.29 | 2.43 | 11.58 |
| TOTAL | | 361.83 | 371.02 | 319.55 | 327.5 | 1,379.90 |

| STATE: | NORTH CAROLINA | | | | | |
|---|---|---|---|---|---|---|
| ZIP CODE | | QUARTER 1 | QUARTER 2 | QUARTER 3 | QUARTER 4 | TOTAL GRAMS |
| | 270 | 0.43 | 0.43 | 1.43 | 1 | 3.29 |
| | 271 | 13.14 | 15.71 | 18.28 | 16.28 | 63.41 |
| | 272 | 2.29 | 1.43 | 1.71 | 3.14 | 8.57 |
| | 273 | 0.71 | 0.71 | 0.14 | 0.29 | 1.85 |
| | 274 | 0.29 | 1.43 | 0.71 | 0.57 | 3 |
| | 275 | 20.93 | 7.43 | 15.71 | 8.14 | 52.21 |
| | 276 | 2.14 | 0.86 | 2.29 | 2.29 | 7.58 |
| | 277 | 5.29 | 2.04 | 2.43 | 3.04 | 12.8 |
| | 278 | 15.71 | 18.14 | 11 | 11.57 | 56.42 |
| | 279 | 1.86 | 1 | 2 | 2.86 | 7.72 |
| | 280 | 18.14 | 14.43 | 17.43 | 16.28 | 66.28 |
| | 281 | 1.71 | 1 | 1.57 | 0.93 | 5.21 |
| | 282 | 4 | 8.82 | 8.29 | 6.14 | 27.25 |

| | | | | | | |
|---|---|---|---|---|---|---|
| | 283 | 1.43 | 2.14 | 0.86 | 1.43 | 5.86 |
| | 284 | 11.32 | 7.86 | 11.14 | 10.71 | 41.03 |
| | 285 | 0.86 | 5 | 2.14 | 1.29 | 9.29 |
| | 286 | 25.43 | 37.68 | 39.1 | 43.57 | 145.78 |
| | 287 | 8.14 | 12.71 | 12.57 | 12 | 45.42 |
| | 288 | 1.71 | 2.86 | 3.14 | 1 | 8.71 |
| TOTAL | | 135.53 | 141.68 | 151.94 | 142.53 | 571.68 |

STATE: NORTH DAKOTA

| ZIP CODE | | QUARTER 1 | QUARTER 2 | QUARTER 3 | QUARTER 4 | TOTAL GRAMS |
|---|---|---|---|---|---|---|
| | 581 | 1.86 | 3.29 | 1.14 | 2.29 | 8.58 |
| | 582 | 7.29 | 6.29 | 9 | 5.29 | 27.87 |
| | 583 | 0 | 0 | 0.29 | 0 | 0.29 |
| | 584 | 0 | 1.71 | 1.57 | 1.43 | 4.71 |
| | 585 | 16.25 | 50.82 | 18.11 | 33.82 | 119 |
| | 586 | 1.14 | 0 | 2.86 | 0.86 | 4.86 |
| | 587 | 6.43 | 4.29 | 4.57 | 0.57 | 15.86 |
| | 588 | 0.29 | 0 | 0.29 | 0.29 | 0.87 |
| TOTAL | | 33.26 | 66.4 | 37.83 | 44.55 | 182.04 |

STATE: OHIO

| ZIP CODE | | QUARTER 1 | QUARTER 2 | QUARTER 3 | QUARTER 4 | TOTAL GRAMS |
|---|---|---|---|---|---|---|
| | 430 | 3.43 | 5.43 | 5.14 | 3.29 | 17.29 |
| | 431 | 1 | 0.86 | 1.29 | 0.29 | 3.44 |
| | 432 | 9.86 | 11.57 | 13.18 | 23.93 | 58.54 |
| | 433 | 3.71 | 5.86 | 2.29 | 4.86 | 16.72 |
| | 435 | 4.43 | 5 | 6.43 | 7.57 | 23.43 |
| | 436 | 4.29 | 2.43 | 2 | 3.14 | 11.86 |
| | 437 | 1.71 | 2.14 | 1.43 | 1.14 | 6.42 |
| | 439 | 4 | 6.29 | 1.43 | 2.57 | 14.29 |
| | 440 | 18.57 | 20 | 20.71 | 17 | 76.28 |
| | 441 | 21.14 | 29.57 | 24.71 | 17.86 | 93.28 |
| | 442 | 11.86 | 18.14 | 13.14 | 11.86 | 55 |
| | 443 | 4.14 | 2.57 | 4.71 | 3.57 | 14.99 |
| | 444 | 1.14 | 3 | 1.86 | 3 | 9 |

| | | | | | | |
|---|---|---|---|---|---|---|
| | 445 | 13.57 | 14.57 | 11.86 | 13.43 | 53.43 |
| | 446 | 1 | 0.43 | 1.57 | 1.71 | 4.71 |
| | 447 | 1.14 | 2.29 | 0.86 | 1.71 | 6 |
| | 448 | 4.14 | 10 | 4.57 | 4.57 | 23.28 |
| | 449 | 0 | 0 | 1.71 | 0 | 1.71 |
| | 450 | 11.71 | 10.86 | 13 | 12.57 | 48.14 |
| | 451 | 0.29 | 0.57 | 0.14 | 0.29 | 1.29 |
| | 452 | 14.14 | 20.32 | 17.86 | 21.32 | 73.64 |
| | 453 | 7 | 6 | 4.57 | 3.43 | 21 |
| | 454 | 17.64 | 7.14 | 6.57 | 9.29 | 40.64 |
| | 455 | 2.86 | 6.29 | 0.57 | 6.14 | 15.86 |
| | 456 | 8.14 | 8.14 | 5.14 | 6 | 27.42 |
| | 457 | 0 | 0 | 0.71 | 0.14 | 0.85 |
| | 458 | 4.57 | 2.57 | 2.57 | 2.57 | 12.28 |
| TOTAL | | 175.48 | 202.04 | 170.02 | 183.25 | 730.79 |

STATE: OKLAHOMA

| ZIP CODE | | QUARTER 1 | QUARTER 2 | QUARTER 3 | QUARTER 4 | TOTAL GRAMS |
|---|---|---|---|---|---|---|
| | 730 | 2.86 | 2.57 | 2.86 | 2.43 | 10.72 |
| | 731 | 9.86 | 7.14 | 9.14 | 9.43 | 35.57 |
| | 735 | 0.57 | 2.29 | 0.71 | 0.43 | 4 |
| | 737 | 0.29 | 0.71 | 0 | 0 | 1 |
| | 740 | 3.57 | 1 | 1 | 3.86 | 9.43 |
| | 741 | 13.43 | 19.43 | 13.71 | 21.71 | 68.28 |
| | 743 | 0 | 0 | 0 | 0.29 | 0.29 |
| | 744 | 0.71 | 0.86 | 0.71 | 1.43 | 3.71 |
| TOTAL | | 31.29 | 34 | 28.13 | 39.58 | 133 |

STATE: OREGON

| ZIP CODE | | QUARTER 1 | QUARTER 2 | QUARTER 3 | QUARTER 4 | TOTAL GRAMS |
|---|---|---|---|---|---|---|
| | 970 | 10.86 | 7.43 | 8.29 | 7.86 | 34.44 |
| | 971 | 5.14 | 5.14 | 4.29 | 5.43 | 20 |
| | 972 | 35.32 | 34.03 | 33.28 | 38.75 | 141.38 |
| | 973 | 7.43 | 11.04 | 6.71 | 6.71 | 31.89 |

| | | | | | | |
|---|---|---|---|---|---|---|
| | 974 | 8.57 | 9.14 | 7.86 | 7.57 | 33.14 |
| | 975 | 27.57 | 24.57 | 35.64 | 15.36 | 103.14 |
| | 977 | 3.14 | 0.86 | 0.29 | 0.29 | 4.58 |
| | 978 | 3.29 | 2.14 | 3.14 | 2.29 | 10.86 |
| | 979 | 2.14 | 1.57 | 0.86 | 0.57 | 5.14 |
| TOTAL | | 103.46 | 95.92 | 100.36 | 84.83 | 384.57 |

STATE: PENNSYLVANIA

| ZIP CODE | QUARTER 1 | QUARTER 2 | QUARTER 3 | QUARTER 4 | TOTAL GRAMS |
|---|---|---|---|---|---|
| 150 | 4.57 | 6 | 5.43 | 6.86 | 22.86 |
| 151 | 1.29 | 1.71 | 1.14 | 2.71 | 6.85 |
| 152 | 10.75 | 10.43 | 8 | 11 | 40.18 |
| 154 | 2 | 0.43 | 1.57 | 0.57 | 4.57 |
| 155 | 0.71 | 0 | 0.86 | 1.43 | 3 |
| 156 | 3.57 | 7.14 | 2.14 | 5 | 17.85 |
| 157 | 0.86 | 4.29 | 3 | 1.57 | 9.72 |
| 162 | 1.71 | 2.43 | 1.14 | 1.86 | 7.14 |
| 163 | 0 | 0.14 | 0.14 | 0 | 0.28 |
| 165 | 2.29 | 1.71 | 2 | 2.71 | 8.71 |
| 166 | 0.14 | 0.29 | 0 | 0 | 0.43 |
| 167 | 0.71 | 0 | 0.57 | 0 | 1.28 |
| 168 | 8.93 | 10.64 | 10.64 | 5.61 | 35.82 |
| 169 | 1.43 | 1.71 | 2 | 2.14 | 7.28 |
| 170 | 6.14 | 11.43 | 4.14 | 2.57 | 24.28 |
| 171 | 5.61 | 2.86 | 1.43 | 4.71 | 14.61 |
| 172 | 2.14 | 1.14 | 1 | 2.14 | 6.42 |
| 173 | 0.29 | 0 | 0 | 0.14 | 0.43 |
| 174 | 9.86 | 5.57 | 13.89 | 16.18 | 45.5 |
| 175 | 2.43 | 3.14 | 3.43 | 1.43 | 10.43 |
| 176 | 17 | 13.57 | 16.71 | 17.14 | 64.42 |
| 177 | 14.28 | 19.28 | 8.86 | 14.71 | 57.13 |
| 178 | 10.57 | 18.18 | 7.29 | 7.86 | 43.9 |
| 179 | 9.29 | 12.43 | 6.29 | 8.57 | 36.58 |
| 180 | 6 | 14.14 | 8.14 | 11.71 | 39.99 |
| 181 | 10.57 | 14.57 | 9.79 | 12.71 | 47.64 |
| 182 | 0.29 | 0.57 | 0.57 | 0.29 | 1.72 |

| | | | | | | |
|---|---|---|---|---|---|---|
| | 183 | 3.43 | 3.57 | 4.86 | 7.29 | 19.15 |
| | 184 | 0.43 | 0 | 0.14 | 0.29 | 0.86 |
| | 185 | 3.04 | 2.29 | 3.14 | 2.18 | 10.65 |
| | 186 | 0.86 | 4.57 | 2.43 | 1.57 | 9.43 |
| | 187 | 5.36 | 5.14 | 5.11 | 2.43 | 18.04 |
| | 188 | 3.71 | 1.86 | 1 | 1.14 | 7.71 |
| | 189 | 13.71 | 16.71 | 14 | 11.18 | 55.6 |
| | 190 | 70.71 | 56 | 61.71 | 63.85 | 252.27 |
| | 191 | 118.89 | 99.64 | 90.85 | 97.49 | 406.87 |
| | 193 | 29.14 | 26.43 | 19.28 | 31 | 105.85 |
| | 194 | 28.43 | 28.71 | 30.43 | 15.43 | 103 |
| | 196 | 1.57 | 1.43 | 3.29 | 2 | 8.29 |
| TOTAL | | 412.71 | 410.15 | 356.41 | 377.47 | 1,556.74 |

STATE: RHODE ISLAND

| ZIP CODE | | QUARTER 1 | QUARTER 2 | QUARTER 3 | QUARTER 4 | TOTAL GRAMS |
|---|---|---|---|---|---|---|
| | 28 | 14.43 | 12.14 | 7.71 | 12.86 | 47.14 |
| | 29 | 10.86 | 4.29 | 9.71 | 4 | 28.86 |
| TOTAL | | 25.29 | 16.43 | 17.42 | 16.86 | 76 |

STATE: SOUTH CAROLINA

| ZIP CODE | | QUARTER 1 | QUARTER 2 | QUARTER 3 | QUARTER 4 | TOTAL GRAMS |
|---|---|---|---|---|---|---|
| | 290 | 0 | 2.86 | 0.71 | 0 | 3.57 |
| | 291 | 3.71 | 2.29 | 3.86 | 3.57 | 13.43 |
| | 292 | 2.32 | 2.29 | 3.57 | 3.14 | 11.32 |
| | 293 | 0.71 | 0.71 | 0.57 | 0.57 | 2.56 |
| | 294 | 4.86 | 12 | 2.86 | 1.71 | 21.43 |
| | 295 | 7.14 | 4.71 | 9.14 | 8.71 | 29.7 |
| | 296 | 10.14 | 5.86 | 6.57 | 9 | 31.57 |
| | 297 | 1.71 | 0.57 | 0.71 | 1.43 | 4.42 |
| | 298 | 0 | 0.29 | 0 | 0 | 0.29 |
| TOTAL | | 30.59 | 31.58 | 27.99 | 28.13 | 118.29 |

STATE: SOUTH DAKOTA

| ZIP CODE | QUARTER 1 | QUARTER 2 | QUARTER 3 | QUARTER 4 | TOTAL GRAMS |
|---|---|---|---|---|---|
| 570 | 6.79 | 7.79 | 11.25 | 5.68 | 31.51 |
| 571 | 24.46 | 6.57 | 7.64 | 33.11 | 71.78 |
| 572 | 1 | 0.29 | 0.14 | 1.43 | 2.86 |
| 573 | 0.29 | 0.57 | 0.43 | 0.43 | 1.72 |
| 574 | 0.14 | 0.14 | 0.29 | 0.29 | 0.86 |
| 575 | 0 | 0 | 0.86 | 0.86 | 1.72 |
| 577 | 11.57 | 9.43 | 6.86 | 10.57 | 38.43 |
| TOTAL | 44.25 | 24.79 | 27.47 | 52.37 | 148.88 |

STATE: TENNESSEE

| ZIP CODE | QUARTER 1 | QUARTER 2 | QUARTER 3 | QUARTER 4 | TOTAL GRAMS |
|---|---|---|---|---|---|
| 370 | 21 | 17.43 | 23.14 | 22.14 | 83.71 |
| 371 | 14.93 | 10.71 | 11.14 | 14.43 | 51.21 |
| 372 | 21.87 | 20.71 | 15.43 | 13.71 | 71.72 |
| 373 | 4.29 | 5.14 | 5.86 | 5.57 | 20.86 |
| 374 | 9.43 | 15.78 | 6.14 | 9 | 40.35 |
| 376 | 17.86 | 15.57 | 24 | 22.14 | 79.57 |
| 377 | 3 | 2.86 | 3.86 | 3.29 | 13.01 |
| 378 | 7.71 | 8.14 | 5.57 | 6.71 | 28.13 |
| 379 | 35 | 31.14 | 32.71 | 31.14 | 129.99 |
| 380 | 2.71 | 1.29 | 1.86 | 1.43 | 7.29 |
| 381 | 31.25 | 19.14 | 19.64 | 24.86 | 94.89 |
| 382 | 0.57 | 0 | 0.14 | 0.43 | 1.14 |
| 383 | 9.29 | 10.71 | 7.86 | 11.57 | 39.43 |
| 384 | 1.29 | 0.57 | 0.86 | 1.14 | 3.86 |
| 385 | 0.86 | 0.93 | 1.5 | 1.57 | 4.86 |
| TOTAL | 181.06 | 160.12 | 159.71 | 169.13 | 670.02 |

STATE: TEXAS

| ZIP CODE | QUARTER 1 | QUARTER 2 | QUARTER 3 | QUARTER 4 | TOTAL GRAMS |
|---|---|---|---|---|---|
| 750 | 9.04 | 12.71 | 10.57 | 17.57 | 49.89 |
| 751 | 1 | 2.43 | 0.43 | 0.71 | 4.57 |
| 752 | 14.18 | 15 | 21.57 | 16.86 | 67.61 |
| 753 | 1.29 | 2.29 | 2.14 | 1.14 | 6.86 |

| | | | | | |
|---|---|---|---|---|---|
| 754 | 0 | 0 | 0.71 | 0.43 | 1.14 |
| 756 | 1.43 | 1.86 | 1.86 | 2.43 | 7.58 |
| 757 | 1.14 | 1.14 | 0.86 | 1.86 | 5 |
| 758 | 0.14 | 0.29 | 0 | 0 | 0.43 |
| 759 | 1 | 2 | 3.43 | 3 | 9.43 |
| 760 | 33.57 | 28.86 | 26 | 35.71 | 124.14 |
| 761 | 48.43 | 45.28 | 43.43 | 54.35 | 191.49 |
| 762 | 8.46 | 7.18 | 1.29 | 8.46 | 25.39 |
| 763 | 0 | 0.14 | 0 | 1.14 | 1.28 |
| 764 | 0 | 0 | 0 | 0.14 | 0.14 |
| 765 | 2.29 | 2.14 | 4.29 | 2.14 | 10.86 |
| 767 | 0.29 | 0 | 0 | 0 | 0.29 |
| 768 | 0.57 | 0 | 0 | 0 | 0.57 |
| 769 | 5.57 | 7.43 | 4 | 4.71 | 21.71 |
| 770 | 39.85 | 71.53 | 47.35 | 70.17 | 228.9 |
| 773 | 14.57 | 27.03 | 10.29 | 22 | 73.89 |
| 774 | 3.57 | 3.29 | 5.14 | 4.43 | 16.43 |
| 775 | 13.57 | 11.14 | 11.43 | 11.14 | 47.28 |
| 776 | 4.57 | 6.43 | 4 | 7.43 | 22.43 |
| 777 | 1.43 | 0.29 | 0.86 | 0.57 | 3.15 |
| 779 | 3.29 | 3.29 | 6.43 | 5.71 | 18.72 |
| 780 | 0.43 | 0.43 | 1.29 | 0.43 | 2.58 |
| 781 | 0.43 | 0.57 | 0.43 | 1.14 | 2.57 |
| 782 | 13.43 | 13.28 | 15 | 16.86 | 58.57 |
| 784 | 3.14 | 2.86 | 3.86 | 4.29 | 14.15 |
| 785 | 4.14 | 7.57 | 3.86 | 4.14 | 19.71 |
| 786 | 12.28 | 18.14 | 13.43 | 20.28 | 64.13 |
| 787 | 48 | 42.64 | 41.71 | 52.57 | 184.92 |
| 788 | 0 | 0.14 | 0.14 | 0 | 0.28 |
| 789 | 0 | 0 | 0.29 | 0.14 | 0.43 |
| 790 | 0 | 0 | 0.14 | 0 | 0.14 |
| 791 | 4.29 | 2.29 | 3.71 | 2 | 12.29 |
| 794 | 0.43 | 0.86 | 0.57 | 1.29 | 3.15 |
| 795 | 0.14 | 0.43 | 0.29 | 0.29 | 1.15 |
| 796 | 5.89 | 4.89 | 0.43 | 0 | 11.21 |
| 797 | 4.57 | 7.57 | 8 | 6.57 | 26.71 |
| 799 | 26.28 | 31.28 | 27.43 | 27.57 | 112.56 |

| | | | | | |
|---|---|---|---|---|---|
| TOTAL | 332.7 | 384.7 | 326.66 | 409.67 | 1,453.73 |

STATE: UTAH

| ZIP CODE | QUARTER 1 | QUARTER 2 | QUARTER 3 | QUARTER 4 | TOTAL GRAMS |
|---|---|---|---|---|---|
| 840 | 26.86 | 23.43 | 28.28 | 24.86 | 103.43 |
| 841 | 19.71 | 27.03 | 16 | 25.5 | 88.24 |
| 843 | 7.29 | 7.14 | 8.86 | 14.14 | 37.43 |
| 844 | 44.14 | 35.57 | 33.07 | 28.71 | 141.49 |
| 845 | 0.71 | 0.14 | 0 | 0.14 | 0.99 |
| 846 | 3 | 2.71 | 1.57 | 3.71 | 10.99 |
| 847 | 1.57 | 2.57 | 2.14 | 2.14 | 8.42 |
| TOTAL | 103.28 | 98.59 | 89.92 | 99.2 | 390.99 |

STATE: VERMONT

| ZIP CODE | QUARTER 1 | QUARTER 2 | QUARTER 3 | QUARTER 4 | TOTAL GRAMS |
|---|---|---|---|---|---|
| 50 | 0.43 | 0.86 | 0.57 | 0 | 1.86 |
| 52 | 0 | 0.57 | 0.57 | 0.29 | 1.43 |
| 53 | 0.29 | 0.29 | 0 | 0.14 | 0.72 |
| 54 | 8.21 | 10.18 | 3 | 7.07 | 28.46 |
| 56 | 1.43 | 1.29 | 2.14 | 0.86 | 5.72 |
| 58 | 0 | 0.29 | 0 | 0 | 0.29 |
| TOTAL | 10.36 | 13.48 | 6.28 | 8.36 | 38.48 |

STATE: VIRGINIA

| ZIP CODE | QUARTER 1 | QUARTER 2 | QUARTER 3 | QUARTER 4 | TOTAL GRAMS |
|---|---|---|---|---|---|
| 201 | 11.43 | 14.14 | 14.71 | 13.14 | 53.42 |
| 220 | 31.43 | 32.57 | 29.57 | 26 | 119.57 |
| 221 | 6.14 | 11.71 | 8.36 | 8.64 | 34.85 |
| 222 | 0.86 | 1.14 | 0 | 1.29 | 3.29 |
| 223 | 5.43 | 2.14 | 2.86 | 3.14 | 13.57 |
| 224 | 8.57 | 5.57 | 6.29 | 7 | 27.43 |
| 225 | 0.29 | 0.71 | 0.57 | 1 | 2.57 |
| 226 | 0.43 | 0.86 | 0.71 | 2 | 4 |
| 227 | 0 | 0 | 0.57 | 0 | 0.57 |

| | | | | | | |
|---|---|---|---|---|---|---|
| | 228 | 1.79 | 0 | 1 | 0 | 2.79 |
| | 229 | 2.71 | 2 | 2.57 | 0.86 | 8.14 |
| | 230 | 0.57 | 0 | 0 | 0.29 | 0.86 |
| | 231 | 10.71 | 10.57 | 9.14 | 11.43 | 41.85 |
| | 232 | 23.71 | 19.46 | 17.28 | 18.86 | 79.31 |
| | 233 | 1.43 | 0 | 2.43 | 1.29 | 5.15 |
| | 234 | 8.86 | 8.29 | 8.86 | 7.14 | 33.15 |
| | 235 | 8.14 | 9.29 | 8.71 | 5 | 31.14 |
| | 236 | 19.57 | 16.86 | 14 | 22.43 | 72.86 |
| | 237 | 0.86 | 0.43 | 3 | 0.86 | 5.15 |
| | 238 | 2.14 | 2.14 | 2.14 | 0.71 | 7.13 |
| | 239 | 1.71 | 2.29 | 1.86 | 0.71 | 6.57 |
| | 240 | 1 | 4.14 | 2 | 4.86 | 12 |
| | 241 | 9.21 | 2 | 13.53 | 1.71 | 26.45 |
| | 242 | 2 | 3.43 | 1.29 | 2 | 8.72 |
| | 243 | 0.57 | 0 | 0 | 1.71 | 2.28 |
| | 244 | 0.29 | 0.86 | 2.14 | 0.57 | 3.86 |
| | 245 | 2 | 2.57 | 1.29 | 1.57 | 7.43 |
| | 246 | 0 | 0 | 0 | 0.43 | 0.43 |
| TOTAL | | 161.85 | 153.17 | 154.88 | 144.64 | 614.54 |

| STATE: | WASHINGTON | | | | | |
|---|---|---|---|---|---|---|
| ZIP CODE | | QUARTER 1 | QUARTER 2 | QUARTER 3 | QUARTER 4 | TOTAL GRAMS |
| | 980 | 30.43 | 12.14 | 31.86 | 23.14 | 97.57 |
| | 981 | 43.75 | 48.32 | 49.71 | 42 | 183.78 |
| | 982 | 3.57 | 4.14 | 5.71 | 2.86 | 16.28 |
| | 983 | 3.57 | 3.57 | 4.71 | 4.86 | 16.71 |
| | 984 | 9.57 | 8 | 6.43 | 6.29 | 30.29 |
| | 985 | 1 | 4.29 | 1.43 | 1.86 | 8.58 |
| | 986 | 3 | 3.43 | 4.14 | 4.89 | 15.46 |
| | 988 | 0.57 | 1 | 1.29 | 0 | 2.86 |
| | 989 | 1.14 | 0.29 | 2 | 1 | 4.43 |
| | 991 | 0 | 0.14 | 0.57 | 0.29 | 1 |
| | 992 | 1.29 | 0.86 | 1.43 | 3.71 | 7.29 |
| | 993 | 15.86 | 16.43 | 14.71 | 19.57 | 66.57 |
| | 994 | 0.71 | 1.57 | 0.43 | 0.57 | 3.28 |

| | | | | | |
|---|---|---|---|---|---|
| TOTAL | 114.46 | 104.18 | 124.42 | 111.04 | 454.1 |

STATE: WEST VIRGINIA

| ZIP CODE | QUARTER 1 | QUARTER 2 | QUARTER 3 | QUARTER 4 | TOTAL GRAMS |
|---|---|---|---|---|---|
| 247 | 4.29 | 0.57 | 6.43 | 6.57 | 17.86 |
| 249 | 0.29 | 0.29 | 0.29 | 0.14 | 1.01 |
| 251 | 0.89 | 0 | 0.89 | 0.89 | 2.67 |
| 252 | 0.14 | 0.14 | 0.29 | 0 | 0.57 |
| 253 | 7.36 | 9.79 | 11.71 | 10.21 | 39.07 |
| 254 | 4.14 | 3.71 | 1.71 | 1.71 | 11.27 |
| 255 | 0.14 | 0.14 | 0.43 | 0.57 | 1.28 |
| 256 | 0.57 | 1.14 | 1.57 | 1.14 | 4.42 |
| 257 | 2.71 | 3.57 | 3.14 | 2.43 | 11.85 |
| 258 | 2 | 3.43 | 2.86 | 4.57 | 12.86 |
| 260 | 5 | 6.14 | 5.14 | 3.43 | 19.71 |
| 261 | 0.86 | 0.29 | 0.57 | 0.57 | 2.29 |
| 262 | 0.43 | 0 | 0.29 | 0.29 | 1.01 |
| 263 | 10.86 | 10 | 8.14 | 10.29 | 39.29 |
| 264 | 1.71 | 0 | 1.29 | 1.14 | 4.14 |
| 265 | 11.14 | 9.14 | 5.14 | 7.07 | 32.49 |
| TOTAL | 52.53 | 48.35 | 49.89 | 51.02 | 201.79 |

STATE: WISCONSIN

| ZIP CODE | QUARTER 1 | QUARTER 2 | QUARTER 3 | QUARTER 4 | TOTAL GRAMS |
|---|---|---|---|---|---|
| 530 | 14.14 | 13 | 11.57 | 16.43 | 55.14 |
| 531 | 15.28 | 15.43 | 11.57 | 14.71 | 56.99 |
| 532 | 29.86 | 33.57 | 27.86 | 24.71 | 116 |
| 534 | 5.14 | 4.86 | 4.29 | 3.57 | 17.86 |
| 535 | 30.64 | 28.93 | 22 | 27.43 | 109 |
| 537 | 24.71 | 21.86 | 17.71 | 18.71 | 82.99 |
| 538 | 0.29 | 1.79 | 1.07 | 0.29 | 3.44 |
| 539 | 3.29 | 3.29 | 2.21 | 1.71 | 10.5 |
| 540 | 1.14 | 0.57 | 1.71 | 0.43 | 3.85 |
| 541 | 0.71 | 2.43 | 1 | 0.43 | 4.57 |

| | | | | | | |
|---|---|---|---|---|---|---|
| | 542 | 0.71 | 2.71 | 0.86 | 0.29 | 4.57 |
| | 543 | 5.29 | 6 | 3.14 | 5.14 | 19.57 |
| | 544 | 2.21 | 1.07 | 2.07 | 3.07 | 8.42 |
| | 545 | 0.57 | 3 | 1 | 0.29 | 4.86 |
| | 546 | 8 | 3.29 | 2.29 | 3.79 | 17.37 |
| | 547 | 0.57 | 0.57 | 1 | 0.86 | 3 |
| | 548 | 3.43 | 2.86 | 3.43 | 4 | 13.72 |
| | 549 | 18.43 | 13 | 15.86 | 25.71 | 73 |
| TOTAL | | 164.41 | 158.23 | 130.64 | 151.57 | 604.85 |

STATE: WYOMING

| ZIP CODE | | QUARTER 1 | QUARTER 2 | QUARTER 3 | QUARTER 4 | TOTAL GRAMS |
|---|---|---|---|---|---|---|
| | 820 | 2.71 | 0.71 | 1 | 1 | 5.42 |
| | 822 | 1 | 0.43 | 0.71 | 0.71 | 2.85 |
| | 823 | 0 | 0.71 | 0 | 0 | 0.71 |
| | 824 | 1 | 1 | 2.57 | 1.14 | 5.71 |
| | 825 | 1 | 0.29 | 0.57 | 0.57 | 2.43 |
| | 826 | 1.71 | 2.86 | 2.43 | 2 | 9 |
| | 827 | 0.57 | 0.29 | 1.14 | 1 | 3 |
| | 829 | 9.71 | 5.71 | 9.71 | 10.43 | 35.56 |
| | 830 | 0 | 0.71 | 0.29 | 0.29 | 1.29 |
| | 831 | 0.29 | 1.14 | 1.86 | 0 | 3.29 |
| TOTAL | | 17.99 | 13.85 | 20.28 | 17.14 | 69.26 |

DRUG CODE: 9050 DRUG NAME: CODEINE

STATE: ALABAMA

| ZIP CODE | | QUARTER 1 | QUARTER 2 | QUARTER 3 | QUARTER 4 | TOTAL GRAMS |
|---|---|---|---|---|---|---|
| | 350 | 5,975.76 | 6,003.04 | 6,076.96 | 6,075.81 | 24,131.57 |
| | 351 | 3,877.13 | 4,024.98 | 3,676.91 | 3,565.01 | 15,144.03 |
| | 352 | 5,756.60 | 5,523.13 | 5,030.43 | 5,490.17 | 21,800.33 |
| | 354 | 1,570.92 | 1,741.17 | 1,883.04 | 1,650.39 | 6,845.52 |
| | 355 | 1,720.35 | 1,820.70 | 1,807.32 | 1,967.35 | 7,315.72 |
| | 356 | 4,999.03 | 4,919.23 | 4,855.32 | 5,287.13 | 20,060.71 |
| | 357 | 908.4 | 824.4 | 732.81 | 654.16 | 3,119.77 |

| | | | | | | |
|---|---|---|---|---|---|---|
| | 358 | 1,112.08 | 1,022.20 | 1,020.80 | 961.24 | 4,116.32 |
| | 359 | 2,416.57 | 2,135.72 | 2,138.62 | 2,222.78 | 8,913.69 |
| | 360 | 2,924.28 | 3,235.34 | 3,663.03 | 3,253.05 | 13,075.70 |
| | 361 | 2,636.54 | 2,650.83 | 2,772.04 | 2,570.84 | 10,630.25 |
| | 362 | 1,447.57 | 1,311.93 | 1,308 | 1,325.52 | 5,393.02 |
| | 363 | 1,907.49 | 1,742.39 | 1,727.66 | 1,607.06 | 6,984.60 |
| | 364 | 2,046.63 | 2,023.63 | 2,034.67 | 1,769.46 | 7,874.39 |
| | 365 | 3,851.73 | 4,096.88 | 3,910.02 | 4,126.01 | 15,984.64 |
| | 366 | 3,946.26 | 4,146.93 | 3,978.03 | 4,642.92 | 16,714.14 |
| | 367 | 2,382.47 | 2,465.96 | 2,498.47 | 2,578.22 | 9,925.12 |
| | 368 | 2,269.32 | 2,349.16 | 2,162.22 | 2,251.43 | 9,032.13 |
| TOTAL | | 52,097.22 | 52,426.60 | 51,821.14 | 52,546.65 | 208,891.61 |

| STATE: | ALASKA | | | | | |
|---|---|---|---|---|---|---|
| ZIP CODE | | QUARTER 1 | QUARTER 2 | QUARTER 3 | QUARTER 4 | TOTAL GRAMS |
| | 995 | 3,134.65 | 2,932.57 | 2,701.47 | 2,562.46 | 11,331.15 |
| | 996 | 1,713.78 | 1,410.44 | 1,388.38 | 1,249.32 | 5,761.92 |
| | 997 | 1,720.54 | 1,841.48 | 1,999.09 | 1,699.89 | 7,261 |
| | 998 | 561.98 | 577.47 | 596.83 | 460.43 | 2,196.71 |
| | 999 | 285.47 | 310.32 | 354.72 | 296.15 | 1,246.66 |
| TOTAL | | 7,416.42 | 7,072.28 | 7,040.49 | 6,268.25 | 27,797.44 |

| STATE: | AMERICAN SAMOA | | | | | |
|---|---|---|---|---|---|---|
| ZIP CODE | | QUARTER 1 | QUARTER 2 | QUARTER 3 | QUARTER 4 | TOTAL GRAMS |
| | 967 | 353.62 | 340.36 | 353.62 | 313.83 | 1,361.43 |
| TOTAL | | 353.62 | 340.36 | 353.62 | 313.83 | 1,361.43 |

| STATE: | ARIZONA | | | | | |
|---|---|---|---|---|---|---|
| ZIP CODE | | QUARTER 1 | QUARTER 2 | QUARTER 3 | QUARTER 4 | TOTAL GRAMS |
| | 850 | 19,008.98 | 19,381.91 | 17,940.22 | 19,124.91 | 75,456.02 |
| | 851 | 3,162.34 | 3,093.95 | 2,903.30 | 2,860.94 | 12,020.53 |
| | 852 | 22,323.96 | 21,462.29 | 20,792.20 | 21,321.80 | 85,900.25 |
| | 853 | 11,431.36 | 9,911.33 | 9,636.58 | 10,321.75 | 41,301.02 |

| | | | | | | |
|---|---|---|---|---|---|---|
| | 855 | 1,088.18 | 949.94 | 822.69 | 940.55 | 3,801.36 |
| | 856 | 2,682.89 | 2,341.01 | 2,051.99 | 2,204.85 | 9,280.74 |
| | 857 | 9,916.04 | 9,806.41 | 8,810.65 | 9,121.90 | 37,655 |
| | 859 | 722.61 | 708.25 | 777.62 | 544.12 | 2,752.60 |
| | 860 | 1,689.62 | 1,290.28 | 1,186.89 | 1,326.14 | 5,492.93 |
| | 863 | 2,011.20 | 2,197.91 | 1,976.38 | 1,929.31 | 8,114.80 |
| | 864 | 2,762.71 | 2,671.56 | 2,443.66 | 2,403.57 | 10,281.50 |
| | 865 | 1,596.42 | 1,239.66 | 1,257.13 | 998.58 | 5,091.79 |
| TOTAL | | 78,396.31 | 75,054.50 | 70,599.31 | 73,098.42 | 297,148.54 |

STATE: ARKANSAS

| ZIP CODE | | QUARTER 1 | QUARTER 2 | QUARTER 3 | QUARTER 4 | TOTAL GRAMS |
|---|---|---|---|---|---|---|
| | 716 | 3,902.25 | 3,806.18 | 3,723.69 | 3,616.62 | 15,048.74 |
| | 717 | 3,642.76 | 3,808.18 | 3,533.46 | 3,322.25 | 14,306.65 |
| | 718 | 5,824.34 | 6,179.61 | 5,636.11 | 5,699.21 | 23,339.27 |
| | 719 | 5,692.48 | 5,379.38 | 5,036.35 | 5,298.39 | 21,406.60 |
| | 720 | 7,222.83 | 7,255.82 | 7,094.29 | 6,501.98 | 28,074.92 |
| | 721 | 7,657.61 | 7,632.73 | 7,200.12 | 7,171.16 | 29,661.62 |
| | 722 | 3,185.99 | 3,137.37 | 3,013.70 | 2,935.61 | 12,272.67 |
| | 723 | 9,387.58 | 9,809.48 | 9,445.86 | 8,674.73 | 37,317.65 |
| | 724 | 8,212.81 | 8,749.40 | 8,350.94 | 8,239.97 | 33,553.12 |
| | 725 | 4,421.96 | 4,704.25 | 4,410.94 | 4,900.53 | 18,437.68 |
| | 726 | 2,671.03 | 2,667.57 | 2,300.33 | 2,280.43 | 9,919.36 |
| | 727 | 3,786.49 | 4,123.72 | 3,986.87 | 3,623.23 | 15,520.31 |
| | 728 | 3,038.45 | 3,404.84 | 2,970.56 | 2,938.90 | 12,352.75 |
| | 729 | 5,473.68 | 5,805.74 | 5,270.61 | 5,047.71 | 21,597.74 |
| TOTAL | | 74,120.26 | 76,464.27 | 71,973.83 | 70,250.72 | 292,809.08 |

STATE: CALIFORNIA

| ZIP CODE | | QUARTER 1 | QUARTER 2 | QUARTER 3 | QUARTER 4 | TOTAL GRAMS |
|---|---|---|---|---|---|---|
| | 900 | 30,148.97 | 28,696.47 | 27,334.41 | 26,234.99 | 112,414.84 |
| | 902 | 25,766.03 | 25,738.62 | 23,395.66 | 22,119.55 | 97,019.86 |
| | 903 | 4,073.66 | 3,669.17 | 2,940.90 | 2,981.40 | 13,665.13 |
| | 904 | 1,313.23 | 1,203.86 | 1,172.17 | 1,161.61 | 4,850.87 |
| | 905 | 2,622.93 | 2,219.84 | 2,224.59 | 2,042.06 | 9,109.42 |

| | | | | | |
|---|---|---|---|---|---|
| 906 | 7,837.51 | 7,338.25 | 7,190.78 | 7,165.73 | 29,532.27 |
| 907 | 10,168.72 | 9,495.84 | 9,228.53 | 9,060.07 | 37,953.16 |
| 908 | 7,907.95 | 7,490.81 | 7,287.29 | 7,235.34 | 29,921.39 |
| 910 | 3,321.37 | 3,312.24 | 3,248.78 | 3,021.34 | 12,903.73 |
| 911 | 2,321.17 | 2,352 | 2,347.10 | 2,100.96 | 9,121.23 |
| 912 | 3,093.20 | 3,276.50 | 2,917.85 | 3,105.55 | 12,393.10 |
| 913 | 14,554.66 | 14,582.33 | 13,619.31 | 13,207.91 | 55,964.21 |
| 914 | 5,168.25 | 5,027.59 | 5,137.24 | 5,023.38 | 20,356.46 |
| 915 | 1,683.56 | 1,597.48 | 1,393.46 | 1,634.39 | 6,308.89 |
| 916 | 1,288.94 | 1,364.90 | 1,138.24 | 1,170.60 | 4,962.68 |
| 917 | 25,096.23 | 23,783.46 | 22,695.06 | 22,273.04 | 93,847.79 |
| 918 | 1,110.33 | 1,010.04 | 927.84 | 957.78 | 4,005.99 |
| 919 | 4,814.05 | 4,864.45 | 4,215.41 | 4,179.69 | 18,073.60 |
| 920 | 19,324.34 | 18,618.63 | 18,320.60 | 18,348.74 | 74,612.31 |
| 921 | 12,067.04 | 12,095.28 | 11,972.97 | 11,138.94 | 47,274.23 |
| 922 | 11,026.58 | 9,822.50 | 9,660.26 | 9,665.96 | 40,175.30 |
| 923 | 20,355.11 | 20,127.95 | 19,367.92 | 18,299.94 | 78,150.92 |
| 924 | 3,622.42 | 3,445.17 | 3,341.34 | 3,126.47 | 13,535.40 |
| 925 | 19,227.82 | 18,453.56 | 18,377.10 | 17,495.72 | 73,554.20 |
| 926 | 16,448.85 | 11,695.35 | 14,199.70 | 11,352.49 | 53,696.39 |
| 927 | 3,927.80 | 3,875.59 | 3,772.31 | 3,686.55 | 15,262.25 |
| 928 | 13,266.11 | 12,775.49 | 12,590.30 | 12,055.55 | 50,687.45 |
| 930 | 7,890.09 | 8,180.60 | 7,613.37 | 6,991.53 | 30,675.59 |
| 931 | 1,886.71 | 1,885.75 | 1,753.98 | 1,569.10 | 7,095.54 |
| 932 | 11,491.74 | 11,141.65 | 10,453.39 | 10,914.04 | 44,000.82 |
| 933 | 8,534.78 | 7,905.45 | 7,256.22 | 7,403.37 | 31,099.82 |
| 934 | 5,906.51 | 6,815.27 | 6,120.62 | 5,867.68 | 24,710.08 |
| 935 | 11,256.11 | 10,887.73 | 10,814.40 | 10,476.22 | 43,434.46 |
| 936 | 9,488.88 | 8,553.50 | 7,803.51 | 8,361.05 | 34,206.94 |
| 937 | 10,434.14 | 9,918.06 | 10,319.10 | 10,484.31 | 41,155.61 |
| 939 | 4,168.94 | 4,103.58 | 3,772.48 | 3,730.63 | 15,775.63 |
| 940 | 6,193.60 | 6,059.95 | 6,356.87 | 6,588.28 | 25,198.70 |
| 941 | 9,409.34 | 8,454.05 | 8,440.85 | 8,768.22 | 35,072.46 |
| 943 | 737.19 | 658.64 | 773.73 | 521.24 | 2,690.80 |
| 944 | 959.03 | 827.08 | 909.96 | 953.36 | 3,649.43 |
| 945 | 38,692.95 | 38,201.08 | 30,926.13 | 29,035.79 | 136,855.95 |
| 946 | 4,213.23 | 3,783.25 | 4,238.72 | 4,226.55 | 16,461.75 |

| | | | | | | |
|---|---|---|---|---|---|---|
| | 947 | 1,074.49 | 1,036.38 | 1,060.67 | 954.59 | 4,126.13 |
| | 948 | 1,630.63 | 1,460.45 | 1,455.01 | 1,563.55 | 6,109.64 |
| | 949 | 3,550.91 | 3,222.91 | 3,683.80 | 3,695.98 | 14,153.60 |
| | 950 | 6,874.31 | 5,938.49 | 6,320.74 | 6,738.95 | 25,872.49 |
| | 951 | 7,380.29 | 6,894.71 | 6,153.15 | 6,607.01 | 27,035.16 |
| | 952 | 11,606.54 | 11,141.63 | 11,153.50 | 10,825.02 | 44,726.69 |
| | 953 | 16,860.59 | 15,765.01 | 16,620.04 | 16,752.54 | 65,998.18 |
| | 954 | 5,222.15 | 5,617.78 | 5,123.28 | 4,878.89 | 20,842.10 |
| | 955 | 3,402.32 | 3,650.15 | 3,295.16 | 2,850.31 | 13,197.94 |
| | 956 | 16,212.25 | 16,080.79 | 15,134.54 | 15,850.34 | 63,277.92 |
| | 957 | 2,535.98 | 2,553.30 | 2,521.11 | 2,333.11 | 9,943.50 |
| | 958 | 9,057.49 | 7,816.57 | 7,740.04 | 8,173.90 | 32,788 |
| | 959 | 7,966.17 | 8,161.52 | 7,475.68 | 7,826.04 | 31,429.41 |
| | 960 | 3,712.11 | 3,790.12 | 3,746.16 | 3,643.54 | 14,891.93 |
| | 961 | 1,536.29 | 1,509.86 | 1,493.99 | 1,350.88 | 5,891.02 |
| TOTAL | | 501,442.59 | 479,948.68 | 460,547.32 | 449,781.77 | 1,891,720.36 |

STATE: COLORADO

| ZIP CODE | | QUARTER 1 | QUARTER 2 | QUARTER 3 | QUARTER 4 | TOTAL GRAMS |
|---|---|---|---|---|---|---|
| | 800 | 4,652.42 | 4,407.04 | 4,460.78 | 4,295.93 | 17,816.17 |
| | 801 | 2,967.21 | 3,262.62 | 2,893.44 | 3,100.23 | 12,223.50 |
| | 802 | 4,862.67 | 4,639.22 | 4,456.39 | 4,401.44 | 18,359.72 |
| | 803 | 444.77 | 392.12 | 313.65 | 404.61 | 1,555.15 |
| | 804 | 685.09 | 691.3 | 708.7 | 565.8 | 2,650.89 |
| | 805 | 2,291.46 | 2,383.52 | 2,368.05 | 2,335.82 | 9,378.85 |
| | 806 | 1,161 | 984.56 | 960.71 | 937.48 | 4,043.75 |
| | 807 | 431.52 | 340.54 | 397.92 | 380.31 | 1,550.29 |
| | 808 | 686.33 | 684.38 | 764.49 | 718.45 | 2,853.65 |
| | 809 | 2,702.68 | 2,533.25 | 2,483.24 | 2,465.28 | 10,184.45 |
| | 810 | 2,115.64 | 1,898.10 | 2,079.40 | 1,862.59 | 7,955.73 |
| | 811 | 392.47 | 504.45 | 475.16 | 367.61 | 1,739.69 |
| | 812 | 799.04 | 713.45 | 763.81 | 764.1 | 3,040.40 |
| | 813 | 314.94 | 404.63 | 279.67 | 302.97 | 1,302.21 |
| | 814 | 547.66 | 593.61 | 647.29 | 577.2 | 2,365.76 |
| | 815 | 1,033.62 | 1,041.46 | 952.24 | 1,019.45 | 4,046.77 |
| | 816 | 464.67 | 417.34 | 419.5 | 500.35 | 1,801.86 |

| | | | | | | |
|---|---|---|---|---|---|---|
| TOTAL | | 26,553.19 | 25,891.59 | 25,424.44 | 24,999.62 | 102,868.84 |

| STATE: | CONNECTICUT | | | | | |
|---|---|---|---|---|---|---|
| ZIP CODE | | QUARTER 1 | QUARTER 2 | QUARTER 3 | QUARTER 4 | TOTAL GRAMS |
| | 060 | 5,466.79 | 5,414.37 | 4,936.10 | 4,654.62 | 20,471.88 |
| | 061 | 1,733.74 | 1,664.91 | 1,493.23 | 1,521.94 | 6,413.82 |
| | 062 | 789.64 | 792.4 | 697 | 722.6 | 3,001.64 |
| | 063 | 1,575.22 | 1,795.79 | 1,653.09 | 1,663.42 | 6,687.52 |
| | 064 | 5,416.14 | 5,695.85 | 5,613 | 5,235.61 | 21,960.60 |
| | 065 | 1,592.85 | 1,620.79 | 1,641.50 | 1,651.45 | 6,506.59 |
| | 066 | 1,511.07 | 1,632.54 | 1,408.33 | 1,557.76 | 6,109.70 |
| | 067 | 2,473.49 | 2,612.77 | 2,416.54 | 2,462.17 | 9,964.97 |
| | 068 | 3,272.84 | 2,997.87 | 3,075.85 | 2,974.36 | 12,320.92 |
| | 069 | 710.43 | 752.13 | 744.23 | 671.71 | 2,878.50 |
| TOTAL | | 24,542.21 | 24,979.42 | 23,678.87 | 23,115.64 | 96,316.14 |

| STATE: | DELAWARE | | | | | |
|---|---|---|---|---|---|---|
| ZIP CODE | | QUARTER 1 | QUARTER 2 | QUARTER 3 | QUARTER 4 | TOTAL GRAMS |
| | 197 | 3,789.77 | 3,538.12 | 3,173.74 | 3,161.97 | 13,663.60 |
| | 198 | 2,595.97 | 2,548.28 | 2,222.49 | 2,242.47 | 9,609.21 |
| | 199 | 3,520.69 | 3,437.45 | 3,325.35 | 3,430.78 | 13,714.27 |
| TOTAL | | 9,906.43 | 9,523.85 | 8,721.58 | 8,835.22 | 36,987.08 |

| STATE: | DISTRICT OF COLUMBIA | | | | | |
|---|---|---|---|---|---|---|
| ZIP CODE | | QUARTER 1 | QUARTER 2 | QUARTER 3 | QUARTER 4 | TOTAL GRAMS |
| | 200 | 12,414.44 | 12,277.52 | 11,652.91 | 10,877.95 | 47,222.82 |
| | 203 | 22.47 | 7.18 | 39.78 | 0 | 69.43 |
| | 204 | 309.39 | 292.83 | 311.62 | 207.75 | 1,121.59 |
| | 205 | 0 | 0 | 2.21 | 4.42 | 6.63 |
| TOTAL | | 12,746.30 | 12,577.53 | 12,006.52 | 11,090.12 | 48,420.47 |

| STATE: | FLORIDA | | | | | |
|---|---|---|---|---|---|---|
| ZIP CODE | | QUARTER 1 | QUARTER 2 | QUARTER 3 | QUARTER 4 | TOTAL GRAMS |

| | | | | | | |
|---|---|---|---|---|---|---|
| | 320 | 5,345.94 | 5,280.65 | 4,952.03 | 5,109.42 | 20,688.04 |
| | 321 | 6,451 | 6,177.37 | 5,388.62 | 5,676.65 | 23,693.64 |
| | 322 | 6,914.87 | 6,564.70 | 6,988.39 | 6,622.72 | 27,090.68 |
| | 323 | 3,922.88 | 3,839.42 | 3,464.92 | 3,712.41 | 14,939.63 |
| | 324 | 3,598.55 | 3,496.16 | 3,127.18 | 3,273.06 | 13,494.95 |
| | 325 | 4,472.48 | 4,043.95 | 3,931.68 | 4,093.73 | 16,541.84 |
| | 326 | 2,571.05 | 2,489.47 | 2,140.62 | 2,577.05 | 9,778.19 |
| | 327 | 7,832.51 | 7,664.85 | 7,367.62 | 7,763.98 | 30,628.96 |
| | 328 | 6,424.90 | 5,816.85 | 6,055.86 | 5,587.21 | 23,884.82 |
| | 329 | 5,357.06 | 5,192.75 | 5,062.79 | 5,155.39 | 20,767.99 |
| | 330 | 14,359.39 | 13,443.96 | 12,806.51 | 12,977.03 | 53,586.89 |
| | 331 | 16,426.32 | 16,874.26 | 15,196.94 | 15,488.15 | 63,985.67 |
| | 333 | 6,681.66 | 6,848.63 | 6,625.89 | 7,062.15 | 27,218.33 |
| | 334 | 13,063.62 | 12,000.27 | 11,834.92 | 12,144.81 | 49,043.62 |
| | 335 | 8,892.98 | 8,577.59 | 7,529.19 | 8,652.85 | 33,652.61 |
| | 336 | 9,313.41 | 8,827.56 | 8,520.66 | 8,637.44 | 35,299.07 |
| | 337 | 9,893.78 | 9,812.72 | 9,512.71 | 9,279.27 | 38,498.48 |
| | 338 | 9,271.87 | 8,392.51 | 7,904.54 | 8,075.30 | 33,644.22 |
| | 339 | 8,457.80 | 7,194.42 | 6,571.35 | 7,285.99 | 29,509.56 |
| | 341 | 2,962.29 | 2,317.99 | 1,627.26 | 2,464.54 | 9,372.08 |
| | 342 | 6,284.53 | 5,973.71 | 5,365.55 | 5,161.04 | 22,784.83 |
| | 344 | 6,020.24 | 5,409.67 | 5,647.63 | 5,839.43 | 22,916.97 |
| | 346 | 7,321.36 | 6,792.32 | 6,594.70 | 7,381.78 | 28,090.16 |
| | 347 | 5,315.87 | 4,700.84 | 4,725.61 | 4,693.20 | 19,435.52 |
| | 349 | 3,530.14 | 3,104.28 | 2,781.33 | 3,424.89 | 12,840.64 |
| TOTAL | | 180,686.50 | 170,836.90 | 161,724.50 | 168,139.49 | 681,387.39 |

| STATE: | | GEORGIA | | | | |
|---|---|---|---|---|---|---|
| ZIP CODE | | QUARTER 1 | QUARTER 2 | QUARTER 3 | QUARTER 4 | TOTAL GRAMS |
| | 300 | 15,962.23 | 16,583.80 | 15,201.65 | 15,874.79 | 63,622.47 |
| | 301 | 9,646.30 | 9,949.21 | 9,103.91 | 9,518.33 | 38,217.75 |
| | 302 | 12,084.63 | 12,883.20 | 12,038.11 | 11,739.74 | 48,745.68 |
| | 303 | 8,441.01 | 8,915.81 | 7,580.27 | 8,036.22 | 32,973.31 |
| | 304 | 3,389.95 | 3,185.22 | 3,162.60 | 3,003.94 | 12,741.71 |
| | 305 | 7,123.35 | 7,073.04 | 6,810.80 | 6,533.38 | 27,540.57 |
| | 306 | 6,244.81 | 6,208.75 | 5,817.44 | 5,986.09 | 24,257.09 |

| | | | | | | |
|---|---|---|---|---|---|---|
| | 307 | 2,800.40 | 2,768.39 | 2,763.83 | 2,844.03 | 11,176.65 |
| | 308 | 2,306.82 | 2,448.46 | 2,354.82 | 2,338.70 | 9,448.80 |
| | 309 | 3,988.76 | 4,087.09 | 4,059.80 | 3,877.09 | 16,012.74 |
| | 310 | 6,022.29 | 6,227.66 | 5,808.39 | 5,894.29 | 23,952.63 |
| | 312 | 2,215.63 | 2,026.08 | 1,964.11 | 2,097.06 | 8,302.88 |
| | 313 | 2,058.04 | 2,050.53 | 1,865.12 | 1,853.74 | 7,827.43 |
| | 314 | 1,807.39 | 1,633.35 | 1,620.23 | 1,790.74 | 6,851.71 |
| | 315 | 6,756.97 | 6,990.69 | 6,664.59 | 6,255.02 | 26,667.27 |
| | 316 | 2,445.80 | 2,722.42 | 2,715.67 | 2,569.21 | 10,453.10 |
| | 317 | 7,026.53 | 7,175.62 | 6,549.76 | 6,539.71 | 27,291.62 |
| | 318 | 560.18 | 562.54 | 419.92 | 454.09 | 1,996.73 |
| | 319 | 3,381.72 | 3,470.18 | 3,361.47 | 2,924.04 | 13,137.41 |
| | 398 | 2,094.46 | 2,312.87 | 2,302.25 | 2,337.75 | 9,047.33 |
| TOTAL | | 106,357.27 | 109,274.91 | 102,164.74 | 102,467.96 | 420,264.88 |

STATE: GUAM

| ZIP CODE | | QUARTER 1 | QUARTER 2 | QUARTER 3 | QUARTER 4 | TOTAL GRAMS |
|---|---|---|---|---|---|---|
| | 969 | 1,289.01 | 1,191.33 | 1,196.49 | 1,088.84 | 4,765.67 |
| TOTAL | | 1,289.01 | 1,191.33 | 1,196.49 | 1,088.84 | 4,765.67 |

STATE: HAWAII

| ZIP CODE | | QUARTER 1 | QUARTER 2 | QUARTER 3 | QUARTER 4 | TOTAL GRAMS |
|---|---|---|---|---|---|---|
| | 967 | 8,744.06 | 9,259.56 | 8,840.53 | 8,456.65 | 35,300.80 |
| | 968 | 5,063.47 | 4,920.04 | 4,925.94 | 4,352.50 | 19,261.95 |
| TOTAL | | 13,807.53 | 14,179.60 | 13,766.47 | 12,809.15 | 54,562.75 |

STATE: IDAHO

| ZIP CODE | | QUARTER 1 | QUARTER 2 | QUARTER 3 | QUARTER 4 | TOTAL GRAMS |
|---|---|---|---|---|---|---|
| | 832 | 1,659.68 | 1,529.84 | 1,542.77 | 1,692 | 6,424.29 |
| | 833 | 1,243.51 | 1,528.35 | 1,388.48 | 1,275.29 | 5,435.63 |
| | 834 | 1,756.58 | 1,729.11 | 1,627.53 | 1,429.56 | 6,542.78 |
| | 835 | 1,086.03 | 1,182.29 | 946.08 | 1,103.90 | 4,318.30 |
| | 836 | 3,495.99 | 3,609.01 | 3,790.75 | 3,582.61 | 14,478.36 |
| | 837 | 2,055.40 | 1,897.73 | 1,842.18 | 1,912.99 | 7,708.30 |

| | | | | | | |
|---|---|---|---|---|---|---|
| | 838 | 2,482.56 | 2,512.54 | 2,457.99 | 2,282.93 | 9,736.02 |
| TOTAL | | 13,779.75 | 13,988.87 | 13,595.78 | 13,279.28 | 54,643.68 |

| STATE: | ILLINOIS | | | | |
|---|---|---|---|---|---|
| ZIP CODE | QUARTER 1 | QUARTER 2 | QUARTER 3 | QUARTER 4 | TOTAL GRAMS |
| 600 | 21,191.02 | 21,771.86 | 20,165.68 | 21,747.71 | 84,876.27 |
| 601 | 14,888.81 | 14,534.54 | 14,093.75 | 13,976.37 | 57,493.47 |
| 602 | 530.8 | 518.28 | 502.06 | 498.47 | 2,049.61 |
| 603 | 1,176.95 | 1,264.92 | 1,203.93 | 1,343.73 | 4,989.53 |
| 604 | 19,862.19 | 19,549.56 | 19,223.15 | 18,583.27 | 77,218.17 |
| 605 | 9,891.28 | 9,695.02 | 9,542.42 | 9,820.39 | 38,949.11 |
| 606 | 34,165.52 | 32,410.14 | 31,870.33 | 31,041.27 | 129,487.26 |
| 607 | 1,676.56 | 1,620.31 | 1,547.73 | 1,549.46 | 6,394.06 |
| 608 | 1,971.78 | 2,232.91 | 1,945.48 | 2,214.41 | 8,364.58 |
| 609 | 2,685.54 | 2,677.82 | 2,586.32 | 2,325.34 | 10,275.02 |
| 610 | 3,201.62 | 3,299.18 | 2,942.13 | 3,216.78 | 12,659.71 |
| 611 | 5,250.41 | 4,977.82 | 4,638.31 | 4,069.75 | 18,936.29 |
| 612 | 2,003.84 | 1,998.79 | 1,982.20 | 2,082.23 | 8,067.06 |
| 613 | 1,714.70 | 1,761.93 | 1,579.47 | 1,752.97 | 6,809.07 |
| 614 | 2,130.32 | 1,845.69 | 2,069.71 | 1,979.51 | 8,025.23 |
| 615 | 2,393.06 | 2,398.13 | 2,579.32 | 2,148.57 | 9,519.08 |
| 616 | 3,088.37 | 3,056.60 | 3,131.32 | 2,884.33 | 12,160.62 |
| 617 | 3,597.72 | 3,318.92 | 3,298.43 | 3,359.73 | 13,574.80 |
| 618 | 4,122.82 | 4,080.06 | 3,642.13 | 3,868.06 | 15,713.07 |
| 619 | 1,520.90 | 1,304.49 | 1,282.22 | 1,039.29 | 5,146.90 |
| 620 | 5,134.29 | 5,377.80 | 5,119.11 | 4,667.71 | 20,298.91 |
| 622 | 6,743.83 | 6,249.01 | 6,399.84 | 5,942.10 | 25,334.78 |
| 623 | 1,531.65 | 1,371.81 | 1,311.50 | 1,399.71 | 5,614.67 |
| 624 | 1,698.63 | 1,459.11 | 1,601.66 | 1,442.53 | 6,201.93 |
| 625 | 2,674.34 | 2,728.67 | 2,621.78 | 2,447.63 | 10,472.42 |
| 626 | 2,193.83 | 1,976.29 | 2,073.88 | 1,969.40 | 8,213.40 |
| 627 | 3,180.44 | 3,190.75 | 3,230.25 | 3,082.18 | 12,683.62 |
| 628 | 2,560.83 | 2,663.53 | 2,347.85 | 2,267.37 | 9,839.58 |
| 629 | 4,513.91 | 4,647.52 | 4,532.69 | 4,215.82 | 17,909.94 |
| TOTAL | 167,295.96 | 163,981.46 | 159,064.65 | 156,936.09 | 647,278.16 |

RETAIL DRUG DISTRIBUTION BY ZIP CODE WITHIN STATE BY GRAMS WT

STATE: INDIANA

| ZIP CODE | QUARTER 1 | QUARTER 2 | QUARTER 3 | QUARTER 4 | TOTAL GRAMS |
|---|---|---|---|---|---|
| 460 | 2,739.17 | 2,536.90 | 2,133.61 | 2,137.22 | 9,546.90 |
| 461 | 3,574.03 | 3,674.31 | 3,710.16 | 3,486.61 | 14,445.11 |
| 462 | 7,393.83 | 7,051.29 | 6,306.52 | 5,793.56 | 26,545.20 |
| 463 | 8,951.06 | 9,254.13 | 8,184.06 | 8,994.84 | 35,384.09 |
| 464 | 2,296.27 | 2,292.07 | 2,189.97 | 2,105.09 | 8,883.40 |
| 465 | 3,077.89 | 2,856.62 | 2,805.47 | 2,749.53 | 11,489.51 |
| 466 | 1,122.37 | 1,119.77 | 1,009.05 | 977.37 | 4,228.56 |
| 467 | 1,909.47 | 1,878.29 | 1,604.04 | 1,916.98 | 7,308.78 |
| 468 | 2,356.42 | 2,045.89 | 1,830 | 2,037.43 | 8,269.74 |
| 469 | 2,759.42 | 2,586.92 | 2,022.77 | 2,027.91 | 9,397.02 |
| 470 | 748.23 | 596.18 | 856.77 | 627.47 | 2,828.65 |
| 471 | 2,085.65 | 3,125.57 | 3,644.73 | 3,952.03 | 12,807.98 |
| 472 | 1,419.97 | 1,306.98 | 1,219.97 | 1,183.10 | 5,130.02 |
| 473 | 1,390.34 | 1,556.96 | 1,426.57 | 1,759.60 | 6,133.47 |
| 474 | 1,630.88 | 1,786.57 | 1,429.96 | 1,514.13 | 6,361.54 |
| 475 | 1,163.80 | 1,104.06 | 958.31 | 969.94 | 4,196.11 |
| 476 | 913.73 | 877.49 | 744.9 | 784.38 | 3,320.50 |
| 477 | 2,417.64 | 2,284.44 | 1,817.23 | 2,030.35 | 8,549.66 |
| 478 | 2,296.70 | 1,979.35 | 1,758.19 | 1,927.76 | 7,962 |
| 479 | 1,891.32 | 2,103.64 | 1,832.57 | 1,849.63 | 7,677.16 |
| TOTAL | 52,138.19 | 52,017.43 | 47,484.85 | 48,824.93 | 200,465.40 |

STATE: IOWA

| ZIP CODE | QUARTER 1 | QUARTER 2 | QUARTER 3 | QUARTER 4 | TOTAL GRAMS |
|---|---|---|---|---|---|
| 500 | 1,302.11 | 1,265.88 | 1,196.75 | 1,362.57 | 5,127.31 |
| 501 | 1,092.07 | 1,112.94 | 1,095.99 | 1,034.85 | 4,335.85 |
| 502 | 1,145.47 | 1,072.03 | 1,063.62 | 1,141.13 | 4,422.25 |
| 503 | 2,152.90 | 2,186.62 | 2,224.83 | 2,009.37 | 8,573.72 |
| 504 | 520.48 | 512.93 | 339.43 | 496.71 | 1,869.55 |
| 505 | 705.14 | 701.83 | 685.42 | 774.63 | 2,867.02 |
| 506 | 776.07 | 772.3 | 779.42 | 788.08 | 3,115.87 |
| 507 | 442.1 | 546.43 | 572.03 | 572.39 | 2,132.95 |
| 508 | 197.8 | 182.33 | 240.9 | 270 | 891.03 |

| | | | | | |
|---|---|---|---|---|---|
| | 510 | 464.65 | 443.84 | 477.93 | 441.46 | 1,827.88 |
| | 511 | 686.03 | 720.84 | 720.47 | 746.64 | 2,873.98 |
| | 512 | 141.45 | 251.25 | 122.66 | 167.97 | 683.33 |
| | 513 | 368.35 | 452.69 | 391.7 | 350.11 | 1,562.85 |
| | 514 | 370.73 | 317.88 | 366.31 | 300.37 | 1,355.29 |
| | 515 | 1,010.61 | 1,009.50 | 1,018.62 | 840.12 | 3,878.85 |
| | 516 | 265.8 | 276.63 | 264.11 | 197.61 | 1,004.15 |
| | 520 | 1,156.85 | 1,225.76 | 1,209.91 | 1,081 | 4,673.52 |
| | 521 | 329.85 | 321.01 | 298.36 | 218.8 | 1,168.02 |
| | 522 | 1,192.15 | 1,221.97 | 1,111.55 | 1,207.77 | 4,733.44 |
| | 523 | 618.07 | 603.9 | 538.24 | 476.26 | 2,236.47 |
| | 524 | 902.63 | 792.13 | 730.43 | 837.43 | 3,262.62 |
| | 525 | 957.13 | 987.06 | 847.64 | 846.25 | 3,638.08 |
| | 526 | 784.83 | 910.82 | 955.6 | 909.21 | 3,560.46 |
| | 527 | 1,262.57 | 1,374.12 | 1,436.01 | 1,536.01 | 5,608.71 |
| | 528 | 1,216.12 | 1,141.44 | 1,037.07 | 1,098.37 | 4,493 |
| TOTAL | | 20,061.96 | 20,404.13 | 19,725 | 19,705.11 | 79,896.20 |

STATE: KANSAS

| ZIP CODE | QUARTER 1 | QUARTER 2 | QUARTER 3 | QUARTER 4 | TOTAL GRAMS |
|---|---|---|---|---|---|
| 660 | 3,108.69 | 3,002.13 | 3,030.36 | 2,741.66 | 11,882.84 |
| 661 | 1,040.15 | 1,144.76 | 1,109.37 | 951.02 | 4,245.30 |
| 662 | 7,853.72 | 7,633.27 | 6,261.70 | 7,405.88 | 29,154.57 |
| 664 | 590.64 | 608.34 | 510.05 | 603.49 | 2,312.52 |
| 665 | 895.4 | 955.35 | 855.88 | 785.69 | 3,492.32 |
| 666 | 1,244.55 | 1,050.01 | 1,132.49 | 1,014.62 | 4,441.67 |
| 667 | 1,080.53 | 1,262.88 | 1,158.72 | 1,104.74 | 4,606.87 |
| 668 | 287.35 | 312.98 | 398.69 | 397.4 | 1,396.42 |
| 669 | 245.32 | 309.41 | 276.63 | 350.3 | 1,181.66 |
| 670 | 1,793.11 | 1,590.64 | 1,743.38 | 1,572.27 | 6,699.40 |
| 671 | 1,022.35 | 1,189.75 | 1,075.46 | 1,165.28 | 4,452.84 |
| 672 | 3,939.71 | 3,943.97 | 3,872.13 | 4,289.35 | 16,045.16 |
| 673 | 984.7 | 758.97 | 732.27 | 693.24 | 3,169.18 |
| 674 | 1,599.25 | 1,522.19 | 1,421.79 | 1,539.30 | 6,082.53 |
| 675 | 861.94 | 768.58 | 825.37 | 847.66 | 3,303.55 |
| 676 | 377.16 | 338.59 | 372.6 | 311.42 | 1,399.77 |

| | | | | | | |
|---|---|---|---|---|---|---|
| | 677 | 151.48 | 216.59 | 227.64 | 269.63 | 865.34 |
| | 678 | 1,033.71 | 1,039.91 | 1,035.03 | 815.4 | 3,924.05 |
| | 679 | 887.45 | 791.22 | 748.48 | 712.76 | 3,139.91 |
| TOTAL | | 28,997.21 | 28,439.54 | 26,788.04 | 27,571.11 | 111,795.90 |

| STATE: | KENTUCKY | | | | | |
|---|---|---|---|---|---|---|
| ZIP CODE | | QUARTER 1 | QUARTER 2 | QUARTER 3 | QUARTER 4 | TOTAL GRAMS |
| | 400 | 1,924.94 | 2,065.90 | 1,906.13 | 1,758.84 | 7,655.81 |
| | 401 | 861.3 | 1,144.27 | 1,079.19 | 994.34 | 4,079.10 |
| | 402 | 6,340.10 | 6,603.84 | 6,305.52 | 5,716.66 | 24,966.12 |
| | 403 | 2,482.44 | 2,467.89 | 2,521.47 | 2,486.39 | 9,958.19 |
| | 404 | 1,158.11 | 1,062.12 | 1,166.52 | 1,055.67 | 4,442.42 |
| | 405 | 1,368.86 | 1,400.97 | 1,310.24 | 1,388.78 | 5,468.85 |
| | 406 | 167.97 | 164.74 | 269.63 | 144.76 | 747.1 |
| | 407 | 1,931.25 | 1,929.60 | 1,830.14 | 1,759.05 | 7,450.04 |
| | 408 | 870.78 | 804.48 | 766.9 | 771.32 | 3,213.48 |
| | 409 | 2,700.74 | 2,433.50 | 2,283.58 | 2,252.27 | 9,670.09 |
| | 410 | 3,863.59 | 3,748.74 | 3,671.56 | 3,723.15 | 15,007.04 |
| | 411 | 1,358.45 | 1,257.85 | 1,169.82 | 1,401.41 | 5,187.53 |
| | 412 | 1,664.21 | 1,841.01 | 1,657.58 | 1,891.85 | 7,054.65 |
| | 413 | 1,640.62 | 1,604.53 | 1,635.47 | 1,405.62 | 6,286.24 |
| | 414 | 496.34 | 421.39 | 451.84 | 494.73 | 1,864.30 |
| | 415 | 2,368.12 | 2,411.22 | 2,592.45 | 2,273.01 | 9,644.80 |
| | 416 | 1,005.37 | 1,149.25 | 1,068.58 | 1,010.02 | 4,233.22 |
| | 417 | 614.02 | 717.87 | 555.44 | 683.26 | 2,570.59 |
| | 418 | 592.31 | 537.2 | 651.98 | 627.67 | 2,409.16 |
| | 420 | 2,349.97 | 2,161.57 | 2,236.99 | 2,213.69 | 8,962.22 |
| | 421 | 2,748.49 | 2,416.51 | 2,550.04 | 2,267.88 | 9,982.92 |
| | 422 | 1,505.17 | 1,434.45 | 1,486.10 | 1,278.63 | 5,704.35 |
| | 423 | 1,938.16 | 2,114.27 | 2,118.30 | 1,895.71 | 8,066.44 |
| | 424 | 1,895.92 | 1,647.54 | 1,631.30 | 1,596.18 | 6,770.94 |
| | 425 | 554.92 | 645.35 | 676.29 | 638.72 | 2,515.28 |
| | 426 | 1,375.05 | 1,323.08 | 1,121.04 | 1,179.42 | 4,998.59 |
| | 427 | 2,899.26 | 2,725.14 | 2,858.95 | 2,735 | 11,218.35 |
| TOTAL | | 48,676.46 | 48,234.28 | 47,573.05 | 45,644.03 | 190,127.82 |

STATE: LOUISIANA

| ZIP CODE | QUARTER 1 | QUARTER 2 | QUARTER 3 | QUARTER 4 | TOTAL GRAMS |
|---|---|---|---|---|---|
| 700 | 5,179.85 | 5,327.24 | 4,714.36 | 4,598.45 | 19,819.90 |
| 701 | 3,109.93 | 2,716.33 | 2,572.81 | 2,606.77 | 11,005.84 |
| 703 | 2,127.28 | 2,155.12 | 1,879.20 | 1,889.22 | 8,050.82 |
| 704 | 3,529.33 | 3,643.46 | 3,428.85 | 3,240 | 13,841.64 |
| 705 | 8,744.65 | 8,918.03 | 8,213.23 | 8,003.93 | 33,879.84 |
| 706 | 3,344.70 | 3,280.18 | 3,398.77 | 3,283.10 | 13,306.75 |
| 707 | 2,037.77 | 2,295.14 | 1,866.43 | 2,056.99 | 8,256.33 |
| 708 | 2,479.22 | 2,295.19 | 2,187.66 | 2,045.64 | 9,007.71 |
| 710 | 1,404.95 | 1,724.40 | 1,370.09 | 1,341.56 | 5,841 |
| 711 | 3,319.79 | 3,534.40 | 3,237.69 | 2,816 | 12,907.88 |
| 712 | 4,728.96 | 4,573.94 | 4,435.80 | 4,255.58 | 17,994.28 |
| 713 | 3,220.26 | 3,261.29 | 2,937.88 | 2,766.80 | 12,186.23 |
| 714 | 2,047.07 | 1,868.57 | 1,842.04 | 1,939.76 | 7,697.44 |
| TOTAL | 45,273.76 | 45,593.29 | 42,084.81 | 40,843.80 | 173,795.66 |

STATE: MAINE

| ZIP CODE | QUARTER 1 | QUARTER 2 | QUARTER 3 | QUARTER 4 | TOTAL GRAMS |
|---|---|---|---|---|---|
| 39 | 247.53 | 143.66 | 121.74 | 130.58 | 643.51 |
| 40 | 2,136.48 | 2,055.36 | 1,906.17 | 1,921.95 | 8,019.96 |
| 41 | 445.56 | 496.78 | 417.22 | 370.71 | 1,730.27 |
| 42 | 1,771.62 | 1,655.05 | 1,374.09 | 1,644.81 | 6,445.57 |
| 43 | 877.78 | 926.42 | 883.28 | 916.69 | 3,604.17 |
| 44 | 1,558.23 | 1,633.88 | 1,394.06 | 1,563.34 | 6,149.51 |
| 45 | 283.44 | 306.74 | 323.22 | 116.55 | 1,029.95 |
| 46 | 699.65 | 870.74 | 752.02 | 817.13 | 3,139.54 |
| 47 | 1,260.28 | 1,400.06 | 1,122.98 | 1,148.40 | 4,931.72 |
| 48 | 268.24 | 278.08 | 256.15 | 240.14 | 1,042.61 |
| 49 | 1,281.09 | 1,241.02 | 1,224.54 | 1,072.67 | 4,819.32 |
| TOTAL | 10,829.90 | 11,007.79 | 9,775.47 | 9,942.97 | 41,556.13 |

STATE: MARYLAND

| ZIP CODE | QUARTER 1 | QUARTER 2 | QUARTER 3 | QUARTER 4 | TOTAL GRAMS |
|---|---|---|---|---|---|

| | | | | | | |
|---|---|---|---|---|---|---|
| | 206 | 4,491.58 | 4,371.72 | 4,075.03 | 4,235.10 | 17,173.43 |
| | 207 | 15,659.40 | 14,382.17 | 13,811.62 | 12,926.23 | 56,779.42 |
| | 208 | 6,243.04 | 5,920.59 | 6,022.69 | 5,678.36 | 23,864.68 |
| | 209 | 2,614.14 | 2,548.08 | 2,280.93 | 2,150.08 | 9,593.23 |
| | 210 | 6,652.94 | 6,458.44 | 5,669.74 | 5,971.24 | 24,752.36 |
| | 211 | 3,367.17 | 3,446.25 | 2,822.72 | 3,044.81 | 12,680.95 |
| | 212 | 11,429.56 | 11,227.11 | 10,336.40 | 9,794.88 | 42,787.95 |
| | 214 | 1,174.75 | 1,149.89 | 1,062.75 | 1,155.62 | 4,543.01 |
| | 215 | 1,014.66 | 990.66 | 961.95 | 903.12 | 3,870.39 |
| | 216 | 1,546.19 | 1,590.08 | 1,502 | 1,325.87 | 5,964.14 |
| | 217 | 5,252.35 | 4,874.35 | 4,436.23 | 5,129.23 | 19,692.16 |
| | 218 | 1,619.97 | 1,524 | 1,294.13 | 1,431.43 | 5,869.53 |
| | 219 | 1,095.18 | 1,107.77 | 1,079.14 | 1,074.16 | 4,356.25 |
| TOTAL | | 62,160.93 | 59,591.11 | 55,355.33 | 54,820.13 | 231,927.50 |

| STATE: | MASSACHUSETTS | | | | | |
|---|---|---|---|---|---|---|
| ZIP CODE | | QUARTER 1 | QUARTER 2 | QUARTER 3 | QUARTER 4 | TOTAL GRAMS |
| | 010 | 3,316.18 | 3,078.84 | 2,988.53 | 3,044.63 | 12,428.18 |
| | 011 | 1,197.72 | 1,258.30 | 1,169.52 | 1,142.63 | 4,768.17 |
| | 012 | 474.95 | 617.57 | 482.95 | 628.44 | 2,203.91 |
| | 013 | 628.5 | 562.19 | 629.17 | 523.48 | 2,343.34 |
| | 014 | 2,017.63 | 1,703.16 | 1,777.94 | 1,698.65 | 7,197.38 |
| | 015 | 2,962.44 | 2,925.39 | 2,947.90 | 2,841.28 | 11,677.01 |
| | 016 | 2,104.31 | 2,183.74 | 1,905.45 | 2,027.65 | 8,221.15 |
| | 017 | 2,829.77 | 2,653.27 | 2,341.77 | 2,572.11 | 10,396.92 |
| | 018 | 8,881.03 | 8,370.17 | 7,925.13 | 7,504.41 | 32,680.74 |
| | 019 | 3,285.01 | 2,895.21 | 3,146.43 | 2,938.05 | 12,264.70 |
| | 020 | 2,385.99 | 2,258.59 | 2,073.21 | 2,108.45 | 8,826.24 |
| | 021 | 8,128.77 | 7,582.01 | 7,316.55 | 7,408.80 | 30,436.13 |
| | 022 | 212.85 | 193.32 | 160.3 | 250.07 | 816.54 |
| | 023 | 2,692.09 | 2,737.51 | 2,661.52 | 2,862.75 | 10,953.87 |
| | 024 | 1,777.67 | 1,553.21 | 1,583.18 | 1,540.82 | 6,454.88 |
| | 025 | 719.7 | 824 | 894.4 | 775.24 | 3,213.34 |
| | 026 | 1,077.53 | 1,237.90 | 1,256.09 | 1,164.09 | 4,735.61 |
| | 027 | 3,968.99 | 4,098.90 | 3,873.28 | 3,670.79 | 15,611.96 |
| TOTAL | | 48,661.13 | 46,733.28 | 45,133.32 | 44,702.34 | 185,230.07 |

STATE: MICHIGAN

| ZIP CODE | QUARTER 1 | QUARTER 2 | QUARTER 3 | QUARTER 4 | TOTAL GRAMS |
|---|---|---|---|---|---|
| 480 | 36,198.17 | 36,064.11 | 34,190.63 | 32,880.12 | 139,333.03 |
| 481 | 54,923.43 | 53,274.98 | 52,388.73 | 49,324 | 209,911.14 |
| 482 | 55,634.31 | 52,423.66 | 50,127.91 | 46,584.23 | 204,770.11 |
| 483 | 17,753.25 | 18,850.11 | 18,561.04 | 18,491.20 | 73,655.60 |
| 484 | 9,508.69 | 9,880.58 | 9,735.13 | 9,482.42 | 38,606.82 |
| 485 | 9,588.87 | 10,460.94 | 9,639.44 | 8,120.08 | 37,809.33 |
| 486 | 13,136.89 | 13,358.88 | 12,638.50 | 12,238.48 | 51,372.75 |
| 487 | 6,712.88 | 6,757.66 | 6,988.39 | 6,475.27 | 26,934.20 |
| 488 | 10,476.09 | 10,980.74 | 11,218.03 | 10,345.98 | 43,020.84 |
| 489 | 3,496.55 | 3,664.48 | 3,839.48 | 3,492.28 | 14,492.79 |
| 490 | 12,602.41 | 11,890.26 | 11,943.26 | 11,073.94 | 47,509.87 |
| 491 | 1,090.67 | 1,072.07 | 975.1 | 892.31 | 4,030.15 |
| 492 | 6,588.58 | 6,457.53 | 5,809.07 | 5,620.99 | 24,476.17 |
| 493 | 3,073.31 | 2,819.31 | 2,673.66 | 2,752.42 | 11,318.70 |
| 494 | 8,559.94 | 8,282.64 | 8,220.50 | 7,880.76 | 32,943.84 |
| 495 | 4,638.66 | 4,545.02 | 4,049.81 | 4,388.92 | 17,622.41 |
| 496 | 3,717.73 | 3,926.96 | 3,583.69 | 3,388.45 | 14,616.83 |
| 497 | 4,126.17 | 4,221.14 | 4,261.51 | 4,093.81 | 16,702.63 |
| 498 | 2,775.92 | 2,702.79 | 2,820.60 | 2,640.77 | 10,940.08 |
| 499 | 1,273.47 | 1,330.09 | 1,264.89 | 1,339.07 | 5,207.52 |
| TOTAL | 265,875.99 | 262,963.95 | 254,929.37 | 241,505.50 | 1,025,274.81 |

STATE: MINNESOTA

| ZIP CODE | QUARTER 1 | QUARTER 2 | QUARTER 3 | QUARTER 4 | TOTAL GRAMS |
|---|---|---|---|---|---|
| 550 | 3,913.32 | 3,663.07 | 3,867.32 | 3,690.28 | 15,133.99 |
| 551 | 6,456.85 | 6,113.22 | 5,907.36 | 5,872.51 | 24,349.94 |
| 553 | 5,820.03 | 5,872.72 | 5,319.57 | 5,789.02 | 22,801.34 |
| 554 | 7,537.42 | 6,847.31 | 7,173.64 | 6,566.23 | 28,124.60 |
| 556 | 121.28 | 184.86 | 142.05 | 137.75 | 585.94 |
| 557 | 1,753.96 | 1,898.05 | 1,976 | 1,847.78 | 7,475.79 |
| 558 | 1,004.66 | 766.47 | 736.56 | 736.3 | 3,243.99 |
| 559 | 1,570.42 | 1,373.86 | 1,341.42 | 1,270.06 | 5,555.76 |

| | | | | | | |
|---|---|---|---|---|---|---|
| | 560 | 1,587.20 | 1,601 | 1,644.79 | 1,540.70 | 6,373.69 |
| | 561 | 565.38 | 504.63 | 729.1 | 617.05 | 2,416.16 |
| | 562 | 1,633.36 | 1,549.33 | 1,716.66 | 1,429.92 | 6,329.27 |
| | 563 | 2,686.26 | 2,538.61 | 2,440.48 | 2,351.72 | 10,017.07 |
| | 564 | 1,206.77 | 1,315.85 | 1,193.24 | 1,303.54 | 5,019.40 |
| | 565 | 1,394.46 | 1,377.85 | 1,333.09 | 1,384.54 | 5,489.94 |
| | 566 | 823.78 | 885.93 | 786.97 | 852.4 | 3,349.08 |
| | 567 | 839.84 | 636.67 | 692.96 | 763.15 | 2,932.62 |
| TOTAL | | 38,914.99 | 37,129.43 | 37,001.21 | 36,152.95 | 149,198.58 |

| STATE: | MISSISSIPPI | | | | | |
|---|---|---|---|---|---|---|
| ZIP CODE | | QUARTER 1 | QUARTER 2 | QUARTER 3 | QUARTER 4 | TOTAL GRAMS |
| | 386 | 3,407.14 | 3,110.90 | 3,138.67 | 2,910.95 | 12,567.66 |
| | 387 | 1,571.38 | 1,529.39 | 1,421.09 | 1,600.11 | 6,121.97 |
| | 388 | 3,137.07 | 3,169.72 | 2,626.01 | 2,995.88 | 11,928.68 |
| | 389 | 1,256.44 | 1,196.77 | 1,326.06 | 1,364.74 | 5,144.01 |
| | 390 | 2,367.33 | 2,450.53 | 2,587.96 | 2,288.69 | 9,694.51 |
| | 391 | 2,934.44 | 3,084.61 | 3,205.24 | 2,753.25 | 11,977.54 |
| | 392 | 1,578.51 | 1,580.44 | 1,631.28 | 1,472 | 6,262.23 |
| | 393 | 2,451.65 | 2,552.07 | 2,574.79 | 2,726.24 | 10,304.75 |
| | 394 | 3,440.50 | 3,384.19 | 3,180.63 | 3,215.25 | 13,220.57 |
| | 395 | 2,975.02 | 2,705.84 | 2,966.04 | 2,596.73 | 11,243.63 |
| | 396 | 1,104.61 | 1,000.85 | 916.2 | 798.97 | 3,820.63 |
| | 397 | 1,843.50 | 1,804.34 | 1,742.57 | 1,936.76 | 7,327.17 |
| TOTAL | | 28,067.59 | 27,569.65 | 27,316.54 | 26,659.57 | 109,613.35 |

| STATE: | MISSOURI | | | | | |
|---|---|---|---|---|---|---|
| ZIP CODE | | QUARTER 1 | QUARTER 2 | QUARTER 3 | QUARTER 4 | TOTAL GRAMS |
| | 630 | 7,013.04 | 7,415.04 | 7,286.34 | 7,398.78 | 29,113.20 |
| | 631 | 19,049.55 | 19,023.57 | 17,722.22 | 17,993.63 | 73,788.97 |
| | 633 | 3,785.93 | 3,825.03 | 3,558.02 | 3,610.19 | 14,779.17 |
| | 634 | 623.98 | 818.91 | 745.61 | 678.58 | 2,867.08 |
| | 635 | 821.05 | 912.77 | 844.26 | 803.37 | 3,381.45 |
| | 636 | 1,922.85 | 1,916.59 | 2,048.53 | 1,925.01 | 7,812.98 |
| | 637 | 1,312.32 | 1,164.62 | 1,147.89 | 1,103.91 | 4,728.74 |

| | | | | | | |
|---|---|---|---|---|---|---|
| | 638 | 2,331.19 | 2,546.04 | 2,614.08 | 2,466.62 | 9,957.93 |
| | 639 | 1,347.42 | 1,455.44 | 1,433.34 | 1,385.91 | 5,622.11 |
| | 640 | 5,482.72 | 6,028.61 | 5,313.70 | 5,578.80 | 22,403.83 |
| | 641 | 5,638.29 | 5,723.65 | 5,360.75 | 5,619.47 | 22,342.16 |
| | 644 | 601.99 | 632.45 | 622.51 | 782.92 | 2,639.87 |
| | 645 | 664.24 | 798.28 | 665.56 | 609.1 | 2,737.18 |
| | 646 | 701.1 | 732.91 | 755.32 | 676.5 | 2,865.83 |
| | 647 | 1,645.51 | 1,491.91 | 1,541.53 | 1,669.52 | 6,348.47 |
| | 648 | 1,976.29 | 1,587.93 | 1,908.23 | 1,924.97 | 7,397.42 |
| | 650 | 1,247.41 | 1,400.22 | 1,108.74 | 1,209.29 | 4,965.66 |
| | 651 | 663.31 | 829.41 | 918.13 | 753.69 | 3,164.54 |
| | 652 | 2,051.77 | 2,023.30 | 1,922.09 | 2,026.16 | 8,023.32 |
| | 653 | 1,168.65 | 1,044.14 | 970.03 | 1,118.12 | 4,300.94 |
| | 654 | 722.7 | 796.36 | 797.83 | 845.17 | 3,162.06 |
| | 655 | 1,279.75 | 1,273.72 | 1,176.37 | 1,296.11 | 5,025.95 |
| | 656 | 1,562.61 | 1,505.14 | 1,510.29 | 1,482.60 | 6,060.64 |
| | 657 | 2,784.95 | 3,026.42 | 2,989.29 | 2,802.93 | 11,603.59 |
| | 658 | 2,254.39 | 2,116.57 | 2,062.86 | 2,271.49 | 8,705.31 |
| TOTAL | | 68,653.01 | 70,089.03 | 67,023.52 | 68,032.84 | 273,798.40 |

STATE: MONTANA

| ZIP CODE | | QUARTER 1 | QUARTER 2 | QUARTER 3 | QUARTER 4 | TOTAL GRAMS |
|---|---|---|---|---|---|---|
| | 590 | 672.85 | 722.86 | 592.1 | 665.22 | 2,653.03 |
| | 591 | 1,133.24 | 1,177.67 | 1,075.75 | 1,069.61 | 4,456.27 |
| | 592 | 483.39 | 483 | 415.27 | 356.89 | 1,738.55 |
| | 593 | 442.2 | 457.67 | 351.59 | 426.73 | 1,678.19 |
| | 594 | 1,284.58 | 1,179.14 | 1,299.67 | 1,124.40 | 4,887.79 |
| | 595 | 303.32 | 274.05 | 209.96 | 311.62 | 1,098.95 |
| | 596 | 1,048.92 | 934.79 | 826.32 | 919.81 | 3,729.84 |
| | 597 | 1,315.44 | 1,522.55 | 1,469.20 | 1,317.09 | 5,624.28 |
| | 598 | 2,174.07 | 2,357.77 | 2,256.96 | 2,298.64 | 9,087.44 |
| | 599 | 1,416.36 | 1,399.85 | 1,396.99 | 1,413.39 | 5,626.59 |
| TOTAL | | 10,274.37 | 10,509.35 | 9,893.81 | 9,903.40 | 40,580.93 |

STATE: NEBRASKA

| ZIP CODE | QUARTER 1 | QUARTER 2 | QUARTER 3 | QUARTER 4 | TOTAL GRAMS |
|---|---|---|---|---|---|
| 680 | 1,160.89 | 1,257.25 | 1,097.66 | 1,132.90 | 4,648.70 |
| 681 | 4,455.03 | 4,157.17 | 4,189.23 | 4,296.68 | 17,098.11 |
| 683 | 318.59 | 231.31 | 349.9 | 306.63 | 1,206.43 |
| 684 | 200.47 | 302.28 | 239.57 | 181.74 | 924.06 |
| 685 | 1,969.44 | 2,077.69 | 1,768.72 | 1,669.77 | 7,485.62 |
| 686 | 198.26 | 159.67 | 228.19 | 138.95 | 725.07 |
| 687 | 775.13 | 771.91 | 898.94 | 840.21 | 3,286.19 |
| 688 | 1,065.95 | 954.66 | 1,023.81 | 1,119.40 | 4,163.82 |
| 689 | 426.73 | 407.95 | 349.47 | 400.02 | 1,584.17 |
| 690 | 152.5 | 179.02 | 99.45 | 123.77 | 554.74 |
| 691 | 871.67 | 674.8 | 850.68 | 634.83 | 3,031.98 |
| 692 | 59.67 | 35.36 | 53.04 | 17.68 | 165.75 |
| 693 | 520.09 | 555.34 | 493.82 | 521.38 | 2,090.63 |
| TOTAL | 12,174.42 | 11,764.41 | 11,642.48 | 11,383.96 | 46,965.27 |

STATE: NEVADA

| ZIP CODE | QUARTER 1 | QUARTER 2 | QUARTER 3 | QUARTER 4 | TOTAL GRAMS |
|---|---|---|---|---|---|
| 890 | 6,289.11 | 6,265.73 | 6,168.22 | 6,247.43 | 24,970.49 |
| 891 | 14,734.96 | 15,092.23 | 13,891.57 | 14,236.35 | 57,955.11 |
| 893 | 43.1 | 70.72 | 101.09 | 113.82 | 328.73 |
| 894 | 2,570.95 | 2,698.96 | 2,516.43 | 2,652.06 | 10,438.40 |
| 895 | 2,297.74 | 2,462.78 | 2,148.06 | 2,286.37 | 9,194.95 |
| 897 | 741.96 | 615.67 | 651.19 | 840.33 | 2,849.15 |
| 898 | 348.09 | 245.32 | 343.67 | 275.16 | 1,212.24 |
| TOTAL | 27,025.91 | 27,451.41 | 25,820.23 | 26,651.52 | 106,949.07 |

STATE: NEW HAMPSHIRE

| ZIP CODE | QUARTER 1 | QUARTER 2 | QUARTER 3 | QUARTER 4 | TOTAL GRAMS |
|---|---|---|---|---|---|
| 30 | 2,207.62 | 1,973.64 | 1,935.31 | 1,968.27 | 8,084.84 |
| 31 | 1,387.53 | 1,389.69 | 1,221.35 | 1,400.10 | 5,398.67 |
| 32 | 935.92 | 777.71 | 826.05 | 962.14 | 3,501.82 |
| 33 | 502.49 | 514.38 | 457.93 | 560.05 | 2,034.85 |
| 34 | 519.63 | 414.65 | 407.75 | 446.25 | 1,788.28 |
| 35 | 380.12 | 356.46 | 282.13 | 342.09 | 1,360.80 |

| | | | | | | |
|---|---|---|---|---|---|---|
| | 36 | 48.62 | 30.94 | 26.52 | 48.62 | 154.7 |
| | 37 | 521.56 | 508.12 | 533.71 | 538.15 | 2,101.54 |
| | 38 | 1,377.74 | 1,578.77 | 1,636.60 | 1,328.75 | 5,921.86 |
| TOTAL | | 7,881.23 | 7,544.36 | 7,327.35 | 7,594.42 | 30,347.36 |

| STATE: | NEW JERSEY | | | | |
|---|---|---|---|---|---|
| ZIP CODE | QUARTER 1 | QUARTER 2 | QUARTER 3 | QUARTER 4 | TOTAL GRAMS |
| 70 | 13,590 | 12,766.01 | 12,498.90 | 11,965.54 | 50,820.45 |
| 71 | 2,381.90 | 2,689.18 | 2,693.57 | 2,342.86 | 10,107.51 |
| 72 | 1,329.54 | 1,191.91 | 1,034.35 | 897.78 | 4,453.58 |
| 73 | 2,271.65 | 2,295.12 | 2,379.03 | 2,330.81 | 9,276.61 |
| 74 | 4,606.01 | 4,464.60 | 4,348.25 | 4,336.10 | 17,754.96 |
| 75 | 1,626.20 | 1,695.79 | 1,696.86 | 1,649.41 | 6,668.26 |
| 76 | 4,026.16 | 4,045.96 | 3,565.72 | 3,604.30 | 15,242.14 |
| 77 | 6,298.29 | 5,698.95 | 5,823.52 | 5,664.61 | 23,485.37 |
| 78 | 3,147.58 | 2,963.36 | 2,546.34 | 2,746.29 | 11,403.57 |
| 79 | 1,456.68 | 1,329.09 | 1,355.91 | 1,441.57 | 5,583.25 |
| 80 | 14,716.88 | 14,422.60 | 13,359.83 | 13,522.96 | 56,022.27 |
| 81 | 2,468.88 | 2,443.82 | 2,194.10 | 2,221.46 | 9,328.26 |
| 82 | 2,696.63 | 2,454.98 | 2,901.68 | 2,579.54 | 10,632.83 |
| 83 | 3,357.83 | 3,117.53 | 3,019.36 | 2,419.92 | 11,914.64 |
| 84 | 448.92 | 452.67 | 531.79 | 452.14 | 1,885.52 |
| 85 | 1,951.31 | 2,019.60 | 1,815.46 | 1,636.08 | 7,422.45 |
| 86 | 3,130.49 | 2,393.99 | 2,303.76 | 2,151.83 | 9,980.07 |
| 87 | 4,413.63 | 4,286.85 | 4,119 | 4,105.97 | 16,925.45 |
| 88 | 7,672.89 | 7,415.51 | 6,821.14 | 7,352.60 | 29,262.14 |
| 89 | 601.19 | 683.5 | 747.26 | 612.57 | 2,644.52 |
| TOTAL | 82,192.66 | 78,831.02 | 75,755.83 | 74,034.34 | 310,813.85 |

| STATE: | NEW MEXICO | | | | |
|---|---|---|---|---|---|
| ZIP CODE | QUARTER 1 | QUARTER 2 | QUARTER 3 | QUARTER 4 | TOTAL GRAMS |
| 870 | 2,107.80 | 1,993.95 | 2,016.63 | 2,061.46 | 8,179.84 |
| 871 | 5,975.27 | 5,675.43 | 5,769.59 | 5,796.10 | 23,216.39 |
| 873 | 1,720.83 | 1,816.99 | 1,682.70 | 1,582.90 | 6,803.42 |
| 874 | 1,294.38 | 1,238.94 | 1,217.95 | 1,082.21 | 4,833.48 |

| | | | | | | |
|---|---|---|---|---|---|---|
| | 875 | 1,141.47 | 1,103.44 | 1,059.55 | 1,082.05 | 4,386.51 |
| | 877 | 323.76 | 418.8 | 371.83 | 373.23 | 1,487.62 |
| | 878 | 72.93 | 24.31 | 53.04 | 77.54 | 227.82 |
| | 879 | 120.54 | 119.35 | 108.29 | 97.24 | 445.42 |
| | 880 | 1,962.83 | 1,998.36 | 2,230.70 | 2,518.33 | 8,710.22 |
| | 881 | 1,054.02 | 1,092.69 | 1,085.95 | 1,151.46 | 4,384.12 |
| | 882 | 2,936.56 | 2,759.46 | 2,881.79 | 2,663.75 | 11,241.56 |
| | 883 | 978.83 | 885.78 | 948.28 | 899.42 | 3,712.31 |
| | 884 | 135.99 | 120.52 | 101.66 | 200.09 | 558.26 |
| TOTAL | | 19,825.21 | 19,248.02 | 19,527.96 | 19,585.78 | 78,186.97 |

STATE: NEW YORK

| ZIP CODE | QUARTER 1 | QUARTER 2 | QUARTER 3 | QUARTER 4 | TOTAL GRAMS |
|---|---|---|---|---|---|
| 063 | 2.21 | 0 | 0 | 2.21 | 4.42 |
| 100 | 15,198.52 | 14,410.61 | 13,904.56 | 13,232.12 | 56,745.81 |
| 101 | 696.23 | 545.74 | 496.26 | 721.35 | 2,459.58 |
| 102 | 22.19 | 33.7 | 30.11 | 26.52 | 112.52 |
| 103 | 2,931.70 | 2,929.95 | 2,801.76 | 2,508.06 | 11,171.47 |
| 104 | 11,645.37 | 11,830.33 | 10,454.68 | 10,346.74 | 44,277.12 |
| 105 | 3,338.41 | 3,321.95 | 3,162.41 | 3,174.97 | 12,997.74 |
| 106 | 618.82 | 603.72 | 480.14 | 637.42 | 2,340.10 |
| 107 | 1,575.04 | 1,752.87 | 1,510.86 | 1,434.16 | 6,272.93 |
| 108 | 595.98 | 469.36 | 442.29 | 346.41 | 1,854.04 |
| 109 | 4,070.31 | 3,931.41 | 4,045.01 | 3,938.15 | 15,984.88 |
| 110 | 1,465.08 | 1,573.48 | 1,469.33 | 1,265.72 | 5,773.61 |
| 111 | 1,253.68 | 1,594.21 | 1,052.66 | 1,178.54 | 5,079.09 |
| 112 | 18,536.39 | 17,496.51 | 17,328.61 | 16,632.79 | 69,994.30 |
| 113 | 7,288.47 | 6,944.95 | 6,543.07 | 6,481.63 | 27,258.12 |
| 114 | 3,758.07 | 3,700.71 | 3,326.65 | 3,025.84 | 13,811.27 |
| 115 | 4,713.18 | 4,515.22 | 4,220.44 | 4,261.48 | 17,710.32 |
| 116 | 913.88 | 998.13 | 801.53 | 790.12 | 3,503.66 |
| 117 | 8,455.74 | 8,534.14 | 7,854.47 | 7,479.90 | 32,324.25 |
| 118 | 502.37 | 572.03 | 671.92 | 540.86 | 2,287.18 |
| 119 | 1,194.35 | 1,191.55 | 1,201.10 | 1,159.14 | 4,746.14 |
| 120 | 1,579.85 | 1,633.85 | 1,613.83 | 1,643.61 | 6,471.14 |
| 121 | 912.66 | 562.46 | 690.93 | 770.49 | 2,936.54 |

| | | | | | | |
|---|---|---|---|---|---|---|
| | 122 | 700.53 | 512.28 | 660.56 | 654.76 | 2,528.13 |
| | 123 | 761.91 | 796.63 | 753.81 | 754.01 | 3,066.36 |
| | 124 | 961.46 | 1,074.73 | 1,206.30 | 1,072.78 | 4,315.27 |
| | 125 | 2,716.24 | 2,703.02 | 2,853.63 | 2,664.12 | 10,937.01 |
| | 126 | 507.86 | 487.78 | 512.1 | 510.07 | 2,017.81 |
| | 127 | 1,171.15 | 1,077.03 | 1,284.78 | 1,287.18 | 4,820.14 |
| | 128 | 1,477.22 | 1,649.31 | 1,489.09 | 1,714.42 | 6,330.04 |
| | 129 | 1,547.96 | 1,586.16 | 1,447.31 | 1,530.38 | 6,111.81 |
| | 130 | 3,095.77 | 3,172.97 | 3,181.60 | 3,083.15 | 12,533.49 |
| | 131 | 1,217.45 | 1,213.40 | 1,142.49 | 1,088.71 | 4,662.05 |
| | 132 | 2,092.45 | 1,834.27 | 1,778.16 | 1,807.57 | 7,512.45 |
| | 133 | 796.17 | 790.74 | 670.38 | 748.37 | 3,005.66 |
| | 134 | 909.33 | 1,066.44 | 938.06 | 957.21 | 3,871.04 |
| | 135 | 577.47 | 456.84 | 566.51 | 521.02 | 2,121.84 |
| | 136 | 3,257.87 | 3,428.42 | 3,605.89 | 3,585.96 | 13,878.14 |
| | 137 | 914.45 | 948.27 | 825.24 | 871.54 | 3,559.50 |
| | 138 | 1,270.77 | 1,491.43 | 1,146.08 | 1,162.58 | 5,070.86 |
| | 139 | 965.66 | 935.94 | 924.36 | 959.03 | 3,784.99 |
| | 140 | 4,988.26 | 5,124.57 | 4,601.93 | 4,820.45 | 19,535.21 |
| | 141 | 2,490.38 | 2,571.86 | 2,271.22 | 2,304.62 | 9,638.08 |
| | 142 | 10,084.59 | 9,999.99 | 9,760.50 | 9,781.47 | 39,626.55 |
| | 143 | 817.34 | 747.63 | 673.69 | 765.12 | 3,003.78 |
| | 144 | 2,485.89 | 2,763.92 | 2,632.44 | 2,492.94 | 10,375.19 |
| | 145 | 2,101.49 | 1,954.10 | 2,181.36 | 1,704.88 | 7,941.83 |
| | 146 | 4,361.39 | 4,172.21 | 3,868.90 | 4,100.23 | 16,502.73 |
| | 147 | 1,183.29 | 1,204.47 | 1,088.24 | 1,198.47 | 4,674.47 |
| | 148 | 2,202.59 | 2,236.63 | 2,006.62 | 1,985.60 | 8,431.44 |
| | 149 | 478.84 | 422.03 | 451.77 | 425.25 | 1,777.89 |
| TOTAL | | 147,404.28 | 145,569.95 | 138,625.64 | 136,150.12 | 567,749.99 |

STATE: NORTH CAROLINA

| ZIP CODE | | QUARTER 1 | QUARTER 2 | QUARTER 3 | QUARTER 4 | TOTAL GRAMS |
|---|---|---|---|---|---|---|
| | 270 | 1,127.90 | 1,353.98 | 918.62 | 1,143.79 | 4,544.29 |
| | 271 | 1,203.97 | 947.78 | 1,249.77 | 1,255.82 | 4,657.34 |
| | 272 | 2,713.53 | 2,404.64 | 2,217.99 | 2,327.80 | 9,663.96 |

| ZIP CODE | QUARTER 1 | QUARTER 2 | QUARTER 3 | QUARTER 4 | TOTAL GRAMS |
|---|---|---|---|---|---|
| 273 | 1,918.36 | 1,707.72 | 1,630.12 | 1,733.03 | 6,989.23 |
| 274 | 2,042.22 | 1,760.23 | 1,905.68 | 1,948.80 | 7,656.93 |
| 275 | 5,758.38 | 5,632.59 | 5,123.46 | 5,202.48 | 21,716.91 |
| 276 | 1,768.64 | 1,857.17 | 1,520.57 | 1,794.48 | 6,940.86 |
| 277 | 1,562.28 | 1,444.81 | 1,285.99 | 1,381.73 | 5,674.81 |
| 278 | 3,173.32 | 2,919.69 | 2,748.63 | 3,047.89 | 11,889.53 |
| 279 | 1,455.86 | 1,437.90 | 1,263.87 | 1,281.23 | 5,438.86 |
| 280 | 2,799.41 | 3,054.79 | 2,471.49 | 3,093.89 | 11,419.58 |
| 281 | 2,463.57 | 2,085.65 | 2,176.63 | 2,138.70 | 8,864.55 |
| 282 | 2,585.52 | 2,359.61 | 2,280.72 | 1,937.06 | 9,162.91 |
| 283 | 7,871.30 | 7,492 | 7,138.18 | 7,037.69 | 29,539.17 |
| 284 | 2,414.38 | 2,210.11 | 2,322.58 | 2,228.76 | 9,175.83 |
| 285 | 3,475.05 | 3,162.12 | 3,289.84 | 3,066.25 | 12,993.26 |
| 286 | 3,425.27 | 3,304.69 | 3,102.86 | 3,052.55 | 12,885.37 |
| 287 | 2,125.19 | 2,303.96 | 2,053.63 | 2,022.64 | 8,505.42 |
| 288 | 867.81 | 821.98 | 719.48 | 699.34 | 3,108.61 |
| 289 | 760.43 | 733.74 | 729.67 | 565.87 | 2,789.71 |
| TOTAL | 51,512.39 | 48,995.16 | 46,149.78 | 46,959.80 | 193,617.13 |

STATE: NORTH DAKOTA

| ZIP CODE | QUARTER 1 | QUARTER 2 | QUARTER 3 | QUARTER 4 | TOTAL GRAMS |
|---|---|---|---|---|---|
| 580 | 446.15 | 500.76 | 462.98 | 410.49 | 1,820.38 |
| 581 | 987.24 | 917.45 | 1,052.79 | 795.94 | 3,753.42 |
| 582 | 868.34 | 889.27 | 947.3 | 788.43 | 3,493.34 |
| 583 | 517.16 | 499.48 | 490.64 | 461.91 | 1,969.19 |
| 584 | 235.74 | 268.15 | 230.36 | 248.4 | 982.65 |
| 585 | 1,153.01 | 1,087.87 | 1,109.21 | 1,135.55 | 4,485.64 |
| 586 | 253.88 | 296.7 | 305.81 | 225.78 | 1,082.17 |
| 587 | 635.6 | 582.9 | 567.97 | 561.34 | 2,347.81 |
| 588 | 655.42 | 775.98 | 694.86 | 717.53 | 2,843.79 |
| TOTAL | 5,752.54 | 5,818.56 | 5,861.92 | 5,345.37 | 22,778.39 |

STATE: OHIO

| ZIP CODE | QUARTER 1 | QUARTER 2 | QUARTER 3 | QUARTER 4 | TOTAL GRAMS |
|---|---|---|---|---|---|
| 430 | 5,209.81 | 4,880.36 | 4,493.28 | 4,604.19 | 19,187.64 |

| | | | | | | |
|---|---|---|---|---|---|---|
| | 431 | 3,422.74 | 3,238.31 | 3,032.04 | 3,286.78 | 12,979.87 |
| | 432 | 15,748.78 | 13,875.46 | 12,949.03 | 13,682.15 | 56,255.42 |
| | 433 | 919.08 | 1,113.81 | 978.77 | 837.5 | 3,849.16 |
| | 434 | 1,390.73 | 1,315.21 | 1,411.53 | 1,151.25 | 5,268.72 |
| | 435 | 3,515.92 | 3,165.43 | 3,054.70 | 3,396.04 | 13,132.09 |
| | 436 | 6,822.65 | 6,374.10 | 5,813.38 | 6,238.69 | 25,248.82 |
| | 437 | 1,884.64 | 1,756.99 | 1,778.35 | 1,583.89 | 7,003.87 |
| | 438 | 439.07 | 495.79 | 358.22 | 280.23 | 1,573.31 |
| | 439 | 2,087.34 | 1,994.34 | 1,993.13 | 1,948.29 | 8,023.10 |
| | 440 | 5,845.97 | 5,138.25 | 5,433.32 | 4,919.72 | 21,337.26 |
| | 441 | 11,236.97 | 10,843.86 | 10,135.68 | 9,512.74 | 41,729.25 |
| | 442 | 2,932.42 | 2,630.82 | 2,794.03 | 2,546.33 | 10,903.60 |
| | 443 | 1,617.49 | 1,563.75 | 1,365.99 | 1,298.07 | 5,845.30 |
| | 444 | 2,732.60 | 2,768 | 2,588.67 | 2,548.27 | 10,637.54 |
| | 445 | 1,861.33 | 1,838.12 | 1,686.93 | 1,637.18 | 7,023.56 |
| | 446 | 2,654.56 | 2,519.26 | 2,327.10 | 2,225.24 | 9,726.16 |
| | 447 | 2,031.96 | 1,646.55 | 1,920.90 | 1,802.90 | 7,402.31 |
| | 448 | 2,645.79 | 2,334.77 | 2,439.60 | 2,336.20 | 9,756.36 |
| | 449 | 1,208 | 1,292.90 | 1,120.88 | 891.42 | 4,513.20 |
| | 450 | 4,297.01 | 4,602.57 | 4,224.61 | 4,040.38 | 17,164.57 |
| | 451 | 2,154.42 | 2,254.31 | 1,831.41 | 2,094.80 | 8,334.94 |
| | 452 | 6,918.75 | 7,421.54 | 6,192.78 | 6,048.95 | 26,582.02 |
| | 453 | 3,894.22 | 3,364.05 | 3,589.46 | 3,126.51 | 13,974.24 |
| | 454 | 5,919.52 | 4,911.97 | 4,705.80 | 4,368.57 | 19,905.86 |
| | 455 | 845.16 | 886.38 | 800.67 | 892.11 | 3,424.32 |
| | 456 | 4,117.79 | 3,798.32 | 3,325.74 | 3,098.01 | 14,339.86 |
| | 457 | 1,490.41 | 1,644.48 | 1,524.22 | 1,480.87 | 6,139.98 |
| | 458 | 3,182.78 | 3,055.32 | 2,833.45 | 2,672.11 | 11,743.66 |
| TOTAL | | 109,027.91 | 102,725.02 | 96,703.67 | 94,549.39 | 403,005.99 |

| STATE: | OKLAHOMA | | | | |
|---|---|---|---|---|---|
| ZIP CODE | QUARTER 1 | QUARTER 2 | QUARTER 3 | QUARTER 4 | TOTAL GRAMS |
| 730 | 14,040.17 | 14,482.89 | 14,144.40 | 14,172.20 | 56,839.66 |
| 731 | 12,337.46 | 12,966.56 | 12,313.42 | 12,941.30 | 50,558.74 |
| 734 | 3,671.12 | 3,655.40 | 3,752.27 | 3,907.08 | 14,985.87 |
| 735 | 9,351.30 | 10,001.02 | 9,938.78 | 9,351.93 | 38,643.03 |

| | | | | | | |
|---|---|---|---|---|---|---|
| | 736 | 2,841.51 | 2,306.54 | 1,961.61 | 2,228.81 | 9,338.47 |
| | 737 | 1,871.78 | 2,115.42 | 2,035.95 | 1,851.94 | 7,875.09 |
| | 738 | 635.4 | 681.26 | 485.09 | 600.02 | 2,401.77 |
| | 739 | 470.93 | 395.79 | 530.61 | 449.01 | 1,846.34 |
| | 740 | 13,033.28 | 13,100.71 | 12,257.46 | 11,881.99 | 50,273.44 |
| | 741 | 5,009.89 | 4,729.62 | 4,693.13 | 4,510.83 | 18,943.47 |
| | 743 | 3,710.58 | 4,090.50 | 3,849.83 | 3,839.95 | 15,490.86 |
| | 744 | 6,392.52 | 6,281.72 | 6,514.06 | 6,644.36 | 25,832.66 |
| | 745 | 1,970 | 2,057.60 | 1,945.99 | 2,075.04 | 8,048.63 |
| | 746 | 1,768.08 | 1,591.27 | 1,673.33 | 1,811.10 | 6,843.78 |
| | 747 | 6,699.45 | 6,185.05 | 6,344.58 | 6,156.62 | 25,385.70 |
| | 748 | 9,206.61 | 8,603.52 | 8,847.02 | 8,427.79 | 35,084.94 |
| | 749 | 3,642.41 | 3,617.93 | 3,346.09 | 2,919.54 | 13,525.97 |
| TOTAL | | 96,652.49 | 96,862.80 | 94,633.62 | 93,769.51 | 381,918.42 |

STATE: OREGON

| ZIP CODE | | QUARTER 1 | QUARTER 2 | QUARTER 3 | QUARTER 4 | TOTAL GRAMS |
|---|---|---|---|---|---|---|
| | 970 | 6,574.76 | 6,529.98 | 6,330.91 | 6,652.94 | 26,088.59 |
| | 971 | 2,390.98 | 2,172.07 | 2,285.26 | 1,982.82 | 8,831.13 |
| | 972 | 8,438.78 | 7,809.75 | 7,412.76 | 7,739.90 | 31,401.19 |
| | 973 | 4,876.79 | 4,436.04 | 4,784.49 | 4,712.57 | 18,809.89 |
| | 974 | 5,100.73 | 5,191.59 | 5,212.67 | 4,993.72 | 20,498.71 |
| | 975 | 3,337.90 | 3,443.43 | 3,333.90 | 3,505.49 | 13,620.72 |
| | 976 | 500.28 | 543.39 | 493.44 | 519.04 | 2,056.15 |
| | 977 | 1,793.90 | 1,602.41 | 1,650.22 | 1,598.66 | 6,645.19 |
| | 978 | 1,661.71 | 1,701.80 | 1,634.85 | 1,500.43 | 6,498.79 |
| | 979 | 450.86 | 279.72 | 340.54 | 338.77 | 1,409.89 |
| TOTAL | | 35,126.69 | 33,710.18 | 33,479.04 | 33,544.34 | 135,860.25 |

STATE: PENNSYLVANIA

| ZIP CODE | | QUARTER 1 | QUARTER 2 | QUARTER 3 | QUARTER 4 | TOTAL GRAMS |
|---|---|---|---|---|---|---|
| | 150 | 3,646.82 | 3,570.22 | 3,592.82 | 3,328.87 | 14,138.73 |
| | 151 | 3,082.74 | 2,975.23 | 2,745.46 | 2,952.76 | 11,756.19 |
| | 152 | 5,142.40 | 5,405.54 | 5,390.28 | 4,942.20 | 20,880.42 |
| | 153 | 1,639.24 | 1,370.46 | 1,599.25 | 1,349.56 | 5,958.51 |

| | | | | | |
|---|---|---|---|---|---|
| 154 | 1,108.91 | 1,159.87 | 1,161.94 | 1,138.65 | 4,569.37 |
| 155 | 2,516.73 | 2,941.50 | 3,024.24 | 2,439.19 | 10,921.66 |
| 156 | 2,158.67 | 1,860.76 | 1,969.94 | 2,219.93 | 8,209.30 |
| 157 | 1,621.64 | 1,460.17 | 1,444.76 | 1,657.24 | 6,183.81 |
| 158 | 712.2 | 800.97 | 783.01 | 699.95 | 2,996.13 |
| 159 | 1,382.10 | 1,268.71 | 1,444.61 | 1,460.73 | 5,556.15 |
| 160 | 1,574.27 | 1,817.19 | 1,540.97 | 1,615 | 6,547.43 |
| 161 | 1,873.16 | 1,632.47 | 1,786.98 | 1,542.10 | 6,834.71 |
| 162 | 1,331.95 | 1,493.98 | 1,302.79 | 1,040.44 | 5,169.16 |
| 163 | 1,205.19 | 1,471.11 | 1,193.22 | 1,343.04 | 5,212.56 |
| 164 | 756.03 | 618.39 | 615.38 | 611.01 | 2,600.81 |
| 165 | 1,621.20 | 1,851.09 | 1,935.90 | 1,467.41 | 6,875.60 |
| 166 | 1,555.27 | 1,622.44 | 1,637.91 | 1,636.80 | 6,452.42 |
| 167 | 585.68 | 638.72 | 491.75 | 624.54 | 2,340.69 |
| 168 | 821.3 | 833.81 | 738.92 | 675.57 | 3,069.60 |
| 169 | 400.76 | 392.29 | 351.5 | 374.42 | 1,518.97 |
| 170 | 5,212.70 | 4,955.12 | 4,676.13 | 4,676.73 | 19,520.68 |
| 171 | 1,228.37 | 1,363.75 | 1,176.49 | 1,193.34 | 4,961.95 |
| 172 | 1,487.05 | 1,524.58 | 1,456.66 | 1,480.05 | 5,948.34 |
| 173 | 2,657.46 | 2,584.60 | 2,522.35 | 2,273.87 | 10,038.28 |
| 174 | 1,626.69 | 1,683.27 | 1,687.03 | 1,431.44 | 6,428.43 |
| 175 | 1,822.62 | 1,582.29 | 1,428.93 | 1,460.93 | 6,294.77 |
| 176 | 1,265.22 | 1,314.22 | 1,364.71 | 1,185.62 | 5,129.77 |
| 177 | 1,411.95 | 1,244.24 | 1,140.97 | 946.46 | 4,743.62 |
| 178 | 1,874.64 | 1,983.98 | 1,820.66 | 1,726.74 | 7,406.02 |
| 179 | 1,803.52 | 1,690.67 | 1,665.48 | 1,526.53 | 6,686.20 |
| 180 | 3,116.34 | 3,084.21 | 3,070.93 | 3,204.27 | 12,475.75 |
| 181 | 1,550.38 | 1,418 | 1,490.70 | 1,563.70 | 6,022.78 |
| 182 | 1,220.13 | 1,425.36 | 1,212.48 | 1,198.03 | 5,056 |
| 183 | 1,470.83 | 1,785.28 | 1,801.60 | 1,658.96 | 6,716.67 |
| 184 | 1,984.19 | 1,839.46 | 1,988.88 | 1,955.66 | 7,768.19 |
| 185 | 1,759.72 | 1,693.09 | 1,692.43 | 1,744.26 | 6,889.50 |
| 186 | 2,667.94 | 2,126.83 | 2,305.44 | 2,230.58 | 9,330.79 |
| 187 | 4,328.51 | 4,513.90 | 3,801.57 | 4,934.54 | 17,578.52 |
| 188 | 824.82 | 934.96 | 904.48 | 674.08 | 3,338.34 |
| 189 | 3,334.80 | 5,736.55 | 5,650.37 | 4,831.81 | 19,553.53 |
| 190 | 14,994.89 | 15,616.63 | 14,596.06 | 14,531.29 | 59,738.87 |

| | | | | | |
|---|---|---|---|---|---|
| | 191 | 30,199.52 | 30,144.69 | 26,804.23 | 26,529.11 | 113,677.55 |
| | 193 | 3,696.62 | 3,235.49 | 3,200.40 | 3,092.83 | 13,225.34 |
| | 194 | 6,417.29 | 11,029.68 | 15,755.26 | 8,240.19 | 41,442.42 |
| | 195 | 1,671.79 | 1,640.48 | 1,613.41 | 1,582.82 | 6,508.50 |
| | 196 | 2,635.80 | 2,574.92 | 2,423.73 | 2,557.41 | 10,191.86 |
| TOTAL | | 137,000.05 | 143,911.17 | 142,003.01 | 131,550.66 | 554,464.89 |

STATE: PUERTO RICO

| ZIP CODE | QUARTER 1 | QUARTER 2 | QUARTER 3 | QUARTER 4 | TOTAL GRAMS |
|---|---|---|---|---|---|
| 6 | 5,153.95 | 5,120.80 | 4,074.32 | 4,751.72 | 19,100.79 |
| 7 | 5,054.50 | 5,084.34 | 4,277.65 | 4,665.52 | 19,082.01 |
| 9 | 3,375.93 | 3,269.84 | 3,277.58 | 3,119.56 | 13,042.91 |
| TOTAL | 13,584.38 | 13,474.98 | 11,629.55 | 12,536.80 | 51,225.71 |

STATE: RHODE ISLAND

| ZIP CODE | QUARTER 1 | QUARTER 2 | QUARTER 3 | QUARTER 4 | TOTAL GRAMS |
|---|---|---|---|---|---|
| 28 | 4,857.47 | 4,671.41 | 4,158.87 | 4,380.09 | 18,067.84 |
| 29 | 3,149.74 | 2,702.36 | 2,715.05 | 2,481.75 | 11,048.90 |
| TOTAL | 8,007.21 | 7,373.77 | 6,873.92 | 6,861.84 | 29,116.74 |

STATE: SOUTH CAROLINA

| ZIP CODE | QUARTER 1 | QUARTER 2 | QUARTER 3 | QUARTER 4 | TOTAL GRAMS |
|---|---|---|---|---|---|
| 290 | 2,985.41 | 2,834.10 | 2,670.11 | 2,803.59 | 11,293.21 |
| 291 | 3,480.59 | 3,217.44 | 3,188.20 | 3,100.16 | 12,986.39 |
| 292 | 3,691.28 | 3,559.99 | 3,564.18 | 3,503.07 | 14,318.52 |
| 293 | 3,579.41 | 3,752.27 | 3,144.28 | 3,833.95 | 14,309.91 |
| 294 | 42,395.43 | 33,184.63 | 38,047.10 | 49,310.49 | 162,937.65 |
| 295 | 8,902.40 | 9,031.58 | 9,847.89 | 9,537.18 | 37,319.05 |
| 296 | 7,912.62 | 7,287.29 | 6,764.53 | 7,238.89 | 29,203.33 |
| 297 | 1,859.63 | 1,692.21 | 1,802.37 | 1,747.39 | 7,101.60 |
| 298 | 2,338.74 | 2,282.04 | 2,431.92 | 2,328.63 | 9,381.33 |
| 299 | 1,389.81 | 1,451.66 | 1,110.92 | 1,232.62 | 5,185.01 |
| TOTAL | 78,535.32 | 68,293.21 | 72,571.50 | 84,635.97 | 304,036 |

STATE: SOUTH DAKOTA

| ZIP CODE | QUARTER 1 | QUARTER 2 | QUARTER 3 | QUARTER 4 | TOTAL GRAMS |
|---|---|---|---|---|---|
| 570 | 911.7 | 1,227.33 | 1,007.82 | 1,009.05 | 4,155.90 |
| 571 | 2,215.46 | 2,092.65 | 1,831.10 | 2,253.07 | 8,392.28 |
| 572 | 529.67 | 458.04 | 506.84 | 499.47 | 1,994.02 |
| 573 | 550.25 | 626.11 | 568.06 | 527.1 | 2,271.52 |
| 574 | 642.55 | 513.85 | 543.03 | 608.51 | 2,307.94 |
| 575 | 377.91 | 334.09 | 283.62 | 303.73 | 1,299.35 |
| 576 | 273.79 | 232.42 | 234.27 | 243.11 | 983.59 |
| 577 | 1,748.87 | 1,592.02 | 1,760.97 | 1,378.70 | 6,480.56 |
| TOTAL | 7,250.20 | 7,076.51 | 6,735.71 | 6,822.74 | 27,885.16 |

STATE: TENNESSEE

| ZIP CODE | QUARTER 1 | QUARTER 2 | QUARTER 3 | QUARTER 4 | TOTAL GRAMS |
|---|---|---|---|---|---|
| 370 | 5,210.26 | 5,068.46 | 4,893.34 | 4,839.50 | 20,011.56 |
| 371 | 51,881.42 | 46,969.21 | 45,136.53 | 33,433.89 | 177,421.05 |
| 372 | 2,620.83 | 2,486.06 | 2,574.22 | 2,193.29 | 9,874.40 |
| 373 | 3,537.29 | 3,751.89 | 3,541.53 | 3,717.88 | 14,548.59 |
| 374 | 1,463.07 | 1,496.69 | 1,295.30 | 1,604.17 | 5,859.23 |
| 376 | 2,266.36 | 2,715.60 | 2,531.61 | 2,636.62 | 10,150.19 |
| 377 | 2,726.36 | 2,829.47 | 2,580.31 | 2,816.17 | 10,952.31 |
| 378 | 4,328.99 | 4,171.50 | 4,131.63 | 4,375.97 | 17,008.09 |
| 379 | 2,487.23 | 2,611.39 | 2,329.71 | 2,572.90 | 10,001.23 |
| 380 | 2,705.61 | 2,736.85 | 2,551.79 | 2,386.48 | 10,380.73 |
| 381 | 5,512.49 | 4,992.02 | 5,133.52 | 5,350.35 | 20,988.38 |
| 382 | 869.92 | 1,004.10 | 863.76 | 866.35 | 3,604.13 |
| 383 | 3,925.14 | 3,683.86 | 3,755.37 | 3,553.25 | 14,917.62 |
| 384 | 1,240.69 | 1,012.68 | 1,085.34 | 1,042.61 | 4,381.32 |
| 385 | 2,745.67 | 2,872.66 | 3,004.35 | 2,762.63 | 11,385.31 |
| TOTAL | 93,521.33 | 88,402.44 | 85,408.31 | 74,152.06 | 341,484.14 |

STATE: TEXAS

| ZIP CODE | QUARTER 1 | QUARTER 2 | QUARTER 3 | QUARTER 4 | TOTAL GRAMS |
|---|---|---|---|---|---|

| | | | | | | |
|---|---|---|---|---|---|---|
| 750 | 64,478.68 | 69,663.32 | | 66,539.25 | 65,850.21 | 266,531.46 |
| 751 | 32,321.07 | 33,252.23 | | 31,161.21 | 30,844.29 | 127,578.80 |
| 752 | 39,190.30 | 39,028.19 | | 37,114.83 | 37,224.18 | 152,557.50 |
| 753 | 509.89 | 581.26 | | 515.5 | 561.73 | 2,168.38 |
| 754 | 21,681.99 | 21,187.31 | | 20,838.50 | 19,568.85 | 83,276.65 |
| 755 | 8,946.84 | 8,912.23 | | 9,045.55 | 8,885.99 | 35,790.61 |
| 756 | 24,762.61 | 25,592.98 | | 23,440.42 | 24,230.82 | 98,026.83 |
| 757 | 25,579.02 | 26,665.45 | | 24,861.18 | 23,329.68 | 100,435.33 |
| 758 | 4,906.72 | 5,015.82 | | 4,620.36 | 4,794.19 | 19,337.09 |
| 759 | 19,223.39 | 18,851.07 | | 18,865.92 | 18,471.44 | 75,411.82 |
| 760 | 49,200.89 | 51,867.07 | | 48,410.95 | 48,977.34 | 198,456.25 |
| 761 | 40,275.58 | 39,252.59 | | 38,632.02 | 38,232.84 | 156,393.03 |
| 762 | 17,622.08 | 18,139.59 | | 17,871.09 | 17,416.31 | 71,049.07 |
| 763 | 11,621.55 | 12,165.04 | | 12,088.86 | 11,817.16 | 47,692.61 |
| 764 | 6,685.65 | 7,333.41 | | 7,230.89 | 7,279.35 | 28,529.30 |
| 765 | 17,348.50 | 17,303.69 | | 16,389.26 | 16,226.88 | 67,268.33 |
| 766 | 5,840.07 | 5,680.14 | | 5,207.56 | 5,300.66 | 22,028.43 |
| 767 | 9,162.90 | 9,376.04 | | 8,429.41 | 8,827.69 | 35,796.04 |
| 768 | 7,696.67 | 8,426.01 | | 8,135.38 | 7,908.82 | 32,166.88 |
| 769 | 7,376.21 | 7,768.50 | | 7,066.98 | 7,002.70 | 29,214.39 |
| 770 | 91,240.52 | 91,893.43 | | 85,391.02 | 87,480 | 356,004.97 |
| 772 | 2.82 | 23.4 | | 6.15 | 0 | 32.37 |
| 773 | 45,813.57 | 45,338.67 | | 44,197.74 | 44,372.11 | 179,722.09 |
| 774 | 33,842.02 | 34,438.30 | | 33,524.90 | 35,218.73 | 137,023.95 |
| 775 | 48,644.48 | 50,264.47 | | 47,412.71 | 48,484.61 | 194,806.27 |
| 776 | 16,260.67 | 17,457.66 | | 16,176.90 | 16,209.19 | 66,104.42 |
| 777 | 9,277.49 | 9,153.70 | | 9,200.32 | 9,031.93 | 36,663.44 |
| 778 | 12,631.07 | 12,175.74 | | 12,507.10 | 11,982.04 | 49,295.95 |
| 779 | 5,444.36 | 5,677.73 | | 5,564.81 | 5,730.60 | 22,417.50 |
| 780 | 15,777.25 | 15,864.59 | | 16,065.88 | 15,535.89 | 63,243.61 |
| 781 | 13,879.43 | 13,942.06 | | 13,935.72 | 13,800.93 | 55,558.14 |
| 782 | 77,855.15 | 77,217.98 | | 75,551.18 | 74,897.30 | 305,521.61 |
| 783 | 14,059.50 | 14,454.11 | | 13,194.79 | 12,748.45 | 54,456.85 |
| 784 | 17,964.41 | 18,205.50 | | 17,521.18 | 17,350.21 | 71,041.30 |
| 785 | 32,124.97 | 31,213.64 | | 31,663.61 | 31,991.50 | 126,993.72 |
| 786 | 22,975.65 | 22,697.05 | | 22,464.75 | 22,221.48 | 90,358.93 |
| 787 | 20,140.76 | 18,817.29 | | 17,772.70 | 17,605.44 | 74,336.19 |

| | | | | | | |
|---|---|---|---|---|---|---|
| | 788 | 5,843.50 | 5,681.24 | 5,651.23 | 5,382.87 | 22,558.84 |
| | 789 | 1,892.95 | 1,792.76 | 1,852.17 | 1,954.66 | 7,492.54 |
| | 790 | 8,083.69 | 8,265.34 | 8,359.18 | 8,150.56 | 32,858.77 |
| | 791 | 14,778.46 | 15,612.88 | 15,547.72 | 15,459.88 | 61,398.94 |
| | 792 | 1,467.83 | 1,363.07 | 1,324.03 | 1,276.88 | 5,431.81 |
| | 793 | 6,166.67 | 5,825.48 | 6,155.66 | 5,671.80 | 23,819.61 |
| | 794 | 13,897.34 | 14,785.79 | 13,646.50 | 14,338.36 | 56,667.99 |
| | 795 | 3,416.65 | 3,381.45 | 3,060.05 | 3,518.24 | 13,376.39 |
| | 796 | 9,311.47 | 9,651.31 | 9,332.18 | 9,744.90 | 38,039.86 |
| | 797 | 23,069.44 | 23,360.57 | 23,783.14 | 24,285.85 | 94,499 |
| | 798 | 1,238.52 | 1,268.70 | 1,186.74 | 1,353.69 | 5,047.65 |
| | 799 | 28,403.85 | 28,857.65 | 28,557.78 | 27,258.23 | 113,077.51 |
| TOTAL | | 1,009,935.10 | 1,024,743.50 | 987,072.96 | 985,807.46 | 4,007,559.02 |

| STATE: | | UTAH | | | | |
|---|---|---|---|---|---|---|
| ZIP CODE | | QUARTER 1 | QUARTER 2 | QUARTER 3 | QUARTER 4 | TOTAL GRAMS |
| | 840 | 7,191.31 | 7,205.94 | 6,836.25 | 6,944.59 | 28,178.09 |
| | 841 | 3,358.34 | 3,471.10 | 3,621.58 | 3,577.64 | 14,028.66 |
| | 843 | 829.49 | 866.44 | 773.43 | 721.21 | 3,190.57 |
| | 844 | 910.6 | 918.84 | 921.72 | 970.79 | 3,721.95 |
| | 845 | 569.83 | 514.86 | 467.44 | 478.67 | 2,030.80 |
| | 846 | 1,930.39 | 1,810.98 | 2,038.06 | 1,787.76 | 7,567.19 |
| | 847 | 1,665.52 | 1,794.99 | 1,719.87 | 1,474.01 | 6,654.39 |
| TOTAL | | 16,455.48 | 16,583.15 | 16,378.35 | 15,954.67 | 65,371.65 |

| STATE: | | VERMONT | | | | |
|---|---|---|---|---|---|---|
| ZIP CODE | | QUARTER 1 | QUARTER 2 | QUARTER 3 | QUARTER 4 | TOTAL GRAMS |
| | 50 | 302.79 | 289.07 | 241.74 | 247.46 | 1,081.06 |
| | 51 | 150.65 | 87.1 | 117.68 | 44.57 | 400 |
| | 52 | 238.03 | 233.61 | 177.9 | 214 | 863.54 |
| | 53 | 184.35 | 199.82 | 175.25 | 181.4 | 740.82 |
| | 54 | 1,258.42 | 1,218.47 | 1,172.44 | 1,139.99 | 4,789.32 |
| | 56 | 560.68 | 551.38 | 521 | 475.42 | 2,108.48 |
| | 57 | 385.19 | 493.76 | 342.92 | 396.88 | 1,618.75 |
| | 58 | 485.44 | 385.53 | 282.11 | 360.86 | 1,513.94 |

| | | | | | |
|---|---|---|---|---|---|
| TOTAL | 3,565.55 | 3,458.74 | 3,031.04 | 3,060.58 | 13,115.91 |

STATE: VIRGIN ISLANDS

| ZIP CODE | QUARTER 1 | QUARTER 2 | QUARTER 3 | QUARTER 4 | TOTAL GRAMS |
|---|---|---|---|---|---|
| 8 | 950.85 | 652.13 | 565.79 | 830.26 | 2,999.03 |
| TOTAL | 950.85 | 652.13 | 565.79 | 830.26 | 2,999.03 |

STATE: VIRGINIA

| ZIP CODE | QUARTER 1 | QUARTER 2 | QUARTER 3 | QUARTER 4 | TOTAL GRAMS |
|---|---|---|---|---|---|
| 201 | 6,001.67 | 5,713.16 | 5,164.22 | 5,423.98 | 22,303.03 |
| 220 | 4,002.45 | 3,962.17 | 3,741.03 | 3,598.69 | 15,304.34 |
| 221 | 3,277.93 | 3,345.37 | 2,804.46 | 2,827.82 | 12,255.58 |
| 222 | 1,322.43 | 1,198.66 | 1,086.10 | 1,145.71 | 4,752.90 |
| 223 | 2,211.05 | 2,142.26 | 1,844.78 | 1,980.65 | 8,178.74 |
| 224 | 2,394.71 | 2,220.78 | 2,027.15 | 2,280.45 | 8,923.09 |
| 225 | 1,317.48 | 1,212.98 | 1,181.65 | 1,142.45 | 4,854.56 |
| 226 | 1,203.12 | 1,276.07 | 923.49 | 1,231.93 | 4,634.61 |
| 227 | 352.31 | 376.45 | 309.95 | 354.81 | 1,393.52 |
| 228 | 1,034.11 | 1,078.32 | 974.73 | 1,168.59 | 4,255.75 |
| 229 | 1,599.47 | 1,471.87 | 1,245.25 | 1,327.91 | 5,644.50 |
| 230 | 1,008.48 | 1,097.05 | 1,192.64 | 1,059.84 | 4,358.01 |
| 231 | 2,221.30 | 2,137.11 | 1,689.45 | 1,834.42 | 7,882.28 |
| 232 | 4,871.96 | 4,410.78 | 4,237.37 | 4,615.59 | 18,135.70 |
| 233 | 1,609.96 | 1,496.41 | 1,524.67 | 1,308.81 | 5,939.85 |
| 234 | 3,983.35 | 3,902.43 | 3,776.90 | 3,405.72 | 15,068.40 |
| 235 | 1,503.09 | 1,551.63 | 1,524.40 | 1,467.47 | 6,046.59 |
| 236 | 2,876.27 | 3,247.50 | 2,881.52 | 2,488.35 | 11,493.64 |
| 237 | 1,039.37 | 1,000.29 | 872.99 | 798.46 | 3,711.11 |
| 238 | 2,626.25 | 2,457.18 | 2,157.72 | 1,943.02 | 9,184.17 |
| 239 | 823.73 | 682.92 | 485.85 | 539.39 | 2,531.89 |
| 240 | 1,861.41 | 1,947.30 | 1,784.33 | 1,563.97 | 7,157.01 |
| 241 | 1,727.85 | 1,626.62 | 1,373.36 | 1,320.58 | 6,048.41 |
| 242 | 1,683.35 | 1,735.30 | 1,422.49 | 1,557.16 | 6,398.30 |
| 243 | 1,670.07 | 1,326.33 | 1,134.26 | 1,179.24 | 5,309.90 |

| | | | | | | |
|---|---|---|---|---|---|---|
| | 244 | 675.79 | 638.25 | 632.39 | 527.13 | 2,473.56 |
| | 245 | 2,296.19 | 2,014.67 | 1,736.44 | 1,855.29 | 7,902.59 |
| | 246 | 2,093.81 | 1,762.75 | 1,673.52 | 1,904.23 | 7,434.31 |
| TOTAL | | 59,288.96 | 57,032.61 | 51,403.11 | 51,851.66 | 219,576.34 |

STATE: WASHINGTON

| ZIP CODE | | QUARTER 1 | QUARTER 2 | QUARTER 3 | QUARTER 4 | TOTAL GRAMS |
|---|---|---|---|---|---|---|
| | 980 | 12,032.25 | 11,700.46 | 11,382.08 | 11,093.76 | 46,208.55 |
| | 981 | 7,681.65 | 7,162.01 | 6,936.36 | 6,843.51 | 28,623.53 |
| | 982 | 8,203.77 | 8,370.82 | 8,085.45 | 7,941.25 | 32,601.29 |
| | 983 | 6,833.41 | 6,884.66 | 6,580.23 | 6,548.64 | 26,846.94 |
| | 984 | 4,080.94 | 4,382.17 | 3,968.91 | 4,109.82 | 16,541.84 |
| | 985 | 4,237.98 | 3,960.19 | 3,890.36 | 3,775.50 | 15,864.03 |
| | 986 | 4,614.31 | 4,769.20 | 4,748.93 | 4,523.80 | 18,656.24 |
| | 988 | 1,671.18 | 1,740.63 | 1,354.44 | 1,526.75 | 6,293 |
| | 989 | 2,380.75 | 2,309.82 | 2,288.79 | 2,128.50 | 9,107.86 |
| | 990 | 1,194.69 | 1,310.55 | 1,110.37 | 1,144.62 | 4,760.23 |
| | 991 | 1,200.13 | 1,155.35 | 1,216.05 | 1,209.07 | 4,780.60 |
| | 992 | 5,960.09 | 6,068.32 | 5,340.39 | 5,488.85 | 22,857.65 |
| | 993 | 2,708.63 | 2,603.72 | 2,443.26 | 2,441.44 | 10,197.05 |
| | 994 | 571.68 | 641.66 | 640.19 | 557.13 | 2,410.66 |
| TOTAL | | 63,371.46 | 63,059.56 | 59,985.81 | 59,332.64 | 245,749.47 |

STATE: WEST VIRGINIA

| ZIP CODE | | QUARTER 1 | QUARTER 2 | QUARTER 3 | QUARTER 4 | TOTAL GRAMS |
|---|---|---|---|---|---|---|
| | 247 | 829.66 | 922.6 | 902.21 | 917.43 | 3,571.90 |
| | 248 | 1,646.52 | 1,693.96 | 1,364.66 | 2,095.09 | 6,800.23 |
| | 249 | 426.39 | 456.5 | 472.92 | 344.05 | 1,699.86 |
| | 250 | 1,071.74 | 1,169.14 | 1,159.49 | 1,269.92 | 4,670.29 |
| | 251 | 779.02 | 904.76 | 679.42 | 880.17 | 3,243.37 |
| | 252 | 1,098.26 | 1,045.76 | 1,077.80 | 1,284.53 | 4,506.35 |
| | 253 | 2,523.46 | 2,015.75 | 2,211.54 | 2,089.48 | 8,840.23 |
| | 254 | 1,542.23 | 1,495.62 | 1,356.66 | 1,364.33 | 5,758.84 |
| | 255 | 2,393.92 | 2,404.81 | 2,191.11 | 2,535.80 | 9,525.64 |
| | 256 | 3,713.41 | 3,585.16 | 3,708.55 | 4,092.71 | 15,099.83 |

| | | | | | |
|---|---|---|---|---|---|
| 257 | 1,459.17 | 1,345.22 | 1,254.64 | 1,286.32 | 5,345.35 |
| 258 | 2,174.51 | 1,853.53 | 1,993.11 | 1,833.20 | 7,854.35 |
| 259 | 632.99 | 702.06 | 657.96 | 654.18 | 2,647.19 |
| 260 | 1,529.74 | 1,651.11 | 1,417.40 | 1,345.65 | 5,943.90 |
| 261 | 1,624.33 | 1,719.89 | 1,392.18 | 1,628.20 | 6,364.60 |
| 262 | 1,593.16 | 1,797.07 | 1,907.12 | 1,779.50 | 7,076.85 |
| 263 | 1,351.52 | 1,353.15 | 1,276.54 | 1,304.88 | 5,286.09 |
| 264 | 436.68 | 517.35 | 437.69 | 451.97 | 1,843.69 |
| 265 | 2,375.60 | 2,547.44 | 2,816.25 | 2,917.33 | 10,656.62 |
| 266 | 962.59 | 1,033.22 | 1,078.53 | 1,034.33 | 4,108.67 |
| 267 | 600.76 | 632.6 | 633.55 | 615.32 | 2,482.23 |
| 268 | 319.8 | 286.21 | 271.13 | 257.87 | 1,135.01 |
| TOTAL | 31,085.46 | 31,132.91 | 30,260.46 | 31,982.26 | 124,461.09 |

STATE: WISCONSIN

| ZIP CODE | QUARTER 1 | QUARTER 2 | QUARTER 3 | QUARTER 4 | TOTAL GRAMS |
|---|---|---|---|---|---|
| 530 | 5,889.73 | 4,812.05 | 4,764.63 | 4,877.16 | 20,343.57 |
| 531 | 5,702.19 | 5,434.90 | 4,970.32 | 5,694.41 | 21,801.82 |
| 532 | 9,914.30 | 9,653.27 | 9,136.12 | 8,991.22 | 37,694.91 |
| 534 | 1,593.24 | 1,437.29 | 1,347.59 | 1,413.44 | 5,791.56 |
| 535 | 3,794.93 | 3,696.92 | 3,586.86 | 3,407.43 | 14,486.14 |
| 537 | 2,153.22 | 2,048.01 | 2,088.97 | 2,199.75 | 8,489.95 |
| 538 | 434.89 | 577.51 | 427.59 | 467.9 | 1,907.89 |
| 539 | 2,065.90 | 1,699.93 | 1,566.10 | 1,572.47 | 6,904.40 |
| 540 | 811.53 | 768.89 | 863.38 | 1,011.49 | 3,455.29 |
| 541 | 1,565.21 | 1,497.40 | 1,397.11 | 1,392.04 | 5,851.76 |
| 542 | 1,134.71 | 1,005.85 | 994.66 | 893.52 | 4,028.74 |
| 543 | 2,142.79 | 2,203.40 | 1,974.10 | 1,885.63 | 8,205.92 |
| 544 | 3,351.09 | 3,167.39 | 3,243.12 | 3,236.61 | 12,998.21 |
| 545 | 1,073.58 | 1,049.80 | 1,020.52 | 899.99 | 4,043.89 |
| 546 | 1,400.17 | 1,406.60 | 1,505.15 | 1,326.43 | 5,638.35 |
| 547 | 1,774.07 | 1,605.44 | 1,414.05 | 1,430.89 | 6,224.45 |
| 548 | 1,827.01 | 1,832.60 | 1,773.13 | 1,740.57 | 7,173.31 |
| 549 | 4,071.02 | 3,868.16 | 3,845.30 | 3,729.57 | 15,514.05 |
| TOTAL | 50,699.58 | 47,765.41 | 45,918.70 | 46,170.52 | 190,554.21 |

STATE: WYOMING

| ZIP CODE | QUARTER 1 | QUARTER 2 | QUARTER 3 | QUARTER 4 | TOTAL GRAMS |
|---|---|---|---|---|---|
| 820 | 1,139.37 | 995.42 | 902.13 | 916.8 | 3,953.72 |
| 822 | 137.21 | 221.01 | 152.5 | 181.41 | 692.13 |
| 823 | 108.52 | 17.68 | 55.25 | 48.62 | 230.07 |
| 824 | 610.14 | 590.07 | 651.02 | 636.28 | 2,487.51 |
| 825 | 416.82 | 400.03 | 408.87 | 359.14 | 1,584.86 |
| 826 | 585.3 | 587.33 | 671.78 | 638.25 | 2,482.66 |
| 827 | 389.71 | 272.75 | 302.77 | 341.81 | 1,307.04 |
| 828 | 285.96 | 248.94 | 396.1 | 268.46 | 1,199.46 |
| 829 | 552.53 | 459.7 | 444.41 | 643.32 | 2,099.96 |
| 830 | 83.54 | 139.37 | 77.35 | 158.24 | 458.5 |
| 831 | 57.46 | 98.8 | 65.65 | 48.62 | 270.53 |
| TOTAL | 4,366.56 | 4,031.10 | 4,127.83 | 4,240.95 | 16,766.44 |

DRUG CODE: 9056 DRUG NAME: ETORPHINE

STATE: WYOMING

| ZIP CODE | QUARTER 1 | QUARTER 2 | QUARTER 3 | QUARTER 4 | TOTAL GRAMS |
|---|---|---|---|---|---|
| 820 | 28 | 0 | 0 | 29 | 57 |
| TOTAL | 28 | 0 | 0 | 29 | 57 |

DRUG CODE: 9064 DRUG NAME: BUPRENORPHINE

STATE: ALABAMA

| ZIP CODE | QUARTER 1 | QUARTER 2 | QUARTER 3 | QUARTER 4 | TOTAL GRAMS |
|---|---|---|---|---|---|
| 350 | 3,590.18 | 3,842.02 | 3,726.56 | 3,801.61 | 14,960.37 |
| 351 | 2,096.48 | 2,163.26 | 2,186.44 | 2,308.21 | 8,754.39 |
| 352 | 3,008.61 | 3,090.22 | 3,186.79 | 3,188.10 | 12,473.72 |
| 354 | 339.53 | 371.63 | 408.04 | 431.07 | 1,550.27 |
| 355 | 1,598.66 | 1,711.08 | 1,757.30 | 1,846.13 | 6,913.17 |
| 356 | 2,748.27 | 2,849.65 | 2,932.20 | 2,935.10 | 11,465.22 |
| 357 | 828.11 | 851.29 | 819.45 | 861.35 | 3,360.20 |
| 358 | 1,125.79 | 1,194.48 | 1,190.28 | 1,211.59 | 4,722.14 |

| | | | | | | |
|---|---|---|---|---|---|---|
| | 359 | 2,373.75 | 2,628.52 | 2,781.55 | 2,982.75 | 10,766.57 |
| | 360 | 454.1 | 531.31 | 496.02 | 580.7 | 2,062.13 |
| | 361 | 321.54 | 345.49 | 317.34 | 356.63 | 1,341 |
| | 362 | 871.59 | 907.42 | 940.08 | 958.82 | 3,677.91 |
| | 363 | 666.87 | 685.02 | 639 | 732.59 | 2,723.48 |
| | 364 | 348.48 | 373.87 | 349.8 | 369.08 | 1,441.23 |
| | 365 | 1,580.38 | 1,725.77 | 1,816.30 | 1,810.42 | 6,932.87 |
| | 366 | 1,059.32 | 1,121.95 | 1,124.14 | 1,284.65 | 4,590.06 |
| | 367 | 189.59 | 204.02 | 202.61 | 229.15 | 825.37 |
| | 368 | 502.59 | 533.69 | 513.42 | 530.55 | 2,080.25 |
| | 369 | 31.31 | 29.86 | 26.68 | 26.9 | 114.75 |
| TOTAL | | 23,735.15 | 25,160.55 | 25,414 | 26,445.40 | 100,755.10 |

| STATE: | ALASKA | | | | | |
|---|---|---|---|---|---|---|
| ZIP CODE | | QUARTER 1 | QUARTER 2 | QUARTER 3 | QUARTER 4 | TOTAL GRAMS |
| | 995 | 982.76 | 1,078.78 | 1,089.94 | 1,127.90 | 4,279.38 |
| | 996 | 645.28 | 664.79 | 698.87 | 680.02 | 2,688.96 |
| | 997 | 174.46 | 192.36 | 212.05 | 236.12 | 814.99 |
| | 998 | 226.98 | 260.68 | 273.64 | 282.72 | 1,044.02 |
| | 999 | 148.91 | 154.95 | 156.9 | 170.07 | 630.83 |
| TOTAL | | 2,178.39 | 2,351.56 | 2,431.40 | 2,496.83 | 9,458.18 |

| STATE: | AMERICAN SAMOA | | | | | |
|---|---|---|---|---|---|---|
| ZIP CODE | | QUARTER 1 | QUARTER 2 | QUARTER 3 | QUARTER 4 | TOTAL GRAMS |
| | 967 | 0 | 0 | 0 | 0 | 0 |
| TOTAL | | 0 | 0 | 0 | 0 | 0 |

| STATE: | ARIZONA | | | | | |
|---|---|---|---|---|---|---|
| ZIP CODE | | QUARTER 1 | QUARTER 2 | QUARTER 3 | QUARTER 4 | TOTAL GRAMS |
| | 850 | 2,082.32 | 2,180.18 | 2,338.50 | 2,505.57 | 9,106.57 |
| | 851 | 547.64 | 564.59 | 610.7 | 623.33 | 2,346.26 |
| | 852 | 2,595.04 | 2,716.44 | 2,721.24 | 2,705.33 | 10,738.05 |
| | 853 | 1,353.54 | 1,437.59 | 1,484.45 | 1,550.12 | 5,825.70 |
| | 855 | 86 | 92.35 | 88.13 | 104.33 | 370.81 |

| | | | | | |
|---|---|---|---|---|---|
| | 856 | 101 | 93.16 | 93.62 | 110.01 | 397.79 |
| | 857 | 1,205.72 | 1,273.08 | 1,291.99 | 1,337.67 | 5,108.46 |
| | 859 | 120.32 | 140.16 | 135.96 | 156.5 | 552.94 |
| | 860 | 103.55 | 109.87 | 116.76 | 115.11 | 445.29 |
| | 863 | 544.1 | 521.07 | 524.33 | 559.82 | 2,149.32 |
| | 864 | 167.02 | 150.88 | 145.14 | 141.62 | 604.66 |
| | 865 | 11.16 | 9 | 16.62 | 17.64 | 54.42 |
| TOTAL | | 8,917.41 | 9,288.37 | 9,567.44 | 9,927.05 | 37,700.27 |

STATE: ARKANSAS

| ZIP CODE | | QUARTER 1 | QUARTER 2 | QUARTER 3 | QUARTER 4 | TOTAL GRAMS |
|---|---|---|---|---|---|---|
| | 716 | 93.28 | 103.97 | 105.66 | 114.9 | 417.81 |
| | 717 | 118.91 | 107.03 | 145.45 | 162.46 | 533.85 |
| | 718 | 174.81 | 204.83 | 201.05 | 205.21 | 785.9 |
| | 719 | 887.88 | 910.48 | 877.99 | 923.76 | 3,600.11 |
| | 720 | 595.46 | 648.65 | 688.57 | 714.07 | 2,646.75 |
| | 721 | 1,059.22 | 1,056.68 | 1,069.42 | 1,096.39 | 4,281.71 |
| | 722 | 593.67 | 614.78 | 612.88 | 646.75 | 2,468.08 |
| | 723 | 141.7 | 154.46 | 155.5 | 153.89 | 605.55 |
| | 724 | 688.37 | 703.64 | 737.47 | 772.93 | 2,902.41 |
| | 725 | 172.04 | 201.45 | 207.44 | 215.49 | 796.42 |
| | 726 | 225.96 | 233.33 | 264.74 | 248.3 | 972.33 |
| | 727 | 680.61 | 749.11 | 826.9 | 844.22 | 3,100.84 |
| | 728 | 120.34 | 117.28 | 131.71 | 137.09 | 506.42 |
| | 729 | 324.6 | 374.15 | 346.13 | 393.35 | 1,438.23 |
| TOTAL | | 5,876.85 | 6,179.84 | 6,370.91 | 6,628.81 | 25,056.41 |

STATE: CALIFORNIA

| ZIP CODE | | QUARTER 1 | QUARTER 2 | QUARTER 3 | QUARTER 4 | TOTAL GRAMS |
|---|---|---|---|---|---|---|
| | 900 | 888.59 | 974.69 | 1,015.17 | 979.87 | 3,858.32 |
| | 902 | 797.75 | 812.14 | 845.81 | 861.63 | 3,317.33 |
| | 903 | 27.52 | 31.63 | 25.08 | 25.42 | 109.65 |
| | 904 | 199.95 | 201.34 | 216.46 | 231.17 | 848.92 |
| | 905 | 162.08 | 172.46 | 187.53 | 196.17 | 718.24 |
| | 906 | 270.61 | 277.63 | 279.64 | 282.46 | 1,110.34 |

| | | | | | |
|---|---|---|---|---|---|
| 907 | 599.29 | 634.98 | 690.11 | 657.4 | 2,581.78 |
| 908 | 291.3 | 302.28 | 322.65 | 347.22 | 1,263.45 |
| 910 | 108.66 | 107.59 | 110.47 | 117.92 | 444.64 |
| 911 | 137.54 | 161.26 | 158.89 | 168.39 | 626.08 |
| 912 | 223.98 | 239.17 | 255.47 | 265.9 | 984.52 |
| 913 | 1,098.09 | 1,150.85 | 1,185.68 | 1,270.97 | 4,705.59 |
| 914 | 456.76 | 475.47 | 459.07 | 494.51 | 1,885.81 |
| 915 | 189.93 | 195.66 | 229.34 | 218.48 | 833.41 |
| 916 | 114.89 | 123.14 | 118.92 | 124.82 | 481.77 |
| 917 | 858.43 | 860.94 | 911.8 | 827.66 | 3,458.83 |
| 918 | 7.91 | 8.93 | 6.67 | 42.7 | 66.21 |
| 919 | 223.4 | 222.63 | 238.79 | 230.85 | 915.67 |
| 920 | 1,319.78 | 1,357.54 | 1,409.58 | 1,390.84 | 5,477.74 |
| 921 | 1,169.23 | 1,132.86 | 1,196.62 | 1,209.33 | 4,708.04 |
| 922 | 338.09 | 327.76 | 324.16 | 353 | 1,343.01 |
| 923 | 397.05 | 445.42 | 455.39 | 483.88 | 1,781.74 |
| 924 | 51.29 | 51.9 | 59.6 | 55.5 | 218.29 |
| 925 | 1,302.99 | 1,363.35 | 1,406.09 | 1,376.55 | 5,448.98 |
| 926 | 1,824.73 | 1,792.88 | 1,825.53 | 1,801.09 | 7,244.23 |
| 927 | 285.54 | 268.12 | 286.76 | 298.98 | 1,139.40 |
| 928 | 772.02 | 816.66 | 820.15 | 826.97 | 3,235.80 |
| 930 | 633.82 | 639.01 | 678.8 | 682.18 | 2,633.81 |
| 931 | 428.19 | 448.55 | 499.6 | 455.01 | 1,831.35 |
| 932 | 311.05 | 332.45 | 355.86 | 373.52 | 1,372.88 |
| 933 | 321.11 | 291.52 | 234.61 | 266.65 | 1,113.89 |
| 934 | 803.76 | 959.97 | 1,038.43 | 925.06 | 3,727.22 |
| 935 | 390.18 | 405.32 | 452.9 | 435.72 | 1,684.12 |
| 936 | 201.84 | 181.69 | 177.71 | 193.26 | 754.5 |
| 937 | 306.73 | 315.92 | 325.98 | 313.64 | 1,262.27 |
| 939 | 605.2 | 637.09 | 672.94 | 662.49 | 2,577.72 |
| 940 | 645.96 | 691.03 | 683.11 | 667.86 | 2,687.96 |
| 941 | 962.68 | 1,025.64 | 1,057.64 | 1,209.09 | 4,255.05 |
| 943 | 99.06 | 106.61 | 115.48 | 112.53 | 433.68 |
| 944 | 116.06 | 126.34 | 124.28 | 138.45 | 505.13 |
| 945 | 2,502.84 | 2,585.55 | 2,778.73 | 2,796.08 | 10,663.20 |
| 946 | 330.42 | 328.59 | 378.39 | 395.24 | 1,432.64 |
| 947 | 106.47 | 120.3 | 112.43 | 115.92 | 455.12 |

| | | | | | | |
|---|---|---|---|---|---|---|
| | 948 | 116.23 | 112.68 | 115.17 | 142.56 | 486.64 |
| | 949 | 814.79 | 831.89 | 896.05 | 913.68 | 3,456.41 |
| | 950 | 948.95 | 948.31 | 1,092.95 | 1,080.44 | 4,070.65 |
| | 951 | 584.17 | 608.18 | 643.56 | 682.27 | 2,518.18 |
| | 952 | 334.59 | 323.35 | 353.79 | 346.58 | 1,358.31 |
| | 953 | 983.36 | 956.29 | 1,022.94 | 1,038.77 | 4,001.36 |
| | 954 | 2,041.69 | 2,102.16 | 2,210.73 | 2,257.73 | 8,612.31 |
| | 955 | 1,191.25 | 1,225.70 | 1,247.48 | 1,324.02 | 4,988.45 |
| | 956 | 1,780 | 1,892.22 | 2,102.31 | 2,174.63 | 7,949.16 |
| | 957 | 268.96 | 272.54 | 287 | 306.03 | 1,134.53 |
| | 958 | 2,063.31 | 2,049.25 | 2,083.84 | 2,128.46 | 8,324.86 |
| | 959 | 1,226.82 | 1,271.57 | 1,387.07 | 1,358.43 | 5,243.89 |
| | 960 | 828.5 | 839.18 | 972.12 | 1,034.72 | 3,674.52 |
| | 961 | 118.49 | 136.45 | 154.38 | 140.92 | 550.24 |
| TOTAL | | 36,183.88 | 37,274.63 | 39,297.71 | 39,811.62 | 152,567.84 |

STATE: COLORADO

| ZIP CODE | | QUARTER 1 | QUARTER 2 | QUARTER 3 | QUARTER 4 | TOTAL GRAMS |
|---|---|---|---|---|---|---|
| | 800 | 1,167.88 | 1,205.95 | 1,262.95 | 1,234.04 | 4,870.82 |
| | 801 | 771.77 | 793.61 | 838.22 | 847.46 | 3,251.06 |
| | 802 | 1,548.32 | 1,597.83 | 1,688.52 | 1,759.52 | 6,594.19 |
| | 803 | 130.95 | 141.42 | 135.89 | 148.63 | 556.89 |
| | 804 | 133.56 | 120.27 | 143.26 | 141.21 | 538.3 |
| | 805 | 895.4 | 949.36 | 998.61 | 985.48 | 3,828.85 |
| | 806 | 404.28 | 440.61 | 450.95 | 482.38 | 1,778.22 |
| | 807 | 50.61 | 54.08 | 71.95 | 62.05 | 238.69 |
| | 808 | 71.75 | 86.18 | 92.13 | 112.8 | 362.86 |
| | 809 | 666.85 | 688.25 | 731.51 | 791.07 | 2,877.68 |
| | 810 | 504.7 | 470.8 | 468.27 | 506.86 | 1,950.63 |
| | 811 | 80.37 | 83.27 | 78.38 | 75.9 | 317.92 |
| | 812 | 104.34 | 101.65 | 97.97 | 108.25 | 412.21 |
| | 813 | 106.08 | 138.91 | 147.96 | 111.98 | 504.93 |
| | 814 | 47.47 | 57.89 | 55.95 | 49.59 | 210.9 |
| | 815 | 226.26 | 193.49 | 252.05 | 194.41 | 866.21 |
| | 816 | 118.99 | 122.09 | 103.21 | 108.64 | 452.93 |
| TOTAL | | 7,029.58 | 7,245.66 | 7,617.78 | 7,720.27 | 29,613.29 |

STATE: CONNECTICUT

| ZIP CODE | QUARTER 1 | QUARTER 2 | QUARTER 3 | QUARTER 4 | TOTAL GRAMS |
|---|---|---|---|---|---|
| 60 | 2,925.17 | 3,064.05 | 3,179.72 | 3,215.22 | 12,384.16 |
| 61 | 1,236.08 | 1,234.83 | 1,283.57 | 1,411.31 | 5,165.79 |
| 62 | 742.87 | 785.76 | 781.23 | 786.5 | 3,096.36 |
| 63 | 1,326.77 | 1,361.95 | 1,388.46 | 1,424.21 | 5,501.39 |
| 64 | 3,284.84 | 3,422.05 | 3,493.47 | 3,595.60 | 13,795.96 |
| 65 | 2,155.96 | 2,346.40 | 2,289.13 | 2,266.26 | 9,057.75 |
| 66 | 295.3 | 308.31 | 302.28 | 320.33 | 1,226.22 |
| 67 | 1,377.86 | 1,427.09 | 1,453.54 | 1,440.02 | 5,698.51 |
| 68 | 619.56 | 643.83 | 668.22 | 717.49 | 2,649.10 |
| 69 | 180.05 | 177.93 | 183.01 | 176.54 | 717.53 |
| TOTAL | 14,144.46 | 14,772.20 | 15,022.63 | 15,353.48 | 59,292.77 |

STATE: DELAWARE

| ZIP CODE | QUARTER 1 | QUARTER 2 | QUARTER 3 | QUARTER 4 | TOTAL GRAMS |
|---|---|---|---|---|---|
| 197 | 1,056.27 | 1,039.01 | 1,118.95 | 1,125.69 | 4,339.92 |
| 198 | 859.87 | 976.49 | 924.51 | 969 | 3,729.87 |
| 199 | 1,467.50 | 1,618.31 | 1,556.06 | 1,804.25 | 6,446.12 |
| TOTAL | 3,383.64 | 3,633.81 | 3,599.52 | 3,898.94 | 14,515.91 |

STATE: DISTRICT OF COLUMBIA

| ZIP CODE | QUARTER 1 | QUARTER 2 | QUARTER 3 | QUARTER 4 | TOTAL GRAMS |
|---|---|---|---|---|---|
| 200 | 2,102.20 | 2,244.83 | 2,439.47 | 2,581.05 | 9,367.55 |
| 204 | 24.36 | 20.22 | 23.6 | 18.6 | 86.78 |
| TOTAL | 2,126.56 | 2,265.05 | 2,463.07 | 2,599.65 | 9,454.33 |

STATE: FLORIDA

| ZIP CODE | QUARTER 1 | QUARTER 2 | QUARTER 3 | QUARTER 4 | TOTAL GRAMS |
|---|---|---|---|---|---|
| 320 | 2,938.69 | 2,962.34 | 3,025.39 | 3,083.04 | 12,009.46 |
| 321 | 2,191.94 | 2,255.64 | 2,268.02 | 2,354.40 | 9,070 |
| 322 | 3,623.26 | 3,719.08 | 3,854.84 | 3,892.53 | 15,089.71 |

| | | | | | | |
|---|---|---|---|---|---|---|
| | 323 | 660.3 | 682.59 | 711.28 | 695.85 | 2,750.02 |
| | 324 | 787.04 | 862.35 | 819.25 | 863.27 | 3,331.91 |
| | 325 | 1,761.12 | 1,835.73 | 2,009.74 | 2,123.82 | 7,730.41 |
| | 326 | 2,264.34 | 2,421.03 | 2,557.30 | 2,679.70 | 9,922.37 |
| | 327 | 2,510.96 | 2,566.94 | 2,383.40 | 2,634.33 | 10,095.63 |
| | 328 | 1,072.02 | 1,067.88 | 1,139.39 | 1,044.17 | 4,323.46 |
| | 329 | 2,811.95 | 2,809.95 | 2,783.95 | 2,881.07 | 11,286.92 |
| | 330 | 1,455.96 | 1,518.36 | 1,516.84 | 1,577.96 | 6,069.12 |
| | 331 | 808.19 | 837.77 | 846.82 | 858.68 | 3,351.46 |
| | 333 | 1,078.52 | 1,132.48 | 1,194.80 | 1,271.79 | 4,677.59 |
| | 334 | 3,662.76 | 3,769.31 | 3,811.03 | 3,786.07 | 15,029.17 |
| | 335 | 1,357.27 | 1,489.20 | 1,372.42 | 1,495.99 | 5,714.88 |
| | 336 | 1,241.45 | 1,303.20 | 1,429.48 | 1,377.56 | 5,351.69 |
| | 337 | 2,329.36 | 2,393.44 | 2,525.01 | 2,426.90 | 9,674.71 |
| | 338 | 804.01 | 856.22 | 865.63 | 933.85 | 3,459.71 |
| | 339 | 2,370.95 | 2,440.90 | 1,971.09 | 2,524.41 | 9,307.35 |
| | 341 | 873.52 | 854.79 | 892.39 | 924.55 | 3,545.25 |
| | 342 | 3,333.35 | 3,422.85 | 3,155.27 | 3,497.20 | 13,408.67 |
| | 344 | 1,256.23 | 1,281.51 | 1,031.64 | 1,422.50 | 4,991.88 |
| | 346 | 1,815.96 | 1,868.71 | 1,957.55 | 2,038.90 | 7,681.12 |
| | 347 | 1,054.74 | 1,093.81 | 962.55 | 1,101.32 | 4,212.42 |
| | 349 | 1,298.53 | 1,390.52 | 1,441.70 | 1,437.05 | 5,567.80 |
| TOTAL | | 45,362.42 | 46,836.60 | 46,526.78 | 48,926.91 | 187,652.71 |

STATE: GEORGIA

| ZIP CODE | | QUARTER 1 | QUARTER 2 | QUARTER 3 | QUARTER 4 | TOTAL GRAMS |
|---|---|---|---|---|---|---|
| | 300 | 2,341.68 | 2,422.71 | 2,440.27 | 2,517.73 | 9,722.39 |
| | 301 | 2,124.29 | 2,198.30 | 2,256.07 | 2,234.54 | 8,813.20 |
| | 302 | 1,322.68 | 1,391.55 | 1,418.59 | 1,430.65 | 5,563.47 |
| | 303 | 691.53 | 703.86 | 688.11 | 687.37 | 2,770.87 |
| | 304 | 739.76 | 759.55 | 737 | 769.86 | 3,006.17 |
| | 305 | 867.12 | 922.66 | 945.97 | 954.59 | 3,690.34 |
| | 306 | 742.37 | 797.53 | 843.82 | 865.75 | 3,249.47 |
| | 307 | 456.45 | 541.25 | 535.48 | 571.99 | 2,105.17 |
| | 308 | 126.52 | 129.46 | 125.6 | 123.15 | 504.73 |

| | | | | | | |
|---|---|---|---|---|---|---|
| | 309 | 266.54 | 281.24 | 319.26 | 330.43 | 1,197.47 |
| | 310 | 994.95 | 1,061.15 | 940.57 | 1,090.71 | 4,087.38 |
| | 312 | 382.35 | 413.79 | 396.32 | 402.97 | 1,595.43 |
| | 313 | 744.46 | 745.7 | 795.05 | 748.76 | 3,033.97 |
| | 314 | 775.63 | 823.84 | 847.24 | 880.42 | 3,327.13 |
| | 315 | 1,497.87 | 1,653.83 | 1,648.88 | 1,762.60 | 6,563.18 |
| | 316 | 142.4 | 160.42 | 160.76 | 163.33 | 626.91 |
| | 317 | 334.62 | 375.28 | 432.9 | 421.88 | 1,564.68 |
| | 318 | 12.59 | 14.86 | 14.87 | 18.05 | 60.37 |
| | 319 | 255.17 | 289.14 | 271.44 | 287.73 | 1,103.48 |
| | 398 | 58.1 | 62.84 | 78.4 | 86.31 | 285.65 |
| TOTAL | | 14,877.08 | 15,748.96 | 15,896.60 | 16,348.82 | 62,871.46 |

| STATE: | GUAM | | | | | |
|---|---|---|---|---|---|---|
| ZIP CODE | | QUARTER 1 | QUARTER 2 | QUARTER 3 | QUARTER 4 | TOTAL GRAMS |
| | 969 | 0.82 | 1.99 | 0 | 1.68 | 4.49 |
| TOTAL | | 0.82 | 1.99 | 0 | 1.68 | 4.49 |

| STATE: | HAWAII | | | | | |
|---|---|---|---|---|---|---|
| ZIP CODE | | QUARTER 1 | QUARTER 2 | QUARTER 3 | QUARTER 4 | TOTAL GRAMS |
| | 967 | 1,404.90 | 1,493.29 | 1,537.86 | 1,579.39 | 6,015.44 |
| | 968 | 456.04 | 493.91 | 474.99 | 438.95 | 1,863.89 |
| TOTAL | | 1,860.94 | 1,987.20 | 2,012.85 | 2,018.34 | 7,879.33 |

| STATE: | IDAHO | | | | | |
|---|---|---|---|---|---|---|
| ZIP CODE | | QUARTER 1 | QUARTER 2 | QUARTER 3 | QUARTER 4 | TOTAL GRAMS |
| | 832 | 311.54 | 346.17 | 359.26 | 372.14 | 1,389.11 |
| | 833 | 183.16 | 189.12 | 212.5 | 225.84 | 810.62 |
| | 834 | 217.7 | 215.03 | 241.79 | 218.37 | 892.89 |
| | 835 | 271.15 | 288.49 | 310.75 | 357.57 | 1,227.96 |
| | 836 | 451.79 | 489.19 | 497.88 | 600.8 | 2,039.66 |
| | 837 | 527 | 565.53 | 609.55 | 659.69 | 2,361.77 |
| | 838 | 1,291.52 | 1,361.71 | 1,400.21 | 1,440.57 | 5,494.01 |
| TOTAL | | 3,253.86 | 3,455.24 | 3,631.94 | 3,874.98 | 14,216.02 |

STATE: ILLINOIS

| ZIP CODE | QUARTER 1 | QUARTER 2 | QUARTER 3 | QUARTER 4 | TOTAL GRAMS |
|---|---|---|---|---|---|
| 600 | 2,056.66 | 2,059.97 | 2,086.86 | 2,172.58 | 8,376.07 |
| 601 | 1,082.04 | 1,133.27 | 1,137.76 | 1,107.65 | 4,460.72 |
| 602 | 28.96 | 32.39 | 34.49 | 32.59 | 128.43 |
| 603 | 115.81 | 166.3 | 179.36 | 232.28 | 693.75 |
| 604 | 1,857.06 | 1,872.27 | 1,922.98 | 1,973.91 | 7,626.22 |
| 605 | 895.95 | 926.66 | 935.19 | 984.86 | 3,742.66 |
| 606 | 1,584.50 | 1,796.30 | 1,875.74 | 1,868.72 | 7,125.26 |
| 607 | 105.92 | 111.9 | 94.69 | 102.98 | 415.49 |
| 608 | 74.76 | 79.43 | 71.46 | 72.09 | 297.74 |
| 609 | 147.47 | 162.68 | 176.39 | 179.72 | 666.26 |
| 610 | 175.19 | 183.53 | 186.55 | 187.66 | 732.93 |
| 611 | 542.69 | 579.71 | 596.21 | 608.13 | 2,326.74 |
| 612 | 185.92 | 173.51 | 182.16 | 173.77 | 715.36 |
| 613 | 243.39 | 242.5 | 265.87 | 273.36 | 1,025.12 |
| 614 | 54.22 | 61.03 | 54.73 | 62.57 | 232.55 |
| 615 | 182.05 | 192.39 | 191.36 | 207.13 | 772.93 |
| 616 | 327.13 | 382.42 | 382.38 | 386.73 | 1,478.66 |
| 617 | 115.17 | 139.73 | 135.62 | 116.93 | 507.45 |
| 618 | 727.19 | 865.14 | 806.88 | 864.77 | 3,263.98 |
| 619 | 81.33 | 83.78 | 94.74 | 88.41 | 348.26 |
| 620 | 312.54 | 372.33 | 379.49 | 366.49 | 1,430.85 |
| 622 | 267.27 | 281.47 | 290.03 | 288.58 | 1,127.35 |
| 623 | 77.8 | 100.2 | 134.08 | 148.54 | 460.62 |
| 624 | 144.5 | 165.44 | 151.54 | 144.1 | 605.58 |
| 625 | 274.56 | 274.14 | 302.38 | 325 | 1,176.08 |
| 626 | 146.74 | 175.76 | 161.17 | 170.85 | 654.52 |
| 627 | 313.16 | 357.66 | 338.12 | 336.49 | 1,345.43 |
| 628 | 300.33 | 323.27 | 355.38 | 360.11 | 1,339.09 |
| 629 | 1,014.92 | 1,084.76 | 1,145.32 | 1,206.79 | 4,451.79 |
| TOTAL | 13,435.23 | 14,379.94 | 14,668.93 | 15,043.79 | 57,527.89 |

STATE: INDIANA

| ZIP CODE | QUARTER 1 | QUARTER 2 | QUARTER 3 | QUARTER 4 | TOTAL GRAMS |
|---|---|---|---|---|---|
| 460 | 1,757.70 | 2,070.04 | 2,248.45 | 2,317.27 | 8,393.46 |
| 461 | 2,025.71 | 2,184.69 | 2,170.19 | 2,129.13 | 8,509.72 |
| 462 | 3,168.30 | 3,349.31 | 3,410.18 | 3,710.12 | 13,637.91 |
| 463 | 2,048.64 | 2,162.23 | 2,188.26 | 2,222.63 | 8,621.76 |
| 464 | 150.52 | 151.34 | 147.33 | 183.29 | 632.48 |
| 465 | 483.75 | 511.74 | 522.6 | 569.2 | 2,087.29 |
| 466 | 163.36 | 187.96 | 224.08 | 243.86 | 819.26 |
| 467 | 109.21 | 133.77 | 149.52 | 193.7 | 586.2 |
| 468 | 248.13 | 263.21 | 368.62 | 422.19 | 1,302.15 |
| 469 | 621.48 | 676.61 | 710.58 | 797 | 2,805.67 |
| 470 | 619.97 | 699.06 | 729.57 | 673.55 | 2,722.15 |
| 471 | 1,636.25 | 1,874.52 | 1,856.67 | 1,786.90 | 7,154.34 |
| 472 | 1,083.18 | 1,293.64 | 1,284.24 | 1,394.20 | 5,055.26 |
| 473 | 1,554.61 | 1,756.46 | 1,783.03 | 1,853.72 | 6,947.82 |
| 474 | 814.69 | 957.59 | 1,136.73 | 1,267.41 | 4,176.42 |
| 475 | 188.4 | 181.59 | 181.35 | 193.9 | 745.24 |
| 476 | 51.55 | 69.16 | 78.05 | 89.23 | 287.99 |
| 477 | 337.4 | 442.26 | 455.66 | 562.27 | 1,797.59 |
| 478 | 374.24 | 418.45 | 443.98 | 464.91 | 1,701.58 |
| 479 | 366.64 | 391.53 | 448.04 | 454.19 | 1,660.40 |
| TOTAL | 17,803.73 | 19,775.16 | 20,537.13 | 21,528.67 | 79,644.69 |

STATE: IOWA

| ZIP CODE | QUARTER 1 | QUARTER 2 | QUARTER 3 | QUARTER 4 | TOTAL GRAMS |
|---|---|---|---|---|---|
| 500 | 52.04 | 66.76 | 79.56 | 91.07 | 289.43 |
| 501 | 64.17 | 70.37 | 71.55 | 68.33 | 274.42 |
| 502 | 43.09 | 71.56 | 71.3 | 80.91 | 266.86 |
| 503 | 333.21 | 288.14 | 321.81 | 346.17 | 1,289.33 |
| 504 | 130.96 | 125.35 | 128 | 159.79 | 544.1 |
| 505 | 35.72 | 39.79 | 42.97 | 54.2 | 172.68 |
| 506 | 19.29 | 25.49 | 23.48 | 19.42 | 87.68 |
| 507 | 31 | 28.67 | 31.29 | 44.01 | 134.97 |
| 508 | 8.43 | 9.54 | 10.58 | 8.22 | 36.77 |
| 510 | 91.81 | 109.81 | 96.86 | 81.79 | 380.27 |
| 511 | 114.92 | 130.83 | 136.67 | 148.8 | 531.22 |

| | | | | | | |
|---|---|---|---|---|---|---|
| | 512 | 25.61 | 32.82 | 30.02 | 37.3 | 125.75 |
| | 513 | 15.7 | 16.63 | 19 | 23.53 | 74.86 |
| | 514 | 8.61 | 7.07 | 6.48 | 9.88 | 32.04 |
| | 515 | 61.85 | 61.3 | 63.56 | 71.3 | 258.01 |
| | 516 | 8.72 | 7.72 | 5.02 | 4.64 | 26.1 |
| | 520 | 66.46 | 79.06 | 63.29 | 59.36 | 268.17 |
| | 521 | 4.63 | 3.57 | 5.2 | 3.04 | 16.44 |
| | 522 | 98.91 | 96.01 | 105.21 | 107.47 | 407.6 |
| | 523 | 17.83 | 11.79 | 13.76 | 12.74 | 56.12 |
| | 524 | 32.53 | 40.31 | 51.15 | 47.59 | 171.58 |
| | 525 | 37.66 | 45.71 | 36.16 | 39.32 | 158.85 |
| | 526 | 49.42 | 42.28 | 46.08 | 49.62 | 187.4 |
| | 527 | 161.77 | 165.34 | 172.89 | 163.67 | 663.67 |
| | 528 | 71.09 | 62.28 | 67.21 | 75.42 | 276 |
| TOTAL | | 1,585.43 | 1,638.20 | 1,699.10 | 1,807.59 | 6,730.32 |

| STATE: | KANSAS | | | | | |
|---|---|---|---|---|---|---|
| ZIP CODE | | QUARTER 1 | QUARTER 2 | QUARTER 3 | QUARTER 4 | TOTAL GRAMS |
| | 660 | 526.04 | 537.35 | 584.88 | 539.6 | 2,187.87 |
| | 661 | 62.5 | 57.23 | 76.11 | 96.57 | 292.41 |
| | 662 | 642.47 | 613.23 | 627.25 | 618.65 | 2,501.60 |
| | 664 | 32.88 | 27.47 | 43.59 | 32.62 | 136.56 |
| | 665 | 45.47 | 48.31 | 44.78 | 47.31 | 185.87 |
| | 666 | 138.2 | 149.88 | 154.02 | 145.27 | 587.37 |
| | 667 | 148.07 | 156.19 | 148.59 | 159.3 | 612.15 |
| | 668 | 10.83 | 12.31 | 13.38 | 10.36 | 46.88 |
| | 669 | 4.64 | 3.08 | 1.64 | 2.91 | 12.27 |
| | 670 | 87.11 | 90.83 | 99.77 | 98.36 | 376.07 |
| | 671 | 53.7 | 48.04 | 49.77 | 56.13 | 207.64 |
| | 672 | 472.5 | 492.43 | 526.05 | 479.94 | 1,970.92 |
| | 673 | 22.95 | 28.95 | 29.09 | 30.9 | 111.89 |
| | 674 | 57.35 | 63.36 | 75.87 | 79.45 | 276.03 |
| | 675 | 56.49 | 56.05 | 58.08 | 53.19 | 223.81 |
| | 676 | 57.89 | 47.97 | 49.14 | 35.07 | 190.07 |
| | 677 | 18.28 | 16.34 | 12.1 | 6.66 | 53.38 |
| | 678 | 42.99 | 43.61 | 46.53 | 48.29 | 181.42 |

| | | | | | | |
|---|---|---|---|---|---|---|
| | 679 | 13.64 | 14.51 | 20.09 | 21.07 | 69.31 |
| TOTAL | | 2,494 | 2,507.14 | 2,660.73 | 2,561.65 | 10,223.52 |

| STATE: | KENTUCKY | | | | | |
|---|---|---|---|---|---|---|
| ZIP CODE | | QUARTER 1 | QUARTER 2 | QUARTER 3 | QUARTER 4 | TOTAL GRAMS |
| | 400 | 633.5 | 676.67 | 660.04 | 679.56 | 2,649.77 |
| | 401 | 150.6 | 164.21 | 163.43 | 204.26 | 682.5 |
| | 402 | 2,569.75 | 2,825.60 | 2,757.71 | 2,970.40 | 11,123.46 |
| | 403 | 3,967.59 | 4,047.72 | 3,933.18 | 4,080.91 | 16,029.40 |
| | 404 | 2,205.57 | 2,082.21 | 2,167.10 | 2,223.18 | 8,678.06 |
| | 405 | 2,420.38 | 2,495.06 | 2,581.75 | 2,477.94 | 9,975.13 |
| | 406 | 421.8 | 490.91 | 534.05 | 496.97 | 1,943.73 |
| | 407 | 1,881.45 | 1,993.23 | 1,951.98 | 1,789.16 | 7,615.82 |
| | 408 | 343.38 | 356.34 | 348.45 | 350.16 | 1,398.33 |
| | 409 | 1,497.38 | 1,529.47 | 1,553.31 | 1,474.05 | 6,054.21 |
| | 410 | 1,943.60 | 1,952.36 | 1,992.98 | 1,988.83 | 7,877.77 |
| | 411 | 1,720.43 | 1,727.58 | 1,683.50 | 1,633.02 | 6,764.53 |
| | 412 | 1,128.82 | 1,016.63 | 989.11 | 928.35 | 4,062.91 |
| | 413 | 801.22 | 881.43 | 879.9 | 847.98 | 3,410.53 |
| | 414 | 605.43 | 664.73 | 635.05 | 661.52 | 2,566.73 |
| | 415 | 2,513.85 | 2,509.59 | 2,521.09 | 2,609.73 | 10,154.26 |
| | 416 | 1,743.96 | 1,898.41 | 1,962.99 | 1,905.59 | 7,510.95 |
| | 417 | 1,158.63 | 1,210.07 | 1,200.07 | 1,206.15 | 4,774.92 |
| | 418 | 428.71 | 426.03 | 446.37 | 427.39 | 1,728.50 |
| | 420 | 948.76 | 1,094.63 | 1,158.88 | 1,245.17 | 4,447.44 |
| | 421 | 880.69 | 900.52 | 876.63 | 1,041.86 | 3,699.70 |
| | 422 | 243.13 | 254.38 | 242.75 | 243.8 | 984.06 |
| | 423 | 633.68 | 634.34 | 624.89 | 603.24 | 2,496.15 |
| | 424 | 313.02 | 334.9 | 338.22 | 338.34 | 1,324.48 |
| | 425 | 638.01 | 659.06 | 559.11 | 680.63 | 2,536.81 |
| | 426 | 887.5 | 788.75 | 775.33 | 757.23 | 3,208.81 |
| | 427 | 1,067.49 | 1,135.39 | 1,107.50 | 1,073.27 | 4,383.65 |
| TOTAL | | 33,748.33 | 34,750.22 | 34,645.37 | 34,938.69 | 138,082.61 |

STATE: LOUISIANA

| ZIP CODE | QUARTER 1 | QUARTER 2 | QUARTER 3 | QUARTER 4 | TOTAL GRAMS |
|---|---|---|---|---|---|
| 700 | 3,747.24 | 3,841.59 | 3,791.32 | 3,855.43 | 15,235.58 |
| 701 | 840.18 | 912.17 | 941.37 | 923.26 | 3,616.98 |
| 703 | 1,000.15 | 1,026.18 | 1,031.13 | 1,078.13 | 4,135.59 |
| 704 | 3,161.41 | 3,367.36 | 3,392.93 | 3,437.19 | 13,358.89 |
| 705 | 2,375.89 | 2,498.72 | 2,496.86 | 2,454.58 | 9,826.05 |
| 706 | 1,103.66 | 1,163.91 | 1,200.92 | 1,228.32 | 4,696.81 |
| 707 | 1,695.56 | 1,710.52 | 1,704.56 | 1,743.44 | 6,854.08 |
| 708 | 790.98 | 812.35 | 903.87 | 928 | 3,435.20 |
| 710 | 397.68 | 421.28 | 453.31 | 277.29 | 1,549.56 |
| 711 | 1,170.92 | 1,254.93 | 1,756.59 | 1,619.34 | 5,801.78 |
| 712 | 1,025.20 | 1,089.61 | 1,125.05 | 1,192.43 | 4,432.29 |
| 713 | 422.05 | 494.83 | 555.3 | 530.19 | 2,002.37 |
| 714 | 323.75 | 360.09 | 329.78 | 355.2 | 1,368.82 |
| TOTAL | 18,054.67 | 18,953.54 | 19,682.99 | 19,622.80 | 76,314 |

STATE: MAINE

| ZIP CODE | QUARTER 1 | QUARTER 2 | QUARTER 3 | QUARTER 4 | TOTAL GRAMS |
|---|---|---|---|---|---|
| 39 | 60.31 | 50.43 | 65.4 | 64.07 | 240.21 |
| 40 | 2,080.86 | 2,254.33 | 2,336.51 | 2,369.53 | 9,041.23 |
| 41 | 954.3 | 1,038.21 | 1,019.96 | 1,010.78 | 4,023.25 |
| 42 | 1,268.94 | 1,367.69 | 1,424.18 | 1,497.23 | 5,558.04 |
| 43 | 684.32 | 771.05 | 798.29 | 797.38 | 3,051.04 |
| 44 | 1,019.93 | 1,035.60 | 1,076.11 | 1,065.52 | 4,197.16 |
| 45 | 331.7 | 373.59 | 352.04 | 383.19 | 1,440.52 |
| 46 | 498.37 | 585.37 | 557.02 | 643.78 | 2,284.54 |
| 47 | 404.79 | 404.17 | 353.34 | 476.24 | 1,638.54 |
| 48 | 660.96 | 698.3 | 748.38 | 715.17 | 2,822.81 |
| 49 | 1,007.41 | 1,094.34 | 1,142.41 | 1,178.88 | 4,423.04 |
| TOTAL | 8,971.89 | 9,673.08 | 9,873.64 | 10,201.77 | 38,720.38 |

STATE: MARYLAND

| ZIP CODE | QUARTER 1 | QUARTER 2 | QUARTER 3 | QUARTER 4 | TOTAL GRAMS |
|---|---|---|---|---|---|
| 206 | 1,469.67 | 1,530.06 | 1,642.12 | 1,721.58 | 6,363.43 |
| 207 | 1,012.23 | 1,001.59 | 945.43 | 1,024.04 | 3,983.29 |

| | | | | | | |
|---|---|---|---|---|---|---|
| | 208 | 1,070.75 | 1,107.72 | 1,082.61 | 993.33 | 4,254.41 |
| | 209 | 371.2 | 426.75 | 461.22 | 425.26 | 1,684.43 |
| | 210 | 3,112.32 | 3,254.68 | 3,242.65 | 3,368.21 | 12,977.86 |
| | 211 | 1,541.36 | 1,643.58 | 1,458.87 | 1,607.15 | 6,250.96 |
| | 212 | 9,853.75 | 10,290.05 | 10,700.69 | 11,278.59 | 42,123.08 |
| | 214 | 283.29 | 368.55 | 445.83 | 504.87 | 1,602.54 |
| | 215 | 988.1 | 1,060.60 | 1,020.37 | 1,179.93 | 4,249 |
| | 216 | 743.62 | 751.98 | 747.91 | 752.22 | 2,995.73 |
| | 217 | 1,800.43 | 2,042.64 | 2,047.48 | 2,120.25 | 8,010.80 |
| | 218 | 783.49 | 666.73 | 722.2 | 745.74 | 2,918.16 |
| | 219 | 1,094.86 | 1,121.67 | 1,155.78 | 1,276.76 | 4,649.07 |
| TOTAL | | 24,125.07 | 25,266.60 | 25,673.16 | 26,997.93 | 102,062.76 |

STATE: MASSACHUSETTS

| ZIP CODE | | QUARTER 1 | QUARTER 2 | QUARTER 3 | QUARTER 4 | TOTAL GRAMS |
|---|---|---|---|---|---|---|
| | 10 | 3,355.85 | 3,502.12 | 3,457.57 | 3,673.98 | 13,989.52 |
| | 11 | 1,250.64 | 1,288.46 | 1,226.45 | 1,281.12 | 5,046.67 |
| | 12 | 1,145.41 | 1,228.69 | 1,201.43 | 1,250.32 | 4,825.85 |
| | 13 | 852.87 | 918.88 | 913.18 | 958.57 | 3,643.50 |
| | 14 | 1,030.71 | 1,086.07 | 1,052.92 | 1,146.57 | 4,316.27 |
| | 15 | 1,404.40 | 1,414.52 | 1,389.58 | 1,434.63 | 5,643.13 |
| | 16 | 2,316.50 | 2,438.45 | 2,291.32 | 2,165.17 | 9,211.44 |
| | 17 | 1,376.40 | 1,498.34 | 1,265.70 | 1,417.62 | 5,558.06 |
| | 18 | 3,250.59 | 3,462.23 | 3,359.08 | 3,406.36 | 13,478.26 |
| | 19 | 2,638.99 | 2,904.16 | 2,700.20 | 2,927.67 | 11,171.02 |
| | 20 | 1,572.51 | 1,665.54 | 1,558.46 | 1,632.85 | 6,429.36 |
| | 21 | 8,408.79 | 9,024.98 | 8,648.17 | 8,982.41 | 35,064.35 |
| | 22 | 89.08 | 84.99 | 79.33 | 87.54 | 340.94 |
| | 23 | 2,783.86 | 3,058.29 | 2,992.61 | 3,061.08 | 11,895.84 |
| | 24 | 1,050.49 | 1,142.48 | 1,021.21 | 1,048.30 | 4,262.48 |
| | 25 | 941.81 | 1,022.98 | 1,027.65 | 1,017.58 | 4,010.02 |
| | 26 | 1,269.48 | 1,320.72 | 1,289.70 | 1,368.83 | 5,248.73 |
| | 27 | 5,067.11 | 5,477.60 | 5,281.21 | 5,351.97 | 21,177.89 |
| TOTAL | | 39,805.49 | 42,539.50 | 40,755.77 | 42,212.57 | 165,313.33 |

STATE: MICHIGAN

| ZIP CODE | QUARTER 1 | QUARTER 2 | QUARTER 3 | QUARTER 4 | TOTAL GRAMS |
|---|---|---|---|---|---|
| 480 | 3,763.09 | 4,064.16 | 4,115.63 | 4,229.83 | 16,172.71 |
| 481 | 7,314.58 | 7,522.53 | 7,297.38 | 7,857.04 | 29,991.53 |
| 482 | 866.01 | 923.44 | 946.96 | 995.12 | 3,731.53 |
| 483 | 2,814.45 | 2,943.16 | 2,989.52 | 3,100.54 | 11,847.67 |
| 484 | 870.78 | 869.05 | 897.4 | 932.36 | 3,569.59 |
| 485 | 524.6 | 554.67 | 546.36 | 579.7 | 2,205.33 |
| 486 | 937.15 | 959.58 | 999.49 | 1,010.86 | 3,907.08 |
| 487 | 620.14 | 615.5 | 633.87 | 645.29 | 2,514.80 |
| 488 | 1,002.55 | 1,086.41 | 1,106.97 | 1,192.70 | 4,388.63 |
| 489 | 212.48 | 226.39 | 216.96 | 214.36 | 870.19 |
| 490 | 1,245.58 | 1,339.44 | 1,410.67 | 1,443.95 | 5,439.64 |
| 491 | 95.38 | 95.95 | 98.46 | 94.09 | 383.88 |
| 492 | 715.75 | 756.47 | 781.7 | 802.93 | 3,056.85 |
| 493 | 271.93 | 327.47 | 388.12 | 383.07 | 1,370.59 |
| 494 | 680.42 | 725.22 | 752.21 | 799.98 | 2,957.83 |
| 495 | 783.79 | 846.32 | 842.96 | 960.43 | 3,433.50 |
| 496 | 1,124.77 | 1,148.40 | 1,182.64 | 1,245.63 | 4,701.44 |
| 497 | 1,029.11 | 1,098.86 | 1,110.01 | 1,174.54 | 4,412.52 |
| 498 | 740.9 | 877.81 | 826.93 | 869.08 | 3,314.72 |
| 499 | 97.42 | 83.67 | 92.11 | 101.34 | 374.54 |
| TOTAL | 25,710.88 | 27,064.50 | 27,236.35 | 28,632.84 | 108,644.57 |

STATE: MINNESOTA

| ZIP CODE | QUARTER 1 | QUARTER 2 | QUARTER 3 | QUARTER 4 | TOTAL GRAMS |
|---|---|---|---|---|---|
| 550 | 869.67 | 889.52 | 905.35 | 948.12 | 3,612.66 |
| 551 | 1,238.48 | 1,354.29 | 1,435.65 | 1,417.28 | 5,445.70 |
| 553 | 858.4 | 933.26 | 982.85 | 1,007.94 | 3,782.45 |
| 554 | 1,828.70 | 1,929.99 | 2,036.31 | 2,054.70 | 7,849.70 |
| 556 | 11.8 | 11.53 | 11.29 | 17.32 | 51.94 |
| 557 | 180.3 | 188.8 | 177.78 | 197.3 | 744.18 |
| 558 | 318.85 | 330.84 | 340.5 | 364.8 | 1,354.99 |
| 559 | 223.82 | 227.43 | 226.59 | 227.37 | 905.21 |
| 560 | 182.76 | 206.73 | 220.71 | 238.68 | 848.88 |
| 561 | 5.63 | 8.17 | 10.2 | 11.08 | 35.08 |

| | | | | | | |
|---|---|---|---|---|---|---|
| | 562 | 42.68 | 56.41 | 53.73 | 58.58 | 211.4 |
| | 563 | 296.3 | 294.17 | 300.84 | 320.93 | 1,212.24 |
| | 564 | 126.53 | 139.99 | 147.37 | 145.72 | 559.61 |
| | 565 | 254.66 | 256.64 | 282.08 | 336.42 | 1,129.80 |
| | 566 | 497.17 | 536.89 | 525.89 | 592.58 | 2,152.53 |
| | 567 | 63.69 | 80.23 | 90.99 | 83.45 | 318.36 |
| TOTAL | | 6,999.44 | 7,444.89 | 7,748.13 | 8,022.27 | 30,214.73 |

| STATE: | MISSISSIPPI | | | | | |
|---|---|---|---|---|---|---|
| ZIP CODE | | QUARTER 1 | QUARTER 2 | QUARTER 3 | QUARTER 4 | TOTAL GRAMS |
| | 386 | 1,199.56 | 1,232.65 | 1,272.76 | 1,269.65 | 4,974.62 |
| | 387 | 72.74 | 89.75 | 88.75 | 95.34 | 346.58 |
| | 388 | 1,572.48 | 1,663.10 | 1,602.68 | 1,548.35 | 6,386.61 |
| | 389 | 133.78 | 139.74 | 139.45 | 131.82 | 544.79 |
| | 390 | 620.92 | 628.51 | 623.44 | 640.26 | 2,513.13 |
| | 391 | 702.92 | 694.32 | 727.54 | 710.61 | 2,835.39 |
| | 392 | 533.79 | 518.73 | 559.78 | 518.37 | 2,130.67 |
| | 393 | 470.53 | 484.86 | 476.79 | 492.74 | 1,924.92 |
| | 394 | 1,706.85 | 1,720.82 | 1,701.03 | 1,706.46 | 6,835.16 |
| | 395 | 2,239.04 | 2,392.86 | 2,366.57 | 2,505.14 | 9,503.61 |
| | 396 | 274.72 | 267.57 | 248.61 | 250 | 1,040.90 |
| | 397 | 162.43 | 162.94 | 154.93 | 155.47 | 635.77 |
| TOTAL | | 9,689.76 | 9,995.85 | 9,962.33 | 10,024.21 | 39,672.15 |

| STATE: | MISSOURI | | | | | |
|---|---|---|---|---|---|---|
| ZIP CODE | | QUARTER 1 | QUARTER 2 | QUARTER 3 | QUARTER 4 | TOTAL GRAMS |
| | 630 | 971.07 | 973.65 | 1,019.08 | 1,036.50 | 4,000.30 |
| | 631 | 1,493.22 | 1,520.94 | 1,606.36 | 1,907.76 | 6,528.28 |
| | 633 | 512.37 | 557.1 | 572.95 | 626.44 | 2,268.86 |
| | 634 | 60.8 | 61 | 65.06 | 77.14 | 264 |
| | 635 | 19.33 | 25.95 | 24.85 | 30.54 | 100.67 |
| | 636 | 420.61 | 454.89 | 527.16 | 654.52 | 2,057.18 |
| | 637 | 95.55 | 120.84 | 100.34 | 156.2 | 472.93 |
| | 638 | 165.58 | 164.99 | 174.01 | 198.83 | 703.41 |
| | 639 | 121.6 | 168.58 | 173.06 | 201.79 | 665.03 |

| | | | | | |
|---|---|---|---|---|---|
| 640 | 666 | 680 | 717.09 | 724.52 | 2,787.61 |
| 641 | 619.13 | 597.05 | 658.25 | 663.64 | 2,538.07 |
| 644 | 56.75 | 61.72 | 59.17 | 57.59 | 235.23 |
| 645 | 236.02 | 242.47 | 253.65 | 252.74 | 984.88 |
| 646 | 20.97 | 23.66 | 31.43 | 26.43 | 102.49 |
| 647 | 82.31 | 93.63 | 94.8 | 107.82 | 378.56 |
| 648 | 178.46 | 182.54 | 208.45 | 229.43 | 798.88 |
| 650 | 138.78 | 168.96 | 181.7 | 180.43 | 669.87 |
| 651 | 109.28 | 115.16 | 117.27 | 117.73 | 459.44 |
| 652 | 448.27 | 486.94 | 522.08 | 585.12 | 2,042.41 |
| 653 | 78.34 | 90.95 | 112.1 | 139.47 | 420.86 |
| 654 | 109.86 | 124.16 | 142.86 | 159.33 | 536.21 |
| 655 | 150.76 | 176.91 | 183.81 | 199.92 | 711.4 |
| 656 | 211.36 | 194.95 | 212.77 | 218.85 | 837.93 |
| 657 | 301.81 | 348.59 | 379.48 | 420.47 | 1,450.35 |
| 658 | 1,362.62 | 1,572.75 | 1,566.27 | 1,710.17 | 6,211.81 |
| TOTAL | 8,630.85 | 9,208.38 | 9,704.05 | 10,683.38 | 38,226.66 |

STATE: MONTANA

| ZIP CODE | QUARTER 1 | QUARTER 2 | QUARTER 3 | QUARTER 4 | TOTAL GRAMS |
|---|---|---|---|---|---|
| 590 | 45.91 | 52.39 | 73.82 | 86.64 | 258.76 |
| 591 | 233.1 | 274.19 | 301.87 | 329.41 | 1,138.57 |
| 592 | 66.67 | 79.12 | 86.01 | 92.4 | 324.2 |
| 593 | 30.25 | 29.96 | 34.77 | 35.31 | 130.29 |
| 594 | 610.89 | 617.49 | 663.74 | 679.32 | 2,571.44 |
| 595 | 22.51 | 28.38 | 21.95 | 26.6 | 99.44 |
| 596 | 239.54 | 256.63 | 281.31 | 277.81 | 1,055.29 |
| 597 | 239.54 | 252.08 | 248.65 | 265.97 | 1,006.24 |
| 598 | 517.16 | 536.88 | 570.79 | 603.79 | 2,228.62 |
| 599 | 214.79 | 188.82 | 213.01 | 222.69 | 839.31 |
| TOTAL | 2,220.36 | 2,315.94 | 2,495.92 | 2,619.94 | 9,652.16 |

STATE: NEBRASKA

| ZIP CODE | QUARTER 1 | QUARTER 2 | QUARTER 3 | QUARTER 4 | TOTAL GRAMS |
|---|---|---|---|---|---|
| 680 | 69.86 | 68.47 | 67.84 | 77.19 | 283.36 |

| | | | | | | |
|---|---|---|---|---|---|---|
| | 681 | 500.32 | 538.6 | 565.76 | 556.4 | 2,161.08 |
| | 683 | 44.14 | 40.27 | 35.6 | 35.87 | 155.88 |
| | 684 | 22.12 | 16.8 | 24.23 | 19.28 | 82.43 |
| | 685 | 282.59 | 292.41 | 291.43 | 285.4 | 1,151.83 |
| | 686 | 30.07 | 29.81 | 28.97 | 31.96 | 120.81 |
| | 687 | 57.28 | 61.92 | 63.98 | 65.53 | 248.71 |
| | 688 | 69.72 | 74.64 | 87.54 | 98.78 | 330.68 |
| | 689 | 26.78 | 20.39 | 23.18 | 27.22 | 97.57 |
| | 690 | 1.82 | 3.54 | 3.75 | 5.2 | 14.31 |
| | 691 | 133.65 | 138.16 | 147.6 | 145.88 | 565.29 |
| | 692 | 1.32 | 0.4 | 0.52 | 1.18 | 3.42 |
| | 693 | 24.86 | 23.1 | 23.16 | 22.33 | 93.45 |
| TOTAL | | 1,264.53 | 1,308.51 | 1,363.56 | 1,372.22 | 5,308.82 |

| STATE: | NEVADA | | | | | |
|---|---|---|---|---|---|---|
| ZIP CODE | | QUARTER 1 | QUARTER 2 | QUARTER 3 | QUARTER 4 | TOTAL GRAMS |
| | 890 | 698.46 | 718.9 | 730.33 | 672.3 | 2,819.99 |
| | 891 | 1,511.85 | 1,700.85 | 1,607.98 | 1,679.90 | 6,500.58 |
| | 893 | 12.16 | 13 | 15.7 | 9.8 | 50.66 |
| | 894 | 276.97 | 307.25 | 322.35 | 324.73 | 1,231.30 |
| | 895 | 568.74 | 572.52 | 599.54 | 580.73 | 2,321.53 |
| | 897 | 104.65 | 100.72 | 110.37 | 131.86 | 447.6 |
| | 898 | 65.96 | 63.31 | 74.96 | 69.32 | 273.55 |
| TOTAL | | 3,238.79 | 3,476.55 | 3,461.23 | 3,468.64 | 13,645.21 |

| STATE: | NEW HAMPSHIRE | | | | | |
|---|---|---|---|---|---|---|
| ZIP CODE | | QUARTER 1 | QUARTER 2 | QUARTER 3 | QUARTER 4 | TOTAL GRAMS |
| | 030 | 1,225.17 | 1,262.65 | 1,307.97 | 1,293.36 | 5,089.15 |
| | 031 | 1,171.92 | 1,214.56 | 1,233.18 | 1,224.56 | 4,844.22 |
| | 032 | 1,100.97 | 1,182.68 | 1,258.64 | 1,275.86 | 4,818.15 |
| | 033 | 732.74 | 821.49 | 884.52 | 869.76 | 3,308.51 |
| | 034 | 320.91 | 325.95 | 305.01 | 320.16 | 1,272.03 |
| | 035 | 329.29 | 322.69 | 322.03 | 290.94 | 1,264.95 |
| | 036 | 9.9 | 11.8 | 10.32 | 9.6 | 41.62 |
| | 037 | 1,151.18 | 1,189.73 | 1,182.09 | 1,141.71 | 4,664.71 |

| | | | | | | |
|---|---|---|---|---|---|---|
| | 038 | 1,628.51 | 1,727.09 | 1,709.70 | 1,724.62 | 6,789.92 |
| TOTAL | | 7,670.59 | 8,058.64 | 8,213.46 | 8,150.57 | 32,093.26 |

STATE: NEW JERSEY

| ZIP CODE | | QUARTER 1 | QUARTER 2 | QUARTER 3 | QUARTER 4 | TOTAL GRAMS |
|---|---|---|---|---|---|---|
| | 070 | 2,748.49 | 2,841.99 | 2,833.38 | 2,929.92 | 11,353.78 |
| | 071 | 918.78 | 895.06 | 898.32 | 900.99 | 3,613.15 |
| | 072 | 237.79 | 241.53 | 228.85 | 228.18 | 936.35 |
| | 073 | 163.96 | 162.25 | 166.02 | 166.28 | 658.51 |
| | 074 | 1,142.80 | 1,200.50 | 1,180.73 | 1,173.57 | 4,697.60 |
| | 075 | 285.92 | 255.78 | 260.5 | 266.03 | 1,068.23 |
| | 076 | 774.7 | 829.61 | 833.13 | 848.84 | 3,286.28 |
| | 077 | 1,989.24 | 2,115.79 | 2,089.15 | 2,082.89 | 8,277.07 |
| | 078 | 748.19 | 852.24 | 872.25 | 839.01 | 3,311.69 |
| | 079 | 355.89 | 367.03 | 365.27 | 352.64 | 1,440.83 |
| | 080 | 6,913.98 | 7,289.78 | 6,757.97 | 7,260.17 | 28,221.90 |
| | 081 | 725.13 | 833.13 | 849.85 | 886.58 | 3,294.69 |
| | 082 | 1,738.13 | 1,871.58 | 1,869.34 | 1,842.11 | 7,321.16 |
| | 083 | 965.14 | 1,055.30 | 996.24 | 1,040.67 | 4,057.35 |
| | 084 | 223.26 | 234.34 | 237.72 | 251.48 | 946.8 |
| | 085 | 544.89 | 589.02 | 575.93 | 594.52 | 2,304.36 |
| | 086 | 737.23 | 743.25 | 709.36 | 730 | 2,919.84 |
| | 087 | 2,438.20 | 2,565.54 | 2,517.35 | 2,592.46 | 10,113.55 |
| | 088 | 1,710.24 | 1,813.31 | 1,736.38 | 1,788.81 | 7,048.74 |
| | 089 | 185.27 | 164.77 | 169.28 | 161.61 | 680.93 |
| TOTAL | | 25,547.23 | 26,921.80 | 26,147.02 | 26,936.76 | 105,552.81 |

STATE: NEW MEXICO

| ZIP CODE | | QUARTER 1 | QUARTER 2 | QUARTER 3 | QUARTER 4 | TOTAL GRAMS |
|---|---|---|---|---|---|---|
| | 870 | 352.15 | 391.12 | 395.09 | 405.03 | 1,543.39 |
| | 871 | 3,307.59 | 3,483.25 | 3,593.10 | 3,693.80 | 14,077.74 |
| | 873 | 100.93 | 117.55 | 136.67 | 116.24 | 471.39 |
| | 874 | 298.3 | 419.2 | 418.89 | 330.13 | 1,466.52 |
| | 875 | 1,706.49 | 1,715.30 | 1,909.64 | 2,086.75 | 7,418.18 |
| | 877 | 156.02 | 179.98 | 199.17 | 198.93 | 734.1 |

| | | | | | | |
|---|---|---|---|---|---|---|
| | 878 | 50.97 | 54.5 | 60.53 | 66.75 | 232.75 |
| | 879 | 12.9 | 16.56 | 11.34 | 13 | 53.8 |
| | 880 | 364.08 | 362.93 | 349.76 | 372.4 | 1,449.17 |
| | 881 | 22.89 | 26.88 | 35.54 | 47.6 | 132.91 |
| | 882 | 358.86 | 365.86 | 384.53 | 388.56 | 1,497.81 |
| | 883 | 89.24 | 97.28 | 100.45 | 113.07 | 400.04 |
| | 884 | 5.5 | 3.54 | 4.02 | 4.74 | 17.8 |
| TOTAL | | 6,825.92 | 7,233.95 | 7,598.73 | 7,837 | 29,495.60 |

| STATE: | NEW YORK | | | | | |
|---|---|---|---|---|---|---|
| ZIP CODE | | QUARTER 1 | QUARTER 2 | QUARTER 3 | QUARTER 4 | TOTAL GRAMS |
| | 100 | 2,591.90 | 2,494.75 | 2,612.07 | 2,725 | 10,423.72 |
| | 101 | 95.14 | 93.36 | 70.85 | 68.81 | 328.16 |
| | 102 | 0.7 | 3.3 | 3.53 | 1.74 | 9.27 |
| | 103 | 1,744.13 | 1,811.78 | 1,797.04 | 1,865 | 7,217.95 |
| | 104 | 2,295.54 | 2,311.36 | 2,491.87 | 2,493.44 | 9,592.21 |
| | 105 | 1,030.18 | 1,078.82 | 1,074.92 | 1,113.84 | 4,297.76 |
| | 106 | 234.68 | 219.81 | 229.13 | 243.8 | 927.42 |
| | 107 | 348.99 | 363.92 | 387.05 | 389.35 | 1,489.31 |
| | 108 | 102.41 | 99.57 | 98.29 | 110.52 | 410.79 |
| | 109 | 1,383.19 | 1,397.52 | 1,414.29 | 1,492.48 | 5,687.48 |
| | 110 | 480.92 | 526.07 | 531.74 | 567.91 | 2,106.64 |
| | 111 | 249.9 | 253.64 | 277.48 | 279.47 | 1,060.49 |
| | 112 | 2,919.72 | 2,939.97 | 2,876.76 | 2,964.02 | 11,700.47 |
| | 113 | 1,113.91 | 1,115.39 | 1,086.89 | 1,081.46 | 4,397.65 |
| | 114 | 488.97 | 515.4 | 529.32 | 515.5 | 2,049.19 |
| | 115 | 1,260.28 | 1,399.72 | 1,380.04 | 1,444.35 | 5,484.39 |
| | 116 | 98.64 | 106.64 | 104.13 | 118.08 | 427.49 |
| | 117 | 5,730.90 | 5,875.66 | 5,906.63 | 6,121.60 | 23,634.79 |
| | 118 | 470.49 | 519.35 | 486.5 | 531.57 | 2,007.91 |
| | 119 | 1,143.35 | 1,190.61 | 1,190.69 | 1,207.88 | 4,732.53 |
| | 120 | 1,575.11 | 1,752.77 | 1,743.70 | 1,915.45 | 6,987.03 |
| | 121 | 828.96 | 869.37 | 896.34 | 930.03 | 3,524.70 |
| | 122 | 616.47 | 633.66 | 675.19 | 758.58 | 2,683.90 |
| | 123 | 600.43 | 649.35 | 648.67 | 684.86 | 2,583.31 |
| | 124 | 741.23 | 782.99 | 842.02 | 891.54 | 3,257.78 |

| | | | | | | |
|---|---|---|---|---|---|---|
| | 125 | 1,268.07 | 1,355.59 | 1,400.70 | 1,545.14 | 5,569.50 |
| | 126 | 282.09 | 324.18 | 318.35 | 319.8 | 1,244.42 |
| | 127 | 787.86 | 827.66 | 844.21 | 834.48 | 3,294.21 |
| | 128 | 1,297.11 | 1,442.06 | 1,515.68 | 1,530.12 | 5,784.97 |
| | 129 | 964.29 | 1,097.27 | 1,251.91 | 1,335.78 | 4,649.25 |
| | 130 | 981.74 | 1,049.82 | 1,087.46 | 1,112.31 | 4,231.33 |
| | 131 | 332.17 | 346.6 | 343.04 | 364.16 | 1,385.97 |
| | 132 | 876.26 | 910.81 | 927.86 | 1,032.47 | 3,747.40 |
| | 133 | 313.77 | 316.84 | 269.57 | 325.32 | 1,225.50 |
| | 134 | 640.29 | 636.95 | 620.14 | 727.9 | 2,625.28 |
| | 135 | 348.97 | 355.03 | 367.14 | 396.44 | 1,467.58 |
| | 136 | 824.09 | 856.29 | 804.26 | 941.2 | 3,425.84 |
| | 137 | 274.6 | 325.15 | 335.72 | 363.4 | 1,298.87 |
| | 138 | 652.56 | 744.01 | 769.87 | 801.97 | 2,968.41 |
| | 139 | 332.87 | 351.16 | 363.41 | 412.97 | 1,460.41 |
| | 140 | 2,142.17 | 2,244.12 | 2,262.44 | 2,455.53 | 9,104.26 |
| | 141 | 956.52 | 1,014.60 | 1,025.88 | 1,013.12 | 4,010.12 |
| | 142 | 3,107.29 | 3,319.83 | 3,321.17 | 3,515.09 | 13,263.38 |
| | 143 | 657.42 | 687.56 | 680.05 | 823.68 | 2,848.71 |
| | 144 | 853.79 | 868.34 | 875.65 | 914.52 | 3,512.30 |
| | 145 | 612.11 | 637.06 | 662.48 | 692.17 | 2,603.82 |
| | 146 | 2,067.97 | 2,099.29 | 2,159.70 | 2,247.84 | 8,574.80 |
| | 147 | 721.9 | 790.45 | 763.28 | 792.74 | 3,068.37 |
| | 148 | 832.44 | 948.04 | 980.05 | 1,195.15 | 3,955.68 |
| | 149 | 213.6 | 251.13 | 267.02 | 283.16 | 1,014.91 |
| TOTAL | | 50,488.09 | 52,804.62 | 53,572.18 | 56,492.74 | 213,357.63 |

STATE: NORTH CAROLINA

| ZIP CODE | | QUARTER 1 | QUARTER 2 | QUARTER 3 | QUARTER 4 | TOTAL GRAMS |
|---|---|---|---|---|---|---|
| | 270 | 570.24 | 597.14 | 614.8 | 647.98 | 2,430.16 |
| | 271 | 597.07 | 645.91 | 657.33 | 714.32 | 2,614.63 |
| | 272 | 1,235.66 | 1,270.70 | 1,245.30 | 1,474.48 | 5,226.14 |
| | 273 | 912.87 | 989.92 | 928.51 | 1,043.23 | 3,874.53 |
| | 274 | 664.28 | 668.94 | 688.03 | 680.41 | 2,701.66 |
| | 275 | 1,947.11 | 2,078.72 | 2,182.74 | 2,325.21 | 8,533.78 |
| | 276 | 1,076.26 | 1,048.23 | 1,077.71 | 1,135.93 | 4,338.13 |

| | | | | | | |
|---|---|---|---|---|---|---|
| | 277 | 1,534.63 | 1,647.57 | 1,443.65 | 1,631.72 | 6,257.57 |
| | 278 | 873.03 | 983.75 | 1,080.28 | 1,215.77 | 4,152.83 |
| | 279 | 472.96 | 490.54 | 490.54 | 486.53 | 1,940.57 |
| | 280 | 2,614.36 | 2,812.75 | 2,419.40 | 2,993.74 | 10,840.25 |
| | 281 | 1,746.46 | 1,925.11 | 1,768.78 | 2,211.83 | 7,652.18 |
| | 282 | 1,445.13 | 1,638.39 | 1,540.10 | 1,573.98 | 6,197.60 |
| | 283 | 2,290.01 | 2,596.70 | 2,527.54 | 2,883.36 | 10,297.61 |
| | 284 | 3,163.23 | 3,525.43 | 3,366.97 | 3,524.62 | 13,580.25 |
| | 285 | 1,184.63 | 1,177.88 | 1,112.38 | 1,208.44 | 4,683.33 |
| | 286 | 4,411.92 | 4,834.07 | 4,793.25 | 4,976.57 | 19,015.81 |
| | 287 | 2,128.76 | 2,328.73 | 2,275.51 | 2,448.30 | 9,181.30 |
| | 288 | 1,320.56 | 1,629.09 | 1,684.93 | 1,916.62 | 6,551.20 |
| | 289 | 176.05 | 185.06 | 194.67 | 216.79 | 772.57 |
| TOTAL | | 30,365.22 | 33,074.63 | 32,092.42 | 35,309.83 | 130,842.10 |

STATE: NORTH DAKOTA

| ZIP CODE | | QUARTER 1 | QUARTER 2 | QUARTER 3 | QUARTER 4 | TOTAL GRAMS |
|---|---|---|---|---|---|---|
| | 580 | 3.48 | 6.95 | 12.42 | 15.03 | 37.88 |
| | 581 | 269.53 | 268.66 | 269.24 | 267.17 | 1,074.60 |
| | 582 | 42.48 | 49.06 | 50.84 | 48.47 | 190.85 |
| | 583 | 41.76 | 46.56 | 59.94 | 59.22 | 207.48 |
| | 584 | 12.72 | 13.87 | 14.97 | 18.43 | 59.99 |
| | 585 | 341.98 | 343.82 | 380.1 | 376.15 | 1,442.05 |
| | 586 | 39.68 | 44.2 | 44.04 | 50.78 | 178.7 |
| | 587 | 35.3 | 41.9 | 42.93 | 44.74 | 164.87 |
| | 588 | 65.02 | 68.11 | 69.51 | 76.28 | 278.92 |
| TOTAL | | 851.95 | 883.13 | 943.99 | 956.27 | 3,635.34 |

STATE: OHIO

| ZIP CODE | | QUARTER 1 | QUARTER 2 | QUARTER 3 | QUARTER 4 | TOTAL GRAMS |
|---|---|---|---|---|---|---|
| | 430 | 3,070.56 | 3,398.57 | 3,246.23 | 3,329.23 | 13,044.59 |
| | 431 | 2,469.57 | 2,587.06 | 2,556.69 | 2,595.27 | 10,208.59 |
| | 432 | 5,064.78 | 5,419.57 | 5,552.02 | 5,444.05 | 21,480.42 |
| | 433 | 895.7 | 1,032.89 | 1,059.67 | 1,104.02 | 4,092.28 |
| | 434 | 249.41 | 292.03 | 310.22 | 326.39 | 1,178.05 |

| | | | | | | |
|---|---|---|---|---|---|---|
| | 435 | 719.05 | 684.59 | 670.34 | 675.45 | 2,749.43 |
| | 436 | 1,886.33 | 1,890.31 | 1,962.89 | 2,082.99 | 7,822.52 |
| | 437 | 1,308.55 | 1,368.39 | 1,333.28 | 1,374.08 | 5,384.30 |
| | 438 | 65.25 | 74.73 | 74.33 | 75.49 | 289.8 |
| | 439 | 1,758.14 | 1,777.51 | 1,662.78 | 1,832.94 | 7,031.37 |
| | 440 | 2,209.35 | 2,324 | 2,206.10 | 2,252.94 | 8,992.39 |
| | 441 | 2,976.97 | 3,170.22 | 3,054.01 | 3,145.46 | 12,346.66 |
| | 442 | 1,446.99 | 1,571.72 | 1,540.37 | 1,568.91 | 6,127.99 |
| | 443 | 806.78 | 775 | 780.06 | 812.92 | 3,174.76 |
| | 444 | 1,305.95 | 1,270.52 | 1,301.29 | 1,347.34 | 5,225.10 |
| | 445 | 1,442.92 | 1,553.19 | 1,443.77 | 1,478.66 | 5,918.54 |
| | 446 | 800.69 | 808.2 | 808.87 | 800.1 | 3,217.86 |
| | 447 | 760.29 | 770.04 | 828.24 | 777.5 | 3,136.07 |
| | 448 | 1,070.57 | 1,121.44 | 1,104.79 | 1,135.25 | 4,432.05 |
| | 449 | 866.77 | 933.09 | 901.91 | 934.18 | 3,635.95 |
| | 450 | 2,833.07 | 2,953.07 | 2,857.63 | 2,964.34 | 11,608.11 |
| | 451 | 2,128.80 | 2,333.70 | 2,213.87 | 2,356.36 | 9,032.73 |
| | 452 | 4,811.19 | 4,883.85 | 4,836.58 | 4,804.93 | 19,336.55 |
| | 453 | 1,051.22 | 1,113.06 | 1,074 | 1,019.14 | 4,257.42 |
| | 454 | 2,041.08 | 2,099.37 | 2,209.13 | 2,164.88 | 8,514.46 |
| | 455 | 796.89 | 793.4 | 808.69 | 759.82 | 3,158.80 |
| | 456 | 4,328.32 | 4,557.93 | 4,709.42 | 4,751.30 | 18,346.97 |
| | 457 | 1,125.37 | 1,123.23 | 1,102.28 | 1,104.32 | 4,455.20 |
| | 458 | 467.18 | 473.54 | 456.83 | 502.37 | 1,899.92 |
| TOTAL | | 50,757.74 | 53,154.22 | 52,666.29 | 53,520.63 | 210,098.88 |

STATE: OKLAHOMA

| ZIP CODE | | QUARTER 1 | QUARTER 2 | QUARTER 3 | QUARTER 4 | TOTAL GRAMS |
|---|---|---|---|---|---|---|
| | 730 | 1,615.82 | 1,652.38 | 1,783.55 | 1,826.46 | 6,878.21 |
| | 731 | 2,315.92 | 2,289.22 | 2,584.22 | 2,629.68 | 9,819.04 |
| | 734 | 95.62 | 97.33 | 104.44 | 123.65 | 421.04 |
| | 735 | 546.3 | 536.11 | 615.41 | 552.25 | 2,250.07 |
| | 736 | 87.91 | 99.85 | 117.71 | 133.11 | 438.58 |
| | 737 | 242.03 | 227.69 | 221.79 | 203.2 | 894.71 |
| | 738 | 58.07 | 48.04 | 74.42 | 75.45 | 255.98 |
| | 739 | 18.05 | 15.78 | 13.53 | 9.16 | 56.52 |

| | | | | | | |
|---|---|---|---|---|---|---|
| | 740 | 1,792.75 | 1,875.67 | 2,108.06 | 2,136.49 | 7,912.97 |
| | 741 | 1,368.98 | 1,451.85 | 1,550.46 | 1,544.42 | 5,915.71 |
| | 743 | 161.81 | 172.91 | 177.49 | 208.47 | 720.68 |
| | 744 | 622.04 | 673.69 | 791.79 | 836.57 | 2,924.09 |
| | 745 | 187.96 | 189.77 | 201.42 | 206.77 | 785.92 |
| | 746 | 177.67 | 172.15 | 193.17 | 181.1 | 724.09 |
| | 747 | 80.97 | 84.38 | 97.58 | 114.51 | 377.44 |
| | 748 | 620.86 | 601.88 | 669.89 | 671.64 | 2,564.27 |
| | 749 | 183.04 | 173.62 | 209.51 | 215.4 | 781.57 |
| TOTAL | | 10,175.80 | 10,362.32 | 11,514.44 | 11,668.33 | 43,720.89 |

STATE: OREGON

| ZIP CODE | | QUARTER 1 | QUARTER 2 | QUARTER 3 | QUARTER 4 | TOTAL GRAMS |
|---|---|---|---|---|---|---|
| | 970 | 1,643.70 | 1,782.82 | 1,870.38 | 1,900.19 | 7,197.09 |
| | 971 | 597.89 | 657.04 | 712.04 | 765.07 | 2,732.04 |
| | 972 | 2,939.62 | 3,084.58 | 2,858.62 | 3,094.96 | 11,977.78 |
| | 973 | 1,322.93 | 1,415.18 | 1,443.87 | 1,558.09 | 5,740.07 |
| | 974 | 1,261.64 | 1,361.76 | 1,494 | 1,600.29 | 5,717.69 |
| | 975 | 970.43 | 936.89 | 961.06 | 1,021 | 3,889.38 |
| | 976 | 33.88 | 39.24 | 37.68 | 43.51 | 154.31 |
| | 977 | 365.17 | 358.51 | 383.53 | 391.61 | 1,498.82 |
| | 978 | 554.23 | 607.44 | 628.31 | 666.48 | 2,456.46 |
| | 979 | 35.14 | 35 | 47.84 | 54.85 | 172.83 |
| TOTAL | | 9,724.63 | 10,278.46 | 10,437.33 | 11,096.05 | 41,536.47 |

STATE: PENNSYLVANIA

| ZIP CODE | | QUARTER 1 | QUARTER 2 | QUARTER 3 | QUARTER 4 | TOTAL GRAMS |
|---|---|---|---|---|---|---|
| | 150 | 3,273.95 | 3,334.42 | 3,275.91 | 3,448.23 | 13,332.51 |
| | 151 | 2,310.30 | 2,483.84 | 2,198.21 | 2,557.78 | 9,550.13 |
| | 152 | 5,856.44 | 6,141.96 | 5,989.90 | 6,224.55 | 24,212.85 |
| | 153 | 2,118.19 | 2,116.92 | 1,924.19 | 2,096.19 | 8,255.49 |
| | 154 | 1,256.61 | 1,250.12 | 1,120.36 | 1,209.14 | 4,836.23 |
| | 155 | 360.21 | 377.67 | 410.69 | 426.5 | 1,575.07 |
| | 156 | 2,647.99 | 2,936.76 | 3,150.14 | 3,277.60 | 12,012.49 |
| | 157 | 681.48 | 677.44 | 708.91 | 700.12 | 2,767.95 |

| | | | | | |
|---|---|---|---|---|---|
| 158 | 638.24 | 651.74 | 619.01 | 685 | 2,593.99 |
| 159 | 1,065.55 | 1,108.12 | 1,094.82 | 1,166.39 | 4,434.88 |
| 160 | 1,347.29 | 1,354.15 | 1,375.58 | 1,401.78 | 5,478.80 |
| 161 | 983.72 | 1,077.98 | 961.12 | 1,227.56 | 4,250.38 |
| 162 | 747.15 | 790.32 | 790.35 | 824.49 | 3,152.31 |
| 163 | 641.28 | 675.47 | 657.92 | 678.78 | 2,653.45 |
| 164 | 263.2 | 294.34 | 292.55 | 286.4 | 1,136.49 |
| 165 | 1,657.13 | 1,733.32 | 1,758.90 | 1,785.21 | 6,934.56 |
| 166 | 1,611.61 | 1,683.76 | 1,708.23 | 1,847.96 | 6,851.56 |
| 167 | 118.49 | 121.81 | 137.02 | 131.84 | 509.16 |
| 168 | 530.12 | 560.54 | 553.32 | 629 | 2,272.98 |
| 169 | 125.5 | 146.92 | 146.66 | 129.97 | 549.05 |
| 170 | 1,549.04 | 1,626.10 | 1,626.59 | 1,762.77 | 6,564.50 |
| 171 | 339.98 | 387.24 | 421.18 | 500.95 | 1,649.35 |
| 172 | 796.38 | 871.98 | 876.05 | 857.84 | 3,402.25 |
| 173 | 987.13 | 1,009 | 1,001.93 | 1,055.26 | 4,053.32 |
| 174 | 426.07 | 488.93 | 506.91 | 555.99 | 1,977.90 |
| 175 | 329.2 | 322.98 | 345.34 | 337.22 | 1,334.74 |
| 176 | 410.39 | 427.95 | 445.57 | 494.1 | 1,778.01 |
| 177 | 1,381.54 | 1,460.43 | 1,581.32 | 1,525.96 | 5,949.25 |
| 178 | 715.17 | 799.89 | 736.66 | 840.95 | 3,092.67 |
| 179 | 415.15 | 457.97 | 446.43 | 477.54 | 1,797.09 |
| 180 | 800.38 | 810.34 | 834.09 | 861.11 | 3,305.92 |
| 181 | 391.89 | 437.18 | 440.74 | 444.1 | 1,713.91 |
| 182 | 343.61 | 424.72 | 407.75 | 460.32 | 1,636.40 |
| 183 | 447.44 | 481.85 | 455.49 | 490.33 | 1,875.11 |
| 184 | 825.52 | 831.54 | 779.67 | 1,041.15 | 3,477.88 |
| 185 | 1,812.77 | 1,909.81 | 1,999.99 | 2,137.48 | 7,860.05 |
| 186 | 795.39 | 839.46 | 840.12 | 1,006.73 | 3,481.70 |
| 187 | 1,947.14 | 1,981.89 | 1,529.73 | 2,064.57 | 7,523.33 |
| 188 | 269.38 | 307.34 | 312.53 | 348.48 | 1,237.73 |
| 189 | 1,018.29 | 1,065.04 | 1,087.58 | 1,105.39 | 4,276.30 |
| 190 | 5,167.84 | 5,287.97 | 5,200.06 | 5,388.57 | 21,044.44 |
| 191 | 7,286.14 | 7,780.70 | 7,932.56 | 8,027.11 | 31,026.51 |
| 193 | 939.5 | 947.41 | 992.1 | 959.89 | 3,838.90 |
| 194 | 1,863.71 | 1,880.86 | 1,743.32 | 1,888.21 | 7,376.10 |
| 195 | 258.95 | 301.72 | 293.86 | 293.42 | 1,147.95 |

| | | | | | |
|---|---|---|---|---|---|
| 196 | 579.35 | 587.43 | 480.55 | 658.8 | 2,306.13 |
| TOTAL | 60,331.80 | 63,245.33 | 62,191.91 | 66,318.73 | 252,087.77 |

STATE: PUERTO RICO

| ZIP CODE | QUARTER 1 | QUARTER 2 | QUARTER 3 | QUARTER 4 | TOTAL GRAMS |
|---|---|---|---|---|---|
| 6 | 562.15 | 602.75 | 478.21 | 536.9 | 2,180.01 |
| 7 | 1,371.81 | 1,426.03 | 1,204.97 | 1,603.31 | 5,606.12 |
| 9 | 558.29 | 541.95 | 536.47 | 533.47 | 2,170.18 |
| TOTAL | 2,492.25 | 2,570.73 | 2,219.65 | 2,673.68 | 9,956.31 |

STATE: RHODE ISLAND

| ZIP CODE | QUARTER 1 | QUARTER 2 | QUARTER 3 | QUARTER 4 | TOTAL GRAMS |
|---|---|---|---|---|---|
| 28 | 3,874.27 | 4,236.70 | 4,077.32 | 4,330.03 | 16,518.32 |
| 29 | 3,204.45 | 3,464.66 | 3,412.42 | 3,497.44 | 13,578.97 |
| TOTAL | 7,078.72 | 7,701.36 | 7,489.74 | 7,827.47 | 30,097.29 |

STATE: SOUTH CAROLINA

| ZIP CODE | QUARTER 1 | QUARTER 2 | QUARTER 3 | QUARTER 4 | TOTAL GRAMS |
|---|---|---|---|---|---|
| 290 | 286.52 | 322.11 | 338.73 | 335.23 | 1,282.59 |
| 291 | 353.97 | 384.19 | 379.25 | 450.32 | 1,567.73 |
| 292 | 426.48 | 462.85 | 498.03 | 486.57 | 1,873.93 |
| 293 | 1,353.48 | 1,422.68 | 1,364.99 | 1,497.74 | 5,638.89 |
| 294 | 1,701.99 | 1,774.57 | 1,803.70 | 1,876.42 | 7,156.68 |
| 295 | 1,498.76 | 1,688.03 | 1,842.33 | 2,074.04 | 7,103.16 |
| 296 | 3,029.11 | 3,167.19 | 3,309.04 | 3,463.44 | 12,968.78 |
| 297 | 558.41 | 623.49 | 643.08 | 656.64 | 2,481.62 |
| 298 | 176.31 | 175.69 | 160.92 | 172.37 | 685.29 |
| 299 | 459.54 | 479.86 | 455.28 | 485.41 | 1,880.09 |
| TOTAL | 9,844.57 | 10,500.66 | 10,795.35 | 11,498.18 | 42,638.76 |

STATE: SOUTH DAKOTA

| ZIP CODE | QUARTER 1 | QUARTER 2 | QUARTER 3 | QUARTER 4 | TOTAL GRAMS |
|---|---|---|---|---|---|
| 570 | 33.39 | 39.13 | 43.39 | 48.64 | 164.55 |

| | | | | | | |
|---|---|---|---|---|---|---|
| | 571 | 124.68 | 152.66 | 140.2 | 161.33 | 578.87 |
| | 572 | 49.45 | 49.76 | 46.24 | 49.92 | 195.37 |
| | 573 | 17.6 | 17.36 | 11.29 | 16.57 | 62.82 |
| | 574 | 82.41 | 93.9 | 83.28 | 74.78 | 334.37 |
| | 575 | 10.68 | 9.43 | 11.33 | 11.19 | 42.63 |
| | 576 | 6.08 | 5.94 | 6.98 | 9.69 | 28.69 |
| | 577 | 86.32 | 96.97 | 106.85 | 100.34 | 390.48 |
| TOTAL | | 410.61 | 465.15 | 449.56 | 472.46 | 1,797.78 |

STATE: TENNESSEE

| ZIP CODE | | QUARTER 1 | QUARTER 2 | QUARTER 3 | QUARTER 4 | TOTAL GRAMS |
|---|---|---|---|---|---|---|
| | 370 | 2,884.86 | 3,172.89 | 3,156.95 | 3,273.82 | 12,488.52 |
| | 371 | 3,302.32 | 3,480.67 | 3,153.46 | 3,813.40 | 13,749.85 |
| | 372 | 2,049.44 | 2,264.30 | 2,349.99 | 2,446.73 | 9,110.46 |
| | 373 | 1,319.10 | 1,417.45 | 1,419.52 | 1,551.45 | 5,707.52 |
| | 374 | 630.6 | 662.64 | 639.58 | 743.46 | 2,676.28 |
| | 376 | 7,171.82 | 7,494.14 | 7,325.21 | 7,280.22 | 29,271.39 |
| | 377 | 2,976.56 | 3,119.40 | 3,179.06 | 3,239.65 | 12,514.67 |
| | 378 | 4,533.97 | 4,803.61 | 4,846.49 | 4,962.37 | 19,146.44 |
| | 379 | 2,202.27 | 2,305.71 | 2,300.76 | 2,340 | 9,148.74 |
| | 380 | 771.23 | 820.87 | 857.25 | 870.38 | 3,319.73 |
| | 381 | 1,165.33 | 1,185.50 | 1,114.79 | 1,148.74 | 4,614.36 |
| | 382 | 474.08 | 490.85 | 486.48 | 491.92 | 1,943.33 |
| | 383 | 1,108.41 | 1,237.53 | 1,310.61 | 1,338.68 | 4,995.23 |
| | 384 | 841.11 | 938.73 | 867.38 | 986.89 | 3,634.11 |
| | 385 | 1,640.60 | 1,759.50 | 1,731.29 | 1,833.71 | 6,965.10 |
| TOTAL | | 33,071.70 | 35,153.79 | 34,738.82 | 36,321.42 | 139,285.73 |

STATE: TEXAS

| ZIP CODE | | QUARTER 1 | QUARTER 2 | QUARTER 3 | QUARTER 4 | TOTAL GRAMS |
|---|---|---|---|---|---|---|
| | 750 | 2,520.21 | 2,640.63 | 2,766.52 | 2,692.39 | 10,619.75 |
| | 751 | 755.64 | 795.43 | 808.37 | 805.11 | 3,164.55 |
| | 752 | 1,000.52 | 1,052.78 | 1,029.91 | 1,033.15 | 4,116.36 |
| | 753 | 1.92 | 2.8 | 1.34 | 3.33 | 9.39 |
| | 754 | 455.69 | 457.15 | 470.25 | 495.02 | 1,878.11 |

| | | | | | |
|---|---|---|---|---|---|
| 755 | 126.18 | 128.3 | 135.2 | 148.47 | 538.15 |
| 756 | 516.32 | 531.73 | 516.49 | 512.89 | 2,077.43 |
| 757 | 441.05 | 435.42 | 419.4 | 411.03 | 1,706.90 |
| 758 | 62.51 | 67.11 | 65.17 | 57.13 | 251.92 |
| 759 | 337.04 | 335.15 | 342.17 | 347.11 | 1,361.47 |
| 760 | 1,906.93 | 2,004.82 | 2,012.97 | 2,015.48 | 7,940.20 |
| 761 | 979.58 | 1,034.58 | 1,079.97 | 1,022.22 | 4,116.35 |
| 762 | 550.47 | 547.67 | 585.97 | 569.97 | 2,254.08 |
| 763 | 277.15 | 296.73 | 317.99 | 310.62 | 1,202.49 |
| 764 | 97.63 | 103.07 | 106.4 | 119.82 | 426.92 |
| 765 | 291.11 | 296.22 | 328.99 | 299.89 | 1,216.21 |
| 766 | 65.91 | 56.28 | 67.6 | 69.4 | 259.19 |
| 767 | 122.6 | 132.51 | 145.09 | 145.19 | 545.39 |
| 768 | 39.55 | 46.65 | 49.15 | 43.47 | 178.82 |
| 769 | 65.87 | 66.11 | 67.5 | 68.9 | 268.38 |
| 770 | 2,341.22 | 2,423.47 | 2,348.38 | 2,489.70 | 9,602.77 |
| 772 | 0 | 0 | 0 | 0 | 0 |
| 773 | 2,169.62 | 2,196.14 | 2,242.45 | 2,311.98 | 8,920.19 |
| 774 | 954.89 | 943.46 | 937.3 | 954.71 | 3,790.36 |
| 775 | 2,272.12 | 2,264.79 | 2,249.86 | 2,234.19 | 9,020.96 |
| 776 | 885.08 | 897.96 | 881.85 | 868.73 | 3,533.62 |
| 777 | 299.21 | 298.94 | 291.16 | 302.89 | 1,192.20 |
| 778 | 293.88 | 300.74 | 305.76 | 316.03 | 1,216.41 |
| 779 | 140.66 | 138.28 | 142.01 | 150.64 | 571.59 |
| 780 | 367.77 | 354.24 | 406.37 | 403.11 | 1,531.49 |
| 781 | 310.79 | 310.45 | 319.54 | 317.76 | 1,258.54 |
| 782 | 950.65 | 973.97 | 1,023.77 | 1,012.99 | 3,961.38 |
| 783 | 154.13 | 155.21 | 154.4 | 151.49 | 615.23 |
| 784 | 302.93 | 363.16 | 366.87 | 345.9 | 1,378.86 |
| 785 | 118.99 | 112.29 | 112 | 99.74 | 443.02 |
| 786 | 857.54 | 926.92 | 992.29 | 1,012.31 | 3,789.06 |
| 787 | 1,249.24 | 1,297.97 | 1,361.17 | 1,327.19 | 5,235.57 |
| 788 | 32.11 | 27.46 | 28.59 | 29.2 | 117.36 |
| 789 | 30.5 | 24.42 | 25.89 | 27.71 | 108.52 |
| 790 | 34.35 | 26.23 | 25.49 | 24.82 | 110.89 |
| 791 | 104.69 | 91.62 | 113.87 | 113.06 | 423.24 |
| 792 | 5.26 | 3.64 | 5.1 | 3.52 | 17.52 |

| | | | | | |
|---|---|---|---|---|---|
| 793 | 36.08 | 42.96 | 48.16 | 46.37 | 173.57 |
| 794 | 149.39 | 165.49 | 198.66 | 192.52 | 706.06 |
| 795 | 29.42 | 32.76 | 29.13 | 31.84 | 123.15 |
| 796 | 185.85 | 201.13 | 210.84 | 225.85 | 823.67 |
| 797 | 454.12 | 470.29 | 505.19 | 543.44 | 1,973.04 |
| 798 | 9.64 | 8.32 | 6.4 | 3.92 | 28.28 |
| 799 | 126.76 | 128.34 | 146.12 | 136.48 | 537.7 |
| TOTAL | 25,480.77 | 26,211.79 | 26,795.07 | 26,848.68 | 105,336.31 |

STATE: UTAH

| ZIP CODE | QUARTER 1 | QUARTER 2 | QUARTER 3 | QUARTER 4 | TOTAL GRAMS |
|---|---|---|---|---|---|
| 840 | 4,360.55 | 4,399.34 | 4,481.35 | 4,467.78 | 17,709.02 |
| 841 | 2,293.53 | 2,364.75 | 2,335.24 | 2,426.57 | 9,420.09 |
| 843 | 370.53 | 422.33 | 426.06 | 392.22 | 1,611.14 |
| 844 | 776.28 | 786.27 | 825.81 | 808.77 | 3,197.13 |
| 845 | 192.22 | 219.24 | 232.3 | 249.95 | 893.71 |
| 846 | 1,006.33 | 1,053.10 | 1,081.86 | 1,166.27 | 4,307.56 |
| 847 | 298.53 | 348.39 | 380.03 | 396.83 | 1,423.78 |
| TOTAL | 9,297.97 | 9,593.42 | 9,762.65 | 9,908.39 | 38,562.43 |

STATE: VERMONT

| ZIP CODE | QUARTER 1 | QUARTER 2 | QUARTER 3 | QUARTER 4 | TOTAL GRAMS |
|---|---|---|---|---|---|
| 50 | 657.9 | 683.03 | 696.57 | 716.77 | 2,754.27 |
| 51 | 243 | 267.65 | 240.48 | 251.53 | 1,002.66 |
| 52 | 362.04 | 392.04 | 380.13 | 389.97 | 1,524.18 |
| 53 | 370.08 | 361.18 | 361.72 | 350.89 | 1,443.87 |
| 54 | 2,209.37 | 2,447.99 | 2,481.21 | 2,579.33 | 9,717.90 |
| 56 | 775.96 | 800.44 | 811.66 | 776.17 | 3,164.23 |
| 57 | 937.97 | 1,046.55 | 970.29 | 1,037.81 | 3,992.62 |
| 58 | 414.72 | 402.58 | 404.09 | 397.97 | 1,619.36 |
| TOTAL | 5,971.04 | 6,401.46 | 6,346.15 | 6,500.44 | 25,219.09 |

STATE: VIRGIN ISLANDS

| ZIP CODE | QUARTER 1 | QUARTER 2 | QUARTER 3 | QUARTER 4 | TOTAL GRAMS |
|---|---|---|---|---|---|
| 8 | 19.68 | 17.54 | 12.98 | 12.51 | 62.71 |
| TOTAL | 19.68 | 17.54 | 12.98 | 12.51 | 62.71 |

STATE: VIRGINIA

| ZIP CODE | QUARTER 1 | QUARTER 2 | QUARTER 3 | QUARTER 4 | TOTAL GRAMS |
|---|---|---|---|---|---|
| 201 | 1,466.26 | 1,323.28 | 1,318.85 | 1,319.08 | 5,427.47 |
| 220 | 819.8 | 802.88 | 794 | 779.97 | 3,196.65 |
| 221 | 594.13 | 581.07 | 552.74 | 528.52 | 2,256.46 |
| 222 | 631.52 | 895.03 | 892.43 | 897.54 | 3,316.52 |
| 223 | 210.13 | 197.15 | 199.15 | 196.61 | 803.04 |
| 224 | 493.36 | 504.5 | 499.14 | 498.51 | 1,995.51 |
| 225 | 238.74 | 242.92 | 218.71 | 214.86 | 915.23 |
| 226 | 1,094.79 | 1,086.22 | 1,047.48 | 1,074.26 | 4,302.75 |
| 227 | 149.62 | 167 | 186.06 | 216.57 | 719.25 |
| 228 | 202.96 | 201.89 | 187.32 | 189.68 | 781.85 |
| 229 | 378.22 | 411.72 | 418.13 | 439.67 | 1,647.74 |
| 230 | 347.44 | 308.93 | 315.14 | 310.11 | 1,281.62 |
| 231 | 945.72 | 996.77 | 764.39 | 883.58 | 3,590.46 |
| 232 | 1,030.43 | 1,058.64 | 1,176.82 | 1,240.62 | 4,506.51 |
| 233 | 293.25 | 275.52 | 182.55 | 277.64 | 1,028.96 |
| 234 | 639.47 | 666.37 | 696.25 | 689.53 | 2,691.62 |
| 235 | 118.64 | 140.8 | 145.25 | 150.75 | 555.44 |
| 236 | 632.23 | 705.97 | 701.77 | 737.36 | 2,777.33 |
| 237 | 99.54 | 109.59 | 103 | 103.5 | 415.63 |
| 238 | 425.9 | 407.68 | 387.44 | 353.74 | 1,574.76 |
| 239 | 139.86 | 133.49 | 125.55 | 137.81 | 536.71 |
| 240 | 1,904.04 | 1,907.62 | 1,583.15 | 1,858.23 | 7,253.04 |
| 241 | 779.3 | 847.06 | 807.39 | 783.67 | 3,217.42 |
| 242 | 3,485.64 | 3,377.34 | 3,271.49 | 3,368.65 | 13,503.12 |
| 243 | 1,103.66 | 850.19 | 771.9 | 819.27 | 3,545.02 |
| 244 | 162 | 140.42 | 131.23 | 140.82 | 574.47 |
| 245 | 776.54 | 784.4 | 719.49 | 729.72 | 3,010.15 |
| 246 | 1,049.14 | 988.73 | 993.82 | 1,002.24 | 4,033.93 |
| TOTAL | 20,212.33 | 20,113.18 | 19,190.64 | 19,942.51 | 79,458.66 |

STATE: WASHINGTON

| ZIP CODE | QUARTER 1 | QUARTER 2 | QUARTER 3 | QUARTER 4 | TOTAL GRAMS |
|---|---|---|---|---|---|
| 980 | 3,298.56 | 3,453.70 | 3,562.07 | 3,546.25 | 13,860.58 |
| 981 | 2,049.22 | 2,210.67 | 2,262.52 | 2,384.85 | 8,907.26 |
| 982 | 3,465.57 | 3,607.62 | 3,638.03 | 3,826.76 | 14,537.98 |
| 983 | 1,608.61 | 1,682.57 | 1,706.29 | 1,826.01 | 6,823.48 |
| 984 | 1,169.92 | 1,207.34 | 1,165.21 | 1,192.17 | 4,734.64 |
| 985 | 1,481.49 | 1,576.20 | 1,628.48 | 1,758.19 | 6,444.36 |
| 986 | 1,106.09 | 1,155.05 | 1,230.90 | 1,389.34 | 4,881.38 |
| 988 | 466.68 | 520.46 | 568.94 | 652.64 | 2,208.72 |
| 989 | 587.1 | 583.95 | 651.57 | 688.69 | 2,511.31 |
| 990 | 102.84 | 113.56 | 126.15 | 137.61 | 480.16 |
| 991 | 275.68 | 285.78 | 302.79 | 264.8 | 1,129.05 |
| 992 | 1,461.97 | 1,619.71 | 1,659.31 | 1,722.71 | 6,463.70 |
| 993 | 733.84 | 787.39 | 879.26 | 922.88 | 3,323.37 |
| 994 | 174.86 | 163.41 | 172.24 | 182.98 | 693.49 |
| TOTAL | 17,982.43 | 18,967.41 | 19,553.76 | 20,495.88 | 76,999.48 |

STATE: WEST VIRGINIA

| ZIP CODE | QUARTER 1 | QUARTER 2 | QUARTER 3 | QUARTER 4 | TOTAL GRAMS |
|---|---|---|---|---|---|
| 247 | 204.8 | 237.41 | 274.15 | 279.73 | 996.09 |
| 248 | 473.35 | 539.9 | 498.78 | 453.87 | 1,965.90 |
| 249 | 127.24 | 115.85 | 123.62 | 119.67 | 486.38 |
| 250 | 806.91 | 816.14 | 803.95 | 788.88 | 3,215.88 |
| 251 | 270.19 | 276.74 | 253.77 | 246.18 | 1,046.88 |
| 252 | 313.04 | 321.05 | 269.61 | 240.35 | 1,144.05 |
| 253 | 1,357.24 | 1,383.11 | 1,411.59 | 1,367.10 | 5,519.04 |
| 254 | 1,030.58 | 1,114.96 | 1,142.18 | 1,182.67 | 4,470.39 |
| 255 | 1,313.74 | 1,292.51 | 1,243.56 | 1,374.95 | 5,224.76 |
| 256 | 1,323.27 | 1,426.53 | 1,324.66 | 863.84 | 4,938.30 |
| 257 | 940.68 | 931.49 | 1,042.62 | 950.94 | 3,865.73 |
| 258 | 1,519.26 | 1,794.82 | 1,944.92 | 2,192.29 | 7,451.29 |
| 259 | 272.38 | 276.3 | 333.41 | 322.86 | 1,204.95 |

| | | | | | | |
|---|---|---|---|---|---|---|
| | 260 | 1,540.47 | 1,568.35 | 1,794 | 1,732.11 | 6,634.93 |
| | 261 | 556.55 | 583.3 | 543.13 | 576.1 | 2,259.08 |
| | 262 | 484.44 | 480.83 | 554.99 | 594.3 | 2,114.56 |
| | 263 | 776.61 | 739.78 | 792.48 | 804.35 | 3,113.22 |
| | 264 | 151.72 | 153.26 | 145.12 | 144.89 | 594.99 |
| | 265 | 1,226.44 | 1,235.83 | 1,253.28 | 1,244.32 | 4,959.87 |
| | 266 | 102.23 | 90.99 | 93.19 | 94.25 | 380.66 |
| | 267 | 91.49 | 111.07 | 117.72 | 226.75 | 547.03 |
| | 268 | 15.43 | 17.82 | 11.99 | 11.41 | 56.65 |
| TOTAL | | 14,898.06 | 15,508.04 | 15,972.72 | 15,811.81 | 62,190.63 |

STATE: WISCONSIN

| ZIP CODE | | QUARTER 1 | QUARTER 2 | QUARTER 3 | QUARTER 4 | TOTAL GRAMS |
|---|---|---|---|---|---|---|
| | 530 | 1,101.49 | 1,180.04 | 1,128.75 | 1,185.44 | 4,595.72 |
| | 531 | 1,241.02 | 1,283.62 | 1,314.97 | 1,391.14 | 5,230.75 |
| | 532 | 2,452.05 | 2,421.57 | 2,448.45 | 2,515.23 | 9,837.30 |
| | 534 | 199.89 | 195.97 | 209.08 | 217.09 | 822.03 |
| | 535 | 830.73 | 888.82 | 935.34 | 967.73 | 3,622.62 |
| | 537 | 726.53 | 759.81 | 782.09 | 866.44 | 3,134.87 |
| | 538 | 22.21 | 21.67 | 20.76 | 29.57 | 94.21 |
| | 539 | 325.43 | 340.42 | 361.72 | 410.01 | 1,437.58 |
| | 540 | 81.62 | 83.02 | 93.41 | 91.07 | 349.12 |
| | 541 | 255.63 | 263.14 | 271.33 | 276.59 | 1,066.69 |
| | 542 | 101.92 | 102.96 | 114.89 | 124.21 | 443.98 |
| | 543 | 213.77 | 234.84 | 242.08 | 242.34 | 933.03 |
| | 544 | 443.98 | 455.56 | 427.4 | 442.39 | 1,769.33 |
| | 545 | 213.95 | 238.26 | 252.29 | 286.32 | 990.82 |
| | 546 | 356.55 | 346.39 | 326.56 | 328.8 | 1,358.30 |
| | 547 | 181.54 | 183.6 | 168.72 | 179.6 | 713.46 |
| | 548 | 164.71 | 179.68 | 204.92 | 191.84 | 741.15 |
| | 549 | 970.09 | 1,002.66 | 970.04 | 1,020.58 | 3,963.37 |
| TOTAL | | 9,883.11 | 10,182.03 | 10,272.80 | 10,766.39 | 41,104.33 |

STATE: WYOMING

| ZIP CODE | | QUARTER 1 | QUARTER 2 | QUARTER 3 | QUARTER 4 | TOTAL GRAMS |
|---|---|---|---|---|---|---|

| | | | | | | |
|---|---|---|---|---|---|---|
| | 820 | 134.05 | 373.69 | 336.5 | 458.03 | 1,302.27 |
| | 822 | 7.2 | 8.57 | 10.36 | 12.36 | 38.49 |
| | 823 | 5.64 | 8.98 | 13.7 | 8.87 | 37.19 |
| | 824 | 111.25 | 101.48 | 123.69 | 110.71 | 447.13 |
| | 825 | 71.46 | 67.82 | 60.71 | 72.69 | 272.68 |
| | 826 | 340.34 | 343.41 | 417.87 | 404.18 | 1,505.80 |
| | 827 | 83.67 | 83.57 | 80.17 | 80.95 | 328.36 |
| | 828 | 40.07 | 47.7 | 56.61 | 56.34 | 200.72 |
| | 829 | 155.52 | 162.84 | 168.52 | 200.67 | 687.55 |
| | 830 | 39.38 | 41.34 | 32.94 | 35.11 | 148.77 |
| | 831 | 13.96 | 17.42 | 15.96 | 16.6 | 63.94 |
| TOTAL | | 1,002.54 | 1,256.82 | 1,317.03 | 1,456.51 | 5,032.90 |

DRUG CODE: 9120 DRUG NAME: DIHYDROCODEINE

STATE: ALABAMA

| ZIP CODE | | QUARTER 1 | QUARTER 2 | QUARTER 3 | QUARTER 4 | TOTAL GRAMS |
|---|---|---|---|---|---|---|
| | 350 | 117.02 | 122.73 | 136.51 | 128.71 | 504.97 |
| | 351 | 24.99 | 36.85 | 30.5 | 29.43 | 121.77 |
| | 352 | 40.8 | 56.72 | 49.78 | 56.13 | 203.43 |
| | 354 | 129.51 | 111.25 | 99.5 | 76.91 | 417.17 |
| | 355 | 45.18 | 56.02 | 39.15 | 41.23 | 181.58 |
| | 356 | 244.29 | 252.57 | 281.94 | 298.29 | 1,077.09 |
| | 357 | 18.91 | 24.78 | 30.87 | 16.56 | 91.12 |
| | 358 | 14.95 | 5.87 | 13.03 | 17.72 | 51.57 |
| | 359 | 26.28 | 14.95 | 9.19 | 14.31 | 64.73 |
| | 360 | 8.55 | 20.3 | 7.48 | 7.48 | 43.81 |
| | 361 | 14.95 | 21.36 | 12.82 | 9.61 | 58.74 |
| | 362 | 14.21 | 15.33 | 19.01 | 22.7 | 71.25 |
| | 363 | 10.68 | 4.27 | 3.2 | 3.2 | 21.35 |
| | 364 | 1.07 | 2.14 | 1.07 | 1.07 | 5.35 |
| | 365 | 23.63 | 16.2 | 18.96 | 17.53 | 76.32 |
| | 366 | 37.99 | 26.44 | 19.9 | 29.91 | 114.24 |
| | 367 | 3.2 | 2.14 | 6.41 | 5.34 | 17.09 |
| | 368 | 7.48 | 7.48 | 6.41 | 5.34 | 26.71 |
| | 369 | 0 | 1.07 | 0 | 0 | 1.07 |

| | | | | | | |
|---|---|---|---|---|---|---|
| TOTAL | | 783.69 | 798.47 | 785.73 | 781.47 | 3,149.36 |

STATE: ALASKA

| ZIP CODE | | QUARTER 1 | QUARTER 2 | QUARTER 3 | QUARTER 4 | TOTAL GRAMS |
|---|---|---|---|---|---|---|
| | 996 | 8.55 | 3.2 | 0 | 2.14 | 13.89 |
| TOTAL | | 8.55 | 3.2 | 0 | 2.14 | 13.89 |

STATE: ARIZONA

| ZIP CODE | | QUARTER 1 | QUARTER 2 | QUARTER 3 | QUARTER 4 | TOTAL GRAMS |
|---|---|---|---|---|---|---|
| | 850 | 14.31 | 9.61 | 5.34 | 0 | 29.26 |
| | 852 | 53.41 | 75.84 | 69.43 | 81.18 | 279.86 |
| | 853 | 1.17 | 0 | 0 | 2.14 | 3.31 |
| | 856 | 0 | 1.07 | 1.07 | 0 | 2.14 |
| | 863 | 0.21 | 0 | 0 | 0 | 0.21 |
| | 864 | 4.7 | 0 | 0 | 0 | 4.7 |
| TOTAL | | 73.8 | 86.52 | 75.84 | 83.32 | 319.48 |

STATE: ARKANSAS

| ZIP CODE | | QUARTER 1 | QUARTER 2 | QUARTER 3 | QUARTER 4 | TOTAL GRAMS |
|---|---|---|---|---|---|---|
| | 716 | 1.07 | 7.48 | 5.66 | 0 | 14.21 |
| | 717 | 1.07 | 2.14 | 3.2 | 0 | 6.41 |
| | 718 | 13.03 | 9.61 | 13.89 | 4.27 | 40.8 |
| | 719 | 8.28 | 8.28 | 33.7 | 35.25 | 85.51 |
| | 720 | 12.5 | 26.76 | 13.65 | 8.55 | 61.46 |
| | 721 | 4.27 | 10.68 | 1.07 | 3.2 | 19.22 |
| | 722 | 11.54 | 8.6 | 4.01 | 2.14 | 26.29 |
| | 723 | 2.24 | 19.49 | 37.12 | 32.47 | 91.32 |
| | 724 | 18.16 | 20.03 | 25.8 | 19.28 | 83.27 |
| | 726 | 0 | 1.07 | 0 | 0 | 1.07 |
| | 727 | 5.53 | 11.88 | 10.68 | 10.68 | 38.77 |
| | 728 | 3.2 | 0 | 0 | 2.14 | 5.34 |
| | 729 | 24.78 | 26.33 | 26.7 | 17.09 | 94.9 |
| TOTAL | | 105.67 | 152.35 | 175.48 | 135.07 | 568.57 |

RETAIL DRUG DISTRIBUTION BY ZIP CODE WITHIN STATE BY GRAMS WT

STATE: CALIFORNIA

| ZIP CODE | QUARTER 1 | QUARTER 2 | QUARTER 3 | QUARTER 4 | TOTAL GRAMS |
|---|---|---|---|---|---|
| 900 | 7.48 | 8.55 | 2.14 | 0 | 18.17 |
| 902 | 17.09 | 0 | 1.07 | 0 | 18.16 |
| 903 | 1.07 | 0 | 0 | 0 | 1.07 |
| 913 | 6.41 | 160.86 | 37.39 | 274.3 | 478.96 |
| 914 | 0 | 0 | 1.07 | 0 | 1.07 |
| 917 | 53.73 | 14.95 | 10.68 | 0 | 79.36 |
| 920 | 17.09 | 22.43 | 3.2 | 9.61 | 52.33 |
| 921 | 1.07 | 0 | 1.07 | 0.21 | 2.35 |
| 925 | 1.07 | 1.07 | 0 | 0 | 2.14 |
| 927 | 4.27 | 3.2 | 0 | 0 | 7.47 |
| 928 | 21.26 | 3.2 | 2.14 | 3.2 | 29.8 |
| 930 | 33.97 | 0.64 | 0 | 3.2 | 37.81 |
| 931 | 2.14 | 1.07 | 0 | 2.14 | 5.35 |
| 934 | 21.36 | 5.34 | 1.07 | 2.14 | 29.91 |
| 935 | 1.07 | 0 | 0 | 0 | 1.07 |
| 936 | 0 | 0 | 1.07 | 1.07 | 2.14 |
| 937 | 0 | 0 | 0.13 | 0 | 0.13 |
| 941 | 12.82 | 14.95 | 1.07 | 3.2 | 32.04 |
| 945 | 1.07 | 1.07 | 2.14 | 1.07 | 5.35 |
| 949 | 0 | 0 | 2.14 | 0 | 2.14 |
| 950 | 2.14 | 1.07 | 1.07 | 0 | 4.28 |
| 951 | 1.07 | 4.27 | 1.07 | 2.14 | 8.55 |
| 952 | 0 | 0 | 1.07 | 0 | 1.07 |
| 953 | 0 | 1.07 | 1.07 | 0 | 2.14 |
| 954 | 1.07 | 2.14 | 0 | 0 | 3.21 |
| 955 | 2.14 | 2.14 | 3.2 | 0 | 7.48 |
| 956 | 2.14 | 1.07 | 0 | 0 | 3.21 |
| 957 | 0 | 1.07 | 0 | 0 | 1.07 |
| 958 | 2.14 | 0 | 0 | 0 | 2.14 |
| 959 | 0 | 2.14 | 0 | 0 | 2.14 |
| 960 | 1.07 | 0 | 0 | 0 | 1.07 |
| TOTAL | 214.74 | 252.3 | 73.86 | 302.28 | 843.18 |

STATE: COLORADO

| ZIP CODE | QUARTER 1 | QUARTER 2 | QUARTER 3 | QUARTER 4 | TOTAL GRAMS |
|---|---|---|---|---|---|
| 806 | 0 | 2.14 | 3.2 | 2.14 | 7.48 |
| 808 | 0 | 0 | 2.14 | 0 | 2.14 |
| TOTAL | 0 | 2.14 | 5.34 | 2.14 | 9.62 |

STATE: CONNECTICUT

| ZIP CODE | QUARTER 1 | QUARTER 2 | QUARTER 3 | QUARTER 4 | TOTAL GRAMS |
|---|---|---|---|---|---|
| 64 | 1.07 | 0 | 1.07 | 0 | 2.14 |
| 68 | 0.13 | 0.13 | 1.09 | 0 | 1.35 |
| 69 | 0 | 0 | 0.27 | 0 | 0.27 |
| TOTAL | 1.2 | 0.13 | 2.43 | 0 | 3.76 |

STATE: DELAWARE

| ZIP CODE | QUARTER 1 | QUARTER 2 | QUARTER 3 | QUARTER 4 | TOTAL GRAMS |
|---|---|---|---|---|---|
| 197 | 3.67 | 0.43 | 3.63 | 2.94 | 10.67 |
| 198 | 3.14 | 4.57 | 4.64 | 4.7 | 17.05 |
| 199 | 6.32 | 8.25 | 4.17 | 8.06 | 26.8 |
| TOTAL | 13.13 | 13.25 | 12.44 | 15.7 | 54.52 |

STATE: FLORIDA

| ZIP CODE | QUARTER 1 | QUARTER 2 | QUARTER 3 | QUARTER 4 | TOTAL GRAMS |
|---|---|---|---|---|---|
| 321 | 2.14 | 1.07 | 2.14 | 13.89 | 19.24 |
| 323 | 3.2 | 3.2 | 4.27 | 0 | 10.67 |
| 324 | 1.07 | 1.07 | 0 | 0 | 2.14 |
| 325 | 10.09 | 7.48 | 11.47 | 8.83 | 37.87 |
| 326 | 0 | 1.07 | 0 | 0 | 1.07 |
| 327 | 1.17 | 5.71 | 7.23 | 9.63 | 23.74 |
| 328 | 40.27 | 47.27 | 147.92 | 70.88 | 306.34 |
| 329 | 0 | 1.07 | 0 | 1.07 | 2.14 |
| 330 | 19.22 | 1.76 | 2.56 | 3.2 | 26.74 |
| 331 | 29.91 | 6.14 | 2.35 | 6.2 | 44.6 |
| 333 | 3.85 | 3.26 | 1.5 | 0.64 | 9.25 |
| 334 | 3.69 | 0.96 | 0 | 5.13 | 9.78 |

| | | | | | | |
|---|---|---|---|---|---|---|
| | 335 | 14.95 | 21.36 | 9.61 | 18.16 | 64.08 |
| | 336 | 11.75 | 1.07 | 1.17 | 12.18 | 26.17 |
| | 337 | 1.07 | 2.35 | 5.34 | 22.38 | 31.14 |
| | 338 | 3.2 | 23.01 | 11.46 | 9.99 | 47.66 |
| | 339 | 0 | 0 | 0 | 3.52 | 3.52 |
| | 341 | 2.14 | 4.27 | 1.07 | 2.67 | 10.15 |
| | 342 | 1.07 | 1.07 | 0 | 2.35 | 4.49 |
| | 344 | 0 | 0 | 2.14 | 0 | 2.14 |
| | 346 | 2.14 | 3.2 | 4.01 | 14.85 | 24.2 |
| | 347 | 2.56 | 4.49 | 22.32 | 10.34 | 39.71 |
| | 349 | 5.34 | 6.41 | 2.14 | 3.2 | 17.09 |
| TOTAL | | 158.83 | 147.29 | 238.7 | 219.11 | 763.93 |

| STATE: | GEORGIA | | | | | |
|---|---|---|---|---|---|---|
| ZIP CODE | | QUARTER 1 | QUARTER 2 | QUARTER 3 | QUARTER 4 | TOTAL GRAMS |
| | 300 | 36 | 26.06 | 25.38 | 63.19 | 150.63 |
| | 301 | 5.66 | 4.27 | 19.55 | 111.4 | 140.88 |
| | 302 | 54.76 | 98.27 | 70.56 | 59.13 | 282.72 |
| | 303 | 9.99 | 3.63 | 6.38 | 17.85 | 37.85 |
| | 304 | 94.93 | 43.53 | 42.73 | 30.98 | 212.17 |
| | 305 | 9.29 | 11.75 | 15.87 | 26.11 | 63.02 |
| | 306 | 9.61 | 9.61 | 5.34 | 6.41 | 30.97 |
| | 307 | 2.14 | 3.2 | 5.34 | 3.2 | 13.88 |
| | 310 | 41.02 | 21.37 | 68.74 | 66.6 | 197.73 |
| | 312 | 9.99 | 20.87 | 22.7 | 24.35 | 77.91 |
| | 313 | 1.98 | 21.2 | 11.75 | 10.68 | 45.61 |
| | 314 | 24.79 | 37.01 | 33.11 | 19.23 | 114.14 |
| | 315 | 2.14 | 0 | 0 | 0 | 2.14 |
| | 316 | 0.16 | 0 | 1.07 | 4.27 | 5.5 |
| | 317 | 121.66 | 241.51 | 170.42 | 224.1 | 757.69 |
| | 318 | 1.07 | 1.07 | 0 | 1.07 | 3.21 |
| | 319 | 3.2 | 3.2 | 2.14 | 0 | 8.54 |
| | 398 | 0 | 3.2 | 8.55 | 41.66 | 53.41 |
| TOTAL | | 428.39 | 549.75 | 509.63 | 710.23 | 2,198 |

RETAIL DRUG DISTRIBUTION BY ZIP CODE WITHIN STATE BY GRAMS WT

STATE: HAWAII

| ZIP CODE | QUARTER 1 | QUARTER 2 | QUARTER 3 | QUARTER 4 | TOTAL GRAMS |
|---|---|---|---|---|---|
| 967 | 1.07 | 0 | 0 | 0 | 1.07 |
| TOTAL | 1.07 | 0 | 0 | 0 | 1.07 |

STATE: IDAHO

| ZIP CODE | QUARTER 1 | QUARTER 2 | QUARTER 3 | QUARTER 4 | TOTAL GRAMS |
|---|---|---|---|---|---|
| 836 | 12.82 | 10.68 | 5.34 | 9.61 | 38.45 |
| 837 | 10.68 | 29.91 | 26.7 | 3.2 | 70.49 |
| TOTAL | 23.5 | 40.59 | 32.04 | 12.81 | 108.94 |

STATE: ILLINOIS

| ZIP CODE | QUARTER 1 | QUARTER 2 | QUARTER 3 | QUARTER 4 | TOTAL GRAMS |
|---|---|---|---|---|---|
| 600 | 27.77 | 45.29 | 47.21 | 38.45 | 158.72 |
| 601 | 9.61 | 74.13 | 60.24 | 48.71 | 192.69 |
| 603 | 0 | 0 | 0 | 0.09 | 0.09 |
| 604 | 12.82 | 16.02 | 26.28 | 11.96 | 67.08 |
| 605 | 1.07 | 3.2 | 0 | 2.14 | 6.41 |
| 606 | 49.99 | 67.29 | 51.27 | 20.3 | 188.85 |
| 607 | 12.82 | 0 | 0 | 0 | 12.82 |
| 613 | 6.41 | 4.27 | 0.64 | 0 | 11.32 |
| 614 | 1.07 | 0 | 0 | 0 | 1.07 |
| 619 | 1.07 | 0 | 0 | 0 | 1.07 |
| 624 | 0 | 0 | 4.27 | 0 | 4.27 |
| 627 | 2.14 | 0 | 0 | 12.82 | 14.96 |
| 629 | 2.14 | 4.27 | 2.14 | 3.2 | 11.75 |
| TOTAL | 126.91 | 214.47 | 192.05 | 137.67 | 671.1 |

STATE: INDIANA

| ZIP CODE | QUARTER 1 | QUARTER 2 | QUARTER 3 | QUARTER 4 | TOTAL GRAMS |
|---|---|---|---|---|---|
| 460 | 1.07 | 0 | 0 | 0 | 1.07 |
| 468 | 0 | 0 | 1.07 | 0 | 1.07 |
| 473 | 0 | 0 | 1.07 | 0 | 1.07 |
| 476 | 0 | 0 | 1.07 | 1.07 | 2.14 |

| | | | | | | |
|---|---|---|---|---|---|---|
| | 477 | 2.14 | 0 | 0 | 0 | 2.14 |
| TOTAL | | 3.21 | 0 | 3.21 | 1.07 | 7.49 |

| STATE: | IOWA | | | | |
|---|---|---|---|---|---|
| ZIP CODE | QUARTER 1 | QUARTER 2 | QUARTER 3 | QUARTER 4 | TOTAL GRAMS |
| 500 | 0 | 0 | 0 | 1.07 | 1.07 |
| 528 | 0 | 1.07 | 0 | 0 | 1.07 |
| TOTAL | 0 | 1.07 | 0 | 1.07 | 2.14 |

| STATE: | KANSAS | | | | |
|---|---|---|---|---|---|
| ZIP CODE | QUARTER 1 | QUARTER 2 | QUARTER 3 | QUARTER 4 | TOTAL GRAMS |
| 661 | 0 | 0 | 0 | 2.14 | 2.14 |
| 662 | 1.07 | 10.68 | 0 | 4.27 | 16.02 |
| TOTAL | 1.07 | 10.68 | 0 | 6.41 | 18.16 |

| STATE: | KENTUCKY | | | | |
|---|---|---|---|---|---|
| ZIP CODE | QUARTER 1 | QUARTER 2 | QUARTER 3 | QUARTER 4 | TOTAL GRAMS |
| 401 | 8.55 | 6.41 | 3.2 | 5.34 | 23.5 |
| 402 | 5.34 | 0 | 0 | 4.27 | 9.61 |
| 405 | 1.07 | 0 | 1.07 | 0 | 2.14 |
| 406 | 0 | 1.07 | 0 | 0 | 1.07 |
| 410 | 1.07 | 2.19 | 0 | 0 | 3.26 |
| 421 | 0 | 0 | 3.2 | 2.14 | 5.34 |
| 422 | 6.41 | 6.41 | 6.41 | 6.41 | 25.64 |
| 427 | 8.55 | 3.2 | 0 | 3.2 | 14.95 |
| TOTAL | 30.99 | 19.28 | 13.88 | 21.36 | 85.51 |

| STATE: | LOUISIANA | | | | |
|---|---|---|---|---|---|
| ZIP CODE | QUARTER 1 | QUARTER 2 | QUARTER 3 | QUARTER 4 | TOTAL GRAMS |
| 700 | 73.66 | 85.24 | 65.37 | 59.28 | 283.55 |
| 701 | 7.58 | 7.05 | 2.14 | 5.66 | 22.43 |
| 703 | 34.61 | 30.76 | 35.04 | 26.81 | 127.22 |

| | | | | | | |
|---|---|---|---|---|---|---|
| | 704 | 24.05 | 47.31 | 36.13 | 49.78 | 157.27 |
| | 705 | 391.27 | 539.16 | 438.99 | 270.5 | 1,639.92 |
| | 706 | 7.58 | 2.14 | 2.33 | 4.49 | 16.54 |
| | 707 | 7.99 | 21.41 | 6.05 | 6.02 | 41.47 |
| | 708 | 35.23 | 19.72 | 18.13 | 13.12 | 86.2 |
| | 710 | 3.2 | 8.71 | 5.34 | 2.14 | 19.39 |
| | 711 | 12.39 | 15.06 | 11.34 | 12.48 | 51.27 |
| | 712 | 21.26 | 19.77 | 23.37 | 31.07 | 95.47 |
| | 713 | 0 | 4.27 | 0 | 2.14 | 6.41 |
| | 714 | 2.14 | 2.14 | 0 | 1.07 | 5.35 |
| TOTAL | | 620.96 | 802.74 | 644.23 | 484.56 | 2,552.49 |

STATE: MAINE

| ZIP CODE | | QUARTER 1 | QUARTER 2 | QUARTER 3 | QUARTER 4 | TOTAL GRAMS |
|---|---|---|---|---|---|---|
| | 043 | 0 | 0 | 1.07 | 0 | 1.07 |
| TOTAL | | 0 | 0 | 1.07 | 0 | 1.07 |

STATE: MARYLAND

| ZIP CODE | | QUARTER 1 | QUARTER 2 | QUARTER 3 | QUARTER 4 | TOTAL GRAMS |
|---|---|---|---|---|---|---|
| | 208 | 0 | 4.81 | 0 | 3.2 | 8.01 |
| | 210 | 2.14 | 1.07 | 0 | 1.71 | 4.92 |
| | 211 | 3.2 | 1.07 | 7.48 | 2.14 | 13.89 |
| | 212 | 1.07 | 2.14 | 1.07 | 2.14 | 6.42 |
| | 214 | 0 | 0 | 0.21 | 0.75 | 0.96 |
| | 217 | 1.07 | 1.07 | 0 | 0 | 2.14 |
| | 219 | 0 | 0 | 1.55 | 0.21 | 1.76 |
| TOTAL | | 7.48 | 10.16 | 10.31 | 10.15 | 38.1 |

STATE: MASSACHUSETTS

| ZIP CODE | | QUARTER 1 | QUARTER 2 | QUARTER 3 | QUARTER 4 | TOTAL GRAMS |
|---|---|---|---|---|---|---|
| | 18 | 21.36 | 16.02 | 33.11 | 55.54 | 126.03 |
| TOTAL | | 21.36 | 16.02 | 33.11 | 55.54 | 126.03 |

STATE: MICHIGAN

| ZIP CODE | QUARTER 1 | QUARTER 2 | QUARTER 3 | QUARTER 4 | TOTAL GRAMS |
|---|---|---|---|---|---|
| 480 | 0.16 | 0 | 2.14 | 1.07 | 3.37 |
| 481 | 0 | 0 | 1.07 | 0 | 1.07 |
| 482 | 1.07 | 0 | 0 | 0 | 1.07 |
| 488 | 0 | 0 | 1.07 | 0 | 1.07 |
| 489 | 33.01 | 0 | 0 | 0 | 33.01 |
| 494 | 0 | 0 | 1.07 | 0 | 1.07 |
| 496 | 0 | 0 | 0 | 0.21 | 0.21 |
| TOTAL | 34.24 | 0 | 5.35 | 1.28 | 40.87 |

STATE: MINNESOTA

| ZIP CODE | QUARTER 1 | QUARTER 2 | QUARTER 3 | QUARTER 4 | TOTAL GRAMS |
|---|---|---|---|---|---|
| 551 | 0 | 2.14 | 0 | 2.14 | 4.28 |
| 565 | 0 | 2.14 | 1.07 | 0 | 3.21 |
| 566 | 1.71 | 0 | 0.64 | 0 | 2.35 |
| TOTAL | 1.71 | 4.28 | 1.71 | 2.14 | 9.84 |

STATE: MISSISSIPPI

| ZIP CODE | QUARTER 1 | QUARTER 2 | QUARTER 3 | QUARTER 4 | TOTAL GRAMS |
|---|---|---|---|---|---|
| 386 | 39.58 | 49.78 | 50.1 | 29.53 | 168.99 |
| 387 | 20.03 | 34.02 | 21.68 | 16.02 | 91.75 |
| 388 | 28.2 | 34.93 | 32.15 | 14.95 | 110.23 |
| 389 | 34.23 | 30.5 | 35.2 | 11.75 | 111.68 |
| 390 | 63.34 | 79.74 | 69.48 | 78.94 | 291.5 |
| 391 | 42.25 | 48.28 | 37.65 | 79.58 | 207.76 |
| 392 | 72.47 | 94.16 | 46.3 | 41.93 | 254.86 |
| 393 | 4.27 | 27.51 | 18.21 | 14.69 | 64.68 |
| 394 | 20.94 | 34.61 | 16.29 | 19.23 | 91.07 |
| 395 | 12.18 | 12.82 | 11.75 | 8.55 | 45.3 |
| 396 | 8.55 | 6.41 | 8.55 | 13.89 | 37.4 |
| 397 | 21.52 | 38.77 | 32.95 | 37.55 | 130.79 |
| TOTAL | 367.56 | 491.53 | 380.31 | 366.61 | 1,606.01 |

STATE: MISSOURI

| ZIP CODE | QUARTER 1 | QUARTER 2 | QUARTER 3 | QUARTER 4 | TOTAL GRAMS |
|---|---|---|---|---|---|
| 630 | 34.18 | 28.84 | 37.39 | 25.64 | 126.05 |
| 631 | 2.14 | 7.48 | 8.55 | 5.34 | 23.51 |
| 633 | 5.34 | 4.27 | 4.27 | 4.27 | 18.15 |
| 641 | 4.27 | 0 | 4.27 | 0 | 8.54 |
| 646 | 0 | 2.14 | 0 | 1.07 | 3.21 |
| 656 | 0 | 0 | 0 | 5.34 | 5.34 |
| TOTAL | 45.93 | 42.73 | 54.48 | 41.66 | 184.8 |

STATE: MONTANA

| ZIP CODE | QUARTER 1 | QUARTER 2 | QUARTER 3 | QUARTER 4 | TOTAL GRAMS |
|---|---|---|---|---|---|
| 594 | 1.07 | 0 | 0 | 0 | 1.07 |
| TOTAL | 1.07 | 0 | 0 | 0 | 1.07 |

STATE: NEBRASKA

| ZIP CODE | QUARTER 1 | QUARTER 2 | QUARTER 3 | QUARTER 4 | TOTAL GRAMS |
|---|---|---|---|---|---|
| 681 | 1.07 | 0 | 0.64 | 2.78 | 4.49 |
| 685 | 1.07 | 1.07 | 3.2 | 0 | 5.34 |
| 686 | 1.07 | 1.07 | 0 | 0 | 2.14 |
| TOTAL | 3.21 | 2.14 | 3.84 | 2.78 | 11.97 |

STATE: NEVADA

| ZIP CODE | QUARTER 1 | QUARTER 2 | QUARTER 3 | QUARTER 4 | TOTAL GRAMS |
|---|---|---|---|---|---|
| 890 | 0 | 2.14 | 0 | 0 | 2.14 |
| 891 | 43.37 | 28.84 | 1.07 | 0 | 73.28 |
| TOTAL | 43.37 | 30.98 | 1.07 | 0 | 75.42 |

STATE: NEW JERSEY

| ZIP CODE | QUARTER 1 | QUARTER 2 | QUARTER 3 | QUARTER 4 | TOTAL GRAMS |
|---|---|---|---|---|---|
| 70 | 9.3 | 5.68 | 13.25 | 18.68 | 46.91 |
| 71 | 229.65 | 256.36 | 149.54 | 122.84 | 758.39 |
| 72 | 0 | 0 | 2.14 | 0 | 2.14 |
| 73 | 0.51 | 0.38 | 2.24 | 7.9 | 11.03 |
| 74 | 2.86 | 3.12 | 2.73 | 3.39 | 12.1 |
| 76 | 0 | 0 | 3.01 | 4.44 | 7.45 |
| 77 | 2.16 | 1.07 | 1.6 | 6.09 | 10.92 |
| 79 | 4.72 | 0.85 | 0.51 | 3.48 | 9.56 |
| 80 | 54.36 | 71.43 | 14.25 | 8.91 | 148.95 |
| 81 | 0 | 0 | 0.45 | 0 | 0.45 |
| 82 | 30.02 | 22.88 | 38.03 | 94.85 | 185.78 |
| 83 | 1.07 | 0 | 0 | 2.14 | 3.21 |
| 84 | 0 | 0 | 0 | 1.07 | 1.07 |
| 85 | 1.67 | 2.48 | 0.98 | 0.26 | 5.39 |
| 86 | 3.85 | 0.45 | 1.58 | 3.2 | 9.08 |
| 87 | 15.7 | 4.49 | 7.48 | 6.11 | 33.78 |
| 88 | 0 | 0 | 0 | 2.14 | 2.14 |
| 89 | 0 | 0.19 | 0 | 0.21 | 0.4 |
| TOTAL | 355.87 | 369.38 | 237.79 | 285.71 | 1,248.75 |

STATE: NEW MEXICO

| ZIP CODE | QUARTER 1 | QUARTER 2 | QUARTER 3 | QUARTER 4 | TOTAL GRAMS |
|---|---|---|---|---|---|
| 870 | 2.14 | 1.07 | 0 | 0 | 3.21 |
| 871 | 2.14 | 5.34 | 12.82 | 8.55 | 28.85 |
| 874 | 1.07 | 0 | 0 | 0 | 1.07 |
| 875 | 13.89 | 4.27 | 10.68 | 7.48 | 36.32 |
| 877 | 0 | 1.07 | 0 | 0 | 1.07 |
| 880 | 116.43 | 76.91 | 76.91 | 1.07 | 271.32 |
| 882 | 0 | 2.14 | 0 | 0 | 2.14 |
| TOTAL | 135.67 | 90.8 | 100.41 | 17.1 | 343.98 |

STATE: NEW YORK

| ZIP CODE | QUARTER 1 | QUARTER 2 | QUARTER 3 | QUARTER 4 | TOTAL GRAMS |
|---|---|---|---|---|---|
| 100 | 9.61 | 65.16 | 3.2 | 0 | 77.97 |

| | | | | | |
|---|---|---|---|---|---|
| 103 | 0 | 0 | 0 | 0.53 | 0.53 |
| 104 | 1.07 | 3.2 | 0 | 1.07 | 5.34 |
| 109 | 0 | 0 | 0 | 1.07 | 1.07 |
| 110 | 1.07 | 2.14 | 2.14 | 1.07 | 6.42 |
| 111 | 0 | 0 | 2.14 | 1.07 | 3.21 |
| 112 | 34.18 | 42.73 | 21.36 | 49.14 | 147.41 |
| 113 | 22.43 | 37.39 | 47 | 51.27 | 158.09 |
| 114 | 2.14 | 0 | 1.07 | 3.2 | 6.41 |
| 115 | 1.07 | 2.14 | 19.23 | 6.41 | 28.85 |
| 117 | 23.5 | 9.61 | 8.55 | 9.61 | 51.27 |
| 118 | 0 | 0 | 6.41 | 6.41 | 12.82 |
| 125 | 1.07 | 0 | 0 | 1.07 | 2.14 |
| 130 | 5.34 | 0 | 0 | 0 | 5.34 |
| 136 | 0 | 5.34 | 5.34 | 3.2 | 13.88 |
| 141 | 5.34 | 0 | 0 | 0 | 5.34 |
| 143 | 2.14 | 0 | 0 | 1.07 | 3.21 |
| TOTAL | 108.96 | 167.71 | 116.44 | 136.19 | 529.3 |

STATE: NORTH CAROLINA

| ZIP CODE | QUARTER 1 | QUARTER 2 | QUARTER 3 | QUARTER 4 | TOTAL GRAMS |
|---|---|---|---|---|---|
| 271 | 0 | 0 | 1.07 | 1.07 | 2.14 |
| 272 | 1.07 | 2.14 | 1.07 | 1.07 | 5.35 |
| 273 | 0 | 1.07 | 0 | 0 | 1.07 |
| 275 | 2.14 | 0 | 1.07 | 0 | 3.21 |
| 276 | 0 | 2.14 | 0 | 0 | 2.14 |
| 278 | 3.2 | 2.14 | 0 | 0 | 5.34 |
| 281 | 0 | 0 | 1.07 | 3.2 | 4.27 |
| 282 | 0 | 0 | 0 | 1.07 | 1.07 |
| 283 | 1.07 | 1.07 | 0 | 0 | 2.14 |
| 284 | 1.07 | 0 | 0 | 0 | 1.07 |
| 285 | 0 | 0 | 0 | 1.07 | 1.07 |
| 286 | 5.34 | 1.07 | 1.07 | 1.07 | 8.55 |
| 289 | 0 | 0 | 1.07 | 0 | 1.07 |
| TOTAL | 13.89 | 9.63 | 6.42 | 8.55 | 38.49 |

STATE: OHIO

| ZIP CODE | QUARTER 1 | QUARTER 2 | QUARTER 3 | QUARTER 4 | TOTAL GRAMS |
|---|---|---|---|---|---|
| 430 | 5.34 | 9.61 | 4.27 | 5.34 | 24.56 |
| 432 | 33.11 | 8.55 | 20.3 | 8.55 | 70.51 |
| 440 | 1.07 | 0 | 0 | 0 | 1.07 |
| 441 | 0 | 2.14 | 2.14 | 0 | 4.28 |
| 450 | 3.2 | 2.4 | 0 | 1.6 | 7.2 |
| 451 | 0 | 0 | 1.6 | 0 | 1.6 |
| 452 | 1.87 | 8.01 | 4.01 | 4.01 | 17.9 |
| 453 | 1.28 | 0 | 3.74 | 1.07 | 6.09 |
| 454 | 0 | 0.96 | 0.8 | 0 | 1.76 |
| 455 | 0 | 0 | 1.07 | 0 | 1.07 |
| 456 | 2.14 | 2.14 | 0 | 0 | 4.28 |
| TOTAL | 48.01 | 33.81 | 37.93 | 20.57 | 140.32 |

STATE: OKLAHOMA

| ZIP CODE | QUARTER 1 | QUARTER 2 | QUARTER 3 | QUARTER 4 | TOTAL GRAMS |
|---|---|---|---|---|---|
| 730 | 55.44 | 62.97 | 60.05 | 49.03 | 227.49 |
| 731 | 97.95 | 90.63 | 103.61 | 75.2 | 367.39 |
| 734 | 22 | 26.7 | 15.27 | 19.23 | 83.2 |
| 735 | 25.42 | 19.23 | 11.75 | 20.3 | 76.7 |
| 736 | 0 | 2.14 | 1.07 | 0 | 3.21 |
| 737 | 0 | 3.2 | 1.07 | 2.14 | 6.41 |
| 738 | 1.07 | 0 | 0 | 0 | 1.07 |
| 740 | 15.06 | 15.25 | 18.69 | 20.46 | 69.46 |
| 741 | 7.07 | 11.39 | 20.66 | 26.4 | 65.52 |
| 743 | 18.16 | 17.09 | 16.02 | 12.82 | 64.09 |
| 744 | 16.54 | 20.58 | 32.78 | 24.03 | 93.93 |
| 745 | 0 | 0 | 1.07 | 1.07 | 2.14 |
| 746 | 0 | 5.34 | 1.07 | 0 | 6.41 |
| 747 | 2.24 | 4.27 | 5.34 | 1.07 | 12.92 |
| 748 | 11.75 | 10.68 | 5.34 | 7.48 | 35.25 |
| 749 | 0.11 | 9.38 | 1.5 | 3.2 | 14.19 |
| TOTAL | 272.81 | 298.85 | 295.29 | 262.43 | 1,129.38 |

DATE RANGE: 01/01/2017 TO 12/31/2017 | ARCOS 3 - REPORT 1 | Run Date: 07/03/2018

RETAIL DRUG DISTRIBUTION BY ZIP CODE WITHIN STATE BY GRAMS WT

STATE: PENNSYLVANIA

| ZIP CODE | QUARTER 1 | QUARTER 2 | QUARTER 3 | QUARTER 4 | TOTAL GRAMS |
|---|---|---|---|---|---|
| 157 | 2.14 | 0 | 1.07 | 0 | 3.21 |
| 170 | 1.07 | 0 | 0 | 0 | 1.07 |
| 180 | 0 | 1.28 | 0.98 | 5.34 | 7.6 |
| 181 | 0 | 0 | 1.15 | 0.51 | 1.66 |
| 182 | 4.27 | 1.07 | 5.34 | 2.14 | 12.82 |
| 184 | 0 | 0 | 0 | 2.14 | 2.14 |
| 186 | 2.14 | 0 | 0 | 0 | 2.14 |
| 187 | 1.07 | 1.07 | 0 | 0 | 2.14 |
| 189 | 0 | 1.07 | 0 | 0 | 1.07 |
| 190 | 73.88 | 25.02 | 43.04 | 25.68 | 167.62 |
| 191 | 115.97 | 10.87 | 7.97 | 6.71 | 141.52 |
| 193 | 5.04 | 1.74 | 3.59 | 5.34 | 15.71 |
| 194 | 0 | 0 | 1.39 | 2.48 | 3.87 |
| TOTAL | 205.58 | 42.12 | 64.53 | 50.34 | 362.57 |

STATE: RHODE ISLAND

| ZIP CODE | QUARTER 1 | QUARTER 2 | QUARTER 3 | QUARTER 4 | TOTAL GRAMS |
|---|---|---|---|---|---|
| 28 | 0 | 1.07 | 0 | 2.14 | 3.21 |
| TOTAL | 0 | 1.07 | 0 | 2.14 | 3.21 |

STATE: SOUTH CAROLINA

| ZIP CODE | QUARTER 1 | QUARTER 2 | QUARTER 3 | QUARTER 4 | TOTAL GRAMS |
|---|---|---|---|---|---|
| 290 | 0 | 0 | 0 | 1.07 | 1.07 |
| 291 | 1.07 | 1.07 | 1.07 | 1.07 | 4.28 |
| 293 | 2.14 | 2.14 | 1.07 | 2.14 | 7.49 |
| 294 | 3.2 | 0 | 0 | 2.14 | 5.34 |
| 295 | 6.41 | 7.48 | 4.27 | 7.48 | 25.64 |
| 296 | 0 | 0 | 1.07 | 0 | 1.07 |
| 297 | 1.07 | 2.14 | 2.14 | 0 | 5.35 |
| 298 | 0 | 1.07 | 1.07 | 0 | 2.14 |
| TOTAL | 13.89 | 13.9 | 10.69 | 13.9 | 52.38 |

STATE: SOUTH DAKOTA

| ZIP CODE | QUARTER 1 | QUARTER 2 | QUARTER 3 | QUARTER 4 | TOTAL GRAMS |
|---|---|---|---|---|---|
| 572 | 0 | 0 | 0 | 1.07 | 1.07 |
| TOTAL | 0 | 0 | 0 | 1.07 | 1.07 |

STATE: TENNESSEE

| ZIP CODE | QUARTER 1 | QUARTER 2 | QUARTER 3 | QUARTER 4 | TOTAL GRAMS |
|---|---|---|---|---|---|
| 370 | 258.79 | 210.13 | 167.7 | 96.37 | 732.99 |
| 371 | 15.89 | 28.75 | 18.16 | 9.82 | 72.62 |
| 372 | 50.8 | 42.3 | 43.79 | 48.22 | 185.11 |
| 373 | 3.2 | 2.14 | 4.27 | 3.2 | 12.81 |
| 374 | 9.61 | 4.27 | 0 | 3.2 | 17.08 |
| 377 | 15.6 | 92.08 | 59.6 | 11.32 | 178.6 |
| 378 | 48.87 | 25.85 | 32.21 | 30.55 | 137.48 |
| 379 | 36.17 | 23.23 | 54.12 | 92.45 | 205.97 |
| 380 | 21.15 | 19.97 | 24.35 | 14.53 | 80 |
| 381 | 14.47 | 26.33 | 25.69 | 23.07 | 89.56 |
| 382 | 0.51 | 2.95 | 2.14 | 2.35 | 7.95 |
| 383 | 24.67 | 32.21 | 72.53 | 64.62 | 194.03 |
| 384 | 6.41 | 5.34 | 3.2 | 2.14 | 17.09 |
| 385 | 1.23 | 1.07 | 1.07 | 2.35 | 5.72 |
| TOTAL | 507.37 | 516.62 | 508.83 | 404.19 | 1,937.01 |

STATE: TEXAS

| ZIP CODE | QUARTER 1 | QUARTER 2 | QUARTER 3 | QUARTER 4 | TOTAL GRAMS |
|---|---|---|---|---|---|
| 750 | 353.99 | 307.77 | 273.43 | 256.91 | 1,192.10 |
| 751 | 83.15 | 75.43 | 83.57 | 68.45 | 310.6 |
| 752 | 227.08 | 236.8 | 197.4 | 142.5 | 803.78 |
| 754 | 42.26 | 39.52 | 42.94 | 31.28 | 156 |
| 755 | 26.17 | 28.84 | 32.04 | 25.32 | 112.37 |
| 756 | 7.69 | 12.07 | 6.73 | 11.06 | 37.55 |
| 757 | 8.55 | 5.34 | 3.2 | 2.14 | 19.23 |
| 758 | 2.14 | 3.2 | 7.48 | 7.48 | 20.3 |

| | | | | | | |
|---|---|---|---|---|---|---|
| 759 | 47.69 | 52.06 | | 25.85 | 18.39 | 143.99 |
| 760 | 134.5 | 171.15 | | 86.92 | 97.54 | 490.11 |
| 761 | 96.74 | 134.43 | | 156.87 | 295.26 | 683.3 |
| 762 | 96.24 | 82.84 | | 100.78 | 63.18 | 343.04 |
| 763 | 116.96 | 114.1 | | 144.33 | 135.28 | 510.67 |
| 764 | 13.89 | 7.48 | | 6.41 | 3.2 | 30.98 |
| 765 | 13.89 | 20.3 | | 14.95 | 17.09 | 66.23 |
| 766 | 1.07 | 5.34 | | 4.27 | 2.14 | 12.82 |
| 767 | 0 | 0 | | 0 | 2.14 | 2.14 |
| 768 | 0 | 0 | | 1.07 | 5.34 | 6.41 |
| 769 | 1.39 | 0.21 | | 0.21 | 0 | 1.81 |
| 770 | 500.63 | 619.89 | | 436.15 | 503.27 | 2,059.94 |
| 773 | 96.13 | 120.56 | | 268.59 | 312.91 | 798.19 |
| 774 | 99.86 | 79.89 | | 64.89 | 159.63 | 404.27 |
| 775 | 119.15 | 141.29 | | 95.71 | 116.8 | 472.95 |
| 776 | 23.71 | 15.38 | | 7.48 | 26.06 | 72.63 |
| 777 | 16.66 | 14.32 | | 4.7 | 15.49 | 51.17 |
| 778 | 5.34 | 6.41 | | 1.07 | 3.2 | 16.02 |
| 779 | 8.94 | 4.7 | | 4.49 | 12.82 | 30.95 |
| 780 | 29 | 28.84 | | 25.64 | 18.16 | 101.64 |
| 781 | 50.42 | 48.28 | | 72.74 | 59.25 | 230.69 |
| 782 | 108.85 | 102.49 | | 55.9 | 50.04 | 317.28 |
| 783 | 84.65 | 86.52 | | 113.01 | 89.47 | 373.65 |
| 784 | 164.98 | 160.01 | | 182.23 | 108.03 | 615.25 |
| 785 | 61.1 | 51.7 | | 129.46 | 163.11 | 405.37 |
| 786 | 11.75 | 7.48 | | 1.07 | 4.27 | 24.57 |
| 787 | 7.48 | 6.41 | | 4.27 | 9.61 | 27.77 |
| 788 | 1.07 | 1.07 | | 0 | 0 | 2.14 |
| 789 | 1.17 | 0 | | 2.14 | 0 | 3.31 |
| 790 | 1.07 | 0 | | 0 | 0 | 1.07 |
| 791 | 0 | 0 | | 2.14 | 0 | 2.14 |
| 792 | 1.07 | 1.07 | | 0 | 0 | 2.14 |
| 793 | 2.14 | 0 | | 0 | 0 | 2.14 |
| 794 | 6.41 | 3.2 | | 4.27 | 1.07 | 14.95 |
| 796 | 1.07 | 1.07 | | 0 | 0 | 2.14 |
| 797 | 6.41 | 10.68 | | 13.89 | 13.89 | 44.87 |
| 799 | 5.34 | 0 | | 1.07 | 2.14 | 8.55 |

| | | | | | | |
|---|---|---|---|---|---|---|
| TOTAL | | 2,687.80 | 2,808.14 | 2,679.36 | 2,853.92 | 11,029.22 |

| STATE: | UTAH | | | | | |
|---|---|---|---|---|---|---|
| ZIP CODE | | QUARTER 1 | QUARTER 2 | QUARTER 3 | QUARTER 4 | TOTAL GRAMS |
| | 841 | 0 | 0 | 1.07 | 0 | 1.07 |
| TOTAL | | 0 | 0 | 1.07 | 0 | 1.07 |

| STATE: | VERMONT | | | | | |
|---|---|---|---|---|---|---|
| ZIP CODE | | QUARTER 1 | QUARTER 2 | QUARTER 3 | QUARTER 4 | TOTAL GRAMS |
| | 57 | 0 | 1.07 | 0 | 0 | 1.07 |
| TOTAL | | 0 | 1.07 | 0 | 0 | 1.07 |

| STATE: | VIRGINIA | | | | | |
|---|---|---|---|---|---|---|
| ZIP CODE | | QUARTER 1 | QUARTER 2 | QUARTER 3 | QUARTER 4 | TOTAL GRAMS |
| | 221 | 0 | 0 | 0 | 1.07 | 1.07 |
| | 226 | 3.2 | 0 | 0 | 1.07 | 4.27 |
| | 228 | 0 | 2.14 | 2.14 | 2.14 | 6.42 |
| | 229 | 2.14 | 0 | 0 | 0 | 2.14 |
| | 230 | 0 | 1.07 | 0 | 0 | 1.07 |
| | 232 | 3.2 | 1.07 | 0 | 0 | 4.27 |
| | 234 | 0 | 0 | 1.07 | 0 | 1.07 |
| | 235 | 1.07 | 0 | 0 | 3.2 | 4.27 |
| | 236 | 1.07 | 0 | 0 | 0 | 1.07 |
| | 245 | 0 | 0 | 0 | 1.07 | 1.07 |
| TOTAL | | 10.68 | 4.28 | 3.21 | 8.55 | 26.72 |

| STATE: | WEST VIRGINIA | | | | | |
|---|---|---|---|---|---|---|
| ZIP CODE | | QUARTER 1 | QUARTER 2 | QUARTER 3 | QUARTER 4 | TOTAL GRAMS |
| | 260 | 5.34 | 2.14 | 0 | 0 | 7.48 |
| | 265 | 0 | 0.21 | 0 | 0 | 0.21 |
| TOTAL | | 5.34 | 2.35 | 0 | 0 | 7.69 |

STATE: WISCONSIN

| ZIP CODE | QUARTER 1 | QUARTER 2 | QUARTER 3 | QUARTER 4 | TOTAL GRAMS |
|---|---|---|---|---|---|
| 531 | 4.27 | 4.27 | 5.34 | 2.14 | 16.02 |
| 532 | 0 | 3.2 | 0 | 0 | 3.2 |
| 535 | 0 | 0 | 1.07 | 1.07 | 2.14 |
| TOTAL | 4.27 | 7.47 | 6.41 | 3.21 | 21.36 |

DRUG CODE: 9143 DRUG NAME: OXYCODONE

STATE: ALABAMA

| ZIP CODE | QUARTER 1 | QUARTER 2 | QUARTER 3 | QUARTER 4 | TOTAL GRAMS |
|---|---|---|---|---|---|
| 350 | 29,789.33 | 28,630.79 | 27,401.23 | 26,192.96 | 112,014.31 |
| 351 | 16,285.31 | 15,074.68 | 15,191.16 | 15,139.27 | 61,690.42 |
| 352 | 23,345.29 | 22,157.51 | 21,566.74 | 21,852.12 | 88,921.66 |
| 354 | 8,763.44 | 8,645.65 | 8,065.72 | 7,928.77 | 33,403.58 |
| 355 | 13,165.55 | 13,076.66 | 12,564.58 | 12,249.29 | 51,056.08 |
| 356 | 21,930.62 | 20,682.05 | 19,727.50 | 19,458.45 | 81,798.62 |
| 357 | 11,061.92 | 10,580.64 | 9,965.46 | 10,010.97 | 41,618.99 |
| 358 | 16,204.15 | 15,565.50 | 14,520.18 | 13,776.42 | 60,066.25 |
| 359 | 20,925.80 | 19,394.30 | 18,500.69 | 19,613.59 | 78,434.38 |
| 360 | 8,578.75 | 8,338.33 | 8,006.03 | 7,660.58 | 32,583.69 |
| 361 | 6,094.88 | 6,037.76 | 5,132.88 | 5,000.56 | 22,266.08 |
| 362 | 11,114.85 | 10,171.87 | 9,759.19 | 9,684.88 | 40,730.79 |
| 363 | 9,438.82 | 9,599.10 | 8,988.36 | 9,118.79 | 37,145.07 |
| 364 | 5,113.84 | 5,105.06 | 4,216.31 | 5,014.52 | 19,449.73 |
| 365 | 19,848.44 | 18,335.25 | 16,846.91 | 16,820.27 | 71,850.87 |
| 366 | 13,808.94 | 13,688.42 | 13,548.65 | 12,848.94 | 53,894.95 |
| 367 | 2,648.15 | 2,403.07 | 2,254.35 | 2,256.19 | 9,561.76 |
| 368 | 6,287.84 | 5,984.71 | 5,914.59 | 5,987.05 | 24,174.19 |
| 369 | 628.22 | 614.77 | 667.44 | 669.26 | 2,579.69 |
| TOTAL | 245,034.14 | 234,086.12 | 222,837.97 | 221,282.88 | 923,241.11 |

STATE: ALASKA

| ZIP CODE | QUARTER 1 | QUARTER 2 | QUARTER 3 | QUARTER 4 | TOTAL GRAMS |
|---|---|---|---|---|---|
| 995 | 19,826.95 | 19,533.82 | 18,778.82 | 18,691.95 | 76,831.54 |
| 996 | 10,652.08 | 10,396.10 | 9,833.01 | 9,890.24 | 40,771.43 |
| 997 | 3,553.02 | 3,457.86 | 3,472.52 | 2,912.13 | 13,395.53 |
| 998 | 1,269.50 | 1,201.51 | 1,168.81 | 1,100.88 | 4,740.70 |
| 999 | 608.41 | 517.5 | 490.7 | 439.8 | 2,056.41 |
| TOTAL | 35,909.96 | 35,106.79 | 33,743.86 | 33,035 | 137,795.61 |

STATE: AMERICAN SAMOA

| ZIP CODE | QUARTER 1 | QUARTER 2 | QUARTER 3 | QUARTER 4 | TOTAL GRAMS |
|---|---|---|---|---|---|
| 967 | 14.34 | 12.1 | 10.76 | 21.52 | 58.72 |
| TOTAL | 14.34 | 12.1 | 10.76 | 21.52 | 58.72 |

STATE: ARIZONA

| ZIP CODE | QUARTER 1 | QUARTER 2 | QUARTER 3 | QUARTER 4 | TOTAL GRAMS |
|---|---|---|---|---|---|
| 850 | 97,820.04 | 91,456.11 | 86,068.50 | 84,010.42 | 359,355.07 |
| 851 | 22,355.40 | 21,921.31 | 20,871.70 | 21,204.52 | 86,352.93 |
| 852 | 113,162.34 | 106,998.55 | 97,836.47 | 94,617.24 | 412,614.60 |
| 853 | 84,090.25 | 77,518.58 | 73,761.02 | 72,757.88 | 308,127.73 |
| 855 | 8,685.15 | 8,069.04 | 7,334.34 | 7,071.28 | 31,159.81 |
| 856 | 14,364.38 | 13,734.35 | 13,008.98 | 13,248.12 | 54,355.83 |
| 857 | 68,536.84 | 65,986.49 | 64,002.85 | 62,804.54 | 261,330.72 |
| 859 | 6,984.83 | 6,810.24 | 6,569.16 | 6,922.18 | 27,286.41 |
| 860 | 7,220.14 | 6,809.27 | 6,365.51 | 6,087.90 | 26,482.82 |
| 863 | 13,960.82 | 13,913.49 | 13,292.38 | 12,756.95 | 53,923.64 |
| 864 | 21,839.49 | 19,401.73 | 18,907.39 | 18,138.50 | 78,287.11 |
| 865 | 291.36 | 300.33 | 257.74 | 275.67 | 1,125.10 |
| TOTAL | 459,311.04 | 432,919.49 | 408,276.04 | 399,895.20 | 1,700,401.77 |

STATE: ARKANSAS

| ZIP CODE | QUARTER 1 | QUARTER 2 | QUARTER 3 | QUARTER 4 | TOTAL GRAMS |
|---|---|---|---|---|---|
| 716 | 4,639.27 | 4,369.22 | 4,408.93 | 4,342.09 | 17,759.51 |
| 717 | 3,103.90 | 3,038 | 2,871.51 | 2,701.07 | 11,714.48 |
| 718 | 3,322.80 | 3,343.09 | 3,362.69 | 3,129.13 | 13,157.71 |

| | | | | | | |
|---|---|---|---|---|---|---|
| | 719 | 6,269.45 | 5,669.43 | 5,477.76 | 5,160.95 | 22,577.59 |
| | 720 | 17,888.14 | 17,430.40 | 16,813.54 | 16,735.65 | 68,867.73 |
| | 721 | 16,785.75 | 16,633.40 | 16,097.51 | 15,106.30 | 64,622.96 |
| | 722 | 12,595.52 | 13,017.89 | 12,411.20 | 12,223.51 | 50,248.12 |
| | 723 | 4,947.87 | 5,010.23 | 5,137.27 | 4,876.14 | 19,971.51 |
| | 724 | 6,407.17 | 6,058.76 | 5,839.41 | 5,795.42 | 24,100.76 |
| | 725 | 5,816.53 | 5,705.79 | 5,634.29 | 5,649.07 | 22,805.68 |
| | 726 | 8,148.99 | 7,439.84 | 7,321.47 | 7,259.90 | 30,170.20 |
| | 727 | 33,789.07 | 32,423.69 | 31,182.23 | 29,617.50 | 127,012.49 |
| | 728 | 5,768.08 | 5,831.11 | 5,750.73 | 5,487.01 | 22,836.93 |
| | 729 | 21,021.09 | 21,216.58 | 20,291.65 | 19,612.66 | 82,141.98 |
| TOTAL | | 150,503.63 | 147,187.43 | 142,600.19 | 137,696.40 | 577,987.65 |

STATE: CALIFORNIA

| ZIP CODE | | QUARTER 1 | QUARTER 2 | QUARTER 3 | QUARTER 4 | TOTAL GRAMS |
|---|---|---|---|---|---|---|
| | 900 | 26,993.80 | 26,621.11 | 26,204.41 | 24,265.40 | 104,084.72 |
| | 902 | 16,748.92 | 16,630.90 | 16,084.64 | 16,141.05 | 65,605.51 |
| | 903 | 1,239.70 | 1,215.35 | 1,427.24 | 1,289.18 | 5,171.47 |
| | 904 | 4,921.01 | 4,891.87 | 4,920.79 | 5,013.74 | 19,747.41 |
| | 905 | 3,907.79 | 3,989.17 | 3,730.84 | 3,652.29 | 15,280.09 |
| | 906 | 8,684.15 | 7,956.90 | 7,794.22 | 7,567.49 | 32,002.76 |
| | 907 | 8,977.89 | 9,030.32 | 9,139.76 | 8,468.77 | 35,616.74 |
| | 908 | 7,058.19 | 6,469.63 | 6,226.69 | 5,797.22 | 25,551.73 |
| | 910 | 5,140.67 | 5,262.59 | 4,997 | 4,324.78 | 19,725.04 |
| | 911 | 2,886.67 | 2,997.48 | 2,908.41 | 2,853.87 | 11,646.43 |
| | 912 | 3,628.52 | 3,329.35 | 3,397.49 | 3,270.24 | 13,625.60 |
| | 913 | 33,102.07 | 31,982.62 | 31,331.52 | 30,635.12 | 127,051.33 |
| | 914 | 8,467.49 | 8,808.70 | 8,449.28 | 8,980.74 | 34,706.21 |
| | 915 | 3,834.17 | 3,514.81 | 3,573.46 | 3,336.86 | 14,259.30 |
| | 916 | 2,230.08 | 1,935.75 | 1,828.81 | 1,849.84 | 7,844.48 |
| | 917 | 28,285.33 | 27,739.72 | 27,270.29 | 25,868.10 | 109,163.44 |
| | 918 | 438.84 | 498.23 | 500.02 | 465.26 | 1,902.35 |
| | 919 | 12,859.10 | 13,044.15 | 12,319.32 | 11,860.38 | 50,082.95 |
| | 920 | 55,312.42 | 53,778.20 | 52,443.25 | 50,287.18 | 211,821.05 |
| | 921 | 37,417.13 | 35,167.06 | 34,229.01 | 32,502.15 | 139,315.35 |

| | | | | | |
|---|---|---|---|---|---|
| 922 | 31,436.40 | 30,022.88 | 28,844.25 | 28,144.60 | 118,448.13 |
| 923 | 33,315.16 | 32,816.02 | 32,349.35 | 31,074.27 | 129,554.80 |
| 924 | 5,114.22 | 5,342.18 | 5,155.20 | 5,250.74 | 20,862.34 |
| 925 | 32,543.54 | 32,174.82 | 30,555.56 | 30,247.56 | 125,521.48 |
| 926 | 30,259.09 | 29,399.79 | 28,648.98 | 27,962.58 | 116,270.44 |
| 927 | 6,204.33 | 6,470.96 | 5,636.40 | 5,283.70 | 23,595.39 |
| 928 | 25,069.36 | 24,333.50 | 23,312.08 | 22,888.31 | 95,603.25 |
| 930 | 18,642.50 | 18,232.61 | 17,398.98 | 16,609.46 | 70,883.55 |
| 931 | 3,157.93 | 3,187.06 | 2,811.16 | 2,702.27 | 11,858.42 |
| 932 | 10,671.63 | 10,167.02 | 10,411.43 | 10,163.43 | 41,413.51 |
| 933 | 14,513.41 | 14,103.70 | 13,872.18 | 13,607.71 | 56,097 |
| 934 | 16,433.35 | 15,534.56 | 15,137.79 | 14,297.66 | 61,403.36 |
| 935 | 23,227.78 | 22,448.56 | 21,424.35 | 21,623.23 | 88,723.92 |
| 936 | 12,680.05 | 12,788.21 | 12,073.27 | 11,524.62 | 49,066.15 |
| 937 | 14,594.84 | 14,564.68 | 14,324.16 | 14,084.09 | 57,567.77 |
| 939 | 7,977.98 | 7,689.27 | 7,332.84 | 6,714.69 | 29,714.78 |
| 940 | 14,276.82 | 13,738.46 | 13,880.60 | 13,584.93 | 55,480.81 |
| 941 | 24,862.07 | 23,735.45 | 22,912.49 | 22,875.33 | 94,385.34 |
| 943 | 3,969.55 | 3,640.80 | 3,265.98 | 3,133.95 | 14,010.28 |
| 944 | 2,429.69 | 2,222.52 | 2,237.07 | 2,137.19 | 9,026.47 |
| 945 | 58,725.70 | 56,371.42 | 54,772.63 | 52,056.46 | 221,926.21 |
| 946 | 7,132.42 | 6,661.65 | 5,853.84 | 5,594.49 | 25,242.40 |
| 947 | 1,589.05 | 1,476.90 | 1,615.77 | 1,626.81 | 6,308.53 |
| 948 | 1,936.39 | 1,887.64 | 1,927.05 | 1,808.61 | 7,559.69 |
| 949 | 10,955.98 | 11,087.04 | 10,685.77 | 10,517.30 | 43,246.09 |
| 950 | 15,054.45 | 14,610.97 | 13,478.07 | 13,231.12 | 56,374.61 |
| 951 | 10,308.65 | 9,738.87 | 9,707.74 | 9,359.97 | 39,115.23 |
| 952 | 11,613.35 | 11,275.85 | 10,741.41 | 9,807.05 | 43,437.66 |
| 953 | 40,167.97 | 39,429.06 | 38,433.67 | 36,833.37 | 154,864.07 |
| 954 | 26,075.09 | 25,407.62 | 23,536.43 | 21,771.02 | 96,790.16 |
| 955 | 6,822.33 | 6,869.11 | 6,415.89 | 6,194.81 | 26,302.14 |
| 956 | 37,813.87 | 35,526.49 | 34,344.40 | 33,785.19 | 141,469.95 |
| 957 | 6,866.11 | 6,481.29 | 6,115.23 | 6,040.75 | 25,503.38 |
| 958 | 23,831.90 | 23,954.98 | 22,443.25 | 22,241.99 | 92,472.12 |
| 959 | 25,443.40 | 24,583.66 | 22,906.14 | 21,960.28 | 94,893.48 |
| 960 | 13,807.70 | 12,680.93 | 12,282 | 11,633.83 | 50,404.46 |
| 961 | 3,660.75 | 3,378.68 | 3,181.69 | 3,009.83 | 13,230.95 |

| | | | | | | |
|---|---|---|---|---|---|---|
| TOTAL | | 905,318.75 | 878,899.12 | 848,801.55 | 819,812.86 | 3,452,832.28 |

| | | | | | | |
|---|---|---|---|---|---|---|
| STATE: | COLORADO | | | | | |
| ZIP CODE | | QUARTER 1 | QUARTER 2 | QUARTER 3 | QUARTER 4 | TOTAL GRAMS |
| | 800 | 38,570.68 | 37,557.23 | 35,810.85 | 34,826.69 | 146,765.45 |
| | 801 | 27,148.98 | 26,568.62 | 25,370.52 | 26,258.44 | 105,346.56 |
| | 802 | 46,177.90 | 45,987.03 | 43,966.09 | 42,189.81 | 178,320.83 |
| | 803 | 4,598.36 | 4,332 | 3,799.29 | 3,594.25 | 16,323.90 |
| | 804 | 6,529.08 | 6,437.87 | 6,200.12 | 6,364.97 | 25,532.04 |
| | 805 | 21,112.47 | 20,517.25 | 19,098.91 | 18,702.60 | 79,431.23 |
| | 806 | 10,740.80 | 10,522.02 | 9,931.54 | 10,006.05 | 41,200.41 |
| | 807 | 2,246.90 | 2,210.84 | 2,078.13 | 2,133.73 | 8,669.60 |
| | 808 | 5,486.77 | 5,413.05 | 5,078.76 | 5,113.01 | 21,091.59 |
| | 809 | 29,035.05 | 28,312.13 | 27,690.80 | 26,687.81 | 111,725.79 |
| | 810 | 20,698.20 | 20,621.65 | 19,622.63 | 19,252.05 | 80,194.53 |
| | 811 | 3,159.88 | 2,381.55 | 2,456.04 | 2,261.31 | 10,258.78 |
| | 812 | 3,693.77 | 3,567.30 | 3,371.32 | 3,117.88 | 13,750.27 |
| | 813 | 1,686.21 | 1,574.64 | 1,671.60 | 1,479.31 | 6,411.76 |
| | 814 | 2,545.96 | 2,511.92 | 2,406.60 | 2,332.44 | 9,796.92 |
| | 815 | 6,025.55 | 5,953.22 | 5,234.63 | 5,167.96 | 22,381.36 |
| | 816 | 4,097.32 | 3,784.93 | 3,756.83 | 3,766.91 | 15,405.99 |
| TOTAL | | 233,553.88 | 228,253.25 | 217,544.66 | 213,255.22 | 892,607.01 |

| | | | | | | |
|---|---|---|---|---|---|---|
| STATE: | CONNECTICUT | | | | | |
| ZIP CODE | | QUARTER 1 | QUARTER 2 | QUARTER 3 | QUARTER 4 | TOTAL GRAMS |
| | 60 | 32,191.37 | 31,126.43 | 29,398.55 | 27,515.35 | 120,231.70 |
| | 61 | 9,039.01 | 9,125.32 | 8,860.27 | 8,246.86 | 35,271.46 |
| | 62 | 5,263.50 | 4,846.78 | 4,689.37 | 4,366.11 | 19,165.76 |
| | 63 | 14,415.65 | 14,504.69 | 14,152.20 | 12,964.90 | 56,037.44 |
| | 64 | 47,393.93 | 45,703.96 | 42,786.99 | 42,604.68 | 178,489.56 |
| | 65 | 17,426.61 | 17,028.21 | 15,973.40 | 16,801.97 | 67,230.19 |
| | 66 | 10,895.05 | 10,798 | 9,996.72 | 10,031.78 | 41,721.55 |
| | 67 | 19,876.66 | 18,682.01 | 17,136.34 | 17,067.11 | 72,762.12 |
| | 68 | 16,381.99 | 16,107.02 | 14,990.50 | 14,945.10 | 62,424.61 |

| | | | | | | |
|---|---|---|---|---|---|---|
| | 69 | 2,530.28 | 2,657.73 | 2,459.04 | 2,479.25 | 10,126.30 |
| TOTAL | | 175,414.05 | 170,580.15 | 160,443.38 | 157,023.11 | 663,460.69 |

STATE: DELAWARE

| ZIP CODE | | QUARTER 1 | QUARTER 2 | QUARTER 3 | QUARTER 4 | TOTAL GRAMS |
|---|---|---|---|---|---|---|
| | 197 | 25,785.21 | 24,403.34 | 23,442.69 | 23,666.38 | 97,297.62 |
| | 198 | 15,735.70 | 14,870.84 | 14,636.04 | 14,135.64 | 59,378.22 |
| | 199 | 30,538.56 | 29,348.89 | 28,676.51 | 28,567.77 | 117,131.73 |
| TOTAL | | 72,059.47 | 68,623.07 | 66,755.24 | 66,369.79 | 273,807.57 |

STATE: DISTRICT OF COLUMBIA

| ZIP CODE | | QUARTER 1 | QUARTER 2 | QUARTER 3 | QUARTER 4 | TOTAL GRAMS |
|---|---|---|---|---|---|---|
| | 200 | 13,862.86 | 13,929.59 | 13,387.86 | 12,537.71 | 53,718.02 |
| | 203 | 63.2 | 41.24 | 25.55 | 8.97 | 138.96 |
| | 204 | 1,185.80 | 1,072.12 | 1,145.64 | 893.18 | 4,296.74 |
| | 205 | 1.34 | 0 | 0.9 | 0 | 2.24 |
| TOTAL | | 15,113.20 | 15,042.95 | 14,559.95 | 13,439.86 | 58,155.96 |

STATE: FLORIDA

| ZIP CODE | | QUARTER 1 | QUARTER 2 | QUARTER 3 | QUARTER 4 | TOTAL GRAMS |
|---|---|---|---|---|---|---|
| | 320 | 35,269.44 | 34,882.46 | 33,540.32 | 33,680.45 | 137,372.67 |
| | 321 | 31,127.34 | 30,124.09 | 28,608.90 | 30,509.14 | 120,369.47 |
| | 322 | 64,163.16 | 63,726.14 | 62,909.21 | 63,131.08 | 253,929.59 |
| | 323 | 13,168.98 | 13,219.87 | 12,676.56 | 12,944 | 52,009.41 |
| | 324 | 16,257.04 | 15,952.46 | 15,148.72 | 16,083.35 | 63,441.57 |
| | 325 | 31,504.49 | 31,077.23 | 30,160.78 | 29,796.30 | 122,538.80 |
| | 326 | 24,188.49 | 24,080.41 | 23,411.47 | 23,542.77 | 95,223.14 |
| | 327 | 45,952.58 | 46,277.32 | 44,703.74 | 45,627.35 | 182,560.99 |
| | 328 | 30,000.83 | 29,648.36 | 28,773.59 | 29,418.26 | 117,841.04 |
| | 329 | 55,015.84 | 53,381.09 | 52,434.69 | 53,606.99 | 214,438.61 |
| | 330 | 66,921.29 | 68,438.85 | 64,967.67 | 69,053.38 | 269,381.19 |
| | 331 | 55,407.98 | 54,837.02 | 54,580.02 | 58,444.26 | 223,269.28 |
| | 333 | 46,263.16 | 46,158.93 | 46,793.70 | 48,575.53 | 187,791.32 |

| | | | | | | |
|---|---|---|---|---|---|---|
| | 334 | 83,210.52 | 83,015 | 81,632.12 | 83,222.19 | 331,079.83 |
| | 335 | 41,636.02 | 40,917.26 | 39,846.72 | 42,199.85 | 164,599.85 |
| | 336 | 58,394.15 | 58,661.86 | 57,432.21 | 57,647.53 | 232,135.75 |
| | 337 | 52,622.73 | 51,830.13 | 51,305.33 | 52,131.76 | 207,889.95 |
| | 338 | 34,394.80 | 32,752.11 | 30,438.30 | 34,076.06 | 131,661.27 |
| | 339 | 46,502.15 | 45,075.71 | 41,729.06 | 45,745.04 | 179,051.96 |
| | 341 | 18,546.75 | 17,719.56 | 16,932.75 | 18,733.41 | 71,932.47 |
| | 342 | 51,537.84 | 50,297.84 | 46,560.36 | 50,189.95 | 198,585.99 |
| | 344 | 32,343.35 | 32,857.87 | 31,223.73 | 33,192.30 | 129,617.25 |
| | 346 | 50,400.99 | 50,727.21 | 48,980.95 | 50,739.24 | 200,848.39 |
| | 347 | 25,826.64 | 24,562.46 | 22,987.68 | 25,165.82 | 98,542.60 |
| | 349 | 29,980.31 | 29,141.47 | 28,585.19 | 30,250.12 | 117,957.09 |
| TOTAL | | 1,040,636.87 | 1,029,362.71 | 996,363.77 | 1,037,706.13 | 4,104,069.48 |

STATE: GEORGIA

| ZIP CODE | QUARTER 1 | QUARTER 2 | QUARTER 3 | QUARTER 4 | TOTAL GRAMS |
|---|---|---|---|---|---|
| 300 | 80,080.25 | 79,569.02 | 78,361.46 | 79,018.01 | 317,028.74 |
| 301 | 69,406.49 | 69,440.67 | 68,052.62 | 67,732.63 | 274,632.41 |
| 302 | 41,502.82 | 40,988.79 | 40,872.38 | 40,828.31 | 164,192.30 |
| 303 | 28,428.48 | 28,780.22 | 27,315.74 | 26,961.14 | 111,485.58 |
| 304 | 10,286.27 | 10,115.76 | 9,743.80 | 9,892.50 | 40,038.33 |
| 305 | 39,437.12 | 39,528.43 | 38,077.03 | 38,350.24 | 155,392.82 |
| 306 | 27,952.08 | 27,980.56 | 27,710.74 | 28,471.92 | 112,115.30 |
| 307 | 19,549.39 | 18,804.03 | 18,194.75 | 17,358.53 | 73,906.70 |
| 308 | 8,159.09 | 7,703.45 | 7,278.83 | 7,632.80 | 30,774.17 |
| 309 | 17,309.37 | 16,089.57 | 15,658.87 | 15,175.38 | 64,233.19 |
| 310 | 26,029.56 | 26,346.63 | 24,964.64 | 25,268.61 | 102,609.44 |
| 312 | 12,528.42 | 12,247 | 12,360.09 | 12,005.78 | 49,141.29 |
| 313 | 12,640.05 | 12,204.21 | 10,834.05 | 9,128.58 | 44,806.89 |
| 314 | 10,812.66 | 10,448.39 | 10,158.20 | 9,976.52 | 41,395.77 |
| 315 | 20,312.54 | 20,516.42 | 19,901.87 | 18,427.73 | 79,158.56 |
| 316 | 6,727.58 | 6,486.91 | 6,330.60 | 6,966.73 | 26,511.82 |
| 317 | 12,958.99 | 12,804.33 | 12,130.63 | 12,773.92 | 50,667.87 |
| 318 | 906.79 | 916.35 | 943.75 | 913.35 | 3,680.24 |
| 319 | 7,764.38 | 7,774.27 | 7,968 | 6,495.50 | 30,002.15 |
| 398 | 2,731.15 | 2,784.03 | 2,666.05 | 2,670.47 | 10,851.70 |

| | | | | | | |
|---|---|---|---|---|---|---|
| TOTAL | | 455,523.48 | 451,529.04 | 439,524.10 | 436,048.65 | 1,782,625.27 |

| STATE: | GUAM | | | | | |
|---|---|---|---|---|---|---|
| ZIP CODE | | QUARTER 1 | QUARTER 2 | QUARTER 3 | QUARTER 4 | TOTAL GRAMS |
| | 969 | 697.48 | 908.57 | 980.23 | 835.09 | 3,421.37 |
| TOTAL | | 697.48 | 908.57 | 980.23 | 835.09 | 3,421.37 |

| STATE: | HAWAII | | | | | |
|---|---|---|---|---|---|---|
| ZIP CODE | | QUARTER 1 | QUARTER 2 | QUARTER 3 | QUARTER 4 | TOTAL GRAMS |
| | 967 | 32,036.17 | 30,560.72 | 29,798.67 | 28,221.53 | 120,617.09 |
| | 968 | 13,571.25 | 13,465.52 | 13,419.34 | 12,357.53 | 52,813.64 |
| TOTAL | | 45,607.42 | 44,026.24 | 43,218.01 | 40,579.06 | 173,430.73 |

| STATE: | IDAHO | | | | | |
|---|---|---|---|---|---|---|
| ZIP CODE | | QUARTER 1 | QUARTER 2 | QUARTER 3 | QUARTER 4 | TOTAL GRAMS |
| | 832 | 6,491.97 | 6,386.24 | 6,269.95 | 6,374.24 | 25,522.40 |
| | 833 | 7,171.66 | 7,015.35 | 6,983.06 | 6,665.87 | 27,835.94 |
| | 834 | 5,983.88 | 5,886.70 | 5,672.78 | 5,650.20 | 23,193.56 |
| | 835 | 2,009.05 | 1,884.76 | 1,898.35 | 1,811.66 | 7,603.82 |
| | 836 | 18,276.40 | 18,288.65 | 18,011.76 | 17,843.22 | 72,420.03 |
| | 837 | 13,485.84 | 13,207.94 | 13,360.08 | 12,893.64 | 52,947.50 |
| | 838 | 8,040.48 | 7,958.41 | 8,062.23 | 7,768.88 | 31,830 |
| TOTAL | | 61,459.28 | 60,628.05 | 60,258.21 | 59,007.71 | 241,353.25 |

| STATE: | ILLINOIS | | | | | |
|---|---|---|---|---|---|---|
| ZIP CODE | | QUARTER 1 | QUARTER 2 | QUARTER 3 | QUARTER 4 | TOTAL GRAMS |
| | 600 | 26,360.75 | 25,865.97 | 24,851.11 | 24,022.09 | 101,099.92 |
| | 601 | 16,626.99 | 15,895.41 | 15,013.04 | 14,851.68 | 62,387.12 |
| | 602 | 571.52 | 542.41 | 560.62 | 489.87 | 2,164.42 |
| | 603 | 558.12 | 643.06 | 529.7 | 530.84 | 2,261.72 |
| | 604 | 23,206.59 | 22,321.68 | 20,626.17 | 20,900.79 | 87,055.23 |
| | 605 | 11,072.87 | 10,790.88 | 10,411.04 | 10,156.61 | 42,431.40 |
| | 606 | 13,574.04 | 13,406.78 | 13,201.32 | 13,172.86 | 53,355 |

| | | | | | | |
|---|---|---|---|---|---|---|
| | 607 | 1,449.38 | 1,548.21 | 1,535.02 | 1,352.66 | 5,885.27 |
| | 608 | 928.19 | 704.52 | 720.21 | 660.05 | 3,012.97 |
| | 609 | 3,004.17 | 3,071.56 | 2,839.21 | 2,827.71 | 11,742.65 |
| | 610 | 3,279.84 | 3,135 | 3,063.84 | 2,988.92 | 12,467.60 |
| | 611 | 3,633.97 | 3,546.97 | 3,380.87 | 3,592.14 | 14,153.95 |
| | 612 | 1,886.04 | 1,743.77 | 1,692.92 | 1,564.62 | 6,887.35 |
| | 613 | 1,827.81 | 1,750.30 | 1,670.05 | 1,852.98 | 7,101.14 |
| | 614 | 1,417.37 | 1,328.93 | 1,301.05 | 1,252.16 | 5,299.51 |
| | 615 | 2,579.64 | 2,412.10 | 2,282.22 | 2,126.37 | 9,400.33 |
| | 616 | 3,082.88 | 2,951.32 | 2,784.23 | 2,738.38 | 11,556.81 |
| | 617 | 2,427.10 | 2,404.80 | 2,200.31 | 2,194.68 | 9,226.89 |
| | 618 | 3,845.95 | 3,807.11 | 3,970.23 | 3,746.82 | 15,370.11 |
| | 619 | 932.94 | 887.98 | 894.89 | 863.05 | 3,578.86 |
| | 620 | 9,149.70 | 8,689.01 | 8,244.65 | 8,613.02 | 34,696.38 |
| | 622 | 10,153.35 | 9,767.73 | 9,584.40 | 9,247.10 | 38,752.58 |
| | 623 | 1,979.92 | 2,022.96 | 1,892.53 | 1,847.66 | 7,743.07 |
| | 624 | 2,198.94 | 2,117.99 | 2,092.19 | 2,080.39 | 8,489.51 |
| | 625 | 1,999.93 | 2,018.34 | 1,860.13 | 2,049.83 | 7,928.23 |
| | 626 | 2,298.32 | 2,009.36 | 2,059.80 | 2,020.17 | 8,387.65 |
| | 627 | 1,761.53 | 1,599.13 | 1,661.56 | 1,578.35 | 6,600.57 |
| | 628 | 4,923.32 | 4,854.62 | 4,703.47 | 4,688.46 | 19,169.87 |
| | 629 | 8,917.27 | 8,825.05 | 8,548.41 | 8,116.66 | 34,407.39 |
| TOTAL | | 165,648.44 | 160,662.95 | 154,175.19 | 152,126.92 | 632,613.50 |

| STATE: | | INDIANA | | | | |
|---|---|---|---|---|---|---|
| ZIP CODE | | QUARTER 1 | QUARTER 2 | QUARTER 3 | QUARTER 4 | TOTAL GRAMS |
| | 460 | 13,315.83 | 13,099.48 | 12,494.55 | 12,372.35 | 51,282.21 |
| | 461 | 18,720.37 | 17,834.81 | 16,892.37 | 16,813.58 | 70,261.13 |
| | 462 | 42,099.76 | 41,528.52 | 39,828.32 | 40,819.78 | 164,276.38 |
| | 463 | 19,237.20 | 18,461.57 | 17,314.72 | 16,622.48 | 71,635.97 |
| | 464 | 2,024.39 | 1,983.79 | 1,953.72 | 1,811.60 | 7,773.50 |
| | 465 | 18,557.13 | 18,358.04 | 17,015.58 | 17,174.43 | 71,105.18 |
| | 466 | 7,357.54 | 7,643.49 | 7,033.52 | 6,791.08 | 28,825.63 |
| | 467 | 8,447.81 | 8,280.04 | 7,913.89 | 7,891.43 | 32,533.17 |
| | 468 | 11,366.91 | 11,045.37 | 10,738.41 | 10,284.46 | 43,435.15 |
| | 469 | 10,457.80 | 10,030.13 | 9,416.02 | 9,095.82 | 38,999.77 |

| | | | | | | |
|---|---|---|---|---|---|---|
| | 470 | 7,073.84 | 7,527.54 | 7,116.96 | 7,517.28 | 29,235.62 |
| | 471 | 17,913.99 | 18,491.43 | 17,161.91 | 15,510.01 | 69,077.34 |
| | 472 | 7,931.82 | 7,433.46 | 7,080.63 | 7,118.56 | 29,564.47 |
| | 473 | 13,280.37 | 12,507.16 | 12,447.26 | 12,117.49 | 50,352.28 |
| | 474 | 8,854.62 | 8,243.54 | 7,828.03 | 7,698.81 | 32,625 |
| | 475 | 5,720.92 | 5,480.60 | 5,106.10 | 5,115.27 | 21,422.89 |
| | 476 | 4,435.86 | 4,164.25 | 4,135.75 | 4,084.79 | 16,820.65 |
| | 477 | 11,913.88 | 11,917.13 | 11,270.88 | 10,939.22 | 46,041.11 |
| | 478 | 3,483.13 | 3,231.17 | 3,122.84 | 3,139.84 | 12,976.98 |
| | 479 | 9,384.11 | 9,204.57 | 8,908.27 | 8,566.86 | 36,063.81 |
| TOTAL | | 241,577.28 | 236,466.09 | 224,779.73 | 221,485.14 | 924,308.24 |

STATE: IOWA

| ZIP CODE | | QUARTER 1 | QUARTER 2 | QUARTER 3 | QUARTER 4 | TOTAL GRAMS |
|---|---|---|---|---|---|---|
| | 500 | 4,770.32 | 4,623.21 | 4,406.12 | 4,458.01 | 18,257.66 |
| | 501 | 3,412.74 | 3,233.23 | 3,118.26 | 3,158.08 | 12,922.31 |
| | 502 | 3,723.85 | 3,555.71 | 3,396.58 | 3,368.84 | 14,044.98 |
| | 503 | 10,967.18 | 10,505.18 | 10,434.83 | 10,417.04 | 42,324.23 |
| | 504 | 1,973.51 | 2,017.63 | 1,671.60 | 1,797.29 | 7,460.03 |
| | 505 | 1,636.16 | 1,671.89 | 1,517.86 | 1,553.70 | 6,379.61 |
| | 506 | 1,602.07 | 1,557.71 | 1,467.93 | 1,537.24 | 6,164.95 |
| | 507 | 1,672.53 | 1,572.84 | 1,412.77 | 1,428.76 | 6,086.90 |
| | 508 | 439.24 | 446.59 | 448.61 | 477.26 | 1,811.70 |
| | 510 | 698.88 | 791.28 | 721.97 | 718.93 | 2,931.06 |
| | 511 | 1,341.75 | 1,393.76 | 1,452.01 | 1,538.17 | 5,725.69 |
| | 512 | 466.27 | 443.77 | 381.06 | 378.59 | 1,669.69 |
| | 513 | 538.02 | 506.87 | 529.5 | 450.75 | 2,025.14 |
| | 514 | 820.36 | 805.69 | 690.32 | 798.96 | 3,115.33 |
| | 515 | 3,874.49 | 3,697.65 | 3,240.46 | 3,153.45 | 13,966.05 |
| | 516 | 448.34 | 477.7 | 445.96 | 485.68 | 1,857.68 |
| | 520 | 1,461.70 | 1,272.96 | 1,223.90 | 1,210 | 5,168.56 |
| | 521 | 670.51 | 572.77 | 489.98 | 561.88 | 2,295.14 |
| | 522 | 2,950.48 | 2,897.73 | 2,741.67 | 2,850.71 | 11,440.59 |
| | 523 | 1,418.76 | 1,340.31 | 1,232.13 | 1,265.41 | 5,256.61 |
| | 524 | 2,777.37 | 2,817.30 | 2,624.32 | 2,703.25 | 10,922.24 |

| | | | | | | |
|---|---|---|---|---|---|---|
| | 525 | 2,127.35 | 2,034.37 | 2,125.55 | 2,181.47 | 8,468.74 |
| | 526 | 2,138.82 | 2,179.48 | 2,077.84 | 2,077.01 | 8,473.15 |
| | 527 | 1,984.65 | 1,843.90 | 1,642.41 | 1,739.73 | 7,210.69 |
| | 528 | 2,131.40 | 1,973.20 | 1,807.62 | 1,932.74 | 7,844.96 |
| TOTAL | | 56,046.75 | 54,232.73 | 51,301.26 | 52,242.95 | 213,823.69 |

STATE: KANSAS

| ZIP CODE | | QUARTER 1 | QUARTER 2 | QUARTER 3 | QUARTER 4 | TOTAL GRAMS |
|---|---|---|---|---|---|---|
| | 660 | 17,228.91 | 16,851.95 | 16,985.87 | 16,859.87 | 67,926.60 |
| | 661 | 8,673.22 | 8,280.74 | 7,956.36 | 7,912.14 | 32,822.46 |
| | 662 | 15,430.25 | 15,260.52 | 14,592.71 | 14,924.02 | 60,207.50 |
| | 664 | 2,799.12 | 2,480.30 | 2,528.76 | 2,167.01 | 9,975.19 |
| | 665 | 2,861.62 | 3,100.41 | 2,967.35 | 2,957.42 | 11,886.80 |
| | 666 | 6,873.50 | 6,899.27 | 6,157.09 | 6,027.50 | 25,957.36 |
| | 667 | 5,461.66 | 5,634.48 | 5,474.84 | 5,278.83 | 21,849.81 |
| | 668 | 1,962.11 | 1,961.08 | 1,864.62 | 1,665.74 | 7,453.55 |
| | 669 | 710.97 | 718.5 | 673.5 | 683.08 | 2,786.05 |
| | 670 | 8,974.39 | 9,052.07 | 8,803.44 | 8,551.90 | 35,381.80 |
| | 671 | 3,963.87 | 4,030.08 | 4,241.60 | 3,709.89 | 15,945.44 |
| | 672 | 26,000.47 | 26,366.39 | 25,877.83 | 25,731.05 | 103,975.74 |
| | 673 | 2,409.44 | 2,470.61 | 2,423.96 | 2,473.06 | 9,777.07 |
| | 674 | 5,312.98 | 5,386.19 | 5,561.29 | 5,252.69 | 21,513.15 |
| | 675 | 6,159.25 | 5,719.70 | 5,472.82 | 5,381.25 | 22,733.02 |
| | 676 | 2,681.48 | 2,518.58 | 2,699.08 | 2,561.94 | 10,461.08 |
| | 677 | 1,337.07 | 1,436.78 | 1,315.84 | 1,481.32 | 5,571.01 |
| | 678 | 4,459.73 | 4,454.85 | 4,495.91 | 4,386.01 | 17,796.50 |
| | 679 | 1,174.36 | 1,097.95 | 1,129.32 | 1,075.68 | 4,477.31 |
| TOTAL | | 124,474.40 | 123,720.45 | 121,222.19 | 119,080.40 | 488,497.44 |

STATE: KENTUCKY

| ZIP CODE | | QUARTER 1 | QUARTER 2 | QUARTER 3 | QUARTER 4 | TOTAL GRAMS |
|---|---|---|---|---|---|---|
| | 400 | 8,649.42 | 8,614.20 | 7,947.11 | 7,958.64 | 33,169.37 |
| | 401 | 4,439.89 | 4,446.65 | 3,894.01 | 4,027.82 | 16,808.37 |
| | 402 | 37,021.50 | 35,923.74 | 34,662.27 | 34,697.89 | 142,305.40 |

| | | | | | | |
|---|---|---|---|---|---|---|
| | 403 | 14,545.63 | 14,587.12 | 13,572.05 | 13,871.78 | 56,576.58 |
| | 404 | 7,733.55 | 7,784.02 | 7,406.87 | 7,385.60 | 30,310.04 |
| | 405 | 12,563.98 | 11,801.45 | 11,741.84 | 11,224.34 | 47,331.61 |
| | 406 | 1,458.56 | 1,486.45 | 1,396.63 | 1,432.16 | 5,773.80 |
| | 407 | 6,405.01 | 6,663.52 | 6,455.96 | 6,442.13 | 25,966.62 |
| | 408 | 1,238.25 | 1,227.26 | 1,227.54 | 1,194.59 | 4,887.64 |
| | 409 | 3,682.60 | 3,426.06 | 3,620.12 | 3,626.72 | 14,355.50 |
| | 410 | 30,478.38 | 29,580.49 | 28,051.07 | 28,321.90 | 116,431.84 |
| | 411 | 5,953.06 | 5,392.14 | 4,912.27 | 4,668.70 | 20,926.17 |
| | 412 | 2,077.70 | 2,139.77 | 2,001.17 | 2,163.90 | 8,382.54 |
| | 413 | 1,561.59 | 1,556.86 | 1,563.36 | 1,732.44 | 6,414.25 |
| | 414 | 830.61 | 841.59 | 779.06 | 814.3 | 3,265.56 |
| | 415 | 3,556.24 | 3,820.35 | 3,619.84 | 3,704.06 | 14,700.49 |
| | 416 | 3,097.12 | 2,984.87 | 3,052.90 | 3,105.23 | 12,240.12 |
| | 417 | 4,419.37 | 4,375.70 | 4,262.38 | 4,233.68 | 17,291.13 |
| | 418 | 1,798.42 | 1,809.05 | 1,719.98 | 1,760.79 | 7,088.24 |
| | 420 | 12,616.49 | 12,449.32 | 12,104.29 | 12,281.52 | 49,451.62 |
| | 421 | 10,380.44 | 10,070.76 | 10,282.41 | 10,115.61 | 40,849.22 |
| | 422 | 6,382.14 | 6,331.68 | 6,508.51 | 5,941.24 | 25,163.57 |
| | 423 | 7,385.45 | 7,311.09 | 6,934.57 | 6,847.28 | 28,478.39 |
| | 424 | 8,606.37 | 8,442.92 | 8,028.99 | 8,014.94 | 33,093.22 |
| | 425 | 2,578.86 | 2,743.65 | 2,550.40 | 2,965.87 | 10,838.78 |
| | 426 | 3,655.86 | 3,432.18 | 3,719.71 | 3,505.38 | 14,313.13 |
| | 427 | 7,574.28 | 7,494.38 | 7,368.14 | 6,831.81 | 29,268.61 |
| TOTAL | | 210,690.77 | 206,737.27 | 199,383.45 | 198,870.32 | 815,681.81 |

| STATE: | LOUISIANA | | | | | |
|---|---|---|---|---|---|---|
| ZIP CODE | | QUARTER 1 | QUARTER 2 | QUARTER 3 | QUARTER 4 | TOTAL GRAMS |
| | 700 | 38,533.60 | 36,003.49 | 32,969.74 | 32,039.88 | 139,546.71 |
| | 701 | 13,990 | 13,980.22 | 13,321.14 | 13,061.44 | 54,352.80 |
| | 703 | 9,696.27 | 9,410.14 | 8,980.66 | 8,463.08 | 36,550.15 |
| | 704 | 30,016.34 | 28,386.23 | 27,701.56 | 26,441.66 | 112,545.79 |
| | 705 | 22,011.24 | 22,002.08 | 21,198.66 | 20,931.98 | 86,143.96 |
| | 706 | 6,649.97 | 6,452.10 | 6,348.63 | 6,196.58 | 25,647.28 |
| | 707 | 22,502.06 | 21,617.31 | 20,774.82 | 20,556.03 | 85,450.22 |
| | 708 | 15,788.16 | 15,163.43 | 14,810 | 14,419.72 | 60,181.31 |

| | | | | | | |
|---|---|---|---|---|---|---|
| | 710 | 3,812.50 | 3,797.31 | 3,735.77 | 3,924.29 | 15,269.87 |
| | 711 | 14,858.35 | 14,602.02 | 14,397.98 | 14,179.93 | 58,038.28 |
| | 712 | 7,177.17 | 6,901.86 | 6,678.16 | 6,335.40 | 27,092.59 |
| | 713 | 12,403.35 | 11,902.59 | 11,465.91 | 11,112.82 | 46,884.67 |
| | 714 | 6,239.56 | 5,827.53 | 5,360.35 | 5,107.95 | 22,535.39 |
| TOTAL | | 203,678.57 | 196,046.31 | 187,743.38 | 182,770.76 | 770,239.02 |

STATE: MAINE

| ZIP CODE | | QUARTER 1 | QUARTER 2 | QUARTER 3 | QUARTER 4 | TOTAL GRAMS |
|---|---|---|---|---|---|---|
| | 039 | 985.48 | 771.39 | 810.93 | 748.04 | 3,315.84 |
| | 040 | 14,683.11 | 13,702.52 | 13,602.34 | 13,284.42 | 55,272.39 |
| | 041 | 3,859.76 | 3,459.40 | 3,316.78 | 3,288.32 | 13,924.26 |
| | 042 | 6,133.49 | 5,935.37 | 5,886.86 | 5,972.56 | 23,928.28 |
| | 043 | 7,122.56 | 6,913.49 | 6,843.73 | 6,752.96 | 27,632.74 |
| | 044 | 5,695.31 | 5,295.34 | 4,909.85 | 4,918.43 | 20,818.93 |
| | 045 | 1,475.63 | 1,514.93 | 1,537.58 | 1,387.80 | 5,915.94 |
| | 046 | 2,941.05 | 2,593.72 | 2,453.17 | 2,331.77 | 10,319.71 |
| | 047 | 1,896.54 | 1,868.34 | 1,635.47 | 1,643.89 | 7,044.24 |
| | 048 | 1,182.02 | 1,197.56 | 1,206.84 | 1,184.54 | 4,770.96 |
| | 049 | 8,186.17 | 7,787.83 | 7,854.97 | 7,535.41 | 31,364.38 |
| TOTAL | | 54,161.12 | 51,039.89 | 50,058.52 | 49,048.14 | 204,307.67 |

STATE: MARYLAND

| ZIP CODE | | QUARTER 1 | QUARTER 2 | QUARTER 3 | QUARTER 4 | TOTAL GRAMS |
|---|---|---|---|---|---|---|
| | 206 | 25,821.83 | 25,131.23 | 23,476.59 | 22,999.35 | 97,429 |
| | 207 | 29,375.27 | 29,545.13 | 28,383.13 | 27,183.54 | 114,487.07 |
| | 208 | 24,323.84 | 23,986.81 | 22,431.08 | 21,199.46 | 91,941.19 |
| | 209 | 4,718.57 | 4,738.53 | 4,186.81 | 4,345.44 | 17,989.35 |
| | 210 | 55,543.20 | 54,174.71 | 50,978.92 | 49,398.14 | 210,094.97 |
| | 211 | 26,669.07 | 25,241.95 | 23,251.80 | 22,255.71 | 97,418.53 |
| | 212 | 105,152.54 | 101,300.49 | 95,119.43 | 95,180.40 | 396,752.86 |
| | 214 | 4,750.10 | 4,473.73 | 4,076.89 | 4,144.33 | 17,445.05 |
| | 215 | 9,038.34 | 8,522.51 | 7,944.14 | 7,508.14 | 33,013.13 |
| | 216 | 10,523.70 | 10,395.78 | 9,910.97 | 9,797.71 | 40,628.16 |
| | 217 | 29,801.14 | 28,455.47 | 27,305.82 | 27,091.67 | 112,654.10 |

| | | | | | | |
|---|---|---|---|---|---|---|
| | 218 | 9,715.03 | 9,040.24 | 9,087.01 | 8,504.23 | 36,346.51 |
| | 219 | 7,961.29 | 7,407.72 | 6,788.86 | 7,019.70 | 29,177.57 |
| TOTAL | | 343,393.92 | 332,414.30 | 312,941.45 | 306,627.82 | 1,295,377.49 |

STATE: MASSACHUSETTS

| ZIP CODE | | QUARTER 1 | QUARTER 2 | QUARTER 3 | QUARTER 4 | TOTAL GRAMS |
|---|---|---|---|---|---|---|
| | 010 | 26,189.68 | 25,598.40 | 24,110.76 | 23,949.31 | 99,848.15 |
| | 011 | 10,387.39 | 10,243.14 | 9,694.26 | 9,186.94 | 39,511.73 |
| | 012 | 4,270.86 | 4,184.70 | 4,037.03 | 4,005.21 | 16,497.80 |
| | 013 | 3,961.91 | 4,075.64 | 4,012.18 | 3,819.60 | 15,869.33 |
| | 014 | 8,323.48 | 7,855.42 | 7,545.98 | 7,425.12 | 31,150 |
| | 015 | 11,702.07 | 11,091.49 | 11,096.50 | 10,900.60 | 44,790.66 |
| | 016 | 10,023.36 | 9,384.89 | 8,945.29 | 8,674.25 | 37,027.79 |
| | 017 | 9,706.18 | 9,423.24 | 8,795.97 | 8,711.65 | 36,637.04 |
| | 018 | 33,128.24 | 31,667.10 | 29,581.90 | 28,614.60 | 122,991.84 |
| | 019 | 16,796.53 | 16,627.35 | 15,327.50 | 15,276.29 | 64,027.67 |
| | 020 | 11,428.71 | 11,212.91 | 10,773.33 | 10,427.67 | 43,842.62 |
| | 021 | 37,608.22 | 35,967.49 | 34,866.89 | 34,124.48 | 142,567.08 |
| | 022 | 3,088.19 | 2,567.84 | 2,484.22 | 2,406.88 | 10,547.13 |
| | 023 | 19,686.92 | 18,674.98 | 17,620.23 | 16,919.09 | 72,901.22 |
| | 024 | 5,840.91 | 5,761.96 | 5,193.07 | 5,468.09 | 22,264.03 |
| | 025 | 4,828.31 | 4,988.12 | 4,622.47 | 4,733.18 | 19,172.08 |
| | 026 | 6,794.48 | 6,674.81 | 6,577.90 | 6,401.11 | 26,448.30 |
| | 027 | 29,359.63 | 28,622.17 | 27,891.50 | 27,369.65 | 113,242.95 |
| TOTAL | | 253,125.07 | 244,621.65 | 233,176.98 | 228,413.72 | 959,337.42 |

STATE: MICHIGAN

| ZIP CODE | | QUARTER 1 | QUARTER 2 | QUARTER 3 | QUARTER 4 | TOTAL GRAMS |
|---|---|---|---|---|---|---|
| | 480 | 52,416.25 | 50,719.33 | 50,184.96 | 47,948.08 | 201,268.62 |
| | 481 | 58,417.69 | 57,343.20 | 54,639.13 | 51,916.98 | 222,317 |
| | 482 | 43,868.87 | 41,311.03 | 39,713.05 | 39,263.97 | 164,156.92 |
| | 483 | 37,979.52 | 36,402.70 | 35,060.27 | 32,945.43 | 142,387.92 |
| | 484 | 9,029.08 | 8,132.97 | 7,922.50 | 7,920.70 | 33,005.25 |
| | 485 | 4,466.75 | 4,406.50 | 4,112.56 | 3,758.48 | 16,744.29 |
| | 486 | 10,574.83 | 9,379.09 | 8,685.53 | 7,971.73 | 36,611.18 |

| | | | | | | |
|---|---|---|---|---|---|---|
| | 487 | 5,130.57 | 4,831.10 | 4,319.07 | 4,063.28 | 18,344.02 |
| | 488 | 11,524.10 | 10,966.65 | 11,003.61 | 10,656.94 | 44,151.30 |
| | 489 | 5,133.75 | 4,884.56 | 4,955.39 | 4,782.45 | 19,756.15 |
| | 490 | 19,435.95 | 19,007.19 | 17,937.46 | 16,819.03 | 73,199.63 |
| | 491 | 2,193.24 | 2,098.76 | 1,929.89 | 1,967.45 | 8,189.34 |
| | 492 | 10,575.87 | 10,736.14 | 10,423.95 | 9,968.72 | 41,704.68 |
| | 493 | 3,483.09 | 3,234.70 | 3,359.27 | 2,941 | 13,018.06 |
| | 494 | 7,822.81 | 7,571.99 | 7,509.68 | 6,862.22 | 29,766.70 |
| | 495 | 7,942.63 | 8,083.84 | 7,509.54 | 7,329.60 | 30,865.61 |
| | 496 | 9,399.12 | 9,495.37 | 9,244.21 | 8,645.36 | 36,784.06 |
| | 497 | 8,630.99 | 8,560.76 | 7,875.61 | 7,622.70 | 32,690.06 |
| | 498 | 4,505.46 | 4,495.35 | 4,499.06 | 4,247.47 | 17,747.34 |
| | 499 | 1,625.36 | 1,486.65 | 1,524.89 | 1,484.69 | 6,121.59 |
| TOTAL | | 314,155.93 | 303,147.88 | 292,409.63 | 279,116.28 | 1,188,829.72 |

STATE: MINNESOTA

| ZIP CODE | | QUARTER 1 | QUARTER 2 | QUARTER 3 | QUARTER 4 | TOTAL GRAMS |
|---|---|---|---|---|---|---|
| | 550 | 16,667.86 | 16,265.54 | 15,954.72 | 15,629.56 | 64,517.68 |
| | 551 | 27,029.38 | 25,667.01 | 24,960.50 | 24,793.80 | 102,450.69 |
| | 553 | 23,565.45 | 22,558.32 | 21,865.78 | 22,128.72 | 90,118.27 |
| | 554 | 40,706.29 | 39,099.53 | 37,755.15 | 36,727.89 | 154,288.86 |
| | 556 | 228.21 | 221.36 | 191.54 | 193.82 | 834.93 |
| | 557 | 4,982.29 | 4,862.26 | 4,634.63 | 4,448.92 | 18,928.10 |
| | 558 | 2,778.09 | 2,842.26 | 2,618.99 | 2,563.14 | 10,802.48 |
| | 559 | 7,704.30 | 7,357.46 | 6,925.91 | 6,838.13 | 28,825.80 |
| | 560 | 4,540.52 | 4,489.69 | 4,425.57 | 4,348.38 | 17,804.16 |
| | 561 | 1,271.56 | 1,216.12 | 1,228.92 | 1,149.33 | 4,865.93 |
| | 562 | 2,562.78 | 2,384.27 | 2,304.84 | 2,205.68 | 9,457.57 |
| | 563 | 7,953.36 | 7,517.03 | 7,219 | 6,941.15 | 29,630.54 |
| | 564 | 3,679.94 | 3,713.70 | 3,399.84 | 3,434.30 | 14,227.78 |
| | 565 | 3,185.66 | 3,108.63 | 2,981.83 | 2,838.27 | 12,114.39 |
| | 566 | 1,398.86 | 1,388.52 | 1,395.21 | 1,276.85 | 5,459.44 |
| | 567 | 1,596.48 | 1,539.18 | 1,600.57 | 1,570.82 | 6,307.05 |
| TOTAL | | 149,851.03 | 144,230.88 | 139,463 | 137,088.76 | 570,633.67 |

STATE: MISSISSIPPI

| ZIP CODE | QUARTER 1 | QUARTER 2 | QUARTER 3 | QUARTER 4 | TOTAL GRAMS |
|---|---|---|---|---|---|
| 386 | 14,208.29 | 14,261.18 | 13,976.26 | 14,032.87 | 56,478.60 |
| 387 | 1,811.02 | 1,867.41 | 1,725.63 | 1,658.39 | 7,062.45 |
| 388 | 9,210.33 | 9,120.89 | 8,912.44 | 9,092.61 | 36,336.27 |
| 389 | 2,091.32 | 1,908.41 | 1,784.61 | 1,648.28 | 7,432.62 |
| 390 | 6,321.69 | 6,055.35 | 5,795.75 | 5,522.18 | 23,694.97 |
| 391 | 5,427.66 | 5,403.04 | 5,170.79 | 4,823.58 | 20,825.07 |
| 392 | 6,455.44 | 6,521.21 | 6,388.40 | 6,348.37 | 25,713.42 |
| 393 | 6,996.80 | 6,720.41 | 6,449.34 | 6,274.22 | 26,440.77 |
| 394 | 22,046.67 | 21,442.10 | 20,830.99 | 20,471.75 | 84,791.51 |
| 395 | 25,294.90 | 24,584.22 | 23,162.13 | 22,638.94 | 95,680.19 |
| 396 | 5,572.93 | 5,361.92 | 4,889.05 | 4,605.72 | 20,429.62 |
| 397 | 3,889.32 | 3,698.54 | 3,429.91 | 3,351.27 | 14,369.04 |
| TOTAL | 109,326.37 | 106,944.68 | 102,515.30 | 100,468.18 | 419,254.53 |

STATE: MISSOURI

| ZIP CODE | QUARTER 1 | QUARTER 2 | QUARTER 3 | QUARTER 4 | TOTAL GRAMS |
|---|---|---|---|---|---|
| 630 | 36,287.42 | 35,671.94 | 33,260.63 | 33,345.88 | 138,565.87 |
| 631 | 60,759.28 | 61,572.24 | 57,727.85 | 56,255.09 | 236,314.46 |
| 633 | 19,261.95 | 18,624.91 | 17,146.60 | 16,786.93 | 71,820.39 |
| 634 | 1,510.20 | 1,529.41 | 1,468.74 | 1,507.30 | 6,015.65 |
| 635 | 2,477.34 | 2,435.49 | 2,307.16 | 2,144.52 | 9,364.51 |
| 636 | 11,110.04 | 11,730.02 | 11,099.30 | 10,727.11 | 44,666.47 |
| 637 | 5,964.25 | 5,707.20 | 5,439.53 | 5,070.70 | 22,181.68 |
| 638 | 6,042.73 | 6,092.27 | 5,525.70 | 5,260.27 | 22,920.97 |
| 639 | 2,932.55 | 2,695.46 | 2,618.09 | 2,453.86 | 10,699.96 |
| 640 | 26,240.93 | 25,743.97 | 24,292.37 | 23,930.74 | 100,208.01 |
| 641 | 24,549.59 | 23,746.44 | 21,889.70 | 21,665.54 | 91,851.27 |
| 644 | 2,409.55 | 2,361.25 | 2,198.42 | 2,205.90 | 9,175.12 |
| 645 | 5,770.73 | 5,506.17 | 5,336.99 | 5,224.11 | 21,838 |
| 646 | 2,757.94 | 2,501.93 | 2,568.60 | 2,511.15 | 10,339.62 |
| 647 | 6,943.85 | 6,071.86 | 5,752.66 | 5,498.21 | 24,266.58 |
| 648 | 11,961.12 | 12,018.72 | 11,716.36 | 10,912.48 | 46,608.68 |
| 650 | 4,607.86 | 4,476.34 | 4,307.12 | 4,119.07 | 17,510.39 |
| 651 | 2,155.38 | 2,138.05 | 2,135.59 | 1,950.92 | 8,379.94 |

| | | | | | | |
|---|---|---|---|---|---|---|
| | 652 | 11,757.79 | 11,490.69 | 10,790.22 | 10,091.48 | 44,130.18 |
| | 653 | 4,021.94 | 3,810.20 | 3,772.37 | 3,660.53 | 15,265.04 |
| | 654 | 6,443.26 | 6,233.24 | 6,403.12 | 5,732.16 | 24,811.78 |
| | 655 | 7,951.19 | 7,827.92 | 7,499.60 | 7,319.77 | 30,598.48 |
| | 656 | 12,107.07 | 11,724.25 | 11,451.36 | 10,773.13 | 46,055.81 |
| | 657 | 15,851.87 | 15,825.72 | 15,200.28 | 14,707.70 | 61,585.57 |
| | 658 | 17,951.99 | 17,948.33 | 17,058.52 | 16,535.82 | 69,494.66 |
| TOTAL | | 309,827.82 | 305,484.02 | 288,966.88 | 280,390.37 | 1,184,669.09 |

STATE: MONTANA

| ZIP CODE | | QUARTER 1 | QUARTER 2 | QUARTER 3 | QUARTER 4 | TOTAL GRAMS |
|---|---|---|---|---|---|---|
| | 590 | 2,886.99 | 2,585.77 | 2,460.84 | 2,458.30 | 10,391.90 |
| | 591 | 8,951.09 | 8,767.71 | 8,342.23 | 8,297.62 | 34,358.65 |
| | 592 | 1,239.39 | 1,298.54 | 1,203.73 | 1,240 | 4,981.66 |
| | 593 | 1,762.09 | 1,763.34 | 1,639.48 | 1,524.29 | 6,689.20 |
| | 594 | 6,001.02 | 4,480.11 | 4,259.79 | 3,812.09 | 18,553.01 |
| | 595 | 779.33 | 744.99 | 728.27 | 658.12 | 2,910.71 |
| | 596 | 3,418.48 | 3,242.28 | 3,130.37 | 3,085.79 | 12,876.92 |
| | 597 | 4,913.41 | 4,624.50 | 4,464.04 | 4,246.65 | 18,248.60 |
| | 598 | 5,992.14 | 5,921.35 | 5,822.76 | 5,805.93 | 23,542.18 |
| | 599 | 5,065.52 | 4,947.65 | 5,143.07 | 4,642.14 | 19,798.38 |
| TOTAL | | 41,009.46 | 38,376.24 | 37,194.58 | 35,770.93 | 152,351.21 |

STATE: NEBRASKA

| ZIP CODE | | QUARTER 1 | QUARTER 2 | QUARTER 3 | QUARTER 4 | TOTAL GRAMS |
|---|---|---|---|---|---|---|
| | 680 | 4,165.81 | 4,351.39 | 4,283.95 | 4,316.66 | 17,117.81 |
| | 681 | 20,041.16 | 19,773.61 | 18,673.98 | 18,279.55 | 76,768.30 |
| | 683 | 1,227.07 | 1,148.31 | 1,148.49 | 1,137.70 | 4,661.57 |
| | 684 | 1,058.81 | 1,023.69 | 1,055.20 | 991.95 | 4,129.65 |
| | 685 | 5,304.44 | 5,133.43 | 4,981.63 | 4,913.86 | 20,333.36 |
| | 686 | 622.37 | 535.52 | 589.14 | 592.66 | 2,339.69 |
| | 687 | 1,377.16 | 1,342.37 | 1,277.04 | 1,329.18 | 5,325.75 |
| | 688 | 3,043.84 | 2,988.53 | 2,948.92 | 3,036.92 | 12,018.21 |
| | 689 | 1,180.57 | 1,247.14 | 1,299.86 | 1,294.08 | 5,021.65 |
| | 690 | 655.57 | 628.22 | 647.18 | 576.6 | 2,507.57 |

| | | | | | | |
|---|---|---|---|---|---|---|
| | 691 | 1,978.34 | 1,950.92 | 1,915.31 | 1,838.06 | 7,682.63 |
| | 692 | 247.12 | 249.45 | 196.6 | 180.24 | 873.41 |
| | 693 | 1,974.28 | 1,836.44 | 1,891.35 | 1,767.10 | 7,469.17 |
| TOTAL | | 42,876.54 | 42,209.02 | 40,908.65 | 40,254.56 | 166,248.77 |

STATE: NEVADA

| ZIP CODE | | QUARTER 1 | QUARTER 2 | QUARTER 3 | QUARTER 4 | TOTAL GRAMS |
|---|---|---|---|---|---|---|
| | 890 | 42,370.52 | 41,910.82 | 40,430.87 | 40,381.88 | 165,094.09 |
| | 891 | 101,461.06 | 99,119.49 | 95,610.06 | 95,544.66 | 391,735.27 |
| | 893 | 756.24 | 680.89 | 672.29 | 592.14 | 2,701.56 |
| | 894 | 15,187.45 | 14,364.02 | 14,338.63 | 13,742.67 | 57,632.77 |
| | 895 | 19,045.36 | 18,837.44 | 17,589.26 | 16,620.12 | 72,092.18 |
| | 897 | 5,608.37 | 5,334 | 5,299.82 | 5,141.22 | 21,383.41 |
| | 898 | 1,874.45 | 1,710.43 | 1,604.96 | 1,461.88 | 6,651.72 |
| TOTAL | | 186,303.45 | 181,957.09 | 175,545.89 | 173,484.57 | 717,291 |

STATE: NEW HAMPSHIRE

| ZIP CODE | | QUARTER 1 | QUARTER 2 | QUARTER 3 | QUARTER 4 | TOTAL GRAMS |
|---|---|---|---|---|---|---|
| | 030 | 13,930.44 | 13,514.63 | 13,532.16 | 13,297.71 | 54,274.94 |
| | 031 | 8,562.43 | 8,248.17 | 8,438.10 | 7,692.22 | 32,940.92 |
| | 032 | 7,451.16 | 7,207.10 | 7,354.49 | 6,606.25 | 28,619 |
| | 033 | 4,916.34 | 4,521.65 | 4,468.01 | 4,281.23 | 18,187.23 |
| | 034 | 3,004.28 | 2,873.60 | 2,750.86 | 2,642.50 | 11,271.24 |
| | 035 | 3,016.68 | 2,905.63 | 2,671.02 | 2,298.88 | 10,892.21 |
| | 036 | 283.74 | 277.02 | 244.52 | 258.64 | 1,063.92 |
| | 037 | 4,287.29 | 4,123.68 | 3,789.20 | 3,500.98 | 15,701.15 |
| | 038 | 20,190.60 | 19,148.54 | 18,817.93 | 18,273.19 | 76,430.26 |
| TOTAL | | 65,642.96 | 62,820.02 | 62,066.29 | 58,851.60 | 249,380.87 |

STATE: NEW JERSEY

| ZIP CODE | | QUARTER 1 | QUARTER 2 | QUARTER 3 | QUARTER 4 | TOTAL GRAMS |
|---|---|---|---|---|---|---|
| | 70 | 78,851.21 | 74,380.20 | 69,480.16 | 68,000.57 | 290,712.14 |
| | 71 | 19,028.20 | 17,593.31 | 16,876.31 | 16,213.01 | 69,710.83 |
| | 72 | 6,638.33 | 5,985.78 | 5,914.77 | 5,710.58 | 24,249.46 |

| | | | | | | |
|---|---|---|---|---|---|---|
| | 73 | 10,785.60 | 10,462.77 | 9,945.12 | 9,886.16 | 41,079.65 |
| | 74 | 23,754.38 | 22,525.25 | 20,248.01 | 20,437.60 | 86,965.24 |
| | 75 | 9,490.75 | 8,949.73 | 8,480.43 | 8,084.52 | 35,005.43 |
| | 76 | 26,283.55 | 25,822.77 | 24,270.89 | 23,703.10 | 100,080.31 |
| | 77 | 36,250.42 | 34,643.45 | 32,556.91 | 32,029.56 | 135,480.34 |
| | 78 | 15,260.45 | 14,188.37 | 13,245.05 | 13,024.91 | 55,718.78 |
| | 79 | 7,484.91 | 6,882.86 | 6,753.75 | 6,586.21 | 27,707.73 |
| | 80 | 124,104.61 | 117,228.59 | 109,325.20 | 102,258.85 | 452,917.25 |
| | 81 | 16,621.21 | 15,620.06 | 15,014.75 | 14,391.33 | 61,647.35 |
| | 82 | 24,875.10 | 24,404.33 | 23,377.82 | 22,076.67 | 94,733.92 |
| | 83 | 16,828.39 | 16,076.50 | 15,015.76 | 14,507.17 | 62,427.82 |
| | 84 | 5,142.40 | 4,273.71 | 3,257.89 | 2,934.84 | 15,608.84 |
| | 85 | 14,825.16 | 13,959.99 | 12,527.90 | 11,474.71 | 52,787.76 |
| | 86 | 16,425.66 | 15,879.80 | 15,112.44 | 14,002.85 | 61,420.75 |
| | 87 | 30,068.69 | 27,924.06 | 26,905.19 | 25,832.28 | 110,730.22 |
| | 88 | 51,698.42 | 50,142.92 | 47,460.61 | 45,879.29 | 195,181.24 |
| | 89 | 4,680.69 | 4,362.27 | 4,251.05 | 3,853.18 | 17,147.19 |
| TOTAL | | 539,098.13 | 511,306.72 | 480,020.01 | 460,887.39 | 1,991,312.25 |

STATE: NEW MEXICO

| ZIP CODE | | QUARTER 1 | QUARTER 2 | QUARTER 3 | QUARTER 4 | TOTAL GRAMS |
|---|---|---|---|---|---|---|
| | 870 | 9,655.53 | 9,214.38 | 8,597.10 | 8,362.10 | 35,829.11 |
| | 871 | 47,244.76 | 43,432.47 | 39,977.81 | 38,496.37 | 169,151.41 |
| | 873 | 1,245.42 | 1,167.83 | 1,203.33 | 1,123.22 | 4,739.80 |
| | 874 | 2,734.89 | 2,775.88 | 2,574.61 | 2,410.96 | 10,496.34 |
| | 875 | 15,016.79 | 14,917.84 | 14,073.96 | 14,107.82 | 58,116.41 |
| | 877 | 1,803.98 | 1,815.28 | 1,821.64 | 1,821.63 | 7,262.53 |
| | 878 | 621.23 | 712.76 | 647.72 | 669.82 | 2,651.53 |
| | 879 | 513.83 | 499.98 | 516.06 | 485.1 | 2,014.97 |
| | 880 | 7,105.75 | 6,122.42 | 5,535.47 | 5,468.84 | 24,232.48 |
| | 881 | 1,305.84 | 1,292.53 | 1,143.58 | 1,008.92 | 4,750.87 |
| | 882 | 7,924.30 | 7,313.11 | 7,003.66 | 6,759.70 | 29,000.77 |
| | 883 | 3,171.83 | 3,013.94 | 2,940.49 | 2,740.14 | 11,866.40 |
| | 884 | 327.67 | 339.44 | 348.99 | 261.33 | 1,277.43 |
| TOTAL | | 98,671.82 | 92,617.86 | 86,384.42 | 83,715.95 | 361,390.05 |

RETAIL DRUG DISTRIBUTION BY ZIP CODE WITHIN STATE BY GRAMS WT

STATE: NEW YORK

| ZIP CODE | QUARTER 1 | QUARTER 2 | QUARTER 3 | QUARTER 4 | TOTAL GRAMS |
|---|---|---|---|---|---|
| 063 | 0 | 0 | 0 | 0.45 | 0.45 |
| 100 | 77,204.18 | 76,143.45 | 70,683.21 | 67,014.58 | 291,045.42 |
| 101 | 1,829.54 | 1,831.62 | 1,539.97 | 1,488.20 | 6,689.33 |
| 102 | 98.59 | 93.22 | 85.14 | 72.61 | 349.56 |
| 103 | 32,355.45 | 30,662.21 | 27,694.33 | 26,528.91 | 117,240.90 |
| 104 | 62,754.50 | 57,101.82 | 55,730.50 | 51,852.75 | 227,439.57 |
| 105 | 20,227.89 | 19,731.19 | 18,896.03 | 18,200.26 | 77,055.37 |
| 106 | 3,697.93 | 3,640.74 | 3,293.49 | 3,310.15 | 13,942.31 |
| 107 | 7,916.96 | 7,928.60 | 7,673.20 | 7,518.15 | 31,036.91 |
| 108 | 2,632.12 | 2,437.06 | 2,435.75 | 2,189.50 | 9,694.43 |
| 109 | 29,196.34 | 27,796.26 | 26,397.48 | 26,195.63 | 109,585.71 |
| 110 | 12,560.34 | 12,328.66 | 11,607.53 | 11,536.59 | 48,033.12 |
| 111 | 8,886.17 | 8,323.99 | 7,794.28 | 7,560.92 | 32,565.36 |
| 112 | 77,825.08 | 71,466.62 | 66,816.97 | 63,758.67 | 279,867.34 |
| 113 | 29,638.30 | 28,152.88 | 26,424.95 | 25,899.93 | 110,116.06 |
| 114 | 16,290.41 | 15,409.91 | 14,351.97 | 13,589.34 | 59,641.63 |
| 115 | 30,756.47 | 30,719.17 | 30,366.53 | 29,192.45 | 121,034.62 |
| 116 | 3,789.06 | 3,402.88 | 3,247.94 | 3,361.01 | 13,800.89 |
| 117 | 96,542.96 | 92,503.94 | 87,241.37 | 83,956.01 | 360,244.28 |
| 118 | 3,284.82 | 3,397.16 | 3,404.44 | 3,154.91 | 13,241.33 |
| 119 | 9,826.08 | 9,692.41 | 9,491.75 | 9,117.46 | 38,127.70 |
| 120 | 10,436.71 | 9,938.94 | 10,195.89 | 10,065.46 | 40,637 |
| 121 | 6,170.22 | 5,988.12 | 5,688.19 | 5,394.28 | 23,240.81 |
| 122 | 5,949.64 | 5,638.54 | 5,391.40 | 5,546.62 | 22,526.20 |
| 123 | 3,120.86 | 2,819.68 | 2,723.40 | 2,663.90 | 11,327.84 |
| 124 | 14,049.28 | 13,870.29 | 13,564.78 | 12,943.28 | 54,427.63 |
| 125 | 23,446.30 | 23,129.30 | 22,313.40 | 21,981.17 | 90,870.17 |
| 126 | 6,908.67 | 6,372.23 | 6,431.77 | 6,502.58 | 26,215.25 |
| 127 | 11,883.33 | 11,429.82 | 10,634.96 | 9,957.69 | 43,905.80 |
| 128 | 9,337.45 | 8,860.87 | 9,129.69 | 8,808.89 | 36,136.90 |
| 129 | 6,343.63 | 6,358.55 | 6,445.18 | 6,194.84 | 25,342.20 |
| 130 | 12,207.75 | 12,291.85 | 11,333.47 | 11,119.02 | 46,952.09 |
| 131 | 3,669.51 | 3,690.01 | 3,262.21 | 3,249.10 | 13,870.83 |
| 132 | 8,594.83 | 8,307.16 | 7,848.22 | 7,816.90 | 32,567.11 |

| | | | | | | |
|---|---|---|---|---|---|---|
| | 133 | 4,068.04 | 4,052.81 | 3,398.70 | 3,489.46 | 15,009.01 |
| | 134 | 5,108.72 | 5,051.87 | 4,531.82 | 4,399.36 | 19,091.77 |
| | 135 | 2,137.46 | 2,021.83 | 1,925.27 | 1,879.44 | 7,964 |
| | 136 | 6,927.98 | 6,986.33 | 6,944.65 | 6,704.14 | 27,563.10 |
| | 137 | 4,789.22 | 4,701.20 | 4,405 | 4,542.96 | 18,438.38 |
| | 138 | 9,247.50 | 8,949.97 | 8,196.66 | 8,068.45 | 34,462.58 |
| | 139 | 5,503.17 | 5,466.41 | 5,122.32 | 5,167.71 | 21,259.61 |
| | 140 | 13,919.51 | 13,257.77 | 12,978.31 | 12,233.72 | 52,389.31 |
| | 141 | 6,488.68 | 5,977.69 | 5,912.30 | 5,593.30 | 23,971.97 |
| | 142 | 18,938.94 | 18,268.71 | 17,659.26 | 17,196.61 | 72,063.52 |
| | 143 | 2,244.06 | 2,442.89 | 2,430.76 | 2,347.86 | 9,465.57 |
| | 144 | 9,066.35 | 8,987.92 | 8,654.65 | 8,304.02 | 35,012.94 |
| | 145 | 8,255.74 | 8,011.42 | 7,651.54 | 7,458.45 | 31,377.15 |
| | 146 | 20,564.93 | 19,731.45 | 19,242.94 | 18,069.49 | 77,608.81 |
| | 147 | 4,296.73 | 4,141.70 | 3,870.57 | 3,870.63 | 16,179.63 |
| | 148 | 12,043.57 | 11,892.80 | 11,704.78 | 11,219.73 | 46,860.88 |
| | 149 | 3,729.68 | 3,593.13 | 3,663.54 | 3,239.40 | 14,225.75 |
| TOTAL | | 786,761.65 | 754,995.05 | 718,432.46 | 691,526.94 | 2,951,716.10 |

STATE: NORTH CAROLINA

| ZIP CODE | | QUARTER 1 | QUARTER 2 | QUARTER 3 | QUARTER 4 | TOTAL GRAMS |
|---|---|---|---|---|---|---|
| | 270 | 18,168.81 | 17,605.88 | 16,093.71 | 15,563.17 | 67,431.57 |
| | 271 | 13,544.05 | 12,081.44 | 11,932.04 | 10,956.87 | 48,514.40 |
| | 272 | 38,269.14 | 35,223.99 | 33,148.33 | 32,934.10 | 139,575.56 |
| | 273 | 24,449.29 | 23,245.70 | 22,118.11 | 22,077.93 | 91,891.03 |
| | 274 | 13,194.96 | 12,875.09 | 12,619.11 | 12,550.35 | 51,239.51 |
| | 275 | 52,405.49 | 49,997.22 | 48,367.65 | 47,897.55 | 198,667.91 |
| | 276 | 14,869.93 | 14,219.32 | 13,401 | 13,406.40 | 55,896.65 |
| | 277 | 15,864.05 | 15,446.46 | 14,820.18 | 14,511.73 | 60,642.42 |
| | 278 | 32,224.58 | 30,840.63 | 30,529.41 | 28,984.89 | 122,579.51 |
| | 279 | 13,186.56 | 12,771.76 | 11,753.61 | 12,122.95 | 49,834.88 |
| | 280 | 39,100.50 | 38,197.05 | 35,881.72 | 36,128.66 | 149,307.93 |
| | 281 | 30,987.46 | 30,046.27 | 27,754.71 | 27,984.39 | 116,772.83 |
| | 282 | 23,924.83 | 23,336.77 | 22,286.78 | 21,683.89 | 91,232.27 |
| | 283 | 78,779.50 | 75,405.11 | 72,533.51 | 70,320.87 | 297,038.99 |
| | 284 | 45,740.23 | 43,504.09 | 39,643.50 | 38,521.68 | 167,409.50 |

| | | | | | |
|---|---|---|---|---|---|
| 285 | 37,774.26 | 35,467.49 | 33,299.42 | 31,852.13 | 138,393.30 |
| 286 | 57,781.99 | 54,386.38 | 50,658.17 | 50,305.07 | 213,131.61 |
| 287 | 31,586.89 | 29,835.37 | 28,297.56 | 27,279.33 | 116,999.15 |
| 288 | 12,932.77 | 12,293.70 | 11,797.11 | 11,368.98 | 48,392.56 |
| 289 | 3,483.38 | 3,524.59 | 3,401.61 | 3,712.31 | 14,121.89 |
| TOTAL | 598,268.67 | 570,304.31 | 540,337.24 | 530,163.25 | 2,239,073.47 |

STATE: NORTH DAKOTA

| ZIP CODE | QUARTER 1 | QUARTER 2 | QUARTER 3 | QUARTER 4 | TOTAL GRAMS |
|---|---|---|---|---|---|
| 580 | 705.68 | 618.32 | 597.38 | 582.73 | 2,504.11 |
| 581 | 2,779.59 | 2,585.66 | 2,481.94 | 2,346.40 | 10,193.59 |
| 582 | 2,289.79 | 2,275.32 | 2,120.97 | 2,172.02 | 8,858.10 |
| 583 | 1,344.12 | 1,359.86 | 1,269.58 | 1,195.53 | 5,169.09 |
| 584 | 715.05 | 703.54 | 665.44 | 674.4 | 2,758.43 |
| 585 | 4,174.69 | 4,130.19 | 4,105.11 | 4,095.67 | 16,505.66 |
| 586 | 902.98 | 964.1 | 854.27 | 880.91 | 3,602.26 |
| 587 | 1,333.79 | 1,370.57 | 1,241.83 | 1,232.13 | 5,178.32 |
| 588 | 917.75 | 954.14 | 948.5 | 999.78 | 3,820.17 |
| TOTAL | 15,163.44 | 14,961.70 | 14,285.02 | 14,179.57 | 58,589.73 |

STATE: OHIO

| ZIP CODE | QUARTER 1 | QUARTER 2 | QUARTER 3 | QUARTER 4 | TOTAL GRAMS |
|---|---|---|---|---|---|
| 430 | 29,678.50 | 29,004.58 | 26,845.58 | 26,372.21 | 111,900.87 |
| 431 | 23,448.15 | 22,428.50 | 21,427.52 | 20,261.10 | 87,565.27 |
| 432 | 54,229.77 | 51,608.22 | 47,983.98 | 46,266.83 | 200,088.80 |
| 433 | 6,929.56 | 6,833.81 | 6,606.64 | 6,291.18 | 26,661.19 |
| 434 | 6,033.21 | 5,904.90 | 5,789.48 | 5,657.71 | 23,385.30 |
| 435 | 9,050.29 | 8,488.03 | 8,514.05 | 8,167 | 34,219.37 |
| 436 | 23,547.40 | 22,851.15 | 21,840.15 | 21,137.33 | 89,376.03 |
| 437 | 10,885.24 | 10,303.77 | 10,132.78 | 9,778.25 | 41,100.04 |
| 438 | 1,656.59 | 1,564.18 | 1,580.90 | 1,544.74 | 6,346.41 |
| 439 | 10,657.26 | 10,195.25 | 10,056.56 | 9,674.10 | 40,583.17 |
| 440 | 27,253.31 | 25,630.53 | 24,746.32 | 22,829.54 | 100,459.70 |
| 441 | 40,262.33 | 38,043.88 | 36,270.46 | 34,060.12 | 148,636.79 |
| 442 | 19,618.39 | 18,413.04 | 17,187.15 | 16,034.99 | 71,253.57 |

| | | | | | | |
|---|---|---|---|---|---|---|
| | 443 | 13,223.89 | 12,638.58 | 11,813 | 11,405.88 | 49,081.35 |
| | 444 | 19,176.19 | 18,078.44 | 17,389.70 | 16,356.12 | 71,000.45 |
| | 445 | 9,688.15 | 9,349.88 | 8,439.32 | 8,135.74 | 35,613.09 |
| | 446 | 13,324.14 | 12,728.30 | 12,310.14 | 11,669.25 | 50,031.83 |
| | 447 | 8,205.65 | 7,718.76 | 7,298.23 | 7,119.16 | 30,341.80 |
| | 448 | 12,877.23 | 12,536.80 | 12,012.74 | 11,765.55 | 49,192.32 |
| | 449 | 3,730.62 | 3,550.63 | 3,176.77 | 2,961.41 | 13,419.43 |
| | 450 | 34,339.33 | 32,841.02 | 31,532 | 29,333.70 | 128,046.05 |
| | 451 | 14,510.76 | 14,507.16 | 13,908.45 | 13,717.79 | 56,644.16 |
| | 452 | 43,950.39 | 42,654.03 | 40,095.31 | 38,945.84 | 165,645.57 |
| | 453 | 14,234.73 | 13,780.22 | 13,281.25 | 12,756.44 | 54,052.64 |
| | 454 | 19,993.23 | 19,254.10 | 17,804.75 | 16,418.10 | 73,470.18 |
| | 455 | 6,103.41 | 6,060.18 | 5,741.64 | 5,586.06 | 23,491.29 |
| | 456 | 23,607.33 | 22,643.85 | 21,987.60 | 21,332.49 | 89,571.27 |
| | 457 | 7,751.60 | 7,260.95 | 6,576.95 | 6,044.83 | 27,634.33 |
| | 458 | 7,335.08 | 7,183.54 | 6,685.61 | 6,513.70 | 27,717.93 |
| TOTAL | | 515,301.73 | 494,056.28 | 469,035.03 | 448,137.16 | 1,926,530.20 |

STATE: OKLAHOMA

| ZIP CODE | | QUARTER 1 | QUARTER 2 | QUARTER 3 | QUARTER 4 | TOTAL GRAMS |
|---|---|---|---|---|---|---|
| | 730 | 39,146.44 | 38,792.21 | 37,123.61 | 37,134.31 | 152,196.57 |
| | 731 | 46,385.65 | 45,847.62 | 44,192.54 | 43,299.95 | 179,725.76 |
| | 734 | 4,903.76 | 4,944.54 | 4,787.20 | 4,668.02 | 19,303.52 |
| | 735 | 12,796.79 | 12,280.95 | 12,972.89 | 10,570.52 | 48,621.15 |
| | 736 | 4,430.24 | 3,687.75 | 3,260.60 | 2,832.81 | 14,211.40 |
| | 737 | 3,925.40 | 4,043.51 | 3,764.04 | 3,903.58 | 15,636.53 |
| | 738 | 796.7 | 917.16 | 800.23 | 803.52 | 3,317.61 |
| | 739 | 176.94 | 206.85 | 165.54 | 182.39 | 731.72 |
| | 740 | 37,066.05 | 35,859.43 | 35,092.16 | 34,597.18 | 142,614.82 |
| | 741 | 26,458.92 | 26,294.61 | 25,343.52 | 25,187.42 | 103,284.47 |
| | 743 | 6,280.44 | 5,940.09 | 5,915.46 | 5,898.41 | 24,034.40 |
| | 744 | 17,665.50 | 17,841.47 | 17,314.60 | 16,919.26 | 69,740.83 |
| | 745 | 4,672.80 | 4,891.09 | 4,423.78 | 3,996.40 | 17,984.07 |
| | 746 | 2,705.55 | 2,848.57 | 2,723.96 | 2,913.88 | 11,191.96 |
| | 747 | 4,082.26 | 3,804.58 | 3,529.35 | 3,456.56 | 14,872.75 |
| | 748 | 10,425.70 | 10,430.50 | 9,955.32 | 9,743.69 | 40,555.21 |

| | | | | | |
|---|---|---|---|---|---|
| 749 | 9,943.93 | 9,540.78 | 9,589.51 | 9,645.57 | 38,719.79 |
| TOTAL | 231,863.07 | 228,171.71 | 220,954.31 | 215,753.47 | 896,742.56 |

STATE: OREGON

| ZIP CODE | QUARTER 1 | QUARTER 2 | QUARTER 3 | QUARTER 4 | TOTAL GRAMS |
|---|---|---|---|---|---|
| 970 | 30,469.79 | 29,390.80 | 27,997.52 | 27,313.68 | 115,171.79 |
| 971 | 13,835.21 | 13,189.84 | 13,149.81 | 12,401.90 | 52,576.76 |
| 972 | 37,211.17 | 36,495.59 | 34,998.85 | 33,408.76 | 142,114.37 |
| 973 | 25,997.34 | 23,999.33 | 22,298.88 | 21,355.99 | 93,651.54 |
| 974 | 34,283.25 | 33,445.26 | 32,062.72 | 31,217.41 | 131,008.64 |
| 975 | 21,029.75 | 20,506.77 | 19,791.53 | 19,822.38 | 81,150.43 |
| 976 | 2,876.24 | 2,698.51 | 2,589.32 | 2,501.27 | 10,665.34 |
| 977 | 7,988.64 | 7,960.38 | 7,753.55 | 7,515.54 | 31,218.11 |
| 978 | 7,419.44 | 7,116.10 | 6,883.58 | 6,472.93 | 27,892.05 |
| 979 | 2,054.26 | 1,985.94 | 1,997.63 | 1,850.74 | 7,888.57 |
| TOTAL | 183,165.09 | 176,788.52 | 169,523.39 | 163,860.60 | 693,337.60 |

STATE: PENNSYLVANIA

| ZIP CODE | QUARTER 1 | QUARTER 2 | QUARTER 3 | QUARTER 4 | TOTAL GRAMS |
|---|---|---|---|---|---|
| 150 | 24,916.59 | 24,867.01 | 25,582.02 | 25,196.18 | 100,561.80 |
| 151 | 13,311.85 | 13,037.64 | 12,732.63 | 12,138.87 | 51,220.99 |
| 152 | 36,353.98 | 35,064.94 | 33,545.27 | 32,516.34 | 137,480.53 |
| 153 | 7,079.42 | 6,677.38 | 6,386.68 | 6,339.41 | 26,482.89 |
| 154 | 10,521.12 | 10,325.17 | 10,199.95 | 9,731.12 | 40,777.36 |
| 155 | 4,066.14 | 3,919.91 | 3,657.40 | 3,421.74 | 15,065.19 |
| 156 | 16,002.49 | 15,439.11 | 14,752 | 14,266.45 | 60,460.05 |
| 157 | 6,270.52 | 6,070.93 | 5,896.86 | 5,516.81 | 23,755.12 |
| 158 | 4,835.18 | 4,832.72 | 4,519.36 | 4,470.07 | 18,657.33 |
| 159 | 14,946.72 | 15,067.83 | 14,251.10 | 13,309.81 | 57,575.46 |
| 160 | 7,955.94 | 7,791.51 | 7,620.41 | 7,747 | 31,114.86 |
| 161 | 11,311.67 | 11,210.45 | 9,670.95 | 10,610.95 | 42,804.02 |
| 162 | 4,469.04 | 3,956.14 | 3,884.93 | 3,609.41 | 15,919.52 |
| 163 | 7,838.68 | 7,604.45 | 7,199.98 | 7,401.70 | 30,044.81 |
| 164 | 2,828.39 | 2,673.27 | 2,660.14 | 2,453.18 | 10,614.98 |
| 165 | 5,969.44 | 5,880.98 | 5,471.49 | 5,331.61 | 22,653.52 |

| | | | | | |
|---|---|---|---|---|---|
| 166 | 11,735.04 | 11,521.39 | 11,123.52 | 10,845.29 | 45,225.24 |
| 167 | 1,887.51 | 1,853.20 | 1,820.31 | 1,736.91 | 7,297.93 |
| 168 | 4,884.92 | 4,744.37 | 4,540.26 | 4,235.64 | 18,405.19 |
| 169 | 2,481.85 | 2,395.83 | 2,308.39 | 2,130.20 | 9,316.27 |
| 170 | 32,201.79 | 31,305.38 | 30,144 | 29,208.82 | 122,859.99 |
| 171 | 9,652.15 | 9,176.62 | 9,050.76 | 8,502.81 | 36,382.34 |
| 172 | 9,545.12 | 9,550.70 | 9,238.81 | 8,844.82 | 37,179.45 |
| 173 | 14,535.88 | 14,208.24 | 13,129.80 | 12,714.64 | 54,588.56 |
| 174 | 8,327.64 | 7,765.44 | 7,648.77 | 7,522.84 | 31,264.69 |
| 175 | 12,260 | 11,420.62 | 11,355.32 | 10,411.43 | 45,447.37 |
| 176 | 9,924.35 | 9,806.10 | 8,949.45 | 8,610.02 | 37,289.92 |
| 177 | 8,562.25 | 8,261.21 | 7,935.41 | 7,541.59 | 32,300.46 |
| 178 | 11,945.96 | 11,447.32 | 9,900.49 | 10,015.71 | 43,309.48 |
| 179 | 7,757.32 | 7,370.16 | 6,869.92 | 6,590.28 | 28,587.68 |
| 180 | 18,708.13 | 17,783.29 | 17,106.92 | 16,941.81 | 70,540.15 |
| 181 | 11,328.21 | 10,737.90 | 10,093.71 | 10,261.32 | 42,421.14 |
| 182 | 8,251.92 | 7,698.71 | 7,399.37 | 7,523.35 | 30,873.35 |
| 183 | 11,548.34 | 11,097.79 | 10,376.62 | 10,754.90 | 43,777.65 |
| 184 | 12,220.91 | 11,927.73 | 11,215.88 | 10,503.36 | 45,867.88 |
| 185 | 16,999.60 | 16,731.73 | 15,435.18 | 13,996.33 | 63,162.84 |
| 186 | 10,153.30 | 9,610.34 | 9,449.62 | 9,172.02 | 38,385.28 |
| 187 | 38,521.95 | 35,610.28 | 33,096.47 | 32,582.21 | 139,810.91 |
| 188 | 3,744.05 | 3,655.39 | 3,501.41 | 3,322.13 | 14,222.98 |
| 189 | 17,653.42 | 17,624.21 | 17,257.87 | 16,299.66 | 68,835.16 |
| 190 | 107,524.97 | 103,206.59 | 100,147.18 | 107,582.87 | 418,461.61 |
| 191 | 137,800.49 | 137,841.83 | 134,510.17 | 126,821.12 | 536,973.61 |
| 193 | 20,186.52 | 19,423.79 | 18,859.36 | 17,554.22 | 76,023.89 |
| 194 | 31,193.12 | 29,569.81 | 28,646.13 | 27,605.95 | 117,015.01 |
| 195 | 7,001.55 | 6,499.84 | 5,951.78 | 5,943.80 | 25,396.97 |
| 196 | 9,426.70 | 9,321.31 | 8,562.53 | 8,345.58 | 35,656.12 |
| TOTAL | 786,642.13 | 763,586.56 | 733,656.58 | 718,182.28 | 3,002,067.55 |

STATE: PUERTO RICO

| ZIP CODE | QUARTER 1 | QUARTER 2 | QUARTER 3 | QUARTER 4 | TOTAL GRAMS |
|---|---|---|---|---|---|
| 006 | 4,725.45 | 3,492.41 | 1,980.59 | 1,404.37 | 11,602.82 |
| 007 | 6,550.55 | 4,160.25 | 2,626.34 | 2,020.85 | 15,357.99 |

| | | | | | |
|---|---|---|---|---|---|
| 009 | 8,495.46 | 6,416.25 | 4,785.29 | 4,068.71 | 23,765.71 |
| TOTAL | 19,771.46 | 14,068.91 | 9,392.22 | 7,493.93 | 50,726.52 |

STATE: RHODE ISLAND

| ZIP CODE | QUARTER 1 | QUARTER 2 | QUARTER 3 | QUARTER 4 | TOTAL GRAMS |
|---|---|---|---|---|---|
| 028 | 25,473.70 | 24,222.69 | 23,219.07 | 22,857.21 | 95,772.67 |
| 029 | 18,176.99 | 16,482.99 | 16,386.35 | 15,703.10 | 66,749.43 |
| TOTAL | 43,650.69 | 40,705.68 | 39,605.42 | 38,560.31 | 162,522.10 |

STATE: SOUTH CAROLINA

| ZIP CODE | QUARTER 1 | QUARTER 2 | QUARTER 3 | QUARTER 4 | TOTAL GRAMS |
|---|---|---|---|---|---|
| 290 | 13,162.54 | 12,821.07 | 12,570.79 | 12,529.60 | 51,084 |
| 291 | 16,345.46 | 15,840.78 | 15,016.72 | 15,239.37 | 62,442.33 |
| 292 | 10,870.95 | 10,619.18 | 10,320.23 | 10,124.23 | 41,934.59 |
| 293 | 26,654.68 | 26,134.40 | 25,492.18 | 25,153.23 | 103,434.49 |
| 294 | 42,647.99 | 42,030.83 | 39,471.98 | 39,879.57 | 164,030.37 |
| 295 | 59,792.13 | 57,267.92 | 52,628.40 | 51,209.12 | 220,897.57 |
| 296 | 47,112.97 | 45,670.94 | 42,666.28 | 42,287.04 | 177,737.23 |
| 297 | 11,631.66 | 11,292.36 | 10,325.66 | 9,941.28 | 43,190.96 |
| 298 | 10,495.40 | 10,284.97 | 9,821.67 | 9,777.12 | 40,379.16 |
| 299 | 7,570.75 | 7,568.84 | 7,316.26 | 7,250.75 | 29,706.60 |
| TOTAL | 246,284.53 | 239,531.29 | 225,630.17 | 223,391.31 | 934,837.30 |

STATE: SOUTH DAKOTA

| ZIP CODE | QUARTER 1 | QUARTER 2 | QUARTER 3 | QUARTER 4 | TOTAL GRAMS |
|---|---|---|---|---|---|
| 570 | 2,998.60 | 2,790.54 | 2,704.13 | 2,708.60 | 11,201.87 |
| 571 | 5,516.38 | 5,768.93 | 5,725.05 | 5,270.65 | 22,281.01 |
| 572 | 822.02 | 748.13 | 734.23 | 727.15 | 3,031.53 |
| 573 | 1,000.14 | 929.15 | 905.4 | 910.08 | 3,744.77 |
| 574 | 1,047.96 | 1,017.44 | 973.11 | 910.17 | 3,948.68 |
| 575 | 879.53 | 887.34 | 831.63 | 807.66 | 3,406.16 |
| 576 | 154.2 | 152.67 | 155.23 | 193.11 | 655.21 |
| 577 | 5,591.22 | 5,232.10 | 5,110.97 | 4,741.85 | 20,676.14 |
| TOTAL | 18,010.05 | 17,526.30 | 17,139.75 | 16,269.27 | 68,945.37 |

STATE: TENNESSEE

| ZIP CODE | QUARTER 1 | QUARTER 2 | QUARTER 3 | QUARTER 4 | TOTAL GRAMS |
|---|---|---|---|---|---|
| 370 | 63,885.86 | 63,236.57 | 62,396.84 | 60,891.69 | 250,410.96 |
| 371 | 49,483.97 | 48,285.85 | 46,633.58 | 46,757.12 | 191,160.52 |
| 372 | 33,703.11 | 32,072.82 | 30,803.81 | 30,765.81 | 127,345.55 |
| 373 | 45,866.63 | 44,624.10 | 41,065.48 | 42,471.03 | 174,027.24 |
| 374 | 17,260.83 | 17,494.28 | 16,226.37 | 16,769.22 | 67,750.70 |
| 376 | 39,728.99 | 38,859.83 | 36,700.70 | 36,430.30 | 151,719.82 |
| 377 | 35,831.44 | 35,494.33 | 34,992.14 | 35,605.49 | 141,923.40 |
| 378 | 49,287.50 | 48,402.14 | 47,344.62 | 46,686.71 | 191,720.97 |
| 379 | 25,531.39 | 25,227.94 | 24,159.49 | 24,849.47 | 99,768.29 |
| 380 | 15,567.88 | 15,502.58 | 15,501.81 | 14,833.75 | 61,406.02 |
| 381 | 29,086.64 | 29,667.69 | 27,922.10 | 40,966.77 | 127,643.20 |
| 382 | 8,652.62 | 8,321.64 | 8,184.48 | 7,887.22 | 33,045.96 |
| 383 | 15,631.65 | 15,185.28 | 14,866.76 | 14,939.23 | 60,622.92 |
| 384 | 13,982.26 | 13,687.40 | 12,104.64 | 13,714.64 | 53,488.94 |
| 385 | 30,725.14 | 30,545.70 | 28,735.77 | 29,101.83 | 119,108.44 |
| TOTAL | 474,225.91 | 466,608.15 | 447,638.59 | 462,670.28 | 1,851,142.93 |

STATE: TEXAS

| ZIP CODE | QUARTER 1 | QUARTER 2 | QUARTER 3 | QUARTER 4 | TOTAL GRAMS |
|---|---|---|---|---|---|
| 750 | 40,875.46 | 38,175.18 | 36,841.41 | 34,897.72 | 150,789.77 |
| 751 | 8,261.45 | 8,384.37 | 8,441.16 | 8,192.91 | 33,279.89 |
| 752 | 11,811.03 | 11,842.91 | 11,086.74 | 10,813.05 | 45,553.73 |
| 753 | 151.33 | 146.8 | 161.41 | 198.49 | 658.03 |
| 754 | 5,998.30 | 5,717.10 | 5,002.86 | 4,992.12 | 21,710.38 |
| 755 | 1,698.51 | 1,675.95 | 1,581.19 | 1,519.42 | 6,475.07 |
| 756 | 2,841.62 | 2,884.66 | 2,731.55 | 2,778.95 | 11,236.78 |
| 757 | 4,546.62 | 4,862.76 | 4,704.05 | 4,723.26 | 18,836.69 |
| 758 | 789.78 | 834.51 | 807.89 | 810.08 | 3,242.26 |
| 759 | 1,917.97 | 1,845.51 | 1,843.65 | 1,897.08 | 7,504.21 |
| 760 | 26,485.03 | 26,387.92 | 27,048.24 | 25,650.65 | 105,571.84 |
| 761 | 19,248.89 | 19,719.85 | 18,489.23 | 18,103.16 | 75,561.13 |
| 762 | 8,658.72 | 8,750.71 | 8,464.36 | 8,158.74 | 34,032.53 |

| | | | | | | |
|---|---|---|---|---|---|---|
| | 763 | 3,086.53 | 2,808.30 | 2,789.12 | 2,505.02 | 11,188.97 |
| | 764 | 1,650.91 | 1,559.59 | 1,439.45 | 1,329.47 | 5,979.42 |
| | 765 | 6,528.05 | 6,616.14 | 6,292.10 | 5,839.85 | 25,276.14 |
| | 766 | 1,471.16 | 1,390.79 | 1,529.76 | 1,432.52 | 5,824.23 |
| | 767 | 1,962.41 | 2,286 | 2,329.62 | 2,236.66 | 8,814.69 |
| | 768 | 1,120.58 | 1,123.69 | 1,144.47 | 1,097.46 | 4,486.20 |
| | 769 | 1,327.66 | 1,132.36 | 1,135.26 | 979.4 | 4,574.68 |
| | 770 | 42,322.23 | 45,503.17 | 47,109.99 | 44,392.21 | 179,327.60 |
| | 773 | 13,243.92 | 12,834.87 | 12,740.81 | 12,765.14 | 51,584.74 |
| | 774 | 13,572.93 | 14,266.76 | 13,757.26 | 14,722.37 | 56,319.32 |
| | 775 | 12,916.09 | 13,451.87 | 13,097.25 | 12,845.42 | 52,310.63 |
| | 776 | 2,636.47 | 2,454.76 | 2,378.40 | 2,420.61 | 9,890.24 |
| | 777 | 1,035.10 | 986.94 | 1,130.19 | 1,052.94 | 4,205.17 |
| | 778 | 1,398.65 | 1,445.11 | 1,423.79 | 1,200.37 | 5,467.92 |
| | 779 | 3,422.67 | 3,441.71 | 3,485.10 | 3,441.98 | 13,791.46 |
| | 780 | 3,270.99 | 3,306.69 | 3,462.72 | 3,502.89 | 13,543.29 |
| | 781 | 3,980.04 | 3,939.33 | 3,975.24 | 4,016.96 | 15,911.57 |
| | 782 | 22,753.23 | 22,527.69 | 23,174.98 | 22,706.56 | 91,162.46 |
| | 783 | 1,383.75 | 1,272.50 | 1,358.91 | 1,150.23 | 5,165.39 |
| | 784 | 2,147.29 | 1,925.96 | 1,933.29 | 1,871.89 | 7,878.43 |
| | 785 | 3,820.03 | 3,655.26 | 3,480.78 | 3,239.72 | 14,195.79 |
| | 786 | 19,535.93 | 18,366.42 | 18,548.09 | 17,607.63 | 74,058.07 |
| | 787 | 18,053.11 | 18,130.72 | 15,818.81 | 14,769.60 | 66,772.24 |
| | 788 | 587.07 | 542.02 | 648.29 | 563.02 | 2,340.40 |
| | 789 | 703.04 | 686.81 | 720.46 | 682.09 | 2,792.40 |
| | 790 | 1,628.64 | 1,521.08 | 1,361.80 | 1,282.89 | 5,794.41 |
| | 791 | 2,987.36 | 3,044.83 | 2,946.17 | 3,041.65 | 12,020.01 |
| | 792 | 287.78 | 339.33 | 340.22 | 327 | 1,294.33 |
| | 793 | 632.7 | 635.26 | 677.62 | 636.92 | 2,582.50 |
| | 794 | 2,503.47 | 2,681.18 | 2,807.79 | 2,641.16 | 10,633.60 |
| | 795 | 732.13 | 692.28 | 710.07 | 806.4 | 2,940.88 |
| | 796 | 1,801.40 | 1,806.07 | 1,655.12 | 1,676.56 | 6,939.15 |
| | 797 | 4,106.40 | 3,797.66 | 3,872.51 | 3,744.18 | 15,520.75 |
| | 798 | 158.59 | 140.97 | 138.96 | 148.28 | 586.8 |
| | 799 | 4,874.85 | 4,737.96 | 4,953.52 | 4,123.76 | 18,690.09 |
| TOTAL | | 336,927.87 | 336,280.31 | 331,571.66 | 319,536.44 | 1,324,316.28 |

STATE: UTAH

| ZIP CODE | QUARTER 1 | QUARTER 2 | QUARTER 3 | QUARTER 4 | TOTAL GRAMS |
|---|---|---|---|---|---|
| 840 | 67,735.11 | 67,573.79 | 65,698.53 | 65,184.47 | 266,191.90 |
| 841 | 46,991.41 | 45,113.68 | 43,152.80 | 42,176.80 | 177,434.69 |
| 843 | 5,747.05 | 5,655.67 | 5,508.04 | 5,282.25 | 22,193.01 |
| 844 | 14,517.55 | 14,261.57 | 13,839.33 | 13,528.56 | 56,147.01 |
| 845 | 6,179.66 | 5,338.43 | 5,056.67 | 4,777.65 | 21,352.41 |
| 846 | 13,779.56 | 13,315.86 | 12,422.94 | 12,456.24 | 51,974.60 |
| 847 | 19,350.13 | 18,879.74 | 19,145.59 | 18,318.16 | 75,693.62 |
| TOTAL | 174,300.47 | 170,138.74 | 164,823.90 | 161,724.13 | 670,987.24 |

STATE: VERMONT

| ZIP CODE | QUARTER 1 | QUARTER 2 | QUARTER 3 | QUARTER 4 | TOTAL GRAMS |
|---|---|---|---|---|---|
| 050 | 2,342.51 | 2,414.93 | 1,969.77 | 1,814.32 | 8,541.53 |
| 051 | 1,703.80 | 1,735.89 | 1,510.42 | 1,718.72 | 6,668.83 |
| 052 | 1,263.29 | 1,224.44 | 1,178.96 | 1,238.16 | 4,904.85 |
| 053 | 1,918.77 | 1,867.42 | 1,785.14 | 1,640.91 | 7,212.24 |
| 054 | 10,054.72 | 9,742.22 | 9,640.24 | 9,151.69 | 38,588.87 |
| 056 | 2,315.85 | 2,257.23 | 2,134.58 | 2,007.08 | 8,714.74 |
| 057 | 2,385.85 | 2,203.97 | 2,124.58 | 1,905.64 | 8,620.04 |
| 058 | 1,866.09 | 1,945 | 1,815.40 | 1,866.43 | 7,492.92 |
| TOTAL | 23,850.88 | 23,391.10 | 22,159.09 | 21,342.95 | 90,744.02 |

STATE: VIRGIN ISLANDS

| ZIP CODE | QUARTER 1 | QUARTER 2 | QUARTER 3 | QUARTER 4 | TOTAL GRAMS |
|---|---|---|---|---|---|
| 008 | 584.77 | 578.06 | 432.34 | 506.93 | 2,102.10 |
| TOTAL | 584.77 | 578.06 | 432.34 | 506.93 | 2,102.10 |

STATE: VIRGINIA

| ZIP CODE | QUARTER 1 | QUARTER 2 | QUARTER 3 | QUARTER 4 | TOTAL GRAMS |
|---|---|---|---|---|---|
| 201 | 21,817.77 | 20,877.44 | 19,470.45 | 19,192.28 | 81,357.94 |
| 220 | 13,367.71 | 12,117.07 | 11,522.34 | 10,679.09 | 47,686.21 |
| 221 | 11,106.97 | 10,564.94 | 10,072.94 | 10,696.50 | 42,441.35 |

| | | | | | | |
|---|---|---|---|---|---|---|
| | 222 | 2,928.97 | 3,053.87 | 2,862.79 | 2,932.78 | 11,778.41 |
| | 223 | 7,248.69 | 6,421.68 | 5,942.29 | 5,693.18 | 25,305.84 |
| | 224 | 10,542.84 | 9,318.71 | 9,040.26 | 8,980.31 | 37,882.12 |
| | 225 | 3,751.72 | 3,629.99 | 3,407.15 | 3,320.57 | 14,109.43 |
| | 226 | 10,069.90 | 9,398.52 | 9,313.21 | 9,225.51 | 38,007.14 |
| | 227 | 2,245.95 | 2,103.52 | 1,825.99 | 1,907.80 | 8,083.26 |
| | 228 | 6,301.25 | 5,905.82 | 4,900.46 | 5,172.70 | 22,280.23 |
| | 229 | 9,821.11 | 8,960.18 | 8,589.54 | 8,774.26 | 36,145.09 |
| | 230 | 7,329.62 | 6,831.66 | 6,803.17 | 6,894.04 | 27,858.49 |
| | 231 | 13,933.31 | 12,323.08 | 10,961.85 | 11,507.76 | 48,726 |
| | 232 | 28,631.24 | 25,701.38 | 24,491.94 | 23,937.01 | 102,761.57 |
| | 233 | 9,499.76 | 9,263.25 | 8,754.90 | 8,568.96 | 36,086.87 |
| | 234 | 21,283.48 | 20,341.01 | 19,342.52 | 18,318.32 | 79,285.33 |
| | 235 | 8,471.43 | 8,062.31 | 7,046.70 | 7,298.12 | 30,878.56 |
| | 236 | 13,573.72 | 12,473.05 | 11,714.57 | 10,649.96 | 48,411.30 |
| | 237 | 6,410.70 | 6,181.71 | 5,487.59 | 5,521.28 | 23,601.28 |
| | 238 | 16,809.52 | 16,179.94 | 15,955.01 | 15,894.80 | 64,839.27 |
| | 239 | 2,899.30 | 2,726.97 | 2,679.26 | 2,624.66 | 10,930.19 |
| | 240 | 19,303.26 | 17,989.80 | 17,441.07 | 16,416.57 | 71,150.70 |
| | 241 | 16,903.43 | 16,434.74 | 15,316.71 | 15,240.48 | 63,895.36 |
| | 242 | 22,391.68 | 21,400.10 | 21,165.93 | 20,613.90 | 85,571.61 |
| | 243 | 9,507.97 | 8,351.44 | 8,096.26 | 8,009.44 | 33,965.11 |
| | 244 | 5,035.88 | 4,540.20 | 4,353.98 | 4,407.72 | 18,337.78 |
| | 245 | 14,866.66 | 13,719.73 | 12,315.15 | 12,333.25 | 53,234.79 |
| | 246 | 5,409.61 | 5,491.65 | 5,517.40 | 5,128.15 | 21,546.81 |
| TOTAL | | 321,463.45 | 300,363.76 | 284,391.43 | 279,939.40 | 1,186,158.04 |

| STATE: | WASHINGTON | | | | | |
|---|---|---|---|---|---|---|
| ZIP CODE | | QUARTER 1 | QUARTER 2 | QUARTER 3 | QUARTER 4 | TOTAL GRAMS |
| | 980 | 53,132.24 | 51,229.98 | 48,844.58 | 46,873.82 | 200,080.62 |
| | 981 | 33,150.84 | 32,881.72 | 31,072.18 | 29,485.17 | 126,589.91 |
| | 982 | 39,907.59 | 39,322.24 | 38,116.92 | 36,441.43 | 153,788.18 |
| | 983 | 35,691.52 | 35,189.09 | 33,642.29 | 32,605.45 | 137,128.35 |
| | 984 | 25,468.66 | 24,512.63 | 23,623.71 | 22,582.64 | 96,187.64 |
| | 985 | 20,092.93 | 19,296.15 | 18,934.52 | 18,434.39 | 76,757.99 |
| | 986 | 30,882.21 | 29,525 | 28,460.72 | 27,278.05 | 116,145.98 |

| | | | | | | |
|---|---|---|---|---|---|---|
| | 988 | 9,550.40 | 9,714.72 | 8,693.03 | 8,608.05 | 36,566.20 |
| | 989 | 12,464.93 | 12,103.81 | 11,621.86 | 10,528.14 | 46,718.74 |
| | 990 | 3,059.57 | 3,036.77 | 3,100.34 | 2,991.94 | 12,188.62 |
| | 991 | 5,020.76 | 5,058.13 | 5,066.22 | 4,747.35 | 19,892.46 |
| | 992 | 18,645.81 | 17,841.46 | 17,474.15 | 16,641.08 | 70,602.50 |
| | 993 | 17,572.14 | 16,675.07 | 15,831.13 | 14,902.34 | 64,980.68 |
| | 994 | 1,087.14 | 1,193.24 | 1,107.29 | 1,199.26 | 4,586.93 |
| TOTAL | | 305,726.74 | 297,580.01 | 285,588.94 | 273,319.11 | 1,162,214.80 |

STATE: WEST VIRGINIA

| ZIP CODE | | QUARTER 1 | QUARTER 2 | QUARTER 3 | QUARTER 4 | TOTAL GRAMS |
|---|---|---|---|---|---|---|
| | 247 | 2,829.34 | 2,817.62 | 2,694.29 | 2,584.73 | 10,925.98 |
| | 248 | 1,623.82 | 1,650.71 | 1,355.65 | 1,501.30 | 6,131.48 |
| | 249 | 2,225.68 | 2,165.60 | 2,169.63 | 1,992.71 | 8,553.62 |
| | 250 | 2,702.87 | 2,423.51 | 2,395.94 | 2,154.15 | 9,676.47 |
| | 251 | 1,643.87 | 1,557.88 | 1,558.57 | 1,483.59 | 6,243.91 |
| | 252 | 2,049.64 | 2,004.98 | 1,961.48 | 1,804.81 | 7,820.91 |
| | 253 | 7,666.52 | 7,265.54 | 7,094.64 | 6,780.23 | 28,806.93 |
| | 254 | 15,201.43 | 14,342.61 | 13,923.83 | 13,093.44 | 56,561.31 |
| | 255 | 5,584.43 | 5,129.66 | 4,996.73 | 4,840 | 20,550.82 |
| | 256 | 1,545.55 | 1,451.78 | 1,397.83 | 1,389.72 | 5,784.88 |
| | 257 | 4,721.42 | 4,290.60 | 3,690.80 | 3,767.17 | 16,469.99 |
| | 258 | 4,026.54 | 4,138.99 | 3,656.31 | 3,743.22 | 15,565.06 |
| | 259 | 2,506.52 | 2,187.28 | 1,927.79 | 1,602.94 | 8,224.53 |
| | 260 | 9,907.40 | 9,522.23 | 8,959.14 | 8,782.04 | 37,170.81 |
| | 261 | 3,435.90 | 3,423.39 | 3,367.58 | 3,073.25 | 13,300.12 |
| | 262 | 2,510.99 | 2,463.06 | 2,280.36 | 2,388.97 | 9,643.38 |
| | 263 | 4,940.67 | 4,861.64 | 4,813.59 | 4,458.05 | 19,073.95 |
| | 264 | 1,503.39 | 1,446.51 | 1,407.44 | 1,377.14 | 5,734.48 |
| | 265 | 7,376.20 | 7,040.81 | 7,074.71 | 6,951.45 | 28,443.17 |
| | 266 | 1,534.45 | 1,432.45 | 1,513.98 | 1,350.80 | 5,831.68 |
| | 267 | 2,576.72 | 2,461.03 | 2,272.74 | 2,191.27 | 9,501.76 |
| | 268 | 846.08 | 744.28 | 692.98 | 609.69 | 2,893.03 |
| TOTAL | | 88,959.43 | 84,822.16 | 81,206.01 | 77,920.67 | 332,908.27 |

DATE RANGE: 01/01/2017 TO 12/31/2017 ARCOS 3 - REPORT 1 Run Date: 07/03/2018

RETAIL DRUG DISTRIBUTION BY ZIP CODE WITHIN STATE BY GRAMS WT

STATE: WISCONSIN

| ZIP CODE | QUARTER 1 | QUARTER 2 | QUARTER 3 | QUARTER 4 | TOTAL GRAMS |
|---|---|---|---|---|---|
| 530 | 26,162.27 | 22,936.63 | 21,219.90 | 21,582.30 | 91,901.10 |
| 531 | 29,816.23 | 27,354.12 | 25,977.01 | 25,969.70 | 109,117.06 |
| 532 | 76,809.30 | 71,136.87 | 68,121.45 | 64,317.94 | 280,385.56 |
| 534 | 6,297.20 | 5,276.34 | 5,367.77 | 5,268.45 | 22,209.76 |
| 535 | 18,649.09 | 17,573.05 | 16,603.78 | 15,990.75 | 68,816.67 |
| 537 | 14,501.84 | 13,269.81 | 12,831.89 | 12,062.96 | 52,666.50 |
| 538 | 1,154.56 | 1,057.20 | 1,066.75 | 1,084.37 | 4,362.88 |
| 539 | 7,465.56 | 6,668.61 | 6,364.09 | 6,205.45 | 26,703.71 |
| 540 | 3,418.96 | 3,348.03 | 3,111.93 | 3,066.80 | 12,945.72 |
| 541 | 3,451.78 | 3,347.46 | 3,257.54 | 3,093 | 13,149.78 |
| 542 | 2,423.04 | 2,209.23 | 2,116.45 | 1,989.93 | 8,738.65 |
| 543 | 4,086.25 | 3,922.99 | 3,731.45 | 3,660.13 | 15,400.82 |
| 544 | 8,019.65 | 7,479.05 | 7,070.05 | 6,887.42 | 29,456.17 |
| 545 | 3,059.15 | 2,961.99 | 2,927.27 | 2,508.80 | 11,457.21 |
| 546 | 7,257.62 | 6,851.56 | 6,462.72 | 6,246.35 | 26,818.25 |
| 547 | 5,091.97 | 4,484.38 | 3,907 | 3,886.93 | 17,370.28 |
| 548 | 4,797.55 | 4,741.31 | 4,575.11 | 4,277.96 | 18,391.93 |
| 549 | 13,225.32 | 11,933.53 | 11,445.93 | 11,223.12 | 47,827.90 |
| TOTAL | 235,687.34 | 216,552.16 | 206,158.09 | 199,322.36 | 857,719.95 |

STATE: WYOMING

| ZIP CODE | QUARTER 1 | QUARTER 2 | QUARTER 3 | QUARTER 4 | TOTAL GRAMS |
|---|---|---|---|---|---|
| 820 | 5,526.71 | 5,358.94 | 5,121.86 | 4,890.28 | 20,897.79 |
| 822 | 614.41 | 569.48 | 513.02 | 654.18 | 2,351.09 |
| 823 | 641.21 | 495.47 | 339.58 | 313.48 | 1,789.74 |
| 824 | 3,925.50 | 3,135.73 | 3,038.16 | 3,161.67 | 13,261.06 |
| 825 | 2,129.22 | 2,264.81 | 2,089.14 | 2,258.36 | 8,741.53 |
| 826 | 4,620.78 | 4,460.65 | 4,692.65 | 4,497.31 | 18,271.39 |
| 827 | 1,289.97 | 1,295.06 | 1,199.64 | 1,203.93 | 4,988.60 |
| 828 | 1,756.60 | 1,482.81 | 1,626.21 | 1,489.99 | 6,355.61 |
| 829 | 5,158.25 | 5,438.17 | 5,390.26 | 5,556.53 | 21,543.21 |
| 830 | 321.93 | 253.4 | 296.02 | 273.97 | 1,145.32 |
| 831 | 801.41 | 649.96 | 699.76 | 672.33 | 2,823.46 |

| | | | | | |
|---|---|---|---|---|---|
| TOTAL | 26,785.99 | 25,404.48 | 25,006.30 | 24,972.03 | 102,168.80 |

DRUG CODE: 9150 DRUG NAME: HYDROMORPHONE

STATE: ALABAMA

| ZIP CODE | QUARTER 1 | QUARTER 2 | QUARTER 3 | QUARTER 4 | TOTAL GRAMS |
|---|---|---|---|---|---|
| 350 | 260.35 | 212.42 | 215.45 | 198.34 | 886.56 |
| 351 | 122.63 | 105.25 | 124.37 | 114.09 | 466.34 |
| 352 | 483.38 | 449.03 | 513.06 | 468.15 | 1,913.62 |
| 354 | 128.15 | 128.3 | 130.16 | 115.75 | 502.36 |
| 355 | 55.59 | 50.52 | 58.21 | 55.2 | 219.52 |
| 356 | 203.65 | 173.87 | 189.44 | 192.63 | 759.59 |
| 357 | 121.21 | 117.39 | 112.12 | 105.85 | 456.57 |
| 358 | 267.24 | 281.07 | 251.07 | 277.75 | 1,077.13 |
| 359 | 140.83 | 129.39 | 112.91 | 134.45 | 517.58 |
| 360 | 186.84 | 165.22 | 139.62 | 145.92 | 637.6 |
| 361 | 185.34 | 172.66 | 159.06 | 160.57 | 677.63 |
| 362 | 105.09 | 85.37 | 94.96 | 83.99 | 369.41 |
| 363 | 251.81 | 260.36 | 269.1 | 221.05 | 1,002.32 |
| 364 | 70.43 | 73.95 | 53.78 | 68.61 | 266.77 |
| 365 | 495.76 | 306.67 | 290.21 | 294.46 | 1,387.10 |
| 366 | 290.66 | 309.7 | 364.29 | 333.57 | 1,298.22 |
| 367 | 21.88 | 25.87 | 21.46 | 17.57 | 86.78 |
| 368 | 125.61 | 119.5 | 114.06 | 120.94 | 480.11 |
| 369 | 12.32 | 7.76 | 7.94 | 4.08 | 32.1 |
| TOTAL | 3,528.77 | 3,174.30 | 3,221.27 | 3,112.97 | 13,037.31 |

STATE: ALASKA

| ZIP CODE | QUARTER 1 | QUARTER 2 | QUARTER 3 | QUARTER 4 | TOTAL GRAMS |
|---|---|---|---|---|---|
| 995 | 644.34 | 582.77 | 513.87 | 530.38 | 2,271.36 |
| 996 | 325.09 | 315.47 | 316.22 | 322.56 | 1,279.34 |
| 997 | 47.24 | 41.49 | 40.84 | 50.77 | 180.34 |
| 998 | 28.65 | 29.28 | 29 | 40.66 | 127.59 |
| 999 | 19.93 | 7.88 | 6.61 | 4.18 | 38.6 |
| TOTAL | 1,065.25 | 976.89 | 906.54 | 948.55 | 3,897.23 |

| STATE: | | AMERICAN SAMOA | | | | |
|---|---|---|---|---|---|---|
| ZIP CODE | | QUARTER 1 | QUARTER 2 | QUARTER 3 | QUARTER 4 | TOTAL GRAMS |
| | 967 | 0.89 | 0.09 | 0 | 0 | 0.98 |
| TOTAL | | 0.89 | 0.09 | 0 | 0 | 0.98 |

| STATE: | | ARIZONA | | | | |
|---|---|---|---|---|---|---|
| ZIP CODE | | QUARTER 1 | QUARTER 2 | QUARTER 3 | QUARTER 4 | TOTAL GRAMS |
| | 850 | 1,329.80 | 1,330.03 | 1,212.49 | 1,083.72 | 4,956.04 |
| | 851 | 204.71 | 210.29 | 210.25 | 191.71 | 816.96 |
| | 852 | 1,698.32 | 1,733.01 | 1,723.09 | 1,699.02 | 6,853.44 |
| | 853 | 1,228.47 | 1,139.67 | 1,085.26 | 1,085.61 | 4,539.01 |
| | 855 | 103.79 | 87.46 | 86.61 | 82.04 | 359.9 |
| | 856 | 199.05 | 200.3 | 168.71 | 166.41 | 734.47 |
| | 857 | 1,137.55 | 1,038.75 | 1,024.55 | 988.53 | 4,189.38 |
| | 859 | 114.25 | 141.59 | 113.77 | 115.14 | 484.75 |
| | 860 | 152.95 | 119.82 | 123.31 | 117.91 | 513.99 |
| | 863 | 286.46 | 301.98 | 293.79 | 250.1 | 1,132.33 |
| | 864 | 632.32 | 592.95 | 550.5 | 447.19 | 2,222.96 |
| | 865 | 1.42 | 1.76 | 3.03 | 2.09 | 8.3 |
| TOTAL | | 7,089.09 | 6,897.61 | 6,595.36 | 6,229.47 | 26,811.53 |

| STATE: | | ARKANSAS | | | | |
|---|---|---|---|---|---|---|
| ZIP CODE | | QUARTER 1 | QUARTER 2 | QUARTER 3 | QUARTER 4 | TOTAL GRAMS |
| | 716 | 69.51 | 71.64 | 71.84 | 67.68 | 280.67 |
| | 717 | 52.55 | 44.11 | 42.77 | 26.4 | 165.83 |
| | 718 | 92.47 | 105.6 | 91.96 | 84.99 | 375.02 |
| | 719 | 178.26 | 150.06 | 183.52 | 162.36 | 674.2 |
| | 720 | 283.86 | 269.66 | 289.71 | 260.42 | 1,103.65 |
| | 721 | 283.88 | 300.27 | 255.16 | 268.97 | 1,108.28 |
| | 722 | 533.27 | 554.98 | 529.54 | 577.25 | 2,195.04 |
| | 723 | 74.45 | 55.64 | 54.05 | 43.2 | 227.34 |

| | | | | | | |
|---|---|---|---|---|---|---|
| | 724 | 201.66 | 233.68 | 253.02 | 186.75 | 875.11 |
| | 725 | 145.07 | 150.91 | 142.25 | 131.89 | 570.12 |
| | 726 | 157.37 | 133.19 | 143.25 | 149.67 | 583.48 |
| | 727 | 802.5 | 756.72 | 735.83 | 737.67 | 3,032.72 |
| | 728 | 97.04 | 81.42 | 88.24 | 77.97 | 344.67 |
| | 729 | 404.95 | 379.35 | 339.39 | 394.8 | 1,518.49 |
| TOTAL | | 3,376.84 | 3,287.23 | 3,220.53 | 3,170.02 | 13,054.62 |

| STATE: | CALIFORNIA | | | | | |
|---|---|---|---|---|---|---|
| ZIP CODE | | QUARTER 1 | QUARTER 2 | QUARTER 3 | QUARTER 4 | TOTAL GRAMS |
| | 900 | 1,559.16 | 1,579.69 | 1,526.33 | 1,527.63 | 6,192.81 |
| | 902 | 633.8 | 581.84 | 557.35 | 547.96 | 2,320.95 |
| | 903 | 41.76 | 36.89 | 36.08 | 37.31 | 152.04 |
| | 904 | 242.32 | 266.25 | 271.4 | 264.49 | 1,044.46 |
| | 905 | 508.86 | 572.23 | 527.46 | 521.32 | 2,129.87 |
| | 906 | 416.91 | 420.35 | 425.94 | 340.54 | 1,603.74 |
| | 907 | 483.09 | 474.26 | 464.27 | 402.62 | 1,824.24 |
| | 908 | 1,247.23 | 1,327.37 | 1,251.67 | 688.17 | 4,514.44 |
| | 910 | 485.42 | 473.92 | 424.21 | 433.56 | 1,817.11 |
| | 911 | 190.28 | 171.12 | 162.84 | 168.98 | 693.22 |
| | 912 | 188.91 | 211.38 | 200.12 | 172.26 | 772.67 |
| | 913 | 1,394.70 | 1,362.90 | 1,314.74 | 1,229.25 | 5,301.59 |
| | 914 | 356.94 | 425.63 | 562.05 | 479.87 | 1,824.49 |
| | 915 | 241.83 | 304.88 | 212.36 | 227.95 | 987.02 |
| | 916 | 91.96 | 83.12 | 73.27 | 71.01 | 319.36 |
| | 917 | 1,728.44 | 1,782.08 | 1,720.16 | 1,719.17 | 6,949.85 |
| | 918 | 23.87 | 24.38 | 13.88 | 20.55 | 82.68 |
| | 919 | 424.13 | 404.32 | 384.78 | 443.02 | 1,656.25 |
| | 920 | 1,972.59 | 1,899.56 | 1,753.74 | 1,918.56 | 7,544.45 |
| | 921 | 1,493.63 | 1,384.92 | 1,401.14 | 1,398.20 | 5,677.89 |
| | 922 | 979.94 | 933.74 | 886.11 | 850.95 | 3,650.74 |
| | 923 | 1,446.19 | 1,336.23 | 1,374.46 | 1,278.24 | 5,435.12 |
| | 924 | 210.38 | 188.97 | 168.31 | 181.65 | 749.31 |
| | 925 | 1,110.41 | 1,334.89 | 1,139.32 | 1,206.68 | 4,791.30 |
| | 926 | 930.25 | 874.25 | 831.14 | 765.76 | 3,401.40 |
| | 927 | 315.64 | 270.05 | 297.05 | 292.22 | 1,174.96 |

| | | | | | | |
|---|---|---|---|---|---|---|
| | 928 | 1,529.34 | 1,443.81 | 1,356.17 | 1,537.52 | 5,866.84 |
| | 930 | 696.01 | 653.99 | 595.52 | 543.41 | 2,488.93 |
| | 931 | 167.2 | 163.42 | 139.43 | 121.16 | 591.21 |
| | 932 | 610.95 | 527.33 | 537.82 | 529.2 | 2,205.30 |
| | 933 | 844.71 | 959.61 | 881.54 | 774.94 | 3,460.80 |
| | 934 | 913.34 | 820.14 | 785.9 | 783.74 | 3,303.12 |
| | 935 | 1,313.01 | 1,325.84 | 1,260.16 | 1,190.65 | 5,089.66 |
| | 936 | 298.79 | 337.52 | 357.17 | 353.37 | 1,346.85 |
| | 937 | 413.15 | 391.77 | 405.44 | 417.52 | 1,627.88 |
| | 939 | 299.06 | 262.43 | 264.94 | 340.99 | 1,167.42 |
| | 940 | 591.62 | 544.45 | 514.74 | 510.76 | 2,161.57 |
| | 941 | 1,015.92 | 887.65 | 878.42 | 870.66 | 3,652.65 |
| | 943 | 284.92 | 286.5 | 258.26 | 315.94 | 1,145.62 |
| | 944 | 105.31 | 65.29 | 76.22 | 68.04 | 314.86 |
| | 945 | 2,900.20 | 2,912.43 | 2,793.48 | 2,623.47 | 11,229.58 |
| | 946 | 455.88 | 386.46 | 346.43 | 429.23 | 1,618 |
| | 947 | 166.66 | 192.98 | 192.04 | 186.87 | 738.55 |
| | 948 | 106.18 | 82.74 | 61.7 | 82.53 | 333.15 |
| | 949 | 515.56 | 550.84 | 537.99 | 465.82 | 2,070.21 |
| | 950 | 516.47 | 490.25 | 477.66 | 469.99 | 1,954.37 |
| | 951 | 393.06 | 391.31 | 351.28 | 383.46 | 1,519.11 |
| | 952 | 499.44 | 516.97 | 482.19 | 407.41 | 1,906.01 |
| | 953 | 1,046.53 | 1,062.50 | 950.7 | 913.09 | 3,972.82 |
| | 954 | 988.4 | 936.35 | 788.41 | 678.13 | 3,391.29 |
| | 955 | 287.73 | 284.08 | 270.4 | 255.83 | 1,098.04 |
| | 956 | 1,770.13 | 1,692.63 | 1,689.83 | 1,663.10 | 6,815.69 |
| | 957 | 207.13 | 177.98 | 187.41 | 172.62 | 745.14 |
| | 958 | 1,090.77 | 1,165.27 | 1,220.14 | 1,053.27 | 4,529.45 |
| | 959 | 1,113.08 | 1,040.89 | 920.28 | 930.69 | 4,004.94 |
| | 960 | 544.62 | 508.25 | 499.81 | 477.77 | 2,030.45 |
| | 961 | 73.87 | 82.43 | 63.37 | 81.69 | 301.36 |
| TOTAL | | 40,477.68 | 39,869.33 | 38,125.03 | 36,820.79 | 155,292.83 |

| STATE: | | COLORADO | | | | |
|---|---|---|---|---|---|---|
| ZIP CODE | | QUARTER 1 | QUARTER 2 | QUARTER 3 | QUARTER 4 | TOTAL GRAMS |
| | 800 | 1,367.40 | 1,239.98 | 1,251.22 | 1,242.79 | 5,101.39 |

| | | | | | | |
|---|---|---|---|---|---|---|
| | 801 | 1,332.56 | 1,174.25 | 1,118.03 | 1,185.57 | 4,810.41 |
| | 802 | 1,299.39 | 1,125.26 | 1,097.39 | 1,169.59 | 4,691.63 |
| | 803 | 128 | 123.02 | 101.7 | 92.66 | 445.38 |
| | 804 | 267.61 | 254.76 | 214.03 | 241.15 | 977.55 |
| | 805 | 668.79 | 641.19 | 613.62 | 565.55 | 2,489.15 |
| | 806 | 383.11 | 354.8 | 349.21 | 341.71 | 1,428.83 |
| | 807 | 71.34 | 82.37 | 71.65 | 69.5 | 294.86 |
| | 808 | 144.42 | 127.9 | 137.58 | 117.94 | 527.84 |
| | 809 | 781.27 | 771.45 | 739.53 | 729.36 | 3,021.61 |
| | 810 | 383.8 | 367.56 | 350.92 | 343.92 | 1,446.20 |
| | 811 | 48.98 | 39.88 | 32.97 | 37.08 | 158.91 |
| | 812 | 100.55 | 94.07 | 92.53 | 79.33 | 366.48 |
| | 813 | 61.76 | 49 | 66.75 | 67.54 | 245.05 |
| | 814 | 68.24 | 60.52 | 56.18 | 55.58 | 240.52 |
| | 815 | 160.78 | 139.48 | 129.83 | 157.66 | 587.75 |
| | 816 | 102.79 | 92.16 | 83.85 | 86.24 | 365.04 |
| TOTAL | | 7,370.79 | 6,737.65 | 6,506.99 | 6,583.17 | 27,198.60 |

STATE: CONNECTICUT

| ZIP CODE | | QUARTER 1 | QUARTER 2 | QUARTER 3 | QUARTER 4 | TOTAL GRAMS |
|---|---|---|---|---|---|---|
| | 60 | 1,282.91 | 1,361.76 | 1,117.67 | 1,133.15 | 4,895.49 |
| | 61 | 628.43 | 538.06 | 525.46 | 537.19 | 2,229.14 |
| | 62 | 171.82 | 156.99 | 164.8 | 166.05 | 659.66 |
| | 63 | 327.21 | 330.98 | 306.43 | 304.68 | 1,269.30 |
| | 64 | 1,761.83 | 1,605.27 | 1,745.38 | 1,504.06 | 6,616.54 |
| | 65 | 517.28 | 509.36 | 480.39 | 569.9 | 2,076.93 |
| | 66 | 152.16 | 169.61 | 173.32 | 180.44 | 675.53 |
| | 67 | 449.23 | 404.06 | 422.89 | 423.1 | 1,699.28 |
| | 68 | 1,276.21 | 1,260.48 | 1,161.11 | 1,321.56 | 5,019.36 |
| | 69 | 104.33 | 112.08 | 107.72 | 110.39 | 434.52 |
| TOTAL | | 6,671.41 | 6,448.65 | 6,205.17 | 6,250.52 | 25,575.75 |

STATE: DELAWARE

| ZIP CODE | | QUARTER 1 | QUARTER 2 | QUARTER 3 | QUARTER 4 | TOTAL GRAMS |
|---|---|---|---|---|---|---|
| | 197 | 992.32 | 874.23 | 908.66 | 948.74 | 3,723.95 |

| | | | | | | |
|---|---|---|---|---|---|---|
| | 198 | 411.83 | 351.81 | 333.36 | 347.32 | 1,444.32 |
| | 199 | 890.67 | 853.5 | 797.2 | 813.58 | 3,354.95 |
| TOTAL | | 2,294.82 | 2,079.54 | 2,039.22 | 2,109.64 | 8,523.22 |

| STATE: | DISTRICT OF COLUMBIA | | | | | |
|---|---|---|---|---|---|---|
| ZIP CODE | | QUARTER 1 | QUARTER 2 | QUARTER 3 | QUARTER 4 | TOTAL GRAMS |
| | 200 | 580.45 | 523.3 | 513.86 | 487.21 | 2,104.82 |
| | 203 | 0 | 0 | 0 | 0.35 | 0.35 |
| | 204 | 16.22 | 16.49 | 15.38 | 14.05 | 62.14 |
| TOTAL | | 596.67 | 539.79 | 529.24 | 501.61 | 2,167.31 |

| STATE: | FLORIDA | | | | | |
|---|---|---|---|---|---|---|
| ZIP CODE | | QUARTER 1 | QUARTER 2 | QUARTER 3 | QUARTER 4 | TOTAL GRAMS |
| | 320 | 882.89 | 848.4 | 820.82 | 802.72 | 3,354.83 |
| | 321 | 1,119.11 | 1,276.59 | 1,269.49 | 1,155.20 | 4,820.39 |
| | 322 | 2,189.15 | 1,913.21 | 1,795.38 | 1,780.68 | 7,678.42 |
| | 323 | 431.56 | 402.09 | 381.48 | 393.41 | 1,608.54 |
| | 324 | 353.3 | 333.75 | 279.57 | 326.78 | 1,293.40 |
| | 325 | 934.98 | 758.36 | 737.75 | 749.42 | 3,180.51 |
| | 326 | 571.01 | 539 | 538.9 | 576.23 | 2,225.14 |
| | 327 | 3,390.33 | 3,024.18 | 2,844.10 | 2,760.33 | 12,018.94 |
| | 328 | 1,963.33 | 2,010.66 | 1,906.87 | 1,853.32 | 7,734.18 |
| | 329 | 2,164.34 | 2,056.64 | 2,155.42 | 1,943.14 | 8,319.54 |
| | 330 | 2,109.01 | 2,160.89 | 1,744.29 | 1,819.60 | 7,833.79 |
| | 331 | 1,655.60 | 1,702.91 | 1,522.71 | 1,549.37 | 6,430.59 |
| | 333 | 1,454.54 | 1,360.85 | 1,450.17 | 1,330.48 | 5,596.04 |
| | 334 | 2,423.37 | 2,313.04 | 2,109.64 | 2,158.96 | 9,005.01 |
| | 335 | 2,312.59 | 2,121.06 | 2,021.16 | 2,029.20 | 8,484.01 |
| | 336 | 8,321.09 | 6,505.75 | 6,811.94 | 7,566.74 | 29,205.52 |
| | 337 | 3,479.81 | 3,300.87 | 3,064.39 | 3,163.79 | 13,008.86 |
| | 338 | 1,301.84 | 1,281.29 | 1,086.58 | 1,122.05 | 4,791.76 |
| | 339 | 2,357.40 | 2,243.87 | 1,914.39 | 2,168.41 | 8,684.07 |
| | 341 | 711.78 | 675.97 | 702.19 | 719.62 | 2,809.56 |
| | 342 | 2,683.41 | 2,589.77 | 2,358.89 | 2,521.07 | 10,153.14 |
| | 344 | 1,116.36 | 1,140.89 | 1,075.84 | 1,042.36 | 4,375.45 |

| | | | | | | |
|---|---|---|---|---|---|---|
| | 346 | 3,481.86 | 3,414.81 | 3,287.55 | 3,407.62 | 13,591.84 |
| | 347 | 1,487.03 | 1,496.15 | 1,380.26 | 1,434.66 | 5,798.10 |
| | 349 | 1,262.49 | 1,184.15 | 1,134.74 | 1,143.92 | 4,725.30 |
| TOTAL | | 50,158.18 | 46,655.15 | 44,394.52 | 45,519.08 | 186,726.93 |

| STATE: | GEORGIA | | | | | |
|---|---|---|---|---|---|---|
| ZIP CODE | | QUARTER 1 | QUARTER 2 | QUARTER 3 | QUARTER 4 | TOTAL GRAMS |
| | 300 | 2,011.78 | 1,779.86 | 1,876.30 | 1,746.26 | 7,414.20 |
| | 301 | 994.68 | 933.97 | 949.29 | 956.51 | 3,834.45 |
| | 302 | 669.76 | 620.41 | 576.49 | 580.48 | 2,447.14 |
| | 303 | 902.95 | 807.22 | 827.13 | 804.73 | 3,342.03 |
| | 304 | 142.58 | 156.72 | 142.29 | 115.78 | 557.37 |
| | 305 | 570.17 | 581.96 | 583.53 | 571.96 | 2,307.62 |
| | 306 | 471.1 | 481.73 | 443.98 | 464.13 | 1,860.94 |
| | 307 | 210.09 | 184.88 | 182.84 | 180.72 | 758.53 |
| | 308 | 127.76 | 121.89 | 123.39 | 121.4 | 494.44 |
| | 309 | 279.94 | 391.37 | 350.06 | 319.33 | 1,340.70 |
| | 310 | 278.93 | 263.81 | 252.64 | 269.11 | 1,064.49 |
| | 312 | 156.74 | 163.98 | 221.67 | 191.57 | 733.96 |
| | 313 | 367.77 | 345.84 | 271.84 | 139.49 | 1,124.94 |
| | 314 | 262.65 | 230.66 | 271.56 | 206.99 | 971.86 |
| | 315 | 544.02 | 513.45 | 557.52 | 398.86 | 2,013.85 |
| | 316 | 146.48 | 166.94 | 154.41 | 136.93 | 604.76 |
| | 317 | 426.29 | 394.92 | 434.02 | 433.08 | 1,688.31 |
| | 318 | 6.75 | 6.14 | 8.5 | 6.63 | 28.02 |
| | 319 | 93.27 | 88.95 | 114.16 | 76.94 | 373.32 |
| | 398 | 109.25 | 86.31 | 69.13 | 72.85 | 337.54 |
| TOTAL | | 8,772.96 | 8,321.01 | 8,410.75 | 7,793.75 | 33,298.47 |

| STATE: | GUAM | | | | | |
|---|---|---|---|---|---|---|
| ZIP CODE | | QUARTER 1 | QUARTER 2 | QUARTER 3 | QUARTER 4 | TOTAL GRAMS |
| | 969 | 31.75 | 26.32 | 23.41 | 33.03 | 114.51 |
| TOTAL | | 31.75 | 26.32 | 23.41 | 33.03 | 114.51 |

STATE: HAWAII

| ZIP CODE | QUARTER 1 | QUARTER 2 | QUARTER 3 | QUARTER 4 | TOTAL GRAMS |
|---|---|---|---|---|---|
| 967 | 367.65 | 353.54 | 328.35 | 329.99 | 1,379.53 |
| 968 | 324.99 | 352.68 | 314.52 | 299.5 | 1,291.69 |
| TOTAL | 692.64 | 706.22 | 642.87 | 629.49 | 2,671.22 |

STATE: IDAHO

| ZIP CODE | QUARTER 1 | QUARTER 2 | QUARTER 3 | QUARTER 4 | TOTAL GRAMS |
|---|---|---|---|---|---|
| 832 | 115.23 | 101.81 | 102.12 | 112.63 | 431.79 |
| 833 | 196.49 | 194.28 | 164.83 | 190.29 | 745.89 |
| 834 | 82.98 | 94.3 | 91.46 | 83.97 | 352.71 |
| 835 | 99.86 | 94.02 | 87.2 | 91.28 | 372.36 |
| 836 | 610.39 | 598 | 614 | 613.84 | 2,436.23 |
| 837 | 569.55 | 484.89 | 519.03 | 613.82 | 2,187.29 |
| 838 | 268.16 | 247.61 | 260.75 | 223.03 | 999.55 |
| TOTAL | 1,942.66 | 1,814.91 | 1,839.39 | 1,928.86 | 7,525.82 |

STATE: ILLINOIS

| ZIP CODE | QUARTER 1 | QUARTER 2 | QUARTER 3 | QUARTER 4 | TOTAL GRAMS |
|---|---|---|---|---|---|
| 600 | 1,781.86 | 1,769.08 | 1,563.94 | 1,687.40 | 6,802.28 |
| 601 | 1,306.38 | 1,139.24 | 1,199.82 | 1,264.79 | 4,910.23 |
| 602 | 66.45 | 67.71 | 78.31 | 80.41 | 292.88 |
| 603 | 94.2 | 93.94 | 95.51 | 98.75 | 382.4 |
| 604 | 1,426.01 | 1,399.19 | 1,329.55 | 1,257.06 | 5,411.81 |
| 605 | 790.11 | 773.46 | 733.52 | 754.04 | 3,051.13 |
| 606 | 1,701.99 | 1,659.04 | 1,509.14 | 1,483.21 | 6,353.38 |
| 607 | 57.57 | 56.69 | 62.99 | 70.88 | 248.13 |
| 608 | 141.45 | 138.07 | 136.55 | 113.04 | 529.11 |
| 609 | 179.48 | 146.04 | 124.59 | 115.98 | 566.09 |
| 610 | 79.76 | 73.63 | 72.16 | 61.7 | 287.25 |
| 611 | 192.44 | 176.86 | 206.32 | 192.29 | 767.91 |
| 612 | 81.51 | 57.18 | 74.39 | 76.77 | 289.85 |
| 613 | 63.59 | 61.63 | 62.85 | 67.33 | 255.4 |
| 614 | 51.79 | 48.18 | 54.54 | 46.45 | 200.96 |
| 615 | 99.47 | 93.02 | 88.02 | 88.06 | 368.57 |

| | | | | | | |
|---|---|---|---|---|---|---|
| | 616 | 187.27 | 215.07 | 209.37 | 199.61 | 811.32 |
| | 617 | 193.83 | 218.55 | 232.41 | 187.96 | 832.75 |
| | 618 | 300.04 | 303.55 | 254.3 | 247.07 | 1,104.96 |
| | 619 | 61.93 | 58.75 | 98.55 | 36.73 | 255.96 |
| | 620 | 199.94 | 169.07 | 170.66 | 176.18 | 715.85 |
| | 622 | 248.95 | 247.56 | 234.57 | 212.6 | 943.68 |
| | 623 | 73.73 | 59.41 | 65.63 | 81.26 | 280.03 |
| | 624 | 101.95 | 82.99 | 90.23 | 85.02 | 360.19 |
| | 625 | 230.25 | 248.4 | 225.78 | 216.27 | 920.7 |
| | 626 | 87.07 | 85.48 | 88.98 | 84.11 | 345.64 |
| | 627 | 230.43 | 147.9 | 135.28 | 168.62 | 682.23 |
| | 628 | 103.91 | 91.64 | 89.27 | 100.07 | 384.89 |
| | 629 | 159.39 | 154.48 | 123.54 | 112.16 | 549.57 |
| TOTAL | | 10,292.75 | 9,835.81 | 9,410.77 | 9,365.82 | 38,905.15 |

| STATE: | INDIANA | | | | | |
|---|---|---|---|---|---|---|
| ZIP CODE | | QUARTER 1 | QUARTER 2 | QUARTER 3 | QUARTER 4 | TOTAL GRAMS |
| | 460 | 300.21 | 365.72 | 378.25 | 404.08 | 1,448.26 |
| | 461 | 340.24 | 294.96 | 294.96 | 346.71 | 1,276.87 |
| | 462 | 1,374.39 | 1,454.17 | 1,522.89 | 1,204.27 | 5,555.72 |
| | 463 | 560.25 | 495.42 | 488.26 | 451.19 | 1,995.12 |
| | 464 | 48.2 | 46.51 | 31.53 | 32.84 | 159.08 |
| | 465 | 390.03 | 386.17 | 364.53 | 367.94 | 1,508.67 |
| | 466 | 156.9 | 166.04 | 144.23 | 161.24 | 628.41 |
| | 467 | 330.09 | 276.22 | 281.17 | 226.72 | 1,114.20 |
| | 468 | 442.14 | 431.57 | 356.47 | 415.28 | 1,645.46 |
| | 469 | 169.45 | 147.35 | 133.51 | 138.89 | 589.2 |
| | 470 | 40.02 | 46.94 | 31.7 | 40.44 | 159.1 |
| | 471 | 256.93 | 293.89 | 225.32 | 178.65 | 954.79 |
| | 472 | 191.74 | 162.49 | 172.7 | 186.28 | 713.21 |
| | 473 | 302.85 | 333.38 | 286.54 | 329.39 | 1,252.16 |
| | 474 | 189.55 | 174.36 | 166.23 | 168.73 | 698.87 |
| | 475 | 241.97 | 267.27 | 327.39 | 323.46 | 1,160.09 |
| | 476 | 69.97 | 74.78 | 80.96 | 115.29 | 341 |
| | 477 | 277.27 | 286.31 | 272.76 | 328.65 | 1,164.99 |
| | 478 | 175.58 | 151.97 | 131.61 | 117.17 | 576.33 |

| | | | | | | |
|---|---|---|---|---|---|---|
| | 479 | 203.92 | 229.61 | 228.91 | 224.93 | 887.37 |
| TOTAL | | 6,061.70 | 6,085.13 | 5,919.92 | 5,762.15 | 23,828.90 |

| STATE: | IOWA | | | | | |
|---|---|---|---|---|---|---|
| ZIP CODE | | QUARTER 1 | QUARTER 2 | QUARTER 3 | QUARTER 4 | TOTAL GRAMS |
| | 500 | 295.54 | 304.11 | 249.16 | 210.21 | 1,059.02 |
| | 501 | 201.65 | 178.19 | 180.98 | 171.81 | 732.63 |
| | 502 | 172.18 | 169.44 | 164.98 | 152.86 | 659.46 |
| | 503 | 726.23 | 641.46 | 641.79 | 632.58 | 2,642.06 |
| | 504 | 71.62 | 65.71 | 71.19 | 55.65 | 264.17 |
| | 505 | 44.97 | 47.66 | 50.89 | 62.46 | 205.98 |
| | 506 | 96.23 | 72.95 | 84.57 | 84.84 | 338.59 |
| | 507 | 59.37 | 61.62 | 55.86 | 54.21 | 231.06 |
| | 508 | 19.96 | 16.38 | 20.71 | 13.93 | 70.98 |
| | 510 | 15.86 | 12.83 | 15.77 | 16.3 | 60.76 |
| | 511 | 52.36 | 59.11 | 55.58 | 49.04 | 216.09 |
| | 512 | 10.84 | 8.32 | 6.18 | 9.57 | 34.91 |
| | 513 | 40.02 | 31 | 31.07 | 32.9 | 134.99 |
| | 514 | 30.16 | 32.57 | 29.38 | 27.77 | 119.88 |
| | 515 | 67.08 | 62.5 | 58.39 | 55.46 | 243.43 |
| | 516 | 7.57 | 10.14 | 7.98 | 14.57 | 40.26 |
| | 520 | 75.4 | 77.45 | 68.4 | 55.08 | 276.33 |
| | 521 | 16.92 | 10.64 | 23.39 | 15.1 | 66.05 |
| | 522 | 402.34 | 393.27 | 361.18 | 345.55 | 1,502.34 |
| | 523 | 70.13 | 70.53 | 67.89 | 67.35 | 275.9 |
| | 524 | 128.35 | 143.92 | 155.84 | 145.63 | 573.74 |
| | 525 | 216.71 | 207.52 | 197.42 | 299.52 | 921.17 |
| | 526 | 118.66 | 114.92 | 93.8 | 98.5 | 425.88 |
| | 527 | 132.3 | 113.46 | 96.45 | 98.39 | 440.6 |
| | 528 | 104.44 | 80.43 | 84.21 | 86.8 | 355.88 |
| TOTAL | | 3,176.89 | 2,986.13 | 2,873.06 | 2,856.08 | 11,892.16 |

| STATE: | KANSAS | | | | | |
|---|---|---|---|---|---|---|
| ZIP CODE | | QUARTER 1 | QUARTER 2 | QUARTER 3 | QUARTER 4 | TOTAL GRAMS |
| | 660 | 415.79 | 419.48 | 408.37 | 401.6 | 1,645.24 |

| | | | | | | |
|---|---|---|---|---|---|---|
| | 661 | 228.77 | 181.89 | 256.29 | 156.46 | 823.41 |
| | 662 | 773.47 | 1,014.46 | 706.89 | 593.13 | 3,087.95 |
| | 664 | 111.19 | 88.4 | 87.1 | 98.7 | 385.39 |
| | 665 | 143.97 | 141.96 | 136.84 | 115.08 | 537.85 |
| | 666 | 110.57 | 95.17 | 104.48 | 104.61 | 414.83 |
| | 667 | 122.37 | 113.93 | 130.9 | 98.91 | 466.11 |
| | 668 | 38.65 | 34.39 | 35.42 | 30.51 | 138.97 |
| | 669 | 14.74 | 15.84 | 15.01 | 18.42 | 64.01 |
| | 670 | 154.18 | 158.18 | 156.41 | 133.66 | 602.43 |
| | 671 | 117.94 | 121.81 | 119.63 | 120.08 | 479.46 |
| | 672 | 694.32 | 710.57 | 742.69 | 629.6 | 2,777.18 |
| | 673 | 28.21 | 33.56 | 34.99 | 33.11 | 129.87 |
| | 674 | 175.46 | 162.37 | 190.18 | 148.19 | 676.2 |
| | 675 | 236.27 | 216.36 | 223.17 | 229.19 | 904.99 |
| | 676 | 76.42 | 76.22 | 84.36 | 92.12 | 329.12 |
| | 677 | 28.22 | 22.98 | 27.7 | 19.39 | 98.29 |
| | 678 | 72.61 | 77.09 | 69.12 | 75.71 | 294.53 |
| | 679 | 32.36 | 26.51 | 32.74 | 30.35 | 121.96 |
| TOTAL | | 3,575.51 | 3,711.17 | 3,562.29 | 3,128.82 | 13,977.79 |

STATE: KENTUCKY

| ZIP CODE | | QUARTER 1 | QUARTER 2 | QUARTER 3 | QUARTER 4 | TOTAL GRAMS |
|---|---|---|---|---|---|---|
| | 400 | 112.74 | 104.01 | 85.54 | 89.63 | 391.92 |
| | 401 | 45.55 | 53.87 | 47.18 | 38.86 | 185.46 |
| | 402 | 833.93 | 787.39 | 756.11 | 934.05 | 3,311.48 |
| | 403 | 83.54 | 70.55 | 86.94 | 75.98 | 317.01 |
| | 404 | 33.36 | 70.85 | 54.67 | 33.99 | 192.87 |
| | 405 | 354.19 | 220.03 | 271.61 | 485.57 | 1,331.40 |
| | 406 | 29.52 | 24.23 | 31.48 | 23.68 | 108.91 |
| | 407 | 21.76 | 15.56 | 23.71 | 23.16 | 84.19 |
| | 408 | 5.15 | 6.39 | 4.84 | 5.78 | 22.16 |
| | 409 | 17.85 | 17.99 | 33.31 | 12.38 | 81.53 |
| | 410 | 154.27 | 130.9 | 146.58 | 131.63 | 563.38 |
| | 411 | 77.84 | 47.34 | 71.27 | 72.84 | 269.29 |
| | 412 | 7.78 | 5.87 | 5.09 | 11.99 | 30.73 |
| | 413 | 6.34 | 5.6 | 4.45 | 9.09 | 25.48 |

| | | | | | | |
|---|---|---|---|---|---|---|
| | 414 | 0.4 | 0.04 | 0.8 | 0.68 | 1.92 |
| | 415 | 58.12 | 41.89 | 55.95 | 145.76 | 301.72 |
| | 416 | 6.47 | 9 | 4.94 | 4.68 | 25.09 |
| | 417 | 57.35 | 39.27 | 56.97 | 42.65 | 196.24 |
| | 418 | 5.57 | 2.42 | 2.89 | 2.13 | 13.01 |
| | 420 | 266.8 | 284.63 | 265.55 | 243.2 | 1,060.18 |
| | 421 | 182.95 | 168.31 | 211.99 | 371.62 | 934.87 |
| | 422 | 141.64 | 144.34 | 143.96 | 136.15 | 566.09 |
| | 423 | 159.63 | 129.31 | 143.31 | 128.23 | 560.48 |
| | 424 | 169.45 | 152.04 | 160.26 | 215.04 | 696.79 |
| | 425 | 30.97 | 26.18 | 52.77 | 16.45 | 126.37 |
| | 426 | 6.44 | 3.9 | 6.31 | 3.9 | 20.55 |
| | 427 | 100.5 | 85.55 | 93.22 | 94.97 | 374.24 |
| TOTAL | | 2,970.11 | 2,647.46 | 2,821.70 | 3,354.09 | 11,793.36 |

STATE: LOUISIANA

| ZIP CODE | | QUARTER 1 | QUARTER 2 | QUARTER 3 | QUARTER 4 | TOTAL GRAMS |
|---|---|---|---|---|---|---|
| | 700 | 261.11 | 341.27 | 260.48 | 228.83 | 1,091.69 |
| | 701 | 198.22 | 207.24 | 211.53 | 211.19 | 828.18 |
| | 703 | 127.6 | 125.2 | 109.67 | 104.84 | 467.31 |
| | 704 | 504.63 | 447.75 | 420.99 | 418.75 | 1,792.12 |
| | 705 | 989.45 | 880.12 | 781.67 | 928.03 | 3,579.27 |
| | 706 | 189.03 | 196.12 | 139.3 | 144.31 | 668.76 |
| | 707 | 354.84 | 234.52 | 314.21 | 334.96 | 1,238.53 |
| | 708 | 305.08 | 265.85 | 286.43 | 242.88 | 1,100.24 |
| | 710 | 32.81 | 30.83 | 33.88 | 32.21 | 129.73 |
| | 711 | 373.2 | 169.65 | 185.81 | 157.03 | 885.69 |
| | 712 | 128.39 | 116.7 | 113.34 | 90.13 | 448.56 |
| | 713 | 177.73 | 164.85 | 169.94 | 151.27 | 663.79 |
| | 714 | 72.36 | 75.08 | 68.73 | 63.15 | 279.32 |
| TOTAL | | 3,714.45 | 3,255.18 | 3,095.98 | 3,107.58 | 13,173.19 |

STATE: MAINE

| ZIP CODE | | QUARTER 1 | QUARTER 2 | QUARTER 3 | QUARTER 4 | TOTAL GRAMS |
|---|---|---|---|---|---|---|
| | 39 | 16.52 | 12.95 | 15.74 | 11.38 | 56.59 |

| | | | | | | |
|---|---|---|---|---|---|---|
| | 40 | 384.74 | 338.97 | 323.06 | 325.24 | 1,372.01 |
| | 41 | 233.82 | 316.76 | 262.28 | 358.92 | 1,171.78 |
| | 42 | 115.84 | 122.3 | 101.52 | 141.39 | 481.05 |
| | 43 | 303.81 | 639.6 | 130.88 | 356.66 | 1,430.95 |
| | 44 | 224.39 | 193.88 | 187.59 | 237.1 | 842.96 |
| | 45 | 62.7 | 56.49 | 51.74 | 55.35 | 226.28 |
| | 46 | 59.23 | 92.61 | 83.5 | 70.07 | 305.41 |
| | 47 | 32.21 | 43.84 | 23.39 | 21.85 | 121.29 |
| | 48 | 42.84 | 31.21 | 32.9 | 27.06 | 134.01 |
| | 49 | 135.72 | 156.02 | 109.74 | 142.59 | 544.07 |
| TOTAL | | 1,611.82 | 2,004.63 | 1,322.34 | 1,747.61 | 6,686.40 |

STATE: MARYLAND

| ZIP CODE | | QUARTER 1 | QUARTER 2 | QUARTER 3 | QUARTER 4 | TOTAL GRAMS |
|---|---|---|---|---|---|---|
| | 206 | 470.11 | 433.17 | 415.4 | 422.41 | 1,741.09 |
| | 207 | 785.41 | 730.91 | 722.06 | 634.52 | 2,872.90 |
| | 208 | 1,224.84 | 1,129.47 | 1,032.47 | 1,049.94 | 4,436.72 |
| | 209 | 231.9 | 229.08 | 212.34 | 253.89 | 927.21 |
| | 210 | 1,455.49 | 1,307.21 | 1,294.24 | 1,333.21 | 5,390.15 |
| | 211 | 578.82 | 549.51 | 505.3 | 496.7 | 2,130.33 |
| | 212 | 2,320.61 | 2,288.78 | 2,128.84 | 2,018.49 | 8,756.72 |
| | 214 | 147.44 | 143.34 | 125.91 | 129.13 | 545.82 |
| | 215 | 100.95 | 84.43 | 82.68 | 86.89 | 354.95 |
| | 216 | 149.64 | 130.36 | 120.11 | 103.92 | 504.03 |
| | 217 | 772.2 | 787.69 | 719.17 | 708.13 | 2,987.19 |
| | 218 | 268.54 | 260.47 | 300.89 | 277.38 | 1,107.28 |
| | 219 | 428.01 | 415.12 | 414.23 | 402.65 | 1,660.01 |
| TOTAL | | 8,933.96 | 8,489.54 | 8,073.64 | 7,917.26 | 33,414.40 |

STATE: MASSACHUSETTS

| ZIP CODE | | QUARTER 1 | QUARTER 2 | QUARTER 3 | QUARTER 4 | TOTAL GRAMS |
|---|---|---|---|---|---|---|
| | 10 | 509.51 | 511.36 | 508.25 | 513.79 | 2,042.91 |
| | 11 | 412.92 | 406.77 | 348.47 | 338.41 | 1,506.57 |
| | 12 | 113.35 | 103.12 | 105.15 | 121.01 | 442.63 |
| | 13 | 54.04 | 49.62 | 60.16 | 62.92 | 226.74 |

| | | | | | | |
|---|---|---|---|---|---|---|
| | 14 | 91.85 | 90.27 | 88.84 | 70.54 | 341.5 |
| | 15 | 174.49 | 193.47 | 169.66 | 189.42 | 727.04 |
| | 16 | 176.96 | 196.08 | 193.2 | 225.02 | 791.26 |
| | 17 | 435.88 | 401.4 | 324.05 | 380.11 | 1,541.44 |
| | 18 | 1,157.77 | 1,296.18 | 1,017.24 | 1,223.12 | 4,694.31 |
| | 19 | 426.82 | 414.89 | 392.7 | 381.03 | 1,615.44 |
| | 20 | 634.23 | 535.18 | 423.11 | 475 | 2,067.52 |
| | 21 | 1,690.06 | 1,566.64 | 1,647.80 | 1,667.75 | 6,572.25 |
| | 22 | 324.13 | 345.9 | 262.97 | 316.15 | 1,249.15 |
| | 23 | 502.56 | 410.08 | 360.5 | 370.12 | 1,643.26 |
| | 24 | 248.79 | 233.1 | 223.65 | 252.37 | 957.91 |
| | 25 | 110.24 | 103.49 | 91.73 | 82.4 | 387.86 |
| | 26 | 163.2 | 151.43 | 159.33 | 157.88 | 631.84 |
| | 27 | 565.98 | 515.95 | 490.33 | 462.28 | 2,034.54 |
| TOTAL | | 7,792.78 | 7,524.93 | 6,867.14 | 7,289.32 | 29,474.17 |

| STATE: | MICHIGAN | | | | | |
|---|---|---|---|---|---|---|
| ZIP CODE | | QUARTER 1 | QUARTER 2 | QUARTER 3 | QUARTER 4 | TOTAL GRAMS |
| | 480 | 1,394.70 | 1,361.35 | 1,167.81 | 1,173.37 | 5,097.23 |
| | 481 | 1,262.06 | 1,290 | 1,142.65 | 1,192.29 | 4,887 |
| | 482 | 850.6 | 842.31 | 875.15 | 893.11 | 3,461.17 |
| | 483 | 1,686.54 | 1,033.86 | 1,499.93 | 1,356.37 | 5,576.70 |
| | 484 | 382.97 | 396.3 | 336.17 | 360.88 | 1,476.32 |
| | 485 | 211.33 | 206.12 | 213.61 | 279.58 | 910.64 |
| | 486 | 460.79 | 415.44 | 403.47 | 395.69 | 1,675.39 |
| | 487 | 136.65 | 135.25 | 116.45 | 119.34 | 507.69 |
| | 488 | 347.14 | 341.38 | 342.37 | 305.3 | 1,336.19 |
| | 489 | 191.13 | 217.05 | 187.23 | 194.3 | 789.71 |
| | 490 | 591.35 | 546.58 | 533.9 | 534.95 | 2,206.78 |
| | 491 | 48.67 | 38.87 | 43.58 | 37.12 | 168.24 |
| | 492 | 158.34 | 173.7 | 168.86 | 174.87 | 675.77 |
| | 493 | 150.8 | 121.75 | 121.88 | 108.87 | 503.3 |
| | 494 | 334.14 | 307.27 | 373.66 | 363.75 | 1,378.82 |
| | 495 | 433.9 | 454.89 | 463.13 | 417.71 | 1,769.63 |
| | 496 | 407.88 | 424.95 | 440.5 | 395.76 | 1,669.09 |
| | 497 | 250.55 | 261.1 | 232.47 | 234.99 | 979.11 |

| | | | | | | |
|---|---|---|---|---|---|---|
| | 498 | 228.07 | 226.11 | 233.57 | 209.02 | 896.77 |
| | 499 | 54.69 | 45.77 | 41.76 | 44.26 | 186.48 |
| TOTAL | | 9,582.30 | 8,840.05 | 8,938.15 | 8,791.53 | 36,152.03 |

STATE: MINNESOTA

| ZIP CODE | | QUARTER 1 | QUARTER 2 | QUARTER 3 | QUARTER 4 | TOTAL GRAMS |
|---|---|---|---|---|---|---|
| | 550 | 551.85 | 518.61 | 531.02 | 522.07 | 2,123.55 |
| | 551 | 1,087.22 | 1,251.14 | 1,127.06 | 1,231.72 | 4,697.14 |
| | 553 | 672.75 | 657.66 | 643.64 | 701.32 | 2,675.37 |
| | 554 | 1,917.34 | 1,953.57 | 2,035.15 | 1,847.81 | 7,753.87 |
| | 556 | 5.92 | 4.17 | 15.01 | 7.08 | 32.18 |
| | 557 | 153.25 | 172.04 | 163.58 | 166.54 | 655.41 |
| | 558 | 167.93 | 176.81 | 187.17 | 177.15 | 709.06 |
| | 559 | 569.03 | 537.88 | 545.37 | 451.31 | 2,103.59 |
| | 560 | 147.33 | 130.21 | 135.57 | 124.89 | 538 |
| | 561 | 13.13 | 15.17 | 14.51 | 17.26 | 60.07 |
| | 562 | 84.17 | 57.88 | 53.84 | 57.55 | 253.44 |
| | 563 | 207.91 | 194.07 | 164.15 | 197.1 | 763.23 |
| | 564 | 132.4 | 113.64 | 133.75 | 118.51 | 498.3 |
| | 565 | 163.79 | 178.99 | 151.49 | 184.27 | 678.54 |
| | 566 | 43.59 | 46.98 | 40.75 | 38.49 | 169.81 |
| | 567 | 19.82 | 26.03 | 17.56 | 16.36 | 79.77 |
| TOTAL | | 5,937.43 | 6,034.85 | 5,959.62 | 5,859.43 | 23,791.33 |

STATE: MISSISSIPPI

| ZIP CODE | | QUARTER 1 | QUARTER 2 | QUARTER 3 | QUARTER 4 | TOTAL GRAMS |
|---|---|---|---|---|---|---|
| | 386 | 169.17 | 178.62 | 157.17 | 142.31 | 647.27 |
| | 387 | 16.16 | 14.75 | 33.94 | 13.28 | 78.13 |
| | 388 | 168.27 | 154.43 | 128.28 | 171.45 | 622.43 |
| | 389 | 24.45 | 31.23 | 28.82 | 23.58 | 108.08 |
| | 390 | 110 | 104.15 | 97.8 | 81.11 | 393.06 |
| | 391 | 3,129.62 | 4,550.10 | 4,149.02 | 4,605.48 | 16,434.22 |
| | 392 | 314.93 | 276.8 | 291.62 | 382.25 | 1,265.60 |
| | 393 | 92.58 | 76.87 | 70.33 | 84.86 | 324.64 |
| | 394 | 368.55 | 358.75 | 302.2 | 351.61 | 1,381.11 |

| | | | | | | |
|---|---|---|---|---|---|---|
| | 395 | 504.6 | 362.67 | 385.85 | 501.92 | 1,755.04 |
| | 396 | 90.09 | 71.33 | 57.36 | 33.89 | 252.67 |
| | 397 | 39.43 | 41.42 | 33.93 | 26.63 | 141.41 |
| TOTAL | | 5,027.85 | 6,221.12 | 5,736.32 | 6,418.37 | 23,403.66 |

STATE: MISSOURI

| ZIP CODE | | QUARTER 1 | QUARTER 2 | QUARTER 3 | QUARTER 4 | TOTAL GRAMS |
|---|---|---|---|---|---|---|
| | 630 | 841.58 | 596.1 | 663.95 | 771.06 | 2,872.69 |
| | 631 | 1,205.10 | 1,089.48 | 1,039.50 | 1,006.96 | 4,341.04 |
| | 633 | 282.93 | 264.2 | 260.75 | 229.37 | 1,037.25 |
| | 634 | 31.32 | 27.23 | 42.5 | 47.69 | 148.74 |
| | 635 | 76.16 | 67.02 | 70.27 | 66.41 | 279.86 |
| | 636 | 183.07 | 194.19 | 168.48 | 159.85 | 705.59 |
| | 637 | 106.44 | 109.93 | 106.98 | 85.9 | 409.25 |
| | 638 | 58.72 | 58.22 | 54.56 | 56.66 | 228.16 |
| | 639 | 49.91 | 49.97 | 63.3 | 52.16 | 215.34 |
| | 640 | 515.04 | 461.3 | 433.38 | 433.12 | 1,842.84 |
| | 641 | 472.79 | 454.74 | 407.03 | 436.63 | 1,771.19 |
| | 644 | 129.41 | 115.51 | 115.73 | 95.33 | 455.98 |
| | 645 | 532.43 | 499.68 | 468.23 | 437.24 | 1,937.58 |
| | 646 | 143.51 | 119.05 | 126.09 | 119.19 | 507.84 |
| | 647 | 123.04 | 83.42 | 87.78 | 68.19 | 362.43 |
| | 648 | 261.5 | 270.41 | 263.24 | 239.41 | 1,034.56 |
| | 650 | 93.26 | 100.2 | 103.4 | 104.91 | 401.77 |
| | 651 | 52.07 | 41.76 | 35.88 | 37.8 | 167.51 |
| | 652 | 312.75 | 284.53 | 321.54 | 438.88 | 1,357.70 |
| | 653 | 62.5 | 55.86 | 51.83 | 54.61 | 224.8 |
| | 654 | 38.3 | 33.55 | 40.12 | 24.21 | 136.18 |
| | 655 | 88.85 | 69.02 | 68.19 | 58.8 | 284.86 |
| | 656 | 334.94 | 311.79 | 316.55 | 309.1 | 1,272.38 |
| | 657 | 537.83 | 510.05 | 430.64 | 466.73 | 1,945.25 |
| | 658 | 466.96 | 497.87 | 447.54 | 448.98 | 1,861.35 |
| TOTAL | | 7,000.41 | 6,365.08 | 6,187.46 | 6,249.19 | 25,802.14 |

STATE: MONTANA

| ZIP CODE | QUARTER 1 | QUARTER 2 | QUARTER 3 | QUARTER 4 | TOTAL GRAMS |
|---|---|---|---|---|---|
| 590 | 104.01 | 75.01 | 104.68 | 91.22 | 374.92 |
| 591 | 538.52 | 503.24 | 487.99 | 462.56 | 1,992.31 |
| 592 | 42 | 29.77 | 38.23 | 28.79 | 138.79 |
| 593 | 64.55 | 62.59 | 57.85 | 60.18 | 245.17 |
| 594 | 153.44 | 124.91 | 110.26 | 116.53 | 505.14 |
| 595 | 14.81 | 9.65 | 10.89 | 10.64 | 45.99 |
| 596 | 107.67 | 122.46 | 122.46 | 129.94 | 482.53 |
| 597 | 171.08 | 217.78 | 320.6 | 223.04 | 932.5 |
| 598 | 291.37 | 311.51 | 334.33 | 282.63 | 1,219.84 |
| 599 | 193.7 | 156.29 | 145.93 | 131.38 | 627.3 |
| TOTAL | 1,681.15 | 1,613.21 | 1,733.22 | 1,536.91 | 6,564.49 |

STATE: NEBRASKA

| ZIP CODE | QUARTER 1 | QUARTER 2 | QUARTER 3 | QUARTER 4 | TOTAL GRAMS |
|---|---|---|---|---|---|
| 680 | 224.83 | 226.13 | 184.5 | 172.16 | 807.62 |
| 681 | 595.75 | 590.08 | 510.17 | 550.41 | 2,246.41 |
| 683 | 25.63 | 30.89 | 37.15 | 33.86 | 127.53 |
| 684 | 22.71 | 24.07 | 19.97 | 25.32 | 92.07 |
| 685 | 155.04 | 172.59 | 150.46 | 122.65 | 600.74 |
| 686 | 22.61 | 28.55 | 27.03 | 30.21 | 108.4 |
| 687 | 45.94 | 50.53 | 44.77 | 38.64 | 179.88 |
| 688 | 103.48 | 117.35 | 102.36 | 105.46 | 428.65 |
| 689 | 44.83 | 24.39 | 28.78 | 48.7 | 146.7 |
| 690 | 22.7 | 19.52 | 13.72 | 15.06 | 71 |
| 691 | 58.34 | 52.58 | 55.5 | 42.45 | 208.87 |
| 692 | 5.52 | 4.61 | 1.72 | 1.29 | 13.14 |
| 693 | 49.88 | 34.71 | 46.87 | 44.28 | 175.74 |
| TOTAL | 1,377.26 | 1,376 | 1,223 | 1,230.49 | 5,206.75 |

STATE: NEVADA

| ZIP CODE | QUARTER 1 | QUARTER 2 | QUARTER 3 | QUARTER 4 | TOTAL GRAMS |
|---|---|---|---|---|---|
| 890 | 712.29 | 724.86 | 691.74 | 667.88 | 2,796.77 |
| 891 | 1,803.81 | 1,646.75 | 1,536.08 | 1,563.61 | 6,550.25 |
| 893 | 0.83 | 2.62 | 1.17 | 1.21 | 5.83 |

| | | | | | | |
|---|---|---|---|---|---|---|
| | 894 | 207.46 | 186.44 | 197.01 | 184.22 | 775.13 |
| | 895 | 297.95 | 278.4 | 264.81 | 238.79 | 1,079.95 |
| | 897 | 121.3 | 140.49 | 110.65 | 101.98 | 474.42 |
| | 898 | 30.17 | 28.11 | 31.17 | 24.68 | 114.13 |
| TOTAL | | 3,173.81 | 3,007.67 | 2,832.63 | 2,782.37 | 11,796.48 |

STATE: NEW HAMPSHIRE

| ZIP CODE | | QUARTER 1 | QUARTER 2 | QUARTER 3 | QUARTER 4 | TOTAL GRAMS |
|---|---|---|---|---|---|---|
| | 30 | 397.98 | 374.82 | 347.76 | 350.81 | 1,471.37 |
| | 31 | 341.98 | 383.39 | 418.24 | 250.77 | 1,394.38 |
| | 32 | 196.15 | 166.64 | 164.71 | 154.34 | 681.84 |
| | 33 | 205.2 | 184.7 | 189.62 | 228.63 | 808.15 |
| | 34 | 95.55 | 102.05 | 83.07 | 79.03 | 359.7 |
| | 35 | 97.5 | 109.71 | 95.78 | 88.27 | 391.26 |
| | 36 | 10.6 | 9.68 | 6.38 | 5.66 | 32.32 |
| | 37 | 285.75 | 248.04 | 280.34 | 275.03 | 1,089.16 |
| | 38 | 428.68 | 418.43 | 391.58 | 396.86 | 1,635.55 |
| TOTAL | | 2,059.39 | 1,997.46 | 1,977.48 | 1,829.40 | 7,863.73 |

STATE: NEW JERSEY

| ZIP CODE | | QUARTER 1 | QUARTER 2 | QUARTER 3 | QUARTER 4 | TOTAL GRAMS |
|---|---|---|---|---|---|---|
| | 70 | 1,640.81 | 1,409.10 | 1,309.19 | 1,376.06 | 5,735.16 |
| | 71 | 260.31 | 225.94 | 217.08 | 187.09 | 890.42 |
| | 72 | 86.4 | 71.24 | 90.01 | 83.49 | 331.14 |
| | 73 | 76.1 | 74.68 | 68.03 | 61.14 | 279.95 |
| | 74 | 889.95 | 763.06 | 714.67 | 1,038.55 | 3,406.23 |
| | 75 | 138.66 | 188.17 | 183.88 | 116.55 | 627.26 |
| | 76 | 890.81 | 886.89 | 837.01 | 804.41 | 3,419.12 |
| | 77 | 786 | 766.85 | 665.46 | 705.96 | 2,924.27 |
| | 78 | 403.2 | 374.21 | 339.54 | 325.33 | 1,442.28 |
| | 79 | 407.31 | 413.19 | 392.23 | 345.87 | 1,558.60 |
| | 80 | 1,303.01 | 1,195.46 | 1,060.11 | 1,022.55 | 4,581.13 |
| | 81 | 154.6 | 170.91 | 215.82 | 201.63 | 742.96 |
| | 82 | 268.61 | 202.77 | 232.78 | 151.33 | 855.49 |
| | 83 | 152.72 | 139.49 | 135.03 | 114.66 | 541.9 |

| | | | | | | |
|---|---|---|---|---|---|---|
| | 84 | 25.03 | 22.78 | 30.83 | 21.68 | 100.32 |
| | 85 | 262.24 | 231.85 | 206.33 | 204.76 | 905.18 |
| | 86 | 162.06 | 153.83 | 138.09 | 134.33 | 588.31 |
| | 87 | 381.32 | 383.26 | 358.97 | 384.93 | 1,508.48 |
| | 88 | 1,099.20 | 1,197.78 | 1,093.14 | 1,014.57 | 4,404.69 |
| | 89 | 186.77 | 168.43 | 146.97 | 152.57 | 654.74 |
| TOTAL | | 9,575.11 | 9,039.89 | 8,435.17 | 8,447.46 | 35,497.63 |

STATE: NEW MEXICO

| ZIP CODE | | QUARTER 1 | QUARTER 2 | QUARTER 3 | QUARTER 4 | TOTAL GRAMS |
|---|---|---|---|---|---|---|
| | 870 | 76.66 | 79.26 | 67.57 | 74.62 | 298.11 |
| | 871 | 685.98 | 710.28 | 666.21 | 625.68 | 2,688.15 |
| | 873 | 12.28 | 11.58 | 6.78 | 16.26 | 46.9 |
| | 874 | 86 | 84.71 | 65.23 | 67.53 | 303.47 |
| | 875 | 137.5 | 147.62 | 131.83 | 141.89 | 558.84 |
| | 877 | 34.13 | 24.2 | 27.93 | 32.91 | 119.17 |
| | 878 | 2.54 | 4.61 | 2.84 | 2.68 | 12.67 |
| | 879 | 6.03 | 5.69 | 9.52 | 13.77 | 35.01 |
| | 880 | 231.02 | 182.47 | 125.52 | 113.55 | 652.56 |
| | 881 | 24.11 | 16.67 | 17.55 | 10.3 | 68.63 |
| | 882 | 130.82 | 115.92 | 116.78 | 109.21 | 472.73 |
| | 883 | 37.19 | 33.75 | 31.64 | 32.79 | 135.37 |
| | 884 | 1.1 | 0.97 | 0.88 | 1.07 | 4.02 |
| TOTAL | | 1,465.36 | 1,417.73 | 1,270.28 | 1,242.26 | 5,395.63 |

STATE: NEW YORK

| ZIP CODE | | QUARTER 1 | QUARTER 2 | QUARTER 3 | QUARTER 4 | TOTAL GRAMS |
|---|---|---|---|---|---|---|
| | 100 | 3,308.21 | 3,150.09 | 2,855.72 | 3,215.91 | 12,529.93 |
| | 101 | 41.54 | 49.99 | 51.6 | 41 | 184.13 |
| | 102 | 4.26 | 1.63 | 2.31 | 5.85 | 14.05 |
| | 103 | 526.9 | 499.71 | 757.16 | 765.5 | 2,549.27 |
| | 104 | 815.48 | 719.63 | 903.91 | 689.1 | 3,128.12 |
| | 105 | 517.43 | 489.69 | 472.78 | 620.71 | 2,100.61 |
| | 106 | 120.92 | 133.83 | 132.34 | 128.59 | 515.68 |
| | 107 | 149.07 | 127.87 | 129.71 | 115.46 | 522.11 |

| | | | | | |
|---|---|---|---|---|---|
| 108 | 73.6 | 64.74 | 51.52 | 50.09 | 239.95 |
| 109 | 636.84 | 568.2 | 530.46 | 551.86 | 2,287.36 |
| 110 | 1,137.63 | 1,073.47 | 964.92 | 1,019.83 | 4,195.85 |
| 111 | 163.35 | 168.85 | 180.02 | 151.46 | 663.68 |
| 112 | 1,422.53 | 1,523.89 | 1,260.08 | 1,300.59 | 5,507.09 |
| 113 | 612.13 | 586.04 | 504.16 | 524.97 | 2,227.30 |
| 114 | 289.09 | 275.74 | 261.62 | 232.27 | 1,058.72 |
| 115 | 852.78 | 925.78 | 889.68 | 800.08 | 3,468.32 |
| 116 | 48.27 | 66.1 | 46.63 | 57.03 | 218.03 |
| 117 | 2,665.24 | 2,587.71 | 2,412.94 | 2,344.79 | 10,010.68 |
| 118 | 200.14 | 223.61 | 225.45 | 146.52 | 795.72 |
| 119 | 169.12 | 166.65 | 184.9 | 196.45 | 717.12 |
| 120 | 214.4 | 198.73 | 256.96 | 195.62 | 865.71 |
| 121 | 84.96 | 68.88 | 92.04 | 87.51 | 333.39 |
| 122 | 362.97 | 346.1 | 386.05 | 267.2 | 1,362.32 |
| 123 | 73.14 | 76.05 | 66.51 | 60.79 | 276.49 |
| 124 | 234.02 | 241.91 | 233.57 | 209.62 | 919.12 |
| 125 | 365.72 | 354.71 | 365.84 | 354.74 | 1,441.01 |
| 126 | 144.95 | 133.95 | 147.47 | 158.33 | 584.7 |
| 127 | 189.53 | 139.88 | 145.43 | 134.33 | 609.17 |
| 128 | 254.86 | 234.02 | 231.72 | 211.01 | 931.61 |
| 129 | 344.59 | 336.72 | 356.77 | 351.23 | 1,389.31 |
| 130 | 198.7 | 177.04 | 180.28 | 169.32 | 725.34 |
| 131 | 93.63 | 107.95 | 104.97 | 104 | 410.55 |
| 132 | 159.54 | 164.54 | 149.91 | 140.4 | 614.39 |
| 133 | 105.82 | 123.69 | 81.05 | 84.5 | 395.06 |
| 134 | 91.99 | 96.55 | 74.89 | 86.34 | 349.77 |
| 135 | 76 | 66.88 | 60.03 | 63.79 | 266.7 |
| 136 | 153.62 | 125.13 | 127.49 | 130.34 | 536.58 |
| 137 | 80.15 | 75.91 | 68 | 65.5 | 289.56 |
| 138 | 153.77 | 146.03 | 137.95 | 142.93 | 580.68 |
| 139 | 90.55 | 64.3 | 53.98 | 54.23 | 263.06 |
| 140 | 305.39 | 307.63 | 290.89 | 284.33 | 1,188.24 |
| 141 | 154.32 | 162.18 | 139.76 | 122.19 | 578.45 |
| 142 | 585.48 | 534.84 | 544.42 | 541.6 | 2,206.34 |
| 143 | 54.02 | 42.84 | 30.56 | 27.8 | 155.22 |
| 144 | 349.43 | 330.48 | 317.88 | 291.21 | 1,289 |

| | | | | | | |
|---|---|---|---|---|---|---|
| | 145 | 283.09 | 286.3 | 273.5 | 213.02 | 1,055.91 |
| | 146 | 918.47 | 1,003.42 | 980.03 | 1,008.71 | 3,910.63 |
| | 147 | 71.15 | 81.19 | 79.65 | 76.88 | 308.87 |
| | 148 | 270.51 | 281.27 | 258.39 | 248.45 | 1,058.62 |
| | 149 | 49.3 | 55.89 | 56.3 | 70.43 | 231.92 |
| TOTAL | | 20,268.60 | 19,768.23 | 19,110.20 | 18,914.41 | 78,061.44 |

STATE: NORTH CAROLINA

| ZIP CODE | | QUARTER 1 | QUARTER 2 | QUARTER 3 | QUARTER 4 | TOTAL GRAMS |
|---|---|---|---|---|---|---|
| | 270 | 338.92 | 285.32 | 271.98 | 281.01 | 1,177.23 |
| | 271 | 652.11 | 525.66 | 517.36 | 452.94 | 2,148.07 |
| | 272 | 693.53 | 736.76 | 678.92 | 622.57 | 2,731.78 |
| | 273 | 399.93 | 397.34 | 391.37 | 394.18 | 1,582.82 |
| | 274 | 310.83 | 262.98 | 312.01 | 307.6 | 1,193.42 |
| | 275 | 1,642.09 | 1,229.42 | 1,319 | 1,266.10 | 5,456.61 |
| | 276 | 447.09 | 438.75 | 427.58 | 407.76 | 1,721.18 |
| | 277 | 501.97 | 556.43 | 450.63 | 609.25 | 2,118.28 |
| | 278 | 631.04 | 448.33 | 434.43 | 379.16 | 1,892.96 |
| | 279 | 280.96 | 274.83 | 264.13 | 281.76 | 1,101.68 |
| | 280 | 743.86 | 709.08 | 726.01 | 678.81 | 2,857.76 |
| | 281 | 481.97 | 476.96 | 470.48 | 457.39 | 1,886.80 |
| | 282 | 1,001.88 | 784.09 | 979.41 | 636.71 | 3,402.09 |
| | 283 | 758.04 | 715.14 | 705.46 | 652.42 | 2,831.06 |
| | 284 | 578.6 | 549.08 | 542.3 | 521.7 | 2,191.68 |
| | 285 | 476.01 | 479.01 | 474.86 | 466.92 | 1,896.80 |
| | 286 | 869.17 | 1,079.53 | 866.9 | 817.85 | 3,633.45 |
| | 287 | 469.6 | 464.52 | 432.12 | 351.34 | 1,717.58 |
| | 288 | 291.4 | 265.75 | 304.61 | 301.99 | 1,163.75 |
| | 289 | 86.08 | 85.63 | 84.13 | 76.66 | 332.5 |
| TOTAL | | 11,655.08 | 10,764.61 | 10,653.69 | 9,964.12 | 43,037.50 |

STATE: NORTH DAKOTA

| ZIP CODE | | QUARTER 1 | QUARTER 2 | QUARTER 3 | QUARTER 4 | TOTAL GRAMS |
|---|---|---|---|---|---|---|
| | 580 | 30.11 | 34.79 | 32.04 | 33.81 | 130.75 |
| | 581 | 155.08 | 177.89 | 149.89 | 144.36 | 627.22 |

| | | | | | | |
|---|---|---|---|---|---|---|
| | 582 | 57.09 | 49.44 | 71.49 | 65.35 | 243.37 |
| | 583 | 13.44 | 9.39 | 10.44 | 11.13 | 44.4 |
| | 584 | 16.79 | 25.25 | 11.04 | 10.21 | 63.29 |
| | 585 | 263.89 | 270.18 | 241.02 | 257.04 | 1,032.13 |
| | 586 | 27.86 | 31.37 | 30.49 | 28.93 | 118.65 |
| | 587 | 54.08 | 55.71 | 55.79 | 60.4 | 225.98 |
| | 588 | 26.07 | 24.3 | 28.26 | 30.91 | 109.54 |
| TOTAL | | 644.41 | 678.32 | 630.46 | 642.14 | 2,595.33 |

STATE: OHIO

| ZIP CODE | | QUARTER 1 | QUARTER 2 | QUARTER 3 | QUARTER 4 | TOTAL GRAMS |
|---|---|---|---|---|---|---|
| | 430 | 844.28 | 742.12 | 633.46 | 769.69 | 2,989.55 |
| | 431 | 331.45 | 299.97 | 237.31 | 208.33 | 1,077.06 |
| | 432 | 1,381.49 | 1,305.49 | 1,412.07 | 1,425.03 | 5,524.08 |
| | 433 | 111.92 | 128.73 | 145.01 | 173.32 | 558.98 |
| | 434 | 74.5 | 80.34 | 64.41 | 52.81 | 272.06 |
| | 435 | 268.87 | 330.76 | 262.77 | 369.7 | 1,232.10 |
| | 436 | 269.81 | 237.38 | 255.4 | 263.19 | 1,025.78 |
| | 437 | 152.14 | 155.91 | 187.89 | 180.24 | 676.18 |
| | 438 | 37.28 | 20.79 | 19.73 | 25.54 | 103.34 |
| | 439 | 114.17 | 119.04 | 97.57 | 115.47 | 446.25 |
| | 440 | 379.39 | 383.08 | 380.11 | 335.24 | 1,477.82 |
| | 441 | 1,120.33 | 1,091.76 | 1,114.05 | 1,042.53 | 4,368.67 |
| | 442 | 401.29 | 350.45 | 392.24 | 375.32 | 1,519.30 |
| | 443 | 338.92 | 342.36 | 356.15 | 272.62 | 1,310.05 |
| | 444 | 128.66 | 129.12 | 131.27 | 122.94 | 511.99 |
| | 445 | 80.07 | 103.31 | 86.08 | 101.67 | 371.13 |
| | 446 | 435.92 | 423.07 | 396 | 378.66 | 1,633.65 |
| | 447 | 354.84 | 342.21 | 385.57 | 378.3 | 1,460.92 |
| | 448 | 236.1 | 210.13 | 227.76 | 263.09 | 937.08 |
| | 449 | 32.76 | 40.94 | 34.95 | 56.65 | 165.3 |
| | 450 | 572.16 | 504.06 | 611.54 | 492.67 | 2,180.43 |
| | 451 | 164.92 | 190.15 | 165.4 | 188.75 | 709.22 |
| | 452 | 608.31 | 617.52 | 566.49 | 626.52 | 2,418.84 |
| | 453 | 194.04 | 154.37 | 168.25 | 154.36 | 671.02 |
| | 454 | 328.95 | 344.68 | 359.71 | 292.43 | 1,325.77 |

| | | | | | | |
|---|---|---|---|---|---|---|
| | 455 | 68.63 | 64.82 | 54.02 | 63.04 | 250.51 |
| | 456 | 226.93 | 206.66 | 183.25 | 234.87 | 851.71 |
| | 457 | 93.04 | 82.33 | 82.88 | 69.71 | 327.96 |
| | 458 | 246.09 | 298.97 | 225.87 | 215.02 | 985.95 |
| TOTAL | | 9,597.26 | 9,300.52 | 9,237.21 | 9,247.71 | 37,382.70 |

STATE: OKLAHOMA

| ZIP CODE | | QUARTER 1 | QUARTER 2 | QUARTER 3 | QUARTER 4 | TOTAL GRAMS |
|---|---|---|---|---|---|---|
| | 730 | 701.48 | 734.33 | 634.89 | 586.31 | 2,657.01 |
| | 731 | 899.09 | 884.01 | 877.37 | 835.3 | 3,495.77 |
| | 734 | 78.97 | 95.71 | 81.66 | 85.15 | 341.49 |
| | 735 | 229.85 | 250.06 | 249.79 | 202.69 | 932.39 |
| | 736 | 17.61 | 20.47 | 22.73 | 21.9 | 82.71 |
| | 737 | 55.75 | 55.96 | 59.92 | 63.95 | 235.58 |
| | 738 | 12.03 | 12.61 | 15.24 | 15.2 | 55.08 |
| | 739 | 13.94 | 24.22 | 6.88 | 6.7 | 51.74 |
| | 740 | 432.5 | 454.68 | 400.29 | 413.91 | 1,701.38 |
| | 741 | 519.41 | 496.8 | 537.35 | 528.9 | 2,082.46 |
| | 743 | 77.19 | 81.12 | 94.87 | 100.7 | 353.88 |
| | 744 | 154.68 | 184.87 | 170.94 | 173.02 | 683.51 |
| | 745 | 114.55 | 101.42 | 99.96 | 89.19 | 405.12 |
| | 746 | 23.48 | 22.49 | 17.12 | 23.45 | 86.54 |
| | 747 | 127.14 | 132.19 | 107.66 | 102.19 | 469.18 |
| | 748 | 110.34 | 117.88 | 99.59 | 102.07 | 429.88 |
| | 749 | 98.09 | 109.07 | 159.94 | 169.09 | 536.19 |
| TOTAL | | 3,666.10 | 3,777.89 | 3,636.20 | 3,519.72 | 14,599.91 |

STATE: OREGON

| ZIP CODE | | QUARTER 1 | QUARTER 2 | QUARTER 3 | QUARTER 4 | TOTAL GRAMS |
|---|---|---|---|---|---|---|
| | 970 | 812.85 | 767 | 758.81 | 691 | 3,029.66 |
| | 971 | 340.36 | 352.32 | 345.31 | 327.45 | 1,365.44 |
| | 972 | 1,292.72 | 1,281.10 | 1,266.82 | 1,274.37 | 5,115.01 |
| | 973 | 706.93 | 667.39 | 725.1 | 651.53 | 2,750.95 |
| | 974 | 903.27 | 910.16 | 859.06 | 769.25 | 3,441.74 |
| | 975 | 561.29 | 548.91 | 480.36 | 438.2 | 2,028.76 |

| | | | | | | |
|---|---|---|---|---|---|---|
| | 976 | 40.72 | 49.07 | 43.16 | 30.63 | 163.58 |
| | 977 | 199.24 | 187.59 | 175.1 | 149.38 | 711.31 |
| | 978 | 163.77 | 157.79 | 153.11 | 130.97 | 605.64 |
| | 979 | 34.98 | 28.28 | 34.66 | 35.82 | 133.74 |
| TOTAL | | 5,056.13 | 4,949.61 | 4,841.49 | 4,498.60 | 19,345.83 |

STATE: PENNSYLVANIA

| ZIP CODE | | QUARTER 1 | QUARTER 2 | QUARTER 3 | QUARTER 4 | TOTAL GRAMS |
|---|---|---|---|---|---|---|
| | 150 | 390.21 | 352.32 | 356.29 | 351 | 1,449.82 |
| | 151 | 133.41 | 148.1 | 121.21 | 115.41 | 518.13 |
| | 152 | 887.45 | 862.3 | 819.87 | 797.42 | 3,367.04 |
| | 153 | 122.15 | 107.01 | 101.84 | 106.49 | 437.49 |
| | 154 | 81.49 | 71.65 | 76.69 | 90.14 | 319.97 |
| | 155 | 81.36 | 76.1 | 73.28 | 73.36 | 304.1 |
| | 156 | 232.21 | 174.68 | 180.53 | 172.95 | 760.37 |
| | 157 | 60.93 | 62.21 | 68.39 | 57.34 | 248.87 |
| | 158 | 71.52 | 79.8 | 78.21 | 62.4 | 291.93 |
| | 159 | 96.79 | 106.13 | 126.86 | 119.18 | 448.96 |
| | 160 | 100.32 | 86.95 | 80.46 | 77.65 | 345.38 |
| | 161 | 175.11 | 179.34 | 142.23 | 157.91 | 654.59 |
| | 162 | 39.8 | 46.28 | 48.5 | 36.69 | 171.27 |
| | 163 | 372.15 | 327 | 252.27 | 241.17 | 1,192.59 |
| | 164 | 86.11 | 79.4 | 75.76 | 85.1 | 326.37 |
| | 165 | 119.39 | 120.5 | 118.47 | 114.11 | 472.47 |
| | 166 | 170.87 | 170.69 | 160.33 | 164.1 | 665.99 |
| | 167 | 18.26 | 24.13 | 28.66 | 19.03 | 90.08 |
| | 168 | 91.26 | 99.5 | 94.61 | 76.1 | 361.47 |
| | 169 | 34.74 | 35.38 | 24.32 | 34.68 | 129.12 |
| | 170 | 657.18 | 658.05 | 646.18 | 608.98 | 2,570.39 |
| | 171 | 193.58 | 201.14 | 187.55 | 153.71 | 735.98 |
| | 172 | 155.86 | 152.15 | 134.1 | 144.39 | 586.5 |
| | 173 | 255.24 | 243.75 | 217.4 | 211.52 | 927.91 |
| | 174 | 163.25 | 146.33 | 131.84 | 130.57 | 571.99 |
| | 175 | 225.31 | 219.65 | 236.08 | 216 | 897.04 |
| | 176 | 332.05 | 298.5 | 330.28 | 390.75 | 1,351.58 |
| | 177 | 137.3 | 127.6 | 87.36 | 94.11 | 446.37 |

| | | | | | | |
|---|---|---|---|---|---|---|
| | 178 | 329.19 | 350.81 | 292.52 | 275.46 | 1,247.98 |
| | 179 | 31.7 | 26.79 | 31.87 | 36.51 | 126.87 |
| | 180 | 433.52 | 431.85 | 421.9 | 402.33 | 1,689.60 |
| | 181 | 322.55 | 309.84 | 332.06 | 286.85 | 1,251.30 |
| | 182 | 52.38 | 63.96 | 64.02 | 61.8 | 242.16 |
| | 183 | 198.51 | 164.62 | 160.63 | 174.17 | 697.93 |
| | 184 | 169.85 | 162.18 | 180.69 | 174.06 | 686.78 |
| | 185 | 165.2 | 163.49 | 166.98 | 230.53 | 726.2 |
| | 186 | 84.09 | 78.8 | 82.33 | 80.98 | 326.2 |
| | 187 | 884.01 | 823.34 | 733.58 | 732.09 | 3,173.02 |
| | 188 | 58.32 | 57.53 | 50.57 | 39.89 | 206.31 |
| | 189 | 366.91 | 353.92 | 377.35 | 330.75 | 1,428.93 |
| | 190 | 5,983.58 | 3,071.16 | 2,892.68 | 3,162.96 | 15,110.38 |
| | 191 | 1,741.44 | 1,723.58 | 1,598.22 | 1,795.38 | 6,858.62 |
| | 193 | 417.76 | 587.04 | 421.56 | 419.42 | 1,845.78 |
| | 194 | 769.37 | 803.32 | 866.04 | 777.22 | 3,215.95 |
| | 195 | 91.27 | 87.31 | 91.19 | 74.34 | 344.11 |
| | 196 | 171.25 | 172.09 | 166.5 | 169.74 | 679.58 |
| TOTAL | | 17,756.20 | 14,688.27 | 13,930.26 | 14,126.74 | 60,501.47 |

STATE: PUERTO RICO

| ZIP CODE | | QUARTER 1 | QUARTER 2 | QUARTER 3 | QUARTER 4 | TOTAL GRAMS |
|---|---|---|---|---|---|---|
| | 6 | 15.6 | 13.83 | 9.93 | 8.16 | 47.52 |
| | 7 | 6.56 | 6.03 | 4.61 | 6.92 | 24.12 |
| | 9 | 53.53 | 45.22 | 66.39 | 59.86 | 225 |
| TOTAL | | 75.69 | 65.08 | 80.93 | 74.94 | 296.64 |

STATE: RHODE ISLAND

| ZIP CODE | | QUARTER 1 | QUARTER 2 | QUARTER 3 | QUARTER 4 | TOTAL GRAMS |
|---|---|---|---|---|---|---|
| | 28 | 536.93 | 583.51 | 486.75 | 460 | 2,067.19 |
| | 29 | 349.93 | 451.24 | 324.34 | 340.26 | 1,465.77 |
| TOTAL | | 886.86 | 1,034.75 | 811.09 | 800.26 | 3,532.96 |

STATE: SOUTH CAROLINA

| ZIP CODE | QUARTER 1 | QUARTER 2 | QUARTER 3 | QUARTER 4 | TOTAL GRAMS |
|---|---|---|---|---|---|
| 290 | 410.68 | 386.48 | 393.16 | 420.51 | 1,610.83 |
| 291 | 570.59 | 598.58 | 611.77 | 702.09 | 2,483.03 |
| 292 | 590.39 | 646.16 | 570.24 | 573.1 | 2,379.89 |
| 293 | 294.99 | 273.85 | 259.75 | 243.82 | 1,072.41 |
| 294 | 999.07 | 938.89 | 974.01 | 924.83 | 3,836.80 |
| 295 | 1,164.13 | 1,118.95 | 1,024.25 | 1,032.75 | 4,340.08 |
| 296 | 980.69 | 910.95 | 855.66 | 795.57 | 3,542.87 |
| 297 | 215.17 | 228.06 | 230.42 | 184.16 | 857.81 |
| 298 | 261.66 | 243.19 | 229.68 | 252.78 | 987.31 |
| 299 | 514.74 | 484.86 | 361.06 | 504.76 | 1,865.42 |
| TOTAL | 6,002.11 | 5,829.97 | 5,510 | 5,634.37 | 22,976.45 |

STATE: SOUTH DAKOTA

| ZIP CODE | QUARTER 1 | QUARTER 2 | QUARTER 3 | QUARTER 4 | TOTAL GRAMS |
|---|---|---|---|---|---|
| 570 | 69.21 | 59.95 | 70.77 | 59.73 | 259.66 |
| 571 | 253.37 | 259.71 | 258.41 | 393.17 | 1,164.66 |
| 572 | 21.7 | 24.22 | 15.94 | 19.76 | 81.62 |
| 573 | 23.37 | 31.26 | 21.89 | 15.57 | 92.09 |
| 574 | 23.11 | 20.79 | 20.12 | 18.99 | 83.01 |
| 575 | 19.7 | 18.59 | 20.69 | 20.99 | 79.97 |
| 576 | 4.33 | 9.11 | 5.85 | 7.97 | 27.26 |
| 577 | 104.89 | 101.95 | 99.17 | 105.42 | 411.43 |
| TOTAL | 519.68 | 525.58 | 512.84 | 641.6 | 2,199.70 |

STATE: TENNESSEE

| ZIP CODE | QUARTER 1 | QUARTER 2 | QUARTER 3 | QUARTER 4 | TOTAL GRAMS |
|---|---|---|---|---|---|
| 370 | 542.39 | 536.39 | 528.1 | 574.97 | 2,181.85 |
| 371 | 427.81 | 467.68 | 479.97 | 436.67 | 1,812.13 |
| 372 | 881.3 | 771.78 | 906.42 | 913.86 | 3,473.36 |
| 373 | 470.87 | 490.35 | 472.61 | 526.11 | 1,959.94 |
| 374 | 359.19 | 335.87 | 300.61 | 293.08 | 1,288.75 |
| 376 | 355.06 | 320.16 | 314.94 | 362.43 | 1,352.59 |
| 377 | 102.21 | 124.93 | 114.96 | 114.67 | 456.77 |
| 378 | 237.85 | 234.52 | 237.33 | 254.28 | 963.98 |

| | | | | | | |
|---|---|---|---|---|---|---|
| | 379 | 389.91 | 445.21 | 518.28 | 250.77 | 1,604.17 |
| | 380 | 215.82 | 260.72 | 216.9 | 214.22 | 907.66 |
| | 381 | 3,745.82 | 1,764.20 | 1,926.92 | 1,509.92 | 8,946.86 |
| | 382 | 63.2 | 63.49 | 59.82 | 51.49 | 238 |
| | 383 | 114.11 | 96.27 | 120.25 | 107.13 | 437.76 |
| | 384 | 104.86 | 86.38 | 62.21 | 62.44 | 315.89 |
| | 385 | 192.21 | 184.22 | 190.08 | 150.15 | 716.66 |
| TOTAL | | 8,202.61 | 6,182.17 | 6,449.40 | 5,822.19 | 26,656.37 |

STATE: TEXAS

| ZIP CODE | | QUARTER 1 | QUARTER 2 | QUARTER 3 | QUARTER 4 | TOTAL GRAMS |
|---|---|---|---|---|---|---|
| | 750 | 2,790.08 | 2,630.88 | 2,559.04 | 2,294.31 | 10,274.31 |
| | 751 | 473.63 | 495.77 | 478.28 | 489.99 | 1,937.67 |
| | 752 | 1,422.38 | 1,567.63 | 1,489.86 | 1,383.46 | 5,863.33 |
| | 753 | 121.08 | 112.34 | 104.73 | 127.66 | 465.81 |
| | 754 | 532.17 | 508.36 | 455.36 | 504.64 | 2,000.53 |
| | 755 | 85.34 | 70.3 | 72.47 | 74.74 | 302.85 |
| | 756 | 606.42 | 699.42 | 596.36 | 977.79 | 2,879.99 |
| | 757 | 273.68 | 297.02 | 399.72 | 288.82 | 1,259.24 |
| | 758 | 54.54 | 47.7 | 50.8 | 35.66 | 188.7 |
| | 759 | 183.51 | 179.21 | 190.23 | 171.33 | 724.28 |
| | 760 | 1,143.44 | 1,124.78 | 1,010.42 | 950.03 | 4,228.67 |
| | 761 | 1,170.20 | 1,117.91 | 1,187.56 | 1,136 | 4,611.67 |
| | 762 | 647.7 | 658.57 | 661.68 | 579.96 | 2,547.91 |
| | 763 | 282.36 | 270.47 | 261.15 | 253.02 | 1,067 |
| | 764 | 115.44 | 112.64 | 104.16 | 95.35 | 427.59 |
| | 765 | 418.99 | 438.95 | 390.6 | 411.94 | 1,660.48 |
| | 766 | 59.76 | 46.9 | 50.33 | 56.56 | 213.55 |
| | 767 | 90.89 | 83.48 | 95.36 | 92.18 | 361.91 |
| | 768 | 51.93 | 55.97 | 82.62 | 52.99 | 243.51 |
| | 769 | 73.09 | 75.07 | 52.27 | 50.02 | 250.45 |
| | 770 | 2,963.47 | 2,710.64 | 2,615.67 | 2,231.79 | 10,521.57 |
| | 773 | 1,025.19 | 1,004.30 | 974.6 | 989.24 | 3,993.33 |
| | 774 | 834.34 | 740.28 | 738.86 | 716.6 | 3,030.08 |
| | 775 | 1,083.33 | 1,272.79 | 1,298.80 | 1,006.54 | 4,661.46 |
| | 776 | 489.7 | 502.59 | 458.1 | 455.37 | 1,905.76 |

| | | | | | | |
|---|---|---|---|---|---|---|
| | 777 | 244.5 | 248.52 | 248.86 | 247.38 | 989.26 |
| | 778 | 127.81 | 129.04 | 117.43 | 102.08 | 476.36 |
| | 779 | 457.63 | 440.6 | 444.09 | 425.62 | 1,767.94 |
| | 780 | 239.13 | 252.45 | 202.35 | 227.29 | 921.22 |
| | 781 | 308.71 | 302.99 | 305.34 | 287.48 | 1,204.52 |
| | 782 | 1,940.07 | 1,957.60 | 1,897.03 | 1,865.52 | 7,660.22 |
| | 783 | 133.26 | 127.9 | 110.54 | 112.95 | 484.65 |
| | 784 | 278.83 | 290.89 | 257.16 | 239.53 | 1,066.41 |
| | 785 | 345.56 | 292.97 | 321.78 | 309.34 | 1,269.65 |
| | 786 | 969.74 | 877.32 | 924.9 | 855.12 | 3,627.08 |
| | 787 | 1,063.01 | 939.21 | 974.34 | 819.48 | 3,796.04 |
| | 788 | 33.65 | 35.27 | 37.3 | 36.98 | 143.2 |
| | 789 | 47.96 | 40.88 | 47.76 | 38.83 | 175.43 |
| | 790 | 70.96 | 72.23 | 78.97 | 83.23 | 305.39 |
| | 791 | 188.36 | 229.45 | 206.71 | 200.39 | 824.91 |
| | 792 | 5.45 | 7.89 | 6.56 | 6.74 | 26.64 |
| | 793 | 71.46 | 85.06 | 83.24 | 86.68 | 326.44 |
| | 794 | 399.62 | 373.65 | 345.83 | 369.29 | 1,488.39 |
| | 795 | 41.36 | 39.58 | 37.71 | 32.88 | 151.53 |
| | 796 | 85.52 | 83.7 | 87.07 | 72.67 | 328.96 |
| | 797 | 227.51 | 228.22 | 211.17 | 201.4 | 868.3 |
| | 798 | 13.33 | 11.66 | 2.71 | 16.18 | 43.88 |
| | 799 | 459.82 | 482.69 | 460.81 | 438.45 | 1,841.77 |
| TOTAL | | 24,745.91 | 24,373.74 | 23,788.69 | 22,501.50 | 95,409.84 |

STATE: UTAH

| ZIP CODE | | QUARTER 1 | QUARTER 2 | QUARTER 3 | QUARTER 4 | TOTAL GRAMS |
|---|---|---|---|---|---|---|
| | 840 | 865.82 | 963.13 | 1,083.41 | 1,017.76 | 3,930.12 |
| | 841 | 893.37 | 853.42 | 895.82 | 1,110.09 | 3,752.70 |
| | 843 | 121.45 | 132.64 | 136.91 | 148.79 | 539.79 |
| | 844 | 192.81 | 198.05 | 197.83 | 210.86 | 799.55 |
| | 845 | 20.82 | 20.09 | 23.21 | 31.81 | 95.93 |
| | 846 | 214.33 | 211.63 | 213.27 | 209.71 | 848.94 |
| | 847 | 249.95 | 232.06 | 232.35 | 224.06 | 938.42 |
| TOTAL | | 2,558.55 | 2,611.02 | 2,782.80 | 2,953.08 | 10,905.45 |

STATE: VERMONT

| ZIP CODE | QUARTER 1 | QUARTER 2 | QUARTER 3 | QUARTER 4 | TOTAL GRAMS |
|---|---|---|---|---|---|
| 50 | 61.31 | 63.75 | 49.15 | 35.85 | 210.06 |
| 51 | 53.77 | 48.21 | 55.88 | 50.23 | 208.09 |
| 52 | 41.21 | 52.07 | 42.47 | 39.8 | 175.55 |
| 53 | 29.3 | 34.3 | 40.7 | 31.38 | 135.68 |
| 54 | 697.23 | 725.84 | 626.43 | 575.13 | 2,624.63 |
| 56 | 128.01 | 117.49 | 117.12 | 115.18 | 477.8 |
| 57 | 159.03 | 164.8 | 206.26 | 254.88 | 784.97 |
| 58 | 68.97 | 63.37 | 58.74 | 64.94 | 256.02 |
| TOTAL | 1,238.83 | 1,269.83 | 1,196.75 | 1,167.39 | 4,872.80 |

STATE: VIRGIN ISLANDS

| ZIP CODE | QUARTER 1 | QUARTER 2 | QUARTER 3 | QUARTER 4 | TOTAL GRAMS |
|---|---|---|---|---|---|
| 8 | 26.6 | 18.71 | 13.94 | 14.9 | 74.15 |
| TOTAL | 26.6 | 18.71 | 13.94 | 14.9 | 74.15 |

STATE: VIRGINIA

| ZIP CODE | QUARTER 1 | QUARTER 2 | QUARTER 3 | QUARTER 4 | TOTAL GRAMS |
|---|---|---|---|---|---|
| 201 | 1,179.88 | 1,114.83 | 987.19 | 1,059.46 | 4,341.36 |
| 220 | 665.64 | 628.21 | 564.35 | 466.73 | 2,324.93 |
| 221 | 463.54 | 396.36 | 355.68 | 359.41 | 1,574.99 |
| 222 | 170.99 | 161.75 | 144.73 | 126.12 | 603.59 |
| 223 | 239.73 | 218.81 | 198.18 | 204.43 | 861.15 |
| 224 | 398.47 | 377.25 | 344.61 | 337.83 | 1,458.16 |
| 225 | 134.93 | 129.96 | 109.21 | 125.92 | 500.02 |
| 226 | 554.78 | 507.49 | 482.29 | 456.65 | 2,001.21 |
| 227 | 51.34 | 64.69 | 56.42 | 46.31 | 218.76 |
| 228 | 338.81 | 325.08 | 289.61 | 289.14 | 1,242.64 |
| 229 | 693.1 | 563.96 | 450.04 | 503.12 | 2,210.22 |
| 230 | 349.07 | 340.23 | 332.67 | 277.26 | 1,299.23 |
| 231 | 648.31 | 554.58 | 488.2 | 560.82 | 2,251.91 |
| 232 | 1,371.12 | 1,145.07 | 1,166.87 | 1,136.24 | 4,819.30 |

| | | | | | | |
|---|---|---|---|---|---|---|
| | 233 | 536.91 | 444.66 | 415.73 | 430.7 | 1,828 |
| | 234 | 1,035.16 | 946.61 | 869.48 | 810.85 | 3,662.10 |
| | 235 | 555.05 | 575 | 530.17 | 516.73 | 2,176.95 |
| | 236 | 670.31 | 556.18 | 508.55 | 483.28 | 2,218.32 |
| | 237 | 283.49 | 233.52 | 225.96 | 251.38 | 994.35 |
| | 238 | 307.91 | 307.35 | 253.37 | 242.89 | 1,111.52 |
| | 239 | 65.27 | 74.68 | 74.77 | 65.4 | 280.12 |
| | 240 | 565.04 | 501.21 | 504.78 | 559.43 | 2,130.46 |
| | 241 | 398.71 | 352.12 | 345.3 | 362.35 | 1,458.48 |
| | 242 | 186.27 | 200.02 | 190.63 | 175.45 | 752.37 |
| | 243 | 124.55 | 112.86 | 105.55 | 107.06 | 450.02 |
| | 244 | 176.23 | 159.11 | 139.51 | 154.04 | 628.89 |
| | 245 | 326.14 | 339.57 | 314.45 | 319.21 | 1,299.37 |
| | 246 | 65.51 | 55.15 | 70.52 | 54.73 | 245.91 |
| TOTAL | | 12,556.26 | 11,386.31 | 10,518.82 | 10,482.94 | 44,944.33 |

STATE: WASHINGTON

| ZIP CODE | | QUARTER 1 | QUARTER 2 | QUARTER 3 | QUARTER 4 | TOTAL GRAMS |
|---|---|---|---|---|---|---|
| | 980 | 1,842.58 | 1,681.19 | 1,762.27 | 1,680.73 | 6,966.77 |
| | 981 | 1,872.75 | 2,086.69 | 1,996.36 | 1,542.90 | 7,498.70 |
| | 982 | 1,457.67 | 1,370.11 | 1,298.68 | 1,315.70 | 5,442.16 |
| | 983 | 945.21 | 897.78 | 857.89 | 887.8 | 3,588.68 |
| | 984 | 573.93 | 595.3 | 497.22 | 524.45 | 2,190.90 |
| | 985 | 838.31 | 785.7 | 789.03 | 694.35 | 3,107.39 |
| | 986 | 1,149.97 | 1,226.58 | 1,065.46 | 1,040.50 | 4,482.51 |
| | 988 | 192.23 | 287.6 | 267.57 | 171.11 | 918.51 |
| | 989 | 292.29 | 319.52 | 254.99 | 300.51 | 1,167.31 |
| | 990 | 91.3 | 86.67 | 107.03 | 93.31 | 378.31 |
| | 991 | 124.34 | 124.11 | 123.41 | 125.13 | 496.99 |
| | 992 | 1,004.03 | 999.88 | 907.53 | 926.36 | 3,837.80 |
| | 993 | 708.83 | 570.51 | 572.48 | 478.97 | 2,330.79 |
| | 994 | 85.09 | 80.85 | 163.52 | 90.54 | 420 |
| TOTAL | | 11,178.53 | 11,112.49 | 10,663.44 | 9,872.36 | 42,826.82 |

STATE: WEST VIRGINIA

| ZIP CODE | QUARTER 1 | QUARTER 2 | QUARTER 3 | QUARTER 4 | TOTAL GRAMS |
|---|---|---|---|---|---|
| 247 | 43.51 | 44.94 | 56.75 | 42.88 | 188.08 |
| 248 | 9.75 | 4.79 | 6.21 | 8.87 | 29.62 |
| 249 | 23.81 | 27.62 | 30.07 | 24.42 | 105.92 |
| 250 | 5.39 | 2.06 | 5.25 | 4.04 | 16.74 |
| 251 | 8.64 | 96.88 | 13.8 | 4.97 | 124.29 |
| 252 | 9.92 | 5.03 | 6.54 | 5.79 | 27.28 |
| 253 | 227.17 | 131.45 | 113.85 | 93.09 | 565.56 |
| 254 | 409.09 | 382.61 | 379.35 | 354.26 | 1,525.31 |
| 255 | 44.64 | 43.65 | 50.37 | 42.93 | 181.59 |
| 256 | 11.98 | 9.3 | 9.19 | 7.66 | 38.13 |
| 257 | 84.85 | 75.63 | 116.42 | 84.07 | 360.97 |
| 258 | 132.65 | 99.09 | 92.9 | 92.92 | 417.56 |
| 259 | 17.02 | 21.14 | 8.47 | 12.77 | 59.4 |
| 260 | 92.57 | 91 | 73.95 | 82.58 | 340.1 |
| 261 | 59.81 | 62.3 | 70.89 | 49.95 | 242.95 |
| 262 | 20.78 | 20.97 | 20.94 | 21.53 | 84.22 |
| 263 | 95.4 | 81.98 | 87.48 | 85.73 | 350.59 |
| 264 | 14.68 | 14.89 | 16.18 | 13.4 | 59.15 |
| 265 | 273.49 | 253.19 | 229.38 | 261.13 | 1,017.19 |
| 266 | 18.34 | 11.95 | 10.27 | 10.44 | 51 |
| 267 | 50.45 | 48.43 | 32.42 | 39.65 | 170.95 |
| 268 | 31.87 | 22.08 | 22.92 | 20.67 | 97.54 |
| TOTAL | 1,685.81 | 1,550.98 | 1,453.60 | 1,363.75 | 6,054.14 |

STATE: WISCONSIN

| ZIP CODE | QUARTER 1 | QUARTER 2 | QUARTER 3 | QUARTER 4 | TOTAL GRAMS |
|---|---|---|---|---|---|
| 530 | 274.29 | 267.32 | 239.11 | 305.14 | 1,085.86 |
| 531 | 470.37 | 356.99 | 400.71 | 464.98 | 1,693.05 |
| 532 | 797.47 | 719.04 | 707.53 | 757 | 2,981.04 |
| 534 | 79.92 | 69.22 | 68.2 | 60.55 | 277.89 |
| 535 | 274.31 | 255.53 | 226.33 | 247.19 | 1,003.36 |
| 537 | 268.28 | 313.9 | 242.54 | 266.49 | 1,091.21 |
| 538 | 47.19 | 34.47 | 39.64 | 26.02 | 147.32 |
| 539 | 77.56 | 85.05 | 77.2 | 70.39 | 310.2 |
| 540 | 88.37 | 75.2 | 81.84 | 82.47 | 327.88 |

| | | | | | | |
|---|---|---|---|---|---|---|
| | 541 | 46.74 | 61.12 | 53.31 | 59.31 | 220.48 |
| | 542 | 45.28 | 32.13 | 39.34 | 36.8 | 153.55 |
| | 543 | 139.75 | 118.87 | 190.65 | 177.05 | 626.32 |
| | 544 | 191.68 | 158.53 | 164.4 | 148.16 | 662.77 |
| | 545 | 52.19 | 54.71 | 57.23 | 49.69 | 213.82 |
| | 546 | 261.13 | 235.27 | 228.83 | 251.78 | 977.01 |
| | 547 | 239.09 | 136.77 | 136.17 | 140.82 | 652.85 |
| | 548 | 127.68 | 144.18 | 140.61 | 129.36 | 541.83 |
| | 549 | 373.16 | 392.98 | 605.66 | 523.95 | 1,895.75 |
| TOTAL | | 3,854.46 | 3,511.28 | 3,699.30 | 3,797.15 | 14,862.19 |

STATE: WYOMING

| ZIP CODE | | QUARTER 1 | QUARTER 2 | QUARTER 3 | QUARTER 4 | TOTAL GRAMS |
|---|---|---|---|---|---|---|
| | 820 | 137.22 | 137.71 | 144.85 | 131.46 | 551.24 |
| | 822 | 6.53 | 8.59 | 7.33 | 7.94 | 30.39 |
| | 823 | 4.88 | 5.54 | 7.1 | 5.78 | 23.3 |
| | 824 | 128.69 | 103.19 | 97.3 | 103.73 | 432.91 |
| | 825 | 34.79 | 39.08 | 30.47 | 31.05 | 135.39 |
| | 826 | 82.87 | 99.16 | 85.59 | 75.19 | 342.81 |
| | 827 | 48.59 | 32.45 | 34.39 | 32.71 | 148.14 |
| | 828 | 39.4 | 38.19 | 35.99 | 41.86 | 155.44 |
| | 829 | 131.23 | 151.66 | 133.21 | 91.82 | 507.92 |
| | 830 | 4.44 | 7.4 | 13.05 | 4.75 | 29.64 |
| | 831 | 33.52 | 19.46 | 15.31 | 13.86 | 82.15 |
| TOTAL | | 652.16 | 642.43 | 604.59 | 540.15 | 2,439.33 |

DRUG CODE: 9193 DRUG NAME: HYDROCODONE

STATE: ALABAMA

| ZIP CODE | | QUARTER 1 | QUARTER 2 | QUARTER 3 | QUARTER 4 | TOTAL GRAMS |
|---|---|---|---|---|---|---|
| | 350 | 29,344.31 | 27,829.50 | 26,898.56 | 26,569.78 | 110,642.15 |
| | 351 | 14,695.22 | 13,917.38 | 13,821.14 | 13,662.31 | 56,096.05 |
| | 352 | 25,633.79 | 24,760.75 | 23,410.37 | 24,340.35 | 98,145.26 |
| | 354 | 11,497.65 | 11,163.93 | 10,693.11 | 10,306.43 | 43,661.12 |
| | 355 | 12,138.34 | 11,803.67 | 11,306.35 | 10,900.13 | 46,148.49 |

| | | | | | | |
|---|---|---|---|---|---|---|
| | 356 | 24,658.52 | 23,643.96 | 22,919.27 | 22,350.84 | 93,572.59 |
| | 357 | 10,113.79 | 9,832.34 | 9,149.85 | 8,986.52 | 38,082.50 |
| | 358 | 11,780.59 | 11,160.83 | 10,407.75 | 10,315.37 | 43,664.54 |
| | 359 | 20,667.83 | 20,312.67 | 18,761.27 | 19,752.69 | 79,494.46 |
| | 360 | 12,206.03 | 11,467.85 | 10,674.47 | 10,747.34 | 45,095.69 |
| | 361 | 10,305.45 | 9,672.11 | 8,326.79 | 8,153.56 | 36,457.91 |
| | 362 | 10,144.90 | 9,571.85 | 8,958.21 | 9,364.30 | 38,039.26 |
| | 363 | 9,866.70 | 9,428.59 | 8,832.58 | 9,070.86 | 37,198.73 |
| | 364 | 6,321.70 | 5,683.23 | 5,158.94 | 5,263.09 | 22,426.96 |
| | 365 | 20,532.22 | 19,196.48 | 18,128.04 | 18,151.79 | 76,008.53 |
| | 366 | 15,977.46 | 15,247.07 | 14,557.72 | 14,102.03 | 59,884.28 |
| | 367 | 3,661.87 | 3,468.80 | 3,276.98 | 3,306.86 | 13,714.51 |
| | 368 | 7,628.04 | 7,295.50 | 7,120.14 | 7,319.83 | 29,363.51 |
| | 369 | 1,170.69 | 1,055.88 | 938.02 | 1,034.81 | 4,199.40 |
| TOTAL | | 258,345.10 | 246,512.39 | 233,339.56 | 233,698.89 | 971,895.94 |

STATE: ALASKA

| ZIP CODE | | QUARTER 1 | QUARTER 2 | QUARTER 3 | QUARTER 4 | TOTAL GRAMS |
|---|---|---|---|---|---|---|
| | 995 | 5,551.47 | 5,430.07 | 5,015.08 | 4,810.26 | 20,806.88 |
| | 996 | 3,904.79 | 3,680.86 | 3,525.24 | 3,498.60 | 14,609.49 |
| | 997 | 1,257.48 | 1,231.23 | 1,192.98 | 1,030.33 | 4,712.02 |
| | 998 | 651.8 | 669.98 | 555.19 | 507.26 | 2,384.23 |
| | 999 | 581.71 | 480.62 | 486.78 | 468.37 | 2,017.48 |
| TOTAL | | 11,947.25 | 11,492.76 | 10,775.27 | 10,314.82 | 44,530.10 |

STATE: AMERICAN SAMOA

| ZIP CODE | | QUARTER 1 | QUARTER 2 | QUARTER 3 | QUARTER 4 | TOTAL GRAMS |
|---|---|---|---|---|---|---|
| | 967 | 4 | 0.27 | 3.63 | 0 | 7.9 |
| TOTAL | | 4 | 0.27 | 3.63 | 0 | 7.9 |

STATE: ARIZONA

| ZIP CODE | | QUARTER 1 | QUARTER 2 | QUARTER 3 | QUARTER 4 | TOTAL GRAMS |
|---|---|---|---|---|---|---|
| | 850 | 17,021.07 | 15,620.90 | 14,700.79 | 14,539.48 | 61,882.24 |
| | 851 | 5,787.11 | 5,455.15 | 5,070.71 | 5,435.75 | 21,748.72 |

| | | | | | | |
|---|---|---|---|---|---|---|
| | 852 | 28,070.11 | 26,336.32 | 24,240.36 | 24,063.61 | 102,710.40 |
| | 853 | 19,702.89 | 17,740.07 | 16,451.85 | 16,952.48 | 70,847.29 |
| | 855 | 2,180.12 | 2,160.36 | 1,957.86 | 1,941.09 | 8,239.43 |
| | 856 | 3,962.37 | 3,715.85 | 3,455.43 | 3,572.48 | 14,706.13 |
| | 857 | 14,278.23 | 13,148.04 | 12,724.28 | 12,370.04 | 52,520.59 |
| | 859 | 2,307.71 | 2,281.46 | 2,650.44 | 2,291.21 | 9,530.82 |
| | 860 | 2,282.09 | 2,163.90 | 2,076.41 | 1,886.69 | 8,409.09 |
| | 863 | 7,350.04 | 7,199.94 | 6,761.37 | 6,417.34 | 27,728.69 |
| | 864 | 9,037.17 | 8,583.58 | 8,160.19 | 8,245.16 | 34,026.10 |
| | 865 | 121.38 | 116.48 | 112.72 | 103.55 | 454.13 |
| TOTAL | | 112,100.29 | 104,522.05 | 98,362.41 | 97,818.88 | 412,803.63 |

| STATE: | ARKANSAS | | | | | |
|---|---|---|---|---|---|---|
| ZIP CODE | | QUARTER 1 | QUARTER 2 | QUARTER 3 | QUARTER 4 | TOTAL GRAMS |
| | 716 | 6,493.54 | 6,262.25 | 6,005.43 | 5,838.70 | 24,599.92 |
| | 717 | 5,492.46 | 5,460.89 | 5,317.51 | 5,294.07 | 21,564.93 |
| | 718 | 7,303.04 | 7,783.65 | 7,676.21 | 7,526.94 | 30,289.84 |
| | 719 | 11,287.64 | 10,359.91 | 9,964.36 | 9,636.03 | 41,247.94 |
| | 720 | 13,216.10 | 12,832.43 | 12,322.13 | 12,216.47 | 50,587.13 |
| | 721 | 16,287.95 | 15,909.94 | 15,339.27 | 15,064.50 | 62,601.66 |
| | 722 | 8,251.23 | 8,197.79 | 7,767.20 | 7,580.57 | 31,796.79 |
| | 723 | 8,269.52 | 7,981.45 | 7,794.55 | 7,740.12 | 31,785.64 |
| | 724 | 12,925.14 | 12,161.88 | 11,679.62 | 12,031.15 | 48,797.79 |
| | 725 | 6,931.25 | 7,033.24 | 6,513.41 | 6,683.10 | 27,161 |
| | 726 | 7,028.41 | 6,783.18 | 6,379.04 | 6,291.81 | 26,482.44 |
| | 727 | 18,120.04 | 17,545.19 | 16,025.13 | 16,187.05 | 67,877.41 |
| | 728 | 4,865.99 | 4,815.17 | 4,370.08 | 4,262.03 | 18,313.27 |
| | 729 | 13,733.40 | 13,342.73 | 12,779.12 | 12,716.07 | 52,571.32 |
| TOTAL | | 140,205.71 | 136,469.70 | 129,933.06 | 129,068.61 | 535,677.08 |

| STATE: | CALIFORNIA | | | | | |
|---|---|---|---|---|---|---|
| ZIP CODE | | QUARTER 1 | QUARTER 2 | QUARTER 3 | QUARTER 4 | TOTAL GRAMS |
| | 900 | 21,580.72 | 20,478.08 | 20,288.49 | 19,988.06 | 82,335.35 |
| | 902 | 17,272.59 | 17,114.49 | 16,641.44 | 16,676.92 | 67,705.44 |
| | 903 | 1,946.11 | 1,908.91 | 1,890.75 | 1,730.79 | 7,476.56 |

| | | | | | |
|---|---|---|---|---|---|
| 904 | 2,041.65 | 2,107.74 | 1,941.92 | 1,922.62 | 8,013.93 |
| 905 | 4,481.23 | 4,328.77 | 4,086.15 | 3,974.36 | 16,870.51 |
| 906 | 13,138.02 | 12,947.35 | 12,427.52 | 12,212.19 | 50,725.08 |
| 907 | 12,632.74 | 12,746.89 | 12,335.58 | 12,151.75 | 49,866.96 |
| 908 | 8,672.60 | 8,269.21 | 8,193.41 | 7,988.61 | 33,123.83 |
| 910 | 5,331.80 | 5,296.80 | 4,961.32 | 4,119.97 | 19,709.89 |
| 911 | 2,667.88 | 2,672.83 | 2,585.16 | 2,497.19 | 10,423.06 |
| 912 | 3,399.73 | 3,410.14 | 3,242.93 | 3,365.08 | 13,417.88 |
| 913 | 25,958.38 | 24,798.39 | 24,293.65 | 24,128.39 | 99,178.81 |
| 914 | 6,423.80 | 6,459.39 | 6,328.04 | 6,313.88 | 25,525.11 |
| 915 | 2,257.93 | 2,206.90 | 2,064.63 | 2,045.49 | 8,574.95 |
| 916 | 1,643.76 | 1,534.77 | 1,410.43 | 1,349.98 | 5,938.94 |
| 917 | 35,276.80 | 35,129.81 | 35,323.98 | 35,034.17 | 140,764.76 |
| 918 | 998.49 | 1,055.56 | 952.64 | 944.57 | 3,951.26 |
| 919 | 9,263.04 | 9,096.06 | 8,732.80 | 8,248.19 | 35,340.09 |
| 920 | 31,199.64 | 29,962.99 | 28,928.61 | 28,538.99 | 118,630.23 |
| 921 | 22,421.93 | 21,506.49 | 20,368.49 | 19,436.91 | 83,733.82 |
| 922 | 22,120.41 | 20,978.38 | 19,989.93 | 19,982.50 | 83,071.22 |
| 923 | 48,085.79 | 47,287.38 | 45,549.25 | 45,198.10 | 186,120.52 |
| 924 | 8,192.42 | 8,184.35 | 7,766.72 | 7,888.52 | 32,032.01 |
| 925 | 41,924.62 | 41,394.63 | 39,201 | 39,134.17 | 161,654.42 |
| 926 | 25,053.23 | 24,461.43 | 23,325.26 | 23,087.35 | 95,927.27 |
| 927 | 7,482.87 | 7,158.26 | 6,897.73 | 6,737.34 | 28,276.20 |
| 928 | 26,129.59 | 25,088.42 | 24,461.29 | 24,036.41 | 99,715.71 |
| 930 | 17,389.37 | 17,006.04 | 16,451.11 | 15,857.83 | 66,704.35 |
| 931 | 3,261.87 | 3,026.77 | 2,964.89 | 2,742.81 | 11,996.34 |
| 932 | 24,789.16 | 24,315.63 | 22,909.82 | 22,918.90 | 94,933.51 |
| 933 | 20,426.17 | 20,642.16 | 19,886.09 | 19,297 | 80,251.42 |
| 934 | 15,954.52 | 15,027.46 | 14,785.94 | 14,888.89 | 60,656.81 |
| 935 | 18,799.39 | 18,374.08 | 17,682.73 | 17,863.27 | 72,719.47 |
| 936 | 19,093.31 | 19,117.64 | 17,679.47 | 17,740.08 | 73,630.50 |
| 937 | 20,043.43 | 20,014.31 | 19,397.44 | 19,193.29 | 78,648.47 |
| 939 | 8,883.85 | 8,808.31 | 8,397.65 | 7,910.91 | 34,000.72 |
| 940 | 10,269.59 | 10,070.66 | 9,598.16 | 9,582.29 | 39,520.70 |
| 941 | 9,538.64 | 9,361.96 | 8,908.90 | 8,642.46 | 36,451.96 |
| 943 | 2,057.70 | 1,953.03 | 1,853.68 | 1,886.07 | 7,750.48 |
| 944 | 1,656.44 | 1,665.72 | 1,620.67 | 1,514.62 | 6,457.45 |

| | | | | | |
|---|---|---|---|---|---|
| 945 | 64,880.16 | 63,287.83 | 60,561.28 | 59,621.35 | 248,350.62 |
| 946 | 10,197.64 | 10,185.98 | 9,507.46 | 9,457.56 | 39,348.64 |
| 947 | 1,539.93 | 1,485.09 | 1,507.91 | 1,554.10 | 6,087.03 |
| 948 | 3,607.03 | 3,418.46 | 3,392.85 | 3,318.01 | 13,736.35 |
| 949 | 8,780.21 | 8,683.42 | 8,674.04 | 8,496.81 | 34,634.48 |
| 950 | 14,408.06 | 13,871.82 | 12,790.56 | 12,713.78 | 53,784.22 |
| 951 | 9,643.54 | 9,251.13 | 8,985.43 | 8,614.59 | 36,494.69 |
| 952 | 20,070.11 | 19,476.85 | 18,444.55 | 18,199.75 | 76,191.26 |
| 953 | 51,419.99 | 50,607.84 | 48,561.38 | 47,837.07 | 198,426.28 |
| 954 | 18,489.88 | 18,423.59 | 17,412.64 | 16,331.88 | 70,657.99 |
| 955 | 8,684.31 | 8,700.51 | 8,179.74 | 7,683.91 | 33,248.47 |
| 956 | 56,251.65 | 54,163 | 51,941.69 | 50,438.90 | 212,795.24 |
| 957 | 10,165.33 | 10,059.87 | 9,359.69 | 8,992.29 | 38,577.18 |
| 958 | 34,147.39 | 33,927.26 | 31,814.90 | 31,515.39 | 131,404.94 |
| 959 | 34,120.01 | 33,768.86 | 31,994.75 | 31,118.89 | 131,002.51 |
| 960 | 22,604.81 | 22,363.33 | 21,371.17 | 20,374.65 | 86,713.96 |
| 961 | 3,555.44 | 3,343.37 | 3,191.39 | 2,989.63 | 13,079.83 |
| TOTAL | 952,326.70 | 931,986.40 | 894,017.10 | 878,029.48 | 3,656,359.68 |

STATE: COLORADO

| ZIP CODE | QUARTER 1 | QUARTER 2 | QUARTER 3 | QUARTER 4 | TOTAL GRAMS |
|---|---|---|---|---|---|
| 800 | 9,797.46 | 9,517.71 | 8,953.15 | 8,708.99 | 36,977.31 |
| 801 | 7,999.55 | 7,403.15 | 7,143.27 | 7,255.66 | 29,801.63 |
| 802 | 11,126.67 | 10,598.99 | 10,352.98 | 9,939.68 | 42,018.32 |
| 803 | 1,034.45 | 988.98 | 934.96 | 921.09 | 3,879.48 |
| 804 | 2,398.97 | 2,096.46 | 2,037.63 | 2,081.04 | 8,614.10 |
| 805 | 7,052.47 | 6,881.36 | 6,701.63 | 6,583.94 | 27,219.40 |
| 806 | 2,920.80 | 2,849.99 | 2,761.60 | 2,735.73 | 11,268.12 |
| 807 | 965.12 | 909.68 | 913.13 | 875.01 | 3,662.94 |
| 808 | 1,839.70 | 1,765.30 | 1,769.29 | 1,595.46 | 6,969.75 |
| 809 | 8,160.80 | 7,735.91 | 7,636 | 7,524.20 | 31,056.91 |
| 810 | 6,049.63 | 5,875.10 | 5,583.64 | 5,611.19 | 23,119.56 |
| 811 | 842.69 | 751.4 | 769.63 | 694.62 | 3,058.34 |
| 812 | 1,516.39 | 1,513 | 1,410.91 | 1,415.37 | 5,855.67 |
| 813 | 1,310.25 | 1,205.88 | 1,142.49 | 1,100.17 | 4,758.79 |

| | | | | | |
|---|---|---|---|---|---|
| 814 | 1,272.56 | 1,239.70 | 1,145.80 | 1,144.77 | 4,802.83 |
| 815 | 2,795.21 | 2,730.79 | 2,563.61 | 2,606.54 | 10,696.15 |
| 816 | 1,746.82 | 1,508.73 | 1,497.10 | 1,406.65 | 6,159.30 |
| TOTAL | 68,829.54 | 65,572.13 | 63,316.82 | 62,200.11 | 259,918.60 |

STATE: CONNECTICUT

| ZIP CODE | QUARTER 1 | QUARTER 2 | QUARTER 3 | QUARTER 4 | TOTAL GRAMS |
|---|---|---|---|---|---|
| 60 | 4,797.48 | 4,471.66 | 4,203.80 | 4,146 | 17,618.94 |
| 61 | 1,348.01 | 1,285.53 | 1,189.92 | 1,117.10 | 4,940.56 |
| 62 | 1,066.65 | 970.42 | 912.27 | 921.1 | 3,870.44 |
| 63 | 2,303.32 | 2,274.90 | 2,154.67 | 2,115.53 | 8,848.42 |
| 64 | 5,054.55 | 4,774.77 | 4,504.97 | 4,503.51 | 18,837.80 |
| 65 | 1,370.86 | 1,300.69 | 1,146.75 | 1,222.61 | 5,040.91 |
| 66 | 948.98 | 903.91 | 845.43 | 810.03 | 3,508.35 |
| 67 | 2,722.20 | 2,532.95 | 2,413.41 | 2,400.98 | 10,069.54 |
| 68 | 1,788.05 | 1,655.40 | 1,576.43 | 1,505.33 | 6,525.21 |
| 69 | 299.26 | 278.49 | 261.99 | 259.48 | 1,099.22 |
| TOTAL | 21,699.36 | 20,448.72 | 19,209.64 | 19,001.67 | 80,359.39 |

STATE: DELAWARE

| ZIP CODE | QUARTER 1 | QUARTER 2 | QUARTER 3 | QUARTER 4 | TOTAL GRAMS |
|---|---|---|---|---|---|
| 197 | 1,840 | 1,721.29 | 1,710.75 | 1,650.80 | 6,922.84 |
| 198 | 1,220.17 | 1,174.65 | 1,094.77 | 1,085.35 | 4,574.94 |
| 199 | 3,329.53 | 3,020.86 | 3,020.59 | 3,091.27 | 12,462.25 |
| TOTAL | 6,389.70 | 5,916.80 | 5,826.11 | 5,827.42 | 23,960.03 |

STATE: DISTRICT OF COLUMBIA

| ZIP CODE | QUARTER 1 | QUARTER 2 | QUARTER 3 | QUARTER 4 | TOTAL GRAMS |
|---|---|---|---|---|---|
| 200 | 1,451.26 | 1,357.39 | 1,324.12 | 1,197.17 | 5,329.94 |
| 203 | 5.45 | 11.81 | 3.03 | 3.63 | 23.92 |
| 204 | 115.03 | 90.81 | 116.84 | 88.39 | 411.07 |
| 205 | 0 | 0.61 | 0.91 | 0 | 1.52 |
| TOTAL | 1,571.74 | 1,460.62 | 1,444.90 | 1,289.19 | 5,766.45 |

STATE: FLORIDA

| ZIP CODE | QUARTER 1 | QUARTER 2 | QUARTER 3 | QUARTER 4 | TOTAL GRAMS |
|---|---|---|---|---|---|
| 320 | 20,649.85 | 19,923.35 | 18,958.93 | 19,313.33 | 78,845.46 |
| 321 | 22,475.36 | 21,964.07 | 20,633.25 | 21,616.78 | 86,689.46 |
| 322 | 37,175.53 | 35,211.06 | 33,668.57 | 33,520.14 | 139,575.30 |
| 323 | 8,275.61 | 7,864.37 | 7,744.25 | 7,960.83 | 31,845.06 |
| 324 | 18,205.54 | 17,950.60 | 16,208.94 | 17,270.74 | 69,635.82 |
| 325 | 28,324.24 | 27,196.22 | 25,910.79 | 26,089.03 | 107,520.28 |
| 326 | 7,101.61 | 6,430.10 | 6,432.89 | 6,769.47 | 26,734.07 |
| 327 | 20,947.10 | 20,054.27 | 18,639.37 | 19,716.82 | 79,357.56 |
| 328 | 9,516.36 | 9,350.47 | 8,907.01 | 9,033.84 | 36,807.68 |
| 329 | 17,500.61 | 16,329.83 | 15,243.16 | 16,168.03 | 65,241.63 |
| 330 | 10,474.58 | 9,990.95 | 8,841.68 | 9,327.10 | 38,634.31 |
| 331 | 5,375.18 | 5,002.67 | 4,703.31 | 4,683.41 | 19,764.57 |
| 333 | 7,028.91 | 6,559.42 | 6,277.48 | 6,563.48 | 26,429.29 |
| 334 | 15,607.02 | 14,458.94 | 13,519.02 | 14,107.70 | 57,692.68 |
| 335 | 13,744.03 | 13,117.84 | 12,126.82 | 12,711.77 | 51,700.46 |
| 336 | 14,028.98 | 13,519.51 | 12,858.84 | 12,629.15 | 53,036.48 |
| 337 | 20,392.27 | 19,317.05 | 17,763.27 | 18,557.44 | 76,030.03 |
| 338 | 13,966.27 | 12,953.61 | 12,021.83 | 12,604.52 | 51,546.23 |
| 339 | 16,386.76 | 14,801.92 | 13,053.74 | 14,662.51 | 58,904.93 |
| 341 | 5,412.10 | 4,129.46 | 3,610.27 | 4,288.56 | 17,440.39 |
| 342 | 19,813.29 | 17,712.21 | 16,295.88 | 17,616.98 | 71,438.36 |
| 344 | 14,754.91 | 14,196.47 | 12,941.83 | 14,061.38 | 55,954.59 |
| 346 | 19,404.10 | 18,670.32 | 17,172.61 | 18,017.66 | 73,264.69 |
| 347 | 9,637.20 | 9,171.07 | 8,398.95 | 8,897.03 | 36,104.25 |
| 349 | 7,982.90 | 7,531.88 | 6,829.12 | 7,481.09 | 29,824.99 |
| TOTAL | 384,180.31 | 363,407.66 | 338,761.81 | 353,668.79 | 1,440,018.57 |

STATE: GEORGIA

| ZIP CODE | QUARTER 1 | QUARTER 2 | QUARTER 3 | QUARTER 4 | TOTAL GRAMS |
|---|---|---|---|---|---|
| 300 | 33,372.96 | 31,953.63 | 30,285.87 | 30,226.25 | 125,838.71 |
| 301 | 35,915.68 | 34,524.22 | 33,070.40 | 33,180.35 | 136,690.65 |
| 302 | 21,057.96 | 20,336.16 | 19,805.39 | 19,174.46 | 80,373.97 |

| | | | | | | |
|---|---|---|---|---|---|---|
| | 303 | 7,499.78 | 6,922.72 | 6,516.58 | 6,277.04 | 27,216.12 |
| | 304 | 7,027.69 | 6,908.16 | 6,570.94 | 6,802.68 | 27,309.47 |
| | 305 | 21,785.66 | 21,315.45 | 20,436.70 | 20,165.67 | 83,703.48 |
| | 306 | 15,862.16 | 15,172.09 | 14,633.10 | 14,809.98 | 60,477.33 |
| | 307 | 13,468.66 | 12,984.96 | 12,499.64 | 12,162.39 | 51,115.65 |
| | 308 | 3,273.05 | 3,083.79 | 2,821.76 | 2,871.17 | 12,049.77 |
| | 309 | 6,591.07 | 5,977.77 | 5,764.36 | 5,711.26 | 24,044.46 |
| | 310 | 23,568.60 | 23,077.65 | 21,568.10 | 22,218.88 | 90,433.23 |
| | 312 | 8,552.01 | 8,103.40 | 7,841.63 | 7,927.50 | 32,424.54 |
| | 313 | 5,541.28 | 5,379.35 | 4,916.04 | 4,580.19 | 20,416.86 |
| | 314 | 5,352.73 | 5,268.78 | 4,916.03 | 4,921.57 | 20,459.11 |
| | 315 | 17,552.69 | 17,053.39 | 16,111.55 | 16,328.62 | 67,046.25 |
| | 316 | 9,177.81 | 9,014.62 | 7,948.59 | 8,513.34 | 34,654.36 |
| | 317 | 11,728.09 | 11,482.79 | 10,770.99 | 10,945.99 | 44,927.86 |
| | 318 | 1,293.74 | 1,203.97 | 1,179.40 | 1,230.60 | 4,907.71 |
| | 319 | 6,665.73 | 6,803.74 | 6,555.41 | 5,881.90 | 25,906.78 |
| | 398 | 3,751.39 | 3,579.35 | 3,354.59 | 3,376.14 | 14,061.47 |
| TOTAL | | 259,038.74 | 250,145.99 | 237,567.07 | 237,305.98 | 984,057.78 |

STATE: GUAM

| ZIP CODE | | QUARTER 1 | QUARTER 2 | QUARTER 3 | QUARTER 4 | TOTAL GRAMS |
|---|---|---|---|---|---|---|
| | 969 | 148.02 | 196.51 | 149.38 | 136.06 | 629.97 |
| TOTAL | | 148.02 | 196.51 | 149.38 | 136.06 | 629.97 |

STATE: HAWAII

| ZIP CODE | | QUARTER 1 | QUARTER 2 | QUARTER 3 | QUARTER 4 | TOTAL GRAMS |
|---|---|---|---|---|---|---|
| | 967 | 13,770.54 | 13,278.33 | 12,607.75 | 12,196.82 | 51,853.44 |
| | 968 | 4,842.77 | 4,451.68 | 4,491.23 | 4,246.40 | 18,032.08 |
| TOTAL | | 18,613.31 | 17,730.01 | 17,098.98 | 16,443.22 | 69,885.52 |

STATE: IDAHO

| ZIP CODE | | QUARTER 1 | QUARTER 2 | QUARTER 3 | QUARTER 4 | TOTAL GRAMS |
|---|---|---|---|---|---|---|
| | 832 | 5,794.76 | 5,738.47 | 5,484.93 | 5,535.61 | 22,553.77 |
| | 833 | 6,851.19 | 6,888.44 | 6,591.69 | 6,741.64 | 27,072.96 |

| | | | | | | |
|---|---|---|---|---|---|---|
| | 834 | 4,773.54 | 4,808.36 | 4,700.02 | 4,677.92 | 18,959.84 |
| | 835 | 3,664.57 | 3,604.08 | 3,535.69 | 3,336.74 | 14,141.08 |
| | 836 | 16,783.40 | 16,530.03 | 16,022.10 | 16,100.89 | 65,436.42 |
| | 837 | 10,576.70 | 10,581.39 | 9,866.77 | 10,004.65 | 41,029.51 |
| | 838 | 10,102.26 | 9,718.67 | 9,611.32 | 9,433.21 | 38,865.46 |
| TOTAL | | 58,546.42 | 57,869.44 | 55,812.52 | 55,830.66 | 228,059.04 |

| STATE: | ILLINOIS | | | | | |
|---|---|---|---|---|---|---|
| ZIP CODE | | QUARTER 1 | QUARTER 2 | QUARTER 3 | QUARTER 4 | TOTAL GRAMS |
| | 600 | 37,900.19 | 37,090.51 | 35,814.16 | 35,750.66 | 146,555.52 |
| | 601 | 32,640.73 | 31,756.10 | 30,383.44 | 30,978.71 | 125,758.98 |
| | 602 | 1,091.14 | 1,138.15 | 1,002.96 | 1,016.32 | 4,248.57 |
| | 603 | 1,545.82 | 1,516.38 | 1,433.30 | 1,467.89 | 5,963.39 |
| | 604 | 41,012.33 | 40,315.63 | 38,217.68 | 38,053.30 | 157,598.94 |
| | 605 | 19,887.48 | 19,149.29 | 18,389.12 | 18,640.88 | 76,066.77 |
| | 606 | 34,966.73 | 33,699.31 | 32,549.74 | 31,531.23 | 132,747.01 |
| | 607 | 3,343.47 | 3,374.05 | 3,180.65 | 3,217.50 | 13,115.67 |
| | 608 | 2,498.81 | 2,519.17 | 2,445.67 | 2,445.55 | 9,909.20 |
| | 609 | 6,403.54 | 6,103.85 | 5,917.71 | 5,742.55 | 24,167.65 |
| | 610 | 6,857.51 | 6,773.20 | 6,538.94 | 6,565.73 | 26,735.38 |
| | 611 | 10,615.44 | 10,481.12 | 10,121.54 | 10,061.35 | 41,279.45 |
| | 612 | 3,658.86 | 3,555.14 | 3,407.03 | 3,393.93 | 14,014.96 |
| | 613 | 4,489.99 | 4,386.57 | 4,205.84 | 4,307.35 | 17,389.75 |
| | 614 | 4,422.84 | 4,365.67 | 4,153.53 | 4,235.17 | 17,177.21 |
| | 615 | 5,889.65 | 5,723.70 | 5,272.16 | 5,462.99 | 22,348.50 |
| | 616 | 8,352.17 | 8,266.06 | 7,986.48 | 7,787.78 | 32,392.49 |
| | 617 | 5,168.55 | 4,995.24 | 4,784.08 | 4,829.91 | 19,777.78 |
| | 618 | 8,919.57 | 8,866.47 | 8,654.73 | 8,421.10 | 34,861.87 |
| | 619 | 2,862.94 | 2,819.48 | 2,590.50 | 2,560.53 | 10,833.45 |
| | 620 | 12,817.02 | 12,567.24 | 12,120.09 | 12,006.15 | 49,510.50 |
| | 622 | 11,954.44 | 11,709.72 | 11,216.59 | 11,026.73 | 45,907.48 |
| | 623 | 4,146.39 | 4,044.72 | 3,880.87 | 3,868 | 15,939.98 |
| | 624 | 4,520.33 | 4,495.57 | 4,409.68 | 4,335.94 | 17,761.52 |
| | 625 | 5,539.99 | 5,566.28 | 5,276.71 | 5,496.03 | 21,879.01 |
| | 626 | 5,159.48 | 4,999.27 | 4,886.49 | 4,715.83 | 19,761.07 |
| | 627 | 6,844.36 | 6,493.27 | 6,261.48 | 6,384.35 | 25,983.46 |

| | | | | | | |
|---|---|---|---|---|---|---|
| | 628 | 8,409.17 | 8,115.44 | 7,957.85 | 7,739.87 | 32,222.33 |
| | 629 | 18,127.12 | 17,885.69 | 17,045.21 | 16,228.41 | 69,286.43 |
| TOTAL | | 320,046.06 | 312,772.29 | 300,104.23 | 298,271.74 | 1,231,194.32 |
| STATE: | | INDIANA | | | | |
| ZIP CODE | | QUARTER 1 | QUARTER 2 | QUARTER 3 | QUARTER 4 | TOTAL GRAMS |
| | 460 | 14,741.47 | 14,288.14 | 13,669.87 | 13,418.05 | 56,117.53 |
| | 461 | 19,487.03 | 19,279.83 | 18,019.29 | 17,680.28 | 74,466.43 |
| | 462 | 32,380.04 | 31,590.99 | 30,817.06 | 30,984.58 | 125,772.67 |
| | 463 | 29,135.87 | 29,122.72 | 27,474.38 | 26,640.69 | 112,373.66 |
| | 464 | 3,989.07 | 3,831.97 | 3,355 | 3,058.47 | 14,234.51 |
| | 465 | 10,926.15 | 10,941.04 | 9,979.94 | 9,886.14 | 41,733.27 |
| | 466 | 3,825.44 | 3,848.01 | 3,471.22 | 3,583.51 | 14,728.18 |
| | 467 | 6,369.07 | 6,240.12 | 5,934.47 | 6,070.80 | 24,614.46 |
| | 468 | 8,607.72 | 8,269.67 | 7,984.43 | 7,694.70 | 32,556.52 |
| | 469 | 13,637.14 | 13,260.08 | 12,488.09 | 12,351.38 | 51,736.69 |
| | 470 | 3,630.98 | 3,587.91 | 3,353.24 | 3,233.05 | 13,805.18 |
| | 471 | 16,539.84 | 16,635.78 | 15,874.03 | 15,053.93 | 64,103.58 |
| | 472 | 7,807.75 | 7,664.63 | 6,984.50 | 6,986.28 | 29,443.16 |
| | 473 | 14,765.83 | 14,117.28 | 13,368.52 | 13,061.62 | 55,313.25 |
| | 474 | 10,310.07 | 9,878.60 | 9,199.79 | 8,975.35 | 38,363.81 |
| | 475 | 7,585.39 | 7,254.21 | 6,822.03 | 6,875.70 | 28,537.33 |
| | 476 | 5,214.38 | 5,081.09 | 4,778.01 | 4,771.17 | 19,844.65 |
| | 477 | 11,983.78 | 12,208.34 | 11,343.88 | 11,407.45 | 46,943.45 |
| | 478 | 5,860.06 | 5,639.99 | 5,058.17 | 4,879.20 | 21,437.42 |
| | 479 | 10,108.21 | 10,022.30 | 9,401.42 | 9,250.13 | 38,782.06 |
| TOTAL | | 236,905.29 | 232,762.70 | 219,377.34 | 215,862.48 | 904,907.81 |
| STATE: | | IOWA | | | | |
| ZIP CODE | | QUARTER 1 | QUARTER 2 | QUARTER 3 | QUARTER 4 | TOTAL GRAMS |
| | 500 | 4,421.90 | 4,405.68 | 4,207.71 | 4,251.43 | 17,286.72 |
| | 501 | 3,835.93 | 3,897.94 | 3,810.33 | 3,665.70 | 15,209.90 |
| | 502 | 4,061.78 | 3,957.64 | 3,860.62 | 3,874.95 | 15,754.99 |
| | 503 | 9,720.81 | 9,465.81 | 9,148.95 | 9,000.05 | 37,335.62 |
| | 504 | 2,108.74 | 2,125.06 | 2,026.28 | 1,857.80 | 8,117.88 |

| | | | | | | |
|---|---|---|---|---|---|---|
| | 505 | 2,104.53 | 2,058.34 | 1,983.74 | 2,036.09 | 8,182.70 |
| | 506 | 1,815.34 | 1,738.03 | 1,747.81 | 1,741.15 | 7,042.33 |
| | 507 | 1,654.54 | 1,627.45 | 1,552.90 | 1,508.94 | 6,343.83 |
| | 508 | 692.07 | 726.99 | 694.95 | 695.72 | 2,809.73 |
| | 510 | 1,042.50 | 998.9 | 966.67 | 994.2 | 4,002.27 |
| | 511 | 2,109.05 | 2,173.38 | 2,109.24 | 2,167.89 | 8,559.56 |
| | 512 | 552.12 | 441.08 | 468.97 | 474.17 | 1,936.34 |
| | 513 | 763.91 | 767.45 | 751.97 | 761.68 | 3,045.01 |
| | 514 | 809.59 | 796 | 764.55 | 740.23 | 3,110.37 |
| | 515 | 2,918.09 | 2,845.89 | 2,562.34 | 2,476.23 | 10,802.55 |
| | 516 | 527.63 | 520.77 | 522.31 | 484.49 | 2,055.20 |
| | 520 | 1,961.07 | 1,862.74 | 1,764.64 | 1,814.34 | 7,402.79 |
| | 521 | 597.89 | 566.42 | 578.79 | 547.21 | 2,290.31 |
| | 522 | 2,842.39 | 2,714.45 | 2,616.95 | 2,441.26 | 10,615.05 |
| | 523 | 1,589.74 | 1,628.95 | 1,458.16 | 1,432.55 | 6,109.40 |
| | 524 | 2,637.83 | 2,606.57 | 2,537.04 | 2,531.94 | 10,313.38 |
| | 525 | 3,141.74 | 3,224.14 | 3,116.46 | 3,040.20 | 12,522.54 |
| | 526 | 2,325.70 | 2,372.13 | 2,247.64 | 2,230.69 | 9,176.16 |
| | 527 | 2,286.38 | 2,211.73 | 2,100.33 | 2,074.54 | 8,672.98 |
| | 528 | 2,873.26 | 2,843.03 | 2,602.25 | 2,661.18 | 10,979.72 |
| TOTAL | | 59,394.53 | 58,576.57 | 56,201.60 | 55,504.63 | 229,677.33 |

| STATE: | KANSAS | | | | | |
|---|---|---|---|---|---|---|
| ZIP CODE | | QUARTER 1 | QUARTER 2 | QUARTER 3 | QUARTER 4 | TOTAL GRAMS |
| | 660 | 11,096.83 | 10,743.54 | 10,412.69 | 10,268.35 | 42,521.41 |
| | 661 | 3,494.39 | 3,351.26 | 3,285.21 | 3,156.38 | 13,287.24 |
| | 662 | 8,409.98 | 8,122.74 | 7,900 | 7,991.65 | 32,424.37 |
| | 664 | 1,430.21 | 1,365.60 | 1,439.12 | 1,286.86 | 5,521.79 |
| | 665 | 1,901.50 | 1,968.64 | 1,871.79 | 1,859.29 | 7,601.22 |
| | 666 | 4,935.94 | 4,720.32 | 4,442.43 | 4,352.97 | 18,451.66 |
| | 667 | 5,171.84 | 5,081.44 | 4,887.79 | 4,797.65 | 19,938.72 |
| | 668 | 1,328.37 | 1,359.96 | 1,245.13 | 1,244.90 | 5,178.36 |
| | 669 | 700.06 | 667.71 | 633.55 | 629.77 | 2,631.09 |
| | 670 | 6,503.79 | 6,496.69 | 6,223.97 | 6,140.79 | 25,365.24 |
| | 671 | 3,597.32 | 3,591.19 | 3,476.27 | 3,434.29 | 14,099.07 |
| | 672 | 18,991.36 | 18,545.74 | 18,073.25 | 17,967.34 | 73,577.69 |

| | | | | | | |
|---|---|---|---|---|---|---|
| | 673 | 2,715.13 | 2,784.40 | 2,562 | 2,615.68 | 10,677.21 |
| | 674 | 4,613.78 | 4,506.69 | 4,340.62 | 4,327.27 | 17,788.36 |
| | 675 | 3,760.94 | 3,550.57 | 3,456.73 | 3,541.71 | 14,309.95 |
| | 676 | 1,447.21 | 1,512.72 | 1,345.13 | 1,402.15 | 5,707.21 |
| | 677 | 700.27 | 666.03 | 631.47 | 683.14 | 2,680.91 |
| | 678 | 2,360.49 | 2,311.09 | 2,181.08 | 2,235.21 | 9,087.87 |
| | 679 | 1,000 | 896.16 | 978.85 | 946.76 | 3,821.77 |
| TOTAL | | 84,159.41 | 82,242.49 | 79,387.08 | 78,882.16 | 324,671.14 |

STATE: KENTUCKY

| ZIP CODE | | QUARTER 1 | QUARTER 2 | QUARTER 3 | QUARTER 4 | TOTAL GRAMS |
|---|---|---|---|---|---|---|
| | 400 | 8,351.37 | 8,031.57 | 7,762.56 | 7,559.08 | 31,704.58 |
| | 401 | 4,644.27 | 4,779.75 | 4,378.23 | 4,271.67 | 18,073.92 |
| | 402 | 37,502.32 | 36,424.01 | 34,345.92 | 34,259.33 | 142,531.58 |
| | 403 | 11,585.29 | 11,370.04 | 10,884.07 | 10,972.03 | 44,811.43 |
| | 404 | 7,413.70 | 7,284.19 | 7,076.17 | 7,048.29 | 28,822.35 |
| | 405 | 8,217.53 | 7,710.55 | 7,616.50 | 7,404 | 30,948.58 |
| | 406 | 2,062.41 | 1,982.94 | 2,036.58 | 1,904.04 | 7,985.97 |
| | 407 | 6,449.63 | 6,334.40 | 5,988.20 | 6,177.44 | 24,949.67 |
| | 408 | 1,834.91 | 1,811.53 | 1,689.23 | 1,738.27 | 7,073.94 |
| | 409 | 8,473.60 | 7,874.82 | 7,768.18 | 7,638.70 | 31,755.30 |
| | 410 | 10,595.25 | 10,335.12 | 9,739.31 | 9,753.40 | 40,423.08 |
| | 411 | 4,028.28 | 3,991.80 | 3,704.90 | 3,684.80 | 15,409.78 |
| | 412 | 3,019.61 | 3,139.34 | 3,059.16 | 3,007.91 | 12,226.02 |
| | 413 | 3,008.07 | 2,970.24 | 2,860.47 | 2,873.75 | 11,712.53 |
| | 414 | 1,335.07 | 1,345.17 | 1,233.64 | 1,256.67 | 5,170.55 |
| | 415 | 6,987.40 | 6,905.67 | 6,622.04 | 6,168.27 | 26,683.38 |
| | 416 | 4,684.41 | 4,577.86 | 4,357.54 | 4,246.01 | 17,865.82 |
| | 417 | 4,922.05 | 4,967.18 | 4,674.57 | 4,477.64 | 19,041.44 |
| | 418 | 2,077.49 | 2,100.97 | 2,014.84 | 2,009.66 | 8,202.96 |
| | 420 | 14,948.36 | 14,958.03 | 13,706.43 | 13,900.14 | 57,512.96 |
| | 421 | 13,270.72 | 12,758.67 | 12,378.97 | 12,548.66 | 50,957.02 |
| | 422 | 6,643.05 | 6,481.36 | 6,570.17 | 6,406.05 | 26,100.63 |
| | 423 | 10,112.97 | 9,779.19 | 9,399.33 | 9,324.34 | 38,615.83 |
| | 424 | 10,233.91 | 10,182.88 | 9,485.36 | 9,577.46 | 39,479.61 |
| | 425 | 2,953.44 | 2,896.86 | 2,476.29 | 2,805.82 | 11,132.41 |

| | | | | | | |
|---|---|---|---|---|---|---|
| | 426 | 3,840.56 | 3,702.96 | 3,707.56 | 3,539.19 | 14,790.27 |
| | 427 | 9,252.66 | 8,899.87 | 8,827.36 | 8,607.30 | 35,587.19 |
| TOTAL | | 208,448.33 | 203,596.97 | 194,363.58 | 193,159.92 | 799,568.80 |

STATE: LOUISIANA

| ZIP CODE | | QUARTER 1 | QUARTER 2 | QUARTER 3 | QUARTER 4 | TOTAL GRAMS |
|---|---|---|---|---|---|---|
| | 700 | 18,544.42 | 17,428.49 | 16,264.09 | 15,448.49 | 67,685.49 |
| | 701 | 8,214.66 | 7,521.16 | 7,066.42 | 6,650.72 | 29,452.96 |
| | 703 | 11,962.20 | 11,223.48 | 10,747.81 | 10,291.33 | 44,224.82 |
| | 704 | 17,373.97 | 16,943.32 | 16,151.73 | 15,345.52 | 65,814.54 |
| | 705 | 28,543.57 | 28,223.46 | 27,074.87 | 27,011.58 | 110,853.48 |
| | 706 | 8,030.29 | 7,709.25 | 7,356.24 | 7,518.69 | 30,614.47 |
| | 707 | 14,989.79 | 14,436.07 | 13,598.75 | 13,478.08 | 56,502.69 |
| | 708 | 12,826.86 | 12,223.38 | 11,375.20 | 11,188.28 | 47,613.72 |
| | 710 | 4,891.87 | 4,748.42 | 4,396.50 | 4,663.25 | 18,700.04 |
| | 711 | 15,459.03 | 14,668.60 | 14,472.57 | 13,784.04 | 58,384.24 |
| | 712 | 12,404.31 | 11,791.04 | 11,238.62 | 11,387.67 | 46,821.64 |
| | 713 | 14,310.70 | 13,794.98 | 13,096.53 | 12,980.05 | 54,182.26 |
| | 714 | 5,683.59 | 5,354.70 | 5,122.95 | 5,068.57 | 21,229.81 |
| TOTAL | | 173,235.26 | 166,066.35 | 157,962.28 | 154,816.27 | 652,080.16 |

STATE: MAINE

| ZIP CODE | | QUARTER 1 | QUARTER 2 | QUARTER 3 | QUARTER 4 | TOTAL GRAMS |
|---|---|---|---|---|---|---|
| | 039 | 206.01 | 176.16 | 187.7 | 168.98 | 738.85 |
| | 040 | 4,458.16 | 4,340.63 | 4,238.10 | 4,217.05 | 17,253.94 |
| | 041 | 933.08 | 896.07 | 944.24 | 888.95 | 3,662.34 |
| | 042 | 2,318.57 | 2,140.15 | 2,126.46 | 2,096.30 | 8,681.48 |
| | 043 | 2,056.34 | 1,949.49 | 1,947.26 | 1,918.33 | 7,871.42 |
| | 044 | 2,178.78 | 2,112.83 | 1,964.59 | 1,904.81 | 8,161.01 |
| | 045 | 535.5 | 547.48 | 546.81 | 527.09 | 2,156.88 |
| | 046 | 1,134.32 | 1,111.83 | 1,123.21 | 1,074.11 | 4,443.47 |
| | 047 | 1,159.03 | 1,216.75 | 1,057.06 | 1,102.86 | 4,535.70 |
| | 048 | 506.29 | 506.68 | 470.31 | 460.86 | 1,944.14 |
| | 049 | 2,290.38 | 2,298.41 | 2,233.27 | 2,188.52 | 9,010.58 |
| TOTAL | | 17,776.46 | 17,296.48 | 16,839.01 | 16,547.86 | 68,459.81 |

STATE: MARYLAND

| ZIP CODE | QUARTER 1 | QUARTER 2 | QUARTER 3 | QUARTER 4 | TOTAL GRAMS |
|---|---|---|---|---|---|
| 206 | 2,830.68 | 2,704.79 | 2,622.43 | 2,612.61 | 10,770.51 |
| 207 | 3,448.50 | 3,176.04 | 2,965.91 | 2,952.57 | 12,543.02 |
| 208 | 2,770.76 | 2,660.79 | 2,412.13 | 2,362.52 | 10,206.20 |
| 209 | 631.64 | 621.91 | 522.38 | 573.83 | 2,349.76 |
| 210 | 6,249.38 | 5,771.44 | 5,505.30 | 5,337.94 | 22,864.06 |
| 211 | 3,172.10 | 2,951.50 | 2,735.13 | 2,711.01 | 11,569.74 |
| 212 | 6,899.30 | 6,584.51 | 6,203.10 | 6,085.62 | 25,772.53 |
| 214 | 765.41 | 666.17 | 656.45 | 645.81 | 2,733.84 |
| 215 | 2,738.43 | 2,606.34 | 2,319.51 | 2,222.01 | 9,886.29 |
| 216 | 1,544.72 | 1,416.56 | 1,381.62 | 1,395.26 | 5,738.16 |
| 217 | 4,185.26 | 3,841.69 | 3,667.83 | 3,772.03 | 15,466.81 |
| 218 | 1,594.96 | 1,462.40 | 1,418.40 | 1,364.71 | 5,840.47 |
| 219 | 737.95 | 667.33 | 622.05 | 699.28 | 2,726.61 |
| TOTAL | 37,569.09 | 35,131.47 | 33,032.24 | 32,735.20 | 138,468 |

STATE: MASSACHUSETTS

| ZIP CODE | QUARTER 1 | QUARTER 2 | QUARTER 3 | QUARTER 4 | TOTAL GRAMS |
|---|---|---|---|---|---|
| 10 | 2,376.37 | 2,275.55 | 2,167.47 | 2,133.28 | 8,952.67 |
| 11 | 511.22 | 492.76 | 434.9 | 445.18 | 1,884.06 |
| 12 | 662.01 | 618.1 | 609.19 | 620.36 | 2,509.66 |
| 13 | 347.1 | 376.6 | 326.93 | 360.96 | 1,411.59 |
| 14 | 1,354.33 | 1,319.28 | 1,234.50 | 1,191.26 | 5,099.37 |
| 15 | 2,127.24 | 1,997.57 | 1,929.01 | 1,958.85 | 8,012.67 |
| 16 | 1,383.50 | 1,257.86 | 1,157.11 | 1,176.61 | 4,975.08 |
| 17 | 1,482.51 | 1,507.47 | 1,407.08 | 1,465.24 | 5,862.30 |
| 18 | 5,873.91 | 5,724.77 | 5,389.40 | 5,195.75 | 22,183.83 |
| 19 | 2,806.97 | 2,683.04 | 2,581.58 | 2,637.80 | 10,709.39 |
| 20 | 1,503.05 | 1,422.37 | 1,289.39 | 1,368.31 | 5,583.12 |
| 21 | 3,459.66 | 3,343.40 | 3,142.23 | 3,169.78 | 13,115.07 |
| 22 | 76.03 | 79.76 | 66.31 | 47.05 | 269.15 |
| 23 | 2,340.12 | 2,222.81 | 2,089.71 | 2,015.21 | 8,667.85 |
| 24 | 839.19 | 816.24 | 734.61 | 804.55 | 3,194.59 |

| | | | | | | |
|---|---|---|---|---|---|---|
| | 25 | 556.8 | 554.97 | 542.97 | 502.45 | 2,157.19 |
| | 26 | 918.48 | 927.7 | 867.04 | 880.35 | 3,593.57 |
| | 27 | 3,187.97 | 3,178.45 | 3,067.80 | 3,055.81 | 12,490.03 |
| TOTAL | | 31,806.46 | 30,798.70 | 29,037.23 | 29,028.80 | 120,671.19 |

| STATE: | MICHIGAN | | | | | |
|---|---|---|---|---|---|---|
| ZIP CODE | | QUARTER 1 | QUARTER 2 | QUARTER 3 | QUARTER 4 | TOTAL GRAMS |
| | 480 | 56,093.96 | 55,010.89 | 52,498.72 | 50,420.03 | 214,023.60 |
| | 481 | 70,871.60 | 70,217.16 | 66,229.98 | 63,235.94 | 270,554.68 |
| | 482 | 33,235.82 | 31,645.41 | 29,010.32 | 28,430.71 | 122,322.26 |
| | 483 | 36,008.97 | 35,133.24 | 33,460.12 | 32,189.87 | 136,792.20 |
| | 484 | 19,743.36 | 19,211.93 | 18,893.52 | 18,089.68 | 75,938.49 |
| | 485 | 12,003.08 | 12,363.10 | 11,638.44 | 11,343.53 | 47,348.15 |
| | 486 | 21,822.11 | 21,602.97 | 20,416.14 | 19,377.99 | 83,219.21 |
| | 487 | 11,617.51 | 11,394.02 | 10,735.93 | 10,247.15 | 43,994.61 |
| | 488 | 21,321.29 | 21,295.74 | 20,422.94 | 19,568.11 | 82,608.08 |
| | 489 | 8,698.19 | 8,413.71 | 8,152.08 | 7,730.22 | 32,994.20 |
| | 490 | 30,459.37 | 29,218.03 | 28,076.98 | 26,397.79 | 114,152.17 |
| | 491 | 2,807.12 | 2,730.15 | 2,538.53 | 2,547.45 | 10,623.25 |
| | 492 | 12,896.87 | 12,646.08 | 12,307.41 | 11,800.04 | 49,650.40 |
| | 493 | 9,066.96 | 9,007.97 | 8,627.32 | 8,229.54 | 34,931.79 |
| | 494 | 23,016.32 | 22,767.61 | 21,878.30 | 21,347.02 | 89,009.25 |
| | 495 | 16,837.35 | 17,116.10 | 16,181.81 | 14,984.45 | 65,119.71 |
| | 496 | 11,327.44 | 11,211.63 | 11,086.23 | 10,465.64 | 44,090.94 |
| | 497 | 9,557.11 | 9,606 | 9,212.32 | 8,647.78 | 37,023.21 |
| | 498 | 8,336.75 | 7,782.80 | 7,763.71 | 7,245.98 | 31,129.24 |
| | 499 | 2,543.92 | 2,448.25 | 2,424.95 | 2,241.64 | 9,658.76 |
| TOTAL | | 418,265.10 | 410,822.79 | 391,555.75 | 374,540.56 | 1,595,184.20 |

| STATE: | MINNESOTA | | | | | |
|---|---|---|---|---|---|---|
| ZIP CODE | | QUARTER 1 | QUARTER 2 | QUARTER 3 | QUARTER 4 | TOTAL GRAMS |
| | 550 | 4,915.67 | 4,615.60 | 4,551.39 | 4,351.63 | 18,434.29 |
| | 551 | 5,254.95 | 4,973.58 | 4,940.46 | 4,696.29 | 19,865.28 |
| | 553 | 6,475.11 | 6,150.15 | 6,069.36 | 6,018.10 | 24,712.72 |
| | 554 | 8,444.13 | 7,992.94 | 7,840.39 | 7,509.79 | 31,787.25 |

| | ZIP CODE | QUARTER 1 | QUARTER 2 | QUARTER 3 | QUARTER 4 | TOTAL GRAMS |
|---|---|---|---|---|---|---|
| | 556 | 251.01 | 221.72 | 231.9 | 226.79 | 931.42 |
| | 557 | 3,491.56 | 3,441.81 | 3,251.74 | 3,208.29 | 13,393.40 |
| | 558 | 2,483.59 | 2,390.16 | 2,281.30 | 2,322.08 | 9,477.13 |
| | 559 | 2,246.37 | 2,119.37 | 2,028.23 | 1,947.29 | 8,341.26 |
| | 560 | 2,008.87 | 1,941.30 | 1,889.79 | 1,784.40 | 7,624.36 |
| | 561 | 865.75 | 855.26 | 793.99 | 774.69 | 3,289.69 |
| | 562 | 1,670.30 | 1,592.72 | 1,516.07 | 1,453.65 | 6,232.74 |
| | 563 | 3,995.42 | 3,797.31 | 3,595.25 | 3,418.38 | 14,806.36 |
| | 564 | 1,859.98 | 1,783.66 | 1,760.07 | 1,642.92 | 7,046.63 |
| | 565 | 2,132.47 | 2,118.10 | 2,010.43 | 1,913.20 | 8,174.20 |
| | 566 | 1,255.91 | 1,264.14 | 1,170.73 | 1,114.40 | 4,805.18 |
| | 567 | 979.49 | 911.75 | 886.59 | 882.02 | 3,659.85 |
| TOTAL | | 48,330.58 | 46,169.57 | 44,817.69 | 43,263.92 | 182,581.76 |

STATE: MISSISSIPPI

| ZIP CODE | | QUARTER 1 | QUARTER 2 | QUARTER 3 | QUARTER 4 | TOTAL GRAMS |
|---|---|---|---|---|---|---|
| | 386 | 11,128.94 | 10,809.02 | 9,995.51 | 9,770.03 | 41,703.50 |
| | 387 | 3,432.66 | 3,411.10 | 3,091.30 | 2,760.28 | 12,695.34 |
| | 388 | 14,433.56 | 13,391.42 | 12,642 | 12,325.72 | 52,792.70 |
| | 389 | 3,841.81 | 3,740.99 | 3,386.15 | 3,131.03 | 14,099.98 |
| | 390 | 8,641.47 | 8,134.69 | 7,386.96 | 6,835.79 | 30,998.91 |
| | 391 | 9,096 | 8,653.34 | 7,673.54 | 7,226.58 | 32,649.46 |
| | 392 | 9,105.27 | 8,483.68 | 7,915.20 | 7,208.25 | 32,712.40 |
| | 393 | 12,567.11 | 12,128.84 | 10,904.71 | 10,418.54 | 46,019.20 |
| | 394 | 21,722.44 | 20,899.51 | 20,000.89 | 18,756.32 | 81,379.16 |
| | 395 | 20,822.15 | 20,209.44 | 18,998.61 | 18,445.80 | 78,476 |
| | 396 | 5,615.41 | 5,652.33 | 5,197.67 | 4,866.39 | 21,331.80 |
| | 397 | 5,929.40 | 5,665.97 | 5,168.65 | 4,924.72 | 21,688.74 |
| TOTAL | | 126,336.22 | 121,180.33 | 112,361.19 | 106,669.45 | 466,547.19 |

STATE: MISSOURI

| ZIP CODE | | QUARTER 1 | QUARTER 2 | QUARTER 3 | QUARTER 4 | TOTAL GRAMS |
|---|---|---|---|---|---|---|
| | 630 | 17,359.94 | 17,091.28 | 16,032.73 | 16,129.65 | 66,613.60 |
| | 631 | 23,122.31 | 22,945.94 | 21,832.41 | 21,109.06 | 89,009.72 |
| | 633 | 10,168.16 | 9,962.32 | 9,369.39 | 9,555.07 | 39,054.94 |

| | | | | | | |
|---|---|---|---|---|---|---|
| | 634 | 2,022.42 | 2,035.47 | 1,942.23 | 1,989.94 | 7,990.06 |
| | 635 | 1,812.09 | 1,922.58 | 1,694.75 | 1,761.15 | 7,190.57 |
| | 636 | 7,138.90 | 7,431.98 | 7,084.29 | 6,673.83 | 28,329 |
| | 637 | 5,630.49 | 5,486.28 | 5,165.56 | 4,836.75 | 21,119.08 |
| | 638 | 10,157.33 | 10,158.72 | 9,725.19 | 8,965.29 | 39,006.53 |
| | 639 | 7,011.09 | 6,895.69 | 6,161.85 | 5,724.65 | 25,793.28 |
| | 640 | 17,359.72 | 16,891.33 | 16,463.96 | 16,430.83 | 67,145.84 |
| | 641 | 13,832.45 | 13,278.93 | 12,565.66 | 12,268.28 | 51,945.32 |
| | 644 | 2,207.23 | 2,156.58 | 2,034.43 | 2,112.07 | 8,510.31 |
| | 645 | 4,603.96 | 4,285.94 | 4,102.60 | 3,989.40 | 16,981.90 |
| | 646 | 1,600.33 | 1,569.42 | 1,463.74 | 1,475.14 | 6,108.63 |
| | 647 | 4,762.61 | 4,649.25 | 4,295.49 | 4,226.62 | 17,933.97 |
| | 648 | 7,596.89 | 7,513.25 | 6,975.42 | 6,783.51 | 28,869.07 |
| | 650 | 3,384.67 | 3,421.32 | 3,200.86 | 3,236.03 | 13,242.88 |
| | 651 | 1,678.47 | 1,689.12 | 1,553.15 | 1,582.43 | 6,503.17 |
| | 652 | 4,938.45 | 4,837.03 | 4,480.79 | 4,612.69 | 18,868.96 |
| | 653 | 2,381.22 | 2,421.17 | 2,338.72 | 2,238.97 | 9,380.08 |
| | 654 | 2,301.24 | 2,259.96 | 2,217.35 | 2,088.36 | 8,866.91 |
| | 655 | 3,852.24 | 3,860.55 | 3,709.59 | 3,622.63 | 15,045.01 |
| | 656 | 7,484.38 | 7,564.45 | 7,225.30 | 6,885.87 | 29,160 |
| | 657 | 11,156.93 | 11,401.03 | 10,621.06 | 10,680.05 | 43,859.07 |
| | 658 | 10,826.02 | 10,451.66 | 10,212.45 | 9,930.92 | 41,421.05 |
| TOTAL | | 184,389.54 | 182,181.25 | 172,468.97 | 168,909.19 | 707,948.95 |

| STATE: | | MONTANA | | | | |
|---|---|---|---|---|---|---|
| ZIP CODE | | QUARTER 1 | QUARTER 2 | QUARTER 3 | QUARTER 4 | TOTAL GRAMS |
| | 590 | 1,181.70 | 1,099.71 | 1,172.37 | 1,179.63 | 4,633.41 |
| | 591 | 3,589.13 | 3,502.31 | 3,317.32 | 3,351.29 | 13,760.05 |
| | 592 | 562.04 | 584.45 | 529.13 | 526.54 | 2,202.16 |
| | 593 | 758.77 | 743.83 | 695.63 | 719.45 | 2,917.68 |
| | 594 | 3,308.42 | 3,122.36 | 3,062.37 | 2,915.30 | 12,408.45 |
| | 595 | 703.06 | 697.75 | 641.52 | 647.16 | 2,689.49 |
| | 596 | 3,073.86 | 3,132.19 | 2,993.37 | 2,881.94 | 12,081.36 |
| | 597 | 3,161.03 | 3,037.79 | 2,974.76 | 2,859.44 | 12,033.02 |
| | 598 | 6,310.87 | 6,240.01 | 5,979.36 | 5,898.65 | 24,428.89 |
| | 599 | 4,140.72 | 3,929.42 | 3,923.14 | 3,730.65 | 15,723.93 |

| | | | | | | |
|---|---|---|---|---|---|---|
| TOTAL | | 26,789.60 | 26,089.82 | 25,288.97 | 24,710.05 | 102,878.44 |

| STATE: | NEBRASKA | | | | | |
|---|---|---|---|---|---|---|
| ZIP CODE | | QUARTER 1 | QUARTER 2 | QUARTER 3 | QUARTER 4 | TOTAL GRAMS |
| | 680 | 3,665.17 | 3,578.74 | 3,546.07 | 3,564.78 | 14,354.76 |
| | 681 | 12,042.97 | 11,373.09 | 10,902.91 | 10,700 | 45,018.97 |
| | 683 | 1,459.05 | 1,422.41 | 1,310.90 | 1,318.59 | 5,510.95 |
| | 684 | 1,002.09 | 959.5 | 990.46 | 935.55 | 3,887.60 |
| | 685 | 5,259.74 | 4,996.51 | 5,027.32 | 5,075.85 | 20,359.42 |
| | 686 | 871.8 | 859.36 | 798.69 | 835.49 | 3,365.34 |
| | 687 | 1,833.90 | 1,855.08 | 1,873.67 | 1,870.44 | 7,433.09 |
| | 688 | 3,066.76 | 3,000.04 | 2,872.62 | 2,922.26 | 11,861.68 |
| | 689 | 1,202.89 | 1,162.31 | 1,184.53 | 1,197.57 | 4,747.30 |
| | 690 | 478.57 | 371.25 | 429.54 | 404.01 | 1,683.37 |
| | 691 | 1,609.40 | 1,550.54 | 1,551.54 | 1,479.17 | 6,190.65 |
| | 692 | 199.97 | 161.94 | 201 | 185.3 | 748.21 |
| | 693 | 1,294.52 | 1,192.78 | 1,183.43 | 1,139.34 | 4,810.07 |
| TOTAL | | 33,986.83 | 32,483.55 | 31,872.68 | 31,628.35 | 129,971.41 |

| STATE: | NEVADA | | | | | |
|---|---|---|---|---|---|---|
| ZIP CODE | | QUARTER 1 | QUARTER 2 | QUARTER 3 | QUARTER 4 | TOTAL GRAMS |
| | 890 | 25,124.22 | 24,470.08 | 23,033.28 | 23,304.71 | 95,932.29 |
| | 891 | 50,948.63 | 48,958.70 | 46,495.43 | 46,602.11 | 193,004.87 |
| | 893 | 514.61 | 508.02 | 523.66 | 477.75 | 2,024.04 |
| | 894 | 8,730.40 | 8,685.98 | 8,386.01 | 8,387.17 | 34,189.56 |
| | 895 | 9,722.05 | 9,847.69 | 8,962.19 | 8,700.54 | 37,232.47 |
| | 897 | 2,688.96 | 2,713.02 | 2,543.41 | 2,655.69 | 10,601.08 |
| | 898 | 1,183.35 | 1,167.45 | 1,088.72 | 1,081.33 | 4,520.85 |
| TOTAL | | 98,912.22 | 96,350.94 | 91,032.70 | 91,209.30 | 377,505.16 |

| STATE: | NEW HAMPSHIRE | | | | | |
|---|---|---|---|---|---|---|
| ZIP CODE | | QUARTER 1 | QUARTER 2 | QUARTER 3 | QUARTER 4 | TOTAL GRAMS |
| | 030 | 2,239.69 | 2,104.99 | 2,114.37 | 2,156.70 | 8,615.75 |
| | 031 | 1,364.90 | 1,271.36 | 1,222.39 | 1,251.14 | 5,109.79 |

| | | | | | | |
|---|---|---|---|---|---|---|
| | 032 | 1,159.60 | 1,132.46 | 1,088.21 | 1,054.79 | 4,435.06 |
| | 033 | 706.84 | 662.4 | 656.58 | 639.77 | 2,665.59 |
| | 034 | 569.62 | 539.67 | 536.01 | 511.19 | 2,156.49 |
| | 035 | 371.66 | 366.18 | 335.42 | 334.49 | 1,407.75 |
| | 036 | 36.62 | 39.53 | 32.66 | 34.01 | 142.82 |
| | 037 | 489.28 | 463.5 | 419.83 | 434.66 | 1,807.27 |
| | 038 | 2,553.48 | 2,452.86 | 2,347.01 | 2,429.49 | 9,782.84 |
| TOTAL | | 9,491.69 | 9,032.95 | 8,752.48 | 8,846.24 | 36,123.36 |

STATE: NEW JERSEY

| ZIP CODE | | QUARTER 1 | QUARTER 2 | QUARTER 3 | QUARTER 4 | TOTAL GRAMS |
|---|---|---|---|---|---|---|
| | 070 | 4,449.83 | 3,961.61 | 3,702.72 | 3,830.34 | 15,944.50 |
| | 071 | 413.45 | 345.12 | 347.26 | 325.54 | 1,431.37 |
| | 072 | 252.4 | 203.28 | 194.92 | 200.09 | 850.69 |
| | 073 | 381.35 | 396.23 | 345.55 | 333.87 | 1,457 |
| | 074 | 2,312.89 | 2,023.49 | 1,960.77 | 1,979.70 | 8,276.85 |
| | 075 | 294.46 | 233.11 | 276.55 | 235.25 | 1,039.37 |
| | 076 | 1,476.23 | 1,343.83 | 1,209.21 | 1,242.55 | 5,271.82 |
| | 077 | 3,344.62 | 2,954.21 | 2,846.86 | 2,859.34 | 12,005.03 |
| | 078 | 2,128.38 | 1,894 | 1,677.31 | 1,693.47 | 7,393.16 |
| | 079 | 878.58 | 717.29 | 653.73 | 667.9 | 2,917.50 |
| | 080 | 8,911.86 | 8,250.34 | 7,758.61 | 7,483.90 | 32,404.71 |
| | 081 | 639.79 | 569.03 | 566.3 | 566.05 | 2,341.17 |
| | 082 | 2,234.85 | 2,197.01 | 2,122.53 | 2,025.84 | 8,580.23 |
| | 083 | 1,954.15 | 1,759.69 | 1,734.80 | 1,717.60 | 7,166.24 |
| | 084 | 318.14 | 306.75 | 269.22 | 288.82 | 1,182.93 |
| | 085 | 1,085.19 | 922.5 | 841.25 | 902.97 | 3,751.91 |
| | 086 | 1,199.80 | 1,075.77 | 1,070.81 | 1,047.46 | 4,393.84 |
| | 087 | 2,717.88 | 2,436.13 | 2,393.83 | 2,316.40 | 9,864.24 |
| | 088 | 3,076.45 | 2,794.99 | 2,606.79 | 2,644.73 | 11,122.96 |
| | 089 | 211.22 | 186.04 | 179.3 | 155.93 | 732.49 |
| TOTAL | | 38,281.52 | 34,570.42 | 32,758.32 | 32,517.75 | 138,128.01 |

STATE: NEW MEXICO

| ZIP CODE | | QUARTER 1 | QUARTER 2 | QUARTER 3 | QUARTER 4 | TOTAL GRAMS |
|---|---|---|---|---|---|---|

| | | | | | | |
|---|---|---|---|---|---|---|
| | 870 | 2,775.40 | 2,636.57 | 2,443.32 | 2,549.02 | 10,404.31 |
| | 871 | 13,189.75 | 12,717.66 | 11,699.75 | 11,499.31 | 49,106.47 |
| | 873 | 656.86 | 640.61 | 598.04 | 622.54 | 2,518.05 |
| | 874 | 2,018.86 | 1,998.34 | 1,693.14 | 1,713.07 | 7,423.41 |
| | 875 | 4,213.33 | 3,944.16 | 3,891.40 | 3,858.67 | 15,907.56 |
| | 877 | 966.78 | 926.92 | 915.13 | 896.48 | 3,705.31 |
| | 878 | 235.44 | 215.05 | 226.56 | 226.73 | 903.78 |
| | 879 | 272.4 | 254.75 | 250.39 | 226.81 | 1,004.35 |
| | 880 | 3,153.94 | 3,017.12 | 2,897.98 | 2,883.10 | 11,952.14 |
| | 881 | 1,299.96 | 1,311.60 | 1,363.67 | 1,258.96 | 5,234.19 |
| | 882 | 6,141.67 | 6,032.85 | 5,823.73 | 5,768.02 | 23,766.27 |
| | 883 | 1,657.80 | 1,651.92 | 1,605.11 | 1,571.27 | 6,486.10 |
| | 884 | 325.53 | 277.77 | 247.42 | 281 | 1,131.72 |
| TOTAL | | 36,907.72 | 35,625.32 | 33,655.64 | 33,354.98 | 139,543.66 |

STATE: NEW YORK

| ZIP CODE | QUARTER 1 | QUARTER 2 | QUARTER 3 | QUARTER 4 | TOTAL GRAMS |
|---|---|---|---|---|---|
| 063 | 0.3 | 0 | 0.24 | 0 | 0.54 |
| 100 | 4,602.45 | 4,398.72 | 3,935.58 | 3,972.72 | 16,909.47 |
| 101 | 263.74 | 197.74 | 159.85 | 184.82 | 806.15 |
| 102 | 21.19 | 11.39 | 18.65 | 12.89 | 64.12 |
| 103 | 2,774.50 | 2,655.08 | 2,497.62 | 2,442.32 | 10,369.52 |
| 104 | 1,744.89 | 1,663.22 | 1,545.35 | 1,386.51 | 6,339.97 |
| 105 | 2,767.93 | 2,629.94 | 2,429.42 | 2,367.39 | 10,194.68 |
| 106 | 379.15 | 325.24 | 337.46 | 339.82 | 1,381.67 |
| 107 | 921.33 | 916.24 | 800.7 | 768.21 | 3,406.48 |
| 108 | 235.39 | 261.52 | 225.61 | 226.5 | 949.02 |
| 109 | 2,899.76 | 2,670.90 | 2,532.76 | 2,448.89 | 10,552.31 |
| 110 | 1,045.39 | 1,002.83 | 971.96 | 934.7 | 3,954.88 |
| 111 | 577.75 | 577.19 | 493.37 | 471.77 | 2,120.08 |
| 112 | 4,185.59 | 4,055.95 | 3,791.95 | 3,696.89 | 15,730.38 |
| 113 | 2,195.62 | 2,006.83 | 1,871.55 | 1,819.28 | 7,893.28 |
| 114 | 1,088.06 | 994.6 | 986.34 | 903.02 | 3,972.02 |
| 115 | 3,100.95 | 2,994.10 | 2,904.21 | 2,922.40 | 11,921.66 |
| 116 | 295.42 | 315.29 | 282.23 | 263.37 | 1,156.31 |
| 117 | 13,342.46 | 13,047.80 | 12,482.72 | 12,288.39 | 51,161.37 |
| 118 | 402.67 | 433.48 | 412.07 | 390.46 | 1,638.68 |

| | | | | | | |
|---|---|---|---|---|---|---|
| | 119 | 2,146.24 | 2,170.31 | 2,043.74 | 1,975.69 | 8,335.98 |
| | 120 | 7,162.78 | 6,834.95 | 6,142.49 | 6,300.20 | 26,440.42 |
| | 121 | 3,956.37 | 3,591.10 | 3,530.21 | 3,439.05 | 14,516.73 |
| | 122 | 3,174.55 | 2,955.57 | 2,829.72 | 2,664.06 | 11,623.90 |
| | 123 | 3,015.35 | 2,769.36 | 2,706.46 | 2,638.91 | 11,130.08 |
| | 124 | 2,766.53 | 2,643.16 | 2,535.36 | 2,343.68 | 10,288.73 |
| | 125 | 4,710.48 | 4,616.56 | 4,392.65 | 4,272 | 17,991.69 |
| | 126 | 769.56 | 792.6 | 702.59 | 645.54 | 2,910.29 |
| | 127 | 1,078.50 | 1,043.09 | 1,019.36 | 950.26 | 4,091.21 |
| | 128 | 4,841.85 | 4,594.74 | 4,408.47 | 4,237.53 | 18,082.59 |
| | 129 | 2,944.77 | 2,889.35 | 2,807.53 | 2,677.56 | 11,319.21 |
| | 130 | 5,751.61 | 5,585.06 | 5,192.55 | 5,227.97 | 21,757.19 |
| | 131 | 2,001.43 | 1,951.90 | 1,846.33 | 1,861.38 | 7,661.04 |
| | 132 | 4,402.65 | 4,195.91 | 4,023.27 | 3,832.46 | 16,454.29 |
| | 133 | 2,778.51 | 2,581.27 | 2,303.23 | 2,396.23 | 10,059.24 |
| | 134 | 4,157.52 | 3,949.13 | 3,526.76 | 3,480.04 | 15,113.45 |
| | 135 | 1,928.08 | 1,894.33 | 1,803.48 | 1,730.43 | 7,356.32 |
| | 136 | 3,786.01 | 3,785.61 | 3,581.09 | 3,581.49 | 14,734.20 |
| | 137 | 1,760.60 | 1,759.51 | 1,630.18 | 1,611.32 | 6,761.61 |
| | 138 | 3,932.03 | 3,818.66 | 3,487.41 | 3,648.51 | 14,886.61 |
| | 139 | 1,668.56 | 1,618.68 | 1,531.62 | 1,574.25 | 6,393.11 |
| | 140 | 12,157.98 | 12,034.55 | 11,643.75 | 11,278.17 | 47,114.45 |
| | 141 | 6,097.82 | 5,982.77 | 5,827.30 | 5,527.15 | 23,435.04 |
| | 142 | 22,619.75 | 21,787.09 | 20,905.22 | 20,712.62 | 86,024.68 |
| | 143 | 2,792.38 | 2,766.89 | 2,707.92 | 2,717.62 | 10,984.81 |
| | 144 | 4,547.97 | 4,482.29 | 4,180.67 | 4,193.66 | 17,404.59 |
| | 145 | 3,636.45 | 3,608.06 | 3,444.64 | 3,320 | 14,009.15 |
| | 146 | 6,503.43 | 6,260.35 | 5,844.29 | 5,822.77 | 24,430.84 |
| | 147 | 4,859.18 | 4,762.09 | 4,528 | 4,384.84 | 18,534.11 |
| | 148 | 4,420.50 | 4,319.59 | 4,219.84 | 4,049.20 | 17,009.13 |
| | 149 | 961.59 | 987.21 | 923.21 | 900.56 | 3,772.57 |
| TOTAL | | 180,175.57 | 174,189.80 | 164,948.98 | 161,835.50 | 681,149.85 |

STATE: NORTH CAROLINA

| ZIP CODE | | QUARTER 1 | QUARTER 2 | QUARTER 3 | QUARTER 4 | TOTAL GRAMS |
|---|---|---|---|---|---|---|
| | 270 | 8,113.48 | 7,525.78 | 7,157.36 | 7,008.86 | 29,805.48 |

| | | | | | | |
|---|---|---|---|---|---|---|
| | 271 | 5,844.27 | 5,249.90 | 5,032.64 | 4,853.45 | 20,980.26 |
| | 272 | 14,878.18 | 13,183.19 | 12,252.28 | 12,064.28 | 52,377.93 |
| | 273 | 8,221.94 | 7,527.60 | 6,926.06 | 6,888.90 | 29,564.50 |
| | 274 | 4,968.94 | 4,500.63 | 4,169.30 | 4,226.40 | 17,865.27 |
| | 275 | 15,937.70 | 14,465.21 | 13,139.91 | 13,617.80 | 57,160.62 |
| | 276 | 5,292.75 | 4,678.24 | 4,337.59 | 4,601.56 | 18,910.14 |
| | 277 | 4,073.16 | 3,637.50 | 3,265.98 | 3,518.79 | 14,495.43 |
| | 278 | 10,238.36 | 9,341.77 | 8,740.25 | 8,934.59 | 37,254.97 |
| | 279 | 3,089.40 | 3,020.34 | 2,690.95 | 2,948.94 | 11,749.63 |
| | 280 | 23,075.42 | 21,619.68 | 19,816.07 | 20,198.50 | 84,709.67 |
| | 281 | 16,594.08 | 15,220.92 | 14,114.52 | 14,398.58 | 60,328.10 |
| | 282 | 11,791.57 | 10,837.14 | 9,926.61 | 9,937.74 | 42,493.06 |
| | 283 | 18,721.37 | 17,222.11 | 16,214.37 | 15,908.57 | 68,066.42 |
| | 284 | 12,052.10 | 11,658.20 | 10,458.23 | 10,137.46 | 44,305.99 |
| | 285 | 9,205.81 | 8,125.78 | 7,686.33 | 7,359.40 | 32,377.32 |
| | 286 | 20,740.49 | 19,410.81 | 17,862.10 | 17,787.95 | 75,801.35 |
| | 287 | 11,663.13 | 11,030.55 | 10,203.47 | 9,998.34 | 42,895.49 |
| | 288 | 6,019.59 | 5,626.55 | 5,403.81 | 4,916.64 | 21,966.59 |
| | 289 | 1,545.56 | 1,601.65 | 1,464.77 | 1,600.98 | 6,212.96 |
| TOTAL | | 212,067.30 | 195,483.55 | 180,862.60 | 180,907.73 | 769,321.18 |

STATE: NORTH DAKOTA

| ZIP CODE | | QUARTER 1 | QUARTER 2 | QUARTER 3 | QUARTER 4 | TOTAL GRAMS |
|---|---|---|---|---|---|---|
| | 580 | 613.56 | 592.52 | 540.42 | 548.02 | 2,294.52 |
| | 581 | 2,044.75 | 1,962.19 | 1,797.88 | 1,777.13 | 7,581.95 |
| | 582 | 1,277.49 | 1,252.97 | 1,192.96 | 1,119.43 | 4,842.85 |
| | 583 | 703.28 | 696.49 | 640.14 | 665.95 | 2,705.86 |
| | 584 | 608.45 | 584.08 | 588.07 | 583.31 | 2,363.91 |
| | 585 | 3,630.17 | 3,394.41 | 3,260.32 | 3,248.84 | 13,533.74 |
| | 586 | 648.87 | 676.99 | 637.85 | 656.27 | 2,619.98 |
| | 587 | 803.25 | 775.49 | 755.55 | 724.79 | 3,059.08 |
| | 588 | 626.17 | 625.87 | 635.74 | 603.67 | 2,491.45 |
| TOTAL | | 10,955.99 | 10,561.01 | 10,048.93 | 9,927.41 | 41,493.34 |

STATE: OHIO

| ZIP CODE | QUARTER 1 | QUARTER 2 | QUARTER 3 | QUARTER 4 | TOTAL GRAMS |
|---|---|---|---|---|---|
| 430 | 10,540.25 | 10,058.94 | 9,443.27 | 9,293.30 | 39,335.76 |
| 431 | 7,740.24 | 7,595.81 | 7,065.10 | 6,835.41 | 29,236.56 |
| 432 | 18,045.52 | 16,908.09 | 15,977.54 | 15,472.46 | 66,403.61 |
| 433 | 2,995.62 | 2,965.66 | 2,802.68 | 2,796.33 | 11,560.29 |
| 434 | 1,901.17 | 1,850.88 | 1,783.38 | 1,764.44 | 7,299.87 |
| 435 | 4,049.52 | 3,868.29 | 3,823.38 | 3,688.02 | 15,429.21 |
| 436 | 7,261.81 | 6,906.12 | 6,860.29 | 6,263.87 | 27,292.09 |
| 437 | 4,265.02 | 4,028.52 | 3,798.21 | 3,686.30 | 15,778.05 |
| 438 | 609.61 | 544.52 | 567.18 | 519.24 | 2,240.55 |
| 439 | 3,679.53 | 3,701.13 | 3,416.98 | 3,257.22 | 14,054.86 |
| 440 | 9,514.25 | 8,963.96 | 8,404.20 | 8,065.64 | 34,948.05 |
| 441 | 11,171.03 | 10,340.80 | 9,810.41 | 9,622.33 | 40,944.57 |
| 442 | 7,155.18 | 6,821.73 | 6,472.89 | 6,033.04 | 26,482.84 |
| 443 | 3,660.03 | 3,336.98 | 3,135.52 | 3,009.23 | 13,141.76 |
| 444 | 8,435.62 | 7,954.89 | 7,533.53 | 7,433.39 | 31,357.43 |
| 445 | 5,249.71 | 5,056.93 | 4,672.15 | 4,559.66 | 19,538.45 |
| 446 | 6,602.58 | 6,267.09 | 6,035.66 | 5,642.54 | 24,547.87 |
| 447 | 3,624.92 | 3,394.51 | 3,306.69 | 3,223.84 | 13,549.96 |
| 448 | 3,851.20 | 3,689.88 | 3,470.94 | 3,312.52 | 14,324.54 |
| 449 | 997.76 | 952.58 | 879.92 | 817.45 | 3,647.71 |
| 450 | 16,618.14 | 15,691.92 | 14,933.69 | 14,752.45 | 61,996.20 |
| 451 | 5,547.45 | 5,215.36 | 5,083.49 | 4,959.92 | 20,806.22 |
| 452 | 12,979.39 | 12,418.57 | 11,880.80 | 11,131.72 | 48,410.48 |
| 453 | 9,825.97 | 9,576.55 | 9,224.29 | 9,106.67 | 37,733.48 |
| 454 | 12,025.73 | 11,475.35 | 11,156.42 | 10,525.59 | 45,183.09 |
| 455 | 2,686.23 | 2,601.19 | 2,536.86 | 2,471.02 | 10,295.30 |
| 456 | 9,509.21 | 9,020.01 | 8,434.53 | 8,185.50 | 35,149.25 |
| 457 | 3,652.44 | 3,528.98 | 3,240.88 | 3,044.10 | 13,466.40 |
| 458 | 4,930.23 | 4,748.81 | 4,541.08 | 4,526.26 | 18,746.38 |
| TOTAL | 199,125.36 | 189,484.05 | 180,291.96 | 173,999.46 | 742,900.83 |

STATE: OKLAHOMA

| ZIP CODE | QUARTER 1 | QUARTER 2 | QUARTER 3 | QUARTER 4 | TOTAL GRAMS |
|---|---|---|---|---|---|
| 730 | 26,792.76 | 26,400.88 | 25,420.03 | 25,429.27 | 104,042.94 |
| 731 | 37,140.25 | 36,797.98 | 34,804.11 | 35,133.97 | 143,876.31 |

| | | | | | | |
|---|---|---|---|---|---|---|
| | 734 | 5,673.16 | 5,821.18 | 5,303.51 | 5,243.62 | 22,041.47 |
| | 735 | 10,253.59 | 10,174.68 | 9,945.99 | 9,613.53 | 39,987.79 |
| | 736 | 1,869.62 | 1,887.01 | 1,819.96 | 1,673.68 | 7,250.27 |
| | 737 | 4,319.66 | 4,351.66 | 4,067.29 | 3,948.55 | 16,687.16 |
| | 738 | 743.63 | 728.84 | 712.82 | 688.75 | 2,874.04 |
| | 739 | 387.22 | 389.58 | 375.19 | 379.36 | 1,531.35 |
| | 740 | 30,201.09 | 30,048.99 | 28,181.92 | 28,320.64 | 116,752.64 |
| | 741 | 21,209.69 | 20,890.74 | 20,157.82 | 19,930.73 | 82,188.98 |
| | 743 | 5,144.20 | 5,164.26 | 4,965.11 | 4,921.49 | 20,195.06 |
| | 744 | 15,462.39 | 15,234.24 | 15,024.80 | 14,339.73 | 60,061.16 |
| | 745 | 4,944.42 | 4,968.66 | 4,607.52 | 4,460.17 | 18,980.77 |
| | 746 | 1,958.32 | 2,017.70 | 1,957.90 | 2,011.27 | 7,945.19 |
| | 747 | 6,733.63 | 6,675.37 | 6,347.39 | 6,038.04 | 25,794.43 |
| | 748 | 9,433.24 | 8,901.25 | 8,792.22 | 8,426.12 | 35,552.83 |
| | 749 | 5,191.90 | 5,142.45 | 4,780.98 | 4,973.19 | 20,088.52 |
| TOTAL | | 187,458.77 | 185,595.47 | 177,264.56 | 175,532.11 | 725,850.91 |

STATE: OREGON

| ZIP CODE | | QUARTER 1 | QUARTER 2 | QUARTER 3 | QUARTER 4 | TOTAL GRAMS |
|---|---|---|---|---|---|---|
| | 970 | 15,081.98 | 14,377.68 | 13,768.78 | 13,861.38 | 57,089.82 |
| | 971 | 6,412.78 | 6,366.98 | 6,151.53 | 6,078.08 | 25,009.37 |
| | 972 | 18,410.92 | 17,624.12 | 17,016.35 | 16,556.51 | 69,607.90 |
| | 973 | 12,887.21 | 12,130.36 | 11,553.47 | 11,205.85 | 47,776.89 |
| | 974 | 19,535.40 | 19,037.15 | 18,196.47 | 17,797.57 | 74,566.59 |
| | 975 | 11,936.11 | 11,307.62 | 11,301.07 | 11,287.21 | 45,832.01 |
| | 976 | 2,879.89 | 2,702.76 | 2,742.63 | 2,473.81 | 10,799.09 |
| | 977 | 5,435.04 | 5,383.50 | 5,230.18 | 4,952.14 | 21,000.86 |
| | 978 | 4,691.58 | 4,442.60 | 4,337.52 | 4,169.58 | 17,641.28 |
| | 979 | 1,954.63 | 1,875.05 | 1,788.03 | 1,759.57 | 7,377.28 |
| TOTAL | | 99,225.54 | 95,247.82 | 92,086.03 | 90,141.70 | 376,701.09 |

STATE: PENNSYLVANIA

| ZIP CODE | | QUARTER 1 | QUARTER 2 | QUARTER 3 | QUARTER 4 | TOTAL GRAMS |
|---|---|---|---|---|---|---|
| | 150 | 8,993.72 | 8,826.88 | 8,804.69 | 8,160.14 | 34,785.43 |
| | 151 | 6,577.79 | 6,094.37 | 5,948.01 | 5,718.48 | 24,338.65 |

| | | | | | | |
|---|---|---|---|---|---|---|
| 152 | 12,672.58 | 12,150.82 | | 11,409.36 | 11,011.34 | 47,244.10 |
| 153 | 3,965.32 | 3,232.35 | | 3,720.07 | 3,044.86 | 13,962.60 |
| 154 | 4,853.08 | 4,634.73 | | 4,416.30 | 4,336.13 | 18,240.24 |
| 155 | 1,698.22 | 1,617.60 | | 1,586.51 | 1,269.62 | 6,171.95 |
| 156 | 6,100.67 | 5,926.48 | | 5,578.05 | 5,219.10 | 22,824.30 |
| 157 | 2,512.76 | 2,479.88 | | 2,478.24 | 2,198.70 | 9,669.58 |
| 158 | 2,143.29 | 2,130.87 | | 2,035.48 | 1,969.47 | 8,279.11 |
| 159 | 2,727.22 | 2,663.98 | | 2,460.94 | 2,393.80 | 10,245.94 |
| 160 | 2,962.81 | 2,781.20 | | 2,818.78 | 2,673.83 | 11,236.62 |
| 161 | 4,232.01 | 4,127.86 | | 3,918.30 | 3,883.17 | 16,161.34 |
| 162 | 1,717.23 | 1,650.49 | | 1,565.29 | 1,534.28 | 6,467.29 |
| 163 | 3,598.14 | 3,457.19 | | 3,259.23 | 3,170.34 | 13,484.90 |
| 164 | 2,269.62 | 2,300.20 | | 2,159.64 | 2,140.33 | 8,869.79 |
| 165 | 7,891.93 | 7,845.61 | | 7,244.02 | 7,102.53 | 30,084.09 |
| 166 | 5,575.51 | 5,447.91 | | 5,201.41 | 5,076.88 | 21,301.71 |
| 167 | 1,361.79 | 1,396.81 | | 1,321.14 | 1,317.89 | 5,397.63 |
| 168 | 1,570.99 | 1,532.62 | | 1,485.13 | 1,373.44 | 5,962.18 |
| 169 | 993.68 | 931.53 | | 865.66 | 802.78 | 3,593.65 |
| 170 | 8,553.44 | 8,229.80 | | 7,784.32 | 7,565.29 | 32,132.85 |
| 171 | 2,055.12 | 1,998.47 | | 1,843.21 | 1,741.49 | 7,638.29 |
| 172 | 1,552.14 | 1,493.79 | | 1,424.68 | 1,380.73 | 5,851.34 |
| 173 | 2,784.76 | 2,733.33 | | 2,478.29 | 2,377.71 | 10,374.09 |
| 174 | 1,644.13 | 1,571.81 | | 1,424.11 | 1,448.42 | 6,088.47 |
| 175 | 2,974.35 | 2,834.75 | | 2,685.18 | 2,547.40 | 11,041.68 |
| 176 | 2,210.89 | 2,096.12 | | 2,012.80 | 1,952.27 | 8,272.08 |
| 177 | 3,157.30 | 3,084.67 | | 2,931.25 | 2,743.98 | 11,917.20 |
| 178 | 2,106.78 | 2,068.05 | | 1,933.94 | 1,848.25 | 7,957.02 |
| 179 | 1,887.55 | 1,791.20 | | 1,744.12 | 1,654.81 | 7,077.68 |
| 180 | 3,309.71 | 3,099.13 | | 2,982.09 | 2,965.64 | 12,356.57 |
| 181 | 1,445.58 | 1,303.64 | | 1,290.40 | 1,262.75 | 5,302.37 |
| 182 | 2,232.15 | 2,117.55 | | 1,878.36 | 1,894.65 | 8,122.71 |
| 183 | 1,696.10 | 1,628.55 | | 1,553.44 | 1,514.68 | 6,392.77 |
| 184 | 3,124.98 | 3,002.11 | | 2,983.79 | 2,797.60 | 11,908.48 |
| 185 | 4,297.65 | 4,138.49 | | 3,933.27 | 3,684.45 | 16,053.86 |
| 186 | 3,223.84 | 3,137.77 | | 3,040.72 | 2,820.79 | 12,223.12 |
| 187 | 8,655.75 | 7,994.97 | | 7,442.98 | 7,759.83 | 31,853.53 |
| 188 | 1,178.25 | 1,106.35 | | 1,079.11 | 1,008.77 | 4,372.48 |

| | | | | | | |
|---|---|---|---|---|---|---|
| | 189 | 2,574.08 | 2,477.28 | 2,379.14 | 2,305.41 | 9,735.91 |
| | 190 | 12,740.83 | 9,994.78 | 14,061.52 | 8,867.46 | 45,664.59 |
| | 191 | 7,453.96 | 7,235.37 | 6,780.14 | 6,438.20 | 27,907.67 |
| | 193 | 2,731.91 | 2,653.46 | 2,575.21 | 2,398.64 | 10,359.22 |
| | 194 | 4,352.72 | 4,240.98 | 3,965.47 | 3,901.12 | 16,460.29 |
| | 195 | 1,786.32 | 1,748.89 | 1,613.25 | 1,578.67 | 6,727.13 |
| | 196 | 2,056.41 | 2,088.23 | 1,952.22 | 1,879.23 | 7,976.09 |
| TOTAL | | 176,205.06 | 167,098.92 | 164,049.26 | 152,735.35 | 660,088.59 |

STATE: PUERTO RICO

| ZIP CODE | QUARTER 1 | QUARTER 2 | QUARTER 3 | QUARTER 4 | TOTAL GRAMS |
|---|---|---|---|---|---|
| 6 | 412.28 | 269.35 | 151.46 | 128.61 | 961.7 |
| 7 | 322.57 | 194.56 | 123.65 | 126.64 | 767.42 |
| 9 | 362.58 | 196.86 | 125 | 133.12 | 817.56 |
| TOTAL | 1,097.43 | 660.77 | 400.11 | 388.37 | 2,546.68 |

STATE: RHODE ISLAND

| ZIP CODE | QUARTER 1 | QUARTER 2 | QUARTER 3 | QUARTER 4 | TOTAL GRAMS |
|---|---|---|---|---|---|
| 28 | 6,660.98 | 6,195.48 | 5,926.90 | 5,882.04 | 24,665.40 |
| 29 | 4,538.74 | 4,307.82 | 4,081.98 | 3,893.71 | 16,822.25 |
| TOTAL | 11,199.72 | 10,503.30 | 10,008.88 | 9,775.75 | 41,487.65 |

STATE: SOUTH CAROLINA

| ZIP CODE | QUARTER 1 | QUARTER 2 | QUARTER 3 | QUARTER 4 | TOTAL GRAMS |
|---|---|---|---|---|---|
| 290 | 8,193.78 | 7,653.58 | 7,185.13 | 7,285.05 | 30,317.54 |
| 291 | 8,969.60 | 8,505.71 | 7,942.34 | 8,084.60 | 33,502.25 |
| 292 | 6,511.33 | 6,059.46 | 5,606.93 | 5,600.62 | 23,778.34 |
| 293 | 23,463.65 | 22,250.66 | 20,832.74 | 21,174 | 87,721.05 |
| 294 | 21,054.27 | 20,053.32 | 18,794.73 | 18,754.35 | 78,656.67 |
| 295 | 24,589.63 | 23,755.82 | 21,863.42 | 22,092.52 | 92,301.39 |
| 296 | 40,235.04 | 38,481.96 | 36,233.35 | 35,927.91 | 150,878.26 |
| 297 | 7,410.19 | 6,958.48 | 6,265.46 | 6,303.06 | 26,937.19 |
| 298 | 3,822.91 | 3,774.80 | 3,275.55 | 3,394.38 | 14,267.64 |
| 299 | 3,232.14 | 3,053.67 | 2,856.09 | 3,017.81 | 12,159.71 |

| | | | | | | |
|---|---|---|---|---|---|---|
| TOTAL | | 147,482.54 | 140,547.46 | 130,855.74 | 131,634.30 | 550,520.04 |

| STATE: | SOUTH DAKOTA | | | | | |
|---|---|---|---|---|---|---|
| ZIP CODE | | QUARTER 1 | QUARTER 2 | QUARTER 3 | QUARTER 4 | TOTAL GRAMS |
| | 570 | 2,430.12 | 2,320.17 | 2,264.49 | 2,306.09 | 9,320.87 |
| | 571 | 5,237.68 | 5,021.36 | 4,739.96 | 4,679.75 | 19,678.75 |
| | 572 | 1,114.16 | 1,010.66 | 993.19 | 982.65 | 4,100.66 |
| | 573 | 1,314.37 | 1,243.36 | 1,212.07 | 1,207.87 | 4,977.67 |
| | 574 | 967.48 | 896.4 | 864.71 | 846.73 | 3,575.32 |
| | 575 | 1,157.83 | 1,088.54 | 1,074.26 | 966.87 | 4,287.50 |
| | 576 | 258.8 | 232.83 | 260.4 | 240.38 | 992.41 |
| | 577 | 3,320.41 | 3,104.26 | 3,009.74 | 2,839.42 | 12,273.83 |
| TOTAL | | 15,800.85 | 14,917.58 | 14,418.82 | 14,069.76 | 59,207.01 |

| STATE: | TENNESSEE | | | | | |
|---|---|---|---|---|---|---|
| ZIP CODE | | QUARTER 1 | QUARTER 2 | QUARTER 3 | QUARTER 4 | TOTAL GRAMS |
| | 370 | 30,618.73 | 29,775.60 | 28,006.25 | 27,636.95 | 116,037.53 |
| | 371 | 27,668.25 | 28,204.42 | 28,643.27 | 27,950.13 | 112,466.07 |
| | 372 | 16,863.90 | 14,714.70 | 12,338.70 | 11,961.62 | 55,878.92 |
| | 373 | 28,201.72 | 27,574.46 | 24,997.05 | 25,809.77 | 106,583 |
| | 374 | 9,884 | 10,152.62 | 9,455.03 | 9,884.62 | 39,376.27 |
| | 376 | 18,935.99 | 18,000.37 | 17,055.08 | 16,630.09 | 70,621.53 |
| | 377 | 14,294.32 | 14,084.50 | 13,447.26 | 13,069.84 | 54,895.92 |
| | 378 | 22,561.43 | 22,310.21 | 21,607.34 | 21,061.91 | 87,540.89 |
| | 379 | 11,457.40 | 10,989.35 | 10,420.91 | 10,401.05 | 43,268.71 |
| | 380 | 13,816.38 | 12,931.84 | 12,808.76 | 12,876.34 | 52,433.32 |
| | 381 | 19,631.05 | 17,405.53 | 21,421.25 | 14,351.03 | 72,808.86 |
| | 382 | 8,038.66 | 7,966.36 | 7,581.36 | 7,632.53 | 31,218.91 |
| | 383 | 16,439.02 | 15,952.75 | 15,361.70 | 15,419.93 | 63,173.40 |
| | 384 | 9,163.44 | 8,710.18 | 8,222.89 | 8,309.60 | 34,406.11 |
| | 385 | 8,358.12 | 8,355.28 | 7,720.53 | 7,704.67 | 32,138.60 |
| TOTAL | | 255,932.41 | 247,128.17 | 239,087.38 | 230,700.08 | 972,848.04 |

RETAIL DRUG DISTRIBUTION BY ZIP CODE WITHIN STATE BY GRAMS WT

STATE: TEXAS

| ZIP CODE | QUARTER 1 | QUARTER 2 | QUARTER 3 | QUARTER 4 | TOTAL GRAMS |
|---|---|---|---|---|---|
| 750 | 55,559.04 | 53,461.62 | 51,433.46 | 51,043.94 | 211,498.06 |
| 751 | 23,398.39 | 22,971.93 | 22,310.22 | 22,003.90 | 90,684.44 |
| 752 | 29,700.30 | 28,932.92 | 27,784.43 | 27,135.94 | 113,553.59 |
| 753 | 294.75 | 289.4 | 312.66 | 276.46 | 1,173.27 |
| 754 | 16,735.83 | 15,424.19 | 14,385.50 | 14,374.01 | 60,919.53 |
| 755 | 5,656.96 | 5,434.55 | 5,039.93 | 4,778.96 | 20,910.40 |
| 756 | 11,123.32 | 10,865.18 | 10,990.50 | 10,858.23 | 43,837.23 |
| 757 | 14,938.15 | 15,216.44 | 14,562.42 | 14,382.62 | 59,099.63 |
| 758 | 3,001.44 | 3,004.71 | 2,789.04 | 2,817.67 | 11,612.86 |
| 759 | 9,045.73 | 9,015.40 | 8,954.77 | 8,693.12 | 35,709.02 |
| 760 | 40,087.65 | 40,453.55 | 38,679.82 | 39,162.24 | 158,383.26 |
| 761 | 34,689.57 | 34,880.40 | 33,092.22 | 33,624.08 | 136,286.27 |
| 762 | 14,841.44 | 14,941.23 | 14,409.58 | 14,562.55 | 58,754.80 |
| 763 | 7,366.78 | 7,131.29 | 6,798.11 | 7,035.85 | 28,332.03 |
| 764 | 4,747.24 | 4,549.29 | 4,539.76 | 4,609.94 | 18,446.23 |
| 765 | 14,944.78 | 14,569.67 | 13,809.71 | 13,071.77 | 56,395.93 |
| 766 | 3,707.41 | 3,702.66 | 3,719.16 | 3,842.88 | 14,972.11 |
| 767 | 10,534.61 | 10,455.72 | 9,836.01 | 9,717.57 | 40,543.91 |
| 768 | 3,547.11 | 3,578.98 | 3,469.46 | 3,420.06 | 14,015.61 |
| 769 | 4,143.08 | 4,123.64 | 3,855.62 | 3,915.78 | 16,038.12 |
| 770 | 58,043.34 | 57,885.07 | 54,654.30 | 54,025.33 | 224,608.04 |
| 773 | 31,417.52 | 31,112 | 29,859.82 | 30,380.15 | 122,769.49 |
| 774 | 19,302.19 | 18,929.47 | 18,441.93 | 18,862.49 | 75,536.08 |
| 775 | 34,649.20 | 34,991.46 | 33,727.20 | 33,523.62 | 136,891.48 |
| 776 | 9,765.42 | 9,130.25 | 9,217.40 | 8,931.46 | 37,044.53 |
| 777 | 4,484.55 | 4,245.83 | 4,327.35 | 4,199.30 | 17,257.03 |
| 778 | 7,211.62 | 6,934 | 6,690.85 | 6,198.54 | 27,035.01 |
| 779 | 7,989.75 | 7,714.04 | 7,092.70 | 7,005.66 | 29,802.15 |
| 780 | 9,816.92 | 9,792.97 | 9,520.18 | 9,578.09 | 38,708.16 |
| 781 | 11,248.39 | 10,792.30 | 10,831.59 | 10,542.35 | 43,414.63 |
| 782 | 46,421.96 | 45,652.80 | 45,341.01 | 44,797.30 | 182,213.07 |
| 783 | 6,659.44 | 6,494.46 | 6,156.31 | 5,855.88 | 25,166.09 |
| 784 | 12,496.64 | 11,868.47 | 11,839.36 | 11,116.91 | 47,321.38 |
| 785 | 8,681.35 | 8,513.34 | 8,202.97 | 8,041.77 | 33,439.43 |
| 786 | 23,913.36 | 23,513.81 | 22,633.62 | 21,927.10 | 91,987.89 |

| | | | | | |
|---|---|---|---|---|---|
| 787 | 23,066.78 | 21,755.73 | 20,304.56 | 19,186.37 | 84,313.44 |
| 788 | 3,054.37 | 2,973.87 | 2,823.40 | 2,765.13 | 11,616.77 |
| 789 | 1,696.71 | 1,693.18 | 1,598.63 | 1,668.81 | 6,657.33 |
| 790 | 4,562.67 | 4,500.21 | 4,142.85 | 4,271.19 | 17,476.92 |
| 791 | 9,304.12 | 8,682.70 | 9,059.80 | 7,775.22 | 34,821.84 |
| 792 | 955.4 | 974.73 | 898.91 | 862.4 | 3,691.44 |
| 793 | 3,813.29 | 3,794.61 | 3,616.74 | 3,683.77 | 14,908.41 |
| 794 | 11,123.90 | 10,874.54 | 10,959.75 | 11,040.32 | 43,998.51 |
| 795 | 3,170.61 | 2,898.66 | 2,818.53 | 2,781.22 | 11,669.02 |
| 796 | 6,104.75 | 5,947.16 | 5,887.28 | 5,731.21 | 23,670.40 |
| 797 | 12,262.44 | 11,732.08 | 11,115.57 | 11,322.98 | 46,433.07 |
| 798 | 371.58 | 367.61 | 309.84 | 326.07 | 1,375.10 |
| 799 | 9,044.83 | 8,427.34 | 8,176.56 | 7,667.56 | 33,316.29 |
| TOTAL | 688,696.68 | 675,195.46 | 651,021.39 | 643,395.77 | 2,658,309.30 |

STATE: UTAH

| ZIP CODE | QUARTER 1 | QUARTER 2 | QUARTER 3 | QUARTER 4 | TOTAL GRAMS |
|---|---|---|---|---|---|
| 840 | 22,819.99 | 22,058.92 | 21,419.53 | 21,190.29 | 87,488.73 |
| 841 | 15,468.33 | 14,916.11 | 14,062.54 | 13,975.70 | 58,422.68 |
| 843 | 2,830.70 | 2,851.76 | 2,758.98 | 2,678.62 | 11,120.06 |
| 844 | 4,530.34 | 4,412.86 | 4,205.19 | 4,345.59 | 17,493.98 |
| 845 | 1,952.37 | 1,790.57 | 1,668.05 | 1,657.57 | 7,068.56 |
| 846 | 5,789.31 | 5,419 | 5,164.49 | 5,146.53 | 21,519.33 |
| 847 | 7,995.91 | 7,858.36 | 7,611.09 | 7,298.83 | 30,764.19 |
| TOTAL | 61,386.95 | 59,307.58 | 56,889.87 | 56,293.13 | 233,877.53 |

STATE: VERMONT

| ZIP CODE | QUARTER 1 | QUARTER 2 | QUARTER 3 | QUARTER 4 | TOTAL GRAMS |
|---|---|---|---|---|---|
| 50 | 344.84 | 410.72 | 324.77 | 331.41 | 1,411.74 |
| 51 | 224.02 | 198.07 | 177.49 | 175.64 | 775.22 |
| 52 | 262.84 | 247.66 | 247.37 | 250.54 | 1,008.41 |
| 53 | 305.74 | 292.52 | 308.26 | 300.8 | 1,207.32 |
| 54 | 2,049.66 | 1,961.26 | 1,831.75 | 1,803.31 | 7,645.98 |
| 56 | 844.51 | 796.68 | 748.04 | 723.49 | 3,112.72 |

| | | | | | | |
|---|---|---|---|---|---|---|
| | 57 | 520.63 | 422.41 | 387.6 | 406.58 | 1,737.22 |
| | 58 | 582.61 | 580.79 | 539.96 | 535.62 | 2,238.98 |
| TOTAL | | 5,134.85 | 4,910.11 | 4,565.24 | 4,527.39 | 19,137.59 |

STATE: VIRGIN ISLANDS

| ZIP CODE | | QUARTER 1 | QUARTER 2 | QUARTER 3 | QUARTER 4 | TOTAL GRAMS |
|---|---|---|---|---|---|---|
| | 8 | 363.62 | 340.97 | 223.69 | 230.32 | 1,158.60 |
| TOTAL | | 363.62 | 340.97 | 223.69 | 230.32 | 1,158.60 |

STATE: VIRGINIA

| ZIP CODE | | QUARTER 1 | QUARTER 2 | QUARTER 3 | QUARTER 4 | TOTAL GRAMS |
|---|---|---|---|---|---|---|
| | 201 | 4,850.46 | 4,297.86 | 3,964.53 | 4,081.77 | 17,194.62 |
| | 220 | 2,140.16 | 1,896.44 | 1,779.37 | 1,840.30 | 7,656.27 |
| | 221 | 1,906.61 | 1,657.60 | 1,514.64 | 1,714.83 | 6,793.68 |
| | 222 | 602.35 | 576.31 | 515.94 | 485.26 | 2,179.86 |
| | 223 | 1,073.81 | 941.49 | 881.68 | 930.24 | 3,827.22 |
| | 224 | 2,905.74 | 2,683.07 | 2,398.97 | 2,504.55 | 10,492.33 |
| | 225 | 1,233.24 | 1,118.62 | 1,074.48 | 1,071.05 | 4,497.39 |
| | 226 | 2,869.59 | 2,609.63 | 2,398.17 | 2,475.51 | 10,352.90 |
| | 227 | 612.77 | 506.28 | 479.1 | 502.58 | 2,100.73 |
| | 228 | 2,432.79 | 2,162.94 | 1,906.65 | 2,006.83 | 8,509.21 |
| | 229 | 2,723.80 | 2,502.07 | 2,308.28 | 2,223.40 | 9,757.55 |
| | 230 | 2,520.54 | 2,326.92 | 2,176.42 | 2,054.23 | 9,078.11 |
| | 231 | 4,633.65 | 4,072.08 | 3,490.16 | 3,736.86 | 15,932.75 |
| | 232 | 7,380.02 | 6,543.85 | 5,999.15 | 5,649.05 | 25,572.07 |
| | 233 | 3,188.37 | 2,856.29 | 2,646.58 | 2,634.84 | 11,326.08 |
| | 234 | 6,975.14 | 6,166.48 | 5,666.63 | 5,805.01 | 24,613.26 |
| | 235 | 2,703.16 | 2,260.74 | 2,082.24 | 2,175.82 | 9,221.96 |
| | 236 | 4,769.86 | 4,045.31 | 3,696.24 | 3,517.05 | 16,028.46 |
| | 237 | 2,505.85 | 2,338.27 | 1,983.13 | 1,913.75 | 8,741 |
| | 238 | 3,734.82 | 3,258.04 | 2,905.81 | 2,924.48 | 12,823.15 |
| | 239 | 1,352.48 | 1,216.65 | 1,109.39 | 1,107.46 | 4,785.98 |
| | 240 | 9,522.81 | 8,823.80 | 8,445.46 | 8,088.04 | 34,880.11 |
| | 241 | 9,661.55 | 8,921.62 | 8,259.94 | 8,145.30 | 34,988.41 |
| | 242 | 13,820.84 | 12,993.96 | 12,508.54 | 12,069.64 | 51,392.98 |

| | | | | | | |
|---|---|---|---|---|---|---|
| | 243 | 5,182.88 | 4,580.11 | 4,393.09 | 4,283.30 | 18,439.38 |
| | 244 | 2,901.22 | 2,599.92 | 2,461.37 | 2,492 | 10,454.51 |
| | 245 | 7,882.63 | 7,176.87 | 6,603.81 | 6,664.62 | 28,327.93 |
| | 246 | 4,307.69 | 4,298.55 | 3,888.83 | 3,876.47 | 16,371.54 |
| TOTAL | | 116,394.83 | 105,431.77 | 97,538.60 | 96,974.24 | 416,339.44 |

STATE: WASHINGTON

| ZIP CODE | | QUARTER 1 | QUARTER 2 | QUARTER 3 | QUARTER 4 | TOTAL GRAMS |
|---|---|---|---|---|---|---|
| | 980 | 17,936.93 | 16,886.77 | 16,150.02 | 16,047.80 | 67,021.52 |
| | 981 | 8,673.97 | 8,417.28 | 7,918.72 | 7,699.30 | 32,709.27 |
| | 982 | 15,792.98 | 15,200.57 | 14,717.36 | 14,331.62 | 60,042.53 |
| | 983 | 13,054.89 | 13,026.25 | 12,406.13 | 12,083.97 | 50,571.24 |
| | 984 | 7,138.38 | 6,857.09 | 6,558.59 | 6,329.88 | 26,883.94 |
| | 985 | 8,845.69 | 8,814.84 | 8,393.77 | 8,083.82 | 34,138.12 |
| | 986 | 12,980.03 | 12,771.64 | 12,537.87 | 12,051.48 | 50,341.02 |
| | 988 | 4,890.39 | 4,752.45 | 4,413.51 | 4,410.95 | 18,467.30 |
| | 989 | 5,983.87 | 5,988.35 | 5,654.84 | 5,290.25 | 22,917.31 |
| | 990 | 3,457.29 | 3,520.34 | 3,453.82 | 3,506.08 | 13,937.53 |
| | 991 | 4,907.60 | 4,557.63 | 4,451.80 | 4,328.86 | 18,245.89 |
| | 992 | 19,988.26 | 19,132.18 | 19,022.28 | 17,753.64 | 75,896.36 |
| | 993 | 11,578.19 | 11,278.27 | 10,564.97 | 10,334.79 | 43,756.22 |
| | 994 | 2,237.12 | 2,257.53 | 1,976.21 | 2,069.30 | 8,540.16 |
| TOTAL | | 137,465.59 | 133,461.19 | 128,219.89 | 124,321.74 | 523,468.41 |

STATE: WEST VIRGINIA

| ZIP CODE | | QUARTER 1 | QUARTER 2 | QUARTER 3 | QUARTER 4 | TOTAL GRAMS |
|---|---|---|---|---|---|---|
| | 247 | 2,309.02 | 2,243.84 | 1,981.78 | 1,877.48 | 8,412.12 |
| | 248 | 1,510.49 | 1,441.75 | 1,321.21 | 1,236.17 | 5,509.62 |
| | 249 | 1,250.47 | 1,132.21 | 1,109.02 | 1,097.97 | 4,589.67 |
| | 250 | 2,748.54 | 2,656.92 | 2,500.23 | 2,421.36 | 10,327.05 |
| | 251 | 1,883.82 | 1,812.45 | 1,857.16 | 1,821.77 | 7,375.20 |
| | 252 | 1,723.42 | 1,515.67 | 1,466.02 | 1,454.95 | 6,160.06 |
| | 253 | 6,034.27 | 5,811.22 | 5,638.47 | 5,459.56 | 22,943.52 |
| | 254 | 2,393.80 | 2,369.11 | 2,263.02 | 2,186.82 | 9,212.75 |
| | 255 | 5,507.91 | 5,216.76 | 5,124.46 | 4,980.14 | 20,829.27 |

| | | | | | | |
|---|---|---|---|---|---|---|
| | 256 | 3,209.75 | 3,060.86 | 2,948.88 | 2,658.91 | 11,878.40 |
| | 257 | 4,887.81 | 4,680.82 | 4,183.72 | 3,828.56 | 17,580.91 |
| | 258 | 4,324.28 | 4,147.35 | 3,863.11 | 3,870.24 | 16,204.98 |
| | 259 | 1,566.52 | 1,393.94 | 1,240.80 | 1,262.90 | 5,464.16 |
| | 260 | 3,716.06 | 3,558.09 | 3,441.39 | 3,433.37 | 14,148.91 |
| | 261 | 3,990.86 | 3,918.05 | 3,701.03 | 3,518.80 | 15,128.74 |
| | 262 | 2,304.31 | 2,278.15 | 2,167.97 | 2,129.01 | 8,879.44 |
| | 263 | 5,091.60 | 5,102.25 | 5,001.51 | 4,643.26 | 19,838.62 |
| | 264 | 1,253.79 | 1,305.38 | 1,246.44 | 1,200.79 | 5,006.40 |
| | 265 | 4,447.62 | 4,320.03 | 4,043.32 | 3,833.96 | 16,644.93 |
| | 266 | 1,293.48 | 1,172.67 | 1,135.95 | 1,144.06 | 4,746.16 |
| | 267 | 868.65 | 823.05 | 762.72 | 771.09 | 3,225.51 |
| | 268 | 429.89 | 374.99 | 354.3 | 335.98 | 1,495.16 |
| TOTAL | | 62,746.36 | 60,335.56 | 57,352.51 | 55,167.15 | 235,601.58 |

| STATE: | WISCONSIN | | | | | |
|---|---|---|---|---|---|---|
| ZIP CODE | | QUARTER 1 | QUARTER 2 | QUARTER 3 | QUARTER 4 | TOTAL GRAMS |
| | 530 | 7,508.67 | 7,130.27 | 6,946.78 | 7,180.39 | 28,766.11 |
| | 531 | 10,812.57 | 10,119.37 | 9,726.25 | 10,005.97 | 40,664.16 |
| | 532 | 11,228.78 | 10,677.71 | 10,358.64 | 10,202.34 | 42,467.47 |
| | 534 | 2,694.32 | 2,408.72 | 2,419.73 | 2,416.92 | 9,939.69 |
| | 535 | 8,196.46 | 7,869.04 | 7,479.48 | 7,480.68 | 31,025.66 |
| | 537 | 4,083.30 | 3,760.12 | 3,566.23 | 3,532.01 | 14,941.66 |
| | 538 | 753.45 | 750.1 | 726.33 | 718.9 | 2,948.78 |
| | 539 | 4,072.41 | 3,920.30 | 3,720.80 | 3,796.51 | 15,510.02 |
| | 540 | 887.25 | 861.66 | 895.11 | 828.45 | 3,472.47 |
| | 541 | 2,639.90 | 2,533.42 | 2,516.37 | 2,460.22 | 10,149.91 |
| | 542 | 1,476.08 | 1,357.90 | 1,373.10 | 1,327 | 5,534.08 |
| | 543 | 2,848.41 | 2,628.79 | 2,614.77 | 2,587.07 | 10,679.04 |
| | 544 | 5,607.78 | 5,263.89 | 4,956.55 | 5,002.69 | 20,830.91 |
| | 545 | 1,527.70 | 1,436.53 | 1,540.24 | 1,325.96 | 5,830.43 |
| | 546 | 4,732.97 | 4,499.88 | 4,266.56 | 4,300.39 | 17,799.80 |
| | 547 | 3,238.96 | 2,973.83 | 2,786.85 | 2,791.75 | 11,791.39 |
| | 548 | 3,276.96 | 3,135.93 | 3,030.41 | 2,932.55 | 12,375.85 |
| | 549 | 6,952.63 | 6,650.10 | 6,606.64 | 6,596.08 | 26,805.45 |
| TOTAL | | 82,538.60 | 77,977.56 | 75,530.84 | 75,485.88 | 311,532.88 |

STATE: WYOMING

| ZIP CODE | QUARTER 1 | QUARTER 2 | QUARTER 3 | QUARTER 4 | TOTAL GRAMS |
|---|---|---|---|---|---|
| 820 | 2,417.31 | 2,326.17 | 2,182.18 | 2,120.60 | 9,046.26 |
| 822 | 450.03 | 454.33 | 433.27 | 469.19 | 1,806.82 |
| 823 | 561.41 | 378.06 | 205.4 | 206.21 | 1,351.08 |
| 824 | 1,264.57 | 1,153.51 | 1,186.42 | 1,186.31 | 4,790.81 |
| 825 | 783.63 | 787.44 | 713.63 | 740.74 | 3,025.44 |
| 826 | 2,274.77 | 2,204.36 | 2,075.86 | 2,109.29 | 8,664.28 |
| 827 | 847.26 | 802.93 | 772.29 | 759.14 | 3,181.62 |
| 828 | 1,156.13 | 1,242.40 | 1,169.19 | 1,150.49 | 4,718.21 |
| 829 | 1,745.09 | 1,689.25 | 1,712.04 | 1,677.10 | 6,823.48 |
| 830 | 234.18 | 211.55 | 237.34 | 205.04 | 888.11 |
| 831 | 450.04 | 443.33 | 424.52 | 395.44 | 1,713.33 |
| TOTAL | 12,184.42 | 11,693.33 | 11,112.14 | 11,019.55 | 46,009.44 |

DRUG CODE: 9220L DRUG NAME: LEVORPHANOL

STATE: ALABAMA

| ZIP CODE | QUARTER 1 | QUARTER 2 | QUARTER 3 | QUARTER 4 | TOTAL GRAMS |
|---|---|---|---|---|---|
| 350 | 1.51 | 0.58 | 1.04 | 0.46 | 3.59 |
| 351 | 0 | 0.12 | 0.12 | 0.12 | 0.36 |
| 352 | 1.04 | 0.93 | 1.28 | 1.28 | 4.53 |
| 354 | 0.93 | 0.46 | 0.81 | 1.04 | 3.24 |
| 357 | 0.12 | 0 | 0 | 0 | 0.12 |
| 359 | 0.23 | 0.35 | 0.46 | 0.35 | 1.39 |
| 361 | 1.04 | 0.93 | 0.58 | 0.35 | 2.9 |
| 364 | 0.81 | 0.58 | 0.46 | 0.46 | 2.31 |
| 365 | 0.23 | 0.35 | 0.35 | 0.23 | 1.16 |
| 366 | 0 | 0 | 0.23 | 0.23 | 0.46 |
| TOTAL | 5.91 | 4.3 | 5.33 | 4.52 | 20.06 |

STATE: ARIZONA

| ZIP CODE | QUARTER 1 | QUARTER 2 | QUARTER 3 | QUARTER 4 | TOTAL GRAMS |
|---|---|---|---|---|---|
| 850 | 7.2 | 11.26 | 14.28 | 23.68 | 56.42 |
| 851 | 0.12 | 1.16 | 1.16 | 0.81 | 3.25 |
| 852 | 10.68 | 9.63 | 13.35 | 4.29 | 37.95 |
| 853 | 0.23 | 0 | 0.58 | 1.16 | 1.97 |
| 860 | 0.23 | 0 | 0 | 0 | 0.23 |
| 863 | 0 | 0 | 0 | 0.12 | 0.12 |
| TOTAL | 18.46 | 22.05 | 29.37 | 30.06 | 99.94 |

STATE: ARKANSAS

| ZIP CODE | QUARTER 1 | QUARTER 2 | QUARTER 3 | QUARTER 4 | TOTAL GRAMS |
|---|---|---|---|---|---|
| 716 | 0 | 0 | 0.35 | 0.12 | 0.47 |
| 719 | 2.79 | 1.74 | 0.58 | 0.58 | 5.69 |
| 720 | 0 | 0.23 | 0.58 | 0.12 | 0.93 |
| 727 | 0 | 1.39 | 0.46 | 0.12 | 1.97 |
| TOTAL | 2.79 | 3.36 | 1.97 | 0.94 | 9.06 |

STATE: CALIFORNIA

| ZIP CODE | QUARTER 1 | QUARTER 2 | QUARTER 3 | QUARTER 4 | TOTAL GRAMS |
|---|---|---|---|---|---|
| 900 | 3.02 | 3.37 | 3.25 | 2.32 | 11.96 |
| 902 | 2.32 | 2.44 | 3.13 | 2.09 | 9.98 |
| 904 | 14.74 | 19.85 | 19.97 | 23.33 | 77.89 |
| 905 | 0 | 0.12 | 0.12 | 1.16 | 1.4 |
| 907 | 1.04 | 1.04 | 0.7 | 1.28 | 4.06 |
| 908 | 0 | 0.12 | 0.46 | 0.12 | 0.7 |
| 910 | 0.46 | 0.12 | 0.35 | 0.23 | 1.16 |
| 911 | 0.7 | 0.35 | 0 | 0 | 1.05 |
| 912 | 0 | 0 | 0 | 0.12 | 0.12 |
| 913 | 3.25 | 2.79 | 1.86 | 4.64 | 12.54 |
| 914 | 0.35 | 0.35 | 0.46 | 0.23 | 1.39 |
| 915 | 0 | 0 | 0 | 0.35 | 0.35 |
| 917 | 43.76 | 38.77 | 38.31 | 21.71 | 142.55 |
| 919 | 1.28 | 1.04 | 0.93 | 0.7 | 3.95 |
| 920 | 3.02 | 2.09 | 3.25 | 6.04 | 14.4 |
| 921 | 2.21 | 1.39 | 0.58 | 0.7 | 4.88 |

| | | | | | | |
|---|---|---|---|---|---|---|
| | 922 | 0.46 | 0.12 | 0.46 | 0.35 | 1.39 |
| | 923 | 2.32 | 1.28 | 1.51 | 1.86 | 6.97 |
| | 924 | 0.58 | 1.04 | 1.63 | 1.74 | 4.99 |
| | 925 | 0.12 | 0.81 | 1.63 | 1.63 | 4.19 |
| | 926 | 5.11 | 4.29 | 4.88 | 3.6 | 17.88 |
| | 927 | 0 | 0 | 0 | 0.23 | 0.23 |
| | 928 | 0 | 0.12 | 0.58 | 0.58 | 1.28 |
| | 930 | 0.12 | 1.39 | 0.23 | 0 | 1.74 |
| | 931 | 0.12 | 1.74 | 0.12 | 0 | 1.98 |
| | 932 | 0 | 0.7 | 0.7 | 0.35 | 1.75 |
| | 934 | 0.35 | 0 | 0.35 | 0 | 0.7 |
| | 935 | 0.7 | 1.28 | 0.81 | 0.58 | 3.37 |
| | 936 | 0.12 | 0.23 | 0.23 | 0.7 | 1.28 |
| | 937 | 0.23 | 1.16 | 0.35 | 0.46 | 2.2 |
| | 939 | 0.58 | 0.7 | 0.81 | 0.23 | 2.32 |
| | 940 | 0.93 | 0.7 | 0.46 | 1.28 | 3.37 |
| | 941 | 1.63 | 1.86 | 1.51 | 3.71 | 8.71 |
| | 943 | 0 | 0.12 | 0 | 0.23 | 0.35 |
| | 945 | 2.79 | 1.86 | 2.32 | 1.63 | 8.6 |
| | 949 | 0.81 | 0.23 | 0.12 | 0.23 | 1.39 |
| | 950 | 2.32 | 2.32 | 3.13 | 2.9 | 10.67 |
| | 951 | 0 | 0 | 0.46 | 0.7 | 1.16 |
| | 953 | 0.35 | 0.23 | 0.12 | 0.12 | 0.82 |
| | 954 | 1.04 | 1.63 | 1.04 | 1.51 | 5.22 |
| | 956 | 0.58 | 0.58 | 0.23 | 0.46 | 1.85 |
| | 957 | 0 | 0.12 | 0 | 0 | 0.12 |
| | 958 | 12.42 | 3.6 | 1.16 | 1.39 | 18.57 |
| | 959 | 0 | 0.23 | 0.7 | 0.23 | 1.16 |
| TOTAL | | 109.83 | 102.18 | 98.91 | 91.72 | 402.64 |

STATE: COLORADO

| ZIP CODE | | QUARTER 1 | QUARTER 2 | QUARTER 3 | QUARTER 4 | TOTAL GRAMS |
|---|---|---|---|---|---|---|
| | 800 | 0.46 | 0.7 | 1.63 | 1.74 | 4.53 |
| | 801 | 0.23 | 0.23 | 1.51 | 4.06 | 6.03 |
| | 802 | 2.44 | 8.01 | 6.96 | 8.94 | 26.35 |
| | 803 | 0 | 0.23 | 0 | 0 | 0.23 |

| | | | | | | |
|---|---|---|---|---|---|---|
| | 804 | 0 | 0.23 | 0 | 0 | 0.23 |
| | 805 | 0.7 | 1.39 | 1.39 | 1.28 | 4.76 |
| | 806 | 1.63 | 1.63 | 1.74 | 1.39 | 6.39 |
| | 807 | 0 | 0 | 0.12 | 0.23 | 0.35 |
| | 808 | 0.12 | 0 | 0.12 | 0 | 0.24 |
| | 809 | 0.46 | 0.46 | 0.23 | 0.46 | 1.61 |
| | 815 | 0 | 0 | 0.12 | 0 | 0.12 |
| TOTAL | | 6.04 | 12.88 | 13.82 | 18.1 | 50.84 |

| STATE: | CONNECTICUT | | | | |
|---|---|---|---|---|---|
| ZIP CODE | QUARTER 1 | QUARTER 2 | QUARTER 3 | QUARTER 4 | TOTAL GRAMS |
| 60 | 0 | 0.7 | 0.7 | 0.81 | 2.21 |
| 61 | 0.23 | 0.23 | 0.23 | 0.46 | 1.15 |
| 63 | 0.23 | 0 | 0 | 0 | 0.23 |
| 64 | 0.58 | 1.16 | 2.21 | 3.6 | 7.55 |
| 65 | 0.12 | 0.58 | 0.93 | 2.44 | 4.07 |
| 66 | 0.35 | 0.7 | 0.58 | 1.51 | 3.14 |
| 67 | 0.12 | 0 | 0 | 0 | 0.12 |
| 68 | 0 | 0 | 0 | 0.12 | 0.12 |
| 69 | 0 | 0.35 | 0.46 | 0.81 | 1.62 |
| TOTAL | 1.63 | 3.72 | 5.11 | 9.75 | 20.21 |

| STATE: | DELAWARE | | | | |
|---|---|---|---|---|---|
| ZIP CODE | QUARTER 1 | QUARTER 2 | QUARTER 3 | QUARTER 4 | TOTAL GRAMS |
| 197 | 0.12 | 0 | 0 | 0 | 0.12 |
| 199 | 0.81 | 0 | 0.58 | 1.04 | 2.43 |
| TOTAL | 0.93 | 0 | 0.58 | 1.04 | 2.55 |

| STATE: | FLORIDA | | | | |
|---|---|---|---|---|---|
| ZIP CODE | QUARTER 1 | QUARTER 2 | QUARTER 3 | QUARTER 4 | TOTAL GRAMS |
| 320 | 0 | 0.12 | 0 | 0 | 0.12 |
| 321 | 0 | 0.23 | 0.12 | 0 | 0.35 |
| 322 | 0 | 0 | 0.23 | 0.35 | 0.58 |
| 324 | 0 | 0.23 | 0.35 | 0.58 | 1.16 |

| | | | | | | |
|---|---|---|---|---|---|---|
| | 325 | 0.46 | 0.81 | 0.93 | 0.81 | 3.01 |
| | 326 | 0.23 | 0 | 0.12 | 1.28 | 1.63 |
| | 327 | 21.01 | 29.95 | 27.98 | 31.34 | 110.28 |
| | 328 | 0.35 | 0 | 0 | 0 | 0.35 |
| | 330 | 0.23 | 0.35 | 0.23 | 0 | 0.81 |
| | 331 | 0 | 0 | 0 | 0.12 | 0.12 |
| | 333 | 0 | 0 | 0 | 0.23 | 0.23 |
| | 334 | 0 | 0 | 0 | 0.12 | 0.12 |
| | 335 | 0 | 0.23 | 0.7 | 1.39 | 2.32 |
| | 336 | 0 | 0.12 | 0 | 0.35 | 0.47 |
| | 337 | 0 | 0.7 | 0.12 | 0 | 0.82 |
| | 338 | 0.12 | 0.46 | 0 | 0 | 0.58 |
| | 339 | 0.23 | 0 | 0.46 | 0.46 | 1.15 |
| | 341 | 0.12 | 0.12 | 0 | 0 | 0.24 |
| | 342 | 3.48 | 2.55 | 2.9 | 2.55 | 11.48 |
| | 344 | 0 | 0 | 0.23 | 0 | 0.23 |
| TOTAL | | 26.23 | 35.87 | 34.37 | 39.58 | 136.05 |

| STATE: | GEORGIA | | | | |
|---|---|---|---|---|---|
| ZIP CODE | QUARTER 1 | QUARTER 2 | QUARTER 3 | QUARTER 4 | TOTAL GRAMS |
| 300 | 0 | 0.23 | 0.58 | 0.7 | 1.51 |
| 301 | 0.23 | 0.35 | 0.46 | 0.81 | 1.85 |
| 303 | 0.46 | 0.81 | 0.93 | 1.51 | 3.71 |
| 305 | 0 | 0.23 | 0.35 | 0 | 0.58 |
| 306 | 0 | 0.23 | 0.46 | 0.23 | 0.92 |
| 308 | 0 | 0 | 0.12 | 0.23 | 0.35 |
| 309 | 0 | 0.23 | 0 | 0 | 0.23 |
| 314 | 0 | 0.46 | 0 | 0 | 0.46 |
| TOTAL | 0.69 | 2.54 | 2.9 | 3.48 | 9.61 |

| STATE: | IDAHO | | | | |
|---|---|---|---|---|---|
| ZIP CODE | QUARTER 1 | QUARTER 2 | QUARTER 3 | QUARTER 4 | TOTAL GRAMS |
| 838 | 0 | 0 | 0.12 | 0.12 | 0.24 |
| TOTAL | 0 | 0 | 0.12 | 0.12 | 0.24 |

DATE RANGE: 01/01/2017 TO 12/31/2017 ARCOS 3 - REPORT 1 Run Date: 07/03/2018

RETAIL DRUG DISTRIBUTION BY ZIP CODE WITHIN STATE BY GRAMS WT

STATE: ILLINOIS

| ZIP CODE | QUARTER 1 | QUARTER 2 | QUARTER 3 | QUARTER 4 | TOTAL GRAMS |
|---|---|---|---|---|---|
| 600 | 0.7 | 0.46 | 1.39 | 1.16 | 3.71 |
| 601 | 2.32 | 5.11 | 3.6 | 4.64 | 15.67 |
| 604 | 0.81 | 0.93 | 1.28 | 1.28 | 4.3 |
| 605 | 0.93 | 1.28 | 1.86 | 1.74 | 5.81 |
| 606 | 0.81 | 0.81 | 1.63 | 2.55 | 5.8 |
| 607 | 0 | 0.46 | 0 | 0 | 0.46 |
| 609 | 0 | 0.35 | 0.23 | 0.58 | 1.16 |
| 613 | 0 | 0 | 0.12 | 0.12 | 0.24 |
| 618 | 0.35 | 0 | 0 | 0 | 0.35 |
| 622 | 1.86 | 2.67 | 3.95 | 2.79 | 11.27 |
| 627 | 0.58 | 0.81 | 0.58 | 0.7 | 2.67 |
| 628 | 0.35 | 0.23 | 0.23 | 0.23 | 1.04 |
| 629 | 0 | 0 | 0 | 0.23 | 0.23 |
| TOTAL | 8.71 | 13.11 | 14.87 | 16.02 | 52.71 |

STATE: INDIANA

| ZIP CODE | QUARTER 1 | QUARTER 2 | QUARTER 3 | QUARTER 4 | TOTAL GRAMS |
|---|---|---|---|---|---|
| 460 | 0.46 | 0.7 | 0.7 | 0.58 | 2.44 |
| 461 | 0.35 | 0 | 0 | 0 | 0.35 |
| 462 | 1.16 | 1.63 | 1.74 | 1.39 | 5.92 |
| 465 | 0.35 | 0.58 | 0 | 0 | 0.93 |
| 467 | 11.49 | 8.94 | 9.52 | 9.63 | 39.58 |
| 468 | 1.51 | 2.21 | 1.63 | 0.81 | 6.16 |
| 469 | 4.53 | 5.46 | 3.71 | 4.88 | 18.58 |
| 470 | 0 | 0 | 0 | 0.12 | 0.12 |
| 471 | 1.97 | 1.16 | 2.55 | 0 | 5.68 |
| 472 | 0.23 | 0.58 | 0.93 | 0.23 | 1.97 |
| 473 | 2.9 | 2.32 | 3.13 | 3.48 | 11.83 |
| 479 | 0.93 | 0.93 | 0.93 | 1.04 | 3.83 |
| TOTAL | 25.88 | 24.51 | 24.84 | 22.16 | 97.39 |

STATE: IOWA

| ZIP CODE | QUARTER 1 | QUARTER 2 | QUARTER 3 | QUARTER 4 | TOTAL GRAMS |
|---|---|---|---|---|---|
| 500 | 0 | 0 | 0.35 | 0.23 | 0.58 |
| 501 | 0.12 | 0 | 0 | 0 | 0.12 |
| 503 | 0.46 | 0.35 | 0.46 | 0.35 | 1.62 |
| 504 | 0 | 0 | 0.12 | 0 | 0.12 |
| 506 | 1.74 | 0.81 | 0.81 | 0.7 | 4.06 |
| 507 | 0.7 | 1.04 | 0.35 | 0 | 2.09 |
| 514 | 0 | 0 | 0 | 0.35 | 0.35 |
| 515 | 0.58 | 1.28 | 0.81 | 1.86 | 4.53 |
| 516 | 0 | 0.23 | 0.23 | 0.23 | 0.69 |
| 520 | 0.58 | 0 | 0 | 0 | 0.58 |
| 524 | 0.23 | 0.23 | 0.23 | 0 | 0.69 |
| TOTAL | 4.41 | 3.94 | 3.36 | 3.72 | 15.43 |

STATE: KANSAS

| ZIP CODE | QUARTER 1 | QUARTER 2 | QUARTER 3 | QUARTER 4 | TOTAL GRAMS |
|---|---|---|---|---|---|
| 660 | 3.13 | 3.13 | 3.13 | 3.02 | 12.41 |
| 662 | 8.13 | 6.85 | 3.71 | 4.18 | 22.87 |
| 665 | 0 | 0 | 0.35 | 0 | 0.35 |
| 677 | 0 | 0 | 0 | 0.23 | 0.23 |
| TOTAL | 11.26 | 9.98 | 7.19 | 7.43 | 35.86 |

STATE: KENTUCKY

| ZIP CODE | QUARTER 1 | QUARTER 2 | QUARTER 3 | QUARTER 4 | TOTAL GRAMS |
|---|---|---|---|---|---|
| 410 | 0 | 0 | 0.23 | 0.12 | 0.35 |
| TOTAL | 0 | 0 | 0.23 | 0.12 | 0.35 |

STATE: LOUISIANA

| ZIP CODE | QUARTER 1 | QUARTER 2 | QUARTER 3 | QUARTER 4 | TOTAL GRAMS |
|---|---|---|---|---|---|
| 700 | 0.58 | 0.12 | 0.12 | 0.23 | 1.05 |
| 701 | 0.35 | 0 | 0 | 0 | 0.35 |
| 703 | 0.58 | 1.16 | 0.93 | 0.12 | 2.79 |
| 704 | 0.23 | 0 | 0.23 | 0.35 | 0.81 |
| 705 | 14.63 | 17.53 | 18.69 | 17.99 | 68.84 |

| | | | | | | |
|---|---|---|---|---|---|---|
| | 707 | 0.12 | 0.35 | 0.81 | 0.81 | 2.09 |
| | 708 | 0 | 0.23 | 0 | 0 | 0.23 |
| | 711 | 0 | 0.12 | 0 | 0 | 0.12 |
| TOTAL | | 16.49 | 19.51 | 20.78 | 19.5 | 76.28 |

STATE: MARYLAND

| ZIP CODE | | QUARTER 1 | QUARTER 2 | QUARTER 3 | QUARTER 4 | TOTAL GRAMS |
|---|---|---|---|---|---|---|
| | 206 | 0 | 0 | 0 | 0.58 | 0.58 |
| | 207 | 0 | 0 | 0 | 0.58 | 0.58 |
| | 210 | 0.81 | 1.97 | 1.16 | 1.63 | 5.57 |
| | 211 | 1.28 | 0.81 | 0.93 | 1.16 | 4.18 |
| | 212 | 0.7 | 2.32 | 0.93 | 2.21 | 6.16 |
| | 214 | 0 | 0.58 | 0.81 | 2.21 | 3.6 |
| | 215 | 0 | 0.12 | 0 | 0 | 0.12 |
| | 217 | 0.23 | 0 | 0 | 0 | 0.23 |
| | 218 | 0.12 | 0.23 | 0 | 0 | 0.35 |
| TOTAL | | 3.14 | 6.03 | 3.83 | 8.37 | 21.37 |

STATE: MASSACHUSETTS

| ZIP CODE | | QUARTER 1 | QUARTER 2 | QUARTER 3 | QUARTER 4 | TOTAL GRAMS |
|---|---|---|---|---|---|---|
| | 10 | 6.27 | 8.36 | 7.89 | 8.01 | 30.53 |
| | 11 | 3.02 | 1.74 | 2.67 | 1.74 | 9.17 |
| | 13 | 0.7 | 0.35 | 0.35 | 0.58 | 1.98 |
| | 14 | 0 | 0 | 0.12 | 0 | 0.12 |
| | 15 | 0.35 | 0 | 0 | 0 | 0.35 |
| | 16 | 0.12 | 0 | 0 | 0 | 0.12 |
| | 18 | 5.8 | 5.92 | 6.15 | 6.73 | 24.6 |
| | 20 | 0.35 | 0.35 | 0.46 | 0.35 | 1.51 |
| | 21 | 1.04 | 1.04 | 0.7 | 1.51 | 4.29 |
| | 23 | 3.02 | 1.86 | 1.63 | 1.16 | 7.67 |
| | 27 | 0.12 | 0 | 0.23 | 0 | 0.35 |
| TOTAL | | 20.79 | 19.62 | 20.2 | 20.08 | 80.69 |

STATE: MICHIGAN

| ZIP CODE | QUARTER 1 | QUARTER 2 | QUARTER 3 | QUARTER 4 | TOTAL GRAMS |
|---|---|---|---|---|---|
| 480 | 0.12 | 0.23 | 0 | 0 | 0.35 |
| 483 | 0 | 0.12 | 0 | 0 | 0.12 |
| 486 | 0 | 0 | 0.12 | 0 | 0.12 |
| 496 | 0 | 0 | 0.23 | 0 | 0.23 |
| 497 | 0.23 | 0 | 0 | 0 | 0.23 |
| TOTAL | 0.35 | 0.35 | 0.35 | 0 | 1.05 |

STATE: MINNESOTA

| ZIP CODE | QUARTER 1 | QUARTER 2 | QUARTER 3 | QUARTER 4 | TOTAL GRAMS |
|---|---|---|---|---|---|
| 551 | 0.35 | 0.46 | 0.81 | 1.16 | 2.78 |
| TOTAL | 0.35 | 0.46 | 0.81 | 1.16 | 2.78 |

STATE: MISSISSIPPI

| ZIP CODE | QUARTER 1 | QUARTER 2 | QUARTER 3 | QUARTER 4 | TOTAL GRAMS |
|---|---|---|---|---|---|
| 394 | 0.23 | 0 | 0 | 0 | 0.23 |
| TOTAL | 0.23 | 0 | 0 | 0 | 0.23 |

STATE: MISSOURI

| ZIP CODE | QUARTER 1 | QUARTER 2 | QUARTER 3 | QUARTER 4 | TOTAL GRAMS |
|---|---|---|---|---|---|
| 630 | 0.35 | 0.23 | 1.74 | 0.46 | 2.78 |
| 631 | 11.49 | 15.55 | 21.13 | 23.22 | 71.39 |
| 633 | 1.51 | 2.67 | 2.09 | 2.09 | 8.36 |
| 636 | 0 | 0.23 | 0.35 | 0.12 | 0.7 |
| 637 | 0 | 0 | 0.58 | 0.7 | 1.28 |
| 640 | 0 | 0 | 0 | 0.35 | 0.35 |
| 641 | 0 | 0.46 | 0 | 0 | 0.46 |
| 657 | 0 | 0 | 0.35 | 0.35 | 0.7 |
| 658 | 0.46 | 0.46 | 0 | 0 | 0.92 |
| TOTAL | 13.81 | 19.6 | 26.24 | 27.29 | 86.94 |

DATE RANGE: 01/01/2017 TO 12/31/2017 | ARCOS 3 - REPORT 1 | Run Date: 07/03/2018

RETAIL DRUG DISTRIBUTION BY ZIP CODE WITHIN STATE BY GRAMS WT

STATE: NEBRASKA

| ZIP CODE | QUARTER 1 | QUARTER 2 | QUARTER 3 | QUARTER 4 | TOTAL GRAMS |
|---|---|---|---|---|---|
| 680 | 1.04 | 0.81 | 1.16 | 0.81 | 3.82 |
| 681 | 3.02 | 4.06 | 4.06 | 7.2 | 18.34 |
| 685 | 0 | 0.23 | 0.23 | 1.16 | 1.62 |
| TOTAL | 4.06 | 5.1 | 5.45 | 9.17 | 23.78 |

STATE: NEVADA

| ZIP CODE | QUARTER 1 | QUARTER 2 | QUARTER 3 | QUARTER 4 | TOTAL GRAMS |
|---|---|---|---|---|---|
| 890 | 0 | 0 | 0 | 0.12 | 0.12 |
| 891 | 2.55 | 2.32 | 4.29 | 2.67 | 11.83 |
| 894 | 0.35 | 0.23 | 0 | 0 | 0.58 |
| 895 | 0 | 0.35 | 0.58 | 0.7 | 1.63 |
| TOTAL | 2.9 | 2.9 | 4.87 | 3.49 | 14.16 |

STATE: NEW HAMPSHIRE

| ZIP CODE | QUARTER 1 | QUARTER 2 | QUARTER 3 | QUARTER 4 | TOTAL GRAMS |
|---|---|---|---|---|---|
| 30 | 0 | 0.12 | 0 | 0 | 0.12 |
| 38 | 1.16 | 1.74 | 1.63 | 0.7 | 5.23 |
| TOTAL | 1.16 | 1.86 | 1.63 | 0.7 | 5.35 |

STATE: NEW JERSEY

| ZIP CODE | QUARTER 1 | QUARTER 2 | QUARTER 3 | QUARTER 4 | TOTAL GRAMS |
|---|---|---|---|---|---|
| 70 | 311.67 | 399.55 | 405.7 | 496.82 | 1,613.74 |
| 74 | 0 | 0.12 | 0.12 | 0 | 0.24 |
| 77 | 0.46 | 0.7 | 0.46 | 0.35 | 1.97 |
| 78 | 0.46 | 0.46 | 0 | 0 | 0.92 |
| 79 | 0.23 | 0 | 0.12 | 0 | 0.35 |
| 80 | 0.46 | 0 | 0.35 | 0.23 | 1.04 |
| 82 | 0.23 | 0 | 0 | 0.46 | 0.69 |
| 85 | 0.23 | 0 | 0 | 0.12 | 0.35 |
| 86 | 0.12 | 0 | 0 | 0 | 0.12 |

| | | | | | | |
|---|---|---|---|---|---|---|
| | 87 | 0 | 0 | 0.23 | 0.35 | 0.58 |
| | 88 | 0 | 0 | 0 | 0.23 | 0.23 |
| TOTAL | | 313.86 | 400.83 | 406.98 | 498.56 | 1,620.23 |

STATE: NEW YORK

| ZIP CODE | QUARTER 1 | QUARTER 2 | QUARTER 3 | QUARTER 4 | TOTAL GRAMS |
|---|---|---|---|---|---|
| 100 | 1.04 | 1.39 | 1.63 | 0.81 | 4.87 |
| 106 | 0.23 | 0.35 | 0.35 | 0.7 | 1.63 |
| 110 | 0.35 | 0 | 0.35 | 0 | 0.7 |
| 111 | 0 | 0 | 0 | 0.23 | 0.23 |
| 112 | 0 | 0.12 | 0.12 | 0.12 | 0.36 |
| 113 | 0.35 | 0.23 | 0.12 | 0.35 | 1.05 |
| 114 | 0.12 | 0 | 0.35 | 0.23 | 0.7 |
| 115 | 1.63 | 1.39 | 1.86 | 1.63 | 6.51 |
| 117 | 8.71 | 15.32 | 11.26 | 11.03 | 46.32 |
| 118 | 0.58 | 0 | 0.46 | 4.88 | 5.92 |
| 119 | 0 | 0.23 | 0.46 | 0.93 | 1.62 |
| 120 | 1.16 | 1.28 | 1.28 | 1.74 | 5.46 |
| 121 | 2.55 | 2.09 | 2.55 | 2.32 | 9.51 |
| 122 | 2.9 | 3.48 | 2.21 | 2.21 | 10.8 |
| 123 | 0.23 | 0.12 | 0.35 | 0.23 | 0.93 |
| 125 | 0 | 0 | 0.12 | 0.35 | 0.47 |
| 128 | 0 | 0 | 0.7 | 1.04 | 1.74 |
| 129 | 0.12 | 0.23 | 0.58 | 0.46 | 1.39 |
| 130 | 0.58 | 0.58 | 0.58 | 0 | 1.74 |
| 132 | 0 | 0 | 0 | 0.7 | 0.7 |
| 133 | 0.46 | 0.12 | 0.46 | 0 | 1.04 |
| 134 | 1.16 | 0.58 | 1.16 | 1.51 | 4.41 |
| 135 | 0.23 | 0.12 | 0.46 | 0.23 | 1.04 |
| 136 | 0.23 | 0 | 0 | 0 | 0.23 |
| 137 | 0.7 | 1.04 | 1.16 | 3.37 | 6.27 |
| 138 | 1.97 | 1.63 | 4.64 | 4.64 | 12.88 |
| 139 | 1.51 | 2.79 | 3.37 | 4.64 | 12.31 |
| 140 | 0.58 | 0.93 | 1.63 | 1.39 | 4.53 |
| 141 | 0.46 | 0.46 | 0.46 | 0.93 | 2.31 |
| 142 | 3.13 | 2.9 | 1.97 | 1.39 | 9.39 |

| | | | | | | |
|---|---|---|---|---|---|---|
| | 145 | 0.23 | 0 | 0 | 0 | 0.23 |
| | 146 | 0.58 | 0 | 0 | 0 | 0.58 |
| | 148 | 1.39 | 1.28 | 0.35 | 0.23 | 3.25 |
| TOTAL | | 33.18 | 38.66 | 40.99 | 48.29 | 161.12 |

STATE: NORTH CAROLINA

| ZIP CODE | | QUARTER 1 | QUARTER 2 | QUARTER 3 | QUARTER 4 | TOTAL GRAMS |
|---|---|---|---|---|---|---|
| | 270 | 1.04 | 0.35 | 0.7 | 0.7 | 2.79 |
| | 271 | 0.46 | 1.04 | 1.04 | 1.39 | 3.93 |
| | 272 | 0.35 | 0.23 | 0.35 | 1.63 | 2.56 |
| | 273 | 0 | 0.35 | 0.46 | 0.58 | 1.39 |
| | 274 | 0 | 0.12 | 0.12 | 0.23 | 0.47 |
| | 275 | 15.67 | 8.59 | 9.75 | 10.22 | 44.23 |
| | 276 | 0.93 | 1.97 | 2.44 | 1.39 | 6.73 |
| | 277 | 0.23 | 1.16 | 0 | 0.12 | 1.51 |
| | 278 | 5.69 | 5.57 | 6.38 | 5.57 | 23.21 |
| | 279 | 0.12 | 0 | 0.12 | 0.12 | 0.36 |
| | 280 | 1.97 | 2.21 | 2.32 | 4.29 | 10.79 |
| | 281 | 1.04 | 1.51 | 3.37 | 1.74 | 7.66 |
| | 282 | 2.9 | 2.09 | 2.09 | 0.58 | 7.66 |
| | 283 | 10.33 | 12.3 | 24.96 | 60.59 | 108.18 |
| | 284 | 2.67 | 2.55 | 2.09 | 1.51 | 8.82 |
| | 285 | 4.99 | 5.92 | 4.88 | 6.04 | 21.83 |
| | 286 | 1.63 | 1.97 | 3.48 | 4.64 | 11.72 |
| | 287 | 0.35 | 0 | 0 | 0.23 | 0.58 |
| | 288 | 0.46 | 0.58 | 0.12 | 0.12 | 1.28 |
| TOTAL | | 50.83 | 48.51 | 64.67 | 101.69 | 265.7 |

STATE: OHIO

| ZIP CODE | | QUARTER 1 | QUARTER 2 | QUARTER 3 | QUARTER 4 | TOTAL GRAMS |
|---|---|---|---|---|---|---|
| | 430 | 0.58 | 0.46 | 0 | 0 | 1.04 |
| | 432 | 7.2 | 12.54 | 10.45 | 7.66 | 37.85 |
| | 439 | 0.12 | 0.35 | 0.58 | 0.46 | 1.51 |
| | 440 | 0 | 0.46 | 0.12 | 0.35 | 0.93 |

| | | | | | | |
|---|---|---|---|---|---|---|
| | 441 | 0 | 0.23 | 0 | 0 | 0.23 |
| | 444 | 0 | 0.35 | 0.35 | 0.58 | 1.28 |
| | 445 | 0 | 0.23 | 0.23 | 0 | 0.46 |
| | 450 | 0 | 0 | 0 | 0.12 | 0.12 |
| | 458 | 0.12 | 0 | 0 | 0 | 0.12 |
| TOTAL | | 8.02 | 14.62 | 11.73 | 9.17 | 43.54 |

STATE: OREGON

| ZIP CODE | | QUARTER 1 | QUARTER 2 | QUARTER 3 | QUARTER 4 | TOTAL GRAMS |
|---|---|---|---|---|---|---|
| | 970 | 0.35 | 0.46 | 0.35 | 0.35 | 1.51 |
| | 971 | 0 | 0 | 0.12 | 0.12 | 0.24 |
| | 972 | 0.12 | 0.12 | 0.23 | 0.35 | 0.82 |
| | 974 | 2.09 | 2.21 | 1.74 | 2.21 | 8.25 |
| | 977 | 0.12 | 0 | 0 | 0 | 0.12 |
| | 978 | 0.81 | 0.7 | 0.46 | 0.7 | 2.67 |
| TOTAL | | 3.49 | 3.49 | 2.9 | 3.73 | 13.61 |

STATE: PENNSYLVANIA

| ZIP CODE | | QUARTER 1 | QUARTER 2 | QUARTER 3 | QUARTER 4 | TOTAL GRAMS |
|---|---|---|---|---|---|---|
| | 150 | 0 | 0.35 | 0.12 | 0.23 | 0.7 |
| | 151 | 0 | 0.58 | 0 | 0 | 0.58 |
| | 152 | 0 | 0.23 | 0 | 0 | 0.23 |
| | 153 | 0.58 | 0.12 | 0 | 0 | 0.7 |
| | 154 | 0 | 0 | 0.23 | 0.12 | 0.35 |
| | 156 | 0 | 0 | 0 | 0.35 | 0.35 |
| | 159 | 0.12 | 0 | 0.12 | 0 | 0.24 |
| | 160 | 0 | 0 | 0.23 | 0 | 0.23 |
| | 161 | 0 | 1.28 | 1.39 | 1.39 | 4.06 |
| | 163 | 0.46 | 0 | 0 | 0 | 0.46 |
| | 164 | 0 | 0 | 0 | 0.12 | 0.12 |
| | 170 | 1.04 | 0.81 | 0.81 | 0.23 | 2.89 |
| | 177 | 0.35 | 0.7 | 0.7 | 0.58 | 2.33 |
| | 178 | 2.09 | 2.44 | 1.97 | 1.51 | 8.01 |
| | 179 | 0.23 | 0 | 0 | 0.12 | 0.35 |

| | | | | | |
|---|---|---|---|---|---|
| 180 | 0.35 | 0.35 | 0.35 | 0.23 | 1.28 |
| 183 | 0.35 | 0.81 | 1.16 | 0.7 | 3.02 |
| 186 | 0.12 | 0 | 0 | 0 | 0.12 |
| 187 | 0.23 | 0.93 | 1.74 | 1.16 | 4.06 |
| 188 | 0.23 | 0.23 | 0.23 | 0.23 | 0.92 |
| 189 | 0.7 | 0 | 0.46 | 0.81 | 1.97 |
| 190 | 1.86 | 4.06 | 2.55 | 1.86 | 10.33 |
| 191 | 1.74 | 2.21 | 1.86 | 2.21 | 8.02 |
| 193 | 0.12 | 0.12 | 0.35 | 0.81 | 1.4 |
| 194 | 0 | 0 | 0.7 | 0.23 | 0.93 |
| TOTAL | 10.57 | 15.22 | 14.97 | 12.89 | 53.65 |

STATE: SOUTH CAROLINA

| ZIP CODE | QUARTER 1 | QUARTER 2 | QUARTER 3 | QUARTER 4 | TOTAL GRAMS |
|---|---|---|---|---|---|
| 290 | 0 | 0.23 | 0.46 | 0.23 | 0.92 |
| 291 | 0 | 0.35 | 0 | 0.12 | 0.47 |
| 292 | 0.12 | 0.12 | 0.46 | 0.46 | 1.16 |
| 294 | 0.23 | 0.35 | 0.35 | 0.23 | 1.16 |
| 295 | 0.12 | 0 | 0.23 | 0.23 | 0.58 |
| 296 | 0.35 | 0.7 | 0.93 | 1.74 | 3.72 |
| 297 | 0 | 0.12 | 0.35 | 0.35 | 0.82 |
| 298 | 0 | 1.51 | 0.46 | 0.23 | 2.2 |
| TOTAL | 0.82 | 3.38 | 3.24 | 3.59 | 11.03 |

STATE: TENNESSEE

| ZIP CODE | QUARTER 1 | QUARTER 2 | QUARTER 3 | QUARTER 4 | TOTAL GRAMS |
|---|---|---|---|---|---|
| 378 | 0.12 | 0.12 | 0.23 | 0 | 0.47 |
| 381 | 0 | 0.12 | 0 | 0 | 0.12 |
| 383 | 0 | 0 | 0.12 | 0 | 0.12 |
| TOTAL | 0.12 | 0.24 | 0.35 | 0 | 0.71 |

STATE: TEXAS

| ZIP CODE | QUARTER 1 | QUARTER 2 | QUARTER 3 | QUARTER 4 | TOTAL GRAMS |
|---|---|---|---|---|---|
| 750 | 25.89 | 35.06 | 42.49 | 39.24 | 142.68 |

| | | | | | | |
|---|---|---|---|---|---|---|
| | 751 | 0.58 | 0.58 | 2.55 | 2.55 | 6.26 |
| | 752 | 1.51 | 1.63 | 1.74 | 1.74 | 6.62 |
| | 754 | 0.58 | 0.35 | 0 | 0 | 0.93 |
| | 757 | 0.35 | 0.58 | 0.46 | 0.23 | 1.62 |
| | 760 | 4.41 | 5.46 | 1.97 | 4.64 | 16.48 |
| | 761 | 0.12 | 0.35 | 0.81 | 1.16 | 2.44 |
| | 762 | 0 | 0 | 2.32 | 3.02 | 5.34 |
| | 763 | 0 | 0 | 0.23 | 0.35 | 0.58 |
| | 764 | 0.35 | 0 | 0 | 0 | 0.35 |
| | 765 | 1.39 | 1.04 | 1.16 | 0.81 | 4.4 |
| | 769 | 0 | 0 | 0.35 | 0 | 0.35 |
| | 770 | 1.04 | 0.93 | 0.46 | 0.35 | 2.78 |
| | 773 | 0.23 | 0 | 0.23 | 0.12 | 0.58 |
| | 774 | 0.23 | 0 | 0.12 | 0.46 | 0.81 |
| | 775 | 0.23 | 0.23 | 0.23 | 0 | 0.69 |
| | 776 | 0.12 | 0.23 | 0.12 | 0.12 | 0.59 |
| | 778 | 0 | 0 | 0.46 | 0.23 | 0.69 |
| | 780 | 0 | 0.23 | 0 | 0 | 0.23 |
| | 781 | 0.46 | 0.12 | 0.12 | 0.12 | 0.82 |
| | 782 | 0 | 0.35 | 0.58 | 1.86 | 2.79 |
| | 783 | 0 | 0.12 | 0.35 | 0.46 | 0.93 |
| | 784 | 0 | 0 | 0.12 | 0.35 | 0.47 |
| | 786 | 0.58 | 0.93 | 0.7 | 0.35 | 2.56 |
| | 787 | 0.93 | 5.22 | 3.71 | 7.89 | 17.75 |
| | 791 | 0 | 0 | 0.12 | 0 | 0.12 |
| | 797 | 0 | 0.23 | 0 | 0 | 0.23 |
| | 799 | 0.23 | 0 | 0 | 0 | 0.23 |
| TOTAL | | 39.23 | 53.64 | 61.4 | 66.05 | 220.32 |

| STATE: | VERMONT | | | | |
|---|---|---|---|---|---|
| ZIP CODE | QUARTER 1 | QUARTER 2 | QUARTER 3 | QUARTER 4 | TOTAL GRAMS |
| 54 | 0.23 | 0.46 | 0.46 | 0.46 | 1.61 |
| TOTAL | 0.23 | 0.46 | 0.46 | 0.46 | 1.61 |

STATE: VIRGINIA

| ZIP CODE | QUARTER 1 | QUARTER 2 | QUARTER 3 | QUARTER 4 | TOTAL GRAMS |
|---|---|---|---|---|---|
| 201 | 0 | 0 | 0.12 | 0 | 0.12 |
| 220 | 0 | 0 | 0.23 | 1.16 | 1.39 |
| 221 | 0 | 0 | 0.12 | 1.04 | 1.16 |
| 222 | 0.12 | 0.58 | 1.16 | 4.18 | 6.04 |
| 223 | 0 | 0 | 0.93 | 2.44 | 3.37 |
| 224 | 0 | 0 | 0.23 | 1.16 | 1.39 |
| 229 | 0 | 0.12 | 0 | 0 | 0.12 |
| 230 | 0.35 | 0.12 | 0 | 0 | 0.47 |
| 231 | 1.04 | 0.46 | 0.46 | 0.46 | 2.42 |
| 232 | 0.81 | 0.35 | 0.35 | 0 | 1.51 |
| 233 | 0.12 | 0 | 0 | 0.58 | 0.7 |
| 234 | 0.93 | 1.63 | 2.79 | 0.93 | 6.28 |
| 236 | 1.28 | 0.81 | 0.93 | 1.04 | 4.06 |
| 237 | 0.12 | 0.93 | 1.39 | 1.97 | 4.41 |
| 238 | 0.7 | 0.58 | 2.21 | 1.04 | 4.53 |
| 243 | 0.35 | 0.23 | 0.23 | 0.23 | 1.04 |
| TOTAL | 5.82 | 5.81 | 11.15 | 16.23 | 39.01 |

STATE: WASHINGTON

| ZIP CODE | QUARTER 1 | QUARTER 2 | QUARTER 3 | QUARTER 4 | TOTAL GRAMS |
|---|---|---|---|---|---|
| 980 | 4.18 | 5.11 | 8.13 | 10.33 | 27.75 |
| 981 | 0.46 | 0.7 | 0.46 | 1.16 | 2.78 |
| 982 | 8.13 | 8.13 | 9.4 | 8.36 | 34.02 |
| 983 | 2.09 | 2.79 | 3.6 | 2.9 | 11.38 |
| 984 | 0.46 | 0.46 | 0.93 | 1.74 | 3.59 |
| 985 | 6.85 | 6.85 | 8.13 | 5.69 | 27.52 |
| 986 | 0.35 | 0.23 | 0.23 | 0.35 | 1.16 |
| 988 | 0.12 | 0.12 | 0 | 0.35 | 0.59 |
| 990 | 0 | 0 | 0 | 0.23 | 0.23 |
| 992 | 0 | 0 | 0 | 0.12 | 0.12 |
| 993 | 0 | 0.12 | 0 | 0 | 0.12 |
| TOTAL | 22.64 | 24.51 | 30.88 | 31.23 | 109.26 |

STATE: WISCONSIN

| ZIP CODE | QUARTER 1 | QUARTER 2 | QUARTER 3 | QUARTER 4 | TOTAL GRAMS |
|---|---|---|---|---|---|
| 530 | 0.23 | 0.23 | 0.81 | 0.23 | 1.5 |
| 532 | 0 | 0.12 | 0 | 0 | 0.12 |
| 535 | 0.81 | 1.74 | 0.35 | 1.28 | 4.18 |
| 537 | 0.35 | 0.81 | 0.93 | 1.04 | 3.13 |
| 539 | 0 | 0.12 | 1.63 | 1.74 | 3.49 |
| 544 | 0.35 | 0.7 | 1.28 | 0.7 | 3.03 |
| 545 | 0 | 0.12 | 0 | 0 | 0.12 |
| TOTAL | 1.74 | 3.84 | 5 | 4.99 | 15.57 |

DRUG CODE: 9230 DRUG NAME: MEPERIDINE (PETHIDINE)

STATE: ALABAMA

| ZIP CODE | QUARTER 1 | QUARTER 2 | QUARTER 3 | QUARTER 4 | TOTAL GRAMS |
|---|---|---|---|---|---|
| 350 | 480.98 | 646.94 | 574.75 | 568.33 | 2,271 |
| 351 | 372.78 | 318.21 | 323.54 | 226.05 | 1,240.58 |
| 352 | 802.98 | 776.11 | 690.93 | 629.16 | 2,899.18 |
| 354 | 620.94 | 476.06 | 468.65 | 435.64 | 2,001.29 |
| 355 | 225.17 | 200.03 | 220.49 | 134.97 | 780.66 |
| 356 | 525.06 | 500.48 | 518.67 | 572.12 | 2,116.33 |
| 357 | 284.33 | 270.69 | 254.04 | 222.23 | 1,031.29 |
| 358 | 620.73 | 537.17 | 470.39 | 441.52 | 2,069.81 |
| 359 | 448.17 | 463.51 | 363.89 | 313.02 | 1,588.59 |
| 360 | 327.9 | 266.24 | 330.89 | 339.12 | 1,264.15 |
| 361 | 293.15 | 277.18 | 191.51 | 253.5 | 1,015.34 |
| 362 | 434.44 | 392.61 | 392.07 | 489.89 | 1,709.01 |
| 363 | 514.05 | 419.54 | 407.6 | 518.85 | 1,860.04 |
| 364 | 306.11 | 238.05 | 233.34 | 234.22 | 1,011.72 |
| 365 | 705.26 | 533.9 | 479.33 | 427.8 | 2,146.29 |
| 366 | 548.74 | 608.96 | 542.79 | 844.13 | 2,544.62 |
| 367 | 177.24 | 138.35 | 106.32 | 174.63 | 596.54 |
| 368 | 311.02 | 305.83 | 286.51 | 316.14 | 1,219.50 |
| 369 | 10.89 | 22.44 | 7.19 | 8.72 | 49.24 |

| | | | | | |
|---|---|---|---|---|---|
| TOTAL | 8,009.94 | 7,392.30 | 6,862.90 | 7,150.04 | 29,415.18 |

STATE: ALASKA

| ZIP CODE | QUARTER 1 | QUARTER 2 | QUARTER 3 | QUARTER 4 | TOTAL GRAMS |
|---|---|---|---|---|---|
| 995 | 519.74 | 468.43 | 373 | 423.98 | 1,785.15 |
| 996 | 201.75 | 161.23 | 194.02 | 139 | 696 |
| 997 | 111.12 | 70.16 | 73.42 | 67.98 | 322.68 |
| 998 | 16.12 | 3.05 | 20.04 | 1.74 | 40.95 |
| 999 | 1.42 | 1.31 | 1.96 | 1.53 | 6.22 |
| TOTAL | 850.15 | 704.18 | 662.44 | 634.23 | 2,851 |

STATE: ARIZONA

| ZIP CODE | QUARTER 1 | QUARTER 2 | QUARTER 3 | QUARTER 4 | TOTAL GRAMS |
|---|---|---|---|---|---|
| 850 | 1,061.92 | 916.14 | 933.29 | 947.91 | 3,859.26 |
| 851 | 226.48 | 117.76 | 60.02 | 107.85 | 512.11 |
| 852 | 1,671.54 | 1,652.80 | 1,559.64 | 1,689.95 | 6,573.93 |
| 853 | 720.51 | 648.18 | 563.64 | 769.32 | 2,701.65 |
| 855 | 142.49 | 164.5 | 96.52 | 130.07 | 533.58 |
| 856 | 181.27 | 222.67 | 175.72 | 152.51 | 732.17 |
| 857 | 1,071.62 | 783.48 | 689.57 | 692.52 | 3,237.19 |
| 859 | 13.73 | 20.7 | 14.16 | 14.16 | 62.75 |
| 860 | 64.49 | 61.44 | 38.56 | 31.16 | 195.65 |
| 863 | 89.98 | 108.94 | 86.28 | 92.71 | 377.91 |
| 864 | 200.66 | 223.32 | 149.79 | 131.71 | 705.48 |
| 865 | 6.1 | 1.31 | 6.54 | 0 | 13.95 |
| TOTAL | 5,450.79 | 4,921.24 | 4,373.73 | 4,759.87 | 19,505.63 |

STATE: ARKANSAS

| ZIP CODE | QUARTER 1 | QUARTER 2 | QUARTER 3 | QUARTER 4 | TOTAL GRAMS |
|---|---|---|---|---|---|
| 716 | 627.22 | 632.88 | 497.19 | 390.87 | 2,148.16 |
| 717 | 282.89 | 307.42 | 220.93 | 229.73 | 1,040.97 |
| 718 | 75.69 | 68.7 | 70.48 | 45.97 | 260.84 |
| 719 | 490.33 | 436.29 | 382.7 | 328.01 | 1,637.33 |
| 720 | 1,392.66 | 1,157.31 | 1,108.48 | 1,008.30 | 4,666.75 |

| | | | | | | |
|---|---|---|---|---|---|---|
| | 721 | 1,439.70 | 1,342.48 | 1,204.46 | 1,092.32 | 5,078.96 |
| | 722 | 1,086.65 | 812.43 | 992.2 | 752.43 | 3,643.71 |
| | 723 | 447.95 | 352.09 | 270.82 | 283.78 | 1,354.64 |
| | 724 | 600.25 | 637.26 | 492.72 | 508.67 | 2,238.90 |
| | 725 | 197.83 | 229.57 | 205.13 | 168.42 | 800.95 |
| | 726 | 194.13 | 200.01 | 216.24 | 118.31 | 728.69 |
| | 727 | 1,056.91 | 1,186.87 | 933.05 | 1,011.70 | 4,188.53 |
| | 728 | 187.92 | 194.56 | 208.4 | 113.84 | 704.72 |
| | 729 | 268.97 | 281.39 | 282.58 | 273.32 | 1,106.26 |
| TOTAL | | 8,349.10 | 7,839.26 | 7,085.38 | 6,325.67 | 29,599.41 |

STATE: CALIFORNIA

| ZIP CODE | | QUARTER 1 | QUARTER 2 | QUARTER 3 | QUARTER 4 | TOTAL GRAMS |
|---|---|---|---|---|---|---|
| | 900 | 777.81 | 803.63 | 749.27 | 896.01 | 3,226.72 |
| | 902 | 431.07 | 521.37 | 446.43 | 336.83 | 1,735.70 |
| | 903 | 3.81 | 4.36 | 13.62 | 5.99 | 27.78 |
| | 904 | 106.32 | 120.48 | 130.51 | 121.14 | 478.45 |
| | 905 | 275.18 | 199.25 | 184.65 | 229.75 | 888.83 |
| | 906 | 203.06 | 120.38 | 138.9 | 76.8 | 539.14 |
| | 907 | 79.74 | 84.64 | 86.5 | 98.04 | 348.92 |
| | 908 | 147.94 | 97.5 | 97.72 | 72.55 | 415.71 |
| | 910 | 68.3 | 57.19 | 103.71 | 71.9 | 301.1 |
| | 911 | 96.52 | 93.03 | 90.42 | 103.49 | 383.46 |
| | 912 | 153.17 | 187.26 | 173.1 | 183.12 | 696.65 |
| | 913 | 909.85 | 797.64 | 706.24 | 698.94 | 3,112.67 |
| | 914 | 209.7 | 222.78 | 280.62 | 183.34 | 896.44 |
| | 915 | 139.66 | 159.05 | 150.33 | 106.98 | 556.02 |
| | 916 | 8.72 | 15.25 | 9.8 | 7.63 | 41.4 |
| | 917 | 740.01 | 888.28 | 917.47 | 849.49 | 3,395.25 |
| | 918 | 50.11 | 49.46 | 49.89 | 53.16 | 202.62 |
| | 919 | 61.33 | 87.69 | 91.07 | 60.35 | 300.44 |
| | 920 | 883.59 | 1,022.81 | 882.61 | 911.48 | 3,700.49 |
| | 921 | 649.05 | 661.36 | 650.36 | 691.97 | 2,652.74 |
| | 922 | 242.71 | 277.68 | 327.68 | 216.13 | 1,064.20 |
| | 923 | 376.71 | 322.78 | 382.26 | 693.17 | 1,774.92 |
| | 924 | 36.39 | 23.53 | 27.89 | 35.95 | 123.76 |

| | | | | | | |
|---|---|---|---|---|---|---|
| | 925 | 767.36 | 844.81 | 568.11 | 561.14 | 2,741.42 |
| | 926 | 577.59 | 403.4 | 555.25 | 441.52 | 1,977.76 |
| | 927 | 261.45 | 208.72 | 295.22 | 268.2 | 1,033.59 |
| | 928 | 495.56 | 536.84 | 543.92 | 641.32 | 2,217.64 |
| | 930 | 231.27 | 190.64 | 177.13 | 155.89 | 754.93 |
| | 931 | 217.88 | 205.02 | 159.05 | 144.89 | 726.84 |
| | 932 | 380.19 | 278.92 | 317.44 | 211.34 | 1,187.89 |
| | 933 | 816.16 | 699.27 | 599.37 | 592.29 | 2,707.09 |
| | 934 | 258.07 | 272.34 | 202.95 | 284.98 | 1,018.34 |
| | 935 | 114.71 | 109.92 | 120.16 | 145.21 | 490 |
| | 936 | 158.74 | 101.79 | 98.41 | 143.14 | 502.08 |
| | 937 | 374.96 | 495.99 | 362.11 | 563.1 | 1,796.16 |
| | 939 | 169.18 | 165.69 | 171.58 | 195.22 | 701.67 |
| | 940 | 182.36 | 193.69 | 177.57 | 212.97 | 766.59 |
| | 941 | 540.77 | 304.81 | 487.6 | 545.23 | 1,878.41 |
| | 943 | 161.01 | 187.26 | 205.13 | 181.93 | 735.33 |
| | 944 | 20.26 | 5.66 | 14.92 | 19.06 | 59.9 |
| | 945 | 885.66 | 1,001.24 | 866.16 | 763.65 | 3,516.71 |
| | 946 | 17.32 | 11.22 | 27.02 | 17.97 | 73.53 |
| | 947 | 2.72 | 4.36 | 11.77 | 5.45 | 24.3 |
| | 948 | 0 | 10.89 | 0.22 | 0 | 11.11 |
| | 949 | 49.57 | 52.73 | 49.24 | 74.84 | 226.38 |
| | 950 | 225.94 | 138.13 | 150.77 | 92.27 | 607.11 |
| | 951 | 119.29 | 105.78 | 112.53 | 90.96 | 428.56 |
| | 952 | 128.98 | 122.23 | 98.81 | 51.53 | 401.55 |
| | 953 | 655.15 | 790.89 | 547.41 | 746.22 | 2,739.67 |
| | 954 | 239.12 | 166.78 | 217.11 | 183.45 | 806.46 |
| | 955 | 50.55 | 65.14 | 64.71 | 13.07 | 193.47 |
| | 956 | 494.79 | 650.57 | 534.01 | 583.25 | 2,262.62 |
| | 957 | 56.65 | 26.58 | 18.52 | 14.16 | 115.91 |
| | 958 | 308.73 | 265.15 | 259.27 | 327.47 | 1,160.62 |
| | 959 | 264.65 | 296.4 | 240.86 | 267.22 | 1,069.13 |
| | 960 | 84.3 | 76.56 | 167.11 | 105.23 | 433.2 |
| | 961 | 13.07 | 67.32 | 36.39 | 26.15 | 142.93 |
| TOTAL | | 15,974.76 | 15,874.14 | 15,148.88 | 15,374.53 | 62,372.31 |

RETAIL DRUG DISTRIBUTION BY ZIP CODE WITHIN STATE BY GRAMS WT

STATE: COLORADO

| ZIP CODE | QUARTER 1 | QUARTER 2 | QUARTER 3 | QUARTER 4 | TOTAL GRAMS |
|---|---|---|---|---|---|
| 800 | 237.48 | 147.39 | 134.76 | 93.25 | 612.88 |
| 801 | 306.77 | 260.73 | 290.86 | 261.23 | 1,119.59 |
| 802 | 359.49 | 295.98 | 204.58 | 239.66 | 1,099.71 |
| 803 | 46.3 | 39.87 | 40.74 | 29.41 | 156.32 |
| 804 | 56.54 | 65.69 | 61.77 | 70.48 | 254.48 |
| 805 | 191.73 | 187.59 | 231.93 | 126.59 | 737.84 |
| 806 | 104.36 | 150.66 | 108.28 | 61.01 | 424.31 |
| 807 | 36.82 | 20.26 | 33.77 | 21.79 | 112.64 |
| 808 | 83.66 | 81.27 | 69.72 | 71.25 | 305.9 |
| 809 | 285.09 | 186.28 | 175.83 | 178.55 | 825.75 |
| 810 | 97.28 | 126.8 | 110.46 | 99.24 | 433.78 |
| 811 | 18.52 | 16.34 | 18.52 | 13.07 | 66.45 |
| 812 | 26.04 | 40.96 | 37.47 | 36.71 | 141.18 |
| 813 | 40.52 | 22.22 | 35.95 | 20.26 | 118.95 |
| 814 | 100 | 98.26 | 57.95 | 66.45 | 322.66 |
| 815 | 74.3 | 64.27 | 47.93 | 39.54 | 226.04 |
| 816 | 106.43 | 86.39 | 69.18 | 58.5 | 320.5 |
| TOTAL | 2,171.33 | 1,890.96 | 1,729.70 | 1,486.99 | 7,278.98 |

STATE: CONNECTICUT

| ZIP CODE | QUARTER 1 | QUARTER 2 | QUARTER 3 | QUARTER 4 | TOTAL GRAMS |
|---|---|---|---|---|---|
| 60 | 38.13 | 68.85 | 43.47 | 64.27 | 214.72 |
| 61 | 82.68 | 65.25 | 69.18 | 64.82 | 281.93 |
| 62 | 19.61 | 5.66 | 36.39 | 22.88 | 84.54 |
| 63 | 21.79 | 44.45 | 94.99 | 56.65 | 217.88 |
| 64 | 382.59 | 329.43 | 298.71 | 359.38 | 1,370.11 |
| 65 | 78.76 | 118.52 | 65.36 | 76.47 | 339.11 |
| 66 | 104.58 | 89.98 | 113.4 | 89.33 | 397.29 |
| 67 | 157.52 | 98.04 | 99.35 | 102.84 | 457.75 |
| 68 | 209.27 | 211.34 | 166.35 | 220.05 | 807.01 |
| 69 | 9.15 | 20.7 | 49.24 | 13.07 | 92.16 |
| TOTAL | 1,104.08 | 1,052.22 | 1,036.44 | 1,069.76 | 4,262.50 |

RETAIL DRUG DISTRIBUTION BY ZIP CODE WITHIN STATE BY GRAMS WT

STATE: DELAWARE

| ZIP CODE | QUARTER 1 | QUARTER 2 | QUARTER 3 | QUARTER 4 | TOTAL GRAMS |
|---|---|---|---|---|---|
| 197 | 78.44 | 74.4 | 86.5 | 58.83 | 298.17 |
| 198 | 42.05 | 26.36 | 25.82 | 14.6 | 108.83 |
| 199 | 127.89 | 173.76 | 103.49 | 58.61 | 463.75 |
| TOTAL | 248.38 | 274.52 | 215.81 | 132.04 | 870.75 |

STATE: DISTRICT OF COLUMBIA

| ZIP CODE | QUARTER 1 | QUARTER 2 | QUARTER 3 | QUARTER 4 | TOTAL GRAMS |
|---|---|---|---|---|---|
| 200 | 215.48 | 220.05 | 168.96 | 147.94 | 752.43 |
| 204 | 2.18 | 0 | 0 | 0 | 2.18 |
| TOTAL | 217.66 | 220.05 | 168.96 | 147.94 | 754.61 |

STATE: FLORIDA

| ZIP CODE | QUARTER 1 | QUARTER 2 | QUARTER 3 | QUARTER 4 | TOTAL GRAMS |
|---|---|---|---|---|---|
| 320 | 409.5 | 447.19 | 334.44 | 406.34 | 1,597.47 |
| 321 | 331.5 | 306.99 | 191.4 | 201.21 | 1,031.10 |
| 322 | 690.34 | 730.86 | 535.65 | 499.37 | 2,456.22 |
| 323 | 510.59 | 407.1 | 394.68 | 413.96 | 1,726.33 |
| 324 | 221.91 | 185.74 | 123.1 | 216.89 | 747.64 |
| 325 | 754.61 | 697.31 | 594.8 | 586.41 | 2,633.13 |
| 326 | 194.13 | 179.64 | 230.29 | 134.54 | 738.6 |
| 327 | 334 | 288.9 | 257.2 | 212.97 | 1,093.07 |
| 328 | 168.85 | 129.31 | 114.17 | 133.78 | 546.11 |
| 329 | 568.76 | 505.69 | 430.09 | 398.71 | 1,903.25 |
| 330 | 624.65 | 582.05 | 478.13 | 479.65 | 2,164.48 |
| 331 | 641.1 | 757.99 | 605.26 | 623.67 | 2,628.02 |
| 333 | 493.6 | 523.66 | 492.18 | 727.92 | 2,237.36 |
| 334 | 495.45 | 561.9 | 470.07 | 636.52 | 2,163.94 |
| 335 | 386.29 | 296.42 | 252.95 | 361.45 | 1,297.11 |
| 336 | 482.7 | 430.96 | 285.63 | 414.51 | 1,613.80 |
| 337 | 196.96 | 155.02 | 200.23 | 131.81 | 684.02 |
| 338 | 406.01 | 365.38 | 354.81 | 390 | 1,516.20 |
| 339 | 495.77 | 408.84 | 319.4 | 352.19 | 1,576.20 |
| 341 | 90.96 | 99.35 | 60.13 | 97.39 | 347.83 |

| | | | | | | |
|---|---|---|---|---|---|---|
| | 342 | 234.76 | 256.44 | 263.74 | 193.8 | 948.74 |
| | 344 | 347.07 | 356.33 | 249.9 | 241.3 | 1,194.60 |
| | 346 | 359.93 | 326.92 | 167.44 | 291.84 | 1,146.13 |
| | 347 | 183.12 | 181.16 | 141.73 | 179.09 | 685.1 |
| | 349 | 329.32 | 329.86 | 261.01 | 217.88 | 1,138.07 |
| TOTAL | | 9,951.88 | 9,511.01 | 7,808.43 | 8,543.20 | 35,814.52 |

STATE: GEORGIA

| ZIP CODE | | QUARTER 1 | QUARTER 2 | QUARTER 3 | QUARTER 4 | TOTAL GRAMS |
|---|---|---|---|---|---|---|
| | 300 | 1,078.26 | 1,011.38 | 1,107.89 | 1,031.42 | 4,228.95 |
| | 301 | 1,124.67 | 1,060.18 | 982.73 | 906.36 | 4,073.94 |
| | 302 | 878.36 | 703.24 | 632.49 | 552.2 | 2,766.29 |
| | 303 | 753.63 | 477.26 | 552.42 | 585.1 | 2,368.41 |
| | 304 | 287.92 | 233.98 | 327.25 | 518.54 | 1,367.69 |
| | 305 | 679.27 | 633.69 | 525.51 | 501.66 | 2,340.13 |
| | 306 | 457.1 | 452.07 | 404.05 | 455.9 | 1,769.12 |
| | 307 | 785.35 | 705.92 | 635.65 | 602.32 | 2,729.24 |
| | 308 | 183.67 | 240.97 | 244.35 | 250.67 | 919.66 |
| | 309 | 543.27 | 489.35 | 421.37 | 491.74 | 1,945.73 |
| | 310 | 814.83 | 721.91 | 637.46 | 720.93 | 2,895.13 |
| | 312 | 339.56 | 239.66 | 269.08 | 191.29 | 1,039.59 |
| | 313 | 277.25 | 259.92 | 229.31 | 276.92 | 1,043.40 |
| | 314 | 287.6 | 299.03 | 314.83 | 361.13 | 1,262.59 |
| | 315 | 668.88 | 665.17 | 595.13 | 573.58 | 2,502.76 |
| | 316 | 889.56 | 869.1 | 803.85 | 708.77 | 3,271.28 |
| | 317 | 784.89 | 796.68 | 685.41 | 690.03 | 2,957.01 |
| | 318 | 53.38 | 70.05 | 47.93 | 66.45 | 237.81 |
| | 319 | 401.54 | 299.45 | 197.98 | 339.49 | 1,238.46 |
| | 398 | 209.62 | 214.59 | 100.22 | 109.63 | 634.06 |
| TOTAL | | 11,498.61 | 10,443.60 | 9,714.91 | 9,934.13 | 41,591.25 |

STATE: GUAM

| ZIP CODE | | QUARTER 1 | QUARTER 2 | QUARTER 3 | QUARTER 4 | TOTAL GRAMS |
|---|---|---|---|---|---|---|
| | 969 | 30.61 | 34.64 | 66.23 | 52.18 | 183.66 |
| TOTAL | | 30.61 | 34.64 | 66.23 | 52.18 | 183.66 |

STATE: HAWAII

| ZIP CODE | QUARTER 1 | QUARTER 2 | QUARTER 3 | QUARTER 4 | TOTAL GRAMS |
|---|---|---|---|---|---|
| 967 | 239.77 | 223.98 | 157.52 | 146.52 | 767.79 |
| 968 | 142.49 | 170.38 | 148.37 | 128.22 | 589.46 |
| TOTAL | 382.26 | 394.36 | 305.89 | 274.74 | 1,357.25 |

STATE: IDAHO

| ZIP CODE | QUARTER 1 | QUARTER 2 | QUARTER 3 | QUARTER 4 | TOTAL GRAMS |
|---|---|---|---|---|---|
| 832 | 117.11 | 132.69 | 110.57 | 76.8 | 437.17 |
| 833 | 98.78 | 139.7 | 49.07 | 116.61 | 404.16 |
| 834 | 138.35 | 173.1 | 120.05 | 116.02 | 547.52 |
| 835 | 33.77 | 36.82 | 42.16 | 54.03 | 166.78 |
| 836 | 136.61 | 129.64 | 236.18 | 153.82 | 656.25 |
| 837 | 226.59 | 236.5 | 124.73 | 229.53 | 817.35 |
| 838 | 96.85 | 76.78 | 58 | 69.18 | 300.81 |
| TOTAL | 848.06 | 925.23 | 740.76 | 815.99 | 3,330.04 |

STATE: ILLINOIS

| ZIP CODE | QUARTER 1 | QUARTER 2 | QUARTER 3 | QUARTER 4 | TOTAL GRAMS |
|---|---|---|---|---|---|
| 600 | 217.55 | 243.15 | 212.75 | 186.17 | 859.62 |
| 601 | 317.33 | 505.47 | 529.98 | 306.66 | 1,659.44 |
| 602 | 7.84 | 0.44 | 15.25 | 16.34 | 39.87 |
| 603 | 0.44 | 1.09 | 15.25 | 1.09 | 17.87 |
| 604 | 286.72 | 259.49 | 344.35 | 267.99 | 1,158.55 |
| 605 | 192.38 | 175.28 | 202.51 | 107.63 | 677.8 |
| 606 | 169.4 | 197.83 | 163.95 | 92.71 | 623.89 |
| 607 | 1.53 | 4.36 | 0 | 9.15 | 15.04 |
| 608 | 44.45 | 44.01 | 61.44 | 35.3 | 185.2 |
| 609 | 0.22 | 2.29 | 1.09 | 5.45 | 9.05 |
| 610 | 100.44 | 130.29 | 90.85 | 145.76 | 467.34 |
| 611 | 34.42 | 69.5 | 74.51 | 65.91 | 244.34 |
| 612 | 7.41 | 15.03 | 29.85 | 17.97 | 70.26 |
| 613 | 48.59 | 66.89 | 55.34 | 56.21 | 227.03 |

| | | | | | | |
|---|---|---|---|---|---|---|
| | 614 | 77.35 | 79.52 | 51.2 | 69.72 | 277.79 |
| | 615 | 58.06 | 55.12 | 27.56 | 55.99 | 196.73 |
| | 616 | 162.32 | 177.13 | 155.56 | 152.73 | 647.74 |
| | 617 | 39.44 | 43.4 | 19.5 | 32.46 | 134.8 |
| | 618 | 152.73 | 211.12 | 173.54 | 142.05 | 679.44 |
| | 619 | 63.95 | 47.17 | 56.54 | 57.74 | 225.4 |
| | 620 | 27.89 | 27.45 | 33.33 | 7.41 | 96.08 |
| | 622 | 337.38 | 278.01 | 312.11 | 215.26 | 1,142.76 |
| | 623 | 23.42 | 14.64 | 37.47 | 34.86 | 110.39 |
| | 624 | 16.67 | 11.16 | 24.51 | 30.94 | 83.28 |
| | 625 | 9.04 | 24.62 | 15.03 | 28.32 | 77.01 |
| | 626 | 167.48 | 149.24 | 178 | 88.24 | 582.96 |
| | 627 | 67 | 104.8 | 61.66 | 66.02 | 299.48 |
| | 628 | 131.6 | 87.72 | 66.56 | 58.61 | 344.49 |
| | 629 | 227.9 | 248.03 | 191.73 | 189.44 | 857.1 |
| TOTAL | | 2,990.95 | 3,274.25 | 3,201.42 | 2,544.13 | 12,010.75 |

| STATE: | INDIANA | | | | | |
|---|---|---|---|---|---|---|
| ZIP CODE | | QUARTER 1 | QUARTER 2 | QUARTER 3 | QUARTER 4 | TOTAL GRAMS |
| | 460 | 283.89 | 273.43 | 258.94 | 308.51 | 1,124.77 |
| | 461 | 326.16 | 508.08 | 367.56 | 309.16 | 1,510.96 |
| | 462 | 1,102.23 | 1,169.99 | 984.36 | 729.12 | 3,985.70 |
| | 463 | 279.42 | 231.71 | 269.29 | 161.66 | 942.08 |
| | 464 | 23.75 | 32.03 | 46.41 | 17.43 | 119.62 |
| | 465 | 41.94 | 41.18 | 35.19 | 70.81 | 189.12 |
| | 466 | 39.22 | 25.06 | 30.94 | 18.52 | 113.74 |
| | 467 | 105.89 | 46.84 | 52.73 | 46.63 | 252.09 |
| | 468 | 150.12 | 142.82 | 150.55 | 119.5 | 562.99 |
| | 469 | 90.74 | 96.24 | 146.67 | 91.29 | 424.94 |
| | 470 | 121.4 | 97.39 | 87.54 | 77.78 | 384.11 |
| | 471 | 227.64 | 165.78 | 160.01 | 183.23 | 736.66 |
| | 472 | 121.57 | 137.91 | 137.26 | 117.65 | 514.39 |
| | 473 | 140.36 | 155.78 | 132.9 | 109.81 | 538.85 |
| | 474 | 111.77 | 72.23 | 73.86 | 46.73 | 304.59 |
| | 475 | 35.62 | 46.56 | 53.38 | 36.6 | 172.16 |
| | 476 | 66.89 | 48.7 | 48.48 | 187.37 | 351.44 |

| | | | | | | |
|---|---|---|---|---|---|---|
| | 477 | 130.07 | 71.03 | 74.84 | 221.47 | 497.41 |
| | 478 | 115.47 | 77.56 | 80.83 | 114.38 | 388.24 |
| | 479 | 75.6 | 84.64 | 68.96 | 82.57 | 311.77 |
| TOTAL | | 3,589.75 | 3,524.96 | 3,260.70 | 3,050.22 | 13,425.63 |

STATE: IOWA

| ZIP CODE | | QUARTER 1 | QUARTER 2 | QUARTER 3 | QUARTER 4 | TOTAL GRAMS |
|---|---|---|---|---|---|---|
| | 500 | 97.83 | 74.73 | 130.94 | 74.84 | 378.34 |
| | 501 | 25.93 | 27.67 | 34.21 | 26.15 | 113.96 |
| | 502 | 216.79 | 194.45 | 186.07 | 160.79 | 758.1 |
| | 503 | 376.92 | 395.66 | 414.62 | 432.92 | 1,620.12 |
| | 504 | 4.36 | 7.84 | 8.17 | 4.36 | 24.73 |
| | 505 | 73.97 | 76.69 | 71.46 | 110.9 | 333.02 |
| | 506 | 64.93 | 41.72 | 48.15 | 44.12 | 198.92 |
| | 507 | 282.8 | 305.9 | 298.92 | 378.88 | 1,266.50 |
| | 508 | 22.88 | 12.83 | 11.77 | 0.54 | 48.02 |
| | 510 | 12.64 | 13.51 | 15.03 | 27.45 | 68.63 |
| | 511 | 74.4 | 55.88 | 63.62 | 73.64 | 267.54 |
| | 512 | 5.66 | 4.14 | 6.32 | 5.45 | 21.57 |
| | 513 | 13.73 | 11.77 | 18.74 | 25.06 | 69.3 |
| | 514 | 12.64 | 11.11 | 13.07 | 9.59 | 46.41 |
| | 515 | 46.52 | 21.77 | 38.02 | 29.3 | 135.61 |
| | 516 | 5.45 | 5.45 | 2.18 | 0.87 | 13.95 |
| | 520 | 52.51 | 41.83 | 56.65 | 40.42 | 191.41 |
| | 521 | 0 | 0.44 | 3.7 | 0 | 4.14 |
| | 522 | 70.16 | 111.55 | 142.93 | 107.63 | 432.27 |
| | 523 | 29.63 | 40.57 | 31.81 | 50.55 | 152.56 |
| | 524 | 236.83 | 236.18 | 210.79 | 197.83 | 881.63 |
| | 525 | 106.76 | 104.19 | 88.57 | 97.39 | 396.91 |
| | 526 | 15.47 | 22.7 | 24.23 | 11.98 | 74.38 |
| | 527 | 42.16 | 33.77 | 44.99 | 23.97 | 144.89 |
| | 528 | 36.39 | 41.61 | 26.25 | 33.77 | 138.02 |
| TOTAL | | 1,927.36 | 1,893.96 | 1,991.21 | 1,968.40 | 7,780.93 |

STATE: KANSAS

| ZIP CODE | QUARTER 1 | QUARTER 2 | QUARTER 3 | QUARTER 4 | TOTAL GRAMS |
|---|---|---|---|---|---|
| 660 | 200.77 | 304.92 | 220.38 | 304.48 | 1,030.55 |
| 661 | 72.33 | 47.93 | 58.17 | 50.66 | 229.09 |
| 662 | 209.81 | 171.84 | 177.68 | 242.39 | 801.72 |
| 664 | 47.04 | 77.02 | 20.94 | 28.3 | 173.3 |
| 665 | 51.85 | 34.42 | 45.32 | 41.4 | 172.99 |
| 666 | 74.51 | 63.18 | 56.65 | 16.78 | 211.12 |
| 667 | 78.87 | 61.24 | 75.71 | 53.6 | 269.42 |
| 668 | 51.75 | 52.18 | 46.84 | 60.68 | 211.45 |
| 669 | 8.72 | 11.33 | 16.56 | 16.45 | 53.06 |
| 670 | 157.2 | 114.8 | 118.09 | 92.92 | 483.01 |
| 671 | 41.51 | 37.47 | 66.34 | 31.59 | 176.91 |
| 672 | 334.55 | 306.64 | 320.43 | 306.55 | 1,268.17 |
| 673 | 69.94 | 63.18 | 44.66 | 44.66 | 222.44 |
| 674 | 89.98 | 85.67 | 75.17 | 79.63 | 330.45 |
| 675 | 170.16 | 133.38 | 167.22 | 136.72 | 607.48 |
| 676 | 66.23 | 70.31 | 73.1 | 51.42 | 261.06 |
| 677 | 16.56 | 11.33 | 22.44 | 31.16 | 81.49 |
| 678 | 47 | 87.56 | 51.85 | 53.6 | 240.01 |
| 679 | 71.9 | 58.43 | 54.69 | 62.31 | 247.33 |
| TOTAL | 1,860.68 | 1,792.83 | 1,712.24 | 1,705.30 | 7,071.05 |

STATE: KENTUCKY

| ZIP CODE | QUARTER 1 | QUARTER 2 | QUARTER 3 | QUARTER 4 | TOTAL GRAMS |
|---|---|---|---|---|---|
| 400 | 42.27 | 61.88 | 52.73 | 52.12 | 209 |
| 401 | 47.28 | 30.5 | 34.86 | 5.45 | 118.09 |
| 402 | 331.37 | 305.92 | 411.13 | 323.54 | 1,371.96 |
| 403 | 63.73 | 69.18 | 72.55 | 93.47 | 298.93 |
| 404 | 40.96 | 46.63 | 53.82 | 52.29 | 193.7 |
| 405 | 516.36 | 44.56 | 119.18 | 46.63 | 726.73 |
| 406 | 14.16 | 9.8 | 0.22 | 9.04 | 33.22 |
| 407 | 53.82 | 55.12 | 49.24 | 41.29 | 199.47 |
| 408 | 41.07 | 37.47 | 42.05 | 46.19 | 166.78 |
| 409 | 85.19 | 78.65 | 69.18 | 57.74 | 290.76 |
| 410 | 140.09 | 155.56 | 53.27 | 99.9 | 448.82 |
| 411 | 54.47 | 51.2 | 43.03 | 56.1 | 204.8 |

| | | | | | | |
|---|---|---|---|---|---|---|
| | 412 | 1.09 | 0 | 1.09 | 0 | 2.18 |
| | 413 | 18.52 | 15.25 | 17.43 | 23.97 | 75.17 |
| | 414 | 0 | 0 | 0.44 | 0 | 0.44 |
| | 415 | 40.74 | 35.3 | 27.13 | 22.33 | 125.5 |
| | 416 | 4.9 | 6.75 | 23.64 | 13.07 | 48.36 |
| | 417 | 2.51 | 5.45 | 5.01 | 1.63 | 14.6 |
| | 418 | 45.75 | 36.49 | 50.11 | 47.39 | 179.74 |
| | 420 | 286.46 | 308.99 | 288.58 | 310.91 | 1,194.94 |
| | 421 | 374.92 | 246.42 | 311.02 | 300.12 | 1,232.48 |
| | 422 | 177.89 | 140.42 | 183.67 | 137.48 | 639.46 |
| | 423 | 123.32 | 158.66 | 148.16 | 114.71 | 544.85 |
| | 424 | 176.57 | 177.55 | 147.83 | 139.88 | 641.83 |
| | 425 | 40.74 | 39.44 | 56.32 | 36.28 | 172.78 |
| | 426 | 38.35 | 26.84 | 54.47 | 12.31 | 131.97 |
| | 427 | 117.98 | 155.93 | 153.38 | 83.66 | 510.95 |
| TOTAL | | 2,880.51 | 2,299.96 | 2,469.54 | 2,127.50 | 9,777.51 |

STATE: LOUISIANA

| ZIP CODE | | QUARTER 1 | QUARTER 2 | QUARTER 3 | QUARTER 4 | TOTAL GRAMS |
|---|---|---|---|---|---|---|
| | 700 | 983.05 | 821.93 | 804.83 | 634.78 | 3,244.59 |
| | 701 | 156.87 | 187.81 | 153.6 | 131.81 | 630.09 |
| | 703 | 400.32 | 407.6 | 307.42 | 352.3 | 1,467.64 |
| | 704 | 839.8 | 788.77 | 633.14 | 456.99 | 2,718.70 |
| | 705 | 1,609.88 | 1,586.28 | 1,483.27 | 1,522.92 | 6,202.35 |
| | 706 | 650.23 | 646.24 | 524.69 | 499.06 | 2,320.22 |
| | 707 | 454.27 | 399.8 | 363.31 | 393.81 | 1,611.19 |
| | 708 | 549.68 | 562.47 | 507.37 | 562.1 | 2,181.62 |
| | 710 | 244.02 | 207.09 | 233.45 | 265.26 | 949.82 |
| | 711 | 995.8 | 792.96 | 690.12 | 650.36 | 3,129.24 |
| | 712 | 530.96 | 476.58 | 424.81 | 593.93 | 2,026.28 |
| | 713 | 477.47 | 446.32 | 415.38 | 330.41 | 1,669.58 |
| | 714 | 218.53 | 246.55 | 173.43 | 229.42 | 867.93 |
| TOTAL | | 8,110.88 | 7,570.40 | 6,714.82 | 6,623.15 | 29,019.25 |

STATE: MAINE

| ZIP CODE | QUARTER 1 | QUARTER 2 | QUARTER 3 | QUARTER 4 | TOTAL GRAMS |
|---|---|---|---|---|---|
| 39 | 13.51 | 11.11 | 6.75 | 11.11 | 42.48 |
| 40 | 95.65 | 115.91 | 87.26 | 73.21 | 372.03 |
| 41 | 19.61 | 3.92 | 25.82 | 15.25 | 64.6 |
| 42 | 11.33 | 18.08 | 15.47 | 10.24 | 55.12 |
| 43 | 9.59 | 10.46 | 19.83 | 4.36 | 44.24 |
| 44 | 146.85 | 96.52 | 99.79 | 85.41 | 428.57 |
| 45 | 9.15 | 0 | 1.31 | 0 | 10.46 |
| 46 | 63.84 | 42.05 | 55.78 | 37.47 | 199.14 |
| 47 | 66.13 | 66.02 | 21.68 | 77.35 | 231.18 |
| 48 | 0 | 4.36 | 4.36 | 0 | 8.72 |
| 49 | 29.74 | 40.96 | 21.02 | 29.96 | 121.68 |
| TOTAL | 465.4 | 409.39 | 359.07 | 344.36 | 1,578.22 |

STATE: MARYLAND

| ZIP CODE | QUARTER 1 | QUARTER 2 | QUARTER 3 | QUARTER 4 | TOTAL GRAMS |
|---|---|---|---|---|---|
| 206 | 109.16 | 117.11 | 99.57 | 81.7 | 407.54 |
| 207 | 317.55 | 267.99 | 292.39 | 205.57 | 1,083.50 |
| 208 | 329.97 | 322.67 | 336.51 | 341.19 | 1,330.34 |
| 209 | 60.35 | 39 | 77.78 | 57.63 | 234.76 |
| 210 | 551.44 | 577.91 | 261.45 | 376.16 | 1,766.96 |
| 211 | 174.08 | 136.61 | 110.35 | 149.68 | 570.72 |
| 212 | 590.44 | 454.38 | 439.02 | 349.47 | 1,833.31 |
| 214 | 131.92 | 119.07 | 64.6 | 78.87 | 394.46 |
| 215 | 65.8 | 63.62 | 97.93 | 107.85 | 335.2 |
| 216 | 179.42 | 121.57 | 97.83 | 102.62 | 501.44 |
| 217 | 116.35 | 126.8 | 96.95 | 83.88 | 423.98 |
| 218 | 100.11 | 109.16 | 134.86 | 90.64 | 434.77 |
| 219 | 24.08 | 18.52 | 6.1 | 4.36 | 53.06 |
| TOTAL | 2,750.67 | 2,474.41 | 2,115.34 | 2,029.62 | 9,370.04 |

STATE: MASSACHUSETTS

| ZIP CODE | QUARTER 1 | QUARTER 2 | QUARTER 3 | QUARTER 4 | TOTAL GRAMS |
|---|---|---|---|---|---|
| 010 | 296.96 | 293.26 | 319.19 | 276.92 | 1,186.33 |
| 011 | 38.56 | 93.69 | 23.31 | 64.16 | 219.72 |

| | | | | | | |
|---|---|---|---|---|---|---|
| | 012 | 27.02 | 31.16 | 1.74 | 35.95 | 95.87 |
| | 013 | 17.87 | 37.04 | 9.7 | 17.43 | 82.04 |
| | 014 | 105.02 | 118.42 | 93.25 | 61.01 | 377.7 |
| | 015 | 67.54 | 53.16 | 86.28 | 60.35 | 267.33 |
| | 016 | 33.66 | 30.94 | 30.61 | 11.44 | 106.65 |
| | 017 | 73.42 | 126.48 | 106.32 | 127.35 | 433.57 |
| | 018 | 161.66 | 149.68 | 131.6 | 139.88 | 582.82 |
| | 019 | 198.92 | 188.68 | 245.33 | 116.56 | 749.49 |
| | 020 | 114.17 | 116.02 | 121.36 | 71.79 | 423.34 |
| | 021 | 374.85 | 342.22 | 367.77 | 231.82 | 1,316.66 |
| | 022 | 39.65 | 40.31 | 62.75 | 12.09 | 154.8 |
| | 023 | 242.49 | 220.49 | 192.82 | 174.74 | 830.54 |
| | 024 | 102.07 | 123.97 | 64.93 | 168.85 | 459.82 |
| | 025 | 83.88 | 100.22 | 85.84 | 56.21 | 326.15 |
| | 026 | 53.6 | 26.15 | 46.84 | 34.86 | 161.45 |
| | 27 | 237.48 | 158.83 | 179.64 | 104.8 | 680.75 |
| TOTAL | | 2,268.82 | 2,250.72 | 2,169.28 | 1,766.21 | 8,455.03 |

| STATE: | MICHIGAN | | | | | |
|---|---|---|---|---|---|---|
| ZIP CODE | | QUARTER 1 | QUARTER 2 | QUARTER 3 | QUARTER 4 | TOTAL GRAMS |
| | 480 | 414.29 | 419.3 | 369.84 | 280.51 | 1,483.94 |
| | 481 | 301.54 | 279.21 | 240.32 | 213.84 | 1,034.91 |
| | 482 | 70.59 | 26.8 | 100.33 | 53.82 | 251.54 |
| | 483 | 79.42 | 152.29 | 102.95 | 83.45 | 418.11 |
| | 484 | 110.68 | 98.7 | 107.52 | 128.44 | 445.34 |
| | 485 | 29.41 | 45.43 | 32.68 | 72.99 | 180.51 |
| | 486 | 267.81 | 325.09 | 270.17 | 188.94 | 1,052.01 |
| | 487 | 39.87 | 45.97 | 103.27 | 52.29 | 241.4 |
| | 488 | 197.07 | 170.71 | 169.62 | 164.71 | 702.11 |
| | 489 | 218.09 | 178.55 | 142.82 | 181.38 | 720.84 |
| | 490 | 325.29 | 292.39 | 267.44 | 172.88 | 1,058 |
| | 491 | 2.18 | 8.06 | 2.18 | 3.27 | 15.69 |
| | 492 | 158.83 | 124.62 | 119.61 | 118.96 | 522.02 |
| | 493 | 40.09 | 87.8 | 64.71 | 21.79 | 214.39 |
| | 494 | 111.99 | 63.29 | 92.92 | 93.47 | 361.67 |
| | 495 | 265.15 | 147.07 | 107.3 | 117.33 | 636.85 |

| | | | | | | |
|---|---|---|---|---|---|---|
| | 496 | 75.93 | 74.62 | 94.12 | 45.75 | 290.42 |
| | 497 | 220.93 | 209.6 | 172.34 | 183.02 | 785.89 |
| | 498 | 76.26 | 114.82 | 66.23 | 57.74 | 315.05 |
| | 499 | 50.76 | 41.94 | 38.78 | 66.23 | 197.71 |
| TOTAL | | 3,056.18 | 2,906.26 | 2,665.15 | 2,300.81 | 10,928.40 |

STATE: MINNESOTA

| ZIP CODE | | QUARTER 1 | QUARTER 2 | QUARTER 3 | QUARTER 4 | TOTAL GRAMS |
|---|---|---|---|---|---|---|
| | 550 | 50.33 | 59.92 | 44.23 | 60.79 | 215.27 |
| | 551 | 26.8 | 28.98 | 34.75 | 20.48 | 111.01 |
| | 553 | 90.53 | 38.89 | 61.11 | 89.22 | 279.75 |
| | 554 | 81.27 | 111.66 | 90.2 | 57.63 | 340.76 |
| | 556 | 4.36 | 0 | 4.79 | 0 | 9.15 |
| | 557 | 67.76 | 74.3 | 41.83 | 42.49 | 226.38 |
| | 558 | 98.26 | 68.19 | 49.68 | 78.22 | 294.35 |
| | 559 | 136.39 | 88.46 | 117.76 | 100.55 | 443.16 |
| | 560 | 16.56 | 22.44 | 20.7 | 18.3 | 78 |
| | 561 | 5.45 | 4.36 | 6.32 | 2.07 | 18.2 |
| | 562 | 18.41 | 22.66 | 31.92 | 6.75 | 79.74 |
| | 563 | 50.76 | 40.74 | 31.37 | 43.47 | 166.34 |
| | 564 | 59.7 | 43.62 | 38.78 | 25.03 | 167.13 |
| | 565 | 58.83 | 25.27 | 28.19 | 8.72 | 121.01 |
| | 566 | 33.55 | 5.01 | 11.77 | 16.89 | 67.22 |
| | 567 | 36.71 | 16.56 | 38.56 | 24.62 | 116.45 |
| TOTAL | | 835.67 | 651.06 | 651.96 | 595.23 | 2,733.92 |

STATE: MISSISSIPPI

| ZIP CODE | | QUARTER 1 | QUARTER 2 | QUARTER 3 | QUARTER 4 | TOTAL GRAMS |
|---|---|---|---|---|---|---|
| | 386 | 493.6 | 608.87 | 468.98 | 462 | 2,033.45 |
| | 387 | 292.8 | 336.18 | 298.58 | 300.93 | 1,228.49 |
| | 388 | 448.6 | 396.71 | 280.62 | 372.89 | 1,498.82 |
| | 389 | 253.87 | 236.94 | 210.84 | 203.63 | 905.28 |
| | 390 | 305.46 | 313.2 | 281.49 | 294.68 | 1,194.83 |
| | 391 | 346.09 | 322.02 | 266.46 | 281.28 | 1,215.85 |
| | 392 | 436.08 | 412.33 | 395.44 | 309.93 | 1,553.78 |

| | | | | | | |
|---|---|---|---|---|---|---|
| | 393 | 365.4 | 319.4 | 366.27 | 414.62 | 1,465.69 |
| | 394 | 461.9 | 482.79 | 389.17 | 365.64 | 1,699.50 |
| | 395 | 907.67 | 769.69 | 731.95 | 781.95 | 3,191.26 |
| | 396 | 130.73 | 187.26 | 78.54 | 97.93 | 494.46 |
| | 397 | 228.29 | 234.78 | 222.6 | 147.48 | 833.15 |
| TOTAL | | 4,670.49 | 4,620.17 | 3,990.94 | 4,032.96 | 17,314.56 |

| STATE: | MISSOURI | | | | | |
|---|---|---|---|---|---|---|
| ZIP CODE | | QUARTER 1 | QUARTER 2 | QUARTER 3 | QUARTER 4 | TOTAL GRAMS |
| | 630 | 214.5 | 217.96 | 206.44 | 265.15 | 904.05 |
| | 631 | 1,307.18 | 912.46 | 1,028.48 | 511.57 | 3,759.69 |
| | 633 | 133.45 | 77.89 | 72.33 | 66.45 | 350.12 |
| | 634 | 7.19 | 15.47 | 17.43 | 16.67 | 56.76 |
| | 635 | 108.39 | 117.48 | 61.22 | 69.72 | 356.81 |
| | 636 | 84.86 | 87.15 | 94.56 | 79.74 | 346.31 |
| | 637 | 59.31 | 73.86 | 31.16 | 25.06 | 189.39 |
| | 638 | 99.92 | 130.07 | 129.16 | 108.94 | 468.09 |
| | 639 | 78 | 54.9 | 48.26 | 61.55 | 242.71 |
| | 640 | 359.47 | 267.11 | 230.01 | 263.08 | 1,119.67 |
| | 641 | 268.2 | 235.41 | 216.89 | 294.35 | 1,014.85 |
| | 644 | 34.42 | 41.4 | 29.85 | 20.15 | 125.82 |
| | 645 | 28.76 | 32.03 | 15.9 | 31.92 | 108.61 |
| | 646 | 27.89 | 19.61 | 7.84 | 18.52 | 73.86 |
| | 647 | 76.47 | 52.29 | 74.27 | 61.88 | 264.91 |
| | 648 | 217.11 | 272.45 | 133.67 | 141.07 | 764.3 |
| | 650 | 48.15 | 26.36 | 40.96 | 66.78 | 182.25 |
| | 651 | 89.33 | 79.74 | 100.22 | 73.31 | 342.6 |
| | 652 | 350.26 | 303.94 | 225.06 | 230.51 | 1,109.77 |
| | 653 | 68.63 | 88.11 | 63.84 | 100.22 | 320.8 |
| | 654 | 63.4 | 88.24 | 78.22 | 44.66 | 274.52 |
| | 655 | 215.91 | 196.52 | 124.19 | 160.57 | 697.19 |
| | 656 | 108.94 | 87.15 | 94.34 | 101.31 | 391.74 |
| | 657 | 364.72 | 237.92 | 235.2 | 184.1 | 1,021.94 |
| | 658 | 213.63 | 197.39 | 166.78 | 145.87 | 723.67 |
| TOTAL | | 4,628.09 | 3,912.91 | 3,526.28 | 3,143.15 | 15,210.43 |

DATE RANGE: 01/01/2017 TO 12/31/2017 | ARCOS 3 - REPORT 1 | Run Date: 07/03/2018

RETAIL DRUG DISTRIBUTION BY ZIP CODE WITHIN STATE BY GRAMS WT

STATE: MONTANA

| ZIP CODE | QUARTER 1 | QUARTER 2 | QUARTER 3 | QUARTER 4 | TOTAL GRAMS |
|---|---|---|---|---|---|
| 590 | 10.02 | 6.75 | 25.06 | 8.93 | 50.76 |
| 591 | 48.8 | 49.13 | 57.95 | 50.98 | 206.86 |
| 592 | 12.2 | 28.32 | 19.61 | 13.07 | 73.2 |
| 593 | 27.45 | 44.99 | 34.21 | 5.66 | 112.31 |
| 594 | 160.84 | 177.66 | 137.04 | 158.18 | 633.72 |
| 595 | 7.84 | 1.53 | 1.53 | 6.54 | 17.44 |
| 596 | 95.43 | 112.42 | 96.74 | 86.71 | 391.3 |
| 597 | 55.34 | 66.89 | 58.28 | 37.26 | 217.77 |
| 598 | 87.8 | 90.2 | 58.28 | 90.85 | 327.13 |
| 599 | 151.21 | 196.74 | 168.42 | 139.44 | 655.81 |
| TOTAL | 656.93 | 774.63 | 657.12 | 597.62 | 2,686.30 |

STATE: NEBRASKA

| ZIP CODE | QUARTER 1 | QUARTER 2 | QUARTER 3 | QUARTER 4 | TOTAL GRAMS |
|---|---|---|---|---|---|
| 680 | 47.17 | 57.95 | 63.62 | 26.47 | 195.21 |
| 681 | 220.27 | 247.07 | 134.65 | 156.43 | 758.42 |
| 683 | 14.38 | 17.43 | 12.2 | 12.31 | 56.32 |
| 684 | 26.36 | 11.59 | 26.36 | 3.05 | 67.36 |
| 685 | 164.76 | 163.91 | 151.64 | 128.66 | 608.97 |
| 686 | 20.26 | 21.57 | 23.53 | 29.2 | 94.56 |
| 687 | 34.97 | 52.94 | 42.92 | 23.42 | 154.25 |
| 688 | 114.71 | 80.85 | 89.22 | 43.9 | 328.68 |
| 689 | 61.22 | 43.9 | 39.33 | 40.63 | 185.08 |
| 690 | 19.39 | 14.05 | 14.27 | 19.06 | 66.77 |
| 691 | 31.37 | 25.27 | 45.54 | 39.11 | 141.29 |
| 692 | 17.56 | 15.25 | 2.61 | 10.89 | 46.31 |
| 693 | 29.63 | 21.79 | 23.97 | 6.32 | 81.71 |
| TOTAL | 802.05 | 773.57 | 669.86 | 539.45 | 2,784.93 |

STATE: NEVADA

| ZIP CODE | QUARTER 1 | QUARTER 2 | QUARTER 3 | QUARTER 4 | TOTAL GRAMS |
|---|---|---|---|---|---|

| | | | | | | |
|---|---|---|---|---|---|---|
| | 890 | 290.54 | 214.28 | 228.01 | 222.01 | 954.84 |
| | 891 | 988.28 | 700.03 | 772.58 | 1,063.67 | 3,524.56 |
| | 893 | 10.89 | 19.61 | 3.27 | 9.26 | 43.03 |
| | 894 | 73.97 | 48.15 | 49.78 | 64.27 | 236.17 |
| | 895 | 194.34 | 174.08 | 132.25 | 130.51 | 631.18 |
| | 897 | 42.7 | 57.39 | 43.4 | 30.5 | 173.99 |
| | 898 | 32.14 | 25.06 | 23.31 | 26.58 | 107.09 |
| TOTAL | | 1,632.86 | 1,238.60 | 1,252.60 | 1,546.80 | 5,670.86 |

| STATE: | NEW HAMPSHIRE | | | | | |
|---|---|---|---|---|---|---|
| ZIP CODE | | QUARTER 1 | QUARTER 2 | QUARTER 3 | QUARTER 4 | TOTAL GRAMS |
| | 030 | 81.05 | 41.51 | 42.05 | 47.61 | 212.22 |
| | 031 | 63.4 | 84.1 | 62.53 | 64.71 | 274.74 |
| | 032 | 115.91 | 110.46 | 98.37 | 106.98 | 431.72 |
| | 033 | 195 | 185.19 | 209.6 | 114.38 | 704.17 |
| | 034 | 22.44 | 31.59 | 23.97 | 28.32 | 106.32 |
| | 035 | 8.72 | 11.33 | 15.03 | 4.36 | 39.44 |
| | 037 | 29.41 | 25.82 | 40.96 | 20.7 | 116.89 |
| | 038 | 160.25 | 120.92 | 120.05 | 63.18 | 464.4 |
| TOTAL | | 676.18 | 610.92 | 612.56 | 450.24 | 2,349.90 |

| STATE: | NEW JERSEY | | | | | |
|---|---|---|---|---|---|---|
| ZIP CODE | | QUARTER 1 | QUARTER 2 | QUARTER 3 | QUARTER 4 | TOTAL GRAMS |
| | 70 | 297.73 | 258.51 | 224.52 | 210.69 | 991.45 |
| | 71 | 29.85 | 35.08 | 14.05 | 16.12 | 95.1 |
| | 72 | 2.18 | 4.36 | 1.09 | 1.09 | 8.72 |
| | 73 | 119.29 | 56.21 | 62.97 | 45.75 | 284.22 |
| | 74 | 122.88 | 163.62 | 106 | 30.28 | 422.78 |
| | 75 | 27.23 | 30.39 | 19.39 | 34.32 | 111.33 |
| | 76 | 202.62 | 128.55 | 148.48 | 154.69 | 634.34 |
| | 77 | 203.93 | 275.5 | 185.85 | 202.73 | 868.01 |
| | 78 | 58.17 | 45.86 | 47.39 | 16.56 | 167.98 |
| | 79 | 65.04 | 57.95 | 84.54 | 63.95 | 271.48 |
| | 80 | 328.34 | 285.85 | 247.18 | 154.8 | 1,016.17 |
| | 81 | 23.09 | 9.7 | 0 | 9.26 | 42.05 |

| | | | | | | |
|---|---|---|---|---|---|---|
| | 82 | 29.63 | 57.74 | 35.95 | 23.42 | 146.74 |
| | 83 | 26.25 | 25.27 | 27.56 | 2.72 | 81.8 |
| | 84 | 0 | 5.45 | 0 | 0.87 | 6.32 |
| | 85 | 49.57 | 31.16 | 37.91 | 51.75 | 170.39 |
| | 86 | 17.87 | 41.51 | 23.09 | 20.48 | 102.95 |
| | 87 | 106.43 | 90.64 | 102.62 | 67.65 | 367.34 |
| | 88 | 423.98 | 539.35 | 451.65 | 387.27 | 1,802.25 |
| | 89 | 52.51 | 37.69 | 12.42 | 8.72 | 111.34 |
| TOTAL | | 2,186.59 | 2,180.39 | 1,832.66 | 1,503.12 | 7,702.76 |

| STATE: | NEW MEXICO | | | | | |
|---|---|---|---|---|---|---|
| ZIP CODE | | QUARTER 1 | QUARTER 2 | QUARTER 3 | QUARTER 4 | TOTAL GRAMS |
| | 870 | 185.63 | 52.29 | 75.17 | 54.9 | 367.99 |
| | 871 | 709.18 | 634.67 | 521.59 | 503.07 | 2,368.51 |
| | 873 | 7.41 | 4.58 | 7.19 | 0.44 | 19.62 |
| | 874 | 52.51 | 32.57 | 40.09 | 44.66 | 169.83 |
| | 875 | 260.47 | 175.17 | 220.27 | 196.09 | 852 |
| | 877 | 17.54 | 23.53 | 42.7 | 14.82 | 98.59 |
| | 878 | 0 | 9.59 | 1.74 | 6.1 | 17.43 |
| | 879 | 0 | 0 | 0.44 | 5.23 | 5.67 |
| | 880 | 132.36 | 260.58 | 173.21 | 239.12 | 805.27 |
| | 881 | 84.75 | 62.79 | 88.46 | 57.08 | 293.08 |
| | 882 | 183.02 | 130.83 | 126.26 | 98.92 | 539.03 |
| | 883 | 34.86 | 23.42 | 16.89 | 17.43 | 92.6 |
| | 884 | 13.07 | 6.54 | 0.87 | 0.44 | 20.92 |
| TOTAL | | 1,680.80 | 1,416.56 | 1,314.88 | 1,238.30 | 5,650.54 |

| STATE: | NEW YORK | | | | | |
|---|---|---|---|---|---|---|
| ZIP CODE | | QUARTER 1 | QUARTER 2 | QUARTER 3 | QUARTER 4 | TOTAL GRAMS |
| | 100 | 709.73 | 665.28 | 566.58 | 563.32 | 2,504.91 |
| | 101 | 24.4 | 47.28 | 51.85 | 39.22 | 162.75 |
| | 103 | 157.09 | 132.47 | 104.14 | 86.17 | 479.87 |
| | 104 | 144.45 | 164.82 | 150.33 | 133.34 | 592.94 |
| | 105 | 240.53 | 222.67 | 218.53 | 179.53 | 861.26 |
| | 106 | 8.06 | 26.15 | 0 | 0 | 34.21 |

| | | | | | | |
|---|---|---|---|---|---|---|
| 107 | 74.62 | 62.31 | | 38.67 | 61.22 | 236.82 |
| 108 | 9.59 | 14.38 | | 3.7 | 12.31 | 39.98 |
| 109 | 98.26 | 40.31 | | 33.33 | 63.07 | 234.97 |
| 110 | 123.54 | 154.69 | | 152.95 | 116.78 | 547.96 |
| 111 | 11.98 | 11.98 | | 29.41 | 8.17 | 61.54 |
| 112 | 200.99 | 197.94 | | 205.67 | 220.82 | 825.42 |
| 113 | 106.11 | 63.62 | | 99.68 | 122.23 | 391.64 |
| 114 | 35.95 | 28.98 | | 44.88 | 23.97 | 133.78 |
| 115 | 65.04 | 125.82 | | 104.36 | 95.65 | 390.87 |
| 116 | 9.8 | 22.88 | | 8.72 | 0 | 41.4 |
| 117 | 412.98 | 339.89 | | 276.37 | 246.63 | 1,275.87 |
| 118 | 113.84 | 78.87 | | 79.52 | 69.72 | 341.95 |
| 119 | 21.79 | 11.11 | | 19.72 | 9.26 | 61.88 |
| 120 | 129.64 | 105.02 | | 75.6 | 91.29 | 401.55 |
| 121 | 65.8 | 61.66 | | 29.63 | 45.97 | 203.06 |
| 122 | 31.37 | 52.07 | | 28.21 | 17.65 | 129.3 |
| 123 | 82.14 | 99.79 | | 121.57 | 68.19 | 371.69 |
| 124 | 113.95 | 143.8 | | 106.54 | 86.61 | 450.9 |
| 125 | 108.39 | 58.28 | | 65.47 | 57.85 | 289.99 |
| 126 | 60.68 | 114.93 | | 105.12 | 125.28 | 406.01 |
| 127 | 31.7 | 8.72 | | 29.41 | 4.36 | 74.19 |
| 128 | 121.79 | 79.31 | | 83.88 | 80.07 | 365.05 |
| 129 | 118.96 | 20.59 | | 75.06 | 120.05 | 334.66 |
| 130 | 256.11 | 194.8 | | 178.22 | 210.25 | 839.38 |
| 131 | 31.16 | 33.66 | | 61.01 | 17.43 | 143.26 |
| 132 | 89.44 | 98.59 | | 71.46 | 87.37 | 346.86 |
| 133 | 69.2 | 65.49 | | 82.66 | 56.12 | 273.47 |
| 134 | 56.84 | 39.98 | | 27.78 | 40.2 | 164.8 |
| 135 | 40.85 | 37.37 | | 35.73 | 39.22 | 153.17 |
| 136 | 26.47 | 15.08 | | 6.75 | 7.34 | 55.64 |
| 137 | 8.72 | 12.85 | | 3.92 | 12.64 | 38.13 |
| 138 | 51.42 | 63.88 | | 72.99 | 54.47 | 242.76 |
| 139 | 10.78 | 10.13 | | 12.2 | 8.72 | 41.83 |
| 140 | 84.64 | 42.05 | | 53.71 | 38.24 | 218.64 |
| 141 | 97.17 | 59.7 | | 52.51 | 70.81 | 280.19 |
| 142 | 370.61 | 282.47 | | 254.91 | 282.47 | 1,190.46 |
| 143 | 101.31 | 100.77 | | 119.83 | 135.08 | 456.99 |

| | | | | | | |
|---|---|---|---|---|---|---|
| | 144 | 31.92 | 19.5 | 13.18 | 35.84 | 100.44 |
| | 145 | 31.16 | 66.89 | 54.9 | 34.42 | 187.37 |
| | 146 | 92.49 | 111.16 | 84.32 | 51.2 | 339.17 |
| | 147 | 53.38 | 77.65 | 15.95 | 59.48 | 206.46 |
| | 148 | 155.78 | 148.16 | 164.71 | 183.78 | 652.43 |
| | 149 | 30.72 | 27.23 | 26.15 | 27.23 | 111.33 |
| TOTAL | | 5,123.34 | 4,663.03 | 4,301.79 | 4,201.04 | 18,289.20 |

STATE: NORTH CAROLINA

| ZIP CODE | | QUARTER 1 | QUARTER 2 | QUARTER 3 | QUARTER 4 | TOTAL GRAMS |
|---|---|---|---|---|---|---|
| | 270 | 273.22 | 252.95 | 216.13 | 161.23 | 903.53 |
| | 271 | 233.02 | 188.14 | 180.62 | 73.86 | 675.64 |
| | 272 | 328.12 | 253.39 | 274.74 | 293.7 | 1,149.95 |
| | 273 | 277.57 | 209.81 | 160.79 | 153.6 | 801.77 |
| | 274 | 207.96 | 253.06 | 151.31 | 138.02 | 750.35 |
| | 275 | 749.05 | 656.89 | 632.93 | 478.67 | 2,517.54 |
| | 276 | 203.93 | 187.7 | 178.98 | 174.41 | 745.02 |
| | 277 | 49.78 | 27.45 | 23.53 | 62.42 | 163.18 |
| | 278 | 439.89 | 378.27 | 320.71 | 419.95 | 1,558.82 |
| | 279 | 277.79 | 208.51 | 202.3 | 198.81 | 887.41 |
| | 280 | 628.79 | 571.05 | 401.54 | 434.77 | 2,036.15 |
| | 281 | 723.67 | 609.83 | 564.41 | 644.15 | 2,542.06 |
| | 282 | 606.04 | 677.68 | 565.86 | 493.97 | 2,343.55 |
| | 283 | 410.48 | 378.64 | 308.08 | 402.85 | 1,500.05 |
| | 284 | 366.57 | 306.77 | 264.72 | 251.65 | 1,189.71 |
| | 285 | 325.72 | 301.32 | 218.09 | 250.23 | 1,095.36 |
| | 286 | 702.97 | 587.06 | 420.17 | 555.47 | 2,265.67 |
| | 287 | 314.39 | 364.29 | 284.33 | 215.37 | 1,178.38 |
| | 288 | 67.65 | 44.23 | 35.95 | 44.12 | 191.95 |
| | 289 | 4.36 | 5.45 | 26.15 | 9.8 | 45.76 |
| TOTAL | | 7,190.97 | 6,462.49 | 5,431.34 | 5,457.05 | 24,541.85 |

STATE: NORTH DAKOTA

| ZIP CODE | | QUARTER 1 | QUARTER 2 | QUARTER 3 | QUARTER 4 | TOTAL GRAMS |
|---|---|---|---|---|---|---|

| | | | | | | |
|---|---|---|---|---|---|---|
| | 580 | 11.98 | 14.38 | 16.56 | 13.07 | 55.99 |
| | 581 | 41.83 | 27.45 | 35.3 | 24.4 | 128.98 |
| | 582 | 48.37 | 37.26 | 33.55 | 9.26 | 128.44 |
| | 583 | 23.97 | 19.61 | 23.09 | 23.53 | 90.2 |
| | 584 | 6.97 | 17.21 | 24.18 | 10.13 | 58.49 |
| | 585 | 50.55 | 79.31 | 59.26 | 100.22 | 289.34 |
| | 586 | 11.11 | 12.42 | 11.98 | 9.8 | 45.31 |
| | 587 | 37.47 | 23.31 | 43.36 | 27.78 | 131.92 |
| | 588 | 5.23 | 6.65 | 6.43 | 4.79 | 23.1 |
| TOTAL | | 237.48 | 237.6 | 253.71 | 222.98 | 951.77 |

STATE: OHIO

| ZIP CODE | | QUARTER 1 | QUARTER 2 | QUARTER 3 | QUARTER 4 | TOTAL GRAMS |
|---|---|---|---|---|---|---|
| | 430 | 166.13 | 171.47 | 93.9 | 159.48 | 590.98 |
| | 431 | 272.78 | 256.87 | 267.77 | 157.31 | 954.73 |
| | 432 | 524.75 | 381.06 | 639.14 | 688.59 | 2,233.54 |
| | 433 | 28.32 | 28.02 | 22.66 | 25.06 | 104.06 |
| | 434 | 136.17 | 119.4 | 105.23 | 94.78 | 455.58 |
| | 435 | 102.62 | 70.59 | 61.33 | 60.35 | 294.89 |
| | 436 | 90.53 | 128.11 | 144.02 | 93.8 | 456.46 |
| | 437 | 211.56 | 205.02 | 188.24 | 171.47 | 776.29 |
| | 438 | 0.44 | 0.87 | 2.18 | 4.36 | 7.85 |
| | 439 | 93.14 | 132.03 | 88.46 | 77.35 | 390.98 |
| | 440 | 293.26 | 220.16 | 245.44 | 176.04 | 934.9 |
| | 441 | 873.03 | 989.81 | 970.31 | 565.6 | 3,398.75 |
| | 442 | 173.86 | 121.36 | 86.82 | 73.1 | 455.14 |
| | 443 | 64.06 | 67.11 | 73.64 | 45.21 | 250.02 |
| | 444 | 96.74 | 70.37 | 108.28 | 33.55 | 308.94 |
| | 445 | 79.85 | 89 | 55.34 | 29.52 | 253.71 |
| | 446 | 107.78 | 114.38 | 134.97 | 83.66 | 440.79 |
| | 447 | 44.01 | 48.59 | 76.15 | 38.35 | 207.1 |
| | 448 | 186.72 | 177.02 | 164.28 | 184.98 | 713 |
| | 449 | 50.55 | 21.24 | 49.13 | 6.43 | 127.35 |
| | 450 | 245.55 | 298.38 | 332.26 | 231.71 | 1,107.90 |
| | 451 | 126.8 | 113.19 | 86.71 | 128.98 | 455.68 |
| | 452 | 671.86 | 647.79 | 774.07 | 563.48 | 2,657.20 |

| | | | | | | |
|---|---|---|---|---|---|---|
| | 453 | 116.89 | 167.76 | 136.72 | 166.67 | 588.04 |
| | 454 | 256.66 | 215.48 | 272.67 | 173.54 | 918.35 |
| | 455 | 26.36 | 27.23 | 31.59 | 11.98 | 97.16 |
| | 456 | 39.11 | 28.21 | 43.62 | 17.97 | 128.91 |
| | 457 | 37.15 | 26.8 | 39.33 | 23.97 | 127.25 |
| | 458 | 145.24 | 153.82 | 122.03 | 124.3 | 545.39 |
| TOTAL | | 5,261.92 | 5,091.14 | 5,416.29 | 4,211.59 | 19,980.94 |

STATE: OKLAHOMA

| ZIP CODE | | QUARTER 1 | QUARTER 2 | QUARTER 3 | QUARTER 4 | TOTAL GRAMS |
|---|---|---|---|---|---|---|
| | 730 | 1,342.11 | 1,221.82 | 1,224.57 | 1,033.53 | 4,822.03 |
| | 731 | 1,901.94 | 1,750.02 | 1,689.82 | 1,760.32 | 7,102.10 |
| | 734 | 64.27 | 49.89 | 61.88 | 47.39 | 223.43 |
| | 735 | 719.34 | 801.8 | 575.32 | 691.08 | 2,787.54 |
| | 736 | 87.69 | 52.62 | 77.45 | 82.79 | 300.55 |
| | 737 | 134.04 | 144.23 | 128.76 | 119.18 | 526.21 |
| | 738 | 20.59 | 10.46 | 6.1 | 15.36 | 52.51 |
| | 739 | 9.8 | 28.32 | 13.29 | 26.8 | 78.21 |
| | 740 | 1,203.87 | 1,132.56 | 1,097.48 | 1,197.44 | 4,631.35 |
| | 741 | 694.15 | 785.88 | 591.42 | 671.6 | 2,743.05 |
| | 743 | 148.16 | 125.17 | 161.12 | 134.43 | 568.88 |
| | 744 | 409.04 | 445.12 | 359.06 | 298.49 | 1,511.71 |
| | 745 | 189.05 | 211.23 | 185.85 | 131.71 | 717.84 |
| | 746 | 33.33 | 33.01 | 49.35 | 33.23 | 148.92 |
| | 747 | 181.38 | 147.28 | 178.11 | 186.28 | 693.05 |
| | 748 | 337.71 | 271.52 | 226.81 | 235.85 | 1,071.89 |
| | 749 | 178.88 | 174.13 | 181.49 | 141.62 | 676.12 |
| TOTAL | | 7,655.35 | 7,385.06 | 6,807.88 | 6,807.10 | 28,655.39 |

STATE: OREGON

| ZIP CODE | | QUARTER 1 | QUARTER 2 | QUARTER 3 | QUARTER 4 | TOTAL GRAMS |
|---|---|---|---|---|---|---|
| | 970 | 371.04 | 313.85 | 223.32 | 199.9 | 1,108.11 |
| | 971 | 146.52 | 154.36 | 212.43 | 110.9 | 624.21 |

| | | | | | | |
|---|---|---|---|---|---|---|
| | 972 | 303.28 | 213.08 | 242.49 | 269.62 | 1,028.47 |
| | 973 | 330.52 | 261.34 | 263.74 | 194.56 | 1,050.16 |
| | 974 | 242.82 | 229.47 | 238.36 | 216.13 | 926.78 |
| | 975 | 168.85 | 162.53 | 182.58 | 165.15 | 679.11 |
| | 976 | 57.74 | 33.33 | 69.07 | 42.92 | 203.06 |
| | 977 | 45.54 | 38.78 | 51.53 | 44.45 | 180.3 |
| | 978 | 81.49 | 51.68 | 88.13 | 62.31 | 283.61 |
| | 979 | 1.74 | 5.88 | 14.16 | 9.8 | 31.58 |
| TOTAL | | 1,749.54 | 1,464.30 | 1,585.81 | 1,315.74 | 6,115.39 |

STATE: PENNSYLVANIA

| ZIP CODE | | QUARTER 1 | QUARTER 2 | QUARTER 3 | QUARTER 4 | TOTAL GRAMS |
|---|---|---|---|---|---|---|
| | 150 | 128.55 | 125.5 | 96.95 | 96.95 | 447.95 |
| | 151 | 96.74 | 92.6 | 84.1 | 63.62 | 337.06 |
| | 152 | 197.94 | 163.41 | 144.45 | 247.29 | 753.09 |
| | 153 | 54.03 | 54.03 | 43.58 | 47.17 | 198.81 |
| | 154 | 7.84 | 11.98 | 8.06 | 13.51 | 41.39 |
| | 155 | 10.89 | 10.33 | 8.72 | 3.81 | 33.75 |
| | 156 | 10.24 | 41.83 | 40.09 | 27.23 | 119.39 |
| | 157 | 11.98 | 14.86 | 7.84 | 24.95 | 59.63 |
| | 158 | 108.94 | 69.72 | 142.71 | 128.55 | 449.92 |
| | 159 | 66.34 | 65.36 | 59.26 | 47.93 | 238.89 |
| | 160 | 20.92 | 14.16 | 30.28 | 28.98 | 94.34 |
| | 161 | 61.11 | 51.64 | 36.6 | 26.04 | 175.39 |
| | 162 | 9.15 | 0.22 | 5.01 | 6.1 | 20.48 |
| | 163 | 46.19 | 46.19 | 40.52 | 23.97 | 156.87 |
| | 164 | 4.36 | 39.22 | 35.4 | 17.43 | 96.41 |
| | 165 | 65.04 | 44.23 | 42.7 | 54.8 | 206.77 |
| | 166 | 109.37 | 90.85 | 70.81 | 70.81 | 341.84 |
| | 167 | 4.79 | 4.36 | 0 | 4.36 | 13.51 |
| | 168 | 45.32 | 22.44 | 35.73 | 26.8 | 130.29 |
| | 169 | 0.87 | 0.44 | 3.05 | 1.53 | 5.89 |
| | 170 | 155.34 | 203.71 | 137.81 | 176.26 | 673.12 |
| | 171 | 93.9 | 30.5 | 46.63 | 20.04 | 191.07 |
| | 172 | 15.25 | 22.88 | 35.73 | 11.11 | 84.97 |

| | | | | | | |
|---|---|---|---|---|---|---|
| | 173 | 186.61 | 199.68 | 237.27 | 218.96 | 842.52 |
| | 174 | 54.69 | 86.71 | 43.14 | 127.57 | 312.11 |
| | 175 | 32.03 | 34.21 | 28.76 | 50.22 | 145.22 |
| | 176 | 64.16 | 47.5 | 48.37 | 57.08 | 217.11 |
| | 177 | 69.61 | 79.96 | 49.13 | 68.63 | 267.33 |
| | 178 | 65.47 | 42.81 | 35.73 | 38.78 | 182.79 |
| | 179 | 25.06 | 14.16 | 23.42 | 22.33 | 84.97 |
| | 180 | 29.96 | 31.16 | 37.37 | 53.92 | 152.41 |
| | 181 | 109.7 | 93.03 | 125.71 | 96.19 | 424.63 |
| | 182 | 61.44 | 47.28 | 33.77 | 13.62 | 156.11 |
| | 183 | 8.06 | 18.52 | 0 | 4.36 | 30.94 |
| | 184 | 60.13 | 41.18 | 39.22 | 37.04 | 177.57 |
| | 185 | 94.12 | 49.68 | 82.36 | 62.31 | 288.47 |
| | 186 | 71.35 | 39.44 | 26.58 | 36.49 | 173.86 |
| | 187 | 316.9 | 274.41 | 347.29 | 247.83 | 1,186.43 |
| | 188 | 99.83 | 83.32 | 65.8 | 124.52 | 373.47 |
| | 189 | 61.88 | 69.07 | 34.53 | 52.83 | 218.31 |
| | 190 | 490.11 | 415.6 | 409.39 | 350.02 | 1,665.12 |
| | 191 | 328.01 | 259.71 | 283.24 | 212.97 | 1,083.93 |
| | 193 | 223.43 | 180.84 | 77.35 | 116.89 | 598.51 |
| | 194 | 79.42 | 100.77 | 131.6 | 59.7 | 371.49 |
| | 195 | 33.12 | 27.45 | 34.64 | 31.81 | 127.02 |
| | 196 | 53.82 | 60.79 | 74.62 | 58.83 | 248.06 |
| TOTAL | | 3,944.01 | 3,517.74 | 3,425.32 | 3,312.14 | 14,199.21 |

STATE: PUERTO RICO

| ZIP CODE | | QUARTER 1 | QUARTER 2 | QUARTER 3 | QUARTER 4 | TOTAL GRAMS |
|---|---|---|---|---|---|---|
| | 6 | 1,536.35 | 1,894.42 | 1,320.65 | 937.84 | 5,689.26 |
| | 7 | 1,905.43 | 1,213.56 | 1,072.05 | 543.49 | 4,734.53 |
| | 9 | 2,211.21 | 1,823.72 | 1,726.01 | 1,187.09 | 6,948.03 |
| TOTAL | | 5,652.99 | 4,931.70 | 4,118.71 | 2,668.42 | 17,371.82 |

STATE: RHODE ISLAND

| ZIP CODE | | QUARTER 1 | QUARTER 2 | QUARTER 3 | QUARTER 4 | TOTAL GRAMS |
|---|---|---|---|---|---|---|
| | 28 | 89.76 | 80.61 | 76.04 | 92.38 | 338.79 |

| ZIP CODE | QUARTER 1 | QUARTER 2 | QUARTER 3 | QUARTER 4 | TOTAL GRAMS |
|---|---|---|---|---|---|
| 29 | 163.19 | 184 | 180.73 | 140.09 | 668.01 |
| TOTAL | 252.95 | 264.61 | 256.77 | 232.47 | 1,006.80 |

STATE: SOUTH CAROLINA

| ZIP CODE | QUARTER 1 | QUARTER 2 | QUARTER 3 | QUARTER 4 | TOTAL GRAMS |
|---|---|---|---|---|---|
| 290 | 854.07 | 732.06 | 654.71 | 598.07 | 2,838.91 |
| 291 | 602.64 | 521.48 | 419.63 | 456.12 | 1,999.87 |
| 292 | 998.41 | 773.89 | 887.51 | 791.98 | 3,451.79 |
| 293 | 468.76 | 571.81 | 389.34 | 337.71 | 1,767.62 |
| 294 | 991.55 | 1,075.10 | 815.18 | 780.65 | 3,662.48 |
| 295 | 933.59 | 623.45 | 637.94 | 652.86 | 2,847.84 |
| 296 | 1,047.87 | 1,009.52 | 892.85 | 887.4 | 3,837.64 |
| 297 | 306.33 | 307.86 | 245.76 | 192.06 | 1,052.01 |
| 298 | 371.91 | 343.37 | 367.34 | 294.13 | 1,376.75 |
| 299 | 205.35 | 160.57 | 214.5 | 101.86 | 682.28 |
| TOTAL | 6,780.48 | 6,119.11 | 5,524.76 | 5,092.84 | 23,517.19 |

STATE: SOUTH DAKOTA

| ZIP CODE | QUARTER 1 | QUARTER 2 | QUARTER 3 | QUARTER 4 | TOTAL GRAMS |
|---|---|---|---|---|---|
| 570 | 159.27 | 142.05 | 136.39 | 117.54 | 555.25 |
| 571 | 161.23 | 138.79 | 153.71 | 157.31 | 611.04 |
| 572 | 44.01 | 55.88 | 36.17 | 37.26 | 173.32 |
| 573 | 22.01 | 32.25 | 48.37 | 15.14 | 117.77 |
| 574 | 28.43 | 28.98 | 26.58 | 11.11 | 95.1 |
| 575 | 16.12 | 10.24 | 8.28 | 0.44 | 35.08 |
| 576 | 6.54 | 0.87 | 1.74 | 0 | 9.15 |
| 577 | 113.3 | 149.68 | 118.52 | 98.04 | 479.54 |
| TOTAL | 550.91 | 558.74 | 529.76 | 436.84 | 2,076.25 |

STATE: TENNESSEE

| ZIP CODE | QUARTER 1 | QUARTER 2 | QUARTER 3 | QUARTER 4 | TOTAL GRAMS |
|---|---|---|---|---|---|
| 370 | 1,583.41 | 1,413.68 | 1,046.56 | 968.35 | 5,012 |
| 371 | 1,263.89 | 1,202.56 | 1,090.57 | 921.94 | 4,478.96 |

| | | | | | | |
|---|---|---|---|---|---|---|
| | 372 | 823.02 | 704.06 | 622.69 | 489.67 | 2,639.44 |
| | 373 | 885.01 | 759.43 | 635.58 | 713.43 | 2,993.45 |
| | 374 | 426.16 | 369.89 | 259.92 | 288.03 | 1,344 |
| | 376 | 318.64 | 286.94 | 196.41 | 255.24 | 1,057.23 |
| | 377 | 249.14 | 268.42 | 187.15 | 180.29 | 885 |
| | 378 | 395.51 | 356.81 | 313.83 | 327.79 | 1,393.94 |
| | 379 | 354.37 | 244.52 | 212.36 | 277.51 | 1,088.76 |
| | 380 | 381.72 | 371.52 | 389.67 | 261.34 | 1,404.25 |
| | 381 | 840.45 | 710.16 | 668.11 | 623.23 | 2,841.95 |
| | 382 | 239.88 | 185.63 | 219.73 | 187.07 | 832.31 |
| | 383 | 453.51 | 482.27 | 416.47 | 388.36 | 1,740.61 |
| | 384 | 322.89 | 288.36 | 219.4 | 266.9 | 1,097.55 |
| | 385 | 212.97 | 204.19 | 262.76 | 202.73 | 882.65 |
| TOTAL | | 8,750.57 | 7,848.44 | 6,741.21 | 6,351.88 | 29,692.10 |

| STATE: | TEXAS | | | | | |
|---|---|---|---|---|---|---|
| ZIP CODE | | QUARTER 1 | QUARTER 2 | QUARTER 3 | QUARTER 4 | TOTAL GRAMS |
| | 750 | 808.77 | 756.53 | 769.8 | 684.5 | 3,019.60 |
| | 751 | 140.68 | 170.75 | 164.06 | 145.43 | 620.92 |
| | 752 | 401.41 | 418.78 | 437.93 | 378.01 | 1,636.13 |
| | 753 | 7.63 | 18.63 | 13.29 | 0 | 39.55 |
| | 754 | 100.03 | 103.38 | 119.4 | 128.98 | 451.79 |
| | 755 | 75.17 | 66.45 | 93.69 | 47.82 | 283.13 |
| | 756 | 353.48 | 214.59 | 233.37 | 158.74 | 960.18 |
| | 757 | 191.51 | 213.54 | 188.27 | 201.58 | 794.9 |
| | 758 | 21.57 | 16.25 | 20.52 | 35.34 | 93.68 |
| | 759 | 158.29 | 165.3 | 151.53 | 138.5 | 613.62 |
| | 760 | 397.01 | 498.56 | 508.74 | 308.95 | 1,713.26 |
| | 761 | 323 | 412.87 | 570.94 | 447.19 | 1,754 |
| | 762 | 152.51 | 180.62 | 97.17 | 237.37 | 667.67 |
| | 763 | 113.84 | 105.54 | 81.16 | 114.38 | 414.92 |
| | 764 | 52.09 | 63.88 | 64.71 | 51.75 | 232.43 |
| | 765 | 123.1 | 114.86 | 129.31 | 136.93 | 504.2 |
| | 766 | 50.33 | 67.8 | 56.04 | 32.46 | 206.63 |
| | 767 | 57.63 | 89.55 | 109.48 | 86.5 | 343.16 |

| | | | | | | |
|---|---|---|---|---|---|---|
| | 768 | 52.29 | 46.08 | 45.75 | 53.92 | 198.04 |
| | 769 | 82.25 | 66.02 | 106.98 | 67.43 | 322.68 |
| | 770 | 2,074 | 1,701.17 | 1,846.93 | 2,476.69 | 8,098.79 |
| | 773 | 379.09 | 397.56 | 396.3 | 487.6 | 1,660.55 |
| | 774 | 347.73 | 264.76 | 359.1 | 315.16 | 1,286.75 |
| | 775 | 616.26 | 427.58 | 413.85 | 437.06 | 1,894.75 |
| | 776 | 134.1 | 145.43 | 151.75 | 200.45 | 631.73 |
| | 777 | 136.93 | 75.6 | 97.06 | 192.82 | 502.41 |
| | 778 | 97.61 | 148.7 | 137.26 | 113.84 | 497.41 |
| | 779 | 53.49 | 53.92 | 53.71 | 50.66 | 211.78 |
| | 780 | 152.08 | 190.53 | 194.13 | 186.83 | 723.57 |
| | 781 | 55.34 | 60.29 | 66.13 | 78.22 | 259.98 |
| | 782 | 896.77 | 774.44 | 832.39 | 809.1 | 3,312.70 |
| | 783 | 30.39 | 37.26 | 13.51 | 31.59 | 112.75 |
| | 784 | 277.88 | 225.06 | 258.4 | 250.34 | 1,011.68 |
| | 785 | 906.69 | 918.82 | 1,012.84 | 874.33 | 3,712.68 |
| | 786 | 202.19 | 168.09 | 204.58 | 208.4 | 783.26 |
| | 787 | 273.87 | 210.9 | 332.7 | 289.12 | 1,106.59 |
| | 788 | 36.1 | 52.4 | 64.16 | 78.87 | 231.53 |
| | 789 | 17.43 | 20.7 | 6.1 | 10.46 | 54.69 |
| | 790 | 38.67 | 36.25 | 30.5 | 34.32 | 139.74 |
| | 791 | 189.01 | 209.81 | 185.19 | 150.12 | 734.13 |
| | 792 | 15.8 | 19.06 | 20.15 | 4.9 | 59.91 |
| | 793 | 83.38 | 70.83 | 95.21 | 147.28 | 396.7 |
| | 794 | 196.96 | 156.5 | 144.56 | 161.99 | 660.01 |
| | 795 | 72.01 | 65.36 | 24.4 | 29.74 | 191.51 |
| | 796 | 103.16 | 67.65 | 176.15 | 63.18 | 410.14 |
| | 797 | 475.51 | 357.32 | 372.02 | 545.99 | 1,750.84 |
| | 798 | 2.18 | 1.31 | 11.44 | 0 | 14.93 |
| | 799 | 252.84 | 164.06 | 327.36 | 264.06 | 1,008.32 |
| TOTAL | | 11,780.06 | 10,811.34 | 11,790.02 | 11,948.90 | 46,330.32 |

STATE: UTAH

| ZIP CODE | | QUARTER 1 | QUARTER 2 | QUARTER 3 | QUARTER 4 | TOTAL GRAMS |
|---|---|---|---|---|---|---|
| | 840 | 772.45 | 642.23 | 683.26 | 697.51 | 2,795.45 |
| | 841 | 478.78 | 329.1 | 416.47 | 420.5 | 1,644.85 |

| | | | | | | |
|---|---|---|---|---|---|---|
| | 843 | 84.32 | 71.14 | 103.6 | 107.3 | 366.36 |
| | 844 | 202.62 | 133.12 | 91.94 | 188.68 | 616.36 |
| | 845 | 127.57 | 67.21 | 62.09 | 78.44 | 335.31 |
| | 846 | 167.76 | 183.06 | 197.83 | 250.88 | 799.53 |
| | 847 | 177.24 | 135.41 | 233.32 | 91.73 | 637.7 |
| TOTAL | | 2,010.74 | 1,561.27 | 1,788.51 | 1,835.04 | 7,195.56 |

STATE: VERMONT

| ZIP CODE | | QUARTER 1 | QUARTER 2 | QUARTER 3 | QUARTER 4 | TOTAL GRAMS |
|---|---|---|---|---|---|---|
| | 50 | 7.63 | 1.09 | 4.36 | 0 | 13.08 |
| | 51 | 0.44 | 0.44 | 0.44 | 0 | 1.32 |
| | 52 | 0.22 | 0.44 | 1.96 | 1.09 | 3.71 |
| | 53 | 2.18 | 0 | 0 | 0 | 2.18 |
| | 54 | 231.49 | 234.22 | 200.23 | 319.4 | 985.34 |
| | 56 | 19.61 | 9.8 | 11.98 | 14.49 | 55.88 |
| | 57 | 52.73 | 61.01 | 46.19 | 46.41 | 206.34 |
| | 58 | 9.59 | 8.06 | 0.44 | 1.96 | 20.05 |
| TOTAL | | 323.89 | 315.06 | 265.6 | 383.35 | 1,287.90 |

STATE: VIRGIN ISLANDS

| ZIP CODE | | QUARTER 1 | QUARTER 2 | QUARTER 3 | QUARTER 4 | TOTAL GRAMS |
|---|---|---|---|---|---|---|
| | 8 | 41.94 | 67.65 | 41.94 | 102.4 | 253.93 |
| TOTAL | | 41.94 | 67.65 | 41.94 | 102.4 | 253.93 |

STATE: VIRGINIA

| ZIP CODE | | QUARTER 1 | QUARTER 2 | QUARTER 3 | QUARTER 4 | TOTAL GRAMS |
|---|---|---|---|---|---|---|
| | 201 | 344.24 | 382.59 | 273 | 290.75 | 1,290.58 |
| | 220 | 210.14 | 230.51 | 243.58 | 198.48 | 882.71 |
| | 221 | 113.08 | 84.1 | 99.68 | 112.97 | 409.83 |
| | 222 | 29.85 | 30.94 | 60.79 | 46.08 | 167.66 |
| | 223 | 189.55 | 159.27 | 247.94 | 174.08 | 770.84 |
| | 224 | 280.41 | 172.34 | 191.73 | 174.3 | 818.78 |
| | 225 | 111.12 | 112.21 | 31.7 | 47.06 | 302.09 |
| | 226 | 189.66 | 161.23 | 166.46 | 158.18 | 675.53 |

| | | | | | | |
|---|---|---|---|---|---|---|
| | 227 | 54.47 | 10.89 | 12.2 | 19.17 | 96.73 |
| | 228 | 127.24 | 118.74 | 77.35 | 75.17 | 398.5 |
| | 229 | 304.37 | 270.71 | 250.45 | 305.35 | 1,130.88 |
| | 230 | 252.95 | 181.71 | 141.62 | 194.02 | 770.3 |
| | 231 | 642.73 | 606.67 | 443.48 | 435.42 | 2,128.30 |
| | 232 | 884.68 | 807.16 | 708.42 | 662.56 | 3,062.82 |
| | 233 | 298.92 | 175.72 | 206.55 | 129.42 | 810.61 |
| | 234 | 810.5 | 733.91 | 532.16 | 432.92 | 2,509.49 |
| | 235 | 250.99 | 206.55 | 168.64 | 165.26 | 791.44 |
| | 236 | 290.65 | 247.72 | 371.26 | 238.36 | 1,147.99 |
| | 237 | 93.25 | 67.32 | 92.27 | 35.51 | 288.35 |
| | 238 | 297.29 | 165.8 | 234.22 | 264.06 | 961.37 |
| | 239 | 89.33 | 72.16 | 112.25 | 99.79 | 373.53 |
| | 240 | 242.17 | 202.73 | 164.71 | 158.4 | 768.01 |
| | 241 | 187.15 | 137.48 | 108.61 | 66.89 | 500.13 |
| | 242 | 271.04 | 245.33 | 247.51 | 189.55 | 953.43 |
| | 243 | 164.71 | 94.99 | 109.59 | 54.25 | 423.54 |
| | 244 | 72.88 | 91.73 | 35.73 | 77.89 | 278.23 |
| | 245 | 167.11 | 157.26 | 126.37 | 102.29 | 553.03 |
| | 246 | 8.72 | 4.36 | 1.31 | 0 | 14.39 |
| TOTAL | | 6,979.20 | 5,932.13 | 5,459.58 | 4,908.18 | 23,279.09 |

| STATE: | WASHINGTON | | | | |
|---|---|---|---|---|---|
| ZIP CODE | QUARTER 1 | QUARTER 2 | QUARTER 3 | QUARTER 4 | TOTAL GRAMS |
| 980 | 723.13 | 582.92 | 594.25 | 494.14 | 2,394.44 |
| 981 | 436.84 | 450.78 | 377.14 | 407.86 | 1,672.62 |
| 982 | 605.69 | 518 | 556.89 | 597.96 | 2,278.54 |
| 983 | 373.76 | 338.58 | 370.5 | 308.29 | 1,391.13 |
| 984 | 194.89 | 185.52 | 139.33 | 90.31 | 610.05 |
| 985 | 258.4 | 190.75 | 167.76 | 208.29 | 825.2 |
| 986 | 235.31 | 173.43 | 175.61 | 200.77 | 785.12 |
| 988 | 163.52 | 162.1 | 178.22 | 151.97 | 655.81 |
| 989 | 209.27 | 146.63 | 119.18 | 83.45 | 558.53 |
| 990 | 17.43 | 26.15 | 27.23 | 33.33 | 104.14 |
| 991 | 166.78 | 144.12 | 137.7 | 163.62 | 612.22 |
| 992 | 159.59 | 183.34 | 210.9 | 146.96 | 700.79 |

| | | | | | | |
|---|---|---|---|---|---|---|
| | 993 | 194.34 | 236.15 | 189.33 | 214.72 | 834.54 |
| | 994 | 34.42 | 32.68 | 20.92 | 32.25 | 120.27 |
| TOTAL | | 3,773.37 | 3,371.15 | 3,264.96 | 3,133.92 | 13,543.40 |

STATE: WEST VIRGINIA

| ZIP CODE | | QUARTER 1 | QUARTER 2 | QUARTER 3 | QUARTER 4 | TOTAL GRAMS |
|---|---|---|---|---|---|---|
| | 247 | 40.09 | 31.59 | 37.04 | 39.44 | 148.16 |
| | 248 | 19.61 | 13.07 | 10.89 | 8.72 | 52.29 |
| | 249 | 39.54 | 23.86 | 20.92 | 34.86 | 119.18 |
| | 250 | 4.36 | 5.45 | 8.72 | 4.36 | 22.89 |
| | 251 | 2.51 | 1.63 | 2.94 | 2.18 | 9.26 |
| | 252 | 6.32 | 1.74 | 1.96 | 1.31 | 11.33 |
| | 253 | 90.64 | 73.64 | 54.25 | 43.68 | 262.21 |
| | 254 | 93.9 | 54.47 | 61.44 | 49.02 | 258.83 |
| | 255 | 29.2 | 26.15 | 34.32 | 4.79 | 94.46 |
| | 256 | 14.16 | 20.7 | 6.43 | 37.04 | 78.33 |
| | 257 | 53.49 | 71.57 | 72.23 | 35.4 | 232.69 |
| | 258 | 15.47 | 12.31 | 13.84 | 12.2 | 53.82 |
| | 259 | 9.26 | 11.98 | 10.89 | 15.25 | 47.38 |
| | 260 | 87.15 | 62.53 | 39.33 | 85.95 | 274.96 |
| | 261 | 107.74 | 128.11 | 110.35 | 73.21 | 419.41 |
| | 262 | 3.49 | 1.74 | 0.87 | 0 | 6.1 |
| | 263 | 48.15 | 57.08 | 57.19 | 38.89 | 201.31 |
| | 264 | 3.27 | 6.54 | 2.18 | 3.27 | 15.26 |
| | 265 | 42.05 | 37.26 | 27.89 | 7.52 | 114.72 |
| | 266 | 14.27 | 2.18 | 5.88 | 1.31 | 23.64 |
| | 267 | 5.01 | 11.11 | 0.44 | 10.24 | 26.8 |
| | 268 | 1.96 | 7.73 | 6.21 | 1.53 | 17.43 |
| TOTAL | | 731.64 | 662.44 | 586.21 | 510.17 | 2,490.46 |

STATE: WISCONSIN

| ZIP CODE | | QUARTER 1 | QUARTER 2 | QUARTER 3 | QUARTER 4 | TOTAL GRAMS |
|---|---|---|---|---|---|---|
| | 530 | 230.51 | 222.34 | 188.57 | 207.74 | 849.16 |
| | 531 | 307.09 | 287.92 | 343.15 | 282.26 | 1,220.42 |
| | 532 | 140.53 | 152.29 | 106.11 | 121.14 | 520.07 |

| | | | | | | |
|---|---|---|---|---|---|---|
| | 534 | 34.86 | 32.25 | 0 | 24.4 | 91.51 |
| | 535 | 95.21 | 115.04 | 96.3 | 71.9 | 378.45 |
| | 537 | 104.58 | 115.91 | 85.84 | 81.92 | 388.25 |
| | 538 | 4.36 | 0 | 2.61 | 0 | 6.97 |
| | 539 | 11.44 | 12.42 | 11.33 | 23.42 | 58.61 |
| | 540 | 6.97 | 10.89 | 13.29 | 0.54 | 31.69 |
| | 541 | 22.66 | 17.43 | 22.01 | 40.52 | 102.62 |
| | 542 | 46.3 | 31.59 | 46.19 | 40.31 | 164.39 |
| | 543 | 173.1 | 187.81 | 162.1 | 167.76 | 690.77 |
| | 544 | 125.06 | 69.07 | 93.03 | 95.54 | 382.7 |
| | 545 | 11.77 | 14.6 | 11.55 | 13.07 | 50.99 |
| | 546 | 37.47 | 33.99 | 47.06 | 18.3 | 136.82 |
| | 547 | 38.56 | 21.57 | 29.52 | 30.07 | 119.72 |
| | 548 | 31.37 | 31.81 | 46.84 | 27.02 | 137.04 |
| | 549 | 136.72 | 140.53 | 161.12 | 84.43 | 522.8 |
| TOTAL | | 1,558.56 | 1,497.46 | 1,466.62 | 1,330.34 | 5,852.98 |

STATE: WYOMING

| ZIP CODE | | QUARTER 1 | QUARTER 2 | QUARTER 3 | QUARTER 4 | TOTAL GRAMS |
|---|---|---|---|---|---|---|
| | 820 | 33.55 | 38.65 | 15.69 | 28.32 | 116.21 |
| | 822 | 20.7 | 42.92 | 40.31 | 64.27 | 168.2 |
| | 823 | 13.07 | 0 | 0 | 17.43 | 30.5 |
| | 824 | 33.99 | 18.78 | 9.37 | 30.07 | 92.21 |
| | 825 | 44.66 | 36.82 | 24.95 | 27.23 | 133.66 |
| | 826 | 81.59 | 39.44 | 64.38 | 58.72 | 244.13 |
| | 827 | 26.15 | 28.76 | 16.34 | 10.35 | 81.6 |
| | 828 | 39.65 | 39.91 | 35.95 | 58.83 | 174.34 |
| | 829 | 17.43 | 23.09 | 19.17 | 32.68 | 92.37 |
| | 830 | 1.09 | 2.18 | 1.09 | 1.09 | 5.45 |
| | 831 | 66.45 | 47.71 | 27.02 | 21.79 | 162.97 |
| TOTAL | | 378.33 | 318.26 | 254.27 | 350.78 | 1,301.64 |

DRUG CODE: 9250B DRUG NAME: METHADONE

STATE: ALABAMA

| ZIP CODE | QUARTER 1 | QUARTER 2 | QUARTER 3 | QUARTER 4 | TOTAL GRAMS |
|---|---|---|---|---|---|
| 350 | 10,609.82 | 13,421.81 | 10,981.26 | 11,844.09 | 46,856.98 |
| 351 | 1,380.24 | 1,339.72 | 1,291.51 | 1,274.97 | 5,286.44 |
| 352 | 13,820.45 | 11,566.47 | 13,443.66 | 12,995.64 | 51,826.22 |
| 354 | 5,013.74 | 4,096.73 | 3,974.27 | 3,931.51 | 17,016.25 |
| 355 | 6,822.48 | 5,457.18 | 6,274.39 | 5,687.04 | 24,241.09 |
| 356 | 994.57 | 1,680.40 | 2,380.89 | 3,059.65 | 8,115.51 |
| 357 | 988.31 | 935.54 | 901.56 | 880.54 | 3,705.95 |
| 358 | 11,544.83 | 13,033.91 | 11,835.33 | 13,310.01 | 49,724.08 |
| 359 | 1,328.01 | 1,906.41 | 2,564.16 | 3,311.96 | 9,110.54 |
| 360 | 721.24 | 686.63 | 550.95 | 529.75 | 2,488.57 |
| 361 | 2,372.84 | 1,491.86 | 1,450.27 | 1,356.63 | 6,671.60 |
| 362 | 3,726.52 | 4,654.46 | 3,282.45 | 4,014.78 | 15,678.21 |
| 363 | 2,243.16 | 2,207.83 | 2,620.59 | 2,219.01 | 9,290.59 |
| 364 | 326.46 | 354.63 | 226.73 | 292.92 | 1,200.74 |
| 365 | 3,874.54 | 4,020.78 | 3,338.35 | 3,022.62 | 14,256.29 |
| 366 | 8,926.20 | 12,380.91 | 8,393.32 | 11,506.01 | 41,206.44 |
| 367 | 629.21 | 592.09 | 598.8 | 467.32 | 2,287.42 |
| 368 | 927.94 | 875.71 | 791.37 | 777.41 | 3,372.43 |
| 369 | 29.52 | 23.25 | 25.94 | 8.05 | 86.76 |
| TOTAL | 76,280.08 | 80,726.32 | 74,925.80 | 80,489.91 | 312,422.11 |

STATE: ALASKA

| ZIP CODE | QUARTER 1 | QUARTER 2 | QUARTER 3 | QUARTER 4 | TOTAL GRAMS |
|---|---|---|---|---|---|
| 995 | 4,423.61 | 5,561.11 | 5,290.29 | 4,757.85 | 20,032.86 |
| 996 | 2,395.74 | 1,852.39 | 1,803.11 | 1,712.33 | 7,763.57 |
| 997 | 754.87 | 789.93 | 1,081.96 | 764.26 | 3,391.02 |
| 998 | 190.95 | 178.88 | 180.85 | 153.39 | 704.07 |
| 999 | 44.72 | 52.77 | 45.17 | 52.77 | 195.43 |
| TOTAL | 7,809.89 | 8,435.08 | 8,401.38 | 7,440.60 | 32,086.95 |

STATE: ARIZONA

| ZIP CODE | QUARTER 1 | QUARTER 2 | QUARTER 3 | QUARTER 4 | TOTAL GRAMS |
|---|---|---|---|---|---|
| 850 | 28,493.80 | 26,415.34 | 26,562.29 | 25,498.23 | 106,969.66 |
| 851 | 718.02 | 578.14 | 588.16 | 529.84 | 2,414.16 |

| | | | | | | |
|---|---|---|---|---|---|---|
| | 852 | 18,036.38 | 18,385.02 | 19,068.52 | 17,992.64 | 73,482.56 |
| | 853 | 6,364.22 | 6,025.39 | 6,947.97 | 5,464.16 | 24,801.74 |
| | 855 | 634.13 | 609.98 | 554.08 | 560.34 | 2,358.53 |
| | 856 | 579.75 | 524.12 | 474.93 | 443.26 | 2,022.06 |
| | 857 | 25,579.21 | 27,596.62 | 23,946.93 | 27,061.41 | 104,184.17 |
| | 859 | 344.25 | 374.22 | 352.93 | 325.83 | 1,397.23 |
| | 860 | 1,045.11 | 1,005.31 | 1,022.84 | 976.77 | 4,050.03 |
| | 863 | 1,869.30 | 2,122.86 | 1,518.69 | 1,860.35 | 7,371.20 |
| | 864 | 1,518.69 | 1,447.14 | 1,392.13 | 2,299.95 | 6,657.91 |
| | 865 | 7.6 | 3.58 | 8.94 | 4.02 | 24.14 |
| TOTAL | | 85,190.46 | 85,087.72 | 82,438.41 | 83,016.80 | 335,733.39 |

DRUG CODE:9250BDRUG NAME:METHADONE

STATE: ARKANSAS

| ZIP CODE | QUARTER 1 | QUARTER 2 | QUARTER 3 | QUARTER 4 | TOTAL GRAMS |
|---|---|---|---|---|---|
| 716 | 264.83 | 234.33 | 249.98 | 197.22 | 946.36 |
| 717 | 497.38 | 435.57 | 413.21 | 376.1 | 1,722.26 |
| 718 | 1,718.59 | 1,686.21 | 1,668.95 | 1,282.57 | 6,356.32 |
| 719 | 634.13 | 401.23 | 364.02 | 373.86 | 1,773.24 |
| 720 | 842.08 | 789.31 | 696.38 | 654.52 | 2,982.29 |
| 721 | 2,970.75 | 2,895.71 | 2,204.70 | 3,181.20 | 11,252.36 |
| 722 | 602.24 | 1,000.65 | 1,386.59 | 490.22 | 3,479.70 |
| 723 | 228.97 | 214.21 | 219.13 | 489.68 | 1,151.99 |
| 724 | 2,528.47 | 2,524.89 | 2,498.95 | 2,500.30 | 10,052.61 |
| 725 | 301.86 | 306.33 | 275.92 | 253.56 | 1,137.67 |
| 726 | 560.52 | 517.41 | 493.26 | 453.01 | 2,024.20 |
| 727 | 4,165.40 | 4,126.94 | 3,690.38 | 3,602.55 | 15,585.27 |
| 728 | 256.69 | 255.8 | 227.62 | 196.77 | 936.88 |
| 729 | 1,306.27 | 1,219.51 | 1,179.71 | 1,134.99 | 4,840.48 |
| TOTAL | 16,878.18 | 16,608.10 | 15,568.80 | 15,186.55 | 64,241.63 |

STATE: CALIFORNIA

| ZIP CODE | QUARTER 1 | QUARTER 2 | QUARTER 3 | QUARTER 4 | TOTAL GRAMS |
|---|---|---|---|---|---|
| 900 | 16,395.98 | 16,881.41 | 16,238.52 | 13,911.13 | 63,427.04 |
| 902 | 3,219.21 | 4,213.43 | 3,267.33 | 3,421.62 | 14,121.59 |

| | | | | | |
|---|---|---|---|---|---|
| 903 | 1,398.84 | 1,433.72 | 1,060.31 | 1,398.84 | 5,291.71 |
| 904 | 284.15 | 271.54 | 231.74 | 234.78 | 1,022.21 |
| 905 | 403.37 | 376.9 | 315.99 | 346.94 | 1,443.20 |
| 906 | 5,602.34 | 5,850.27 | 6,615.88 | 5,814.49 | 23,882.98 |
| 907 | 5,154.07 | 4,259.22 | 4,730.12 | 4,909.27 | 19,052.68 |
| 908 | 2,886.59 | 1,659.02 | 2,978.98 | 1,233.91 | 8,758.50 |
| 910 | 551.22 | 629.03 | 565.98 | 495.32 | 2,241.55 |
| 911 | 932.5 | 587.26 | 598.8 | 1,227.39 | 3,345.95 |
| 912 | 1,316.91 | 1,651.15 | 1,288.83 | 1,656.07 | 5,912.96 |
| 913 | 6,042.12 | 6,032.55 | 6,121.90 | 5,723.53 | 23,920.10 |
| 914 | 3,105.36 | 3,034.34 | 3,373.95 | 2,917.35 | 12,431 |
| 915 | 289.34 | 272.43 | 228.34 | 234.87 | 1,024.98 |
| 916 | 916.31 | 700.76 | 880.54 | 961.03 | 3,458.64 |
| 917 | 8,152.72 | 8,908.49 | 9,275.29 | 7,619.04 | 33,955.54 |
| 918 | 35.33 | 40.25 | 36.67 | 33.54 | 145.79 |
| 919 | 2,773.98 | 3,113.41 | 2,857.79 | 2,878.63 | 11,623.81 |
| 920 | 13,497.48 | 14,296.09 | 12,552.81 | 14,259.78 | 54,606.16 |
| 921 | 11,052.46 | 12,496.38 | 11,033.77 | 9,241.30 | 43,823.91 |
| 922 | 6,854.41 | 5,001.40 | 7,389.35 | 5,988.46 | 25,233.62 |
| 923 | 8,831.40 | 7,579.31 | 7,592 | 8,199.63 | 32,202.34 |
| 924 | 243.9 | 226.28 | 189.52 | 222.26 | 881.96 |
| 925 | 7,438.46 | 7,664.29 | 6,983.83 | 6,951.46 | 29,038.04 |
| 926 | 2,292.81 | 2,272.15 | 2,136.92 | 2,116 | 8,817.88 |
| 927 | 3,443.62 | 3,510.43 | 3,532.34 | 3,232.63 | 13,719.02 |
| 928 | 5,234.04 | 5,376.60 | 5,033.37 | 4,783.99 | 20,428 |
| 930 | 10,055.83 | 10,110.21 | 10,051.62 | 9,208.21 | 39,425.87 |
| 931 | 2,736.15 | 2,700.82 | 2,691.25 | 2,668.62 | 10,796.84 |
| 932 | 8,628.10 | 9,885.09 | 8,316.40 | 8,784.80 | 35,614.39 |
| 933 | 9,598.16 | 9,488.78 | 9,443.16 | 9,162.68 | 37,692.78 |
| 934 | 6,362.49 | 6,113.76 | 6,101.24 | 5,995.61 | 24,573.10 |
| 935 | 3,352.93 | 4,601.51 | 3,847.53 | 4,129.71 | 15,931.68 |
| 936 | 2,417.21 | 2,299.59 | 2,112.22 | 2,106.41 | 8,935.43 |
| 937 | 16,716.51 | 16,152.15 | 18,068.76 | 17,468.97 | 68,406.39 |
| 939 | 2,514.70 | 2,424 | 2,518.99 | 2,403.25 | 9,860.94 |
| 940 | 5,173.48 | 4,646.94 | 4,561.08 | 3,968.01 | 18,349.51 |
| 941 | 20,405.52 | 24,888.98 | 20,099.41 | 22,050.68 | 87,444.59 |
| 943 | 469.01 | 484.74 | 433.34 | 447.4 | 1,834.49 |

| | | | | | | |
|---|---|---|---|---|---|---|
| | 944 | 333.16 | 311.79 | 325.02 | 322.34 | 1,292.31 |
| | 945 | 23,719.67 | 22,914.17 | 23,232.93 | 22,388.89 | 92,255.66 |
| | 946 | 8,429.09 | 10,536.39 | 9,023.42 | 9,273.94 | 37,262.84 |
| | 947 | 1,476.03 | 1,076.95 | 1,427.02 | 1,109.32 | 5,089.32 |
| | 948 | 3,940.91 | 2,732.03 | 2,742.95 | 3,861.93 | 13,277.82 |
| | 949 | 2,513.26 | 3,318.76 | 3,249.99 | 3,269.22 | 12,351.23 |
| | 950 | 7,918.31 | 6,828.48 | 7,453.58 | 6,484.68 | 28,685.05 |
| | 951 | 2,859.40 | 2,822.28 | 2,815.39 | 3,603.90 | 12,100.97 |
| | 952 | 16,733.33 | 15,186.73 | 15,576.78 | 15,121.44 | 62,618.28 |
| | 953 | 18,343.52 | 16,506.15 | 15,841.26 | 16,814.09 | 67,505.02 |
| | 954 | 6,735.10 | 7,803.55 | 6,592.18 | 6,506.58 | 27,637.41 |
| | 955 | 1,365.93 | 1,384.35 | 1,252.16 | 1,200.37 | 5,202.81 |
| | 956 | 14,877.27 | 15,412.75 | 13,860.70 | 14,658.50 | 58,809.22 |
| | 957 | 789.93 | 724.73 | 684.75 | 613.83 | 2,813.24 |
| | 958 | 18,937.54 | 20,041.04 | 18,910.18 | 18,881.52 | 76,770.28 |
| | 959 | 12,943.22 | 12,100.43 | 10,703.20 | 11,451.54 | 47,198.39 |
| | 960 | 3,400.60 | 3,789.84 | 3,454.17 | 3,228.43 | 13,873.04 |
| | 961 | 351.05 | 368.94 | 303.38 | 341.21 | 1,364.58 |
| TOTAL | | 352,446.33 | 355,994.04 | 342,804.93 | 337,539.34 | 1,388,784.64 |

| STATE: | COLORADO | | | | |
|---|---|---|---|---|---|
| ZIP CODE | QUARTER 1 | QUARTER 2 | QUARTER 3 | QUARTER 4 | TOTAL GRAMS |
| 800 | 8,402.27 | 10,014.20 | 9,668.39 | 8,318.02 | 36,402.88 |
| 801 | 2,157.92 | 1,983.33 | 2,411.39 | 2,968.25 | 9,520.89 |
| 802 | 19,193.65 | 17,657.78 | 18,201.76 | 18,051.94 | 73,105.13 |
| 803 | 248.64 | 448.54 | 448 | 762.03 | 1,907.21 |
| 804 | 314.02 | 280.84 | 305.44 | 338.53 | 1,238.83 |
| 805 | 3,163.22 | 3,061.35 | 3,453.99 | 3,001.79 | 12,680.35 |
| 806 | 427.52 | 468.4 | 452.3 | 391.66 | 1,739.88 |
| 807 | 84.07 | 63.06 | 38.46 | 54.11 | 239.7 |
| 808 | 290.41 | 255.8 | 225.39 | 184.69 | 956.29 |
| 809 | 3,319.92 | 3,961.39 | 3,911.39 | 3,699.15 | 14,891.85 |
| 810 | 1,897.29 | 2,347.35 | 2,496.18 | 2,362.47 | 9,103.29 |
| 811 | 373.41 | 367.6 | 421.71 | 226.28 | 1,389 |
| 812 | 186.93 | 173.96 | 145.34 | 164.84 | 671.07 |
| 813 | 531.72 | 743.69 | 517.86 | 733.86 | 2,527.13 |

| | | | | | | |
|---|---|---|---|---|---|---|
| | 814 | 107.33 | 105.99 | 109.56 | 123.43 | 446.31 |
| | 815 | 1,261.82 | 1,234.09 | 2,055.15 | 1,174.97 | 5,726.03 |
| | 816 | 140.42 | 126.56 | 107.33 | 98.38 | 472.69 |
| TOTAL | | 42,100.56 | 43,293.93 | 44,969.64 | 42,654.40 | 173,018.53 |

STATE: CONNECTICUT

| ZIP CODE | | QUARTER 1 | QUARTER 2 | QUARTER 3 | QUARTER 4 | TOTAL GRAMS |
|---|---|---|---|---|---|---|
| | 60 | 16,305.99 | 10,906.85 | 14,142.88 | 17,324.26 | 58,679.98 |
| | 61 | 12,760.58 | 16,133.23 | 12,998.49 | 10,982.83 | 52,875.13 |
| | 62 | 3,741.72 | 5,074.38 | 4,888.79 | 3,939.38 | 17,644.27 |
| | 63 | 7,013.53 | 6,960.58 | 10,209.22 | 10,131.58 | 34,314.91 |
| | 64 | 15,192.55 | 13,017.10 | 8,436.79 | 15,123.77 | 51,770.21 |
| | 65 | 21,776.58 | 35,617.96 | 13,649.84 | 37,163.93 | 108,208.31 |
| | 66 | 23,560.87 | 17,712.52 | 19,532.44 | 12,828.11 | 73,633.94 |
| | 67 | 10,324.69 | 11,818.60 | 12,729.73 | 9,989.82 | 44,862.84 |
| | 68 | 6,658.63 | 6,702.37 | 6,650.04 | 6,655.41 | 26,666.45 |
| | 69 | 1,923.85 | 1,967.77 | 1,931.46 | 1,925.64 | 7,748.72 |
| TOTAL | | 119,258.99 | 125,911.36 | 105,169.68 | 126,064.73 | 476,404.76 |

STATE: DELAWARE

| ZIP CODE | | QUARTER 1 | QUARTER 2 | QUARTER 3 | QUARTER 4 | TOTAL GRAMS |
|---|---|---|---|---|---|---|
| | 197 | 11,191.54 | 12,796.54 | 11,738.64 | 11,544.74 | 47,271.46 |
| | 198 | 6,679.23 | 6,409.18 | 6,609.44 | 5,316.76 | 25,014.61 |
| | 199 | 7,389.26 | 11,480.97 | 8,053.18 | 10,783.87 | 37,707.28 |
| TOTAL | | 25,260.03 | 30,686.69 | 26,401.26 | 27,645.37 | 109,993.35 |

STATE: DISTRICT OF COLUMBIA

| ZIP CODE | | QUARTER 1 | QUARTER 2 | QUARTER 3 | QUARTER 4 | TOTAL GRAMS |
|---|---|---|---|---|---|---|
| | 200 | 8,566.52 | 10,074.43 | 8,568.98 | 10,207.66 | 37,417.59 |
| | 204 | 647.55 | 607.3 | 660.96 | 575.99 | 2,491.80 |
| TOTAL | | 9,214.07 | 10,681.73 | 9,229.94 | 10,783.65 | 39,909.39 |

STATE: FLORIDA

| ZIP CODE | QUARTER 1 | QUARTER 2 | QUARTER 3 | QUARTER 4 | TOTAL GRAMS |
|---|---|---|---|---|---|
| 320 | 7,731.37 | 6,395.59 | 8,383.66 | 7,324.96 | 29,835.58 |
| 321 | 7,984.85 | 8,031.89 | 10,629.05 | 7,877.25 | 34,523.04 |
| 322 | 18,868.44 | 22,222.44 | 22,126.54 | 19,866.39 | 83,083.81 |
| 323 | 3,937.15 | 4,044.48 | 5,588.21 | 2,951.70 | 16,521.54 |
| 324 | 875.62 | 2,712.27 | 6,284.68 | 4,457.69 | 14,330.26 |
| 325 | 11,833.99 | 15,307.03 | 14,260.49 | 11,659.22 | 53,060.73 |
| 326 | 2,352.18 | 3,411.60 | 5,025.01 | 4,633.35 | 15,422.14 |
| 327 | 2,518.27 | 2,517.29 | 2,217.66 | 2,364.88 | 9,618.10 |
| 328 | 8,218.82 | 12,742.43 | 10,423.96 | 14,484.36 | 45,869.57 |
| 329 | 3,072.26 | 5,732.66 | 7,167.27 | 6,817.12 | 22,789.31 |
| 330 | 6,420.61 | 5,419.93 | 6,221.27 | 6,173.91 | 24,235.72 |
| 331 | 2,466.01 | 2,744.51 | 2,913.42 | 2,159.69 | 10,283.63 |
| 333 | 2,820.76 | 3,319.39 | 3,368.13 | 2,807.70 | 12,315.98 |
| 334 | 8,570.14 | 8,863.06 | 9,442.45 | 10,138.02 | 37,013.67 |
| 335 | 2,585.89 | 2,485.36 | 2,305.94 | 2,291.10 | 9,668.29 |
| 336 | 13,615.99 | 13,287.85 | 12,424.70 | 11,783.01 | 51,111.55 |
| 337 | 15,571.19 | 12,719.17 | 21,129.13 | 18,913.79 | 68,333.28 |
| 338 | 2,321.50 | 2,233.76 | 2,551.37 | 2,060.79 | 9,167.42 |
| 339 | 6,204.01 | 7,124.07 | 17,075.80 | 15,038.89 | 45,442.77 |
| 341 | 4,280.33 | 3,332.89 | 4,182.57 | 3,301.32 | 15,097.11 |
| 342 | 10,071.21 | 8,834.53 | 15,914.24 | 16,644.07 | 51,464.05 |
| 344 | 9,121.45 | 9,192.55 | 10,974.91 | 8,838.28 | 38,127.19 |
| 346 | 13,017.63 | 11,353.07 | 17,077.67 | 16,234.08 | 57,682.45 |
| 347 | 4,590.96 | 4,430.86 | 4,338.11 | 4,291.78 | 17,651.71 |
| 349 | 2,726.67 | 2,430.62 | 2,458.26 | 2,448.51 | 10,064.06 |
| TOTAL | 171,777.30 | 180,889.30 | 224,484.50 | 205,561.86 | 782,712.96 |

STATE: GEORGIA

| ZIP CODE | QUARTER 1 | QUARTER 2 | QUARTER 3 | QUARTER 4 | TOTAL GRAMS |
|---|---|---|---|---|---|
| 300 | 15,765.62 | 17,397.34 | 15,499.16 | 15,686.35 | 64,348.47 |
| 301 | 12,828.02 | 13,277.90 | 13,231.14 | 13,074.90 | 52,411.96 |
| 302 | 7,416.99 | 7,951.48 | 8,675.23 | 8,221.68 | 32,265.38 |
| 303 | 5,052.47 | 4,805.66 | 4,526.65 | 5,244.14 | 19,628.92 |
| 304 | 1,470.84 | 1,696.23 | 2,516.66 | 841.72 | 6,525.45 |

| | | | | | | |
|---|---|---|---|---|---|---|
| | 305 | 14,433.96 | 12,951.74 | 12,198.70 | 14,256.20 | 53,840.60 |
| | 306 | 5,823.44 | 5,731.49 | 5,995.25 | 5,690.35 | 23,240.53 |
| | 307 | 23,711.44 | 29,392.04 | 23,675.22 | 25,995.38 | 102,774.08 |
| | 308 | 437.81 | 398.9 | 951.46 | 1,772.25 | 3,560.42 |
| | 309 | 2,166.56 | 3,542.59 | 2,709.99 | 3,391.92 | 11,811.06 |
| | 310 | 5,347.80 | 5,188.77 | 5,357.19 | 3,875.08 | 19,768.84 |
| | 312 | 2,305.76 | 2,310.32 | 2,891.77 | 2,370.34 | 9,878.19 |
| | 313 | 496.66 | 484.59 | 416.88 | 396.67 | 1,794.80 |
| | 314 | 3,786.62 | 5,111.05 | 4,067.73 | 4,345 | 17,310.40 |
| | 315 | 1,989.15 | 2,380 | 3,783.76 | 3,158.13 | 11,311.04 |
| | 316 | 457.93 | 1,006.38 | 2,141.19 | 1,586.22 | 5,191.72 |
| | 317 | 3,141.40 | 3,293.45 | 3,126.82 | 3,129.51 | 12,691.18 |
| | 318 | 135.5 | 138.63 | 132.37 | 128.61 | 535.11 |
| | 319 | 3,211.70 | 3,313.30 | 3,145.25 | 2,643.67 | 12,313.92 |
| | 398 | 988.67 | 1,119.79 | 1,351.44 | 936.88 | 4,396.78 |
| TOTAL | | 110,968.34 | 121,491.65 | 116,393.86 | 116,745 | 465,598.85 |

| STATE: | GUAM | | | | |
|---|---|---|---|---|---|
| ZIP CODE | QUARTER 1 | QUARTER 2 | QUARTER 3 | QUARTER 4 | TOTAL GRAMS |
| 969 | 58.14 | 38.46 | 42.48 | 32.2 | 171.28 |
| TOTAL | 58.14 | 38.46 | 42.48 | 32.2 | 171.28 |

| STATE: | HAWAII | | | | |
|---|---|---|---|---|---|
| ZIP CODE | QUARTER 1 | QUARTER 2 | QUARTER 3 | QUARTER 4 | TOTAL GRAMS |
| 967 | 3,975.34 | 4,450.18 | 3,932.95 | 3,504.08 | 15,862.55 |
| 968 | 3,884.11 | 4,531.03 | 3,678.49 | 4,339.54 | 16,433.17 |
| TOTAL | 7,859.45 | 8,981.21 | 7,611.44 | 7,843.62 | 32,295.72 |

| STATE: | IDAHO | | | | |
|---|---|---|---|---|---|
| ZIP CODE | QUARTER 1 | QUARTER 2 | QUARTER 3 | QUARTER 4 | TOTAL GRAMS |
| 832 | 955.04 | 904.78 | 842.08 | 828.3 | 3,530.20 |
| 833 | 536.64 | 490.13 | 484.32 | 408.74 | 1,919.83 |
| 834 | 529.57 | 515.71 | 464.91 | 441.21 | 1,951.40 |
| 835 | 413.66 | 381.46 | 372.96 | 343 | 1,511.08 |

| | | | | | | |
|---|---|---|---|---|---|---|
| | 836 | 2,744.20 | 2,294.49 | 2,743.04 | 2,135.38 | 9,917.11 |
| | 837 | 5,576.67 | 6,199.89 | 5,707.26 | 6,161.34 | 23,645.16 |
| | 838 | 1,304.04 | 1,265.67 | 1,125.60 | 1,062.73 | 4,758.04 |
| TOTAL | | 12,059.82 | 12,052.13 | 11,740.17 | 11,380.70 | 47,232.82 |

STATE: ILLINOIS

| ZIP CODE | | QUARTER 1 | QUARTER 2 | QUARTER 3 | QUARTER 4 | TOTAL GRAMS |
|---|---|---|---|---|---|---|
| | 600 | 9,701.65 | 8,971.37 | 8,586.42 | 8,529.71 | 35,789.15 |
| | 601 | 8,246.55 | 7,947.10 | 6,897.52 | 7,476.02 | 30,567.19 |
| | 602 | 4,530.31 | 4,704.54 | 4,463.15 | 4,642.03 | 18,340.03 |
| | 603 | 52.77 | 42.04 | 52.32 | 42.48 | 189.61 |
| | 604 | 6,586.54 | 6,555.06 | 6,720.25 | 5,657.80 | 25,519.65 |
| | 605 | 6,743.69 | 6,418.57 | 5,601.98 | 6,377.97 | 25,142.21 |
| | 606 | 52,807.25 | 56,306.15 | 54,356.09 | 53,041.77 | 216,511.26 |
| | 607 | 588.27 | 1,375.77 | 1,243.39 | 1,129.63 | 4,337.06 |
| | 608 | 4,141.07 | 3,857.10 | 4,068.18 | 3,654.07 | 15,720.42 |
| | 609 | 748.79 | 981.96 | 927.4 | 838.05 | 3,496.20 |
| | 610 | 867.12 | 1,279.44 | 1,084.55 | 768.74 | 3,999.85 |
| | 611 | 2,447.44 | 2,380.45 | 3,094.18 | 3,077.99 | 11,000.06 |
| | 612 | 1,321.92 | 1,487.83 | 1,109.95 | 1,097.43 | 5,017.13 |
| | 613 | 589.68 | 580.29 | 566.42 | 571.97 | 2,308.36 |
| | 614 | 200.79 | 212.87 | 184.69 | 156.52 | 754.87 |
| | 615 | 292.02 | 277.71 | 237.55 | 186.93 | 994.21 |
| | 616 | 8,132.86 | 10,165.42 | 7,530.85 | 9,597.72 | 35,426.85 |
| | 617 | 2,270.34 | 1,368.52 | 1,325.05 | 1,825.20 | 6,789.11 |
| | 618 | 2,508.34 | 2,516.84 | 1,880.74 | 2,475.25 | 9,381.17 |
| | 619 | 75.58 | 69.32 | 65.38 | 57.69 | 267.97 |
| | 620 | 2,295.48 | 2,269.45 | 2,215.52 | 2,790.98 | 9,571.43 |
| | 622 | 1,563.86 | 1,023.55 | 1,530.14 | 1,523.16 | 5,640.71 |
| | 623 | 430.12 | 425.64 | 404.27 | 414.55 | 1,674.58 |
| | 624 | 192.3 | 174.86 | 167.25 | 160.54 | 694.95 |
| | 625 | 1,607.95 | 1,956.95 | 1,551.78 | 1,962.76 | 7,079.44 |
| | 626 | 170.83 | 207.05 | 220.47 | 187.38 | 785.73 |
| | 627 | 1,247.69 | 2,340.20 | 2,433.22 | 3,062.87 | 9,083.98 |
| | 628 | 200.79 | 226.28 | 210.18 | 207.5 | 844.75 |
| | 629 | 660.96 | 673.75 | 596.74 | 564.72 | 2,496.17 |

| | | | | | |
|---|---|---|---|---|---|
| TOTAL | 121,222.96 | 126,796.08 | 119,325.63 | 122,079.43 | 489,424.10 |

STATE: INDIANA

| ZIP CODE | QUARTER 1 | QUARTER 2 | QUARTER 3 | QUARTER 4 | TOTAL GRAMS |
|---|---|---|---|---|---|
| 460 | 703.45 | 594.95 | 630.28 | 652.11 | 2,580.79 |
| 461 | 1,418.70 | 1,353.67 | 1,278.54 | 1,187.23 | 5,238.14 |
| 462 | 18,333.80 | 17,855.23 | 17,699.38 | 17,479.62 | 71,368.03 |
| 463 | 2,195.87 | 2,226.52 | 1,768.23 | 2,132.70 | 8,323.32 |
| 464 | 5,469.70 | 6,147.21 | 5,576.58 | 6,482.61 | 23,676.10 |
| 465 | 2,320.88 | 2,272.13 | 2,069.82 | 1,954.26 | 8,617.09 |
| 466 | 3,204.19 | 3,676.88 | 3,701.92 | 3,701.03 | 14,284.02 |
| 467 | 1,125.33 | 1,101.90 | 1,043.23 | 985.45 | 4,255.91 |
| 468 | 6,659.70 | 6,584.39 | 6,492 | 5,692.41 | 25,428.50 |
| 469 | 7,656.96 | 7,518.33 | 7,541.04 | 7,869.74 | 30,586.07 |
| 470 | 7,499.28 | 7,452.14 | 7,503.21 | 6,100.17 | 28,554.80 |
| 471 | 13,610.17 | 13,571.45 | 12,310.88 | 12,233.51 | 51,726.01 |
| 472 | 553.19 | 489.68 | 483.96 | 505.78 | 2,032.61 |
| 473 | 7,100.37 | 6,050.44 | 5,586.15 | 6,773.56 | 25,510.52 |
| 474 | 958.35 | 845.21 | 842.97 | 721.78 | 3,368.31 |
| 475 | 327.35 | 279.95 | 275.92 | 279.95 | 1,163.17 |
| 476 | 294.7 | 272.34 | 284.87 | 287.55 | 1,139.46 |
| 477 | 3,587.98 | 3,322.87 | 3,563.29 | 2,987.12 | 13,461.26 |
| 478 | 322.88 | 261.61 | 180.67 | 174.41 | 939.57 |
| 479 | 1,221.48 | 1,172.29 | 1,161.83 | 1,125.78 | 4,681.38 |
| TOTAL | 84,564.33 | 83,049.19 | 79,994.77 | 79,326.77 | 326,935.06 |

STATE: IOWA

| ZIP CODE | QUARTER 1 | QUARTER 2 | QUARTER 3 | QUARTER 4 | TOTAL GRAMS |
|---|---|---|---|---|---|
| 500 | 262.51 | 267.87 | 216.53 | 232.1 | 979.01 |
| 501 | 210.01 | 188.81 | 166.36 | 173.33 | 738.51 |
| 502 | 240.15 | 232.45 | 224.49 | 220.2 | 917.29 |
| 503 | 3,961.92 | 5,381.16 | 4,639.70 | 4,938.61 | 18,921.39 |
| 504 | 157.41 | 138.18 | 157.06 | 147.58 | 600.23 |

| | | | | | | |
|---|---|---|---|---|---|---|
| | 505 | 194.08 | 216.89 | 200.79 | 209.74 | 821.5 |
| | 506 | 803.89 | 454.89 | 775.44 | 1,132.58 | 3,166.80 |
| | 507 | 64.84 | 80.67 | 60.37 | 67.88 | 273.76 |
| | 508 | 92.57 | 96.15 | 82.28 | 80.5 | 351.5 |
| | 510 | 191.4 | 186.93 | 192.3 | 190.06 | 760.69 |
| | 511 | 413.93 | 850.13 | 398.28 | 851.02 | 2,513.36 |
| | 512 | 21.02 | 15.65 | 12.07 | 9.84 | 58.58 |
| | 513 | 45.17 | 43.38 | 51.88 | 28.17 | 168.6 |
| | 514 | 69.32 | 39 | 49.1 | 44.99 | 202.41 |
| | 515 | 381.91 | 355.52 | 315.28 | 266.98 | 1,319.69 |
| | 516 | 34.43 | 42.48 | 35.33 | 35.78 | 148.02 |
| | 520 | 122.71 | 119.04 | 128.79 | 104.47 | 475.01 |
| | 521 | 14.31 | 29.52 | 12.52 | 16.99 | 73.34 |
| | 522 | 253.47 | 219.66 | 231.11 | 214.75 | 918.99 |
| | 523 | 1,150.91 | 845.48 | 1,188.21 | 1,028.92 | 4,213.52 |
| | 524 | 1,241.07 | 1,338.92 | 1,268.53 | 1,575.22 | 5,423.74 |
| | 525 | 219.58 | 211.08 | 199 | 179.77 | 809.43 |
| | 526 | 177.72 | 182.01 | 154.46 | 168.5 | 682.69 |
| | 527 | 162.78 | 166.81 | 134.16 | 144.71 | 608.46 |
| | 528 | 1,812.50 | 1,788.53 | 1,762.42 | 1,761.16 | 7,124.61 |
| TOTAL | | 12,299.61 | 13,491.21 | 12,656.46 | 13,823.85 | 52,271.13 |

| STATE: | KANSAS | | | | | |
|---|---|---|---|---|---|---|
| ZIP CODE | | QUARTER 1 | QUARTER 2 | QUARTER 3 | QUARTER 4 | TOTAL GRAMS |
| | 660 | 1,665.73 | 2,030.38 | 2,469.89 | 1,555.72 | 7,721.72 |
| | 661 | 3,824.45 | 3,498.18 | 3,554.79 | 3,673.75 | 14,551.17 |
| | 662 | 1,585.50 | 1,820.64 | 1,760.27 | 1,537.38 | 6,703.79 |
| | 664 | 112.52 | 118.51 | 117.61 | 121.64 | 470.28 |
| | 665 | 146.5 | 123.87 | 120.3 | 138.36 | 529.03 |
| | 666 | 1,993.80 | 2,246.46 | 1,858.83 | 1,736.48 | 7,835.57 |
| | 667 | 402.48 | 414.55 | 349.71 | 382.36 | 1,549.10 |
| | 668 | 143.55 | 113.59 | 85.42 | 86.76 | 429.32 |
| | 669 | 98.38 | 99.28 | 106.88 | 92.12 | 396.66 |
| | 670 | 704.79 | 697.63 | 636.81 | 647.99 | 2,687.22 |
| | 671 | 324.76 | 307.94 | 313.31 | 304.1 | 1,250.11 |
| | 672 | 10,567.25 | 12,019.30 | 10,145.89 | 11,495.10 | 44,227.54 |

| | | | | | | |
|---|---|---|---|---|---|---|
| | 673 | 148.02 | 166.81 | 114.93 | 130.14 | 559.9 |
| | 674 | 370.73 | 319.3 | 317.06 | 276.82 | 1,283.91 |
| | 675 | 232.99 | 217.79 | 190.06 | 204.37 | 845.21 |
| | 676 | 101.96 | 106.88 | 84.97 | 98.83 | 392.64 |
| | 677 | 46.51 | 41.59 | 42.48 | 43.83 | 174.41 |
| | 678 | 186.48 | 185.59 | 167.7 | 185.59 | 725.36 |
| | 679 | 31.3 | 25.94 | 28.17 | 27.28 | 112.69 |
| TOTAL | | 22,687.70 | 24,554.23 | 22,465.08 | 22,738.62 | 92,445.63 |

STATE: KENTUCKY

| ZIP CODE | | QUARTER 1 | QUARTER 2 | QUARTER 3 | QUARTER 4 | TOTAL GRAMS |
|---|---|---|---|---|---|---|
| | 400 | 465.98 | 462.85 | 451.14 | 421.26 | 1,801.23 |
| | 401 | 216.89 | 200.35 | 176.2 | 181.56 | 775 |
| | 402 | 9,927.66 | 8,085.38 | 10,393.46 | 9,366.87 | 37,773.37 |
| | 403 | 2,572.47 | 2,027.16 | 2,314.44 | 2,014.64 | 8,928.71 |
| | 404 | 708.81 | 666.78 | 1,002.18 | 644.68 | 3,022.45 |
| | 405 | 4,390.39 | 4,611.98 | 4,700.23 | 4,372.73 | 18,075.33 |
| | 406 | 1,892.10 | 1,863.48 | 1,892.10 | 1,866.17 | 7,513.85 |
| | 407 | 1,840.68 | 1,852.75 | 1,827.71 | 1,812.95 | 7,334.09 |
| | 408 | 86.31 | 100.62 | 78.71 | 81.39 | 347.03 |
| | 409 | 193.64 | 173.07 | 165.91 | 162.33 | 694.95 |
| | 410 | 9,801.91 | 9,602.64 | 11,154.24 | 9,815.59 | 40,374.38 |
| | 411 | 3,729.20 | 4,082.85 | 4,079.72 | 4,708.12 | 16,599.89 |
| | 412 | 1,249.48 | 1,247.69 | 1,254.84 | 1,245 | 4,997.01 |
| | 413 | 197.66 | 196.77 | 193.64 | 192.74 | 780.81 |
| | 414 | 99.73 | 93.02 | 87.2 | 86.76 | 366.71 |
| | 415 | 1,218.89 | 1,068 | 1,614.66 | 1,214.95 | 5,116.50 |
| | 416 | 170.83 | 173.07 | 186.93 | 151.15 | 681.98 |
| | 417 | 2,659.95 | 3,363.84 | 1,922.51 | 2,582.13 | 10,528.43 |
| | 418 | 37.12 | 39.35 | 39.8 | 30.86 | 147.13 |
| | 420 | 2,211.85 | 2,079.57 | 2,275.44 | 2,258.09 | 8,824.95 |
| | 421 | 2,266.05 | 2,332.60 | 2,859.84 | 2,511.48 | 9,969.97 |
| | 422 | 1,759.28 | 1,728.43 | 1,749 | 1,716.35 | 6,953.06 |
| | 423 | 1,007.45 | 1,138.57 | 1,225.78 | 1,332.66 | 4,704.46 |
| | 424 | 436.47 | 455.25 | 436.91 | 418.58 | 1,747.21 |
| | 425 | 148.92 | 157.41 | 131.48 | 430.65 | 868.46 |

| | | | | | | |
|---|---|---|---|---|---|---|
| | 426 | 95.7 | 84.52 | 82.28 | 87.65 | 350.15 |
| | 427 | 1,723.06 | 1,713.67 | 1,700.25 | 1,706.96 | 6,843.94 |
| TOTAL | | 51,108.48 | 49,601.67 | 53,996.60 | 51,414.30 | 206,121.05 |

STATE: LOUISIANA

| ZIP CODE | | QUARTER 1 | QUARTER 2 | QUARTER 3 | QUARTER 4 | TOTAL GRAMS |
|---|---|---|---|---|---|---|
| | 700 | 6,325.91 | 9,776.24 | 7,480.94 | 5,929.87 | 29,512.96 |
| | 701 | 4,211.73 | 4,857.75 | 5,251.92 | 3,894.31 | 18,215.71 |
| | 703 | 193.64 | 154.73 | 179.77 | 182.46 | 710.6 |
| | 704 | 4,071.76 | 3,968.27 | 3,936.70 | 3,924.72 | 15,901.45 |
| | 705 | 3,895.56 | 4,651.77 | 4,547.85 | 3,603.90 | 16,699.08 |
| | 706 | 1,576.38 | 1,983.15 | 1,488.28 | 1,566.09 | 6,613.90 |
| | 707 | 560.61 | 475.11 | 474.3 | 409.01 | 1,919.03 |
| | 708 | 4,143.76 | 3,611.32 | 4,207.70 | 3,017.88 | 14,980.66 |
| | 710 | 495.94 | 486.11 | 473.41 | 461.33 | 1,916.79 |
| | 711 | 4,220.41 | 4,079 | 4,040.24 | 3,932.03 | 16,271.68 |
| | 712 | 1,668.95 | 1,042.69 | 997.26 | 976.68 | 4,685.58 |
| | 713 | 3,941.17 | 1,772.70 | 3,732.33 | 3,868.73 | 13,314.93 |
| | 714 | 526.8 | 521.88 | 425.73 | 419.92 | 1,894.33 |
| TOTAL | | 35,832.62 | 37,380.72 | 37,236.43 | 32,186.93 | 142,636.70 |

STATE: MAINE

| ZIP CODE | | QUARTER 1 | QUARTER 2 | QUARTER 3 | QUARTER 4 | TOTAL GRAMS |
|---|---|---|---|---|---|---|
| | 39 | 83.18 | 67.08 | 76.02 | 70.66 | 296.94 |
| | 40 | 5,002.56 | 4,862.50 | 3,783.13 | 4,031.96 | 17,680.15 |
| | 41 | 5,482.40 | 5,859.66 | 5,903.93 | 6,472.06 | 23,718.05 |
| | 42 | 2,671.93 | 2,002.47 | 2,018.21 | 2,543.05 | 9,235.66 |
| | 43 | 1,065.77 | 783.76 | 729.92 | 707.02 | 3,286.47 |
| | 44 | 8,960.81 | 10,398.47 | 9,441.29 | 10,346.06 | 39,146.63 |
| | 45 | 124.77 | 108.67 | 101.96 | 100.17 | 435.57 |
| | 46 | 1,524.50 | 1,776.73 | 1,441.77 | 1,411.18 | 6,154.18 |
| | 47 | 105.09 | 96.6 | 92.12 | 100.62 | 394.43 |
| | 48 | 1,580.40 | 1,360.38 | 1,467.17 | 1,420.93 | 5,828.88 |
| | 49 | 5,745.45 | 5,486.07 | 4,437.12 | 4,379.70 | 20,048.34 |
| TOTAL | | 32,346.86 | 32,802.39 | 29,492.64 | 31,583.41 | 126,225.30 |

STATE: MARYLAND

| ZIP CODE | QUARTER 1 | QUARTER 2 | QUARTER 3 | QUARTER 4 | TOTAL GRAMS |
|---|---|---|---|---|---|
| 206 | 5,229.29 | 5,098.17 | 4,553.21 | 5,150.67 | 20,031.34 |
| 207 | 3,619.19 | 3,715.07 | 2,741.07 | 3,265.72 | 13,341.05 |
| 208 | 3,585.29 | 2,846.43 | 3,441.38 | 2,855.91 | 12,729.01 |
| 209 | 852.81 | 857.28 | 913.63 | 834.92 | 3,458.64 |
| 210 | 32,397.23 | 33,278.48 | 36,911.62 | 37,150.42 | 139,737.75 |
| 211 | 5,313.90 | 6,369.92 | 5,091.37 | 6,980.34 | 23,755.53 |
| 212 | 107,563.30 | 105,697.31 | 99,834.74 | 99,997.96 | 413,093.31 |
| 214 | 2,356.57 | 1,554.11 | 2,636.15 | 2,088.33 | 8,635.16 |
| 215 | 3,606.85 | 3,594.15 | 4,073.81 | 3,627.24 | 14,902.05 |
| 216 | 3,641.82 | 3,573.04 | 4,374.69 | 3,218.41 | 14,807.96 |
| 217 | 14,725.31 | 14,354.32 | 12,154.81 | 13,552.13 | 54,786.57 |
| 218 | 3,191.49 | 3,990.01 | 3,516.96 | 4,362.44 | 15,060.90 |
| 219 | 6,687.79 | 8,029.65 | 7,194.37 | 8,022.77 | 29,934.58 |
| TOTAL | 192,770.84 | 192,957.94 | 187,437.81 | 191,107.26 | 764,273.85 |

STATE: MASSACHUSETTS

| ZIP CODE | QUARTER 1 | QUARTER 2 | QUARTER 3 | QUARTER 4 | TOTAL GRAMS |
|---|---|---|---|---|---|
| 10 | 11,037.79 | 11,804.83 | 11,436.87 | 12,194.61 | 46,474.10 |
| 11 | 8,142.35 | 7,396.51 | 6,865.15 | 7,790.40 | 30,194.41 |
| 12 | 4,799.26 | 5,892.75 | 4,350.36 | 5,581.95 | 20,624.32 |
| 13 | 1,368.16 | 1,465.12 | 1,575.75 | 1,323.09 | 5,732.12 |
| 14 | 7,034.90 | 6,232.63 | 7,398.30 | 6,552.82 | 27,218.65 |
| 15 | 3,243.09 | 3,386.74 | 4,461.36 | 4,036.70 | 15,127.89 |
| 16 | 10,495.19 | 11,041.44 | 9,271.66 | 9,620.07 | 40,428.36 |
| 17 | 4,123.01 | 3,824.90 | 3,982.14 | 3,983.66 | 15,913.71 |
| 18 | 18,859.77 | 17,046.37 | 18,295.67 | 15,601.11 | 69,802.92 |
| 19 | 15,753.69 | 14,694.63 | 13,901.48 | 16,272.89 | 60,622.69 |
| 20 | 716.15 | 722.85 | 572.95 | 541.83 | 2,553.78 |
| 21 | 24,936.99 | 24,458.31 | 24,038.65 | 22,727.19 | 96,161.14 |
| 22 | 295.56 | 325.35 | 283.52 | 307.67 | 1,212.10 |

| | | | | | |
|---|---|---|---|---|---|
| 23 | 9,834.29 | 7,797.02 | 10,000.11 | 8,941.85 | 36,573.27 |
| 24 | 1,341.06 | 1,739.88 | 1,326.57 | 1,712.95 | 6,120.46 |
| 25 | 3,144.62 | 3,764.53 | 2,754.57 | 3,575.90 | 13,239.62 |
| 26 | 3,296.31 | 3,250.16 | 2,538.49 | 3,393.35 | 12,478.31 |
| 27 | 18,745.82 | 21,045.95 | 18,615.86 | 20,489.18 | 78,896.81 |
| TOTAL | 147,168.01 | 145,889.97 | 141,669.46 | 144,647.22 | 579,374.66 |

STATE: MICHIGAN

| ZIP CODE | QUARTER 1 | QUARTER 2 | QUARTER 3 | QUARTER 4 | TOTAL GRAMS |
|---|---|---|---|---|---|
| 480 | 15,733.30 | 18,922.64 | 16,045.07 | 16,499.22 | 67,200.23 |
| 481 | 15,833.47 | 16,624.93 | 13,891.64 | 13,954.88 | 60,304.92 |
| 482 | 15,225.66 | 14,681.99 | 14,813.63 | 15,073.25 | 59,794.53 |
| 483 | 11,465.58 | 11,538.12 | 10,348.74 | 11,766.37 | 45,118.81 |
| 484 | 1,329.53 | 1,276.31 | 1,250.82 | 1,191.34 | 5,048 |
| 485 | 10,130.33 | 6,494.24 | 11,878.97 | 8,887.21 | 37,390.75 |
| 486 | 5,276.33 | 5,028.59 | 4,979.93 | 4,822.60 | 20,107.45 |
| 487 | 738.77 | 651.57 | 591.82 | 578.86 | 2,561.02 |
| 488 | 5,623.18 | 4,212.53 | 7,308.32 | 5,832.02 | 22,976.05 |
| 489 | 7,294.57 | 9,350.16 | 7,273.62 | 8,369.45 | 32,287.80 |
| 490 | 12,351.04 | 10,472.89 | 11,027.24 | 10,713.12 | 44,564.29 |
| 491 | 464.1 | 477.7 | 441.21 | 452.12 | 1,835.13 |
| 492 | 4,798.63 | 5,527.84 | 5,418.28 | 4,675.48 | 20,420.23 |
| 493 | 1,245.90 | 1,212.63 | 1,156.46 | 1,071.49 | 4,686.48 |
| 494 | 8,564.60 | 8,174.10 | 8,870.12 | 9,720.61 | 35,329.43 |
| 495 | 9,944.58 | 10,594.92 | 9,661.77 | 10,189.44 | 40,390.71 |
| 496 | 2,947.76 | 3,021.55 | 2,735.43 | 2,637.85 | 11,342.59 |
| 497 | 4,075.69 | 5,534.90 | 5,231.79 | 3,730.27 | 18,572.65 |
| 498 | 839.39 | 843.87 | 826.43 | 852.36 | 3,362.05 |
| 499 | 219.58 | 173.96 | 160.54 | 164.93 | 719.01 |
| TOTAL | 134,101.99 | 134,815.44 | 133,911.83 | 131,182.87 | 534,012.13 |

STATE: MINNESOTA

| ZIP CODE | QUARTER 1 | QUARTER 2 | QUARTER 3 | QUARTER 4 | TOTAL GRAMS |
|---|---|---|---|---|---|
| 550 | 1,114.42 | 1,100.11 | 1,048.33 | 996.81 | 4,259.67 |
| 551 | 6,716.06 | 8,344.84 | 11,926.15 | 13,240.61 | 40,227.66 |

| | | | | | | |
|---|---|---|---|---|---|---|
| | 553 | 2,459.69 | 2,448.87 | 2,430.53 | 3,151.24 | 10,490.33 |
| | 554 | 23,732.28 | 27,980.88 | 24,288.63 | 29,024.60 | 105,026.39 |
| | 556 | 28.17 | 17.89 | 38.01 | 27.28 | 111.35 |
| | 557 | 401.23 | 368.05 | 368.49 | 359.82 | 1,497.59 |
| | 558 | 2,652.52 | 3,756.12 | 2,848.04 | 4,057.45 | 13,314.13 |
| | 559 | 908.17 | 1,455.01 | 1,535.86 | 1,581.39 | 5,480.43 |
| | 560 | 195.34 | 211.35 | 224.67 | 245.78 | 877.14 |
| | 561 | 32.65 | 31.75 | 29.52 | 30.86 | 124.78 |
| | 562 | 118.96 | 113.41 | 110.01 | 111.62 | 454 |
| | 563 | 1,743.54 | 2,360.41 | 1,907.04 | 2,366.85 | 8,377.84 |
| | 564 | 3,973.46 | 3,952.80 | 3,917.92 | 3,882.32 | 15,726.50 |
| | 565 | 225.57 | 198.38 | 200.44 | 184.25 | 808.64 |
| | 566 | 132.01 | 186.84 | 116.81 | 126.11 | 561.77 |
| | 567 | 117.61 | 124.77 | 118.06 | 112.52 | 472.96 |
| TOTAL | | 44,551.68 | 52,651.48 | 51,108.51 | 59,499.51 | 207,811.18 |

| STATE: | MISSISSIPPI | | | | | |
|---|---|---|---|---|---|---|
| ZIP CODE | | QUARTER 1 | QUARTER 2 | QUARTER 3 | QUARTER 4 | TOTAL GRAMS |
| | 386 | 681.98 | 552.74 | 773.66 | 849.23 | 2,857.61 |
| | 387 | 379.67 | 360.44 | 339.42 | 327.35 | 1,406.88 |
| | 388 | 235.94 | 194.98 | 199.81 | 174.94 | 805.67 |
| | 389 | 103.3 | 109.56 | 96.6 | 106.88 | 416.34 |
| | 390 | 225.84 | 208.4 | 227.62 | 224.94 | 886.8 |
| | 391 | 212.87 | 199.45 | 217.34 | 217.79 | 847.45 |
| | 392 | 1,011.92 | 1,606.07 | 1,447.77 | 1,444.46 | 5,510.22 |
| | 393 | 911.39 | 732.07 | 1,239.64 | 1,119.34 | 4,002.44 |
| | 394 | 680.64 | 598.8 | 572.42 | 461.24 | 2,313.10 |
| | 395 | 1,334 | 1,189.10 | 1,915.36 | 2,011.86 | 6,450.32 |
| | 396 | 162.33 | 168.15 | 147.58 | 146.68 | 624.74 |
| | 397 | 145.97 | 142.66 | 129.69 | 103.75 | 522.07 |
| TOTAL | | 6,085.85 | 6,062.42 | 7,306.91 | 7,188.46 | 26,643.64 |

| STATE: | MISSOURI | | | | | |
|---|---|---|---|---|---|---|
| ZIP CODE | | QUARTER 1 | QUARTER 2 | QUARTER 3 | QUARTER 4 | TOTAL GRAMS |
| | 630 | 5,187.61 | 4,646.94 | 4,376.21 | 4,863.39 | 19,074.15 |

| | | | | | | |
|---|---|---|---|---|---|---|
| | 631 | 13,106.06 | 9,687.49 | 12,865.09 | 13,005.83 | 48,664.47 |
| | 633 | 1,254.13 | 1,143.31 | 1,074.62 | 1,036.16 | 4,508.22 |
| | 634 | 278.16 | 278.16 | 255.8 | 259.38 | 1,071.50 |
| | 635 | 67.97 | 68.87 | 61.27 | 58.58 | 256.69 |
| | 636 | 330.57 | 336.03 | 289.25 | 270.02 | 1,225.87 |
| | 637 | 2,069.37 | 1,128.11 | 2,096.12 | 2,059.27 | 7,352.87 |
| | 638 | 138.72 | 142.66 | 145.07 | 146.68 | 573.13 |
| | 639 | 1,047.61 | 839.39 | 942.25 | 435.13 | 3,264.38 |
| | 640 | 1,198.50 | 1,131.42 | 1,066.04 | 993.23 | 4,389.19 |
| | 641 | 4,834.95 | 4,894.60 | 4,965.62 | 5,179.38 | 19,874.55 |
| | 644 | 118.96 | 121.19 | 104.64 | 105.99 | 450.78 |
| | 645 | 2,269.54 | 1,622.44 | 2,301.74 | 2,354.96 | 8,548.68 |
| | 646 | 175.75 | 161.44 | 157.41 | 137.74 | 632.34 |
| | 647 | 199.9 | 186.48 | 177.54 | 171.72 | 735.64 |
| | 648 | 1,627.45 | 2,160.33 | 1,619.31 | 2,773.71 | 8,180.80 |
| | 650 | 156.52 | 155.18 | 153.39 | 157.41 | 622.5 |
| | 651 | 146.68 | 146.68 | 129.24 | 125.22 | 547.82 |
| | 652 | 1,551.34 | 1,439.36 | 1,391.51 | 1,447.59 | 5,829.80 |
| | 653 | 92.12 | 93.91 | 84.97 | 96.15 | 367.15 |
| | 654 | 176.2 | 167.7 | 172.62 | 149.81 | 666.33 |
| | 655 | 262.06 | 230.76 | 211.08 | 212.42 | 916.32 |
| | 656 | 568.39 | 559.63 | 505.25 | 515.17 | 2,148.44 |
| | 657 | 1,701.24 | 1,681.47 | 1,707.86 | 1,563.86 | 6,654.43 |
| | 658 | 5,307.28 | 4,470.48 | 4,950.24 | 4,697.12 | 19,425.12 |
| TOTAL | | 43,867.08 | 37,494.03 | 41,804.14 | 42,815.92 | 165,981.17 |

STATE: MONTANA

| ZIP CODE | QUARTER 1 | QUARTER 2 | QUARTER 3 | QUARTER 4 | TOTAL GRAMS |
|---|---|---|---|---|---|
| 590 | 90.33 | 83.63 | 75.58 | 77.81 | 327.35 |
| 591 | 1,842.46 | 818.64 | 1,389.09 | 1,396.25 | 5,446.44 |
| 592 | 18.34 | 21.02 | 22.81 | 19.23 | 81.4 |
| 593 | 51.88 | 40.25 | 21.47 | 28.62 | 142.22 |
| 594 | 279.5 | 269.66 | 210.63 | 177.54 | 937.33 |
| 595 | 39.35 | 42.04 | 42.04 | 36.22 | 159.65 |
| 596 | 355.97 | 322.43 | 344.79 | 267.34 | 1,290.53 |
| 597 | 755.59 | 770.7 | 1,017.83 | 968.19 | 3,512.31 |

| | | | | | | |
|---|---|---|---|---|---|---|
| | 598 | 2,275 | 2,722.73 | 1,829.32 | 2,213.82 | 9,040.87 |
| | 599 | 1,971.97 | 2,393.86 | 1,543.73 | 2,373.29 | 8,282.85 |
| TOTAL | | 7,680.39 | 7,484.96 | 6,497.29 | 7,558.31 | 29,220.95 |

STATE: NEBRASKA

| ZIP CODE | | QUARTER 1 | QUARTER 2 | QUARTER 3 | QUARTER 4 | TOTAL GRAMS |
|---|---|---|---|---|---|---|
| | 680 | 476.45 | 394.88 | 356.42 | 350.6 | 1,578.35 |
| | 681 | 5,913.95 | 4,972.51 | 5,452.98 | 5,053.90 | 21,393.34 |
| | 683 | 78.26 | 80.94 | 73.79 | 55.01 | 288 |
| | 684 | 41.14 | 44.27 | 35.33 | 41.14 | 161.88 |
| | 685 | 1,374.55 | 1,626.02 | 1,314.64 | 1,644.94 | 5,960.15 |
| | 686 | 72.45 | 59.03 | 66.63 | 62.16 | 260.27 |
| | 687 | 169.94 | 168.15 | 140.87 | 162.33 | 641.29 |
| | 688 | 275.92 | 254.9 | 231.2 | 253.29 | 1,015.31 |
| | 689 | 53.84 | 52.32 | 46.78 | 51.07 | 204.01 |
| | 690 | 46.51 | 41.59 | 33.09 | 29.96 | 151.15 |
| | 691 | 137.11 | 127.72 | 130.58 | 112.25 | 507.66 |
| | 692 | 28.62 | 21.47 | 22.36 | 12.07 | 84.52 |
| | 693 | 131.3 | 115.02 | 116.27 | 116.16 | 478.75 |
| TOTAL | | 8,800.04 | 7,958.82 | 8,020.94 | 7,944.88 | 32,724.68 |

STATE: NEVADA

| ZIP CODE | | QUARTER 1 | QUARTER 2 | QUARTER 3 | QUARTER 4 | TOTAL GRAMS |
|---|---|---|---|---|---|---|
| | 890 | 11,961.08 | 10,356.62 | 10,495.43 | 9,406.58 | 42,219.71 |
| | 891 | 15,228.13 | 16,566.61 | 14,693.06 | 16,342.97 | 62,830.77 |
| | 893 | 35.78 | 30.41 | 33.09 | 33.09 | 132.37 |
| | 894 | 4,290.53 | 5,499.58 | 3,983.75 | 3,275.74 | 17,049.60 |
| | 895 | 4,389.36 | 4,428.17 | 4,183.91 | 4,131.86 | 17,133.30 |
| | 897 | 995.56 | 1,006.11 | 1,573.25 | 1,487.83 | 5,062.75 |
| | 898 | 77.37 | 60.37 | 67.08 | 62.7 | 267.52 |
| TOTAL | | 36,977.81 | 37,947.87 | 35,029.57 | 34,740.77 | 144,696.02 |

STATE: NEW HAMPSHIRE

DATE RANGE: 01/01/2017 TO 12/31/2017 ARCOS 3 - REPORT 1 Run Date: 07/03/2018

RETAIL DRUG DISTRIBUTION BY ZIP CODE WITHIN STATE BY GRAMS WT

| ZIP CODE | QUARTER 1 | QUARTER 2 | QUARTER 3 | QUARTER 4 | TOTAL GRAMS |
|---|---|---|---|---|---|
| 30 | 4,070.68 | 3,500.50 | 4,020.51 | 4,014.51 | 15,606.20 |
| 31 | 9,281.10 | 10,069.33 | 7,602.31 | 10,051.45 | 37,004.19 |
| 32 | 642.36 | 638.6 | 591.2 | 564.37 | 2,436.53 |
| 33 | 5,270.34 | 4,805.61 | 4,134.81 | 4,187.31 | 18,398.07 |
| 34 | 2,010.16 | 2,007.93 | 3,847.71 | 2,001.67 | 9,867.47 |
| 35 | 286.21 | 294.7 | 241.94 | 207.95 | 1,030.80 |
| 36 | 32.2 | 34.43 | 31.75 | 29.52 | 127.9 |
| 37 | 2,532.49 | 3,157.23 | 3,136.75 | 2,612.10 | 11,438.57 |
| 38 | 6,791 | 6,725.98 | 6,973.01 | 6,933.93 | 27,423.92 |
| TOTAL | 30,916.54 | 31,234.31 | 30,579.99 | 30,602.81 | 123,333.65 |

STATE: NEW JERSEY

| ZIP CODE | QUARTER 1 | QUARTER 2 | QUARTER 3 | QUARTER 4 | TOTAL GRAMS |
|---|---|---|---|---|---|
| 70 | 9,863.44 | 8,346.63 | 7,664.83 | 8,847.58 | 34,722.48 |
| 71 | 9,470.18 | 12,585.43 | 12,807.92 | 11,333.55 | 46,197.08 |
| 72 | 1,851.86 | 2,744.02 | 1,839.60 | 1,886.29 | 8,321.77 |
| 73 | 3,879.01 | 5,720.58 | 5,314.08 | 4,420.12 | 19,333.79 |
| 74 | 795.12 | 762.03 | 746.82 | 675.72 | 2,979.69 |
| 75 | 2,860.29 | 4,679.59 | 1,015.32 | 3,190.77 | 11,745.97 |
| 76 | 2,698.49 | 2,625.24 | 2,177.51 | 2,593.58 | 10,094.82 |
| 77 | 8,340.01 | 8,896.69 | 8,814.49 | 9,720.34 | 35,771.53 |
| 78 | 4,694.71 | 435.57 | 1,314.50 | 2,568.54 | 9,013.32 |
| 79 | 240.86 | 232.99 | 210.36 | 237.55 | 921.76 |
| 80 | 3,893.41 | 3,728.13 | 5,372.03 | 3,751.20 | 16,744.77 |
| 81 | 25,520.36 | 25,484.23 | 18,716.48 | 25,190.06 | 94,911.13 |
| 82 | 7,352.15 | 9,324.66 | 7,959 | 9,190.94 | 33,826.75 |
| 83 | 4,554.73 | 5,543.94 | 4,272.55 | 5,576.67 | 19,947.89 |
| 84 | 135.23 | 135.14 | 102.95 | 103.75 | 477.07 |
| 85 | 437.99 | 439.96 | 437.36 | 411.42 | 1,726.73 |
| 86 | 6,334.05 | 5,615.40 | 4,460.37 | 4,822.16 | 21,231.98 |
| 87 | 8,965.73 | 10,352.68 | 11,456.37 | 10,557.05 | 41,331.83 |
| 88 | 8,545.37 | 10,613.40 | 9,392.63 | 9,551.83 | 38,103.23 |
| 89 | 2,255.59 | 2,953.40 | 4,353.31 | 2,595.10 | 12,157.40 |
| TOTAL | 112,688.58 | 121,219.71 | 108,428.48 | 117,224.22 | 459,560.99 |

STATE: NEW MEXICO

| ZIP CODE | QUARTER 1 | QUARTER 2 | QUARTER 3 | QUARTER 4 | TOTAL GRAMS |
|---|---|---|---|---|---|
| 870 | 1,284.81 | 1,121.58 | 1,834.41 | 1,261.55 | 5,502.35 |
| 871 | 31,745.39 | 31,155.80 | 31,549.69 | 33,368.90 | 127,819.78 |
| 873 | 54.11 | 74.24 | 72.89 | 51.43 | 252.67 |
| 874 | 644.86 | 653.81 | 1,215.49 | 1,207.44 | 3,721.60 |
| 875 | 5,800.09 | 5,958.05 | 6,111.61 | 6,562.66 | 24,432.41 |
| 877 | 56.79 | 55.01 | 46.51 | 61.71 | 220.02 |
| 878 | 8.94 | 8.05 | 9.84 | 12.52 | 39.35 |
| 879 | 13.86 | 17.44 | 11.18 | 6.26 | 48.74 |
| 880 | 2,105.06 | 2,018.66 | 1,840.05 | 2,350.48 | 8,314.25 |
| 881 | 23.7 | 26.38 | 16.99 | 22.09 | 89.16 |
| 882 | 1,089.74 | 1,352.24 | 1,341.60 | 1,331.76 | 5,115.34 |
| 883 | 178.88 | 206.16 | 190.06 | 217.79 | 792.89 |
| 884 | 16.1 | 10.29 | 11.63 | 9.84 | 47.86 |
| TOTAL | 43,022.33 | 42,657.71 | 44,251.95 | 46,464.43 | 176,396.42 |

STATE: NEW YORK

| ZIP CODE | QUARTER 1 | QUARTER 2 | QUARTER 3 | QUARTER 4 | TOTAL GRAMS |
|---|---|---|---|---|---|
| 100 | 82,282.73 | 87,971.78 | 87,815.14 | 83,258.85 | 341,328.50 |
| 101 | 1,136.25 | 1,773.24 | 1,324.16 | 1,197.60 | 5,431.25 |
| 102 | 16.1 | 16.1 | 17.89 | 19.23 | 69.32 |
| 103 | 11,747.32 | 13,064.68 | 11,512.45 | 12,800.74 | 49,125.19 |
| 104 | 59,137.28 | 66,725.73 | 58,511.20 | 61,302 | 245,676.21 |
| 105 | 4,903.64 | 4,910.52 | 4,881.37 | 5,366.94 | 20,062.47 |
| 106 | 135.77 | 140.87 | 105.54 | 140.15 | 522.33 |
| 107 | 4,836.56 | 4,850.60 | 5,689.82 | 4,824.93 | 20,201.91 |
| 108 | 1,389 | 1,659.11 | 1,294.20 | 1,826.36 | 6,168.67 |
| 109 | 2,985.86 | 3,232.45 | 3,002.50 | 3,260 | 12,480.81 |
| 110 | 3,343.42 | 3,433.74 | 3,480.32 | 3,136.12 | 13,393.60 |
| 111 | 221.81 | 213.76 | 168.15 | 170.38 | 774.1 |
| 112 | 33,143.86 | 36,813.68 | 32,411.89 | 34,826.15 | 137,195.58 |
| 113 | 4,305.28 | 4,835.48 | 5,747.68 | 5,048.71 | 19,937.15 |
| 114 | 7,467.88 | 7,301.88 | 6,884.47 | 7,634.33 | 29,288.56 |

| | | | | | | | |
|---|---|---|---|---|---|---|---|
| | 115 | 2,774.52 | 4,866.97 | | 4,646.68 | 4,840.15 | 17,128.32 |
| | 116 | 131.92 | 110.46 | | 134.34 | 145.34 | 522.06 |
| | 117 | 10,452.23 | 10,303.40 | | 9,204.54 | 10,069.87 | 40,030.04 |
| | 118 | 146.68 | 128.35 | | 119.85 | 116.72 | 511.6 |
| | 119 | 2,556.82 | 3,292.73 | | 2,526.68 | 2,549.04 | 10,925.27 |
| | 120 | 1,408.23 | 1,193.58 | | 1,697.66 | 1,333.10 | 5,632.57 |
| | 121 | 435.13 | 548.27 | | 502.65 | 630.1 | 2,116.15 |
| | 122 | 5,775.95 | 5,665.49 | | 6,812.82 | 6,934.37 | 25,188.63 |
| | 123 | 433.78 | 352.84 | | 239.25 | 372.07 | 1,397.94 |
| | 124 | 3,436.64 | 3,174.94 | | 3,673.21 | 2,259.25 | 12,544.04 |
| | 125 | 4,387.03 | 5,295.74 | | 5,608.78 | 6,331.46 | 21,623.01 |
| | 126 | 2,093.79 | 2,123.57 | | 2,404.15 | 3,087.56 | 9,709.07 |
| | 127 | 435.13 | 435.13 | | 434.23 | 408.92 | 1,713.41 |
| | 128 | 703.62 | 710.69 | | 740.12 | 705.32 | 2,859.75 |
| | 129 | 838.05 | 984.73 | | 1,015.86 | 1,326.93 | 4,165.57 |
| | 130 | 1,675.21 | 1,715.46 | | 1,739.16 | 1,819.48 | 6,949.31 |
| | 131 | 273.24 | 270.11 | | 287.1 | 494.16 | 1,324.61 |
| | 132 | 6,109.65 | 6,233.79 | | 8,135.02 | 7,709.73 | 28,188.19 |
| | 133 | 227.89 | 223.6 | | 142.21 | 214.75 | 808.45 |
| | 134 | 1,052.98 | 690.03 | | 1,103.69 | 1,463.24 | 4,309.94 |
| | 135 | 176.82 | 171.28 | | 173.16 | 149.72 | 670.98 |
| | 136 | 629.21 | 722.68 | | 716.41 | 712.03 | 2,780.33 |
| | 137 | 219.58 | 207.05 | | 226.28 | 229.86 | 882.77 |
| | 138 | 587.26 | 525.55 | | 538.88 | 512.04 | 2,163.73 |
| | 139 | 1,116.21 | 1,476.65 | | 1,182.40 | 1,271.84 | 5,047.10 |
| | 140 | 1,494.01 | 1,444.72 | | 1,420.49 | 1,242.86 | 5,602.08 |
| | 141 | 631.63 | 594.42 | | 560.88 | 490.13 | 2,277.06 |
| | 142 | 13,062.44 | 16,605.94 | | 14,265.98 | 15,300.86 | 59,235.22 |
| | 143 | 1,456.62 | 1,481.04 | | 1,465.03 | 1,477.01 | 5,879.70 |
| | 144 | 682.87 | 675.36 | | 663.82 | 660.87 | 2,682.92 |
| | 145 | 394.61 | 391.75 | | 384.77 | 397.56 | 1,568.69 |
| | 146 | 6,732.95 | 4,891.74 | | 6,679.56 | 6,709.25 | 25,013.50 |
| | 147 | 604.97 | 544.78 | | 537.62 | 529.13 | 2,216.50 |
| | 148 | 778.49 | 739.49 | | 761.94 | 712.03 | 2,991.95 |
| | 149 | 265.19 | 284.42 | | 280.39 | 244.62 | 1,074.62 |
| TOTAL | | 291,234.11 | 316,020.37 | | 303,872.39 | 308,263.86 | 1,219,390.73 |

STATE: NORTH CAROLINA

| ZIP CODE | QUARTER 1 | QUARTER 2 | QUARTER 3 | QUARTER 4 | TOTAL GRAMS |
|---|---|---|---|---|---|
| 270 | 606.4 | 591.65 | 571.52 | 541.11 | 2,310.68 |
| 271 | 4,072.20 | 4,095.90 | 4,352.15 | 4,113.79 | 16,634.04 |
| 272 | 6,979.45 | 7,134.60 | 7,140.42 | 7,505.36 | 28,759.83 |
| 273 | 2,828.09 | 4,493.02 | 4,134.36 | 2,644.74 | 14,100.21 |
| 274 | 6,268.58 | 8,416.75 | 7,823.32 | 4,967.94 | 27,476.59 |
| 275 | 6,675.65 | 7,600.68 | 7,559.38 | 7,616.38 | 29,452.09 |
| 276 | 5,271.06 | 5,475.96 | 5,078.31 | 5,090.12 | 20,915.45 |
| 277 | 6,643.78 | 6,878.38 | 7,777.34 | 6,768.37 | 28,067.87 |
| 278 | 5,860.20 | 6,024.23 | 5,173.21 | 6,336.82 | 23,394.46 |
| 279 | 575.99 | 854.6 | 864.44 | 500.42 | 2,795.45 |
| 280 | 9,638.05 | 8,865.38 | 8,146.46 | 8,676.22 | 35,326.11 |
| 281 | 5,853.58 | 6,846.63 | 6,371.62 | 6,552.46 | 25,624.29 |
| 282 | 7,126.94 | 6,683.23 | 7,731.73 | 7,159.58 | 28,701.48 |
| 283 | 10,672.52 | 10,303 | 11,277.94 | 10,675.69 | 42,929.15 |
| 284 | 6,273.67 | 5,852.41 | 5,963.77 | 6,590.35 | 24,680.20 |
| 285 | 5,494.93 | 5,188.86 | 5,445.64 | 5,363.45 | 21,492.88 |
| 286 | 14,935.94 | 13,120.40 | 13,062.98 | 14,005.95 | 55,125.27 |
| 287 | 9,438.25 | 10,075.86 | 8,707.88 | 8,562.18 | 36,784.17 |
| 288 | 13,967.67 | 15,416.33 | 14,575.14 | 14,849.28 | 58,808.42 |
| 289 | 251.77 | 703.27 | 819.27 | 1,343.39 | 3,117.70 |
| TOTAL | 129,434.72 | 134,621.14 | 132,576.88 | 129,863.60 | 526,496.34 |

STATE: NORTH DAKOTA

| ZIP CODE | QUARTER 1 | QUARTER 2 | QUARTER 3 | QUARTER 4 | TOTAL GRAMS |
|---|---|---|---|---|---|
| 580 | 63.95 | 77.19 | 76.83 | 65.74 | 283.71 |
| 581 | 246.59 | 507.84 | 930.98 | 1,092.06 | 2,777.47 |
| 582 | 219.13 | 249.09 | 179.15 | 197.57 | 844.94 |
| 583 | 36.67 | 32.2 | 40.25 | 31.3 | 140.42 |
| 584 | 38.91 | 38.46 | 28.17 | 25.49 | 131.03 |
| 585 | 535.48 | 232.01 | 510.7 | 532.62 | 1,810.81 |
| 586 | 43.38 | 47.85 | 33.99 | 28.17 | 153.39 |
| 587 | 341.66 | 620.53 | 328.42 | 691.37 | 1,981.98 |
| 588 | 64.4 | 66.19 | 64.4 | 53.66 | 248.65 |

| | | | | | |
|---|---|---|---|---|---|
| TOTAL | 1,590.17 | 1,871.36 | 2,192.89 | 2,717.98 | 8,372.40 |

STATE: OHIO

| ZIP CODE | QUARTER 1 | QUARTER 2 | QUARTER 3 | QUARTER 4 | TOTAL GRAMS |
|---|---|---|---|---|---|
| 430 | 1,663.85 | 1,616.99 | 1,520.30 | 1,424.96 | 6,226.10 |
| 431 | 917.65 | 899.05 | 862.02 | 738.33 | 3,417.05 |
| 432 | 16,054.08 | 14,802.60 | 15,143.41 | 14,643.11 | 60,643.20 |
| 433 | 696.65 | 670.98 | 620.45 | 634.4 | 2,622.48 |
| 434 | 304.54 | 289.34 | 287.55 | 270.56 | 1,151.99 |
| 435 | 419.03 | 341.66 | 279.14 | 243.9 | 1,283.73 |
| 436 | 4,114.60 | 4,894.87 | 5,330.80 | 4,688.53 | 19,028.80 |
| 437 | 400.24 | 372.16 | 343.63 | 345.95 | 1,461.98 |
| 438 | 113.68 | 78.26 | 86.76 | 96.15 | 374.85 |
| 439 | 518.57 | 497.82 | 474.93 | 423.95 | 1,915.27 |
| 440 | 1,616 | 1,561.44 | 1,549.10 | 1,412.26 | 6,138.80 |
| 441 | 10,599.71 | 9,463.83 | 9,100.70 | 8,217.84 | 37,382.08 |
| 442 | 919.89 | 940.01 | 764.98 | 775.36 | 3,400.24 |
| 443 | 4,254.30 | 4,269.24 | 4,228.19 | 4,491.59 | 17,243.32 |
| 444 | 1,629.60 | 1,377.38 | 1,486.94 | 1,512.88 | 6,006.80 |
| 445 | 4,183.29 | 4,076.94 | 4,021.67 | 3,945.74 | 16,227.64 |
| 446 | 620.09 | 607.48 | 639.59 | 596.12 | 2,463.28 |
| 447 | 1,824.31 | 1,880.12 | 1,789.96 | 1,796.85 | 7,291.24 |
| 448 | 777.41 | 750.94 | 739.13 | 684.39 | 2,951.87 |
| 449 | 171.99 | 141.32 | 144.45 | 115.38 | 573.14 |
| 450 | 3,200.61 | 3,560.25 | 3,428.24 | 3,419.55 | 13,608.65 |
| 451 | 852.81 | 823.65 | 808.98 | 834.48 | 3,319.92 |
| 452 | 13,487.78 | 12,911.02 | 11,859.89 | 12,214.52 | 50,473.21 |
| 453 | 930.53 | 867.12 | 840.83 | 765.16 | 3,403.64 |
| 454 | 5,156.48 | 4,774.49 | 4,351.43 | 4,673.24 | 18,955.64 |
| 455 | 152.05 | 140.87 | 152.05 | 147.22 | 592.19 |
| 456 | 854.87 | 862.56 | 799.59 | 802.19 | 3,319.21 |
| 457 | 449.79 | 436.02 | 374.31 | 406.95 | 1,667.07 |
| 458 | 1,285.25 | 868.91 | 810.24 | 1,205.47 | 4,169.87 |
| TOTAL | 78,169.65 | 74,777.32 | 72,839.26 | 71,527.03 | 297,313.26 |

STATE: OKLAHOMA

| ZIP CODE | QUARTER 1 | QUARTER 2 | QUARTER 3 | QUARTER 4 | TOTAL GRAMS |
|---|---|---|---|---|---|
| 730 | 1,266.92 | 1,177.12 | 1,142.69 | 1,007.72 | 4,594.45 |
| 731 | 10,146.52 | 13,001.98 | 11,001.57 | 10,561.70 | 44,711.77 |
| 734 | 2,655.12 | 2,945.80 | 2,196.29 | 1,900.06 | 9,697.27 |
| 735 | 1,315.66 | 1,690.42 | 1,124.71 | 1,546.86 | 5,677.65 |
| 736 | 58.58 | 67.53 | 66.19 | 73.79 | 266.09 |
| 737 | 153.75 | 150.26 | 154.28 | 147.13 | 605.42 |
| 738 | 81.84 | 79.6 | 56.35 | 53.22 | 271.01 |
| 739 | 21.91 | 32.65 | 14.31 | 18.34 | 87.21 |
| 740 | 2,871.02 | 3,113.05 | 2,649.66 | 2,763.70 | 11,397.43 |
| 741 | 12,567.30 | 12,631.16 | 11,594.55 | 13,129.17 | 49,922.18 |
| 743 | 1,334.89 | 1,304.93 | 766.5 | 1,303.14 | 4,709.46 |
| 744 | 758 | 687.79 | 656.04 | 621.61 | 2,723.44 |
| 745 | 260.27 | 512.49 | 233.44 | 451.22 | 1,457.42 |
| 746 | 208.57 | 743.69 | 738.77 | 742.35 | 2,433.38 |
| 747 | 292.47 | 268.32 | 191.4 | 192.56 | 944.75 |
| 748 | 330.48 | 321.09 | 264.74 | 270.56 | 1,186.87 |
| 749 | 3,829.37 | 4,486.76 | 3,768.55 | 4,234.98 | 16,319.66 |
| TOTAL | 38,152.67 | 43,214.64 | 36,620.04 | 39,018.11 | 157,005.46 |

STATE: OREGON

| ZIP CODE | QUARTER 1 | QUARTER 2 | QUARTER 3 | QUARTER 4 | TOTAL GRAMS |
|---|---|---|---|---|---|
| 970 | 2,637.76 | 2,422.75 | 2,278.22 | 2,215.88 | 9,554.61 |
| 971 | 1,548.39 | 1,520.75 | 1,279.35 | 1,255.74 | 5,604.23 |
| 972 | 34,531.08 | 35,210.05 | 34,679.53 | 33,844.31 | 138,264.97 |
| 973 | 8,679.35 | 9,242.18 | 8,896.06 | 8,956.16 | 35,773.75 |
| 974 | 7,145.63 | 7,043.22 | 7,523.69 | 6,408.20 | 28,120.74 |
| 975 | 6,625.72 | 7,101.09 | 6,410.79 | 6,658.54 | 26,796.14 |
| 976 | 324.67 | 270.56 | 296.05 | 270.56 | 1,161.84 |
| 977 | 1,522.09 | 1,359.31 | 1,323.18 | 1,624.41 | 5,828.99 |
| 978 | 570.18 | 487.98 | 490.4 | 470.01 | 2,018.57 |
| 979 | 148.47 | 167.16 | 169.67 | 146.68 | 631.98 |
| TOTAL | 63,733.34 | 64,825.05 | 63,346.94 | 61,850.49 | 253,755.82 |

RETAIL DRUG DISTRIBUTION BY ZIP CODE WITHIN STATE BY GRAMS WT

STATE: PENNSYLVANIA

| ZIP CODE | QUARTER 1 | QUARTER 2 | QUARTER 3 | QUARTER 4 | TOTAL GRAMS |
|---|---|---|---|---|---|
| 150 | 3,229.68 | 3,785.01 | 2,958.59 | 4,212.27 | 14,185.55 |
| 151 | 623.84 | 595.31 | 559.27 | 461.78 | 2,240.20 |
| 152 | 22,096.69 | 22,753.89 | 20,935.67 | 22,610.88 | 88,397.13 |
| 153 | 3,883.66 | 4,373.88 | 4,142.86 | 4,317.27 | 16,717.67 |
| 154 | 3,144.71 | 2,585.98 | 2,632.67 | 2,974.33 | 11,337.69 |
| 155 | 101.78 | 87.2 | 83.18 | 71.55 | 343.71 |
| 156 | 8,081.17 | 8,734.35 | 7,789.33 | 8,529.71 | 33,134.56 |
| 157 | 1,159.23 | 820.16 | 989.21 | 1,033.03 | 4,001.63 |
| 158 | 148.2 | 139.97 | 156.79 | 148.83 | 593.79 |
| 159 | 3,189.16 | 3,827.23 | 3,870.07 | 3,121.37 | 14,007.83 |
| 160 | 4,408.68 | 5,105.50 | 4,208.60 | 4,697.21 | 18,419.99 |
| 161 | 5,211.85 | 7,960.16 | 4,706.42 | 7,687.64 | 25,566.07 |
| 162 | 232.99 | 224.05 | 206.61 | 245.69 | 909.34 |
| 163 | 2,264.08 | 1,641.58 | 2,185.02 | 2,136.27 | 8,226.95 |
| 164 | 114.48 | 113.14 | 122.53 | 83.63 | 433.78 |
| 165 | 3,346.84 | 3,877.05 | 3,748.52 | 3,832.06 | 14,804.47 |
| 166 | 6,683.31 | 6,475.01 | 6,157.14 | 5,875.85 | 25,191.31 |
| 167 | 199 | 207.77 | 220.29 | 161.44 | 788.5 |
| 168 | 3,171.10 | 3,153.21 | 2,879.52 | 3,015.02 | 12,218.85 |
| 169 | 136.31 | 124.32 | 109.56 | 112.25 | 482.44 |
| 170 | 6,890.54 | 5,896.11 | 8,226.19 | 6,533.39 | 27,546.23 |
| 171 | 3,938.49 | 3,984.10 | 4,801.14 | 4,791.30 | 17,515.03 |
| 172 | 560.79 | 553.01 | 520.09 | 500.86 | 2,134.75 |
| 173 | 1,630.49 | 1,535.51 | 1,421.20 | 1,934.14 | 6,521.34 |
| 174 | 3,588.33 | 3,544.06 | 3,460.34 | 3,372.34 | 13,965.07 |
| 175 | 511.6 | 470.45 | 425.38 | 365.81 | 1,773.24 |
| 176 | 6,003.03 | 4,193.66 | 5,210.51 | 5,040.26 | 20,447.46 |
| 177 | 3,771.68 | 4,607.50 | 3,812.38 | 4,538.72 | 16,730.28 |
| 178 | 1,420.66 | 1,394.37 | 1,249.92 | 1,288.20 | 5,353.15 |
| 179 | 178.43 | 183.35 | 178.43 | 175.75 | 715.96 |
| 180 | 5,353.07 | 5,232.69 | 5,270.34 | 950.21 | 16,806.31 |
| 181 | 2,009.45 | 2,458.44 | 2,098.98 | 2,079.66 | 8,646.53 |
| 182 | 196.14 | 195.87 | 187.38 | 197.22 | 776.61 |
| 183 | 3,384.41 | 3,378.60 | 3,371.44 | 3,341.48 | 13,475.93 |

| | | | | | | |
|---|---|---|---|---|---|---|
| | 184 | 355.17 | 317.42 | 301.59 | 287.37 | 1,261.55 |
| | 185 | 1,903.91 | 2,743.04 | 2,733.38 | 2,265.96 | 9,646.29 |
| | 186 | 578.77 | 537.53 | 531.72 | 492.46 | 2,140.48 |
| | 187 | 5,935.83 | 5,633.83 | 6,224.17 | 11,503.86 | 29,297.69 |
| | 188 | 243.72 | 213.76 | 194.53 | 185.59 | 837.6 |
| | 189 | 6,099.81 | 5,467.91 | 5,927.10 | 6,208.92 | 23,703.74 |
| | 190 | 14,773.01 | 13,989.85 | 14,426.76 | 14,453.95 | 57,643.57 |
| | 191 | 36,717.74 | 42,567.48 | 37,857.65 | 42,779.02 | 159,921.89 |
| | 193 | 4,723.68 | 3,885.18 | 4,701.77 | 4,708.39 | 18,019.02 |
| | 194 | 9,209.55 | 6,797.53 | 6,108.13 | 6,681.17 | 28,796.38 |
| | 195 | 169.49 | 161.44 | 146.23 | 166.27 | 643.43 |
| | 196 | 2,527.13 | 2,473.91 | 4,927.89 | 3,057.60 | 12,986.53 |
| TOTAL | | 194,101.68 | 199,001.37 | 192,976.49 | 203,227.98 | 789,307.52 |

STATE: PUERTO RICO

| ZIP CODE | QUARTER 1 | QUARTER 2 | QUARTER 3 | QUARTER 4 | TOTAL GRAMS |
|---|---|---|---|---|---|
| 6 | 1,404.48 | 1,510.64 | 1,620.21 | 1,082.22 | 5,617.55 |
| 7 | 5,194.32 | 4,495.08 | 5,393.05 | 3,605.95 | 18,688.40 |
| 9 | 15,791.53 | 12,306.50 | 17,053.97 | 14,570.67 | 59,722.67 |
| TOTAL | 22,390.33 | 18,312.22 | 24,067.23 | 19,258.84 | 84,028.62 |

STATE: RHODE ISLAND

| ZIP CODE | QUARTER 1 | QUARTER 2 | QUARTER 3 | QUARTER 4 | TOTAL GRAMS |
|---|---|---|---|---|---|
| 28 | 19,899.42 | 20,458.51 | 23,998.27 | 21,608.26 | 85,964.46 |
| 29 | 20,782.46 | 22,638.96 | 21,511.54 | 26,211.55 | 91,144.51 |
| TOTAL | 40,681.88 | 43,097.47 | 45,509.81 | 47,819.81 | 177,108.97 |

STATE: SOUTH CAROLINA

| ZIP CODE | QUARTER 1 | QUARTER 2 | QUARTER 3 | QUARTER 4 | TOTAL GRAMS |
|---|---|---|---|---|---|
| 290 | 911.39 | 842.08 | 855.05 | 879.64 | 3,488.16 |
| 291 | 3,182.72 | 3,184.69 | 3,078.79 | 2,064.54 | 11,510.74 |
| 292 | 2,529.81 | 3,192.38 | 3,074.77 | 2,494.57 | 11,291.53 |
| 293 | 5,214.17 | 5,051.03 | 5,090.21 | 5,842.85 | 21,198.26 |

| | | | | | | |
|---|---|---|---|---|---|---|
| | 294 | 6,824.81 | 6,779.73 | 8,225.80 | 9,373.22 | 31,203.56 |
| | 295 | 18,339.14 | 16,621.53 | 14,123.56 | 16,216.72 | 65,300.95 |
| | 296 | 12,477.60 | 13,115.39 | 13,519.84 | 12,143.36 | 51,256.19 |
| | 297 | 4,573.96 | 4,558.76 | 5,574.80 | 4,300.19 | 19,007.71 |
| | 298 | 2,697.96 | 1,597.85 | 1,702.04 | 1,017.38 | 7,015.23 |
| | 299 | 932.86 | 1,033.03 | 1,043.76 | 1,096.09 | 4,105.74 |
| TOTAL | | 57,684.42 | 55,976.47 | 56,288.62 | 55,428.56 | 225,378.07 |

STATE: SOUTH DAKOTA

| ZIP CODE | | QUARTER 1 | QUARTER 2 | QUARTER 3 | QUARTER 4 | TOTAL GRAMS |
|---|---|---|---|---|---|---|
| | 570 | 433.78 | 427.17 | 388.8 | 395.68 | 1,645.43 |
| | 571 | 1,283.46 | 1,184.63 | 1,331.67 | 1,242.59 | 5,042.35 |
| | 572 | 37.12 | 34.88 | 35.78 | 22.81 | 130.59 |
| | 573 | 105.99 | 99.73 | 85.42 | 85.86 | 377 |
| | 574 | 31.3 | 30.86 | 36.67 | 23.7 | 122.53 |
| | 575 | 30.86 | 29.07 | 34.43 | 21.91 | 116.27 |
| | 576 | 8.94 | 9.84 | 12.07 | 11.63 | 42.48 |
| | 577 | 333.52 | 330.93 | 306.87 | 309.91 | 1,281.23 |
| TOTAL | | 2,264.97 | 2,147.11 | 2,231.71 | 2,114.09 | 8,757.88 |

STATE: TENNESSEE

| ZIP CODE | | QUARTER 1 | QUARTER 2 | QUARTER 3 | QUARTER 4 | TOTAL GRAMS |
|---|---|---|---|---|---|---|
| | 370 | 1,763.22 | 1,622.26 | 1,530.94 | 1,403.85 | 6,320.27 |
| | 371 | 1,403.13 | 1,291.60 | 1,232.13 | 1,129.36 | 5,056.22 |
| | 372 | 4,866.79 | 4,737.61 | 5,553.63 | 4,816.63 | 19,974.66 |
| | 373 | 1,115.41 | 1,092.33 | 983.84 | 891.63 | 4,083.21 |
| | 374 | 8,530.07 | 8,159.43 | 7,164.23 | 7,428.80 | 31,282.53 |
| | 376 | 695.66 | 722.32 | 644.42 | 1,000.83 | 3,063.23 |
| | 377 | 192.74 | 209.74 | 190.06 | 167.25 | 759.79 |
| | 378 | 231.11 | 249.09 | 249.98 | 216.44 | 946.62 |
| | 379 | 10,298.70 | 11,238.49 | 9,120.38 | 10,774.39 | 41,431.96 |
| | 380 | 3,009.66 | 4,019.61 | 2,615.85 | 3,502.47 | 13,147.59 |
| | 381 | 9,782.77 | 8,704.66 | 9,697.35 | 8,917.62 | 37,102.40 |
| | 382 | 3,077.18 | 2,844.64 | 3,729.20 | 2,639.82 | 12,290.84 |
| | 383 | 2,561.29 | 2,792.32 | 3,322.16 | 4,114.51 | 12,790.28 |

| | | | | | | |
|---|---|---|---|---|---|---|
| | 384 | 1,485.78 | 1,063.26 | 1,460.02 | 1,035 | 5,044.06 |
| | 385 | 312.15 | 295.15 | 206.16 | 196.32 | 1,009.78 |
| TOTAL | | 49,325.66 | 49,042.51 | 47,700.35 | 48,234.92 | 194,303.44 |

STATE: TEXAS

| ZIP CODE | | QUARTER 1 | QUARTER 2 | QUARTER 3 | QUARTER 4 | TOTAL GRAMS |
|---|---|---|---|---|---|---|
| | 750 | 9,240.23 | 8,408.25 | 6,388.43 | 7,480.05 | 31,516.96 |
| | 751 | 1,213.70 | 1,191.34 | 1,149.30 | 947.62 | 4,501.96 |
| | 752 | 9,448.98 | 9,213.22 | 9,084.02 | 8,581.10 | 36,327.32 |
| | 753 | 11.54 | 22.99 | 29.07 | 12.52 | 76.12 |
| | 754 | 1,735.58 | 1,472.18 | 1,103.51 | 1,093.85 | 5,405.12 |
| | 755 | 300.52 | 321.98 | 338.98 | 265.19 | 1,226.67 |
| | 756 | 2,873.08 | 4,099.30 | 3,472.96 | 3,280.12 | 13,725.46 |
| | 757 | 5,225 | 4,975.28 | 5,563.70 | 4,446.06 | 20,210.04 |
| | 758 | 263.04 | 248.64 | 267.43 | 245.07 | 1,024.18 |
| | 759 | 1,421.83 | 1,558.67 | 1,351.98 | 1,980.20 | 6,312.68 |
| | 760 | 4,728.96 | 3,229.95 | 2,426.15 | 3,826.96 | 14,212.02 |
| | 761 | 9,836.43 | 11,236.17 | 11,454.58 | 9,929.09 | 42,456.27 |
| | 762 | 2,153 | 2,565.14 | 2,052.20 | 2,415.77 | 9,186.11 |
| | 763 | 711.94 | 665.17 | 655.15 | 691.55 | 2,723.81 |
| | 764 | 245.16 | 239.61 | 241.49 | 237.02 | 963.28 |
| | 765 | 994.3 | 934.65 | 492.9 | 874.54 | 3,296.39 |
| | 766 | 265.19 | 226.73 | 198.56 | 202.58 | 893.06 |
| | 767 | 2,286.09 | 2,258.36 | 2,251.20 | 1,622.89 | 8,418.54 |
| | 768 | 319.3 | 349.8 | 290.68 | 302.4 | 1,262.18 |
| | 769 | 1,439.27 | 1,045.64 | 1,047.88 | 1,055.75 | 4,588.54 |
| | 770 | 13,625.52 | 13,699.35 | 13,062.99 | 15,412.26 | 55,800.12 |
| | 772 | 429.31 | 429.31 | 429.31 | 429.31 | 1,717.24 |
| | 773 | 4,330.15 | 3,338.96 | 4,896.69 | 3,713.49 | 16,279.29 |
| | 774 | 1,106.46 | 1,087.14 | 987.6 | 905.67 | 4,086.87 |
| | 775 | 4,705.17 | 4,938.79 | 5,779.97 | 4,425.40 | 19,849.33 |
| | 776 | 999.49 | 1,578.17 | 980.44 | 2,238.15 | 5,796.25 |
| | 777 | 228.07 | 778.58 | 1,088.75 | 1,364.68 | 3,460.08 |
| | 778 | 243.55 | 233.98 | 268.32 | 242.2 | 988.05 |
| | 779 | 1,646.14 | 825.26 | 1,199.84 | 1,579.06 | 5,250.30 |

| | | | | | | |
|---|---|---|---|---|---|---|
| | 780 | 2,065.53 | 2,069.91 | 1,992.72 | 3,030.50 | 9,158.66 |
| | 781 | 1,951.40 | 2,124.74 | 1,975.10 | 1,788.35 | 7,839.59 |
| | 782 | 27,039.05 | 31,506.04 | 26,557.87 | 29,896.48 | 114,999.44 |
| | 783 | 353.74 | 348.82 | 358.65 | 274.13 | 1,335.34 |
| | 784 | 5,459.86 | 7,262.08 | 6,541.55 | 6,411.95 | 25,675.44 |
| | 785 | 1,865.72 | 1,278.99 | 1,853.20 | 1,637.38 | 6,635.29 |
| | 786 | 2,702.07 | 2,027.07 | 1,755.35 | 1,809.10 | 8,293.59 |
| | 787 | 7,505.54 | 8,483.65 | 6,646.47 | 8,047.72 | 30,683.38 |
| | 788 | 107.33 | 98.38 | 80.85 | 68.42 | 354.98 |
| | 789 | 50.98 | 47.85 | 52.77 | 50.53 | 202.13 |
| | 790 | 277.26 | 243.28 | 267.87 | 241.04 | 1,029.45 |
| | 791 | 1,362.53 | 1,240.09 | 942.25 | 1,301.17 | 4,846.04 |
| | 792 | 46.96 | 51.88 | 41.14 | 38.91 | 178.89 |
| | 793 | 143.55 | 145.34 | 139.53 | 104.2 | 532.62 |
| | 794 | 784.75 | 1,946.04 | 1,870.55 | 1,940.67 | 6,542.01 |
| | 795 | 197.22 | 177.54 | 201.24 | 160.54 | 736.54 |
| | 796 | 548.71 | 518.75 | 462.94 | 428.86 | 1,959.26 |
| | 797 | 2,409.07 | 2,390.28 | 1,954.35 | 2,372.75 | 9,126.45 |
| | 798 | 17.89 | 6.26 | 8.94 | 7.16 | 40.25 |
| | 799 | 2,520.69 | 2,753.95 | 2,393.68 | 2,626.50 | 10,294.82 |
| TOTAL | | 139,436.85 | 145,893.55 | 134,651.10 | 142,036.91 | 562,018.41 |

STATE: UTAH

| ZIP CODE | | QUARTER 1 | QUARTER 2 | QUARTER 3 | QUARTER 4 | TOTAL GRAMS |
|---|---|---|---|---|---|---|
| | 840 | 7,583.08 | 7,605.53 | 7,248.40 | 7,816.43 | 30,253.44 |
| | 841 | 11,550.46 | 8,802.15 | 14,723.17 | 11,021.60 | 46,097.38 |
| | 843 | 314.38 | 359.55 | 331.82 | 295.15 | 1,300.90 |
| | 844 | 2,604.94 | 2,821.38 | 2,499.40 | 2,510.40 | 10,436.12 |
| | 845 | 205.71 | 229.59 | 168.59 | 200.26 | 804.15 |
| | 846 | 1,977.70 | 1,409.57 | 1,346.34 | 1,838.98 | 6,572.59 |
| | 847 | 2,647.25 | 2,640.81 | 2,218.29 | 2,715.49 | 10,221.84 |
| TOTAL | | 26,883.52 | 23,868.58 | 28,536.01 | 26,398.31 | 105,686.42 |

STATE: VERMONT

| ZIP CODE | QUARTER 1 | QUARTER 2 | QUARTER 3 | QUARTER 4 | TOTAL GRAMS |
|---|---|---|---|---|---|
| 050 | 508.47 | 534.4 | 447.65 | 474.48 | 1,965 |
| 051 | 281.29 | 254.46 | 242.83 | 232.99 | 1,011.57 |
| 052 | 163.68 | 144 | 166.36 | 152.5 | 626.54 |
| 053 | 2,069.64 | 2,080.37 | 1,765.10 | 2,036.55 | 7,951.66 |
| 054 | 7,153.86 | 5,220.17 | 7,567.52 | 6,688.32 | 26,629.87 |
| 056 | 2,217.22 | 2,151.03 | 2,403.25 | 2,255.23 | 9,026.73 |
| 057 | 2,299.59 | 2,324.99 | 3,535.38 | 2,478.92 | 10,638.88 |
| 058 | 4,158.60 | 4,342.49 | 4,038.04 | 3,609.89 | 16,149.02 |
| TOTAL | 18,852.35 | 17,051.91 | 20,166.13 | 17,928.88 | 73,999.27 |

STATE: VIRGIN ISLANDS

| ZIP CODE | QUARTER 1 | QUARTER 2 | QUARTER 3 | QUARTER 4 | TOTAL GRAMS |
|---|---|---|---|---|---|
| 008 | 28.17 | 386.38 | 23.25 | 19.68 | 457.48 |
| TOTAL | 28.17 | 386.38 | 23.25 | 19.68 | 457.48 |

STATE: VIRGINIA

| ZIP CODE | QUARTER 1 | QUARTER 2 | QUARTER 3 | QUARTER 4 | TOTAL GRAMS |
|---|---|---|---|---|---|
| 201 | 2,783.28 | 2,753.05 | 3,045.34 | 3,045.97 | 11,627.64 |
| 220 | 1,771.81 | 1,704.37 | 1,534.07 | 1,924.39 | 6,934.64 |
| 221 | 504.89 | 502.21 | 443.98 | 491.2 | 1,942.28 |
| 222 | 114.93 | 135.5 | 116.09 | 94.18 | 460.7 |
| 223 | 708.72 | 661.86 | 663.38 | 650.68 | 2,684.64 |
| 224 | 4,187.40 | 4,086.87 | 4,422.18 | 3,461.78 | 16,158.23 |
| 225 | 217.79 | 186.48 | 167.25 | 164.66 | 736.18 |
| 226 | 3,863.45 | 4,289.27 | 4,517.97 | 4,992.72 | 17,663.41 |
| 227 | 231.2 | 222.71 | 183.8 | 154.28 | 791.99 |
| 228 | 2,416.85 | 2,367.92 | 4,059.59 | 2,311.76 | 11,156.12 |
| 229 | 6,379.84 | 4,146.53 | 3,964.92 | 3,969.53 | 18,460.82 |
| 230 | 2,377.76 | 2,006.59 | 2,907.96 | 3,089.70 | 10,382.01 |
| 231 | 1,665.28 | 1,462.97 | 1,179.45 | 1,229.62 | 5,537.32 |
| 232 | 16,984.03 | 16,757.30 | 13,568.32 | 18,476.60 | 65,786.25 |
| 233 | 3,587.26 | 3,137.38 | 3,578.58 | 2,749.48 | 13,052.70 |
| 234 | 4,746.04 | 5,230.54 | 5,397.97 | 5,930.14 | 21,304.69 |

| ZIP CODE | QUARTER 1 | QUARTER 2 | QUARTER 3 | QUARTER 4 | TOTAL GRAMS |
|---|---|---|---|---|---|
| 235 | 2,087.23 | 1,485.95 | 1,462.05 | 1,972.24 | 7,007.47 |
| 236 | 5,386.88 | 5,212.21 | 5,778.72 | 4,899.88 | 21,277.69 |
| 237 | 857.01 | 875.62 | 882.77 | 870.88 | 3,486.28 |
| 238 | 476.72 | 656.49 | 1,510.28 | 1,720.38 | 4,363.87 |
| 239 | 148.47 | 126.56 | 134.16 | 169.49 | 578.68 |
| 240 | 4,793.27 | 4,350.36 | 3,785.15 | 4,756.55 | 17,685.33 |
| 241 | 1,130.07 | 1,035.89 | 989.74 | 953.79 | 4,109.49 |
| 242 | 665.08 | 625.9 | 663.29 | 648.71 | 2,602.98 |
| 243 | 4,840.22 | 4,403.04 | 4,932.26 | 5,056.94 | 19,232.46 |
| 244 | 500.42 | 464.19 | 427.97 | 611.14 | 2,003.72 |
| 245 | 4,025.43 | 3,680.10 | 3,999.49 | 4,157.80 | 15,862.82 |
| 246 | 4,214.41 | 2,834.98 | 4,288.92 | 2,844.19 | 14,182.50 |
| TOTAL | 81,665.74 | 75,402.84 | 78,605.65 | 81,398.68 | 317,072.91 |

STATE: WASHINGTON

| ZIP CODE | QUARTER 1 | QUARTER 2 | QUARTER 3 | QUARTER 4 | TOTAL GRAMS |
|---|---|---|---|---|---|
| 980 | 14,454.58 | 14,739.35 | 17,563.87 | 16,352.40 | 63,110.20 |
| 981 | 13,608.21 | 18,574.81 | 21,134.85 | 25,951.37 | 79,269.24 |
| 982 | 15,833.03 | 15,361.14 | 15,253.90 | 15,147.65 | 61,595.72 |
| 983 | 4,037.41 | 3,907.81 | 3,645.57 | 3,422.69 | 15,013.48 |
| 984 | 12,160.35 | 12,338.25 | 11,868.60 | 12,559.79 | 48,926.99 |
| 985 | 1,308.06 | 3,920.33 | 8,466.75 | 10,398.47 | 24,093.61 |
| 986 | 7,238.47 | 8,043.16 | 8,188.77 | 7,375.13 | 30,845.53 |
| 988 | 793.69 | 739.4 | 687.61 | 664.18 | 2,884.88 |
| 989 | 1,928.86 | 1,964.55 | 2,198.79 | 2,153.72 | 8,245.92 |
| 990 | 319.75 | 316.53 | 323.33 | 272.79 | 1,232.40 |
| 991 | 624.65 | 637.71 | 622.41 | 563.11 | 2,447.88 |
| 992 | 11,189.12 | 11,513.16 | 12,848.59 | 12,161.60 | 47,712.47 |
| 993 | 1,353.50 | 1,121.94 | 986.88 | 925.26 | 4,387.58 |
| 994 | 286.66 | 299.62 | 246.68 | 241.76 | 1,074.72 |
| TOTAL | 85,136.34 | 93,477.76 | 104,036.60 | 108,189.92 | 390,840.62 |

STATE: WEST VIRGINIA

| ZIP CODE | QUARTER 1 | QUARTER 2 | QUARTER 3 | QUARTER 4 | TOTAL GRAMS |
|---|---|---|---|---|---|
| 247 | 70.21 | 64.84 | 57.24 | 46.06 | 238.35 |

| | | | | | | |
|---|---|---|---|---|---|---|
| | 248 | 21.47 | 25.49 | 18.34 | 18.34 | 83.64 |
| | 249 | 48.74 | 50.98 | 45.61 | 36.67 | 182 |
| | 250 | 34.88 | 29.52 | 28.62 | 24.15 | 117.17 |
| | 251 | 58.58 | 56.35 | 67.53 | 27.28 | 209.74 |
| | 252 | 50.53 | 50.53 | 49.64 | 51.43 | 202.13 |
| | 253 | 2,309.25 | 1,947.20 | 2,109.89 | 2,280.72 | 8,647.06 |
| | 254 | 4,140.45 | 3,946.54 | 3,754.24 | 3,368.31 | 15,209.54 |
| | 255 | 92.39 | 97.49 | 100.17 | 90.78 | 380.83 |
| | 256 | 2,697.51 | 2,797.68 | 2,692.59 | 2,701.09 | 10,888.87 |
| | 257 | 4,512.61 | 5,300.57 | 4,775.29 | 4,794.70 | 19,383.17 |
| | 258 | 3,257.40 | 3,448.81 | 3,259.19 | 3,042.75 | 13,008.15 |
| | 259 | 76.02 | 54.56 | 46.51 | 34.43 | 211.52 |
| | 260 | 1,321.48 | 1,352.78 | 819.63 | 1,694.26 | 5,188.15 |
| | 261 | 1,177.03 | 1,222.20 | 1,321.92 | 1,116.21 | 4,837.36 |
| | 262 | 135.5 | 117.17 | 109.56 | 101.96 | 464.19 |
| | 263 | 1,558.04 | 1,104.14 | 1,624.95 | 1,110.84 | 5,397.97 |
| | 264 | 84.52 | 72.89 | 67.08 | 70.66 | 295.15 |
| | 265 | 1,619.94 | 1,553.93 | 1,826.01 | 1,748.64 | 6,748.52 |
| | 266 | 17.89 | 17.89 | 18.78 | 13.86 | 68.42 |
| | 267 | 175.75 | 131.92 | 139.53 | 109.56 | 556.76 |
| | 268 | 75.13 | 65.11 | 70.66 | 40.7 | 251.6 |
| TOTAL | | 23,535.32 | 23,508.59 | 23,002.98 | 22,523.40 | 92,570.29 |

| STATE: | WISCONSIN | | | | |
|---|---|---|---|---|---|
| ZIP CODE | QUARTER 1 | QUARTER 2 | QUARTER 3 | QUARTER 4 | TOTAL GRAMS |
| 530 | 3,757.64 | 4,025.96 | 3,600.94 | 3,579.30 | 14,963.84 |
| 531 | 5,003.99 | 5,144.59 | 4,745.69 | 5,253.44 | 20,147.71 |
| 532 | 15,835.35 | 17,381.77 | 16,239.40 | 14,491.16 | 63,947.68 |
| 534 | 3,442.10 | 3,307.04 | 3,386.82 | 3,288.89 | 13,424.85 |
| 535 | 3,763.01 | 4,627.63 | 3,428.41 | 4,242.23 | 16,061.28 |
| 537 | 9,730.98 | 9,821.85 | 10,061.11 | 10,199.92 | 39,813.86 |
| 538 | 125.22 | 118.96 | 124.32 | 107.78 | 476.28 |
| 539 | 726.7 | 648.89 | 557.66 | 480.29 | 2,413.54 |
| 540 | 252.49 | 230.67 | 228.88 | 228.07 | 940.11 |
| 541 | 260.27 | 248.2 | 256.25 | 212.6 | 977.32 |
| 542 | 113.59 | 105.54 | 93.46 | 92.57 | 405.16 |

| | | | | | | |
|---|---|---|---|---|---|---|
| | 543 | 4,549.37 | 4,412.43 | 4,456.88 | 4,197.96 | 17,616.64 |
| | 544 | 2,599.84 | 2,876.75 | 2,470.24 | 2,508.79 | 10,455.62 |
| | 545 | 77.81 | 50.98 | 54.56 | 43.2 | 226.55 |
| | 546 | 3,541.20 | 4,321.20 | 3,415.80 | 3,374.30 | 14,652.50 |
| | 547 | 2,265.34 | 2,604.67 | 2,124.47 | 2,824.87 | 9,819.35 |
| | 548 | 375.65 | 342.11 | 379.23 | 354.09 | 1,451.08 |
| | 549 | 9,293.35 | 8,601.27 | 7,907.66 | 8,551.81 | 34,354.09 |
| TOTAL | | 65,713.90 | 68,870.51 | 63,531.78 | 64,031.27 | 262,147.46 |

STATE: WYOMING

| ZIP CODE | | QUARTER 1 | QUARTER 2 | QUARTER 3 | QUARTER 4 | TOTAL GRAMS |
|---|---|---|---|---|---|---|
| | 820 | 259.29 | 225.39 | 208.84 | 167.7 | 861.22 |
| | 822 | 44.72 | 45.61 | 45.61 | 52.77 | 188.71 |
| | 823 | 16.99 | 14.31 | 13.42 | 21.47 | 66.19 |
| | 824 | 85.42 | 55.9 | 66.63 | 74.24 | 282.19 |
| | 825 | 117.7 | 129.51 | 116.99 | 129.69 | 493.89 |
| | 826 | 267.43 | 264.3 | 277.71 | 257.14 | 1,066.58 |
| | 827 | 45.17 | 40.7 | 41.14 | 42.04 | 169.05 |
| | 828 | 88.99 | 84.07 | 84.07 | 80.5 | 337.63 |
| | 829 | 244.62 | 229.86 | 250.43 | 235.67 | 960.58 |
| | 830 | 52.77 | 48.3 | 50.09 | 51.43 | 202.59 |
| | 831 | 54.56 | 26.83 | 27.73 | 18.78 | 127.9 |
| TOTAL | | 1,277.66 | 1,164.78 | 1,182.66 | 1,131.43 | 4,756.53 |

DRUG CODE: 9300 DRUG NAME: MORPHINE

STATE: ALABAMA

| ZIP CODE | | QUARTER 1 | QUARTER 2 | QUARTER 3 | QUARTER 4 | TOTAL GRAMS |
|---|---|---|---|---|---|---|
| | 350 | 8,176.02 | 7,756.76 | 7,988.34 | 7,393.18 | 31,314.30 |
| | 351 | 4,093.96 | 3,918.21 | 4,133.97 | 4,200.48 | 16,346.62 |
| | 352 | 7,176.48 | 6,685.96 | 6,572.60 | 6,260.92 | 26,695.96 |
| | 354 | 4,504.70 | 4,581.33 | 4,271.30 | 3,951.20 | 17,308.53 |
| | 355 | 2,582.03 | 2,463.72 | 2,303.07 | 2,134.03 | 9,482.85 |
| | 356 | 5,221.12 | 4,801.92 | 4,664.44 | 4,519.04 | 19,206.52 |
| | 357 | 2,754.65 | 2,885.37 | 2,435.41 | 2,431.54 | 10,506.97 |

| | | | | | |
|---|---|---|---|---|---|
| 358 | 4,408 | 4,470.34 | 4,199 | 4,022.14 | 17,099.48 |
| 359 | 6,089.01 | 5,608.58 | 5,499.96 | 5,536.26 | 22,733.81 |
| 360 | 4,154.50 | 3,917.14 | 3,455.85 | 3,376.01 | 14,903.50 |
| 361 | 3,543.60 | 3,265.73 | 2,653.15 | 2,786.78 | 12,249.26 |
| 362 | 2,542.25 | 2,334.49 | 2,436.61 | 2,376.90 | 9,690.25 |
| 363 | 2,221.85 | 2,151.11 | 2,189.08 | 2,047.10 | 8,609.14 |
| 364 | 1,129.08 | 1,163.94 | 952.62 | 1,168.07 | 4,413.71 |
| 365 | 6,325.72 | 5,819.16 | 5,452.36 | 5,102.54 | 22,699.78 |
| 366 | 5,092.95 | 4,961.25 | 4,416.34 | 4,490.30 | 18,960.84 |
| 367 | 1,107.13 | 1,000.64 | 848.24 | 860.16 | 3,816.17 |
| 368 | 4,049.21 | 3,808.42 | 3,651.97 | 3,715.91 | 15,225.51 |
| 369 | 213.48 | 157.74 | 218.29 | 214.12 | 803.63 |
| TOTAL | 75,385.74 | 71,751.81 | 68,342.60 | 66,586.68 | 282,066.83 |

STATE: ALASKA

| ZIP CODE | QUARTER 1 | QUARTER 2 | QUARTER 3 | QUARTER 4 | TOTAL GRAMS |
|---|---|---|---|---|---|
| 995 | 5,452.38 | 5,448.40 | 5,435.14 | 4,921.72 | 21,257.64 |
| 996 | 4,216.18 | 4,084.01 | 3,852.07 | 4,031.78 | 16,184.04 |
| 997 | 1,638.48 | 1,470.38 | 1,330.72 | 1,102.91 | 5,542.49 |
| 998 | 466.8 | 469.13 | 432.04 | 410.19 | 1,778.16 |
| 999 | 301.24 | 203.16 | 228.55 | 237.34 | 970.29 |
| TOTAL | 12,075.08 | 11,675.08 | 11,278.52 | 10,703.94 | 45,732.62 |

STATE: AMERICAN SAMOA

| ZIP CODE | QUARTER 1 | QUARTER 2 | QUARTER 3 | QUARTER 4 | TOTAL GRAMS |
|---|---|---|---|---|---|
| 967 | 30.08 | 27.45 | 3.01 | 27.07 | 87.61 |
| TOTAL | 30.08 | 27.45 | 3.01 | 27.07 | 87.61 |

STATE: ARIZONA

| ZIP CODE | QUARTER 1 | QUARTER 2 | QUARTER 3 | QUARTER 4 | TOTAL GRAMS |
|---|---|---|---|---|---|
| 850 | 32,152.70 | 29,936.31 | 26,364.22 | 24,195.34 | 112,648.57 |
| 851 | 6,430.73 | 6,351.86 | 5,692.29 | 5,742.91 | 24,217.79 |
| 852 | 37,644.31 | 35,032.94 | 32,958.76 | 31,202.74 | 136,838.75 |
| 853 | 27,318.40 | 23,762.18 | 21,809.48 | 21,181.64 | 94,071.70 |

| | | | | | | |
|---|---|---|---|---|---|---|
| | 855 | 4,663.32 | 3,968.82 | 3,518.20 | 3,244.25 | 15,394.59 |
| | 856 | 7,176.40 | 6,614.23 | 6,239.01 | 6,134.90 | 26,164.54 |
| | 857 | 38,277.62 | 35,929.03 | 35,670.81 | 32,631.70 | 142,509.16 |
| | 859 | 3,596.53 | 3,935.47 | 3,648.40 | 3,176.93 | 14,357.33 |
| | 860 | 3,573.27 | 3,320.93 | 3,396.87 | 2,750.79 | 13,041.86 |
| | 863 | 8,522.27 | 8,342.11 | 7,986.55 | 6,685.96 | 31,536.89 |
| | 864 | 10,116.14 | 9,236.91 | 9,035.32 | 7,900.91 | 36,289.28 |
| | 865 | 277.74 | 189.4 | 303.16 | 239.2 | 1,009.50 |
| TOTAL | | 179,749.43 | 166,620.19 | 156,623.07 | 145,087.27 | 648,079.96 |

STATE: ARKANSAS

| ZIP CODE | | QUARTER 1 | QUARTER 2 | QUARTER 3 | QUARTER 4 | TOTAL GRAMS |
|---|---|---|---|---|---|---|
| | 716 | 1,649.65 | 1,482.77 | 1,547.40 | 1,363.99 | 6,043.81 |
| | 717 | 1,603.24 | 1,416.31 | 1,555.89 | 1,319.54 | 5,894.98 |
| | 718 | 1,254.79 | 1,240.75 | 1,080.49 | 959.66 | 4,535.69 |
| | 719 | 2,989.33 | 2,445.90 | 2,706.37 | 2,548.21 | 10,689.81 |
| | 720 | 5,208.93 | 5,060.41 | 4,853.04 | 4,693.89 | 19,816.27 |
| | 721 | 6,349.29 | 5,942.78 | 5,504.03 | 5,367.65 | 23,163.75 |
| | 722 | 3,345.42 | 3,125.90 | 3,148.74 | 2,871.44 | 12,491.50 |
| | 723 | 1,251.66 | 1,173.63 | 1,196.61 | 1,127.67 | 4,749.57 |
| | 724 | 3,523.78 | 3,400.49 | 3,147.20 | 2,988.13 | 13,059.60 |
| | 725 | 2,296.26 | 1,936.20 | 1,749.51 | 1,664.27 | 7,646.24 |
| | 726 | 3,342.96 | 3,100.44 | 3,048.95 | 3,004.61 | 12,496.96 |
| | 727 | 9,346.27 | 8,950.26 | 8,820.17 | 8,111.52 | 35,228.22 |
| | 728 | 2,126.56 | 1,812.10 | 1,697.09 | 1,649.32 | 7,285.07 |
| | 729 | 4,856.61 | 4,557.07 | 4,385.78 | 3,989.51 | 17,788.97 |
| TOTAL | | 49,144.75 | 45,645.01 | 44,441.27 | 41,659.41 | 180,890.44 |

STATE: CALIFORNIA

| ZIP CODE | | QUARTER 1 | QUARTER 2 | QUARTER 3 | QUARTER 4 | TOTAL GRAMS |
|---|---|---|---|---|---|---|
| | 900 | 8,064.93 | 7,456.93 | 7,578.16 | 7,260.52 | 30,360.54 |
| | 902 | 5,463.89 | 5,167.52 | 5,231.23 | 5,014.27 | 20,876.91 |
| | 903 | 275.38 | 246.09 | 303.64 | 278.86 | 1,103.97 |
| | 904 | 1,316.64 | 1,434.96 | 1,272.36 | 1,183.86 | 5,207.82 |
| | 905 | 2,341.02 | 2,391.42 | 2,148.24 | 2,352.42 | 9,233.10 |

| | | | | | |
|---|---|---|---|---|---|
| 906 | 5,682.48 | 5,781.22 | 6,391.30 | 6,234.38 | 24,089.38 |
| 907 | 4,591.58 | 4,277.50 | 4,616.65 | 4,194.51 | 17,680.24 |
| 908 | 4,740.51 | 5,482.04 | 4,476.01 | 4,653.61 | 19,352.17 |
| 910 | 3,112.39 | 2,937.12 | 2,953.56 | 2,286.27 | 11,289.34 |
| 911 | 1,435.79 | 1,411.69 | 1,667.23 | 1,394.36 | 5,909.07 |
| 912 | 2,648.48 | 3,216.07 | 3,134.86 | 3,032.32 | 12,031.73 |
| 913 | 12,750.22 | 11,768.79 | 11,681.24 | 11,613.29 | 47,813.54 |
| 914 | 4,354.14 | 4,468.54 | 4,124.41 | 4,210.09 | 17,157.18 |
| 915 | 948.26 | 961.19 | 898.31 | 1,019.52 | 3,827.28 |
| 916 | 776.42 | 633.79 | 384.91 | 384.12 | 2,179.24 |
| 917 | 17,250.58 | 16,094.77 | 16,075.04 | 15,358.72 | 64,779.11 |
| 918 | 206.85 | 223.4 | 237.71 | 221.76 | 889.72 |
| 919 | 4,980.97 | 4,967.20 | 4,457.35 | 4,624.95 | 19,030.47 |
| 920 | 23,986.14 | 22,990.47 | 22,539.13 | 21,662.53 | 91,178.27 |
| 921 | 15,166.29 | 14,840.81 | 13,961.43 | 13,558.64 | 57,527.17 |
| 922 | 10,764.29 | 10,210.17 | 9,889.56 | 9,321.40 | 40,185.42 |
| 923 | 20,324.08 | 18,867.53 | 18,812.01 | 18,393.85 | 76,397.47 |
| 924 | 2,403.70 | 2,236.07 | 2,199.20 | 2,200.91 | 9,039.88 |
| 925 | 14,657.01 | 14,860.51 | 13,282.57 | 13,586.95 | 56,387.04 |
| 926 | 11,756.33 | 12,276.33 | 11,765.42 | 11,551.72 | 47,349.80 |
| 927 | 3,161.94 | 2,870.99 | 3,028.22 | 3,147.86 | 12,209.01 |
| 928 | 12,322.60 | 12,211.79 | 11,824.34 | 11,341.65 | 47,700.38 |
| 930 | 8,056.68 | 7,856.83 | 7,159.04 | 6,535.63 | 29,608.18 |
| 931 | 1,185.11 | 1,103.14 | 1,120.51 | 947.64 | 4,356.40 |
| 932 | 6,777.29 | 6,637.05 | 6,789.52 | 6,652.05 | 26,855.91 |
| 933 | 9,861.82 | 9,053.80 | 8,460.45 | 7,885.73 | 35,261.80 |
| 934 | 9,469.56 | 8,692.59 | 8,212.79 | 7,799.11 | 34,174.05 |
| 935 | 11,080.55 | 10,831.63 | 10,458.13 | 10,261.80 | 42,632.11 |
| 936 | 7,192.30 | 7,114.40 | 7,044.20 | 6,247.52 | 27,598.42 |
| 937 | 9,502 | 9,395.77 | 9,157.04 | 9,026.20 | 37,081.01 |
| 939 | 4,325.23 | 3,611.46 | 3,728.90 | 3,362.71 | 15,028.30 |
| 940 | 7,278.79 | 6,919.52 | 6,502.32 | 6,917.82 | 27,618.45 |
| 941 | 14,051.30 | 12,715.17 | 12,727.82 | 11,969.22 | 51,463.51 |
| 943 | 1,746.56 | 1,440.69 | 1,432.64 | 1,485.84 | 6,105.73 |
| 944 | 1,121.93 | 1,237.75 | 1,159.69 | 1,269.20 | 4,788.57 |
| 945 | 37,730.78 | 36,404.03 | 36,057.11 | 34,998.07 | 145,189.99 |
| 946 | 4,118.03 | 3,749.21 | 3,736.31 | 3,370.85 | 14,974.40 |

| | | | | | | |
|---|---|---|---|---|---|---|
| | 947 | 847.68 | 745.71 | 754.33 | 651.06 | 2,998.78 |
| | 948 | 1,305.43 | 1,212.73 | 1,187.87 | 1,262.96 | 4,968.99 |
| | 949 | 6,336.77 | 6,467.40 | 6,207.75 | 5,667.29 | 24,679.21 |
| | 950 | 6,724.77 | 6,403.54 | 5,986.23 | 5,982.02 | 25,096.56 |
| | 951 | 3,460.45 | 3,494.10 | 3,585.82 | 3,217.53 | 13,757.90 |
| | 952 | 8,754.27 | 8,573.19 | 7,684.52 | 7,562.12 | 32,574.10 |
| | 953 | 22,261.14 | 21,614.40 | 21,000.20 | 20,131.88 | 85,007.62 |
| | 954 | 13,966.47 | 12,918.62 | 11,757.75 | 11,423.99 | 50,066.83 |
| | 955 | 5,524.29 | 5,573.32 | 5,082.01 | 5,063.23 | 21,242.85 |
| | 956 | 27,246.97 | 25,427.01 | 25,422.74 | 23,464.57 | 101,561.29 |
| | 957 | 3,610.91 | 3,029.28 | 3,139.89 | 2,880.16 | 12,660.24 |
| | 958 | 15,076.92 | 14,483.82 | 14,119.48 | 13,854.81 | 57,535.03 |
| | 959 | 12,776.15 | 12,277.54 | 11,209.45 | 11,000.07 | 47,263.21 |
| | 960 | 9,490.07 | 8,683.90 | 8,455.80 | 8,700.19 | 35,329.96 |
| | 961 | 1,949.28 | 1,841.94 | 1,689.03 | 1,752.13 | 7,232.38 |
| TOTAL | | 468,312.41 | 449,794.45 | 435,963.43 | 421,426.74 | 1,775,497.03 |

| STATE: | COLORADO | | | | | |
|---|---|---|---|---|---|---|
| ZIP CODE | | QUARTER 1 | QUARTER 2 | QUARTER 3 | QUARTER 4 | TOTAL GRAMS |
| | 800 | 13,710.54 | 13,320.65 | 12,436.75 | 12,030.89 | 51,498.83 |
| | 801 | 9,641.81 | 9,156.84 | 9,146.45 | 9,182.89 | 37,127.99 |
| | 802 | 16,378.95 | 15,303.73 | 14,876.84 | 14,639.03 | 61,198.55 |
| | 803 | 1,310.49 | 1,258.07 | 1,083.44 | 959.99 | 4,611.99 |
| | 804 | 3,171.81 | 3,001.82 | 3,138.51 | 3,056.56 | 12,368.70 |
| | 805 | 8,276.11 | 7,741.88 | 7,882.83 | 6,988.67 | 30,889.49 |
| | 806 | 3,524.95 | 3,277.01 | 3,239.71 | 3,193.04 | 13,234.71 |
| | 807 | 1,342.39 | 1,204.60 | 1,112.05 | 1,043.73 | 4,702.77 |
| | 808 | 1,865.41 | 1,467.54 | 1,577.65 | 1,522.67 | 6,433.27 |
| | 809 | 9,275.34 | 9,345.93 | 8,681.17 | 8,707.73 | 36,010.17 |
| | 810 | 5,040.34 | 4,848.88 | 4,684.75 | 4,400.90 | 18,974.87 |
| | 811 | 712.03 | 706.7 | 684.6 | 618.61 | 2,721.94 |
| | 812 | 1,433.25 | 1,521.35 | 1,335.37 | 1,213.39 | 5,503.36 |
| | 813 | 541.24 | 783.11 | 584.74 | 423.33 | 2,332.42 |
| | 814 | 694.78 | 742.08 | 629.3 | 660.6 | 2,726.76 |
| | 815 | 2,664.12 | 2,493.91 | 2,373.06 | 2,355.55 | 9,886.64 |
| | 816 | 1,367.34 | 1,297.86 | 1,321.13 | 1,251.10 | 5,237.43 |

| | | | | | | |
|---|---|---|---|---|---|---|
| TOTAL | | 80,950.90 | 77,471.96 | 74,788.35 | 72,248.68 | 305,459.89 |

| STATE: | CONNECTICUT | | | | | |
|---|---|---|---|---|---|---|
| ZIP CODE | | QUARTER 1 | QUARTER 2 | QUARTER 3 | QUARTER 4 | TOTAL GRAMS |
| | 60 | 7,175.55 | 6,946.82 | 6,399.93 | 5,830 | 26,352.30 |
| | 61 | 1,856.28 | 1,777.63 | 1,833.40 | 1,738.59 | 7,205.90 |
| | 62 | 1,949.64 | 1,617.45 | 1,422.20 | 1,399.17 | 6,388.46 |
| | 63 | 2,278.86 | 2,123.39 | 2,256.59 | 2,029.13 | 8,687.97 |
| | 64 | 10,393.61 | 10,050.17 | 9,861.29 | 9,471.36 | 39,776.43 |
| | 65 | 4,139.66 | 3,949.74 | 4,385.82 | 4,445.68 | 16,920.90 |
| | 66 | 1,454.50 | 1,521.78 | 1,460.60 | 1,467.26 | 5,904.14 |
| | 67 | 3,150.09 | 3,197.39 | 3,080.14 | 2,965.88 | 12,393.50 |
| | 68 | 4,299.77 | 4,172.73 | 4,160.37 | 4,088.44 | 16,721.31 |
| | 69 | 264.49 | 294.41 | 271.35 | 273.76 | 1,104.01 |
| TOTAL | | 36,962.45 | 35,651.51 | 35,131.69 | 33,709.27 | 141,454.92 |

| STATE: | DELAWARE | | | | | |
|---|---|---|---|---|---|---|
| ZIP CODE | | QUARTER 1 | QUARTER 2 | QUARTER 3 | QUARTER 4 | TOTAL GRAMS |
| | 197 | 9,115.47 | 8,719.68 | 8,430.16 | 8,474.06 | 34,739.37 |
| | 198 | 4,704.89 | 4,847.05 | 4,570.01 | 4,371.43 | 18,493.38 |
| | 199 | 8,666.09 | 8,326.71 | 8,333.98 | 8,127.98 | 33,454.76 |
| TOTAL | | 22,486.45 | 21,893.44 | 21,334.15 | 20,973.47 | 86,687.51 |

| STATE: | DISTRICT OF COLUMBIA | | | | | |
|---|---|---|---|---|---|---|
| ZIP CODE | | QUARTER 1 | QUARTER 2 | QUARTER 3 | QUARTER 4 | TOTAL GRAMS |
| | 200 | 2,622.88 | 2,907.60 | 2,641.99 | 2,567.63 | 10,740.10 |
| | 203 | 16.92 | 32.71 | 40.67 | 54.2 | 144.5 |
| | 204 | 1,073.71 | 911.91 | 1,107.58 | 596.02 | 3,689.22 |
| | 205 | 0.08 | 1.28 | 0.08 | 0 | 1.44 |
| TOTAL | | 3,713.59 | 3,853.50 | 3,790.32 | 3,217.85 | 14,575.26 |

STATE: FLORIDA

| ZIP CODE | | QUARTER 1 | QUARTER 2 | QUARTER 3 | QUARTER 4 | TOTAL GRAMS |
|---|---|---|---|---|---|---|
| | 320 | 14,035.33 | 13,355.13 | 12,834.44 | 12,948.34 | 53,173.24 |
| | 321 | 15,771.35 | 15,660.74 | 14,624.40 | 15,064.51 | 61,121 |
| | 322 | 23,174.29 | 22,283.59 | 22,573.71 | 22,009.65 | 90,041.24 |
| | 323 | 4,976.97 | 4,880.32 | 4,721.91 | 4,823.84 | 19,403.04 |
| | 324 | 4,303.71 | 4,168.10 | 3,952.38 | 4,314.66 | 16,738.85 |
| | 325 | 9,480.47 | 8,748.39 | 8,494.18 | 8,097.10 | 34,820.14 |
| | 326 | 9,673.65 | 8,880.23 | 8,866.09 | 8,627.71 | 36,047.68 |
| | 327 | 21,176.86 | 20,303.30 | 19,160.51 | 19,829.54 | 80,470.21 |
| | 328 | 11,941.18 | 11,926.42 | 11,152.19 | 10,591.29 | 45,611.08 |
| | 329 | 19,382.72 | 18,127.10 | 18,046.06 | 18,023.90 | 73,579.78 |
| | 330 | 22,853.30 | 22,171.40 | 20,325.91 | 19,233.42 | 84,584.03 |
| | 331 | 8,471.72 | 8,118.44 | 7,276.74 | 7,570.62 | 31,437.52 |
| | 333 | 17,067.60 | 16,599.88 | 17,624.11 | 18,076.62 | 69,368.21 |
| | 334 | 29,241.82 | 28,773.17 | 27,936.34 | 26,532.31 | 112,483.64 |
| | 335 | 16,863.73 | 16,239.17 | 16,286.04 | 16,484.60 | 65,873.54 |
| | 336 | 28,110.58 | 24,175.56 | 26,469.11 | 23,630.39 | 102,385.64 |
| | 337 | 27,571.06 | 26,453.03 | 25,296.06 | 25,465.30 | 104,785.45 |
| | 338 | 9,066.62 | 8,460.86 | 7,447.48 | 8,270.45 | 33,245.41 |
| | 339 | 23,100.09 | 21,721.25 | 19,727.18 | 22,329.68 | 86,878.20 |
| | 341 | 6,624.12 | 6,431.60 | 6,076.97 | 6,366.87 | 25,499.56 |
| | 342 | 22,260.45 | 21,674.36 | 20,197.79 | 20,671.25 | 84,803.85 |
| | 344 | 14,006.83 | 13,667.17 | 13,274.87 | 13,531.12 | 54,479.99 |
| | 346 | 28,166.53 | 27,711.09 | 27,074.68 | 26,272.06 | 109,224.36 |
| | 347 | 10,574.55 | 9,887.26 | 9,332.71 | 9,788.19 | 39,582.71 |
| | 349 | 14,792.57 | 14,581 | 14,633.15 | 14,014.32 | 58,021.04 |
| TOTAL | | 412,688.10 | 394,998.56 | 383,405.01 | 382,567.74 | 1,573,659.41 |

STATE: GEORGIA

| ZIP CODE | | QUARTER 1 | QUARTER 2 | QUARTER 3 | QUARTER 4 | TOTAL GRAMS |
|---|---|---|---|---|---|---|
| | 300 | 22,767.52 | 22,961.67 | 22,508.57 | 22,047.89 | 90,285.65 |
| | 301 | 17,029.36 | 15,886.85 | 16,213.56 | 15,954.40 | 65,084.17 |
| | 302 | 10,890.93 | 10,692.96 | 10,806.21 | 10,201.52 | 42,591.62 |
| | 303 | 6,568.50 | 6,098.39 | 6,124.18 | 6,196.11 | 24,987.18 |
| | 304 | 2,764.53 | 2,637.19 | 2,580.37 | 2,524.73 | 10,506.82 |
| | 305 | 8,627.12 | 8,449.56 | 8,179.28 | 8,639.70 | 33,895.66 |

| | | | | | | |
|---|---|---|---|---|---|---|
| | 306 | 4,742.38 | 4,513.21 | 4,235.83 | 4,373.71 | 17,865.13 |
| | 307 | 3,998.90 | 3,615.26 | 3,460.35 | 3,561.75 | 14,636.26 |
| | 308 | 2,326.70 | 2,109.04 | 1,957.83 | 1,963.88 | 8,357.45 |
| | 309 | 4,792.39 | 4,461.01 | 4,477.29 | 3,745.24 | 17,475.93 |
| | 310 | 7,338.01 | 6,906.58 | 6,789.87 | 6,462.87 | 27,497.33 |
| | 312 | 2,628.18 | 2,816.29 | 2,996.10 | 2,652.01 | 11,092.58 |
| | 313 | 3,083.29 | 2,974.09 | 2,886.57 | 2,654.74 | 11,598.69 |
| | 314 | 2,636.95 | 2,555.65 | 2,548.93 | 2,527.42 | 10,268.95 |
| | 315 | 4,438.25 | 4,042.19 | 4,012.03 | 4,163.40 | 16,655.87 |
| | 316 | 3,070.51 | 2,950.98 | 2,642.47 | 2,886.97 | 11,550.93 |
| | 317 | 3,661.04 | 3,077.02 | 3,028.81 | 2,904.37 | 12,671.24 |
| | 318 | 402.49 | 446.79 | 427.87 | 472.45 | 1,749.60 |
| | 319 | 4,073 | 4,184.46 | 4,315.58 | 3,621.10 | 16,194.14 |
| | 398 | 991.14 | 923.35 | 831.56 | 843.8 | 3,589.85 |
| TOTAL | | 116,831.19 | 112,302.54 | 111,023.26 | 108,398.06 | 448,555.05 |

STATE: GUAM

| ZIP CODE | | QUARTER 1 | QUARTER 2 | QUARTER 3 | QUARTER 4 | TOTAL GRAMS |
|---|---|---|---|---|---|---|
| | 969 | 1,270.56 | 829.71 | 1,031 | 866.98 | 3,998.25 |
| TOTAL | | 1,270.56 | 829.71 | 1,031 | 866.98 | 3,998.25 |

STATE: HAWAII

| ZIP CODE | | QUARTER 1 | QUARTER 2 | QUARTER 3 | QUARTER 4 | TOTAL GRAMS |
|---|---|---|---|---|---|---|
| | 967 | 15,740.49 | 14,905.20 | 13,576.07 | 12,646.22 | 56,867.98 |
| | 968 | 7,669.92 | 7,583.10 | 7,611.82 | 7,426.18 | 30,291.02 |
| TOTAL | | 23,410.41 | 22,488.30 | 21,187.89 | 20,072.40 | 87,159 |

STATE: IDAHO

| ZIP CODE | | QUARTER 1 | QUARTER 2 | QUARTER 3 | QUARTER 4 | TOTAL GRAMS |
|---|---|---|---|---|---|---|
| | 832 | 2,995.31 | 2,850.85 | 2,656.47 | 2,703.77 | 11,206.40 |
| | 833 | 3,316.24 | 3,411.31 | 3,092.94 | 2,975.01 | 12,795.50 |
| | 834 | 2,047.67 | 1,883.06 | 1,863.59 | 1,695.81 | 7,490.13 |
| | 835 | 1,917.52 | 1,613.52 | 1,554.08 | 1,566.91 | 6,652.03 |
| | 836 | 7,807.21 | 7,976.24 | 7,502.53 | 7,455.93 | 30,741.91 |

| | | | | | | |
|---|---|---|---|---|---|---|
| | 837 | 6,583.21 | 6,276.56 | 5,844.89 | 6,136.10 | 24,840.76 |
| | 838 | 5,109.48 | 4,544.28 | 4,676.68 | 4,469.64 | 18,800.08 |
| TOTAL | | 29,776.64 | 28,555.82 | 27,191.18 | 27,003.17 | 112,526.81 |

STATE: ILLINOIS

| ZIP CODE | | QUARTER 1 | QUARTER 2 | QUARTER 3 | QUARTER 4 | TOTAL GRAMS |
|---|---|---|---|---|---|---|
| | 600 | 18,871.30 | 18,261.08 | 18,066.47 | 17,949.63 | 73,148.48 |
| | 601 | 10,465.10 | 9,739.91 | 9,293.99 | 8,878.58 | 38,377.58 |
| | 602 | 368.79 | 410.77 | 371.34 | 318.77 | 1,469.67 |
| | 603 | 568.68 | 580.83 | 557.42 | 562.42 | 2,269.35 |
| | 604 | 11,573.92 | 11,404.12 | 10,675.79 | 10,415.23 | 44,069.06 |
| | 605 | 6,248.43 | 5,767.33 | 5,480.02 | 5,513.21 | 23,008.99 |
| | 606 | 11,263.59 | 10,917.92 | 10,065.11 | 10,368.86 | 42,615.48 |
| | 607 | 660.12 | 664.61 | 597.78 | 625.15 | 2,547.66 |
| | 608 | 981.06 | 1,005.11 | 887.16 | 678.68 | 3,552.01 |
| | 609 | 2,041.98 | 1,853.10 | 1,624.23 | 1,590.54 | 7,109.85 |
| | 610 | 3,074.26 | 2,906.93 | 2,976.76 | 2,998.63 | 11,956.58 |
| | 611 | 3,953.51 | 3,949.74 | 3,652.68 | 3,604.28 | 15,160.21 |
| | 612 | 1,441.35 | 1,125.94 | 1,192.25 | 1,329.83 | 5,089.37 |
| | 613 | 1,367.96 | 1,401.40 | 1,267.15 | 1,206.31 | 5,242.82 |
| | 614 | 996.48 | 938.98 | 1,048.74 | 1,023.91 | 4,008.11 |
| | 615 | 1,687.68 | 1,542.16 | 1,740.26 | 1,992.61 | 6,962.71 |
| | 616 | 3,562.91 | 3,372.01 | 3,169.51 | 2,924.29 | 13,028.72 |
| | 617 | 1,600 | 1,533.02 | 1,792.58 | 1,636.54 | 6,562.14 |
| | 618 | 2,737.10 | 2,701.42 | 2,775.64 | 2,705.63 | 10,919.79 |
| | 619 | 1,028.03 | 844.76 | 885.52 | 995.92 | 3,754.23 |
| | 620 | 3,894.12 | 3,598.65 | 3,573.59 | 3,458.76 | 14,525.12 |
| | 622 | 4,398.70 | 3,973.77 | 4,164.48 | 3,875.83 | 16,412.78 |
| | 623 | 1,254.82 | 1,284.69 | 1,136.71 | 1,033.51 | 4,709.73 |
| | 624 | 1,334.59 | 1,379.80 | 1,364.66 | 1,173.56 | 5,252.61 |
| | 625 | 2,273.61 | 2,220.74 | 2,036.43 | 2,092.32 | 8,623.10 |
| | 626 | 1,806.66 | 1,678.62 | 1,630.85 | 1,430.36 | 6,546.49 |
| | 627 | 1,688.83 | 1,613.78 | 1,623.92 | 1,592.62 | 6,519.15 |
| | 628 | 2,104.76 | 2,040.89 | 2,037.97 | 1,850.46 | 8,034.08 |
| | 629 | 4,763.42 | 4,508.25 | 4,395.60 | 4,211.30 | 17,878.57 |
| TOTAL | | 108,011.76 | 103,220.33 | 100,084.61 | 98,037.74 | 409,354.44 |

STATE: INDIANA

| ZIP CODE | QUARTER 1 | QUARTER 2 | QUARTER 3 | QUARTER 4 | TOTAL GRAMS |
|---|---|---|---|---|---|
| 460 | 5,273.67 | 5,046.54 | 5,047.55 | 4,970.48 | 20,338.24 |
| 461 | 7,438.45 | 7,121.69 | 6,802.03 | 6,775.35 | 28,137.52 |
| 462 | 17,519.90 | 16,373.78 | 15,796.61 | 16,480.37 | 66,170.66 |
| 463 | 7,647.31 | 7,123.45 | 7,308.28 | 6,653.44 | 28,732.48 |
| 464 | 718.51 | 658.44 | 555.87 | 643.64 | 2,576.46 |
| 465 | 6,470.81 | 6,750.03 | 5,532.87 | 5,692 | 24,445.71 |
| 466 | 2,221.37 | 2,227.31 | 1,908.38 | 1,867.53 | 8,224.59 |
| 467 | 3,479.30 | 2,900.21 | 2,967.08 | 2,909.05 | 12,255.64 |
| 468 | 5,105.73 | 4,475.13 | 4,057.30 | 4,278.34 | 17,916.50 |
| 469 | 4,856.74 | 4,463.65 | 4,147.33 | 4,138.84 | 17,606.56 |
| 470 | 1,779.50 | 1,781.16 | 1,693.86 | 1,544.94 | 6,799.46 |
| 471 | 5,647.46 | 5,589.94 | 4,788.37 | 4,359.01 | 20,384.78 |
| 472 | 2,718.52 | 2,742.42 | 2,644.06 | 2,527.02 | 10,632.02 |
| 473 | 5,807.78 | 5,165.65 | 5,201.36 | 4,919.72 | 21,094.51 |
| 474 | 2,051.39 | 2,005.51 | 1,791.98 | 1,634.67 | 7,483.55 |
| 475 | 2,158.09 | 2,036.70 | 1,954.40 | 1,814.97 | 7,964.16 |
| 476 | 1,327.74 | 1,253.82 | 1,373.92 | 1,273.14 | 5,228.62 |
| 477 | 3,866.42 | 3,902.46 | 4,100.78 | 3,745.35 | 15,615.01 |
| 478 | 2,481.29 | 2,041.23 | 1,866.81 | 1,683.26 | 8,072.59 |
| 479 | 3,582.37 | 3,713.75 | 3,482.90 | 3,331.86 | 14,110.88 |
| TOTAL | 92,152.35 | 87,372.87 | 83,021.74 | 81,242.98 | 343,789.94 |

STATE: IOWA

| ZIP CODE | QUARTER 1 | QUARTER 2 | QUARTER 3 | QUARTER 4 | TOTAL GRAMS |
|---|---|---|---|---|---|
| 500 | 2,421.35 | 2,488.67 | 2,318.14 | 2,342.75 | 9,570.91 |
| 501 | 2,461.16 | 2,364.55 | 2,335.67 | 2,511.60 | 9,672.98 |
| 502 | 2,231.40 | 2,262.40 | 2,151.80 | 2,019.93 | 8,665.53 |
| 503 | 5,631.33 | 5,308.19 | 5,035.26 | 5,060.42 | 21,035.20 |
| 504 | 1,346.64 | 1,416.26 | 1,306.60 | 1,315.36 | 5,384.86 |
| 505 | 1,598.35 | 1,583.56 | 1,597.77 | 1,502.87 | 6,282.55 |
| 506 | 1,175.51 | 1,060.15 | 1,076.91 | 1,044.08 | 4,356.65 |
| 507 | 879.17 | 820.73 | 820.88 | 813.2 | 3,333.98 |

| | | | | | | |
|---|---|---|---|---|---|---|
| | 508 | 432.11 | 410.12 | 517.42 | 439.57 | 1,799.22 |
| | 510 | 601.84 | 569.46 | 606.05 | 553.63 | 2,330.98 |
| | 511 | 1,023.58 | 958.44 | 1,043.30 | 1,030.58 | 4,055.90 |
| | 512 | 228.01 | 248.4 | 270.99 | 232.45 | 979.85 |
| | 513 | 414.47 | 392.25 | 404.22 | 372.75 | 1,583.69 |
| | 514 | 614.59 | 521.46 | 525.4 | 642.37 | 2,303.82 |
| | 515 | 1,316.59 | 1,281.42 | 1,225.26 | 1,251.77 | 5,075.04 |
| | 516 | 182.37 | 219.46 | 177.89 | 165.28 | 745 |
| | 520 | 1,418.47 | 1,485.87 | 1,452.30 | 1,569.88 | 5,926.52 |
| | 521 | 439.18 | 376.18 | 332.69 | 305.24 | 1,453.29 |
| | 522 | 2,165.28 | 2,215.08 | 2,110.37 | 2,057.67 | 8,548.40 |
| | 523 | 865.76 | 772.06 | 771.69 | 662.47 | 3,071.98 |
| | 524 | 1,651.97 | 1,600.78 | 1,487.94 | 1,419.79 | 6,160.48 |
| | 525 | 1,740.17 | 1,652 | 1,533.43 | 1,614.53 | 6,540.13 |
| | 526 | 1,149.15 | 1,190.11 | 1,128.21 | 1,117.20 | 4,584.67 |
| | 527 | 1,243.33 | 1,194.46 | 964.58 | 1,122.31 | 4,524.68 |
| | 528 | 1,637.48 | 1,534.25 | 1,414.15 | 1,402.75 | 5,988.63 |
| TOTAL | | 34,869.26 | 33,926.31 | 32,608.92 | 32,570.45 | 133,974.94 |

STATE: KANSAS

| ZIP CODE | | QUARTER 1 | QUARTER 2 | QUARTER 3 | QUARTER 4 | TOTAL GRAMS |
|---|---|---|---|---|---|---|
| | 660 | 6,997.18 | 6,529.31 | 6,205.62 | 6,395.23 | 26,127.34 |
| | 661 | 1,912.58 | 1,837.69 | 1,474.09 | 1,754.82 | 6,979.18 |
| | 662 | 6,527.68 | 6,427.24 | 6,219.42 | 6,129.25 | 25,303.59 |
| | 664 | 978.26 | 811.56 | 778.98 | 608.28 | 3,177.08 |
| | 665 | 1,350.77 | 1,227.17 | 1,250.95 | 1,129.53 | 4,958.42 |
| | 666 | 3,381.01 | 3,000.60 | 3,087.61 | 2,818.89 | 12,288.11 |
| | 667 | 2,982.36 | 2,728.87 | 2,890.97 | 2,566.90 | 11,169.10 |
| | 668 | 788.48 | 802.88 | 811.64 | 718.4 | 3,121.40 |
| | 669 | 685.25 | 662.02 | 693.44 | 610.67 | 2,651.38 |
| | 670 | 2,593.04 | 2,601.66 | 2,639.31 | 2,394.07 | 10,228.08 |
| | 671 | 1,678.11 | 1,543.28 | 1,664.50 | 1,546.40 | 6,432.29 |
| | 672 | 8,635.07 | 8,270.40 | 8,539.45 | 7,894.55 | 33,339.47 |
| | 673 | 1,564.17 | 1,422.07 | 1,319.26 | 1,351.57 | 5,657.07 |
| | 674 | 2,323.97 | 2,146.79 | 2,175.90 | 2,088.42 | 8,735.08 |
| | 675 | 1,342.61 | 1,250.14 | 1,380.73 | 1,343.63 | 5,317.11 |

| | | | | | | |
|---|---|---|---|---|---|---|
| | 676 | 1,018.73 | 1,057.51 | 969.86 | 1,178.92 | 4,225.02 |
| | 677 | 706.56 | 684.15 | 698.47 | 703.03 | 2,792.21 |
| | 678 | 1,005.65 | 938.92 | 848.35 | 877.44 | 3,670.36 |
| | 679 | 371.94 | 376.82 | 385.84 | 367.13 | 1,501.73 |
| TOTAL | | 46,843.42 | 44,319.08 | 44,034.39 | 42,477.13 | 177,674.02 |

| STATE: | KENTUCKY | | | | | |
|---|---|---|---|---|---|---|
| ZIP CODE | | QUARTER 1 | QUARTER 2 | QUARTER 3 | QUARTER 4 | TOTAL GRAMS |
| | 400 | 1,728.65 | 1,499.23 | 1,367.80 | 1,349.19 | 5,944.87 |
| | 401 | 1,296.87 | 1,197.98 | 1,062.61 | 1,093.36 | 4,650.82 |
| | 402 | 8,509.39 | 8,030.36 | 7,287.26 | 7,417.66 | 31,244.67 |
| | 403 | 3,268.79 | 3,041.99 | 2,700.02 | 2,767.60 | 11,778.40 |
| | 404 | 1,554.85 | 1,485.23 | 1,433.69 | 1,454.06 | 5,927.83 |
| | 405 | 4,185.35 | 3,569.69 | 3,775.72 | 3,688.60 | 15,219.36 |
| | 406 | 352.01 | 313.43 | 336.32 | 282.88 | 1,284.64 |
| | 407 | 1,198.67 | 1,196.24 | 1,142.36 | 1,140.53 | 4,677.80 |
| | 408 | 304.58 | 336.52 | 349.53 | 301.75 | 1,292.38 |
| | 409 | 625.57 | 589.48 | 648.18 | 517.93 | 2,381.16 |
| | 410 | 6,059.10 | 5,852.09 | 5,756.35 | 5,886.01 | 23,553.55 |
| | 411 | 1,394.44 | 1,270.22 | 1,216.98 | 1,168.05 | 5,049.69 |
| | 412 | 355.44 | 273.33 | 294.25 | 318.52 | 1,241.54 |
| | 413 | 435.12 | 394.36 | 363.19 | 382.68 | 1,575.35 |
| | 414 | 198.3 | 208.19 | 230.92 | 155.09 | 792.5 |
| | 415 | 579.37 | 550.47 | 482.52 | 448.72 | 2,061.08 |
| | 416 | 311.41 | 307.85 | 386.66 | 313.19 | 1,319.11 |
| | 417 | 1,176.96 | 1,252.41 | 898.39 | 1,066.47 | 4,394.23 |
| | 418 | 198.33 | 176.12 | 168.24 | 171.63 | 714.32 |
| | 420 | 3,014.78 | 2,674.92 | 2,692.52 | 2,880.35 | 11,262.57 |
| | 421 | 3,178.15 | 3,263.89 | 2,959.36 | 3,032.49 | 12,433.89 |
| | 422 | 1,854.51 | 1,462.82 | 1,753.74 | 1,395.35 | 6,466.42 |
| | 423 | 2,169.34 | 1,977.39 | 1,970.33 | 1,806.82 | 7,923.88 |
| | 424 | 2,407.14 | 2,245.96 | 2,146.14 | 2,175.99 | 8,975.23 |
| | 425 | 732.94 | 753.21 | 583.77 | 684.63 | 2,754.55 |
| | 426 | 598.32 | 543.58 | 510.75 | 471.98 | 2,124.63 |
| | 427 | 2,211.09 | 2,127.50 | 2,108.81 | 1,950.87 | 8,398.27 |

| | | | | | |
|---|---|---|---|---|---|
| TOTAL | 49,899.47 | 46,594.46 | 44,626.41 | 44,322.40 | 185,442.74 |

STATE: LOUISIANA

| ZIP CODE | QUARTER 1 | QUARTER 2 | QUARTER 3 | QUARTER 4 | TOTAL GRAMS |
|---|---|---|---|---|---|
| 700 | 7,751.27 | 7,287.18 | 6,702.92 | 6,304.83 | 28,046.20 |
| 701 | 4,226.16 | 3,660.21 | 3,408.41 | 3,427.69 | 14,722.47 |
| 703 | 1,877.47 | 1,952.14 | 1,871.95 | 1,764.30 | 7,465.86 |
| 704 | 6,828.24 | 6,337.92 | 6,177.39 | 5,814.16 | 25,157.71 |
| 705 | 5,803.39 | 6,047.76 | 5,769.26 | 5,675.93 | 23,296.34 |
| 706 | 1,668.29 | 1,625.88 | 1,713.76 | 1,666.55 | 6,674.48 |
| 707 | 6,108.44 | 5,758.81 | 5,476.36 | 5,454.08 | 22,797.69 |
| 708 | 5,084.26 | 5,001.20 | 4,647.85 | 4,547.92 | 19,281.23 |
| 710 | 1,653.56 | 1,683.56 | 1,779.62 | 1,698.83 | 6,815.57 |
| 711 | 6,044.06 | 6,182.41 | 6,087 | 6,136.33 | 24,449.80 |
| 712 | 2,119.05 | 1,863.59 | 2,069.64 | 1,931.93 | 7,984.21 |
| 713 | 4,864.61 | 4,862.69 | 4,640.79 | 4,183.18 | 18,551.27 |
| 714 | 2,267.07 | 2,094.65 | 2,213 | 1,943.60 | 8,518.32 |
| TOTAL | 56,295.87 | 54,358 | 52,557.95 | 50,549.33 | 213,761.15 |

STATE: MAINE

| ZIP CODE | QUARTER 1 | QUARTER 2 | QUARTER 3 | QUARTER 4 | TOTAL GRAMS |
|---|---|---|---|---|---|
| 039 | 407.58 | 339.24 | 372.62 | 377.27 | 1,496.71 |
| 040 | 4,129.27 | 3,958.48 | 3,915.93 | 3,811.32 | 15,815 |
| 041 | 883.75 | 884.16 | 920.44 | 838.07 | 3,526.42 |
| 042 | 2,017.59 | 1,944.85 | 1,720.32 | 1,667.30 | 7,350.06 |
| 043 | 2,925.81 | 3,141.26 | 3,082.67 | 2,757.42 | 11,907.16 |
| 044 | 1,646.28 | 1,626.28 | 1,505.19 | 1,386.99 | 6,164.74 |
| 045 | 329.64 | 321.16 | 391.5 | 315.24 | 1,357.54 |
| 046 | 756.09 | 636.18 | 607.4 | 614.36 | 2,614.03 |
| 047 | 576.62 | 570.53 | 545.34 | 564.01 | 2,256.50 |
| 048 | 572.59 | 765.84 | 683.17 | 627.7 | 2,649.30 |
| 049 | 2,800.70 | 2,670.82 | 2,675.94 | 2,648.97 | 10,796.43 |
| TOTAL | 17,045.92 | 16,858.80 | 16,420.52 | 15,608.65 | 65,933.89 |

DATE RANGE: 01/01/2017 TO 12/31/2017 ARCOS 3 - REPORT 1 Run Date: 07/03/2018

RETAIL DRUG DISTRIBUTION BY ZIP CODE WITHIN STATE BY GRAMS WT

STATE: MARYLAND

| ZIP CODE | QUARTER 1 | QUARTER 2 | QUARTER 3 | QUARTER 4 | TOTAL GRAMS |
|---|---|---|---|---|---|
| 206 | 5,538.27 | 5,293.99 | 4,840.72 | 4,611.28 | 20,284.26 |
| 207 | 6,865.47 | 6,352.22 | 6,374.18 | 5,462.38 | 25,054.25 |
| 208 | 6,480.01 | 6,662.92 | 5,419.03 | 5,709.06 | 24,271.02 |
| 209 | 993.89 | 996.72 | 976.44 | 864.68 | 3,831.73 |
| 210 | 13,778.89 | 13,126.19 | 12,152.66 | 11,480.73 | 50,538.47 |
| 211 | 6,016.72 | 5,850 | 5,404.14 | 5,417.25 | 22,688.11 |
| 212 | 20,814.18 | 19,802.55 | 18,169.17 | 17,911.32 | 76,697.22 |
| 214 | 1,355.99 | 1,131.71 | 996.54 | 910.8 | 4,395.04 |
| 215 | 2,271.84 | 2,126.52 | 1,871.49 | 1,840.45 | 8,110.30 |
| 216 | 2,721.37 | 2,550.70 | 2,606.07 | 2,407.84 | 10,285.98 |
| 217 | 8,855.82 | 8,117.73 | 7,646.18 | 7,615.59 | 32,235.32 |
| 218 | 2,298.52 | 2,302.23 | 2,136.50 | 2,047.92 | 8,785.17 |
| 219 | 4,097.89 | 4,217.25 | 3,939.16 | 3,712.15 | 15,966.45 |
| TOTAL | 82,088.86 | 78,530.73 | 72,532.28 | 69,991.45 | 303,143.32 |

STATE: MASSACHUSETTS

| ZIP CODE | QUARTER 1 | QUARTER 2 | QUARTER 3 | QUARTER 4 | TOTAL GRAMS |
|---|---|---|---|---|---|
| 10 | 7,995.51 | 7,470.58 | 7,113.38 | 7,042.93 | 29,622.40 |
| 11 | 3,107.95 | 3,332.28 | 2,828.15 | 2,896.29 | 12,164.67 |
| 12 | 1,276.98 | 1,130.84 | 1,127.70 | 1,345.17 | 4,880.69 |
| 13 | 1,561.79 | 1,548.07 | 1,391.78 | 1,263.10 | 5,764.74 |
| 14 | 2,528.57 | 2,478.50 | 2,408.19 | 2,203.42 | 9,618.68 |
| 15 | 3,303.17 | 3,325.90 | 3,242.70 | 3,453.36 | 13,325.13 |
| 16 | 2,324.76 | 2,205.89 | 2,070.50 | 2,134.37 | 8,735.52 |
| 17 | 3,764.77 | 3,418.37 | 3,262.79 | 3,084.94 | 13,530.87 |
| 18 | 9,896.63 | 9,745.45 | 9,051.93 | 9,186.31 | 37,880.32 |
| 19 | 4,309.47 | 3,881.81 | 3,887.92 | 3,883.44 | 15,962.64 |
| 20 | 2,492.38 | 2,301.10 | 2,373.86 | 2,062.97 | 9,230.31 |
| 21 | 8,018.84 | 7,353.47 | 7,195.18 | 6,944.31 | 29,511.80 |
| 22 | 903.69 | 1,062.44 | 660.25 | 1,038.14 | 3,664.52 |
| 23 | 5,384.77 | 5,030.93 | 4,554.35 | 4,489.87 | 19,459.92 |
| 24 | 1,318.02 | 1,243.74 | 1,221.60 | 1,279.29 | 5,062.65 |
| 25 | 1,009.29 | 1,165.59 | 1,115.70 | 1,089.40 | 4,379.98 |
| 26 | 1,737.57 | 1,502.01 | 1,575.20 | 1,508.39 | 6,323.17 |

| | | | | | | |
|---|---|---|---|---|---|---|
| | 27 | 8,634.53 | 8,310.78 | 8,066.35 | 8,068.61 | 33,080.27 |
| TOTAL | | 69,568.69 | 66,507.75 | 63,147.53 | 62,974.31 | 262,198.28 |

STATE: MICHIGAN

| ZIP CODE | | QUARTER 1 | QUARTER 2 | QUARTER 3 | QUARTER 4 | TOTAL GRAMS |
|---|---|---|---|---|---|---|
| | 480 | 23,053.47 | 22,165.51 | 20,520.78 | 19,122.98 | 84,862.74 |
| | 481 | 23,096.70 | 22,929.40 | 22,076.78 | 20,720.12 | 88,823 |
| | 482 | 13,220.06 | 12,065.09 | 11,928.30 | 11,187.74 | 48,401.19 |
| | 483 | 18,910.41 | 17,848.03 | 16,507.76 | 14,940.67 | 68,206.87 |
| | 484 | 7,518.49 | 7,071.13 | 7,406.61 | 7,054.66 | 29,050.89 |
| | 485 | 4,970.76 | 4,729.47 | 4,284.49 | 4,259.72 | 18,244.44 |
| | 486 | 14,066.62 | 13,294.49 | 12,448.68 | 11,782.70 | 51,592.49 |
| | 487 | 6,812.49 | 6,944.29 | 6,194.67 | 5,851.11 | 25,802.56 |
| | 488 | 10,541.11 | 9,727.73 | 9,973.22 | 9,318.14 | 39,560.20 |
| | 489 | 4,535.84 | 4,497.10 | 4,348.07 | 4,014.89 | 17,395.90 |
| | 490 | 17,698.16 | 16,601.79 | 16,038.33 | 15,084.90 | 65,423.18 |
| | 491 | 1,623.49 | 1,614.43 | 1,805.05 | 1,660.02 | 6,702.99 |
| | 492 | 7,015.92 | 6,877.85 | 6,763.37 | 6,338.75 | 26,995.89 |
| | 493 | 3,993.35 | 3,795.65 | 3,638.41 | 3,541.44 | 14,968.85 |
| | 494 | 10,628.61 | 9,982.45 | 9,888.84 | 9,935.21 | 40,435.11 |
| | 495 | 8,547.98 | 8,861.18 | 8,253.62 | 7,513.52 | 33,176.30 |
| | 496 | 9,953.22 | 9,488.33 | 9,414.17 | 8,981.95 | 37,837.67 |
| | 497 | 8,485.85 | 8,386.96 | 8,588.93 | 7,731.11 | 33,192.85 |
| | 498 | 4,743.67 | 4,471.02 | 4,513.80 | 3,784.16 | 17,512.65 |
| | 499 | 1,432.61 | 1,336.33 | 1,384.36 | 1,231.10 | 5,384.40 |
| TOTAL | | 200,848.81 | 192,688.23 | 185,978.24 | 174,054.89 | 753,570.17 |

STATE: MINNESOTA

| ZIP CODE | | QUARTER 1 | QUARTER 2 | QUARTER 3 | QUARTER 4 | TOTAL GRAMS |
|---|---|---|---|---|---|---|
| | 550 | 5,270.62 | 4,867.29 | 4,823.79 | 4,766.50 | 19,728.20 |
| | 551 | 7,813.69 | 7,052.19 | 6,620.09 | 6,082.58 | 27,568.55 |
| | 553 | 6,596.13 | 6,177.43 | 6,057.87 | 6,078.06 | 24,909.49 |
| | 554 | 11,468.31 | 11,253.96 | 10,573.85 | 10,418.40 | 43,714.52 |
| | 556 | 208.99 | 180.47 | 223.43 | 182.65 | 795.54 |
| | 557 | 1,762.71 | 1,711.97 | 1,615.39 | 1,621.27 | 6,711.34 |

| | | | | | | |
|---|---|---|---|---|---|---|
| | 558 | 1,633.54 | 1,503.66 | 1,583.19 | 1,507.17 | 6,227.56 |
| | 559 | 2,136.14 | 2,211.76 | 2,135.96 | 1,938.57 | 8,422.43 |
| | 560 | 2,012.08 | 1,774.63 | 1,781.57 | 1,611.37 | 7,179.65 |
| | 561 | 789.36 | 800.8 | 716.61 | 660.18 | 2,966.95 |
| | 562 | 1,145.30 | 1,117.59 | 1,089.54 | 1,059.77 | 4,412.20 |
| | 563 | 3,882.67 | 3,715.35 | 3,452.94 | 3,332.09 | 14,383.05 |
| | 564 | 1,728.36 | 1,672.80 | 1,533.53 | 1,513.13 | 6,447.82 |
| | 565 | 1,529.48 | 1,373.49 | 1,470.39 | 1,394.60 | 5,767.96 |
| | 566 | 829.05 | 731.88 | 859.34 | 684.26 | 3,104.53 |
| | 567 | 742.21 | 799.26 | 839.67 | 751.29 | 3,132.43 |
| TOTAL | | 49,548.64 | 46,944.53 | 45,377.16 | 43,601.89 | 185,472.22 |

STATE: MISSISSIPPI

| ZIP CODE | | QUARTER 1 | QUARTER 2 | QUARTER 3 | QUARTER 4 | TOTAL GRAMS |
|---|---|---|---|---|---|---|
| | 386 | 3,932.82 | 3,936.91 | 3,467.78 | 3,792.70 | 15,130.21 |
| | 387 | 506 | 478.92 | 408.96 | 376.39 | 1,770.27 |
| | 388 | 4,376.44 | 4,474.06 | 4,079.97 | 4,091.81 | 17,022.28 |
| | 389 | 853.69 | 814.2 | 710.42 | 763.59 | 3,141.90 |
| | 390 | 3,245.38 | 3,046.41 | 3,005.78 | 2,701.44 | 11,999.01 |
| | 391 | 5,345.69 | 6,127.83 | 6,188.03 | 6,801.66 | 24,463.21 |
| | 392 | 3,546.66 | 3,490.03 | 3,378.70 | 3,300.55 | 13,715.94 |
| | 393 | 3,769.77 | 3,480.25 | 3,198.93 | 2,840.69 | 13,289.64 |
| | 394 | 5,957.48 | 5,422.07 | 5,513.65 | 5,381.93 | 22,275.13 |
| | 395 | 6,394.19 | 5,866.08 | 5,784.95 | 5,993.68 | 24,038.90 |
| | 396 | 2,134.43 | 2,009.51 | 1,765.59 | 1,666.89 | 7,576.42 |
| | 397 | 1,271.21 | 1,399.25 | 1,341.68 | 1,376.32 | 5,388.46 |
| TOTAL | | 41,333.76 | 40,545.52 | 38,844.44 | 39,087.65 | 159,811.37 |

STATE: MISSOURI

| ZIP CODE | | QUARTER 1 | QUARTER 2 | QUARTER 3 | QUARTER 4 | TOTAL GRAMS |
|---|---|---|---|---|---|---|
| | 630 | 10,797.13 | 10,030.35 | 9,656.81 | 9,744.02 | 40,228.31 |
| | 631 | 16,737.88 | 16,075.05 | 15,566.29 | 15,491.72 | 63,870.94 |
| | 633 | 5,935.67 | 5,768.22 | 5,520.16 | 5,125.16 | 22,349.21 |
| | 634 | 565.99 | 616.98 | 660.67 | 642.31 | 2,485.95 |
| | 635 | 1,492.97 | 1,215.57 | 1,248.69 | 1,170.09 | 5,127.32 |

| | | | | | | |
|---|---|---|---|---|---|---|
| | 636 | 3,359.81 | 3,193.73 | 3,336.25 | 3,131.54 | 13,021.33 |
| | 637 | 1,590.37 | 1,488.78 | 1,559.27 | 1,562.74 | 6,201.16 |
| | 638 | 1,675.25 | 1,482.40 | 1,402.89 | 1,301.06 | 5,861.60 |
| | 639 | 1,940.90 | 1,731.36 | 1,412.12 | 1,271.25 | 6,355.63 |
| | 640 | 7,726.61 | 7,270.13 | 7,339.49 | 6,955.04 | 29,291.27 |
| | 641 | 7,581.28 | 6,994.76 | 6,557.63 | 6,358.16 | 27,491.83 |
| | 644 | 988.91 | 841.48 | 816.81 | 831.32 | 3,478.52 |
| | 645 | 1,516.99 | 1,437.20 | 1,380.24 | 1,307.28 | 5,641.71 |
| | 646 | 952.71 | 813.12 | 871.35 | 796.73 | 3,433.91 |
| | 647 | 2,781.60 | 2,605.08 | 2,336.80 | 2,341.79 | 10,065.27 |
| | 648 | 3,883.25 | 3,708.67 | 3,698.94 | 3,408.75 | 14,699.61 |
| | 650 | 1,821.90 | 1,793.28 | 1,579.74 | 1,708.81 | 6,903.73 |
| | 651 | 1,215.95 | 1,282.53 | 1,120.65 | 1,011.20 | 4,630.33 |
| | 652 | 4,430.99 | 4,495.69 | 4,635.18 | 4,486.52 | 18,048.38 |
| | 653 | 1,183.91 | 1,149.75 | 1,062.57 | 996.48 | 4,392.71 |
| | 654 | 1,531.72 | 1,508.33 | 1,519.19 | 1,258.36 | 5,817.60 |
| | 655 | 2,568.88 | 2,487.33 | 2,314.27 | 1,994.70 | 9,365.18 |
| | 656 | 4,325.66 | 4,367.79 | 4,281.52 | 4,082.01 | 17,056.98 |
| | 657 | 7,411.28 | 7,113.90 | 7,274.42 | 7,161.95 | 28,961.55 |
| | 658 | 8,505.57 | 7,607.24 | 7,603.84 | 7,436.90 | 31,153.55 |
| TOTAL | | 102,523.18 | 97,078.72 | 94,755.79 | 91,575.89 | 385,933.58 |

| STATE: | MONTANA | | | | | |
|---|---|---|---|---|---|---|
| ZIP CODE | | QUARTER 1 | QUARTER 2 | QUARTER 3 | QUARTER 4 | TOTAL GRAMS |
| | 590 | 1,285.03 | 1,073.50 | 1,195.62 | 1,093.99 | 4,648.14 |
| | 591 | 3,827.06 | 3,352.43 | 3,275.68 | 3,436.95 | 13,892.12 |
| | 592 | 666.03 | 564.11 | 543.46 | 534.03 | 2,307.63 |
| | 593 | 725.4 | 674.94 | 666.03 | 538.2 | 2,604.57 |
| | 594 | 2,702.35 | 2,022.27 | 1,904.90 | 1,888.46 | 8,517.98 |
| | 595 | 392.12 | 313.28 | 281.5 | 277.25 | 1,264.15 |
| | 596 | 2,254.05 | 2,197.20 | 2,132.72 | 1,993.51 | 8,577.48 |
| | 597 | 1,903.21 | 1,879.28 | 2,003.36 | 1,698.64 | 7,484.49 |
| | 598 | 2,834.32 | 2,759.30 | 2,571.61 | 2,587.41 | 10,752.64 |
| | 599 | 2,477.17 | 2,397.75 | 2,428.90 | 2,288.93 | 9,592.75 |
| TOTAL | | 19,066.74 | 17,234.06 | 17,003.78 | 16,337.37 | 69,641.95 |

STATE: NEBRASKA

| ZIP CODE | QUARTER 1 | QUARTER 2 | QUARTER 3 | QUARTER 4 | TOTAL GRAMS |
|---|---|---|---|---|---|
| 680 | 2,035.45 | 2,252.57 | 2,279.04 | 2,110.03 | 8,677.09 |
| 681 | 6,799.80 | 6,531.56 | 6,561.65 | 6,304.67 | 26,197.68 |
| 683 | 664.06 | 669.78 | 695.68 | 667.59 | 2,697.11 |
| 684 | 664.82 | 603.01 | 546.3 | 532.51 | 2,346.64 |
| 685 | 2,918.29 | 2,611.28 | 2,729.72 | 2,634.62 | 10,893.91 |
| 686 | 452.75 | 464.19 | 461.03 | 410.91 | 1,788.88 |
| 687 | 971.9 | 978.91 | 845.42 | 914.97 | 3,711.20 |
| 688 | 2,121.21 | 1,903.50 | 1,808.18 | 1,881.88 | 7,714.77 |
| 689 | 644.38 | 646.63 | 634.29 | 645.71 | 2,571.01 |
| 690 | 417.41 | 342.52 | 414.27 | 326.53 | 1,500.73 |
| 691 | 932.52 | 782.67 | 827.31 | 701.62 | 3,244.12 |
| 692 | 55.68 | 99.22 | 43.66 | 61.18 | 259.74 |
| 693 | 1,092.51 | 965.23 | 1,092.52 | 939.95 | 4,090.21 |
| TOTAL | 19,770.78 | 18,851.07 | 18,939.07 | 18,132.17 | 75,693.09 |

STATE: NEVADA

| ZIP CODE | QUARTER 1 | QUARTER 2 | QUARTER 3 | QUARTER 4 | TOTAL GRAMS |
|---|---|---|---|---|---|
| 890 | 15,732.38 | 16,074.10 | 15,291.48 | 14,266.85 | 61,364.81 |
| 891 | 32,568.86 | 31,710.61 | 30,451.07 | 29,849.01 | 124,579.55 |
| 893 | 197.31 | 152.87 | 131.3 | 103.1 | 584.58 |
| 894 | 4,692.47 | 4,424.03 | 4,428.70 | 4,031.66 | 17,576.86 |
| 895 | 7,167.54 | 6,293.31 | 6,190.44 | 5,194.55 | 24,845.84 |
| 897 | 1,810.63 | 1,674.21 | 1,667.71 | 1,666.75 | 6,819.30 |
| 898 | 465.58 | 465.2 | 401.77 | 410.88 | 1,743.43 |
| TOTAL | 62,634.77 | 60,794.33 | 58,562.47 | 55,522.80 | 237,514.37 |

STATE: NEW HAMPSHIRE

| ZIP CODE | QUARTER 1 | QUARTER 2 | QUARTER 3 | QUARTER 4 | TOTAL GRAMS |
|---|---|---|---|---|---|
| 30 | 4,710.48 | 4,597.81 | 4,595.49 | 4,651.82 | 18,555.60 |
| 31 | 2,753.72 | 2,694.15 | 2,598.74 | 2,782.96 | 10,829.57 |
| 32 | 1,964.32 | 1,897.14 | 1,847.82 | 1,834.67 | 7,543.95 |
| 33 | 1,950.08 | 1,869.13 | 1,916.42 | 1,897.06 | 7,632.69 |

| | | | | | | |
|---|---|---|---|---|---|---|
| | 34 | 1,067.14 | 945 | 930.64 | 998.49 | 3,941.27 |
| | 35 | 731.87 | 700.34 | 641.25 | 632.25 | 2,705.71 |
| | 36 | 136.86 | 177.47 | 155.06 | 205.85 | 675.24 |
| | 37 | 1,086.71 | 1,014.67 | 1,032.15 | 1,074.18 | 4,207.71 |
| | 38 | 4,181.73 | 3,945.97 | 4,184.29 | 4,073.73 | 16,385.72 |
| TOTAL | | 18,582.91 | 17,841.68 | 17,901.86 | 18,151.01 | 72,477.46 |

STATE: NEW JERSEY

| ZIP CODE | | QUARTER 1 | QUARTER 2 | QUARTER 3 | QUARTER 4 | TOTAL GRAMS |
|---|---|---|---|---|---|---|
| | 70 | 13,467.31 | 11,266.54 | 10,555.02 | 10,186.11 | 45,474.98 |
| | 71 | 1,842.55 | 1,612.15 | 1,524.95 | 1,624.05 | 6,603.70 |
| | 72 | 751.66 | 652.5 | 621.27 | 554.98 | 2,580.41 |
| | 73 | 1,037.49 | 1,138.23 | 1,056.88 | 905.63 | 4,138.23 |
| | 74 | 5,168.37 | 4,874.83 | 4,575.95 | 4,920.53 | 19,539.68 |
| | 75 | 935.17 | 756.21 | 771.92 | 710.45 | 3,173.75 |
| | 76 | 6,606.63 | 5,712.04 | 5,573.98 | 5,487.99 | 23,380.64 |
| | 77 | 6,274.32 | 6,072.04 | 5,566.72 | 6,047.04 | 23,960.12 |
| | 78 | 3,376.13 | 3,084.22 | 2,709.05 | 2,620.69 | 11,790.09 |
| | 79 | 2,424.64 | 1,902.40 | 2,028.63 | 2,144.07 | 8,499.74 |
| | 80 | 21,015.28 | 19,916.83 | 18,519.36 | 17,280.63 | 76,732.10 |
| | 81 | 1,716 | 1,498.08 | 1,531.19 | 1,570.48 | 6,315.75 |
| | 82 | 3,716.60 | 3,689.32 | 3,357.87 | 3,104.93 | 13,868.72 |
| | 83 | 2,241.25 | 2,088.27 | 2,215.21 | 2,090.68 | 8,635.41 |
| | 84 | 418.68 | 452.3 | 415.7 | 420.23 | 1,706.91 |
| | 85 | 2,678.05 | 2,511.50 | 2,162.91 | 1,901.41 | 9,253.87 |
| | 86 | 2,550.95 | 2,334.04 | 2,312.91 | 2,482.45 | 9,680.35 |
| | 87 | 5,876.81 | 5,895.21 | 5,765.23 | 5,431.29 | 22,968.54 |
| | 88 | 10,179.19 | 10,135.43 | 9,390.18 | 9,483.69 | 39,188.49 |
| | 89 | 813.7 | 616.19 | 665.8 | 654.73 | 2,750.42 |
| TOTAL | | 93,090.78 | 86,208.33 | 81,320.73 | 79,622.06 | 340,241.90 |

STATE: NEW MEXICO

| ZIP CODE | | QUARTER 1 | QUARTER 2 | QUARTER 3 | QUARTER 4 | TOTAL GRAMS |
|---|---|---|---|---|---|---|
| | 870 | 2,229.12 | 1,996.98 | 1,830.10 | 1,835.45 | 7,891.65 |
| | 871 | 13,629.86 | 12,466.40 | 11,940.91 | 10,612.92 | 48,650.09 |

| | | | | | | |
|---|---|---|---|---|---|---|
| | 873 | 1,056.03 | 975.8 | 858.91 | 915.4 | 3,806.14 |
| | 874 | 1,070.26 | 991 | 931.12 | 834.59 | 3,826.97 |
| | 875 | 3,078.79 | 2,855.88 | 2,905.19 | 2,643.96 | 11,483.82 |
| | 877 | 381.84 | 378.75 | 406.6 | 381.78 | 1,548.97 |
| | 878 | 128.44 | 122.39 | 113.39 | 89.1 | 453.32 |
| | 879 | 231.93 | 227.59 | 254.25 | 266.64 | 980.41 |
| | 880 | 2,735.27 | 2,277.94 | 2,333.43 | 2,189.04 | 9,535.68 |
| | 881 | 701.29 | 576.16 | 559.9 | 485.36 | 2,322.71 |
| | 882 | 2,605.33 | 2,362.40 | 2,446.33 | 2,319.61 | 9,733.67 |
| | 883 | 630.6 | 663.94 | 669.36 | 555.13 | 2,519.03 |
| | 884 | 100.21 | 98.19 | 107.13 | 95.04 | 400.57 |
| TOTAL | | 28,578.97 | 25,993.42 | 25,356.62 | 23,224.02 | 103,153.03 |

| STATE: | NEW YORK | | | | | |
|---|---|---|---|---|---|---|
| ZIP CODE | | QUARTER 1 | QUARTER 2 | QUARTER 3 | QUARTER 4 | TOTAL GRAMS |
| | 63 | 0 | 0.03 | 0 | 0 | 0.03 |
| | 100 | 13,616.40 | 12,937.31 | 12,632.55 | 12,544.28 | 51,730.54 |
| | 101 | 188.7 | 234.46 | 182.13 | 179.27 | 784.56 |
| | 102 | 6.77 | 6.77 | 5.64 | 2.26 | 21.44 |
| | 103 | 6,813.91 | 6,466.89 | 5,808.94 | 5,624.06 | 24,713.80 |
| | 104 | 7,043 | 6,314.03 | 6,576.06 | 6,687.82 | 26,620.91 |
| | 105 | 2,529.11 | 2,605.13 | 2,350.82 | 2,669.36 | 10,154.42 |
| | 106 | 370.3 | 328.51 | 355.91 | 511.71 | 1,566.43 |
| | 107 | 920.63 | 934.45 | 852.32 | 888.52 | 3,595.92 |
| | 108 | 363.58 | 348.34 | 364.43 | 259.65 | 1,336 |
| | 109 | 3,649.90 | 3,097.10 | 3,095.37 | 3,076.06 | 12,918.43 |
| | 110 | 3,981.08 | 4,080.31 | 3,431.70 | 3,894.57 | 15,387.66 |
| | 111 | 853.77 | 871.75 | 741.62 | 722.18 | 3,189.32 |
| | 112 | 9,054.29 | 8,755.67 | 8,233.89 | 8,015.14 | 34,058.99 |
| | 113 | 3,631.55 | 3,108.64 | 3,105.08 | 2,858.86 | 12,704.13 |
| | 114 | 2,053.32 | 1,810.81 | 1,884.87 | 1,637.18 | 7,386.18 |
| | 115 | 4,947.47 | 5,152.59 | 5,157.24 | 4,971.40 | 20,228.70 |
| | 116 | 554.13 | 547.28 | 551.75 | 480.91 | 2,134.07 |
| | 117 | 19,670.12 | 18,380.83 | 18,036.93 | 17,473.38 | 73,561.26 |
| | 118 | 776.96 | 624.6 | 506.75 | 463.25 | 2,371.56 |
| | 119 | 1,892.93 | 1,830.21 | 1,765.74 | 2,115.28 | 7,604.16 |

| | | | | | |
|---|---|---|---|---|---|
| 120 | 3,671.88 | 3,674.77 | 3,574.13 | 3,634.69 | 14,555.47 |
| 121 | 1,867.71 | 1,720.02 | 1,670.59 | 1,565.73 | 6,824.05 |
| 122 | 3,153.45 | 3,068.39 | 2,612.95 | 2,555.80 | 11,390.59 |
| 123 | 1,000.98 | 853.08 | 778.08 | 912.33 | 3,544.47 |
| 124 | 2,626.16 | 2,388.88 | 2,337.51 | 2,436.79 | 9,789.34 |
| 125 | 3,561.40 | 3,482.28 | 3,315.74 | 3,503.64 | 13,863.06 |
| 126 | 790.51 | 932.78 | 835.39 | 737.3 | 3,295.98 |
| 127 | 1,841.88 | 1,727.38 | 1,587.98 | 1,248.25 | 6,405.49 |
| 128 | 3,105.44 | 2,746.92 | 2,896.45 | 2,810.79 | 11,559.60 |
| 129 | 2,313.39 | 2,297.37 | 2,263.51 | 2,280.60 | 9,154.87 |
| 130 | 4,159.98 | 4,263.78 | 4,178.66 | 4,219.30 | 16,821.72 |
| 131 | 1,445.23 | 1,433.03 | 1,309.25 | 1,435.04 | 5,622.55 |
| 132 | 4,285.62 | 4,282.34 | 3,777.38 | 3,840.87 | 16,186.21 |
| 133 | 1,537.57 | 1,571.52 | 1,259.73 | 1,260.27 | 5,629.09 |
| 134 | 1,899.24 | 1,934.21 | 1,620.86 | 1,911.17 | 7,365.48 |
| 135 | 831.49 | 709.92 | 732.49 | 689.3 | 2,963.20 |
| 136 | 2,516.45 | 2,553.39 | 2,339.77 | 2,204.75 | 9,614.36 |
| 137 | 1,004.89 | 1,033.17 | 908.1 | 1,031.43 | 3,977.59 |
| 138 | 3,146.20 | 3,167.33 | 3,022.21 | 3,168.26 | 12,504 |
| 139 | 813.71 | 780.36 | 750.6 | 876.08 | 3,220.75 |
| 140 | 5,153.34 | 5,123.84 | 5,005.90 | 4,929.81 | 20,212.89 |
| 141 | 2,425.34 | 2,364.81 | 2,326.08 | 2,206.95 | 9,323.18 |
| 142 | 6,791.82 | 6,496.12 | 6,551.67 | 6,326.72 | 26,166.33 |
| 143 | 1,060.37 | 919.05 | 862.43 | 871.28 | 3,713.13 |
| 144 | 3,399.24 | 3,223.28 | 3,051.83 | 2,943.22 | 12,617.57 |
| 145 | 2,881.93 | 2,748.70 | 2,617.61 | 2,387.12 | 10,635.36 |
| 146 | 6,392.88 | 5,874.73 | 5,853.08 | 5,737.43 | 23,858.12 |
| 147 | 2,241.06 | 2,172.21 | 2,220.39 | 2,157.16 | 8,790.82 |
| 148 | 3,959.14 | 3,792.69 | 4,032.58 | 3,904 | 15,688.41 |
| 149 | 866.26 | 823.01 | 798.74 | 907.57 | 3,395.58 |
| TOTAL | 163,662.48 | 156,595.07 | 150,731.43 | 149,768.79 | 620,757.77 |

STATE: NORTH CAROLINA

| ZIP CODE | QUARTER 1 | QUARTER 2 | QUARTER 3 | QUARTER 4 | TOTAL GRAMS |
|---|---|---|---|---|---|
| 270 | 4,445.92 | 3,580.23 | 3,718.77 | 3,584.54 | 15,329.46 |
| 271 | 4,239.95 | 3,675.98 | 3,390.51 | 3,205.27 | 14,511.71 |

| | | | | | |
|---|---|---|---|---|---|
| 272 | 8,309.99 | 6,955.88 | 6,938.56 | 6,439.15 | 28,643.58 |
| 273 | 4,665.06 | 4,466.54 | 4,227.28 | 4,412.05 | 17,770.93 |
| 274 | 2,996.69 | 2,800.01 | 3,035.34 | 2,973.13 | 11,805.17 |
| 275 | 11,087.16 | 10,742.11 | 10,447.90 | 10,586.08 | 42,863.25 |
| 276 | 4,091.66 | 3,751.43 | 3,711.93 | 3,471.36 | 15,026.38 |
| 277 | 5,664.24 | 6,023.69 | 5,302.35 | 5,400.88 | 22,391.16 |
| 278 | 7,201.14 | 6,795.63 | 6,941.72 | 6,711.64 | 27,650.13 |
| 279 | 2,463.33 | 2,432.16 | 2,228.05 | 2,110.03 | 9,233.57 |
| 280 | 9,639.18 | 9,367.39 | 8,787.89 | 8,664.95 | 36,459.41 |
| 281 | 7,938.23 | 7,813.64 | 7,435.49 | 7,526.41 | 30,713.77 |
| 282 | 5,949.73 | 5,494.37 | 5,443.15 | 5,281.63 | 22,168.88 |
| 283 | 9,221.91 | 8,468.29 | 8,759.74 | 7,673.90 | 34,123.84 |
| 284 | 7,565.64 | 7,512.71 | 6,876.97 | 6,814.05 | 28,769.37 |
| 285 | 6,953.11 | 6,635.54 | 6,517.20 | 6,260.39 | 26,366.24 |
| 286 | 11,329.07 | 11,200.42 | 11,058.55 | 10,549.73 | 44,137.77 |
| 287 | 8,264.10 | 7,784.48 | 7,428.80 | 7,127.95 | 30,605.33 |
| 288 | 3,870.79 | 3,712.17 | 3,439.19 | 3,240.48 | 14,262.63 |
| 289 | 777.78 | 835.19 | 776.33 | 918.15 | 3,307.45 |
| TOTAL | 126,674.68 | 120,047.86 | 116,465.72 | 112,951.77 | 476,140.03 |

STATE: NORTH DAKOTA

| ZIP CODE | QUARTER 1 | QUARTER 2 | QUARTER 3 | QUARTER 4 | TOTAL GRAMS |
|---|---|---|---|---|---|
| 580 | 477.07 | 358.42 | 331.28 | 309.57 | 1,476.34 |
| 581 | 1,520.55 | 1,507.54 | 1,422.25 | 1,238.30 | 5,688.64 |
| 582 | 1,014.08 | 921.47 | 902.6 | 877.76 | 3,715.91 |
| 583 | 370.16 | 366.55 | 343.72 | 303.02 | 1,383.45 |
| 584 | 358.85 | 346.51 | 303.49 | 324.88 | 1,333.73 |
| 585 | 2,099.74 | 2,046.90 | 1,870.14 | 2,098.58 | 8,115.36 |
| 586 | 245.38 | 232.79 | 249.69 | 237.96 | 965.82 |
| 587 | 354.34 | 402.76 | 284.67 | 475.93 | 1,517.70 |
| 588 | 246.23 | 238.68 | 243.72 | 206.25 | 934.88 |
| TOTAL | 6,686.40 | 6,421.62 | 5,951.56 | 6,072.25 | 25,131.83 |

STATE: OHIO

| ZIP CODE | QUARTER 1 | QUARTER 2 | QUARTER 3 | QUARTER 4 | TOTAL GRAMS |
|---|---|---|---|---|---|
| 430 | 7,127.95 | 7,034.01 | 6,464.67 | 6,716.43 | 27,343.06 |
| 431 | 4,074.31 | 4,103.57 | 3,836.28 | 3,745.74 | 15,759.90 |
| 432 | 14,366.67 | 13,559.23 | 12,860.81 | 12,308.79 | 53,095.50 |
| 433 | 1,325.01 | 1,367.96 | 1,410.63 | 1,315.68 | 5,419.28 |
| 434 | 1,194.30 | 1,125.83 | 1,275.84 | 1,183.27 | 4,779.24 |
| 435 | 3,329.65 | 2,889.54 | 3,299.64 | 3,304.92 | 12,823.75 |
| 436 | 5,476.96 | 5,396.84 | 5,416.98 | 5,146.33 | 21,437.11 |
| 437 | 2,133.52 | 2,149.85 | 2,199.35 | 1,938.84 | 8,421.56 |
| 438 | 385.02 | 397.81 | 340.79 | 282.41 | 1,406.03 |
| 439 | 2,351.35 | 2,261.57 | 2,320.88 | 2,188.37 | 9,122.17 |
| 440 | 5,932.81 | 5,966.13 | 5,933.46 | 5,269.15 | 23,101.55 |
| 441 | 9,310.26 | 8,630.81 | 8,592.26 | 7,894.42 | 34,427.75 |
| 442 | 5,665.53 | 5,441.51 | 5,222.48 | 4,956.94 | 21,286.46 |
| 443 | 2,727.17 | 2,726.33 | 2,547.58 | 2,558.05 | 10,559.13 |
| 444 | 4,109.54 | 3,867.39 | 3,639.05 | 3,611.28 | 15,227.26 |
| 445 | 2,702.36 | 2,632.46 | 2,472.76 | 2,260.62 | 10,068.20 |
| 446 | 3,684.05 | 3,613.97 | 3,619.55 | 3,298.49 | 14,216.06 |
| 447 | 2,610.40 | 2,403.22 | 2,568.32 | 2,477.47 | 10,059.41 |
| 448 | 2,771.58 | 2,587.73 | 2,683.68 | 2,652.47 | 10,695.46 |
| 449 | 728.74 | 832.17 | 692 | 825.1 | 3,078.01 |
| 450 | 9,982.38 | 9,401.92 | 9,226.88 | 8,337.78 | 36,948.96 |
| 451 | 2,902.77 | 2,938.52 | 3,178.31 | 3,016.16 | 12,035.76 |
| 452 | 9,230.48 | 9,094.45 | 8,741.22 | 8,632.98 | 35,699.13 |
| 453 | 4,591.17 | 4,660.42 | 4,515.75 | 4,396.61 | 18,163.95 |
| 454 | 6,070.66 | 5,725.47 | 5,571.64 | 5,274.30 | 22,642.07 |
| 455 | 991.49 | 937.41 | 979.93 | 900.53 | 3,809.36 |
| 456 | 5,550.28 | 5,554.64 | 5,494.90 | 5,155.63 | 21,755.45 |
| 457 | 1,242.34 | 1,251.40 | 1,185.85 | 1,264.48 | 4,944.07 |
| 458 | 2,390.53 | 2,495.58 | 2,208.76 | 2,301.55 | 9,396.42 |
| TOTAL | 124,959.28 | 121,047.74 | 118,500.25 | 113,214.79 | 477,722.06 |

STATE: OKLAHOMA

| ZIP CODE | QUARTER 1 | QUARTER 2 | QUARTER 3 | QUARTER 4 | TOTAL GRAMS |
|---|---|---|---|---|---|
| 730 | 17,235.74 | 16,392.74 | 15,277.57 | 14,798.15 | 63,704.20 |

| | | | | | | |
|---|---|---|---|---|---|---|
| | 731 | 20,942.62 | 20,073.42 | 19,314.73 | 18,992.60 | 79,323.37 |
| | 734 | 2,369.94 | 2,336.35 | 2,283.66 | 2,012.55 | 9,002.50 |
| | 735 | 6,120.95 | 6,196.38 | 5,485.66 | 4,977.48 | 22,780.47 |
| | 736 | 1,709.45 | 1,393.87 | 1,069.69 | 952.66 | 5,125.67 |
| | 737 | 1,563.66 | 1,536.14 | 1,333.26 | 1,320.32 | 5,753.38 |
| | 738 | 468.07 | 370.91 | 326.66 | 281.05 | 1,446.69 |
| | 739 | 96.33 | 76.83 | 100.8 | 78.15 | 352.11 |
| | 740 | 17,481.89 | 15,879.10 | 16,375.39 | 15,779.90 | 65,516.28 |
| | 741 | 10,575.96 | 9,669.08 | 9,872.31 | 9,462.33 | 39,579.68 |
| | 743 | 2,386 | 2,249.23 | 2,285.01 | 2,218.89 | 9,139.13 |
| | 744 | 5,871.74 | 6,090.54 | 6,158.58 | 5,293.87 | 23,414.73 |
| | 745 | 1,865.04 | 1,823.99 | 1,779.29 | 1,660.24 | 7,128.56 |
| | 746 | 1,056.75 | 1,142.20 | 1,063.91 | 1,016.76 | 4,279.62 |
| | 747 | 1,500.61 | 1,525.65 | 1,071.18 | 1,028.67 | 5,126.11 |
| | 748 | 4,044.07 | 4,042.30 | 3,492.71 | 3,490.35 | 15,069.43 |
| | 749 | 1,694.16 | 1,793.66 | 1,835.82 | 1,683.31 | 7,006.95 |
| TOTAL | | 96,982.98 | 92,592.39 | 89,126.23 | 85,047.28 | 363,748.88 |

STATE: OREGON

| ZIP CODE | | QUARTER 1 | QUARTER 2 | QUARTER 3 | QUARTER 4 | TOTAL GRAMS |
|---|---|---|---|---|---|---|
| | 970 | 12,202.73 | 11,303.25 | 11,048.23 | 10,343.37 | 44,897.58 |
| | 971 | 6,198.12 | 5,720.45 | 5,753.47 | 5,535.70 | 23,207.74 |
| | 972 | 18,411.24 | 17,025.82 | 16,205.71 | 15,701.90 | 67,344.67 |
| | 973 | 9,268.99 | 8,693.51 | 8,100.97 | 7,773.70 | 33,837.17 |
| | 974 | 15,254.40 | 14,575.34 | 13,468.40 | 13,048.58 | 56,346.72 |
| | 975 | 7,843.50 | 7,240.49 | 6,759.54 | 6,374.90 | 28,218.43 |
| | 976 | 1,462.17 | 1,246.94 | 1,255.37 | 1,269.92 | 5,234.40 |
| | 977 | 3,029.32 | 3,050.64 | 3,185.02 | 3,283.20 | 12,548.18 |
| | 978 | 2,913.91 | 2,749.76 | 2,614.22 | 2,390.59 | 10,668.48 |
| | 979 | 649.64 | 471.52 | 451.91 | 442.6 | 2,015.67 |
| TOTAL | | 77,234.02 | 72,077.72 | 68,842.84 | 66,164.46 | 284,319.04 |

STATE: PENNSYLVANIA

| ZIP CODE | | QUARTER 1 | QUARTER 2 | QUARTER 3 | QUARTER 4 | TOTAL GRAMS |
|---|---|---|---|---|---|---|
| | 150 | 12,447.91 | 12,439.22 | 12,417.48 | 11,547 | 48,851.61 |

| | | | | | |
|---|---|---|---|---|---|
| 151 | 3,125.44 | 3,167.01 | 3,189.04 | 3,040.39 | 12,521.88 |
| 152 | 10,318.72 | 9,759.32 | 9,282.72 | 9,046.01 | 38,406.77 |
| 153 | 2,456.65 | 2,350.31 | 2,516.89 | 2,312.55 | 9,636.40 |
| 154 | 2,381.88 | 2,231.03 | 2,140.05 | 2,116.70 | 8,869.66 |
| 155 | 1,493.26 | 1,523.54 | 1,649.93 | 1,307.77 | 5,974.50 |
| 156 | 5,557.68 | 5,267.46 | 5,533.76 | 5,000.46 | 21,359.36 |
| 157 | 2,926.17 | 2,668.65 | 2,892.37 | 2,906.21 | 11,393.40 |
| 158 | 2,095.86 | 1,959.68 | 2,012.04 | 1,934.76 | 8,002.34 |
| 159 | 3,035.57 | 2,733.82 | 2,871.23 | 2,635.10 | 11,275.72 |
| 160 | 3,271.12 | 3,139.04 | 3,130.73 | 3,019.85 | 12,560.74 |
| 161 | 4,549.22 | 4,580.96 | 4,451.07 | 4,353.97 | 17,935.22 |
| 162 | 1,519.78 | 1,429.26 | 1,309.96 | 1,409 | 5,668 |
| 163 | 4,816.17 | 4,811.19 | 4,516.53 | 4,606.46 | 18,750.35 |
| 164 | 1,517.42 | 1,526.53 | 1,554.94 | 1,489.84 | 6,088.73 |
| 165 | 4,608.94 | 4,459.91 | 4,343.12 | 3,944.21 | 17,356.18 |
| 166 | 4,835 | 4,711.45 | 4,473.73 | 4,219.62 | 18,239.80 |
| 167 | 2,049.26 | 2,121.42 | 2,085.60 | 1,877.37 | 8,133.65 |
| 168 | 1,299.57 | 1,269.63 | 1,378.73 | 1,175.69 | 5,123.62 |
| 169 | 816.27 | 790.5 | 709.83 | 740.23 | 3,056.83 |
| 170 | 12,510.90 | 11,770.98 | 11,242.12 | 11,365.27 | 46,889.27 |
| 171 | 2,816.79 | 2,796.03 | 2,456.36 | 2,411.98 | 10,481.16 |
| 172 | 2,677.39 | 2,510.31 | 2,385.96 | 2,556.91 | 10,130.57 |
| 173 | 4,662.26 | 4,565.13 | 4,224.05 | 4,358.92 | 17,810.36 |
| 174 | 3,172.78 | 2,755.17 | 2,921.02 | 2,751.49 | 11,600.46 |
| 175 | 3,945.12 | 3,754.36 | 3,608.10 | 3,559.20 | 14,866.78 |
| 176 | 3,929.77 | 3,308.81 | 3,191.43 | 3,431.08 | 13,861.09 |
| 177 | 2,231.90 | 2,116.76 | 2,155.51 | 2,069.80 | 8,573.97 |
| 178 | 3,349.48 | 3,513.39 | 3,132.20 | 3,130.93 | 13,126 |
| 179 | 1,593.49 | 1,513.49 | 1,410.67 | 1,307.48 | 5,825.13 |
| 180 | 4,593.61 | 4,510.79 | 4,332.77 | 4,156.21 | 17,593.38 |
| 181 | 4,506.10 | 4,027.10 | 3,626.15 | 3,766.94 | 15,926.29 |
| 182 | 2,409.50 | 2,196.24 | 1,848.34 | 2,025.79 | 8,479.87 |
| 183 | 2,861.23 | 2,451.06 | 2,481.82 | 2,502.85 | 10,296.96 |
| 184 | 3,218.77 | 3,171.34 | 3,141.68 | 2,948.85 | 12,480.64 |
| 185 | 4,056.42 | 3,937.81 | 3,635.40 | 3,199.90 | 14,829.53 |
| 186 | 2,746.42 | 2,625.93 | 2,589.11 | 2,405.34 | 10,366.80 |
| 187 | 10,656.35 | 9,601.92 | 8,829 | 9,043.97 | 38,131.24 |

| | | | | | | |
|---|---|---|---|---|---|---|
| | 188 | 1,022.82 | 939.11 | 890.11 | 944.16 | 3,796.20 |
| | 189 | 4,082.87 | 4,164.90 | 4,135.58 | 4,118.10 | 16,501.45 |
| | 190 | 70,396.01 | 44,439.27 | 42,353.42 | 43,523.03 | 200,711.73 |
| | 191 | 17,181.26 | 16,987.51 | 15,949.18 | 14,957.02 | 65,074.97 |
| | 193 | 6,089.08 | 5,918.47 | 5,432.69 | 5,258.88 | 22,699.12 |
| | 194 | 9,685.86 | 9,400.86 | 9,004.22 | 9,110.66 | 37,201.60 |
| | 195 | 1,764.61 | 1,771 | 1,631.28 | 1,618.91 | 6,785.80 |
| | 196 | 2,335.52 | 2,210.62 | 2,249.47 | 2,189.39 | 8,985 |
| TOTAL | | 263,618.20 | 229,898.29 | 221,317.39 | 217,396.25 | 932,230.13 |

STATE: PUERTO RICO

| ZIP CODE | | QUARTER 1 | QUARTER 2 | QUARTER 3 | QUARTER 4 | TOTAL GRAMS |
|---|---|---|---|---|---|---|
| | 006 | 600.31 | 476.31 | 336.96 | 211.71 | 1,625.29 |
| | 007 | 554.52 | 498.21 | 379.52 | 323.53 | 1,755.78 |
| | 009 | 1,433.64 | 1,266.76 | 1,148.94 | 1,169.34 | 5,018.68 |
| TOTAL | | 2,588.47 | 2,241.28 | 1,865.42 | 1,704.58 | 8,399.75 |

STATE: RHODE ISLAND

| ZIP CODE | | QUARTER 1 | QUARTER 2 | QUARTER 3 | QUARTER 4 | TOTAL GRAMS |
|---|---|---|---|---|---|---|
| | 28 | 7,506.91 | 7,560.77 | 7,617.95 | 6,893.52 | 29,579.15 |
| | 29 | 4,563.02 | 4,129.10 | 4,404.60 | 3,952.64 | 17,049.36 |
| TOTAL | | 12,069.93 | 11,689.87 | 12,022.55 | 10,846.16 | 46,628.51 |

STATE: SOUTH CAROLINA

| ZIP CODE | | QUARTER 1 | QUARTER 2 | QUARTER 3 | QUARTER 4 | TOTAL GRAMS |
|---|---|---|---|---|---|---|
| | 290 | 5,781.25 | 5,351.58 | 5,359.14 | 5,369.54 | 21,861.51 |
| | 291 | 4,701.48 | 4,591.87 | 4,680.92 | 4,751.28 | 18,725.55 |
| | 292 | 5,377.13 | 5,269.33 | 5,036.19 | 4,800.47 | 20,483.12 |
| | 293 | 6,491.23 | 6,171.77 | 5,840.98 | 6,080.76 | 24,584.74 |
| | 294 | 13,784.97 | 12,639.51 | 12,433.86 | 12,494.56 | 51,352.90 |
| | 295 | 9,963.45 | 9,545.18 | 9,919.70 | 9,602.27 | 39,030.60 |
| | 296 | 14,805.11 | 13,702.48 | 13,156.17 | 13,352.99 | 55,016.75 |
| | 297 | 2,856.81 | 2,803.12 | 2,494.53 | 2,706.85 | 10,861.31 |
| | 298 | 2,133.54 | 2,073.86 | 2,151.44 | 2,122.38 | 8,481.22 |

| | | | | | |
|---|---|---|---|---|---|
| | 299 | 2,831.83 | 2,654.44 | 2,810.18 | 2,751.89 | 11,048.34 |
| TOTAL | | 68,726.80 | 64,803.14 | 63,883.11 | 64,032.99 | 261,446.04 |

STATE: SOUTH DAKOTA

| ZIP CODE | | QUARTER 1 | QUARTER 2 | QUARTER 3 | QUARTER 4 | TOTAL GRAMS |
|---|---|---|---|---|---|---|
| | 570 | 1,449.32 | 1,501.47 | 1,361.52 | 1,323 | 5,635.31 |
| | 571 | 3,126.60 | 2,988.51 | 2,899.56 | 2,880.58 | 11,895.25 |
| | 572 | 437.31 | 353.16 | 414.3 | 361.81 | 1,566.58 |
| | 573 | 453.07 | 465.36 | 444.03 | 440.08 | 1,802.54 |
| | 574 | 443.66 | 464.62 | 455.33 | 452.91 | 1,816.52 |
| | 575 | 529 | 525.5 | 493.96 | 463.79 | 2,012.25 |
| | 576 | 63.24 | 54.99 | 68.9 | 114.28 | 301.41 |
| | 577 | 2,602.91 | 2,391.35 | 2,538.40 | 2,238.54 | 9,771.20 |
| TOTAL | | 9,105.11 | 8,744.96 | 8,676 | 8,274.99 | 34,801.06 |

STATE: TENNESSEE

| ZIP CODE | | QUARTER 1 | QUARTER 2 | QUARTER 3 | QUARTER 4 | TOTAL GRAMS |
|---|---|---|---|---|---|---|
| | 370 | 19,042.14 | 18,276.75 | 17,669.22 | 16,258.52 | 71,246.63 |
| | 371 | 18,054.71 | 17,363.53 | 15,936.06 | 15,298.49 | 66,652.79 |
| | 372 | 10,484.43 | 9,961.24 | 9,402.92 | 9,153.50 | 39,002.09 |
| | 373 | 13,951.34 | 12,816.93 | 12,243.84 | 11,276.38 | 50,288.49 |
| | 374 | 4,879.80 | 4,693.90 | 4,154.51 | 4,180.79 | 17,909 |
| | 376 | 10,657.22 | 9,783.13 | 9,101.16 | 9,086.23 | 38,627.74 |
| | 377 | 8,368.06 | 8,063.48 | 8,145.82 | 8,135.56 | 32,712.92 |
| | 378 | 13,558.83 | 12,409.19 | 12,148.86 | 11,440.79 | 49,557.67 |
| | 379 | 8,748.22 | 7,954.01 | 7,607.27 | 7,943.75 | 32,253.25 |
| | 380 | 5,009.62 | 4,656.88 | 4,527.02 | 4,392.67 | 18,586.19 |
| | 381 | 47,822.96 | 28,703.92 | 31,943.81 | 30,975.40 | 139,446.09 |
| | 382 | 3,160.55 | 3,061.87 | 3,037.74 | 2,920.03 | 12,180.19 |
| | 383 | 7,963.42 | 7,553.60 | 7,305.21 | 6,629.42 | 29,451.65 |
| | 384 | 4,859.43 | 4,686.64 | 4,208.87 | 3,983.99 | 17,738.93 |
| | 385 | 9,163.37 | 9,097.75 | 8,262.96 | 7,845.21 | 34,369.29 |
| TOTAL | | 185,724.10 | 159,082.82 | 155,695.27 | 149,520.73 | 650,022.92 |

STATE: TEXAS

| ZIP CODE | QUARTER 1 | QUARTER 2 | QUARTER 3 | QUARTER 4 | TOTAL GRAMS |
|---|---|---|---|---|---|
| 750 | 18,438.42 | 17,632.79 | 16,894.20 | 15,880.02 | 68,845.43 |
| 751 | 5,271.46 | 5,262.56 | 5,379.93 | 5,360.24 | 21,274.19 |
| 752 | 8,658.89 | 8,198.85 | 7,273.91 | 8,037.59 | 32,169.24 |
| 753 | 198.81 | 176.94 | 213.45 | 215.31 | 804.51 |
| 754 | 5,372.34 | 4,716.14 | 3,992.07 | 3,806.91 | 17,887.46 |
| 755 | 1,239.54 | 1,273.81 | 1,254.11 | 1,165.76 | 4,933.22 |
| 756 | 3,648.39 | 3,421.81 | 3,647.53 | 3,678.86 | 14,396.59 |
| 757 | 3,599.52 | 3,627.09 | 3,564.03 | 3,497.91 | 14,288.55 |
| 758 | 794.19 | 834.57 | 900.83 | 781.04 | 3,310.63 |
| 759 | 2,337.04 | 2,598.65 | 2,416.81 | 2,501.03 | 9,853.53 |
| 760 | 12,376.69 | 11,809.52 | 11,944.51 | 11,770.14 | 47,900.86 |
| 761 | 13,402.45 | 12,224.12 | 13,084.61 | 12,445.30 | 51,156.48 |
| 762 | 4,357.05 | 4,559.79 | 4,459.36 | 4,106.58 | 17,482.78 |
| 763 | 3,094.83 | 2,921.43 | 2,798.09 | 2,520.83 | 11,335.18 |
| 764 | 1,449.35 | 1,383.95 | 1,365.26 | 1,280.61 | 5,479.17 |
| 765 | 4,442.74 | 3,875.35 | 4,003.10 | 3,589.63 | 15,910.82 |
| 766 | 969.91 | 921.17 | 1,103.68 | 1,088.84 | 4,083.60 |
| 767 | 3,165.27 | 3,467.78 | 3,184.68 | 3,275.90 | 13,093.63 |
| 768 | 1,311.91 | 1,319.40 | 1,063.54 | 1,139.24 | 4,834.09 |
| 769 | 1,610.36 | 1,506.23 | 1,402.57 | 1,289.82 | 5,808.98 |
| 770 | 13,028.10 | 13,321.46 | 12,527.10 | 12,169.05 | 51,045.71 |
| 773 | 8,567.87 | 8,121.93 | 8,343.41 | 8,388.02 | 33,421.23 |
| 774 | 6,085.44 | 5,876.38 | 5,686.82 | 5,922 | 23,570.64 |
| 775 | 11,122.52 | 11,470.50 | 10,627.64 | 11,430.16 | 44,650.82 |
| 776 | 2,358.84 | 2,548.90 | 2,438.02 | 2,397.92 | 9,743.68 |
| 777 | 1,489.22 | 1,437.27 | 1,391.01 | 1,740.75 | 6,058.25 |
| 778 | 2,145.91 | 2,040.09 | 2,093.93 | 2,069.15 | 8,349.08 |
| 779 | 2,455.36 | 2,504.12 | 2,233.39 | 2,198.50 | 9,391.37 |
| 780 | 2,745.48 | 2,682.71 | 2,639.58 | 2,528.15 | 10,595.92 |
| 781 | 3,263.53 | 2,823.53 | 3,040.12 | 2,860.98 | 11,988.16 |
| 782 | 16,672.17 | 15,903.67 | 15,981.30 | 15,531.88 | 64,089.02 |
| 783 | 1,893.27 | 1,740.40 | 1,699.21 | 1,362.73 | 6,695.61 |
| 784 | 3,274.29 | 2,934.40 | 2,782.92 | 2,963.56 | 11,955.17 |
| 785 | 2,720.24 | 2,580 | 2,628.88 | 2,655.81 | 10,584.93 |

| | | | | | |
|---|---|---|---|---|---|
| 786 | 10,021.53 | 9,608.55 | 9,233.39 | 8,787.84 | 37,651.31 |
| 787 | 8,910.53 | 8,722.53 | 8,055.20 | 7,995.02 | 33,683.28 |
| 788 | 595.41 | 557.93 | 507.19 | 471.52 | 2,132.05 |
| 789 | 545.36 | 506.26 | 463.58 | 510.05 | 2,025.25 |
| 790 | 1,670.73 | 1,552.88 | 1,586.14 | 1,348.76 | 6,158.51 |
| 791 | 4,620.76 | 4,228.31 | 4,400.60 | 3,607.73 | 16,857.40 |
| 792 | 329.86 | 318.1 | 334.05 | 248.48 | 1,230.49 |
| 793 | 855.48 | 730.43 | 822.5 | 740.81 | 3,149.22 |
| 794 | 3,312.32 | 3,286.38 | 3,436.28 | 3,347.32 | 13,382.30 |
| 795 | 845.21 | 814.31 | 673.09 | 681.57 | 3,014.18 |
| 796 | 2,214.53 | 2,252.70 | 2,073.09 | 2,019.92 | 8,560.24 |
| 797 | 3,099.53 | 3,075.76 | 2,960.56 | 2,838.74 | 11,974.59 |
| 798 | 85.2 | 99.99 | 86 | 106.11 | 377.3 |
| 799 | 3,964.11 | 3,472.91 | 3,254.65 | 3,261.09 | 13,952.76 |
| TOTAL | 214,631.96 | 206,944.35 | 201,945.92 | 197,615.18 | 821,137.41 |

STATE: UTAH

| ZIP CODE | QUARTER 1 | QUARTER 2 | QUARTER 3 | QUARTER 4 | TOTAL GRAMS |
|---|---|---|---|---|---|
| 840 | 19,924.87 | 19,305.17 | 19,044.81 | 18,045.81 | 76,320.66 |
| 841 | 14,382.27 | 13,846.15 | 13,912.09 | 13,429.56 | 55,570.07 |
| 843 | 2,104.47 | 2,072.12 | 2,026.22 | 1,700.27 | 7,903.08 |
| 844 | 4,461.20 | 4,337.24 | 4,437.38 | 4,231.60 | 17,467.42 |
| 845 | 1,494.47 | 1,177.27 | 1,123.73 | 1,247.87 | 5,043.34 |
| 846 | 3,409.93 | 3,203.40 | 2,923.33 | 2,960.82 | 12,497.48 |
| 847 | 5,445.76 | 5,458.97 | 5,578.24 | 5,267.77 | 21,750.74 |
| TOTAL | 51,222.97 | 49,400.32 | 49,045.80 | 46,883.70 | 196,552.79 |

STATE: VERMONT

| ZIP CODE | QUARTER 1 | QUARTER 2 | QUARTER 3 | QUARTER 4 | TOTAL GRAMS |
|---|---|---|---|---|---|
| 50 | 1,354.24 | 1,305.89 | 984.97 | 1,130.19 | 4,775.29 |
| 51 | 549.29 | 455.19 | 569.93 | 564.44 | 2,138.85 |
| 52 | 511.19 | 495.74 | 484.99 | 417.59 | 1,909.51 |
| 53 | 659.88 | 659.88 | 665.52 | 710.73 | 2,696.01 |
| 54 | 4,480 | 4,106.84 | 4,204.79 | 4,024.79 | 16,816.42 |
| 56 | 896.44 | 825.89 | 853.71 | 747.99 | 3,324.03 |

| | | | | | | |
|---|---|---|---|---|---|---|
| | 57 | 1,445.13 | 1,520.48 | 1,217.72 | 1,388.21 | 5,571.54 |
| | 58 | 850.04 | 821.2 | 777.17 | 848.5 | 3,296.91 |
| TOTAL | | 10,746.21 | 10,191.11 | 9,758.80 | 9,832.44 | 40,528.56 |

| STATE: | VIRGIN ISLANDS | | | | |
|---|---|---|---|---|---|
| ZIP CODE | QUARTER 1 | QUARTER 2 | QUARTER 3 | QUARTER 4 | TOTAL GRAMS |
| 8 | 139.62 | 152.01 | 119.42 | 113.85 | 524.9 |
| TOTAL | 139.62 | 152.01 | 119.42 | 113.85 | 524.9 |

| STATE: | VIRGINIA | | | | |
|---|---|---|---|---|---|
| ZIP CODE | QUARTER 1 | QUARTER 2 | QUARTER 3 | QUARTER 4 | TOTAL GRAMS |
| 201 | 6,366.14 | 6,674.81 | 5,939.50 | 5,924.56 | 24,905.01 |
| 220 | 3,310.22 | 2,659.15 | 2,706.90 | 2,658.35 | 11,334.62 |
| 221 | 3,148.23 | 2,954.19 | 2,739.39 | 2,916.87 | 11,758.68 |
| 222 | 684.02 | 715.65 | 607.32 | 625.24 | 2,632.23 |
| 223 | 1,351.47 | 1,365.09 | 1,404.24 | 1,447.90 | 5,568.70 |
| 224 | 2,985.89 | 2,647.03 | 2,689.44 | 2,639.02 | 10,961.38 |
| 225 | 799.79 | 969.72 | 903.27 | 862.38 | 3,535.16 |
| 226 | 3,061.63 | 2,912.86 | 2,955.79 | 3,032.31 | 11,962.59 |
| 227 | 581.45 | 489.87 | 439.98 | 519 | 2,030.30 |
| 228 | 2,272.35 | 2,119.35 | 1,866.54 | 1,957.01 | 8,215.25 |
| 229 | 2,641.46 | 2,438.74 | 2,410.38 | 2,331.03 | 9,821.61 |
| 230 | 2,272.12 | 2,049.26 | 2,002.65 | 1,972.45 | 8,296.48 |
| 231 | 3,297.50 | 3,019.17 | 2,678.68 | 3,034.87 | 12,030.22 |
| 232 | 8,702.01 | 8,032.56 | 7,551.38 | 6,779.42 | 31,065.37 |
| 233 | 2,417.73 | 2,403.66 | 2,407.33 | 2,408.02 | 9,636.74 |
| 234 | 5,629.76 | 5,268.29 | 5,390.56 | 5,316.98 | 21,605.59 |
| 235 | 2,192.86 | 2,318.76 | 1,794.30 | 2,198.99 | 8,504.91 |
| 236 | 4,390.54 | 3,490.52 | 3,882.64 | 3,358.48 | 15,122.18 |
| 237 | 2,009.73 | 1,793.98 | 1,823.40 | 1,906.99 | 7,534.10 |
| 238 | 3,573.51 | 3,130.19 | 3,169.23 | 2,921.56 | 12,794.49 |
| 239 | 705.26 | 693.1 | 640.97 | 673.84 | 2,713.17 |
| 240 | 4,259.56 | 3,931.96 | 3,945.56 | 3,884.25 | 16,021.33 |
| 241 | 5,039.05 | 4,744.65 | 4,449.34 | 4,250.59 | 18,483.63 |
| 242 | 4,351.80 | 4,192.70 | 4,132.62 | 3,855.86 | 16,532.98 |

| | | | | | | |
|---|---|---|---|---|---|---|
| | 243 | 2,402.21 | 2,346.05 | 2,239.68 | 1,955.22 | 8,943.16 |
| | 244 | 1,403.44 | 1,293.12 | 1,234.84 | 1,265.37 | 5,196.77 |
| | 245 | 3,446.34 | 3,295.36 | 2,948.07 | 2,643.05 | 12,332.82 |
| | 246 | 1,162.40 | 1,113.33 | 1,137.61 | 915.97 | 4,329.31 |
| TOTAL | | 84,458.47 | 79,063.12 | 76,091.61 | 74,255.58 | 313,868.78 |

| STATE: | WASHINGTON | | | | | |
|---|---|---|---|---|---|---|
| ZIP CODE | | QUARTER 1 | QUARTER 2 | QUARTER 3 | QUARTER 4 | TOTAL GRAMS |
| | 980 | 16,067.26 | 15,160.26 | 15,063.45 | 13,793.83 | 60,084.80 |
| | 981 | 12,077.40 | 11,677.89 | 11,124.39 | 10,437.54 | 45,317.22 |
| | 982 | 13,445.88 | 12,510.22 | 12,020.14 | 11,328.36 | 49,304.60 |
| | 983 | 12,934 | 12,671.59 | 12,059.41 | 11,653.15 | 49,318.15 |
| | 984 | 8,316.48 | 8,009.63 | 7,557.49 | 7,034.92 | 30,918.52 |
| | 985 | 6,857.76 | 6,557.71 | 6,685.11 | 6,249.67 | 26,350.25 |
| | 986 | 14,874.43 | 14,272.62 | 14,067.99 | 13,143.31 | 56,358.35 |
| | 988 | 3,163.04 | 3,131.83 | 2,970.17 | 3,116.90 | 12,381.94 |
| | 989 | 3,976.17 | 3,659.23 | 3,537.08 | 3,329.33 | 14,501.81 |
| | 990 | 1,559.48 | 1,603.99 | 1,623.34 | 1,585.33 | 6,372.14 |
| | 991 | 2,622.49 | 2,719.84 | 2,469.79 | 2,506.03 | 10,318.15 |
| | 992 | 11,458.35 | 11,046.08 | 10,885.25 | 10,342.86 | 43,732.54 |
| | 993 | 6,835.10 | 6,250.95 | 5,707.59 | 5,588.88 | 24,382.52 |
| | 994 | 978.16 | 1,117.06 | 1,111.65 | 1,130.82 | 4,337.69 |
| TOTAL | | 115,166 | 110,388.90 | 106,882.85 | 101,240.93 | 433,678.68 |

| STATE: | WEST VIRGINIA | | | | | |
|---|---|---|---|---|---|---|
| ZIP CODE | | QUARTER 1 | QUARTER 2 | QUARTER 3 | QUARTER 4 | TOTAL GRAMS |
| | 247 | 914.8 | 631.72 | 708.44 | 609.5 | 2,864.46 |
| | 248 | 233.24 | 208.35 | 223.09 | 247.96 | 912.64 |
| | 249 | 547.89 | 449.66 | 449.47 | 382.4 | 1,829.42 |
| | 250 | 615.81 | 550.48 | 640.48 | 695.52 | 2,502.29 |
| | 251 | 404.87 | 730.37 | 304.16 | 316.32 | 1,755.72 |
| | 252 | 511.15 | 489.58 | 525.03 | 550.92 | 2,076.68 |
| | 253 | 1,576.35 | 1,449.40 | 1,452.77 | 1,326.02 | 5,804.54 |
| | 254 | 4,142.38 | 3,899.04 | 4,024.28 | 3,442.03 | 15,507.73 |
| | 255 | 1,000.15 | 1,062.94 | 1,090.76 | 1,062.17 | 4,216.02 |

| | | | | | | |
|---|---|---|---|---|---|---|
| | 256 | 350.25 | 351.67 | 394.77 | 384.03 | 1,480.72 |
| | 257 | 1,627.77 | 1,370.29 | 1,220.52 | 1,245.70 | 5,464.28 |
| | 258 | 1,301.62 | 1,284.80 | 1,205.37 | 1,228.08 | 5,019.87 |
| | 259 | 369.38 | 356.58 | 303.04 | 323.09 | 1,352.09 |
| | 260 | 2,243.97 | 2,144.18 | 2,075.97 | 2,027.37 | 8,491.49 |
| | 261 | 917.58 | 844.69 | 903.85 | 983.56 | 3,649.68 |
| | 262 | 773.6 | 755.88 | 818.61 | 802.67 | 3,150.76 |
| | 263 | 1,962.09 | 1,706.73 | 1,855.34 | 1,611.54 | 7,135.70 |
| | 264 | 454.82 | 457.26 | 411.7 | 418.84 | 1,742.62 |
| | 265 | 1,762.32 | 1,597.13 | 1,553.60 | 1,500.54 | 6,413.59 |
| | 266 | 445.65 | 517.49 | 491.88 | 452.21 | 1,907.23 |
| | 267 | 815.05 | 689.6 | 691.09 | 647.06 | 2,842.80 |
| | 268 | 418.28 | 328.83 | 295.28 | 283.85 | 1,326.24 |
| TOTAL | | 23,389.02 | 21,876.67 | 21,639.50 | 20,541.38 | 87,446.57 |

| STATE: | WISCONSIN | | | | | |
|---|---|---|---|---|---|---|
| ZIP CODE | | QUARTER 1 | QUARTER 2 | QUARTER 3 | QUARTER 4 | TOTAL GRAMS |
| | 530 | 9,113.16 | 8,168.73 | 8,152.01 | 8,130.65 | 33,564.55 |
| | 531 | 9,257.27 | 8,482.98 | 7,920.98 | 8,240.84 | 33,902.07 |
| | 532 | 15,639.95 | 14,617.87 | 13,755.32 | 13,352.53 | 57,365.67 |
| | 534 | 1,676.11 | 1,639.84 | 1,628.76 | 1,568.21 | 6,512.92 |
| | 535 | 8,636.71 | 8,171.29 | 8,164.43 | 7,687.07 | 32,659.50 |
| | 537 | 7,537.16 | 7,026.98 | 6,655.77 | 6,647.56 | 27,867.47 |
| | 538 | 770.99 | 643.98 | 641.14 | 625.82 | 2,681.93 |
| | 539 | 4,372.42 | 3,976.84 | 3,690.11 | 3,693.06 | 15,732.43 |
| | 540 | 1,139.22 | 1,056.18 | 982.04 | 952.45 | 4,129.89 |
| | 541 | 1,661.83 | 1,600.04 | 1,578.97 | 1,323.14 | 6,163.98 |
| | 542 | 1,193.97 | 1,213.51 | 1,115.75 | 1,076.07 | 4,599.30 |
| | 543 | 2,402.07 | 2,076.28 | 2,137.54 | 1,903.04 | 8,518.93 |
| | 544 | 3,851.21 | 3,310.25 | 3,256.49 | 3,168.42 | 13,586.37 |
| | 545 | 1,073.91 | 1,147.61 | 1,052.41 | 848.76 | 4,122.69 |
| | 546 | 3,590.24 | 3,492.01 | 3,537.34 | 3,449.66 | 14,069.25 |
| | 547 | 3,264.96 | 2,721.25 | 2,680.43 | 2,441.55 | 11,108.19 |
| | 548 | 2,374.80 | 2,436.48 | 2,095.61 | 1,982.10 | 8,888.99 |
| | 549 | 6,135.10 | 5,766.73 | 5,600.45 | 5,446.19 | 22,948.47 |
| TOTAL | | 83,691.08 | 77,548.85 | 74,645.55 | 72,537.12 | 308,422.60 |

STATE: WYOMING

| ZIP CODE | QUARTER 1 | QUARTER 2 | QUARTER 3 | QUARTER 4 | TOTAL GRAMS |
|---|---|---|---|---|---|
| 820 | 2,043.76 | 1,899.02 | 1,565.61 | 1,498.31 | 7,006.70 |
| 822 | 255.24 | 315.21 | 260.64 | 256.57 | 1,087.66 |
| 823 | 139.09 | 156.81 | 138.36 | 138.5 | 572.76 |
| 824 | 1,039.50 | 909.25 | 917.08 | 848.71 | 3,714.54 |
| 825 | 828.49 | 731.24 | 685.91 | 678.28 | 2,923.92 |
| 826 | 1,388.49 | 1,299.63 | 1,316.52 | 1,182.09 | 5,186.73 |
| 827 | 362.05 | 439.99 | 316.98 | 375.25 | 1,494.27 |
| 828 | 1,069.10 | 1,205.53 | 1,229.55 | 1,028.59 | 4,532.77 |
| 829 | 1,786.23 | 2,053.30 | 2,176.87 | 1,979.29 | 7,995.69 |
| 830 | 52.92 | 81.1 | 80.78 | 85.57 | 300.37 |
| 831 | 256.45 | 243.9 | 311.8 | 239.93 | 1,052.08 |
| TOTAL | 9,221.32 | 9,334.98 | 9,000.10 | 8,311.09 | 35,867.49 |

DRUG CODE: 9333 DRUG NAME: THEBAINE

STATE: TEXAS

| ZIP CODE | QUARTER 1 | QUARTER 2 | QUARTER 3 | QUARTER 4 | TOTAL GRAMS |
|---|---|---|---|---|---|
| 782 | 0 | 0 | 25 | 0 | 25 |
| TOTAL | 0 | 0 | 25 | 0 | 25 |

DRUG CODE: 9630 DRUG NAME: OPIUM TINCTURE (LAUDANUM)

STATE: CALIFORNIA

| ZIP CODE | QUARTER 1 | QUARTER 2 | QUARTER 3 | QUARTER 4 | TOTAL GRAMS |
|---|---|---|---|---|---|
| 907 | 1.18 | 0 | 0 | 0 | 1.18 |
| 951 | 4.73 | 0 | 0 | 0 | 4.73 |
| TOTAL | 5.91 | 0 | 0 | 0 | 5.91 |

STATE: CONNECTICUT

| ZIP CODE | QUARTER 1 | QUARTER 2 | QUARTER 3 | QUARTER 4 | TOTAL GRAMS |
|---|---|---|---|---|---|
| 64 | 0 | 0 | 4.73 | 0 | 4.73 |
| TOTAL | 0 | 0 | 4.73 | 0 | 4.73 |

STATE: DELAWARE

| ZIP CODE | QUARTER 1 | QUARTER 2 | QUARTER 3 | QUARTER 4 | TOTAL GRAMS |
|---|---|---|---|---|---|
| 197 | 0 | 1.18 | 0 | 0 | 1.18 |
| TOTAL | 0 | 1.18 | 0 | 0 | 1.18 |

STATE: FLORIDA

| ZIP CODE | QUARTER 1 | QUARTER 2 | QUARTER 3 | QUARTER 4 | TOTAL GRAMS |
|---|---|---|---|---|---|
| 321 | 9.46 | 4.73 | 0 | 0 | 14.19 |
| TOTAL | 9.46 | 4.73 | 0 | 0 | 14.19 |

STATE: ILLINOIS

| ZIP CODE | QUARTER 1 | QUARTER 2 | QUARTER 3 | QUARTER 4 | TOTAL GRAMS |
|---|---|---|---|---|---|
| 616 | 4.73 | 4.73 | 0 | 0 | 9.46 |
| 618 | 1.18 | 1.18 | 0 | 0 | 2.36 |
| 626 | 0 | 4.73 | 0 | 0 | 4.73 |
| TOTAL | 5.91 | 10.64 | 0 | 0 | 16.55 |

STATE: INDIANA

| ZIP CODE | QUARTER 1 | QUARTER 2 | QUARTER 3 | QUARTER 4 | TOTAL GRAMS |
|---|---|---|---|---|---|
| 460 | 1.18 | 0 | 0 | 0 | 1.18 |
| 461 | 18.92 | 0 | 0 | 0 | 18.92 |
| 462 | 1.18 | 9.46 | 0 | 0 | 10.64 |
| 463 | 0 | 4.73 | 0 | 0 | 4.73 |
| TOTAL | 21.28 | 14.19 | 0 | 0 | 35.47 |

STATE: KANSAS

| ZIP CODE | QUARTER 1 | QUARTER 2 | QUARTER 3 | QUARTER 4 | TOTAL GRAMS |
|---|---|---|---|---|---|

| | | | | | |
|---|---|---|---|---|---|
| 660 | 4.73 | 0 | 0 | 0 | 4.73 |
| 673 | 1.18 | 0 | 0 | 0 | 1.18 |
| TOTAL | 5.91 | 0 | 0 | 0 | 5.91 |

STATE: KENTUCKY

| ZIP CODE | QUARTER 1 | QUARTER 2 | QUARTER 3 | QUARTER 4 | TOTAL GRAMS |
|---|---|---|---|---|---|
| 410 | 2.36 | 0 | 0 | 0 | 2.36 |
| TOTAL | 2.36 | 0 | 0 | 0 | 2.36 |

STATE: MARYLAND

| ZIP CODE | QUARTER 1 | QUARTER 2 | QUARTER 3 | QUARTER 4 | TOTAL GRAMS |
|---|---|---|---|---|---|
| 212 | 0 | 2.36 | 0 | 0 | 2.36 |
| 214 | 0 | 1.18 | 0 | 0 | 1.18 |
| 217 | 4.72 | 4.73 | 0 | 0 | 9.45 |
| TOTAL | 4.72 | 8.27 | 0 | 0 | 12.99 |

STATE: MICHIGAN

| ZIP CODE | QUARTER 1 | QUARTER 2 | QUARTER 3 | QUARTER 4 | TOTAL GRAMS |
|---|---|---|---|---|---|
| 480 | 9.46 | 0 | 0 | 0 | 9.46 |
| 481 | 7.08 | 4.72 | 0 | 0 | 11.8 |
| 483 | 4.73 | 0 | 0 | 0 | 4.73 |
| 489 | 4.73 | 0 | 0 | 0 | 4.73 |
| TOTAL | 26 | 4.72 | 0 | 0 | 30.72 |

STATE: NEW HAMPSHIRE

| ZIP CODE | QUARTER 1 | QUARTER 2 | QUARTER 3 | QUARTER 4 | TOTAL GRAMS |
|---|---|---|---|---|---|
| 33 | 4.73 | 0 | 0 | 0 | 4.73 |
| TOTAL | 4.73 | 0 | 0 | 0 | 4.73 |

STATE: NEW JERSEY

| ZIP CODE | QUARTER 1 | QUARTER 2 | QUARTER 3 | QUARTER 4 | TOTAL GRAMS |
|---|---|---|---|---|---|
| 74 | 0 | 1.18 | 0 | 0 | 1.18 |
| TOTAL | 0 | 1.18 | 0 | 0 | 1.18 |

STATE: NEW YORK

| ZIP CODE | QUARTER 1 | QUARTER 2 | QUARTER 3 | QUARTER 4 | TOTAL GRAMS |
|---|---|---|---|---|---|
| 141 | 4.73 | 0 | 0 | 0 | 4.73 |
| TOTAL | 4.73 | 0 | 0 | 0 | 4.73 |

STATE: OHIO

| ZIP CODE | QUARTER 1 | QUARTER 2 | QUARTER 3 | QUARTER 4 | TOTAL GRAMS |
|---|---|---|---|---|---|
| 430 | 5.9 | 2.36 | 0 | 0 | 8.26 |
| 431 | 1.18 | 0 | 0 | 0 | 1.18 |
| 432 | 18.91 | 1.18 | 0 | 0 | 20.09 |
| 440 | 9.44 | 1.18 | 0 | 0 | 10.62 |
| 442 | 4.72 | 0 | 0 | 0 | 4.72 |
| 444 | 4.72 | 0 | 0 | 0 | 4.72 |
| 450 | 2.36 | 0 | 0 | 0 | 2.36 |
| 452 | 1.18 | 0 | 0 | 0 | 1.18 |
| 453 | 13 | 0 | 0 | 0 | 13 |
| 456 | 9.46 | 4.73 | 0 | 0 | 14.19 |
| 457 | 1.18 | 0 | 0 | 0 | 1.18 |
| 458 | 1.18 | 0 | 0 | 0 | 1.18 |
| TOTAL | 73.23 | 9.45 | 0 | 0 | 82.68 |

STATE: OREGON

| ZIP CODE | QUARTER 1 | QUARTER 2 | QUARTER 3 | QUARTER 4 | TOTAL GRAMS |
|---|---|---|---|---|---|
| 970 | 1.18 | 0 | 0 | 0 | 1.18 |
| 971 | 1.18 | 0 | 0 | 0 | 1.18 |
| TOTAL | 2.36 | 0 | 0 | 0 | 2.36 |

STATE: PENNSYLVANIA

| ZIP CODE | QUARTER 1 | QUARTER 2 | QUARTER 3 | QUARTER 4 | TOTAL GRAMS |
|---|---|---|---|---|---|

| | | | | | | |
|---|---|---|---|---|---|---|
| | 152 | 8.26 | 9.44 | 0 | 0 | 17.7 |
| | 180 | 0 | 1.18 | 0 | 0 | 1.18 |
| | 190 | 2.36 | 0 | 0 | 0 | 2.36 |
| TOTAL | | 10.62 | 10.62 | 0 | 0 | 21.24 |

STATE: TEXAS

| ZIP CODE | QUARTER 1 | QUARTER 2 | QUARTER 3 | QUARTER 4 | TOTAL GRAMS |
|---|---|---|---|---|---|
| 750 | 1.18 | 2.36 | 0 | 0 | 3.54 |
| 760 | 4.72 | 1.18 | 0 | 0 | 5.9 |
| 761 | 1.18 | 1.18 | 0 | 0 | 2.36 |
| 790 | 2.36 | 0 | 0 | 0 | 2.36 |
| TOTAL | 9.44 | 4.72 | 0 | 0 | 14.16 |

STATE: VIRGINIA

| ZIP CODE | QUARTER 1 | QUARTER 2 | QUARTER 3 | QUARTER 4 | TOTAL GRAMS |
|---|---|---|---|---|---|
| 201 | 8.26 | 0 | 0 | 0 | 8.26 |
| 223 | 2.36 | 0 | 0 | 0 | 2.36 |
| TOTAL | 10.62 | 0 | 0 | 0 | 10.62 |

STATE: WASHINGTON

| ZIP CODE | QUARTER 1 | QUARTER 2 | QUARTER 3 | QUARTER 4 | TOTAL GRAMS |
|---|---|---|---|---|---|
| 982 | 1.18 | 0 | 0 | 0 | 1.18 |
| 984 | 2.36 | 0 | 0 | 0 | 2.36 |
| TOTAL | 3.54 | 0 | 0 | 0 | 3.54 |

STATE: WEST VIRGINIA

| ZIP CODE | QUARTER 1 | QUARTER 2 | QUARTER 3 | QUARTER 4 | TOTAL GRAMS |
|---|---|---|---|---|---|
| 253 | 2.36 | 1.18 | 0 | 0 | 3.54 |
| 254 | 1.18 | 1.18 | 0 | 0 | 2.36 |
| TOTAL | 3.54 | 2.36 | 0 | 0 | 5.9 |

STATE: WISCONSIN

| ZIP CODE | QUARTER 1 | QUARTER 2 | QUARTER 3 | QUARTER 4 | TOTAL GRAMS |
|---|---|---|---|---|---|
| 537 | 2.36 | 1.18 | 0 | 0 | 3.54 |
| 543 | 1.18 | 0 | 0 | 0 | 1.18 |
| 546 | 0 | 4.73 | 0 | 0 | 4.73 |
| 547 | 0 | 4.73 | 0 | 0 | 4.73 |
| TOTAL | 3.54 | 10.64 | 0 | 0 | 14.18 |

DRUG CODE: 9639 DRUG NAME: OPIUM POWDERED

STATE: ALABAMA

| ZIP CODE | QUARTER 1 | QUARTER 2 | QUARTER 3 | QUARTER 4 | TOTAL GRAMS |
|---|---|---|---|---|---|
| 350 | 4.68 | 6.84 | 5.4 | 5.4 | 22.32 |
| 351 | 1.44 | 4.32 | 0 | 0 | 5.76 |
| 352 | 37.44 | 27.36 | 27.72 | 15.48 | 108 |
| 354 | 2.52 | 7.2 | 7.56 | 10.08 | 27.36 |
| 355 | 0 | 0.72 | 0.72 | 0 | 1.44 |
| 356 | 5.76 | 9.72 | 3.96 | 4.32 | 23.76 |
| 357 | 0.72 | 4.68 | 1.44 | 0.72 | 7.56 |
| 358 | 6.48 | 11.52 | 6.84 | 9.72 | 34.56 |
| 359 | 1.08 | 2.88 | 2.88 | 3.96 | 10.8 |
| 361 | 4.68 | 5.04 | 10.08 | 7.2 | 27 |
| 362 | 0.72 | 0.36 | 1.44 | 1.8 | 4.32 |
| 363 | 0 | 0 | 0.72 | 2.16 | 2.88 |
| 364 | 0 | 0.72 | 2.52 | 1.8 | 5.04 |
| 365 | 14.04 | 5.4 | 6.12 | 9 | 34.56 |
| 366 | 2.52 | 7.92 | 8.28 | 6.48 | 25.2 |
| 367 | 0 | 0 | 0 | 0.36 | 0.36 |
| 368 | 0 | 0 | 0.72 | 0 | 0.72 |
| TOTAL | 82.08 | 94.68 | 86.4 | 78.48 | 341.64 |

STATE: ALASKA

| ZIP CODE | QUARTER 1 | QUARTER 2 | QUARTER 3 | QUARTER 4 | TOTAL GRAMS |
|---|---|---|---|---|---|
| 995 | 21.6 | 11.52 | 12.6 | 11.88 | 57.6 |
| 996 | 5.76 | 4.32 | 4.32 | 8.64 | 23.04 |
| 997 | 0.72 | 5.76 | 1.8 | 2.52 | 10.8 |

| | | | | | | |
|---|---|---|---|---|---|---|
| | 998 | 1.44 | 3.6 | 0.72 | 4.32 | 10.08 |
| | 999 | 0 | 0 | 0.36 | 0.72 | 1.08 |
| TOTAL | | 29.52 | 25.2 | 19.8 | 28.08 | 102.6 |

STATE: ARIZONA

| ZIP CODE | | QUARTER 1 | QUARTER 2 | QUARTER 3 | QUARTER 4 | TOTAL GRAMS |
|---|---|---|---|---|---|---|
| | 850 | 111.24 | 97.92 | 106.2 | 102.24 | 417.6 |
| | 851 | 2.16 | 2.52 | 0 | 4.32 | 9 |
| | 852 | 89.28 | 84.6 | 95.4 | 113.4 | 382.68 |
| | 853 | 17.28 | 37.44 | 42.12 | 45 | 141.84 |
| | 855 | 0.72 | 0 | 0.36 | 7.56 | 8.64 |
| | 856 | 0.72 | 0.36 | 3.96 | 1.44 | 6.48 |
| | 857 | 29.88 | 26.28 | 24.12 | 46.44 | 126.72 |
| | 859 | 0.36 | 0 | 0.36 | 0 | 0.72 |
| | 860 | 1.44 | 0.72 | 2.16 | 2.16 | 6.48 |
| | 863 | 21.96 | 18.36 | 17.28 | 10.8 | 68.4 |
| | 864 | 2.52 | 5.4 | 0.36 | 4.32 | 12.6 |
| TOTAL | | 277.56 | 273.6 | 292.32 | 337.68 | 1,181.16 |

STATE: ARKANSAS

| ZIP CODE | | QUARTER 1 | QUARTER 2 | QUARTER 3 | QUARTER 4 | TOTAL GRAMS |
|---|---|---|---|---|---|---|
| | 716 | 4.32 | 4.32 | 4.32 | 3.6 | 16.56 |
| | 717 | 0.36 | 0 | 0 | 0.72 | 1.08 |
| | 719 | 3.6 | 1.44 | 5.4 | 3.96 | 14.4 |
| | 720 | 1.08 | 6.48 | 6.12 | 3.24 | 16.92 |
| | 721 | 28.44 | 34.2 | 23.04 | 37.8 | 123.48 |
| | 722 | 20.88 | 15.48 | 21.24 | 19.08 | 76.68 |
| | 723 | 1.44 | 4.68 | 0 | 0.36 | 6.48 |
| | 724 | 10.08 | 15.12 | 14.76 | 13.68 | 53.64 |
| | 725 | 2.88 | 1.8 | 1.8 | 1.44 | 7.92 |
| | 726 | 0.36 | 0.72 | 1.08 | 1.44 | 3.6 |
| | 727 | 7.92 | 11.88 | 8.28 | 7.2 | 35.28 |
| | 728 | 0 | 0 | 0.36 | 0.36 | 0.72 |
| | 729 | 2.16 | 0.72 | 0.72 | 0.72 | 4.32 |
| TOTAL | | 83.52 | 96.84 | 87.12 | 93.6 | 361.08 |

STATE: CALIFORNIA

| ZIP CODE | QUARTER 1 | QUARTER 2 | QUARTER 3 | QUARTER 4 | TOTAL GRAMS |
|---|---|---|---|---|---|
| 900 | 11.16 | 25.92 | 13.32 | 20.16 | 70.56 |
| 902 | 1.08 | 3.24 | 3.24 | 7.92 | 15.48 |
| 903 | 0 | 0.36 | 0.36 | 0.36 | 1.08 |
| 904 | 8.64 | 1.44 | 5.04 | 7.92 | 23.04 |
| 905 | 2.52 | 0 | 1.44 | 0.36 | 4.32 |
| 906 | 0.36 | 1.8 | 0.36 | 1.8 | 4.32 |
| 907 | 8.64 | 12.24 | 9.72 | 9 | 39.6 |
| 908 | 3.24 | 6.48 | 5.4 | 7.2 | 22.32 |
| 910 | 1.44 | 5.4 | 10.44 | 4.68 | 21.96 |
| 911 | 2.88 | 5.04 | 2.52 | 2.88 | 13.32 |
| 912 | 2.16 | 5.04 | 1.44 | 2.16 | 10.8 |
| 913 | 5.76 | 9.36 | 3.96 | 11.16 | 30.24 |
| 914 | 0.72 | 0.72 | 0 | 0.72 | 2.16 |
| 915 | 0.72 | 2.16 | 1.08 | 0.36 | 4.32 |
| 917 | 15.84 | 16.56 | 8.64 | 11.16 | 52.2 |
| 918 | 0 | 0.72 | 0 | 0 | 0.72 |
| 919 | 2.16 | 5.04 | 13.68 | 0.36 | 21.24 |
| 920 | 42.48 | 22.32 | 41.76 | 36.36 | 142.92 |
| 921 | 23.4 | 43.2 | 38.88 | 34.92 | 140.4 |
| 922 | 26.64 | 21.24 | 23.4 | 17.64 | 88.92 |
| 923 | 9.36 | 10.08 | 6.84 | 3.24 | 29.52 |
| 925 | 6.84 | 7.2 | 12.6 | 9 | 35.64 |
| 926 | 11.88 | 7.56 | 17.64 | 12.96 | 50.04 |
| 927 | 2.88 | 5.04 | 4.68 | 7.56 | 20.16 |
| 928 | 8.28 | 8.28 | 11.88 | 9 | 37.44 |
| 930 | 5.76 | 7.2 | 2.88 | 3.96 | 19.8 |
| 931 | 0.72 | 3.24 | 5.4 | 0 | 9.36 |
| 932 | 4.68 | 5.4 | 6.84 | 5.4 | 22.32 |
| 933 | 5.4 | 3.24 | 3.24 | 1.08 | 12.96 |
| 934 | 6.12 | 10.8 | 7.92 | 7.92 | 32.76 |
| 935 | 0.36 | 0.72 | 1.8 | 5.76 | 8.64 |
| 936 | 0.72 | 1.08 | 1.44 | 1.44 | 4.68 |
| 937 | 5.4 | 8.64 | 22.68 | 12.6 | 49.32 |

| | | | | | | |
|---|---|---|---|---|---|---|
| | 939 | 1.8 | 1.8 | 2.16 | 1.44 | 7.2 |
| | 940 | 12.24 | 6.84 | 9.72 | 4.32 | 33.12 |
| | 941 | 30.96 | 27.72 | 31.68 | 25.92 | 116.28 |
| | 943 | 2.16 | 9.36 | 9.72 | 7.92 | 29.16 |
| | 944 | 0 | 0.72 | 0 | 0 | 0.72 |
| | 945 | 23.76 | 27.72 | 23.4 | 24.12 | 99 |
| | 946 | 6.84 | 3.96 | 3.96 | 6.48 | 21.24 |
| | 947 | 1.08 | 0 | 1.08 | 0.36 | 2.52 |
| | 948 | 0 | 0 | 7.2 | 1.44 | 8.64 |
| | 949 | 2.88 | 1.8 | 2.88 | 3.24 | 10.8 |
| | 950 | 12.6 | 9.36 | 8.28 | 9.72 | 39.96 |
| | 951 | 3.96 | 4.68 | 6.12 | 3.96 | 18.72 |
| | 952 | 11.52 | 6.12 | 4.32 | 11.16 | 33.12 |
| | 953 | 19.08 | 15.84 | 10.44 | 13.68 | 59.04 |
| | 954 | 7.56 | 6.48 | 5.76 | 9.72 | 29.52 |
| | 955 | 2.88 | 0 | 0.72 | 0.72 | 4.32 |
| | 956 | 14.04 | 21.24 | 26.64 | 21.6 | 83.52 |
| | 957 | 5.04 | 1.08 | 2.16 | 0 | 8.28 |
| | 958 | 12.24 | 29.88 | 12.96 | 28.44 | 83.52 |
| | 959 | 11.16 | 26.28 | 18.36 | 16.92 | 72.72 |
| | 960 | 2.52 | 2.52 | 4.32 | 5.4 | 14.76 |
| | 961 | 0.36 | 0.36 | 2.16 | 0 | 2.88 |
| TOTAL | | 412.92 | 470.52 | 484.56 | 453.6 | 1,821.60 |

| STATE: | COLORADO | | | | |
|---|---|---|---|---|---|
| ZIP CODE | QUARTER 1 | QUARTER 2 | QUARTER 3 | QUARTER 4 | TOTAL GRAMS |
| 800 | 37.08 | 41.4 | 46.08 | 28.08 | 152.64 |
| 801 | 21.24 | 27.36 | 36.36 | 55.08 | 140.04 |
| 802 | 43.56 | 59.4 | 70.92 | 66.24 | 240.12 |
| 803 | 1.44 | 0.72 | 2.88 | 2.16 | 7.2 |
| 804 | 8.64 | 11.52 | 19.8 | 15.84 | 55.8 |
| 805 | 44.64 | 53.28 | 41.4 | 41.04 | 180.36 |
| 806 | 0.72 | 8.28 | 3.6 | 3.96 | 16.56 |
| 807 | 2.16 | 4.68 | 0.72 | 2.52 | 10.08 |
| 808 | 0 | 0 | 0 | 0.72 | 0.72 |
| 809 | 48.6 | 33.48 | 45.72 | 32.4 | 160.2 |

| | | | | | | |
|---|---|---|---|---|---|---|
| | 810 | 5.4 | 3.6 | 3.6 | 4.68 | 17.28 |
| | 811 | 0.36 | 0 | 0 | 0 | 0.36 |
| | 812 | 0.72 | 0.36 | 0.72 | 1.8 | 3.6 |
| | 813 | 0 | 1.44 | 2.52 | 3.6 | 7.56 |
| | 814 | 5.04 | 7.2 | 6.84 | 9.72 | 28.8 |
| | 815 | 12.24 | 6.84 | 6.48 | 7.92 | 33.48 |
| | 816 | 0 | 1.08 | 0.72 | 3.6 | 5.4 |
| TOTAL | | 231.84 | 260.64 | 288.36 | 279.36 | 1,060.20 |

STATE: CONNECTICUT

| ZIP CODE | | QUARTER 1 | QUARTER 2 | QUARTER 3 | QUARTER 4 | TOTAL GRAMS |
|---|---|---|---|---|---|---|
| | 60 | 24.48 | 11.16 | 9 | 12.24 | 56.88 |
| | 61 | 3.96 | 18.36 | 7.92 | 7.56 | 37.8 |
| | 62 | 0 | 0.72 | 0 | 0.72 | 1.44 |
| | 63 | 10.08 | 11.16 | 12.96 | 21.96 | 56.16 |
| | 64 | 42.48 | 37.08 | 30.96 | 8.64 | 119.16 |
| | 65 | 11.52 | 15.48 | 7.2 | 9 | 43.2 |
| | 66 | 6.12 | 13.68 | 7.2 | 2.16 | 29.16 |
| | 67 | 13.68 | 11.88 | 6.12 | 5.04 | 36.72 |
| | 68 | 2.16 | 1.44 | 0.36 | 0 | 3.96 |
| | 69 | 0 | 0 | 0 | 0.72 | 0.72 |
| TOTAL | | 114.48 | 120.96 | 81.72 | 68.04 | 385.2 |

STATE: DELAWARE

| ZIP CODE | | QUARTER 1 | QUARTER 2 | QUARTER 3 | QUARTER 4 | TOTAL GRAMS |
|---|---|---|---|---|---|---|
| | 197 | 1.08 | 2.88 | 6.12 | 6.12 | 16.2 |
| | 198 | 7.92 | 2.52 | 3.96 | 3.96 | 18.36 |
| | 199 | 8.64 | 9.36 | 6.12 | 1.44 | 25.56 |
| TOTAL | | 17.64 | 14.76 | 16.2 | 11.52 | 60.12 |

STATE: DISTRICT OF COLUMBIA

| ZIP CODE | | QUARTER 1 | QUARTER 2 | QUARTER 3 | QUARTER 4 | TOTAL GRAMS |
|---|---|---|---|---|---|---|
| | 200 | 11.88 | 10.8 | 18.36 | 14.04 | 55.08 |
| | 203 | 0 | 1.44 | 0 | 0 | 1.44 |

| | | | | | |
|---|---|---|---|---|---|
| 204 | 0.36 | 0.72 | 0 | 1.44 | 2.52 |
| TOTAL | 12.24 | 12.96 | 18.36 | 15.48 | 59.04 |

STATE: FLORIDA

| ZIP CODE | QUARTER 1 | QUARTER 2 | QUARTER 3 | QUARTER 4 | TOTAL GRAMS |
|---|---|---|---|---|---|
| 320 | 6.12 | 5.76 | 27.36 | 17.28 | 56.52 |
| 321 | 17.64 | 24.48 | 15.48 | 16.56 | 74.16 |
| 322 | 47.88 | 42.12 | 26.28 | 31.32 | 147.6 |
| 323 | 0.72 | 2.16 | 7.56 | 14.76 | 25.2 |
| 324 | 7.92 | 7.2 | 6.48 | 6.84 | 28.44 |
| 325 | 57.96 | 33.48 | 37.8 | 43.92 | 173.16 |
| 326 | 10.8 | 9.72 | 5.76 | 6.12 | 32.4 |
| 327 | 24.48 | 27.72 | 30.24 | 31.68 | 114.12 |
| 328 | 25.92 | 29.88 | 28.44 | 26.28 | 110.52 |
| 329 | 46.08 | 32.04 | 28.08 | 36.72 | 142.92 |
| 330 | 13.68 | 12.96 | 14.4 | 7.2 | 48.24 |
| 331 | 19.8 | 23.04 | 20.52 | 19.44 | 82.8 |
| 333 | 8.64 | 12.96 | 6.12 | 16.56 | 44.28 |
| 334 | 56.52 | 44.28 | 43.92 | 32.76 | 177.48 |
| 335 | 5.76 | 9.72 | 7.2 | 10.8 | 33.48 |
| 336 | 26.28 | 32.76 | 23.76 | 40.68 | 123.48 |
| 337 | 18.36 | 21.24 | 22.32 | 19.44 | 81.36 |
| 338 | 6.48 | 8.28 | 7.56 | 16.92 | 39.24 |
| 339 | 28.44 | 33.84 | 20.16 | 32.04 | 114.48 |
| 341 | 24.84 | 15.12 | 13.68 | 26.64 | 80.28 |
| 342 | 20.16 | 19.8 | 10.8 | 38.16 | 88.92 |
| 344 | 0.36 | 0.36 | 0.72 | 1.44 | 2.88 |
| 346 | 20.52 | 10.08 | 15.12 | 18.36 | 64.08 |
| 347 | 28.8 | 33.48 | 27.36 | 37.44 | 127.08 |
| 349 | 2.88 | 5.76 | 5.4 | 4.32 | 18.36 |
| TOTAL | 527.04 | 498.24 | 452.52 | 553.68 | 2,031.48 |

STATE: GEORGIA

| ZIP CODE | QUARTER 1 | QUARTER 2 | QUARTER 3 | QUARTER 4 | TOTAL GRAMS |
|---|---|---|---|---|---|

| | | | | | | |
|---|---|---|---|---|---|---|
| | 300 | 21.96 | 31.68 | 23.76 | 27 | 104.4 |
| | 301 | 31.68 | 34.92 | 42.12 | 28.08 | 136.8 |
| | 302 | 17.28 | 11.52 | 15.48 | 21.24 | 65.52 |
| | 303 | 21.24 | 28.44 | 20.52 | 19.8 | 90 |
| | 304 | 3.24 | 3.6 | 1.08 | 2.52 | 10.44 |
| | 305 | 6.12 | 7.2 | 10.08 | 9.72 | 33.12 |
| | 306 | 9.36 | 14.4 | 10.08 | 13.32 | 47.16 |
| | 307 | 9 | 12.24 | 6.84 | 7.92 | 36 |
| | 308 | 0 | 0.36 | 0 | 0 | 0.36 |
| | 309 | 24.84 | 15.12 | 12.6 | 23.04 | 75.6 |
| | 310 | 1.8 | 9.36 | 2.16 | 4.32 | 17.64 |
| | 312 | 5.4 | 12.96 | 8.28 | 3.96 | 30.6 |
| | 313 | 1.44 | 0 | 1.44 | 0.36 | 3.24 |
| | 314 | 7.2 | 11.16 | 12.24 | 5.76 | 36.36 |
| | 315 | 0.72 | 6.12 | 5.04 | 8.64 | 20.52 |
| | 316 | 5.76 | 2.16 | 3.24 | 1.8 | 12.96 |
| | 317 | 4.32 | 9 | 5.76 | 6.84 | 25.92 |
| | 319 | 2.88 | 0.36 | 4.32 | 0 | 7.56 |
| | 398 | 0 | 0 | 0 | 1.44 | 1.44 |
| TOTAL | | 174.24 | 210.6 | 185.04 | 185.76 | 755.64 |

STATE: GUAM

| ZIP CODE | | QUARTER 1 | QUARTER 2 | QUARTER 3 | QUARTER 4 | TOTAL GRAMS |
|---|---|---|---|---|---|---|
| | 969 | 0.36 | 0 | 0 | 0 | 0.36 |
| TOTAL | | 0.36 | 0 | 0 | 0 | 0.36 |

STATE: HAWAII

| ZIP CODE | | QUARTER 1 | QUARTER 2 | QUARTER 3 | QUARTER 4 | TOTAL GRAMS |
|---|---|---|---|---|---|---|
| | 967 | 8.28 | 11.88 | 16.2 | 11.52 | 47.88 |
| | 968 | 16.56 | 15.84 | 18 | 16.92 | 67.32 |
| TOTAL | | 24.84 | 27.72 | 34.2 | 28.44 | 115.2 |

STATE: IDAHO

| ZIP CODE | | QUARTER 1 | QUARTER 2 | QUARTER 3 | QUARTER 4 | TOTAL GRAMS |
|---|---|---|---|---|---|---|
| | 832 | 3.6 | 4.32 | 2.16 | 4.68 | 14.76 |
| | 833 | 2.16 | 3.96 | 1.8 | 3.24 | 11.16 |
| | 834 | 5.04 | 5.76 | 4.32 | 1.08 | 16.2 |
| | 835 | 0.36 | 0.36 | 0 | 0 | 0.72 |
| | 836 | 18.36 | 14.4 | 21.96 | 23.04 | 77.76 |
| | 837 | 6.48 | 9.36 | 9.36 | 7.2 | 32.4 |
| | 838 | 9.72 | 12.96 | 9.36 | 10.44 | 42.48 |
| TOTAL | | 45.72 | 51.12 | 48.96 | 49.68 | 195.48 |

STATE: ILLINOIS

| ZIP CODE | | QUARTER 1 | QUARTER 2 | QUARTER 3 | QUARTER 4 | TOTAL GRAMS |
|---|---|---|---|---|---|---|
| | 600 | 32.4 | 31.32 | 23.04 | 23.4 | 110.16 |
| | 601 | 35.28 | 33.84 | 28.8 | 32.04 | 129.96 |
| | 602 | 2.52 | 3.24 | 0 | 0.36 | 6.12 |
| | 603 | 1.44 | 0 | 2.52 | 0.36 | 4.32 |
| | 604 | 20.88 | 23.76 | 25.92 | 24.84 | 95.4 |
| | 605 | 10.8 | 8.28 | 7.2 | 5.76 | 32.04 |
| | 606 | 14.4 | 19.44 | 9.72 | 17.28 | 60.84 |
| | 608 | 1.44 | 0 | 2.88 | 1.08 | 5.4 |
| | 609 | 0 | 0 | 0 | 3.96 | 3.96 |
| | 610 | 1.08 | 2.52 | 3.24 | 3.6 | 10.44 |
| | 611 | 11.16 | 17.64 | 13.68 | 16.92 | 59.4 |
| | 612 | 0.36 | 4.68 | 3.6 | 0.72 | 9.36 |
| | 613 | 1.08 | 2.16 | 1.44 | 5.76 | 10.44 |
| | 614 | 2.16 | 2.16 | 2.88 | 6.84 | 14.04 |
| | 615 | 0.36 | 0 | 0 | 1.8 | 2.16 |
| | 616 | 5.4 | 18 | 10.08 | 18 | 51.48 |
| | 617 | 2.52 | 2.52 | 3.24 | 2.88 | 11.16 |
| | 618 | 2.16 | 3.24 | 2.16 | 2.88 | 10.44 |
| | 619 | 0.72 | 0 | 0 | 1.08 | 1.8 |
| | 620 | 0.36 | 5.76 | 2.88 | 4.32 | 13.32 |
| | 622 | 3.96 | 0.72 | 3.96 | 1.08 | 9.72 |
| | 623 | 1.08 | 3.96 | 5.04 | 4.32 | 14.4 |
| | 624 | 1.8 | 0.72 | 1.44 | 0 | 3.96 |
| | 625 | 1.08 | 0.72 | 1.8 | 1.08 | 4.68 |

| | | | | | | |
|---|---|---|---|---|---|---|
| | 626 | 0 | 1.8 | 1.08 | 0.72 | 3.6 |
| | 627 | 0.72 | 3.24 | 10.08 | 8.28 | 22.32 |
| | 628 | 0.36 | 0 | 0 | 0 | 0.36 |
| | 629 | 2.88 | 4.32 | 5.4 | 3.6 | 16.2 |
| TOTAL | | 158.4 | 194.04 | 172.08 | 192.96 | 717.48 |

| STATE: | INDIANA | | | | | |
|---|---|---|---|---|---|---|
| ZIP CODE | | QUARTER 1 | QUARTER 2 | QUARTER 3 | QUARTER 4 | TOTAL GRAMS |
| | 460 | 21.96 | 30.24 | 28.08 | 33.12 | 113.4 |
| | 461 | 26.28 | 25.92 | 30.6 | 14.04 | 96.84 |
| | 462 | 72 | 82.8 | 76.68 | 98.28 | 329.76 |
| | 463 | 9.36 | 5.76 | 3.24 | 7.2 | 25.56 |
| | 464 | 0.36 | 0.36 | 1.08 | 1.44 | 3.24 |
| | 465 | 12.6 | 14.76 | 10.44 | 14.4 | 52.2 |
| | 466 | 4.32 | 3.6 | 3.96 | 6.48 | 18.36 |
| | 467 | 0.72 | 1.44 | 0.36 | 6.48 | 9 |
| | 468 | 8.64 | 14.4 | 11.16 | 17.64 | 51.84 |
| | 469 | 5.04 | 3.6 | 3.6 | 6.48 | 18.72 |
| | 470 | 1.44 | 1.08 | 1.08 | 0.72 | 4.32 |
| | 471 | 13.32 | 12.96 | 11.52 | 13.32 | 51.12 |
| | 472 | 6.84 | 7.2 | 8.64 | 3.24 | 25.92 |
| | 473 | 9 | 15.12 | 12.6 | 12.6 | 49.32 |
| | 474 | 4.68 | 2.52 | 9.36 | 6.48 | 23.04 |
| | 475 | 7.2 | 11.52 | 6.84 | 7.92 | 33.48 |
| | 476 | 3.96 | 2.52 | 1.44 | 1.44 | 9.36 |
| | 477 | 8.64 | 12.6 | 10.8 | 12.6 | 44.64 |
| | 478 | 2.16 | 3.24 | 1.44 | 2.88 | 9.72 |
| | 479 | 4.32 | 2.16 | 1.8 | 3.6 | 11.88 |
| TOTAL | | 222.84 | 253.8 | 234.72 | 270.36 | 981.72 |

| STATE: | IOWA | | | | | |
|---|---|---|---|---|---|---|
| ZIP CODE | | QUARTER 1 | QUARTER 2 | QUARTER 3 | QUARTER 4 | TOTAL GRAMS |
| | 500 | 4.32 | 5.04 | 2.16 | 6.84 | 18.36 |
| | 501 | 19.44 | 6.12 | 3.24 | 1.8 | 30.6 |

| | | | | | | |
|---|---|---|---|---|---|---|
| | 502 | 1.08 | 1.8 | 3.96 | 2.88 | 9.72 |
| | 503 | 3.96 | 3.6 | 5.04 | 12.24 | 24.84 |
| | 504 | 0 | 4.32 | 1.44 | 0.72 | 6.48 |
| | 505 | 0.36 | 1.08 | 2.88 | 1.44 | 5.76 |
| | 506 | 2.52 | 0.72 | 10.44 | 1.44 | 15.12 |
| | 507 | 6.84 | 12.6 | 10.8 | 9 | 39.24 |
| | 508 | 0 | 0.36 | 0 | 0.36 | 0.72 |
| | 510 | 2.52 | 0.72 | 1.08 | 1.8 | 6.12 |
| | 511 | 4.68 | 7.2 | 4.32 | 4.32 | 20.52 |
| | 512 | 0.36 | 1.08 | 0.72 | 0 | 2.16 |
| | 513 | 2.52 | 2.16 | 1.8 | 2.88 | 9.36 |
| | 514 | 3.24 | 1.08 | 1.44 | 0.36 | 6.12 |
| | 515 | 6.48 | 15.12 | 8.64 | 6.48 | 36.72 |
| | 520 | 1.08 | 2.52 | 1.08 | 1.08 | 5.76 |
| | 521 | 0 | 0.72 | 1.44 | 0 | 2.16 |
| | 522 | 29.88 | 8.64 | 19.44 | 14.4 | 72.36 |
| | 523 | 0.72 | 1.08 | 0.72 | 1.44 | 3.96 |
| | 524 | 5.76 | 6.48 | 5.4 | 9.72 | 27.36 |
| | 525 | 0.36 | 1.44 | 0 | 1.44 | 3.24 |
| | 526 | 0.72 | 3.96 | 0 | 0 | 4.68 |
| | 527 | 0.72 | 0 | 0.72 | 0 | 1.44 |
| | 528 | 14.76 | 12.96 | 10.08 | 11.88 | 49.68 |
| TOTAL | | 112.32 | 100.8 | 96.84 | 92.52 | 402.48 |

| STATE: | KANSAS | | | | |
|---|---|---|---|---|---|
| ZIP CODE | QUARTER 1 | QUARTER 2 | QUARTER 3 | QUARTER 4 | TOTAL GRAMS |
| 660 | 14.4 | 30.96 | 24.84 | 28.08 | 98.28 |
| 661 | 17.28 | 9.72 | 6.84 | 18.72 | 52.56 |
| 662 | 60.84 | 73.08 | 68.04 | 69.12 | 271.08 |
| 664 | 1.8 | 0 | 2.52 | 0 | 4.32 |
| 665 | 13.32 | 14.76 | 16.92 | 16.2 | 61.2 |
| 666 | 2.88 | 5.76 | 5.04 | 6.48 | 20.16 |
| 667 | 2.88 | 1.8 | 0.72 | 1.8 | 7.2 |
| 668 | 0.36 | 1.08 | 1.08 | 0.36 | 2.88 |
| 669 | 0 | 0.36 | 1.44 | 0 | 1.8 |

| | | | | | | |
|---|---|---|---|---|---|---|
| | 670 | 3.96 | 9.36 | 3.6 | 2.88 | 19.8 |
| | 671 | 1.08 | 0 | 0.72 | 0.72 | 2.52 |
| | 672 | 26.28 | 33.12 | 22.32 | 24.12 | 105.84 |
| | 673 | 0 | 0 | 1.44 | 2.16 | 3.6 |
| | 674 | 9.36 | 6.12 | 10.08 | 9.72 | 35.28 |
| | 675 | 3.6 | 0.36 | 3.6 | 0.72 | 8.28 |
| | 676 | 6.12 | 3.6 | 6.48 | 1.44 | 17.64 |
| | 677 | 0 | 0 | 0 | 1.44 | 1.44 |
| | 678 | 1.44 | 1.8 | 1.8 | 1.08 | 6.12 |
| TOTAL | | 165.6 | 191.88 | 177.48 | 185.04 | 720 |

| STATE: | KENTUCKY | | | | |
|---|---|---|---|---|---|
| ZIP CODE | QUARTER 1 | QUARTER 2 | QUARTER 3 | QUARTER 4 | TOTAL GRAMS |
| 400 | 2.16 | 5.4 | 8.28 | 1.8 | 17.64 |
| 401 | 2.16 | 0 | 1.08 | 0.36 | 3.6 |
| 402 | 53.28 | 39.6 | 36 | 40.32 | 169.2 |
| 403 | 5.76 | 1.44 | 6.48 | 2.88 | 16.56 |
| 404 | 30.6 | 28.44 | 7.2 | 6.12 | 72.36 |
| 405 | 43.92 | 30.96 | 32.4 | 37.44 | 144.72 |
| 406 | 0 | 0.72 | 0 | 2.16 | 2.88 |
| 407 | 0 | 21.96 | 4.32 | 24.12 | 50.4 |
| 408 | 2.16 | 0 | 0 | 0 | 2.16 |
| 409 | 0 | 0.72 | 0 | 0.72 | 1.44 |
| 410 | 6.48 | 11.52 | 6.84 | 11.88 | 36.72 |
| 411 | 1.44 | 2.16 | 1.44 | 3.96 | 9 |
| 412 | 0 | 1.44 | 0 | 0.72 | 2.16 |
| 413 | 0.72 | 0 | 1.08 | 0.36 | 2.16 |
| 414 | 0 | 0.36 | 0 | 0.36 | 0.72 |
| 415 | 0.36 | 1.08 | 0.36 | 1.44 | 3.24 |
| 416 | 0 | 0.36 | 0 | 0.36 | 0.72 |
| 417 | 1.44 | 1.8 | 1.08 | 1.08 | 5.4 |
| 418 | 0 | 0 | 0 | 0.36 | 0.36 |
| 420 | 16.92 | 9 | 17.28 | 16.92 | 60.12 |
| 421 | 11.52 | 12.24 | 6.84 | 7.2 | 37.8 |
| 422 | 1.44 | 5.04 | 4.32 | 3.6 | 14.4 |
| 423 | 1.44 | 0.72 | 4.32 | 2.52 | 9 |

| | | | | | | |
|---|---|---|---|---|---|---|
| | 424 | 8.64 | 2.52 | 2.52 | 2.52 | 16.2 |
| | 425 | 1.44 | 1.08 | 3.6 | 3.96 | 10.08 |
| | 426 | 0.72 | 0 | 0 | 0 | 0.72 |
| | 427 | 4.32 | 1.08 | 0.72 | 0.72 | 6.84 |
| TOTAL | | 196.92 | 179.64 | 146.16 | 173.88 | 696.6 |

| STATE: | LOUISIANA | | | | |
|---|---|---|---|---|---|
| ZIP CODE | QUARTER 1 | QUARTER 2 | QUARTER 3 | QUARTER 4 | TOTAL GRAMS |
| 700 | 18.72 | 21.96 | 17.28 | 10.44 | 68.4 |
| 701 | 15.84 | 11.52 | 8.28 | 14.4 | 50.04 |
| 703 | 7.92 | 10.8 | 2.16 | 4.32 | 25.2 |
| 704 | 5.76 | 8.64 | 7.92 | 6.48 | 28.8 |
| 705 | 20.88 | 17.28 | 20.16 | 28.8 | 87.12 |
| 706 | 5.4 | 8.64 | 7.56 | 6.48 | 28.08 |
| 707 | 6.48 | 0 | 1.44 | 1.44 | 9.36 |
| 708 | 4.68 | 16.56 | 21.24 | 12.6 | 55.08 |
| 710 | 7.92 | 0 | 0.36 | 0 | 8.28 |
| 711 | 10.8 | 7.2 | 9 | 7.92 | 34.92 |
| 712 | 0 | 4.68 | 1.44 | 6.48 | 12.6 |
| 713 | 1.08 | 5.04 | 1.44 | 5.04 | 12.6 |
| 714 | 0 | 0.72 | 0 | 0 | 0.72 |
| TOTAL | 105.48 | 113.04 | 98.28 | 104.4 | 421.2 |

| STATE: | MAINE | | | | |
|---|---|---|---|---|---|
| ZIP CODE | QUARTER 1 | QUARTER 2 | QUARTER 3 | QUARTER 4 | TOTAL GRAMS |
| 39 | 0.36 | 0.36 | 0 | 1.08 | 1.8 |
| 40 | 3.24 | 3.96 | 14.04 | 4.68 | 25.92 |
| 41 | 10.44 | 10.44 | 4.32 | 10.8 | 36 |
| 42 | 9.36 | 8.28 | 11.16 | 9.72 | 38.52 |
| 43 | 1.44 | 3.6 | 7.2 | 7.92 | 20.16 |
| 44 | 10.44 | 12.24 | 7.56 | 16.2 | 46.44 |
| 45 | 1.44 | 0 | 0 | 0 | 1.44 |
| 46 | 1.44 | 4.32 | 1.44 | 1.8 | 9 |
| 47 | 1.08 | 1.8 | 0.72 | 1.44 | 5.04 |
| 48 | 0 | 0.36 | 6.84 | 3.24 | 10.44 |

| | | | | | | |
|---|---|---|---|---|---|---|
| | 49 | 0 | 2.52 | 0.72 | 1.08 | 4.32 |
| TOTAL | | 39.24 | 47.88 | 54 | 57.96 | 199.08 |

STATE: MARYLAND

| ZIP CODE | | QUARTER 1 | QUARTER 2 | QUARTER 3 | QUARTER 4 | TOTAL GRAMS |
|---|---|---|---|---|---|---|
| | 206 | 2.88 | 4.32 | 2.52 | 3.6 | 13.32 |
| | 207 | 6.84 | 4.32 | 4.32 | 5.76 | 21.24 |
| | 208 | 18 | 19.08 | 14.76 | 34.56 | 86.4 |
| | 209 | 0 | 1.08 | 0.72 | 0.72 | 2.52 |
| | 210 | 11.88 | 11.52 | 10.44 | 8.28 | 42.12 |
| | 211 | 2.16 | 6.84 | 4.32 | 3.6 | 16.92 |
| | 212 | 11.52 | 28.8 | 18 | 33.48 | 91.8 |
| | 214 | 1.08 | 0.72 | 1.44 | 0.72 | 3.96 |
| | 215 | 1.44 | 0.72 | 2.16 | 2.52 | 6.84 |
| | 216 | 3.6 | 10.44 | 5.76 | 3.96 | 23.76 |
| | 217 | 2.16 | 6.84 | 2.88 | 2.88 | 14.76 |
| | 218 | 1.44 | 2.16 | 2.52 | 3.96 | 10.08 |
| | 219 | 0 | 0.72 | 0.72 | 0 | 1.44 |
| TOTAL | | 63 | 97.56 | 70.56 | 104.04 | 335.16 |

STATE: MASSACHUSETTS

| ZIP CODE | | QUARTER 1 | QUARTER 2 | QUARTER 3 | QUARTER 4 | TOTAL GRAMS |
|---|---|---|---|---|---|---|
| | 10 | 23.04 | 43.92 | 28.44 | 42.48 | 137.88 |
| | 11 | 22.68 | 26.64 | 25.56 | 18.72 | 93.6 |
| | 12 | 1.44 | 0 | 0 | 0 | 1.44 |
| | 13 | 0 | 0.72 | 0 | 0.36 | 1.08 |
| | 14 | 2.52 | 1.8 | 5.04 | 5.04 | 14.4 |
| | 15 | 0 | 8.28 | 10.8 | 6.12 | 25.2 |
| | 16 | 0.72 | 7.56 | 6.84 | 3.6 | 18.72 |
| | 17 | 6.12 | 1.8 | 5.76 | 15.84 | 29.52 |
| | 18 | 14.76 | 9.72 | 7.2 | 1.44 | 33.12 |
| | 19 | 7.92 | 10.8 | 10.44 | 19.8 | 48.96 |
| | 20 | 9.72 | 16.2 | 12.96 | 14.4 | 53.28 |
| | 21 | 42.48 | 43.92 | 38.16 | 54.36 | 178.92 |
| | 22 | 3.96 | 2.16 | 1.44 | 2.52 | 10.08 |

| | | | | | |
|---|---|---|---|---|---|
| 23 | 7.56 | 10.44 | 6.48 | 7.56 | 32.04 |
| 24 | 16.2 | 17.28 | 2.88 | 16.92 | 53.28 |
| 25 | 4.68 | 5.04 | 6.12 | 12.24 | 28.08 |
| 26 | 1.44 | 0.72 | 0.72 | 3.6 | 6.48 |
| 27 | 7.56 | 7.56 | 8.28 | 13.68 | 37.08 |
| TOTAL | 172.8 | 214.56 | 177.12 | 238.68 | 803.16 |

STATE: MICHIGAN

| ZIP CODE | QUARTER 1 | QUARTER 2 | QUARTER 3 | QUARTER 4 | TOTAL GRAMS |
|---|---|---|---|---|---|
| 480 | 44.28 | 43.92 | 26.64 | 46.44 | 161.28 |
| 481 | 74.52 | 63 | 73.44 | 60.84 | 271.8 |
| 482 | 18 | 32.04 | 30.6 | 27 | 107.64 |
| 483 | 37.44 | 36.72 | 30.96 | 48.6 | 153.72 |
| 484 | 5.04 | 5.4 | 8.28 | 5.76 | 24.48 |
| 485 | 2.88 | 5.4 | 5.04 | 7.2 | 20.52 |
| 486 | 10.44 | 17.28 | 11.88 | 16.92 | 56.52 |
| 487 | 6.48 | 2.88 | 6.48 | 2.88 | 18.72 |
| 488 | 5.04 | 13.68 | 38.16 | 26.28 | 83.16 |
| 489 | 9.72 | 12.24 | 9.36 | 13.68 | 45 |
| 490 | 32.4 | 31.68 | 29.16 | 28.8 | 122.04 |
| 491 | 0.72 | 0.72 | 2.16 | 5.76 | 9.36 |
| 492 | 20.16 | 18.72 | 13.32 | 22.32 | 74.52 |
| 493 | 1.08 | 0 | 1.44 | 1.08 | 3.6 |
| 494 | 10.08 | 10.44 | 10.08 | 11.52 | 42.12 |
| 495 | 61.56 | 50.76 | 41.04 | 51.84 | 205.2 |
| 496 | 17.64 | 8.28 | 18.36 | 20.16 | 64.44 |
| 497 | 9 | 10.08 | 14.04 | 13.68 | 46.8 |
| 498 | 7.92 | 5.76 | 5.4 | 11.16 | 30.24 |
| 499 | 0 | 0.72 | 0 | 0.36 | 1.08 |
| TOTAL | 374.4 | 369.72 | 375.84 | 422.28 | 1,542.24 |

STATE: MINNESOTA

| ZIP CODE | QUARTER 1 | QUARTER 2 | QUARTER 3 | QUARTER 4 | TOTAL GRAMS |
|---|---|---|---|---|---|
| 550 | 24.48 | 31.32 | 18.72 | 21.96 | 96.48 |
| 551 | 34.56 | 35.64 | 28.8 | 54.72 | 153.72 |

| | | | | | | |
|---|---|---|---|---|---|---|
| | 553 | 43.92 | 43.2 | 47.16 | 41.76 | 176.04 |
| | 554 | 142.2 | 129.96 | 132.48 | 135.36 | 540 |
| | 556 | 0.72 | 0 | 0 | 0 | 0.72 |
| | 557 | 5.04 | 1.08 | 2.88 | 1.8 | 10.8 |
| | 558 | 6.48 | 7.92 | 5.76 | 15.84 | 36 |
| | 559 | 55.08 | 61.2 | 61.92 | 57.6 | 235.8 |
| | 560 | 14.4 | 13.32 | 10.44 | 12.24 | 50.4 |
| | 561 | 0.72 | 2.16 | 0 | 1.44 | 4.32 |
| | 562 | 19.08 | 5.76 | 4.32 | 2.88 | 32.04 |
| | 563 | 33.84 | 27 | 30.6 | 21.24 | 112.68 |
| | 564 | 15.12 | 16.2 | 17.64 | 18.36 | 67.32 |
| | 565 | 5.04 | 5.04 | 2.52 | 2.88 | 15.48 |
| | 566 | 0.72 | 0 | 0.72 | 0.72 | 2.16 |
| | 567 | 0 | 1.44 | 0 | 0.36 | 1.8 |
| TOTAL | | 401.4 | 381.24 | 363.96 | 389.16 | 1,535.76 |

STATE: MISSISSIPPI

| ZIP CODE | | QUARTER 1 | QUARTER 2 | QUARTER 3 | QUARTER 4 | TOTAL GRAMS |
|---|---|---|---|---|---|---|
| | 386 | 7.56 | 3.96 | 7.92 | 5.4 | 24.84 |
| | 387 | 0.36 | 0 | 2.52 | 0.72 | 3.6 |
| | 388 | 3.96 | 6.84 | 9 | 6.84 | 26.64 |
| | 389 | 0 | 0 | 0.72 | 0 | 0.72 |
| | 391 | 0 | 0.36 | 1.08 | 0 | 1.44 |
| | 392 | 2.52 | 10.44 | 7.92 | 2.88 | 23.76 |
| | 393 | 4.32 | 1.8 | 0.72 | 1.08 | 7.92 |
| | 394 | 2.16 | 5.04 | 0 | 0.36 | 7.56 |
| | 395 | 8.28 | 7.2 | 4.32 | 4.32 | 24.12 |
| | 396 | 0 | 0.72 | 0 | 1.08 | 1.8 |
| | 397 | 3.96 | 3.6 | 2.16 | 2.52 | 12.24 |
| TOTAL | | 33.12 | 39.96 | 36.36 | 25.2 | 134.64 |

STATE: MISSOURI

| ZIP CODE | | QUARTER 1 | QUARTER 2 | QUARTER 3 | QUARTER 4 | TOTAL GRAMS |
|---|---|---|---|---|---|---|
| | 630 | 3.6 | 7.92 | 3.24 | 5.76 | 20.52 |
| | 631 | 24.48 | 31.32 | 32.04 | 42.12 | 129.96 |

| | | | | | | |
|---|---|---|---|---|---|---|
| | 633 | 3.6 | 4.68 | 7.2 | 3.6 | 19.08 |
| | 634 | 0.72 | 0 | 1.08 | 0.72 | 2.52 |
| | 635 | 2.52 | 0 | 0 | 0.72 | 3.24 |
| | 636 | 0 | 0 | 0.72 | 0 | 0.72 |
| | 637 | 6.12 | 7.92 | 4.32 | 3.6 | 21.96 |
| | 638 | 0 | 0 | 2.52 | 2.16 | 4.68 |
| | 639 | 0.72 | 1.44 | 0.72 | 0 | 2.88 |
| | 640 | 27.36 | 23.04 | 24.12 | 22.68 | 97.2 |
| | 641 | 35.64 | 30.24 | 27 | 34.56 | 127.44 |
| | 644 | 0 | 1.08 | 0 | 1.8 | 2.88 |
| | 645 | 6.48 | 4.32 | 8.28 | 6.48 | 25.56 |
| | 646 | 0 | 0 | 0 | 1.44 | 1.44 |
| | 647 | 12.24 | 11.88 | 11.16 | 7.2 | 42.48 |
| | 648 | 0.72 | 4.68 | 4.68 | 3.96 | 14.04 |
| | 650 | 0 | 0.72 | 0 | 0.72 | 1.44 |
| | 651 | 1.08 | 0.36 | 0.72 | 2.52 | 4.68 |
| | 652 | 11.16 | 8.28 | 13.32 | 4.68 | 37.44 |
| | 653 | 3.6 | 1.44 | 2.88 | 1.44 | 9.36 |
| | 654 | 4.32 | 0.72 | 4.32 | 0.72 | 10.08 |
| | 655 | 3.6 | 7.92 | 5.04 | 15.12 | 31.68 |
| | 656 | 1.08 | 0 | 0 | 0 | 1.08 |
| | 657 | 0 | 0 | 0 | 1.8 | 1.8 |
| | 658 | 20.88 | 15.48 | 12.96 | 10.8 | 60.12 |
| TOTAL | | 169.92 | 163.44 | 166.32 | 174.6 | 674.28 |

STATE: MONTANA

| ZIP CODE | | QUARTER 1 | QUARTER 2 | QUARTER 3 | QUARTER 4 | TOTAL GRAMS |
|---|---|---|---|---|---|---|
| | 590 | 0 | 0 | 0 | 0.36 | 0.36 |
| | 591 | 16.2 | 18.36 | 20.16 | 21.6 | 76.32 |
| | 592 | 0 | 0 | 0 | 1.8 | 1.8 |
| | 594 | 9.72 | 10.44 | 7.2 | 7.56 | 34.92 |
| | 595 | 0 | 1.44 | 2.16 | 0.72 | 4.32 |
| | 596 | 6.48 | 5.76 | 1.44 | 0 | 13.68 |
| | 597 | 3.6 | 1.8 | 1.08 | 2.16 | 8.64 |
| | 598 | 12.96 | 11.16 | 11.52 | 11.88 | 47.52 |
| | 599 | 4.32 | 3.96 | 4.32 | 3.96 | 16.56 |

| | | | | | |
|---|---|---|---|---|---|
| TOTAL | 53.28 | 52.92 | 47.88 | 50.04 | 204.12 |

STATE: NEBRASKA

| ZIP CODE | QUARTER 1 | QUARTER 2 | QUARTER 3 | QUARTER 4 | TOTAL GRAMS |
|---|---|---|---|---|---|
| 680 | 12.24 | 15.48 | 13.32 | 11.52 | 52.56 |
| 681 | 42.48 | 54.36 | 50.04 | 54 | 200.88 |
| 683 | 2.52 | 0 | 0 | 0 | 2.52 |
| 684 | 0 | 0 | 0 | 1.08 | 1.08 |
| 685 | 9.36 | 5.76 | 8.64 | 11.88 | 35.64 |
| 686 | 0 | 0.72 | 0.72 | 0.72 | 2.16 |
| 687 | 1.8 | 6.48 | 5.4 | 1.8 | 15.48 |
| 688 | 16.2 | 11.88 | 6.48 | 14.4 | 48.96 |
| 689 | 3.96 | 6.84 | 3.6 | 5.4 | 19.8 |
| 690 | 0 | 0 | 0.72 | 0 | 0.72 |
| 691 | 0.36 | 1.8 | 1.08 | 1.8 | 5.04 |
| 692 | 0.72 | 0 | 0 | 0 | 0.72 |
| 693 | 2.16 | 2.88 | 0.36 | 2.88 | 8.28 |
| TOTAL | 91.8 | 106.2 | 90.36 | 105.48 | 393.84 |

STATE: NEVADA

| ZIP CODE | QUARTER 1 | QUARTER 2 | QUARTER 3 | QUARTER 4 | TOTAL GRAMS |
|---|---|---|---|---|---|
| 890 | 11.52 | 3.24 | 3.96 | 9.36 | 28.08 |
| 891 | 6.48 | 11.88 | 16.92 | 19.08 | 54.36 |
| 894 | 2.88 | 1.08 | 2.88 | 2.88 | 9.72 |
| 895 | 8.28 | 8.64 | 9 | 7.2 | 33.12 |
| 897 | 0.72 | 2.88 | 1.8 | 2.16 | 7.56 |
| 898 | 0.72 | 0.36 | 0.36 | 0 | 1.44 |
| TOTAL | 30.6 | 28.08 | 34.92 | 40.68 | 134.28 |

STATE: NEW HAMPSHIRE

| ZIP CODE | QUARTER 1 | QUARTER 2 | QUARTER 3 | QUARTER 4 | TOTAL GRAMS |
|---|---|---|---|---|---|
| 30 | 16.92 | 11.88 | 14.76 | 18.72 | 62.28 |
| 31 | 1.44 | 3.6 | 0.72 | 2.16 | 7.92 |

| | | | | | | |
|---|---|---|---|---|---|---|
| | 32 | 0 | 0.72 | 0 | 0.36 | 1.08 |
| | 33 | 8.28 | 10.44 | 9.36 | 8.28 | 36.36 |
| | 34 | 0 | 4.32 | 0 | 1.44 | 5.76 |
| | 35 | 0.72 | 0.72 | 1.44 | 0.72 | 3.6 |
| | 37 | 2.52 | 5.04 | 8.28 | 7.92 | 23.76 |
| | 38 | 0.36 | 0.72 | 2.88 | 1.8 | 5.76 |
| TOTAL | | 30.24 | 37.44 | 37.44 | 41.4 | 146.52 |

STATE: NEW JERSEY

| ZIP CODE | | QUARTER 1 | QUARTER 2 | QUARTER 3 | QUARTER 4 | TOTAL GRAMS |
|---|---|---|---|---|---|---|
| | 70 | 5.76 | 6.12 | 1.08 | 6.12 | 19.08 |
| | 74 | 2.52 | 3.96 | 5.76 | 3.96 | 16.2 |
| | 76 | 1.44 | 8.28 | 3.6 | 6.84 | 20.16 |
| | 77 | 6.48 | 4.68 | 2.88 | 7.56 | 21.6 |
| | 78 | 2.52 | 2.52 | 4.68 | 2.16 | 11.88 |
| | 79 | 0 | 1.44 | 0.72 | 0 | 2.16 |
| | 80 | 47.52 | 69.48 | 57.24 | 47.16 | 221.4 |
| | 81 | 1.44 | 0.72 | 16.56 | 3.24 | 21.96 |
| | 82 | 1.08 | 5.04 | 0 | 0.72 | 6.84 |
| | 83 | 1.44 | 2.88 | 3.96 | 2.88 | 11.16 |
| | 84 | 0 | 0 | 0.36 | 0 | 0.36 |
| | 85 | 4.32 | 3.24 | 3.6 | 2.88 | 14.04 |
| | 86 | 2.16 | 4.32 | 1.08 | 2.52 | 10.08 |
| | 87 | 3.24 | 2.16 | 3.24 | 2.52 | 11.16 |
| | 88 | 4.68 | 12.24 | 9 | 15.12 | 41.04 |
| | 89 | 2.52 | 6.48 | 3.96 | 1.08 | 14.04 |
| TOTAL | | 87.12 | 133.56 | 117.72 | 104.76 | 443.16 |

STATE: NEW MEXICO

| ZIP CODE | | QUARTER 1 | QUARTER 2 | QUARTER 3 | QUARTER 4 | TOTAL GRAMS |
|---|---|---|---|---|---|---|
| | 871 | 16.56 | 22.68 | 10.08 | 9.72 | 59.04 |
| | 874 | 1.44 | 5.76 | 0 | 0 | 7.2 |
| | 875 | 9.36 | 7.2 | 6.12 | 8.64 | 31.32 |
| | 880 | 7.56 | 6.48 | 7.92 | 7.2 | 29.16 |
| | 881 | 0 | 0.36 | 0 | 1.8 | 2.16 |

| | | | | | | |
|---|---|---|---|---|---|---|
| | 882 | 2.16 | 6.48 | 5.04 | 0.72 | 14.4 |
| | 883 | 0.72 | 0.36 | 0 | 0.36 | 1.44 |
| TOTAL | | 37.8 | 49.32 | 29.16 | 28.44 | 144.72 |

STATE: NEW YORK

| ZIP CODE | | QUARTER 1 | QUARTER 2 | QUARTER 3 | QUARTER 4 | TOTAL GRAMS |
|---|---|---|---|---|---|---|
| | 100 | 27 | 54.36 | 22.32 | 35.64 | 139.32 |
| | 103 | 0 | 0.36 | 0 | 0.36 | 0.72 |
| | 104 | 0.72 | 4.32 | 0.72 | 6.12 | 11.88 |
| | 105 | 3.6 | 1.8 | 0.72 | 8.64 | 14.76 |
| | 108 | 0 | 0 | 1.08 | 0 | 1.08 |
| | 109 | 0.72 | 0 | 0 | 0.72 | 1.44 |
| | 110 | 16.2 | 9 | 8.64 | 8.28 | 42.12 |
| | 111 | 0 | 0 | 1.8 | 0 | 1.8 |
| | 112 | 2.52 | 3.6 | 3.6 | 3.96 | 13.68 |
| | 113 | 0 | 0.36 | 0 | 0 | 0.36 |
| | 114 | 0 | 0.36 | 0 | 0 | 0.36 |
| | 115 | 2.52 | 1.8 | 0.36 | 3.24 | 7.92 |
| | 117 | 15.48 | 10.08 | 15.48 | 14.4 | 55.44 |
| | 118 | 0.72 | 1.44 | 0 | 0 | 2.16 |
| | 119 | 0.72 | 0 | 0 | 5.04 | 5.76 |
| | 120 | 7.56 | 3.6 | 4.68 | 6.48 | 22.32 |
| | 121 | 0.72 | 7.92 | 1.44 | 0.72 | 10.8 |
| | 122 | 0.72 | 2.88 | 6.12 | 8.64 | 18.36 |
| | 123 | 15.84 | 5.4 | 8.28 | 8.64 | 38.16 |
| | 124 | 0 | 2.52 | 1.44 | 2.52 | 6.48 |
| | 125 | 2.16 | 0 | 0 | 0 | 2.16 |
| | 128 | 0.36 | 5.76 | 7.2 | 3.6 | 16.92 |
| | 129 | 15.84 | 7.92 | 6.84 | 13.68 | 44.28 |
| | 130 | 1.44 | 6.84 | 10.08 | 4.68 | 23.04 |
| | 131 | 0 | 0.36 | 0 | 0.36 | 0.72 |
| | 132 | 7.2 | 13.68 | 4.32 | 12.6 | 37.8 |
| | 133 | 1.44 | 0.72 | 3.6 | 1.44 | 7.2 |
| | 134 | 5.04 | 5.76 | 5.04 | 7.56 | 23.4 |
| | 135 | 5.4 | 2.88 | 4.68 | 2.16 | 15.12 |
| | 136 | 7.92 | 3.96 | 3.96 | 9.72 | 25.56 |

| | | | | | | |
|---|---|---|---|---|---|---|
| | 137 | 0.72 | 1.08 | 0.72 | 0 | 2.52 |
| | 138 | 0.72 | 0 | 0.72 | 0.36 | 1.8 |
| | 139 | 2.88 | 1.44 | 2.16 | 2.52 | 9 |
| | 140 | 1.8 | 0.36 | 9 | 5.76 | 16.92 |
| | 141 | 0 | 0 | 3.6 | 1.08 | 4.68 |
| | 142 | 34.92 | 37.44 | 37.8 | 38.52 | 148.68 |
| | 143 | 0 | 0 | 0.72 | 0 | 0.72 |
| | 144 | 3.24 | 1.44 | 0 | 1.44 | 6.12 |
| | 145 | 3.6 | 2.16 | 2.88 | 3.6 | 12.24 |
| | 146 | 17.28 | 12.6 | 16.2 | 13.32 | 59.4 |
| | 147 | 0.36 | 0 | 0.72 | 2.16 | 3.24 |
| | 148 | 0.72 | 3.24 | 4.32 | 1.8 | 10.08 |
| | 149 | 0.72 | 0 | 0.72 | 1.44 | 2.88 |
| TOTAL | | 208.8 | 217.44 | 201.96 | 241.2 | 869.4 |

STATE: NORTH CAROLINA

| ZIP CODE | | QUARTER 1 | QUARTER 2 | QUARTER 3 | QUARTER 4 | TOTAL GRAMS |
|---|---|---|---|---|---|---|
| | 270 | 0.72 | 3.24 | 1.44 | 0.72 | 6.12 |
| | 271 | 15.12 | 13.32 | 20.52 | 14.4 | 63.36 |
| | 272 | 22.32 | 22.32 | 17.28 | 21.24 | 83.16 |
| | 273 | 6.48 | 6.12 | 5.76 | 6.84 | 25.2 |
| | 274 | 8.64 | 6.48 | 7.56 | 5.4 | 28.08 |
| | 275 | 21.96 | 20.88 | 19.8 | 29.88 | 92.52 |
| | 276 | 24.48 | 14.76 | 9 | 19.8 | 68.04 |
| | 277 | 10.08 | 9.36 | 10.8 | 11.52 | 41.76 |
| | 278 | 9.36 | 7.92 | 7.56 | 7.56 | 32.4 |
| | 279 | 5.76 | 7.2 | 3.24 | 5.4 | 21.6 |
| | 280 | 10.44 | 12.24 | 16.2 | 5.76 | 44.64 |
| | 281 | 19.08 | 24.84 | 17.28 | 18 | 79.2 |
| | 282 | 22.32 | 14.76 | 21.6 | 24.12 | 82.8 |
| | 283 | 10.44 | 6.84 | 12.24 | 16.92 | 46.44 |
| | 284 | 19.8 | 19.8 | 19.44 | 22.32 | 81.36 |
| | 285 | 4.68 | 3.24 | 7.2 | 5.04 | 20.16 |
| | 286 | 17.64 | 11.16 | 8.64 | 10.44 | 47.88 |
| | 287 | 8.28 | 7.92 | 8.64 | 12.6 | 37.44 |
| | 288 | 0.72 | 3.96 | 5.76 | 2.16 | 12.6 |

| | | | | | | |
|---|---|---|---|---|---|---|
| | 289 | 0 | 2.16 | 0 | 0.72 | 2.88 |
| TOTAL | | 238.32 | 218.52 | 219.96 | 240.84 | 917.64 |

STATE: NORTH DAKOTA

| ZIP CODE | | QUARTER 1 | QUARTER 2 | QUARTER 3 | QUARTER 4 | TOTAL GRAMS |
|---|---|---|---|---|---|---|
| | 580 | 0.72 | 0.72 | 0.36 | 0 | 1.8 |
| | 581 | 5.04 | 14.76 | 15.84 | 8.64 | 44.28 |
| | 582 | 1.44 | 2.16 | 3.6 | 5.04 | 12.24 |
| | 583 | 0 | 0 | 0 | 1.44 | 1.44 |
| | 584 | 1.08 | 1.44 | 1.8 | 2.16 | 6.48 |
| | 585 | 14.76 | 21.6 | 22.32 | 25.2 | 83.88 |
| | 586 | 3.6 | 0.72 | 0 | 1.08 | 5.4 |
| | 587 | 2.88 | 0.72 | 1.08 | 1.8 | 6.48 |
| | 588 | 1.44 | 0 | 0.72 | 0.72 | 2.88 |
| TOTAL | | 30.96 | 42.12 | 45.72 | 46.08 | 164.88 |

STATE: OHIO

| ZIP CODE | | QUARTER 1 | QUARTER 2 | QUARTER 3 | QUARTER 4 | TOTAL GRAMS |
|---|---|---|---|---|---|---|
| | 430 | 15.48 | 14.04 | 19.8 | 19.8 | 69.12 |
| | 431 | 3.24 | 2.52 | 5.4 | 6.12 | 17.28 |
| | 432 | 44.28 | 55.44 | 27 | 31.68 | 158.4 |
| | 433 | 5.04 | 2.52 | 3.6 | 6.48 | 17.64 |
| | 434 | 6.84 | 3.24 | 1.44 | 1.8 | 13.32 |
| | 435 | 18 | 6.12 | 6.48 | 6.48 | 37.08 |
| | 436 | 22.32 | 10.44 | 18.36 | 24.48 | 75.6 |
| | 437 | 2.88 | 0.36 | 2.52 | 3.6 | 9.36 |
| | 438 | 0 | 0.36 | 0 | 0 | 0.36 |
| | 439 | 0 | 0 | 1.44 | 1.08 | 2.52 |
| | 440 | 11.16 | 14.4 | 22.32 | 18.36 | 66.24 |
| | 441 | 62.64 | 71.28 | 62.28 | 66.96 | 263.16 |
| | 442 | 12.6 | 14.4 | 15.48 | 10.08 | 52.56 |
| | 443 | 15.12 | 15.84 | 12.96 | 13.32 | 57.24 |
| | 444 | 1.8 | 14.4 | 15.12 | 16.56 | 47.88 |
| | 445 | 9.72 | 12.24 | 11.52 | 10.8 | 44.28 |

| | | | | | | |
|---|---|---|---|---|---|---|
| | 446 | 3.96 | 4.32 | 3.96 | 2.16 | 14.4 |
| | 447 | 8.64 | 8.64 | 7.92 | 7.2 | 32.4 |
| | 448 | 7.2 | 1.8 | 3.24 | 7.2 | 19.44 |
| | 449 | 6.84 | 2.16 | 2.16 | 0 | 11.16 |
| | 450 | 5.76 | 7.56 | 10.44 | 11.88 | 35.64 |
| | 451 | 19.44 | 26.28 | 19.44 | 22.68 | 87.84 |
| | 452 | 36.72 | 35.64 | 43.2 | 41.4 | 156.96 |
| | 453 | 6.84 | 6.48 | 10.08 | 11.16 | 34.56 |
| | 454 | 26.64 | 29.52 | 19.8 | 29.52 | 105.48 |
| | 455 | 2.88 | 0.72 | 3.96 | 1.44 | 9 |
| | 456 | 5.4 | 6.12 | 16.92 | 4.68 | 33.12 |
| | 457 | 0.72 | 2.52 | 0.36 | 2.52 | 6.12 |
| | 458 | 9.72 | 10.8 | 9 | 6.84 | 36.36 |
| TOTAL | | 371.88 | 380.16 | 376.2 | 386.28 | 1,514.52 |

| STATE: | OKLAHOMA | | | | | |
|---|---|---|---|---|---|---|
| ZIP CODE | | QUARTER 1 | QUARTER 2 | QUARTER 3 | QUARTER 4 | TOTAL GRAMS |
| | 730 | 3.96 | 3.24 | 7.92 | 1.8 | 16.92 |
| | 731 | 31.68 | 30.24 | 19.44 | 19.08 | 100.44 |
| | 734 | 0 | 1.44 | 0 | 0 | 1.44 |
| | 735 | 12.6 | 3.24 | 4.68 | 2.52 | 23.04 |
| | 736 | 0 | 0.36 | 0.36 | 0.36 | 1.08 |
| | 737 | 0.72 | 1.08 | 2.88 | 0.72 | 5.4 |
| | 738 | 1.8 | 3.6 | 2.88 | 0 | 8.28 |
| | 740 | 0.72 | 8.28 | 2.52 | 3.96 | 15.48 |
| | 741 | 16.2 | 19.8 | 26.64 | 18.36 | 81 |
| | 743 | 0 | 3.6 | 4.68 | 2.16 | 10.44 |
| | 744 | 2.52 | 3.6 | 8.64 | 0.36 | 15.12 |
| | 745 | 0 | 0 | 6.48 | 1.08 | 7.56 |
| | 746 | 1.44 | 0 | 3.96 | 0.36 | 5.76 |
| | 747 | 2.88 | 2.88 | 2.16 | 2.88 | 10.8 |
| | 748 | 0.72 | 2.16 | 2.16 | 1.44 | 6.48 |
| | 749 | 1.44 | 0 | 0 | 0 | 1.44 |
| TOTAL | | 76.68 | 83.52 | 95.4 | 55.08 | 310.68 |

RETAIL DRUG DISTRIBUTION BY ZIP CODE WITHIN STATE BY GRAMS WT

STATE: OREGON

| ZIP CODE | QUARTER 1 | QUARTER 2 | QUARTER 3 | QUARTER 4 | TOTAL GRAMS |
|---|---|---|---|---|---|
| 970 | 19.8 | 25.56 | 15.48 | 18 | 78.84 |
| 971 | 6.48 | 12.6 | 10.08 | 16.92 | 46.08 |
| 972 | 38.52 | 50.4 | 64.8 | 60.48 | 214.2 |
| 973 | 22.32 | 40.68 | 15.12 | 14.04 | 92.16 |
| 974 | 36.36 | 32.76 | 49.32 | 48.6 | 167.04 |
| 975 | 37.44 | 36 | 39.24 | 44.28 | 156.96 |
| 976 | 1.8 | 3.96 | 0.72 | 1.8 | 8.28 |
| 977 | 3.96 | 10.8 | 6.48 | 4.68 | 25.92 |
| 978 | 1.44 | 1.8 | 1.44 | 1.8 | 6.48 |
| 979 | 0.36 | 0.72 | 0.36 | 0.72 | 2.16 |
| TOTAL | 168.48 | 215.28 | 203.04 | 211.32 | 798.12 |

STATE: PENNSYLVANIA

| ZIP CODE | QUARTER 1 | QUARTER 2 | QUARTER 3 | QUARTER 4 | TOTAL GRAMS |
|---|---|---|---|---|---|
| 150 | 6.48 | 2.16 | 4.68 | 2.88 | 16.2 |
| 151 | 1.44 | 0.36 | 2.52 | 2.52 | 6.84 |
| 152 | 9.72 | 8.28 | 15.84 | 10.08 | 43.92 |
| 153 | 2.52 | 7.2 | 3.96 | 5.04 | 18.72 |
| 154 | 0.36 | 0.72 | 0 | 0.36 | 1.44 |
| 155 | 1.08 | 1.44 | 1.44 | 2.16 | 6.12 |
| 156 | 1.8 | 1.44 | 2.52 | 11.88 | 17.64 |
| 157 | 2.88 | 0 | 1.8 | 3.6 | 8.28 |
| 158 | 2.88 | 3.24 | 2.16 | 3.96 | 12.24 |
| 159 | 0 | 0.36 | 0.36 | 0 | 0.72 |
| 160 | 0 | 3.24 | 1.08 | 2.16 | 6.48 |
| 161 | 1.8 | 0 | 0.72 | 5.4 | 7.92 |
| 162 | 0.72 | 2.52 | 1.44 | 1.08 | 5.76 |
| 163 | 1.08 | 1.08 | 2.52 | 1.44 | 6.12 |
| 164 | 0 | 4.32 | 0 | 0 | 4.32 |
| 165 | 2.88 | 2.52 | 4.32 | 10.08 | 19.8 |
| 166 | 0.36 | 2.16 | 1.08 | 1.08 | 4.68 |
| 167 | 1.08 | 0.36 | 0 | 0.36 | 1.8 |
| 168 | 2.88 | 7.2 | 6.48 | 8.64 | 25.2 |
| 169 | 0 | 0 | 0 | 0.36 | 0.36 |

| | | | | | | |
|---|---|---|---|---|---|---|
| | 170 | 15.84 | 24.84 | 32.4 | 19.44 | 92.52 |
| | 171 | 5.4 | 6.48 | 6.12 | 2.88 | 20.88 |
| | 172 | 11.16 | 12.24 | 12.24 | 12.24 | 47.88 |
| | 173 | 5.4 | 5.4 | 6.48 | 4.32 | 21.6 |
| | 174 | 8.64 | 5.76 | 1.44 | 5.04 | 20.88 |
| | 175 | 0.72 | 0 | 1.8 | 1.8 | 4.32 |
| | 176 | 5.04 | 7.56 | 2.16 | 5.4 | 20.16 |
| | 177 | 1.44 | 1.44 | 1.44 | 16.56 | 20.88 |
| | 178 | 6.84 | 13.68 | 15.12 | 18 | 53.64 |
| | 179 | 8.64 | 8.28 | 8.28 | 9.72 | 34.92 |
| | 180 | 23.04 | 26.28 | 20.16 | 20.88 | 90.36 |
| | 181 | 5.4 | 4.32 | 4.32 | 6.84 | 20.88 |
| | 182 | 4.32 | 1.44 | 1.44 | 2.88 | 10.08 |
| | 183 | 1.44 | 1.8 | 0 | 2.16 | 5.4 |
| | 184 | 0 | 0.72 | 3.24 | 0 | 3.96 |
| | 185 | 6.84 | 5.04 | 8.64 | 11.16 | 31.68 |
| | 186 | 3.24 | 7.56 | 5.76 | 3.96 | 20.52 |
| | 187 | 39.96 | 20.16 | 12.6 | 42.84 | 115.56 |
| | 188 | 2.16 | 1.08 | 2.16 | 2.88 | 8.28 |
| | 189 | 1.8 | 2.52 | 10.8 | 2.16 | 17.28 |
| | 190 | 26.64 | 21.96 | 56.88 | 47.88 | 153.36 |
| | 191 | 47.52 | 43.2 | 33.84 | 47.52 | 172.08 |
| | 193 | 12.24 | 9.36 | 19.08 | 14.76 | 55.44 |
| | 194 | 10.8 | 7.56 | 10.8 | 3.96 | 33.12 |
| | 195 | 2.16 | 0.72 | 4.68 | 1.8 | 9.36 |
| | 196 | 0 | 0 | 0 | 0.72 | 0.72 |
| TOTAL | | 296.64 | 288 | 334.8 | 380.88 | 1,300.32 |

STATE: RHODE ISLAND

| ZIP CODE | | QUARTER 1 | QUARTER 2 | QUARTER 3 | QUARTER 4 | TOTAL GRAMS |
|---|---|---|---|---|---|---|
| | 028 | 12.24 | 16.56 | 17.64 | 12.24 | 58.68 |
| | 029 | 20.52 | 17.64 | 27.36 | 13.32 | 78.84 |
| TOTAL | | 32.76 | 34.2 | 45 | 25.56 | 137.52 |

STATE: SOUTH CAROLINA

| ZIP CODE | QUARTER 1 | QUARTER 2 | QUARTER 3 | QUARTER 4 | TOTAL GRAMS |
|---|---|---|---|---|---|
| 290 | 4.32 | 3.24 | 3.24 | 3.24 | 14.04 |
| 291 | 6.84 | 8.64 | 7.2 | 8.64 | 31.32 |
| 292 | 2.88 | 9.36 | 9.36 | 3.24 | 24.84 |
| 293 | 10.44 | 9 | 9 | 6.48 | 34.92 |
| 294 | 21.96 | 25.92 | 21.96 | 22.68 | 92.52 |
| 295 | 20.16 | 32.4 | 24.84 | 32.76 | 110.16 |
| 296 | 30.96 | 27.72 | 21.96 | 25.2 | 105.84 |
| 297 | 7.92 | 9.72 | 4.68 | 14.4 | 36.72 |
| 298 | 8.64 | 7.92 | 7.92 | 11.88 | 36.36 |
| 299 | 13.68 | 20.88 | 17.28 | 16.56 | 68.4 |
| TOTAL | 127.8 | 154.8 | 127.44 | 145.08 | 555.12 |

STATE: SOUTH DAKOTA

| ZIP CODE | QUARTER 1 | QUARTER 2 | QUARTER 3 | QUARTER 4 | TOTAL GRAMS |
|---|---|---|---|---|---|
| 570 | 5.04 | 3.96 | 4.68 | 9.36 | 23.04 |
| 571 | 48.96 | 36 | 17.64 | 27.72 | 130.32 |
| 572 | 1.08 | 1.08 | 0.72 | 0.72 | 3.6 |
| 573 | 2.52 | 1.08 | 1.44 | 3.6 | 8.64 |
| 574 | 1.8 | 1.08 | 0.72 | 0.72 | 4.32 |
| 575 | 2.16 | 1.08 | 0.72 | 0.36 | 4.32 |
| 577 | 2.52 | 2.52 | 1.08 | 0.36 | 6.48 |
| TOTAL | 64.08 | 46.8 | 27 | 42.84 | 180.72 |

STATE: TENNESSEE

| ZIP CODE | QUARTER 1 | QUARTER 2 | QUARTER 3 | QUARTER 4 | TOTAL GRAMS |
|---|---|---|---|---|---|
| 370 | 12.24 | 13.32 | 20.88 | 11.88 | 58.32 |
| 371 | 5.04 | 3.96 | 3.96 | 2.52 | 15.48 |
| 372 | 28.8 | 28.8 | 31.32 | 58.68 | 147.6 |
| 373 | 4.32 | 3.6 | 7.56 | 4.68 | 20.16 |
| 374 | 6.48 | 6.12 | 12.6 | 10.8 | 36 |
| 376 | 17.64 | 23.4 | 22.68 | 17.64 | 81.36 |
| 377 | 10.08 | 9 | 8.28 | 11.16 | 38.52 |
| 378 | 6.84 | 9.36 | 11.52 | 11.52 | 39.24 |
| 379 | 16.56 | 18.36 | 14.76 | 20.52 | 70.2 |

| | | | | | | |
|---|---|---|---|---|---|---|
| | 380 | 2.52 | 2.88 | 1.08 | 1.8 | 8.28 |
| | 381 | 42.12 | 53.64 | 38.52 | 35.64 | 169.92 |
| | 382 | 0.72 | 2.52 | 1.8 | 1.8 | 6.84 |
| | 383 | 12.96 | 11.88 | 6.48 | 9 | 40.32 |
| | 384 | 1.44 | 1.8 | 2.88 | 4.32 | 10.44 |
| | 385 | 3.24 | 3.96 | 6.48 | 3.6 | 17.28 |
| TOTAL | | 171 | 192.6 | 190.8 | 205.56 | 759.96 |

STATE: TEXAS

| ZIP CODE | | QUARTER 1 | QUARTER 2 | QUARTER 3 | QUARTER 4 | TOTAL GRAMS |
|---|---|---|---|---|---|---|
| | 750 | 50.4 | 41.4 | 43.92 | 56.16 | 191.88 |
| | 751 | 1.44 | 9.72 | 7.56 | 3.6 | 22.32 |
| | 752 | 61.2 | 63.36 | 68.4 | 76.68 | 269.64 |
| | 753 | 10.08 | 20.52 | 9.36 | 12.96 | 52.92 |
| | 754 | 3.24 | 1.8 | 2.16 | 3.6 | 10.8 |
| | 755 | 3.96 | 5.04 | 5.4 | 5.76 | 20.16 |
| | 756 | 9.36 | 6.84 | 10.8 | 13.32 | 40.32 |
| | 757 | 24.84 | 21.6 | 30.96 | 18.72 | 96.12 |
| | 758 | 0 | 0.72 | 0 | 0 | 0.72 |
| | 759 | 10.08 | 12.96 | 10.44 | 12.96 | 46.44 |
| | 760 | 32.76 | 21.24 | 39.6 | 34.2 | 127.8 |
| | 761 | 25.92 | 28.8 | 24.48 | 27.72 | 106.92 |
| | 762 | 9.72 | 8.28 | 8.28 | 14.4 | 40.68 |
| | 763 | 6.12 | 3.6 | 1.44 | 3.6 | 14.76 |
| | 764 | 1.44 | 0 | 0 | 0.36 | 1.8 |
| | 765 | 6.84 | 14.4 | 11.88 | 12.24 | 45.36 |
| | 766 | 0 | 0.72 | 1.8 | 2.16 | 4.68 |
| | 767 | 5.04 | 4.32 | 3.6 | 6.48 | 19.44 |
| | 768 | 0.72 | 0.72 | 1.44 | 0 | 2.88 |
| | 769 | 2.52 | 3.6 | 1.08 | 1.8 | 9 |
| | 770 | 55.44 | 54 | 59.04 | 51.12 | 219.6 |
| | 773 | 5.04 | 6.12 | 1.8 | 4.32 | 17.28 |
| | 774 | 6.48 | 5.04 | 7.56 | 10.8 | 29.88 |
| | 775 | 26.64 | 24.84 | 27.36 | 21.96 | 100.8 |
| | 776 | 1.08 | 1.44 | 0.72 | 0 | 3.24 |
| | 777 | 1.08 | 0.72 | 1.8 | 2.16 | 5.76 |

| | | | | | |
|---|---|---|---|---|---|
| | 778 | 4.32 | 10.44 | 2.16 | 5.4 | 22.32 |
| | 779 | 5.4 | 1.08 | 3.6 | 3.24 | 13.32 |
| | 780 | 1.44 | 0 | 0.36 | 0.72 | 2.52 |
| | 781 | 4.32 | 3.24 | 2.16 | 1.44 | 11.16 |
| | 782 | 40.32 | 36.72 | 33.12 | 47.88 | 158.04 |
| | 784 | 1.8 | 3.6 | 0.36 | 1.8 | 7.56 |
| | 785 | 3.24 | 3.24 | 1.8 | 2.16 | 10.44 |
| | 786 | 17.28 | 20.88 | 18.36 | 21.6 | 78.12 |
| | 787 | 30.24 | 34.92 | 30.6 | 36 | 131.76 |
| | 788 | 0.72 | 0.72 | 0 | 1.8 | 3.24 |
| | 789 | 0 | 0.36 | 0 | 1.08 | 1.44 |
| | 790 | 2.88 | 2.16 | 2.88 | 2.88 | 10.8 |
| | 791 | 14.76 | 10.8 | 18.72 | 16.2 | 60.48 |
| | 792 | 0 | 0 | 0 | 0.72 | 0.72 |
| | 794 | 9 | 7.56 | 5.76 | 18.36 | 40.68 |
| | 796 | 1.08 | 1.08 | 1.8 | 2.52 | 6.48 |
| | 797 | 0 | 1.44 | 2.16 | 3.6 | 7.2 |
| | 799 | 5.4 | 4.32 | 8.64 | 2.88 | 21.24 |
| TOTAL | | 503.64 | 504.36 | 513.36 | 567.36 | 2,088.72 |

| STATE: | UTAH | | | | |
|---|---|---|---|---|---|
| ZIP CODE | QUARTER 1 | QUARTER 2 | QUARTER 3 | QUARTER 4 | TOTAL GRAMS |
| 840 | 23.04 | 29.88 | 28.44 | 23.4 | 104.76 |
| 841 | 29.52 | 23.4 | 20.52 | 28.44 | 101.88 |
| 843 | 1.08 | 0.36 | 0.36 | 0.72 | 2.52 |
| 844 | 2.16 | 1.44 | 2.16 | 2.16 | 7.92 |
| 846 | 4.68 | 2.88 | 8.28 | 5.4 | 21.24 |
| 847 | 7.2 | 6.12 | 2.88 | 6.12 | 22.32 |
| TOTAL | 67.68 | 64.08 | 62.64 | 66.24 | 260.64 |

| STATE: | VERMONT | | | | |
|---|---|---|---|---|---|
| ZIP CODE | QUARTER 1 | QUARTER 2 | QUARTER 3 | QUARTER 4 | TOTAL GRAMS |
| 50 | 5.4 | 1.8 | 1.08 | 1.8 | 10.08 |
| 51 | 9 | 0 | 0 | 0 | 9 |
| 53 | 0 | 0.36 | 0.36 | 0.36 | 1.08 |

| | | | | | |
|---|---|---|---|---|---|
| 54 | 14.4 | 21.6 | 25.2 | 30.24 | 91.44 |
| 56 | 4.32 | 9 | 5.04 | 2.88 | 21.24 |
| 57 | 7.92 | 7.2 | 5.04 | 8.28 | 28.44 |
| 58 | 0.36 | 0 | 0.72 | 0.36 | 1.44 |
| TOTAL | 41.4 | 39.96 | 37.44 | 43.92 | 162.72 |

STATE: VIRGIN ISLANDS

| ZIP CODE | QUARTER 1 | QUARTER 2 | QUARTER 3 | QUARTER 4 | TOTAL GRAMS |
|---|---|---|---|---|---|
| 8 | 0 | 0 | 1.8 | 0 | 1.8 |
| TOTAL | 0 | 0 | 1.8 | 0 | 1.8 |

STATE: VIRGINIA

| ZIP CODE | QUARTER 1 | QUARTER 2 | QUARTER 3 | QUARTER 4 | TOTAL GRAMS |
|---|---|---|---|---|---|
| 201 | 14.04 | 18 | 28.08 | 18.72 | 78.84 |
| 220 | 21.24 | 10.08 | 12.24 | 20.88 | 64.44 |
| 221 | 5.04 | 9 | 7.92 | 6.84 | 28.8 |
| 222 | 2.52 | 2.88 | 1.44 | 4.68 | 11.52 |
| 223 | 0.72 | 2.52 | 3.6 | 3.24 | 10.08 |
| 224 | 1.8 | 1.8 | 2.16 | 1.44 | 7.2 |
| 225 | 1.08 | 0.72 | 1.44 | 0 | 3.24 |
| 226 | 1.44 | 1.44 | 2.16 | 1.44 | 6.48 |
| 227 | 0 | 0.36 | 0 | 0 | 0.36 |
| 228 | 3.6 | 1.8 | 3.6 | 7.2 | 16.2 |
| 229 | 19.08 | 26.28 | 32.4 | 16.56 | 94.32 |
| 230 | 5.04 | 0 | 5.04 | 0 | 10.08 |
| 231 | 9 | 5.4 | 11.88 | 8.28 | 34.56 |
| 232 | 23.4 | 30.24 | 16.92 | 18.72 | 89.28 |
| 233 | 5.04 | 3.6 | 10.8 | 5.4 | 24.84 |
| 234 | 9.72 | 9.36 | 11.52 | 11.88 | 42.48 |
| 235 | 5.76 | 3.6 | 8.28 | 8.28 | 25.92 |
| 236 | 3.6 | 9.72 | 4.68 | 4.68 | 22.68 |
| 237 | 9.72 | 10.8 | 10.44 | 9.36 | 40.32 |
| 238 | 0 | 2.16 | 1.8 | 0 | 3.96 |
| 239 | 0.36 | 0 | 0.72 | 0.72 | 1.8 |
| 240 | 4.32 | 10.44 | 5.04 | 11.16 | 30.96 |

| | | | | | | |
|---|---|---|---|---|---|---|
| | 241 | 1.8 | 6.12 | 3.96 | 1.08 | 12.96 |
| | 242 | 14.76 | 1.8 | 1.8 | 2.52 | 20.88 |
| | 243 | 0 | 0.72 | 0.36 | 0.36 | 1.44 |
| | 244 | 0 | 0 | 0 | 0.36 | 0.36 |
| | 245 | 7.56 | 6.48 | 11.16 | 10.44 | 35.64 |
| | 246 | 0 | 2.88 | 0 | 0.36 | 3.24 |
| TOTAL | | 170.64 | 178.2 | 199.44 | 174.6 | 722.88 |

STATE: WASHINGTON

| ZIP CODE | | QUARTER 1 | QUARTER 2 | QUARTER 3 | QUARTER 4 | TOTAL GRAMS |
|---|---|---|---|---|---|---|
| | 980 | 31.68 | 25.2 | 34.92 | 37.08 | 128.88 |
| | 981 | 77.76 | 84.24 | 118.44 | 111.24 | 391.68 |
| | 982 | 22.32 | 33.12 | 30.24 | 20.16 | 105.84 |
| | 983 | 34.56 | 35.28 | 31.68 | 43.92 | 145.44 |
| | 984 | 21.24 | 41.76 | 26.28 | 26.64 | 115.92 |
| | 985 | 14.4 | 16.56 | 10.8 | 13.32 | 55.08 |
| | 986 | 16.2 | 19.44 | 23.76 | 17.64 | 77.04 |
| | 988 | 33.48 | 32.04 | 18.72 | 28.08 | 112.32 |
| | 989 | 1.44 | 0.72 | 1.8 | 3.24 | 7.2 |
| | 990 | 0 | 0.36 | 0 | 0.72 | 1.08 |
| | 991 | 3.6 | 2.88 | 4.32 | 3.24 | 14.04 |
| | 992 | 17.28 | 17.64 | 10.08 | 16.56 | 61.56 |
| | 993 | 7.92 | 11.16 | 12.96 | 7.2 | 39.24 |
| | 994 | 0 | 0.72 | 1.44 | 0 | 2.16 |
| TOTAL | | 281.88 | 321.12 | 325.44 | 329.04 | 1,257.48 |

STATE: WEST VIRGINIA

| ZIP CODE | | QUARTER 1 | QUARTER 2 | QUARTER 3 | QUARTER 4 | TOTAL GRAMS |
|---|---|---|---|---|---|---|
| | 247 | 1.44 | 1.8 | 1.44 | 0.72 | 5.4 |
| | 249 | 7.2 | 6.48 | 6.48 | 6.48 | 26.64 |
| | 250 | 0 | 1.08 | 0.72 | 0.36 | 2.16 |
| | 252 | 2.88 | 2.16 | 1.44 | 2.16 | 8.64 |
| | 253 | 22.32 | 33.48 | 27.36 | 33.84 | 117 |
| | 254 | 1.08 | 0.72 | 0 | 1.44 | 3.24 |

| | | | | | | |
|---|---|---|---|---|---|---|
| | 255 | 5.76 | 3.6 | 2.88 | 3.24 | 15.48 |
| | 256 | 0 | 0 | 2.52 | 2.16 | 4.68 |
| | 257 | 17.28 | 18 | 19.08 | 18.72 | 73.08 |
| | 258 | 2.16 | 7.2 | 4.32 | 5.04 | 18.72 |
| | 259 | 11.16 | 10.8 | 10.8 | 10.8 | 43.56 |
| | 260 | 1.8 | 3.6 | 1.08 | 2.52 | 9 |
| | 261 | 0.72 | 1.44 | 0 | 2.16 | 4.32 |
| | 262 | 2.52 | 1.44 | 1.08 | 2.52 | 7.56 |
| | 263 | 1.44 | 11.16 | 3.96 | 0.72 | 17.28 |
| | 264 | 0 | 0 | 1.44 | 0 | 1.44 |
| | 265 | 3.24 | 7.92 | 2.16 | 5.4 | 18.72 |
| | 266 | 3.6 | 0 | 0.36 | 0 | 3.96 |
| TOTAL | | 84.6 | 110.88 | 87.12 | 98.28 | 380.88 |

STATE: WISCONSIN

| ZIP CODE | | QUARTER 1 | QUARTER 2 | QUARTER 3 | QUARTER 4 | TOTAL GRAMS |
|---|---|---|---|---|---|---|
| | 530 | 21.96 | 27.72 | 15.12 | 21.24 | 86.04 |
| | 531 | 6.84 | 12.6 | 25.92 | 13.68 | 59.04 |
| | 532 | 34.56 | 47.88 | 38.52 | 33.48 | 154.44 |
| | 534 | 1.44 | 1.44 | 2.88 | 1.44 | 7.2 |
| | 535 | 15.84 | 7.92 | 14.04 | 17.28 | 55.08 |
| | 537 | 30.6 | 50.76 | 52.92 | 38.16 | 172.44 |
| | 538 | 0 | 0.36 | 0.36 | 2.52 | 3.24 |
| | 539 | 3.96 | 6.84 | 12.96 | 7.56 | 31.32 |
| | 540 | 2.16 | 4.32 | 2.16 | 2.88 | 11.52 |
| | 541 | 1.08 | 0 | 2.88 | 9.36 | 13.32 |
| | 542 | 2.52 | 2.16 | 1.44 | 3.96 | 10.08 |
| | 543 | 6.84 | 13.68 | 10.8 | 12.24 | 43.56 |
| | 544 | 7.92 | 6.12 | 7.2 | 10.08 | 31.32 |
| | 545 | 6.84 | 5.04 | 5.04 | 2.52 | 19.44 |
| | 546 | 9 | 13.68 | 9.72 | 9.36 | 41.76 |
| | 547 | 20.16 | 7.92 | 11.16 | 7.2 | 46.44 |
| | 548 | 0.72 | 2.52 | 6.84 | 3.24 | 13.32 |
| | 549 | 10.8 | 12.96 | 19.08 | 12.24 | 55.08 |
| TOTAL | | 183.24 | 223.92 | 239.04 | 208.44 | 854.64 |

STATE: WYOMING

| ZIP CODE | QUARTER 1 | QUARTER 2 | QUARTER 3 | QUARTER 4 | TOTAL GRAMS |
|---|---|---|---|---|---|
| 820 | 11.52 | 4.32 | 2.16 | 5.76 | 23.76 |
| 822 | 1.8 | 0 | 0 | 0 | 1.8 |
| 824 | 0 | 0.36 | 1.08 | 0 | 1.44 |
| 825 | 0 | 1.44 | 0 | 0.72 | 2.16 |
| 826 | 9.36 | 15.48 | 15.12 | 18.36 | 58.32 |
| 827 | 0.72 | 1.44 | 0 | 0.36 | 2.52 |
| 828 | 1.44 | 0.72 | 1.44 | 1.44 | 5.04 |
| 829 | 0.72 | 0.36 | 0 | 0.72 | 1.8 |
| 830 | 0 | 1.44 | 0 | 0 | 1.44 |
| TOTAL | 25.56 | 25.56 | 19.8 | 27.36 | 98.28 |

DRUG CODE:9652DRUG NAME:OXYMORPHONE

STATE: ALABAMA

| ZIP CODE | QUARTER 1 | QUARTER 2 | QUARTER 3 | QUARTER 4 | TOTAL GRAMS |
|---|---|---|---|---|---|
| 350 | 766.22 | 643.61 | 345.64 | 209.38 | 1,964.85 |
| 351 | 398.99 | 381.24 | 241.13 | 163.3 | 1,184.66 |
| 352 | 386.81 | 329.5 | 155.23 | 92.87 | 964.41 |
| 354 | 195.41 | 206.48 | 136.94 | 107.32 | 646.15 |
| 355 | 256.43 | 234.35 | 175.88 | 105.13 | 771.79 |
| 356 | 539.77 | 458.09 | 264.24 | 208.17 | 1,470.27 |
| 357 | 453.77 | 398.77 | 252.69 | 163.43 | 1,268.66 |
| 358 | 775.86 | 644.54 | 374.64 | 236.14 | 2,031.18 |
| 359 | 998.04 | 887.28 | 554.66 | 451.67 | 2,891.65 |
| 360 | 226.33 | 162.32 | 83.9 | 56.65 | 529.2 |
| 361 | 120.21 | 98.76 | 73.33 | 36.49 | 328.79 |
| 362 | 507.87 | 368.66 | 221.24 | 215 | 1,312.77 |
| 363 | 143.54 | 148.45 | 78.24 | 82.25 | 452.48 |
| 364 | 30.96 | 46.84 | 35.86 | 15.61 | 129.27 |
| 365 | 799.95 | 748.34 | 416.21 | 290.2 | 2,254.70 |
| 366 | 685.89 | 652.94 | 410.27 | 259.69 | 2,008.79 |
| 367 | 105.36 | 108.88 | 84.39 | 51.74 | 350.37 |
| 368 | 254.34 | 233.42 | 185.47 | 169.86 | 843.09 |

| | | | | | | |
|---|---|---|---|---|---|---|
| | 369 | 28.9 | 29.84 | 18.64 | 23.37 | 100.75 |
| TOTAL | | 7,674.65 | 6,782.31 | 4,108.60 | 2,938.27 | 21,503.83 |

| STATE: | ALASKA | | | | | |
|---|---|---|---|---|---|---|
| ZIP CODE | | QUARTER 1 | QUARTER 2 | QUARTER 3 | QUARTER 4 | TOTAL GRAMS |
| | 995 | 104.82 | 115.66 | 104.64 | 68.96 | 394.08 |
| | 996 | 158.53 | 152.37 | 126.5 | 105.71 | 543.11 |
| | 997 | 19.8 | 16.41 | 11.06 | 7.14 | 54.41 |
| | 998 | 0 | 0.27 | 0 | 0 | 0.27 |
| | 999 | 4.28 | 2.14 | 4.28 | 2.14 | 12.84 |
| TOTAL | | 287.43 | 286.85 | 246.48 | 183.95 | 1,004.71 |

| STATE: | ARIZONA | | | | | |
|---|---|---|---|---|---|---|
| ZIP CODE | | QUARTER 1 | QUARTER 2 | QUARTER 3 | QUARTER 4 | TOTAL GRAMS |
| | 850 | 1,407.92 | 1,351.17 | 918.38 | 554.62 | 4,232.09 |
| | 851 | 411.12 | 359.07 | 217.4 | 133.28 | 1,120.87 |
| | 852 | 2,613.78 | 2,276.95 | 1,355.14 | 778.4 | 7,024.27 |
| | 853 | 1,484.68 | 1,308.44 | 810.43 | 566.93 | 4,170.48 |
| | 855 | 182.66 | 187.43 | 147.2 | 75.29 | 592.58 |
| | 856 | 282.08 | 220.44 | 157.68 | 85.95 | 746.15 |
| | 857 | 1,284.95 | 1,100.94 | 762.28 | 359.36 | 3,507.53 |
| | 859 | 106.25 | 85.78 | 49.6 | 13.07 | 254.7 |
| | 860 | 105.09 | 63.03 | 60.44 | 31.4 | 259.96 |
| | 863 | 330.57 | 277.93 | 192.25 | 127.39 | 928.14 |
| | 864 | 393.46 | 264.46 | 173.47 | 82.7 | 914.09 |
| TOTAL | | 8,602.56 | 7,495.64 | 4,844.27 | 2,808.39 | 23,750.86 |

| STATE: | ARKANSAS | | | | | |
|---|---|---|---|---|---|---|
| ZIP CODE | | QUARTER 1 | QUARTER 2 | QUARTER 3 | QUARTER 4 | TOTAL GRAMS |
| | 716 | 51.12 | 60.26 | 50.4 | 33.99 | 195.77 |
| | 717 | 52.63 | 64.37 | 39.39 | 19.85 | 176.24 |
| | 718 | 25.87 | 30.96 | 18.2 | 5 | 80.03 |
| | 719 | 112.58 | 86.49 | 65.3 | 36.84 | 301.21 |
| | 720 | 335.25 | 359.2 | 270.22 | 157.81 | 1,122.48 |

| | | | | | | |
|---|---|---|---|---|---|---|
| | 721 | 270.93 | 252.69 | 170.3 | 154.78 | 848.7 |
| | 722 | 177.17 | 133.64 | 102.5 | 66.37 | 479.68 |
| | 723 | 56.02 | 51.03 | 33.19 | 26.85 | 167.09 |
| | 724 | 98.67 | 83.1 | 64.5 | 23.19 | 269.46 |
| | 725 | 90.33 | 69.94 | 60.4 | 33.1 | 253.77 |
| | 726 | 56.34 | 37.56 | 33.45 | 12.27 | 139.62 |
| | 727 | 452.74 | 494.45 | 326.95 | 187.52 | 1,461.66 |
| | 728 | 62.63 | 51.25 | 30.33 | 18.64 | 162.85 |
| | 729 | 436.73 | 410.81 | 286.72 | 80.2 | 1,214.46 |
| TOTAL | | 2,279.01 | 2,185.75 | 1,551.85 | 856.41 | 6,873.02 |

STATE: CALIFORNIA

| ZIP CODE | | QUARTER 1 | QUARTER 2 | QUARTER 3 | QUARTER 4 | TOTAL GRAMS |
|---|---|---|---|---|---|---|
| | 900 | 538.39 | 594.28 | 510.47 | 428.3 | 2,071.44 |
| | 902 | 425.89 | 383.69 | 252.29 | 199.16 | 1,261.03 |
| | 903 | 134.89 | 28.37 | 41.22 | 44.07 | 248.55 |
| | 904 | 57.9 | 71.1 | 68.87 | 63.07 | 260.94 |
| | 905 | 15.61 | 25.78 | 32.83 | 15.88 | 90.1 |
| | 906 | 104.78 | 91.98 | 69.49 | 51.03 | 317.28 |
| | 907 | 175.92 | 166.2 | 137.78 | 78.86 | 558.76 |
| | 908 | 122.66 | 83.14 | 79.13 | 74.22 | 359.15 |
| | 910 | 68.25 | 90.37 | 48.35 | 20.7 | 227.67 |
| | 911 | 132.03 | 130.43 | 101.88 | 114.55 | 478.89 |
| | 912 | 24.98 | 26.85 | 31.58 | 25.78 | 109.19 |
| | 913 | 573.34 | 427.81 | 356.66 | 315.18 | 1,672.99 |
| | 914 | 132.48 | 134.66 | 152.46 | 136.13 | 555.73 |
| | 915 | 35.06 | 36.22 | 20.25 | 8.47 | 100 |
| | 916 | 39.07 | 25.42 | 29.66 | 25.07 | 119.22 |
| | 917 | 608.38 | 483.61 | 255.77 | 234 | 1,581.76 |
| | 918 | 21.41 | 9.28 | 18.91 | 3.57 | 53.17 |
| | 919 | 226.95 | 216.74 | 170.88 | 80.96 | 695.53 |
| | 920 | 1,629.17 | 1,605.20 | 1,000.13 | 996.39 | 5,230.89 |
| | 921 | 678.71 | 697.85 | 712.3 | 372.36 | 2,461.22 |
| | 922 | 360.72 | 324.5 | 241.85 | 174.41 | 1,101.48 |
| | 923 | 419.11 | 287.48 | 227.49 | 142.07 | 1,076.15 |
| | 924 | 112.44 | 101.61 | 63.43 | 59.41 | 336.89 |

| | | | | | | |
|---|---|---|---|---|---|---|
| | 925 | 282.13 | 315.89 | 158.17 | 98.76 | 854.95 |
| | 926 | 777.32 | 705.98 | 526.53 | 420.71 | 2,430.54 |
| | 927 | 113.07 | 128.6 | 81.63 | 44.07 | 367.37 |
| | 928 | 539.99 | 483.61 | 518.89 | 368.53 | 1,911.02 |
| | 930 | 383.12 | 376.73 | 262.19 | 241.76 | 1,263.80 |
| | 931 | 24.27 | 28.55 | 21.41 | 2.23 | 76.46 |
| | 932 | 91.67 | 117.8 | 82.88 | 54.6 | 346.95 |
| | 933 | 92.89 | 113.64 | 44.96 | 17.75 | 269.24 |
| | 934 | 219.55 | 201.35 | 189.04 | 131.67 | 741.61 |
| | 935 | 364.07 | 352.29 | 171.06 | 92.56 | 979.98 |
| | 936 | 471.56 | 409.56 | 339.53 | 255.77 | 1,476.42 |
| | 937 | 446.59 | 524.06 | 377.72 | 311.16 | 1,659.53 |
| | 939 | 133.06 | 104.29 | 60.13 | 42.64 | 340.12 |
| | 940 | 415.91 | 412.72 | 309.02 | 190.64 | 1,328.29 |
| | 941 | 314.65 | 249.07 | 185.33 | 110.98 | 860.03 |
| | 943 | 21.41 | 31.22 | 10.53 | 7.58 | 70.74 |
| | 944 | 117.25 | 74.13 | 67.44 | 49.6 | 308.42 |
| | 945 | 2,027.44 | 1,719.56 | 1,268.61 | 660.38 | 5,675.99 |
| | 946 | 39.79 | 24.18 | 11.69 | 0 | 75.66 |
| | 947 | 30.87 | 16.68 | 14.5 | 6.42 | 68.47 |
| | 948 | 35.77 | 22.84 | 16.15 | 3.48 | 78.24 |
| | 949 | 148.86 | 157.9 | 78.15 | 74.22 | 459.13 |
| | 950 | 285.63 | 307.43 | 217.98 | 112.14 | 923.18 |
| | 951 | 113.3 | 114.23 | 59.64 | 32.96 | 320.13 |
| | 952 | 199.96 | 157.37 | 143.18 | 70.83 | 571.34 |
| | 953 | 564.62 | 520.76 | 353.36 | 153.44 | 1,592.18 |
| | 954 | 301.35 | 287.69 | 224.54 | 164.41 | 977.99 |
| | 955 | 34.97 | 36.58 | 40.95 | 27.48 | 139.98 |
| | 956 | 648.24 | 610.62 | 362.82 | 214.24 | 1,835.92 |
| | 957 | 112.14 | 100.09 | 87.78 | 51.03 | 351.04 |
| | 958 | 377.53 | 344.32 | 177.48 | 98.22 | 997.55 |
| | 959 | 390.07 | 311.74 | 235.96 | 188.68 | 1,126.45 |
| | 960 | 259.82 | 191.8 | 69.9 | 48.44 | 569.96 |
| | 961 | 73.46 | 63.96 | 43.04 | 17.84 | 198.3 |
| TOTAL | | 17,090.47 | 15,659.81 | 11,435.87 | 8,028.86 | 52,215.01 |

RETAIL DRUG DISTRIBUTION BY ZIP CODE WITHIN STATE BY GRAMS WT

STATE: COLORADO

| ZIP CODE | QUARTER 1 | QUARTER 2 | QUARTER 3 | QUARTER 4 | TOTAL GRAMS |
|---|---|---|---|---|---|
| 800 | 654.76 | 573.13 | 372.18 | 226.64 | 1,826.71 |
| 801 | 714.88 | 663.39 | 543.51 | 388.33 | 2,310.11 |
| 802 | 839.56 | 776.44 | 522.41 | 412.33 | 2,550.74 |
| 803 | 139.02 | 102.59 | 76.72 | 45.32 | 363.65 |
| 804 | 130.07 | 103.93 | 87.38 | 21.41 | 342.79 |
| 805 | 574.22 | 449.71 | 408.27 | 271.42 | 1,703.62 |
| 806 | 222.77 | 172.71 | 101.79 | 75.11 | 572.38 |
| 807 | 42.29 | 39.07 | 27.74 | 12.98 | 122.08 |
| 808 | 165.22 | 145.37 | 127.66 | 56.29 | 494.54 |
| 809 | 467.28 | 450.56 | 355.9 | 206.61 | 1,480.35 |
| 810 | 217.53 | 168.29 | 119.01 | 49.33 | 554.16 |
| 811 | 9.63 | 8.03 | 6.78 | 6.78 | 31.22 |
| 812 | 40.77 | 29.17 | 35.33 | 27.12 | 132.39 |
| 813 | 7.4 | 7.58 | 1.78 | 0 | 16.76 |
| 814 | 1.61 | 2.41 | 2.68 | 2.68 | 9.38 |
| 815 | 87.52 | 80.38 | 54.95 | 19.36 | 242.21 |
| 816 | 26.67 | 21.41 | 11.69 | 9.28 | 69.05 |
| TOTAL | 4,341.20 | 3,794.17 | 2,855.78 | 1,830.99 | 12,822.14 |

STATE: CONNECTICUT

| ZIP CODE | QUARTER 1 | QUARTER 2 | QUARTER 3 | QUARTER 4 | TOTAL GRAMS |
|---|---|---|---|---|---|
| 60 | 463.59 | 409.47 | 285.61 | 226.64 | 1,385.31 |
| 61 | 120.61 | 118.92 | 72.62 | 32.92 | 345.07 |
| 62 | 77.7 | 63.7 | 44.52 | 27.83 | 213.75 |
| 63 | 256.21 | 251.57 | 141.93 | 78.68 | 728.39 |
| 64 | 1,167.82 | 1,086.35 | 935.9 | 525.85 | 3,715.92 |
| 65 | 451.32 | 386.42 | 292.81 | 214.95 | 1,345.50 |
| 66 | 222.85 | 219.32 | 163.08 | 140.33 | 745.58 |
| 67 | 275.88 | 251.97 | 127.93 | 100.72 | 756.5 |
| 68 | 369.55 | 341.72 | 218.07 | 167.18 | 1,096.52 |
| 69 | 50.31 | 31.94 | 37.83 | 42.29 | 162.37 |
| TOTAL | 3,455.84 | 3,161.38 | 2,320.30 | 1,557.39 | 10,494.91 |

STATE: DELAWARE

| ZIP CODE | QUARTER 1 | QUARTER 2 | QUARTER 3 | QUARTER 4 | TOTAL GRAMS |
|---|---|---|---|---|---|
| 197 | 875.86 | 898.61 | 838.75 | 670.86 | 3,284.08 |
| 198 | 518.89 | 480.04 | 329.63 | 323.3 | 1,651.86 |
| 199 | 759.99 | 809.05 | 675.28 | 545.39 | 2,789.71 |
| TOTAL | 2,154.74 | 2,187.70 | 1,843.66 | 1,539.55 | 7,725.65 |

STATE: DISTRICT OF COLUMBIA

| ZIP CODE | QUARTER 1 | QUARTER 2 | QUARTER 3 | QUARTER 4 | TOTAL GRAMS |
|---|---|---|---|---|---|
| 200 | 120.14 | 126.32 | 101.43 | 130.83 | 478.72 |
| 204 | 0 | 0.54 | 0 | 0 | 0.54 |
| TOTAL | 120.14 | 126.86 | 101.43 | 130.83 | 479.26 |

STATE: FLORIDA

| ZIP CODE | QUARTER 1 | QUARTER 2 | QUARTER 3 | QUARTER 4 | TOTAL GRAMS |
|---|---|---|---|---|---|
| 320 | 1,001.25 | 907.53 | 662.25 | 566.22 | 3,137.25 |
| 321 | 575.06 | 557.43 | 307.28 | 205.9 | 1,645.67 |
| 322 | 2,044.56 | 1,938.40 | 1,375.75 | 970.25 | 6,328.96 |
| 323 | 400.91 | 393.59 | 365.81 | 288.91 | 1,449.22 |
| 324 | 521.66 | 482.31 | 398.28 | 266.96 | 1,669.21 |
| 325 | 271.33 | 235.34 | 176.46 | 101.7 | 784.83 |
| 326 | 588.3 | 583.83 | 433.43 | 340.69 | 1,946.25 |
| 327 | 1,451.05 | 1,318.30 | 947.77 | 676.43 | 4,393.55 |
| 328 | 716.62 | 682.01 | 518.85 | 338.51 | 2,255.99 |
| 329 | 2,066.77 | 1,905.48 | 1,470.89 | 1,009.19 | 6,452.33 |
| 330 | 1,633.77 | 1,642.09 | 1,086.80 | 553.9 | 4,916.56 |
| 331 | 672.06 | 668.32 | 498.15 | 247.83 | 2,086.36 |
| 333 | 883.54 | 902.31 | 871.14 | 660.56 | 3,317.55 |
| 334 | 2,195.67 | 2,235.57 | 1,730.54 | 1,358.18 | 7,519.96 |
| 335 | 1,142.78 | 1,129.89 | 774.79 | 404.26 | 3,451.72 |
| 336 | 1,261.43 | 1,164.28 | 805.03 | 477.54 | 3,708.28 |
| 337 | 1,631.78 | 1,505.98 | 1,133.68 | 587.89 | 4,859.33 |
| 338 | 465.54 | 415.5 | 260.45 | 203.22 | 1,344.71 |
| 339 | 1,206.74 | 1,132.74 | 669.48 | 415 | 3,423.96 |
| 341 | 519.48 | 451.18 | 220.53 | 107.23 | 1,298.42 |

| | | | | | | |
|---|---|---|---|---|---|---|
| | 342 | 1,783.71 | 1,655.56 | 1,047.50 | 826.17 | 5,312.94 |
| | 344 | 632.94 | 560.33 | 399.17 | 202.24 | 1,794.68 |
| | 346 | 2,444.67 | 2,331.55 | 1,814.09 | 1,127.97 | 7,718.28 |
| | 347 | 701.01 | 640.35 | 474.86 | 340.51 | 2,156.73 |
| | 349 | 1,225.57 | 1,077.48 | 963.11 | 638.48 | 3,904.64 |
| TOTAL | | 28,038.20 | 26,517.35 | 19,406.09 | 12,915.74 | 86,877.38 |

STATE: GEORGIA

| ZIP CODE | | QUARTER 1 | QUARTER 2 | QUARTER 3 | QUARTER 4 | TOTAL GRAMS |
|---|---|---|---|---|---|---|
| | 300 | 1,323.54 | 1,310.15 | 864.22 | 649.85 | 4,147.76 |
| | 301 | 1,379.01 | 1,353.85 | 1,073.20 | 704.8 | 4,510.86 |
| | 302 | 844.27 | 724.39 | 441.63 | 324.86 | 2,335.15 |
| | 303 | 364.16 | 310 | 246.67 | 171.73 | 1,092.56 |
| | 304 | 184.04 | 187.79 | 87.96 | 62.27 | 522.06 |
| | 305 | 578.71 | 609.57 | 402.29 | 273.35 | 1,863.92 |
| | 306 | 367.01 | 330.21 | 243.28 | 158.44 | 1,098.94 |
| | 307 | 327.45 | 298.59 | 242.83 | 139.26 | 1,008.13 |
| | 308 | 78.86 | 69.32 | 65.57 | 43.89 | 257.64 |
| | 309 | 126.99 | 129.93 | 72.3 | 63.07 | 392.29 |
| | 310 | 517.24 | 452.79 | 326.95 | 196.98 | 1,493.96 |
| | 312 | 178.69 | 151.66 | 80.56 | 53.97 | 464.88 |
| | 313 | 447.43 | 473.21 | 262.9 | 121.86 | 1,305.40 |
| | 314 | 350.95 | 395.2 | 140.24 | 53.17 | 939.56 |
| | 315 | 259.29 | 249.79 | 150.63 | 102.86 | 762.57 |
| | 316 | 627.37 | 602.44 | 236.54 | 145.68 | 1,612.03 |
| | 317 | 249.25 | 240.29 | 223.87 | 142.2 | 855.61 |
| | 318 | 16.86 | 26.32 | 6.07 | 7.32 | 56.57 |
| | 319 | 285.25 | 294.08 | 189.3 | 162.54 | 931.17 |
| | 398 | 58.43 | 40.32 | 46.08 | 24.89 | 169.72 |
| TOTAL | | 8,564.80 | 8,249.90 | 5,403.09 | 3,602.99 | 25,820.78 |

STATE: HAWAII

| ZIP CODE | | QUARTER 1 | QUARTER 2 | QUARTER 3 | QUARTER 4 | TOTAL GRAMS |
|---|---|---|---|---|---|---|
| | 967 | 461.53 | 481.2 | 262.81 | 241.85 | 1,447.39 |

| | | | | | | |
|---|---|---|---|---|---|---|
| | 968 | 216.6 | 218.56 | 132.03 | 131.67 | 698.86 |
| TOTAL | | 678.13 | 699.76 | 394.84 | 373.52 | 2,146.25 |

STATE: IDAHO

| ZIP CODE | | QUARTER 1 | QUARTER 2 | QUARTER 3 | QUARTER 4 | TOTAL GRAMS |
|---|---|---|---|---|---|---|
| | 832 | 88.5 | 80.16 | 62.05 | 47.91 | 278.62 |
| | 833 | 186.05 | 184.26 | 161.56 | 115.88 | 647.75 |
| | 834 | 72.35 | 84.84 | 76.19 | 55.04 | 288.42 |
| | 835 | 34.3 | 31.22 | 22.66 | 28.73 | 116.91 |
| | 836 | 647.31 | 562.74 | 414.65 | 326.33 | 1,951.03 |
| | 837 | 453.91 | 456.5 | 341.85 | 266.11 | 1,518.37 |
| | 838 | 104.61 | 92.38 | 95.9 | 51.12 | 344.01 |
| TOTAL | | 1,587.03 | 1,492.10 | 1,174.86 | 891.12 | 5,145.11 |

STATE: ILLINOIS

| ZIP CODE | | QUARTER 1 | QUARTER 2 | QUARTER 3 | QUARTER 4 | TOTAL GRAMS |
|---|---|---|---|---|---|---|
| | 600 | 1,299.79 | 1,254.87 | 990.1 | 655.78 | 4,200.54 |
| | 601 | 507.78 | 473.88 | 403.59 | 289.66 | 1,674.91 |
| | 602 | 52.19 | 43.71 | 41.35 | 21.28 | 158.53 |
| | 603 | 49.96 | 41.48 | 38.18 | 27.21 | 156.83 |
| | 604 | 789.37 | 716.8 | 490.3 | 323.92 | 2,320.39 |
| | 605 | 540.61 | 480.4 | 319.15 | 235.29 | 1,575.45 |
| | 606 | 447.99 | 447.74 | 334.98 | 255.01 | 1,485.72 |
| | 607 | 33.54 | 20.34 | 15.7 | 29.08 | 98.66 |
| | 608 | 30.51 | 19.98 | 19.54 | 11.78 | 81.81 |
| | 609 | 89.03 | 47.82 | 33.72 | 16.95 | 187.52 |
| | 610 | 46.6 | 52.14 | 38.18 | 30.51 | 167.43 |
| | 611 | 127.66 | 128.02 | 126.86 | 40.86 | 423.4 |
| | 612 | 40.1 | 43.94 | 46.43 | 16.28 | 146.75 |
| | 613 | 67.18 | 70.12 | 58.34 | 46.21 | 241.85 |
| | 614 | 14.51 | 13.78 | 26.23 | 1.07 | 55.59 |
| | 615 | 10.19 | 12.6 | 9.1 | 6.6 | 38.49 |
| | 616 | 31.95 | 42.5 | 21.59 | 19.8 | 115.84 |
| | 617 | 117.85 | 95.54 | 52.54 | 25.16 | 291.09 |
| | 618 | 264.15 | 200.32 | 107.54 | 30.33 | 602.34 |

| | | | | | | |
|---|---|---|---|---|---|---|
| | 619 | 57.54 | 50.27 | 23.51 | 4.55 | 135.87 |
| | 620 | 154.51 | 185.65 | 71.9 | 39.88 | 451.94 |
| | 622 | 169.41 | 152.82 | 108.35 | 76.27 | 506.85 |
| | 623 | 30.96 | 24.71 | 18.91 | 18.47 | 93.05 |
| | 624 | 43.09 | 35.42 | 33.01 | 20.7 | 132.22 |
| | 625 | 82.61 | 81.98 | 32.12 | 10.35 | 207.06 |
| | 626 | 55.22 | 65.75 | 47.99 | 21.68 | 190.64 |
| | 627 | 13.02 | 26.76 | 33.81 | 20.61 | 94.2 |
| | 628 | 30.51 | 22.39 | 7.14 | 7.32 | 67.36 |
| | 629 | 106.47 | 132.92 | 88.5 | 67.35 | 395.24 |
| TOTAL | | 5,304.30 | 4,984.65 | 3,638.66 | 2,369.96 | 16,297.57 |

| STATE: | INDIANA | | | | | |
|---|---|---|---|---|---|---|
| ZIP CODE | | QUARTER 1 | QUARTER 2 | QUARTER 3 | QUARTER 4 | TOTAL GRAMS |
| | 460 | 385.88 | 341.92 | 274.72 | 137.65 | 1,140.17 |
| | 461 | 640.88 | 466.57 | 337.12 | 259.51 | 1,704.08 |
| | 462 | 1,091.20 | 871.85 | 654.8 | 574.11 | 3,191.96 |
| | 463 | 551.06 | 507.56 | 359.61 | 283.29 | 1,701.52 |
| | 464 | 2.68 | 0 | 0 | 0 | 2.68 |
| | 465 | 537.76 | 528.88 | 376.29 | 254.07 | 1,697 |
| | 466 | 102.5 | 92.51 | 44.16 | 34.17 | 273.34 |
| | 467 | 181.68 | 166.87 | 85.37 | 43.09 | 477.01 |
| | 468 | 243.45 | 196.84 | 114.23 | 60.75 | 615.27 |
| | 469 | 289.65 | 248 | 142.78 | 79.13 | 759.56 |
| | 470 | 151.57 | 116.33 | 94.38 | 69.94 | 432.22 |
| | 471 | 446.18 | 374.77 | 407.42 | 121.59 | 1,349.96 |
| | 472 | 158.88 | 138.54 | 93.94 | 45.68 | 437.04 |
| | 473 | 286.5 | 254.43 | 184.93 | 115.88 | 841.74 |
| | 474 | 227.08 | 191.04 | 140.51 | 89.12 | 647.75 |
| | 475 | 176.9 | 177.44 | 114.37 | 90.37 | 559.08 |
| | 476 | 144.97 | 126.05 | 101.57 | 75.11 | 447.7 |
| | 477 | 447.92 | 392.35 | 295.95 | 178.87 | 1,315.09 |
| | 478 | 408.05 | 369.86 | 243.28 | 215.53 | 1,236.72 |
| | 479 | 200.39 | 213.7 | 149.2 | 74.85 | 638.14 |
| TOTAL | | 6,675.18 | 5,775.51 | 4,214.63 | 2,802.71 | 19,468.03 |

DATE RANGE: 01/01/2017 TO 12/31/2017 | ARCOS 3 - REPORT 1 | Run Date: 07/03/2018

RETAIL DRUG DISTRIBUTION BY ZIP CODE WITHIN STATE BY GRAMS WT

STATE: IOWA

| ZIP CODE | QUARTER 1 | QUARTER 2 | QUARTER 3 | QUARTER 4 | TOTAL GRAMS |
|---|---|---|---|---|---|
| 500 | 128.74 | 125.8 | 91.53 | 63.96 | 410.03 |
| 501 | 100.54 | 119.59 | 77.35 | 51.92 | 349.4 |
| 502 | 169.37 | 146.57 | 120.08 | 102.06 | 538.08 |
| 503 | 439.76 | 359.38 | 262.41 | 144.79 | 1,206.34 |
| 504 | 54.51 | 43.53 | 40.14 | 30.87 | 169.05 |
| 505 | 21.5 | 19.98 | 11.42 | 13.74 | 66.64 |
| 506 | 20.07 | 19.67 | 11.11 | 2.01 | 52.86 |
| 507 | 10.71 | 4.95 | 1.07 | 0 | 16.73 |
| 508 | 3.39 | 2.68 | 3.12 | 2.68 | 11.87 |
| 510 | 21.41 | 25.96 | 29.17 | 16.24 | 92.78 |
| 511 | 19.98 | 21.9 | 7.85 | 9.86 | 59.59 |
| 512 | 1.07 | 0.54 | 0.8 | 0.27 | 2.68 |
| 513 | 3.48 | 3.39 | 2.14 | 1.96 | 10.97 |
| 514 | 6.96 | 7.49 | 2.14 | 0.27 | 16.86 |
| 515 | 110.13 | 128.91 | 66.19 | 44.43 | 349.66 |
| 516 | 6.51 | 2.5 | 2.59 | 0 | 11.6 |
| 520 | 32.03 | 28.46 | 46.03 | 3.21 | 109.73 |
| 521 | 0 | 1.07 | 0 | 0 | 1.07 |
| 522 | 13.56 | 14.32 | 7.14 | 2.85 | 37.87 |
| 523 | 23.73 | 25.16 | 27.57 | 23.46 | 99.92 |
| 524 | 21.23 | 27.92 | 27.97 | 12.31 | 89.43 |
| 525 | 50.63 | 35.15 | 21.86 | 19.09 | 126.73 |
| 526 | 15.52 | 12.58 | 2.41 | 3.21 | 33.72 |
| 527 | 36.95 | 29.8 | 14.63 | 1.69 | 83.07 |
| 528 | 64.05 | 43 | 34.52 | 19 | 160.57 |
| TOTAL | 1,375.83 | 1,250.30 | 911.24 | 569.88 | 4,107.25 |

STATE: KANSAS

| ZIP CODE | QUARTER 1 | QUARTER 2 | QUARTER 3 | QUARTER 4 | TOTAL GRAMS |
|---|---|---|---|---|---|
| 660 | 708.15 | 570.94 | 347.38 | 293.59 | 1,920.06 |
| 661 | 107.68 | 94.65 | 88.59 | 103.93 | 394.85 |
| 662 | 490.93 | 437.58 | 314.96 | 195.33 | 1,438.80 |

| | | | | | | |
|---|---|---|---|---|---|---|
| | 664 | 49.15 | 40.14 | 44.78 | 32.03 | 166.1 |
| | 665 | 18.02 | 18.38 | 20.16 | 14.1 | 70.66 |
| | 666 | 140.51 | 131.18 | 90.1 | 39.07 | 400.86 |
| | 667 | 44.07 | 39.97 | 38.09 | 21.05 | 143.18 |
| | 668 | 25.78 | 23.82 | 21.95 | 13.38 | 84.93 |
| | 669 | 18.56 | 17.75 | 9.9 | 6.87 | 53.08 |
| | 670 | 147.64 | 195.82 | 129.18 | 88.14 | 560.78 |
| | 671 | 52.9 | 57.36 | 37.91 | 37.47 | 185.64 |
| | 672 | 384 | 407.29 | 292.61 | 114.95 | 1,198.85 |
| | 673 | 41.66 | 34.7 | 23.55 | 16.5 | 116.41 |
| | 674 | 50.05 | 35.42 | 27.74 | 19.45 | 132.66 |
| | 675 | 89.21 | 97.51 | 72.71 | 69.49 | 328.92 |
| | 676 | 32.38 | 29.17 | 19.27 | 20.16 | 100.98 |
| | 677 | 9.55 | 12.67 | 10.35 | 2.5 | 35.07 |
| | 678 | 29.8 | 39.43 | 33.45 | 24.53 | 127.21 |
| | 679 | 18.2 | 23.64 | 8.3 | 1.87 | 52.01 |
| TOTAL | | 2,458.24 | 2,307.42 | 1,630.98 | 1,114.41 | 7,511.05 |

| STATE: | KENTUCKY | | | | |
|---|---|---|---|---|---|
| ZIP CODE | QUARTER 1 | QUARTER 2 | QUARTER 3 | QUARTER 4 | TOTAL GRAMS |
| 400 | 107.59 | 93.63 | 49.02 | 26.36 | 276.6 |
| 401 | 94.74 | 111.16 | 77.43 | 74.94 | 358.27 |
| 402 | 610.06 | 530.93 | 355.64 | 244.3 | 1,740.93 |
| 403 | 319.28 | 339.58 | 279.99 | 254.83 | 1,193.68 |
| 404 | 240.47 | 181.77 | 145.01 | 108.43 | 675.68 |
| 405 | 217 | 203.53 | 173.65 | 164.64 | 758.82 |
| 406 | 31.76 | 15.79 | 13.02 | 6.82 | 67.39 |
| 407 | 363.49 | 359.65 | 315.27 | 259.42 | 1,297.83 |
| 408 | 51.79 | 47.64 | 20.79 | 14.27 | 134.49 |
| 409 | 180.96 | 166.06 | 110.89 | 95.72 | 553.63 |
| 410 | 562.47 | 513.54 | 356.57 | 189.71 | 1,622.29 |
| 411 | 140.06 | 146.71 | 95.01 | 79.49 | 461.27 |
| 412 | 13.2 | 13.56 | 7.49 | 7.49 | 41.74 |
| 413 | 19.36 | 25.34 | 18.29 | 6.42 | 69.41 |
| 414 | 6.96 | 7.49 | 1.61 | 0.54 | 16.6 |
| 415 | 229.05 | 258.89 | 121.95 | 88.41 | 698.3 |

| | | | | | |
|---|---|---|---|---|---|
| 416 | 74.31 | 78.15 | 64.63 | 79.04 | 296.13 |
| 417 | 58.12 | 62.58 | 29.71 | 13.78 | 164.19 |
| 418 | 7.49 | 2.85 | 2.85 | 1.78 | 14.97 |
| 420 | 197.42 | 185.65 | 124.27 | 54.51 | 561.85 |
| 421 | 241.67 | 240.02 | 187.79 | 130.96 | 800.44 |
| 422 | 207.9 | 251.17 | 197.82 | 154.33 | 811.22 |
| 423 | 120.83 | 136.22 | 77.61 | 38.27 | 372.93 |
| 424 | 125.96 | 115.97 | 69.32 | 52.28 | 363.53 |
| 425 | 75.52 | 83.46 | 59.19 | 55.31 | 273.48 |
| 426 | 34.17 | 40.77 | 28.37 | 14.81 | 118.12 |
| 427 | 222.49 | 207.59 | 150.14 | 139.12 | 719.34 |
| TOTAL | 4,554.12 | 4,419.70 | 3,133.33 | 2,355.98 | 14,463.13 |

STATE: LOUISIANA

| ZIP CODE | QUARTER 1 | QUARTER 2 | QUARTER 3 | QUARTER 4 | TOTAL GRAMS |
|---|---|---|---|---|---|
| 700 | 1,110.98 | 978.41 | 690.57 | 633.39 | 3,413.35 |
| 701 | 154.15 | 163.34 | 188.86 | 164.68 | 671.03 |
| 703 | 350.77 | 326.82 | 221.69 | 209.55 | 1,108.83 |
| 704 | 1,352.20 | 1,186.09 | 839.24 | 684.91 | 4,062.44 |
| 705 | 649.94 | 662.34 | 534.19 | 445.51 | 2,291.98 |
| 706 | 86.09 | 64.5 | 47.91 | 23.02 | 221.52 |
| 707 | 1,973.73 | 1,797.40 | 1,381.60 | 1,211.56 | 6,364.29 |
| 708 | 1,183.77 | 1,180.34 | 950.76 | 803.2 | 4,118.07 |
| 710 | 232.12 | 227.26 | 135.42 | 77.97 | 672.77 |
| 711 | 792.54 | 791.16 | 551.09 | 344.13 | 2,478.92 |
| 712 | 68.96 | 59.77 | 71.19 | 27.57 | 227.49 |
| 713 | 156.07 | 141.75 | 98.84 | 60.57 | 457.23 |
| 714 | 94.12 | 84.88 | 44.96 | 23.28 | 247.24 |
| TOTAL | 8,205.44 | 7,664.06 | 5,756.32 | 4,709.34 | 26,335.16 |

STATE: MAINE

| ZIP CODE | QUARTER 1 | QUARTER 2 | QUARTER 3 | QUARTER 4 | TOTAL GRAMS |
|---|---|---|---|---|---|
| 39 | 5.35 | 1.96 | 0.89 | 1.07 | 9.27 |
| 40 | 285.25 | 241.76 | 190.86 | 120.88 | 838.75 |
| 41 | 12.12 | 12.04 | 12.4 | 12.22 | 48.78 |

| | | | | | |
|---|---|---|---|---|---|
| 42 | 210.98 | 196.53 | 151.17 | 113.43 | 672.11 |
| 43 | 310.45 | 237.74 | 262.86 | 225.43 | 1,036.48 |
| 44 | 28.55 | 28.9 | 21.68 | 10.97 | 90.1 |
| 45 | 62.18 | 57.81 | 64.23 | 40.59 | 224.81 |
| 46 | 3.75 | 3.48 | 4.1 | 3.03 | 14.36 |
| 47 | 21.68 | 14.18 | 4.55 | 1.07 | 41.48 |
| 48 | 11.51 | 12.49 | 5.08 | 6.42 | 35.5 |
| 49 | 238.28 | 198.98 | 139.97 | 109.59 | 686.82 |
| TOTAL | 1,190.10 | 1,005.87 | 857.79 | 644.7 | 3,698.46 |

STATE: MARYLAND

| ZIP CODE | QUARTER 1 | QUARTER 2 | QUARTER 3 | QUARTER 4 | TOTAL GRAMS |
|---|---|---|---|---|---|
| 206 | 1,196.44 | 1,246.53 | 954.06 | 901.24 | 4,298.27 |
| 207 | 813.82 | 757.71 | 662.92 | 583.97 | 2,818.42 |
| 208 | 874.08 | 775.73 | 617.82 | 465.1 | 2,732.73 |
| 209 | 106.43 | 107.32 | 85.91 | 39.88 | 339.54 |
| 210 | 2,294.44 | 2,114.81 | 1,505.02 | 1,268.70 | 7,182.97 |
| 211 | 961.42 | 929.57 | 719.52 | 647.62 | 3,258.13 |
| 212 | 1,347.29 | 1,297.96 | 932.74 | 770.57 | 4,348.56 |
| 214 | 265.22 | 252.87 | 189.39 | 166.29 | 873.77 |
| 215 | 419.02 | 342.39 | 293.41 | 249.65 | 1,304.47 |
| 216 | 306.44 | 289.49 | 238.99 | 214.59 | 1,049.51 |
| 217 | 2,173.91 | 1,994.47 | 1,652.75 | 1,362.50 | 7,183.63 |
| 218 | 211.87 | 183.95 | 102.41 | 69.14 | 567.37 |
| 219 | 316.07 | 308.93 | 242.83 | 224.94 | 1,092.77 |
| TOTAL | 11,286.45 | 10,601.73 | 8,197.77 | 6,964.19 | 37,050.14 |

STATE: MASSACHUSETTS

| ZIP CODE | QUARTER 1 | QUARTER 2 | QUARTER 3 | QUARTER 4 | TOTAL GRAMS |
|---|---|---|---|---|---|
| 10 | 119.72 | 110.62 | 84.21 | 41.39 | 355.94 |
| 11 | 11.6 | 16.24 | 10.08 | 1.78 | 39.7 |
| 12 | 31.05 | 36.93 | 33.72 | 19.98 | 121.68 |
| 13 | 25.92 | 36.62 | 28.37 | 22.03 | 112.94 |
| 14 | 34.44 | 21.59 | 12.58 | 11.24 | 79.85 |
| 15 | 131.7 | 110.44 | 104.73 | 78.77 | 425.64 |

| | | | | | | |
|---|---|---|---|---|---|---|
| | 16 | 35.15 | 31.4 | 41.13 | 12.04 | 119.72 |
| | 17 | 91.57 | 103.57 | 64.23 | 10.71 | 270.08 |
| | 18 | 1,214.15 | 1,114.01 | 658.24 | 275.44 | 3,261.84 |
| | 19 | 77.97 | 91.57 | 92.29 | 80.65 | 342.48 |
| | 20 | 68.44 | 62.54 | 82.56 | 59.59 | 273.13 |
| | 21 | 119.43 | 99.99 | 111.29 | 109.1 | 439.81 |
| | 22 | 106.52 | 103.57 | 21.59 | 36.75 | 268.43 |
| | 23 | 71.33 | 98.09 | 78.77 | 47.82 | 296.01 |
| | 24 | 94.29 | 117.13 | 77.08 | 30.6 | 319.1 |
| | 25 | 22.21 | 30.26 | 19.89 | 22.21 | 94.57 |
| | 26 | 42.73 | 40.23 | 36.75 | 28.64 | 148.35 |
| | 27 | 564.85 | 505.64 | 411.08 | 250.32 | 1,731.89 |
| TOTAL | | 2,863.07 | 2,730.44 | 1,968.59 | 1,139.06 | 8,701.16 |

STATE: MICHIGAN

| ZIP CODE | QUARTER 1 | QUARTER 2 | QUARTER 3 | QUARTER 4 | TOTAL GRAMS |
|---|---|---|---|---|---|
| 480 | 4,027.62 | 4,084.88 | 3,818.99 | 3,537.09 | 15,468.58 |
| 481 | 2,854.81 | 3,454.70 | 2,964.81 | 2,769.35 | 12,043.67 |
| 482 | 8,564.16 | 8,751.14 | 7,608.54 | 7,069.62 | 31,993.46 |
| 483 | 2,168.25 | 1,834.47 | 1,612.87 | 1,351.89 | 6,967.48 |
| 484 | 275.93 | 236.94 | 181.81 | 150.5 | 845.18 |
| 485 | 292.92 | 243.59 | 202.46 | 125.47 | 864.44 |
| 486 | 197.87 | 176.9 | 126.86 | 91.62 | 593.25 |
| 487 | 128.28 | 120.17 | 64.19 | 44.84 | 357.48 |
| 488 | 214.36 | 220.3 | 179.4 | 151.48 | 765.54 |
| 489 | 130.87 | 116.29 | 87.43 | 71.19 | 405.78 |
| 490 | 438.56 | 441.86 | 369.6 | 263.08 | 1,513.10 |
| 491 | 54.6 | 52.99 | 50.49 | 47.28 | 205.36 |
| 492 | 177.71 | 198.14 | 152.46 | 108.75 | 637.06 |
| 493 | 45.27 | 64.77 | 51.65 | 35.24 | 196.93 |
| 494 | 208.93 | 224.54 | 189.08 | 106.61 | 729.16 |
| 495 | 164.15 | 118.74 | 81.54 | 61.02 | 425.45 |
| 496 | 192.65 | 175.16 | 146.48 | 122.04 | 636.33 |
| 497 | 227.22 | 291.4 | 216.16 | 170.52 | 905.3 |
| 498 | 17.71 | 24.35 | 22.44 | 5.44 | 69.94 |
| 499 | 15.52 | 33.45 | 2.32 | 21.23 | 72.52 |

| | | | | | |
|---|---|---|---|---|---|
| TOTAL | 20,397.39 | 20,864.78 | 18,129.58 | 16,304.26 | 75,696.01 |

STATE: MINNESOTA

| ZIP CODE | QUARTER 1 | QUARTER 2 | QUARTER 3 | QUARTER 4 | TOTAL GRAMS |
|---|---|---|---|---|---|
| 550 | 76.02 | 85.11 | 60.22 | 40.77 | 262.12 |
| 551 | 112 | 110.95 | 77.7 | 65.3 | 365.95 |
| 553 | 86.49 | 104.02 | 87.25 | 70.83 | 348.59 |
| 554 | 62.49 | 63.7 | 61.55 | 28.32 | 216.06 |
| 557 | 29.35 | 27.3 | 33.01 | 14.99 | 104.65 |
| 558 | 14.45 | 20.34 | 14.63 | 6.42 | 55.84 |
| 559 | 8.94 | 12.77 | 12.4 | 6.96 | 41.07 |
| 560 | 16.5 | 16.33 | 15.52 | 8.3 | 56.65 |
| 561 | 2.68 | 6.42 | 0 | 0 | 9.1 |
| 562 | 10.17 | 8.39 | 5.71 | 3.21 | 27.48 |
| 563 | 44.25 | 28.55 | 14.81 | 8.56 | 96.17 |
| 564 | 14.76 | 12.31 | 3.35 | 0.67 | 31.09 |
| 565 | 7.14 | 8.92 | 7.14 | 6.24 | 29.44 |
| 566 | 3.57 | 1.07 | 0 | 0 | 4.64 |
| 567 | 12.49 | 13.2 | 10.88 | 12.67 | 49.24 |
| TOTAL | 501.3 | 519.38 | 404.17 | 273.24 | 1,698.09 |

STATE: MISSISSIPPI

| ZIP CODE | QUARTER 1 | QUARTER 2 | QUARTER 3 | QUARTER 4 | TOTAL GRAMS |
|---|---|---|---|---|---|
| 386 | 224.27 | 236.27 | 161.96 | 127.57 | 750.07 |
| 387 | 20.12 | 28.32 | 22.57 | 15.7 | 86.71 |
| 388 | 495.76 | 524.02 | 414.65 | 291.58 | 1,726.01 |
| 389 | 61.55 | 43.62 | 39.52 | 21.95 | 166.64 |
| 390 | 209.51 | 198.27 | 160.04 | 76.63 | 644.45 |
| 391 | 171.02 | 107.41 | 65.57 | 46.39 | 390.39 |
| 392 | 260.23 | 220.57 | 142.87 | 93.76 | 717.43 |
| 393 | 174 | 172.04 | 113.43 | 54.28 | 513.75 |
| 394 | 374.77 | 358.4 | 224.85 | 147.82 | 1,105.84 |
| 395 | 1,720.68 | 1,527.23 | 1,245.68 | 1,046.61 | 5,540.20 |
| 396 | 98.4 | 118.56 | 76.32 | 55.67 | 348.95 |
| 397 | 76.63 | 54.64 | 71.41 | 55.13 | 257.81 |

| | | | | | |
|---|---|---|---|---|---|
| TOTAL | 3,886.94 | 3,589.35 | 2,738.87 | 2,033.09 | 12,248.25 |

STATE: MISSOURI

| ZIP CODE | QUARTER 1 | QUARTER 2 | QUARTER 3 | QUARTER 4 | TOTAL GRAMS |
|---|---|---|---|---|---|
| 630 | 705.38 | 666.18 | 442.93 | 253.71 | 2,068.20 |
| 631 | 1,274.55 | 1,061.96 | 811.1 | 611.18 | 3,758.79 |
| 633 | 461.48 | 437.22 | 285.47 | 143.98 | 1,328.15 |
| 634 | 87.92 | 80.56 | 67.09 | 41.66 | 277.23 |
| 635 | 12.94 | 20.61 | 14.99 | 6.69 | 55.23 |
| 636 | 202.6 | 195.77 | 132.57 | 71.01 | 601.95 |
| 637 | 133.1 | 91.4 | 55.71 | 25.69 | 305.9 |
| 638 | 27.3 | 24.44 | 15.34 | 4.91 | 71.99 |
| 639 | 3.66 | 5.35 | 0.45 | 1.34 | 10.8 |
| 640 | 526.34 | 496.1 | 300.19 | 197.24 | 1,519.87 |
| 641 | 523.66 | 428.83 | 249.61 | 197.69 | 1,399.79 |
| 644 | 22.12 | 16.33 | 9.63 | 3.21 | 51.29 |
| 645 | 216.51 | 231.32 | 143.09 | 88.59 | 679.51 |
| 646 | 94.56 | 84.84 | 85.02 | 28.19 | 292.61 |
| 647 | 133.46 | 157.37 | 107.68 | 50.31 | 448.82 |
| 648 | 155.31 | 151.92 | 105.49 | 28.81 | 441.53 |
| 650 | 103.26 | 108.12 | 85.46 | 59.68 | 356.52 |
| 651 | 96.84 | 89.25 | 89.17 | 53.79 | 329.05 |
| 652 | 121.59 | 98.62 | 55.13 | 49.78 | 325.12 |
| 653 | 43.98 | 37.16 | 23.91 | 17.66 | 122.71 |
| 654 | 62.27 | 58.74 | 52.77 | 17.75 | 191.53 |
| 655 | 99.56 | 101.21 | 66.46 | 38.32 | 305.55 |
| 656 | 113.07 | 120.17 | 108.88 | 30.06 | 372.18 |
| 657 | 306.08 | 304.92 | 206.21 | 166.33 | 983.54 |
| 658 | 326.11 | 262.41 | 207.37 | 134.35 | 930.24 |
| TOTAL | 5,853.65 | 5,330.80 | 3,721.72 | 2,321.93 | 17,228.10 |

STATE: MONTANA

| ZIP CODE | QUARTER 1 | QUARTER 2 | QUARTER 3 | QUARTER 4 | TOTAL GRAMS |
|---|---|---|---|---|---|
| 590 | 6.24 | 4.73 | 1.61 | 0 | 12.58 |
| 591 | 48.82 | 66.28 | 42.82 | 31.22 | 189.14 |
| 592 | 0.89 | 0 | 0 | 0 | 0.89 |

| | | | | | | |
|---|---|---|---|---|---|---|
| | 593 | 3.03 | 1.34 | 0 | 0 | 4.37 |
| | 594 | 111.42 | 39.74 | 40.55 | 24.09 | 215.8 |
| | 596 | 17.8 | 4.28 | 6.42 | 2.14 | 30.64 |
| | 597 | 35.83 | 27.12 | 19.98 | 5.53 | 88.46 |
| | 598 | 42.46 | 32.47 | 42.37 | 24 | 141.3 |
| | 599 | 121.5 | 120.57 | 136.22 | 71.28 | 449.57 |
| TOTAL | | 387.99 | 296.53 | 289.97 | 158.26 | 1,132.75 |

STATE: NEBRASKA

| ZIP CODE | | QUARTER 1 | QUARTER 2 | QUARTER 3 | QUARTER 4 | TOTAL GRAMS |
|---|---|---|---|---|---|---|
| | 680 | 92.42 | 108.39 | 63.07 | 24.44 | 288.32 |
| | 681 | 816.9 | 753.78 | 542.22 | 293.37 | 2,406.27 |
| | 683 | 12.04 | 9.28 | 9.99 | 15.25 | 46.56 |
| | 684 | 9.1 | 21.68 | 19.54 | 18.47 | 68.79 |
| | 685 | 277.09 | 252.06 | 172.93 | 120.52 | 822.6 |
| | 686 | 47.1 | 53.84 | 47.24 | 45.99 | 194.17 |
| | 687 | 132.25 | 135.64 | 88.36 | 73.2 | 429.45 |
| | 688 | 43.53 | 55.09 | 31.45 | 7.36 | 137.43 |
| | 689 | 25.92 | 29.53 | 5.17 | 3.39 | 64.01 |
| | 690 | 4.64 | 0 | 0 | 0 | 4.64 |
| | 691 | 37.91 | 24.18 | 19.8 | 8.03 | 89.92 |
| | 692 | 2.68 | 5.26 | 4.46 | 0 | 12.4 |
| | 693 | 65.61 | 63.16 | 53.84 | 20.16 | 202.77 |
| TOTAL | | 1,567.19 | 1,511.89 | 1,058.07 | 630.18 | 4,767.33 |

STATE: NEVADA

| ZIP CODE | | QUARTER 1 | QUARTER 2 | QUARTER 3 | QUARTER 4 | TOTAL GRAMS |
|---|---|---|---|---|---|---|
| | 890 | 1,360.41 | 1,243.50 | 983.14 | 590.48 | 4,177.53 |
| | 891 | 3,349.13 | 2,936.47 | 2,063.25 | 1,253.27 | 9,602.12 |
| | 893 | 2.99 | 0 | 0 | 1.78 | 4.77 |
| | 894 | 257.24 | 226.99 | 193.9 | 113.97 | 792.1 |
| | 895 | 344.48 | 319.91 | 192.34 | 82.74 | 939.47 |
| | 897 | 114.55 | 108.21 | 78.5 | 95.45 | 396.71 |

| | | | | | |
|---|---|---|---|---|---|
| | 898 | 5.35 | 4.28 | 3.75 | 4.28 | 17.66 |
| TOTAL | | 5,434.15 | 4,839.36 | 3,514.88 | 2,141.97 | 15,930.36 |

STATE: NEW HAMPSHIRE

| ZIP CODE | QUARTER 1 | QUARTER 2 | QUARTER 3 | QUARTER 4 | TOTAL GRAMS |
|---|---|---|---|---|---|
| 30 | 246.26 | 206.88 | 202.77 | 137.65 | 793.56 |
| 31 | 106.35 | 108.48 | 68.33 | 34.61 | 317.77 |
| 32 | 145.01 | 142.25 | 113.52 | 85.55 | 486.33 |
| 33 | 149.38 | 142.74 | 110.35 | 101.92 | 504.39 |
| 34 | 20.16 | 17.93 | 13.65 | 9.72 | 61.46 |
| 35 | 19.72 | 25.25 | 22.3 | 11.78 | 79.05 |
| 37 | 33.01 | 13.2 | 20.96 | 8.03 | 75.2 |
| 38 | 375.87 | 336.1 | 274.32 | 187.52 | 1,173.81 |
| TOTAL | 1,095.76 | 992.83 | 826.2 | 576.78 | 3,491.57 |

STATE: NEW JERSEY

| ZIP CODE | QUARTER 1 | QUARTER 2 | QUARTER 3 | QUARTER 4 | TOTAL GRAMS |
|---|---|---|---|---|---|
| 70 | 1,258.71 | 1,185.69 | 842.41 | 741.07 | 4,027.88 |
| 71 | 89.57 | 107.85 | 80.02 | 44.16 | 321.6 |
| 72 | 47.91 | 48.98 | 40.41 | 23.55 | 160.85 |
| 73 | 55.67 | 58.61 | 38.18 | 31.58 | 184.04 |
| 74 | 389.04 | 335.22 | 245.42 | 216.25 | 1,185.93 |
| 75 | 87.25 | 87.96 | 83.05 | 61.73 | 319.99 |
| 76 | 417.45 | 470.63 | 320.4 | 226.06 | 1,434.54 |
| 77 | 732.55 | 648.2 | 488.83 | 412.95 | 2,282.53 |
| 78 | 271.15 | 247.69 | 167.09 | 126.41 | 812.34 |
| 79 | 173.34 | 136.31 | 127.84 | 70.74 | 508.23 |
| 80 | 2,088.02 | 1,928.01 | 1,183.24 | 826.31 | 6,025.58 |
| 81 | 133.15 | 101.74 | 102.5 | 79.04 | 416.43 |
| 82 | 189.08 | 183.15 | 97.68 | 62.63 | 532.54 |
| 83 | 212.99 | 193.99 | 113.92 | 57.81 | 578.71 |
| 84 | 20.96 | 24.18 | 21.95 | 16.68 | 83.77 |
| 85 | 304.56 | 236.23 | 198.31 | 179.94 | 919.04 |
| 86 | 167.71 | 135.15 | 96.35 | 45.76 | 444.97 |
| 87 | 632.72 | 593.16 | 373.25 | 315.27 | 1,914.40 |

| | | | | | | |
|---|---|---|---|---|---|---|
| | 88 | 1,060.57 | 1,031.09 | 778.71 | 497.79 | 3,368.16 |
| | 89 | 59.86 | 55.85 | 23.37 | 33.45 | 172.53 |
| TOTAL | | 8,392.26 | 7,809.69 | 5,422.93 | 4,069.18 | 25,694.06 |

STATE: NEW MEXICO

| ZIP CODE | | QUARTER 1 | QUARTER 2 | QUARTER 3 | QUARTER 4 | TOTAL GRAMS |
|---|---|---|---|---|---|---|
| | 870 | 101.34 | 91.75 | 94.52 | 74.8 | 362.41 |
| | 871 | 671.8 | 518.82 | 435.43 | 347.07 | 1,973.12 |
| | 873 | 45.1 | 30.24 | 33.72 | 33.86 | 142.92 |
| | 874 | 12.76 | 13.56 | 14.81 | 14.81 | 55.94 |
| | 875 | 165.66 | 175.57 | 89.66 | 71.1 | 501.99 |
| | 877 | 5.44 | 12.22 | 4.19 | 9.37 | 31.22 |
| | 879 | 15.61 | 23.11 | 8.03 | 4.46 | 51.21 |
| | 880 | 137.12 | 118.47 | 68.83 | 22.21 | 346.63 |
| | 881 | 1.28 | 1.47 | 0 | 0 | 2.75 |
| | 882 | 57.94 | 55 | 36.53 | 50.85 | 200.32 |
| | 883 | 34.7 | 36.13 | 23.91 | 24.09 | 118.83 |
| | 884 | 3.21 | 3.21 | 4.28 | 0 | 10.7 |
| TOTAL | | 1,251.96 | 1,079.55 | 813.91 | 652.62 | 3,798.04 |

STATE: NEW YORK

| ZIP CODE | | QUARTER 1 | QUARTER 2 | QUARTER 3 | QUARTER 4 | TOTAL GRAMS |
|---|---|---|---|---|---|---|
| | 100 | 1,718.54 | 1,695.18 | 1,458.58 | 1,246.53 | 6,118.83 |
| | 101 | 64.5 | 59.41 | 46.57 | 36.58 | 207.06 |
| | 102 | 1.61 | 4.55 | 3.48 | 0 | 9.64 |
| | 103 | 1,155.54 | 1,136.22 | 797.94 | 664.26 | 3,753.96 |
| | 104 | 783 | 742.32 | 575.36 | 407.82 | 2,508.50 |
| | 105 | 453.72 | 456.67 | 338.91 | 272.71 | 1,522.01 |
| | 106 | 106.78 | 97.42 | 50.85 | 32.56 | 287.61 |
| | 107 | 215.75 | 194.03 | 108.3 | 67.44 | 585.52 |
| | 108 | 71.19 | 50.14 | 47.73 | 36.31 | 205.37 |
| | 109 | 926.71 | 882.96 | 720.46 | 555.24 | 3,085.37 |
| | 110 | 237.39 | 238.77 | 222 | 169.1 | 867.26 |
| | 111 | 216.91 | 196.35 | 158.17 | 191.09 | 762.52 |
| | 112 | 2,134.72 | 2,105.98 | 1,823.41 | 1,433.16 | 7,497.27 |

| | | | | | |
|---|---|---|---|---|---|
| 113 | 547.04 | 442.39 | 335.12 | 299.3 | 1,623.85 |
| 114 | 222.67 | 223.65 | 194.3 | 178.87 | 819.49 |
| 115 | 634.77 | 618.85 | 468.49 | 358.94 | 2,081.05 |
| 116 | 91.26 | 77.88 | 76.63 | 62.45 | 308.22 |
| 117 | 2,400.46 | 2,287.08 | 1,467.19 | 1,173.42 | 7,328.15 |
| 118 | 39.61 | 47.01 | 26.32 | 22.66 | 135.6 |
| 119 | 195.01 | 223.92 | 151.66 | 111.24 | 681.83 |
| 120 | 213.21 | 233.73 | 154.69 | 125.7 | 727.33 |
| 121 | 134.26 | 120.7 | 97.82 | 38.63 | 391.41 |
| 122 | 55.13 | 66.46 | 55.04 | 21.14 | 197.77 |
| 123 | 47.73 | 55.49 | 34.35 | 21.14 | 158.71 |
| 124 | 374.73 | 386.37 | 318.21 | 269.95 | 1,349.26 |
| 125 | 633.97 | 662.96 | 529.24 | 390.74 | 2,216.91 |
| 126 | 182.43 | 179.49 | 172.18 | 176.19 | 710.29 |
| 127 | 338.15 | 269.59 | 268.03 | 152.86 | 1,028.63 |
| 128 | 261.57 | 227.84 | 197.78 | 127.04 | 814.23 |
| 129 | 483.56 | 488.96 | 410.81 | 292.21 | 1,675.54 |
| 130 | 605.33 | 545.56 | 466.12 | 332.22 | 1,949.23 |
| 131 | 153.13 | 134.13 | 88.85 | 81.09 | 457.2 |
| 132 | 217.49 | 198.58 | 110 | 68.25 | 594.32 |
| 133 | 152.46 | 166.06 | 122.49 | 95.37 | 536.38 |
| 134 | 209.78 | 194.79 | 119.18 | 47.64 | 571.39 |
| 135 | 59.73 | 47.91 | 33.63 | 17.08 | 158.35 |
| 136 | 270.13 | 219.46 | 204.51 | 137.43 | 831.53 |
| 137 | 146.44 | 177.17 | 135.06 | 108.93 | 567.6 |
| 138 | 345.6 | 370.94 | 277.8 | 210.98 | 1,205.32 |
| 139 | 232.39 | 214.59 | 202.69 | 179.04 | 828.71 |
| 140 | 1,145.46 | 995.76 | 812.69 | 593.29 | 3,547.20 |
| 141 | 661.36 | 621.35 | 483.52 | 301.35 | 2,067.58 |
| 142 | 1,249.79 | 1,117.62 | 854.45 | 676.93 | 3,898.79 |
| 143 | 338.2 | 275.61 | 225.97 | 239.04 | 1,078.82 |
| 144 | 183.77 | 146.62 | 141.22 | 113.48 | 585.09 |
| 145 | 146.46 | 131.41 | 118.11 | 74.67 | 470.65 |
| 146 | 143.18 | 131.58 | 76.85 | 96.66 | 448.27 |
| 147 | 182.79 | 169.77 | 143.98 | 95.32 | 591.86 |
| 148 | 451.67 | 424.1 | 392.7 | 281.28 | 1,549.75 |
| 149 | 66.33 | 70.39 | 52.46 | 27.48 | 216.66 |

| | | | | | | |
|---|---|---|---|---|---|---|
| TOTAL | | 21,903.41 | 20,825.77 | 16,371.90 | 12,712.81 | 71,813.89 |

STATE: NORTH CAROLINA

| ZIP CODE | | QUARTER 1 | QUARTER 2 | QUARTER 3 | QUARTER 4 | TOTAL GRAMS |
|---|---|---|---|---|---|---|
| | 270 | 1,313.04 | 1,167.31 | 650.61 | 327.94 | 3,458.90 |
| | 271 | 946.19 | 619.61 | 506.45 | 303.05 | 2,375.30 |
| | 272 | 2,240.37 | 1,982.16 | 1,178.95 | 685.49 | 6,086.97 |
| | 273 | 1,042.33 | 850.22 | 637.36 | 306.61 | 2,836.52 |
| | 274 | 543.78 | 517.86 | 428.61 | 300.41 | 1,790.66 |
| | 275 | 2,269.77 | 2,044.47 | 1,384.54 | 851.15 | 6,549.93 |
| | 276 | 574.02 | 524.06 | 353.67 | 281.37 | 1,733.12 |
| | 277 | 400.51 | 439.93 | 267.99 | 200.9 | 1,309.33 |
| | 278 | 2,305.81 | 2,331.55 | 1,957.18 | 1,703.42 | 8,297.96 |
| | 279 | 414.38 | 334.36 | 304.92 | 264.24 | 1,317.90 |
| | 280 | 2,603.90 | 2,390.59 | 1,718.45 | 1,426.60 | 8,139.54 |
| | 281 | 1,732.02 | 1,602.27 | 1,156.34 | 852.62 | 5,343.25 |
| | 282 | 948.15 | 921.58 | 781.79 | 633.3 | 3,284.82 |
| | 283 | 3,004.55 | 2,604.31 | 1,439.63 | 803.43 | 7,851.92 |
| | 284 | 1,774.16 | 1,590.88 | 940.45 | 606.32 | 4,911.81 |
| | 285 | 1,469.24 | 1,394.71 | 1,094.70 | 944.47 | 4,903.12 |
| | 286 | 3,207.80 | 2,965.88 | 2,013.78 | 1,320.66 | 9,508.12 |
| | 287 | 671.89 | 614.34 | 375.62 | 206.34 | 1,868.19 |
| | 288 | 139.57 | 142.6 | 73.69 | 29.71 | 385.57 |
| | 289 | 292.07 | 298.59 | 231.23 | 181.54 | 1,003.43 |
| TOTAL | | 27,893.55 | 25,337.28 | 17,495.96 | 12,229.57 | 82,956.36 |

STATE: NORTH DAKOTA

| ZIP CODE | | QUARTER 1 | QUARTER 2 | QUARTER 3 | QUARTER 4 | TOTAL GRAMS |
|---|---|---|---|---|---|---|
| | 580 | 0.89 | 0.89 | 1.78 | 0.89 | 4.45 |
| | 581 | 11.6 | 6.42 | 5.44 | 4.55 | 28.01 |
| | 582 | 6.6 | 7.76 | 5.71 | 0.89 | 20.96 |
| | 583 | 2.68 | 3.48 | 14.45 | 8.03 | 28.64 |
| | 584 | 8.56 | 6.24 | 8.56 | 3.12 | 26.48 |
| | 585 | 83.32 | 85.73 | 57.01 | 42.82 | 268.88 |
| | 586 | 4.28 | 3.21 | 6.96 | 4.64 | 19.09 |

| | | | | | | |
|---|---|---|---|---|---|---|
| | 587 | 32.65 | 33.81 | 34.61 | 32.92 | 133.99 |
| | 588 | 23.64 | 33.19 | 25.16 | 17.13 | 99.12 |
| TOTAL | | 174.22 | 180.73 | 159.68 | 114.99 | 629.62 |

| STATE: | OHIO | | | | | |
|---|---|---|---|---|---|---|
| ZIP CODE | | QUARTER 1 | QUARTER 2 | QUARTER 3 | QUARTER 4 | TOTAL GRAMS |
| | 430 | 575.63 | 592.59 | 465.05 | 303.31 | 1,936.58 |
| | 431 | 433.78 | 387.04 | 302.42 | 157.81 | 1,281.05 |
| | 432 | 1,315.21 | 1,167.94 | 780.72 | 520.86 | 3,784.73 |
| | 433 | 152.82 | 107.19 | 76.63 | 54.37 | 391.01 |
| | 434 | 270.48 | 257.42 | 198.8 | 196.17 | 922.87 |
| | 435 | 326.39 | 270.71 | 253.62 | 156.47 | 1,007.19 |
| | 436 | 632.28 | 572.55 | 436.33 | 320.17 | 1,961.33 |
| | 437 | 120.52 | 125.25 | 77.52 | 66.64 | 389.93 |
| | 438 | 42.73 | 42.91 | 44.07 | 39.7 | 169.41 |
| | 439 | 473.93 | 454.52 | 389.94 | 263.08 | 1,581.47 |
| | 440 | 479.1 | 516.21 | 354.43 | 194.25 | 1,543.99 |
| | 441 | 390.12 | 388.2 | 301.93 | 130.43 | 1,210.68 |
| | 442 | 401.4 | 382.18 | 294.17 | 173.65 | 1,251.40 |
| | 443 | 152.5 | 126.32 | 99.65 | 54.15 | 432.62 |
| | 444 | 371.6 | 373.66 | 287.57 | 172.89 | 1,205.72 |
| | 445 | 216.47 | 196.08 | 180.83 | 157.5 | 750.88 |
| | 446 | 353.18 | 311.66 | 279.67 | 200.77 | 1,145.28 |
| | 447 | 263.44 | 269.24 | 215.67 | 135.24 | 883.59 |
| | 448 | 477.72 | 509.52 | 437.04 | 363.8 | 1,788.08 |
| | 449 | 60.13 | 14.9 | 44.16 | 24.44 | 143.63 |
| | 450 | 1,093.67 | 969.31 | 708.91 | 527.05 | 3,298.94 |
| | 451 | 467.86 | 394.32 | 320.31 | 223.56 | 1,406.05 |
| | 452 | 686.18 | 610.51 | 436.37 | 264.78 | 1,997.84 |
| | 453 | 364.33 | 320.71 | 224.54 | 188.95 | 1,098.53 |
| | 454 | 408.32 | 394 | 286.27 | 162.45 | 1,251.04 |
| | 455 | 124.05 | 104.55 | 69.41 | 22.66 | 320.67 |
| | 456 | 1,319.51 | 1,349.12 | 1,045.10 | 910.7 | 4,624.43 |
| | 457 | 167.67 | 184.49 | 118.74 | 55.67 | 526.57 |
| | 458 | 79.08 | 67.93 | 64.37 | 32.47 | 243.85 |

| | | | | | |
|---|---|---|---|---|---|
| TOTAL | 12,220.10 | 11,461.03 | 8,794.24 | 6,073.99 | 38,549.36 |

STATE: OKLAHOMA

| ZIP CODE | QUARTER 1 | QUARTER 2 | QUARTER 3 | QUARTER 4 | TOTAL GRAMS |
|---|---|---|---|---|---|
| 730 | 1,859.54 | 1,711.05 | 1,429.05 | 1,205.41 | 6,205.05 |
| 731 | 1,517.21 | 1,380.53 | 959.59 | 607.07 | 4,464.40 |
| 734 | 86.71 | 102.41 | 83.77 | 49.07 | 321.96 |
| 735 | 286.45 | 277.67 | 257.68 | 117.18 | 938.98 |
| 736 | 88.76 | 79.26 | 59.28 | 31.4 | 258.7 |
| 737 | 60.04 | 52.81 | 26.94 | 24.53 | 164.32 |
| 738 | 20.88 | 41.66 | 26.05 | 15.52 | 104.11 |
| 739 | 0 | 0.27 | 0 | 0 | 0.27 |
| 740 | 916.94 | 882.2 | 673.76 | 369.73 | 2,842.63 |
| 741 | 785 | 744.28 | 475.94 | 256.88 | 2,262.10 |
| 743 | 224.68 | 171.73 | 114.55 | 64.05 | 575.01 |
| 744 | 295.29 | 279.94 | 199.3 | 124.27 | 898.8 |
| 745 | 185.47 | 203.71 | 149.43 | 131.58 | 670.19 |
| 746 | 62.8 | 51.92 | 71.23 | 50.67 | 236.62 |
| 747 | 69.14 | 78.33 | 35.55 | 41.04 | 224.06 |
| 748 | 355.28 | 349.44 | 213.93 | 161.74 | 1,080.39 |
| 749 | 169.68 | 180.16 | 111.51 | 51.92 | 513.27 |
| TOTAL | 6,983.87 | 6,587.37 | 4,887.56 | 3,302.06 | 21,760.86 |

STATE: OREGON

| ZIP CODE | QUARTER 1 | QUARTER 2 | QUARTER 3 | QUARTER 4 | TOTAL GRAMS |
|---|---|---|---|---|---|
| 970 | 369.92 | 362.12 | 292.52 | 178.78 | 1,203.34 |
| 971 | 66.25 | 81.27 | 59.41 | 47.82 | 254.75 |
| 972 | 375.04 | 368.91 | 287.79 | 164.32 | 1,196.06 |
| 973 | 332.95 | 245.42 | 198.4 | 129.35 | 906.12 |
| 974 | 489.19 | 451.63 | 453.37 | 339.44 | 1,733.63 |
| 975 | 133.01 | 137.29 | 90.5 | 63.79 | 424.59 |
| 976 | 11.06 | 12.76 | 8.39 | 13.11 | 45.32 |
| 977 | 123.67 | 111.82 | 105 | 76.27 | 416.76 |
| 978 | 75.68 | 72.08 | 52.63 | 68.16 | 268.55 |
| 979 | 55.85 | 33.54 | 28.01 | 14.18 | 131.58 |

| | | | | | |
|---|---|---|---|---|---|
| TOTAL | 2,032.62 | 1,876.84 | 1,576.02 | 1,095.22 | 6,580.70 |

STATE: PENNSYLVANIA

| ZIP CODE | QUARTER 1 | QUARTER 2 | QUARTER 3 | QUARTER 4 | TOTAL GRAMS |
|---|---|---|---|---|---|
| 150 | 1,986.13 | 1,952.27 | 1,323.61 | 1,003.97 | 6,265.98 |
| 151 | 1,012.93 | 960.52 | 835.14 | 672.64 | 3,481.23 |
| 152 | 2,743.83 | 2,629.46 | 1,911.06 | 1,616.35 | 8,900.70 |
| 153 | 701.1 | 658.64 | 545.52 | 464.92 | 2,370.18 |
| 154 | 1,373.39 | 1,356.39 | 1,127.57 | 991.79 | 4,849.14 |
| 155 | 447.74 | 444.53 | 407.87 | 268.61 | 1,568.75 |
| 156 | 1,326.24 | 1,417.46 | 1,186.54 | 951.83 | 4,882.07 |
| 157 | 663.54 | 562.34 | 473.93 | 359.61 | 2,059.42 |
| 158 | 499.93 | 537.58 | 464.87 | 403.5 | 1,905.88 |
| 159 | 1,591.06 | 1,574.02 | 1,366.88 | 1,100.41 | 5,632.37 |
| 160 | 631.79 | 635.49 | 502.39 | 472.55 | 2,242.22 |
| 161 | 1,587.27 | 1,568.67 | 1,381.64 | 1,206.07 | 5,743.65 |
| 162 | 463.31 | 494.31 | 418.8 | 375.13 | 1,751.55 |
| 163 | 606.27 | 605.96 | 566.53 | 495.43 | 2,274.19 |
| 164 | 163.88 | 149.87 | 142.78 | 106.78 | 563.31 |
| 165 | 325.39 | 319.68 | 254.07 | 198.8 | 1,097.94 |
| 166 | 859.58 | 829.25 | 662.65 | 615.73 | 2,967.21 |
| 167 | 163.79 | 234.58 | 210.45 | 193.23 | 802.05 |
| 168 | 153.42 | 128.33 | 99.42 | 66.06 | 447.23 |
| 169 | 20.34 | 19.27 | 14.45 | 18.73 | 72.79 |
| 170 | 895.36 | 927.11 | 781.3 | 601.41 | 3,205.18 |
| 171 | 232.26 | 236.41 | 214.33 | 163.61 | 846.61 |
| 172 | 480.44 | 497.21 | 362.5 | 294.97 | 1,635.12 |
| 173 | 587.76 | 562.42 | 410.1 | 364.16 | 1,924.44 |
| 174 | 340.83 | 355.1 | 326.91 | 242.87 | 1,265.71 |
| 175 | 298.5 | 301.26 | 229.72 | 167.36 | 996.84 |
| 176 | 168.12 | 189.3 | 153.84 | 146.48 | 657.74 |
| 177 | 64.68 | 64.77 | 57.63 | 35.59 | 222.67 |
| 178 | 98.04 | 100.58 | 57.09 | 23.46 | 279.17 |
| 179 | 40.77 | 40.77 | 25.6 | 18.56 | 125.7 |
| 180 | 223.43 | 250.19 | 155.05 | 103.31 | 731.98 |
| 181 | 140.28 | 139.21 | 87.07 | 81.76 | 448.32 |

| | | | | | | |
|---|---|---|---|---|---|---|
| | 182 | 32.78 | 31.8 | 38.49 | 104.96 | 208.03 |
| | 183 | 112.81 | 116.29 | 109.55 | 35.55 | 374.2 |
| | 184 | 184.58 | 191.27 | 153.26 | 77.79 | 606.9 |
| | 185 | 146.08 | 148.71 | 85.02 | 57.27 | 437.08 |
| | 186 | 143.72 | 141.09 | 146.39 | 109.55 | 540.75 |
| | 187 | 1,447.57 | 1,289.40 | 862.71 | 585.31 | 4,184.99 |
| | 188 | 35.37 | 33.81 | 39.88 | 24.8 | 133.86 |
| | 189 | 219.99 | 256.12 | 214.95 | 157.14 | 848.2 |
| | 190 | 1,874.88 | 1,710.24 | 1,362.46 | 929.66 | 5,877.24 |
| | 191 | 657.75 | 593.42 | 441.01 | 414.17 | 2,106.35 |
| | 193 | 509.39 | 420.4 | 337.79 | 228.73 | 1,496.31 |
| | 194 | 386.41 | 353 | 265.31 | 208.39 | 1,213.11 |
| | 195 | 109.01 | 86.4 | 77.52 | 48.53 | 321.46 |
| | 196 | 175.03 | 116.2 | 116.06 | 93.67 | 500.96 |
| TOTAL | | 26,926.77 | 26,231.10 | 21,007.71 | 16,901.20 | 91,066.78 |

STATE: RHODE ISLAND

| ZIP CODE | QUARTER 1 | QUARTER 2 | QUARTER 3 | QUARTER 4 | TOTAL GRAMS |
|---|---|---|---|---|---|
| 28 | 306.86 | 280.57 | 185.65 | 70.03 | 843.11 |
| 29 | 270 | 206.01 | 178.73 | 104.15 | 758.89 |
| TOTAL | 576.86 | 486.58 | 364.38 | 174.18 | 1,602 |

STATE: SOUTH CAROLINA

| ZIP CODE | QUARTER 1 | QUARTER 2 | QUARTER 3 | QUARTER 4 | TOTAL GRAMS |
|---|---|---|---|---|---|
| 290 | 927.92 | 842.63 | 517.64 | 393.42 | 2,681.61 |
| 291 | 710.51 | 732.99 | 512.6 | 379.32 | 2,335.42 |
| 292 | 910.57 | 958.16 | 805.25 | 656.81 | 3,330.79 |
| 293 | 363.4 | 306.75 | 136.67 | 68.42 | 875.24 |
| 294 | 1,687.14 | 1,642.71 | 1,091.17 | 751.82 | 5,172.84 |
| 295 | 1,004.01 | 939.69 | 735.58 | 479.15 | 3,158.43 |
| 296 | 1,175.69 | 1,187.39 | 703.38 | 487.98 | 3,554.44 |
| 297 | 495.34 | 405.28 | 264.24 | 161.87 | 1,326.73 |
| 298 | 178.69 | 199.92 | 101.07 | 94.74 | 574.42 |
| 299 | 313.8 | 287.84 | 239.17 | 151.92 | 992.73 |
| TOTAL | 7,767.07 | 7,503.36 | 5,106.77 | 3,625.45 | 24,002.65 |

RETAIL DRUG DISTRIBUTION BY ZIP CODE WITHIN STATE BY GRAMS WT

STATE: SOUTH DAKOTA

| ZIP CODE | QUARTER 1 | QUARTER 2 | QUARTER 3 | QUARTER 4 | TOTAL GRAMS |
|---|---|---|---|---|---|
| 570 | 78.46 | 62.45 | 43.09 | 43 | 227 |
| 571 | 135.85 | 133.37 | 111.87 | 63.43 | 444.52 |
| 572 | 6.42 | 7.49 | 4.28 | 4.46 | 22.65 |
| 573 | 5.62 | 5.62 | 1.34 | 1.78 | 14.36 |
| 574 | 24.44 | 7.85 | 9.28 | 6.96 | 48.53 |
| 575 | 11.15 | 7.32 | 13.47 | 6.24 | 38.18 |
| 576 | 6.42 | 3.57 | 0 | 1.43 | 11.42 |
| 577 | 363.38 | 305.9 | 235.78 | 200.45 | 1,105.51 |
| TOTAL | 631.74 | 533.57 | 419.11 | 327.75 | 1,912.17 |

STATE: TENNESSEE

| ZIP CODE | QUARTER 1 | QUARTER 2 | QUARTER 3 | QUARTER 4 | TOTAL GRAMS |
|---|---|---|---|---|---|
| 370 | 3,408.09 | 3,262.05 | 2,451.01 | 2,019.09 | 11,140.24 |
| 371 | 2,166.20 | 2,011.46 | 1,496.27 | 1,179.13 | 6,853.06 |
| 372 | 1,636.95 | 1,348.50 | 1,013.65 | 933.76 | 4,932.86 |
| 373 | 1,688.66 | 1,649.27 | 1,165.31 | 856.59 | 5,359.83 |
| 374 | 849.86 | 865.6 | 706.19 | 490.92 | 2,912.57 |
| 376 | 1,539.10 | 1,375.80 | 1,065.57 | 651.59 | 4,632.06 |
| 377 | 4,124.89 | 4,025.29 | 3,516.03 | 3,304.43 | 14,970.64 |
| 378 | 4,791.38 | 4,384.49 | 3,651.32 | 3,202.24 | 16,029.43 |
| 379 | 1,798.16 | 1,612.60 | 1,328.20 | 1,064.05 | 5,803.01 |
| 380 | 203.84 | 169.9 | 161.2 | 89.52 | 624.46 |
| 381 | 197.73 | 191.62 | 107.1 | 134.17 | 630.62 |
| 382 | 264.33 | 229.49 | 178.2 | 123.87 | 795.89 |
| 383 | 534.41 | 464.47 | 353.54 | 292.88 | 1,645.30 |
| 384 | 464.52 | 393.19 | 285.47 | 177.44 | 1,320.62 |
| 385 | 1,521.79 | 1,362.77 | 974.22 | 724.79 | 4,583.57 |
| TOTAL | 25,189.91 | 23,346.50 | 18,453.28 | 15,244.47 | 82,234.16 |

STATE: TEXAS

| ZIP CODE | QUARTER 1 | QUARTER 2 | QUARTER 3 | QUARTER 4 | TOTAL GRAMS |
|---|---|---|---|---|---|

| | | | | | | |
|---|---|---|---|---|---|---|
| 750 | 2,068.56 | 1,811.72 | | 1,232.57 | 884.96 | 5,997.81 |
| 751 | 295.96 | 298.9 | | 182.61 | 105.98 | 883.45 |
| 752 | 448.52 | 423.21 | | 329.15 | 206.3 | 1,407.18 |
| 754 | 245.51 | 207.86 | | 83.59 | 49.42 | 586.38 |
| 755 | 9.37 | 11.46 | | 8.43 | 0 | 29.26 |
| 756 | 29.97 | 33.81 | | 25.42 | 5.08 | 94.28 |
| 757 | 59.86 | 72.04 | | 63.25 | 39.16 | 234.31 |
| 758 | 62.27 | 50.49 | | 49.51 | 22.03 | 184.3 |
| 759 | 88.81 | 76.05 | | 54.06 | 34.52 | 253.44 |
| 760 | 928.23 | 927.74 | | 704.8 | 432.49 | 2,993.26 |
| 761 | 752.62 | 738.17 | | 543.07 | 238.28 | 2,272.14 |
| 762 | 274.41 | 353.67 | | 170.17 | 119.45 | 917.7 |
| 763 | 276.77 | 277.76 | | 210.09 | 194.88 | 959.5 |
| 764 | 74 | 46.17 | | 36.93 | 27.48 | 184.58 |
| 765 | 153.89 | 147.46 | | 85.24 | 49.42 | 436.01 |
| 766 | 22.08 | 24.35 | | 5.8 | 1.61 | 53.84 |
| 767 | 18.69 | 36.04 | | 24 | 21.5 | 100.23 |
| 768 | 48.89 | 34.35 | | 14.45 | 22.75 | 120.44 |
| 769 | 93.22 | 72.08 | | 57.63 | 57.09 | 280.02 |
| 770 | 816.76 | 823.32 | | 560.06 | 372.45 | 2,572.59 |
| 773 | 601.41 | 550.11 | | 338.15 | 219.55 | 1,709.22 |
| 774 | 805.57 | 691.07 | | 406.26 | 265.18 | 2,168.08 |
| 775 | 849.73 | 757.66 | | 589.68 | 394.98 | 2,592.05 |
| 776 | 328.96 | 285.83 | | 168.83 | 135.42 | 919.04 |
| 777 | 125.12 | 162.45 | | 93.67 | 85.64 | 466.88 |
| 778 | 142.91 | 123.96 | | 98.98 | 45.85 | 411.7 |
| 779 | 145.01 | 124.85 | | 81.14 | 55.18 | 406.18 |
| 780 | 231.59 | 206.79 | | 154.15 | 77.43 | 669.96 |
| 781 | 268.88 | 268.34 | | 184.84 | 83.05 | 805.11 |
| 782 | 1,745.63 | 1,592.28 | | 1,149.34 | 664.48 | 5,151.73 |
| 783 | 73.82 | 64.77 | | 31.58 | 20.07 | 190.24 |
| 784 | 225.52 | 212.72 | | 135.24 | 80.11 | 653.59 |
| 785 | 42.29 | 57.9 | | 27.03 | 5.53 | 132.75 |
| 786 | 1,208.55 | 1,097.91 | | 779.7 | 612.29 | 3,698.45 |
| 787 | 1,083.81 | 957.45 | | 678.84 | 438.33 | 3,158.43 |
| 788 | 48.84 | 37.11 | | 31.49 | 13.56 | 131 |
| 789 | 33.63 | 39.07 | | 21.81 | 14.45 | 108.96 |

| | | | | | | |
|---|---|---|---|---|---|---|
| | 790 | 84.93 | 64.81 | 62.76 | 16.15 | 228.65 |
| | 791 | 182.17 | 248.05 | 162.99 | 105.45 | 698.66 |
| | 792 | 1.16 | 1.87 | 0 | 0 | 3.03 |
| | 793 | 33.63 | 26.05 | 18.02 | 4.73 | 82.43 |
| | 794 | 67.44 | 59.9 | 37.6 | 24.58 | 189.52 |
| | 795 | 0.54 | 3.66 | 1.61 | 1.25 | 7.06 |
| | 796 | 43.94 | 33.19 | 30.06 | 12.13 | 119.32 |
| | 797 | 152.64 | 156.83 | 80.38 | 32.12 | 421.97 |
| | 798 | 12.85 | 0 | 0 | 0 | 12.85 |
| | 799 | 173.78 | 140.33 | 90.19 | 66.37 | 470.67 |
| TOTAL | | 15,482.74 | 14,431.61 | 9,895.17 | 6,358.73 | 46,168.25 |

STATE: UTAH

| ZIP CODE | | QUARTER 1 | QUARTER 2 | QUARTER 3 | QUARTER 4 | TOTAL GRAMS |
|---|---|---|---|---|---|---|
| | 840 | 2,406.80 | 2,277.58 | 1,909.36 | 1,693.79 | 8,287.53 |
| | 841 | 873.28 | 771.94 | 653.02 | 523.66 | 2,821.90 |
| | 843 | 167.54 | 154.87 | 117.44 | 89.34 | 529.19 |
| | 844 | 515.05 | 525.13 | 418.57 | 368.93 | 1,827.68 |
| | 845 | 82.12 | 74.53 | 73.06 | 64.01 | 293.72 |
| | 846 | 380.17 | 375.31 | 320.13 | 261.7 | 1,337.31 |
| | 847 | 391.36 | 333.82 | 345.91 | 231.01 | 1,302.10 |
| TOTAL | | 4,816.32 | 4,513.18 | 3,837.49 | 3,232.44 | 16,399.43 |

STATE: VERMONT

| ZIP CODE | | QUARTER 1 | QUARTER 2 | QUARTER 3 | QUARTER 4 | TOTAL GRAMS |
|---|---|---|---|---|---|---|
| | 50 | 8.83 | 16.06 | 13.11 | 10.88 | 48.88 |
| | 51 | 11.33 | 4.01 | 1.07 | 0 | 16.41 |
| | 52 | 27.57 | 17.84 | 17.84 | 22.3 | 85.55 |
| | 53 | 5.89 | 6.42 | 5.62 | 0 | 17.93 |
| | 54 | 212.5 | 216.42 | 174.14 | 115.17 | 718.23 |
| | 56 | 28.37 | 25.59 | 18.2 | 9.46 | 81.62 |
| | 57 | 65.08 | 35.51 | 42.73 | 20.43 | 163.75 |
| | 58 | 3.12 | 3.57 | 2.68 | 3.12 | 12.49 |
| TOTAL | | 362.69 | 325.42 | 275.39 | 181.36 | 1,144.86 |

STATE: VIRGINIA

| ZIP CODE | QUARTER 1 | QUARTER 2 | QUARTER 3 | QUARTER 4 | TOTAL GRAMS |
|---|---|---|---|---|---|
| 201 | 952.9 | 902.14 | 732.99 | 618.58 | 3,206.61 |
| 220 | 744.74 | 704.76 | 537.13 | 494.13 | 2,480.76 |
| 221 | 742.6 | 714.3 | 623.71 | 585.89 | 2,666.50 |
| 222 | 168.92 | 155.76 | 140.51 | 148.89 | 614.08 |
| 223 | 219.77 | 193.99 | 138.05 | 185.33 | 737.14 |
| 224 | 1,145.90 | 948.17 | 711.54 | 674.43 | 3,480.04 |
| 225 | 150.32 | 188.59 | 135.02 | 108.48 | 582.41 |
| 226 | 472.01 | 475.62 | 411.61 | 338.28 | 1,697.52 |
| 227 | 58.03 | 51.34 | 49.11 | 46.03 | 204.51 |
| 228 | 63.79 | 71.37 | 40.99 | 29.35 | 205.5 |
| 229 | 169.23 | 177.8 | 128.15 | 112.58 | 587.76 |
| 230 | 266.96 | 300.24 | 312.64 | 333.07 | 1,212.91 |
| 231 | 377.09 | 353.27 | 272.05 | 135.06 | 1,137.47 |
| 232 | 557.7 | 455.64 | 276.96 | 205 | 1,495.30 |
| 233 | 381.32 | 429.37 | 302.29 | 250.59 | 1,363.57 |
| 234 | 872.91 | 798.93 | 624.02 | 498.15 | 2,794.01 |
| 235 | 251.24 | 313.26 | 188.63 | 162.36 | 915.49 |
| 236 | 266.29 | 251.75 | 189.75 | 123.56 | 831.35 |
| 237 | 173.07 | 216.51 | 129.35 | 102.15 | 621.08 |
| 238 | 428.88 | 381.11 | 293.33 | 264.11 | 1,367.43 |
| 239 | 76.1 | 86.89 | 79.17 | 59.41 | 301.57 |
| 240 | 587 | 457.25 | 379.77 | 259.65 | 1,683.67 |
| 241 | 279.81 | 280.07 | 165.26 | 149.61 | 874.75 |
| 242 | 504.57 | 451.4 | 387.71 | 271.24 | 1,614.92 |
| 243 | 667.16 | 233.28 | 155.58 | 124 | 1,180.02 |
| 244 | 90.28 | 63.96 | 22.57 | 31.4 | 208.21 |
| 245 | 454.61 | 457.78 | 327.13 | 242.65 | 1,482.17 |
| 246 | 263.44 | 260.58 | 144.16 | 97.42 | 765.6 |
| TOTAL | 11,386.64 | 10,375.13 | 7,899.18 | 6,651.40 | 36,312.35 |

STATE: WASHINGTON

| ZIP CODE | QUARTER 1 | QUARTER 2 | QUARTER 3 | QUARTER 4 | TOTAL GRAMS |
|---|---|---|---|---|---|
| 980 | 835.1 | 714.68 | 572.77 | 399.08 | 2,521.63 |

| | | | | | | |
|---|---|---|---|---|---|---|
| | 981 | 217.93 | 215.52 | 190.64 | 126.23 | 750.32 |
| | 982 | 439.06 | 403.45 | 329.54 | 306.88 | 1,478.93 |
| | 983 | 287.15 | 306.66 | 229.58 | 164.77 | 988.16 |
| | 984 | 184.97 | 171.02 | 125.65 | 95.19 | 576.83 |
| | 985 | 303.89 | 263.3 | 179.89 | 143 | 890.08 |
| | 986 | 501.83 | 483.58 | 400.2 | 310.09 | 1,695.70 |
| | 988 | 57.72 | 42.73 | 30.87 | 20.96 | 152.28 |
| | 989 | 112.05 | 94.29 | 70.3 | 52.63 | 329.27 |
| | 990 | 42.15 | 21.01 | 25.2 | 20.16 | 108.52 |
| | 991 | 31.6 | 19.89 | 18.91 | 4.64 | 75.04 |
| | 992 | 345.78 | 298.59 | 236.5 | 145.01 | 1,025.88 |
| | 993 | 258.17 | 214.91 | 171.91 | 113.65 | 758.64 |
| | 994 | 16.59 | 17.13 | 12.31 | 8.03 | 54.06 |
| TOTAL | | 3,633.99 | 3,266.76 | 2,594.27 | 1,910.32 | 11,405.34 |

STATE: WEST VIRGINIA

| ZIP CODE | | QUARTER 1 | QUARTER 2 | QUARTER 3 | QUARTER 4 | TOTAL GRAMS |
|---|---|---|---|---|---|---|
| | 247 | 58.88 | 83.77 | 55.13 | 37.33 | 235.11 |
| | 248 | 137.34 | 129.18 | 119.05 | 72.35 | 457.92 |
| | 249 | 63.07 | 66.73 | 42.64 | 28.73 | 201.17 |
| | 250 | 121.91 | 123.47 | 98.49 | 16.24 | 360.11 |
| | 251 | 107.68 | 114.28 | 35.95 | 18.91 | 276.82 |
| | 252 | 43.27 | 31.58 | 18.2 | 6.87 | 99.92 |
| | 253 | 206.57 | 225.12 | 209.24 | 112.58 | 753.51 |
| | 254 | 915.61 | 901.2 | 873.46 | 726.66 | 3,416.93 |
| | 255 | 242.87 | 241.71 | 206.92 | 131.45 | 822.95 |
| | 256 | 78.01 | 86.8 | 47.28 | 49.42 | 261.51 |
| | 257 | 138.68 | 85.15 | 68.78 | 58.88 | 351.49 |
| | 258 | 88.41 | 126.68 | 59.68 | 16.59 | 291.36 |
| | 259 | 46.75 | 49.78 | 38.18 | 20.79 | 155.5 |
| | 260 | 366.3 | 356.97 | 297.78 | 194.7 | 1,215.75 |
| | 261 | 42.73 | 34.17 | 36.58 | 9.19 | 122.67 |
| | 262 | 39.21 | 41.04 | 38.14 | 15.52 | 133.91 |
| | 263 | 39.21 | 28.77 | 21.54 | 20.88 | 110.4 |
| | 264 | 18.56 | 17.49 | 14.27 | 6.07 | 56.39 |
| | 265 | 58.61 | 79.49 | 46.3 | 39.79 | 224.19 |

| | | | | | | |
|---|---|---|---|---|---|---|
| | 266 | 23.51 | 20.88 | 10.17 | 8.39 | 62.95 |
| | 267 | 88.85 | 67.8 | 61.11 | 57.9 | 275.66 |
| | 268 | 3.48 | 17.4 | 11.24 | 5.35 | 37.47 |
| TOTAL | | 2,929.51 | 2,929.46 | 2,410.13 | 1,654.59 | 9,923.69 |

STATE: WISCONSIN

| ZIP CODE | | QUARTER 1 | QUARTER 2 | QUARTER 3 | QUARTER 4 | TOTAL GRAMS |
|---|---|---|---|---|---|---|
| | 530 | 423.22 | 385.58 | 255.14 | 156.92 | 1,220.86 |
| | 531 | 685.72 | 575.78 | 433.29 | 288.15 | 1,982.94 |
| | 532 | 522.4 | 431.52 | 302.33 | 155.76 | 1,412.01 |
| | 534 | 124.89 | 100.81 | 93.14 | 91.53 | 410.37 |
| | 535 | 132.83 | 122.04 | 81.63 | 59.86 | 396.36 |
| | 537 | 70.85 | 54.87 | 34.88 | 15.88 | 176.48 |
| | 538 | 8.13 | 3.48 | 1.61 | 3.21 | 16.43 |
| | 539 | 66.73 | 43.71 | 30.55 | 18.29 | 159.28 |
| | 540 | 26.85 | 26.23 | 19.63 | 14.27 | 86.98 |
| | 541 | 97.48 | 77.26 | 40.01 | 24.09 | 238.84 |
| | 542 | 39.21 | 37.96 | 19.98 | 14.81 | 111.96 |
| | 543 | 69.99 | 89.03 | 72.48 | 38 | 269.5 |
| | 544 | 132.66 | 126.14 | 122.98 | 95.41 | 477.19 |
| | 545 | 25.25 | 34.44 | 18.73 | 10.44 | 88.86 |
| | 546 | 1.87 | 0 | 0.54 | 4.01 | 6.42 |
| | 547 | 31.05 | 20.34 | 9.9 | 11.33 | 72.62 |
| | 548 | 24 | 35.06 | 20.79 | 18.64 | 98.49 |
| | 549 | 441.42 | 375.44 | 286.23 | 182.7 | 1,285.79 |
| TOTAL | | 2,924.55 | 2,539.69 | 1,843.84 | 1,203.30 | 8,511.38 |

STATE: WYOMING

| ZIP CODE | | QUARTER 1 | QUARTER 2 | QUARTER 3 | QUARTER 4 | TOTAL GRAMS |
|---|---|---|---|---|---|---|
| | 820 | 40.14 | 38.72 | 26.23 | 25.78 | 130.87 |
| | 822 | 12.31 | 18.02 | 5.89 | 0 | 36.22 |
| | 823 | 0.89 | 0 | 1.78 | 2.68 | 5.35 |
| | 824 | 6.16 | 3.21 | 5.89 | 2.68 | 17.94 |
| | 825 | 30.96 | 51.3 | 43.31 | 44.2 | 169.77 |

| | | | | | | |
|---|---|---|---|---|---|---|
| | 826 | 45.68 | 47.28 | 55.76 | 47.55 | 196.27 |
| | 827 | 24.8 | 26.67 | 23.11 | 29.44 | 104.02 |
| | 828 | 56.92 | 53.35 | 17.57 | 12.67 | 140.51 |
| | 829 | 73.06 | 68.87 | 47.1 | 20.96 | 209.99 |
| | 830 | 4.28 | 6.78 | 10.71 | 0 | 21.77 |
| | 831 | 27.3 | 28.64 | 18.02 | 13.38 | 87.34 |
| TOTAL | | 322.5 | 342.84 | 255.37 | 199.34 | 1,120.05 |

DRUG CODE:9668DRUG NAME:NOROXYMORPHONE

STATE: ARIZONA

| ZIP CODE | | QUARTER 1 | QUARTER 2 | QUARTER 3 | QUARTER 4 | TOTAL GRAMS |
|---|---|---|---|---|---|---|
| | 857 | 0 | 0 | 0.1 | 0 | 0.1 |
| TOTAL | | 0 | 0 | 0.1 | 0 | 0.1 |

STATE: ARKANSAS

| ZIP CODE | | QUARTER 1 | QUARTER 2 | QUARTER 3 | QUARTER 4 | TOTAL GRAMS |
|---|---|---|---|---|---|---|
| | 722 | 7.22 | 7.13 | 11.64 | 7.34 | 33.33 |
| | 727 | 7.41 | 10.08 | 15.21 | 12.04 | 44.74 |
| TOTAL | | 14.63 | 17.21 | 26.85 | 19.38 | 78.07 |

STATE: CALIFORNIA

| ZIP CODE | | QUARTER 1 | QUARTER 2 | QUARTER 3 | QUARTER 4 | TOTAL GRAMS |
|---|---|---|---|---|---|---|
| | 902 | 6.65 | 10.23 | 3.17 | 15.49 | 35.54 |
| | 905 | 5.68 | 4.37 | 3.49 | 2.37 | 15.91 |
| | 906 | 4.23 | 6.22 | 5.84 | 7.83 | 24.12 |
| | 907 | 2.71 | 4.78 | 5.23 | 3.79 | 16.51 |
| | 908 | 1.06 | 0 | 4 | 4.91 | 9.97 |
| | 912 | 2.25 | 2.25 | 7.61 | 0.62 | 12.73 |
| | 914 | 0 | 0 | 0.1 | 0 | 0.1 |
| | 919 | 3.74 | 4.01 | 5.2 | 2.59 | 15.54 |
| | 920 | 3.73 | 4.07 | 4.84 | 6.74 | 19.38 |
| | 921 | 2.04 | 2.36 | 2.78 | 1.12 | 8.3 |
| | 923 | 2.69 | 7.11 | 0.1 | 4.87 | 14.77 |
| | 925 | 0 | 1.84 | 5.84 | 0 | 7.68 |

| | | | | | | |
|---|---|---|---|---|---|---|
| | 927 | 0 | 0 | 0 | 0.1 | 0.1 |
| | 928 | 17.07 | 20.41 | 15.85 | 13.24 | 66.57 |
| | 943 | 0 | 0 | 1.56 | 0 | 1.56 |
| | 946 | 0.75 | 0.87 | 4.23 | 4.97 | 10.82 |
| TOTAL | | 52.6 | 68.52 | 69.84 | 68.64 | 259.6 |

STATE: FLORIDA

| ZIP CODE | | QUARTER 1 | QUARTER 2 | QUARTER 3 | QUARTER 4 | TOTAL GRAMS |
|---|---|---|---|---|---|---|
| | 322 | 0 | 0.1 | 1.56 | 0.08 | 1.74 |
| | 327 | 0.04 | 0 | 0 | 0 | 0.04 |
| | 328 | 0 | 0.29 | 1.06 | 0.06 | 1.41 |
| | 329 | 1.45 | 1.81 | 0 | 0 | 3.26 |
| | 330 | 0 | 0.25 | 0 | 0 | 0.25 |
| | 331 | 2.42 | 2.67 | 1.93 | 4.91 | 11.93 |
| | 333 | 8.7 | 7.15 | 8.96 | 10.9 | 35.71 |
| | 334 | 0 | 1.56 | 0 | 1.06 | 2.62 |
| | 336 | 0 | 0 | 0 | 1.06 | 1.06 |
| TOTAL | | 12.61 | 13.83 | 13.51 | 18.07 | 58.02 |

STATE: GEORGIA

| ZIP CODE | | QUARTER 1 | QUARTER 2 | QUARTER 3 | QUARTER 4 | TOTAL GRAMS |
|---|---|---|---|---|---|---|
| | 300 | 3.06 | 0.37 | 2.59 | 0.12 | 6.14 |
| | 303 | 0.12 | 1.75 | 4.95 | 3.63 | 10.45 |
| | 309 | 0 | 1.06 | 1.76 | 1.05 | 3.87 |
| TOTAL | | 3.18 | 3.18 | 9.3 | 4.8 | 20.46 |

STATE: ILLINOIS

| ZIP CODE | | QUARTER 1 | QUARTER 2 | QUARTER 3 | QUARTER 4 | TOTAL GRAMS |
|---|---|---|---|---|---|---|
| | 601 | 0.32 | 0 | 0.1 | 0 | 0.42 |
| | 606 | 4.1 | 9.76 | 14.99 | 8.75 | 37.6 |
| TOTAL | | 4.42 | 9.76 | 15.09 | 8.75 | 38.02 |

STATE: KANSAS

| ZIP CODE | QUARTER 1 | QUARTER 2 | QUARTER 3 | QUARTER 4 | TOTAL GRAMS |
|---|---|---|---|---|---|
| 662 | 2.75 | 0 | 0 | 0 | 2.75 |
| TOTAL | 2.75 | 0 | 0 | 0 | 2.75 |

STATE: MARYLAND

| ZIP CODE | QUARTER 1 | QUARTER 2 | QUARTER 3 | QUARTER 4 | TOTAL GRAMS |
|---|---|---|---|---|---|
| 212 | 0 | 0.1 | 0 | 0.1 | 0.2 |
| TOTAL | 0 | 0.1 | 0 | 0.1 | 0.2 |

STATE: MASSACHUSETTS

| ZIP CODE | QUARTER 1 | QUARTER 2 | QUARTER 3 | QUARTER 4 | TOTAL GRAMS |
|---|---|---|---|---|---|
| 24 | 1.14 | 1.9 | 0 | 0 | 3.04 |
| TOTAL | 1.14 | 1.9 | 0 | 0 | 3.04 |

STATE: MICHIGAN

| ZIP CODE | QUARTER 1 | QUARTER 2 | QUARTER 3 | QUARTER 4 | TOTAL GRAMS |
|---|---|---|---|---|---|
| 490 | 0 | 0 | 1.56 | 0 | 1.56 |
| 494 | 0 | 0 | 1.56 | 0 | 1.56 |
| 495 | 0 | 0 | 0 | 1.17 | 1.17 |
| TOTAL | 0 | 0 | 3.12 | 1.17 | 4.29 |

STATE: MINNESOTA

| ZIP CODE | QUARTER 1 | QUARTER 2 | QUARTER 3 | QUARTER 4 | TOTAL GRAMS |
|---|---|---|---|---|---|
| TOTAL | 0 | 0.5 | 0 | 0 | 0.5 |

STATE: MISSISSIPPI

| ZIP CODE | QUARTER 1 | QUARTER 2 | QUARTER 3 | QUARTER 4 | TOTAL GRAMS |
|---|---|---|---|---|---|
| 392 | 2.4 | 1.57 | 2.22 | 0 | 6.19 |
| TOTAL | 2.4 | 1.57 | 2.22 | 0 | 6.19 |

STATE: MISSOURI

| ZIP CODE | QUARTER 1 | QUARTER 2 | QUARTER 3 | QUARTER 4 | TOTAL GRAMS |
|---|---|---|---|---|---|
| 631 | 7.33 | 13 | 9.03 | 5.8 | 35.16 |
| 633 | 0.37 | 0.1 | 0 | 0 | 0.47 |
| 641 | 0 | 0 | 1.56 | 0 | 1.56 |
| TOTAL | 7.7 | 13.1 | 10.59 | 5.8 | 37.19 |

STATE: NEVADA

| ZIP CODE | QUARTER 1 | QUARTER 2 | QUARTER 3 | QUARTER 4 | TOTAL GRAMS |
|---|---|---|---|---|---|
| 891 | 6.64 | 2.87 | 7.45 | 8.31 | 25.27 |
| TOTAL | 6.64 | 2.87 | 7.45 | 8.31 | 25.27 |

STATE: NEW JERSEY

| ZIP CODE | QUARTER 1 | QUARTER 2 | QUARTER 3 | QUARTER 4 | TOTAL GRAMS |
|---|---|---|---|---|---|
| 80 | 7.12 | 12.03 | 1.56 | 3 | 23.71 |
| TOTAL | 7.12 | 12.03 | 1.56 | 3 | 23.71 |

STATE: NEW YORK

| ZIP CODE | QUARTER 1 | QUARTER 2 | QUARTER 3 | QUARTER 4 | TOTAL GRAMS |
|---|---|---|---|---|---|
| 101 | 0.41 | 0 | 0 | 0 | 0.41 |
| 105 | 0 | 0 | 0.1 | 0.14 | 0.24 |
| 112 | 0.87 | 2 | 1.75 | 2.87 | 7.49 |
| 114 | 0.25 | 1.54 | 1.86 | 0 | 3.65 |
| 146 | 1.12 | 2.15 | 5.59 | 1.47 | 10.33 |
| TOTAL | 2.65 | 5.69 | 9.3 | 4.48 | 22.12 |

STATE: NORTH CAROLINA

| ZIP CODE | QUARTER 1 | QUARTER 2 | QUARTER 3 | QUARTER 4 | TOTAL GRAMS |
|---|---|---|---|---|---|
| 277 | 0 | 0 | 0 | 1.56 | 1.56 |
| 282 | 0 | 0 | 2.12 | 0 | 2.12 |
| TOTAL | 0 | 0 | 2.12 | 1.56 | 3.68 |

STATE: OHIO

| ZIP CODE | QUARTER 1 | QUARTER 2 | QUARTER 3 | QUARTER 4 | TOTAL GRAMS |
|---|---|---|---|---|---|
| 447 | 0.42 | 0.46 | 1.82 | 2.88 | 5.58 |
| 452 | 0 | 2.22 | 0 | 0 | 2.22 |
| 454 | 5.47 | 6.68 | 4.37 | 0 | 16.52 |
| TOTAL | 5.89 | 9.36 | 6.19 | 2.88 | 24.32 |

STATE: OKLAHOMA

| ZIP CODE | QUARTER 1 | QUARTER 2 | QUARTER 3 | QUARTER 4 | TOTAL GRAMS |
|---|---|---|---|---|---|
| 731 | 0 | 0 | 0.1 | 0.55 | 0.65 |
| TOTAL | 0 | 0 | 0.1 | 0.55 | 0.65 |

STATE: OREGON

| ZIP CODE | QUARTER 1 | QUARTER 2 | QUARTER 3 | QUARTER 4 | TOTAL GRAMS |
|---|---|---|---|---|---|
| 972 | 0.15 | 0.04 | 0 | 0 | 0.19 |
| 974 | 0 | 0 | 1.81 | 0 | 1.81 |
| TOTAL | 0.15 | 0.04 | 1.81 | 0 | 2 |

STATE: PENNSYLVANIA

| ZIP CODE | QUARTER 1 | QUARTER 2 | QUARTER 3 | QUARTER 4 | TOTAL GRAMS |
|---|---|---|---|---|---|
| 181 | 0 | 0.51 | 0.47 | 0 | 0.98 |
| TOTAL | 0 | 0.51 | 0.47 | 0 | 0.98 |

STATE: PUERTO RICO

| ZIP CODE | QUARTER 1 | QUARTER 2 | QUARTER 3 | QUARTER 4 | TOTAL GRAMS |
|---|---|---|---|---|---|
| 9 | 1.86 | 4.44 | 11.94 | 6.83 | 25.07 |
| TOTAL | 1.86 | 4.44 | 11.94 | 6.83 | 25.07 |

STATE: RHODE ISLAND

| ZIP CODE | QUARTER 1 | QUARTER 2 | QUARTER 3 | QUARTER 4 | TOTAL GRAMS |
|---|---|---|---|---|---|
| 28 | 0 | 0 | 0.1 | 0 | 0.1 |
| TOTAL | 0 | 0 | 0.1 | 0 | 0.1 |

STATE: TENNESSEE

| ZIP CODE | QUARTER 1 | QUARTER 2 | QUARTER 3 | QUARTER 4 | TOTAL GRAMS |
|---|---|---|---|---|---|
| 381 | 0.13 | 0 | 0 | 0 | 0.13 |
| TOTAL | 0.13 | 0 | 0 | 0 | 0.13 |

STATE: TEXAS

| ZIP CODE | QUARTER 1 | QUARTER 2 | QUARTER 3 | QUARTER 4 | TOTAL GRAMS |
|---|---|---|---|---|---|
| 750 | 10.17 | 8.49 | 14.83 | 11.11 | 44.6 |
| 751 | 5.58 | 11.06 | 5.59 | 3.78 | 26.01 |
| 753 | 0 | 0 | 0 | 0.01 | 0.01 |
| 761 | 0 | 0 | 3.12 | 0 | 3.12 |
| 763 | 0.15 | 0.24 | 0 | 0 | 0.39 |
| 770 | 0 | 0 | 2.22 | 2.46 | 4.68 |
| 782 | 0 | 0 | 1.56 | 1.06 | 2.62 |
| 787 | 5.87 | 9.8 | 3.97 | 7.77 | 27.41 |
| TOTAL | 21.77 | 29.59 | 31.29 | 26.19 | 108.84 |

STATE: VERMONT

| ZIP CODE | QUARTER 1 | QUARTER 2 | QUARTER 3 | QUARTER 4 | TOTAL GRAMS |
|---|---|---|---|---|---|
| 50 | 0 | 0 | 0.1 | 0 | 0.1 |
| TOTAL | 0 | 0 | 0.1 | 0 | 0.1 |

STATE: WASHINGTON

| ZIP CODE | QUARTER 1 | QUARTER 2 | QUARTER 3 | QUARTER 4 | TOTAL GRAMS |
|---|---|---|---|---|---|
| 980 | 0.41 | 0.58 | 3.11 | 3.94 | 8.04 |
| 992 | 0.24 | 0.1 | 0.1 | 0 | 0.44 |
| TOTAL | 0.65 | 0.68 | 3.21 | 3.94 | 8.48 |

DATE RANGE: 01/01/2017 TO 12/31/2017 — ARCOS 3 - REPORT 1 — Run Date: 07/03/2018

RETAIL DRUG DISTRIBUTION BY ZIP CODE WITHIN STATE BY GRAMS WT

DRUG CODE:9737DRUG NAME:ALFENTANIL

STATE: ALABAMA

| ZIP CODE | QUARTER 1 | QUARTER 2 | QUARTER 3 | QUARTER 4 | TOTAL GRAMS |
|---|---|---|---|---|---|
| 357 | 0 | 0.13 | 0 | 0 | 0.13 |
| 363 | 0.03 | 0 | 0.03 | 0.03 | 0.09 |
| 368 | 0.01 | 0 | 0.02 | 0 | 0.03 |
| TOTAL | 0.04 | 0.13 | 0.05 | 0.03 | 0.25 |

STATE: ALASKA

| ZIP CODE | QUARTER 1 | QUARTER 2 | QUARTER 3 | QUARTER 4 | TOTAL GRAMS |
|---|---|---|---|---|---|
| 995 | 0.74 | 0.74 | 0.5 | 0.86 | 2.84 |
| 996 | 0.01 | 0.01 | 0 | 0.03 | 0.05 |
| TOTAL | 0.75 | 0.75 | 0.5 | 0.89 | 2.89 |

STATE: ARIZONA

| ZIP CODE | QUARTER 1 | QUARTER 2 | QUARTER 3 | QUARTER 4 | TOTAL GRAMS |
|---|---|---|---|---|---|
| 850 | 0.08 | 0.08 | 0.12 | 0.11 | 0.39 |
| 851 | 0.02 | 0.06 | 0 | 0.05 | 0.13 |
| 852 | 0.18 | 0.09 | 0.04 | 0.21 | 0.52 |
| 853 | 0 | 0 | 0.06 | 0.06 | 0.12 |
| 855 | 0 | 0.04 | 0.03 | 0 | 0.07 |
| 857 | 0.84 | 0.59 | 0.44 | 0.55 | 2.42 |
| 860 | 0 | 0.1 | 0 | 0 | 0.1 |
| TOTAL | 1.12 | 0.96 | 0.69 | 0.98 | 3.75 |

STATE: ARKANSAS

| ZIP CODE | QUARTER 1 | QUARTER 2 | QUARTER 3 | QUARTER 4 | TOTAL GRAMS |
|---|---|---|---|---|---|
| 718 | 0.01 | 0 | 0.02 | 0.01 | 0.04 |
| 726 | 0 | 0 | 0.02 | 0.02 | 0.04 |
| TOTAL | 0.01 | 0 | 0.04 | 0.03 | 0.08 |

STATE: CALIFORNIA

| ZIP CODE | QUARTER 1 | QUARTER 2 | QUARTER 3 | QUARTER 4 | TOTAL GRAMS |
|---|---|---|---|---|---|
| 900 | 1.84 | 1.9 | 1.39 | 2.83 | 7.96 |
| 902 | 0.39 | 0.65 | 0.08 | 0.35 | 1.47 |
| 904 | 0.1 | 0.29 | 0.05 | 0.1 | 0.54 |
| 905 | 0 | 0.05 | 0 | 0 | 0.05 |
| 906 | 0.15 | 0.25 | 0.32 | 0.28 | 1 |
| 910 | 0 | 0 | 0 | 0.01 | 0.01 |
| 912 | 0 | 0 | 0.01 | 0 | 0.01 |
| 913 | 0.44 | 0.28 | 0.18 | 0.4 | 1.3 |
| 914 | 0.08 | 0.1 | 0.05 | 0.15 | 0.38 |
| 915 | 0.05 | 0.06 | 0.02 | 0 | 0.13 |
| 917 | 1.11 | 0.6 | 1.46 | 0.75 | 3.92 |
| 919 | 0.26 | 0.13 | 0.15 | 0.12 | 0.66 |
| 920 | 0.94 | 0.37 | 0.59 | 1.31 | 3.21 |
| 921 | 1.38 | 0.71 | 0.9 | 0.99 | 3.98 |
| 922 | 0.52 | 0.76 | 0.6 | 0.49 | 2.37 |
| 923 | 1.02 | 1.47 | 1.79 | 1.95 | 6.23 |
| 925 | 0.26 | 0.22 | 0.21 | 0.24 | 0.93 |
| 926 | 0.5 | 0.43 | 0.4 | 0.25 | 1.58 |
| 927 | 0.04 | 0 | 0.18 | 0.04 | 0.26 |

DRUG CODE:9737DRUG NAME:ALFENTANIL

STATE:CALIFORNIA

| ZIP CODE | QUARTER 1 | QUARTER 2 | QUARTER 3 | QUARTER 4 | TOTAL GRAMS |
|---|---|---|---|---|---|
| 928 | 0.55 | 0.41 | 0 | 0 | 0.96 |
| 930 | 0.18 | 0.24 | 0.21 | 0.21 | 0.84 |
| 931 | 0.26 | 0.36 | 0.2 | 0.29 | 1.11 |
| 932 | 0.11 | 0 | 0.1 | 0.03 | 0.24 |
| 934 | 0.05 | 0.09 | 0 | 0.04 | 0.18 |
| 935 | 0.03 | 0.05 | 0.13 | 0.21 | 0.42 |
| 936 | 0.03 | 0 | 0.07 | 0.07 | 0.17 |
| 937 | 1.41 | 1.34 | 0.96 | 0.9 | 4.61 |
| 939 | 0.03 | 0.04 | 0 | 0.02 | 0.09 |
| 940 | 0.46 | 0.28 | 0.58 | 0.44 | 1.76 |

| | | | | | | |
|---|---|---|---|---|---|---|
| | 941 | 0.08 | 0.32 | 0.01 | 0.16 | 0.57 |
| | 943 | 1.07 | 0.56 | 0.47 | 0.76 | 2.86 |
| | 944 | 0.02 | 0.05 | 0.07 | 0.1 | 0.24 |
| | 945 | 0.9 | 1.27 | 1.28 | 0.84 | 4.29 |
| | 946 | 0.21 | 0.23 | 0.49 | 0.15 | 1.08 |
| | 948 | 0.03 | 0 | 0.02 | 0 | 0.05 |
| | 949 | 0 | 0 | 0.03 | 0.08 | 0.11 |
| | 950 | 0.01 | 0.02 | 0.03 | 0.05 | 0.11 |
| | 951 | 0.58 | 0.55 | 0.45 | 0.5 | 2.08 |
| | 952 | 0.29 | 0.04 | 0.13 | 0.38 | 0.84 |
| | 953 | 0.62 | 0.65 | 0.63 | 0.53 | 2.43 |
| | 954 | 0.19 | 0.2 | 0.15 | 0.34 | 0.88 |
| | 956 | 0.06 | 0 | 0.04 | 0.04 | 0.14 |
| | 958 | 0.86 | 0.13 | 1.6 | 0.3 | 2.89 |
| | 959 | 0.24 | 0.22 | 0.24 | 0.22 | 0.92 |
| | 960 | 0.03 | 0.01 | 0.07 | 0.14 | 0.25 |
| TOTAL | | 17.38 | 15.33 | 16.34 | 17.06 | 66.11 |

STATE: COLORADO

| ZIP CODE | | QUARTER 1 | QUARTER 2 | QUARTER 3 | QUARTER 4 | TOTAL GRAMS |
|---|---|---|---|---|---|---|
| | 800 | 0.89 | 0.4 | 0.47 | 0.8 | 2.56 |
| | 801 | 0.03 | 0.09 | 0.04 | 0.07 | 0.23 |
| | 805 | 0.09 | 0.06 | 0 | 0.01 | 0.16 |
| | 808 | 0.06 | 0.08 | 0.13 | 0 | 0.27 |
| | 811 | 0 | 0 | 0.03 | 0 | 0.03 |
| | 812 | 0 | 0 | 0 | 0.01 | 0.01 |
| | 813 | 0.08 | 0.08 | 0.12 | 0.1 | 0.38 |
| | 814 | 0.08 | 0.01 | 0 | 0 | 0.09 |
| | 815 | 0 | 0 | 0 | 0.07 | 0.07 |
| | 816 | 0.04 | 0 | 0 | 0.02 | 0.06 |
| TOTAL | | 1.27 | 0.72 | 0.79 | 1.08 | 3.86 |

STATE: CONNECTICUT

| ZIP CODE | QUARTER 1 | QUARTER 2 | QUARTER 3 | QUARTER 4 | TOTAL GRAMS |
|---|---|---|---|---|---|
| 64 | 0.1 | 0 | 0 | 0 | 0.1 |
| 65 | 0.98 | 0.77 | 0.79 | 0.95 | 3.49 |
| 68 | 0.44 | 0.43 | 0.25 | 0.6 | 1.72 |
| TOTAL | 1.52 | 1.2 | 1.04 | 1.55 | 5.31 |

STATE: FLORIDA

| ZIP CODE | QUARTER 1 | QUARTER 2 | QUARTER 3 | QUARTER 4 | TOTAL GRAMS |
|---|---|---|---|---|---|
| 321 | 0.03 | 0.02 | 0 | 0 | 0.05 |
| 322 | 0.1 | 0.21 | 0.37 | 0.37 | 1.05 |
| 324 | 0.59 | 0.38 | 0.72 | 0.07 | 1.76 |
| 325 | 0 | 0.33 | 0 | 0.03 | 0.36 |
| 326 | 2.5 | 2.4 | 2.35 | 4.1 | 11.35 |
| 327 | 0 | 0.09 | 0 | 0 | 0.09 |
| 328 | 0.15 | 0.16 | 0.15 | 0.24 | 0.7 |
| 329 | 0.4 | 0.57 | 0.48 | 0.52 | 1.97 |
| 330 | 0 | 0.18 | 0 | 0.1 | 0.28 |
| 331 | 0.1 | 0.2 | 0.13 | 0.1 | 0.53 |
| 333 | 0 | 0 | 0 | 0.01 | 0.01 |
| 334 | 0.64 | 0.53 | 0.55 | 0.43 | 2.15 |
| 336 | 0.03 | 0.02 | 0 | 0 | 0.05 |
| 337 | 0 | 0.2 | 0 | 0 | 0.2 |
| 339 | 0 | 0 | 0.01 | 0.1 | 0.11 |
| 342 | 0.01 | 0.03 | 0.01 | 0.03 | 0.08 |
| 344 | 0 | 0.09 | 0.02 | 0 | 0.11 |
| 346 | 0 | 0.05 | 0 | 0 | 0.05 |
| 349 | 0 | 0.02 | 0 | 0.01 | 0.03 |
| TOTAL | 4.55 | 5.48 | 4.79 | 6.11 | 20.93 |

STATE: GEORGIA

| ZIP CODE | QUARTER 1 | QUARTER 2 | QUARTER 3 | QUARTER 4 | TOTAL GRAMS |
|---|---|---|---|---|---|
| 301 | 0.02 | 0.01 | 0.03 | 0.02 | 0.08 |
| 303 | 0.62 | 0.73 | 2.13 | 0.4 | 3.88 |
| 305 | 0 | 0 | 0.01 | 0.01 | 0.02 |

| | | | | | | |
|---|---|---|---|---|---|---|
| | 308 | 0.06 | 0.04 | 0.06 | 0.08 | 0.24 |
| | 309 | 1.34 | 1.19 | 1.69 | 1.21 | 5.43 |
| | 312 | 0.16 | 0.22 | 0.12 | 0.24 | 0.74 |
| | 314 | 0 | 0 | 0 | 0.05 | 0.05 |
| | 315 | 0.3 | 0.15 | 0.2 | 0.07 | 0.72 |
| | 317 | 0.42 | 0.41 | 0.4 | 0.49 | 1.72 |
| | 319 | 0 | 0.04 | 0.02 | 0.1 | 0.16 |
| | 398 | 0 | 0 | 0 | 0.02 | 0.02 |
| TOTAL | | 2.92 | 2.79 | 4.66 | 2.69 | 13.06 |

STATE: GUAM

| ZIP CODE | | QUARTER 1 | QUARTER 2 | QUARTER 3 | QUARTER 4 | TOTAL GRAMS |
|---|---|---|---|---|---|---|
| | 969 | 0 | 0 | 0.05 | 0.03 | 0.08 |
| TOTAL | | 0 | 0 | 0.05 | 0.03 | 0.08 |

STATE: HAWAII

| ZIP CODE | | QUARTER 1 | QUARTER 2 | QUARTER 3 | QUARTER 4 | TOTAL GRAMS |
|---|---|---|---|---|---|---|
| | 967 | 0.23 | 0.28 | 0.25 | 0.38 | 1.14 |
| | 968 | 0.22 | 0.22 | 0.37 | 0.46 | 1.27 |
| TOTAL | | 0.45 | 0.5 | 0.62 | 0.84 | 2.41 |

STATE: IDAHO

| ZIP CODE | | QUARTER 1 | QUARTER 2 | QUARTER 3 | QUARTER 4 | TOTAL GRAMS |
|---|---|---|---|---|---|---|
| | 833 | 0.1 | 0.15 | 0.13 | 0.08 | 0.46 |
| | 836 | 0 | 0 | 0.05 | 0.08 | 0.13 |
| | 837 | 1.36 | 1.24 | 0.97 | 1.31 | 4.88 |
| TOTAL | | 1.46 | 1.39 | 1.15 | 1.47 | 5.47 |

STATE: ILLINOIS

| ZIP CODE | | QUARTER 1 | QUARTER 2 | QUARTER 3 | QUARTER 4 | TOTAL GRAMS |
|---|---|---|---|---|---|---|
| | 600 | 0.16 | 0.18 | 0.19 | 0.21 | 0.74 |
| | 601 | 1.03 | 0.88 | 1.05 | 1.69 | 4.65 |
| | 604 | 0 | 0.63 | 0.03 | 0.37 | 1.03 |

| | | | | | | |
|---|---|---|---|---|---|---|
| | 605 | 0.05 | 0 | 0.1 | 0.08 | 0.23 |
| | 606 | 0.5 | 0.8 | 0.77 | 0.81 | 2.88 |
| | 608 | 0.06 | 0.05 | 0.04 | 0.05 | 0.2 |
| | 610 | 0.52 | 0.44 | 0.32 | 0.27 | 1.55 |
| | 611 | 0.97 | 0.97 | 0.65 | 0.96 | 3.55 |
| | 612 | 0.06 | 0 | 0.03 | 0.05 | 0.14 |
| | 622 | 0 | 0.01 | 0 | 0.01 | 0.02 |
| | 624 | 0.04 | 0.06 | 0.04 | 0.05 | 0.19 |
| | 627 | 0.11 | 0.06 | 0.08 | 0.37 | 0.62 |
| | 628 | 0.23 | 0.12 | 0.07 | 0.18 | 0.6 |
| | 629 | 0.05 | 0.04 | 0.01 | 0.06 | 0.16 |
| TOTAL | | 3.78 | 4.24 | 3.38 | 5.16 | 16.56 |

STATE: INDIANA

| ZIP CODE | | QUARTER 1 | QUARTER 2 | QUARTER 3 | QUARTER 4 | TOTAL GRAMS |
|---|---|---|---|---|---|---|
| | 460 | 0.75 | 1.08 | 1 | 0.96 | 3.79 |
| | 461 | 0.03 | 0 | 0 | 0 | 0.03 |
| | 462 | 0.47 | 0.59 | 0.63 | 0.68 | 2.37 |
| | 463 | 0.04 | 0 | 0.02 | 0 | 0.06 |
| | 464 | 0.01 | 0 | 0.01 | 0.01 | 0.03 |
| | 465 | 0 | 0.03 | 0 | 0.02 | 0.05 |
| | 466 | 0.67 | 0.67 | 0.52 | 0.5 | 2.36 |
| | 467 | 0 | 0 | 0.01 | 0.01 | 0.02 |
| | 469 | 0.02 | 0.02 | 0.02 | 0.01 | 0.07 |
| | 470 | 0 | 0 | 0.02 | 0.01 | 0.03 |
| | 471 | 0 | 0.45 | 0.01 | 0.01 | 0.47 |
| | 472 | 0.08 | 0.1 | 0.05 | 0 | 0.23 |
| | 473 | 0.03 | 0.05 | 0.13 | 0.1 | 0.31 |
| | 475 | 0.02 | 0.01 | 0 | 0 | 0.03 |
| | 477 | 0.05 | 0.1 | 0.1 | 0.03 | 0.28 |
| | 479 | 0.07 | 0.08 | 0.02 | 0.01 | 0.18 |
| TOTAL | | 2.24 | 3.18 | 2.54 | 2.35 | 10.31 |

STATE: IOWA

| ZIP CODE | | QUARTER 1 | QUARTER 2 | QUARTER 3 | QUARTER 4 | TOTAL GRAMS |
|---|---|---|---|---|---|---|

| | | | | | | |
|---|---|---|---|---|---|---|
| | 500 | 0 | 0.03 | 0.04 | 0.03 | 0.1 |
| | 503 | 0.02 | 0 | 0 | 0 | 0.02 |
| | 505 | 0 | 0.04 | 0 | 0.02 | 0.06 |
| | 506 | 0 | 0 | 0.01 | 0 | 0.01 |
| | 507 | 0.01 | 0.02 | 0.04 | 0.01 | 0.08 |
| | 520 | 0 | 0 | 0 | 0.09 | 0.09 |
| | 522 | 1.41 | 0.85 | 0.83 | 0.86 | 3.95 |
| | 523 | 0 | 0 | 0.01 | 0.01 | 0.02 |
| | 524 | 0.21 | 0.15 | 0.13 | 0.21 | 0.7 |
| | 526 | 0 | 0.2 | 0 | 0 | 0.2 |
| | 527 | 0.13 | 0.02 | 0.06 | 0.11 | 0.32 |
| | 528 | 0.27 | 0.19 | 0.28 | 0.41 | 1.15 |
| TOTAL | | 2.05 | 1.5 | 1.4 | 1.75 | 6.7 |

STATE: KANSAS

| ZIP CODE | | QUARTER 1 | QUARTER 2 | QUARTER 3 | QUARTER 4 | TOTAL GRAMS |
|---|---|---|---|---|---|---|
| | 660 | 0 | 0 | 0 | 0.02 | 0.02 |
| | 662 | 0 | 0.02 | 0.01 | 0.02 | 0.05 |
| | 667 | 0 | 0 | 0 | 0.04 | 0.04 |
| | 669 | 0.1 | 0.05 | 0 | 0 | 0.15 |
| | 671 | 0.02 | 0.02 | 0.02 | 0.02 | 0.08 |
| | 674 | 0.99 | 0.88 | 0.69 | 0.98 | 3.54 |
| | 675 | 0.27 | 0.2 | 0.24 | 0.18 | 0.89 |
| | 676 | 0 | 0 | 0.01 | 0.01 | 0.02 |
| | 678 | 0.24 | 0.34 | 0.29 | 0.33 | 1.2 |
| TOTAL | | 1.62 | 1.51 | 1.26 | 1.6 | 5.99 |

STATE: KENTUCKY

| ZIP CODE | | QUARTER 1 | QUARTER 2 | QUARTER 3 | QUARTER 4 | TOTAL GRAMS |
|---|---|---|---|---|---|---|
| | 402 | 0.15 | 0.44 | 0.2 | 0.3 | 1.09 |
| | 404 | 0.03 | 0.03 | 0.04 | 0.06 | 0.16 |
| | 408 | 0.01 | 0 | 0.01 | 0 | 0.02 |
| | 410 | 4.75 | 2.5 | 2.5 | 2.5 | 12.25 |
| | 411 | 0.04 | 0.13 | 0.04 | 0.06 | 0.27 |
| | 423 | 0.02 | 0.81 | 0.88 | 0.93 | 2.64 |

| | | | | | | |
|---|---|---|---|---|---|---|
| TOTAL | | 5 | 3.91 | 3.67 | 3.85 | 16.43 |

| STATE: | LOUISIANA | | | | | |
|---|---|---|---|---|---|---|
| ZIP CODE | | QUARTER 1 | QUARTER 2 | QUARTER 3 | QUARTER 4 | TOTAL GRAMS |
| | 700 | 0 | 0 | 0 | 0.02 | 0.02 |
| | 701 | 0.2 | 0.28 | 0.15 | 0.28 | 0.91 |
| | 704 | 0.69 | 0.8 | 0.65 | 0.87 | 3.01 |
| | 705 | 0.3 | 0.13 | 0.3 | 0.21 | 0.94 |
| | 708 | 0.1 | 0.18 | 0.1 | 0.17 | 0.55 |
| | 710 | 0.03 | 0 | 0.04 | 0 | 0.07 |
| | 711 | 0.95 | 0.84 | 1.36 | 0.93 | 4.08 |
| | 712 | 0.04 | 0.05 | 0.02 | 0.01 | 0.12 |
| | 713 | 0.13 | 0.01 | 0.18 | 0.02 | 0.34 |
| | 714 | 0 | 0 | 0.04 | 0 | 0.04 |
| TOTAL | | 2.44 | 2.29 | 2.84 | 2.51 | 10.08 |

| STATE: | MAINE | | | | | |
|---|---|---|---|---|---|---|
| ZIP CODE | | QUARTER 1 | QUARTER 2 | QUARTER 3 | QUARTER 4 | TOTAL GRAMS |
| | 39 | 0.13 | 0.16 | 0.08 | 0.14 | 0.51 |
| | 40 | 0.12 | 0.21 | 0.22 | 0.22 | 0.77 |
| | 41 | 0.26 | 0.16 | 0.23 | 0.06 | 0.71 |
| | 42 | 0.01 | 0 | 0 | 0 | 0.01 |
| | 44 | 0.06 | 0.23 | 0.09 | 0.12 | 0.5 |
| | 46 | 0.02 | 0.03 | 0.01 | 0.03 | 0.09 |
| | 48 | 0.02 | 0 | 0 | 0.08 | 0.1 |
| TOTAL | | 0.62 | 0.79 | 0.63 | 0.65 | 2.69 |

| STATE: | MARYLAND | | | | | |
|---|---|---|---|---|---|---|
| ZIP CODE | | QUARTER 1 | QUARTER 2 | QUARTER 3 | QUARTER 4 | TOTAL GRAMS |
| | 207 | 0.02 | 0 | 0 | 0.02 | 0.04 |
| | 210 | 0.22 | 0.19 | 0.22 | 0.07 | 0.7 |
| | 212 | 0.66 | 0.05 | 0.63 | 0.82 | 2.16 |
| | 217 | 0 | 0 | 0 | 0.01 | 0.01 |
| TOTAL | | 0.9 | 0.24 | 0.85 | 0.92 | 2.91 |

STATE: MASSACHUSETTS

| ZIP CODE | QUARTER 1 | QUARTER 2 | QUARTER 3 | QUARTER 4 | TOTAL GRAMS |
|---|---|---|---|---|---|
| 18 | 0 | 0.01 | 0 | 0 | 0.01 |
| 21 | 0.2 | 0.13 | 0.25 | 0.18 | 0.76 |
| 24 | 1.15 | 0.59 | 0.4 | 0.55 | 2.69 |
| TOTAL | 1.35 | 0.73 | 0.65 | 0.73 | 3.46 |

STATE: MICHIGAN

| ZIP CODE | QUARTER 1 | QUARTER 2 | QUARTER 3 | QUARTER 4 | TOTAL GRAMS |
|---|---|---|---|---|---|
| 481 | 0.23 | 0.24 | 0.2 | 0.15 | 0.82 |
| 483 | 0.18 | 0.16 | 0.2 | 0.28 | 0.82 |
| 485 | 0.04 | 0 | 0.02 | 0.02 | 0.08 |
| 486 | 0.03 | 0 | 0.01 | 0 | 0.04 |
| 488 | 0.26 | 0.67 | 0.34 | 0.52 | 1.79 |
| 489 | 0.05 | 0.05 | 0 | 0.05 | 0.15 |
| 490 | 0.08 | 0.1 | 0.18 | 0.1 | 0.46 |
| 492 | 0.04 | 0.03 | 0 | 0.04 | 0.11 |
| 494 | 0 | 0 | 0 | 0.02 | 0.02 |
| 497 | 0 | 0.03 | 0 | 0.03 | 0.06 |
| 498 | 0 | 0.04 | 0.1 | 0 | 0.14 |
| TOTAL | 0.91 | 1.32 | 1.05 | 1.21 | 4.49 |

STATE: MINNESOTA

| ZIP CODE | QUARTER 1 | QUARTER 2 | QUARTER 3 | QUARTER 4 | TOTAL GRAMS |
|---|---|---|---|---|---|
| 550 | 0.08 | 0.09 | 0.07 | 0.12 | 0.36 |
| 551 | 1.03 | 0.53 | 0.66 | 0.93 | 3.15 |
| 553 | 0.01 | 0.17 | 0 | 0.02 | 0.2 |
| 554 | 1.92 | 2.36 | 2.11 | 2.49 | 8.88 |
| 557 | 0.01 | 0 | 0 | 0 | 0.01 |
| 558 | 0 | 0.01 | 0 | 0 | 0.01 |
| 559 | 0.1 | 0.2 | 0.23 | 0.33 | 0.86 |
| 560 | 0 | 0 | 0 | 0.01 | 0.01 |
| 561 | 0 | 0 | 0.05 | 0 | 0.05 |

| | | | | | | |
|---|---|---|---|---|---|---|
| | 562 | 0.1 | 0.05 | 0.11 | 0.12 | 0.38 |
| | 563 | 0 | 0 | 0 | 0.03 | 0.03 |
| | 565 | 0 | 0 | 0.01 | 0 | 0.01 |
| TOTAL | | 3.25 | 3.41 | 3.24 | 4.05 | 13.95 |

| STATE: | MISSISSIPPI | | | | | |
|---|---|---|---|---|---|---|
| ZIP CODE | | QUARTER 1 | QUARTER 2 | QUARTER 3 | QUARTER 4 | TOTAL GRAMS |
| | 387 | 0 | 0.01 | 0 | 0 | 0.01 |
| | 389 | 0 | 0.01 | 0 | 0 | 0.01 |
| | 390 | 0.02 | 0.03 | 0.02 | 0 | 0.07 |
| | 393 | 0.02 | 0.04 | 0.03 | 0.02 | 0.11 |
| | 394 | 0 | 0.02 | 0.06 | 0.03 | 0.11 |
| | 395 | 0 | 0.01 | 0 | 0 | 0.01 |
| | 396 | 0 | 0 | 0.04 | 0.02 | 0.06 |
| | 397 | 0.03 | 0 | 0.02 | 0 | 0.05 |
| TOTAL | | 0.07 | 0.12 | 0.17 | 0.07 | 0.43 |

| STATE: | MISSOURI | | | | | |
|---|---|---|---|---|---|---|
| ZIP CODE | | QUARTER 1 | QUARTER 2 | QUARTER 3 | QUARTER 4 | TOTAL GRAMS |
| | 631 | 0 | 0.06 | 0.04 | 0.04 | 0.14 |
| | 636 | 0.38 | 0.4 | 0.33 | 0.41 | 1.52 |
| | 640 | 0.07 | 0.08 | 0.01 | 0.09 | 0.25 |
| | 641 | 0.05 | 0.09 | 0 | 0.1 | 0.24 |
| | 646 | 0.09 | 0.05 | 0 | 0 | 0.14 |
| | 652 | 0.03 | 0.01 | 0.03 | 0.03 | 0.1 |
| | 654 | 0.24 | 0.17 | 0.22 | 0.2 | 0.83 |
| | 657 | 0.35 | 0.14 | 0.27 | 0.35 | 1.11 |
| TOTAL | | 1.21 | 1 | 0.9 | 1.22 | 4.33 |

| STATE: | MONTANA | | | | | |
|---|---|---|---|---|---|---|
| ZIP CODE | | QUARTER 1 | QUARTER 2 | QUARTER 3 | QUARTER 4 | TOTAL GRAMS |
| | 590 | 0.12 | 0.06 | 0.09 | 0.1 | 0.37 |
| | 591 | 0 | 0.02 | 0 | 0.01 | 0.03 |
| | 592 | 0.01 | 0 | 0.03 | 0 | 0.04 |

| | | | | | | |
|---|---|---|---|---|---|---|
| | 596 | 0.05 | 0 | 0.02 | 0 | 0.07 |
| | 597 | 0.12 | 0.15 | 0.14 | 0.17 | 0.58 |
| | 598 | 0.06 | 0.04 | 0.05 | 0.06 | 0.21 |
| TOTAL | | 0.36 | 0.27 | 0.33 | 0.34 | 1.3 |

| STATE: | NEBRASKA | | | | | |
|---|---|---|---|---|---|---|
| ZIP CODE | | QUARTER 1 | QUARTER 2 | QUARTER 3 | QUARTER 4 | TOTAL GRAMS |
| | 680 | 0 | 0 | 0 | 0.01 | 0.01 |
| | 681 | 0.08 | 0.1 | 0.23 | 0 | 0.41 |
| | 683 | 0.12 | 0.02 | 0.03 | 0.09 | 0.26 |
| | 684 | 0.03 | 0.04 | 0.05 | 0.03 | 0.15 |
| | 685 | 0.01 | 0 | 0 | 0 | 0.01 |
| | 686 | 0 | 0.01 | 0.01 | 0 | 0.02 |
| | 687 | 0.1 | 0.01 | 0.33 | 0.19 | 0.63 |
| | 688 | 0.09 | 0.13 | 0.12 | 0.19 | 0.53 |
| | 689 | 0.35 | 0.35 | 0.24 | 0.38 | 1.32 |
| | 691 | 0.04 | 0.04 | 0.05 | 0.04 | 0.17 |
| | 693 | 0.03 | 0 | 0.02 | 0.01 | 0.06 |
| TOTAL | | 0.85 | 0.7 | 1.08 | 0.94 | 3.57 |

| STATE: | NEVADA | | | | | |
|---|---|---|---|---|---|---|
| ZIP CODE | | QUARTER 1 | QUARTER 2 | QUARTER 3 | QUARTER 4 | TOTAL GRAMS |
| | 890 | 0.01 | 0 | 0.03 | 0.01 | 0.05 |
| | 891 | 0.81 | 1.05 | 1.16 | 1.61 | 4.63 |
| | 894 | 0.05 | 0.05 | 0.07 | 0.1 | 0.27 |
| | 895 | 0.38 | 0.44 | 0.46 | 0.36 | 1.64 |
| | 898 | 0.03 | 0.05 | 0.03 | 0.28 | 0.39 |
| TOTAL | | 1.28 | 1.59 | 1.75 | 2.36 | 6.98 |

| STATE: | NEW HAMPSHIRE | | | | | |
|---|---|---|---|---|---|---|
| ZIP CODE | | QUARTER 1 | QUARTER 2 | QUARTER 3 | QUARTER 4 | TOTAL GRAMS |
| | 30 | 0.09 | 0 | 0 | 0.03 | 0.12 |
| | 32 | 0.03 | 0.02 | 0.01 | 0.02 | 0.08 |
| | 37 | 0 | 0 | 0 | 0.01 | 0.01 |

| | | | | | |
|---|---|---|---|---|---|
| 38 | 0.36 | 0.4 | 0.43 | 0.55 | 1.74 |
| TOTAL | 0.48 | 0.42 | 0.44 | 0.61 | 1.95 |

STATE: NEW JERSEY

| ZIP CODE | QUARTER 1 | QUARTER 2 | QUARTER 3 | QUARTER 4 | TOTAL GRAMS |
|---|---|---|---|---|---|
| 70 | 0.02 | 0.03 | 0.02 | 0.13 | 0.2 |
| 74 | 0.02 | 0.01 | 0.06 | 0.02 | 0.11 |
| 75 | 0 | 0.05 | 0 | 0.02 | 0.07 |
| 77 | 0.07 | 0.02 | 0.04 | 0.07 | 0.2 |
| 88 | 0.08 | 0.16 | 0.11 | 0.11 | 0.46 |
| 89 | 0.01 | 0 | 0 | 0 | 0.01 |
| TOTAL | 0.2 | 0.27 | 0.23 | 0.35 | 1.05 |

STATE: NEW MEXICO

| ZIP CODE | QUARTER 1 | QUARTER 2 | QUARTER 3 | QUARTER 4 | TOTAL GRAMS |
|---|---|---|---|---|---|
| 871 | 0.87 | 0.83 | 1.14 | 1.13 | 3.97 |
| 874 | 0 | 0.05 | 0 | 0.02 | 0.07 |
| 875 | 0 | 0 | 0 | 0.06 | 0.06 |
| TOTAL | 0.87 | 0.88 | 1.14 | 1.21 | 4.1 |

STATE: NEW YORK

| ZIP CODE | QUARTER 1 | QUARTER 2 | QUARTER 3 | QUARTER 4 | TOTAL GRAMS |
|---|---|---|---|---|---|
| 100 | 0.1 | 0.1 | 0.29 | 2.35 | 2.84 |
| 104 | 0 | 0 | 0.08 | 0.01 | 0.09 |
| 105 | 0.03 | 0.05 | 0.14 | 0.1 | 0.32 |
| 106 | 0.03 | 0 | 0 | 0 | 0.03 |
| 112 | 0.62 | 0.73 | 0.78 | 1.09 | 3.22 |
| 113 | 0 | 0 | 0 | 0.01 | 0.01 |
| 117 | 0.1 | 0 | 0.05 | 0 | 0.15 |
| 122 | 0.15 | 0.1 | 0.06 | 0.16 | 0.47 |
| 123 | 0.05 | 0.02 | 0.04 | 0.05 | 0.16 |
| 129 | 0.02 | 0 | 0 | 0.03 | 0.05 |
| 135 | 0 | 0.02 | 0 | 0.03 | 0.05 |
| 136 | 0.02 | 0 | 0 | 0 | 0.02 |

| | | | | | |
|---|---|---|---|---|---|
| 138 | 0.02 | 0 | 0.04 | 0.01 | 0.07 |
| 142 | 0.07 | 0.1 | 0.06 | 0.14 | 0.37 |
| 146 | 0.73 | 0 | 0 | 0.07 | 0.8 |
| 148 | 0.19 | 0.1 | 0.12 | 0.12 | 0.53 |
| TOTAL | 2.13 | 1.22 | 1.66 | 4.17 | 9.18 |

STATE: NORTH CAROLINA

| ZIP CODE | QUARTER 1 | QUARTER 2 | QUARTER 3 | QUARTER 4 | TOTAL GRAMS |
|---|---|---|---|---|---|
| 270 | 0.14 | 0.15 | 0.13 | 0.21 | 0.63 |
| 271 | 0.57 | 0.8 | 0.58 | 0.65 | 2.6 |
| 272 | 0.1 | 0.07 | 0.13 | 0.05 | 0.35 |
| 273 | 0.02 | 0.01 | 0.04 | 0.05 | 0.12 |
| 274 | 0.43 | 0.8 | 0.4 | 0.67 | 2.3 |
| 276 | 0.14 | 0.08 | 0.14 | 0.11 | 0.47 |
| 277 | 0 | 0.02 | 0 | 0.04 | 0.06 |
| 278 | 0 | 0 | 0 | 0.03 | 0.03 |
| 280 | 0.02 | 0 | 0.01 | 0.04 | 0.07 |
| 281 | 0.05 | 0.08 | 0.06 | 0.12 | 0.31 |
| 282 | 0.87 | 0.87 | 0.99 | 1.07 | 3.8 |
| 283 | 0.11 | 0.1 | 0.07 | 0.2 | 0.48 |
| 286 | 0.33 | 0.4 | 0.37 | 0.26 | 1.36 |
| TOTAL | 2.78 | 3.38 | 2.92 | 3.5 | 12.58 |

STATE: NORTH DAKOTA

| ZIP CODE | QUARTER 1 | QUARTER 2 | QUARTER 3 | QUARTER 4 | TOTAL GRAMS |
|---|---|---|---|---|---|
| 582 | 0.03 | 0.06 | 0.05 | 0.06 | 0.2 |
| 584 | 0.08 | 0.07 | 0.04 | 0.05 | 0.24 |
| 585 | 0 | 0.14 | 0 | 0 | 0.14 |
| 586 | 0 | 0.01 | 0 | 0 | 0.01 |
| TOTAL | 0.11 | 0.28 | 0.09 | 0.11 | 0.59 |

STATE: OHIO

| ZIP CODE | QUARTER 1 | QUARTER 2 | QUARTER 3 | QUARTER 4 | TOTAL GRAMS |
|---|---|---|---|---|---|
| 430 | 0 | 0 | 0.13 | 0.05 | 0.18 |

| | | | | | | |
|---|---|---|---|---|---|---|
| | 432 | 0 | 0 | 0 | 0.24 | 0.24 |
| | 433 | 0.01 | 0.02 | 0.03 | 0.04 | 0.1 |
| | 435 | 0.02 | 0.02 | 0.03 | 0.05 | 0.12 |
| | 436 | 0 | 0 | 0 | 0.15 | 0.15 |
| | 437 | 0 | 0 | 0.01 | 0 | 0.01 |
| | 440 | 0.15 | 0.15 | 0.18 | 0.25 | 0.73 |
| | 441 | 2.67 | 4.37 | 1.28 | 2.59 | 10.91 |
| | 442 | 0.05 | 0.09 | 0.1 | 0.04 | 0.28 |
| | 443 | 0 | 0.04 | 0.06 | 0.04 | 0.14 |
| | 444 | 0.28 | 0.29 | 0.29 | 0.12 | 0.98 |
| | 445 | 0.78 | 0.46 | 0.76 | 0.66 | 2.66 |
| | 448 | 0.04 | 0.05 | 0 | 0.06 | 0.15 |
| | 449 | 0.03 | 0 | 0 | 0 | 0.03 |
| | 452 | 0 | 0 | 0 | 0.05 | 0.05 |
| | 454 | 0.46 | 0.65 | 0.64 | 0.64 | 2.39 |
| TOTAL | | 4.49 | 6.14 | 3.51 | 4.98 | 19.12 |

STATE: OKLAHOMA

| ZIP CODE | | QUARTER 1 | QUARTER 2 | QUARTER 3 | QUARTER 4 | TOTAL GRAMS |
|---|---|---|---|---|---|---|
| | 731 | 0 | 0.04 | 0 | 0.03 | 0.07 |
| | 740 | 0 | 0.04 | 0 | 0.01 | 0.05 |
| | 741 | 0.22 | 0.12 | 0.14 | 0 | 0.48 |
| | 744 | 0.31 | 0.48 | 0.4 | 0.43 | 1.62 |
| | 748 | 0.3 | 0.33 | 0.3 | 0.37 | 1.3 |
| TOTAL | | 0.83 | 1.01 | 0.84 | 0.84 | 3.52 |

STATE: OREGON

| ZIP CODE | | QUARTER 1 | QUARTER 2 | QUARTER 3 | QUARTER 4 | TOTAL GRAMS |
|---|---|---|---|---|---|---|
| | 970 | 0.66 | 0.58 | 0.77 | 0.98 | 2.99 |
| | 971 | 0.05 | 0.05 | 0.09 | 0.11 | 0.3 |
| | 972 | 2.53 | 2.53 | 4.08 | 2.77 | 11.91 |
| | 973 | 0 | 0 | 0.03 | 0.01 | 0.04 |
| | 974 | 0 | 0.01 | 0 | 0.01 | 0.02 |
| | 975 | 0 | 0.2 | 0 | 0 | 0.2 |

| | | | | | | |
|---|---|---|---|---|---|---|
| | 976 | 0.57 | 0.93 | 0.67 | 0.96 | 3.13 |
| | 977 | 0.03 | 0.02 | 0.03 | 0.03 | 0.11 |
| | 978 | 0.01 | 0 | 0 | 0 | 0.01 |
| TOTAL | | 3.85 | 4.32 | 5.67 | 4.87 | 18.71 |

STATE: PENNSYLVANIA

| ZIP CODE | QUARTER 1 | QUARTER 2 | QUARTER 3 | QUARTER 4 | TOTAL GRAMS |
|---|---|---|---|---|---|
| 152 | 0.02 | 0 | 0 | 0 | 0.02 |
| 161 | 0.03 | 0 | 0 | 0 | 0.03 |
| 163 | 0.09 | 0.04 | 0.03 | 0.05 | 0.21 |
| 164 | 0.02 | 0 | 0 | 0 | 0.02 |
| 165 | 0.11 | 0.11 | 0.05 | 0.04 | 0.31 |
| 170 | 0.1 | 0.09 | 0.1 | 0.22 | 0.51 |
| 171 | 0.25 | 0 | 0.1 | 0.1 | 0.45 |
| 173 | 0 | 0.01 | 0 | 0 | 0.01 |
| 174 | 0.08 | 0.05 | 0.07 | 0.05 | 0.25 |
| 181 | 0 | 0.12 | 0 | 0 | 0.12 |
| 185 | 0 | 0.01 | 0 | 0 | 0.01 |
| 190 | 0 | 0 | 0.02 | 0 | 0.02 |
| 191 | 0 | 0.05 | 0 | 0 | 0.05 |
| TOTAL | 0.7 | 0.48 | 0.37 | 0.46 | 2.01 |

STATE: RHODE ISLAND

| ZIP CODE | QUARTER 1 | QUARTER 2 | QUARTER 3 | QUARTER 4 | TOTAL GRAMS |
|---|---|---|---|---|---|
| 28 | 0.05 | 0.08 | 0.13 | 0 | 0.26 |
| TOTAL | 0.05 | 0.08 | 0.13 | 0 | 0.26 |

STATE: SOUTH CAROLINA

| ZIP CODE | QUARTER 1 | QUARTER 2 | QUARTER 3 | QUARTER 4 | TOTAL GRAMS |
|---|---|---|---|---|---|
| 293 | 0.12 | 0.04 | 0.05 | 0.13 | 0.34 |
| 294 | 0.01 | 0.01 | 0 | 0 | 0.02 |
| TOTAL | 0.13 | 0.05 | 0.05 | 0.13 | 0.36 |

DATE RANGE: 01/01/2017 TO 12/31/2017 ARCOS 3 - REPORT 1 Run Date: 07/03/2018

RETAIL DRUG DISTRIBUTION BY ZIP CODE WITHIN STATE BY GRAMS WT

STATE: SOUTH DAKOTA

| ZIP CODE | QUARTER 1 | QUARTER 2 | QUARTER 3 | QUARTER 4 | TOTAL GRAMS |
|---|---|---|---|---|---|
| 570 | 0.05 | 0 | 0.02 | 0.04 | 0.11 |
| 572 | 0.06 | 0.06 | 0.08 | 0.02 | 0.22 |
| TOTAL | 0.11 | 0.06 | 0.1 | 0.06 | 0.33 |

STATE: TENNESSEE

| ZIP CODE | QUARTER 1 | QUARTER 2 | QUARTER 3 | QUARTER 4 | TOTAL GRAMS |
|---|---|---|---|---|---|
| 371 | 0.92 | 1.01 | 1.08 | 1.11 | 4.12 |
| 372 | 0 | 0 | 0 | 0.35 | 0.35 |
| 376 | 0 | 0.02 | 0.01 | 0 | 0.03 |
| 379 | 0 | 0 | 0 | 0.05 | 0.05 |
| 382 | 0.03 | 0.05 | 0.07 | 0.02 | 0.17 |
| 384 | 0.02 | 0 | 0 | 0.03 | 0.05 |
| TOTAL | 0.97 | 1.08 | 1.16 | 1.56 | 4.77 |

STATE: TEXAS

| ZIP CODE | QUARTER 1 | QUARTER 2 | QUARTER 3 | QUARTER 4 | TOTAL GRAMS |
|---|---|---|---|---|---|
| 750 | 0 | 0.09 | 0.31 | 0 | 0.4 |
| 751 | 0.12 | 0.09 | 0.12 | 0.18 | 0.51 |
| 752 | 0.25 | 0.14 | 0 | 0.18 | 0.57 |
| 755 | 0 | 0.01 | 0 | 0 | 0.01 |
| 756 | 0 | 0 | 0.02 | 0 | 0.02 |
| 757 | 0 | 0 | 0.01 | 0 | 0.01 |
| 760 | 0 | 0.25 | 0 | 0 | 0.25 |
| 761 | 0 | 0.01 | 0 | 0.01 | 0.02 |
| 770 | 0.36 | 0.5 | 0.44 | 0.76 | 2.06 |
| 773 | 0.05 | 0 | 0 | 0.04 | 0.09 |
| 774 | 0.04 | 0.05 | 0 | 0 | 0.09 |
| 775 | 0.09 | 0.15 | 0.2 | 0.2 | 0.64 |
| 777 | 0.02 | 0 | 0.03 | 0.01 | 0.06 |
| 779 | 0 | 0 | 0.02 | 0 | 0.02 |

| | | | | | | |
|---|---|---|---|---|---|---|
| | 780 | 0.02 | 0.06 | 0.07 | 0.05 | 0.2 |
| | 782 | 1.66 | 1.65 | 1.65 | 1.82 | 6.78 |
| | 785 | 0.02 | 0 | 0 | 0 | 0.02 |
| | 787 | 0.01 | 0.03 | 0 | 0.02 | 0.06 |
| | 788 | 0.01 | 0.04 | 0.02 | 0 | 0.07 |
| | 794 | 0 | 0 | 0.03 | 0.09 | 0.12 |
| | 797 | 0.01 | 0.01 | 0.01 | 0 | 0.03 |
| | 799 | 2.4 | 2.99 | 2.29 | 2.36 | 10.04 |
| TOTAL | | 5.06 | 6.07 | 5.22 | 5.72 | 22.07 |

STATE: UTAH

| ZIP CODE | | QUARTER 1 | QUARTER 2 | QUARTER 3 | QUARTER 4 | TOTAL GRAMS |
|---|---|---|---|---|---|---|
| | 841 | 0 | 0.03 | 0.15 | 1.18 | 1.36 |
| | 844 | 0.44 | 0.31 | 0.31 | 0.58 | 1.64 |
| | 845 | 0 | 0 | 0.02 | 0 | 0.02 |
| | 847 | 0.03 | 0.01 | 0.04 | 0.01 | 0.09 |
| TOTAL | | 0.47 | 0.35 | 0.52 | 1.77 | 3.11 |

STATE: VERMONT

| ZIP CODE | | QUARTER 1 | QUARTER 2 | QUARTER 3 | QUARTER 4 | TOTAL GRAMS |
|---|---|---|---|---|---|---|
| | 56 | 0.01 | 0 | 0.01 | 0 | 0.02 |
| | 57 | 0 | 0 | 0 | 0.02 | 0.02 |
| TOTAL | | 0.01 | 0 | 0.01 | 0.02 | 0.04 |

STATE: VIRGINIA

| ZIP CODE | | QUARTER 1 | QUARTER 2 | QUARTER 3 | QUARTER 4 | TOTAL GRAMS |
|---|---|---|---|---|---|---|
| | 201 | 0.02 | 0 | 0.04 | 0 | 0.06 |
| | 229 | 0.06 | 0.03 | 0.03 | 0.03 | 0.15 |
| | 232 | 0.43 | 0.41 | 0.35 | 0.46 | 1.65 |
| | 234 | 0.13 | 0.16 | 0.17 | 0.15 | 0.61 |
| | 236 | 0.03 | 0.02 | 0 | 0.01 | 0.06 |
| | 237 | 0 | 0 | 0 | 0.01 | 0.01 |
| | 240 | 0.08 | 0.06 | 0.09 | 0.08 | 0.31 |
| | 241 | 0.02 | 0 | 0 | 0 | 0.02 |

RETAIL DRUG DISTRIBUTION BY ZIP CODE WITHIN STATE BY GRAMS WT

| | | | | | | |
|---|---|---|---|---|---|---|
| | 242 | 0.1 | 0.45 | 0 | 0.2 | 0.75 |
| TOTAL | | 0.87 | 1.13 | 0.68 | 0.94 | 3.62 |

STATE: WASHINGTON

| ZIP CODE | | QUARTER 1 | QUARTER 2 | QUARTER 3 | QUARTER 4 | TOTAL GRAMS |
|---|---|---|---|---|---|---|
| | 980 | 0.06 | 0.07 | 0.1 | 0.12 | 0.35 |
| | 981 | 1.95 | 2.37 | 2.64 | 2.27 | 9.23 |
| | 982 | 0.2 | 0.2 | 0.12 | 0.3 | 0.82 |
| | 983 | 0.17 | 0.43 | 0.21 | 0.19 | 1 |
| | 984 | 0.09 | 0.14 | 0.18 | 0.19 | 0.6 |
| | 985 | 0.01 | 0.01 | 0 | 0 | 0.02 |
| | 986 | 0 | 0.01 | 0.01 | 0 | 0.02 |
| | 988 | 0 | 0 | 0.04 | 0 | 0.04 |
| | 989 | 0.01 | 0.03 | 0.06 | 0 | 0.1 |
| | 991 | 0 | 0.01 | 0.01 | 0.06 | 0.08 |
| | 992 | 0.03 | 0.05 | 0.04 | 0.08 | 0.2 |
| | 993 | 0.01 | 0.05 | 0.07 | 0.07 | 0.2 |
| TOTAL | | 2.53 | 3.37 | 3.48 | 3.28 | 12.66 |

STATE: WISCONSIN

| ZIP CODE | | QUARTER 1 | QUARTER 2 | QUARTER 3 | QUARTER 4 | TOTAL GRAMS |
|---|---|---|---|---|---|---|
| | 530 | 0.55 | 0.53 | 0.55 | 0.51 | 2.14 |
| | 531 | 0.01 | 0.02 | 0.04 | 0.01 | 0.08 |
| | 532 | 2.4 | 2.96 | 2.89 | 3.15 | 11.4 |
| | 535 | 0.3 | 0.26 | 0.26 | 0.35 | 1.17 |
| | 537 | 0.62 | 0.68 | 0.66 | 0.89 | 2.85 |
| | 538 | 0.1 | 0.23 | 0.15 | 0.28 | 0.76 |
| | 539 | 0.01 | 0.01 | 0 | 0.02 | 0.04 |
| | 542 | 0 | 0 | 0 | 0.06 | 0.06 |
| | 544 | 0.01 | 0 | 0.01 | 0.02 | 0.04 |
| | 546 | 0.05 | 0.06 | 0.03 | 0.07 | 0.21 |
| | 547 | 0.08 | 0.07 | 0.23 | 0.16 | 0.54 |
| TOTAL | | 4.13 | 4.82 | 4.82 | 5.52 | 19.29 |

STATE: WYOMING

| ZIP CODE | QUARTER 1 | QUARTER 2 | QUARTER 3 | QUARTER 4 | TOTAL GRAMS |
|---|---|---|---|---|---|
| 823 | 0 | 0 | 0 | 0.06 | 0.06 |
| 826 | 0.01 | 0 | 0 | 0.03 | 0.04 |
| 828 | 0 | 0 | 0.02 | 0 | 0.02 |
| TOTAL | 0.01 | 0 | 0.02 | 0.09 | 0.12 |

DRUG CODE: 9739 DRUG NAME: REMIFENTANIL

STATE: ALABAMA

| ZIP CODE | QUARTER 1 | QUARTER 2 | QUARTER 3 | QUARTER 4 | TOTAL GRAMS |
|---|---|---|---|---|---|
| 350 | 0 | 0.01 | 0 | 0 | 0.01 |
| 352 | 1.86 | 2.36 | 2.2 | 1.87 | 8.29 |
| 354 | 0.02 | 0 | 0.03 | 0.01 | 0.06 |
| 356 | 0.04 | 0.06 | 0.04 | 0.1 | 0.24 |
| 357 | 0 | 0 | 0.01 | 0 | 0.01 |
| 358 | 0.05 | 0.04 | 0.06 | 0 | 0.15 |
| 361 | 0.54 | 0.51 | 0.5 | 0.45 | 2 |
| 363 | 0.06 | 0.08 | 0.06 | 0.08 | 0.28 |
| 366 | 0.59 | 0.34 | 0.54 | 0.63 | 2.1 |
| 368 | 0 | 0.01 | 0 | 0 | 0.01 |
| TOTAL | 3.16 | 3.41 | 3.44 | 3.14 | 13.15 |

STATE: ALASKA

| ZIP CODE | QUARTER 1 | QUARTER 2 | QUARTER 3 | QUARTER 4 | TOTAL GRAMS |
|---|---|---|---|---|---|
| 995 | 2.34 | 2.68 | 2.37 | 2.7 | 10.09 |
| 996 | 0.16 | 0.07 | 0.22 | 0.27 | 0.72 |
| 997 | 0.23 | 0.26 | 0.19 | 0.17 | 0.85 |
| 999 | 0 | 0 | 0.02 | 0 | 0.02 |
| TOTAL | 2.73 | 3.01 | 2.8 | 3.14 | 11.68 |

RETAIL DRUG DISTRIBUTION BY ZIP CODE WITHIN STATE BY GRAMS WT

STATE: ARIZONA

| ZIP CODE | QUARTER 1 | QUARTER 2 | QUARTER 3 | QUARTER 4 | TOTAL GRAMS |
|---|---|---|---|---|---|
| 850 | 1.82 | 1.59 | 1.59 | 1.88 | 6.88 |
| 851 | 0 | 0 | 0 | 0.03 | 0.03 |
| 852 | 0.84 | 0.69 | 0.6 | 0.77 | 2.9 |
| 853 | 0.16 | 0.1 | 0.24 | 0.4 | 0.9 |
| 855 | 0.03 | 0 | 0.01 | 0.05 | 0.09 |
| 857 | 1.03 | 1.15 | 0.83 | 0.89 | 3.9 |
| 859 | 0.04 | 0.03 | 0.06 | 0 | 0.13 |
| 860 | 0.22 | 0.18 | 0.3 | 0.14 | 0.84 |
| 863 | 0.01 | 0 | 0 | 0 | 0.01 |
| 864 | 0.01 | 0 | 0 | 0 | 0.01 |
| TOTAL | 4.16 | 3.74 | 3.63 | 4.16 | 15.69 |

STATE: ARKANSAS

| ZIP CODE | QUARTER 1 | QUARTER 2 | QUARTER 3 | QUARTER 4 | TOTAL GRAMS |
|---|---|---|---|---|---|
| 720 | 0.06 | 0.06 | 0.13 | 0.05 | 0.3 |
| 722 | 1.43 | 1.45 | 1.59 | 1.37 | 5.84 |
| 724 | 0.04 | 0 | 0 | 0.02 | 0.06 |
| 725 | 0 | 0 | 0 | 0.01 | 0.01 |
| 727 | 0.01 | 0 | 0.04 | 0.01 | 0.06 |
| 728 | 0 | 0 | 0.06 | 0 | 0.06 |
| 729 | 0.03 | 0.01 | 0 | 0.02 | 0.06 |
| TOTAL | 1.57 | 1.52 | 1.82 | 1.48 | 6.39 |

STATE: CALIFORNIA

| ZIP CODE | QUARTER 1 | QUARTER 2 | QUARTER 3 | QUARTER 4 | TOTAL GRAMS |
|---|---|---|---|---|---|
| 900 | 4.41 | 4.45 | 4.03 | 4.21 | 17.1 |
| 902 | 0.53 | 0.22 | 0.35 | 0.28 | 1.38 |
| 904 | 0.68 | 0.68 | 0.83 | 0.96 | 3.15 |
| 905 | 0.45 | 0.33 | 0.38 | 0.34 | 1.5 |
| 906 | 0 | 0 | 0.01 | 0 | 0.01 |
| 907 | 0.11 | 0.09 | 0.26 | 0.14 | 0.6 |
| 908 | 0.28 | 0.3 | 0.32 | 0.4 | 1.3 |
| 910 | 0.04 | 0.04 | 0.03 | 0.02 | 0.13 |

| | | | | | |
|---|---|---|---|---|---|
| 911 | 0.28 | 0.26 | 0.14 | 0.24 | 0.92 |
| 912 | 0 | 0 | 0.06 | 0.04 | 0.1 |
| 913 | 0.15 | 0.04 | 0.15 | 0.15 | 0.49 |
| 917 | 0.07 | 0.03 | 0.11 | 0.11 | 0.32 |
| 919 | 0.19 | 0.21 | 0.21 | 0.15 | 0.76 |
| 920 | 0.78 | 0.91 | 0.97 | 1.11 | 3.77 |
| 921 | 0.92 | 0.89 | 1.27 | 0.89 | 3.97 |
| 922 | 0.24 | 0.25 | 0.19 | 0.27 | 0.95 |
| 923 | 0.96 | 1.31 | 0.94 | 0.96 | 4.17 |
| 924 | 0.26 | 0.24 | 0.3 | 0.12 | 0.92 |
| 925 | 0.26 | 0.19 | 0.28 | 0.2 | 0.93 |
| 926 | 1.6 | 1.94 | 1.46 | 2.13 | 7.13 |
| 927 | 0.21 | 0.21 | 0.21 | 0.23 | 0.86 |
| 928 | 1.84 | 1.87 | 1.97 | 1.78 | 7.46 |
| 930 | 0.33 | 0.49 | 0.3 | 0.44 | 1.56 |
| 931 | 0.45 | 0.64 | 0.37 | 0.43 | 1.89 |
| 932 | 0.32 | 0.08 | 0.15 | 0.21 | 0.76 |
| 933 | 0.1 | 0.15 | 0.05 | 0.1 | 0.4 |
| 934 | 0.05 | 0.01 | 0.05 | 0.08 | 0.19 |
| 935 | 0.02 | 0.01 | 0 | 0.01 | 0.04 |
| 936 | 0.31 | 0.43 | 0.27 | 0.24 | 1.25 |
| 937 | 1.13 | 1.56 | 0.61 | 0.99 | 4.29 |
| 939 | 0.04 | 0.04 | 0.01 | 0.04 | 0.13 |
| 940 | 1.82 | 1.44 | 1.5 | 1.45 | 6.21 |
| 941 | 3.11 | 4.85 | 1.56 | 3.03 | 12.55 |
| 943 | 5.32 | 4.32 | 5.43 | 4.89 | 19.96 |
| 944 | 0 | 0.03 | 0.02 | 0.07 | 0.12 |
| 945 | 0.77 | 0.81 | 1.49 | 0.54 | 3.61 |
| 946 | 0.1 | 0.12 | 0.09 | 0.18 | 0.49 |
| 948 | 0 | 0 | 0.02 | 0 | 0.02 |
| 949 | 0.1 | 0.05 | 0.09 | 0.1 | 0.34 |
| 950 | 0.35 | 0.31 | 0.22 | 0.24 | 1.12 |
| 951 | 1.51 | 1.2 | 1.27 | 1.26 | 5.24 |
| 952 | 0.09 | 0.09 | 0.1 | 0.14 | 0.42 |
| 953 | 0.06 | 0.08 | 0.03 | 0.05 | 0.22 |
| 954 | 0.09 | 0.1 | 0.14 | 0.19 | 0.52 |
| 955 | 0.09 | 0.09 | 0.05 | 0 | 0.23 |

| | | | | | | |
|---|---|---|---|---|---|---|
| | 956 | 0.76 | 0.59 | 0.91 | 1.08 | 3.34 |
| | 957 | 0.01 | 0 | 0.01 | 0 | 0.02 |
| | 958 | 1.84 | 2.31 | 1.82 | 3.65 | 9.62 |
| | 959 | 0 | 0.05 | 0.02 | 0.02 | 0.09 |
| | 960 | 0.18 | 0.25 | 0.2 | 0.18 | 0.81 |
| TOTAL | | 33.21 | 34.56 | 31.25 | 34.34 | 133.36 |

| STATE: | COLORADO | | | | | |
|---|---|---|---|---|---|---|
| ZIP CODE | | QUARTER 1 | QUARTER 2 | QUARTER 3 | QUARTER 4 | TOTAL GRAMS |
| | 800 | 2.53 | 2.6 | 3.03 | 3.56 | 11.72 |
| | 801 | 0.44 | 0.53 | 0.31 | 0.51 | 1.79 |
| | 802 | 1.05 | 1.03 | 0.99 | 0.95 | 4.02 |
| | 803 | 0.27 | 0.3 | 0.26 | 0.17 | 1 |
| | 804 | 0.23 | 0.15 | 0.04 | 0.04 | 0.46 |
| | 805 | 0.03 | 0.04 | 0.2 | 0.11 | 0.38 |
| | 806 | 0.1 | 0.12 | 0.1 | 0.04 | 0.36 |
| | 807 | 0.01 | 0.01 | 0.01 | 0 | 0.03 |
| | 808 | 0 | 0.21 | 0 | 0 | 0.21 |
| | 809 | 0.18 | 0.27 | 0.19 | 0.23 | 0.87 |
| | 810 | 0.38 | 0.44 | 0.28 | 0.63 | 1.73 |
| | 812 | 0 | 0 | 0 | 0.01 | 0.01 |
| | 813 | 0.39 | 0.33 | 0.29 | 0.4 | 1.41 |
| | 814 | 0.01 | 0.01 | 0.03 | 0.02 | 0.07 |
| | 815 | 0.26 | 0.42 | 0.36 | 0.4 | 1.44 |
| | 816 | 0.08 | 0.07 | 0.1 | 0.09 | 0.34 |
| TOTAL | | 5.96 | 6.53 | 6.19 | 7.16 | 25.84 |

| STATE: | CONNECTICUT | | | | | |
|---|---|---|---|---|---|---|
| ZIP CODE | | QUARTER 1 | QUARTER 2 | QUARTER 3 | QUARTER 4 | TOTAL GRAMS |
| | 60 | 0.12 | 0.11 | 0.1 | 0.17 | 0.5 |
| | 61 | 1.01 | 0.38 | 0.46 | 0.56 | 2.41 |
| | 62 | 0 | 0.02 | 0.04 | 0.08 | 0.14 |
| | 63 | 0.23 | 0.2 | 0.18 | 0.25 | 0.86 |
| | 64 | 0.43 | 0.56 | 0.6 | 0.76 | 2.35 |

| | | | | | | |
|---|---|---|---|---|---|---|
| | 65 | 5.93 | 5.01 | 5.19 | 5.32 | 21.45 |
| | 66 | 1.31 | 0.71 | 0.5 | 0.69 | 3.21 |
| | 67 | 0.14 | 0.09 | 0.1 | 0.06 | 0.39 |
| | 68 | 0.36 | 0.31 | 0.38 | 0.52 | 1.57 |
| | 69 | 0.28 | 0.33 | 0.27 | 0.3 | 1.18 |
| TOTAL | | 9.81 | 7.72 | 7.82 | 8.71 | 34.06 |

| STATE: | DELAWARE | | | | | |
|---|---|---|---|---|---|---|
| ZIP CODE | | QUARTER 1 | QUARTER 2 | QUARTER 3 | QUARTER 4 | TOTAL GRAMS |
| | 197 | 0.6 | 0.72 | 0.74 | 0.9 | 2.96 |
| | 198 | 1.75 | 3.01 | 1.83 | 1.44 | 8.03 |
| | 199 | 0.32 | 0.29 | 0.24 | 0.2 | 1.05 |
| TOTAL | | 2.67 | 4.02 | 2.81 | 2.54 | 12.04 |

| STATE: | DISTRICT OF COLUMBIA | | | | | |
|---|---|---|---|---|---|---|
| ZIP CODE | | QUARTER 1 | QUARTER 2 | QUARTER 3 | QUARTER 4 | TOTAL GRAMS |
| | 200 | 0.83 | 0.86 | 0.52 | 0.56 | 2.77 |
| | 204 | 0.05 | 0 | 0 | 0.02 | 0.07 |
| TOTAL | | 0.88 | 0.86 | 0.52 | 0.58 | 2.84 |

| STATE: | FLORIDA | | | | | |
|---|---|---|---|---|---|---|
| ZIP CODE | | QUARTER 1 | QUARTER 2 | QUARTER 3 | QUARTER 4 | TOTAL GRAMS |
| | 320 | 0.01 | 0 | 0.02 | 0.02 | 0.05 |
| | 321 | 0.11 | 0.05 | 0.03 | 0.04 | 0.23 |
| | 322 | 1.59 | 2 | 1.68 | 1.9 | 7.17 |
| | 323 | 0.08 | 0 | 0.04 | 0.02 | 0.14 |
| | 325 | 0.92 | 0.92 | 0.69 | 1 | 3.53 |
| | 326 | 1.32 | 1.87 | 1.41 | 1.97 | 6.57 |
| | 327 | 0.83 | 0.75 | 0.83 | 0.87 | 3.28 |
| | 328 | 2.4 | 2.77 | 2.47 | 2.9 | 10.54 |
| | 329 | 0.11 | 0.2 | 0.21 | 0.08 | 0.6 |
| | 330 | 0.45 | 0.48 | 0.53 | 0.62 | 2.08 |
| | 331 | 4.96 | 4.61 | 4.85 | 4.31 | 18.73 |

| | | | | | | |
|---|---|---|---|---|---|---|
| | 333 | 0.78 | 1.06 | 0.9 | 1.07 | 3.81 |
| | 334 | 3.63 | 3.77 | 3.35 | 4.41 | 15.16 |
| | 335 | 0.19 | 0.15 | 0.16 | 0.52 | 1.02 |
| | 336 | 4.68 | 4.56 | 4.63 | 6.1 | 19.97 |
| | 337 | 0.82 | 0.77 | 0.79 | 0.83 | 3.21 |
| | 338 | 0.27 | 0.44 | 0.35 | 0.26 | 1.32 |
| | 339 | 0.29 | 0.24 | 0.23 | 0.16 | 0.92 |
| | 341 | 0.13 | 0.08 | 0.14 | 0.03 | 0.38 |
| | 342 | 0.4 | 0.35 | 0.41 | 0.49 | 1.65 |
| | 344 | 0.1 | 0.14 | 0.06 | 0.06 | 0.36 |
| | 346 | 0.5 | 0.38 | 0.33 | 0.39 | 1.6 |
| | 347 | 0.79 | 0.77 | 0.68 | 0.94 | 3.18 |
| | 349 | 0.39 | 0.56 | 0.44 | 0.56 | 1.95 |
| TOTAL | | 25.75 | 26.92 | 25.23 | 29.55 | 107.45 |

| STATE: | GEORGIA | | | | | |
|---|---|---|---|---|---|---|
| ZIP CODE | | QUARTER 1 | QUARTER 2 | QUARTER 3 | QUARTER 4 | TOTAL GRAMS |
| | 300 | 0.92 | 0.67 | 0.97 | 1.02 | 3.58 |
| | 301 | 0.1 | 0.18 | 0.1 | 0.13 | 0.51 |
| | 302 | 0.4 | 0.22 | 0.31 | 0.18 | 1.11 |
| | 303 | 0.77 | 1.06 | 0.9 | 0.69 | 3.42 |
| | 305 | 0.05 | 0.06 | 0.04 | 0.06 | 0.21 |
| | 306 | 0.08 | 0.15 | 0.06 | 0.09 | 0.38 |
| | 307 | 0.06 | 0.04 | 0.05 | 0.06 | 0.21 |
| | 309 | 1.38 | 1.52 | 1.32 | 1.85 | 6.07 |
| | 310 | 0.01 | 0 | 0 | 0 | 0.01 |
| | 312 | 0.57 | 0.51 | 0.58 | 0.51 | 2.17 |
| | 313 | 0 | 0.18 | 0 | 0 | 0.18 |
| | 314 | 0.09 | 0.16 | 0.11 | 0.05 | 0.41 |
| | 315 | 0.06 | 0.04 | 0.06 | 0 | 0.16 |
| | 316 | 0 | 0.03 | 0.04 | 0.02 | 0.09 |
| | 317 | 0.15 | 0.2 | 0.08 | 0.13 | 0.56 |
| | 319 | 0 | 0.04 | 0.03 | 0 | 0.07 |
| | 398 | 0.01 | 0.03 | 0.02 | 0 | 0.06 |
| TOTAL | | 4.65 | 5.09 | 4.67 | 4.79 | 19.2 |

| STATE: | GUAM | | | | |
|---|---|---|---|---|---|
| ZIP CODE | QUARTER 1 | QUARTER 2 | QUARTER 3 | QUARTER 4 | TOTAL GRAMS |
| 969 | 0.02 | 0.04 | 0 | 0.25 | 0.31 |
| TOTAL | 0.02 | 0.04 | 0 | 0.25 | 0.31 |

| STATE: | HAWAII | | | | |
|---|---|---|---|---|---|
| ZIP CODE | QUARTER 1 | QUARTER 2 | QUARTER 3 | QUARTER 4 | TOTAL GRAMS |
| 967 | 0.01 | 0.06 | 0.02 | 0.06 | 0.15 |
| 968 | 1.28 | 1.07 | 1.32 | 1.32 | 4.99 |
| TOTAL | 1.29 | 1.13 | 1.34 | 1.38 | 5.14 |

| STATE: | IDAHO | | | | |
|---|---|---|---|---|---|
| ZIP CODE | QUARTER 1 | QUARTER 2 | QUARTER 3 | QUARTER 4 | TOTAL GRAMS |
| 832 | 0.07 | 0.09 | 0.11 | 0.06 | 0.33 |
| 833 | 0.49 | 0.29 | 0.23 | 0.13 | 1.14 |
| 834 | 0.67 | 0.66 | 0.53 | 0.78 | 2.64 |
| 836 | 0.14 | 0.18 | 0.11 | 0.23 | 0.66 |
| 837 | 0.82 | 0.9 | 1.12 | 0.83 | 3.67 |
| 838 | 0.06 | 0.07 | 0.02 | 0.11 | 0.26 |
| TOTAL | 2.25 | 2.19 | 2.12 | 2.14 | 8.7 |

| STATE: | ILLINOIS | | | | |
|---|---|---|---|---|---|
| ZIP CODE | QUARTER 1 | QUARTER 2 | QUARTER 3 | QUARTER 4 | TOTAL GRAMS |
| 600 | 1.29 | 0.93 | 0.94 | 1 | 4.16 |
| 601 | 2.78 | 2.26 | 2.62 | 2.63 | 10.29 |
| 602 | 0.46 | 0.55 | 0.36 | 0.68 | 2.05 |
| 603 | 0 | 0 | 0.05 | 0.2 | 0.25 |
| 604 | 1.13 | 1.4 | 1.29 | 1.54 | 5.36 |
| 605 | 0.7 | 0.87 | 0.88 | 1.27 | 3.72 |
| 606 | 6.17 | 5.68 | 5.9 | 5.97 | 23.72 |
| 607 | 0.14 | 0.14 | 0.12 | 0.14 | 0.54 |

| | | | | | | |
|---|---|---|---|---|---|---|
| | 610 | 0 | 0 | 0.02 | 0.01 | 0.03 |
| | 611 | 0.79 | 1.07 | 0.71 | 1.13 | 3.7 |
| | 612 | 0.13 | 0.14 | 0.18 | 0.12 | 0.57 |
| | 614 | 0.22 | 0.2 | 0.22 | 0.29 | 0.93 |
| | 616 | 0.34 | 0.39 | 0.41 | 0.55 | 1.69 |
| | 617 | 0.07 | 0.14 | 0.11 | 0.16 | 0.48 |
| | 618 | 0.02 | 0.08 | 0.04 | 0.1 | 0.24 |
| | 619 | 0 | 0.04 | 0 | 0.02 | 0.06 |
| | 620 | 0 | 0 | 0.01 | 0.03 | 0.04 |
| | 622 | 0.21 | 0.24 | 0.2 | 0.21 | 0.86 |
| | 625 | 0 | 0.02 | 0.02 | 0.06 | 0.1 |
| | 627 | 0.24 | 0.21 | 0.39 | 0.66 | 1.5 |
| | 629 | 0.12 | 0.12 | 0.11 | 0.12 | 0.47 |
| TOTAL | | 14.81 | 14.48 | 14.58 | 16.89 | 60.76 |

| STATE: | INDIANA | | | | | |
|---|---|---|---|---|---|---|
| ZIP CODE | | QUARTER 1 | QUARTER 2 | QUARTER 3 | QUARTER 4 | TOTAL GRAMS |
| | 460 | 1.1 | 0.98 | 0.93 | 1.16 | 4.17 |
| | 461 | 0.08 | 0.12 | 0.08 | 0.08 | 0.36 |
| | 462 | 3.55 | 3.48 | 3.38 | 3.24 | 13.65 |
| | 463 | 0.49 | 0.45 | 0.48 | 0.52 | 1.94 |
| | 464 | 0.1 | 0.09 | 0.06 | 0.12 | 0.37 |
| | 465 | 0.52 | 0.43 | 0.57 | 0.76 | 2.28 |
| | 466 | 0.08 | 0.14 | 0.14 | 0.15 | 0.51 |
| | 468 | 0.38 | 0.34 | 0.5 | 0.36 | 1.58 |
| | 469 | 0.04 | 0.04 | 0.02 | 0.01 | 0.11 |
| | 471 | 0.1 | 0.15 | 0.1 | 0.05 | 0.4 |
| | 472 | 0.2 | 0.24 | 0.25 | 0.5 | 1.19 |
| | 473 | 0.69 | 0.6 | 0.51 | 0.55 | 2.35 |
| | 474 | 0.17 | 0.16 | 0.12 | 0.14 | 0.59 |
| | 475 | 0.11 | 0.12 | 0.16 | 0.05 | 0.44 |
| | 476 | 0.06 | 0.03 | 0.07 | 0.06 | 0.22 |
| | 477 | 0.17 | 0.12 | 0.27 | 0.08 | 0.64 |
| | 478 | 0.16 | 0.15 | 0.21 | 0.13 | 0.65 |
| | 479 | 0.1 | 0.11 | 0.12 | 0.15 | 0.48 |

| | | | | | |
|---|---|---|---|---|---|
| TOTAL | 8.1 | 7.75 | 7.97 | 8.11 | 31.93 |

STATE: IOWA

| ZIP CODE | QUARTER 1 | QUARTER 2 | QUARTER 3 | QUARTER 4 | TOTAL GRAMS |
|---|---|---|---|---|---|
| 500 | 0.22 | 0.22 | 0.16 | 0.18 | 0.78 |
| 501 | 0 | 0.06 | 0 | 0 | 0.06 |
| 502 | 0.02 | 0.02 | 0 | 0.02 | 0.06 |
| 503 | 0.01 | 0.04 | 0.02 | 0.02 | 0.09 |
| 504 | 0 | 0.01 | 0 | 0 | 0.01 |
| 505 | 0.05 | 0.05 | 0.04 | 0.03 | 0.17 |
| 507 | 0.14 | 0.12 | 0.2 | 0.18 | 0.64 |
| 510 | 0 | 0 | 0 | 0.04 | 0.04 |
| 512 | 0 | 0.02 | 0 | 0.04 | 0.06 |
| 520 | 0.15 | 0.1 | 0.12 | 0.13 | 0.5 |
| 522 | 2.6 | 2.47 | 2.19 | 1.42 | 8.68 |
| 524 | 0.05 | 0 | 0.01 | 0.06 | 0.12 |
| 527 | 0.03 | 0.03 | 0.04 | 0.02 | 0.12 |
| 528 | 0.3 | 0.1 | 0.55 | 0.3 | 1.25 |
| TOTAL | 3.57 | 3.24 | 3.33 | 2.44 | 12.58 |

STATE: KANSAS

| ZIP CODE | QUARTER 1 | QUARTER 2 | QUARTER 3 | QUARTER 4 | TOTAL GRAMS |
|---|---|---|---|---|---|
| 660 | 0.5 | 0.39 | 0.23 | 0.57 | 1.69 |
| 661 | 0.85 | 0.88 | 0.74 | 0.92 | 3.39 |
| 662 | 0.21 | 0.45 | 0.38 | 0.22 | 1.26 |
| 664 | 0 | 0.18 | 0 | 0 | 0.18 |
| 665 | 0.05 | 0.03 | 0.03 | 0.04 | 0.15 |
| 666 | 0.23 | 0.31 | 0.4 | 0.2 | 1.14 |
| 670 | 0.04 | 0.02 | 0.04 | 0.06 | 0.16 |
| 672 | 0.17 | 0.22 | 0.21 | 0.29 | 0.89 |
| 674 | 0.03 | 0.05 | 0.02 | 0.03 | 0.13 |
| 675 | 0.02 | 0 | 0.06 | 0 | 0.08 |
| TOTAL | 2.1 | 2.53 | 2.11 | 2.33 | 9.07 |

STATE: KENTUCKY

| ZIP CODE | QUARTER 1 | QUARTER 2 | QUARTER 3 | QUARTER 4 | TOTAL GRAMS |
|---|---|---|---|---|---|
| 402 | 1.71 | 1.21 | 1.38 | 1.48 | 5.78 |
| 403 | 0.02 | 0 | 0.07 | 0.01 | 0.1 |
| 404 | 0.35 | 0.15 | 0.29 | 0.33 | 1.12 |
| 405 | 1.32 | 1.38 | 1.74 | 1.72 | 6.16 |
| 407 | 0.02 | 0 | 0 | 0 | 0.02 |
| 410 | 0.12 | 0.1 | 0.08 | 0 | 0.3 |
| 411 | 0.1 | 0.13 | 0.18 | 0.11 | 0.52 |
| 415 | 0.02 | 0 | 0.02 | 0.02 | 0.06 |
| 420 | 0.36 | 0.26 | 0.36 | 0.26 | 1.24 |
| 421 | 0.97 | 0.9 | 1.09 | 0.67 | 3.63 |
| 422 | 0.03 | 0.1 | 0.01 | 0 | 0.14 |
| 423 | 0 | 0.03 | 0 | 0.01 | 0.04 |
| 424 | 0 | 0 | 0.02 | 0 | 0.02 |
| 425 | 0 | 0.01 | 0 | 0 | 0.01 |
| 427 | 0 | 0 | 0.02 | 0 | 0.02 |
| TOTAL | 5.02 | 4.27 | 5.26 | 4.61 | 19.16 |

STATE: LOUISIANA

| ZIP CODE | QUARTER 1 | QUARTER 2 | QUARTER 3 | QUARTER 4 | TOTAL GRAMS |
|---|---|---|---|---|---|
| 700 | 0.37 | 0.22 | 0.25 | 0.29 | 1.13 |
| 701 | 2.34 | 2.88 | 2.33 | 2.18 | 9.73 |
| 703 | 0.08 | 0.02 | 0.01 | 0.08 | 0.19 |
| 704 | 0.19 | 0.18 | 0.31 | 0.28 | 0.96 |
| 705 | 0.47 | 0.41 | 0.52 | 0.63 | 2.03 |
| 706 | 0 | 0 | 0.01 | 0 | 0.01 |
| 708 | 0.45 | 0.52 | 0.76 | 1.24 | 2.97 |
| 711 | 0.49 | 0.33 | 0.31 | 0.26 | 1.39 |
| 712 | 0.02 | 0 | 0 | 0 | 0.02 |
| 713 | 0 | 0.06 | 0 | 0 | 0.06 |
| 714 | 0 | 0.08 | 0 | 0 | 0.08 |
| TOTAL | 4.41 | 4.7 | 4.5 | 4.96 | 18.57 |

STATE: MAINE

| ZIP CODE | QUARTER 1 | QUARTER 2 | QUARTER 3 | QUARTER 4 | TOTAL GRAMS |
|---|---|---|---|---|---|
| 40 | 0.02 | 0.06 | 0.01 | 0.01 | 0.1 |
| 41 | 0.66 | 0.3 | 0.34 | 0.33 | 1.63 |
| 42 | 0.13 | 0.23 | 0.14 | 0.22 | 0.72 |
| 43 | 0.09 | 0.03 | 0.1 | 0.07 | 0.29 |
| 44 | 0.41 | 0.51 | 0.43 | 0.26 | 1.61 |
| 46 | 0.02 | 0.02 | 0.04 | 0.01 | 0.09 |
| 47 | 0.02 | 0 | 0 | 0.02 | 0.04 |
| 49 | 0.02 | 0.05 | 0.05 | 0.01 | 0.13 |
| TOTAL | 1.37 | 1.2 | 1.11 | 0.93 | 4.61 |

STATE: MARYLAND

| ZIP CODE | QUARTER 1 | QUARTER 2 | QUARTER 3 | QUARTER 4 | TOTAL GRAMS |
|---|---|---|---|---|---|
| 207 | 0.09 | 0.2 | 0.04 | 0.06 | 0.39 |
| 208 | 1.39 | 0.88 | 0.86 | 1.12 | 4.25 |
| 209 | 0.04 | 0 | 0.02 | 0.02 | 0.08 |
| 210 | 0.21 | 0.19 | 0.15 | 0.19 | 0.74 |
| 211 | 0.05 | 0 | 0.06 | 0 | 0.11 |
| 212 | 5.05 | 5.41 | 4.4 | 5.48 | 20.34 |
| 214 | 0.02 | 0.02 | 0.02 | 0.04 | 0.1 |
| 216 | 0.02 | 0 | 0.04 | 0 | 0.06 |
| 217 | 0.08 | 0.05 | 0.03 | 0.05 | 0.21 |
| 218 | 0.24 | 0.38 | 0.26 | 0.24 | 1.12 |
| TOTAL | 7.19 | 7.13 | 5.88 | 7.2 | 27.4 |

STATE: MASSACHUSETTS

| ZIP CODE | QUARTER 1 | QUARTER 2 | QUARTER 3 | QUARTER 4 | TOTAL GRAMS |
|---|---|---|---|---|---|
| 10 | 0.18 | 0.11 | 0.1 | 0 | 0.39 |
| 11 | 0.2 | 0.21 | 0.22 | 0.17 | 0.8 |
| 12 | 0.08 | 0.07 | 0.02 | 0.06 | 0.23 |
| 14 | 0.07 | 0.11 | 0.08 | 0.09 | 0.35 |
| 15 | 0.02 | 0.03 | 0.01 | 0 | 0.06 |

| | | | | | |
|---|---|---|---|---|---|
| 16 | 1.09 | 1.03 | 1.06 | 1.42 | 4.6 |
| 17 | 0.23 | 0.26 | 0.21 | 0.45 | 1.15 |
| 18 | 0.96 | 1.05 | 0.92 | 1.21 | 4.14 |
| 19 | 0.47 | 0.58 | 0.41 | 0.56 | 2.02 |
| 20 | 0.35 | 0.6 | 0.32 | 0.42 | 1.69 |
| 21 | 15.32 | 17.75 | 16.7 | 17.31 | 67.08 |
| 22 | 1.09 | 1.17 | 1.02 | 0.74 | 4.02 |
| 23 | 0.14 | 0.28 | 0.14 | 0.23 | 0.79 |
| 24 | 0.33 | 0.4 | 0.17 | 0.28 | 1.18 |
| 25 | 0.04 | 0.03 | 0.05 | 0.03 | 0.15 |
| 26 | 0.09 | 0.13 | 0.11 | 0.06 | 0.39 |
| 27 | 0.02 | 0.09 | 0.08 | 0.04 | 0.23 |
| TOTAL | 20.68 | 23.9 | 21.62 | 23.07 | 89.27 |

STATE: MICHIGAN

| ZIP CODE | QUARTER 1 | QUARTER 2 | QUARTER 3 | QUARTER 4 | TOTAL GRAMS |
|---|---|---|---|---|---|
| 480 | 0.79 | 1.31 | 0.57 | 1.09 | 3.76 |
| 481 | 3.68 | 4.27 | 4.24 | 3.66 | 15.85 |
| 482 | 2.61 | 2.91 | 2.39 | 2.6 | 10.51 |
| 483 | 0.97 | 0.84 | 1.21 | 0.81 | 3.83 |
| 485 | 0.02 | 0.08 | 0.04 | 0.1 | 0.24 |
| 486 | 0.34 | 0.33 | 0.29 | 0.35 | 1.31 |
| 487 | 0.29 | 0.31 | 0.28 | 0.18 | 1.06 |
| 489 | 0.17 | 0.15 | 0.16 | 0.14 | 0.62 |
| 490 | 0.16 | 0.25 | 0.09 | 0.23 | 0.73 |
| 492 | 0.12 | 0.11 | 0.08 | 0.09 | 0.4 |
| 494 | 0.04 | 0.11 | 0.05 | 0.05 | 0.25 |
| 495 | 0.26 | 0.23 | 0.2 | 0.12 | 0.81 |
| 497 | 0.03 | 0.03 | 0.06 | 0.05 | 0.17 |
| 498 | 0.03 | 0.02 | 0.02 | 0.02 | 0.09 |
| TOTAL | 9.51 | 10.95 | 9.68 | 9.49 | 39.63 |

STATE: MINNESOTA

| ZIP CODE | QUARTER 1 | QUARTER 2 | QUARTER 3 | QUARTER 4 | TOTAL GRAMS |
|---|---|---|---|---|---|
| 550 | 0 | 0.01 | 0.01 | 0 | 0.02 |

| | | | | | |
|---|---|---|---|---|---|
| | 551 | 0.79 | 0.86 | 0.87 | 0.83 | 3.35 |
| | 553 | 0.05 | 0.06 | 0.04 | 0.06 | 0.21 |
| | 554 | 3.37 | 3.62 | 3.84 | 3.85 | 14.68 |
| | 558 | 0.24 | 0.76 | 0.15 | 0.34 | 1.49 |
| | 559 | 2.44 | 2.45 | 2.87 | 4.57 | 12.33 |
| | 562 | 0 | 0.02 | 0 | 0 | 0.02 |
| | 563 | 0.12 | 0.11 | 0.13 | 0.2 | 0.56 |
| | 564 | 0.01 | 0.01 | 0.01 | 0 | 0.03 |
| TOTAL | | 7.02 | 7.9 | 7.92 | 9.85 | 32.69 |

STATE: MISSISSIPPI

| ZIP CODE | QUARTER 1 | QUARTER 2 | QUARTER 3 | QUARTER 4 | TOTAL GRAMS |
|---|---|---|---|---|---|
| 392 | 0.37 | 0.41 | 0.23 | 0.25 | 1.26 |
| 394 | 0 | 0.12 | 0 | 0 | 0.12 |
| 395 | 0.18 | 0.07 | 0.04 | 0.06 | 0.35 |
| 396 | 0.01 | 0 | 0 | 0.01 | 0.02 |
| 397 | 0 | 0.03 | 0 | 0 | 0.03 |
| TOTAL | 0.56 | 0.63 | 0.27 | 0.32 | 1.78 |

STATE: MISSOURI

| ZIP CODE | QUARTER 1 | QUARTER 2 | QUARTER 3 | QUARTER 4 | TOTAL GRAMS |
|---|---|---|---|---|---|
| 630 | 0.07 | 0.06 | 0.09 | 0.06 | 0.28 |
| 631 | 2.09 | 2.25 | 2.36 | 2.62 | 9.32 |
| 633 | 0 | 0.02 | 0.01 | 0 | 0.03 |
| 637 | 0.26 | 0.17 | 0.25 | 0.28 | 0.96 |
| 638 | 0 | 0 | 0.02 | 0.04 | 0.06 |
| 640 | 0.07 | 0.06 | 0.06 | 0.08 | 0.27 |
| 641 | 1.44 | 1.48 | 1.78 | 1.34 | 6.04 |
| 645 | 0.05 | 0.1 | 0.04 | 0.05 | 0.24 |
| 648 | 0.04 | 0.08 | 0.14 | 0.12 | 0.38 |
| 650 | 0.17 | 0.1 | 0.15 | 0.17 | 0.59 |
| 651 | 0.06 | 0.06 | 0.04 | 0.12 | 0.28 |
| 652 | 0.55 | 0.65 | 0.68 | 0.44 | 2.32 |
| 654 | 0.06 | 0.06 | 0.06 | 0.06 | 0.24 |

| | | | | | | |
|---|---|---|---|---|---|---|
| | 656 | 0.06 | 0.06 | 0.04 | 0.06 | 0.22 |
| | 658 | 0.1 | 0.16 | 0.14 | 0.14 | 0.54 |
| TOTAL | | 5.02 | 5.31 | 5.86 | 5.58 | 21.77 |
| STATE: | MONTANA | | | | | |
| ZIP CODE | QUARTER 1 | QUARTER 2 | QUARTER 3 | QUARTER 4 | TOTAL GRAMS | |
| | 591 | 0.03 | 0.12 | 0.09 | 0.05 | 0.29 |
| | 592 | 0.02 | 0 | 0.03 | 0 | 0.05 |
| | 593 | 0.01 | 0 | 0.01 | 0 | 0.02 |
| | 594 | 0.02 | 0.26 | 0 | 0.04 | 0.32 |
| | 596 | 0.1 | 0.08 | 0.08 | 0.04 | 0.3 |
| | 597 | 0.04 | 0.08 | 0.11 | 0.14 | 0.37 |
| | 598 | 1.22 | 1.25 | 1.38 | 1.69 | 5.54 |
| | 599 | 0.01 | 0.04 | 0.02 | 0.03 | 0.1 |
| TOTAL | | 1.45 | 1.83 | 1.72 | 1.99 | 6.99 |
| STATE: | NEBRASKA | | | | | |
| ZIP CODE | QUARTER 1 | QUARTER 2 | QUARTER 3 | QUARTER 4 | TOTAL GRAMS | |
| | 680 | 0.05 | 0.05 | 0.02 | 0.05 | 0.17 |
| | 681 | 0.85 | 0.67 | 0.47 | 0.68 | 2.67 |
| | 685 | 0.01 | 0.06 | 0.14 | 0.04 | 0.25 |
| | 686 | 0.01 | 0 | 0.01 | 0.01 | 0.03 |
| | 687 | 0 | 0 | 0.01 | 0 | 0.01 |
| | 688 | 0.07 | 0.1 | 0.11 | 0.19 | 0.47 |
| | 689 | 0.01 | 0.03 | 0.03 | 0.14 | 0.21 |
| | 691 | 0.01 | 0 | 0 | 0.02 | 0.03 |
| | 693 | 0.13 | 0.06 | 0.12 | 0.14 | 0.45 |
| TOTAL | | 1.14 | 0.97 | 0.91 | 1.27 | 4.29 |
| STATE: | NEVADA | | | | | |
| ZIP CODE | QUARTER 1 | QUARTER 2 | QUARTER 3 | QUARTER 4 | TOTAL GRAMS | |
| | 890 | 0.03 | 0.04 | 0.06 | 0.06 | 0.19 |
| | 891 | 0.27 | 0.24 | 0.43 | 0.24 | 1.18 |
| | 894 | 0.02 | 0.03 | 0.02 | 0.02 | 0.09 |
| | 895 | 0.42 | 0.43 | 0.58 | 0.62 | 2.05 |

| | | | | | | |
|---|---|---|---|---|---|---|
| | 897 | 0.15 | 0 | 0 | 0.02 | 0.17 |
| | 898 | 0 | 0 | 0.02 | 0 | 0.02 |
| TOTAL | | 0.89 | 0.74 | 1.11 | 0.96 | 3.7 |

STATE: NEW HAMPSHIRE

| ZIP CODE | | QUARTER 1 | QUARTER 2 | QUARTER 3 | QUARTER 4 | TOTAL GRAMS |
|---|---|---|---|---|---|---|
| | 30 | 0.1 | 0.14 | 0.1 | 0.12 | 0.46 |
| | 31 | 0.29 | 0.32 | 0.43 | 0.68 | 1.72 |
| | 32 | 0.02 | 0 | 0.01 | 0.01 | 0.04 |
| | 33 | 0.08 | 0.13 | 0.1 | 0.16 | 0.47 |
| | 34 | 0 | 0.01 | 0.03 | 0 | 0.04 |
| | 35 | 0 | 0.01 | 0 | 0 | 0.01 |
| | 37 | 0.37 | 0.54 | 0.47 | 0.97 | 2.35 |
| | 38 | 0.29 | 0.37 | 0.29 | 0.48 | 1.43 |
| TOTAL | | 1.15 | 1.52 | 1.43 | 2.42 | 6.52 |

STATE: NEW JERSEY

| ZIP CODE | | QUARTER 1 | QUARTER 2 | QUARTER 3 | QUARTER 4 | TOTAL GRAMS |
|---|---|---|---|---|---|---|
| | 070 | 0.76 | 1.06 | 0.9 | 0.76 | 3.48 |
| | 071 | 1.93 | 1.12 | 1.31 | 1.96 | 6.32 |
| | 072 | 0.1 | 0.02 | 0.08 | 0.16 | 0.36 |
| | 073 | 0.1 | 0.08 | 0.12 | 0.12 | 0.42 |
| | 074 | 0.44 | 0.42 | 0.38 | 0.38 | 1.62 |
| | 075 | 0.48 | 0.77 | 0.64 | 0.62 | 2.51 |
| | 076 | 1.03 | 1.16 | 1.19 | 1.33 | 4.71 |
| | 077 | 0.73 | 1.11 | 0.89 | 1.37 | 4.1 |
| | 078 | 0 | 0.02 | 0.04 | 0.04 | 0.1 |
| | 079 | 1.22 | 0.98 | 0.88 | 1.16 | 4.24 |
| | 080 | 1.51 | 1.68 | 1.23 | 1.53 | 5.95 |
| | 081 | 1.14 | 0.81 | 0.81 | 0.97 | 3.73 |
| | 082 | 0.58 | 0.54 | 0.73 | 0.4 | 2.25 |
| | 084 | 0.2 | 0.3 | 0.42 | 0.18 | 1.1 |
| | 085 | 0.16 | 0.26 | 0.27 | 0.29 | 0.98 |
| | 086 | 0.17 | 0.3 | 0.26 | 0.31 | 1.04 |
| | 087 | 0.02 | 0.25 | 0.08 | 0.1 | 0.45 |

| | | | | | |
|---|---|---|---|---|---|
| | 088 | 0.64 | 0.54 | 0.7 | 0.73 | 2.61 |
| | 089 | 1.58 | 1.59 | 2.01 | 1.76 | 6.94 |
| TOTAL | | 12.79 | 13.01 | 12.94 | 14.17 | 52.91 |

STATE: NEW MEXICO

| ZIP CODE | | QUARTER 1 | QUARTER 2 | QUARTER 3 | QUARTER 4 | TOTAL GRAMS |
|---|---|---|---|---|---|---|
| | 871 | 0.43 | 0.69 | 0.73 | 0.49 | 2.34 |
| | 874 | 0.06 | 0.06 | 0.05 | 0.02 | 0.19 |
| | 875 | 0.14 | 0.13 | 0.18 | 0.24 | 0.69 |
| | 880 | 0.31 | 0.2 | 0.26 | 0.23 | 1 |
| | 882 | 0.02 | 0 | 0.01 | 0.01 | 0.04 |
| | 884 | 0.04 | 0.06 | 0.04 | 0.04 | 0.18 |
| TOTAL | | 1 | 1.14 | 1.27 | 1.03 | 4.44 |

STATE: NEW YORK

| ZIP CODE | | QUARTER 1 | QUARTER 2 | QUARTER 3 | QUARTER 4 | TOTAL GRAMS |
|---|---|---|---|---|---|---|
| | 100 | 19.87 | 20.51 | 20.33 | 21.26 | 81.97 |
| | 101 | 0.02 | 0.04 | 0 | 0 | 0.06 |
| | 103 | 0.2 | 0.31 | 0.2 | 0.22 | 0.93 |
| | 104 | 1.53 | 1.61 | 1.5 | 0.85 | 5.49 |
| | 105 | 0.96 | 0.96 | 1.18 | 1.57 | 4.67 |
| | 106 | 0.28 | 0.3 | 0.22 | 0.4 | 1.2 |
| | 107 | 0 | 0 | 0 | 0.02 | 0.02 |
| | 109 | 0.08 | 0.05 | 0.1 | 0.03 | 0.26 |
| | 110 | 3.58 | 3.71 | 4.42 | 3.72 | 15.43 |
| | 112 | 2.37 | 2.27 | 3.18 | 2.52 | 10.34 |
| | 113 | 0.16 | 0.16 | 0.3 | 0.29 | 0.91 |
| | 114 | 0.13 | 0.21 | 0.13 | 0.17 | 0.64 |
| | 115 | 1.03 | 0.83 | 1.09 | 0.82 | 3.77 |
| | 116 | 0 | 0 | 0.02 | 0 | 0.02 |
| | 117 | 0.78 | 0.76 | 0.78 | 0.91 | 3.23 |
| | 119 | 0.01 | 0.03 | 0 | 0 | 0.04 |
| | 120 | 0 | 0 | 0 | 0.04 | 0.04 |
| | 121 | 0.21 | 0.24 | 0.19 | 0.03 | 0.67 |
| | 122 | 1.06 | 1.02 | 1.24 | 0.76 | 4.08 |

| | | | | | | |
|---|---|---|---|---|---|---|
| | 123 | 0.06 | 0.11 | 0.08 | 0.09 | 0.34 |
| | 124 | 0.02 | 0.04 | 0 | 0.03 | 0.09 |
| | 125 | 0.14 | 0.03 | 0.09 | 0.22 | 0.48 |
| | 126 | 0.19 | 0.31 | 0.32 | 0.52 | 1.34 |
| | 127 | 0.04 | 0.02 | 0.03 | 0.04 | 0.13 |
| | 128 | 0.04 | 0.06 | 0.06 | 0.03 | 0.19 |
| | 129 | 0 | 0.02 | 0 | 0 | 0.02 |
| | 132 | 1.25 | 1.35 | 1.27 | 1.4 | 5.27 |
| | 133 | 0.17 | 0.14 | 0.14 | 0.17 | 0.62 |
| | 134 | 0.01 | 0 | 0.02 | 0.02 | 0.05 |
| | 135 | 0.13 | 0.16 | 0.22 | 0.03 | 0.54 |
| | 136 | 0.07 | 0.05 | 0.09 | 0.06 | 0.27 |
| | 137 | 0.3 | 0.3 | 0.34 | 0.24 | 1.18 |
| | 138 | 0.02 | 0.03 | 0 | 0.02 | 0.07 |
| | 139 | 0.04 | 0.02 | 0.02 | 0 | 0.08 |
| | 140 | 0.02 | 0.01 | 0.03 | 0.01 | 0.07 |
| | 142 | 1.34 | 0.95 | 1.72 | 2.83 | 6.84 |
| | 144 | 0.02 | 0.06 | 0.05 | 0.02 | 0.15 |
| | 146 | 0.52 | 1 | 0.88 | 0.88 | 3.28 |
| | 147 | 0 | 0 | 0.01 | 0.01 | 0.02 |
| | 148 | 0.04 | 0.05 | 0.06 | 0.08 | 0.23 |
| TOTAL | | 36.69 | 37.72 | 40.31 | 40.31 | 155.03 |

| STATE: | NORTH CAROLINA | | | | | |
|---|---|---|---|---|---|---|
| ZIP CODE | | QUARTER 1 | QUARTER 2 | QUARTER 3 | QUARTER 4 | TOTAL GRAMS |
| | 270 | 0 | 0.01 | 0 | 0.02 | 0.03 |
| | 271 | 1.38 | 1.88 | 1.51 | 2.1 | 6.87 |
| | 272 | 0.72 | 0.57 | 0.45 | 0.68 | 2.42 |
| | 273 | 0.02 | 0 | 0.04 | 0 | 0.06 |
| | 274 | 0.14 | 0.31 | 0.28 | 0.12 | 0.85 |
| | 275 | 1.73 | 1.8 | 1.9 | 1.47 | 6.9 |
| | 276 | 2.09 | 2.01 | 1.3 | 1.31 | 6.71 |
| | 277 | 2.86 | 2.91 | 3.48 | 3.52 | 12.77 |
| | 278 | 0.44 | 0.52 | 0.67 | 0.74 | 2.37 |
| | 280 | 1.15 | 1.05 | 1.01 | 1.08 | 4.29 |
| | 281 | 0.21 | 0.18 | 0.29 | 0.21 | 0.89 |

| | | | | | |
|---|---|---|---|---|---|
| 282 | 1.13 | 1.48 | 1.04 | 1.85 | 5.5 |
| 283 | 0.47 | 0.5 | 0.43 | 0.45 | 1.85 |
| 284 | 0.01 | 0 | 0.02 | 0 | 0.03 |
| 285 | 0.05 | 0.09 | 0.08 | 0.09 | 0.31 |
| 286 | 0.91 | 0.94 | 0.85 | 0.97 | 3.67 |
| 287 | 0.21 | 0.2 | 0.16 | 0.17 | 0.74 |
| 288 | 0.35 | 0.4 | 0.33 | 0.4 | 1.48 |
| TOTAL | 13.87 | 14.85 | 13.84 | 15.18 | 57.74 |

STATE: NORTH DAKOTA

| ZIP CODE | QUARTER 1 | QUARTER 2 | QUARTER 3 | QUARTER 4 | TOTAL GRAMS |
|---|---|---|---|---|---|
| 581 | 0.44 | 0.18 | 0.27 | 0.23 | 1.12 |
| 585 | 0.13 | 0.1 | 0.06 | 0.12 | 0.41 |
| 587 | 0 | 0.02 | 0.02 | 0.04 | 0.08 |
| 588 | 0.01 | 0.02 | 0.03 | 0 | 0.06 |
| TOTAL | 0.58 | 0.32 | 0.38 | 0.39 | 1.67 |

STATE: OHIO

| ZIP CODE | QUARTER 1 | QUARTER 2 | QUARTER 3 | QUARTER 4 | TOTAL GRAMS |
|---|---|---|---|---|---|
| 430 | 0.36 | 0.09 | 0.22 | 0.21 | 0.88 |
| 432 | 2.36 | 3.06 | 3.11 | 4.36 | 12.89 |
| 433 | 0.05 | 0.06 | 0 | 0.1 | 0.21 |
| 434 | 0 | 0 | 0.01 | 0 | 0.01 |
| 435 | 0 | 0.03 | 0 | 0 | 0.03 |
| 436 | 0.17 | 0.26 | 0.31 | 0.33 | 1.07 |
| 437 | 0.79 | 0.82 | 0.24 | 0.3 | 2.15 |
| 439 | 0.02 | 0 | 0 | 0 | 0.02 |
| 440 | 0.24 | 0.23 | 0.24 | 0.23 | 0.94 |
| 441 | 6.86 | 7.59 | 4.52 | 4.99 | 23.96 |
| 442 | 0.08 | 0.1 | 0.03 | 0.03 | 0.24 |
| 443 | 0.78 | 0.96 | 0.65 | 1.11 | 3.5 |
| 444 | 0.1 | 0.09 | 0.13 | 0.09 | 0.41 |
| 445 | 2.38 | 1.34 | 1.55 | 1.92 | 7.19 |
| 446 | 0.06 | 0.03 | 0.03 | 0.03 | 0.15 |

| | ZIP CODE | QUARTER 1 | QUARTER 2 | QUARTER 3 | QUARTER 4 | TOTAL GRAMS |
|---|---|---|---|---|---|---|
| | 447 | 0.07 | 0.03 | 0.1 | 0.09 | 0.29 |
| | 448 | 0.05 | 0.01 | 0.07 | 0.08 | 0.21 |
| | 449 | 0.05 | 0.07 | 0.04 | 0.07 | 0.23 |
| | 450 | 0.39 | 0.29 | 0.44 | 0.5 | 1.62 |
| | 452 | 2.89 | 3.37 | 2.98 | 3.6 | 12.84 |
| | 453 | 0 | 0 | 0.02 | 0 | 0.02 |
| | 454 | 1.03 | 1.1 | 0.84 | 1.55 | 4.52 |
| | 455 | 0.01 | 0.01 | 0 | 0 | 0.02 |
| | 456 | 0.24 | 0.24 | 0.16 | 0.54 | 1.18 |
| | 457 | 0.23 | 0.18 | 0.2 | 0.21 | 0.82 |
| | 458 | 0.06 | 0.04 | 0.04 | 0.1 | 0.24 |
| TOTAL | | 19.27 | 20 | 15.93 | 20.44 | 75.64 |

STATE: OKLAHOMA

| ZIP CODE | | QUARTER 1 | QUARTER 2 | QUARTER 3 | QUARTER 4 | TOTAL GRAMS |
|---|---|---|---|---|---|---|
| | 730 | 0.04 | 0.1 | 0 | 0.06 | 0.2 |
| | 731 | 1.96 | 1.92 | 1.98 | 1.96 | 7.82 |
| | 734 | 0.02 | 0 | 0.08 | 0.08 | 0.18 |
| | 735 | 0.05 | 0.05 | 0.14 | 0.02 | 0.26 |
| | 740 | 0.01 | 0.04 | 0.01 | 0.01 | 0.07 |
| | 741 | 0.45 | 0.75 | 0.39 | 0.44 | 2.03 |
| | 743 | 0 | 0 | 0.02 | 0 | 0.02 |
| | 744 | 0.04 | 0 | 0.02 | 0.03 | 0.09 |
| | 745 | 0.02 | 0.02 | 0.01 | 0.01 | 0.06 |
| TOTAL | | 2.59 | 2.88 | 2.65 | 2.61 | 10.73 |

STATE: OREGON

| ZIP CODE | | QUARTER 1 | QUARTER 2 | QUARTER 3 | QUARTER 4 | TOTAL GRAMS |
|---|---|---|---|---|---|---|
| | 970 | 0.4 | 0.27 | 0.42 | 0.34 | 1.43 |
| | 971 | 0.11 | 0.1 | 0.12 | 0.1 | 0.43 |
| | 972 | 0.36 | 0.52 | 0.56 | 0.46 | 1.9 |
| | 973 | 0.12 | 0.11 | 0.09 | 0.03 | 0.35 |
| | 974 | 0.88 | 1.12 | 0.87 | 1.17 | 4.04 |
| | 975 | 0.32 | 0.6 | 0.08 | 0.38 | 1.38 |
| | 977 | 0.5 | 0.53 | 0.38 | 0.43 | 1.84 |

| | | | | | | |
|---|---|---|---|---|---|---|
| | 978 | 0 | 0.01 | 0.01 | 0 | 0.02 |
| | 979 | 0 | 0.01 | 0 | 0 | 0.01 |
| TOTAL | | 2.69 | 3.27 | 2.53 | 2.91 | 11.4 |

| STATE: | PENNSYLVANIA | | | | |
|---|---|---|---|---|---|
| ZIP CODE | QUARTER 1 | QUARTER 2 | QUARTER 3 | QUARTER 4 | TOTAL GRAMS |
| 150 | 0.36 | 0.51 | 0.33 | 0.39 | 1.59 |
| 151 | 0.5 | 0.36 | 0.54 | 0.55 | 1.95 |
| 152 | 7.65 | 8.28 | 8.12 | 9.07 | 33.12 |
| 153 | 0.07 | 0.07 | 0.04 | 0.05 | 0.23 |
| 155 | 0.02 | 0.01 | 0.01 | 0.02 | 0.06 |
| 156 | 0.08 | 0.1 | 0.06 | 0.15 | 0.39 |
| 158 | 0.08 | 0.07 | 0.13 | 0.15 | 0.43 |
| 159 | 0.12 | 0.09 | 0.07 | 0.09 | 0.37 |
| 160 | 0.29 | 0.47 | 0.31 | 0.54 | 1.61 |
| 161 | 0 | 0.01 | 0.01 | 0 | 0.02 |
| 162 | 0.01 | 0.03 | 0.03 | 0.01 | 0.08 |
| 165 | 0.46 | 0.45 | 0.26 | 0.55 | 1.72 |
| 166 | 0.02 | 0.05 | 0.06 | 0.28 | 0.41 |
| 168 | 0.04 | 0.12 | 0.03 | 0.04 | 0.23 |
| 170 | 0.98 | 0.99 | 1.01 | 0.54 | 3.52 |
| 171 | 0.07 | 0.04 | 0.15 | 0.03 | 0.29 |
| 172 | 0 | 0 | 0.02 | 0 | 0.02 |
| 174 | 0.04 | 0.1 | 0 | 0.02 | 0.16 |
| 175 | 0.1 | 0.1 | 0.1 | 0 | 0.3 |
| 176 | 0.01 | 0.02 | 0.01 | 0.03 | 0.07 |
| 178 | 0.91 | 1.36 | 1.06 | 1.12 | 4.45 |
| 179 | 0.04 | 0 | 0.02 | 0.02 | 0.08 |
| 180 | 1.17 | 1.38 | 0.81 | 1.13 | 4.49 |
| 181 | 0.92 | 1.12 | 0.9 | 1.07 | 4.01 |
| 182 | 0 | 0.01 | 0.01 | 0 | 0.02 |
| 183 | 0 | 0.31 | 0.26 | 0.16 | 0.73 |
| 184 | 0.08 | 0.06 | 0.05 | 0.09 | 0.28 |
| 185 | 0.5 | 0.37 | 0.5 | 0.45 | 1.82 |
| 186 | 0.01 | 0.03 | 0.02 | 0 | 0.06 |
| 187 | 0.52 | 0.52 | 0.56 | 0.48 | 2.08 |

| | | | | | | |
|---|---|---|---|---|---|---|
| | 188 | 0.05 | 0 | 0.05 | 0.11 | 0.21 |
| | 189 | 0.06 | 0 | 0 | 0 | 0.06 |
| | 190 | 1.68 | 1.62 | 1.66 | 1.7 | 6.66 |
| | 191 | 18.36 | 16.11 | 16.97 | 16.31 | 67.75 |
| | 193 | 0.59 | 0.58 | 0.61 | 0.86 | 2.64 |
| | 194 | 0.3 | 0.19 | 0.23 | 0.24 | 0.96 |
| | 196 | 0.1 | 0.06 | 0.09 | 0.12 | 0.37 |
| TOTAL | | 36.19 | 35.59 | 35.09 | 36.37 | 143.24 |

STATE: RHODE ISLAND

| ZIP CODE | | QUARTER 1 | QUARTER 2 | QUARTER 3 | QUARTER 4 | TOTAL GRAMS |
|---|---|---|---|---|---|---|
| | 28 | 0.13 | 0.01 | 0.1 | 0.16 | 0.4 |
| | 29 | 0.55 | 0.5 | 0.66 | 1.06 | 2.77 |
| TOTAL | | 0.68 | 0.51 | 0.76 | 1.22 | 3.17 |

STATE: SOUTH CAROLINA

| ZIP CODE | | QUARTER 1 | QUARTER 2 | QUARTER 3 | QUARTER 4 | TOTAL GRAMS |
|---|---|---|---|---|---|---|
| | 290 | 0 | 0.03 | 0 | 0.01 | 0.04 |
| | 291 | 0.93 | 0.64 | 0.87 | 1.09 | 3.53 |
| | 292 | 0.61 | 0.57 | 0.61 | 0.61 | 2.4 |
| | 293 | 0.3 | 0.18 | 0.2 | 0.26 | 0.94 |
| | 294 | 1.68 | 1.55 | 1.69 | 1.48 | 6.4 |
| | 295 | 0.19 | 0.18 | 0.14 | 0.16 | 0.67 |
| | 296 | 1.2 | 1.12 | 1.02 | 0.84 | 4.18 |
| | 297 | 0.16 | 0.07 | 0.08 | 0.09 | 0.4 |
| | 299 | 0.01 | 0.02 | 0.03 | 0.03 | 0.09 |
| TOTAL | | 5.08 | 4.36 | 4.64 | 4.57 | 18.65 |

STATE: SOUTH DAKOTA

| ZIP CODE | | QUARTER 1 | QUARTER 2 | QUARTER 3 | QUARTER 4 | TOTAL GRAMS |
|---|---|---|---|---|---|---|
| | 571 | 0.08 | 0.1 | 0.11 | 0.02 | 0.31 |
| | 572 | 0.01 | 0 | 0 | 0.01 | 0.02 |
| | 573 | 0.02 | 0 | 0 | 0.01 | 0.03 |
| | 574 | 0 | 0.04 | 0 | 0 | 0.04 |

| | | | | | |
|---|---|---|---|---|---|
| 575 | 0 | 0 | 0 | 0.02 | 0.02 |
| 577 | 0.36 | 0.52 | 0.33 | 0.26 | 1.47 |
| TOTAL | 0.47 | 0.66 | 0.44 | 0.32 | 1.89 |

STATE: TENNESSEE

| ZIP CODE | QUARTER 1 | QUARTER 2 | QUARTER 3 | QUARTER 4 | TOTAL GRAMS |
|---|---|---|---|---|---|
| 370 | 0.32 | 0.31 | 0.31 | 0.26 | 1.2 |
| 371 | 0.07 | 0.04 | 0.07 | 0.05 | 0.23 |
| 372 | 3.08 | 3.61 | 4.28 | 4.19 | 15.16 |
| 373 | 0 | 0.01 | 0.01 | 0 | 0.02 |
| 374 | 0.07 | 0.23 | 0.18 | 0.24 | 0.72 |
| 376 | 0.06 | 0.13 | 0.01 | 0.1 | 0.3 |
| 378 | 0.18 | 0.09 | 0.71 | 0.07 | 1.05 |
| 379 | 0.33 | 0.37 | 0.41 | 0.5 | 1.61 |
| 380 | 0 | 0 | 0.04 | 0.02 | 0.06 |
| 381 | 0.41 | 0.46 | 0.46 | 0.76 | 2.09 |
| 382 | 0.03 | 0 | 0.01 | 0 | 0.04 |
| 383 | 0.02 | 0.04 | 0.02 | 0.16 | 0.24 |
| 384 | 0.09 | 0.11 | 0.16 | 0.12 | 0.48 |
| 385 | 0 | 0.02 | 0.02 | 0 | 0.04 |
| TOTAL | 4.66 | 5.42 | 6.69 | 6.47 | 23.24 |

STATE: TEXAS

| ZIP CODE | QUARTER 1 | QUARTER 2 | QUARTER 3 | QUARTER 4 | TOTAL GRAMS |
|---|---|---|---|---|---|
| 750 | 0.68 | 0.57 | 0.76 | 0.86 | 2.87 |
| 751 | 0.15 | 0.12 | 0.06 | 0.15 | 0.48 |
| 752 | 2.99 | 3.06 | 3.24 | 2.36 | 11.65 |
| 753 | 3.26 | 3.62 | 3.33 | 3.4 | 13.61 |
| 754 | 0.08 | 0.07 | 0.05 | 0.07 | 0.27 |
| 756 | 0 | 0.04 | 0 | 0.02 | 0.06 |
| 757 | 0.28 | 0.41 | 0.22 | 0.5 | 1.41 |
| 759 | 0.01 | 0.01 | 0 | 0 | 0.02 |
| 760 | 0.53 | 0.79 | 0.77 | 0.44 | 2.53 |
| 761 | 3.66 | 3.63 | 4.28 | 4.88 | 16.45 |
| 762 | 0.11 | 0.13 | 0.2 | 0.24 | 0.68 |

| | | | | | | |
|---|---|---|---|---|---|---|
| | 763 | 0.02 | 0 | 0.03 | 0 | 0.05 |
| | 765 | 0.57 | 0.39 | 0.48 | 0.51 | 1.95 |
| | 767 | 0.05 | 0.07 | 0.17 | 0.15 | 0.44 |
| | 769 | 0.02 | 0.04 | 0 | 0 | 0.06 |
| | 770 | 6.25 | 4.12 | 5.07 | 4.94 | 20.38 |
| | 773 | 0.23 | 0.69 | 0.32 | 0.17 | 1.41 |
| | 774 | 0.43 | 0.33 | 0.59 | 0.33 | 1.68 |
| | 775 | 0.99 | 1.16 | 1.04 | 1.82 | 5.01 |
| | 778 | 0.13 | 0.15 | 0.09 | 0.11 | 0.48 |
| | 779 | 0.01 | 0 | 0.01 | 0.03 | 0.05 |
| | 781 | 0.02 | 0 | 0.05 | 0.1 | 0.17 |
| | 782 | 2.12 | 2.79 | 1.64 | 3.66 | 10.21 |
| | 784 | 0.7 | 0.69 | 0.58 | 0.63 | 2.6 |
| | 785 | 0.02 | 0.04 | 0.03 | 0.04 | 0.13 |
| | 786 | 0.21 | 0.21 | 0.26 | 0.24 | 0.92 |
| | 787 | 0.86 | 1.11 | 1.03 | 1.1 | 4.1 |
| | 791 | 0.06 | 0.02 | 0.04 | 0.04 | 0.16 |
| | 794 | 1.09 | 1.73 | 1.39 | 2.11 | 6.32 |
| | 796 | 0.09 | 0.08 | 0.15 | 0.08 | 0.4 |
| | 797 | 0.2 | 0.2 | 0.26 | 0.16 | 0.82 |
| | 798 | 0 | 0.01 | 0 | 0 | 0.01 |
| | 799 | 0.97 | 1.14 | 1.06 | 1.31 | 4.48 |
| TOTAL | | 26.79 | 27.42 | 27.2 | 30.45 | 111.86 |

STATE: UTAH

| ZIP CODE | | QUARTER 1 | QUARTER 2 | QUARTER 3 | QUARTER 4 | TOTAL GRAMS |
|---|---|---|---|---|---|---|
| | 840 | 1.81 | 2.02 | 1.77 | 2.15 | 7.75 |
| | 841 | 11.67 | 7.75 | 9.33 | 17.54 | 46.29 |
| | 843 | 0.07 | 0.08 | 0.01 | 0.05 | 0.21 |
| | 844 | 0.34 | 0.23 | 0.39 | 0.72 | 1.68 |
| | 846 | 1.99 | 1.97 | 2.21 | 2.61 | 8.78 |
| | 847 | 1.94 | 2.03 | 1.67 | 1.43 | 7.07 |
| TOTAL | | 17.82 | 14.08 | 15.38 | 24.5 | 71.78 |

STATE: VERMONT

| ZIP CODE | QUARTER 1 | QUARTER 2 | QUARTER 3 | QUARTER 4 | TOTAL GRAMS |
|---|---|---|---|---|---|
| 50 | 0.18 | 0.28 | 0.1 | 0.18 | 0.74 |
| 54 | 0.49 | 0.62 | 0.41 | 0.54 | 2.06 |
| 57 | 0.01 | 0.01 | 0.01 | 0 | 0.03 |
| TOTAL | 0.68 | 0.91 | 0.52 | 0.72 | 2.83 |

STATE: VIRGINIA

| ZIP CODE | QUARTER 1 | QUARTER 2 | QUARTER 3 | QUARTER 4 | TOTAL GRAMS |
|---|---|---|---|---|---|
| 201 | 0.08 | 0.08 | 0.06 | 0.12 | 0.34 |
| 220 | 0.44 | 0.32 | 0.4 | 0.48 | 1.64 |
| 221 | 0.03 | 0.02 | 0.01 | 0.01 | 0.07 |
| 222 | 0.43 | 0.35 | 0.39 | 0.48 | 1.65 |
| 224 | 0 | 0.02 | 0 | 0 | 0.02 |
| 226 | 0.2 | 0.24 | 0.1 | 0.22 | 0.76 |
| 228 | 0.01 | 0.01 | 0 | 0.01 | 0.03 |
| 229 | 0.51 | 0.34 | 0.36 | 0.8 | 2.01 |
| 231 | 0.01 | 0 | 0 | 0 | 0.01 |
| 232 | 1.98 | 2.23 | 2.05 | 2.35 | 8.61 |
| 233 | 0.1 | 0.1 | 0.12 | 0.06 | 0.38 |
| 234 | 0.14 | 0.25 | 0.21 | 0.24 | 0.84 |
| 235 | 0.87 | 1.67 | 0.73 | 1.15 | 4.42 |
| 236 | 0.43 | 0.51 | 0.44 | 0.63 | 2.01 |
| 237 | 0.12 | 0.24 | 0.24 | 0.12 | 0.72 |
| 238 | 0.15 | 0.1 | 0.05 | 0.06 | 0.36 |
| 239 | 0 | 0 | 0 | 0.01 | 0.01 |
| 240 | 0.44 | 0.45 | 0.48 | 0.28 | 1.65 |
| 241 | 0.15 | 0.22 | 0.26 | 0.27 | 0.9 |
| 244 | 0 | 0.01 | 0.01 | 0 | 0.02 |
| 245 | 0.18 | 0.23 | 0.13 | 0.18 | 0.72 |
| TOTAL | 6.27 | 7.39 | 6.04 | 7.47 | 27.17 |

STATE: WASHINGTON

| ZIP CODE | QUARTER 1 | QUARTER 2 | QUARTER 3 | QUARTER 4 | TOTAL GRAMS |
|---|---|---|---|---|---|
| 980 | 0.5 | 0.49 | 0.79 | 0.64 | 2.42 |
| 981 | 8.53 | 9.04 | 10.59 | 7.74 | 35.9 |

| | | | | | |
|---|---|---|---|---|---|
| 982 | 0.41 | 0.27 | 0.52 | 0.67 | 1.87 |
| 983 | 0.08 | 0.1 | 0.12 | 0.09 | 0.39 |
| 984 | 1.11 | 0.66 | 1.03 | 0.48 | 3.28 |
| 985 | 0.27 | 0.41 | 0.34 | 0.23 | 1.25 |
| 986 | 0.02 | 0.11 | 0.09 | 0.03 | 0.25 |
| 988 | 0.08 | 0.06 | 0.16 | 0.09 | 0.39 |
| 989 | 0.37 | 0.27 | 0.24 | 0.18 | 1.06 |
| 991 | 0 | 0 | 0.03 | 0 | 0.03 |
| 992 | 0.32 | 0.12 | 0.16 | 0.13 | 0.73 |
| 993 | 1.02 | 0.55 | 0.57 | 0.3 | 2.44 |
| TOTAL | 12.71 | 12.08 | 14.64 | 10.58 | 50.01 |

STATE: WEST VIRGINIA

| ZIP CODE | QUARTER 1 | QUARTER 2 | QUARTER 3 | QUARTER 4 | TOTAL GRAMS |
|---|---|---|---|---|---|
| 247 | 0.01 | 0.01 | 0.01 | 0 | 0.03 |
| 249 | 0.01 | 0 | 0 | 0.01 | 0.02 |
| 253 | 2.2 | 2.83 | 1.81 | 1.16 | 8 |
| 254 | 0.01 | 0 | 0.03 | 0.04 | 0.08 |
| 255 | 0.16 | 0.13 | 0.11 | 0.15 | 0.55 |
| 257 | 0.67 | 0.74 | 0.65 | 0.93 | 2.99 |
| 260 | 0 | 0 | 0.03 | 0.06 | 0.09 |
| 261 | 0.17 | 0.08 | 0.16 | 0.14 | 0.55 |
| 263 | 0.08 | 0.13 | 0.14 | 0.23 | 0.58 |
| 265 | 2.06 | 2.1 | 2.01 | 1.69 | 7.86 |
| TOTAL | 5.37 | 6.02 | 4.95 | 4.41 | 20.75 |

STATE: WISCONSIN

| ZIP CODE | QUARTER 1 | QUARTER 2 | QUARTER 3 | QUARTER 4 | TOTAL GRAMS |
|---|---|---|---|---|---|
| 530 | 0.25 | 0.16 | 0.32 | 0.35 | 1.08 |
| 531 | 0.11 | 0.14 | 0.13 | 0.06 | 0.44 |
| 532 | 2.68 | 4.16 | 1.81 | 3.02 | 11.67 |
| 534 | 0.22 | 0.19 | 0.17 | 0.11 | 0.69 |
| 535 | 0.39 | 0.39 | 0.28 | 0.17 | 1.23 |
| 537 | 1.28 | 1.24 | 1.35 | 1.75 | 5.62 |

| | | | | | | |
|---|---|---|---|---|---|---|
| | 539 | 0.02 | 0 | 0.01 | 0 | 0.03 |
| | 540 | 0 | 0.01 | 0.01 | 0 | 0.02 |
| | 543 | 0.09 | 0.16 | 0.14 | 0.42 | 0.81 |
| | 544 | 0.45 | 0.7 | 0.49 | 0.66 | 2.3 |
| | 545 | 0 | 0 | 0.02 | 0.02 | 0.04 |
| | 546 | 0.12 | 0.05 | 0.11 | 0.46 | 0.74 |
| | 547 | 0.01 | 0.03 | 0 | 0.02 | 0.06 |
| | 548 | 0 | 0 | 0.08 | 0.04 | 0.12 |
| | 549 | 0.43 | 0.33 | 0.4 | 0.72 | 1.88 |
| TOTAL | | 6.05 | 7.56 | 5.32 | 7.8 | 26.73 |

STATE: WYOMING

| ZIP CODE | | QUARTER 1 | QUARTER 2 | QUARTER 3 | QUARTER 4 | TOTAL GRAMS |
|---|---|---|---|---|---|---|
| | 820 | 0.03 | 0.16 | 0.07 | 0.11 | 0.37 |
| | 822 | 0 | 0 | 0.01 | 0 | 0.01 |
| | 823 | 0 | 0.02 | 0.02 | 0 | 0.04 |
| | 824 | 0.01 | 0.03 | 0.03 | 0.02 | 0.09 |
| | 826 | 0.24 | 0.05 | 0.06 | 0.1 | 0.45 |
| | 827 | 0.04 | 0.02 | 0.06 | 0.05 | 0.17 |
| | 829 | 0 | 0 | 0.02 | 0 | 0.02 |
| | 830 | 0.08 | 0.08 | 0.1 | 0.12 | 0.38 |
| | 831 | 0.1 | 0.03 | 0.06 | 0.06 | 0.25 |
| TOTAL | | 0.5 | 0.39 | 0.43 | 0.46 | 1.78 |

DRUG CODE: 9740 DRUG NAME: SUFENTANIL BASE

STATE: ALABAMA

| ZIP CODE | | QUARTER 1 | QUARTER 2 | QUARTER 3 | QUARTER 4 | TOTAL GRAMS |
|---|---|---|---|---|---|---|
| | 352 | 0.04 | 0.03 | 0.03 | 0.04 | 0.14 |
| | 354 | 0 | 0 | 0 | 0 | 0 |
| | 356 | 0.01 | 0.02 | 0.01 | 0.03 | 0.07 |
| | 358 | 0.71 | 1.37 | 1.4 | 1.64 | 5.12 |
| | 359 | 0.02 | 0.02 | 0.01 | 0.02 | 0.07 |
| | 360 | 0.01 | 0.01 | 0 | 0 | 0.02 |
| | 361 | 0.02 | 0.02 | 0.05 | 0.02 | 0.11 |

| | | | | | |
|---|---|---|---|---|---|
| 363 | 0.06 | 0.09 | 0.09 | 0.08 | 0.32 |
| 364 | 0 | 0 | 0 | 0 | 0 |
| 365 | 0.02 | 0.05 | 0.01 | 0.01 | 0.09 |
| 366 | 0.14 | 0.16 | 0.14 | 0.16 | 0.6 |
| 367 | 0 | 0 | 0 | 0 | 0 |
| TOTAL | 1.03 | 1.77 | 1.74 | 2 | 6.54 |

STATE: ALASKA

| ZIP CODE | QUARTER 1 | QUARTER 2 | QUARTER 3 | QUARTER 4 | TOTAL GRAMS |
|---|---|---|---|---|---|
| 995 | 0 | 0.01 | 0.01 | 0.01 | 0.03 |
| 996 | 0 | 0 | 0 | 0 | 0 |
| 997 | 0 | 0 | 0 | 0 | 0 |
| 998 | 0 | 0 | 0 | 0 | 0 |
| TOTAL | 0 | 0.01 | 0.01 | 0.01 | 0.03 |

STATE: ARIZONA

| ZIP CODE | QUARTER 1 | QUARTER 2 | QUARTER 3 | QUARTER 4 | TOTAL GRAMS |
|---|---|---|---|---|---|
| 850 | 0.04 | 0.04 | 0.03 | 0.07 | 0.18 |
| 852 | 0 | 0.01 | 0.02 | 0.02 | 0.05 |
| 853 | 0 | 0 | 0.01 | 0.01 | 0.02 |
| 857 | 0.02 | 0.02 | 0.01 | 0.03 | 0.08 |
| 859 | 0 | 0 | 0 | 0 | 0 |
| 860 | 0 | 0 | 0 | 0 | 0 |
| 863 | 0 | 0 | 0 | 0 | 0 |
| TOTAL | 0.06 | 0.07 | 0.07 | 0.13 | 0.33 |

STATE: ARKANSAS

| ZIP CODE | QUARTER 1 | QUARTER 2 | QUARTER 3 | QUARTER 4 | TOTAL GRAMS |
|---|---|---|---|---|---|
| 716 | 0 | 0 | 0 | 0 | 0 |
| 717 | 0.01 | 0.01 | 0.01 | 0.01 | 0.04 |
| 720 | 0 | 0 | 0 | 0 | 0 |
| 722 | 0.06 | 0.06 | 0.09 | 0.06 | 0.27 |
| 723 | 0 | 0 | 0 | 0 | 0 |

| | | | | | | |
|---|---|---|---|---|---|---|
| | 724 | 0.03 | 0.02 | 0.02 | 0.03 | 0.1 |
| | 725 | 0 | 0 | 0 | 0 | 0 |
| | 726 | 0 | 0 | 0 | 0 | 0 |
| | 727 | 0.04 | 0.01 | 0.03 | 0.04 | 0.12 |
| | 729 | 0.01 | 0 | 0.01 | 0.01 | 0.03 |
| TOTAL | | 0.15 | 0.1 | 0.16 | 0.15 | 0.56 |

STATE: CALIFORNIA

| ZIP CODE | QUARTER 1 | QUARTER 2 | QUARTER 3 | QUARTER 4 | TOTAL GRAMS |
|---|---|---|---|---|---|
| 900 | 0.11 | 0.13 | 0.21 | 0.17 | 0.62 |
| 902 | 0 | 0.02 | 0.05 | 0.01 | 0.08 |
| 904 | 0.01 | 0 | 0 | 0.01 | 0.02 |
| 905 | 0.01 | 0.01 | 0 | 0.01 | 0.03 |
| 906 | 0 | 0.01 | 0 | 0 | 0.01 |
| 907 | 0.01 | 0.01 | 0.01 | 0 | 0.03 |
| 908 | 7.35 | 10.69 | 9.36 | 7.36 | 34.76 |
| 910 | 0 | 0.02 | 0 | 0.01 | 0.03 |
| 911 | 0 | 0 | 0 | 0 | 0 |
| 913 | 0.01 | 0.01 | 0.01 | 0.01 | 0.04 |
| 917 | 0 | 0.03 | 0.02 | 0.03 | 0.08 |
| 919 | 0 | 0 | 0 | 0 | 0 |
| 920 | 0.01 | 0.02 | 0.01 | 0.04 | 0.08 |
| 921 | 0.06 | 0.01 | 0.06 | 0.02 | 0.15 |
| 922 | 0.01 | 0.01 | 0.02 | 0.01 | 0.05 |
| 923 | 0.02 | 0.04 | 0.01 | 0.04 | 0.11 |
| 925 | 1.47 | 1.2 | 0.85 | 1.46 | 4.98 |
| 926 | 0.01 | 0.01 | 0.03 | 0.03 | 0.08 |
| 927 | 0 | 0 | 0 | 0.01 | 0.01 |
| 928 | 0.02 | 0.01 | 0.02 | 0.01 | 0.06 |
| 930 | 0 | 0 | 0.01 | 0.03 | 0.04 |
| 931 | 0.01 | 0.01 | 0 | 0.01 | 0.03 |
| 932 | 0 | 0.01 | 0 | 0.01 | 0.02 |
| 933 | 0 | 0.07 | 0 | 0 | 0.07 |
| 934 | 0.03 | 0.01 | 0.01 | 0.02 | 0.07 |
| 935 | 0 | 0 | 0.01 | 0.01 | 0.02 |
| 936 | 0 | 0 | 0 | 0 | 0 |

| | | | | | | |
|---|---|---|---|---|---|---|
| | 937 | 0.01 | 0.01 | 0.01 | 0.03 | 0.06 |
| | 939 | 0.01 | 0 | 0 | 0 | 0.01 |
| | 940 | 0 | 0 | 0.01 | 0 | 0.01 |
| | 941 | 0.1 | 0.12 | 0.11 | 0.14 | 0.47 |
| | 943 | 0.11 | 0.12 | 0.14 | 0.14 | 0.51 |
| | 945 | 0.02 | 0.02 | 0.04 | 0.03 | 0.11 |
| | 946 | 0 | 0 | 0.01 | 0.14 | 0.15 |
| | 948 | 0 | 0 | 0 | 0 | 0 |
| | 949 | 0 | 0 | 0 | 0 | 0 |
| | 950 | 0.01 | 0 | 0.01 | 0.02 | 0.04 |
| | 951 | 0.12 | 0.03 | 0.03 | 0.02 | 0.2 |
| | 952 | 0.01 | 0.01 | 0.01 | 0.01 | 0.04 |
| | 953 | 0.01 | 0.01 | 0.03 | 0.02 | 0.07 |
| | 954 | 0 | 0 | 0 | 0 | 0 |
| | 955 | 0.03 | 0.04 | 0.03 | 0.03 | 0.13 |
| | 956 | 0.01 | 0.01 | 0.01 | 0.07 | 0.1 |
| | 958 | 0.07 | 0.06 | 0.09 | 0.06 | 0.28 |
| | 959 | 0 | 0 | 0 | 0 | 0 |
| | 960 | 0.02 | 0.01 | 0.01 | 0.01 | 0.05 |
| | 961 | 0 | 0 | 0 | 0 | 0 |
| TOTAL | | 9.67 | 12.77 | 11.23 | 10.03 | 43.7 |

| STATE: | COLORADO | | | | |
|---|---|---|---|---|---|
| ZIP CODE | QUARTER 1 | QUARTER 2 | QUARTER 3 | QUARTER 4 | TOTAL GRAMS |
| 800 | 0.06 | 0.06 | 0.07 | 0.06 | 0.25 |
| 801 | 0 | 0.01 | 0 | 0.01 | 0.02 |
| 802 | 0.02 | 0.02 | 0.06 | 0.11 | 0.21 |
| 803 | 0 | 0 | 0 | 0.01 | 0.01 |
| 805 | 0 | 0.01 | 0.01 | 0 | 0.02 |
| 806 | 0.01 | 0.01 | 0.01 | 0.01 | 0.04 |
| 807 | 0 | 0 | 0 | 0 | 0 |
| 808 | 0 | 0 | 0 | 0 | 0 |
| 809 | 0.07 | 0.04 | 0.02 | 0.03 | 0.16 |
| 810 | 0 | 0 | 0 | 0.02 | 0.02 |
| 811 | 0 | 0 | 0 | 0 | 0 |
| 812 | 0 | 0 | 0 | 0 | 0 |

| | ZIP CODE | QUARTER 1 | QUARTER 2 | QUARTER 3 | QUARTER 4 | TOTAL GRAMS |
|---|---|---|---|---|---|---|
| | 813 | 0 | 0.01 | 0 | 0 | 0.01 |
| | 814 | 0 | 0 | 0 | 0 | 0 |
| | 815 | 0.02 | 0.01 | 0.02 | 0.02 | 0.07 |
| | 816 | 0 | 0 | 0 | 0 | 0 |
| TOTAL | | 0.18 | 0.17 | 0.19 | 0.27 | 0.81 |

STATE: CONNECTICUT

| ZIP CODE | QUARTER 1 | QUARTER 2 | QUARTER 3 | QUARTER 4 | TOTAL GRAMS |
|---|---|---|---|---|---|
| 60 | 0 | 0 | 0 | 0.01 | 0.01 |
| 61 | 0.05 | 0.01 | 0.07 | 0.02 | 0.15 |
| 63 | 0 | 0 | 0 | 0.01 | 0.01 |
| 64 | 0 | 0 | 0 | 0 | 0 |
| 65 | 0.08 | 0.11 | 0.1 | 0.1 | 0.39 |
| 67 | 0 | 0 | 0 | 0 | 0 |
| 68 | 0.27 | 0.34 | 0.4 | 0.8 | 1.81 |
| TOTAL | 0.4 | 0.46 | 0.57 | 0.94 | 2.37 |

STATE: DELAWARE

| ZIP CODE | QUARTER 1 | QUARTER 2 | QUARTER 3 | QUARTER 4 | TOTAL GRAMS |
|---|---|---|---|---|---|
| 198 | 0.01 | 0.01 | 0.01 | 0.01 | 0.04 |
| 199 | 0 | 0 | 0 | 0 | 0 |
| TOTAL | 0.01 | 0.01 | 0.01 | 0.01 | 0.04 |

STATE: DISTRICT OF COLUMBIA

| ZIP CODE | QUARTER 1 | QUARTER 2 | QUARTER 3 | QUARTER 4 | TOTAL GRAMS |
|---|---|---|---|---|---|
| 200 | 0.01 | 0 | 0.01 | 0 | 0.02 |
| TOTAL | 0.01 | 0 | 0.01 | 0 | 0.02 |

STATE: FLORIDA

| ZIP CODE | QUARTER 1 | QUARTER 2 | QUARTER 3 | QUARTER 4 | TOTAL GRAMS |
|---|---|---|---|---|---|
| 321 | 0 | 0.02 | 0.01 | 0.01 | 0.04 |
| 322 | 0.03 | 0.05 | 0.09 | 0.02 | 0.19 |
| 323 | 0.02 | 0.01 | 0.02 | 0.04 | 0.09 |

| | | | | | | |
|---|---|---|---|---|---|---|
| | 324 | 0.05 | 0.04 | 0.05 | 0.04 | 0.18 |
| | 325 | 0.02 | 0.01 | 0.01 | 0.01 | 0.05 |
| | 326 | 0.07 | 0.06 | 0.04 | 0.21 | 0.38 |
| | 327 | 0 | 0 | 0 | 0 | 0 |
| | 328 | 0.08 | 0.08 | 0.08 | 0.08 | 0.32 |
| | 329 | 1.35 | 1.35 | 1.35 | 1.35 | 5.4 |
| | 330 | 0 | 0 | 0 | 0 | 0 |
| | 331 | 0.08 | 0.14 | 0.1 | 0.11 | 0.43 |
| | 333 | 0 | 0 | 0 | 0.01 | 0.01 |
| | 334 | 0.01 | 0.01 | 0.01 | 0.01 | 0.04 |
| | 335 | 0 | 0 | 0 | 0.01 | 0.01 |
| | 336 | 2.47 | 0.15 | 1.45 | 0.94 | 5.01 |
| | 337 | 0.01 | 0.02 | 0.01 | 0.01 | 0.05 |
| | 339 | 0 | 0 | 0 | 0 | 0 |
| | 341 | 0 | 0 | 0 | 0 | 0 |
| | 342 | 0 | 0 | 0.03 | 0.04 | 0.07 |
| | 344 | 0.03 | 0.02 | 0.01 | 0.04 | 0.1 |
| | 346 | 0.02 | 0.02 | 0.01 | 0.03 | 0.08 |
| | 347 | 0.01 | 0.03 | 0.05 | 0.08 | 0.17 |
| | 349 | 0 | 0 | 0 | 0 | 0 |
| TOTAL | | 4.25 | 2.01 | 3.32 | 3.04 | 12.62 |

STATE: GEORGIA

| ZIP CODE | | QUARTER 1 | QUARTER 2 | QUARTER 3 | QUARTER 4 | TOTAL GRAMS |
|---|---|---|---|---|---|---|
| | 300 | 0.04 | 0.03 | 0.03 | 0.01 | 0.11 |
| | 301 | 0.08 | 0.08 | 0.02 | 0.03 | 0.21 |
| | 302 | 0 | 0 | 0.08 | 0.01 | 0.09 |
| | 303 | 0.01 | 0.02 | 0.19 | 0.01 | 0.23 |
| | 304 | 0 | 0 | 0 | 0.04 | 0.04 |
| | 305 | 0.04 | 0.04 | 0.04 | 0.05 | 0.17 |
| | 306 | 0.03 | 0.02 | 0.02 | 0.02 | 0.09 |
| | 309 | 0.04 | 0.03 | 0.03 | 0.05 | 0.15 |
| | 310 | 0.01 | 0.02 | 0.03 | 0.01 | 0.07 |
| | 312 | 0.07 | 0.06 | 0.07 | 0.07 | 0.27 |
| | 313 | 0 | 0 | 0 | 0 | 0 |
| | 314 | 0 | 0.02 | 0 | 0.3 | 0.32 |

| | | | | | | |
|---|---|---|---|---|---|---|
| | 317 | 0 | 0 | 0 | 0 | 0 |
| | 319 | 0 | 0 | 0 | 0.01 | 0.01 |
| TOTAL | | 0.32 | 0.32 | 0.51 | 0.61 | 1.76 |

| STATE: | HAWAII | | | | | |
|---|---|---|---|---|---|---|
| ZIP CODE | | QUARTER 1 | QUARTER 2 | QUARTER 3 | QUARTER 4 | TOTAL GRAMS |
| | 967 | 0 | 0 | 0 | 0 | 0 |
| | 968 | 0.01 | 0 | 0.01 | 0.01 | 0.03 |
| TOTAL | | 0.01 | 0 | 0.01 | 0.01 | 0.03 |

| STATE: | IDAHO | | | | | |
|---|---|---|---|---|---|---|
| ZIP CODE | | QUARTER 1 | QUARTER 2 | QUARTER 3 | QUARTER 4 | TOTAL GRAMS |
| | 832 | 0 | 0 | 0 | 0 | 0 |
| | 833 | 0 | 0 | 0.01 | 0 | 0.01 |
| | 834 | 0.01 | 0.02 | 0.02 | 0.01 | 0.06 |
| | 836 | 0 | 0.01 | 0.01 | 0.01 | 0.03 |
| | 837 | 0.02 | 0.03 | 0.02 | 0.02 | 0.09 |
| | 838 | 0 | 0 | 0 | 0 | 0 |
| TOTAL | | 0.03 | 0.06 | 0.06 | 0.04 | 0.19 |

| STATE: | ILLINOIS | | | | | |
|---|---|---|---|---|---|---|
| ZIP CODE | | QUARTER 1 | QUARTER 2 | QUARTER 3 | QUARTER 4 | TOTAL GRAMS |
| | 600 | 0.08 | 0.04 | 0.07 | 0.04 | 0.23 |
| | 601 | 0.03 | 0.05 | 0.05 | 0.06 | 0.19 |
| | 602 | 0.03 | 0.04 | 0.01 | 0.01 | 0.09 |
| | 603 | 0.01 | 0.01 | 0 | 0 | 0.02 |
| | 604 | 0.06 | 0.07 | 0.06 | 0.06 | 0.25 |
| | 605 | 0.02 | 0.06 | 0.01 | 0.01 | 0.1 |
| | 606 | 0.11 | 0.11 | 0.11 | 0.15 | 0.48 |
| | 609 | 0 | 0 | 0 | 0 | 0 |
| | 610 | 0 | 0 | 0 | 0 | 0 |
| | 611 | 0.01 | 0 | 0.01 | 0.01 | 0.03 |
| | 612 | 0.01 | 0.01 | 0 | 0.01 | 0.03 |
| | 613 | 0 | 0 | 0 | 0 | 0 |

| | | | | | |
|---|---|---|---|---|---|
| 614 | 0 | 0 | 0 | 0 | 0 |
| 615 | 0 | 0 | 0 | 0 | 0 |
| 616 | 0.01 | 0.01 | 0.01 | 0.02 | 0.05 |
| 618 | 0.02 | 0.02 | 0.02 | 0.01 | 0.07 |
| 619 | 0 | 0 | 0 | 0 | 0 |
| 620 | 0 | 0 | 0 | 0 | 0 |
| 622 | 0.14 | 0.16 | 0.16 | 0.12 | 0.58 |
| 623 | 0 | 0.01 | 0.01 | 0 | 0.02 |
| 624 | 0 | 0 | 0 | 0 | 0 |
| 625 | 0 | 0 | 0 | 0.01 | 0.01 |
| 626 | 0 | 0 | 0 | 0 | 0 |
| 627 | 0.01 | 0 | 0.01 | 0.02 | 0.04 |
| 628 | 0 | 0 | 0 | 0 | 0 |
| 629 | 0 | 0.01 | 0.03 | 0.01 | 0.05 |
| TOTAL | 0.54 | 0.6 | 0.56 | 0.54 | 2.24 |

STATE: INDIANA

| ZIP CODE | QUARTER 1 | QUARTER 2 | QUARTER 3 | QUARTER 4 | TOTAL GRAMS |
|---|---|---|---|---|---|
| 460 | 0 | 0 | 0 | 0.05 | 0.05 |
| 461 | 0 | 0 | 0 | 0 | 0 |
| 462 | 0.24 | 0.3 | 0.19 | 0.1 | 0.83 |
| 463 | 0.01 | 0.02 | 0.01 | 0.01 | 0.05 |
| 464 | 0 | 0 | 0 | 0 | 0 |
| 465 | 0.01 | 0 | 0 | 0 | 0.01 |
| 466 | 0 | 0 | 0.01 | 0.01 | 0.02 |
| 467 | 0 | 0 | 0 | 0 | 0 |
| 468 | 0.02 | 0.01 | 0 | 0.02 | 0.05 |
| 469 | 0 | 0 | 0 | 0 | 0 |
| 471 | 0 | 0 | 0 | 0 | 0 |
| 472 | 0 | 0 | 0 | 0.01 | 0.01 |
| 473 | 0 | 0 | 0 | 0 | 0 |
| 474 | 0 | 0 | 0.01 | 0 | 0.01 |
| 475 | 0 | 0 | 0 | 0 | 0 |
| 476 | 0 | 0 | 0 | 0 | 0 |
| 477 | 0 | 0.01 | 0 | 0.01 | 0.02 |

| | | | | | | |
|---|---|---|---|---|---|---|
| | 478 | 0.02 | 0.01 | 0.02 | 0.01 | 0.06 |
| | 479 | 0.01 | 0 | 0 | 0 | 0.01 |
| TOTAL | | 0.31 | 0.35 | 0.24 | 0.22 | 1.12 |

STATE: IOWA

| ZIP CODE | | QUARTER 1 | QUARTER 2 | QUARTER 3 | QUARTER 4 | TOTAL GRAMS |
|---|---|---|---|---|---|---|
| | 500 | 0 | 0 | 0 | 0 | 0 |
| | 501 | 0 | 0 | 0 | 0 | 0 |
| | 502 | 0 | 0 | 0 | 0 | 0 |
| | 503 | 0.03 | 0.01 | 0.01 | 0.03 | 0.08 |
| | 505 | 0 | 0 | 0 | 0 | 0 |
| | 506 | 0 | 0 | 0 | 0 | 0 |
| | 507 | 0 | 0 | 0 | 0 | 0 |
| | 511 | 0 | 0 | 0 | 0.02 | 0.02 |
| | 513 | 0 | 0 | 0 | 0 | 0 |
| | 515 | 0 | 0 | 0 | 0 | 0 |
| | 520 | 0 | 0 | 0 | 0 | 0 |
| | 523 | 0 | 0 | 0 | 0 | 0 |
| | 524 | 0.01 | 0.01 | 0.01 | 0 | 0.03 |
| | 526 | 0 | 0 | 0 | 0 | 0 |
| | 527 | 0 | 0 | 0 | 0 | 0 |
| | 528 | 0.02 | 0.01 | 0.01 | 0.01 | 0.05 |
| TOTAL | | 0.06 | 0.03 | 0.03 | 0.06 | 0.18 |

STATE: KANSAS

| ZIP CODE | | QUARTER 1 | QUARTER 2 | QUARTER 3 | QUARTER 4 | TOTAL GRAMS |
|---|---|---|---|---|---|---|
| | 660 | 0 | 0.01 | 0.01 | 0.04 | 0.06 |
| | 661 | 0 | 0.01 | 0.02 | 0.03 | 0.06 |
| | 662 | 0.09 | 0.1 | 0.11 | 0.05 | 0.35 |
| | 664 | 0 | 0 | 0 | 0.01 | 0.01 |
| | 665 | 0 | 0 | 0 | 0 | 0 |
| | 666 | 0.02 | 0.04 | 0.03 | 0.03 | 0.12 |
| | 667 | 0.01 | 0.01 | 0.01 | 0.01 | 0.04 |
| | 668 | 0 | 0 | 0.01 | 0 | 0.01 |

| | | | | | | |
|---|---|---|---|---|---|---|
| | 669 | 0 | 0 | 0 | 0 | 0 |
| | 670 | 0.01 | 0.02 | 0 | 0.01 | 0.04 |
| | 671 | 0.01 | 0.01 | 0.01 | 0.01 | 0.04 |
| | 672 | 0.08 | 0.05 | 0.1 | 0.07 | 0.3 |
| | 673 | 0 | 0 | 0 | 0 | 0 |
| | 674 | 0.01 | 0.02 | 0.02 | 0.03 | 0.08 |
| | 675 | 0.01 | 0 | 0.01 | 0.01 | 0.03 |
| | 676 | 0 | 0 | 0.01 | 0 | 0.01 |
| | 678 | 0 | 0 | 0 | 0 | 0 |
| | 679 | 0 | 0 | 0 | 0 | 0 |
| TOTAL | | 0.24 | 0.27 | 0.34 | 0.3 | 1.15 |

STATE: KENTUCKY

| ZIP CODE | | QUARTER 1 | QUARTER 2 | QUARTER 3 | QUARTER 4 | TOTAL GRAMS |
|---|---|---|---|---|---|---|
| | 400 | 0.01 | 0.01 | 0.01 | 0.01 | 0.04 |
| | 402 | 0.04 | 0.06 | 0.04 | 0.04 | 0.18 |
| | 403 | 0 | 0 | 0 | 0 | 0 |
| | 404 | 0 | 0.01 | 0 | 0 | 0.01 |
| | 405 | 0.08 | 0.13 | 0.13 | 0.13 | 0.47 |
| | 407 | 0 | 0 | 0 | 0 | 0 |
| | 411 | 0.01 | 0.03 | 0.02 | 0.01 | 0.07 |
| | 415 | 0 | 0 | 0 | 0 | 0 |
| | 420 | 0.14 | 0.07 | 0.14 | 0.14 | 0.49 |
| | 421 | 0.03 | 0.03 | 0.04 | 0.04 | 0.14 |
| | 422 | 0 | 0.04 | 0.02 | 0 | 0.06 |
| | 423 | 0 | 0.01 | 0 | 0 | 0.01 |
| | 424 | 0 | 0 | 0 | 0.01 | 0.01 |
| | 427 | 0 | 0 | 0 | 0 | 0 |
| TOTAL | | 0.31 | 0.39 | 0.4 | 0.38 | 1.48 |

STATE: LOUISIANA

| ZIP CODE | | QUARTER 1 | QUARTER 2 | QUARTER 3 | QUARTER 4 | TOTAL GRAMS |
|---|---|---|---|---|---|---|
| | 700 | 0.01 | 0.02 | 0.01 | 0.05 | 0.09 |
| | 701 | 0.06 | 0.06 | 0.08 | 0 | 0.2 |
| | 703 | 0 | 0 | 0 | 0.01 | 0.01 |

| | | | | | | |
|---|---|---|---|---|---|---|
| | 704 | 0.03 | 0.01 | 0.01 | 0.03 | 0.08 |
| | 705 | 0.03 | 1.7 | 0.7 | 0.69 | 3.12 |
| | 706 | 0.01 | 0.01 | 0.01 | 0 | 0.03 |
| | 707 | 0.07 | 0.07 | 0.13 | 0.07 | 0.34 |
| | 708 | 0.02 | 0.02 | 0.02 | 0.05 | 0.11 |
| | 711 | 0.09 | 0.09 | 0.1 | 0.09 | 0.37 |
| | 712 | 0.03 | 0.05 | 0.02 | 0.06 | 0.16 |
| | 713 | 0 | 0 | 0 | 0.11 | 0.11 |
| | 714 | 0 | 0 | 0 | 0 | 0 |
| TOTAL | | 0.35 | 2.03 | 1.08 | 1.16 | 4.62 |

STATE: MAINE

| ZIP CODE | | QUARTER 1 | QUARTER 2 | QUARTER 3 | QUARTER 4 | TOTAL GRAMS |
|---|---|---|---|---|---|---|
| | 40 | 0 | 0 | 0 | 0 | 0 |
| | 41 | 0 | 0 | 0 | 0 | 0 |
| | 42 | 0 | 0 | 0 | 0 | 0 |
| | 43 | 0.01 | 0 | 0.01 | 0.01 | 0.03 |
| | 44 | 0 | 0 | 0 | 0.01 | 0.01 |
| | 46 | 0 | 0 | 0 | 0 | 0 |
| | 47 | 0 | 0 | 0 | 0 | 0 |
| | 48 | 0 | 0 | 0 | 0 | 0 |
| TOTAL | | 0.01 | 0 | 0.01 | 0.02 | 0.04 |

STATE: MARYLAND

| ZIP CODE | | QUARTER 1 | QUARTER 2 | QUARTER 3 | QUARTER 4 | TOTAL GRAMS |
|---|---|---|---|---|---|---|
| | 207 | 0 | 0.01 | 0 | 0 | 0.01 |
| | 208 | 0.02 | 0.01 | 0.01 | 0.01 | 0.05 |
| | 210 | 0 | 0.01 | 0 | 0.01 | 0.02 |
| | 212 | 0.01 | 0.03 | 0.04 | 0.02 | 0.1 |
| | 215 | 0.01 | 0.01 | 0.01 | 0 | 0.03 |
| | 216 | 0 | 0 | 0 | 0 | 0 |
| | 217 | 0 | 0 | 0 | 0 | 0 |
| | 218 | 0 | 0 | 0.01 | 0 | 0.01 |
| TOTAL | | 0.04 | 0.07 | 0.07 | 0.04 | 0.22 |

STATE: MASSACHUSETTS

| ZIP CODE | QUARTER 1 | QUARTER 2 | QUARTER 3 | QUARTER 4 | TOTAL GRAMS |
|---|---|---|---|---|---|
| 10 | 0.03 | 0.04 | 0.03 | 0.04 | 0.14 |
| 11 | 0 | 0 | 0 | 0 | 0 |
| 12 | 0 | 0 | 0 | 0 | 0 |
| 16 | 0.01 | 0 | 0.01 | 0.01 | 0.03 |
| 18 | 0 | 0 | 0 | 0.01 | 0.01 |
| 20 | 0 | 0 | 0 | 0 | 0 |
| 21 | 0.05 | 0.06 | 0.07 | 0.09 | 0.27 |
| 22 | 0 | 0 | 0 | 0.03 | 0.03 |
| 25 | 0 | 0 | 0 | 0 | 0 |
| 27 | 0 | 0.02 | 0.01 | 0.01 | 0.04 |
| TOTAL | 0.09 | 0.12 | 0.12 | 0.19 | 0.52 |

STATE: MICHIGAN

| ZIP CODE | QUARTER 1 | QUARTER 2 | QUARTER 3 | QUARTER 4 | TOTAL GRAMS |
|---|---|---|---|---|---|
| 480 | 0.12 | 0.12 | 0.04 | 0.17 | 0.45 |
| 481 | 0.06 | 0.06 | 0.05 | 0.12 | 0.29 |
| 482 | 0.04 | 0.06 | 0.05 | 0.06 | 0.21 |
| 483 | 0.2 | 0.01 | 0.01 | 0.49 | 0.71 |
| 484 | 0 | 0.01 | 0.02 | 0.02 | 0.05 |
| 485 | 0 | 0 | 0 | 0 | 0 |
| 486 | 0.01 | 0.01 | 0 | 0.01 | 0.03 |
| 487 | 0 | 0 | 0 | 0.02 | 0.02 |
| 488 | 0 | 0.01 | 0.02 | 0.02 | 0.05 |
| 489 | 0.01 | 0.01 | 0 | 0 | 0.02 |
| 490 | 0 | 0.01 | 0 | 0.01 | 0.02 |
| 492 | 0.01 | 0.01 | 0.01 | 0.02 | 0.05 |
| 494 | 0.01 | 0 | 0 | 0.01 | 0.02 |
| 495 | 0.01 | 0 | 0.05 | 0.01 | 0.07 |
| 496 | 0.06 | 0.07 | 0.06 | 0.06 | 0.25 |
| 497 | 0 | 0 | 0 | 0.01 | 0.01 |
| 498 | 0 | 0.01 | 0.01 | 0 | 0.02 |

| | | | | | | |
|---|---|---|---|---|---|---|
| | 499 | 0 | 0 | 0 | 0 | 0 |
| TOTAL | | 0.53 | 0.39 | 0.32 | 1.03 | 2.27 |

STATE: MINNESOTA

| ZIP CODE | | QUARTER 1 | QUARTER 2 | QUARTER 3 | QUARTER 4 | TOTAL GRAMS |
|---|---|---|---|---|---|---|
| | 551 | 0 | 0 | 0 | 0.09 | 0.09 |
| | 553 | 0 | 0 | 0 | 0 | 0 |
| | 554 | 0.01 | 0.02 | 0.01 | 0.01 | 0.05 |
| | 557 | 0 | 0 | 0 | 0 | 0 |
| | 558 | 0 | 0 | 0 | 0.05 | 0.05 |
| | 559 | 0.11 | 0.09 | 0.08 | 0.12 | 0.4 |
| | 561 | 0 | 0 | 0 | 0 | 0 |
| | 562 | 0.01 | 0.01 | 0.02 | 0.01 | 0.05 |
| | 563 | 0.02 | 0.03 | 0.03 | 0.03 | 0.11 |
| | 564 | 0 | 0 | 0 | 0.01 | 0.01 |
| | 565 | 0 | 0 | 0 | 0 | 0 |
| | 566 | 0 | 0 | 0 | 0 | 0 |
| | 567 | 0 | 0 | 0 | 0 | 0 |
| TOTAL | | 0.15 | 0.15 | 0.14 | 0.32 | 0.76 |

STATE: MISSISSIPPI

| ZIP CODE | | QUARTER 1 | QUARTER 2 | QUARTER 3 | QUARTER 4 | TOTAL GRAMS |
|---|---|---|---|---|---|---|
| | 386 | 0.02 | 0.02 | 0.02 | 0.02 | 0.08 |
| | 390 | 0 | 0 | 0 | 0 | 0 |
| | 391 | 14.66 | 10.36 | 12.36 | 14.38 | 51.76 |
| | 392 | 0.01 | 0.03 | 0.07 | 0.02 | 0.13 |
| | 393 | 0.03 | 0.02 | 0 | 0.02 | 0.07 |
| | 394 | 0.01 | 0 | 0 | 0 | 0.01 |
| | 395 | 0.01 | 0.01 | 0.01 | 0.02 | 0.05 |
| | 396 | 0 | 0 | 0 | 0 | 0 |
| | 397 | 0 | 0 | 0 | 0 | 0 |
| TOTAL | | 14.74 | 10.44 | 12.46 | 14.46 | 52.1 |

STATE: MISSOURI

| ZIP CODE | QUARTER 1 | QUARTER 2 | QUARTER 3 | QUARTER 4 | TOTAL GRAMS |
|---|---|---|---|---|---|
| 630 | 0.01 | 0.02 | 0 | 0 | 0.03 |
| 631 | 0.09 | 0.08 | 0.1 | 0.08 | 0.35 |
| 633 | 0 | 0 | 0 | 0 | 0 |
| 635 | 0 | 0 | 0 | 0 | 0 |
| 636 | 0.01 | 0.01 | 0.01 | 0.01 | 0.04 |
| 637 | 0.04 | 0.05 | 0.08 | 0.07 | 0.24 |
| 639 | 0.03 | 0.04 | 0.04 | 0.03 | 0.14 |
| 640 | 0 | 0 | 0 | 0 | 0 |
| 641 | 0.04 | 0.05 | 0.03 | 0.04 | 0.16 |
| 645 | 0.02 | 0 | 0.02 | 0.01 | 0.05 |
| 647 | 0 | 0 | 0 | 0 | 0 |
| 648 | 0.02 | 0.02 | 0.02 | 0.01 | 0.07 |
| 652 | 0.04 | 0.04 | 0.05 | 0.03 | 0.16 |
| 653 | 0.01 | 0 | 0 | 0 | 0.01 |
| 654 | 0 | 0 | 0 | 0 | 0 |
| 657 | 0 | 0 | 0 | 0 | 0 |
| 658 | 0 | 0 | 0 | 0 | 0 |
| TOTAL | 0.31 | 0.31 | 0.35 | 0.28 | 1.25 |

STATE: MONTANA

| ZIP CODE | QUARTER 1 | QUARTER 2 | QUARTER 3 | QUARTER 4 | TOTAL GRAMS |
|---|---|---|---|---|---|
| 590 | 0 | 0 | 0 | 0 | 0 |
| 591 | 0.01 | 0.02 | 0.01 | 0.02 | 0.06 |
| 592 | 0 | 0 | 0 | 0 | 0 |
| 594 | 0 | 0 | 0 | 0 | 0 |
| 596 | 0 | 0 | 0 | 0 | 0 |
| 597 | 0.01 | 0.01 | 0.01 | 0.01 | 0.04 |
| 598 | 0.01 | 0.01 | 0.01 | 0.02 | 0.05 |
| 599 | 0.01 | 0.01 | 0 | 0 | 0.02 |
| TOTAL | 0.04 | 0.05 | 0.03 | 0.05 | 0.17 |

STATE: NEBRASKA

| ZIP CODE | QUARTER 1 | QUARTER 2 | QUARTER 3 | QUARTER 4 | TOTAL GRAMS |
|---|---|---|---|---|---|

| | | | | | |
|---|---|---|---|---|---|
| 680 | 0 | 0 | 0 | 0 | 0 |
| 681 | 0.06 | 0.06 | 0.05 | 0.11 | 0.28 |
| 683 | 0 | 0 | 0 | 0 | 0 |
| 684 | 0 | 0 | 0 | 0 | 0 |
| 685 | 0.01 | 0.01 | 0.02 | 0.02 | 0.06 |
| 686 | 0 | 0 | 0 | 0 | 0 |
| 687 | 0 | 0.01 | 0 | 0 | 0.01 |
| 688 | 0 | 0 | 0 | 0 | 0 |
| 689 | 0 | 0 | 0 | 0 | 0 |
| 690 | 0 | 0 | 0 | 0 | 0 |
| 691 | 0 | 0 | 0 | 0 | 0 |
| TOTAL | 0.07 | 0.08 | 0.07 | 0.13 | 0.35 |

STATE: NEVADA

| ZIP CODE | QUARTER 1 | QUARTER 2 | QUARTER 3 | QUARTER 4 | TOTAL GRAMS |
|---|---|---|---|---|---|
| 890 | 0.01 | 0.01 | 0.01 | 0.02 | 0.05 |
| 891 | 0.04 | 0.02 | 0.09 | 0.05 | 0.2 |
| 894 | 0.01 | 0 | 0 | 0.01 | 0.02 |
| 895 | 0.01 | 0.02 | 0.01 | 0.02 | 0.06 |
| 897 | 0 | 0 | 0 | 0 | 0 |
| 898 | 0.01 | 0.01 | 0.01 | 0.02 | 0.05 |
| TOTAL | 0.08 | 0.06 | 0.12 | 0.12 | 0.38 |

STATE: NEW HAMPSHIRE

| ZIP CODE | QUARTER 1 | QUARTER 2 | QUARTER 3 | QUARTER 4 | TOTAL GRAMS |
|---|---|---|---|---|---|
| 32 | 0 | 0 | 0 | 0 | 0 |
| 33 | 0 | 0 | 0 | 0 | 0 |
| 34 | 0 | 0 | 0 | 0 | 0 |
| 35 | 0 | 0 | 0 | 0 | 0 |
| 37 | 0 | 0 | 0.01 | 0 | 0.01 |
| 38 | 0.01 | 0.01 | 0.01 | 0.01 | 0.04 |
| TOTAL | 0.01 | 0.01 | 0.02 | 0.01 | 0.05 |

STATE: NEW JERSEY

| ZIP CODE | QUARTER 1 | QUARTER 2 | QUARTER 3 | QUARTER 4 | TOTAL GRAMS |
|---|---|---|---|---|---|
| 70 | 0.01 | 0.01 | 0.01 | 0.01 | 0.04 |
| 71 | 0.01 | 0.01 | 0.01 | 0.01 | 0.04 |
| 73 | 0 | 0 | 0 | 0 | 0 |
| 74 | 0.71 | 0.73 | 0.04 | 1.38 | 2.86 |
| 75 | 0.03 | 0.04 | 0.03 | 0.03 | 0.13 |
| 76 | 0.04 | 0.03 | 0 | 0 | 0.07 |
| 77 | 0.03 | 0.04 | 0.03 | 0.03 | 0.13 |
| 79 | 0 | 0 | 0 | 0 | 0 |
| 80 | 0 | 0 | 0 | 0 | 0 |
| 82 | 0.02 | 0.02 | 0.01 | 0.01 | 0.06 |
| 84 | 0 | 0 | 0 | 0 | 0 |
| 85 | 0 | 0 | 0 | 0 | 0 |
| 86 | 0 | 0 | 0 | 0 | 0 |
| 88 | 0.01 | 0 | 0 | 0 | 0.01 |
| 89 | 0.1 | 0.12 | 0.1 | 0.08 | 0.4 |
| TOTAL | 0.96 | 1 | 0.23 | 1.55 | 3.74 |

STATE: NEW MEXICO

| ZIP CODE | QUARTER 1 | QUARTER 2 | QUARTER 3 | QUARTER 4 | TOTAL GRAMS |
|---|---|---|---|---|---|
| 871 | 0.05 | 0.03 | 0.02 | 0.08 | 0.18 |
| 873 | 0 | 0 | 0 | 0 | 0 |
| 874 | 0 | 0 | 0 | 0 | 0 |
| 875 | 0 | 0 | 0 | 0 | 0 |
| 880 | 0 | 0.01 | 0 | 0 | 0.01 |
| 881 | 0 | 0 | 0 | 0 | 0 |
| 882 | 0 | 0 | 0 | 0 | 0 |
| TOTAL | 0.05 | 0.04 | 0.02 | 0.08 | 0.19 |

STATE: NEW YORK

| ZIP CODE | QUARTER 1 | QUARTER 2 | QUARTER 3 | QUARTER 4 | TOTAL GRAMS |
|---|---|---|---|---|---|
| 100 | 1.18 | 1.19 | 1.4 | 0.7 | 4.47 |
| 103 | 0 | 0 | 0 | 0 | 0 |

| | | | | | | |
|---|---|---|---|---|---|---|
| | 104 | 0.03 | 0.01 | 0.01 | 0.03 | 0.08 |
| | 105 | 0 | 0 | 0 | 0 | 0 |
| | 110 | 0.05 | 0.03 | 0.05 | 0.03 | 0.16 |
| | 112 | 0 | 0 | 0.01 | 0.01 | 0.02 |
| | 113 | 0.05 | 0.06 | 0.06 | 0.06 | 0.23 |
| | 114 | 0.13 | 0 | 0.13 | 0 | 0.26 |
| | 117 | 0.03 | 0.03 | 0.01 | 0.05 | 0.12 |
| | 119 | 0 | 0 | 0 | 0 | 0 |
| | 122 | 0.08 | 0.05 | 0.1 | 0.2 | 0.43 |
| | 123 | 0 | 0 | 0 | 0 | 0 |
| | 124 | 0 | 0 | 0 | 0 | 0 |
| | 125 | 0 | 0 | 0 | 0 | 0 |
| | 126 | 0 | 0.01 | 0 | 0.01 | 0.02 |
| | 127 | 0 | 0 | 0 | 0 | 0 |
| | 128 | 0.02 | 0.02 | 0.02 | 0.02 | 0.08 |
| | 129 | 0 | 0 | 0 | 0 | 0 |
| | 133 | 0.01 | 0.01 | 0.01 | 0.01 | 0.04 |
| | 135 | 0 | 0 | 0 | 0 | 0 |
| | 137 | 0.01 | 0 | 0.01 | 0.01 | 0.03 |
| | 139 | 0 | 0 | 0 | 0 | 0 |
| | 140 | 0 | 0 | 0 | 0 | 0 |
| | 142 | 0.09 | 0.08 | 0.1 | 0.15 | 0.42 |
| | 144 | 0 | 0 | 0 | 0 | 0 |
| | 146 | 0.1 | 0.08 | 0.11 | 0.08 | 0.37 |
| TOTAL | | 1.78 | 1.57 | 2.02 | 1.36 | 6.73 |

STATE: NORTH CAROLINA

| ZIP CODE | | QUARTER 1 | QUARTER 2 | QUARTER 3 | QUARTER 4 | TOTAL GRAMS |
|---|---|---|---|---|---|---|
| | 270 | 0 | 0 | 0.01 | 0.01 | 0.02 |
| | 271 | 0.07 | 0.16 | 0.18 | 0.07 | 0.48 |
| | 272 | 0.02 | 0.01 | 0.02 | 0.02 | 0.07 |
| | 273 | 0.01 | 0 | 0 | 0 | 0.01 |
| | 274 | 0.01 | 0.02 | 0.07 | 0 | 0.1 |
| | 275 | 0 | 0 | 0.01 | 0.03 | 0.04 |
| | 276 | 0 | 0.01 | 0.01 | 0.03 | 0.05 |

| | | | | | | |
|---|---|---|---|---|---|---|
| | 277 | 0.02 | 0.05 | 0.05 | 0.08 | 0.2 |
| | 278 | 0.02 | 0.01 | 0.01 | 0.01 | 0.05 |
| | 279 | 0 | 0 | 0 | 0 | 0 |
| | 280 | 0 | 0 | 0 | 0.01 | 0.01 |
| | 282 | 0.01 | 0.03 | 0.01 | 0.01 | 0.06 |
| | 283 | 0.03 | 0.03 | 0.02 | 0.03 | 0.11 |
| | 284 | 0.01 | 0.02 | 0.04 | 0.03 | 0.1 |
| | 285 | 0.01 | 0.01 | 0 | 0.01 | 0.03 |
| | 286 | 0 | 0 | 0.01 | 0.02 | 0.03 |
| | 287 | 0.01 | 0 | 0 | 0.02 | 0.03 |
| | 288 | 0.07 | 0.06 | 0.08 | 0.07 | 0.28 |
| | 289 | 0 | 0 | 0 | 0 | 0 |
| TOTAL | | 0.29 | 0.41 | 0.52 | 0.45 | 1.67 |

| STATE: | NORTH DAKOTA | | | | |
|---|---|---|---|---|---|
| ZIP CODE | QUARTER 1 | QUARTER 2 | QUARTER 3 | QUARTER 4 | TOTAL GRAMS |
| 581 | 0 | 0 | 0 | 0.01 | 0.01 |
| 582 | 0.02 | 0.02 | 0.02 | 0.02 | 0.08 |
| 583 | 0 | 0 | 0 | 0 | 0 |
| 585 | 0 | 0.04 | 0 | 0 | 0.04 |
| 587 | 0 | 0 | 0 | 0 | 0 |
| 588 | 0 | 0 | 0 | 0 | 0 |
| TOTAL | 0.02 | 0.06 | 0.02 | 0.03 | 0.13 |

| STATE: | OHIO | | | | |
|---|---|---|---|---|---|
| ZIP CODE | QUARTER 1 | QUARTER 2 | QUARTER 3 | QUARTER 4 | TOTAL GRAMS |
| 430 | 0 | 0.02 | 0.01 | 0.04 | 0.07 |
| 431 | 0 | 0 | 0 | 0.01 | 0.01 |
| 432 | 0.05 | 0.04 | 0.08 | 0.13 | 0.3 |
| 433 | 0 | 0 | 0 | 0 | 0 |
| 434 | 0 | 0 | 0 | 0.02 | 0.02 |
| 435 | 0.01 | 0.07 | 0.01 | 0.08 | 0.17 |
| 436 | 0.07 | 0.1 | 0.09 | 0.1 | 0.36 |
| 437 | 0.01 | 0.03 | 0.01 | 0.01 | 0.06 |
| 440 | 0 | 0.07 | 0.02 | 0.01 | 0.1 |

| | | | | | | |
|---|---|---|---|---|---|---|
| | 441 | 0.05 | 0.09 | 0.07 | 0.02 | 0.23 |
| | 442 | 0 | 0.01 | 0 | 0.01 | 0.02 |
| | 443 | 0 | 0 | 0 | 0 | 0 |
| | 445 | 0.01 | 0 | 0.01 | 0 | 0.02 |
| | 446 | 0 | 0 | 0 | 0 | 0 |
| | 448 | 0.01 | 0.01 | 0 | 0.01 | 0.03 |
| | 449 | 0.02 | 0.01 | 0.01 | 0.05 | 0.09 |
| | 450 | 0 | 0.01 | 0 | 0 | 0.01 |
| | 451 | 0 | 0 | 0 | 0 | 0 |
| | 452 | 0.03 | 0.03 | 0.02 | 0.1 | 0.18 |
| | 453 | 0 | 0 | 0 | 0 | 0 |
| | 454 | 0.02 | 0.03 | 0.06 | 0.07 | 0.18 |
| | 455 | 0 | 0 | 0 | 0 | 0 |
| | 457 | 0.01 | 0.01 | 0.01 | 0.01 | 0.04 |
| | 458 | 0.02 | 0.01 | 0.01 | 0.04 | 0.08 |
| TOTAL | | 0.31 | 0.54 | 0.41 | 0.71 | 1.97 |

STATE: OKLAHOMA

| ZIP CODE | | QUARTER 1 | QUARTER 2 | QUARTER 3 | QUARTER 4 | TOTAL GRAMS |
|---|---|---|---|---|---|---|
| | 730 | 0.03 | 0.03 | 0.03 | 0.03 | 0.12 |
| | 731 | 0.11 | 0.09 | 0.07 | 0.07 | 0.34 |
| | 735 | 0.01 | 0.01 | 0.01 | 0.01 | 0.04 |
| | 737 | 0 | 0 | 0 | 0 | 0 |
| | 738 | 0 | 0 | 0 | 0 | 0 |
| | 741 | 0.04 | 0.04 | 0.04 | 0.05 | 0.17 |
| | 743 | 0 | 0 | 0 | 0 | 0 |
| | 744 | 0 | 0 | 0 | 0 | 0 |
| | 748 | 0 | 0 | 0 | 0 | 0 |
| TOTAL | | 0.19 | 0.17 | 0.15 | 0.16 | 0.67 |

STATE: OREGON

| ZIP CODE | | QUARTER 1 | QUARTER 2 | QUARTER 3 | QUARTER 4 | TOTAL GRAMS |
|---|---|---|---|---|---|---|
| | 970 | 0.03 | 0.03 | 0.03 | 0.04 | 0.13 |
| | 971 | 0.04 | 0 | 0.01 | 0.01 | 0.06 |
| | 972 | 0.2 | 0.2 | 0.25 | 0.29 | 0.94 |

| | | | | | | |
|---|---|---|---|---|---|---|
| | 973 | 0.01 | 0.01 | 0.01 | 0.01 | 0.04 |
| | 974 | 0 | 0 | 0 | 0 | 0 |
| | 975 | 0 | 0 | 0 | 0.02 | 0.02 |
| | 977 | 0 | 0 | 0.01 | 0.01 | 0.02 |
| | 978 | 0 | 0.01 | 0 | 0 | 0.01 |
| TOTAL | | 0.28 | 0.25 | 0.31 | 0.38 | 1.22 |

STATE: PENNSYLVANIA

| ZIP CODE | | QUARTER 1 | QUARTER 2 | QUARTER 3 | QUARTER 4 | TOTAL GRAMS |
|---|---|---|---|---|---|---|
| | 150 | 0.01 | 0.01 | 0.01 | 0.02 | 0.05 |
| | 151 | 0 | 0 | 0 | 0 | 0 |
| | 152 | 0 | 0.01 | 0.01 | 0.01 | 0.03 |
| | 154 | 0 | 0 | 0 | 0 | 0 |
| | 158 | 0 | 0.01 | 0 | 0 | 0.01 |
| | 159 | 0 | 0 | 0.01 | 0.01 | 0.02 |
| | 163 | 0 | 0 | 0 | 0 | 0 |
| | 165 | 0 | 0.02 | 0.01 | 0.01 | 0.04 |
| | 166 | 0.01 | 0.01 | 0 | 0.01 | 0.03 |
| | 167 | 0 | 0 | 0 | 0 | 0 |
| | 169 | 0.01 | 0 | 0 | 0 | 0.01 |
| | 170 | 0.02 | 0.01 | 0 | 0 | 0.03 |
| | 171 | 0.01 | 0 | 0 | 0.01 | 0.02 |
| | 174 | 0 | 0 | 0 | 0.01 | 0.01 |
| | 175 | 0.01 | 0.03 | 0.04 | 0.03 | 0.11 |
| | 176 | 0 | 0 | 0 | 0 | 0 |
| | 177 | 0 | 0 | 0 | 0 | 0 |
| | 178 | 0.01 | 0.01 | 0.01 | 0.02 | 0.05 |
| | 180 | 0 | 0 | 0 | 0.01 | 0.01 |
| | 181 | 0.01 | 0.01 | 0.03 | 0.14 | 0.19 |
| | 185 | 0.02 | 0.01 | 0.02 | 0.01 | 0.06 |
| | 187 | 0.02 | 0.03 | 0.02 | 0.04 | 0.11 |
| | 188 | 0 | 0 | 0 | 0.03 | 0.03 |
| | 190 | 0.34 | 1.67 | 4.02 | 0.7 | 6.73 |
| | 191 | 0.1 | 0.14 | 0.13 | 0.2 | 0.57 |
| | 196 | 0 | 0.01 | 0 | 0 | 0.01 |
| TOTAL | | 0.57 | 1.98 | 4.31 | 1.26 | 8.12 |

STATE: PUERTO RICO

| ZIP CODE | QUARTER 1 | QUARTER 2 | QUARTER 3 | QUARTER 4 | TOTAL GRAMS |
|---|---|---|---|---|---|
| 6 | 0 | 0 | 0 | 0.06 | 0.06 |
| 7 | 0.03 | 0.01 | 0.03 | 0.02 | 0.09 |
| 9 | 0.18 | 0.15 | 0.21 | 0.08 | 0.62 |
| TOTAL | 0.21 | 0.16 | 0.24 | 0.16 | 0.77 |

STATE: RHODE ISLAND

| ZIP CODE | QUARTER 1 | QUARTER 2 | QUARTER 3 | QUARTER 4 | TOTAL GRAMS |
|---|---|---|---|---|---|
| 28 | 0 | 0.01 | 0 | 0 | 0.01 |
| TOTAL | 0 | 0.01 | 0 | 0 | 0.01 |

STATE: SOUTH CAROLINA

| ZIP CODE | QUARTER 1 | QUARTER 2 | QUARTER 3 | QUARTER 4 | TOTAL GRAMS |
|---|---|---|---|---|---|
| 290 | 0 | 0 | 0 | 0 | 0 |
| 291 | 0 | 0.01 | 0 | 0 | 0.01 |
| 292 | 0.02 | 0.01 | 0.04 | 0.09 | 0.16 |
| 293 | 0.01 | 0.01 | 0.02 | 0.01 | 0.05 |
| 294 | 0.04 | 0.06 | 0.06 | 0.07 | 0.23 |
| 295 | 0.01 | 0.01 | 0.01 | 0.03 | 0.06 |
| 296 | 0.04 | 0.04 | 0.03 | 0.03 | 0.14 |
| 299 | 0.67 | 0.67 | 0.67 | 0 | 2.01 |
| TOTAL | 0.79 | 0.81 | 0.83 | 0.23 | 2.66 |

STATE: SOUTH DAKOTA

| ZIP CODE | QUARTER 1 | QUARTER 2 | QUARTER 3 | QUARTER 4 | TOTAL GRAMS |
|---|---|---|---|---|---|
| 570 | 0.01 | 0.01 | 0.01 | 0.02 | 0.05 |
| 571 | 0 | 0 | 0 | 0.05 | 0.05 |
| 575 | 0 | 0 | 0 | 0 | 0 |
| 577 | 0.01 | 0.02 | 0.02 | 0.02 | 0.07 |
| TOTAL | 0.02 | 0.03 | 0.03 | 0.09 | 0.17 |

STATE: TENNESSEE

| ZIP CODE | QUARTER 1 | QUARTER 2 | QUARTER 3 | QUARTER 4 | TOTAL GRAMS |
|---|---|---|---|---|---|
| 370 | 0.43 | 0.25 | 0.33 | 0.3 | 1.31 |
| 371 | 0.05 | 0.05 | 0.06 | 0.05 | 0.21 |
| 372 | 0.04 | 0.05 | 0.05 | 0.1 | 0.24 |
| 373 | 0 | 0.01 | 0.01 | 0 | 0.02 |
| 376 | 0.02 | 0.02 | 0.01 | 0.02 | 0.07 |
| 377 | 0 | 0.02 | 0.01 | 0 | 0.03 |
| 378 | 0.02 | 0.01 | 0.01 | 0.02 | 0.06 |
| 379 | 0.02 | 0.01 | 0.02 | 0.03 | 0.08 |
| 380 | 0 | 0 | 0 | 0 | 0 |
| 381 | 0.05 | 0.06 | 0.09 | 0.14 | 0.34 |
| 382 | 0 | 0 | 0 | 0 | 0 |
| 383 | 0.01 | 0.03 | 0.02 | 0.03 | 0.09 |
| 384 | 0.02 | 0.01 | 0.02 | 0.02 | 0.07 |
| 385 | 0.01 | 0.01 | 0.01 | 0.01 | 0.04 |
| TOTAL | 0.67 | 0.53 | 0.64 | 0.72 | 2.56 |

STATE: TEXAS

| ZIP CODE | QUARTER 1 | QUARTER 2 | QUARTER 3 | QUARTER 4 | TOTAL GRAMS |
|---|---|---|---|---|---|
| 750 | 0.04 | 0.1 | 0.05 | 0.05 | 0.24 |
| 751 | 0 | 0 | 0 | 0 | 0 |
| 752 | 0.05 | 0.06 | 0.06 | 0.08 | 0.25 |
| 753 | 0.01 | 0.01 | 0.02 | 0.02 | 0.06 |
| 754 | 0 | 0.01 | 0 | 0.01 | 0.02 |
| 755 | 0 | 0 | 0 | 0 | 0 |
| 756 | 0.18 | 0.19 | 0.06 | 0.19 | 0.62 |
| 757 | 0.02 | 0.03 | 0.02 | 0.04 | 0.11 |
| 759 | 0.01 | 0 | 0 | 0 | 0.01 |
| 760 | 0.01 | 0.03 | 0.01 | 0.02 | 0.07 |
| 761 | 0.13 | 0.15 | 0.13 | 0.18 | 0.59 |
| 762 | 0.01 | 0.01 | 0.01 | 0.01 | 0.04 |
| 763 | 0 | 0.01 | 0.02 | 0.02 | 0.05 |
| 765 | 0.04 | 0.05 | 0.05 | 0.04 | 0.18 |

| | | | | | | |
|---|---|---|---|---|---|---|
| | 767 | 0.01 | 0 | 0.01 | 0 | 0.02 |
| | 769 | 0 | 0 | 0 | 0 | 0 |
| | 770 | 1.06 | 0.28 | 0.23 | 1.23 | 2.8 |
| | 773 | 0 | 0.03 | 0 | 0.03 | 0.06 |
| | 774 | 0.01 | 0.01 | 0.01 | 0.01 | 0.04 |
| | 775 | 0 | 0.01 | 0.02 | 0.01 | 0.04 |
| | 777 | 0 | 0 | 0 | 0 | 0 |
| | 778 | 0.01 | 0.01 | 0.01 | 0.01 | 0.04 |
| | 779 | 0 | 0 | 0 | 0 | 0 |
| | 781 | 0 | 0 | 0 | 0 | 0 |
| | 782 | 0.06 | 0.13 | 0.03 | 0.06 | 0.28 |
| | 784 | 0.02 | 0.03 | 0.03 | 0.03 | 0.11 |
| | 785 | 0.08 | 0.09 | 0.1 | 0.08 | 0.35 |
| | 786 | 0 | 0 | 0 | 0 | 0 |
| | 787 | 0.01 | 0.02 | 0.01 | 0.02 | 0.06 |
| | 788 | 0 | 0 | 0 | 0 | 0 |
| | 789 | 0 | 0 | 0 | 0 | 0 |
| | 790 | 0 | 0 | 0 | 0 | 0 |
| | 791 | 0 | 0 | 0.03 | 0.01 | 0.04 |
| | 794 | 0.08 | 0.06 | 0.07 | 0.09 | 0.3 |
| | 796 | 0.03 | 0.02 | 0.03 | 0 | 0.08 |
| | 797 | 0 | 0 | 0 | 0.01 | 0.01 |
| | 799 | 0.03 | 0.03 | 0.03 | 0.08 | 0.17 |
| TOTAL | | 1.9 | 1.37 | 1.04 | 2.33 | 6.64 |

STATE: UTAH

| ZIP CODE | | QUARTER 1 | QUARTER 2 | QUARTER 3 | QUARTER 4 | TOTAL GRAMS |
|---|---|---|---|---|---|---|
| | 840 | 0.01 | 0.01 | 0.02 | 0.01 | 0.05 |
| | 841 | 0.21 | 0.17 | 0.14 | 0.19 | 0.71 |
| | 843 | 0 | 0 | 0 | 0 | 0 |
| | 844 | 0.01 | 0.01 | 0.01 | 0.03 | 0.06 |
| | 846 | 0 | 0 | 0.01 | 0.01 | 0.02 |
| | 847 | 0.02 | 0.02 | 0.02 | 0.01 | 0.07 |
| TOTAL | | 0.25 | 0.21 | 0.2 | 0.25 | 0.91 |

STATE: VERMONT

| ZIP CODE | QUARTER 1 | QUARTER 2 | QUARTER 3 | QUARTER 4 | TOTAL GRAMS |
|---|---|---|---|---|---|
| 54 | 0.01 | 0.01 | 0 | 0.01 | 0.03 |
| 57 | 0 | 0 | 0 | 0 | 0 |
| TOTAL | 0.01 | 0.01 | 0 | 0.01 | 0.03 |

STATE: VIRGINIA

| ZIP CODE | QUARTER 1 | QUARTER 2 | QUARTER 3 | QUARTER 4 | TOTAL GRAMS |
|---|---|---|---|---|---|
| 201 | 0 | 0 | 0 | 0 | 0 |
| 220 | 0.07 | 0.06 | 0.05 | 0.06 | 0.24 |
| 222 | 0.02 | 0.02 | 0.03 | 0.03 | 0.1 |
| 223 | 0.01 | 0.01 | 0.01 | 0.01 | 0.04 |
| 224 | 0.03 | 0.03 | 0.03 | 0.02 | 0.11 |
| 225 | 0.02 | 0.02 | 0.01 | 0.02 | 0.07 |
| 226 | 0 | 0 | 0 | 0.01 | 0.01 |
| 228 | 0 | 0.01 | 0 | 0 | 0.01 |
| 229 | 0.06 | 0.05 | 0.06 | 0.06 | 0.23 |
| 231 | 0.02 | 0.01 | 0.01 | 0.02 | 0.06 |
| 232 | 0.09 | 0.05 | 0.06 | 0.12 | 0.32 |
| 233 | 0 | 0 | 0 | 0 | 0 |
| 234 | 0 | 0 | 0 | 0 | 0 |
| 235 | 0 | 0 | 0 | 0 | 0 |
| 236 | 0 | 0 | 0 | 0.01 | 0.01 |
| 237 | 0 | 0 | 0.01 | 0 | 0.01 |
| 240 | 0.03 | 0.04 | 0.04 | 0.03 | 0.14 |
| 241 | 0 | 0 | 0 | 0 | 0 |
| 242 | 0 | 0.01 | 0 | 0 | 0.01 |
| 243 | 0 | 0 | 0 | 0 | 0 |
| 245 | 0.05 | 0.06 | 0.05 | 0.06 | 0.22 |
| TOTAL | 0.4 | 0.37 | 0.36 | 0.45 | 1.58 |

STATE: WASHINGTON

| ZIP CODE | QUARTER 1 | QUARTER 2 | QUARTER 3 | QUARTER 4 | TOTAL GRAMS |
|---|---|---|---|---|---|
| 980 | 0.02 | 0.02 | 0.03 | 0.02 | 0.09 |
| 981 | 0.13 | 0.11 | 0.2 | 0.16 | 0.6 |

| | | | | | | |
|---|---|---|---|---|---|---|
| | 982 | 0.02 | 0 | 0 | 0 | 0.02 |
| | 983 | 0 | 0 | 0.01 | 0 | 0.01 |
| | 984 | 0.02 | 0.03 | 0.03 | 0.03 | 0.11 |
| | 985 | 0 | 0 | 0 | 0 | 0 |
| | 986 | 0 | 0 | 0 | 0 | 0 |
| | 988 | 0 | 0 | 0 | 0 | 0 |
| | 989 | 0 | 0 | 0 | 0 | 0 |
| | 991 | 0 | 0 | 0 | 0 | 0 |
| | 992 | 0.04 | 0.04 | 0.05 | 0.05 | 0.18 |
| | 993 | 0 | 0 | 0 | 0 | 0 |
| TOTAL | | 0.23 | 0.2 | 0.32 | 0.26 | 1.01 |

STATE: WEST VIRGINIA

| ZIP CODE | | QUARTER 1 | QUARTER 2 | QUARTER 3 | QUARTER 4 | TOTAL GRAMS |
|---|---|---|---|---|---|---|
| | 253 | 0.02 | 0.02 | 0.03 | 0.01 | 0.08 |
| | 254 | 0 | 0 | 0 | 0 | 0 |
| | 256 | 0 | 0 | 0 | 0 | 0 |
| | 257 | 0 | 0 | 0.03 | 0.01 | 0.04 |
| | 258 | 0 | 0 | 0 | 0 | 0 |
| | 261 | 0 | 0.01 | 0.01 | 0.01 | 0.03 |
| | 265 | 0.03 | 0.02 | 0.02 | 0.03 | 0.1 |
| TOTAL | | 0.05 | 0.05 | 0.09 | 0.06 | 0.25 |

STATE: WISCONSIN

| ZIP CODE | | QUARTER 1 | QUARTER 2 | QUARTER 3 | QUARTER 4 | TOTAL GRAMS |
|---|---|---|---|---|---|---|
| | 530 | 0 | 0.01 | 0.01 | 0.01 | 0.03 |
| | 531 | 0 | 0 | 0 | 0 | 0 |
| | 532 | 0.72 | 0.73 | 0.05 | 0.74 | 2.24 |
| | 535 | 0 | 0 | 0 | 0 | 0 |
| | 537 | 0.02 | 0.03 | 0.01 | 0.06 | 0.12 |
| | 539 | 0 | 0 | 0 | 0 | 0 |
| | 540 | 0 | 0 | 0 | 0 | 0 |
| | 541 | 0 | 0 | 0 | 0 | 0 |
| | 542 | 0 | 0 | 0 | 0 | 0 |
| | 543 | 0 | 0.01 | 0.01 | 0.01 | 0.03 |

| | | | | | | |
|---|---|---|---|---|---|---|
| | 544 | 0 | 0 | 0 | 0 | 0 |
| | 546 | 0.01 | 0 | 0 | 0.01 | 0.02 |
| | 547 | 0 | 0 | 0 | 0 | 0 |
| | 549 | 0.01 | 0.01 | 0.01 | 0.02 | 0.05 |
| TOTAL | | 0.76 | 0.79 | 0.09 | 0.85 | 2.49 |

STATE: WYOMING

| ZIP CODE | | QUARTER 1 | QUARTER 2 | QUARTER 3 | QUARTER 4 | TOTAL GRAMS |
|---|---|---|---|---|---|---|
| | 820 | 0 | 0 | 0.02 | 0.02 | 0.04 |
| | 822 | 0 | 0 | 0 | 0 | 0 |
| | 824 | 0 | 0 | 0 | 0 | 0 |
| | 826 | 0 | 0 | 0 | 0 | 0 |
| | 827 | 0 | 0 | 0 | 0 | 0 |
| | 829 | 0 | 0 | 0 | 0.01 | 0.01 |
| | 831 | 0 | 0 | 0 | 0 | 0 |
| TOTAL | | 0 | 0 | 0.02 | 0.03 | 0.05 |

DRUG CODE: 9780 DRUG NAME: TAPENTADOL
STATE: ALABAMA

| ZIP CODE | | QUARTER 1 | QUARTER 2 | QUARTER 3 | QUARTER 4 | TOTAL GRAMS |
|---|---|---|---|---|---|---|
| | 350 | 7,809.50 | 8,395 | 7,573 | 7,725.50 | 31,503 |
| | 351 | 4,541.50 | 4,358.50 | 4,476 | 3,995 | 17,371 |
| | 352 | 4,557 | 4,194.50 | 3,996 | 3,764.50 | 16,512 |
| | 354 | 4,533 | 3,703 | 3,478.50 | 3,340 | 15,054.50 |
| | 355 | 3,180.50 | 3,158.50 | 2,805.50 | 2,903.50 | 12,048 |
| | 356 | 4,295.50 | 4,050 | 3,746.50 | 3,941 | 16,033 |
| | 357 | 2,122.50 | 1,964 | 1,787 | 1,654.50 | 7,528 |
| | 358 | 2,662.50 | 2,480 | 2,523 | 2,276 | 9,941.50 |
| | 359 | 2,571 | 2,574.50 | 2,664 | 2,292.50 | 10,102 |
| | 360 | 5,150 | 4,357 | 4,398.50 | 3,933 | 17,838.50 |
| | 361 | 2,585 | 2,579.50 | 2,208 | 2,663 | 10,035.50 |
| | 362 | 1,844.50 | 1,874 | 1,778 | 1,682.50 | 7,179 |
| | 363 | 1,577 | 1,681 | 1,191 | 1,238 | 5,687 |
| | 364 | 633 | 593.5 | 454.5 | 394 | 2,075 |
| | 365 | 4,978.50 | 4,369.50 | 4,062.50 | 3,766 | 17,176.50 |

| | | | | | | |
|---|---|---|---|---|---|---|
| | 366 | 2,880.50 | 2,805.50 | 2,918 | 2,491 | 11,095 |
| | 367 | 598 | 684 | 476 | 514 | 2,272 |
| | 368 | 2,288 | 2,291.50 | 1,797.50 | 2,009.50 | 8,386.50 |
| | 369 | 223 | 181 | 141 | 178 | 723 |
| TOTAL | | 59,030.50 | 56,294.50 | 52,474.50 | 50,761.50 | 218,561 |
| STATE: | ALASKA | | | | | |
| ZIP CODE | QUARTER 1 | | QUARTER 2 | QUARTER 3 | QUARTER 4 | TOTAL GRAMS |
| | 995 | 1,081 | 1,159.50 | 1,281 | 1,312.50 | 4,834 |
| | 996 | 809.5 | 809.5 | 851.5 | 857 | 3,327.50 |
| | 997 | 144 | 122.5 | 117.5 | 165.5 | 549.5 |
| | 998 | 51 | 61 | 54 | 69 | 235 |
| | 999 | 10 | 15 | 15 | 5 | 45 |
| TOTAL | | 2,095.50 | 2,167.50 | 2,319 | 2,409 | 8,991 |
| STATE: | ARIZONA | | | | | |
| ZIP CODE | QUARTER 1 | | QUARTER 2 | QUARTER 3 | QUARTER 4 | TOTAL GRAMS |
| | 850 | 6,252.50 | 6,126 | 4,716.50 | 7,425 | 24,520 |
| | 851 | 1,262.50 | 1,079 | 971 | 984.5 | 4,297 |
| | 852 | 16,105.50 | 15,137 | 13,440.50 | 13,806 | 58,489 |
| | 853 | 7,090 | 6,465.50 | 5,916 | 5,940.50 | 25,412 |
| | 855 | 289 | 436 | 401 | 415.5 | 1,541.50 |
| | 856 | 942.5 | 660.5 | 809 | 877.5 | 3,289.50 |
| | 857 | 4,136 | 4,039.50 | 4,323.50 | 4,779 | 17,278 |
| | 859 | 128.5 | 116.5 | 53.5 | 43.5 | 342 |
| | 860 | 313 | 192.5 | 207.5 | 273.5 | 986.5 |
| | 863 | 977 | 1,119 | 1,040.50 | 880.5 | 4,017 |
| | 864 | 689 | 374.5 | 359 | 528 | 1,950.50 |
| TOTAL | | 38,185.50 | 35,746 | 32,238 | 35,953.50 | 142,123 |
| STATE: | ARKANSAS | | | | | |
| ZIP CODE | QUARTER 1 | | QUARTER 2 | QUARTER 3 | QUARTER 4 | TOTAL GRAMS |
| | 716 | 727.5 | 584.5 | 645 | 662.5 | 2,619.50 |
| | 717 | 1,231.50 | 1,142.50 | 944 | 902.5 | 4,220.50 |
| | 718 | 334 | 302.5 | 298.5 | 281 | 1,216 |

| | | | | | | |
|---|---|---|---|---|---|---|
| | 719 | 1,110.50 | 1,146.50 | 1,193 | 995.5 | 4,445.50 |
| | 720 | 4,088 | 3,769.50 | 3,434 | 3,923.50 | 15,215 |
| | 721 | 2,841.50 | 2,765 | 2,320 | 2,513.50 | 10,440 |
| | 722 | 1,230 | 1,403.50 | 1,511 | 1,294 | 5,438.50 |
| | 723 | 450 | 330.5 | 285 | 277.5 | 1,343 |
| | 724 | 1,618 | 1,736.50 | 1,305.50 | 1,614 | 6,274 |
| | 725 | 990.5 | 886 | 775 | 754.5 | 3,406 |
| | 726 | 1,203.50 | 1,159.50 | 1,037.50 | 1,068.50 | 4,469 |
| | 727 | 4,352 | 4,204 | 3,240 | 3,177.50 | 14,973.50 |
| | 728 | 581 | 437 | 326 | 418.5 | 1,762.50 |
| | 729 | 2,535 | 2,523 | 1,743.50 | 1,310 | 8,111.50 |
| TOTAL | | 23,293 | 22,390.50 | 19,058 | 19,193 | 83,934.50 |

STATE: CALIFORNIA

| ZIP CODE | QUARTER 1 | QUARTER 2 | QUARTER 3 | QUARTER 4 | TOTAL GRAMS |
|---|---|---|---|---|---|
| 900 | 2,382 | 2,434 | 2,524 | 2,532.50 | 9,872.50 |
| 902 | 2,613.50 | 2,635.50 | 2,356 | 2,313 | 9,918 |
| 903 | 117.5 | 82 | 47.5 | 79 | 326 |
| 904 | 1,429.50 | 1,831.50 | 1,507.50 | 1,770.50 | 6,539 |
| 905 | 228.5 | 270.5 | 252 | 353 | 1,104 |
| 906 | 1,211 | 1,234.50 | 806 | 880.5 | 4,132 |
| 907 | 1,487.50 | 1,362 | 1,355 | 1,219.50 | 5,424 |
| 908 | 913 | 959.5 | 761 | 801 | 3,434.50 |
| 910 | 647 | 775.5 | 617 | 661.5 | 2,701 |
| 911 | 453 | 549.5 | 568 | 598.5 | 2,169 |
| 912 | 287 | 255.5 | 223.5 | 266.5 | 1,032.50 |
| 913 | 4,384 | 4,256 | 4,734.50 | 4,576.50 | 17,951 |
| 914 | 1,241.50 | 1,265 | 1,092 | 1,349 | 4,947.50 |
| 915 | 196 | 158.5 | 150 | 213 | 717.5 |
| 916 | 153 | 167.5 | 95 | 98 | 513.5 |
| 917 | 9,358 | 9,152 | 8,375 | 6,664.50 | 33,549.50 |
| 918 | 50 | 75 | 72.5 | 68.5 | 266 |
| 919 | 1,236 | 1,268.50 | 1,143.50 | 1,310.50 | 4,958.50 |
| 920 | 8,760.50 | 8,735 | 8,613.50 | 9,041 | 35,150 |
| 921 | 3,449 | 3,430 | 3,090 | 3,305 | 13,274 |
| 922 | 2,856 | 2,775 | 2,411 | 2,274.50 | 10,316.50 |

| | | | | | | |
|---|---|---|---|---|---|---|
| | 923 | 3,837 | 4,020.50 | 3,651.50 | 3,443 | 14,952 |
| | 924 | 1,222 | 1,000 | 926 | 875 | 4,023 |
| | 925 | 7,171.50 | 7,174 | 5,863 | 7,242.50 | 27,451 |
| | 926 | 6,783.50 | 6,645.50 | 6,334.95 | 6,122.90 | 25,886.85 |
| | 927 | 1,062.50 | 1,102 | 989 | 839 | 3,992.50 |
| | 928 | 4,010.50 | 4,328 | 4,126 | 4,026 | 16,490.50 |
| | 930 | 2,651 | 2,376 | 2,261 | 2,670 | 9,958 |
| | 931 | 498 | 447.5 | 376.5 | 424 | 1,746 |
| | 932 | 4,046.50 | 3,330.50 | 3,675.50 | 3,138 | 14,190.50 |
| | 933 | 2,316 | 2,231 | 1,929.50 | 1,780.50 | 8,257 |
| | 934 | 2,514.50 | 2,718 | 2,193 | 2,388.50 | 9,814 |
| | 935 | 1,753.50 | 2,120 | 1,987 | 1,895.50 | 7,756 |
| | 936 | 2,769.50 | 2,827 | 2,284 | 2,255 | 10,135.50 |
| | 937 | 2,580 | 2,753.50 | 2,936 | 3,147 | 11,416.50 |
| | 939 | 1,308 | 1,378.50 | 1,371.50 | 1,372 | 5,430 |
| | 940 | 1,432 | 1,124.50 | 1,027 | 1,283.50 | 4,867 |
| | 941 | 693.5 | 504.5 | 646 | 572.5 | 2,416.50 |
| | 943 | 57 | 84 | 39 | 36 | 216 |
| | 944 | 203.5 | 170 | 151 | 254.5 | 779 |
| | 945 | 4,368 | 4,281.50 | 3,764.50 | 4,224 | 16,638 |
| | 946 | 321 | 358.5 | 249.5 | 269.5 | 1,198.50 |
| | 947 | 223 | 184 | 142 | 134.5 | 683.5 |
| | 948 | 74 | 81 | 94 | 74.5 | 323.5 |
| | 949 | 327.5 | 320.5 | 406 | 404.5 | 1,458.50 |
| | 950 | 1,017 | 985 | 915.5 | 968 | 3,885.50 |
| | 951 | 415 | 427.5 | 570.5 | 507 | 1,920 |
| | 952 | 710.5 | 612 | 452.5 | 466.5 | 2,241.50 |
| | 953 | 2,013.50 | 2,190 | 2,036 | 2,121 | 8,360.50 |
| | 954 | 920 | 787 | 708 | 805 | 3,220 |
| | 955 | 113.5 | 67 | 21 | 29 | 230.5 |
| | 956 | 3,064.50 | 3,202.50 | 2,751.50 | 2,487 | 11,505.50 |
| | 957 | 1,199 | 938 | 972.5 | 1,024.50 | 4,134 |
| | 958 | 1,812 | 1,776.50 | 1,177 | 1,156.50 | 5,922 |
| | 959 | 2,337.50 | 2,680.50 | 2,502.50 | 2,174 | 9,694.50 |
| | 960 | 1,220 | 1,497.50 | 1,262.50 | 1,207.50 | 5,187.50 |
| | 961 | 58 | 35 | 56 | 50 | 199 |
| TOTAL | | 110,557.50 | 110,431.50 | 101,643.45 | 102,244.40 | 424,876.85 |

STATE: COLORADO

| ZIP CODE | QUARTER 1 | QUARTER 2 | QUARTER 3 | QUARTER 4 | TOTAL GRAMS |
|---|---|---|---|---|---|
| 800 | 4,166 | 3,932.50 | 3,648 | 3,642.50 | 15,389 |
| 801 | 5,037 | 5,290 | 5,296.50 | 5,723 | 21,346.50 |
| 802 | 3,580.50 | 4,007.50 | 3,580.50 | 3,841.50 | 15,010 |
| 803 | 631 | 751.5 | 718 | 935 | 3,035.50 |
| 804 | 688 | 691.5 | 582.5 | 691 | 2,653 |
| 805 | 3,324.50 | 3,393 | 3,273.50 | 3,223 | 13,214 |
| 806 | 1,791.50 | 1,833 | 1,777 | 1,499 | 6,900.50 |
| 807 | 471.5 | 380.5 | 313 | 223 | 1,388 |
| 808 | 578 | 585.5 | 597 | 633.5 | 2,394 |
| 809 | 3,853 | 4,300 | 3,889 | 3,800.50 | 15,842.50 |
| 810 | 1,553.50 | 1,699.50 | 2,085.50 | 2,327 | 7,665.50 |
| 811 | 76 | 118.5 | 74 | 125.5 | 394 |
| 812 | 534.5 | 788 | 567 | 646.5 | 2,536 |
| 813 | 374 | 323 | 253 | 295 | 1,245 |
| 814 | 81 | 60 | 52 | 103 | 296 |
| 815 | 526 | 553 | 543 | 578 | 2,200 |
| 816 | 201 | 182 | 223.5 | 136.5 | 743 |
| TOTAL | 27,467 | 28,889 | 27,473 | 28,423.50 | 112,252.50 |

STATE: CONNECTICUT

| ZIP CODE | QUARTER 1 | QUARTER 2 | QUARTER 3 | QUARTER 4 | TOTAL GRAMS |
|---|---|---|---|---|---|
| 60 | 1,872 | 1,787.50 | 1,646 | 1,531.50 | 6,837 |
| 61 | 358.5 | 309 | 316 | 344.5 | 1,328 |
| 62 | 649 | 592.5 | 497.5 | 656 | 2,395 |
| 63 | 1,882.50 | 1,725.50 | 1,433.50 | 1,722 | 6,763.50 |
| 64 | 3,541 | 3,338.50 | 3,172.50 | 2,964 | 13,016 |
| 65 | 1,025.50 | 1,000 | 874 | 686 | 3,585.50 |
| 66 | 659.5 | 533.5 | 374 | 239 | 1,806 |
| 67 | 997 | 941.5 | 934.5 | 938 | 3,811 |
| 68 | 1,460.50 | 1,402.50 | 1,441 | 1,316 | 5,620 |
| 69 | 212 | 265 | 249 | 161 | 887 |

| | | | | | | |
|---|---|---|---|---|---|---|
| TOTAL | | 12,657.50 | 11,895.50 | 10,938 | 10,558 | 46,049 |

STATE: DELAWARE

| ZIP CODE | | QUARTER 1 | QUARTER 2 | QUARTER 3 | QUARTER 4 | TOTAL GRAMS |
|---|---|---|---|---|---|---|
| | 197 | 2,605.50 | 2,595 | 2,524.50 | 2,594.50 | 10,319.50 |
| | 198 | 1,655.50 | 1,720.50 | 1,803 | 1,758.50 | 6,937.50 |
| | 199 | 3,378 | 3,116 | 3,076.50 | 2,846.50 | 12,417 |
| TOTAL | | 7,639 | 7,431.50 | 7,404 | 7,199.50 | 29,674 |

STATE: DISTRICT OF COLUMBIA

| ZIP CODE | | QUARTER 1 | QUARTER 2 | QUARTER 3 | QUARTER 4 | TOTAL GRAMS |
|---|---|---|---|---|---|---|
| | 200 | 1,209.50 | 1,086 | 886 | 788 | 3,969.50 |
| | 204 | 0 | 25 | 30 | 72 | 127 |
| TOTAL | | 1,209.50 | 1,111 | 916 | 860 | 4,096.50 |

STATE: FLORIDA

| ZIP CODE | | QUARTER 1 | QUARTER 2 | QUARTER 3 | QUARTER 4 | TOTAL GRAMS |
|---|---|---|---|---|---|---|
| | 320 | 3,779.50 | 3,972.50 | 3,438 | 3,344 | 14,534 |
| | 321 | 2,795 | 2,594 | 2,745 | 2,560 | 10,694 |
| | 322 | 6,649.50 | 6,364.50 | 5,903 | 5,787 | 24,704 |
| | 323 | 3,292.50 | 3,009 | 3,065 | 2,616 | 11,982.50 |
| | 324 | 2,042 | 2,018.50 | 1,638.50 | 1,688 | 7,387 |
| | 325 | 5,403 | 4,308.50 | 4,169 | 3,755.50 | 17,636 |
| | 326 | 1,921 | 1,947 | 1,927.50 | 2,045.50 | 7,841 |
| | 327 | 5,627.50 | 5,609.50 | 5,196 | 5,130.50 | 21,563.50 |
| | 328 | 3,020 | 3,038 | 2,670.50 | 2,410 | 11,138.50 |
| | 329 | 4,048.50 | 3,896 | 4,077 | 3,919 | 15,940.50 |
| | 330 | 3,525 | 3,590 | 3,017.50 | 2,826 | 12,958.50 |
| | 331 | 705.5 | 809.5 | 549.5 | 435 | 2,499.50 |
| | 333 | 2,257 | 2,182 | 1,957 | 1,964 | 8,360 |
| | 334 | 8,529 | 7,831 | 7,094 | 6,969.50 | 30,423.50 |
| | 335 | 6,598.50 | 6,366.50 | 5,714.50 | 5,807.50 | 24,487 |
| | 336 | 5,076 | 4,424 | 3,698.50 | 3,505 | 16,703.50 |
| | 337 | 4,223 | 3,591 | 3,027.50 | 3,220 | 14,061.50 |

| | | | | | |
|---|---|---|---|---|---|
| 338 | 3,440.50 | 3,406 | 2,965.50 | 3,307.50 | 13,119.50 |
| 339 | 11,730.50 | 10,822.50 | 9,221 | 9,619 | 41,393 |
| 341 | 4,764.50 | 4,260.50 | 3,752.50 | 4,181.50 | 16,959 |
| 342 | 7,013 | 6,186.50 | 5,217.50 | 5,049.50 | 23,466.50 |
| 344 | 2,571.50 | 2,672.50 | 2,512 | 2,486 | 10,242 |
| 346 | 4,371.50 | 4,191.50 | 3,507 | 3,764 | 15,834 |
| 347 | 3,168 | 3,267.50 | 2,768.50 | 2,713.50 | 11,917.50 |
| 349 | 2,872.50 | 2,664 | 2,418.50 | 2,456 | 10,411 |
| TOTAL | 109,424.50 | 103,022.50 | 92,250.50 | 91,559.50 | 396,257 |

STATE: GEORGIA

| ZIP CODE | QUARTER 1 | QUARTER 2 | QUARTER 3 | QUARTER 4 | TOTAL GRAMS |
|---|---|---|---|---|---|
| 300 | 12,561.50 | 12,113 | 11,174.50 | 11,510.50 | 47,359.50 |
| 301 | 11,351 | 11,542.50 | 9,731 | 10,329.50 | 42,954 |
| 302 | 5,267.50 | 5,018 | 4,854 | 5,016 | 20,155.50 |
| 303 | 3,060.50 | 3,202.50 | 2,852 | 3,061 | 12,176 |
| 304 | 3,874.50 | 4,076 | 3,700.50 | 3,903.50 | 15,554.50 |
| 305 | 4,335 | 4,161.50 | 3,270.50 | 3,396.50 | 15,163.50 |
| 306 | 2,207 | 2,010 | 1,673.50 | 1,691.50 | 7,582 |
| 307 | 1,413 | 1,718.50 | 1,443.50 | 1,428 | 6,003 |
| 308 | 996.5 | 750.5 | 652 | 582.5 | 2,981.50 |
| 309 | 880 | 962.5 | 1,030.50 | 1,014.50 | 3,887.50 |
| 310 | 3,665 | 4,018 | 3,811.50 | 3,432 | 14,926.50 |
| 312 | 1,108.50 | 1,323.50 | 1,274.50 | 1,153.50 | 4,860 |
| 313 | 4,835 | 4,832 | 4,354.50 | 3,987 | 18,008.50 |
| 314 | 2,576 | 2,367.50 | 2,139 | 2,265.50 | 9,348 |
| 315 | 7,692 | 7,777 | 6,746.50 | 6,566.50 | 28,782 |
| 316 | 1,471.50 | 1,462.50 | 1,643 | 1,533 | 6,110 |
| 317 | 1,765 | 1,756 | 1,602.50 | 1,459 | 6,582.50 |
| 318 | 222.5 | 215 | 222.5 | 200.5 | 860.5 |
| 319 | 1,355.50 | 1,176 | 1,015 | 1,021.50 | 4,568 |
| 398 | 477.5 | 415.5 | 443.5 | 431 | 1,767.50 |
| TOTAL | 71,115 | 70,898 | 63,634.50 | 63,983 | 269,630.50 |

STATE: HAWAII

| ZIP CODE | QUARTER 1 | QUARTER 2 | QUARTER 3 | QUARTER 4 | TOTAL GRAMS |
|---|---|---|---|---|---|
| 967 | 1,342 | 1,322 | 1,255 | 1,734 | 5,653 |
| 968 | 1,523 | 1,962 | 1,873.50 | 1,582.50 | 6,941 |
| TOTAL | 2,865 | 3,284 | 3,128.50 | 3,316.50 | 12,594 |

STATE: IDAHO

| ZIP CODE | QUARTER 1 | QUARTER 2 | QUARTER 3 | QUARTER 4 | TOTAL GRAMS |
|---|---|---|---|---|---|
| 832 | 1,172 | 1,167.50 | 1,129 | 1,606.50 | 5,075 |
| 833 | 345 | 404 | 564 | 462.5 | 1,775.50 |
| 834 | 946 | 859.5 | 705 | 774 | 3,284.50 |
| 835 | 274 | 94 | 137 | 92 | 597 |
| 836 | 1,885 | 1,569 | 1,544 | 1,701.50 | 6,699.50 |
| 837 | 1,061.50 | 923.5 | 949.5 | 1,042.50 | 3,977 |
| 838 | 742.5 | 640 | 680.5 | 655 | 2,718 |
| TOTAL | 6,426 | 5,657.50 | 5,709 | 6,334 | 24,126.50 |

STATE: ILLINOIS

| ZIP CODE | QUARTER 1 | QUARTER 2 | QUARTER 3 | QUARTER 4 | TOTAL GRAMS |
|---|---|---|---|---|---|
| 600 | 6,719 | 6,416.50 | 5,879.50 | 5,615.50 | 24,630.50 |
| 601 | 4,228.50 | 3,674.50 | 3,104.50 | 3,057.50 | 14,065 |
| 602 | 268.5 | 190.5 | 239 | 376 | 1,074 |
| 603 | 515.5 | 653.5 | 388.5 | 354 | 1,911.50 |
| 604 | 5,311 | 4,635.50 | 4,202 | 4,293 | 18,441.50 |
| 605 | 4,186.50 | 4,054.50 | 3,943 | 3,680.50 | 15,864.50 |
| 606 | 3,274 | 3,077 | 2,633 | 2,785 | 11,769 |
| 607 | 532 | 448 | 415.5 | 442.5 | 1,838 |
| 608 | 45.5 | 35 | 93.5 | 136.5 | 310.5 |
| 609 | 718.5 | 717 | 707.5 | 786.5 | 2,929.50 |
| 610 | 1,012.50 | 1,026 | 914.5 | 852.5 | 3,805.50 |
| 611 | 1,665.50 | 1,290.50 | 1,295.50 | 1,301.50 | 5,553 |
| 612 | 229.5 | 239 | 145.5 | 178.5 | 792.5 |
| 613 | 301 | 227.5 | 167 | 132 | 827.5 |
| 614 | 195 | 134 | 195 | 149 | 673 |

| | | | | | | |
|---|---|---|---|---|---|---|
| | 615 | 440.5 | 390 | 495 | 422 | 1,747.50 |
| | 616 | 339.5 | 292 | 324 | 309.5 | 1,265 |
| | 617 | 1,089 | 785.5 | 1,006 | 904.5 | 3,785 |
| | 618 | 1,767 | 1,859 | 2,233.50 | 1,841.50 | 7,701 |
| | 619 | 456 | 399.5 | 373.5 | 245 | 1,474 |
| | 620 | 1,743.50 | 1,641 | 1,380.50 | 1,136 | 5,901 |
| | 622 | 3,626.50 | 3,506 | 3,050 | 3,259 | 13,441.50 |
| | 623 | 291.5 | 401 | 347.5 | 364 | 1,404 |
| | 624 | 397 | 352.5 | 237.5 | 342 | 1,329 |
| | 625 | 698.5 | 738 | 937.5 | 1,093 | 3,467 |
| | 626 | 216 | 198 | 238.5 | 133 | 785.5 |
| | 627 | 273 | 243 | 237 | 150 | 903 |
| | 628 | 444 | 547 | 476 | 464 | 1,931 |
| | 629 | 465 | 568.5 | 508.5 | 536.5 | 2,078.50 |
| TOTAL | | 41,449.50 | 38,740 | 36,168.50 | 35,340.50 | 151,698.50 |

| STATE: | INDIANA | | | | | |
|---|---|---|---|---|---|---|
| ZIP CODE | | QUARTER 1 | QUARTER 2 | QUARTER 3 | QUARTER 4 | TOTAL GRAMS |
| | 460 | 1,683.50 | 1,803.50 | 1,697 | 1,366 | 6,550 |
| | 461 | 2,821.50 | 2,665 | 2,303.50 | 2,775.50 | 10,565.50 |
| | 462 | 2,754 | 2,497.50 | 2,546.50 | 2,549.50 | 10,347.50 |
| | 463 | 3,725 | 4,494 | 4,499 | 4,662.50 | 17,380.50 |
| | 464 | 193 | 262 | 368 | 423 | 1,246 |
| | 465 | 2,394.50 | 2,155.50 | 2,617.50 | 2,359 | 9,526.50 |
| | 466 | 661.5 | 735.5 | 811.5 | 636.5 | 2,845 |
| | 467 | 3,304 | 3,176 | 3,096.50 | 3,038.50 | 12,615 |
| | 468 | 3,764 | 3,939 | 3,965.50 | 4,081.50 | 15,750 |
| | 469 | 1,531.50 | 1,676 | 1,414 | 1,536 | 6,157.50 |
| | 470 | 351 | 451.5 | 364.5 | 334.5 | 1,501.50 |
| | 471 | 1,667.50 | 1,418.50 | 1,088 | 896 | 5,070 |
| | 472 | 787 | 672.5 | 684 | 675 | 2,818.50 |
| | 473 | 1,059.50 | 929 | 884.5 | 680 | 3,553 |
| | 474 | 670 | 660.5 | 727 | 753 | 2,810.50 |
| | 475 | 869 | 728 | 563 | 620 | 2,780 |
| | 476 | 1,065.50 | 1,031 | 939 | 928.5 | 3,964 |
| | 477 | 1,544 | 1,410.50 | 1,260 | 1,276 | 5,490.50 |

| | | | | | | |
|---|---|---|---|---|---|---|
| | 478 | 6,315 | 5,986.50 | 5,323.50 | 5,881.50 | 23,506.50 |
| | 479 | 1,261.50 | 1,207.50 | 1,395 | 1,089.50 | 4,953.50 |
| TOTAL | | 38,422.50 | 37,899.50 | 36,547.50 | 36,562 | 149,431.50 |

| STATE: | IOWA | | | | | |
|---|---|---|---|---|---|---|
| ZIP CODE | | QUARTER 1 | QUARTER 2 | QUARTER 3 | QUARTER 4 | TOTAL GRAMS |
| | 500 | 1,768 | 1,754 | 1,308.50 | 1,614 | 6,444.50 |
| | 501 | 1,288 | 1,225.50 | 943.5 | 1,115 | 4,572 |
| | 502 | 1,793.50 | 1,908.50 | 1,549.50 | 1,620.50 | 6,872 |
| | 503 | 3,509 | 2,918.50 | 2,287 | 2,439 | 11,153.50 |
| | 504 | 105 | 86 | 101 | 20 | 312 |
| | 505 | 251 | 349.5 | 223 | 276.5 | 1,100 |
| | 506 | 467.5 | 409 | 417 | 437.5 | 1,731 |
| | 507 | 394 | 346 | 282.5 | 325.5 | 1,348 |
| | 508 | 280.5 | 254 | 176 | 254 | 964.5 |
| | 510 | 175 | 240 | 201 | 198 | 814 |
| | 511 | 134 | 97 | 203.5 | 376 | 810.5 |
| | 512 | 165.5 | 102.5 | 73.5 | 110 | 451.5 |
| | 513 | 78 | 55 | 50 | 124 | 307 |
| | 514 | 15 | 15 | 26 | 15 | 71 |
| | 515 | 646.5 | 384.5 | 398 | 544.5 | 1,973.50 |
| | 516 | 273 | 289 | 221 | 257.5 | 1,040.50 |
| | 520 | 430 | 414 | 389 | 384 | 1,617 |
| | 521 | 21 | 15 | 20 | 62.5 | 118.5 |
| | 522 | 159 | 134 | 122 | 159 | 574 |
| | 523 | 484.5 | 431.5 | 425 | 387.5 | 1,728.50 |
| | 524 | 632 | 757 | 599 | 584 | 2,572 |
| | 525 | 354 | 398.5 | 346 | 398.5 | 1,497 |
| | 526 | 65 | 35 | 86.5 | 89.5 | 276 |
| | 527 | 200.5 | 111 | 124.5 | 125 | 561 |
| | 528 | 283.5 | 191 | 136 | 181 | 791.5 |
| TOTAL | | 13,973 | 12,921 | 10,709 | 12,098 | 49,701 |

| STATE: | KANSAS | | | | | |
|---|---|---|---|---|---|---|
| ZIP CODE | | QUARTER 1 | QUARTER 2 | QUARTER 3 | QUARTER 4 | TOTAL GRAMS |

| | | | | | | |
|---|---|---|---|---|---|---|
| | 660 | 2,189 | 2,079 | 1,955 | 2,091.50 | 8,314.50 |
| | 661 | 197 | 265 | 326.5 | 301.5 | 1,090 |
| | 662 | 2,446 | 2,188 | 1,964 | 2,040.50 | 8,638.50 |
| | 664 | 277 | 331 | 312.5 | 307.5 | 1,228 |
| | 665 | 292 | 256.5 | 261.5 | 229 | 1,039 |
| | 666 | 1,184 | 951 | 857.5 | 976.5 | 3,969 |
| | 667 | 534.5 | 470.5 | 398.5 | 337.5 | 1,741 |
| | 668 | 73 | 107 | 107 | 142.5 | 429.5 |
| | 669 | 24 | 64.5 | 21 | 0 | 109.5 |
| | 670 | 273 | 155 | 171 | 148 | 747 |
| | 671 | 56 | 113 | 108 | 120 | 397 |
| | 672 | 828 | 689 | 867.5 | 608 | 2,992.50 |
| | 673 | 254 | 252.5 | 189.5 | 209 | 905 |
| | 674 | 337 | 321 | 297.5 | 384 | 1,339.50 |
| | 675 | 159.5 | 185 | 105 | 184 | 633.5 |
| | 676 | 195.5 | 188.5 | 206.5 | 204 | 794.5 |
| | 677 | 152 | 102 | 94 | 70 | 418 |
| | 678 | 47 | 52 | 46 | 67 | 212 |
| | 679 | 10 | 19 | 0 | 15 | 44 |
| TOTAL | | 9,528.50 | 8,789.50 | 8,288.50 | 8,435.50 | 35,042 |

STATE: KENTUCKY

| ZIP CODE | | QUARTER 1 | QUARTER 2 | QUARTER 3 | QUARTER 4 | TOTAL GRAMS |
|---|---|---|---|---|---|---|
| | 400 | 625.5 | 672.5 | 640 | 549 | 2,487 |
| | 401 | 378 | 363.5 | 251.5 | 252 | 1,245 |
| | 402 | 2,717 | 3,000.50 | 3,121.50 | 2,624 | 11,463 |
| | 403 | 733.5 | 702 | 870 | 795.5 | 3,101 |
| | 404 | 294 | 317.5 | 404 | 462 | 1,477.50 |
| | 405 | 640 | 627 | 750.5 | 606 | 2,623.50 |
| | 406 | 53 | 57.5 | 100 | 76 | 286.5 |
| | 407 | 158 | 214 | 155 | 114.5 | 641.5 |
| | 408 | 20 | 15 | 0 | 0 | 35 |
| | 409 | 111.5 | 85 | 52 | 40.5 | 289 |
| | 410 | 2,308 | 2,328.50 | 2,263.50 | 2,136.50 | 9,036.50 |
| | 411 | 283.5 | 295.5 | 224.5 | 278.5 | 1,082 |
| | 412 | 106 | 90 | 46.5 | 71 | 313.5 |

| | | | | | | |
|---|---|---|---|---|---|---|
| | 413 | 35 | 15 | 15 | 22.5 | 87.5 |
| | 415 | 292 | 233 | 171 | 148 | 844 |
| | 416 | 175 | 239 | 254 | 262 | 930 |
| | 417 | 66.5 | 65 | 40.5 | 63.5 | 235.5 |
| | 418 | 54 | 46.5 | 61.5 | 59 | 221 |
| | 420 | 1,578 | 1,658 | 1,665.50 | 1,832 | 6,733.50 |
| | 421 | 533.5 | 426.5 | 443 | 432 | 1,835 |
| | 422 | 832.5 | 908 | 1,132.50 | 821.5 | 3,694.50 |
| | 423 | 1,577.50 | 1,368.50 | 1,327.50 | 1,417.50 | 5,691 |
| | 424 | 822 | 752 | 716.5 | 689.5 | 2,980 |
| | 425 | 20 | 20 | 39 | 56 | 135 |
| | 426 | 23 | 34 | 33 | 45 | 135 |
| | 427 | 724.5 | 619.5 | 552 | 635 | 2,531 |
| TOTAL | | 15,161.50 | 15,153.50 | 15,330 | 14,489 | 60,134 |

STATE: LOUISIANA

| ZIP CODE | | QUARTER 1 | QUARTER 2 | QUARTER 3 | QUARTER 4 | TOTAL GRAMS |
|---|---|---|---|---|---|---|
| | 700 | 1,605.50 | 1,811 | 1,753.50 | 1,808 | 6,978 |
| | 701 | 152 | 254 | 176 | 152.5 | 734.5 |
| | 703 | 543 | 547 | 415 | 660.5 | 2,165.50 |
| | 704 | 3,250 | 2,769.50 | 2,636.50 | 2,937.50 | 11,593.50 |
| | 705 | 1,811.50 | 1,677 | 2,019 | 1,893.50 | 7,401 |
| | 706 | 332.5 | 394 | 409 | 579.5 | 1,715 |
| | 707 | 2,747 | 2,664 | 2,868 | 3,255.50 | 11,534.50 |
| | 708 | 1,568.50 | 1,456.50 | 1,508.50 | 1,662.50 | 6,196 |
| | 710 | 300.5 | 298 | 302.5 | 338.5 | 1,239.50 |
| | 711 | 1,161.50 | 1,226 | 1,003.50 | 1,093.50 | 4,484.50 |
| | 712 | 411.5 | 491.5 | 434.5 | 287 | 1,624.50 |
| | 713 | 984 | 931 | 737 | 831.5 | 3,483.50 |
| | 714 | 436 | 504.5 | 334 | 366 | 1,640.50 |
| TOTAL | | 15,303.50 | 15,024 | 14,597 | 15,866 | 60,790.50 |

STATE: MAINE

| ZIP CODE | | QUARTER 1 | QUARTER 2 | QUARTER 3 | QUARTER 4 | TOTAL GRAMS |
|---|---|---|---|---|---|---|
| | 39 | 34 | 20 | 5 | 23 | 82 |

| | | | | | | |
|---|---|---|---|---|---|---|
| | 40 | 525.5 | 492 | 396 | 391 | 1,804.50 |
| | 41 | 10 | 74 | 80 | 33 | 197 |
| | 42 | 409 | 360 | 301 | 287 | 1,357 |
| | 43 | 54 | 66.5 | 76 | 69 | 265.5 |
| | 44 | 55 | 84 | 72 | 47 | 258 |
| | 45 | 98.5 | 125 | 89 | 100.5 | 413 |
| | 46 | 0 | 6 | 12 | 7.5 | 25.5 |
| | 47 | 56.5 | 37 | 139 | 226 | 458.5 |
| | 48 | 30 | 44 | 51 | 9 | 134 |
| | 49 | 176 | 179 | 121 | 164.5 | 640.5 |
| TOTAL | | 1,448.50 | 1,487.50 | 1,342 | 1,357.50 | 5,635.50 |

STATE: MARYLAND

| ZIP CODE | | QUARTER 1 | QUARTER 2 | QUARTER 3 | QUARTER 4 | TOTAL GRAMS |
|---|---|---|---|---|---|---|
| | 206 | 2,958.50 | 2,995 | 2,773 | 2,697 | 11,423.50 |
| | 207 | 3,070.50 | 2,965 | 2,681 | 2,577.50 | 11,294 |
| | 208 | 3,777.50 | 3,583.50 | 3,077 | 2,915.50 | 13,353.50 |
| | 209 | 568 | 435 | 388 | 431.5 | 1,822.50 |
| | 210 | 7,398 | 7,430.50 | 6,779 | 6,361.50 | 27,969 |
| | 211 | 2,971 | 2,914 | 2,696.50 | 2,750 | 11,331.50 |
| | 212 | 4,245.50 | 4,565.50 | 4,113.50 | 3,967 | 16,891.50 |
| | 214 | 996 | 1,033 | 934 | 897.5 | 3,860.50 |
| | 215 | 598 | 678 | 643 | 598 | 2,517 |
| | 216 | 1,438.50 | 1,452 | 1,383.50 | 1,613.50 | 5,887.50 |
| | 217 | 5,132 | 4,866 | 4,684 | 4,465 | 19,147 |
| | 218 | 681.5 | 684.5 | 585 | 784 | 2,735 |
| | 219 | 469.5 | 378.5 | 435.5 | 488.5 | 1,772 |
| TOTAL | | 34,304.50 | 33,980.50 | 31,173 | 30,546.50 | 130,004.50 |

STATE: MASSACHUSETTS

| ZIP CODE | | QUARTER 1 | QUARTER 2 | QUARTER 3 | QUARTER 4 | TOTAL GRAMS |
|---|---|---|---|---|---|---|
| | 10 | 1,200.50 | 1,015 | 992 | 1,068.50 | 4,276 |
| | 11 | 469.5 | 490.5 | 439 | 372 | 1,771 |
| | 12 | 109 | 120 | 92 | 75 | 396 |
| | 13 | 129.5 | 92 | 161 | 107 | 489.5 |

| | | | | | | |
|---|---|---|---|---|---|---|
| | 14 | 387 | 446.5 | 354.5 | 360 | 1,548 |
| | 15 | 622 | 571 | 498 | 576 | 2,267 |
| | 16 | 169 | 252 | 242 | 278.5 | 941.5 |
| | 17 | 464 | 528 | 534.5 | 565.5 | 2,092 |
| | 18 | 9,896 | 10,406 | 10,198.50 | 9,475 | 39,975.50 |
| | 19 | 931 | 1,046 | 978 | 1,313 | 4,268 |
| | 20 | 372 | 375 | 289 | 462 | 1,498 |
| | 21 | 701 | 617.5 | 842 | 880.5 | 3,041 |
| | 22 | 76 | 48 | 39.5 | 61.5 | 225 |
| | 23 | 925 | 911 | 735.5 | 907.5 | 3,479 |
| | 24 | 396.5 | 341 | 317 | 280 | 1,334.50 |
| | 25 | 172 | 197 | 237.5 | 172 | 778.5 |
| | 26 | 264 | 253 | 261.5 | 137.5 | 916 |
| | 27 | 1,954.50 | 1,885 | 2,084 | 1,856 | 7,779.50 |
| TOTAL | | 19,238.50 | 19,594.50 | 19,295.50 | 18,947.50 | 77,076 |

| STATE: | MICHIGAN | | | | | |
|---|---|---|---|---|---|---|
| ZIP CODE | | QUARTER 1 | QUARTER 2 | QUARTER 3 | QUARTER 4 | TOTAL GRAMS |
| | 480 | 2,647.50 | 2,571 | 2,628.50 | 2,799 | 10,646 |
| | 481 | 3,505 | 3,584.50 | 3,482 | 3,712.50 | 14,284 |
| | 482 | 331.5 | 269 | 310 | 368.5 | 1,279 |
| | 483 | 2,302 | 2,279 | 2,520.50 | 2,548.50 | 9,650 |
| | 484 | 555.5 | 537 | 690 | 628 | 2,410.50 |
| | 485 | 391.5 | 384.5 | 451.5 | 639 | 1,866.50 |
| | 486 | 762 | 682 | 765.5 | 780.5 | 2,990 |
| | 487 | 307.5 | 243 | 359.5 | 351.5 | 1,261.50 |
| | 488 | 1,752 | 1,741.50 | 1,597.50 | 1,708 | 6,799 |
| | 489 | 579 | 487 | 426 | 388.5 | 1,880.50 |
| | 490 | 2,591 | 2,331 | 2,334.50 | 2,326 | 9,582.50 |
| | 491 | 447 | 427.5 | 462.5 | 428 | 1,765 |
| | 492 | 723 | 676 | 646 | 638 | 2,683 |
| | 493 | 888.5 | 1,019.50 | 1,026.50 | 999.5 | 3,934 |
| | 494 | 1,025.50 | 977.5 | 889 | 729 | 3,621 |
| | 495 | 1,551 | 1,319.50 | 1,246.50 | 1,260.50 | 5,377.50 |
| | 496 | 741 | 738 | 780 | 680.5 | 2,939.50 |

| | | | | | | |
|---|---|---|---|---|---|---|
| | 497 | 442 | 630.5 | 700 | 552 | 2,324.50 |
| | 498 | 241 | 253 | 221 | 255 | 970 |
| | 499 | 75 | 100 | 95 | 105 | 375 |
| TOTAL | | 21,858.50 | 21,251 | 21,632 | 21,897.50 | 86,639 |

| STATE: | MINNESOTA | | | | | |
|---|---|---|---|---|---|---|
| ZIP CODE | | QUARTER 1 | QUARTER 2 | QUARTER 3 | QUARTER 4 | TOTAL GRAMS |
| | 550 | 689 | 632.5 | 592 | 660 | 2,573.50 |
| | 551 | 849 | 925 | 853.5 | 925.5 | 3,553 |
| | 553 | 444.5 | 480 | 529.5 | 571 | 2,025 |
| | 554 | 567.5 | 579.5 | 620 | 876 | 2,643 |
| | 556 | 5 | 10 | 0 | 15 | 30 |
| | 557 | 34 | 15 | 43 | 18 | 110 |
| | 558 | 52 | 51 | 47.5 | 39 | 189.5 |
| | 559 | 199 | 149.5 | 130 | 179.5 | 658 |
| | 560 | 222 | 171.5 | 260 | 168 | 821.5 |
| | 561 | 276.5 | 210 | 275 | 273.5 | 1,035 |
| | 562 | 224.5 | 138 | 176 | 62.5 | 601 |
| | 563 | 207 | 180.5 | 178 | 236 | 801.5 |
| | 564 | 81 | 119.5 | 169 | 250.5 | 620 |
| | 565 | 116 | 240 | 215.5 | 92.5 | 664 |
| | 566 | 89 | 99.5 | 75.5 | 102 | 366 |
| | 567 | 260.5 | 225.5 | 240.5 | 184 | 910.5 |
| TOTAL | | 4,316.50 | 4,227 | 4,405 | 4,653 | 17,601.50 |

| STATE: | MISSISSIPPI | | | | | |
|---|---|---|---|---|---|---|
| ZIP CODE | | QUARTER 1 | QUARTER 2 | QUARTER 3 | QUARTER 4 | TOTAL GRAMS |
| | 386 | 1,132.50 | 1,197 | 1,317 | 1,284.50 | 4,931 |
| | 387 | 261 | 264 | 334 | 247 | 1,106 |
| | 388 | 1,206 | 1,296.50 | 1,085 | 1,111.50 | 4,699 |
| | 389 | 193 | 216 | 201 | 167 | 777 |
| | 390 | 1,096 | 1,039.50 | 1,140 | 1,036 | 4,311.50 |
| | 391 | 1,048.50 | 883.5 | 788 | 762.5 | 3,482.50 |
| | 392 | 976 | 1,116.50 | 1,122.50 | 907 | 4,122 |

| | | | | | | |
|---|---|---|---|---|---|---|
| | 393 | 822 | 764.5 | 788 | 731.5 | 3,106 |
| | 394 | 1,667.50 | 1,601 | 1,498 | 1,563.50 | 6,330 |
| | 395 | 3,792 | 3,130 | 2,690 | 2,184.50 | 11,796.50 |
| | 396 | 345.5 | 323.5 | 306.5 | 230.5 | 1,206 |
| | 397 | 601.5 | 692.5 | 564.5 | 559.5 | 2,418 |
| TOTAL | | 13,141.50 | 12,524.50 | 11,834.50 | 10,785 | 48,285.50 |

STATE: MISSOURI

| ZIP CODE | | QUARTER 1 | QUARTER 2 | QUARTER 3 | QUARTER 4 | TOTAL GRAMS |
|---|---|---|---|---|---|---|
| | 630 | 3,539.50 | 3,075.50 | 2,771.50 | 2,892.50 | 12,279 |
| | 631 | 5,592.50 | 5,155.50 | 5,233.50 | 5,311.50 | 21,293 |
| | 633 | 3,076 | 2,922.50 | 3,024.50 | 2,793 | 11,816 |
| | 634 | 229.5 | 212.5 | 265 | 141.5 | 848.5 |
| | 635 | 191 | 228.5 | 316 | 294 | 1,029.50 |
| | 636 | 337 | 400 | 368.5 | 467.5 | 1,573 |
| | 637 | 1,067 | 912 | 1,019 | 1,089.50 | 4,087.50 |
| | 638 | 428.5 | 473 | 465 | 492.5 | 1,859 |
| | 639 | 324 | 268.5 | 125 | 194 | 911.5 |
| | 640 | 2,304.50 | 2,185 | 1,800 | 1,848.50 | 8,138 |
| | 641 | 1,618 | 1,845.50 | 1,328 | 1,267.50 | 6,059 |
| | 644 | 151 | 193.5 | 173.5 | 121.5 | 639.5 |
| | 645 | 749 | 810.5 | 641.5 | 647.5 | 2,848.50 |
| | 646 | 92 | 106.5 | 74 | 131 | 403.5 |
| | 647 | 504 | 518.5 | 400 | 470 | 1,892.50 |
| | 648 | 988.5 | 1,021 | 778.5 | 737.5 | 3,525.50 |
| | 650 | 288.5 | 434 | 497.5 | 452 | 1,672 |
| | 651 | 660 | 658 | 460 | 533 | 2,311 |
| | 652 | 413.5 | 422.5 | 443.5 | 516 | 1,795.50 |
| | 653 | 487 | 495 | 344.5 | 289.5 | 1,616 |
| | 654 | 257 | 232.5 | 287 | 391 | 1,167.50 |
| | 655 | 251.5 | 350.5 | 232.5 | 344.5 | 1,179 |
| | 656 | 664 | 589.5 | 447.5 | 473 | 2,174 |
| | 657 | 690.5 | 694 | 636.5 | 569 | 2,590 |
| | 658 | 947 | 1,028 | 953.5 | 929 | 3,857.50 |
| TOTAL | | 25,851 | 25,232.50 | 23,086 | 23,396.50 | 97,566 |

STATE: MONTANA

| ZIP CODE | QUARTER 1 | QUARTER 2 | QUARTER 3 | QUARTER 4 | TOTAL GRAMS |
|---|---|---|---|---|---|
| 590 | 201.5 | 158 | 199.5 | 202 | 761 |
| 591 | 471.5 | 380.5 | 400 | 336 | 1,588 |
| 592 | 12 | 3 | 20 | 0 | 35 |
| 593 | 104 | 88 | 98 | 44.5 | 334.5 |
| 594 | 481 | 272 | 232 | 236 | 1,221 |
| 595 | 179.5 | 76 | 77.5 | 66 | 399 |
| 596 | 93 | 98 | 135 | 130 | 456 |
| 597 | 385 | 359 | 344 | 437 | 1,525 |
| 598 | 179 | 174 | 146.5 | 264.5 | 764 |
| 599 | 95 | 60 | 106 | 90 | 351 |
| TOTAL | 2,201.50 | 1,668.50 | 1,758.50 | 1,806 | 7,434.50 |

STATE: NEBRASKA

| ZIP CODE | QUARTER 1 | QUARTER 2 | QUARTER 3 | QUARTER 4 | TOTAL GRAMS |
|---|---|---|---|---|---|
| 680 | 989.5 | 853 | 806.5 | 997 | 3,646 |
| 681 | 4,385 | 3,804 | 4,105 | 4,103.50 | 16,397.50 |
| 683 | 259 | 201 | 152 | 194 | 806 |
| 684 | 179 | 223 | 141 | 142 | 685 |
| 685 | 1,517.50 | 1,519 | 1,705.50 | 1,597 | 6,339 |
| 686 | 424 | 377 | 374 | 362 | 1,537 |
| 687 | 654.5 | 425.5 | 388 | 408 | 1,876 |
| 688 | 1,334 | 1,205 | 967.5 | 1,283.50 | 4,790 |
| 689 | 216 | 177 | 184 | 269.5 | 846.5 |
| 690 | 18 | 37 | 41.5 | 62 | 158.5 |
| 691 | 385 | 467 | 361.5 | 341 | 1,554.50 |
| 692 | 0 | 0 | 0 | 20 | 20 |
| 693 | 84.5 | 53.5 | 5 | 21 | 164 |
| TOTAL | 10,446 | 9,342 | 9,231.50 | 9,800.50 | 38,820 |

STATE: NEVADA

| ZIP CODE | QUARTER 1 | QUARTER 2 | QUARTER 3 | QUARTER 4 | TOTAL GRAMS |
|---|---|---|---|---|---|
| 890 | 3,445.50 | 3,745 | 3,968 | 3,206 | 14,364.50 |

| | | | | | | |
|---|---|---|---|---|---|---|
| | 891 | 7,168.50 | 6,763.50 | 5,958.50 | 7,332.50 | 27,223 |
| | 893 | 36 | 18 | 9 | 9 | 72 |
| | 894 | 841 | 587 | 654.5 | 630 | 2,712.50 |
| | 895 | 713 | 720.5 | 630.5 | 726 | 2,790 |
| | 897 | 239.5 | 162 | 249 | 317 | 967.5 |
| | 898 | 150 | 168.5 | 190.5 | 167 | 676 |
| TOTAL | | 12,593.50 | 12,164.50 | 11,660 | 12,387.50 | 48,805.50 |

STATE: NEW HAMPSHIRE

| ZIP CODE | | QUARTER 1 | QUARTER 2 | QUARTER 3 | QUARTER 4 | TOTAL GRAMS |
|---|---|---|---|---|---|---|
| | 30 | 2,109.50 | 2,181 | 1,905 | 1,762 | 7,957.50 |
| | 31 | 567.5 | 722.5 | 676.5 | 812 | 2,778.50 |
| | 32 | 1,189 | 1,064.50 | 1,080.50 | 823 | 4,157 |
| | 33 | 675 | 626.5 | 572.5 | 571.5 | 2,445.50 |
| | 34 | 137 | 79.5 | 81.5 | 57 | 355 |
| | 35 | 290 | 296 | 354 | 406 | 1,346 |
| | 37 | 120 | 147.5 | 61 | 195.5 | 524 |
| | 38 | 2,485.50 | 2,618 | 2,333.50 | 2,415 | 9,852 |
| TOTAL | | 7,573.50 | 7,735.50 | 7,064.50 | 7,042 | 29,415.50 |

STATE: NEW JERSEY

| ZIP CODE | | QUARTER 1 | QUARTER 2 | QUARTER 3 | QUARTER 4 | TOTAL GRAMS |
|---|---|---|---|---|---|---|
| | 70 | 7,081 | 6,715 | 6,146.50 | 7,015.50 | 26,958 |
| | 71 | 449.5 | 513 | 502 | 529 | 1,993.50 |
| | 72 | 432.5 | 441 | 416 | 289 | 1,578.50 |
| | 73 | 343 | 413.5 | 393.5 | 310 | 1,460 |
| | 74 | 2,285.50 | 2,303 | 2,136.50 | 2,261.50 | 8,986.50 |
| | 75 | 258.5 | 272.5 | 157 | 278 | 966 |
| | 76 | 2,404 | 2,270 | 2,201.50 | 2,259.50 | 9,135 |
| | 77 | 4,607.50 | 4,323.50 | 4,572.50 | 4,271 | 17,774.50 |
| | 78 | 1,829.50 | 2,016.50 | 1,907 | 1,918.50 | 7,671.50 |
| | 79 | 1,213 | 987.5 | 1,039 | 1,197.50 | 4,437 |
| | 80 | 10,429.50 | 10,605.50 | 10,337 | 8,739 | 40,111 |
| | 81 | 572.5 | 726.5 | 657 | 665.5 | 2,621.50 |

| | | | | | |
|---|---|---|---|---|---|
| 82 | 1,964 | 2,151.50 | 2,052 | 2,313.50 | 8,481 |
| 83 | 1,866 | 1,877.50 | 1,978 | 1,951.50 | 7,673 |
| 84 | 45 | 44 | 36.5 | 18 | 143.5 |
| 85 | 1,975.50 | 2,002.50 | 1,920 | 1,689.50 | 7,587.50 |
| 86 | 1,942.50 | 1,825.50 | 1,656.50 | 1,596.50 | 7,021 |
| 87 | 3,574.50 | 3,420.50 | 3,022.50 | 3,279.50 | 13,297 |
| 88 | 4,931.50 | 4,746.50 | 4,628.50 | 5,066.50 | 19,373 |
| 89 | 564.5 | 421 | 489 | 482.5 | 1,957 |
| TOTAL | 48,769.50 | 48,076.50 | 46,248.50 | 46,131.50 | 189,226 |

STATE: NEW MEXICO

| ZIP CODE | QUARTER 1 | QUARTER 2 | QUARTER 3 | QUARTER 4 | TOTAL GRAMS |
|---|---|---|---|---|---|
| 870 | 360 | 251.5 | 245.5 | 319.5 | 1,176.50 |
| 871 | 1,381.50 | 1,204 | 1,442 | 1,402.50 | 5,430 |
| 873 | 0 | 12 | 0 | 6 | 18 |
| 874 | 227 | 186 | 143 | 153 | 709 |
| 875 | 312 | 254 | 238.5 | 257 | 1,061.50 |
| 877 | 73.5 | 64 | 34 | 40 | 211.5 |
| 878 | 30 | 80 | 10 | 39 | 159 |
| 879 | 0 | 0 | 24 | 16 | 40 |
| 880 | 632 | 540 | 546 | 525 | 2,243 |
| 881 | 59 | 46 | 52 | 72 | 229 |
| 882 | 372 | 370 | 276 | 226 | 1,244 |
| 883 | 80 | 86 | 41 | 17 | 224 |
| 884 | 9 | 36 | 24 | 12 | 81 |
| TOTAL | 3,536 | 3,129.50 | 3,076 | 3,085 | 12,826.50 |

STATE: NEW YORK

| ZIP CODE | QUARTER 1 | QUARTER 2 | QUARTER 3 | QUARTER 4 | TOTAL GRAMS |
|---|---|---|---|---|---|
| 100 | 4,403.50 | 4,129 | 3,811 | 3,728 | 16,071.50 |
| 101 | 120.5 | 78 | 31.5 | 43 | 273 |
| 102 | 0 | 10 | 5 | 0 | 15 |
| 103 | 2,623 | 2,571 | 2,715.50 | 2,472.50 | 10,382 |
| 104 | 1,690 | 1,704 | 1,677 | 1,506.50 | 6,577.50 |
| 105 | 1,451 | 1,669 | 1,416 | 1,522.50 | 6,058.50 |

| | | | | | |
|---|---|---|---|---|---|
| 106 | 87.5 | 154.5 | 181.5 | 130 | 553.5 |
| 107 | 451 | 533 | 467.5 | 433 | 1,884.50 |
| 108 | 289 | 183 | 250 | 160.5 | 882.5 |
| 109 | 1,871 | 1,732 | 1,919 | 1,779 | 7,301 |
| 110 | 2,201 | 1,873.50 | 1,768 | 1,656 | 7,498.50 |
| 111 | 1,284.50 | 1,105 | 1,011 | 1,207.50 | 4,608 |
| 112 | 4,285.50 | 4,124.50 | 4,043.50 | 4,066 | 16,519.50 |
| 113 | 3,291 | 3,082.50 | 2,834.50 | 2,562 | 11,770 |
| 114 | 991 | 954 | 930 | 1,035.50 | 3,910.50 |
| 115 | 4,159.50 | 4,258 | 4,074.50 | 4,248.50 | 16,740.50 |
| 116 | 218.5 | 166 | 220.5 | 99 | 704 |
| 117 | 13,772 | 13,884.50 | 13,368.50 | 13,258 | 54,283 |
| 118 | 485.5 | 591.5 | 757 | 667.5 | 2,501.50 |
| 119 | 1,392 | 1,636.50 | 1,918 | 1,586 | 6,532.50 |
| 120 | 1,057 | 670.5 | 843.5 | 732.5 | 3,303.50 |
| 121 | 272.5 | 317 | 413.5 | 309 | 1,312 |
| 122 | 1,069 | 1,053.50 | 933 | 1,155 | 4,210.50 |
| 123 | 342 | 373 | 226 | 244 | 1,185 |
| 124 | 588.5 | 719.5 | 506 | 552 | 2,366 |
| 125 | 1,862 | 1,873 | 2,072.50 | 1,857 | 7,664.50 |
| 126 | 342 | 241.5 | 148 | 171 | 902.5 |
| 127 | 511.5 | 461 | 501.5 | 435.5 | 1,909.50 |
| 128 | 1,364 | 1,258 | 1,259.50 | 1,234.50 | 5,116 |
| 129 | 826 | 873.5 | 740 | 965 | 3,404.50 |
| 130 | 5,373.50 | 5,063 | 5,086 | 4,884 | 20,406.50 |
| 131 | 1,321 | 1,306.50 | 1,194 | 1,349 | 5,170.50 |
| 132 | 2,131.50 | 2,251.50 | 1,930 | 2,096.50 | 8,409.50 |
| 133 | 2,111 | 2,170 | 1,939 | 1,891.50 | 8,111.50 |
| 134 | 2,034 | 2,071.50 | 1,768.50 | 2,010.50 | 7,884.50 |
| 135 | 1,105 | 1,173.50 | 860 | 833 | 3,971.50 |
| 136 | 3,567 | 3,426 | 3,310.50 | 3,303.50 | 13,607 |
| 137 | 639.5 | 797.5 | 652 | 876.5 | 2,965.50 |
| 138 | 873.5 | 974 | 964 | 996 | 3,807.50 |
| 139 | 1,066.50 | 987.5 | 1,077 | 1,111 | 4,242 |
| 140 | 6,302 | 5,782.50 | 5,785.50 | 5,491.50 | 23,361.50 |
| 141 | 2,907 | 2,747 | 2,712 | 2,260 | 10,626 |
| 142 | 7,074.50 | 6,786 | 6,427 | 6,052 | 26,339.50 |

| | | | | | | |
|---|---|---|---|---|---|---|
| | 143 | 1,471 | 1,336.50 | 1,032 | 1,032.50 | 4,872 |
| | 144 | 1,695 | 1,766 | 1,619 | 1,656.50 | 6,736.50 |
| | 145 | 1,610 | 1,674.50 | 1,550 | 1,677 | 6,511.50 |
| | 146 | 2,578.50 | 2,528 | 2,449.50 | 2,333.50 | 9,889.50 |
| | 147 | 665 | 621.5 | 689.5 | 491.5 | 2,467.50 |
| | 148 | 1,952 | 1,751 | 1,777.50 | 1,732.50 | 7,213 |
| | 149 | 209 | 202 | 215.5 | 224 | 850.5 |
| TOTAL | | 99,988 | 97,696 | 94,081.50 | 92,119 | 383,884.50 |

STATE: NORTH CAROLINA

| ZIP CODE | | QUARTER 1 | QUARTER 2 | QUARTER 3 | QUARTER 4 | TOTAL GRAMS |
|---|---|---|---|---|---|---|
| | 270 | 5,233.50 | 5,134.50 | 4,281.50 | 4,532 | 19,181.50 |
| | 271 | 4,528.50 | 4,752 | 4,410 | 4,712.50 | 18,403 |
| | 272 | 7,529.50 | 7,364 | 6,842 | 7,030.50 | 28,766 |
| | 273 | 4,621 | 4,362.50 | 3,884 | 4,271.50 | 17,139 |
| | 274 | 2,953.50 | 2,904 | 2,667 | 2,669 | 11,193.50 |
| | 275 | 11,443 | 10,910 | 10,599 | 10,444.50 | 43,396.50 |
| | 276 | 3,468 | 3,877.50 | 3,854.50 | 4,273.50 | 15,473.50 |
| | 277 | 2,048.50 | 1,972.50 | 2,088 | 2,025 | 8,134 |
| | 278 | 4,172 | 4,050 | 3,644 | 3,702 | 15,568 |
| | 279 | 1,631 | 1,778.50 | 1,570.50 | 1,498.50 | 6,478.50 |
| | 280 | 7,874.50 | 7,316 | 6,743.50 | 6,558 | 28,492 |
| | 281 | 6,155.50 | 5,789.50 | 5,521.50 | 5,779 | 23,245.50 |
| | 282 | 4,554 | 4,412.50 | 4,013 | 4,025.50 | 17,005 |
| | 283 | 8,298 | 8,329.50 | 7,439.50 | 5,918.50 | 29,985.50 |
| | 284 | 5,925.50 | 6,336 | 6,576.50 | 5,876 | 24,714 |
| | 285 | 8,084.50 | 7,818 | 7,643 | 6,858.50 | 30,404 |
| | 286 | 8,736.50 | 8,867 | 7,893 | 7,938.50 | 33,435 |
| | 287 | 2,478.50 | 2,674.50 | 2,630.50 | 2,888 | 10,671.50 |
| | 288 | 980.5 | 1,048 | 1,016 | 914.5 | 3,959 |
| | 289 | 231 | 219.5 | 159.5 | 272.5 | 882.5 |
| TOTAL | | 100,947 | 99,916 | 93,476.50 | 92,188 | 386,527.50 |

STATE: NORTH DAKOTA

| ZIP CODE | | QUARTER 1 | QUARTER 2 | QUARTER 3 | QUARTER 4 | TOTAL GRAMS |
|---|---|---|---|---|---|---|

| | | | | | |
|---|---|---|---|---|---|
| 580 | 244 | 200 | 156 | 221.5 | 821.5 |
| 581 | 124.5 | 207 | 224.5 | 87.5 | 643.5 |
| 582 | 396.5 | 375.5 | 266 | 301.5 | 1,339.50 |
| 583 | 20 | 25 | 30 | 0 | 75 |
| 584 | 85.5 | 86.5 | 115 | 101.5 | 388.5 |
| 585 | 1,169 | 1,094.50 | 1,070.50 | 1,104 | 4,438 |
| 586 | 154.5 | 135.5 | 179.5 | 212 | 681.5 |
| 587 | 339 | 363 | 242 | 362 | 1,306 |
| 588 | 15 | 24 | 15 | 28 | 82 |
| TOTAL | 2,548 | 2,511 | 2,298.50 | 2,418 | 9,775.50 |

STATE: OHIO

| ZIP CODE | QUARTER 1 | QUARTER 2 | QUARTER 3 | QUARTER 4 | TOTAL GRAMS |
|---|---|---|---|---|---|
| 430 | 4,167.50 | 4,078.50 | 3,599 | 3,237 | 15,082 |
| 431 | 2,571 | 2,435 | 2,488 | 2,098 | 9,592 |
| 432 | 7,283.78 | 6,236.45 | 6,015.45 | 6,307.83 | 25,843.51 |
| 433 | 496.5 | 511 | 617 | 595.5 | 2,220 |
| 434 | 1,086 | 1,238 | 1,158 | 1,251 | 4,733 |
| 435 | 3,316 | 3,048.50 | 3,040.50 | 3,077 | 12,482 |
| 436 | 5,741 | 5,615.50 | 4,987 | 5,467.50 | 21,811 |
| 437 | 577 | 472 | 420.5 | 345.5 | 1,815 |
| 438 | 361 | 286 | 246.5 | 200.5 | 1,094 |
| 439 | 1,199 | 1,274 | 1,400.50 | 1,216 | 5,089.50 |
| 440 | 6,304 | 6,008.50 | 5,029.50 | 5,216.50 | 22,558.50 |
| 441 | 4,576.50 | 4,399 | 3,537 | 3,560 | 16,072.50 |
| 442 | 2,680 | 2,421.50 | 2,208 | 2,226.50 | 9,536 |
| 443 | 873 | 646.5 | 675.5 | 613 | 2,808 |
| 444 | 1,430 | 1,235.50 | 1,258.50 | 1,225.50 | 5,149.50 |
| 445 | 986 | 791.5 | 770.5 | 712 | 3,260 |
| 446 | 2,789 | 3,053.50 | 2,547 | 2,766.50 | 11,156 |
| 447 | 1,301.50 | 1,297.50 | 1,268.50 | 1,284.50 | 5,152 |
| 448 | 955 | 749.5 | 785.5 | 821 | 3,311 |
| 449 | 292.5 | 310.5 | 268 | 214.5 | 1,085.50 |
| 450 | 4,018 | 4,080 | 3,590.50 | 3,438.50 | 15,127 |
| 451 | 1,540 | 1,368 | 1,477.50 | 1,133 | 5,518.50 |
| 452 | 3,534 | 3,525 | 3,053.50 | 2,876.50 | 12,989 |

| | | | | | | |
|---|---|---|---|---|---|---|
| | 453 | 1,950 | 1,955 | 1,770.50 | 1,566.50 | 7,242 |
| | 454 | 2,150.50 | 2,198.50 | 2,090 | 1,626.50 | 8,065.50 |
| | 455 | 426.5 | 283 | 341.5 | 364.5 | 1,415.50 |
| | 456 | 2,024.50 | 1,883 | 1,796 | 1,604 | 7,307.50 |
| | 457 | 409 | 402.5 | 227 | 348 | 1,386.50 |
| | 458 | 1,148 | 872 | 898 | 938.5 | 3,856.50 |
| TOTAL | | 66,186.78 | 62,675.45 | 57,564.95 | 56,331.83 | 242,759.01 |

STATE: OKLAHOMA

| ZIP CODE | | QUARTER 1 | QUARTER 2 | QUARTER 3 | QUARTER 4 | TOTAL GRAMS |
|---|---|---|---|---|---|---|
| | 730 | 5,304 | 5,036.50 | 4,989.50 | 4,578.50 | 19,908.50 |
| | 731 | 3,764.50 | 3,811.50 | 3,446.50 | 3,139 | 14,161.50 |
| | 734 | 1,539 | 1,373.50 | 1,022 | 948.5 | 4,883 |
| | 735 | 619 | 745.5 | 801 | 901.5 | 3,067 |
| | 736 | 36.5 | 68.5 | 67 | 175.5 | 347.5 |
| | 737 | 344.5 | 413 | 216.5 | 272.5 | 1,246.50 |
| | 738 | 99.5 | 81.5 | 55 | 69 | 305 |
| | 739 | 6 | 44 | 10 | 23 | 83 |
| | 740 | 6,300.50 | 5,871 | 5,172 | 5,478 | 22,821.50 |
| | 741 | 3,189 | 3,556 | 3,692.50 | 3,742.50 | 14,180 |
| | 743 | 587 | 636 | 515.5 | 514 | 2,252.50 |
| | 744 | 1,206.50 | 1,118 | 990 | 993.5 | 4,308 |
| | 745 | 258.5 | 371.5 | 275 | 274.5 | 1,179.50 |
| | 746 | 183 | 223 | 217 | 308.5 | 931.5 |
| | 747 | 69.5 | 87 | 177 | 152 | 485.5 |
| | 748 | 1,112 | 1,064 | 996.5 | 844 | 4,016.50 |
| | 749 | 191.5 | 218 | 197 | 134 | 740.5 |
| TOTAL | | 24,810.50 | 24,718.50 | 22,840 | 22,548.50 | 94,917.50 |

STATE: OREGON

| ZIP CODE | | QUARTER 1 | QUARTER 2 | QUARTER 3 | QUARTER 4 | TOTAL GRAMS |
|---|---|---|---|---|---|---|
| | 970 | 1,555 | 1,416.50 | 1,665.50 | 1,693.50 | 6,330.50 |
| | 971 | 325 | 340 | 423.5 | 242 | 1,330.50 |
| | 972 | 778.5 | 671.5 | 741 | 853 | 3,044 |
| | 973 | 1,189 | 1,062.50 | 1,193 | 1,257.50 | 4,702 |

| | | | | | | |
|---|---|---|---|---|---|---|
| | 974 | 1,147 | 1,504 | 1,417 | 1,225 | 5,293 |
| | 975 | 1,382 | 1,506.50 | 1,304.50 | 1,329 | 5,522 |
| | 976 | 184 | 209 | 157 | 194 | 744 |
| | 977 | 471 | 511 | 454 | 524.5 | 1,960.50 |
| | 978 | 274 | 154 | 133.5 | 141 | 702.5 |
| | 979 | 28 | 66 | 53 | 39 | 186 |
| TOTAL | | 7,333.50 | 7,441 | 7,542 | 7,498.50 | 29,815 |

| STATE: | PENNSYLVANIA | | | | | |
|---|---|---|---|---|---|---|
| ZIP CODE | | QUARTER 1 | QUARTER 2 | QUARTER 3 | QUARTER 4 | TOTAL GRAMS |
| | 150 | 1,814 | 1,968 | 1,990 | 2,113.50 | 7,885.50 |
| | 151 | 790 | 770.5 | 942 | 916 | 3,418.50 |
| | 152 | 835 | 640 | 393 | 468.5 | 2,336.50 |
| | 153 | 594 | 570.5 | 579 | 675.5 | 2,419 |
| | 154 | 204 | 203.5 | 211 | 239 | 857.5 |
| | 155 | 602 | 615 | 510.5 | 461 | 2,188.50 |
| | 156 | 1,071.50 | 773 | 570 | 525 | 2,939.50 |
| | 157 | 302.5 | 277 | 194 | 225 | 998.5 |
| | 158 | 374 | 218 | 184 | 186.5 | 962.5 |
| | 159 | 938 | 1,200.50 | 1,118 | 901.5 | 4,158 |
| | 160 | 272 | 327.5 | 345.5 | 327.5 | 1,272.50 |
| | 161 | 780 | 809 | 653.5 | 722 | 2,964.50 |
| | 162 | 442.5 | 432 | 413.5 | 399.5 | 1,687.50 |
| | 163 | 640.5 | 692.5 | 635.5 | 574 | 2,542.50 |
| | 164 | 230 | 242 | 250 | 292 | 1,014 |
| | 165 | 641 | 804.5 | 848 | 779 | 3,072.50 |
| | 166 | 1,068 | 1,004.50 | 794.5 | 933.5 | 3,800.50 |
| | 167 | 15 | 84 | 107.5 | 62.5 | 269 |
| | 168 | 908 | 994.5 | 770.5 | 711.5 | 3,384.50 |
| | 169 | 145 | 105 | 110 | 106 | 466 |
| | 170 | 3,615.50 | 4,087.50 | 3,046.50 | 3,184.50 | 13,934 |
| | 171 | 1,042.50 | 1,025.50 | 919.5 | 997 | 3,984.50 |
| | 172 | 925.5 | 974.5 | 889.5 | 936 | 3,725.50 |
| | 173 | 2,764.50 | 3,272.50 | 2,925.50 | 3,035 | 11,997.50 |
| | 174 | 1,350 | 1,357 | 1,579 | 1,678 | 5,964 |
| | 175 | 381.5 | 360 | 303.5 | 349 | 1,394 |

| | | | | | | |
|---|---|---|---|---|---|---|
| | 176 | 634 | 620 | 472 | 534.5 | 2,260.50 |
| | 177 | 356.5 | 211 | 308 | 280 | 1,155.50 |
| | 178 | 356 | 318.5 | 371.5 | 189.5 | 1,235.50 |
| | 179 | 398.5 | 427.5 | 483 | 536.5 | 1,845.50 |
| | 180 | 6,051.50 | 5,891.50 | 5,422 | 6,085.50 | 23,450.50 |
| | 181 | 3,189.50 | 3,226 | 2,782.50 | 2,973.50 | 12,171.50 |
| | 182 | 556 | 812 | 869 | 1,613 | 3,850 |
| | 183 | 3,303 | 3,221.50 | 3,208.50 | 2,974.50 | 12,707.50 |
| | 184 | 1,312.50 | 1,416.50 | 1,479 | 1,387.50 | 5,595.50 |
| | 185 | 776 | 703 | 723 | 654.5 | 2,856.50 |
| | 186 | 357 | 342 | 344.5 | 276.5 | 1,320 |
| | 187 | 6,500.50 | 6,727.50 | 5,943.50 | 5,984 | 25,155.50 |
| | 188 | 228.5 | 161.5 | 137 | 122.5 | 649.5 |
| | 189 | 2,101.50 | 2,080.50 | 1,824 | 1,890 | 7,896 |
| | 190 | 8,142.50 | 8,186 | 8,302 | 7,762 | 32,392.50 |
| | 191 | 2,237.50 | 2,661.50 | 2,232 | 2,058 | 9,189 |
| | 193 | 2,125.50 | 2,239 | 2,107 | 2,385.50 | 8,857 |
| | 194 | 3,212 | 2,881 | 3,048 | 2,616 | 11,757 |
| | 195 | 1,544 | 1,578.50 | 1,281.50 | 1,353 | 5,757 |
| | 196 | 1,352.50 | 1,159.50 | 1,171.50 | 1,048.50 | 4,732 |
| TOTAL | | 67,481.50 | 68,673 | 63,793 | 64,523.50 | 264,471 |

STATE: PUERTO RICO

| ZIP CODE | | QUARTER 1 | QUARTER 2 | QUARTER 3 | QUARTER 4 | TOTAL GRAMS |
|---|---|---|---|---|---|---|
| | 006 | 156 | 124.5 | 112.5 | 58.5 | 451.5 |
| | 007 | 133.5 | 158.5 | 123.5 | 113.5 | 529 |
| | 009 | 615 | 707 | 593 | 478.5 | 2,393.50 |
| TOTAL | | 904.5 | 990 | 829 | 650.5 | 3,374 |

STATE: RHODE ISLAND

| ZIP CODE | | QUARTER 1 | QUARTER 2 | QUARTER 3 | QUARTER 4 | TOTAL GRAMS |
|---|---|---|---|---|---|---|
| | 028 | 1,442 | 1,623 | 1,481 | 1,439 | 5,985 |
| | 029 | 625.5 | 543.5 | 579 | 657 | 2,405 |
| TOTAL | | 2,067.50 | 2,166.50 | 2,060 | 2,096 | 8,390 |

| STATE: | SOUTH CAROLINA | | | | |
|---|---|---|---|---|---|
| ZIP CODE | QUARTER 1 | QUARTER 2 | QUARTER 3 | QUARTER 4 | TOTAL GRAMS |
| 290 | 3,348.50 | 2,850 | 2,571.50 | 2,934.50 | 11,704.50 |
| 291 | 2,520 | 2,924.50 | 2,689 | 2,769 | 10,902.50 |
| 292 | 3,389.50 | 3,332.50 | 3,168.50 | 3,071.50 | 12,962 |
| 293 | 3,999.50 | 3,653.50 | 3,465.50 | 3,961.50 | 15,080 |
| 294 | 5,064 | 5,414.50 | 5,151.50 | 4,808 | 20,438 |
| 295 | 6,286 | 6,371 | 6,189 | 5,670.50 | 24,516.50 |
| 296 | 12,128 | 12,487.50 | 12,023 | 12,770 | 49,408.50 |
| 297 | 2,545.50 | 2,663.50 | 2,730 | 2,288.50 | 10,227.50 |
| 298 | 2,045 | 2,105.50 | 1,960.50 | 2,263.50 | 8,374.50 |
| 299 | 2,946 | 2,753.50 | 2,503.50 | 2,193 | 10,396 |
| TOTAL | 44,272 | 44,556 | 42,452 | 42,730 | 174,010 |

| STATE: | SOUTH DAKOTA | | | | |
|---|---|---|---|---|---|
| ZIP CODE | QUARTER 1 | QUARTER 2 | QUARTER 3 | QUARTER 4 | TOTAL GRAMS |
| 570 | 716.5 | 620.5 | 531 | 702 | 2,570 |
| 571 | 1,737 | 1,680 | 1,367 | 1,272 | 6,056 |
| 572 | 300.5 | 294 | 256 | 242.5 | 1,093 |
| 573 | 344.5 | 279 | 327.5 | 324 | 1,275 |
| 574 | 598 | 579 | 539.5 | 545 | 2,261.50 |
| 575 | 135 | 137.5 | 115 | 130 | 517.5 |
| 576 | 136.5 | 155 | 135 | 125 | 551.5 |
| 577 | 1,243.50 | 1,099 | 1,050 | 1,143 | 4,535.50 |
| TOTAL | 5,211.50 | 4,844 | 4,321 | 4,483.50 | 18,860 |

| STATE: | TENNESSEE | | | | |
|---|---|---|---|---|---|
| ZIP CODE | QUARTER 1 | QUARTER 2 | QUARTER 3 | QUARTER 4 | TOTAL GRAMS |
| 370 | 6,622 | 6,688.50 | 6,384 | 6,319.50 | 26,014 |
| 371 | 5,878.50 | 5,655.50 | 5,556 | 5,212.50 | 22,302.50 |
| 372 | 1,787.50 | 1,542 | 1,661 | 1,872 | 6,862.50 |
| 373 | 3,434.50 | 3,449 | 3,421 | 3,928.50 | 14,233 |
| 374 | 2,098 | 1,909.50 | 2,028.50 | 2,278 | 8,314 |
| 376 | 2,095.50 | 2,029 | 1,852.50 | 2,115.50 | 8,092.50 |

| | | | | | |
|---|---|---|---|---|---|
| 377 | 888.5 | 815.5 | 742.5 | 740 | 3,186.50 |
| 378 | 1,542.50 | 1,449.50 | 1,470.50 | 1,492 | 5,954.50 |
| 379 | 1,327.50 | 1,233.50 | 1,153.50 | 999.5 | 4,714 |
| 380 | 1,407 | 1,139 | 1,464 | 1,531 | 5,541 |
| 381 | 1,434 | 1,038 | 1,270 | 1,482.50 | 5,224.50 |
| 382 | 428.5 | 351 | 558.5 | 753.5 | 2,091.50 |
| 383 | 1,658.50 | 1,312 | 1,398.50 | 1,814 | 6,183 |
| 384 | 1,150.50 | 1,082.50 | 993.5 | 915.5 | 4,142 |
| 385 | 1,130 | 1,094 | 1,134 | 1,001.50 | 4,359.50 |
| TOTAL | 32,883 | 30,788.50 | 31,088 | 32,455.50 | 127,215 |

STATE: TEXAS

| ZIP CODE | QUARTER 1 | QUARTER 2 | QUARTER 3 | QUARTER 4 | TOTAL GRAMS |
|---|---|---|---|---|---|
| 750 | 14,377.50 | 13,407.50 | 12,285 | 11,905 | 51,975 |
| 751 | 4,062 | 3,855.50 | 3,585.50 | 3,490 | 14,993 |
| 752 | 4,757.50 | 4,634.50 | 4,361 | 4,147.50 | 17,900.50 |
| 754 | 1,223.50 | 1,201 | 936 | 1,169 | 4,529.50 |
| 755 | 131 | 141 | 164.5 | 54 | 490.5 |
| 756 | 597.5 | 784 | 869.5 | 697.5 | 2,948.50 |
| 757 | 1,038 | 964.5 | 1,487 | 1,112.50 | 4,602 |
| 758 | 643 | 570 | 509.5 | 693 | 2,415.50 |
| 759 | 299 | 263.5 | 152.5 | 259 | 974 |
| 760 | 7,256 | 7,196.50 | 6,300 | 6,623.50 | 27,376 |
| 761 | 4,570.50 | 4,254 | 3,807 | 3,791.50 | 16,423 |
| 762 | 1,730 | 2,219 | 1,964 | 2,258 | 8,171 |
| 763 | 1,023.50 | 1,137.50 | 1,050.50 | 1,121.50 | 4,333 |
| 764 | 424.5 | 515.5 | 337 | 323 | 1,600 |
| 765 | 811 | 1,096 | 1,361.50 | 959.5 | 4,228 |
| 766 | 193 | 183 | 111.5 | 146 | 633.5 |
| 767 | 352 | 470.5 | 356.5 | 381 | 1,560 |
| 768 | 69 | 65.5 | 93 | 79 | 306.5 |
| 769 | 813 | 657.5 | 677 | 707 | 2,854.50 |
| 770 | 5,517 | 5,661 | 5,163.50 | 5,496 | 21,837.50 |
| 773 | 4,940.50 | 4,809.50 | 4,429.50 | 5,078.50 | 19,258 |
| 774 | 4,684.50 | 4,794.50 | 4,601 | 4,727.50 | 18,807.50 |

| | | | | | | |
|---|---|---|---|---|---|---|
| | 775 | 5,849.50 | 5,893.50 | 5,622 | 5,258 | 22,623 |
| | 776 | 1,006 | 1,052 | 785 | 874.5 | 3,717.50 |
| | 777 | 614.5 | 462 | 333 | 346.5 | 1,756 |
| | 778 | 755.5 | 884 | 743.5 | 857 | 3,240 |
| | 779 | 321.5 | 319 | 186.5 | 213 | 1,040 |
| | 780 | 1,484 | 1,584.50 | 1,387 | 1,579.50 | 6,035 |
| | 781 | 2,181 | 2,341 | 1,970 | 2,195.50 | 8,687.50 |
| | 782 | 11,111 | 10,877 | 9,507.50 | 10,130.50 | 41,626 |
| | 783 | 785 | 697 | 473.5 | 571 | 2,526.50 |
| | 784 | 1,992 | 1,833 | 1,341 | 1,603.50 | 6,769.50 |
| | 785 | 744.5 | 609.5 | 486 | 534 | 2,374 |
| | 786 | 4,145 | 4,189 | 3,512 | 4,076.50 | 15,922.50 |
| | 787 | 4,889.50 | 4,879.50 | 4,171 | 5,255.50 | 19,195.50 |
| | 788 | 262 | 344 | 250 | 176.5 | 1,032.50 |
| | 790 | 633 | 473.5 | 433 | 419.5 | 1,959 |
| | 791 | 1,063.50 | 1,341 | 1,080 | 1,399.50 | 4,884 |
| | 792 | 33 | 9 | 0 | 5 | 47 |
| | 793 | 134 | 117 | 148 | 90 | 489 |
| | 794 | 916.5 | 992 | 917 | 1,419 | 4,244.50 |
| | 795 | 50.5 | 24 | 27 | 28 | 129.5 |
| | 796 | 107.5 | 130.5 | 119 | 100.5 | 457.5 |
| | 797 | 310 | 401 | 282.5 | 299 | 1,292.50 |
| | 798 | 9 | 6 | 0 | 3 | 18 |
| | 799 | 2,898 | 2,637.50 | 2,562.50 | 2,792.50 | 10,890.50 |
| TOTAL | | 102,077.50 | 101,164 | 91,158.50 | 95,703 | 390,103 |

| STATE: | UTAH | | | | |
|---|---|---|---|---|---|
| ZIP CODE | QUARTER 1 | QUARTER 2 | QUARTER 3 | QUARTER 4 | TOTAL GRAMS |
| 840 | 7,117.50 | 6,217.50 | 6,253.50 | 5,415.50 | 25,004 |
| 841 | 2,503 | 2,223 | 1,935 | 1,852 | 8,513 |
| 843 | 487.5 | 590.5 | 575 | 557.5 | 2,210.50 |
| 844 | 1,130.50 | 834.5 | 1,075 | 1,068 | 4,108 |
| 845 | 787 | 733 | 707 | 732.5 | 2,959.50 |
| 846 | 1,196 | 1,139 | 988.5 | 1,122.50 | 4,446 |
| 847 | 2,375.50 | 1,846.50 | 1,926 | 1,501.50 | 7,649.50 |
| TOTAL | 15,597 | 13,584 | 13,460 | 12,249.50 | 54,890.50 |

STATE: VERMONT

| ZIP CODE | QUARTER 1 | QUARTER 2 | QUARTER 3 | QUARTER 4 | TOTAL GRAMS |
|---|---|---|---|---|---|
| 50 | 35 | 25 | 25 | 20 | 105 |
| 52 | 140 | 129 | 120 | 175 | 564 |
| 53 | 0 | 0 | 0 | 9 | 9 |
| 54 | 606.5 | 420 | 300.5 | 343.5 | 1,670.50 |
| 56 | 171 | 139 | 197 | 155 | 662 |
| 57 | 311 | 289.5 | 284 | 355.5 | 1,240 |
| 58 | 78 | 93.5 | 112 | 86 | 369.5 |
| TOTAL | 1,341.50 | 1,096 | 1,038.50 | 1,144 | 4,620 |

STATE: VIRGIN ISLANDS

| ZIP CODE | QUARTER 1 | QUARTER 2 | QUARTER 3 | QUARTER 4 | TOTAL GRAMS |
|---|---|---|---|---|---|
| 8 | 9 | 6 | 3 | 0 | 18 |
| TOTAL | 9 | 6 | 3 | 0 | 18 |

STATE: VIRGINIA

| ZIP CODE | QUARTER 1 | QUARTER 2 | QUARTER 3 | QUARTER 4 | TOTAL GRAMS |
|---|---|---|---|---|---|
| 201 | 5,539 | 5,244 | 4,669 | 4,521 | 19,973 |
| 220 | 2,477 | 2,309.50 | 2,319 | 1,590 | 8,695.50 |
| 221 | 2,005.50 | 2,000 | 1,757 | 1,628.50 | 7,391 |
| 222 | 1,183.50 | 1,177.50 | 981.5 | 1,008 | 4,350.50 |
| 223 | 1,585.50 | 1,505.25 | 1,513.50 | 1,245 | 5,849.25 |
| 224 | 2,019 | 1,793.50 | 1,489.50 | 1,503.50 | 6,805.50 |
| 225 | 701 | 705.5 | 663.5 | 584.5 | 2,654.50 |
| 226 | 1,585.50 | 1,456.50 | 1,491.50 | 1,280.50 | 5,814 |
| 227 | 209.5 | 151 | 219 | 140 | 719.5 |
| 228 | 642.5 | 530 | 648.5 | 668 | 2,489 |
| 229 | 478 | 499.5 | 440 | 428.5 | 1,846 |
| 230 | 648 | 553.5 | 394 | 406.5 | 2,002 |
| 231 | 1,868.50 | 1,793 | 1,152.50 | 1,537 | 6,351 |
| 232 | 2,126.50 | 2,008 | 1,851 | 1,898.50 | 7,884 |
| 233 | 2,507.50 | 2,093 | 2,089.50 | 2,089.50 | 8,779.50 |

| | | | | | | |
|---|---|---|---|---|---|---|
| | 234 | 5,754.50 | 5,702.50 | 5,108.50 | 5,564.50 | 22,130 |
| | 235 | 1,299 | 1,386 | 1,090 | 1,428.50 | 5,203.50 |
| | 236 | 2,296.50 | 2,345.50 | 1,917 | 1,872 | 8,431 |
| | 237 | 942 | 819.5 | 791 | 761 | 3,313.50 |
| | 238 | 1,740.50 | 1,487.50 | 1,245 | 992 | 5,465 |
| | 239 | 322 | 339.5 | 383 | 325.5 | 1,370 |
| | 240 | 1,784 | 1,502.25 | 1,243.50 | 1,162.50 | 5,692.25 |
| | 241 | 1,800 | 1,473 | 1,255.50 | 1,488 | 6,016.50 |
| | 242 | 736 | 717 | 775 | 793 | 3,021 |
| | 243 | 1,090.50 | 1,060.50 | 992.5 | 793 | 3,936.50 |
| | 244 | 468 | 398.5 | 396.5 | 407.5 | 1,670.50 |
| | 245 | 1,886.50 | 1,972 | 1,454 | 1,280 | 6,592.50 |
| | 246 | 299 | 295 | 274 | 253 | 1,121 |
| TOTAL | | 45,995 | 43,318.50 | 38,604.50 | 37,649.50 | 165,567.50 |

STATE: WASHINGTON

| ZIP CODE | | QUARTER 1 | QUARTER 2 | QUARTER 3 | QUARTER 4 | TOTAL GRAMS |
|---|---|---|---|---|---|---|
| | 980 | 3,341.50 | 3,535 | 2,659 | 2,339 | 11,874.50 |
| | 981 | 1,253.50 | 1,155.50 | 1,119.50 | 971.5 | 4,500 |
| | 982 | 1,584 | 1,709.50 | 1,574 | 1,176.50 | 6,044 |
| | 983 | 2,675.50 | 2,532 | 2,770 | 2,933 | 10,910.50 |
| | 984 | 926.5 | 1,009 | 922.5 | 910 | 3,768 |
| | 985 | 902 | 1,020.50 | 1,031.50 | 953 | 3,907 |
| | 986 | 1,574.50 | 1,588.50 | 1,690 | 1,688.50 | 6,541.50 |
| | 988 | 493.5 | 539 | 454.5 | 511.5 | 1,998.50 |
| | 989 | 536.5 | 541 | 375 | 467 | 1,919.50 |
| | 990 | 545 | 416.5 | 432.5 | 415.5 | 1,809.50 |
| | 991 | 228 | 233 | 142 | 183 | 786 |
| | 992 | 2,938.50 | 3,021 | 2,630.50 | 2,926 | 11,516 |
| | 993 | 1,266 | 1,022.50 | 1,088.50 | 1,118.50 | 4,495.50 |
| | 994 | 56 | 103 | 55 | 66 | 280 |
| TOTAL | | 18,321 | 18,426 | 16,944.50 | 16,659 | 70,350.50 |

STATE: WEST VIRGINIA

| ZIP CODE | | QUARTER 1 | QUARTER 2 | QUARTER 3 | QUARTER 4 | TOTAL GRAMS |
|---|---|---|---|---|---|---|

| | | | | | | |
|---|---|---|---|---|---|---|
| | 247 | 208 | 203 | 271.5 | 224.5 | 907 |
| | 248 | 3 | 16 | 0 | 5 | 24 |
| | 249 | 22 | 6 | 0 | 15 | 43 |
| | 250 | 148.5 | 88 | 112 | 64.5 | 413 |
| | 251 | 125 | 157 | 121 | 37 | 440 |
| | 252 | 42 | 56 | 55 | 38 | 191 |
| | 253 | 354.5 | 342.5 | 347.5 | 280.5 | 1,325 |
| | 254 | 1,860 | 1,897.50 | 1,421 | 1,314.50 | 6,493 |
| | 255 | 422.5 | 583 | 514 | 486.5 | 2,006 |
| | 256 | 71 | 79 | 57 | 83 | 290 |
| | 257 | 647 | 774.5 | 750 | 784.5 | 2,956 |
| | 258 | 86 | 92 | 98 | 100.5 | 376.5 |
| | 259 | 33 | 48 | 37 | 17 | 135 |
| | 260 | 1,359 | 1,297 | 1,258.50 | 1,215 | 5,129.50 |
| | 261 | 304.5 | 304.5 | 277.5 | 334.5 | 1,221 |
| | 262 | 81.5 | 106 | 97.5 | 57 | 342 |
| | 263 | 109 | 109.5 | 93 | 75 | 386.5 |
| | 264 | 24 | 41 | 34 | 10 | 109 |
| | 265 | 220.5 | 184 | 229.5 | 238 | 872 |
| | 266 | 30 | 33 | 34.5 | 12 | 109.5 |
| | 267 | 69.5 | 61.5 | 114 | 84 | 329 |
| | 268 | 62.5 | 114 | 135 | 135 | 446.5 |
| TOTAL | | 6,283 | 6,593 | 6,057.50 | 5,611 | 24,544.50 |

STATE: WISCONSIN

| ZIP CODE | | QUARTER 1 | QUARTER 2 | QUARTER 3 | QUARTER 4 | TOTAL GRAMS |
|---|---|---|---|---|---|---|
| | 530 | 2,731 | 2,884.50 | 3,090.50 | 2,613.50 | 11,319.50 |
| | 531 | 2,503.50 | 2,518 | 2,405 | 2,244.50 | 9,671 |
| | 532 | 3,199 | 3,008.50 | 2,473.50 | 2,585 | 11,266 |
| | 534 | 907 | 853.5 | 776 | 859 | 3,395.50 |
| | 535 | 720.5 | 813 | 886.5 | 871.5 | 3,291.50 |
| | 537 | 284 | 301.5 | 193.5 | 207 | 986 |
| | 538 | 134 | 143 | 64.5 | 66 | 407.5 |
| | 539 | 309 | 244.5 | 262.5 | 234 | 1,050 |
| | 540 | 479 | 652 | 444 | 511.5 | 2,086.50 |
| | 541 | 271 | 483 | 578.5 | 334 | 1,666.50 |

| | | | | | | |
|---|---|---|---|---|---|---|
| | 542 | 269 | 276 | 288 | 297.5 | 1,130.50 |
| | 543 | 448.5 | 437 | 469 | 453.5 | 1,808 |
| | 544 | 965.5 | 788 | 819 | 797 | 3,369.50 |
| | 545 | 297 | 286 | 249 | 289 | 1,121 |
| | 546 | 90 | 68 | 67 | 64 | 289 |
| | 547 | 197 | 165 | 132 | 230.5 | 724.5 |
| | 548 | 258.5 | 286 | 281.5 | 186 | 1,012 |
| | 549 | 1,435 | 1,205.50 | 1,085 | 1,155 | 4,880.50 |
| TOTAL | | 15,498.50 | 15,413 | 14,565 | 13,998.50 | 59,475 |

STATE: WYOMING

| ZIP CODE | | QUARTER 1 | QUARTER 2 | QUARTER 3 | QUARTER 4 | TOTAL GRAMS |
|---|---|---|---|---|---|---|
| | 820 | 1,147 | 1,082 | 916.5 | 1,048.50 | 4,194 |
| | 822 | 77 | 69 | 56 | 49 | 251 |
| | 823 | 25 | 35 | 50 | 30 | 140 |
| | 824 | 127 | 70 | 68 | 68 | 333 |
| | 825 | 15 | 10 | 0 | 0 | 25 |
| | 826 | 210 | 230 | 212 | 217 | 869 |
| | 827 | 181 | 454.5 | 375.5 | 380 | 1,391 |
| | 828 | 65 | 55 | 65 | 75 | 260 |
| | 829 | 83 | 68 | 54 | 74 | 279 |
| | 830 | 3 | 0 | 0 | 6 | 9 |
| | 831 | 15 | 0 | 0 | 0 | 15 |
| TOTAL | | 1,948 | 2,073.50 | 1,797 | 1,947.50 | 7,766 |

DRUG CODE: 9801 DRUG NAME: FENTANYL BASE

STATE: ALABAMA

| ZIP CODE | | QUARTER 1 | QUARTER 2 | QUARTER 3 | QUARTER 4 | TOTAL GRAMS |
|---|---|---|---|---|---|---|
| | 350 | 183.09 | 179.05 | 164.93 | 153.55 | 680.62 |
| | 351 | 104.62 | 93.67 | 87.06 | 84.21 | 369.56 |
| | 352 | 151.23 | 140.34 | 135.95 | 136.18 | 563.7 |
| | 354 | 91.79 | 84.11 | 81.66 | 81 | 338.56 |
| | 355 | 67.2 | 67.12 | 61.21 | 53.88 | 249.41 |
| | 356 | 112.54 | 105.82 | 97.51 | 88.92 | 404.79 |

| | | | | | | |
|---|---|---|---|---|---|---|
| | 357 | 39.84 | 37.84 | 32.21 | 32.85 | 142.74 |
| | 358 | 73.42 | 78.99 | 73.7 | 73.11 | 299.22 |
| | 359 | 132.22 | 125.71 | 113.69 | 108.61 | 480.23 |
| | 360 | 94.63 | 91.15 | 87.05 | 79.54 | 352.37 |
| | 361 | 94.86 | 93.28 | 81.86 | 80.37 | 350.37 |
| | 362 | 50.44 | 46.03 | 43.03 | 45.52 | 185.02 |
| | 363 | 91.04 | 87.83 | 83.13 | 82.05 | 344.05 |
| | 364 | 35.73 | 37.69 | 26.45 | 29.06 | 128.93 |
| | 365 | 113.02 | 101.23 | 88.32 | 83.77 | 386.34 |
| | 366 | 83.89 | 76.17 | 66.88 | 70.4 | 297.34 |
| | 367 | 31.04 | 28.27 | 29.98 | 28.77 | 118.06 |
| | 368 | 61.98 | 61.74 | 60.62 | 57.56 | 241.9 |
| | 369 | 7.98 | 7.26 | 8.25 | 6.41 | 29.9 |
| TOTAL | | 1,620.56 | 1,543.30 | 1,423.49 | 1,375.76 | 5,963.11 |

STATE: ALASKA

| ZIP CODE | | QUARTER 1 | QUARTER 2 | QUARTER 3 | QUARTER 4 | TOTAL GRAMS |
|---|---|---|---|---|---|---|
| | 995 | 102.59 | 103.2 | 103.31 | 95.22 | 404.32 |
| | 996 | 85.4 | 88.68 | 83.89 | 84.71 | 342.68 |
| | 997 | 28.67 | 26.14 | 26.66 | 27.55 | 109.02 |
| | 998 | 12.63 | 12.26 | 11.14 | 13.04 | 49.07 |
| | 999 | 10.59 | 9.12 | 8.05 | 7.75 | 35.51 |
| TOTAL | | 239.88 | 239.4 | 233.05 | 228.27 | 940.6 |

STATE: AMERICAN SAMOA

| ZIP CODE | | QUARTER 1 | QUARTER 2 | QUARTER 3 | QUARTER 4 | TOTAL GRAMS |
|---|---|---|---|---|---|---|
| | 967 | 0.24 | 0.23 | 0 | 0 | 0.47 |
| TOTAL | | 0.24 | 0.23 | 0 | 0 | 0.47 |

STATE: ARIZONA

| ZIP CODE | | QUARTER 1 | QUARTER 2 | QUARTER 3 | QUARTER 4 | TOTAL GRAMS |
|---|---|---|---|---|---|---|
| | 850 | 360.01 | 323.86 | 291.63 | 263.42 | 1,238.92 |

| | | | | | | |
|---|---|---|---|---|---|---|
| | 851 | 63.73 | 59.21 | 51.62 | 57.45 | 232.01 |
| | 852 | 619.79 | 584.4 | 525.34 | 510.61 | 2,240.14 |
| | 853 | 320.47 | 275.23 | 243.81 | 253.44 | 1,092.95 |
| | 855 | 25.74 | 22.15 | 19.49 | 18.93 | 86.31 |
| | 856 | 71.65 | 61.72 | 54.07 | 58.18 | 245.62 |
| | 857 | 299.53 | 275.58 | 257.61 | 251.61 | 1,084.33 |
| | 859 | 18.54 | 16.08 | 17.81 | 14.57 | 67 |
| | 860 | 27.83 | 28.31 | 26.49 | 24.3 | 106.93 |
| | 863 | 81.23 | 75.69 | 65.58 | 67.94 | 290.44 |
| | 864 | 87.08 | 78.77 | 71.39 | 63.3 | 300.54 |
| | 865 | 2.33 | 2.41 | 1.3 | 2.14 | 8.18 |
| TOTAL | | 1,977.93 | 1,803.41 | 1,626.14 | 1,585.89 | 6,993.37 |

STATE: ARKANSAS

| ZIP CODE | | QUARTER 1 | QUARTER 2 | QUARTER 3 | QUARTER 4 | TOTAL GRAMS |
|---|---|---|---|---|---|---|
| | 716 | 46.01 | 46.3 | 41.12 | 38.4 | 171.83 |
| | 717 | 31.77 | 28.83 | 28.61 | 27.45 | 116.66 |
| | 718 | 25.48 | 24.33 | 24.09 | 21.45 | 95.35 |
| | 719 | 75.5 | 71.71 | 71.53 | 69.23 | 287.97 |
| | 720 | 132.52 | 123.47 | 106.68 | 96.34 | 459.01 |
| | 721 | 148.54 | 145.26 | 131.39 | 121.16 | 546.35 |
| | 722 | 100.79 | 95.21 | 87.49 | 87.13 | 370.62 |
| | 723 | 29.81 | 29.64 | 24.06 | 24.56 | 108.07 |
| | 724 | 60.07 | 61.92 | 55.27 | 52.66 | 229.92 |
| | 725 | 48.45 | 52.6 | 46.09 | 44.29 | 191.43 |
| | 726 | 50 | 51.01 | 44.84 | 42.25 | 188.1 |
| | 727 | 160.88 | 152.33 | 139.53 | 137.85 | 590.59 |
| | 728 | 40.82 | 36.97 | 37.34 | 32.7 | 147.83 |
| | 729 | 59.81 | 56.81 | 48.85 | 46.02 | 211.49 |
| TOTAL | | 1,010.45 | 976.39 | 886.89 | 841.49 | 3,715.22 |

STATE: CALIFORNIA

| ZIP CODE | | QUARTER 1 | QUARTER 2 | QUARTER 3 | QUARTER 4 | TOTAL GRAMS |
|---|---|---|---|---|---|---|
| | 900 | 224.37 | 247.9 | 211.91 | 189.1 | 873.28 |

| | | | | | |
|---|---|---|---|---|---|
| 902 | 91.03 | 82.54 | 82.01 | 77.13 | 332.71 |
| 903 | 5.88 | 4.41 | 5.18 | 5 | 20.47 |
| 904 | 59.69 | 56.83 | 55.35 | 66.52 | 238.39 |
| 905 | 39.52 | 38.04 | 35.79 | 34.5 | 147.85 |
| 906 | 88.56 | 89.16 | 88.28 | 77.76 | 343.76 |
| 907 | 71.63 | 72.59 | 70.53 | 67.67 | 282.42 |
| 908 | 154.55 | 132.21 | 164.02 | 112.48 | 563.26 |
| 910 | 60.06 | 60.18 | 64.12 | 46.62 | 230.98 |
| 911 | 25.27 | 26.6 | 23.36 | 23.19 | 98.42 |
| 912 | 36.69 | 39.64 | 38.4 | 35.18 | 149.91 |
| 913 | 206.06 | 206.11 | 197.35 | 198.67 | 808.19 |
| 914 | 84.64 | 83.18 | 79.01 | 76.19 | 323.02 |
| 915 | 18.55 | 18.76 | 14.61 | 13.67 | 65.59 |
| 916 | 11.61 | 11.32 | 7.07 | 7.18 | 37.18 |
| 917 | 297.66 | 270.2 | 287.31 | 274.72 | 1,129.89 |
| 918 | 6.36 | 7.45 | 6.75 | 5.2 | 25.76 |
| 919 | 57.26 | 53.22 | 49.9 | 50.56 | 210.94 |
| 920 | 371.83 | 361.18 | 351.73 | 341.21 | 1,425.95 |
| 921 | 393.49 | 413.51 | 371.79 | 354.21 | 1,533 |
| 922 | 217.92 | 213.6 | 197.58 | 197.61 | 826.71 |
| 923 | 296.42 | 275.54 | 250.97 | 243.34 | 1,066.27 |
| 924 | 64.79 | 54.54 | 50.32 | 38.46 | 208.11 |
| 925 | 284.16 | 272.05 | 256.97 | 248.36 | 1,061.54 |
| 926 | 286.86 | 273.93 | 268.41 | 256.65 | 1,085.85 |
| 927 | 60.46 | 53.71 | 50.75 | 53.52 | 218.44 |
| 928 | 242.33 | 241.13 | 222.1 | 210.48 | 916.04 |
| 930 | 138.57 | 134.39 | 124.8 | 117.29 | 515.05 |
| 931 | 24.75 | 25.81 | 24.94 | 25.7 | 101.2 |
| 932 | 116.75 | 120.8 | 113.44 | 107.18 | 458.17 |
| 933 | 162.66 | 161.07 | 137.31 | 164.9 | 625.94 |
| 934 | 125.22 | 132.95 | 115.05 | 98.54 | 471.76 |
| 935 | 105.22 | 102.05 | 90.72 | 86.95 | 384.94 |
| 936 | 75.51 | 73.97 | 70.91 | 69.5 | 289.89 |
| 937 | 106.7 | 104.43 | 101.31 | 101.73 | 414.17 |
| 939 | 36.51 | 35.93 | 34.87 | 34.85 | 142.16 |
| 940 | 123.67 | 115.88 | 111.62 | 110.56 | 461.73 |
| 941 | 138.53 | 130.88 | 126.35 | 128.14 | 523.9 |

| | | | | | | |
|---|---|---|---|---|---|---|
| | 943 | 35.7 | 31.98 | 29.2 | 30.94 | 127.82 |
| | 944 | 19.38 | 15.59 | 16.24 | 15.57 | 66.78 |
| | 945 | 498.62 | 464.86 | 451.83 | 414.19 | 1,829.50 |
| | 946 | 49.12 | 48.82 | 45.85 | 44.44 | 188.23 |
| | 947 | 14.22 | 14.85 | 16.1 | 16.91 | 62.08 |
| | 948 | 16.3 | 14.28 | 14.05 | 12.43 | 57.06 |
| | 949 | 64.18 | 64.95 | 61.68 | 56.88 | 247.69 |
| | 950 | 123.54 | 122.7 | 115.87 | 118.08 | 480.19 |
| | 951 | 65.33 | 65.82 | 61.6 | 63.03 | 255.78 |
| | 952 | 128.13 | 123.93 | 107.04 | 107.13 | 466.23 |
| | 953 | 281 | 267.87 | 262.41 | 253.76 | 1,065.04 |
| | 954 | 142.83 | 129.89 | 116.04 | 107.07 | 495.83 |
| | 955 | 54.42 | 55.69 | 51.22 | 47.86 | 209.19 |
| | 956 | 362.6 | 336.83 | 332.77 | 313.92 | 1,346.12 |
| | 957 | 58.23 | 54.55 | 51.64 | 47.64 | 212.06 |
| | 958 | 208.56 | 198.79 | 194.04 | 183.91 | 785.3 |
| | 959 | 205.1 | 192.43 | 173.03 | 169.1 | 739.66 |
| | 960 | 136.69 | 122.26 | 122.23 | 112.51 | 493.69 |
| | 961 | 18.94 | 19.14 | 18.9 | 17.23 | 74.21 |
| TOTAL | | 7,394.63 | 7,142.92 | 6,794.63 | 6,483.12 | 27,815.30 |

STATE: COLORADO

| ZIP CODE | | QUARTER 1 | QUARTER 2 | QUARTER 3 | QUARTER 4 | TOTAL GRAMS |
|---|---|---|---|---|---|---|
| | 800 | 219.67 | 203.56 | 194.97 | 183.67 | 801.87 |
| | 801 | 176.95 | 153.9 | 157.14 | 147.05 | 635.04 |
| | 802 | 214.14 | 207.77 | 198.46 | 192.8 | 813.17 |
| | 803 | 33.56 | 33.85 | 29.52 | 29.92 | 126.85 |
| | 804 | 54.42 | 51.68 | 43.78 | 49.66 | 199.54 |
| | 805 | 158.79 | 144.97 | 134.02 | 126.47 | 564.25 |
| | 806 | 56.71 | 52.29 | 49.62 | 50.98 | 209.6 |
| | 807 | 28.44 | 27.72 | 24.42 | 23.56 | 104.14 |
| | 808 | 32.09 | 27.69 | 25.03 | 24.41 | 109.22 |
| | 809 | 149.79 | 147.48 | 144.78 | 134.73 | 576.78 |
| | 810 | 80.5 | 78.48 | 77.77 | 72 | 308.75 |
| | 811 | 15.97 | 13.71 | 13.35 | 11.1 | 54.13 |
| | 812 | 25.5 | 23.76 | 22.72 | 22.01 | 93.99 |

| | | | | | |
|---|---|---|---|---|---|
| 813 | 17.05 | 15.76 | 16.66 | 12.62 | 62.09 |
| 814 | 18.23 | 16.12 | 15.36 | 12.52 | 62.23 |
| 815 | 61.73 | 53.84 | 51.4 | 48.42 | 215.39 |
| 816 | 18.7 | 19.52 | 17.63 | 17.69 | 73.54 |
| TOTAL | 1,362.24 | 1,272.10 | 1,216.63 | 1,159.61 | 5,010.58 |

STATE: CONNECTICUT

| ZIP CODE | QUARTER 1 | QUARTER 2 | QUARTER 3 | QUARTER 4 | TOTAL GRAMS |
|---|---|---|---|---|---|
| 60 | 205.2 | 195.33 | 181.51 | 157.68 | 739.72 |
| 61 | 49.14 | 49.73 | 47.39 | 48.4 | 194.66 |
| 62 | 40.23 | 36.82 | 36.95 | 35.27 | 149.27 |
| 63 | 75.45 | 75.46 | 72.77 | 64.9 | 288.58 |
| 64 | 252.95 | 243.68 | 233.53 | 232.98 | 963.14 |
| 65 | 82.74 | 79.11 | 76.54 | 85.7 | 324.09 |
| 66 | 40.59 | 42.53 | 36.43 | 35.27 | 154.82 |
| 67 | 91.72 | 85.49 | 84.43 | 75.58 | 337.22 |
| 68 | 152.96 | 164.25 | 141.34 | 130.97 | 589.52 |
| 69 | 15.21 | 16.88 | 13.31 | 14.19 | 59.59 |
| TOTAL | 1,006.19 | 989.28 | 924.2 | 880.94 | 3,800.61 |

STATE: DELAWARE

| ZIP CODE | QUARTER 1 | QUARTER 2 | QUARTER 3 | QUARTER 4 | TOTAL GRAMS |
|---|---|---|---|---|---|
| 197 | 135.29 | 130.61 | 129.89 | 127.83 | 523.62 |
| 198 | 70.56 | 66.83 | 62.69 | 62.36 | 262.44 |
| 199 | 152.42 | 152.77 | 132.18 | 137.17 | 574.54 |
| TOTAL | 358.27 | 350.21 | 324.76 | 327.36 | 1,360.60 |

STATE: DISTRICT OF COLUMBIA

| ZIP CODE | QUARTER 1 | QUARTER 2 | QUARTER 3 | QUARTER 4 | TOTAL GRAMS |
|---|---|---|---|---|---|
| 200 | 84.93 | 83.59 | 74.69 | 71.06 | 314.27 |
| 203 | 0 | 0.08 | 0.04 | 0.1 | 0.22 |
| 204 | 7.48 | 6.47 | 5.63 | 4.68 | 24.26 |
| 205 | 0 | 0.02 | 0 | 0 | 0.02 |

| | | | | | | |
|---|---|---|---|---|---|---|
| TOTAL | | 92.41 | 90.16 | 80.36 | 75.84 | 338.77 |

STATE: FLORIDA

| ZIP CODE | | QUARTER 1 | QUARTER 2 | QUARTER 3 | QUARTER 4 | TOTAL GRAMS |
|---|---|---|---|---|---|---|
| | 320 | 237.27 | 225.69 | 208.58 | 207.69 | 879.23 |
| | 321 | 246.03 | 218.29 | 202.7 | 214.69 | 881.71 |
| | 322 | 477.67 | 429.32 | 411.35 | 401.18 | 1,719.52 |
| | 323 | 90.04 | 90.31 | 84.78 | 89.45 | 354.58 |
| | 324 | 130.74 | 119.95 | 114.62 | 117.25 | 482.56 |
| | 325 | 160.26 | 159.85 | 148.43 | 134.6 | 603.14 |
| | 326 | 124.35 | 120.61 | 121.94 | 124.14 | 491.04 |
| | 327 | 353.14 | 334.41 | 308.43 | 311.79 | 1,307.77 |
| | 328 | 188.91 | 174.31 | 161.86 | 162.84 | 687.92 |
| | 329 | 358.79 | 346.69 | 310.73 | 327.09 | 1,343.30 |
| | 330 | 296.99 | 286.48 | 252.92 | 272.62 | 1,109.01 |
| | 331 | 226.44 | 215.64 | 195.3 | 205.22 | 842.6 |
| | 333 | 230.05 | 210.82 | 212.65 | 219.91 | 873.43 |
| | 334 | 511.29 | 469.04 | 449.59 | 455.44 | 1,885.36 |
| | 335 | 220.14 | 198.78 | 193.99 | 198.48 | 811.39 |
| | 336 | 463.13 | 401.91 | 387.6 | 332.09 | 1,584.73 |
| | 337 | 314.37 | 291.14 | 288.08 | 293.05 | 1,186.64 |
| | 338 | 172.81 | 157.77 | 145.74 | 159.74 | 636.06 |
| | 339 | 304.82 | 265.05 | 229.04 | 261.3 | 1,060.21 |
| | 341 | 98.47 | 88.85 | 77.82 | 85.29 | 350.43 |
| | 342 | 344.2 | 313.6 | 274.22 | 306.7 | 1,238.72 |
| | 344 | 169.8 | 159.31 | 142.81 | 152.84 | 624.76 |
| | 346 | 314.66 | 296.06 | 276.57 | 287.31 | 1,174.60 |
| | 347 | 151.69 | 135.67 | 113.01 | 131.52 | 531.89 |
| | 349 | 185.54 | 174.26 | 159.99 | 172.34 | 692.13 |
| TOTAL | | 6,371.60 | 5,883.81 | 5,472.75 | 5,624.57 | 23,352.73 |

STATE: GEORGIA

| ZIP CODE | | QUARTER 1 | QUARTER 2 | QUARTER 3 | QUARTER 4 | TOTAL GRAMS |
|---|---|---|---|---|---|---|
| | 300 | 564.82 | 529.47 | 509.22 | 516.16 | 2,119.67 |
| | 301 | 375.86 | 362.64 | 341.16 | 333.97 | 1,413.63 |

| | | | | | | |
|---|---|---|---|---|---|---|
| | 302 | 241.18 | 234.09 | 217 | 210.82 | 903.09 |
| | 303 | 181.88 | 171.1 | 151.32 | 170.68 | 674.98 |
| | 304 | 90.98 | 95.5 | 87.91 | 77.43 | 351.82 |
| | 305 | 228.24 | 217.26 | 200.13 | 197.29 | 842.92 |
| | 306 | 111.67 | 107.81 | 104.99 | 104.08 | 428.55 |
| | 307 | 71.69 | 66.32 | 61.95 | 55.17 | 255.13 |
| | 308 | 60.28 | 54.27 | 53.3 | 52.35 | 220.2 |
| | 309 | 119.42 | 108.61 | 95.95 | 92.98 | 416.96 |
| | 310 | 199.92 | 201.16 | 186.63 | 175.43 | 763.14 |
| | 312 | 77.16 | 70.42 | 66.24 | 65.76 | 279.58 |
| | 313 | 87.34 | 79.96 | 66.84 | 52.73 | 286.87 |
| | 314 | 76.1 | 71.09 | 70.44 | 64.45 | 282.08 |
| | 315 | 141.91 | 137.17 | 129.89 | 117.33 | 526.3 |
| | 316 | 68.8 | 69.74 | 62.69 | 69.51 | 270.74 |
| | 317 | 129.92 | 119.79 | 120.68 | 118.75 | 489.14 |
| | 318 | 8.57 | 9.01 | 7.26 | 7.02 | 31.86 |
| | 319 | 62.72 | 56.19 | 57.27 | 51.56 | 227.74 |
| | 398 | 34.56 | 30.21 | 28.63 | 30.11 | 123.51 |
| TOTAL | | 2,933.02 | 2,791.81 | 2,619.50 | 2,563.58 | 10,907.91 |

STATE: GUAM

| ZIP CODE | | QUARTER 1 | QUARTER 2 | QUARTER 3 | QUARTER 4 | TOTAL GRAMS |
|---|---|---|---|---|---|---|
| TOTAL | | 12.2 | 10.02 | 9.17 | 9.95 | 41.34 |

STATE: HAWAII

| ZIP CODE | | QUARTER 1 | QUARTER 2 | QUARTER 3 | QUARTER 4 | TOTAL GRAMS |
|---|---|---|---|---|---|---|
| | 967 | 120.58 | 114.18 | 104.19 | 99.5 | 438.45 |
| | 968 | 77.6 | 79.59 | 71.01 | 75.65 | 303.85 |
| TOTAL | | 198.18 | 193.77 | 175.2 | 175.15 | 742.3 |

STATE: IDAHO

| ZIP CODE | | QUARTER 1 | QUARTER 2 | QUARTER 3 | QUARTER 4 | TOTAL GRAMS |
|---|---|---|---|---|---|---|
| | 832 | 54.32 | 57.56 | 56.59 | 55.36 | 223.83 |
| | 833 | 60.46 | 62.31 | 61.1 | 55.82 | 239.69 |

| | | | | | | |
|---|---|---|---|---|---|---|
| | 834 | 55.44 | 62.69 | 55.01 | 55.05 | 228.19 |
| | 835 | 39.37 | 38.3 | 39.48 | 38.05 | 155.2 |
| | 836 | 169.69 | 167.1 | 162.68 | 154.66 | 654.13 |
| | 837 | 125.17 | 121.74 | 112.07 | 120.2 | 479.18 |
| | 838 | 90.1 | 79.52 | 78.72 | 76.34 | 324.68 |
| TOTAL | | 594.55 | 589.22 | 565.65 | 555.48 | 2,304.90 |

STATE: ILLINOIS

| ZIP CODE | QUARTER 1 | QUARTER 2 | QUARTER 3 | QUARTER 4 | TOTAL GRAMS |
|---|---|---|---|---|---|
| 600 | 431.51 | 418.01 | 410.04 | 394.44 | 1,654 |
| 601 | 268.12 | 254.87 | 243.67 | 255.42 | 1,022.08 |
| 602 | 13.31 | 11.22 | 10.01 | 11.52 | 46.06 |
| 603 | 12.14 | 11.6 | 10.53 | 12.68 | 46.95 |
| 604 | 366.07 | 336.89 | 328.39 | 318.02 | 1,349.37 |
| 605 | 207.42 | 197.57 | 189.15 | 185.17 | 779.31 |
| 606 | 282.69 | 279.56 | 258.43 | 256.44 | 1,077.12 |
| 607 | 22.87 | 22.34 | 23.32 | 26.63 | 95.16 |
| 608 | 20.19 | 20.87 | 17.7 | 23.82 | 82.58 |
| 609 | 47.34 | 46.46 | 43.6 | 44.08 | 181.48 |
| 610 | 109.39 | 106.62 | 99.03 | 97.17 | 412.21 |
| 611 | 117.88 | 119.07 | 114.16 | 104.03 | 455.14 |
| 612 | 36.72 | 36.82 | 32.91 | 31.12 | 137.57 |
| 613 | 45.07 | 41.41 | 38.49 | 37.12 | 162.09 |
| 614 | 50.23 | 46.97 | 46.27 | 44.18 | 187.65 |
| 615 | 66.93 | 67.72 | 65.63 | 65.84 | 266.12 |
| 616 | 142.56 | 136.23 | 128.83 | 121.75 | 529.37 |
| 617 | 74.61 | 71.11 | 70.71 | 67.34 | 283.77 |
| 618 | 73.12 | 68.77 | 66.51 | 65.2 | 273.6 |
| 619 | 33.74 | 34.23 | 33.5 | 33.36 | 134.83 |
| 620 | 114.4 | 108.87 | 103.78 | 100.76 | 427.81 |
| 622 | 116.42 | 105.74 | 104.47 | 92.57 | 419.2 |
| 623 | 45.57 | 46.67 | 47.07 | 43.6 | 182.91 |
| 624 | 45.01 | 44.61 | 43.76 | 44.51 | 177.89 |
| 625 | 93.01 | 83.02 | 91.43 | 82.57 | 350.03 |
| 626 | 59.78 | 58.7 | 56.63 | 53.8 | 228.91 |
| 627 | 73.89 | 68.2 | 66.39 | 69.44 | 277.92 |

| | | | | | | |
|---|---|---|---|---|---|---|
| | 628 | 54.52 | 49.99 | 49.01 | 48.08 | 201.6 |
| | 629 | 99.21 | 96.23 | 87.5 | 83.49 | 366.43 |
| TOTAL | | 3,123.72 | 2,990.37 | 2,880.92 | 2,814.15 | 11,809.16 |

STATE: INDIANA

| ZIP CODE | | QUARTER 1 | QUARTER 2 | QUARTER 3 | QUARTER 4 | TOTAL GRAMS |
|---|---|---|---|---|---|---|
| | 460 | 138.04 | 126.7 | 118.05 | 117.89 | 500.68 |
| | 461 | 151.66 | 139.09 | 132.53 | 130.77 | 554.05 |
| | 462 | 362.62 | 344.36 | 317.92 | 349.14 | 1,374.04 |
| | 463 | 193.36 | 194.14 | 179.99 | 175.11 | 742.6 |
| | 464 | 15.91 | 14.96 | 12.77 | 15.14 | 58.78 |
| | 465 | 144.27 | 136.05 | 126.4 | 130.52 | 537.24 |
| | 466 | 49.36 | 48.32 | 41.2 | 41.84 | 180.72 |
| | 467 | 62.89 | 60.79 | 59.31 | 56.56 | 239.55 |
| | 468 | 120.63 | 118.63 | 113.88 | 107.97 | 461.11 |
| | 469 | 106.14 | 96.13 | 88.12 | 82.75 | 373.14 |
| | 470 | 48.92 | 47.25 | 43.63 | 44.39 | 184.19 |
| | 471 | 133.7 | 131.54 | 114.39 | 90.54 | 470.17 |
| | 472 | 57.27 | 51.55 | 47.28 | 48.23 | 204.33 |
| | 473 | 104.73 | 98.17 | 100.97 | 96 | 399.87 |
| | 474 | 75.85 | 72.1 | 64.25 | 58.92 | 271.12 |
| | 475 | 63.23 | 52.61 | 51.14 | 49.32 | 216.3 |
| | 476 | 32.83 | 33.07 | 33.09 | 21.65 | 120.64 |
| | 477 | 53.91 | 52.2 | 47.94 | 50.78 | 204.83 |
| | 478 | 42.78 | 40.18 | 36.73 | 34.35 | 154.04 |
| | 479 | 82.68 | 64.23 | 63.02 | 66.23 | 276.16 |
| TOTAL | | 2,040.78 | 1,922.07 | 1,792.61 | 1,768.10 | 7,523.56 |

STATE: IOWA

| ZIP CODE | | QUARTER 1 | QUARTER 2 | QUARTER 3 | QUARTER 4 | TOTAL GRAMS |
|---|---|---|---|---|---|---|
| | 500 | 65.07 | 69.27 | 66.76 | 68.94 | 270.04 |
| | 501 | 81.13 | 75.41 | 77.69 | 77.4 | 311.63 |
| | 502 | 69.66 | 70.79 | 63.56 | 64.6 | 268.61 |

| | | | | | | |
|---|---|---|---|---|---|---|
| | 503 | 109.25 | 95.58 | 94.78 | 93.42 | 393.03 |
| | 504 | 43.88 | 44.45 | 39.04 | 39.83 | 167.2 |
| | 505 | 43.21 | 40.9 | 41.52 | 41.98 | 167.61 |
| | 506 | 50.72 | 50.81 | 45.89 | 46.06 | 193.48 |
| | 507 | 20.54 | 17.54 | 18.66 | 18.09 | 74.83 |
| | 508 | 13.95 | 12.5 | 11.75 | 11.2 | 49.4 |
| | 510 | 12.55 | 12.56 | 12.28 | 11.43 | 48.82 |
| | 511 | 24.81 | 28.39 | 24.64 | 23.48 | 101.32 |
| | 512 | 13.49 | 11.76 | 10.33 | 8.39 | 43.97 |
| | 513 | 13.07 | 11.41 | 12.49 | 11.33 | 48.3 |
| | 514 | 26.07 | 24.11 | 25.84 | 24.5 | 100.52 |
| | 515 | 37.82 | 36.89 | 33.99 | 30.94 | 139.64 |
| | 516 | 7.3 | 6.84 | 6.27 | 7.47 | 27.88 |
| | 520 | 39.26 | 38.83 | 39.11 | 37.55 | 154.75 |
| | 521 | 16.62 | 16.49 | 15.33 | 15 | 63.44 |
| | 522 | 32.26 | 32.61 | 30.88 | 30.06 | 125.81 |
| | 523 | 25.71 | 24.61 | 24.37 | 22.89 | 97.58 |
| | 524 | 29.56 | 34.52 | 32.31 | 30.81 | 127.2 |
| | 525 | 46.21 | 42.52 | 41.47 | 40.26 | 170.46 |
| | 526 | 29.02 | 29.07 | 26.21 | 27.1 | 111.4 |
| | 527 | 23.13 | 25.36 | 20.98 | 22.41 | 91.88 |
| | 528 | 33.76 | 33.7 | 29.75 | 30.55 | 127.76 |
| TOTAL | | 908.05 | 886.92 | 845.9 | 835.69 | 3,476.56 |

| STATE: | KANSAS | | | | |
|---|---|---|---|---|---|
| ZIP CODE | | QUARTER 1 | QUARTER 2 | QUARTER 3 | QUARTER 4 | TOTAL GRAMS |

| ZIP CODE | | QUARTER 1 | QUARTER 2 | QUARTER 3 | QUARTER 4 | TOTAL GRAMS |
|---|---|---|---|---|---|---|
| | 660 | 166.93 | 158.86 | 156.72 | 144.37 | 626.88 |
| | 661 | 52.53 | 43 | 46.27 | 50.54 | 192.34 |
| | 662 | 176.47 | 175.61 | 158.41 | 152.26 | 662.75 |
| | 664 | 27.96 | 28.35 | 28.14 | 25.03 | 109.48 |
| | 665 | 44.05 | 44.45 | 42.16 | 43.85 | 174.51 |
| | 666 | 67.02 | 68.87 | 64.9 | 62.24 | 263.03 |
| | 667 | 65.09 | 65.8 | 62.78 | 143.68 | 337.35 |
| | 668 | 11.22 | 12.06 | 12 | 10.84 | 46.12 |
| | 669 | 18.19 | 18.73 | 19.85 | 17.63 | 74.4 |
| | 670 | 58.92 | 56.12 | 57.45 | 55.02 | 227.51 |

| | | | | | | |
|---|---|---|---|---|---|---|
| | 671 | 37.74 | 39.31 | 35.12 | 37.65 | 149.82 |
| | 672 | 148.47 | 145.19 | 139.2 | 131.05 | 563.91 |
| | 673 | 31.31 | 28.92 | 26.88 | 26.47 | 113.58 |
| | 674 | 66.86 | 72.03 | 70.85 | 68.15 | 277.89 |
| | 675 | 38.02 | 36.2 | 33.96 | 29.93 | 138.11 |
| | 676 | 36.28 | 37.2 | 36.49 | 36.05 | 146.02 |
| | 677 | 20.88 | 22.01 | 18.77 | 20.09 | 81.75 |
| | 678 | 51.21 | 51.56 | 49.8 | 49.01 | 201.58 |
| | 679 | 20.88 | 17.21 | 18.84 | 17.99 | 74.92 |
| TOTAL | | 1,140.03 | 1,121.48 | 1,078.59 | 1,121.85 | 4,461.95 |

STATE: KENTUCKY

| ZIP CODE | | QUARTER 1 | QUARTER 2 | QUARTER 3 | QUARTER 4 | TOTAL GRAMS |
|---|---|---|---|---|---|---|
| | 400 | 54 | 48.32 | 46.65 | 45.78 | 194.75 |
| | 401 | 22.14 | 22.59 | 18.66 | 19.42 | 82.81 |
| | 402 | 230.43 | 212.89 | 195.78 | 194.17 | 833.27 |
| | 403 | 79.4 | 83.83 | 74.43 | 74.34 | 312 |
| | 404 | 46.75 | 45.83 | 43.44 | 39.89 | 175.91 |
| | 405 | 104.54 | 96.79 | 89.5 | 90.64 | 381.47 |
| | 406 | 7.49 | 7.62 | 7.78 | 8.05 | 30.94 |
| | 407 | 37.84 | 33.23 | 34.34 | 33.41 | 138.82 |
| | 408 | 7.42 | 7.39 | 6.37 | 5.72 | 26.9 |
| | 409 | 16.59 | 14.29 | 13.55 | 11.76 | 56.19 |
| | 410 | 148.4 | 135.32 | 128.42 | 121.27 | 533.41 |
| | 411 | 33 | 34.03 | 30.66 | 27.52 | 125.21 |
| | 412 | 14.99 | 13.52 | 11.05 | 12.07 | 51.63 |
| | 413 | 13.08 | 10.93 | 10.43 | 9.01 | 43.45 |
| | 414 | 3.06 | 2.87 | 2.73 | 2.56 | 11.22 |
| | 415 | 22.64 | 21.05 | 17.16 | 16.88 | 77.73 |
| | 416 | 13.59 | 13.5 | 13.14 | 11.71 | 51.94 |
| | 417 | 26.58 | 28.54 | 31.63 | 31.19 | 117.94 |
| | 418 | 3.4 | 3.22 | 2.59 | 3.27 | 12.48 |
| | 420 | 56.82 | 55.33 | 51.8 | 52.9 | 216.85 |
| | 421 | 71.36 | 69.04 | 66.63 | 63.73 | 270.76 |
| | 422 | 28.32 | 27.56 | 28.57 | 24.37 | 108.82 |

| | | | | | |
|---|---|---|---|---|---|
| 423 | 40.17 | 41.68 | 36.97 | 37.43 | 156.25 |
| 424 | 55.55 | 50.22 | 49.02 | 45.64 | 200.43 |
| 425 | 24.78 | 26.75 | 20.47 | 18.38 | 90.38 |
| 426 | 17.5 | 15.67 | 15.42 | 16.01 | 64.6 |
| 427 | 52.98 | 52.92 | 51.96 | 43.05 | 200.91 |
| TOTAL | 1,232.82 | 1,174.93 | 1,099.15 | 1,060.17 | 4,567.07 |

STATE: LOUISIANA

| ZIP CODE | QUARTER 1 | QUARTER 2 | QUARTER 3 | QUARTER 4 | TOTAL GRAMS |
|---|---|---|---|---|---|
| 700 | 109.48 | 105.46 | 96.79 | 90.14 | 401.87 |
| 701 | 77.43 | 71.64 | 67.28 | 65.66 | 282.01 |
| 703 | 87.32 | 76.91 | 74.53 | 66.5 | 305.26 |
| 704 | 131.98 | 116.79 | 104.33 | 100.58 | 453.68 |
| 705 | 225.83 | 229.68 | 226.14 | 196.43 | 878.08 |
| 706 | 45.96 | 42.08 | 48.82 | 44.25 | 181.11 |
| 707 | 141.59 | 119.55 | 114.61 | 114.61 | 490.36 |
| 708 | 103.09 | 98.05 | 89.59 | 92.03 | 382.76 |
| 710 | 32.1 | 32.93 | 31.25 | 30.97 | 127.25 |
| 711 | 92.15 | 88.32 | 80.39 | 77.86 | 338.72 |
| 712 | 104.66 | 99.96 | 96.2 | 91.01 | 391.83 |
| 713 | 105.37 | 97.24 | 88.1 | 79.33 | 370.04 |
| 714 | 44.4 | 37.87 | 35.8 | 34.56 | 152.63 |
| TOTAL | 1,301.36 | 1,216.48 | 1,153.83 | 1,083.93 | 4,755.60 |

STATE: MAINE

| ZIP CODE | QUARTER 1 | QUARTER 2 | QUARTER 3 | QUARTER 4 | TOTAL GRAMS |
|---|---|---|---|---|---|
| 39 | 5.46 | 3.84 | 3.47 | 2.85 | 15.62 |
| 40 | 97.03 | 79.55 | 76.03 | 77.85 | 330.46 |
| 41 | 26.78 | 31.69 | 32.34 | 31.04 | 121.85 |
| 42 | 67.6 | 38.48 | 34.33 | 33.9 | 174.31 |
| 43 | 46.79 | 47.9 | 39.92 | 41.37 | 175.98 |
| 44 | 46.51 | 42.62 | 38.9 | 35.93 | 163.96 |
| 45 | 6.13 | 5.04 | 5.31 | 5.56 | 22.04 |
| 46 | 14.76 | 12.48 | 11.25 | 12.95 | 51.44 |

| | | | | | | |
|---|---|---|---|---|---|---|
| | 47 | 24.25 | 22.78 | 21 | 19.17 | 87.2 |
| | 48 | 8.99 | 8.31 | 7.09 | 7.59 | 31.98 |
| | 49 | 43.97 | 38.02 | 34.24 | 31.74 | 147.97 |
| TOTAL | | 388.27 | 330.71 | 303.88 | 299.95 | 1,322.81 |

| STATE: | | MARYLAND | | | | |
|---|---|---|---|---|---|---|
| ZIP CODE | | QUARTER 1 | QUARTER 2 | QUARTER 3 | QUARTER 4 | TOTAL GRAMS |
| | 206 | 94.46 | 86.64 | 80.06 | 75.64 | 336.8 |
| | 207 | 133.08 | 121.36 | 128.15 | 101.07 | 483.66 |
| | 208 | 175.25 | 149.78 | 152.35 | 134.74 | 612.12 |
| | 209 | 38.7 | 39.33 | 30.68 | 35.15 | 143.86 |
| | 210 | 222.22 | 209.85 | 194.83 | 180.67 | 807.57 |
| | 211 | 125.24 | 109.24 | 98.51 | 94.79 | 427.78 |
| | 212 | 342.69 | 313.52 | 299.63 | 277.65 | 1,233.49 |
| | 214 | 30.93 | 26.49 | 27.32 | 24.8 | 109.54 |
| | 215 | 57.13 | 54.06 | 46.38 | 37.16 | 194.73 |
| | 216 | 45.06 | 43.16 | 39.38 | 37.17 | 164.77 |
| | 217 | 192.33 | 180.71 | 164.63 | 163.13 | 700.8 |
| | 218 | 56.69 | 48.44 | 48.72 | 43.88 | 197.73 |
| | 219 | 39.02 | 34.25 | 34.36 | 31.93 | 139.56 |
| TOTAL | | 1,552.80 | 1,416.83 | 1,345 | 1,237.78 | 5,552.41 |

| STATE: | | MASSACHUSETTS | | | | |
|---|---|---|---|---|---|---|
| ZIP CODE | | QUARTER 1 | QUARTER 2 | QUARTER 3 | QUARTER 4 | TOTAL GRAMS |
| | 10 | 144.74 | 139.05 | 138.74 | 132.93 | 555.46 |
| | 11 | 47.36 | 47.4 | 42.91 | 39.12 | 176.79 |
| | 12 | 29.63 | 26.66 | 27.26 | 24.69 | 108.24 |
| | 13 | 14.48 | 15.01 | 17.13 | 15.14 | 61.76 |
| | 14 | 38.46 | 35.84 | 34.12 | 32.01 | 140.43 |
| | 15 | 63.48 | 63.93 | 57.27 | 59.49 | 244.17 |
| | 16 | 51.48 | 47.15 | 41.83 | 40.73 | 181.19 |
| | 17 | 71.26 | 69.72 | 61.87 | 59.64 | 262.49 |
| | 18 | 212.18 | 208.92 | 195.04 | 175.29 | 791.43 |
| | 19 | 86.34 | 82.18 | 79.13 | 85.69 | 333.34 |
| | 20 | 58.74 | 53.02 | 48.94 | 50.54 | 211.24 |

| | | | | | | |
|---|---|---|---|---|---|---|
| | 21 | 181.34 | 172.93 | 166.13 | 174.99 | 695.39 |
| | 22 | 27.6 | 26.87 | 19.91 | 22.96 | 97.34 |
| | 23 | 108.33 | 103.44 | 92.34 | 91.17 | 395.28 |
| | 24 | 45.21 | 41.2 | 35.99 | 37.2 | 159.6 |
| | 25 | 25.75 | 24.4 | 24.43 | 21.34 | 95.92 |
| | 26 | 24.81 | 22.02 | 24.66 | 23.63 | 95.12 |
| | 27 | 158.66 | 157.6 | 158.71 | 148.88 | 623.85 |
| TOTAL | | 1,389.85 | 1,337.34 | 1,266.41 | 1,235.44 | 5,229.04 |

STATE: MICHIGAN

| ZIP CODE | | QUARTER 1 | QUARTER 2 | QUARTER 3 | QUARTER 4 | TOTAL GRAMS |
|---|---|---|---|---|---|---|
| | 480 | 386.97 | 363.73 | 338.98 | 318.51 | 1,408.19 |
| | 481 | 443.02 | 433.49 | 407.1 | 396.45 | 1,680.06 |
| | 482 | 97.97 | 97.42 | 92.93 | 82.83 | 371.15 |
| | 483 | 331.26 | 323.66 | 296.55 | 278.47 | 1,229.94 |
| | 484 | 133.23 | 127.94 | 121.55 | 113.77 | 496.49 |
| | 485 | 69.62 | 67.06 | 61.85 | 67.4 | 265.93 |
| | 486 | 212.9 | 194.43 | 195.1 | 188.1 | 790.53 |
| | 487 | 100.52 | 96.07 | 85.93 | 83.17 | 365.69 |
| | 488 | 175.85 | 174.56 | 157.76 | 149.18 | 657.35 |
| | 489 | 61.73 | 60.54 | 57.46 | 54.69 | 234.42 |
| | 490 | 348.21 | 333.4 | 316.83 | 299.96 | 1,298.40 |
| | 491 | 33.77 | 34.11 | 33.04 | 33.66 | 134.58 |
| | 492 | 110.89 | 112.27 | 100.35 | 95.2 | 418.71 |
| | 493 | 68.07 | 66.64 | 66.63 | 58.97 | 260.31 |
| | 494 | 184.55 | 175.87 | 173.56 | 172.39 | 706.37 |
| | 495 | 182.83 | 181.21 | 176.11 | 166.09 | 706.24 |
| | 496 | 145.1 | 138.8 | 136.1 | 130.54 | 550.54 |
| | 497 | 102.53 | 104.49 | 95.46 | 92.32 | 394.8 |
| | 498 | 79.68 | 75.11 | 79.65 | 74.58 | 309.02 |
| | 499 | 26.79 | 24.83 | 23.94 | 21.7 | 97.26 |
| TOTAL | | 3,295.49 | 3,185.63 | 3,016.88 | 2,877.98 | 12,375.98 |

STATE: MINNESOTA

| ZIP CODE | | QUARTER 1 | QUARTER 2 | QUARTER 3 | QUARTER 4 | TOTAL GRAMS |
|---|---|---|---|---|---|---|

| | | | | | | |
|---|---|---|---|---|---|---|
| | 550 | 108.68 | 103.7 | 100.47 | 93.99 | 406.84 |
| | 551 | 150.84 | 149.08 | 144.17 | 140.68 | 584.77 |
| | 553 | 137 | 126.67 | 118.33 | 120.16 | 502.16 |
| | 554 | 225.96 | 252.74 | 216.13 | 212.78 | 907.61 |
| | 556 | 2.51 | 2.16 | 2.48 | 2.58 | 9.73 |
| | 557 | 39.89 | 37.56 | 36.04 | 33.27 | 146.76 |
| | 558 | 29.56 | 30.23 | 30.5 | 24.66 | 114.95 |
| | 559 | 65.36 | 62.94 | 62.12 | 64.59 | 255.01 |
| | 560 | 46.07 | 38.49 | 41.81 | 38.18 | 164.55 |
| | 561 | 20.97 | 19.55 | 17.41 | 19.26 | 77.19 |
| | 562 | 45.61 | 41.92 | 40.04 | 36.35 | 163.92 |
| | 563 | 79.63 | 76.01 | 72.7 | 66.72 | 295.06 |
| | 564 | 39.1 | 38.32 | 34.17 | 34.71 | 146.3 |
| | 565 | 41.07 | 41.31 | 34.25 | 32.36 | 148.99 |
| | 566 | 18.74 | 18.94 | 18.21 | 15.74 | 71.63 |
| | 567 | 29.42 | 29.58 | 32.03 | 29.16 | 120.19 |
| TOTAL | | 1,080.41 | 1,069.20 | 1,000.86 | 965.19 | 4,115.66 |

STATE: MISSISSIPPI

| ZIP CODE | | QUARTER 1 | QUARTER 2 | QUARTER 3 | QUARTER 4 | TOTAL GRAMS |
|---|---|---|---|---|---|---|
| | 386 | 136.15 | 127.89 | 119.44 | 110.3 | 493.78 |
| | 387 | 26.71 | 24.59 | 21.82 | 22.14 | 95.26 |
| | 388 | 106.48 | 107.46 | 103.25 | 103.17 | 420.36 |
| | 389 | 30.35 | 30.84 | 26.11 | 25.76 | 113.06 |
| | 390 | 80.26 | 77.34 | 75.7 | 65.42 | 298.72 |
| | 391 | 275.01 | 362.24 | 331.9 | 316.27 | 1,285.42 |
| | 392 | 107.11 | 96.51 | 98.79 | 92.62 | 395.03 |
| | 393 | 105.82 | 102.26 | 94.97 | 86.06 | 389.11 |
| | 394 | 166.77 | 165.19 | 154.39 | 140.64 | 626.99 |
| | 395 | 122.41 | 116.15 | 101.27 | 95.45 | 435.28 |
| | 396 | 61.44 | 67.11 | 54.37 | 52.16 | 235.08 |
| | 397 | 34.57 | 34.12 | 32.14 | 33.7 | 134.53 |
| TOTAL | | 1,253.08 | 1,311.70 | 1,214.15 | 1,143.69 | 4,922.62 |

STATE: MISSOURI

| ZIP CODE | QUARTER 1 | QUARTER 2 | QUARTER 3 | QUARTER 4 | TOTAL GRAMS |
|---|---|---|---|---|---|
| 630 | 206 | 203.98 | 183.46 | 186.94 | 780.38 |
| 631 | 373.02 | 360.81 | 352.99 | 830.57 | 1,917.39 |
| 633 | 125.9 | 124.08 | 113.65 | 109.79 | 473.42 |
| 634 | 28.88 | 29.74 | 28.61 | 26.93 | 114.16 |
| 635 | 34.88 | 34.05 | 32.24 | 31.91 | 133.08 |
| 636 | 52.67 | 50.58 | 48.77 | 52.48 | 204.5 |
| 637 | 43.36 | 44.75 | 39.55 | 36.19 | 163.85 |
| 638 | 46.71 | 43.16 | 44.66 | 38.88 | 173.41 |
| 639 | 22.24 | 21.55 | 15.89 | 16.08 | 75.76 |
| 640 | 217.31 | 210.12 | 209.16 | 197.2 | 833.79 |
| 641 | 198.92 | 173.61 | 171.33 | 169.18 | 713.04 |
| 644 | 32.79 | 30.48 | 30.82 | 30.54 | 124.63 |
| 645 | 46.89 | 50.53 | 44.42 | 40.36 | 182.2 |
| 646 | 29.47 | 28.1 | 28.14 | 27.91 | 113.62 |
| 647 | 67.21 | 66.86 | 60.66 | 55.83 | 250.56 |
| 648 | 79.9 | 77.68 | 72.2 | 73.67 | 303.45 |
| 650 | 45.02 | 47.15 | 47.2 | 45.41 | 184.78 |
| 651 | 29.52 | 27.25 | 28.61 | 24.7 | 110.08 |
| 652 | 81.02 | 82.19 | 81.26 | 77.5 | 321.97 |
| 653 | 33.11 | 35 | 30.95 | 29.9 | 128.96 |
| 654 | 41.78 | 39.42 | 37.62 | 33.69 | 152.51 |
| 655 | 43.46 | 46.34 | 40.04 | 36.55 | 166.39 |
| 656 | 88.56 | 80.97 | 83.33 | 76.24 | 329.1 |
| 657 | 131.53 | 122.28 | 119.49 | 117.35 | 490.65 |
| 658 | 158.64 | 168.98 | 157.76 | 158.01 | 643.39 |
| TOTAL | 2,258.79 | 2,199.66 | 2,102.81 | 2,523.81 | 9,085.07 |

STATE: MONTANA

| ZIP CODE | QUARTER 1 | QUARTER 2 | QUARTER 3 | QUARTER 4 | TOTAL GRAMS |
|---|---|---|---|---|---|
| 590 | 19.94 | 18.87 | 19.61 | 19.72 | 78.14 |
| 591 | 71.31 | 68.4 | 59.45 | 61.23 | 260.39 |
| 592 | 14.23 | 13.27 | 12.26 | 12.67 | 52.43 |
| 593 | 16.64 | 15.19 | 13.66 | 13.02 | 58.51 |
| 594 | 52.52 | 45.09 | 42.41 | 42.39 | 182.41 |
| 595 | 11.65 | 12.26 | 11.87 | 10.78 | 46.56 |

| | | | | | |
|---|---|---|---|---|---|
| | 596 | 35.48 | 33.43 | 29.88 | 29.64 | 128.43 |
| | 597 | 34.74 | 33.07 | 30.68 | 31.29 | 129.78 |
| | 598 | 58.95 | 58.65 | 58.18 | 54.24 | 230.02 |
| | 599 | 38.52 | 38.39 | 37.93 | 36.59 | 151.43 |
| TOTAL | | 353.98 | 336.62 | 315.93 | 311.57 | 1,318.10 |

STATE: NEBRASKA

| ZIP CODE | | QUARTER 1 | QUARTER 2 | QUARTER 3 | QUARTER 4 | TOTAL GRAMS |
|---|---|---|---|---|---|---|
| | 680 | 68.26 | 68.08 | 67.89 | 64.42 | 268.65 |
| | 681 | 160.68 | 143.96 | 133.93 | 133.02 | 571.59 |
| | 683 | 25.4 | 28.11 | 26.24 | 25.39 | 105.14 |
| | 684 | 17.35 | 16.93 | 16.52 | 15.84 | 66.64 |
| | 685 | 57.08 | 55.99 | 54.25 | 54.36 | 221.68 |
| | 686 | 14.99 | 13.39 | 13.64 | 13.26 | 55.28 |
| | 687 | 39.89 | 35.5 | 34.87 | 33.19 | 143.45 |
| | 688 | 55.37 | 61.41 | 58.34 | 59.02 | 234.14 |
| | 689 | 35.25 | 28.77 | 26.4 | 25.66 | 116.08 |
| | 690 | 16.84 | 16.98 | 18.37 | 17.25 | 69.44 |
| | 691 | 33.51 | 33.6 | 31.42 | 30.09 | 128.62 |
| | 692 | 2.92 | 3.44 | 3.08 | 2.49 | 11.93 |
| | 693 | 34.18 | 34.01 | 35.31 | 32.9 | 136.4 |
| TOTAL | | 561.72 | 540.17 | 520.26 | 506.89 | 2,129.04 |

STATE: NEVADA

| ZIP CODE | | QUARTER 1 | QUARTER 2 | QUARTER 3 | QUARTER 4 | TOTAL GRAMS |
|---|---|---|---|---|---|---|
| | 890 | 168.55 | 152.54 | 140.95 | 136.12 | 598.16 |
| | 891 | 331.16 | 297.6 | 281.47 | 277.66 | 1,187.89 |
| | 893 | 1.53 | 1.56 | 1.54 | 2.01 | 6.64 |
| | 894 | 48.69 | 43.34 | 42.13 | 38.52 | 172.68 |
| | 895 | 76.54 | 75.27 | 69.07 | 59.89 | 280.77 |
| | 897 | 18.52 | 18.13 | 14.21 | 15.05 | 65.91 |
| | 898 | 11.34 | 11.22 | 10.16 | 9.14 | 41.86 |
| TOTAL | | 656.33 | 599.66 | 559.53 | 538.39 | 2,353.91 |

STATE: NEW HAMPSHIRE

| ZIP CODE | QUARTER 1 | QUARTER 2 | QUARTER 3 | QUARTER 4 | TOTAL GRAMS |
|---|---|---|---|---|---|
| 30 | 101.43 | 100.7 | 94.56 | 95.32 | 392.01 |
| 31 | 48.73 | 46.12 | 43.84 | 43.65 | 182.34 |
| 32 | 29.52 | 25.54 | 27.39 | 23.93 | 106.38 |
| 33 | 45.43 | 44.74 | 41.53 | 39.13 | 170.83 |
| 34 | 9.9 | 10.13 | 10.02 | 10.11 | 40.16 |
| 35 | 17.81 | 15.17 | 15.05 | 12.09 | 60.12 |
| 36 | 1.96 | 1.88 | 1.31 | 1.43 | 6.58 |
| 37 | 30.1 | 26.45 | 26.28 | 23.86 | 106.69 |
| 38 | 87.19 | 80.47 | 80.46 | 77.89 | 326.01 |
| TOTAL | 372.07 | 351.2 | 340.44 | 327.41 | 1,391.12 |

STATE: NEW JERSEY

| ZIP CODE | QUARTER 1 | QUARTER 2 | QUARTER 3 | QUARTER 4 | TOTAL GRAMS |
|---|---|---|---|---|---|
| 70 | 447.33 | 453.38 | 389.93 | 365.68 | 1,656.32 |
| 71 | 40.98 | 36.04 | 34.88 | 33.53 | 145.43 |
| 72 | 13.98 | 14.43 | 12.93 | 11.84 | 53.18 |
| 73 | 18.12 | 18.11 | 14.71 | 13.16 | 64.1 |
| 74 | 172.49 | 141.06 | 135.43 | 171.06 | 620.04 |
| 75 | 30.83 | 30.67 | 26.8 | 25.7 | 114 |
| 76 | 214.03 | 204 | 200.92 | 191.74 | 810.69 |
| 77 | 189.47 | 183.26 | 168.88 | 166.52 | 708.13 |
| 78 | 74.62 | 74.43 | 63.96 | 60.15 | 273.16 |
| 79 | 75.19 | 61.89 | 66.69 | 67.92 | 271.69 |
| 80 | 438.07 | 417.65 | 362.95 | 336.23 | 1,554.90 |
| 81 | 36.41 | 36.98 | 32.21 | 26.46 | 132.06 |
| 82 | 82.45 | 74.9 | 70.71 | 57.27 | 285.33 |
| 83 | 39.67 | 35.35 | 32.61 | 31.75 | 139.38 |
| 84 | 11.84 | 10.82 | 10.18 | 9.88 | 42.72 |
| 85 | 70.49 | 71.52 | 63.01 | 58.48 | 263.5 |
| 86 | 74.72 | 73.13 | 66.39 | 65.32 | 279.56 |
| 87 | 141.2 | 140.5 | 129.44 | 126.44 | 537.58 |
| 88 | 298.52 | 293.63 | 263.58 | 264.72 | 1,120.45 |
| 89 | 26.01 | 27.51 | 25.19 | 27.82 | 106.53 |

| | | | | | | |
|---|---|---|---|---|---|---|
| TOTAL | | 2,496.42 | 2,399.26 | 2,171.40 | 2,111.67 | 9,178.75 |

STATE: NEW MEXICO

| ZIP CODE | | QUARTER 1 | QUARTER 2 | QUARTER 3 | QUARTER 4 | TOTAL GRAMS |
|---|---|---|---|---|---|---|
| | 870 | 29.17 | 28.86 | 24.93 | 23.54 | 106.5 |
| | 871 | 220.32 | 206.23 | 182.89 | 170.12 | 779.56 |
| | 873 | 8.38 | 7.17 | 5.68 | 6.5 | 27.73 |
| | 874 | 29.88 | 26.89 | 26.69 | 21.6 | 105.06 |
| | 875 | 55.74 | 48.96 | 47.12 | 46.34 | 198.16 |
| | 877 | 7.46 | 8.34 | 7.68 | 7.45 | 30.93 |
| | 878 | 3.09 | 1.65 | 1.41 | 1.61 | 7.76 |
| | 879 | 3.72 | 4.34 | 4.04 | 4.54 | 16.64 |
| | 880 | 33.21 | 28.73 | 28.07 | 29.23 | 119.24 |
| | 881 | 12.31 | 12.15 | 11.4 | 11.62 | 47.48 |
| | 882 | 59.85 | 52.02 | 49.56 | 48.99 | 210.42 |
| | 883 | 18.95 | 17.17 | 17.93 | 17.55 | 71.6 |
| | 884 | 4.08 | 3.61 | 2.66 | 3.61 | 13.96 |
| TOTAL | | 486.16 | 446.12 | 410.06 | 392.7 | 1,735.04 |

STATE: NEW YORK

| ZIP CODE | | QUARTER 1 | QUARTER 2 | QUARTER 3 | QUARTER 4 | TOTAL GRAMS |
|---|---|---|---|---|---|---|
| | 100 | 436.67 | 415.32 | 376.28 | 367.11 | 1,595.38 |
| | 101 | 9.7 | 10.37 | 9.66 | 8.94 | 38.67 |
| | 102 | 0.28 | 0.73 | 0.61 | 0.41 | 2.03 |
| | 103 | 149.1 | 152.17 | 131.65 | 136.38 | 569.3 |
| | 104 | 161.72 | 164.77 | 147.28 | 143.07 | 616.84 |
| | 105 | 122.88 | 115.28 | 107.95 | 103.03 | 449.14 |
| | 106 | 18.19 | 20.66 | 18.55 | 22.15 | 79.55 |
| | 107 | 42.99 | 36.22 | 28.61 | 28.05 | 135.87 |
| | 108 | 13.44 | 13.96 | 11.6 | 10.91 | 49.91 |
| | 109 | 152.7 | 128.23 | 113.42 | 109.44 | 503.79 |
| | 110 | 95.99 | 94.5 | 84.31 | 82.3 | 357.1 |
| | 111 | 23.5 | 20.91 | 17.97 | 19.51 | 81.89 |
| | 112 | 260.58 | 258.06 | 236.34 | 220.57 | 975.55 |
| | 113 | 113.65 | 115.15 | 100.04 | 101.08 | 429.92 |

| | | | | | | |
|---|---|---|---|---|---|---|
| | 114 | 46.6 | 46.2 | 50.75 | 62.21 | 205.76 |
| | 115 | 164.76 | 158.61 | 160.45 | 152.34 | 636.16 |
| | 116 | 10.59 | 11.69 | 10.24 | 12.02 | 44.54 |
| | 117 | 512.34 | 492.43 | 454.55 | 437 | 1,896.32 |
| | 118 | 20.72 | 26.03 | 22.12 | 17.29 | 86.16 |
| | 119 | 65.96 | 65.89 | 55.32 | 58.69 | 245.86 |
| | 120 | 95.41 | 100.67 | 98.87 | 95.88 | 390.83 |
| | 121 | 43.88 | 43.14 | 40.43 | 38.49 | 165.94 |
| | 122 | 88.06 | 74.19 | 69.84 | 73.37 | 305.46 |
| | 123 | 31.46 | 28.15 | 27.35 | 26.25 | 113.21 |
| | 124 | 52.49 | 52.29 | 53.89 | 47.96 | 206.63 |
| | 125 | 103.63 | 95.21 | 90.89 | 92.33 | 382.06 |
| | 126 | 30.76 | 29.51 | 25.26 | 26.88 | 112.41 |
| | 127 | 44.54 | 37.41 | 36.84 | 34.68 | 153.47 |
| | 128 | 98.66 | 90.74 | 89.07 | 86.48 | 364.95 |
| | 129 | 57.28 | 52.19 | 54.99 | 48.53 | 212.99 |
| | 130 | 103.41 | 96.67 | 92.13 | 89.22 | 381.43 |
| | 131 | 38.23 | 32.58 | 30.53 | 28.58 | 129.92 |
| | 132 | 122.28 | 122.76 | 106.47 | 109.58 | 461.09 |
| | 133 | 48.1 | 47.52 | 37.9 | 38.12 | 171.64 |
| | 134 | 73.79 | 71.24 | 59.71 | 56.2 | 260.94 |
| | 135 | 30.19 | 27.12 | 28.37 | 25.2 | 110.88 |
| | 136 | 73.64 | 69.54 | 63.84 | 63.68 | 270.7 |
| | 137 | 31.12 | 32.25 | 29.63 | 28.18 | 121.18 |
| | 138 | 90.98 | 86.18 | 81.34 | 77.66 | 336.16 |
| | 139 | 34.8 | 34.24 | 32.15 | 32.2 | 133.39 |
| | 140 | 122.06 | 112.19 | 106.71 | 99.11 | 440.07 |
| | 141 | 68.59 | 67.14 | 59.48 | 56.62 | 251.83 |
| | 142 | 200.2 | 184.18 | 160.29 | 159.98 | 704.65 |
| | 143 | 26.22 | 24.78 | 20.01 | 18.69 | 89.7 |
| | 144 | 80.05 | 80.79 | 76.71 | 75.81 | 313.36 |
| | 145 | 78.88 | 74.42 | 68.87 | 61.26 | 283.43 |
| | 146 | 159.23 | 151.57 | 147.21 | 136.21 | 594.22 |
| | 147 | 66.42 | 63.75 | 56.55 | 51.64 | 238.36 |
| | 148 | 115.68 | 109.35 | 100.31 | 98.78 | 424.12 |
| | 149 | 28.77 | 30 | 28.72 | 25.16 | 112.65 |
| TOTAL | | 4,661.17 | 4,468.95 | 4,112.06 | 3,995.23 | 17,237.41 |

STATE: NORTH CAROLINA

| ZIP CODE | QUARTER 1 | QUARTER 2 | QUARTER 3 | QUARTER 4 | TOTAL GRAMS |
|---|---|---|---|---|---|
| 270 | 86.49 | 76.91 | 69.24 | 63.73 | 296.37 |
| 271 | 112.55 | 100.78 | 96.95 | 89.6 | 399.88 |
| 272 | 195.89 | 181.49 | 162.56 | 169.41 | 709.35 |
| 273 | 116.11 | 100.51 | 97.38 | 94.36 | 408.36 |
| 274 | 61.79 | 59.43 | 55.68 | 59.5 | 236.4 |
| 275 | 272.58 | 270.76 | 245.93 | 248.97 | 1,038.24 |
| 276 | 130.15 | 122.65 | 110.24 | 115.29 | 478.33 |
| 277 | 96.92 | 107.5 | 80.91 | 82.08 | 367.41 |
| 278 | 208.97 | 191.01 | 180.53 | 180.48 | 760.99 |
| 279 | 66.96 | 63.36 | 60.49 | 63.45 | 254.26 |
| 280 | 237.68 | 213.97 | 199.96 | 199.38 | 850.99 |
| 281 | 182.9 | 172.18 | 153.52 | 150.19 | 658.79 |
| 282 | 147.39 | 136.47 | 128.33 | 122.79 | 534.98 |
| 283 | 256.31 | 234.53 | 201.6 | 182.84 | 875.28 |
| 284 | 197.07 | 181.71 | 164.92 | 165.86 | 709.56 |
| 285 | 195.07 | 195.53 | 173.84 | 167.22 | 731.66 |
| 286 | 276.79 | 265.3 | 244.7 | 237.15 | 1,023.94 |
| 287 | 165.54 | 152.81 | 139.01 | 127.51 | 584.87 |
| 288 | 70.86 | 69.29 | 60.45 | 65.87 | 266.47 |
| 289 | 20.53 | 20.37 | 18.81 | 20.39 | 80.1 |
| TOTAL | 3,098.55 | 2,916.56 | 2,645.05 | 2,606.07 | 11,266.23 |

STATE: NORTH DAKOTA

| ZIP CODE | QUARTER 1 | QUARTER 2 | QUARTER 3 | QUARTER 4 | TOTAL GRAMS |
|---|---|---|---|---|---|
| 580 | 14.38 | 13.68 | 12.46 | 13.03 | 53.55 |
| 581 | 50.3 | 50.94 | 50.25 | 44.33 | 195.82 |
| 582 | 43.34 | 39.56 | 39.29 | 40.84 | 163.03 |
| 583 | 20.64 | 17.88 | 16.64 | 16.32 | 71.48 |
| 584 | 18.43 | 16.39 | 14.92 | 14.26 | 64 |
| 585 | 57.25 | 58.63 | 54.26 | 57.8 | 227.94 |
| 586 | 17.61 | 17.13 | 16.38 | 15.22 | 66.34 |
| 587 | 16.92 | 17.13 | 17.45 | 17.12 | 68.62 |

| | | | | | | |
|---|---|---|---|---|---|---|
| | 588 | 9.28 | 9.07 | 9.09 | 9.64 | 37.08 |
| TOTAL | | 248.15 | 240.41 | 230.74 | 228.56 | 947.86 |

| STATE: | OHIO | | | | |
|---|---|---|---|---|---|
| ZIP CODE | QUARTER 1 | QUARTER 2 | QUARTER 3 | QUARTER 4 | TOTAL GRAMS |
| 430 | 160.12 | 157.31 | 141.19 | 138.6 | 597.22 |
| 431 | 80.83 | 80.39 | 76.24 | 75.19 | 312.65 |
| 432 | 357.46 | 355.27 | 318.39 | 305.76 | 1,336.88 |
| 433 | 37.23 | 33.96 | 30.84 | 28.91 | 130.94 |
| 434 | 32.95 | 33.51 | 32.2 | 29.9 | 128.56 |
| 435 | 86.5 | 87.78 | 83.79 | 78.01 | 336.08 |
| 436 | 159.65 | 153.18 | 152.31 | 136.88 | 602.02 |
| 437 | 39.93 | 37.87 | 36.56 | 33.15 | 147.51 |
| 438 | 4.36 | 3.93 | 3.77 | 3.43 | 15.49 |
| 439 | 39.94 | 38.48 | 39.01 | 34.71 | 152.14 |
| 440 | 128.89 | 122.69 | 111.81 | 102.31 | 465.7 |
| 441 | 183.32 | 180.11 | 173.17 | 162.84 | 699.44 |
| 442 | 93.91 | 82.58 | 75.02 | 69.08 | 320.59 |
| 443 | 39.23 | 37.46 | 33.41 | 34.66 | 144.76 |
| 444 | 57.01 | 53.79 | 49.45 | 46.72 | 206.97 |
| 445 | 63.87 | 61.08 | 61.62 | 58.29 | 244.86 |
| 446 | 94.32 | 81.02 | 72.13 | 64.7 | 312.17 |
| 447 | 55.12 | 52.32 | 47.81 | 48.28 | 203.53 |
| 448 | 63.02 | 53.85 | 53.06 | 50.91 | 220.84 |
| 449 | 23.15 | 20.14 | 16.76 | 16.63 | 76.68 |
| 450 | 180.9 | 174.46 | 168.47 | 150.59 | 674.42 |
| 451 | 53.84 | 52.66 | 51.12 | 49.17 | 206.79 |
| 452 | 182.99 | 178.21 | 173.79 | 167.45 | 702.44 |
| 453 | 108 | 106.23 | 97.79 | 93 | 405.02 |
| 454 | 119.33 | 113.75 | 103.21 | 106.93 | 443.22 |
| 455 | 27.91 | 25.99 | 22.65 | 22.01 | 98.56 |
| 456 | 90.58 | 85.53 | 86.27 | 83.97 | 346.35 |
| 457 | 29.57 | 27.64 | 26.28 | 26.01 | 109.5 |
| 458 | 57.87 | 53.31 | 48.82 | 48.89 | 208.89 |
| TOTAL | 2,651.80 | 2,544.50 | 2,386.94 | 2,266.98 | 9,850.22 |

STATE: OKLAHOMA

| ZIP CODE | QUARTER 1 | QUARTER 2 | QUARTER 3 | QUARTER 4 | TOTAL GRAMS |
|---|---|---|---|---|---|
| 730 | 293.33 | 277.28 | 265.35 | 239.87 | 1,075.83 |
| 731 | 354.13 | 301.43 | 278.95 | 265.6 | 1,200.11 |
| 734 | 45.65 | 44.01 | 43.22 | 37.7 | 170.58 |
| 735 | 121.95 | 115.92 | 115.34 | 104.29 | 457.5 |
| 736 | 45.86 | 39.73 | 34.8 | 28.93 | 149.32 |
| 737 | 50.42 | 48.38 | 43.98 | 42.05 | 184.83 |
| 738 | 12.24 | 13.09 | 11.21 | 11.18 | 47.72 |
| 739 | 3.82 | 2.35 | 3.31 | 2.64 | 12.12 |
| 740 | 304.14 | 294.16 | 285.07 | 268.76 | 1,152.13 |
| 741 | 239.33 | 213.72 | 213.95 | 214.87 | 881.87 |
| 743 | 59.09 | 56.41 | 55.36 | 53.41 | 224.27 |
| 744 | 107.48 | 113.9 | 97.73 | 90.42 | 409.53 |
| 745 | 45.51 | 44.91 | 37.72 | 39.4 | 167.54 |
| 746 | 24.7 | 26.96 | 25.24 | 24.26 | 101.16 |
| 747 | 41.18 | 35.49 | 30.88 | 29.21 | 136.76 |
| 748 | 71.77 | 74.66 | 65.84 | 66.8 | 279.07 |
| 749 | 26.85 | 27.9 | 25.62 | 24.44 | 104.81 |
| TOTAL | 1,847.45 | 1,730.30 | 1,633.57 | 1,543.83 | 6,755.15 |

STATE: OREGON

| ZIP CODE | QUARTER 1 | QUARTER 2 | QUARTER 3 | QUARTER 4 | TOTAL GRAMS |
|---|---|---|---|---|---|
| 970 | 153.12 | 140.76 | 132.16 | 122.52 | 548.56 |
| 971 | 78.97 | 73.6 | 74.55 | 72.48 | 299.6 |
| 972 | 221.65 | 218.72 | 189.72 | 197.72 | 827.81 |
| 973 | 135.65 | 130.97 | 123.52 | 116.85 | 506.99 |
| 974 | 183.49 | 169.68 | 160.28 | 151.66 | 665.11 |
| 975 | 93.57 | 83.8 | 76.53 | 75.47 | 329.37 |
| 976 | 15.82 | 13.94 | 14.65 | 12.74 | 57.15 |
| 977 | 54.04 | 58.54 | 54.89 | 51.83 | 219.3 |
| 978 | 53.33 | 48.72 | 45.32 | 43.61 | 190.98 |
| 979 | 16.87 | 15.97 | 15.2 | 12.73 | 60.77 |
| TOTAL | 1,006.51 | 954.7 | 886.82 | 857.61 | 3,705.64 |

DATE RANGE: 01/01/2017 TO 12/31/2017

ARCOS 3 - REPORT 1

Run Date: 07/03/2018

RETAIL DRUG DISTRIBUTION BY ZIP CODE WITHIN STATE BY GRAMS WT

STATE: PENNSYLVANIA

| ZIP CODE | QUARTER 1 | QUARTER 2 | QUARTER 3 | QUARTER 4 | TOTAL GRAMS |
|---|---|---|---|---|---|
| 150 | 251.59 | 253.32 | 235.81 | 232.61 | 973.33 |
| 151 | 109.41 | 114.42 | 110.31 | 104 | 438.14 |
| 152 | 266.37 | 259.24 | 245.08 | 225.74 | 996.43 |
| 153 | 48.07 | 48.42 | 42.31 | 39.53 | 178.33 |
| 154 | 54.69 | 51.24 | 49.69 | 44.21 | 199.83 |
| 155 | 48.75 | 43.39 | 42.77 | 38.72 | 173.63 |
| 156 | 124.94 | 124.03 | 108.77 | 106.86 | 464.6 |
| 157 | 88.48 | 90.43 | 83.76 | 74.87 | 337.54 |
| 158 | 68.81 | 64.25 | 61.73 | 58.27 | 253.06 |
| 159 | 101.25 | 96.46 | 95.21 | 81.57 | 374.49 |
| 160 | 79.63 | 76.91 | 79.57 | 76.18 | 312.29 |
| 161 | 87.98 | 83.09 | 78.03 | 71.8 | 320.9 |
| 162 | 46 | 43.77 | 40.32 | 34.81 | 164.9 |
| 163 | 97.44 | 96.34 | 92.83 | 91.73 | 378.34 |
| 164 | 26.62 | 25.87 | 25.3 | 27.07 | 104.86 |
| 165 | 79.96 | 74.59 | 70.59 | 65.35 | 290.49 |
| 166 | 115.02 | 118.2 | 114.3 | 102 | 449.52 |
| 167 | 19.25 | 20.31 | 18.74 | 16.46 | 74.76 |
| 168 | 46.01 | 49.03 | 45.74 | 38.29 | 179.07 |
| 169 | 25.41 | 24.46 | 24.47 | 20.25 | 94.59 |
| 170 | 235.36 | 214.16 | 200.38 | 193.33 | 843.23 |
| 171 | 65.55 | 58.57 | 55.55 | 53.14 | 232.81 |
| 172 | 51.86 | 51.4 | 53.19 | 48.94 | 205.39 |
| 173 | 114.78 | 109.48 | 104.13 | 95.33 | 423.72 |
| 174 | 71.77 | 69.32 | 67.36 | 64.89 | 273.34 |
| 175 | 69.31 | 61.58 | 59.09 | 53.87 | 243.85 |
| 176 | 58.68 | 56.19 | 53.98 | 55.82 | 224.67 |
| 177 | 90.38 | 87.61 | 77.75 | 77.75 | 333.49 |
| 178 | 118.8 | 111.23 | 104.79 | 99.07 | 433.89 |
| 179 | 38.08 | 40.57 | 38.53 | 35.59 | 152.77 |
| 180 | 159.42 | 154.39 | 138.59 | 130.74 | 583.14 |
| 181 | 111.66 | 100.23 | 92.39 | 94.87 | 399.15 |
| 182 | 69.38 | 67.75 | 58.96 | 59.38 | 255.47 |
| 183 | 58.21 | 53.62 | 54.42 | 53.8 | 220.05 |

| | | | | | | |
|---|---|---|---|---|---|---|
| | 184 | 73.65 | 71.72 | 67.95 | 63.49 | 276.81 |
| | 185 | 113.94 | 106.43 | 95.83 | 90.2 | 406.4 |
| | 186 | 85.69 | 79.56 | 81.16 | 72.35 | 318.76 |
| | 187 | 323.01 | 330.8 | 271.93 | 282.3 | 1,208.04 |
| | 188 | 35.82 | 36.27 | 32.69 | 32.76 | 137.54 |
| | 189 | 123.01 | 111.68 | 104.4 | 100.87 | 439.96 |
| | 190 | 712.99 | 690.26 | 641.1 | 629.52 | 2,673.87 |
| | 191 | 383.57 | 385.12 | 350.67 | 328.11 | 1,447.47 |
| | 193 | 120.35 | 114.28 | 98.79 | 100.79 | 434.21 |
| | 194 | 200.91 | 195.81 | 186.31 | 172.24 | 755.27 |
| | 195 | 50.99 | 45.42 | 46.51 | 41.45 | 184.37 |
| | 196 | 51.51 | 49.43 | 49.29 | 49.58 | 199.81 |
| TOTAL | | 5,374.36 | 5,210.65 | 4,851.07 | 4,630.50 | 20,066.58 |

STATE: PUERTO RICO

| ZIP CODE | | QUARTER 1 | QUARTER 2 | QUARTER 3 | QUARTER 4 | TOTAL GRAMS |
|---|---|---|---|---|---|---|
| | 006 | 51.42 | 30.57 | 20.77 | 16.64 | 119.4 |
| | 007 | 66.56 | 53.6 | 44.01 | 30.04 | 194.21 |
| | 009 | 78.89 | 66.84 | 61.79 | 57.12 | 264.64 |
| TOTAL | | 196.87 | 151.01 | 126.57 | 103.8 | 578.25 |

STATE: RHODE ISLAND

| ZIP CODE | | QUARTER 1 | QUARTER 2 | QUARTER 3 | QUARTER 4 | TOTAL GRAMS |
|---|---|---|---|---|---|---|
| | 028 | 121.94 | 110.33 | 108.55 | 107.6 | 448.42 |
| | 029 | 75.86 | 76.45 | 69.2 | 65.04 | 286.55 |
| TOTAL | | 197.8 | 186.78 | 177.75 | 172.64 | 734.97 |

STATE: SOUTH CAROLINA

| ZIP CODE | | QUARTER 1 | QUARTER 2 | QUARTER 3 | QUARTER 4 | TOTAL GRAMS |
|---|---|---|---|---|---|---|
| | 290 | 90.49 | 89.98 | 83.93 | 79.21 | 343.61 |
| | 291 | 102.8 | 99.33 | 88.82 | 97.39 | 388.34 |
| | 292 | 100.55 | 106.37 | 94.49 | 84.77 | 386.18 |
| | 293 | 125.96 | 121.88 | 108.72 | 109.04 | 465.6 |
| | 294 | 208.32 | 195.79 | 197 | 191.31 | 792.42 |

| | | | | | |
|---|---|---|---|---|---|
| 295 | 268.23 | 245.39 | 225.57 | 223.89 | 963.08 |
| 296 | 297.73 | 281.29 | 255.04 | 252.68 | 1,086.74 |
| 297 | 68.91 | 64.53 | 56.43 | 54.24 | 244.11 |
| 298 | 79.91 | 74.76 | 72.97 | 72.26 | 299.9 |
| 299 | 81.12 | 67.09 | 76.14 | 67.2 | 291.55 |
| TOTAL | 1,424.02 | 1,346.41 | 1,259.11 | 1,231.99 | 5,261.53 |

STATE: SOUTH DAKOTA

| ZIP CODE | QUARTER 1 | QUARTER 2 | QUARTER 3 | QUARTER 4 | TOTAL GRAMS |
|---|---|---|---|---|---|
| 570 | 48.87 | 47.82 | 47.43 | 45.81 | 189.93 |
| 571 | 145.72 | 127.19 | 125.62 | 122.05 | 520.58 |
| 572 | 19.67 | 16.45 | 17.17 | 17.36 | 70.65 |
| 573 | 23.06 | 21.77 | 23.2 | 23.36 | 91.39 |
| 574 | 21.45 | 22.06 | 19.55 | 18.98 | 82.04 |
| 575 | 19.45 | 19.04 | 15.09 | 13.91 | 67.49 |
| 576 | 2.84 | 3.6 | 3.84 | 4.45 | 14.73 |
| 577 | 44.26 | 43.95 | 43.89 | 35.32 | 167.42 |
| TOTAL | 325.32 | 301.88 | 295.79 | 281.24 | 1,204.23 |

STATE: TENNESSEE

| ZIP CODE | QUARTER 1 | QUARTER 2 | QUARTER 3 | QUARTER 4 | TOTAL GRAMS |
|---|---|---|---|---|---|
| 370 | 247.61 | 232.23 | 214.12 | 209.84 | 903.8 |
| 371 | 191.73 | 189.67 | 178.3 | 159.66 | 719.36 |
| 372 | 165 | 154.22 | 145.57 | 154.79 | 619.58 |
| 373 | 168.32 | 154.04 | 129.02 | 128.67 | 580.05 |
| 374 | 69.72 | 66.22 | 61.89 | 54.09 | 251.92 |
| 376 | 137.37 | 130.07 | 119.53 | 118.31 | 505.28 |
| 377 | 78.44 | 69.6 | 67.09 | 62.19 | 277.32 |
| 378 | 143.12 | 133.46 | 122.46 | 115.59 | 514.63 |
| 379 | 106.09 | 98.58 | 85.9 | 86.38 | 376.95 |
| 380 | 90.94 | 86.52 | 84.73 | 81.25 | 343.44 |
| 381 | 423.65 | 399.61 | 399.17 | 390.3 | 1,612.73 |
| 382 | 30.8 | 28.2 | 28.68 | 28.28 | 115.96 |

| | | | | | | |
|---|---|---|---|---|---|---|
| | 383 | 85.34 | 77.17 | 79.3 | 72.23 | 314.04 |
| | 384 | 52.2 | 50.26 | 43.06 | 44.03 | 189.55 |
| | 385 | 103.59 | 95.3 | 86.83 | 79.08 | 364.8 |
| TOTAL | | 2,093.92 | 1,965.15 | 1,845.65 | 1,784.69 | 7,689.41 |

STATE: TEXAS

| ZIP CODE | QUARTER 1 | QUARTER 2 | QUARTER 3 | QUARTER 4 | TOTAL GRAMS |
|---|---|---|---|---|---|
| 750 | 576.37 | 539.43 | 491.48 | 469.62 | 2,076.90 |
| 751 | 144.66 | 145.4 | 142.25 | 135.74 | 568.05 |
| 752 | 249.37 | 233.6 | 210.63 | 206.09 | 899.69 |
| 753 | 11.25 | 10.54 | 11.21 | 12.21 | 45.21 |
| 754 | 170.97 | 159.67 | 151.96 | 138.91 | 621.51 |
| 755 | 21.84 | 21.85 | 22.57 | 21.6 | 87.86 |
| 756 | 105.82 | 103.32 | 89.4 | 112.79 | 411.33 |
| 757 | 133.27 | 137.65 | 130.31 | 128.83 | 530.06 |
| 758 | 23.69 | 20.33 | 21.89 | 22.29 | 88.2 |
| 759 | 61.24 | 58 | 56.15 | 55.39 | 230.78 |
| 760 | 359.76 | 357.89 | 323.8 | 311.86 | 1,353.31 |
| 761 | 351.14 | 343.81 | 315.16 | 311.26 | 1,321.37 |
| 762 | 140.53 | 137.49 | 133.96 | 130.54 | 542.52 |
| 763 | 91.76 | 87 | 80.5 | 78.92 | 338.18 |
| 764 | 57.12 | 51.68 | 52.62 | 51.76 | 213.18 |
| 765 | 101.64 | 90.77 | 88.02 | 89.61 | 370.04 |
| 766 | 27.2 | 27.62 | 24.71 | 26.96 | 106.49 |
| 767 | 48.93 | 44.09 | 44.02 | 44.6 | 181.64 |
| 768 | 43.28 | 46.25 | 44.79 | 44.42 | 178.74 |
| 769 | 28.86 | 29.77 | 27.53 | 27.82 | 113.98 |
| 770 | 364.8 | 368.36 | 318.67 | 340.65 | 1,392.48 |
| 772 | 1.19 | 0.04 | 0.02 | 0.03 | 1.28 |
| 773 | 262.62 | 254.27 | 244.47 | 251 | 1,012.36 |
| 774 | 192.63 | 182.74 | 171.78 | 167.42 | 714.57 |
| 775 | 241.92 | 229.62 | 229.64 | 216.16 | 917.34 |
| 776 | 64.96 | 63.93 | 57.11 | 56.97 | 242.97 |
| 777 | 30.08 | 30 | 26.51 | 26.22 | 112.81 |
| 778 | 67.44 | 63.27 | 59.8 | 56.82 | 247.33 |
| 779 | 61.85 | 60.8 | 58.69 | 55.29 | 236.63 |

| | | | | | | |
|---|---|---|---|---|---|---|
| | 780 | 79.32 | 77.5 | 76.35 | 75.93 | 309.1 |
| | 781 | 89.44 | 88.39 | 82.61 | 81.21 | 341.65 |
| | 782 | 456.38 | 435.38 | 413.03 | 409.71 | 1,714.50 |
| | 783 | 53.14 | 48.72 | 45.3 | 41.16 | 188.32 |
| | 784 | 108.83 | 103.99 | 97.75 | 90.76 | 401.33 |
| | 785 | 134.84 | 124.63 | 129.59 | 126.74 | 515.8 |
| | 786 | 256.34 | 247.26 | 235.27 | 223.82 | 962.69 |
| | 787 | 244.34 | 232.25 | 208.19 | 198.57 | 883.35 |
| | 788 | 25.96 | 17.88 | 16.91 | 16.42 | 77.17 |
| | 789 | 12.12 | 10.79 | 12.86 | 11.68 | 47.45 |
| | 790 | 66.39 | 64.53 | 60.68 | 57 | 248.6 |
| | 791 | 120.98 | 117.66 | 111.34 | 108.83 | 458.81 |
| | 792 | 14.1 | 10.03 | 10.49 | 9.51 | 44.13 |
| | 793 | 38.6 | 36.91 | 35.02 | 33.3 | 143.83 |
| | 794 | 103.85 | 102.23 | 101.71 | 97.35 | 405.14 |
| | 795 | 35.24 | 34.51 | 33.52 | 32.41 | 135.68 |
| | 796 | 61.15 | 68.63 | 59.84 | 62.71 | 252.33 |
| | 797 | 54.35 | 58.43 | 55.79 | 54.35 | 222.92 |
| | 798 | 5.58 | 4.55 | 4.83 | 4.22 | 19.18 |
| | 799 | 131.52 | 129.84 | 122.3 | 113.69 | 497.35 |
| TOTAL | | 6,128.66 | 5,913.30 | 5,543.03 | 5,441.15 | 23,026.14 |

STATE: UTAH

| ZIP CODE | | QUARTER 1 | QUARTER 2 | QUARTER 3 | QUARTER 4 | TOTAL GRAMS |
|---|---|---|---|---|---|---|
| | 840 | 299.67 | 286.67 | 277.62 | 273.55 | 1,137.51 |
| | 841 | 220.92 | 205.9 | 204.15 | 199.43 | 830.4 |
| | 843 | 44.9 | 45.7 | 44.69 | 44.82 | 180.11 |
| | 844 | 64.82 | 63.1 | 61.26 | 58.68 | 247.86 |
| | 845 | 34.01 | 29.87 | 26.53 | 26.59 | 117 |
| | 846 | 81.88 | 78.67 | 78.36 | 72.09 | 311 |
| | 847 | 116.63 | 111.09 | 113.32 | 110.65 | 451.69 |
| TOTAL | | 862.83 | 821 | 805.93 | 785.81 | 3,275.57 |

STATE: VERMONT

| ZIP CODE | QUARTER 1 | QUARTER 2 | QUARTER 3 | QUARTER 4 | TOTAL GRAMS |
|---|---|---|---|---|---|

| | | | | | | |
|---|---|---|---|---|---|---|
| | 050 | 11.58 | 11.81 | 10.25 | 10.6 | 44.24 |
| | 051 | 9.72 | 6.45 | 6.45 | 5.44 | 28.06 |
| | 052 | 12.94 | 12.03 | 12 | 11.9 | 48.87 |
| | 053 | 5.89 | 4.67 | 5.84 | 3.67 | 20.07 |
| | 054 | 81.4 | 85.44 | 73.96 | 70.66 | 311.46 |
| | 056 | 27.93 | 25.8 | 24.17 | 19.88 | 97.78 |
| | 057 | 34.71 | 33.34 | 30.95 | 29.07 | 128.07 |
| | 058 | 18.58 | 17.18 | 16.53 | 17.21 | 69.5 |
| TOTAL | | 202.75 | 196.72 | 180.15 | 168.43 | 748.05 |

STATE: VIRGIN ISLANDS

| ZIP CODE | | QUARTER 1 | QUARTER 2 | QUARTER 3 | QUARTER 4 | TOTAL GRAMS |
|---|---|---|---|---|---|---|
| | 008 | 4.01 | 3.46 | 3.02 | 3.47 | 13.96 |
| TOTAL | | 4.01 | 3.46 | 3.02 | 3.47 | 13.96 |

STATE: VIRGINIA

| ZIP CODE | | QUARTER 1 | QUARTER 2 | QUARTER 3 | QUARTER 4 | TOTAL GRAMS |
|---|---|---|---|---|---|---|
| | 201 | 133.66 | 123.51 | 110.19 | 110.43 | 477.79 |
| | 220 | 73.94 | 60.76 | 65.1 | 52.49 | 252.29 |
| | 221 | 48.44 | 46.16 | 37.47 | 37.08 | 169.15 |
| | 222 | 15.77 | 14.94 | 13.93 | 9.98 | 54.62 |
| | 223 | 33.28 | 34.89 | 31.17 | 26.8 | 126.14 |
| | 224 | 48.85 | 44.37 | 41.39 | 40.47 | 175.08 |
| | 225 | 17.22 | 15.09 | 14.88 | 14.53 | 61.72 |
| | 226 | 56.15 | 59.32 | 55.49 | 54.71 | 225.67 |
| | 227 | 15.52 | 13 | 10.37 | 9.24 | 48.13 |
| | 228 | 47.2 | 49.31 | 43.79 | 46.28 | 186.58 |
| | 229 | 77.27 | 70.98 | 67.12 | 60.72 | 276.09 |
| | 230 | 64.5 | 65.77 | 59.34 | 56.69 | 246.3 |
| | 231 | 92.85 | 81.59 | 65.84 | 74.79 | 315.07 |
| | 232 | 179.99 | 156.32 | 148.69 | 133.47 | 618.47 |
| | 233 | 73.52 | 65.59 | 65.21 | 66.23 | 270.55 |
| | 234 | 158.37 | 149.43 | 139.51 | 134.04 | 581.35 |
| | 235 | 56.02 | 50.92 | 46.39 | 46.6 | 199.93 |

| | | | | | | |
|---|---|---|---|---|---|---|
| | 236 | 94.98 | 82.84 | 79.22 | 69.92 | 326.96 |
| | 237 | 50.14 | 43.66 | 41.14 | 41.64 | 176.58 |
| | 238 | 69.8 | 68.58 | 57.19 | 55.16 | 250.73 |
| | 239 | 20.28 | 17.29 | 16.59 | 14.99 | 69.15 |
| | 240 | 110.39 | 101.7 | 99.36 | 90.58 | 402.03 |
| | 241 | 88.43 | 83.55 | 71.34 | 67.81 | 311.13 |
| | 242 | 93.76 | 83.82 | 76.19 | 71.23 | 325 |
| | 243 | 43.54 | 41.29 | 35.45 | 32.93 | 153.21 |
| | 244 | 46.79 | 48.95 | 43.03 | 44.93 | 183.7 |
| | 245 | 86.43 | 83.55 | 85.13 | 82.48 | 337.59 |
| | 246 | 24.21 | 24.47 | 22.81 | 21.01 | 92.5 |
| TOTAL | | 1,921.30 | 1,781.65 | 1,643.33 | 1,567.23 | 6,913.51 |

STATE: WASHINGTON

| ZIP CODE | | QUARTER 1 | QUARTER 2 | QUARTER 3 | QUARTER 4 | TOTAL GRAMS |
|---|---|---|---|---|---|---|
| | 980 | 209.69 | 199.24 | 185.64 | 171.11 | 765.68 |
| | 981 | 170.15 | 183.94 | 151.42 | 155.94 | 661.45 |
| | 982 | 172.51 | 166.4 | 154.99 | 144.61 | 638.51 |
| | 983 | 155.33 | 155.78 | 150.46 | 143.13 | 604.7 |
| | 984 | 98.32 | 89.05 | 85.71 | 91.24 | 364.32 |
| | 985 | 78.43 | 71.39 | 72.02 | 70.08 | 291.92 |
| | 986 | 193.24 | 181.21 | 167.36 | 163.72 | 705.53 |
| | 988 | 35.33 | 37.15 | 35.41 | 36.66 | 144.55 |
| | 989 | 74.31 | 73.48 | 69.88 | 65.77 | 283.44 |
| | 990 | 22.88 | 23.89 | 24.77 | 21.79 | 93.33 |
| | 991 | 37.69 | 34.95 | 34.16 | 32.95 | 139.75 |
| | 992 | 178.35 | 175.27 | 172.79 | 161.79 | 688.2 |
| | 993 | 164.34 | 142.31 | 133 | 122.93 | 562.58 |
| | 994 | 24.02 | 22.49 | 21.33 | 20.6 | 88.44 |
| TOTAL | | 1,614.59 | 1,556.55 | 1,458.94 | 1,402.32 | 6,032.40 |

STATE: WEST VIRGINIA

| ZIP CODE | | QUARTER 1 | QUARTER 2 | QUARTER 3 | QUARTER 4 | TOTAL GRAMS |
|---|---|---|---|---|---|---|
| | 247 | 14.41 | 14.96 | 12.98 | 14.45 | 56.8 |
| | 248 | 5.45 | 4.83 | 4.24 | 5.08 | 19.6 |

| | | | | | | |
|---|---|---|---|---|---|---|
| | 249 | 13.41 | 9.07 | 8.3 | 7.6 | 38.38 |
| | 250 | 14.82 | 14.57 | 14.45 | 14.42 | 58.26 |
| | 251 | 10 | 8.48 | 7.69 | 9.28 | 35.45 |
| | 252 | 15.01 | 15.79 | 15.27 | 13.64 | 59.71 |
| | 253 | 71.92 | 68.34 | 67.97 | 59.32 | 267.55 |
| | 254 | 73.44 | 68.75 | 65.18 | 62 | 269.37 |
| | 255 | 29.38 | 28.98 | 27.8 | 25.93 | 112.09 |
| | 256 | 10.74 | 9.06 | 10.1 | 9.26 | 39.16 |
| | 257 | 51.38 | 49 | 43.16 | 40.16 | 183.7 |
| | 258 | 19.3 | 20.18 | 20 | 18.33 | 77.81 |
| | 259 | 12.42 | 11.41 | 9.97 | 8.7 | 42.5 |
| | 260 | 39.42 | 38.99 | 37.82 | 35.36 | 151.59 |
| | 261 | 29.05 | 29.59 | 27.97 | 26.35 | 112.96 |
| | 262 | 19.83 | 20.51 | 21.29 | 20.46 | 82.09 |
| | 263 | 35.61 | 36.27 | 31.09 | 29.34 | 132.31 |
| | 264 | 11.41 | 11.11 | 10.54 | 9.27 | 42.33 |
| | 265 | 63.25 | 61.59 | 57.84 | 56.41 | 239.09 |
| | 266 | 8.83 | 7.7 | 7.78 | 7.19 | 31.5 |
| | 267 | 19.99 | 20.62 | 20.18 | 17.92 | 78.71 |
| | 268 | 8.56 | 9.34 | 9.2 | 9.39 | 36.49 |
| TOTAL | | 577.63 | 559.14 | 530.82 | 499.86 | 2,167.45 |

| STATE: | WISCONSIN | | | | |
|---|---|---|---|---|---|
| ZIP CODE | QUARTER 1 | QUARTER 2 | QUARTER 3 | QUARTER 4 | TOTAL GRAMS |
| 530 | 161.07 | 147.86 | 142.48 | 136.22 | 587.63 |
| 531 | 155.35 | 155.87 | 132.47 | 126.47 | 570.16 |
| 532 | 250.59 | 226.51 | 210.82 | 205.07 | 892.99 |
| 534 | 29.09 | 25.75 | 22.52 | 23.28 | 100.64 |
| 535 | 159.85 | 150.08 | 133.45 | 127.08 | 570.46 |
| 537 | 118.99 | 100.96 | 90.54 | 89.73 | 400.22 |
| 538 | 22.37 | 22.36 | 22.7 | 21.28 | 88.71 |
| 539 | 74.56 | 69.83 | 62.03 | 54.01 | 260.43 |
| 540 | 25.81 | 24.3 | 24.02 | 21.38 | 95.51 |
| 541 | 38.93 | 38.54 | 34.89 | 32.63 | 144.99 |
| 542 | 24.76 | 23.2 | 21.94 | 22.2 | 92.1 |
| 543 | 57.1 | 59.22 | 54.02 | 52.81 | 223.15 |

| | | | | | | |
|---|---|---|---|---|---|---|
| | 544 | 82.56 | 71.01 | 66.39 | 65.49 | 285.45 |
| | 545 | 16.09 | 16.27 | 17.51 | 14.08 | 63.95 |
| | 546 | 96.7 | 98 | 89.84 | 87.62 | 372.16 |
| | 547 | 82.05 | 77.48 | 66.67 | 67.19 | 293.39 |
| | 548 | 41.87 | 40.56 | 33.12 | 35.44 | 150.99 |
| | 549 | 199.73 | 176.56 | 165.58 | 163.32 | 705.19 |
| TOTAL | | 1,637.47 | 1,524.36 | 1,390.99 | 1,345.30 | 5,898.12 |

STATE: WYOMING

| ZIP CODE | | QUARTER 1 | QUARTER 2 | QUARTER 3 | QUARTER 4 | TOTAL GRAMS |
|---|---|---|---|---|---|---|
| | 820 | 36.39 | 29.53 | 32.62 | 29.42 | 127.96 |
| | 822 | 7.57 | 7.14 | 5.7 | 5.81 | 26.22 |
| | 823 | 1.36 | 1.36 | 1.43 | 1.68 | 5.83 |
| | 824 | 17.39 | 16.99 | 18.9 | 17.93 | 71.21 |
| | 825 | 8.28 | 6.88 | 7.66 | 5.73 | 28.55 |
| | 826 | 31.74 | 29.68 | 27.33 | 25.68 | 114.43 |
| | 827 | 6.13 | 4.53 | 6.97 | 5.45 | 23.08 |
| | 828 | 11.27 | 9.59 | 10.36 | 10.06 | 41.28 |
| | 829 | 36.07 | 34.32 | 36.63 | 33.35 | 140.37 |
| | 830 | 6.07 | 5.04 | 6.32 | 5.16 | 22.59 |
| | 831 | 6 | 4.45 | 4.1 | 4.94 | 19.49 |
| TOTAL | | 168.27 | 149.51 | 158.02 | 145.21 | 621.01 |

DRUG CODE: 9809 DRUG NAME: OPIUM COMBINATION PRODUCT (C-III)

STATE: ALABAMA

| ZIP CODE | | QUARTER 1 | QUARTER 2 | QUARTER 3 | QUARTER 4 | TOTAL GRAMS |
|---|---|---|---|---|---|---|
| | 350 | 0 | 0.76 | 0.76 | 0.38 | 1.9 |
| | 351 | 0.38 | 0.57 | 2.08 | 0.19 | 3.22 |
| | 352 | 3.03 | 3.59 | 1.7 | 3.03 | 11.35 |
| | 354 | 0.19 | 0 | 0.19 | 0 | 0.38 |
| | 355 | 0 | 0 | 0.19 | 0 | 0.19 |
| | 356 | 0 | 0.19 | 0 | 0.19 | 0.38 |
| | 357 | 1.32 | 0.95 | 0.95 | 0.95 | 4.17 |
| | 358 | 1.51 | 2.27 | 1.7 | 1.14 | 6.62 |

| | | | | | |
|---|---|---|---|---|---|
| 359 | 3.78 | 3.41 | 3.41 | 3.22 | 13.82 |
| 361 | 0 | 0.19 | 0 | 0 | 0.19 |
| 362 | 1.32 | 0.19 | 0.19 | 0.38 | 2.08 |
| 363 | 0 | 0 | 0 | 0.19 | 0.19 |
| 364 | 0 | 0 | 0.38 | 0.19 | 0.57 |
| 365 | 0.38 | 0.38 | 0.19 | 0.38 | 1.33 |
| 366 | 0 | 0.19 | 0 | 0.38 | 0.57 |
| 368 | 0 | 0 | 0 | 0.19 | 0.19 |
| TOTAL | 11.91 | 12.69 | 11.74 | 10.81 | 47.15 |

STATE: ALASKA

| ZIP CODE | QUARTER 1 | QUARTER 2 | QUARTER 3 | QUARTER 4 | TOTAL GRAMS |
|---|---|---|---|---|---|
| 997 | 0 | 0 | 0.19 | 0 | 0.19 |
| TOTAL | 0 | 0 | 0.19 | 0 | 0.19 |

STATE: ARIZONA

| ZIP CODE | QUARTER 1 | QUARTER 2 | QUARTER 3 | QUARTER 4 | TOTAL GRAMS |
|---|---|---|---|---|---|
| 850 | 1.51 | 0.95 | 0.38 | 1.89 | 4.73 |
| 851 | 0.19 | 0 | 0 | 0 | 0.19 |
| 852 | 10.03 | 4.16 | 6.24 | 5.49 | 25.92 |
| 853 | 0.19 | 0.19 | 0 | 0.19 | 0.57 |
| 857 | 0.19 | 0.19 | 0.57 | 0.76 | 1.71 |
| 864 | 0 | 0 | 0 | 0.19 | 0.19 |
| TOTAL | 12.11 | 5.49 | 7.19 | 8.52 | 33.31 |

STATE: ARKANSAS

| ZIP CODE | QUARTER 1 | QUARTER 2 | QUARTER 3 | QUARTER 4 | TOTAL GRAMS |
|---|---|---|---|---|---|
| 719 | 0 | 0 | 0.57 | 0 | 0.57 |
| 720 | 0.76 | 0.95 | 0 | 0.38 | 2.09 |
| 721 | 0 | 0 | 0 | 0.19 | 0.19 |
| 722 | 0 | 0 | 0.19 | 0.19 | 0.38 |
| 727 | 0.57 | 1.32 | 1.89 | 1.32 | 5.1 |
| 729 | 0 | 0 | 0.19 | 0 | 0.19 |
| TOTAL | 1.33 | 2.27 | 2.84 | 2.08 | 8.52 |

STATE: CALIFORNIA

| ZIP CODE | QUARTER 1 | QUARTER 2 | QUARTER 3 | QUARTER 4 | TOTAL GRAMS |
|---|---|---|---|---|---|
| 900 | 3.59 | 1.89 | 3.03 | 3.03 | 11.54 |
| 902 | 0 | 0 | 0 | 0.19 | 0.19 |
| 903 | 0 | 0.38 | 0 | 0 | 0.38 |
| 904 | 0 | 0 | 0.19 | 0 | 0.19 |
| 905 | 0.38 | 0 | 0 | 0 | 0.38 |
| 906 | 0 | 0.19 | 0 | 0 | 0.19 |
| 907 | 0 | 0.19 | 0 | 0 | 0.19 |
| 908 | 0.57 | 1.14 | 0.38 | 0.57 | 2.66 |
| 910 | 0.19 | 0 | 0 | 0 | 0.19 |
| 913 | 0.19 | 0 | 0 | 1.32 | 1.51 |
| 914 | 0.19 | 0.57 | 0.19 | 0 | 0.95 |
| 915 | 0.38 | 0.19 | 0.38 | 0.38 | 1.33 |
| 917 | 0 | 0.19 | 0.19 | 0.38 | 0.76 |
| 920 | 3.41 | 1.14 | 1.89 | 2.84 | 9.28 |
| 921 | 1.7 | 0.57 | 1.7 | 1.32 | 5.29 |
| 922 | 0.19 | 0.19 | 0 | 0.19 | 0.57 |
| 925 | 0.38 | 0.57 | 0.76 | 0.57 | 2.28 |
| 926 | 0 | 0.19 | 0.19 | 0 | 0.38 |
| 928 | 0.38 | 0.38 | 0.19 | 0.38 | 1.33 |
| 930 | 0 | 0.19 | 0 | 0.38 | 0.57 |
| 932 | 0.57 | 0 | 0 | 0 | 0.57 |
| 933 | 0.76 | 0.76 | 0.76 | 0.76 | 3.04 |
| 934 | 0.57 | 0.76 | 2.08 | 2.84 | 6.25 |
| 935 | 0.38 | 0.19 | 0 | 0 | 0.57 |
| 936 | 0 | 0.19 | 0 | 0 | 0.19 |
| 939 | 0.19 | 0 | 0 | 0.19 | 0.38 |
| 940 | 0 | 0.19 | 0.19 | 1.32 | 1.7 |
| 941 | 0 | 0 | 0.57 | 0.19 | 0.76 |
| 943 | 0.19 | 0 | 0 | 0.19 | 0.38 |
| 944 | 0 | 0 | 0 | 0.19 | 0.19 |
| 945 | 0 | 1.32 | 0.38 | 0 | 1.7 |
| 947 | 0 | 0.19 | 0 | 0 | 0.19 |
| 948 | 0.19 | 0 | 0 | 0 | 0.19 |

| | | | | | |
|---|---|---|---|---|---|
| 949 | 0.19 | 0.38 | 0 | 0.38 | 0.95 |
| 950 | 0.76 | 0.38 | 1.14 | 0.38 | 2.66 |
| 951 | 0 | 0.19 | 0.38 | 0.19 | 0.76 |
| 953 | 1.51 | 2.46 | 1.7 | 2.27 | 7.94 |
| 954 | 0 | 0.76 | 0.95 | 0.76 | 2.47 |
| 955 | 0.19 | 0 | 0.57 | 0.76 | 1.52 |
| 956 | 0.19 | 0.57 | 0 | 0 | 0.76 |
| 957 | 0.38 | 0 | 0 | 0 | 0.38 |
| 958 | 0 | 0.57 | 0.19 | 0.38 | 1.14 |
| 959 | 0.57 | 1.14 | 0 | 0.19 | 1.9 |
| TOTAL | 18.19 | 18.02 | 18 | 22.54 | 76.75 |

STATE: COLORADO

| ZIP CODE | QUARTER 1 | QUARTER 2 | QUARTER 3 | QUARTER 4 | TOTAL GRAMS |
|---|---|---|---|---|---|
| 800 | 0 | 0.19 | 0 | 0 | 0.19 |
| 801 | 0.19 | 0 | 0.19 | 0.38 | 0.76 |
| 802 | 1.14 | 1.32 | 0.95 | 0.76 | 4.17 |
| 804 | 0 | 0.19 | 0 | 0 | 0.19 |
| 805 | 0.57 | 0.19 | 0.95 | 0.57 | 2.28 |
| 809 | 0.19 | 0.38 | 0.19 | 0.57 | 1.33 |
| 810 | 1.14 | 0.76 | 0 | 0 | 1.9 |
| 811 | 0 | 0 | 0 | 0.19 | 0.19 |
| TOTAL | 3.23 | 3.03 | 2.28 | 2.47 | 11.01 |

STATE: CONNECTICUT

| ZIP CODE | QUARTER 1 | QUARTER 2 | QUARTER 3 | QUARTER 4 | TOTAL GRAMS |
|---|---|---|---|---|---|
| 60 | 1.7 | 0.95 | 1.14 | 2.27 | 6.06 |
| 61 | 0.19 | 0.38 | 0.38 | 0.57 | 1.52 |
| 62 | 0.57 | 0.19 | 0.38 | 0.19 | 1.33 |
| 64 | 1.89 | 0.95 | 0.38 | 0.38 | 3.6 |
| 66 | 0 | 0 | 0.19 | 0.19 | 0.38 |
| 68 | 0.76 | 1.14 | 1.14 | 0.57 | 3.61 |
| 69 | 0 | 0.19 | 2.84 | 0 | 3.03 |
| TOTAL | 5.11 | 3.8 | 6.45 | 4.17 | 19.53 |

STATE: DELAWARE

| ZIP CODE | QUARTER 1 | QUARTER 2 | QUARTER 3 | QUARTER 4 | TOTAL GRAMS |
|---|---|---|---|---|---|
| 197 | 0 | 0.57 | 0 | 0.19 | 0.76 |
| 199 | 0.19 | 0.19 | 0.19 | 0.76 | 1.33 |
| TOTAL | 0.19 | 0.76 | 0.19 | 0.95 | 2.09 |

STATE: DISTRICT OF COLUMBIA

| ZIP CODE | QUARTER 1 | QUARTER 2 | QUARTER 3 | QUARTER 4 | TOTAL GRAMS |
|---|---|---|---|---|---|
| 200 | 0.19 | 0.38 | 0 | 0.57 | 1.14 |
| 204 | 0 | 0 | 0.38 | 0 | 0.38 |
| TOTAL | 0.19 | 0.38 | 0.38 | 0.57 | 1.52 |

STATE: FLORIDA

| ZIP CODE | QUARTER 1 | QUARTER 2 | QUARTER 3 | QUARTER 4 | TOTAL GRAMS |
|---|---|---|---|---|---|
| 320 | 1.32 | 0.57 | 0.76 | 0.19 | 2.84 |
| 321 | 0 | 0.76 | 0.19 | 0.76 | 1.71 |
| 322 | 1.7 | 0.95 | 0 | 0.76 | 3.41 |
| 323 | 0.38 | 0.57 | 0.57 | 0 | 1.52 |
| 324 | 0.76 | 0.57 | 0.38 | 0.38 | 2.09 |
| 325 | 0.38 | 0.38 | 1.14 | 0.76 | 2.66 |
| 326 | 3.97 | 4.35 | 2.46 | 5.49 | 16.27 |
| 327 | 0.38 | 0.95 | 0.76 | 0.38 | 2.47 |
| 328 | 1.7 | 1.7 | 0.95 | 1.14 | 5.49 |
| 329 | 4.54 | 2.84 | 4.73 | 3.22 | 15.33 |
| 330 | 2.08 | 2.08 | 1.32 | 2.46 | 7.94 |
| 331 | 0.95 | 4.73 | 2.65 | 2.84 | 11.17 |
| 333 | 2.08 | 4.16 | 1.89 | 1.89 | 10.02 |
| 334 | 6.24 | 4.35 | 3.97 | 3.59 | 18.15 |
| 335 | 0.95 | 0.57 | 0.19 | 0.19 | 1.9 |
| 336 | 1.14 | 1.32 | 2.27 | 0.95 | 5.68 |
| 337 | 1.14 | 0.38 | 0.57 | 0.38 | 2.47 |
| 338 | 0.76 | 0.19 | 0.57 | 0.57 | 2.09 |

| | | | | | | |
|---|---|---|---|---|---|---|
| | 339 | 1.14 | 0.19 | 1.14 | 1.51 | 3.98 |
| | 341 | 0 | 0.38 | 0 | 0 | 0.38 |
| | 342 | 2.65 | 3.03 | 2.84 | 1.51 | 10.03 |
| | 344 | 0.19 | 0.19 | 0.19 | 0 | 0.57 |
| | 346 | 0.57 | 0.76 | 0.19 | 0 | 1.52 |
| | 347 | 0 | 0.57 | 0 | 0.19 | 0.76 |
| | 349 | 1.51 | 1.51 | 2.08 | 2.27 | 7.37 |
| TOTAL | | 36.53 | 38.05 | 31.81 | 31.43 | 137.82 |

STATE: GEORGIA

| ZIP CODE | | QUARTER 1 | QUARTER 2 | QUARTER 3 | QUARTER 4 | TOTAL GRAMS |
|---|---|---|---|---|---|---|
| | 300 | 1.51 | 0.38 | 0 | 0.38 | 2.27 |
| | 301 | 0.57 | 0.38 | 0.57 | 0.95 | 2.47 |
| | 302 | 0.76 | 0.95 | 0.57 | 0.19 | 2.47 |
| | 303 | 0.57 | 0.76 | 1.32 | 1.32 | 3.97 |
| | 305 | 0.76 | 1.7 | 1.32 | 1.51 | 5.29 |
| | 306 | 0.38 | 0.38 | 0.38 | 0.38 | 1.52 |
| | 307 | 0.19 | 0 | 0 | 0.19 | 0.38 |
| | 308 | 0 | 1.32 | 0.76 | 1.14 | 3.22 |
| | 309 | 0.57 | 0.57 | 0.19 | 0 | 1.33 |
| | 312 | 0 | 0.38 | 0.38 | 0 | 0.76 |
| | 314 | 0.19 | 0.38 | 0.38 | 0 | 0.95 |
| | 315 | 0.19 | 0.95 | 0.95 | 0.76 | 2.85 |
| | 316 | 0.19 | 0.19 | 0.57 | 0.38 | 1.33 |
| | 317 | 0.19 | 0 | 0 | 0 | 0.19 |
| | 319 | 0 | 0.38 | 0.19 | 0 | 0.57 |
| TOTAL | | 6.07 | 8.72 | 7.58 | 7.2 | 29.57 |

STATE: HAWAII

| ZIP CODE | | QUARTER 1 | QUARTER 2 | QUARTER 3 | QUARTER 4 | TOTAL GRAMS |
|---|---|---|---|---|---|---|
| | 968 | 1.89 | 1.51 | 1.32 | 0 | 4.72 |
| TOTAL | | 1.89 | 1.51 | 1.32 | 0 | 4.72 |

STATE: IDAHO

| ZIP CODE | QUARTER 1 | QUARTER 2 | QUARTER 3 | QUARTER 4 | TOTAL GRAMS |
|---|---|---|---|---|---|
| 832 | 0.76 | 1.14 | 0.38 | 0.57 | 2.85 |
| 833 | 0.19 | 0.19 | 0.38 | 0.38 | 1.14 |
| 834 | 0.19 | 0 | 0.19 | 0 | 0.38 |
| 836 | 0.19 | 0 | 0 | 0 | 0.19 |
| 838 | 2.65 | 2.46 | 2.65 | 2.84 | 10.6 |
| TOTAL | 3.98 | 3.79 | 3.6 | 3.79 | 15.16 |

STATE: ILLINOIS

| ZIP CODE | QUARTER 1 | QUARTER 2 | QUARTER 3 | QUARTER 4 | TOTAL GRAMS |
|---|---|---|---|---|---|
| 600 | 0.19 | 1.14 | 2.08 | 1.51 | 4.92 |
| 601 | 0.38 | 0.19 | 0.19 | 0.57 | 1.33 |
| 602 | 0 | 0 | 0 | 0.19 | 0.19 |
| 603 | 0.57 | 0.57 | 0.38 | 0.38 | 1.9 |
| 604 | 0 | 0.19 | 0 | 0 | 0.19 |
| 605 | 0 | 0 | 0.57 | 0 | 0.57 |
| 606 | 2.08 | 0.95 | 1.14 | 0.95 | 5.12 |
| 609 | 0.19 | 0.95 | 0.38 | 0 | 1.52 |
| 610 | 0 | 0 | 0 | 0.19 | 0.19 |
| 612 | 0 | 0.19 | 0 | 0 | 0.19 |
| 613 | 0 | 0.19 | 0 | 0 | 0.19 |
| 617 | 0.57 | 0 | 0.19 | 0 | 0.76 |
| 618 | 0.38 | 0.57 | 0.57 | 0.19 | 1.71 |
| 622 | 0.95 | 0.76 | 0.57 | 0.76 | 3.04 |
| 626 | 0.19 | 0 | 0 | 0 | 0.19 |
| 627 | 0.38 | 0.38 | 0.57 | 0.38 | 1.71 |
| 628 | 0.19 | 0 | 0 | 0 | 0.19 |
| TOTAL | 6.07 | 6.08 | 6.64 | 5.12 | 23.91 |

STATE: INDIANA

| ZIP CODE | QUARTER 1 | QUARTER 2 | QUARTER 3 | QUARTER 4 | TOTAL GRAMS |
|---|---|---|---|---|---|
| 460 | 0 | 0.19 | 0.19 | 0 | 0.38 |
| 462 | 0.76 | 0.57 | 0.38 | 2.65 | 4.36 |
| 463 | 0 | 0.57 | 0 | 0 | 0.57 |

| | | | | | | |
|---|---|---|---|---|---|---|
| | 465 | 0 | 0.19 | 0 | 0.38 | 0.57 |
| | 467 | 0.19 | 0 | 0 | 0 | 0.19 |
| | 468 | 0.19 | 0 | 0 | 0 | 0.19 |
| | 471 | 0 | 0 | 0.19 | 0 | 0.19 |
| | 472 | 3.22 | 1.89 | 3.78 | 0.76 | 9.65 |
| | 473 | 0 | 0 | 0 | 0.19 | 0.19 |
| | 474 | 2.27 | 3.03 | 3.78 | 2.84 | 11.92 |
| | 475 | 0.19 | 0.38 | 0 | 0.38 | 0.95 |
| TOTAL | | 6.82 | 6.82 | 8.32 | 7.2 | 29.16 |

STATE: IOWA

| ZIP CODE | | QUARTER 1 | QUARTER 2 | QUARTER 3 | QUARTER 4 | TOTAL GRAMS |
|---|---|---|---|---|---|---|
| | 503 | 0.38 | 0 | 0 | 0 | 0.38 |
| | 513 | 0.95 | 0.76 | 0.57 | 0.76 | 3.04 |
| | 522 | 1.51 | 1.14 | 1.14 | 1.32 | 5.11 |
| | 525 | 0.19 | 0 | 0 | 0 | 0.19 |
| | 528 | 0.19 | 0 | 0 | 0 | 0.19 |
| TOTAL | | 3.22 | 1.9 | 1.71 | 2.08 | 8.91 |

STATE: KANSAS

| ZIP CODE | | QUARTER 1 | QUARTER 2 | QUARTER 3 | QUARTER 4 | TOTAL GRAMS |
|---|---|---|---|---|---|---|
| | 662 | 0.19 | 0 | 0.19 | 0.38 | 0.76 |
| | 666 | 0 | 0 | 0 | 0.57 | 0.57 |
| | 668 | 0.19 | 0 | 0 | 0 | 0.19 |
| | 670 | 0 | 0.19 | 0 | 0 | 0.19 |
| | 671 | 0 | 0.19 | 0 | 0.19 | 0.38 |
| | 672 | 0 | 0.19 | 0 | 0.19 | 0.38 |
| | 674 | 0.19 | 0 | 0 | 0 | 0.19 |
| | 679 | 0.19 | 0 | 0 | 0 | 0.19 |
| TOTAL | | 0.76 | 0.57 | 0.19 | 1.33 | 2.85 |

STATE: KENTUCKY

| ZIP CODE | | QUARTER 1 | QUARTER 2 | QUARTER 3 | QUARTER 4 | TOTAL GRAMS |
|---|---|---|---|---|---|---|

| | | | | | | |
|---|---|---|---|---|---|---|
| | 400 | 0 | 0 | 0 | 0.38 | 0.38 |
| | 401 | 0 | 0 | 0.19 | 0 | 0.19 |
| | 402 | 0.76 | 0.95 | 0.57 | 0.57 | 2.85 |
| | 404 | 0.57 | 0.95 | 0.95 | 0.57 | 3.04 |
| | 405 | 2.27 | 1.51 | 1.51 | 0.94 | 6.23 |
| | 406 | 2.65 | 3.03 | 1.7 | 1.89 | 9.27 |
| | 407 | 0.76 | 1.32 | 0.76 | 0.76 | 3.6 |
| | 410 | 0.19 | 0.19 | 0.19 | 0.38 | 0.95 |
| | 413 | 0 | 0 | 0.19 | 0 | 0.19 |
| | 414 | 0 | 0.19 | 0 | 0 | 0.19 |
| | 415 | 0 | 0 | 0 | 0.38 | 0.38 |
| | 418 | 0.19 | 0 | 0.19 | 0.38 | 0.76 |
| | 420 | 1.89 | 2.08 | 0.76 | 0.76 | 5.49 |
| | 421 | 0 | 0 | 0 | 0.19 | 0.19 |
| | 422 | 0.57 | 0 | 0.38 | 0 | 0.95 |
| | 425 | 0.38 | 0.57 | 0 | 0.57 | 1.52 |
| | 427 | 0.38 | 0.19 | 0.38 | 0 | 0.95 |
| TOTAL | | 10.61 | 10.98 | 7.77 | 7.77 | 37.13 |

STATE: LOUISIANA

| ZIP CODE | | QUARTER 1 | QUARTER 2 | QUARTER 3 | QUARTER 4 | TOTAL GRAMS |
|---|---|---|---|---|---|---|
| | 700 | 4.16 | 3.97 | 4.54 | 3.03 | 15.7 |
| | 701 | 1.32 | 0.57 | 2.27 | 3.03 | 7.19 |
| | 703 | 0.38 | 0.19 | 0.19 | 0 | 0.76 |
| | 704 | 1.14 | 3.03 | 1.89 | 1.14 | 7.2 |
| | 705 | 0 | 0 | 0 | 0.38 | 0.38 |
| | 706 | 0 | 0 | 0 | 0.19 | 0.19 |
| | 707 | 0 | 0.19 | 0.19 | 0 | 0.38 |
| | 708 | 0 | 0.19 | 0.19 | 0.19 | 0.57 |
| | 711 | 0.19 | 0.19 | 0 | 0.19 | 0.57 |
| | 712 | 0 | 0.38 | 0 | 0 | 0.38 |
| TOTAL | | 7.19 | 8.71 | 9.27 | 8.15 | 33.32 |

STATE: MAINE

| ZIP CODE | QUARTER 1 | QUARTER 2 | QUARTER 3 | QUARTER 4 | TOTAL GRAMS |
|---|---|---|---|---|---|
| 40 | 0.38 | 0.38 | 0.57 | 1.51 | 2.84 |
| 41 | 0 | 0 | 0.38 | 0 | 0.38 |
| 42 | 0 | 0.38 | 0.57 | 0.19 | 1.14 |
| 43 | 0.38 | 0.38 | 0.38 | 0.57 | 1.71 |
| 44 | 0 | 0 | 0 | 0.57 | 0.57 |
| 48 | 0 | 0 | 0.19 | 0 | 0.19 |
| TOTAL | 0.76 | 1.14 | 2.09 | 2.84 | 6.83 |

STATE: MARYLAND

| ZIP CODE | QUARTER 1 | QUARTER 2 | QUARTER 3 | QUARTER 4 | TOTAL GRAMS |
|---|---|---|---|---|---|
| 206 | 1.14 | 1.32 | 0.57 | 0.57 | 3.6 |
| 208 | 0.38 | 0.19 | 0 | 0.38 | 0.95 |
| 209 | 0 | 0.19 | 0 | 0 | 0.19 |
| 210 | 1.14 | 0.38 | 0.19 | 0 | 1.71 |
| 211 | 0 | 0 | 0 | 0.19 | 0.19 |
| 212 | 2.27 | 3.03 | 3.97 | 2.84 | 12.11 |
| 214 | 0.19 | 0.19 | 0 | 0 | 0.38 |
| 216 | 3.22 | 2.46 | 3.59 | 3.03 | 12.3 |
| 217 | 0 | 0.19 | 0.38 | 0.95 | 1.52 |
| 218 | 0 | 0.76 | 0.19 | 0 | 0.95 |
| 219 | 0.19 | 0 | 0 | 0 | 0.19 |
| TOTAL | 8.53 | 8.71 | 8.89 | 7.96 | 34.09 |

STATE: MASSACHUSETTS

| ZIP CODE | QUARTER 1 | QUARTER 2 | QUARTER 3 | QUARTER 4 | TOTAL GRAMS |
|---|---|---|---|---|---|
| 10 | 0.19 | 0.57 | 0 | 0 | 0.76 |
| 12 | 0 | 0.19 | 0.19 | 0.57 | 0.95 |
| 15 | 0.38 | 0 | 0 | 0 | 0.38 |
| 17 | 0.38 | 0.38 | 0 | 0 | 0.76 |
| 18 | 0 | 0 | 0.19 | 0 | 0.19 |
| 19 | 0.76 | 0.57 | 0.38 | 0.38 | 2.09 |
| 21 | 0 | 0.57 | 0.38 | 0 | 0.95 |
| 24 | 0.19 | 0 | 0.19 | 0 | 0.38 |
| 27 | 0.19 | 0.19 | 0 | 0 | 0.38 |

| | | | | | |
|---|---|---|---|---|---|
| TOTAL | 2.09 | 2.47 | 1.33 | 0.95 | 6.84 |

STATE: MICHIGAN

| ZIP CODE | QUARTER 1 | QUARTER 2 | QUARTER 3 | QUARTER 4 | TOTAL GRAMS |
|---|---|---|---|---|---|
| 480 | 0.38 | 0.76 | 1.89 | 0.75 | 3.78 |
| 481 | 0.38 | 0.57 | 0.38 | 0 | 1.33 |
| 482 | 0 | 0.19 | 0.57 | 0 | 0.76 |
| 483 | 1.32 | 0.76 | 2.08 | 1.7 | 5.86 |
| 484 | 0 | 0 | 0 | 0.19 | 0.19 |
| 487 | 0.19 | 0 | 0 | 0 | 0.19 |
| 490 | 0 | 0.38 | 0.76 | 0 | 1.14 |
| 494 | 0.19 | 0.38 | 0.38 | 0 | 0.95 |
| 495 | 0.95 | 0.38 | 1.32 | 0.38 | 3.03 |
| 496 | 0.57 | 0.57 | 0.57 | 0.57 | 2.28 |
| 497 | 0 | 0.19 | 0 | 0 | 0.19 |
| 499 | 0.76 | 0.76 | 0.57 | 0.95 | 3.04 |
| TOTAL | 4.74 | 4.94 | 8.52 | 4.54 | 22.74 |

STATE: MINNESOTA

| ZIP CODE | QUARTER 1 | QUARTER 2 | QUARTER 3 | QUARTER 4 | TOTAL GRAMS |
|---|---|---|---|---|---|
| 550 | 0.38 | 0.57 | 0.38 | 0.38 | 1.71 |
| 551 | 0 | 0.38 | 0 | 0 | 0.38 |
| 553 | 0 | 0.19 | 0 | 0.76 | 0.95 |
| 554 | 0.19 | 0.19 | 0.38 | 0 | 0.76 |
| 557 | 0 | 0 | 0 | 0.19 | 0.19 |
| 558 | 0 | 0 | 0 | 0.19 | 0.19 |
| 559 | 0 | 0.57 | 0.76 | 0.95 | 2.28 |
| 560 | 0 | 0 | 0.38 | 0 | 0.38 |
| 561 | 0.95 | 0 | 0 | 0.38 | 1.33 |
| 563 | 0.19 | 0.38 | 0.19 | 0.19 | 0.95 |
| TOTAL | 1.71 | 2.28 | 2.09 | 3.04 | 9.12 |

STATE: MISSISSIPPI

| ZIP CODE | QUARTER 1 | QUARTER 2 | QUARTER 3 | QUARTER 4 | TOTAL GRAMS |
|---|---|---|---|---|---|
| 386 | 0.19 | 0 | 0 | 0 | 0.19 |
| 388 | 0.95 | 0.38 | 1.51 | 0 | 2.84 |
| 389 | 0.38 | 0 | 0 | 0 | 0.38 |
| 393 | 0.19 | 0.19 | 0 | 0 | 0.38 |
| 394 | 2.08 | 0.76 | 0.57 | 0.38 | 3.79 |
| 395 | 0.38 | 0.57 | 0.57 | 0.57 | 2.09 |
| 396 | 0.38 | 0.38 | 0.38 | 0 | 1.14 |
| 397 | 0 | 0.19 | 0 | 0.76 | 0.95 |
| TOTAL | 4.55 | 2.47 | 3.03 | 1.71 | 11.76 |

STATE: MISSOURI

| ZIP CODE | QUARTER 1 | QUARTER 2 | QUARTER 3 | QUARTER 4 | TOTAL GRAMS |
|---|---|---|---|---|---|
| 630 | 0.95 | 0 | 0 | 0.19 | 1.14 |
| 631 | 4.16 | 1.51 | 1.89 | 7.38 | 14.94 |
| 633 | 0 | 0 | 0 | 0.19 | 0.19 |
| 640 | 1.14 | 0.76 | 0.38 | 0 | 2.28 |
| 641 | 0.76 | 0.19 | 0 | 0.19 | 1.14 |
| 645 | 0.19 | 0.19 | 0 | 0 | 0.38 |
| 646 | 0 | 0.19 | 0 | 0 | 0.19 |
| 648 | 0 | 0 | 0.19 | 0 | 0.19 |
| 651 | 0.95 | 0.76 | 0 | 0 | 1.71 |
| 652 | 1.7 | 1.51 | 1.51 | 1.32 | 6.04 |
| 658 | 0.38 | 0 | 0 | 0 | 0.38 |
| TOTAL | 10.23 | 5.11 | 3.97 | 9.27 | 28.58 |

STATE: MONTANA

| ZIP CODE | QUARTER 1 | QUARTER 2 | QUARTER 3 | QUARTER 4 | TOTAL GRAMS |
|---|---|---|---|---|---|
| 594 | 0 | 0 | 0 | 0.19 | 0.19 |
| 597 | 0.19 | 0 | 0 | 0 | 0.19 |
| 598 | 0.76 | 0.19 | 0.95 | 0.76 | 2.66 |
| 599 | 0 | 0 | 0.38 | 0 | 0.38 |
| TOTAL | 0.95 | 0.19 | 1.33 | 0.95 | 3.42 |

STATE: NEBRASKA

| ZIP CODE | QUARTER 1 | QUARTER 2 | QUARTER 3 | QUARTER 4 | TOTAL GRAMS |
|---|---|---|---|---|---|
| 680 | 0 | 0 | 0.19 | 0 | 0.19 |
| 681 | 0.95 | 0.57 | 0.38 | 0 | 1.9 |
| 684 | 0.19 | 0 | 0 | 0.57 | 0.76 |
| 693 | 0 | 0 | 0.19 | 0 | 0.19 |
| TOTAL | 1.14 | 0.57 | 0.76 | 0.57 | 3.04 |

STATE: NEVADA

| ZIP CODE | QUARTER 1 | QUARTER 2 | QUARTER 3 | QUARTER 4 | TOTAL GRAMS |
|---|---|---|---|---|---|
| 890 | 0 | 0.57 | 0.19 | 0 | 0.76 |
| 891 | 0.57 | 0.19 | 1.51 | 0.38 | 2.65 |
| 895 | 0.19 | 0.19 | 0 | 0 | 0.38 |
| 897 | 0 | 0.38 | 0 | 0 | 0.38 |
| TOTAL | 0.76 | 1.33 | 1.7 | 0.38 | 4.17 |

STATE: NEW HAMPSHIRE

| ZIP CODE | QUARTER 1 | QUARTER 2 | QUARTER 3 | QUARTER 4 | TOTAL GRAMS |
|---|---|---|---|---|---|
| 30 | 0 | 0.76 | 0.57 | 0.57 | 1.9 |
| 32 | 0 | 0 | 0.38 | 0 | 0.38 |
| 34 | 0 | 0.19 | 0.19 | 0 | 0.38 |
| 35 | 0.76 | 0.19 | 0 | 0.38 | 1.33 |
| 37 | 0 | 0 | 0 | 0.38 | 0.38 |
| 38 | 0.95 | 0.76 | 0.76 | 2.08 | 4.55 |
| TOTAL | 1.71 | 1.9 | 1.9 | 3.41 | 8.92 |

STATE: NEW JERSEY

| ZIP CODE | QUARTER 1 | QUARTER 2 | QUARTER 3 | QUARTER 4 | TOTAL GRAMS |
|---|---|---|---|---|---|
| 70 | 0.57 | 0.76 | 0.76 | 0.19 | 2.28 |
| 72 | 0 | 0.38 | 0 | 0 | 0.38 |
| 74 | 0.19 | 0.76 | 0.19 | 0 | 1.14 |
| 76 | 0.76 | 0.57 | 0 | 0 | 1.33 |
| 77 | 0.19 | 0.38 | 0.57 | 0 | 1.14 |

| | | | | | | |
|---|---|---|---|---|---|---|
| | 78 | 0.19 | 0.19 | 0 | 0 | 0.38 |
| | 79 | 0.19 | 0 | 0 | 0 | 0.19 |
| | 80 | 1.89 | 1.51 | 0.38 | 1.32 | 5.1 |
| | 81 | 0 | 0 | 0.38 | 0 | 0.38 |
| | 85 | 0.95 | 0 | 0 | 0 | 0.95 |
| | 87 | 0.38 | 1.14 | 2.08 | 1.89 | 5.49 |
| | 88 | 0.38 | 0 | 0.95 | 0.76 | 2.09 |
| TOTAL | | 5.69 | 5.69 | 5.31 | 4.16 | 20.85 |

| STATE: | NEW MEXICO | | | | |
|---|---|---|---|---|---|
| ZIP CODE | QUARTER 1 | QUARTER 2 | QUARTER 3 | QUARTER 4 | TOTAL GRAMS |
| 871 | 1.32 | 1.32 | 0.19 | 0.19 | 3.02 |
| 875 | 0.57 | 0.57 | 0.76 | 1.32 | 3.22 |
| 877 | 0 | 0 | 0 | 0.19 | 0.19 |
| 882 | 0 | 0 | 0 | 0.19 | 0.19 |
| TOTAL | 1.89 | 1.89 | 0.95 | 1.89 | 6.62 |

| STATE: | NEW YORK | | | | |
|---|---|---|---|---|---|
| ZIP CODE | QUARTER 1 | QUARTER 2 | QUARTER 3 | QUARTER 4 | TOTAL GRAMS |
| 100 | 0.76 | 0.57 | 1.32 | 1.14 | 3.79 |
| 103 | 0 | 0 | 0.19 | 0 | 0.19 |
| 104 | 0 | 0.38 | 0.19 | 0 | 0.57 |
| 105 | 0.76 | 0.57 | 0.19 | 0.95 | 2.47 |
| 109 | 0 | 0 | 0 | 0.38 | 0.38 |
| 110 | 0 | 0.19 | 0 | 0 | 0.19 |
| 112 | 0.38 | 0.38 | 0 | 0.19 | 0.95 |
| 113 | 0.76 | 0.19 | 0.38 | 0 | 1.33 |
| 114 | 0.19 | 0.19 | 0 | 0 | 0.38 |
| 115 | 3.03 | 5.11 | 2.84 | 2.46 | 13.44 |
| 117 | 0.19 | 0 | 0.38 | 1.51 | 2.08 |
| 118 | 0 | 0.38 | 0.57 | 0.76 | 1.71 |
| 119 | 0.38 | 0 | 0 | 0 | 0.38 |
| 123 | 0 | 0 | 0 | 0.19 | 0.19 |
| 124 | 0 | 0 | 0 | 0.19 | 0.19 |

| | | | | | |
|---|---|---|---|---|---|
| 125 | 0.19 | 0 | 0 | 0.19 | 0.38 |
| 127 | 0 | 0 | 0.38 | 0 | 0.38 |
| 128 | 0 | 0 | 0.19 | 0 | 0.19 |
| 129 | 0 | 0 | 0 | 0.38 | 0.38 |
| 130 | 0.19 | 0 | 0.19 | 0 | 0.38 |
| 132 | 0 | 0 | 0.19 | 0 | 0.19 |
| 133 | 0 | 0 | 0.19 | 0 | 0.19 |
| 134 | 0 | 0 | 0.19 | 0.38 | 0.57 |
| 138 | 0.19 | 0 | 0 | 0 | 0.19 |
| 140 | 0.57 | 0.19 | 0.19 | 0.57 | 1.52 |
| 141 | 0 | 0.19 | 0 | 0.38 | 0.57 |
| 142 | 0.19 | 0.76 | 0.38 | 0.19 | 1.52 |
| 144 | 0.19 | 0.76 | 0 | 0.19 | 1.14 |
| 145 | 0 | 0 | 0.19 | 0 | 0.19 |
| 146 | 0.19 | 0.38 | 0.19 | 0.19 | 0.95 |
| 148 | 0.19 | 0.19 | 0.19 | 0.57 | 1.14 |
| TOTAL | 8.35 | 10.43 | 8.53 | 10.81 | 38.12 |

STATE: NORTH CAROLINA

| ZIP CODE | QUARTER 1 | QUARTER 2 | QUARTER 3 | QUARTER 4 | TOTAL GRAMS |
|---|---|---|---|---|---|
| 270 | 0.57 | 0.76 | 0.38 | 0.57 | 2.28 |
| 271 | 0.76 | 0.19 | 0.19 | 0.19 | 1.33 |
| 272 | 0.57 | 0.95 | 1.89 | 1.7 | 5.11 |
| 273 | 0.19 | 0.38 | 0 | 0 | 0.57 |
| 274 | 0.95 | 1.51 | 0.95 | 0.95 | 4.36 |
| 275 | 0.57 | 0.76 | 0.76 | 0.19 | 2.28 |
| 276 | 0 | 0 | 0 | 0.38 | 0.38 |
| 278 | 0.76 | 0 | 0.38 | 0.19 | 1.33 |
| 279 | 0 | 0 | 0 | 0.19 | 0.19 |
| 280 | 1.51 | 1.14 | 1.32 | 0.57 | 4.54 |
| 281 | 0.19 | 0.38 | 0.76 | 0.57 | 1.9 |
| 282 | 0.57 | 0.38 | 0.19 | 0.19 | 1.33 |
| 283 | 0.19 | 0.19 | 0.19 | 0.19 | 0.76 |
| 284 | 0.57 | 1.32 | 0.76 | 1.32 | 3.97 |
| 285 | 0.95 | 0.95 | 0.57 | 0.76 | 3.23 |
| 286 | 0.76 | 1.51 | 1.7 | 1.14 | 5.11 |

| | | | | | | |
|---|---|---|---|---|---|---|
| | 287 | 0.57 | 1.32 | 0.57 | 0.95 | 3.41 |
| | 288 | 0.95 | 0.19 | 0.57 | 0 | 1.71 |
| | 289 | 1.51 | 2.84 | 3.97 | 1.89 | 10.21 |
| TOTAL | | 12.14 | 14.77 | 15.15 | 11.94 | 54 |

STATE: NORTH DAKOTA

| ZIP CODE | | QUARTER 1 | QUARTER 2 | QUARTER 3 | QUARTER 4 | TOTAL GRAMS |
|---|---|---|---|---|---|---|
| | 585 | 0 | 0.38 | 0.57 | 0.57 | 1.52 |
| | 588 | 0 | 0 | 0.19 | 0.95 | 1.14 |
| TOTAL | | 0 | 0.38 | 0.76 | 1.52 | 2.66 |

STATE: OHIO

| ZIP CODE | | QUARTER 1 | QUARTER 2 | QUARTER 3 | QUARTER 4 | TOTAL GRAMS |
|---|---|---|---|---|---|---|
| | 430 | 0 | 0.76 | 0.57 | 0.19 | 1.52 |
| | 431 | 0 | 0.38 | 0 | 0 | 0.38 |
| | 432 | 8.89 | 6.62 | 3.22 | 10.6 | 29.33 |
| | 436 | 0 | 0 | 0.38 | 0 | 0.38 |
| | 440 | 0.76 | 0.19 | 0.19 | 0 | 1.14 |
| | 441 | 0.76 | 1.32 | 2.44 | 0.76 | 5.28 |
| | 442 | 1.89 | 0.19 | 1.14 | 0.95 | 4.17 |
| | 443 | 0 | 0 | 0 | 0.19 | 0.19 |
| | 444 | 0 | 0 | 0 | 0.19 | 0.19 |
| | 445 | 0 | 0 | 0 | 0.19 | 0.19 |
| | 446 | 0 | 0.19 | 0 | 0.38 | 0.57 |
| | 447 | 1.14 | 0.38 | 0.57 | 0.19 | 2.28 |
| | 448 | 0 | 0.19 | 0 | 0 | 0.19 |
| | 449 | 0.38 | 0.38 | 0.38 | 0.38 | 1.52 |
| | 450 | 0.19 | 1.89 | 0.38 | 1.7 | 4.16 |
| | 451 | 0 | 0.19 | 0.19 | 0 | 0.38 |
| | 452 | 0.19 | 0.19 | 0.95 | 0.76 | 2.09 |
| | 453 | 0.38 | 0.19 | 0 | 0 | 0.57 |
| | 454 | 0 | 0.76 | 0.57 | 0.76 | 2.09 |
| | 456 | 0.19 | 0 | 0 | 0.57 | 0.76 |
| | 457 | 0.19 | 0 | 0.19 | 0.38 | 0.76 |
| | 458 | 0 | 0 | 0 | 0.19 | 0.19 |

| | | | | | |
|---|---|---|---|---|---|
| TOTAL | 14.96 | 13.82 | 11.17 | 18.38 | 58.33 |

STATE: OKLAHOMA

| ZIP CODE | QUARTER 1 | QUARTER 2 | QUARTER 3 | QUARTER 4 | TOTAL GRAMS |
|---|---|---|---|---|---|
| 730 | 0.19 | 0.38 | 0.19 | 0 | 0.76 |
| 731 | 0 | 0.19 | 0 | 0.19 | 0.38 |
| 734 | 0.19 | 0.19 | 0.38 | 0 | 0.76 |
| 735 | 0 | 0 | 0.19 | 0 | 0.19 |
| 740 | 0.76 | 0 | 0 | 1.14 | 1.9 |
| 741 | 0.95 | 1.32 | 2.46 | 2.27 | 7 |
| 744 | 0.38 | 0 | 0 | 0.76 | 1.14 |
| 745 | 0.76 | 0 | 0 | 0 | 0.76 |
| TOTAL | 3.23 | 2.08 | 3.22 | 4.36 | 12.89 |

STATE: OREGON

| ZIP CODE | QUARTER 1 | QUARTER 2 | QUARTER 3 | QUARTER 4 | TOTAL GRAMS |
|---|---|---|---|---|---|
| 970 | 0 | 0.38 | 0.19 | 0.57 | 1.14 |
| 971 | 0 | 0 | 0.19 | 0 | 0.19 |
| 972 | 0 | 0 | 0 | 0.19 | 0.19 |
| 973 | 0.19 | 0 | 0.38 | 0 | 0.57 |
| 975 | 0 | 0.19 | 0.19 | 1.14 | 1.52 |
| TOTAL | 0.19 | 0.57 | 0.95 | 1.9 | 3.61 |

STATE: PENNSYLVANIA

| ZIP CODE | QUARTER 1 | QUARTER 2 | QUARTER 3 | QUARTER 4 | TOTAL GRAMS |
|---|---|---|---|---|---|
| 150 | 0 | 0.76 | 0.57 | 0.76 | 2.09 |
| 151 | 0.76 | 0.57 | 1.51 | 0.95 | 3.79 |
| 152 | 5.87 | 4.16 | 7.38 | 3.78 | 21.19 |
| 153 | 0.19 | 0.18 | 0.57 | 0.38 | 1.32 |
| 154 | 0 | 0 | 0 | 0.57 | 0.57 |
| 155 | 0 | 0 | 0.19 | 0 | 0.19 |
| 156 | 1.32 | 1.14 | 1.14 | 0.76 | 4.36 |
| 158 | 0 | 0 | 0.19 | 0 | 0.19 |
| 160 | 0 | 0.19 | 0.76 | 0 | 0.95 |

| | | | | | | |
|---|---|---|---|---|---|---|
| | 161 | 0.76 | 0.38 | 0.38 | 0.57 | 2.09 |
| | 162 | 0.19 | 0.19 | 0.19 | 0.38 | 0.95 |
| | 163 | 0.19 | 0.76 | 0 | 0 | 0.95 |
| | 165 | 0.38 | 0.57 | 0.38 | 0.38 | 1.71 |
| | 166 | 0 | 0 | 0 | 0.38 | 0.38 |
| | 170 | 0.38 | 0 | 0 | 0.57 | 0.95 |
| | 171 | 0.19 | 0 | 0 | 0 | 0.19 |
| | 172 | 0 | 0.57 | 0 | 0 | 0.57 |
| | 173 | 0.38 | 0.38 | 0 | 0 | 0.76 |
| | 174 | 0.19 | 0 | 0.19 | 0.38 | 0.76 |
| | 177 | 0.38 | 0.19 | 0.19 | 0.38 | 1.14 |
| | 178 | 1.14 | 1.32 | 1.14 | 1.32 | 4.92 |
| | 179 | 0 | 0 | 0 | 0.38 | 0.38 |
| | 180 | 0 | 0 | 0 | 0.19 | 0.19 |
| | 181 | 0.57 | 0.38 | 0.19 | 0 | 1.14 |
| | 182 | 5.87 | 4.54 | 6.43 | 5.3 | 22.14 |
| | 184 | 0 | 0 | 0.38 | 0.38 | 0.76 |
| | 186 | 0.38 | 0 | 0.19 | 0 | 0.57 |
| | 187 | 8.89 | 17.6 | 0.19 | 16.08 | 42.76 |
| | 189 | 0.38 | 0.57 | 0.57 | 0 | 1.52 |
| | 190 | 0.95 | 1.32 | 0.38 | 0.76 | 3.41 |
| | 191 | 0.76 | 0.76 | 1.14 | 0.76 | 3.42 |
| | 193 | 0.19 | 0.19 | 0.19 | 0.19 | 0.76 |
| | 194 | 1.14 | 1.89 | 2.84 | 2.46 | 8.33 |
| | 195 | 0 | 0.38 | 0 | 0 | 0.38 |
| TOTAL | | 31.45 | 38.99 | 27.28 | 38.06 | 135.78 |

| STATE: | RHODE ISLAND | | | | |
|---|---|---|---|---|---|
| ZIP CODE | QUARTER 1 | QUARTER 2 | QUARTER 3 | QUARTER 4 | TOTAL GRAMS |
| 28 | 0 | 0.19 | 0.19 | 0 | 0.38 |
| TOTAL | 0 | 0.19 | 0.19 | 0 | 0.38 |

| STATE: | SOUTH CAROLINA | | | | |
|---|---|---|---|---|---|
| ZIP CODE | QUARTER 1 | QUARTER 2 | QUARTER 3 | QUARTER 4 | TOTAL GRAMS |
| 290 | 0 | 0.76 | 0.57 | 0.95 | 2.28 |

| | | | | | | |
|---|---|---|---|---|---|---|
| | 291 | 0.19 | 0 | 0.57 | 0.19 | 0.95 |
| | 292 | 0.57 | 0.95 | 0.76 | 0.57 | 2.85 |
| | 293 | 3.22 | 4.35 | 3.97 | 4.35 | 15.89 |
| | 294 | 1.32 | 1.51 | 1.32 | 0.19 | 4.34 |
| | 295 | 0.95 | 1.32 | 1.51 | 1.32 | 5.1 |
| | 296 | 1.32 | 1.51 | 1.32 | 1.51 | 5.66 |
| | 297 | 0.38 | 0.19 | 0.19 | 0.38 | 1.14 |
| | 298 | 0.38 | 0 | 0 | 0 | 0.38 |
| | 299 | 0 | 0.19 | 0.57 | 0.76 | 1.52 |
| TOTAL | | 8.33 | 10.78 | 10.78 | 10.22 | 40.11 |

STATE: SOUTH DAKOTA

| ZIP CODE | | QUARTER 1 | QUARTER 2 | QUARTER 3 | QUARTER 4 | TOTAL GRAMS |
|---|---|---|---|---|---|---|
| | 571 | 1.14 | 0.57 | 0.57 | 0 | 2.28 |
| | 577 | 0.19 | 0 | 0 | 0.19 | 0.38 |
| TOTAL | | 1.33 | 0.57 | 0.57 | 0.19 | 2.66 |

STATE: TENNESSEE

| ZIP CODE | | QUARTER 1 | QUARTER 2 | QUARTER 3 | QUARTER 4 | TOTAL GRAMS |
|---|---|---|---|---|---|---|
| | 370 | 0.38 | 0.57 | 0.19 | 0.95 | 2.09 |
| | 371 | 0.76 | 0.76 | 1.32 | 1.14 | 3.98 |
| | 372 | 0.19 | 0.19 | 0.19 | 0.38 | 0.95 |
| | 373 | 0 | 0 | 0.38 | 0 | 0.38 |
| | 374 | 0.95 | 0.76 | 0 | 0 | 1.71 |
| | 376 | 0.19 | 0.19 | 0.38 | 0.57 | 1.33 |
| | 377 | 0.38 | 0.38 | 0.19 | 0.38 | 1.33 |
| | 378 | 0.95 | 1.14 | 1.7 | 1.7 | 5.49 |
| | 379 | 0.38 | 0.57 | 0.38 | 0.76 | 2.09 |
| | 380 | 0.38 | 0 | 0.38 | 0.95 | 1.71 |
| | 381 | 0.57 | 0.38 | 0.57 | 0.76 | 2.28 |
| | 382 | 0.19 | 0.19 | 0 | 0.19 | 0.57 |
| | 383 | 0.19 | 0.38 | 0 | 0 | 0.57 |
| | 384 | 0.19 | 0 | 0 | 0.38 | 0.57 |
| TOTAL | | 5.7 | 5.51 | 5.68 | 8.16 | 25.05 |

STATE: TEXAS

| ZIP CODE | QUARTER 1 | QUARTER 2 | QUARTER 3 | QUARTER 4 | TOTAL GRAMS |
|---|---|---|---|---|---|
| 750 | 0.76 | 0.57 | 0.76 | 0.38 | 2.47 |
| 752 | 0.76 | 0.76 | 0.76 | 0.95 | 3.23 |
| 753 | 0 | 0 | 0.19 | 0 | 0.19 |
| 754 | 0 | 0 | 0 | 0.19 | 0.19 |
| 756 | 0 | 0 | 0.19 | 0 | 0.19 |
| 757 | 0.19 | 0.76 | 1.89 | 1.51 | 4.35 |
| 760 | 0.38 | 0.76 | 0.38 | 0.19 | 1.71 |
| 761 | 0.76 | 0.95 | 0.19 | 0.38 | 2.28 |
| 762 | 0 | 0 | 0 | 0.38 | 0.38 |
| 764 | 0.19 | 0 | 0 | 0 | 0.19 |
| 765 | 0.76 | 0.76 | 0 | 0 | 1.52 |
| 766 | 0.57 | 0.38 | 0.38 | 0.76 | 2.09 |
| 768 | 0 | 1.32 | 0.76 | 0.76 | 2.84 |
| 769 | 1.51 | 3.03 | 2.46 | 2.27 | 9.27 |
| 770 | 0 | 0.19 | 0.57 | 0.38 | 1.14 |
| 773 | 0.76 | 0.19 | 0 | 0 | 0.95 |
| 774 | 0.38 | 0.76 | 1.14 | 0.57 | 2.85 |
| 775 | 0.76 | 0 | 0.95 | 0.57 | 2.28 |
| 777 | 0 | 0 | 0 | 0.19 | 0.19 |
| 780 | 0.19 | 0 | 0 | 0 | 0.19 |
| 782 | 0.38 | 0.76 | 0.76 | 0.57 | 2.47 |
| 786 | 0.38 | 0.76 | 0.38 | 0.95 | 2.47 |
| 787 | 2.65 | 1.51 | 0.76 | 1.32 | 6.24 |
| 790 | 0 | 0.38 | 0 | 0 | 0.38 |
| 793 | 0 | 0 | 0 | 0.19 | 0.19 |
| 796 | 0 | 0.38 | 0.57 | 1.7 | 2.65 |
| 799 | 0 | 0 | 0 | 0.19 | 0.19 |
| TOTAL | 11.38 | 14.22 | 13.09 | 14.4 | 53.09 |

STATE: UTAH

| ZIP CODE | QUARTER 1 | QUARTER 2 | QUARTER 3 | QUARTER 4 | TOTAL GRAMS |
|---|---|---|---|---|---|
| 840 | 5.68 | 4.35 | 3.97 | 6.24 | 20.24 |
| 841 | 2.27 | 2.08 | 2.46 | 1.7 | 8.51 |

| | | | | | |
|---|---|---|---|---|---|
| 843 | 0.19 | 0.19 | 0.19 | 0.38 | 0.95 |
| 844 | 1.7 | 2.27 | 2.27 | 2.84 | 9.08 |
| 846 | 2.84 | 3.59 | 1.51 | 0.76 | 8.7 |
| 847 | 0.38 | 0.19 | 0.57 | 0.95 | 2.09 |
| TOTAL | 13.06 | 12.67 | 10.97 | 12.87 | 49.57 |

STATE: VERMONT

| ZIP CODE | QUARTER 1 | QUARTER 2 | QUARTER 3 | QUARTER 4 | TOTAL GRAMS |
|---|---|---|---|---|---|
| 50 | 0 | 0.38 | 0.19 | 0.38 | 0.95 |
| TOTAL | 0 | 0.38 | 0.19 | 0.38 | 0.95 |

STATE: VIRGINIA

| ZIP CODE | QUARTER 1 | QUARTER 2 | QUARTER 3 | QUARTER 4 | TOTAL GRAMS |
|---|---|---|---|---|---|
| 201 | 0 | 0 | 0.57 | 0 | 0.57 |
| 221 | 0.95 | 0.76 | 0.19 | 0.19 | 2.09 |
| 224 | 0.95 | 0.19 | 0.38 | 0 | 1.52 |
| 226 | 0 | 0.19 | 0 | 0.19 | 0.38 |
| 228 | 0.38 | 0.57 | 0 | 0.19 | 1.14 |
| 229 | 0.76 | 0.19 | 0.76 | 0.57 | 2.28 |
| 230 | 0.38 | 0.19 | 0.19 | 0 | 0.76 |
| 231 | 0.57 | 1.32 | 0.38 | 0.19 | 2.46 |
| 232 | 0.19 | 0.19 | 0.76 | 0.95 | 2.09 |
| 234 | 0.57 | 0.38 | 0.19 | 0 | 1.14 |
| 235 | 1.14 | 0.19 | 0 | 0.19 | 1.52 |
| 236 | 0 | 0 | 0.19 | 0 | 0.19 |
| 238 | 0 | 0 | 0 | 0.19 | 0.19 |
| 239 | 0 | 0 | 0 | 0.19 | 0.19 |
| 240 | 0.95 | 0 | 0.19 | 0.19 | 1.33 |
| 242 | 0.76 | 0.57 | 0.38 | 0.38 | 2.09 |
| 243 | 0 | 0.38 | 0.38 | 0.19 | 0.95 |
| 244 | 0.19 | 0.38 | 0.76 | 1.14 | 2.47 |
| 245 | 0.19 | 0 | 0.19 | 0 | 0.38 |
| 246 | 0 | 0 | 0.19 | 0 | 0.19 |
| TOTAL | 7.98 | 5.5 | 5.7 | 4.75 | 23.93 |

DATE RANGE: 01/01/2017 TO 12/31/2017 | ARCOS 3 - REPORT 1 | Run Date: 07/03/2018

RETAIL DRUG DISTRIBUTION BY ZIP CODE WITHIN STATE BY GRAMS WT

STATE: WASHINGTON

| ZIP CODE | QUARTER 1 | QUARTER 2 | QUARTER 3 | QUARTER 4 | TOTAL GRAMS |
|---|---|---|---|---|---|
| 980 | 0 | 0 | 0.38 | 0.19 | 0.57 |
| 981 | 0 | 0.57 | 0 | 0 | 0.57 |
| 982 | 0.38 | 0.76 | 1.14 | 0.19 | 2.47 |
| 983 | 0.38 | 0 | 0.19 | 0.38 | 0.95 |
| 984 | 0 | 0 | 0.19 | 0 | 0.19 |
| 985 | 0 | 0.19 | 0 | 0 | 0.19 |
| 986 | 0 | 0 | 0 | 0.19 | 0.19 |
| 988 | 0 | 0 | 0.19 | 0 | 0.19 |
| 991 | 0.38 | 0.19 | 0.38 | 0.57 | 1.52 |
| 992 | 0.57 | 0.19 | 0.56 | 0.57 | 1.89 |
| 993 | 0.38 | 0.19 | 1.14 | 0 | 1.71 |
| TOTAL | 2.09 | 2.09 | 4.17 | 2.09 | 10.44 |

STATE: WEST VIRGINIA

| ZIP CODE | QUARTER 1 | QUARTER 2 | QUARTER 3 | QUARTER 4 | TOTAL GRAMS |
|---|---|---|---|---|---|
| 247 | 0.57 | 0.38 | 0.95 | 0.57 | 2.47 |
| 251 | 0.19 | 0.38 | 0 | 0.19 | 0.76 |
| 253 | 0.19 | 0 | 0.19 | 0.19 | 0.57 |
| 255 | 0.95 | 0.76 | 0.57 | 1.14 | 3.42 |
| 257 | 0.38 | 0.19 | 0 | 0 | 0.57 |
| 258 | 0.19 | 0 | 0 | 0.19 | 0.38 |
| 260 | 0 | 0 | 0 | 0.19 | 0.19 |
| 261 | 0 | 0 | 0 | 0.19 | 0.19 |
| 263 | 0 | 0.19 | 0.19 | 0 | 0.38 |
| 265 | 0.19 | 0 | 0.57 | 0 | 0.76 |
| TOTAL | 2.66 | 1.9 | 2.47 | 2.66 | 9.69 |

STATE: WISCONSIN

| ZIP CODE | QUARTER 1 | QUARTER 2 | QUARTER 3 | QUARTER 4 | TOTAL GRAMS |
|---|---|---|---|---|---|
| 532 | 0 | 1.7 | 1.89 | 1.7 | 5.29 |
| 535 | 0 | 0 | 0 | 0.19 | 0.19 |
| 537 | 2.08 | 0 | 0.19 | 0.57 | 2.84 |

| | | | | | | |
|---|---|---|---|---|---|---|
| | 541 | 0.19 | 0 | 0.19 | 0 | 0.38 |
| | 544 | 2.84 | 3.03 | 2.84 | 4.54 | 13.25 |
| | 545 | 0.19 | 0 | 0 | 0 | 0.19 |
| | 546 | 0.19 | 0 | 0 | 0 | 0.19 |
| | 548 | 0 | 0.19 | 0.19 | 0 | 0.38 |
| | 549 | 0.19 | 0 | 0.19 | 0 | 0.38 |
| TOTAL | | 5.68 | 4.92 | 5.49 | 7 | 23.09 |

STATE: WYOMING

| ZIP CODE | | QUARTER 1 | QUARTER 2 | QUARTER 3 | QUARTER 4 | TOTAL GRAMS |
|---|---|---|---|---|---|---|
| | 820 | 0 | 0.38 | 0.57 | 1.51 | 2.46 |
| | 826 | 0 | 0.19 | 0.19 | 0 | 0.38 |
| | 829 | 0 | 0 | 0 | 0.19 | 0.19 |
| TOTAL | | 0 | 0.57 | 0.76 | 1.7 | 3.03 |

DRUG CODE:1100 DRUG NAME:AMPHETAMINE

| STATE | QUARTER 1 | QUARTER 2 | QUARTER 3 | QUARTER 4 | TOTAL GRAMS |
|---|---|---|---|---|---|
| ALABAMA | 129,522.52 | 129,197.88 | 129,399.68 | 133,902.59 | 522,022.67 |
| ALASKA | 8,017.63 | 8,035.54 | 8,190.14 | 8,091.96 | 32,335.27 |
| AMERICAN SAMOA | 1.25 | 1.25 | 0 | 0 | 2.5 |
| ARIZONA | 81,311.77 | 80,184.72 | 80,199.21 | 85,874.24 | 327,569.94 |
| ARKANSAS | 50,604.83 | 50,551.75 | 50,903.79 | 52,311.33 | 204,371.70 |
| CALIFORNIA | 295,548.59 | 301,541.39 | 296,156.77 | 304,886.80 | 1,198,133.55 |
| COLORADO | 77,961.82 | 78,762.94 | 80,330.49 | 82,398.93 | 319,454.18 |
| CONNECTICUT | 51,291.82 | 52,762.24 | 52,806.74 | 54,411.74 | 211,272.54 |
| DELAWARE | 21,634.53 | 21,501.74 | 21,199.97 | 21,865.65 | 86,201.89 |
| DISTRICT OF COLUMBIA | 11,565.87 | 11,640.51 | 11,504.66 | 11,955.82 | 46,666.86 |
| FLORIDA | 296,894.70 | 298,005.58 | 291,272.31 | 315,032.71 | 1,201,205.30 |
| GEORGIA | 186,484.74 | 184,712.71 | 186,327.72 | 192,738.54 | 750,263.71 |
| GUAM | 411.77 | 559.5 | 574.21 | 358.63 | 1,904.11 |
| HAWAII | 9,569.19 | 9,834.42 | 9,939.96 | 9,576.50 | 38,920.07 |
| IDAHO | 24,608.37 | 25,240.99 | 25,335.53 | 26,579.68 | 101,764.57 |
| ILLINOIS | 188,728.87 | 188,378.21 | 189,659.18 | 197,200.90 | 763,967.16 |
| INDIANA | 127,718.22 | 128,736.61 | 129,954 | 133,581.79 | 519,990.62 |
| IOWA | 48,435.44 | 49,394.50 | 49,817.41 | 52,122.76 | 199,770.11 |
| KANSAS | 58,640.79 | 59,886.78 | 60,660.15 | 62,570.96 | 241,758.68 |
| KENTUCKY | 68,115.50 | 66,555.89 | 67,282.59 | 69,993.68 | 271,947.66 |
| LOUISIANA | 152,840.05 | 150,402.28 | 152,820.35 | 161,545.77 | 617,608.45 |
| MAINE | 21,419.93 | 22,777.34 | 23,454.54 | 23,020.49 | 90,672.30 |
| MARYLAND | 90,180.32 | 91,107.65 | 91,040.76 | 93,934.66 | 366,263.39 |
| MASSACHUSETTS | 174,995.37 | 179,718.58 | 175,691.40 | 181,453.66 | 711,859.01 |
| MICHIGAN | 238,888.13 | 239,018.63 | 239,220.46 | 244,814.10 | 961,941.32 |
| MINNESOTA | 106,640.37 | 107,219.41 | 106,743.34 | 111,048.37 | 431,651.49 |
| MISSISSIPPI | 62,603.84 | 60,446.21 | 60,437.34 | 62,550.25 | 246,037.64 |
| MISSOURI | 131,873.12 | 132,162.08 | 133,905.97 | 138,655.85 | 536,597.02 |
| MONTANA | 13,605.53 | 13,878.40 | 14,159.86 | 14,352.84 | 55,996.63 |
| NEBRASKA | 25,323.08 | 25,225.28 | 26,017.59 | 26,750.34 | 103,316.29 |
| NEVADA | 25,495.19 | 25,984.38 | 25,970.40 | 26,697.57 | 104,147.54 |
| NEW HAMPSHIRE | 37,268.50 | 38,741.66 | 38,476.01 | 39,382.95 | 153,869.12 |
| NEW JERSEY | 101,969.53 | 103,664.89 | 103,491.02 | 107,553.26 | 416,678.70 |
| NEW MEXICO | 16,802.06 | 16,226.35 | 15,622.66 | 15,009.26 | 63,660.33 |
| NEW YORK | 228,378.91 | 235,698.16 | 233,426.04 | 244,230.77 | 941,733.88 |

| | | | | | |
|---|---|---|---|---|---|
| NORTH CAROLINA | 200,834.82 | 201,804.95 | 201,294.03 | 208,597.67 | 812,531.47 |
| NORTH DAKOTA | 8,808.92 | 8,886.90 | 9,070.85 | 9,197.64 | 35,964.31 |
| OHIO | 207,941.78 | 208,201.07 | 209,912.76 | 220,964.23 | 847,019.84 |
| OKLAHOMA | 52,284.73 | 52,570.95 | 52,516.13 | 54,232.93 | 211,604.74 |
| OREGON | 57,185.60 | 59,272.39 | 58,568.95 | 59,537.23 | 234,564.17 |
| PENNSYLVANIA | 208,503.68 | 211,247.55 | 208,839.24 | 215,744.51 | 844,334.98 |
| PUERTO RICO | 9,076.62 | 8,442.37 | 7,845.57 | 7,750.67 | 33,115.23 |
| RHODE ISLAND | 35,384.78 | 36,481.19 | 36,854.42 | 37,818.74 | 146,539.13 |
| SOUTH CAROLINA | 136,661.36 | 134,954.13 | 133,936.64 | 139,640.78 | 545,192.91 |
| SOUTH DAKOTA | 15,983.46 | 14,915.82 | 14,655.77 | 15,257.17 | 60,812.22 |
| TENNESSEE | 129,020.72 | 128,485.65 | 128,719.73 | 134,212.64 | 520,438.74 |
| TEXAS | 407,471.01 | 411,025.34 | 407,717.31 | 429,646.03 | 1,655,859.69 |
| UTAH | 79,212.88 | 81,029.06 | 81,748.72 | 85,695.73 | 327,686.39 |
| VERMONT | 9,800.02 | 9,909.35 | 9,781.02 | 10,199.01 | 39,689.40 |
| VIRGIN ISLANDS | 368.82 | 364.19 | 251.15 | 233.08 | 1,217.24 |
| VIRGINIA | 177,158.94 | 178,362.49 | 176,595.37 | 181,966.29 | 714,083.09 |
| WASHINGTON | 107,196.40 | 109,746.86 | 108,248.01 | 110,209.82 | 435,401.09 |
| WEST VIRGINIA | 23,855.35 | 23,267.05 | 23,223.58 | 23,626.77 | 93,972.75 |
| WISCONSIN | 129,986.53 | 126,614.29 | 125,467.50 | 128,764.70 | 510,833.02 |
| WYOMING | 6,609.86 | 6,232.11 | 6,410.92 | 6,483.05 | 25,735.94 |
| TOTAL | 5,168,698.43 | 5,199,570.16 | 5,183,649.92 | 5,386,234.04 | 20,938,152.55 |

DRUG CODE:1105B DRUG NAME:DL-METHAMPHETAMINE RACEMIC BASE

| STATE | QUARTER 1 | QUARTER 2 | QUARTER 3 | QUARTER 4 | TOTAL GRAMS |
|---|---|---|---|---|---|
| ALABAMA | 22.5 | 24.51 | 27.73 | 21.3 | 96.04 |
| ALASKA | 0.8 | 4.82 | 4.02 | 1.21 | 10.85 |
| ARIZONA | 34.16 | 32.15 | 28.53 | 39.38 | 134.22 |
| ARKANSAS | 5.22 | 7.23 | 6.83 | 6.03 | 25.31 |
| CALIFORNIA | 320.68 | 311.03 | 271.25 | 249.95 | 1,152.91 |
| COLORADO | 49.03 | 54.25 | 49.83 | 57.46 | 210.57 |
| CONNECTICUT | 20.49 | 24.51 | 20.09 | 24.51 | 89.6 |
| DELAWARE | 2.41 | 3.62 | 4.82 | 4.42 | 15.27 |
| DISTRICT OF COLUMBIA | 4.02 | 2.01 | 0 | 0.8 | 6.83 |
| FLORIDA | 28.13 | 39.38 | 36.57 | 31.75 | 135.83 |
| GEORGIA | 32.15 | 24.51 | 17.68 | 20.49 | 94.83 |
| HAWAII | 5.22 | 6.83 | 4.02 | 4.42 | 20.49 |

| | | | | | |
|---|---|---|---|---|---|
| IDAHO | 7.64 | 6.83 | 10.45 | 10.05 | 34.97 |
| ILLINOIS | 63.89 | 58.27 | 63.49 | 62.69 | 248.34 |
| INDIANA | 29.74 | 26.12 | 29.74 | 18.08 | 103.68 |
| IOWA | 14.87 | 18.89 | 18.89 | 21.3 | 73.95 |
| KANSAS | 10.85 | 12.46 | 8.04 | 10.45 | 41.8 |
| KENTUCKY | 10.85 | 10.05 | 9.64 | 9.24 | 39.78 |
| LOUISIANA | 14.06 | 14.87 | 14.06 | 12.06 | 55.05 |
| MAINE | 3.21 | 6.83 | 10.45 | 12.86 | 33.35 |
| MARYLAND | 41.39 | 44.61 | 38.18 | 40.19 | 164.37 |
| MASSACHUSETTS | 44.61 | 41.39 | 43 | 39.38 | 168.38 |
| MICHIGAN | 22.91 | 22.5 | 21.7 | 24.51 | 91.62 |
| MINNESOTA | 78.36 | 87.2 | 63.89 | 59.88 | 289.33 |
| MISSISSIPPI | 8.44 | 8.04 | 8.84 | 7.64 | 32.96 |
| MISSOURI | 49.03 | 62.29 | 16.07 | 62.69 | 190.08 |
| MONTANA | 1.21 | 0.8 | 0.4 | 0.4 | 2.81 |
| NEBRASKA | 6.03 | 8.84 | 8.84 | 9.24 | 32.95 |
| NEVADA | 23.71 | 29.34 | 22.1 | 22.5 | 97.65 |
| NEW HAMPSHIRE | 8.04 | 7.64 | 6.83 | 3.62 | 26.13 |
| NEW JERSEY | 38.18 | 30.54 | 31.75 | 37.77 | 138.24 |
| NEW MEXICO | 27.33 | 20.9 | 12.86 | 4.82 | 65.91 |
| NEW YORK | 74.34 | 80.77 | 76.75 | 93.63 | 325.49 |
| NORTH CAROLINA | 47.82 | 56.66 | 42.19 | 42.6 | 189.27 |
| NORTH DAKOTA | 2.81 | 7.64 | 6.03 | 7.23 | 23.71 |
| OHIO | 77.56 | 80.77 | 48.62 | 93.23 | 300.18 |
| OKLAHOMA | 23.71 | 16.48 | 20.49 | 16.48 | 77.16 |
| OREGON | 44.2 | 32.95 | 19.29 | 31.34 | 127.78 |
| PENNSYLVANIA | 118.55 | 134.22 | 116.13 | 120.15 | 489.05 |
| RHODE ISLAND | 4.82 | 5.63 | 2.81 | 3.62 | 16.88 |
| SOUTH CAROLINA | 18.49 | 12.06 | 16.48 | 13.26 | 60.29 |
| SOUTH DAKOTA | 15.67 | 22.1 | 17.28 | 14.47 | 69.52 |
| TENNESSEE | 10.45 | 12.46 | 12.46 | 15.27 | 50.64 |
| TEXAS | 116.54 | 116.13 | 91.22 | 102.87 | 426.76 |
| UTAH | 25.32 | 23.31 | 26.52 | 28.93 | 104.08 |
| VERMONT | 0.8 | 2.01 | 2.01 | 2.41 | 7.23 |
| VIRGINIA | 51.03 | 40.59 | 39.78 | 43.8 | 175.2 |
| WASHINGTON | 90.82 | 95.64 | 91.22 | 103.68 | 381.36 |
| WEST VIRGINIA | 3.21 | 4.42 | 5.63 | 3.21 | 16.47 |

| | | | | | |
|---|---|---|---|---|---|
| WISCONSIN | 61.48 | 62.29 | 42.19 | 31.34 | 197.3 |
| WYOMING | 6.03 | 7.23 | 4.82 | 3.21 | 21.29 |
| TOTAL | 1,822.81 | 1,866.62 | 1,592.51 | 1,701.82 | 6,983.76 |

DRUG CODE:1105D DRUG NAME:D-METHAMPHETAMINE

| STATE | QUARTER 1 | QUARTER 2 | QUARTER 3 | QUARTER 4 | TOTAL GRAMS |
|---|---|---|---|---|---|
| ALABAMA | 0 | 0.8 | 6.83 | 7.23 | 14.86 |
| ALASKA | 0 | 0 | 0.8 | 0 | 0.8 |
| ARIZONA | 0 | 4.42 | 4.02 | 11.25 | 19.69 |
| CALIFORNIA | 0 | 23.06 | 61.14 | 58.65 | 142.85 |
| CONNECTICUT | 0 | 0 | 3.62 | 4.42 | 8.04 |
| DISTRICT OF COLUMBIA | 0 | 0.8 | 2.81 | 1.61 | 5.22 |
| FLORIDA | 0 | 4.82 | 4.42 | 3.21 | 12.45 |
| GEORGIA | 0 | 1.61 | 4.82 | 4.82 | 11.25 |
| HAWAII | 0 | 0 | 1.61 | 1.61 | 3.22 |
| IDAHO | 0 | 1.21 | 5.22 | 4.02 | 10.45 |
| ILLINOIS | 0 | 0 | 3.62 | 2.41 | 6.03 |
| KANSAS | 0 | 0.8 | 0.8 | 2.81 | 4.41 |
| KENTUCKY | 0 | 0.4 | 1.61 | 1.61 | 3.62 |
| LOUISIANA | 0 | 0 | 2.01 | 2.41 | 4.42 |
| MARYLAND | 0 | 0 | 1.21 | 3.62 | 4.83 |
| MASSACHUSETTS | 0 | 0.8 | 3.62 | 2.41 | 6.83 |
| MINNESOTA | 0 | 4.42 | 25.71 | 22.9 | 53.03 |
| MISSISSIPPI | 0 | 0 | 1.61 | 0.8 | 2.41 |
| MISSOURI | 0 | 3.21 | 12.05 | 16.87 | 32.13 |
| NEBRASKA | 0 | 0 | 0 | 0.8 | 0.8 |
| NEVADA | 0 | 0 | 0.4 | 0 | 0.4 |
| NEW JERSEY | 0 | 0.8 | 2.81 | 2.01 | 5.62 |
| NEW YORK | 4.02 | 0 | 6.83 | 6.91 | 17.76 |
| NORTH CAROLINA | 0 | 2.01 | 9.64 | 10.04 | 21.69 |
| NORTH DAKOTA | 0 | 0 | 5.62 | 0.4 | 6.02 |
| OHIO | 0 | 0.4 | 8.44 | 12.45 | 21.29 |
| OKLAHOMA | 0 | 0.4 | 4.02 | 2.81 | 7.23 |
| OREGON | 0 | 9.24 | 14.06 | 14.46 | 37.76 |
| PENNSYLVANIA | 0 | 3.21 | 11.65 | 11.25 | 26.11 |
| RHODE ISLAND | 0 | 0 | 1.61 | 2.01 | 3.62 |

| | | | | | |
|---|---|---|---|---|---|
| SOUTH CAROLINA | 0 | 2.01 | 4.02 | 2.81 | 8.84 |
| TENNESSEE | 0 | 0 | 0 | 0.4 | 0.4 |
| TEXAS | 0 | 4.42 | 18.08 | 17.27 | 39.77 |
| WASHINGTON | 0 | 0 | 0.8 | 1.61 | 2.41 |
| WEST VIRGINIA | 0 | 0 | 2.41 | 1.21 | 3.62 |
| WISCONSIN | 0 | 1.61 | 13.66 | 12.85 | 28.12 |
| TOTAL | 4.02 | 70.45 | 251.58 | 251.95 | 578 |

DRUG CODE:1205 DRUG NAME:LISDEXAMFETAMINE

| STATE | QUARTER 1 | QUARTER 2 | QUARTER 3 | QUARTER 4 | TOTAL GRAMS |
|---|---|---|---|---|---|
| ALABAMA | 73,124.45 | 69,008.38 | 69,770.43 | 73,421.78 | 285,325.04 |
| ALASKA | 2,366.74 | 2,413.57 | 2,534.39 | 2,551.16 | 9,865.86 |
| ARIZONA | 35,652.58 | 34,515.46 | 35,081.42 | 38,130.90 | 143,380.36 |
| ARKANSAS | 31,934.82 | 28,732.73 | 29,007.32 | 31,528.74 | 121,203.61 |
| CALIFORNIA | 111,261.70 | 111,775.06 | 108,794.14 | 114,459.16 | 446,290.06 |
| COLORADO | 27,319.27 | 27,395.58 | 27,495.59 | 28,822.33 | 111,032.77 |
| CONNECTICUT | 35,946.26 | 36,527.83 | 34,903.37 | 37,374.17 | 144,751.63 |
| DELAWARE | 14,390.07 | 14,231.09 | 13,743.17 | 14,852.55 | 57,216.88 |
| DISTRICT OF COLUMBIA | 5,127.17 | 5,166.48 | 5,261.29 | 5,454.95 | 21,009.89 |
| FLORIDA | 143,974.07 | 135,084.05 | 128,667.31 | 141,074.99 | 548,800.42 |
| GEORGIA | 105,611.35 | 98,907.13 | 101,109.11 | 103,946.40 | 409,573.99 |
| GUAM | 37.58 | 27.17 | 12.14 | 13.87 | 90.76 |
| HAWAII | 1,348.71 | 1,304.77 | 1,331.94 | 1,372.99 | 5,358.41 |
| IDAHO | 8,492.29 | 8,748.97 | 8,782.50 | 9,373.31 | 35,397.07 |
| ILLINOIS | 71,184.34 | 69,483 | 69,469.12 | 73,303.66 | 283,440.12 |
| INDIANA | 86,359.47 | 83,479.37 | 83,966.13 | 87,564.23 | 341,369.20 |
| IOWA | 35,187.21 | 34,025.81 | 33,274.86 | 35,151.95 | 137,639.83 |
| KANSAS | 31,604.15 | 31,149.76 | 31,014.49 | 33,384.12 | 127,152.52 |
| KENTUCKY | 43,977.79 | 41,332.99 | 41,486.74 | 42,833.74 | 169,631.26 |
| LOUISIANA | 98,011.07 | 88,695.57 | 89,750.60 | 94,902.51 | 371,359.75 |
| MAINE | 16,286.23 | 16,819.82 | 16,980.53 | 17,223.91 | 67,310.49 |
| MARYLAND | 56,770.58 | 55,794.17 | 53,445.92 | 57,445.85 | 223,456.52 |
| MASSACHUSETTS | 35,567.60 | 36,220.86 | 34,222.94 | 36,886.25 | 142,897.65 |
| MICHIGAN | 60,273.80 | 58,677.73 | 57,238.14 | 61,662.92 | 237,852.59 |
| MINNESOTA | 31,804.17 | 32,248.73 | 31,807.64 | 30,819.09 | 126,679.63 |
| MISSISSIPPI | 40,741.02 | 37,115.18 | 37,862.08 | 40,202.81 | 155,921.09 |

| | | | | | |
|---|---|---|---|---|---|
| MISSOURI | 67,909.41 | 64,820.04 | 65,065.16 | 68,418.14 | 266,212.75 |
| MONTANA | 5,844.59 | 5,535.31 | 5,568.84 | 5,961.37 | 22,910.11 |
| NEBRASKA | 16,416.88 | 15,918.56 | 16,075.92 | 17,354.48 | 65,765.84 |
| NEVADA | 7,799.73 | 7,551.72 | 7,207.06 | 7,719.95 | 30,278.46 |
| NEW HAMPSHIRE | 13,951.29 | 14,138.59 | 13,564.54 | 13,983.66 | 55,638.08 |
| NEW JERSEY | 49,063.35 | 49,824.70 | 47,456.23 | 49,616.59 | 195,960.87 |
| NEW MEXICO | 4,711.52 | 4,368.70 | 4,101.62 | 4,077.92 | 17,259.76 |
| NEW YORK | 83,860.89 | 84,888.75 | 81,943.94 | 87,724.90 | 338,418.48 |
| NORTH CAROLINA | 104,400.84 | 101,389.49 | 97,896.61 | 106,885.02 | 410,571.96 |
| NORTH DAKOTA | 5,931.88 | 5,516.23 | 5,592.54 | 5,851.53 | 22,892.18 |
| OHIO | 115,341.01 | 109,211.18 | 108,377.10 | 113,584.68 | 446,513.97 |
| OKLAHOMA | 37,115.11 | 35,305.15 | 35,866.35 | 38,390.49 | 146,677.10 |
| OREGON | 14,860.64 | 15,086.10 | 14,691.83 | 15,479.18 | 60,117.75 |
| PENNSYLVANIA | 86,193.55 | 85,037.93 | 83,572.45 | 88,797.32 | 343,601.25 |
| PUERTO RICO | 2,120.47 | 1,865.53 | 1,730.25 | 1,889.23 | 7,605.48 |
| RHODE ISLAND | 7,438.99 | 7,675.43 | 7,171.91 | 7,825.16 | 30,111.49 |
| SOUTH CAROLINA | 62,814.61 | 59,107.26 | 58,061.36 | 64,448.06 | 244,431.29 |
| SOUTH DAKOTA | 11,455.05 | 10,899.50 | 10,515.06 | 10,965.98 | 43,835.59 |
| TENNESSEE | 72,415.12 | 68,092.09 | 68,641.40 | 71,450.56 | 280,599.17 |
| TEXAS | 282,570.08 | 270,317.25 | 258,324.68 | 277,763.06 | 1,088,975.07 |
| UTAH | 21,398.37 | 21,111.63 | 21,113.37 | 22,982.37 | 86,605.74 |
| VERMONT | 6,875.34 | 6,792.68 | 6,786.89 | 7,249.37 | 27,704.28 |
| VIRGIN ISLANDS | 78.62 | 68.22 | 47.98 | 72.84 | 267.66 |
| VIRGINIA | 69,037.86 | 68,163.77 | 65,077.30 | 68,702.91 | 270,981.84 |
| WASHINGTON | 33,478.93 | 34,401 | 33,707.74 | 34,724.65 | 136,312.32 |
| WEST VIRGINIA | 20,520.82 | 19,364.62 | 18,863.40 | 19,838.66 | 78,587.50 |
| WISCONSIN | 65,047.23 | 65,814.37 | 65,424.73 | 71,265.28 | 267,551.61 |
| WYOMING | 2,701.46 | 2,558.67 | 2,568.50 | 2,621.68 | 10,450.31 |
| TOTAL | 2,475,708.13 | 2,393,705.73 | 2,352,028.07 | 2,499,403.38 | 9,720,845.31 |

DRUG CODE:1724 DRUG NAME:METHYLPHENIDATE (DL;D;L;ISOMERS)

| STATE | QUARTER 1 | QUARTER 2 | QUARTER 3 | QUARTER 4 | TOTAL GRAMS |
|---|---|---|---|---|---|
| ALABAMA | 76,441.86 | 69,939.23 | 69,273.14 | 75,215.07 | 290,869.30 |
| ALASKA | 8,308.25 | 7,938.66 | 7,812.94 | 8,079.10 | 32,138.95 |
| AMERICAN SAMOA | 2.59 | 6.05 | 4.32 | 4.76 | 17.72 |
| ARIZONA | 76,843.41 | 71,741.10 | 71,852.92 | 77,682.89 | 298,120.32 |

| | | | | | |
|---|---|---|---|---|---|
| ARKANSAS | 42,738.26 | 38,885.47 | 37,985.81 | 40,491.31 | 160,100.85 |
| CALIFORNIA | 280,017.65 | 271,790.79 | 259,368.75 | 276,806.18 | 1,087,983.37 |
| COLORADO | 62,961.72 | 60,411.10 | 59,560.82 | 63,461.83 | 246,395.47 |
| CONNECTICUT | 55,654.28 | 55,007.99 | 50,532.25 | 55,121.41 | 216,315.93 |
| DELAWARE | 19,422.76 | 18,908.96 | 17,653.36 | 19,110.67 | 75,095.75 |
| DISTRICT OF COLUMBIA | 8,149.44 | 8,276.15 | 7,546.97 | 8,227.44 | 32,200 |
| FLORIDA | 222,746.10 | 205,780.52 | 184,896.75 | 209,350.28 | 822,773.65 |
| GEORGIA | 175,509.10 | 154,308.03 | 156,407.40 | 167,480.79 | 653,705.32 |
| GUAM | 562.07 | 394.32 | 633.74 | 324.62 | 1,914.75 |
| HAWAII | 10,893.55 | 10,932.98 | 10,782.82 | 11,026.95 | 43,636.30 |
| IDAHO | 27,982.04 | 27,772.05 | 25,796.99 | 28,036.03 | 109,587.11 |
| ILLINOIS | 183,062.06 | 171,136.73 | 164,334.65 | 177,772.06 | 696,305.50 |
| INDIANA | 128,505.04 | 119,020.53 | 116,461.55 | 121,311.99 | 485,299.11 |
| IOWA | 85,301.45 | 81,314.10 | 77,115.85 | 82,345.02 | 326,076.42 |
| KANSAS | 52,329.49 | 49,623.14 | 47,814.40 | 50,486.24 | 200,253.27 |
| KENTUCKY | 79,957.38 | 71,563.96 | 72,425.05 | 76,221.48 | 300,167.87 |
| LOUISIANA | 93,211.93 | 78,871.78 | 79,234.69 | 87,401.96 | 338,720.36 |
| MAINE | 35,708.93 | 35,725.10 | 34,610.63 | 35,180.12 | 141,224.78 |
| MARYLAND | 104,696.27 | 101,052.01 | 93,114.43 | 101,335.63 | 400,198.34 |
| MASSACHUSETTS | 149,723.22 | 149,480.86 | 136,831.79 | 148,614.43 | 584,650.30 |
| MICHIGAN | 196,629.98 | 188,328.66 | 177,232.93 | 192,684.13 | 754,875.70 |
| MINNESOTA | 103,381.49 | 100,563.87 | 94,501.53 | 104,893.88 | 403,340.77 |
| MISSISSIPPI | 46,464.57 | 40,093.82 | 41,116.20 | 43,645.13 | 171,319.72 |
| MISSOURI | 135,184.09 | 126,144.06 | 128,490.32 | 136,556.50 | 526,374.97 |
| MONTANA | 18,347.86 | 17,175.60 | 16,958.78 | 17,493.61 | 69,975.85 |
| NEBRASKA | 33,026.53 | 30,615.36 | 29,742.71 | 31,755.77 | 125,140.37 |
| NEVADA | 15,406.98 | 14,798.82 | 13,977.47 | 15,058.02 | 59,241.29 |
| NEW HAMPSHIRE | 34,410.12 | 34,736.63 | 32,581.78 | 33,911.45 | 135,639.98 |
| NEW JERSEY | 96,262.44 | 93,216.84 | 84,140.32 | 93,160.74 | 366,780.34 |
| NEW MEXICO | 20,020.32 | 18,861.11 | 18,673.30 | 18,715.70 | 76,270.43 |
| NEW YORK | 234,957.87 | 233,012.46 | 209,800.05 | 232,310.15 | 910,080.53 |
| NORTH CAROLINA | 205,316.64 | 189,744.23 | 175,482.80 | 191,636.06 | 762,179.73 |
| NORTH DAKOTA | 15,693.46 | 14,870.97 | 14,200.42 | 15,363.74 | 60,128.59 |
| OHIO | 241,069.37 | 223,831.04 | 221,728.35 | 240,266.34 | 926,895.10 |
| OKLAHOMA | 51,387.79 | 46,391.81 | 46,687.87 | 49,045.13 | 193,512.60 |
| OREGON | 62,327.98 | 63,350.18 | 58,177.20 | 62,048.85 | 245,904.21 |
| PENNSYLVANIA | 229,798.99 | 224,625.98 | 214,395.79 | 230,994 | 899,814.76 |

| | | | | | |
|---|---|---|---|---|---|
| PUERTO RICO | 22,532.32 | 17,863.66 | 14,050.16 | 13,418.65 | 67,864.79 |
| RHODE ISLAND | 19,040.20 | 19,549.72 | 17,663.33 | 19,046.02 | 75,299.27 |
| SOUTH CAROLINA | 95,241.64 | 84,944.70 | 83,046.40 | 90,539.45 | 353,772.19 |
| SOUTH DAKOTA | 24,985.21 | 20,180.07 | 22,968.82 | 24,851.08 | 92,985.18 |
| TENNESSEE | 88,695.13 | 80,906.13 | 80,894.85 | 85,146.69 | 335,642.80 |
| TEXAS | 359,410.70 | 333,662.03 | 305,197.30 | 338,505.30 | 1,336,775.33 |
| UTAH | 47,750.91 | 45,267.45 | 43,816.53 | 47,326.10 | 184,160.99 |
| VERMONT | 19,251.52 | 19,375.66 | 18,661.95 | 19,638.19 | 76,927.32 |
| VIRGIN ISLANDS | 210 | 180.55 | 99.13 | 107.51 | 597.19 |
| VIRGINIA | 140,383.28 | 134,905.93 | 125,005.22 | 135,489.52 | 535,783.95 |
| WASHINGTON | 113,786.25 | 114,638.48 | 105,145.17 | 113,286.62 | 446,856.52 |
| WEST VIRGINIA | 33,556.50 | 30,899.27 | 30,340.53 | 31,580.48 | 126,376.78 |
| WISCONSIN | 107,110.53 | 103,041.93 | 97,436.49 | 106,075.78 | 413,664.73 |
| WYOMING | 8,742.70 | 7,947.27 | 7,794.85 | 8,091.15 | 32,575.97 |
| TOTAL | 4,801,112.22 | 4,533,605.90 | 4,312,060.57 | 4,663,730 | 18,310,508.69 |

DRUG CODE:2012 DRUG NAME:GAMMA HYDROXYBUTYRIC ACID(FDA APPROVED-CSA

| STATE | QUARTER 1 | QUARTER 2 | QUARTER 3 | QUARTER 4 | TOTAL GRAMS |
|---|---|---|---|---|---|
| CALIFORNIA | 1,560.38 | 1,337.47 | 1,337.47 | 3,789.50 | 8,024.82 |
| ILLINOIS | 520.13 | 743.04 | 668.74 | 743.04 | 2,674.95 |
| LOUISIANA | 297.22 | 148.61 | 0 | 0 | 445.83 |
| MASSACHUSETTS | 0 | 1,560.38 | 297.22 | 817.34 | 2,674.94 |
| MICHIGAN | 594.43 | 0 | 297.22 | 445.82 | 1,337.47 |
| MISSOURI | 9,221,200.70 | 5,490,842.69 | 8,990,338.18 | 6,709,576.90 | 30,411,958.47 |
| NEW YORK | 0 | 0 | 2,972.16 | 4,978.37 | 7,950.53 |
| NORTH CAROLINA | 222.91 | 1,411.78 | 2,006.21 | 1,337.47 | 4,978.37 |
| OHIO | 1,188.86 | 2,080.51 | 1,931.90 | 2,229.12 | 7,430.39 |
| SOUTH CAROLINA | 965.95 | 2,749.25 | 371.52 | 3,492.29 | 7,579.01 |
| TENNESSEE | 594.43 | 594.43 | 594.43 | 594.43 | 2,377.72 |
| TEXAS | 4,161.02 | 2,972.16 | 2,229.12 | 1,783.30 | 11,145.60 |
| VIRGINIA | 0 | 2,229.12 | 1,634.69 | 2,229.12 | 6,092.93 |
| WASHINGTON | 371.52 | 817.34 | 594.43 | 445.82 | 2,229.11 |
| TOTAL | 9,231,677.55 | 5,507,486.78 | 9,005,273.29 | 6,732,462.52 | 30,476,900.14 |

DRUG CODE:2100 DRUG NAME:BARBITURIC ACID DERIVIATIVE OR SALT [PER 21CFR 1

RETAIL DRUG DISTRIBUTION BY STATE WITHIN DRUG CODE BY GRAMS WT

| STATE | QUARTER 1 | QUARTER 2 | QUARTER 3 | QUARTER 4 | TOTAL GRAMS |
|---|---|---|---|---|---|
| ALABAMA | 1,865 | 1,415 | 1,420 | 1,445 | 6,145 |
| ALASKA | 225 | 175 | 165 | 150 | 715 |
| ARIZONA | 5,820 | 5,380 | 4,530 | 4,520 | 20,250 |
| ARKANSAS | 1,640 | 1,335 | 1,265 | 1,085 | 5,325 |
| CALIFORNIA | 15,045 | 14,100 | 16,110 | 13,928.50 | 59,183.50 |
| COLORADO | 1,975 | 1,840 | 1,440 | 1,330 | 6,585 |
| CONNECTICUT | 2,160 | 2,085 | 2,450 | 2,015 | 8,710 |
| DELAWARE | 480 | 380 | 490 | 490 | 1,840 |
| DISTRICT OF COLUMBIA | 280 | 180 | 285 | 225 | 970 |
| FLORIDA | 20,135 | 16,140 | 17,445 | 16,125 | 69,845 |
| GEORGIA | 6,285 | 5,975 | 4,775 | 3,990 | 21,025 |
| GUAM | 0 | 10 | 0 | 0 | 10 |
| HAWAII | 770 | 930 | 940 | 740 | 3,380 |
| IDAHO | 640 | 645 | 445 | 435 | 2,165 |
| ILLINOIS | 7,355 | 7,290 | 7,975 | 7,615 | 30,235 |
| INDIANA | 3,425 | 3,300 | 3,315 | 3,120 | 13,160 |
| IOWA | 720 | 675 | 650 | 735 | 2,780 |
| KANSAS | 1,305 | 1,490 | 1,030 | 995 | 4,820 |
| KENTUCKY | 2,060 | 1,995 | 1,739 | 1,465 | 7,259 |
| LOUISIANA | 3,370 | 3,320 | 3,785 | 2,950 | 13,425 |
| MAINE | 480 | 375 | 405 | 430 | 1,690 |
| MARYLAND | 2,610 | 2,300 | 2,520 | 2,110 | 9,540 |
| MASSACHUSETTS | 2,625 | 2,555 | 3,185 | 2,455 | 10,820 |
| MICHIGAN | 6,140 | 7,125 | 7,090 | 6,555 | 26,910 |
| MINNESOTA | 1,390 | 1,320 | 1,345 | 1,260 | 5,315 |
| MISSISSIPPI | 1,200 | 1,050 | 1,080 | 690 | 4,020 |
| MISSOURI | 4,180 | 4,515 | 3,910 | 3,215 | 15,820 |
| MONTANA | 490 | 540 | 590 | 375 | 1,995 |
| NEBRASKA | 715 | 705 | 800 | 750 | 2,970 |
| NEVADA | 5,020 | 4,835 | 4,195 | 4,225 | 18,275 |
| NEW HAMPSHIRE | 485 | 510 | 475 | 525 | 1,995 |
| NEW JERSEY | 4,580 | 4,205 | 5,065 | 4,260 | 18,110 |
| NEW MEXICO | 840 | 835 | 860 | 790 | 3,325 |
| NEW YORK | 4,720 | 4,390 | 4,410 | 4,065 | 17,585 |
| NORTH CAROLINA | 3,875 | 3,715 | 4,085 | 3,590 | 15,265 |
| NORTH DAKOTA | 170 | 170 | 125 | 100 | 565 |

| | | | | | |
|---|---|---|---|---|---|
| OHIO | 5,420 | 5,220 | 3,425 | 2,730 | 16,795 |
| OKLAHOMA | 2,665 | 2,700 | 3,055 | 2,835 | 11,255 |
| OREGON | 1,205 | 1,115 | 830 | 715 | 3,865 |
| PENNSYLVANIA | 5,740 | 5,845 | 5,560 | 4,530 | 21,675 |
| PUERTO RICO | 135 | 150 | 165 | 155 | 605 |
| RHODE ISLAND | 635 | 600 | 690 | 595 | 2,520 |
| SOUTH CAROLINA | 2,620 | 2,205 | 2,375 | 2,195 | 9,395 |
| SOUTH DAKOTA | 270 | 220 | 225 | 190 | 905 |
| TENNESSEE | 2,860 | 2,915 | 2,880 | 2,650 | 11,305 |
| TEXAS | 12,555 | 12,640 | 13,294.90 | 10,815 | 49,304.90 |
| UTAH | 1,630 | 1,710 | 1,125 | 1,220 | 5,685 |
| VERMONT | 100 | 60 | 85 | 40 | 285 |
| VIRGIN ISLANDS | 20 | 45 | 25 | 15 | 105 |
| VIRGINIA | 3,670 | 3,965 | 3,145 | 3,120 | 13,900 |
| WASHINGTON | 2,710 | 2,700 | 2,130 | 1,700 | 9,240 |
| WEST VIRGINIA | 665 | 550 | 500 | 405 | 2,120 |
| WISCONSIN | 1,620 | 1,450 | 1,750 | 1,540 | 6,360 |
| WYOMING | 430 | 395 | 350 | 350 | 1,525 |
| TOTAL | 160,025 | 152,290 | 152,003.90 | 134,553.50 | 598,872.40 |

DRUG CODE:2125 DRUG NAME:AMOBARBITAL (SCHEDULE 2)

| STATE | QUARTER 1 | QUARTER 2 | QUARTER 3 | QUARTER 4 | TOTAL GRAMS |
|---|---|---|---|---|---|
| ARIZONA | 12.76 | 11.39 | 12.76 | 8.2 | 45.11 |
| ARKANSAS | 0 | 0.46 | 0.46 | 0 | 0.92 |
| CALIFORNIA | 27.34 | 20.96 | 30.08 | 22.33 | 100.71 |
| COLORADO | 27.8 | 15.95 | 14.58 | 10.03 | 68.36 |
| CONNECTICUT | 0 | 2.28 | 0 | 1.37 | 3.65 |
| DISTRICT OF COLUMBIA | 0.46 | 0 | 1.82 | 4.56 | 6.84 |
| FLORIDA | 4.1 | 2.28 | 8.66 | 8.2 | 23.24 |
| GEORGIA | 1.82 | 0.91 | 10.48 | 13.22 | 26.43 |
| ILLINOIS | 4.56 | 0.91 | 6.84 | 3.65 | 15.96 |
| INDIANA | 0 | 0 | 2.28 | 1.82 | 4.1 |
| IOWA | 0 | 0 | 0.46 | 0 | 0.46 |
| KANSAS | 1.82 | 5.01 | 0.46 | 6.38 | 13.67 |
| KENTUCKY | 4.56 | 3.65 | 3.65 | 7.29 | 19.15 |
| MAINE | 0 | 0 | 0.46 | 2.28 | 2.74 |

| | | | | | |
|---|---|---|---|---|---|
| MARYLAND | 7.29 | 3.19 | 6.38 | 10.48 | 27.34 |
| MASSACHUSETTS | 9.57 | 1.82 | 0.91 | 7.75 | 20.05 |
| MICHIGAN | 5.92 | 4.1 | 5.92 | 2.73 | 18.67 |
| MINNESOTA | 5.01 | 3.19 | 7.29 | 7.75 | 23.24 |
| MISSISSIPPI | 1.82 | 0 | 1.37 | 1.37 | 4.56 |
| MISSOURI | 0.46 | 0.91 | 2.73 | 2.73 | 6.83 |
| NEBRASKA | 2.73 | 0 | 0 | 0.91 | 3.64 |
| NEW HAMPSHIRE | 4.1 | 1.82 | 2.73 | 3.65 | 12.3 |
| NEW JERSEY | 3.65 | 3.19 | 7.29 | 16.41 | 30.54 |
| NEW YORK | 54.68 | 21.87 | 26.89 | 49.22 | 152.66 |
| NORTH CAROLINA | 16.41 | 5.01 | 5.47 | 11.39 | 38.28 |
| OHIO | 3.19 | 3.65 | 5.47 | 5.47 | 17.78 |
| OREGON | 1.37 | 1.37 | 1.37 | 2.73 | 6.84 |
| PENNSYLVANIA | 13.67 | 2.73 | 4.1 | 8.66 | 29.16 |
| RHODE ISLAND | 3.65 | 0 | 0 | 0 | 3.65 |
| SOUTH CAROLINA | 1.37 | 4.1 | 5.47 | 4.1 | 15.04 |
| TENNESSEE | 4.1 | 5.01 | 5.01 | 1.82 | 15.94 |
| TEXAS | 25.52 | 13.67 | 27.8 | 16.41 | 83.4 |
| UTAH | 1.82 | 13.67 | 14.58 | 4.1 | 34.17 |
| VERMONT | 0 | 0 | 0.91 | 0 | 0.91 |
| VIRGINIA | 3.65 | 2.73 | 2.73 | 2.28 | 11.39 |
| WASHINGTON | 4.56 | 0 | 11.39 | 20.96 | 36.91 |
| WEST VIRGINIA | 0 | 0 | 0 | 1.82 | 1.82 |
| WISCONSIN | 6.38 | 3.19 | 7.29 | 10.03 | 26.89 |
| TOTAL | 266.14 | 159.02 | 246.09 | 282.1 | 953.35 |

DRUG CODE:2165 DRUG NAME:BUTALBITAL

| STATE | QUARTER 1 | QUARTER 2 | QUARTER 3 | QUARTER 4 | TOTAL GRAMS |
|---|---|---|---|---|---|
| ALABAMA | 4,415 | 4,745 | 4,615 | 4,025 | 17,800 |
| ALASKA | 180 | 265 | 275 | 205 | 925 |
| AMERICAN SAMOA | 10 | 0 | 0 | 0 | 10 |
| ARIZONA | 4,710 | 4,705 | 5,495 | 5,625 | 20,535 |
| ARKANSAS | 3,295 | 3,435 | 3,520 | 3,515 | 13,765 |
| CALIFORNIA | 24,630 | 24,940 | 25,060 | 23,680 | 98,310 |
| COLORADO | 1,715 | 1,835 | 1,985 | 2,275 | 7,810 |
| CONNECTICUT | 3,255 | 3,040 | 3,350 | 2,655 | 12,300 |

| | | | | | |
|---|---|---|---|---|---|
| DELAWARE | 560 | 550 | 495 | 505 | 2,110 |
| DISTRICT OF COLUMBIA | 315 | 350 | 395 | 340 | 1,400 |
| FLORIDA | 14,835 | 16,625 | 17,215 | 17,045 | 65,720 |
| GEORGIA | 9,170 | 9,660 | 10,605 | 10,365 | 39,800 |
| GUAM | 25 | 5 | 5 | 5 | 40 |
| HAWAII | 1,110 | 1,075 | 1,045 | 930 | 4,160 |
| IDAHO | 1,180 | 1,240 | 1,430 | 1,395 | 5,245 |
| ILLINOIS | 7,300 | 7,740 | 7,450 | 7,170 | 29,660 |
| INDIANA | 3,810 | 3,680 | 4,190 | 3,915 | 15,595 |
| IOWA | 1,230 | 1,290 | 1,335 | 1,245 | 5,100 |
| KANSAS | 2,220 | 1,875 | 2,580 | 2,130 | 8,805 |
| KENTUCKY | 2,385 | 2,410 | 2,830 | 2,740 | 10,365 |
| LOUISIANA | 4,150 | 4,050 | 3,835 | 3,715 | 15,750 |
| MAINE | 750 | 635 | 610 | 525 | 2,520 |
| MARYLAND | 4,850 | 4,490 | 4,895 | 4,755 | 18,990 |
| MASSACHUSETTS | 2,530 | 2,405 | 2,350 | 2,015 | 9,300 |
| MICHIGAN | 9,265 | 8,495 | 8,940 | 8,820 | 35,520 |
| MINNESOTA | 1,535 | 1,640 | 1,595 | 1,935 | 6,705 |
| MISSISSIPPI | 2,130 | 2,130 | 2,185 | 2,100 | 8,545 |
| MISSOURI | 5,405 | 5,725 | 6,105 | 6,515 | 23,750 |
| MONTANA | 735 | 770 | 880 | 695 | 3,080 |
| NEBRASKA | 1,050 | 1,080 | 1,105 | 1,200 | 4,435 |
| NEVADA | 3,435 | 3,420 | 4,025 | 3,935 | 14,815 |
| NEW HAMPSHIRE | 625 | 615 | 710 | 715 | 2,665 |
| NEW JERSEY | 6,570 | 6,190 | 6,305 | 5,605 | 24,670 |
| NEW MEXICO | 820 | 1,010 | 995 | 875 | 3,700 |
| NEW YORK | 5,965 | 6,075 | 5,765 | 4,970 | 22,775 |
| NORTH CAROLINA | 5,140 | 5,030 | 5,105 | 5,115 | 20,390 |
| NORTH DAKOTA | 310 | 335 | 305 | 335 | 1,285 |
| OHIO | 5,915 | 5,455 | 7,495 | 7,765 | 26,630 |
| OKLAHOMA | 5,445 | 5,275 | 4,825 | 4,445 | 19,990 |
| OREGON | 2,370 | 2,075 | 2,390 | 2,550 | 9,385 |
| PENNSYLVANIA | 12,020 | 11,445 | 12,105 | 11,230 | 46,800 |
| RHODE ISLAND | 375 | 415 | 360 | 320 | 1,470 |
| SOUTH CAROLINA | 3,295 | 3,405 | 3,430 | 3,350 | 13,480 |
| SOUTH DAKOTA | 660 | 635 | 650 | 650 | 2,595 |
| TENNESSEE | 3,530 | 3,625 | 4,025 | 4,085 | 15,265 |

| | | | | | |
|---|---|---|---|---|---|
| TEXAS | 18,785 | 18,150 | 18,445 | 19,415 | 74,795 |
| UTAH | 3,055 | 2,735 | 3,430 | 3,500 | 12,720 |
| VERMONT | 150 | 85 | 95 | 140 | 470 |
| VIRGIN ISLANDS | 0 | 0 | 20 | 25 | 45 |
| VIRGINIA | 5,545 | 5,155 | 5,165 | 5,005 | 20,870 |
| WASHINGTON | 3,765 | 4,230 | 4,445 | 4,965 | 17,405 |
| WEST VIRGINIA | 1,215 | 1,240 | 1,355 | 1,260 | 5,070 |
| WISCONSIN | 1,670 | 1,745 | 1,940 | 1,640 | 6,995 |
| WYOMING | 340 | 375 | 470 | 445 | 1,630 |
| TOTAL | 209,750 | 209,605 | 220,230 | 214,380 | 853,965 |

DRUG CODE:2270 DRUG NAME:PENTOBARBITAL (SCHEDULE 2)

| STATE | QUARTER 1 | QUARTER 2 | QUARTER 3 | QUARTER 4 | TOTAL GRAMS |
|---|---|---|---|---|---|
| ALABAMA | 11,145.13 | 14,500.33 | 22,368.20 | 17,579.89 | 65,593.55 |
| ALASKA | 5,687.14 | 4,996.29 | 6,220.31 | 5,887.64 | 22,791.38 |
| ARIZONA | 44,885.44 | 44,597.77 | 56,444.25 | 69,138.71 | 215,066.17 |
| ARKANSAS | 7,552.68 | 13,890.83 | 8,916.40 | 6,067.28 | 36,427.19 |
| CALIFORNIA | 173,418.97 | 173,257.94 | 189,476.46 | 172,640.31 | 708,793.68 |
| COLORADO | 47,273.77 | 38,582.56 | 33,020.93 | 60,578.29 | 179,455.55 |
| CONNECTICUT | 14,888.95 | 14,529.60 | 21,462.52 | 21,570.99 | 72,452.06 |
| DELAWARE | 787.45 | 1,976.09 | 1,709.50 | 2,675.85 | 7,148.89 |
| DISTRICT OF COLUMBIA | 195.95 | 9.11 | 871.3 | 1,476.47 | 2,552.83 |
| FLORIDA | 65,115.01 | 76,163.21 | 70,442.04 | 72,885.89 | 284,606.15 |
| GEORGIA | 37,024.55 | 33,886.33 | 38,688.78 | 44,214.69 | 153,814.35 |
| GUAM | 0 | 3,215.42 | 0 | 3,215.42 | 6,430.84 |
| HAWAII | 4,631.28 | 1,230.39 | 17,224.55 | 4,857.76 | 27,943.98 |
| IDAHO | 16,141.35 | 20,099.10 | 28,465.10 | 25,743.86 | 90,449.41 |
| ILLINOIS | 56,158.15 | 60,069.08 | 59,541.08 | 101,493.52 | 277,261.83 |
| INDIANA | 41,277.70 | 38,050.43 | 43,439.08 | 57,017.45 | 179,784.66 |
| IOWA | 11,284.04 | 16,575.81 | 14,194.51 | 29,448.79 | 71,503.15 |
| KANSAS | 21,721.40 | 14,447.51 | 21,334.96 | 32,986.30 | 90,490.17 |
| KENTUCKY | 20,477 | 22,887.26 | 35,235.21 | 32,743.94 | 111,343.41 |
| LOUISIANA | 24,820.76 | 21,484.63 | 27,264.59 | 26,765.45 | 100,335.43 |
| MAINE | 21,326.21 | 17,117 | 19,406.17 | 23,031.16 | 80,880.54 |
| MARYLAND | 23,025.95 | 14,663.50 | 23,838.64 | 27,402.48 | 88,930.57 |
| MASSACHUSETTS | 21,722.39 | 26,302.18 | 31,710.33 | 33,023.51 | 112,758.41 |

| | | | | | |
|---|---|---|---|---|---|
| MICHIGAN | 89,535.30 | 78,113.67 | 110,086.02 | 107,206.29 | 384,941.28 |
| MINNESOTA | 26,227.77 | 36,097.90 | 31,190.18 | 51,363.94 | 144,879.79 |
| MISSISSIPPI | 11,126.08 | 5,525.17 | 6,281.72 | 9,590.44 | 32,523.41 |
| MISSOURI | 36,952.86 | 26,105.75 | 47,154.64 | 50,814.53 | 161,027.78 |
| MONTANA | 12,442.79 | 10,492.04 | 21,509.95 | 29,680.65 | 74,125.43 |
| NEBRASKA | 12,527.65 | 19,068.77 | 20,501.94 | 30,369.58 | 82,467.94 |
| NEVADA | 12,224.69 | 11,557.92 | 18,280.86 | 19,692.25 | 61,755.72 |
| NEW HAMPSHIRE | 11,871.16 | 16,646.96 | 16,792.07 | 16,615.07 | 61,925.26 |
| NEW JERSEY | 18,830.01 | 15,715.07 | 26,327.21 | 21,045.85 | 81,918.14 |
| NEW MEXICO | 13,888.18 | 18,120.85 | 26,506.79 | 21,787.28 | 80,303.10 |
| NEW YORK | 56,153.83 | 76,688.34 | 83,875.93 | 89,677.25 | 306,395.35 |
| NORTH CAROLINA | 51,212.01 | 55,633.78 | 60,744.81 | 74,943.77 | 242,534.37 |
| NORTH DAKOTA | 8,726.66 | 6,497.83 | 4,790.77 | 8,352.98 | 28,368.24 |
| OHIO | 59,844.72 | 65,744.16 | 78,226.93 | 91,305.99 | 295,121.80 |
| OKLAHOMA | 13,683.85 | 21,513.21 | 14,100.72 | 26,814.93 | 76,112.71 |
| OREGON | 28,973.41 | 35,321.39 | 43,992.82 | 46,399.10 | 154,686.72 |
| PENNSYLVANIA | 53,524.62 | 63,201.28 | 87,707.74 | 64,757.20 | 269,190.84 |
| PUERTO RICO | 0 | 0 | 0.2 | 0 | 0.2 |
| RHODE ISLAND | 3,207.67 | 6,945.87 | 7,290.47 | 7,118.40 | 24,562.41 |
| SOUTH CAROLINA | 12,579.37 | 12,854.39 | 13,920.90 | 16,360.09 | 55,714.75 |
| SOUTH DAKOTA | 5,781.01 | 5,426.02 | 6,676.01 | 7,109.83 | 24,992.87 |
| TENNESSEE | 26,638.02 | 41,338.06 | 35,204.43 | 39,413 | 142,593.51 |
| TEXAS | 105,413.74 | 107,956.51 | 142,377.96 | 140,844.93 | 496,593.14 |
| UTAH | 9,644.89 | 11,271.74 | 13,304.40 | 22,693.86 | 56,914.89 |
| VERMONT | 9,795.63 | 9,364.52 | 17,431.72 | 17,770.38 | 54,362.25 |
| VIRGINIA | 42,855.88 | 47,223.42 | 47,155 | 53,755.62 | 190,989.92 |
| WASHINGTON | 55,558.36 | 53,612.17 | 57,017.68 | 74,631.70 | 240,819.91 |
| WEST VIRGINIA | 6,970.84 | 12,573.12 | 14,259.29 | 8,298.29 | 42,101.54 |
| WISCONSIN | 40,971.17 | 43,561.15 | 52,120.04 | 72,768.15 | 209,420.51 |
| WYOMING | 4,531.94 | 12,623.80 | 9,096.68 | 14,217.84 | 40,470.26 |
| TOTAL | 1,512,255.38 | 1,599,327.23 | 1,885,200.79 | 2,107,844.84 | 7,104,628.24 |

DRUG CODE:2315 DRUG NAME:SECOBARBITAL (SCHEDULE 2)

| STATE | QUARTER 1 | QUARTER 2 | QUARTER 3 | QUARTER 4 | TOTAL GRAMS |
|---|---|---|---|---|---|
| ARIZONA | 9.16 | 0 | 0 | 9.16 | 18.32 |
| CALIFORNIA | 897.19 | 988.74 | 1,162.69 | 1,327.48 | 4,376.10 |

| | | | | | |
|---|---|---|---|---|---|
| COLORADO | 109.86 | 91.55 | 109.86 | 64.09 | 375.36 |
| CONNECTICUT | 36.62 | 27.47 | 9.16 | 18.31 | 91.56 |
| DELAWARE | 0 | 9.16 | 9.16 | 9.16 | 27.48 |
| FLORIDA | 100.71 | 73.24 | 100.71 | 91.55 | 366.21 |
| GEORGIA | 36.62 | 9.16 | 0 | 0 | 45.78 |
| HAWAII | 9.16 | 0 | 0 | 0 | 9.16 |
| ILLINOIS | 45.78 | 54.93 | 36.62 | 27.47 | 164.8 |
| KANSAS | 36.62 | 18.31 | 18.31 | 27.47 | 100.71 |
| MARYLAND | 64.09 | 64.09 | 45.78 | 45.78 | 219.74 |
| MASSACHUSETTS | 54.93 | 45.78 | 36.62 | 54.93 | 192.26 |
| MICHIGAN | 0 | 0 | 0 | 18.31 | 18.31 |
| MINNESOTA | 9.16 | 0 | 0 | 9.16 | 18.32 |
| MISSOURI | 27.47 | 18.31 | 36.62 | 18.31 | 100.71 |
| NEVADA | 9.16 | 0 | 0 | 0 | 9.16 |
| NEW HAMPSHIRE | 18.31 | 9.16 | 0 | 0 | 27.47 |
| NEW JERSEY | 73.24 | 54.93 | 54.93 | 45.78 | 228.88 |
| NEW YORK | 466.91 | 402.82 | 292.96 | 338.74 | 1,501.43 |
| NORTH CAROLINA | 0 | 9.16 | 0 | 0 | 9.16 |
| OHIO | 54.93 | 18.31 | 27.47 | 36.62 | 137.33 |
| OKLAHOMA | 0 | 0 | 27.47 | 9.16 | 36.63 |
| OREGON | 256.34 | 173.95 | 219.72 | 302.12 | 952.13 |
| PENNSYLVANIA | 36.62 | 73.24 | 82.4 | 73.24 | 265.5 |
| RHODE ISLAND | 9.16 | 9.16 | 0 | 18.31 | 36.63 |
| SOUTH CAROLINA | 0 | 0 | 18.31 | 18.31 | 36.62 |
| SOUTH DAKOTA | 0 | 0 | 0 | 9.16 | 9.16 |
| TENNESSEE | 0 | 9.16 | 0 | 0 | 9.16 |
| TEXAS | 9.16 | 0 | 9.16 | 27.47 | 45.79 |
| VERMONT | 54.93 | 27.47 | 0 | 54.93 | 137.33 |
| VIRGINIA | 27.47 | 27.47 | 27.47 | 27.47 | 109.88 |
| WASHINGTON | 228.88 | 173.95 | 155.64 | 164.79 | 723.26 |
| WISCONSIN | 9.16 | 0 | 0 | 0 | 9.16 |
| TOTAL | 2,691.64 | 2,389.52 | 2,481.06 | 2,847.28 | 10,409.50 |

DRUG CODE:7365 DRUG NAME:DRONABINOL IN AN ORAL SOLUTION IN FDA APPROVE

| STATE | QUARTER 1 | QUARTER 2 | QUARTER 3 | QUARTER 4 | TOTAL GRAMS |
|---|---|---|---|---|---|
| ALABAMA | 0 | 0 | 1.5 | 2.7 | 4.2 |

| | | | | | |
|---|---|---|---|---|---|
| ARIZONA | 0 | 0 | 14.4 | 7.2 | 21.6 |
| ARKANSAS | 0 | 0 | 0.6 | 0.6 | 1.2 |
| CALIFORNIA | 0 | 0 | 15.15 | 28.65 | 43.8 |
| COLORADO | 0 | 0 | 3.6 | 0.15 | 3.75 |
| CONNECTICUT | 0 | 0 | 1.2 | 0.45 | 1.65 |
| DELAWARE | 0 | 0 | 0.15 | 0 | 0.15 |
| DISTRICT OF COLUMBIA | 0 | 0 | 0 | 0.75 | 0.75 |
| FLORIDA | 0 | 0 | 13.95 | 22.65 | 36.6 |
| GEORGIA | 0 | 0 | 1.35 | 3.3 | 4.65 |
| ILLINOIS | 0 | 0 | 0.3 | 0.75 | 1.05 |
| INDIANA | 0 | 0 | 1.65 | 0 | 1.65 |
| KANSAS | 0 | 0 | 0 | 0.6 | 0.6 |
| KENTUCKY | 0 | 0 | 0.75 | 0.75 | 1.5 |
| LOUISIANA | 0 | 0 | 0.15 | 0.15 | 0.3 |
| MAINE | 0 | 0 | 0.3 | 0 | 0.3 |
| MARYLAND | 0 | 0 | 0.15 | 1.8 | 1.95 |
| MASSACHUSETTS | 0 | 0 | 0.75 | 3 | 3.75 |
| MICHIGAN | 0 | 0 | 1.05 | 1.35 | 2.4 |
| MINNESOTA | 0 | 0 | 0 | 2.85 | 2.85 |
| MISSISSIPPI | 0 | 0 | 0.6 | 0.15 | 0.75 |
| MISSOURI | 0 | 0 | 4.35 | 4.2 | 8.55 |
| MONTANA | 0 | 0 | 0 | 0.3 | 0.3 |
| NEVADA | 0 | 0 | 4.95 | 6.6 | 11.55 |
| NEW HAMPSHIRE | 0 | 0 | 0.75 | 1.05 | 1.8 |
| NEW JERSEY | 0 | 0 | 70.8 | 72.75 | 143.55 |
| NEW MEXICO | 0 | 0 | 0.15 | 0 | 0.15 |
| NEW YORK | 0 | 0 | 0.75 | 1.2 | 1.95 |
| NORTH CAROLINA | 0 | 0 | 0.45 | 0.6 | 1.05 |
| OHIO | 0 | 0 | 0.45 | 8.25 | 8.7 |
| PENNSYLVANIA | 0 | 0 | 14.4 | 1.95 | 16.35 |
| SOUTH CAROLINA | 0 | 0 | 0.3 | 0 | 0.3 |
| TENNESSEE | 0 | 0 | 1.5 | 2.1 | 3.6 |
| TEXAS | 0 | 0 | 6.6 | 5.1 | 11.7 |
| UTAH | 0 | 0 | 0.45 | 0.15 | 0.6 |
| VIRGINIA | 0 | 0 | 0 | 0.75 | 0.75 |
| WISCONSIN | 0 | 0 | 0.15 | 0 | 0.15 |
| TOTAL | 0 | 0 | 163.65 | 182.85 | 346.5 |

DRUG CODE:7379 DRUG NAME:NABILONE

| STATE | QUARTER 1 | QUARTER 2 | QUARTER 3 | QUARTER 4 | TOTAL GRAMS |
|---|---|---|---|---|---|
| ARIZONA | 0 | 0 | 0.15 | 0.05 | 0.2 |
| CALIFORNIA | 0 | 0.25 | 0.05 | 0.35 | 0.65 |
| COLORADO | 0.15 | 0.05 | 0 | 0.15 | 0.35 |
| CONNECTICUT | 0.1 | 0.15 | 0.1 | 0 | 0.35 |
| FLORIDA | 1.05 | 0.45 | 0.45 | 0.2 | 2.15 |
| HAWAII | 0.45 | 0 | 0.15 | 0 | 0.6 |
| ILLINOIS | 0.25 | 0.05 | 0.1 | 0.1 | 0.5 |
| IOWA | 0.05 | 0.05 | 0 | 0 | 0.1 |
| KENTUCKY | 0.15 | 0.2 | 0 | 0 | 0.35 |
| MAINE | 0.1 | 0.1 | 0.1 | 0 | 0.3 |
| MARYLAND | 1.05 | 1.1 | 1.05 | 0.9 | 4.1 |
| MINNESOTA | 0.1 | 0 | 0 | 0 | 0.1 |
| MISSOURI | 0.2 | 0.1 | 0.1 | 0.15 | 0.55 |
| NEBRASKA | 0.15 | 0 | 0 | 0.15 | 0.3 |
| NEW HAMPSHIRE | 0 | 0 | 0 | 0.05 | 0.05 |
| NEW JERSEY | 0.1 | 0.15 | 0.1 | 0 | 0.35 |
| NEW MEXICO | 0 | 0 | 0.1 | 0 | 0.1 |
| NEW YORK | 0.2 | 0.15 | 0.05 | 0 | 0.4 |
| NORTH CAROLINA | 0.2 | 0.15 | 0.15 | 0.25 | 0.75 |
| OHIO | 0.1 | 0.1 | 0.1 | 0.7 | 1 |
| OREGON | 0 | 0 | 0 | 0.05 | 0.05 |
| PENNSYLVANIA | 0.05 | 0.05 | 0.15 | 0.1 | 0.35 |
| VIRGINIA | 0.2 | 0.35 | 0.2 | 0 | 0.75 |
| WASHINGTON | 0 | 0 | 0 | 0.05 | 0.05 |
| TOTAL | 4.65 | 3.45 | 3.1 | 3.25 | 14.45 |

DRUG CODE:9041L DRUG NAME:COCAINE

| STATE | QUARTER 1 | QUARTER 2 | QUARTER 3 | QUARTER 4 | TOTAL GRAMS |
|---|---|---|---|---|---|
| ALABAMA | 62.42 | 65.85 | 49.85 | 50.57 | 228.69 |
| ALASKA | 8.57 | 19.78 | 9.57 | 10 | 47.92 |
| ARIZONA | 110.49 | 109.67 | 99.57 | 99.85 | 419.58 |
| ARKANSAS | 56.32 | 40.93 | 38.18 | 40.18 | 175.61 |

| | | | | | |
|---|---|---|---|---|---|
| CALIFORNIA | 927.62 | 879.09 | 835.77 | 828.09 | 3,470.57 |
| COLORADO | 82.1 | 109.17 | 71.21 | 80.57 | 343.05 |
| CONNECTICUT | 39.07 | 32.28 | 50.14 | 29.43 | 150.92 |
| DELAWARE | 6.57 | 18.86 | 14.86 | 11.57 | 51.86 |
| DISTRICT OF COLUMBIA | 22.53 | 21.46 | 28.14 | 20.75 | 92.88 |
| FLORIDA | 289.16 | 306.77 | 294.66 | 271.52 | 1,162.11 |
| GEORGIA | 154.45 | 171.17 | 147.13 | 184.13 | 656.88 |
| GUAM | 0 | 0 | 0.36 | 0 | 0.36 |
| HAWAII | 28.28 | 23.86 | 24.86 | 20.43 | 97.43 |
| IDAHO | 64.6 | 37.43 | 46.85 | 67.67 | 216.55 |
| ILLINOIS | 326.62 | 356.83 | 272.7 | 333.98 | 1,290.13 |
| INDIANA | 251.13 | 195.74 | 153.13 | 200.92 | 800.92 |
| IOWA | 105.42 | 102.85 | 86.1 | 98.64 | 393.01 |
| KANSAS | 63.53 | 84.89 | 82.32 | 97.89 | 328.63 |
| KENTUCKY | 109.89 | 117.89 | 88.49 | 118.14 | 434.41 |
| LOUISIANA | 92.32 | 96.71 | 86.71 | 113.71 | 389.45 |
| MAINE | 22.43 | 24.93 | 22.86 | 17.21 | 87.43 |
| MARYLAND | 140.78 | 146.6 | 119.53 | 110.67 | 517.58 |
| MASSACHUSETTS | 228.41 | 202.7 | 180.56 | 203.77 | 815.44 |
| MICHIGAN | 367.55 | 422.15 | 336.59 | 381.58 | 1,507.87 |
| MINNESOTA | 211.52 | 233.59 | 199.24 | 211.17 | 855.52 |
| MISSISSIPPI | 79.67 | 101.53 | 75.25 | 82.53 | 338.98 |
| MISSOURI | 164.2 | 180.88 | 159.92 | 154.56 | 659.56 |
| MONTANA | 48.85 | 55 | 41.71 | 66.03 | 211.59 |
| NEBRASKA | 21.57 | 31 | 37.03 | 29.57 | 119.17 |
| NEVADA | 44.6 | 52.1 | 43.93 | 39.32 | 179.95 |
| NEW HAMPSHIRE | 38.71 | 54.89 | 40.57 | 56.14 | 190.31 |
| NEW JERSEY | 176.02 | 168.7 | 148.88 | 152.13 | 645.73 |
| NEW MEXICO | 76.35 | 73.71 | 73.78 | 66.57 | 290.41 |
| NEW YORK | 361.8 | 371.01 | 319.55 | 327.51 | 1,379.87 |
| NORTH CAROLINA | 135.53 | 141.67 | 151.95 | 142.53 | 571.68 |
| NORTH DAKOTA | 33.25 | 66.39 | 37.82 | 44.53 | 181.99 |
| OHIO | 175.49 | 202.02 | 170.02 | 183.24 | 730.77 |
| OKLAHOMA | 31.28 | 34 | 28.14 | 39.57 | 132.99 |
| OREGON | 103.46 | 95.92 | 100.35 | 84.82 | 384.55 |
| PENNSYLVANIA | 412.69 | 410.15 | 356.41 | 377.48 | 1,556.73 |
| RHODE ISLAND | 25.28 | 16.43 | 17.43 | 16.86 | 76 |

| | | | | | |
|---|---|---|---|---|---|
| SOUTH CAROLINA | 30.61 | 31.57 | 28 | 28.14 | 118.32 |
| SOUTH DAKOTA | 44.25 | 24.78 | 27.46 | 52.35 | 148.84 |
| TENNESSEE | 181.04 | 160.13 | 159.7 | 169.13 | 670 |
| TEXAS | 332.69 | 384.69 | 326.62 | 409.69 | 1,453.69 |
| UTAH | 103.28 | 98.6 | 89.92 | 99.21 | 391.01 |
| VERMONT | 10.36 | 13.46 | 6.29 | 8.36 | 38.47 |
| VIRGINIA | 161.85 | 153.17 | 154.88 | 144.63 | 614.53 |
| WASHINGTON | 114.46 | 104.17 | 124.42 | 111.03 | 454.08 |
| WEST VIRGINIA | 52.53 | 48.35 | 49.89 | 51.03 | 201.8 |
| WISCONSIN | 164.42 | 158.2 | 130.63 | 151.56 | 604.81 |
| WYOMING | 18 | 13.86 | 20.28 | 17.14 | 69.28 |
| TOTAL | 6,914.02 | 7,067.58 | 6,260.21 | 6,708.10 | 26,949.91 |

DRUG CODE:9050 DRUG NAME:CODEINE

| STATE | QUARTER 1 | QUARTER 2 | QUARTER 3 | QUARTER 4 | TOTAL GRAMS |
|---|---|---|---|---|---|
| ALABAMA | 52,097.22 | 52,426.61 | 51,821.15 | 52,546.65 | 208,891.63 |
| ALASKA | 7,416.42 | 7,072.30 | 7,040.49 | 6,268.26 | 27,797.47 |
| AMERICAN SAMOA | 353.62 | 340.36 | 353.62 | 313.83 | 1,361.43 |
| ARIZONA | 78,396.32 | 75,054.50 | 70,599.31 | 73,098.41 | 297,148.54 |
| ARKANSAS | 74,120.24 | 76,464.28 | 71,973.79 | 70,250.71 | 292,809.02 |
| CALIFORNIA | 501,442.60 | 479,948.70 | 460,547.34 | 449,781.78 | 1,891,720.42 |
| COLORADO | 26,553.20 | 25,891.58 | 25,424.45 | 24,999.62 | 102,868.85 |
| CONNECTICUT | 24,542.20 | 24,979.42 | 23,678.88 | 23,115.66 | 96,316.16 |
| DELAWARE | 9,906.43 | 9,523.85 | 8,721.58 | 8,835.22 | 36,987.08 |
| DISTRICT OF COLUMBIA | 12,746.29 | 12,577.52 | 12,006.52 | 11,090.12 | 48,420.45 |
| FLORIDA | 180,686.49 | 170,836.92 | 161,724.52 | 168,139.52 | 681,387.45 |
| GEORGIA | 106,357.27 | 109,274.90 | 102,164.73 | 102,467.93 | 420,264.83 |
| GUAM | 1,289.01 | 1,191.33 | 1,196.49 | 1,088.84 | 4,765.67 |
| HAWAII | 13,807.53 | 14,179.60 | 13,766.47 | 12,809.15 | 54,562.75 |
| IDAHO | 13,779.75 | 13,988.86 | 13,595.78 | 13,279.28 | 54,643.67 |
| ILLINOIS | 167,295.96 | 163,981.42 | 159,064.67 | 156,936.08 | 647,278.13 |
| INDIANA | 52,138.19 | 52,017.41 | 47,484.82 | 48,824.93 | 200,465.35 |
| IOWA | 20,061.94 | 20,404.12 | 19,724.99 | 19,705.10 | 79,896.15 |
| KANSAS | 28,997.24 | 28,439.53 | 26,788.02 | 27,571.11 | 111,795.90 |
| KENTUCKY | 48,676.44 | 48,234.28 | 47,573.05 | 45,644.01 | 190,127.78 |
| LOUISIANA | 45,273.75 | 45,593.29 | 42,084.80 | 40,843.81 | 173,795.65 |

| | | | | | |
|---|---|---|---|---|---|
| MAINE | 10,829.90 | 11,007.78 | 9,775.47 | 9,942.97 | 41,556.12 |
| MARYLAND | 62,160.93 | 59,591.10 | 55,355.33 | 54,820.13 | 231,927.49 |
| MASSACHUSETTS | 48,661.15 | 46,733.29 | 45,133.33 | 44,702.33 | 185,230.10 |
| MICHIGAN | 265,875.99 | 262,963.95 | 254,929.37 | 241,505.52 | 1,025,274.83 |
| MINNESOTA | 38,915.01 | 37,129.43 | 37,001.21 | 36,152.95 | 149,198.60 |
| MISSISSIPPI | 28,067.59 | 27,569.65 | 27,316.55 | 26,659.58 | 109,613.37 |
| MISSOURI | 68,652.99 | 70,089.01 | 67,023.53 | 68,032.84 | 273,798.37 |
| MONTANA | 10,274.37 | 10,509.37 | 9,893.81 | 9,903.40 | 40,580.95 |
| NEBRASKA | 12,174.42 | 11,764.41 | 11,642.48 | 11,383.96 | 46,965.27 |
| NEVADA | 27,025.90 | 27,451.42 | 25,820.24 | 26,651.52 | 106,949.08 |
| NEW HAMPSHIRE | 7,881.23 | 7,544.36 | 7,327.35 | 7,594.42 | 30,347.36 |
| NEW JERSEY | 82,192.68 | 78,830.99 | 75,755.85 | 74,034.34 | 310,813.86 |
| NEW MEXICO | 19,825.21 | 19,248.02 | 19,527.98 | 19,585.79 | 78,187 |
| NEW YORK | 147,404.26 | 145,569.93 | 138,625.64 | 136,150.10 | 567,749.93 |
| NORTH CAROLINA | 51,512.39 | 48,995.17 | 46,149.79 | 46,959.82 | 193,617.17 |
| NORTH DAKOTA | 5,752.54 | 5,818.55 | 5,861.92 | 5,345.38 | 22,778.39 |
| OHIO | 109,027.91 | 102,725.01 | 96,703.69 | 94,549.37 | 403,005.98 |
| OKLAHOMA | 96,652.50 | 96,862.79 | 94,633.62 | 93,769.52 | 381,918.43 |
| OREGON | 35,126.69 | 33,710.18 | 33,479.03 | 33,544.33 | 135,860.23 |
| PENNSYLVANIA | 137,000.07 | 143,911.20 | 142,003.02 | 131,550.66 | 554,464.95 |
| PUERTO RICO | 13,584.38 | 13,474.98 | 11,629.55 | 12,536.79 | 51,225.70 |
| RHODE ISLAND | 8,007.21 | 7,373.78 | 6,873.92 | 6,861.83 | 29,116.74 |
| SOUTH CAROLINA | 78,535.31 | 68,293.22 | 72,571.49 | 84,635.97 | 304,035.99 |
| SOUTH DAKOTA | 7,250.21 | 7,076.52 | 6,735.71 | 6,822.74 | 27,885.18 |
| TENNESSEE | 93,521.32 | 88,402.47 | 85,408.29 | 74,152.05 | 341,484.13 |
| TEXAS | 1,009,935.10 | 1,024,743.49 | 987,072.94 | 985,807.44 | 4,007,558.97 |
| UTAH | 16,455.49 | 16,583.15 | 16,378.36 | 15,954.67 | 65,371.67 |
| VERMONT | 3,565.55 | 3,458.76 | 3,031.04 | 3,060.57 | 13,115.92 |
| VIRGIN ISLANDS | 950.85 | 652.13 | 565.79 | 830.26 | 2,999.03 |
| VIRGINIA | 59,288.95 | 57,032.64 | 51,403.11 | 51,851.65 | 219,576.35 |
| WASHINGTON | 63,371.45 | 63,059.58 | 59,985.80 | 59,332.65 | 245,749.48 |
| WEST VIRGINIA | 31,085.46 | 31,132.91 | 30,260.45 | 31,982.24 | 124,461.06 |
| WISCONSIN | 50,699.55 | 47,765.40 | 45,918.70 | 46,170.52 | 190,554.17 |
| WYOMING | 4,366.56 | 4,031.10 | 4,127.83 | 4,240.97 | 16,766.46 |
| TOTAL | 4,171,569.23 | 4,113,526.52 | 3,953,257.66 | 3,914,689.30 | 16,153,042.71 |

DRUG CODE:9056 DRUG NAME:ETORPHINE

| STATE | QUARTER 1 | QUARTER 2 | QUARTER 3 | QUARTER 4 | TOTAL GRAMS |
|---|---|---|---|---|---|
| WYOMING | 28 | 0 | 0 | 29 | 57 |
| TOTAL | 28 | 0 | 0 | 29 | 57 |

DRUG CODE:9064 DRUG NAME:BUPRENORPHINE

| STATE | QUARTER 1 | QUARTER 2 | QUARTER 3 | QUARTER 4 | TOTAL GRAMS |
|---|---|---|---|---|---|
| ALABAMA | 23,735.15 | 25,160.56 | 25,414 | 26,445.37 | 100,755.08 |
| ALASKA | 2,178.38 | 2,351.56 | 2,431.40 | 2,496.82 | 9,458.16 |
| AMERICAN SAMOA | 0 | 0 | 0 | 0 | 0 |
| ARIZONA | 8,917.41 | 9,288.38 | 9,567.44 | 9,927.03 | 37,700.26 |
| ARKANSAS | 5,876.84 | 6,179.83 | 6,370.91 | 6,628.80 | 25,056.38 |
| CALIFORNIA | 36,183.89 | 37,274.64 | 39,297.69 | 39,811.60 | 152,567.82 |
| COLORADO | 7,029.58 | 7,245.65 | 7,617.76 | 7,720.27 | 29,613.26 |
| CONNECTICUT | 14,144.45 | 14,772.20 | 15,022.64 | 15,353.47 | 59,292.76 |
| DELAWARE | 3,383.64 | 3,633.81 | 3,599.52 | 3,898.94 | 14,515.91 |
| DISTRICT OF COLUMBIA | 2,126.56 | 2,265.05 | 2,463.07 | 2,599.65 | 9,454.33 |
| FLORIDA | 45,362.39 | 46,836.59 | 46,526.79 | 48,926.92 | 187,652.69 |
| GEORGIA | 14,877.07 | 15,748.96 | 15,896.57 | 16,348.80 | 62,871.40 |
| GUAM | 0.82 | 1.99 | 0 | 1.68 | 4.49 |
| HAWAII | 1,860.94 | 1,987.21 | 2,012.85 | 2,018.34 | 7,879.34 |
| IDAHO | 3,253.87 | 3,455.24 | 3,631.94 | 3,874.99 | 14,216.04 |
| ILLINOIS | 13,435.24 | 14,379.93 | 14,668.92 | 15,043.80 | 57,527.89 |
| INDIANA | 17,803.76 | 19,775.16 | 20,537.14 | 21,528.68 | 79,644.74 |
| IOWA | 1,585.44 | 1,638.21 | 1,699.13 | 1,807.58 | 6,730.36 |
| KANSAS | 2,494.01 | 2,507.15 | 2,660.73 | 2,561.66 | 10,223.55 |
| KENTUCKY | 33,748.36 | 34,750.20 | 34,645.38 | 34,938.66 | 138,082.60 |
| LOUISIANA | 18,054.67 | 18,953.53 | 19,682.99 | 19,622.81 | 76,314 |
| MAINE | 8,971.92 | 9,673.06 | 9,873.65 | 10,201.77 | 38,720.40 |
| MARYLAND | 24,125.10 | 25,266.60 | 25,673.14 | 26,997.92 | 102,062.76 |
| MASSACHUSETTS | 39,805.50 | 42,539.51 | 40,755.78 | 42,212.58 | 165,313.37 |
| MICHIGAN | 25,710.86 | 27,064.52 | 27,236.37 | 28,632.85 | 108,644.60 |
| MINNESOTA | 6,999.44 | 7,444.88 | 7,748.16 | 8,022.26 | 30,214.74 |
| MISSISSIPPI | 9,689.76 | 9,995.82 | 9,962.33 | 10,024.20 | 39,672.11 |
| MISSOURI | 8,630.87 | 9,208.38 | 9,704.05 | 10,683.37 | 38,226.67 |
| MONTANA | 2,220.35 | 2,315.94 | 2,495.93 | 2,619.94 | 9,652.16 |

| | | | | | |
|---|---|---|---|---|---|
| NEBRASKA | 1,264.53 | 1,308.52 | 1,363.58 | 1,372.23 | 5,308.86 |
| NEVADA | 3,238.79 | 3,476.56 | 3,461.23 | 3,468.65 | 13,645.23 |
| NEW HAMPSHIRE | 7,670.59 | 8,058.64 | 8,213.44 | 8,150.59 | 32,093.26 |
| NEW JERSEY | 25,547.24 | 26,921.80 | 26,147.03 | 26,936.73 | 105,552.80 |
| NEW MEXICO | 6,825.93 | 7,233.94 | 7,598.72 | 7,837.01 | 29,495.60 |
| NEW YORK | 50,488.07 | 52,804.61 | 53,572.20 | 56,492.76 | 213,357.64 |
| NORTH CAROLINA | 30,365.22 | 33,074.64 | 32,092.39 | 35,309.84 | 130,842.09 |
| NORTH DAKOTA | 851.95 | 883.13 | 943.98 | 956.29 | 3,635.35 |
| OHIO | 50,757.75 | 53,154.23 | 52,666.30 | 53,520.61 | 210,098.89 |
| OKLAHOMA | 10,175.80 | 10,362.32 | 11,514.45 | 11,668.33 | 43,720.90 |
| OREGON | 9,724.64 | 10,278.46 | 10,437.33 | 11,096.05 | 41,536.48 |
| PENNSYLVANIA | 60,331.80 | 63,245.33 | 62,191.87 | 66,318.71 | 252,087.71 |
| PUERTO RICO | 2,492.26 | 2,570.74 | 2,219.65 | 2,673.68 | 9,956.33 |
| RHODE ISLAND | 7,078.72 | 7,701.37 | 7,489.74 | 7,827.47 | 30,097.30 |
| SOUTH CAROLINA | 9,844.56 | 10,500.67 | 10,795.35 | 11,498.19 | 42,638.77 |
| SOUTH DAKOTA | 410.6 | 465.16 | 449.56 | 472.47 | 1,797.79 |
| TENNESSEE | 33,071.70 | 35,153.80 | 34,738.83 | 36,321.42 | 139,285.75 |
| TEXAS | 25,480.77 | 26,211.78 | 26,795.05 | 26,848.72 | 105,336.32 |
| UTAH | 9,297.97 | 9,593.42 | 9,762.65 | 9,908.39 | 38,562.43 |
| VERMONT | 5,971.04 | 6,401.46 | 6,346.14 | 6,500.42 | 25,219.06 |
| VIRGIN ISLANDS | 19.68 | 17.54 | 12.98 | 12.51 | 62.71 |
| VIRGINIA | 20,212.32 | 20,113.19 | 19,190.64 | 19,942.50 | 79,458.65 |
| WASHINGTON | 17,982.44 | 18,967.40 | 19,553.76 | 20,495.88 | 76,999.48 |
| WEST VIRGINIA | 14,898.04 | 15,508.03 | 15,972.70 | 15,811.79 | 62,190.56 |
| WISCONSIN | 9,883.10 | 10,182.04 | 10,272.80 | 10,766.41 | 41,104.35 |
| WYOMING | 1,002.54 | 1,256.80 | 1,317.04 | 1,456.51 | 5,032.89 |
| TOTAL | 797,094.32 | 837,160.14 | 842,315.69 | 874,614.92 | 3,351,185.07 |

DRUG CODE:9120 DRUG NAME:DIHYDROCODEINE

| STATE | QUARTER 1 | QUARTER 2 | QUARTER 3 | QUARTER 4 | TOTAL GRAMS |
|---|---|---|---|---|---|
| ALABAMA | 783.7 | 798.47 | 785.72 | 781.48 | 3,149.37 |
| ALASKA | 8.55 | 3.2 | 0 | 2.14 | 13.89 |
| ARIZONA | 73.81 | 86.52 | 75.84 | 83.32 | 319.49 |
| ARKANSAS | 105.67 | 152.34 | 175.48 | 135.07 | 568.56 |
| CALIFORNIA | 214.7 | 252.3 | 73.83 | 302.29 | 843.12 |
| COLORADO | 0 | 2.14 | 5.34 | 2.14 | 9.62 |

| | | | | | |
|---|---|---|---|---|---|
| CONNECTICUT | 1.2 | 0.13 | 2.42 | 0 | 3.75 |
| DELAWARE | 13.14 | 13.25 | 12.43 | 15.7 | 54.52 |
| FLORIDA | 158.82 | 147.29 | 238.7 | 219.11 | 763.92 |
| GEORGIA | 428.39 | 549.78 | 509.62 | 710.23 | 2,198.02 |
| HAWAII | 1.07 | 0 | 0 | 0 | 1.07 |
| IDAHO | 23.5 | 40.59 | 32.04 | 12.82 | 108.95 |
| ILLINOIS | 126.9 | 214.49 | 192.06 | 137.66 | 671.11 |
| INDIANA | 3.2 | 0 | 3.2 | 1.07 | 7.47 |
| IOWA | 0 | 1.07 | 0 | 1.07 | 2.14 |
| KANSAS | 1.07 | 10.68 | 0 | 6.41 | 18.16 |
| KENTUCKY | 30.98 | 19.28 | 13.89 | 21.36 | 85.51 |
| LOUISIANA | 620.96 | 802.73 | 644.22 | 484.55 | 2,552.46 |
| MAINE | 0 | 0 | 1.07 | 0 | 1.07 |
| MARYLAND | 7.48 | 10.15 | 10.31 | 10.15 | 38.09 |
| MASSACHUSETTS | 21.36 | 16.02 | 33.11 | 55.54 | 126.03 |
| MICHIGAN | 34.23 | 0 | 5.34 | 1.28 | 40.85 |
| MINNESOTA | 1.71 | 4.27 | 1.71 | 2.14 | 9.83 |
| MISSISSIPPI | 367.55 | 491.51 | 380.32 | 366.59 | 1,605.97 |
| MISSOURI | 45.93 | 42.73 | 54.48 | 41.66 | 184.8 |
| MONTANA | 1.07 | 0 | 0 | 0 | 1.07 |
| NEBRASKA | 3.2 | 2.14 | 3.85 | 2.78 | 11.97 |
| NEVADA | 43.37 | 30.98 | 1.07 | 0 | 75.42 |
| NEW JERSEY | 355.87 | 369.38 | 237.79 | 285.71 | 1,248.75 |
| NEW MEXICO | 135.66 | 90.79 | 100.41 | 17.09 | 343.95 |
| NEW YORK | 108.95 | 167.7 | 116.43 | 136.19 | 529.27 |
| NORTH CAROLINA | 13.89 | 9.61 | 6.41 | 8.55 | 38.46 |
| OHIO | 48.01 | 33.81 | 37.92 | 20.56 | 140.3 |
| OKLAHOMA | 272.81 | 298.86 | 295.29 | 262.41 | 1,129.37 |
| PENNSYLVANIA | 205.58 | 42.12 | 64.53 | 50.33 | 362.56 |
| RHODE ISLAND | 0 | 1.07 | 0 | 2.14 | 3.21 |
| SOUTH CAROLINA | 13.89 | 13.89 | 10.68 | 13.89 | 52.35 |
| SOUTH DAKOTA | 0 | 0 | 0 | 1.07 | 1.07 |
| TENNESSEE | 507.39 | 516.62 | 508.84 | 404.19 | 1,937.04 |
| TEXAS | 2,687.78 | 2,808.14 | 2,679.34 | 2,853.92 | 11,029.18 |
| UTAH | 0 | 0 | 1.07 | 0 | 1.07 |
| VERMONT | 0 | 1.07 | 0 | 0 | 1.07 |
| VIRGINIA | 10.68 | 4.27 | 3.2 | 8.55 | 26.7 |

| | | | | | |
|---|---|---|---|---|---|
| WEST VIRGINIA | 5.34 | 2.35 | 0 | 0 | 7.69 |
| WISCONSIN | 4.27 | 7.48 | 6.41 | 3.2 | 21.36 |
| TOTAL | 7,491.68 | 8,059.22 | 7,324.37 | 7,464.36 | 30,339.63 |

DRUG CODE:9143 DRUG NAME:OXYCODONE

| STATE | QUARTER 1 | QUARTER 2 | QUARTER 3 | QUARTER 4 | TOTAL GRAMS |
|---|---|---|---|---|---|
| ALABAMA | 245,034.14 | 234,086.14 | 222,837.98 | 221,282.88 | 923,241.14 |
| ALASKA | 35,909.96 | 35,106.79 | 33,743.86 | 33,035.01 | 137,795.62 |
| AMERICAN SAMOA | 14.34 | 12.1 | 10.76 | 21.52 | 58.72 |
| ARIZONA | 459,311.04 | 432,919.49 | 408,276.05 | 399,895.18 | 1,700,401.76 |
| ARKANSAS | 150,503.65 | 147,187.43 | 142,600.16 | 137,696.38 | 577,987.62 |
| CALIFORNIA | 905,318.77 | 878,899.08 | 848,801.57 | 819,812.84 | 3,452,832.26 |
| COLORADO | 233,553.88 | 228,253.27 | 217,544.66 | 213,255.23 | 892,607.04 |
| CONNECTICUT | 175,414.04 | 170,580.13 | 160,443.37 | 157,023.12 | 663,460.66 |
| DELAWARE | 72,059.47 | 68,623.07 | 66,755.23 | 66,369.80 | 273,807.57 |
| DISTRICT OF COLUMBIA | 15,113.21 | 15,042.95 | 14,559.95 | 13,439.86 | 58,155.97 |
| FLORIDA | 1,040,636.86 | 1,029,362.72 | 996,363.76 | 1,037,706.14 | 4,104,069.48 |
| GEORGIA | 455,523.47 | 451,529.04 | 439,524.11 | 436,048.64 | 1,782,625.26 |
| GUAM | 697.48 | 908.57 | 980.23 | 835.09 | 3,421.37 |
| HAWAII | 45,607.42 | 44,026.24 | 43,218.01 | 40,579.06 | 173,430.73 |
| IDAHO | 61,459.28 | 60,628.06 | 60,258.22 | 59,007.70 | 241,353.26 |
| ILLINOIS | 165,648.45 | 160,662.93 | 154,175.18 | 152,126.92 | 632,613.48 |
| INDIANA | 241,577.26 | 236,466.08 | 224,779.73 | 221,485.13 | 924,308.20 |
| IOWA | 56,046.72 | 54,232.73 | 51,301.26 | 52,242.96 | 213,823.67 |
| KANSAS | 124,474.40 | 123,720.45 | 121,222.15 | 119,080.42 | 488,497.42 |
| KENTUCKY | 210,690.77 | 206,737.28 | 199,383.42 | 198,870.30 | 815,681.77 |
| LOUISIANA | 203,678.58 | 196,046.31 | 187,743.37 | 182,770.76 | 770,239.02 |
| MAINE | 54,161.13 | 51,039.90 | 50,058.51 | 49,048.14 | 204,307.68 |
| MARYLAND | 343,393.91 | 332,414.30 | 312,941.46 | 306,627.83 | 1,295,377.50 |
| MASSACHUSETTS | 253,125.08 | 244,621.64 | 233,176.98 | 228,413.72 | 959,337.42 |
| MICHIGAN | 314,155.93 | 303,147.88 | 292,409.66 | 279,116.29 | 1,188,829.76 |
| MINNESOTA | 149,851.01 | 144,230.88 | 139,462.99 | 137,088.75 | 570,633.63 |
| MISSISSIPPI | 109,326.39 | 106,944.69 | 102,515.30 | 100,468.18 | 419,254.56 |
| MISSOURI | 309,827.79 | 305,484.02 | 288,966.90 | 280,390.36 | 1,184,669.07 |
| MONTANA | 41,009.46 | 38,376.24 | 37,194.59 | 35,770.94 | 152,351.23 |
| NEBRASKA | 42,876.54 | 42,209.02 | 40,908.67 | 40,254.56 | 166,248.79 |

| | | | | | |
|---|---|---|---|---|---|
| NEVADA | 186,303.46 | 181,957.10 | 175,545.89 | 173,484.57 | 717,291.02 |
| NEW HAMPSHIRE | 65,642.96 | 62,820.01 | 62,066.27 | 58,851.59 | 249,380.83 |
| NEW JERSEY | 539,098.15 | 511,306.71 | 480,020.01 | 460,887.38 | 1,991,312.25 |
| NEW MEXICO | 98,671.82 | 92,617.85 | 86,384.43 | 83,715.95 | 361,390.05 |
| NEW YORK | 786,761.65 | 754,995.06 | 718,432.45 | 691,526.96 | 2,951,716.12 |
| NORTH CAROLINA | 598,268.67 | 570,304.32 | 540,337.25 | 530,163.25 | 2,239,073.49 |
| NORTH DAKOTA | 15,163.44 | 14,961.70 | 14,285.01 | 14,179.56 | 58,589.71 |
| OHIO | 515,301.72 | 494,056.29 | 469,035.01 | 448,137.18 | 1,926,530.20 |
| OKLAHOMA | 231,863.07 | 228,171.70 | 220,954.32 | 215,753.47 | 896,742.56 |
| OREGON | 183,165.09 | 176,788.51 | 169,523.38 | 163,860.60 | 693,337.58 |
| PENNSYLVANIA | 786,642.17 | 763,586.57 | 733,656.61 | 718,182.30 | 3,002,067.65 |
| PUERTO RICO | 19,771.46 | 14,068.91 | 9,392.23 | 7,493.92 | 50,726.52 |
| RHODE ISLAND | 43,650.69 | 40,705.67 | 39,605.42 | 38,560.31 | 162,522.09 |
| SOUTH CAROLINA | 246,284.54 | 239,531.29 | 225,630.18 | 223,391.32 | 934,837.33 |
| SOUTH DAKOTA | 18,010.05 | 17,526.31 | 17,139.74 | 16,269.27 | 68,945.37 |
| TENNESSEE | 474,225.91 | 466,608.14 | 447,638.60 | 462,670.28 | 1,851,142.93 |
| TEXAS | 336,927.85 | 336,280.34 | 331,571.65 | 319,536.42 | 1,324,316.26 |
| UTAH | 174,300.46 | 170,138.74 | 164,823.91 | 161,724.13 | 670,987.24 |
| VERMONT | 23,850.89 | 23,391.11 | 22,159.10 | 21,342.96 | 90,744.06 |
| VIRGIN ISLANDS | 584.77 | 578.06 | 432.34 | 506.93 | 2,102.10 |
| VIRGINIA | 321,463.45 | 300,363.76 | 284,391.42 | 279,939.38 | 1,186,158.01 |
| WASHINGTON | 305,726.74 | 297,580.02 | 285,588.93 | 273,319.11 | 1,162,214.80 |
| WEST VIRGINIA | 88,959.43 | 84,822.16 | 81,205.99 | 77,920.68 | 332,908.26 |
| WISCONSIN | 235,687.35 | 216,552.16 | 206,158.06 | 199,322.36 | 857,719.93 |
| WYOMING | 26,786 | 25,404.48 | 25,006.32 | 24,972.03 | 102,168.83 |
| TOTAL | 12,839,112.22 | 12,428,616.40 | 11,903,142.61 | 11,685,475.62 | 48,856,346.85 |

DRUG CODE:9150 DRUG NAME:HYDROMORPHONE

| STATE | QUARTER 1 | QUARTER 2 | QUARTER 3 | QUARTER 4 | TOTAL GRAMS |
|---|---|---|---|---|---|
| ALABAMA | 3,528.78 | 3,174.30 | 3,221.27 | 3,112.98 | 13,037.33 |
| ALASKA | 1,065.25 | 976.89 | 906.53 | 948.56 | 3,897.23 |
| AMERICAN SAMOA | 0.89 | 0.09 | 0 | 0 | 0.98 |
| ARIZONA | 7,089.09 | 6,897.62 | 6,595.39 | 6,229.48 | 26,811.58 |
| ARKANSAS | 3,376.84 | 3,287.22 | 3,220.53 | 3,170.03 | 13,054.62 |
| CALIFORNIA | 40,477.70 | 39,869.34 | 38,125 | 36,820.78 | 155,292.82 |
| COLORADO | 7,370.82 | 6,737.63 | 6,506.99 | 6,583.16 | 27,198.60 |

| | | | | | |
|---|---|---|---|---|---|
| CONNECTICUT | 6,671.40 | 6,448.66 | 6,205.18 | 6,250.52 | 25,575.76 |
| DELAWARE | 2,294.82 | 2,079.53 | 2,039.22 | 2,109.63 | 8,523.20 |
| DISTRICT OF COLUMBIA | 596.68 | 539.79 | 529.24 | 501.62 | 2,167.33 |
| FLORIDA | 50,158.18 | 46,655.17 | 44,394.53 | 45,519.06 | 186,726.94 |
| GEORGIA | 8,772.96 | 8,321.02 | 8,410.75 | 7,793.73 | 33,298.46 |
| GUAM | 31.75 | 26.32 | 23.41 | 33.03 | 114.51 |
| HAWAII | 692.64 | 706.22 | 642.87 | 629.49 | 2,671.22 |
| IDAHO | 1,942.65 | 1,814.90 | 1,839.38 | 1,928.86 | 7,525.79 |
| ILLINOIS | 10,292.76 | 9,835.82 | 9,410.79 | 9,365.83 | 38,905.20 |
| INDIANA | 6,061.70 | 6,085.13 | 5,919.92 | 5,762.15 | 23,828.90 |
| IOWA | 3,176.90 | 2,986.13 | 2,873.08 | 2,856.05 | 11,892.16 |
| KANSAS | 3,575.53 | 3,711.19 | 3,562.32 | 3,128.83 | 13,977.87 |
| KENTUCKY | 2,970.10 | 2,647.44 | 2,821.70 | 3,354.10 | 11,793.34 |
| LOUISIANA | 3,714.46 | 3,255.18 | 3,095.98 | 3,107.57 | 13,173.19 |
| MAINE | 1,611.81 | 2,004.63 | 1,322.33 | 1,747.62 | 6,686.39 |
| MARYLAND | 8,933.97 | 8,489.56 | 8,073.63 | 7,917.26 | 33,414.42 |
| MASSACHUSETTS | 7,792.78 | 7,524.92 | 6,867.12 | 7,289.33 | 29,474.15 |
| MICHIGAN | 9,582.29 | 8,840.05 | 8,938.15 | 8,791.55 | 36,152.04 |
| MINNESOTA | 5,937.44 | 6,034.87 | 5,959.61 | 5,859.45 | 23,791.37 |
| MISSISSIPPI | 5,027.87 | 6,221.14 | 5,736.32 | 6,418.37 | 23,403.70 |
| MISSOURI | 7,000.39 | 6,365.07 | 6,187.46 | 6,249.20 | 25,802.12 |
| MONTANA | 1,681.14 | 1,613.19 | 1,733.22 | 1,536.91 | 6,564.46 |
| NEBRASKA | 1,377.27 | 1,375.99 | 1,223.01 | 1,230.50 | 5,206.77 |
| NEVADA | 3,173.81 | 3,007.67 | 2,832.63 | 2,782.37 | 11,796.48 |
| NEW HAMPSHIRE | 2,059.38 | 1,997.46 | 1,977.49 | 1,829.39 | 7,863.72 |
| NEW JERSEY | 9,575.10 | 9,039.88 | 8,435.17 | 8,447.42 | 35,497.57 |
| NEW MEXICO | 1,465.35 | 1,417.73 | 1,270.28 | 1,242.25 | 5,395.61 |
| NEW YORK | 20,268.56 | 19,768.22 | 19,110.20 | 18,914.43 | 78,061.41 |
| NORTH CAROLINA | 11,655.08 | 10,764.60 | 10,653.70 | 9,964.11 | 43,037.49 |
| NORTH DAKOTA | 644.43 | 678.33 | 630.46 | 642.14 | 2,595.36 |
| OHIO | 9,597.27 | 9,300.50 | 9,237.22 | 9,247.73 | 37,382.72 |
| OKLAHOMA | 3,666.11 | 3,777.89 | 3,636.19 | 3,519.73 | 14,599.92 |
| OREGON | 5,056.14 | 4,949.63 | 4,841.48 | 4,498.62 | 19,345.87 |
| PENNSYLVANIA | 17,756.18 | 14,688.28 | 13,930.26 | 14,126.71 | 60,501.43 |
| PUERTO RICO | 75.7 | 65.08 | 80.93 | 74.93 | 296.64 |
| RHODE ISLAND | 886.87 | 1,034.75 | 811.1 | 800.25 | 3,532.97 |
| SOUTH CAROLINA | 6,002.10 | 5,829.97 | 5,510 | 5,634.37 | 22,976.44 |

| | | | | | |
|---|---|---|---|---|---|
| SOUTH DAKOTA | 519.68 | 525.6 | 512.84 | 641.6 | 2,199.72 |
| TENNESSEE | 8,202.61 | 6,182.19 | 6,449.40 | 5,822.20 | 26,656.40 |
| TEXAS | 24,745.91 | 24,373.73 | 23,788.71 | 22,501.46 | 95,409.81 |
| UTAH | 2,558.55 | 2,611.03 | 2,782.81 | 2,953.08 | 10,905.47 |
| VERMONT | 1,238.83 | 1,269.84 | 1,196.76 | 1,167.38 | 4,872.81 |
| VIRGIN ISLANDS | 26.6 | 18.71 | 13.94 | 14.9 | 74.15 |
| VIRGINIA | 12,556.24 | 11,386.29 | 10,518.79 | 10,482.93 | 44,944.25 |
| WASHINGTON | 11,178.55 | 11,112.49 | 10,663.43 | 9,872.34 | 42,826.81 |
| WEST VIRGINIA | 1,685.85 | 1,550.98 | 1,453.60 | 1,363.74 | 6,054.17 |
| WISCONSIN | 3,854.46 | 3,511.27 | 3,699.29 | 3,797.14 | 14,862.16 |
| WYOMING | 652.17 | 642.44 | 604.6 | 540.15 | 2,439.36 |
| TOTAL | 371,908.39 | 354,029.57 | 341,026.21 | 337,127.02 | 1,404,091.19 |

DRUG CODE:9193 DRUG NAME:HYDROCODONE

| STATE | QUARTER 1 | QUARTER 2 | QUARTER 3 | QUARTER 4 | TOTAL GRAMS |
|---|---|---|---|---|---|
| ALABAMA | 258,345.10 | 246,512.38 | 233,339.55 | 233,698.90 | 971,895.93 |
| ALASKA | 11,947.24 | 11,492.75 | 10,775.27 | 10,314.83 | 44,530.09 |
| AMERICAN SAMOA | 4 | 0.27 | 3.63 | 0 | 7.9 |
| ARIZONA | 112,100.29 | 104,522.05 | 98,362.40 | 97,818.88 | 412,803.62 |
| ARKANSAS | 140,205.70 | 136,469.71 | 129,933.06 | 129,068.61 | 535,677.08 |
| CALIFORNIA | 952,326.68 | 931,986.40 | 894,017.09 | 878,029.49 | 3,656,359.66 |
| COLORADO | 68,829.55 | 65,572.12 | 63,316.81 | 62,200.10 | 259,918.58 |
| CONNECTICUT | 21,699.36 | 20,448.70 | 19,209.66 | 19,001.68 | 80,359.40 |
| DELAWARE | 6,389.70 | 5,916.80 | 5,826.12 | 5,827.43 | 23,960.05 |
| DISTRICT OF COLUMBIA | 1,571.73 | 1,460.61 | 1,444.90 | 1,289.19 | 5,766.43 |
| FLORIDA | 384,180.30 | 363,407.66 | 338,761.82 | 353,668.78 | 1,440,018.56 |
| GEORGIA | 259,038.74 | 250,146 | 237,567.07 | 237,305.97 | 984,057.78 |
| GUAM | 148.02 | 196.51 | 149.38 | 136.06 | 629.97 |
| HAWAII | 18,613.32 | 17,730.01 | 17,098.98 | 16,443.22 | 69,885.53 |
| IDAHO | 58,546.42 | 57,869.44 | 55,812.53 | 55,830.66 | 228,059.05 |
| ILLINOIS | 320,046.07 | 312,772.26 | 300,104.23 | 298,271.74 | 1,231,194.30 |
| INDIANA | 236,905.30 | 232,762.71 | 219,377.34 | 215,862.49 | 904,907.84 |
| IOWA | 59,394.52 | 58,576.56 | 56,201.60 | 55,504.63 | 229,677.31 |
| KANSAS | 84,159.41 | 82,242.50 | 79,387.08 | 78,882.16 | 324,671.15 |
| KENTUCKY | 208,448.35 | 203,596.96 | 194,363.59 | 193,159.90 | 799,568.80 |
| LOUISIANA | 173,235.24 | 166,066.34 | 157,962.30 | 154,816.27 | 652,080.15 |

| | | | | | |
|---|---|---|---|---|---|
| MAINE | 17,776.45 | 17,296.48 | 16,839.01 | 16,547.86 | 68,459.80 |
| MARYLAND | 37,569.09 | 35,131.47 | 33,032.25 | 32,735.19 | 138,468 |
| MASSACHUSETTS | 31,806.48 | 30,798.70 | 29,037.25 | 29,028.79 | 120,671.22 |
| MICHIGAN | 418,265.12 | 410,822.78 | 391,555.73 | 374,540.57 | 1,595,184.20 |
| MINNESOTA | 48,330.57 | 46,169.58 | 44,817.69 | 43,263.92 | 182,581.76 |
| MISSISSIPPI | 126,336.22 | 121,180.34 | 112,361.19 | 106,669.46 | 466,547.21 |
| MISSOURI | 184,389.53 | 182,181.22 | 172,468.98 | 168,909.19 | 707,948.92 |
| MONTANA | 26,789.61 | 26,089.84 | 25,288.98 | 24,710.06 | 102,878.49 |
| NEBRASKA | 33,986.82 | 32,483.56 | 31,872.68 | 31,628.37 | 129,971.43 |
| NEVADA | 98,912.23 | 96,350.94 | 91,032.69 | 91,209.30 | 377,505.16 |
| NEW HAMPSHIRE | 9,491.69 | 9,032.94 | 8,752.47 | 8,846.23 | 36,123.33 |
| NEW JERSEY | 38,281.54 | 34,570.40 | 32,758.32 | 32,517.77 | 138,128.03 |
| NEW MEXICO | 36,907.72 | 35,625.33 | 33,655.64 | 33,354.99 | 139,543.68 |
| NEW YORK | 180,175.57 | 174,189.76 | 164,949 | 161,835.48 | 681,149.81 |
| NORTH CAROLINA | 212,067.30 | 195,483.54 | 180,862.60 | 180,907.74 | 769,321.18 |
| NORTH DAKOTA | 10,955.98 | 10,561.01 | 10,048.93 | 9,927.41 | 41,493.33 |
| OHIO | 199,125.37 | 189,484.03 | 180,291.94 | 173,999.46 | 742,900.80 |
| OKLAHOMA | 187,458.77 | 185,595.49 | 177,264.58 | 175,532.11 | 725,850.95 |
| OREGON | 99,225.54 | 95,247.81 | 92,086.04 | 90,141.72 | 376,701.11 |
| PENNSYLVANIA | 176,205.06 | 167,098.93 | 164,049.26 | 152,735.32 | 660,088.57 |
| PUERTO RICO | 1,097.43 | 660.78 | 400.12 | 388.37 | 2,546.70 |
| RHODE ISLAND | 11,199.72 | 10,503.30 | 10,008.87 | 9,775.76 | 41,487.65 |
| SOUTH CAROLINA | 147,482.52 | 140,547.45 | 130,855.75 | 131,634.30 | 550,520.02 |
| SOUTH DAKOTA | 15,800.85 | 14,917.59 | 14,418.82 | 14,069.75 | 59,207.01 |
| TENNESSEE | 255,932.40 | 247,128.17 | 239,087.40 | 230,700.09 | 972,848.06 |
| TEXAS | 688,696.66 | 675,195.44 | 651,021.39 | 643,395.78 | 2,658,309.27 |
| UTAH | 61,386.94 | 59,307.58 | 56,889.89 | 56,293.13 | 233,877.54 |
| VERMONT | 5,134.85 | 4,910.12 | 4,565.24 | 4,527.41 | 19,137.62 |
| VIRGIN ISLANDS | 363.62 | 340.97 | 223.69 | 230.32 | 1,158.60 |
| VIRGINIA | 116,394.84 | 105,431.76 | 97,538.61 | 96,974.23 | 416,339.44 |
| WASHINGTON | 137,465.59 | 133,461.20 | 128,219.88 | 124,321.75 | 523,468.42 |
| WEST VIRGINIA | 62,746.35 | 60,335.58 | 57,352.51 | 55,167.17 | 235,601.61 |
| WISCONSIN | 82,538.61 | 77,977.55 | 75,530.83 | 75,485.88 | 311,532.87 |
| WYOMING | 12,184.40 | 11,693.32 | 11,112.16 | 11,019.53 | 46,009.41 |
| TOTAL | 7,148,616.48 | 6,907,553.70 | 6,583,264.80 | 6,490,155.40 | 27,129,590.38 |

RETAIL DRUG DISTRIBUTION BY STATE WITHIN DRUG CODE BY GRAMS WT

DRUG CODE:9220L DRUG NAME:LEVORPHANOL

| STATE | QUARTER 1 | QUARTER 2 | QUARTER 3 | QUARTER 4 | TOTAL GRAMS |
|---|---|---|---|---|---|
| ALABAMA | 5.92 | 4.29 | 5.34 | 4.53 | 20.08 |
| ARIZONA | 18.46 | 22.06 | 29.37 | 30.06 | 99.95 |
| ARKANSAS | 2.79 | 3.37 | 1.97 | 0.93 | 9.06 |
| CALIFORNIA | 109.81 | 102.15 | 98.9 | 91.7 | 402.56 |
| COLORADO | 6.04 | 12.88 | 13.81 | 18.11 | 50.84 |
| CONNECTICUT | 1.63 | 3.71 | 5.11 | 9.75 | 20.2 |
| DELAWARE | 0.93 | 0 | 0.58 | 1.04 | 2.55 |
| FLORIDA | 26.23 | 35.87 | 34.36 | 39.58 | 136.04 |
| GEORGIA | 0.7 | 2.55 | 2.9 | 3.48 | 9.63 |
| IDAHO | 0 | 0 | 0.12 | 0.12 | 0.24 |
| ILLINOIS | 8.71 | 13.12 | 14.86 | 16.02 | 52.71 |
| INDIANA | 25.89 | 24.49 | 24.84 | 22.17 | 97.39 |
| IOWA | 4.41 | 3.95 | 3.37 | 3.71 | 15.44 |
| KANSAS | 11.26 | 9.98 | 7.2 | 7.43 | 35.87 |
| KENTUCKY | 0 | 0 | 0.23 | 0.12 | 0.35 |
| LOUISIANA | 16.48 | 19.5 | 20.78 | 19.5 | 76.26 |
| MARYLAND | 3.13 | 6.04 | 3.83 | 8.36 | 21.36 |
| MASSACHUSETTS | 20.78 | 19.62 | 20.2 | 20.08 | 80.68 |
| MICHIGAN | 0.35 | 0.35 | 0.35 | 0 | 1.05 |
| MINNESOTA | 0.35 | 0.46 | 0.81 | 1.16 | 2.78 |
| MISSISSIPPI | 0.23 | 0 | 0 | 0 | 0.23 |
| MISSOURI | 13.81 | 19.62 | 26.23 | 27.28 | 86.94 |
| NEBRASKA | 4.06 | 5.11 | 5.46 | 9.17 | 23.8 |
| NEVADA | 2.9 | 2.9 | 4.88 | 3.48 | 14.16 |
| NEW HAMPSHIRE | 1.16 | 1.86 | 1.63 | 0.7 | 5.35 |
| NEW JERSEY | 313.88 | 400.82 | 406.98 | 498.56 | 1,620.24 |
| NEW YORK | 33.2 | 38.65 | 40.98 | 48.29 | 161.12 |
| NORTH CAROLINA | 50.84 | 48.52 | 64.66 | 101.69 | 265.71 |
| OHIO | 8.01 | 14.63 | 11.72 | 9.17 | 43.53 |
| OREGON | 3.48 | 3.48 | 2.9 | 3.71 | 13.57 |
| PENNSYLVANIA | 10.56 | 15.21 | 14.97 | 12.88 | 53.62 |
| SOUTH CAROLINA | 0.81 | 3.37 | 3.25 | 3.6 | 11.03 |
| TENNESSEE | 0.12 | 0.23 | 0.35 | 0 | 0.7 |
| TEXAS | 39.24 | 53.63 | 61.41 | 66.05 | 220.33 |
| VERMONT | 0.23 | 0.46 | 0.46 | 0.46 | 1.61 |

| | | | | | |
|---|---|---|---|---|---|
| VIRGINIA | 5.8 | 5.8 | 11.14 | 16.25 | 38.99 |
| WASHINGTON | 22.64 | 24.49 | 30.88 | 31.23 | 109.24 |
| WISCONSIN | 1.74 | 3.83 | 4.99 | 4.99 | 15.55 |
| TOTAL | 776.58 | 927 | 981.82 | 1,135.36 | 3,820.76 |

DRUG CODE:9230 DRUG NAME:MEPERIDINE (PETHIDINE)

| STATE | QUARTER 1 | QUARTER 2 | QUARTER 3 | QUARTER 4 | TOTAL GRAMS |
|---|---|---|---|---|---|
| ALABAMA | 8,009.96 | 7,392.30 | 6,862.91 | 7,150.03 | 29,415.20 |
| ALASKA | 850.15 | 704.17 | 662.45 | 634.23 | 2,851 |
| ARIZONA | 5,450.80 | 4,921.23 | 4,373.73 | 4,759.85 | 19,505.61 |
| ARKANSAS | 8,349.08 | 7,839.27 | 7,085.38 | 6,325.67 | 29,599.40 |
| CALIFORNIA | 15,974.75 | 15,874.20 | 15,148.89 | 15,374.57 | 62,372.41 |
| COLORADO | 2,171.34 | 1,890.98 | 1,729.71 | 1,487 | 7,279.03 |
| CONNECTICUT | 1,104.08 | 1,052.23 | 1,036.43 | 1,069.77 | 4,262.51 |
| DELAWARE | 248.38 | 274.52 | 215.81 | 132.03 | 870.74 |
| DISTRICT OF COLUMBIA | 217.66 | 220.05 | 168.96 | 147.94 | 754.61 |
| FLORIDA | 9,951.88 | 9,511.01 | 7,808.42 | 8,543.21 | 35,814.52 |
| GEORGIA | 11,498.61 | 10,443.58 | 9,714.92 | 9,934.14 | 41,591.25 |
| GUAM | 30.61 | 34.64 | 66.23 | 52.18 | 183.66 |
| HAWAII | 382.26 | 394.35 | 305.9 | 274.74 | 1,357.25 |
| IDAHO | 848.06 | 925.23 | 740.75 | 815.99 | 3,330.03 |
| ILLINOIS | 2,990.92 | 3,274.25 | 3,201.46 | 2,544.13 | 12,010.76 |
| INDIANA | 3,589.75 | 3,524.96 | 3,260.70 | 3,050.25 | 13,425.66 |
| IOWA | 1,927.32 | 1,893.97 | 1,991.20 | 1,968.39 | 7,780.88 |
| KANSAS | 1,860.67 | 1,792.85 | 1,712.24 | 1,705.29 | 7,071.05 |
| KENTUCKY | 2,880.50 | 2,299.95 | 2,469.50 | 2,127.48 | 9,777.43 |
| LOUISIANA | 8,110.88 | 7,570.39 | 6,714.82 | 6,623.16 | 29,019.25 |
| MAINE | 465.38 | 409.39 | 359.06 | 344.35 | 1,578.18 |
| MARYLAND | 2,750.67 | 2,474.41 | 2,115.35 | 2,029.61 | 9,370.04 |
| MASSACHUSETTS | 2,268.84 | 2,250.69 | 2,169.27 | 1,766.20 | 8,455 |
| MICHIGAN | 3,056.18 | 2,906.26 | 2,665.16 | 2,300.80 | 10,928.40 |
| MINNESOTA | 835.66 | 651.05 | 651.97 | 595.21 | 2,733.89 |
| MISSISSIPPI | 4,670.48 | 4,620.17 | 3,990.95 | 4,032.95 | 17,314.55 |
| MISSOURI | 4,628.10 | 3,912.93 | 3,526.29 | 3,143.17 | 15,210.49 |
| MONTANA | 656.94 | 774.63 | 657.11 | 597.63 | 2,686.31 |
| NEBRASKA | 802.06 | 773.59 | 669.86 | 539.46 | 2,784.97 |

| | | | | | |
|---|---|---|---|---|---|
| NEVADA | 1,632.86 | 1,238.60 | 1,252.61 | 1,546.80 | 5,670.87 |
| NEW HAMPSHIRE | 676.18 | 610.92 | 612.56 | 450.24 | 2,349.90 |
| NEW JERSEY | 2,186.59 | 2,180.38 | 1,832.66 | 1,503.12 | 7,702.75 |
| NEW MEXICO | 1,680.80 | 1,416.56 | 1,314.88 | 1,238.29 | 5,650.53 |
| NEW YORK | 5,123.33 | 4,663 | 4,301.85 | 4,201.02 | 18,289.20 |
| NORTH CAROLINA | 7,190.99 | 6,462.50 | 5,431.34 | 5,457.05 | 24,541.88 |
| NORTH DAKOTA | 237.48 | 237.59 | 253.72 | 223 | 951.79 |
| OHIO | 5,261.90 | 5,091.15 | 5,416.29 | 4,211.58 | 19,980.92 |
| OKLAHOMA | 7,655.34 | 7,385.05 | 6,807.87 | 6,807.09 | 28,655.35 |
| OREGON | 1,749.54 | 1,464.32 | 1,585.80 | 1,315.75 | 6,115.41 |
| PENNSYLVANIA | 3,944.02 | 3,517.70 | 3,425.32 | 3,312.14 | 14,199.18 |
| PUERTO RICO | 5,652.98 | 4,931.71 | 4,118.71 | 2,668.42 | 17,371.82 |
| RHODE ISLAND | 252.95 | 264.61 | 256.77 | 232.47 | 1,006.80 |
| SOUTH CAROLINA | 6,780.49 | 6,119.13 | 5,524.77 | 5,092.83 | 23,517.22 |
| SOUTH DAKOTA | 550.9 | 558.74 | 529.76 | 436.84 | 2,076.24 |
| TENNESSEE | 8,750.58 | 7,848.45 | 6,741.23 | 6,351.88 | 29,692.14 |
| TEXAS | 11,780.05 | 10,811.35 | 11,790.01 | 11,948.92 | 46,330.33 |
| UTAH | 2,010.75 | 1,561.27 | 1,788.51 | 1,835.03 | 7,195.56 |
| VERMONT | 323.87 | 315.05 | 265.59 | 383.35 | 1,287.86 |
| VIRGIN ISLANDS | 41.94 | 67.65 | 41.94 | 102.4 | 253.93 |
| VIRGINIA | 6,979.19 | 5,932.13 | 5,459.56 | 4,908.18 | 23,279.06 |
| WASHINGTON | 3,773.38 | 3,371.16 | 3,264.97 | 3,133.91 | 13,543.42 |
| WEST VIRGINIA | 731.62 | 662.45 | 586.19 | 510.15 | 2,490.41 |
| WISCONSIN | 1,558.57 | 1,497.45 | 1,466.63 | 1,330.34 | 5,852.99 |
| WYOMING | 378.34 | 318.27 | 254.26 | 350.78 | 1,301.65 |
| TOTAL | 193,486.61 | 179,134.44 | 166,399.23 | 159,621.01 | 698,641.29 |

DRUG CODE:9250B DRUG NAME:METHADONE

| STATE | QUARTER 1 | QUARTER 2 | QUARTER 3 | QUARTER 4 | TOTAL GRAMS |
|---|---|---|---|---|---|
| ALABAMA | 76,280.07 | 80,726.33 | 74,925.79 | 80,489.93 | 312,422.12 |
| ALASKA | 7,809.90 | 8,435.09 | 8,401.37 | 7,440.60 | 32,086.96 |
| ARIZONA | 85,190.46 | 85,087.72 | 82,438.41 | 83,016.82 | 335,733.41 |
| ARKANSAS | 16,878.18 | 16,608.11 | 15,568.82 | 15,186.55 | 64,241.66 |
| CALIFORNIA | 352,446.36 | 355,994.06 | 342,804.92 | 337,539.34 | 1,388,784.68 |
| COLORADO | 42,100.58 | 43,293.93 | 44,969.64 | 42,654.39 | 173,018.54 |
| CONNECTICUT | 119,258.98 | 125,911.35 | 105,169.68 | 126,064.74 | 476,404.75 |

| | | | | | |
|---|---|---|---|---|---|
| DELAWARE | 25,260.03 | 30,686.69 | 26,401.26 | 27,645.37 | 109,993.35 |
| DISTRICT OF COLUMBIA | 9,214.07 | 10,681.73 | 9,229.94 | 10,783.66 | 39,909.40 |
| FLORIDA | 171,777.30 | 180,889.29 | 224,484.51 | 205,561.84 | 782,712.94 |
| GEORGIA | 110,968.35 | 121,491.67 | 116,393.86 | 116,744.99 | 465,598.87 |
| GUAM | 58.14 | 38.46 | 42.48 | 32.2 | 171.28 |
| HAWAII | 7,859.45 | 8,981.21 | 7,611.43 | 7,843.62 | 32,295.71 |
| IDAHO | 12,059.82 | 12,052.13 | 11,740.16 | 11,380.70 | 47,232.81 |
| ILLINOIS | 121,222.97 | 126,796.08 | 119,325.66 | 122,079.43 | 489,424.14 |
| INDIANA | 84,564.33 | 83,049.21 | 79,994.79 | 79,326.75 | 326,935.08 |
| IOWA | 12,299.61 | 13,491.22 | 12,656.48 | 13,823.85 | 52,271.16 |
| KANSAS | 22,687.71 | 24,554.23 | 22,465.09 | 22,738.60 | 92,445.63 |
| KENTUCKY | 51,108.47 | 49,601.65 | 53,996.61 | 51,414.32 | 206,121.05 |
| LOUISIANA | 35,832.62 | 37,380.73 | 37,236.43 | 32,186.93 | 142,636.71 |
| MAINE | 32,346.87 | 32,802.39 | 29,492.66 | 31,583.41 | 126,225.33 |
| MARYLAND | 192,770.82 | 192,957.93 | 187,437.82 | 191,107.27 | 764,273.84 |
| MASSACHUSETTS | 147,168.01 | 145,889.95 | 141,669.46 | 144,647.23 | 579,374.65 |
| MICHIGAN | 134,102.02 | 134,815.42 | 133,911.84 | 131,182.87 | 534,012.15 |
| MINNESOTA | 44,551.68 | 52,651.48 | 51,108.51 | 59,499.49 | 207,811.16 |
| MISSISSIPPI | 6,085.86 | 6,062.42 | 7,306.89 | 7,188.47 | 26,643.64 |
| MISSOURI | 43,867.07 | 37,494.03 | 41,804.11 | 42,815.91 | 165,981.12 |
| MONTANA | 7,680.39 | 7,484.97 | 6,497.28 | 7,558.31 | 29,220.95 |
| NEBRASKA | 8,800.04 | 7,958.82 | 8,020.94 | 7,944.89 | 32,724.69 |
| NEVADA | 36,977.79 | 37,947.87 | 35,029.57 | 34,740.78 | 144,696.01 |
| NEW HAMPSHIRE | 30,916.55 | 31,234.33 | 30,579.98 | 30,602.79 | 123,333.65 |
| NEW JERSEY | 112,688.59 | 121,219.70 | 108,428.50 | 117,224.25 | 459,561.04 |
| NEW MEXICO | 43,022.34 | 42,657.69 | 44,251.96 | 46,464.44 | 176,396.43 |
| NEW YORK | 291,234.12 | 316,020.36 | 303,872.37 | 308,263.87 | 1,219,390.72 |
| NORTH CAROLINA | 129,434.71 | 134,621.14 | 132,576.89 | 129,863.61 | 526,496.35 |
| NORTH DAKOTA | 1,590.15 | 1,871.35 | 2,192.89 | 2,717.99 | 8,372.38 |
| OHIO | 78,169.68 | 74,777.31 | 72,839.25 | 71,527 | 297,313.24 |
| OKLAHOMA | 38,152.69 | 43,214.64 | 36,620.05 | 39,018.11 | 157,005.49 |
| OREGON | 63,733.33 | 64,825.05 | 63,346.93 | 61,850.48 | 253,755.79 |
| PENNSYLVANIA | 194,101.72 | 199,001.41 | 192,976.48 | 203,227.96 | 789,307.57 |
| PUERTO RICO | 22,390.32 | 18,312.21 | 24,067.23 | 19,258.85 | 84,028.61 |
| RHODE ISLAND | 40,681.87 | 43,097.47 | 45,509.81 | 47,819.81 | 177,108.96 |
| SOUTH CAROLINA | 57,684.42 | 55,976.47 | 56,288.62 | 55,428.56 | 225,378.07 |
| SOUTH DAKOTA | 2,264.98 | 2,147.10 | 2,231.71 | 2,114.09 | 8,757.88 |

| | | | | | |
|---|---|---|---|---|---|
| TENNESSEE | 49,325.67 | 49,042.52 | 47,700.36 | 48,234.92 | 194,303.47 |
| TEXAS | 139,436.83 | 145,893.54 | 134,651.11 | 142,036.93 | 562,018.41 |
| UTAH | 26,883.52 | 23,868.58 | 28,536.01 | 26,398.31 | 105,686.42 |
| VERMONT | 18,852.34 | 17,051.91 | 20,166.13 | 17,928.87 | 73,999.25 |
| VIRGIN ISLANDS | 28.17 | 386.38 | 23.25 | 19.68 | 457.48 |
| VIRGINIA | 81,665.76 | 75,402.83 | 78,605.66 | 81,398.67 | 317,072.92 |
| WASHINGTON | 85,136.33 | 93,477.77 | 104,036.61 | 108,189.93 | 390,840.64 |
| WEST VIRGINIA | 23,535.33 | 23,508.59 | 23,002.98 | 22,523.41 | 92,570.31 |
| WISCONSIN | 65,713.89 | 68,870.50 | 63,531.78 | 64,031.26 | 262,147.43 |
| WYOMING | 1,277.65 | 1,164.78 | 1,182.67 | 1,131.42 | 4,756.52 |
| TOTAL | 3,617,148.91 | 3,719,459.80 | 3,655,355.64 | 3,699,498.47 | 14,691,462.82 |

DRUG CODE:9300 DRUG NAME:MORPHINE

| STATE | QUARTER 1 | QUARTER 2 | QUARTER 3 | QUARTER 4 | TOTAL GRAMS |
|---|---|---|---|---|---|
| ALABAMA | 75,385.73 | 71,751.81 | 68,342.60 | 66,586.68 | 282,066.82 |
| ALASKA | 12,075.08 | 11,675.08 | 11,278.53 | 10,703.94 | 45,732.63 |
| AMERICAN SAMOA | 30.08 | 27.45 | 3.01 | 27.07 | 87.61 |
| ARIZONA | 179,749.43 | 166,620.20 | 156,623.07 | 145,087.27 | 648,079.97 |
| ARKANSAS | 49,144.75 | 45,645 | 44,441.27 | 41,659.42 | 180,890.44 |
| CALIFORNIA | 468,312.40 | 449,794.43 | 435,963.44 | 421,426.73 | 1,775,497 |
| COLORADO | 80,950.89 | 77,471.94 | 74,788.34 | 72,248.67 | 305,459.84 |
| CONNECTICUT | 36,962.44 | 35,651.52 | 35,131.70 | 33,709.26 | 141,454.92 |
| DELAWARE | 22,486.45 | 21,893.44 | 21,334.15 | 20,973.47 | 86,687.51 |
| DISTRICT OF COLUMBIA | 3,713.59 | 3,853.50 | 3,790.31 | 3,217.86 | 14,575.26 |
| FLORIDA | 412,688.12 | 394,998.55 | 383,405.02 | 382,567.73 | 1,573,659.42 |
| GEORGIA | 116,831.19 | 112,302.56 | 111,023.27 | 108,398.04 | 448,555.06 |
| GUAM | 1,270.56 | 829.71 | 1,031 | 866.98 | 3,998.25 |
| HAWAII | 23,410.41 | 22,488.31 | 21,187.89 | 20,072.40 | 87,159.01 |
| IDAHO | 29,776.64 | 28,555.83 | 27,191.18 | 27,003.18 | 112,526.83 |
| ILLINOIS | 108,011.75 | 103,220.32 | 100,084.61 | 98,037.74 | 409,354.42 |
| INDIANA | 92,152.33 | 87,372.85 | 83,021.72 | 81,242.99 | 343,789.89 |
| IOWA | 34,869.28 | 33,926.30 | 32,608.91 | 32,570.43 | 133,974.92 |
| KANSAS | 46,843.42 | 44,319.08 | 44,034.39 | 42,477.13 | 177,674.02 |
| KENTUCKY | 49,899.50 | 46,594.48 | 44,626.41 | 44,322.40 | 185,442.79 |
| LOUISIANA | 56,295.87 | 54,358.01 | 52,557.96 | 50,549.33 | 213,761.17 |
| MAINE | 17,045.94 | 16,858.81 | 16,420.51 | 15,608.65 | 65,933.91 |

| | | | | | |
|---|---|---|---|---|---|
| MARYLAND | 82,088.87 | 78,530.74 | 72,532.28 | 69,991.46 | 303,143.35 |
| MASSACHUSETTS | 69,568.66 | 66,507.75 | 63,147.53 | 62,974.31 | 262,198.25 |
| MICHIGAN | 200,848.83 | 192,688.22 | 185,978.26 | 174,054.87 | 753,570.18 |
| MINNESOTA | 49,548.65 | 46,944.54 | 45,377.16 | 43,601.89 | 185,472.24 |
| MISSISSIPPI | 41,333.75 | 40,545.50 | 38,844.42 | 39,087.65 | 159,811.32 |
| MISSOURI | 102,523.17 | 97,078.72 | 94,755.80 | 91,575.87 | 385,933.56 |
| MONTANA | 19,066.75 | 17,234.06 | 17,003.77 | 16,337.37 | 69,641.95 |
| NEBRASKA | 19,770.78 | 18,851.07 | 18,939.08 | 18,132.17 | 75,693.10 |
| NEVADA | 62,634.76 | 60,794.33 | 58,562.47 | 55,522.81 | 237,514.37 |
| NEW HAMPSHIRE | 18,582.91 | 17,841.69 | 17,901.86 | 18,151.01 | 72,477.47 |
| NEW JERSEY | 93,090.78 | 86,208.33 | 81,320.72 | 79,622.09 | 340,241.92 |
| NEW MEXICO | 28,578.98 | 25,993.41 | 25,356.63 | 23,224.01 | 103,153.03 |
| NEW YORK | 163,662.50 | 156,595.10 | 150,731.44 | 149,768.80 | 620,757.84 |
| NORTH CAROLINA | 126,674.67 | 120,047.86 | 116,465.73 | 112,951.75 | 476,140.01 |
| NORTH DAKOTA | 6,686.39 | 6,421.62 | 5,951.58 | 6,072.25 | 25,131.84 |
| OHIO | 124,959.28 | 121,047.73 | 118,500.26 | 113,214.79 | 477,722.06 |
| OKLAHOMA | 96,982.97 | 92,592.40 | 89,126.22 | 85,047.29 | 363,748.88 |
| OREGON | 77,234.02 | 72,077.72 | 68,842.86 | 66,164.46 | 284,319.06 |
| PENNSYLVANIA | 263,618.17 | 229,898.27 | 221,317.41 | 217,396.23 | 932,230.08 |
| PUERTO RICO | 2,588.47 | 2,241.27 | 1,865.42 | 1,704.59 | 8,399.75 |
| RHODE ISLAND | 12,069.92 | 11,689.87 | 12,022.55 | 10,846.17 | 46,628.51 |
| SOUTH CAROLINA | 68,726.78 | 64,803.14 | 63,883.11 | 64,033 | 261,446.03 |
| SOUTH DAKOTA | 9,105.11 | 8,744.97 | 8,676 | 8,274.99 | 34,801.07 |
| TENNESSEE | 185,724.11 | 159,082.83 | 155,695.27 | 149,520.74 | 650,022.95 |
| TEXAS | 214,631.96 | 206,944.37 | 201,945.89 | 197,615.22 | 821,137.44 |
| UTAH | 51,222.98 | 49,400.32 | 49,045.82 | 46,883.70 | 196,552.82 |
| VERMONT | 10,746.22 | 10,191.11 | 9,758.79 | 9,832.44 | 40,528.56 |
| VIRGIN ISLANDS | 139.62 | 152.01 | 119.42 | 113.85 | 524.9 |
| VIRGINIA | 84,458.48 | 79,063.13 | 76,091.63 | 74,255.58 | 313,868.82 |
| WASHINGTON | 115,166.01 | 110,388.89 | 106,882.84 | 101,240.93 | 433,678.67 |
| WEST VIRGINIA | 23,389.02 | 21,876.66 | 21,639.48 | 20,541.36 | 87,446.52 |
| WISCONSIN | 83,691.07 | 77,548.84 | 74,645.54 | 72,537.13 | 308,422.58 |
| WYOMING | 9,221.32 | 9,334.98 | 9,000.08 | 8,311.06 | 35,867.44 |
| TOTAL | 4,436,241.81 | 4,189,570.63 | 4,050,816.61 | 3,927,959.21 | 16,604,588.26 |

DRUG CODE:9333 DRUG NAME:THEBAINE

| STATE | QUARTER 1 | QUARTER 2 | QUARTER 3 | QUARTER 4 | TOTAL GRAMS |
|---|---|---|---|---|---|
| TEXAS | 0 | 0 | 25 | 0 | 25 |
| TOTAL | 0 | 0 | 25 | 0 | 25 |

DRUG CODE:9630 DRUG NAME:OPIUM TINCTURE (LAUDANUM)

| STATE | QUARTER 1 | QUARTER 2 | QUARTER 3 | QUARTER 4 | TOTAL GRAMS |
|---|---|---|---|---|---|
| CALIFORNIA | 5.91 | 0 | 0 | 0 | 5.91 |
| CONNECTICUT | 0 | 0 | 4.73 | 0 | 4.73 |
| DELAWARE | 0 | 1.18 | 0 | 0 | 1.18 |
| FLORIDA | 9.46 | 4.73 | 0 | 0 | 14.19 |
| ILLINOIS | 5.91 | 10.64 | 0 | 0 | 16.55 |
| INDIANA | 21.28 | 14.19 | 0 | 0 | 35.47 |
| KANSAS | 5.91 | 0 | 0 | 0 | 5.91 |
| KENTUCKY | 2.36 | 0 | 0 | 0 | 2.36 |
| MARYLAND | 4.72 | 8.27 | 0 | 0 | 12.99 |
| MICHIGAN | 26 | 4.72 | 0 | 0 | 30.72 |
| NEW HAMPSHIRE | 4.73 | 0 | 0 | 0 | 4.73 |
| NEW JERSEY | 0 | 1.18 | 0 | 0 | 1.18 |
| NEW YORK | 4.73 | 0 | 0 | 0 | 4.73 |
| OHIO | 73.23 | 9.45 | 0 | 0 | 82.68 |
| OREGON | 2.36 | 0 | 0 | 0 | 2.36 |
| PENNSYLVANIA | 10.62 | 10.62 | 0 | 0 | 21.24 |
| TEXAS | 9.44 | 4.72 | 0 | 0 | 14.16 |
| VIRGINIA | 10.62 | 0 | 0 | 0 | 10.62 |
| WASHINGTON | 3.54 | 0 | 0 | 0 | 3.54 |
| WEST VIRGINIA | 3.54 | 2.36 | 0 | 0 | 5.9 |
| WISCONSIN | 3.54 | 10.64 | 0 | 0 | 14.18 |
| TOTAL | 207.9 | 82.7 | 4.73 | 0 | 295.33 |

DRUG CODE:9639 DRUG NAME:OPIUM POWDERED

| STATE | QUARTER 1 | QUARTER 2 | QUARTER 3 | QUARTER 4 | TOTAL GRAMS |
|---|---|---|---|---|---|
| ALABAMA | 82.08 | 94.68 | 86.4 | 78.48 | 341.64 |
| ALASKA | 29.52 | 25.2 | 19.8 | 28.08 | 102.6 |
| ARIZONA | 277.56 | 273.6 | 292.32 | 337.68 | 1,181.16 |
| ARKANSAS | 83.52 | 96.84 | 87.12 | 93.6 | 361.08 |

| | | | | | |
|---|---|---|---|---|---|
| CALIFORNIA | 412.92 | 470.52 | 484.56 | 453.6 | 1,821.60 |
| COLORADO | 231.84 | 260.64 | 288.36 | 279.36 | 1,060.20 |
| CONNECTICUT | 114.48 | 120.96 | 81.72 | 68.04 | 385.2 |
| DELAWARE | 17.64 | 14.76 | 16.2 | 11.52 | 60.12 |
| DISTRICT OF COLUMBIA | 12.24 | 12.96 | 18.36 | 15.48 | 59.04 |
| FLORIDA | 527.04 | 498.24 | 452.52 | 553.68 | 2,031.48 |
| GEORGIA | 174.24 | 210.6 | 185.04 | 185.76 | 755.64 |
| GUAM | 0.36 | 0 | 0 | 0 | 0.36 |
| HAWAII | 24.84 | 27.72 | 34.2 | 28.44 | 115.2 |
| IDAHO | 45.72 | 51.12 | 48.96 | 49.68 | 195.48 |
| ILLINOIS | 158.4 | 194.04 | 172.08 | 192.96 | 717.48 |
| INDIANA | 222.84 | 253.8 | 234.72 | 270.36 | 981.72 |
| IOWA | 112.32 | 100.8 | 96.84 | 92.52 | 402.48 |
| KANSAS | 165.6 | 191.88 | 177.48 | 185.04 | 720 |
| KENTUCKY | 196.92 | 179.64 | 146.16 | 173.88 | 696.6 |
| LOUISIANA | 105.48 | 113.04 | 98.28 | 104.4 | 421.2 |
| MAINE | 39.24 | 47.88 | 54 | 57.96 | 199.08 |
| MARYLAND | 63 | 97.56 | 70.56 | 104.04 | 335.16 |
| MASSACHUSETTS | 172.8 | 214.56 | 177.12 | 238.68 | 803.16 |
| MICHIGAN | 374.4 | 369.72 | 375.84 | 422.28 | 1,542.24 |
| MINNESOTA | 401.4 | 381.24 | 363.96 | 389.16 | 1,535.76 |
| MISSISSIPPI | 33.12 | 39.96 | 36.36 | 25.2 | 134.64 |
| MISSOURI | 169.92 | 163.44 | 166.32 | 174.6 | 674.28 |
| MONTANA | 53.28 | 52.92 | 47.88 | 50.04 | 204.12 |
| NEBRASKA | 91.8 | 106.2 | 90.36 | 105.48 | 393.84 |
| NEVADA | 30.6 | 28.08 | 34.92 | 40.68 | 134.28 |
| NEW HAMPSHIRE | 30.24 | 37.44 | 37.44 | 41.4 | 146.52 |
| NEW JERSEY | 87.12 | 133.56 | 117.72 | 104.76 | 443.16 |
| NEW MEXICO | 37.8 | 49.32 | 29.16 | 28.44 | 144.72 |
| NEW YORK | 208.8 | 217.44 | 201.96 | 241.2 | 869.4 |
| NORTH CAROLINA | 238.32 | 218.52 | 219.96 | 240.84 | 917.64 |
| NORTH DAKOTA | 30.96 | 42.12 | 45.72 | 46.08 | 164.88 |
| OHIO | 371.88 | 380.16 | 376.2 | 386.28 | 1,514.52 |
| OKLAHOMA | 76.68 | 83.52 | 95.4 | 55.08 | 310.68 |
| OREGON | 168.48 | 215.28 | 203.04 | 211.32 | 798.12 |
| PENNSYLVANIA | 296.64 | 288 | 334.8 | 380.88 | 1,300.32 |
| RHODE ISLAND | 32.76 | 34.2 | 45 | 25.56 | 137.52 |

| | | | | | |
|---|---|---|---|---|---|
| SOUTH CAROLINA | 127.8 | 154.8 | 127.44 | 145.08 | 555.12 |
| SOUTH DAKOTA | 64.08 | 46.8 | 27 | 42.84 | 180.72 |
| TENNESSEE | 171 | 192.6 | 190.8 | 205.56 | 759.96 |
| TEXAS | 503.64 | 504.36 | 513.36 | 567.36 | 2,088.72 |
| UTAH | 67.68 | 64.08 | 62.64 | 66.24 | 260.64 |
| VERMONT | 41.4 | 39.96 | 37.44 | 43.92 | 162.72 |
| VIRGIN ISLANDS | 0 | 0 | 1.8 | 0 | 1.8 |
| VIRGINIA | 170.64 | 178.2 | 199.44 | 174.6 | 722.88 |
| WASHINGTON | 281.88 | 321.12 | 325.44 | 329.04 | 1,257.48 |
| WEST VIRGINIA | 84.6 | 110.88 | 87.12 | 98.28 | 380.88 |
| WISCONSIN | 183.24 | 223.92 | 239.04 | 208.44 | 854.64 |
| WYOMING | 25.56 | 25.56 | 19.8 | 27.36 | 98.28 |
| TOTAL | 7,726.32 | 8,254.44 | 7,976.16 | 8,481.24 | 32,438.16 |

DRUG CODE:9652 DRUG NAME:OXYMORPHONE

| STATE | QUARTER 1 | QUARTER 2 | QUARTER 3 | QUARTER 4 | TOTAL GRAMS |
|---|---|---|---|---|---|
| ALABAMA | 7,674.65 | 6,782.29 | 4,108.61 | 2,938.27 | 21,503.82 |
| ALASKA | 287.43 | 286.85 | 246.49 | 183.95 | 1,004.72 |
| ARIZONA | 8,602.56 | 7,495.65 | 4,844.27 | 2,808.40 | 23,750.88 |
| ARKANSAS | 2,279 | 2,185.73 | 1,551.85 | 856.42 | 6,873 |
| CALIFORNIA | 17,090.45 | 15,659.83 | 11,435.85 | 8,028.86 | 52,214.99 |
| COLORADO | 4,341.19 | 3,794.17 | 2,855.79 | 1,830.99 | 12,822.14 |
| CONNECTICUT | 3,455.85 | 3,161.39 | 2,320.28 | 1,557.38 | 10,494.90 |
| DELAWARE | 2,154.74 | 2,187.70 | 1,843.66 | 1,539.54 | 7,725.64 |
| DISTRICT OF COLUMBIA | 120.14 | 126.86 | 101.43 | 130.83 | 479.26 |
| FLORIDA | 28,038.19 | 26,517.35 | 19,406.08 | 12,915.73 | 86,877.35 |
| GEORGIA | 8,564.78 | 8,249.88 | 5,403.09 | 3,602.98 | 25,820.73 |
| HAWAII | 678.13 | 699.76 | 394.84 | 373.52 | 2,146.25 |
| IDAHO | 1,587.02 | 1,492.09 | 1,174.85 | 891.12 | 5,145.08 |
| ILLINOIS | 5,304.30 | 4,984.65 | 3,638.65 | 2,369.95 | 16,297.55 |
| INDIANA | 6,675.18 | 5,775.53 | 4,214.64 | 2,802.71 | 19,468.06 |
| IOWA | 1,375.82 | 1,250.29 | 911.24 | 569.87 | 4,107.22 |
| KANSAS | 2,458.24 | 2,307.42 | 1,630.98 | 1,114.41 | 7,511.05 |
| KENTUCKY | 4,554.13 | 4,419.69 | 3,133.32 | 2,355.99 | 14,463.13 |
| LOUISIANA | 8,205.45 | 7,664.08 | 5,756.32 | 4,709.35 | 26,335.20 |
| MAINE | 1,190.10 | 1,005.89 | 857.8 | 644.72 | 3,698.51 |

| | | | | | |
|---|---|---|---|---|---|
| MARYLAND | 11,286.45 | 10,601.73 | 8,197.77 | 6,964.20 | 37,050.15 |
| MASSACHUSETTS | 2,863.05 | 2,730.45 | 1,968.60 | 1,139.08 | 8,701.18 |
| MICHIGAN | 20,397.38 | 20,864.79 | 18,129.57 | 16,304.25 | 75,695.99 |
| MINNESOTA | 501.31 | 519.37 | 404.17 | 273.25 | 1,698.10 |
| MISSISSIPPI | 3,886.94 | 3,589.36 | 2,738.88 | 2,033.10 | 12,248.28 |
| MISSOURI | 5,853.66 | 5,330.79 | 3,721.71 | 2,321.96 | 17,228.12 |
| MONTANA | 388 | 296.53 | 289.98 | 158.26 | 1,132.77 |
| NEBRASKA | 1,567.20 | 1,511.89 | 1,058.08 | 630.18 | 4,767.35 |
| NEVADA | 5,434.15 | 4,839.37 | 3,514.87 | 2,141.98 | 15,930.37 |
| NEW HAMPSHIRE | 1,095.76 | 992.83 | 826.22 | 576.79 | 3,491.60 |
| NEW JERSEY | 8,392.25 | 7,809.68 | 5,422.94 | 4,069.18 | 25,694.05 |
| NEW MEXICO | 1,251.97 | 1,079.55 | 813.91 | 652.62 | 3,798.05 |
| NEW YORK | 21,903.43 | 20,825.78 | 16,371.89 | 12,712.78 | 71,813.88 |
| NORTH CAROLINA | 27,893.54 | 25,337.27 | 17,495.95 | 12,229.58 | 82,956.34 |
| NORTH DAKOTA | 174.23 | 180.74 | 159.69 | 114.99 | 629.65 |
| OHIO | 12,220.12 | 11,461 | 8,794.23 | 6,074.01 | 38,549.36 |
| OKLAHOMA | 6,983.86 | 6,587.36 | 4,887.55 | 3,302.06 | 21,760.83 |
| OREGON | 2,032.61 | 1,876.84 | 1,576.03 | 1,095.23 | 6,580.71 |
| PENNSYLVANIA | 26,926.77 | 26,231.13 | 21,007.71 | 16,901.20 | 91,066.81 |
| RHODE ISLAND | 576.86 | 486.59 | 364.38 | 174.18 | 1,602.01 |
| SOUTH CAROLINA | 7,767.06 | 7,503.36 | 5,106.78 | 3,625.45 | 24,002.65 |
| SOUTH DAKOTA | 631.75 | 533.57 | 419.11 | 327.76 | 1,912.19 |
| TENNESSEE | 25,189.91 | 23,346.52 | 18,453.28 | 15,244.47 | 82,234.18 |
| TEXAS | 15,482.71 | 14,431.61 | 9,895.18 | 6,358.75 | 46,168.25 |
| UTAH | 4,816.31 | 4,513.19 | 3,837.50 | 3,232.44 | 16,399.44 |
| VERMONT | 362.68 | 325.42 | 275.39 | 181.36 | 1,144.85 |
| VIRGINIA | 11,386.63 | 10,375.13 | 7,899.21 | 6,651.41 | 36,312.38 |
| WASHINGTON | 3,634.01 | 3,266.76 | 2,594.27 | 1,910.34 | 11,405.38 |
| WEST VIRGINIA | 2,929.48 | 2,929.43 | 2,410.14 | 1,654.58 | 9,923.63 |
| WISCONSIN | 2,924.54 | 2,539.68 | 1,843.84 | 1,203.31 | 8,511.37 |
| WYOMING | 322.49 | 342.83 | 255.36 | 199.34 | 1,120.02 |
| TOTAL | 351,714.46 | 329,307.65 | 246,564.23 | 182,683.08 | 1,110,269.42 |

DRUG CODE:9668 DRUG NAME:NOROXYMORPHONE

| STATE | QUARTER 1 | QUARTER 2 | QUARTER 3 | QUARTER 4 | TOTAL GRAMS |
|---|---|---|---|---|---|
| ARIZONA | 0 | 0 | 0.1 | 0 | 0.1 |

| | | | | | |
|---|---|---|---|---|---|
| ARKANSAS | 14.63 | 17.21 | 26.85 | 19.38 | 78.07 |
| CALIFORNIA | 52.59 | 68.53 | 69.85 | 68.65 | 259.62 |
| FLORIDA | 12.61 | 13.83 | 13.51 | 18.08 | 58.03 |
| GEORGIA | 3.18 | 3.18 | 9.3 | 4.8 | 20.46 |
| ILLINOIS | 4.42 | 9.76 | 15.09 | 8.75 | 38.02 |
| KANSAS | 2.75 | 0 | 0 | 0 | 2.75 |
| MARYLAND | 0 | 0.1 | 0 | 0.1 | 0.2 |
| MASSACHUSETTS | 1.14 | 1.9 | 0 | 0 | 3.04 |
| MICHIGAN | 0 | 0 | 3.12 | 1.17 | 4.29 |
| MINNESOTA | 0 | 0.5 | 0 | 0 | 0.5 |
| MISSISSIPPI | 2.4 | 1.57 | 2.22 | 0 | 6.19 |
| MISSOURI | 7.71 | 13.1 | 10.59 | 5.8 | 37.2 |
| NEVADA | 6.64 | 2.87 | 7.45 | 8.31 | 25.27 |
| NEW JERSEY | 7.12 | 12.03 | 1.56 | 3 | 23.71 |
| NEW YORK | 2.65 | 5.69 | 9.3 | 4.49 | 22.13 |
| NORTH CAROLINA | 0 | 0 | 2.12 | 1.56 | 3.68 |
| OHIO | 5.89 | 9.36 | 6.19 | 2.88 | 24.32 |
| OKLAHOMA | 0 | 0 | 0.1 | 0.55 | 0.65 |
| OREGON | 0.15 | 0.04 | 1.81 | 0 | 2 |
| PENNSYLVANIA | 0 | 0.51 | 0.47 | 0 | 0.98 |
| PUERTO RICO | 1.86 | 4.44 | 11.94 | 6.83 | 25.07 |
| RHODE ISLAND | 0 | 0 | 0.1 | 0 | 0.1 |
| TENNESSEE | 0.13 | 0 | 0 | 0 | 0.13 |
| TEXAS | 21.77 | 29.59 | 31.28 | 26.19 | 108.83 |
| VERMONT | 0 | 0 | 0.1 | 0 | 0.1 |
| WASHINGTON | 0.65 | 0.68 | 3.21 | 3.94 | 8.48 |
| TOTAL | 148.29 | 194.89 | 226.26 | 184.48 | 753.92 |

DRUG CODE:9737 DRUG NAME:ALFENTANIL

| STATE | QUARTER 1 | QUARTER 2 | QUARTER 3 | QUARTER 4 | TOTAL GRAMS |
|---|---|---|---|---|---|
| ALABAMA | 0.04 | 0.13 | 0.05 | 0.03 | 0.25 |
| ALASKA | 0.75 | 0.75 | 0.5 | 0.89 | 2.89 |
| ARIZONA | 1.12 | 0.96 | 0.69 | 0.97 | 3.74 |
| ARKANSAS | 0.01 | 0 | 0.04 | 0.03 | 0.08 |
| CALIFORNIA | 17.35 | 15.3 | 16.28 | 17 | 65.93 |
| COLORADO | 1.27 | 0.72 | 0.79 | 1.08 | 3.86 |

| | | | | | |
|---|---|---|---|---|---|
| CONNECTICUT | 1.52 | 1.2 | 1.03 | 1.55 | 5.3 |
| FLORIDA | 4.55 | 5.48 | 4.78 | 6.1 | 20.91 |
| GEORGIA | 2.92 | 2.79 | 4.66 | 2.69 | 13.06 |
| GUAM | 0 | 0 | 0.05 | 0.03 | 0.08 |
| HAWAII | 0.45 | 0.5 | 0.62 | 0.84 | 2.41 |
| IDAHO | 1.46 | 1.39 | 1.15 | 1.46 | 5.46 |
| ILLINOIS | 3.76 | 4.23 | 3.37 | 5.14 | 16.5 |
| INDIANA | 2.24 | 3.18 | 2.54 | 2.34 | 10.3 |
| IOWA | 2.05 | 1.5 | 1.4 | 1.74 | 6.69 |
| KANSAS | 1.62 | 1.51 | 1.26 | 1.6 | 5.99 |
| KENTUCKY | 5 | 3.91 | 3.67 | 3.85 | 16.43 |
| LOUISIANA | 2.44 | 2.28 | 2.84 | 2.5 | 10.06 |
| MAINE | 0.62 | 0.79 | 0.63 | 0.65 | 2.69 |
| MARYLAND | 0.89 | 0.24 | 0.85 | 0.92 | 2.9 |
| MASSACHUSETTS | 1.35 | 0.73 | 0.65 | 0.73 | 3.46 |
| MICHIGAN | 0.91 | 1.32 | 1.05 | 1.21 | 4.49 |
| MINNESOTA | 3.24 | 3.41 | 3.23 | 4.04 | 13.92 |
| MISSISSIPPI | 0.07 | 0.12 | 0.17 | 0.07 | 0.43 |
| MISSOURI | 1.21 | 1 | 0.9 | 1.22 | 4.33 |
| MONTANA | 0.36 | 0.27 | 0.33 | 0.34 | 1.3 |
| NEBRASKA | 0.84 | 0.7 | 1.08 | 0.94 | 3.56 |
| NEVADA | 1.27 | 1.59 | 1.74 | 2.36 | 6.96 |
| NEW HAMPSHIRE | 0.48 | 0.42 | 0.44 | 0.61 | 1.95 |
| NEW JERSEY | 0.2 | 0.27 | 0.23 | 0.35 | 1.05 |
| NEW MEXICO | 0.87 | 0.88 | 1.14 | 1.21 | 4.1 |
| NEW YORK | 2.12 | 1.22 | 1.66 | 4.16 | 9.16 |
| NORTH CAROLINA | 2.78 | 3.38 | 2.92 | 3.5 | 12.58 |
| NORTH DAKOTA | 0.11 | 0.28 | 0.09 | 0.11 | 0.59 |
| OHIO | 4.49 | 6.14 | 3.5 | 4.97 | 19.1 |
| OKLAHOMA | 0.83 | 1.01 | 0.84 | 0.84 | 3.52 |
| OREGON | 3.84 | 4.32 | 5.66 | 4.86 | 18.68 |
| PENNSYLVANIA | 0.7 | 0.48 | 0.37 | 0.46 | 2.01 |
| RHODE ISLAND | 0.05 | 0.08 | 0.13 | 0 | 0.26 |
| SOUTH CAROLINA | 0.13 | 0.05 | 0.05 | 0.13 | 0.36 |
| SOUTH DAKOTA | 0.11 | 0.06 | 0.1 | 0.06 | 0.33 |
| TENNESSEE | 0.97 | 1.08 | 1.16 | 1.56 | 4.77 |
| TEXAS | 5.06 | 6.06 | 5.21 | 5.72 | 22.05 |

| | | | | | |
|---|---|---|---|---|---|
| UTAH | 0.47 | 0.35 | 0.52 | 1.76 | 3.1 |
| VERMONT | 0.01 | 0 | 0.01 | 0.02 | 0.04 |
| VIRGINIA | 0.87 | 1.13 | 0.68 | 0.94 | 3.62 |
| WASHINGTON | 2.53 | 3.37 | 3.48 | 3.28 | 12.66 |
| WISCONSIN | 4.13 | 4.82 | 4.81 | 5.52 | 19.28 |
| WYOMING | 0.01 | 0 | 0.02 | 0.09 | 0.12 |
| TOTAL | 90.07 | 91.4 | 89.37 | 102.47 | 373.31 |

DRUG CODE:9739 DRUG NAME:REMIFENTANIL

| STATE | QUARTER 1 | QUARTER 2 | QUARTER 3 | QUARTER 4 | TOTAL GRAMS |
|---|---|---|---|---|---|
| ALABAMA | 3.16 | 3.41 | 3.44 | 3.14 | 13.15 |
| ALASKA | 2.73 | 3.01 | 2.8 | 3.14 | 11.68 |
| ARIZONA | 4.16 | 3.74 | 3.63 | 4.16 | 15.69 |
| ARKANSAS | 1.57 | 1.52 | 1.82 | 1.48 | 6.39 |
| CALIFORNIA | 33.2 | 34.56 | 31.25 | 34.34 | 133.35 |
| COLORADO | 5.96 | 6.53 | 6.18 | 7.17 | 25.84 |
| CONNECTICUT | 9.81 | 7.72 | 7.82 | 8.71 | 34.06 |
| DELAWARE | 2.67 | 4.02 | 2.81 | 2.54 | 12.04 |
| DISTRICT OF COLUMBIA | 0.88 | 0.86 | 0.52 | 0.58 | 2.84 |
| FLORIDA | 25.75 | 26.92 | 25.23 | 29.55 | 107.45 |
| GEORGIA | 4.65 | 5.09 | 4.67 | 4.79 | 19.2 |
| GUAM | 0.02 | 0.04 | 0 | 0.25 | 0.31 |
| HAWAII | 1.29 | 1.13 | 1.34 | 1.38 | 5.14 |
| IDAHO | 2.25 | 2.19 | 2.12 | 2.14 | 8.7 |
| ILLINOIS | 14.81 | 14.48 | 14.58 | 16.89 | 60.76 |
| INDIANA | 8.1 | 7.75 | 7.97 | 8.11 | 31.93 |
| IOWA | 3.57 | 3.24 | 3.33 | 2.44 | 12.58 |
| KANSAS | 2.1 | 2.53 | 2.11 | 2.33 | 9.07 |
| KENTUCKY | 5.02 | 4.27 | 5.26 | 4.61 | 19.16 |
| LOUISIANA | 4.41 | 4.7 | 4.5 | 4.96 | 18.57 |
| MAINE | 1.37 | 1.2 | 1.11 | 0.93 | 4.61 |
| MARYLAND | 7.19 | 7.13 | 5.88 | 7.2 | 27.4 |
| MASSACHUSETTS | 20.68 | 23.9 | 21.61 | 23.06 | 89.25 |
| MICHIGAN | 9.51 | 10.95 | 9.68 | 9.49 | 39.63 |
| MINNESOTA | 7.02 | 7.9 | 7.92 | 9.85 | 32.69 |
| MISSISSIPPI | 0.56 | 0.63 | 0.27 | 0.32 | 1.78 |

| | | | | | |
|---|---|---|---|---|---|
| MISSOURI | 5.02 | 5.31 | 5.86 | 5.58 | 21.77 |
| MONTANA | 1.45 | 1.83 | 1.72 | 1.99 | 6.99 |
| NEBRASKA | 1.14 | 0.97 | 0.91 | 1.27 | 4.29 |
| NEVADA | 0.89 | 0.74 | 1.11 | 0.96 | 3.7 |
| NEW HAMPSHIRE | 1.15 | 1.52 | 1.43 | 2.42 | 6.52 |
| NEW JERSEY | 12.79 | 13.01 | 12.94 | 14.17 | 52.91 |
| NEW MEXICO | 1 | 1.14 | 1.27 | 1.03 | 4.44 |
| NEW YORK | 36.7 | 37.72 | 40.31 | 40.31 | 155.04 |
| NORTH CAROLINA | 13.88 | 14.85 | 13.84 | 15.18 | 57.75 |
| NORTH DAKOTA | 0.58 | 0.32 | 0.38 | 0.39 | 1.67 |
| OHIO | 19.27 | 20 | 15.93 | 20.44 | 75.64 |
| OKLAHOMA | 2.59 | 2.88 | 2.65 | 2.61 | 10.73 |
| OREGON | 2.69 | 3.27 | 2.53 | 2.91 | 11.4 |
| PENNSYLVANIA | 36.19 | 35.59 | 35.09 | 36.37 | 143.24 |
| RHODE ISLAND | 0.68 | 0.51 | 0.76 | 1.22 | 3.17 |
| SOUTH CAROLINA | 5.08 | 4.36 | 4.64 | 4.57 | 18.65 |
| SOUTH DAKOTA | 0.47 | 0.66 | 0.44 | 0.32 | 1.89 |
| TENNESSEE | 4.66 | 5.42 | 6.69 | 6.47 | 23.24 |
| TEXAS | 26.79 | 27.42 | 27.2 | 30.45 | 111.86 |
| UTAH | 17.82 | 14.08 | 15.38 | 24.5 | 71.78 |
| VERMONT | 0.68 | 0.91 | 0.52 | 0.72 | 2.83 |
| VIRGINIA | 6.27 | 7.39 | 6.04 | 7.47 | 27.17 |
| WASHINGTON | 12.71 | 12.08 | 14.64 | 10.58 | 50.01 |
| WEST VIRGINIA | 5.37 | 6.02 | 4.95 | 4.41 | 20.75 |
| WISCONSIN | 6.05 | 7.56 | 5.32 | 7.8 | 26.73 |
| WYOMING | 0.5 | 0.39 | 0.43 | 0.46 | 1.78 |
| TOTAL | 404.86 | 415.37 | 400.83 | 438.16 | 1,659.22 |

DRUG CODE:9740 DRUG NAME:SUFENTANIL BASE

| STATE | QUARTER 1 | QUARTER 2 | QUARTER 3 | QUARTER 4 | TOTAL GRAMS |
|---|---|---|---|---|---|
| ALABAMA | 1.04 | 1.76 | 1.76 | 2 | 6.56 |
| ALASKA | 0.01 | 0.01 | 0.01 | 0.01 | 0.04 |
| ARIZONA | 0.07 | 0.07 | 0.06 | 0.14 | 0.34 |
| ARKANSAS | 0.14 | 0.1 | 0.17 | 0.15 | 0.56 |
| CALIFORNIA | 9.68 | 12.78 | 11.23 | 10.05 | 43.74 |
| COLORADO | 0.19 | 0.16 | 0.21 | 0.29 | 0.85 |

| | | | | | |
|---|---|---|---|---|---|
| CONNECTICUT | 0.41 | 0.46 | 0.58 | 0.94 | 2.39 |
| DELAWARE | 0.01 | 0.01 | 0.01 | 0.01 | 0.04 |
| DISTRICT OF COLUMBIA | 0.01 | 0 | 0.01 | 0 | 0.02 |
| FLORIDA | 4.25 | 2.01 | 3.33 | 3.06 | 12.65 |
| GEORGIA | 0.32 | 0.34 | 0.52 | 0.61 | 1.79 |
| HAWAII | 0.01 | 0 | 0.01 | 0.02 | 0.04 |
| IDAHO | 0.04 | 0.05 | 0.04 | 0.04 | 0.17 |
| ILLINOIS | 0.55 | 0.61 | 0.58 | 0.54 | 2.28 |
| INDIANA | 0.31 | 0.37 | 0.25 | 0.22 | 1.15 |
| IOWA | 0.06 | 0.04 | 0.04 | 0.07 | 0.21 |
| KANSAS | 0.23 | 0.28 | 0.35 | 0.29 | 1.15 |
| KENTUCKY | 0.32 | 0.4 | 0.4 | 0.39 | 1.51 |
| LOUISIANA | 0.35 | 2.02 | 1.09 | 1.17 | 4.63 |
| MAINE | 0.01 | 0.01 | 0.02 | 0.03 | 0.07 |
| MARYLAND | 0.04 | 0.06 | 0.07 | 0.05 | 0.22 |
| MASSACHUSETTS | 0.09 | 0.12 | 0.12 | 0.19 | 0.52 |
| MICHIGAN | 0.54 | 0.37 | 0.32 | 1.02 | 2.25 |
| MINNESOTA | 0.16 | 0.16 | 0.15 | 0.31 | 0.78 |
| MISSISSIPPI | 14.74 | 10.44 | 12.48 | 14.46 | 52.12 |
| MISSOURI | 0.29 | 0.32 | 0.36 | 0.29 | 1.26 |
| MONTANA | 0.04 | 0.04 | 0.03 | 0.06 | 0.17 |
| NEBRASKA | 0.08 | 0.09 | 0.08 | 0.15 | 0.4 |
| NEVADA | 0.07 | 0.05 | 0.13 | 0.12 | 0.37 |
| NEW HAMPSHIRE | 0.01 | 0.02 | 0.02 | 0.02 | 0.07 |
| NEW JERSEY | 0.95 | 0.99 | 0.23 | 1.56 | 3.73 |
| NEW MEXICO | 0.05 | 0.04 | 0.02 | 0.08 | 0.19 |
| NEW YORK | 1.78 | 1.57 | 2.02 | 1.37 | 6.74 |
| NORTH CAROLINA | 0.27 | 0.42 | 0.52 | 0.45 | 1.66 |
| NORTH DAKOTA | 0.02 | 0.06 | 0.02 | 0.03 | 0.13 |
| OHIO | 0.31 | 0.54 | 0.41 | 0.7 | 1.96 |
| OKLAHOMA | 0.18 | 0.17 | 0.16 | 0.17 | 0.68 |
| OREGON | 0.28 | 0.26 | 0.31 | 0.39 | 1.24 |
| PENNSYLVANIA | 0.59 | 1.97 | 4.31 | 1.25 | 8.12 |
| PUERTO RICO | 0.21 | 0.16 | 0.23 | 0.15 | 0.75 |
| RHODE ISLAND | 0 | 0.01 | 0 | 0 | 0.01 |
| SOUTH CAROLINA | 0.79 | 0.81 | 0.82 | 0.23 | 2.65 |
| SOUTH DAKOTA | 0.03 | 0.03 | 0.04 | 0.09 | 0.19 |

| | | | | | |
|---|---|---|---|---|---|
| TENNESSEE | 0.67 | 0.54 | 0.64 | 0.71 | 2.56 |
| TEXAS | 1.91 | 1.38 | 1.06 | 2.31 | 6.66 |
| UTAH | 0.26 | 0.21 | 0.21 | 0.24 | 0.92 |
| VERMONT | 0.01 | 0.01 | 0 | 0.01 | 0.03 |
| VIRGINIA | 0.41 | 0.36 | 0.34 | 0.44 | 1.55 |
| WASHINGTON | 0.24 | 0.21 | 0.32 | 0.27 | 1.04 |
| WEST VIRGINIA | 0.04 | 0.06 | 0.09 | 0.06 | 0.25 |
| WISCONSIN | 0.77 | 0.8 | 0.09 | 0.87 | 2.53 |
| WYOMING | 0 | 0 | 0.03 | 0.03 | 0.06 |
| TOTAL | 43.84 | 43.75 | 46.3 | 48.11 | 182 |

DRUG CODE:9780 DRUG NAME:TAPENTADOL

| STATE | QUARTER 1 | QUARTER 2 | QUARTER 3 | QUARTER 4 | TOTAL GRAMS |
|---|---|---|---|---|---|
| ALABAMA | 59,030.50 | 56,294.50 | 52,474.50 | 50,761.50 | 218,561 |
| ALASKA | 2,095.50 | 2,167.50 | 2,319 | 2,409 | 8,991 |
| ARIZONA | 38,185.50 | 35,746 | 32,238 | 35,953.50 | 142,123 |
| ARKANSAS | 23,293 | 22,390.50 | 19,058 | 19,193 | 83,934.50 |
| CALIFORNIA | 110,557.50 | 110,431.50 | 101,643.45 | 102,244.40 | 424,876.85 |
| COLORADO | 27,467 | 28,889 | 27,473 | 28,423.50 | 112,252.50 |
| CONNECTICUT | 12,657.50 | 11,895.50 | 10,938 | 10,558 | 46,049 |
| DELAWARE | 7,639 | 7,431.50 | 7,404 | 7,199.50 | 29,674 |
| DISTRICT OF COLUMBIA | 1,209.50 | 1,111 | 916 | 860 | 4,096.50 |
| FLORIDA | 109,424.50 | 103,022.50 | 92,250.50 | 91,559.50 | 396,257 |
| GEORGIA | 71,115 | 70,898 | 63,634.50 | 63,983 | 269,630.50 |
| HAWAII | 2,865 | 3,284 | 3,128.50 | 3,316.50 | 12,594 |
| IDAHO | 6,426 | 5,657.50 | 5,709 | 6,334 | 24,126.50 |
| ILLINOIS | 41,449.50 | 38,740 | 36,168.50 | 35,340.50 | 151,698.50 |
| INDIANA | 38,422.50 | 37,899.50 | 36,547.50 | 36,562 | 149,431.50 |
| IOWA | 13,973 | 12,921 | 10,709 | 12,098 | 49,701 |
| KANSAS | 9,528.50 | 8,789.50 | 8,288.50 | 8,435.50 | 35,042 |
| KENTUCKY | 15,161.50 | 15,153.50 | 15,330 | 14,489 | 60,134 |
| LOUISIANA | 15,303.50 | 15,024 | 14,597 | 15,866 | 60,790.50 |
| MAINE | 1,448.50 | 1,487.50 | 1,342 | 1,357.50 | 5,635.50 |
| MARYLAND | 34,304.50 | 33,980.50 | 31,173 | 30,546.50 | 130,004.50 |
| MASSACHUSETTS | 19,238.50 | 19,594.50 | 19,295.50 | 18,947.50 | 77,076 |
| MICHIGAN | 21,858.50 | 21,251 | 21,632 | 21,897.50 | 86,639 |

| | | | | | |
|---|---|---|---|---|---|
| MINNESOTA | 4,316.50 | 4,227 | 4,405 | 4,653 | 17,601.50 |
| MISSISSIPPI | 13,141.50 | 12,524.50 | 11,834.50 | 10,785 | 48,285.50 |
| MISSOURI | 25,851 | 25,232.50 | 23,086 | 23,396.50 | 97,566 |
| MONTANA | 2,201.50 | 1,668.50 | 1,758.50 | 1,806 | 7,434.50 |
| NEBRASKA | 10,446 | 9,342 | 9,231.50 | 9,800.50 | 38,820 |
| NEVADA | 12,593.50 | 12,164.50 | 11,660 | 12,387.50 | 48,805.50 |
| NEW HAMPSHIRE | 7,573.50 | 7,735.50 | 7,064.50 | 7,042 | 29,415.50 |
| NEW JERSEY | 48,769.50 | 48,076.50 | 46,248.50 | 46,131.50 | 189,226 |
| NEW MEXICO | 3,536 | 3,129.50 | 3,076 | 3,085 | 12,826.50 |
| NEW YORK | 99,988 | 97,696 | 94,081.50 | 92,119 | 383,884.50 |
| NORTH CAROLINA | 100,947 | 99,916 | 93,476.50 | 92,188 | 386,527.50 |
| NORTH DAKOTA | 2,548 | 2,511 | 2,298.50 | 2,418 | 9,775.50 |
| OHIO | 66,186.78 | 62,675.45 | 57,564.95 | 56,331.83 | 242,759.01 |
| OKLAHOMA | 24,810.50 | 24,718.50 | 22,840 | 22,548.50 | 94,917.50 |
| OREGON | 7,333.50 | 7,441 | 7,542 | 7,498.50 | 29,815 |
| PENNSYLVANIA | 67,481.50 | 68,673 | 63,793 | 64,523.50 | 264,471 |
| PUERTO RICO | 904.5 | 990 | 829 | 650.5 | 3,374 |
| RHODE ISLAND | 2,067.50 | 2,166.50 | 2,060 | 2,096 | 8,390 |
| SOUTH CAROLINA | 44,272 | 44,556 | 42,452 | 42,730 | 174,010 |
| SOUTH DAKOTA | 5,211.50 | 4,844 | 4,321 | 4,483.50 | 18,860 |
| TENNESSEE | 32,883 | 30,788.50 | 31,088 | 32,455.50 | 127,215 |
| TEXAS | 102,077.50 | 101,164 | 91,158.50 | 95,703 | 390,103 |
| UTAH | 15,597 | 13,584 | 13,460 | 12,249.50 | 54,890.50 |
| VERMONT | 1,341.50 | 1,096 | 1,038.50 | 1,144 | 4,620 |
| VIRGIN ISLANDS | 9 | 6 | 3 | 0 | 18 |
| VIRGINIA | 45,995 | 43,318.50 | 38,604.50 | 37,649.50 | 165,567.50 |
| WASHINGTON | 18,321 | 18,426 | 16,944.50 | 16,659 | 70,350.50 |
| WEST VIRGINIA | 6,283 | 6,593 | 6,057.50 | 5,611 | 24,544.50 |
| WISCONSIN | 15,498.50 | 15,413 | 14,565 | 13,998.50 | 59,475 |
| WYOMING | 1,948 | 2,073.50 | 1,797 | 1,947.50 | 7,766 |
| TOTAL | 1,472,787.28 | 1,436,810.45 | 1,338,608.90 | 1,342,427.23 | 5,590,633.86 |

DRUG CODE:9801 DRUG NAME:FENTANYL BASE

| STATE | QUARTER 1 | QUARTER 2 | QUARTER 3 | QUARTER 4 | TOTAL GRAMS |
|---|---|---|---|---|---|
| ALABAMA | 1,620.56 | 1,543.32 | 1,423.48 | 1,375.76 | 5,963.12 |
| ALASKA | 239.88 | 239.41 | 233.05 | 228.27 | 940.61 |

| | | | | | |
|---|---|---|---|---|---|
| AMERICAN SAMOA | 0.24 | 0.23 | 0 | 0 | 0.47 |
| ARIZONA | 1,977.94 | 1,803.41 | 1,626.13 | 1,585.90 | 6,993.38 |
| ARKANSAS | 1,010.45 | 976.38 | 886.88 | 841.48 | 3,715.19 |
| CALIFORNIA | 7,394.64 | 7,142.92 | 6,794.64 | 6,483.09 | 27,815.29 |
| COLORADO | 1,362.25 | 1,272.08 | 1,216.64 | 1,159.62 | 5,010.59 |
| CONNECTICUT | 1,006.18 | 989.28 | 924.2 | 880.94 | 3,800.60 |
| DELAWARE | 358.26 | 350.22 | 324.76 | 327.37 | 1,360.61 |
| DISTRICT OF COLUMBIA | 92.41 | 90.16 | 80.36 | 75.83 | 338.76 |
| FLORIDA | 6,371.59 | 5,883.81 | 5,472.76 | 5,624.55 | 23,352.71 |
| GEORGIA | 2,933.02 | 2,791.81 | 2,619.49 | 2,563.56 | 10,907.88 |
| GUAM | 12.2 | 10.02 | 9.17 | 9.95 | 41.34 |
| HAWAII | 198.18 | 193.78 | 175.2 | 175.15 | 742.31 |
| IDAHO | 594.55 | 589.21 | 565.65 | 555.48 | 2,304.89 |
| ILLINOIS | 3,123.72 | 2,990.38 | 2,880.90 | 2,814.15 | 11,809.15 |
| INDIANA | 2,040.79 | 1,922.07 | 1,792.60 | 1,768.11 | 7,523.57 |
| IOWA | 908.05 | 886.91 | 845.88 | 835.7 | 3,476.54 |
| KANSAS | 1,140.04 | 1,121.48 | 1,078.58 | 1,121.86 | 4,461.96 |
| KENTUCKY | 1,232.80 | 1,174.90 | 1,099.14 | 1,060.15 | 4,566.99 |
| LOUISIANA | 1,301.35 | 1,216.49 | 1,153.84 | 1,083.95 | 4,755.63 |
| MAINE | 388.28 | 330.7 | 303.9 | 299.95 | 1,322.83 |
| MARYLAND | 1,552.80 | 1,416.82 | 1,344.99 | 1,237.78 | 5,552.39 |
| MASSACHUSETTS | 1,389.85 | 1,337.33 | 1,266.41 | 1,235.44 | 5,229.03 |
| MICHIGAN | 3,295.49 | 3,185.63 | 3,016.89 | 2,877.98 | 12,375.99 |
| MINNESOTA | 1,080.41 | 1,069.20 | 1,000.89 | 965.18 | 4,115.68 |
| MISSISSIPPI | 1,253.09 | 1,311.71 | 1,214.16 | 1,143.68 | 4,922.64 |
| MISSOURI | 2,258.79 | 2,199.65 | 2,102.82 | 2,523.82 | 9,085.08 |
| MONTANA | 353.98 | 336.62 | 315.94 | 311.55 | 1,318.09 |
| NEBRASKA | 561.72 | 540.17 | 520.25 | 506.88 | 2,129.02 |
| NEVADA | 656.33 | 599.66 | 559.53 | 538.39 | 2,353.91 |
| NEW HAMPSHIRE | 372.07 | 351.19 | 340.42 | 327.42 | 1,391.10 |
| NEW JERSEY | 2,496.43 | 2,399.25 | 2,171.39 | 2,111.68 | 9,178.75 |
| NEW MEXICO | 486.16 | 446.12 | 410.05 | 392.71 | 1,735.04 |
| NEW YORK | 4,661.14 | 4,468.97 | 4,112.05 | 3,995.21 | 17,237.37 |
| NORTH CAROLINA | 3,098.54 | 2,916.55 | 2,645.07 | 2,606.06 | 11,266.22 |
| NORTH DAKOTA | 248.16 | 240.41 | 230.75 | 228.56 | 947.88 |
| OHIO | 2,651.77 | 2,544.49 | 2,386.96 | 2,266.99 | 9,850.21 |
| OKLAHOMA | 1,847.46 | 1,730.28 | 1,633.57 | 1,543.84 | 6,755.15 |

| | | | | | |
|---|---|---|---|---|---|
| OREGON | 1,006.51 | 954.7 | 886.81 | 857.63 | 3,705.65 |
| PENNSYLVANIA | 5,374.35 | 5,210.59 | 4,851.08 | 4,630.51 | 20,066.53 |
| PUERTO RICO | 196.87 | 151.01 | 126.57 | 103.8 | 578.25 |
| RHODE ISLAND | 197.8 | 186.79 | 177.75 | 172.64 | 734.98 |
| SOUTH CAROLINA | 1,424.02 | 1,346.42 | 1,259.11 | 1,231.99 | 5,261.54 |
| SOUTH DAKOTA | 325.31 | 301.89 | 295.79 | 281.25 | 1,204.24 |
| TENNESSEE | 2,093.91 | 1,965.15 | 1,845.64 | 1,784.70 | 7,689.40 |
| TEXAS | 6,128.66 | 5,913.30 | 5,543.02 | 5,441.14 | 23,026.12 |
| UTAH | 862.83 | 820.99 | 805.92 | 785.81 | 3,275.55 |
| VERMONT | 202.75 | 196.73 | 180.15 | 168.44 | 748.07 |
| VIRGIN ISLANDS | 4.01 | 3.46 | 3.02 | 3.47 | 13.96 |
| VIRGINIA | 1,921.32 | 1,781.64 | 1,643.32 | 1,567.23 | 6,913.51 |
| WASHINGTON | 1,614.60 | 1,556.54 | 1,458.94 | 1,402.32 | 6,032.40 |
| WEST VIRGINIA | 577.65 | 559.14 | 530.84 | 499.87 | 2,167.50 |
| WISCONSIN | 1,637.47 | 1,524.37 | 1,390.97 | 1,345.32 | 5,898.13 |
| WYOMING | 168.25 | 149.5 | 158.02 | 145.2 | 620.97 |
| TOTAL | 87,307.88 | 83,235.24 | 77,936.34 | 76,105.31 | 324,584.77 |

DRUG CODE:9809 DRUG NAME:OPIUM COMBINATION PRODUCT (C-III)

| STATE | QUARTER 1 | QUARTER 2 | QUARTER 3 | QUARTER 4 | TOTAL GRAMS |
|---|---|---|---|---|---|
| ALABAMA | 11.92 | 12.68 | 11.73 | 10.78 | 47.11 |
| ALASKA | 0 | 0 | 0.19 | 0 | 0.19 |
| ARIZONA | 12.11 | 5.49 | 7.19 | 8.51 | 33.3 |
| ARKANSAS | 1.32 | 2.27 | 2.84 | 2.08 | 8.51 |
| CALIFORNIA | 18.16 | 17.97 | 17.97 | 22.51 | 76.61 |
| COLORADO | 3.22 | 3.03 | 2.27 | 2.46 | 10.98 |
| CONNECTICUT | 5.11 | 3.78 | 6.43 | 4.16 | 19.48 |
| DELAWARE | 0.19 | 0.76 | 0.19 | 0.95 | 2.09 |
| DISTRICT OF COLUMBIA | 0.19 | 0.38 | 0.38 | 0.57 | 1.52 |
| FLORIDA | 36.52 | 38.03 | 31.79 | 31.41 | 137.75 |
| GEORGIA | 6.05 | 8.7 | 7.57 | 7.19 | 29.51 |
| HAWAII | 1.89 | 1.51 | 1.32 | 0 | 4.72 |
| IDAHO | 3.97 | 3.78 | 3.59 | 3.78 | 15.12 |
| ILLINOIS | 6.05 | 6.05 | 6.62 | 5.11 | 23.83 |
| INDIANA | 6.81 | 6.81 | 8.32 | 7.19 | 29.13 |
| IOWA | 3.22 | 1.89 | 1.7 | 2.08 | 8.89 |

| | | | | | |
|---|---|---|---|---|---|
| KANSAS | 0.76 | 0.57 | 0.19 | 1.32 | 2.84 |
| KENTUCKY | 10.59 | 10.97 | 7.76 | 7.75 | 37.07 |
| LOUISIANA | 7.19 | 8.7 | 9.27 | 8.14 | 33.3 |
| MAINE | 0.76 | 1.14 | 2.08 | 2.84 | 6.82 |
| MARYLAND | 8.51 | 8.7 | 8.89 | 7.95 | 34.05 |
| MASSACHUSETTS | 2.08 | 2.46 | 1.32 | 0.95 | 6.81 |
| MICHIGAN | 4.73 | 4.92 | 8.51 | 4.54 | 22.7 |
| MINNESOTA | 1.7 | 2.27 | 2.08 | 3.03 | 9.08 |
| MISSISSIPPI | 4.54 | 2.46 | 3.03 | 1.7 | 11.73 |
| MISSOURI | 10.22 | 5.11 | 3.97 | 9.27 | 28.57 |
| MONTANA | 0.95 | 0.19 | 1.32 | 0.95 | 3.41 |
| NEBRASKA | 1.14 | 0.57 | 0.76 | 0.57 | 3.04 |
| NEVADA | 0.76 | 1.32 | 1.7 | 0.38 | 4.16 |
| NEW HAMPSHIRE | 1.7 | 1.89 | 1.89 | 3.41 | 8.89 |
| NEW JERSEY | 5.68 | 5.68 | 5.3 | 4.16 | 20.82 |
| NEW MEXICO | 1.89 | 1.89 | 0.95 | 1.89 | 6.62 |
| NEW YORK | 8.32 | 10.41 | 8.51 | 10.78 | 38.02 |
| NORTH CAROLINA | 12.11 | 14.76 | 15.14 | 11.92 | 53.93 |
| NORTH DAKOTA | 0 | 0.38 | 0.76 | 1.51 | 2.65 |
| OHIO | 14.95 | 13.81 | 11.14 | 18.35 | 58.25 |
| OKLAHOMA | 3.22 | 2.08 | 3.22 | 4.35 | 12.87 |
| OREGON | 0.19 | 0.57 | 0.95 | 1.89 | 3.6 |
| PENNSYLVANIA | 31.41 | 38.97 | 27.24 | 38.03 | 135.65 |
| RHODE ISLAND | 0 | 0.19 | 0.19 | 0 | 0.38 |
| SOUTH CAROLINA | 8.32 | 10.78 | 10.78 | 10.22 | 40.1 |
| SOUTH DAKOTA | 1.32 | 0.57 | 0.57 | 0.19 | 2.65 |
| TENNESSEE | 5.68 | 5.49 | 5.68 | 8.14 | 24.99 |
| TEXAS | 11.35 | 14.19 | 13.06 | 14.38 | 52.98 |
| UTAH | 13.05 | 12.68 | 10.97 | 12.87 | 49.57 |
| VERMONT | 0 | 0.38 | 0.19 | 0.38 | 0.95 |
| VIRGINIA | 7.95 | 5.49 | 5.68 | 4.73 | 23.85 |
| WASHINGTON | 2.08 | 2.08 | 4.16 | 2.08 | 10.4 |
| WEST VIRGINIA | 2.65 | 1.89 | 2.46 | 2.65 | 9.65 |
| WISCONSIN | 5.68 | 4.92 | 5.49 | 7 | 23.09 |
| WYOMING | 0 | 0.57 | 0.76 | 1.7 | 3.03 |
| TOTAL | 308.21 | 312.18 | 296.07 | 318.8 | 1,235.26 |

DRUG CODE:1100 DRUG NAME: AMPHETAMINE

| STATE | 1ST QUARTER | 2ND QUARTER | 3RD QUARTER | 4TH QUARTER | TOTAL GRAMS |
|---|---|---|---|---|---|
| ALABAMA | 2,709.83 | 2,703.03 | 2,707.26 | 2,801.46 | 10,921.58 |
| ALASKA | 1,128.88 | 1,131.40 | 1,153.17 | 1,139.34 | 4,552.78 |
| AMERICAN SAMOA | 1.91 | 1.91 | 0 | 0 | 3.81 |
| ARIZONA | 1,272.08 | 1,254.45 | 1,254.68 | 1,343.46 | 5,124.67 |
| ARKANSAS | 1,735.47 | 1,733.65 | 1,745.72 | 1,793.99 | 7,008.83 |
| CALIFORNIA | 793.33 | 809.42 | 794.97 | 818.4 | 3,216.12 |
| COLORADO | 1,550.18 | 1,566.11 | 1,597.28 | 1,638.41 | 6,351.99 |
| CONNECTICUT | 1,435.10 | 1,476.24 | 1,477.48 | 1,522.39 | 5,911.21 |
| DELAWARE | 2,409.37 | 2,394.58 | 2,360.97 | 2,435.11 | 9,600.03 |
| DISTRICT OF COLUMBIA | 1,922.13 | 1,934.53 | 1,911.95 | 1,986.93 | 7,755.54 |
| FLORIDA | 1,579.12 | 1,585.03 | 1,549.21 | 1,675.59 | 6,388.94 |
| GEORGIA | 1,924.97 | 1,906.68 | 1,923.35 | 1,989.53 | 7,744.54 |
| GUAM | 230.78 | 313.57 | 321.81 | 200.99 | 1,067.15 |
| HAWAII | 703.46 | 722.96 | 730.72 | 704 | 2,861.14 |
| IDAHO | 1,569.83 | 1,610.19 | 1,616.22 | 1,695.58 | 6,491.82 |
| ILLINOIS | 1,470.92 | 1,468.19 | 1,478.17 | 1,536.95 | 5,954.24 |
| INDIANA | 1,969.80 | 1,985.51 | 2,004.29 | 2,060.24 | 8,019.84 |
| IOWA | 1,589.95 | 1,621.43 | 1,635.31 | 1,710.99 | 6,557.68 |
| KANSAS | 2,055.32 | 2,098.99 | 2,126.10 | 2,193.07 | 8,473.49 |
| KENTUCKY | 1,569.71 | 1,533.77 | 1,550.52 | 1,612.99 | 6,266.99 |
| LOUISIANA | 3,371.44 | 3,317.67 | 3,371.01 | 3,563.48 | 13,623.60 |
| MAINE | 1,612.51 | 1,714.70 | 1,765.68 | 1,733 | 6,825.88 |
| MARYLAND | 1,561.96 | 1,578.02 | 1,576.86 | 1,626.98 | 6,343.81 |
| MASSACHUSETTS | 2,672.65 | 2,744.79 | 2,683.28 | 2,771.29 | 10,872.01 |
| MICHIGAN | 2,417.01 | 2,418.33 | 2,420.37 | 2,476.96 | 9,732.66 |
| MINNESOTA | 2,010.59 | 2,021.51 | 2,012.53 | 2,093.70 | 8,138.34 |
| MISSISSIPPI | 2,109.79 | 2,037.08 | 2,036.78 | 2,107.99 | 8,291.64 |
| MISSOURI | 2,201.95 | 2,206.77 | 2,235.89 | 2,315.20 | 8,959.82 |
| MONTANA | 1,375.11 | 1,402.69 | 1,431.13 | 1,450.64 | 5,659.57 |
| NEBRASKA | 1,386.55 | 1,381.19 | 1,424.57 | 1,464.70 | 5,657.01 |
| NEVADA | 944.07 | 962.19 | 961.67 | 988.6 | 3,856.53 |
| NEW HAMPSHIRE | 2,830.94 | 2,942.84 | 2,922.67 | 2,991.56 | 11,688.01 |
| NEW JERSEY | 1,159.81 | 1,179.10 | 1,177.12 | 1,223.32 | 4,739.35 |
| NEW MEXICO | 815.96 | 788 | 758.68 | 728.9 | 3,091.54 |

DRUG CODE:1100 DRUG NAME: AMPHETAMINE

| STATE | 1ST QUARTER | 2ND QUARTER | 3RD QUARTER | 4TH QUARTER | TOTAL GRAMS |
|---|---|---|---|---|---|
| NEW YORK | 1,178.54 | 1,216.31 | 1,204.59 | 1,260.34 | 4,859.78 |
| NORTH CAROLINA | 2,106.18 | 2,116.36 | 2,111 | 2,187.59 | 8,521.14 |
| NORTH DAKOTA | 1,309.70 | 1,321.29 | 1,348.64 | 1,367.49 | 5,347.13 |
| OHIO | 1,802.47 | 1,804.72 | 1,819.55 | 1,915.35 | 7,342.08 |
| OKLAHOMA | 1,393.76 | 1,401.39 | 1,399.93 | 1,445.69 | 5,640.76 |
| OREGON | 1,492.68 | 1,547.15 | 1,528.79 | 1,554.06 | 6,122.67 |
| PENNSYLVANIA | 1,641.45 | 1,663.06 | 1,644.10 | 1,698.46 | 6,647.06 |
| PUERTO RICO | 243.62 | 226.59 | 210.57 | 208.03 | 888.81 |
| RHODE ISLAND | 3,361.76 | 3,465.93 | 3,501.38 | 3,593 | 13,922.07 |
| SOUTH CAROLINA | 2,954.61 | 2,917.70 | 2,895.70 | 3,019.02 | 11,787.03 |
| SOUTH DAKOTA | 1,963.14 | 1,832 | 1,800.07 | 1,873.93 | 7,469.14 |
| TENNESSEE | 2,033.07 | 2,024.64 | 2,028.33 | 2,114.88 | 8,200.92 |
| TEXAS | 1,620.45 | 1,634.58 | 1,621.43 | 1,708.64 | 6,585.10 |
| UTAH | 2,866 | 2,931.71 | 2,957.75 | 3,100.55 | 11,856.01 |
| VERMONT | 1,566.15 | 1,583.62 | 1,563.11 | 1,629.91 | 6,342.78 |
| VIRGIN ISLANDS | 335.82 | 331.61 | 228.68 | 212.23 | 1,108.34 |
| VIRGINIA | 2,214.20 | 2,229.25 | 2,207.16 | 2,274.29 | 8,924.90 |
| WASHINGTON | 1,594.11 | 1,632.04 | 1,609.75 | 1,638.92 | 6,474.81 |
| WEST VIRGINIA | 1,287.39 | 1,255.65 | 1,253.30 | 1,275.06 | 5,071.40 |

| | | | | | |
|---|---|---|---|---|---|
| WISCONSIN | 2,285.68 | 2,226.39 | 2,206.22 | 2,264.20 | 8,982.49 |
| WYOMING | 1,172.74 | 1,105.72 | 1,137.44 | 1,150.24 | 4,566.14 |
| U.S. TOTAL | 1,652.28 | 1,662.14 | 1,657.05 | 1,721.82 | 6,693.29 |

DRUG CODE:1105B DRUG NAME: DL-METHAMPHETAMINE RACEMIC BASE

| STATE | 1ST QUARTER | 2ND QUARTER | 3RD QUARTER | 4TH QUARTER | TOTAL GRAMS |
|---|---|---|---|---|---|
| ALABAMA | 0.47 | 0.51 | 0.58 | 0.45 | 2.01 |
| ALASKA | 0.11 | 0.68 | 0.57 | 0.17 | 1.53 |
| ARIZONA | 0.53 | 0.5 | 0.45 | 0.62 | 2.1 |
| ARKANSAS | 0.18 | 0.25 | 0.23 | 0.21 | 0.87 |
| CALIFORNIA | 0.86 | 0.83 | 0.73 | 0.67 | 3.09 |
| COLORADO | 0.97 | 1.08 | 0.99 | 1.14 | 4.19 |
| CONNECTICUT | 0.57 | 0.69 | 0.56 | 0.69 | 2.51 |
| DELAWARE | 0.27 | 0.4 | 0.54 | 0.49 | 1.7 |
| DISTRICT OF COLUMBIA | 0.67 | 0.33 | 0 | 0.13 | 1.14 |
| FLORIDA | 0.15 | 0.21 | 0.19 | 0.17 | 0.72 |
| GEORGIA | 0.33 | 0.25 | 0.18 | 0.21 | 0.98 |
| HAWAII | 0.38 | 0.5 | 0.3 | 0.32 | 1.51 |
| IDAHO | 0.49 | 0.44 | 0.67 | 0.64 | 2.23 |
| ILLINOIS | 0.5 | 0.45 | 0.49 | 0.49 | 1.94 |
| INDIANA | 0.46 | 0.4 | 0.46 | 0.28 | 1.6 |
| IOWA | 0.49 | 0.62 | 0.62 | 0.7 | 2.43 |
| KANSAS | 0.38 | 0.44 | 0.28 | 0.37 | 1.46 |
| KENTUCKY | 0.25 | 0.23 | 0.22 | 0.21 | 0.92 |
| LOUISIANA | 0.31 | 0.33 | 0.31 | 0.27 | 1.21 |
| MAINE | 0.24 | 0.51 | 0.79 | 0.97 | 2.51 |
| MARYLAND | 0.72 | 0.77 | 0.66 | 0.7 | 2.85 |
| MASSACHUSETTS | 0.68 | 0.63 | 0.66 | 0.6 | 2.57 |
| MICHIGAN | 0.23 | 0.23 | 0.22 | 0.25 | 0.93 |
| MINNESOTA | 1.48 | 1.64 | 1.2 | 1.13 | 5.46 |
| MISSISSIPPI | 0.28 | 0.27 | 0.3 | 0.26 | 1.11 |
| MISSOURI | 0.82 | 1.04 | 0.27 | 1.05 | 3.17 |
| MONTANA | 0.12 | 0.08 | 0.04 | 0.04 | 0.28 |
| NEBRASKA | 0.33 | 0.48 | 0.48 | 0.51 | 1.8 |
| NEVADA | 0.88 | 1.09 | 0.82 | 0.83 | 3.62 |
| NEW HAMPSHIRE | 0.61 | 0.58 | 0.52 | 0.27 | 1.98 |
| NEW JERSEY | 0.43 | 0.35 | 0.36 | 0.43 | 1.57 |
| NEW MEXICO | 1.33 | 1.01 | 0.62 | 0.23 | 3.2 |
| NEW YORK | 0.38 | 0.42 | 0.4 | 0.48 | 1.68 |
| NORTH CAROLINA | 0.5 | 0.59 | 0.44 | 0.45 | 1.98 |
| NORTH DAKOTA | 0.42 | 1.14 | 0.9 | 1.08 | 3.53 |
| OHIO | 0.67 | 0.7 | 0.42 | 0.81 | 2.6 |
| OKLAHOMA | 0.63 | 0.44 | 0.55 | 0.44 | 2.06 |
| OREGON | 1.15 | 0.86 | 0.5 | 0.82 | 3.34 |
| PENNSYLVANIA | 0.93 | 1.06 | 0.91 | 0.95 | 3.85 |
| RHODE ISLAND | 0.46 | 0.53 | 0.27 | 0.34 | 1.6 |
| SOUTH CAROLINA | 0.4 | 0.26 | 0.36 | 0.29 | 1.3 |
| SOUTH DAKOTA | 1.92 | 2.71 | 2.12 | 1.78 | 8.54 |
| TENNESSEE | 0.16 | 0.2 | 0.2 | 0.24 | 0.8 |
| TEXAS | 0.46 | 0.46 | 0.36 | 0.41 | 1.7 |
| UTAH | 0.92 | 0.84 | 0.96 | 1.05 | 3.77 |
| VERMONT | 0.13 | 0.32 | 0.32 | 0.39 | 1.16 |
| VIRGINIA | 0.64 | 0.51 | 0.5 | 0.55 | 2.19 |
| WASHINGTON | 1.35 | 1.42 | 1.36 | 1.54 | 5.67 |
| WEST VIRGINIA | 0.17 | 0.24 | 0.3 | 0.17 | 0.89 |
| WISCONSIN | 1.08 | 1.1 | 0.74 | 0.55 | 3.47 |
| WYOMING | 1.07 | 1.28 | 0.86 | 0.57 | 3.78 |
| U.S. TOTAL | 0.58 | 0.6 | 0.51 | 0.54 | 2.23 |

DRUG CODE:1105D DRUG NAME: D-METHAMPHETAMINE

| STATE | 1ST QUARTER | 2ND QUARTER | 3RD QUARTER | 4TH QUARTER | TOTAL GRAMS |
|---|---|---|---|---|---|
| ALABAMA | 0 | 0.02 | 0.14 | 0.15 | 0.31 |
| ALASKA | 0 | 0 | 0.11 | 0 | 0.11 |
| ARIZONA | 0 | 0.07 | 0.06 | 0.18 | 0.31 |
| CALIFORNIA | 0 | 0.06 | 0.16 | 0.16 | 0.38 |
| CONNECTICUT | 0 | 0 | 0.1 | 0.12 | 0.22 |
| DISTRICT OF COLUMBIA | 0 | 0.13 | 0.47 | 0.27 | 0.87 |
| FLORIDA | 0 | 0.03 | 0.02 | 0.02 | 0.07 |
| GEORGIA | 0 | 0.02 | 0.05 | 0.05 | 0.12 |
| HAWAII | 0 | 0 | 0.12 | 0.12 | 0.24 |
| IDAHO | 0 | 0.08 | 0.33 | 0.26 | 0.67 |
| ILLINOIS | 0 | 0 | 0.03 | 0.02 | 0.05 |
| KANSAS | 0 | 0.03 | 0.03 | 0.1 | 0.15 |
| KENTUCKY | 0 | 0.01 | 0.04 | 0.04 | 0.08 |
| LOUISIANA | 0 | 0 | 0.04 | 0.05 | 0.1 |
| MARYLAND | 0 | 0 | 0.02 | 0.06 | 0.08 |
| MASSACHUSETTS | 0 | 0.01 | 0.06 | 0.04 | 0.1 |
| MINNESOTA | 0 | 0.08 | 0.48 | 0.43 | 1 |
| MISSISSIPPI | 0 | 0 | 0.05 | 0.03 | 0.08 |
| MISSOURI | 0 | 0.05 | 0.2 | 0.28 | 0.54 |
| NEBRASKA | 0 | 0 | 0 | 0.04 | 0.04 |
| NEVADA | 0 | 0 | 0.01 | 0 | 0.01 |
| NEW JERSEY | 0 | 0.01 | 0.03 | 0.02 | 0.06 |
| NEW YORK | 0.02 | 0 | 0.04 | 0.04 | 0.09 |
| NORTH CAROLINA | 0 | 0.02 | 0.1 | 0.11 | 0.23 |
| NORTH DAKOTA | 0 | 0 | 0.84 | 0.06 | 0.9 |
| OHIO | 0 | 0 | 0.07 | 0.11 | 0.18 |
| OKLAHOMA | 0 | 0.01 | 0.11 | 0.07 | 0.19 |
| OREGON | 0 | 0.24 | 0.37 | 0.38 | 0.99 |
| PENNSYLVANIA | 0 | 0.03 | 0.09 | 0.09 | 0.21 |
| RHODE ISLAND | 0 | 0 | 0.15 | 0.19 | 0.34 |
| SOUTH CAROLINA | 0 | 0.04 | 0.09 | 0.06 | 0.19 |
| TENNESSEE | 0 | 0 | 0 | 0.01 | 0.01 |
| TEXAS | 0 | 0.02 | 0.07 | 0.07 | 0.16 |
| WASHINGTON | 0 | 0 | 0.01 | 0.02 | 0.04 |
| WEST VIRGINIA | 0 | 0 | 0.13 | 0.07 | 0.2 |
| WISCONSIN | 0 | 0.03 | 0.24 | 0.23 | 0.49 |
| U.S. TOTAL | 0 | 0.02 | 0.08 | 0.08 | 0.18 |

DRUG CODE:1205 DRUG NAME: LISDEXAMFETAMINE

| STATE | 1ST QUARTER | 2ND QUARTER | 3RD QUARTER | 4TH QUARTER | TOTAL GRAMS |
|---|---|---|---|---|---|
| ALABAMA | 1,529.88 | 1,443.77 | 1,459.71 | 1,536.11 | 5,969.47 |
| ALASKA | 333.24 | 339.83 | 356.84 | 359.2 | 1,389.11 |
| ARIZONA | 557.77 | 539.98 | 548.83 | 596.54 | 2,243.12 |
| ARKANSAS | 1,095.19 | 985.37 | 994.79 | 1,081.26 | 4,156.62 |
| CALIFORNIA | 298.66 | 300.04 | 292.03 | 307.24 | 1,197.97 |
| COLORADO | 543.21 | 544.73 | 546.72 | 573.1 | 2,207.76 |
| CONNECTICUT | 1,005.74 | 1,022.02 | 976.56 | 1,045.70 | 4,050.02 |
| DELAWARE | 1,602.57 | 1,584.87 | 1,530.53 | 1,654.08 | 6,372.06 |
| DISTRICT OF COLUMBIA | 852.08 | 858.61 | 874.37 | 906.56 | 3,491.62 |
| FLORIDA | 765.77 | 718.48 | 684.35 | 750.35 | 2,918.95 |
| GEORGIA | 1,090.16 | 1,020.96 | 1,043.69 | 1,072.98 | 4,227.79 |
| GUAM | 21.06 | 15.23 | 6.8 | 7.78 | 50.87 |
| HAWAII | 99.15 | 95.92 | 97.92 | 100.93 | 393.91 |

| | | | | | |
|---|---|---|---|---|---|
| IDAHO | 541.74 | 558.12 | 560.26 | 597.95 | 2,258.07 |
| ILLINOIS | 554.8 | 541.54 | 541.43 | 571.32 | 2,209.09 |
| INDIANA | 1,331.93 | 1,287.51 | 1,295.01 | 1,350.51 | 5,264.95 |
| IOWA | 1,155.06 | 1,116.94 | 1,092.28 | 1,153.90 | 4,518.18 |
| KANSAS | 1,107.71 | 1,091.78 | 1,087.04 | 1,170.09 | 4,456.62 |
| KENTUCKY | 1,013.46 | 952.51 | 956.06 | 987.1 | 3,909.12 |
| LOUISIANA | 2,161.99 | 1,956.50 | 1,979.78 | 2,093.42 | 8,191.69 |
| MAINE | 1,226.04 | 1,266.21 | 1,278.31 | 1,296.63 | 5,067.18 |
| MARYLAND | 983.29 | 966.38 | 925.7 | 994.98 | 3,870.35 |
| MASSACHUSETTS | 543.21 | 553.19 | 522.68 | 563.35 | 2,182.43 |
| MICHIGAN | 609.83 | 593.69 | 579.12 | 623.89 | 2,406.53 |
| MINNESOTA | 599.63 | 608.02 | 599.7 | 581.06 | 2,388.41 |
| MISSISSIPPI | 1,373 | 1,250.81 | 1,275.98 | 1,354.86 | 5,254.65 |
| MISSOURI | 1,133.92 | 1,082.33 | 1,086.42 | 1,142.41 | 4,445.08 |
| MONTANA | 590.71 | 559.45 | 562.84 | 602.51 | 2,315.52 |
| NEBRASKA | 898.89 | 871.61 | 880.23 | 950.23 | 3,600.96 |
| NEVADA | 288.82 | 279.64 | 266.87 | 285.87 | 1,121.20 |
| NEW HAMPSHIRE | 1,059.75 | 1,073.98 | 1,030.37 | 1,062.21 | 4,226.31 |
| NEW JERSEY | 558.05 | 566.71 | 539.77 | 564.34 | 2,228.88 |
| NEW MEXICO | 228.81 | 212.16 | 199.19 | 198.04 | 838.19 |
| NEW YORK | 432.76 | 438.07 | 422.87 | 452.7 | 1,746.40 |
| NORTH CAROLINA | 1,094.87 | 1,063.29 | 1,026.66 | 1,120.92 | 4,305.73 |
| NORTH DAKOTA | 881.95 | 820.15 | 831.49 | 870 | 3,403.58 |
| OHIO | 999.79 | 946.66 | 939.43 | 984.57 | 3,870.44 |
| OKLAHOMA | 989.38 | 941.13 | 956.09 | 1,023.38 | 3,909.98 |
| OREGON | 387.9 | 393.78 | 383.49 | 404.04 | 1,569.21 |
| PENNSYLVANIA | 678.56 | 669.46 | 657.93 | 699.06 | 2,705.01 |
| PUERTO RICO | 56.91 | 50.07 | 46.44 | 50.71 | 204.13 |
| RHODE ISLAND | 706.75 | 729.21 | 681.37 | 743.44 | 2,860.77 |
| SOUTH CAROLINA | 1,358.05 | 1,277.89 | 1,255.28 | 1,393.36 | 5,284.58 |
| SOUTH DAKOTA | 1,406.94 | 1,338.71 | 1,291.49 | 1,346.87 | 5,384.02 |
| TENNESSEE | 1,141.10 | 1,072.97 | 1,081.63 | 1,125.90 | 4,421.60 |
| TEXAS | 1,123.74 | 1,075.01 | 1,027.32 | 1,104.62 | 4,330.69 |
| UTAH | 774.21 | 763.84 | 763.9 | 831.52 | 3,133.48 |
| VERMONT | 1,098.75 | 1,085.54 | 1,084.62 | 1,158.53 | 4,427.44 |
| VIRGIN ISLANDS | 71.59 | 62.11 | 43.69 | 66.32 | 243.72 |
| VIRGINIA | 862.86 | 851.94 | 813.36 | 858.68 | 3,386.84 |
| WASHINGTON | 497.86 | 511.57 | 501.26 | 516.39 | 2,027.09 |
| WEST VIRGINIA | 1,107.44 | 1,045.04 | 1,018 | 1,070.63 | 4,241.11 |
| WISCONSIN | 1,143.79 | 1,157.28 | 1,150.43 | 1,253.13 | 4,704.63 |
| WYOMING | 479.3 | 453.97 | 455.71 | 465.15 | 1,854.12 |
| U.S. TOTAL | 791.4 | 765.19 | 751.87 | 798.98 | 3,107.44 |

DRUG CODE:1724 DRUG NAME: METHYLPHENIDATE (DL;D;L;ISOMERS)

| STATE | 1ST QUARTER | 2ND QUARTER | 3RD QUARTER | 4TH QUARTER | TOTAL GRAMS |
|---|---|---|---|---|---|
| ALABAMA | 1,599.29 | 1,463.24 | 1,449.31 | 1,573.62 | 6,085.47 |
| ALASKA | 1,169.80 | 1,117.76 | 1,100.06 | 1,137.53 | 4,525.14 |
| AMERICAN SAMOA | 3.95 | 9.22 | 6.59 | 7.25 | 27.01 |
| ARIZONA | 1,202.18 | 1,122.35 | 1,124.10 | 1,215.31 | 4,663.95 |
| ARKANSAS | 1,465.69 | 1,333.56 | 1,302.70 | 1,388.63 | 5,490.58 |
| CALIFORNIA | 751.65 | 729.56 | 696.22 | 743.02 | 2,920.45 |
| COLORADO | 1,251.92 | 1,201.21 | 1,184.30 | 1,261.87 | 4,899.30 |
| CONNECTICUT | 1,557.16 | 1,539.07 | 1,413.85 | 1,542.25 | 6,052.32 |
| DELAWARE | 2,163.05 | 2,105.83 | 1,966 | 2,128.29 | 8,363.17 |
| DISTRICT OF COLUMBIA | 1,354.35 | 1,375.41 | 1,254.23 | 1,367.31 | 5,351.30 |
| FLORIDA | 1,184.74 | 1,094.50 | 983.42 | 1,113.49 | 4,376.15 |
| GEORGIA | 1,811.68 | 1,592.83 | 1,614.50 | 1,728.81 | 6,747.82 |
| GUAM | 315.01 | 221 | 355.18 | 181.93 | 1,073.11 |
| HAWAII | 800.82 | 803.72 | 792.68 | 810.63 | 3,207.84 |

| | | | | | |
|---|---|---|---|---|---|
| IDAHO | 1,785.04 | 1,771.65 | 1,645.65 | 1,788.49 | 6,990.84 |
| ILLINOIS | 1,426.76 | 1,333.81 | 1,280.80 | 1,385.53 | 5,426.90 |
| INDIANA | 1,981.94 | 1,835.66 | 1,796.19 | 1,871 | 7,484.79 |
| IOWA | 2,800.12 | 2,669.23 | 2,531.41 | 2,703.07 | 10,703.82 |
| KANSAS | 1,834.12 | 1,739.26 | 1,675.86 | 1,769.51 | 7,018.75 |
| KENTUCKY | 1,842.60 | 1,649.18 | 1,669.02 | 1,756.51 | 6,917.32 |
| LOUISIANA | 2,056.13 | 1,739.80 | 1,747.81 | 1,927.97 | 7,471.71 |
| MAINE | 2,688.19 | 2,689.41 | 2,605.51 | 2,648.39 | 10,631.51 |
| MARYLAND | 1,813.38 | 1,750.26 | 1,612.78 | 1,755.17 | 6,931.58 |
| MASSACHUSETTS | 2,286.68 | 2,282.98 | 2,089.79 | 2,269.74 | 8,929.19 |
| MICHIGAN | 1,989.45 | 1,905.46 | 1,793.19 | 1,949.53 | 7,637.63 |
| MINNESOTA | 1,949.15 | 1,896.03 | 1,781.73 | 1,977.67 | 7,604.57 |
| MISSISSIPPI | 1,565.89 | 1,351.19 | 1,385.64 | 1,470.87 | 5,773.60 |
| MISSOURI | 2,257.23 | 2,106.29 | 2,145.46 | 2,280.15 | 8,789.14 |
| MONTANA | 1,854.41 | 1,735.93 | 1,714.02 | 1,768.08 | 7,072.45 |
| NEBRASKA | 1,808.34 | 1,676.32 | 1,628.54 | 1,738.76 | 6,851.97 |
| NEVADA | 570.51 | 547.99 | 517.58 | 557.59 | 2,193.67 |
| NEW HAMPSHIRE | 2,613.82 | 2,638.62 | 2,474.94 | 2,575.94 | 10,303.31 |
| NEW JERSEY | 1,094.90 | 1,060.26 | 957.02 | 1,059.62 | 4,171.80 |
| NEW MEXICO | 972.25 | 915.95 | 906.83 | 908.89 | 3,703.92 |
| NEW YORK | 1,212.49 | 1,202.45 | 1,082.67 | 1,198.83 | 4,696.44 |
| NORTH CAROLINA | 2,153.19 | 1,989.88 | 1,840.31 | 2,009.72 | 7,993.09 |
| NORTH DAKOTA | 2,333.28 | 2,211 | 2,111.30 | 2,284.26 | 8,939.85 |
| OHIO | 2,089.62 | 1,940.20 | 1,921.97 | 2,082.66 | 8,034.45 |
| OKLAHOMA | 1,369.85 | 1,236.67 | 1,244.56 | 1,307.40 | 5,158.48 |
| OREGON | 1,626.91 | 1,653.59 | 1,518.56 | 1,619.62 | 6,418.68 |
| PENNSYLVANIA | 1,809.10 | 1,768.38 | 1,687.84 | 1,818.51 | 7,083.83 |
| PUERTO RICO | 604.77 | 479.46 | 377.11 | 360.16 | 1,821.49 |
| RHODE ISLAND | 1,808.93 | 1,857.34 | 1,678.12 | 1,809.48 | 7,153.87 |
| SOUTH CAROLINA | 2,059.12 | 1,836.50 | 1,795.46 | 1,957.46 | 7,648.53 |
| SOUTH DAKOTA | 3,068.76 | 2,478.58 | 2,821.10 | 3,052.28 | 11,420.71 |
| TENNESSEE | 1,397.63 | 1,274.89 | 1,274.72 | 1,341.72 | 5,288.96 |
| TEXAS | 1,429.32 | 1,326.92 | 1,213.72 | 1,346.18 | 5,316.15 |
| UTAH | 1,727.67 | 1,637.82 | 1,585.32 | 1,712.30 | 6,663.12 |
| VERMONT | 3,076.60 | 3,096.43 | 2,982.38 | 3,138.39 | 12,293.80 |
| VIRGIN ISLANDS | 191.22 | 164.4 | 90.26 | 97.89 | 543.76 |
| VIRGINIA | 1,754.57 | 1,686.11 | 1,562.37 | 1,693.40 | 6,696.44 |
| WASHINGTON | 1,692.10 | 1,704.78 | 1,563.60 | 1,684.67 | 6,645.16 |
| WEST VIRGINIA | 1,810.93 | 1,667.53 | 1,637.38 | 1,704.29 | 6,820.14 |
| WISCONSIN | 1,883.43 | 1,811.89 | 1,713.32 | 1,865.24 | 7,273.88 |
| WYOMING | 1,551.15 | 1,410.03 | 1,382.98 | 1,435.55 | 5,779.72 |
| U.S. TOTAL | 1,534.78 | 1,449.28 | 1,378.44 | 1,490.88 | 5,853.38 |

DRUG CODE:2012 DRUG NAME: GAMMA HYDROXYBUTYRIC ACID(FDA APPROVED-CSA III)

| STATE | 1ST QUARTER | 2ND QUARTER | 3RD QUARTER | 4TH QUARTER | TOTAL GRAMS |
|---|---|---|---|---|---|
| CALIFORNIA | 4.19 | 3.59 | 3.59 | 10.17 | 21.54 |
| ILLINOIS | 4.05 | 5.79 | 5.21 | 5.79 | 20.85 |
| LOUISIANA | 6.56 | 3.28 | 0 | 0 | 9.83 |
| MASSACHUSETTS | 0 | 23.83 | 4.54 | 12.48 | 40.85 |
| MICHIGAN | 6.01 | 0 | 3.01 | 4.51 | 13.53 |
| MISSOURI | 153,970.83 | 91,683.25 | 150,116.01 | 112,033.04 | 507,803.13 |
| NEW YORK | 0 | 0 | 15.34 | 25.69 | 41.03 |
| NORTH CAROLINA | 2.34 | 14.81 | 21.04 | 14.03 | 52.21 |
| OHIO | 10.31 | 18.03 | 16.75 | 19.32 | 64.41 |
| SOUTH CAROLINA | 20.88 | 59.44 | 8.03 | 75.5 | 163.86 |
| TENNESSEE | 9.37 | 9.37 | 9.37 | 9.37 | 37.47 |
| TEXAS | 16.55 | 11.82 | 8.86 | 7.09 | 44.32 |
| VIRGINIA | 0 | 27.86 | 20.43 | 27.86 | 76.15 |
| WASHINGTON | 5.52 | 12.15 | 8.84 | 6.63 | 33.15 |

| | | | | | |
|---|---|---|---|---|---|
| U.S. TOTAL | 2,951.07 | 1,760.56 | 2,878.69 | 2,152.15 | 9,742.47 |

DRUG CODE:2100 DRUG NAME: BARBITURIC ACID DERIVIATIVE OR SALT [PER 21CFR 1308.13(C)(3)]

| STATE | 1ST QUARTER | 2ND QUARTER | 3RD QUARTER | 4TH QUARTER | TOTAL GRAMS |
|---|---|---|---|---|---|
| ALABAMA | 39.02 | 29.6 | 29.71 | 30.23 | 128.56 |
| ALASKA | 31.68 | 24.64 | 23.23 | 21.12 | 100.67 |
| ARIZONA | 91.05 | 84.17 | 70.87 | 70.71 | 316.8 |
| ARKANSAS | 56.24 | 45.78 | 43.38 | 37.21 | 182.62 |
| CALIFORNIA | 40.38 | 37.85 | 43.24 | 37.39 | 158.87 |
| COLORADO | 39.27 | 36.59 | 28.63 | 26.45 | 130.94 |
| CONNECTICUT | 60.43 | 58.34 | 68.55 | 56.38 | 243.7 |
| DELAWARE | 53.46 | 42.32 | 54.57 | 54.57 | 204.91 |
| DISTRICT OF COLUMBIA | 46.53 | 29.91 | 47.36 | 37.39 | 161.2 |
| FLORIDA | 107.09 | 85.85 | 92.79 | 85.77 | 371.49 |
| GEORGIA | 64.88 | 61.68 | 49.29 | 41.19 | 217.03 |
| GUAM | 0 | 5.6 | 0 | 0 | 5.6 |
| HAWAII | 56.61 | 68.37 | 69.1 | 54.4 | 248.47 |
| IDAHO | 40.83 | 41.15 | 28.39 | 27.75 | 138.11 |
| ILLINOIS | 57.32 | 56.82 | 62.16 | 59.35 | 235.65 |
| INDIANA | 52.82 | 50.9 | 51.13 | 48.12 | 202.97 |
| IOWA | 23.63 | 22.16 | 21.34 | 24.13 | 91.26 |
| KANSAS | 45.74 | 52.22 | 36.1 | 34.87 | 168.94 |
| KENTUCKY | 47.47 | 45.97 | 40.07 | 33.76 | 167.28 |
| LOUISIANA | 74.34 | 73.23 | 83.49 | 65.07 | 296.14 |
| MAINE | 36.13 | 28.23 | 30.49 | 32.37 | 127.22 |
| MARYLAND | 45.21 | 39.84 | 43.65 | 36.55 | 165.24 |
| MASSACHUSETTS | 40.09 | 39.02 | 48.64 | 37.49 | 165.25 |
| MICHIGAN | 62.12 | 72.09 | 71.73 | 66.32 | 272.27 |
| MINNESOTA | 26.21 | 24.89 | 25.36 | 23.76 | 100.21 |
| MISSISSIPPI | 40.44 | 35.39 | 36.4 | 23.25 | 135.48 |
| MISSOURI | 69.8 | 75.39 | 65.29 | 53.68 | 264.15 |
| MONTANA | 49.52 | 54.58 | 59.63 | 37.9 | 201.63 |
| NEBRASKA | 39.15 | 38.6 | 43.8 | 41.07 | 162.62 |
| NEVADA | 185.89 | 179.04 | 155.34 | 156.45 | 676.71 |
| NEW HAMPSHIRE | 36.84 | 38.74 | 36.08 | 39.88 | 151.54 |
| NEW JERSEY | 52.09 | 47.83 | 57.61 | 48.45 | 205.99 |
| NEW MEXICO | 40.79 | 40.55 | 41.76 | 38.36 | 161.47 |
| NEW YORK | 24.36 | 22.65 | 22.76 | 20.98 | 90.75 |
| NORTH CAROLINA | 40.64 | 38.96 | 42.84 | 37.65 | 160.09 |
| NORTH DAKOTA | 25.28 | 25.28 | 18.58 | 14.87 | 84 |
| OHIO | 46.98 | 45.25 | 29.69 | 23.66 | 145.58 |
| OKLAHOMA | 71.04 | 71.97 | 81.44 | 75.57 | 300.03 |
| OREGON | 31.45 | 29.1 | 21.66 | 18.66 | 100.89 |
| PENNSYLVANIA | 45.19 | 46.02 | 43.77 | 35.66 | 170.64 |
| PUERTO RICO | 3.62 | 4.03 | 4.43 | 4.16 | 16.24 |
| RHODE ISLAND | 60.33 | 57 | 65.55 | 56.53 | 239.41 |
| SOUTH CAROLINA | 56.64 | 47.67 | 51.35 | 47.46 | 203.12 |
| SOUTH DAKOTA | 33.16 | 27.02 | 27.64 | 23.34 | 111.15 |
| TENNESSEE | 45.07 | 45.93 | 45.38 | 41.76 | 178.14 |
| TEXAS | 49.93 | 50.27 | 52.87 | 43.01 | 196.08 |
| UTAH | 58.97 | 61.87 | 40.7 | 44.14 | 205.69 |
| VERMONT | 15.98 | 9.59 | 13.58 | 6.39 | 45.55 |
| VIRGIN ISLANDS | 18.21 | 40.97 | 22.76 | 13.66 | 95.61 |
| VIRGINIA | 45.87 | 49.56 | 39.31 | 39 | 173.73 |
| WASHINGTON | 40.3 | 40.15 | 31.68 | 25.28 | 137.41 |
| WEST VIRGINIA | 35.89 | 29.68 | 26.98 | 21.86 | 114.41 |
| WISCONSIN | 28.49 | 25.5 | 30.77 | 27.08 | 111.83 |
| WYOMING | 76.29 | 70.08 | 62.1 | 62.1 | 270.57 |

| | | | | | |
|---|---|---|---|---|---|
| U.S. TOTAL | 51.15 | 48.68 | 48.59 | 43.01 | 191.43 |

DRUG CODE:2125 DRUG NAME: AMOBARBITAL (SCHEDULE 2)

| STATE | 1ST QUARTER | 2ND QUARTER | 3RD QUARTER | 4TH QUARTER | TOTAL GRAMS |
|---|---|---|---|---|---|
| ARIZONA | 0.2 | 0.18 | 0.2 | 0.13 | 0.71 |
| ARKANSAS | 0 | 0.02 | 0.02 | 0 | 0.03 |
| CALIFORNIA | 0.07 | 0.06 | 0.08 | 0.06 | 0.27 |
| COLORADO | 0.55 | 0.32 | 0.29 | 0.2 | 1.36 |
| CONNECTICUT | 0 | 0.06 | 0 | 0.04 | 0.1 |
| DISTRICT OF COLUMBIA | 0.08 | 0 | 0.3 | 0.76 | 1.14 |
| FLORIDA | 0.02 | 0.01 | 0.05 | 0.04 | 0.12 |
| GEORGIA | 0.02 | 0.01 | 0.11 | 0.14 | 0.27 |
| ILLINOIS | 0.04 | 0.01 | 0.05 | 0.03 | 0.12 |
| INDIANA | 0 | 0 | 0.04 | 0.03 | 0.06 |
| IOWA | 0 | 0 | 0.01 | 0 | 0.01 |
| KANSAS | 0.06 | 0.18 | 0.02 | 0.22 | 0.48 |
| KENTUCKY | 0.11 | 0.08 | 0.08 | 0.17 | 0.44 |
| MAINE | 0 | 0 | 0.03 | 0.17 | 0.21 |
| MARYLAND | 0.13 | 0.06 | 0.11 | 0.18 | 0.47 |
| MASSACHUSETTS | 0.15 | 0.03 | 0.01 | 0.12 | 0.31 |
| MICHIGAN | 0.06 | 0.04 | 0.06 | 0.03 | 0.19 |
| MINNESOTA | 0.09 | 0.06 | 0.14 | 0.15 | 0.44 |
| MISSISSIPPI | 0.06 | 0 | 0.05 | 0.05 | 0.15 |
| MISSOURI | 0.01 | 0.02 | 0.05 | 0.05 | 0.11 |
| NEBRASKA | 0.15 | 0 | 0 | 0.05 | 0.2 |
| NEW HAMPSHIRE | 0.31 | 0.14 | 0.21 | 0.28 | 0.93 |
| NEW JERSEY | 0.04 | 0.04 | 0.08 | 0.19 | 0.35 |
| NEW YORK | 0.28 | 0.11 | 0.14 | 0.25 | 0.79 |
| NORTH CAROLINA | 0.17 | 0.05 | 0.06 | 0.12 | 0.4 |
| OHIO | 0.03 | 0.03 | 0.05 | 0.05 | 0.15 |
| OREGON | 0.04 | 0.04 | 0.04 | 0.07 | 0.18 |
| PENNSYLVANIA | 0.11 | 0.02 | 0.03 | 0.07 | 0.23 |
| RHODE ISLAND | 0.35 | 0 | 0 | 0 | 0.35 |
| SOUTH CAROLINA | 0.03 | 0.09 | 0.12 | 0.09 | 0.33 |
| TENNESSEE | 0.06 | 0.08 | 0.08 | 0.03 | 0.25 |
| TEXAS | 0.1 | 0.05 | 0.11 | 0.07 | 0.33 |
| UTAH | 0.07 | 0.49 | 0.53 | 0.15 | 1.24 |
| VERMONT | 0 | 0 | 0.15 | 0 | 0.15 |
| VIRGINIA | 0.05 | 0.03 | 0.03 | 0.03 | 0.14 |
| WASHINGTON | 0.07 | 0 | 0.17 | 0.31 | 0.55 |
| WEST VIRGINIA | 0 | 0 | 0 | 0.1 | 0.1 |
| WISCONSIN | 0.11 | 0.06 | 0.13 | 0.18 | 0.47 |
| U.S. TOTAL | 0.09 | 0.05 | 0.08 | 0.09 | 0.31 |

DRUG CODE:2165 DRUG NAME: BUTALBITAL

| STATE | 1ST QUARTER | 2ND QUARTER | 3RD QUARTER | 4TH QUARTER | TOTAL GRAMS |
|---|---|---|---|---|---|
| ALABAMA | 92.37 | 99.27 | 96.55 | 84.21 | 372.41 |
| ALASKA | 25.34 | 37.31 | 38.72 | 28.86 | 130.24 |
| AMERICAN SAMOA | 15.24 | 0 | 0 | 0 | 15.24 |
| ARIZONA | 73.69 | 73.61 | 85.97 | 88 | 321.26 |
| ARKANSAS | 113 | 117.8 | 120.72 | 120.55 | 472.06 |
| CALIFORNIA | 66.11 | 66.95 | 67.27 | 63.56 | 263.89 |
| COLORADO | 34.1 | 36.49 | 39.47 | 45.24 | 155.29 |
| CONNECTICUT | 91.07 | 85.06 | 93.73 | 74.28 | 344.14 |
| DELAWARE | 62.37 | 61.25 | 55.13 | 56.24 | 234.98 |
| DISTRICT OF COLUMBIA | 52.35 | 58.17 | 65.64 | 56.5 | 232.67 |

| | | | | | |
|---|---|---|---|---|---|
| FLORIDA | 78.9 | 88.42 | 91.56 | 90.66 | 349.55 |
| GEORGIA | 94.66 | 99.71 | 109.47 | 106.99 | 410.83 |
| GUAM | 14.01 | 2.8 | 2.8 | 2.8 | 22.42 |
| HAWAII | 81.6 | 79.03 | 76.82 | 68.37 | 305.81 |
| IDAHO | 75.28 | 79.1 | 91.22 | 88.99 | 334.59 |
| ILLINOIS | 56.9 | 60.32 | 58.06 | 55.88 | 231.17 |
| INDIANA | 58.76 | 56.76 | 64.62 | 60.38 | 240.52 |
| IOWA | 40.38 | 42.35 | 43.82 | 40.87 | 167.41 |
| KANSAS | 77.81 | 65.72 | 90.43 | 74.66 | 308.61 |
| KENTUCKY | 54.96 | 55.54 | 65.22 | 63.14 | 238.86 |
| LOUISIANA | 91.54 | 89.34 | 84.59 | 81.95 | 347.42 |
| MAINE | 56.46 | 47.8 | 45.92 | 39.52 | 189.71 |
| MARYLAND | 84 | 77.77 | 84.78 | 82.36 | 328.91 |
| MASSACHUSETTS | 38.64 | 36.73 | 35.89 | 30.77 | 142.04 |
| MICHIGAN | 93.74 | 85.95 | 90.45 | 89.24 | 359.38 |
| MINNESOTA | 28.94 | 30.92 | 30.07 | 36.48 | 126.42 |
| MISSISSIPPI | 71.78 | 71.78 | 73.64 | 70.77 | 287.97 |
| MISSOURI | 90.25 | 95.59 | 101.94 | 108.78 | 396.57 |
| MONTANA | 74.29 | 77.82 | 88.94 | 70.24 | 311.3 |
| NEBRASKA | 57.49 | 59.13 | 60.5 | 65.71 | 242.84 |
| NEVADA | 127.2 | 126.64 | 149.04 | 145.71 | 548.59 |
| NEW HAMPSHIRE | 47.48 | 46.72 | 53.93 | 54.31 | 202.44 |
| NEW JERSEY | 74.73 | 70.41 | 71.71 | 63.75 | 280.6 |
| NEW MEXICO | 39.82 | 49.05 | 48.32 | 42.49 | 179.68 |
| NEW YORK | 30.78 | 31.35 | 29.75 | 25.65 | 117.53 |
| NORTH CAROLINA | 53.9 | 52.75 | 53.54 | 53.64 | 213.83 |
| NORTH DAKOTA | 46.09 | 49.81 | 45.35 | 49.81 | 191.05 |
| OHIO | 51.27 | 47.28 | 64.97 | 67.31 | 230.83 |
| OKLAHOMA | 145.15 | 140.62 | 128.62 | 118.49 | 532.87 |
| OREGON | 61.86 | 54.16 | 62.38 | 66.56 | 244.97 |
| PENNSYLVANIA | 94.63 | 90.1 | 95.3 | 88.41 | 368.43 |
| RHODE ISLAND | 35.63 | 39.43 | 34.2 | 30.4 | 139.66 |
| SOUTH CAROLINA | 71.24 | 73.62 | 74.16 | 72.43 | 291.44 |
| SOUTH DAKOTA | 81.06 | 77.99 | 79.83 | 79.83 | 318.73 |
| TENNESSEE | 55.62 | 57.12 | 63.42 | 64.37 | 240.54 |
| TEXAS | 74.71 | 72.18 | 73.35 | 77.21 | 297.45 |
| UTAH | 110.53 | 98.95 | 124.1 | 126.63 | 460.22 |
| VERMONT | 23.97 | 13.58 | 15.18 | 22.37 | 75.11 |
| VIRGIN ISLANDS | 0 | 0 | 18.21 | 22.76 | 40.97 |
| VIRGINIA | 69.3 | 64.43 | 64.55 | 62.55 | 260.84 |
| WASHINGTON | 55.99 | 62.9 | 66.1 | 73.83 | 258.83 |
| WEST VIRGINIA | 65.57 | 66.92 | 73.12 | 68 | 273.61 |
| WISCONSIN | 29.37 | 30.68 | 34.11 | 28.84 | 123 |
| WYOMING | 60.32 | 66.53 | 83.39 | 78.95 | 289.2 |
| U.S. TOTAL | 67.05 | 67 | 70.4 | 68.53 | 272.98 |

DRUG CODE:2270 DRUG NAME: PENTOBARBITAL (SCHEDULE 2)

| STATE | 1ST QUARTER | 2ND QUARTER | 3RD QUARTER | 4TH QUARTER | TOTAL GRAMS |
|---|---|---|---|---|---|
| ALABAMA | 233.17 | 303.37 | 467.98 | 367.8 | 1,372.33 |
| ALASKA | 800.74 | 703.47 | 875.81 | 828.98 | 3,209.01 |
| ARIZONA | 702.21 | 697.71 | 883.04 | 1,081.64 | 3,364.61 |
| ARKANSAS | 259.02 | 476.38 | 305.78 | 208.07 | 1,249.25 |
| CALIFORNIA | 465.5 | 465.07 | 508.61 | 463.41 | 1,902.60 |
| COLORADO | 939.99 | 767.17 | 656.58 | 1,204.53 | 3,568.28 |
| CONNECTICUT | 416.58 | 406.53 | 600.5 | 603.54 | 2,027.14 |
| DELAWARE | 87.7 | 220.07 | 190.38 | 298 | 796.15 |
| DISTRICT OF COLUMBIA | 32.56 | 1.51 | 144.8 | 245.37 | 424.25 |
| FLORIDA | 346.33 | 405.1 | 374.67 | 387.66 | 1,513.76 |
| GEORGIA | 382.18 | 349.79 | 399.36 | 456.4 | 1,587.74 |

| | | | | | |
|---|---|---|---|---|---|
| GUAM | 0 | 1,802.06 | 0 | 1,802.06 | 3,604.12 |
| HAWAII | 340.46 | 90.45 | 1,266.23 | 357.11 | 2,054.25 |
| IDAHO | 1,029.70 | 1,282.17 | 1,815.86 | 1,642.27 | 5,770 |
| ILLINOIS | 437.69 | 468.17 | 464.05 | 791.03 | 2,160.94 |
| INDIANA | 636.63 | 586.85 | 669.96 | 879.38 | 2,772.83 |
| IOWA | 370.41 | 544.12 | 465.95 | 966.69 | 2,347.17 |
| KANSAS | 761.32 | 506.38 | 747.78 | 1,156.15 | 3,171.62 |
| KENTUCKY | 471.89 | 527.43 | 811.99 | 754.58 | 2,565.89 |
| LOUISIANA | 547.51 | 473.92 | 601.42 | 590.41 | 2,213.26 |
| MAINE | 1,605.45 | 1,288.58 | 1,460.91 | 1,733.80 | 6,088.75 |
| MARYLAND | 398.82 | 253.98 | 412.89 | 474.62 | 1,540.31 |
| MASSACHUSETTS | 331.76 | 401.71 | 484.3 | 504.36 | 1,722.13 |
| MICHIGAN | 905.89 | 790.33 | 1,113.82 | 1,084.68 | 3,894.73 |
| MINNESOTA | 494.5 | 680.59 | 588.06 | 968.41 | 2,731.56 |
| MISSISSIPPI | 374.96 | 186.2 | 211.7 | 323.2 | 1,096.06 |
| MISSOURI | 617.02 | 435.9 | 787.36 | 848.47 | 2,688.76 |
| MONTANA | 1,257.59 | 1,060.43 | 2,174.01 | 2,999.82 | 7,491.84 |
| NEBRASKA | 685.94 | 1,044.10 | 1,122.57 | 1,662.86 | 4,515.47 |
| NEVADA | 452.67 | 427.98 | 676.93 | 729.19 | 2,286.78 |
| NEW HAMPSHIRE | 901.74 | 1,264.51 | 1,275.54 | 1,262.09 | 4,703.89 |
| NEW JERSEY | 214.17 | 178.75 | 299.45 | 239.38 | 931.75 |
| NEW MEXICO | 674.45 | 880 | 1,287.25 | 1,058.06 | 3,899.76 |
| NEW YORK | 289.78 | 395.75 | 432.84 | 462.78 | 1,581.14 |
| NORTH CAROLINA | 537.07 | 583.44 | 637.04 | 785.95 | 2,543.49 |
| NORTH DAKOTA | 1,297.47 | 966.09 | 712.29 | 1,241.91 | 4,217.75 |
| OHIO | 518.74 | 569.88 | 678.08 | 791.45 | 2,558.16 |
| OKLAHOMA | 364.77 | 573.48 | 375.88 | 714.81 | 2,028.94 |
| OREGON | 756.27 | 921.97 | 1,148.32 | 1,211.13 | 4,037.69 |
| PENNSYLVANIA | 421.37 | 497.55 | 690.48 | 509.8 | 2,119.22 |
| PUERTO RICO | 0 | 0 | 0.01 | 0 | 0.01 |
| RHODE ISLAND | 304.75 | 659.9 | 692.64 | 676.29 | 2,333.57 |
| SOUTH CAROLINA | 271.96 | 277.91 | 300.97 | 353.7 | 1,204.55 |
| SOUTH DAKOTA | 710.04 | 666.44 | 819.97 | 873.25 | 3,069.70 |
| TENNESSEE | 419.75 | 651.39 | 554.74 | 621.06 | 2,246.95 |
| TEXAS | 419.21 | 429.33 | 566.22 | 560.12 | 1,974.87 |
| UTAH | 348.96 | 407.82 | 481.37 | 821.09 | 2,059.24 |
| VERMONT | 1,565.44 | 1,496.55 | 2,785.77 | 2,839.89 | 8,687.66 |
| VIRGINIA | 535.63 | 590.22 | 589.36 | 671.86 | 2,387.07 |
| WASHINGTON | 826.2 | 797.26 | 847.9 | 1,109.84 | 3,581.21 |
| WEST VIRGINIA | 376.19 | 678.53 | 769.53 | 447.83 | 2,272.08 |
| WISCONSIN | 720.44 | 765.98 | 916.48 | 1,279.56 | 3,682.45 |
| WYOMING | 804.07 | 2,239.75 | 1,613.96 | 2,522.57 | 7,180.34 |
| U.S. TOTAL | 483.42 | 511.25 | 602.64 | 673.81 | 2,271.12 |

DRUG CODE:2315 DRUG NAME: SECOBARBITAL (SCHEDULE 2)

| STATE | 1ST QUARTER | 2ND QUARTER | 3RD QUARTER | 4TH QUARTER | TOTAL GRAMS |
|---|---|---|---|---|---|
| ARIZONA | 0.14 | 0 | 0 | 0.14 | 0.29 |
| CALIFORNIA | 2.41 | 2.65 | 3.12 | 3.56 | 11.75 |
| COLORADO | 2.18 | 1.82 | 2.18 | 1.27 | 7.46 |
| CONNECTICUT | 1.02 | 0.77 | 0.26 | 0.51 | 2.56 |
| DELAWARE | 0 | 1.02 | 1.02 | 1.02 | 3.06 |
| FLORIDA | 0.54 | 0.39 | 0.54 | 0.49 | 1.95 |
| GEORGIA | 0.38 | 0.09 | 0 | 0 | 0.47 |
| HAWAII | 0.67 | 0 | 0 | 0 | 0.67 |
| ILLINOIS | 0.36 | 0.43 | 0.29 | 0.21 | 1.28 |
| KANSAS | 1.28 | 0.64 | 0.64 | 0.96 | 3.53 |
| MARYLAND | 1.11 | 1.11 | 0.79 | 0.79 | 3.81 |
| MASSACHUSETTS | 0.84 | 0.7 | 0.56 | 0.84 | 2.94 |
| MICHIGAN | 0 | 0 | 0 | 0.19 | 0.19 |

| | | | | | |
|---|---|---|---|---|---|
| MINNESOTA | 0.17 | 0 | 0 | 0.17 | 0.35 |
| MISSOURI | 0.46 | 0.31 | 0.61 | 0.31 | 1.68 |
| NEVADA | 0.34 | 0 | 0 | 0 | 0.34 |
| NEW HAMPSHIRE | 1.39 | 0.7 | 0 | 0 | 2.09 |
| NEW JERSEY | 0.83 | 0.62 | 0.62 | 0.52 | 2.6 |
| NEW YORK | 2.41 | 2.08 | 1.51 | 1.75 | 7.75 |
| NORTH CAROLINA | 0 | 0.1 | 0 | 0 | 0.1 |
| OHIO | 0.48 | 0.16 | 0.24 | 0.32 | 1.19 |
| OKLAHOMA | 0 | 0 | 0.73 | 0.24 | 0.98 |
| OREGON | 6.69 | 4.54 | 5.74 | 7.89 | 24.85 |
| PENNSYLVANIA | 0.29 | 0.58 | 0.65 | 0.58 | 2.09 |
| RHODE ISLAND | 0.87 | 0.87 | 0 | 1.74 | 3.48 |
| SOUTH CAROLINA | 0 | 0 | 0.4 | 0.4 | 0.79 |
| SOUTH DAKOTA | 0 | 0 | 0 | 1.12 | 1.12 |
| TENNESSEE | 0 | 0.14 | 0 | 0 | 0.14 |
| TEXAS | 0.04 | 0 | 0.04 | 0.11 | 0.18 |
| VERMONT | 8.78 | 4.39 | 0 | 8.78 | 21.95 |
| VIRGINIA | 0.34 | 0.34 | 0.34 | 0.34 | 1.37 |
| WASHINGTON | 3.4 | 2.59 | 2.31 | 2.45 | 10.76 |
| WISCONSIN | 0.16 | 0 | 0 | 0 | 0.16 |
| U.S. TOTAL | 0.86 | 0.76 | 0.79 | 0.91 | 3.32 |

DRUG CODE:7365 DRUG NAME: DRONABINOL IN AN ORAL SOLUTION IN FDA APPROVED
DRUG PRODUCT (SYNDROS - CII)

| STATE | 1ST QUARTER | 2ND QUARTER | 3RD QUARTER | 4TH QUARTER | TOTAL GRAMS |
|---|---|---|---|---|---|
| ALABAMA | 0 | 0 | 0.03 | 0.06 | 0.09 |
| ARIZONA | 0 | 0 | 0.23 | 0.11 | 0.34 |
| ARKANSAS | 0 | 0 | 0.02 | 0.02 | 0.04 |
| CALIFORNIA | 0 | 0 | 0.04 | 0.08 | 0.12 |
| COLORADO | 0 | 0 | 0.07 | 0 | 0.07 |
| CONNECTICUT | 0 | 0 | 0.03 | 0.01 | 0.05 |
| DELAWARE | 0 | 0 | 0.02 | 0 | 0.02 |
| DISTRICT OF COLUMBIA | 0 | 0 | 0 | 0.12 | 0.12 |
| FLORIDA | 0 | 0 | 0.07 | 0.12 | 0.19 |
| GEORGIA | 0 | 0 | 0.01 | 0.03 | 0.05 |
| ILLINOIS | 0 | 0 | 0 | 0.01 | 0.01 |
| INDIANA | 0 | 0 | 0.03 | 0 | 0.03 |
| KANSAS | 0 | 0 | 0 | 0.02 | 0.02 |
| KENTUCKY | 0 | 0 | 0.02 | 0.02 | 0.03 |
| LOUISIANA | 0 | 0 | 0 | 0 | 0.01 |
| MAINE | 0 | 0 | 0.02 | 0 | 0.02 |
| MARYLAND | 0 | 0 | 0 | 0.03 | 0.03 |
| MASSACHUSETTS | 0 | 0 | 0.01 | 0.05 | 0.06 |
| MICHIGAN | 0 | 0 | 0.01 | 0.01 | 0.02 |
| MINNESOTA | 0 | 0 | 0 | 0.05 | 0.05 |
| MISSISSIPPI | 0 | 0 | 0.02 | 0.01 | 0.03 |
| MISSOURI | 0 | 0 | 0.07 | 0.07 | 0.14 |
| MONTANA | 0 | 0 | 0 | 0.03 | 0.03 |
| NEVADA | 0 | 0 | 0.18 | 0.24 | 0.43 |
| NEW HAMPSHIRE | 0 | 0 | 0.06 | 0.08 | 0.14 |
| NEW JERSEY | 0 | 0 | 0.81 | 0.83 | 1.63 |
| NEW MEXICO | 0 | 0 | 0.01 | 0 | 0.01 |
| NEW YORK | 0 | 0 | 0 | 0.01 | 0.01 |
| NORTH CAROLINA | 0 | 0 | 0 | 0.01 | 0.01 |
| OHIO | 0 | 0 | 0 | 0.07 | 0.08 |
| PENNSYLVANIA | 0 | 0 | 0.11 | 0.02 | 0.13 |
| SOUTH CAROLINA | 0 | 0 | 0.01 | 0 | 0.01 |
| TENNESSEE | 0 | 0 | 0.02 | 0.03 | 0.06 |
| TEXAS | 0 | 0 | 0.03 | 0.02 | 0.05 |

| | | | | | |
|---|---|---|---|---|---|
| UTAH | 0 | 0 | 0.02 | 0.01 | 0.02 |
| VIRGINIA | 0 | 0 | 0 | 0.01 | 0.01 |
| WISCONSIN | 0 | 0 | 0 | 0 | 0 |
| U.S. TOTAL | 0 | 0 | 0.05 | 0.06 | 0.11 |

DRUG CODE:7379 DRUG NAME: NABILONE

| STATE | 1ST QUARTER | 2ND QUARTER | 3RD QUARTER | 4TH QUARTER | TOTAL GRAMS |
|---|---|---|---|---|---|
| ARIZONA | 0 | 0 | 0 | 0 | 0 |
| CALIFORNIA | 0 | 0 | 0 | 0 | 0 |
| COLORADO | 0 | 0 | 0 | 0 | 0.01 |
| CONNECTICUT | 0 | 0 | 0 | 0 | 0.01 |
| FLORIDA | 0.01 | 0 | 0 | 0 | 0.01 |
| HAWAII | 0.03 | 0 | 0.01 | 0 | 0.04 |
| ILLINOIS | 0 | 0 | 0 | 0 | 0 |
| IOWA | 0 | 0 | 0 | 0 | 0 |
| KENTUCKY | 0 | 0 | 0 | 0 | 0.01 |
| MAINE | 0.01 | 0.01 | 0.01 | 0 | 0.02 |
| MARYLAND | 0.02 | 0.02 | 0.02 | 0.02 | 0.07 |
| MINNESOTA | 0 | 0 | 0 | 0 | 0 |
| MISSOURI | 0 | 0 | 0 | 0 | 0.01 |
| NEBRASKA | 0.01 | 0 | 0 | 0.01 | 0.02 |
| NEW HAMPSHIRE | 0 | 0 | 0 | 0 | 0 |
| NEW JERSEY | 0 | 0 | 0 | 0 | 0 |
| NEW MEXICO | 0 | 0 | 0 | 0 | 0 |
| NEW YORK | 0 | 0 | 0 | 0 | 0 |
| NORTH CAROLINA | 0 | 0 | 0 | 0 | 0.01 |
| OHIO | 0 | 0 | 0 | 0.01 | 0.01 |
| OREGON | 0 | 0 | 0 | 0 | 0 |
| PENNSYLVANIA | 0 | 0 | 0 | 0 | 0 |
| VIRGINIA | 0 | 0 | 0 | 0 | 0.01 |
| WASHINGTON | 0 | 0 | 0 | 0 | 0 |
| U.S. TOTAL | 0 | 0 | 0 | 0 | 0 |

DRUG CODE:9041L DRUG NAME: COCAINE

| STATE | 1ST QUARTER | 2ND QUARTER | 3RD QUARTER | 4TH QUARTER | TOTAL GRAMS |
|---|---|---|---|---|---|
| ALABAMA | 1.31 | 1.38 | 1.04 | 1.06 | 4.78 |
| ALASKA | 1.21 | 2.79 | 1.35 | 1.41 | 6.75 |
| ARIZONA | 1.73 | 1.72 | 1.56 | 1.56 | 6.56 |
| ARKANSAS | 1.93 | 1.4 | 1.31 | 1.38 | 6.02 |
| CALIFORNIA | 2.49 | 2.36 | 2.24 | 2.22 | 9.32 |
| COLORADO | 1.63 | 2.17 | 1.42 | 1.6 | 6.82 |
| CONNECTICUT | 1.09 | 0.9 | 1.4 | 0.82 | 4.22 |
| DELAWARE | 0.73 | 2.1 | 1.65 | 1.29 | 5.77 |
| DISTRICT OF COLUMBIA | 3.74 | 3.57 | 4.68 | 3.45 | 15.44 |
| FLORIDA | 1.54 | 1.63 | 1.57 | 1.44 | 6.18 |
| GEORGIA | 1.59 | 1.77 | 1.52 | 1.9 | 6.78 |
| GUAM | 0 | 0 | 0.2 | 0 | 0.2 |
| HAWAII | 2.08 | 1.75 | 1.83 | 1.5 | 7.16 |
| IDAHO | 4.12 | 2.39 | 2.99 | 4.32 | 13.81 |
| ILLINOIS | 2.55 | 2.78 | 2.13 | 2.6 | 10.06 |
| INDIANA | 3.87 | 3.02 | 2.36 | 3.1 | 12.35 |
| IOWA | 3.46 | 3.38 | 2.83 | 3.24 | 12.9 |
| KANSAS | 2.23 | 2.98 | 2.89 | 3.43 | 11.52 |
| KENTUCKY | 2.53 | 2.72 | 2.04 | 2.72 | 10.01 |
| LOUISIANA | 2.04 | 2.13 | 1.91 | 2.51 | 8.59 |
| MAINE | 1.69 | 1.88 | 1.72 | 1.3 | 6.58 |

| | | | | | |
|---|---|---|---|---|---|
| MARYLAND | 2.44 | 2.54 | 2.07 | 1.92 | 8.96 |
| MASSACHUSETTS | 3.49 | 3.1 | 2.76 | 3.11 | 12.45 |
| MICHIGAN | 3.72 | 4.27 | 3.41 | 3.86 | 15.26 |
| MINNESOTA | 3.99 | 4.4 | 3.76 | 3.98 | 16.13 |
| MISSISSIPPI | 2.69 | 3.42 | 2.54 | 2.78 | 11.42 |
| MISSOURI | 2.74 | 3.02 | 2.67 | 2.58 | 11.01 |
| MONTANA | 4.94 | 5.56 | 4.22 | 6.67 | 21.39 |
| NEBRASKA | 1.18 | 1.7 | 2.03 | 1.62 | 6.53 |
| NEVADA | 1.65 | 1.93 | 1.63 | 1.46 | 6.66 |
| NEW HAMPSHIRE | 2.94 | 4.17 | 3.08 | 4.26 | 14.46 |
| NEW JERSEY | 2 | 1.92 | 1.69 | 1.73 | 7.34 |
| NEW MEXICO | 3.71 | 3.58 | 3.58 | 3.23 | 14.1 |
| NEW YORK | 1.87 | 1.91 | 1.65 | 1.69 | 7.12 |
| NORTH CAROLINA | 1.42 | 1.49 | 1.59 | 1.49 | 6 |
| NORTH DAKOTA | 4.94 | 9.87 | 5.62 | 6.62 | 27.06 |
| OHIO | 1.52 | 1.75 | 1.47 | 1.59 | 6.33 |
| OKLAHOMA | 0.83 | 0.91 | 0.75 | 1.05 | 3.55 |
| OREGON | 2.7 | 2.5 | 2.62 | 2.21 | 10.04 |
| PENNSYLVANIA | 3.25 | 3.23 | 2.81 | 2.97 | 12.26 |
| RHODE ISLAND | 2.4 | 1.56 | 1.66 | 1.6 | 7.22 |
| SOUTH CAROLINA | 0.66 | 0.68 | 0.61 | 0.61 | 2.56 |
| SOUTH DAKOTA | 5.43 | 3.04 | 3.37 | 6.43 | 18.28 |
| TENNESSEE | 2.85 | 2.52 | 2.52 | 2.67 | 10.56 |
| TEXAS | 1.32 | 1.53 | 1.3 | 1.63 | 5.78 |
| UTAH | 3.74 | 3.57 | 3.25 | 3.59 | 14.15 |
| VERMONT | 1.66 | 2.15 | 1 | 1.34 | 6.15 |
| VIRGINIA | 2.02 | 1.91 | 1.94 | 1.81 | 7.68 |
| WASHINGTON | 1.7 | 1.55 | 1.85 | 1.65 | 6.75 |
| WEST VIRGINIA | 2.83 | 2.61 | 2.69 | 2.75 | 10.89 |
| WISCONSIN | 2.89 | 2.78 | 2.3 | 2.67 | 10.64 |
| WYOMING | 3.19 | 2.46 | 3.6 | 3.04 | 12.29 |
| U.S. TOTAL | 2.21 | 2.26 | 2 | 2.14 | 8.61 |

DRUG CODE:9050 DRUG NAME: CODEINE

| STATE | 1ST QUARTER | 2ND QUARTER | 3RD QUARTER | 4TH QUARTER | TOTAL GRAMS |
|---|---|---|---|---|---|
| ALABAMA | 1,089.96 | 1,096.85 | 1,084.18 | 1,099.36 | 4,370.36 |
| ALASKA | 1,044.23 | 995.77 | 991.3 | 882.57 | 3,913.86 |
| AMERICAN SAMOA | 538.82 | 518.61 | 538.82 | 478.2 | 2,074.45 |
| ARIZONA | 1,226.47 | 1,174.19 | 1,104.49 | 1,143.59 | 4,648.74 |
| ARKANSAS | 2,541.92 | 2,622.31 | 2,468.31 | 2,409.21 | 10,041.74 |
| CALIFORNIA | 1,346.01 | 1,288.32 | 1,236.24 | 1,207.34 | 5,077.90 |
| COLORADO | 527.98 | 514.83 | 505.54 | 497.09 | 2,045.43 |
| CONNECTICUT | 686.67 | 698.9 | 662.51 | 646.76 | 2,694.84 |
| DELAWARE | 1,103.25 | 1,060.64 | 971.29 | 983.95 | 4,119.13 |
| DISTRICT OF COLUMBIA | 2,118.30 | 2,090.25 | 1,995.36 | 1,843.06 | 8,046.97 |
| FLORIDA | 961.03 | 908.64 | 860.18 | 894.3 | 3,624.15 |
| GEORGIA | 1,097.86 | 1,127.98 | 1,054.59 | 1,057.72 | 4,338.15 |
| GUAM | 722.42 | 667.68 | 670.56 | 610.23 | 2,670.89 |
| HAWAII | 1,015.03 | 1,042.39 | 1,012.02 | 941.64 | 4,011.08 |
| IDAHO | 879.05 | 892.38 | 867.31 | 847.12 | 3,485.86 |
| ILLINOIS | 1,303.88 | 1,278.05 | 1,239.73 | 1,223.14 | 5,044.79 |
| INDIANA | 804.13 | 802.27 | 732.36 | 753.03 | 3,091.79 |
| IOWA | 658.56 | 669.79 | 647.49 | 646.84 | 2,622.68 |
| KANSAS | 1,016.33 | 996.79 | 938.9 | 966.35 | 3,918.38 |
| KENTUCKY | 1,121.74 | 1,111.55 | 1,096.31 | 1,051.86 | 4,381.46 |
| LOUISIANA | 998.68 | 1,005.73 | 928.33 | 900.96 | 3,833.69 |
| MAINE | 815.28 | 828.67 | 735.9 | 748.51 | 3,128.38 |
| MARYLAND | 1,076.65 | 1,032.14 | 958.77 | 949.5 | 4,017.07 |
| MASSACHUSETTS | 743.19 | 713.74 | 689.31 | 682.73 | 2,828.96 |

| | | | | | |
|---|---|---|---|---|---|
| MICHIGAN | 2,690.06 | 2,660.60 | 2,579.31 | 2,443.49 | 10,373.45 |
| MINNESOTA | 733.7 | 700.04 | 697.62 | 681.63 | 2,812.98 |
| MISSISSIPPI | 945.9 | 929.12 | 920.59 | 898.45 | 3,694.05 |
| MISSOURI | 1,146.33 | 1,170.31 | 1,119.12 | 1,135.98 | 4,571.74 |
| MONTANA | 1,038.43 | 1,062.18 | 999.97 | 1,000.93 | 4,101.51 |
| NEBRASKA | 666.6 | 644.15 | 637.48 | 623.32 | 2,571.55 |
| NEVADA | 1,000.76 | 1,016.51 | 956.11 | 986.89 | 3,960.27 |
| NEW HAMPSHIRE | 598.66 | 573.08 | 556.59 | 576.88 | 2,305.21 |
| NEW JERSEY | 934.87 | 896.63 | 861.66 | 842.08 | 3,535.23 |
| NEW MEXICO | 962.77 | 934.74 | 948.34 | 951.15 | 3,797 |
| NEW YORK | 760.67 | 751.21 | 715.37 | 702.6 | 2,929.85 |
| NORTH CAROLINA | 540.22 | 513.82 | 483.98 | 492.47 | 2,030.49 |
| NORTH DAKOTA | 855.28 | 865.09 | 871.54 | 794.74 | 3,386.66 |
| OHIO | 945.07 | 890.43 | 838.24 | 819.57 | 3,493.31 |
| OKLAHOMA | 2,576.47 | 2,582.08 | 2,522.65 | 2,499.62 | 10,180.82 |
| OREGON | 916.89 | 879.91 | 873.88 | 875.59 | 3,546.27 |
| PENNSYLVANIA | 1,078.54 | 1,132.95 | 1,117.92 | 1,035.64 | 4,365.05 |
| PUERTO RICO | 364.6 | 361.67 | 312.14 | 336.49 | 1,374.90 |
| RHODE ISLAND | 760.73 | 700.55 | 653.06 | 651.91 | 2,766.26 |
| SOUTH CAROLINA | 1,697.93 | 1,476.49 | 1,568.99 | 1,829.82 | 6,573.23 |
| SOUTH DAKOTA | 890.49 | 869.16 | 827.3 | 837.99 | 3,424.94 |
| TENNESSEE | 1,473.68 | 1,393.02 | 1,345.84 | 1,168.47 | 5,381 |
| TEXAS | 4,016.36 | 4,075.25 | 3,925.44 | 3,920.40 | 15,937.44 |
| UTAH | 595.38 | 599.99 | 592.58 | 577.26 | 2,365.21 |
| VERMONT | 569.81 | 552.75 | 484.39 | 489.11 | 2,096.06 |
| VIRGIN ISLANDS | 865.79 | 593.79 | 515.17 | 755.99 | 2,730.73 |
| VIRGINIA | 741.02 | 712.82 | 642.46 | 648.06 | 2,744.35 |
| WASHINGTON | 942.39 | 937.75 | 892.04 | 882.33 | 3,654.52 |
| WEST VIRGINIA | 1,677.58 | 1,680.14 | 1,633.06 | 1,725.98 | 6,716.75 |
| WISCONSIN | 891.5 | 839.91 | 807.43 | 811.86 | 3,350.71 |
| WYOMING | 774.73 | 715.21 | 732.37 | 752.44 | 2,974.75 |
| U.S. TOTAL | 1,333.63 | 1,315.04 | 1,263.89 | 1,251.50 | 5,164.06 |

DRUG CODE:9056 DRUG NAME: ETORPHINE

| STATE | 1ST QUARTER | 2ND QUARTER | 3RD QUARTER | 4TH QUARTER | TOTAL GRAMS |
|---|---|---|---|---|---|
| WYOMING | 4.97 | 0 | 0 | 5.15 | 10.11 |
| U.S. TOTAL | 0.01 | 0 | 0 | 0.01 | 0.02 |

DRUG CODE:9064 DRUG NAME: BUPRENORPHINE

| STATE | 1ST QUARTER | 2ND QUARTER | 3RD QUARTER | 4TH QUARTER | TOTAL GRAMS |
|---|---|---|---|---|---|
| ALABAMA | 496.58 | 526.4 | 531.7 | 553.28 | 2,107.96 |
| ALASKA | 306.71 | 331.1 | 342.34 | 351.55 | 1,331.70 |
| AMERICAN SAMOA | 0 | 0 | 0 | 0 | 0.01 |
| ARIZONA | 139.51 | 145.31 | 149.68 | 155.3 | 589.8 |
| ARKANSAS | 201.54 | 211.93 | 218.49 | 227.33 | 859.3 |
| CALIFORNIA | 97.13 | 100.06 | 105.49 | 106.87 | 409.53 |
| COLORADO | 139.78 | 144.07 | 151.47 | 153.51 | 588.83 |
| CONNECTICUT | 395.75 | 413.31 | 420.32 | 429.58 | 1,658.96 |
| DELAWARE | 376.82 | 404.69 | 400.87 | 434.21 | 1,616.59 |
| DISTRICT OF COLUMBIA | 353.41 | 376.43 | 409.34 | 432.03 | 1,571.21 |
| FLORIDA | 241.27 | 249.11 | 247.47 | 260.23 | 998.08 |
| GEORGIA | 153.57 | 162.57 | 164.09 | 168.76 | 648.98 |
| GUAM | 0.46 | 1.11 | 0 | 0.94 | 2.52 |
| HAWAII | 136.8 | 146.09 | 147.97 | 148.37 | 579.23 |
| IDAHO | 207.57 | 220.42 | 231.69 | 247.2 | 906.88 |
| ILLINOIS | 104.71 | 112.07 | 114.33 | 117.25 | 448.36 |

| | | | | | |
|---|---|---|---|---|---|
| INDIANA | 274.59 | 304.99 | 316.75 | 332.04 | 1,228.36 |
| IOWA | 52.04 | 53.78 | 55.78 | 59.34 | 220.93 |
| KANSAS | 87.41 | 87.87 | 93.26 | 89.78 | 358.33 |
| KENTUCKY | 777.73 | 800.81 | 798.4 | 805.16 | 3,182.09 |
| LOUISIANA | 398.26 | 418.09 | 434.18 | 432.85 | 1,683.38 |
| MAINE | 675.41 | 728.2 | 743.3 | 768 | 2,914.90 |
| MARYLAND | 417.86 | 437.63 | 444.67 | 467.61 | 1,767.76 |
| MASSACHUSETTS | 607.94 | 649.69 | 622.45 | 644.7 | 2,524.78 |
| MICHIGAN | 260.14 | 273.83 | 275.57 | 289.7 | 1,099.24 |
| MINNESOTA | 131.97 | 140.37 | 146.08 | 151.25 | 569.67 |
| MISSISSIPPI | 326.55 | 336.87 | 335.74 | 337.82 | 1,336.98 |
| MISSOURI | 144.11 | 153.76 | 162.03 | 178.39 | 638.29 |
| MONTANA | 224.41 | 234.07 | 252.26 | 264.8 | 975.54 |
| NEBRASKA | 69.24 | 71.65 | 74.66 | 75.14 | 290.68 |
| NEVADA | 119.93 | 128.74 | 128.17 | 128.44 | 505.28 |
| NEW HAMPSHIRE | 582.66 | 612.14 | 623.9 | 619.12 | 2,437.83 |
| NEW JERSEY | 290.58 | 306.21 | 297.4 | 306.38 | 1,200.57 |
| NEW MEXICO | 331.49 | 351.3 | 369.02 | 380.59 | 1,432.40 |
| NEW YORK | 260.54 | 272.5 | 276.46 | 291.53 | 1,101.02 |
| NORTH CAROLINA | 318.44 | 346.86 | 336.56 | 370.3 | 1,372.16 |
| NORTH DAKOTA | 126.67 | 131.3 | 140.35 | 142.18 | 540.5 |
| OHIO | 439.98 | 460.75 | 456.52 | 463.92 | 1,821.17 |
| OKLAHOMA | 271.26 | 276.23 | 306.94 | 311.04 | 1,165.47 |
| OREGON | 253.84 | 268.29 | 272.44 | 289.63 | 1,084.20 |
| PENNSYLVANIA | 474.96 | 497.9 | 489.61 | 522.1 | 1,984.57 |
| PUERTO RICO | 66.89 | 69 | 59.58 | 71.76 | 267.23 |
| RHODE ISLAND | 672.52 | 731.67 | 711.57 | 743.66 | 2,859.42 |
| SOUTH CAROLINA | 212.84 | 227.02 | 233.39 | 248.59 | 921.85 |
| SOUTH DAKOTA | 50.43 | 57.13 | 55.22 | 58.03 | 220.81 |
| TENNESSEE | 521.13 | 553.94 | 547.4 | 572.34 | 2,194.82 |
| TEXAS | 101.33 | 104.24 | 106.56 | 106.77 | 418.91 |
| UTAH | 336.41 | 347.1 | 353.22 | 358.49 | 1,395.23 |
| VERMONT | 954.23 | 1,023.02 | 1,014.18 | 1,038.84 | 4,030.27 |
| VIRGIN ISLANDS | 17.92 | 15.97 | 11.82 | 11.4 | 57.11 |
| VIRGINIA | 252.62 | 251.38 | 239.85 | 249.25 | 993.11 |
| WASHINGTON | 267.42 | 282.06 | 290.78 | 304.79 | 1,145.05 |
| WEST VIRGINIA | 804 | 836.92 | 861.99 | 853.31 | 3,356.22 |
| WISCONSIN | 173.78 | 179.04 | 180.64 | 189.32 | 722.78 |
| WYOMING | 177.87 | 222.99 | 233.67 | 258.42 | 892.95 |
| U.S. TOTAL | 254.81 | 267.61 | 269.26 | 279.59 | 1,071.27 |

DRUG CODE:9120 DRUG NAME: DIHYDROCODEINE

| STATE | 1ST QUARTER | 2ND QUARTER | 3RD QUARTER | 4TH QUARTER | TOTAL GRAMS |
|---|---|---|---|---|---|
| ALABAMA | 16.4 | 16.71 | 16.44 | 16.35 | 65.89 |
| ALASKA | 1.2 | 0.45 | 0 | 0.3 | 1.96 |
| ARIZONA | 1.15 | 1.35 | 1.19 | 1.3 | 5 |
| ARKANSAS | 3.62 | 5.22 | 6.02 | 4.63 | 19.5 |
| CALIFORNIA | 0.58 | 0.68 | 0.2 | 0.81 | 2.26 |
| COLORADO | 0 | 0.04 | 0.11 | 0.04 | 0.19 |
| CONNECTICUT | 0.03 | 0 | 0.07 | 0 | 0.1 |
| DELAWARE | 1.46 | 1.48 | 1.38 | 1.75 | 6.07 |
| FLORIDA | 0.84 | 0.78 | 1.27 | 1.17 | 4.06 |
| GEORGIA | 4.42 | 5.68 | 5.26 | 7.33 | 22.69 |
| HAWAII | 0.08 | 0 | 0 | 0 | 0.08 |
| IDAHO | 1.5 | 2.59 | 2.04 | 0.82 | 6.95 |
| ILLINOIS | 0.99 | 1.67 | 1.5 | 1.07 | 5.23 |
| INDIANA | 0.05 | 0 | 0.05 | 0.02 | 0.12 |
| IOWA | 0 | 0.04 | 0 | 0.04 | 0.07 |
| KANSAS | 0.04 | 0.37 | 0 | 0.22 | 0.64 |

| | | | | | |
|---|---|---|---|---|---|
| KENTUCKY | 0.71 | 0.44 | 0.32 | 0.49 | 1.97 |
| LOUISIANA | 13.7 | 17.71 | 14.21 | 10.69 | 56.3 |
| MAINE | 0 | 0 | 0.08 | 0 | 0.08 |
| MARYLAND | 0.13 | 0.18 | 0.18 | 0.18 | 0.66 |
| MASSACHUSETTS | 0.33 | 0.24 | 0.51 | 0.85 | 1.93 |
| MICHIGAN | 0.35 | 0 | 0.05 | 0.01 | 0.41 |
| MINNESOTA | 0.03 | 0.08 | 0.03 | 0.04 | 0.19 |
| MISSISSIPPI | 12.39 | 16.56 | 12.82 | 12.35 | 54.12 |
| MISSOURI | 0.77 | 0.71 | 0.91 | 0.7 | 3.09 |
| MONTANA | 0.11 | 0 | 0 | 0 | 0.11 |
| NEBRASKA | 0.18 | 0.12 | 0.21 | 0.15 | 0.66 |
| NEVADA | 1.61 | 1.15 | 0.04 | 0 | 2.79 |
| NEW JERSEY | 4.05 | 4.2 | 2.7 | 3.25 | 14.2 |
| NEW MEXICO | 6.59 | 4.41 | 4.88 | 0.83 | 16.7 |
| NEW YORK | 0.56 | 0.87 | 0.6 | 0.7 | 2.73 |
| NORTH CAROLINA | 0.15 | 0.1 | 0.07 | 0.09 | 0.4 |
| OHIO | 0.42 | 0.29 | 0.33 | 0.18 | 1.22 |
| OKLAHOMA | 7.27 | 7.97 | 7.87 | 7 | 30.11 |
| PENNSYLVANIA | 1.62 | 0.33 | 0.51 | 0.4 | 2.85 |
| RHODE ISLAND | 0 | 0.1 | 0 | 0.2 | 0.3 |
| SOUTH CAROLINA | 0.3 | 0.3 | 0.23 | 0.3 | 1.13 |
| SOUTH DAKOTA | 0 | 0 | 0 | 0.13 | 0.13 |
| TENNESSEE | 8 | 8.14 | 8.02 | 6.37 | 30.52 |
| TEXAS | 10.69 | 11.17 | 10.66 | 11.35 | 43.86 |
| UTAH | 0 | 0 | 0.04 | 0 | 0.04 |
| VERMONT | 0 | 0.17 | 0 | 0 | 0.17 |
| VIRGINIA | 0.13 | 0.05 | 0.04 | 0.11 | 0.33 |
| WEST VIRGINIA | 0.29 | 0.13 | 0 | 0 | 0.42 |
| WISCONSIN | 0.08 | 0.13 | 0.11 | 0.06 | 0.38 |
| U.S. TOTAL | 2.39 | 2.58 | 2.34 | 2.39 | 9.7 |

DRUG CODE:9143 DRUG NAME: OXYCODONE

| STATE | 1ST QUARTER | 2ND QUARTER | 3RD QUARTER | 4TH QUARTER | TOTAL GRAMS |
|---|---|---|---|---|---|
| ALABAMA | 5,126.52 | 4,897.47 | 4,662.14 | 4,629.60 | 19,315.74 |
| ALASKA | 5,056.10 | 4,943.01 | 4,751.11 | 4,651.30 | 19,401.52 |
| AMERICAN SAMOA | 21.86 | 18.44 | 16.39 | 32.78 | 89.48 |
| ARIZONA | 7,185.70 | 6,772.82 | 6,387.28 | 6,256.17 | 26,601.96 |
| ARKANSAS | 5,161.45 | 5,047.72 | 4,890.40 | 4,722.23 | 19,821.81 |
| CALIFORNIA | 2,430.13 | 2,359.21 | 2,278.42 | 2,200.61 | 9,268.36 |
| COLORADO | 4,643.96 | 4,538.56 | 4,325.63 | 4,240.34 | 17,748.50 |
| CONNECTICUT | 4,907.93 | 4,772.68 | 4,489.06 | 4,393.36 | 18,563.03 |
| DELAWARE | 8,025.03 | 7,642.33 | 7,434.31 | 7,391.39 | 30,493.06 |
| DISTRICT OF COLUMBIA | 2,511.66 | 2,499.98 | 2,419.71 | 2,233.56 | 9,664.91 |
| FLORIDA | 5,534.92 | 5,474.95 | 5,299.44 | 5,519.33 | 21,828.64 |
| GEORGIA | 4,702.10 | 4,660.87 | 4,536.95 | 4,501.08 | 18,401 |
| GUAM | 390.9 | 509.2 | 549.37 | 468.02 | 1,917.49 |
| HAWAII | 3,352.74 | 3,236.51 | 3,177.09 | 2,983.09 | 12,749.44 |
| IDAHO | 3,920.64 | 3,867.62 | 3,844.02 | 3,764.25 | 15,396.53 |
| ILLINOIS | 1,291.04 | 1,252.18 | 1,201.62 | 1,185.65 | 4,930.49 |
| INDIANA | 3,725.86 | 3,647.03 | 3,466.79 | 3,415.98 | 14,255.65 |
| IOWA | 1,839.80 | 1,780.25 | 1,684.02 | 1,714.93 | 7,019 |
| KANSAS | 4,362.75 | 4,336.32 | 4,248.76 | 4,173.69 | 17,121.53 |
| KENTUCKY | 4,855.33 | 4,764.23 | 4,594.76 | 4,582.93 | 18,797.25 |
| LOUISIANA | 4,492.87 | 4,324.51 | 4,141.36 | 4,031.67 | 16,990.42 |
| MAINE | 4,077.29 | 3,842.32 | 3,768.44 | 3,692.38 | 15,380.43 |
| MARYLAND | 5,947.71 | 5,757.54 | 5,420.26 | 5,310.90 | 22,436.40 |
| MASSACHUSETTS | 3,865.90 | 3,736.03 | 3,561.24 | 3,488.50 | 14,651.68 |
| MICHIGAN | 3,178.54 | 3,067.17 | 2,958.52 | 2,824.02 | 12,028.26 |
| MINNESOTA | 2,825.29 | 2,719.32 | 2,629.43 | 2,584.67 | 10,758.70 |

| | | | | | |
|---|---|---|---|---|---|
| MISSISSIPPI | 3,684.38 | 3,604.11 | 3,454.84 | 3,385.85 | 14,129.17 |
| MISSOURI | 5,173.34 | 5,100.81 | 4,825.02 | 4,681.81 | 19,780.99 |
| MONTANA | 4,144.82 | 3,878.68 | 3,759.25 | 3,615.36 | 15,398.11 |
| NEBRASKA | 2,347.67 | 2,311.12 | 2,239.93 | 2,204.11 | 9,102.83 |
| NEVADA | 6,898.72 | 6,737.78 | 6,500.37 | 6,424.04 | 26,560.91 |
| NEW HAMPSHIRE | 4,986.29 | 4,771.85 | 4,714.60 | 4,470.41 | 18,943.15 |
| NEW JERSEY | 6,131.76 | 5,815.66 | 5,459.80 | 5,242.19 | 22,649.41 |
| NEW MEXICO | 4,791.80 | 4,497.80 | 4,195.09 | 4,065.50 | 17,550.20 |
| NEW YORK | 4,060.06 | 3,896.12 | 3,707.44 | 3,568.60 | 15,232.23 |
| NORTH CAROLINA | 6,274.13 | 5,980.86 | 5,666.60 | 5,559.90 | 23,481.49 |
| NORTH DAKOTA | 2,254.48 | 2,224.49 | 2,123.88 | 2,108.20 | 8,711.05 |
| OHIO | 4,466.71 | 4,282.55 | 4,065.66 | 3,884.51 | 16,699.43 |
| OKLAHOMA | 6,180.79 | 6,082.39 | 5,889.99 | 5,751.35 | 23,904.52 |
| OREGON | 4,781.04 | 4,614.59 | 4,424.96 | 4,277.15 | 18,097.73 |
| PENNSYLVANIA | 6,192.87 | 6,011.37 | 5,775.74 | 5,653.92 | 23,633.90 |
| PUERTO RICO | 530.66 | 377.61 | 252.09 | 201.14 | 1,361.50 |
| RHODE ISLAND | 4,147.07 | 3,867.28 | 3,762.75 | 3,663.45 | 15,440.55 |
| SOUTH CAROLINA | 5,324.65 | 5,178.65 | 4,878.11 | 4,829.70 | 20,211.11 |
| SOUTH DAKOTA | 2,212.05 | 2,152.63 | 2,105.15 | 1,998.24 | 8,468.07 |
| TENNESSEE | 7,472.71 | 7,352.67 | 7,053.75 | 7,290.62 | 29,169.75 |
| TEXAS | 1,339.91 | 1,337.33 | 1,318.61 | 1,270.75 | 5,266.60 |
| UTAH | 6,306.36 | 6,155.78 | 5,963.49 | 5,851.33 | 24,276.96 |
| VERMONT | 3,811.62 | 3,738.15 | 3,541.26 | 3,410.83 | 14,501.86 |
| VIRGIN ISLANDS | 532.46 | 526.35 | 393.66 | 461.58 | 1,914.04 |
| VIRGINIA | 4,017.78 | 3,754.07 | 3,554.44 | 3,498.79 | 14,825.08 |
| WASHINGTON | 4,546.43 | 4,425.28 | 4,246.97 | 4,064.50 | 17,283.19 |
| WEST VIRGINIA | 4,800.85 | 4,577.57 | 4,382.42 | 4,205.12 | 17,965.97 |
| WISCONSIN | 4,144.33 | 3,807.85 | 3,625.08 | 3,504.89 | 15,082.15 |
| WYOMING | 4,752.44 | 4,507.33 | 4,436.69 | 4,430.60 | 18,127.06 |
| U.S. TOTAL | 4,104.53 | 3,973.31 | 3,805.28 | 3,735.78 | 15,618.90 |

DRUG CODE:9150 DRUG NAME: HYDROMORPHONE

| STATE | 1ST QUARTER | 2ND QUARTER | 3RD QUARTER | 4TH QUARTER | TOTAL GRAMS |
|---|---|---|---|---|---|
| ALABAMA | 73.83 | 66.41 | 67.39 | 65.13 | 272.76 |
| ALASKA | 149.99 | 137.55 | 127.64 | 133.56 | 548.73 |
| AMERICAN SAMOA | 1.35 | 0.14 | 0 | 0 | 1.49 |
| ARIZONA | 110.91 | 107.91 | 103.18 | 97.46 | 419.45 |
| ARKANSAS | 115.81 | 112.73 | 110.45 | 108.71 | 447.7 |
| CALIFORNIA | 108.65 | 107.02 | 102.34 | 98.84 | 416.85 |
| COLORADO | 146.56 | 133.97 | 129.38 | 130.9 | 540.81 |
| CONNECTICUT | 186.66 | 180.43 | 173.62 | 174.88 | 715.59 |
| DELAWARE | 255.57 | 231.59 | 227.1 | 234.94 | 949.2 |
| DISTRICT OF COLUMBIA | 99.16 | 89.71 | 87.95 | 83.36 | 360.19 |
| FLORIDA | 266.78 | 248.15 | 236.12 | 242.11 | 993.16 |
| GEORGIA | 90.56 | 85.89 | 86.82 | 80.45 | 343.72 |
| GUAM | 17.79 | 14.75 | 13.12 | 18.51 | 64.18 |
| HAWAII | 50.92 | 51.92 | 47.26 | 46.28 | 196.37 |
| IDAHO | 123.93 | 115.78 | 117.34 | 123.05 | 480.09 |
| ILLINOIS | 80.22 | 76.66 | 73.35 | 73 | 303.22 |
| INDIANA | 93.49 | 93.85 | 91.3 | 88.87 | 367.51 |
| IOWA | 104.29 | 98.02 | 94.31 | 93.75 | 390.37 |
| KANSAS | 125.32 | 130.07 | 124.86 | 109.66 | 489.92 |
| KENTUCKY | 68.45 | 61.01 | 65.03 | 77.29 | 271.78 |
| LOUISIANA | 81.94 | 71.8 | 68.29 | 68.55 | 290.58 |
| MAINE | 121.34 | 150.91 | 99.55 | 131.56 | 503.36 |
| MARYLAND | 154.74 | 147.04 | 139.84 | 137.13 | 578.75 |
| MASSACHUSETTS | 119.02 | 114.93 | 104.88 | 111.33 | 450.15 |
| MICHIGAN | 96.95 | 89.44 | 90.43 | 88.95 | 365.78 |
| MINNESOTA | 111.94 | 113.78 | 112.36 | 110.47 | 448.56 |

| | | | | | |
|---|---|---|---|---|---|
| MISSISSIPPI | 169.44 | 209.66 | 193.32 | 216.3 | 788.72 |
| MISSOURI | 116.89 | 106.28 | 103.32 | 104.35 | 430.83 |
| MONTANA | 169.91 | 163.04 | 175.18 | 155.33 | 663.47 |
| NEBRASKA | 75.41 | 75.34 | 66.97 | 67.37 | 285.09 |
| NEVADA | 117.52 | 111.37 | 104.89 | 103.03 | 436.82 |
| NEW HAMPSHIRE | 156.43 | 151.73 | 150.21 | 138.96 | 597.33 |
| NEW JERSEY | 108.91 | 102.82 | 95.94 | 96.08 | 403.75 |
| NEW MEXICO | 71.16 | 68.85 | 61.69 | 60.33 | 262.03 |
| NEW YORK | 104.6 | 102.01 | 98.62 | 97.61 | 402.83 |
| NORTH CAROLINA | 122.23 | 112.89 | 111.73 | 104.5 | 451.34 |
| NORTH DAKOTA | 95.81 | 100.85 | 93.74 | 95.47 | 385.87 |
| OHIO | 83.19 | 80.62 | 80.07 | 80.16 | 324.04 |
| OKLAHOMA | 97.73 | 100.71 | 96.93 | 93.83 | 389.19 |
| OREGON | 131.98 | 129.2 | 126.37 | 117.42 | 504.97 |
| PENNSYLVANIA | 139.79 | 115.63 | 109.67 | 111.21 | 476.3 |
| PUERTO RICO | 2.03 | 1.75 | 2.17 | 2.01 | 7.96 |
| RHODE ISLAND | 84.26 | 98.31 | 77.06 | 76.03 | 335.65 |
| SOUTH CAROLINA | 129.76 | 126.04 | 119.13 | 121.81 | 496.75 |
| SOUTH DAKOTA | 63.83 | 64.56 | 62.99 | 78.8 | 270.17 |
| TENNESSEE | 129.25 | 97.42 | 101.63 | 91.74 | 420.04 |
| TEXAS | 98.41 | 96.93 | 94.6 | 89.48 | 379.43 |
| UTAH | 92.57 | 94.47 | 100.68 | 106.85 | 394.57 |
| VERMONT | 197.98 | 202.93 | 191.25 | 186.56 | 778.73 |
| VIRGIN ISLANDS | 24.22 | 17.03 | 12.69 | 13.57 | 67.51 |
| VIRGINIA | 156.93 | 142.31 | 131.47 | 131.02 | 561.73 |
| WASHINGTON | 166.24 | 165.25 | 158.57 | 146.81 | 636.87 |
| WEST VIRGINIA | 90.98 | 83.7 | 78.45 | 73.6 | 326.72 |
| WISCONSIN | 67.78 | 61.74 | 65.05 | 66.77 | 261.34 |
| WYOMING | 115.71 | 113.98 | 107.27 | 95.83 | 432.8 |
| U.S. TOTAL | 118.89 | 113.17 | 109.02 | 107.77 | 448.85 |

DRUG CODE:9193 DRUG NAME: HYDROCODONE

| STATE | 1ST QUARTER | 2ND QUARTER | 3RD QUARTER | 4TH QUARTER | TOTAL GRAMS |
|---|---|---|---|---|---|
| ALABAMA | 5,405.01 | 5,157.45 | 4,881.85 | 4,889.37 | 20,333.67 |
| ALASKA | 1,682.16 | 1,618.17 | 1,517.15 | 1,452.32 | 6,269.80 |
| AMERICAN SAMOA | 6.09 | 0.42 | 5.53 | 0 | 12.04 |
| ARIZONA | 1,753.75 | 1,635.20 | 1,538.83 | 1,530.33 | 6,458.11 |
| ARKANSAS | 4,808.29 | 4,680.16 | 4,455.99 | 4,426.35 | 18,370.79 |
| CALIFORNIA | 2,556.31 | 2,501.71 | 2,399.79 | 2,356.88 | 9,814.69 |
| COLORADO | 1,368.60 | 1,303.83 | 1,258.98 | 1,236.78 | 5,168.19 |
| CONNECTICUT | 607.13 | 572.14 | 537.47 | 531.65 | 2,248.38 |
| DELAWARE | 711.6 | 658.94 | 648.84 | 648.98 | 2,668.35 |
| DISTRICT OF COLUMBIA | 261.21 | 242.74 | 240.13 | 214.25 | 958.32 |
| FLORIDA | 2,043.37 | 1,932.88 | 1,801.80 | 1,881.09 | 7,659.14 |
| GEORGIA | 2,673.91 | 2,582.11 | 2,452.27 | 2,449.57 | 10,157.86 |
| GUAM | 82.96 | 110.13 | 83.72 | 76.26 | 353.07 |
| HAWAII | 1,368.32 | 1,303.39 | 1,257 | 1,208.79 | 5,137.50 |
| IDAHO | 3,734.82 | 3,691.64 | 3,560.42 | 3,561.58 | 14,548.46 |
| ILLINOIS | 2,494.39 | 2,437.70 | 2,338.97 | 2,324.68 | 9,595.74 |
| INDIANA | 3,653.80 | 3,589.91 | 3,383.47 | 3,329.26 | 13,956.44 |
| IOWA | 1,949.69 | 1,922.84 | 1,844.88 | 1,822 | 7,539.41 |
| KANSAS | 2,949.73 | 2,882.55 | 2,782.47 | 2,764.77 | 11,379.52 |
| KENTUCKY | 4,803.66 | 4,691.86 | 4,479.08 | 4,451.34 | 18,425.93 |
| LOUISIANA | 3,821.33 | 3,663.20 | 3,484.43 | 3,415.04 | 14,384 |
| MAINE | 1,338.22 | 1,302.09 | 1,267.65 | 1,245.74 | 5,153.70 |
| MARYLAND | 650.71 | 608.49 | 572.13 | 566.99 | 2,398.32 |
| MASSACHUSETTS | 485.77 | 470.38 | 443.48 | 443.35 | 1,842.98 |
| MICHIGAN | 4,231.89 | 4,156.59 | 3,961.66 | 3,789.50 | 16,139.64 |
| MINNESOTA | 911.22 | 870.48 | 844.99 | 815.7 | 3,442.39 |

| | | | | | |
|---|---|---|---|---|---|
| MISSISSIPPI | 4,257.62 | 4,083.86 | 3,786.65 | 3,594.84 | 15,722.97 |
| MISSOURI | 3,078.84 | 3,041.97 | 2,879.80 | 2,820.36 | 11,820.96 |
| MONTANA | 2,707.62 | 2,636.90 | 2,555.95 | 2,497.44 | 10,397.91 |
| NEBRASKA | 1,860.92 | 1,778.61 | 1,745.17 | 1,731.79 | 7,116.49 |
| NEVADA | 3,662.67 | 3,567.83 | 3,370.89 | 3,377.43 | 13,978.82 |
| NEW HAMPSHIRE | 721 | 686.15 | 664.84 | 671.97 | 2,743.95 |
| NEW JERSEY | 435.42 | 393.21 | 372.6 | 369.86 | 1,571.08 |
| NEW MEXICO | 1,792.35 | 1,730.07 | 1,634.42 | 1,619.82 | 6,776.67 |
| NEW YORK | 929.79 | 898.9 | 851.21 | 835.15 | 3,515.05 |
| NORTH CAROLINA | 2,223.98 | 2,050.06 | 1,896.73 | 1,897.21 | 8,067.98 |
| NORTH DAKOTA | 1,628.92 | 1,570.20 | 1,494.06 | 1,475.99 | 6,169.18 |
| OHIO | 1,726.05 | 1,642.47 | 1,562.80 | 1,508.25 | 6,439.57 |
| OKLAHOMA | 4,997.10 | 4,947.43 | 4,725.35 | 4,679.17 | 19,349.05 |
| OREGON | 2,590.02 | 2,486.19 | 2,403.66 | 2,352.91 | 9,832.78 |
| PENNSYLVANIA | 1,387.18 | 1,315.49 | 1,291.48 | 1,202.42 | 5,196.57 |
| PUERTO RICO | 29.45 | 17.74 | 10.74 | 10.42 | 68.35 |
| RHODE ISLAND | 1,064.04 | 997.87 | 950.9 | 928.75 | 3,941.57 |
| SOUTH CAROLINA | 3,188.56 | 3,038.62 | 2,829.09 | 2,845.92 | 11,902.20 |
| SOUTH DAKOTA | 1,940.71 | 1,832.22 | 1,770.96 | 1,728.09 | 7,271.98 |
| TENNESSEE | 4,032.91 | 3,894.17 | 3,767.47 | 3,635.30 | 15,329.84 |
| TEXAS | 2,738.84 | 2,685.15 | 2,589.01 | 2,558.69 | 10,571.68 |
| UTAH | 2,221.04 | 2,145.81 | 2,058.33 | 2,036.74 | 8,461.91 |
| VERMONT | 820.6 | 784.69 | 729.57 | 723.53 | 3,058.39 |
| VIRGIN ISLANDS | 331.09 | 310.47 | 203.67 | 209.71 | 1,054.95 |
| VIRGINIA | 1,454.75 | 1,317.73 | 1,219.08 | 1,212.02 | 5,203.58 |
| WASHINGTON | 2,044.24 | 1,984.69 | 1,906.75 | 1,848.78 | 7,784.45 |
| WEST VIRGINIA | 3,386.21 | 3,256.11 | 3,095.13 | 2,977.19 | 12,714.64 |
| WISCONSIN | 1,451.36 | 1,371.16 | 1,328.13 | 1,327.34 | 5,478 |
| WYOMING | 2,161.79 | 2,074.66 | 1,971.55 | 1,955.11 | 8,163.11 |
| U.S. TOTAL | 2,285.19 | 2,208.13 | 2,104.46 | 2,074.70 | 8,672.48 |

DRUG CODE:9220L DRUG NAME: LEVORPHANOL

| STATE | 1ST QUARTER | 2ND QUARTER | 3RD QUARTER | 4TH QUARTER | TOTAL GRAMS |
|---|---|---|---|---|---|
| ALABAMA | 0.12 | 0.09 | 0.11 | 0.09 | 0.42 |
| ARIZONA | 0.29 | 0.35 | 0.46 | 0.47 | 1.56 |
| ARKANSAS | 0.1 | 0.12 | 0.07 | 0.03 | 0.31 |
| CALIFORNIA | 0.29 | 0.27 | 0.27 | 0.25 | 1.08 |
| COLORADO | 0.12 | 0.26 | 0.27 | 0.36 | 1.01 |
| CONNECTICUT | 0.05 | 0.1 | 0.14 | 0.27 | 0.57 |
| DELAWARE | 0.1 | 0 | 0.06 | 0.12 | 0.28 |
| FLORIDA | 0.14 | 0.19 | 0.18 | 0.21 | 0.72 |
| GEORGIA | 0.01 | 0.03 | 0.03 | 0.04 | 0.1 |
| IDAHO | 0 | 0 | 0.01 | 0.01 | 0.01 |
| ILLINOIS | 0.07 | 0.1 | 0.12 | 0.12 | 0.41 |
| INDIANA | 0.4 | 0.38 | 0.38 | 0.34 | 1.5 |
| IOWA | 0.14 | 0.13 | 0.11 | 0.12 | 0.51 |
| KANSAS | 0.39 | 0.35 | 0.25 | 0.26 | 1.26 |
| KENTUCKY | 0 | 0 | 0.01 | 0 | 0.01 |
| LOUISIANA | 0.36 | 0.43 | 0.46 | 0.43 | 1.68 |
| MARYLAND | 0.05 | 0.1 | 0.07 | 0.14 | 0.37 |
| MASSACHUSETTS | 0.32 | 0.3 | 0.31 | 0.31 | 1.23 |
| MICHIGAN | 0 | 0 | 0 | 0 | 0.01 |
| MINNESOTA | 0.01 | 0.01 | 0.02 | 0.02 | 0.05 |
| MISSISSIPPI | 0.01 | 0 | 0 | 0 | 0.01 |
| MISSOURI | 0.23 | 0.33 | 0.44 | 0.46 | 1.45 |
| NEBRASKA | 0.22 | 0.28 | 0.3 | 0.5 | 1.3 |
| NEVADA | 0.11 | 0.11 | 0.18 | 0.13 | 0.52 |
| NEW HAMPSHIRE | 0.09 | 0.14 | 0.12 | 0.05 | 0.41 |
| NEW JERSEY | 3.57 | 4.56 | 4.63 | 5.67 | 18.43 |

| | | | | | |
|---|---|---|---|---|---|
| NEW YORK | 0.17 | 0.2 | 0.21 | 0.25 | 0.83 |
| NORTH CAROLINA | 0.53 | 0.51 | 0.68 | 1.07 | 2.79 |
| OHIO | 0.07 | 0.13 | 0.1 | 0.08 | 0.38 |
| OREGON | 0.09 | 0.09 | 0.08 | 0.1 | 0.35 |
| PENNSYLVANIA | 0.08 | 0.12 | 0.12 | 0.1 | 0.42 |
| SOUTH CAROLINA | 0.02 | 0.07 | 0.07 | 0.08 | 0.24 |
| TENNESSEE | 0 | 0 | 0.01 | 0 | 0.01 |
| TEXAS | 0.16 | 0.21 | 0.24 | 0.26 | 0.88 |
| VERMONT | 0.04 | 0.07 | 0.07 | 0.07 | 0.26 |
| VIRGINIA | 0.07 | 0.07 | 0.14 | 0.2 | 0.49 |
| WASHINGTON | 0.34 | 0.36 | 0.46 | 0.46 | 1.62 |
| WISCONSIN | 0.03 | 0.07 | 0.09 | 0.09 | 0.27 |
| U.S. TOTAL | 0.25 | 0.3 | 0.31 | 0.36 | 1.22 |

DRUG CODE:9230 DRUG NAME: MEPERIDINE (PETHIDINE)

| STATE | 1ST QUARTER | 2ND QUARTER | 3RD QUARTER | 4TH QUARTER | TOTAL GRAMS |
|---|---|---|---|---|---|
| ALABAMA | 167.58 | 154.66 | 143.58 | 149.59 | 615.41 |
| ALASKA | 119.7 | 99.15 | 93.27 | 89.3 | 401.42 |
| ARIZONA | 85.28 | 76.99 | 68.42 | 74.47 | 305.16 |
| ARKANSAS | 286.33 | 268.84 | 242.99 | 216.94 | 1,015.10 |
| CALIFORNIA | 42.88 | 42.61 | 40.66 | 41.27 | 167.42 |
| COLORADO | 43.17 | 37.6 | 34.39 | 29.57 | 144.74 |
| CONNECTICUT | 30.89 | 29.44 | 29 | 29.93 | 119.26 |
| DELAWARE | 27.66 | 30.57 | 24.03 | 14.7 | 96.97 |
| DISTRICT OF COLUMBIA | 36.17 | 36.57 | 28.08 | 24.59 | 125.41 |
| FLORIDA | 52.93 | 50.59 | 41.53 | 45.44 | 190.49 |
| GEORGIA | 118.69 | 107.8 | 100.28 | 102.54 | 429.32 |
| GUAM | 17.16 | 19.41 | 37.12 | 29.24 | 102.94 |
| HAWAII | 28.1 | 28.99 | 22.49 | 20.2 | 99.78 |
| IDAHO | 54.1 | 59.02 | 47.25 | 52.05 | 212.43 |
| ILLINOIS | 23.31 | 25.52 | 24.95 | 19.83 | 93.61 |
| INDIANA | 55.36 | 54.37 | 50.29 | 47.04 | 207.06 |
| IOWA | 63.27 | 62.17 | 65.36 | 64.61 | 255.42 |
| KANSAS | 65.22 | 62.84 | 60.01 | 59.77 | 247.84 |
| KENTUCKY | 66.38 | 53 | 56.91 | 49.03 | 225.32 |
| LOUISIANA | 178.91 | 166.99 | 148.12 | 146.1 | 640.13 |
| MAINE | 35.03 | 30.82 | 27.03 | 25.92 | 118.81 |
| MARYLAND | 47.64 | 42.86 | 36.64 | 35.15 | 162.29 |
| MASSACHUSETTS | 34.65 | 34.37 | 33.13 | 26.97 | 129.13 |
| MICHIGAN | 30.92 | 29.4 | 26.97 | 23.28 | 110.57 |
| MINNESOTA | 15.76 | 12.27 | 12.29 | 11.22 | 51.54 |
| MISSISSIPPI | 157.4 | 155.7 | 134.5 | 135.91 | 583.51 |
| MISSOURI | 77.28 | 65.34 | 58.88 | 52.48 | 253.98 |
| MONTANA | 66.4 | 78.29 | 66.41 | 60.4 | 271.51 |
| NEBRASKA | 43.92 | 42.36 | 36.68 | 29.54 | 152.49 |
| NEVADA | 60.46 | 45.86 | 46.38 | 57.28 | 209.99 |
| NEW HAMPSHIRE | 51.36 | 46.41 | 46.53 | 34.2 | 178.5 |
| NEW JERSEY | 24.87 | 24.8 | 20.84 | 17.1 | 87.61 |
| NEW MEXICO | 81.62 | 68.79 | 63.85 | 60.14 | 274.41 |
| NEW YORK | 26.44 | 24.06 | 22.2 | 21.68 | 94.38 |
| NORTH CAROLINA | 75.41 | 67.77 | 56.96 | 57.23 | 257.37 |
| NORTH DAKOTA | 35.31 | 35.32 | 37.72 | 33.15 | 141.51 |
| OHIO | 45.61 | 44.13 | 46.95 | 36.51 | 173.2 |
| OKLAHOMA | 204.07 | 196.86 | 181.48 | 181.46 | 763.87 |
| OREGON | 45.67 | 38.22 | 41.39 | 34.34 | 159.63 |
| PENNSYLVANIA | 31.05 | 27.69 | 26.97 | 26.07 | 111.78 |
| PUERTO RICO | 151.73 | 132.37 | 110.55 | 71.62 | 466.26 |
| RHODE ISLAND | 24.03 | 25.14 | 24.39 | 22.09 | 95.65 |
| SOUTH CAROLINA | 146.59 | 132.3 | 119.44 | 110.11 | 508.44 |

| | | | | | |
|---|---|---|---|---|---|
| SOUTH DAKOTA | 67.66 | 68.63 | 65.07 | 53.65 | 255.01 |
| TENNESSEE | 137.89 | 123.67 | 106.23 | 100.09 | 467.88 |
| TEXAS | 46.85 | 43 | 46.89 | 47.52 | 184.25 |
| UTAH | 72.75 | 56.49 | 64.71 | 66.39 | 260.34 |
| VERMONT | 51.76 | 50.35 | 42.44 | 61.26 | 205.81 |
| VIRGIN ISLANDS | 38.19 | 61.6 | 38.19 | 93.24 | 231.22 |
| VIRGINIA | 87.23 | 74.14 | 68.24 | 61.34 | 290.95 |
| WASHINGTON | 56.11 | 50.13 | 48.55 | 46.6 | 201.4 |
| WEST VIRGINIA | 39.48 | 35.75 | 31.63 | 27.53 | 134.4 |
| WISCONSIN | 27.41 | 26.33 | 25.79 | 23.39 | 102.92 |
| WYOMING | 67.13 | 56.47 | 45.11 | 62.24 | 230.94 |
| U.S. TOTAL | 61.86 | 57.26 | 53.2 | 51.03 | 223.35 |

DRUG CODE:9250B DRUG NAME: METHADONE

| STATE | 1ST QUARTER | 2ND QUARTER | 3RD QUARTER | 4TH QUARTER | TOTAL GRAMS |
|---|---|---|---|---|---|
| ALABAMA | 1,595.91 | 1,688.93 | 1,567.57 | 1,683.98 | 6,536.39 |
| ALASKA | 1,099.63 | 1,187.65 | 1,182.91 | 1,047.63 | 4,517.82 |
| ARIZONA | 1,332.76 | 1,331.16 | 1,289.71 | 1,298.76 | 5,252.39 |
| ARKANSAS | 578.83 | 569.57 | 533.93 | 520.82 | 2,203.14 |
| CALIFORNIA | 946.06 | 955.59 | 920.18 | 906.05 | 3,727.89 |
| COLORADO | 837.12 | 860.85 | 894.17 | 848.14 | 3,440.28 |
| CONNECTICUT | 3,336.76 | 3,522.89 | 2,942.55 | 3,527.18 | 13,329.37 |
| DELAWARE | 2,813.13 | 3,417.48 | 2,940.22 | 3,078.77 | 12,249.60 |
| DISTRICT OF COLUMBIA | 1,531.28 | 1,775.19 | 1,533.92 | 1,792.13 | 6,632.52 |
| FLORIDA | 913.65 | 962.11 | 1,193.98 | 1,093.34 | 4,163.08 |
| GEORGIA | 1,145.46 | 1,254.09 | 1,201.47 | 1,205.09 | 4,806.11 |
| GUAM | 32.58 | 21.55 | 23.81 | 18.05 | 95.99 |
| HAWAII | 577.77 | 660.24 | 559.54 | 576.61 | 2,374.16 |
| IDAHO | 769.33 | 768.84 | 748.93 | 726 | 3,013.10 |
| ILLINOIS | 944.79 | 988.23 | 930.01 | 951.47 | 3,814.50 |
| INDIANA | 1,304.24 | 1,280.87 | 1,233.76 | 1,223.46 | 5,042.34 |
| IOWA | 403.75 | 442.86 | 415.46 | 453.78 | 1,715.86 |
| KANSAS | 795.19 | 860.61 | 787.39 | 796.97 | 3,240.16 |
| KENTUCKY | 1,177.79 | 1,143.06 | 1,244.34 | 1,184.83 | 4,750.03 |
| LOUISIANA | 790.42 | 824.57 | 821.38 | 710 | 3,146.37 |
| MAINE | 2,435.10 | 2,469.39 | 2,220.23 | 2,377.62 | 9,502.34 |
| MARYLAND | 3,338.86 | 3,342.10 | 3,246.49 | 3,310.05 | 13,237.50 |
| MASSACHUSETTS | 2,247.65 | 2,228.13 | 2,163.68 | 2,209.15 | 8,848.62 |
| MICHIGAN | 1,356.81 | 1,364.03 | 1,354.88 | 1,327.27 | 5,402.99 |
| MINNESOTA | 839.98 | 992.69 | 963.6 | 1,121.80 | 3,918.06 |
| MISSISSIPPI | 205.1 | 204.31 | 246.25 | 242.26 | 897.91 |
| MISSOURI | 732.47 | 626.06 | 698.02 | 714.92 | 2,771.47 |
| MONTANA | 776.26 | 756.5 | 656.68 | 763.92 | 2,953.36 |
| NEBRASKA | 481.84 | 435.78 | 439.18 | 435.02 | 1,791.82 |
| NEVADA | 1,369.27 | 1,405.19 | 1,297.13 | 1,286.43 | 5,358.02 |
| NEW HAMPSHIRE | 2,348.44 | 2,372.58 | 2,322.88 | 2,324.61 | 9,368.51 |
| NEW JERSEY | 1,281.73 | 1,378.77 | 1,233.28 | 1,333.32 | 5,227.10 |
| NEW MEXICO | 2,089.30 | 2,071.59 | 2,149.01 | 2,256.45 | 8,566.35 |
| NEW YORK | 1,502.90 | 1,630.81 | 1,568.12 | 1,590.78 | 6,292.62 |
| NORTH CAROLINA | 1,357.40 | 1,411.79 | 1,390.35 | 1,361.90 | 5,521.44 |
| NORTH DAKOTA | 236.42 | 278.23 | 326.04 | 404.11 | 1,244.80 |
| OHIO | 677.59 | 648.18 | 631.38 | 620.01 | 2,577.15 |
| OKLAHOMA | 1,017.04 | 1,151.98 | 976.18 | 1,040.11 | 4,185.30 |
| OREGON | 1,663.59 | 1,692.09 | 1,653.50 | 1,614.44 | 6,623.62 |
| PENNSYLVANIA | 1,528.07 | 1,566.65 | 1,519.22 | 1,599.92 | 6,213.86 |
| PUERTO RICO | 600.96 | 491.5 | 645.96 | 516.91 | 2,255.32 |
| RHODE ISLAND | 3,865.02 | 4,094.51 | 4,323.70 | 4,543.16 | 16,826.38 |
| SOUTH CAROLINA | 1,247.13 | 1,210.21 | 1,216.96 | 1,198.36 | 4,872.66 |
| SOUTH DAKOTA | 278.19 | 263.71 | 274.1 | 259.66 | 1,075.67 |

| | | | | | |
|---|---|---|---|---|---|
| TENNESSEE | 777.26 | 772.8 | 751.65 | 760.07 | 3,061.78 |
| TEXAS | 554.52 | 580.2 | 535.49 | 564.86 | 2,235.06 |
| UTAH | 972.67 | 863.59 | 1,032.46 | 955.12 | 3,823.84 |
| VERMONT | 3,012.80 | 2,725.08 | 3,222.76 | 2,865.22 | 11,825.86 |
| VIRGIN ISLANDS | 25.65 | 351.81 | 21.17 | 17.92 | 416.56 |
| VIRGINIA | 1,020.69 | 942.41 | 982.45 | 1,017.35 | 3,962.90 |
| WASHINGTON | 1,266.05 | 1,390.10 | 1,547.12 | 1,608.88 | 5,812.15 |
| WEST VIRGINIA | 1,270.12 | 1,268.68 | 1,241.40 | 1,215.51 | 4,995.72 |
| WISCONSIN | 1,155.51 | 1,211.02 | 1,117.14 | 1,125.93 | 4,609.60 |
| WYOMING | 226.68 | 206.66 | 209.83 | 200.74 | 843.91 |
| U.S. TOTAL | 1,156.30 | 1,189 | 1,168.51 | 1,182.62 | 4,696.43 |

DRUG CODE:9300 DRUG NAME: MORPHINE

| STATE | 1ST QUARTER | 2ND QUARTER | 3RD QUARTER | 4TH QUARTER | TOTAL GRAMS |
|---|---|---|---|---|---|
| ALABAMA | 1,577.19 | 1,501.17 | 1,429.84 | 1,393.10 | 5,901.31 |
| ALASKA | 1,700.16 | 1,643.84 | 1,588.01 | 1,507.11 | 6,439.12 |
| AMERICAN SAMOA | 45.83 | 41.82 | 4.58 | 41.25 | 133.49 |
| ARIZONA | 2,812.09 | 2,606.69 | 2,450.29 | 2,269.82 | 10,138.90 |
| ARKANSAS | 1,685.40 | 1,565.37 | 1,524.09 | 1,428.69 | 6,203.55 |
| CALIFORNIA | 1,257.08 | 1,207.37 | 1,170.25 | 1,131.23 | 4,765.93 |
| COLORADO | 1,609.62 | 1,540.44 | 1,487.08 | 1,436.58 | 6,073.73 |
| CONNECTICUT | 1,034.18 | 997.5 | 982.95 | 943.15 | 3,957.78 |
| DELAWARE | 2,504.24 | 2,438.20 | 2,375.92 | 2,335.75 | 9,654.11 |
| DISTRICT OF COLUMBIA | 617.16 | 640.41 | 629.91 | 534.77 | 2,422.25 |
| FLORIDA | 2,195 | 2,100.91 | 2,039.25 | 2,034.79 | 8,369.95 |
| GEORGIA | 1,205.98 | 1,159.23 | 1,146.03 | 1,118.93 | 4,630.17 |
| GUAM | 712.08 | 465.01 | 577.82 | 485.89 | 2,240.80 |
| HAWAII | 1,720.97 | 1,653.19 | 1,557.59 | 1,475.59 | 6,407.33 |
| IDAHO | 1,899.53 | 1,821.65 | 1,734.59 | 1,722.60 | 7,178.37 |
| ILLINOIS | 841.83 | 804.48 | 780.04 | 764.09 | 3,190.45 |
| INDIANA | 1,421.27 | 1,347.56 | 1,280.45 | 1,253.01 | 5,302.29 |
| IOWA | 1,144.62 | 1,113.67 | 1,070.42 | 1,069.16 | 4,397.88 |
| KANSAS | 1,641.83 | 1,553.36 | 1,543.38 | 1,488.80 | 6,227.36 |
| KENTUCKY | 1,149.93 | 1,073.76 | 1,028.41 | 1,021.40 | 4,273.50 |
| LOUISIANA | 1,241.81 | 1,199.06 | 1,159.36 | 1,115.05 | 4,715.28 |
| MAINE | 1,283.23 | 1,269.14 | 1,236.15 | 1,175.03 | 4,963.55 |
| MARYLAND | 1,421.81 | 1,360.18 | 1,256.29 | 1,212.28 | 5,250.55 |
| MASSACHUSETTS | 1,062.50 | 1,015.75 | 964.43 | 961.79 | 4,004.48 |
| MICHIGAN | 2,032.13 | 1,949.57 | 1,881.68 | 1,761.04 | 7,624.42 |
| MINNESOTA | 934.19 | 885.09 | 855.54 | 822.07 | 3,496.89 |
| MISSISSIPPI | 1,392.98 | 1,366.41 | 1,309.08 | 1,317.28 | 5,385.75 |
| MISSOURI | 1,711.88 | 1,620.97 | 1,582.18 | 1,529.09 | 6,444.12 |
| MONTANA | 1,927.07 | 1,741.84 | 1,718.57 | 1,651.21 | 7,038.70 |
| NEBRASKA | 1,082.53 | 1,032.18 | 1,037 | 992.81 | 4,144.52 |
| NEVADA | 2,319.33 | 2,251.18 | 2,168.54 | 2,055.98 | 8,795.03 |
| NEW HAMPSHIRE | 1,411.57 | 1,355.27 | 1,359.84 | 1,378.76 | 5,505.44 |
| NEW JERSEY | 1,058.83 | 980.54 | 924.95 | 905.63 | 3,869.95 |
| NEW MEXICO | 1,387.88 | 1,262.32 | 1,231.39 | 1,127.83 | 5,009.42 |
| NEW YORK | 844.57 | 808.1 | 777.84 | 772.88 | 3,203.40 |
| NORTH CAROLINA | 1,328.46 | 1,258.96 | 1,221.39 | 1,184.54 | 4,993.35 |
| NORTH DAKOTA | 994.12 | 954.76 | 884.87 | 902.81 | 3,736.57 |
| OHIO | 1,083.16 | 1,049.26 | 1,027.18 | 981.36 | 4,140.96 |
| OKLAHOMA | 2,585.28 | 2,468.24 | 2,375.84 | 2,267.11 | 9,696.48 |
| OREGON | 2,015.99 | 1,881.40 | 1,796.96 | 1,727.05 | 7,421.39 |
| PENNSYLVANIA | 2,075.34 | 1,809.88 | 1,742.33 | 1,711.46 | 7,339.02 |
| PUERTO RICO | 69.47 | 60.16 | 50.07 | 45.75 | 225.45 |
| RHODE ISLAND | 1,146.71 | 1,110.61 | 1,142.21 | 1,030.45 | 4,429.98 |
| SOUTH CAROLINA | 1,485.87 | 1,401.04 | 1,381.15 | 1,384.39 | 5,652.44 |
| SOUTH DAKOTA | 1,118.32 | 1,074.08 | 1,065.61 | 1,016.36 | 4,274.37 |

| | | | | | |
|---|---|---|---|---|---|
| TENNESSEE | 2,926.58 | 2,506.78 | 2,453.40 | 2,356.10 | 10,242.86 |
| TEXAS | 853.56 | 822.99 | 803.11 | 785.89 | 3,265.54 |
| UTAH | 1,853.30 | 1,787.35 | 1,774.52 | 1,696.30 | 7,111.47 |
| VERMONT | 1,717.36 | 1,628.65 | 1,559.56 | 1,571.33 | 6,476.89 |
| VIRGIN ISLANDS | 127.13 | 138.41 | 108.73 | 103.66 | 477.93 |
| VIRGINIA | 1,055.60 | 988.16 | 951.02 | 928.08 | 3,922.86 |
| WASHINGTON | 1,712.62 | 1,641.58 | 1,589.44 | 1,505.54 | 6,449.19 |
| WEST VIRGINIA | 1,262.23 | 1,180.61 | 1,167.81 | 1,108.55 | 4,719.20 |
| WISCONSIN | 1,471.62 | 1,363.62 | 1,312.57 | 1,275.49 | 5,423.30 |
| WYOMING | 1,636.07 | 1,656.24 | 1,596.82 | 1,474.57 | 6,363.70 |
| U.S. TOTAL | 1,418.27 | 1,339.43 | 1,295.02 | 1,255.83 | 5,308.55 |

DRUG CODE:9333 DRUG NAME: THEBAINE

| STATE | 1ST QUARTER | 2ND QUARTER | 3RD QUARTER | 4TH QUARTER | TOTAL GRAMS |
|---|---|---|---|---|---|
| TEXAS | 0 | 0 | 0.1 | 0 | 0.1 |
| U.S. TOTAL | 0 | 0 | 0.01 | 0 | 0.01 |

DRUG CODE:9630 DRUG NAME: OPIUM TINCTURE (LAUDANUM)

| STATE | 1ST QUARTER | 2ND QUARTER | 3RD QUARTER | 4TH QUARTER | TOTAL GRAMS |
|---|---|---|---|---|---|
| CALIFORNIA | 0.02 | 0 | 0 | 0 | 0.02 |
| CONNECTICUT | 0 | 0 | 0.13 | 0 | 0.13 |
| DELAWARE | 0 | 0.13 | 0 | 0 | 0.13 |
| FLORIDA | 0.05 | 0.03 | 0 | 0 | 0.08 |
| ILLINOIS | 0.05 | 0.08 | 0 | 0 | 0.13 |
| INDIANA | 0.33 | 0.22 | 0 | 0 | 0.55 |
| KANSAS | 0.21 | 0 | 0 | 0 | 0.21 |
| KENTUCKY | 0.05 | 0 | 0 | 0 | 0.05 |
| MARYLAND | 0.08 | 0.14 | 0 | 0 | 0.22 |
| MICHIGAN | 0.26 | 0.05 | 0 | 0 | 0.31 |
| NEW HAMPSHIRE | 0.36 | 0 | 0 | 0 | 0.36 |
| NEW JERSEY | 0 | 0.01 | 0 | 0 | 0.01 |
| NEW YORK | 0.02 | 0 | 0 | 0 | 0.02 |
| OHIO | 0.63 | 0.08 | 0 | 0 | 0.72 |
| OREGON | 0.06 | 0 | 0 | 0 | 0.06 |
| PENNSYLVANIA | 0.08 | 0.08 | 0 | 0 | 0.17 |
| TEXAS | 0.04 | 0.02 | 0 | 0 | 0.06 |
| VIRGINIA | 0.13 | 0 | 0 | 0 | 0.13 |
| WASHINGTON | 0.05 | 0 | 0 | 0 | 0.05 |
| WEST VIRGINIA | 0.19 | 0.13 | 0 | 0 | 0.32 |
| WISCONSIN | 0.06 | 0.19 | 0 | 0 | 0.25 |
| U.S. TOTAL | 0.07 | 0.03 | 0 | 0 | 0.1 |

DRUG CODE:9639 DRUG NAME: OPIUM POWDERED

| STATE | 1ST QUARTER | 2ND QUARTER | 3RD QUARTER | 4TH QUARTER | TOTAL GRAMS |
|---|---|---|---|---|---|
| ALABAMA | 1.72 | 1.98 | 1.81 | 1.64 | 7.15 |
| ALASKA | 4.16 | 3.55 | 2.79 | 3.95 | 14.45 |
| ARIZONA | 4.34 | 4.28 | 4.57 | 5.28 | 18.48 |
| ARKANSAS | 2.86 | 3.32 | 2.99 | 3.21 | 12.38 |
| CALIFORNIA | 1.11 | 1.26 | 1.3 | 1.22 | 4.89 |
| COLORADO | 4.61 | 5.18 | 5.73 | 5.55 | 21.08 |
| CONNECTICUT | 3.2 | 3.38 | 2.29 | 1.9 | 10.78 |
| DELAWARE | 1.96 | 1.64 | 1.8 | 1.28 | 6.7 |
| DISTRICT OF COLUMBIA | 2.03 | 2.15 | 3.05 | 2.57 | 9.81 |

| | | | | | |
|---|---|---|---|---|---|
| FLORIDA | 2.8 | 2.65 | 2.41 | 2.94 | 10.8 |
| GEORGIA | 1.8 | 2.17 | 1.91 | 1.92 | 7.8 |
| GUAM | 0.2 | 0 | 0 | 0 | 0.2 |
| HAWAII | 1.83 | 2.04 | 2.51 | 2.09 | 8.47 |
| IDAHO | 2.92 | 3.26 | 3.12 | 3.17 | 12.47 |
| ILLINOIS | 1.23 | 1.51 | 1.34 | 1.5 | 5.59 |
| INDIANA | 3.44 | 3.91 | 3.62 | 4.17 | 15.14 |
| IOWA | 3.69 | 3.31 | 3.18 | 3.04 | 13.21 |
| KANSAS | 5.8 | 6.73 | 6.22 | 6.49 | 25.24 |
| KENTUCKY | 4.54 | 4.14 | 3.37 | 4.01 | 16.05 |
| LOUISIANA | 2.33 | 2.49 | 2.17 | 2.3 | 9.29 |
| MAINE | 2.95 | 3.6 | 4.07 | 4.36 | 14.99 |
| MARYLAND | 1.09 | 1.69 | 1.22 | 1.8 | 5.81 |
| MASSACHUSETTS | 2.64 | 3.28 | 2.71 | 3.65 | 12.27 |
| MICHIGAN | 3.79 | 3.74 | 3.8 | 4.27 | 15.6 |
| MINNESOTA | 7.57 | 7.19 | 6.86 | 7.34 | 28.96 |
| MISSISSIPPI | 1.12 | 1.35 | 1.23 | 0.85 | 4.54 |
| MISSOURI | 2.84 | 2.73 | 2.78 | 2.92 | 11.26 |
| MONTANA | 5.39 | 5.35 | 4.84 | 5.06 | 20.63 |
| NEBRASKA | 5.03 | 5.81 | 4.95 | 5.78 | 21.56 |
| NEVADA | 1.13 | 1.04 | 1.29 | 1.51 | 4.97 |
| NEW HAMPSHIRE | 2.3 | 2.84 | 2.84 | 3.14 | 11.13 |
| NEW JERSEY | 0.99 | 1.52 | 1.34 | 1.19 | 5.04 |
| NEW MEXICO | 1.84 | 2.4 | 1.42 | 1.38 | 7.03 |
| NEW YORK | 1.08 | 1.12 | 1.04 | 1.24 | 4.49 |
| NORTH CAROLINA | 2.5 | 2.29 | 2.31 | 2.53 | 9.62 |
| NORTH DAKOTA | 4.6 | 6.26 | 6.8 | 6.85 | 24.51 |
| OHIO | 3.22 | 3.3 | 3.26 | 3.35 | 13.13 |
| OKLAHOMA | 2.04 | 2.23 | 2.54 | 1.47 | 8.28 |
| OREGON | 4.4 | 5.62 | 5.3 | 5.52 | 20.83 |
| PENNSYLVANIA | 2.34 | 2.27 | 2.64 | 3 | 10.24 |
| RHODE ISLAND | 3.11 | 3.25 | 4.28 | 2.43 | 13.07 |
| SOUTH CAROLINA | 2.76 | 3.35 | 2.76 | 3.14 | 12 |
| SOUTH DAKOTA | 7.87 | 5.75 | 3.32 | 5.26 | 22.2 |
| TENNESSEE | 2.69 | 3.03 | 3.01 | 3.24 | 11.98 |
| TEXAS | 2 | 2.01 | 2.04 | 2.26 | 8.31 |
| UTAH | 2.45 | 2.32 | 2.27 | 2.4 | 9.43 |
| VERMONT | 6.62 | 6.39 | 5.98 | 7.02 | 26 |
| VIRGIN ISLANDS | 0 | 0 | 1.64 | 0 | 1.64 |
| VIRGINIA | 2.13 | 2.23 | 2.49 | 2.18 | 9.03 |
| WASHINGTON | 4.19 | 4.78 | 4.84 | 4.89 | 18.7 |
| WEST VIRGINIA | 4.57 | 5.98 | 4.7 | 5.3 | 20.55 |
| WISCONSIN | 3.22 | 3.94 | 4.2 | 3.67 | 15.03 |
| WYOMING | 4.53 | 4.53 | 3.51 | 4.85 | 17.44 |
| U.S. TOTAL | 2.47 | 2.64 | 2.55 | 2.71 | 10.37 |

DRUG CODE:9652 DRUG NAME: OXYMORPHONE

| STATE | 1ST QUARTER | 2ND QUARTER | 3RD QUARTER | 4TH QUARTER | TOTAL GRAMS |
|---|---|---|---|---|---|
| ALABAMA | 160.57 | 141.9 | 85.96 | 61.47 | 449.9 |
| ALASKA | 40.47 | 40.39 | 34.71 | 25.9 | 141.46 |
| ARIZONA | 134.58 | 117.27 | 75.79 | 43.94 | 371.57 |
| ARKANSAS | 78.16 | 74.96 | 53.22 | 29.37 | 235.71 |
| CALIFORNIA | 45.88 | 42.04 | 30.7 | 21.55 | 140.16 |
| COLORADO | 86.32 | 75.44 | 56.78 | 36.41 | 254.95 |
| CONNECTICUT | 96.69 | 88.45 | 64.92 | 43.57 | 293.64 |
| DELAWARE | 239.97 | 243.64 | 205.32 | 171.45 | 860.38 |
| DISTRICT OF COLUMBIA | 19.97 | 21.08 | 16.86 | 21.74 | 79.65 |
| FLORIDA | 149.13 | 141.04 | 103.22 | 68.7 | 462.08 |
| GEORGIA | 88.41 | 85.16 | 55.77 | 37.19 | 266.53 |

| | | | | | |
|---|---|---|---|---|---|
| HAWAII | 49.85 | 51.44 | 29.03 | 27.46 | 157.78 |
| IDAHO | 101.24 | 95.18 | 74.95 | 56.85 | 328.22 |
| ILLINOIS | 41.34 | 38.85 | 28.36 | 18.47 | 127.02 |
| INDIANA | 102.95 | 89.08 | 65 | 43.23 | 300.26 |
| IOWA | 45.16 | 41.04 | 29.91 | 18.71 | 134.82 |
| KANSAS | 86.16 | 80.87 | 57.16 | 39.06 | 263.26 |
| KENTUCKY | 104.95 | 101.85 | 72.21 | 54.29 | 333.3 |
| LOUISIANA | 181 | 169.06 | 126.98 | 103.88 | 580.92 |
| MAINE | 89.59 | 75.72 | 64.58 | 48.54 | 278.43 |
| MARYLAND | 195.49 | 183.63 | 141.99 | 120.62 | 641.72 |
| MASSACHUSETTS | 43.73 | 41.7 | 30.07 | 17.4 | 132.89 |
| MICHIGAN | 206.38 | 211.1 | 183.43 | 164.96 | 765.87 |
| MINNESOTA | 9.45 | 9.79 | 7.62 | 5.15 | 32.02 |
| MISSISSIPPI | 130.99 | 120.96 | 92.3 | 68.52 | 412.78 |
| MISSOURI | 97.74 | 89.01 | 62.14 | 38.77 | 287.67 |
| MONTANA | 39.22 | 29.97 | 29.31 | 16 | 114.49 |
| NEBRASKA | 85.81 | 82.78 | 57.93 | 34.51 | 261.03 |
| NEVADA | 201.22 | 179.2 | 130.15 | 79.32 | 589.89 |
| NEW HAMPSHIRE | 83.23 | 75.42 | 62.76 | 43.81 | 265.22 |
| NEW JERSEY | 95.45 | 88.83 | 61.68 | 46.28 | 292.25 |
| NEW MEXICO | 60.8 | 52.43 | 39.53 | 31.69 | 184.44 |
| NEW YORK | 113.03 | 107.47 | 84.49 | 65.6 | 370.59 |
| NORTH CAROLINA | 292.52 | 265.72 | 183.48 | 128.25 | 869.98 |
| NORTH DAKOTA | 25.9 | 26.87 | 23.74 | 17.1 | 93.61 |
| OHIO | 105.93 | 99.35 | 76.23 | 52.65 | 334.15 |
| OKLAHOMA | 186.17 | 175.6 | 130.29 | 88.02 | 580.08 |
| OREGON | 53.06 | 48.99 | 41.14 | 28.59 | 171.77 |
| PENNSYLVANIA | 211.98 | 206.51 | 165.38 | 133.06 | 716.93 |
| RHODE ISLAND | 54.8 | 46.23 | 34.62 | 16.55 | 152.2 |
| SOUTH CAROLINA | 167.92 | 162.22 | 110.41 | 78.38 | 518.94 |
| SOUTH DAKOTA | 77.59 | 65.53 | 51.48 | 40.26 | 234.86 |
| TENNESSEE | 396.93 | 367.89 | 290.78 | 240.22 | 1,295.82 |
| TEXAS | 61.57 | 57.39 | 39.35 | 25.29 | 183.6 |
| UTAH | 174.26 | 163.29 | 138.84 | 116.95 | 593.35 |
| VERMONT | 57.96 | 52.01 | 44.01 | 28.98 | 182.96 |
| VIRGINIA | 142.31 | 129.67 | 98.73 | 83.13 | 453.85 |
| WASHINGTON | 54.04 | 48.58 | 38.58 | 28.41 | 169.61 |
| WEST VIRGINIA | 158.09 | 158.09 | 130.07 | 89.29 | 535.55 |
| WISCONSIN | 51.43 | 44.66 | 32.42 | 21.16 | 149.66 |
| WYOMING | 57.22 | 60.83 | 45.31 | 35.37 | 198.72 |
| U.S. TOTAL | 112.43 | 105.27 | 78.82 | 58.4 | 354.92 |

DRUG CODE:9668 DRUG NAME: NOROXYMORPHONE

| STATE | 1ST QUARTER | 2ND QUARTER | 3RD QUARTER | 4TH QUARTER | TOTAL GRAMS |
|---|---|---|---|---|---|
| ARIZONA | 0 | 0 | 0 | 0 | 0 |
| ARKANSAS | 0.5 | 0.59 | 0.92 | 0.66 | 2.68 |
| CALIFORNIA | 0.14 | 0.18 | 0.19 | 0.18 | 0.7 |
| FLORIDA | 0.07 | 0.07 | 0.07 | 0.1 | 0.31 |
| GEORGIA | 0.03 | 0.03 | 0.1 | 0.05 | 0.21 |
| ILLINOIS | 0.03 | 0.08 | 0.12 | 0.07 | 0.3 |
| KANSAS | 0.1 | 0 | 0 | 0 | 0.1 |
| MARYLAND | 0 | 0 | 0 | 0 | 0 |
| MASSACHUSETTS | 0.02 | 0.03 | 0 | 0 | 0.05 |
| MICHIGAN | 0 | 0 | 0.03 | 0.01 | 0.04 |
| MINNESOTA | 0 | 0.01 | 0 | 0 | 0.01 |
| MISSISSIPPI | 0.08 | 0.05 | 0.07 | 0 | 0.21 |
| MISSOURI | 0.13 | 0.22 | 0.18 | 0.1 | 0.62 |
| NEVADA | 0.25 | 0.11 | 0.28 | 0.31 | 0.94 |
| NEW JERSEY | 0.08 | 0.14 | 0.02 | 0.03 | 0.27 |

| | | | | | |
|---|---|---|---|---|---|
| NEW YORK | 0.01 | 0.03 | 0.05 | 0.02 | 0.11 |
| NORTH CAROLINA | 0 | 0 | 0.02 | 0.02 | 0.04 |
| OHIO | 0.05 | 0.08 | 0.05 | 0.02 | 0.21 |
| OKLAHOMA | 0 | 0 | 0 | 0.01 | 0.02 |
| OREGON | 0 | 0 | 0.05 | 0 | 0.05 |
| PENNSYLVANIA | 0 | 0 | 0 | 0 | 0.01 |
| PUERTO RICO | 0.05 | 0.12 | 0.32 | 0.18 | 0.67 |
| RHODE ISLAND | 0 | 0 | 0.01 | 0 | 0.01 |
| TENNESSEE | 0 | 0 | 0 | 0 | 0 |
| TEXAS | 0.09 | 0.12 | 0.12 | 0.1 | 0.43 |
| VERMONT | 0 | 0 | 0.02 | 0 | 0.02 |
| WASHINGTON | 0.01 | 0.01 | 0.05 | 0.06 | 0.13 |
| U.S. TOTAL | 0.05 | 0.06 | 0.07 | 0.06 | 0.24 |

DRUG CODE:9737 DRUG NAME: ALFENTANIL

| STATE | 1ST QUARTER | 2ND QUARTER | 3RD QUARTER | 4TH QUARTER | TOTAL GRAMS |
|---|---|---|---|---|---|
| ALABAMA | 0 | 0 | 0 | 0 | 0.01 |
| ALASKA | 0.11 | 0.11 | 0.07 | 0.13 | 0.41 |
| ARIZONA | 0.02 | 0.02 | 0.01 | 0.02 | 0.06 |
| ARKANSAS | 0 | 0 | 0 | 0 | 0 |
| CALIFORNIA | 0.05 | 0.04 | 0.04 | 0.05 | 0.18 |
| COLORADO | 0.03 | 0.01 | 0.02 | 0.02 | 0.08 |
| CONNECTICUT | 0.04 | 0.03 | 0.03 | 0.04 | 0.15 |
| FLORIDA | 0.02 | 0.03 | 0.03 | 0.03 | 0.11 |
| GEORGIA | 0.03 | 0.03 | 0.05 | 0.03 | 0.13 |
| GUAM | 0 | 0 | 0.03 | 0.02 | 0.04 |
| HAWAII | 0.03 | 0.04 | 0.05 | 0.06 | 0.18 |
| IDAHO | 0.09 | 0.09 | 0.07 | 0.09 | 0.35 |
| ILLINOIS | 0.03 | 0.03 | 0.03 | 0.04 | 0.13 |
| INDIANA | 0.03 | 0.05 | 0.04 | 0.04 | 0.16 |
| IOWA | 0.07 | 0.05 | 0.05 | 0.06 | 0.22 |
| KANSAS | 0.06 | 0.05 | 0.04 | 0.06 | 0.21 |
| KENTUCKY | 0.12 | 0.09 | 0.08 | 0.09 | 0.38 |
| LOUISIANA | 0.05 | 0.05 | 0.06 | 0.06 | 0.22 |
| MAINE | 0.05 | 0.06 | 0.05 | 0.05 | 0.2 |
| MARYLAND | 0.02 | 0 | 0.01 | 0.02 | 0.05 |
| MASSACHUSETTS | 0.02 | 0.01 | 0.01 | 0.01 | 0.05 |
| MICHIGAN | 0.01 | 0.01 | 0.01 | 0.01 | 0.05 |
| MINNESOTA | 0.06 | 0.06 | 0.06 | 0.08 | 0.26 |
| MISSISSIPPI | 0 | 0 | 0.01 | 0 | 0.01 |
| MISSOURI | 0.02 | 0.02 | 0.02 | 0.02 | 0.07 |
| MONTANA | 0.04 | 0.03 | 0.03 | 0.03 | 0.13 |
| NEBRASKA | 0.05 | 0.04 | 0.06 | 0.05 | 0.19 |
| NEVADA | 0.05 | 0.06 | 0.06 | 0.09 | 0.26 |
| NEW HAMPSHIRE | 0.04 | 0.03 | 0.03 | 0.05 | 0.15 |
| NEW JERSEY | 0 | 0 | 0 | 0 | 0.01 |
| NEW MEXICO | 0.04 | 0.04 | 0.06 | 0.06 | 0.2 |
| NEW YORK | 0.01 | 0.01 | 0.01 | 0.02 | 0.05 |
| NORTH CAROLINA | 0.03 | 0.04 | 0.03 | 0.04 | 0.13 |
| NORTH DAKOTA | 0.02 | 0.04 | 0.01 | 0.02 | 0.09 |
| OHIO | 0.04 | 0.05 | 0.03 | 0.04 | 0.17 |
| OKLAHOMA | 0.02 | 0.03 | 0.02 | 0.02 | 0.09 |
| OREGON | 0.1 | 0.11 | 0.15 | 0.13 | 0.49 |
| PENNSYLVANIA | 0.01 | 0 | 0 | 0 | 0.02 |
| RHODE ISLAND | 0 | 0.01 | 0.01 | 0 | 0.02 |
| SOUTH CAROLINA | 0 | 0 | 0 | 0 | 0.01 |
| SOUTH DAKOTA | 0.01 | 0.01 | 0.01 | 0.01 | 0.04 |
| TENNESSEE | 0.02 | 0.02 | 0.02 | 0.02 | 0.08 |
| TEXAS | 0.02 | 0.02 | 0.02 | 0.02 | 0.09 |

| | | | | | |
|---|---|---|---|---|---|
| UTAH | 0.02 | 0.01 | 0.02 | 0.06 | 0.11 |
| VERMONT | 0 | 0 | 0 | 0 | 0.01 |
| VIRGINIA | 0.01 | 0.01 | 0.01 | 0.01 | 0.05 |
| WASHINGTON | 0.04 | 0.05 | 0.05 | 0.05 | 0.19 |
| WISCONSIN | 0.07 | 0.08 | 0.08 | 0.1 | 0.34 |
| WYOMING | 0 | 0 | 0 | 0.02 | 0.02 |
| U.S. TOTAL | 0.03 | 0.03 | 0.03 | 0.03 | 0.12 |

DRUG CODE:9739 DRUG NAME: REMIFENTANIL

| STATE | 1ST QUARTER | 2ND QUARTER | 3RD QUARTER | 4TH QUARTER | TOTAL GRAMS |
|---|---|---|---|---|---|
| ALABAMA | 0.07 | 0.07 | 0.07 | 0.07 | 0.28 |
| ALASKA | 0.38 | 0.42 | 0.39 | 0.44 | 1.64 |
| ARIZONA | 0.07 | 0.06 | 0.06 | 0.07 | 0.25 |
| ARKANSAS | 0.05 | 0.05 | 0.06 | 0.05 | 0.22 |
| CALIFORNIA | 0.09 | 0.09 | 0.08 | 0.09 | 0.36 |
| COLORADO | 0.12 | 0.13 | 0.12 | 0.14 | 0.51 |
| CONNECTICUT | 0.27 | 0.22 | 0.22 | 0.24 | 0.95 |
| DELAWARE | 0.3 | 0.45 | 0.31 | 0.28 | 1.34 |
| DISTRICT OF COLUMBIA | 0.15 | 0.14 | 0.09 | 0.1 | 0.47 |
| FLORIDA | 0.14 | 0.14 | 0.13 | 0.16 | 0.57 |
| GEORGIA | 0.05 | 0.05 | 0.05 | 0.05 | 0.2 |
| GUAM | 0.01 | 0.02 | 0 | 0.14 | 0.17 |
| HAWAII | 0.09 | 0.08 | 0.1 | 0.1 | 0.38 |
| IDAHO | 0.14 | 0.14 | 0.14 | 0.14 | 0.55 |
| ILLINOIS | 0.12 | 0.11 | 0.11 | 0.13 | 0.47 |
| INDIANA | 0.12 | 0.12 | 0.12 | 0.13 | 0.49 |
| IOWA | 0.12 | 0.11 | 0.11 | 0.08 | 0.41 |
| KANSAS | 0.07 | 0.09 | 0.07 | 0.08 | 0.32 |
| KENTUCKY | 0.12 | 0.1 | 0.12 | 0.11 | 0.44 |
| LOUISIANA | 0.1 | 0.1 | 0.1 | 0.11 | 0.41 |
| MAINE | 0.1 | 0.09 | 0.08 | 0.07 | 0.35 |
| MARYLAND | 0.12 | 0.12 | 0.1 | 0.12 | 0.47 |
| MASSACHUSETTS | 0.32 | 0.37 | 0.33 | 0.35 | 1.36 |
| MICHIGAN | 0.1 | 0.11 | 0.1 | 0.1 | 0.4 |
| MINNESOTA | 0.13 | 0.15 | 0.15 | 0.19 | 0.62 |
| MISSISSIPPI | 0.02 | 0.02 | 0.01 | 0.01 | 0.06 |
| MISSOURI | 0.08 | 0.09 | 0.1 | 0.09 | 0.36 |
| MONTANA | 0.15 | 0.18 | 0.17 | 0.2 | 0.71 |
| NEBRASKA | 0.06 | 0.05 | 0.05 | 0.07 | 0.23 |
| NEVADA | 0.03 | 0.03 | 0.04 | 0.04 | 0.14 |
| NEW HAMPSHIRE | 0.09 | 0.12 | 0.11 | 0.18 | 0.5 |
| NEW JERSEY | 0.15 | 0.15 | 0.15 | 0.16 | 0.6 |
| NEW MEXICO | 0.05 | 0.06 | 0.06 | 0.05 | 0.22 |
| NEW YORK | 0.19 | 0.19 | 0.21 | 0.21 | 0.8 |
| NORTH CAROLINA | 0.15 | 0.16 | 0.15 | 0.16 | 0.61 |
| NORTH DAKOTA | 0.09 | 0.05 | 0.06 | 0.06 | 0.25 |
| OHIO | 0.17 | 0.17 | 0.14 | 0.18 | 0.66 |
| OKLAHOMA | 0.07 | 0.08 | 0.07 | 0.07 | 0.29 |
| OREGON | 0.07 | 0.09 | 0.07 | 0.08 | 0.3 |
| PENNSYLVANIA | 0.28 | 0.28 | 0.28 | 0.29 | 1.13 |
| RHODE ISLAND | 0.06 | 0.05 | 0.07 | 0.12 | 0.3 |
| SOUTH CAROLINA | 0.11 | 0.09 | 0.1 | 0.1 | 0.4 |
| SOUTH DAKOTA | 0.06 | 0.08 | 0.05 | 0.04 | 0.23 |
| TENNESSEE | 0.07 | 0.09 | 0.11 | 0.1 | 0.37 |
| TEXAS | 0.11 | 0.11 | 0.11 | 0.12 | 0.44 |
| UTAH | 0.64 | 0.51 | 0.56 | 0.89 | 2.6 |
| VERMONT | 0.11 | 0.15 | 0.08 | 0.12 | 0.45 |
| VIRGINIA | 0.08 | 0.09 | 0.08 | 0.09 | 0.34 |
| WASHINGTON | 0.19 | 0.18 | 0.22 | 0.16 | 0.74 |

| | | | | | |
|---|---|---|---|---|---|
| WEST VIRGINIA | 0.29 | 0.32 | 0.27 | 0.24 | 1.12 |
| WISCONSIN | 0.11 | 0.13 | 0.09 | 0.14 | 0.47 |
| WYOMING | 0.09 | 0.07 | 0.08 | 0.08 | 0.32 |
| U.S. TOTAL | 0.13 | 0.13 | 0.13 | 0.14 | 0.53 |

DRUG CODE:9740 DRUG NAME: SUFENTANIL BASE

| STATE | 1ST QUARTER | 2ND QUARTER | 3RD QUARTER | 4TH QUARTER | TOTAL GRAMS |
|---|---|---|---|---|---|
| ALABAMA | 0.02 | 0.04 | 0.04 | 0.04 | 0.14 |
| ALASKA | 0 | 0 | 0 | 0 | 0.01 |
| ARIZONA | 0 | 0 | 0 | 0 | 0.01 |
| ARKANSAS | 0 | 0 | 0.01 | 0 | 0.02 |
| CALIFORNIA | 0.03 | 0.03 | 0.03 | 0.03 | 0.12 |
| COLORADO | 0 | 0 | 0 | 0.01 | 0.02 |
| CONNECTICUT | 0.01 | 0.01 | 0.02 | 0.03 | 0.07 |
| DELAWARE | 0 | 0 | 0 | 0 | 0 |
| DISTRICT OF COLUMBIA | 0 | 0 | 0 | 0 | 0 |
| FLORIDA | 0.02 | 0.01 | 0.02 | 0.02 | 0.07 |
| GEORGIA | 0 | 0 | 0.01 | 0.01 | 0.02 |
| HAWAII | 0 | 0 | 0 | 0 | 0 |
| IDAHO | 0 | 0 | 0 | 0 | 0.01 |
| ILLINOIS | 0 | 0 | 0 | 0 | 0.02 |
| INDIANA | 0 | 0.01 | 0 | 0 | 0.02 |
| IOWA | 0 | 0 | 0 | 0 | 0.01 |
| KANSAS | 0.01 | 0.01 | 0.01 | 0.01 | 0.04 |
| KENTUCKY | 0.01 | 0.01 | 0.01 | 0.01 | 0.03 |
| LOUISIANA | 0.01 | 0.04 | 0.02 | 0.03 | 0.1 |
| MAINE | 0 | 0 | 0 | 0 | 0.01 |
| MARYLAND | 0 | 0 | 0 | 0 | 0 |
| MASSACHUSETTS | 0 | 0 | 0 | 0 | 0.01 |
| MICHIGAN | 0.01 | 0 | 0 | 0.01 | 0.02 |
| MINNESOTA | 0 | 0 | 0 | 0.01 | 0.01 |
| MISSISSIPPI | 0.5 | 0.35 | 0.42 | 0.49 | 1.76 |
| MISSOURI | 0 | 0.01 | 0.01 | 0 | 0.02 |
| MONTANA | 0 | 0 | 0 | 0.01 | 0.02 |
| NEBRASKA | 0 | 0 | 0 | 0.01 | 0.02 |
| NEVADA | 0 | 0 | 0 | 0 | 0.01 |
| NEW HAMPSHIRE | 0 | 0 | 0 | 0 | 0 |
| NEW JERSEY | 0.01 | 0.01 | 0 | 0.02 | 0.04 |
| NEW MEXICO | 0 | 0 | 0 | 0 | 0.01 |
| NEW YORK | 0.01 | 0.01 | 0.01 | 0.01 | 0.03 |
| NORTH CAROLINA | 0 | 0 | 0.01 | 0 | 0.02 |
| NORTH DAKOTA | 0 | 0.01 | 0 | 0 | 0.02 |
| OHIO | 0 | 0 | 0 | 0.01 | 0.02 |
| OKLAHOMA | 0 | 0 | 0 | 0 | 0.02 |
| OREGON | 0.01 | 0.01 | 0.01 | 0.01 | 0.03 |
| PENNSYLVANIA | 0 | 0.02 | 0.03 | 0.01 | 0.06 |
| PUERTO RICO | 0.01 | 0 | 0.01 | 0 | 0.02 |
| RHODE ISLAND | 0 | 0 | 0 | 0 | 0 |
| SOUTH CAROLINA | 0.02 | 0.02 | 0.02 | 0 | 0.06 |
| SOUTH DAKOTA | 0 | 0 | 0 | 0.01 | 0.02 |
| TENNESSEE | 0.01 | 0.01 | 0.01 | 0.01 | 0.04 |
| TEXAS | 0.01 | 0.01 | 0 | 0.01 | 0.03 |
| UTAH | 0.01 | 0.01 | 0.01 | 0.01 | 0.03 |
| VERMONT | 0 | 0 | 0 | 0 | 0 |
| VIRGINIA | 0.01 | 0 | 0 | 0.01 | 0.02 |
| WASHINGTON | 0 | 0 | 0 | 0 | 0.02 |
| WEST VIRGINIA | 0 | 0 | 0.01 | 0 | 0.01 |
| WISCONSIN | 0.01 | 0.01 | 0 | 0.02 | 0.04 |
| WYOMING | 0 | 0 | 0 | 0.01 | 0.01 |

| | | | | | |
|---|---|---|---|---|---|
| U.S. TOTAL | 0.01 | 0.01 | 0.01 | 0.02 | 0.05 |

DRUG CODE:9780 DRUG NAME: TAPENTADOL

| STATE | 1ST QUARTER | 2ND QUARTER | 3RD QUARTER | 4TH QUARTER | TOTAL GRAMS |
|---|---|---|---|---|---|
| ALABAMA | 1,235.02 | 1,177.77 | 1,097.85 | 1,062.01 | 4,572.66 |
| ALASKA | 295.04 | 305.18 | 326.51 | 339.19 | 1,265.93 |
| ARIZONA | 597.39 | 559.23 | 504.35 | 562.48 | 2,223.45 |
| ARKANSAS | 798.82 | 767.87 | 653.58 | 658.21 | 2,878.49 |
| CALIFORNIA | 296.77 | 296.43 | 272.84 | 274.45 | 1,140.49 |
| COLORADO | 546.15 | 574.43 | 546.27 | 565.17 | 2,232.02 |
| CONNECTICUT | 354.15 | 332.83 | 306.04 | 295.4 | 1,288.41 |
| DELAWARE | 850.73 | 827.62 | 824.56 | 801.78 | 3,304.70 |
| DISTRICT OF COLUMBIA | 201.01 | 184.64 | 152.23 | 142.92 | 680.79 |
| FLORIDA | 582 | 547.95 | 490.66 | 486.98 | 2,107.60 |
| GEORGIA | 734.08 | 731.84 | 656.86 | 660.46 | 2,783.24 |
| HAWAII | 210.62 | 241.42 | 229.99 | 243.81 | 925.82 |
| IDAHO | 409.93 | 360.91 | 364.19 | 404.06 | 1,539.09 |
| ILLINOIS | 323.05 | 301.93 | 281.89 | 275.44 | 1,182.32 |
| INDIANA | 592.59 | 584.53 | 563.67 | 563.9 | 2,304.69 |
| IOWA | 458.68 | 424.15 | 351.53 | 397.13 | 1,631.49 |
| KANSAS | 333.97 | 308.07 | 290.51 | 295.66 | 1,228.20 |
| KENTUCKY | 349.39 | 349.21 | 353.28 | 333.9 | 1,385.78 |
| LOUISIANA | 337.57 | 331.41 | 321.99 | 349.98 | 1,340.96 |
| MAINE | 109.04 | 111.98 | 101.03 | 102.19 | 424.24 |
| MARYLAND | 594.17 | 588.55 | 539.93 | 529.08 | 2,251.72 |
| MASSACHUSETTS | 293.82 | 299.26 | 294.69 | 289.38 | 1,177.16 |
| MICHIGAN | 221.16 | 215.01 | 218.87 | 221.55 | 876.59 |
| MINNESOTA | 81.38 | 79.7 | 83.05 | 87.73 | 331.86 |
| MISSISSIPPI | 442.88 | 422.08 | 398.83 | 363.46 | 1,627.26 |
| MISSOURI | 431.65 | 421.32 | 385.48 | 390.66 | 1,629.11 |
| MONTANA | 222.51 | 168.64 | 177.73 | 182.53 | 751.4 |
| NEBRASKA | 571.96 | 511.51 | 505.46 | 536.62 | 2,125.56 |
| NEVADA | 466.33 | 450.45 | 431.76 | 458.7 | 1,807.24 |
| NEW HAMPSHIRE | 575.29 | 587.59 | 536.62 | 534.92 | 2,234.42 |
| NEW JERSEY | 554.71 | 546.83 | 526.04 | 524.7 | 2,152.28 |
| NEW MEXICO | 171.72 | 151.98 | 149.38 | 149.82 | 622.89 |
| NEW YORK | 515.98 | 504.16 | 485.5 | 475.38 | 1,981.02 |
| NORTH CAROLINA | 1,058.65 | 1,047.83 | 980.3 | 966.79 | 4,053.57 |
| NORTH DAKOTA | 378.83 | 373.33 | 341.74 | 359.51 | 1,453.41 |
| OHIO | 573.72 | 543.28 | 498.98 | 488.29 | 2,104.27 |
| OKLAHOMA | 661.38 | 658.92 | 608.85 | 601.08 | 2,530.22 |
| OREGON | 191.42 | 194.23 | 196.86 | 195.73 | 778.24 |
| PENNSYLVANIA | 531.25 | 540.63 | 502.21 | 507.96 | 2,082.06 |
| PUERTO RICO | 24.28 | 26.57 | 22.25 | 17.46 | 90.56 |
| RHODE ISLAND | 196.42 | 205.83 | 195.71 | 199.13 | 797.1 |
| SOUTH CAROLINA | 957.16 | 963.3 | 917.81 | 923.82 | 3,762.08 |
| SOUTH DAKOTA | 640.09 | 594.95 | 530.72 | 550.68 | 2,316.44 |
| TENNESSEE | 518.16 | 485.16 | 489.88 | 511.42 | 2,004.62 |
| TEXAS | 405.95 | 402.31 | 362.52 | 380.6 | 1,551.38 |
| UTAH | 564.31 | 491.48 | 487 | 443.2 | 1,985.99 |
| VERMONT | 214.39 | 175.15 | 165.96 | 182.82 | 738.32 |
| VIRGIN ISLANDS | 8.19 | 5.46 | 2.73 | 0 | 16.39 |
| VIRGINIA | 574.86 | 541.41 | 482.49 | 470.56 | 2,069.33 |
| WASHINGTON | 272.45 | 274.01 | 251.98 | 247.73 | 1,046.18 |
| WEST VIRGINIA | 339.07 | 355.8 | 326.9 | 302.81 | 1,324.59 |
| WISCONSIN | 272.53 | 271.02 | 256.11 | 246.15 | 1,045.81 |
| WYOMING | 345.62 | 367.89 | 318.83 | 345.53 | 1,377.86 |
| U.S. TOTAL | 470.8 | 459.3 | 427.91 | 429.13 | 1,787.14 |

DRUG CODE:9801 DRUG NAME: FENTANYL BASE

| STATE | 1ST QUARTER | 2ND QUARTER | 3RD QUARTER | 4TH QUARTER | TOTAL GRAMS |
|---|---|---|---|---|---|
| ALABAMA | 33.9 | 32.29 | 29.78 | 28.78 | 124.76 |
| ALASKA | 33.78 | 33.71 | 32.81 | 32.14 | 132.44 |
| AMERICAN SAMOA | 0.36 | 0.35 | 0 | 0 | 0.71 |
| ARIZONA | 30.94 | 28.21 | 25.44 | 24.81 | 109.41 |
| ARKANSAS | 34.65 | 33.48 | 30.42 | 28.86 | 127.41 |
| CALIFORNIA | 19.85 | 19.17 | 18.24 | 17.4 | 74.66 |
| COLORADO | 27.09 | 25.29 | 24.19 | 23.06 | 99.63 |
| CONNECTICUT | 28.15 | 27.68 | 25.86 | 24.65 | 106.34 |
| DELAWARE | 39.9 | 39 | 36.17 | 36.46 | 151.53 |
| DISTRICT OF COLUMBIA | 15.36 | 14.98 | 13.35 | 12.6 | 56.3 |
| FLORIDA | 33.89 | 31.29 | 29.11 | 29.92 | 124.21 |
| GEORGIA | 30.28 | 28.82 | 27.04 | 26.46 | 112.6 |
| GUAM | 6.84 | 5.61 | 5.14 | 5.57 | 23.16 |
| HAWAII | 14.57 | 14.25 | 12.88 | 12.88 | 54.57 |
| IDAHO | 37.93 | 37.59 | 36.08 | 35.44 | 147.04 |
| ILLINOIS | 24.35 | 23.31 | 22.45 | 21.93 | 92.04 |
| INDIANA | 31.48 | 29.64 | 27.65 | 27.27 | 116.04 |
| IOWA | 29.81 | 29.11 | 27.77 | 27.43 | 114.12 |
| KANSAS | 39.96 | 39.31 | 37.8 | 39.32 | 156.39 |
| KENTUCKY | 28.41 | 27.08 | 25.33 | 24.43 | 105.25 |
| LOUISIANA | 28.71 | 26.83 | 25.45 | 23.91 | 104.9 |
| MAINE | 29.23 | 24.9 | 22.88 | 22.58 | 99.58 |
| MARYLAND | 26.9 | 24.54 | 23.3 | 21.44 | 96.17 |
| MASSACHUSETTS | 21.23 | 20.42 | 19.34 | 18.87 | 79.86 |
| MICHIGAN | 33.34 | 32.23 | 30.52 | 29.12 | 125.22 |
| MINNESOTA | 20.37 | 20.16 | 18.87 | 18.2 | 77.6 |
| MISSISSIPPI | 42.23 | 44.21 | 40.92 | 38.54 | 165.9 |
| MISSOURI | 37.72 | 36.73 | 35.11 | 42.14 | 151.7 |
| MONTANA | 35.78 | 34.02 | 31.93 | 31.49 | 133.22 |
| NEBRASKA | 30.76 | 29.58 | 28.49 | 27.75 | 116.57 |
| NEVADA | 24.3 | 22.21 | 20.72 | 19.94 | 87.16 |
| NEW HAMPSHIRE | 28.26 | 26.68 | 25.86 | 24.87 | 105.67 |
| NEW JERSEY | 28.39 | 27.29 | 24.7 | 24.02 | 104.4 |
| NEW MEXICO | 23.61 | 21.66 | 19.91 | 19.07 | 84.26 |
| NEW YORK | 24.05 | 23.06 | 21.22 | 20.62 | 88.95 |
| NORTH CAROLINA | 32.49 | 30.59 | 27.74 | 27.33 | 118.15 |
| NORTH DAKOTA | 36.9 | 35.74 | 34.31 | 33.98 | 140.93 |
| OHIO | 22.99 | 22.06 | 20.69 | 19.65 | 85.38 |
| OKLAHOMA | 49.25 | 46.12 | 43.55 | 41.15 | 180.07 |
| OREGON | 26.27 | 24.92 | 23.15 | 22.39 | 96.73 |
| PENNSYLVANIA | 42.31 | 41.02 | 38.19 | 36.45 | 157.97 |
| PUERTO RICO | 5.28 | 4.05 | 3.4 | 2.79 | 15.52 |
| RHODE ISLAND | 18.79 | 17.75 | 16.89 | 16.4 | 69.83 |
| SOUTH CAROLINA | 30.79 | 29.11 | 27.22 | 26.64 | 113.75 |
| SOUTH DAKOTA | 39.96 | 37.08 | 36.33 | 34.54 | 147.91 |
| TENNESSEE | 33 | 30.97 | 29.08 | 28.12 | 121.17 |
| TEXAS | 24.37 | 23.52 | 22.04 | 21.64 | 91.57 |
| UTAH | 31.22 | 29.7 | 29.16 | 28.43 | 118.51 |
| VERMONT | 32.4 | 31.44 | 28.79 | 26.92 | 119.55 |
| VIRGIN ISLANDS | 3.65 | 3.15 | 2.75 | 3.16 | 12.71 |
| VIRGINIA | 24.01 | 22.27 | 20.54 | 19.59 | 86.41 |
| WASHINGTON | 24.01 | 23.15 | 21.7 | 20.85 | 89.71 |
| WEST VIRGINIA | 31.17 | 30.17 | 28.65 | 26.98 | 116.97 |
| WISCONSIN | 28.79 | 26.8 | 24.46 | 23.66 | 103.71 |
| WYOMING | 29.85 | 26.53 | 28.04 | 25.76 | 110.18 |
| U.S. TOTAL | 27.91 | 26.61 | 24.92 | 24.33 | 103.77 |

DRUG CODE:9809 DRUG NAME: OPIUM COMBINATION PRODUCT (C-III)

| STATE | 1ST QUARTER | 2ND QUARTER | 3RD QUARTER | 4TH QUARTER | TOTAL GRAMS |
|---|---|---|---|---|---|
| ALABAMA | 0.25 | 0.27 | 0.25 | 0.23 | 0.99 |
| ALASKA | 0 | 0 | 0.03 | 0 | 0.03 |
| ARIZONA | 0.19 | 0.09 | 0.11 | 0.13 | 0.52 |
| ARKANSAS | 0.05 | 0.08 | 0.1 | 0.07 | 0.29 |
| CALIFORNIA | 0.05 | 0.05 | 0.05 | 0.06 | 0.21 |
| COLORADO | 0.06 | 0.06 | 0.05 | 0.05 | 0.22 |
| CONNECTICUT | 0.14 | 0.11 | 0.18 | 0.12 | 0.55 |
| DELAWARE | 0.02 | 0.08 | 0.02 | 0.11 | 0.23 |
| DISTRICT OF COLUMBIA | 0.03 | 0.06 | 0.06 | 0.09 | 0.25 |
| FLORIDA | 0.19 | 0.2 | 0.17 | 0.17 | 0.73 |
| GEORGIA | 0.06 | 0.09 | 0.08 | 0.07 | 0.3 |
| HAWAII | 0.14 | 0.11 | 0.1 | 0 | 0.35 |
| IDAHO | 0.25 | 0.24 | 0.23 | 0.24 | 0.97 |
| ILLINOIS | 0.05 | 0.05 | 0.05 | 0.04 | 0.19 |
| INDIANA | 0.11 | 0.11 | 0.13 | 0.11 | 0.45 |
| IOWA | 0.11 | 0.06 | 0.06 | 0.07 | 0.29 |
| KANSAS | 0.03 | 0.02 | 0.01 | 0.05 | 0.1 |
| KENTUCKY | 0.24 | 0.25 | 0.18 | 0.18 | 0.85 |
| LOUISIANA | 0.16 | 0.19 | 0.2 | 0.18 | 0.73 |
| MAINE | 0.06 | 0.09 | 0.16 | 0.21 | 0.51 |
| MARYLAND | 0.15 | 0.15 | 0.15 | 0.14 | 0.59 |
| MASSACHUSETTS | 0.03 | 0.04 | 0.02 | 0.01 | 0.1 |
| MICHIGAN | 0.05 | 0.05 | 0.09 | 0.05 | 0.23 |
| MINNESOTA | 0.03 | 0.04 | 0.04 | 0.06 | 0.17 |
| MISSISSIPPI | 0.15 | 0.08 | 0.1 | 0.06 | 0.4 |
| MISSOURI | 0.17 | 0.09 | 0.07 | 0.15 | 0.48 |
| MONTANA | 0.1 | 0.02 | 0.13 | 0.1 | 0.34 |
| NEBRASKA | 0.06 | 0.03 | 0.04 | 0.03 | 0.17 |
| NEVADA | 0.03 | 0.05 | 0.06 | 0.01 | 0.15 |
| NEW HAMPSHIRE | 0.13 | 0.14 | 0.14 | 0.26 | 0.68 |
| NEW JERSEY | 0.06 | 0.06 | 0.06 | 0.05 | 0.24 |
| NEW MEXICO | 0.09 | 0.09 | 0.05 | 0.09 | 0.32 |
| NEW YORK | 0.04 | 0.05 | 0.04 | 0.06 | 0.2 |
| NORTH CAROLINA | 0.13 | 0.15 | 0.16 | 0.13 | 0.57 |
| NORTH DAKOTA | 0 | 0.06 | 0.11 | 0.23 | 0.39 |
| OHIO | 0.13 | 0.12 | 0.1 | 0.16 | 0.5 |
| OKLAHOMA | 0.09 | 0.06 | 0.09 | 0.12 | 0.34 |
| OREGON | 0 | 0.01 | 0.02 | 0.05 | 0.09 |
| PENNSYLVANIA | 0.25 | 0.31 | 0.21 | 0.3 | 1.07 |
| RHODE ISLAND | 0 | 0.02 | 0.02 | 0 | 0.04 |
| SOUTH CAROLINA | 0.18 | 0.23 | 0.23 | 0.22 | 0.87 |
| SOUTH DAKOTA | 0.16 | 0.07 | 0.07 | 0.02 | 0.33 |
| TENNESSEE | 0.09 | 0.09 | 0.09 | 0.13 | 0.39 |
| TEXAS | 0.05 | 0.06 | 0.05 | 0.06 | 0.21 |
| UTAH | 0.47 | 0.46 | 0.4 | 0.47 | 1.79 |
| VERMONT | 0 | 0.06 | 0.03 | 0.06 | 0.15 |
| VIRGINIA | 0.1 | 0.07 | 0.07 | 0.06 | 0.3 |
| WASHINGTON | 0.03 | 0.03 | 0.06 | 0.03 | 0.15 |
| WEST VIRGINIA | 0.14 | 0.1 | 0.13 | 0.14 | 0.52 |
| WISCONSIN | 0.1 | 0.09 | 0.1 | 0.12 | 0.41 |
| WYOMING | 0 | 0.1 | 0.13 | 0.3 | 0.54 |
| U.S. TOTAL | 0.1 | 0.1 | 0.09 | 0.1 | 0.39 |

**DRUG CODE:1100** **DRUG NAME:AMPHETAMINE**

| RANK | STATE | 2010 POP | TOTAL GRAMS | GRAMS/100K POP |
|---|---|---|---|---|
| 1 | RHODE ISLAND | 1,052,567 | 146,539.12 | 13,922.07 |
| 2 | LOUISIANA | 4,533,372 | 617,608.44 | 13,623.60 |
| 3 | UTAH | 2,763,885 | 327,686.38 | 11,856.01 |
| 4 | SOUTH CAROLINA | 4,625,364 | 545,192.92 | 11,787.03 |
| 5 | NEW HAMPSHIRE | 1,316,470 | 153,869.12 | 11,688.01 |
| 6 | ALABAMA | 4,779,736 | 522,022.67 | 10,921.58 |
| 7 | MASSACHUSETTS | 6,547,629 | 711,859.01 | 10,872.01 |
| 8 | MICHIGAN | 9,883,640 | 961,941.32 | 9,732.66 |
| 9 | DELAWARE | 897,934 | 86,201.90 | 9,600.03 |
| 10 | WISCONSIN | 5,686,986 | 510,833.02 | 8,982.49 |
| 11 | MISSOURI | 5,988,927 | 536,597.02 | 8,959.82 |
| 12 | VIRGINIA | 8,001,024 | 714,083.09 | 8,924.90 |
| 13 | NORTH CAROLINA | 9,535,483 | 812,531.47 | 8,521.14 |
| 14 | KANSAS | 2,853,118 | 241,758.68 | 8,473.49 |
| 15 | MISSISSIPPI | 2,967,297 | 246,037.64 | 8,291.64 |
| 16 | TENNESSEE | 6,346,105 | 520,438.74 | 8,200.92 |
| 17 | MINNESOTA | 5,303,925 | 431,651.49 | 8,138.34 |
| 18 | INDIANA | 6,483,802 | 519,990.62 | 8,019.84 |
| 19 | DISTRICT OF COLUMBIA | 601,723 | 46,666.86 | 7,755.54 |
| 20 | GEORGIA | 9,687,653 | 750,263.70 | 7,744.54 |
| 21 | SOUTH DAKOTA | 814,180 | 60,812.21 | 7,469.14 |
| 22 | OHIO | 11,536,504 | 847,019.84 | 7,342.08 |
| 23 | ARKANSAS | 2,915,918 | 204,371.71 | 7,008.83 |
| 24 | MAINE | 1,328,361 | 90,672.30 | 6,825.88 |
| 25 | PENNSYLVANIA | 12,702,379 | 844,334.98 | 6,647.06 |
| 26 | TEXAS | 25,145,561 | 1,655,859.69 | 6,585.10 |
| 27 | IOWA | 3,046,355 | 199,770.10 | 6,557.68 |
| 28 | IDAHO | 1,567,582 | 101,764.56 | 6,491.82 |
| 29 | WASHINGTON | 6,724,540 | 435,401.09 | 6,474.81 |
| 30 | FLORIDA | 18,801,310 | 1,201,205.30 | 6,388.94 |
| 31 | COLORADO | 5,029,196 | 319,454.18 | 6,351.99 |
| 32 | MARYLAND | 5,773,552 | 366,263.40 | 6,343.81 |
| 33 | VERMONT | 625,741 | 39,689.40 | 6,342.78 |
| 34 | KENTUCKY | 4,339,367 | 271,947.67 | 6,266.99 |
| 35 | OREGON | 3,831,074 | 234,564.17 | 6,122.67 |
| 36 | ILLINOIS | 12,830,632 | 763,967.16 | 5,954.24 |
| 37 | CONNECTICUT | 3,574,097 | 211,272.54 | 5,911.21 |
| 38 | MONTANA | 989,415 | 55,996.63 | 5,659.57 |
| 39 | NEBRASKA | 1,826,341 | 103,316.29 | 5,657.01 |
| 40 | OKLAHOMA | 3,751,351 | 211,604.73 | 5,640.76 |
| 41 | NORTH DAKOTA | 672,591 | 35,964.31 | 5,347.13 |
| 42 | ARIZONA | 6,392,017 | 327,569.95 | 5,124.67 |
| 43 | WEST VIRGINIA | 1,852,994 | 93,972.75 | 5,071.40 |
| 44 | NEW YORK | 19,378,102 | 941,733.87 | 4,859.78 |
| 45 | NEW JERSEY | 8,791,894 | 416,678.70 | 4,739.35 |
| 46 | WYOMING | 563,626 | 25,735.95 | 4,566.14 |
| 47 | ALASKA | 710,231 | 32,335.27 | 4,552.78 |
| 48 | NEVADA | 2,700,551 | 104,147.55 | 3,856.53 |
| 49 | CALIFORNIA | 37,253,956 | 1,198,133.55 | 3,216.12 |
| 50 | NEW MEXICO | 2,059,179 | 63,660.33 | 3,091.54 |
| 51 | HAWAII | 1,360,301 | 38,920.07 | 2,861.14 |
| 52 | VIRGIN ISLANDS | 109,825 | 1,217.23 | 1,108.34 |
| 53 | GUAM | 178,430 | 1,904.11 | 1,067.15 |
| 54 | PUERTO RICO | 3,725,789 | 33,115.23 | 888.81 |
| 55 | AMERICAN SAMOA | 65,628 | 2.5 | 3.81 |
| **U.S. GRAMS / PER 100K:** | | 312,825,210 | 20,938,152.53 | 6,693.24 |

**DRUG CODE:1105B** **DRUG NAME:DL-METHAMPHETAMINE RACEMIC BASE**

CUMULATIVE DISTRIBUTION BY STATE IN GRAMS PER 100,000 POPULATION

| RANK | STATE | 2010 POP | TOTAL GRAMS | GRAMS/100K POP |
|---|---|---|---|---|
| 1 | SOUTH DAKOTA | 814,180 | 69.52 | 8.54 |
| 2 | WASHINGTON | 6,724,540 | 381.36 | 5.67 |
| 3 | MINNESOTA | 5,303,925 | 289.33 | 5.46 |
| 4 | COLORADO | 5,029,196 | 210.57 | 4.19 |
| 5 | PENNSYLVANIA | 12,702,379 | 489.05 | 3.85 |
| 6 | WYOMING | 563,626 | 21.3 | 3.78 |
| 7 | UTAH | 2,763,885 | 104.08 | 3.77 |
| 8 | NEVADA | 2,700,551 | 97.65 | 3.62 |
| 9 | NORTH DAKOTA | 672,591 | 23.71 | 3.53 |
| 10 | WISCONSIN | 5,686,986 | 197.31 | 3.47 |
| 11 | OREGON | 3,831,074 | 127.79 | 3.34 |
| 12 | NEW MEXICO | 2,059,179 | 65.9 | 3.2 |
| 13 | MISSOURI | 5,988,927 | 190.08 | 3.17 |
| 14 | CALIFORNIA | 37,253,956 | 1,152.91 | 3.09 |
| 15 | MARYLAND | 5,773,552 | 164.36 | 2.85 |
| 16 | OHIO | 11,536,504 | 300.18 | 2.6 |
| 17 | MASSACHUSETTS | 6,547,629 | 168.38 | 2.57 |
| 18 | MAINE | 1,328,361 | 33.35 | 2.51 |
| 19 | CONNECTICUT | 3,574,097 | 89.61 | 2.51 |
| 20 | IOWA | 3,046,355 | 73.94 | 2.43 |
| 21 | IDAHO | 1,567,582 | 34.96 | 2.23 |
| 22 | VIRGINIA | 8,001,024 | 175.21 | 2.19 |
| 23 | ARIZONA | 6,392,017 | 134.22 | 2.1 |
| 24 | OKLAHOMA | 3,751,351 | 77.16 | 2.06 |
| 25 | ALABAMA | 4,779,736 | 96.04 | 2.01 |
| 26 | NEW HAMPSHIRE | 1,316,470 | 26.12 | 1.98 |
| 27 | NORTH CAROLINA | 9,535,483 | 189.27 | 1.98 |
| 28 | ILLINOIS | 12,830,632 | 248.34 | 1.94 |
| 29 | NEBRASKA | 1,826,341 | 32.95 | 1.8 |
| 30 | DELAWARE | 897,934 | 15.27 | 1.7 |
| 31 | TEXAS | 25,145,561 | 426.76 | 1.7 |
| 32 | NEW YORK | 19,378,102 | 325.5 | 1.68 |
| 33 | RHODE ISLAND | 1,052,567 | 16.88 | 1.6 |
| 34 | INDIANA | 6,483,802 | 103.68 | 1.6 |
| 35 | NEW JERSEY | 8,791,894 | 138.24 | 1.57 |
| 36 | ALASKA | 710,231 | 10.85 | 1.53 |
| 37 | HAWAII | 1,360,301 | 20.49 | 1.51 |
| 38 | KANSAS | 2,853,118 | 41.79 | 1.46 |
| 39 | SOUTH CAROLINA | 4,625,364 | 60.28 | 1.3 |
| 40 | LOUISIANA | 4,533,372 | 55.05 | 1.21 |
| 41 | VERMONT | 625,741 | 7.23 | 1.16 |
| 42 | DISTRICT OF COLUMBIA | 601,723 | 6.83 | 1.14 |
| 43 | MISSISSIPPI | 2,967,297 | 32.95 | 1.11 |
| 44 | GEORGIA | 9,687,653 | 94.84 | 0.98 |
| 45 | MICHIGAN | 9,883,640 | 91.62 | 0.93 |
| 46 | KENTUCKY | 4,339,367 | 39.78 | 0.92 |
| 47 | WEST VIRGINIA | 1,852,994 | 16.48 | 0.89 |
| 48 | ARKANSAS | 2,915,918 | 25.32 | 0.87 |
| 49 | TENNESSEE | 6,346,105 | 50.63 | 0.8 |
| 50 | FLORIDA | 18,801,310 | 135.83 | 0.72 |
| 51 | MONTANA | 989,415 | 2.81 | 0.28 |
| **U.S. GRAMS / PER 100K:** | | 312,825,210 | 6,983.75 | 2.23 |

**DRUG CODE:1105D** **DRUG NAME:D-METHAMPHETAMINE**

| RANK | STATE | 2010 POP | TOTAL GRAMS | GRAMS/100K POP |
|---|---|---|---|---|
| 1 | MINNESOTA | 5,303,925 | 53.02 | 1 |
| 2 | OREGON | 3,831,074 | 37.76 | 0.99 |
| 3 | NORTH DAKOTA | 672,591 | 6.03 | 0.9 |
| 4 | DISTRICT OF COLUMBIA | 601,723 | 5.22 | 0.87 |
| 5 | IDAHO | 1,567,582 | 10.44 | 0.67 |

| | | | | |
|---|---|---|---|---|
| 6 | MISSOURI | 5,988,927 | 32.14 | 0.54 |
| 7 | WISCONSIN | 5,686,986 | 28.12 | 0.49 |
| 8 | CALIFORNIA | 37,253,956 | 142.84 | 0.38 |
| 9 | RHODE ISLAND | 1,052,567 | 3.62 | 0.34 |
| 10 | ARIZONA | 6,392,017 | 19.68 | 0.31 |
| 11 | ALABAMA | 4,779,736 | 14.86 | 0.31 |
| 12 | HAWAII | 1,360,301 | 3.21 | 0.24 |
| 13 | NORTH CAROLINA | 9,535,483 | 21.69 | 0.23 |
| 14 | CONNECTICUT | 3,574,097 | 8.03 | 0.22 |
| 15 | PENNSYLVANIA | 12,702,379 | 26.11 | 0.21 |
| 16 | WEST VIRGINIA | 1,852,994 | 3.62 | 0.2 |
| 17 | SOUTH CAROLINA | 4,625,364 | 8.84 | 0.19 |
| 18 | OKLAHOMA | 3,751,351 | 7.23 | 0.19 |
| 19 | OHIO | 11,536,504 | 21.29 | 0.18 |
| 20 | TEXAS | 25,145,561 | 39.77 | 0.16 |
| 21 | KANSAS | 2,853,118 | 4.42 | 0.15 |
| 22 | GEORGIA | 9,687,653 | 11.25 | 0.12 |
| 23 | ALASKA | 710,231 | 0.8 | 0.11 |
| 24 | LOUISIANA | 4,533,372 | 4.42 | 0.1 |
| 25 | MASSACHUSETTS | 6,547,629 | 6.83 | 0.1 |
| 26 | NEW YORK | 19,378,102 | 17.76 | 0.09 |
| 27 | MISSISSIPPI | 2,967,297 | 2.41 | 0.08 |
| 28 | KENTUCKY | 4,339,367 | 3.62 | 0.08 |
| 29 | MARYLAND | 5,773,552 | 4.82 | 0.08 |
| 30 | FLORIDA | 18,801,310 | 12.45 | 0.07 |
| 31 | NEW JERSEY | 8,791,894 | 5.62 | 0.06 |
| 32 | ILLINOIS | 12,830,632 | 6.03 | 0.05 |
| 33 | WASHINGTON | 6,724,540 | 2.41 | 0.04 |
| 34 | NEBRASKA | 1,826,341 | 0.8 | 0.04 |
| 35 | NEVADA | 2,700,551 | 0.4 | 0.01 |
| 36 | TENNESSEE | 6,346,105 | 0.4 | 0.01 |
| **U.S. GRAMS / PER 100K:** | | 312,825,210 | 577.97 | 0.18 |

**DRUG CODE:1205** **DRUG NAME:LISDEXAMFETAMINE**

| **RANK** | **STATE** | **2010 POP** | **TOTAL GRAMS** | **GRAMS/100K POP** |
|---|---|---|---|---|
| 1 | LOUISIANA | 4,533,372 | 371,359.76 | 8,191.69 |
| 2 | DELAWARE | 897,934 | 57,216.87 | 6,372.06 |
| 3 | ALABAMA | 4,779,736 | 285,325.03 | 5,969.47 |
| 4 | SOUTH DAKOTA | 814,180 | 43,835.59 | 5,384.02 |
| 5 | SOUTH CAROLINA | 4,625,364 | 244,431.29 | 5,284.58 |
| 6 | INDIANA | 6,483,802 | 341,369.21 | 5,264.95 |
| 7 | MISSISSIPPI | 2,967,297 | 155,921.09 | 5,254.65 |
| 8 | MAINE | 1,328,361 | 67,310.50 | 5,067.18 |
| 9 | WISCONSIN | 5,686,986 | 267,551.62 | 4,704.63 |
| 10 | IOWA | 3,046,355 | 137,639.83 | 4,518.18 |
| 11 | KANSAS | 2,853,118 | 127,152.52 | 4,456.62 |
| 12 | MISSOURI | 5,988,927 | 266,212.74 | 4,445.08 |
| 13 | VERMONT | 625,741 | 27,704.29 | 4,427.44 |
| 14 | TENNESSEE | 6,346,105 | 280,599.16 | 4,421.60 |
| 15 | TEXAS | 25,145,561 | 1,088,975.06 | 4,330.69 |
| 16 | NORTH CAROLINA | 9,535,483 | 410,571.97 | 4,305.73 |
| 17 | WEST VIRGINIA | 1,852,994 | 78,587.49 | 4,241.11 |
| 18 | GEORGIA | 9,687,653 | 409,573.99 | 4,227.79 |
| 19 | NEW HAMPSHIRE | 1,316,470 | 55,638.08 | 4,226.31 |
| 20 | ARKANSAS | 2,915,918 | 121,203.61 | 4,156.62 |
| 21 | CONNECTICUT | 3,574,097 | 144,751.62 | 4,050.02 |
| 22 | OKLAHOMA | 3,751,351 | 146,677.10 | 3,909.98 |
| 23 | KENTUCKY | 4,339,367 | 169,631.26 | 3,909.12 |
| 24 | OHIO | 11,536,504 | 446,513.98 | 3,870.44 |
| 25 | MARYLAND | 5,773,552 | 223,456.52 | 3,870.35 |
| 26 | NEBRASKA | 1,826,341 | 65,765.84 | 3,600.96 |

| | | | | |
|---|---|---|---|---|
| 27 | DISTRICT OF COLUMBIA | 601,723 | 21,009.89 | 3,491.62 |
| 28 | NORTH DAKOTA | 672,591 | 22,892.18 | 3,403.58 |
| 29 | VIRGINIA | 8,001,024 | 270,981.83 | 3,386.84 |
| 30 | UTAH | 2,763,885 | 86,605.74 | 3,133.48 |
| 31 | FLORIDA | 18,801,310 | 548,800.42 | 2,918.95 |
| 32 | RHODE ISLAND | 1,052,567 | 30,111.49 | 2,860.77 |
| 33 | PENNSYLVANIA | 12,702,379 | 343,601.25 | 2,705.01 |
| 34 | MICHIGAN | 9,883,640 | 237,852.60 | 2,406.53 |
| 35 | MINNESOTA | 5,303,925 | 126,679.63 | 2,388.41 |
| 36 | MONTANA | 989,415 | 22,910.10 | 2,315.52 |
| 37 | IDAHO | 1,567,582 | 35,397.06 | 2,258.07 |
| 38 | ARIZONA | 6,392,017 | 143,380.36 | 2,243.12 |
| 39 | NEW JERSEY | 8,791,894 | 195,960.87 | 2,228.88 |
| 40 | ILLINOIS | 12,830,632 | 283,440.12 | 2,209.09 |
| 41 | COLORADO | 5,029,196 | 111,032.78 | 2,207.76 |
| 42 | MASSACHUSETTS | 6,547,629 | 142,897.65 | 2,182.43 |
| 43 | WASHINGTON | 6,724,540 | 136,312.31 | 2,027.09 |
| 44 | WYOMING | 563,626 | 10,450.31 | 1,854.12 |
| 45 | NEW YORK | 19,378,102 | 338,418.49 | 1,746.40 |
| 46 | OREGON | 3,831,074 | 60,117.75 | 1,569.21 |
| 47 | ALASKA | 710,231 | 9,865.85 | 1,389.11 |
| 48 | CALIFORNIA | 37,253,956 | 446,290.06 | 1,197.97 |
| 49 | NEVADA | 2,700,551 | 30,278.45 | 1,121.20 |
| 50 | NEW MEXICO | 2,059,179 | 17,259.75 | 838.19 |
| 51 | HAWAII | 1,360,301 | 5,358.41 | 393.91 |
| 52 | VIRGIN ISLANDS | 109,825 | 267.66 | 243.72 |
| 53 | PUERTO RICO | 3,725,789 | 7,605.48 | 204.13 |
| 54 | GUAM | 178,430 | 90.76 | 50.87 |
| **U.S. GRAMS / PER 100K:** | | 312,825,210 | 9,720,845.25 | 3,107.44 |

**DRUG CODE:1724** **DRUG NAME:METHYLPHENIDATE (DL;D;L;ISOMERS)**

| **RANK** | **STATE** | **2010 POP** | **TOTAL GRAMS** | **GRAMS/100K POP** |
|---|---|---|---|---|
| 1 | VERMONT | 625,741 | 76,927.33 | 12,293.80 |
| 2 | SOUTH DAKOTA | 814,180 | 92,985.17 | 11,420.71 |
| 3 | IOWA | 3,046,355 | 326,076.42 | 10,703.82 |
| 4 | MAINE | 1,328,361 | 141,224.78 | 10,631.51 |
| 5 | NEW HAMPSHIRE | 1,316,470 | 135,639.97 | 10,303.31 |
| 6 | NORTH DAKOTA | 672,591 | 60,128.60 | 8,939.85 |
| 7 | MASSACHUSETTS | 6,547,629 | 584,650.30 | 8,929.19 |
| 8 | MISSOURI | 5,988,927 | 526,374.97 | 8,789.14 |
| 9 | DELAWARE | 897,934 | 75,095.75 | 8,363.17 |
| 10 | OHIO | 11,536,504 | 926,895.09 | 8,034.45 |
| 11 | NORTH CAROLINA | 9,535,483 | 762,179.72 | 7,993.09 |
| 12 | SOUTH CAROLINA | 4,625,364 | 353,772.19 | 7,648.53 |
| 13 | MICHIGAN | 9,883,640 | 754,875.69 | 7,637.63 |
| 14 | MINNESOTA | 5,303,925 | 403,340.78 | 7,604.57 |
| 15 | INDIANA | 6,483,802 | 485,299.10 | 7,484.79 |
| 16 | LOUISIANA | 4,533,372 | 338,720.36 | 7,471.71 |
| 17 | WISCONSIN | 5,686,986 | 413,664.73 | 7,273.88 |
| 18 | RHODE ISLAND | 1,052,567 | 75,299.27 | 7,153.87 |
| 19 | PENNSYLVANIA | 12,702,379 | 899,814.76 | 7,083.83 |
| 20 | MONTANA | 989,415 | 69,975.85 | 7,072.45 |
| 21 | KANSAS | 2,853,118 | 200,253.28 | 7,018.75 |
| 22 | IDAHO | 1,567,582 | 109,587.11 | 6,990.84 |
| 23 | MARYLAND | 5,773,552 | 400,198.33 | 6,931.58 |
| 24 | KENTUCKY | 4,339,367 | 300,167.86 | 6,917.32 |
| 25 | NEBRASKA | 1,826,341 | 125,140.37 | 6,851.97 |
| 26 | WEST VIRGINIA | 1,852,994 | 126,376.79 | 6,820.14 |
| 27 | GEORGIA | 9,687,653 | 653,705.32 | 6,747.82 |
| 28 | VIRGINIA | 8,001,024 | 535,783.95 | 6,696.44 |
| 29 | UTAH | 2,763,885 | 184,160.98 | 6,663.12 |

| | | | | |
|---|---|---|---|---|
| 30 | WASHINGTON | 6,724,540 | 446,856.52 | 6,645.16 |
| 31 | OREGON | 3,831,074 | 245,904.22 | 6,418.68 |
| 32 | ALABAMA | 4,779,736 | 290,869.30 | 6,085.47 |
| 33 | CONNECTICUT | 3,574,097 | 216,315.94 | 6,052.32 |
| 34 | WYOMING | 563,626 | 32,575.98 | 5,779.72 |
| 35 | MISSISSIPPI | 2,967,297 | 171,319.72 | 5,773.60 |
| 36 | ARKANSAS | 2,915,918 | 160,100.85 | 5,490.58 |
| 37 | ILLINOIS | 12,830,632 | 696,305.50 | 5,426.90 |
| 38 | DISTRICT OF COLUMBIA | 601,723 | 32,200 | 5,351.30 |
| 39 | TEXAS | 25,145,561 | 1,336,775.33 | 5,316.15 |
| 40 | TENNESSEE | 6,346,105 | 335,642.81 | 5,288.96 |
| 41 | OKLAHOMA | 3,751,351 | 193,512.61 | 5,158.48 |
| 42 | COLORADO | 5,029,196 | 246,395.47 | 4,899.30 |
| 43 | NEW YORK | 19,378,102 | 910,080.54 | 4,696.44 |
| 44 | ARIZONA | 6,392,017 | 298,120.31 | 4,663.95 |
| 45 | ALASKA | 710,231 | 32,138.95 | 4,525.14 |
| 46 | FLORIDA | 18,801,310 | 822,773.66 | 4,376.15 |
| 47 | NEW JERSEY | 8,791,894 | 366,780.33 | 4,171.80 |
| 48 | NEW MEXICO | 2,059,179 | 76,270.43 | 3,703.92 |
| 49 | HAWAII | 1,360,301 | 43,636.31 | 3,207.84 |
| 50 | CALIFORNIA | 37,253,956 | 1,087,983.37 | 2,920.45 |
| 51 | NEVADA | 2,700,551 | 59,241.28 | 2,193.67 |
| 52 | PUERTO RICO | 3,725,789 | 67,864.79 | 1,821.49 |
| 53 | GUAM | 178,430 | 1,914.75 | 1,073.11 |
| 54 | VIRGIN ISLANDS | 109,825 | 597.19 | 543.76 |
| 55 | AMERICAN SAMOA | 65,628 | 17.73 | 27.01 |
| **U.S. GRAMS / PER 100K:** | | 312,825,210 | 18,310,508.74 | 5,853.27 |

**DRUG CODE:2100 1308.13(C)(3)]** **DRUG NAME:BARBITURIC ACID DERIVIATIVE OR SALT [PER 21CFR**

| **RANK** | **STATE** | **2010 POP** | **TOTAL GRAMS** | **GRAMS/100K POP** |
|---|---|---|---|---|
| 1 | NEVADA | 2,700,551 | 18,275 | 676.71 |
| 2 | FLORIDA | 18,801,310 | 69,845 | 371.49 |
| 3 | ARIZONA | 6,392,017 | 20,250 | 316.8 |
| 4 | OKLAHOMA | 3,751,351 | 11,255 | 300.03 |
| 5 | LOUISIANA | 4,533,372 | 13,425 | 296.14 |
| 6 | MICHIGAN | 9,883,640 | 26,910 | 272.27 |
| 7 | WYOMING | 563,626 | 1,525 | 270.57 |
| 8 | MISSOURI | 5,988,927 | 15,820 | 264.15 |
| 9 | HAWAII | 1,360,301 | 3,380 | 248.47 |
| 10 | CONNECTICUT | 3,574,097 | 8,710 | 243.7 |
| 11 | RHODE ISLAND | 1,052,567 | 2,520 | 239.41 |
| 12 | ILLINOIS | 12,830,632 | 30,235 | 235.65 |
| 13 | GEORGIA | 9,687,653 | 21,025 | 217.03 |
| 14 | NEW JERSEY | 8,791,894 | 18,110 | 205.99 |
| 15 | UTAH | 2,763,885 | 5,685 | 205.69 |
| 16 | DELAWARE | 897,934 | 1,840 | 204.91 |
| 17 | SOUTH CAROLINA | 4,625,364 | 9,395 | 203.12 |
| 18 | INDIANA | 6,483,802 | 13,160 | 202.97 |
| 19 | MONTANA | 989,415 | 1,995 | 201.63 |
| 20 | TEXAS | 25,145,561 | 49,304.90 | 196.08 |
| 21 | ARKANSAS | 2,915,918 | 5,325 | 182.62 |
| 22 | TENNESSEE | 6,346,105 | 11,305 | 178.14 |
| 23 | VIRGINIA | 8,001,024 | 13,900 | 173.73 |
| 24 | PENNSYLVANIA | 12,702,379 | 21,675 | 170.64 |
| 25 | KANSAS | 2,853,118 | 4,820 | 168.94 |
| 26 | KENTUCKY | 4,339,367 | 7,259 | 167.28 |
| 27 | MASSACHUSETTS | 6,547,629 | 10,820 | 165.25 |
| 28 | MARYLAND | 5,773,552 | 9,540 | 165.24 |
| 29 | NEBRASKA | 1,826,341 | 2,970 | 162.62 |
| 30 | NEW MEXICO | 2,059,179 | 3,325 | 161.47 |
| 31 | DISTRICT OF COLUMBIA | 601,723 | 970 | 161.2 |

| | | | | |
|---|---|---|---|---|
| 32 | NORTH CAROLINA | 9,535,483 | 15,265 | 160.09 |
| 33 | CALIFORNIA | 37,253,956 | 59,183.50 | 158.87 |
| 34 | NEW HAMPSHIRE | 1,316,470 | 1,995 | 151.54 |
| 35 | OHIO | 11,536,504 | 16,795 | 145.58 |
| 36 | IDAHO | 1,567,582 | 2,165 | 138.11 |
| 37 | WASHINGTON | 6,724,540 | 9,240 | 137.41 |
| 38 | MISSISSIPPI | 2,967,297 | 4,020 | 135.48 |
| 39 | COLORADO | 5,029,196 | 6,585 | 130.94 |
| 40 | ALABAMA | 4,779,736 | 6,145 | 128.56 |
| 41 | MAINE | 1,328,361 | 1,690 | 127.22 |
| 42 | WEST VIRGINIA | 1,852,994 | 2,120 | 114.41 |
| 43 | WISCONSIN | 5,686,986 | 6,360 | 111.83 |
| 44 | SOUTH DAKOTA | 814,180 | 905 | 111.15 |
| 45 | OREGON | 3,831,074 | 3,865 | 100.89 |
| 46 | ALASKA | 710,231 | 715 | 100.67 |
| 47 | MINNESOTA | 5,303,925 | 5,315 | 100.21 |
| 48 | VIRGIN ISLANDS | 109,825 | 105 | 95.61 |
| 49 | IOWA | 3,046,355 | 2,780 | 91.26 |
| 50 | NEW YORK | 19,378,102 | 17,585 | 90.75 |
| 51 | NORTH DAKOTA | 672,591 | 565 | 84 |
| 52 | VERMONT | 625,741 | 285 | 45.55 |
| 53 | PUERTO RICO | 3,725,789 | 605 | 16.24 |
| 54 | GUAM | 178,430 | 10 | 5.6 |
| **U.S. GRAMS / PER 100K:** | | 312,825,210 | 598,872.40 | 191.44 |

**DRUG CODE:2125** **DRUG NAME:AMOBARBITAL (SCHEDULE 2)**

| RANK | STATE | 2010 POP | TOTAL GRAMS | GRAMS/100K POP |
|---|---|---|---|---|
| 1 | COLORADO | 5,029,196 | 68.36 | 1.36 |
| 2 | UTAH | 2,763,885 | 34.18 | 1.24 |
| 3 | DISTRICT OF COLUMBIA | 601,723 | 6.84 | 1.14 |
| 4 | NEW HAMPSHIRE | 1,316,470 | 12.3 | 0.93 |
| 5 | NEW YORK | 19,378,102 | 152.66 | 0.79 |
| 6 | ARIZONA | 6,392,017 | 45.11 | 0.71 |
| 7 | WASHINGTON | 6,724,540 | 36.91 | 0.55 |
| 8 | KANSAS | 2,853,118 | 13.67 | 0.48 |
| 9 | WISCONSIN | 5,686,986 | 26.89 | 0.47 |
| 10 | MARYLAND | 5,773,552 | 27.34 | 0.47 |
| 11 | KENTUCKY | 4,339,367 | 19.14 | 0.44 |
| 12 | MINNESOTA | 5,303,925 | 23.24 | 0.44 |
| 13 | NORTH CAROLINA | 9,535,483 | 38.28 | 0.4 |
| 14 | NEW JERSEY | 8,791,894 | 30.53 | 0.35 |
| 15 | RHODE ISLAND | 1,052,567 | 3.65 | 0.35 |
| 16 | TEXAS | 25,145,561 | 83.39 | 0.33 |
| 17 | SOUTH CAROLINA | 4,625,364 | 15.04 | 0.33 |
| 18 | MASSACHUSETTS | 6,547,629 | 20.05 | 0.31 |
| 19 | CALIFORNIA | 37,253,956 | 100.71 | 0.27 |
| 20 | GEORGIA | 9,687,653 | 26.43 | 0.27 |
| 21 | TENNESSEE | 6,346,105 | 15.95 | 0.25 |
| 22 | PENNSYLVANIA | 12,702,379 | 29.16 | 0.23 |
| 23 | MAINE | 1,328,361 | 2.73 | 0.21 |
| 24 | NEBRASKA | 1,826,341 | 3.65 | 0.2 |
| 25 | MICHIGAN | 9,883,640 | 18.68 | 0.19 |
| 26 | OREGON | 3,831,074 | 6.84 | 0.18 |
| 27 | MISSISSIPPI | 2,967,297 | 4.56 | 0.15 |
| 28 | VERMONT | 625,741 | 0.91 | 0.15 |
| 29 | OHIO | 11,536,504 | 17.77 | 0.15 |
| 30 | VIRGINIA | 8,001,024 | 11.39 | 0.14 |
| 31 | FLORIDA | 18,801,310 | 23.24 | 0.12 |
| 32 | ILLINOIS | 12,830,632 | 15.95 | 0.12 |
| 33 | MISSOURI | 5,988,927 | 6.84 | 0.11 |
| 34 | WEST VIRGINIA | 1,852,994 | 1.82 | 0.1 |

| | | | | |
|---|---|---|---|---|
| 35 | CONNECTICUT | 3,574,097 | 3.65 | 0.1 |
| 36 | INDIANA | 6,483,802 | 4.1 | 0.06 |
| 37 | ARKANSAS | 2,915,918 | 0.91 | 0.03 |
| 38 | IOWA | 3,046,355 | 0.46 | 0.01 |
| **U.S. GRAMS / PER 100K:** | | 312,825,210 | 953.32 | 0.3 |

**DRUG CODE:2165** **DRUG NAME:BUTALBITAL**

| **RANK** | **STATE** | **2010 POP** | **TOTAL GRAMS** | **GRAMS/100K POP** |
|---|---|---|---|---|
| 1 | NEVADA | 2,700,551 | 14,815 | 548.59 |
| 2 | OKLAHOMA | 3,751,351 | 19,990 | 532.87 |
| 3 | ARKANSAS | 2,915,918 | 13,765 | 472.06 |
| 4 | UTAH | 2,763,885 | 12,720 | 460.22 |
| 5 | GEORGIA | 9,687,653 | 39,800 | 410.83 |
| 6 | MISSOURI | 5,988,927 | 23,750 | 396.57 |
| 7 | ALABAMA | 4,779,736 | 17,800 | 372.41 |
| 8 | PENNSYLVANIA | 12,702,379 | 46,800 | 368.43 |
| 9 | MICHIGAN | 9,883,640 | 35,520 | 359.38 |
| 10 | FLORIDA | 18,801,310 | 65,720 | 349.55 |
| 11 | LOUISIANA | 4,533,372 | 15,750 | 347.42 |
| 12 | CONNECTICUT | 3,574,097 | 12,300 | 344.14 |
| 13 | IDAHO | 1,567,582 | 5,245 | 334.59 |
| 14 | MARYLAND | 5,773,552 | 18,990 | 328.91 |
| 15 | ARIZONA | 6,392,017 | 20,535 | 321.26 |
| 16 | SOUTH DAKOTA | 814,180 | 2,595 | 318.73 |
| 17 | MONTANA | 989,415 | 3,080 | 311.3 |
| 18 | KANSAS | 2,853,118 | 8,805 | 308.61 |
| 19 | HAWAII | 1,360,301 | 4,160 | 305.81 |
| 20 | TEXAS | 25,145,561 | 74,795 | 297.45 |
| 21 | SOUTH CAROLINA | 4,625,364 | 13,480 | 291.44 |
| 22 | WYOMING | 563,626 | 1,630 | 289.2 |
| 23 | MISSISSIPPI | 2,967,297 | 8,545 | 287.97 |
| 24 | NEW JERSEY | 8,791,894 | 24,670 | 280.6 |
| 25 | WEST VIRGINIA | 1,852,994 | 5,070 | 273.61 |
| 26 | CALIFORNIA | 37,253,956 | 98,310 | 263.89 |
| 27 | VIRGINIA | 8,001,024 | 20,870 | 260.84 |
| 28 | WASHINGTON | 6,724,540 | 17,405 | 258.83 |
| 29 | OREGON | 3,831,074 | 9,385 | 244.97 |
| 30 | NEBRASKA | 1,826,341 | 4,435 | 242.84 |
| 31 | TENNESSEE | 6,346,105 | 15,265 | 240.54 |
| 32 | INDIANA | 6,483,802 | 15,595 | 240.52 |
| 33 | KENTUCKY | 4,339,367 | 10,365 | 238.86 |
| 34 | DELAWARE | 897,934 | 2,110 | 234.98 |
| 35 | DISTRICT OF COLUMBIA | 601,723 | 1,400 | 232.67 |
| 36 | ILLINOIS | 12,830,632 | 29,660 | 231.17 |
| 37 | OHIO | 11,536,504 | 26,630 | 230.83 |
| 38 | NORTH CAROLINA | 9,535,483 | 20,390 | 213.83 |
| 39 | NEW HAMPSHIRE | 1,316,470 | 2,665 | 202.44 |
| 40 | NORTH DAKOTA | 672,591 | 1,285 | 191.05 |
| 41 | MAINE | 1,328,361 | 2,520 | 189.71 |
| 42 | NEW MEXICO | 2,059,179 | 3,700 | 179.68 |
| 43 | IOWA | 3,046,355 | 5,100 | 167.41 |
| 44 | COLORADO | 5,029,196 | 7,810 | 155.29 |
| 45 | MASSACHUSETTS | 6,547,629 | 9,300 | 142.04 |
| 46 | RHODE ISLAND | 1,052,567 | 1,470 | 139.66 |
| 47 | ALASKA | 710,231 | 925 | 130.24 |
| 48 | MINNESOTA | 5,303,925 | 6,705 | 126.42 |
| 49 | WISCONSIN | 5,686,986 | 6,995 | 123 |
| 50 | NEW YORK | 19,378,102 | 22,775 | 117.53 |
| 51 | VERMONT | 625,741 | 470 | 75.11 |
| 52 | VIRGIN ISLANDS | 109,825 | 45 | 40.97 |
| 53 | GUAM | 178,430 | 40 | 22.42 |

| | | | | |
|---|---|---|---|---|
| 54 | AMERICAN SAMOA | 65,628 | 10 | 15.24 |
| **U.S. GRAMS / PER 100K:** | | 312,825,210 | 853,965 | 272.98 |

**DRUG CODE:2270** **DRUG NAME:PENTOBARBITAL (SCHEDULE 2)**

| RANK | STATE | 2010 POP | TOTAL GRAMS | GRAMS/100K POP |
|---|---|---|---|---|
| 1 | VERMONT | 625,741 | 54,362.24 | 8,687.66 |
| 2 | MONTANA | 989,415 | 74,125.43 | 7,491.84 |
| 3 | WYOMING | 563,626 | 40,470.26 | 7,180.34 |
| 4 | MAINE | 1,328,361 | 80,880.53 | 6,088.75 |
| 5 | IDAHO | 1,567,582 | 90,449.41 | 5,770 |
| 6 | NEW HAMPSHIRE | 1,316,470 | 61,925.27 | 4,703.89 |
| 7 | NEBRASKA | 1,826,341 | 82,467.93 | 4,515.47 |
| 8 | NORTH DAKOTA | 672,591 | 28,368.24 | 4,217.75 |
| 9 | OREGON | 3,831,074 | 154,686.71 | 4,037.69 |
| 10 | NEW MEXICO | 2,059,179 | 80,303.10 | 3,899.76 |
| 11 | MICHIGAN | 9,883,640 | 384,941.28 | 3,894.73 |
| 12 | WISCONSIN | 5,686,986 | 209,420.50 | 3,682.45 |
| 13 | GUAM | 178,430 | 6,430.84 | 3,604.12 |
| 14 | WASHINGTON | 6,724,540 | 240,819.92 | 3,581.21 |
| 15 | COLORADO | 5,029,196 | 179,455.55 | 3,568.28 |
| 16 | ARIZONA | 6,392,017 | 215,066.18 | 3,364.61 |
| 17 | ALASKA | 710,231 | 22,791.38 | 3,209.01 |
| 18 | KANSAS | 2,853,118 | 90,490.17 | 3,171.62 |
| 19 | SOUTH DAKOTA | 814,180 | 24,992.87 | 3,069.70 |
| 20 | INDIANA | 6,483,802 | 179,784.67 | 2,772.83 |
| 21 | MINNESOTA | 5,303,925 | 144,879.78 | 2,731.56 |
| 22 | MISSOURI | 5,988,927 | 161,027.78 | 2,688.76 |
| 23 | KENTUCKY | 4,339,367 | 111,343.42 | 2,565.89 |
| 24 | OHIO | 11,536,504 | 295,121.79 | 2,558.16 |
| 25 | NORTH CAROLINA | 9,535,483 | 242,534.37 | 2,543.49 |
| 26 | VIRGINIA | 8,001,024 | 190,989.91 | 2,387.07 |
| 27 | IOWA | 3,046,355 | 71,503.15 | 2,347.17 |
| 28 | RHODE ISLAND | 1,052,567 | 24,562.40 | 2,333.57 |
| 29 | NEVADA | 2,700,551 | 61,755.73 | 2,286.78 |
| 30 | WEST VIRGINIA | 1,852,994 | 42,101.54 | 2,272.08 |
| 31 | TENNESSEE | 6,346,105 | 142,593.51 | 2,246.95 |
| 32 | LOUISIANA | 4,533,372 | 100,335.42 | 2,213.26 |
| 33 | ILLINOIS | 12,830,632 | 277,261.83 | 2,160.94 |
| 34 | PENNSYLVANIA | 12,702,379 | 269,190.85 | 2,119.22 |
| 35 | UTAH | 2,763,885 | 56,914.89 | 2,059.24 |
| 36 | HAWAII | 1,360,301 | 27,943.98 | 2,054.25 |
| 37 | OKLAHOMA | 3,751,351 | 76,112.70 | 2,028.94 |
| 38 | CONNECTICUT | 3,574,097 | 72,452.06 | 2,027.14 |
| 39 | TEXAS | 25,145,561 | 496,593.15 | 1,974.87 |
| 40 | CALIFORNIA | 37,253,956 | 708,793.68 | 1,902.60 |
| 41 | MASSACHUSETTS | 6,547,629 | 112,758.41 | 1,722.13 |
| 42 | GEORGIA | 9,687,653 | 153,814.35 | 1,587.74 |
| 43 | NEW YORK | 19,378,102 | 306,395.35 | 1,581.14 |
| 44 | MARYLAND | 5,773,552 | 88,930.58 | 1,540.31 |
| 45 | FLORIDA | 18,801,310 | 284,606.14 | 1,513.76 |
| 46 | ALABAMA | 4,779,736 | 65,593.55 | 1,372.33 |
| 47 | ARKANSAS | 2,915,918 | 36,427.18 | 1,249.25 |
| 48 | SOUTH CAROLINA | 4,625,364 | 55,714.74 | 1,204.55 |
| 49 | MISSISSIPPI | 2,967,297 | 32,523.41 | 1,096.06 |
| 50 | NEW JERSEY | 8,791,894 | 81,918.15 | 931.75 |
| 51 | DELAWARE | 897,934 | 7,148.89 | 796.15 |
| 52 | DISTRICT OF COLUMBIA | 601,723 | 2,552.83 | 424.25 |
| 53 | PUERTO RICO | 3,725,789 | 0.2 | 0.01 |
| U.S. GRAMS / PER 100K: | | 312,825,210 | 7,104,628.21 | 2,271.12 |

**DRUG CODE:2315** **DRUG NAME:SECOBARBITAL (SCHEDULE 2)**

CUMULATIVE DISTRIBUTION BY STATE IN GRAMS PER 100,000 POPULATION

| RANK | STATE | 2010 POP | TOTAL GRAMS | GRAMS/100K POP |
|---|---|---|---|---|
| 1 | OREGON | 3,831,074 | 952.12 | 24.85 |
| 2 | VERMONT | 625,741 | 137.33 | 21.95 |
| 3 | CALIFORNIA | 37,253,956 | 4,376.09 | 11.75 |
| 4 | WASHINGTON | 6,724,540 | 723.25 | 10.76 |
| 5 | NEW YORK | 19,378,102 | 1,501.42 | 7.75 |
| 6 | COLORADO | 5,029,196 | 375.36 | 7.46 |
| 7 | MARYLAND | 5,773,552 | 219.72 | 3.81 |
| 8 | KANSAS | 2,853,118 | 100.71 | 3.53 |
| 9 | RHODE ISLAND | 1,052,567 | 36.62 | 3.48 |
| 10 | DELAWARE | 897,934 | 27.47 | 3.06 |
| 11 | MASSACHUSETTS | 6,547,629 | 192.26 | 2.94 |
| 12 | NEW JERSEY | 8,791,894 | 228.88 | 2.6 |
| 13 | CONNECTICUT | 3,574,097 | 91.55 | 2.56 |
| 14 | PENNSYLVANIA | 12,702,379 | 265.5 | 2.09 |
| 15 | NEW HAMPSHIRE | 1,316,470 | 27.47 | 2.09 |
| 16 | FLORIDA | 18,801,310 | 366.2 | 1.95 |
| 17 | MISSOURI | 5,988,927 | 100.71 | 1.68 |
| 18 | VIRGINIA | 8,001,024 | 109.86 | 1.37 |
| 19 | ILLINOIS | 12,830,632 | 164.79 | 1.28 |
| 20 | OHIO | 11,536,504 | 137.33 | 1.19 |
| 21 | SOUTH DAKOTA | 814,180 | 9.16 | 1.12 |
| 22 | OKLAHOMA | 3,751,351 | 36.62 | 0.98 |
| 23 | SOUTH CAROLINA | 4,625,364 | 36.62 | 0.79 |
| 24 | HAWAII | 1,360,301 | 9.16 | 0.67 |
| 25 | GEORGIA | 9,687,653 | 45.78 | 0.47 |
| 26 | MINNESOTA | 5,303,925 | 18.31 | 0.35 |
| 27 | NEVADA | 2,700,551 | 9.16 | 0.34 |
| 28 | ARIZONA | 6,392,017 | 18.31 | 0.29 |
| 29 | MICHIGAN | 9,883,640 | 18.31 | 0.19 |
| 30 | TEXAS | 25,145,561 | 45.78 | 0.18 |
| 31 | WISCONSIN | 5,686,986 | 9.16 | 0.16 |
| 32 | TENNESSEE | 6,346,105 | 9.16 | 0.14 |
| 33 | NORTH CAROLINA | 9,535,483 | 9.16 | 0.1 |
| **U.S. GRAMS / PER 100K:** | | 312,825,210 | 10,409.24 | 3.33 |

**DRUG CODE:7365 DRUG PRODUCT (SYNI DRUG NAME:DRONABINOL IN AN ORAL SOLUTION IN FDA APPROVED**

| RANK | STATE | 2010 POP | TOTAL GRAMS | GRAMS/100K POP |
|---|---|---|---|---|
| 1 | NEW JERSEY | 8,791,894 | 143.55 | 1.63 |
| 2 | NEVADA | 2,700,551 | 11.55 | 0.43 |
| 3 | ARIZONA | 6,392,017 | 21.6 | 0.34 |
| 4 | FLORIDA | 18,801,310 | 36.6 | 0.19 |
| 5 | MISSOURI | 5,988,927 | 8.55 | 0.14 |
| 6 | NEW HAMPSHIRE | 1,316,470 | 1.8 | 0.14 |
| 7 | PENNSYLVANIA | 12,702,379 | 16.35 | 0.13 |
| 8 | CALIFORNIA | 37,253,956 | 43.8 | 0.12 |
| 9 | DISTRICT OF COLUMBIA | 601,723 | 0.75 | 0.12 |
| 10 | ALABAMA | 4,779,736 | 4.2 | 0.09 |
| 11 | OHIO | 11,536,504 | 8.7 | 0.08 |
| 12 | COLORADO | 5,029,196 | 3.75 | 0.07 |
| 13 | MASSACHUSETTS | 6,547,629 | 3.75 | 0.06 |
| 14 | TENNESSEE | 6,346,105 | 3.6 | 0.06 |
| 15 | MINNESOTA | 5,303,925 | 2.85 | 0.05 |
| 16 | CONNECTICUT | 3,574,097 | 1.65 | 0.05 |
| 17 | TEXAS | 25,145,561 | 11.7 | 0.05 |
| 18 | GEORGIA | 9,687,653 | 4.65 | 0.05 |
| 19 | ARKANSAS | 2,915,918 | 1.2 | 0.04 |
| 20 | MISSISSIPPI | 2,967,297 | 0.75 | 0.03 |
| 21 | KENTUCKY | 4,339,367 | 1.5 | 0.03 |
| 22 | MARYLAND | 5,773,552 | 1.95 | 0.03 |
| 23 | MONTANA | 989,415 | 0.3 | 0.03 |

| | | | | |
|---|---|---|---|---|
| 24 | INDIANA | 6,483,802 | 1.65 | 0.03 |
| 25 | KANSAS | 2,853,118 | 0.6 | 0.02 |
| 26 | DELAWARE | 897,934 | 0.15 | 0.02 |
| 27 | MAINE | 1,328,361 | 0.3 | 0.02 |
| 28 | UTAH | 2,763,885 | 0.6 | 0.02 |
| 29 | MICHIGAN | 9,883,640 | 2.4 | 0.02 |
| 30 | NEW MEXICO | 2,059,179 | 0.15 | 0.01 |
| 31 | SOUTH CAROLINA | 4,625,364 | 0.3 | 0.01 |
| 32 | ILLINOIS | 12,830,632 | 1.05 | 0.01 |
| 33 | VIRGINIA | 8,001,024 | 0.75 | 0.01 |
| 34 | LOUISIANA | 4,533,372 | 0.3 | 0.01 |
| 35 | NEW YORK | 19,378,102 | 1.95 | 0.01 |
| 36 | NORTH CAROLINA | 9,535,483 | 1.05 | 0.01 |
| 37 | WISCONSIN | 5,686,986 | 0.15 | 0 |
| **U.S. GRAMS / PER 100K:** | | 312,825,210 | 346.5 | 0.11 |

| **DRUG CODE:7379** | **DRUG NAME:NABILONE** | | | |
|---|---|---|---|---|
| **RANK** | **STATE** | **2010 POP** | **TOTAL GRAMS** | **GRAMS/100K POP** |
| 1 | MARYLAND | 5,773,552 | 4.1 | 0.07 |
| 2 | HAWAII | 1,360,301 | 0.6 | 0.04 |
| 3 | NEBRASKA | 1,826,341 | 0.3 | 0.02 |
| 4 | MAINE | 1,328,361 | 0.3 | 0.02 |
| 5 | MISSOURI | 5,988,927 | 0.55 | 0.01 |
| 6 | COLORADO | 5,029,196 | 0.35 | 0.01 |
| 7 | NORTH CAROLINA | 9,535,483 | 0.75 | 0.01 |
| 8 | KENTUCKY | 4,339,367 | 0.35 | 0.01 |
| 9 | VIRGINIA | 8,001,024 | 0.75 | 0.01 |
| 10 | OHIO | 11,536,504 | 1 | 0.01 |
| 11 | CONNECTICUT | 3,574,097 | 0.35 | 0.01 |
| 12 | FLORIDA | 18,801,310 | 2.15 | 0.01 |
| 13 | PENNSYLVANIA | 12,702,379 | 0.35 | 0 |
| 14 | ARIZONA | 6,392,017 | 0.2 | 0 |
| 15 | MINNESOTA | 5,303,925 | 0.1 | 0 |
| 16 | NEW YORK | 19,378,102 | 0.4 | 0 |
| 17 | NEW JERSEY | 8,791,894 | 0.35 | 0 |
| 18 | NEW MEXICO | 2,059,179 | 0.1 | 0 |
| 19 | IOWA | 3,046,355 | 0.1 | 0 |
| 20 | OREGON | 3,831,074 | 0.05 | 0 |
| 21 | CALIFORNIA | 37,253,956 | 0.65 | 0 |
| 22 | ILLINOIS | 12,830,632 | 0.5 | 0 |
| 23 | NEW HAMPSHIRE | 1,316,470 | 0.05 | 0 |
| 24 | WASHINGTON | 6,724,540 | 0.05 | 0 |
| **U.S. GRAMS / PER 100K:** | | 312,825,210 | 14.45 | 0 |

| **DRUG CODE:9041L** | **DRUG NAME:COCAINE** | | | |
|---|---|---|---|---|
| **RANK** | **STATE** | **2010 POP** | **TOTAL GRAMS** | **GRAMS/100K POP** |
| 1 | NORTH DAKOTA | 672,591 | 181.99 | 27.06 |
| 2 | MONTANA | 989,415 | 211.59 | 21.39 |
| 3 | SOUTH DAKOTA | 814,180 | 148.85 | 18.28 |
| 4 | MINNESOTA | 5,303,925 | 855.52 | 16.13 |
| 5 | DISTRICT OF COLUMBIA | 601,723 | 92.89 | 15.44 |
| 6 | MICHIGAN | 9,883,640 | 1,507.87 | 15.26 |
| 7 | NEW HAMPSHIRE | 1,316,470 | 190.31 | 14.46 |
| 8 | UTAH | 2,763,885 | 391.01 | 14.15 |
| 9 | NEW MEXICO | 2,059,179 | 290.41 | 14.1 |
| 10 | IDAHO | 1,567,582 | 216.56 | 13.81 |
| 11 | IOWA | 3,046,355 | 393.01 | 12.9 |
| 12 | MASSACHUSETTS | 6,547,629 | 815.45 | 12.45 |
| 13 | INDIANA | 6,483,802 | 800.91 | 12.35 |
| 14 | WYOMING | 563,626 | 69.28 | 12.29 |

| | | | | |
|---|---|---|---|---|
| 15 | PENNSYLVANIA | 12,702,379 | 1,556.72 | 12.26 |
| 16 | KANSAS | 2,853,118 | 328.62 | 11.52 |
| 17 | MISSISSIPPI | 2,967,297 | 338.98 | 11.42 |
| 18 | MISSOURI | 5,988,927 | 659.56 | 11.01 |
| 19 | WEST VIRGINIA | 1,852,994 | 201.81 | 10.89 |
| 20 | WISCONSIN | 5,686,986 | 604.82 | 10.64 |
| 21 | TENNESSEE | 6,346,105 | 670.01 | 10.56 |
| 22 | ILLINOIS | 12,830,632 | 1,290.13 | 10.06 |
| 23 | OREGON | 3,831,074 | 384.55 | 10.04 |
| 24 | KENTUCKY | 4,339,367 | 434.4 | 10.01 |
| 25 | CALIFORNIA | 37,253,956 | 3,470.56 | 9.32 |
| 26 | MARYLAND | 5,773,552 | 517.57 | 8.96 |
| 27 | LOUISIANA | 4,533,372 | 389.44 | 8.59 |
| 28 | VIRGINIA | 8,001,024 | 614.53 | 7.68 |
| 29 | NEW JERSEY | 8,791,894 | 645.74 | 7.34 |
| 30 | RHODE ISLAND | 1,052,567 | 76 | 7.22 |
| 31 | HAWAII | 1,360,301 | 97.42 | 7.16 |
| 32 | NEW YORK | 19,378,102 | 1,379.88 | 7.12 |
| 33 | COLORADO | 5,029,196 | 343.05 | 6.82 |
| 34 | GEORGIA | 9,687,653 | 656.89 | 6.78 |
| 35 | WASHINGTON | 6,724,540 | 454.08 | 6.75 |
| 36 | ALASKA | 710,231 | 47.93 | 6.75 |
| 37 | NEVADA | 2,700,551 | 179.95 | 6.66 |
| 38 | MAINE | 1,328,361 | 87.42 | 6.58 |
| 39 | ARIZONA | 6,392,017 | 419.58 | 6.56 |
| 40 | NEBRASKA | 1,826,341 | 119.17 | 6.53 |
| 41 | OHIO | 11,536,504 | 730.77 | 6.33 |
| 42 | FLORIDA | 18,801,310 | 1,162.10 | 6.18 |
| 43 | VERMONT | 625,741 | 38.46 | 6.15 |
| 44 | ARKANSAS | 2,915,918 | 175.6 | 6.02 |
| 45 | NORTH CAROLINA | 9,535,483 | 571.68 | 6 |
| 46 | TEXAS | 25,145,561 | 1,453.69 | 5.78 |
| 47 | DELAWARE | 897,934 | 51.85 | 5.77 |
| 48 | ALABAMA | 4,779,736 | 228.7 | 4.78 |
| 49 | CONNECTICUT | 3,574,097 | 150.92 | 4.22 |
| 50 | OKLAHOMA | 3,751,351 | 132.99 | 3.55 |
| 51 | SOUTH CAROLINA | 4,625,364 | 118.31 | 2.56 |
| 52 | GUAM | 178,430 | 0.36 | 0.2 |
| **U.S. GRAMS / PER 100K:** | | 312,825,210 | 26,949.90 | 8.62 |

| **DRUG CODE:9050** | **DRUG NAME:CODEINE** | | | |
|---|---|---|---|---|
| **RANK** | **STATE** | **2010 POP** | **TOTAL GRAMS** | **GRAMS/100K POP** |
| 1 | TEXAS | 25,145,561 | 4,007,558.98 | 15,937.44 |
| 2 | MICHIGAN | 9,883,640 | 1,025,274.84 | 10,373.45 |
| 3 | OKLAHOMA | 3,751,351 | 381,918.42 | 10,180.82 |
| 4 | ARKANSAS | 2,915,918 | 292,809.02 | 10,041.74 |
| 5 | DISTRICT OF COLUMBIA | 601,723 | 48,420.46 | 8,046.97 |
| 6 | WEST VIRGINIA | 1,852,994 | 124,461.06 | 6,716.75 |
| 7 | SOUTH CAROLINA | 4,625,364 | 304,035.99 | 6,573.23 |
| 8 | TENNESSEE | 6,346,105 | 341,484.13 | 5,381 |
| 9 | CALIFORNIA | 37,253,956 | 1,891,720.41 | 5,077.90 |
| 10 | ILLINOIS | 12,830,632 | 647,278.14 | 5,044.79 |
| 11 | ARIZONA | 6,392,017 | 297,148.54 | 4,648.74 |
| 12 | MISSOURI | 5,988,927 | 273,798.37 | 4,571.74 |
| 13 | KENTUCKY | 4,339,367 | 190,127.79 | 4,381.46 |
| 14 | ALABAMA | 4,779,736 | 208,891.64 | 4,370.36 |
| 15 | PENNSYLVANIA | 12,702,379 | 554,464.95 | 4,365.05 |
| 16 | GEORGIA | 9,687,653 | 420,264.84 | 4,338.15 |
| 17 | DELAWARE | 897,934 | 36,987.08 | 4,119.13 |
| 18 | MONTANA | 989,415 | 40,580.95 | 4,101.51 |
| 19 | MARYLAND | 5,773,552 | 231,927.48 | 4,017.07 |

| | | | | |
|---|---|---|---|---|
| 20 | HAWAII | 1,360,301 | 54,562.75 | 4,011.08 |
| 21 | NEVADA | 2,700,551 | 106,949.08 | 3,960.27 |
| 22 | KANSAS | 2,853,118 | 111,795.90 | 3,918.38 |
| 23 | ALASKA | 710,231 | 27,797.47 | 3,913.86 |
| 24 | LOUISIANA | 4,533,372 | 173,795.65 | 3,833.69 |
| 25 | NEW MEXICO | 2,059,179 | 78,187 | 3,797 |
| 26 | MISSISSIPPI | 2,967,297 | 109,613.37 | 3,694.05 |
| 27 | WASHINGTON | 6,724,540 | 245,749.48 | 3,654.52 |
| 28 | FLORIDA | 18,801,310 | 681,387.45 | 3,624.15 |
| 29 | OREGON | 3,831,074 | 135,860.23 | 3,546.27 |
| 30 | NEW JERSEY | 8,791,894 | 310,813.87 | 3,535.23 |
| 31 | OHIO | 11,536,504 | 403,005.98 | 3,493.31 |
| 32 | IDAHO | 1,567,582 | 54,643.68 | 3,485.86 |
| 33 | SOUTH DAKOTA | 814,180 | 27,885.17 | 3,424.94 |
| 34 | NORTH DAKOTA | 672,591 | 22,778.39 | 3,386.66 |
| 35 | WISCONSIN | 5,686,986 | 190,554.17 | 3,350.71 |
| 36 | MAINE | 1,328,361 | 41,556.12 | 3,128.38 |
| 37 | INDIANA | 6,483,802 | 200,465.35 | 3,091.79 |
| 38 | WYOMING | 563,626 | 16,766.46 | 2,974.75 |
| 39 | NEW YORK | 19,378,102 | 567,749.93 | 2,929.85 |
| 40 | MASSACHUSETTS | 6,547,629 | 185,230.09 | 2,828.96 |
| 41 | MINNESOTA | 5,303,925 | 149,198.60 | 2,812.98 |
| 42 | RHODE ISLAND | 1,052,567 | 29,116.74 | 2,766.26 |
| 43 | VIRGINIA | 8,001,024 | 219,576.35 | 2,744.35 |
| 44 | VIRGIN ISLANDS | 109,825 | 2,999.03 | 2,730.73 |
| 45 | CONNECTICUT | 3,574,097 | 96,316.16 | 2,694.84 |
| 46 | GUAM | 178,430 | 4,765.67 | 2,670.89 |
| 47 | IOWA | 3,046,355 | 79,896.15 | 2,622.68 |
| 48 | NEBRASKA | 1,826,341 | 46,965.27 | 2,571.55 |
| 49 | UTAH | 2,763,885 | 65,371.67 | 2,365.21 |
| 50 | NEW HAMPSHIRE | 1,316,470 | 30,347.37 | 2,305.21 |
| 51 | VERMONT | 625,741 | 13,115.92 | 2,096.06 |
| 52 | AMERICAN SAMOA | 65,628 | 1,361.42 | 2,074.45 |
| 53 | COLORADO | 5,029,196 | 102,868.85 | 2,045.43 |
| 54 | NORTH CAROLINA | 9,535,483 | 193,617.17 | 2,030.49 |
| 55 | PUERTO RICO | 3,725,789 | 51,225.70 | 1,374.90 |
| **U.S. GRAMS / PER 100K:** | | 312,825,210 | 16,153,042.74 | 5,163.60 |

| **DRUG CODE:9056** | **DRUG NAME:ETORPHINE** | | | |
|---|---|---|---|---|
| **RANK** | **STATE** | **2010 POP** | **TOTAL GRAMS** | **GRAMS/100K POP** |
| 1 | WYOMING | 563,626 | 57 | 10.11 |
| **U.S. GRAMS / PER 100K:** | | 312,825,210 | 57 | 0.02 |

| **DRUG CODE:9064** | **DRUG NAME:BUPRENORPHINE** | | | |
|---|---|---|---|---|
| **RANK** | **STATE** | **2010 POP** | **TOTAL GRAMS** | **GRAMS/100K POP** |
| 1 | VERMONT | 625,741 | 25,219.07 | 4,030.27 |
| 2 | WEST VIRGINIA | 1,852,994 | 62,190.56 | 3,356.22 |
| 3 | KENTUCKY | 4,339,367 | 138,082.60 | 3,182.09 |
| 4 | MAINE | 1,328,361 | 38,720.40 | 2,914.90 |
| 5 | RHODE ISLAND | 1,052,567 | 30,097.29 | 2,859.42 |
| 6 | MASSACHUSETTS | 6,547,629 | 165,313.37 | 2,524.78 |
| 7 | NEW HAMPSHIRE | 1,316,470 | 32,093.27 | 2,437.83 |
| 8 | TENNESSEE | 6,346,105 | 139,285.76 | 2,194.82 |
| 9 | ALABAMA | 4,779,736 | 100,755.08 | 2,107.96 |
| 10 | PENNSYLVANIA | 12,702,379 | 252,087.71 | 1,984.57 |
| 11 | OHIO | 11,536,504 | 210,098.89 | 1,821.17 |
| 12 | MARYLAND | 5,773,552 | 102,062.77 | 1,767.76 |
| 13 | LOUISIANA | 4,533,372 | 76,313.99 | 1,683.38 |
| 14 | CONNECTICUT | 3,574,097 | 59,292.76 | 1,658.96 |
| 15 | DELAWARE | 897,934 | 14,515.91 | 1,616.59 |

| | | | | |
|---|---|---|---|---|
| 16 | DISTRICT OF COLUMBIA | 601,723 | 9,454.34 | 1,571.21 |
| 17 | NEW MEXICO | 2,059,179 | 29,495.60 | 1,432.40 |
| 18 | UTAH | 2,763,885 | 38,562.44 | 1,395.23 |
| 19 | NORTH CAROLINA | 9,535,483 | 130,842.10 | 1,372.16 |
| 20 | MISSISSIPPI | 2,967,297 | 39,672.12 | 1,336.98 |
| 21 | ALASKA | 710,231 | 9,458.16 | 1,331.70 |
| 22 | INDIANA | 6,483,802 | 79,644.73 | 1,228.36 |
| 23 | NEW JERSEY | 8,791,894 | 105,552.79 | 1,200.57 |
| 24 | OKLAHOMA | 3,751,351 | 43,720.90 | 1,165.47 |
| 25 | WASHINGTON | 6,724,540 | 76,999.48 | 1,145.05 |
| 26 | NEW YORK | 19,378,102 | 213,357.64 | 1,101.02 |
| 27 | MICHIGAN | 9,883,640 | 108,644.60 | 1,099.24 |
| 28 | OREGON | 3,831,074 | 41,536.48 | 1,084.20 |
| 29 | FLORIDA | 18,801,310 | 187,652.68 | 998.08 |
| 30 | VIRGINIA | 8,001,024 | 79,458.64 | 993.11 |
| 31 | MONTANA | 989,415 | 9,652.16 | 975.54 |
| 32 | SOUTH CAROLINA | 4,625,364 | 42,638.77 | 921.85 |
| 33 | IDAHO | 1,567,582 | 14,216.04 | 906.88 |
| 34 | WYOMING | 563,626 | 5,032.89 | 892.95 |
| 35 | ARKANSAS | 2,915,918 | 25,056.38 | 859.3 |
| 36 | WISCONSIN | 5,686,986 | 41,104.34 | 722.78 |
| 37 | GEORGIA | 9,687,653 | 62,871.40 | 648.98 |
| 38 | MISSOURI | 5,988,927 | 38,226.66 | 638.29 |
| 39 | ARIZONA | 6,392,017 | 37,700.26 | 589.8 |
| 40 | COLORADO | 5,029,196 | 29,613.27 | 588.83 |
| 41 | HAWAII | 1,360,301 | 7,879.33 | 579.23 |
| 42 | MINNESOTA | 5,303,925 | 30,214.73 | 569.67 |
| 43 | NORTH DAKOTA | 672,591 | 3,635.35 | 540.5 |
| 44 | NEVADA | 2,700,551 | 13,645.22 | 505.28 |
| 45 | ILLINOIS | 12,830,632 | 57,527.89 | 448.36 |
| 46 | TEXAS | 25,145,561 | 105,336.32 | 418.91 |
| 47 | CALIFORNIA | 37,253,956 | 152,567.81 | 409.53 |
| 48 | KANSAS | 2,853,118 | 10,223.55 | 358.33 |
| 49 | NEBRASKA | 1,826,341 | 5,308.86 | 290.68 |
| 50 | PUERTO RICO | 3,725,789 | 9,956.33 | 267.23 |
| 51 | IOWA | 3,046,355 | 6,730.35 | 220.93 |
| 52 | SOUTH DAKOTA | 814,180 | 1,797.80 | 220.81 |
| 53 | VIRGIN ISLANDS | 109,825 | 62.72 | 57.11 |
| 54 | GUAM | 178,430 | 4.49 | 2.52 |
| 55 | AMERICAN SAMOA | 65,628 | 0.01 | 0.01 |
| **U.S. GRAMS / PER 100K:** | | 312,825,210 | 3,351,185.07 | 1,071.26 |

**DRUG CODE:9120** **DRUG NAME:DIHYDROCODEINE**

| **RANK** | **STATE** | **2010 POP** | **TOTAL GRAMS** | **GRAMS/100K POP** |
|---|---|---|---|---|
| 1 | ALABAMA | 4,779,736 | 3,149.36 | 65.89 |
| 2 | LOUISIANA | 4,533,372 | 2,552.46 | 56.3 |
| 3 | MISSISSIPPI | 2,967,297 | 1,605.98 | 54.12 |
| 4 | TEXAS | 25,145,561 | 11,029.18 | 43.86 |
| 5 | TENNESSEE | 6,346,105 | 1,937.03 | 30.52 |
| 6 | OKLAHOMA | 3,751,351 | 1,129.38 | 30.11 |
| 7 | GEORGIA | 9,687,653 | 2,198.02 | 22.69 |
| 8 | ARKANSAS | 2,915,918 | 568.56 | 19.5 |
| 9 | NEW MEXICO | 2,059,179 | 343.95 | 16.7 |
| 10 | NEW JERSEY | 8,791,894 | 1,248.75 | 14.2 |
| 11 | IDAHO | 1,567,582 | 108.95 | 6.95 |
| 12 | DELAWARE | 897,934 | 54.52 | 6.07 |
| 13 | ILLINOIS | 12,830,632 | 671.1 | 5.23 |
| 14 | ARIZONA | 6,392,017 | 319.49 | 5 |
| 15 | FLORIDA | 18,801,310 | 763.93 | 4.06 |
| 16 | MISSOURI | 5,988,927 | 184.79 | 3.09 |
| 17 | PENNSYLVANIA | 12,702,379 | 362.55 | 2.85 |

| | | | | |
|---|---|---|---|---|
| 18 | NEVADA | 2,700,551 | 75.41 | 2.79 |
| 19 | NEW YORK | 19,378,102 | 529.27 | 2.73 |
| 20 | CALIFORNIA | 37,253,956 | 843.12 | 2.26 |
| 21 | KENTUCKY | 4,339,367 | 85.51 | 1.97 |
| 22 | ALASKA | 710,231 | 13.89 | 1.96 |
| 23 | MASSACHUSETTS | 6,547,629 | 126.04 | 1.93 |
| 24 | OHIO | 11,536,504 | 140.3 | 1.22 |
| 25 | SOUTH CAROLINA | 4,625,364 | 52.34 | 1.13 |
| 26 | MARYLAND | 5,773,552 | 38.08 | 0.66 |
| 27 | NEBRASKA | 1,826,341 | 11.96 | 0.66 |
| 28 | KANSAS | 2,853,118 | 18.16 | 0.64 |
| 29 | WEST VIRGINIA | 1,852,994 | 7.69 | 0.42 |
| 30 | MICHIGAN | 9,883,640 | 40.86 | 0.41 |
| 31 | NORTH CAROLINA | 9,535,483 | 38.45 | 0.4 |
| 32 | WISCONSIN | 5,686,986 | 21.36 | 0.38 |
| 33 | VIRGINIA | 8,001,024 | 26.7 | 0.33 |
| 34 | RHODE ISLAND | 1,052,567 | 3.2 | 0.3 |
| 35 | COLORADO | 5,029,196 | 9.61 | 0.19 |
| 36 | MINNESOTA | 5,303,925 | 9.83 | 0.19 |
| 37 | VERMONT | 625,741 | 1.07 | 0.17 |
| 38 | SOUTH DAKOTA | 814,180 | 1.07 | 0.13 |
| 39 | INDIANA | 6,483,802 | 7.48 | 0.12 |
| 40 | MONTANA | 989,415 | 1.07 | 0.11 |
| 41 | CONNECTICUT | 3,574,097 | 3.75 | 0.1 |
| 42 | HAWAII | 1,360,301 | 1.07 | 0.08 |
| 43 | MAINE | 1,328,361 | 1.07 | 0.08 |
| 44 | IOWA | 3,046,355 | 2.14 | 0.07 |
| 45 | UTAH | 2,763,885 | 1.07 | 0.04 |
| **U.S. GRAMS / PER 100K:** | | 312,825,210 | 30,339.58 | 9.7 |

**DRUG CODE:9143** **DRUG NAME:OXYCODONE**

| **RANK** | **STATE** | **2010 POP** | **TOTAL GRAMS** | **GRAMS/100K POP** |
|---|---|---|---|---|
| 1 | DELAWARE | 897,934 | 273,807.57 | 30,493.06 |
| 2 | TENNESSEE | 6,346,105 | 1,851,142.93 | 29,169.75 |
| 3 | ARIZONA | 6,392,017 | 1,700,401.77 | 26,601.96 |
| 4 | NEVADA | 2,700,551 | 717,291.03 | 26,560.91 |
| 5 | UTAH | 2,763,885 | 670,987.24 | 24,276.96 |
| 6 | OKLAHOMA | 3,751,351 | 896,742.56 | 23,904.52 |
| 7 | PENNSYLVANIA | 12,702,379 | 3,002,067.64 | 23,633.90 |
| 8 | NORTH CAROLINA | 9,535,483 | 2,239,073.49 | 23,481.49 |
| 9 | NEW JERSEY | 8,791,894 | 1,991,312.25 | 22,649.41 |
| 10 | MARYLAND | 5,773,552 | 1,295,377.50 | 22,436.40 |
| 11 | FLORIDA | 18,801,310 | 4,104,069.47 | 21,828.64 |
| 12 | SOUTH CAROLINA | 4,625,364 | 934,837.33 | 20,211.11 |
| 13 | ARKANSAS | 2,915,918 | 577,987.63 | 19,821.81 |
| 14 | MISSOURI | 5,988,927 | 1,184,669.07 | 19,780.99 |
| 15 | ALASKA | 710,231 | 137,795.62 | 19,401.52 |
| 16 | ALABAMA | 4,779,736 | 923,241.15 | 19,315.74 |
| 17 | NEW HAMPSHIRE | 1,316,470 | 249,380.84 | 18,943.15 |
| 18 | KENTUCKY | 4,339,367 | 815,681.77 | 18,797.25 |
| 19 | CONNECTICUT | 3,574,097 | 663,460.65 | 18,563.03 |
| 20 | GEORGIA | 9,687,653 | 1,782,625.27 | 18,401 |
| 21 | WYOMING | 563,626 | 102,168.83 | 18,127.06 |
| 22 | OREGON | 3,831,074 | 693,337.57 | 18,097.73 |
| 23 | WEST VIRGINIA | 1,852,994 | 332,908.26 | 17,965.97 |
| 24 | COLORADO | 5,029,196 | 892,607.05 | 17,748.50 |
| 25 | NEW MEXICO | 2,059,179 | 361,390.05 | 17,550.20 |
| 26 | WASHINGTON | 6,724,540 | 1,162,214.79 | 17,283.19 |
| 27 | KANSAS | 2,853,118 | 488,497.43 | 17,121.53 |
| 28 | LOUISIANA | 4,533,372 | 770,239.02 | 16,990.42 |
| 29 | OHIO | 11,536,504 | 1,926,530.19 | 16,699.43 |

| | | | | |
|---|---|---|---|---|
| 30 | RHODE ISLAND | 1,052,567 | 162,522.08 | 15,440.55 |
| 31 | MONTANA | 989,415 | 152,351.22 | 15,398.11 |
| 32 | IDAHO | 1,567,582 | 241,353.25 | 15,396.53 |
| 33 | MAINE | 1,328,361 | 204,307.68 | 15,380.43 |
| 34 | NEW YORK | 19,378,102 | 2,951,716.13 | 15,232.23 |
| 35 | WISCONSIN | 5,686,986 | 857,719.93 | 15,082.15 |
| 36 | VIRGINIA | 8,001,024 | 1,186,158.02 | 14,825.08 |
| 37 | MASSACHUSETTS | 6,547,629 | 959,337.42 | 14,651.68 |
| 38 | VERMONT | 625,741 | 90,744.06 | 14,501.86 |
| 39 | INDIANA | 6,483,802 | 924,308.19 | 14,255.65 |
| 40 | MISSISSIPPI | 2,967,297 | 419,254.56 | 14,129.17 |
| 41 | HAWAII | 1,360,301 | 173,430.73 | 12,749.44 |
| 42 | MICHIGAN | 9,883,640 | 1,188,829.75 | 12,028.26 |
| 43 | MINNESOTA | 5,303,925 | 570,633.64 | 10,758.70 |
| 44 | DISTRICT OF COLUMBIA | 601,723 | 58,155.97 | 9,664.91 |
| 45 | CALIFORNIA | 37,253,956 | 3,452,832.26 | 9,268.36 |
| 46 | NEBRASKA | 1,826,341 | 166,248.79 | 9,102.83 |
| 47 | NORTH DAKOTA | 672,591 | 58,589.72 | 8,711.05 |
| 48 | SOUTH DAKOTA | 814,180 | 68,945.36 | 8,468.07 |
| 49 | IOWA | 3,046,355 | 213,823.68 | 7,019 |
| 50 | TEXAS | 25,145,561 | 1,324,316.26 | 5,266.60 |
| 51 | ILLINOIS | 12,830,632 | 632,613.48 | 4,930.49 |
| 52 | GUAM | 178,430 | 3,421.37 | 1,917.49 |
| 53 | VIRGIN ISLANDS | 109,825 | 2,102.10 | 1,914.04 |
| 54 | PUERTO RICO | 3,725,789 | 50,726.52 | 1,361.50 |
| 55 | AMERICAN SAMOA | 65,628 | 58.72 | 89.48 |
| **U.S. GRAMS / PER 100K:** | | 312,825,210 | 48,856,346.90 | 15,617.78 |

| **DRUG CODE:9150** | **DRUG NAME:HYDROMORPHONE** | | | |
|---|---|---|---|---|
| **RANK** | **STATE** | **2010 POP** | **TOTAL GRAMS** | **GRAMS/100K POP** |
| 1 | FLORIDA | 18,801,310 | 186,726.94 | 993.16 |
| 2 | DELAWARE | 897,934 | 8,523.20 | 949.2 |
| 3 | MISSISSIPPI | 2,967,297 | 23,403.69 | 788.72 |
| 4 | VERMONT | 625,741 | 4,872.81 | 778.73 |
| 5 | CONNECTICUT | 3,574,097 | 25,575.75 | 715.59 |
| 6 | MONTANA | 989,415 | 6,564.46 | 663.47 |
| 7 | WASHINGTON | 6,724,540 | 42,826.80 | 636.87 |
| 8 | NEW HAMPSHIRE | 1,316,470 | 7,863.71 | 597.33 |
| 9 | MARYLAND | 5,773,552 | 33,414.42 | 578.75 |
| 10 | VIRGINIA | 8,001,024 | 44,944.26 | 561.73 |
| 11 | ALASKA | 710,231 | 3,897.24 | 548.73 |
| 12 | COLORADO | 5,029,196 | 27,198.61 | 540.81 |
| 13 | OREGON | 3,831,074 | 19,345.86 | 504.97 |
| 14 | MAINE | 1,328,361 | 6,686.40 | 503.36 |
| 15 | SOUTH CAROLINA | 4,625,364 | 22,976.43 | 496.75 |
| 16 | KANSAS | 2,853,118 | 13,977.87 | 489.92 |
| 17 | IDAHO | 1,567,582 | 7,525.79 | 480.09 |
| 18 | PENNSYLVANIA | 12,702,379 | 60,501.44 | 476.3 |
| 19 | NORTH CAROLINA | 9,535,483 | 43,037.48 | 451.34 |
| 20 | MASSACHUSETTS | 6,547,629 | 29,474.16 | 450.15 |
| 21 | MINNESOTA | 5,303,925 | 23,791.36 | 448.56 |
| 22 | ARKANSAS | 2,915,918 | 13,054.62 | 447.7 |
| 23 | NEVADA | 2,700,551 | 11,796.48 | 436.82 |
| 24 | WYOMING | 563,626 | 2,439.36 | 432.8 |
| 25 | MISSOURI | 5,988,927 | 25,802.13 | 430.83 |
| 26 | TENNESSEE | 6,346,105 | 26,656.40 | 420.04 |
| 27 | ARIZONA | 6,392,017 | 26,811.58 | 419.45 |
| 28 | CALIFORNIA | 37,253,956 | 155,292.82 | 416.85 |
| 29 | NEW JERSEY | 8,791,894 | 35,497.58 | 403.75 |
| 30 | NEW YORK | 19,378,102 | 78,061.42 | 402.83 |
| 31 | UTAH | 2,763,885 | 10,905.47 | 394.57 |

| | | | | |
|---|---|---|---|---|
| 32 | IOWA | 3,046,355 | 11,892.16 | 390.37 |
| 33 | OKLAHOMA | 3,751,351 | 14,599.92 | 389.19 |
| 34 | NORTH DAKOTA | 672,591 | 2,595.36 | 385.87 |
| 35 | TEXAS | 25,145,561 | 95,409.81 | 379.43 |
| 36 | INDIANA | 6,483,802 | 23,828.90 | 367.51 |
| 37 | MICHIGAN | 9,883,640 | 36,152.05 | 365.78 |
| 38 | DISTRICT OF COLUMBIA | 601,723 | 2,167.32 | 360.19 |
| 39 | GEORGIA | 9,687,653 | 33,298.47 | 343.72 |
| 40 | RHODE ISLAND | 1,052,567 | 3,532.96 | 335.65 |
| 41 | WEST VIRGINIA | 1,852,994 | 6,054.18 | 326.72 |
| 42 | OHIO | 11,536,504 | 37,382.73 | 324.04 |
| 43 | ILLINOIS | 12,830,632 | 38,905.20 | 303.22 |
| 44 | LOUISIANA | 4,533,372 | 13,173.20 | 290.58 |
| 45 | NEBRASKA | 1,826,341 | 5,206.77 | 285.09 |
| 46 | ALABAMA | 4,779,736 | 13,037.33 | 272.76 |
| 47 | KENTUCKY | 4,339,367 | 11,793.33 | 271.78 |
| 48 | SOUTH DAKOTA | 814,180 | 2,199.71 | 270.17 |
| 49 | NEW MEXICO | 2,059,179 | 5,395.61 | 262.03 |
| 50 | WISCONSIN | 5,686,986 | 14,862.16 | 261.34 |
| 51 | HAWAII | 1,360,301 | 2,671.22 | 196.37 |
| 52 | VIRGIN ISLANDS | 109,825 | 74.15 | 67.51 |
| 53 | GUAM | 178,430 | 114.51 | 64.18 |
| 54 | PUERTO RICO | 3,725,789 | 296.63 | 7.96 |
| 55 | AMERICAN SAMOA | 65,628 | 0.98 | 1.49 |
| **U.S. GRAMS / PER 100K:** | | 312,825,210 | 1,404,091.19 | 448.84 |

**DRUG CODE:9193** **DRUG NAME:HYDROCODONE**

| **RANK** | **STATE** | **2010 POP** | **TOTAL GRAMS** | **GRAMS/100K POP** |
|---|---|---|---|---|
| 1 | ALABAMA | 4,779,736 | 971,895.93 | 20,333.67 |
| 2 | OKLAHOMA | 3,751,351 | 725,850.95 | 19,349.05 |
| 3 | KENTUCKY | 4,339,367 | 799,568.80 | 18,425.93 |
| 4 | ARKANSAS | 2,915,918 | 535,677.07 | 18,370.79 |
| 5 | MICHIGAN | 9,883,640 | 1,595,184.20 | 16,139.64 |
| 6 | MISSISSIPPI | 2,967,297 | 466,547.22 | 15,722.97 |
| 7 | TENNESSEE | 6,346,105 | 972,848.06 | 15,329.84 |
| 8 | IDAHO | 1,567,582 | 228,059.05 | 14,548.46 |
| 9 | LOUISIANA | 4,533,372 | 652,080.15 | 14,384 |
| 10 | NEVADA | 2,700,551 | 377,505.15 | 13,978.82 |
| 11 | INDIANA | 6,483,802 | 904,907.84 | 13,956.44 |
| 12 | WEST VIRGINIA | 1,852,994 | 235,601.60 | 12,714.64 |
| 13 | SOUTH CAROLINA | 4,625,364 | 550,520.03 | 11,902.20 |
| 14 | MISSOURI | 5,988,927 | 707,948.92 | 11,820.96 |
| 15 | KANSAS | 2,853,118 | 324,671.16 | 11,379.52 |
| 16 | TEXAS | 25,145,561 | 2,658,309.26 | 10,571.68 |
| 17 | MONTANA | 989,415 | 102,878.49 | 10,397.91 |
| 18 | GEORGIA | 9,687,653 | 984,057.78 | 10,157.86 |
| 19 | OREGON | 3,831,074 | 376,701.12 | 9,832.78 |
| 20 | CALIFORNIA | 37,253,956 | 3,656,359.66 | 9,814.69 |
| 21 | ILLINOIS | 12,830,632 | 1,231,194.30 | 9,595.74 |
| 22 | UTAH | 2,763,885 | 233,877.54 | 8,461.91 |
| 23 | WYOMING | 563,626 | 46,009.42 | 8,163.11 |
| 24 | NORTH CAROLINA | 9,535,483 | 769,321.19 | 8,067.98 |
| 25 | WASHINGTON | 6,724,540 | 523,468.42 | 7,784.45 |
| 26 | FLORIDA | 18,801,310 | 1,440,018.56 | 7,659.14 |
| 27 | IOWA | 3,046,355 | 229,677.31 | 7,539.41 |
| 28 | SOUTH DAKOTA | 814,180 | 59,207.01 | 7,271.98 |
| 29 | NEBRASKA | 1,826,341 | 129,971.43 | 7,116.49 |
| 30 | NEW MEXICO | 2,059,179 | 139,543.68 | 6,776.67 |
| 31 | ARIZONA | 6,392,017 | 412,803.61 | 6,458.11 |
| 32 | OHIO | 11,536,504 | 742,900.80 | 6,439.57 |
| 33 | ALASKA | 710,231 | 44,530.10 | 6,269.80 |

| | | | | |
|---|---|---|---|---|
| 34 | NORTH DAKOTA | 672,591 | 41,493.33 | 6,169.18 |
| 35 | WISCONSIN | 5,686,986 | 311,532.88 | 5,478 |
| 36 | VIRGINIA | 8,001,024 | 416,339.44 | 5,203.58 |
| 37 | PENNSYLVANIA | 12,702,379 | 660,088.57 | 5,196.57 |
| 38 | COLORADO | 5,029,196 | 259,918.58 | 5,168.19 |
| 39 | MAINE | 1,328,361 | 68,459.79 | 5,153.70 |
| 40 | HAWAII | 1,360,301 | 69,885.53 | 5,137.50 |
| 41 | RHODE ISLAND | 1,052,567 | 41,487.65 | 3,941.57 |
| 42 | NEW YORK | 19,378,102 | 681,149.81 | 3,515.05 |
| 43 | MINNESOTA | 5,303,925 | 182,581.75 | 3,442.39 |
| 44 | VERMONT | 625,741 | 19,137.62 | 3,058.39 |
| 45 | NEW HAMPSHIRE | 1,316,470 | 36,123.33 | 2,743.95 |
| 46 | DELAWARE | 897,934 | 23,960.06 | 2,668.35 |
| 47 | MARYLAND | 5,773,552 | 138,467.99 | 2,398.32 |
| 48 | CONNECTICUT | 3,574,097 | 80,359.40 | 2,248.38 |
| 49 | MASSACHUSETTS | 6,547,629 | 120,671.22 | 1,842.98 |
| 50 | NEW JERSEY | 8,791,894 | 138,128.04 | 1,571.08 |
| 51 | VIRGIN ISLANDS | 109,825 | 1,158.60 | 1,054.95 |
| 52 | DISTRICT OF COLUMBIA | 601,723 | 5,766.43 | 958.32 |
| 53 | GUAM | 178,430 | 629.98 | 353.07 |
| 54 | PUERTO RICO | 3,725,789 | 2,546.70 | 68.35 |
| 55 | AMERICAN SAMOA | 65,628 | 7.9 | 12.04 |
| **U.S. GRAMS / PER 100K:** | | 312,825,210 | 27,129,590.40 | 8,672.44 |

**DRUG CODE:9220L** **DRUG NAME:LEVORPHANOL**

| RANK | STATE | 2010 POP | TOTAL GRAMS | GRAMS/100K POP |
|---|---|---|---|---|
| 1 | NEW JERSEY | 8,791,894 | 1,620.24 | 18.43 |
| 2 | NORTH CAROLINA | 9,535,483 | 265.71 | 2.79 |
| 3 | LOUISIANA | 4,533,372 | 76.26 | 1.68 |
| 4 | WASHINGTON | 6,724,540 | 109.23 | 1.62 |
| 5 | ARIZONA | 6,392,017 | 99.94 | 1.56 |
| 6 | INDIANA | 6,483,802 | 97.39 | 1.5 |
| 7 | MISSOURI | 5,988,927 | 86.94 | 1.45 |
| 8 | NEBRASKA | 1,826,341 | 23.8 | 1.3 |
| 9 | KANSAS | 2,853,118 | 35.87 | 1.26 |
| 10 | MASSACHUSETTS | 6,547,629 | 80.68 | 1.23 |
| 11 | CALIFORNIA | 37,253,956 | 402.57 | 1.08 |
| 12 | COLORADO | 5,029,196 | 50.84 | 1.01 |
| 13 | TEXAS | 25,145,561 | 220.32 | 0.88 |
| 14 | NEW YORK | 19,378,102 | 161.12 | 0.83 |
| 15 | FLORIDA | 18,801,310 | 136.05 | 0.72 |
| 16 | CONNECTICUT | 3,574,097 | 20.2 | 0.57 |
| 17 | NEVADA | 2,700,551 | 14.16 | 0.52 |
| 18 | IOWA | 3,046,355 | 15.44 | 0.51 |
| 19 | VIRGINIA | 8,001,024 | 39 | 0.49 |
| 20 | PENNSYLVANIA | 12,702,379 | 53.63 | 0.42 |
| 21 | ALABAMA | 4,779,736 | 20.08 | 0.42 |
| 22 | NEW HAMPSHIRE | 1,316,470 | 5.34 | 0.41 |
| 23 | ILLINOIS | 12,830,632 | 52.7 | 0.41 |
| 24 | OHIO | 11,536,504 | 43.53 | 0.38 |
| 25 | MARYLAND | 5,773,552 | 21.36 | 0.37 |
| 26 | OREGON | 3,831,074 | 13.58 | 0.35 |
| 27 | ARKANSAS | 2,915,918 | 9.05 | 0.31 |
| 28 | DELAWARE | 897,934 | 2.55 | 0.28 |
| 29 | WISCONSIN | 5,686,986 | 15.55 | 0.27 |
| 30 | VERMONT | 625,741 | 1.63 | 0.26 |
| 31 | SOUTH CAROLINA | 4,625,364 | 11.03 | 0.24 |
| 32 | GEORGIA | 9,687,653 | 9.63 | 0.1 |
| 33 | MINNESOTA | 5,303,925 | 2.79 | 0.05 |
| 34 | KENTUCKY | 4,339,367 | 0.35 | 0.01 |
| 35 | IDAHO | 1,567,582 | 0.23 | 0.01 |

| | | | | |
|---|---|---|---|---|
| 36 | MISSISSIPPI | 2,967,297 | 0.23 | 0.01 |
| 37 | MICHIGAN | 9,883,640 | 1.04 | 0.01 |
| 38 | TENNESSEE | 6,346,105 | 0.7 | 0.01 |
| **U.S. GRAMS / PER 100K:** | | 312,825,210 | 3,820.77 | 1.22 |

**DRUG CODE:9230** **DRUG NAME:MEPERIDINE (PETHIDINE)**

| RANK | STATE | 2010 POP | TOTAL GRAMS | GRAMS/100K POP |
|---|---|---|---|---|
| 1 | ARKANSAS | 2,915,918 | 29,599.41 | 1,015.10 |
| 2 | OKLAHOMA | 3,751,351 | 28,655.36 | 763.87 |
| 3 | LOUISIANA | 4,533,372 | 29,019.25 | 640.13 |
| 4 | ALABAMA | 4,779,736 | 29,415.19 | 615.41 |
| 5 | MISSISSIPPI | 2,967,297 | 17,314.55 | 583.51 |
| 6 | SOUTH CAROLINA | 4,625,364 | 23,517.21 | 508.44 |
| 7 | TENNESSEE | 6,346,105 | 29,692.14 | 467.88 |
| 8 | PUERTO RICO | 3,725,789 | 17,371.83 | 466.26 |
| 9 | GEORGIA | 9,687,653 | 41,591.25 | 429.32 |
| 10 | ALASKA | 710,231 | 2,851 | 401.42 |
| 11 | ARIZONA | 6,392,017 | 19,505.61 | 305.16 |
| 12 | VIRGINIA | 8,001,024 | 23,279.05 | 290.95 |
| 13 | NEW MEXICO | 2,059,179 | 5,650.52 | 274.41 |
| 14 | MONTANA | 989,415 | 2,686.31 | 271.51 |
| 15 | UTAH | 2,763,885 | 7,195.56 | 260.34 |
| 16 | NORTH CAROLINA | 9,535,483 | 24,541.88 | 257.37 |
| 17 | IOWA | 3,046,355 | 7,780.88 | 255.42 |
| 18 | SOUTH DAKOTA | 814,180 | 2,076.24 | 255.01 |
| 19 | MISSOURI | 5,988,927 | 15,210.49 | 253.98 |
| 20 | KANSAS | 2,853,118 | 7,071.05 | 247.84 |
| 21 | VIRGIN ISLANDS | 109,825 | 253.93 | 231.22 |
| 22 | WYOMING | 563,626 | 1,301.65 | 230.94 |
| 23 | KENTUCKY | 4,339,367 | 9,777.45 | 225.32 |
| 24 | IDAHO | 1,567,582 | 3,330.02 | 212.43 |
| 25 | NEVADA | 2,700,551 | 5,670.87 | 209.99 |
| 26 | INDIANA | 6,483,802 | 13,425.65 | 207.06 |
| 27 | VERMONT | 625,741 | 1,287.86 | 205.81 |
| 28 | WASHINGTON | 6,724,540 | 13,543.42 | 201.4 |
| 29 | FLORIDA | 18,801,310 | 35,814.51 | 190.49 |
| 30 | TEXAS | 25,145,561 | 46,330.33 | 184.25 |
| 31 | NEW HAMPSHIRE | 1,316,470 | 2,349.89 | 178.5 |
| 32 | OHIO | 11,536,504 | 19,980.92 | 173.2 |
| 33 | CALIFORNIA | 37,253,956 | 62,372.41 | 167.42 |
| 34 | MARYLAND | 5,773,552 | 9,370.04 | 162.29 |
| 35 | OREGON | 3,831,074 | 6,115.40 | 159.63 |
| 36 | NEBRASKA | 1,826,341 | 2,784.97 | 152.49 |
| 37 | COLORADO | 5,029,196 | 7,279.03 | 144.74 |
| 38 | NORTH DAKOTA | 672,591 | 951.79 | 141.51 |
| 39 | WEST VIRGINIA | 1,852,994 | 2,490.42 | 134.4 |
| 40 | MASSACHUSETTS | 6,547,629 | 8,455.01 | 129.13 |
| 41 | DISTRICT OF COLUMBIA | 601,723 | 754.61 | 125.41 |
| 42 | CONNECTICUT | 3,574,097 | 4,262.51 | 119.26 |
| 43 | MAINE | 1,328,361 | 1,578.18 | 118.81 |
| 44 | PENNSYLVANIA | 12,702,379 | 14,199.18 | 111.78 |
| 45 | MICHIGAN | 9,883,640 | 10,928.39 | 110.57 |
| 46 | GUAM | 178,430 | 183.67 | 102.94 |
| 47 | WISCONSIN | 5,686,986 | 5,852.99 | 102.92 |
| 48 | HAWAII | 1,360,301 | 1,357.25 | 99.78 |
| 49 | DELAWARE | 897,934 | 870.74 | 96.97 |
| 50 | RHODE ISLAND | 1,052,567 | 1,006.80 | 95.65 |
| 51 | NEW YORK | 19,378,102 | 18,289.21 | 94.38 |
| 52 | ILLINOIS | 12,830,632 | 12,010.75 | 93.61 |
| 53 | NEW JERSEY | 8,791,894 | 7,702.75 | 87.61 |
| 54 | MINNESOTA | 5,303,925 | 2,733.90 | 51.54 |

| | | | | |
|---|---|---|---|---|
| **U.S. GRAMS / PER 100K:** | | 312,825,210 | 698,641.25 | 223.33 |

| **DRUG CODE:9250B** | **DRUG NAME:METHADONE** | | | |
|---|---|---|---|---|
| **RANK** | **STATE** | **2010 POP** | **TOTAL GRAMS** | **GRAMS/100K POP** |
| 1 | RHODE ISLAND | 1,052,567 | 177,108.96 | 16,826.38 |
| 2 | CONNECTICUT | 3,574,097 | 476,404.75 | 13,329.37 |
| 3 | MARYLAND | 5,773,552 | 764,273.85 | 13,237.50 |
| 4 | DELAWARE | 897,934 | 109,993.34 | 12,249.60 |
| 5 | VERMONT | 625,741 | 73,999.26 | 11,825.86 |
| 6 | MAINE | 1,328,361 | 126,225.33 | 9,502.34 |
| 7 | NEW HAMPSHIRE | 1,316,470 | 123,333.65 | 9,368.51 |
| 8 | MASSACHUSETTS | 6,547,629 | 579,374.66 | 8,848.62 |
| 9 | NEW MEXICO | 2,059,179 | 176,396.43 | 8,566.35 |
| 10 | DISTRICT OF COLUMBIA | 601,723 | 39,909.39 | 6,632.52 |
| 11 | OREGON | 3,831,074 | 253,755.79 | 6,623.62 |
| 12 | ALABAMA | 4,779,736 | 312,422.11 | 6,536.39 |
| 13 | NEW YORK | 19,378,102 | 1,219,390.72 | 6,292.62 |
| 14 | PENNSYLVANIA | 12,702,379 | 789,307.56 | 6,213.86 |
| 15 | WASHINGTON | 6,724,540 | 390,840.64 | 5,812.15 |
| 16 | NORTH CAROLINA | 9,535,483 | 526,496.35 | 5,521.44 |
| 17 | MICHIGAN | 9,883,640 | 534,012.15 | 5,402.99 |
| 18 | NEVADA | 2,700,551 | 144,696.01 | 5,358.02 |
| 19 | ARIZONA | 6,392,017 | 335,733.41 | 5,252.39 |
| 20 | NEW JERSEY | 8,791,894 | 459,561.04 | 5,227.10 |
| 21 | INDIANA | 6,483,802 | 326,935.08 | 5,042.34 |
| 22 | WEST VIRGINIA | 1,852,994 | 92,570.31 | 4,995.72 |
| 23 | SOUTH CAROLINA | 4,625,364 | 225,378.07 | 4,872.66 |
| 24 | GEORGIA | 9,687,653 | 465,598.88 | 4,806.11 |
| 25 | KENTUCKY | 4,339,367 | 206,121.05 | 4,750.03 |
| 26 | WISCONSIN | 5,686,986 | 262,147.43 | 4,609.60 |
| 27 | ALASKA | 710,231 | 32,086.96 | 4,517.82 |
| 28 | OKLAHOMA | 3,751,351 | 157,005.48 | 4,185.30 |
| 29 | FLORIDA | 18,801,310 | 782,712.94 | 4,163.08 |
| 30 | VIRGINIA | 8,001,024 | 317,072.93 | 3,962.90 |
| 31 | MINNESOTA | 5,303,925 | 207,811.17 | 3,918.06 |
| 32 | UTAH | 2,763,885 | 105,686.42 | 3,823.84 |
| 33 | ILLINOIS | 12,830,632 | 489,424.13 | 3,814.50 |
| 34 | CALIFORNIA | 37,253,956 | 1,388,784.67 | 3,727.89 |
| 35 | COLORADO | 5,029,196 | 173,018.55 | 3,440.28 |
| 36 | KANSAS | 2,853,118 | 92,445.63 | 3,240.16 |
| 37 | LOUISIANA | 4,533,372 | 142,636.71 | 3,146.37 |
| 38 | TENNESSEE | 6,346,105 | 194,303.46 | 3,061.78 |
| 39 | IDAHO | 1,567,582 | 47,232.82 | 3,013.10 |
| 40 | MONTANA | 989,415 | 29,220.94 | 2,953.36 |
| 41 | MISSOURI | 5,988,927 | 165,981.13 | 2,771.47 |
| 42 | OHIO | 11,536,504 | 297,313.24 | 2,577.15 |
| 43 | HAWAII | 1,360,301 | 32,295.71 | 2,374.16 |
| 44 | PUERTO RICO | 3,725,789 | 84,028.61 | 2,255.32 |
| 45 | TEXAS | 25,145,561 | 562,018.41 | 2,235.06 |
| 46 | ARKANSAS | 2,915,918 | 64,241.67 | 2,203.14 |
| 47 | NEBRASKA | 1,826,341 | 32,724.69 | 1,791.82 |
| 48 | IOWA | 3,046,355 | 52,271.15 | 1,715.86 |
| 49 | NORTH DAKOTA | 672,591 | 8,372.39 | 1,244.80 |
| 50 | SOUTH DAKOTA | 814,180 | 8,757.88 | 1,075.67 |
| 51 | MISSISSIPPI | 2,967,297 | 26,643.64 | 897.91 |
| 52 | WYOMING | 563,626 | 4,756.51 | 843.91 |
| 53 | VIRGIN ISLANDS | 109,825 | 457.49 | 416.56 |
| 54 | GUAM | 178,430 | 171.28 | 95.99 |
| **U.S. GRAMS / PER 100K:** | | 312,825,210 | 14,691,462.79 | 4,696.38 |

**DRUG CODE:9300** **DRUG NAME:MORPHINE**

| RANK | STATE | 2010 POP | TOTAL GRAMS | GRAMS/100K POP |
|---|---|---|---|---|
| 1 | TENNESSEE | 6,346,105 | 650,022.95 | 10,242.86 |
| 2 | ARIZONA | 6,392,017 | 648,079.96 | 10,138.90 |
| 3 | OKLAHOMA | 3,751,351 | 363,748.88 | 9,696.48 |
| 4 | DELAWARE | 897,934 | 86,687.52 | 9,654.11 |
| 5 | NEVADA | 2,700,551 | 237,514.37 | 8,795.03 |
| 6 | FLORIDA | 18,801,310 | 1,573,659.42 | 8,369.95 |
| 7 | MICHIGAN | 9,883,640 | 753,570.18 | 7,624.42 |
| 8 | OREGON | 3,831,074 | 284,319.06 | 7,421.39 |
| 9 | PENNSYLVANIA | 12,702,379 | 932,230.09 | 7,339.02 |
| 10 | IDAHO | 1,567,582 | 112,526.83 | 7,178.37 |
| 11 | UTAH | 2,763,885 | 196,552.81 | 7,111.47 |
| 12 | MONTANA | 989,415 | 69,641.95 | 7,038.70 |
| 13 | VERMONT | 625,741 | 40,528.56 | 6,476.89 |
| 14 | WASHINGTON | 6,724,540 | 433,678.66 | 6,449.19 |
| 15 | MISSOURI | 5,988,927 | 385,933.56 | 6,444.12 |
| 16 | ALASKA | 710,231 | 45,732.63 | 6,439.12 |
| 17 | HAWAII | 1,360,301 | 87,159 | 6,407.33 |
| 18 | WYOMING | 563,626 | 35,867.45 | 6,363.70 |
| 19 | KANSAS | 2,853,118 | 177,674.01 | 6,227.36 |
| 20 | ARKANSAS | 2,915,918 | 180,890.44 | 6,203.55 |
| 21 | COLORADO | 5,029,196 | 305,459.85 | 6,073.73 |
| 22 | ALABAMA | 4,779,736 | 282,066.83 | 5,901.31 |
| 23 | SOUTH CAROLINA | 4,625,364 | 261,446.04 | 5,652.44 |
| 24 | NEW HAMPSHIRE | 1,316,470 | 72,477.48 | 5,505.44 |
| 25 | WISCONSIN | 5,686,986 | 308,422.58 | 5,423.30 |
| 26 | MISSISSIPPI | 2,967,297 | 159,811.32 | 5,385.75 |
| 27 | INDIANA | 6,483,802 | 343,789.89 | 5,302.29 |
| 28 | MARYLAND | 5,773,552 | 303,143.35 | 5,250.55 |
| 29 | NEW MEXICO | 2,059,179 | 103,153.03 | 5,009.42 |
| 30 | NORTH CAROLINA | 9,535,483 | 476,140 | 4,993.35 |
| 31 | MAINE | 1,328,361 | 65,933.90 | 4,963.55 |
| 32 | CALIFORNIA | 37,253,956 | 1,775,497 | 4,765.93 |
| 33 | WEST VIRGINIA | 1,852,994 | 87,446.52 | 4,719.20 |
| 34 | LOUISIANA | 4,533,372 | 213,761.16 | 4,715.28 |
| 35 | GEORGIA | 9,687,653 | 448,555.06 | 4,630.17 |
| 36 | RHODE ISLAND | 1,052,567 | 46,628.51 | 4,429.98 |
| 37 | IOWA | 3,046,355 | 133,974.93 | 4,397.88 |
| 38 | SOUTH DAKOTA | 814,180 | 34,801.07 | 4,274.37 |
| 39 | KENTUCKY | 4,339,367 | 185,442.79 | 4,273.50 |
| 40 | NEBRASKA | 1,826,341 | 75,693.09 | 4,144.52 |
| 41 | OHIO | 11,536,504 | 477,722.06 | 4,140.96 |
| 42 | MASSACHUSETTS | 6,547,629 | 262,198.26 | 4,004.48 |
| 43 | CONNECTICUT | 3,574,097 | 141,454.92 | 3,957.78 |
| 44 | VIRGINIA | 8,001,024 | 313,868.81 | 3,922.86 |
| 45 | NEW JERSEY | 8,791,894 | 340,241.92 | 3,869.95 |
| 46 | NORTH DAKOTA | 672,591 | 25,131.85 | 3,736.57 |
| 47 | MINNESOTA | 5,303,925 | 185,472.24 | 3,496.89 |
| 48 | TEXAS | 25,145,561 | 821,137.44 | 3,265.54 |
| 49 | NEW YORK | 19,378,102 | 620,757.85 | 3,203.40 |
| 50 | ILLINOIS | 12,830,632 | 409,354.42 | 3,190.45 |
| 51 | DISTRICT OF COLUMBIA | 601,723 | 14,575.26 | 2,422.25 |
| 52 | GUAM | 178,430 | 3,998.25 | 2,240.80 |
| 53 | VIRGIN ISLANDS | 109,825 | 524.89 | 477.93 |
| 54 | PUERTO RICO | 3,725,789 | 8,399.74 | 225.45 |
| 55 | AMERICAN SAMOA | 65,628 | 87.61 | 133.49 |
| **U.S. GRAMS / PER 100K:** | | 312,825,210 | 16,604,588.25 | 5,307.94 |

**DRUG CODE:9333** **DRUG NAME:THEBAINE**

| RANK | STATE | 2010 POP | TOTAL GRAMS | GRAMS/100K POP |
|---|---|---|---|---|

| | | | | |
|---|---|---|---|---|
| 1 | TEXAS | 25,145,561 | 25 | 0.1 |
| **U.S. GRAMS / PER 100K:** | | 312,825,210 | 25 | 0.01 |

**DRUG CODE:9630** **DRUG NAME:OPIUM TINCTURE (LAUDANUM)**

| RANK | STATE | 2010 POP | TOTAL GRAMS | GRAMS/100K POP |
|---|---|---|---|---|
| 1 | OHIO | 11,536,504 | 82.68 | 0.72 |
| 2 | INDIANA | 6,483,802 | 35.47 | 0.55 |
| 3 | NEW HAMPSHIRE | 1,316,470 | 4.73 | 0.36 |
| 4 | WEST VIRGINIA | 1,852,994 | 5.9 | 0.32 |
| 5 | MICHIGAN | 9,883,640 | 30.72 | 0.31 |
| 6 | WISCONSIN | 5,686,986 | 14.18 | 0.25 |
| 7 | MARYLAND | 5,773,552 | 12.99 | 0.22 |
| 8 | KANSAS | 2,853,118 | 5.91 | 0.21 |
| 9 | PENNSYLVANIA | 12,702,379 | 21.24 | 0.17 |
| 10 | DELAWARE | 897,934 | 1.18 | 0.13 |
| 11 | CONNECTICUT | 3,574,097 | 4.73 | 0.13 |
| 12 | VIRGINIA | 8,001,024 | 10.62 | 0.13 |
| 13 | ILLINOIS | 12,830,632 | 16.55 | 0.13 |
| 14 | FLORIDA | 18,801,310 | 14.19 | 0.08 |
| 15 | TEXAS | 25,145,561 | 14.16 | 0.06 |
| 16 | OREGON | 3,831,074 | 2.36 | 0.06 |
| 17 | KENTUCKY | 4,339,367 | 2.36 | 0.05 |
| 18 | WASHINGTON | 6,724,540 | 3.54 | 0.05 |
| 19 | CALIFORNIA | 37,253,956 | 5.91 | 0.02 |
| 20 | NEW YORK | 19,378,102 | 4.73 | 0.02 |
| 21 | NEW JERSEY | 8,791,894 | 1.18 | 0.01 |
| **U.S. GRAMS / PER 100K:** | | 312,825,210 | 295.33 | 0.09 |

**DRUG CODE:9639** **DRUG NAME:OPIUM POWDERED**

| RANK | STATE | 2010 POP | TOTAL GRAMS | GRAMS/100K POP |
|---|---|---|---|---|
| 1 | MINNESOTA | 5,303,925 | 1,535.76 | 28.96 |
| 2 | VERMONT | 625,741 | 162.72 | 26 |
| 3 | KANSAS | 2,853,118 | 720 | 25.24 |
| 4 | NORTH DAKOTA | 672,591 | 164.88 | 24.51 |
| 5 | SOUTH DAKOTA | 814,180 | 180.72 | 22.2 |
| 6 | NEBRASKA | 1,826,341 | 393.84 | 21.56 |
| 7 | COLORADO | 5,029,196 | 1,060.20 | 21.08 |
| 8 | OREGON | 3,831,074 | 798.12 | 20.83 |
| 9 | MONTANA | 989,415 | 204.12 | 20.63 |
| 10 | WEST VIRGINIA | 1,852,994 | 380.88 | 20.55 |
| 11 | WASHINGTON | 6,724,540 | 1,257.48 | 18.7 |
| 12 | ARIZONA | 6,392,017 | 1,181.16 | 18.48 |
| 13 | WYOMING | 563,626 | 98.28 | 17.44 |
| 14 | KENTUCKY | 4,339,367 | 696.6 | 16.05 |
| 15 | MICHIGAN | 9,883,640 | 1,542.24 | 15.6 |
| 16 | INDIANA | 6,483,802 | 981.72 | 15.14 |
| 17 | WISCONSIN | 5,686,986 | 854.64 | 15.03 |
| 18 | MAINE | 1,328,361 | 199.08 | 14.99 |
| 19 | ALASKA | 710,231 | 102.6 | 14.45 |
| 20 | IOWA | 3,046,355 | 402.48 | 13.21 |
| 21 | OHIO | 11,536,504 | 1,514.52 | 13.13 |
| 22 | RHODE ISLAND | 1,052,567 | 137.52 | 13.07 |
| 23 | IDAHO | 1,567,582 | 195.48 | 12.47 |
| 24 | ARKANSAS | 2,915,918 | 361.08 | 12.38 |
| 25 | MASSACHUSETTS | 6,547,629 | 803.16 | 12.27 |
| 26 | SOUTH CAROLINA | 4,625,364 | 555.12 | 12 |
| 27 | TENNESSEE | 6,346,105 | 759.96 | 11.98 |
| 28 | MISSOURI | 5,988,927 | 674.28 | 11.26 |
| 29 | NEW HAMPSHIRE | 1,316,470 | 146.52 | 11.13 |
| 30 | FLORIDA | 18,801,310 | 2,031.48 | 10.8 |

| | | | | |
|---|---|---|---|---|
| 31 | CONNECTICUT | 3,574,097 | 385.2 | 10.78 |
| 32 | PENNSYLVANIA | 12,702,379 | 1,300.32 | 10.24 |
| 33 | DISTRICT OF COLUMBIA | 601,723 | 59.04 | 9.81 |
| 34 | NORTH CAROLINA | 9,535,483 | 917.64 | 9.62 |
| 35 | UTAH | 2,763,885 | 260.64 | 9.43 |
| 36 | LOUISIANA | 4,533,372 | 421.2 | 9.29 |
| 37 | VIRGINIA | 8,001,024 | 722.88 | 9.03 |
| 38 | HAWAII | 1,360,301 | 115.2 | 8.47 |
| 39 | TEXAS | 25,145,561 | 2,088.72 | 8.31 |
| 40 | OKLAHOMA | 3,751,351 | 310.68 | 8.28 |
| 41 | GEORGIA | 9,687,653 | 755.64 | 7.8 |
| 42 | ALABAMA | 4,779,736 | 341.64 | 7.15 |
| 43 | NEW MEXICO | 2,059,179 | 144.72 | 7.03 |
| 44 | DELAWARE | 897,934 | 60.12 | 6.7 |
| 45 | MARYLAND | 5,773,552 | 335.16 | 5.81 |
| 46 | ILLINOIS | 12,830,632 | 717.48 | 5.59 |
| 47 | NEW JERSEY | 8,791,894 | 443.16 | 5.04 |
| 48 | NEVADA | 2,700,551 | 134.28 | 4.97 |
| 49 | CALIFORNIA | 37,253,956 | 1,821.60 | 4.89 |
| 50 | MISSISSIPPI | 2,967,297 | 134.64 | 4.54 |
| 51 | NEW YORK | 19,378,102 | 869.4 | 4.49 |
| 52 | VIRGIN ISLANDS | 109,825 | 1.8 | 1.64 |
| 53 | GUAM | 178,430 | 0.36 | 0.2 |
| **U.S. GRAMS / PER 100K:** | | 312,825,210 | 32,438.16 | 10.37 |

**DRUG CODE:9652** **DRUG NAME:OXYMORPHONE**

| RANK | STATE | 2010 POP | TOTAL GRAMS | GRAMS/100K POP |
|---|---|---|---|---|
| 1 | TENNESSEE | 6,346,105 | 82,234.18 | 1,295.82 |
| 2 | NORTH CAROLINA | 9,535,483 | 82,956.34 | 869.98 |
| 3 | DELAWARE | 897,934 | 7,725.64 | 860.38 |
| 4 | MICHIGAN | 9,883,640 | 75,695.99 | 765.87 |
| 5 | PENNSYLVANIA | 12,702,379 | 91,066.81 | 716.93 |
| 6 | MARYLAND | 5,773,552 | 37,050.14 | 641.72 |
| 7 | UTAH | 2,763,885 | 16,399.44 | 593.35 |
| 8 | NEVADA | 2,700,551 | 15,930.36 | 589.89 |
| 9 | LOUISIANA | 4,533,372 | 26,335.19 | 580.92 |
| 10 | OKLAHOMA | 3,751,351 | 21,760.84 | 580.08 |
| 11 | WEST VIRGINIA | 1,852,994 | 9,923.63 | 535.55 |
| 12 | SOUTH CAROLINA | 4,625,364 | 24,002.65 | 518.94 |
| 13 | FLORIDA | 18,801,310 | 86,877.35 | 462.08 |
| 14 | VIRGINIA | 8,001,024 | 36,312.38 | 453.85 |
| 15 | ALABAMA | 4,779,736 | 21,503.81 | 449.9 |
| 16 | MISSISSIPPI | 2,967,297 | 12,248.28 | 412.78 |
| 17 | ARIZONA | 6,392,017 | 23,750.89 | 371.57 |
| 18 | NEW YORK | 19,378,102 | 71,813.88 | 370.59 |
| 19 | OHIO | 11,536,504 | 38,549.36 | 334.15 |
| 20 | KENTUCKY | 4,339,367 | 14,463.13 | 333.3 |
| 21 | IDAHO | 1,567,582 | 5,145.08 | 328.22 |
| 22 | INDIANA | 6,483,802 | 19,468.06 | 300.26 |
| 23 | CONNECTICUT | 3,574,097 | 10,494.91 | 293.64 |
| 24 | NEW JERSEY | 8,791,894 | 25,694.05 | 292.25 |
| 25 | MISSOURI | 5,988,927 | 17,228.11 | 287.67 |
| 26 | MAINE | 1,328,361 | 3,698.50 | 278.43 |
| 27 | GEORGIA | 9,687,653 | 25,820.74 | 266.53 |
| 28 | NEW HAMPSHIRE | 1,316,470 | 3,491.59 | 265.22 |
| 29 | KANSAS | 2,853,118 | 7,511.04 | 263.26 |
| 30 | NEBRASKA | 1,826,341 | 4,767.34 | 261.03 |
| 31 | COLORADO | 5,029,196 | 12,822.14 | 254.95 |
| 32 | ARKANSAS | 2,915,918 | 6,873.01 | 235.71 |
| 33 | SOUTH DAKOTA | 814,180 | 1,912.19 | 234.86 |
| 34 | WYOMING | 563,626 | 1,120.03 | 198.72 |

| | | | | |
|---|---|---|---|---|
| 35 | NEW MEXICO | 2,059,179 | 3,798.04 | 184.44 |
| 36 | TEXAS | 25,145,561 | 46,168.26 | 183.6 |
| 37 | VERMONT | 625,741 | 1,144.86 | 182.96 |
| 38 | OREGON | 3,831,074 | 6,580.72 | 171.77 |
| 39 | WASHINGTON | 6,724,540 | 11,405.38 | 169.61 |
| 40 | HAWAII | 1,360,301 | 2,146.26 | 157.78 |
| 41 | RHODE ISLAND | 1,052,567 | 1,602.01 | 152.2 |
| 42 | WISCONSIN | 5,686,986 | 8,511.37 | 149.66 |
| 43 | ALASKA | 710,231 | 1,004.73 | 141.46 |
| 44 | CALIFORNIA | 37,253,956 | 52,214.98 | 140.16 |
| 45 | IOWA | 3,046,355 | 4,107.22 | 134.82 |
| 46 | MASSACHUSETTS | 6,547,629 | 8,701.18 | 132.89 |
| 47 | ILLINOIS | 12,830,632 | 16,297.56 | 127.02 |
| 48 | MONTANA | 989,415 | 1,132.77 | 114.49 |
| 49 | NORTH DAKOTA | 672,591 | 629.64 | 93.61 |
| 50 | DISTRICT OF COLUMBIA | 601,723 | 479.25 | 79.65 |
| 51 | MINNESOTA | 5,303,925 | 1,698.09 | 32.02 |
| **U.S. GRAMS / PER 100K:** | | 312,825,210 | 1,110,269.42 | 354.92 |

**DRUG CODE:9668** **DRUG NAME:NOROXYMORPHONE**

| **RANK** | **STATE** | **2010 POP** | **TOTAL GRAMS** | **GRAMS/100K POP** |
|---|---|---|---|---|
| 1 | ARKANSAS | 2,915,918 | 78.06 | 2.68 |
| 2 | NEVADA | 2,700,551 | 25.26 | 0.94 |
| 3 | CALIFORNIA | 37,253,956 | 259.61 | 0.7 |
| 4 | PUERTO RICO | 3,725,789 | 25.07 | 0.67 |
| 5 | MISSOURI | 5,988,927 | 37.2 | 0.62 |
| 6 | TEXAS | 25,145,561 | 108.84 | 0.43 |
| 7 | FLORIDA | 18,801,310 | 58.03 | 0.31 |
| 8 | ILLINOIS | 12,830,632 | 38.02 | 0.3 |
| 9 | NEW JERSEY | 8,791,894 | 23.7 | 0.27 |
| 10 | MISSISSIPPI | 2,967,297 | 6.19 | 0.21 |
| 11 | OHIO | 11,536,504 | 24.31 | 0.21 |
| 12 | GEORGIA | 9,687,653 | 20.46 | 0.21 |
| 13 | WASHINGTON | 6,724,540 | 8.47 | 0.13 |
| 14 | NEW YORK | 19,378,102 | 22.13 | 0.11 |
| 15 | KANSAS | 2,853,118 | 2.75 | 0.1 |
| 16 | OREGON | 3,831,074 | 2 | 0.05 |
| 17 | MASSACHUSETTS | 6,547,629 | 3.03 | 0.05 |
| 18 | MICHIGAN | 9,883,640 | 4.29 | 0.04 |
| 19 | NORTH CAROLINA | 9,535,483 | 3.68 | 0.04 |
| 20 | OKLAHOMA | 3,751,351 | 0.64 | 0.02 |
| 21 | VERMONT | 625,741 | 0.1 | 0.02 |
| 22 | RHODE ISLAND | 1,052,567 | 0.1 | 0.01 |
| 23 | PENNSYLVANIA | 12,702,379 | 0.99 | 0.01 |
| 24 | MINNESOTA | 5,303,925 | 0.5 | 0.01 |
| 25 | TENNESSEE | 6,346,105 | 0.13 | 0 |
| 26 | ARIZONA | 6,392,017 | 0.1 | 0 |
| 27 | MARYLAND | 5,773,552 | 0.2 | 0 |
| **U.S. GRAMS / PER 100K:** | | 312,825,210 | 753.86 | 0.24 |

**DRUG CODE:9737** **DRUG NAME:ALFENTANIL**

| **RANK** | **STATE** | **2010 POP** | **TOTAL GRAMS** | **GRAMS/100K POP** |
|---|---|---|---|---|
| 1 | OREGON | 3,831,074 | 18.67 | 0.49 |
| 2 | ALASKA | 710,231 | 2.89 | 0.41 |
| 3 | KENTUCKY | 4,339,367 | 16.43 | 0.38 |
| 4 | IDAHO | 1,567,582 | 5.45 | 0.35 |
| 5 | WISCONSIN | 5,686,986 | 19.27 | 0.34 |
| 6 | NEVADA | 2,700,551 | 6.95 | 0.26 |
| 7 | MINNESOTA | 5,303,925 | 13.91 | 0.26 |
| 8 | IOWA | 3,046,355 | 6.68 | 0.22 |

| | | | | |
|---|---|---|---|---|
| 9 | LOUISIANA | 4,533,372 | 10.05 | 0.22 |
| 10 | KANSAS | 2,853,118 | 5.98 | 0.21 |
| 11 | NEW MEXICO | 2,059,179 | 4.1 | 0.2 |
| 12 | MAINE | 1,328,361 | 2.69 | 0.2 |
| 13 | NEBRASKA | 1,826,341 | 3.56 | 0.19 |
| 14 | WASHINGTON | 6,724,540 | 12.65 | 0.19 |
| 15 | HAWAII | 1,360,301 | 2.4 | 0.18 |
| 16 | CALIFORNIA | 37,253,956 | 65.92 | 0.18 |
| 17 | OHIO | 11,536,504 | 19.09 | 0.17 |
| 18 | INDIANA | 6,483,802 | 10.29 | 0.16 |
| 19 | CONNECTICUT | 3,574,097 | 5.29 | 0.15 |
| 20 | NEW HAMPSHIRE | 1,316,470 | 1.95 | 0.15 |
| 21 | GEORGIA | 9,687,653 | 13.05 | 0.13 |
| 22 | MONTANA | 989,415 | 1.3 | 0.13 |
| 23 | NORTH CAROLINA | 9,535,483 | 12.57 | 0.13 |
| 24 | ILLINOIS | 12,830,632 | 16.5 | 0.13 |
| 25 | UTAH | 2,763,885 | 3.09 | 0.11 |
| 26 | FLORIDA | 18,801,310 | 20.91 | 0.11 |
| 27 | OKLAHOMA | 3,751,351 | 3.52 | 0.09 |
| 28 | NORTH DAKOTA | 672,591 | 0.59 | 0.09 |
| 29 | TEXAS | 25,145,561 | 22.04 | 0.09 |
| 30 | TENNESSEE | 6,346,105 | 4.77 | 0.08 |
| 31 | COLORADO | 5,029,196 | 3.86 | 0.08 |
| 32 | MISSOURI | 5,988,927 | 4.33 | 0.07 |
| 33 | ARIZONA | 6,392,017 | 3.73 | 0.06 |
| 34 | NEW YORK | 19,378,102 | 9.15 | 0.05 |
| 35 | MICHIGAN | 9,883,640 | 4.48 | 0.05 |
| 36 | VIRGINIA | 8,001,024 | 3.62 | 0.05 |
| 37 | MARYLAND | 5,773,552 | 2.89 | 0.05 |
| 38 | MASSACHUSETTS | 6,547,629 | 3.45 | 0.05 |
| 39 | SOUTH DAKOTA | 814,180 | 0.33 | 0.04 |
| 40 | GUAM | 178,430 | 0.08 | 0.04 |
| 41 | PENNSYLVANIA | 12,702,379 | 2.01 | 0.02 |
| 42 | RHODE ISLAND | 1,052,567 | 0.26 | 0.02 |
| 43 | WYOMING | 563,626 | 0.12 | 0.02 |
| 44 | VERMONT | 625,741 | 0.04 | 0.01 |
| 45 | SOUTH CAROLINA | 4,625,364 | 0.35 | 0.01 |
| 46 | ALABAMA | 4,779,736 | 0.25 | 0.01 |
| 47 | MISSISSIPPI | 2,967,297 | 0.43 | 0.01 |
| 48 | NEW JERSEY | 8,791,894 | 1.05 | 0.01 |
| 49 | ARKANSAS | 2,915,918 | 0.08 | 0 |
| **U.S. GRAMS / PER 100K:** | | 312,825,210 | 372.99 | 0.12 |

**DRUG CODE:9739** **DRUG NAME:REMIFENTANIL**

| **RANK** | **STATE** | **2010 POP** | **TOTAL GRAMS** | **GRAMS/100K POP** |
|---|---|---|---|---|
| 1 | UTAH | 2,763,885 | 71.78 | 2.6 |
| 2 | ALASKA | 710,231 | 11.68 | 1.64 |
| 3 | MASSACHUSETTS | 6,547,629 | 89.26 | 1.36 |
| 4 | DELAWARE | 897,934 | 12.04 | 1.34 |
| 5 | PENNSYLVANIA | 12,702,379 | 143.24 | 1.13 |
| 6 | WEST VIRGINIA | 1,852,994 | 20.75 | 1.12 |
| 7 | CONNECTICUT | 3,574,097 | 34.06 | 0.95 |
| 8 | NEW YORK | 19,378,102 | 155.04 | 0.8 |
| 9 | WASHINGTON | 6,724,540 | 50.01 | 0.74 |
| 10 | MONTANA | 989,415 | 6.99 | 0.71 |
| 11 | OHIO | 11,536,504 | 75.63 | 0.66 |
| 12 | MINNESOTA | 5,303,925 | 32.7 | 0.62 |
| 13 | NORTH CAROLINA | 9,535,483 | 57.75 | 0.61 |
| 14 | NEW JERSEY | 8,791,894 | 52.91 | 0.6 |
| 15 | FLORIDA | 18,801,310 | 107.44 | 0.57 |
| 16 | IDAHO | 1,567,582 | 8.69 | 0.55 |

| | | | | |
|---|---|---|---|---|
| 17 | COLORADO | 5,029,196 | 25.83 | 0.51 |
| 18 | NEW HAMPSHIRE | 1,316,470 | 6.52 | 0.5 |
| 19 | INDIANA | 6,483,802 | 31.93 | 0.49 |
| 20 | DISTRICT OF COLUMBIA | 601,723 | 2.84 | 0.47 |
| 21 | MARYLAND | 5,773,552 | 27.4 | 0.47 |
| 22 | ILLINOIS | 12,830,632 | 60.76 | 0.47 |
| 23 | WISCONSIN | 5,686,986 | 26.73 | 0.47 |
| 24 | VERMONT | 625,741 | 2.83 | 0.45 |
| 25 | TEXAS | 25,145,561 | 111.86 | 0.44 |
| 26 | KENTUCKY | 4,339,367 | 19.16 | 0.44 |
| 27 | LOUISIANA | 4,533,372 | 18.57 | 0.41 |
| 28 | IOWA | 3,046,355 | 12.58 | 0.41 |
| 29 | MICHIGAN | 9,883,640 | 39.63 | 0.4 |
| 30 | SOUTH CAROLINA | 4,625,364 | 18.65 | 0.4 |
| 31 | HAWAII | 1,360,301 | 5.14 | 0.38 |
| 32 | TENNESSEE | 6,346,105 | 23.24 | 0.37 |
| 33 | MISSOURI | 5,988,927 | 21.77 | 0.36 |
| 34 | CALIFORNIA | 37,253,956 | 133.36 | 0.36 |
| 35 | MAINE | 1,328,361 | 4.61 | 0.35 |
| 36 | VIRGINIA | 8,001,024 | 27.17 | 0.34 |
| 37 | WYOMING | 563,626 | 1.78 | 0.32 |
| 38 | KANSAS | 2,853,118 | 9.07 | 0.32 |
| 39 | OREGON | 3,831,074 | 11.4 | 0.3 |
| 40 | RHODE ISLAND | 1,052,567 | 3.17 | 0.3 |
| 41 | OKLAHOMA | 3,751,351 | 10.74 | 0.29 |
| 42 | ALABAMA | 4,779,736 | 13.15 | 0.28 |
| 43 | ARIZONA | 6,392,017 | 15.69 | 0.25 |
| 44 | NORTH DAKOTA | 672,591 | 1.67 | 0.25 |
| 45 | SOUTH DAKOTA | 814,180 | 1.89 | 0.23 |
| 46 | NEBRASKA | 1,826,341 | 4.29 | 0.23 |
| 47 | ARKANSAS | 2,915,918 | 6.4 | 0.22 |
| 48 | NEW MEXICO | 2,059,179 | 4.44 | 0.22 |
| 49 | GEORGIA | 9,687,653 | 19.21 | 0.2 |
| 50 | GUAM | 178,430 | 0.31 | 0.17 |
| 51 | NEVADA | 2,700,551 | 3.7 | 0.14 |
| 52 | MISSISSIPPI | 2,967,297 | 1.78 | 0.06 |
| U.S. GRAMS / PER 100K: | | 312,825,210 | 1,659.21 | 0.53 |

**DRUG CODE:9740** **DRUG NAME:SUFENTANIL BASE**

| RANK | STATE | 2010 POP | TOTAL GRAMS | GRAMS/100K POP |
|---|---|---|---|---|
| 1 | MISSISSIPPI | 2,967,297 | 52.13 | 1.76 |
| 2 | ALABAMA | 4,779,736 | 6.56 | 0.14 |
| 3 | CALIFORNIA | 37,253,956 | 43.74 | 0.12 |
| 4 | LOUISIANA | 4,533,372 | 4.63 | 0.1 |
| 5 | CONNECTICUT | 3,574,097 | 2.39 | 0.07 |
| 6 | FLORIDA | 18,801,310 | 12.66 | 0.07 |
| 7 | PENNSYLVANIA | 12,702,379 | 8.12 | 0.06 |
| 8 | SOUTH CAROLINA | 4,625,364 | 2.64 | 0.06 |
| 9 | WISCONSIN | 5,686,986 | 2.53 | 0.04 |
| 10 | NEW JERSEY | 8,791,894 | 3.74 | 0.04 |
| 11 | TENNESSEE | 6,346,105 | 2.57 | 0.04 |
| 12 | KANSAS | 2,853,118 | 1.16 | 0.04 |
| 13 | TEXAS | 25,145,561 | 6.66 | 0.03 |
| 14 | KENTUCKY | 4,339,367 | 1.51 | 0.03 |
| 15 | NEW YORK | 19,378,102 | 6.74 | 0.03 |
| 16 | UTAH | 2,763,885 | 0.92 | 0.03 |
| 17 | OREGON | 3,831,074 | 1.23 | 0.03 |
| 18 | MICHIGAN | 9,883,640 | 2.25 | 0.02 |
| 19 | NEBRASKA | 1,826,341 | 0.4 | 0.02 |
| 20 | NORTH DAKOTA | 672,591 | 0.13 | 0.02 |
| 21 | WASHINGTON | 6,724,540 | 1.04 | 0.02 |

| | | | | |
|---|---|---|---|---|
| 22 | OKLAHOMA | 3,751,351 | 0.69 | 0.02 |
| 23 | MONTANA | 989,415 | 0.17 | 0.02 |
| 24 | PUERTO RICO | 3,725,789 | 0.75 | 0.02 |
| 25 | GEORGIA | 9,687,653 | 1.79 | 0.02 |
| 26 | MISSOURI | 5,988,927 | 1.26 | 0.02 |
| 27 | ILLINOIS | 12,830,632 | 2.29 | 0.02 |
| 28 | OHIO | 11,536,504 | 1.95 | 0.02 |
| 29 | VIRGINIA | 8,001,024 | 1.54 | 0.02 |
| 30 | COLORADO | 5,029,196 | 0.84 | 0.02 |
| 31 | INDIANA | 6,483,802 | 1.14 | 0.02 |
| 32 | NORTH CAROLINA | 9,535,483 | 1.67 | 0.02 |
| 33 | SOUTH DAKOTA | 814,180 | 0.18 | 0.02 |
| 34 | ARKANSAS | 2,915,918 | 0.56 | 0.02 |
| 35 | MASSACHUSETTS | 6,547,629 | 0.52 | 0.01 |
| 36 | IOWA | 3,046,355 | 0.21 | 0.01 |
| 37 | WEST VIRGINIA | 1,852,994 | 0.26 | 0.01 |
| 38 | ARIZONA | 6,392,017 | 0.34 | 0.01 |
| 39 | MINNESOTA | 5,303,925 | 0.77 | 0.01 |
| 40 | IDAHO | 1,567,582 | 0.18 | 0.01 |
| 41 | NEVADA | 2,700,551 | 0.37 | 0.01 |
| 42 | MAINE | 1,328,361 | 0.08 | 0.01 |
| 43 | ALASKA | 710,231 | 0.04 | 0.01 |
| 44 | NEW MEXICO | 2,059,179 | 0.19 | 0.01 |
| 45 | WYOMING | 563,626 | 0.07 | 0.01 |
| 46 | RHODE ISLAND | 1,052,567 | 0.01 | 0 |
| 47 | HAWAII | 1,360,301 | 0.03 | 0 |
| 48 | NEW HAMPSHIRE | 1,316,470 | 0.06 | 0 |
| 49 | DISTRICT OF COLUMBIA | 601,723 | 0.03 | 0 |
| 50 | DELAWARE | 897,934 | 0.03 | 0 |
| 51 | VERMONT | 625,741 | 0.03 | 0 |
| 52 | MARYLAND | 5,773,552 | 0.22 | 0 |
| **U.S. GRAMS / PER 100K:** | | 312,825,210 | 182.02 | 0.06 |

**DRUG CODE:9780** **DRUG NAME:TAPENTADOL**

| **RANK** | **STATE** | **2010 POP** | **TOTAL GRAMS** | **GRAMS/100K POP** |
|---|---|---|---|---|
| 1 | ALABAMA | 4,779,736 | 218,561 | 4,572.66 |
| 2 | NORTH CAROLINA | 9,535,483 | 386,527.50 | 4,053.57 |
| 3 | SOUTH CAROLINA | 4,625,364 | 174,010 | 3,762.08 |
| 4 | DELAWARE | 897,934 | 29,674 | 3,304.70 |
| 5 | ARKANSAS | 2,915,918 | 83,934.50 | 2,878.49 |
| 6 | GEORGIA | 9,687,653 | 269,630.50 | 2,783.24 |
| 7 | OKLAHOMA | 3,751,351 | 94,917.50 | 2,530.22 |
| 8 | SOUTH DAKOTA | 814,180 | 18,860 | 2,316.44 |
| 9 | INDIANA | 6,483,802 | 149,431.50 | 2,304.69 |
| 10 | MARYLAND | 5,773,552 | 130,004.50 | 2,251.72 |
| 11 | NEW HAMPSHIRE | 1,316,470 | 29,415.50 | 2,234.42 |
| 12 | COLORADO | 5,029,196 | 112,252.50 | 2,232.02 |
| 13 | ARIZONA | 6,392,017 | 142,123 | 2,223.45 |
| 14 | NEW JERSEY | 8,791,894 | 189,226 | 2,152.28 |
| 15 | NEBRASKA | 1,826,341 | 38,820 | 2,125.56 |
| 16 | FLORIDA | 18,801,310 | 396,257 | 2,107.60 |
| 17 | OHIO | 11,536,504 | 242,759 | 2,104.27 |
| 18 | PENNSYLVANIA | 12,702,379 | 264,471 | 2,082.06 |
| 19 | VIRGINIA | 8,001,024 | 165,567.50 | 2,069.33 |
| 20 | TENNESSEE | 6,346,105 | 127,215 | 2,004.62 |
| 21 | UTAH | 2,763,885 | 54,890.50 | 1,985.99 |
| 22 | NEW YORK | 19,378,102 | 383,884.50 | 1,981.02 |
| 23 | NEVADA | 2,700,551 | 48,805.50 | 1,807.24 |
| 24 | IOWA | 3,046,355 | 49,701 | 1,631.49 |
| 25 | MISSOURI | 5,988,927 | 97,566 | 1,629.11 |
| 26 | MISSISSIPPI | 2,967,297 | 48,285.50 | 1,627.26 |

| | | | | |
|---|---|---|---|---|
| 27 | TEXAS | 25,145,561 | 390,103 | 1,551.38 |
| 28 | IDAHO | 1,567,582 | 24,126.50 | 1,539.09 |
| 29 | NORTH DAKOTA | 672,591 | 9,775.50 | 1,453.41 |
| 30 | KENTUCKY | 4,339,367 | 60,134 | 1,385.78 |
| 31 | WYOMING | 563,626 | 7,766 | 1,377.86 |
| 32 | LOUISIANA | 4,533,372 | 60,790.50 | 1,340.96 |
| 33 | WEST VIRGINIA | 1,852,994 | 24,544.50 | 1,324.59 |
| 34 | CONNECTICUT | 3,574,097 | 46,049 | 1,288.41 |
| 35 | ALASKA | 710,231 | 8,991 | 1,265.93 |
| 36 | KANSAS | 2,853,118 | 35,042 | 1,228.20 |
| 37 | ILLINOIS | 12,830,632 | 151,698.50 | 1,182.32 |
| 38 | MASSACHUSETTS | 6,547,629 | 77,076 | 1,177.16 |
| 39 | CALIFORNIA | 37,253,956 | 424,876.85 | 1,140.49 |
| 40 | WASHINGTON | 6,724,540 | 70,350.50 | 1,046.18 |
| 41 | WISCONSIN | 5,686,986 | 59,475 | 1,045.81 |
| 42 | HAWAII | 1,360,301 | 12,594 | 925.82 |
| 43 | MICHIGAN | 9,883,640 | 86,639 | 876.59 |
| 44 | RHODE ISLAND | 1,052,567 | 8,390 | 797.1 |
| 45 | OREGON | 3,831,074 | 29,815 | 778.24 |
| 46 | MONTANA | 989,415 | 7,434.50 | 751.4 |
| 47 | VERMONT | 625,741 | 4,620 | 738.32 |
| 48 | DISTRICT OF COLUMBIA | 601,723 | 4,096.50 | 680.79 |
| 49 | NEW MEXICO | 2,059,179 | 12,826.50 | 622.89 |
| 50 | MAINE | 1,328,361 | 5,635.50 | 424.24 |
| 51 | MINNESOTA | 5,303,925 | 17,601.50 | 331.86 |
| 52 | PUERTO RICO | 3,725,789 | 3,374 | 90.56 |
| 53 | VIRGIN ISLANDS | 109,825 | 18 | 16.39 |
| **U.S. GRAMS / PER 100K:** | | 312,825,210 | 5,590,633.85 | 1,787.14 |

**DRUG CODE:9801** **DRUG NAME:FENTANYL BASE**

| **RANK** | **STATE** | **2010 POP** | **TOTAL GRAMS** | **GRAMS/100K POP** |
|---|---|---|---|---|
| 1 | OKLAHOMA | 3,751,351 | 6,755.14 | 180.07 |
| 2 | MISSISSIPPI | 2,967,297 | 4,922.64 | 165.9 |
| 3 | PENNSYLVANIA | 12,702,379 | 20,066.53 | 157.97 |
| 4 | KANSAS | 2,853,118 | 4,461.96 | 156.39 |
| 5 | MISSOURI | 5,988,927 | 9,085.08 | 151.7 |
| 6 | DELAWARE | 897,934 | 1,360.60 | 151.53 |
| 7 | SOUTH DAKOTA | 814,180 | 1,204.24 | 147.91 |
| 8 | IDAHO | 1,567,582 | 2,304.90 | 147.04 |
| 9 | NORTH DAKOTA | 672,591 | 947.88 | 140.93 |
| 10 | MONTANA | 989,415 | 1,318.09 | 133.22 |
| 11 | ALASKA | 710,231 | 940.62 | 132.44 |
| 12 | ARKANSAS | 2,915,918 | 3,715.20 | 127.41 |
| 13 | MICHIGAN | 9,883,640 | 12,375.98 | 125.22 |
| 14 | ALABAMA | 4,779,736 | 5,963.13 | 124.76 |
| 15 | FLORIDA | 18,801,310 | 23,352.70 | 124.21 |
| 16 | TENNESSEE | 6,346,105 | 7,689.41 | 121.17 |
| 17 | VERMONT | 625,741 | 748.07 | 119.55 |
| 18 | UTAH | 2,763,885 | 3,275.55 | 118.51 |
| 19 | NORTH CAROLINA | 9,535,483 | 11,266.22 | 118.15 |
| 20 | WEST VIRGINIA | 1,852,994 | 2,167.50 | 116.97 |
| 21 | NEBRASKA | 1,826,341 | 2,129.02 | 116.57 |
| 22 | INDIANA | 6,483,802 | 7,523.57 | 116.04 |
| 23 | IOWA | 3,046,355 | 3,476.55 | 114.12 |
| 24 | SOUTH CAROLINA | 4,625,364 | 5,261.53 | 113.75 |
| 25 | GEORGIA | 9,687,653 | 10,907.88 | 112.6 |
| 26 | WYOMING | 563,626 | 620.98 | 110.18 |
| 27 | ARIZONA | 6,392,017 | 6,993.38 | 109.41 |
| 28 | CONNECTICUT | 3,574,097 | 3,800.61 | 106.34 |
| 29 | NEW HAMPSHIRE | 1,316,470 | 1,391.09 | 105.67 |
| 30 | KENTUCKY | 4,339,367 | 4,567 | 105.25 |

| | | | | |
|---|---|---|---|---|
| 31 | LOUISIANA | 4,533,372 | 4,755.62 | 104.9 |
| 32 | NEW JERSEY | 8,791,894 | 9,178.74 | 104.4 |
| 33 | WISCONSIN | 5,686,986 | 5,898.14 | 103.71 |
| 34 | COLORADO | 5,029,196 | 5,010.59 | 99.63 |
| 35 | MAINE | 1,328,361 | 1,322.82 | 99.58 |
| 36 | OREGON | 3,831,074 | 3,705.65 | 96.73 |
| 37 | MARYLAND | 5,773,552 | 5,552.39 | 96.17 |
| 38 | ILLINOIS | 12,830,632 | 11,809.15 | 92.04 |
| 39 | TEXAS | 25,145,561 | 23,026.12 | 91.57 |
| 40 | WASHINGTON | 6,724,540 | 6,032.40 | 89.71 |
| 41 | NEW YORK | 19,378,102 | 17,237.37 | 88.95 |
| 42 | NEVADA | 2,700,551 | 2,353.92 | 87.16 |
| 43 | VIRGINIA | 8,001,024 | 6,913.51 | 86.41 |
| 44 | OHIO | 11,536,504 | 9,850.21 | 85.38 |
| 45 | NEW MEXICO | 2,059,179 | 1,735.05 | 84.26 |
| 46 | MASSACHUSETTS | 6,547,629 | 5,229.02 | 79.86 |
| 47 | MINNESOTA | 5,303,925 | 4,115.67 | 77.6 |
| 48 | CALIFORNIA | 37,253,956 | 27,815.29 | 74.66 |
| 49 | RHODE ISLAND | 1,052,567 | 734.98 | 69.83 |
| 50 | DISTRICT OF COLUMBIA | 601,723 | 338.77 | 56.3 |
| 51 | HAWAII | 1,360,301 | 742.32 | 54.57 |
| 52 | GUAM | 178,430 | 41.33 | 23.16 |
| 53 | PUERTO RICO | 3,725,789 | 578.26 | 15.52 |
| 54 | VIRGIN ISLANDS | 109,825 | 13.96 | 12.71 |
| 55 | AMERICAN SAMOA | 65,628 | 0.47 | 0.71 |
| **U.S. GRAMS / PER 100K:** | | 312,825,210 | 324,584.80 | 103.76 |

**DRUG CODE:9809** **DRUG NAME:OPIUM COMBINATION PRODUCT (C-III)**

| RANK | STATE | 2010 POP | TOTAL GRAMS | GRAMS/100K POP |
|---|---|---|---|---|
| 1 | UTAH | 2,763,885 | 49.57 | 1.79 |
| 2 | PENNSYLVANIA | 12,702,379 | 135.65 | 1.07 |
| 3 | ALABAMA | 4,779,736 | 47.11 | 0.99 |
| 4 | IDAHO | 1,567,582 | 15.14 | 0.97 |
| 5 | SOUTH CAROLINA | 4,625,364 | 40.11 | 0.87 |
| 6 | KENTUCKY | 4,339,367 | 37.07 | 0.85 |
| 7 | LOUISIANA | 4,533,372 | 33.3 | 0.73 |
| 8 | FLORIDA | 18,801,310 | 137.74 | 0.73 |
| 9 | NEW HAMPSHIRE | 1,316,470 | 8.89 | 0.68 |
| 10 | MARYLAND | 5,773,552 | 34.06 | 0.59 |
| 11 | NORTH CAROLINA | 9,535,483 | 53.92 | 0.57 |
| 12 | CONNECTICUT | 3,574,097 | 19.49 | 0.55 |
| 13 | WYOMING | 563,626 | 3.03 | 0.54 |
| 14 | ARIZONA | 6,392,017 | 33.3 | 0.52 |
| 15 | WEST VIRGINIA | 1,852,994 | 9.65 | 0.52 |
| 16 | MAINE | 1,328,361 | 6.81 | 0.51 |
| 17 | OHIO | 11,536,504 | 58.25 | 0.5 |
| 18 | MISSOURI | 5,988,927 | 28.57 | 0.48 |
| 19 | INDIANA | 6,483,802 | 29.14 | 0.45 |
| 20 | WISCONSIN | 5,686,986 | 23.08 | 0.41 |
| 21 | MISSISSIPPI | 2,967,297 | 11.73 | 0.4 |
| 22 | NORTH DAKOTA | 672,591 | 2.65 | 0.39 |
| 23 | TENNESSEE | 6,346,105 | 24.97 | 0.39 |
| 24 | HAWAII | 1,360,301 | 4.73 | 0.35 |
| 25 | MONTANA | 989,415 | 3.41 | 0.34 |
| 26 | OKLAHOMA | 3,751,351 | 12.87 | 0.34 |
| 27 | SOUTH DAKOTA | 814,180 | 2.65 | 0.33 |
| 28 | NEW MEXICO | 2,059,179 | 6.62 | 0.32 |
| 29 | GEORGIA | 9,687,653 | 29.52 | 0.3 |
| 30 | VIRGINIA | 8,001,024 | 23.84 | 0.3 |
| 31 | ARKANSAS | 2,915,918 | 8.51 | 0.29 |
| 32 | IOWA | 3,046,355 | 8.89 | 0.29 |

CUMULATIVE DISTRIBUTION BY STATE IN GRAMS PER 100,000 POPULATION

| | | | | |
|---|---|---|---|---|
| 33 | DISTRICT OF COLUMBIA | 601,723 | 1.51 | 0.25 |
| 34 | NEW JERSEY | 8,791,894 | 20.81 | 0.24 |
| 35 | MICHIGAN | 9,883,640 | 22.7 | 0.23 |
| 36 | DELAWARE | 897,934 | 2.08 | 0.23 |
| 37 | COLORADO | 5,029,196 | 10.97 | 0.22 |
| 38 | TEXAS | 25,145,561 | 52.97 | 0.21 |
| 39 | CALIFORNIA | 37,253,956 | 76.63 | 0.21 |
| 40 | NEW YORK | 19,378,102 | 38.03 | 0.2 |
| 41 | ILLINOIS | 12,830,632 | 23.84 | 0.19 |
| 42 | NEBRASKA | 1,826,341 | 3.03 | 0.17 |
| 43 | MINNESOTA | 5,303,925 | 9.08 | 0.17 |
| 44 | NEVADA | 2,700,551 | 4.16 | 0.15 |
| 45 | WASHINGTON | 6,724,540 | 10.4 | 0.15 |
| 46 | VERMONT | 625,741 | 0.95 | 0.15 |
| 47 | MASSACHUSETTS | 6,547,629 | 6.81 | 0.1 |
| 48 | KANSAS | 2,853,118 | 2.84 | 0.1 |
| 49 | OREGON | 3,831,074 | 3.59 | 0.09 |
| 50 | RHODE ISLAND | 1,052,567 | 0.38 | 0.04 |
| 51 | ALASKA | 710,231 | 0.19 | 0.03 |
| **U.S. GRAMS / PER 100K:** | | 312,825,210 | 1,235.23 | 0.39 |

STATE:ALABAMA BUSINESS ACTIVITY:A - PHARMACIES

| DRUG NAME | DRUG CODE | BUYERS | TOTAL GRAMS | AVG GRAMS |
|---|---|---|---|---|
| AMPHETAMINE | 1100 | 1,371 | 513,195.41 | 374.32 |
| DL-METHAMPHETAMINE RACEMIC BASE | 1105B | 32 | 96.04 | 3 |
| D-METHAMPHETAMINE | 1105D | 4 | 14.86 | 3.72 |
| LISDEXAMFETAMINE | 1205 | 1,328 | 282,505.33 | 212.73 |
| METHYLPHENIDATE (DL;D;L;ISOMERS) | 1724 | 1,352 | 283,489.85 | 209.68 |
| BARBITURIC ACID DERIVIATIVE OR SALT [PER 21C | 2100 | 292 | 6,130 | 20.99 |
| BUTALBITAL | 2165 | 610 | 17,770 | 29.13 |
| DRONABINOL IN AN ORAL SOLUTION IN FDA APPR | 7365 | 9 | 4.2 | 0.47 |
| CODEINE | 9050 | 1,371 | 201,385.95 | 146.89 |
| BUPRENORPHINE | 9064 | 1,340 | 99,420.27 | 74.19 |
| DIHYDROCODEINE | 9120 | 519 | 3,017.55 | 5.81 |
| OXYCODONE | 9143 | 1,379 | 888,450.18 | 644.27 |
| HYDROMORPHONE | 9150 | 1,137 | 9,924.86 | 8.73 |
| HYDROCODONE | 9193 | 1,394 | 924,556.65 | 663.24 |
| LEVORPHANOL | 9220L | 23 | 20.08 | 0.87 |
| MEPERIDINE (PETHIDINE) | 9230 | 832 | 17,119.96 | 20.58 |
| METHADONE | 9250B | 1,145 | 72,727.60 | 63.52 |
| MORPHINE | 9300 | 1,341 | 255,863.42 | 190.8 |
| OPIUM POWDERED | 9639 | 58 | 82.44 | 1.42 |
| OXYMORPHONE | 9652 | 874 | 21,416.15 | 24.5 |
| TAPENTADOL | 9780 | 1,134 | 217,710.50 | 191.98 |
| FENTANYL BASE | 9801 | 1,308 | 5,465.95 | 4.18 |
| OPIUM COMBINATION PRODUCT (C-III) | 9809 | 55 | 43.71 | 0.79 |

STATE:ALABAMA BUSINESS ACTIVITY:B - HOSPITALS

| DRUG NAME | DRUG CODE | BUYERS | TOTAL GRAMS | AVG GRAMS |
|---|---|---|---|---|
| AMPHETAMINE | 1100 | 62 | 8,827.26 | 142.38 |
| LISDEXAMFETAMINE | 1205 | 28 | 2,801.47 | 100.05 |
| METHYLPHENIDATE (DL;D;L;ISOMERS) | 1724 | 77 | 7,301.88 | 94.83 |
| BARBITURIC ACID DERIVIATIVE OR SALT [PER 21C | 2100 | 1 | 15 | 15 |
| BUTALBITAL | 2165 | 4 | 30 | 7.5 |
| PENTOBARBITAL (SCHEDULE 2) | 2270 | 3 | 290.28 | 96.76 |

| | | | | |
|---|---|---|---|---|
| COCAINE | 9041L | 41 | 137.42 | 3.35 |
| CODEINE | 9050 | 116 | 5,082.31 | 43.81 |
| BUPRENORPHINE | 9064 | 95 | 1,009.87 | 10.63 |
| DIHYDROCODEINE | 9120 | 3 | 4.27 | 1.42 |
| OXYCODONE | 9143 | 139 | 33,735.34 | 242.7 |
| HYDROMORPHONE | 9150 | 130 | 2,910.62 | 22.39 |
| HYDROCODONE | 9193 | 144 | 45,670.59 | 317.16 |
| MEPERIDINE (PETHIDINE) | 9230 | 119 | 10,291.54 | 86.48 |
| METHADONE | 9250B | 101 | 3,654.64 | 36.18 |
| MORPHINE | 9300 | 144 | 25,226.60 | 175.18 |
| OPIUM POWDERED | 9639 | 51 | 254.88 | 5 |
| OXYMORPHONE | 9652 | 18 | 86.13 | 4.79 |
| ALFENTANIL | 9737 | 3 | 0.25 | 0.08 |
| REMIFENTANIL | 9739 | 18 | 11.41 | 0.63 |
| SUFENTANIL BASE | 9740 | 31 | 1.55 | 0.05 |
| TAPENTADOL | 9780 | 31 | 850.5 | 27.44 |
| FENTANYL BASE | 9801 | 133 | 457.5 | 3.44 |
| OPIUM COMBINATION PRODUCT (C-III) | 9809 | 4 | 3.41 | 0.85 |

STATE:ALABAMA BUSINESS ACTIVITY:C - PRACTITIONERS

| DRUG NAME | DRUG CODE | BUYERS | TOTAL GRAMS | AVG GRAMS |
|---|---|---|---|---|
| LISDEXAMFETAMINE | 1205 | 1 | 18.22 | 18.22 |
| METHYLPHENIDATE (DL;D;L;ISOMERS) | 1724 | 1 | 77.56 | 77.56 |
| PENTOBARBITAL (SCHEDULE 2) | 2270 | 72 | 42,999.03 | 597.21 |
| COCAINE | 9041L | 8 | 91.28 | 11.41 |
| CODEINE | 9050 | 220 | 2,389.75 | 10.86 |
| BUPRENORPHINE | 9064 | 257 | 73.53 | 0.29 |
| DIHYDROCODEINE | 9120 | 120 | 124.33 | 1.04 |
| OXYCODONE | 9143 | 39 | 1,055.64 | 27.07 |
| HYDROMORPHONE | 9150 | 119 | 201.85 | 1.7 |
| HYDROCODONE | 9193 | 132 | 1,668.70 | 12.64 |
| MEPERIDINE (PETHIDINE) | 9230 | 114 | 2,003.69 | 17.58 |
| METHADONE | 9250B | 3 | 4.13 | 1.38 |
| MORPHINE | 9300 | 142 | 976.81 | 6.88 |
| OPIUM POWDERED | 9639 | 2 | 4.32 | 2.16 |

| | | | | |
|---|---|---|---|---|
| OXYMORPHONE | 9652 | 1 | 1.52 | 1.52 |
| REMIFENTANIL | 9739 | 1 | 1.74 | 1.74 |
| SUFENTANIL BASE | 9740 | 7 | 5.01 | 0.72 |
| FENTANYL BASE | 9801 | 231 | 39.68 | 0.17 |

STATE:ALABAMA BUSINESS ACTIVITY:D - TEACHING INSTITUTIONS

| DRUG NAME | DRUG CODE | BUYERS | TOTAL GRAMS | AVG GRAMS |
|---|---|---|---|---|
| BUPRENORPHINE | 9064 | 1 | 0.01 | 0.01 |

STATE:ALABAMA BUSINESS ACTIVITY:M - MID-LEVEL PRACTITIONERS

| DRUG NAME | DRUG CODE | BUYERS | TOTAL GRAMS | AVG GRAMS |
|---|---|---|---|---|
| PENTOBARBITAL (SCHEDULE 2) | 2270 | 21 | 22,304.24 | 1,062.11 |
| CODEINE | 9050 | 14 | 33.64 | 2.4 |
| BUPRENORPHINE | 9064 | 1 | 0.12 | 0.12 |
| DIHYDROCODEINE | 9120 | 10 | 3.2 | 0.32 |

STATE:ALABAMA BUSINESS ACTIVITY:N-U NARCOTIC TREATMENT PROGRAMS

| DRUG NAME | DRUG CODE | BUYERS | TOTAL GRAMS | AVG GRAMS |
|---|---|---|---|---|
| BUPRENORPHINE | 9064 | 8 | 251.28 | 31.41 |
| METHADONE | 9250B | 22 | 236,035.74 | 10,728.90 |

STATE:ALASKA BUSINESS ACTIVITY:A - PHARMACIES

| DRUG NAME | DRUG CODE | BUYERS | TOTAL GRAMS | AVG GRAMS |
|---|---|---|---|---|
| AMPHETAMINE | 1100 | 107 | 25,907.13 | 242.12 |
| DL-METHAMPHETAMINE RACEMIC BASE | 1105B | 5 | 10.85 | 2.17 |
| D-METHAMPHETAMINE | 1105D | 1 | 0.8 | 0.8 |
| LISDEXAMFETAMINE | 1205 | 96 | 8,931.65 | 93.04 |
| METHYLPHENIDATE (DL;D;L;ISOMERS) | 1724 | 106 | 24,505.89 | 231.19 |
| BARBITURIC ACID DERIVIATIVE OR SALT [PER 21C | 2100 | 26 | 715 | 27.5 |

| | | | | |
|---|---|---|---|---|
| BUTALBITAL | 2165 | 36 | 910 | 25.28 |
| COCAINE | 9041L | 1 | 0.29 | 0.29 |
| CODEINE | 9050 | 110 | 19,955.10 | 181.41 |
| BUPRENORPHINE | 9064 | 104 | 8,772.51 | 84.35 |
| DIHYDROCODEINE | 9120 | 1 | 13.89 | 13.89 |
| OXYCODONE | 9143 | 111 | 123,965.53 | 1,116.81 |
| HYDROMORPHONE | 9150 | 95 | 3,237.94 | 34.08 |
| HYDROCODONE | 9193 | 110 | 39,318.08 | 357.44 |
| MEPERIDINE (PETHIDINE) | 9230 | 58 | 2,378.32 | 41.01 |
| METHADONE | 9250B | 96 | 15,470.26 | 161.15 |
| MORPHINE | 9300 | 109 | 38,003.31 | 348.65 |
| OPIUM POWDERED | 9639 | 12 | 21.96 | 1.83 |
| OXYMORPHONE | 9652 | 34 | 991.17 | 29.15 |
| ALFENTANIL | 9737 | 3 | 0.14 | 0.05 |
| REMIFENTANIL | 9739 | 4 | 0.22 | 0.06 |
| TAPENTADOL | 9780 | 58 | 8,259 | 142.4 |
| FENTANYL BASE | 9801 | 106 | 750.65 | 7.08 |
| OPIUM COMBINATION PRODUCT (C-III) | 9809 | 1 | 0.19 | 0.19 |

STATE:ALASKA BUSINESS ACTIVITY:B - HOSPITALS

| DRUG NAME | DRUG CODE | BUYERS | TOTAL GRAMS | AVG GRAMS |
|---|---|---|---|---|
| AMPHETAMINE | 1100 | 27 | 6,428.14 | 238.08 |
| LISDEXAMFETAMINE | 1205 | 17 | 934.21 | 54.95 |
| METHYLPHENIDATE (DL;D;L;ISOMERS) | 1724 | 31 | 7,633.06 | 246.23 |
| BUTALBITAL | 2165 | 1 | 15 | 15 |
| PENTOBARBITAL (SCHEDULE 2) | 2270 | 3 | 2,086.65 | 695.55 |
| COCAINE | 9041L | 11 | 38.07 | 3.46 |
| CODEINE | 9050 | 38 | 7,751.98 | 204 |
| BUPRENORPHINE | 9064 | 33 | 654.25 | 19.83 |
| OXYCODONE | 9143 | 47 | 13,725.79 | 292.04 |
| HYDROMORPHONE | 9150 | 45 | 622.59 | 13.84 |
| HYDROCODONE | 9193 | 50 | 5,041.10 | 100.82 |
| MEPERIDINE (PETHIDINE) | 9230 | 37 | 431.83 | 11.67 |
| METHADONE | 9250B | 25 | 1,254.66 | 50.19 |
| MORPHINE | 9300 | 51 | 7,650.67 | 150.01 |

| | | | | |
|---|---|---|---|---|
| OPIUM POWDERED | 9639 | 17 | 80.28 | 4.72 |
| OXYMORPHONE | 9652 | 2 | 5 | 2.5 |
| ALFENTANIL | 9737 | 4 | 2.45 | 0.61 |
| REMIFENTANIL | 9739 | 18 | 11.03 | 0.61 |
| SUFENTANIL BASE | 9740 | 6 | 0.04 | 0.01 |
| TAPENTADOL | 9780 | 6 | 732 | 122 |
| FENTANYL BASE | 9801 | 51 | 178.82 | 3.51 |

STATE:ALASKA BUSINESS ACTIVITY:C - PRACTITIONERS

| DRUG NAME | DRUG CODE | BUYERS | TOTAL GRAMS | AVG GRAMS |
|---|---|---|---|---|
| PENTOBARBITAL (SCHEDULE 2) | 2270 | 32 | 19,282.95 | 602.59 |
| COCAINE | 9041L | 2 | 9.57 | 4.79 |
| CODEINE | 9050 | 10 | 73.82 | 7.38 |
| BUPRENORPHINE | 9064 | 51 | 5.01 | 0.1 |
| OXYCODONE | 9143 | 8 | 104.31 | 13.04 |
| HYDROMORPHONE | 9150 | 53 | 36.69 | 0.69 |
| HYDROCODONE | 9193 | 43 | 164.86 | 3.83 |
| MEPERIDINE (PETHIDINE) | 9230 | 15 | 40.85 | 2.72 |
| METHADONE | 9250B | 3 | 14.13 | 4.71 |
| MORPHINE | 9300 | 35 | 77.71 | 2.22 |
| OPIUM POWDERED | 9639 | 1 | 0.36 | 0.36 |
| OXYMORPHONE | 9652 | 1 | 8.56 | 8.56 |
| ALFENTANIL | 9737 | 1 | 0.3 | 0.3 |
| REMIFENTANIL | 9739 | 6 | 0.43 | 0.07 |
| FENTANYL BASE | 9801 | 72 | 11.14 | 0.15 |

STATE:ALASKA BUSINESS ACTIVITY:M - MID-LEVEL PRACTITIONERS

| DRUG NAME | DRUG CODE | BUYERS | TOTAL GRAMS | AVG GRAMS |
|---|---|---|---|---|
| PENTOBARBITAL (SCHEDULE 2) | 2270 | 4 | 1,421.78 | 355.45 |
| CODEINE | 9050 | 3 | 16.58 | 5.53 |
| HYDROMORPHONE | 9150 | 1 | 0.02 | 0.02 |
| HYDROCODONE | 9193 | 5 | 6.05 | 1.21 |
| MORPHINE | 9300 | 5 | 0.94 | 0.19 |

| | | | | |
|---|---|---|---|---|
| FENTANYL BASE | 9801 | 3 | 0.01 | 0 |

STATE:ALASKA BUSINESS ACTIVITY:N-U NARCOTIC TREATMENT PROGRAMS

| DRUG NAME | DRUG CODE | BUYERS | TOTAL GRAMS | AVG GRAMS |
|---|---|---|---|---|
| BUPRENORPHINE | 9064 | 2 | 26.4 | 13.2 |
| METHADONE | 9250B | 4 | 15,347.90 | 3,836.98 |

STATE:AMERICAN SAMOA BUSINESS ACTIVITY:B - HOSPITALS

| DRUG NAME | DRUG CODE | BUYERS | TOTAL GRAMS | AVG GRAMS |
|---|---|---|---|---|
| AMPHETAMINE | 1100 | 1 | 2.5 | 2.5 |
| METHYLPHENIDATE (DL;D;L;ISOMERS) | 1724 | 1 | 17.73 | 17.73 |
| BUTALBITAL | 2165 | 1 | 10 | 10 |
| CODEINE | 9050 | 1 | 1,361.42 | 1,361.42 |
| OXYCODONE | 9143 | 1 | 58.72 | 58.72 |
| HYDROMORPHONE | 9150 | 1 | 0.98 | 0.98 |
| HYDROCODONE | 9193 | 1 | 7.9 | 7.9 |
| MORPHINE | 9300 | 1 | 87.61 | 87.61 |
| FENTANYL BASE | 9801 | 1 | 0.47 | 0.47 |

STATE:AMERICAN SAMOA BUSINESS ACTIVITY:C - PRACTITIONERS

| DRUG NAME | DRUG CODE | BUYERS | TOTAL GRAMS | AVG GRAMS |
|---|---|---|---|---|
| BUPRENORPHINE | 9064 | 1 | 0.01 | 0.01 |

STATE:ARIZONA BUSINESS ACTIVITY:A - PHARMACIES

| DRUG NAME | DRUG CODE | BUYERS | TOTAL GRAMS | AVG GRAMS |
|---|---|---|---|---|
| AMPHETAMINE | 1100 | 1,112 | 319,806.38 | 287.6 |
| DL-METHAMPHETAMINE RACEMIC BASE | 1105B | 28 | 134.22 | 4.79 |
| D-METHAMPHETAMINE | 1105D | 5 | 19.68 | 3.94 |
| LISDEXAMFETAMINE | 1205 | 1,052 | 141,762.84 | 134.76 |

| | | | | |
|---|---|---|---|---|
| METHYLPHENIDATE (DL;D;L;ISOMERS) | 1724 | 1,097 | 282,504.72 | 257.52 |
| BARBITURIC ACID DERIVIATIVE OR SALT [PER 21C | 2100 | 511 | 20,245 | 39.62 |
| BUTALBITAL | 2165 | 486 | 20,450 | 42.08 |
| SECOBARBITAL (SCHEDULE 2) | 2315 | 2 | 18.31 | 9.16 |
| DRONABINOL IN AN ORAL SOLUTION IN FDA APPR | 7365 | 4 | 21.6 | 5.4 |
| NABILONE | 7379 | 1 | 0.2 | 0.2 |
| COCAINE | 9041L | 2 | 4.61 | 2.3 |
| CODEINE | 9050 | 1,112 | 270,203.88 | 242.99 |
| BUPRENORPHINE | 9064 | 1,059 | 34,656.01 | 32.73 |
| DIHYDROCODEINE | 9120 | 25 | 316.18 | 12.65 |
| OXYCODONE | 9143 | 1,133 | 1,631,548.23 | 1,440.02 |
| HYDROMORPHONE | 9150 | 1,042 | 23,513.97 | 22.57 |
| HYDROCODONE | 9193 | 1,139 | 390,210.36 | 342.59 |
| LEVORPHANOL | 9220L | 52 | 99.94 | 1.92 |
| MEPERIDINE (PETHIDINE) | 9230 | 520 | 11,101.71 | 21.35 |
| METHADONE | 9250B | 943 | 50,743.96 | 53.81 |
| MORPHINE | 9300 | 1,121 | 594,891.33 | 530.68 |
| OPIUM POWDERED | 9639 | 110 | 597.24 | 5.43 |
| OXYMORPHONE | 9652 | 882 | 23,704.75 | 26.88 |
| REMIFENTANIL | 9739 | 1 | 0.03 | 0.03 |
| TAPENTADOL | 9780 | 837 | 141,441.50 | 168.99 |
| FENTANYL BASE | 9801 | 1,056 | 6,367.25 | 6.03 |
| OPIUM COMBINATION PRODUCT (C-III) | 9809 | 26 | 31.22 | 1.2 |

STATE:ARIZONA BUSINESS ACTIVITY:B - HOSPITALS

| DRUG NAME | DRUG CODE | BUYERS | TOTAL GRAMS | AVG GRAMS |
|---|---|---|---|---|
| AMPHETAMINE | 1100 | 96 | 7,763.57 | 80.87 |
| LISDEXAMFETAMINE | 1205 | 37 | 1,617.52 | 43.72 |
| METHYLPHENIDATE (DL;D;L;ISOMERS) | 1724 | 110 | 15,615.60 | 141.96 |
| BARBITURIC ACID DERIVIATIVE OR SALT [PER 21C | 2100 | 1 | 5 | 5 |
| AMOBARBITAL (SCHEDULE 2) | 2125 | 5 | 45.11 | 9.02 |
| BUTALBITAL | 2165 | 6 | 85 | 14.17 |
| PENTOBARBITAL (SCHEDULE 2) | 2270 | 17 | 1,262.74 | 74.28 |
| COCAINE | 9041L | 62 | 293.98 | 4.74 |
| CODEINE | 9050 | 129 | 22,187.70 | 172 |

| | | | | |
|---|---|---|---|---|
| BUPRENORPHINE | 9064 | 68 | 2,541.31 | 37.37 |
| OXYCODONE | 9143 | 160 | 68,431.55 | 427.7 |
| HYDROMORPHONE | 9150 | 126 | 3,079.61 | 24.44 |
| HYDROCODONE | 9193 | 148 | 21,067.88 | 142.35 |
| MEPERIDINE (PETHIDINE) | 9230 | 90 | 5,314.95 | 59.06 |
| METHADONE | 9250B | 108 | 4,880.90 | 45.19 |
| MORPHINE | 9300 | 155 | 52,777.89 | 340.5 |
| OPIUM POWDERED | 9639 | 57 | 569.16 | 9.99 |
| OXYMORPHONE | 9652 | 11 | 44.07 | 4.01 |
| ALFENTANIL | 9737 | 9 | 1.65 | 0.18 |
| REMIFENTANIL | 9739 | 35 | 13.65 | 0.39 |
| SUFENTANIL BASE | 9740 | 33 | 0.31 | 0.01 |
| TAPENTADOL | 9780 | 13 | 681.5 | 52.42 |
| FENTANYL BASE | 9801 | 138 | 566.69 | 4.11 |
| OPIUM COMBINATION PRODUCT (C-III) | 9809 | 3 | 2.08 | 0.69 |

STATE:ARIZONA BUSINESS ACTIVITY:C - PRACTITIONERS

| DRUG NAME | DRUG CODE | BUYERS | TOTAL GRAMS | AVG GRAMS |
|---|---|---|---|---|
| PENTOBARBITAL (SCHEDULE 2) | 2270 | 194 | 213,625.71 | 1,101.16 |
| COCAINE | 9041L | 28 | 120.99 | 4.32 |
| CODEINE | 9050 | 154 | 4,483.84 | 29.12 |
| BUPRENORPHINE | 9064 | 591 | 84.31 | 0.14 |
| DIHYDROCODEINE | 9120 | 5 | 1.6 | 0.32 |
| OXYCODONE | 9143 | 102 | 421.55 | 4.13 |
| HYDROMORPHONE | 9150 | 441 | 217.96 | 0.49 |
| HYDROCODONE | 9193 | 320 | 1,481.80 | 4.63 |
| MEPERIDINE (PETHIDINE) | 9230 | 284 | 3,088.94 | 10.88 |
| METHADONE | 9250B | 15 | 69.67 | 4.64 |
| MORPHINE | 9300 | 288 | 410.53 | 1.43 |
| OPIUM POWDERED | 9639 | 8 | 14.76 | 1.85 |
| OXYMORPHONE | 9652 | 13 | 2.07 | 0.16 |
| NOROXYMORPHONE | 9668 | 1 | 0.1 | 0.1 |
| ALFENTANIL | 9737 | 20 | 2.09 | 0.1 |
| REMIFENTANIL | 9739 | 27 | 2.01 | 0.07 |
| SUFENTANIL BASE | 9740 | 9 | 0.03 | 0 |

| | | | | |
|---|---|---|---|---|
| FENTANYL BASE | 9801 | 712 | 59.43 | 0.08 |

STATE:ARIZONA BUSINESS ACTIVITY:M - MID-LEVEL PRACTITIONERS

| DRUG NAME | DRUG CODE | BUYERS | TOTAL GRAMS | AVG GRAMS |
|---|---|---|---|---|
| PENTOBARBITAL (SCHEDULE 2) | 2270 | 1 | 177.72 | 177.72 |
| CODEINE | 9050 | 73 | 273.12 | 3.74 |
| BUPRENORPHINE | 9064 | 1 | 0.48 | 0.48 |
| DIHYDROCODEINE | 9120 | 5 | 1.71 | 0.34 |
| OXYCODONE | 9143 | 1 | 0.45 | 0.45 |
| HYDROMORPHONE | 9150 | 1 | 0.04 | 0.04 |
| HYDROCODONE | 9193 | 2 | 43.57 | 21.79 |
| MORPHINE | 9300 | 2 | 0.22 | 0.11 |
| FENTANYL BASE | 9801 | 1 | 0 | 0 |

STATE:ARIZONA BUSINESS ACTIVITY:N-U NARCOTIC TREATMENT PROGRAMS

| DRUG NAME | DRUG CODE | BUYERS | TOTAL GRAMS | AVG GRAMS |
|---|---|---|---|---|
| BUPRENORPHINE | 9064 | 12 | 418.16 | 34.85 |
| METHADONE | 9250B | 40 | 280,038.88 | 7,000.97 |

STATE:ARKANSAS BUSINESS ACTIVITY:A - PHARMACIES

| DRUG NAME | DRUG CODE | BUYERS | TOTAL GRAMS | AVG GRAMS |
|---|---|---|---|---|
| AMPHETAMINE | 1100 | 787 | 200,355.92 | 254.58 |
| DL-METHAMPHETAMINE RACEMIC BASE | 1105B | 10 | 25.32 | 2.53 |
| LISDEXAMFETAMINE | 1205 | 777 | 120,313.02 | 154.84 |
| METHYLPHENIDATE (DL;D;L;ISOMERS) | 1724 | 784 | 156,082.16 | 199.08 |
| BARBITURIC ACID DERIVIATIVE OR SALT [PER 21C | 2100 | 194 | 5,320 | 27.42 |
| BUTALBITAL | 2165 | 355 | 13,525 | 38.1 |
| DRONABINOL IN AN ORAL SOLUTION IN FDA APPR | 7365 | 3 | 1.2 | 0.4 |
| COCAINE | 9041L | 1 | 4.46 | 4.46 |
| CODEINE | 9050 | 789 | 287,273.84 | 364.1 |
| BUPRENORPHINE | 9064 | 737 | 24,053.25 | 32.64 |

| | | | | |
|---|---|---|---|---|
| DIHYDROCODEINE | 9120 | 128 | 527.67 | 4.12 |
| OXYCODONE | 9143 | 795 | 551,165.83 | 693.29 |
| HYDROMORPHONE | 9150 | 746 | 11,018.03 | 14.77 |
| HYDROCODONE | 9193 | 799 | 508,544.91 | 636.48 |
| LEVORPHANOL | 9220L | 10 | 9.05 | 0.91 |
| MEPERIDINE (PETHIDINE) | 9230 | 655 | 23,073.51 | 35.23 |
| METHADONE | 9250B | 713 | 32,064.87 | 44.97 |
| MORPHINE | 9300 | 799 | 163,540.84 | 204.68 |
| OPIUM POWDERED | 9639 | 26 | 59.04 | 2.27 |
| OXYMORPHONE | 9652 | 471 | 6,819.57 | 14.48 |
| REMIFENTANIL | 9739 | 1 | 0.18 | 0.18 |
| TAPENTADOL | 9780 | 649 | 82,731 | 127.47 |
| FENTANYL BASE | 9801 | 772 | 3,277.14 | 4.24 |
| OPIUM COMBINATION PRODUCT (C-III) | 9809 | 8 | 8.14 | 1.02 |

STATE:ARKANSAS BUSINESS ACTIVITY:B - HOSPITALS

| DRUG NAME | DRUG CODE | BUYERS | TOTAL GRAMS | AVG GRAMS |
|---|---|---|---|---|
| AMPHETAMINE | 1100 | 50 | 4,015.79 | 80.32 |
| LISDEXAMFETAMINE | 1205 | 26 | 877.56 | 33.75 |
| METHYLPHENIDATE (DL;D;L;ISOMERS) | 1724 | 70 | 4,015.23 | 57.36 |
| BARBITURIC ACID DERIVIATIVE OR SALT [PER 21C | 2100 | 1 | 5 | 5 |
| AMOBARBITAL (SCHEDULE 2) | 2125 | 1 | 0.91 | 0.91 |
| BUTALBITAL | 2165 | 5 | 240 | 48 |
| PENTOBARBITAL (SCHEDULE 2) | 2270 | 7 | 262.48 | 37.5 |
| COCAINE | 9041L | 30 | 146.99 | 4.9 |
| CODEINE | 9050 | 91 | 4,979.88 | 54.72 |
| BUPRENORPHINE | 9064 | 43 | 970.56 | 22.57 |
| DIHYDROCODEINE | 9120 | 1 | 1.07 | 1.07 |
| OXYCODONE | 9143 | 122 | 26,792.67 | 219.61 |
| HYDROMORPHONE | 9150 | 106 | 2,012.45 | 18.99 |
| HYDROCODONE | 9193 | 124 | 26,950.06 | 217.34 |
| MEPERIDINE (PETHIDINE) | 9230 | 101 | 5,933.72 | 58.75 |
| METHADONE | 9250B | 76 | 2,933.50 | 38.6 |
| MORPHINE | 9300 | 119 | 17,241.88 | 144.89 |
| OPIUM POWDERED | 9639 | 37 | 298.08 | 8.06 |

| | | | | |
|---|---|---|---|---|
| OXYMORPHONE | 9652 | 8 | 53.44 | 6.68 |
| ALFENTANIL | 9737 | 2 | 0.08 | 0.04 |
| REMIFENTANIL | 9739 | 13 | 6.08 | 0.47 |
| SUFENTANIL BASE | 9740 | 21 | 0.56 | 0.03 |
| TAPENTADOL | 9780 | 28 | 1,203.50 | 42.98 |
| FENTANYL BASE | 9801 | 127 | 424.22 | 3.34 |
| OPIUM COMBINATION PRODUCT (C-III) | 9809 | 1 | 0.38 | 0.38 |

STATE:ARKANSAS BUSINESS ACTIVITY:C - PRACTITIONERS

| DRUG NAME | DRUG CODE | BUYERS | TOTAL GRAMS | AVG GRAMS |
|---|---|---|---|---|
| LISDEXAMFETAMINE | 1205 | 1 | 13.04 | 13.04 |
| METHYLPHENIDATE (DL;D;L;ISOMERS) | 1724 | 1 | 3.46 | 3.46 |
| PENTOBARBITAL (SCHEDULE 2) | 2270 | 47 | 24,167.03 | 514.19 |
| COCAINE | 9041L | 3 | 24.14 | 8.05 |
| CODEINE | 9050 | 40 | 541.65 | 13.54 |
| BUPRENORPHINE | 9064 | 133 | 6.65 | 0.05 |
| DIHYDROCODEINE | 9120 | 25 | 18.17 | 0.73 |
| OXYCODONE | 9143 | 6 | 29.14 | 4.86 |
| HYDROMORPHONE | 9150 | 58 | 24.14 | 0.42 |
| HYDROCODONE | 9193 | 74 | 182.1 | 2.46 |
| MEPERIDINE (PETHIDINE) | 9230 | 43 | 592.18 | 13.77 |
| MORPHINE | 9300 | 86 | 107.72 | 1.25 |
| OPIUM POWDERED | 9639 | 3 | 3.96 | 1.32 |
| NOROXYMORPHONE | 9668 | 2 | 78.06 | 39.03 |
| REMIFENTANIL | 9739 | 3 | 0.14 | 0.05 |
| FENTANYL BASE | 9801 | 144 | 13.84 | 0.1 |

STATE:ARKANSAS BUSINESS ACTIVITY:D - TEACHING INSTITUTIONS

| DRUG NAME | DRUG CODE | BUYERS | TOTAL GRAMS | AVG GRAMS |
|---|---|---|---|---|
| BUPRENORPHINE | 9064 | 1 | 0 | 0 |

STATE:ARKANSAS BUSINESS ACTIVITY:M - MID-LEVEL PRACTITIONERS

| DRUG NAME | DRUG CODE | BUYERS | TOTAL GRAMS | AVG GRAMS |
|---|---|---|---|---|
| PENTOBARBITAL (SCHEDULE 2) | 2270 | 11 | 11,997.67 | 1,090.70 |
| CODEINE | 9050 | 7 | 13.65 | 1.95 |
| DIHYDROCODEINE | 9120 | 17 | 21.65 | 1.27 |

STATE:ARKANSAS BUSINESS ACTIVITY:N-U NARCOTIC TREATMENT PROGRAMS

| DRUG NAME | DRUG CODE | BUYERS | TOTAL GRAMS | AVG GRAMS |
|---|---|---|---|---|
| BUPRENORPHINE | 9064 | 1 | 25.92 | 25.92 |
| METHADONE | 9250B | 7 | 29,243.30 | 4,177.61 |

STATE:CALIFORNIA BUSINESS ACTIVITY:A - PHARMACIES

| DRUG NAME | DRUG CODE | BUYERS | TOTAL GRAMS | AVG GRAMS |
|---|---|---|---|---|
| AMPHETAMINE | 1100 | 5,552 | 1,154,151.07 | 207.88 |
| DL-METHAMPHETAMINE RACEMIC BASE | 1105B | 301 | 1,129.20 | 3.75 |
| D-METHAMPHETAMINE | 1105D | 30 | 142.6 | 4.75 |
| LISDEXAMFETAMINE | 1205 | 4,855 | 438,739.17 | 90.37 |
| METHYLPHENIDATE (DL;D;L;ISOMERS) | 1724 | 5,466 | 1,040,896.98 | 190.43 |
| BARBITURIC ACID DERIVIATIVE OR SALT [PER 21C | 2100 | 1,616 | 58,865 | 36.43 |
| BUTALBITAL | 2165 | 2,342 | 97,400 | 41.59 |
| PENTOBARBITAL (SCHEDULE 2) | 2270 | 1 | 2.73 | 2.73 |
| SECOBARBITAL (SCHEDULE 2) | 2315 | 64 | 4,275.39 | 66.8 |
| DRONABINOL IN AN ORAL SOLUTION IN FDA APPR | 7365 | 25 | 43.65 | 1.75 |
| NABILONE | 7379 | 1 | 0.65 | 0.65 |
| COCAINE | 9041L | 38 | 364.94 | 9.6 |
| CODEINE | 9050 | 6,208 | 1,769,207.32 | 284.99 |
| BUPRENORPHINE | 9064 | 5,225 | 145,614.02 | 27.87 |
| DIHYDROCODEINE | 9120 | 53 | 740.23 | 13.97 |
| OXYCODONE | 9143 | 5,891 | 3,289,504.71 | 558.39 |
| HYDROMORPHONE | 9150 | 5,195 | 130,572.18 | 25.13 |
| HYDROCODONE | 9193 | 6,179 | 3,461,545.80 | 560.21 |
| LEVORPHANOL | 9220L | 216 | 398.04 | 1.84 |
| MEPERIDINE (PETHIDINE) | 9230 | 1,002 | 26,944.17 | 26.89 |

| | | | | |
|---|---|---|---|---|
| METHADONE | 9250B | 4,745 | 344,813.47 | 72.67 |
| MORPHINE | 9300 | 5,510 | 1,613,860.54 | 292.9 |
| OPIUM TINCTURE (LAUDANUM) | 9630 | 2 | 5.91 | 2.96 |
| OPIUM POWDERED | 9639 | 226 | 410.4 | 1.82 |
| OXYMORPHONE | 9652 | 2,360 | 51,574.47 | 21.85 |
| NOROXYMORPHONE | 9668 | 1 | 1.49 | 1.49 |
| ALFENTANIL | 9737 | 15 | 5.17 | 0.34 |
| REMIFENTANIL | 9739 | 14 | 1.27 | 0.09 |
| SUFENTANIL BASE | 9740 | 9 | 39.64 | 4.4 |
| TAPENTADOL | 9780 | 3,440 | 420,533 | 122.25 |
| FENTANYL BASE | 9801 | 5,083 | 24,570.52 | 4.83 |
| OPIUM COMBINATION PRODUCT (C-III) | 9809 | 99 | 71.52 | 0.72 |

STATE:CALIFORNIA BUSINESS ACTIVITY:B - HOSPITALS

| DRUG NAME | DRUG CODE | BUYERS | TOTAL GRAMS | AVG GRAMS |
|---|---|---|---|---|
| AMPHETAMINE | 1100 | 266 | 43,132.35 | 162.15 |
| DL-METHAMPHETAMINE RACEMIC BASE | 1105B | 2 | 23.71 | 11.85 |
| LISDEXAMFETAMINE | 1205 | 98 | 7,500.27 | 76.53 |
| METHYLPHENIDATE (DL;D;L;ISOMERS) | 1724 | 401 | 47,016.35 | 117.25 |
| GAMMA HYDROXYBUTYRIC ACID(FDA APPROVED | 2012 | 1 | 2,080.51 | 2,080.51 |
| BARBITURIC ACID DERIVIATIVE OR SALT [PER 21C | 2100 | 22 | 240 | 10.91 |
| AMOBARBITAL (SCHEDULE 2) | 2125 | 21 | 100.71 | 4.8 |
| BUTALBITAL | 2165 | 50 | 910 | 18.2 |
| PENTOBARBITAL (SCHEDULE 2) | 2270 | 78 | 13,302.79 | 170.55 |
| SECOBARBITAL (SCHEDULE 2) | 2315 | 3 | 100.71 | 33.57 |
| DRONABINOL IN AN ORAL SOLUTION IN FDA APPR | 7365 | 1 | 0.15 | 0.15 |
| COCAINE | 9041L | 401 | 2,855.96 | 7.12 |
| CODEINE | 9050 | 578 | 85,565.98 | 148.04 |
| BUPRENORPHINE | 9064 | 322 | 5,402.04 | 16.78 |
| OXYCODONE | 9143 | 647 | 162,276.17 | 250.81 |
| HYDROMORPHONE | 9150 | 672 | 23,609.25 | 35.13 |
| HYDROCODONE | 9193 | 729 | 182,538.46 | 250.4 |
| LEVORPHANOL | 9220L | 5 | 4.53 | 0.91 |
| MEPERIDINE (PETHIDINE) | 9230 | 643 | 26,660.41 | 41.46 |
| METHADONE | 9250B | 504 | 47,308.37 | 93.87 |

| | | | | |
|---|---|---|---|---|
| MORPHINE | 9300 | 750 | 159,934.38 | 213.25 |
| OPIUM POWDERED | 9639 | 303 | 1,401.12 | 4.62 |
| OXYMORPHONE | 9652 | 61 | 632.48 | 10.37 |
| NOROXYMORPHONE | 9668 | 2 | 7.04 | 3.52 |
| ALFENTANIL | 9737 | 142 | 51.43 | 0.36 |
| REMIFENTANIL | 9739 | 196 | 128.36 | 0.65 |
| SUFENTANIL BASE | 9740 | 153 | 3.96 | 0.03 |
| TAPENTADOL | 9780 | 73 | 4,343.85 | 59.5 |
| FENTANYL BASE | 9801 | 844 | 2,942.75 | 3.49 |
| OPIUM COMBINATION PRODUCT (C-III) | 9809 | 10 | 5.11 | 0.51 |

STATE:CALIFORNIA BUSINESS ACTIVITY:C - PRACTITIONERS

| DRUG NAME | DRUG CODE | BUYERS | TOTAL GRAMS | AVG GRAMS |
|---|---|---|---|---|
| AMPHETAMINE | 1100 | 7 | 850.13 | 121.45 |
| D-METHAMPHETAMINE | 1105D | 2 | 0.24 | 0.12 |
| LISDEXAMFETAMINE | 1205 | 6 | 50.63 | 8.44 |
| METHYLPHENIDATE (DL;D;L;ISOMERS) | 1724 | 3 | 70.04 | 23.35 |
| GAMMA HYDROXYBUTYRIC ACID(FDA APPROVED | 2012 | 3 | 5,944.32 | 1,981.44 |
| BARBITURIC ACID DERIVIATIVE OR SALT [PER 21C | 2100 | 3 | 78.5 | 26.17 |
| PENTOBARBITAL (SCHEDULE 2) | 2270 | 613 | 689,267.84 | 1,124.42 |
| COCAINE | 9041L | 104 | 249.66 | 2.4 |
| CODEINE | 9050 | 1,163 | 36,282.66 | 31.2 |
| BUPRENORPHINE | 9064 | 2,697 | 1,066.33 | 0.4 |
| DIHYDROCODEINE | 9120 | 12 | 102.89 | 8.57 |
| OXYCODONE | 9143 | 268 | 1,051.39 | 3.92 |
| HYDROMORPHONE | 9150 | 1,743 | 1,110.82 | 0.64 |
| HYDROCODONE | 9193 | 2,072 | 12,275.39 | 5.92 |
| MEPERIDINE (PETHIDINE) | 9230 | 846 | 8,767.83 | 10.36 |
| METHADONE | 9250B | 141 | 345.06 | 2.45 |
| MORPHINE | 9300 | 1,049 | 1,699.63 | 1.62 |
| OPIUM POWDERED | 9639 | 10 | 10.08 | 1.01 |
| OXYMORPHONE | 9652 | 52 | 8.03 | 0.15 |
| NOROXYMORPHONE | 9668 | 22 | 251.08 | 11.41 |
| ALFENTANIL | 9737 | 59 | 9.32 | 0.16 |
| REMIFENTANIL | 9739 | 63 | 3.72 | 0.06 |

| | | | | |
|---|---|---|---|---|
| SUFENTANIL BASE | 9740 | 19 | 0.14 | 0.01 |
| FENTANYL BASE | 9801 | 2,379 | 302.02 | 0.13 |

STATE:CALIFORNIA BUSINESS ACTIVITY:D - TEACHING INSTITUTIONS

| DRUG NAME | DRUG CODE | BUYERS | TOTAL GRAMS | AVG GRAMS |
|---|---|---|---|---|
| PENTOBARBITAL (SCHEDULE 2) | 2270 | 2 | 622.03 | 311.02 |
| BUPRENORPHINE | 9064 | 7 | 0.04 | 0.01 |
| HYDROMORPHONE | 9150 | 4 | 0.57 | 0.14 |
| MORPHINE | 9300 | 1 | 2.44 | 2.44 |

STATE:CALIFORNIA BUSINESS ACTIVITY:M - MID-LEVEL PRACTITIONERS

| DRUG NAME | DRUG CODE | BUYERS | TOTAL GRAMS | AVG GRAMS |
|---|---|---|---|---|
| PENTOBARBITAL (SCHEDULE 2) | 2270 | 4 | 5,598.27 | 1,399.57 |
| CODEINE | 9050 | 133 | 664.45 | 5 |

STATE:CALIFORNIA BUSINESS ACTIVITY:N-U NARCOTIC TREATMENT PROGRAMS

| DRUG NAME | DRUG CODE | BUYERS | TOTAL GRAMS | AVG GRAMS |
|---|---|---|---|---|
| BUPRENORPHINE | 9064 | 52 | 485.38 | 9.33 |
| METHADONE | 9250B | 151 | 996,317.77 | 6,598.13 |

STATE:COLORADO BUSINESS ACTIVITY:A - PHARMACIES

| DRUG NAME | DRUG CODE | BUYERS | TOTAL GRAMS | AVG GRAMS |
|---|---|---|---|---|
| AMPHETAMINE | 1100 | 819 | 273,180.24 | 333.55 |
| DL-METHAMPHETAMINE RACEMIC BASE | 1105B | 83 | 205.75 | 2.48 |
| LISDEXAMFETAMINE | 1205 | 785 | 104,788.14 | 133.49 |
| METHYLPHENIDATE (DL;D;L;ISOMERS) | 1724 | 823 | 203,662.76 | 247.46 |
| BARBITURIC ACID DERIVIATIVE OR SALT [PER 21C | 2100 | 243 | 6,480 | 26.67 |
| BUTALBITAL | 2165 | 291 | 7,390 | 25.4 |
| SECOBARBITAL (SCHEDULE 2) | 2315 | 14 | 228.88 | 16.35 |

| | | | | |
|---|---|---|---|---|
| DRONABINOL IN AN ORAL SOLUTION IN FDA APPR | 7365 | 2 | 3.6 | 1.8 |
| NABILONE | 7379 | 2 | 0.35 | 0.18 |
| COCAINE | 9041L | 5 | 4.36 | 0.87 |
| CODEINE | 9050 | 804 | 86,742.09 | 107.89 |
| BUPRENORPHINE | 9064 | 798 | 25,709.27 | 32.22 |
| DIHYDROCODEINE | 9120 | 2 | 9.61 | 4.81 |
| OXYCODONE | 9143 | 837 | 766,152.43 | 915.36 |
| HYDROMORPHONE | 9150 | 809 | 20,978.29 | 25.93 |
| HYDROCODONE | 9193 | 840 | 221,473.23 | 263.66 |
| LEVORPHANOL | 9220L | 54 | 50.73 | 0.94 |
| MEPERIDINE (PETHIDINE) | 9230 | 263 | 5,760.18 | 21.9 |
| METHADONE | 9250B | 700 | 31,599.42 | 45.14 |
| MORPHINE | 9300 | 834 | 241,902.95 | 290.05 |
| OPIUM POWDERED | 9639 | 82 | 298.8 | 3.64 |
| OXYMORPHONE | 9652 | 532 | 12,244.12 | 23.02 |
| SUFENTANIL BASE | 9740 | 1 | 0.07 | 0.07 |
| TAPENTADOL | 9780 | 675 | 107,492 | 159.25 |
| FENTANYL BASE | 9801 | 802 | 3,994.35 | 4.98 |
| OPIUM COMBINATION PRODUCT (C-III) | 9809 | 25 | 9.46 | 0.38 |

STATE:COLORADO BUSINESS ACTIVITY:B - HOSPITALS

| DRUG NAME | DRUG CODE | BUYERS | TOTAL GRAMS | AVG GRAMS |
|---|---|---|---|---|
| AMPHETAMINE | 1100 | 114 | 46,273.94 | 405.91 |
| DL-METHAMPHETAMINE RACEMIC BASE | 1105B | 3 | 4.82 | 1.61 |
| LISDEXAMFETAMINE | 1205 | 70 | 6,244.64 | 89.21 |
| METHYLPHENIDATE (DL;D;L;ISOMERS) | 1724 | 128 | 42,732.71 | 333.85 |
| BARBITURIC ACID DERIVIATIVE OR SALT [PER 21C | 2100 | 5 | 105 | 21 |
| AMOBARBITAL (SCHEDULE 2) | 2125 | 4 | 68.36 | 17.09 |
| BUTALBITAL | 2165 | 29 | 420 | 14.48 |
| PENTOBARBITAL (SCHEDULE 2) | 2270 | 16 | 1,273.68 | 79.61 |
| SECOBARBITAL (SCHEDULE 2) | 2315 | 2 | 146.48 | 73.24 |
| DRONABINOL IN AN ORAL SOLUTION IN FDA APPR | 7365 | 1 | 0.15 | 0.15 |
| COCAINE | 9041L | 53 | 230.99 | 4.36 |
| CODEINE | 9050 | 147 | 14,835.10 | 100.92 |
| BUPRENORPHINE | 9064 | 104 | 3,421.99 | 32.9 |

| | | | | |
|---|---|---|---|---|
| OXYCODONE | 9143 | 200 | 125,647.41 | 628.24 |
| HYDROMORPHONE | 9150 | 194 | 5,888.80 | 30.35 |
| HYDROCODONE | 9193 | 210 | 37,304.42 | 177.64 |
| LEVORPHANOL | 9220L | 1 | 0.12 | 0.12 |
| MEPERIDINE (PETHIDINE) | 9230 | 102 | 1,099.88 | 10.78 |
| METHADONE | 9250B | 126 | 9,089.61 | 72.14 |
| MORPHINE | 9300 | 204 | 62,931.40 | 308.49 |
| OPIUM POWDERED | 9639 | 73 | 654.12 | 8.96 |
| OXYMORPHONE | 9652 | 49 | 576.74 | 11.77 |
| ALFENTANIL | 9737 | 10 | 3.45 | 0.35 |
| REMIFENTANIL | 9739 | 58 | 24.68 | 0.43 |
| SUFENTANIL BASE | 9740 | 41 | 0.71 | 0.02 |
| TAPENTADOL | 9780 | 57 | 4,757.50 | 83.46 |
| FENTANYL BASE | 9801 | 201 | 934.83 | 4.65 |
| OPIUM COMBINATION PRODUCT (C-III) | 9809 | 3 | 1.51 | 0.5 |

STATE:COLORADO BUSINESS ACTIVITY:C - PRACTITIONERS

| DRUG NAME | DRUG CODE | BUYERS | TOTAL GRAMS | AVG GRAMS |
|---|---|---|---|---|
| PENTOBARBITAL (SCHEDULE 2) | 2270 | 203 | 154,095.84 | 759.09 |
| COCAINE | 9041L | 19 | 107.71 | 5.67 |
| CODEINE | 9050 | 96 | 1,198.31 | 12.48 |
| BUPRENORPHINE | 9064 | 726 | 55.22 | 0.08 |
| OXYCODONE | 9143 | 124 | 806.31 | 6.5 |
| HYDROMORPHONE | 9150 | 541 | 330.75 | 0.61 |
| HYDROCODONE | 9193 | 392 | 1,140.92 | 2.91 |
| MEPERIDINE (PETHIDINE) | 9230 | 139 | 418.97 | 3.01 |
| METHADONE | 9250B | 32 | 38.82 | 1.21 |
| MORPHINE | 9300 | 352 | 614.79 | 1.75 |
| OPIUM POWDERED | 9639 | 18 | 107.28 | 5.96 |
| OXYMORPHONE | 9652 | 11 | 1.28 | 0.12 |
| ALFENTANIL | 9737 | 5 | 0.41 | 0.08 |
| REMIFENTANIL | 9739 | 20 | 1.15 | 0.06 |
| SUFENTANIL BASE | 9740 | 10 | 0.06 | 0.01 |
| TAPENTADOL | 9780 | 1 | 3 | 3 |
| FENTANYL BASE | 9801 | 583 | 79.92 | 0.14 |

STATE:COLORADO BUSINESS ACTIVITY:D - TEACHING INSTITUTIONS

| DRUG NAME | DRUG CODE | BUYERS | TOTAL GRAMS | AVG GRAMS |
|---|---|---|---|---|
| MORPHINE | 9300 | 1 | 0.19 | 0.19 |

STATE:COLORADO BUSINESS ACTIVITY:M - MID-LEVEL PRACTITIONERS

| DRUG NAME | DRUG CODE | BUYERS | TOTAL GRAMS | AVG GRAMS |
|---|---|---|---|---|
| PENTOBARBITAL (SCHEDULE 2) | 2270 | 17 | 24,086.02 | 1,416.82 |
| CODEINE | 9050 | 39 | 93.35 | 2.39 |
| OXYCODONE | 9143 | 1 | 0.9 | 0.9 |
| HYDROMORPHONE | 9150 | 7 | 0.76 | 0.11 |
| MORPHINE | 9300 | 30 | 10.51 | 0.35 |
| FENTANYL BASE | 9801 | 49 | 1.49 | 0.03 |

STATE:COLORADO BUSINESS ACTIVITY:N-U NARCOTIC TREATMENT PROGRAMS

| DRUG NAME | DRUG CODE | BUYERS | TOTAL GRAMS | AVG GRAMS |
|---|---|---|---|---|
| BUPRENORPHINE | 9064 | 12 | 426.79 | 35.57 |
| METHADONE | 9250B | 23 | 132,290.70 | 5,751.77 |

STATE:CONNECTICUT BUSINESS ACTIVITY:A - PHARMACIES

| DRUG NAME | DRUG CODE | BUYERS | TOTAL GRAMS | AVG GRAMS |
|---|---|---|---|---|
| AMPHETAMINE | 1100 | 728 | 206,885.22 | 284.18 |
| DL-METHAMPHETAMINE RACEMIC BASE | 1105B | 23 | 84.39 | 3.67 |
| D-METHAMPHETAMINE | 1105D | 1 | 0.4 | 0.4 |
| LISDEXAMFETAMINE | 1205 | 714 | 142,992.46 | 200.27 |
| METHYLPHENIDATE (DL;D;L;ISOMERS) | 1724 | 728 | 212,034.03 | 291.26 |
| BARBITURIC ACID DERIVIATIVE OR SALT [PER 21C | 2100 | 244 | 8,705 | 35.68 |
| BUTALBITAL | 2165 | 301 | 12,250 | 40.7 |
| SECOBARBITAL (SCHEDULE 2) | 2315 | 5 | 91.55 | 18.31 |

| | | | | |
|---|---|---|---|---|
| DRONABINOL IN AN ORAL SOLUTION IN FDA APPR | 7365 | 3 | 1.5 | 0.5 |
| NABILONE | 7379 | 3 | 0.35 | 0.12 |
| CODEINE | 9050 | 717 | 91,857.14 | 128.11 |
| BUPRENORPHINE | 9064 | 721 | 56,869.95 | 78.88 |
| DIHYDROCODEINE | 9120 | 1 | 2.14 | 2.14 |
| OXYCODONE | 9143 | 736 | 642,640.49 | 873.15 |
| HYDROMORPHONE | 9150 | 704 | 21,634.27 | 30.73 |
| HYDROCODONE | 9193 | 736 | 77,719.32 | 105.6 |
| LEVORPHANOL | 9220L | 52 | 20.08 | 0.39 |
| MEPERIDINE (PETHIDINE) | 9230 | 115 | 3,549.73 | 30.87 |
| METHADONE | 9250B | 548 | 22,552.56 | 41.15 |
| MORPHINE | 9300 | 699 | 128,704.46 | 184.13 |
| OPIUM TINCTURE (LAUDANUM) | 9630 | 1 | 4.73 | 4.73 |
| OPIUM POWDERED | 9639 | 65 | 267.84 | 4.12 |
| OXYMORPHONE | 9652 | 428 | 10,428.20 | 24.36 |
| SUFENTANIL BASE | 9740 | 1 | 1.8 | 1.8 |
| TAPENTADOL | 9780 | 494 | 45,647.50 | 92.4 |
| FENTANYL BASE | 9801 | 684 | 3,444.83 | 5.04 |
| OPIUM COMBINATION PRODUCT (C-III) | 9809 | 25 | 16.27 | 0.65 |

STATE:CONNECTICUT BUSINESS ACTIVITY:B - HOSPITALS

| DRUG NAME | DRUG CODE | BUYERS | TOTAL GRAMS | AVG GRAMS |
|---|---|---|---|---|
| AMPHETAMINE | 1100 | 40 | 4,376.06 | 109.4 |
| DL-METHAMPHETAMINE RACEMIC BASE | 1105B | 1 | 5.22 | 5.22 |
| D-METHAMPHETAMINE | 1105D | 1 | 7.63 | 7.63 |
| LISDEXAMFETAMINE | 1205 | 20 | 1,759.16 | 87.96 |
| METHYLPHENIDATE (DL;D;L;ISOMERS) | 1724 | 53 | 4,281.91 | 80.79 |
| BARBITURIC ACID DERIVIATIVE OR SALT [PER 21C | 2100 | 1 | 5 | 5 |
| AMOBARBITAL (SCHEDULE 2) | 2125 | 2 | 3.65 | 1.82 |
| BUTALBITAL | 2165 | 4 | 50 | 12.5 |
| PENTOBARBITAL (SCHEDULE 2) | 2270 | 6 | 498.99 | 83.17 |
| DRONABINOL IN AN ORAL SOLUTION IN FDA APPR | 7365 | 1 | 0.15 | 0.15 |
| COCAINE | 9041L | 25 | 128.35 | 5.13 |
| CODEINE | 9050 | 49 | 4,123.28 | 84.15 |
| BUPRENORPHINE | 9064 | 52 | 2,306.85 | 44.36 |

| | | | | |
|---|---|---|---|---|
| OXYCODONE | 9143 | 71 | 19,490.04 | 274.51 |
| HYDROMORPHONE | 9150 | 62 | 3,809.77 | 61.45 |
| HYDROCODONE | 9193 | 55 | 2,076.54 | 37.76 |
| LEVORPHANOL | 9220L | 1 | 0.12 | 0.12 |
| MEPERIDINE (PETHIDINE) | 9230 | 47 | 415.27 | 8.84 |
| METHADONE | 9250B | 49 | 4,550.39 | 92.87 |
| MORPHINE | 9300 | 62 | 12,266.95 | 197.85 |
| OPIUM POWDERED | 9639 | 32 | 116.64 | 3.65 |
| OXYMORPHONE | 9652 | 7 | 66.54 | 9.51 |
| ALFENTANIL | 9737 | 4 | 3.93 | 0.98 |
| REMIFENTANIL | 9739 | 24 | 33.4 | 1.39 |
| SUFENTANIL BASE | 9740 | 10 | 0.59 | 0.06 |
| TAPENTADOL | 9780 | 6 | 401.5 | 66.92 |
| FENTANYL BASE | 9801 | 64 | 323.04 | 5.05 |
| OPIUM COMBINATION PRODUCT (C-III) | 9809 | 3 | 3.22 | 1.07 |

STATE:CONNECTICUT BUSINESS ACTIVITY:C - PRACTITIONERS

| DRUG NAME | DRUG CODE | BUYERS | TOTAL GRAMS | AVG GRAMS |
|---|---|---|---|---|
| AMPHETAMINE | 1100 | 1 | 11.26 | 11.26 |
| PENTOBARBITAL (SCHEDULE 2) | 2270 | 85 | 71,953.07 | 846.51 |
| COCAINE | 9041L | 3 | 22.57 | 7.52 |
| CODEINE | 9050 | 75 | 325.73 | 4.34 |
| BUPRENORPHINE | 9064 | 321 | 23.26 | 0.07 |
| DIHYDROCODEINE | 9120 | 5 | 1.61 | 0.32 |
| OXYCODONE | 9143 | 56 | 1,330.12 | 23.75 |
| HYDROMORPHONE | 9150 | 173 | 131.72 | 0.76 |
| HYDROCODONE | 9193 | 105 | 563.54 | 5.37 |
| MEPERIDINE (PETHIDINE) | 9230 | 31 | 297.51 | 9.6 |
| METHADONE | 9250B | 20 | 72.85 | 3.64 |
| MORPHINE | 9300 | 118 | 483.51 | 4.1 |
| OPIUM POWDERED | 9639 | 2 | 0.72 | 0.36 |
| OXYMORPHONE | 9652 | 3 | 0.16 | 0.05 |
| ALFENTANIL | 9737 | 3 | 1.36 | 0.45 |
| REMIFENTANIL | 9739 | 8 | 0.66 | 0.08 |
| SUFENTANIL BASE | 9740 | 2 | 0 | 0 |

| | | | | |
|---|---|---|---|---|
| FENTANYL BASE | 9801 | 245 | 32.6 | 0.13 |

STATE:CONNECTICUT BUSINESS ACTIVITY:M - MID-LEVEL PRACTITIONERS

| DRUG NAME | DRUG CODE | BUYERS | TOTAL GRAMS | AVG GRAMS |
|---|---|---|---|---|
| CODEINE | 9050 | 7 | 10.01 | 1.43 |
| FENTANYL BASE | 9801 | 1 | 0.14 | 0.14 |

STATE:CONNECTICUT BUSINESS ACTIVITY:N-U NARCOTIC TREATMENT PROGRAMS

| DRUG NAME | DRUG CODE | BUYERS | TOTAL GRAMS | AVG GRAMS |
|---|---|---|---|---|
| BUPRENORPHINE | 9064 | 4 | 92.7 | 23.18 |
| METHADONE | 9250B | 41 | 449,228.95 | 10,956.80 |

STATE:DELAWARE BUSINESS ACTIVITY:A - PHARMACIES

| DRUG NAME | DRUG CODE | BUYERS | TOTAL GRAMS | AVG GRAMS |
|---|---|---|---|---|
| AMPHETAMINE | 1100 | 209 | 84,389.92 | 403.78 |
| DL-METHAMPHETAMINE RACEMIC BASE | 1105B | 3 | 15.27 | 5.09 |
| LISDEXAMFETAMINE | 1205 | 206 | 56,716.81 | 275.32 |
| METHYLPHENIDATE (DL;D;L;ISOMERS) | 1724 | 207 | 73,396.46 | 354.57 |
| BARBITURIC ACID DERIVIATIVE OR SALT [PER 21C | 2100 | 60 | 1,840 | 30.67 |
| BUTALBITAL | 2165 | 67 | 2,095 | 31.27 |
| SECOBARBITAL (SCHEDULE 2) | 2315 | 1 | 27.47 | 27.47 |
| DRONABINOL IN AN ORAL SOLUTION IN FDA APPR | 7365 | 1 | 0.15 | 0.15 |
| CODEINE | 9050 | 210 | 35,516.15 | 169.12 |
| BUPRENORPHINE | 9064 | 201 | 11,393.49 | 56.68 |
| DIHYDROCODEINE | 9120 | 16 | 29.91 | 1.87 |
| OXYCODONE | 9143 | 211 | 264,822.16 | 1,255.08 |
| HYDROMORPHONE | 9150 | 204 | 7,852.12 | 38.49 |
| HYDROCODONE | 9193 | 211 | 22,889.35 | 108.48 |
| LEVORPHANOL | 9220L | 4 | 2.55 | 0.64 |
| MEPERIDINE (PETHIDINE) | 9230 | 31 | 614.41 | 19.82 |
| METHADONE | 9250B | 168 | 11,380.97 | 67.74 |

| | | | | |
|---|---|---|---|---|
| MORPHINE | 9300 | 205 | 80,773.28 | 394.02 |
| OPIUM POWDERED | 9639 | 10 | 27.72 | 2.77 |
| OXYMORPHONE | 9652 | 151 | 7,688.79 | 50.92 |
| TAPENTADOL | 9780 | 175 | 29,534.50 | 168.77 |
| FENTANYL BASE | 9801 | 203 | 1,260.05 | 6.21 |
| OPIUM COMBINATION PRODUCT (C-III) | 9809 | 4 | 2.08 | 0.52 |

STATE:DELAWARE BUSINESS ACTIVITY:B - HOSPITALS

| DRUG NAME | DRUG CODE | BUYERS | TOTAL GRAMS | AVG GRAMS |
|---|---|---|---|---|
| AMPHETAMINE | 1100 | 14 | 1,811.98 | 129.43 |
| LISDEXAMFETAMINE | 1205 | 7 | 500.06 | 71.44 |
| METHYLPHENIDATE (DL;D;L;ISOMERS) | 1724 | 15 | 1,689.96 | 112.66 |
| BUTALBITAL | 2165 | 1 | 15 | 15 |
| PENTOBARBITAL (SCHEDULE 2) | 2270 | 2 | 402.84 | 201.42 |
| COCAINE | 9041L | 11 | 51.85 | 4.71 |
| CODEINE | 9050 | 19 | 1,255.38 | 66.07 |
| BUPRENORPHINE | 9064 | 18 | 811.51 | 45.08 |
| OXYCODONE | 9143 | 27 | 8,980.44 | 332.61 |
| HYDROMORPHONE | 9150 | 27 | 655.98 | 24.3 |
| HYDROCODONE | 9193 | 24 | 1,018.62 | 42.44 |
| MEPERIDINE (PETHIDINE) | 9230 | 16 | 252.63 | 15.79 |
| METHADONE | 9250B | 17 | 1,555.72 | 91.51 |
| MORPHINE | 9300 | 27 | 5,820.77 | 215.58 |
| OPIUM TINCTURE (LAUDANUM) | 9630 | 1 | 1.18 | 1.18 |
| OPIUM POWDERED | 9639 | 7 | 32.4 | 4.63 |
| OXYMORPHONE | 9652 | 5 | 36.84 | 7.37 |
| REMIFENTANIL | 9739 | 11 | 12.04 | 1.09 |
| SUFENTANIL BASE | 9740 | 3 | 0.03 | 0.01 |
| TAPENTADOL | 9780 | 4 | 139.5 | 34.88 |
| FENTANYL BASE | 9801 | 35 | 95.4 | 2.73 |

STATE:DELAWARE BUSINESS ACTIVITY:C - PRACTITIONERS

| DRUG NAME | DRUG CODE | BUYERS | TOTAL GRAMS | AVG GRAMS |
|---|---|---|---|---|

| | | | | |
|---|---|---|---|---|
| METHYLPHENIDATE (DL;D;L;ISOMERS) | 1724 | 1 | 9.34 | 9.34 |
| PENTOBARBITAL (SCHEDULE 2) | 2270 | 11 | 6,746.05 | 613.28 |
| CODEINE | 9050 | 26 | 209.22 | 8.05 |
| BUPRENORPHINE | 9064 | 57 | 3.79 | 0.07 |
| DIHYDROCODEINE | 9120 | 55 | 22.6 | 0.41 |
| OXYCODONE | 9143 | 5 | 3.85 | 0.77 |
| HYDROMORPHONE | 9150 | 32 | 14.27 | 0.45 |
| HYDROCODONE | 9193 | 22 | 52.08 | 2.37 |
| MEPERIDINE (PETHIDINE) | 9230 | 2 | 3.7 | 1.85 |
| METHADONE | 9250B | 4 | 1.99 | 0.5 |
| MORPHINE | 9300 | 12 | 50.5 | 4.21 |
| OXYMORPHONE | 9652 | 1 | 0.01 | 0.01 |
| FENTANYL BASE | 9801 | 54 | 4.35 | 0.08 |

STATE:DELAWARE BUSINESS ACTIVITY:M - MID-LEVEL PRACTITIONERS

| DRUG NAME | DRUG CODE | BUYERS | TOTAL GRAMS | AVG GRAMS |
|---|---|---|---|---|
| CODEINE | 9050 | 4 | 6.33 | 1.58 |
| DIHYDROCODEINE | 9120 | 7 | 2.01 | 0.29 |
| OXYCODONE | 9143 | 1 | 1.12 | 1.12 |
| HYDROMORPHONE | 9150 | 1 | 0.83 | 0.83 |
| METHADONE | 9250B | 1 | 0.63 | 0.63 |
| MORPHINE | 9300 | 1 | 42.98 | 42.98 |
| FENTANYL BASE | 9801 | 1 | 0.8 | 0.8 |

STATE:DELAWARE BUSINESS ACTIVITY:N-U NARCOTIC TREATMENT PROGRAMS

| DRUG NAME | DRUG CODE | BUYERS | TOTAL GRAMS | AVG GRAMS |
|---|---|---|---|---|
| BUPRENORPHINE | 9064 | 13 | 2,307.12 | 177.47 |
| METHADONE | 9250B | 12 | 97,054.03 | 8,087.84 |

STATE:DISTRICT OF COLUMBIA BUSINESS ACTIVITY:A - PHARMACIES

| DRUG NAME | DRUG CODE | BUYERS | TOTAL GRAMS | AVG GRAMS |
|---|---|---|---|---|

| | | | | |
|---|---|---|---|---|
| AMPHETAMINE | 1100 | 140 | 43,950.05 | 313.93 |
| DL-METHAMPHETAMINE RACEMIC BASE | 1105B | 6 | 6.83 | 1.14 |
| D-METHAMPHETAMINE | 1105D | 1 | 5.22 | 5.22 |
| LISDEXAMFETAMINE | 1205 | 129 | 20,641.06 | 160.01 |
| METHYLPHENIDATE (DL;D;L;ISOMERS) | 1724 | 141 | 28,623.51 | 203 |
| BARBITURIC ACID DERIVIATIVE OR SALT [PER 21C | 2100 | 44 | 970 | 22.05 |
| BUTALBITAL | 2165 | 57 | 1,390 | 24.39 |
| DRONABINOL IN AN ORAL SOLUTION IN FDA APPR | 7365 | 2 | 0.75 | 0.38 |
| CODEINE | 9050 | 150 | 46,132.75 | 307.55 |
| BUPRENORPHINE | 9064 | 130 | 9,305.34 | 71.58 |
| OXYCODONE | 9143 | 146 | 49,582.71 | 339.61 |
| HYDROMORPHONE | 9150 | 119 | 1,139.38 | 9.57 |
| HYDROCODONE | 9193 | 140 | 5,194.16 | 37.1 |
| MEPERIDINE (PETHIDINE) | 9230 | 16 | 472.03 | 29.5 |
| METHADONE | 9250B | 46 | 1,225.69 | 26.65 |
| MORPHINE | 9300 | 108 | 9,062.96 | 83.92 |
| OPIUM POWDERED | 9639 | 6 | 6.48 | 1.08 |
| OXYMORPHONE | 9652 | 28 | 458.81 | 16.39 |
| TAPENTADOL | 9780 | 51 | 3,937 | 77.2 |
| FENTANYL BASE | 9801 | 95 | 193.66 | 2.04 |
| OPIUM COMBINATION PRODUCT (C-III) | 9809 | 3 | 0.95 | 0.32 |

STATE:DISTRICT OF COLUMBIA BUSINESS ACTIVITY:B - HOSPITALS

| DRUG NAME | DRUG CODE | BUYERS | TOTAL GRAMS | AVG GRAMS |
|---|---|---|---|---|
| AMPHETAMINE | 1100 | 11 | 2,715.55 | 246.87 |
| LISDEXAMFETAMINE | 1205 | 4 | 368.83 | 92.21 |
| METHYLPHENIDATE (DL;D;L;ISOMERS) | 1724 | 16 | 3,576.50 | 223.53 |
| AMOBARBITAL (SCHEDULE 2) | 2125 | 3 | 6.84 | 2.28 |
| BUTALBITAL | 2165 | 1 | 10 | 10 |
| PENTOBARBITAL (SCHEDULE 2) | 2270 | 6 | 2,108.52 | 351.42 |
| COCAINE | 9041L | 6 | 92.89 | 15.48 |
| CODEINE | 9050 | 16 | 2,193.04 | 137.06 |
| BUPRENORPHINE | 9064 | 22 | 142.4 | 6.47 |
| OXYCODONE | 9143 | 20 | 8,564.29 | 428.21 |
| HYDROMORPHONE | 9150 | 24 | 1,026.06 | 42.75 |

| | | | | |
|---|---|---|---|---|
| HYDROCODONE | 9193 | 21 | 559.86 | 26.66 |
| MEPERIDINE (PETHIDINE) | 9230 | 10 | 252.74 | 25.27 |
| METHADONE | 9250B | 16 | 1,190.46 | 74.4 |
| MORPHINE | 9300 | 22 | 5,509.52 | 250.43 |
| OPIUM POWDERED | 9639 | 7 | 52.56 | 7.51 |
| OXYMORPHONE | 9652 | 4 | 20.45 | 5.11 |
| REMIFENTANIL | 9739 | 6 | 2.84 | 0.47 |
| SUFENTANIL BASE | 9740 | 3 | 0.03 | 0.01 |
| TAPENTADOL | 9780 | 3 | 159.5 | 53.17 |
| FENTANYL BASE | 9801 | 22 | 144.75 | 6.58 |
| OPIUM COMBINATION PRODUCT (C-III) | 9809 | 2 | 0.57 | 0.28 |

STATE:DISTRICT OF COLUMBIA BUSINESS ACTIVITY:C - PRACTITIONERS

| DRUG NAME | DRUG CODE | BUYERS | TOTAL GRAMS | AVG GRAMS |
|---|---|---|---|---|
| AMPHETAMINE | 1100 | 1 | 1.25 | 1.25 |
| PENTOBARBITAL (SCHEDULE 2) | 2270 | 3 | 444.31 | 148.1 |
| CODEINE | 9050 | 14 | 92.5 | 6.61 |
| BUPRENORPHINE | 9064 | 14 | 0.6 | 0.04 |
| OXYCODONE | 9143 | 5 | 8.97 | 1.79 |
| HYDROMORPHONE | 9150 | 3 | 1.88 | 0.63 |
| HYDROCODONE | 9193 | 6 | 12.41 | 2.07 |
| MEPERIDINE (PETHIDINE) | 9230 | 4 | 29.85 | 7.46 |
| MORPHINE | 9300 | 7 | 2.78 | 0.4 |
| FENTANYL BASE | 9801 | 14 | 0.31 | 0.02 |

STATE:DISTRICT OF COLUMBIA BUSINESS ACTIVITY:D - TEACHING INSTITUTIONS

| DRUG NAME | DRUG CODE | BUYERS | TOTAL GRAMS | AVG GRAMS |
|---|---|---|---|---|
| BUPRENORPHINE | 9064 | 1 | 0 | 0 |

STATE:DISTRICT OF COLUMBIA BUSINESS ACTIVITY:M - MID-LEVEL PRACTITIONERS

| DRUG NAME | DRUG CODE | BUYERS | TOTAL GRAMS | AVG GRAMS |
|---|---|---|---|---|

| | | | | |
|---|---|---|---|---|
| CODEINE | 9050 | 1 | 2.17 | 2.17 |
| FENTANYL BASE | 9801 | 1 | 0.06 | 0.06 |

STATE:DISTRICT OF COLUMBIA BUSINESS ACTIVITY:N-U NARCOTIC TREATMENT PROGRAMS

| DRUG NAME | DRUG CODE | BUYERS | TOTAL GRAMS | AVG GRAMS |
|---|---|---|---|---|
| BUPRENORPHINE | 9064 | 1 | 6 | 6 |
| METHADONE | 9250B | 5 | 37,493.25 | 7,498.65 |

STATE:FLORIDA BUSINESS ACTIVITY:A - PHARMACIES

| DRUG NAME | DRUG CODE | BUYERS | TOTAL GRAMS | AVG GRAMS |
|---|---|---|---|---|
| AMPHETAMINE | 1100 | 4,348 | 1,171,199.22 | 269.37 |
| DL-METHAMPHETAMINE RACEMIC BASE | 1105B | 77 | 130.2 | 1.69 |
| D-METHAMPHETAMINE | 1105D | 3 | 8.44 | 2.81 |
| LISDEXAMFETAMINE | 1205 | 3,969 | 541,680.86 | 136.48 |
| METHYLPHENIDATE (DL;D;L;ISOMERS) | 1724 | 4,198 | 791,875.97 | 188.63 |
| BARBITURIC ACID DERIVIATIVE OR SALT [PER 21C | 2100 | 1,908 | 69,705 | 36.53 |
| BUTALBITAL | 2165 | 1,769 | 65,295 | 36.91 |
| SECOBARBITAL (SCHEDULE 2) | 2315 | 14 | 366.2 | 26.16 |
| DRONABINOL IN AN ORAL SOLUTION IN FDA APPR | 7365 | 24 | 36.6 | 1.53 |
| NABILONE | 7379 | 12 | 2.15 | 0.18 |
| COCAINE | 9041L | 11 | 7.93 | 0.72 |
| CODEINE | 9050 | 4,405 | 656,120.95 | 148.95 |
| BUPRENORPHINE | 9064 | 3,988 | 178,910.18 | 44.86 |
| DIHYDROCODEINE | 9120 | 164 | 711.39 | 4.34 |
| OXYCODONE | 9143 | 4,526 | 3,961,522.67 | 875.28 |
| HYDROMORPHONE | 9150 | 4,091 | 171,984.53 | 42.04 |
| HYDROCODONE | 9193 | 4,474 | 1,376,414.11 | 307.65 |
| LEVORPHANOL | 9220L | 54 | 133.61 | 2.47 |
| MEPERIDINE (PETHIDINE) | 9230 | 1,353 | 23,971.15 | 17.72 |
| METHADONE | 9250B | 3,351 | 217,131.70 | 64.8 |
| MORPHINE | 9300 | 4,238 | 1,467,377.71 | 346.24 |
| OPIUM TINCTURE (LAUDANUM) | 9630 | 1 | 14.19 | 14.19 |

| | | | | |
|---|---|---|---|---|
| OPIUM POWDERED | 9639 | 209 | 538.56 | 2.58 |
| OXYMORPHONE | 9652 | 2,672 | 86,554.40 | 32.39 |
| NOROXYMORPHONE | 9668 | 1 | 1.56 | 1.56 |
| REMIFENTANIL | 9739 | 3 | 0.16 | 0.05 |
| SUFENTANIL BASE | 9740 | 5 | 9.79 | 1.96 |
| TAPENTADOL | 9780 | 2,883 | 390,040 | 135.29 |
| FENTANYL BASE | 9801 | 4,103 | 21,059.95 | 5.13 |
| OPIUM COMBINATION PRODUCT (C-III) | 9809 | 181 | 123.74 | 0.68 |

STATE:FLORIDA BUSINESS ACTIVITY:B - HOSPITALS

| DRUG NAME | DRUG CODE | BUYERS | TOTAL GRAMS | AVG GRAMS |
|---|---|---|---|---|
| AMPHETAMINE | 1100 | 189 | 29,995.22 | 158.7 |
| DL-METHAMPHETAMINE RACEMIC BASE | 1105B | 1 | 5.63 | 5.63 |
| LISDEXAMFETAMINE | 1205 | 87 | 7,035.48 | 80.87 |
| METHYLPHENIDATE (DL;D;L;ISOMERS) | 1724 | 278 | 30,350.09 | 109.17 |
| BARBITURIC ACID DERIVIATIVE OR SALT [PER 21C | 2100 | 12 | 140 | 11.67 |
| AMOBARBITAL (SCHEDULE 2) | 2125 | 4 | 23.24 | 5.81 |
| BUTALBITAL | 2165 | 21 | 355 | 16.9 |
| PENTOBARBITAL (SCHEDULE 2) | 2270 | 44 | 3,986.01 | 90.59 |
| COCAINE | 9041L | 241 | 1,103.64 | 4.58 |
| CODEINE | 9050 | 396 | 23,330.35 | 58.92 |
| BUPRENORPHINE | 9064 | 258 | 6,440.64 | 24.96 |
| OXYCODONE | 9143 | 574 | 139,094.80 | 242.33 |
| HYDROMORPHONE | 9150 | 591 | 14,398.63 | 24.36 |
| HYDROCODONE | 9193 | 576 | 60,189.11 | 104.49 |
| LEVORPHANOL | 9220L | 1 | 2.44 | 2.44 |
| MEPERIDINE (PETHIDINE) | 9230 | 499 | 9,182.78 | 18.4 |
| METHADONE | 9250B | 323 | 10,558.81 | 32.69 |
| MORPHINE | 9300 | 606 | 105,102.40 | 173.44 |
| OPIUM POWDERED | 9639 | 248 | 1,492.92 | 6.02 |
| OXYMORPHONE | 9652 | 30 | 319.06 | 10.64 |
| NOROXYMORPHONE | 9668 | 1 | 2.62 | 2.62 |
| ALFENTANIL | 9737 | 30 | 19.3 | 0.64 |
| REMIFENTANIL | 9739 | 173 | 106.67 | 0.62 |
| SUFENTANIL BASE | 9740 | 93 | 2.72 | 0.03 |

| | | | | |
|---|---|---|---|---|
| TAPENTADOL | 9780 | 90 | 6,217 | 69.08 |
| FENTANYL BASE | 9801 | 788 | 2,210.05 | 2.8 |
| OPIUM COMBINATION PRODUCT (C-III) | 9809 | 17 | 14 | 0.82 |

STATE:FLORIDA BUSINESS ACTIVITY:C - PRACTITIONERS

| DRUG NAME | DRUG CODE | BUYERS | TOTAL GRAMS | AVG GRAMS |
|---|---|---|---|---|
| AMPHETAMINE | 1100 | 7 | 10.85 | 1.55 |
| D-METHAMPHETAMINE | 1105D | 1 | 4.02 | 4.02 |
| LISDEXAMFETAMINE | 1205 | 7 | 84.08 | 12.01 |
| METHYLPHENIDATE (DL;D;L;ISOMERS) | 1724 | 7 | 547.59 | 78.23 |
| BUTALBITAL | 2165 | 1 | 70 | 70 |
| PENTOBARBITAL (SCHEDULE 2) | 2270 | 326 | 178,462.22 | 547.43 |
| COCAINE | 9041L | 23 | 50.53 | 2.2 |
| CODEINE | 9050 | 285 | 1,933.98 | 6.79 |
| BUPRENORPHINE | 9064 | 1,924 | 612.77 | 0.32 |
| DIHYDROCODEINE | 9120 | 123 | 51.4 | 0.42 |
| OXYCODONE | 9143 | 103 | 3,452 | 33.51 |
| HYDROMORPHONE | 9150 | 741 | 343.79 | 0.46 |
| HYDROCODONE | 9193 | 898 | 3,415.34 | 3.8 |
| MEPERIDINE (PETHIDINE) | 9230 | 291 | 2,660.58 | 9.14 |
| METHADONE | 9250B | 21 | 54.38 | 2.59 |
| MORPHINE | 9300 | 484 | 1,179.30 | 2.44 |
| OXYMORPHONE | 9652 | 8 | 3.9 | 0.49 |
| NOROXYMORPHONE | 9668 | 12 | 53.85 | 4.49 |
| ALFENTANIL | 9737 | 6 | 1.61 | 0.27 |
| REMIFENTANIL | 9739 | 14 | 0.61 | 0.04 |
| SUFENTANIL BASE | 9740 | 12 | 0.14 | 0.01 |
| FENTANYL BASE | 9801 | 1,225 | 82.71 | 0.07 |

STATE:FLORIDA BUSINESS ACTIVITY:M - MID-LEVEL PRACTITIONERS

| DRUG NAME | DRUG CODE | BUYERS | TOTAL GRAMS | AVG GRAMS |
|---|---|---|---|---|
| PENTOBARBITAL (SCHEDULE 2) | 2270 | 51 | 102,157.91 | 2,003.10 |
| CODEINE | 9050 | 2 | 2.17 | 1.08 |

| | | | | |
|---|---|---|---|---|
| BUPRENORPHINE | 9064 | 1 | 0.01 | 0.01 |
| DIHYDROCODEINE | 9120 | 3 | 1.13 | 0.38 |

STATE:FLORIDA BUSINESS ACTIVITY:N-U NARCOTIC TREATMENT PROGRAMS

| DRUG NAME | DRUG CODE | BUYERS | TOTAL GRAMS | AVG GRAMS |
|---|---|---|---|---|
| BUPRENORPHINE | 9064 | 53 | 1,689.09 | 31.87 |
| METHADONE | 9250B | 57 | 554,968.04 | 9,736.28 |

STATE:GEORGIA BUSINESS ACTIVITY:A - PHARMACIES

| DRUG NAME | DRUG CODE | BUYERS | TOTAL GRAMS | AVG GRAMS |
|---|---|---|---|---|
| AMPHETAMINE | 1100 | 2,239 | 731,893.92 | 326.88 |
| DL-METHAMPHETAMINE RACEMIC BASE | 1105B | 35 | 90.82 | 2.59 |
| D-METHAMPHETAMINE | 1105D | 1 | 4.82 | 4.82 |
| LISDEXAMFETAMINE | 1205 | 2,159 | 403,897.08 | 187.08 |
| METHYLPHENIDATE (DL;D;L;ISOMERS) | 1724 | 2,231 | 632,076.38 | 283.32 |
| BARBITURIC ACID DERIVIATIVE OR SALT [PER 21C | 2100 | 756 | 20,990 | 27.76 |
| BUTALBITAL | 2165 | 1,167 | 39,635 | 33.96 |
| SECOBARBITAL (SCHEDULE 2) | 2315 | 3 | 45.78 | 15.26 |
| DRONABINOL IN AN ORAL SOLUTION IN FDA APPR | 7365 | 8 | 4.35 | 0.54 |
| COCAINE | 9041L | 4 | 13.03 | 3.26 |
| CODEINE | 9050 | 2,245 | 402,112.23 | 179.11 |
| BUPRENORPHINE | 9064 | 2,097 | 59,649.42 | 28.45 |
| DIHYDROCODEINE | 9120 | 324 | 2,046.59 | 6.32 |
| OXYCODONE | 9143 | 2,262 | 1,715,676.50 | 758.48 |
| HYDROMORPHONE | 9150 | 2,016 | 28,005.87 | 13.89 |
| HYDROCODONE | 9193 | 2,280 | 932,795.26 | 409.12 |
| LEVORPHANOL | 9220L | 16 | 9.63 | 0.6 |
| MEPERIDINE (PETHIDINE) | 9230 | 1,329 | 31,934.59 | 24.03 |
| METHADONE | 9250B | 1,866 | 97,497.74 | 52.25 |
| MORPHINE | 9300 | 2,182 | 410,549.64 | 188.15 |
| OPIUM POWDERED | 9639 | 136 | 189.36 | 1.39 |
| OXYMORPHONE | 9652 | 1,260 | 25,730.22 | 20.42 |
| REMIFENTANIL | 9739 | 1 | 0.02 | 0.02 |

| | | | | |
|---|---|---|---|---|
| SUFENTANIL BASE | 9740 | 6 | 0.19 | 0.03 |
| TAPENTADOL | 9780 | 1,728 | 266,584 | 154.27 |
| FENTANYL BASE | 9801 | 2,152 | 9,589.62 | 4.46 |
| OPIUM COMBINATION PRODUCT (C-III) | 9809 | 62 | 28.19 | 0.45 |

STATE:GEORGIA BUSINESS ACTIVITY:B - HOSPITALS

| DRUG NAME | DRUG CODE | BUYERS | TOTAL GRAMS | AVG GRAMS |
|---|---|---|---|---|
| AMPHETAMINE | 1100 | 100 | 18,290.65 | 182.91 |
| DL-METHAMPHETAMINE RACEMIC BASE | 1105B | 1 | 4.02 | 4.02 |
| D-METHAMPHETAMINE | 1105D | 1 | 6.43 | 6.43 |
| LISDEXAMFETAMINE | 1205 | 55 | 5,669.43 | 103.08 |
| METHYLPHENIDATE (DL;D;L;ISOMERS) | 1724 | 145 | 21,622.98 | 149.12 |
| BARBITURIC ACID DERIVIATIVE OR SALT [PER 21C | 2100 | 5 | 30 | 6 |
| AMOBARBITAL (SCHEDULE 2) | 2125 | 5 | 26.43 | 5.29 |
| BUTALBITAL | 2165 | 12 | 165 | 13.75 |
| PENTOBARBITAL (SCHEDULE 2) | 2270 | 20 | 1,446.39 | 72.32 |
| DRONABINOL IN AN ORAL SOLUTION IN FDA APPR | 7365 | 1 | 0.3 | 0.3 |
| COCAINE | 9041L | 83 | 584.14 | 7.04 |
| CODEINE | 9050 | 183 | 13,325.50 | 72.82 |
| BUPRENORPHINE | 9064 | 107 | 1,662.74 | 15.54 |
| DIHYDROCODEINE | 9120 | 2 | 9.61 | 4.81 |
| OXYCODONE | 9143 | 228 | 63,289.58 | 277.59 |
| HYDROMORPHONE | 9150 | 203 | 5,072.56 | 24.99 |
| HYDROCODONE | 9193 | 227 | 46,932.24 | 206.75 |
| MEPERIDINE (PETHIDINE) | 9230 | 174 | 8,144.67 | 46.81 |
| METHADONE | 9250B | 162 | 3,924.45 | 24.22 |
| MORPHINE | 9300 | 225 | 36,615.88 | 162.74 |
| OPIUM POWDERED | 9639 | 90 | 535.68 | 5.95 |
| OXYMORPHONE | 9652 | 16 | 88.85 | 5.55 |
| NOROXYMORPHONE | 9668 | 2 | 5.42 | 2.71 |
| ALFENTANIL | 9737 | 12 | 9.41 | 0.78 |
| REMIFENTANIL | 9739 | 52 | 18.66 | 0.36 |
| SUFENTANIL BASE | 9740 | 38 | 1.43 | 0.04 |
| TAPENTADOL | 9780 | 54 | 3,041.50 | 56.32 |
| FENTANYL BASE | 9801 | 210 | 1,223.01 | 5.82 |

| | | | | |
|---|---|---|---|---|
| OPIUM COMBINATION PRODUCT (C-III) | 9809 | 4 | 1.32 | 0.33 |

STATE:GEORGIA BUSINESS ACTIVITY:C - PRACTITIONERS

| DRUG NAME | DRUG CODE | BUYERS | TOTAL GRAMS | AVG GRAMS |
|---|---|---|---|---|
| AMPHETAMINE | 1100 | 8 | 79.13 | 9.89 |
| LISDEXAMFETAMINE | 1205 | 1 | 7.49 | 7.49 |
| METHYLPHENIDATE (DL;D;L;ISOMERS) | 1724 | 2 | 5.96 | 2.98 |
| BARBITURIC ACID DERIVIATIVE OR SALT [PER 21C | 2100 | 1 | 5 | 5 |
| PENTOBARBITAL (SCHEDULE 2) | 2270 | 183 | 152,367.96 | 832.61 |
| COCAINE | 9041L | 26 | 59.71 | 2.3 |
| CODEINE | 9050 | 334 | 4,755.64 | 14.24 |
| BUPRENORPHINE | 9064 | 791 | 88.05 | 0.11 |
| DIHYDROCODEINE | 9120 | 207 | 107.3 | 0.52 |
| OXYCODONE | 9143 | 114 | 3,659.19 | 32.1 |
| HYDROMORPHONE | 9150 | 380 | 220.04 | 0.58 |
| HYDROCODONE | 9193 | 469 | 4,330.28 | 9.23 |
| MEPERIDINE (PETHIDINE) | 9230 | 244 | 1,511.99 | 6.2 |
| METHADONE | 9250B | 10 | 59.3 | 5.93 |
| MORPHINE | 9300 | 326 | 1,389.54 | 4.26 |
| OPIUM POWDERED | 9639 | 11 | 30.6 | 2.78 |
| OXYMORPHONE | 9652 | 6 | 1.67 | 0.28 |
| NOROXYMORPHONE | 9668 | 4 | 15.04 | 3.76 |
| ALFENTANIL | 9737 | 8 | 3.64 | 0.46 |
| REMIFENTANIL | 9739 | 6 | 0.53 | 0.09 |
| SUFENTANIL BASE | 9740 | 8 | 0.17 | 0.02 |
| TAPENTADOL | 9780 | 1 | 5 | 5 |
| FENTANYL BASE | 9801 | 807 | 95.25 | 0.12 |

STATE:GEORGIA BUSINESS ACTIVITY:D - TEACHING INSTITUTIONS

| DRUG NAME | DRUG CODE | BUYERS | TOTAL GRAMS | AVG GRAMS |
|---|---|---|---|---|
| BUPRENORPHINE | 9064 | 1 | 1.95 | 1.95 |

STATISTICAL SUMMARY FOR RETAIL DRUG PURCHASES BY GRAMS WT

STATE:GEORGIA BUSINESS ACTIVITY:M - MID-LEVEL PRACTITIONERS

| DRUG NAME | DRUG CODE | BUYERS | TOTAL GRAMS | AVG GRAMS |
|---|---|---|---|---|
| CODEINE | 9050 | 54 | 71.47 | 1.32 |
| DIHYDROCODEINE | 9120 | 48 | 34.51 | 0.72 |

STATE:GEORGIA BUSINESS ACTIVITY:N-U NARCOTIC TREATMENT PROGRAMS

| DRUG NAME | DRUG CODE | BUYERS | TOTAL GRAMS | AVG GRAMS |
|---|---|---|---|---|
| BUPRENORPHINE | 9064 | 32 | 1,469.25 | 45.91 |
| METHADONE | 9250B | 73 | 364,117.40 | 4,987.91 |

STATE:GUAM BUSINESS ACTIVITY:A - PHARMACIES

| DRUG NAME | DRUG CODE | BUYERS | TOTAL GRAMS | AVG GRAMS |
|---|---|---|---|---|
| AMPHETAMINE | 1100 | 9 | 1,083.51 | 120.39 |
| LISDEXAMFETAMINE | 1205 | 4 | 30.06 | 7.52 |
| METHYLPHENIDATE (DL;D;L;ISOMERS) | 1724 | 9 | 667.16 | 74.13 |
| BARBITURIC ACID DERIVIATIVE OR SALT [PER 21C | 2100 | 1 | 10 | 10 |
| BUTALBITAL | 2165 | 2 | 40 | 20 |
| CODEINE | 9050 | 17 | 3,972.65 | 233.69 |
| BUPRENORPHINE | 9064 | 3 | 3.84 | 1.28 |
| OXYCODONE | 9143 | 11 | 2,489.10 | 226.28 |
| HYDROMORPHONE | 9150 | 6 | 83.16 | 13.86 |
| HYDROCODONE | 9193 | 11 | 385.19 | 35.02 |
| MEPERIDINE (PETHIDINE) | 9230 | 3 | 57.52 | 19.17 |
| METHADONE | 9250B | 3 | 150.26 | 50.09 |
| MORPHINE | 9300 | 8 | 3,329.71 | 416.21 |
| FENTANYL BASE | 9801 | 7 | 22.44 | 3.21 |

STATE:GUAM BUSINESS ACTIVITY:B - HOSPITALS

| DRUG NAME | DRUG CODE | BUYERS | TOTAL GRAMS | AVG GRAMS |
|---|---|---|---|---|
| AMPHETAMINE | 1100 | 4 | 820.6 | 205.15 |

| | | | | |
|---|---|---|---|---|
| LISDEXAMFETAMINE | 1205 | 3 | 60.7 | 20.23 |
| METHYLPHENIDATE (DL;D;L;ISOMERS) | 1724 | 6 | 1,247.59 | 207.93 |
| COCAINE | 9041L | 1 | 0.36 | 0.36 |
| CODEINE | 9050 | 6 | 775.34 | 129.22 |
| BUPRENORPHINE | 9064 | 2 | 0.48 | 0.24 |
| OXYCODONE | 9143 | 7 | 932.27 | 133.18 |
| HYDROMORPHONE | 9150 | 8 | 31.28 | 3.91 |
| HYDROCODONE | 9193 | 5 | 231.17 | 46.23 |
| MEPERIDINE (PETHIDINE) | 9230 | 6 | 77.24 | 12.87 |
| METHADONE | 9250B | 3 | 21.02 | 7.01 |
| MORPHINE | 9300 | 7 | 668.1 | 95.44 |
| OPIUM POWDERED | 9639 | 1 | 0.36 | 0.36 |
| REMIFENTANIL | 9739 | 2 | 0.31 | 0.16 |
| FENTANYL BASE | 9801 | 7 | 18.65 | 2.66 |

STATE:GUAM BUSINESS ACTIVITY:C - PRACTITIONERS

| DRUG NAME | DRUG CODE | BUYERS | TOTAL GRAMS | AVG GRAMS |
|---|---|---|---|---|
| CODEINE | 9050 | 4 | 17.68 | 4.42 |
| BUPRENORPHINE | 9064 | 2 | 0.17 | 0.08 |
| HYDROMORPHONE | 9150 | 1 | 0.07 | 0.07 |
| HYDROCODONE | 9193 | 4 | 13.62 | 3.41 |
| MEPERIDINE (PETHIDINE) | 9230 | 3 | 48.91 | 16.3 |
| MORPHINE | 9300 | 3 | 0.44 | 0.15 |
| ALFENTANIL | 9737 | 1 | 0.08 | 0.08 |
| FENTANYL BASE | 9801 | 4 | 0.24 | 0.06 |

STATE:GUAM BUSINESS ACTIVITY:M - MID-LEVEL PRACTITIONERS

| DRUG NAME | DRUG CODE | BUYERS | TOTAL GRAMS | AVG GRAMS |
|---|---|---|---|---|
| PENTOBARBITAL (SCHEDULE 2) | 2270 | 1 | 6,430.84 | 6,430.84 |

STATE:HAWAII BUSINESS ACTIVITY:A - PHARMACIES

| DRUG NAME | DRUG CODE | BUYERS | TOTAL GRAMS | AVG GRAMS |
|---|---|---|---|---|
| AMPHETAMINE | 1100 | 221 | 30,177.86 | 136.55 |
| DL-METHAMPHETAMINE RACEMIC BASE | 1105B | 8 | 20.49 | 2.56 |
| D-METHAMPHETAMINE | 1105D | 1 | 3.21 | 3.21 |
| LISDEXAMFETAMINE | 1205 | 156 | 4,142.66 | 26.56 |
| METHYLPHENIDATE (DL;D;L;ISOMERS) | 1724 | 222 | 33,018.17 | 148.73 |
| BARBITURIC ACID DERIVIATIVE OR SALT [PER 21C | 2100 | 77 | 3,380 | 43.9 |
| BUTALBITAL | 2165 | 92 | 4,160 | 45.22 |
| SECOBARBITAL (SCHEDULE 2) | 2315 | 1 | 9.16 | 9.16 |
| COCAINE | 9041L | 3 | 12.43 | 4.14 |
| CODEINE | 9050 | 229 | 49,304.87 | 215.31 |
| BUPRENORPHINE | 9064 | 203 | 7,598.52 | 37.43 |
| DIHYDROCODEINE | 9120 | 1 | 1.07 | 1.07 |
| OXYCODONE | 9143 | 231 | 153,634.24 | 665.08 |
| HYDROMORPHONE | 9150 | 162 | 2,063.28 | 12.74 |
| HYDROCODONE | 9193 | 232 | 62,805.34 | 270.71 |
| MEPERIDINE (PETHIDINE) | 9230 | 37 | 897.86 | 24.27 |
| METHADONE | 9250B | 156 | 12,734.74 | 81.63 |
| MORPHINE | 9300 | 211 | 78,654.33 | 372.77 |
| OPIUM POWDERED | 9639 | 8 | 15.48 | 1.94 |
| OXYMORPHONE | 9652 | 79 | 1,679.02 | 21.25 |
| REMIFENTANIL | 9739 | 1 | 0.03 | 0.03 |
| SUFENTANIL BASE | 9740 | 1 | 0 | 0 |
| TAPENTADOL | 9780 | 95 | 11,776 | 123.96 |
| FENTANYL BASE | 9801 | 188 | 571.74 | 3.04 |
| OPIUM COMBINATION PRODUCT (C-III) | 9809 | 1 | 4.73 | 4.73 |

STATE:HAWAII BUSINESS ACTIVITY:B - HOSPITALS

| DRUG NAME | DRUG CODE | BUYERS | TOTAL GRAMS | AVG GRAMS |
|---|---|---|---|---|
| AMPHETAMINE | 1100 | 17 | 8,742.21 | 514.25 |
| LISDEXAMFETAMINE | 1205 | 10 | 1,215.74 | 121.57 |
| METHYLPHENIDATE (DL;D;L;ISOMERS) | 1724 | 23 | 10,618.14 | 461.66 |
| PENTOBARBITAL (SCHEDULE 2) | 2270 | 5 | 601.52 | 120.3 |
| NABILONE | 7379 | 2 | 0.6 | 0.3 |
| COCAINE | 9041L | 14 | 83.28 | 5.95 |

| | | | | |
|---|---|---|---|---|
| CODEINE | 9050 | 37 | 4,838.22 | 130.76 |
| BUPRENORPHINE | 9064 | 20 | 275.83 | 13.79 |
| OXYCODONE | 9143 | 40 | 18,191.54 | 454.79 |
| HYDROMORPHONE | 9150 | 35 | 580.01 | 16.57 |
| HYDROCODONE | 9193 | 40 | 5,322.58 | 133.06 |
| MEPERIDINE (PETHIDINE) | 9230 | 30 | 291.41 | 9.71 |
| METHADONE | 9250B | 29 | 1,138.30 | 39.25 |
| MORPHINE | 9300 | 42 | 8,119.21 | 193.31 |
| OPIUM POWDERED | 9639 | 14 | 97.56 | 6.97 |
| OXYMORPHONE | 9652 | 6 | 462.55 | 77.09 |
| ALFENTANIL | 9737 | 5 | 2.08 | 0.42 |
| REMIFENTANIL | 9739 | 10 | 5.11 | 0.51 |
| SUFENTANIL BASE | 9740 | 5 | 0.03 | 0.01 |
| TAPENTADOL | 9780 | 9 | 818 | 90.89 |
| FENTANYL BASE | 9801 | 47 | 166.95 | 3.55 |

STATE:HAWAII BUSINESS ACTIVITY:C - PRACTITIONERS

| DRUG NAME | DRUG CODE | BUYERS | TOTAL GRAMS | AVG GRAMS |
|---|---|---|---|---|
| PENTOBARBITAL (SCHEDULE 2) | 2270 | 17 | 27,342.46 | 1,608.38 |
| COCAINE | 9041L | 1 | 1.71 | 1.71 |
| CODEINE | 9050 | 9 | 419.65 | 46.63 |
| BUPRENORPHINE | 9064 | 77 | 4.98 | 0.06 |
| OXYCODONE | 9143 | 16 | 1,604.96 | 100.31 |
| HYDROMORPHONE | 9150 | 48 | 27.93 | 0.58 |
| HYDROCODONE | 9193 | 55 | 1,757.61 | 31.96 |
| MEPERIDINE (PETHIDINE) | 9230 | 18 | 167.98 | 9.33 |
| METHADONE | 9250B | 2 | 5.19 | 2.59 |
| MORPHINE | 9300 | 26 | 384.52 | 14.79 |
| OPIUM POWDERED | 9639 | 1 | 2.16 | 2.16 |
| OXYMORPHONE | 9652 | 2 | 4.68 | 2.34 |
| ALFENTANIL | 9737 | 2 | 0.32 | 0.16 |
| FENTANYL BASE | 9801 | 49 | 3.62 | 0.07 |

STATE:HAWAII BUSINESS ACTIVITY:M - MID-LEVEL PRACTITIONERS

| DRUG NAME | DRUG CODE | BUYERS | TOTAL GRAMS | AVG GRAMS |
|---|---|---|---|---|
| MORPHINE | 9300 | 1 | 0.94 | 0.94 |
| FENTANYL BASE | 9801 | 1 | 0.01 | 0.01 |

STATE:HAWAII BUSINESS ACTIVITY:N-U NARCOTIC TREATMENT PROGRAMS

| DRUG NAME | DRUG CODE | BUYERS | TOTAL GRAMS | AVG GRAMS |
|---|---|---|---|---|
| METHADONE | 9250B | 4 | 18,417.48 | 4,604.37 |

STATE:IDAHO BUSINESS ACTIVITY:A - PHARMACIES

| DRUG NAME | DRUG CODE | BUYERS | TOTAL GRAMS | AVG GRAMS |
|---|---|---|---|---|
| AMPHETAMINE | 1100 | 309 | 100,261.34 | 324.47 |
| DL-METHAMPHETAMINE RACEMIC BASE | 1105B | 14 | 34.96 | 2.5 |
| D-METHAMPHETAMINE | 1105D | 1 | 10.44 | 10.44 |
| LISDEXAMFETAMINE | 1205 | 297 | 35,229.99 | 118.62 |
| METHYLPHENIDATE (DL;D;L;ISOMERS) | 1724 | 309 | 107,761.78 | 348.74 |
| BARBITURIC ACID DERIVIATIVE OR SALT [PER 21C | 2100 | 63 | 2,150 | 34.13 |
| BUTALBITAL | 2165 | 136 | 5,245 | 38.57 |
| COCAINE | 9041L | 3 | 64.17 | 21.39 |
| CODEINE | 9050 | 309 | 49,961.71 | 161.69 |
| BUPRENORPHINE | 9064 | 300 | 13,653.30 | 45.51 |
| OXYCODONE | 9143 | 311 | 229,195.06 | 736.96 |
| HYDROMORPHONE | 9150 | 300 | 6,605.30 | 22.02 |
| HYDROCODONE | 9193 | 315 | 217,517.68 | 690.53 |
| LEVORPHANOL | 9220L | 2 | 0.23 | 0.12 |
| MEPERIDINE (PETHIDINE) | 9230 | 108 | 2,233.22 | 20.68 |
| METHADONE | 9250B | 299 | 24,522.39 | 82.01 |
| MORPHINE | 9300 | 315 | 102,543.09 | 325.53 |
| OPIUM POWDERED | 9639 | 26 | 34.92 | 1.34 |
| OXYMORPHONE | 9652 | 211 | 5,131.85 | 24.32 |
| REMIFENTANIL | 9739 | 1 | 0.02 | 0.02 |
| TAPENTADOL | 9780 | 228 | 23,937.50 | 104.99 |
| FENTANYL BASE | 9801 | 305 | 2,056.93 | 6.74 |

| | | | | |
|---|---|---|---|---|
| OPIUM COMBINATION PRODUCT (C-III) | 9809 | 10 | 14.95 | 1.49 |

STATE:IDAHO BUSINESS ACTIVITY:B - HOSPITALS

| DRUG NAME | DRUG CODE | BUYERS | TOTAL GRAMS | AVG GRAMS |
|---|---|---|---|---|
| AMPHETAMINE | 1100 | 25 | 1,503.22 | 60.13 |
| LISDEXAMFETAMINE | 1205 | 10 | 167.07 | 16.71 |
| METHYLPHENIDATE (DL;D;L;ISOMERS) | 1724 | 32 | 1,825.33 | 57.04 |
| BARBITURIC ACID DERIVIATIVE OR SALT [PER 21C | 2100 | 2 | 15 | 7.5 |
| PENTOBARBITAL (SCHEDULE 2) | 2270 | 6 | 201.42 | 33.57 |
| COCAINE | 9041L | 23 | 143.81 | 6.25 |
| CODEINE | 9050 | 44 | 1,995.30 | 45.35 |
| BUPRENORPHINE | 9064 | 35 | 423.07 | 12.09 |
| OXYCODONE | 9143 | 64 | 12,004.85 | 187.58 |
| HYDROMORPHONE | 9150 | 56 | 873.17 | 15.59 |
| HYDROCODONE | 9193 | 66 | 10,088.70 | 152.86 |
| MEPERIDINE (PETHIDINE) | 9230 | 44 | 846.47 | 19.24 |
| METHADONE | 9250B | 46 | 1,411.81 | 30.69 |
| MORPHINE | 9300 | 62 | 9,750.10 | 157.26 |
| OPIUM POWDERED | 9639 | 24 | 160.56 | 6.69 |
| OXYMORPHONE | 9652 | 4 | 12.22 | 3.06 |
| ALFENTANIL | 9737 | 7 | 5.34 | 0.76 |
| REMIFENTANIL | 9739 | 18 | 8.31 | 0.46 |
| SUFENTANIL BASE | 9740 | 20 | 0.17 | 0.01 |
| TAPENTADOL | 9780 | 8 | 189 | 23.63 |
| FENTANYL BASE | 9801 | 61 | 228.26 | 3.74 |
| OPIUM COMBINATION PRODUCT (C-III) | 9809 | 1 | 0.19 | 0.19 |

STATE:IDAHO BUSINESS ACTIVITY:C - PRACTITIONERS

| DRUG NAME | DRUG CODE | BUYERS | TOTAL GRAMS | AVG GRAMS |
|---|---|---|---|---|
| PENTOBARBITAL (SCHEDULE 2) | 2270 | 82 | 77,336.64 | 943.13 |
| COCAINE | 9041L | 2 | 8.57 | 4.29 |
| CODEINE | 9050 | 40 | 2,557.93 | 63.95 |
| BUPRENORPHINE | 9064 | 126 | 15.12 | 0.12 |

| | | | | |
|---|---|---|---|---|
| DIHYDROCODEINE | 9120 | 5 | 108.95 | 21.79 |
| OXYCODONE | 9143 | 17 | 153.34 | 9.02 |
| HYDROMORPHONE | 9150 | 91 | 47.24 | 0.52 |
| HYDROCODONE | 9193 | 105 | 449.37 | 4.28 |
| MEPERIDINE (PETHIDINE) | 9230 | 26 | 243.04 | 9.35 |
| METHADONE | 9250B | 2 | 11.9 | 5.95 |
| MORPHINE | 9300 | 80 | 233.3 | 2.92 |
| OXYMORPHONE | 9652 | 4 | 1.01 | 0.25 |
| ALFENTANIL | 9737 | 1 | 0.11 | 0.11 |
| REMIFENTANIL | 9739 | 4 | 0.36 | 0.09 |
| SUFENTANIL BASE | 9740 | 1 | 0 | 0 |
| FENTANYL BASE | 9801 | 127 | 19.42 | 0.15 |

STATE:IDAHO BUSINESS ACTIVITY:M - MID-LEVEL PRACTITIONERS

| DRUG NAME | DRUG CODE | BUYERS | TOTAL GRAMS | AVG GRAMS |
|---|---|---|---|---|
| PENTOBARBITAL (SCHEDULE 2) | 2270 | 11 | 12,911.35 | 1,173.76 |
| CODEINE | 9050 | 4 | 128.73 | 32.18 |
| HYDROMORPHONE | 9150 | 1 | 0.09 | 0.09 |
| HYDROCODONE | 9193 | 3 | 3.3 | 1.1 |
| MEPERIDINE (PETHIDINE) | 9230 | 3 | 7.3 | 2.43 |
| MORPHINE | 9300 | 3 | 0.34 | 0.11 |
| FENTANYL BASE | 9801 | 5 | 0.3 | 0.06 |

STATE:IDAHO BUSINESS ACTIVITY:N-U NARCOTIC TREATMENT PROGRAMS

| DRUG NAME | DRUG CODE | BUYERS | TOTAL GRAMS | AVG GRAMS |
|---|---|---|---|---|
| BUPRENORPHINE | 9064 | 3 | 124.55 | 41.52 |
| METHADONE | 9250B | 3 | 21,286.72 | 7,095.57 |

STATE:ILLINOIS BUSINESS ACTIVITY:A - PHARMACIES

| DRUG NAME | DRUG CODE | BUYERS | TOTAL GRAMS | AVG GRAMS |
|---|---|---|---|---|
| AMPHETAMINE | 1100 | 2,181 | 755,498.65 | 346.4 |

| | | | | |
|---|---|---|---|---|
| DL-METHAMPHETAMINE RACEMIC BASE | 1105B | 81 | 246.74 | 3.05 |
| D-METHAMPHETAMINE | 1105D | 2 | 5.62 | 2.81 |
| LISDEXAMFETAMINE | 1205 | 1,958 | 281,275.71 | 143.65 |
| METHYLPHENIDATE (DL;D;L;ISOMERS) | 1724 | 2,162 | 686,506.49 | 317.53 |
| BARBITURIC ACID DERIVIATIVE OR SALT [PER 21C | 2100 | 823 | 30,160 | 36.65 |
| BUTALBITAL | 2165 | 753 | 29,480 | 39.15 |
| SECOBARBITAL (SCHEDULE 2) | 2315 | 7 | 164.79 | 23.54 |
| DRONABINOL IN AN ORAL SOLUTION IN FDA APPR | 7365 | 6 | 1.05 | 0.18 |
| NABILONE | 7379 | 3 | 0.5 | 0.17 |
| COCAINE | 9041L | 5 | 42.82 | 8.56 |
| CODEINE | 9050 | 2,244 | 630,936.51 | 281.17 |
| BUPRENORPHINE | 9064 | 1,932 | 54,012.46 | 27.96 |
| DIHYDROCODEINE | 9120 | 16 | 121.77 | 7.61 |
| OXYCODONE | 9143 | 2,137 | 605,547.95 | 283.36 |
| HYDROMORPHONE | 9150 | 1,883 | 31,714.94 | 16.84 |
| HYDROCODONE | 9193 | 2,252 | 1,167,638.05 | 518.49 |
| LEVORPHANOL | 9220L | 48 | 52.7 | 1.1 |
| MEPERIDINE (PETHIDINE) | 9230 | 244 | 6,430.25 | 26.35 |
| METHADONE | 9250B | 1,508 | 48,612.25 | 32.24 |
| MORPHINE | 9300 | 2,041 | 370,205.33 | 181.38 |
| OPIUM TINCTURE (LAUDANUM) | 9630 | 3 | 16.55 | 5.52 |
| OPIUM POWDERED | 9639 | 119 | 174.96 | 1.47 |
| OXYMORPHONE | 9652 | 975 | 15,986.43 | 16.4 |
| ALFENTANIL | 9737 | 1 | 0.05 | 0.05 |
| REMIFENTANIL | 9739 | 1 | 0.01 | 0.01 |
| TAPENTADOL | 9780 | 1,342 | 150,273 | 111.98 |
| FENTANYL BASE | 9801 | 2,038 | 10,531.53 | 5.17 |
| OPIUM COMBINATION PRODUCT (C-III) | 9809 | 37 | 22.33 | 0.6 |

STATE:ILLINOIS BUSINESS ACTIVITY:B - HOSPITALS

| DRUG NAME | DRUG CODE | BUYERS | TOTAL GRAMS | AVG GRAMS |
|---|---|---|---|---|
| AMPHETAMINE | 1100 | 132 | 8,461.82 | 64.1 |
| DL-METHAMPHETAMINE RACEMIC BASE | 1105B | 1 | 1.61 | 1.61 |
| D-METHAMPHETAMINE | 1105D | 1 | 0.4 | 0.4 |
| LISDEXAMFETAMINE | 1205 | 53 | 2,164.41 | 40.84 |

| | | | | |
|---|---|---|---|---|
| METHYLPHENIDATE (DL;D;L;ISOMERS) | 1724 | 188 | 9,748.73 | 51.85 |
| GAMMA HYDROXYBUTYRIC ACID(FDA APPROVED | 2012 | 1 | 2,674.94 | 2,674.94 |
| BARBITURIC ACID DERIVIATIVE OR SALT [PER 21C | 2100 | 9 | 75 | 8.33 |
| AMOBARBITAL (SCHEDULE 2) | 2125 | 9 | 15.95 | 1.77 |
| BUTALBITAL | 2165 | 16 | 180 | 11.25 |
| PENTOBARBITAL (SCHEDULE 2) | 2270 | 27 | 2,103.51 | 77.91 |
| COCAINE | 9041L | 133 | 881.76 | 6.63 |
| CODEINE | 9050 | 233 | 12,116.88 | 52 |
| BUPRENORPHINE | 9064 | 138 | 1,562.62 | 11.32 |
| OXYCODONE | 9143 | 229 | 26,695.81 | 116.58 |
| HYDROMORPHONE | 9150 | 228 | 6,838.18 | 29.99 |
| HYDROCODONE | 9193 | 248 | 59,244.22 | 238.89 |
| MEPERIDINE (PETHIDINE) | 9230 | 184 | 4,303.10 | 23.39 |
| METHADONE | 9250B | 177 | 6,321.13 | 35.71 |
| MORPHINE | 9300 | 235 | 38,564.19 | 164.1 |
| OPIUM POWDERED | 9639 | 129 | 537.12 | 4.16 |
| OXYMORPHONE | 9652 | 27 | 311.1 | 11.52 |
| ALFENTANIL | 9737 | 23 | 8.2 | 0.36 |
| REMIFENTANIL | 9739 | 83 | 60.32 | 0.73 |
| SUFENTANIL BASE | 9740 | 75 | 2.28 | 0.03 |
| TAPENTADOL | 9780 | 47 | 1,420.50 | 30.22 |
| FENTANYL BASE | 9801 | 236 | 1,186.29 | 5.03 |
| OPIUM COMBINATION PRODUCT (C-III) | 9809 | 6 | 1.51 | 0.25 |

STATE:ILLINOIS BUSINESS ACTIVITY:C - PRACTITIONERS

| DRUG NAME | DRUG CODE | BUYERS | TOTAL GRAMS | AVG GRAMS |
|---|---|---|---|---|
| AMPHETAMINE | 1100 | 2 | 6.69 | 3.34 |
| METHYLPHENIDATE (DL;D;L;ISOMERS) | 1724 | 1 | 50.28 | 50.28 |
| PENTOBARBITAL (SCHEDULE 2) | 2270 | 384 | 243,551.43 | 634.25 |
| COCAINE | 9041L | 40 | 365.55 | 9.14 |
| CODEINE | 9050 | 342 | 4,194.42 | 12.26 |
| BUPRENORPHINE | 9064 | 847 | 62.58 | 0.07 |
| DIHYDROCODEINE | 9120 | 15 | 549.33 | 36.62 |
| OXYCODONE | 9143 | 56 | 369.72 | 6.6 |
| HYDROMORPHONE | 9150 | 456 | 340.08 | 0.75 |

| | | | | |
|---|---|---|---|---|
| HYDROCODONE | 9193 | 588 | 4,300.34 | 7.31 |
| MEPERIDINE (PETHIDINE) | 9230 | 148 | 1,277.40 | 8.63 |
| METHADONE | 9250B | 20 | 39.98 | 2 |
| MORPHINE | 9300 | 274 | 564.98 | 2.06 |
| OPIUM POWDERED | 9639 | 8 | 5.4 | 0.68 |
| OXYMORPHONE | 9652 | 1 | 0.03 | 0.03 |
| NOROXYMORPHONE | 9668 | 4 | 38.02 | 9.5 |
| ALFENTANIL | 9737 | 8 | 8.25 | 1.03 |
| REMIFENTANIL | 9739 | 10 | 0.43 | 0.04 |
| SUFENTANIL BASE | 9740 | 2 | 0.01 | 0 |
| TAPENTADOL | 9780 | 1 | 5 | 5 |
| FENTANYL BASE | 9801 | 641 | 88.23 | 0.14 |

STATE:ILLINOIS BUSINESS ACTIVITY:D - TEACHING INSTITUTIONS

| DRUG NAME | DRUG CODE | BUYERS | TOTAL GRAMS | AVG GRAMS |
|---|---|---|---|---|
| PENTOBARBITAL (SCHEDULE 2) | 2270 | 3 | 10,397.71 | 3,465.90 |
| CODEINE | 9050 | 1 | 2.39 | 2.39 |
| BUPRENORPHINE | 9064 | 2 | 0.73 | 0.36 |
| HYDROMORPHONE | 9150 | 1 | 12 | 12 |
| HYDROCODONE | 9193 | 1 | 11.69 | 11.69 |
| METHADONE | 9250B | 1 | 10.73 | 10.73 |
| MORPHINE | 9300 | 1 | 19.93 | 19.93 |
| FENTANYL BASE | 9801 | 2 | 3.1 | 1.55 |

STATE:ILLINOIS BUSINESS ACTIVITY:M - MID-LEVEL PRACTITIONERS

| DRUG NAME | DRUG CODE | BUYERS | TOTAL GRAMS | AVG GRAMS |
|---|---|---|---|---|
| PENTOBARBITAL (SCHEDULE 2) | 2270 | 15 | 21,209.19 | 1,413.95 |
| CODEINE | 9050 | 12 | 27.94 | 2.33 |

STATE:ILLINOIS BUSINESS ACTIVITY:N-U NARCOTIC TREATMENT PROGRAMS

| DRUG NAME | DRUG CODE | BUYERS | TOTAL GRAMS | AVG GRAMS |
|---|---|---|---|---|

| | | | | |
|---|---|---|---|---|
| BUPRENORPHINE | 9064 | 11 | 1,889.50 | 171.77 |
| METHADONE | 9250B | 78 | 434,440.04 | 5,569.74 |

STATE:INDIANA BUSINESS ACTIVITY:A - PHARMACIES

| DRUG NAME | DRUG CODE | BUYERS | TOTAL GRAMS | AVG GRAMS |
|---|---|---|---|---|
| AMPHETAMINE | 1100 | 1,186 | 511,273.61 | 431.09 |
| DL-METHAMPHETAMINE RACEMIC BASE | 1105B | 27 | 103.68 | 3.84 |
| LISDEXAMFETAMINE | 1205 | 1,168 | 337,130.58 | 288.64 |
| METHYLPHENIDATE (DL;D;L;ISOMERS) | 1724 | 1,188 | 477,422.05 | 401.87 |
| BARBITURIC ACID DERIVIATIVE OR SALT [PER 21C | 2100 | 443 | 13,115 | 29.6 |
| BUTALBITAL | 2165 | 460 | 15,355 | 33.38 |
| DRONABINOL IN AN ORAL SOLUTION IN FDA APPR | 7365 | 3 | 1.65 | 0.55 |
| COCAINE | 9041L | 1 | 8.93 | 8.93 |
| CODEINE | 9050 | 1,162 | 195,023.17 | 167.83 |
| BUPRENORPHINE | 9064 | 1,134 | 75,445.03 | 66.53 |
| DIHYDROCODEINE | 9120 | 6 | 7.48 | 1.25 |
| OXYCODONE | 9143 | 1,194 | 886,532.77 | 742.49 |
| HYDROMORPHONE | 9150 | 1,051 | 18,399.72 | 17.51 |
| HYDROCODONE | 9193 | 1,204 | 861,296.73 | 715.36 |
| LEVORPHANOL | 9220L | 40 | 97.28 | 2.43 |
| MEPERIDINE (PETHIDINE) | 9230 | 334 | 5,546.23 | 16.61 |
| METHADONE | 9250B | 1,012 | 66,355.89 | 65.57 |
| MORPHINE | 9300 | 1,168 | 314,519.94 | 269.28 |
| OPIUM TINCTURE (LAUDANUM) | 9630 | 6 | 35.47 | 5.91 |
| OPIUM POWDERED | 9639 | 99 | 274.68 | 2.77 |
| OXYMORPHONE | 9652 | 780 | 19,363.57 | 24.83 |
| REMIFENTANIL | 9739 | 1 | 0.12 | 0.12 |
| TAPENTADOL | 9780 | 891 | 148,182 | 166.31 |
| FENTANYL BASE | 9801 | 1,144 | 6,586.88 | 5.76 |
| OPIUM COMBINATION PRODUCT (C-III) | 9809 | 20 | 20.62 | 1.03 |

STATE:INDIANA BUSINESS ACTIVITY:B - HOSPITALS

| DRUG NAME | DRUG CODE | BUYERS | TOTAL GRAMS | AVG GRAMS |
|---|---|---|---|---|

| | | | | |
|---|---|---|---|---|
| AMPHETAMINE | 1100 | 126 | 8,717.01 | 69.18 |
| LISDEXAMFETAMINE | 1205 | 74 | 4,238.63 | 57.28 |
| METHYLPHENIDATE (DL;D;L;ISOMERS) | 1724 | 146 | 7,870.13 | 53.91 |
| BARBITURIC ACID DERIVIATIVE OR SALT [PER 21C | 2100 | 6 | 45 | 7.5 |
| AMOBARBITAL (SCHEDULE 2) | 2125 | 2 | 4.1 | 2.05 |
| BUTALBITAL | 2165 | 21 | 240 | 11.43 |
| PENTOBARBITAL (SCHEDULE 2) | 2270 | 18 | 10,266.37 | 570.35 |
| COCAINE | 9041L | 107 | 790.84 | 7.39 |
| CODEINE | 9050 | 172 | 4,943.54 | 28.74 |
| BUPRENORPHINE | 9064 | 120 | 1,982.98 | 16.52 |
| OXYCODONE | 9143 | 241 | 37,689.78 | 156.39 |
| HYDROMORPHONE | 9150 | 239 | 5,318.78 | 22.25 |
| HYDROCODONE | 9193 | 261 | 42,959.24 | 164.59 |
| LEVORPHANOL | 9220L | 1 | 0.12 | 0.12 |
| MEPERIDINE (PETHIDINE) | 9230 | 220 | 7,708.61 | 35.04 |
| METHADONE | 9250B | 138 | 2,621.52 | 19 |
| MORPHINE | 9300 | 260 | 29,019.77 | 111.61 |
| OPIUM POWDERED | 9639 | 130 | 707.04 | 5.44 |
| OXYMORPHONE | 9652 | 22 | 104.44 | 4.75 |
| ALFENTANIL | 9737 | 37 | 9.96 | 0.27 |
| REMIFENTANIL | 9739 | 82 | 31.76 | 0.39 |
| SUFENTANIL BASE | 9740 | 50 | 1.12 | 0.02 |
| TAPENTADOL | 9780 | 36 | 1,249.50 | 34.71 |
| FENTANYL BASE | 9801 | 292 | 883.3 | 3.02 |
| OPIUM COMBINATION PRODUCT (C-III) | 9809 | 2 | 8.51 | 4.26 |

STATE:INDIANA BUSINESS ACTIVITY:C - PRACTITIONERS

| DRUG NAME | DRUG CODE | BUYERS | TOTAL GRAMS | AVG GRAMS |
|---|---|---|---|---|
| METHYLPHENIDATE (DL;D;L;ISOMERS) | 1724 | 1 | 6.92 | 6.92 |
| PENTOBARBITAL (SCHEDULE 2) | 2270 | 215 | 153,774.86 | 715.23 |
| COCAINE | 9041L | 3 | 1.14 | 0.38 |
| CODEINE | 9050 | 91 | 456.12 | 5.01 |
| BUPRENORPHINE | 9064 | 438 | 42.05 | 0.1 |
| OXYCODONE | 9143 | 16 | 85.64 | 5.35 |
| HYDROMORPHONE | 9150 | 233 | 110.41 | 0.47 |

| | | | | |
|---|---|---|---|---|
| HYDROCODONE | 9193 | 258 | 651.26 | 2.52 |
| MEPERIDINE (PETHIDINE) | 9230 | 60 | 170.81 | 2.85 |
| METHADONE | 9250B | 4 | 3.76 | 0.94 |
| MORPHINE | 9300 | 143 | 250.19 | 1.75 |
| OXYMORPHONE | 9652 | 2 | 0.05 | 0.03 |
| ALFENTANIL | 9737 | 3 | 0.33 | 0.11 |
| REMIFENTANIL | 9739 | 1 | 0.05 | 0.05 |
| SUFENTANIL BASE | 9740 | 3 | 0.03 | 0.01 |
| FENTANYL BASE | 9801 | 257 | 53.39 | 0.21 |

STATE:INDIANA BUSINESS ACTIVITY:M - MID-LEVEL PRACTITIONERS

| DRUG NAME | DRUG CODE | BUYERS | TOTAL GRAMS | AVG GRAMS |
|---|---|---|---|---|
| PENTOBARBITAL (SCHEDULE 2) | 2270 | 24 | 15,743.43 | 655.98 |
| CODEINE | 9050 | 18 | 42.53 | 2.36 |
| HYDROCODONE | 9193 | 1 | 0.61 | 0.61 |

STATE:INDIANA BUSINESS ACTIVITY:N-U NARCOTIC TREATMENT PROGRAMS

| DRUG NAME | DRUG CODE | BUYERS | TOTAL GRAMS | AVG GRAMS |
|---|---|---|---|---|
| BUPRENORPHINE | 9064 | 9 | 2,174.67 | 241.63 |
| METHADONE | 9250B | 14 | 257,953.90 | 18,425.28 |

STATE:IOWA BUSINESS ACTIVITY:A - PHARMACIES

| DRUG NAME | DRUG CODE | BUYERS | TOTAL GRAMS | AVG GRAMS |
|---|---|---|---|---|
| AMPHETAMINE | 1100 | 688 | 194,438.69 | 282.61 |
| DL-METHAMPHETAMINE RACEMIC BASE | 1105B | 21 | 73.94 | 3.52 |
| LISDEXAMFETAMINE | 1205 | 672 | 135,650.01 | 201.86 |
| METHYLPHENIDATE (DL;D;L;ISOMERS) | 1724 | 694 | 318,706.20 | 459.23 |
| BARBITURIC ACID DERIVIATIVE OR SALT [PER 21C | 2100 | 104 | 2,780 | 26.73 |
| BUTALBITAL | 2165 | 192 | 5,090 | 26.51 |
| NABILONE | 7379 | 1 | 0.1 | 0.1 |
| COCAINE | 9041L | 2 | 0.43 | 0.21 |

| | | | | |
|---|---|---|---|---|
| CODEINE | 9050 | 689 | 76,566.52 | 111.13 |
| BUPRENORPHINE | 9064 | 541 | 5,967.20 | 11.03 |
| DIHYDROCODEINE | 9120 | 2 | 2.14 | 1.07 |
| OXYCODONE | 9143 | 696 | 199,787.63 | 287.05 |
| HYDROMORPHONE | 9150 | 649 | 9,520.96 | 14.67 |
| HYDROCODONE | 9193 | 707 | 214,597.76 | 303.53 |
| LEVORPHANOL | 9220L | 17 | 15.32 | 0.9 |
| MEPERIDINE (PETHIDINE) | 9230 | 123 | 2,008.59 | 16.33 |
| METHADONE | 9250B | 543 | 16,170.48 | 29.78 |
| MORPHINE | 9300 | 691 | 119,127.13 | 172.4 |
| OPIUM POWDERED | 9639 | 75 | 104.76 | 1.4 |
| OXYMORPHONE | 9652 | 265 | 4,048.04 | 15.28 |
| SUFENTANIL BASE | 9740 | 1 | 0.01 | 0.01 |
| TAPENTADOL | 9780 | 436 | 48,801 | 111.93 |
| FENTANYL BASE | 9801 | 678 | 3,141.82 | 4.63 |
| OPIUM COMBINATION PRODUCT (C-III) | 9809 | 6 | 3.97 | 0.66 |

STATE:IOWA BUSINESS ACTIVITY:B - HOSPITALS

| DRUG NAME | DRUG CODE | BUYERS | TOTAL GRAMS | AVG GRAMS |
|---|---|---|---|---|
| AMPHETAMINE | 1100 | 48 | 5,331.42 | 111.07 |
| LISDEXAMFETAMINE | 1205 | 36 | 1,989.82 | 55.27 |
| METHYLPHENIDATE (DL;D;L;ISOMERS) | 1724 | 84 | 7,370.22 | 87.74 |
| AMOBARBITAL (SCHEDULE 2) | 2125 | 1 | 0.46 | 0.46 |
| BUTALBITAL | 2165 | 2 | 10 | 5 |
| PENTOBARBITAL (SCHEDULE 2) | 2270 | 8 | 1,193.48 | 149.18 |
| COCAINE | 9041L | 68 | 354.69 | 5.22 |
| CODEINE | 9050 | 114 | 3,015.04 | 26.45 |
| BUPRENORPHINE | 9064 | 55 | 303.55 | 5.52 |
| OXYCODONE | 9143 | 143 | 13,814.30 | 96.6 |
| HYDROMORPHONE | 9150 | 143 | 2,335.97 | 16.34 |
| HYDROCODONE | 9193 | 149 | 14,670.64 | 98.46 |
| LEVORPHANOL | 9220L | 1 | 0.12 | 0.12 |
| MEPERIDINE (PETHIDINE) | 9230 | 125 | 4,288.46 | 34.31 |
| METHADONE | 9250B | 74 | 1,252.52 | 16.93 |
| MORPHINE | 9300 | 147 | 14,705.59 | 100.04 |

| | | | | |
|---|---|---|---|---|
| OPIUM POWDERED | 9639 | 64 | 296.64 | 4.64 |
| OXYMORPHONE | 9652 | 12 | 59.18 | 4.93 |
| ALFENTANIL | 9737 | 22 | 6.37 | 0.29 |
| REMIFENTANIL | 9739 | 23 | 12.58 | 0.55 |
| SUFENTANIL BASE | 9740 | 27 | 0.19 | 0.01 |
| TAPENTADOL | 9780 | 31 | 900 | 29.03 |
| FENTANYL BASE | 9801 | 151 | 310.3 | 2.05 |
| OPIUM COMBINATION PRODUCT (C-III) | 9809 | 1 | 4.92 | 4.92 |

STATE:IOWA BUSINESS ACTIVITY:C - PRACTITIONERS

| DRUG NAME | DRUG CODE | BUYERS | TOTAL GRAMS | AVG GRAMS |
|---|---|---|---|---|
| PENTOBARBITAL (SCHEDULE 2) | 2270 | 98 | 67,910.41 | 692.96 |
| COCAINE | 9041L | 5 | 37.89 | 7.58 |
| CODEINE | 9050 | 21 | 314.59 | 14.98 |
| BUPRENORPHINE | 9064 | 234 | 11.94 | 0.05 |
| OXYCODONE | 9143 | 9 | 221.75 | 24.64 |
| HYDROMORPHONE | 9150 | 95 | 35.09 | 0.37 |
| HYDROCODONE | 9193 | 103 | 408.91 | 3.97 |
| MEPERIDINE (PETHIDINE) | 9230 | 30 | 1,483.84 | 49.46 |
| METHADONE | 9250B | 1 | 0.54 | 0.54 |
| MORPHINE | 9300 | 72 | 141.84 | 1.97 |
| OPIUM POWDERED | 9639 | 2 | 1.08 | 0.54 |
| ALFENTANIL | 9737 | 1 | 0.31 | 0.31 |
| SUFENTANIL BASE | 9740 | 1 | 0.01 | 0.01 |
| FENTANYL BASE | 9801 | 94 | 24.42 | 0.26 |

STATE:IOWA BUSINESS ACTIVITY:D - TEACHING INSTITUTIONS

| DRUG NAME | DRUG CODE | BUYERS | TOTAL GRAMS | AVG GRAMS |
|---|---|---|---|---|
| HYDROMORPHONE | 9150 | 2 | 0.14 | 0.07 |
| MORPHINE | 9300 | 1 | 0.38 | 0.38 |

STATE:IOWA BUSINESS ACTIVITY:M - MID-LEVEL PRACTITIONERS

| DRUG NAME | DRUG CODE | BUYERS | TOTAL GRAMS | AVG GRAMS |
|---|---|---|---|---|
| PENTOBARBITAL (SCHEDULE 2) | 2270 | 5 | 2,399.26 | 479.85 |
| FENTANYL BASE | 9801 | 1 | 0.01 | 0.01 |

STATE:IOWA BUSINESS ACTIVITY:N-U NARCOTIC TREATMENT PROGRAMS

| DRUG NAME | DRUG CODE | BUYERS | TOTAL GRAMS | AVG GRAMS |
|---|---|---|---|---|
| BUPRENORPHINE | 9064 | 6 | 447.66 | 74.61 |
| METHADONE | 9250B | 8 | 34,847.61 | 4,355.95 |

STATE:KANSAS BUSINESS ACTIVITY:A - PHARMACIES

| DRUG NAME | DRUG CODE | BUYERS | TOTAL GRAMS | AVG GRAMS |
|---|---|---|---|---|
| AMPHETAMINE | 1100 | 623 | 232,401.60 | 373.04 |
| DL-METHAMPHETAMINE RACEMIC BASE | 1105B | 10 | 39.78 | 3.98 |
| D-METHAMPHETAMINE | 1105D | 2 | 1.61 | 0.8 |
| LISDEXAMFETAMINE | 1205 | 609 | 125,099.11 | 205.42 |
| METHYLPHENIDATE (DL;D;L;ISOMERS) | 1724 | 625 | 190,841.84 | 305.35 |
| BARBITURIC ACID DERIVIATIVE OR SALT [PER 21C | 2100 | 142 | 4,815 | 33.91 |
| BUTALBITAL | 2165 | 240 | 8,710 | 36.29 |
| SECOBARBITAL (SCHEDULE 2) | 2315 | 3 | 100.71 | 33.57 |
| DRONABINOL IN AN ORAL SOLUTION IN FDA APPR | 7365 | 1 | 0.6 | 0.6 |
| COCAINE | 9041L | 3 | 19.43 | 6.48 |
| CODEINE | 9050 | 623 | 105,267.54 | 168.97 |
| BUPRENORPHINE | 9064 | 521 | 9,622.72 | 18.47 |
| DIHYDROCODEINE | 9120 | 3 | 18.16 | 6.05 |
| OXYCODONE | 9143 | 631 | 456,787.68 | 723.91 |
| HYDROMORPHONE | 9150 | 585 | 11,704.80 | 20.01 |
| HYDROCODONE | 9193 | 633 | 305,595.23 | 482.77 |
| LEVORPHANOL | 9220L | 11 | 35.87 | 3.26 |
| MEPERIDINE (PETHIDINE) | 9230 | 228 | 4,910.79 | 21.54 |
| METHADONE | 9250B | 529 | 28,409.27 | 53.7 |
| MORPHINE | 9300 | 626 | 159,707.27 | 255.12 |
| OPIUM TINCTURE (LAUDANUM) | 9630 | 2 | 5.91 | 2.96 |

| | | | | |
|---|---|---|---|---|
| OPIUM POWDERED | 9639 | 85 | 317.16 | 3.73 |
| OXYMORPHONE | 9652 | 340 | 7,318.65 | 21.53 |
| SUFENTANIL BASE | 9740 | 4 | 0.34 | 0.08 |
| TAPENTADOL | 9780 | 369 | 33,927.50 | 91.94 |
| FENTANYL BASE | 9801 | 619 | 3,897.10 | 6.3 |
| OPIUM COMBINATION PRODUCT (C-III) | 9809 | 11 | 2.84 | 0.26 |

STATE:KANSAS BUSINESS ACTIVITY:B - HOSPITALS

| DRUG NAME | DRUG CODE | BUYERS | TOTAL GRAMS | AVG GRAMS |
|---|---|---|---|---|
| AMPHETAMINE | 1100 | 55 | 9,356.34 | 170.12 |
| DL-METHAMPHETAMINE RACEMIC BASE | 1105B | 2 | 2.01 | 1 |
| D-METHAMPHETAMINE | 1105D | 1 | 2.81 | 2.81 |
| LISDEXAMFETAMINE | 1205 | 27 | 2,053.41 | 76.05 |
| METHYLPHENIDATE (DL;D;L;ISOMERS) | 1724 | 90 | 9,402.80 | 104.48 |
| BARBITURIC ACID DERIVIATIVE OR SALT [PER 21C | 2100 | 1 | 5 | 5 |
| AMOBARBITAL (SCHEDULE 2) | 2125 | 3 | 13.67 | 4.56 |
| BUTALBITAL | 2165 | 1 | 95 | 95 |
| PENTOBARBITAL (SCHEDULE 2) | 2270 | 8 | 9,444.84 | 1,180.60 |
| COCAINE | 9041L | 52 | 284.55 | 5.47 |
| CODEINE | 9050 | 137 | 5,718 | 41.74 |
| BUPRENORPHINE | 9064 | 40 | 376.44 | 9.41 |
| OXYCODONE | 9143 | 190 | 31,366.19 | 165.09 |
| HYDROMORPHONE | 9150 | 164 | 2,218.75 | 13.53 |
| HYDROCODONE | 9193 | 195 | 18,835.04 | 96.59 |
| MEPERIDINE (PETHIDINE) | 9230 | 149 | 2,073.32 | 13.91 |
| METHADONE | 9250B | 85 | 1,836.20 | 21.6 |
| MORPHINE | 9300 | 195 | 17,866.07 | 91.62 |
| OPIUM POWDERED | 9639 | 76 | 402.84 | 5.3 |
| OXYMORPHONE | 9652 | 13 | 191.58 | 14.74 |
| ALFENTANIL | 9737 | 15 | 5.78 | 0.39 |
| REMIFENTANIL | 9739 | 27 | 8.67 | 0.32 |
| SUFENTANIL BASE | 9740 | 51 | 0.82 | 0.02 |
| TAPENTADOL | 9780 | 31 | 1,114.50 | 35.95 |
| FENTANYL BASE | 9801 | 201 | 555.23 | 2.76 |

STATISTICAL SUMMARY FOR RETAIL DRUG PURCHASES BY GRAMS WT

STATE:KANSAS BUSINESS ACTIVITY:C - PRACTITIONERS

| DRUG NAME | DRUG CODE | BUYERS | TOTAL GRAMS | AVG GRAMS |
|---|---|---|---|---|
| AMPHETAMINE | 1100 | 1 | 0.74 | 0.74 |
| METHYLPHENIDATE (DL;D;L;ISOMERS) | 1724 | 1 | 8.65 | 8.65 |
| PENTOBARBITAL (SCHEDULE 2) | 2270 | 124 | 81,045.33 | 653.59 |
| COCAINE | 9041L | 1 | 24.64 | 24.64 |
| CODEINE | 9050 | 21 | 810.36 | 38.59 |
| BUPRENORPHINE | 9064 | 256 | 42.53 | 0.17 |
| OXYCODONE | 9143 | 12 | 343.56 | 28.63 |
| HYDROMORPHONE | 9150 | 110 | 53.59 | 0.49 |
| HYDROCODONE | 9193 | 95 | 240.89 | 2.54 |
| MEPERIDINE (PETHIDINE) | 9230 | 18 | 86.93 | 4.83 |
| MORPHINE | 9300 | 60 | 93.3 | 1.55 |
| OXYMORPHONE | 9652 | 1 | 0.81 | 0.81 |
| NOROXYMORPHONE | 9668 | 1 | 2.75 | 2.75 |
| ALFENTANIL | 9737 | 1 | 0.2 | 0.2 |
| REMIFENTANIL | 9739 | 1 | 0.4 | 0.4 |
| SUFENTANIL BASE | 9740 | 3 | 0.01 | 0 |
| FENTANYL BASE | 9801 | 106 | 9.2 | 0.09 |

STATE:KANSAS BUSINESS ACTIVITY:D - TEACHING INSTITUTIONS

| DRUG NAME | DRUG CODE | BUYERS | TOTAL GRAMS | AVG GRAMS |
|---|---|---|---|---|
| BUPRENORPHINE | 9064 | 1 | 0 | 0 |

STATE:KANSAS BUSINESS ACTIVITY:M - MID-LEVEL PRACTITIONERS

| DRUG NAME | DRUG CODE | BUYERS | TOTAL GRAMS | AVG GRAMS |
|---|---|---|---|---|
| HYDROMORPHONE | 9150 | 13 | 0.73 | 0.06 |
| MORPHINE | 9300 | 24 | 7.38 | 0.31 |
| FENTANYL BASE | 9801 | 35 | 0.43 | 0.01 |

STATE:KANSAS BUSINESS ACTIVITY:N-U NARCOTIC TREATMENT PROGRAMS

| DRUG NAME | DRUG CODE | BUYERS | TOTAL GRAMS | AVG GRAMS |
|---|---|---|---|---|
| BUPRENORPHINE | 9064 | 6 | 181.86 | 30.31 |
| METHADONE | 9250B | 9 | 62,200.15 | 6,911.13 |

STATE:KENTUCKY BUSINESS ACTIVITY:A - PHARMACIES

| DRUG NAME | DRUG CODE | BUYERS | TOTAL GRAMS | AVG GRAMS |
|---|---|---|---|---|
| AMPHETAMINE | 1100 | 1,110 | 263,851.55 | 237.7 |
| DL-METHAMPHETAMINE RACEMIC BASE | 1105B | 22 | 39.78 | 1.81 |
| D-METHAMPHETAMINE | 1105D | 1 | 3.62 | 3.62 |
| LISDEXAMFETAMINE | 1205 | 1,049 | 167,557.66 | 159.73 |
| METHYLPHENIDATE (DL;D;L;ISOMERS) | 1724 | 1,107 | 291,149.77 | 263.01 |
| BARBITURIC ACID DERIVIATIVE OR SALT [PER 21C | 2100 | 224 | 7,244 | 32.34 |
| BUTALBITAL | 2165 | 358 | 10,350 | 28.91 |
| DRONABINOL IN AN ORAL SOLUTION IN FDA APPR | 7365 | 3 | 1.5 | 0.5 |
| NABILONE | 7379 | 1 | 0.35 | 0.35 |
| COCAINE | 9041L | 2 | 6.18 | 3.09 |
| CODEINE | 9050 | 1,088 | 184,573.48 | 169.64 |
| BUPRENORPHINE | 9064 | 1,023 | 136,014.80 | 132.96 |
| DIHYDROCODEINE | 9120 | 21 | 84.38 | 4.02 |
| OXYCODONE | 9143 | 1,124 | 781,156.07 | 694.98 |
| HYDROMORPHONE | 9150 | 836 | 8,689.11 | 10.39 |
| HYDROCODONE | 9193 | 1,138 | 770,019.86 | 676.64 |
| LEVORPHANOL | 9220L | 1 | 0.35 | 0.35 |
| MEPERIDINE (PETHIDINE) | 9230 | 293 | 4,457.29 | 15.21 |
| METHADONE | 9250B | 912 | 48,033.66 | 52.67 |
| MORPHINE | 9300 | 1,090 | 164,328.23 | 150.76 |
| OPIUM TINCTURE (LAUDANUM) | 9630 | 1 | 2.36 | 2.36 |
| OPIUM POWDERED | 9639 | 107 | 305.64 | 2.86 |
| OXYMORPHONE | 9652 | 655 | 14,388.81 | 21.97 |
| REMIFENTANIL | 9739 | 1 | 0.02 | 0.02 |
| SUFENTANIL BASE | 9740 | 1 | 0 | 0 |
| TAPENTADOL | 9780 | 629 | 57,822 | 91.93 |
| FENTANYL BASE | 9801 | 1,051 | 3,919.27 | 3.73 |

| | | | | |
|---|---|---|---|---|
| OPIUM COMBINATION PRODUCT (C-III) | 9809 | 39 | 34.43 | 0.88 |

STATE:KENTUCKY BUSINESS ACTIVITY:B - HOSPITALS

| DRUG NAME | DRUG CODE | BUYERS | TOTAL GRAMS | AVG GRAMS |
|---|---|---|---|---|
| AMPHETAMINE | 1100 | 51 | 8,096.12 | 158.75 |
| LISDEXAMFETAMINE | 1205 | 26 | 2,073.60 | 79.75 |
| METHYLPHENIDATE (DL;D;L;ISOMERS) | 1724 | 83 | 9,018.09 | 108.65 |
| BARBITURIC ACID DERIVIATIVE OR SALT [PER 21C | 2100 | 3 | 15 | 5 |
| AMOBARBITAL (SCHEDULE 2) | 2125 | 3 | 19.14 | 6.38 |
| BUTALBITAL | 2165 | 3 | 15 | 5 |
| PENTOBARBITAL (SCHEDULE 2) | 2270 | 6 | 1,496.97 | 249.5 |
| COCAINE | 9041L | 71 | 426.08 | 6 |
| CODEINE | 9050 | 129 | 4,860.51 | 37.68 |
| BUPRENORPHINE | 9064 | 83 | 1,205.18 | 14.52 |
| OXYCODONE | 9143 | 142 | 34,425.58 | 242.43 |
| HYDROMORPHONE | 9150 | 129 | 2,830.51 | 21.94 |
| HYDROCODONE | 9193 | 145 | 28,967.44 | 199.78 |
| MEPERIDINE (PETHIDINE) | 9230 | 122 | 4,403.01 | 36.09 |
| METHADONE | 9250B | 92 | 2,853.98 | 31.02 |
| MORPHINE | 9300 | 139 | 20,441.54 | 147.06 |
| OPIUM POWDERED | 9639 | 66 | 374.4 | 5.67 |
| OXYMORPHONE | 9652 | 20 | 74.31 | 3.72 |
| ALFENTANIL | 9737 | 10 | 4.13 | 0.41 |
| REMIFENTANIL | 9739 | 35 | 19 | 0.54 |
| SUFENTANIL BASE | 9740 | 33 | 1.35 | 0.04 |
| TAPENTADOL | 9780 | 20 | 2,312 | 115.6 |
| FENTANYL BASE | 9801 | 139 | 610.82 | 4.39 |
| OPIUM COMBINATION PRODUCT (C-III) | 9809 | 7 | 2.64 | 0.38 |

STATE:KENTUCKY BUSINESS ACTIVITY:C - PRACTITIONERS

| DRUG NAME | DRUG CODE | BUYERS | TOTAL GRAMS | AVG GRAMS |
|---|---|---|---|---|
| PENTOBARBITAL (SCHEDULE 2) | 2270 | 126 | 90,647.63 | 719.43 |
| COCAINE | 9041L | 3 | 2.14 | 0.71 |

| | | | | |
|---|---|---|---|---|
| CODEINE | 9050 | 14 | 682.08 | 48.72 |
| BUPRENORPHINE | 9064 | 282 | 25.32 | 0.09 |
| DIHYDROCODEINE | 9120 | 1 | 1.12 | 1.12 |
| OXYCODONE | 9143 | 13 | 100.12 | 7.7 |
| HYDROMORPHONE | 9150 | 91 | 273.71 | 3.01 |
| HYDROCODONE | 9193 | 119 | 581.51 | 4.89 |
| MEPERIDINE (PETHIDINE) | 9230 | 43 | 481.39 | 11.2 |
| METHADONE | 9250B | 1 | 0.89 | 0.89 |
| MORPHINE | 9300 | 136 | 673.02 | 4.95 |
| OPIUM POWDERED | 9639 | 1 | 16.56 | 16.56 |
| ALFENTANIL | 9737 | 2 | 12.3 | 6.15 |
| REMIFENTANIL | 9739 | 2 | 0.14 | 0.07 |
| SUFENTANIL BASE | 9740 | 2 | 0.16 | 0.08 |
| FENTANYL BASE | 9801 | 260 | 36.91 | 0.14 |

STATE:KENTUCKY BUSINESS ACTIVITY:D - TEACHING INSTITUTIONS

| DRUG NAME | DRUG CODE | BUYERS | TOTAL GRAMS | AVG GRAMS |
|---|---|---|---|---|
| PENTOBARBITAL (SCHEDULE 2) | 2270 | 1 | 88.86 | 88.86 |
| BUPRENORPHINE | 9064 | 1 | 0 | 0 |

STATE:KENTUCKY BUSINESS ACTIVITY:M - MID-LEVEL PRACTITIONERS

| DRUG NAME | DRUG CODE | BUYERS | TOTAL GRAMS | AVG GRAMS |
|---|---|---|---|---|
| PENTOBARBITAL (SCHEDULE 2) | 2270 | 19 | 19,109.95 | 1,005.79 |
| CODEINE | 9050 | 2 | 11.71 | 5.86 |

STATE:KENTUCKY BUSINESS ACTIVITY:N-U NARCOTIC TREATMENT PROGRAMS

| DRUG NAME | DRUG CODE | BUYERS | TOTAL GRAMS | AVG GRAMS |
|---|---|---|---|---|
| BUPRENORPHINE | 9064 | 19 | 837.29 | 44.07 |
| MEPERIDINE (PETHIDINE) | 9230 | 1 | 435.75 | 435.75 |
| METHADONE | 9250B | 26 | 155,232.51 | 5,970.48 |

STATE:LOUISIANA BUSINESS ACTIVITY:A - PHARMACIES

| DRUG NAME | DRUG CODE | BUYERS | TOTAL GRAMS | AVG GRAMS |
|---|---|---|---|---|
| AMPHETAMINE | 1100 | 1,175 | 610,985.05 | 519.99 |
| DL-METHAMPHETAMINE RACEMIC BASE | 1105B | 19 | 55.05 | 2.9 |
| D-METHAMPHETAMINE | 1105D | 1 | 4.42 | 4.42 |
| LISDEXAMFETAMINE | 1205 | 1,155 | 369,286.81 | 319.73 |
| METHYLPHENIDATE (DL;D;L;ISOMERS) | 1724 | 1,178 | 333,860.42 | 283.41 |
| BARBITURIC ACID DERIVIATIVE OR SALT [PER 21C | 2100 | 416 | 13,385 | 32.18 |
| BUTALBITAL | 2165 | 448 | 15,625 | 34.88 |
| DRONABINOL IN AN ORAL SOLUTION IN FDA APPR | 7365 | 1 | 0.3 | 0.3 |
| COCAINE | 9041L | 5 | 91.17 | 18.23 |
| CODEINE | 9050 | 1,180 | 169,554.20 | 143.69 |
| BUPRENORPHINE | 9064 | 1,064 | 75,317.98 | 70.79 |
| DIHYDROCODEINE | 9120 | 212 | 2,426.86 | 11.45 |
| OXYCODONE | 9143 | 1,196 | 740,196.42 | 618.89 |
| HYDROMORPHONE | 9150 | 979 | 10,401.87 | 10.62 |
| HYDROCODONE | 9193 | 1,204 | 614,356.93 | 510.26 |
| LEVORPHANOL | 9220L | 19 | 76.26 | 4.01 |
| MEPERIDINE (PETHIDINE) | 9230 | 845 | 20,806.39 | 24.62 |
| METHADONE | 9250B | 732 | 32,861.15 | 44.89 |
| MORPHINE | 9300 | 1,099 | 194,000.76 | 176.52 |
| OPIUM POWDERED | 9639 | 47 | 136.8 | 2.91 |
| OXYMORPHONE | 9652 | 586 | 26,242.24 | 44.78 |
| ALFENTANIL | 9737 | 4 | 0.98 | 0.24 |
| REMIFENTANIL | 9739 | 3 | 0.18 | 0.06 |
| SUFENTANIL BASE | 9740 | 3 | 3.34 | 1.11 |
| TAPENTADOL | 9780 | 662 | 60,225 | 90.97 |
| FENTANYL BASE | 9801 | 1,098 | 4,292.54 | 3.91 |
| OPIUM COMBINATION PRODUCT (C-III) | 9809 | 46 | 31.79 | 0.69 |

STATE:LOUISIANA BUSINESS ACTIVITY:B - HOSPITALS

| DRUG NAME | DRUG CODE | BUYERS | TOTAL GRAMS | AVG GRAMS |
|---|---|---|---|---|
| AMPHETAMINE | 1100 | 54 | 6,623.39 | 122.66 |

| | | | | |
|---|---|---|---|---|
| LISDEXAMFETAMINE | 1205 | 30 | 2,067.29 | 68.91 |
| METHYLPHENIDATE (DL;D;L;ISOMERS) | 1724 | 101 | 4,859.95 | 48.12 |
| BARBITURIC ACID DERIVIATIVE OR SALT [PER 21C | 2100 | 4 | 40 | 10 |
| BUTALBITAL | 2165 | 7 | 125 | 17.86 |
| PENTOBARBITAL (SCHEDULE 2) | 2270 | 13 | 664.87 | 51.14 |
| COCAINE | 9041L | 55 | 265.7 | 4.83 |
| CODEINE | 9050 | 140 | 3,325.39 | 23.75 |
| BUPRENORPHINE | 9064 | 94 | 674.26 | 7.17 |
| OXYCODONE | 9143 | 195 | 29,874.09 | 153.2 |
| HYDROMORPHONE | 9150 | 185 | 2,727.42 | 14.74 |
| HYDROCODONE | 9193 | 203 | 37,227.27 | 183.39 |
| MEPERIDINE (PETHIDINE) | 9230 | 173 | 7,322.95 | 42.33 |
| METHADONE | 9250B | 112 | 2,520.28 | 22.5 |
| MORPHINE | 9300 | 193 | 19,603.41 | 101.57 |
| OPIUM POWDERED | 9639 | 74 | 261.36 | 3.53 |
| OXYMORPHONE | 9652 | 18 | 92.96 | 5.16 |
| ALFENTANIL | 9737 | 14 | 6.66 | 0.48 |
| REMIFENTANIL | 9739 | 37 | 18.32 | 0.5 |
| SUFENTANIL BASE | 9740 | 40 | 1.23 | 0.03 |
| TAPENTADOL | 9780 | 17 | 560.5 | 32.97 |
| FENTANYL BASE | 9801 | 191 | 435.19 | 2.28 |
| OPIUM COMBINATION PRODUCT (C-III) | 9809 | 2 | 1.51 | 0.76 |

STATE:LOUISIANA BUSINESS ACTIVITY:C - PRACTITIONERS

| DRUG NAME | DRUG CODE | BUYERS | TOTAL GRAMS | AVG GRAMS |
|---|---|---|---|---|
| LISDEXAMFETAMINE | 1205 | 1 | 5.67 | 5.67 |
| GAMMA HYDROXYBUTYRIC ACID(FDA APPROVED | 2012 | 1 | 445.82 | 445.82 |
| PENTOBARBITAL (SCHEDULE 2) | 2270 | 96 | 60,574.80 | 630.99 |
| COCAINE | 9041L | 15 | 32.57 | 2.17 |
| CODEINE | 9050 | 112 | 913.41 | 8.16 |
| BUPRENORPHINE | 9064 | 302 | 11.86 | 0.04 |
| DIHYDROCODEINE | 9120 | 103 | 102.74 | 1 |
| OXYCODONE | 9143 | 16 | 168.51 | 10.53 |
| HYDROMORPHONE | 9150 | 118 | 43.76 | 0.37 |
| HYDROCODONE | 9193 | 215 | 495.95 | 2.31 |

| | | | | |
|---|---|---|---|---|
| MEPERIDINE (PETHIDINE) | 9230 | 123 | 889.91 | 7.24 |
| METHADONE | 9250B | 4 | 9.57 | 2.39 |
| MORPHINE | 9300 | 142 | 156.99 | 1.11 |
| OPIUM POWDERED | 9639 | 4 | 23.04 | 5.76 |
| ALFENTANIL | 9737 | 4 | 2.42 | 0.6 |
| REMIFENTANIL | 9739 | 2 | 0.07 | 0.04 |
| SUFENTANIL BASE | 9740 | 3 | 0.05 | 0.02 |
| TAPENTADOL | 9780 | 1 | 5 | 5 |
| FENTANYL BASE | 9801 | 243 | 27.87 | 0.11 |

STATE:LOUISIANA BUSINESS ACTIVITY:D - TEACHING INSTITUTIONS

| DRUG NAME | DRUG CODE | BUYERS | TOTAL GRAMS | AVG GRAMS |
|---|---|---|---|---|
| HYDROMORPHONE | 9150 | 1 | 0.14 | 0.14 |
| FENTANYL BASE | 9801 | 1 | 0.02 | 0.02 |

STATE:LOUISIANA BUSINESS ACTIVITY:M - MID-LEVEL PRACTITIONERS

| DRUG NAME | DRUG CODE | BUYERS | TOTAL GRAMS | AVG GRAMS |
|---|---|---|---|---|
| PENTOBARBITAL (SCHEDULE 2) | 2270 | 19 | 39,095.76 | 2,057.67 |
| CODEINE | 9050 | 2 | 2.66 | 1.33 |
| DIHYDROCODEINE | 9120 | 26 | 22.87 | 0.88 |

STATE:LOUISIANA BUSINESS ACTIVITY:N-U NARCOTIC TREATMENT PROGRAMS

| DRUG NAME | DRUG CODE | BUYERS | TOTAL GRAMS | AVG GRAMS |
|---|---|---|---|---|
| BUPRENORPHINE | 9064 | 7 | 309.9 | 44.27 |
| METHADONE | 9250B | 11 | 107,245.72 | 9,749.61 |

STATE:MAINE BUSINESS ACTIVITY:A - PHARMACIES

| DRUG NAME | DRUG CODE | BUYERS | TOTAL GRAMS | AVG GRAMS |
|---|---|---|---|---|
| AMPHETAMINE | 1100 | 299 | 89,282.54 | 298.6 |

| | | | | |
|---|---|---|---|---|
| DL-METHAMPHETAMINE RACEMIC BASE | 1105B | 11 | 33.35 | 3.03 |
| LISDEXAMFETAMINE | 1205 | 298 | 66,857.84 | 224.36 |
| METHYLPHENIDATE (DL;D;L;ISOMERS) | 1724 | 304 | 138,784.44 | 456.53 |
| BARBITURIC ACID DERIVIATIVE OR SALT [PER 21C | 2100 | 66 | 1,680 | 25.45 |
| BUTALBITAL | 2165 | 98 | 2,515 | 25.66 |
| DRONABINOL IN AN ORAL SOLUTION IN FDA APPR | 7365 | 1 | 0.3 | 0.3 |
| NABILONE | 7379 | 2 | 0.3 | 0.15 |
| COCAINE | 9041L | 4 | 4.14 | 1.04 |
| CODEINE | 9050 | 301 | 40,255.14 | 133.74 |
| BUPRENORPHINE | 9064 | 305 | 37,705.27 | 123.62 |
| DIHYDROCODEINE | 9120 | 1 | 1.07 | 1.07 |
| OXYCODONE | 9143 | 304 | 193,989.81 | 638.12 |
| HYDROMORPHONE | 9150 | 304 | 5,459.90 | 17.96 |
| HYDROCODONE | 9193 | 305 | 64,434.89 | 211.26 |
| MEPERIDINE (PETHIDINE) | 9230 | 64 | 1,363.03 | 21.3 |
| METHADONE | 9250B | 287 | 18,667.56 | 65.04 |
| MORPHINE | 9300 | 306 | 60,438.30 | 197.51 |
| OPIUM POWDERED | 9639 | 23 | 48.6 | 2.11 |
| OXYMORPHONE | 9652 | 175 | 3,693.12 | 21.1 |
| SUFENTANIL BASE | 9740 | 1 | 0.03 | 0.03 |
| TAPENTADOL | 9780 | 119 | 5,572.50 | 46.83 |
| FENTANYL BASE | 9801 | 301 | 1,132.04 | 3.76 |
| OPIUM COMBINATION PRODUCT (C-III) | 9809 | 11 | 5.11 | 0.46 |

STATE:MAINE BUSINESS ACTIVITY:B - HOSPITALS

| DRUG NAME | DRUG CODE | BUYERS | TOTAL GRAMS | AVG GRAMS |
|---|---|---|---|---|
| AMPHETAMINE | 1100 | 24 | 1,389.76 | 57.91 |
| LISDEXAMFETAMINE | 1205 | 14 | 452.65 | 32.33 |
| METHYLPHENIDATE (DL;D;L;ISOMERS) | 1724 | 39 | 2,440.33 | 62.57 |
| BARBITURIC ACID DERIVIATIVE OR SALT [PER 21C | 2100 | 1 | 10 | 10 |
| AMOBARBITAL (SCHEDULE 2) | 2125 | 1 | 2.73 | 2.73 |
| BUTALBITAL | 2165 | 1 | 5 | 5 |
| PENTOBARBITAL (SCHEDULE 2) | 2270 | 2 | 109.37 | 54.68 |
| COCAINE | 9041L | 24 | 83.28 | 3.47 |
| CODEINE | 9050 | 44 | 1,175.44 | 26.71 |

| | | | | |
|---|---|---|---|---|
| BUPRENORPHINE | 9064 | 38 | 881.61 | 23.2 |
| OXYCODONE | 9143 | 46 | 10,305.09 | 224.02 |
| HYDROMORPHONE | 9150 | 40 | 1,191.32 | 29.78 |
| HYDROCODONE | 9193 | 46 | 3,924.26 | 85.31 |
| MEPERIDINE (PETHIDINE) | 9230 | 37 | 195.11 | 5.27 |
| METHADONE | 9250B | 35 | 1,040.10 | 29.72 |
| MORPHINE | 9300 | 45 | 5,431.99 | 120.71 |
| OPIUM POWDERED | 9639 | 27 | 150.48 | 5.57 |
| OXYMORPHONE | 9652 | 2 | 5.35 | 2.68 |
| ALFENTANIL | 9737 | 9 | 2.28 | 0.25 |
| REMIFENTANIL | 9739 | 13 | 4.59 | 0.35 |
| SUFENTANIL BASE | 9740 | 13 | 0.05 | 0 |
| TAPENTADOL | 9780 | 2 | 63 | 31.5 |
| FENTANYL BASE | 9801 | 43 | 158.88 | 3.69 |
| OPIUM COMBINATION PRODUCT (C-III) | 9809 | 1 | 1.7 | 1.7 |

STATE:MAINE BUSINESS ACTIVITY:C - PRACTITIONERS

| DRUG NAME | DRUG CODE | BUYERS | TOTAL GRAMS | AVG GRAMS |
|---|---|---|---|---|
| PENTOBARBITAL (SCHEDULE 2) | 2270 | 108 | 80,060.54 | 741.3 |
| CODEINE | 9050 | 6 | 121.95 | 20.32 |
| BUPRENORPHINE | 9064 | 158 | 9.72 | 0.06 |
| OXYCODONE | 9143 | 6 | 12.78 | 2.13 |
| HYDROMORPHONE | 9150 | 77 | 35.18 | 0.46 |
| HYDROCODONE | 9193 | 57 | 99.73 | 1.75 |
| MEPERIDINE (PETHIDINE) | 9230 | 3 | 20.04 | 6.68 |
| MORPHINE | 9300 | 38 | 63.61 | 1.67 |
| OXYMORPHONE | 9652 | 1 | 0.04 | 0.04 |
| ALFENTANIL | 9737 | 2 | 0.41 | 0.21 |
| REMIFENTANIL | 9739 | 1 | 0.02 | 0.02 |
| FENTANYL BASE | 9801 | 53 | 31.9 | 0.6 |

STATE:MAINE BUSINESS ACTIVITY:D - TEACHING INSTITUTIONS

| DRUG NAME | DRUG CODE | BUYERS | TOTAL GRAMS | AVG GRAMS |
|---|---|---|---|---|

| | | | | |
|---|---|---|---|---|
| BUPRENORPHINE | 9064 | 1 | 0.03 | 0.03 |

STATE:MAINE BUSINESS ACTIVITY:M - MID-LEVEL PRACTITIONERS

| DRUG NAME | DRUG CODE | BUYERS | TOTAL GRAMS | AVG GRAMS |
|---|---|---|---|---|
| PENTOBARBITAL (SCHEDULE 2) | 2270 | 3 | 710.62 | 236.87 |
| CODEINE | 9050 | 3 | 3.58 | 1.19 |
| HYDROCODONE | 9193 | 1 | 0.91 | 0.91 |

STATE:MAINE BUSINESS ACTIVITY:N-U NARCOTIC TREATMENT PROGRAMS

| DRUG NAME | DRUG CODE | BUYERS | TOTAL GRAMS | AVG GRAMS |
|---|---|---|---|---|
| BUPRENORPHINE | 9064 | 3 | 123.78 | 41.26 |
| METHADONE | 9250B | 10 | 106,517.67 | 10,651.77 |

STATE:MARYLAND BUSINESS ACTIVITY:A - PHARMACIES

| DRUG NAME | DRUG CODE | BUYERS | TOTAL GRAMS | AVG GRAMS |
|---|---|---|---|---|
| AMPHETAMINE | 1100 | 1,233 | 356,309.52 | 288.98 |
| DL-METHAMPHETAMINE RACEMIC BASE | 1105B | 42 | 164.36 | 3.91 |
| D-METHAMPHETAMINE | 1105D | 2 | 4.82 | 2.41 |
| LISDEXAMFETAMINE | 1205 | 1,194 | 221,752.80 | 185.72 |
| METHYLPHENIDATE (DL;D;L;ISOMERS) | 1724 | 1,230 | 385,280.49 | 313.24 |
| BARBITURIC ACID DERIVIATIVE OR SALT [PER 21C | 2100 | 325 | 9,530 | 29.32 |
| BUTALBITAL | 2165 | 561 | 18,965 | 33.81 |
| PENTOBARBITAL (SCHEDULE 2) | 2270 | 1 | 1.82 | 1.82 |
| SECOBARBITAL (SCHEDULE 2) | 2315 | 5 | 219.72 | 43.94 |
| DRONABINOL IN AN ORAL SOLUTION IN FDA APPR | 7365 | 5 | 1.95 | 0.39 |
| NABILONE | 7379 | 10 | 4.1 | 0.41 |
| COCAINE | 9041L | 9 | 144.28 | 16.03 |
| CODEINE | 9050 | 1,244 | 218,426.99 | 175.58 |
| BUPRENORPHINE | 9064 | 1,180 | 94,834.75 | 80.37 |
| DIHYDROCODEINE | 9120 | 12 | 25.64 | 2.14 |
| OXYCODONE | 9143 | 1,264 | 1,241,338.50 | 982.07 |

| | | | | |
|---|---|---|---|---|
| HYDROMORPHONE | 9150 | 1,177 | 27,570.06 | 23.42 |
| HYDROCODONE | 9193 | 1,251 | 132,976.94 | 106.3 |
| LEVORPHANOL | 9220L | 39 | 21.36 | 0.55 |
| MEPERIDINE (PETHIDINE) | 9230 | 327 | 7,490.65 | 22.91 |
| METHADONE | 9250B | 979 | 59,545.93 | 60.82 |
| MORPHINE | 9300 | 1,211 | 281,301.48 | 232.29 |
| OPIUM TINCTURE (LAUDANUM) | 9630 | 5 | 12.99 | 2.6 |
| OPIUM POWDERED | 9639 | 64 | 125.64 | 1.96 |
| OXYMORPHONE | 9652 | 870 | 36,335.77 | 41.77 |
| ALFENTANIL | 9737 | 3 | 0.7 | 0.23 |
| REMIFENTANIL | 9739 | 4 | 0.22 | 0.06 |
| TAPENTADOL | 9780 | 886 | 127,109.50 | 143.46 |
| FENTANYL BASE | 9801 | 1,134 | 4,905.98 | 4.33 |
| OPIUM COMBINATION PRODUCT (C-III) | 9809 | 41 | 31.22 | 0.76 |

STATE:MARYLAND BUSINESS ACTIVITY:B - HOSPITALS

| DRUG NAME | DRUG CODE | BUYERS | TOTAL GRAMS | AVG GRAMS |
|---|---|---|---|---|
| AMPHETAMINE | 1100 | 55 | 9,952.40 | 180.95 |
| LISDEXAMFETAMINE | 1205 | 21 | 1,703.72 | 81.13 |
| METHYLPHENIDATE (DL;D;L;ISOMERS) | 1724 | 70 | 14,917.85 | 213.11 |
| BARBITURIC ACID DERIVIATIVE OR SALT [PER 21C | 2100 | 2 | 10 | 5 |
| AMOBARBITAL (SCHEDULE 2) | 2125 | 6 | 27.34 | 4.56 |
| BUTALBITAL | 2165 | 2 | 25 | 12.5 |
| PENTOBARBITAL (SCHEDULE 2) | 2270 | 12 | 3,301.55 | 275.13 |
| COCAINE | 9041L | 57 | 357.3 | 6.27 |
| CODEINE | 9050 | 84 | 5,102.59 | 60.75 |
| BUPRENORPHINE | 9064 | 71 | 1,898.44 | 26.74 |
| OXYCODONE | 9143 | 128 | 50,967.30 | 398.18 |
| HYDROMORPHONE | 9150 | 125 | 5,666.44 | 45.33 |
| HYDROCODONE | 9193 | 107 | 3,909.45 | 36.54 |
| MEPERIDINE (PETHIDINE) | 9230 | 95 | 1,402.24 | 14.76 |
| METHADONE | 9250B | 67 | 5,577.98 | 83.25 |
| MORPHINE | 9300 | 115 | 20,682.94 | 179.85 |
| OPIUM POWDERED | 9639 | 41 | 208.44 | 5.08 |
| OXYMORPHONE | 9652 | 9 | 262.9 | 29.21 |

| | | | | |
|---|---|---|---|---|
| ALFENTANIL | 9737 | 5 | 2.2 | 0.44 |
| REMIFENTANIL | 9739 | 30 | 26.25 | 0.88 |
| SUFENTANIL BASE | 9740 | 13 | 0.21 | 0.02 |
| TAPENTADOL | 9780 | 20 | 2,430 | 121.5 |
| FENTANYL BASE | 9801 | 193 | 574.45 | 2.98 |
| OPIUM COMBINATION PRODUCT (C-III) | 9809 | 3 | 2.84 | 0.95 |

STATE:MARYLAND BUSINESS ACTIVITY:C - PRACTITIONERS

| DRUG NAME | DRUG CODE | BUYERS | TOTAL GRAMS | AVG GRAMS |
|---|---|---|---|---|
| AMPHETAMINE | 1100 | 2 | 1.47 | 0.73 |
| PENTOBARBITAL (SCHEDULE 2) | 2270 | 96 | 64,319.59 | 670 |
| COCAINE | 9041L | 9 | 16 | 1.78 |
| CODEINE | 9050 | 281 | 8,353.46 | 29.73 |
| BUPRENORPHINE | 9064 | 537 | 614.76 | 1.14 |
| DIHYDROCODEINE | 9120 | 12 | 11.8 | 0.98 |
| OXYCODONE | 9143 | 69 | 3,071.25 | 44.51 |
| HYDROMORPHONE | 9150 | 261 | 177.92 | 0.68 |
| HYDROCODONE | 9193 | 268 | 1,581.30 | 5.9 |
| MEPERIDINE (PETHIDINE) | 9230 | 71 | 477.15 | 6.72 |
| METHADONE | 9250B | 17 | 77.05 | 4.53 |
| MORPHINE | 9300 | 161 | 1,158.89 | 7.2 |
| OPIUM POWDERED | 9639 | 1 | 1.08 | 1.08 |
| OXYMORPHONE | 9652 | 18 | 451.47 | 25.08 |
| NOROXYMORPHONE | 9668 | 1 | 0.2 | 0.2 |
| REMIFENTANIL | 9739 | 6 | 0.93 | 0.16 |
| SUFENTANIL BASE | 9740 | 3 | 0.01 | 0 |
| TAPENTADOL | 9780 | 8 | 465 | 58.13 |
| FENTANYL BASE | 9801 | 367 | 71.95 | 0.2 |

STATE:MARYLAND BUSINESS ACTIVITY:D - TEACHING INSTITUTIONS

| DRUG NAME | DRUG CODE | BUYERS | TOTAL GRAMS | AVG GRAMS |
|---|---|---|---|---|
| MORPHINE | 9300 | 1 | 0.03 | 0.03 |
| FENTANYL BASE | 9801 | 1 | 0.01 | 0.01 |

STATE:MARYLAND BUSINESS ACTIVITY:M - MID-LEVEL PRACTITIONERS

| DRUG NAME | DRUG CODE | BUYERS | TOTAL GRAMS | AVG GRAMS |
|---|---|---|---|---|
| PENTOBARBITAL (SCHEDULE 2) | 2270 | 14 | 21,307.62 | 1,521.97 |
| CODEINE | 9050 | 36 | 44.45 | 1.23 |
| BUPRENORPHINE | 9064 | 5 | 9.53 | 1.91 |
| DIHYDROCODEINE | 9120 | 2 | 0.64 | 0.32 |
| OXYCODONE | 9143 | 1 | 0.45 | 0.45 |
| HYDROCODONE | 9193 | 1 | 0.3 | 0.3 |

STATE:MARYLAND BUSINESS ACTIVITY:N-U NARCOTIC TREATMENT PROGRAMS

| DRUG NAME | DRUG CODE | BUYERS | TOTAL GRAMS | AVG GRAMS |
|---|---|---|---|---|
| BUPRENORPHINE | 9064 | 41 | 4,705.29 | 114.76 |
| METHADONE | 9250B | 81 | 699,072.88 | 8,630.53 |

STATE:MASSACHUSETTS BUSINESS ACTIVITY:A - PHARMACIES

| DRUG NAME | DRUG CODE | BUYERS | TOTAL GRAMS | AVG GRAMS |
|---|---|---|---|---|
| AMPHETAMINE | 1100 | 1,142 | 682,803.50 | 597.9 |
| DL-METHAMPHETAMINE RACEMIC BASE | 1105B | 33 | 168.38 | 5.1 |
| D-METHAMPHETAMINE | 1105D | 4 | 6.83 | 1.71 |
| LISDEXAMFETAMINE | 1205 | 1,083 | 139,220.93 | 128.55 |
| METHYLPHENIDATE (DL;D;L;ISOMERS) | 1724 | 1,139 | 555,629.86 | 487.82 |
| BARBITURIC ACID DERIVIATIVE OR SALT [PER 21C | 2100 | 423 | 10,755 | 25.43 |
| BUTALBITAL | 2165 | 359 | 8,995 | 25.06 |
| SECOBARBITAL (SCHEDULE 2) | 2315 | 8 | 192.26 | 24.03 |
| DRONABINOL IN AN ORAL SOLUTION IN FDA APPR | 7365 | 4 | 1.2 | 0.3 |
| CODEINE | 9050 | 1,128 | 175,363.93 | 155.46 |
| BUPRENORPHINE | 9064 | 1,113 | 155,993.82 | 140.16 |
| DIHYDROCODEINE | 9120 | 2 | 126.04 | 63.02 |
| OXYCODONE | 9143 | 1,150 | 890,076.90 | 773.98 |
| HYDROMORPHONE | 9150 | 1,089 | 20,855.92 | 19.15 |

| | | | | |
|---|---|---|---|---|
| HYDROCODONE | 9193 | 1,140 | 116,077.32 | 101.82 |
| LEVORPHANOL | 9220L | 37 | 80.56 | 2.18 |
| MEPERIDINE (PETHIDINE) | 9230 | 198 | 6,080.24 | 30.71 |
| METHADONE | 9250B | 972 | 65,587.78 | 67.48 |
| MORPHINE | 9300 | 1,098 | 233,599.19 | 212.75 |
| OPIUM POWDERED | 9639 | 125 | 571.32 | 4.57 |
| OXYMORPHONE | 9652 | 282 | 7,966.23 | 28.25 |
| TAPENTADOL | 9780 | 515 | 75,084 | 145.79 |
| FENTANYL BASE | 9801 | 1,035 | 4,472.70 | 4.32 |
| OPIUM COMBINATION PRODUCT (C-III) | 9809 | 18 | 6.43 | 0.36 |

STATE:MASSACHUSETTS BUSINESS ACTIVITY:B - HOSPITALS

| DRUG NAME | DRUG CODE | BUYERS | TOTAL GRAMS | AVG GRAMS |
|---|---|---|---|---|
| AMPHETAMINE | 1100 | 150 | 29,055.51 | 193.7 |
| LISDEXAMFETAMINE | 1205 | 69 | 3,676.72 | 53.29 |
| METHYLPHENIDATE (DL;D;L;ISOMERS) | 1724 | 165 | 29,018.28 | 175.87 |
| GAMMA HYDROXYBUTYRIC ACID(FDA APPROVED | 2012 | 1 | 2,674.94 | 2,674.94 |
| BARBITURIC ACID DERIVIATIVE OR SALT [PER 21C | 2100 | 4 | 65 | 16.25 |
| AMOBARBITAL (SCHEDULE 2) | 2125 | 7 | 20.05 | 2.86 |
| BUTALBITAL | 2165 | 15 | 305 | 20.33 |
| PENTOBARBITAL (SCHEDULE 2) | 2270 | 16 | 10,487.26 | 655.45 |
| DRONABINOL IN AN ORAL SOLUTION IN FDA APPR | 7365 | 1 | 2.55 | 2.55 |
| COCAINE | 9041L | 77 | 784.16 | 10.18 |
| CODEINE | 9050 | 161 | 8,682.23 | 53.93 |
| BUPRENORPHINE | 9064 | 158 | 8,767.77 | 55.49 |
| OXYCODONE | 9143 | 202 | 69,224.93 | 342.7 |
| HYDROMORPHONE | 9150 | 175 | 8,513.83 | 48.65 |
| HYDROCODONE | 9193 | 175 | 4,182.92 | 23.9 |
| LEVORPHANOL | 9220L | 1 | 0.12 | 0.12 |
| MEPERIDINE (PETHIDINE) | 9230 | 122 | 1,984.56 | 16.27 |
| METHADONE | 9250B | 156 | 11,573.04 | 74.19 |
| MORPHINE | 9300 | 195 | 28,470.48 | 146 |
| OPIUM POWDERED | 9639 | 58 | 227.52 | 3.92 |
| OXYMORPHONE | 9652 | 17 | 733.89 | 43.17 |
| ALFENTANIL | 9737 | 3 | 3.45 | 1.15 |

| | | | | |
|---|---|---|---|---|
| REMIFENTANIL | 9739 | 67 | 87.02 | 1.3 |
| SUFENTANIL BASE | 9740 | 18 | 0.52 | 0.03 |
| TAPENTADOL | 9780 | 24 | 1,992 | 83 |
| FENTANYL BASE | 9801 | 185 | 721.87 | 3.9 |
| OPIUM COMBINATION PRODUCT (C-III) | 9809 | 1 | 0.38 | 0.38 |

STATE:MASSACHUSETTS BUSINESS ACTIVITY:C - PRACTITIONERS

| DRUG NAME | DRUG CODE | BUYERS | TOTAL GRAMS | AVG GRAMS |
|---|---|---|---|---|
| METHYLPHENIDATE (DL;D;L;ISOMERS) | 1724 | 2 | 2.16 | 1.08 |
| PENTOBARBITAL (SCHEDULE 2) | 2270 | 187 | 102,271.15 | 546.9 |
| COCAINE | 9041L | 4 | 31.28 | 7.82 |
| CODEINE | 9050 | 42 | 1,178.29 | 28.05 |
| BUPRENORPHINE | 9064 | 529 | 59.77 | 0.11 |
| OXYCODONE | 9143 | 20 | 35.59 | 1.78 |
| HYDROMORPHONE | 9150 | 241 | 104.41 | 0.43 |
| HYDROCODONE | 9193 | 128 | 410.98 | 3.21 |
| MEPERIDINE (PETHIDINE) | 9230 | 37 | 390.21 | 10.55 |
| METHADONE | 9250B | 24 | 93.02 | 3.88 |
| MORPHINE | 9300 | 109 | 128.58 | 1.18 |
| OPIUM POWDERED | 9639 | 2 | 4.32 | 2.16 |
| OXYMORPHONE | 9652 | 10 | 1.05 | 0.11 |
| NOROXYMORPHONE | 9668 | 1 | 3.03 | 3.03 |
| REMIFENTANIL | 9739 | 20 | 2.24 | 0.11 |
| SUFENTANIL BASE | 9740 | 1 | 0 | 0 |
| FENTANYL BASE | 9801 | 305 | 34.45 | 0.11 |

STATE:MASSACHUSETTS BUSINESS ACTIVITY:M - MID-LEVEL PRACTITIONERS

| DRUG NAME | DRUG CODE | BUYERS | TOTAL GRAMS | AVG GRAMS |
|---|---|---|---|---|
| CODEINE | 9050 | 5 | 5.63 | 1.13 |
| BUPRENORPHINE | 9064 | 1 | 0.6 | 0.6 |

STATE:MASSACHUSETTS BUSINESS ACTIVITY:N-U NARCOTIC TREATMENT PROGRAMS

| DRUG NAME | DRUG CODE | BUYERS | TOTAL GRAMS | AVG GRAMS |
|---|---|---|---|---|
| BUPRENORPHINE | 9064 | 33 | 491.4 | 14.89 |
| METHADONE | 9250B | 74 | 502,120.81 | 6,785.42 |

STATE:MICHIGAN BUSINESS ACTIVITY:A - PHARMACIES

| DRUG NAME | DRUG CODE | BUYERS | TOTAL GRAMS | AVG GRAMS |
|---|---|---|---|---|
| AMPHETAMINE | 1100 | 2,409 | 943,474.67 | 391.65 |
| DL-METHAMPHETAMINE RACEMIC BASE | 1105B | 37 | 91.62 | 2.48 |
| LISDEXAMFETAMINE | 1205 | 2,169 | 233,844.86 | 107.81 |
| METHYLPHENIDATE (DL;D;L;ISOMERS) | 1724 | 2,319 | 737,432.11 | 318 |
| BARBITURIC ACID DERIVIATIVE OR SALT [PER 21C | 2100 | 779 | 26,750 | 34.34 |
| BUTALBITAL | 2165 | 1,043 | 35,215 | 33.76 |
| SECOBARBITAL (SCHEDULE 2) | 2315 | 1 | 18.31 | 18.31 |
| DRONABINOL IN AN ORAL SOLUTION IN FDA APPR | 7365 | 8 | 2.4 | 0.3 |
| COCAINE | 9041L | 11 | 19.57 | 1.78 |
| CODEINE | 9050 | 2,472 | 998,622.91 | 403.97 |
| BUPRENORPHINE | 9064 | 2,209 | 105,473.70 | 47.75 |
| DIHYDROCODEINE | 9120 | 5 | 7.48 | 1.5 |
| OXYCODONE | 9143 | 2,441 | 1,127,113.21 | 461.74 |
| HYDROMORPHONE | 9150 | 1,967 | 29,477.48 | 14.99 |
| HYDROCODONE | 9193 | 2,500 | 1,517,211.61 | 606.88 |
| LEVORPHANOL | 9220L | 4 | 0.7 | 0.17 |
| MEPERIDINE (PETHIDINE) | 9230 | 262 | 6,823.30 | 26.04 |
| METHADONE | 9250B | 1,911 | 137,989.55 | 72.21 |
| MORPHINE | 9300 | 2,325 | 694,249.52 | 298.6 |
| OPIUM TINCTURE (LAUDANUM) | 9630 | 6 | 30.72 | 5.12 |
| OPIUM POWDERED | 9639 | 156 | 487.08 | 3.12 |
| OXYMORPHONE | 9652 | 1,198 | 75,289.05 | 62.85 |
| NOROXYMORPHONE | 9668 | 2 | 3.12 | 1.56 |
| REMIFENTANIL | 9739 | 4 | 2.59 | 0.65 |
| SUFENTANIL BASE | 9740 | 3 | 0.72 | 0.24 |
| TAPENTADOL | 9780 | 1,055 | 85,282 | 80.84 |
| FENTANYL BASE | 9801 | 2,205 | 11,127.47 | 5.05 |
| OPIUM COMBINATION PRODUCT (C-III) | 9809 | 27 | 18.92 | 0.7 |

STATE:MICHIGAN BUSINESS ACTIVITY:B - HOSPITALS

| DRUG NAME | DRUG CODE | BUYERS | TOTAL GRAMS | AVG GRAMS |
|---|---|---|---|---|
| AMPHETAMINE | 1100 | 107 | 18,425.14 | 172.2 |
| LISDEXAMFETAMINE | 1205 | 58 | 3,996.41 | 68.9 |
| METHYLPHENIDATE (DL;D;L;ISOMERS) | 1724 | 159 | 17,443.59 | 109.71 |
| GAMMA HYDROXYBUTYRIC ACID(FDA APPROVED | 2012 | 1 | 1,337.47 | 1,337.47 |
| BARBITURIC ACID DERIVIATIVE OR SALT [PER 21C | 2100 | 8 | 160 | 20 |
| AMOBARBITAL (SCHEDULE 2) | 2125 | 8 | 18.68 | 2.34 |
| BUTALBITAL | 2165 | 23 | 255 | 11.09 |
| PENTOBARBITAL (SCHEDULE 2) | 2270 | 26 | 2,524.12 | 97.08 |
| COCAINE | 9041L | 109 | 1,347.66 | 12.36 |
| CODEINE | 9050 | 185 | 23,026.56 | 124.47 |
| BUPRENORPHINE | 9064 | 125 | 2,660.55 | 21.28 |
| OXYCODONE | 9143 | 193 | 60,934.80 | 315.72 |
| HYDROMORPHONE | 9150 | 183 | 6,483.74 | 35.43 |
| HYDROCODONE | 9193 | 198 | 76,031.56 | 384 |
| LEVORPHANOL | 9220L | 1 | 0.35 | 0.35 |
| MEPERIDINE (PETHIDINE) | 9230 | 138 | 2,873.23 | 20.82 |
| METHADONE | 9250B | 151 | 9,337.97 | 61.84 |
| MORPHINE | 9300 | 194 | 58,655.75 | 302.35 |
| OPIUM POWDERED | 9639 | 105 | 1,046.52 | 9.97 |
| OXYMORPHONE | 9652 | 29 | 405.15 | 13.97 |
| ALFENTANIL | 9737 | 12 | 2.4 | 0.2 |
| REMIFENTANIL | 9739 | 54 | 37.01 | 0.69 |
| SUFENTANIL BASE | 9740 | 43 | 1.47 | 0.03 |
| TAPENTADOL | 9780 | 28 | 1,357 | 48.46 |
| FENTANYL BASE | 9801 | 188 | 1,165.48 | 6.2 |
| OPIUM COMBINATION PRODUCT (C-III) | 9809 | 5 | 3.78 | 0.76 |

STATE:MICHIGAN BUSINESS ACTIVITY:C - PRACTITIONERS

| DRUG NAME | DRUG CODE | BUYERS | TOTAL GRAMS | AVG GRAMS |
|---|---|---|---|---|
| AMPHETAMINE | 1100 | 2 | 41.51 | 20.75 |

| | | | | |
|---|---|---|---|---|
| LISDEXAMFETAMINE | 1205 | 2 | 11.33 | 5.67 |
| BUTALBITAL | 2165 | 1 | 50 | 50 |
| PENTOBARBITAL (SCHEDULE 2) | 2270 | 448 | 342,339.26 | 764.15 |
| COCAINE | 9041L | 29 | 140.63 | 4.85 |
| CODEINE | 9050 | 173 | 2,643.64 | 15.28 |
| BUPRENORPHINE | 9064 | 688 | 378.79 | 0.55 |
| DIHYDROCODEINE | 9120 | 1 | 33.01 | 33.01 |
| OXYCODONE | 9143 | 40 | 781.74 | 19.54 |
| HYDROMORPHONE | 9150 | 414 | 188.5 | 0.46 |
| HYDROCODONE | 9193 | 283 | 1,916.02 | 6.77 |
| MEPERIDINE (PETHIDINE) | 9230 | 117 | 1,230.78 | 10.52 |
| METHADONE | 9250B | 8 | 16.73 | 2.09 |
| MORPHINE | 9300 | 305 | 661.53 | 2.17 |
| OPIUM POWDERED | 9639 | 7 | 8.64 | 1.23 |
| OXYMORPHONE | 9652 | 4 | 1.79 | 0.45 |
| NOROXYMORPHONE | 9668 | 1 | 1.17 | 1.17 |
| ALFENTANIL | 9737 | 7 | 2.08 | 0.3 |
| REMIFENTANIL | 9739 | 1 | 0.03 | 0.03 |
| SUFENTANIL BASE | 9740 | 3 | 0.03 | 0.01 |
| FENTANYL BASE | 9801 | 527 | 79.7 | 0.15 |

STATE:MICHIGAN BUSINESS ACTIVITY:D - TEACHING INSTITUTIONS

| DRUG NAME | DRUG CODE | BUYERS | TOTAL GRAMS | AVG GRAMS |
|---|---|---|---|---|
| PENTOBARBITAL (SCHEDULE 2) | 2270 | 1 | 13,329.23 | 13,329.23 |
| CODEINE | 9050 | 1 | 745.17 | 745.17 |
| BUPRENORPHINE | 9064 | 1 | 1.3 | 1.3 |
| HYDROMORPHONE | 9150 | 1 | 2.32 | 2.32 |
| HYDROCODONE | 9193 | 1 | 25 | 25 |
| MEPERIDINE (PETHIDINE) | 9230 | 1 | 1.09 | 1.09 |
| METHADONE | 9250B | 1 | 31.3 | 31.3 |
| MORPHINE | 9300 | 1 | 3.38 | 3.38 |
| SUFENTANIL BASE | 9740 | 1 | 0.02 | 0.02 |
| FENTANYL BASE | 9801 | 2 | 3.33 | 1.67 |

STATE:MICHIGAN BUSINESS ACTIVITY:M - MID-LEVEL PRACTITIONERS

| DRUG NAME | DRUG CODE | BUYERS | TOTAL GRAMS | AVG GRAMS |
|---|---|---|---|---|
| PENTOBARBITAL (SCHEDULE 2) | 2270 | 25 | 26,748.67 | 1,069.95 |
| CODEINE | 9050 | 10 | 236.55 | 23.66 |
| DIHYDROCODEINE | 9120 | 2 | 0.37 | 0.19 |

STATE:MICHIGAN BUSINESS ACTIVITY:N-U NARCOTIC TREATMENT PROGRAMS

| DRUG NAME | DRUG CODE | BUYERS | TOTAL GRAMS | AVG GRAMS |
|---|---|---|---|---|
| BUPRENORPHINE | 9064 | 6 | 130.26 | 21.71 |
| METHADONE | 9250B | 44 | 386,636.60 | 8,787.20 |

STATE:MINNESOTA BUSINESS ACTIVITY:A - PHARMACIES

| DRUG NAME | DRUG CODE | BUYERS | TOTAL GRAMS | AVG GRAMS |
|---|---|---|---|---|
| AMPHETAMINE | 1100 | 1,026 | 425,084.45 | 414.31 |
| DL-METHAMPHETAMINE RACEMIC BASE | 1105B | 47 | 264.02 | 5.62 |
| D-METHAMPHETAMINE | 1105D | 3 | 10.04 | 3.35 |
| LISDEXAMFETAMINE | 1205 | 1,010 | 125,218.77 | 123.98 |
| METHYLPHENIDATE (DL;D;L;ISOMERS) | 1724 | 1,030 | 395,570.50 | 384.05 |
| BARBITURIC ACID DERIVIATIVE OR SALT [PER 21C | 2100 | 221 | 5,285 | 23.91 |
| BUTALBITAL | 2165 | 280 | 6,670 | 23.82 |
| PENTOBARBITAL (SCHEDULE 2) | 2270 | 1 | 802.94 | 802.94 |
| SECOBARBITAL (SCHEDULE 2) | 2315 | 1 | 18.31 | 18.31 |
| DRONABINOL IN AN ORAL SOLUTION IN FDA APPR | 7365 | 2 | 2.55 | 1.28 |
| NABILONE | 7379 | 2 | 0.1 | 0.05 |
| COCAINE | 9041L | 3 | 19.39 | 6.46 |
| CODEINE | 9050 | 1,025 | 142,618.66 | 139.14 |
| BUPRENORPHINE | 9064 | 890 | 27,024.13 | 30.36 |
| DIHYDROCODEINE | 9120 | 2 | 7.48 | 3.74 |
| OXYCODONE | 9143 | 1,041 | 538,155.51 | 516.96 |
| HYDROMORPHONE | 9150 | 995 | 18,867.01 | 18.96 |
| HYDROCODONE | 9193 | 1,046 | 169,756.86 | 162.29 |
| LEVORPHANOL | 9220L | 4 | 2.79 | 0.7 |

| | | | | |
|---|---|---|---|---|
| MEPERIDINE (PETHIDINE) | 9230 | 131 | 1,562.27 | 11.93 |
| METHADONE | 9250B | 863 | 30,269 | 35.07 |
| MORPHINE | 9300 | 1,016 | 167,092.25 | 164.46 |
| OPIUM POWDERED | 9639 | 175 | 597.96 | 3.42 |
| OXYMORPHONE | 9652 | 196 | 1,678.58 | 8.56 |
| NOROXYMORPHONE | 9668 | 1 | 0.5 | 0.5 |
| ALFENTANIL | 9737 | 2 | 0.98 | 0.49 |
| REMIFENTANIL | 9739 | 1 | 0.02 | 0.02 |
| SUFENTANIL BASE | 9740 | 1 | 0.09 | 0.09 |
| TAPENTADOL | 9780 | 340 | 17,409 | 51.2 |
| FENTANYL BASE | 9801 | 987 | 3,563.36 | 3.61 |
| OPIUM COMBINATION PRODUCT (C-III) | 9809 | 17 | 8.51 | 0.5 |

STATE:MINNESOTA BUSINESS ACTIVITY:B - HOSPITALS

| DRUG NAME | DRUG CODE | BUYERS | TOTAL GRAMS | AVG GRAMS |
|---|---|---|---|---|
| AMPHETAMINE | 1100 | 81 | 6,557.53 | 80.96 |
| DL-METHAMPHETAMINE RACEMIC BASE | 1105B | 3 | 25.32 | 8.44 |
| D-METHAMPHETAMINE | 1105D | 2 | 42.98 | 21.49 |
| LISDEXAMFETAMINE | 1205 | 41 | 1,460.86 | 35.63 |
| METHYLPHENIDATE (DL;D;L;ISOMERS) | 1724 | 97 | 7,770.28 | 80.11 |
| BARBITURIC ACID DERIVIATIVE OR SALT [PER 21C | 2100 | 4 | 30 | 7.5 |
| AMOBARBITAL (SCHEDULE 2) | 2125 | 5 | 23.24 | 4.65 |
| BUTALBITAL | 2165 | 5 | 35 | 7 |
| PENTOBARBITAL (SCHEDULE 2) | 2270 | 14 | 13,997.74 | 999.84 |
| DRONABINOL IN AN ORAL SOLUTION IN FDA APPR | 7365 | 1 | 0.3 | 0.3 |
| COCAINE | 9041L | 87 | 670.92 | 7.71 |
| CODEINE | 9050 | 138 | 5,381.46 | 39 |
| BUPRENORPHINE | 9064 | 74 | 1,191.12 | 16.1 |
| OXYCODONE | 9143 | 149 | 32,200.38 | 216.11 |
| HYDROMORPHONE | 9150 | 150 | 4,765.02 | 31.77 |
| HYDROCODONE | 9193 | 148 | 12,228.16 | 82.62 |
| MEPERIDINE (PETHIDINE) | 9230 | 119 | 911.7 | 7.66 |
| METHADONE | 9250B | 97 | 3,250.62 | 33.51 |
| MORPHINE | 9300 | 156 | 18,164.54 | 116.44 |
| OPIUM POWDERED | 9639 | 82 | 923.04 | 11.26 |

| | | | | |
|---|---|---|---|---|
| OXYMORPHONE | 9652 | 9 | 19.46 | 2.16 |
| ALFENTANIL | 9737 | 21 | 10.98 | 0.52 |
| REMIFENTANIL | 9739 | 24 | 31.85 | 1.33 |
| SUFENTANIL BASE | 9740 | 23 | 0.68 | 0.03 |
| TAPENTADOL | 9780 | 11 | 192.5 | 17.5 |
| FENTANYL BASE | 9801 | 151 | 501.92 | 3.32 |
| OPIUM COMBINATION PRODUCT (C-III) | 9809 | 3 | 0.57 | 0.19 |

STATE:MINNESOTA BUSINESS ACTIVITY:C - PRACTITIONERS

| DRUG NAME | DRUG CODE | BUYERS | TOTAL GRAMS | AVG GRAMS |
|---|---|---|---|---|
| AMPHETAMINE | 1100 | 4 | 9.51 | 2.38 |
| PENTOBARBITAL (SCHEDULE 2) | 2270 | 179 | 130,079.10 | 726.7 |
| COCAINE | 9041L | 22 | 165.2 | 7.51 |
| CODEINE | 9050 | 42 | 1,175.30 | 27.98 |
| BUPRENORPHINE | 9064 | 521 | 34.61 | 0.07 |
| OXYCODONE | 9143 | 62 | 277.75 | 4.48 |
| HYDROMORPHONE | 9150 | 360 | 159.33 | 0.44 |
| HYDROCODONE | 9193 | 251 | 596.46 | 2.38 |
| MEPERIDINE (PETHIDINE) | 9230 | 57 | 259.92 | 4.56 |
| METHADONE | 9250B | 2 | 3.4 | 1.7 |
| MORPHINE | 9300 | 194 | 215.45 | 1.11 |
| OPIUM POWDERED | 9639 | 10 | 14.76 | 1.48 |
| OXYMORPHONE | 9652 | 2 | 0.06 | 0.03 |
| ALFENTANIL | 9737 | 7 | 1.95 | 0.28 |
| REMIFENTANIL | 9739 | 7 | 0.83 | 0.12 |
| SUFENTANIL BASE | 9740 | 2 | 0 | 0 |
| FENTANYL BASE | 9801 | 349 | 50.38 | 0.14 |

STATE:MINNESOTA BUSINESS ACTIVITY:M - MID-LEVEL PRACTITIONERS

| DRUG NAME | DRUG CODE | BUYERS | TOTAL GRAMS | AVG GRAMS |
|---|---|---|---|---|
| CODEINE | 9050 | 4 | 23.19 | 5.8 |
| DIHYDROCODEINE | 9120 | 1 | 2.35 | 2.35 |
| HYDROCODONE | 9193 | 1 | 0.27 | 0.27 |

| | | | | |
|---|---|---|---|---|
| FENTANYL BASE | 9801 | 2 | 0.01 | 0.01 |

STATE:MINNESOTA BUSINESS ACTIVITY:N-U NARCOTIC TREATMENT PROGRAMS

| DRUG NAME | DRUG CODE | BUYERS | TOTAL GRAMS | AVG GRAMS |
|---|---|---|---|---|
| BUPRENORPHINE | 9064 | 14 | 1,964.87 | 140.35 |
| METHADONE | 9250B | 18 | 174,288.15 | 9,682.68 |

STATE:MISSISSIPPI BUSINESS ACTIVITY:A - PHARMACIES

| DRUG NAME | DRUG CODE | BUYERS | TOTAL GRAMS | AVG GRAMS |
|---|---|---|---|---|
| AMPHETAMINE | 1100 | 773 | 239,922.92 | 310.38 |
| DL-METHAMPHETAMINE RACEMIC BASE | 1105B | 7 | 32.95 | 4.71 |
| D-METHAMPHETAMINE | 1105D | 1 | 2.41 | 2.41 |
| LISDEXAMFETAMINE | 1205 | 750 | 152,360.57 | 203.15 |
| METHYLPHENIDATE (DL;D;L;ISOMERS) | 1724 | 763 | 166,639.05 | 218.4 |
| BARBITURIC ACID DERIVIATIVE OR SALT [PER 21C | 2100 | 171 | 3,995 | 23.36 |
| BUTALBITAL | 2165 | 255 | 8,435 | 33.08 |
| DRONABINOL IN AN ORAL SOLUTION IN FDA APPR | 7365 | 2 | 0.75 | 0.38 |
| COCAINE | 9041L | 4 | 68.39 | 17.1 |
| CODEINE | 9050 | 769 | 104,788.51 | 136.27 |
| BUPRENORPHINE | 9064 | 709 | 38,754.40 | 54.66 |
| DIHYDROCODEINE | 9120 | 297 | 1,529.61 | 5.15 |
| OXYCODONE | 9143 | 780 | 403,226.73 | 516.96 |
| HYDROMORPHONE | 9150 | 626 | 21,501.04 | 34.35 |
| HYDROCODONE | 9193 | 789 | 441,908.96 | 560.09 |
| LEVORPHANOL | 9220L | 1 | 0.23 | 0.23 |
| MEPERIDINE (PETHIDINE) | 9230 | 510 | 10,989.83 | 21.55 |
| METHADONE | 9250B | 474 | 15,377.06 | 32.44 |
| MORPHINE | 9300 | 742 | 147,472.65 | 198.75 |
| OPIUM POWDERED | 9639 | 33 | 36.36 | 1.1 |
| OXYMORPHONE | 9652 | 416 | 12,163.29 | 29.24 |
| SUFENTANIL BASE | 9740 | 1 | 51.75 | 51.75 |
| TAPENTADOL | 9780 | 515 | 46,908.50 | 91.08 |
| FENTANYL BASE | 9801 | 755 | 4,527.85 | 6 |

| | | | | |
|---|---|---|---|---|
| OPIUM COMBINATION PRODUCT (C-III) | 9809 | 22 | 11.73 | 0.53 |

STATE:MISSISSIPPI BUSINESS ACTIVITY:B - HOSPITALS

| DRUG NAME | DRUG CODE | BUYERS | TOTAL GRAMS | AVG GRAMS |
|---|---|---|---|---|
| AMPHETAMINE | 1100 | 54 | 6,114.73 | 113.24 |
| LISDEXAMFETAMINE | 1205 | 34 | 3,560.52 | 104.72 |
| METHYLPHENIDATE (DL;D;L;ISOMERS) | 1724 | 65 | 4,680.68 | 72.01 |
| BARBITURIC ACID DERIVIATIVE OR SALT [PER 21C | 2100 | 2 | 25 | 12.5 |
| AMOBARBITAL (SCHEDULE 2) | 2125 | 1 | 4.56 | 4.56 |
| BUTALBITAL | 2165 | 12 | 110 | 9.17 |
| PENTOBARBITAL (SCHEDULE 2) | 2270 | 3 | 185.01 | 61.67 |
| COCAINE | 9041L | 26 | 244.73 | 9.41 |
| CODEINE | 9050 | 107 | 4,698.42 | 43.91 |
| BUPRENORPHINE | 9064 | 64 | 894.5 | 13.98 |
| DIHYDROCODEINE | 9120 | 7 | 32.04 | 4.58 |
| OXYCODONE | 9143 | 134 | 15,299.78 | 114.18 |
| HYDROMORPHONE | 9150 | 124 | 1,887.45 | 15.22 |
| HYDROCODONE | 9193 | 146 | 24,535.44 | 168.05 |
| MEPERIDINE (PETHIDINE) | 9230 | 118 | 5,719.35 | 48.47 |
| METHADONE | 9250B | 57 | 766.32 | 13.44 |
| MORPHINE | 9300 | 140 | 12,259.86 | 87.57 |
| OPIUM POWDERED | 9639 | 30 | 96.84 | 3.23 |
| OXYMORPHONE | 9652 | 13 | 84.99 | 6.54 |
| ALFENTANIL | 9737 | 9 | 0.43 | 0.05 |
| REMIFENTANIL | 9739 | 7 | 1.59 | 0.23 |
| SUFENTANIL BASE | 9740 | 18 | 0.35 | 0.02 |
| TAPENTADOL | 9780 | 15 | 1,377 | 91.8 |
| FENTANYL BASE | 9801 | 139 | 385.67 | 2.77 |

STATE:MISSISSIPPI BUSINESS ACTIVITY:C - PRACTITIONERS

| DRUG NAME | DRUG CODE | BUYERS | TOTAL GRAMS | AVG GRAMS |
|---|---|---|---|---|
| PENTOBARBITAL (SCHEDULE 2) | 2270 | 36 | 32,338.39 | 898.29 |
| COCAINE | 9041L | 5 | 25.86 | 5.17 |

| | | | | |
|---|---|---|---|---|
| CODEINE | 9050 | 15 | 116.91 | 7.79 |
| BUPRENORPHINE | 9064 | 111 | 5.21 | 0.05 |
| DIHYDROCODEINE | 9120 | 45 | 30.5 | 0.68 |
| OXYCODONE | 9143 | 10 | 728.05 | 72.81 |
| HYDROMORPHONE | 9150 | 33 | 15.1 | 0.46 |
| HYDROCODONE | 9193 | 41 | 102.82 | 2.51 |
| MEPERIDINE (PETHIDINE) | 9230 | 40 | 605.37 | 15.13 |
| MORPHINE | 9300 | 54 | 78.81 | 1.46 |
| OPIUM POWDERED | 9639 | 1 | 1.44 | 1.44 |
| NOROXYMORPHONE | 9668 | 1 | 6.19 | 6.19 |
| REMIFENTANIL | 9739 | 4 | 0.19 | 0.05 |
| SUFENTANIL BASE | 9740 | 3 | 0.03 | 0.01 |
| FENTANYL BASE | 9801 | 105 | 9.12 | 0.09 |

STATE:MISSISSIPPI BUSINESS ACTIVITY:D - TEACHING INSTITUTIONS

| DRUG NAME | DRUG CODE | BUYERS | TOTAL GRAMS | AVG GRAMS |
|---|---|---|---|---|
| BUPRENORPHINE | 9064 | 1 | 0 | 0 |
| HYDROMORPHONE | 9150 | 1 | 0.11 | 0.11 |

STATE:MISSISSIPPI BUSINESS ACTIVITY:M - MID-LEVEL PRACTITIONERS

| DRUG NAME | DRUG CODE | BUYERS | TOTAL GRAMS | AVG GRAMS |
|---|---|---|---|---|
| CODEINE | 9050 | 9 | 9.53 | 1.06 |
| DIHYDROCODEINE | 9120 | 21 | 13.83 | 0.66 |

STATE:MISSISSIPPI BUSINESS ACTIVITY:N-U NARCOTIC TREATMENT PROGRAMS

| DRUG NAME | DRUG CODE | BUYERS | TOTAL GRAMS | AVG GRAMS |
|---|---|---|---|---|
| BUPRENORPHINE | 9064 | 3 | 18 | 6 |
| METHADONE | 9250B | 5 | 10,500.26 | 2,100.05 |

STATE:MISSOURI BUSINESS ACTIVITY:A - PHARMACIES

| DRUG NAME | DRUG CODE | BUYERS | TOTAL GRAMS | AVG GRAMS |
|---|---|---|---|---|
| AMPHETAMINE | 1100 | 1,315 | 526,983.86 | 400.75 |
| DL-METHAMPHETAMINE RACEMIC BASE | 1105B | 28 | 190.08 | 6.79 |
| D-METHAMPHETAMINE | 1105D | 3 | 32.14 | 10.71 |
| LISDEXAMFETAMINE | 1205 | 1,281 | 263,101.40 | 205.39 |
| METHYLPHENIDATE (DL;D;L;ISOMERS) | 1724 | 1,318 | 515,769.49 | 391.33 |
| GAMMA HYDROXYBUTYRIC ACID(FDA APPROVED 30,411,958.46 | 2012 | 1 | 30,411,958.46 | |
| BARBITURIC ACID DERIVIATIVE OR SALT [PER 21C | 2100 | 378 | 15,805 | 41.81 |
| BUTALBITAL | 2165 | 530 | 23,555 | 44.44 |
| SECOBARBITAL (SCHEDULE 2) | 2315 | 4 | 100.71 | 25.18 |
| DRONABINOL IN AN ORAL SOLUTION IN FDA APPR | 7365 | 8 | 8.55 | 1.07 |
| NABILONE | 7379 | 4 | 0.55 | 0.14 |
| COCAINE | 9041L | 2 | 9.14 | 4.57 |
| CODEINE | 9050 | 1,323 | 263,296.65 | 199.01 |
| BUPRENORPHINE | 9064 | 1,234 | 35,576.31 | 28.83 |
| DIHYDROCODEINE | 9120 | 12 | 65.16 | 5.43 |
| OXYCODONE | 9143 | 1,333 | 1,129,075.33 | 847.02 |
| HYDROMORPHONE | 9150 | 1,191 | 22,113.99 | 18.57 |
| HYDROCODONE | 9193 | 1,337 | 678,034.38 | 507.13 |
| LEVORPHANOL | 9220L | 36 | 86.94 | 2.42 |
| MEPERIDINE (PETHIDINE) | 9230 | 468 | 10,250.47 | 21.9 |
| METHADONE | 9250B | 1,108 | 50,516.25 | 45.59 |
| MORPHINE | 9300 | 1,313 | 351,780.35 | 267.92 |
| OPIUM POWDERED | 9639 | 101 | 322.56 | 3.19 |
| OXYMORPHONE | 9652 | 796 | 17,158.29 | 21.56 |
| REMIFENTANIL | 9739 | 1 | 0.04 | 0.04 |
| TAPENTADOL | 9780 | 907 | 95,896 | 105.73 |
| FENTANYL BASE | 9801 | 1,294 | 7,821.11 | 6.04 |
| OPIUM COMBINATION PRODUCT (C-III) | 9809 | 26 | 27.81 | 1.07 |

STATE:MISSOURI BUSINESS ACTIVITY:B - HOSPITALS

| DRUG NAME | DRUG CODE | BUYERS | TOTAL GRAMS | AVG GRAMS |
|---|---|---|---|---|
| AMPHETAMINE | 1100 | 95 | 9,611.44 | 101.17 |

| | | | | |
|---|---|---|---|---|
| LISDEXAMFETAMINE | 1205 | 50 | 3,111.33 | 62.23 |
| METHYLPHENIDATE (DL;D;L;ISOMERS) | 1724 | 127 | 10,605.48 | 83.51 |
| BARBITURIC ACID DERIVIATIVE OR SALT [PER 21C | 2100 | 2 | 15 | 7.5 |
| AMOBARBITAL (SCHEDULE 2) | 2125 | 4 | 6.84 | 1.71 |
| BUTALBITAL | 2165 | 6 | 195 | 32.5 |
| PENTOBARBITAL (SCHEDULE 2) | 2270 | 17 | 1,238.14 | 72.83 |
| COCAINE | 9041L | 107 | 650.28 | 6.08 |
| CODEINE | 9050 | 156 | 7,419.22 | 47.56 |
| BUPRENORPHINE | 9064 | 91 | 1,138.33 | 12.51 |
| OXYCODONE | 9143 | 221 | 55,217.48 | 249.85 |
| HYDROMORPHONE | 9150 | 182 | 3,445.01 | 18.93 |
| HYDROCODONE | 9193 | 234 | 28,309.72 | 120.98 |
| MEPERIDINE (PETHIDINE) | 9230 | 168 | 4,671.96 | 27.81 |
| METHADONE | 9250B | 114 | 3,886.24 | 34.09 |
| MORPHINE | 9300 | 231 | 33,689.82 | 145.84 |
| OPIUM POWDERED | 9639 | 86 | 351.72 | 4.09 |
| OXYMORPHONE | 9652 | 18 | 68.91 | 3.83 |
| ALFENTANIL | 9737 | 15 | 4.33 | 0.29 |
| REMIFENTANIL | 9739 | 43 | 21.53 | 0.5 |
| SUFENTANIL BASE | 9740 | 45 | 1.22 | 0.03 |
| TAPENTADOL | 9780 | 30 | 1,670 | 55.67 |
| FENTANYL BASE | 9801 | 252 | 1,243.84 | 4.94 |
| OPIUM COMBINATION PRODUCT (C-III) | 9809 | 2 | 0.76 | 0.38 |

STATE:MISSOURI BUSINESS ACTIVITY:C - PRACTITIONERS

| DRUG NAME | DRUG CODE | BUYERS | TOTAL GRAMS | AVG GRAMS |
|---|---|---|---|---|
| AMPHETAMINE | 1100 | 2 | 1.72 | 0.86 |
| PENTOBARBITAL (SCHEDULE 2) | 2270 | 199 | 150,903.50 | 758.31 |
| COCAINE | 9041L | 1 | 0.14 | 0.14 |
| CODEINE | 9050 | 89 | 2,983.04 | 33.52 |
| BUPRENORPHINE | 9064 | 445 | 47.65 | 0.11 |
| DIHYDROCODEINE | 9120 | 2 | 119.63 | 59.82 |
| OXYCODONE | 9143 | 12 | 376.27 | 31.36 |
| HYDROMORPHONE | 9150 | 164 | 234.81 | 1.43 |
| HYDROCODONE | 9193 | 211 | 1,597.36 | 7.57 |

| | | | | |
|---|---|---|---|---|
| MEPERIDINE (PETHIDINE) | 9230 | 29 | 287.18 | 9.9 |
| METHADONE | 9250B | 1 | 1.79 | 1.79 |
| MORPHINE | 9300 | 161 | 440.26 | 2.73 |
| OXYMORPHONE | 9652 | 2 | 0.9 | 0.45 |
| NOROXYMORPHONE | 9668 | 5 | 37.2 | 7.44 |
| REMIFENTANIL | 9739 | 1 | 0.2 | 0.2 |
| SUFENTANIL BASE | 9740 | 3 | 0.04 | 0.01 |
| FENTANYL BASE | 9801 | 174 | 17.84 | 0.1 |

STATE:MISSOURI BUSINESS ACTIVITY:D - TEACHING INSTITUTIONS

| DRUG NAME | DRUG CODE | BUYERS | TOTAL GRAMS | AVG GRAMS |
|---|---|---|---|---|
| PENTOBARBITAL (SCHEDULE 2) | 2270 | 1 | 8,886.15 | 8,886.15 |
| CODEINE | 9050 | 1 | 99.45 | 99.45 |
| BUPRENORPHINE | 9064 | 2 | 0.43 | 0.22 |
| HYDROMORPHONE | 9150 | 2 | 8.24 | 4.12 |
| HYDROCODONE | 9193 | 2 | 7.46 | 3.73 |
| MORPHINE | 9300 | 2 | 5.85 | 2.93 |
| FENTANYL BASE | 9801 | 4 | 1.64 | 0.41 |

STATE:MISSOURI BUSINESS ACTIVITY:M - MID-LEVEL PRACTITIONERS

| DRUG NAME | DRUG CODE | BUYERS | TOTAL GRAMS | AVG GRAMS |
|---|---|---|---|---|
| HYDROMORPHONE | 9150 | 3 | 0.08 | 0.03 |
| MEPERIDINE (PETHIDINE) | 9230 | 1 | 0.87 | 0.87 |
| MORPHINE | 9300 | 39 | 17.28 | 0.44 |
| FENTANYL BASE | 9801 | 51 | 0.65 | 0.01 |

STATE:MISSOURI BUSINESS ACTIVITY:N-U NARCOTIC TREATMENT PROGRAMS

| DRUG NAME | DRUG CODE | BUYERS | TOTAL GRAMS | AVG GRAMS |
|---|---|---|---|---|
| BUPRENORPHINE | 9064 | 15 | 1,463.95 | 97.6 |
| METHADONE | 9250B | 16 | 111,576.85 | 6,973.55 |

STATE:MONTANA BUSINESS ACTIVITY:A - PHARMACIES

| DRUG NAME | DRUG CODE | BUYERS | TOTAL GRAMS | AVG GRAMS |
|---|---|---|---|---|
| AMPHETAMINE | 1100 | 234 | 52,028.33 | 222.34 |
| DL-METHAMPHETAMINE RACEMIC BASE | 1105B | 4 | 2.81 | 0.7 |
| LISDEXAMFETAMINE | 1205 | 231 | 21,587.41 | 93.45 |
| METHYLPHENIDATE (DL;D;L;ISOMERS) | 1724 | 236 | 64,115.64 | 271.68 |
| BARBITURIC ACID DERIVIATIVE OR SALT [PER 21C | 2100 | 53 | 1,935 | 36.51 |
| BUTALBITAL | 2165 | 82 | 2,990 | 36.46 |
| COCAINE | 9041L | 4 | 1.29 | 0.32 |
| CODEINE | 9050 | 234 | 35,922.31 | 153.51 |
| BUPRENORPHINE | 9064 | 222 | 8,319.59 | 37.48 |
| DIHYDROCODEINE | 9120 | 1 | 1.07 | 1.07 |
| OXYCODONE | 9143 | 238 | 137,599.01 | 578.15 |
| HYDROMORPHONE | 9150 | 233 | 5,451.12 | 23.4 |
| HYDROCODONE | 9193 | 240 | 87,270.44 | 363.63 |
| MEPERIDINE (PETHIDINE) | 9230 | 101 | 1,757.16 | 17.4 |
| METHADONE | 9250B | 202 | 7,636.39 | 37.8 |
| MORPHINE | 9300 | 238 | 57,885.39 | 243.22 |
| OPIUM POWDERED | 9639 | 18 | 32.4 | 1.8 |
| OXYMORPHONE | 9652 | 90 | 1,123.38 | 12.48 |
| TAPENTADOL | 9780 | 120 | 7,185.50 | 59.88 |
| FENTANYL BASE | 9801 | 238 | 1,061.84 | 4.46 |
| OPIUM COMBINATION PRODUCT (C-III) | 9809 | 6 | 3.41 | 0.57 |

STATE:MONTANA BUSINESS ACTIVITY:B - HOSPITALS

| DRUG NAME | DRUG CODE | BUYERS | TOTAL GRAMS | AVG GRAMS |
|---|---|---|---|---|
| AMPHETAMINE | 1100 | 38 | 3,968.30 | 104.43 |
| LISDEXAMFETAMINE | 1205 | 25 | 1,322.69 | 52.91 |
| METHYLPHENIDATE (DL;D;L;ISOMERS) | 1724 | 45 | 5,860.21 | 130.23 |
| BARBITURIC ACID DERIVIATIVE OR SALT [PER 21C | 2100 | 3 | 60 | 20 |
| BUTALBITAL | 2165 | 5 | 90 | 18 |
| PENTOBARBITAL (SCHEDULE 2) | 2270 | 2 | 3.65 | 1.82 |
| DRONABINOL IN AN ORAL SOLUTION IN FDA APPR | 7365 | 1 | 0.3 | 0.3 |

| | | | | |
|---|---|---|---|---|
| COCAINE | 9041L | 27 | 209.81 | 7.77 |
| CODEINE | 9050 | 60 | 4,467.96 | 74.47 |
| BUPRENORPHINE | 9064 | 27 | 749.11 | 27.74 |
| OXYCODONE | 9143 | 81 | 14,746.38 | 182.05 |
| HYDROMORPHONE | 9150 | 68 | 1,083.42 | 15.93 |
| HYDROCODONE | 9193 | 83 | 15,433.04 | 185.94 |
| MEPERIDINE (PETHIDINE) | 9230 | 59 | 859.76 | 14.57 |
| METHADONE | 9250B | 34 | 1,005.31 | 29.57 |
| MORPHINE | 9300 | 85 | 11,668.02 | 137.27 |
| OPIUM POWDERED | 9639 | 24 | 171.72 | 7.16 |
| OXYMORPHONE | 9652 | 2 | 9.37 | 4.68 |
| ALFENTANIL | 9737 | 12 | 1.25 | 0.1 |
| REMIFENTANIL | 9739 | 15 | 6.99 | 0.47 |
| SUFENTANIL BASE | 9740 | 18 | 0.16 | 0.01 |
| TAPENTADOL | 9780 | 9 | 249 | 27.67 |
| FENTANYL BASE | 9801 | 81 | 249.14 | 3.08 |

STATE:MONTANA BUSINESS ACTIVITY:C - PRACTITIONERS

| DRUG NAME | DRUG CODE | BUYERS | TOTAL GRAMS | AVG GRAMS |
|---|---|---|---|---|
| PENTOBARBITAL (SCHEDULE 2) | 2270 | 88 | 72,254.79 | 821.08 |
| COCAINE | 9041L | 1 | 0.5 | 0.5 |
| CODEINE | 9050 | 13 | 190.69 | 14.67 |
| BUPRENORPHINE | 9064 | 139 | 5.58 | 0.04 |
| OXYCODONE | 9143 | 4 | 5.83 | 1.46 |
| HYDROMORPHONE | 9150 | 80 | 29.91 | 0.37 |
| HYDROCODONE | 9193 | 75 | 175 | 2.33 |
| MEPERIDINE (PETHIDINE) | 9230 | 8 | 69.39 | 8.67 |
| METHADONE | 9250B | 1 | 8.05 | 8.05 |
| MORPHINE | 9300 | 71 | 88.25 | 1.24 |
| OXYMORPHONE | 9652 | 2 | 0.03 | 0.01 |
| ALFENTANIL | 9737 | 2 | 0.05 | 0.03 |
| SUFENTANIL BASE | 9740 | 1 | 0.01 | 0.01 |
| FENTANYL BASE | 9801 | 78 | 7.1 | 0.09 |

STATE:MONTANA BUSINESS ACTIVITY:D - TEACHING INSTITUTIONS

| DRUG NAME | DRUG CODE | BUYERS | TOTAL GRAMS | AVG GRAMS |
|---|---|---|---|---|
| BUPRENORPHINE | 9064 | 2 | 0.02 | 0.01 |

STATE:MONTANA BUSINESS ACTIVITY:M - MID-LEVEL PRACTITIONERS

| DRUG NAME | DRUG CODE | BUYERS | TOTAL GRAMS | AVG GRAMS |
|---|---|---|---|---|
| PENTOBARBITAL (SCHEDULE 2) | 2270 | 4 | 1,867 | 466.75 |
| MORPHINE | 9300 | 1 | 0.3 | 0.3 |
| FENTANYL BASE | 9801 | 2 | 0.01 | 0 |

STATE:MONTANA BUSINESS ACTIVITY:N-U NARCOTIC TREATMENT PROGRAMS

| DRUG NAME | DRUG CODE | BUYERS | TOTAL GRAMS | AVG GRAMS |
|---|---|---|---|---|
| BUPRENORPHINE | 9064 | 4 | 577.87 | 144.47 |
| METHADONE | 9250B | 4 | 20,571.20 | 5,142.80 |

STATE:NEBRASKA BUSINESS ACTIVITY:A - PHARMACIES

| DRUG NAME | DRUG CODE | BUYERS | TOTAL GRAMS | AVG GRAMS |
|---|---|---|---|---|
| AMPHETAMINE | 1100 | 437 | 99,599.28 | 227.92 |
| DL-METHAMPHETAMINE RACEMIC BASE | 1105B | 12 | 32.95 | 2.75 |
| D-METHAMPHETAMINE | 1105D | 1 | 0.8 | 0.8 |
| LISDEXAMFETAMINE | 1205 | 428 | 65,132.21 | 152.18 |
| METHYLPHENIDATE (DL;D;L;ISOMERS) | 1724 | 447 | 120,734.10 | 270.1 |
| BARBITURIC ACID DERIVIATIVE OR SALT [PER 21C | 2100 | 88 | 2,970 | 33.75 |
| BUTALBITAL | 2165 | 143 | 4,430 | 30.98 |
| NABILONE | 7379 | 2 | 0.3 | 0.15 |
| CODEINE | 9050 | 439 | 43,284.92 | 98.6 |
| BUPRENORPHINE | 9064 | 375 | 4,950.55 | 13.2 |
| DIHYDROCODEINE | 9120 | 5 | 9.61 | 1.92 |
| OXYCODONE | 9143 | 450 | 155,885.36 | 346.41 |
| HYDROMORPHONE | 9150 | 404 | 4,300.13 | 10.64 |

| | | | | |
|---|---|---|---|---|
| HYDROCODONE | 9193 | 455 | 121,696.44 | 267.46 |
| LEVORPHANOL | 9220L | 22 | 23.45 | 1.07 |
| MEPERIDINE (PETHIDINE) | 9230 | 108 | 1,573.17 | 14.57 |
| METHADONE | 9250B | 354 | 11,368.81 | 32.12 |
| MORPHINE | 9300 | 449 | 65,820.14 | 146.59 |
| OPIUM POWDERED | 9639 | 52 | 89.64 | 1.72 |
| OXYMORPHONE | 9652 | 238 | 4,731.21 | 19.88 |
| ALFENTANIL | 9737 | 2 | 0.3 | 0.15 |
| SUFENTANIL BASE | 9740 | 1 | 0 | 0 |
| TAPENTADOL | 9780 | 333 | 37,786.50 | 113.47 |
| FENTANYL BASE | 9801 | 432 | 1,877.69 | 4.35 |
| OPIUM COMBINATION PRODUCT (C-III) | 9809 | 7 | 3.03 | 0.43 |

STATE:NEBRASKA BUSINESS ACTIVITY:B - HOSPITALS

| DRUG NAME | DRUG CODE | BUYERS | TOTAL GRAMS | AVG GRAMS |
|---|---|---|---|---|
| AMPHETAMINE | 1100 | 37 | 3,715.76 | 100.43 |
| LISDEXAMFETAMINE | 1205 | 26 | 622.61 | 23.95 |
| METHYLPHENIDATE (DL;D;L;ISOMERS) | 1724 | 59 | 4,405.40 | 74.67 |
| AMOBARBITAL (SCHEDULE 2) | 2125 | 2 | 3.65 | 1.82 |
| BUTALBITAL | 2165 | 1 | 5 | 5 |
| PENTOBARBITAL (SCHEDULE 2) | 2270 | 3 | 349.07 | 116.36 |
| COCAINE | 9041L | 48 | 117.03 | 2.44 |
| CODEINE | 9050 | 76 | 2,646.88 | 34.83 |
| BUPRENORPHINE | 9064 | 43 | 325.15 | 7.56 |
| DIHYDROCODEINE | 9120 | 1 | 1.07 | 1.07 |
| OXYCODONE | 9143 | 118 | 10,097.63 | 85.57 |
| HYDROMORPHONE | 9150 | 105 | 830.31 | 7.91 |
| HYDROCODONE | 9193 | 128 | 8,021.33 | 62.67 |
| MEPERIDINE (PETHIDINE) | 9230 | 115 | 1,106.67 | 9.62 |
| METHADONE | 9250B | 53 | 1,316.05 | 24.83 |
| MORPHINE | 9300 | 128 | 9,734.93 | 76.05 |
| OPIUM POWDERED | 9639 | 55 | 297 | 5.4 |
| OXYMORPHONE | 9652 | 11 | 36.13 | 3.28 |
| ALFENTANIL | 9737 | 21 | 3.01 | 0.14 |
| REMIFENTANIL | 9739 | 20 | 4.29 | 0.21 |

| | | | | |
|---|---|---|---|---|
| SUFENTANIL BASE | 9740 | 26 | 0.4 | 0.02 |
| TAPENTADOL | 9780 | 37 | 943.5 | 25.5 |
| FENTANYL BASE | 9801 | 132 | 238.88 | 1.81 |

STATE:NEBRASKA BUSINESS ACTIVITY:C - PRACTITIONERS

| DRUG NAME | DRUG CODE | BUYERS | TOTAL GRAMS | AVG GRAMS |
|---|---|---|---|---|
| AMPHETAMINE | 1100 | 1 | 1.25 | 1.25 |
| LISDEXAMFETAMINE | 1205 | 1 | 11.01 | 11.01 |
| METHYLPHENIDATE (DL;D;L;ISOMERS) | 1724 | 1 | 0.86 | 0.86 |
| PENTOBARBITAL (SCHEDULE 2) | 2270 | 120 | 82,118.87 | 684.32 |
| COCAINE | 9041L | 1 | 2.14 | 2.14 |
| CODEINE | 9050 | 34 | 983.7 | 28.93 |
| BUPRENORPHINE | 9064 | 153 | 8.91 | 0.06 |
| DIHYDROCODEINE | 9120 | 1 | 1.28 | 1.28 |
| OXYCODONE | 9143 | 8 | 265.8 | 33.22 |
| HYDROMORPHONE | 9150 | 85 | 75.56 | 0.89 |
| HYDROCODONE | 9193 | 73 | 253.67 | 3.47 |
| LEVORPHANOL | 9220L | 1 | 0.35 | 0.35 |
| MEPERIDINE (PETHIDINE) | 9230 | 23 | 105.12 | 4.57 |
| METHADONE | 9250B | 2 | 5.28 | 2.64 |
| MORPHINE | 9300 | 45 | 130.51 | 2.9 |
| OPIUM POWDERED | 9639 | 2 | 7.2 | 3.6 |
| ALFENTANIL | 9737 | 2 | 0.25 | 0.13 |
| SUFENTANIL BASE | 9740 | 1 | 0 | 0 |
| TAPENTADOL | 9780 | 2 | 90 | 45 |
| FENTANYL BASE | 9801 | 88 | 11.93 | 0.14 |

STATE:NEBRASKA BUSINESS ACTIVITY:D - TEACHING INSTITUTIONS

| DRUG NAME | DRUG CODE | BUYERS | TOTAL GRAMS | AVG GRAMS |
|---|---|---|---|---|
| BUPRENORPHINE | 9064 | 1 | 0 | 0 |

STATE:NEBRASKA BUSINESS ACTIVITY:M - MID-LEVEL PRACTITIONERS

| DRUG NAME | DRUG CODE | BUYERS | TOTAL GRAMS | AVG GRAMS |
|---|---|---|---|---|
| CODEINE | 9050 | 14 | 49.77 | 3.55 |
| HYDROMORPHONE | 9150 | 4 | 0.76 | 0.19 |
| MORPHINE | 9300 | 23 | 7.52 | 0.33 |
| FENTANYL BASE | 9801 | 30 | 0.51 | 0.02 |

STATE:NEBRASKA BUSINESS ACTIVITY:N-U NARCOTIC TREATMENT PROGRAMS

| DRUG NAME | DRUG CODE | BUYERS | TOTAL GRAMS | AVG GRAMS |
|---|---|---|---|---|
| BUPRENORPHINE | 9064 | 1 | 24.24 | 24.24 |
| METHADONE | 9250B | 3 | 20,034.56 | 6,678.19 |

STATE:NEVADA BUSINESS ACTIVITY:A - PHARMACIES

| DRUG NAME | DRUG CODE | BUYERS | TOTAL GRAMS | AVG GRAMS |
|---|---|---|---|---|
| AMPHETAMINE | 1100 | 463 | 100,986.79 | 218.11 |
| DL-METHAMPHETAMINE RACEMIC BASE | 1105B | 28 | 97.65 | 3.49 |
| D-METHAMPHETAMINE | 1105D | 1 | 0.4 | 0.4 |
| LISDEXAMFETAMINE | 1205 | 423 | 29,670.98 | 70.14 |
| METHYLPHENIDATE (DL;D;L;ISOMERS) | 1724 | 444 | 54,626.24 | 123.03 |
| BARBITURIC ACID DERIVIATIVE OR SALT [PER 21C | 2100 | 243 | 18,270 | 75.19 |
| BUTALBITAL | 2165 | 256 | 14,230 | 55.59 |
| SECOBARBITAL (SCHEDULE 2) | 2315 | 1 | 9.16 | 9.16 |
| DRONABINOL IN AN ORAL SOLUTION IN FDA APPR | 7365 | 3 | 11.55 | 3.85 |
| COCAINE | 9041L | 2 | 14.28 | 7.14 |
| CODEINE | 9050 | 466 | 102,752.54 | 220.5 |
| BUPRENORPHINE | 9064 | 440 | 12,534.87 | 28.49 |
| DIHYDROCODEINE | 9120 | 6 | 74.77 | 12.46 |
| OXYCODONE | 9143 | 476 | 670,863.26 | 1,409.38 |
| HYDROMORPHONE | 9150 | 444 | 9,415.59 | 21.21 |
| HYDROCODONE | 9193 | 477 | 350,438.22 | 734.67 |
| LEVORPHANOL | 9220L | 13 | 14.16 | 1.09 |
| MEPERIDINE (PETHIDINE) | 9230 | 161 | 2,866.80 | 17.81 |
| METHADONE | 9250B | 448 | 42,900.07 | 95.76 |

| | | | | |
|---|---|---|---|---|
| MORPHINE | 9300 | 467 | 208,424.09 | 446.3 |
| OPIUM POWDERED | 9639 | 24 | 30.24 | 1.26 |
| OXYMORPHONE | 9652 | 386 | 15,788.61 | 40.9 |
| ALFENTANIL | 9737 | 2 | 0.07 | 0.03 |
| SUFENTANIL BASE | 9740 | 1 | 0.01 | 0.01 |
| TAPENTADOL | 9780 | 350 | 47,776.50 | 136.5 |
| FENTANYL BASE | 9801 | 449 | 1,898.68 | 4.23 |
| OPIUM COMBINATION PRODUCT (C-III) | 9809 | 14 | 4.16 | 0.3 |

STATE:NEVADA BUSINESS ACTIVITY:B - HOSPITALS

| DRUG NAME | DRUG CODE | BUYERS | TOTAL GRAMS | AVG GRAMS |
|---|---|---|---|---|
| AMPHETAMINE | 1100 | 35 | 3,135.11 | 89.57 |
| LISDEXAMFETAMINE | 1205 | 12 | 601.8 | 50.15 |
| METHYLPHENIDATE (DL;D;L;ISOMERS) | 1724 | 50 | 4,397 | 87.94 |
| BARBITURIC ACID DERIVIATIVE OR SALT [PER 21C | 2100 | 1 | 5 | 5 |
| BUTALBITAL | 2165 | 7 | 185 | 26.43 |
| PENTOBARBITAL (SCHEDULE 2) | 2270 | 6 | 437.02 | 72.84 |
| COCAINE | 9041L | 31 | 165.53 | 5.34 |
| CODEINE | 9050 | 53 | 3,778.36 | 71.29 |
| BUPRENORPHINE | 9064 | 30 | 391.4 | 13.05 |
| OXYCODONE | 9143 | 86 | 41,851.91 | 486.65 |
| HYDROMORPHONE | 9150 | 77 | 2,173.90 | 28.23 |
| HYDROCODONE | 9193 | 91 | 25,392.72 | 279.04 |
| MEPERIDINE (PETHIDINE) | 9230 | 77 | 2,055.35 | 26.69 |
| METHADONE | 9250B | 46 | 2,939.69 | 63.91 |
| MORPHINE | 9300 | 92 | 27,846.86 | 302.68 |
| OPIUM POWDERED | 9639 | 29 | 103.68 | 3.58 |
| OXYMORPHONE | 9652 | 8 | 121.59 | 15.2 |
| ALFENTANIL | 9737 | 21 | 6.7 | 0.32 |
| REMIFENTANIL | 9739 | 19 | 3.41 | 0.18 |
| SUFENTANIL BASE | 9740 | 23 | 0.36 | 0.02 |
| TAPENTADOL | 9780 | 13 | 879 | 67.62 |
| FENTANYL BASE | 9801 | 119 | 420.12 | 3.53 |

STATISTICAL SUMMARY FOR RETAIL DRUG PURCHASES BY GRAMS WT

STATE:NEVADA BUSINESS ACTIVITY:C - PRACTITIONERS

| DRUG NAME | DRUG CODE | BUYERS | TOTAL GRAMS | AVG GRAMS |
|---|---|---|---|---|
| AMPHETAMINE | 1100 | 1 | 25.65 | 25.65 |
| LISDEXAMFETAMINE | 1205 | 1 | 5.67 | 5.67 |
| METHYLPHENIDATE (DL;D;L;ISOMERS) | 1724 | 1 | 217.18 | 217.18 |
| BUTALBITAL | 2165 | 1 | 135 | 135 |
| PENTOBARBITAL (SCHEDULE 2) | 2270 | 55 | 57,764.25 | 1,050.26 |
| COCAINE | 9041L | 1 | 0.14 | 0.14 |
| CODEINE | 9050 | 39 | 290.12 | 7.44 |
| BUPRENORPHINE | 9064 | 187 | 343.66 | 1.84 |
| DIHYDROCODEINE | 9120 | 2 | 0.64 | 0.32 |
| OXYCODONE | 9143 | 32 | 2,470.20 | 77.19 |
| HYDROMORPHONE | 9150 | 140 | 176.85 | 1.26 |
| HYDROCODONE | 9193 | 124 | 961.21 | 7.75 |
| MEPERIDINE (PETHIDINE) | 9230 | 61 | 738.71 | 12.11 |
| METHADONE | 9250B | 14 | 209.29 | 14.95 |
| MORPHINE | 9300 | 93 | 610.54 | 6.56 |
| OPIUM POWDERED | 9639 | 1 | 0.36 | 0.36 |
| OXYMORPHONE | 9652 | 6 | 6.24 | 1.04 |
| NOROXYMORPHONE | 9668 | 1 | 25.26 | 25.26 |
| ALFENTANIL | 9737 | 5 | 0.19 | 0.04 |
| REMIFENTANIL | 9739 | 5 | 0.29 | 0.06 |
| SUFENTANIL BASE | 9740 | 1 | 0.01 | 0.01 |
| TAPENTADOL | 9780 | 1 | 40 | 40 |
| FENTANYL BASE | 9801 | 183 | 28.57 | 0.16 |

STATE:NEVADA BUSINESS ACTIVITY:D - TEACHING INSTITUTIONS

| DRUG NAME | DRUG CODE | BUYERS | TOTAL GRAMS | AVG GRAMS |
|---|---|---|---|---|
| MEPERIDINE (PETHIDINE) | 9230 | 1 | 2.18 | 2.18 |
| FENTANYL BASE | 9801 | 1 | 0 | 0 |

STATE:NEVADA BUSINESS ACTIVITY:M - MID-LEVEL PRACTITIONERS

| DRUG NAME | DRUG CODE | BUYERS | TOTAL GRAMS | AVG GRAMS |
|---|---|---|---|---|
| METHYLPHENIDATE (DL;D;L;ISOMERS) | 1724 | 1 | 0.86 | 0.86 |
| BUTALBITAL | 2165 | 4 | 265 | 66.25 |
| PENTOBARBITAL (SCHEDULE 2) | 2270 | 9 | 3,554.46 | 394.94 |
| CODEINE | 9050 | 10 | 128.05 | 12.81 |
| OXYCODONE | 9143 | 4 | 2,105.65 | 526.41 |
| HYDROMORPHONE | 9150 | 4 | 30.14 | 7.54 |
| HYDROCODONE | 9193 | 6 | 713.01 | 118.83 |
| MEPERIDINE (PETHIDINE) | 9230 | 1 | 7.84 | 7.84 |
| METHADONE | 9250B | 4 | 355.52 | 88.88 |
| MORPHINE | 9300 | 5 | 632.88 | 126.58 |
| OXYMORPHONE | 9652 | 3 | 13.92 | 4.64 |
| TAPENTADOL | 9780 | 2 | 110 | 55 |
| FENTANYL BASE | 9801 | 4 | 6.55 | 1.64 |

STATE:NEVADA BUSINESS ACTIVITY:N-U NARCOTIC TREATMENT PROGRAMS

| DRUG NAME | DRUG CODE | BUYERS | TOTAL GRAMS | AVG GRAMS |
|---|---|---|---|---|
| BUPRENORPHINE | 9064 | 7 | 375.29 | 53.61 |
| METHADONE | 9250B | 14 | 98,291.43 | 7,020.82 |

STATE:NEW HAMPSHIRE BUSINESS ACTIVITY:A - PHARMACIES

| DRUG NAME | DRUG CODE | BUYERS | TOTAL GRAMS | AVG GRAMS |
|---|---|---|---|---|
| AMPHETAMINE | 1100 | 266 | 151,006.22 | 567.69 |
| DL-METHAMPHETAMINE RACEMIC BASE | 1105B | 7 | 26.12 | 3.73 |
| LISDEXAMFETAMINE | 1205 | 265 | 55,129.35 | 208.04 |
| METHYLPHENIDATE (DL;D;L;ISOMERS) | 1724 | 266 | 133,225.04 | 500.85 |
| BARBITURIC ACID DERIVIATIVE OR SALT [PER 21C | 2100 | 73 | 1,995 | 27.33 |
| BUTALBITAL | 2165 | 104 | 2,665 | 25.63 |
| SECOBARBITAL (SCHEDULE 2) | 2315 | 3 | 27.47 | 9.16 |
| DRONABINOL IN AN ORAL SOLUTION IN FDA APPR | 7365 | 2 | 1.8 | 0.9 |
| NABILONE | 7379 | 1 | 0.05 | 0.05 |
| COCAINE | 9041L | 1 | 0.89 | 0.89 |
| CODEINE | 9050 | 262 | 29,873.77 | 114.02 |

| | | | | |
|---|---|---|---|---|
| BUPRENORPHINE | 9064 | 266 | 30,886.95 | 116.12 |
| OXYCODONE | 9143 | 265 | 239,173.80 | 902.54 |
| HYDROMORPHONE | 9150 | 262 | 6,852.23 | 26.15 |
| HYDROCODONE | 9193 | 267 | 34,627.25 | 129.69 |
| LEVORPHANOL | 9220L | 5 | 5.34 | 1.07 |
| MEPERIDINE (PETHIDINE) | 9230 | 62 | 1,915.56 | 30.9 |
| METHADONE | 9250B | 249 | 22,414.65 | 90.02 |
| MORPHINE | 9300 | 268 | 67,312.62 | 251.17 |
| OPIUM TINCTURE (LAUDANUM) | 9630 | 1 | 4.73 | 4.73 |
| OPIUM POWDERED | 9639 | 15 | 51.84 | 3.46 |
| OXYMORPHONE | 9652 | 160 | 3,487.98 | 21.8 |
| TAPENTADOL | 9780 | 205 | 29,166.50 | 142.28 |
| FENTANYL BASE | 9801 | 256 | 1,224.90 | 4.78 |
| OPIUM COMBINATION PRODUCT (C-III) | 9809 | 11 | 8.89 | 0.81 |

STATE:NEW HAMPSHIRE BUSINESS ACTIVITY:B - HOSPITALS

| DRUG NAME | DRUG CODE | BUYERS | TOTAL GRAMS | AVG GRAMS |
|---|---|---|---|---|
| AMPHETAMINE | 1100 | 23 | 2,862.90 | 124.47 |
| LISDEXAMFETAMINE | 1205 | 12 | 508.73 | 42.39 |
| METHYLPHENIDATE (DL;D;L;ISOMERS) | 1724 | 35 | 2,414.93 | 69 |
| AMOBARBITAL (SCHEDULE 2) | 2125 | 1 | 12.3 | 12.3 |
| PENTOBARBITAL (SCHEDULE 2) | 2270 | 3 | 359.09 | 119.7 |
| COCAINE | 9041L | 26 | 180.42 | 6.94 |
| CODEINE | 9050 | 33 | 457.54 | 13.86 |
| BUPRENORPHINE | 9064 | 26 | 651.12 | 25.04 |
| OXYCODONE | 9143 | 47 | 10,206.15 | 217.15 |
| HYDROMORPHONE | 9150 | 40 | 972.52 | 24.31 |
| HYDROCODONE | 9193 | 45 | 1,480.23 | 32.89 |
| MEPERIDINE (PETHIDINE) | 9230 | 36 | 323.65 | 8.99 |
| METHADONE | 9250B | 31 | 1,379.61 | 44.5 |
| MORPHINE | 9300 | 47 | 5,109.35 | 108.71 |
| OPIUM POWDERED | 9639 | 17 | 94.68 | 5.57 |
| OXYMORPHONE | 9652 | 1 | 3.57 | 3.57 |
| ALFENTANIL | 9737 | 7 | 1.95 | 0.28 |
| REMIFENTANIL | 9739 | 17 | 6.51 | 0.38 |

| | | | | |
|---|---|---|---|---|
| SUFENTANIL BASE | 9740 | 7 | 0.06 | 0.01 |
| TAPENTADOL | 9780 | 1 | 249 | 249 |
| FENTANYL BASE | 9801 | 47 | 150.54 | 3.2 |

STATE:NEW HAMPSHIRE BUSINESS ACTIVITY:C - PRACTITIONERS

| DRUG NAME | DRUG CODE | BUYERS | TOTAL GRAMS | AVG GRAMS |
|---|---|---|---|---|
| PENTOBARBITAL (SCHEDULE 2) | 2270 | 81 | 61,566.17 | 760.08 |
| COCAINE | 9041L | 1 | 9 | 9 |
| CODEINE | 9050 | 3 | 11.64 | 3.88 |
| BUPRENORPHINE | 9064 | 178 | 11.4 | 0.06 |
| OXYCODONE | 9143 | 2 | 0.9 | 0.45 |
| HYDROMORPHONE | 9150 | 84 | 38.97 | 0.46 |
| HYDROCODONE | 9193 | 14 | 15.55 | 1.11 |
| MEPERIDINE (PETHIDINE) | 9230 | 10 | 110.68 | 11.07 |
| METHADONE | 9250B | 3 | 1.61 | 0.54 |
| MORPHINE | 9300 | 53 | 55.51 | 1.05 |
| OXYMORPHONE | 9652 | 4 | 0.04 | 0.01 |
| REMIFENTANIL | 9739 | 1 | 0.01 | 0.01 |
| SUFENTANIL BASE | 9740 | 1 | 0 | 0 |
| FENTANYL BASE | 9801 | 100 | 15.65 | 0.16 |

STATE:NEW HAMPSHIRE BUSINESS ACTIVITY:M - MID-LEVEL PRACTITIONERS

| DRUG NAME | DRUG CODE | BUYERS | TOTAL GRAMS | AVG GRAMS |
|---|---|---|---|---|
| CODEINE | 9050 | 1 | 4.42 | 4.42 |
| HYDROCODONE | 9193 | 1 | 0.3 | 0.3 |
| FENTANYL BASE | 9801 | 1 | 0.01 | 0.01 |

STATE:NEW HAMPSHIRE BUSINESS ACTIVITY:N-U NARCOTIC TREATMENT PROGRAMS

| DRUG NAME | DRUG CODE | BUYERS | TOTAL GRAMS | AVG GRAMS |
|---|---|---|---|---|
| BUPRENORPHINE | 9064 | 4 | 543.8 | 135.95 |
| METHADONE | 9250B | 8 | 99,537.78 | 12,442.22 |

STATISTICAL SUMMARY FOR RETAIL DRUG PURCHASES BY GRAMS WT

STATE:NEW JERSEY BUSINESS ACTIVITY:A - PHARMACIES

| DRUG NAME | DRUG CODE | BUYERS | TOTAL GRAMS | AVG GRAMS |
|---|---|---|---|---|
| AMPHETAMINE | 1100 | 1,980 | 413,434.81 | 208.81 |
| DL-METHAMPHETAMINE RACEMIC BASE | 1105B | 49 | 138.24 | 2.82 |
| D-METHAMPHETAMINE | 1105D | 4 | 5.62 | 1.41 |
| LISDEXAMFETAMINE | 1205 | 1,807 | 194,671.13 | 107.73 |
| METHYLPHENIDATE (DL;D;L;ISOMERS) | 1724 | 1,948 | 363,224.43 | 186.46 |
| BARBITURIC ACID DERIVIATIVE OR SALT [PER 21C | 2100 | 515 | 18,025 | 35 |
| BUTALBITAL | 2165 | 648 | 24,470 | 37.76 |
| PENTOBARBITAL (SCHEDULE 2) | 2270 | 1 | 11 | 11 |
| SECOBARBITAL (SCHEDULE 2) | 2315 | 11 | 228.88 | 20.81 |
| DRONABINOL IN AN ORAL SOLUTION IN FDA APPR | 7365 | 11 | 143.55 | 13.05 |
| NABILONE | 7379 | 4 | 0.35 | 0.09 |
| COCAINE | 9041L | 7 | 118.31 | 16.9 |
| CODEINE | 9050 | 1,994 | 298,841.60 | 149.87 |
| BUPRENORPHINE | 9064 | 1,831 | 103,668.13 | 56.62 |
| DIHYDROCODEINE | 9120 | 82 | 1,154.68 | 14.08 |
| OXYCODONE | 9143 | 2,030 | 1,954,808.71 | 962.96 |
| HYDROMORPHONE | 9150 | 1,779 | 30,522.62 | 17.16 |
| HYDROCODONE | 9193 | 1,985 | 135,813.57 | 68.42 |
| LEVORPHANOL | 9220L | 26 | 1,620.24 | 62.32 |
| MEPERIDINE (PETHIDINE) | 9230 | 236 | 6,194.08 | 26.25 |
| METHADONE | 9250B | 1,193 | 42,942.47 | 36 |
| MORPHINE | 9300 | 1,842 | 324,762.10 | 176.31 |
| OPIUM TINCTURE (LAUDANUM) | 9630 | 1 | 1.18 | 1.18 |
| OPIUM POWDERED | 9639 | 109 | 320.04 | 2.94 |
| OXYMORPHONE | 9652 | 989 | 25,615.09 | 25.9 |
| ALFENTANIL | 9737 | 2 | 0.47 | 0.24 |
| REMIFENTANIL | 9739 | 5 | 0.4 | 0.08 |
| SUFENTANIL BASE | 9740 | 3 | 2.78 | 0.93 |
| TAPENTADOL | 9780 | 1,405 | 187,283 | 133.3 |
| FENTANYL BASE | 9801 | 1,810 | 8,428.03 | 4.66 |
| OPIUM COMBINATION PRODUCT (C-III) | 9809 | 35 | 20.43 | 0.58 |

STATISTICAL SUMMARY FOR RETAIL DRUG PURCHASES BY GRAMS WT

STATE:NEW JERSEY BUSINESS ACTIVITY:B - HOSPITALS

| DRUG NAME | DRUG CODE | BUYERS | TOTAL GRAMS | AVG GRAMS |
|---|---|---|---|---|
| AMPHETAMINE | 1100 | 52 | 3,242.42 | 62.35 |
| LISDEXAMFETAMINE | 1205 | 23 | 1,289.74 | 56.08 |
| METHYLPHENIDATE (DL;D;L;ISOMERS) | 1724 | 96 | 3,555.90 | 37.04 |
| BARBITURIC ACID DERIVIATIVE OR SALT [PER 21C | 2100 | 2 | 85 | 42.5 |
| AMOBARBITAL (SCHEDULE 2) | 2125 | 10 | 30.53 | 3.05 |
| BUTALBITAL | 2165 | 5 | 200 | 40 |
| PENTOBARBITAL (SCHEDULE 2) | 2270 | 17 | 793.37 | 46.67 |
| COCAINE | 9041L | 62 | 351.73 | 5.67 |
| CODEINE | 9050 | 107 | 4,538.21 | 42.41 |
| BUPRENORPHINE | 9064 | 84 | 824.04 | 9.81 |
| DIHYDROCODEINE | 9120 | 2 | 5.34 | 2.67 |
| OXYCODONE | 9143 | 120 | 36,298.47 | 302.49 |
| HYDROMORPHONE | 9150 | 112 | 4,832.17 | 43.14 |
| HYDROCODONE | 9193 | 89 | 1,402.54 | 15.76 |
| MEPERIDINE (PETHIDINE) | 9230 | 76 | 1,166.72 | 15.35 |
| METHADONE | 9250B | 99 | 3,457.55 | 34.92 |
| MORPHINE | 9300 | 115 | 15,255.99 | 132.66 |
| OPIUM POWDERED | 9639 | 42 | 122.04 | 2.91 |
| OXYMORPHONE | 9652 | 7 | 78.19 | 11.17 |
| ALFENTANIL | 9737 | 5 | 0.38 | 0.08 |
| REMIFENTANIL | 9739 | 54 | 50.87 | 0.94 |
| SUFENTANIL BASE | 9740 | 22 | 0.94 | 0.04 |
| TAPENTADOL | 9780 | 36 | 1,935.50 | 53.76 |
| FENTANYL BASE | 9801 | 108 | 685.01 | 6.34 |
| OPIUM COMBINATION PRODUCT (C-III) | 9809 | 2 | 0.38 | 0.19 |

STATE:NEW JERSEY BUSINESS ACTIVITY:C - PRACTITIONERS

| DRUG NAME | DRUG CODE | BUYERS | TOTAL GRAMS | AVG GRAMS |
|---|---|---|---|---|
| AMPHETAMINE | 1100 | 1 | 1.47 | 1.47 |
| PENTOBARBITAL (SCHEDULE 2) | 2270 | 94 | 74,270.34 | 790.11 |
| COCAINE | 9041L | 38 | 175.7 | 4.62 |

| | | | | |
|---|---|---|---|---|
| CODEINE | 9050 | 549 | 6,748.20 | 12.29 |
| BUPRENORPHINE | 9064 | 550 | 408.22 | 0.74 |
| DIHYDROCODEINE | 9120 | 85 | 79.68 | 0.94 |
| OXYCODONE | 9143 | 112 | 205.08 | 1.83 |
| HYDROMORPHONE | 9150 | 295 | 142.79 | 0.48 |
| HYDROCODONE | 9193 | 258 | 911.93 | 3.53 |
| MEPERIDINE (PETHIDINE) | 9230 | 126 | 341.95 | 2.71 |
| METHADONE | 9250B | 22 | 81.48 | 3.7 |
| MORPHINE | 9300 | 198 | 223.81 | 1.13 |
| OPIUM POWDERED | 9639 | 2 | 1.08 | 0.54 |
| OXYMORPHONE | 9652 | 6 | 0.77 | 0.13 |
| NOROXYMORPHONE | 9668 | 2 | 23.7 | 11.85 |
| ALFENTANIL | 9737 | 2 | 0.2 | 0.1 |
| REMIFENTANIL | 9739 | 29 | 1.64 | 0.06 |
| SUFENTANIL BASE | 9740 | 3 | 0.01 | 0 |
| TAPENTADOL | 9780 | 1 | 7.5 | 7.5 |
| FENTANYL BASE | 9801 | 498 | 65.7 | 0.13 |

STATE:NEW JERSEY BUSINESS ACTIVITY:D - TEACHING INSTITUTIONS

| DRUG NAME | DRUG CODE | BUYERS | TOTAL GRAMS | AVG GRAMS |
|---|---|---|---|---|
| BUPRENORPHINE | 9064 | 4 | 0.02 | 0 |
| MORPHINE | 9300 | 1 | 0.02 | 0.02 |

STATE:NEW JERSEY BUSINESS ACTIVITY:M - MID-LEVEL PRACTITIONERS

| DRUG NAME | DRUG CODE | BUYERS | TOTAL GRAMS | AVG GRAMS |
|---|---|---|---|---|
| PENTOBARBITAL (SCHEDULE 2) | 2270 | 10 | 6,843.43 | 684.34 |
| CODEINE | 9050 | 45 | 685.87 | 15.24 |
| BUPRENORPHINE | 9064 | 1 | 0.12 | 0.12 |
| DIHYDROCODEINE | 9120 | 6 | 9.05 | 1.51 |

STATE:NEW JERSEY BUSINESS ACTIVITY:N-U NARCOTIC TREATMENT PROGRAMS

| DRUG NAME | DRUG CODE | BUYERS | TOTAL GRAMS | AVG GRAMS |
|---|---|---|---|---|
| BUPRENORPHINE | 9064 | 18 | 652.27 | 36.24 |
| METHADONE | 9250B | 31 | 413,079.53 | 13,325.15 |

STATE:NEW MEXICO BUSINESS ACTIVITY:A - PHARMACIES

| DRUG NAME | DRUG CODE | BUYERS | TOTAL GRAMS | AVG GRAMS |
|---|---|---|---|---|
| AMPHETAMINE | 1100 | 345 | 60,983.73 | 176.76 |
| DL-METHAMPHETAMINE RACEMIC BASE | 1105B | 23 | 65.9 | 2.87 |
| LISDEXAMFETAMINE | 1205 | 319 | 16,776.46 | 52.59 |
| METHYLPHENIDATE (DL;D;L;ISOMERS) | 1724 | 350 | 69,650.67 | 199 |
| BARBITURIC ACID DERIVIATIVE OR SALT [PER 21C | 2100 | 114 | 3,305 | 28.99 |
| BUTALBITAL | 2165 | 114 | 3,690 | 32.37 |
| DRONABINOL IN AN ORAL SOLUTION IN FDA APPR | 7365 | 1 | 0.15 | 0.15 |
| NABILONE | 7379 | 1 | 0.1 | 0.1 |
| COCAINE | 9041L | 1 | 0.71 | 0.71 |
| CODEINE | 9050 | 349 | 66,843.22 | 191.53 |
| BUPRENORPHINE | 9064 | 335 | 27,314.03 | 81.53 |
| DIHYDROCODEINE | 9120 | 15 | 343.95 | 22.93 |
| OXYCODONE | 9143 | 354 | 334,781.44 | 945.71 |
| HYDROMORPHONE | 9150 | 317 | 4,463.65 | 14.08 |
| HYDROCODONE | 9193 | 355 | 126,982.61 | 357.7 |
| MEPERIDINE (PETHIDINE) | 9230 | 187 | 4,691.50 | 25.09 |
| METHADONE | 9250B | 283 | 8,166.95 | 28.86 |
| MORPHINE | 9300 | 350 | 86,645.42 | 247.56 |
| OPIUM POWDERED | 9639 | 15 | 29.16 | 1.94 |
| OXYMORPHONE | 9652 | 208 | 3,785.05 | 18.2 |
| TAPENTADOL | 9780 | 183 | 12,742 | 69.63 |
| FENTANYL BASE | 9801 | 334 | 1,394.40 | 4.17 |
| OPIUM COMBINATION PRODUCT (C-III) | 9809 | 6 | 6.62 | 1.1 |

STATE:NEW MEXICO BUSINESS ACTIVITY:B - HOSPITALS

| DRUG NAME | DRUG CODE | BUYERS | TOTAL GRAMS | AVG GRAMS |
|---|---|---|---|---|
| AMPHETAMINE | 1100 | 41 | 2,676.61 | 65.28 |

| | | | | |
|---|---|---|---|---|
| LISDEXAMFETAMINE | 1205 | 9 | 483.29 | 53.7 |
| METHYLPHENIDATE (DL;D;L;ISOMERS) | 1724 | 59 | 6,619.76 | 112.2 |
| BARBITURIC ACID DERIVIATIVE OR SALT [PER 21C | 2100 | 3 | 20 | 6.67 |
| BUTALBITAL | 2165 | 1 | 10 | 10 |
| PENTOBARBITAL (SCHEDULE 2) | 2270 | 5 | 123.95 | 24.79 |
| COCAINE | 9041L | 23 | 289.7 | 12.6 |
| CODEINE | 9050 | 71 | 11,150.29 | 157.05 |
| BUPRENORPHINE | 9064 | 44 | 1,744.13 | 39.64 |
| OXYCODONE | 9143 | 87 | 26,561.48 | 305.3 |
| HYDROMORPHONE | 9150 | 65 | 846.69 | 13.03 |
| HYDROCODONE | 9193 | 94 | 12,433.96 | 132.28 |
| MEPERIDINE (PETHIDINE) | 9230 | 58 | 933.96 | 16.1 |
| METHADONE | 9250B | 47 | 2,442.16 | 51.96 |
| MORPHINE | 9300 | 96 | 16,345.95 | 170.27 |
| OPIUM POWDERED | 9639 | 29 | 115.56 | 3.98 |
| OXYMORPHONE | 9652 | 3 | 12.94 | 4.31 |
| ALFENTANIL | 9737 | 10 | 4.1 | 0.41 |
| REMIFENTANIL | 9739 | 17 | 4.42 | 0.26 |
| SUFENTANIL BASE | 9740 | 12 | 0.19 | 0.02 |
| TAPENTADOL | 9780 | 4 | 84.5 | 21.13 |
| FENTANYL BASE | 9801 | 100 | 324.63 | 3.25 |

STATE:NEW MEXICO BUSINESS ACTIVITY:C - PRACTITIONERS

| DRUG NAME | DRUG CODE | BUYERS | TOTAL GRAMS | AVG GRAMS |
|---|---|---|---|---|
| PENTOBARBITAL (SCHEDULE 2) | 2270 | 77 | 64,790.68 | 841.44 |
| CODEINE | 9050 | 20 | 144.21 | 7.21 |
| BUPRENORPHINE | 9064 | 133 | 115.49 | 0.87 |
| OXYCODONE | 9143 | 5 | 47.14 | 9.43 |
| HYDROMORPHONE | 9150 | 75 | 76.01 | 1.01 |
| HYDROCODONE | 9193 | 48 | 122.47 | 2.55 |
| MEPERIDINE (PETHIDINE) | 9230 | 7 | 25.06 | 3.58 |
| METHADONE | 9250B | 3 | 1.34 | 0.45 |
| MORPHINE | 9300 | 51 | 132.72 | 2.6 |
| OXYMORPHONE | 9652 | 2 | 0.06 | 0.03 |
| REMIFENTANIL | 9739 | 2 | 0.02 | 0.01 |

| | | | | |
|---|---|---|---|---|
| SUFENTANIL BASE | 9740 | 2 | 0 | 0 |
| FENTANYL BASE | 9801 | 95 | 15.03 | 0.16 |

STATE:NEW MEXICO BUSINESS ACTIVITY:D - TEACHING INSTITUTIONS

| DRUG NAME | DRUG CODE | BUYERS | TOTAL GRAMS | AVG GRAMS |
|---|---|---|---|---|
| PENTOBARBITAL (SCHEDULE 2) | 2270 | 1 | 177.72 | 177.72 |
| BUPRENORPHINE | 9064 | 1 | 0.01 | 0.01 |
| FENTANYL BASE | 9801 | 1 | 0.02 | 0.02 |

STATE:NEW MEXICO BUSINESS ACTIVITY:M - MID-LEVEL PRACTITIONERS

| DRUG NAME | DRUG CODE | BUYERS | TOTAL GRAMS | AVG GRAMS |
|---|---|---|---|---|
| PENTOBARBITAL (SCHEDULE 2) | 2270 | 9 | 15,210.75 | 1,690.08 |
| CODEINE | 9050 | 14 | 49.29 | 3.52 |
| BUPRENORPHINE | 9064 | 2 | 0.57 | 0.29 |
| HYDROMORPHONE | 9150 | 7 | 9.26 | 1.32 |
| HYDROCODONE | 9193 | 3 | 4.64 | 1.55 |
| MORPHINE | 9300 | 24 | 28.94 | 1.21 |
| FENTANYL BASE | 9801 | 38 | 0.97 | 0.03 |

STATE:NEW MEXICO BUSINESS ACTIVITY:N-U NARCOTIC TREATMENT PROGRAMS

| DRUG NAME | DRUG CODE | BUYERS | TOTAL GRAMS | AVG GRAMS |
|---|---|---|---|---|
| BUPRENORPHINE | 9064 | 4 | 321.37 | 80.34 |
| METHADONE | 9250B | 16 | 165,785.98 | 10,361.62 |

STATE:NEW YORK BUSINESS ACTIVITY:A - PHARMACIES

| DRUG NAME | DRUG CODE | BUYERS | TOTAL GRAMS | AVG GRAMS |
|---|---|---|---|---|
| AMPHETAMINE | 1100 | 4,608 | 933,140.47 | 202.5 |
| DL-METHAMPHETAMINE RACEMIC BASE | 1105B | 99 | 325.5 | 3.29 |
| D-METHAMPHETAMINE | 1105D | 3 | 5.62 | 1.87 |

| | | | | |
|---|---|---|---|---|
| LISDEXAMFETAMINE | 1205 | 3,780 | 336,801.64 | 89.1 |
| METHYLPHENIDATE (DL;D;L;ISOMERS) | 1724 | 4,509 | 897,756.11 | 199.1 |
| BARBITURIC ACID DERIVIATIVE OR SALT [PER 21C | 2100 | 691 | 17,585 | 25.45 |
| BUTALBITAL | 2165 | 844 | 22,770 | 26.98 |
| SECOBARBITAL (SCHEDULE 2) | 2315 | 36 | 1,501.42 | 41.71 |
| DRONABINOL IN AN ORAL SOLUTION IN FDA APPR | 7365 | 6 | 1.35 | 0.23 |
| NABILONE | 7379 | 4 | 0.3 | 0.08 |
| COCAINE | 9041L | 5 | 43.32 | 8.66 |
| CODEINE | 9050 | 4,829 | 548,795.54 | 113.65 |
| BUPRENORPHINE | 9064 | 3,975 | 205,972.50 | 51.82 |
| DIHYDROCODEINE | 9120 | 52 | 528.74 | 10.17 |
| OXYCODONE | 9143 | 4,940 | 2,855,205.02 | 577.98 |
| HYDROMORPHONE | 9150 | 3,962 | 62,142.71 | 15.68 |
| HYDROCODONE | 9193 | 4,691 | 652,797.84 | 139.16 |
| LEVORPHANOL | 9220L | 175 | 160.07 | 0.91 |
| MEPERIDINE (PETHIDINE) | 9230 | 376 | 12,760.50 | 33.94 |
| METHADONE | 9250B | 3,143 | 138,877.51 | 44.19 |
| MORPHINE | 9300 | 4,107 | 559,554.36 | 136.24 |
| OPIUM TINCTURE (LAUDANUM) | 9630 | 1 | 4.73 | 4.73 |
| OPIUM POWDERED | 9639 | 134 | 352.44 | 2.63 |
| OXYMORPHONE | 9652 | 2,137 | 71,418.43 | 33.42 |
| ALFENTANIL | 9737 | 1 | 0.36 | 0.36 |
| REMIFENTANIL | 9739 | 2 | 0.07 | 0.04 |
| SUFENTANIL BASE | 9740 | 1 | 0.27 | 0.27 |
| TAPENTADOL | 9780 | 2,688 | 376,794 | 140.18 |
| FENTANYL BASE | 9801 | 4,070 | 14,662.30 | 3.6 |
| OPIUM COMBINATION PRODUCT (C-III) | 9809 | 71 | 34.43 | 0.48 |

STATE:NEW YORK BUSINESS ACTIVITY:B - HOSPITALS

| DRUG NAME | DRUG CODE | BUYERS | TOTAL GRAMS | AVG GRAMS |
|---|---|---|---|---|
| AMPHETAMINE | 1100 | 183 | 8,586.93 | 46.92 |
| LISDEXAMFETAMINE | 1205 | 84 | 1,611.64 | 19.19 |
| METHYLPHENIDATE (DL;D;L;ISOMERS) | 1724 | 250 | 12,305.75 | 49.22 |
| AMOBARBITAL (SCHEDULE 2) | 2125 | 24 | 152.66 | 6.36 |
| BUTALBITAL | 2165 | 1 | 5 | 5 |

| | | | | |
|---|---|---|---|---|
| PENTOBARBITAL (SCHEDULE 2) | 2270 | 39 | 11,239.38 | 288.19 |
| DRONABINOL IN AN ORAL SOLUTION IN FDA APPR | 7365 | 2 | 0.6 | 0.3 |
| NABILONE | 7379 | 1 | 0.1 | 0.1 |
| COCAINE | 9041L | 146 | 1,156.68 | 7.92 |
| CODEINE | 9050 | 286 | 15,225.18 | 53.23 |
| BUPRENORPHINE | 9064 | 227 | 4,980.06 | 21.94 |
| OXYCODONE | 9143 | 331 | 95,895.83 | 289.72 |
| HYDROMORPHONE | 9150 | 285 | 15,628.55 | 54.84 |
| HYDROCODONE | 9193 | 253 | 26,950.88 | 106.53 |
| LEVORPHANOL | 9220L | 3 | 1.04 | 0.35 |
| MEPERIDINE (PETHIDINE) | 9230 | 208 | 4,064.81 | 19.54 |
| METHADONE | 9250B | 261 | 25,920.08 | 99.31 |
| MORPHINE | 9300 | 320 | 60,556.56 | 189.24 |
| OPIUM POWDERED | 9639 | 84 | 504 | 6 |
| OXYMORPHONE | 9652 | 47 | 394.54 | 8.39 |
| ALFENTANIL | 9737 | 25 | 7.89 | 0.32 |
| REMIFENTANIL | 9739 | 121 | 152.48 | 1.26 |
| SUFENTANIL BASE | 9740 | 48 | 6.43 | 0.13 |
| TAPENTADOL | 9780 | 86 | 7,090.50 | 82.45 |
| FENTANYL BASE | 9801 | 295 | 2,431.02 | 8.24 |
| OPIUM COMBINATION PRODUCT (C-III) | 9809 | 8 | 3.59 | 0.45 |

STATE:NEW YORK BUSINESS ACTIVITY:C - PRACTITIONERS

| DRUG NAME | DRUG CODE | BUYERS | TOTAL GRAMS | AVG GRAMS |
|---|---|---|---|---|
| AMPHETAMINE | 1100 | 2 | 5.74 | 2.87 |
| D-METHAMPHETAMINE | 1105D | 2 | 8.11 | 4.06 |
| LISDEXAMFETAMINE | 1205 | 1 | 5.2 | 5.2 |
| METHYLPHENIDATE (DL;D;L;ISOMERS) | 1724 | 1 | 18.68 | 18.68 |
| GAMMA HYDROXYBUTYRIC ACID(FDA APPROVED | 2012 | 1 | 7,950.53 | 7,950.53 |
| PENTOBARBITAL (SCHEDULE 2) | 2270 | 327 | 283,493.89 | 866.95 |
| COCAINE | 9041L | 47 | 179.88 | 3.83 |
| CODEINE | 9050 | 130 | 3,662.15 | 28.17 |
| BUPRENORPHINE | 9064 | 1,069 | 162.95 | 0.15 |
| DIHYDROCODEINE | 9120 | 3 | 0.53 | 0.18 |
| OXYCODONE | 9143 | 138 | 615.28 | 4.46 |

| | | | | |
|---|---|---|---|---|
| HYDROMORPHONE | 9150 | 615 | 290.15 | 0.47 |
| HYDROCODONE | 9193 | 430 | 1,401.04 | 3.26 |
| MEPERIDINE (PETHIDINE) | 9230 | 214 | 1,462.81 | 6.84 |
| METHADONE | 9250B | 28 | 93.91 | 3.35 |
| MORPHINE | 9300 | 395 | 646.37 | 1.64 |
| OPIUM POWDERED | 9639 | 2 | 12.96 | 6.48 |
| OXYMORPHONE | 9652 | 8 | 0.91 | 0.11 |
| NOROXYMORPHONE | 9668 | 5 | 22.13 | 4.43 |
| ALFENTANIL | 9737 | 5 | 0.9 | 0.18 |
| REMIFENTANIL | 9739 | 29 | 2.48 | 0.09 |
| SUFENTANIL BASE | 9740 | 8 | 0.04 | 0 |
| FENTANYL BASE | 9801 | 1,102 | 143.75 | 0.13 |

STATE:NEW YORK BUSINESS ACTIVITY:D - TEACHING INSTITUTIONS

| DRUG NAME | DRUG CODE | BUYERS | TOTAL GRAMS | AVG GRAMS |
|---|---|---|---|---|
| AMPHETAMINE | 1100 | 1 | 0.73 | 0.73 |
| D-METHAMPHETAMINE | 1105D | 1 | 4.02 | 4.02 |
| PENTOBARBITAL (SCHEDULE 2) | 2270 | 2 | 182.28 | 91.14 |
| BUPRENORPHINE | 9064 | 7 | 0.15 | 0.02 |
| MORPHINE | 9300 | 1 | 0.56 | 0.56 |
| FENTANYL BASE | 9801 | 1 | 0.01 | 0.01 |

STATE:NEW YORK BUSINESS ACTIVITY:M - MID-LEVEL PRACTITIONERS

| DRUG NAME | DRUG CODE | BUYERS | TOTAL GRAMS | AVG GRAMS |
|---|---|---|---|---|
| PENTOBARBITAL (SCHEDULE 2) | 2270 | 9 | 11,479.79 | 1,275.53 |
| CODEINE | 9050 | 4 | 67.06 | 16.76 |
| BUPRENORPHINE | 9064 | 1 | 0.18 | 0.18 |
| HYDROCODONE | 9193 | 1 | 0.06 | 0.06 |
| MEPERIDINE (PETHIDINE) | 9230 | 1 | 1.09 | 1.09 |
| FENTANYL BASE | 9801 | 1 | 0.28 | 0.28 |

STATE:NEW YORK BUSINESS ACTIVITY:N-U NARCOTIC TREATMENT PROGRAMS

STATISTICAL SUMMARY FOR RETAIL DRUG PURCHASES BY GRAMS WT

| DRUG NAME | DRUG CODE | BUYERS | TOTAL GRAMS | AVG GRAMS |
|---|---|---|---|---|
| BUPRENORPHINE | 9064 | 35 | 2,241.80 | 64.05 |
| METHADONE | 9250B | 107 | 1,054,499.21 | 9,855.13 |

STATE:NORTH CAROLINA BUSINESS ACTIVITY:A - PHARMACIES

| DRUG NAME | DRUG CODE | BUYERS | TOTAL GRAMS | AVG GRAMS |
|---|---|---|---|---|
| AMPHETAMINE | 1100 | 2,182 | 783,256.30 | 358.96 |
| DL-METHAMPHETAMINE RACEMIC BASE | 1105B | 53 | 189.27 | 3.57 |
| D-METHAMPHETAMINE | 1105D | 7 | 21.69 | 3.1 |
| LISDEXAMFETAMINE | 1205 | 2,134 | 400,999.64 | 187.91 |
| METHYLPHENIDATE (DL;D;L;ISOMERS) | 1724 | 2,173 | 729,832.98 | 335.86 |
| BARBITURIC ACID DERIVIATIVE OR SALT [PER 21C | 2100 | 622 | 15,190 | 24.42 |
| AMOBARBITAL (SCHEDULE 2) | 2125 | 1 | 0.46 | 0.46 |
| BUTALBITAL | 2165 | 768 | 20,160 | 26.25 |
| SECOBARBITAL (SCHEDULE 2) | 2315 | 1 | 9.16 | 9.16 |
| DRONABINOL IN AN ORAL SOLUTION IN FDA APPR | 7365 | 4 | 1.05 | 0.26 |
| NABILONE | 7379 | 1 | 0.75 | 0.75 |
| COCAINE | 9041L | 8 | 18.21 | 2.28 |
| CODEINE | 9050 | 2,119 | 183,727.15 | 86.7 |
| BUPRENORPHINE | 9064 | 2,065 | 116,841.36 | 56.58 |
| DIHYDROCODEINE | 9120 | 20 | 38.45 | 1.92 |
| OXYCODONE | 9143 | 2,196 | 2,127,760.19 | 968.93 |
| HYDROMORPHONE | 9150 | 2,081 | 35,580.68 | 17.1 |
| HYDROCODONE | 9193 | 2,202 | 721,205.59 | 327.52 |
| LEVORPHANOL | 9220L | 241 | 265.24 | 1.1 |
| MEPERIDINE (PETHIDINE) | 9230 | 1,072 | 21,046.62 | 19.63 |
| METHADONE | 9250B | 1,938 | 101,110.13 | 52.17 |
| MORPHINE | 9300 | 2,150 | 425,240.22 | 197.79 |
| OPIUM POWDERED | 9639 | 145 | 249.12 | 1.72 |
| OXYMORPHONE | 9652 | 1,733 | 81,925.14 | 47.27 |
| ALFENTANIL | 9737 | 2 | 0.52 | 0.26 |
| REMIFENTANIL | 9739 | 5 | 0.15 | 0.03 |
| TAPENTADOL | 9780 | 1,880 | 373,342.50 | 198.59 |
| FENTANYL BASE | 9801 | 2,099 | 9,921.68 | 4.73 |

| | | | | |
|---|---|---|---|---|
| OPIUM COMBINATION PRODUCT (C-III) | 9809 | 79 | 46.16 | 0.58 |

STATE:NORTH CAROLINA BUSINESS ACTIVITY:B - HOSPITALS

| DRUG NAME | DRUG CODE | BUYERS | TOTAL GRAMS | AVG GRAMS |
|---|---|---|---|---|
| AMPHETAMINE | 1100 | 144 | 29,274.43 | 203.29 |
| LISDEXAMFETAMINE | 1205 | 74 | 9,571.60 | 129.35 |
| METHYLPHENIDATE (DL;D;L;ISOMERS) | 1724 | 166 | 32,330.23 | 194.76 |
| GAMMA HYDROXYBUTYRIC ACID(FDA APPROVED | 2012 | 1 | 2,303.42 | 2,303.42 |
| BARBITURIC ACID DERIVIATIVE OR SALT [PER 21C | 2100 | 5 | 75 | 15 |
| AMOBARBITAL (SCHEDULE 2) | 2125 | 5 | 37.82 | 7.56 |
| BUTALBITAL | 2165 | 8 | 230 | 28.75 |
| PENTOBARBITAL (SCHEDULE 2) | 2270 | 58 | 57,754.21 | 995.76 |
| COCAINE | 9041L | 108 | 545.75 | 5.05 |
| CODEINE | 9050 | 179 | 9,394.60 | 52.48 |
| BUPRENORPHINE | 9064 | 237 | 3,060.72 | 12.91 |
| OXYCODONE | 9143 | 259 | 111,067.62 | 428.83 |
| HYDROMORPHONE | 9150 | 320 | 7,288.89 | 22.78 |
| HYDROCODONE | 9193 | 307 | 47,459.63 | 154.59 |
| LEVORPHANOL | 9220L | 1 | 0.46 | 0.46 |
| MEPERIDINE (PETHIDINE) | 9230 | 194 | 3,049.05 | 15.72 |
| METHADONE | 9250B | 170 | 8,413.99 | 49.49 |
| MORPHINE | 9300 | 319 | 50,321.02 | 157.75 |
| OPIUM POWDERED | 9639 | 99 | 668.16 | 6.75 |
| OXYMORPHONE | 9652 | 82 | 1,030.11 | 12.56 |
| ALFENTANIL | 9737 | 29 | 12.05 | 0.42 |
| REMIFENTANIL | 9739 | 80 | 53.01 | 0.66 |
| SUFENTANIL BASE | 9740 | 61 | 1.64 | 0.03 |
| TAPENTADOL | 9780 | 87 | 13,185 | 151.55 |
| FENTANYL BASE | 9801 | 329 | 1,307.64 | 3.97 |
| OPIUM COMBINATION PRODUCT (C-III) | 9809 | 4 | 7.76 | 1.94 |

STATE:NORTH CAROLINA BUSINESS ACTIVITY:C - PRACTITIONERS

| DRUG NAME | DRUG CODE | BUYERS | TOTAL GRAMS | AVG GRAMS |
|---|---|---|---|---|

| | | | | |
|---|---|---|---|---|
| AMPHETAMINE | 1100 | 1 | 0.74 | 0.74 |
| LISDEXAMFETAMINE | 1205 | 1 | 0.72 | 0.72 |
| METHYLPHENIDATE (DL;D;L;ISOMERS) | 1724 | 1 | 16.52 | 16.52 |
| GAMMA HYDROXYBUTYRIC ACID(FDA APPROVED | 2012 | 1 | 2,674.94 | 2,674.94 |
| PENTOBARBITAL (SCHEDULE 2) | 2270 | 199 | 132,180.20 | 664.22 |
| COCAINE | 9041L | 5 | 7.71 | 1.54 |
| CODEINE | 9050 | 119 | 376.08 | 3.16 |
| BUPRENORPHINE | 9064 | 898 | 57.14 | 0.06 |
| OXYCODONE | 9143 | 24 | 245.68 | 10.24 |
| HYDROMORPHONE | 9150 | 393 | 166.39 | 0.42 |
| HYDROCODONE | 9193 | 279 | 655.97 | 2.35 |
| MEPERIDINE (PETHIDINE) | 9230 | 98 | 446.21 | 4.55 |
| METHADONE | 9250B | 7 | 14.13 | 2.02 |
| MORPHINE | 9300 | 212 | 455.78 | 2.15 |
| OPIUM POWDERED | 9639 | 1 | 0.36 | 0.36 |
| OXYMORPHONE | 9652 | 6 | 1.09 | 0.18 |
| NOROXYMORPHONE | 9668 | 1 | 1.56 | 1.56 |
| REMIFENTANIL | 9739 | 24 | 4.59 | 0.19 |
| SUFENTANIL BASE | 9740 | 2 | 0.03 | 0.01 |
| FENTANYL BASE | 9801 | 415 | 35.33 | 0.09 |

STATE:NORTH CAROLINA BUSINESS ACTIVITY:D - TEACHING INSTITUTIONS

| DRUG NAME | DRUG CODE | BUYERS | TOTAL GRAMS | AVG GRAMS |
|---|---|---|---|---|
| PENTOBARBITAL (SCHEDULE 2) | 2270 | 1 | 266.58 | 266.58 |
| BUPRENORPHINE | 9064 | 3 | 0.03 | 0.01 |
| HYDROMORPHONE | 9150 | 2 | 0.14 | 0.07 |
| METHADONE | 9250B | 1 | 0.89 | 0.89 |
| MORPHINE | 9300 | 1 | 0.19 | 0.19 |
| FENTANYL BASE | 9801 | 1 | 0.03 | 0.03 |

STATE:NORTH CAROLINA BUSINESS ACTIVITY:M - MID-LEVEL PRACTITIONERS

| DRUG NAME | DRUG CODE | BUYERS | TOTAL GRAMS | AVG GRAMS |
|---|---|---|---|---|
| PENTOBARBITAL (SCHEDULE 2) | 2270 | 37 | 52,333.38 | 1,414.42 |

| | | | | |
|---|---|---|---|---|
| CODEINE | 9050 | 70 | 119.34 | 1.7 |
| BUPRENORPHINE | 9064 | 2 | 0.04 | 0.02 |
| HYDROMORPHONE | 9150 | 10 | 1.38 | 0.14 |
| MORPHINE | 9300 | 76 | 122.8 | 1.62 |
| NOROXYMORPHONE | 9668 | 1 | 2.12 | 2.12 |
| FENTANYL BASE | 9801 | 82 | 1.53 | 0.02 |

STATE:NORTH CAROLINA BUSINESS ACTIVITY:N-U NARCOTIC TREATMENT PROGRAMS

| DRUG NAME | DRUG CODE | BUYERS | TOTAL GRAMS | AVG GRAMS |
|---|---|---|---|---|
| BUPRENORPHINE | 9064 | 44 | 10,882.82 | 247.34 |
| METHADONE | 9250B | 62 | 416,957.21 | 6,725.12 |

STATE:NORTH DAKOTA BUSINESS ACTIVITY:A - PHARMACIES

| DRUG NAME | DRUG CODE | BUYERS | TOTAL GRAMS | AVG GRAMS |
|---|---|---|---|---|
| AMPHETAMINE | 1100 | 170 | 33,433.63 | 196.67 |
| DL-METHAMPHETAMINE RACEMIC BASE | 1105B | 3 | 22.5 | 7.5 |
| LISDEXAMFETAMINE | 1205 | 165 | 21,507.05 | 130.35 |
| METHYLPHENIDATE (DL;D;L;ISOMERS) | 1724 | 170 | 54,194.44 | 318.79 |
| BARBITURIC ACID DERIVIATIVE OR SALT [PER 21C | 2100 | 24 | 505 | 21.04 |
| BUTALBITAL | 2165 | 36 | 1,275 | 35.42 |
| CODEINE | 9050 | 171 | 18,917.36 | 110.63 |
| BUPRENORPHINE | 9064 | 131 | 3,480.74 | 26.57 |
| OXYCODONE | 9143 | 174 | 49,037.51 | 281.82 |
| HYDROMORPHONE | 9150 | 156 | 1,848.34 | 11.85 |
| HYDROCODONE | 9193 | 176 | 33,229.56 | 188.8 |
| MEPERIDINE (PETHIDINE) | 9230 | 53 | 649.05 | 12.25 |
| METHADONE | 9250B | 122 | 3,385.04 | 27.75 |
| MORPHINE | 9300 | 171 | 19,115.07 | 111.78 |
| OPIUM POWDERED | 9639 | 24 | 46.8 | 1.95 |
| OXYMORPHONE | 9652 | 44 | 608.06 | 13.82 |
| ALFENTANIL | 9737 | 1 | 0.03 | 0.03 |
| TAPENTADOL | 9780 | 95 | 8,464.50 | 89.1 |
| FENTANYL BASE | 9801 | 167 | 756.58 | 4.53 |

| | | | | |
|---|---|---|---|---|
| OPIUM COMBINATION PRODUCT (C-III) | 9809 | 1 | 1.14 | 1.14 |

STATE:NORTH DAKOTA BUSINESS ACTIVITY:B - HOSPITALS

| DRUG NAME | DRUG CODE | BUYERS | TOTAL GRAMS | AVG GRAMS |
|---|---|---|---|---|
| AMPHETAMINE | 1100 | 21 | 2,528.18 | 120.39 |
| DL-METHAMPHETAMINE RACEMIC BASE | 1105B | 1 | 1.21 | 1.21 |
| D-METHAMPHETAMINE | 1105D | 1 | 2.01 | 2.01 |
| LISDEXAMFETAMINE | 1205 | 15 | 1,385.13 | 92.34 |
| METHYLPHENIDATE (DL;D;L;ISOMERS) | 1724 | 25 | 5,932.29 | 237.29 |
| BARBITURIC ACID DERIVIATIVE OR SALT [PER 21C | 2100 | 2 | 60 | 30 |
| BUTALBITAL | 2165 | 2 | 10 | 5 |
| PENTOBARBITAL (SCHEDULE 2) | 2270 | 3 | 110.28 | 36.76 |
| COCAINE | 9041L | 13 | 181.56 | 13.97 |
| CODEINE | 9050 | 50 | 3,728.53 | 74.57 |
| BUPRENORPHINE | 9064 | 16 | 149.78 | 9.36 |
| OXYCODONE | 9143 | 53 | 9,349.43 | 176.4 |
| HYDROMORPHONE | 9150 | 48 | 732.86 | 15.27 |
| HYDROCODONE | 9193 | 54 | 8,074.94 | 149.54 |
| MEPERIDINE (PETHIDINE) | 9230 | 35 | 255.02 | 7.29 |
| METHADONE | 9250B | 22 | 477.97 | 21.73 |
| MORPHINE | 9300 | 56 | 5,988.08 | 106.93 |
| OPIUM POWDERED | 9639 | 17 | 117.36 | 6.9 |
| OXYMORPHONE | 9652 | 2 | 21.59 | 10.79 |
| ALFENTANIL | 9737 | 6 | 0.56 | 0.09 |
| REMIFENTANIL | 9739 | 8 | 1.67 | 0.21 |
| SUFENTANIL BASE | 9740 | 6 | 0.13 | 0.02 |
| TAPENTADOL | 9780 | 6 | 1,311 | 218.5 |
| FENTANYL BASE | 9801 | 53 | 185.39 | 3.5 |
| OPIUM COMBINATION PRODUCT (C-III) | 9809 | 1 | 1.51 | 1.51 |

STATE:NORTH DAKOTA BUSINESS ACTIVITY:C - PRACTITIONERS

| DRUG NAME | DRUG CODE | BUYERS | TOTAL GRAMS | AVG GRAMS |
|---|---|---|---|---|
| AMPHETAMINE | 1100 | 2 | 2.5 | 1.25 |

| | | | | |
|---|---|---|---|---|
| D-METHAMPHETAMINE | 1105D | 1 | 4.02 | 4.02 |
| METHYLPHENIDATE (DL;D;L;ISOMERS) | 1724 | 1 | 1.87 | 1.87 |
| PENTOBARBITAL (SCHEDULE 2) | 2270 | 34 | 28,257.96 | 831.12 |
| COCAINE | 9041L | 2 | 0.43 | 0.21 |
| CODEINE | 9050 | 9 | 132.5 | 14.72 |
| BUPRENORPHINE | 9064 | 58 | 4.82 | 0.08 |
| OXYCODONE | 9143 | 10 | 202.78 | 20.28 |
| HYDROMORPHONE | 9150 | 37 | 14.16 | 0.38 |
| HYDROCODONE | 9193 | 26 | 188.83 | 7.26 |
| MEPERIDINE (PETHIDINE) | 9230 | 14 | 47.71 | 3.41 |
| METHADONE | 9250B | 1 | 1.61 | 1.61 |
| MORPHINE | 9300 | 30 | 28.69 | 0.96 |
| OPIUM POWDERED | 9639 | 1 | 0.72 | 0.72 |
| FENTANYL BASE | 9801 | 43 | 5.91 | 0.14 |

STATE:NORTH DAKOTA BUSINESS ACTIVITY:N-U NARCOTIC TREATMENT PROGRAMS

| DRUG NAME | DRUG CODE | BUYERS | TOTAL GRAMS | AVG GRAMS |
|---|---|---|---|---|
| METHADONE | 9250B | 3 | 4,507.78 | 1,502.59 |

STATE:OHIO BUSINESS ACTIVITY:A - PHARMACIES

| DRUG NAME | DRUG CODE | BUYERS | TOTAL GRAMS | AVG GRAMS |
|---|---|---|---|---|
| AMPHETAMINE | 1100 | 2,239 | 832,478.90 | 371.81 |
| DL-METHAMPHETAMINE RACEMIC BASE | 1105B | 40 | 296.57 | 7.41 |
| D-METHAMPHETAMINE | 1105D | 6 | 19.28 | 3.21 |
| LISDEXAMFETAMINE | 1205 | 2,184 | 443,038.50 | 202.86 |
| METHYLPHENIDATE (DL;D;L;ISOMERS) | 1724 | 2,237 | 910,677.43 | 407.1 |
| BARBITURIC ACID DERIVIATIVE OR SALT [PER 21C | 2100 | 631 | 16,775 | 26.58 |
| BUTALBITAL | 2165 | 833 | 26,475 | 31.78 |
| SECOBARBITAL (SCHEDULE 2) | 2315 | 3 | 137.33 | 45.78 |
| DRONABINOL IN AN ORAL SOLUTION IN FDA APPR | 7365 | 6 | 8.55 | 1.43 |
| NABILONE | 7379 | 2 | 1 | 0.5 |
| COCAINE | 9041L | 3 | 10.71 | 3.57 |
| CODEINE | 9050 | 2,211 | 392,141.04 | 177.36 |

| | | | | |
|---|---|---|---|---|
| BUPRENORPHINE | 9064 | 2,101 | 196,821.05 | 93.68 |
| DIHYDROCODEINE | 9120 | 12 | 108.95 | 9.08 |
| OXYCODONE | 9143 | 2,251 | 1,830,420.30 | 813.16 |
| HYDROMORPHONE | 9150 | 1,922 | 28,209.03 | 14.68 |
| HYDROCODONE | 9193 | 2,263 | 716,367.69 | 316.56 |
| LEVORPHANOL | 9220L | 18 | 43.53 | 2.42 |
| MEPERIDINE (PETHIDINE) | 9230 | 458 | 9,740.32 | 21.27 |
| METHADONE | 9250B | 1,895 | 85,828.95 | 45.29 |
| MORPHINE | 9300 | 2,180 | 430,821.21 | 197.62 |
| OPIUM TINCTURE (LAUDANUM) | 9630 | 22 | 82.68 | 3.76 |
| OPIUM POWDERED | 9639 | 260 | 671.4 | 2.58 |
| OXYMORPHONE | 9652 | 1,422 | 38,057.25 | 26.76 |
| ALFENTANIL | 9737 | 2 | 0.26 | 0.13 |
| REMIFENTANIL | 9739 | 1 | 0.1 | 0.1 |
| SUFENTANIL BASE | 9740 | 2 | 0.14 | 0.07 |
| TAPENTADOL | 9780 | 1,657 | 239,097 | 144.3 |
| FENTANYL BASE | 9801 | 2,083 | 8,506.36 | 4.08 |
| OPIUM COMBINATION PRODUCT (C-III) | 9809 | 62 | 53.35 | 0.86 |

STATE:OHIO BUSINESS ACTIVITY:B - HOSPITALS

| DRUG NAME | DRUG CODE | BUYERS | TOTAL GRAMS | AVG GRAMS |
|---|---|---|---|---|
| AMPHETAMINE | 1100 | 160 | 14,539.95 | 90.87 |
| DL-METHAMPHETAMINE RACEMIC BASE | 1105B | 2 | 3.62 | 1.81 |
| D-METHAMPHETAMINE | 1105D | 1 | 2.01 | 2.01 |
| LISDEXAMFETAMINE | 1205 | 70 | 3,455.88 | 49.37 |
| METHYLPHENIDATE (DL;D;L;ISOMERS) | 1724 | 217 | 16,217.66 | 74.74 |
| GAMMA HYDROXYBUTYRIC ACID(FDA APPROVED | 2012 | 2 | 7,430.40 | 3,715.20 |
| BARBITURIC ACID DERIVIATIVE OR SALT [PER 21C | 2100 | 4 | 20 | 5 |
| AMOBARBITAL (SCHEDULE 2) | 2125 | 5 | 17.77 | 3.55 |
| BUTALBITAL | 2165 | 12 | 155 | 12.92 |
| PENTOBARBITAL (SCHEDULE 2) | 2270 | 32 | 24,444.20 | 763.88 |
| DRONABINOL IN AN ORAL SOLUTION IN FDA APPR | 7365 | 1 | 0.15 | 0.15 |
| COCAINE | 9041L | 165 | 716.78 | 4.34 |
| CODEINE | 9050 | 274 | 10,508.07 | 38.35 |
| BUPRENORPHINE | 9064 | 244 | 5,909.78 | 24.22 |

| | | | | |
|---|---|---|---|---|
| DIHYDROCODEINE | 9120 | 8 | 31.35 | 3.92 |
| OXYCODONE | 9143 | 388 | 95,344.85 | 245.73 |
| HYDROMORPHONE | 9150 | 383 | 8,997.19 | 23.49 |
| HYDROCODONE | 9193 | 396 | 25,747.75 | 65.02 |
| MEPERIDINE (PETHIDINE) | 9230 | 318 | 9,768.02 | 30.72 |
| METHADONE | 9250B | 244 | 9,047.40 | 37.08 |
| MORPHINE | 9300 | 405 | 46,408.03 | 114.59 |
| OPIUM POWDERED | 9639 | 185 | 842.4 | 4.55 |
| OXYMORPHONE | 9652 | 64 | 491.17 | 7.67 |
| ALFENTANIL | 9737 | 29 | 18.53 | 0.64 |
| REMIFENTANIL | 9739 | 115 | 74.21 | 0.65 |
| SUFENTANIL BASE | 9740 | 76 | 1.82 | 0.02 |
| TAPENTADOL | 9780 | 74 | 3,662 | 49.49 |
| FENTANYL BASE | 9801 | 471 | 1,308.75 | 2.78 |
| OPIUM COMBINATION PRODUCT (C-III) | 9809 | 9 | 4.9 | 0.54 |

STATE:OHIO BUSINESS ACTIVITY:C - PRACTITIONERS

| DRUG NAME | DRUG CODE | BUYERS | TOTAL GRAMS | AVG GRAMS |
|---|---|---|---|---|
| AMPHETAMINE | 1100 | 1 | 0.99 | 0.99 |
| LISDEXAMFETAMINE | 1205 | 2 | 19.6 | 9.8 |
| PENTOBARBITAL (SCHEDULE 2) | 2270 | 294 | 237,887.06 | 809.14 |
| COCAINE | 9041L | 7 | 3.29 | 0.47 |
| CODEINE | 9050 | 64 | 332.8 | 5.2 |
| BUPRENORPHINE | 9064 | 861 | 131.59 | 0.15 |
| OXYCODONE | 9143 | 26 | 765.03 | 29.42 |
| HYDROMORPHONE | 9150 | 339 | 175.71 | 0.52 |
| HYDROCODONE | 9193 | 274 | 785.35 | 2.87 |
| MEPERIDINE (PETHIDINE) | 9230 | 69 | 472.35 | 6.85 |
| METHADONE | 9250B | 6 | 7.33 | 1.22 |
| MORPHINE | 9300 | 184 | 476.14 | 2.59 |
| OPIUM POWDERED | 9639 | 1 | 0.72 | 0.72 |
| OXYMORPHONE | 9652 | 5 | 0.94 | 0.19 |
| NOROXYMORPHONE | 9668 | 4 | 24.31 | 6.08 |
| ALFENTANIL | 9737 | 3 | 0.3 | 0.1 |
| REMIFENTANIL | 9739 | 13 | 1.31 | 0.1 |

| | | | | |
|---|---|---|---|---|
| FENTANYL BASE | 9801 | 361 | 33.83 | 0.09 |

STATE:OHIO BUSINESS ACTIVITY:D - TEACHING INSTITUTIONS

| DRUG NAME | DRUG CODE | BUYERS | TOTAL GRAMS | AVG GRAMS |
|---|---|---|---|---|
| PENTOBARBITAL (SCHEDULE 2) | 2270 | 1 | 0.91 | 0.91 |
| BUPRENORPHINE | 9064 | 6 | 0.17 | 0.03 |
| HYDROMORPHONE | 9150 | 4 | 0.25 | 0.06 |
| MORPHINE | 9300 | 1 | 0.03 | 0.03 |
| REMIFENTANIL | 9739 | 1 | 0.01 | 0.01 |
| FENTANYL BASE | 9801 | 1 | 0.15 | 0.15 |

STATE:OHIO BUSINESS ACTIVITY:M - MID-LEVEL PRACTITIONERS

| DRUG NAME | DRUG CODE | BUYERS | TOTAL GRAMS | AVG GRAMS |
|---|---|---|---|---|
| PENTOBARBITAL (SCHEDULE 2) | 2270 | 29 | 32,789.62 | 1,130.68 |
| CODEINE | 9050 | 2 | 1.98 | 0.99 |
| BUPRENORPHINE | 9064 | 5 | 1.75 | 0.35 |
| HYDROMORPHONE | 9150 | 6 | 0.55 | 0.09 |
| MEPERIDINE (PETHIDINE) | 9230 | 1 | 0.22 | 0.22 |
| MORPHINE | 9300 | 93 | 16.64 | 0.18 |
| FENTANYL BASE | 9801 | 152 | 1.11 | 0.01 |

STATE:OHIO BUSINESS ACTIVITY:N-U NARCOTIC TREATMENT PROGRAMS

| DRUG NAME | DRUG CODE | BUYERS | TOTAL GRAMS | AVG GRAMS |
|---|---|---|---|---|
| CODEINE | 9050 | 1 | 22.1 | 22.1 |
| BUPRENORPHINE | 9064 | 19 | 7,234.55 | 380.77 |
| METHADONE | 9250B | 23 | 202,429.55 | 8,801.28 |

STATE:OKLAHOMA BUSINESS ACTIVITY:A - PHARMACIES

| DRUG NAME | DRUG CODE | BUYERS | TOTAL GRAMS | AVG GRAMS |
|---|---|---|---|---|

| | | | | |
|---|---|---|---|---|
| AMPHETAMINE | 1100 | 895 | 205,219.28 | 229.3 |
| DL-METHAMPHETAMINE RACEMIC BASE | 1105B | 18 | 77.16 | 4.29 |
| D-METHAMPHETAMINE | 1105D | 1 | 7.23 | 7.23 |
| LISDEXAMFETAMINE | 1205 | 869 | 142,821.92 | 164.35 |
| METHYLPHENIDATE (DL;D;L;ISOMERS) | 1724 | 894 | 183,848.76 | 205.65 |
| BARBITURIC ACID DERIVIATIVE OR SALT [PER 21C | 2100 | 255 | 11,250 | 44.12 |
| BUTALBITAL | 2165 | 443 | 19,900 | 44.92 |
| SECOBARBITAL (SCHEDULE 2) | 2315 | 1 | 36.62 | 36.62 |
| COCAINE | 9041L | 2 | 0.86 | 0.43 |
| CODEINE | 9050 | 930 | 343,730.75 | 369.6 |
| BUPRENORPHINE | 9064 | 863 | 42,293.71 | 49.01 |
| DIHYDROCODEINE | 9120 | 239 | 1,064.96 | 4.46 |
| OXYCODONE | 9143 | 909 | 860,854.37 | 947.03 |
| HYDROMORPHONE | 9150 | 801 | 12,337.59 | 15.4 |
| HYDROCODONE | 9193 | 926 | 684,169.84 | 738.84 |
| MEPERIDINE (PETHIDINE) | 9230 | 621 | 22,530.67 | 36.28 |
| METHADONE | 9250B | 751 | 32,580.40 | 43.38 |
| MORPHINE | 9300 | 899 | 333,136.65 | 370.56 |
| OPIUM POWDERED | 9639 | 55 | 139.32 | 2.53 |
| OXYMORPHONE | 9652 | 621 | 21,522.63 | 34.66 |
| REMIFENTANIL | 9739 | 2 | 0.31 | 0.16 |
| SUFENTANIL BASE | 9740 | 1 | 0.06 | 0.06 |
| TAPENTADOL | 9780 | 664 | 93,729.50 | 141.16 |
| FENTANYL BASE | 9801 | 894 | 6,131.44 | 6.86 |
| OPIUM COMBINATION PRODUCT (C-III) | 9809 | 19 | 9.65 | 0.51 |

STATE:OKLAHOMA BUSINESS ACTIVITY:B - HOSPITALS

| DRUG NAME | DRUG CODE | BUYERS | TOTAL GRAMS | AVG GRAMS |
|---|---|---|---|---|
| AMPHETAMINE | 1100 | 70 | 6,385.44 | 91.22 |
| LISDEXAMFETAMINE | 1205 | 38 | 3,822.90 | 100.6 |
| METHYLPHENIDATE (DL;D;L;ISOMERS) | 1724 | 91 | 9,663.84 | 106.2 |
| BARBITURIC ACID DERIVIATIVE OR SALT [PER 21C | 2100 | 1 | 5 | 5 |
| BUTALBITAL | 2165 | 8 | 90 | 11.25 |
| PENTOBARBITAL (SCHEDULE 2) | 2270 | 4 | 211.9 | 52.98 |
| COCAINE | 9041L | 42 | 131.56 | 3.13 |

| | | | | |
|---|---|---|---|---|
| CODEINE | 9050 | 162 | 36,000.58 | 222.23 |
| BUPRENORPHINE | 9064 | 71 | 1,133.98 | 15.97 |
| DIHYDROCODEINE | 9120 | 4 | 9.61 | 2.4 |
| OXYCODONE | 9143 | 201 | 35,343.94 | 175.84 |
| HYDROMORPHONE | 9150 | 173 | 2,217.39 | 12.82 |
| HYDROCODONE | 9193 | 206 | 40,352.32 | 195.89 |
| MEPERIDINE (PETHIDINE) | 9230 | 160 | 5,895.37 | 36.85 |
| METHADONE | 9250B | 79 | 1,763.04 | 22.32 |
| MORPHINE | 9300 | 205 | 29,755.72 | 145.15 |
| OPIUM POWDERED | 9639 | 42 | 168.84 | 4.02 |
| OXYMORPHONE | 9652 | 18 | 236.51 | 13.14 |
| ALFENTANIL | 9737 | 8 | 3.52 | 0.44 |
| REMIFENTANIL | 9739 | 28 | 10.43 | 0.37 |
| SUFENTANIL BASE | 9740 | 35 | 0.62 | 0.02 |
| TAPENTADOL | 9780 | 28 | 1,188 | 42.43 |
| FENTANYL BASE | 9801 | 209 | 561.05 | 2.68 |
| OPIUM COMBINATION PRODUCT (C-III) | 9809 | 6 | 3.22 | 0.54 |

STATE:OKLAHOMA BUSINESS ACTIVITY:C - PRACTITIONERS

| DRUG NAME | DRUG CODE | BUYERS | TOTAL GRAMS | AVG GRAMS |
|---|---|---|---|---|
| LISDEXAMFETAMINE | 1205 | 4 | 32.29 | 8.07 |
| PENTOBARBITAL (SCHEDULE 2) | 2270 | 94 | 68,608.69 | 729.88 |
| COCAINE | 9041L | 1 | 0.57 | 0.57 |
| CODEINE | 9050 | 101 | 2,100.55 | 20.8 |
| BUPRENORPHINE | 9064 | 266 | 51.22 | 0.19 |
| DIHYDROCODEINE | 9120 | 67 | 30.45 | 0.45 |
| OXYCODONE | 9143 | 13 | 544.26 | 41.87 |
| HYDROMORPHONE | 9150 | 86 | 38.36 | 0.45 |
| HYDROCODONE | 9193 | 65 | 1,320.32 | 20.31 |
| MEPERIDINE (PETHIDINE) | 9230 | 44 | 229.31 | 5.21 |
| METHADONE | 9250B | 5 | 44.72 | 8.94 |
| MORPHINE | 9300 | 72 | 800.31 | 11.12 |
| OPIUM POWDERED | 9639 | 2 | 2.52 | 1.26 |
| OXYMORPHONE | 9652 | 3 | 1.7 | 0.57 |
| NOROXYMORPHONE | 9668 | 2 | 0.64 | 0.32 |

| | | | | |
|---|---|---|---|---|
| SUFENTANIL BASE | 9740 | 1 | 0.01 | 0.01 |
| FENTANYL BASE | 9801 | 147 | 58.09 | 0.4 |

STATE:OKLAHOMA BUSINESS ACTIVITY:D - TEACHING INSTITUTIONS

| DRUG NAME | DRUG CODE | BUYERS | TOTAL GRAMS | AVG GRAMS |
|---|---|---|---|---|
| PENTOBARBITAL (SCHEDULE 2) | 2270 | 1 | 88.86 | 88.86 |
| BUPRENORPHINE | 9064 | 1 | 0.25 | 0.25 |
| HYDROMORPHONE | 9150 | 1 | 6.03 | 6.03 |
| HYDROCODONE | 9193 | 1 | 8.47 | 8.47 |
| METHADONE | 9250B | 1 | 27.28 | 27.28 |
| MORPHINE | 9300 | 1 | 44.1 | 44.1 |
| FENTANYL BASE | 9801 | 1 | 0.95 | 0.95 |

STATE:OKLAHOMA BUSINESS ACTIVITY:M - MID-LEVEL PRACTITIONERS

| DRUG NAME | DRUG CODE | BUYERS | TOTAL GRAMS | AVG GRAMS |
|---|---|---|---|---|
| PENTOBARBITAL (SCHEDULE 2) | 2270 | 10 | 7,203.25 | 720.32 |
| CODEINE | 9050 | 42 | 86.53 | 2.06 |
| DIHYDROCODEINE | 9120 | 42 | 24.35 | 0.58 |
| HYDROMORPHONE | 9150 | 12 | 0.55 | 0.05 |
| MORPHINE | 9300 | 41 | 12.1 | 0.3 |
| FENTANYL BASE | 9801 | 65 | 3.62 | 0.06 |

STATE:OKLAHOMA BUSINESS ACTIVITY:N-U NARCOTIC TREATMENT PROGRAMS

| DRUG NAME | DRUG CODE | BUYERS | TOTAL GRAMS | AVG GRAMS |
|---|---|---|---|---|
| BUPRENORPHINE | 9064 | 10 | 241.74 | 24.17 |
| METHADONE | 9250B | 19 | 122,590.04 | 6,452.11 |

STATE:OREGON BUSINESS ACTIVITY:A - PHARMACIES

| DRUG NAME | DRUG CODE | BUYERS | TOTAL GRAMS | AVG GRAMS |
|---|---|---|---|---|

| | | | | |
|---|---|---|---|---|
| AMPHETAMINE | 1100 | 657 | 231,398.37 | 352.2 |
| DL-METHAMPHETAMINE RACEMIC BASE | 1105B | 34 | 126.58 | 3.72 |
| D-METHAMPHETAMINE | 1105D | 4 | 32.94 | 8.23 |
| LISDEXAMFETAMINE | 1205 | 618 | 59,688.25 | 96.58 |
| METHYLPHENIDATE (DL;D;L;ISOMERS) | 1724 | 655 | 241,148.11 | 368.17 |
| BARBITURIC ACID DERIVIATIVE OR SALT [PER 21C | 2100 | 157 | 3,785 | 24.11 |
| BUTALBITAL | 2165 | 304 | 9,355 | 30.77 |
| SECOBARBITAL (SCHEDULE 2) | 2315 | 31 | 952.12 | 30.71 |
| NABILONE | 7379 | 1 | 0.05 | 0.05 |
| COCAINE | 9041L | 6 | 111.42 | 18.57 |
| CODEINE | 9050 | 661 | 127,945.55 | 193.56 |
| BUPRENORPHINE | 9064 | 646 | 35,282.71 | 54.62 |
| OXYCODONE | 9143 | 665 | 657,250.78 | 988.35 |
| HYDROMORPHONE | 9150 | 646 | 16,093.84 | 24.91 |
| HYDROCODONE | 9193 | 671 | 351,923.20 | 524.48 |
| LEVORPHANOL | 9220L | 12 | 13.58 | 1.13 |
| MEPERIDINE (PETHIDINE) | 9230 | 219 | 4,671.24 | 21.33 |
| METHADONE | 9250B | 628 | 60,988.69 | 97.12 |
| MORPHINE | 9300 | 669 | 258,694.62 | 386.69 |
| OPIUM TINCTURE (LAUDANUM) | 9630 | 2 | 2.36 | 1.18 |
| OPIUM POWDERED | 9639 | 53 | 123.48 | 2.33 |
| OXYMORPHONE | 9652 | 367 | 6,573.93 | 17.91 |
| ALFENTANIL | 9737 | 2 | 1.84 | 0.92 |
| REMIFENTANIL | 9739 | 3 | 0.11 | 0.04 |
| SUFENTANIL BASE | 9740 | 1 | 0 | 0 |
| TAPENTADOL | 9780 | 364 | 29,756.50 | 81.75 |
| FENTANYL BASE | 9801 | 636 | 3,314.47 | 5.21 |
| OPIUM COMBINATION PRODUCT (C-III) | 9809 | 12 | 3.59 | 0.3 |

STATE:OREGON BUSINESS ACTIVITY:B - HOSPITALS

| DRUG NAME | DRUG CODE | BUYERS | TOTAL GRAMS | AVG GRAMS |
|---|---|---|---|---|
| AMPHETAMINE | 1100 | 44 | 3,165.81 | 71.95 |
| DL-METHAMPHETAMINE RACEMIC BASE | 1105B | 1 | 1.21 | 1.21 |
| D-METHAMPHETAMINE | 1105D | 1 | 0.8 | 0.8 |
| LISDEXAMFETAMINE | 1205 | 20 | 424.9 | 21.25 |

| | | | | |
|---|---|---|---|---|
| METHYLPHENIDATE (DL;D;L;ISOMERS) | 1724 | 54 | 4,756.11 | 88.08 |
| BARBITURIC ACID DERIVIATIVE OR SALT [PER 21C | 2100 | 3 | 80 | 26.67 |
| AMOBARBITAL (SCHEDULE 2) | 2125 | 2 | 6.84 | 3.42 |
| BUTALBITAL | 2165 | 6 | 30 | 5 |
| PENTOBARBITAL (SCHEDULE 2) | 2270 | 3 | 2,184.63 | 728.21 |
| COCAINE | 9041L | 45 | 240.95 | 5.35 |
| CODEINE | 9050 | 89 | 4,144.42 | 46.57 |
| BUPRENORPHINE | 9064 | 57 | 942.4 | 16.53 |
| OXYCODONE | 9143 | 92 | 35,929.50 | 390.54 |
| HYDROMORPHONE | 9150 | 78 | 3,109.96 | 39.87 |
| HYDROCODONE | 9193 | 103 | 24,069.16 | 233.68 |
| MEPERIDINE (PETHIDINE) | 9230 | 65 | 1,036.85 | 15.95 |
| METHADONE | 9250B | 65 | 4,171.95 | 64.18 |
| MORPHINE | 9300 | 94 | 25,293.63 | 269.08 |
| OPIUM POWDERED | 9639 | 47 | 654.84 | 13.93 |
| OXYMORPHONE | 9652 | 6 | 6.11 | 1.02 |
| ALFENTANIL | 9737 | 16 | 15.14 | 0.95 |
| REMIFENTANIL | 9739 | 30 | 6.72 | 0.22 |
| SUFENTANIL BASE | 9740 | 28 | 1.21 | 0.04 |
| TAPENTADOL | 9780 | 7 | 58.5 | 8.36 |
| FENTANYL BASE | 9801 | 82 | 353.66 | 4.31 |

STATE:OREGON BUSINESS ACTIVITY:C - PRACTITIONERS

| DRUG NAME | DRUG CODE | BUYERS | TOTAL GRAMS | AVG GRAMS |
|---|---|---|---|---|
| LISDEXAMFETAMINE | 1205 | 1 | 4.6 | 4.6 |
| PENTOBARBITAL (SCHEDULE 2) | 2270 | 179 | 144,855.44 | 809.25 |
| COCAINE | 9041L | 16 | 31.28 | 1.96 |
| CODEINE | 9050 | 52 | 3,739.11 | 71.91 |
| BUPRENORPHINE | 9064 | 576 | 97.58 | 0.17 |
| OXYCODONE | 9143 | 60 | 156.84 | 2.61 |
| HYDROMORPHONE | 9150 | 379 | 141.79 | 0.37 |
| HYDROCODONE | 9193 | 286 | 690.89 | 2.42 |
| MEPERIDINE (PETHIDINE) | 9230 | 87 | 405.14 | 4.66 |
| METHADONE | 9250B | 11 | 35.06 | 3.19 |
| MORPHINE | 9300 | 200 | 330.25 | 1.65 |

| | | | | |
|---|---|---|---|---|
| OPIUM POWDERED | 9639 | 6 | 19.8 | 3.3 |
| OXYMORPHONE | 9652 | 12 | 0.68 | 0.06 |
| NOROXYMORPHONE | 9668 | 2 | 2 | 1 |
| ALFENTANIL | 9737 | 4 | 1.69 | 0.42 |
| REMIFENTANIL | 9739 | 37 | 4.57 | 0.12 |
| SUFENTANIL BASE | 9740 | 5 | 0.02 | 0 |
| FENTANYL BASE | 9801 | 362 | 37.5 | 0.1 |

STATE:OREGON BUSINESS ACTIVITY:D - TEACHING INSTITUTIONS

| DRUG NAME | DRUG CODE | BUYERS | TOTAL GRAMS | AVG GRAMS |
|---|---|---|---|---|
| D-METHAMPHETAMINE | 1105D | 1 | 4.02 | 4.02 |
| COCAINE | 9041L | 1 | 0.89 | 0.89 |
| BUPRENORPHINE | 9064 | 1 | 0.04 | 0.04 |
| OXYCODONE | 9143 | 1 | 0.45 | 0.45 |
| FENTANYL BASE | 9801 | 1 | 0.03 | 0.03 |

STATE:OREGON BUSINESS ACTIVITY:M - MID-LEVEL PRACTITIONERS

| DRUG NAME | DRUG CODE | BUYERS | TOTAL GRAMS | AVG GRAMS |
|---|---|---|---|---|
| PENTOBARBITAL (SCHEDULE 2) | 2270 | 9 | 7,646.65 | 849.63 |
| CODEINE | 9050 | 5 | 31.16 | 6.23 |
| BUPRENORPHINE | 9064 | 2 | 2.37 | 1.19 |
| HYDROMORPHONE | 9150 | 5 | 0.27 | 0.05 |
| HYDROCODONE | 9193 | 7 | 17.86 | 2.55 |
| MEPERIDINE (PETHIDINE) | 9230 | 1 | 2.18 | 2.18 |
| MORPHINE | 9300 | 3 | 0.56 | 0.19 |
| FENTANYL BASE | 9801 | 1 | 0 | 0 |

STATE:OREGON BUSINESS ACTIVITY:N-U NARCOTIC TREATMENT PROGRAMS

| DRUG NAME | DRUG CODE | BUYERS | TOTAL GRAMS | AVG GRAMS |
|---|---|---|---|---|
| BUPRENORPHINE | 9064 | 15 | 5,211.37 | 347.42 |
| METHADONE | 9250B | 17 | 188,560.09 | 11,091.77 |

STATISTICAL SUMMARY FOR RETAIL DRUG PURCHASES BY GRAMS WT

STATE:PENNSYLVANIA BUSINESS ACTIVITY:A - PHARMACIES

| DRUG NAME | DRUG CODE | BUYERS | TOTAL GRAMS | AVG GRAMS |
|---|---|---|---|---|
| AMPHETAMINE | 1100 | 2,950 | 832,126.70 | 282.08 |
| DL-METHAMPHETAMINE RACEMIC BASE | 1105B | 92 | 489.05 | 5.32 |
| D-METHAMPHETAMINE | 1105D | 3 | 26.11 | 8.7 |
| LISDEXAMFETAMINE | 1205 | 2,742 | 340,381.23 | 124.14 |
| METHYLPHENIDATE (DL;D;L;ISOMERS) | 1724 | 2,920 | 886,809.58 | 303.7 |
| BARBITURIC ACID DERIVIATIVE OR SALT [PER 21C | 2100 | 634 | 21,590 | 34.05 |
| BUTALBITAL | 2165 | 1,203 | 46,575 | 38.72 |
| PENTOBARBITAL (SCHEDULE 2) | 2270 | 1 | 53.77 | 53.77 |
| SECOBARBITAL (SCHEDULE 2) | 2315 | 9 | 265.5 | 29.5 |
| DRONABINOL IN AN ORAL SOLUTION IN FDA APPR | 7365 | 10 | 16.35 | 1.64 |
| NABILONE | 7379 | 2 | 0.35 | 0.18 |
| COCAINE | 9041L | 8 | 17.25 | 2.16 |
| CODEINE | 9050 | 2,974 | 511,587.23 | 172.02 |
| BUPRENORPHINE | 9064 | 2,780 | 242,773.12 | 87.33 |
| DIHYDROCODEINE | 9120 | 55 | 201.88 | 3.67 |
| OXYCODONE | 9143 | 3,024 | 2,878,775.18 | 951.98 |
| HYDROMORPHONE | 9150 | 2,620 | 49,703.35 | 18.97 |
| HYDROCODONE | 9193 | 3,000 | 631,977.42 | 210.66 |
| LEVORPHANOL | 9220L | 68 | 49.8 | 0.73 |
| MEPERIDINE (PETHIDINE) | 9230 | 463 | 11,654.13 | 25.17 |
| METHADONE | 9250B | 2,292 | 127,217.34 | 55.5 |
| MORPHINE | 9300 | 2,925 | 863,020.28 | 295.05 |
| OPIUM TINCTURE (LAUDANUM) | 9630 | 4 | 21.24 | 5.31 |
| OPIUM POWDERED | 9639 | 174 | 570.96 | 3.28 |
| OXYMORPHONE | 9652 | 1,908 | 89,999.06 | 47.17 |
| ALFENTANIL | 9737 | 1 | 0.02 | 0.02 |
| REMIFENTANIL | 9739 | 4 | 1.55 | 0.39 |
| SUFENTANIL BASE | 9740 | 2 | 6.7 | 3.35 |
| TAPENTADOL | 9780 | 1,815 | 262,376.50 | 144.56 |
| FENTANYL BASE | 9801 | 2,862 | 18,307.32 | 6.4 |
| OPIUM COMBINATION PRODUCT (C-III) | 9809 | 117 | 130.74 | 1.12 |

STATISTICAL SUMMARY FOR RETAIL DRUG PURCHASES BY GRAMS WT

STATE:PENNSYLVANIA BUSINESS ACTIVITY:B - HOSPITALS

| DRUG NAME | DRUG CODE | BUYERS | TOTAL GRAMS | AVG GRAMS |
|---|---|---|---|---|
| AMPHETAMINE | 1100 | 166 | 12,200.01 | 73.49 |
| LISDEXAMFETAMINE | 1205 | 52 | 3,201.52 | 61.57 |
| METHYLPHENIDATE (DL;D;L;ISOMERS) | 1724 | 242 | 12,974.45 | 53.61 |
| BARBITURIC ACID DERIVIATIVE OR SALT [PER 21C | 2100 | 4 | 85 | 21.25 |
| AMOBARBITAL (SCHEDULE 2) | 2125 | 11 | 29.16 | 2.65 |
| BUTALBITAL | 2165 | 13 | 225 | 17.31 |
| PENTOBARBITAL (SCHEDULE 2) | 2270 | 30 | 2,568.33 | 85.61 |
| COCAINE | 9041L | 157 | 1,297.27 | 8.26 |
| CODEINE | 9050 | 281 | 15,831.62 | 56.34 |
| BUPRENORPHINE | 9064 | 198 | 7,082.32 | 35.77 |
| OXYCODONE | 9143 | 331 | 122,487.64 | 370.05 |
| HYDROMORPHONE | 9150 | 301 | 10,530.99 | 34.99 |
| HYDROCODONE | 9193 | 298 | 26,864.39 | 90.15 |
| LEVORPHANOL | 9220L | 1 | 3.83 | 3.83 |
| MEPERIDINE (PETHIDINE) | 9230 | 225 | 2,169.86 | 9.64 |
| METHADONE | 9250B | 242 | 13,768.20 | 56.89 |
| MORPHINE | 9300 | 327 | 68,764 | 210.29 |
| OPIUM POWDERED | 9639 | 159 | 727.92 | 4.58 |
| OXYMORPHONE | 9652 | 78 | 1,067.49 | 13.69 |
| ALFENTANIL | 9737 | 13 | 0.97 | 0.07 |
| REMIFENTANIL | 9739 | 116 | 138.94 | 1.2 |
| SUFENTANIL BASE | 9740 | 44 | 1.37 | 0.03 |
| TAPENTADOL | 9780 | 63 | 2,094.50 | 33.25 |
| FENTANYL BASE | 9801 | 365 | 1,678.74 | 4.6 |
| OPIUM COMBINATION PRODUCT (C-III) | 9809 | 12 | 4.91 | 0.41 |

STATE:PENNSYLVANIA BUSINESS ACTIVITY:C - PRACTITIONERS

| DRUG NAME | DRUG CODE | BUYERS | TOTAL GRAMS | AVG GRAMS |
|---|---|---|---|---|
| AMPHETAMINE | 1100 | 1 | 8.26 | 8.26 |
| LISDEXAMFETAMINE | 1205 | 1 | 18.5 | 18.5 |
| METHYLPHENIDATE (DL;D;L;ISOMERS) | 1724 | 1 | 30.29 | 30.29 |

| | | | | |
|---|---|---|---|---|
| PENTOBARBITAL (SCHEDULE 2) | 2270 | 301 | 262,836.57 | 873.21 |
| COCAINE | 9041L | 24 | 242.2 | 10.09 |
| CODEINE | 9050 | 321 | 26,017.67 | 81.05 |
| BUPRENORPHINE | 9064 | 994 | 1,728.07 | 1.74 |
| DIHYDROCODEINE | 9120 | 55 | 157.49 | 2.86 |
| OXYCODONE | 9143 | 69 | 804.83 | 11.66 |
| HYDROMORPHONE | 9150 | 467 | 263.88 | 0.57 |
| HYDROCODONE | 9193 | 393 | 1,185.12 | 3.02 |
| MEPERIDINE (PETHIDINE) | 9230 | 111 | 375.18 | 3.38 |
| METHADONE | 9250B | 32 | 191.76 | 5.99 |
| MORPHINE | 9300 | 255 | 408.99 | 1.6 |
| OPIUM POWDERED | 9639 | 1 | 1.44 | 1.44 |
| OXYMORPHONE | 9652 | 3 | 0.08 | 0.03 |
| NOROXYMORPHONE | 9668 | 1 | 0.99 | 0.99 |
| ALFENTANIL | 9737 | 6 | 1.02 | 0.17 |
| REMIFENTANIL | 9739 | 26 | 2.63 | 0.1 |
| SUFENTANIL BASE | 9740 | 5 | 0.05 | 0.01 |
| FENTANYL BASE | 9801 | 611 | 77.23 | 0.13 |

STATE:PENNSYLVANIA BUSINESS ACTIVITY:D - TEACHING INSTITUTIONS

| DRUG NAME | DRUG CODE | BUYERS | TOTAL GRAMS | AVG GRAMS |
|---|---|---|---|---|
| METHYLPHENIDATE (DL;D;L;ISOMERS) | 1724 | 1 | 0.43 | 0.43 |
| PENTOBARBITAL (SCHEDULE 2) | 2270 | 1 | 3,732.18 | 3,732.18 |
| CODEINE | 9050 | 1 | 964.06 | 964.06 |
| BUPRENORPHINE | 9064 | 2 | 1.41 | 0.7 |
| HYDROMORPHONE | 9150 | 2 | 3.22 | 1.61 |
| HYDROCODONE | 9193 | 1 | 61.63 | 61.63 |
| METHADONE | 9250B | 1 | 35.06 | 35.06 |
| MORPHINE | 9300 | 2 | 36.81 | 18.41 |
| OXYMORPHONE | 9652 | 1 | 0.18 | 0.18 |
| REMIFENTANIL | 9739 | 2 | 0.12 | 0.06 |
| FENTANYL BASE | 9801 | 4 | 3.23 | 0.81 |

STATE:PENNSYLVANIA BUSINESS ACTIVITY:M - MID-LEVEL PRACTITIONERS

| DRUG NAME | DRUG CODE | BUYERS | TOTAL GRAMS | AVG GRAMS |
|---|---|---|---|---|
| CODEINE | 9050 | 32 | 64.37 | 2.01 |
| DIHYDROCODEINE | 9120 | 10 | 3.18 | 0.32 |

STATE:PENNSYLVANIA BUSINESS ACTIVITY:N-U NARCOTIC TREATMENT PROGRAMS

| DRUG NAME | DRUG CODE | BUYERS | TOTAL GRAMS | AVG GRAMS |
|---|---|---|---|---|
| BUPRENORPHINE | 9064 | 15 | 502.79 | 33.52 |
| METHADONE | 9250B | 85 | 648,095.21 | 7,624.65 |

STATE:PUERTO RICO BUSINESS ACTIVITY:A - PHARMACIES

| DRUG NAME | DRUG CODE | BUYERS | TOTAL GRAMS | AVG GRAMS |
|---|---|---|---|---|
| AMPHETAMINE | 1100 | 675 | 32,974.30 | 48.85 |
| LISDEXAMFETAMINE | 1205 | 306 | 7,605.48 | 24.85 |
| METHYLPHENIDATE (DL;D;L;ISOMERS) | 1724 | 732 | 67,579.14 | 92.32 |
| BARBITURIC ACID DERIVIATIVE OR SALT [PER 21C | 2100 | 29 | 605 | 20.86 |
| CODEINE | 9050 | 1,076 | 46,967.94 | 43.65 |
| BUPRENORPHINE | 9064 | 336 | 9,371.98 | 27.89 |
| OXYCODONE | 9143 | 827 | 44,087.66 | 53.31 |
| HYDROMORPHONE | 9150 | 30 | 187.78 | 6.26 |
| HYDROCODONE | 9193 | 547 | 2,349.42 | 4.3 |
| MEPERIDINE (PETHIDINE) | 9230 | 680 | 5,967.49 | 8.78 |
| METHADONE | 9250B | 15 | 229.59 | 15.31 |
| MORPHINE | 9300 | 152 | 3,974.06 | 26.15 |
| TAPENTADOL | 9780 | 71 | 2,733.50 | 38.5 |
| FENTANYL BASE | 9801 | 665 | 408.72 | 0.61 |

STATE:PUERTO RICO BUSINESS ACTIVITY:B - HOSPITALS

| DRUG NAME | DRUG CODE | BUYERS | TOTAL GRAMS | AVG GRAMS |
|---|---|---|---|---|
| AMPHETAMINE | 1100 | 8 | 140.93 | 17.62 |
| METHYLPHENIDATE (DL;D;L;ISOMERS) | 1724 | 17 | 285.65 | 16.8 |

| | | | | |
|---|---|---|---|---|
| CODEINE | 9050 | 82 | 4,257.76 | 51.92 |
| BUPRENORPHINE | 9064 | 16 | 583.42 | 36.46 |
| OXYCODONE | 9143 | 67 | 6,638.85 | 99.09 |
| HYDROMORPHONE | 9150 | 5 | 108.85 | 21.77 |
| HYDROCODONE | 9193 | 26 | 197.27 | 7.59 |
| MEPERIDINE (PETHIDINE) | 9230 | 95 | 11,355.32 | 119.53 |
| METHADONE | 9250B | 1 | 118.96 | 118.96 |
| MORPHINE | 9300 | 85 | 4,423.44 | 52.04 |
| SUFENTANIL BASE | 9740 | 12 | 0.75 | 0.06 |
| TAPENTADOL | 9780 | 5 | 640.5 | 128.1 |
| FENTANYL BASE | 9801 | 81 | 169.13 | 2.09 |

STATE:PUERTO RICO BUSINESS ACTIVITY:C - PRACTITIONERS

| DRUG NAME | DRUG CODE | BUYERS | TOTAL GRAMS | AVG GRAMS |
|---|---|---|---|---|
| BUPRENORPHINE | 9064 | 14 | 0.92 | 0.07 |
| MEPERIDINE (PETHIDINE) | 9230 | 5 | 49.02 | 9.8 |
| MORPHINE | 9300 | 1 | 2.24 | 2.24 |
| NOROXYMORPHONE | 9668 | 2 | 25.07 | 12.53 |
| FENTANYL BASE | 9801 | 10 | 0.42 | 0.04 |

STATE:PUERTO RICO BUSINESS ACTIVITY:D - TEACHING INSTITUTIONS

| DRUG NAME | DRUG CODE | BUYERS | TOTAL GRAMS | AVG GRAMS |
|---|---|---|---|---|
| PENTOBARBITAL (SCHEDULE 2) | 2270 | 1 | 0.2 | 0.2 |

STATE:PUERTO RICO BUSINESS ACTIVITY:N-U NARCOTIC TREATMENT PROGRAMS

| DRUG NAME | DRUG CODE | BUYERS | TOTAL GRAMS | AVG GRAMS |
|---|---|---|---|---|
| METHADONE | 9250B | 6 | 83,680.06 | 13,946.68 |

STATE:RHODE ISLAND BUSINESS ACTIVITY:A - PHARMACIES

STATISTICAL SUMMARY FOR RETAIL DRUG PURCHASES BY GRAMS WT

| DRUG NAME | DRUG CODE | BUYERS | TOTAL GRAMS | AVG GRAMS |
|---|---|---|---|---|
| AMPHETAMINE | 1100 | 189 | 144,014.30 | 761.98 |
| DL-METHAMPHETAMINE RACEMIC BASE | 1105B | 5 | 16.88 | 3.38 |
| D-METHAMPHETAMINE | 1105D | 1 | 3.62 | 3.62 |
| LISDEXAMFETAMINE | 1205 | 179 | 29,753.07 | 166.22 |
| METHYLPHENIDATE (DL;D;L;ISOMERS) | 1724 | 187 | 72,332.93 | 386.81 |
| BARBITURIC ACID DERIVIATIVE OR SALT [PER 21C | 2100 | 71 | 2,515 | 35.42 |
| BUTALBITAL | 2165 | 72 | 1,420 | 19.72 |
| SECOBARBITAL (SCHEDULE 2) | 2315 | 1 | 36.62 | 36.62 |
| CODEINE | 9050 | 184 | 27,925.75 | 151.77 |
| BUPRENORPHINE | 9064 | 191 | 28,706.51 | 150.3 |
| DIHYDROCODEINE | 9120 | 1 | 3.2 | 3.2 |
| OXYCODONE | 9143 | 190 | 152,626.40 | 803.3 |
| HYDROMORPHONE | 9150 | 167 | 2,902.61 | 17.38 |
| HYDROCODONE | 9193 | 190 | 39,272.47 | 206.7 |
| MEPERIDINE (PETHIDINE) | 9230 | 20 | 634.02 | 31.7 |
| METHADONE | 9250B | 152 | 6,364.46 | 41.87 |
| MORPHINE | 9300 | 187 | 41,001.53 | 219.26 |
| OPIUM POWDERED | 9639 | 20 | 30.24 | 1.51 |
| OXYMORPHONE | 9652 | 77 | 1,601.14 | 20.79 |
| TAPENTADOL | 9780 | 104 | 8,380 | 80.58 |
| FENTANYL BASE | 9801 | 176 | 590.9 | 3.36 |
| OPIUM COMBINATION PRODUCT (C-III) | 9809 | 2 | 0.38 | 0.19 |

STATE:RHODE ISLAND BUSINESS ACTIVITY:B - HOSPITALS

| DRUG NAME | DRUG CODE | BUYERS | TOTAL GRAMS | AVG GRAMS |
|---|---|---|---|---|
| AMPHETAMINE | 1100 | 16 | 2,524.82 | 157.8 |
| LISDEXAMFETAMINE | 1205 | 8 | 358.42 | 44.8 |
| METHYLPHENIDATE (DL;D;L;ISOMERS) | 1724 | 18 | 2,966.34 | 164.8 |
| BARBITURIC ACID DERIVIATIVE OR SALT [PER 21C | 2100 | 1 | 5 | 5 |
| AMOBARBITAL (SCHEDULE 2) | 2125 | 1 | 3.65 | 3.65 |
| BUTALBITAL | 2165 | 2 | 50 | 25 |
| PENTOBARBITAL (SCHEDULE 2) | 2270 | 2 | 37.37 | 18.68 |
| COCAINE | 9041L | 11 | 63.57 | 5.78 |
| CODEINE | 9050 | 18 | 1,190.99 | 66.17 |

| | | | | |
|---|---|---|---|---|
| BUPRENORPHINE | 9064 | 16 | 677.79 | 42.36 |
| OXYCODONE | 9143 | 21 | 9,891.65 | 471.03 |
| HYDROMORPHONE | 9150 | 15 | 614.15 | 40.94 |
| HYDROCODONE | 9193 | 20 | 2,129.09 | 106.45 |
| MEPERIDINE (PETHIDINE) | 9230 | 13 | 304.37 | 23.41 |
| METHADONE | 9250B | 14 | 1,317.59 | 94.11 |
| MORPHINE | 9300 | 19 | 5,552.54 | 292.24 |
| OPIUM POWDERED | 9639 | 11 | 100.8 | 9.16 |
| OXYMORPHONE | 9652 | 1 | 0.27 | 0.27 |
| ALFENTANIL | 9737 | 2 | 0.26 | 0.13 |
| REMIFENTANIL | 9739 | 6 | 2.85 | 0.48 |
| SUFENTANIL BASE | 9740 | 3 | 0.01 | 0 |
| TAPENTADOL | 9780 | 1 | 10 | 10 |
| FENTANYL BASE | 9801 | 18 | 135.22 | 7.51 |

STATE:RHODE ISLAND BUSINESS ACTIVITY:C - PRACTITIONERS

| DRUG NAME | DRUG CODE | BUYERS | TOTAL GRAMS | AVG GRAMS |
|---|---|---|---|---|
| PENTOBARBITAL (SCHEDULE 2) | 2270 | 35 | 24,525.04 | 700.72 |
| COCAINE | 9041L | 2 | 12.43 | 6.21 |
| BUPRENORPHINE | 9064 | 75 | 5.48 | 0.07 |
| OXYCODONE | 9143 | 2 | 4.03 | 2.02 |
| HYDROMORPHONE | 9150 | 44 | 16.2 | 0.37 |
| HYDROCODONE | 9193 | 29 | 86.09 | 2.97 |
| MEPERIDINE (PETHIDINE) | 9230 | 14 | 68.41 | 4.89 |
| METHADONE | 9250B | 3 | 16.28 | 5.43 |
| MORPHINE | 9300 | 13 | 74.44 | 5.73 |
| OPIUM POWDERED | 9639 | 2 | 6.48 | 3.24 |
| OXYMORPHONE | 9652 | 4 | 0.6 | 0.15 |
| NOROXYMORPHONE | 9668 | 1 | 0.1 | 0.1 |
| REMIFENTANIL | 9739 | 1 | 0.32 | 0.32 |
| FENTANYL BASE | 9801 | 48 | 8.86 | 0.18 |

STATE:RHODE ISLAND BUSINESS ACTIVITY:N-U NARCOTIC TREATMENT PROGRAMS

| DRUG NAME | DRUG CODE | BUYERS | TOTAL GRAMS | AVG GRAMS |
|---|---|---|---|---|
| BUPRENORPHINE | 9064 | 13 | 707.51 | 54.42 |
| METHADONE | 9250B | 19 | 169,410.63 | 8,916.35 |

STATE:SOUTH CAROLINA BUSINESS ACTIVITY:A - PHARMACIES

| DRUG NAME | DRUG CODE | BUYERS | TOTAL GRAMS | AVG GRAMS |
|---|---|---|---|---|
| AMPHETAMINE | 1100 | 1,107 | 535,663.09 | 483.89 |
| DL-METHAMPHETAMINE RACEMIC BASE | 1105B | 27 | 56.66 | 2.1 |
| LISDEXAMFETAMINE | 1205 | 1,079 | 242,733.78 | 224.96 |
| METHYLPHENIDATE (DL;D;L;ISOMERS) | 1724 | 1,100 | 343,576.83 | 312.34 |
| BARBITURIC ACID DERIVIATIVE OR SALT [PER 21C | 2100 | 324 | 9,390 | 28.98 |
| BUTALBITAL | 2165 | 443 | 13,475 | 30.42 |
| SECOBARBITAL (SCHEDULE 2) | 2315 | 3 | 36.62 | 12.21 |
| COCAINE | 9041L | 1 | 0.89 | 0.89 |
| CODEINE | 9050 | 1,097 | 298,483.53 | 272.09 |
| BUPRENORPHINE | 9064 | 1,015 | 40,272.55 | 39.68 |
| DIHYDROCODEINE | 9120 | 11 | 52.34 | 4.76 |
| OXYCODONE | 9143 | 1,106 | 904,896.83 | 818.17 |
| HYDROMORPHONE | 9150 | 1,021 | 19,550.41 | 19.15 |
| HYDROCODONE | 9193 | 1,120 | 534,772.16 | 477.48 |
| LEVORPHANOL | 9220L | 23 | 11.03 | 0.48 |
| MEPERIDINE (PETHIDINE) | 9230 | 729 | 19,518.11 | 26.77 |
| METHADONE | 9250B | 882 | 36,890.33 | 41.83 |
| MORPHINE | 9300 | 1,094 | 238,347.35 | 217.87 |
| OPIUM POWDERED | 9639 | 62 | 158.4 | 2.55 |
| OXYMORPHONE | 9652 | 813 | 23,968.81 | 29.48 |
| SUFENTANIL BASE | 9740 | 1 | 2 | 2 |
| TAPENTADOL | 9780 | 943 | 172,597 | 183.03 |
| FENTANYL BASE | 9801 | 1,067 | 4,749.77 | 4.45 |
| OPIUM COMBINATION PRODUCT (C-III) | 9809 | 43 | 37.84 | 0.88 |

STATE:SOUTH CAROLINA BUSINESS ACTIVITY:B - HOSPITALS

| DRUG NAME | DRUG CODE | BUYERS | TOTAL GRAMS | AVG GRAMS |
|---|---|---|---|---|

| | | | | |
|---|---|---|---|---|
| AMPHETAMINE | 1100 | 55 | 9,528.35 | 173.24 |
| DL-METHAMPHETAMINE RACEMIC BASE | 1105B | 1 | 3.62 | 3.62 |
| D-METHAMPHETAMINE | 1105D | 1 | 8.84 | 8.84 |
| LISDEXAMFETAMINE | 1205 | 25 | 1,678.80 | 67.15 |
| METHYLPHENIDATE (DL;D;L;ISOMERS) | 1724 | 80 | 10,195.35 | 127.44 |
| GAMMA HYDROXYBUTYRIC ACID(FDA APPROVED | 2012 | 1 | 5,126.98 | 5,126.98 |
| BARBITURIC ACID DERIVIATIVE OR SALT [PER 21C | 2100 | 1 | 5 | 5 |
| AMOBARBITAL (SCHEDULE 2) | 2125 | 3 | 15.04 | 5.01 |
| BUTALBITAL | 2165 | 1 | 5 | 5 |
| PENTOBARBITAL (SCHEDULE 2) | 2270 | 10 | 3,118.36 | 311.84 |
| DRONABINOL IN AN ORAL SOLUTION IN FDA APPR | 7365 | 1 | 0.3 | 0.3 |
| COCAINE | 9041L | 47 | 113.99 | 2.43 |
| CODEINE | 9050 | 91 | 5,258.73 | 57.79 |
| BUPRENORPHINE | 9064 | 67 | 924.79 | 13.8 |
| OXYCODONE | 9143 | 132 | 29,595.87 | 224.21 |
| HYDROMORPHONE | 9150 | 144 | 3,315.90 | 23.03 |
| HYDROCODONE | 9193 | 139 | 15,502.34 | 111.53 |
| MEPERIDINE (PETHIDINE) | 9230 | 122 | 3,443.84 | 28.23 |
| METHADONE | 9250B | 78 | 2,303.62 | 29.53 |
| MORPHINE | 9300 | 150 | 22,845.74 | 152.3 |
| OPIUM POWDERED | 9639 | 59 | 396.72 | 6.72 |
| OXYMORPHONE | 9652 | 8 | 33.81 | 4.23 |
| ALFENTANIL | 9737 | 2 | 0.35 | 0.18 |
| REMIFENTANIL | 9739 | 30 | 18.43 | 0.61 |
| SUFENTANIL BASE | 9740 | 26 | 0.62 | 0.02 |
| TAPENTADOL | 9780 | 29 | 1,413 | 48.72 |
| FENTANYL BASE | 9801 | 166 | 496.04 | 2.99 |
| OPIUM COMBINATION PRODUCT (C-III) | 9809 | 4 | 2.27 | 0.57 |

STATE:SOUTH CAROLINA BUSINESS ACTIVITY:C - PRACTITIONERS

| DRUG NAME | DRUG CODE | BUYERS | TOTAL GRAMS | AVG GRAMS |
|---|---|---|---|---|
| AMPHETAMINE | 1100 | 2 | 1.48 | 0.74 |
| LISDEXAMFETAMINE | 1205 | 2 | 18.7 | 9.35 |
| GAMMA HYDROXYBUTYRIC ACID(FDA APPROVED | 2012 | 1 | 2,452.03 | 2,452.03 |
| PENTOBARBITAL (SCHEDULE 2) | 2270 | 78 | 52,596.39 | 674.31 |

| | | | | |
|---|---|---|---|---|
| COCAINE | 9041L | 2 | 3.43 | 1.71 |
| CODEINE | 9050 | 26 | 285.94 | 11 |
| BUPRENORPHINE | 9064 | 378 | 58.94 | 0.16 |
| OXYCODONE | 9143 | 9 | 344.63 | 38.29 |
| HYDROMORPHONE | 9150 | 149 | 110.05 | 0.74 |
| HYDROCODONE | 9193 | 79 | 245.53 | 3.11 |
| MEPERIDINE (PETHIDINE) | 9230 | 70 | 555.25 | 7.93 |
| METHADONE | 9250B | 2 | 0.89 | 0.45 |
| MORPHINE | 9300 | 112 | 226.25 | 2.02 |
| OXYMORPHONE | 9652 | 1 | 0.03 | 0.03 |
| REMIFENTANIL | 9739 | 4 | 0.22 | 0.06 |
| SUFENTANIL BASE | 9740 | 1 | 0.02 | 0.02 |
| FENTANYL BASE | 9801 | 228 | 15.01 | 0.07 |

STATE:SOUTH CAROLINA BUSINESS ACTIVITY:D - TEACHING INSTITUTIONS

| DRUG NAME | DRUG CODE | BUYERS | TOTAL GRAMS | AVG GRAMS |
|---|---|---|---|---|
| BUPRENORPHINE | 9064 | 1 | 0.02 | 0.02 |
| HYDROMORPHONE | 9150 | 1 | 0.07 | 0.07 |

STATE:SOUTH CAROLINA BUSINESS ACTIVITY:M - MID-LEVEL PRACTITIONERS

| DRUG NAME | DRUG CODE | BUYERS | TOTAL GRAMS | AVG GRAMS |
|---|---|---|---|---|
| CODEINE | 9050 | 6 | 7.8 | 1.3 |
| MORPHINE | 9300 | 62 | 26.7 | 0.43 |
| FENTANYL BASE | 9801 | 66 | 0.71 | 0.01 |

STATE:SOUTH CAROLINA BUSINESS ACTIVITY:N-U NARCOTIC TREATMENT PROGRAMS

| DRUG NAME | DRUG CODE | BUYERS | TOTAL GRAMS | AVG GRAMS |
|---|---|---|---|---|
| BUPRENORPHINE | 9064 | 14 | 1,382.48 | 98.75 |
| METHADONE | 9250B | 23 | 186,183.22 | 8,094.92 |

STATISTICAL SUMMARY FOR RETAIL DRUG PURCHASES BY GRAMS WT

STATE:SOUTH DAKOTA BUSINESS ACTIVITY:A - PHARMACIES

| DRUG NAME | DRUG CODE | BUYERS | TOTAL GRAMS | AVG GRAMS |
|---|---|---|---|---|
| AMPHETAMINE | 1100 | 192 | 57,976.94 | 301.96 |
| DL-METHAMPHETAMINE RACEMIC BASE | 1105B | 9 | 69.52 | 7.72 |
| LISDEXAMFETAMINE | 1205 | 190 | 43,450 | 228.68 |
| METHYLPHENIDATE (DL;D;L;ISOMERS) | 1724 | 193 | 89,268.30 | 462.53 |
| BARBITURIC ACID DERIVIATIVE OR SALT [PER 21C | 2100 | 31 | 900 | 29.03 |
| BUTALBITAL | 2165 | 77 | 2,590 | 33.64 |
| SECOBARBITAL (SCHEDULE 2) | 2315 | 1 | 9.16 | 9.16 |
| COCAINE | 9041L | 1 | 0.14 | 0.14 |
| CODEINE | 9050 | 197 | 24,547.87 | 124.61 |
| BUPRENORPHINE | 9064 | 140 | 1,566 | 11.19 |
| DIHYDROCODEINE | 9120 | 1 | 1.07 | 1.07 |
| OXYCODONE | 9143 | 195 | 61,411.10 | 314.93 |
| HYDROMORPHONE | 9150 | 167 | 1,509.62 | 9.04 |
| HYDROCODONE | 9193 | 197 | 48,729.17 | 247.36 |
| MEPERIDINE (PETHIDINE) | 9230 | 49 | 1,271.41 | 25.95 |
| METHADONE | 9250B | 145 | 4,672.35 | 32.22 |
| MORPHINE | 9300 | 191 | 26,222.82 | 137.29 |
| OPIUM POWDERED | 9639 | 25 | 111.24 | 4.45 |
| OXYMORPHONE | 9652 | 84 | 1,854.90 | 22.08 |
| SUFENTANIL BASE | 9740 | 1 | 0 | 0 |
| TAPENTADOL | 9780 | 140 | 17,807.50 | 127.2 |
| FENTANYL BASE | 9801 | 194 | 1,019.97 | 5.26 |
| OPIUM COMBINATION PRODUCT (C-III) | 9809 | 4 | 2.65 | 0.66 |

STATE:SOUTH DAKOTA BUSINESS ACTIVITY:B - HOSPITALS

| DRUG NAME | DRUG CODE | BUYERS | TOTAL GRAMS | AVG GRAMS |
|---|---|---|---|---|
| AMPHETAMINE | 1100 | 23 | 2,833.81 | 123.21 |
| LISDEXAMFETAMINE | 1205 | 17 | 385.59 | 22.68 |
| METHYLPHENIDATE (DL;D;L;ISOMERS) | 1724 | 36 | 3,716.87 | 103.25 |
| BARBITURIC ACID DERIVIATIVE OR SALT [PER 21C | 2100 | 1 | 5 | 5 |
| BUTALBITAL | 2165 | 1 | 5 | 5 |
| PENTOBARBITAL (SCHEDULE 2) | 2270 | 1 | 11.39 | 11.39 |

| | | | | |
|---|---|---|---|---|
| COCAINE | 9041L | 21 | 148.7 | 7.08 |
| CODEINE | 9050 | 45 | 3,253.32 | 72.3 |
| BUPRENORPHINE | 9064 | 19 | 230.21 | 12.12 |
| OXYCODONE | 9143 | 73 | 7,346.96 | 100.64 |
| HYDROMORPHONE | 9150 | 63 | 630.87 | 10.01 |
| HYDROCODONE | 9193 | 76 | 10,431.18 | 137.25 |
| MEPERIDINE (PETHIDINE) | 9230 | 56 | 671.6 | 11.99 |
| METHADONE | 9250B | 26 | 522.24 | 20.09 |
| MORPHINE | 9300 | 74 | 8,523.35 | 115.18 |
| OPIUM POWDERED | 9639 | 21 | 65.88 | 3.14 |
| OXYMORPHONE | 9652 | 7 | 57.25 | 8.18 |
| ALFENTANIL | 9737 | 5 | 0.33 | 0.07 |
| REMIFENTANIL | 9739 | 9 | 1.8 | 0.2 |
| SUFENTANIL BASE | 9740 | 7 | 0.18 | 0.03 |
| TAPENTADOL | 9780 | 11 | 1,052.50 | 95.68 |
| FENTANYL BASE | 9801 | 77 | 179.88 | 2.34 |

STATE:SOUTH DAKOTA BUSINESS ACTIVITY:C - PRACTITIONERS

| DRUG NAME | DRUG CODE | BUYERS | TOTAL GRAMS | AVG GRAMS |
|---|---|---|---|---|
| PENTOBARBITAL (SCHEDULE 2) | 2270 | 13 | 24,892.61 | 1,914.82 |
| CODEINE | 9050 | 2 | 83.98 | 41.99 |
| BUPRENORPHINE | 9064 | 61 | 1.57 | 0.03 |
| OXYCODONE | 9143 | 5 | 187.31 | 37.46 |
| HYDROMORPHONE | 9150 | 29 | 59.18 | 2.04 |
| HYDROCODONE | 9193 | 21 | 46.66 | 2.22 |
| MEPERIDINE (PETHIDINE) | 9230 | 11 | 133.23 | 12.11 |
| MORPHINE | 9300 | 25 | 54.3 | 2.17 |
| OPIUM POWDERED | 9639 | 1 | 3.6 | 3.6 |
| OXYMORPHONE | 9652 | 1 | 0.03 | 0.03 |
| REMIFENTANIL | 9739 | 2 | 0.09 | 0.05 |
| FENTANYL BASE | 9801 | 41 | 4.25 | 0.1 |

STATE:SOUTH DAKOTA BUSINESS ACTIVITY:D - TEACHING INSTITUTIONS

| DRUG NAME | DRUG CODE | BUYERS | TOTAL GRAMS | AVG GRAMS |
|---|---|---|---|---|
| AMPHETAMINE | 1100 | 1 | 1.47 | 1.47 |
| PENTOBARBITAL (SCHEDULE 2) | 2270 | 1 | 88.86 | 88.86 |
| BUPRENORPHINE | 9064 | 2 | 0.01 | 0 |

STATE:SOUTH DAKOTA BUSINESS ACTIVITY:M - MID-LEVEL PRACTITIONERS

| DRUG NAME | DRUG CODE | BUYERS | TOTAL GRAMS | AVG GRAMS |
|---|---|---|---|---|
| HYDROMORPHONE | 9150 | 1 | 0.04 | 0.04 |
| MORPHINE | 9300 | 4 | 0.59 | 0.15 |
| FENTANYL BASE | 9801 | 6 | 0.13 | 0.02 |

STATE:SOUTH DAKOTA BUSINESS ACTIVITY:N-U NARCOTIC TREATMENT PROGRAMS

| DRUG NAME | DRUG CODE | BUYERS | TOTAL GRAMS | AVG GRAMS |
|---|---|---|---|---|
| METHADONE | 9250B | 1 | 3,563.29 | 3,563.29 |

STATE:TENNESSEE BUSINESS ACTIVITY:A - PHARMACIES

| DRUG NAME | DRUG CODE | BUYERS | TOTAL GRAMS | AVG GRAMS |
|---|---|---|---|---|
| AMPHETAMINE | 1100 | 1,567 | 514,247.55 | 328.17 |
| DL-METHAMPHETAMINE RACEMIC BASE | 1105B | 11 | 50.63 | 4.6 |
| D-METHAMPHETAMINE | 1105D | 1 | 0.4 | 0.4 |
| LISDEXAMFETAMINE | 1205 | 1,498 | 277,271.79 | 185.09 |
| METHYLPHENIDATE (DL;D;L;ISOMERS) | 1724 | 1,536 | 329,808.61 | 214.72 |
| BARBITURIC ACID DERIVIATIVE OR SALT [PER 21C | 2100 | 468 | 11,265 | 24.07 |
| AMOBARBITAL (SCHEDULE 2) | 2125 | 1 | 0.46 | 0.46 |
| BUTALBITAL | 2165 | 477 | 15,200 | 31.87 |
| SECOBARBITAL (SCHEDULE 2) | 2315 | 1 | 9.16 | 9.16 |
| DRONABINOL IN AN ORAL SOLUTION IN FDA APPR | 7365 | 6 | 3.6 | 0.6 |
| COCAINE | 9041L | 3 | 9.82 | 3.27 |
| CODEINE | 9050 | 1,530 | 333,893.63 | 218.23 |
| BUPRENORPHINE | 9064 | 1,472 | 136,431.14 | 92.68 |
| DIHYDROCODEINE | 9120 | 306 | 1,848.98 | 6.04 |

| | | | | |
|---|---|---|---|---|
| OXYCODONE | 9143 | 1,580 | 1,760,211.38 | 1,114.06 |
| HYDROMORPHONE | 9150 | 1,334 | 21,102.49 | 15.82 |
| HYDROCODONE | 9193 | 1,588 | 921,316.31 | 580.17 |
| LEVORPHANOL | 9220L | 3 | 0.7 | 0.23 |
| MEPERIDINE (PETHIDINE) | 9230 | 990 | 24,803.54 | 25.05 |
| METHADONE | 9250B | 1,052 | 39,775.04 | 37.81 |
| MORPHINE | 9300 | 1,560 | 604,791.01 | 387.69 |
| OPIUM POWDERED | 9639 | 106 | 184.68 | 1.74 |
| OXYMORPHONE | 9652 | 1,202 | 81,770.78 | 68.03 |
| REMIFENTANIL | 9739 | 1 | 0.03 | 0.03 |
| SUFENTANIL BASE | 9740 | 2 | 1.2 | 0.6 |
| TAPENTADOL | 9780 | 1,178 | 125,409.50 | 106.46 |
| FENTANYL BASE | 9801 | 1,516 | 6,831.63 | 4.51 |
| OPIUM COMBINATION PRODUCT (C-III) | 9809 | 53 | 23.84 | 0.45 |

STATE:TENNESSEE BUSINESS ACTIVITY:B - HOSPITALS

| DRUG NAME | DRUG CODE | BUYERS | TOTAL GRAMS | AVG GRAMS |
|---|---|---|---|---|
| AMPHETAMINE | 1100 | 81 | 5,471.92 | 67.55 |
| LISDEXAMFETAMINE | 1205 | 39 | 2,497.97 | 64.05 |
| METHYLPHENIDATE (DL;D;L;ISOMERS) | 1724 | 117 | 5,233.94 | 44.73 |
| GAMMA HYDROXYBUTYRIC ACID(FDA APPROVED | 2012 | 1 | 2,377.73 | 2,377.73 |
| BARBITURIC ACID DERIVIATIVE OR SALT [PER 21C | 2100 | 5 | 40 | 8 |
| AMOBARBITAL (SCHEDULE 2) | 2125 | 3 | 15.49 | 5.16 |
| BUTALBITAL | 2165 | 4 | 65 | 16.25 |
| PENTOBARBITAL (SCHEDULE 2) | 2270 | 13 | 4,271.73 | 328.59 |
| COCAINE | 9041L | 82 | 491.13 | 5.99 |
| CODEINE | 9050 | 144 | 4,363.51 | 30.3 |
| BUPRENORPHINE | 9064 | 123 | 1,816.09 | 14.76 |
| DIHYDROCODEINE | 9120 | 2 | 6.41 | 3.2 |
| OXYCODONE | 9143 | 178 | 90,341.63 | 507.54 |
| HYDROMORPHONE | 9150 | 165 | 5,344 | 32.39 |
| HYDROCODONE | 9193 | 193 | 50,195.15 | 260.08 |
| MEPERIDINE (PETHIDINE) | 9230 | 137 | 3,926.67 | 28.66 |
| METHADONE | 9250B | 116 | 2,476.49 | 21.35 |
| MORPHINE | 9300 | 180 | 44,701.49 | 248.34 |

| | | | | |
|---|---|---|---|---|
| OPIUM POWDERED | 9639 | 87 | 514.08 | 5.91 |
| OXYMORPHONE | 9652 | 37 | 462.2 | 12.49 |
| ALFENTANIL | 9737 | 6 | 2.21 | 0.37 |
| REMIFENTANIL | 9739 | 46 | 21.72 | 0.47 |
| SUFENTANIL BASE | 9740 | 49 | 1.3 | 0.03 |
| TAPENTADOL | 9780 | 42 | 1,800.50 | 42.87 |
| FENTANYL BASE | 9801 | 171 | 809.93 | 4.74 |
| OPIUM COMBINATION PRODUCT (C-III) | 9809 | 3 | 1.14 | 0.38 |

STATE:TENNESSEE BUSINESS ACTIVITY:C - PRACTITIONERS

| DRUG NAME | DRUG CODE | BUYERS | TOTAL GRAMS | AVG GRAMS |
|---|---|---|---|---|
| AMPHETAMINE | 1100 | 8 | 719.27 | 89.91 |
| LISDEXAMFETAMINE | 1205 | 8 | 829.4 | 103.68 |
| METHYLPHENIDATE (DL;D;L;ISOMERS) | 1724 | 3 | 600.25 | 200.08 |
| PENTOBARBITAL (SCHEDULE 2) | 2270 | 168 | 117,425.20 | 698.96 |
| COCAINE | 9041L | 23 | 169.06 | 7.35 |
| CODEINE | 9050 | 56 | 3,121.60 | 55.74 |
| BUPRENORPHINE | 9064 | 493 | 41.09 | 0.08 |
| DIHYDROCODEINE | 9120 | 64 | 46.11 | 0.72 |
| OXYCODONE | 9143 | 78 | 589.92 | 7.56 |
| HYDROMORPHONE | 9150 | 272 | 209.91 | 0.77 |
| HYDROCODONE | 9193 | 267 | 1,335.39 | 5 |
| MEPERIDINE (PETHIDINE) | 9230 | 159 | 939.69 | 5.91 |
| METHADONE | 9250B | 10 | 11.54 | 1.15 |
| MORPHINE | 9300 | 172 | 503.89 | 2.93 |
| OPIUM POWDERED | 9639 | 9 | 61.2 | 6.8 |
| OXYMORPHONE | 9652 | 3 | 1.2 | 0.4 |
| NOROXYMORPHONE | 9668 | 1 | 0.13 | 0.13 |
| ALFENTANIL | 9737 | 2 | 2.56 | 1.28 |
| REMIFENTANIL | 9739 | 9 | 1.49 | 0.17 |
| SUFENTANIL BASE | 9740 | 4 | 0.06 | 0.01 |
| TAPENTADOL | 9780 | 1 | 5 | 5 |
| FENTANYL BASE | 9801 | 410 | 46.54 | 0.11 |

STATISTICAL SUMMARY FOR RETAIL DRUG PURCHASES BY GRAMS WT

STATE:TENNESSEE BUSINESS ACTIVITY:D - TEACHING INSTITUTIONS

| DRUG NAME | DRUG CODE | BUYERS | TOTAL GRAMS | AVG GRAMS |
|---|---|---|---|---|
| BUPRENORPHINE | 9064 | 2 | 0.13 | 0.06 |
| FENTANYL BASE | 9801 | 1 | 0 | 0 |

STATE:TENNESSEE BUSINESS ACTIVITY:M - MID-LEVEL PRACTITIONERS

| DRUG NAME | DRUG CODE | BUYERS | TOTAL GRAMS | AVG GRAMS |
|---|---|---|---|---|
| PENTOBARBITAL (SCHEDULE 2) | 2270 | 24 | 20,896.58 | 870.69 |
| CODEINE | 9050 | 13 | 105.39 | 8.11 |
| DIHYDROCODEINE | 9120 | 55 | 35.53 | 0.65 |
| HYDROCODONE | 9193 | 1 | 1.21 | 1.21 |
| MEPERIDINE (PETHIDINE) | 9230 | 10 | 22.22 | 2.22 |
| MORPHINE | 9300 | 62 | 26.55 | 0.43 |
| FENTANYL BASE | 9801 | 78 | 1.3 | 0.02 |

STATE:TENNESSEE BUSINESS ACTIVITY:N-U NARCOTIC TREATMENT PROGRAMS

| DRUG NAME | DRUG CODE | BUYERS | TOTAL GRAMS | AVG GRAMS |
|---|---|---|---|---|
| BUPRENORPHINE | 9064 | 10 | 997.32 | 99.73 |
| METHADONE | 9250B | 13 | 152,040.40 | 11,695.42 |

STATE:TEXAS BUSINESS ACTIVITY:A - PHARMACIES

| DRUG NAME | DRUG CODE | BUYERS | TOTAL GRAMS | AVG GRAMS |
|---|---|---|---|---|
| AMPHETAMINE | 1100 | 4,595 | 1,620,453.17 | 352.66 |
| DL-METHAMPHETAMINE RACEMIC BASE | 1105B | 164 | 423.95 | 2.59 |
| D-METHAMPHETAMINE | 1105D | 13 | 34.14 | 2.63 |
| LISDEXAMFETAMINE | 1205 | 4,431 | 1,079,841.66 | 243.7 |
| METHYLPHENIDATE (DL;D;L;ISOMERS) | 1724 | 4,565 | 1,287,419.38 | 282.02 |
| BARBITURIC ACID DERIVIATIVE OR SALT [PER 21C | 2100 | 1,643 | 49,025 | 29.84 |
| BUTALBITAL | 2165 | 2,065 | 74,475 | 36.07 |
| PENTOBARBITAL (SCHEDULE 2) | 2270 | 3 | 7.75 | 2.58 |

| | | | | |
|---|---|---|---|---|
| SECOBARBITAL (SCHEDULE 2) | 2315 | 3 | 36.62 | 12.21 |
| DRONABINOL IN AN ORAL SOLUTION IN FDA APPR | 7365 | 13 | 11.7 | 0.9 |
| COCAINE | 9041L | 13 | 34.53 | 2.66 |
| CODEINE | 9050 | 4,932 | 3,881,704.12 | 787.04 |
| BUPRENORPHINE | 9064 | 4,200 | 99,861.65 | 23.78 |
| DIHYDROCODEINE | 9120 | 1,565 | 10,415.63 | 6.66 |
| OXYCODONE | 9143 | 4,514 | 1,262,157.71 | 279.61 |
| HYDROMORPHONE | 9150 | 4,014 | 79,229.41 | 19.74 |
| HYDROCODONE | 9193 | 4,956 | 2,501,324.80 | 504.71 |
| LEVORPHANOL | 9220L | 128 | 220.2 | 1.72 |
| MEPERIDINE (PETHIDINE) | 9230 | 1,084 | 18,663.61 | 17.22 |
| METHADONE | 9250B | 3,472 | 141,211.37 | 40.67 |
| MORPHINE | 9300 | 4,342 | 723,947.66 | 166.73 |
| OPIUM TINCTURE (LAUDANUM) | 9630 | 7 | 14.16 | 2.02 |
| OPIUM POWDERED | 9639 | 159 | 280.44 | 1.76 |
| OXYMORPHONE | 9652 | 2,189 | 45,543.49 | 20.81 |
| NOROXYMORPHONE | 9668 | 2 | 5.74 | 2.87 |
| REMIFENTANIL | 9739 | 3 | 0.15 | 0.05 |
| SUFENTANIL BASE | 9740 | 4 | 2.7 | 0.67 |
| TAPENTADOL | 9780 | 3,009 | 384,643.50 | 127.83 |
| FENTANYL BASE | 9801 | 4,359 | 19,817.34 | 4.55 |
| OPIUM COMBINATION PRODUCT (C-III) | 9809 | 84 | 50.89 | 0.61 |

STATE:TEXAS BUSINESS ACTIVITY:B - HOSPITALS

| DRUG NAME | DRUG CODE | BUYERS | TOTAL GRAMS | AVG GRAMS |
|---|---|---|---|---|
| AMPHETAMINE | 1100 | 295 | 35,401.34 | 120 |
| DL-METHAMPHETAMINE RACEMIC BASE | 1105B | 1 | 2.81 | 2.81 |
| D-METHAMPHETAMINE | 1105D | 1 | 5.62 | 5.62 |
| LISDEXAMFETAMINE | 1205 | 123 | 9,090.62 | 73.91 |
| METHYLPHENIDATE (DL;D;L;ISOMERS) | 1724 | 423 | 49,170.99 | 116.24 |
| GAMMA HYDROXYBUTYRIC ACID(FDA APPROVED | 2012 | 1 | 3,269.38 | 3,269.38 |
| BARBITURIC ACID DERIVIATIVE OR SALT [PER 21C | 2100 | 23 | 274.9 | 11.95 |
| AMOBARBITAL (SCHEDULE 2) | 2125 | 15 | 83.39 | 5.56 |
| BUTALBITAL | 2165 | 23 | 320 | 13.91 |
| PENTOBARBITAL (SCHEDULE 2) | 2270 | 51 | 4,768.44 | 93.5 |

| | | | | |
|---|---|---|---|---|
| SECOBARBITAL (SCHEDULE 2) | 2315 | 1 | 9.16 | 9.16 |
| COCAINE | 9041L | 292 | 1,397.87 | 4.79 |
| CODEINE | 9050 | 936 | 120,087.44 | 128.3 |
| BUPRENORPHINE | 9064 | 284 | 3,981.37 | 14.02 |
| DIHYDROCODEINE | 9120 | 7 | 28.84 | 4.12 |
| OXYCODONE | 9143 | 730 | 61,703.86 | 84.53 |
| HYDROMORPHONE | 9150 | 1,121 | 15,755.34 | 14.05 |
| HYDROCODONE | 9193 | 1,203 | 155,005.93 | 128.85 |
| LEVORPHANOL | 9220L | 1 | 0.12 | 0.12 |
| MEPERIDINE (PETHIDINE) | 9230 | 802 | 24,435.38 | 30.47 |
| METHADONE | 9250B | 473 | 11,906.77 | 25.17 |
| MORPHINE | 9300 | 1,285 | 96,275.62 | 74.92 |
| OPIUM POWDERED | 9639 | 322 | 1,803.24 | 5.6 |
| OXYMORPHONE | 9652 | 46 | 619 | 13.46 |
| NOROXYMORPHONE | 9668 | 2 | 4.12 | 2.06 |
| ALFENTANIL | 9737 | 53 | 20.67 | 0.39 |
| REMIFENTANIL | 9739 | 193 | 110.7 | 0.57 |
| SUFENTANIL BASE | 9740 | 197 | 3.9 | 0.02 |
| TAPENTADOL | 9780 | 110 | 5,444.50 | 49.5 |
| FENTANYL BASE | 9801 | 1,283 | 3,075.08 | 2.4 |
| OPIUM COMBINATION PRODUCT (C-III) | 9809 | 6 | 2.08 | 0.35 |

STATE:TEXAS BUSINESS ACTIVITY:C - PRACTITIONERS

| DRUG NAME | DRUG CODE | BUYERS | TOTAL GRAMS | AVG GRAMS |
|---|---|---|---|---|
| AMPHETAMINE | 1100 | 3 | 5.17 | 1.72 |
| LISDEXAMFETAMINE | 1205 | 6 | 42.78 | 7.13 |
| METHYLPHENIDATE (DL;D;L;ISOMERS) | 1724 | 3 | 184.97 | 61.66 |
| GAMMA HYDROXYBUTYRIC ACID(FDA APPROVED | 2012 | 3 | 7,876.22 | 2,625.41 |
| BARBITURIC ACID DERIVIATIVE OR SALT [PER 21C | 2100 | 1 | 5 | 5 |
| PENTOBARBITAL (SCHEDULE 2) | 2270 | 357 | 233,571.43 | 654.26 |
| COCAINE | 9041L | 14 | 21.28 | 1.52 |
| CODEINE | 9050 | 768 | 5,435.66 | 7.08 |
| BUPRENORPHINE | 9064 | 1,877 | 629.55 | 0.34 |
| DIHYDROCODEINE | 9120 | 464 | 461.45 | 0.99 |
| OXYCODONE | 9143 | 33 | 454.69 | 13.78 |

| | | | | |
|---|---|---|---|---|
| HYDROMORPHONE | 9150 | 770 | 411.98 | 0.54 |
| HYDROCODONE | 9193 | 841 | 1,974.04 | 2.35 |
| MEPERIDINE (PETHIDINE) | 9230 | 415 | 3,226.77 | 7.78 |
| METHADONE | 9250B | 32 | 56.88 | 1.78 |
| MORPHINE | 9300 | 514 | 781.74 | 1.52 |
| THEBAINE | 9333 | 1 | 25 | 25 |
| OPIUM POWDERED | 9639 | 2 | 5.04 | 2.52 |
| OXYMORPHONE | 9652 | 7 | 5.59 | 0.8 |
| NOROXYMORPHONE | 9668 | 8 | 98.98 | 12.37 |
| ALFENTANIL | 9737 | 12 | 1.37 | 0.11 |
| REMIFENTANIL | 9739 | 21 | 0.93 | 0.04 |
| SUFENTANIL BASE | 9740 | 7 | 0.06 | 0.01 |
| TAPENTADOL | 9780 | 1 | 15 | 15 |
| FENTANYL BASE | 9801 | 1,341 | 84.08 | 0.06 |

STATE:TEXAS BUSINESS ACTIVITY:D - TEACHING INSTITUTIONS

| DRUG NAME | DRUG CODE | BUYERS | TOTAL GRAMS | AVG GRAMS |
|---|---|---|---|---|
| PENTOBARBITAL (SCHEDULE 2) | 2270 | 2 | 2,843.57 | 1,421.78 |
| BUPRENORPHINE | 9064 | 4 | 0.73 | 0.18 |
| HYDROMORPHONE | 9150 | 1 | 8.44 | 8.44 |
| HYDROCODONE | 9193 | 1 | 3.28 | 3.28 |
| METHADONE | 9250B | 1 | 13.59 | 13.59 |
| MORPHINE | 9300 | 2 | 6.55 | 3.28 |
| OXYMORPHONE | 9652 | 1 | 0.18 | 0.18 |
| REMIFENTANIL | 9739 | 1 | 0.08 | 0.08 |
| FENTANYL BASE | 9801 | 3 | 2.49 | 0.83 |

STATE:TEXAS BUSINESS ACTIVITY:M - MID-LEVEL PRACTITIONERS

| DRUG NAME | DRUG CODE | BUYERS | TOTAL GRAMS | AVG GRAMS |
|---|---|---|---|---|
| PENTOBARBITAL (SCHEDULE 2) | 2270 | 157 | 255,401.97 | 1,626.76 |
| CODEINE | 9050 | 184 | 331.76 | 1.8 |
| DIHYDROCODEINE | 9120 | 174 | 123.27 | 0.71 |
| HYDROMORPHONE | 9150 | 33 | 4.64 | 0.14 |

| | | | | |
|---|---|---|---|---|
| HYDROCODONE | 9193 | 1 | 1.21 | 1.21 |
| MEPERIDINE (PETHIDINE) | 9230 | 4 | 4.58 | 1.14 |
| MORPHINE | 9300 | 376 | 125.86 | 0.33 |
| FENTANYL BASE | 9801 | 634 | 47.13 | 0.07 |

STATE:TEXAS BUSINESS ACTIVITY:N-U NARCOTIC TREATMENT PROGRAMS

| DRUG NAME | DRUG CODE | BUYERS | TOTAL GRAMS | AVG GRAMS |
|---|---|---|---|---|
| BUPRENORPHINE | 9064 | 28 | 863.02 | 30.82 |
| METHADONE | 9250B | 89 | 408,829.79 | 4,593.59 |

STATE:UTAH BUSINESS ACTIVITY:A - PHARMACIES

| DRUG NAME | DRUG CODE | BUYERS | TOTAL GRAMS | AVG GRAMS |
|---|---|---|---|---|
| AMPHETAMINE | 1100 | 549 | 322,909.40 | 588.18 |
| DL-METHAMPHETAMINE RACEMIC BASE | 1105B | 25 | 104.08 | 4.16 |
| LISDEXAMFETAMINE | 1205 | 529 | 85,720.67 | 162.04 |
| METHYLPHENIDATE (DL;D;L;ISOMERS) | 1724 | 543 | 179,635.47 | 330.82 |
| BARBITURIC ACID DERIVIATIVE OR SALT [PER 21C | 2100 | 154 | 5,675 | 36.85 |
| BUTALBITAL | 2165 | 303 | 12,720 | 41.98 |
| DRONABINOL IN AN ORAL SOLUTION IN FDA APPR | 7365 | 1 | 0.45 | 0.45 |
| COCAINE | 9041L | 4 | 4.71 | 1.18 |
| CODEINE | 9050 | 532 | 63,584.26 | 119.52 |
| BUPRENORPHINE | 9064 | 547 | 37,590.94 | 68.72 |
| DIHYDROCODEINE | 9120 | 1 | 1.07 | 1.07 |
| OXYCODONE | 9143 | 549 | 643,887.89 | 1,172.84 |
| HYDROMORPHONE | 9150 | 512 | 9,064.12 | 17.7 |
| HYDROCODONE | 9193 | 551 | 221,703.78 | 402.37 |
| MEPERIDINE (PETHIDINE) | 9230 | 234 | 5,381.29 | 23 |
| METHADONE | 9250B | 503 | 38,100.55 | 75.75 |
| MORPHINE | 9300 | 534 | 181,248.57 | 339.42 |
| OPIUM POWDERED | 9639 | 25 | 36.36 | 1.45 |
| OXYMORPHONE | 9652 | 415 | 16,319.77 | 39.32 |
| REMIFENTANIL | 9739 | 5 | 0.72 | 0.14 |
| SUFENTANIL BASE | 9740 | 3 | 0.08 | 0.03 |

| | | | | |
|---|---|---|---|---|
| TAPENTADOL | 9780 | 400 | 51,783.50 | 129.46 |
| FENTANYL BASE | 9801 | 530 | 2,994.95 | 5.65 |
| OPIUM COMBINATION PRODUCT (C-III) | 9809 | 72 | 49 | 0.68 |

STATE:UTAH BUSINESS ACTIVITY:B - HOSPITALS

| DRUG NAME | DRUG CODE | BUYERS | TOTAL GRAMS | AVG GRAMS |
|---|---|---|---|---|
| AMPHETAMINE | 1100 | 32 | 4,776.99 | 149.28 |
| LISDEXAMFETAMINE | 1205 | 18 | 885.07 | 49.17 |
| METHYLPHENIDATE (DL;D;L;ISOMERS) | 1724 | 43 | 4,496.46 | 104.57 |
| BARBITURIC ACID DERIVIATIVE OR SALT [PER 21C | 2100 | 1 | 10 | 10 |
| AMOBARBITAL (SCHEDULE 2) | 2125 | 3 | 34.18 | 11.39 |
| PENTOBARBITAL (SCHEDULE 2) | 2270 | 3 | 844.87 | 281.62 |
| DRONABINOL IN AN ORAL SOLUTION IN FDA APPR | 7365 | 1 | 0.15 | 0.15 |
| COCAINE | 9041L | 51 | 386.01 | 7.57 |
| CODEINE | 9050 | 37 | 1,538.08 | 41.57 |
| BUPRENORPHINE | 9064 | 41 | 628.1 | 15.32 |
| OXYCODONE | 9143 | 88 | 26,736.46 | 303.82 |
| HYDROMORPHONE | 9150 | 81 | 1,789.16 | 22.09 |
| HYDROCODONE | 9193 | 108 | 11,806.68 | 109.32 |
| MEPERIDINE (PETHIDINE) | 9230 | 77 | 1,649.12 | 21.42 |
| METHADONE | 9250B | 48 | 1,373.53 | 28.62 |
| MORPHINE | 9300 | 100 | 15,208.01 | 152.08 |
| OPIUM POWDERED | 9639 | 34 | 224.28 | 6.6 |
| OXYMORPHONE | 9652 | 8 | 78.86 | 9.86 |
| ALFENTANIL | 9737 | 8 | 3.09 | 0.39 |
| REMIFENTANIL | 9739 | 37 | 70.87 | 1.92 |
| SUFENTANIL BASE | 9740 | 28 | 0.83 | 0.03 |
| TAPENTADOL | 9780 | 22 | 3,107 | 141.23 |
| FENTANYL BASE | 9801 | 105 | 269.31 | 2.56 |
| OPIUM COMBINATION PRODUCT (C-III) | 9809 | 2 | 0.57 | 0.28 |

STATE:UTAH BUSINESS ACTIVITY:C - PRACTITIONERS

| DRUG NAME | DRUG CODE | BUYERS | TOTAL GRAMS | AVG GRAMS |
|---|---|---|---|---|

| | | | | |
|---|---|---|---|---|
| METHYLPHENIDATE (DL;D;L;ISOMERS) | 1724 | 1 | 29.05 | 29.05 |
| PENTOBARBITAL (SCHEDULE 2) | 2270 | 46 | 50,733.77 | 1,102.91 |
| COCAINE | 9041L | 1 | 0.29 | 0.29 |
| CODEINE | 9050 | 7 | 249.33 | 35.62 |
| BUPRENORPHINE | 9064 | 164 | 20.73 | 0.13 |
| OXYCODONE | 9143 | 12 | 362.88 | 30.24 |
| HYDROMORPHONE | 9150 | 89 | 52.06 | 0.58 |
| HYDROCODONE | 9193 | 78 | 367.08 | 4.71 |
| MEPERIDINE (PETHIDINE) | 9230 | 30 | 132.25 | 4.41 |
| METHADONE | 9250B | 10 | 8.85 | 0.89 |
| MORPHINE | 9300 | 95 | 96.23 | 1.01 |
| OXYMORPHONE | 9652 | 1 | 0.81 | 0.81 |
| REMIFENTANIL | 9739 | 2 | 0.15 | 0.08 |
| SUFENTANIL BASE | 9740 | 1 | 0.01 | 0.01 |
| FENTANYL BASE | 9801 | 175 | 10.7 | 0.06 |

STATE:UTAH BUSINESS ACTIVITY:D - TEACHING INSTITUTIONS

| DRUG NAME | DRUG CODE | BUYERS | TOTAL GRAMS | AVG GRAMS |
|---|---|---|---|---|
| BUPRENORPHINE | 9064 | 1 | 0.01 | 0.01 |

STATE:UTAH BUSINESS ACTIVITY:M - MID-LEVEL PRACTITIONERS

| DRUG NAME | DRUG CODE | BUYERS | TOTAL GRAMS | AVG GRAMS |
|---|---|---|---|---|
| PENTOBARBITAL (SCHEDULE 2) | 2270 | 12 | 5,336.25 | 444.69 |
| BUPRENORPHINE | 9064 | 2 | 0.25 | 0.12 |
| HYDROMORPHONE | 9150 | 1 | 0.12 | 0.12 |
| MEPERIDINE (PETHIDINE) | 9230 | 3 | 32.9 | 10.97 |
| REMIFENTANIL | 9739 | 1 | 0.04 | 0.04 |
| FENTANYL BASE | 9801 | 9 | 0.58 | 0.06 |

STATE:UTAH BUSINESS ACTIVITY:N-U NARCOTIC TREATMENT PROGRAMS

| DRUG NAME | DRUG CODE | BUYERS | TOTAL GRAMS | AVG GRAMS |
|---|---|---|---|---|

| | | | | |
|---|---|---|---|---|
| BUPRENORPHINE | 9064 | 12 | 322.42 | 26.87 |
| METHADONE | 9250B | 13 | 66,203.49 | 5,092.58 |

STATE:VERMONT BUSINESS ACTIVITY:A - PHARMACIES

| DRUG NAME | DRUG CODE | BUYERS | TOTAL GRAMS | AVG GRAMS |
|---|---|---|---|---|
| AMPHETAMINE | 1100 | 149 | 38,761.65 | 260.15 |
| DL-METHAMPHETAMINE RACEMIC BASE | 1105B | 2 | 7.23 | 3.62 |
| LISDEXAMFETAMINE | 1205 | 149 | 27,324.47 | 183.39 |
| METHYLPHENIDATE (DL;D;L;ISOMERS) | 1724 | 149 | 74,807.64 | 502.06 |
| BARBITURIC ACID DERIVIATIVE OR SALT [PER 21C | 2100 | 14 | 285 | 20.36 |
| BUTALBITAL | 2165 | 30 | 470 | 15.67 |
| SECOBARBITAL (SCHEDULE 2) | 2315 | 4 | 137.33 | 34.33 |
| COCAINE | 9041L | 1 | 4.46 | 4.46 |
| CODEINE | 9050 | 143 | 12,700.85 | 88.82 |
| BUPRENORPHINE | 9064 | 147 | 20,052.57 | 136.41 |
| DIHYDROCODEINE | 9120 | 1 | 1.07 | 1.07 |
| OXYCODONE | 9143 | 148 | 84,817.39 | 573.09 |
| HYDROMORPHONE | 9150 | 149 | 4,183.76 | 28.08 |
| HYDROCODONE | 9193 | 149 | 18,183.61 | 122.04 |
| LEVORPHANOL | 9220L | 1 | 0.12 | 0.12 |
| MEPERIDINE (PETHIDINE) | 9230 | 24 | 408.52 | 17.02 |
| METHADONE | 9250B | 139 | 13,500.97 | 97.13 |
| MORPHINE | 9300 | 147 | 35,798.07 | 243.52 |
| OPIUM POWDERED | 9639 | 18 | 77.04 | 4.28 |
| OXYMORPHONE | 9652 | 54 | 1,140.55 | 21.12 |
| TAPENTADOL | 9780 | 56 | 4,600 | 82.14 |
| FENTANYL BASE | 9801 | 147 | 689.75 | 4.69 |
| OPIUM COMBINATION PRODUCT (C-III) | 9809 | 1 | 0.57 | 0.57 |

STATE:VERMONT BUSINESS ACTIVITY:B - HOSPITALS

| DRUG NAME | DRUG CODE | BUYERS | TOTAL GRAMS | AVG GRAMS |
|---|---|---|---|---|
| AMPHETAMINE | 1100 | 11 | 927.75 | 84.34 |
| LISDEXAMFETAMINE | 1205 | 7 | 379.81 | 54.26 |

| | | | | |
|---|---|---|---|---|
| METHYLPHENIDATE (DL;D;L;ISOMERS) | 1724 | 15 | 2,119.69 | 141.31 |
| AMOBARBITAL (SCHEDULE 2) | 2125 | 1 | 0.91 | 0.91 |
| PENTOBARBITAL (SCHEDULE 2) | 2270 | 1 | 84.76 | 84.76 |
| COCAINE | 9041L | 8 | 34 | 4.25 |
| CODEINE | 9050 | 13 | 367.69 | 28.28 |
| BUPRENORPHINE | 9064 | 17 | 1,148.38 | 67.55 |
| OXYCODONE | 9143 | 17 | 5,924.88 | 348.52 |
| HYDROMORPHONE | 9150 | 16 | 672.98 | 42.06 |
| HYDROCODONE | 9193 | 17 | 921.13 | 54.18 |
| LEVORPHANOL | 9220L | 1 | 1.51 | 1.51 |
| MEPERIDINE (PETHIDINE) | 9230 | 15 | 878.25 | 58.55 |
| METHADONE | 9250B | 15 | 704.97 | 47 |
| MORPHINE | 9300 | 16 | 4,689.51 | 293.09 |
| OPIUM POWDERED | 9639 | 13 | 85.68 | 6.59 |
| OXYMORPHONE | 9652 | 1 | 4.28 | 4.28 |
| ALFENTANIL | 9737 | 2 | 0.04 | 0.02 |
| REMIFENTANIL | 9739 | 3 | 2.79 | 0.93 |
| SUFENTANIL BASE | 9740 | 2 | 0.03 | 0.01 |
| TAPENTADOL | 9780 | 1 | 20 | 20 |
| FENTANYL BASE | 9801 | 17 | 55.49 | 3.26 |
| OPIUM COMBINATION PRODUCT (C-III) | 9809 | 1 | 0.38 | 0.38 |

STATE:VERMONT BUSINESS ACTIVITY:C - PRACTITIONERS

| DRUG NAME | DRUG CODE | BUYERS | TOTAL GRAMS | AVG GRAMS |
|---|---|---|---|---|
| PENTOBARBITAL (SCHEDULE 2) | 2270 | 74 | 53,566.58 | 723.87 |
| CODEINE | 9050 | 2 | 12.16 | 6.08 |
| BUPRENORPHINE | 9064 | 83 | 3.47 | 0.04 |
| OXYCODONE | 9143 | 1 | 1.79 | 1.79 |
| HYDROMORPHONE | 9150 | 51 | 16.07 | 0.32 |
| HYDROCODONE | 9193 | 20 | 32.88 | 1.64 |
| MEPERIDINE (PETHIDINE) | 9230 | 1 | 1.09 | 1.09 |
| MORPHINE | 9300 | 28 | 40.98 | 1.46 |
| OXYMORPHONE | 9652 | 1 | 0.03 | 0.03 |
| NOROXYMORPHONE | 9668 | 1 | 0.1 | 0.1 |
| REMIFENTANIL | 9739 | 1 | 0.04 | 0.04 |

| | | | | |
|---|---|---|---|---|
| FENTANYL BASE | 9801 | 37 | 2.82 | 0.08 |

STATE:VERMONT BUSINESS ACTIVITY:D - TEACHING INSTITUTIONS

| DRUG NAME | DRUG CODE | BUYERS | TOTAL GRAMS | AVG GRAMS |
|---|---|---|---|---|
| BUPRENORPHINE | 9064 | 2 | 0 | 0 |

STATE:VERMONT BUSINESS ACTIVITY:M - MID-LEVEL PRACTITIONERS

| DRUG NAME | DRUG CODE | BUYERS | TOTAL GRAMS | AVG GRAMS |
|---|---|---|---|---|
| PENTOBARBITAL (SCHEDULE 2) | 2270 | 3 | 710.89 | 236.96 |
| CODEINE | 9050 | 5 | 35.23 | 7.05 |

STATE:VERMONT BUSINESS ACTIVITY:N-U NARCOTIC TREATMENT PROGRAMS

| DRUG NAME | DRUG CODE | BUYERS | TOTAL GRAMS | AVG GRAMS |
|---|---|---|---|---|
| BUPRENORPHINE | 9064 | 10 | 4,014.64 | 401.46 |
| METHADONE | 9250B | 10 | 59,793.32 | 5,979.33 |

STATE:VIRGIN ISLANDS BUSINESS ACTIVITY:A - PHARMACIES

| DRUG NAME | DRUG CODE | BUYERS | TOTAL GRAMS | AVG GRAMS |
|---|---|---|---|---|
| AMPHETAMINE | 1100 | 19 | 1,217.23 | 64.06 |
| LISDEXAMFETAMINE | 1205 | 14 | 267.66 | 19.12 |
| METHYLPHENIDATE (DL;D;L;ISOMERS) | 1724 | 19 | 595.89 | 31.36 |
| BARBITURIC ACID DERIVIATIVE OR SALT [PER 21C | 2100 | 7 | 105 | 15 |
| BUTALBITAL | 2165 | 3 | 45 | 15 |
| CODEINE | 9050 | 21 | 2,939.36 | 139.97 |
| BUPRENORPHINE | 9064 | 11 | 62.59 | 5.69 |
| OXYCODONE | 9143 | 21 | 2,066.24 | 98.39 |
| HYDROMORPHONE | 9150 | 13 | 57.1 | 4.39 |
| HYDROCODONE | 9193 | 21 | 1,126.02 | 53.62 |
| MEPERIDINE (PETHIDINE) | 9230 | 7 | 220.05 | 31.44 |

| | | | | |
|---|---|---|---|---|
| METHADONE | 9250B | 9 | 99.73 | 11.08 |
| MORPHINE | 9300 | 13 | 491.23 | 37.79 |
| TAPENTADOL | 9780 | 2 | 18 | 9 |
| FENTANYL BASE | 9801 | 12 | 12.01 | 1 |

STATE:VIRGIN ISLANDS BUSINESS ACTIVITY:B - HOSPITALS

| DRUG NAME | DRUG CODE | BUYERS | TOTAL GRAMS | AVG GRAMS |
|---|---|---|---|---|
| METHYLPHENIDATE (DL;D;L;ISOMERS) | 1724 | 2 | 1.3 | 0.65 |
| CODEINE | 9050 | 2 | 55.25 | 27.63 |
| OXYCODONE | 9143 | 2 | 35.86 | 17.93 |
| HYDROMORPHONE | 9150 | 2 | 16.95 | 8.48 |
| HYDROCODONE | 9193 | 2 | 28.34 | 14.17 |
| MEPERIDINE (PETHIDINE) | 9230 | 1 | 21.24 | 21.24 |
| MORPHINE | 9300 | 3 | 32.38 | 10.79 |
| OPIUM POWDERED | 9639 | 1 | 1.8 | 1.8 |
| FENTANYL BASE | 9801 | 3 | 1.87 | 0.62 |

STATE:VIRGIN ISLANDS BUSINESS ACTIVITY:C - PRACTITIONERS

| DRUG NAME | DRUG CODE | BUYERS | TOTAL GRAMS | AVG GRAMS |
|---|---|---|---|---|
| CODEINE | 9050 | 2 | 4.42 | 2.21 |
| BUPRENORPHINE | 9064 | 3 | 0.13 | 0.04 |
| HYDROMORPHONE | 9150 | 2 | 0.1 | 0.05 |
| HYDROCODONE | 9193 | 2 | 4.24 | 2.12 |
| MEPERIDINE (PETHIDINE) | 9230 | 2 | 12.64 | 6.32 |
| MORPHINE | 9300 | 1 | 1.28 | 1.28 |
| FENTANYL BASE | 9801 | 3 | 0.07 | 0.02 |

STATE:VIRGIN ISLANDS BUSINESS ACTIVITY:N-U NARCOTIC TREATMENT PROGRAMS

| DRUG NAME | DRUG CODE | BUYERS | TOTAL GRAMS | AVG GRAMS |
|---|---|---|---|---|
| METHADONE | 9250B | 1 | 357.76 | 357.76 |

STATE:VIRGINIA BUSINESS ACTIVITY:A - PHARMACIES

| DRUG NAME | DRUG CODE | BUYERS | TOTAL GRAMS | AVG GRAMS |
|---|---|---|---|---|
| AMPHETAMINE | 1100 | 1,682 | 685,423.29 | 407.5 |
| DL-METHAMPHETAMINE RACEMIC BASE | 1105B | 59 | 175.21 | 2.97 |
| LISDEXAMFETAMINE | 1205 | 1,618 | 263,719.39 | 162.99 |
| METHYLPHENIDATE (DL;D;L;ISOMERS) | 1724 | 1,668 | 501,926.04 | 300.91 |
| BARBITURIC ACID DERIVIATIVE OR SALT [PER 21C | 2100 | 540 | 13,850 | 25.65 |
| BUTALBITAL | 2165 | 725 | 20,780 | 28.66 |
| PENTOBARBITAL (SCHEDULE 2) | 2270 | 1 | 0.91 | 0.91 |
| SECOBARBITAL (SCHEDULE 2) | 2315 | 3 | 109.86 | 36.62 |
| DRONABINOL IN AN ORAL SOLUTION IN FDA APPR | 7365 | 2 | 0.6 | 0.3 |
| NABILONE | 7379 | 4 | 0.75 | 0.19 |
| COCAINE | 9041L | 4 | 47.18 | 11.79 |
| CODEINE | 9050 | 1,645 | 205,635.14 | 125.01 |
| BUPRENORPHINE | 9064 | 1,582 | 75,607.16 | 47.79 |
| DIHYDROCODEINE | 9120 | 13 | 26.7 | 2.05 |
| OXYCODONE | 9143 | 1,704 | 1,116,236.21 | 655.07 |
| HYDROMORPHONE | 9150 | 1,595 | 38,087.17 | 23.88 |
| HYDROCODONE | 9193 | 1,707 | 394,094.50 | 230.87 |
| LEVORPHANOL | 9220L | 51 | 39 | 0.76 |
| MEPERIDINE (PETHIDINE) | 9230 | 744 | 18,804.90 | 25.28 |
| METHADONE | 9250B | 1,312 | 62,956.64 | 47.99 |
| MORPHINE | 9300 | 1,620 | 280,864.81 | 173.37 |
| OPIUM TINCTURE (LAUDANUM) | 9630 | 2 | 10.62 | 5.31 |
| OPIUM POWDERED | 9639 | 109 | 263.16 | 2.41 |
| OXYMORPHONE | 9652 | 1,095 | 36,210.03 | 33.07 |
| REMIFENTANIL | 9739 | 2 | 0.04 | 0.02 |
| TAPENTADOL | 9780 | 1,204 | 163,167.50 | 135.52 |
| FENTANYL BASE | 9801 | 1,556 | 6,046.85 | 3.89 |
| OPIUM COMBINATION PRODUCT (C-III) | 9809 | 58 | 21.19 | 0.37 |

STATE:VIRGINIA BUSINESS ACTIVITY:B - HOSPITALS

| DRUG NAME | DRUG CODE | BUYERS | TOTAL GRAMS | AVG GRAMS |
|---|---|---|---|---|

| | | | | |
|---|---|---|---|---|
| AMPHETAMINE | 1100 | 87 | 28,529.87 | 327.93 |
| LISDEXAMFETAMINE | 1205 | 43 | 7,256.31 | 168.75 |
| METHYLPHENIDATE (DL;D;L;ISOMERS) | 1724 | 121 | 33,798.20 | 279.32 |
| GAMMA HYDROXYBUTYRIC ACID(FDA APPROVED | 2012 | 1 | 6,092.93 | 6,092.93 |
| BARBITURIC ACID DERIVIATIVE OR SALT [PER 21C | 2100 | 4 | 45 | 11.25 |
| AMOBARBITAL (SCHEDULE 2) | 2125 | 4 | 11.39 | 2.85 |
| BUTALBITAL | 2165 | 5 | 90 | 18 |
| PENTOBARBITAL (SCHEDULE 2) | 2270 | 25 | 10,081 | 403.24 |
| DRONABINOL IN AN ORAL SOLUTION IN FDA APPR | 7365 | 1 | 0.15 | 0.15 |
| COCAINE | 9041L | 102 | 481 | 4.72 |
| CODEINE | 9050 | 131 | 7,329.26 | 55.95 |
| BUPRENORPHINE | 9064 | 103 | 1,653.81 | 16.06 |
| OXYCODONE | 9143 | 179 | 69,163.69 | 386.39 |
| HYDROMORPHONE | 9150 | 180 | 6,649.60 | 36.94 |
| HYDROCODONE | 9193 | 177 | 20,517.96 | 115.92 |
| MEPERIDINE (PETHIDINE) | 9230 | 138 | 3,752.22 | 27.19 |
| METHADONE | 9250B | 115 | 3,940.98 | 34.27 |
| MORPHINE | 9300 | 188 | 32,605.56 | 173.43 |
| OPIUM POWDERED | 9639 | 80 | 452.52 | 5.66 |
| OXYMORPHONE | 9652 | 18 | 64.32 | 3.57 |
| ALFENTANIL | 9737 | 16 | 3.56 | 0.22 |
| REMIFENTANIL | 9739 | 51 | 26.39 | 0.52 |
| SUFENTANIL BASE | 9740 | 41 | 1.54 | 0.04 |
| TAPENTADOL | 9780 | 35 | 2,133 | 60.94 |
| FENTANYL BASE | 9801 | 192 | 818.15 | 4.26 |
| OPIUM COMBINATION PRODUCT (C-III) | 9809 | 7 | 2.65 | 0.38 |

STATE:VIRGINIA BUSINESS ACTIVITY:C - PRACTITIONERS

| DRUG NAME | DRUG CODE | BUYERS | TOTAL GRAMS | AVG GRAMS |
|---|---|---|---|---|
| AMPHETAMINE | 1100 | 3 | 129.94 | 43.31 |
| LISDEXAMFETAMINE | 1205 | 1 | 6.13 | 6.13 |
| METHYLPHENIDATE (DL;D;L;ISOMERS) | 1724 | 2 | 59.71 | 29.85 |
| BARBITURIC ACID DERIVIATIVE OR SALT [PER 21C | 2100 | 1 | 5 | 5 |
| PENTOBARBITAL (SCHEDULE 2) | 2270 | 189 | 141,986.67 | 751.25 |
| COCAINE | 9041L | 10 | 84.14 | 8.41 |

| | | | | |
|---|---|---|---|---|
| CODEINE | 9050 | 281 | 6,068.71 | 21.6 |
| BUPRENORPHINE | 9064 | 803 | 56.76 | 0.07 |
| OXYCODONE | 9143 | 38 | 758.12 | 19.95 |
| HYDROMORPHONE | 9150 | 382 | 198.18 | 0.52 |
| HYDROCODONE | 9193 | 237 | 1,720.71 | 7.26 |
| MEPERIDINE (PETHIDINE) | 9230 | 127 | 720.29 | 5.67 |
| METHADONE | 9250B | 15 | 155.98 | 10.4 |
| MORPHINE | 9300 | 211 | 381.66 | 1.81 |
| OPIUM POWDERED | 9639 | 2 | 7.2 | 3.6 |
| OXYMORPHONE | 9652 | 15 | 37.85 | 2.52 |
| ALFENTANIL | 9737 | 1 | 0.06 | 0.06 |
| REMIFENTANIL | 9739 | 8 | 0.7 | 0.09 |
| TAPENTADOL | 9780 | 4 | 267 | 66.75 |
| FENTANYL BASE | 9801 | 473 | 47.52 | 0.1 |

STATE:VIRGINIA BUSINESS ACTIVITY:D - TEACHING INSTITUTIONS

| DRUG NAME | DRUG CODE | BUYERS | TOTAL GRAMS | AVG GRAMS |
|---|---|---|---|---|
| PENTOBARBITAL (SCHEDULE 2) | 2270 | 1 | 7,997.54 | 7,997.54 |
| COCAINE | 9041L | 2 | 2.21 | 1.11 |
| CODEINE | 9050 | 1 | 409.01 | 409.01 |
| BUPRENORPHINE | 9064 | 2 | 0.49 | 0.24 |
| HYDROMORPHONE | 9150 | 1 | 9.22 | 9.22 |
| HYDROCODONE | 9193 | 1 | 6.28 | 6.28 |
| MEPERIDINE (PETHIDINE) | 9230 | 1 | 0.54 | 0.54 |
| METHADONE | 9250B | 2 | 9.48 | 4.74 |
| MORPHINE | 9300 | 1 | 13.35 | 13.35 |
| OXYMORPHONE | 9652 | 1 | 0.18 | 0.18 |
| REMIFENTANIL | 9739 | 1 | 0.04 | 0.04 |
| FENTANYL BASE | 9801 | 1 | 0.79 | 0.79 |

STATE:VIRGINIA BUSINESS ACTIVITY:M - MID-LEVEL PRACTITIONERS

| DRUG NAME | DRUG CODE | BUYERS | TOTAL GRAMS | AVG GRAMS |
|---|---|---|---|---|
| PENTOBARBITAL (SCHEDULE 2) | 2270 | 40 | 30,923.80 | 773.1 |

| | | | | |
|---|---|---|---|---|
| CODEINE | 9050 | 66 | 134.23 | 2.03 |
| HYDROMORPHONE | 9150 | 1 | 0.09 | 0.09 |
| MEPERIDINE (PETHIDINE) | 9230 | 1 | 1.09 | 1.09 |
| MORPHINE | 9300 | 14 | 3.43 | 0.24 |
| FENTANYL BASE | 9801 | 14 | 0.2 | 0.01 |

STATE:VIRGINIA BUSINESS ACTIVITY:N-U NARCOTIC TREATMENT PROGRAMS

| DRUG NAME | DRUG CODE | BUYERS | TOTAL GRAMS | AVG GRAMS |
|---|---|---|---|---|
| BUPRENORPHINE | 9064 | 26 | 2,140.43 | 82.32 |
| METHADONE | 9250B | 39 | 250,009.84 | 6,410.51 |

STATE:WASHINGTON BUSINESS ACTIVITY:A - PHARMACIES

| DRUG NAME | DRUG CODE | BUYERS | TOTAL GRAMS | AVG GRAMS |
|---|---|---|---|---|
| AMPHETAMINE | 1100 | 1,203 | 420,314.25 | 349.39 |
| DL-METHAMPHETAMINE RACEMIC BASE | 1105B | 110 | 381.36 | 3.47 |
| D-METHAMPHETAMINE | 1105D | 2 | 2.41 | 1.21 |
| LISDEXAMFETAMINE | 1205 | 1,124 | 132,656.03 | 118.02 |
| METHYLPHENIDATE (DL;D;L;ISOMERS) | 1724 | 1,205 | 422,540.95 | 350.66 |
| BARBITURIC ACID DERIVIATIVE OR SALT [PER 21C | 2100 | 314 | 9,215 | 29.35 |
| BUTALBITAL | 2165 | 553 | 17,135 | 30.99 |
| SECOBARBITAL (SCHEDULE 2) | 2315 | 21 | 595.08 | 28.34 |
| NABILONE | 7379 | 1 | 0.05 | 0.05 |
| COCAINE | 9041L | 7 | 34.6 | 4.94 |
| CODEINE | 9050 | 1,219 | 229,118.50 | 187.96 |
| BUPRENORPHINE | 9064 | 1,183 | 72,336.58 | 61.15 |
| OXYCODONE | 9143 | 1,222 | 1,073,381.79 | 878.38 |
| HYDROMORPHONE | 9150 | 1,180 | 35,099.43 | 29.75 |
| HYDROCODONE | 9193 | 1,230 | 487,973.82 | 396.73 |
| LEVORPHANOL | 9220L | 104 | 108.53 | 1.04 |
| MEPERIDINE (PETHIDINE) | 9230 | 464 | 10,742.22 | 23.15 |
| METHADONE | 9250B | 1,151 | 102,582.58 | 89.12 |
| MORPHINE | 9300 | 1,212 | 386,631.86 | 319 |
| OPIUM TINCTURE (LAUDANUM) | 9630 | 2 | 3.54 | 1.77 |

| | | | | |
|---|---|---|---|---|
| OPIUM POWDERED | 9639 | 99 | 405 | 4.09 |
| OXYMORPHONE | 9652 | 673 | 11,285.99 | 16.77 |
| REMIFENTANIL | 9739 | 1 | 0.02 | 0.02 |
| TAPENTADOL | 9780 | 743 | 68,635.50 | 92.38 |
| FENTANYL BASE | 9801 | 1,144 | 5,195.40 | 4.54 |
| OPIUM COMBINATION PRODUCT (C-III) | 9809 | 19 | 9.65 | 0.51 |

STATE:WASHINGTON BUSINESS ACTIVITY:B - HOSPITALS

| DRUG NAME | DRUG CODE | BUYERS | TOTAL GRAMS | AVG GRAMS |
|---|---|---|---|---|
| AMPHETAMINE | 1100 | 94 | 15,085.89 | 160.49 |
| LISDEXAMFETAMINE | 1205 | 36 | 3,646.08 | 101.28 |
| METHYLPHENIDATE (DL;D;L;ISOMERS) | 1724 | 113 | 24,292.75 | 214.98 |
| GAMMA HYDROXYBUTYRIC ACID(FDA APPROVED | 2012 | 1 | 2,229.12 | 2,229.12 |
| BARBITURIC ACID DERIVIATIVE OR SALT [PER 21C | 2100 | 4 | 25 | 6.25 |
| AMOBARBITAL (SCHEDULE 2) | 2125 | 8 | 36.91 | 4.61 |
| BUTALBITAL | 2165 | 12 | 270 | 22.5 |
| PENTOBARBITAL (SCHEDULE 2) | 2270 | 12 | 1,935.81 | 161.32 |
| SECOBARBITAL (SCHEDULE 2) | 2315 | 3 | 128.17 | 42.72 |
| COCAINE | 9041L | 62 | 416.62 | 6.72 |
| CODEINE | 9050 | 155 | 9,671.09 | 62.39 |
| BUPRENORPHINE | 9064 | 105 | 2,873.24 | 27.36 |
| OXYCODONE | 9143 | 228 | 88,740.88 | 389.21 |
| HYDROMORPHONE | 9150 | 202 | 7,445.37 | 36.86 |
| HYDROCODONE | 9193 | 218 | 34,311.59 | 157.39 |
| LEVORPHANOL | 9220L | 3 | 0.7 | 0.23 |
| MEPERIDINE (PETHIDINE) | 9230 | 172 | 2,141.04 | 12.45 |
| METHADONE | 9250B | 131 | 10,148.13 | 77.47 |
| MORPHINE | 9300 | 224 | 46,306.44 | 206.73 |
| OPIUM POWDERED | 9639 | 83 | 851.04 | 10.25 |
| OXYMORPHONE | 9652 | 18 | 117.33 | 6.52 |
| ALFENTANIL | 9737 | 19 | 11.27 | 0.59 |
| REMIFENTANIL | 9739 | 60 | 48.39 | 0.81 |
| SUFENTANIL BASE | 9740 | 35 | 1.02 | 0.03 |
| TAPENTADOL | 9780 | 18 | 1,715 | 95.28 |
| FENTANYL BASE | 9801 | 235 | 785.7 | 3.34 |

| | | | | |
|---|---|---|---|---|
| OPIUM COMBINATION PRODUCT (C-III) | 9809 | 2 | 0.75 | 0.38 |

STATE:WASHINGTON BUSINESS ACTIVITY:C - PRACTITIONERS

| DRUG NAME | DRUG CODE | BUYERS | TOTAL GRAMS | AVG GRAMS |
|---|---|---|---|---|
| AMPHETAMINE | 1100 | 1 | 0.95 | 0.95 |
| LISDEXAMFETAMINE | 1205 | 1 | 10.2 | 10.2 |
| METHYLPHENIDATE (DL;D;L;ISOMERS) | 1724 | 1 | 22.83 | 22.83 |
| PENTOBARBITAL (SCHEDULE 2) | 2270 | 218 | 217,366.89 | 997.1 |
| COCAINE | 9041L | 2 | 2.86 | 1.43 |
| CODEINE | 9050 | 86 | 6,927.25 | 80.55 |
| BUPRENORPHINE | 9064 | 770 | 160.01 | 0.21 |
| OXYCODONE | 9143 | 38 | 85.85 | 2.26 |
| HYDROMORPHONE | 9150 | 536 | 280.42 | 0.52 |
| HYDROCODONE | 9193 | 390 | 1,182.41 | 3.03 |
| MEPERIDINE (PETHIDINE) | 9230 | 131 | 652.75 | 4.98 |
| METHADONE | 9250B | 40 | 175.12 | 4.38 |
| MORPHINE | 9300 | 312 | 735.61 | 2.36 |
| OPIUM POWDERED | 9639 | 2 | 1.44 | 0.72 |
| OXYMORPHONE | 9652 | 23 | 2.06 | 0.09 |
| NOROXYMORPHONE | 9668 | 2 | 8.47 | 4.24 |
| ALFENTANIL | 9737 | 8 | 1.38 | 0.17 |
| REMIFENTANIL | 9739 | 43 | 1.6 | 0.04 |
| SUFENTANIL BASE | 9740 | 6 | 0.02 | 0 |
| FENTANYL BASE | 9801 | 642 | 50.97 | 0.08 |

STATE:WASHINGTON BUSINESS ACTIVITY:D - TEACHING INSTITUTIONS

| DRUG NAME | DRUG CODE | BUYERS | TOTAL GRAMS | AVG GRAMS |
|---|---|---|---|---|
| BUPRENORPHINE | 9064 | 1 | 0.01 | 0.01 |

STATE:WASHINGTON BUSINESS ACTIVITY:M - MID-LEVEL PRACTITIONERS

| DRUG NAME | DRUG CODE | BUYERS | TOTAL GRAMS | AVG GRAMS |
|---|---|---|---|---|

| | | | | |
|---|---|---|---|---|
| PENTOBARBITAL (SCHEDULE 2) | 2270 | 22 | 21,514.93 | 977.95 |
| CODEINE | 9050 | 4 | 32.63 | 8.16 |
| BUPRENORPHINE | 9064 | 2 | 1.08 | 0.54 |
| OXYCODONE | 9143 | 2 | 6.28 | 3.14 |
| HYDROMORPHONE | 9150 | 5 | 0.34 | 0.07 |
| HYDROCODONE | 9193 | 1 | 0.61 | 0.61 |
| MEPERIDINE (PETHIDINE) | 9230 | 4 | 7.41 | 1.85 |
| METHADONE | 9250B | 2 | 3.58 | 1.79 |
| MORPHINE | 9300 | 4 | 4.75 | 1.19 |
| FENTANYL BASE | 9801 | 14 | 0.33 | 0.02 |

STATE:WASHINGTON BUSINESS ACTIVITY:N-U NARCOTIC TREATMENT PROGRAMS

| DRUG NAME | DRUG CODE | BUYERS | TOTAL GRAMS | AVG GRAMS |
|---|---|---|---|---|
| PENTOBARBITAL (SCHEDULE 2) | 2270 | 1 | 2.28 | 2.28 |
| BUPRENORPHINE | 9064 | 13 | 1,628.56 | 125.27 |
| HYDROMORPHONE | 9150 | 1 | 1.24 | 1.24 |
| METHADONE | 9250B | 24 | 277,931.22 | 11,580.47 |

STATE:WEST VIRGINIA BUSINESS ACTIVITY:A - PHARMACIES

| DRUG NAME | DRUG CODE | BUYERS | TOTAL GRAMS | AVG GRAMS |
|---|---|---|---|---|
| AMPHETAMINE | 1100 | 557 | 92,437.51 | 165.96 |
| DL-METHAMPHETAMINE RACEMIC BASE | 1105B | 5 | 16.48 | 3.3 |
| D-METHAMPHETAMINE | 1105D | 1 | 3.62 | 3.62 |
| LISDEXAMFETAMINE | 1205 | 547 | 77,857.93 | 142.34 |
| METHYLPHENIDATE (DL;D;L;ISOMERS) | 1724 | 562 | 123,648.70 | 220.02 |
| BARBITURIC ACID DERIVIATIVE OR SALT [PER 21C | 2100 | 82 | 2,105 | 25.67 |
| BUTALBITAL | 2165 | 166 | 5,055 | 30.45 |
| COCAINE | 9041L | 2 | 3.04 | 1.52 |
| CODEINE | 9050 | 558 | 121,528.28 | 217.79 |
| BUPRENORPHINE | 9064 | 492 | 58,295.89 | 118.49 |
| DIHYDROCODEINE | 9120 | 1 | 7.48 | 7.48 |
| OXYCODONE | 9143 | 567 | 314,687.41 | 555 |
| HYDROMORPHONE | 9150 | 403 | 4,667 | 11.58 |

| | | | | |
|---|---|---|---|---|
| HYDROCODONE | 9193 | 565 | 217,071.24 | 384.2 |
| MEPERIDINE (PETHIDINE) | 9230 | 72 | 1,399.96 | 19.44 |
| METHADONE | 9250B | 363 | 15,601.11 | 42.98 |
| MORPHINE | 9300 | 546 | 75,327.60 | 137.96 |
| OPIUM TINCTURE (LAUDANUM) | 9630 | 2 | 5.9 | 2.95 |
| OPIUM POWDERED | 9639 | 34 | 98.28 | 2.89 |
| OXYMORPHONE | 9652 | 281 | 9,891.02 | 35.2 |
| TAPENTADOL | 9780 | 260 | 24,252 | 93.28 |
| FENTANYL BASE | 9801 | 513 | 1,888.46 | 3.68 |
| OPIUM COMBINATION PRODUCT (C-III) | 9809 | 18 | 9.08 | 0.5 |

STATE:WEST VIRGINIA BUSINESS ACTIVITY:B - HOSPITALS

| DRUG NAME | DRUG CODE | BUYERS | TOTAL GRAMS | AVG GRAMS |
|---|---|---|---|---|
| AMPHETAMINE | 1100 | 36 | 1,535.24 | 42.65 |
| LISDEXAMFETAMINE | 1205 | 20 | 729.56 | 36.48 |
| METHYLPHENIDATE (DL;D;L;ISOMERS) | 1724 | 51 | 2,579.06 | 50.57 |
| BARBITURIC ACID DERIVIATIVE OR SALT [PER 21C | 2100 | 2 | 15 | 7.5 |
| AMOBARBITAL (SCHEDULE 2) | 2125 | 1 | 1.82 | 1.82 |
| BUTALBITAL | 2165 | 1 | 15 | 15 |
| PENTOBARBITAL (SCHEDULE 2) | 2270 | 5 | 166.33 | 33.27 |
| COCAINE | 9041L | 39 | 198.77 | 5.1 |
| CODEINE | 9050 | 68 | 2,738.68 | 40.27 |
| BUPRENORPHINE | 9064 | 51 | 2,642.30 | 51.81 |
| OXYCODONE | 9143 | 78 | 18,219.96 | 233.59 |
| HYDROMORPHONE | 9150 | 70 | 1,367.71 | 19.54 |
| HYDROCODONE | 9193 | 78 | 18,331.23 | 235.02 |
| MEPERIDINE (PETHIDINE) | 9230 | 56 | 841.87 | 15.03 |
| METHADONE | 9250B | 47 | 802.1 | 17.07 |
| MORPHINE | 9300 | 75 | 12,067.19 | 160.9 |
| OPIUM POWDERED | 9639 | 33 | 282.6 | 8.56 |
| OXYMORPHONE | 9652 | 10 | 32.61 | 3.26 |
| REMIFENTANIL | 9739 | 24 | 20.53 | 0.86 |
| SUFENTANIL BASE | 9740 | 10 | 0.26 | 0.03 |
| TAPENTADOL | 9780 | 11 | 292.5 | 26.59 |
| FENTANYL BASE | 9801 | 74 | 271.53 | 3.67 |

| | | | | |
|---|---|---|---|---|
| OPIUM COMBINATION PRODUCT (C-III) | 9809 | 2 | 0.57 | 0.28 |

STATE:WEST VIRGINIA BUSINESS ACTIVITY:C - PRACTITIONERS

| DRUG NAME | DRUG CODE | BUYERS | TOTAL GRAMS | AVG GRAMS |
|---|---|---|---|---|
| METHYLPHENIDATE (DL;D;L;ISOMERS) | 1724 | 1 | 149.03 | 149.03 |
| PENTOBARBITAL (SCHEDULE 2) | 2270 | 44 | 34,470.84 | 783.43 |
| CODEINE | 9050 | 46 | 175.04 | 3.81 |
| BUPRENORPHINE | 9064 | 107 | 7.74 | 0.07 |
| DIHYDROCODEINE | 9120 | 1 | 0.21 | 0.21 |
| OXYCODONE | 9143 | 1 | 0.9 | 0.9 |
| HYDROMORPHONE | 9150 | 32 | 19.46 | 0.61 |
| HYDROCODONE | 9193 | 50 | 199.14 | 3.98 |
| MEPERIDINE (PETHIDINE) | 9230 | 10 | 248.6 | 24.86 |
| MORPHINE | 9300 | 28 | 39.94 | 1.43 |
| REMIFENTANIL | 9739 | 1 | 0.22 | 0.22 |
| FENTANYL BASE | 9801 | 52 | 6.95 | 0.13 |

STATE:WEST VIRGINIA BUSINESS ACTIVITY:M - MID-LEVEL PRACTITIONERS

| DRUG NAME | DRUG CODE | BUYERS | TOTAL GRAMS | AVG GRAMS |
|---|---|---|---|---|
| PENTOBARBITAL (SCHEDULE 2) | 2270 | 12 | 7,464.37 | 622.03 |
| CODEINE | 9050 | 8 | 19.06 | 2.38 |
| MORPHINE | 9300 | 41 | 11.8 | 0.29 |
| FENTANYL BASE | 9801 | 44 | 0.55 | 0.01 |

STATE:WEST VIRGINIA BUSINESS ACTIVITY:N-U NARCOTIC TREATMENT PROGRAMS

| DRUG NAME | DRUG CODE | BUYERS | TOTAL GRAMS | AVG GRAMS |
|---|---|---|---|---|
| BUPRENORPHINE | 9064 | 7 | 1,244.64 | 177.81 |
| METHADONE | 9250B | 10 | 76,167.10 | 7,616.71 |

STATE:WISCONSIN BUSINESS ACTIVITY:A - PHARMACIES

STATISTICAL SUMMARY FOR RETAIL DRUG PURCHASES BY GRAMS WT

| DRUG NAME | DRUG CODE | BUYERS | TOTAL GRAMS | AVG GRAMS |
|---|---|---|---|---|
| AMPHETAMINE | 1100 | 1,114 | 499,223.92 | 448.14 |
| DL-METHAMPHETAMINE RACEMIC BASE | 1105B | 50 | 191.68 | 3.83 |
| D-METHAMPHETAMINE | 1105D | 6 | 24.1 | 4.02 |
| LISDEXAMFETAMINE | 1205 | 1,095 | 265,229.39 | 242.22 |
| METHYLPHENIDATE (DL;D;L;ISOMERS) | 1724 | 1,114 | 404,367.21 | 362.99 |
| BARBITURIC ACID DERIVIATIVE OR SALT [PER 21C | 2100 | 266 | 6,325 | 23.78 |
| BUTALBITAL | 2165 | 304 | 6,890 | 22.66 |
| SECOBARBITAL (SCHEDULE 2) | 2315 | 1 | 9.16 | 9.16 |
| DRONABINOL IN AN ORAL SOLUTION IN FDA APPR | 7365 | 1 | 0.15 | 0.15 |
| COCAINE | 9041L | 2 | 0.86 | 0.43 |
| CODEINE | 9050 | 1,101 | 184,148 | 167.26 |
| BUPRENORPHINE | 9064 | 1,047 | 36,539.03 | 34.9 |
| DIHYDROCODEINE | 9120 | 3 | 18.16 | 6.05 |
| OXYCODONE | 9143 | 1,123 | 787,727.19 | 701.45 |
| HYDROMORPHONE | 9150 | 940 | 10,604.79 | 11.28 |
| HYDROCODONE | 9193 | 1,130 | 294,374.34 | 260.51 |
| LEVORPHANOL | 9220L | 12 | 15.55 | 1.3 |
| MEPERIDINE (PETHIDINE) | 9230 | 122 | 2,146.07 | 17.59 |
| METHADONE | 9250B | 913 | 36,011.41 | 39.44 |
| MORPHINE | 9300 | 1,111 | 276,399.77 | 248.78 |
| OPIUM TINCTURE (LAUDANUM) | 9630 | 3 | 10.64 | 3.55 |
| OPIUM POWDERED | 9639 | 114 | 219.24 | 1.92 |
| OXYMORPHONE | 9652 | 552 | 8,341.81 | 15.11 |
| REMIFENTANIL | 9739 | 2 | 0.09 | 0.05 |
| SUFENTANIL BASE | 9740 | 1 | 2 | 2 |
| TAPENTADOL | 9780 | 668 | 57,648.50 | 86.3 |
| FENTANYL BASE | 9801 | 1,076 | 5,173.41 | 4.81 |
| OPIUM COMBINATION PRODUCT (C-III) | 9809 | 20 | 22.14 | 1.11 |

STATE:WISCONSIN BUSINESS ACTIVITY:B - HOSPITALS

| DRUG NAME | DRUG CODE | BUYERS | TOTAL GRAMS | AVG GRAMS |
|---|---|---|---|---|
| AMPHETAMINE | 1100 | 108 | 11,607.85 | 107.48 |
| DL-METHAMPHETAMINE RACEMIC BASE | 1105B | 1 | 5.63 | 5.63 |

| | | | | |
|---|---|---|---|---|
| D-METHAMPHETAMINE | 1105D | 1 | 4.02 | 4.02 |
| LISDEXAMFETAMINE | 1205 | 66 | 2,322.23 | 35.19 |
| METHYLPHENIDATE (DL;D;L;ISOMERS) | 1724 | 127 | 9,297.52 | 73.21 |
| BARBITURIC ACID DERIVIATIVE OR SALT [PER 21C | 2100 | 2 | 35 | 17.5 |
| AMOBARBITAL (SCHEDULE 2) | 2125 | 8 | 26.89 | 3.36 |
| BUTALBITAL | 2165 | 6 | 105 | 17.5 |
| PENTOBARBITAL (SCHEDULE 2) | 2270 | 13 | 11,369.26 | 874.56 |
| COCAINE | 9041L | 91 | 476.54 | 5.24 |
| CODEINE | 9050 | 172 | 5,839.55 | 33.95 |
| BUPRENORPHINE | 9064 | 103 | 2,598.74 | 25.23 |
| OXYCODONE | 9143 | 170 | 69,203.69 | 407.08 |
| HYDROMORPHONE | 9150 | 161 | 4,006.11 | 24.88 |
| HYDROCODONE | 9193 | 171 | 16,505.60 | 96.52 |
| MEPERIDINE (PETHIDINE) | 9230 | 119 | 3,223.24 | 27.09 |
| METHADONE | 9250B | 121 | 3,115.33 | 25.75 |
| MORPHINE | 9300 | 172 | 31,505.05 | 183.17 |
| OPIUM TINCTURE (LAUDANUM) | 9630 | 1 | 3.54 | 3.54 |
| OPIUM POWDERED | 9639 | 110 | 598.32 | 5.44 |
| OXYMORPHONE | 9652 | 24 | 159.53 | 6.65 |
| ALFENTANIL | 9737 | 34 | 15.21 | 0.45 |
| REMIFENTANIL | 9739 | 59 | 25.76 | 0.44 |
| SUFENTANIL BASE | 9740 | 41 | 0.51 | 0.01 |
| TAPENTADOL | 9780 | 31 | 1,826.50 | 58.92 |
| FENTANYL BASE | 9801 | 166 | 675.94 | 4.07 |
| OPIUM COMBINATION PRODUCT (C-III) | 9809 | 2 | 0.95 | 0.47 |

STATE:WISCONSIN BUSINESS ACTIVITY:C - PRACTITIONERS

| DRUG NAME | DRUG CODE | BUYERS | TOTAL GRAMS | AVG GRAMS |
|---|---|---|---|---|
| AMPHETAMINE | 1100 | 1 | 1.25 | 1.25 |
| PENTOBARBITAL (SCHEDULE 2) | 2270 | 245 | 185,498.63 | 757.14 |
| COCAINE | 9041L | 17 | 127.42 | 7.5 |
| CODEINE | 9050 | 56 | 564.39 | 10.08 |
| BUPRENORPHINE | 9064 | 497 | 83.05 | 0.17 |
| DIHYDROCODEINE | 9120 | 1 | 3.2 | 3.2 |
| OXYCODONE | 9143 | 61 | 789.05 | 12.94 |

| | | | | |
|---|---|---|---|---|
| HYDROMORPHONE | 9150 | 408 | 251.27 | 0.62 |
| HYDROCODONE | 9193 | 236 | 652.94 | 2.77 |
| MEPERIDINE (PETHIDINE) | 9230 | 81 | 483.68 | 5.97 |
| METHADONE | 9250B | 13 | 38.73 | 2.98 |
| MORPHINE | 9300 | 231 | 511.93 | 2.22 |
| OPIUM POWDERED | 9639 | 7 | 37.08 | 5.3 |
| OXYMORPHONE | 9652 | 10 | 10.03 | 1 |
| ALFENTANIL | 9737 | 14 | 4.06 | 0.29 |
| REMIFENTANIL | 9739 | 15 | 0.88 | 0.06 |
| SUFENTANIL BASE | 9740 | 3 | 0.01 | 0 |
| FENTANYL BASE | 9801 | 337 | 48.78 | 0.14 |

STATE:WISCONSIN BUSINESS ACTIVITY:M - MID-LEVEL PRACTITIONERS

| DRUG NAME | DRUG CODE | BUYERS | TOTAL GRAMS | AVG GRAMS |
|---|---|---|---|---|
| PENTOBARBITAL (SCHEDULE 2) | 2270 | 21 | 12,552.62 | 597.74 |
| CODEINE | 9050 | 1 | 2.23 | 2.23 |
| MORPHINE | 9300 | 2 | 5.83 | 2.91 |
| FENTANYL BASE | 9801 | 1 | 0.01 | 0.01 |

STATE:WISCONSIN BUSINESS ACTIVITY:N-U NARCOTIC TREATMENT PROGRAMS

| DRUG NAME | DRUG CODE | BUYERS | TOTAL GRAMS | AVG GRAMS |
|---|---|---|---|---|
| BUPRENORPHINE | 9064 | 8 | 1,883.52 | 235.44 |
| METHADONE | 9250B | 19 | 222,981.97 | 11,735.89 |

STATE:WYOMING BUSINESS ACTIVITY:A - PHARMACIES

| DRUG NAME | DRUG CODE | BUYERS | TOTAL GRAMS | AVG GRAMS |
|---|---|---|---|---|
| AMPHETAMINE | 1100 | 119 | 24,045.57 | 202.06 |
| DL-METHAMPHETAMINE RACEMIC BASE | 1105B | 5 | 21.3 | 4.26 |
| LISDEXAMFETAMINE | 1205 | 118 | 10,068.77 | 85.33 |
| METHYLPHENIDATE (DL;D;L;ISOMERS) | 1724 | 119 | 30,396.30 | 255.43 |
| BARBITURIC ACID DERIVIATIVE OR SALT [PER 21C | 2100 | 30 | 1,525 | 50.83 |

| | | | | |
|---|---|---|---|---|
| BUTALBITAL | 2165 | 46 | 1,605 | 34.89 |
| COCAINE | 9041L | 2 | 21.14 | 10.57 |
| CODEINE | 9050 | 119 | 14,898.72 | 125.2 |
| ETORPHINE | 9056 | 1 | 57 | 57 |
| BUPRENORPHINE | 9064 | 120 | 4,718.06 | 39.32 |
| OXYCODONE | 9143 | 121 | 95,190.94 | 786.7 |
| HYDROMORPHONE | 9150 | 120 | 2,194.78 | 18.29 |
| HYDROCODONE | 9193 | 121 | 40,822.05 | 337.37 |
| MEPERIDINE (PETHIDINE) | 9230 | 50 | 1,014.86 | 20.3 |
| METHADONE | 9250B | 103 | 4,216.38 | 40.94 |
| MORPHINE | 9300 | 121 | 28,432.25 | 234.98 |
| OPIUM POWDERED | 9639 | 12 | 9.72 | 0.81 |
| OXYMORPHONE | 9652 | 64 | 1,120.03 | 17.5 |
| REMIFENTANIL | 9739 | 1 | 0.11 | 0.11 |
| TAPENTADOL | 9780 | 63 | 7,696 | 122.16 |
| FENTANYL BASE | 9801 | 119 | 534.41 | 4.49 |
| OPIUM COMBINATION PRODUCT (C-III) | 9809 | 4 | 3.03 | 0.76 |

STATE:WYOMING BUSINESS ACTIVITY:B - HOSPITALS

| DRUG NAME | DRUG CODE | BUYERS | TOTAL GRAMS | AVG GRAMS |
|---|---|---|---|---|
| AMPHETAMINE | 1100 | 15 | 1,690.38 | 112.69 |
| LISDEXAMFETAMINE | 1205 | 10 | 381.55 | 38.15 |
| METHYLPHENIDATE (DL;D;L;ISOMERS) | 1724 | 24 | 2,179.68 | 90.82 |
| BUTALBITAL | 2165 | 2 | 25 | 12.5 |
| PENTOBARBITAL (SCHEDULE 2) | 2270 | 2 | 32.81 | 16.41 |
| COCAINE | 9041L | 22 | 48.14 | 2.19 |
| CODEINE | 9050 | 33 | 1,638.60 | 49.65 |
| BUPRENORPHINE | 9064 | 17 | 312.79 | 18.4 |
| OXYCODONE | 9143 | 43 | 6,967.60 | 162.04 |
| HYDROMORPHONE | 9150 | 42 | 226.07 | 5.38 |
| HYDROCODONE | 9193 | 43 | 5,000.69 | 116.3 |
| MEPERIDINE (PETHIDINE) | 9230 | 38 | 276.77 | 7.28 |
| METHADONE | 9250B | 18 | 540.13 | 30.01 |
| MORPHINE | 9300 | 43 | 7,388.49 | 171.83 |
| OPIUM POWDERED | 9639 | 13 | 88.56 | 6.81 |

| | | | | |
|---|---|---|---|---|
| ALFENTANIL | 9737 | 5 | 0.12 | 0.02 |
| REMIFENTANIL | 9739 | 15 | 1.67 | 0.11 |
| SUFENTANIL BASE | 9740 | 8 | 0.07 | 0.01 |
| TAPENTADOL | 9780 | 5 | 70 | 14 |
| FENTANYL BASE | 9801 | 45 | 84.7 | 1.88 |

STATE:WYOMING BUSINESS ACTIVITY:C - PRACTITIONERS

| DRUG NAME | DRUG CODE | BUYERS | TOTAL GRAMS | AVG GRAMS |
|---|---|---|---|---|
| PENTOBARBITAL (SCHEDULE 2) | 2270 | 38 | 40,259.73 | 1,059.47 |
| CODEINE | 9050 | 13 | 218.35 | 16.8 |
| BUPRENORPHINE | 9064 | 58 | 2.04 | 0.04 |
| OXYCODONE | 9143 | 1 | 9.75 | 9.75 |
| HYDROMORPHONE | 9150 | 40 | 18.51 | 0.46 |
| HYDROCODONE | 9193 | 26 | 186.32 | 7.17 |
| MEPERIDINE (PETHIDINE) | 9230 | 4 | 10.02 | 2.51 |
| MORPHINE | 9300 | 35 | 46.7 | 1.33 |
| FENTANYL BASE | 9801 | 39 | 1.87 | 0.05 |

STATE:WYOMING BUSINESS ACTIVITY:D - TEACHING INSTITUTIONS

| DRUG NAME | DRUG CODE | BUYERS | TOTAL GRAMS | AVG GRAMS |
|---|---|---|---|---|
| BUPRENORPHINE | 9064 | 1 | 0 | 0 |

STATE:WYOMING BUSINESS ACTIVITY:M - MID-LEVEL PRACTITIONERS

| DRUG NAME | DRUG CODE | BUYERS | TOTAL GRAMS | AVG GRAMS |
|---|---|---|---|---|
| PENTOBARBITAL (SCHEDULE 2) | 2270 | 2 | 177.72 | 88.86 |
| CODEINE | 9050 | 1 | 10.8 | 10.8 |
| OXYCODONE | 9143 | 1 | 0.54 | 0.54 |
| HYDROCODONE | 9193 | 1 | 0.36 | 0.36 |

DATE RANGE: 01/01/2017 TO 12/31/2017

ARCOS 3 - REPORT 7

Run Date: 07/05/2018

UNITED STATES SUMMARY FOR RETAIL DRUG PURCHASES BY GRAMS WT

ENTIRE UNITED STATES

BUSINESS ACTIVITY: A - PHARMACIES

| DRUG NAME | DRUG CODE | BUYERS | TOTAL GRAMS | AVG GRAMS |
|---|---|---|---|---|
| AMPHETAMINE | 1100 | 64,254 | 20,407,502.81 | 317.61 |
| DL-METHAMPHETAMINE RACEMIC BASE | 1105B | 1,944 | 6,893.33 | 3.55 |
| D-METHAMPHETAMINE | 1105D | 121 | 469.99 | 3.88 |
| LISDEXAMFETAMINE | 1205 | 59,765 | 9,592,434.05 | 160.5 |
| METHYLPHENIDATE (DL;D;L;ISOMERS) | 1724 | 63,671 | 17,701,935.44 | 278.02 |
| GAMMA HYDROXYBUTYRIC ACID(FDA APPROVED-CSA III) | 2012 | 1 | 30,411,958.46 | 30,411,958.46 |
| BARBITURIC ACID DERIVIATIVE OR SALT [PER 21CFR 1308.13(C)(3)] | 2100 | 18,623 | 596,769 | 32.04 |
| AMOBARBITAL (SCHEDULE 2) | 2125 | 2 | 0.91 | 0.46 |
| BUTALBITAL | 2165 | 24,330 | 847,360 | 34.83 |
| PENTOBARBITAL (SCHEDULE 2) | 2270 | 9 | 880.93 | 97.88 |
| SECOBARBITAL (SCHEDULE 2) | 2315 | 267 | 10,024.73 | 37.55 |
| DRONABINOL IN AN ORAL SOLUTION IN FDA APPROVED DRUG PRODU | 7365 | 191 | 341.25 | 1.79 |
| NABILONE | 7379 | 65 | 13.75 | 0.21 |
| COCAINE | 9041L | 206 | 1,408.73 | 6.84 |
| CODEINE | 9050 | 65,917 | 15,427,479.78 | 234.04 |
| ETORPHINE | 9056 | 1 | 57 | 57 |
| BUPRENORPHINE | 9064 | 59,274 | 3,179,903.93 | 53.65 |
| DIHYDROCODEINE | 9120 | 4,215 | 27,742.25 | 6.58 |
| OXYCODONE | 9143 | 65,710 | 46,519,135.37 | 707.95 |
| HYDROMORPHONE | 9150 | 57,230 | 1,170,251.35 | 20.45 |
| HYDROCODONE | 9193 | 66,146 | 25,666,888.29 | 388.03 |
| LEVORPHANOL | 9220L | 1,642 | 3,804.87 | 2.32 |
| MEPERIDINE (PETHIDINE) | 9230 | 19,735 | 451,854.54 | 22.9 |
| METHADONE | 9250B | 50,318 | 2,740,641.36 | 54.47 |
| MORPHINE | 9300 | 61,951 | 15,050,790.76 | 242.95 |
| OPIUM TINCTURE (LAUDANUM) | 9630 | 74 | 290.61 | 3.93 |
| OPIUM POWDERED | 9639 | 4,044 | 10,634.40 | 2.63 |
| OXYMORPHONE | 9652 | 35,318 | 1,099,737.69 | 31.14 |
| NOROXYMORPHONE | 9668 | 7 | 12.41 | 1.77 |
| ALFENTANIL | 9737 | 43 | 11.88 | 0.28 |
| REMIFENTANIL | 9739 | 79 | 8.98 | 0.11 |
| SUFENTANIL BASE | 9740 | 62 | 125.72 | 2.03 |
| TAPENTADOL | 9780 | 41,713 | 5,496,988 | 131.78 |

BUSINESS ACTIVITY:A - PHARMACIES

| DRUG NAME | DRUG CODE | BUYERS | TOTAL GRAMS | AVG GRAMS |
|---|---|---|---|---|
| FENTANYL BASE | 9801 | 60,607 | 285,984 | 4.72 |
| OPIUM COMBINATION PRODUCT (C-III) | 9809 | 1,639 | 1,136.71 | 0.69 |

ENTIRE UNITED STATES BUSINESS ACTIVITY: B - HOSPITALS

| DRUG NAME | DRUG CODE | BUYERS | TOTAL GRAMS | AVG GRAMS |
|---|---|---|---|---|
| AMPHETAMINE | 1100 | 3,923 | 528,728.62 | 134.78 |
| DL-METHAMPHETAMINE RACEMIC BASE | 1105B | 21 | 90.42 | 4.31 |
| D-METHAMPHETAMINE | 1105D | 12 | 83.55 | 6.96 |
| LISDEXAMFETAMINE | 1205 | 1,877 | 127,215.95 | 67.78 |
| METHYLPHENIDATE (DL;D;L;ISOMERS) | 1724 | 5,376 | 606,458.81 | 112.81 |
| GAMMA HYDROXYBUTYRIC ACID(FDA APPROVED-CSA III) | 2012 | 12 | 37,597.82 | 3133.15 |
| BARBITURIC ACID DERIVIATIVE OR SALT [PER 21CFR 1308.13(C)(3)] | 2100 | 168 | 2,009.90 | 11.96 |
| AMOBARBITAL (SCHEDULE 2) | 2125 | 202 | 952.41 | 4.71 |
| BUTALBITAL | 2165 | 375 | 6,085 | 16.23 |
| PENTOBARBITAL (SCHEDULE 2) | 2270 | 726 | 222,038.76 | 305.84 |
| SECOBARBITAL (SCHEDULE 2) | 2315 | 9 | 384.51 | 42.72 |
| DRONABINOL IN AN ORAL SOLUTION IN FDA APPROVED DRUG PRODUCT (SYNDROS - CII) | 7365 | 13 | 5.25 | 0.4 |
| NABILONE | 7379 | 3 | 0.7 | 0.23 |
| COCAINE | 9041L | 3,696 | 22,874.80 | 6.19 |
| CODEINE | 9050 | 7,298 | 572,528.25 | 78.45 |
| BUPRENORPHINE | 9064 | 4,596 | 97,615.88 | 21.24 |
| DIHYDROCODEINE | 9120 | 37 | 129.62 | 3.5 |
| OXYCODONE | 9143 | 8,760 | 2,304,923.66 | 263.12 |
| HYDROMORPHONE | 9150 | 8,669 | 225,982.09 | 26.07 |
| HYDROCODONE | 9193 | 9,293 | 1,404,591.66 | 151.15 |
| LEVORPHANOL | 9220L | 22 | 15.55 | 0.71 |
| MEPERIDINE (PETHIDINE) | 9230 | 7,155 | 206,659.13 | 28.88 |
| METHADONE | 9250B | 5,536 | 261,684.36 | 47.27 |
| MORPHINE | 9300 | 9,625 | 1,532,432.24 | 159.21 |
| OPIUM TINCTURE (LAUDANUM) | 9630 | 2 | 4.72 | 2.36 |
| OPIUM POWDERED | 9639 | 3,625 | 21,384 | 5.9 |
| OXYMORPHONE | 9652 | 937 | 9,957.38 | 10.63 |
| NOROXYMORPHONE | 9668 | 7 | 19.2 | 2.74 |
| ALFENTANIL | 9737 | 777 | 299.62 | 0.39 |

| | | | | |
|---|---|---|---|---|
| REMIFENTANIL | 9739 | 2,332 | 1,608.88 | 0.69 |
| SUFENTANIL BASE | 9740 | 1,771 | 49.96 | 0.03 |
| TAPENTADOL | 9780 | 1,466 | 92,628.35 | 63.18 |
| FENTANYL BASE | 9801 | 10,114 | 36,371.18 | 3.6 |
| OPIUM COMBINATION PRODUCT (C-III) | 9809 | 152 | 98.52 | 0.65 |

ENTIRE UNITED STATES BUSINESS ACTIVITY: C - PRACTITIONERS

| DRUG NAME | DRUG CODE | BUYERS | TOTAL GRAMS | AVG GRAMS |
|---|---|---|---|---|
| AMPHETAMINE | 1100 | 65 | 1,918.90 | 29.52 |
| D-METHAMPHETAMINE | 1105D | 6 | 16.39 | 2.73 |
| LISDEXAMFETAMINE | 1205 | 49 | 1,195.25 | 24.39 |
| METHYLPHENIDATE (DL;D;L;ISOMERS) | 1724 | 37 | 2,113.19 | 57.11 |
| GAMMA HYDROXYBUTYRIC ACID(FDA APPROVED-CSA III) | 2012 | 10 | 27,343.87 | 2,734.39 |
| BARBITURIC ACID DERIVIATIVE OR SALT [PER 21CFR 1308.13(C)(3)] | 2100 | 6 | 93.5 | 15.58 |
| BUTALBITAL | 2165 | 3 | 255 | 85 |
| PENTOBARBITAL (SCHEDULE 2) | 2270 | 7,610 | 6,005,685.79 | 789.18 |
| COCAINE | 9041L | 571 | 2,663.26 | 4.66 |
| CODEINE | 9050 | 6,512 | 147,102.14 | 22.59 |
| BUPRENORPHINE | 9064 | 24,463 | 7,584.69 | 0.31 |
| DIHYDROCODEINE | 9120 | 1,478 | 2,168.04 | 1.47 |
| OXYCODONE | 9143 | 1,985 | 30,172.04 | 15.2 |
| HYDROMORPHONE | 9150 | 12,946 | 7,754.93 | 0.6 |
| HYDROCODONE | 9193 | 11,928 | 57,192.35 | 4.79 |
| LEVORPHANOL | 9220L | 1 | 0.35 | 0.35 |
| MEPERIDINE (PETHIDINE) | 9230 | 4,768 | 39,600.32 | 8.31 |
| METHADONE | 9250B | 591 | 2,084 | 3.53 |
| MORPHINE | 9300 | 8,490 | 20,123.58 | 2.37 |
| THEBAINE | 9333 | 1 | 25 | 25 |
| OPIUM POWDERED | 9639 | 136 | 419.76 | 3.09 |
| OXYMORPHONE | 9652 | 266 | 559.89 | 2.1 |
| NOROXYMORPHONE | 9668 | 88 | 720.12 | 8.18 |
| ALFENTANIL | 9737 | 207 | 61.49 | 0.3 |
| REMIFENTANIL | 9739 | 481 | 41.06 | 0.09 |
| SUFENTANIL BASE | 9740 | 149 | 6.32 | 0.04 |
| TAPENTADOL | 9780 | 22 | 907.5 | 41.25 |
| FENTANYL BASE | 9801 | 18,026 | 2,144.37 | 0.12 |

ENTIRE UNITED STATES — BUSINESS ACTIVITY: D - TEACHING INSTITUTIONS

| DRUG NAME | DRUG CODE | BUYERS | TOTAL GRAMS | AVG GRAMS |
|---|---|---|---|---|
| AMPHETAMINE | 1100 | 2 | 2.2 | 1.1 |
| D-METHAMPHETAMINE | 1105D | 2 | 8.03 | 4.02 |
| METHYLPHENIDATE (DL;D;L;ISOMERS) | 1724 | 1 | 0.43 | 0.43 |
| PENTOBARBITAL (SCHEDULE 2) | 2270 | 20 | 48,702.68 | 2,435.13 |
| COCAINE | 9041L | 3 | 3.11 | 1.04 |
| CODEINE | 9050 | 5 | 2,220.08 | 444.02 |
| BUPRENORPHINE | 9064 | 64 | 8 | 0.12 |
| OXYCODONE | 9143 | 1 | 0.45 | 0.45 |
| HYDROMORPHONE | 9150 | 24 | 50.89 | 2.12 |
| HYDROCODONE | 9193 | 8 | 123.82 | 15.48 |
| MEPERIDINE (PETHIDINE) | 9230 | 3 | 3.81 | 1.27 |
| METHADONE | 9250B | 8 | 128.35 | 16.04 |
| MORPHINE | 9300 | 18 | 133.82 | 7.43 |
| OXYMORPHONE | 9652 | 3 | 0.54 | 0.18 |
| REMIFENTANIL | 9739 | 5 | 0.25 | 0.05 |
| SUFENTANIL BASE | 9740 | 1 | 0.02 | 0.02 |
| FENTANYL BASE | 9801 | 26 | 15.81 | 0.61 |

ENTIRE UNITED STATES — BUSINESS ACTIVITY: M - MID-LEVEL PRACTITIONERS

| DRUG NAME | DRUG CODE | BUYERS | TOTAL GRAMS | AVG GRAMS |
|---|---|---|---|---|
| METHYLPHENIDATE (DL;D;L;ISOMERS) | 1724 | 1 | 0.86 | 0.86 |
| BUTALBITAL | 2165 | 4 | 265 | 66.25 |
| PENTOBARBITAL (SCHEDULE 2) | 2270 | 664 | 827,317.76 | 1,245.96 |
| CODEINE | 9050 | 967 | 3,690.39 | 3.82 |
| BUPRENORPHINE | 9064 | 26 | 17.09 | 0.66 |
| DIHYDROCODEINE | 9120 | 429 | 299.66 | 0.7 |
| OXYCODONE | 9143 | 11 | 2,115.38 | 192.31 |
| HYDROMORPHONE | 9150 | 116 | 50.69 | 0.44 |
| HYDROCODONE | 9193 | 36 | 794.28 | 22.06 |
| MEPERIDINE (PETHIDINE) | 9230 | 30 | 87.69 | 2.92 |
| METHADONE | 9250B | 7 | 359.73 | 51.39 |
| MORPHINE | 9300 | 936 | 1,107.84 | 1.18 |

| | | | | |
|---|---|---|---|---|
| OXYMORPHONE | 9652 | 3 | 13.92 | 4.64 |
| NOROXYMORPHONE | 9668 | 1 | 2.12 | 2.12 |
| REMIFENTANIL | 9739 | 1 | 0.04 | 0.04 |
| TAPENTADOL | 9780 | 2 | 110 | 55 |
| FENTANYL BASE | 9801 | 1,393 | 69.43 | 0.05 |

ENTIRE UNITED STATES BUSINESS ACTIVITY: N-U NARCOTIC TREATMENT PROGRAMS

| DRUG NAME | DRUG CODE | BUYERS | TOTAL GRAMS | AVG GRAMS |
|---|---|---|---|---|
| PENTOBARBITAL (SCHEDULE 2) | 2270 | 1 | 2.28 | 2.28 |
| CODEINE | 9050 | 1 | 22.1 | 22.1 |
| BUPRENORPHINE | 9064 | 683 | 66,055.48 | 96.71 |
| HYDROMORPHONE | 9150 | 1 | 1.24 | 1.24 |
| MEPERIDINE (PETHIDINE) | 9230 | 1 | 435.75 | 435.75 |
| METHADONE | 9250B | 1,495 | 11,686,565 | 7,817.10 |
