## Supplementary material for "Methadone Distribution Trends from 2017-2019 in the United States": Raw data from 2018

### DEPARTMENT OF JUSTICE DRUG ENFORCEMENT ADMINISTRATION

### DEA DIVERSION CONTROL DIVISION

### ARCOS DRUG RETAIL SUMMARY REPORTS

REPORT 01 by GRAMS - RETAIL DRUG DISTRIBUTION BY ZIP CODE WITHIN STATE

REPORT 02 by GRAMS - RETAIL DRUG DISTRIBUTION BY STATE WITHIN DRUG CODE

REPORT 03 by GRAMS - QUARTERLY DRUG DISTRIBUTION BY STATE PER 100K POPULATION

REPORT 04 by GRAMS - CUMULATIVE DISTRIBUTION BY STATE PER 100K POPULATION

REPORT 05 by GRAMS - STATISTICAL SUMMARY FOR RETAIL DRUG PURCHASES

REPORT 07 by GRAMS - UNITED STATES SUMMARY FOR RETAIL DRUG PURCHASES

DATE RANGE: 01/01/2018 TO 12/31/2018

**DISCLAIMER**: Automated Reports and Consolidated Ordering System (ARCOS) is a data collection system in which manufacturers and distributors report their controlled substances transactions to the Drug Enforcement Administration (DEA). ARCOS provides an acquisition/distribution transactional records of applicable activities to the DEA involving certain controlled substances, in accordance with Title 21, United States Code, Section 827(d)(1), and Title 21, Code of Federal Regulations, Section 1304.33. This information is collected and compiled by DEA in accordance with law for determining quota, distribution trends, internal audits, and other analyses.

Recipients should be aware, prior to use, analysis, studies, or publication of ARCOS data, the information contained in ARCOS generated reports are complete and accurate as to what was provided by DEA registrants to ARCOS in accordance with reporting requirements. This information is susceptible to future updates and corrections, without notice, as new information is obtained. DEA assumes no liability for analysis, conclusions or policy decisions of third parties based on internal interpretation of provided data.

**DRUG: 1100 - AMPHETAMINE**

**STATE: AK - ALASKA**

| REGISTRANT ZIP CODE 3 | 1ST QUARTER | 2ND QUARTER | 3RD QUARTER | 4TH QUARTER | TOTAL GRAMS |
|---|---|---|---|---|---|
| 995 | 4,948.11 | 5,100.40 | 4,978.97 | 5,300.81 | **20,328.28** |
| 996 | 1,473.61 | 1,562.72 | 1,663.91 | 1,567.87 | **6,268.10** |
| 997 | 1,174.32 | 1,197.90 | 1,168.31 | 1,115.16 | **4,655.68** |
| 998 | 443.78 | 482.88 | 501.80 | 506.88 | **1,935.35** |
| 999 | 160.43 | 169.52 | 205.60 | 177.73 | **713.29** |
| **AK - ALASKA - Total** | **8,200.25** | **8,513.41** | **8,518.59** | **8,668.44** | **33,900.69** |

**STATE: AL - ALABAMA**

| REGISTRANT ZIP CODE 3 | 1ST QUARTER | 2ND QUARTER | 3RD QUARTER | 4TH QUARTER | TOTAL GRAMS |
|---|---|---|---|---|---|
| 350 | 18,225.66 | 17,964.26 | 18,406.76 | 18,514.40 | **73,111.08** |
| 351 | 9,178.60 | 9,208.29 | 9,456.49 | 9,401.98 | **37,245.35** |
| 352 | 19,546.01 | 19,641.83 | 20,262.04 | 20,435.80 | **79,885.68** |
| 354 | 8,135.87 | 8,026.41 | 8,170.46 | 8,315.88 | **32,648.62** |
| 355 | 3,508.42 | 3,559.74 | 3,373.07 | 3,586.58 | **14,027.81** |
| 356 | 10,340.95 | 10,035.29 | 10,590.13 | 10,324.00 | **41,290.38** |
| 357 | 4,628.00 | 4,649.74 | 4,687.52 | 4,888.04 | **18,853.30** |
| 358 | 5,860.07 | 5,891.52 | 6,276.07 | 5,952.49 | **23,980.15** |
| 359 | 5,345.57 | 5,194.70 | 5,353.61 | 5,431.57 | **21,325.45** |
| 360 | 6,843.05 | 6,624.82 | 6,757.18 | 6,788.32 | **27,013.36** |
| 361 | 5,668.28 | 5,425.51 | 5,462.06 | 5,358.58 | **21,914.44** |
| 362 | 2,352.69 | 2,435.97 | 2,449.60 | 2,444.72 | **9,682.98** |
| 363 | 5,606.26 | 5,727.53 | 5,754.71 | 5,623.69 | **22,712.19** |
| 364 | 1,281.84 | 1,307.61 | 1,281.14 | 1,260.40 | **5,130.99** |
| 365 | 12,402.62 | 12,724.13 | 12,659.77 | 12,480.66 | **50,267.18** |
| 366 | 8,164.74 | 7,971.68 | 8,133.59 | 8,136.99 | **32,406.99** |
| 367 | 1,636.41 | 1,461.64 | 1,521.36 | 1,504.65 | **6,124.06** |
| 368 | 6,619.67 | 6,545.43 | 6,596.24 | 6,645.50 | **26,406.84** |
| 369 | 131.74 | 143.59 | 142.93 | 157.90 | **576.16** |
| **AL - ALABAMA - Total** | **135,476.45** | **134,539.67** | **137,334.74** | **137,252.15** | **544,603.01** |

**STATE: AR - ARKANSAS**

| REGISTRANT ZIP CODE 3 | 1ST QUARTER | 2ND QUARTER | 3RD QUARTER | 4TH QUARTER | TOTAL GRAMS |
|---|---|---|---|---|---|
| 716 | 2,679.61 | 2,736.23 | 2,848.02 | 2,864.91 | **11,128.77** |
| 717 | 964.59 | 915.68 | 948.19 | 997.76 | **3,826.21** |
| 718 | 960.33 | 958.81 | 939.71 | 928.20 | **3,787.05** |
| 719 | 3,502.38 | 3,456.54 | 3,476.52 | 3,397.64 | **13,833.09** |
| 720 | 8,063.50 | 7,898.68 | 8,245.73 | 8,015.59 | **32,223.50** |
| 721 | 6,311.82 | 6,261.09 | 6,376.39 | 6,232.49 | **25,181.79** |
| 722 | 7,636.18 | 7,426.65 | 7,558.24 | 7,326.02 | **29,947.10** |
| 723 | 1,559.50 | 1,453.38 | 1,590.51 | 1,517.63 | **6,121.02** |
| 724 | 2,911.89 | 2,784.11 | 2,816.20 | 2,868.71 | **11,380.91** |
| 725 | 763.51 | 766.06 | 736.63 | 705.63 | **2,971.83** |

| | | | | | |
|---|---|---|---|---|---|
| 726 | 1,487.26 | 1,579.51 | 1,536.36 | 1,550.74 | **6,153.87** |
| 727 | 12,489.45 | 12,257.38 | 12,758.54 | 13,099.68 | **50,605.06** |
| 728 | 993.53 | 994.00 | 1,040.81 | 1,079.97 | **4,108.31** |
| 729 | 2,001.07 | 1,992.35 | 2,056.07 | 1,954.64 | **8,004.13** |
| **AR - ARKANSAS - Total** | **52,324.62** | **51,480.47** | **52,927.92** | **52,539.62** | **209,272.63** |

**STATE: AS - AMERICAN SAMOA**

| REGISTRANT ZIP CODE 3 | 1ST QUARTER | 2ND QUARTER | 3RD QUARTER | 4TH QUARTER | TOTAL GRAMS |
|---|---|---|---|---|---|
| 967 | 0 | 2.50 | 0 | 0 | **2.50** |
| **AS - AMERICAN SAMOA - Tota** | **0.00** | **2.50** | **0.00** | **0.00** | **2.50** |

**STATE: AZ - ARIZONA**

| REGISTRANT ZIP CODE 3 | 1ST QUARTER | 2ND QUARTER | 3RD QUARTER | 4TH QUARTER | TOTAL GRAMS |
|---|---|---|---|---|---|
| 850 | 17,901.11 | 18,481.09 | 18,786.04 | 19,387.57 | **74,555.80** |
| 851 | 3,573.59 | 3,428.61 | 3,525.47 | 3,774.98 | **14,302.64** |
| 852 | 37,099.25 | 44,040.68 | 51,473.94 | 39,506.43 | **172,120.30** |
| 853 | 11,201.70 | 11,065.18 | 11,055.05 | 11,666.31 | **44,988.24** |
| 855 | 480.74 | 451.64 | 471.75 | 476.05 | **1,880.19** |
| 856 | 1,676.74 | 1,625.59 | 1,714.33 | 1,690.44 | **6,707.11** |
| 857 | 11,159.91 | 10,823.87 | 10,763.84 | 11,136.85 | **43,884.47** |
| 859 | 452.41 | 443.52 | 495.77 | 555.18 | **1,946.87** |
| 860 | 1,277.32 | 1,375.92 | 1,317.83 | 1,410.87 | **5,381.95** |
| 863 | 1,628.61 | 1,637.41 | 1,541.25 | 1,830.06 | **6,637.32** |
| 864 | 983.70 | 940.72 | 854.31 | 977.50 | **3,756.24** |
| 865 | 53.40 | 51.53 | 45.24 | 64.39 | **214.56** |
| **AZ - ARIZONA - Total** | **87,488.48** | **94,365.76** | **102,044.82** | **92,476.65** | **376,375.70** |

**STATE: CA - CALIFORNIA**

| REGISTRANT ZIP CODE 3 | 1ST QUARTER | 2ND QUARTER | 3RD QUARTER | 4TH QUARTER | TOTAL GRAMS |
|---|---|---|---|---|---|
| 900 | 21,606.86 | 23,050.50 | 22,296.73 | 23,029.67 | **89,983.75** |
| 902 | 12,791.67 | 13,168.10 | 13,166.51 | 13,307.87 | **52,434.15** |
| 903 | 234.43 | 209.72 | 224.04 | 315.48 | **983.67** |
| 904 | 4,221.13 | 4,531.51 | 4,238.36 | 4,333.89 | **17,324.89** |
| 905 | 1,790.05 | 1,962.83 | 1,777.60 | 1,867.62 | **7,398.11** |
| 906 | 2,397.55 | 2,463.31 | 2,452.12 | 2,532.10 | **9,845.09** |
| 907 | 3,501.80 | 3,535.94 | 3,720.70 | 3,503.36 | **14,261.80** |
| 908 | 3,008.61 | 3,227.61 | 3,014.44 | 3,321.82 | **12,572.49** |
| 910 | 1,549.76 | 1,660.33 | 1,571.52 | 1,602.71 | **6,384.31** |
| 911 | 2,752.77 | 2,731.66 | 2,760.78 | 2,799.54 | **11,044.75** |
| 912 | 1,529.91 | 1,711.27 | 1,606.01 | 1,644.28 | **6,491.46** |
| 913 | 14,686.88 | 15,155.52 | 15,166.30 | 15,306.72 | **60,315.42** |
| 914 | 4,458.93 | 4,834.46 | 4,747.38 | 4,960.26 | **19,001.03** |
| 915 | 1,751.26 | 1,859.97 | 1,905.56 | 1,826.17 | **7,342.95** |

| | | | | | |
|---|---|---|---|---|---|
| 916 | 1,653.05 | 1,790.55 | 1,719.50 | 1,962.06 | **7,125.16** |
| 917 | 6,968.00 | 7,111.28 | 6,949.71 | 7,196.22 | **28,225.21** |
| 918 | 378.53 | 398.30 | 393.92 | 436.74 | **1,607.50** |
| 919 | 2,747.27 | 2,796.06 | 2,714.30 | 2,910.58 | **11,168.21** |
| 920 | 26,117.41 | 26,794.82 | 26,782.04 | 27,666.83 | **107,361.11** |
| 921 | 17,566.59 | 18,432.72 | 18,263.27 | 18,070.17 | **72,332.76** |
| 922 | 3,754.97 | 3,576.44 | 3,196.86 | 3,522.46 | **14,050.73** |
| 923 | 4,234.96 | 4,412.25 | 4,321.43 | 4,347.21 | **17,315.85** |
| 924 | 319.88 | 328.50 | 268.25 | 302.91 | **1,219.55** |
| 925 | 6,514.48 | 6,706.59 | 6,409.88 | 6,590.49 | **26,221.45** |
| 926 | 21,080.30 | 22,452.04 | 21,963.50 | 21,951.81 | **87,447.65** |
| 927 | 4,586.20 | 4,857.97 | 4,916.19 | 4,892.03 | **19,252.39** |
| 928 | 7,932.98 | 8,419.30 | 8,354.65 | 8,349.14 | **33,056.07** |
| 930 | 5,567.31 | 5,995.91 | 5,823.69 | 5,867.63 | **23,254.54** |
| 931 | 3,366.60 | 3,463.72 | 3,116.67 | 3,386.04 | **13,333.03** |
| 932 | 1,940.85 | 1,905.32 | 1,841.17 | 1,903.46 | **7,590.79** |
| 933 | 1,887.45 | 1,906.65 | 1,910.68 | 1,868.41 | **7,573.18** |
| 934 | 3,281.72 | 3,399.24 | 3,203.04 | 3,260.27 | **13,144.27** |
| 935 | 2,273.66 | 2,326.88 | 2,135.41 | 2,162.52 | **8,898.46** |
| 936 | 2,205.37 | 2,186.87 | 2,079.72 | 2,183.87 | **8,655.83** |
| 937 | 3,395.91 | 3,462.46 | 3,413.21 | 3,628.63 | **13,900.21** |
| 939 | 1,648.95 | 1,670.18 | 1,595.14 | 1,703.45 | **6,617.72** |
| 940 | 5,776.44 | 6,128.43 | 6,156.70 | 6,345.99 | **24,407.56** |
| 941 | 11,756.29 | 12,297.78 | 12,463.68 | 12,753.49 | **49,271.24** |
| 943 | 1,019.10 | 1,106.67 | 1,043.96 | 1,077.77 | **4,247.50** |
| 944 | 1,382.66 | 1,330.86 | 1,352.73 | 1,422.80 | **5,489.06** |
| 945 | 16,564.13 | 16,095.17 | 16,614.63 | 17,331.54 | **66,605.47** |
| 946 | 3,669.62 | 3,817.58 | 3,659.24 | 3,923.86 | **15,070.30** |
| 947 | 1,478.11 | 1,427.02 | 1,445.43 | 1,540.93 | **5,891.49** |
| 948 | 687.50 | 691.27 | 671.73 | 801.53 | **2,852.03** |
| 949 | 5,076.76 | 4,888.19 | 4,840.39 | 4,986.73 | **19,792.07** |
| 950 | 7,164.37 | 7,194.46 | 6,999.46 | 7,500.44 | **28,858.73** |
| 951 | 3,897.99 | 3,951.50 | 4,078.19 | 4,125.51 | **16,053.19** |
| 952 | 2,003.17 | 2,078.25 | 1,915.61 | 2,003.30 | **8,000.34** |
| 953 | 4,988.07 | 4,941.06 | 4,664.05 | 5,085.80 | **19,678.97** |
| 954 | 3,872.37 | 3,914.46 | 3,866.08 | 3,889.18 | **15,542.09** |
| 955 | 907.83 | 872.51 | 961.20 | 933.26 | **3,674.80** |
| 956 | 10,232.86 | 9,825.64 | 10,366.30 | 10,673.11 | **41,097.92** |
| 957 | 2,380.76 | 2,474.96 | 2,453.58 | 2,579.16 | **9,888.47** |
| 958 | 4,974.11 | 5,139.90 | 5,028.77 | 5,238.00 | **20,380.78** |

| | | | | | |
|---|---|---|---|---|---|
| 959 | 4,266.29 | 4,222.45 | 4,093.79 | 4,143.15 | **16,725.67** |
| 960 | 1,796.54 | 1,799.05 | 1,906.94 | 1,884.75 | **7,387.29** |
| 961 | 609.71 | 633.49 | 611.41 | 697.59 | **2,552.20** |
| **CA - CALIFORNIA - Total** | **300,208.74** | **309,327.49** | **305,214.17** | **313,452.33** | **1,228,202.73** |

**STATE: CO - COLORADO**

| REGISTRANT ZIP CODE 3 | 1ST QUARTER | 2ND QUARTER | 3RD QUARTER | 4TH QUARTER | TOTAL GRAMS |
|---|---|---|---|---|---|
| 800 | 12,709.09 | 12,840.49 | 14,243.04 | 14,630.75 | **54,423.38** |
| 801 | 15,227.03 | 15,507.35 | 15,764.69 | 16,219.30 | **62,718.37** |
| 802 | 20,774.59 | 21,767.46 | 21,746.11 | 21,961.45 | **86,249.61** |
| 803 | 3,548.00 | 3,538.72 | 3,358.82 | 3,813.66 | **14,259.19** |
| 804 | 3,233.84 | 3,096.78 | 3,352.55 | 3,371.69 | **13,054.87** |
| 805 | 8,132.50 | 8,254.13 | 8,430.92 | 8,818.43 | **33,635.98** |
| 806 | 2,395.77 | 2,551.37 | 2,564.30 | 2,585.37 | **10,096.82** |
| 807 | 352.68 | 390.70 | 409.20 | 383.47 | **1,536.04** |
| 808 | 1,198.40 | 1,175.95 | 1,280.28 | 1,180.68 | **4,835.31** |
| 809 | 7,784.19 | 8,061.64 | 8,078.07 | 8,093.68 | **32,017.58** |
| 810 | 1,412.43 | 1,504.61 | 1,503.34 | 1,457.77 | **5,878.14** |
| 811 | 269.79 | 247.58 | 324.83 | 311.78 | **1,153.99** |
| 812 | 598.23 | 558.90 | 613.47 | 624.58 | **2,395.19** |
| 813 | 698.89 | 681.89 | 726.91 | 709.17 | **2,816.87** |
| 814 | 573.27 | 593.72 | 667.06 | 677.76 | **2,511.81** |
| 815 | 1,764.52 | 1,753.62 | 1,827.96 | 1,833.23 | **7,179.33** |
| 816 | 2,575.27 | 2,329.40 | 2,617.60 | 2,666.66 | **10,188.93** |
| **CO - COLORADO - Total** | **83,248.50** | **84,854.33** | **87,509.13** | **89,339.44** | **344,951.40** |

**STATE: CT - CONNECTICUT**

| REGISTRANT ZIP CODE 3 | 1ST QUARTER | 2ND QUARTER | 3RD QUARTER | 4TH QUARTER | TOTAL GRAMS |
|---|---|---|---|---|---|
| 060 | 9,499.66 | 9,379.45 | 9,655.00 | 9,581.20 | **38,115.31** |
| 061 | 3,415.48 | 3,431.57 | 3,478.57 | 3,529.40 | **13,855.01** |
| 062 | 2,453.03 | 2,401.36 | 2,455.85 | 2,697.51 | **10,007.76** |
| 063 | 5,603.92 | 5,872.58 | 5,969.45 | 5,962.98 | **23,408.94** |
| 064 | 13,093.00 | 13,150.21 | 13,339.72 | 13,582.32 | **53,165.25** |
| 065 | 4,379.28 | 4,403.76 | 4,389.37 | 4,447.33 | **17,619.74** |
| 066 | 1,543.73 | 1,616.71 | 1,785.29 | 1,852.06 | **6,797.79** |
| 067 | 3,790.94 | 3,740.03 | 3,765.23 | 3,892.46 | **15,188.65** |
| 068 | 9,872.64 | 9,976.99 | 10,586.54 | 10,591.59 | **41,027.76** |
| 069 | 1,977.52 | 1,942.94 | 1,995.81 | 2,114.87 | **8,031.14** |
| **CT - CONNECTICUT - Total** | **55,629.20** | **55,915.61** | **57,420.84** | **58,251.72** | **227,217.37** |

**STATE: DC - DISTRICT OF COLUMBIA**

| REGISTRANT ZIP CODE 3 | 1ST QUARTER | 2ND QUARTER | 3RD QUARTER | 4TH QUARTER | TOTAL GRAMS |
|---|---|---|---|---|---|
| 200 | 11,519.24 | 11,921.60 | 11,849.74 | 11,930.65 | **47,221.23** |

| | | | | | |
|---|---|---|---|---|---|
| 203 | 100.26 | 92.01 | 148.27 | 86.65 | **427.20** |
| 204 | 471.35 | 427.40 | 465.56 | 513.42 | **1,877.72** |
| 205 | 0 | 0.63 | 1.88 | 0 | **2.50** |
| **DC - DISTRICT OF COLUMBIA -** | **12,090.86** | **12,441.63** | **12,465.45** | **12,530.71** | **49,528.65** |

**STATE: DE - DELAWARE**

| REGISTRANT ZIP CODE 3 | 1ST QUARTER | 2ND QUARTER | 3RD QUARTER | 4TH QUARTER | TOTAL GRAMS |
|---|---|---|---|---|---|
| 197 | 8,861.19 | 8,955.25 | 8,727.19 | 9,194.61 | **35,738.23** |
| 198 | 6,545.24 | 6,721.87 | 6,636.39 | 6,755.11 | **26,658.61** |
| 199 | 5,678.63 | 5,584.34 | 5,815.38 | 5,592.05 | **22,670.41** |
| **DE - DELAWARE - Total** | **21,085.06** | **21,261.45** | **21,178.96** | **21,541.78** | **85,067.25** |

**STATE: FL - FLORIDA**

| REGISTRANT ZIP CODE 3 | 1ST QUARTER | 2ND QUARTER | 3RD QUARTER | 4TH QUARTER | TOTAL GRAMS |
|---|---|---|---|---|---|
| 320 | 9,966.58 | 10,078.31 | 10,173.49 | 10,245.91 | **40,464.29** |
| 321 | 7,889.16 | 8,292.18 | 7,928.44 | 8,354.63 | **32,464.42** |
| 322 | 23,160.02 | 23,847.99 | 23,514.09 | 23,368.58 | **93,890.68** |
| 323 | 9,900.64 | 9,888.69 | 9,541.59 | 9,650.44 | **38,981.36** |
| 324 | 8,767.12 | 8,766.69 | 8,490.48 | 7,239.32 | **33,263.61** |
| 325 | 19,391.26 | 19,454.99 | 20,172.83 | 18,745.07 | **77,764.15** |
| 326 | 6,175.93 | 6,034.24 | 6,034.83 | 6,292.93 | **24,537.92** |
| 327 | 18,709.57 | 18,862.59 | 18,996.06 | 19,391.44 | **75,959.66** |
| 328 | 14,083.89 | 13,892.67 | 13,883.33 | 14,147.02 | **56,006.91** |
| 329 | 13,562.89 | 13,876.50 | 14,071.82 | 13,915.00 | **55,426.20** |
| 330 | 15,150.83 | 15,526.42 | 15,264.96 | 15,344.23 | **61,286.44** |
| 331 | 18,145.19 | 18,636.84 | 18,192.93 | 18,842.05 | **73,817.01** |
| 333 | 13,139.48 | 13,459.76 | 12,928.76 | 13,359.77 | **52,887.78** |
| 334 | 25,471.27 | 26,139.65 | 26,066.66 | 26,164.46 | **103,842.04** |
| 335 | 11,691.68 | 11,855.17 | 11,618.42 | 12,242.41 | **47,407.67** |
| 336 | 15,680.06 | 15,951.86 | 15,849.78 | 16,358.52 | **63,840.22** |
| 337 | 14,764.30 | 15,140.32 | 14,929.15 | 15,497.24 | **60,331.00** |
| 338 | 7,589.46 | 7,356.55 | 7,444.58 | 7,652.60 | **30,043.20** |
| 339 | 13,510.54 | 14,025.38 | 13,666.81 | 14,192.08 | **55,394.80** |
| 341 | 7,781.39 | 7,582.74 | 7,574.12 | 7,741.98 | **30,680.23** |
| 342 | 14,556.83 | 14,586.66 | 14,246.42 | 14,484.27 | **57,874.17** |
| 344 | 4,310.23 | 4,236.26 | 4,118.70 | 4,231.06 | **16,896.26** |
| 346 | 9,891.93 | 9,679.42 | 9,865.72 | 10,188.22 | **39,625.29** |
| 347 | 8,642.98 | 8,705.04 | 8,561.01 | 8,950.60 | **34,859.63** |
| 349 | 7,085.93 | 7,441.65 | 7,495.28 | 7,334.40 | **29,357.26** |
| **FL - FLORIDA - Total** | **319,019.15** | **323,318.57** | **320,630.26** | **323,934.22** | **1,286,902.21** |

**STATE: GA - GEORGIA**

| REGISTRANT ZIP CODE 3 | 1ST QUARTER | 2ND QUARTER | 3RD QUARTER | 4TH QUARTER | TOTAL GRAMS |
|---|---|---|---|---|---|

| | | | | | |
|---|---|---|---|---|---|
| 300 | 39,818.27 | 39,372.77 | 40,611.48 | 40,811.60 | **160,614.11** |
| 301 | 23,315.81 | 22,741.69 | 23,298.99 | 23,712.57 | **93,069.07** |
| 302 | 14,299.33 | 14,193.66 | 14,875.82 | 15,188.87 | **58,557.68** |
| 303 | 23,666.66 | 23,798.44 | 24,935.66 | 24,996.88 | **97,397.63** |
| 304 | 7,088.84 | 6,689.99 | 6,879.12 | 6,884.58 | **27,542.52** |
| 305 | 12,492.87 | 12,565.25 | 12,728.91 | 12,972.55 | **50,759.59** |
| 306 | 8,753.25 | 8,188.48 | 8,545.18 | 8,661.07 | **34,147.98** |
| 307 | 6,602.97 | 6,296.43 | 6,434.65 | 6,573.18 | **25,907.22** |
| 308 | 3,704.89 | 3,531.61 | 3,656.05 | 3,765.02 | **14,657.56** |
| 309 | 5,666.63 | 5,529.78 | 5,907.64 | 5,514.14 | **22,618.20** |
| 310 | 7,845.45 | 7,433.82 | 7,468.79 | 7,684.39 | **30,432.45** |
| 312 | 2,825.50 | 2,784.48 | 2,831.34 | 2,736.65 | **11,177.97** |
| 313 | 6,037.64 | 5,785.25 | 6,163.96 | 5,986.89 | **23,973.74** |
| 314 | 6,359.72 | 6,378.09 | 6,505.78 | 6,336.57 | **25,580.17** |
| 315 | 8,606.90 | 8,169.92 | 8,469.97 | 8,284.55 | **33,531.33** |
| 316 | 3,554.58 | 3,369.48 | 3,258.89 | 3,462.39 | **13,645.33** |
| 317 | 6,864.69 | 6,691.24 | 6,866.13 | 6,922.36 | **27,344.41** |
| 318 | 480.50 | 503.03 | 538.39 | 527.17 | **2,049.08** |
| 319 | 6,845.49 | 6,704.97 | 6,849.47 | 6,568.03 | **26,967.95** |
| 398 | 1,579.31 | 1,515.37 | 1,477.65 | 1,534.20 | **6,106.53** |
| **GA - GEORGIA - Total** | **196,409.28** | **192,243.75** | **198,303.85** | **199,123.65** | **786,080.52** |

**STATE: GU - GUAM**

| REGISTRANT ZIP CODE 3 | 1ST QUARTER | 2ND QUARTER | 3RD QUARTER | 4TH QUARTER | TOTAL GRAMS |
|---|---|---|---|---|---|
| 969 | 371.55 | 474.05 | 488.00 | 425.77 | **1,759.37** |
| **GU - GUAM - Total** | **371.55** | **474.05** | **488.00** | **425.77** | **1,759.37** |

**STATE: HI - HAWAII**

| REGISTRANT ZIP CODE 3 | 1ST QUARTER | 2ND QUARTER | 3RD QUARTER | 4TH QUARTER | TOTAL GRAMS |
|---|---|---|---|---|---|
| 967 | 5,112.50 | 5,132.88 | 4,977.85 | 5,215.14 | **20,438.36** |
| 968 | 5,006.63 | 5,185.66 | 5,698.82 | 4,903.88 | **20,795.00** |
| **HI - HAWAII - Total** | **10,119.13** | **10,318.54** | **10,676.67** | **10,119.02** | **41,233.36** |

**STATE: IA - IOWA**

| REGISTRANT ZIP CODE 3 | 1ST QUARTER | 2ND QUARTER | 3RD QUARTER | 4TH QUARTER | TOTAL GRAMS |
|---|---|---|---|---|---|
| 500 | 4,221.10 | 4,176.96 | 4,139.85 | 4,431.86 | **16,969.77** |
| 501 | 1,872.03 | 1,889.57 | 1,969.87 | 1,841.59 | **7,573.06** |
| 502 | 2,992.76 | 3,114.00 | 3,253.32 | 3,274.28 | **12,634.36** |
| 503 | 5,760.47 | 6,008.97 | 5,983.94 | 5,992.14 | **23,745.52** |
| 504 | 839.44 | 847.35 | 901.39 | 902.45 | **3,490.62** |
| 505 | 1,247.74 | 1,288.80 | 1,242.81 | 1,347.18 | **5,126.53** |
| 506 | 1,619.99 | 1,652.26 | 1,604.52 | 1,704.21 | **6,580.99** |
| 507 | 1,201.82 | 1,216.44 | 1,314.10 | 1,302.69 | **5,035.05** |

| | | | | | |
|---|---|---|---|---|---|
| 508 | 162.66 | 150.89 | 152.47 | 148.79 | **614.81** |
| 510 | 1,039.88 | 1,018.41 | 1,042.99 | 1,085.00 | **4,186.28** |
| 511 | 2,792.63 | 2,841.03 | 2,985.74 | 3,053.97 | **11,673.37** |
| 512 | 463.83 | 457.87 | 503.99 | 504.02 | **1,929.70** |
| 513 | 745.69 | 731.67 | 776.94 | 756.06 | **3,010.35** |
| 514 | 371.71 | 388.85 | 401.05 | 386.35 | **1,547.96** |
| 515 | 2,149.65 | 2,185.40 | 2,324.16 | 2,246.71 | **8,905.91** |
| 516 | 211.49 | 207.47 | 224.92 | 197.69 | **841.57** |
| 520 | 3,274.70 | 3,284.59 | 3,400.74 | 3,450.80 | **13,410.82** |
| 521 | 510.07 | 482.27 | 466.16 | 553.58 | **2,012.08** |
| 522 | 4,992.06 | 5,036.52 | 5,122.24 | 5,575.27 | **20,726.09** |
| 523 | 2,546.88 | 2,521.30 | 2,583.62 | 2,580.22 | **10,232.02** |
| 524 | 4,042.53 | 4,125.05 | 4,144.54 | 4,241.81 | **16,553.93** |
| 525 | 1,029.73 | 989.89 | 1,013.85 | 967.93 | **4,001.41** |
| 526 | 1,658.30 | 1,537.64 | 1,657.78 | 1,662.64 | **6,516.36** |
| 527 | 3,206.12 | 3,158.29 | 3,207.79 | 3,274.01 | **12,846.20** |
| 528 | 2,610.66 | 2,630.05 | 2,741.55 | 2,656.29 | **10,638.55** |
| **IA - IOWA - Total** | **51,563.94** | **51,941.54** | **53,160.32** | **54,137.53** | **210,803.33** |

**STATE: ID - IDAHO**

| REGISTRANT ZIP CODE 3 | 1ST QUARTER | 2ND QUARTER | 3RD QUARTER | 4TH QUARTER | **TOTAL GRAMS** |
|---|---|---|---|---|---|
| 832 | 2,254.15 | 2,269.05 | 2,332.15 | 2,544.42 | **9,399.76** |
| 833 | 2,028.26 | 2,056.59 | 2,050.62 | 2,146.31 | **8,281.78** |
| 834 | 5,018.41 | 4,972.78 | 5,044.12 | 5,357.06 | **20,392.37** |
| 835 | 388.05 | 499.68 | 453.65 | 449.47 | **1,790.84** |
| 836 | 7,856.21 | 8,159.17 | 8,151.40 | 8,303.29 | **32,470.07** |
| 837 | 7,477.87 | 7,769.64 | 7,832.30 | 8,187.13 | **31,266.94** |
| 838 | 2,228.28 | 2,232.23 | 2,343.41 | 2,414.83 | **9,218.74** |
| **ID - IDAHO - Total** | **27,251.22** | **27,959.13** | **28,207.65** | **29,402.52** | **112,820.52** |

**STATE: IL - ILLINOIS**

| REGISTRANT ZIP CODE 3 | 1ST QUARTER | 2ND QUARTER | 3RD QUARTER | 4TH QUARTER | **TOTAL GRAMS** |
|---|---|---|---|---|---|
| 600 | 33,517.21 | 34,061.71 | 34,670.26 | 34,889.72 | **137,138.90** |
| 601 | 23,443.97 | 23,673.22 | 24,311.20 | 24,814.18 | **96,242.56** |
| 602 | 1,506.53 | 1,536.39 | 1,499.78 | 1,517.74 | **6,060.44** |
| 603 | 1,356.45 | 1,336.32 | 1,381.23 | 1,448.29 | **5,522.29** |
| 604 | 19,033.52 | 19,164.88 | 19,699.77 | 20,121.25 | **78,019.42** |
| 605 | 18,055.95 | 18,148.10 | 18,612.88 | 19,025.45 | **73,842.38** |
| 606 | 38,567.22 | 39,771.95 | 39,828.97 | 40,752.48 | **158,920.62** |
| 607 | 2,218.83 | 2,306.38 | 2,447.41 | 2,440.93 | **9,413.55** |
| 608 | 710.83 | 692.71 | 767.15 | 741.39 | **2,912.08** |
| 609 | 2,161.30 | 2,029.48 | 2,030.84 | 2,183.87 | **8,405.49** |

| | | | | | |
|---|---|---|---|---|---|
| 610 | 3,426.46 | 3,410.79 | 3,593.16 | 3,714.08 | **14,144.50** |
| 611 | 4,009.81 | 4,049.59 | 4,268.07 | 4,086.44 | **16,413.91** |
| 612 | 3,136.17 | 3,140.87 | 3,090.59 | 3,188.31 | **12,555.94** |
| 613 | 2,493.91 | 2,504.78 | 2,535.28 | 2,782.84 | **10,316.81** |
| 614 | 1,986.08 | 1,904.98 | 1,938.96 | 1,961.26 | **7,791.27** |
| 615 | 3,334.28 | 3,190.59 | 3,278.61 | 3,438.64 | **13,242.11** |
| 616 | 4,253.61 | 4,106.54 | 4,301.04 | 4,382.28 | **17,043.46** |
| 617 | 3,847.50 | 3,532.65 | 3,799.60 | 3,869.53 | **15,049.28** |
| 618 | 3,980.04 | 3,722.83 | 3,720.25 | 4,081.25 | **15,504.37** |
| 619 | 1,262.55 | 1,125.82 | 1,080.57 | 1,164.98 | **4,633.92** |
| 620 | 3,795.51 | 3,769.17 | 3,744.47 | 3,750.20 | **15,059.35** |
| 622 | 5,499.31 | 5,725.97 | 5,695.78 | 5,660.98 | **22,582.04** |
| 623 | 1,923.10 | 1,962.63 | 1,886.10 | 1,968.51 | **7,740.33** |
| 624 | 1,214.04 | 1,074.36 | 1,182.30 | 1,154.79 | **4,625.49** |
| 625 | 1,642.82 | 1,581.14 | 1,558.72 | 1,628.32 | **6,411.00** |
| 626 | 1,617.17 | 1,529.99 | 1,537.90 | 1,617.20 | **6,302.27** |
| 627 | 2,408.10 | 2,363.80 | 2,376.89 | 2,450.59 | **9,599.38** |
| 628 | 2,464.19 | 2,410.05 | 2,482.35 | 2,428.57 | **9,785.15** |
| 629 | 3,945.88 | 3,515.70 | 3,722.01 | 3,664.51 | **14,848.09** |
| **IL - ILLINOIS - Total** | **196,812.32** | **197,343.37** | **201,042.12** | **204,928.58** | **800,126.39** |

**STATE: IN - INDIANA**

| REGISTRANT ZIP CODE 3 | 1ST QUARTER | 2ND QUARTER | 3RD QUARTER | 4TH QUARTER | TOTAL GRAMS |
|---|---|---|---|---|---|
| 460 | 15,666.20 | 16,047.57 | 16,167.36 | 16,207.30 | **64,088.43** |
| 461 | 9,179.26 | 9,206.05 | 9,772.16 | 10,152.71 | **38,310.18** |
| 462 | 16,506.54 | 17,080.95 | 16,880.59 | 16,998.54 | **67,466.62** |
| 463 | 12,478.58 | 12,448.37 | 12,859.72 | 13,042.82 | **50,829.49** |
| 464 | 1,217.05 | 1,203.03 | 1,279.76 | 1,296.58 | **4,996.41** |
| 465 | 17,814.32 | 17,558.38 | 17,938.10 | 17,806.52 | **71,117.32** |
| 466 | 4,936.11 | 4,747.49 | 4,688.38 | 4,788.30 | **19,160.27** |
| 467 | 3,592.13 | 3,412.48 | 3,699.08 | 3,804.17 | **14,507.86** |
| 468 | 6,015.18 | 6,128.25 | 6,241.09 | 6,438.23 | **24,822.76** |
| 469 | 5,673.68 | 5,503.17 | 5,669.20 | 5,720.12 | **22,566.17** |
| 470 | 1,663.69 | 1,633.38 | 1,760.84 | 1,765.39 | **6,823.29** |
| 471 | 7,780.35 | 7,874.28 | 7,221.08 | 6,682.97 | **29,558.68** |
| 472 | 2,357.84 | 2,364.25 | 2,448.86 | 2,518.65 | **9,689.60** |
| 473 | 5,261.09 | 5,022.60 | 5,194.33 | 5,255.84 | **20,733.86** |
| 474 | 3,998.19 | 3,898.90 | 4,090.20 | 4,151.25 | **16,138.55** |
| 475 | 2,332.01 | 2,270.12 | 2,460.14 | 2,539.14 | **9,601.41** |
| 476 | 2,802.09 | 2,792.09 | 2,905.31 | 3,010.72 | **11,510.21** |
| 477 | 6,018.74 | 6,006.68 | 6,087.02 | 6,243.45 | **24,355.90** |

| | | | | | |
|---|---|---|---|---|---|
| 478 | 2,386.83 | 2,267.89 | 2,276.63 | 2,351.54 | **9,282.89** |
| 479 | 7,225.71 | 7,200.80 | 7,417.47 | 7,372.81 | **29,216.79** |
| **IN - INDIANA - Total** | **134,905.57** | **134,666.73** | **137,057.34** | **138,147.04** | **544,776.67** |

**STATE: KS - KANSAS**

| REGISTRANT ZIP CODE 3 | 1ST QUARTER | 2ND QUARTER | 3RD QUARTER | 4TH QUARTER | TOTAL GRAMS |
|---|---|---|---|---|---|
| 660 | 15,047.94 | 14,981.44 | 15,207.27 | 15,468.41 | **60,705.06** |
| 661 | 1,949.41 | 1,936.04 | 1,929.50 | 2,013.89 | **7,828.84** |
| 662 | 19,018.94 | 19,448.39 | 19,979.27 | 19,760.73 | **78,207.33** |
| 664 | 826.71 | 990.71 | 1,021.78 | 789.88 | **3,629.08** |
| 665 | 1,491.33 | 1,558.25 | 1,512.46 | 1,647.48 | **6,209.52** |
| 666 | 5,004.46 | 5,143.78 | 5,129.12 | 5,241.44 | **20,518.80** |
| 667 | 1,705.89 | 1,688.02 | 1,779.33 | 1,748.56 | **6,921.81** |
| 668 | 603.10 | 624.29 | 607.11 | 638.04 | **2,472.55** |
| 669 | 117.06 | 104.38 | 74.03 | 114.42 | **409.89** |
| 670 | 2,620.14 | 2,659.28 | 2,602.52 | 2,639.31 | **10,521.24** |
| 671 | 1,031.69 | 942.08 | 989.04 | 1,037.51 | **4,000.32** |
| 672 | 7,872.45 | 7,927.99 | 7,975.21 | 8,128.50 | **31,904.15** |
| 673 | 838.94 | 804.02 | 795.15 | 775.21 | **3,213.32** |
| 674 | 1,358.08 | 1,264.64 | 1,338.16 | 1,323.36 | **5,284.24** |
| 675 | 1,157.33 | 1,144.44 | 1,169.39 | 1,187.45 | **4,658.61** |
| 676 | 745.98 | 739.58 | 746.90 | 782.51 | **3,014.97** |
| 677 | 346.62 | 383.80 | 346.38 | 369.05 | **1,445.85** |
| 678 | 679.50 | 663.58 | 662.30 | 718.64 | **2,724.03** |
| 679 | 173.81 | 207.41 | 250.32 | 249.96 | **881.49** |
| **KS - KANSAS - Total** | **62,589.40** | **63,212.10** | **64,115.24** | **64,634.36** | **254,551.10** |

**STATE: KY - KENTUCKY**

| REGISTRANT ZIP CODE 3 | 1ST QUARTER | 2ND QUARTER | 3RD QUARTER | 4TH QUARTER | TOTAL GRAMS |
|---|---|---|---|---|---|
| 400 | 4,163.47 | 3,942.54 | 3,977.27 | 4,148.11 | **16,231.40** |
| 401 | 1,879.73 | 1,599.68 | 2,073.30 | 1,354.62 | **6,907.32** |
| 402 | 16,982.26 | 16,794.58 | 17,053.56 | 17,233.90 | **68,064.29** |
| 403 | 4,344.76 | 4,228.94 | 4,223.56 | 4,378.51 | **17,175.76** |
| 404 | 1,839.39 | 1,862.56 | 1,804.24 | 1,843.29 | **7,349.49** |
| 405 | 7,473.50 | 7,351.09 | 7,267.90 | 7,652.41 | **29,744.90** |
| 406 | 762.12 | 742.31 | 799.91 | 771.86 | **3,076.21** |
| 407 | 610.14 | 576.77 | 583.06 | 628.01 | **2,397.98** |
| 408 | 173.00 | 169.35 | 182.21 | 168.77 | **693.34** |
| 409 | 201.26 | 195.73 | 215.71 | 190.82 | **803.51** |
| 410 | 10,231.82 | 9,958.01 | 10,032.01 | 10,482.04 | **40,703.87** |
| 411 | 1,415.63 | 1,415.90 | 1,327.32 | 1,414.79 | **5,573.64** |
| 412 | 358.83 | 339.61 | 361.34 | 361.36 | **1,421.14** |

| | | | | | |
|---|---|---|---|---|---|
| 413 | 117.47 | 133.18 | 150.12 | 144.37 | **545.15** |
| 414 | 137.25 | 136.57 | 127.61 | 133.71 | **535.14** |
| 415 | 580.27 | 509.14 | 579.27 | 608.78 | **2,277.46** |
| 416 | 348.16 | 357.03 | 367.35 | 351.06 | **1,423.61** |
| 417 | 344.21 | 308.93 | 335.64 | 382.80 | **1,371.58** |
| 418 | 100.49 | 97.85 | 88.94 | 106.35 | **393.63** |
| 420 | 4,592.93 | 4,639.80 | 4,663.97 | 4,628.52 | **18,525.21** |
| 421 | 2,984.40 | 3,110.49 | 3,048.01 | 3,054.48 | **12,197.38** |
| 422 | 2,373.05 | 2,096.23 | 2,175.50 | 2,105.33 | **8,750.10** |
| 423 | 3,523.82 | 3,502.09 | 3,476.52 | 3,544.82 | **14,047.26** |
| 424 | 3,516.42 | 3,417.05 | 3,438.64 | 3,578.44 | **13,950.55** |
| 425 | 445.01 | 389.08 | 406.71 | 396.75 | **1,637.56** |
| 426 | 206.07 | 212.47 | 222.94 | 214.89 | **856.36** |
| 427 | 2,491.31 | 2,422.47 | 2,507.14 | 2,433.10 | **9,854.03** |
| **KY - KENTUCKY - Total** | **72,196.77** | **70,509.45** | **71,489.75** | **72,311.88** | **286,507.85** |

**STATE: LA - LOUISIANA**

| REGISTRANT ZIP CODE 3 | 1ST QUARTER | 2ND QUARTER | 3RD QUARTER | 4TH QUARTER | **TOTAL GRAMS** |
|---|---|---|---|---|---|
| 700 | 19,992.53 | 19,911.21 | 19,753.51 | 20,511.11 | **80,168.35** |
| 701 | 10,432.79 | 10,431.94 | 10,468.96 | 11,010.88 | **42,344.57** |
| 703 | 6,201.60 | 6,156.73 | 6,382.06 | 6,343.44 | **25,083.82** |
| 704 | 19,762.00 | 19,992.32 | 20,967.19 | 20,921.97 | **81,643.47** |
| 705 | 32,877.52 | 33,189.24 | 35,535.10 | 34,964.63 | **136,566.49** |
| 706 | 12,063.98 | 12,046.29 | 12,710.46 | 12,776.24 | **49,596.97** |
| 707 | 20,238.10 | 20,033.83 | 21,039.30 | 20,862.16 | **82,173.39** |
| 708 | 16,108.83 | 16,027.83 | 16,662.10 | 16,666.25 | **65,465.01** |
| 710 | 2,082.01 | 2,031.24 | 2,063.50 | 2,134.75 | **8,311.51** |
| 711 | 7,429.89 | 7,473.16 | 7,882.44 | 7,873.56 | **30,659.05** |
| 712 | 6,985.72 | 6,739.95 | 7,160.15 | 7,122.87 | **28,008.69** |
| 713 | 5,851.45 | 5,654.05 | 5,958.59 | 6,054.88 | **23,518.96** |
| 714 | 2,087.12 | 2,002.74 | 2,217.58 | 2,118.30 | **8,425.73** |
| **LA - LOUISIANA - Total** | **162,113.52** | **161,690.52** | **168,800.94** | **169,361.04** | **661,966.01** |

**STATE: MA - MASSACHUSETTS**

| REGISTRANT ZIP CODE 3 | 1ST QUARTER | 2ND QUARTER | 3RD QUARTER | 4TH QUARTER | **TOTAL GRAMS** |
|---|---|---|---|---|---|
| 010 | 9,042.33 | 8,896.92 | 8,579.91 | 9,278.78 | **35,797.95** |
| 011 | 2,195.81 | 2,287.04 | 2,241.46 | 2,487.13 | **9,211.44** |
| 012 | 2,281.88 | 2,396.82 | 2,351.22 | 2,442.06 | **9,471.98** |
| 013 | 2,596.98 | 2,639.84 | 2,625.13 | 2,649.10 | **10,511.05** |
| 014 | 5,251.74 | 5,322.20 | 5,349.09 | 5,637.25 | **21,560.28** |
| 015 | 7,445.40 | 7,549.87 | 7,703.27 | 7,918.54 | **30,617.08** |
| 016 | 5,703.39 | 5,652.15 | 5,645.64 | 5,779.22 | **22,780.40** |

| | | | | | |
|---|---|---|---|---|---|
| 017 | 11,794.57 | 11,466.14 | 11,766.43 | 11,776.26 | **46,803.40** |
| 018 | 20,462.05 | 20,284.98 | 20,631.16 | 20,927.27 | **82,305.46** |
| 019 | 17,773.82 | 17,770.23 | 18,033.36 | 18,017.71 | **71,595.13** |
| 020 | 13,940.32 | 13,916.36 | 14,206.86 | 14,467.55 | **56,531.08** |
| 021 | 34,662.59 | 34,569.47 | 34,661.87 | 35,415.56 | **139,309.49** |
| 022 | 1,579.88 | 1,560.80 | 1,537.59 | 1,701.15 | **6,379.43** |
| 023 | 13,582.65 | 13,820.41 | 13,841.84 | 14,119.96 | **55,364.86** |
| 024 | 10,993.80 | 10,897.60 | 11,295.09 | 11,275.92 | **44,462.41** |
| 025 | 3,934.22 | 4,277.32 | 4,455.33 | 4,332.61 | **16,999.48** |
| 026 | 4,968.70 | 5,098.29 | 5,663.52 | 5,152.29 | **20,882.81** |
| 027 | 12,572.76 | 12,771.97 | 12,958.06 | 13,240.63 | **51,543.42** |
| **MA - MASSACHUSETTS - Total** | **180,782.88** | **181,178.42** | **183,546.84** | **186,618.99** | **732,127.12** |

**STATE: MD - MARYLAND**

| REGISTRANT ZIP CODE 3 | 1ST QUARTER | 2ND QUARTER | 3RD QUARTER | 4TH QUARTER | **TOTAL GRAMS** |
|---|---|---|---|---|---|
| 206 | 4,466.68 | 4,576.48 | 4,713.08 | 4,997.37 | **18,753.60** |
| 207 | 6,133.30 | 6,438.76 | 6,233.88 | 5,926.76 | **24,732.71** |
| 208 | 14,570.10 | 14,712.57 | 14,870.12 | 14,880.17 | **59,032.96** |
| 209 | 2,839.40 | 2,749.42 | 2,742.81 | 2,751.12 | **11,082.74** |
| 210 | 17,728.95 | 18,528.44 | 18,044.63 | 18,658.55 | **72,960.57** |
| 211 | 8,005.08 | 8,246.40 | 8,126.76 | 8,511.83 | **32,890.06** |
| 212 | 16,897.60 | 17,570.00 | 17,028.27 | 17,820.35 | **69,316.23** |
| 214 | 2,851.14 | 2,888.80 | 2,926.58 | 2,944.22 | **11,610.74** |
| 215 | 1,973.43 | 1,934.58 | 1,870.64 | 2,010.20 | **7,788.85** |
| 216 | 3,308.46 | 3,258.28 | 3,241.59 | 3,293.71 | **13,102.03** |
| 217 | 10,933.89 | 11,162.67 | 10,993.73 | 11,213.31 | **44,303.60** |
| 218 | 2,077.58 | 2,169.33 | 2,215.45 | 2,143.21 | **8,605.57** |
| 219 | 2,794.90 | 2,766.25 | 2,856.80 | 2,894.26 | **11,312.22** |
| **MD - MARYLAND - Total** | **94,580.51** | **97,001.98** | **95,864.34** | **98,045.05** | **385,491.88** |

**STATE: ME - MAINE**

| REGISTRANT ZIP CODE 3 | 1ST QUARTER | 2ND QUARTER | 3RD QUARTER | 4TH QUARTER | **TOTAL GRAMS** |
|---|---|---|---|---|---|
| 039 | 913.24 | 982.46 | 972.14 | 994.75 | **3,862.60** |
| 040 | 8,217.43 | 8,234.05 | 8,834.36 | 8,627.82 | **33,913.66** |
| 041 | 4,031.40 | 4,049.99 | 4,240.38 | 4,127.38 | **16,449.15** |
| 042 | 2,067.48 | 2,005.56 | 2,169.44 | 2,280.49 | **8,522.97** |
| 043 | 1,510.69 | 1,591.46 | 1,690.32 | 1,650.88 | **6,443.35** |
| 044 | 2,203.33 | 2,097.82 | 2,161.95 | 2,228.24 | **8,691.33** |
| 045 | 804.58 | 790.07 | 865.89 | 781.08 | **3,241.62** |
| 046 | 716.67 | 736.79 | 791.70 | 779.27 | **3,024.44** |
| 047 | 493.16 | 453.61 | 494.20 | 508.83 | **1,949.80** |
| 048 | 542.91 | 619.06 | 707.51 | 619.98 | **2,489.46** |

| | | | | | |
|---|---|---|---|---|---|
| 049 | 1,991.15 | 1,968.93 | 1,954.09 | 1,904.75 | **7,818.92** |
| **ME - MAINE - Total** | **23,492.06** | **23,529.79** | **24,881.99** | **24,503.48** | **96,407.31** |

**STATE: MI - MICHIGAN**

| REGISTRANT ZIP CODE 3 | 1ST QUARTER | 2ND QUARTER | 3RD QUARTER | 4TH QUARTER | TOTAL GRAMS |
|---|---|---|---|---|---|
| 480 | 39,577.93 | 39,573.13 | 39,388.64 | 39,655.13 | **158,194.83** |
| 481 | 41,234.36 | 41,261.68 | 41,677.74 | 42,121.83 | **166,295.61** |
| 482 | 6,748.60 | 6,690.46 | 6,263.44 | 6,523.13 | **26,225.62** |
| 483 | 32,990.01 | 33,747.10 | 33,056.71 | 33,712.70 | **133,506.53** |
| 484 | 8,879.82 | 8,521.88 | 8,735.51 | 8,698.01 | **34,835.22** |
| 485 | 2,889.71 | 2,869.79 | 2,814.33 | 2,891.80 | **11,465.63** |
| 486 | 5,026.81 | 5,019.86 | 5,092.97 | 5,028.83 | **20,168.47** |
| 487 | 2,901.86 | 2,937.06 | 2,887.12 | 2,994.85 | **11,720.89** |
| 488 | 15,529.69 | 15,048.62 | 14,595.38 | 15,632.96 | **60,806.64** |
| 489 | 4,415.07 | 4,361.44 | 4,307.54 | 4,450.91 | **17,534.96** |
| 490 | 17,737.42 | 17,509.42 | 17,440.17 | 17,771.02 | **70,458.04** |
| 491 | 2,233.52 | 2,224.13 | 2,299.58 | 2,311.48 | **9,068.70** |
| 492 | 4,148.01 | 4,048.81 | 4,017.86 | 4,173.73 | **16,388.42** |
| 493 | 7,071.47 | 7,196.81 | 7,121.63 | 7,385.56 | **28,775.47** |
| 494 | 19,500.42 | 19,420.13 | 19,638.32 | 19,753.75 | **78,312.62** |
| 495 | 19,560.14 | 19,317.35 | 19,472.39 | 19,657.87 | **78,007.75** |
| 496 | 5,791.76 | 5,711.07 | 5,806.79 | 5,751.12 | **23,060.75** |
| 497 | 3,512.59 | 3,545.87 | 3,636.09 | 3,480.73 | **14,175.27** |
| 498 | 3,503.66 | 3,213.23 | 3,471.07 | 3,365.44 | **13,553.40** |
| 499 | 1,604.79 | 1,472.10 | 1,593.62 | 1,602.16 | **6,272.67** |
| **MI - MICHIGAN - Total** | **244,857.64** | **243,689.95** | **243,316.89** | **246,963.00** | **978,827.49** |

**STATE: MN - MINNESOTA**

| REGISTRANT ZIP CODE 3 | 1ST QUARTER | 2ND QUARTER | 3RD QUARTER | 4TH QUARTER | TOTAL GRAMS |
|---|---|---|---|---|---|
| 550 | 12,845.37 | 12,800.42 | 12,665.47 | 13,409.02 | **51,720.28** |
| 551 | 17,709.84 | 18,027.72 | 17,808.72 | 18,396.21 | **71,942.48** |
| 553 | 22,878.90 | 22,958.84 | 23,301.34 | 24,099.77 | **93,238.85** |
| 554 | 26,311.76 | 26,852.70 | 26,618.06 | 27,888.10 | **107,670.62** |
| 556 | 138.50 | 109.20 | 124.83 | 131.63 | **504.16** |
| 557 | 3,186.47 | 3,273.16 | 3,191.78 | 3,355.60 | **13,007.01** |
| 558 | 2,116.78 | 2,062.44 | 2,039.98 | 2,120.95 | **8,340.16** |
| 559 | 5,383.01 | 5,199.52 | 5,194.57 | 5,445.73 | **21,222.84** |
| 560 | 5,410.85 | 5,392.89 | 5,301.11 | 5,484.05 | **21,588.90** |
| 561 | 826.10 | 873.27 | 878.84 | 881.17 | **3,459.38** |
| 562 | 1,723.74 | 1,700.63 | 1,594.73 | 1,799.22 | **6,818.33** |
| 563 | 4,444.02 | 4,266.93 | 4,365.91 | 4,565.13 | **17,641.98** |
| 564 | 2,261.61 | 2,345.41 | 2,511.68 | 2,470.35 | **9,589.05** |

| | | | | | |
|---|---|---|---|---|---|
| 565 | 2,803.29 | 2,845.76 | 2,793.21 | 2,972.14 | **11,414.41** |
| 566 | 1,348.82 | 1,296.44 | 1,243.46 | 1,334.11 | **5,222.84** |
| 567 | 820.46 | 823.52 | 846.67 | 854.09 | **3,344.74** |
| **MN - MINNESOTA - Total** | **110,209.54** | **110,828.85** | **110,480.36** | **115,207.26** | **446,726.01** |

**STATE: MO - MISSOURI**

| REGISTRANT ZIP CODE 3 | 1ST QUARTER | 2ND QUARTER | 3RD QUARTER | 4TH QUARTER | **TOTAL GRAMS** |
|---|---|---|---|---|---|
| 630 | 20,086.85 | 19,759.10 | 19,957.13 | 20,260.49 | **80,063.57** |
| 631 | 39,285.49 | 39,481.88 | 41,948.43 | 54,317.14 | **175,032.94** |
| 633 | 14,443.59 | 14,621.71 | 15,018.41 | 15,060.95 | **59,144.66** |
| 634 | 1,147.59 | 1,148.38 | 1,217.80 | 1,190.28 | **4,704.06** |
| 635 | 1,303.68 | 1,288.99 | 1,240.55 | 1,391.11 | **5,224.33** |
| 636 | 2,102.50 | 2,012.89 | 2,110.84 | 2,016.59 | **8,242.82** |
| 637 | 2,238.37 | 2,052.04 | 2,108.16 | 2,180.76 | **8,579.33** |
| 638 | 1,634.13 | 1,591.61 | 1,604.05 | 1,629.98 | **6,459.77** |
| 639 | 543.69 | 533.64 | 584.39 | 566.97 | **2,228.70** |
| 640 | 14,670.15 | 14,785.01 | 15,137.18 | 15,278.09 | **59,870.43** |
| 641 | 17,566.46 | 18,059.53 | 18,321.52 | 18,155.95 | **72,103.46** |
| 644 | 654.55 | 638.64 | 677.63 | 679.26 | **2,650.08** |
| 645 | 1,176.07 | 1,171.94 | 1,113.41 | 1,219.05 | **4,680.46** |
| 646 | 448.56 | 430.39 | 452.75 | 414.51 | **1,746.21** |
| 647 | 939.74 | 987.81 | 1,076.43 | 1,061.10 | **4,065.07** |
| 648 | 1,481.76 | 1,549.49 | 1,608.89 | 1,677.86 | **6,317.99** |
| 650 | 1,115.67 | 1,192.51 | 1,234.21 | 1,163.36 | **4,705.75** |
| 651 | 1,156.36 | 1,090.06 | 1,178.42 | 1,228.06 | **4,652.90** |
| 652 | 5,444.95 | 5,357.69 | 5,386.87 | 5,528.12 | **21,717.63** |
| 653 | 983.45 | 1,036.15 | 1,047.07 | 1,050.71 | **4,117.38** |
| 654 | 820.73 | 842.16 | 843.14 | 838.30 | **3,344.33** |
| 655 | 691.75 | 672.48 | 624.83 | 654.43 | **2,643.49** |
| 656 | 1,353.66 | 1,325.16 | 1,340.28 | 1,363.71 | **5,382.81** |
| 657 | 1,847.00 | 2,027.22 | 2,146.73 | 2,098.57 | **8,119.51** |
| 658 | 3,597.26 | 3,524.35 | 3,576.62 | 3,756.88 | **14,455.11** |
| **MO - MISSOURI - Total** | **136,734.01** | **137,180.84** | **141,555.73** | **154,782.23** | **570,252.80** |

**STATE: MS - MISSISSIPPI**

| REGISTRANT ZIP CODE 3 | 1ST QUARTER | 2ND QUARTER | 3RD QUARTER | 4TH QUARTER | **TOTAL GRAMS** |
|---|---|---|---|---|---|
| 386 | 9,612.97 | 9,421.73 | 9,816.23 | 9,581.94 | **38,432.87** |
| 387 | 1,567.16 | 1,483.47 | 1,585.80 | 1,531.55 | **6,167.99** |
| 388 | 4,700.08 | 4,747.10 | 4,879.21 | 5,026.47 | **19,352.87** |
| 389 | 1,489.98 | 1,467.13 | 1,503.81 | 1,531.53 | **5,992.45** |
| 390 | 5,112.01 | 5,066.77 | 5,132.09 | 4,955.38 | **20,266.26** |
| 391 | 6,191.63 | 6,094.90 | 6,422.35 | 6,268.42 | **24,977.31** |

| | | | | | |
|---|---|---|---|---|---|
| 392 | 5,214.36 | 5,064.20 | 5,539.15 | 5,305.90 | **21,123.61** |
| 393 | 4,013.25 | 3,870.49 | 3,894.61 | 3,765.56 | **15,543.91** |
| 394 | 8,957.61 | 8,701.69 | 8,741.32 | 8,704.81 | **35,105.43** |
| 395 | 9,053.43 | 9,224.65 | 9,517.65 | 9,175.19 | **36,970.92** |
| 396 | 2,500.84 | 2,417.70 | 2,544.28 | 2,490.12 | **9,952.95** |
| 397 | 3,545.75 | 3,189.30 | 3,346.02 | 3,379.64 | **13,460.71** |
| **MS - MISSISSIPPI - Total** | **61,959.09** | **60,749.14** | **62,922.53** | **61,716.51** | **247,347.27** |

**STATE: MT - MONTANA**

| REGISTRANT ZIP CODE 3 | 1ST QUARTER | 2ND QUARTER | 3RD QUARTER | 4TH QUARTER | TOTAL GRAMS |
|---|---|---|---|---|---|
| 590 | 663.09 | 617.50 | 647.86 | 685.85 | **2,614.31** |
| 591 | 2,412.90 | 2,405.28 | 2,501.45 | 2,593.70 | **9,913.33** |
| 592 | 322.56 | 346.82 | 324.70 | 355.67 | **1,349.76** |
| 593 | 303.78 | 351.93 | 367.30 | 322.23 | **1,345.24** |
| 594 | 1,152.62 | 1,195.65 | 1,086.41 | 1,117.20 | **4,551.88** |
| 595 | 245.07 | 226.24 | 266.69 | 217.97 | **955.96** |
| 596 | 1,137.57 | 1,197.91 | 1,207.71 | 1,142.02 | **4,685.21** |
| 597 | 3,324.98 | 3,484.46 | 3,507.85 | 3,781.12 | **14,098.40** |
| 598 | 3,461.55 | 3,636.78 | 3,638.39 | 3,587.79 | **14,324.51** |
| 599 | 1,294.27 | 1,252.92 | 1,315.73 | 1,386.31 | **5,249.23** |
| **MT - MONTANA - Total** | **14,318.39** | **14,715.49** | **14,864.09** | **15,189.87** | **59,087.84** |

**STATE: NC - NORTH CAROLINA**

| REGISTRANT ZIP CODE 3 | 1ST QUARTER | 2ND QUARTER | 3RD QUARTER | 4TH QUARTER | TOTAL GRAMS |
|---|---|---|---|---|---|
| 270 | 4,112.56 | 3,884.82 | 4,064.91 | 4,313.28 | **16,375.57** |
| 271 | 5,847.66 | 5,612.60 | 5,873.55 | 5,969.36 | **23,303.17** |
| 272 | 11,570.87 | 11,440.27 | 11,916.63 | 12,222.26 | **47,150.03** |
| 273 | 6,193.39 | 5,955.99 | 6,122.81 | 6,536.71 | **24,808.90** |
| 274 | 8,150.29 | 7,916.92 | 8,126.10 | 8,400.88 | **32,594.19** |
| 275 | 24,533.46 | 24,033.13 | 24,433.83 | 26,110.45 | **99,110.87** |
| 276 | 15,418.97 | 15,096.26 | 15,571.65 | 16,100.55 | **62,187.43** |
| 277 | 5,970.36 | 5,972.14 | 5,608.17 | 6,128.40 | **23,679.07** |
| 278 | 9,367.99 | 8,818.44 | 9,103.33 | 9,958.66 | **37,248.42** |
| 279 | 3,295.37 | 3,411.37 | 3,541.47 | 3,639.75 | **13,887.96** |
| 280 | 18,751.10 | 18,398.49 | 18,637.12 | 19,342.89 | **75,129.60** |
| 281 | 12,593.45 | 12,386.92 | 12,822.20 | 13,014.58 | **50,817.14** |
| 282 | 20,525.87 | 20,173.49 | 20,664.97 | 20,842.66 | **82,206.99** |
| 283 | 15,389.52 | 15,593.02 | 15,509.27 | 15,959.15 | **62,450.96** |
| 284 | 17,761.23 | 17,804.45 | 18,176.22 | 19,176.32 | **72,918.21** |
| 285 | 12,158.70 | 11,731.47 | 11,759.85 | 11,609.70 | **47,259.73** |
| 286 | 8,353.28 | 8,182.67 | 8,228.34 | 8,591.45 | **33,355.75** |
| 287 | 7,346.31 | 7,166.57 | 7,581.17 | 7,650.30 | **29,744.36** |

| | | | | | |
|---|---|---|---|---|---|
| 288 | 4,561.56 | 4,475.23 | 4,597.42 | 4,644.39 | **18,278.61** |
| 289 | 387.20 | 494.28 | 460.84 | 461.82 | **1,804.14** |
| **NC - NORTH CAROLINA - Total** | **212,289.15** | **208,548.54** | **212,799.85** | **220,673.55** | **854,311.09** |

**STATE: ND - NORTH DAKOTA**

| REGISTRANT ZIP CODE 3 | 1ST QUARTER | 2ND QUARTER | 3RD QUARTER | 4TH QUARTER | TOTAL GRAMS |
|---|---|---|---|---|---|
| 580 | 714.95 | 710.00 | 698.87 | 779.60 | **2,903.42** |
| 581 | 2,277.92 | 2,267.40 | 2,265.07 | 2,343.83 | **9,154.22** |
| 582 | 1,566.36 | 1,475.85 | 1,495.71 | 1,591.82 | **6,129.74** |
| 583 | 500.36 | 455.98 | 459.85 | 459.15 | **1,875.34** |
| 584 | 272.41 | 279.12 | 296.11 | 286.04 | **1,133.67** |
| 585 | 1,463.35 | 1,470.21 | 1,575.11 | 1,508.92 | **6,017.59** |
| 586 | 365.99 | 411.62 | 419.79 | 430.43 | **1,627.83** |
| 587 | 1,384.10 | 1,427.33 | 1,527.97 | 1,467.01 | **5,806.42** |
| 588 | 654.13 | 685.90 | 748.24 | 770.55 | **2,858.82** |
| **ND - NORTH DAKOTA - Total** | **9,199.58** | **9,183.41** | **9,486.72** | **9,637.35** | **37,507.06** |

**STATE: NE - NEBRASKA**

| REGISTRANT ZIP CODE 3 | 1ST QUARTER | 2ND QUARTER | 3RD QUARTER | 4TH QUARTER | TOTAL GRAMS |
|---|---|---|---|---|---|
| 680 | 2,441.28 | 2,507.91 | 2,561.16 | 2,569.94 | **10,080.30** |
| 681 | 12,777.74 | 13,120.67 | 13,196.68 | 13,602.20 | **52,697.29** |
| 683 | 504.12 | 563.82 | 525.49 | 551.56 | **2,144.99** |
| 684 | 448.17 | 451.49 | 449.29 | 511.62 | **1,860.58** |
| 685 | 5,564.55 | 5,541.57 | 5,576.96 | 5,779.87 | **22,462.95** |
| 686 | 485.67 | 474.44 | 489.69 | 490.43 | **1,940.22** |
| 687 | 1,032.71 | 1,041.98 | 1,105.66 | 1,119.80 | **4,300.15** |
| 688 | 1,588.63 | 1,561.71 | 1,682.99 | 1,667.84 | **6,501.17** |
| 689 | 474.77 | 491.85 | 529.10 | 552.02 | **2,047.74** |
| 690 | 171.31 | 176.53 | 223.29 | 162.47 | **733.61** |
| 691 | 715.68 | 722.72 | 693.57 | 758.24 | **2,890.20** |
| 692 | 37.95 | 42.11 | 34.48 | 44.49 | **159.04** |
| 693 | 517.54 | 543.71 | 553.54 | 585.15 | **2,199.94** |
| **NE - NEBRASKA - Total** | **26,760.12** | **27,240.52** | **27,621.89** | **28,395.64** | **110,018.17** |

**STATE: NH - NEW HAMPSHIRE**

| REGISTRANT ZIP CODE 3 | 1ST QUARTER | 2ND QUARTER | 3RD QUARTER | 4TH QUARTER | TOTAL GRAMS |
|---|---|---|---|---|---|
| 030 | 12,012.72 | 12,268.03 | 12,659.73 | 12,508.25 | **49,448.72** |
| 031 | 6,390.31 | 6,574.51 | 6,615.70 | 6,612.78 | **26,193.30** |
| 032 | 2,880.53 | 3,017.27 | 3,017.30 | 2,908.69 | **11,823.79** |
| 033 | 2,126.19 | 2,180.27 | 2,212.78 | 2,255.74 | **8,774.98** |
| 034 | 1,480.45 | 1,587.96 | 1,588.83 | 1,616.66 | **6,273.90** |
| 035 | 558.59 | 557.23 | 664.44 | 690.51 | **2,470.77** |
| 036 | 93.86 | 105.76 | 113.38 | 121.30 | **434.30** |

| | | | | | |
|---|---|---|---|---|---|
| 037 | 1,789.63 | 1,854.97 | 1,809.70 | 1,709.66 | **7,163.96** |
| 038 | 11,970.22 | 12,090.40 | 12,628.36 | 12,181.19 | **48,870.17** |
| **NH - NEW HAMPSHIRE - Total** | **39,302.49** | **40,236.40** | **41,310.22** | **40,604.80** | **161,453.90** |

**STATE: NJ - NEW JERSEY**

| REGISTRANT ZIP CODE 3 | 1ST QUARTER | 2ND QUARTER | 3RD QUARTER | 4TH QUARTER | TOTAL GRAMS |
|---|---|---|---|---|---|
| 070 | 16,620.66 | 17,468.37 | 16,952.82 | 17,868.44 | **68,910.29** |
| 071 | 1,300.43 | 1,361.80 | 1,254.46 | 1,321.90 | **5,238.58** |
| 072 | 625.84 | 717.01 | 639.97 | 686.92 | **2,669.74** |
| 073 | 1,727.54 | 1,818.64 | 1,897.01 | 1,821.62 | **7,264.81** |
| 074 | 6,312.59 | 6,421.64 | 6,637.47 | 6,899.30 | **26,271.01** |
| 075 | 663.34 | 668.56 | 686.13 | 769.25 | **2,787.28** |
| 076 | 5,468.27 | 5,866.39 | 6,060.20 | 6,062.37 | **23,457.23** |
| 077 | 8,590.22 | 8,758.65 | 9,132.54 | 9,170.38 | **35,651.79** |
| 078 | 5,327.77 | 5,625.38 | 5,542.51 | 5,874.74 | **22,370.40** |
| 079 | 4,704.48 | 4,812.09 | 4,940.56 | 4,965.36 | **19,422.49** |
| 080 | 23,650.55 | 23,734.41 | 23,998.17 | 24,376.79 | **95,759.91** |
| 081 | 1,724.93 | 1,833.84 | 1,788.45 | 1,934.61 | **7,281.83** |
| 082 | 3,912.05 | 4,001.55 | 4,239.59 | 3,809.70 | **15,962.89** |
| 083 | 2,568.31 | 2,596.37 | 2,684.45 | 2,725.00 | **10,574.13** |
| 084 | 310.59 | 416.25 | 399.56 | 401.36 | **1,527.76** |
| 085 | 3,484.42 | 3,620.80 | 3,680.88 | 3,646.46 | **14,432.56** |
| 086 | 2,604.96 | 2,813.49 | 2,712.11 | 2,962.65 | **11,093.21** |
| 087 | 6,059.99 | 6,147.22 | 6,220.03 | 6,224.32 | **24,651.55** |
| 088 | 9,559.72 | 9,620.29 | 9,635.23 | 10,094.22 | **38,909.47** |
| 089 | 966.17 | 1,012.11 | 942.08 | 1,081.99 | **4,002.35** |
| **NJ - NEW JERSEY - Total** | **106,182.82** | **109,314.85** | **110,044.20** | **112,697.38** | **438,239.25** |

**STATE: NM - NEW MEXICO**

| REGISTRANT ZIP CODE 3 | 1ST QUARTER | 2ND QUARTER | 3RD QUARTER | 4TH QUARTER | TOTAL GRAMS |
|---|---|---|---|---|---|
| 870 | 626.74 | 637.16 | 654.17 | 678.84 | **2,596.91** |
| 871 | 6,304.23 | 6,299.13 | 6,235.63 | 6,466.14 | **25,305.13** |
| 873 | 105.58 | 138.16 | 111.17 | 114.95 | **469.85** |
| 874 | 639.74 | 648.00 | 704.68 | 676.16 | **2,668.58** |
| 875 | 1,942.65 | 1,980.89 | 2,100.76 | 1,901.29 | **7,925.58** |
| 877 | 163.73 | 184.19 | 164.10 | 163.94 | **675.96** |
| 878 | 35.53 | 35.65 | 34.75 | 45.55 | **151.49** |
| 879 | 75.81 | 56.95 | 75.28 | 72.65 | **280.69** |
| 880 | 1,933.72 | 2,041.99 | 2,078.25 | 2,113.30 | **8,167.26** |
| 881 | 370.46 | 387.05 | 343.94 | 370.74 | **1,472.20** |
| 882 | 1,852.12 | 1,858.50 | 1,972.30 | 1,983.35 | **7,666.27** |
| 883 | 869.79 | 868.93 | 954.89 | 919.35 | **3,612.96** |

| | | | | | |
|---|---|---|---|---|---|
| 884 | 47.18 | 66.66 | 33.50 | 56.50 | **203.84** |
| **NM - NEW MEXICO - Total** | **14,967.28** | **15,203.27** | **15,463.42** | **15,562.75** | **61,196.72** |

**STATE: NV - NEVADA**

| REGISTRANT ZIP CODE 3 | 1ST QUARTER | 2ND QUARTER | 3RD QUARTER | 4TH QUARTER | TOTAL GRAMS |
|---|---|---|---|---|---|
| 890 | 6,029.64 | 5,883.62 | 6,069.61 | 6,156.98 | **24,139.85** |
| 891 | 14,900.53 | 15,335.23 | 15,553.51 | 15,367.59 | **61,156.87** |
| 893 | 50.24 | 61.64 | 35.51 | 36.57 | **183.96** |
| 894 | 1,659.48 | 1,658.83 | 1,795.76 | 1,817.83 | **6,931.90** |
| 895 | 2,859.54 | 2,782.96 | 2,918.55 | 3,018.25 | **11,579.29** |
| 897 | 524.05 | 526.50 | 542.46 | 555.27 | **2,148.29** |
| 898 | 422.74 | 478.39 | 514.29 | 498.23 | **1,913.65** |
| **NV - NEVADA - Total** | **26,446.23** | **26,727.17** | **27,429.70** | **27,450.72** | **108,053.81** |

**STATE: NY - NEW YORK**

| REGISTRANT ZIP CODE 3 | 1ST QUARTER | 2ND QUARTER | 3RD QUARTER | 4TH QUARTER | TOTAL GRAMS |
|---|---|---|---|---|---|
| 100 | 50,309.45 | 52,983.32 | 51,866.66 | 53,722.18 | **208,881.61** |
| 101 | 2,833.97 | 2,866.72 | 2,817.56 | 2,968.87 | **11,487.12** |
| 102 | 481.68 | 412.27 | 452.74 | 436.34 | **1,783.03** |
| 103 | 3,379.00 | 3,530.09 | 3,559.46 | 3,812.78 | **14,281.33** |
| 104 | 3,983.50 | 4,038.74 | 3,800.04 | 4,049.56 | **15,871.84** |
| 105 | 9,526.14 | 9,720.08 | 9,936.65 | 9,988.15 | **39,171.02** |
| 106 | 1,257.08 | 1,319.53 | 1,301.98 | 1,157.55 | **5,036.15** |
| 107 | 1,775.47 | 1,934.11 | 1,906.00 | 1,982.39 | **7,597.97** |
| 108 | 725.72 | 772.96 | 757.90 | 769.71 | **3,026.29** |
| 109 | 6,561.00 | 6,772.77 | 6,827.93 | 6,840.78 | **27,002.48** |
| 110 | 2,612.80 | 2,680.16 | 2,809.18 | 2,821.24 | **10,923.38** |
| 111 | 2,721.61 | 3,003.90 | 2,931.99 | 3,118.71 | **11,776.22** |
| 112 | 16,399.87 | 17,240.80 | 16,838.42 | 17,191.51 | **67,670.61** |
| 113 | 5,563.75 | 5,632.32 | 5,462.62 | 5,674.00 | **22,332.69** |
| 114 | 1,360.32 | 1,395.37 | 1,293.42 | 1,388.03 | **5,437.14** |
| 115 | 7,541.86 | 7,752.98 | 7,747.57 | 7,943.33 | **30,985.75** |
| 116 | 563.02 | 586.42 | 514.58 | 589.92 | **2,253.95** |
| 117 | 21,434.77 | 21,987.84 | 22,092.59 | 22,614.47 | **88,129.67** |
| 118 | 1,072.75 | 1,166.69 | 1,104.01 | 1,177.86 | **4,521.32** |
| 119 | 3,017.30 | 3,296.27 | 3,472.74 | 3,354.59 | **13,140.90** |
| 120 | 5,328.12 | 5,565.62 | 5,663.33 | 5,753.76 | **22,310.83** |
| 121 | 4,026.68 | 4,108.91 | 4,114.11 | 4,248.52 | **16,498.23** |
| 122 | 2,855.62 | 2,991.15 | 2,988.03 | 3,080.15 | **11,914.95** |
| 123 | 2,295.64 | 2,415.96 | 2,304.32 | 2,410.92 | **9,426.84** |
| 124 | 2,079.55 | 2,012.46 | 2,061.17 | 2,143.95 | **8,297.13** |
| 125 | 5,593.21 | 5,821.81 | 5,865.51 | 5,962.98 | **23,243.52** |

| | | | | | |
|---|---|---|---|---|---|
| 126 | 1,278.01 | 1,310.46 | 1,318.74 | 1,358.41 | **5,265.62** |
| 127 | 1,181.17 | 1,267.77 | 1,301.33 | 1,315.26 | **5,065.54** |
| 128 | 3,415.18 | 3,485.06 | 3,654.17 | 3,646.40 | **14,200.81** |
| 129 | 2,349.18 | 2,402.99 | 2,510.83 | 2,677.42 | **9,940.41** |
| 130 | 5,991.46 | 6,117.28 | 6,141.23 | 6,306.51 | **24,556.48** |
| 131 | 1,981.28 | 1,948.43 | 1,953.29 | 2,090.58 | **7,973.57** |
| 132 | 3,385.42 | 3,319.75 | 3,440.07 | 3,655.35 | **13,800.60** |
| 133 | 1,073.09 | 1,082.09 | 1,045.57 | 1,102.10 | **4,302.84** |
| 134 | 2,009.95 | 2,018.44 | 2,034.85 | 2,091.04 | **8,154.27** |
| 135 | 985.98 | 986.46 | 931.39 | 991.65 | **3,895.47** |
| 136 | 2,783.33 | 2,824.13 | 2,959.17 | 3,092.61 | **11,659.23** |
| 137 | 1,293.83 | 1,276.14 | 1,239.85 | 1,342.26 | **5,152.08** |
| 138 | 1,611.78 | 1,569.82 | 1,537.81 | 1,554.33 | **6,273.75** |
| 139 | 1,062.72 | 1,077.36 | 1,006.00 | 1,014.29 | **4,160.37** |
| 140 | 6,568.28 | 6,527.29 | 6,641.01 | 6,707.00 | **26,443.58** |
| 141 | 2,740.77 | 2,773.72 | 2,811.06 | 2,869.99 | **11,195.55** |
| 142 | 9,387.64 | 9,501.43 | 9,523.19 | 9,818.03 | **38,230.29** |
| 143 | 1,142.36 | 1,298.66 | 1,199.05 | 1,263.84 | **4,903.91** |
| 144 | 5,910.99 | 5,834.20 | 5,728.42 | 6,041.30 | **23,514.90** |
| 145 | 5,376.34 | 5,519.41 | 5,311.78 | 5,459.98 | **21,667.52** |
| 146 | 9,979.43 | 10,083.14 | 10,241.28 | 10,591.68 | **40,895.54** |
| 147 | 1,572.66 | 1,557.70 | 1,579.44 | 1,533.41 | **6,243.20** |
| 148 | 3,686.28 | 3,812.03 | 3,650.12 | 3,793.18 | **14,941.60** |
| 149 | 913.77 | 943.16 | 903.10 | 947.49 | **3,707.52** |
| **NY - NEW YORK - Total** | **242,980.79** | **250,546.17** | **249,153.25** | **256,466.39** | **999,146.60** |

**STATE: OH - OHIO**

| REGISTRANT ZIP CODE 3 | 1ST QUARTER | 2ND QUARTER | 3RD QUARTER | 4TH QUARTER | TOTAL GRAMS |
|---|---|---|---|---|---|
| 430 | 16,045.89 | 16,024.53 | 16,576.14 | 17,118.00 | **65,764.57** |
| 431 | 6,168.77 | 6,258.54 | 6,343.84 | 6,539.92 | **25,311.07** |
| 432 | 29,005.44 | 24,444.74 | 14,384.57 | 14,874.07 | **82,708.81** |
| 433 | 2,389.24 | 2,448.64 | 2,372.89 | 2,415.01 | **9,625.78** |
| 434 | 2,189.13 | 2,179.96 | 2,257.24 | 2,409.81 | **9,036.13** |
| 435 | 5,489.62 | 5,588.02 | 5,653.84 | 5,824.03 | **22,555.51** |
| 436 | 8,696.57 | 8,823.15 | 8,901.44 | 9,349.09 | **35,770.24** |
| 437 | 1,666.85 | 1,691.68 | 1,624.86 | 1,723.65 | **6,707.03** |
| 438 | 234.56 | 267.88 | 260.99 | 231.74 | **995.17** |
| 439 | 2,097.23 | 2,043.07 | 2,059.32 | 2,194.44 | **8,394.07** |
| 440 | 15,328.55 | 15,430.81 | 15,607.83 | 16,048.33 | **62,415.52** |
| 441 | 21,040.63 | 21,095.13 | 21,706.08 | 22,230.16 | **86,071.99** |
| 442 | 8,576.07 | 8,363.29 | 8,511.73 | 8,809.98 | **34,261.07** |

| | | | | | |
|---|---|---|---|---|---|
| 443 | 3,881.06 | 4,060.54 | 3,993.08 | 4,038.44 | **15,973.12** |
| 444 | 4,737.57 | 4,711.36 | 4,582.85 | 4,952.02 | **18,983.80** |
| 445 | 4,862.19 | 5,012.59 | 5,001.28 | 5,129.21 | **20,005.27** |
| 446 | 5,961.67 | 5,931.19 | 5,817.58 | 5,966.91 | **23,677.35** |
| 447 | 5,276.03 | 5,100.24 | 5,263.05 | 5,347.37 | **20,986.70** |
| 448 | 3,500.77 | 3,465.65 | 3,368.02 | 3,460.57 | **13,795.01** |
| 449 | 887.77 | 909.25 | 902.86 | 908.92 | **3,608.79** |
| 450 | 15,396.61 | 15,560.96 | 15,778.41 | 15,982.07 | **62,718.04** |
| 451 | 5,492.36 | 5,568.67 | 5,659.80 | 5,656.90 | **22,377.74** |
| 452 | 21,678.05 | 21,711.42 | 21,940.24 | 22,609.87 | **87,939.59** |
| 453 | 5,093.83 | 5,071.12 | 5,010.22 | 5,418.81 | **20,593.98** |
| 454 | 6,182.26 | 6,159.66 | 6,152.45 | 6,575.47 | **25,069.83** |
| 455 | 1,445.53 | 1,401.37 | 1,400.05 | 1,412.17 | **5,659.12** |
| 456 | 3,858.29 | 3,695.07 | 3,631.13 | 3,839.74 | **15,024.23** |
| 457 | 3,083.11 | 2,838.21 | 2,778.42 | 2,709.56 | **11,409.29** |
| 458 | 3,534.24 | 3,524.16 | 3,501.31 | 3,701.42 | **14,261.13** |
| **OH - OHIO - Total** | **213,799.88** | **209,380.92** | **201,041.51** | **207,477.66** | **831,699.97** |

**STATE: OK - OKLAHOMA**

| REGISTRANT ZIP CODE 3 | 1ST QUARTER | 2ND QUARTER | 3RD QUARTER | 4TH QUARTER | **TOTAL GRAMS** |
|---|---|---|---|---|---|
| 730 | 10,944.45 | 10,982.10 | 11,369.38 | 11,539.53 | **44,835.45** |
| 731 | 9,455.17 | 9,474.07 | 9,878.96 | 9,562.28 | **38,370.48** |
| 734 | 986.49 | 968.79 | 991.67 | 997.33 | **3,944.28** |
| 735 | 2,526.33 | 2,574.63 | 2,684.84 | 2,221.26 | **10,007.05** |
| 736 | 270.40 | 256.44 | 279.74 | 293.29 | **1,099.87** |
| 737 | 1,046.63 | 1,012.95 | 1,159.24 | 1,133.77 | **4,352.59** |
| 738 | 183.62 | 182.60 | 148.10 | 174.02 | **688.34** |
| 739 | 63.86 | 72.52 | 78.16 | 76.31 | **290.85** |
| 740 | 12,408.32 | 12,203.69 | 11,878.66 | 12,225.72 | **48,716.39** |
| 741 | 9,934.51 | 9,755.99 | 9,777.98 | 9,852.38 | **39,320.85** |
| 743 | 948.97 | 1,032.86 | 1,026.06 | 1,095.83 | **4,103.71** |
| 744 | 2,683.85 | 2,421.67 | 2,529.70 | 2,581.29 | **10,216.51** |
| 745 | 504.77 | 504.48 | 451.59 | 477.90 | **1,938.74** |
| 746 | 324.01 | 275.21 | 277.71 | 324.95 | **1,201.89** |
| 747 | 647.32 | 701.87 | 664.82 | 748.19 | **2,762.20** |
| 748 | 1,389.30 | 1,365.54 | 1,391.34 | 1,421.62 | **5,567.80** |
| 749 | 444.90 | 411.95 | 407.59 | 399.05 | **1,663.50** |
| **OK - OKLAHOMA - Total** | **54,762.92** | **54,197.33** | **54,995.55** | **55,124.71** | **219,080.50** |

**STATE: OR - OREGON**

| REGISTRANT ZIP CODE 3 | 1ST QUARTER | 2ND QUARTER | 3RD QUARTER | 4TH QUARTER | **TOTAL GRAMS** |
|---|---|---|---|---|---|
| 970 | 12,954.46 | 13,619.58 | 13,507.51 | 13,577.57 | **53,659.12** |

| | | | | | |
|---|---|---|---|---|---|
| 971 | 5,104.96 | 5,423.74 | 5,189.39 | 5,433.51 | **21,151.59** |
| 972 | 19,003.20 | 19,640.89 | 19,744.20 | 20,147.70 | **78,535.99** |
| 973 | 8,733.99 | 8,871.19 | 8,246.63 | 8,760.21 | **34,612.02** |
| 974 | 7,003.60 | 7,295.85 | 7,204.29 | 7,484.46 | **28,988.20** |
| 975 | 2,375.07 | 2,471.96 | 2,418.91 | 2,559.99 | **9,825.93** |
| 976 | 457.22 | 510.62 | 429.40 | 484.69 | **1,881.92** |
| 977 | 3,706.56 | 4,113.17 | 4,133.17 | 4,163.48 | **16,116.39** |
| 978 | 901.99 | 854.37 | 943.07 | 979.30 | **3,678.74** |
| 979 | 338.88 | 366.06 | 365.94 | 400.59 | **1,471.47** |
| **OR - OREGON - Total** | **60,579.93** | **63,167.43** | **62,182.52** | **63,991.50** | **249,921.37** |

**STATE: PA - PENNSYLVANIA**

| REGISTRANT ZIP CODE 3 | 1ST QUARTER | 2ND QUARTER | 3RD QUARTER | 4TH QUARTER | **TOTAL GRAMS** |
|---|---|---|---|---|---|
| 150 | 6,621.40 | 6,494.96 | 6,572.83 | 6,655.31 | **26,344.50** |
| 151 | 5,445.24 | 5,508.83 | 5,461.86 | 5,590.49 | **22,006.42** |
| 152 | 11,350.88 | 11,313.10 | 11,199.40 | 11,702.09 | **45,565.47** |
| 153 | 2,458.68 | 2,451.16 | 2,457.52 | 2,604.52 | **9,971.88** |
| 154 | 1,906.65 | 1,894.74 | 1,740.17 | 1,940.79 | **7,482.35** |
| 155 | 944.30 | 953.16 | 976.74 | 1,024.41 | **3,898.61** |
| 156 | 4,407.00 | 4,411.98 | 4,341.68 | 4,362.38 | **17,523.04** |
| 157 | 1,396.64 | 1,416.40 | 1,314.66 | 1,285.93 | **5,413.63** |
| 158 | 1,248.78 | 1,285.28 | 1,276.63 | 1,288.02 | **5,098.71** |
| 159 | 1,465.59 | 1,381.22 | 1,363.63 | 1,383.98 | **5,594.43** |
| 160 | 3,137.55 | 3,154.21 | 3,192.87 | 3,347.89 | **12,832.52** |
| 161 | 2,721.80 | 2,663.93 | 2,702.67 | 2,702.92 | **10,791.32** |
| 162 | 968.45 | 980.10 | 977.26 | 934.51 | **3,860.32** |
| 163 | 1,913.04 | 1,839.57 | 1,780.72 | 1,817.49 | **7,350.83** |
| 164 | 1,000.01 | 939.11 | 1,004.54 | 1,047.55 | **3,991.21** |
| 165 | 4,226.55 | 4,167.38 | 4,181.79 | 4,319.92 | **16,895.65** |
| 166 | 2,893.04 | 2,916.83 | 2,812.74 | 2,799.57 | **11,422.17** |
| 167 | 591.45 | 592.79 | 591.82 | 571.13 | **2,347.19** |
| 168 | 2,885.94 | 2,710.57 | 2,761.48 | 2,788.58 | **11,146.56** |
| 169 | 511.20 | 439.92 | 484.52 | 532.78 | **1,968.43** |
| 170 | 11,878.78 | 11,937.95 | 11,731.78 | 11,899.54 | **47,448.05** |
| 171 | 2,990.35 | 3,148.84 | 3,015.22 | 3,115.22 | **12,269.64** |
| 172 | 3,382.14 | 3,239.78 | 3,376.54 | 3,409.18 | **13,407.63** |
| 173 | 5,479.34 | 5,398.02 | 5,680.70 | 5,694.23 | **22,252.30** |
| 174 | 4,071.19 | 4,204.16 | 4,046.41 | 4,128.48 | **16,450.24** |
| 175 | 3,862.69 | 3,862.68 | 3,644.10 | 3,727.30 | **15,096.76** |
| 176 | 4,097.94 | 4,327.06 | 4,225.90 | 4,239.43 | **16,890.33** |
| 177 | 2,224.00 | 2,157.72 | 2,113.76 | 2,256.69 | **8,752.18** |

| | | | | | |
|---|---|---|---|---|---|
| 178 | 2,691.66 | 2,583.46 | 2,616.49 | 2,654.54 | **10,546.15** |
| 179 | 877.24 | 816.24 | 825.59 | 824.15 | **3,343.22** |
| 180 | 5,596.90 | 6,007.09 | 6,017.10 | 5,959.34 | **23,580.42** |
| 181 | 2,773.06 | 3,164.67 | 2,959.65 | 2,785.98 | **11,683.37** |
| 182 | 927.58 | 940.45 | 950.36 | 928.05 | **3,746.45** |
| 183 | 1,462.45 | 1,484.96 | 1,543.92 | 1,463.71 | **5,955.04** |
| 184 | 2,484.13 | 2,535.66 | 2,728.95 | 2,574.24 | **10,322.98** |
| 185 | 3,849.46 | 3,777.76 | 3,996.53 | 3,719.77 | **15,343.51** |
| 186 | 1,928.13 | 1,873.43 | 2,005.67 | 1,894.84 | **7,702.07** |
| 187 | 16,419.90 | 17,304.06 | 18,075.61 | 16,518.03 | **68,317.60** |
| 188 | 607.94 | 614.98 | 614.43 | 617.29 | **2,454.65** |
| 189 | 7,645.13 | 7,615.09 | 7,543.23 | 7,543.83 | **30,347.28** |
| 190 | 22,854.93 | 23,364.99 | 22,846.56 | 23,266.14 | **92,332.62** |
| 191 | 20,525.53 | 21,162.95 | 20,274.20 | 20,897.15 | **82,859.83** |
| 193 | 10,421.40 | 10,328.16 | 10,117.81 | 10,352.20 | **41,219.58** |
| 194 | 11,217.77 | 11,346.04 | 11,056.26 | 11,230.34 | **44,850.42** |
| 195 | 1,898.75 | 1,857.24 | 1,863.39 | 1,915.17 | **7,534.55** |
| 196 | 3,137.42 | 3,073.27 | 3,209.91 | 3,132.74 | **12,553.33** |
| **PA - PENNSYLVANIA - Total** | **213,400.03** | **215,641.98** | **214,275.61** | **215,447.84** | **858,765.46** |

**STATE: PR - PUERTO RICO**

| REGISTRANT ZIP CODE 3 | 1ST QUARTER | 2ND QUARTER | 3RD QUARTER | 4TH QUARTER | TOTAL GRAMS |
|---|---|---|---|---|---|
| 006 | 2,197.55 | 2,084.17 | 2,131.48 | 2,191.07 | **8,604.27** |
| 007 | 2,091.40 | 2,138.57 | 2,151.87 | 2,285.66 | **8,667.50** |
| 009 | 5,281.20 | 4,981.33 | 5,008.74 | 5,312.57 | **20,583.83** |
| **PR - PUERTO RICO - Total** | **9,570.15** | **9,204.07** | **9,292.09** | **9,789.29** | **37,855.60** |

**STATE: RI - RHODE ISLAND**

| REGISTRANT ZIP CODE 3 | 1ST QUARTER | 2ND QUARTER | 3RD QUARTER | 4TH QUARTER | TOTAL GRAMS |
|---|---|---|---|---|---|
| 028 | 24,110.00 | 24,436.82 | 24,991.70 | 24,638.37 | **98,176.89** |
| 029 | 13,982.55 | 13,951.08 | 14,387.98 | 14,017.43 | **56,339.05** |
| **RI - RHODE ISLAND - Total** | **38,092.55** | **38,387.90** | **39,379.68** | **38,655.81** | **154,515.93** |

**STATE: SC - SOUTH CAROLINA**

| REGISTRANT ZIP CODE 3 | 1ST QUARTER | 2ND QUARTER | 3RD QUARTER | 4TH QUARTER | TOTAL GRAMS |
|---|---|---|---|---|---|
| 290 | 12,248.12 | 11,898.83 | 12,107.35 | 12,355.06 | **48,609.35** |
| 291 | 9,925.99 | 9,533.76 | 9,656.97 | 9,796.77 | **38,913.49** |
| 292 | 10,506.55 | 9,924.22 | 10,486.09 | 10,208.35 | **41,125.21** |
| 293 | 8,723.08 | 8,394.19 | 8,626.83 | 8,806.04 | **34,550.13** |
| 294 | 33,840.12 | 33,122.31 | 35,253.86 | 34,658.48 | **136,874.77** |
| 295 | 18,273.45 | 18,075.70 | 18,565.23 | 18,643.35 | **73,557.72** |
| 296 | 28,955.58 | 28,212.96 | 30,040.86 | 29,811.91 | **117,021.31** |
| 297 | 8,160.57 | 8,097.23 | 8,220.48 | 8,404.85 | **32,883.14** |

| | | | | | |
|---|---|---|---|---|---|
| 298 | 4,580.89 | 4,469.43 | 4,446.02 | 4,457.72 | **17,954.06** |
| 299 | 4,075.23 | 3,966.66 | 4,204.97 | 4,021.28 | **16,268.14** |
| **SC - SOUTH CAROLINA - Total** | **139,289.57** | **135,695.31** | **141,608.66** | **141,163.81** | **557,757.34** |

**STATE: SD - SOUTH DAKOTA**

| REGISTRANT ZIP CODE 3 | 1ST QUARTER | 2ND QUARTER | 3RD QUARTER | 4TH QUARTER | TOTAL GRAMS |
|---|---|---|---|---|---|
| 570 | 2,201.03 | 2,082.26 | 2,242.85 | 2,168.13 | **8,694.28** |
| 571 | 8,562.87 | 7,818.21 | 7,837.16 | 8,121.49 | **32,339.73** |
| 572 | 682.49 | 639.48 | 642.93 | 664.19 | **2,629.08** |
| 573 | 619.07 | 634.03 | 648.85 | 623.18 | **2,525.13** |
| 574 | 773.04 | 712.68 | 736.63 | 728.70 | **2,951.06** |
| 575 | 273.00 | 284.24 | 284.56 | 326.98 | **1,168.78** |
| 576 | 51.10 | 63.89 | 47.19 | 40.71 | **202.90** |
| 577 | 2,635.54 | 2,606.70 | 2,763.11 | 2,655.26 | **10,660.60** |
| **SD - SOUTH DAKOTA - Total** | **15,798.14** | **14,841.50** | **15,203.28** | **15,328.63** | **61,171.55** |

**STATE: TN - TENNESSEE**

| REGISTRANT ZIP CODE 3 | 1ST QUARTER | 2ND QUARTER | 3RD QUARTER | 4TH QUARTER | TOTAL GRAMS |
|---|---|---|---|---|---|
| 370 | 20,985.55 | 20,702.56 | 21,530.96 | 20,921.20 | **84,140.27** |
| 371 | 13,296.47 | 13,166.78 | 13,606.57 | 13,708.76 | **53,778.58** |
| 372 | 14,660.40 | 15,040.93 | 15,256.25 | 15,417.49 | **60,375.07** |
| 373 | 12,183.26 | 11,930.30 | 12,261.85 | 12,076.73 | **48,452.14** |
| 374 | 9,116.69 | 8,927.84 | 9,210.06 | 8,895.77 | **36,150.36** |
| 376 | 4,206.60 | 4,187.01 | 4,258.67 | 4,188.22 | **16,840.51** |
| 377 | 4,525.86 | 4,481.97 | 4,589.87 | 4,497.13 | **18,094.83** |
| 378 | 7,027.44 | 6,791.30 | 7,217.27 | 7,043.82 | **28,079.84** |
| 379 | 11,867.60 | 11,716.55 | 11,988.00 | 12,166.94 | **47,739.09** |
| 380 | 10,015.37 | 10,188.37 | 10,463.08 | 10,597.67 | **41,264.50** |
| 381 | 12,169.55 | 12,611.55 | 12,870.08 | 12,879.28 | **50,530.46** |
| 382 | 3,294.80 | 3,230.74 | 3,301.74 | 3,142.49 | **12,969.77** |
| 383 | 4,744.74 | 4,767.02 | 4,917.46 | 4,708.91 | **19,138.13** |
| 384 | 3,393.79 | 3,321.36 | 3,405.85 | 3,381.34 | **13,502.33** |
| 385 | 2,206.12 | 2,238.66 | 2,233.01 | 2,174.76 | **8,852.55** |
| **TN - TENNESSEE - Total** | **133,694.24** | **133,302.95** | **137,110.71** | **135,800.52** | **539,908.42** |

**STATE: TX - TEXAS**

| REGISTRANT ZIP CODE 3 | 1ST QUARTER | 2ND QUARTER | 3RD QUARTER | 4TH QUARTER | TOTAL GRAMS |
|---|---|---|---|---|---|
| 750 | 53,645.99 | 53,634.79 | 56,108.76 | 57,045.92 | **220,435.46** |
| 751 | 9,243.13 | 9,526.36 | 10,015.47 | 10,204.07 | **38,989.03** |
| 752 | 26,851.67 | 28,315.16 | 28,289.79 | 29,437.43 | **112,894.04** |
| 753 | 51.02 | 51.92 | 41.41 | 49.96 | **194.31** |
| 754 | 4,350.55 | 4,495.99 | 4,563.22 | 4,906.28 | **18,316.04** |
| 755 | 1,266.55 | 1,173.26 | 1,326.55 | 1,352.52 | **5,118.88** |

| | | | | | |
|---|---|---|---|---|---|
| 756 | 3,875.56 | 4,001.92 | 4,276.64 | 4,227.43 | **16,381.55** |
| 757 | 5,157.41 | 5,135.48 | 5,242.46 | 5,357.34 | **20,892.70** |
| 758 | 858.88 | 780.19 | 836.57 | 841.78 | **3,317.42** |
| 759 | 2,584.65 | 2,462.03 | 2,624.42 | 2,687.35 | **10,358.46** |
| 760 | 24,994.43 | 24,512.16 | 25,873.51 | 25,748.87 | **101,128.97** |
| 761 | 15,047.06 | 15,292.17 | 15,667.98 | 15,889.51 | **61,896.71** |
| 762 | 12,338.02 | 12,554.05 | 12,995.21 | 13,380.22 | **51,267.50** |
| 763 | 3,616.56 | 3,502.77 | 3,674.42 | 3,539.75 | **14,333.50** |
| 764 | 1,449.87 | 1,478.37 | 1,467.84 | 1,404.38 | **5,800.45** |
| 765 | 6,019.28 | 5,826.93 | 6,321.15 | 6,102.18 | **24,269.55** |
| 766 | 1,170.44 | 1,217.39 | 1,157.16 | 1,219.43 | **4,764.43** |
| 767 | 3,620.63 | 3,399.50 | 3,673.14 | 3,636.67 | **14,329.95** |
| 768 | 818.14 | 864.09 | 832.78 | 868.66 | **3,383.67** |
| 769 | 1,847.49 | 1,761.31 | 1,807.32 | 1,917.45 | **7,333.58** |
| 770 | 40,383.78 | 41,990.26 | 42,049.68 | 42,543.61 | **166,967.33** |
| 772 | 43.18 | 34.29 | 25.19 | 17.83 | **120.49** |
| 773 | 30,148.70 | 30,417.52 | 31,819.89 | 31,869.59 | **124,255.70** |
| 774 | 22,069.75 | 21,641.73 | 22,633.65 | 23,261.12 | **89,606.25** |
| 775 | 24,021.97 | 24,504.46 | 25,293.40 | 25,829.50 | **99,649.33** |
| 776 | 5,016.12 | 4,984.02 | 5,412.02 | 5,252.29 | **20,664.45** |
| 777 | 2,507.28 | 2,460.30 | 2,644.49 | 2,583.77 | **10,195.83** |
| 778 | 7,242.91 | 6,958.61 | 7,244.98 | 7,233.40 | **28,679.91** |
| 779 | 2,211.34 | 2,191.68 | 2,333.92 | 2,327.38 | **9,064.32** |
| 780 | 5,074.83 | 5,022.91 | 5,261.61 | 5,314.56 | **20,673.91** |
| 781 | 6,396.42 | 6,333.43 | 6,997.60 | 6,670.21 | **26,397.67** |
| 782 | 27,883.77 | 27,945.78 | 29,571.06 | 28,941.74 | **114,342.35** |
| 783 | 1,925.16 | 2,046.14 | 2,002.88 | 2,109.94 | **8,084.11** |
| 784 | 5,046.14 | 4,782.55 | 5,011.38 | 5,208.79 | **20,048.85** |
| 785 | 3,357.75 | 3,305.44 | 3,463.12 | 3,538.38 | **13,664.69** |
| 786 | 18,122.31 | 18,424.99 | 19,555.08 | 19,487.57 | **75,589.95** |
| 787 | 29,361.71 | 30,592.17 | 31,792.43 | 32,990.00 | **124,736.31** |
| 788 | 652.60 | 642.77 | 724.59 | 670.21 | **2,690.16** |
| 789 | 725.31 | 734.44 | 819.52 | 769.10 | **3,048.37** |
| 790 | 906.91 | 836.42 | 799.36 | 936.94 | **3,479.62** |
| 791 | 1,942.02 | 2,089.78 | 1,994.88 | 2,232.08 | **8,258.76** |
| 792 | 84.08 | 91.29 | 76.16 | 80.27 | **331.80** |
| 793 | 493.40 | 472.02 | 482.95 | 547.98 | **1,996.34** |
| 794 | 4,770.67 | 4,722.96 | 4,721.57 | 5,011.32 | **19,226.50** |
| 795 | 523.70 | 556.02 | 502.00 | 550.58 | **2,132.30** |
| 796 | 2,314.08 | 2,364.04 | 2,364.09 | 2,499.79 | **9,542.00** |

| | | | | | |
|---|---|---|---|---|---|
| 797 | 6,602.56 | 6,666.97 | 7,053.06 | 7,172.96 | **27,495.55** |
| 798 | 245.15 | 247.71 | 246.16 | 252.04 | **991.06** |
| 799 | 3,256.17 | 3,378.66 | 3,877.51 | 3,335.57 | **13,847.91** |
| **TX - TEXAS - Total** | **432,137.12** | **436,425.19** | **453,570.03** | **459,055.68** | **1,781,188.02** |

**STATE: UT - UTAH**

| REGISTRANT ZIP CODE 3 | 1ST QUARTER | 2ND QUARTER | 3RD QUARTER | 4TH QUARTER | TOTAL GRAMS |
|---|---|---|---|---|---|
| 840 | 44,095.47 | 44,666.27 | 45,665.84 | 47,857.17 | **182,284.76** |
| 841 | 17,205.76 | 17,390.24 | 17,896.57 | 18,674.10 | **71,166.68** |
| 843 | 4,725.93 | 4,600.90 | 4,738.56 | 4,939.11 | **19,004.51** |
| 844 | 4,384.45 | 4,332.32 | 4,618.39 | 4,663.00 | **17,998.16** |
| 845 | 651.88 | 700.90 | 627.35 | 671.93 | **2,652.07** |
| 846 | 9,171.07 | 9,171.45 | 9,628.68 | 9,997.40 | **37,968.61** |
| 847 | 6,743.33 | 6,984.23 | 6,867.50 | 7,019.20 | **27,614.26** |
| **UT - UTAH - Total** | **86,977.90** | **87,846.30** | **90,042.90** | **93,821.93** | **358,689.03** |

**STATE: VA - VIRGINIA**

| REGISTRANT ZIP CODE 3 | 1ST QUARTER | 2ND QUARTER | 3RD QUARTER | 4TH QUARTER | TOTAL GRAMS |
|---|---|---|---|---|---|
| 201 | 18,236.50 | 19,107.10 | 19,234.77 | 19,151.42 | **75,729.79** |
| 220 | 10,613.91 | 11,095.39 | 11,124.69 | 10,295.26 | **43,129.25** |
| 221 | 6,731.15 | 7,502.84 | 7,390.98 | 7,247.29 | **28,872.26** |
| 222 | 4,795.97 | 4,905.54 | 5,092.61 | 4,964.46 | **19,758.58** |
| 223 | 4,233.64 | 4,236.28 | 4,335.79 | 4,375.87 | **17,181.59** |
| 224 | 4,878.77 | 4,810.00 | 4,782.32 | 4,962.54 | **19,433.62** |
| 225 | 2,157.56 | 2,071.48 | 2,126.74 | 2,192.79 | **8,548.56** |
| 226 | 3,380.71 | 3,494.15 | 3,524.11 | 3,456.95 | **13,855.92** |
| 227 | 1,141.79 | 1,163.67 | 1,088.25 | 1,244.37 | **4,638.08** |
| 228 | 3,180.04 | 2,912.94 | 3,038.30 | 3,197.80 | **12,329.08** |
| 229 | 6,526.43 | 6,454.34 | 6,688.01 | 6,674.25 | **26,343.04** |
| 230 | 5,049.23 | 4,793.05 | 5,099.36 | 5,122.89 | **20,064.53** |
| 231 | 12,168.50 | 12,046.09 | 12,158.24 | 12,596.01 | **48,968.84** |
| 232 | 16,538.14 | 16,140.38 | 16,523.91 | 16,719.18 | **65,921.61** |
| 233 | 10,062.38 | 9,720.02 | 9,835.59 | 10,025.44 | **39,643.43** |
| 234 | 24,619.56 | 24,806.07 | 25,090.33 | 25,113.00 | **99,628.96** |
| 235 | 6,147.66 | 6,172.15 | 5,993.34 | 6,034.13 | **24,347.28** |
| 236 | 10,020.47 | 9,718.92 | 10,136.62 | 9,756.15 | **39,632.15** |
| 237 | 4,420.99 | 4,072.40 | 4,328.94 | 3,732.25 | **16,554.57** |
| 238 | 6,857.80 | 6,771.36 | 6,826.14 | 7,008.35 | **27,463.65** |
| 239 | 966.73 | 976.53 | 986.51 | 998.48 | **3,928.25** |
| 240 | 5,389.37 | 5,306.41 | 5,435.56 | 5,633.43 | **21,764.77** |
| 241 | 2,704.17 | 2,668.51 | 2,757.50 | 2,786.14 | **10,916.33** |
| 242 | 1,348.39 | 1,205.97 | 1,274.58 | 1,243.35 | **5,072.29** |

| | | | | | |
|---|---|---|---|---|---|
| 243 | 866.75 | 788.94 | 925.80 | 957.73 | **3,539.22** |
| 244 | 3,365.85 | 3,297.13 | 3,523.36 | 3,438.56 | **13,624.90** |
| 245 | 5,488.87 | 5,083.43 | 5,476.47 | 5,216.35 | **21,265.13** |
| 246 | 417.51 | 451.29 | 454.00 | 422.49 | **1,745.29** |
| **VA - VIRGINIA - Total** | **182,308.82** | **181,772.40** | **185,252.82** | **184,566.91** | **733,900.96** |

**STATE: VI - VIRGIN ISLANDS**

| REGISTRANT ZIP CODE 3 | 1ST QUARTER | 2ND QUARTER | 3RD QUARTER | 4TH QUARTER | TOTAL GRAMS |
|---|---|---|---|---|---|
| 008 | 308.38 | 333.01 | 348.04 | 335.67 | **1,325.10** |
| **VI - VIRGIN ISLANDS - Total** | **308.38** | **333.01** | **348.04** | **335.67** | **1,325.10** |

**STATE: VT - VERMONT**

| REGISTRANT ZIP CODE 3 | 1ST QUARTER | 2ND QUARTER | 3RD QUARTER | 4TH QUARTER | TOTAL GRAMS |
|---|---|---|---|---|---|
| 050 | 590.25 | 616.75 | 613.33 | 588.99 | **2,409.32** |
| 051 | 565.90 | 529.42 | 577.22 | 573.89 | **2,246.44** |
| 052 | 709.74 | 784.54 | 711.61 | 735.53 | **2,941.42** |
| 053 | 1,611.16 | 1,569.25 | 1,603.86 | 1,602.88 | **6,387.16** |
| 054 | 3,441.38 | 3,392.21 | 3,474.89 | 3,561.44 | **13,869.93** |
| 056 | 1,614.90 | 1,624.96 | 1,676.23 | 1,663.87 | **6,579.97** |
| 057 | 906.75 | 845.03 | 817.61 | 832.75 | **3,402.14** |
| 058 | 635.88 | 668.00 | 683.97 | 713.48 | **2,701.33** |
| **VT - VERMONT - Total** | **10,075.97** | **10,030.16** | **10,158.72** | **10,272.84** | **40,537.69** |

**STATE: WA - WASHINGTON**

| REGISTRANT ZIP CODE 3 | 1ST QUARTER | 2ND QUARTER | 3RD QUARTER | 4TH QUARTER | TOTAL GRAMS |
|---|---|---|---|---|---|
| 980 | 27,685.73 | 28,513.26 | 27,960.99 | 29,204.59 | **113,364.58** |
| 981 | 18,087.79 | 18,816.72 | 18,463.78 | 19,544.08 | **74,912.37** |
| 982 | 16,384.84 | 17,085.82 | 17,243.72 | 17,339.03 | **68,053.41** |
| 983 | 10,990.31 | 11,331.77 | 11,164.82 | 11,332.84 | **44,819.74** |
| 984 | 7,476.66 | 7,558.07 | 7,249.48 | 7,429.69 | **29,713.89** |
| 985 | 5,578.38 | 5,787.40 | 5,679.80 | 5,811.71 | **22,857.29** |
| 986 | 8,382.97 | 8,666.22 | 8,521.68 | 8,830.86 | **34,401.74** |
| 988 | 1,938.94 | 2,020.42 | 2,031.06 | 2,058.08 | **8,048.51** |
| 989 | 2,478.99 | 2,502.08 | 2,319.89 | 2,574.02 | **9,874.96** |
| 990 | 1,062.61 | 1,077.38 | 1,036.97 | 1,072.33 | **4,249.29** |
| 991 | 1,087.88 | 1,070.81 | 995.24 | 1,227.54 | **4,381.48** |
| 992 | 5,778.51 | 6,101.36 | 6,000.73 | 6,466.80 | **24,347.40** |
| 993 | 5,548.94 | 5,754.00 | 5,570.85 | 5,813.47 | **22,687.26** |
| 994 | 210.02 | 204.56 | 205.50 | 212.21 | **832.29** |
| **WA - WASHINGTON - Total** | **112,692.56** | **116,489.87** | **114,444.53** | **118,917.24** | **462,544.20** |

**STATE: WI - WISCONSIN**

| REGISTRANT ZIP CODE 3 | 1ST QUARTER | 2ND QUARTER | 3RD QUARTER | 4TH QUARTER | TOTAL GRAMS |
|---|---|---|---|---|---|
| 530 | 14,383.45 | 14,186.24 | 14,436.96 | 14,555.58 | **57,562.24** |

| | | | | | |
|---|---|---|---|---|---|
| 531 | 14,969.87 | 14,735.39 | 14,807.09 | 15,056.24 | **59,568.60** |
| 532 | 13,404.71 | 13,417.42 | 13,296.34 | 13,684.14 | **53,802.62** |
| 534 | 4,122.56 | 4,177.71 | 4,262.56 | 4,163.45 | **16,726.28** |
| 535 | 10,269.50 | 9,934.00 | 10,036.86 | 10,051.42 | **40,291.78** |
| 537 | 8,081.00 | 8,228.30 | 8,106.74 | 8,074.83 | **32,490.87** |
| 538 | 1,044.66 | 1,083.74 | 1,048.65 | 1,120.56 | **4,297.62** |
| 539 | 3,520.95 | 3,563.64 | 3,517.24 | 3,554.22 | **14,156.05** |
| 540 | 2,527.78 | 2,483.90 | 2,497.26 | 2,498.59 | **10,007.53** |
| 541 | 6,892.36 | 6,677.43 | 6,878.48 | 6,834.03 | **27,282.30** |
| 542 | 2,700.56 | 2,679.03 | 2,697.64 | 2,699.32 | **10,776.55** |
| 543 | 8,080.09 | 8,217.37 | 8,195.64 | 8,256.34 | **32,749.44** |
| 544 | 8,097.84 | 8,046.11 | 8,093.88 | 8,154.70 | **32,392.52** |
| 545 | 2,180.39 | 2,135.18 | 2,301.71 | 2,197.08 | **8,814.36** |
| 546 | 4,304.07 | 4,299.77 | 4,198.17 | 4,229.95 | **17,031.95** |
| 547 | 4,343.39 | 4,316.22 | 4,021.07 | 4,215.25 | **16,895.93** |
| 548 | 2,294.28 | 2,259.08 | 2,238.93 | 2,372.35 | **9,164.63** |
| 549 | 15,248.41 | 14,930.21 | 15,248.86 | 15,207.62 | **60,635.11** |
| **WI - WISCONSIN - Total** | **126,465.87** | **125,370.76** | **125,884.09** | **126,925.65** | **504,646.38** |

**STATE: WV - WEST VIRGINIA**

| REGISTRANT ZIP CODE 3 | 1ST QUARTER | 2ND QUARTER | 3RD QUARTER | 4TH QUARTER | **TOTAL GRAMS** |
|---|---|---|---|---|---|
| 247 | 331.03 | 306.94 | 297.02 | 313.30 | **1,248.29** |
| 248 | 198.51 | 181.09 | 207.72 | 227.49 | **814.80** |
| 249 | 421.90 | 388.11 | 408.14 | 402.95 | **1,621.11** |
| 250 | 860.09 | 904.31 | 861.84 | 906.28 | **3,532.52** |
| 251 | 674.29 | 728.95 | 751.33 | 774.93 | **2,929.50** |
| 252 | 456.02 | 417.69 | 417.27 | 442.53 | **1,733.52** |
| 253 | 3,525.09 | 3,356.22 | 3,382.85 | 3,227.71 | **13,491.87** |
| 254 | 2,650.34 | 2,763.13 | 2,758.50 | 2,759.61 | **10,931.57** |
| 255 | 2,312.55 | 2,434.87 | 2,343.32 | 2,322.28 | **9,413.03** |
| 256 | 288.64 | 237.56 | 230.30 | 298.27 | **1,054.76** |
| 257 | 1,771.33 | 1,744.85 | 1,814.95 | 1,910.19 | **7,241.32** |
| 258 | 1,199.14 | 1,107.35 | 1,126.78 | 1,059.17 | **4,492.45** |
| 259 | 244.09 | 235.57 | 251.52 | 228.83 | **960.01** |
| 260 | 1,412.62 | 1,483.29 | 1,412.52 | 1,473.07 | **5,781.50** |
| 261 | 2,271.47 | 2,350.78 | 2,243.85 | 2,295.55 | **9,161.66** |
| 262 | 345.17 | 367.14 | 343.84 | 344.27 | **1,400.42** |
| 263 | 970.15 | 1,051.01 | 1,026.82 | 1,017.22 | **4,065.20** |
| 264 | 257.24 | 234.05 | 234.86 | 237.16 | **963.32** |
| 265 | 2,682.33 | 2,632.71 | 2,759.76 | 2,738.41 | **10,813.20** |
| 266 | 124.18 | 131.53 | 139.45 | 154.76 | **549.93** |

| | | | | | |
|---|---|---|---|---|---|
| 267 | 298.47 | 307.26 | 326.99 | 320.96 | **1,253.69** |
| 268 | 228.74 | 239.23 | 253.78 | 253.22 | **974.97** |
| **WV - WEST VIRGINIA - Total** | **23,523.40** | **23,603.65** | **23,593.42** | **23,708.16** | **94,428.63** |

**STATE: WY - WYOMING**

| REGISTRANT ZIP CODE 3 | 1ST QUARTER | 2ND QUARTER | 3RD QUARTER | 4TH QUARTER | TOTAL GRAMS |
|---|---|---|---|---|---|
| 820 | 1,438.32 | 1,553.17 | 1,430.90 | 1,503.80 | **5,926.20** |
| 822 | 91.19 | 75.16 | 73.46 | 73.92 | **313.73** |
| 823 | 120.24 | 112.87 | 129.20 | 120.49 | **482.80** |
| 824 | 817.03 | 855.02 | 919.18 | 841.58 | **3,432.81** |
| 825 | 342.78 | 322.40 | 286.65 | 358.84 | **1,310.66** |
| 826 | 1,033.02 | 991.88 | 1,053.55 | 1,088.07 | **4,166.51** |
| 827 | 535.39 | 499.85 | 532.30 | 539.37 | **2,106.91** |
| 828 | 612.10 | 548.61 | 567.98 | 531.89 | **2,260.58** |
| 829 | 1,203.95 | 1,166.23 | 1,204.10 | 1,235.09 | **4,809.37** |
| 830 | 541.95 | 530.60 | 593.86 | 623.85 | **2,290.26** |
| 831 | 134.50 | 120.94 | 139.78 | 127.65 | **522.87** |
| **WY - WYOMING - Total** | **6,870.46** | **6,776.74** | **6,930.96** | **7,044.55** | **27,622.71** |

**DRUG: 1105 - METHAMPHETAMINE**

**STATE: AK - ALASKA**

| REGISTRANT ZIP CODE 3 | 1ST QUARTER | 2ND QUARTER | 3RD QUARTER | 4TH QUARTER | TOTAL GRAMS |
|---|---|---|---|---|---|
| 995 | 0.40 | 0 | 0 | 0 | **0.40** |
| 997 | 1.21 | 0.40 | 2.41 | 0 | **4.02** |
| 998 | 0.40 | 1.61 | 1.21 | 0 | **3.21** |
| **AK - ALASKA - Total** | **2.01** | **2.01** | **3.62** | **0.00** | **7.63** |

**STATE: AL - ALABAMA**

| REGISTRANT ZIP CODE 3 | 1ST QUARTER | 2ND QUARTER | 3RD QUARTER | 4TH QUARTER | TOTAL GRAMS |
|---|---|---|---|---|---|
| 350 | 3.62 | 2.41 | 2.81 | 3.21 | **12.05** |
| 351 | 3.62 | 3.21 | 2.41 | 3.62 | **12.86** |
| 352 | 3.62 | 6.03 | 4.82 | 5.62 | **20.09** |
| 354 | 2.01 | 2.01 | 0.80 | 1.61 | **6.43** |
| 355 | 3.21 | 3.21 | 5.63 | 4.42 | **16.47** |
| 356 | 5.62 | 6.43 | 6.83 | 7.23 | **26.11** |
| 357 | 1.21 | 1.61 | 2.01 | 2.41 | **7.23** |
| 359 | 0.40 | 0.80 | 0.40 | 0 | **1.61** |
| 360 | 0 | 0 | 0 | 0.40 | **0.40** |
| 361 | 2.81 | 0 | 0 | 0 | **2.81** |
| 362 | 1.61 | 4.02 | 4.42 | 4.42 | **14.47** |
| 363 | 0 | 0.80 | 0 | 0.40 | **1.21** |
| 365 | 0 | 0.80 | 1.61 | 0.40 | **2.81** |
| 366 | 2.41 | 2.01 | 2.81 | 4.02 | **11.25** |

| | | | | | |
|---|---|---|---|---|---|
| **AL - ALABAMA - Total** | **30.14** | **33.35** | **34.55** | **37.77** | **135.81** |

**STATE: AR - ARKANSAS**

| REGISTRANT ZIP CODE 3 | 1ST QUARTER | 2ND QUARTER | 3RD QUARTER | 4TH QUARTER | TOTAL GRAMS |
|---|---|---|---|---|---|
| 717 | 0 | 0 | 0 | 2.41 | **2.41** |
| 719 | 0 | 0.80 | 0 | 0 | **0.80** |
| 720 | 1.21 | 0.40 | 1.61 | 2.01 | **5.22** |
| 721 | 0.80 | 0.80 | 0.80 | 0.80 | **3.21** |
| 722 | 1.21 | 0.40 | 0 | 2.41 | **4.02** |
| 723 | 0 | 0 | 0 | 1.21 | **1.21** |
| 724 | 0 | 0 | 0 | 0.80 | **0.80** |
| 727 | 4.02 | 2.41 | 0 | 0 | **6.43** |
| 729 | 1.61 | 2.41 | 0 | 0 | **4.02** |
| **AR - ARKANSAS - Total** | **8.84** | **7.23** | **2.41** | **9.64** | **28.13** |

**STATE: AZ - ARIZONA**

| REGISTRANT ZIP CODE 3 | 1ST QUARTER | 2ND QUARTER | 3RD QUARTER | 4TH QUARTER | TOTAL GRAMS |
|---|---|---|---|---|---|
| 850 | 2.41 | 0 | 1.21 | 2.01 | **5.63** |
| 852 | 29.33 | 33.35 | 58.67 | 33.75 | **155.09** |
| 853 | 1.21 | 4.02 | 2.81 | 1.61 | **9.64** |
| 855 | 1.61 | 2.41 | 1.21 | 1.61 | **6.83** |
| 856 | 1.21 | 0 | 0 | 0 | **1.21** |
| 857 | 0.80 | 0.40 | 0 | 0 | **1.21** |
| 863 | 0.40 | 0 | 0 | 0 | **0.40** |
| **AZ - ARIZONA - Total** | **36.97** | **40.18** | **63.89** | **38.97** | **180.01** |

**STATE: CA - CALIFORNIA**

| REGISTRANT ZIP CODE 3 | 1ST QUARTER | 2ND QUARTER | 3RD QUARTER | 4TH QUARTER | TOTAL GRAMS |
|---|---|---|---|---|---|
| 900 | 27.32 | 27.32 | 29.33 | 24.10 | **108.07** |
| 902 | 4.42 | 1.61 | 1.61 | 1.21 | **8.84** |
| 905 | 0 | 0.40 | 0 | 0 | **0.40** |
| 906 | 0.40 | 0 | 0.80 | 0 | **1.21** |
| 907 | 0 | 0.40 | 0 | 0 | **0.40** |
| 908 | 1.21 | 1.61 | 3.21 | 1.61 | **7.63** |
| 910 | 2.81 | 7.23 | 5.22 | 4.82 | **20.09** |
| 911 | 1.61 | 2.81 | 2.41 | 0.40 | **7.23** |
| 912 | 3.62 | 5.62 | 5.62 | 6.83 | **21.70** |
| 913 | 20.89 | 26.92 | 18.48 | 17.28 | **83.57** |
| 914 | 4.02 | 3.62 | 3.21 | 3.62 | **14.47** |
| 917 | 17.68 | 21.69 | 24.91 | 25.31 | **89.58** |
| 918 | 0 | 0 | 0.40 | 0.40 | **0.80** |
| 919 | 5.62 | 2.01 | 4.02 | 2.81 | **14.46** |
| 920 | 38.97 | 34.95 | 36.56 | 41.38 | **151.85** |

| | | | | | |
|---|---|---|---|---|---|
| 921 | 12.46 | 10.85 | 10.85 | 6.83 | **40.99** |
| 922 | 2.81 | 1.21 | 0.40 | 2.41 | **6.83** |
| 923 | 5.22 | 7.23 | 2.81 | 4.02 | **19.28** |
| 925 | 6.83 | 4.82 | 7.63 | 8.03 | **27.32** |
| 926 | 18.48 | 20.09 | 19.29 | 14.87 | **72.72** |
| 927 | 1.61 | 9.24 | 3.21 | 7.23 | **21.29** |
| 928 | 10.45 | 12.45 | 9.24 | 14.87 | **47.01** |
| 930 | 2.41 | 2.01 | 2.81 | 2.81 | **10.05** |
| 931 | 2.01 | 2.01 | 3.62 | 2.01 | **9.64** |
| 934 | 1.21 | 0 | 1.21 | 0.80 | **3.21** |
| 935 | 0.40 | 0.80 | 2.41 | 1.61 | **5.22** |
| 937 | 1.61 | 1.61 | 0.80 | 0 | **4.02** |
| 939 | 1.61 | 2.81 | 1.21 | 0 | **5.63** |
| 940 | 10.85 | 9.64 | 8.44 | 5.22 | **34.15** |
| 941 | 13.66 | 5.63 | 7.23 | 8.04 | **34.55** |
| 943 | 0.40 | 0.40 | 0.80 | 0 | **1.61** |
| 944 | 0 | 0.40 | 0 | 0 | **0.40** |
| 945 | 4.82 | 3.62 | 6.03 | 6.03 | **20.49** |
| 946 | 3.62 | 2.01 | 0 | 0 | **5.63** |
| 947 | 0 | 1.21 | 0 | 0 | **1.21** |
| 948 | 0.40 | 0 | 0 | 0 | **0.40** |
| 949 | 0.80 | 2.41 | 4.42 | 2.41 | **10.04** |
| 950 | 7.63 | 6.83 | 10.85 | 6.83 | **32.14** |
| 951 | 12.46 | 12.86 | 14.06 | 17.68 | **57.05** |
| 953 | 5.62 | 3.62 | 3.62 | 2.41 | **15.27** |
| 954 | 0.80 | 0.40 | 0 | 0 | **1.21** |
| 955 | 1.21 | 0.80 | 0.80 | 1.21 | **4.02** |
| 956 | 4.02 | 2.41 | 5.22 | 4.42 | **16.07** |
| 957 | 2.01 | 0.40 | 0.80 | 0.40 | **3.62** |
| 958 | 4.02 | 4.42 | 5.22 | 3.21 | **16.87** |
| 959 | 4.42 | 4.82 | 4.42 | 6.03 | **19.69** |
| 960 | 0 | 0.80 | 0 | 1.21 | **2.01** |
| **CA - CALIFORNIA - Total** | **272.41** | **274.01** | **273.20** | **260.34** | **1,079.95** |

**STATE: CO - COLORADO**

| REGISTRANT ZIP CODE 3 | 1ST QUARTER | 2ND QUARTER | 3RD QUARTER | 4TH QUARTER | TOTAL GRAMS |
|---|---|---|---|---|---|
| 800 | 11.65 | 13.26 | 14.47 | 14.06 | **53.44** |
| 801 | 16.47 | 19.69 | 19.69 | 11.65 | **67.50** |
| 802 | 14.06 | 16.07 | 21.70 | 20.49 | **72.33** |
| 803 | 1.21 | 1.21 | 0.40 | 2.41 | **5.22** |
| 804 | 3.21 | 0.80 | 2.01 | 4.42 | **10.45** |

| | | | | | |
|---|---|---|---|---|---|
| 805 | 2.41 | 2.81 | 0.40 | 2.41 | **8.04** |
| 806 | 1.21 | 0 | 0.80 | 0 | **2.01** |
| 809 | 2.41 | 0.80 | 0.80 | 0.40 | **4.42** |
| 810 | 0 | 0 | 1.21 | 0.40 | **1.61** |
| 811 | 0 | 1.21 | 0 | 0 | **1.21** |
| 812 | 0.80 | 2.41 | 1.21 | 0 | **4.42** |
| 813 | 1.21 | 0 | 0 | 0 | **1.21** |
| 816 | 2.01 | 0.80 | 0.80 | 0 | **3.62** |
| **CO - COLORADO - Total** | **56.66** | **59.07** | **63.49** | **56.25** | **235.47** |

**STATE: CT - CONNECTICUT**

| REGISTRANT ZIP CODE 3 | 1ST QUARTER | 2ND QUARTER | 3RD QUARTER | 4TH QUARTER | TOTAL GRAMS |
|---|---|---|---|---|---|
| 060 | 7.23 | 6.03 | 8.04 | 9.64 | **30.94** |
| 061 | 3.62 | 1.61 | 1.61 | 0.80 | **7.64** |
| 064 | 0 | 1.21 | 0 | 0.40 | **1.61** |
| 065 | 4.42 | 6.03 | 2.81 | 3.62 | **16.88** |
| 066 | 0.80 | 0.40 | 0.40 | 0.80 | **2.41** |
| 067 | 1.61 | 2.41 | 1.61 | 0.80 | **6.43** |
| 068 | 5.63 | 5.63 | 4.42 | 8.44 | **24.11** |
| 069 | 1.21 | 1.61 | 1.61 | 1.61 | **6.03** |
| **CT - CONNECTICUT - Total** | **24.51** | **24.91** | **20.49** | **26.12** | **96.03** |

**STATE: DC - DISTRICT OF COLUMBIA**

| REGISTRANT ZIP CODE 3 | 1ST QUARTER | 2ND QUARTER | 3RD QUARTER | 4TH QUARTER | TOTAL GRAMS |
|---|---|---|---|---|---|
| 200 | 5.22 | 11.65 | 7.23 | 5.62 | **29.73** |
| **DC - DISTRICT OF COLUMBIA -** | **5.22** | **11.65** | **7.23** | **5.62** | **29.73** |

**STATE: DE - DELAWARE**

| REGISTRANT ZIP CODE 3 | 1ST QUARTER | 2ND QUARTER | 3RD QUARTER | 4TH QUARTER | TOTAL GRAMS |
|---|---|---|---|---|---|
| 197 | 0.40 | 0.80 | 0.80 | 0.80 | **2.81** |
| 198 | 1.21 | 3.21 | 2.41 | 1.21 | **8.04** |
| 199 | 1.61 | 1.61 | 1.21 | 2.41 | **6.83** |
| **DE - DELAWARE - Total** | **3.21** | **5.63** | **4.42** | **4.42** | **17.68** |

**STATE: FL - FLORIDA**

| REGISTRANT ZIP CODE 3 | 1ST QUARTER | 2ND QUARTER | 3RD QUARTER | 4TH QUARTER | TOTAL GRAMS |
|---|---|---|---|---|---|
| 320 | 1.61 | 1.61 | 2.41 | 2.41 | **8.04** |
| 321 | 3.21 | 1.21 | 1.61 | 2.01 | **8.03** |
| 322 | 1.61 | 2.81 | 3.21 | 2.41 | **10.05** |
| 324 | 1.61 | 1.21 | 1.21 | 0.80 | **4.82** |
| 325 | 2.81 | 3.62 | 3.62 | 4.82 | **14.87** |
| 326 | 4.02 | 3.21 | 4.42 | 3.62 | **15.27** |
| 327 | 8.84 | 1.21 | 9.64 | 3.62 | **23.30** |
| 328 | 2.01 | 4.82 | 3.62 | 2.81 | **13.26** |

| | | | | | |
|---|---|---|---|---|---|
| 329 | 3.62 | 4.42 | 3.62 | 4.82 | **16.48** |
| 330 | 0.40 | 0.80 | 2.81 | 0.80 | **4.82** |
| 331 | 4.02 | 2.81 | 2.01 | 4.42 | **13.26** |
| 333 | 2.41 | 0 | 0.80 | 0.80 | **4.02** |
| 334 | 2.41 | 1.21 | 0.80 | 2.01 | **6.43** |
| 335 | 0.40 | 0 | 0.80 | 0.40 | **1.61** |
| 336 | 0 | 0.80 | 0.40 | 0 | **1.21** |
| 337 | 2.01 | 1.61 | 1.61 | 1.61 | **6.83** |
| 338 | 1.61 | 2.81 | 1.61 | 0.40 | **6.43** |
| 339 | 4.02 | 3.62 | 3.21 | 3.62 | **14.47** |
| 341 | 0.40 | 1.61 | 0 | 0 | **2.01** |
| 342 | 1.61 | 4.82 | 3.21 | 3.62 | **13.26** |
| 346 | 1.21 | 2.81 | 2.01 | 2.01 | **8.04** |
| **FL - FLORIDA - Total** | **49.82** | **47.01** | **52.64** | **47.01** | **196.48** |

**STATE: GA - GEORGIA**

| REGISTRANT ZIP CODE 3 | 1ST QUARTER | 2ND QUARTER | 3RD QUARTER | 4TH QUARTER | TOTAL GRAMS |
|---|---|---|---|---|---|
| 300 | 4.42 | 3.62 | 7.23 | 5.63 | **20.90** |
| 301 | 4.42 | 4.82 | 4.82 | 6.03 | **20.09** |
| 302 | 0.40 | 0 | 0 | 2.81 | **3.21** |
| 303 | 4.02 | 3.62 | 4.42 | 3.62 | **15.67** |
| 304 | 2.81 | 0.80 | 1.21 | 1.61 | **6.43** |
| 306 | 0 | 0 | 0 | 0.40 | **0.40** |
| 310 | 3.21 | 2.01 | 2.41 | 2.41 | **10.05** |
| 313 | 0.80 | 0.40 | 0.80 | 0 | **2.01** |
| 315 | 3.62 | 1.61 | 1.61 | 2.81 | **9.64** |
| 316 | 0.40 | 0 | 0 | 0 | **0.40** |
| **GA - GEORGIA - Total** | **24.11** | **16.88** | **22.50** | **25.31** | **88.80** |

**STATE: HI - HAWAII**

| REGISTRANT ZIP CODE 3 | 1ST QUARTER | 2ND QUARTER | 3RD QUARTER | 4TH QUARTER | TOTAL GRAMS |
|---|---|---|---|---|---|
| 967 | 2.01 | 2.01 | 1.61 | 2.81 | **8.44** |
| 968 | 3.21 | 2.81 | 3.21 | 4.42 | **13.66** |
| **HI - HAWAII - Total** | **5.22** | **4.82** | **4.82** | **7.23** | **22.10** |

**STATE: IA - IOWA**

| REGISTRANT ZIP CODE 3 | 1ST QUARTER | 2ND QUARTER | 3RD QUARTER | 4TH QUARTER | TOTAL GRAMS |
|---|---|---|---|---|---|
| 500 | 0.40 | 0 | 0 | 0.40 | **0.80** |
| 501 | 0.40 | 0 | 0 | 0 | **0.40** |
| 502 | 3.62 | 8.04 | 7.23 | 7.63 | **26.52** |
| 503 | 4.42 | 2.01 | 4.42 | 2.01 | **12.86** |
| 504 | 2.41 | 2.41 | 4.02 | 1.21 | **10.05** |
| 505 | 0.80 | 0.80 | 0 | 1.21 | **2.81** |

| | | | | | |
|---|---|---|---|---|---|
| 506 | 0.40 | 0 | 0 | 0 | **0.40** |
| 515 | 2.81 | 2.01 | 2.41 | 0.80 | **8.04** |
| 516 | 0.40 | 0 | 0.80 | 0 | **1.21** |
| 522 | 0.40 | 0 | 0 | 0 | **0.40** |
| 528 | 1.21 | 2.41 | 2.41 | 1.21 | **7.23** |
| **IA - IOWA - Total** | **17.28** | **17.68** | **21.30** | **14.46** | **70.72** |

**STATE: ID - IDAHO**

| REGISTRANT ZIP CODE 3 | 1ST QUARTER | 2ND QUARTER | 3RD QUARTER | 4TH QUARTER | **TOTAL GRAMS** |
|---|---|---|---|---|---|
| 834 | 2.81 | 0.80 | 1.21 | 1.21 | **6.03** |
| 836 | 5.63 | 5.22 | 6.43 | 6.43 | **23.71** |
| 837 | 2.01 | 0.80 | 0.80 | 1.61 | **5.22** |
| 838 | 5.22 | 5.22 | 5.62 | 4.42 | **20.49** |
| **ID - IDAHO - Total** | **15.67** | **12.05** | **14.06** | **13.66** | **55.44** |

**STATE: IL - ILLINOIS**

| REGISTRANT ZIP CODE 3 | 1ST QUARTER | 2ND QUARTER | 3RD QUARTER | 4TH QUARTER | **TOTAL GRAMS** |
|---|---|---|---|---|---|
| 600 | 16.88 | 14.87 | 14.87 | 12.86 | **59.47** |
| 601 | 5.22 | 6.83 | 9.24 | 4.02 | **25.31** |
| 602 | 0 | 0 | 0 | 3.21 | **3.21** |
| 603 | 0.80 | 2.01 | 1.21 | 1.21 | **5.22** |
| 604 | 2.41 | 2.81 | 1.21 | 5.63 | **12.06** |
| 605 | 15.27 | 13.66 | 13.66 | 12.86 | **55.45** |
| 606 | 12.86 | 6.43 | 8.84 | 6.83 | **34.96** |
| 607 | 0.40 | 1.21 | 0.80 | 0 | **2.41** |
| 611 | 0 | 0.40 | 0 | 0 | **0.40** |
| 615 | 1.21 | 0 | 0 | 0 | **1.21** |
| 616 | 0 | 0 | 0.40 | 0 | **0.40** |
| 617 | 0 | 1.21 | 0 | 0 | **1.21** |
| 622 | 0 | 0 | 0.40 | 0 | **0.40** |
| 627 | 0.40 | 2.41 | 1.21 | 2.01 | **6.03** |
| **IL - ILLINOIS - Total** | **55.45** | **51.84** | **51.83** | **48.62** | **207.74** |

**STATE: IN - INDIANA**

| REGISTRANT ZIP CODE 3 | 1ST QUARTER | 2ND QUARTER | 3RD QUARTER | 4TH QUARTER | **TOTAL GRAMS** |
|---|---|---|---|---|---|
| 460 | 9.64 | 6.43 | 6.43 | 5.63 | **28.13** |
| 461 | 0 | 2.01 | 0.40 | 0 | **2.41** |
| 462 | 0 | 2.81 | 0.80 | 1.61 | **5.22** |
| 463 | 1.61 | 1.61 | 2.01 | 2.41 | **7.63** |
| 469 | 11.25 | 10.85 | 10.45 | 10.85 | **43.40** |
| 472 | 0 | 0.40 | 0 | 0 | **0.40** |
| 473 | 0 | 0 | 0.40 | 0 | **0.40** |
| 474 | 2.41 | 0.80 | 2.01 | 2.41 | **7.64** |

| | | | | | |
|---|---|---|---|---|---|
| 477 | 0.80 | 1.21 | 1.21 | 1.21 | **4.42** |
| 479 | 0 | 1.61 | 0.80 | 0.40 | **2.81** |
| **IN - INDIANA - Total** | **25.72** | **27.73** | **24.51** | **24.51** | **102.47** |

**STATE: KS - KANSAS**

| REGISTRANT ZIP CODE 3 | 1ST QUARTER | 2ND QUARTER | 3RD QUARTER | 4TH QUARTER | TOTAL GRAMS |
|---|---|---|---|---|---|
| 660 | 4.42 | 5.22 | 6.43 | 4.02 | **20.09** |
| 662 | 0 | 0 | 0 | 1.61 | **1.61** |
| 665 | 1.61 | 1.21 | 1.61 | 1.61 | **6.03** |
| 670 | 0 | 0 | 0.80 | 1.21 | **2.01** |
| 672 | 8.44 | 9.24 | 6.83 | 7.23 | **31.74** |
| **KS - KANSAS - Total** | **14.47** | **15.67** | **15.67** | **15.67** | **61.48** |

**STATE: KY - KENTUCKY**

| REGISTRANT ZIP CODE 3 | 1ST QUARTER | 2ND QUARTER | 3RD QUARTER | 4TH QUARTER | TOTAL GRAMS |
|---|---|---|---|---|---|
| 402 | 0 | 0 | 0 | 0.80 | **0.80** |
| 403 | 2.01 | 2.41 | 4.02 | 1.61 | **10.05** |
| 405 | 4.02 | 6.43 | 6.83 | 6.03 | **23.30** |
| 406 | 0.80 | 0.40 | 0 | 0 | **1.21** |
| 408 | 2.41 | 1.61 | 2.01 | 1.61 | **7.63** |
| 410 | 0.80 | 0.40 | 0.40 | 0 | **1.61** |
| 416 | 0 | 0.40 | 0.40 | 0 | **0.80** |
| 425 | 0.80 | 0 | 0.40 | 0.40 | **1.61** |
| **KY - KENTUCKY - Total** | **10.85** | **11.65** | **14.06** | **10.45** | **47.01** |

**STATE: LA - LOUISIANA**

| REGISTRANT ZIP CODE 3 | 1ST QUARTER | 2ND QUARTER | 3RD QUARTER | 4TH QUARTER | TOTAL GRAMS |
|---|---|---|---|---|---|
| 701 | 0 | 0 | 0.40 | 0 | **0.40** |
| 703 | 3.21 | 2.01 | 2.01 | 0 | **7.23** |
| 704 | 2.41 | 1.21 | 0.80 | 0 | **4.42** |
| 705 | 0 | 0 | 1.61 | 1.61 | **3.21** |
| 706 | 0.80 | 2.01 | 1.61 | 1.61 | **6.03** |
| 707 | 1.21 | 0.40 | 0 | 0.40 | **2.01** |
| 708 | 2.41 | 4.42 | 3.62 | 5.63 | **16.07** |
| 711 | 2.81 | 3.21 | 3.21 | 3.21 | **12.46** |
| 712 | 0 | 0 | 1.21 | 0 | **1.21** |
| 713 | 2.81 | 1.61 | 1.21 | 1.21 | **6.83** |
| 714 | 0 | 0.40 | 0 | 0 | **0.40** |
| **LA - LOUISIANA - Total** | **15.67** | **15.27** | **15.67** | **13.66** | **60.27** |

**STATE: MA - MASSACHUSETTS**

| REGISTRANT ZIP CODE 3 | 1ST QUARTER | 2ND QUARTER | 3RD QUARTER | 4TH QUARTER | TOTAL GRAMS |
|---|---|---|---|---|---|
| 010 | 15.27 | 8.84 | 3.21 | 2.81 | **30.14** |
| 012 | 1.21 | 0.40 | 0 | 0 | **1.61** |

| | | | | | |
|---|---|---|---|---|---|
| 013 | 0 | 0 | 0 | 2.41 | **2.41** |
| 014 | 0.80 | 2.41 | 1.61 | 2.01 | **6.83** |
| 017 | 3.21 | 3.21 | 3.21 | 4.02 | **13.66** |
| 018 | 4.02 | 4.82 | 3.62 | 3.62 | **16.07** |
| 019 | 6.43 | 8.44 | 6.43 | 2.41 | **23.71** |
| 020 | 0.80 | 0 | 0 | 0 | **0.80** |
| 021 | 3.21 | 4.82 | 5.63 | 4.02 | **17.68** |
| 023 | 0.80 | 0.40 | 0.40 | 0 | **1.61** |
| 024 | 4.42 | 2.01 | 6.43 | 4.42 | **17.28** |
| 026 | 0 | 0 | 0.40 | 0 | **0.40** |
| 027 | 2.81 | 2.41 | 0.80 | 1.21 | **7.23** |
| **MA - MASSACHUSETTS - Total** | **43.00** | **37.77** | **31.74** | **26.92** | **139.43** |

**STATE: MD - MARYLAND**

| REGISTRANT ZIP CODE 3 | 1ST QUARTER | 2ND QUARTER | 3RD QUARTER | 4TH QUARTER | TOTAL GRAMS |
|---|---|---|---|---|---|
| 206 | 0 | 0 | 0.80 | 2.41 | **3.21** |
| 207 | 0 | 0.80 | 0.80 | 0.40 | **2.01** |
| 208 | 6.43 | 8.44 | 6.43 | 10.04 | **31.33** |
| 210 | 6.43 | 8.03 | 8.44 | 8.84 | **31.74** |
| 211 | 9.24 | 7.64 | 12.86 | 11.65 | **41.39** |
| 212 | 11.25 | 7.23 | 8.04 | 10.85 | **37.37** |
| 214 | 1.61 | 0.40 | 0.40 | 0.40 | **2.81** |
| 216 | 0.40 | 0 | 0 | 0 | **0.40** |
| 217 | 1.61 | 0.80 | 0.80 | 0.80 | **4.02** |
| **MD - MARYLAND - Total** | **36.97** | **33.35** | **38.57** | **45.40** | **154.29** |

**STATE: ME - MAINE**

| REGISTRANT ZIP CODE 3 | 1ST QUARTER | 2ND QUARTER | 3RD QUARTER | 4TH QUARTER | TOTAL GRAMS |
|---|---|---|---|---|---|
| 040 | 1.61 | 2.01 | 2.81 | 3.62 | **10.05** |
| 041 | 3.21 | 1.21 | 2.01 | 0 | **6.43** |
| 042 | 4.82 | 5.22 | 8.04 | 6.03 | **24.11** |
| 044 | 0 | 1.21 | 0 | 0 | **1.21** |
| 045 | 0 | 0 | 6.83 | 2.01 | **8.84** |
| 048 | 0.80 | 0.80 | 0.40 | 0.40 | **2.41** |
| **ME - MAINE - Total** | **10.45** | **10.45** | **20.09** | **12.05** | **53.04** |

**STATE: MI - MICHIGAN**

| REGISTRANT ZIP CODE 3 | 1ST QUARTER | 2ND QUARTER | 3RD QUARTER | 4TH QUARTER | TOTAL GRAMS |
|---|---|---|---|---|---|
| 480 | 2.41 | 2.01 | 2.81 | 5.22 | **12.45** |
| 481 | 2.01 | 2.01 | 3.62 | 1.61 | **9.24** |
| 482 | 0 | 0 | 0.80 | 0 | **0.80** |
| 483 | 5.22 | 4.42 | 3.21 | 1.61 | **14.47** |
| 487 | 4.82 | 0 | 0 | 0 | **4.82** |

| | | | | | |
|---|---|---|---|---|---|
| 488 | 2.81 | 0 | 0 | 0.40 | **3.21** |
| 490 | 11.25 | 13.26 | 9.64 | 9.24 | **43.39** |
| 492 | 0 | 0.40 | 1.61 | 0 | **2.01** |
| 493 | 1.61 | 2.01 | 1.61 | 2.01 | **7.23** |
| 494 | 0 | 0.40 | 0.80 | 0.40 | **1.61** |
| 495 | 0 | 0.40 | 0.40 | 0.40 | **1.21** |
| 498 | 0.40 | 0.40 | 0 | 0 | **0.80** |
| **MI - MICHIGAN - Total** | **30.54** | **25.31** | **24.51** | **20.89** | **101.26** |

**STATE: MN - MINNESOTA**

| REGISTRANT ZIP CODE 3 | 1ST QUARTER | 2ND QUARTER | 3RD QUARTER | 4TH QUARTER | **TOTAL GRAMS** |
|---|---|---|---|---|---|
| 550 | 2.01 | 1.61 | 2.81 | 2.41 | **8.84** |
| 551 | 5.63 | 6.03 | 8.04 | 9.64 | **29.33** |
| 553 | 6.83 | 6.43 | 8.44 | 8.44 | **30.14** |
| 554 | 31.74 | 28.13 | 32.15 | 36.57 | **128.59** |
| 557 | 2.41 | 2.81 | 3.21 | 2.81 | **11.25** |
| 558 | 0 | 0 | 0.80 | 0.80 | **1.61** |
| 559 | 20.49 | 16.47 | 14.86 | 10.44 | **62.26** |
| 560 | 1.21 | 2.41 | 0 | 2.41 | **6.03** |
| 561 | 0.40 | 0 | 0 | 0 | **0.40** |
| 562 | 0.40 | 0.80 | 0 | 0 | **1.21** |
| 563 | 1.61 | 2.01 | 0 | 2.01 | **5.63** |
| 565 | 2.81 | 2.41 | 1.21 | 2.41 | **8.84** |
| 566 | 1.61 | 2.01 | 2.41 | 0.80 | **6.83** |
| **MN - MINNESOTA - Total** | **77.14** | **71.12** | **73.93** | **78.75** | **300.95** |

**STATE: MO - MISSOURI**

| REGISTRANT ZIP CODE 3 | 1ST QUARTER | 2ND QUARTER | 3RD QUARTER | 4TH QUARTER | **TOTAL GRAMS** |
|---|---|---|---|---|---|
| 630 | 4.82 | 2.81 | 1.21 | 0.80 | **9.64** |
| 631 | 13.26 | 32.95 | 32.94 | 45.80 | **124.95** |
| 633 | 0 | 1.21 | 0 | 1.61 | **2.81** |
| 634 | 0.40 | 0 | 0 | 0 | **0.40** |
| 636 | 0.40 | 0 | 0.40 | 0 | **0.80** |
| 637 | 0 | 0 | 0.80 | 1.21 | **2.01** |
| 639 | 1.21 | 1.21 | 1.61 | 2.01 | **6.03** |
| 640 | 1.61 | 2.41 | 1.61 | 2.81 | **8.44** |
| 641 | 2.81 | 1.61 | 1.61 | 0 | **6.03** |
| 645 | 2.01 | 0.40 | 0.40 | 1.61 | **4.42** |
| 648 | 0 | 0 | 0 | 0.40 | **0.40** |
| 654 | 0 | 0.40 | 0.40 | 0.40 | **1.21** |
| 655 | 1.21 | 1.61 | 0.80 | 1.61 | **5.22** |
| **MO - MISSOURI - Total** | **27.73** | **44.60** | **41.78** | **58.25** | **172.36** |

**STATE: MS - MISSISSIPPI**

| REGISTRANT ZIP CODE 3 | 1ST QUARTER | 2ND QUARTER | 3RD QUARTER | 4TH QUARTER | TOTAL GRAMS |
|---|---|---|---|---|---|
| 389 | 0 | 0.80 | 0 | 0 | **0.80** |
| 392 | 4.02 | 4.02 | 6.03 | 8.44 | **22.50** |
| 394 | 0.80 | 0 | 0 | 0.80 | **1.61** |
| 396 | 1.61 | 0.80 | 1.61 | 0.40 | **4.42** |
| 397 | 0 | 0.40 | 0.80 | 0.40 | **1.61** |
| **MS - MISSISSIPPI - Total** | **6.43** | **6.03** | **8.44** | **10.04** | **30.94** |

**STATE: MT - MONTANA**

| REGISTRANT ZIP CODE 3 | 1ST QUARTER | 2ND QUARTER | 3RD QUARTER | 4TH QUARTER | TOTAL GRAMS |
|---|---|---|---|---|---|
| 596 | 0 | 0.40 | 0 | 0 | **0.40** |
| **MT - MONTANA - Total** | **0.00** | **0.40** | **0.00** | **0.00** | **0.40** |

**STATE: NC - NORTH CAROLINA**

| REGISTRANT ZIP CODE 3 | 1ST QUARTER | 2ND QUARTER | 3RD QUARTER | 4TH QUARTER | TOTAL GRAMS |
|---|---|---|---|---|---|
| 270 | 0 | 1.61 | 2.01 | 0.80 | **4.42** |
| 272 | 2.01 | 2.01 | 2.41 | 1.21 | **7.64** |
| 273 | 0.40 | 0 | 0 | 0.80 | **1.21** |
| 274 | 2.81 | 2.41 | 1.61 | 2.01 | **8.84** |
| 275 | 12.86 | 13.26 | 15.67 | 10.85 | **52.64** |
| 276 | 9.24 | 6.03 | 3.21 | 3.21 | **21.70** |
| 277 | 2.81 | 2.81 | 1.21 | 0 | **6.83** |
| 278 | 0.40 | 0 | 0 | 0 | **0.40** |
| 280 | 3.62 | 6.83 | 5.63 | 2.01 | **18.08** |
| 281 | 4.42 | 4.02 | 4.82 | 4.42 | **17.68** |
| 282 | 3.21 | 4.82 | 3.21 | 4.02 | **15.27** |
| 283 | 3.21 | 4.42 | 2.41 | 2.41 | **12.46** |
| 284 | 0 | 0.40 | 0.80 | 0 | **1.21** |
| 285 | 0 | 0 | 4.02 | 4.42 | **8.44** |
| 286 | 1.61 | 0.40 | 0 | 0 | **2.01** |
| 287 | 2.41 | 1.21 | 1.21 | 2.01 | **6.83** |
| 288 | 1.21 | 1.61 | 2.01 | 0.80 | **5.62** |
| **NC - NORTH CAROLINA - Total** | **50.23** | **51.83** | **50.23** | **38.97** | **191.26** |

**STATE: ND - NORTH DAKOTA**

| REGISTRANT ZIP CODE 3 | 1ST QUARTER | 2ND QUARTER | 3RD QUARTER | 4TH QUARTER | TOTAL GRAMS |
|---|---|---|---|---|---|
| 581 | 0.80 | 2.01 | 0.40 | 2.01 | **5.22** |
| 584 | 0.80 | 3.62 | 3.62 | 2.01 | **10.04** |
| 587 | 2.01 | 1.61 | 0 | 0.40 | **4.02** |
| **ND - NORTH DAKOTA - Total** | **3.62** | **7.23** | **4.02** | **4.42** | **19.28** |

**STATE: NE - NEBRASKA**

| REGISTRANT ZIP CODE 3 | 1ST QUARTER | 2ND QUARTER | 3RD QUARTER | 4TH QUARTER | TOTAL GRAMS |
|---|---|---|---|---|---|

| | | | | | |
|---|---|---|---|---|---|
| 681 | 1.61 | 2.01 | 4.02 | 1.21 | **8.84** |
| 684 | 1.61 | 1.61 | 0 | 0 | **3.21** |
| 685 | 0.80 | 0.80 | 1.61 | 0 | **3.21** |
| 687 | 0.40 | 0 | 0 | 0 | **0.40** |
| 693 | 0.80 | 1.61 | 2.01 | 0 | **4.42** |
| **NE - NEBRASKA - Total** | **5.22** | **6.03** | **7.63** | **1.21** | **20.09** |

**STATE: NH - NEW HAMPSHIRE**

| REGISTRANT ZIP CODE 3 | 1ST QUARTER | 2ND QUARTER | 3RD QUARTER | 4TH QUARTER | **TOTAL GRAMS** |
|---|---|---|---|---|---|
| 030 | 0.40 | 0 | 0 | 0 | **0.40** |
| 031 | 0.80 | 2.41 | 2.41 | 1.61 | **7.23** |
| 037 | 2.41 | 1.61 | 2.41 | 2.41 | **8.84** |
| 038 | 2.01 | 1.21 | 0 | 0 | **3.21** |
| **NH - NEW HAMPSHIRE - Total** | **5.63** | **5.22** | **4.82** | **4.02** | **19.69** |

**STATE: NJ - NEW JERSEY**

| REGISTRANT ZIP CODE 3 | 1ST QUARTER | 2ND QUARTER | 3RD QUARTER | 4TH QUARTER | **TOTAL GRAMS** |
|---|---|---|---|---|---|
| 070 | 5.22 | 4.82 | 2.41 | 4.42 | **16.88** |
| 073 | 0.80 | 0 | 0 | 0 | **0.80** |
| 074 | 2.01 | 1.21 | 0.40 | 0.40 | **4.02** |
| 076 | 10.85 | 6.43 | 10.05 | 4.42 | **31.74** |
| 077 | 0 | 0 | 0.40 | 0 | **0.40** |
| 078 | 6.43 | 5.63 | 4.82 | 4.02 | **20.90** |
| 079 | 2.01 | 0.80 | 2.01 | 2.01 | **6.83** |
| 080 | 8.44 | 8.04 | 5.63 | 5.22 | **27.32** |
| 081 | 1.21 | 0.80 | 1.21 | 0.40 | **3.62** |
| 082 | 0 | 0 | 0 | 0.40 | **0.40** |
| 083 | 0 | 0 | 0.80 | 0 | **0.80** |
| 085 | 2.01 | 2.01 | 1.61 | 1.61 | **7.23** |
| 086 | 0.40 | 0 | 0 | 0.40 | **0.80** |
| 087 | 0 | 0 | 0 | 0.80 | **0.80** |
| 088 | 4.02 | 2.81 | 4.42 | 5.63 | **16.88** |
| **NJ - NEW JERSEY - Total** | **43.40** | **32.55** | **33.75** | **29.73** | **139.43** |

**STATE: NM - NEW MEXICO**

| REGISTRANT ZIP CODE 3 | 1ST QUARTER | 2ND QUARTER | 3RD QUARTER | 4TH QUARTER | **TOTAL GRAMS** |
|---|---|---|---|---|---|
| 871 | 4.42 | 2.01 | 2.81 | 4.42 | **13.66** |
| 880 | 0 | 0.40 | 3.21 | 3.21 | **6.83** |
| 882 | 0.40 | 0.80 | 0 | 0.80 | **2.01** |
| 883 | 0 | 0 | 0 | 0.40 | **0.40** |
| **NM - NEW MEXICO - Total** | **4.82** | **3.21** | **6.03** | **8.84** | **22.90** |

**STATE: NV - NEVADA**

| REGISTRANT ZIP CODE 3 | 1ST QUARTER | 2ND QUARTER | 3RD QUARTER | 4TH QUARTER | **TOTAL GRAMS** |
|---|---|---|---|---|---|

| | | | | | |
|---|---|---|---|---|---|
| 891 | 16.88 | 22.91 | 15.67 | 12.86 | **68.31** |
| 893 | 0.80 | 0.80 | 0.80 | 0 | **2.41** |
| 894 | 0.40 | 0 | 0.80 | 0 | **1.21** |
| 895 | 1.21 | 0.80 | 3.62 | 1.21 | **6.83** |
| 898 | 0 | 1.21 | 2.01 | 0 | **3.21** |
| **NV - NEVADA - Total** | **19.29** | **25.72** | **22.90** | **14.06** | **81.98** |

**STATE: NY - NEW YORK**

| REGISTRANT ZIP CODE 3 | 1ST QUARTER | 2ND QUARTER | 3RD QUARTER | 4TH QUARTER | **TOTAL GRAMS** |
|---|---|---|---|---|---|
| 100 | 27.32 | 22.50 | 21.70 | 18.48 | **90.01** |
| 101 | 0.40 | 0.40 | 1.21 | 0 | **2.01** |
| 102 | 0.80 | 0 | 0 | 0 | **0.80** |
| 104 | 0 | 0.80 | 0.80 | 0.40 | **2.01** |
| 105 | 0.40 | 4.42 | 1.61 | 2.41 | **8.84** |
| 106 | 10.85 | 9.64 | 12.05 | 6.83 | **39.37** |
| 107 | 0 | 0 | 0 | 2.81 | **2.81** |
| 109 | 5.22 | 2.81 | 2.01 | 2.41 | **12.46** |
| 110 | 0 | 0.40 | 1.61 | 2.81 | **4.82** |
| 111 | 0.40 | 0.80 | 0 | 1.21 | **2.41** |
| 112 | 1.61 | 1.61 | 2.41 | 4.02 | **9.64** |
| 113 | 3.21 | 2.81 | 2.81 | 2.01 | **10.85** |
| 114 | 2.01 | 1.61 | 2.41 | 1.21 | **7.23** |
| 115 | 3.62 | 4.02 | 3.21 | 3.62 | **14.46** |
| 116 | 0 | 1.61 | 0.80 | 1.61 | **4.02** |
| 117 | 11.65 | 10.04 | 10.04 | 12.05 | **43.79** |
| 120 | 1.61 | 1.61 | 1.21 | 2.01 | **6.43** |
| 121 | 0 | 0 | 0.40 | 0 | **0.40** |
| 122 | 0 | 0 | 0.40 | 0.40 | **0.80** |
| 123 | 4.42 | 3.62 | 3.21 | 3.21 | **14.47** |
| 124 | 0.40 | 0 | 0 | 0.40 | **0.80** |
| 127 | 0.80 | 0.40 | 0 | 0 | **1.21** |
| 128 | 1.21 | 1.21 | 1.61 | 0 | **4.02** |
| 130 | 2.01 | 2.41 | 2.01 | 2.01 | **8.44** |
| 133 | 0 | 0 | 0.40 | 0 | **0.40** |
| 138 | 0.40 | 0 | 0 | 0 | **0.40** |
| 139 | 0.80 | 0.80 | 0.40 | 0.80 | **2.81** |
| 140 | 6.03 | 6.03 | 9.24 | 6.43 | **27.73** |
| 141 | 2.81 | 3.21 | 2.81 | 3.21 | **12.06** |
| 142 | 2.81 | 3.21 | 0 | 0 | **6.03** |
| 146 | 2.41 | 0 | 1.21 | 1.61 | **5.22** |
| 148 | 4.42 | 4.42 | 4.02 | 6.03 | **18.89** |

| NY - NEW YORK - Total | 97.64 | 90.41 | 89.60 | 87.99 | 365.64 |
|---|---|---|---|---|---|

**STATE: OH - OHIO**

| REGISTRANT ZIP CODE 3 | 1ST QUARTER | 2ND QUARTER | 3RD QUARTER | 4TH QUARTER | TOTAL GRAMS |
|---|---|---|---|---|---|
| 430 | 0 | 2.81 | 4.02 | 4.42 | **11.25** |
| 431 | 0.80 | 0 | 2.41 | 2.81 | **6.03** |
| 432 | 2.81 | 40.99 | 4.42 | 4.82 | **53.04** |
| 434 | 0 | 0 | 0 | 0.40 | **0.40** |
| 439 | 0 | 0 | 0.40 | 0.40 | **0.80** |
| 440 | 0 | 7.23 | 5.22 | 5.63 | **18.08** |
| 441 | 1.61 | 1.21 | 0.80 | 0.80 | **4.42** |
| 445 | 1.21 | 3.21 | 1.21 | 0 | **5.63** |
| 448 | 1.61 | 0.80 | 1.21 | 1.21 | **4.82** |
| 450 | 4.82 | 7.23 | 5.63 | 4.42 | **22.10** |
| 452 | 0.40 | 2.01 | 0.80 | 0.40 | **3.62** |
| 453 | 0.80 | 0.80 | 1.61 | 1.21 | **4.42** |
| 454 | 4.02 | 4.02 | 5.22 | 5.63 | **18.89** |
| 457 | 2.01 | 1.21 | 0.80 | 0 | **4.02** |
| **OH - OHIO - Total** | **20.09** | **71.53** | **33.75** | **32.15** | **157.52** |

**STATE: OK - OKLAHOMA**

| REGISTRANT ZIP CODE 3 | 1ST QUARTER | 2ND QUARTER | 3RD QUARTER | 4TH QUARTER | TOTAL GRAMS |
|---|---|---|---|---|---|
| 730 | 1.61 | 0.80 | 2.01 | 0.40 | **4.82** |
| 731 | 1.61 | 2.01 | 2.81 | 2.41 | **8.84** |
| 735 | 1.21 | 2.41 | 1.21 | 0 | **4.82** |
| 737 | 1.21 | 0 | 0 | 0 | **1.21** |
| 740 | 4.02 | 5.22 | 6.83 | 6.03 | **22.10** |
| 741 | 2.81 | 0.80 | 0.80 | 0 | **4.42** |
| 743 | 0 | 0 | 0 | 0.40 | **0.40** |
| 745 | 0.40 | 0 | 0 | 0 | **0.40** |
| 747 | 1.61 | 0.40 | 0.80 | 0 | **2.81** |
| 748 | 1.61 | 0.80 | 0.80 | 1.61 | **4.82** |
| **OK - OKLAHOMA - Total** | **16.07** | **12.46** | **15.27** | **10.85** | **54.65** |

**STATE: OR - OREGON**

| REGISTRANT ZIP CODE 3 | 1ST QUARTER | 2ND QUARTER | 3RD QUARTER | 4TH QUARTER | TOTAL GRAMS |
|---|---|---|---|---|---|
| 970 | 7.63 | 6.43 | 11.65 | 11.25 | **36.96** |
| 971 | 0.40 | 0.40 | 0 | 1.61 | **2.41** |
| 972 | 8.44 | 7.63 | 12.86 | 12.05 | **40.98** |
| 973 | 8.44 | 1.61 | 0.80 | 1.61 | **12.46** |
| 974 | 2.01 | 0 | 0.40 | 0.40 | **2.81** |
| 975 | 0.40 | 0.40 | 0.40 | 0.40 | **1.61** |
| 977 | 0.40 | 0 | 0.80 | 0.80 | **2.01** |

| | | | | | |
|---|---|---|---|---|---|
| 978 | 12.05 | 10.85 | 8.03 | 12.05 | **42.98** |
| **OR - OREGON - Total** | **39.78** | **27.32** | **34.95** | **40.17** | **142.23** |

**STATE: PA - PENNSYLVANIA**

| REGISTRANT ZIP CODE 3 | 1ST QUARTER | 2ND QUARTER | 3RD QUARTER | 4TH QUARTER | TOTAL GRAMS |
|---|---|---|---|---|---|
| 150 | 5.22 | 11.25 | 6.03 | 7.23 | **29.73** |
| 151 | 6.43 | 2.01 | 5.62 | 8.44 | **22.50** |
| 152 | 23.30 | 18.89 | 14.06 | 11.65 | **67.90** |
| 156 | 0 | 0 | 0 | 1.21 | **1.21** |
| 160 | 2.01 | 0 | 0 | 0 | **2.01** |
| 168 | 5.22 | 3.62 | 2.41 | 1.21 | **12.46** |
| 169 | 0 | 0 | 0.40 | 0 | **0.40** |
| 170 | 0.80 | 2.81 | 0 | 2.81 | **6.43** |
| 171 | 3.62 | 3.21 | 2.01 | 2.81 | **11.65** |
| 172 | 0 | 0.80 | 0.80 | 1.21 | **2.81** |
| 173 | 2.01 | 0.80 | 0.40 | 0.40 | **3.62** |
| 174 | 1.61 | 0.40 | 0 | 0.40 | **2.41** |
| 177 | 0.40 | 0.40 | 0 | 0 | **0.80** |
| 178 | 2.81 | 3.21 | 2.81 | 2.01 | **10.85** |
| 179 | 0 | 0.80 | 2.41 | 2.81 | **6.03** |
| 180 | 2.81 | 2.81 | 2.81 | 1.61 | **10.04** |
| 181 | 1.61 | 2.01 | 1.61 | 2.01 | **7.23** |
| 182 | 0 | 0.80 | 0.40 | 0 | **1.21** |
| 183 | 0 | 4.02 | 2.81 | 4.02 | **10.85** |
| 184 | 3.62 | 6.03 | 4.42 | 4.42 | **18.48** |
| 186 | 2.41 | 1.61 | 1.61 | 2.01 | **7.63** |
| 187 | 35.36 | 28.53 | 32.15 | 36.57 | **132.60** |
| 189 | 5.63 | 7.23 | 6.43 | 6.43 | **25.72** |
| 190 | 7.64 | 7.63 | 7.23 | 10.04 | **32.55** |
| 191 | 5.22 | 6.03 | 4.42 | 5.63 | **21.30** |
| 193 | 1.21 | 3.62 | 3.21 | 2.41 | **10.45** |
| 194 | 2.81 | 4.02 | 3.62 | 7.63 | **18.08** |
| 195 | 0 | 0.80 | 0 | 0.80 | **1.61** |
| 196 | 0.80 | 0.80 | 0.40 | 0.80 | **2.81** |
| **PA - PENNSYLVANIA - Total** | **122.55** | **124.16** | **108.09** | **126.56** | **481.36** |

**STATE: RI - RHODE ISLAND**

| REGISTRANT ZIP CODE 3 | 1ST QUARTER | 2ND QUARTER | 3RD QUARTER | 4TH QUARTER | TOTAL GRAMS |
|---|---|---|---|---|---|
| 028 | 3.21 | 4.42 | 2.01 | 4.42 | **14.06** |
| 029 | 0.40 | 0 | 0 | 0.40 | **0.80** |
| **RI - RHODE ISLAND - Total** | **3.62** | **4.42** | **2.01** | **4.82** | **14.87** |

**STATE: SC - SOUTH CAROLINA**

| REGISTRANT ZIP CODE 3 | 1ST QUARTER | 2ND QUARTER | 3RD QUARTER | 4TH QUARTER | TOTAL GRAMS |
|---|---|---|---|---|---|
| 290 | 0.40 | 0.80 | 0.80 | 0 | **2.01** |
| 291 | 0 | 0.80 | 0 | 1.21 | **2.01** |
| 292 | 4.82 | 4.02 | 3.21 | 4.82 | **16.87** |
| 293 | 0.80 | 3.62 | 1.61 | 2.81 | **8.84** |
| 294 | 2.81 | 1.61 | 2.01 | 3.62 | **10.05** |
| 295 | 3.21 | 6.03 | 2.81 | 3.62 | **15.67** |
| 296 | 4.02 | 2.41 | 3.62 | 7.23 | **17.28** |
| **SC - SOUTH CAROLINA - Total** | **16.07** | **19.29** | **14.06** | **23.31** | **72.73** |

**STATE: SD - SOUTH DAKOTA**

| REGISTRANT ZIP CODE 3 | 1ST QUARTER | 2ND QUARTER | 3RD QUARTER | 4TH QUARTER | TOTAL GRAMS |
|---|---|---|---|---|---|
| 570 | 0 | 0 | 0 | 0.40 | **0.40** |
| 571 | 10.45 | 6.43 | 5.22 | 7.63 | **29.73** |
| 577 | 0.40 | 0.40 | 0.40 | 0 | **1.21** |
| **SD - SOUTH DAKOTA - Total** | **10.85** | **6.83** | **5.63** | **8.03** | **31.34** |

**STATE: TN - TENNESSEE**

| REGISTRANT ZIP CODE 3 | 1ST QUARTER | 2ND QUARTER | 3RD QUARTER | 4TH QUARTER | TOTAL GRAMS |
|---|---|---|---|---|---|
| 370 | 1.21 | 1.61 | 0.80 | 0.80 | **4.42** |
| 372 | 0.80 | 3.21 | 7.64 | 2.41 | **14.06** |
| 373 | 0.40 | 0 | 0 | 0 | **0.40** |
| 374 | 1.21 | 0 | 0.40 | 0.40 | **2.01** |
| 376 | 0 | 0 | 0.40 | 0 | **0.40** |
| 378 | 1.21 | 2.01 | 0.80 | 1.61 | **5.63** |
| 379 | 1.61 | 0.40 | 1.21 | 0.80 | **4.02** |
| 380 | 0.80 | 0.80 | 2.41 | 0.80 | **4.82** |
| 381 | 7.23 | 4.82 | 4.42 | 6.43 | **22.91** |
| 383 | 0 | 0.40 | 0 | 0 | **0.40** |
| **TN - TENNESSEE - Total** | **14.47** | **13.26** | **18.08** | **13.26** | **59.07** |

**STATE: TX - TEXAS**

| REGISTRANT ZIP CODE 3 | 1ST QUARTER | 2ND QUARTER | 3RD QUARTER | 4TH QUARTER | TOTAL GRAMS |
|---|---|---|---|---|---|
| 750 | 21.70 | 22.91 | 28.53 | 21.30 | **94.43** |
| 751 | 1.61 | 0 | 0.80 | 0 | **2.41** |
| 752 | 18.88 | 22.10 | 14.87 | 16.87 | **72.72** |
| 754 | 2.01 | 2.41 | 2.81 | 2.41 | **9.64** |
| 756 | 1.61 | 3.62 | 4.42 | 0.80 | **10.45** |
| 757 | 1.61 | 2.41 | 3.21 | 4.42 | **11.65** |
| 759 | 1.21 | 0 | 0 | 0 | **1.21** |
| 760 | 8.44 | 2.41 | 2.01 | 2.41 | **15.27** |
| 761 | 1.61 | 2.81 | 1.61 | 3.62 | **9.64** |
| 762 | 4.82 | 2.41 | 0.40 | 1.61 | **9.24** |

| | | | | | |
|---|---|---|---|---|---|
| 764 | 1.21 | 1.21 | 0 | 0.80 | **3.21** |
| 769 | 2.81 | 0.40 | 2.41 | 0 | **5.63** |
| 770 | 12.05 | 13.66 | 11.25 | 10.45 | **47.41** |
| 773 | 3.21 | 2.01 | 1.61 | 2.01 | **8.84** |
| 774 | 2.81 | 3.21 | 2.81 | 2.81 | **11.65** |
| 775 | 0.80 | 0 | 2.01 | 2.01 | **4.82** |
| 778 | 0.40 | 0 | 0 | 0.40 | **0.80** |
| 779 | 0 | 0 | 0 | 0.40 | **0.40** |
| 780 | 0 | 0 | 1.21 | 0.80 | **2.01** |
| 781 | 1.21 | 0.80 | 0 | 0.40 | **2.41** |
| 782 | 1.61 | 1.61 | 4.02 | 4.82 | **12.05** |
| 784 | 0.80 | 1.61 | 1.21 | 4.02 | **7.64** |
| 785 | 1.21 | 1.21 | 2.41 | 2.01 | **6.83** |
| 786 | 2.81 | 2.41 | 1.21 | 2.81 | **9.24** |
| 787 | 6.03 | 12.06 | 8.04 | 10.05 | **36.16** |
| 788 | 3.21 | 4.02 | 3.62 | 3.21 | **14.06** |
| 789 | 0.80 | 0.40 | 1.61 | 1.21 | **4.02** |
| 790 | 1.61 | 2.41 | 1.21 | 1.21 | **6.43** |
| 797 | 0.80 | 0.40 | 0.40 | 2.41 | **4.02** |
| **TX - TEXAS - Total** | **106.88** | **108.49** | **103.66** | **105.27** | **424.31** |

**STATE: UT - UTAH**

| REGISTRANT ZIP CODE 3 | 1ST QUARTER | 2ND QUARTER | 3RD QUARTER | 4TH QUARTER | **TOTAL GRAMS** |
|---|---|---|---|---|---|
| 840 | 10.05 | 6.03 | 4.02 | 6.43 | **26.52** |
| 841 | 4.42 | 4.02 | 2.01 | 1.61 | **12.06** |
| 845 | 0.40 | 0 | 0 | 0 | **0.40** |
| 846 | 6.03 | 6.43 | 7.23 | 4.82 | **24.51** |
| 847 | 8.04 | 7.23 | 8.44 | 7.23 | **30.94** |
| **UT - UTAH - Total** | **28.93** | **23.71** | **21.70** | **20.09** | **94.43** |

**STATE: VA - VIRGINIA**

| REGISTRANT ZIP CODE 3 | 1ST QUARTER | 2ND QUARTER | 3RD QUARTER | 4TH QUARTER | **TOTAL GRAMS** |
|---|---|---|---|---|---|
| 201 | 3.62 | 2.81 | 5.22 | 4.82 | **16.48** |
| 220 | 1.61 | 0.40 | 0 | 0 | **2.01** |
| 221 | 0.80 | 0 | 0 | 0 | **0.80** |
| 222 | 0.80 | 0.40 | 0 | 0 | **1.21** |
| 223 | 2.01 | 2.01 | 2.01 | 2.41 | **8.44** |
| 224 | 2.41 | 0 | 0 | 0.80 | **3.21** |
| 228 | 0 | 0.80 | 2.01 | 0.40 | **3.21** |
| 229 | 1.61 | 0 | 2.01 | 0 | **3.62** |
| 231 | 2.81 | 1.21 | 3.21 | 2.81 | **10.05** |
| 232 | 3.21 | 6.03 | 2.81 | 2.81 | **14.87** |

| | | | | | |
|---|---|---|---|---|---|
| 234 | 2.81 | 3.21 | 4.82 | 2.81 | **13.66** |
| 235 | 2.41 | 2.01 | 1.61 | 2.01 | **8.04** |
| 236 | 0.80 | 0 | 0 | 0 | **0.80** |
| 237 | 0.80 | 1.21 | 1.21 | 0.80 | **4.02** |
| 238 | 2.41 | 1.61 | 2.01 | 2.41 | **8.44** |
| 240 | 4.42 | 6.03 | 4.82 | 4.02 | **19.29** |
| 241 | 2.01 | 0.80 | 0 | 2.01 | **4.82** |
| 242 | 0 | 0.40 | 0.80 | 0.40 | **1.61** |
| 245 | 0.40 | 0 | 0.40 | 0.80 | **1.61** |
| **VA - VIRGINIA - Total** | **34.96** | **28.93** | **32.95** | **29.33** | **126.17** |

**STATE: VT - VERMONT**

| REGISTRANT ZIP CODE 3 | 1ST QUARTER | 2ND QUARTER | 3RD QUARTER | 4TH QUARTER | TOTAL GRAMS |
|---|---|---|---|---|---|
| 054 | 0 | 0 | 0 | 0.40 | **0.40** |
| 056 | 1.21 | 0 | 2.01 | 0.40 | **3.62** |
| 057 | 0.80 | 1.21 | 0.80 | 0 | **2.81** |
| 058 | 0 | 0 | 0 | 1.61 | **1.61** |
| **VT - VERMONT - Total** | **2.01** | **1.21** | **2.81** | **2.41** | **8.44** |

**STATE: WA - WASHINGTON**

| REGISTRANT ZIP CODE 3 | 1ST QUARTER | 2ND QUARTER | 3RD QUARTER | 4TH QUARTER | TOTAL GRAMS |
|---|---|---|---|---|---|
| 980 | 35.76 | 32.55 | 23.30 | 26.11 | **117.73** |
| 981 | 11.25 | 14.06 | 16.07 | 13.26 | **54.65** |
| 982 | 16.07 | 12.86 | 15.27 | 10.04 | **54.24** |
| 983 | 6.43 | 7.23 | 6.03 | 9.24 | **28.93** |
| 984 | 2.41 | 2.81 | 2.41 | 2.41 | **10.05** |
| 985 | 0.40 | 0.40 | 0.40 | 0 | **1.21** |
| 986 | 12.86 | 13.66 | 16.88 | 10.04 | **53.44** |
| 992 | 5.22 | 8.04 | 7.23 | 6.43 | **26.92** |
| 993 | 1.61 | 2.01 | 2.41 | 0 | **6.03** |
| 994 | 0 | 0 | 0 | 0.40 | **0.40** |
| **WA - WASHINGTON - Total** | **92.02** | **93.63** | **90.00** | **77.94** | **353.59** |

**STATE: WI - WISCONSIN**

| REGISTRANT ZIP CODE 3 | 1ST QUARTER | 2ND QUARTER | 3RD QUARTER | 4TH QUARTER | TOTAL GRAMS |
|---|---|---|---|---|---|
| 530 | 2.81 | 0.80 | 0 | 0 | **3.62** |
| 531 | 2.01 | 2.01 | 1.61 | 2.81 | **8.44** |
| 532 | 3.21 | 4.02 | 5.63 | 2.01 | **14.87** |
| 535 | 2.81 | 2.41 | 2.01 | 2.81 | **10.04** |
| 537 | 6.83 | 9.64 | 6.83 | 7.63 | **30.94** |
| 539 | 0.80 | 1.61 | 0.80 | 1.21 | **4.42** |
| 540 | 0 | 0.80 | 1.61 | 0.80 | **3.21** |
| 541 | 0.80 | 0 | 0 | 4.42 | **5.22** |

| | | | | | |
|---|---|---|---|---|---|
| 542 | 6.83 | 6.43 | 7.63 | 5.63 | **26.52** |
| 543 | 2.41 | 3.21 | 3.21 | 1.21 | **10.05** |
| 544 | 0 | 0 | 0.40 | 4.02 | **4.42** |
| 546 | 0 | 1.21 | 0 | 0 | **1.21** |
| 549 | 3.62 | 3.62 | 5.62 | 2.41 | **15.26** |
| **WI - WISCONSIN - Total** | **32.14** | **35.76** | **35.36** | **34.95** | **138.21** |

**STATE: WV - WEST VIRGINIA**

| REGISTRANT ZIP CODE 3 | 1ST QUARTER | 2ND QUARTER | 3RD QUARTER | 4TH QUARTER | TOTAL GRAMS |
|---|---|---|---|---|---|
| 258 | 1.21 | 0.40 | 1.21 | 2.41 | **5.22** |
| 260 | 2.41 | 2.81 | 1.61 | 2.41 | **9.24** |
| 265 | 1.21 | 0.40 | 0.40 | 2.01 | **4.02** |
| **WV - WEST VIRGINIA - Total** | **4.82** | **3.62** | **3.21** | **6.83** | **18.48** |

**STATE: WY - WYOMING**

| REGISTRANT ZIP CODE 3 | 1ST QUARTER | 2ND QUARTER | 3RD QUARTER | 4TH QUARTER | TOTAL GRAMS |
|---|---|---|---|---|---|
| 820 | 4.42 | 2.41 | 2.81 | 2.81 | **12.46** |
| 826 | 0.40 | 1.21 | 2.41 | 1.21 | **5.22** |
| 830 | 0 | 0 | 0 | 0.80 | **0.80** |
| **WY - WYOMING - Total** | **4.82** | **3.62** | **5.22** | **4.82** | **18.48** |

**DRUG: 1205 - LISDEXAMFETAMINE**

**STATE: AK - ALASKA**

| REGISTRANT ZIP CODE 3 | 1ST QUARTER | 2ND QUARTER | 3RD QUARTER | 4TH QUARTER | TOTAL GRAMS |
|---|---|---|---|---|---|
| 995 | 1,268.93 | 1,305.35 | 1,380.50 | 1,413.45 | **5,368.24** |
| 996 | 571.16 | 619.72 | 615.10 | 604.69 | **2,410.68** |
| 997 | 331.25 | 333.56 | 367.67 | 341.66 | **1,374.14** |
| 998 | 246.27 | 219.10 | 217.37 | 207.54 | **890.27** |
| 999 | 145.10 | 152.62 | 159.56 | 127.76 | **585.04** |
| **AK - ALASKA - Total** | **2,562.72** | **2,630.36** | **2,740.19** | **2,695.10** | **10,628.37** |

**STATE: AL - ALABAMA**

| REGISTRANT ZIP CODE 3 | 1ST QUARTER | 2ND QUARTER | 3RD QUARTER | 4TH QUARTER | TOTAL GRAMS |
|---|---|---|---|---|---|
| 350 | 6,905.98 | 6,726.77 | 6,675.90 | 6,889.22 | **27,197.87** |
| 351 | 4,679.14 | 4,276.78 | 4,486.63 | 4,649.66 | **18,092.22** |
| 352 | 9,672.19 | 9,534.03 | 9,601.66 | 10,245.67 | **39,053.55** |
| 354 | 4,588.38 | 4,317.25 | 4,374.48 | 4,660.06 | **17,940.18** |
| 355 | 1,298.99 | 1,244.07 | 1,311.13 | 1,289.16 | **5,143.36** |
| 356 | 4,074.45 | 3,861.13 | 3,997.56 | 4,014.90 | **15,948.04** |
| 357 | 2,373.68 | 2,344.77 | 2,440.74 | 2,486.99 | **9,646.18** |
| 358 | 2,685.27 | 2,653.48 | 2,807.25 | 2,870.84 | **11,016.85** |
| 359 | 2,393.91 | 2,315.29 | 2,344.20 | 2,529.77 | **9,583.16** |
| 360 | 3,818.93 | 3,526.99 | 3,431.02 | 3,551.27 | **14,328.21** |
| 361 | 2,858.13 | 2,795.11 | 2,591.62 | 2,943.11 | **11,187.97** |

| | | | | | |
|---|---|---|---|---|---|
| 362 | 1,870.15 | 1,701.93 | 1,762.63 | 1,782.28 | **7,116.99** |
| 363 | 4,146.31 | 4,097.49 | 4,085.52 | 4,114.34 | **16,443.65** |
| 364 | 1,016.88 | 994.91 | 994.33 | 1,049.25 | **4,055.37** |
| 365 | 7,825.16 | 7,758.68 | 7,839.04 | 8,018.25 | **31,441.12** |
| 366 | 6,264.29 | 6,016.86 | 6,026.69 | 6,170.64 | **24,478.49** |
| 367 | 682.74 | 620.88 | 657.30 | 662.50 | **2,623.42** |
| 368 | 4,809.79 | 4,213.77 | 4,490.10 | 4,486.06 | **17,999.72** |
| 369 | 159.56 | 117.93 | 132.96 | 142.79 | **553.24** |
| **AL - ALABAMA - Total** | **72,123.93** | **69,118.13** | **70,050.78** | **72,556.75** | **283,849.59** |

**STATE: AR - ARKANSAS**

| REGISTRANT ZIP CODE 3 | 1ST QUARTER | 2ND QUARTER | 3RD QUARTER | 4TH QUARTER | TOTAL GRAMS |
|---|---|---|---|---|---|
| 716 | 1,430.22 | 1,384.55 | 1,196.67 | 1,278.18 | **5,289.62** |
| 717 | 672.91 | 627.24 | 628.97 | 625.50 | **2,554.62** |
| 718 | 807.61 | 720.31 | 698.92 | 809.92 | **3,036.76** |
| 719 | 1,874.78 | 1,814.66 | 1,819.86 | 1,844.72 | **7,354.01** |
| 720 | 4,469.87 | 4,259.44 | 4,322.45 | 4,448.48 | **17,500.24** |
| 721 | 3,259.91 | 3,040.81 | 2,952.93 | 3,202.10 | **12,455.74** |
| 722 | 3,599.48 | 3,425.91 | 3,468.48 | 3,643.39 | **14,137.26** |
| 723 | 1,123.83 | 1,083.36 | 1,083.36 | 1,049.25 | **4,339.80** |
| 724 | 2,759.85 | 2,485.25 | 2,531.50 | 2,491.03 | **10,267.63** |
| 725 | 682.16 | 607.01 | 599.49 | 624.93 | **2,513.58** |
| 726 | 819.75 | 752.69 | 750.37 | 755.00 | **3,077.80** |
| 727 | 5,376.33 | 5,263.60 | 5,335.86 | 5,364.19 | **21,339.98** |
| 728 | 1,151.00 | 1,046.94 | 1,007.05 | 1,092.03 | **4,297.02** |
| 729 | 2,628.62 | 2,378.30 | 2,302.57 | 2,479.47 | **9,788.97** |
| **AR - ARKANSAS - Total** | **30,656.30** | **28,890.06** | **28,698.50** | **29,708.18** | **117,953.04** |

**STATE: AZ - ARIZONA**

| REGISTRANT ZIP CODE 3 | 1ST QUARTER | 2ND QUARTER | 3RD QUARTER | 4TH QUARTER | TOTAL GRAMS |
|---|---|---|---|---|---|
| 850 | 6,814.06 | 6,657.40 | 6,758.57 | 7,059.76 | **27,289.79** |
| 851 | 1,377.61 | 1,408.25 | 1,452.19 | 1,591.51 | **5,829.56** |
| 852 | 18,668.01 | 22,284.60 | 29,136.24 | 19,583.72 | **89,672.56** |
| 853 | 4,384.31 | 4,209.72 | 4,299.33 | 4,747.36 | **17,640.72** |
| 855 | 165.91 | 150.88 | 136.43 | 150.88 | **604.11** |
| 856 | 597.76 | 653.25 | 619.72 | 684.47 | **2,555.20** |
| 857 | 4,772.22 | 4,660.06 | 4,530.57 | 4,874.54 | **18,837.39** |
| 859 | 191.93 | 214.48 | 187.30 | 207.54 | **801.25** |
| 860 | 332.41 | 331.83 | 326.05 | 402.36 | **1,392.64** |
| 863 | 807.61 | 728.98 | 768.87 | 809.34 | **3,114.80** |
| 864 | 286.74 | 300.03 | 344.55 | 349.75 | **1,281.07** |
| 865 | 1.73 | 1.16 | 0 | 4.62 | **7.52** |

| AZ - ARIZONA - Total | 38,400.29 | 41,600.65 | 48,559.82 | 40,465.84 | 169,026.61 |
|---|---|---|---|---|---|

**STATE: CA - CALIFORNIA**

| REGISTRANT ZIP CODE 3 | 1ST QUARTER | 2ND QUARTER | 3RD QUARTER | 4TH QUARTER | TOTAL GRAMS |
|---|---|---|---|---|---|
| 900 | 6,494.95 | 6,317.48 | 6,422.69 | 6,694.40 | **25,929.52** |
| 902 | 5,070.52 | 5,146.25 | 5,000.57 | 5,086.12 | **20,303.45** |
| 903 | 101.17 | 105.21 | 106.37 | 86.14 | **398.89** |
| 904 | 1,040.00 | 1,211.70 | 1,174.70 | 1,216.90 | **4,643.30** |
| 905 | 767.14 | 812.23 | 729.56 | 741.70 | **3,050.63** |
| 906 | 1,141.75 | 1,105.33 | 1,040.58 | 1,155.04 | **4,442.70** |
| 907 | 1,524.45 | 1,499.59 | 1,425.02 | 1,640.65 | **6,089.71** |
| 908 | 921.78 | 939.82 | 842.75 | 937.68 | **3,642.03** |
| 910 | 707.02 | 765.40 | 697.77 | 804.14 | **2,974.32** |
| 911 | 1,460.86 | 1,382.24 | 1,515.78 | 1,500.17 | **5,859.04** |
| 912 | 493.70 | 548.04 | 442.25 | 512.77 | **1,996.76** |
| 913 | 5,936.51 | 5,835.92 | 5,931.31 | 5,921.48 | **23,625.21** |
| 914 | 1,277.02 | 1,347.41 | 1,286.27 | 1,366.63 | **5,277.33** |
| 915 | 458.43 | 487.34 | 478.67 | 464.21 | **1,888.65** |
| 916 | 520.29 | 585.62 | 576.94 | 604.69 | **2,287.54** |
| 917 | 2,933.28 | 2,810.14 | 2,752.33 | 3,128.10 | **11,623.86** |
| 918 | 121.40 | 109.84 | 123.14 | 145.68 | **500.06** |
| 919 | 1,010.52 | 889.70 | 968.32 | 905.88 | **3,774.41** |
| 920 | 13,265.08 | 12,974.88 | 13,420.01 | 14,007.94 | **53,667.91** |
| 921 | 4,768.40 | 4,914.63 | 5,009.24 | 5,031.78 | **19,724.05** |
| 922 | 1,320.35 | 1,360.41 | 1,201.49 | 1,437.76 | **5,320.02** |
| 923 | 1,987.31 | 2,051.23 | 1,955.48 | 1,921.69 | **7,915.71** |
| 924 | 166.49 | 164.18 | 153.77 | 142.21 | **626.66** |
| 925 | 2,069.94 | 2,146.86 | 1,916.14 | 2,144.17 | **8,277.12** |
| 926 | 8,163.35 | 8,026.92 | 7,861.58 | 8,306.72 | **32,358.57** |
| 927 | 1,337.15 | 1,292.05 | 1,248.12 | 1,255.06 | **5,132.37** |
| 928 | 2,977.22 | 3,219.44 | 2,923.45 | 3,307.89 | **12,427.99** |
| 930 | 2,906.11 | 3,083.59 | 2,758.12 | 3,080.69 | **11,828.50** |
| 931 | 934.21 | 1,037.69 | 918.60 | 991.44 | **3,881.94** |
| 932 | 1,085.67 | 1,078.16 | 949.82 | 1,143.48 | **4,257.13** |
| 933 | 835.93 | 848.65 | 777.54 | 914.55 | **3,376.68** |
| 934 | 1,929.70 | 2,008.32 | 1,809.45 | 1,951.09 | **7,698.56** |
| 935 | 1,077.58 | 1,045.78 | 1,106.48 | 1,084.52 | **4,314.36** |
| 936 | 1,448.72 | 1,418.08 | 1,290.32 | 1,352.18 | **5,509.29** |
| 937 | 1,999.07 | 1,952.24 | 1,924.49 | 1,923.92 | **7,799.73** |
| 939 | 745.17 | 810.50 | 808.76 | 883.91 | **3,248.34** |
| 940 | 2,321.07 | 2,381.19 | 2,384.08 | 2,410.10 | **9,496.45** |

| | | | | | |
|---|---|---|---|---|---|
| 941 | 3,817.19 | 4,088.44 | 3,955.94 | 4,212.61 | **16,074.19** |
| 943 | 480.98 | 452.65 | 442.25 | 491.96 | **1,867.84** |
| 944 | 517.40 | 543.99 | 538.21 | 532.43 | **2,132.03** |
| 945 | 4,855.46 | 4,871.65 | 5,008.66 | 5,399.45 | **20,135.22** |
| 946 | 1,127.30 | 1,137.12 | 1,111.69 | 1,184.53 | **4,560.63** |
| 947 | 556.71 | 571.16 | 486.76 | 616.25 | **2,230.89** |
| 948 | 103.48 | 124.29 | 89.03 | 90.18 | **406.98** |
| 949 | 1,501.90 | 1,451.03 | 1,448.72 | 1,512.89 | **5,914.54** |
| 950 | 2,310.09 | 2,294.48 | 2,223.95 | 2,414.15 | **9,242.66** |
| 951 | 1,323.85 | 1,306.51 | 1,274.71 | 1,351.60 | **5,256.66** |
| 952 | 1,019.19 | 961.38 | 985.08 | 910.51 | **3,876.16** |
| 953 | 2,341.31 | 2,254.59 | 2,310.09 | 2,454.03 | **9,360.02** |
| 954 | 617.41 | 694.30 | 679.27 | 709.91 | **2,700.88** |
| 955 | 116.78 | 157.82 | 163.02 | 148.57 | **586.19** |
| 956 | 3,310.20 | 3,069.71 | 3,178.97 | 3,525.83 | **13,084.72** |
| 957 | 1,123.83 | 1,027.86 | 1,077.00 | 1,093.77 | **4,322.45** |
| 958 | 1,492.65 | 1,463.75 | 1,469.53 | 1,485.14 | **5,911.07** |
| 959 | 1,438.89 | 1,361.43 | 1,421.55 | 1,346.39 | **5,568.26** |
| 960 | 839.40 | 827.26 | 913.98 | 777.54 | **3,358.18** |
| 961 | 126.60 | 112.15 | 105.21 | 153.20 | **497.17** |
| **CA - CALIFORNIA - Total** | **112,339.92** | **112,485.61** | **110,815.58** | **116,614.45** | **452,255.56** |

**STATE: CO - COLORADO**

| REGISTRANT ZIP CODE 3 | 1ST QUARTER | 2ND QUARTER | 3RD QUARTER | 4TH QUARTER | TOTAL GRAMS |
|---|---|---|---|---|---|
| 800 | 4,179.08 | 4,235.74 | 4,355.98 | 4,595.90 | **17,366.70** |
| 801 | 5,385.00 | 5,398.88 | 5,264.76 | 5,471.72 | **21,520.35** |
| 802 | 6,277.01 | 6,109.94 | 6,076.41 | 6,492.06 | **24,955.42** |
| 803 | 1,650.48 | 1,711.75 | 1,626.20 | 1,883.45 | **6,871.87** |
| 804 | 1,001.85 | 787.95 | 923.23 | 1,007.63 | **3,720.65** |
| 805 | 3,103.24 | 3,121.16 | 3,125.21 | 3,309.62 | **12,659.23** |
| 806 | 1,127.30 | 1,160.82 | 1,142.90 | 1,163.14 | **4,594.16** |
| 807 | 149.15 | 152.04 | 133.54 | 171.12 | **605.85** |
| 808 | 567.69 | 517.40 | 552.09 | 577.52 | **2,214.70** |
| 809 | 3,100.35 | 2,978.37 | 3,140.82 | 2,989.36 | **12,208.89** |
| 810 | 597.76 | 612.79 | 660.77 | 707.59 | **2,578.90** |
| 811 | 106.37 | 134.12 | 105.79 | 108.10 | **454.39** |
| 812 | 113.31 | 128.34 | 132.38 | 149.73 | **523.76** |
| 813 | 175.16 | 147.42 | 198.29 | 209.85 | **730.72** |
| 814 | 121.40 | 124.29 | 121.40 | 149.15 | **516.24** |
| 815 | 503.53 | 469.42 | 432.42 | 534.16 | **1,939.53** |
| 816 | 793.15 | 713.95 | 747.48 | 787.95 | **3,042.54** |

| | | | | | |
|---|---|---|---|---|---|
| **CO - COLORADO - Total** | **28,951.83** | **28,504.38** | **28,739.66** | **30,308.05** | **116,503.91** |

**STATE: CT - CONNECTICUT**

| REGISTRANT ZIP CODE 3 | 1ST QUARTER | 2ND QUARTER | 3RD QUARTER | 4TH QUARTER | TOTAL GRAMS |
|---|---|---|---|---|---|
| 060 | 5,978.71 | 6,207.64 | 5,934.77 | 6,286.84 | **24,407.96** |
| 061 | 2,122.78 | 2,195.62 | 2,007.16 | 2,233.20 | **8,558.77** |
| 062 | 1,785.17 | 1,682.85 | 1,690.36 | 1,882.29 | **7,040.68** |
| 063 | 3,728.17 | 3,773.26 | 3,720.65 | 3,868.65 | **15,090.72** |
| 064 | 7,451.13 | 7,321.64 | 7,252.84 | 7,525.71 | **29,551.32** |
| 065 | 1,870.73 | 1,852.23 | 1,837.20 | 1,957.45 | **7,517.61** |
| 066 | 1,370.10 | 1,449.87 | 1,364.89 | 1,488.61 | **5,673.47** |
| 067 | 3,219.44 | 3,010.17 | 2,910.73 | 2,947.73 | **12,088.07** |
| 068 | 7,521.08 | 7,695.67 | 7,489.86 | 7,834.41 | **30,541.02** |
| 069 | 1,122.09 | 1,004.74 | 1,064.86 | 1,141.75 | **4,333.44** |
| **CT - CONNECTICUT - Total** | **36,169.40** | **36,193.68** | **35,273.35** | **37,166.63** | **144,803.07** |

**STATE: DC - DISTRICT OF COLUMBIA**

| REGISTRANT ZIP CODE 3 | 1ST QUARTER | 2ND QUARTER | 3RD QUARTER | 4TH QUARTER | TOTAL GRAMS |
|---|---|---|---|---|---|
| 200 | 5,458.42 | 5,689.66 | 5,522.01 | 5,756.72 | **22,426.81** |
| 203 | 43.36 | 26.59 | 0 | 8.09 | **78.04** |
| 204 | 30.64 | 19.08 | 32.37 | 21.39 | **103.48** |
| **DC - DISTRICT OF COLUMBIA -** | **5,532.42** | **5,735.33** | **5,554.38** | **5,786.20** | **22,608.33** |

**STATE: DE - DELAWARE**

| REGISTRANT ZIP CODE 3 | 1ST QUARTER | 2ND QUARTER | 3RD QUARTER | 4TH QUARTER | TOTAL GRAMS |
|---|---|---|---|---|---|
| 197 | 5,493.11 | 5,260.71 | 4,969.35 | 5,560.17 | **21,283.33** |
| 198 | 3,476.12 | 3,285.92 | 3,239.09 | 3,380.15 | **13,381.28** |
| 199 | 5,387.31 | 5,258.98 | 5,044.50 | 5,394.25 | **21,085.04** |
| **DE - DELAWARE - Total** | **14,356.54** | **13,805.61** | **13,252.94** | **14,334.57** | **55,749.65** |

**STATE: FL - FLORIDA**

| REGISTRANT ZIP CODE 3 | 1ST QUARTER | 2ND QUARTER | 3RD QUARTER | 4TH QUARTER | TOTAL GRAMS |
|---|---|---|---|---|---|
| 320 | 4,793.61 | 4,571.61 | 4,441.54 | 4,767.01 | **18,573.77** |
| 321 | 2,929.23 | 2,900.91 | 2,751.76 | 3,029.82 | **11,611.72** |
| 322 | 8,570.81 | 8,089.08 | 7,867.91 | 8,742.72 | **33,270.52** |
| 323 | 5,214.46 | 5,089.59 | 4,899.40 | 5,233.54 | **20,436.99** |
| 324 | 3,070.87 | 3,009.01 | 2,910.16 | 2,560.40 | **11,550.44** |
| 325 | 7,729.78 | 7,595.08 | 7,740.76 | 7,856.38 | **30,921.99** |
| 326 | 3,363.96 | 3,083.35 | 2,930.97 | 3,274.94 | **12,653.22** |
| 327 | 9,412.23 | 8,987.34 | 8,578.43 | 9,413.78 | **36,391.79** |
| 328 | 7,813.82 | 7,425.32 | 6,942.08 | 7,836.90 | **30,018.12** |
| 329 | 6,834.88 | 6,722.15 | 6,368.35 | 7,152.25 | **27,077.63** |
| 330 | 6,523.86 | 6,336.55 | 6,011.08 | 6,540.05 | **25,411.54** |
| 331 | 8,351.81 | 7,920.55 | 7,573.11 | 8,383.03 | **32,228.50** |

| | | | | | |
|---|---|---|---|---|---|
| 333 | 3,614.86 | 3,568.61 | 3,400.96 | 3,418.31 | **14,002.74** |
| 334 | 9,073.28 | 9,024.14 | 8,711.97 | 9,296.43 | **36,105.81** |
| 335 | 6,921.01 | 6,748.16 | 6,427.32 | 7,096.18 | **27,192.67** |
| 336 | 6,244.40 | 6,138.06 | 5,992.58 | 6,628.49 | **25,003.55** |
| 337 | 4,751.40 | 4,793.11 | 4,595.90 | 5,060.69 | **19,201.10** |
| 338 | 5,986.80 | 5,465.36 | 5,241.05 | 5,472.87 | **22,166.09** |
| 339 | 4,990.74 | 4,705.73 | 4,577.97 | 5,021.38 | **19,295.82** |
| 341 | 2,316.45 | 2,018.73 | 1,908.89 | 2,204.30 | **8,448.35** |
| 342 | 5,885.64 | 5,575.20 | 5,413.91 | 5,874.65 | **22,749.39** |
| 344 | 2,643.65 | 2,365.59 | 2,308.35 | 2,607.23 | **9,924.82** |
| 346 | 4,158.27 | 3,933.39 | 3,824.71 | 4,372.17 | **16,288.55** |
| 347 | 5,015.02 | 4,873.38 | 4,673.36 | 5,349.16 | **19,910.92** |
| 349 | 2,556.94 | 2,597.98 | 2,630.93 | 2,877.78 | **10,663.63** |
| **FL - FLORIDA - Total** | **138,767.77** | **133,537.99** | **128,723.45** | **140,070.45** | **541,099.66** |

**STATE: GA - GEORGIA**

| REGISTRANT ZIP CODE 3 | 1ST QUARTER | 2ND QUARTER | 3RD QUARTER | 4TH QUARTER | **TOTAL GRAMS** |
|---|---|---|---|---|---|
| 300 | 23,932.76 | 22,753.47 | 22,634.73 | 23,617.12 | **92,938.07** |
| 301 | 11,636.57 | 11,250.98 | 11,286.25 | 11,764.34 | **45,938.14** |
| 302 | 7,607.22 | 7,115.25 | 7,169.60 | 7,328.00 | **29,220.06** |
| 303 | 12,166.69 | 12,228.55 | 12,432.04 | 12,444.76 | **49,272.04** |
| 304 | 2,851.19 | 2,733.83 | 2,722.27 | 2,865.64 | **11,172.94** |
| 305 | 6,478.19 | 5,940.56 | 6,033.05 | 6,227.87 | **24,679.67** |
| 306 | 5,285.57 | 4,836.96 | 4,856.62 | 5,191.34 | **20,170.49** |
| 307 | 2,432.07 | 2,374.83 | 2,278.29 | 2,406.05 | **9,491.25** |
| 308 | 2,044.74 | 1,899.64 | 1,940.68 | 1,985.77 | **7,870.83** |
| 309 | 2,918.25 | 2,755.80 | 2,978.95 | 2,806.68 | **11,459.68** |
| 310 | 5,071.09 | 4,642.72 | 4,761.81 | 4,757.76 | **19,233.39** |
| 312 | 2,064.97 | 2,241.87 | 2,152.84 | 2,247.65 | **8,707.34** |
| 313 | 2,537.28 | 2,300.26 | 2,528.61 | 2,452.88 | **9,819.03** |
| 314 | 3,060.46 | 3,081.27 | 3,007.85 | 3,048.90 | **12,198.49** |
| 315 | 3,385.93 | 2,822.28 | 2,918.25 | 2,921.72 | **12,048.18** |
| 316 | 1,418.66 | 1,304.19 | 1,350.44 | 1,478.78 | **5,552.07** |
| 317 | 2,893.97 | 2,664.46 | 2,826.91 | 2,880.67 | **11,266.01** |
| 318 | 229.51 | 237.60 | 269.39 | 226.62 | **963.11** |
| 319 | 2,160.94 | 1,993.29 | 2,245.92 | 1,973.06 | **8,373.20** |
| 398 | 412.76 | 413.34 | 349.75 | 375.77 | **1,551.62** |
| **GA - GEORGIA - Total** | **100,588.82** | **95,591.18** | **96,744.25** | **99,001.36** | **391,925.61** |

**STATE: GU - GUAM**

| REGISTRANT ZIP CODE 3 | 1ST QUARTER | 2ND QUARTER | 3RD QUARTER | 4TH QUARTER | **TOTAL GRAMS** |
|---|---|---|---|---|---|
| 969 | 9.25 | 18.50 | 8.67 | 7.52 | **43.94** |

| GU - GUAM - Total | 9.25 | 18.50 | 8.67 | 7.52 | 43.94 |
|---|---|---|---|---|---|

STATE: HI - HAWAII

| REGISTRANT ZIP CODE 3 | 1ST QUARTER | 2ND QUARTER | 3RD QUARTER | 4TH QUARTER | TOTAL GRAMS |
|---|---|---|---|---|---|
| 967 | 704.13 | 744.01 | 761.94 | 842.87 | **3,052.95** |
| 968 | 748.06 | 838.25 | 811.07 | 664.82 | **3,062.20** |
| **HI - HAWAII - Total** | **1,452.19** | **1,582.26** | **1,573.01** | **1,507.68** | **6,115.14** |

STATE: IA - IOWA

| REGISTRANT ZIP CODE 3 | 1ST QUARTER | 2ND QUARTER | 3RD QUARTER | 4TH QUARTER | TOTAL GRAMS |
|---|---|---|---|---|---|
| 500 | 2,486.41 | 2,458.08 | 2,364.43 | 2,561.56 | **9,870.48** |
| 501 | 1,418.08 | 1,405.94 | 1,418.66 | 1,425.59 | **5,668.27** |
| 502 | 1,623.30 | 1,549.31 | 1,561.45 | 1,650.48 | **6,384.54** |
| 503 | 2,686.43 | 2,780.08 | 2,587.58 | 2,981.26 | **11,035.35** |
| 504 | 938.26 | 861.37 | 849.81 | 884.49 | **3,533.93** |
| 505 | 1,499.59 | 1,545.84 | 1,365.47 | 1,519.82 | **5,930.73** |
| 506 | 1,465.48 | 1,436.58 | 1,369.52 | 1,420.39 | **5,691.97** |
| 507 | 1,159.09 | 1,190.89 | 1,103.59 | 1,227.88 | **4,681.45** |
| 508 | 169.38 | 193.09 | 198.87 | 167.07 | **728.41** |
| 510 | 1,114.58 | 1,067.75 | 1,052.14 | 1,053.30 | **4,287.77** |
| 511 | 2,655.21 | 2,644.23 | 2,583.53 | 2,715.91 | **10,598.89** |
| 512 | 397.73 | 435.89 | 396.00 | 416.81 | **1,646.43** |
| 513 | 593.71 | 623.19 | 616.25 | 663.08 | **2,496.24** |
| 514 | 369.41 | 348.02 | 314.49 | 323.74 | **1,355.64** |
| 515 | 1,786.33 | 1,842.98 | 1,730.25 | 1,810.03 | **7,169.60** |
| 516 | 184.41 | 201.18 | 210.43 | 186.73 | **782.75** |
| 520 | 1,730.83 | 1,679.38 | 1,632.55 | 1,744.13 | **6,786.89** |
| 521 | 234.71 | 204.07 | 204.65 | 193.09 | **836.51** |
| 522 | 2,409.52 | 2,414.72 | 2,266.15 | 2,415.30 | **9,505.70** |
| 523 | 1,815.81 | 1,745.28 | 1,700.19 | 1,803.67 | **7,064.96** |
| 524 | 2,466.17 | 2,293.90 | 2,266.15 | 2,460.97 | **9,487.20** |
| 525 | 1,184.53 | 1,063.70 | 1,104.17 | 1,080.47 | **4,432.87** |
| 526 | 1,174.70 | 1,116.31 | 1,163.14 | 1,142.90 | **4,597.05** |
| 527 | 1,642.38 | 1,659.73 | 1,609.43 | 1,627.93 | **6,539.47** |
| 528 | 1,431.38 | 1,453.92 | 1,393.80 | 1,453.92 | **5,733.02** |
| **IA - IOWA - Total** | **34,637.44** | **34,215.43** | **33,062.70** | **34,930.54** | **136,846.10** |

STATE: ID - IDAHO

| REGISTRANT ZIP CODE 3 | 1ST QUARTER | 2ND QUARTER | 3RD QUARTER | 4TH QUARTER | TOTAL GRAMS |
|---|---|---|---|---|---|
| 832 | 608.74 | 634.75 | 561.91 | 659.61 | **2,465.02** |
| 833 | 815.12 | 781.01 | 740.55 | 842.87 | **3,179.55** |
| 834 | 1,545.84 | 1,607.70 | 1,586.88 | 1,711.18 | **6,451.60** |
| 835 | 140.48 | 169.96 | 138.17 | 152.62 | **601.22** |

| | | | | | |
|---|---|---|---|---|---|
| 836 | 3,257.59 | 3,188.80 | 3,137.93 | 3,303.26 | **12,887.58** |
| 837 | 2,193.31 | 2,173.66 | 2,165.56 | 2,308.93 | **8,841.46** |
| 838 | 1,090.87 | 1,040.00 | 998.38 | 1,033.64 | **4,162.90** |
| **ID - IDAHO - Total** | **9,651.96** | **9,595.88** | **9,329.38** | **10,012.11** | **38,589.33** |

**STATE: IL - ILLINOIS**

| REGISTRANT ZIP CODE 3 | 1ST QUARTER | 2ND QUARTER | 3RD QUARTER | 4TH QUARTER | **TOTAL GRAMS** |
|---|---|---|---|---|---|
| 600 | 15,571.70 | 16,008.75 | 15,456.66 | 16,055.57 | **63,092.68** |
| 601 | 9,777.41 | 9,827.70 | 9,554.84 | 10,250.29 | **39,410.23** |
| 602 | 813.96 | 768.29 | 717.42 | 815.12 | **3,114.80** |
| 603 | 672.33 | 672.91 | 660.77 | 718.58 | **2,724.59** |
| 604 | 7,234.34 | 7,055.13 | 7,264.40 | 7,610.69 | **29,164.57** |
| 605 | 8,873.84 | 8,696.94 | 8,869.79 | 9,223.59 | **35,664.15** |
| 606 | 10,664.21 | 11,380.48 | 11,191.44 | 11,872.44 | **45,108.56** |
| 607 | 667.13 | 687.36 | 655.57 | 623.77 | **2,633.82** |
| 608 | 150.31 | 178.05 | 178.63 | 166.49 | **673.49** |
| 609 | 1,034.80 | 1,101.28 | 1,015.72 | 1,069.49 | **4,221.29** |
| 610 | 1,000.69 | 934.21 | 900.68 | 937.68 | **3,773.26** |
| 611 | 1,005.32 | 944.62 | 1,026.13 | 960.22 | **3,936.28** |
| 612 | 622.61 | 667.71 | 639.38 | 689.10 | **2,618.79** |
| 613 | 687.94 | 635.91 | 592.55 | 639.96 | **2,556.36** |
| 614 | 765.40 | 709.91 | 671.17 | 774.08 | **2,920.56** |
| 615 | 677.53 | 635.91 | 672.33 | 667.71 | **2,653.48** |
| 616 | 1,162.56 | 1,104.75 | 1,197.25 | 1,253.90 | **4,718.45** |
| 617 | 1,514.04 | 1,351.60 | 1,398.42 | 1,483.40 | **5,747.47** |
| 618 | 1,430.80 | 1,245.81 | 1,347.55 | 1,490.92 | **5,515.07** |
| 619 | 285.58 | 216.21 | 242.22 | 296.57 | **1,040.58** |
| 620 | 1,004.16 | 1,030.75 | 969.47 | 1,103.59 | **4,107.98** |
| 622 | 1,815.23 | 1,819.28 | 1,899.06 | 1,921.60 | **7,455.18** |
| 623 | 799.51 | 898.95 | 776.39 | 793.73 | **3,268.58** |
| 624 | 306.97 | 281.53 | 265.93 | 256.10 | **1,110.53** |
| 625 | 479.24 | 459.59 | 501.21 | 432.42 | **1,872.47** |
| 626 | 348.02 | 376.92 | 315.64 | 387.91 | **1,428.49** |
| 627 | 682.74 | 709.33 | 690.25 | 715.11 | **2,797.43** |
| 628 | 389.06 | 434.15 | 397.15 | 398.31 | **1,618.68** |
| 629 | 1,006.47 | 1,023.82 | 1,041.74 | 1,047.52 | **4,119.54** |
| **IL - ILLINOIS - Total** | **71,443.91** | **71,857.83** | **71,109.77** | **74,655.83** | **289,067.34** |

**STATE: IN - INDIANA**

| REGISTRANT ZIP CODE 3 | 1ST QUARTER | 2ND QUARTER | 3RD QUARTER | 4TH QUARTER | **TOTAL GRAMS** |
|---|---|---|---|---|---|
| 460 | 10,455.52 | 9,806.89 | 10,070.50 | 10,389.61 | **40,722.52** |
| 461 | 6,592.07 | 6,464.89 | 6,904.25 | 6,986.34 | **26,947.55** |

| | | | | | |
|---|---|---|---|---|---|
| 462 | 9,542.70 | 8,877.88 | 8,823.54 | 8,879.04 | **36,123.16** |
| 463 | 8,335.05 | 8,065.65 | 8,158.15 | 8,614.27 | **33,173.11** |
| 464 | 681.00 | 704.13 | 611.63 | 666.55 | **2,663.31** |
| 465 | 8,021.72 | 7,606.64 | 7,420.49 | 8,004.95 | **31,053.80** |
| 466 | 2,430.33 | 2,581.79 | 2,381.19 | 2,447.68 | **9,841.00** |
| 467 | 2,745.40 | 2,543.06 | 2,485.25 | 2,644.81 | **10,418.52** |
| 468 | 3,607.34 | 3,565.72 | 3,465.13 | 3,907.38 | **14,545.57** |
| 469 | 6,466.63 | 6,277.59 | 6,079.30 | 6,050.97 | **24,874.49** |
| 470 | 711.64 | 664.24 | 678.69 | 665.39 | **2,719.96** |
| 471 | 5,227.76 | 4,868.76 | 4,227.07 | 4,219.55 | **18,543.14** |
| 472 | 1,446.98 | 1,494.39 | 1,405.94 | 1,495.54 | **5,842.86** |
| 473 | 3,387.67 | 3,114.80 | 3,244.30 | 3,398.65 | **13,145.42** |
| 474 | 2,832.69 | 2,678.92 | 2,507.22 | 2,870.27 | **10,889.09** |
| 475 | 1,685.74 | 1,515.78 | 1,593.24 | 1,687.47 | **6,482.24** |
| 476 | 2,160.94 | 2,114.69 | 2,288.70 | 2,310.67 | **8,874.99** |
| 477 | 4,933.51 | 4,742.15 | 4,858.93 | 4,822.51 | **19,357.10** |
| 478 | 1,290.90 | 1,177.01 | 1,165.45 | 1,214.01 | **4,847.37** |
| 479 | 3,284.76 | 3,074.34 | 3,132.15 | 3,406.74 | **12,897.99** |
| **IN - INDIANA - Total** | **85,840.33** | **81,939.32** | **81,501.12** | **84,682.40** | **333,963.17** |

**STATE: KS - KANSAS**

| REGISTRANT ZIP CODE 3 | 1ST QUARTER | 2ND QUARTER | 3RD QUARTER | 4TH QUARTER | **TOTAL GRAMS** |
|---|---|---|---|---|---|
| 660 | 7,129.71 | 7,090.40 | 7,040.10 | 7,426.85 | **28,687.06** |
| 661 | 875.82 | 898.37 | 810.50 | 973.52 | **3,558.21** |
| 662 | 7,525.71 | 7,585.83 | 7,643.64 | 7,906.10 | **30,661.27** |
| 664 | 528.38 | 516.82 | 522.02 | 430.68 | **1,997.91** |
| 665 | 864.84 | 870.62 | 860.21 | 938.83 | **3,534.50** |
| 666 | 3,553.00 | 3,548.38 | 3,496.35 | 3,824.71 | **14,422.44** |
| 667 | 1,161.98 | 1,129.03 | 1,128.45 | 1,178.75 | **4,598.21** |
| 668 | 492.54 | 472.31 | 511.62 | 485.03 | **1,961.49** |
| 669 | 102.90 | 127.76 | 85.56 | 129.49 | **445.72** |
| 670 | 1,591.51 | 1,651.05 | 1,570.70 | 1,657.41 | **6,470.67** |
| 671 | 583.30 | 534.74 | 539.37 | 590.82 | **2,248.23** |
| 672 | 3,841.47 | 3,975.02 | 3,728.17 | 4,104.51 | **15,649.17** |
| 673 | 565.38 | 557.29 | 556.13 | 598.33 | **2,277.14** |
| 674 | 918.60 | 904.15 | 885.07 | 933.05 | **3,640.87** |
| 675 | 842.87 | 845.76 | 761.36 | 838.82 | **3,288.81** |
| 676 | 539.37 | 563.65 | 492.54 | 545.15 | **2,140.70** |
| 677 | 123.14 | 146.84 | 114.46 | 112.73 | **497.17** |
| 678 | 329.52 | 326.63 | 298.88 | 320.85 | **1,275.87** |
| 679 | 112.73 | 119.09 | 121.40 | 143.95 | **497.17** |

| KS - KANSAS - Total | 31,682.77 | 31,863.72 | 31,166.53 | 33,139.58 | 127,852.60 |
|---|---|---|---|---|---|

**STATE: KY - KENTUCKY**

| REGISTRANT ZIP CODE 3 | 1ST QUARTER | 2ND QUARTER | 3RD QUARTER | 4TH QUARTER | TOTAL GRAMS |
|---|---|---|---|---|---|
| 400 | 3,186.49 | 3,041.38 | 3,035.60 | 3,217.13 | **12,480.60** |
| 401 | 931.90 | 812.81 | 1,123.83 | 820.32 | **3,688.86** |
| 402 | 10,132.36 | 9,492.40 | 9,549.63 | 9,819.61 | **38,994.00** |
| 403 | 2,426.29 | 2,511.27 | 2,502.59 | 2,616.48 | **10,056.63** |
| 404 | 1,123.83 | 1,015.72 | 976.99 | 1,036.53 | **4,153.07** |
| 405 | 3,922.99 | 3,817.77 | 3,749.79 | 4,107.95 | **15,598.49** |
| 406 | 447.45 | 508.73 | 406.40 | 426.64 | **1,789.22** |
| 407 | 345.70 | 311.60 | 323.16 | 316.80 | **1,297.26** |
| 408 | 100.59 | 100.59 | 65.90 | 93.07 | **360.16** |
| 409 | 115.04 | 131.23 | 109.26 | 115.62 | **471.15** |
| 410 | 4,387.78 | 4,315.52 | 4,066.93 | 4,431.71 | **17,201.94** |
| 411 | 631.29 | 704.13 | 599.49 | 646.89 | **2,581.79** |
| 412 | 249.16 | 204.07 | 217.94 | 198.87 | **870.04** |
| 413 | 50.87 | 67.06 | 51.45 | 56.65 | **226.04** |
| 414 | 96.54 | 86.72 | 69.95 | 82.67 | **335.88** |
| 415 | 479.24 | 373.45 | 426.06 | 433.58 | **1,712.33** |
| 416 | 219.10 | 251.47 | 221.99 | 240.49 | **933.05** |
| 417 | 215.63 | 199.44 | 229.51 | 233.55 | **878.13** |
| 418 | 46.83 | 39.31 | 37.00 | 54.34 | **177.48** |
| 420 | 4,160.01 | 3,884.83 | 3,801.59 | 3,827.60 | **15,674.03** |
| 421 | 1,538.32 | 1,616.95 | 1,574.74 | 1,636.60 | **6,366.62** |
| 422 | 873.51 | 798.93 | 1,131.34 | 668.86 | **3,472.65** |
| 423 | 1,950.51 | 1,848.19 | 1,838.36 | 1,847.03 | **7,484.08** |
| 424 | 2,615.32 | 2,481.78 | 2,408.36 | 2,345.93 | **9,851.40** |
| 425 | 319.11 | 302.92 | 298.88 | 335.30 | **1,256.21** |
| 426 | 111.57 | 120.24 | 115.62 | 143.95 | **491.39** |
| 427 | 1,429.06 | 1,359.11 | 1,298.41 | 1,312.29 | **5,398.88** |
| **KY - KENTUCKY - Total** | **42,106.49** | **40,397.62** | **40,230.79** | **41,066.46** | **163,801.36** |

**STATE: LA - LOUISIANA**

| REGISTRANT ZIP CODE 3 | 1ST QUARTER | 2ND QUARTER | 3RD QUARTER | 4TH QUARTER | TOTAL GRAMS |
|---|---|---|---|---|---|
| 700 | 8,327.53 | 7,815.91 | 8,112.48 | 8,214.80 | **32,470.72** |
| 701 | 4,692.44 | 4,465.82 | 4,286.61 | 4,571.04 | **18,015.91** |
| 703 | 4,992.47 | 4,915.01 | 4,839.85 | 5,051.44 | **19,798.77** |
| 704 | 9,175.03 | 8,759.95 | 8,568.02 | 9,038.59 | **35,541.59** |
| 705 | 17,143.56 | 16,693.79 | 16,598.41 | 17,023.89 | **67,459.65** |
| 706 | 8,944.05 | 8,778.19 | 8,574.38 | 8,966.33 | **35,262.94** |
| 707 | 11,962.05 | 11,451.58 | 11,652.18 | 12,096.74 | **47,162.55** |

| | | | | | |
|---|---|---|---|---|---|
| 708 | 8,714.86 | 8,592.88 | 8,180.69 | 8,485.93 | **33,974.36** |
| 710 | 1,988.09 | 1,956.87 | 2,017.57 | 2,069.02 | **8,031.54** |
| 711 | 6,176.42 | 5,634.16 | 5,895.46 | 5,835.34 | **23,541.39** |
| 712 | 3,889.46 | 3,498.08 | 3,671.51 | 3,906.22 | **14,965.27** |
| 713 | 3,371.48 | 3,272.62 | 3,363.39 | 3,498.66 | **13,506.15** |
| 714 | 1,599.02 | 1,602.49 | 1,654.52 | 1,656.83 | **6,512.87** |
| **LA - LOUISIANA - Total** | **90,976.44** | **87,437.36** | **87,415.08** | **90,414.84** | **356,243.72** |

**STATE: MA - MASSACHUSETTS**

| REGISTRANT ZIP CODE 3 | 1ST QUARTER | 2ND QUARTER | 3RD QUARTER | 4TH QUARTER | TOTAL GRAMS |
|---|---|---|---|---|---|
| 010 | 1,866.11 | 1,930.28 | 1,782.28 | 1,915.82 | **7,494.49** |
| 011 | 631.86 | 700.66 | 661.92 | 749.80 | **2,744.24** |
| 012 | 586.19 | 639.96 | 602.96 | 572.90 | **2,402.01** |
| 013 | 391.37 | 448.61 | 434.73 | 487.34 | **1,762.05** |
| 014 | 1,334.83 | 1,355.64 | 1,357.96 | 1,534.28 | **5,582.71** |
| 015 | 1,592.09 | 1,619.84 | 1,571.85 | 1,678.22 | **6,462.00** |
| 016 | 918.02 | 943.46 | 881.02 | 961.38 | **3,703.89** |
| 017 | 2,731.52 | 2,751.76 | 2,606.07 | 2,745.40 | **10,834.75** |
| 018 | 4,398.18 | 4,374.48 | 4,177.93 | 4,746.78 | **17,697.38** |
| 019 | 3,903.33 | 3,881.94 | 3,747.24 | 4,082.54 | **15,615.06** |
| 020 | 3,166.25 | 3,190.53 | 3,028.67 | 3,394.60 | **12,780.06** |
| 021 | 5,732.44 | 6,153.87 | 5,737.64 | 6,430.78 | **24,054.74** |
| 022 | 347.44 | 366.52 | 345.13 | 482.71 | **1,541.79** |
| 023 | 2,517.63 | 2,580.06 | 2,435.54 | 2,619.95 | **10,153.17** |
| 024 | 2,725.74 | 2,822.86 | 2,749.44 | 2,974.90 | **11,272.95** |
| 025 | 763.09 | 765.40 | 812.81 | 763.67 | **3,104.98** |
| 026 | 884.49 | 835.35 | 981.61 | 902.99 | **3,604.45** |
| 027 | 2,344.77 | 2,406.05 | 2,290.43 | 2,603.18 | **9,644.44** |
| **MA - MASSACHUSETTS - Total** | **36,835.38** | **37,767.27** | **36,205.25** | **39,647.25** | **150,455.15** |

**STATE: MD - MARYLAND**

| REGISTRANT ZIP CODE 3 | 1ST QUARTER | 2ND QUARTER | 3RD QUARTER | 4TH QUARTER | TOTAL GRAMS |
|---|---|---|---|---|---|
| 206 | 3,132.72 | 3,211.35 | 2,945.42 | 3,304.42 | **12,593.91** |
| 207 | 3,967.50 | 3,895.82 | 3,785.98 | 3,855.35 | **15,504.64** |
| 208 | 7,833.26 | 8,035.59 | 7,703.76 | 8,004.95 | **31,577.56** |
| 209 | 1,512.31 | 1,412.88 | 1,364.89 | 1,578.21 | **5,868.29** |
| 210 | 10,180.34 | 10,679.82 | 10,020.79 | 10,871.17 | **41,752.12** |
| 211 | 4,940.44 | 5,058.95 | 4,808.06 | 5,256.66 | **20,064.12** |
| 212 | 9,496.28 | 9,743.79 | 8,845.42 | 9,915.57 | **38,001.06** |
| 214 | 1,563.18 | 1,535.43 | 1,517.51 | 1,625.62 | **6,241.75** |
| 215 | 1,996.18 | 1,957.45 | 1,819.28 | 1,884.61 | **7,657.51** |
| 216 | 2,382.35 | 2,341.31 | 2,225.69 | 2,315.87 | **9,265.21** |

| | | | | | |
|---|---|---|---|---|---|
| 217 | 5,805.28 | 5,751.52 | 5,757.88 | 6,149.25 | **23,463.92** |
| 218 | 2,289.28 | 2,317.60 | 2,082.32 | 2,407.21 | **9,096.40** |
| 219 | 1,396.69 | 1,365.47 | 1,263.73 | 1,322.11 | **5,348.00** |
| **MD - MARYLAND - Total** | **56,495.81** | **57,306.97** | **54,140.71** | **58,491.00** | **226,434.49** |

**STATE: ME - MAINE**

| REGISTRANT ZIP CODE 3 | 1ST QUARTER | 2ND QUARTER | 3RD QUARTER | 4TH QUARTER | TOTAL GRAMS |
|---|---|---|---|---|---|
| 039 | 617.41 | 588.51 | 605.85 | 541.68 | **2,353.45** |
| 040 | 5,637.05 | 5,586.76 | 5,439.34 | 5,586.18 | **22,249.33** |
| 041 | 2,021.62 | 1,977.10 | 2,104.86 | 2,130.88 | **8,234.46** |
| 042 | 1,830.26 | 1,928.54 | 1,839.51 | 1,918.71 | **7,517.03** |
| 043 | 837.09 | 812.23 | 819.75 | 796.62 | **3,265.69** |
| 044 | 1,871.89 | 1,808.30 | 1,751.06 | 1,823.33 | **7,254.58** |
| 045 | 360.73 | 364.78 | 381.55 | 360.73 | **1,467.80** |
| 046 | 826.10 | 839.98 | 723.78 | 771.19 | **3,161.05** |
| 047 | 951.55 | 930.74 | 827.26 | 839.40 | **3,548.96** |
| 048 | 319.69 | 315.64 | 341.66 | 304.08 | **1,281.07** |
| 049 | 1,635.44 | 1,692.68 | 1,603.65 | 1,722.74 | **6,654.51** |
| **ME - MAINE - Total** | **16,908.85** | **16,845.26** | **16,438.27** | **16,795.54** | **66,987.92** |

**STATE: MI - MICHIGAN**

| REGISTRANT ZIP CODE 3 | 1ST QUARTER | 2ND QUARTER | 3RD QUARTER | 4TH QUARTER | TOTAL GRAMS |
|---|---|---|---|---|---|
| 480 | 7,748.85 | 7,669.65 | 7,444.77 | 8,035.01 | **30,898.29** |
| 481 | 7,390.43 | 7,041.26 | 6,869.56 | 7,885.86 | **29,187.11** |
| 482 | 1,646.43 | 1,579.95 | 1,536.59 | 1,680.54 | **6,443.50** |
| 483 | 6,563.17 | 6,979.03 | 6,638.90 | 7,614.16 | **27,795.25** |
| 484 | 1,986.35 | 1,891.54 | 1,856.86 | 1,990.40 | **7,725.15** |
| 485 | 802.40 | 807.61 | 759.05 | 875.24 | **3,244.30** |
| 486 | 2,617.64 | 2,500.86 | 2,375.99 | 2,580.06 | **10,074.55** |
| 487 | 1,098.39 | 1,049.83 | 993.75 | 1,113.42 | **4,255.39** |
| 488 | 4,840.43 | 4,768.17 | 4,441.54 | 4,972.82 | **19,022.96** |
| 489 | 1,217.48 | 1,166.61 | 1,048.67 | 1,203.60 | **4,636.36** |
| 490 | 6,059.64 | 5,694.29 | 5,549.18 | 5,955.59 | **23,258.70** |
| 491 | 901.26 | 839.98 | 802.98 | 905.88 | **3,450.10** |
| 492 | 1,577.06 | 1,493.23 | 1,445.25 | 1,522.72 | **6,038.25** |
| 493 | 1,981.73 | 1,848.76 | 1,853.39 | 2,016.41 | **7,700.29** |
| 494 | 5,408.70 | 5,401.19 | 5,080.92 | 5,601.21 | **21,492.02** |
| 495 | 4,251.35 | 4,144.40 | 4,241.52 | 4,338.06 | **16,975.33** |
| 496 | 2,604.92 | 2,628.62 | 2,521.09 | 2,544.80 | **10,299.43** |
| 497 | 1,406.52 | 1,441.78 | 1,443.52 | 1,500.75 | **5,792.56** |
| 498 | 1,198.40 | 1,235.40 | 1,208.23 | 1,238.29 | **4,880.32** |
| 499 | 442.82 | 376.34 | 397.73 | 416.81 | **1,633.71** |

| MI - MICHIGAN - Total | 61,743.97 | 60,558.49 | 58,509.50 | 63,991.62 | 244,803.58 |
|---|---|---|---|---|---|

**STATE: MN - MINNESOTA**

| REGISTRANT ZIP CODE 3 | 1ST QUARTER | 2ND QUARTER | 3RD QUARTER | 4TH QUARTER | TOTAL GRAMS |
|---|---|---|---|---|---|
| 550 | 2,778.35 | 2,871.42 | 2,860.44 | 2,913.05 | **11,423.26** |
| 551 | 4,129.95 | 4,088.32 | 3,979.06 | 4,275.63 | **16,472.96** |
| 553 | 5,911.65 | 5,868.87 | 5,734.17 | 6,261.98 | **23,776.67** |
| 554 | 5,977.55 | 6,118.61 | 5,811.06 | 6,377.60 | **24,284.82** |
| 556 | 20.81 | 16.76 | 28.33 | 19.08 | **84.98** |
| 557 | 980.46 | 1,018.03 | 969.47 | 1,082.78 | **4,050.75** |
| 558 | 354.38 | 372.87 | 453.81 | 431.26 | **1,612.32** |
| 559 | 1,309.40 | 1,233.09 | 1,221.53 | 1,290.90 | **5,054.91** |
| 560 | 2,981.26 | 2,887.03 | 2,915.94 | 3,083.59 | **11,867.81** |
| 561 | 584.46 | 650.94 | 624.35 | 631.29 | **2,491.03** |
| 562 | 685.05 | 674.64 | 689.67 | 744.01 | **2,793.38** |
| 563 | 1,412.88 | 1,351.02 | 1,377.61 | 1,492.65 | **5,634.16** |
| 564 | 874.09 | 882.76 | 898.37 | 923.80 | **3,579.02** |
| 565 | 1,381.08 | 1,366.63 | 1,355.64 | 1,487.45 | **5,590.81** |
| 566 | 922.65 | 950.97 | 786.22 | 859.06 | **3,518.89** |
| 567 | 550.35 | 517.40 | 476.35 | 481.56 | **2,025.66** |
| **MN - MINNESOTA - Total** | **30,854.35** | **30,869.38** | **30,182.02** | **32,355.68** | **124,261.44** |

**STATE: MO - MISSOURI**

| REGISTRANT ZIP CODE 3 | 1ST QUARTER | 2ND QUARTER | 3RD QUARTER | 4TH QUARTER | TOTAL GRAMS |
|---|---|---|---|---|---|
| 630 | 7,501.43 | 7,195.03 | 7,375.40 | 7,377.71 | **29,449.57** |
| 631 | 20,940.52 | 19,155.75 | 18,950.70 | 30,397.08 | **89,444.04** |
| 633 | 7,301.40 | 7,100.80 | 7,068.43 | 7,502.58 | **28,973.22** |
| 634 | 523.76 | 523.18 | 545.15 | 565.38 | **2,157.47** |
| 635 | 550.93 | 546.88 | 493.12 | 527.23 | **2,118.16** |
| 636 | 830.15 | 799.51 | 787.95 | 831.31 | **3,248.92** |
| 637 | 544.57 | 478.09 | 449.76 | 566.54 | **2,038.96** |
| 638 | 906.46 | 993.75 | 1,077.00 | 1,070.06 | **4,047.28** |
| 639 | 423.75 | 396.00 | 374.61 | 393.69 | **1,588.04** |
| 640 | 5,798.34 | 5,698.33 | 5,497.15 | 6,104.74 | **23,098.56** |
| 641 | 8,314.81 | 8,499.80 | 8,336.78 | 8,655.89 | **33,807.29** |
| 644 | 398.31 | 379.23 | 375.19 | 389.06 | **1,541.79** |
| 645 | 1,178.17 | 1,180.48 | 1,012.83 | 1,122.67 | **4,494.15** |
| 646 | 220.26 | 197.71 | 213.90 | 215.63 | **847.49** |
| 647 | 642.27 | 691.99 | 619.72 | 673.49 | **2,627.46** |
| 648 | 1,112.26 | 1,039.42 | 1,025.55 | 1,059.08 | **4,236.32** |
| 650 | 513.35 | 482.14 | 477.51 | 546.30 | **2,019.30** |
| 651 | 738.81 | 711.64 | 745.17 | 742.86 | **2,938.48** |

| | | | | | |
|---|---|---|---|---|---|
| 652 | 2,192.16 | 2,085.21 | 2,175.39 | 2,348.82 | **8,801.57** |
| 653 | 445.14 | 429.53 | 464.21 | 442.82 | **1,781.70** |
| 654 | 411.03 | 481.56 | 417.97 | 401.78 | **1,712.33** |
| 655 | 601.80 | 566.54 | 576.37 | 600.65 | **2,345.35** |
| 656 | 675.22 | 690.25 | 731.87 | 768.87 | **2,866.22** |
| 657 | 1,185.11 | 1,193.20 | 1,230.20 | 1,205.92 | **4,814.42** |
| 658 | 1,855.12 | 1,753.38 | 1,816.39 | 1,860.33 | **7,285.22** |
| **MO - MISSOURI - Total** | **65,805.12** | **63,269.40** | **62,838.31** | **76,370.48** | **268,283.32** |

**STATE: MS - MISSISSIPPI**

| REGISTRANT ZIP CODE 3 | 1ST QUARTER | 2ND QUARTER | 3RD QUARTER | 4TH QUARTER | TOTAL GRAMS |
|---|---|---|---|---|---|
| 386 | 6,958.01 | 6,383.38 | 6,116.88 | 6,287.42 | **25,745.68** |
| 387 | 556.13 | 563.07 | 537.63 | 513.35 | **2,170.19** |
| 388 | 2,904.37 | 2,678.92 | 2,692.79 | 2,808.99 | **11,085.07** |
| 389 | 826.10 | 833.04 | 753.26 | 791.42 | **3,203.83** |
| 390 | 2,888.19 | 2,602.03 | 2,633.25 | 2,780.66 | **10,904.12** |
| 391 | 3,352.40 | 3,022.31 | 3,191.69 | 3,150.07 | **12,716.47** |
| 392 | 2,883.56 | 2,811.88 | 3,021.15 | 2,893.97 | **11,610.56** |
| 393 | 2,407.21 | 2,287.54 | 2,218.17 | 2,270.20 | **9,183.12** |
| 394 | 6,641.21 | 5,952.12 | 6,353.32 | 6,180.47 | **25,127.12** |
| 395 | 6,064.27 | 5,664.22 | 5,738.22 | 6,001.26 | **23,467.97** |
| 396 | 1,834.31 | 1,634.29 | 1,671.87 | 1,756.27 | **6,896.73** |
| 397 | 2,093.88 | 1,663.19 | 1,868.42 | 1,887.50 | **7,512.99** |
| **MS - MISSISSIPPI - Total** | **39,409.66** | **36,095.99** | **36,796.64** | **37,321.56** | **149,623.84** |

**STATE: MT - MONTANA**

| REGISTRANT ZIP CODE 3 | 1ST QUARTER | 2ND QUARTER | 3RD QUARTER | 4TH QUARTER | TOTAL GRAMS |
|---|---|---|---|---|---|
| 590 | 257.83 | 215.05 | 226.04 | 253.79 | **952.71** |
| 591 | 1,181.64 | 1,166.03 | 1,166.61 | 1,193.20 | **4,707.47** |
| 592 | 125.45 | 135.85 | 109.26 | 142.79 | **513.35** |
| 593 | 230.08 | 234.71 | 215.05 | 237.02 | **916.87** |
| 594 | 750.95 | 709.91 | 682.74 | 691.99 | **2,835.58** |
| 595 | 176.90 | 157.82 | 170.54 | 166.49 | **671.75** |
| 596 | 390.80 | 442.25 | 404.09 | 413.92 | **1,651.05** |
| 597 | 1,197.82 | 1,045.20 | 1,209.96 | 1,317.49 | **4,770.48** |
| 598 | 1,359.11 | 1,332.52 | 1,401.31 | 1,476.47 | **5,569.42** |
| 599 | 486.76 | 513.35 | 510.46 | 539.95 | **2,050.52** |
| **MT - MONTANA - Total** | **6,157.34** | **5,952.70** | **6,096.06** | **6,433.10** | **24,639.20** |

**STATE: NC - NORTH CAROLINA**

| REGISTRANT ZIP CODE 3 | 1ST QUARTER | 2ND QUARTER | 3RD QUARTER | 4TH QUARTER | TOTAL GRAMS |
|---|---|---|---|---|---|
| 270 | 2,358.07 | 2,345.35 | 2,289.28 | 2,580.06 | **9,572.76** |
| 271 | 3,639.72 | 3,396.34 | 3,334.48 | 3,753.03 | **14,123.56** |

| | | | | | |
|---|---|---|---|---|---|
| 272 | 6,098.96 | 6,088.55 | 6,076.41 | 6,483.97 | **24,747.88** |
| 273 | 3,659.37 | 3,450.68 | 3,290.55 | 3,791.76 | **14,192.36** |
| 274 | 3,692.90 | 3,749.56 | 3,596.94 | 3,896.97 | **14,936.37** |
| 275 | 12,426.84 | 12,229.71 | 11,698.43 | 13,680.74 | **50,035.71** |
| 276 | 5,837.08 | 5,600.63 | 5,454.37 | 6,110.52 | **23,002.60** |
| 277 | 2,267.89 | 2,326.85 | 2,134.35 | 2,500.86 | **9,229.94** |
| 278 | 5,283.83 | 4,946.22 | 4,796.50 | 5,615.09 | **20,641.64** |
| 279 | 1,224.42 | 1,216.32 | 1,189.73 | 1,388.02 | **5,018.49** |
| 280 | 10,150.28 | 9,661.79 | 9,357.13 | 10,032.35 | **39,201.54** |
| 281 | 7,517.03 | 6,974.20 | 6,833.14 | 7,153.99 | **28,478.36** |
| 282 | 8,150.05 | 8,191.68 | 7,533.80 | 8,461.65 | **32,337.18** |
| 283 | 8,324.64 | 8,029.23 | 7,228.56 | 8,392.86 | **31,975.29** |
| 284 | 6,526.75 | 6,421.53 | 5,848.06 | 6,472.41 | **25,268.75** |
| 285 | 5,800.66 | 5,716.83 | 5,153.76 | 5,483.28 | **22,154.53** |
| 286 | 8,370.31 | 8,762.84 | 8,240.24 | 8,973.85 | **34,347.23** |
| 287 | 3,955.36 | 3,813.73 | 3,784.82 | 4,116.65 | **15,670.56** |
| 288 | 1,682.27 | 1,705.97 | 1,776.50 | 1,856.28 | **7,021.02** |
| 289 | 274.02 | 291.36 | 265.35 | 293.10 | **1,123.83** |
| **NC - NORTH CAROLINA - Total** | **107,240.44** | **104,919.37** | **99,882.38** | **111,037.40** | **423,079.59** |

**STATE: ND - NORTH DAKOTA**

| REGISTRANT ZIP CODE 3 | 1ST QUARTER | 2ND QUARTER | 3RD QUARTER | 4TH QUARTER | TOTAL GRAMS |
|---|---|---|---|---|---|
| 580 | 666.55 | 554.40 | 535.90 | 630.13 | **2,386.97** |
| 581 | 1,731.99 | 1,585.73 | 1,655.68 | 1,716.38 | **6,689.77** |
| 582 | 1,284.54 | 1,163.14 | 1,193.78 | 1,271.82 | **4,913.27** |
| 583 | 302.92 | 271.13 | 239.33 | 274.02 | **1,087.41** |
| 584 | 200.60 | 193.66 | 180.95 | 198.87 | **774.08** |
| 585 | 545.15 | 548.62 | 541.10 | 659.61 | **2,294.48** |
| 586 | 142.21 | 138.74 | 167.07 | 144.53 | **592.55** |
| 587 | 913.98 | 946.35 | 935.94 | 884.49 | **3,680.76** |
| 588 | 250.90 | 247.43 | 275.75 | 328.94 | **1,103.01** |
| **ND - NORTH DAKOTA - Total** | **6,038.83** | **5,649.19** | **5,725.50** | **6,108.78** | **23,522.31** |

**STATE: NE - NEBRASKA**

| REGISTRANT ZIP CODE 3 | 1ST QUARTER | 2ND QUARTER | 3RD QUARTER | 4TH QUARTER | TOTAL GRAMS |
|---|---|---|---|---|---|
| 680 | 1,644.12 | 1,627.93 | 1,766.67 | 1,763.78 | **6,802.50** |
| 681 | 6,720.41 | 6,605.95 | 6,513.45 | 7,258.62 | **27,098.44** |
| 683 | 518.56 | 505.26 | 488.49 | 496.59 | **2,008.90** |
| 684 | 348.59 | 381.55 | 402.94 | 388.48 | **1,521.56** |
| 685 | 4,115.93 | 4,035.24 | 4,209.31 | 4,444.26 | **16,804.73** |
| 686 | 528.96 | 423.17 | 424.90 | 504.10 | **1,881.14** |
| 687 | 576.37 | 563.07 | 553.82 | 563.65 | **2,256.90** |

| | | | | | |
|---|---|---|---|---|---|
| 688 | 1,307.08 | 1,160.82 | 1,267.20 | 1,359.69 | **5,094.80** |
| 689 | 472.89 | 420.28 | 417.39 | 441.67 | **1,752.22** |
| 690 | 162.45 | 157.24 | 169.38 | 158.98 | **648.05** |
| 691 | 394.26 | 335.88 | 373.45 | 403.51 | **1,507.11** |
| 692 | 21.39 | 30.06 | 29.48 | 50.29 | **131.23** |
| 693 | 508.15 | 489.65 | 447.45 | 497.74 | **1,942.99** |
| **NE - NEBRASKA - Total** | **17,319.15** | **16,736.10** | **17,063.94** | **18,331.38** | **69,450.56** |

**STATE: NH - NEW HAMPSHIRE**

| REGISTRANT ZIP CODE 3 | 1ST QUARTER | 2ND QUARTER | 3RD QUARTER | 4TH QUARTER | TOTAL GRAMS |
|---|---|---|---|---|---|
| 030 | 3,907.38 | 4,081.39 | 3,803.90 | 4,209.15 | **16,001.81** |
| 031 | 2,147.06 | 2,252.86 | 2,230.89 | 2,414.15 | **9,044.95** |
| 032 | 1,181.06 | 1,172.96 | 1,104.17 | 1,192.04 | **4,650.24** |
| 033 | 975.25 | 872.93 | 900.68 | 876.40 | **3,625.27** |
| 034 | 399.47 | 406.40 | 360.16 | 432.42 | **1,598.45** |
| 035 | 310.44 | 282.69 | 300.03 | 289.63 | **1,182.79** |
| 036 | 29.48 | 33.53 | 34.11 | 35.84 | **132.96** |
| 037 | 823.21 | 897.79 | 800.67 | 838.25 | **3,359.92** |
| 038 | 4,068.09 | 4,373.90 | 4,224.75 | 4,275.05 | **16,941.80** |
| **NH - NEW HAMPSHIRE - Total** | **13,841.45** | **14,374.46** | **13,759.36** | **14,562.92** | **56,538.18** |

**STATE: NJ - NEW JERSEY**

| REGISTRANT ZIP CODE 3 | 1ST QUARTER | 2ND QUARTER | 3RD QUARTER | 4TH QUARTER | TOTAL GRAMS |
|---|---|---|---|---|---|
| 070 | 8,480.73 | 8,789.43 | 8,243.71 | 8,967.49 | **34,481.35** |
| 071 | 539.37 | 561.91 | 535.32 | 580.99 | **2,217.59** |
| 072 | 376.34 | 381.55 | 317.96 | 375.19 | **1,451.03** |
| 073 | 603.54 | 616.83 | 671.75 | 686.78 | **2,578.90** |
| 074 | 3,433.91 | 3,429.87 | 3,359.34 | 3,520.05 | **13,743.17** |
| 075 | 413.34 | 377.50 | 341.08 | 412.76 | **1,544.68** |
| 076 | 3,193.42 | 3,341.42 | 3,122.90 | 3,491.15 | **13,148.88** |
| 077 | 3,191.00 | 3,383.07 | 3,269.88 | 3,391.22 | **13,235.17** |
| 078 | 2,904.37 | 2,983.57 | 2,656.95 | 2,928.08 | **11,472.97** |
| 079 | 3,139.66 | 3,120.58 | 3,158.74 | 3,352.98 | **12,771.96** |
| 080 | 8,771.51 | 8,880.77 | 8,299.20 | 8,795.79 | **34,747.28** |
| 081 | 600.07 | 606.43 | 568.85 | 646.89 | **2,422.24** |
| 082 | 1,186.84 | 1,207.07 | 1,297.26 | 1,168.92 | **4,860.09** |
| 083 | 840.56 | 913.40 | 834.78 | 822.06 | **3,410.79** |
| 084 | 83.25 | 94.81 | 122.56 | 92.50 | **393.11** |
| 085 | 2,151.69 | 2,211.23 | 2,060.93 | 2,244.76 | **8,668.61** |
| 086 | 1,463.75 | 1,446.41 | 1,434.84 | 1,450.45 | **5,795.45** |
| 087 | 2,365.01 | 2,303.73 | 2,257.48 | 2,304.31 | **9,230.52** |
| 088 | 4,244.41 | 4,252.50 | 3,954.78 | 4,264.64 | **16,716.34** |

| | | | | | |
|---|---|---|---|---|---|
| 089 | 497.17 | 435.31 | 453.81 | 500.06 | **1,886.34** |
| **NJ - NEW JERSEY - Total** | **48,479.93** | **49,337.40** | **46,962.10** | **49,997.07** | **194,776.49** |

**STATE: NM - NEW MEXICO**

| REGISTRANT ZIP CODE 3 | 1ST QUARTER | 2ND QUARTER | 3RD QUARTER | 4TH QUARTER | TOTAL GRAMS |
|---|---|---|---|---|---|
| 870 | 233.55 | 232.97 | 209.27 | 244.54 | **920.34** |
| 871 | 1,807.72 | 1,728.52 | 1,630.82 | 1,678.80 | **6,845.86** |
| 873 | 17.34 | 31.22 | 23.12 | 32.37 | **104.06** |
| 874 | 126.60 | 131.81 | 139.90 | 127.18 | **525.49** |
| 875 | 431.26 | 357.27 | 416.81 | 464.79 | **1,670.13** |
| 877 | 80.93 | 64.17 | 72.26 | 60.12 | **277.49** |
| 878 | 43.36 | 44.51 | 49.14 | 47.98 | **184.99** |
| 879 | 16.19 | 11.56 | 27.17 | 14.45 | **69.37** |
| 880 | 377.50 | 360.16 | 398.89 | 399.47 | **1,536.01** |
| 881 | 242.80 | 67.06 | 96.54 | 93.07 | **499.48** |
| 882 | 557.87 | 515.09 | 619.15 | 615.10 | **2,307.20** |
| 883 | 183.84 | 132.38 | 167.07 | 172.27 | **655.57** |
| 884 | 26.59 | 13.87 | 14.45 | 27.75 | **82.67** |
| **NM - NEW MEXICO - Total** | **4,145.56** | **3,690.59** | **3,864.60** | **3,977.91** | **15,678.65** |

**STATE: NV - NEVADA**

| REGISTRANT ZIP CODE 3 | 1ST QUARTER | 2ND QUARTER | 3RD QUARTER | 4TH QUARTER | TOTAL GRAMS |
|---|---|---|---|---|---|
| 890 | 1,984.62 | 1,934.32 | 1,809.45 | 1,974.21 | **7,702.60** |
| 891 | 3,949.74 | 3,785.57 | 3,975.65 | 3,803.32 | **15,514.28** |
| 893 | 10.41 | 17.92 | 13.87 | 11.56 | **53.76** |
| 894 | 735.34 | 719.16 | 709.91 | 744.59 | **2,909.00** |
| 895 | 1,027.86 | 1,083.36 | 924.96 | 1,044.05 | **4,080.23** |
| 897 | 211.01 | 234.13 | 206.38 | 213.90 | **865.42** |
| 898 | 76.89 | 101.17 | 83.25 | 94.81 | **356.11** |
| **NV - NEVADA - Total** | **7,995.86** | **7,875.63** | **7,723.47** | **7,886.44** | **31,481.41** |

**STATE: NY - NEW YORK**

| REGISTRANT ZIP CODE 3 | 1ST QUARTER | 2ND QUARTER | 3RD QUARTER | 4TH QUARTER | TOTAL GRAMS |
|---|---|---|---|---|---|
| 100 | 16,697.25 | 17,951.48 | 17,225.65 | 18,767.42 | **70,641.80** |
| 101 | 952.71 | 967.74 | 903.57 | 993.75 | **3,817.77** |
| 102 | 126.03 | 179.79 | 95.96 | 149.15 | **550.93** |
| 103 | 1,139.44 | 1,020.92 | 966.58 | 1,198.40 | **4,325.34** |
| 104 | 1,135.97 | 1,140.59 | 959.07 | 1,133.08 | **4,368.70** |
| 105 | 5,226.60 | 5,298.86 | 4,997.67 | 5,434.14 | **20,957.28** |
| 106 | 513.35 | 596.02 | 471.15 | 601.80 | **2,182.33** |
| 107 | 720.89 | 643.43 | 659.61 | 766.56 | **2,790.49** |
| 108 | 426.64 | 413.92 | 363.62 | 454.96 | **1,659.15** |
| 109 | 2,571.97 | 2,621.68 | 2,429.18 | 2,647.12 | **10,269.95** |

| | | | | | |
|---|---|---|---|---|---|
| 110 | 1,282.80 | 1,302.46 | 1,423.86 | 1,590.35 | **5,599.48** |
| 111 | 759.62 | 699.50 | 736.50 | 781.01 | **2,976.64** |
| 112 | 5,812.80 | 6,136.53 | 5,516.23 | 6,002.99 | **23,468.55** |
| 113 | 1,633.13 | 1,751.64 | 1,624.46 | 1,805.98 | **6,815.22** |
| 114 | 513.35 | 504.10 | 464.21 | 564.23 | **2,045.90** |
| 115 | 4,505.71 | 4,346.16 | 4,228.22 | 4,574.51 | **17,654.60** |
| 116 | 359.00 | 330.67 | 328.94 | 322.58 | **1,341.19** |
| 117 | 8,672.66 | 8,964.60 | 8,156.41 | 9,106.23 | **34,899.90** |
| 118 | 586.19 | 500.63 | 544.57 | 534.74 | **2,166.14** |
| 119 | 998.96 | 1,044.05 | 1,130.76 | 1,004.74 | **4,178.51** |
| 120 | 1,925.65 | 1,938.37 | 1,824.48 | 1,882.29 | **7,570.80** |
| 121 | 1,093.77 | 1,075.84 | 1,089.14 | 1,097.81 | **4,356.56** |
| 122 | 693.72 | 666.55 | 675.22 | 755.00 | **2,790.49** |
| 123 | 773.50 | 803.56 | 711.64 | 771.76 | **3,060.46** |
| 124 | 515.09 | 463.06 | 452.07 | 433.58 | **1,863.79** |
| 125 | 2,292.17 | 2,318.18 | 2,211.23 | 2,281.76 | **9,103.34** |
| 126 | 468.26 | 457.86 | 478.67 | 519.13 | **1,923.92** |
| 127 | 430.68 | 442.25 | 440.51 | 422.01 | **1,735.46** |
| 128 | 1,438.27 | 1,472.97 | 1,425.02 | 1,478.78 | **5,815.04** |
| 129 | 448.61 | 406.40 | 425.48 | 419.12 | **1,699.61** |
| 130 | 1,833.73 | 1,756.85 | 1,683.43 | 1,786.33 | **7,060.34** |
| 131 | 726.67 | 779.86 | 668.28 | 787.95 | **2,962.76** |
| 132 | 1,007.63 | 885.65 | 907.62 | 974.10 | **3,774.99** |
| 133 | 520.29 | 480.98 | 452.07 | 500.63 | **1,953.98** |
| 134 | 711.06 | 664.82 | 650.94 | 669.44 | **2,696.26** |
| 135 | 249.16 | 222.57 | 193.66 | 217.37 | **882.76** |
| 136 | 1,219.21 | 1,243.49 | 1,156.78 | 1,272.98 | **4,892.46** |
| 137 | 646.32 | 550.93 | 534.16 | 565.96 | **2,297.37** |
| 138 | 876.40 | 748.64 | 691.99 | 840.56 | **3,157.58** |
| 139 | 607.01 | 537.05 | 600.65 | 520.87 | **2,265.57** |
| 140 | 1,600.18 | 1,623.30 | 1,445.83 | 1,542.37 | **6,211.68** |
| 141 | 593.13 | 605.27 | 560.18 | 650.36 | **2,408.94** |
| 142 | 2,130.30 | 2,214.12 | 2,032.60 | 2,318.76 | **8,695.78** |
| 143 | 171.70 | 171.70 | 189.62 | 158.40 | **691.41** |
| 144 | 2,201.40 | 2,172.50 | 2,093.88 | 2,256.32 | **8,724.11** |
| 145 | 1,810.61 | 1,839.51 | 1,790.95 | 1,897.32 | **7,338.40** |
| 146 | 2,778.12 | 2,764.76 | 2,850.61 | 2,978.37 | **11,371.86** |
| 147 | 677.53 | 663.08 | 652.67 | 601.22 | **2,594.51** |
| 148 | 1,169.50 | 1,190.89 | 1,068.33 | 1,262.57 | **4,691.28** |
| 149 | 284.43 | 318.53 | 316.80 | 319.11 | **1,238.87** |

| NY - NEW YORK - Total | 86,529.15 | 87,894.32 | 83,500.76 | 90,616.00 | 348,540.24 |
|---|---|---|---|---|---|

**STATE: OH - OHIO**

| REGISTRANT ZIP CODE 3 | 1ST QUARTER | 2ND QUARTER | 3RD QUARTER | 4TH QUARTER | TOTAL GRAMS |
|---|---|---|---|---|---|
| 430 | 9,402.80 | 9,107.39 | 9,151.90 | 9,679.13 | **37,341.21** |
| 431 | 3,749.56 | 3,601.56 | 3,614.28 | 3,775.57 | **14,740.97** |
| 432 | 18,827.56 | 12,695.65 | 6,627.34 | 6,930.26 | **45,080.82** |
| 433 | 904.73 | 879.29 | 863.10 | 905.88 | **3,553.00** |
| 434 | 1,127.30 | 944.04 | 976.41 | 1,111.11 | **4,158.85** |
| 435 | 2,252.86 | 2,163.25 | 2,099.66 | 2,265.57 | **8,781.34** |
| 436 | 2,981.84 | 2,590.47 | 2,625.15 | 2,707.24 | **10,904.70** |
| 437 | 933.63 | 971.21 | 926.69 | 1,017.46 | **3,848.99** |
| 438 | 187.30 | 228.93 | 191.93 | 211.01 | **819.17** |
| 439 | 1,269.51 | 1,138.28 | 1,066.59 | 1,250.43 | **4,724.81** |
| 440 | 9,405.69 | 9,133.40 | 9,039.75 | 9,364.06 | **36,942.90** |
| 441 | 10,527.78 | 10,391.49 | 10,120.22 | 10,597.73 | **41,637.22** |
| 442 | 4,554.85 | 4,737.53 | 4,569.88 | 4,678.56 | **18,540.82** |
| 443 | 1,725.63 | 1,748.17 | 1,721.58 | 1,816.39 | **7,011.77** |
| 444 | 2,317.60 | 2,207.19 | 2,084.05 | 2,151.11 | **8,759.95** |
| 445 | 1,926.23 | 2,017.57 | 1,933.74 | 2,019.88 | **7,897.42** |
| 446 | 3,199.21 | 3,165.68 | 2,973.75 | 3,191.11 | **12,529.74** |
| 447 | 2,687.01 | 2,470.22 | 2,478.89 | 2,692.21 | **10,328.33** |
| 448 | 1,762.05 | 1,700.77 | 1,699.61 | 1,776.50 | **6,938.93** |
| 449 | 707.59 | 695.45 | 642.27 | 712.22 | **2,757.54** |
| 450 | 6,453.59 | 6,307.63 | 6,088.61 | 6,678.85 | **25,528.68** |
| 451 | 2,336.68 | 2,248.23 | 2,223.95 | 2,428.60 | **9,237.46** |
| 452 | 7,619.36 | 7,515.30 | 7,255.73 | 7,870.25 | **30,260.64** |
| 453 | 3,403.85 | 3,257.59 | 3,185.33 | 3,593.47 | **13,440.25** |
| 454 | 3,781.35 | 3,518.89 | 3,544.33 | 3,620.64 | **14,465.22** |
| 455 | 805.87 | 721.47 | 726.67 | 716.27 | **2,970.28** |
| 456 | 1,722.74 | 1,589.78 | 1,567.23 | 1,715.22 | **6,594.96** |
| 457 | 1,565.49 | 1,493.23 | 1,325.58 | 1,449.87 | **5,834.19** |
| 458 | 1,134.23 | 1,210.54 | 1,110.53 | 1,184.53 | **4,639.83** |
| **OH - OHIO - Total** | **109,273.88** | **100,450.21** | **92,434.78** | **98,111.14** | **400,270.01** |

**STATE: OK - OKLAHOMA**

| REGISTRANT ZIP CODE 3 | 1ST QUARTER | 2ND QUARTER | 3RD QUARTER | 4TH QUARTER | TOTAL GRAMS |
|---|---|---|---|---|---|
| 730 | 8,822.96 | 8,658.78 | 8,696.36 | 9,050.73 | **35,228.84** |
| 731 | 7,024.92 | 6,747.71 | 6,663.86 | 6,839.13 | **27,275.63** |
| 734 | 806.45 | 679.85 | 697.77 | 764.25 | **2,948.31** |
| 735 | 1,352.18 | 1,335.41 | 1,324.43 | 1,352.75 | **5,364.77** |
| 736 | 357.84 | 306.39 | 326.63 | 357.27 | **1,348.13** |

| | | | | | |
|---|---|---|---|---|---|
| 737 | 836.51 | 859.06 | 856.74 | 951.55 | **3,503.86** |
| 738 | 94.81 | 93.65 | 98.86 | 92.50 | **379.81** |
| 739 | 70.53 | 54.92 | 60.70 | 64.75 | **250.90** |
| 740 | 7,241.28 | 6,749.32 | 6,734.87 | 7,244.75 | **27,970.21** |
| 741 | 4,505.71 | 4,390.67 | 4,123.01 | 4,432.87 | **17,452.26** |
| 743 | 1,158.51 | 1,155.04 | 1,077.58 | 1,140.59 | **4,531.73** |
| 744 | 1,816.97 | 1,694.99 | 1,761.47 | 1,887.50 | **7,160.92** |
| 745 | 649.78 | 676.38 | 672.91 | 620.30 | **2,619.37** |
| 746 | 461.90 | 371.72 | 357.27 | 389.64 | **1,580.53** |
| 747 | 658.46 | 574.05 | 589.08 | 648.05 | **2,469.64** |
| 748 | 1,596.71 | 1,478.20 | 1,540.06 | 1,668.40 | **6,283.37** |
| 749 | 617.99 | 571.74 | 539.37 | 566.54 | **2,295.64** |
| **OK - OKLAHOMA - Total** | **38,073.52** | **36,397.88** | **36,120.95** | **38,071.56** | **148,663.91** |

**STATE: OR - OREGON**

| REGISTRANT ZIP CODE 3 | 1ST QUARTER | 2ND QUARTER | 3RD QUARTER | 4TH QUARTER | TOTAL GRAMS |
|---|---|---|---|---|---|
| 970 | 3,650.70 | 3,727.10 | 3,564.56 | 3,777.88 | **14,720.25** |
| 971 | 1,116.89 | 1,108.22 | 1,194.93 | 1,128.45 | **4,548.49** |
| 972 | 4,798.81 | 4,899.98 | 4,564.68 | 4,902.87 | **19,166.33** |
| 973 | 1,746.44 | 1,685.16 | 1,649.32 | 1,731.99 | **6,812.91** |
| 974 | 1,779.97 | 1,822.75 | 1,677.07 | 1,832.58 | **7,112.36** |
| 975 | 622.04 | 667.71 | 705.28 | 639.38 | **2,634.40** |
| 976 | 93.65 | 110.42 | 89.61 | 100.01 | **393.69** |
| 977 | 1,186.84 | 1,338.30 | 1,434.84 | 1,269.51 | **5,229.49** |
| 978 | 407.56 | 394.84 | 417.97 | 461.32 | **1,681.69** |
| 979 | 106.37 | 115.62 | 115.62 | 101.17 | **438.78** |
| **OR - OREGON - Total** | **15,509.27** | **15,870.09** | **15,413.88** | **15,945.15** | **62,738.39** |

**STATE: PA - PENNSYLVANIA**

| REGISTRANT ZIP CODE 3 | 1ST QUARTER | 2ND QUARTER | 3RD QUARTER | 4TH QUARTER | TOTAL GRAMS |
|---|---|---|---|---|---|
| 150 | 2,622.84 | 2,824.60 | 2,785.29 | 3,033.87 | **11,266.59** |
| 151 | 2,078.27 | 2,131.45 | 2,032.60 | 2,215.28 | **8,457.60** |
| 152 | 4,411.48 | 4,355.41 | 4,402.23 | 4,797.07 | **17,966.19** |
| 153 | 1,012.25 | 998.96 | 1,038.27 | 1,140.01 | **4,189.49** |
| 154 | 845.76 | 845.76 | 797.78 | 872.93 | **3,362.23** |
| 155 | 339.34 | 289.05 | 286.74 | 344.55 | **1,259.68** |
| 156 | 1,938.37 | 1,881.14 | 1,858.01 | 2,102.55 | **7,780.07** |
| 157 | 1,296.68 | 1,263.15 | 1,155.62 | 1,258.52 | **4,973.97** |
| 158 | 1,142.90 | 1,114.58 | 1,058.50 | 1,087.98 | **4,403.97** |
| 159 | 831.89 | 861.37 | 826.10 | 782.75 | **3,302.11** |
| 160 | 1,473.00 | 1,534.86 | 1,478.78 | 1,533.12 | **6,019.76** |
| 161 | 1,667.24 | 1,667.82 | 1,660.88 | 1,764.36 | **6,760.30** |

| | | | | | |
|---|---|---|---|---|---|
| 162 | 700.66 | 727.25 | 653.83 | 720.31 | **2,802.05** |
| 163 | 1,115.73 | 1,049.25 | 1,022.08 | 1,068.33 | **4,255.39** |
| 164 | 759.05 | 641.11 | 678.69 | 753.26 | **2,832.11** |
| 165 | 2,960.45 | 2,919.98 | 2,786.44 | 2,977.79 | **11,644.67** |
| 166 | 1,466.06 | 1,416.35 | 1,349.29 | 1,492.65 | **5,724.35** |
| 167 | 510.46 | 487.34 | 515.67 | 549.20 | **2,062.66** |
| 168 | 1,507.11 | 1,293.21 | 1,382.82 | 1,374.14 | **5,557.28** |
| 169 | 445.72 | 372.87 | 422.59 | 392.53 | **1,633.71** |
| 170 | 4,606.88 | 4,675.67 | 4,448.48 | 4,634.05 | **18,365.08** |
| 171 | 1,289.74 | 1,327.90 | 1,200.71 | 1,382.24 | **5,200.59** |
| 172 | 1,292.05 | 1,291.48 | 1,325.58 | 1,265.46 | **5,174.57** |
| 173 | 2,126.83 | 2,119.31 | 2,052.83 | 2,114.11 | **8,413.09** |
| 174 | 1,318.07 | 1,317.49 | 1,175.28 | 1,265.46 | **5,076.30** |
| 175 | 1,471.84 | 1,524.45 | 1,398.42 | 1,361.43 | **5,756.14** |
| 176 | 1,592.67 | 1,557.98 | 1,664.93 | 1,551.62 | **6,367.19** |
| 177 | 788.53 | 712.80 | 725.52 | 769.45 | **2,996.29** |
| 178 | 751.53 | 740.55 | 769.45 | 802.40 | **3,063.93** |
| 179 | 315.06 | 290.78 | 302.35 | 273.44 | **1,181.64** |
| 180 | 2,198.51 | 2,335.52 | 2,224.53 | 2,329.74 | **9,088.31** |
| 181 | 915.71 | 862.53 | 838.25 | 952.13 | **3,568.61** |
| 182 | 302.92 | 306.39 | 267.08 | 276.33 | **1,152.73** |
| 183 | 535.90 | 533.59 | 548.62 | 549.77 | **2,167.88** |
| 184 | 500.06 | 583.88 | 605.85 | 578.68 | **2,268.46** |
| 185 | 426.06 | 515.09 | 429.53 | 470.57 | **1,841.25** |
| 186 | 422.01 | 434.73 | 437.04 | 465.95 | **1,759.74** |
| 187 | 11,245.20 | 11,182.77 | 10,490.20 | 10,986.79 | **43,904.96** |
| 188 | 190.19 | 180.37 | 157.82 | 158.40 | **686.78** |
| 189 | 2,367.32 | 2,279.45 | 2,369.05 | 2,317.60 | **9,333.42** |
| 190 | 7,967.37 | 8,074.90 | 7,626.87 | 8,289.95 | **31,959.10** |
| 191 | 6,184.51 | 6,611.73 | 6,091.44 | 7,238.97 | **26,126.65** |
| 193 | 4,634.05 | 4,616.13 | 4,576.24 | 4,716.72 | **18,543.14** |
| 194 | 3,714.87 | 3,710.82 | 3,588.84 | 4,074.45 | **15,088.99** |
| 195 | 935.94 | 915.71 | 849.81 | 897.79 | **3,599.25** |
| 196 | 1,715.22 | 1,710.02 | 1,665.51 | 1,642.96 | **6,733.71** |
| **PA - PENNSYLVANIA - Total** | **88,934.33** | **89,087.52** | **86,022.44** | **91,627.69** | **355,671.98** |

**STATE: PR - PUERTO RICO**

| REGISTRANT ZIP CODE 3 | 1ST QUARTER | 2ND QUARTER | 3RD QUARTER | 4TH QUARTER | **TOTAL GRAMS** |
|---|---|---|---|---|---|
| 006 | 368.83 | 350.33 | 301.77 | 374.03 | **1,394.96** |
| 007 | 283.27 | 256.68 | 247.43 | 315.06 | **1,102.44** |
| 009 | 1,618.10 | 1,435.42 | 1,443.52 | 1,483.40 | **5,980.44** |

| PR - PUERTO RICO - Total | 2,270.20 | 2,042.43 | 1,992.71 | 2,172.50 | 8,477.84 |
|---|---|---|---|---|---|

**STATE: RI - RHODE ISLAND**

| REGISTRANT ZIP CODE 3 | 1ST QUARTER | 2ND QUARTER | 3RD QUARTER | 4TH QUARTER | TOTAL GRAMS |
|---|---|---|---|---|---|
| 028 | 5,137.57 | 5,022.53 | 4,979.18 | 5,175.15 | **20,314.43** |
| 029 | 2,257.48 | 2,380.62 | 2,188.11 | 2,317.60 | **9,143.81** |
| RI - RHODE ISLAND - Total | 7,395.06 | 7,403.15 | 7,167.28 | 7,492.75 | 29,458.24 |

**STATE: SC - SOUTH CAROLINA**

| REGISTRANT ZIP CODE 3 | 1ST QUARTER | 2ND QUARTER | 3RD QUARTER | 4TH QUARTER | TOTAL GRAMS |
|---|---|---|---|---|---|
| 290 | 5,366.50 | 5,156.07 | 5,108.09 | 5,472.29 | **21,102.96** |
| 291 | 3,353.56 | 3,372.06 | 3,111.91 | 3,425.82 | **13,263.35** |
| 292 | 4,790.71 | 4,461.78 | 4,351.36 | 4,672.20 | **18,276.05** |
| 293 | 5,028.89 | 4,909.23 | 4,458.89 | 5,178.04 | **19,575.04** |
| 294 | 13,608.39 | 13,637.84 | 13,218.95 | 13,735.66 | **54,200.83** |
| 295 | 7,559.81 | 7,452.87 | 7,082.30 | 7,600.86 | **29,695.84** |
| 296 | 14,793.58 | 14,065.17 | 13,727.56 | 15,141.02 | **57,727.33** |
| 297 | 3,170.30 | 3,277.25 | 3,072.02 | 3,411.95 | **12,931.52** |
| 298 | 2,271.93 | 2,240.72 | 2,265.57 | 2,453.46 | **9,231.68** |
| 299 | 1,890.97 | 1,784.59 | 1,842.40 | 1,769.56 | **7,287.53** |
| SC - SOUTH CAROLINA - Total | 61,834.65 | 60,357.57 | 58,239.07 | 62,860.86 | 243,292.14 |

**STATE: SD - SOUTH DAKOTA**

| REGISTRANT ZIP CODE 3 | 1ST QUARTER | 2ND QUARTER | 3RD QUARTER | 4TH QUARTER | TOTAL GRAMS |
|---|---|---|---|---|---|
| 570 | 1,677.07 | 1,593.24 | 1,663.77 | 1,813.50 | **6,747.58** |
| 571 | 5,662.49 | 5,482.70 | 5,004.03 | 5,397.72 | **21,546.94** |
| 572 | 455.54 | 447.45 | 476.35 | 445.14 | **1,824.48** |
| 573 | 953.87 | 897.21 | 850.96 | 1,000.11 | **3,702.15** |
| 574 | 528.96 | 459.01 | 522.60 | 520.87 | **2,031.44** |
| 575 | 312.75 | 302.92 | 250.32 | 293.67 | **1,159.67** |
| 576 | 19.08 | 23.12 | 24.28 | 21.39 | **87.87** |
| 577 | 969.47 | 983.93 | 978.15 | 951.55 | **3,883.10** |
| SD - SOUTH DAKOTA - Total | 10,579.23 | 10,189.59 | 9,770.47 | 10,443.95 | 40,983.24 |

**STATE: TN - TENNESSEE**

| REGISTRANT ZIP CODE 3 | 1ST QUARTER | 2ND QUARTER | 3RD QUARTER | 4TH QUARTER | TOTAL GRAMS |
|---|---|---|---|---|---|
| 370 | 11,320.35 | 10,645.13 | 10,572.87 | 10,719.71 | **43,258.07** |
| 371 | 7,338.40 | 7,079.99 | 7,040.68 | 7,139.54 | **28,598.61** |
| 372 | 7,190.41 | 7,121.61 | 7,147.63 | 7,584.09 | **29,043.74** |
| 373 | 5,367.08 | 4,957.79 | 5,074.56 | 5,294.24 | **20,693.67** |
| 374 | 3,176.08 | 3,177.24 | 3,029.24 | 3,155.27 | **12,537.83** |
| 376 | 2,363.27 | 2,240.14 | 2,238.98 | 2,292.74 | **9,135.14** |
| 377 | 2,823.44 | 2,786.44 | 2,634.98 | 2,760.43 | **11,005.29** |
| 378 | 4,250.77 | 4,055.95 | 4,002.19 | 4,220.71 | **16,529.61** |

| | | | | | |
|---|---|---|---|---|---|
| 379 | 5,743.42 | 5,805.86 | 5,887.37 | 6,057.91 | **23,494.56** |
| 380 | 5,338.75 | 5,347.43 | 5,186.71 | 5,400.03 | **21,272.92** |
| 381 | 7,403.64 | 7,074.47 | 7,003.85 | 7,452.26 | **28,934.22** |
| 382 | 848.65 | 801.25 | 765.40 | 781.59 | **3,196.89** |
| 383 | 2,344.20 | 2,059.77 | 2,192.73 | 2,206.61 | **8,803.31** |
| 384 | 1,770.14 | 1,737.77 | 1,653.94 | 1,778.81 | **6,940.67** |
| 385 | 1,671.87 | 1,451.03 | 1,493.23 | 1,470.69 | **6,086.81** |
| **TN - TENNESSEE - Total** | **68,950.48** | **66,341.86** | **65,924.39** | **68,314.63** | **269,531.35** |

**STATE: TX - TEXAS**

| REGISTRANT ZIP CODE 3 | 1ST QUARTER | 2ND QUARTER | 3RD QUARTER | 4TH QUARTER | TOTAL GRAMS |
|---|---|---|---|---|---|
| 750 | 31,035.88 | 30,709.25 | 30,827.76 | 31,460.78 | **124,033.67** |
| 751 | 7,967.95 | 7,535.53 | 7,459.80 | 7,803.77 | **30,767.06** |
| 752 | 15,394.80 | 14,972.21 | 14,704.55 | 15,743.98 | **60,815.54** |
| 753 | 22.55 | 54.34 | 36.42 | 42.20 | **155.51** |
| 754 | 2,641.34 | 2,648.28 | 2,485.83 | 2,724.01 | **10,499.45** |
| 755 | 917.44 | 868.31 | 870.62 | 902.99 | **3,559.36** |
| 756 | 3,663.42 | 3,469.76 | 3,207.88 | 3,438.54 | **13,779.59** |
| 757 | 3,684.81 | 3,722.39 | 3,548.38 | 3,802.16 | **14,757.74** |
| 758 | 785.06 | 709.91 | 700.66 | 750.37 | **2,946.00** |
| 759 | 2,284.65 | 2,159.20 | 2,005.43 | 2,206.61 | **8,655.89** |
| 760 | 13,852.43 | 13,224.62 | 12,997.42 | 13,528.70 | **53,603.17** |
| 761 | 7,176.53 | 7,221.05 | 6,844.70 | 7,195.03 | **28,437.32** |
| 762 | 6,156.19 | 5,931.31 | 5,863.67 | 6,386.27 | **24,337.43** |
| 763 | 1,781.70 | 1,779.39 | 1,675.91 | 1,879.40 | **7,116.41** |
| 764 | 617.99 | 602.38 | 576.37 | 657.88 | **2,454.61** |
| 765 | 3,125.21 | 2,603.18 | 2,802.05 | 2,841.94 | **11,372.38** |
| 766 | 749.22 | 642.27 | 698.92 | 752.69 | **2,843.10** |
| 767 | 2,938.48 | 2,470.22 | 2,545.95 | 2,815.35 | **10,770.00** |
| 768 | 361.31 | 358.42 | 342.81 | 401.78 | **1,464.33** |
| 769 | 1,504.22 | 1,436.58 | 1,364.89 | 1,528.50 | **5,834.19** |
| 770 | 26,565.70 | 26,562.68 | 25,930.82 | 26,783.72 | **105,842.93** |
| 772 | 12.72 | 12.14 | 0 | 0 | **24.86** |
| 773 | 19,784.26 | 19,294.04 | 18,905.81 | 20,236.84 | **78,220.95** |
| 774 | 16,304.73 | 15,122.52 | 15,384.98 | 16,354.45 | **63,166.67** |
| 775 | 12,517.60 | 11,836.60 | 11,679.93 | 12,334.92 | **48,369.05** |
| 776 | 3,285.92 | 3,084.16 | 3,239.09 | 3,507.91 | **13,117.09** |
| 777 | 1,703.08 | 1,554.51 | 1,581.68 | 1,649.32 | **6,488.59** |
| 778 | 5,952.70 | 5,669.43 | 5,642.83 | 5,416.22 | **22,681.18** |
| 779 | 1,956.29 | 1,684.01 | 1,787.49 | 1,774.19 | **7,201.97** |
| 780 | 5,061.27 | 4,830.03 | 4,770.48 | 5,171.68 | **19,833.45** |

| | | | | | |
|---|---|---|---|---|---|
| 781 | 3,866.91 | 3,750.13 | 3,658.79 | 3,938.02 | **15,213.86** |
| 782 | 16,654.54 | 16,233.89 | 15,806.44 | 16,695.56 | **65,390.42** |
| 783 | 2,252.86 | 2,000.23 | 1,903.11 | 2,053.41 | **8,209.60** |
| 784 | 4,025.89 | 3,572.08 | 3,322.92 | 3,683.65 | **14,604.54** |
| 785 | 7,758.68 | 7,113.52 | 6,892.11 | 7,287.53 | **29,051.84** |
| 786 | 9,161.73 | 9,043.80 | 8,759.37 | 9,369.84 | **36,334.74** |
| 787 | 13,976.72 | 13,848.93 | 13,733.86 | 14,812.08 | **56,371.60** |
| 788 | 759.62 | 627.82 | 699.50 | 680.42 | **2,767.36** |
| 789 | 375.19 | 400.05 | 382.12 | 384.44 | **1,541.79** |
| 790 | 591.97 | 574.63 | 520.87 | 567.12 | **2,254.59** |
| 791 | 1,246.38 | 1,215.17 | 1,204.76 | 1,459.70 | **5,126.01** |
| 792 | 97.12 | 57.23 | 72.26 | 100.01 | **326.63** |
| 793 | 348.59 | 336.45 | 350.91 | 337.03 | **1,372.99** |
| 794 | 3,429.87 | 3,300.95 | 3,139.08 | 3,417.15 | **13,287.05** |
| 795 | 357.27 | 400.62 | 366.52 | 378.66 | **1,503.06** |
| 796 | 1,229.62 | 1,311.71 | 1,242.34 | 1,333.68 | **5,117.34** |
| 797 | 3,072.02 | 2,934.44 | 2,614.17 | 3,100.93 | **11,721.56** |
| 798 | 98.86 | 85.56 | 71.68 | 91.92 | **348.02** |
| 799 | 2,901.48 | 2,832.69 | 2,686.43 | 2,835.00 | **11,255.61** |
| **TX - TEXAS - Total** | **272,010.78** | **262,408.59** | **257,910.38** | **272,618.33** | **1,064,948.09** |

**STATE: UT - UTAH**

| REGISTRANT ZIP CODE 3 | 1ST QUARTER | 2ND QUARTER | 3RD QUARTER | 4TH QUARTER | TOTAL GRAMS |
|---|---|---|---|---|---|
| 840 | 13,208.14 | 13,080.34 | 13,129.59 | 14,047.38 | **53,465.45** |
| 841 | 4,164.05 | 4,004.50 | 3,995.25 | 4,373.33 | **16,537.13** |
| 843 | 847.49 | 818.59 | 759.05 | 874.67 | **3,299.79** |
| 844 | 1,082.20 | 1,064.28 | 978.15 | 1,130.76 | **4,255.39** |
| 845 | 123.14 | 128.92 | 116.20 | 147.42 | **515.67** |
| 846 | 2,244.18 | 2,032.02 | 2,082.32 | 2,191.58 | **8,550.10** |
| 847 | 909.35 | 960.80 | 838.25 | 945.19 | **3,653.59** |
| **UT - UTAH - Total** | **22,578.56** | **22,089.45** | **21,898.79** | **23,710.32** | **90,277.12** |

**STATE: VA - VIRGINIA**

| REGISTRANT ZIP CODE 3 | 1ST QUARTER | 2ND QUARTER | 3RD QUARTER | 4TH QUARTER | TOTAL GRAMS |
|---|---|---|---|---|---|
| 201 | 7,012.35 | 7,346.49 | 7,233.77 | 7,582.36 | **29,174.97** |
| 220 | 3,596.36 | 3,559.36 | 3,424.09 | 3,755.92 | **14,335.72** |
| 221 | 2,829.80 | 2,759.27 | 2,929.23 | 2,748.87 | **11,267.17** |
| 222 | 1,997.91 | 2,173.08 | 2,093.30 | 2,221.06 | **8,485.35** |
| 223 | 1,874.20 | 1,881.14 | 1,768.41 | 1,936.64 | **7,460.38** |
| 224 | 2,195.62 | 2,060.93 | 2,082.89 | 2,311.24 | **8,650.69** |
| 225 | 1,021.50 | 1,009.94 | 948.08 | 1,038.27 | **4,017.80** |
| 226 | 1,438.89 | 1,403.05 | 1,477.62 | 1,508.26 | **5,827.83** |

| | | | | | |
|---|---|---|---|---|---|
| 227 | 480.40 | 507.57 | 527.23 | 520.29 | **2,035.49** |
| 228 | 1,026.71 | 1,046.94 | 1,005.89 | 1,128.45 | **4,207.99** |
| 229 | 2,284.07 | 2,315.87 | 2,139.55 | 2,422.24 | **9,161.73** |
| 230 | 1,617.52 | 1,632.55 | 1,574.74 | 1,718.69 | **6,543.51** |
| 231 | 4,529.99 | 4,443.28 | 4,311.47 | 4,638.10 | **17,922.83** |
| 232 | 5,405.81 | 5,263.60 | 4,942.18 | 5,334.13 | **20,945.72** |
| 233 | 3,200.36 | 2,908.42 | 2,835.58 | 2,983.57 | **11,927.94** |
| 234 | 6,746.43 | 6,680.52 | 6,567.22 | 6,853.38 | **26,847.54** |
| 235 | 1,738.92 | 1,566.65 | 1,505.37 | 1,726.78 | **6,537.73** |
| 236 | 3,210.19 | 3,031.56 | 2,877.78 | 3,203.25 | **12,322.78** |
| 237 | 1,179.90 | 1,253.90 | 1,289.16 | 1,148.68 | **4,871.65** |
| 238 | 2,203.62 | 2,130.86 | 1,890.91 | 2,095.87 | **8,321.26** |
| 239 | 549.77 | 490.23 | 455.54 | 537.05 | **2,032.60** |
| 240 | 3,081.27 | 3,044.27 | 2,980.68 | 3,225.22 | **12,331.45** |
| 241 | 1,841.25 | 1,672.44 | 1,747.60 | 1,810.03 | **7,071.32** |
| 242 | 935.94 | 815.70 | 891.43 | 896.06 | **3,539.13** |
| 243 | 946.93 | 841.71 | 882.76 | 902.41 | **3,573.81** |
| 244 | 1,342.35 | 1,382.82 | 1,281.65 | 1,272.98 | **5,279.79** |
| 245 | 3,019.42 | 2,693.95 | 2,729.79 | 2,934.44 | **11,377.59** |
| 246 | 552.66 | 526.07 | 480.98 | 611.05 | **2,170.77** |
| **VA - VIRGINIA - Total** | **67,860.17** | **66,442.17** | **64,874.90** | **69,065.29** | **268,242.53** |

**STATE: VI - VIRGIN ISLANDS**

| REGISTRANT ZIP CODE 3 | 1ST QUARTER | 2ND QUARTER | 3RD QUARTER | 4TH QUARTER | **TOTAL GRAMS** |
|---|---|---|---|---|---|
| 008 | 70.53 | 58.97 | 80.36 | 67.64 | **277.49** |
| **VI - VIRGIN ISLANDS - Total** | **70.53** | **58.97** | **80.36** | **67.64** | **277.49** |

**STATE: VT - VERMONT**

| REGISTRANT ZIP CODE 3 | 1ST QUARTER | 2ND QUARTER | 3RD QUARTER | 4TH QUARTER | **TOTAL GRAMS** |
|---|---|---|---|---|---|
| 050 | 300.61 | 302.35 | 319.69 | 302.35 | **1,224.99** |
| 051 | 275.75 | 256.10 | 259.57 | 261.88 | **1,053.30** |
| 052 | 653.83 | 631.29 | 630.13 | 642.85 | **2,558.09** |
| 053 | 695.45 | 697.77 | 657.30 | 715.69 | **2,766.21** |
| 054 | 2,836.74 | 2,762.16 | 2,733.83 | 2,862.75 | **11,195.48** |
| 056 | 656.72 | 707.02 | 725.52 | 787.37 | **2,876.63** |
| 057 | 1,006.47 | 926.12 | 865.99 | 944.62 | **3,743.20** |
| 058 | 805.87 | 824.37 | 803.56 | 850.96 | **3,284.76** |
| **VT - VERMONT - Total** | **7,231.45** | **7,107.16** | **6,995.59** | **7,368.46** | **28,702.67** |

**STATE: WA - WASHINGTON**

| REGISTRANT ZIP CODE 3 | 1ST QUARTER | 2ND QUARTER | 3RD QUARTER | 4TH QUARTER | **TOTAL GRAMS** |
|---|---|---|---|---|---|
| 980 | 9,673.17 | 9,803.77 | 9,618.89 | 10,442.86 | **39,538.69** |
| 981 | 5,830.72 | 5,987.70 | 5,724.35 | 6,185.67 | **23,728.43** |

| | | | | | |
|---|---|---|---|---|---|
| 982 | 3,660.53 | 3,769.79 | 3,609.66 | 3,630.47 | **14,670.44** |
| 983 | 3,515.43 | 3,599.83 | 3,465.71 | 3,625.84 | **14,206.81** |
| 984 | 2,071.91 | 2,045.90 | 2,007.16 | 2,171.92 | **8,296.89** |
| 985 | 1,925.65 | 1,951.09 | 1,881.72 | 2,034.91 | **7,793.37** |
| 986 | 2,391.02 | 2,527.45 | 2,490.45 | 2,676.60 | **10,085.53** |
| 988 | 289.05 | 345.13 | 330.67 | 381.55 | **1,346.39** |
| 989 | 594.29 | 642.85 | 590.82 | 739.97 | **2,567.92** |
| 990 | 460.17 | 444.56 | 460.17 | 445.72 | **1,810.61** |
| 991 | 360.73 | 298.88 | 343.39 | 408.14 | **1,411.14** |
| 992 | 2,389.87 | 2,436.11 | 2,264.42 | 2,475.42 | **9,565.82** |
| 993 | 1,780.55 | 1,792.69 | 1,759.74 | 1,864.95 | **7,197.92** |
| 994 | 93.65 | 89.03 | 94.23 | 102.32 | **379.23** |
| **WA - WASHINGTON - Total** | **35,036.73** | **35,734.76** | **34,641.37** | **37,186.34** | **142,599.20** |

**STATE: WI - WISCONSIN**

| REGISTRANT ZIP CODE 3 | 1ST QUARTER | 2ND QUARTER | 3RD QUARTER | 4TH QUARTER | TOTAL GRAMS |
|---|---|---|---|---|---|
| 530 | 7,606.06 | 7,637.28 | 7,669.65 | 8,132.71 | **31,045.70** |
| 531 | 8,844.93 | 8,843.77 | 8,817.18 | 9,254.22 | **35,760.11** |
| 532 | 9,519.57 | 9,440.95 | 9,139.76 | 10,400.02 | **38,500.30** |
| 534 | 2,675.45 | 2,564.45 | 2,711.87 | 2,626.89 | **10,578.65** |
| 535 | 5,689.66 | 5,562.48 | 5,463.62 | 5,711.63 | **22,427.39** |
| 537 | 3,174.93 | 3,252.39 | 3,283.03 | 3,560.52 | **13,270.86** |
| 538 | 892.01 | 798.93 | 726.67 | 896.63 | **3,314.25** |
| 539 | 2,127.99 | 2,212.39 | 2,165.56 | 2,260.37 | **8,766.31** |
| 540 | 826.68 | 748.64 | 722.63 | 814.54 | **3,112.49** |
| 541 | 4,054.22 | 3,980.80 | 3,958.83 | 4,179.66 | **16,173.50** |
| 542 | 1,704.82 | 1,700.77 | 1,699.04 | 1,705.97 | **6,810.60** |
| 543 | 5,628.38 | 5,589.07 | 5,472.87 | 5,831.29 | **22,521.62** |
| 544 | 5,433.56 | 5,381.53 | 5,218.51 | 5,574.04 | **21,607.64** |
| 545 | 1,200.14 | 1,254.48 | 1,283.38 | 1,263.73 | **5,001.72** |
| 546 | 2,022.77 | 2,113.53 | 1,894.43 | 2,213.54 | **8,244.28** |
| 547 | 1,526.76 | 1,574.74 | 1,440.63 | 1,559.14 | **6,101.27** |
| 548 | 1,518.67 | 1,556.82 | 1,588.04 | 1,748.17 | **6,411.71** |
| 549 | 7,839.61 | 8,045.42 | 7,890.49 | 8,285.91 | **32,061.43** |
| **WI - WISCONSIN - Total** | **72,286.20** | **72,258.45** | **71,146.19** | **76,018.99** | **291,709.84** |

**STATE: WV - WEST VIRGINIA**

| REGISTRANT ZIP CODE 3 | 1ST QUARTER | 2ND QUARTER | 3RD QUARTER | 4TH QUARTER | TOTAL GRAMS |
|---|---|---|---|---|---|
| 247 | 394.84 | 374.61 | 401.78 | 380.97 | **1,552.20** |
| 248 | 253.21 | 259.57 | 217.94 | 254.36 | **985.08** |
| 249 | 336.45 | 358.42 | 382.12 | 368.25 | **1,445.25** |
| 250 | 711.06 | 702.39 | 672.33 | 724.94 | **2,810.72** |

| | | | | | |
|---|---|---|---|---|---|
| 251 | 563.65 | 494.28 | 520.87 | 605.85 | **2,184.64** |
| 252 | 353.80 | 385.01 | 374.61 | 322.00 | **1,435.42** |
| 253 | 1,908.89 | 1,638.34 | 1,591.51 | 1,663.19 | **6,801.92** |
| 254 | 1,314.60 | 1,314.02 | 1,322.11 | 1,296.10 | **5,246.84** |
| 255 | 1,600.76 | 1,775.92 | 1,660.30 | 1,740.08 | **6,777.07** |
| 256 | 213.32 | 206.96 | 223.72 | 236.44 | **880.45** |
| 257 | 1,479.94 | 1,484.56 | 1,379.92 | 1,682.27 | **6,026.69** |
| 258 | 1,041.74 | 1,032.49 | 970.05 | 1,079.31 | **4,123.59** |
| 259 | 301.77 | 306.97 | 280.38 | 302.35 | **1,191.46** |
| 260 | 1,180.48 | 1,041.16 | 956.18 | 1,006.47 | **4,184.29** |
| 261 | 1,833.16 | 1,970.16 | 1,737.19 | 1,910.04 | **7,450.55** |
| 262 | 648.05 | 608.74 | 587.93 | 652.10 | **2,496.81** |
| 263 | 1,118.05 | 1,195.51 | 1,129.61 | 1,170.65 | **4,613.82** |
| 264 | 346.28 | 325.47 | 304.66 | 322.58 | **1,298.99** |
| 265 | 2,098.50 | 2,040.11 | 2,010.05 | 2,164.98 | **8,313.66** |
| 266 | 243.96 | 228.35 | 213.32 | 253.21 | **938.83** |
| 267 | 736.50 | 691.41 | 629.55 | 715.11 | **2,772.57** |
| 268 | 440.51 | 413.92 | 374.03 | 391.95 | **1,620.41** |
| **WV - WEST VIRGINIA - Total** | **19,119.50** | **18,848.37** | **17,940.18** | **19,243.21** | **75,151.27** |

**STATE: WY - WYOMING**

| REGISTRANT ZIP CODE 3 | 1ST QUARTER | 2ND QUARTER | 3RD QUARTER | 4TH QUARTER | TOTAL GRAMS |
|---|---|---|---|---|---|
| 820 | 599.49 | 628.39 | 560.76 | 598.91 | **2,387.55** |
| 822 | 87.29 | 78.04 | 63.01 | 84.98 | **313.33** |
| 823 | 26.01 | 39.89 | 30.64 | 16.19 | **112.73** |
| 824 | 541.68 | 478.67 | 458.43 | 542.26 | **2,021.04** |
| 825 | 97.70 | 60.70 | 63.59 | 75.73 | **297.72** |
| 826 | 313.33 | 319.11 | 325.47 | 302.92 | **1,260.84** |
| 827 | 303.50 | 318.53 | 290.21 | 325.47 | **1,237.71** |
| 828 | 185.57 | 182.68 | 172.85 | 153.20 | **694.30** |
| 829 | 408.72 | 400.62 | 335.88 | 383.28 | **1,528.50** |
| 830 | 141.63 | 108.10 | 140.48 | 109.84 | **500.06** |
| 831 | 45.67 | 32.95 | 45.09 | 41.05 | **164.76** |
| **WY - WYOMING - Total** | **2,750.60** | **2,647.70** | **2,486.41** | **2,633.82** | **10,518.53** |

**DRUG: 1724 - METHYLPHENIDATE (DL;D;L;ISOMERS)**

**STATE: AK - ALASKA**

| REGISTRANT ZIP CODE 3 | 1ST QUARTER | 2ND QUARTER | 3RD QUARTER | 4TH QUARTER | TOTAL GRAMS |
|---|---|---|---|---|---|
| 995 | 4,424.11 | 4,277.25 | 3,882.77 | 4,670.30 | **17,254.44** |
| 996 | 1,483.63 | 1,452.26 | 1,456.20 | 1,387.64 | **5,779.74** |
| 997 | 1,310.21 | 1,380.23 | 1,158.14 | 1,337.86 | **5,186.44** |
| 998 | 535.80 | 517.57 | 490.66 | 499.55 | **2,043.58** |

| | | | | | |
|---|---|---|---|---|---|
| 999 | 482.94 | 384.28 | 471.71 | 391.02 | **1,729.95** |
| **AK - ALASKA - Total** | **8,236.69** | **8,011.60** | **7,459.48** | **8,286.37** | **31,994.14** |

**STATE: AL - ALABAMA**

| REGISTRANT ZIP CODE 3 | 1ST QUARTER | 2ND QUARTER | 3RD QUARTER | 4TH QUARTER | TOTAL GRAMS |
|---|---|---|---|---|---|
| 350 | 9,003.53 | 8,116.65 | 8,298.63 | 8,539.77 | **33,958.58** |
| 351 | 4,854.67 | 4,325.62 | 4,554.67 | 4,621.48 | **18,356.44** |
| 352 | 8,732.55 | 8,135.19 | 8,059.56 | 8,401.80 | **33,329.10** |
| 354 | 2,760.49 | 2,448.28 | 2,496.24 | 2,747.43 | **10,452.44** |
| 355 | 2,111.26 | 1,968.92 | 1,941.37 | 2,171.47 | **8,193.01** |
| 356 | 4,358.42 | 3,981.39 | 3,841.33 | 4,077.65 | **16,258.79** |
| 357 | 2,746.65 | 2,484.83 | 2,530.77 | 2,633.23 | **10,395.48** |
| 358 | 2,918.80 | 2,785.26 | 3,102.03 | 2,970.89 | **11,776.98** |
| 359 | 3,824.83 | 3,645.33 | 3,561.11 | 3,958.93 | **14,990.19** |
| 360 | 3,714.41 | 3,339.31 | 3,257.38 | 3,641.68 | **13,952.78** |
| 361 | 2,937.42 | 2,848.19 | 2,685.61 | 2,886.18 | **11,357.39** |
| 362 | 1,967.47 | 1,819.17 | 1,893.71 | 2,007.88 | **7,688.22** |
| 363 | 5,922.96 | 5,258.84 | 5,074.32 | 5,332.90 | **21,589.02** |
| 364 | 1,444.96 | 1,266.56 | 1,333.22 | 1,504.21 | **5,548.95** |
| 365 | 6,773.80 | 6,338.17 | 6,430.94 | 6,874.54 | **26,417.44** |
| 366 | 5,093.74 | 4,711.90 | 4,397.24 | 4,785.44 | **18,988.31** |
| 367 | 684.98 | 563.59 | 575.08 | 631.44 | **2,455.10** |
| 368 | 3,919.66 | 3,445.28 | 3,515.39 | 3,872.05 | **14,752.39** |
| 369 | 183.88 | 147.30 | 169.69 | 143.62 | **644.49** |
| **AL - ALABAMA - Total** | **73,954.47** | **67,629.75** | **67,718.28** | **71,802.60** | **281,105.10** |

**STATE: AR - ARKANSAS**

| REGISTRANT ZIP CODE 3 | 1ST QUARTER | 2ND QUARTER | 3RD QUARTER | 4TH QUARTER | TOTAL GRAMS |
|---|---|---|---|---|---|
| 716 | 1,577.23 | 1,507.40 | 1,442.76 | 1,448.82 | **5,976.21** |
| 717 | 955.93 | 877.75 | 830.30 | 819.34 | **3,483.31** |
| 718 | 994.49 | 864.33 | 898.61 | 991.53 | **3,748.97** |
| 719 | 2,260.76 | 2,092.70 | 2,035.97 | 2,149.56 | **8,538.99** |
| 720 | 6,195.60 | 6,015.82 | 6,065.10 | 6,218.48 | **24,494.99** |
| 721 | 4,863.62 | 4,414.89 | 4,192.62 | 4,417.16 | **17,888.30** |
| 722 | 4,165.42 | 3,831.24 | 3,866.99 | 4,049.14 | **15,912.79** |
| 723 | 1,460.20 | 1,262.57 | 1,230.71 | 1,356.00 | **5,309.48** |
| 724 | 3,517.68 | 3,271.23 | 3,265.67 | 3,342.77 | **13,397.35** |
| 725 | 1,085.21 | 1,072.94 | 1,059.09 | 1,119.21 | **4,336.45** |
| 726 | 1,433.29 | 1,210.82 | 1,250.05 | 1,137.83 | **5,031.98** |
| 727 | 6,838.89 | 6,827.22 | 6,725.63 | 6,891.47 | **27,283.20** |
| 728 | 1,409.22 | 1,321.07 | 1,264.81 | 1,310.26 | **5,305.35** |
| 729 | 3,027.94 | 2,813.62 | 2,785.70 | 2,886.26 | **11,513.53** |

| AR - ARKANSAS - Total | 39,785.47 | 37,383.60 | 36,914.01 | 38,137.82 | 152,220.89 |
|---|---|---|---|---|---|

**STATE: AS - AMERICAN SAMOA**

| REGISTRANT ZIP CODE 3 | 1ST QUARTER | 2ND QUARTER | 3RD QUARTER | 4TH QUARTER | TOTAL GRAMS |
|---|---|---|---|---|---|
| 967 | 12.97 | 7.78 | 6.92 | 16.43 | **44.10** |
| AS - AMERICAN SAMOA - Tota | 12.97 | 7.78 | 6.92 | 16.43 | 44.10 |

**STATE: AZ - ARIZONA**

| REGISTRANT ZIP CODE 3 | 1ST QUARTER | 2ND QUARTER | 3RD QUARTER | 4TH QUARTER | TOTAL GRAMS |
|---|---|---|---|---|---|
| 850 | 12,169.45 | 11,702.35 | 11,091.04 | 11,997.56 | **46,960.40** |
| 851 | 4,206.26 | 4,025.42 | 3,976.61 | 4,326.97 | **16,535.27** |
| 852 | 28,155.24 | 35,160.32 | 48,707.86 | 28,335.20 | **140,358.63** |
| 853 | 12,027.27 | 11,519.74 | 10,847.38 | 11,918.82 | **46,313.21** |
| 855 | 811.98 | 718.13 | 694.54 | 760.16 | **2,984.81** |
| 856 | 2,684.15 | 2,616.19 | 2,698.86 | 2,661.79 | **10,660.99** |
| 857 | 11,782.54 | 11,187.09 | 11,572.79 | 11,247.47 | **45,789.89** |
| 859 | 694.38 | 688.03 | 685.56 | 721.89 | **2,789.85** |
| 860 | 1,359.90 | 1,314.00 | 1,168.27 | 1,359.88 | **5,202.05** |
| 863 | 1,949.49 | 1,931.17 | 1,901.67 | 2,002.03 | **7,784.36** |
| 864 | 1,109.88 | 1,030.69 | 987.31 | 1,085.14 | **4,213.01** |
| 865 | 353.92 | 323.19 | 386.95 | 362.87 | **1,426.94** |
| AZ - ARIZONA - Total | 77,304.46 | 82,216.33 | 94,718.85 | 76,779.75 | 331,019.40 |

**STATE: CA - CALIFORNIA**

| REGISTRANT ZIP CODE 3 | 1ST QUARTER | 2ND QUARTER | 3RD QUARTER | 4TH QUARTER | TOTAL GRAMS |
|---|---|---|---|---|---|
| 900 | 9,798.89 | 9,895.45 | 9,038.78 | 9,920.64 | **38,653.75** |
| 902 | 7,413.27 | 7,472.37 | 6,817.48 | 7,364.62 | **29,067.73** |
| 903 | 423.98 | 329.50 | 342.33 | 406.94 | **1,502.74** |
| 904 | 1,208.79 | 1,274.86 | 1,330.70 | 1,429.40 | **5,243.74** |
| 905 | 1,300.77 | 1,376.40 | 1,127.33 | 1,175.97 | **4,980.46** |
| 906 | 3,467.10 | 3,508.99 | 3,107.91 | 3,412.90 | **13,496.91** |
| 907 | 3,443.36 | 3,441.25 | 3,101.43 | 3,310.41 | **13,296.46** |
| 908 | 2,229.94 | 2,249.25 | 2,053.91 | 2,139.32 | **8,672.42** |
| 910 | 1,344.41 | 1,411.89 | 1,349.62 | 1,430.26 | **5,536.18** |
| 911 | 2,226.05 | 2,305.77 | 2,248.29 | 2,255.32 | **9,035.43** |
| 912 | 980.84 | 1,023.81 | 785.97 | 942.61 | **3,733.24** |
| 913 | 11,718.30 | 11,638.01 | 10,658.15 | 11,528.28 | **45,542.75** |
| 914 | 2,795.46 | 2,983.91 | 2,637.97 | 2,934.24 | **11,351.58** |
| 915 | 797.07 | 824.15 | 716.62 | 757.75 | **3,095.59** |
| 916 | 787.79 | 807.39 | 781.54 | 826.73 | **3,203.46** |
| 917 | 8,403.10 | 8,117.39 | 7,741.80 | 8,441.27 | **32,703.56** |
| 918 | 289.11 | 292.29 | 261.31 | 265.70 | **1,108.41** |
| 919 | 2,785.53 | 2,767.85 | 2,733.50 | 2,587.00 | **10,873.88** |

| | | | | | |
|---|---|---|---|---|---|
| 920 | 26,986.05 | 26,235.82 | 25,795.93 | 26,722.73 | **105,740.54** |
| 921 | 11,208.15 | 11,465.05 | 10,722.07 | 10,679.60 | **44,074.87** |
| 922 | 4,088.36 | 4,156.90 | 3,457.20 | 4,247.46 | **15,949.92** |
| 923 | 7,487.93 | 7,304.73 | 6,810.03 | 7,271.70 | **28,874.40** |
| 924 | 848.67 | 855.92 | 682.45 | 798.80 | **3,185.84** |
| 925 | 9,150.29 | 8,794.71 | 8,189.22 | 8,628.99 | **34,763.21** |
| 926 | 11,549.78 | 11,906.83 | 11,034.01 | 11,874.04 | **46,364.66** |
| 927 | 2,859.33 | 2,867.56 | 2,990.10 | 2,852.42 | **11,569.41** |
| 928 | 7,719.83 | 7,906.00 | 7,058.06 | 8,098.41 | **30,782.31** |
| 930 | 5,614.18 | 5,670.36 | 4,985.98 | 5,729.46 | **21,999.99** |
| 931 | 1,605.10 | 1,478.53 | 1,406.47 | 1,628.97 | **6,119.07** |
| 932 | 3,442.17 | 3,113.30 | 2,902.84 | 3,306.34 | **12,764.64** |
| 933 | 2,984.37 | 2,939.25 | 2,481.27 | 2,792.81 | **11,197.71** |
| 934 | 3,197.75 | 3,392.99 | 2,708.59 | 3,211.22 | **12,510.55** |
| 935 | 3,506.35 | 3,446.82 | 3,206.90 | 3,205.36 | **13,365.43** |
| 936 | 3,884.99 | 3,622.34 | 3,382.75 | 3,400.06 | **14,290.14** |
| 937 | 5,224.87 | 4,682.45 | 4,555.74 | 4,934.99 | **19,398.06** |
| 939 | 2,129.88 | 2,103.18 | 1,930.58 | 2,003.26 | **8,166.89** |
| 940 | 5,291.64 | 5,499.05 | 5,125.01 | 5,630.36 | **21,546.06** |
| 941 | 6,285.05 | 6,152.68 | 5,648.76 | 5,962.80 | **24,049.30** |
| 943 | 1,112.08 | 1,072.85 | 1,044.32 | 1,112.27 | **4,341.52** |
| 944 | 1,124.54 | 1,104.57 | 1,060.11 | 1,155.42 | **4,444.64** |
| 945 | 18,487.41 | 16,541.93 | 16,138.82 | 18,147.81 | **69,315.96** |
| 946 | 3,062.19 | 2,835.25 | 2,778.00 | 2,965.23 | **11,640.67** |
| 947 | 859.79 | 856.03 | 757.43 | 887.16 | **3,360.42** |
| 948 | 687.78 | 621.81 | 754.02 | 650.73 | **2,714.34** |
| 949 | 3,932.62 | 3,694.74 | 3,487.50 | 3,727.51 | **14,842.36** |
| 950 | 6,609.70 | 6,510.48 | 5,916.09 | 6,565.37 | **25,601.63** |
| 951 | 3,794.22 | 4,088.69 | 3,587.31 | 4,206.12 | **15,676.35** |
| 952 | 3,000.71 | 2,869.47 | 2,573.84 | 2,642.88 | **11,086.90** |
| 953 | 5,687.24 | 5,580.97 | 4,878.42 | 5,496.08 | **21,642.71** |
| 954 | 3,623.27 | 3,414.42 | 3,251.64 | 3,398.26 | **13,687.59** |
| 955 | 993.52 | 1,027.33 | 960.22 | 935.07 | **3,916.14** |
| 956 | 13,353.31 | 12,135.64 | 12,388.29 | 12,886.85 | **50,764.09** |
| 957 | 3,303.82 | 3,197.15 | 3,085.60 | 3,182.40 | **12,768.96** |
| 958 | 5,591.46 | 5,262.35 | 4,923.44 | 5,708.35 | **21,485.59** |
| 959 | 3,932.63 | 3,694.35 | 3,394.08 | 3,464.27 | **14,485.33** |
| 960 | 2,851.04 | 2,905.32 | 2,669.17 | 2,686.42 | **11,111.95** |
| 961 | 436.81 | 344.51 | 368.07 | 447.74 | **1,597.12** |
| **CA - CALIFORNIA - Total** | **268,922.66** | **263,072.88** | **245,924.94** | **264,375.09** | **1,042,295.57** |

**STATE: CO - COLORADO**

| REGISTRANT ZIP CODE 3 | 1ST QUARTER | 2ND QUARTER | 3RD QUARTER | 4TH QUARTER | TOTAL GRAMS |
|---|---|---|---|---|---|
| 800 | 10,140.27 | 9,918.43 | 9,901.38 | 10,963.58 | **40,923.67** |
| 801 | 11,356.90 | 10,734.76 | 10,884.71 | 11,697.74 | **44,674.11** |
| 802 | 11,253.84 | 10,838.17 | 9,709.11 | 10,635.59 | **42,436.72** |
| 803 | 1,909.11 | 2,007.26 | 1,999.02 | 2,179.21 | **8,094.60** |
| 804 | 1,829.76 | 1,708.43 | 1,758.22 | 1,833.99 | **7,130.40** |
| 805 | 6,719.39 | 6,768.02 | 6,381.02 | 6,885.90 | **26,754.32** |
| 806 | 2,308.88 | 2,200.94 | 2,167.60 | 2,349.63 | **9,027.05** |
| 807 | 475.81 | 497.21 | 444.58 | 440.84 | **1,858.43** |
| 808 | 1,590.03 | 1,425.01 | 1,455.15 | 1,315.59 | **5,785.78** |
| 809 | 8,099.60 | 7,504.18 | 7,417.80 | 7,337.34 | **30,358.92** |
| 810 | 1,928.97 | 1,874.12 | 1,692.49 | 1,935.22 | **7,430.80** |
| 811 | 292.44 | 251.05 | 283.91 | 279.54 | **1,106.95** |
| 812 | 619.56 | 595.85 | 595.32 | 554.50 | **2,365.24** |
| 813 | 485.34 | 489.87 | 449.44 | 497.44 | **1,922.10** |
| 814 | 686.32 | 660.17 | 665.57 | 663.12 | **2,675.18** |
| 815 | 1,991.48 | 1,957.70 | 1,848.04 | 1,940.55 | **7,737.77** |
| 816 | 1,239.96 | 1,116.70 | 1,168.85 | 1,217.08 | **4,742.59** |
| **CO - COLORADO - Total** | **62,927.66** | **60,547.88** | **58,822.20** | **62,726.88** | **245,024.63** |

**STATE: CT - CONNECTICUT**

| REGISTRANT ZIP CODE 3 | 1ST QUARTER | 2ND QUARTER | 3RD QUARTER | 4TH QUARTER | TOTAL GRAMS |
|---|---|---|---|---|---|
| 060 | 12,318.01 | 11,777.04 | 11,009.97 | 11,605.76 | **46,710.78** |
| 061 | 4,656.02 | 4,178.68 | 3,814.80 | 4,328.98 | **16,978.47** |
| 062 | 2,736.75 | 2,543.74 | 2,491.32 | 2,552.62 | **10,324.43** |
| 063 | 5,141.48 | 4,988.39 | 4,384.45 | 4,490.76 | **19,005.07** |
| 064 | 12,881.05 | 12,125.38 | 10,682.88 | 11,741.08 | **47,430.40** |
| 065 | 3,839.17 | 3,516.56 | 3,210.98 | 3,630.50 | **14,197.21** |
| 066 | 1,763.59 | 1,691.31 | 1,328.51 | 1,601.71 | **6,385.12** |
| 067 | 4,909.54 | 4,452.16 | 4,238.35 | 4,625.39 | **18,225.44** |
| 068 | 8,799.36 | 8,667.51 | 8,189.15 | 8,772.89 | **34,428.90** |
| 069 | 1,217.24 | 1,052.71 | 1,029.02 | 1,135.32 | **4,434.29** |
| **CT - CONNECTICUT - Total** | **58,262.20** | **54,993.48** | **50,379.41** | **54,485.01** | **218,120.11** |

**STATE: DC - DISTRICT OF COLUMBIA**

| REGISTRANT ZIP CODE 3 | 1ST QUARTER | 2ND QUARTER | 3RD QUARTER | 4TH QUARTER | TOTAL GRAMS |
|---|---|---|---|---|---|
| 200 | 7,780.28 | 7,260.17 | 6,675.72 | 7,398.21 | **29,114.38** |
| 203 | 190.54 | 189.54 | 241.16 | 207.86 | **829.11** |
| 204 | 520.03 | 312.50 | 417.56 | 436.24 | **1,686.34** |
| 205 | 0 | 0.86 | 6.23 | 4.41 | **11.50** |
| **DC - DISTRICT OF COLUMBIA -** | **8,490.85** | **7,763.08** | **7,340.67** | **8,046.73** | **31,641.33** |

**STATE: DE - DELAWARE**

| REGISTRANT ZIP CODE 3 | 1ST QUARTER | 2ND QUARTER | 3RD QUARTER | 4TH QUARTER | TOTAL GRAMS |
|---|---|---|---|---|---|
| 197 | 7,079.35 | 6,514.14 | 6,210.76 | 6,802.60 | **26,606.85** |
| 198 | 5,004.31 | 4,867.54 | 4,706.85 | 5,027.30 | **19,606.00** |
| 199 | 6,469.47 | 6,085.18 | 5,743.98 | 5,986.96 | **24,285.58** |
| **DE - DELAWARE - Total** | **18,553.13** | **17,466.85** | **16,661.60** | **17,816.85** | **70,498.43** |

**STATE: FL - FLORIDA**

| REGISTRANT ZIP CODE 3 | 1ST QUARTER | 2ND QUARTER | 3RD QUARTER | 4TH QUARTER | TOTAL GRAMS |
|---|---|---|---|---|---|
| 320 | 8,440.59 | 7,815.58 | 7,282.09 | 7,662.01 | **31,200.28** |
| 321 | 5,026.28 | 4,795.17 | 4,506.67 | 5,003.67 | **19,331.78** |
| 322 | 12,939.10 | 12,453.10 | 11,235.99 | 11,834.37 | **48,462.56** |
| 323 | 5,259.75 | 4,761.25 | 4,447.69 | 4,705.92 | **19,174.60** |
| 324 | 6,430.67 | 6,219.77 | 5,633.05 | 4,858.29 | **23,141.78** |
| 325 | 13,347.50 | 12,364.64 | 11,654.31 | 11,706.77 | **49,073.23** |
| 326 | 5,966.40 | 5,531.42 | 5,255.35 | 5,740.43 | **22,493.60** |
| 327 | 12,204.23 | 11,529.09 | 11,150.73 | 11,548.26 | **46,432.31** |
| 328 | 7,942.80 | 7,648.73 | 6,911.97 | 7,680.67 | **30,184.17** |
| 329 | 9,829.64 | 9,185.13 | 9,063.30 | 9,449.37 | **37,527.43** |
| 330 | 10,078.83 | 9,447.47 | 8,491.16 | 9,368.13 | **37,385.60** |
| 331 | 11,460.38 | 10,557.39 | 9,327.90 | 10,173.28 | **41,518.96** |
| 333 | 4,788.38 | 4,541.07 | 4,260.23 | 4,412.22 | **18,001.90** |
| 334 | 13,150.10 | 13,009.70 | 11,374.86 | 12,130.79 | **49,665.45** |
| 335 | 9,892.79 | 9,383.21 | 8,885.00 | 9,650.68 | **37,811.68** |
| 336 | 9,130.78 | 9,130.38 | 8,593.98 | 8,960.91 | **35,816.04** |
| 337 | 6,989.96 | 6,961.68 | 6,461.75 | 6,722.67 | **27,136.05** |
| 338 | 8,758.59 | 7,758.16 | 7,325.34 | 7,613.26 | **31,455.35** |
| 339 | 8,865.04 | 8,459.55 | 8,035.82 | 8,126.30 | **33,486.72** |
| 341 | 3,312.06 | 3,067.44 | 2,616.34 | 2,984.83 | **11,980.68** |
| 342 | 9,980.84 | 8,932.60 | 8,895.16 | 9,133.09 | **36,941.70** |
| 344 | 5,698.79 | 5,206.78 | 5,253.09 | 4,999.15 | **21,157.81** |
| 346 | 7,937.64 | 7,689.39 | 7,441.97 | 7,611.69 | **30,680.68** |
| 347 | 6,998.40 | 6,715.85 | 6,180.83 | 6,555.69 | **26,450.77** |
| 349 | 4,825.32 | 4,816.19 | 4,341.62 | 4,387.41 | **18,370.54** |
| **FL - FLORIDA - Total** | **209,254.86** | **197,980.76** | **184,626.22** | **193,019.84** | **784,881.68** |

**STATE: GA - GEORGIA**

| REGISTRANT ZIP CODE 3 | 1ST QUARTER | 2ND QUARTER | 3RD QUARTER | 4TH QUARTER | TOTAL GRAMS |
|---|---|---|---|---|---|
| 300 | 31,374.14 | 28,646.34 | 28,355.15 | 30,831.41 | **119,207.03** |
| 301 | 19,302.34 | 17,787.80 | 17,653.42 | 19,139.11 | **73,882.68** |
| 302 | 14,720.04 | 12,937.41 | 12,943.28 | 13,568.39 | **54,169.13** |
| 303 | 12,277.70 | 11,943.84 | 11,620.65 | 12,204.58 | **48,046.77** |

| | | | | | |
|---|---|---|---|---|---|
| 304 | 4,442.11 | 3,582.68 | 3,760.40 | 3,890.72 | **15,675.91** |
| 305 | 9,714.86 | 9,350.84 | 9,114.12 | 9,732.95 | **37,912.76** |
| 306 | 6,245.94 | 5,438.36 | 5,579.02 | 6,043.73 | **23,307.04** |
| 307 | 6,497.07 | 6,115.28 | 6,160.21 | 6,395.52 | **25,168.09** |
| 308 | 3,338.34 | 3,108.79 | 3,092.78 | 3,242.48 | **12,782.40** |
| 309 | 4,931.87 | 4,338.74 | 4,555.64 | 4,230.09 | **18,056.34** |
| 310 | 11,858.18 | 10,444.77 | 11,013.71 | 10,864.97 | **44,181.63** |
| 312 | 4,008.09 | 3,593.32 | 3,716.24 | 3,705.16 | **15,022.81** |
| 313 | 5,352.25 | 4,964.88 | 5,233.99 | 4,963.86 | **20,514.97** |
| 314 | 4,000.22 | 3,533.23 | 3,867.85 | 3,762.41 | **15,163.71** |
| 315 | 9,080.05 | 7,690.20 | 8,122.27 | 8,209.12 | **33,101.64** |
| 316 | 4,209.76 | 3,423.61 | 3,733.53 | 3,872.53 | **15,239.43** |
| 317 | 9,282.18 | 7,386.02 | 7,826.22 | 8,248.95 | **32,743.38** |
| 318 | 619.21 | 529.68 | 506.36 | 438.66 | **2,093.91** |
| 319 | 4,992.49 | 4,774.64 | 4,500.44 | 4,709.73 | **18,977.30** |
| 398 | 1,568.75 | 1,340.91 | 1,382.02 | 1,298.85 | **5,590.53** |
| **GA - GEORGIA - Total** | **167,815.59** | **150,931.34** | **152,737.31** | **159,353.22** | **630,837.46** |

**STATE: GU - GUAM**

| REGISTRANT ZIP CODE 3 | 1ST QUARTER | 2ND QUARTER | 3RD QUARTER | 4TH QUARTER | TOTAL GRAMS |
|---|---|---|---|---|---|
| 969 | 404.01 | 477.07 | 675.57 | 428.08 | **1,984.74** |
| **GU - GUAM - Total** | **404.01** | **477.07** | **675.57** | **428.08** | **1,984.74** |

**STATE: HI - HAWAII**

| REGISTRANT ZIP CODE 3 | 1ST QUARTER | 2ND QUARTER | 3RD QUARTER | 4TH QUARTER | TOTAL GRAMS |
|---|---|---|---|---|---|
| 967 | 5,883.53 | 5,420.02 | 5,033.69 | 5,496.97 | **21,834.20** |
| 968 | 5,343.69 | 5,403.46 | 5,980.58 | 5,186.81 | **21,914.53** |
| **HI - HAWAII - Total** | **11,227.21** | **10,823.48** | **11,014.27** | **10,683.77** | **43,748.73** |

**STATE: IA - IOWA**

| REGISTRANT ZIP CODE 3 | 1ST QUARTER | 2ND QUARTER | 3RD QUARTER | 4TH QUARTER | TOTAL GRAMS |
|---|---|---|---|---|---|
| 500 | 6,656.98 | 6,605.07 | 6,179.51 | 6,723.52 | **26,165.08** |
| 501 | 4,040.45 | 3,933.10 | 3,660.82 | 3,791.51 | **15,425.88** |
| 502 | 4,791.16 | 4,667.29 | 4,547.80 | 4,583.68 | **18,589.93** |
| 503 | 7,794.37 | 7,503.44 | 7,035.32 | 7,743.53 | **30,076.67** |
| 504 | 2,015.96 | 2,091.44 | 2,082.78 | 2,074.40 | **8,264.58** |
| 505 | 2,156.98 | 2,237.35 | 1,978.87 | 2,181.66 | **8,554.86** |
| 506 | 3,256.50 | 3,121.27 | 2,946.19 | 3,315.50 | **12,639.45** |
| 507 | 1,907.10 | 1,889.33 | 1,618.68 | 1,753.34 | **7,168.46** |
| 508 | 573.60 | 535.08 | 560.73 | 507.33 | **2,176.74** |
| 510 | 2,207.58 | 2,161.49 | 2,009.39 | 2,034.74 | **8,413.20** |
| 511 | 3,879.95 | 3,858.11 | 3,624.31 | 3,966.14 | **15,328.51** |
| 512 | 1,136.35 | 985.62 | 899.16 | 967.11 | **3,988.25** |

| | | | | | |
|---|---|---|---|---|---|
| 513 | 1,089.07 | 1,037.24 | 960.80 | 981.87 | **4,068.97** |
| 514 | 995.09 | 974.64 | 853.80 | 891.97 | **3,715.50** |
| 515 | 3,375.95 | 3,282.94 | 2,957.73 | 3,139.98 | **12,756.60** |
| 516 | 515.87 | 543.28 | 436.77 | 428.85 | **1,924.77** |
| 520 | 3,827.00 | 3,744.47 | 3,485.46 | 3,707.63 | **14,764.56** |
| 521 | 1,152.15 | 1,196.33 | 1,124.23 | 1,133.73 | **4,606.45** |
| 522 | 6,900.81 | 6,905.52 | 6,428.56 | 6,814.72 | **27,049.62** |
| 523 | 3,969.36 | 3,932.83 | 3,731.59 | 3,779.62 | **15,413.40** |
| 524 | 6,301.96 | 6,109.46 | 5,783.46 | 6,242.34 | **24,437.22** |
| 525 | 2,244.94 | 2,161.57 | 2,036.38 | 2,347.86 | **8,790.76** |
| 526 | 2,978.91 | 2,802.71 | 2,828.15 | 3,000.84 | **11,610.60** |
| 527 | 4,553.70 | 4,406.67 | 4,249.53 | 4,459.26 | **17,669.16** |
| 528 | 3,402.26 | 3,409.13 | 3,276.78 | 3,286.13 | **13,374.29** |
| **IA - IOWA - Total** | **81,724.04** | **80,095.38** | **75,296.80** | **79,857.27** | **316,973.49** |

**STATE: ID - IDAHO**

| REGISTRANT ZIP CODE 3 | 1ST QUARTER | 2ND QUARTER | 3RD QUARTER | 4TH QUARTER | TOTAL GRAMS |
|---|---|---|---|---|---|
| 832 | 2,741.53 | 2,718.87 | 2,639.18 | 2,703.50 | **10,803.09** |
| 833 | 2,899.05 | 2,779.11 | 2,431.86 | 2,650.67 | **10,760.69** |
| 834 | 3,367.18 | 3,357.05 | 3,175.13 | 3,297.19 | **13,196.55** |
| 835 | 810.21 | 825.42 | 699.21 | 721.49 | **3,056.33** |
| 836 | 9,113.35 | 8,755.66 | 8,383.12 | 8,533.93 | **34,786.06** |
| 837 | 7,005.87 | 6,767.37 | 6,332.42 | 6,725.71 | **26,831.36** |
| 838 | 2,754.10 | 2,783.42 | 2,627.27 | 2,868.94 | **11,033.72** |
| **ID - IDAHO - Total** | **28,691.29** | **27,986.90** | **26,288.20** | **27,501.42** | **110,467.80** |

**STATE: IL - ILLINOIS**

| REGISTRANT ZIP CODE 3 | 1ST QUARTER | 2ND QUARTER | 3RD QUARTER | 4TH QUARTER | TOTAL GRAMS |
|---|---|---|---|---|---|
| 600 | 25,859.15 | 25,283.06 | 24,555.02 | 25,582.53 | **101,279.76** |
| 601 | 18,601.06 | 17,679.99 | 16,932.88 | 18,318.02 | **71,531.94** |
| 602 | 1,381.84 | 1,349.34 | 1,156.26 | 1,425.93 | **5,313.37** |
| 603 | 1,397.63 | 1,251.12 | 1,212.53 | 1,247.49 | **5,108.77** |
| 604 | 13,262.06 | 12,441.19 | 11,905.92 | 12,935.55 | **50,544.72** |
| 605 | 15,426.07 | 14,333.47 | 13,837.07 | 15,181.49 | **58,778.10** |
| 606 | 16,613.62 | 16,594.12 | 14,949.57 | 16,290.08 | **64,447.38** |
| 607 | 1,149.49 | 1,270.57 | 1,173.31 | 1,213.94 | **4,807.30** |
| 608 | 639.13 | 579.33 | 581.11 | 596.61 | **2,396.17** |
| 609 | 2,337.73 | 2,377.88 | 2,083.33 | 2,355.13 | **9,154.08** |
| 610 | 3,836.34 | 3,545.55 | 3,575.38 | 3,590.37 | **14,547.65** |
| 611 | 4,010.58 | 3,821.66 | 3,648.62 | 3,816.65 | **15,297.50** |
| 612 | 3,190.85 | 2,873.57 | 2,765.42 | 2,881.30 | **11,711.14** |
| 613 | 2,379.90 | 2,217.36 | 1,996.63 | 2,125.80 | **8,719.70** |

| | | | | | |
|---|---|---|---|---|---|
| 614 | 2,299.36 | 2,010.82 | 1,903.16 | 2,183.74 | **8,397.07** |
| 615 | 4,138.42 | 3,758.91 | 3,741.82 | 4,114.97 | **15,754.13** |
| 616 | 5,198.56 | 4,796.60 | 4,877.93 | 4,982.59 | **19,855.67** |
| 617 | 6,132.45 | 5,579.74 | 5,611.94 | 5,889.13 | **23,213.26** |
| 618 | 6,271.75 | 5,862.96 | 5,568.76 | 5,936.81 | **23,640.28** |
| 619 | 2,310.66 | 2,083.04 | 1,994.22 | 2,123.58 | **8,511.50** |
| 620 | 5,611.93 | 5,139.79 | 4,834.44 | 5,477.58 | **21,063.74** |
| 622 | 7,474.16 | 7,417.08 | 6,935.41 | 7,492.75 | **29,319.40** |
| 623 | 2,117.23 | 2,133.26 | 1,906.95 | 2,191.88 | **8,349.32** |
| 624 | 2,353.81 | 2,048.46 | 1,970.91 | 2,268.69 | **8,641.87** |
| 625 | 2,701.27 | 2,391.69 | 2,532.04 | 2,549.60 | **10,174.60** |
| 626 | 4,040.80 | 3,751.94 | 3,490.07 | 3,830.83 | **15,113.64** |
| 627 | 4,687.07 | 4,336.16 | 4,271.47 | 4,417.59 | **17,712.29** |
| 628 | 3,591.01 | 3,492.44 | 3,199.62 | 3,404.96 | **13,688.04** |
| 629 | 4,315.10 | 3,895.34 | 3,871.30 | 3,925.73 | **16,007.47** |
| **IL - ILLINOIS - Total** | **173,329.01** | **164,316.41** | **157,083.11** | **168,351.34** | **663,079.86** |

**STATE: IN - INDIANA**

| REGISTRANT ZIP CODE 3 | 1ST QUARTER | 2ND QUARTER | 3RD QUARTER | 4TH QUARTER | **TOTAL GRAMS** |
|---|---|---|---|---|---|
| 460 | 13,831.90 | 12,878.41 | 12,721.19 | 13,540.25 | **52,971.75** |
| 461 | 10,709.48 | 10,274.80 | 10,228.64 | 10,613.49 | **41,826.41** |
| 462 | 16,220.97 | 15,397.87 | 14,784.91 | 15,097.43 | **61,501.18** |
| 463 | 9,603.95 | 9,055.77 | 8,476.48 | 9,004.39 | **36,140.59** |
| 464 | 775.13 | 828.68 | 793.76 | 836.99 | **3,234.56** |
| 465 | 8,631.86 | 7,963.55 | 7,429.10 | 8,173.12 | **32,197.63** |
| 466 | 2,669.12 | 2,632.06 | 2,452.65 | 2,505.62 | **10,259.45** |
| 467 | 4,496.38 | 4,442.80 | 4,121.98 | 4,594.21 | **17,655.37** |
| 468 | 6,989.79 | 6,457.41 | 6,121.13 | 6,631.64 | **26,199.97** |
| 469 | 4,921.19 | 4,774.15 | 4,637.08 | 4,871.49 | **19,203.91** |
| 470 | 1,675.41 | 1,485.07 | 1,409.41 | 1,472.48 | **6,042.38** |
| 471 | 6,889.51 | 6,639.78 | 5,666.26 | 5,265.40 | **24,460.95** |
| 472 | 3,804.38 | 3,562.81 | 3,436.61 | 3,701.43 | **14,505.24** |
| 473 | 5,241.52 | 5,031.84 | 4,759.63 | 4,809.47 | **19,842.45** |
| 474 | 3,812.82 | 3,464.31 | 3,396.56 | 3,554.93 | **14,228.61** |
| 475 | 2,862.93 | 2,644.95 | 2,778.76 | 2,672.77 | **10,959.41** |
| 476 | 2,698.15 | 2,580.78 | 2,547.61 | 2,912.00 | **10,738.54** |
| 477 | 5,471.58 | 5,161.44 | 5,024.31 | 5,271.48 | **20,928.80** |
| 478 | 3,353.27 | 3,066.34 | 2,874.42 | 3,035.14 | **12,329.17** |
| 479 | 5,931.12 | 5,478.85 | 5,558.28 | 5,703.12 | **22,671.36** |
| **IN - INDIANA - Total** | **120,590.42** | **113,821.67** | **109,218.78** | **114,266.85** | **457,897.73** |

**STATE: KS - KANSAS**

| REGISTRANT ZIP CODE 3 | 1ST QUARTER | 2ND QUARTER | 3RD QUARTER | 4TH QUARTER | TOTAL GRAMS |
|---|---|---|---|---|---|
| 660 | 9,986.56 | 9,563.84 | 9,457.33 | 10,004.57 | **39,012.31** |
| 661 | 1,228.16 | 1,178.91 | 1,141.83 | 1,239.65 | **4,788.55** |
| 662 | 8,699.90 | 8,217.78 | 8,012.01 | 8,748.74 | **33,678.43** |
| 664 | 1,545.81 | 1,758.14 | 1,543.46 | 1,289.47 | **6,136.88** |
| 665 | 1,763.30 | 1,593.98 | 1,582.74 | 1,804.77 | **6,744.80** |
| 666 | 5,361.12 | 5,438.81 | 5,144.98 | 5,485.42 | **21,430.34** |
| 667 | 1,680.89 | 1,688.81 | 1,735.42 | 1,878.62 | **6,983.74** |
| 668 | 747.87 | 755.52 | 736.73 | 789.37 | **3,029.49** |
| 669 | 291.30 | 294.65 | 267.25 | 299.70 | **1,152.90** |
| 670 | 2,659.78 | 2,805.17 | 2,489.32 | 2,786.79 | **10,741.06** |
| 671 | 1,397.89 | 1,261.25 | 1,210.89 | 1,305.64 | **5,175.67** |
| 672 | 6,228.89 | 6,201.63 | 5,804.80 | 6,373.28 | **24,608.59** |
| 673 | 1,491.47 | 1,530.00 | 1,384.52 | 1,480.47 | **5,886.46** |
| 674 | 1,966.70 | 1,942.37 | 1,809.52 | 1,999.76 | **7,718.34** |
| 675 | 1,756.75 | 1,739.40 | 1,531.66 | 1,751.44 | **6,779.26** |
| 676 | 1,035.04 | 992.06 | 956.07 | 1,108.85 | **4,092.02** |
| 677 | 299.13 | 361.70 | 305.90 | 368.17 | **1,334.90** |
| 678 | 866.55 | 825.93 | 774.54 | 883.25 | **3,350.28** |
| 679 | 148.31 | 162.70 | 148.35 | 155.05 | **614.40** |
| **KS - KANSAS - Total** | **49,155.41** | **48,312.66** | **46,037.33** | **49,753.01** | **193,258.41** |

**STATE: KY - KENTUCKY**

| REGISTRANT ZIP CODE 3 | 1ST QUARTER | 2ND QUARTER | 3RD QUARTER | 4TH QUARTER | TOTAL GRAMS |
|---|---|---|---|---|---|
| 400 | 4,177.28 | 3,933.32 | 3,876.27 | 4,034.78 | **16,021.64** |
| 401 | 2,167.79 | 1,920.96 | 2,389.66 | 1,629.54 | **8,107.94** |
| 402 | 14,013.49 | 12,793.82 | 12,774.23 | 13,248.94 | **52,830.48** |
| 403 | 5,937.74 | 5,786.50 | 5,423.22 | 6,062.57 | **23,210.05** |
| 404 | 2,248.55 | 1,986.29 | 2,009.78 | 2,077.26 | **8,321.86** |
| 405 | 5,427.68 | 5,044.91 | 4,992.86 | 5,213.97 | **20,679.42** |
| 406 | 1,028.52 | 1,018.96 | 1,000.24 | 1,009.56 | **4,057.28** |
| 407 | 1,648.92 | 1,653.14 | 1,607.98 | 1,680.52 | **6,590.56** |
| 408 | 379.25 | 303.43 | 307.84 | 280.23 | **1,270.75** |
| 409 | 667.39 | 599.96 | 571.32 | 675.18 | **2,513.85** |
| 410 | 10,533.40 | 9,687.08 | 9,372.91 | 10,437.80 | **40,031.19** |
| 411 | 2,202.19 | 2,054.05 | 1,902.22 | 2,013.49 | **8,171.94** |
| 412 | 742.85 | 616.03 | 685.69 | 659.23 | **2,703.79** |
| 413 | 363.89 | 453.77 | 375.11 | 429.23 | **1,622.00** |
| 414 | 304.63 | 269.53 | 234.91 | 244.28 | **1,053.35** |
| 415 | 1,286.19 | 1,160.33 | 1,110.44 | 1,180.96 | **4,737.92** |
| 416 | 606.68 | 603.93 | 537.15 | 596.19 | **2,343.95** |

| | | | | | |
|---|---|---|---|---|---|
| 417 | 944.16 | 940.55 | 831.38 | 955.43 | **3,671.52** |
| 418 | 340.35 | 343.11 | 362.74 | 356.21 | **1,402.41** |
| 420 | 4,745.71 | 4,212.88 | 4,310.99 | 4,420.00 | **17,689.58** |
| 421 | 3,102.65 | 2,835.51 | 3,048.03 | 3,027.05 | **12,013.24** |
| 422 | 2,806.37 | 2,493.41 | 2,836.41 | 2,416.54 | **10,552.73** |
| 423 | 2,918.37 | 2,694.81 | 2,639.68 | 2,707.20 | **10,960.06** |
| 424 | 3,692.81 | 3,035.24 | 3,139.45 | 3,188.04 | **13,055.54** |
| 425 | 952.08 | 874.31 | 1,009.27 | 1,014.57 | **3,850.23** |
| 426 | 671.71 | 636.84 | 661.72 | 713.81 | **2,684.09** |
| 427 | 2,858.75 | 2,709.42 | 2,571.50 | 2,607.29 | **10,746.97** |
| **KY - KENTUCKY - Total** | **76,769.40** | **70,662.09** | **70,582.99** | **72,879.85** | **290,894.33** |

**STATE: LA - LOUISIANA**

| REGISTRANT ZIP CODE 3 | 1ST QUARTER | 2ND QUARTER | 3RD QUARTER | 4TH QUARTER | **TOTAL GRAMS** |
|---|---|---|---|---|---|
| 700 | 9,796.91 | 8,821.05 | 8,654.56 | 9,622.56 | **36,895.08** |
| 701 | 4,660.74 | 4,367.03 | 4,135.14 | 4,583.66 | **17,746.57** |
| 703 | 4,698.98 | 4,039.45 | 4,023.06 | 4,598.34 | **17,359.83** |
| 704 | 9,918.88 | 9,103.40 | 8,564.63 | 9,877.34 | **37,464.25** |
| 705 | 13,142.15 | 11,764.77 | 12,243.91 | 13,575.21 | **50,726.03** |
| 706 | 8,144.17 | 7,331.81 | 7,273.50 | 7,631.60 | **30,381.07** |
| 707 | 10,437.53 | 9,499.95 | 9,705.36 | 10,641.30 | **40,284.13** |
| 708 | 7,271.30 | 6,282.10 | 6,549.70 | 7,042.71 | **27,145.82** |
| 710 | 1,949.99 | 1,729.86 | 1,520.58 | 1,705.48 | **6,905.90** |
| 711 | 4,791.99 | 4,504.70 | 4,223.74 | 4,556.28 | **18,076.71** |
| 712 | 5,004.80 | 4,440.19 | 4,240.41 | 4,810.08 | **18,495.48** |
| 713 | 3,559.89 | 3,257.18 | 3,445.28 | 3,741.88 | **14,004.23** |
| 714 | 2,674.96 | 2,509.44 | 2,528.44 | 2,641.14 | **10,353.98** |
| **LA - LOUISIANA - Total** | **86,052.29** | **77,650.93** | **77,108.30** | **85,027.56** | **325,839.09** |

**STATE: MA - MASSACHUSETTS**

| REGISTRANT ZIP CODE 3 | 1ST QUARTER | 2ND QUARTER | 3RD QUARTER | 4TH QUARTER | **TOTAL GRAMS** |
|---|---|---|---|---|---|
| 010 | 10,532.83 | 10,273.71 | 9,179.11 | 10,092.88 | **40,078.53** |
| 011 | 4,260.66 | 4,188.13 | 3,440.59 | 3,955.60 | **15,844.99** |
| 012 | 4,263.71 | 4,336.63 | 3,802.98 | 3,804.67 | **16,207.99** |
| 013 | 2,094.07 | 1,980.55 | 1,925.67 | 2,111.19 | **8,111.49** |
| 014 | 5,990.14 | 5,700.67 | 5,200.64 | 5,779.37 | **22,670.82** |
| 015 | 8,240.36 | 8,049.94 | 7,267.77 | 8,188.98 | **31,747.05** |
| 016 | 4,078.91 | 3,892.23 | 3,436.23 | 4,036.59 | **15,443.96** |
| 017 | 11,002.91 | 10,469.21 | 9,771.91 | 10,348.78 | **41,592.82** |
| 018 | 15,385.08 | 15,275.10 | 13,495.23 | 15,161.09 | **59,316.50** |
| 019 | 11,204.93 | 11,250.14 | 10,225.79 | 11,102.29 | **43,783.16** |
| 020 | 10,013.53 | 9,745.75 | 8,888.54 | 9,766.70 | **38,414.52** |

| | | | | | |
|---|---|---|---|---|---|
| 021 | 21,398.87 | 20,924.19 | 18,697.45 | 20,766.40 | **81,786.90** |
| 022 | 938.69 | 1,022.92 | 845.93 | 1,203.51 | **4,011.05** |
| 023 | 9,244.47 | 8,900.29 | 8,168.05 | 8,778.88 | **35,091.69** |
| 024 | 11,561.70 | 11,742.58 | 10,396.70 | 11,738.66 | **45,439.64** |
| 025 | 3,028.40 | 2,816.87 | 2,838.25 | 2,775.38 | **11,458.91** |
| 026 | 3,775.45 | 3,717.04 | 3,910.46 | 3,515.23 | **14,918.17** |
| 027 | 9,412.01 | 9,376.85 | 7,933.67 | 8,990.13 | **35,712.65** |
| **MA - MASSACHUSETTS - Total** | **146,426.72** | **143,662.81** | **129,424.98** | **142,116.33** | **561,630.85** |

**STATE: MD - MARYLAND**

| REGISTRANT ZIP CODE 3 | 1ST QUARTER | 2ND QUARTER | 3RD QUARTER | 4TH QUARTER | TOTAL GRAMS |
|---|---|---|---|---|---|
| 206 | 3,938.16 | 3,857.87 | 3,476.87 | 3,784.17 | **15,057.08** |
| 207 | 7,030.97 | 6,677.42 | 6,388.07 | 5,958.58 | **26,055.04** |
| 208 | 15,883.16 | 15,330.56 | 14,344.64 | 14,547.27 | **60,105.64** |
| 209 | 2,852.77 | 2,655.70 | 2,438.91 | 2,690.77 | **10,638.15** |
| 210 | 16,916.96 | 17,307.57 | 15,029.54 | 16,730.37 | **65,984.44** |
| 211 | 8,521.24 | 8,365.81 | 7,313.63 | 8,230.79 | **32,431.47** |
| 212 | 20,238.32 | 20,081.82 | 17,456.02 | 19,917.35 | **77,693.51** |
| 214 | 2,021.64 | 1,840.75 | 1,844.69 | 1,917.73 | **7,624.81** |
| 215 | 3,430.44 | 3,395.52 | 3,095.41 | 3,368.15 | **13,289.52** |
| 216 | 3,895.28 | 3,800.21 | 3,439.95 | 3,652.80 | **14,788.25** |
| 217 | 11,796.42 | 11,663.17 | 10,377.56 | 11,579.48 | **45,416.63** |
| 218 | 3,215.68 | 3,156.69 | 2,802.19 | 3,175.78 | **12,350.33** |
| 219 | 1,835.97 | 1,798.88 | 1,711.14 | 1,803.42 | **7,149.41** |
| **MD - MARYLAND - Total** | **101,577.01** | **99,931.99** | **89,718.61** | **97,356.66** | **388,584.27** |

**STATE: ME - MAINE**

| REGISTRANT ZIP CODE 3 | 1ST QUARTER | 2ND QUARTER | 3RD QUARTER | 4TH QUARTER | TOTAL GRAMS |
|---|---|---|---|---|---|
| 039 | 802.97 | 773.87 | 729.33 | 651.96 | **2,958.12** |
| 040 | 7,915.67 | 8,012.40 | 7,194.39 | 7,359.53 | **30,481.99** |
| 041 | 2,801.04 | 2,850.94 | 2,799.73 | 2,817.28 | **11,268.99** |
| 042 | 3,318.29 | 3,078.85 | 2,964.42 | 2,981.28 | **12,342.85** |
| 043 | 2,721.29 | 2,815.75 | 2,740.52 | 2,789.92 | **11,067.48** |
| 044 | 6,494.19 | 6,109.39 | 5,896.15 | 6,110.54 | **24,610.28** |
| 045 | 1,374.44 | 1,389.86 | 1,251.52 | 1,319.76 | **5,335.58** |
| 046 | 2,085.96 | 1,999.11 | 1,933.12 | 1,831.19 | **7,849.37** |
| 047 | 2,130.75 | 2,074.43 | 1,800.27 | 1,726.36 | **7,731.82** |
| 048 | 1,190.46 | 1,311.19 | 1,344.90 | 1,294.57 | **5,141.11** |
| 049 | 5,183.57 | 5,133.34 | 4,856.78 | 4,544.44 | **19,718.13** |
| **ME - MAINE - Total** | **36,018.64** | **35,549.13** | **33,511.13** | **33,426.83** | **138,505.73** |

**STATE: MI - MICHIGAN**

| REGISTRANT ZIP CODE 3 | 1ST QUARTER | 2ND QUARTER | 3RD QUARTER | 4TH QUARTER | TOTAL GRAMS |
|---|---|---|---|---|---|

| | | | | | |
|---|---|---|---|---|---|
| 480 | 23,856.10 | 22,504.76 | 20,175.61 | 22,590.69 | **89,127.16** |
| 481 | 27,205.43 | 26,132.02 | 23,547.15 | 26,160.40 | **103,045.01** |
| 482 | 4,851.83 | 4,484.19 | 4,162.71 | 4,294.79 | **17,793.53** |
| 483 | 19,464.39 | 18,969.19 | 17,361.04 | 18,731.93 | **74,526.55** |
| 484 | 6,370.54 | 6,235.89 | 5,534.22 | 6,259.73 | **24,400.38** |
| 485 | 2,385.50 | 2,243.85 | 1,955.77 | 2,087.72 | **8,672.84** |
| 486 | 7,190.72 | 6,845.46 | 6,456.85 | 6,646.93 | **27,139.95** |
| 487 | 4,321.41 | 4,021.76 | 3,472.29 | 3,852.08 | **15,667.54** |
| 488 | 12,879.43 | 12,229.71 | 11,522.28 | 12,606.71 | **49,238.13** |
| 489 | 3,047.25 | 3,031.34 | 2,758.89 | 2,783.33 | **11,620.81** |
| 490 | 13,449.78 | 13,324.93 | 12,359.83 | 13,205.29 | **52,339.84** |
| 491 | 1,085.63 | 1,007.81 | 943.67 | 999.17 | **4,036.28** |
| 492 | 5,809.56 | 5,321.04 | 4,873.02 | 5,403.82 | **21,407.44** |
| 493 | 6,030.50 | 5,534.76 | 5,184.55 | 5,431.31 | **22,181.12** |
| 494 | 16,666.97 | 16,076.55 | 15,489.71 | 16,159.46 | **64,392.69** |
| 495 | 12,828.16 | 12,204.11 | 11,834.35 | 12,233.97 | **49,100.59** |
| 496 | 7,101.72 | 6,608.77 | 6,262.72 | 6,248.84 | **26,222.05** |
| 497 | 5,289.61 | 5,065.79 | 4,677.90 | 4,922.79 | **19,956.10** |
| 498 | 7,161.21 | 6,131.11 | 6,326.68 | 6,202.49 | **25,821.49** |
| 499 | 1,470.34 | 1,238.25 | 1,323.97 | 1,328.03 | **5,360.59** |
| **MI - MICHIGAN - Total** | **188,466.09** | **179,211.28** | **166,223.22** | **178,149.49** | **712,050.08** |

**STATE: MN - MINNESOTA**

| REGISTRANT ZIP CODE 3 | 1ST QUARTER | 2ND QUARTER | 3RD QUARTER | 4TH QUARTER | **TOTAL GRAMS** |
|---|---|---|---|---|---|
| 550 | 11,512.22 | 11,047.58 | 10,357.00 | 11,666.77 | **44,583.57** |
| 551 | 14,836.77 | 14,578.52 | 13,620.94 | 14,526.79 | **57,563.02** |
| 553 | 21,104.10 | 19,649.41 | 18,639.68 | 20,953.18 | **80,346.37** |
| 554 | 19,593.32 | 19,162.43 | 18,132.48 | 19,734.48 | **76,622.71** |
| 556 | 192.85 | 196.39 | 207.20 | 157.35 | **753.79** |
| 557 | 2,293.44 | 2,173.90 | 2,155.88 | 2,171.13 | **8,794.36** |
| 558 | 2,741.74 | 2,609.56 | 2,503.29 | 2,607.54 | **10,462.13** |
| 559 | 8,521.72 | 8,001.76 | 7,686.81 | 8,358.28 | **32,568.57** |
| 560 | 4,819.53 | 4,524.98 | 4,147.68 | 4,466.26 | **17,958.45** |
| 561 | 1,430.18 | 1,397.19 | 1,336.81 | 1,405.46 | **5,569.64** |
| 562 | 2,400.45 | 2,300.24 | 2,213.75 | 2,340.24 | **9,254.69** |
| 563 | 6,186.71 | 5,765.64 | 5,540.60 | 5,953.00 | **23,445.95** |
| 564 | 2,212.24 | 2,080.79 | 2,062.04 | 2,146.21 | **8,501.28** |
| 565 | 3,749.57 | 3,483.17 | 3,608.49 | 3,613.43 | **14,454.66** |
| 566 | 1,050.90 | 981.88 | 867.42 | 990.39 | **3,890.59** |
| 567 | 1,014.28 | 930.32 | 992.99 | 996.26 | **3,933.85** |
| **MN - MINNESOTA - Total** | **103,660.03** | **98,883.75** | **94,073.07** | **102,086.78** | **398,703.63** |

STATE: MO - MISSOURI

| REGISTRANT ZIP CODE 3 | 1ST QUARTER | 2ND QUARTER | 3RD QUARTER | 4TH QUARTER | TOTAL GRAMS |
|---|---|---|---|---|---|
| 630 | 17,096.97 | 15,563.06 | 15,255.65 | 16,548.06 | **64,463.74** |
| 631 | 36,161.98 | 38,932.22 | 40,946.45 | 62,248.91 | **178,289.56** |
| 633 | 11,543.44 | 10,895.95 | 10,940.55 | 11,641.79 | **45,021.73** |
| 634 | 1,366.10 | 1,228.92 | 1,336.99 | 1,439.83 | **5,371.84** |
| 635 | 779.29 | 717.20 | 731.85 | 831.87 | **3,060.21** |
| 636 | 2,212.62 | 2,121.73 | 2,062.88 | 2,141.07 | **8,538.29** |
| 637 | 2,073.49 | 1,815.20 | 1,894.50 | 2,043.08 | **7,826.27** |
| 638 | 2,235.80 | 2,124.16 | 2,038.85 | 2,233.89 | **8,632.70** |
| 639 | 835.48 | 773.69 | 685.93 | 769.17 | **3,064.28** |
| 640 | 10,044.10 | 9,468.36 | 9,282.39 | 10,074.30 | **38,869.15** |
| 641 | 11,107.40 | 11,100.05 | 10,863.16 | 11,228.48 | **44,299.10** |
| 644 | 962.89 | 931.58 | 904.81 | 956.73 | **3,756.01** |
| 645 | 2,046.36 | 2,084.15 | 1,828.65 | 2,114.02 | **8,073.18** |
| 646 | 637.07 | 507.93 | 569.47 | 537.50 | **2,251.97** |
| 647 | 999.70 | 911.25 | 948.64 | 1,010.91 | **3,870.50** |
| 648 | 3,133.32 | 2,978.87 | 2,865.95 | 3,044.48 | **12,022.62** |
| 650 | 1,383.36 | 1,280.14 | 1,159.38 | 1,291.97 | **5,114.84** |
| 651 | 1,655.17 | 1,402.09 | 1,496.66 | 1,717.58 | **6,271.51** |
| 652 | 3,885.09 | 3,526.20 | 3,636.63 | 4,066.44 | **15,114.35** |
| 653 | 1,139.75 | 1,213.93 | 1,162.38 | 1,120.29 | **4,636.35** |
| 654 | 1,458.63 | 1,358.32 | 1,311.83 | 1,316.70 | **5,445.49** |
| 655 | 1,445.48 | 1,330.88 | 1,288.81 | 1,337.24 | **5,402.42** |
| 656 | 1,652.32 | 1,612.61 | 1,528.46 | 1,698.25 | **6,491.64** |
| 657 | 3,497.32 | 3,383.32 | 3,109.46 | 3,339.88 | **13,329.98** |
| 658 | 4,186.62 | 3,941.26 | 3,809.48 | 4,190.19 | **16,127.55** |
| **MO - MISSOURI - Total** | **123,539.76** | **121,203.09** | **121,659.82** | **148,942.62** | **515,345.28** |

STATE: MS - MISSISSIPPI

| REGISTRANT ZIP CODE 3 | 1ST QUARTER | 2ND QUARTER | 3RD QUARTER | 4TH QUARTER | TOTAL GRAMS |
|---|---|---|---|---|---|
| 386 | 5,939.19 | 5,706.33 | 5,670.34 | 6,092.50 | **23,408.36** |
| 387 | 662.29 | 609.31 | 619.09 | 601.14 | **2,491.83** |
| 388 | 3,195.51 | 3,070.89 | 3,020.85 | 3,316.85 | **12,604.09** |
| 389 | 1,052.44 | 1,065.77 | 1,022.31 | 1,063.05 | **4,203.57** |
| 390 | 3,568.87 | 3,311.87 | 3,264.15 | 3,572.57 | **13,717.45** |
| 391 | 3,420.01 | 3,100.36 | 3,138.82 | 3,510.64 | **13,169.83** |
| 392 | 3,043.10 | 3,121.57 | 3,183.02 | 3,214.39 | **12,562.08** |
| 393 | 2,619.63 | 2,462.20 | 2,412.55 | 2,362.33 | **9,856.71** |
| 394 | 6,418.89 | 6,109.85 | 6,188.71 | 6,648.76 | **25,366.21** |
| 395 | 9,209.73 | 8,031.02 | 8,396.71 | 8,942.33 | **34,579.79** |

| | | | | | |
|---|---|---|---|---|---|
| 396 | 1,871.64 | 1,784.99 | 1,750.39 | 1,733.07 | **7,140.08** |
| 397 | 1,503.87 | 1,309.76 | 1,424.28 | 1,585.22 | **5,823.13** |
| **MS - MISSISSIPPI - Total** | **42,505.17** | **39,683.90** | **40,091.23** | **42,642.84** | **164,923.13** |

**STATE: MT - MONTANA**

| REGISTRANT ZIP CODE 3 | 1ST QUARTER | 2ND QUARTER | 3RD QUARTER | 4TH QUARTER | TOTAL GRAMS |
|---|---|---|---|---|---|
| 590 | 988.44 | 874.99 | 843.18 | 845.81 | **3,552.42** |
| 591 | 4,065.34 | 4,010.45 | 3,716.63 | 4,186.54 | **15,978.95** |
| 592 | 689.08 | 583.03 | 464.40 | 588.06 | **2,324.57** |
| 593 | 779.59 | 719.02 | 636.98 | 706.71 | **2,842.30** |
| 594 | 2,423.23 | 2,294.39 | 2,085.67 | 2,301.13 | **9,104.43** |
| 595 | 395.26 | 351.20 | 322.72 | 274.28 | **1,343.46** |
| 596 | 1,289.26 | 1,337.53 | 1,266.19 | 1,304.28 | **5,197.27** |
| 597 | 2,784.35 | 2,608.10 | 2,532.73 | 2,568.06 | **10,493.24** |
| 598 | 3,058.02 | 2,802.31 | 2,734.28 | 2,822.71 | **11,417.32** |
| 599 | 1,440.35 | 1,364.29 | 1,250.11 | 1,375.19 | **5,429.94** |
| **MT - MONTANA - Total** | **17,912.93** | **16,945.32** | **15,852.89** | **16,972.77** | **67,683.91** |

**STATE: NC - NORTH CAROLINA**

| REGISTRANT ZIP CODE 3 | 1ST QUARTER | 2ND QUARTER | 3RD QUARTER | 4TH QUARTER | TOTAL GRAMS |
|---|---|---|---|---|---|
| 270 | 5,068.71 | 4,319.62 | 4,084.80 | 4,724.99 | **18,198.12** |
| 271 | 5,963.77 | 5,130.60 | 4,653.35 | 5,375.08 | **21,122.80** |
| 272 | 11,213.31 | 10,939.07 | 9,675.20 | 11,081.65 | **42,909.23** |
| 273 | 6,736.85 | 6,741.00 | 5,809.40 | 6,898.82 | **26,186.07** |
| 274 | 5,859.59 | 5,418.23 | 4,795.88 | 5,278.24 | **21,351.94** |
| 275 | 24,835.34 | 23,777.14 | 21,489.87 | 24,728.03 | **94,830.37** |
| 276 | 9,233.29 | 9,042.06 | 8,395.14 | 9,286.46 | **35,956.95** |
| 277 | 5,990.51 | 5,577.99 | 5,095.59 | 5,438.28 | **22,102.37** |
| 278 | 9,148.22 | 8,081.15 | 7,608.55 | 9,052.32 | **33,890.25** |
| 279 | 2,539.47 | 2,327.54 | 2,267.37 | 2,565.10 | **9,699.48** |
| 280 | 18,237.75 | 16,438.16 | 15,099.13 | 16,657.35 | **66,432.39** |
| 281 | 13,341.60 | 12,748.06 | 11,423.43 | 12,764.70 | **50,277.79** |
| 282 | 13,706.56 | 12,825.36 | 11,489.28 | 12,700.04 | **50,721.23** |
| 283 | 16,205.55 | 15,662.88 | 13,460.52 | 14,520.77 | **59,849.72** |
| 284 | 8,608.91 | 7,854.87 | 7,079.33 | 8,035.22 | **31,578.32** |
| 285 | 11,842.15 | 11,234.07 | 10,207.33 | 9,890.58 | **43,174.12** |
| 286 | 11,232.73 | 10,203.89 | 9,108.91 | 10,241.33 | **40,786.86** |
| 287 | 8,942.59 | 8,624.68 | 7,889.27 | 8,680.62 | **34,137.17** |
| 288 | 4,071.83 | 4,008.35 | 3,662.05 | 3,853.28 | **15,595.51** |
| 289 | 550.28 | 667.46 | 541.60 | 458.51 | **2,217.85** |
| **NC - NORTH CAROLINA - Total** | **193,329.00** | **181,622.18** | **163,836.01** | **182,231.37** | **721,018.56** |

**STATE: ND - NORTH DAKOTA**

| REGISTRANT ZIP CODE 3 | 1ST QUARTER | 2ND QUARTER | 3RD QUARTER | 4TH QUARTER | TOTAL GRAMS |
|---|---|---|---|---|---|
| 580 | 1,625.60 | 1,638.62 | 1,410.20 | 1,710.29 | **6,384.72** |
| 581 | 3,776.19 | 3,364.65 | 3,487.31 | 3,604.02 | **14,232.17** |
| 582 | 2,286.89 | 2,289.84 | 2,116.48 | 2,243.87 | **8,937.09** |
| 583 | 1,067.60 | 916.90 | 964.49 | 1,030.80 | **3,979.79** |
| 584 | 708.94 | 642.68 | 571.17 | 634.55 | **2,557.33** |
| 585 | 2,940.67 | 2,963.84 | 2,731.82 | 2,956.49 | **11,592.82** |
| 586 | 854.42 | 670.87 | 685.19 | 809.42 | **3,019.91** |
| 587 | 1,693.22 | 1,708.55 | 1,718.71 | 1,802.94 | **6,923.42** |
| 588 | 708.31 | 621.02 | 612.32 | 690.70 | **2,632.35** |
| **ND - NORTH DAKOTA - Total** | **15,661.85** | **14,816.96** | **14,297.70** | **15,483.09** | **60,259.60** |

**STATE: NE - NEBRASKA**

| REGISTRANT ZIP CODE 3 | 1ST QUARTER | 2ND QUARTER | 3RD QUARTER | 4TH QUARTER | TOTAL GRAMS |
|---|---|---|---|---|---|
| 680 | 3,255.86 | 3,180.47 | 3,206.97 | 3,352.74 | **12,996.03** |
| 681 | 12,477.77 | 12,040.29 | 11,866.83 | 12,468.16 | **48,853.06** |
| 683 | 887.70 | 824.62 | 781.22 | 856.19 | **3,349.73** |
| 684 | 801.22 | 696.62 | 685.33 | 731.01 | **2,914.18** |
| 685 | 5,581.36 | 5,552.03 | 5,538.71 | 5,934.19 | **22,606.29** |
| 686 | 822.48 | 694.09 | 669.34 | 742.79 | **2,928.69** |
| 687 | 1,541.99 | 1,432.78 | 1,445.14 | 1,524.16 | **5,944.06** |
| 688 | 2,488.30 | 2,331.35 | 2,258.91 | 2,303.57 | **9,382.13** |
| 689 | 1,019.87 | 1,005.59 | 944.65 | 980.21 | **3,950.33** |
| 690 | 249.39 | 214.97 | 259.98 | 278.14 | **1,002.47** |
| 691 | 1,150.65 | 1,031.75 | 986.21 | 1,001.10 | **4,169.72** |
| 692 | 42.44 | 45.40 | 39.17 | 55.00 | **182.00** |
| 693 | 859.59 | 826.03 | 782.63 | 882.09 | **3,350.35** |
| **NE - NEBRASKA - Total** | **31,178.63** | **29,875.99** | **29,465.08** | **31,109.35** | **121,629.05** |

**STATE: NH - NEW HAMPSHIRE**

| REGISTRANT ZIP CODE 3 | 1ST QUARTER | 2ND QUARTER | 3RD QUARTER | 4TH QUARTER | TOTAL GRAMS |
|---|---|---|---|---|---|
| 030 | 8,379.23 | 8,764.26 | 7,987.87 | 8,521.85 | **33,653.21** |
| 031 | 5,041.75 | 5,124.50 | 4,678.94 | 4,829.05 | **19,674.24** |
| 032 | 3,165.05 | 3,181.49 | 2,975.09 | 2,979.62 | **12,301.25** |
| 033 | 2,227.34 | 2,283.32 | 2,197.51 | 2,308.69 | **9,016.86** |
| 034 | 1,987.81 | 2,002.38 | 1,939.93 | 1,882.91 | **7,813.03** |
| 035 | 1,920.65 | 1,732.73 | 1,740.31 | 1,701.33 | **7,095.03** |
| 036 | 107.48 | 114.14 | 97.11 | 82.41 | **401.13** |
| 037 | 3,347.06 | 3,550.42 | 2,998.19 | 3,253.14 | **13,148.81** |
| 038 | 7,736.43 | 7,324.42 | 7,234.78 | 7,564.68 | **29,860.30** |
| **NH - NEW HAMPSHIRE - Total** | **33,912.80** | **34,077.66** | **31,849.73** | **33,123.67** | **132,963.86** |

**STATE: NJ - NEW JERSEY**

| REGISTRANT ZIP CODE 3 | 1ST QUARTER | 2ND QUARTER | 3RD QUARTER | 4TH QUARTER | TOTAL GRAMS |
|---|---|---|---|---|---|
| 070 | 15,908.31 | 15,724.63 | 13,826.19 | 15,928.72 | **61,387.85** |
| 071 | 1,902.63 | 1,868.93 | 1,538.42 | 1,943.02 | **7,252.99** |
| 072 | 1,010.07 | 1,030.23 | 829.55 | 952.50 | **3,822.35** |
| 073 | 1,060.99 | 1,125.76 | 890.91 | 940.81 | **4,018.47** |
| 074 | 5,380.62 | 4,952.00 | 4,318.48 | 5,193.59 | **19,844.69** |
| 075 | 904.25 | 845.44 | 684.80 | 787.00 | **3,221.50** |
| 076 | 4,794.76 | 4,619.96 | 4,086.14 | 4,670.40 | **18,171.27** |
| 077 | 6,603.98 | 6,302.13 | 5,849.88 | 6,516.55 | **25,272.54** |
| 078 | 4,652.80 | 4,807.77 | 4,473.65 | 4,416.63 | **18,350.85** |
| 079 | 4,742.89 | 5,015.01 | 4,496.13 | 4,760.87 | **19,014.90** |
| 080 | 17,247.21 | 17,321.94 | 15,677.98 | 17,455.67 | **67,702.81** |
| 081 | 1,558.48 | 1,504.65 | 1,314.49 | 1,502.70 | **5,880.33** |
| 082 | 2,646.41 | 2,576.52 | 2,591.56 | 2,427.52 | **10,242.01** |
| 083 | 2,341.38 | 2,362.66 | 2,072.32 | 2,198.36 | **8,974.72** |
| 084 | 227.01 | 222.28 | 216.86 | 249.90 | **916.04** |
| 085 | 3,970.68 | 3,858.03 | 3,575.11 | 3,826.70 | **15,230.51** |
| 086 | 2,901.81 | 2,770.76 | 2,534.11 | 2,577.16 | **10,783.85** |
| 087 | 4,944.92 | 4,782.65 | 4,624.18 | 4,699.20 | **19,050.95** |
| 088 | 8,731.70 | 8,558.87 | 7,298.81 | 8,455.21 | **33,044.59** |
| 089 | 868.20 | 824.70 | 690.28 | 937.43 | **3,320.62** |
| **NJ - NEW JERSEY - Total** | **92,399.11** | **91,074.92** | **81,589.86** | **90,439.94** | **355,503.83** |

**STATE: NM - NEW MEXICO**

| REGISTRANT ZIP CODE 3 | 1ST QUARTER | 2ND QUARTER | 3RD QUARTER | 4TH QUARTER | TOTAL GRAMS |
|---|---|---|---|---|---|
| 870 | 1,400.03 | 1,297.13 | 1,291.66 | 1,329.27 | **5,318.09** |
| 871 | 8,485.47 | 7,893.06 | 7,519.55 | 7,977.59 | **31,875.67** |
| 873 | 610.56 | 482.38 | 488.47 | 518.82 | **2,100.23** |
| 874 | 1,671.01 | 1,477.94 | 1,447.07 | 1,390.23 | **5,986.25** |
| 875 | 1,905.04 | 1,771.47 | 1,778.47 | 1,911.98 | **7,366.96** |
| 877 | 296.74 | 265.54 | 257.61 | 247.83 | **1,067.72** |
| 878 | 163.31 | 133.36 | 101.19 | 97.74 | **495.59** |
| 879 | 73.67 | 78.95 | 93.22 | 92.70 | **338.54** |
| 880 | 1,746.59 | 1,706.13 | 1,536.00 | 1,711.64 | **6,700.36** |
| 881 | 441.06 | 352.84 | 327.98 | 339.30 | **1,461.17** |
| 882 | 1,855.79 | 1,623.55 | 1,651.83 | 1,764.53 | **6,895.71** |
| 883 | 584.28 | 563.28 | 658.70 | 594.18 | **2,400.44** |
| 884 | 112.44 | 115.87 | 107.81 | 98.93 | **435.05** |
| **NM - NEW MEXICO - Total** | **19,346.00** | **17,761.48** | **17,259.56** | **18,074.75** | **72,441.79** |

**STATE: NV - NEVADA**

| REGISTRANT ZIP CODE 3 | 1ST QUARTER | 2ND QUARTER | 3RD QUARTER | 4TH QUARTER | TOTAL GRAMS |
|---|---|---|---|---|---|

| | | | | | |
|---|---|---|---|---|---|
| 890 | 3,320.04 | 3,206.47 | 3,086.45 | 3,265.81 | **12,878.77** |
| 891 | 6,911.47 | 6,476.71 | 6,756.14 | 6,571.45 | **26,715.77** |
| 893 | 16.08 | 26.29 | 24.56 | 19.20 | **86.13** |
| 894 | 1,611.05 | 1,567.53 | 1,581.43 | 1,693.01 | **6,453.02** |
| 895 | 2,166.94 | 2,127.01 | 2,111.27 | 2,133.02 | **8,538.25** |
| 897 | 576.10 | 457.42 | 485.48 | 414.54 | **1,933.55** |
| 898 | 368.62 | 418.12 | 363.61 | 460.34 | **1,610.68** |
| **NV - NEVADA - Total** | **14,970.31** | **14,279.54** | **14,408.94** | **14,557.37** | **58,216.16** |

**STATE: NY - NEW YORK**

| REGISTRANT ZIP CODE 3 | 1ST QUARTER | 2ND QUARTER | 3RD QUARTER | 4TH QUARTER | TOTAL GRAMS |
|---|---|---|---|---|---|
| 100 | 21,936.58 | 22,564.34 | 19,781.72 | 21,996.60 | **86,279.24** |
| 101 | 1,237.44 | 1,372.55 | 1,017.71 | 1,210.33 | **4,838.02** |
| 102 | 168.62 | 146.35 | 129.29 | 141.91 | **586.16** |
| 103 | 2,589.58 | 2,773.21 | 2,331.07 | 2,689.35 | **10,383.21** |
| 104 | 7,994.66 | 7,726.05 | 6,394.57 | 6,945.77 | **29,061.04** |
| 105 | 10,016.53 | 10,298.45 | 9,054.30 | 10,367.75 | **39,737.02** |
| 106 | 1,048.40 | 1,125.96 | 1,079.81 | 1,058.44 | **4,312.61** |
| 107 | 1,771.65 | 1,841.14 | 1,587.16 | 1,734.37 | **6,934.32** |
| 108 | 915.07 | 938.15 | 830.26 | 840.87 | **3,524.35** |
| 109 | 6,872.54 | 6,735.44 | 6,088.60 | 6,663.55 | **26,360.13** |
| 110 | 2,911.94 | 2,894.48 | 2,861.49 | 3,322.76 | **11,990.68** |
| 111 | 1,195.83 | 1,223.81 | 970.12 | 1,184.26 | **4,574.03** |
| 112 | 12,488.99 | 12,607.32 | 10,363.89 | 11,688.39 | **47,148.58** |
| 113 | 4,951.24 | 5,002.08 | 3,815.26 | 4,626.66 | **18,395.24** |
| 114 | 2,310.73 | 2,444.59 | 1,872.56 | 2,221.36 | **8,849.24** |
| 115 | 8,481.27 | 8,384.02 | 7,233.68 | 8,088.94 | **32,187.92** |
| 116 | 833.50 | 896.32 | 750.30 | 901.51 | **3,381.63** |
| 117 | 19,990.55 | 20,450.54 | 18,019.05 | 19,571.95 | **78,032.09** |
| 118 | 1,286.00 | 1,441.11 | 1,178.91 | 1,483.89 | **5,389.90** |
| 119 | 2,779.37 | 2,797.57 | 2,791.22 | 2,640.01 | **11,008.17** |
| 120 | 7,534.95 | 7,585.76 | 6,936.53 | 7,230.08 | **29,287.32** |
| 121 | 4,764.49 | 4,688.52 | 4,380.33 | 4,995.12 | **18,828.46** |
| 122 | 2,758.76 | 2,918.37 | 2,489.78 | 2,797.02 | **10,963.93** |
| 123 | 3,217.99 | 3,310.41 | 2,955.21 | 3,154.78 | **12,638.40** |
| 124 | 1,835.08 | 1,896.20 | 1,670.87 | 1,639.75 | **7,041.90** |
| 125 | 5,121.71 | 5,131.72 | 4,695.81 | 5,100.54 | **20,049.77** |
| 126 | 1,130.84 | 1,135.03 | 869.84 | 1,099.02 | **4,234.73** |
| 127 | 998.71 | 1,103.08 | 931.48 | 937.54 | **3,970.81** |
| 128 | 5,049.95 | 4,896.24 | 4,749.29 | 4,920.85 | **19,616.32** |
| 129 | 4,100.64 | 4,011.43 | 4,057.84 | 4,287.35 | **16,457.25** |

| | | | | | |
|---|---|---|---|---|---|
| 130 | 6,534.24 | 6,321.82 | 5,946.63 | 6,367.71 | **25,170.40** |
| 131 | 2,256.79 | 2,350.27 | 2,077.62 | 2,157.05 | **8,841.74** |
| 132 | 3,539.86 | 3,395.10 | 3,200.80 | 3,430.05 | **13,565.82** |
| 133 | 2,159.57 | 2,099.98 | 1,796.46 | 1,984.39 | **8,040.39** |
| 134 | 2,899.07 | 2,668.58 | 2,515.18 | 2,634.01 | **10,716.84** |
| 135 | 1,083.84 | 1,015.02 | 993.26 | 1,020.86 | **4,112.98** |
| 136 | 5,889.70 | 5,722.12 | 5,410.50 | 5,799.55 | **22,821.86** |
| 137 | 1,368.22 | 1,316.52 | 1,291.33 | 1,382.62 | **5,358.70** |
| 138 | 2,662.07 | 2,554.28 | 2,369.03 | 2,525.34 | **10,110.72** |
| 139 | 1,093.16 | 1,066.22 | 1,061.94 | 991.35 | **4,212.67** |
| 140 | 5,706.63 | 5,667.00 | 5,249.97 | 5,497.29 | **22,120.88** |
| 141 | 2,676.66 | 2,780.06 | 2,474.89 | 2,687.15 | **10,618.76** |
| 142 | 7,485.45 | 7,380.35 | 6,976.34 | 7,542.43 | **29,384.58** |
| 143 | 624.08 | 697.61 | 583.55 | 684.29 | **2,589.52** |
| 144 | 7,862.87 | 7,598.51 | 6,937.75 | 7,548.05 | **29,947.17** |
| 145 | 8,100.62 | 7,714.88 | 7,273.14 | 7,835.34 | **30,923.98** |
| 146 | 12,708.31 | 12,162.37 | 11,536.01 | 12,351.81 | **48,758.49** |
| 147 | 2,211.40 | 2,107.40 | 2,057.23 | 2,158.56 | **8,534.58** |
| 148 | 5,420.65 | 5,246.44 | 4,939.13 | 5,452.31 | **21,058.53** |
| 149 | 1,194.21 | 1,219.21 | 1,043.84 | 1,259.92 | **4,717.18** |
| **NY - NEW YORK - Total** | **231,770.97** | **231,423.99** | **207,622.54** | **226,850.77** | **897,668.27** |

**STATE: OH - OHIO**

| REGISTRANT ZIP CODE 3 | 1ST QUARTER | 2ND QUARTER | 3RD QUARTER | 4TH QUARTER | TOTAL GRAMS |
|---|---|---|---|---|---|
| 430 | 16,679.70 | 16,209.89 | 15,397.91 | 16,570.54 | **64,858.03** |
| 431 | 8,237.53 | 7,958.01 | 7,679.54 | 8,204.60 | **32,079.68** |
| 432 | 32,137.99 | 24,846.26 | 9,822.82 | 10,787.47 | **77,594.55** |
| 433 | 2,833.01 | 2,694.06 | 2,555.47 | 2,681.19 | **10,763.73** |
| 434 | 2,808.64 | 2,488.33 | 2,541.45 | 2,828.26 | **10,666.68** |
| 435 | 6,322.40 | 5,981.71 | 5,717.37 | 6,526.43 | **24,547.92** |
| 436 | 7,682.73 | 6,968.33 | 6,670.02 | 7,242.74 | **28,563.83** |
| 437 | 2,600.75 | 2,432.28 | 2,339.50 | 2,318.47 | **9,691.00** |
| 438 | 477.21 | 569.66 | 453.67 | 428.04 | **1,928.58** |
| 439 | 2,149.49 | 2,179.08 | 1,978.12 | 1,991.87 | **8,298.56** |
| 440 | 13,561.10 | 13,220.36 | 12,525.15 | 13,660.04 | **52,966.66** |
| 441 | 16,050.05 | 15,226.20 | 14,614.00 | 15,739.84 | **61,630.09** |
| 442 | 8,220.51 | 7,965.92 | 7,588.95 | 8,085.15 | **31,860.54** |
| 443 | 4,052.95 | 3,799.23 | 3,577.69 | 3,769.88 | **15,199.76** |
| 444 | 3,782.78 | 3,514.59 | 3,325.86 | 3,662.90 | **14,286.12** |
| 445 | 2,822.07 | 2,861.51 | 2,504.82 | 2,782.17 | **10,970.58** |
| 446 | 6,187.89 | 5,844.43 | 5,371.45 | 5,642.42 | **23,046.18** |

| | | | | | |
|---|---|---|---|---|---|
| 447 | 4,658.74 | 4,403.69 | 4,254.73 | 4,334.52 | **17,651.69** |
| 448 | 4,728.65 | 4,310.01 | 3,928.73 | 4,386.87 | **17,354.26** |
| 449 | 1,562.36 | 1,455.12 | 1,400.27 | 1,490.52 | **5,908.27** |
| 450 | 15,561.54 | 15,296.03 | 14,331.04 | 16,164.32 | **61,352.93** |
| 451 | 6,460.34 | 6,248.58 | 6,078.31 | 6,502.31 | **25,289.55** |
| 452 | 19,954.29 | 18,499.28 | 17,911.11 | 19,711.50 | **76,076.17** |
| 453 | 8,006.84 | 7,329.55 | 7,392.73 | 8,120.74 | **30,849.86** |
| 454 | 9,085.68 | 8,750.22 | 8,433.76 | 9,264.31 | **35,533.97** |
| 455 | 2,349.41 | 2,187.22 | 2,191.22 | 2,275.80 | **9,003.65** |
| 456 | 6,669.12 | 6,433.14 | 5,978.48 | 6,698.77 | **25,779.51** |
| 457 | 2,496.08 | 2,169.91 | 2,090.62 | 2,332.59 | **9,089.20** |
| 458 | 5,146.69 | 4,769.80 | 4,509.57 | 5,112.21 | **19,538.28** |
| **OH - OHIO - Total** | **223,286.55** | **206,612.44** | **183,164.36** | **199,316.48** | **812,379.83** |

**STATE: OK - OKLAHOMA**

| REGISTRANT ZIP CODE 3 | 1ST QUARTER | 2ND QUARTER | 3RD QUARTER | 4TH QUARTER | **TOTAL GRAMS** |
|---|---|---|---|---|---|
| 730 | 9,725.72 | 9,160.75 | 8,639.90 | 9,505.76 | **37,032.13** |
| 731 | 7,261.92 | 6,750.34 | 6,884.76 | 6,995.19 | **27,892.22** |
| 734 | 728.55 | 624.19 | 623.88 | 673.95 | **2,650.57** |
| 735 | 3,357.34 | 3,107.27 | 2,855.65 | 2,879.75 | **12,200.01** |
| 736 | 793.93 | 707.14 | 718.19 | 795.03 | **3,014.28** |
| 737 | 1,248.78 | 1,184.15 | 1,237.83 | 1,156.99 | **4,827.75** |
| 738 | 211.73 | 185.24 | 190.06 | 187.65 | **774.68** |
| 739 | 56.57 | 50.46 | 47.73 | 52.86 | **207.62** |
| 740 | 10,734.75 | 9,587.78 | 9,358.06 | 9,828.30 | **39,508.90** |
| 741 | 6,165.34 | 5,399.74 | 5,235.34 | 5,423.54 | **22,223.96** |
| 743 | 1,916.10 | 1,637.30 | 1,531.38 | 1,603.19 | **6,687.97** |
| 744 | 3,161.82 | 2,683.50 | 2,794.24 | 2,924.46 | **11,564.01** |
| 745 | 776.46 | 755.82 | 663.40 | 730.27 | **2,925.94** |
| 746 | 740.66 | 667.72 | 622.95 | 717.35 | **2,748.69** |
| 747 | 805.67 | 688.82 | 725.40 | 814.08 | **3,033.97** |
| 748 | 1,721.98 | 1,502.51 | 1,510.55 | 1,712.65 | **6,447.68** |
| 749 | 726.10 | 631.20 | 729.06 | 712.05 | **2,798.41** |
| **OK - OKLAHOMA - Total** | **50,133.42** | **45,323.93** | **44,368.37** | **46,713.07** | **186,538.79** |

**STATE: OR - OREGON**

| REGISTRANT ZIP CODE 3 | 1ST QUARTER | 2ND QUARTER | 3RD QUARTER | 4TH QUARTER | **TOTAL GRAMS** |
|---|---|---|---|---|---|
| 970 | 12,771.24 | 12,558.96 | 11,827.10 | 12,381.80 | **49,539.10** |
| 971 | 5,484.21 | 5,354.89 | 5,292.30 | 5,397.49 | **21,528.89** |
| 972 | 16,040.27 | 16,277.90 | 15,357.42 | 16,299.86 | **63,975.45** |
| 973 | 11,200.88 | 11,109.54 | 10,341.47 | 11,163.48 | **43,815.36** |
| 974 | 7,193.99 | 7,712.05 | 7,065.61 | 7,458.68 | **29,430.34** |

| | | | | | |
|---|---|---|---|---|---|
| 975 | 3,200.86 | 3,507.10 | 3,098.31 | 3,258.30 | **13,064.57** |
| 976 | 625.63 | 699.60 | 592.93 | 588.54 | **2,506.70** |
| 977 | 3,160.48 | 3,345.63 | 3,127.24 | 3,269.29 | **12,902.65** |
| 978 | 1,735.14 | 1,779.38 | 1,623.17 | 1,691.07 | **6,828.77** |
| 979 | 527.76 | 554.34 | 566.25 | 561.53 | **2,209.89** |
| **OR - OREGON - Total** | **61,940.45** | **62,899.40** | **58,891.81** | **62,070.04** | **245,801.71** |

**STATE: PA - PENNSYLVANIA**

| REGISTRANT ZIP CODE 3 | 1ST QUARTER | 2ND QUARTER | 3RD QUARTER | 4TH QUARTER | **TOTAL GRAMS** |
|---|---|---|---|---|---|
| 150 | 7,550.49 | 7,469.57 | 6,937.50 | 7,674.82 | **29,632.39** |
| 151 | 6,721.67 | 6,664.09 | 5,967.99 | 6,051.31 | **25,405.06** |
| 152 | 11,792.52 | 11,646.07 | 10,823.93 | 11,853.27 | **46,115.79** |
| 153 | 3,426.09 | 3,412.84 | 3,203.83 | 3,386.46 | **13,429.22** |
| 154 | 1,466.16 | 1,514.06 | 1,350.25 | 1,470.85 | **5,801.31** |
| 155 | 1,167.80 | 1,136.04 | 1,076.59 | 1,096.50 | **4,476.93** |
| 156 | 5,359.71 | 5,209.86 | 4,604.32 | 5,101.96 | **20,275.85** |
| 157 | 2,358.17 | 2,212.46 | 2,048.08 | 2,142.53 | **8,761.24** |
| 158 | 2,208.94 | 2,032.52 | 2,043.74 | 2,140.54 | **8,425.74** |
| 159 | 2,341.18 | 2,181.19 | 2,049.14 | 2,274.94 | **8,846.45** |
| 160 | 3,914.71 | 3,775.47 | 3,554.81 | 3,739.37 | **14,984.36** |
| 161 | 4,076.79 | 3,865.26 | 3,507.96 | 3,886.52 | **15,336.53** |
| 162 | 1,560.66 | 1,441.02 | 1,350.72 | 1,544.43 | **5,896.83** |
| 163 | 2,316.99 | 2,155.39 | 2,041.49 | 2,130.08 | **8,643.95** |
| 164 | 1,145.14 | 1,168.86 | 1,018.14 | 1,153.84 | **4,485.97** |
| 165 | 4,631.98 | 4,311.69 | 4,070.84 | 4,295.29 | **17,309.79** |
| 166 | 3,659.39 | 3,473.73 | 3,170.34 | 3,415.45 | **13,718.90** |
| 167 | 795.71 | 772.79 | 767.72 | 766.88 | **3,103.10** |
| 168 | 3,547.75 | 3,208.36 | 3,097.94 | 3,270.79 | **13,124.84** |
| 169 | 709.07 | 745.45 | 703.55 | 750.19 | **2,908.25** |
| 170 | 14,819.31 | 14,704.39 | 13,926.97 | 14,202.33 | **57,653.00** |
| 171 | 3,160.47 | 2,945.65 | 2,809.29 | 2,809.39 | **11,724.80** |
| 172 | 3,877.48 | 3,630.92 | 3,532.91 | 3,661.89 | **14,703.20** |
| 173 | 6,272.16 | 6,079.46 | 5,879.48 | 6,111.34 | **24,342.44** |
| 174 | 4,745.18 | 4,783.62 | 4,286.68 | 4,512.26 | **18,327.74** |
| 175 | 5,031.34 | 4,747.46 | 4,414.36 | 4,595.37 | **18,788.54** |
| 176 | 4,404.54 | 4,441.92 | 3,953.93 | 4,359.60 | **17,159.99** |
| 177 | 3,073.96 | 2,879.94 | 2,780.08 | 2,795.99 | **11,529.97** |
| 178 | 3,626.52 | 3,264.60 | 3,094.91 | 3,129.08 | **13,115.10** |
| 179 | 1,370.98 | 1,219.69 | 1,130.66 | 1,301.20 | **5,022.52** |
| 180 | 5,844.80 | 5,753.22 | 5,198.73 | 5,614.09 | **22,410.84** |
| 181 | 2,826.54 | 3,014.62 | 2,734.47 | 2,695.63 | **11,271.26** |

| | | | | | |
|---|---|---|---|---|---|
| 182 | 1,383.16 | 1,332.96 | 1,180.49 | 1,331.62 | **5,228.24** |
| 183 | 1,522.69 | 1,571.89 | 1,378.85 | 1,541.66 | **6,015.09** |
| 184 | 2,191.09 | 2,271.91 | 2,142.25 | 2,204.73 | **8,809.98** |
| 185 | 2,489.90 | 2,542.46 | 2,383.00 | 2,590.64 | **10,006.00** |
| 186 | 2,180.51 | 2,051.76 | 1,980.25 | 2,078.20 | **8,290.71** |
| 187 | 25,512.12 | 24,041.18 | 22,890.97 | 24,516.72 | **96,961.00** |
| 188 | 988.50 | 1,011.50 | 947.11 | 1,121.33 | **4,068.45** |
| 189 | 5,694.99 | 5,772.21 | 5,258.58 | 5,952.43 | **22,678.20** |
| 190 | 17,515.02 | 17,596.52 | 16,601.06 | 18,208.82 | **69,921.42** |
| 191 | 13,291.42 | 13,299.17 | 12,254.10 | 13,453.71 | **52,298.41** |
| 193 | 8,666.57 | 8,389.39 | 7,880.74 | 8,406.12 | **33,342.82** |
| 194 | 8,528.91 | 8,168.16 | 7,722.47 | 8,258.60 | **32,678.15** |
| 195 | 2,509.54 | 2,534.00 | 2,370.73 | 2,637.07 | **10,051.34** |
| 196 | 3,582.37 | 3,430.77 | 3,355.47 | 3,638.03 | **14,006.63** |
| **PA - PENNSYLVANIA - Total** | **225,860.97** | **219,876.07** | **205,477.41** | **219,873.88** | **871,088.33** |

**STATE: PR - PUERTO RICO**

| REGISTRANT ZIP CODE 3 | 1ST QUARTER | 2ND QUARTER | 3RD QUARTER | 4TH QUARTER | TOTAL GRAMS |
|---|---|---|---|---|---|
| 006 | 3,900.56 | 3,767.79 | 3,520.92 | 3,532.39 | **14,721.67** |
| 007 | 4,482.17 | 4,290.90 | 3,888.25 | 4,279.27 | **16,940.59** |
| 009 | 6,819.13 | 6,561.13 | 5,964.13 | 6,383.34 | **25,727.73** |
| **PR - PUERTO RICO - Total** | **15,201.86** | **14,619.82** | **13,373.30** | **14,194.99** | **57,389.98** |

**STATE: RI - RHODE ISLAND**

| REGISTRANT ZIP CODE 3 | 1ST QUARTER | 2ND QUARTER | 3RD QUARTER | 4TH QUARTER | TOTAL GRAMS |
|---|---|---|---|---|---|
| 028 | 12,926.65 | 12,726.46 | 11,517.78 | 12,090.50 | **49,261.39** |
| 029 | 6,332.95 | 5,968.58 | 5,218.21 | 5,756.56 | **23,276.31** |
| **RI - RHODE ISLAND - Total** | **19,259.60** | **18,695.05** | **16,735.99** | **17,847.06** | **72,537.70** |

**STATE: SC - SOUTH CAROLINA**

| REGISTRANT ZIP CODE 3 | 1ST QUARTER | 2ND QUARTER | 3RD QUARTER | 4TH QUARTER | TOTAL GRAMS |
|---|---|---|---|---|---|
| 290 | 6,987.60 | 6,442.73 | 6,261.86 | 6,838.60 | **26,530.79** |
| 291 | 5,609.82 | 5,493.27 | 4,747.60 | 5,212.71 | **21,063.39** |
| 292 | 6,333.92 | 6,053.01 | 5,718.87 | 5,765.50 | **23,871.30** |
| 293 | 7,774.90 | 7,310.96 | 6,951.07 | 7,531.21 | **29,568.14** |
| 294 | 18,745.80 | 17,446.63 | 16,552.44 | 17,729.38 | **70,474.25** |
| 295 | 11,110.91 | 10,496.48 | 9,374.25 | 10,600.01 | **41,581.64** |
| 296 | 19,444.61 | 18,421.65 | 18,180.06 | 19,530.25 | **75,576.57** |
| 297 | 7,530.54 | 7,255.92 | 6,603.19 | 7,488.46 | **28,878.11** |
| 298 | 3,155.73 | 2,882.49 | 2,851.58 | 2,960.96 | **11,850.76** |
| 299 | 2,922.09 | 2,591.48 | 2,480.68 | 2,657.05 | **10,651.30** |
| **SC - SOUTH CAROLINA - Total** | **89,615.93** | **84,394.62** | **79,721.60** | **86,314.11** | **340,046.25** |

**STATE: SD - SOUTH DAKOTA**

| REGISTRANT ZIP CODE 3 | 1ST QUARTER | 2ND QUARTER | 3RD QUARTER | 4TH QUARTER | TOTAL GRAMS |
|---|---|---|---|---|---|
| 570 | 2,680.26 | 2,522.11 | 2,517.40 | 2,767.58 | **10,487.34** |
| 571 | 11,171.75 | 9,380.73 | 9,952.43 | 9,852.18 | **40,357.08** |
| 572 | 1,135.93 | 1,000.16 | 1,039.27 | 1,054.39 | **4,229.75** |
| 573 | 1,724.14 | 1,491.94 | 1,541.29 | 1,671.72 | **6,429.09** |
| 574 | 1,179.18 | 1,208.85 | 1,075.75 | 1,187.86 | **4,651.64** |
| 575 | 972.24 | 682.96 | 815.87 | 926.64 | **3,397.71** |
| 576 | 164.16 | 159.19 | 173.24 | 200.73 | **697.32** |
| 577 | 3,015.43 | 2,831.30 | 2,862.09 | 2,739.12 | **11,447.93** |
| **SD - SOUTH DAKOTA - Total** | **22,043.10** | **19,277.23** | **19,977.33** | **20,400.21** | **81,697.86** |

**STATE: TN - TENNESSEE**

| REGISTRANT ZIP CODE 3 | 1ST QUARTER | 2ND QUARTER | 3RD QUARTER | 4TH QUARTER | TOTAL GRAMS |
|---|---|---|---|---|---|
| 370 | 12,985.63 | 11,574.59 | 11,847.99 | 11,920.34 | **48,328.53** |
| 371 | 8,614.16 | 8,335.22 | 7,951.43 | 8,417.49 | **33,318.31** |
| 372 | 5,592.70 | 5,374.80 | 5,247.89 | 5,518.82 | **21,734.21** |
| 373 | 8,063.61 | 7,464.33 | 7,732.54 | 8,015.59 | **31,276.07** |
| 374 | 3,557.79 | 3,299.29 | 2,992.04 | 3,275.93 | **13,125.05** |
| 376 | 4,887.93 | 4,575.61 | 4,447.26 | 4,482.80 | **18,393.60** |
| 377 | 4,413.98 | 4,216.21 | 4,212.06 | 4,449.83 | **17,292.08** |
| 378 | 6,801.23 | 6,446.54 | 6,622.28 | 6,783.73 | **26,653.79** |
| 379 | 6,078.29 | 5,833.66 | 5,884.89 | 6,133.44 | **23,930.27** |
| 380 | 6,127.57 | 5,817.66 | 5,361.03 | 5,943.30 | **23,249.57** |
| 381 | 7,385.75 | 7,174.36 | 6,917.11 | 7,380.94 | **28,858.15** |
| 382 | 1,546.02 | 1,438.52 | 1,392.82 | 1,478.15 | **5,855.51** |
| 383 | 3,617.90 | 3,353.43 | 3,389.37 | 3,498.49 | **13,859.19** |
| 384 | 2,002.32 | 1,843.95 | 1,802.71 | 1,798.02 | **7,446.99** |
| 385 | 2,033.85 | 1,911.56 | 1,906.24 | 1,991.53 | **7,843.17** |
| **TN - TENNESSEE - Total** | **83,708.72** | **78,659.73** | **77,707.66** | **81,088.40** | **321,164.51** |

**STATE: TX - TEXAS**

| REGISTRANT ZIP CODE 3 | 1ST QUARTER | 2ND QUARTER | 3RD QUARTER | 4TH QUARTER | TOTAL GRAMS |
|---|---|---|---|---|---|
| 750 | 31,472.98 | 30,588.16 | 27,952.43 | 31,189.04 | **121,202.62** |
| 751 | 8,716.65 | 8,418.93 | 7,558.59 | 8,389.10 | **33,083.27** |
| 752 | 11,630.17 | 11,588.42 | 10,042.09 | 11,519.54 | **44,780.22** |
| 753 | 34.24 | 39.34 | 39.08 | 46.61 | **159.28** |
| 754 | 5,299.41 | 4,993.19 | 4,529.64 | 5,131.01 | **19,953.25** |
| 755 | 1,446.92 | 1,244.61 | 1,323.97 | 1,396.91 | **5,412.41** |
| 756 | 4,348.50 | 4,141.31 | 3,718.72 | 4,215.28 | **16,423.81** |
| 757 | 5,357.99 | 5,018.99 | 4,720.77 | 5,233.53 | **20,331.27** |
| 758 | 1,176.45 | 1,127.21 | 1,067.45 | 1,188.69 | **4,559.81** |
| 759 | 4,438.59 | 4,202.76 | 3,915.29 | 4,323.36 | **16,880.00** |

| | | | | | |
|---|---|---|---|---|---|
| 760 | 17,906.20 | 16,977.07 | 16,013.74 | 17,140.55 | **68,037.56** |
| 761 | 10,636.33 | 10,017.63 | 9,107.56 | 10,406.24 | **40,167.75** |
| 762 | 7,975.55 | 7,579.75 | 7,149.69 | 7,907.73 | **30,612.72** |
| 763 | 3,685.54 | 3,311.39 | 3,386.84 | 3,566.04 | **13,949.80** |
| 764 | 1,518.25 | 1,302.60 | 1,200.21 | 1,437.11 | **5,458.17** |
| 765 | 9,935.10 | 9,112.19 | 8,712.06 | 9,404.63 | **37,163.98** |
| 766 | 1,430.94 | 1,347.80 | 1,356.81 | 1,352.13 | **5,487.68** |
| 767 | 4,222.29 | 3,778.05 | 3,533.32 | 4,142.18 | **15,675.84** |
| 768 | 1,342.27 | 1,331.70 | 1,103.20 | 1,371.13 | **5,148.30** |
| 769 | 2,789.78 | 2,359.00 | 2,026.80 | 2,461.04 | **9,636.62** |
| 770 | 23,910.55 | 23,604.70 | 21,214.45 | 24,047.76 | **92,777.47** |
| 772 | 0 | 1.73 | 0 | 2.59 | **4.32** |
| 773 | 17,470.86 | 16,849.65 | 15,525.75 | 17,690.33 | **67,536.60** |
| 774 | 17,772.41 | 16,448.11 | 15,161.64 | 17,681.85 | **67,064.01** |
| 775 | 15,954.91 | 15,592.12 | 14,031.59 | 16,351.45 | **61,930.07** |
| 776 | 3,693.73 | 3,375.51 | 3,009.32 | 3,388.76 | **13,467.32** |
| 777 | 1,496.76 | 1,488.53 | 1,433.22 | 1,582.33 | **6,000.84** |
| 778 | 4,796.52 | 4,327.32 | 4,123.60 | 4,694.82 | **17,942.27** |
| 779 | 2,436.74 | 2,198.32 | 2,010.15 | 2,311.64 | **8,956.84** |
| 780 | 5,645.23 | 5,147.42 | 5,159.25 | 5,951.51 | **21,903.40** |
| 781 | 6,021.24 | 6,035.98 | 5,726.25 | 6,132.63 | **23,916.10** |
| 782 | 25,765.48 | 25,584.48 | 23,082.65 | 25,363.43 | **99,796.04** |
| 783 | 3,062.21 | 2,828.29 | 2,630.96 | 3,004.67 | **11,526.13** |
| 784 | 5,603.86 | 5,362.25 | 4,846.92 | 5,735.89 | **21,548.92** |
| 785 | 11,526.36 | 10,719.89 | 9,494.70 | 10,233.29 | **41,974.24** |
| 786 | 12,741.16 | 12,546.21 | 11,515.43 | 13,113.30 | **49,916.10** |
| 787 | 12,822.38 | 12,596.44 | 11,734.08 | 13,417.45 | **50,570.36** |
| 788 | 1,483.50 | 1,355.22 | 1,207.21 | 1,317.32 | **5,363.24** |
| 789 | 714.96 | 671.25 | 689.69 | 777.00 | **2,852.91** |
| 790 | 1,394.56 | 1,262.68 | 1,101.82 | 1,343.29 | **5,102.36** |
| 791 | 2,318.67 | 2,393.48 | 2,175.76 | 2,425.02 | **9,312.93** |
| 792 | 335.95 | 297.69 | 264.02 | 288.24 | **1,185.90** |
| 793 | 1,000.27 | 1,080.50 | 891.15 | 1,046.82 | **4,018.74** |
| 794 | 5,221.63 | 4,935.90 | 4,501.69 | 4,798.98 | **19,458.20** |
| 795 | 1,054.97 | 1,060.32 | 988.03 | 1,066.70 | **4,170.03** |
| 796 | 3,472.24 | 3,253.42 | 2,991.45 | 3,348.16 | **13,065.27** |
| 797 | 4,504.16 | 4,123.82 | 3,983.48 | 4,363.73 | **16,975.19** |
| 798 | 247.30 | 214.88 | 233.13 | 203.38 | **898.69** |
| 799 | 7,752.46 | 7,665.17 | 7,163.32 | 7,692.07 | **30,273.01** |
| **TX - TEXAS - Total** | **335,585.23** | **321,501.38** | **295,348.98** | **331,196.25** | **1,283,631.84** |

**STATE: UT - UTAH**

| REGISTRANT ZIP CODE 3 | 1ST QUARTER | 2ND QUARTER | 3RD QUARTER | 4TH QUARTER | TOTAL GRAMS |
|---|---|---|---|---|---|
| 840 | 24,446.75 | 23,109.54 | 22,942.92 | 24,535.78 | **95,034.99** |
| 841 | 9,086.95 | 8,761.24 | 8,466.73 | 9,198.50 | **35,513.42** |
| 843 | 2,162.31 | 2,136.84 | 2,028.94 | 2,338.37 | **8,666.45** |
| 844 | 2,881.51 | 2,885.05 | 2,575.00 | 2,952.76 | **11,294.31** |
| 845 | 581.05 | 659.94 | 501.05 | 584.72 | **2,326.76** |
| 846 | 4,827.85 | 4,544.14 | 4,568.53 | 4,959.88 | **18,900.40** |
| 847 | 3,187.35 | 3,024.17 | 2,822.76 | 2,933.23 | **11,967.52** |
| **UT - UTAH - Total** | **47,173.78** | **45,120.91** | **43,905.93** | **47,503.23** | **183,703.86** |

**STATE: VA - VIRGINIA**

| REGISTRANT ZIP CODE 3 | 1ST QUARTER | 2ND QUARTER | 3RD QUARTER | 4TH QUARTER | TOTAL GRAMS |
|---|---|---|---|---|---|
| 201 | 12,238.57 | 12,124.01 | 11,499.94 | 11,953.88 | **47,816.39** |
| 220 | 9,083.47 | 9,387.98 | 8,958.31 | 8,564.46 | **35,994.22** |
| 221 | 5,550.86 | 5,656.51 | 5,934.67 | 5,379.41 | **22,521.46** |
| 222 | 3,063.01 | 2,741.86 | 2,690.51 | 2,724.23 | **11,219.60** |
| 223 | 3,452.92 | 3,424.49 | 2,900.68 | 3,430.24 | **13,208.33** |
| 224 | 4,715.09 | 4,359.31 | 4,195.49 | 4,624.71 | **17,894.61** |
| 225 | 2,389.73 | 2,348.90 | 2,073.65 | 2,278.61 | **9,090.88** |
| 226 | 2,931.38 | 2,797.48 | 2,897.15 | 3,143.94 | **11,769.95** |
| 227 | 926.30 | 947.08 | 841.56 | 914.61 | **3,629.54** |
| 228 | 1,910.31 | 1,976.90 | 1,845.34 | 1,949.91 | **7,682.46** |
| 229 | 6,153.40 | 5,780.26 | 5,885.54 | 6,027.77 | **23,846.97** |
| 230 | 3,685.49 | 3,508.95 | 3,366.37 | 3,703.62 | **14,264.42** |
| 231 | 8,284.98 | 7,919.97 | 7,194.43 | 8,328.14 | **31,727.52** |
| 232 | 9,692.67 | 9,449.04 | 8,471.19 | 8,672.96 | **36,285.86** |
| 233 | 5,961.90 | 5,936.98 | 4,791.06 | 5,514.89 | **22,204.83** |
| 234 | 12,375.21 | 12,102.12 | 10,942.13 | 11,700.85 | **47,120.30** |
| 235 | 3,338.12 | 3,105.64 | 2,990.73 | 3,207.18 | **12,641.67** |
| 236 | 7,684.87 | 7,098.30 | 7,013.40 | 6,991.66 | **28,788.24** |
| 237 | 3,365.06 | 2,975.12 | 3,079.02 | 1,971.93 | **11,391.13** |
| 238 | 4,849.98 | 4,885.39 | 3,994.97 | 4,850.73 | **18,581.07** |
| 239 | 1,274.71 | 1,141.30 | 1,191.02 | 1,300.93 | **4,907.96** |
| 240 | 5,467.42 | 5,301.84 | 5,220.96 | 5,827.86 | **21,818.08** |
| 241 | 3,630.25 | 3,143.29 | 3,280.48 | 3,370.05 | **13,424.07** |
| 242 | 2,315.14 | 2,154.06 | 2,083.53 | 2,122.67 | **8,675.40** |
| 243 | 1,888.60 | 1,786.47 | 1,725.44 | 1,927.64 | **7,328.15** |
| 244 | 2,516.66 | 2,374.64 | 2,310.95 | 2,488.46 | **9,690.71** |
| 245 | 6,149.00 | 5,715.88 | 5,507.01 | 5,727.48 | **23,099.36** |
| 246 | 1,054.84 | 941.00 | 899.21 | 1,037.52 | **3,932.56** |

| VA - VIRGINIA - Total | 135,949.94 | 131,084.77 | 123,784.72 | 129,736.33 | 520,555.76 |
|---|---|---|---|---|---|

**STATE: VI - VIRGIN ISLANDS**

| REGISTRANT ZIP CODE 3 | 1ST QUARTER | 2ND QUARTER | 3RD QUARTER | 4TH QUARTER | TOTAL GRAMS |
|---|---|---|---|---|---|
| 008 | 135.03 | 159.83 | 132.68 | 158.62 | **586.16** |
| VI - VIRGIN ISLANDS - Total | 135.03 | 159.83 | 132.68 | 158.62 | 586.16 |

**STATE: VT - VERMONT**

| REGISTRANT ZIP CODE 3 | 1ST QUARTER | 2ND QUARTER | 3RD QUARTER | 4TH QUARTER | TOTAL GRAMS |
|---|---|---|---|---|---|
| 050 | 1,550.94 | 1,472.29 | 1,342.75 | 1,357.62 | **5,723.61** |
| 051 | 816.19 | 750.60 | 797.12 | 857.48 | **3,221.40** |
| 052 | 1,329.16 | 1,297.38 | 1,241.39 | 1,250.51 | **5,118.45** |
| 053 | 1,168.04 | 1,277.56 | 1,013.72 | 1,195.24 | **4,654.56** |
| 054 | 6,470.35 | 6,435.62 | 6,067.10 | 6,572.34 | **25,545.41** |
| 056 | 3,356.29 | 3,267.41 | 3,315.36 | 3,227.77 | **13,166.83** |
| 057 | 2,407.07 | 2,399.96 | 2,194.78 | 2,508.53 | **9,510.34** |
| 058 | 2,015.42 | 1,988.68 | 1,992.50 | 1,988.19 | **7,984.80** |
| VT - VERMONT - Total | 19,113.46 | 18,889.50 | 17,964.74 | 18,957.69 | 74,925.40 |

**STATE: WA - WASHINGTON**

| REGISTRANT ZIP CODE 3 | 1ST QUARTER | 2ND QUARTER | 3RD QUARTER | 4TH QUARTER | TOTAL GRAMS |
|---|---|---|---|---|---|
| 980 | 24,600.46 | 24,657.67 | 22,386.53 | 24,524.72 | **96,169.38** |
| 981 | 13,358.06 | 13,737.71 | 12,761.44 | 13,816.14 | **53,673.35** |
| 982 | 15,846.53 | 15,543.26 | 14,641.06 | 15,590.95 | **61,621.80** |
| 983 | 12,827.83 | 12,713.81 | 11,927.93 | 12,283.13 | **49,752.70** |
| 984 | 7,945.60 | 7,582.80 | 6,871.99 | 7,366.60 | **29,766.99** |
| 985 | 7,460.74 | 7,666.04 | 7,000.78 | 7,516.81 | **29,644.38** |
| 986 | 9,195.02 | 9,170.53 | 8,369.46 | 9,053.79 | **35,788.80** |
| 988 | 2,559.50 | 2,650.76 | 2,441.32 | 2,607.81 | **10,259.37** |
| 989 | 3,548.72 | 3,421.65 | 3,081.33 | 3,501.78 | **13,553.48** |
| 990 | 1,916.52 | 1,797.36 | 1,718.91 | 1,837.75 | **7,270.54** |
| 991 | 1,304.97 | 1,227.71 | 1,189.06 | 1,283.82 | **5,005.57** |
| 992 | 8,253.55 | 8,309.89 | 7,595.21 | 8,475.73 | **32,634.39** |
| 993 | 5,211.72 | 5,304.56 | 4,853.13 | 5,003.03 | **20,372.44** |
| 994 | 585.39 | 513.96 | 465.08 | 550.00 | **2,114.43** |
| WA - WASHINGTON - Total | 114,614.62 | 114,297.71 | 105,303.23 | 113,412.05 | 447,627.62 |

**STATE: WI - WISCONSIN**

| REGISTRANT ZIP CODE 3 | 1ST QUARTER | 2ND QUARTER | 3RD QUARTER | 4TH QUARTER | TOTAL GRAMS |
|---|---|---|---|---|---|
| 530 | 12,950.08 | 11,941.30 | 11,687.54 | 12,617.38 | **49,196.30** |
| 531 | 11,737.50 | 10,884.26 | 10,503.24 | 11,217.36 | **44,342.36** |
| 532 | 12,365.30 | 11,006.60 | 10,152.96 | 11,626.26 | **45,151.13** |
| 534 | 2,296.92 | 2,333.84 | 2,167.49 | 2,117.47 | **8,915.71** |
| 535 | 10,749.41 | 9,930.34 | 9,685.65 | 10,084.97 | **40,450.37** |

| | | | | | |
|---|---|---|---|---|---|
| 537 | 6,386.68 | 6,084.31 | 6,013.76 | 6,341.21 | **24,825.96** |
| 538 | 1,053.93 | 857.35 | 852.32 | 988.88 | **3,752.48** |
| 539 | 3,726.60 | 3,395.62 | 3,259.13 | 3,301.08 | **13,682.43** |
| 540 | 2,552.15 | 2,414.66 | 2,217.89 | 2,332.60 | **9,517.29** |
| 541 | 4,092.14 | 3,866.50 | 3,809.61 | 3,959.99 | **15,728.23** |
| 542 | 2,303.57 | 2,069.12 | 2,079.46 | 2,007.95 | **8,460.10** |
| 543 | 3,762.34 | 3,712.03 | 3,603.76 | 3,778.28 | **14,856.41** |
| 544 | 7,801.44 | 7,029.62 | 6,717.78 | 7,377.53 | **28,926.36** |
| 545 | 1,757.10 | 1,579.62 | 1,541.40 | 1,518.30 | **6,396.43** |
| 546 | 5,677.44 | 5,234.05 | 4,902.32 | 5,297.49 | **21,111.29** |
| 547 | 5,981.53 | 5,258.52 | 4,781.85 | 5,540.11 | **21,562.01** |
| 548 | 3,146.76 | 2,713.24 | 2,705.69 | 2,728.27 | **11,293.96** |
| 549 | 10,718.34 | 10,164.77 | 9,684.38 | 10,273.20 | **40,840.70** |
| **WI - WISCONSIN - Total** | **109,059.24** | **100,475.74** | **96,366.22** | **103,108.32** | **409,009.53** |

**STATE: WV - WEST VIRGINIA**

| REGISTRANT ZIP CODE 3 | 1ST QUARTER | 2ND QUARTER | 3RD QUARTER | 4TH QUARTER | **TOTAL GRAMS** |
|---|---|---|---|---|---|
| 247 | 1,096.94 | 1,017.81 | 959.08 | 949.59 | **4,023.42** |
| 248 | 467.93 | 338.66 | 391.41 | 320.44 | **1,518.44** |
| 249 | 736.17 | 730.19 | 656.86 | 651.23 | **2,774.44** |
| 250 | 1,152.95 | 1,046.45 | 1,008.37 | 1,078.91 | **4,286.68** |
| 251 | 963.39 | 909.65 | 927.29 | 997.80 | **3,798.13** |
| 252 | 755.38 | 730.72 | 720.74 | 693.07 | **2,899.92** |
| 253 | 3,114.14 | 2,622.49 | 2,548.47 | 2,600.88 | **10,885.98** |
| 254 | 2,934.34 | 2,902.06 | 2,821.58 | 2,549.30 | **11,207.28** |
| 255 | 2,882.04 | 3,021.11 | 2,578.07 | 2,810.83 | **11,292.06** |
| 256 | 778.96 | 663.22 | 644.42 | 796.28 | **2,882.88** |
| 257 | 2,481.98 | 2,261.24 | 2,063.22 | 2,429.19 | **9,235.63** |
| 258 | 2,102.43 | 2,025.17 | 1,743.11 | 1,792.33 | **7,663.03** |
| 259 | 738.30 | 710.22 | 614.25 | 654.42 | **2,717.19** |
| 260 | 2,175.97 | 1,831.60 | 1,744.60 | 1,828.06 | **7,580.24** |
| 261 | 1,985.20 | 2,009.87 | 1,763.96 | 1,874.79 | **7,633.82** |
| 262 | 976.45 | 914.26 | 803.91 | 809.77 | **3,504.39** |
| 263 | 1,489.49 | 1,415.90 | 1,378.71 | 1,375.14 | **5,659.24** |
| 264 | 501.76 | 445.27 | 429.73 | 381.57 | **1,758.33** |
| 265 | 3,064.59 | 3,016.88 | 2,961.26 | 3,083.96 | **12,126.69** |
| 266 | 450.35 | 442.47 | 412.02 | 398.42 | **1,703.26** |
| 267 | 878.03 | 840.34 | 810.16 | 791.71 | **3,320.24** |
| 268 | 610.28 | 644.38 | 491.48 | 525.24 | **2,271.38** |
| **WV - WEST VIRGINIA - Total** | **32,337.10** | **30,539.97** | **28,472.70** | **29,392.91** | **120,742.68** |

**STATE: WY - WYOMING**

| REGISTRANT ZIP CODE 3 | 1ST QUARTER | 2ND QUARTER | 3RD QUARTER | 4TH QUARTER | TOTAL GRAMS |
|---|---|---|---|---|---|
| 820 | 2,211.89 | 2,198.81 | 1,922.74 | 2,203.58 | **8,537.02** |
| 822 | 235.33 | 238.52 | 227.51 | 215.92 | **917.28** |
| 823 | 98.66 | 117.30 | 104.89 | 114.66 | **435.52** |
| 824 | 681.97 | 725.51 | 683.87 | 743.07 | **2,834.43** |
| 825 | 551.24 | 398.13 | 357.91 | 452.95 | **1,760.23** |
| 826 | 1,469.74 | 1,510.41 | 1,337.17 | 1,574.95 | **5,892.28** |
| 827 | 673.59 | 616.41 | 661.46 | 654.87 | **2,606.32** |
| 828 | 558.93 | 557.93 | 526.10 | 581.10 | **2,224.06** |
| 829 | 794.34 | 721.85 | 715.42 | 802.21 | **3,033.82** |
| 830 | 331.96 | 306.28 | 319.97 | 353.41 | **1,311.61** |
| 831 | 209.69 | 155.17 | 171.14 | 199.13 | **735.13** |
| **WY - WYOMING - Total** | **7,817.34** | **7,546.33** | **7,028.17** | **7,895.84** | **30,287.69** |

**DRUG: 9050 - CODEINE**

**STATE: AK - ALASKA**

| REGISTRANT ZIP CODE 3 | 1ST QUARTER | 2ND QUARTER | 3RD QUARTER | 4TH QUARTER | TOTAL GRAMS |
|---|---|---|---|---|---|
| 995 | 2,585.42 | 2,292.93 | 2,508.92 | 2,004.49 | **9,391.76** |
| 996 | 1,291.52 | 1,354.13 | 1,187.58 | 997.96 | **4,831.19** |
| 997 | 1,683.63 | 1,758.01 | 1,759.75 | 1,542.77 | **6,744.15** |
| 998 | 439.06 | 466.31 | 407.71 | 380.68 | **1,693.76** |
| 999 | 246.61 | 221.19 | 231.13 | 209.96 | **908.88** |
| **AK - ALASKA - Total** | **6,246.24** | **6,092.57** | **6,095.09** | **5,135.85** | **23,569.75** |

**STATE: AL - ALABAMA**

| REGISTRANT ZIP CODE 3 | 1ST QUARTER | 2ND QUARTER | 3RD QUARTER | 4TH QUARTER | TOTAL GRAMS |
|---|---|---|---|---|---|
| 350 | 5,700.69 | 5,826.36 | 5,251.00 | 5,510.60 | **22,288.64** |
| 351 | 3,695.73 | 3,390.41 | 3,596.83 | 3,326.91 | **14,009.87** |
| 352 | 5,551.75 | 4,495.23 | 4,742.96 | 4,724.83 | **19,514.76** |
| 354 | 1,812.85 | 1,886.06 | 1,467.66 | 1,721.44 | **6,888.01** |
| 355 | 2,012.28 | 2,014.69 | 1,912.28 | 1,796.99 | **7,736.25** |
| 356 | 6,250.52 | 5,832.80 | 5,719.09 | 5,604.84 | **23,407.25** |
| 357 | 727.30 | 663.59 | 658.69 | 693.14 | **2,742.72** |
| 358 | 1,203.94 | 739.39 | 1,041.80 | 709.38 | **3,694.51** |
| 359 | 2,255.81 | 1,753.27 | 1,947.62 | 1,982.42 | **7,939.13** |
| 360 | 3,465.87 | 3,326.16 | 3,707.81 | 3,594.73 | **14,094.56** |
| 361 | 2,748.70 | 2,323.68 | 2,053.34 | 2,067.73 | **9,193.45** |
| 362 | 1,416.24 | 997.90 | 998.25 | 1,202.29 | **4,614.68** |
| 363 | 1,839.25 | 1,559.77 | 1,676.91 | 1,634.86 | **6,710.78** |
| 364 | 1,811.18 | 1,525.15 | 1,822.86 | 1,642.10 | **6,801.30** |
| 365 | 4,226.68 | 3,609.28 | 3,682.48 | 3,689.20 | **15,207.64** |
| 366 | 4,450.56 | 4,148.35 | 3,989.25 | 4,210.79 | **16,798.96** |

| | | | | | |
|---|---|---|---|---|---|
| 367 | 2,727.77 | 2,337.18 | 2,434.61 | 2,297.40 | **9,796.96** |
| 368 | 2,448.06 | 1,947.21 | 2,014.29 | 2,013.35 | **8,422.91** |
| 369 | 382.35 | 484.01 | 432.07 | 444.23 | **1,742.66** |
| **AL - ALABAMA - Total** | **54,727.52** | **48,860.49** | **49,149.79** | **48,867.23** | **201,605.04** |

**STATE: AR - ARKANSAS**

| REGISTRANT ZIP CODE 3 | 1ST QUARTER | 2ND QUARTER | 3RD QUARTER | 4TH QUARTER | TOTAL GRAMS |
|---|---|---|---|---|---|
| 716 | 3,477.58 | 3,335.02 | 3,427.47 | 3,040.97 | **13,281.04** |
| 717 | 3,025.98 | 3,236.68 | 3,096.88 | 2,830.38 | **12,189.93** |
| 718 | 5,421.05 | 5,430.78 | 5,266.63 | 4,826.94 | **20,945.41** |
| 719 | 4,832.49 | 3,997.88 | 4,383.36 | 4,175.61 | **17,389.33** |
| 720 | 6,102.15 | 5,518.85 | 5,018.02 | 4,618.84 | **21,257.86** |
| 721 | 6,583.06 | 6,114.23 | 5,963.15 | 5,092.14 | **23,752.58** |
| 722 | 3,063.80 | 2,804.81 | 2,551.81 | 2,346.92 | **10,767.34** |
| 723 | 7,524.30 | 6,244.64 | 5,916.44 | 5,632.25 | **25,317.62** |
| 724 | 8,642.39 | 7,366.71 | 7,480.98 | 6,882.04 | **30,372.12** |
| 725 | 5,026.29 | 4,570.29 | 3,769.13 | 3,934.15 | **17,299.86** |
| 726 | 1,924.05 | 2,075.99 | 2,219.10 | 2,059.06 | **8,278.19** |
| 727 | 4,352.27 | 3,443.47 | 3,580.65 | 3,210.08 | **14,586.48** |
| 728 | 2,808.37 | 2,615.84 | 2,546.04 | 2,126.30 | **10,096.54** |
| 729 | 5,103.49 | 4,029.83 | 3,763.24 | 3,316.96 | **16,213.52** |
| **AR - ARKANSAS - Total** | **67,887.27** | **60,785.00** | **58,982.89** | **54,092.64** | **241,747.80** |

**STATE: AS - AMERICAN SAMOA**

| REGISTRANT ZIP CODE 3 | 1ST QUARTER | 2ND QUARTER | 3RD QUARTER | 4TH QUARTER | TOTAL GRAMS |
|---|---|---|---|---|---|
| 967 | 190.07 | 353.62 | 181.23 | 335.94 | **1,060.85** |
| **AS - AMERICAN SAMOA - Tota** | **190.07** | **353.62** | **181.23** | **335.94** | **1,060.85** |

**STATE: AZ - ARIZONA**

| REGISTRANT ZIP CODE 3 | 1ST QUARTER | 2ND QUARTER | 3RD QUARTER | 4TH QUARTER | TOTAL GRAMS |
|---|---|---|---|---|---|
| 850 | 15,027.17 | 13,181.40 | 14,592.50 | 13,455.85 | **56,256.92** |
| 851 | 3,232.52 | 2,261.54 | 2,179.95 | 2,217.55 | **9,891.56** |
| 852 | 20,137.67 | 16,989.11 | 16,143.69 | 15,622.58 | **68,893.05** |
| 853 | 9,861.14 | 8,207.01 | 7,895.83 | 7,458.12 | **33,422.10** |
| 855 | 843.88 | 705.39 | 873.12 | 845.35 | **3,267.74** |
| 856 | 2,209.27 | 2,113.41 | 2,227.38 | 1,795.23 | **8,345.30** |
| 857 | 8,966.82 | 7,383.00 | 7,438.13 | 6,966.75 | **30,754.70** |
| 859 | 571.26 | 455.05 | 683.44 | 663.65 | **2,373.41** |
| 860 | 938.74 | 792.04 | 711.87 | 891.33 | **3,333.99** |
| 863 | 1,997.68 | 1,787.23 | 1,955.11 | 1,793.10 | **7,533.12** |
| 864 | 2,265.87 | 2,479.06 | 2,140.22 | 2,410.71 | **9,295.86** |
| 865 | 1,198.58 | 1,031.55 | 825.82 | 858.79 | **3,914.74** |
| **AZ - ARIZONA - Total** | **67,250.60** | **57,385.77** | **57,667.06** | **54,979.04** | **237,282.47** |

**STATE: CA - CALIFORNIA**

| REGISTRANT ZIP CODE 3 | 1ST QUARTER | 2ND QUARTER | 3RD QUARTER | 4TH QUARTER | TOTAL GRAMS |
|---|---|---|---|---|---|
| 900 | 25,090.51 | 24,081.59 | 23,062.21 | 21,104.13 | **93,338.45** |
| 902 | 22,476.14 | 21,331.42 | 21,442.64 | 20,253.01 | **85,503.21** |
| 903 | 2,644.61 | 2,963.74 | 2,890.81 | 2,731.68 | **11,230.84** |
| 904 | 1,051.15 | 1,114.61 | 1,019.75 | 1,068.01 | **4,253.53** |
| 905 | 2,189.59 | 1,863.35 | 1,846.44 | 1,796.16 | **7,695.54** |
| 906 | 6,766.50 | 6,280.30 | 6,126.49 | 5,835.17 | **25,008.46** |
| 907 | 8,637.61 | 8,633.09 | 8,085.85 | 7,196.74 | **32,553.29** |
| 908 | 6,499.15 | 6,537.96 | 6,187.35 | 5,594.93 | **24,819.38** |
| 910 | 2,731.77 | 2,871.53 | 2,811.65 | 2,632.70 | **11,047.65** |
| 911 | 2,092.52 | 2,113.68 | 2,106.49 | 1,746.54 | **8,059.22** |
| 912 | 2,986.27 | 2,657.75 | 2,827.30 | 2,313.05 | **10,784.38** |
| 913 | 12,708.79 | 12,834.50 | 12,555.66 | 11,038.10 | **49,137.04** |
| 914 | 4,766.04 | 4,482.47 | 4,135.32 | 3,702.56 | **17,086.39** |
| 915 | 1,220.75 | 1,426.54 | 1,441.18 | 1,294.01 | **5,382.49** |
| 916 | 1,245.50 | 1,200.59 | 1,249.88 | 1,217.21 | **4,913.17** |
| 917 | 20,716.23 | 20,128.68 | 19,593.71 | 18,339.33 | **78,777.95** |
| 918 | 871.49 | 835.64 | 693.59 | 718.65 | **3,119.37** |
| 919 | 4,225.10 | 3,615.55 | 3,504.41 | 3,164.42 | **14,509.48** |
| 920 | 19,828.71 | 14,453.49 | 15,061.02 | 14,598.28 | **63,941.50** |
| 921 | 10,512.77 | 10,122.01 | 9,897.64 | 8,829.65 | **39,362.07** |
| 922 | 10,002.73 | 8,784.95 | 8,556.14 | 8,722.73 | **36,066.55** |
| 923 | 17,930.75 | 16,566.55 | 16,897.65 | 15,157.73 | **66,552.68** |
| 924 | 2,981.19 | 2,768.15 | 2,686.38 | 2,483.05 | **10,918.76** |
| 925 | 17,602.26 | 16,175.45 | 16,075.56 | 14,977.88 | **64,831.14** |
| 926 | 10,828.65 | 10,753.08 | 10,827.43 | 10,029.70 | **42,438.86** |
| 927 | 3,365.46 | 3,398.20 | 3,275.82 | 3,145.89 | **13,185.37** |
| 928 | 11,972.05 | 11,267.61 | 11,220.46 | 10,687.97 | **45,148.08** |
| 930 | 6,951.94 | 6,153.10 | 6,004.67 | 6,071.48 | **25,181.18** |
| 931 | 1,561.12 | 1,608.31 | 1,465.91 | 1,322.59 | **5,957.92** |
| 932 | 9,743.73 | 9,006.04 | 8,983.34 | 8,672.87 | **36,405.98** |
| 933 | 7,211.61 | 6,539.75 | 6,588.17 | 6,322.40 | **26,661.93** |
| 934 | 5,451.45 | 5,360.18 | 5,095.43 | 5,144.96 | **21,052.02** |
| 935 | 9,704.12 | 9,070.42 | 9,368.63 | 8,292.96 | **36,436.13** |
| 936 | 7,374.04 | 7,154.16 | 6,997.71 | 6,894.58 | **28,420.49** |
| 937 | 9,678.04 | 9,157.62 | 9,536.31 | 8,514.13 | **36,886.10** |
| 939 | 3,388.66 | 3,805.10 | 3,192.76 | 2,971.51 | **13,358.03** |
| 940 | 6,170.80 | 5,615.97 | 5,368.39 | 4,828.27 | **21,983.43** |
| 941 | 8,135.91 | 7,542.99 | 7,542.08 | 7,225.16 | **30,446.14** |

| | | | | | |
|---|---|---|---|---|---|
| 943 | 578.69 | 603.41 | 573.41 | 612.89 | **2,368.40** |
| 944 | 855.71 | 1,062.36 | 971.99 | 868.04 | **3,758.09** |
| 945 | 28,287.15 | 26,161.83 | 25,770.19 | 24,313.79 | **104,532.96** |
| 946 | 3,925.00 | 3,526.09 | 3,704.69 | 3,405.77 | **14,561.56** |
| 947 | 926.27 | 934.82 | 925.64 | 830.13 | **3,616.87** |
| 948 | 1,395.58 | 1,402.44 | 1,227.96 | 1,060.84 | **5,086.82** |
| 949 | 3,570.87 | 3,248.59 | 3,326.61 | 3,046.45 | **13,192.51** |
| 950 | 5,864.07 | 5,346.86 | 5,217.06 | 5,330.74 | **21,758.73** |
| 951 | 6,436.80 | 5,720.15 | 5,644.61 | 5,023.88 | **22,825.45** |
| 952 | 10,633.09 | 9,379.97 | 9,415.83 | 8,558.65 | **37,987.53** |
| 953 | 15,895.69 | 14,999.74 | 15,648.59 | 14,290.53 | **60,834.55** |
| 954 | 5,097.24 | 4,436.34 | 4,886.85 | 4,522.28 | **18,942.72** |
| 955 | 2,948.57 | 2,892.18 | 2,756.40 | 2,556.04 | **11,153.19** |
| 956 | 13,995.25 | 13,125.89 | 12,988.59 | 11,959.15 | **52,068.88** |
| 957 | 2,230.55 | 2,256.14 | 2,249.95 | 1,976.00 | **8,712.63** |
| 958 | 7,315.15 | 6,938.19 | 7,083.41 | 5,850.99 | **27,187.74** |
| 959 | 7,131.71 | 7,193.71 | 6,635.71 | 5,880.55 | **26,841.68** |
| 960 | 3,282.59 | 3,156.52 | 3,206.75 | 3,221.57 | **12,867.43** |
| 961 | 1,197.58 | 1,061.95 | 1,134.89 | 1,011.12 | **4,405.54** |
| **CA - CALIFORNIA - Total** | **430,883.25** | **403,723.32** | **399,591.36** | **370,959.56** | **1,605,157.49** |

**STATE: CO - COLORADO**

| REGISTRANT ZIP CODE 3 | 1ST QUARTER | 2ND QUARTER | 3RD QUARTER | 4TH QUARTER | **TOTAL GRAMS** |
|---|---|---|---|---|---|
| 800 | 4,357.99 | 3,904.50 | 3,692.57 | 3,141.72 | **15,096.78** |
| 801 | 2,566.36 | 2,475.59 | 2,292.20 | 2,167.25 | **9,501.40** |
| 802 | 4,627.66 | 3,960.40 | 3,513.09 | 3,114.17 | **15,215.31** |
| 803 | 471.40 | 405.88 | 294.82 | 396.67 | **1,568.77** |
| 804 | 553.13 | 471.63 | 465.54 | 410.17 | **1,900.46** |
| 805 | 2,089.65 | 2,010.84 | 2,055.70 | 1,539.60 | **7,695.79** |
| 806 | 957.76 | 815.30 | 730.48 | 692.55 | **3,196.09** |
| 807 | 391.37 | 295.41 | 325.43 | 387.00 | **1,399.22** |
| 808 | 633.06 | 621.91 | 727.18 | 562.82 | **2,544.98** |
| 809 | 2,485.93 | 2,238.47 | 2,216.90 | 1,905.57 | **8,846.87** |
| 810 | 2,131.19 | 1,475.02 | 1,364.17 | 1,570.68 | **6,541.06** |
| 811 | 433.45 | 344.96 | 412.55 | 295.41 | **1,486.37** |
| 812 | 828.09 | 600.16 | 568.49 | 470.70 | **2,467.44** |
| 813 | 237.68 | 170.18 | 246.79 | 127.08 | **781.73** |
| 814 | 516.15 | 414.15 | 469.82 | 300.11 | **1,700.22** |
| 815 | 841.79 | 823.97 | 962.16 | 604.63 | **3,232.55** |
| 816 | 481.03 | 392.25 | 426.45 | 257.30 | **1,557.03** |
| **CO - COLORADO - Total** | **24,603.70** | **21,420.62** | **20,764.32** | **17,943.44** | **84,732.08** |

**STATE: CT - CONNECTICUT**

| REGISTRANT ZIP CODE 3 | 1ST QUARTER | 2ND QUARTER | 3RD QUARTER | 4TH QUARTER | TOTAL GRAMS |
|---|---|---|---|---|---|
| 060 | 4,059.07 | 4,188.15 | 3,878.55 | 3,597.63 | **15,723.39** |
| 061 | 1,230.94 | 1,242.44 | 1,284.10 | 1,255.75 | **5,013.24** |
| 062 | 690.00 | 661.54 | 625.26 | 597.82 | **2,574.62** |
| 063 | 1,599.79 | 1,389.37 | 1,684.77 | 1,350.22 | **6,024.16** |
| 064 | 4,428.58 | 4,820.02 | 3,869.33 | 4,755.31 | **17,873.24** |
| 065 | 1,320.46 | 1,391.99 | 1,421.41 | 1,209.63 | **5,343.48** |
| 066 | 1,327.25 | 1,292.92 | 1,212.78 | 1,269.59 | **5,102.55** |
| 067 | 2,419.51 | 2,303.59 | 2,300.62 | 2,035.09 | **9,058.82** |
| 068 | 2,708.00 | 2,506.92 | 2,260.35 | 2,168.28 | **9,643.55** |
| 069 | 569.32 | 586.96 | 485.94 | 480.96 | **2,123.18** |
| **CT - CONNECTICUT - Total** | **20,352.91** | **20,383.90** | **19,023.12** | **18,720.29** | **78,480.23** |

**STATE: DC - DISTRICT OF COLUMBIA**

| REGISTRANT ZIP CODE 3 | 1ST QUARTER | 2ND QUARTER | 3RD QUARTER | 4TH QUARTER | TOTAL GRAMS |
|---|---|---|---|---|---|
| 200 | 10,707.20 | 10,373.64 | 10,640.26 | 8,761.79 | **40,482.89** |
| 203 | 16.20 | 17.68 | 0 | 0 | **33.88** |
| 204 | 332.24 | 166.30 | 197.25 | 242.73 | **938.53** |
| 205 | 0 | 0 | 0 | 6.63 | **6.63** |
| **DC - DISTRICT OF COLUMBIA -** | **11,055.64** | **10,557.62** | **10,837.50** | **9,011.16** | **41,461.92** |

**STATE: DE - DELAWARE**

| REGISTRANT ZIP CODE 3 | 1ST QUARTER | 2ND QUARTER | 3RD QUARTER | 4TH QUARTER | TOTAL GRAMS |
|---|---|---|---|---|---|
| 197 | 3,230.74 | 3,328.06 | 2,750.04 | 3,037.03 | **12,345.87** |
| 198 | 2,042.51 | 1,780.24 | 2,055.90 | 1,833.72 | **7,712.38** |
| 199 | 3,143.81 | 3,062.59 | 2,832.91 | 2,851.54 | **11,890.85** |
| **DE - DELAWARE - Total** | **8,417.06** | **8,170.89** | **7,638.85** | **7,722.29** | **31,949.10** |

**STATE: FL - FLORIDA**

| REGISTRANT ZIP CODE 3 | 1ST QUARTER | 2ND QUARTER | 3RD QUARTER | 4TH QUARTER | TOTAL GRAMS |
|---|---|---|---|---|---|
| 320 | 5,312.53 | 4,389.92 | 4,833.35 | 4,738.60 | **19,274.39** |
| 321 | 5,942.87 | 5,313.54 | 4,831.68 | 4,967.47 | **21,055.56** |
| 322 | 6,618.68 | 6,375.43 | 6,112.75 | 5,307.84 | **24,414.71** |
| 323 | 3,690.41 | 3,340.58 | 4,220.32 | 3,132.17 | **14,383.47** |
| 324 | 3,337.33 | 3,145.68 | 4,855.98 | 4,438.26 | **15,777.25** |
| 325 | 3,965.33 | 3,307.71 | 3,699.53 | 3,184.44 | **14,157.01** |
| 326 | 2,417.77 | 2,136.02 | 2,124.77 | 2,295.35 | **8,973.92** |
| 327 | 7,601.92 | 6,452.72 | 6,178.22 | 6,051.37 | **26,284.24** |
| 328 | 6,256.30 | 5,102.93 | 4,448.94 | 4,086.69 | **19,894.86** |
| 329 | 5,088.44 | 4,763.52 | 4,268.33 | 4,219.38 | **18,339.67** |
| 330 | 13,677.99 | 12,127.16 | 10,347.48 | 9,270.95 | **45,423.57** |
| 331 | 15,436.37 | 15,200.31 | 12,136.06 | 10,909.04 | **53,681.79** |

| | | | | | |
|---|---|---|---|---|---|
| 333 | 7,278.16 | 6,349.42 | 5,401.55 | 5,028.76 | **24,057.88** |
| 334 | 12,547.28 | 10,946.38 | 9,173.88 | 9,570.54 | **42,238.07** |
| 335 | 8,299.89 | 7,304.70 | 6,543.98 | 6,711.56 | **28,860.12** |
| 336 | 8,104.50 | 7,883.25 | 7,182.87 | 7,567.99 | **30,738.61** |
| 337 | 8,975.74 | 8,040.53 | 7,670.80 | 7,468.95 | **32,156.02** |
| 338 | 8,236.50 | 7,076.26 | 6,616.49 | 7,263.00 | **29,192.26** |
| 339 | 8,387.34 | 7,135.52 | 5,851.87 | 6,421.42 | **27,796.16** |
| 341 | 2,805.63 | 1,760.50 | 1,479.01 | 1,764.83 | **7,809.97** |
| 342 | 6,253.72 | 5,337.87 | 3,754.79 | 4,246.50 | **19,592.88** |
| 344 | 5,491.46 | 5,152.99 | 5,718.12 | 5,253.21 | **21,615.79** |
| 346 | 6,904.53 | 6,332.38 | 6,616.15 | 6,518.65 | **26,371.71** |
| 347 | 5,012.28 | 4,297.71 | 3,847.58 | 3,632.24 | **16,789.81** |
| 349 | 3,205.19 | 3,038.40 | 2,738.61 | 2,635.55 | **11,617.74** |
| **FL - FLORIDA - Total** | **170,848.17** | **152,311.43** | **140,653.09** | **136,684.76** | **600,497.44** |

**STATE: GA - GEORGIA**

| REGISTRANT ZIP CODE 3 | 1ST QUARTER | 2ND QUARTER | 3RD QUARTER | 4TH QUARTER | TOTAL GRAMS |
|---|---|---|---|---|---|
| 300 | 15,265.91 | 14,044.13 | 12,446.06 | 11,677.24 | **53,433.34** |
| 301 | 9,817.43 | 8,765.96 | 8,191.38 | 7,449.08 | **34,223.84** |
| 302 | 11,966.18 | 11,026.16 | 10,139.41 | 8,964.00 | **42,095.75** |
| 303 | 7,478.32 | 6,546.02 | 6,333.99 | 5,561.67 | **25,920.01** |
| 304 | 3,240.94 | 3,161.10 | 2,917.90 | 2,983.00 | **12,302.94** |
| 305 | 7,102.10 | 5,853.06 | 6,218.47 | 5,490.90 | **24,664.52** |
| 306 | 6,235.91 | 5,579.31 | 5,587.31 | 5,394.03 | **22,796.57** |
| 307 | 3,268.74 | 2,472.71 | 2,556.53 | 2,559.48 | **10,857.46** |
| 308 | 2,467.66 | 2,246.55 | 2,339.85 | 2,320.32 | **9,374.37** |
| 309 | 3,392.21 | 3,342.77 | 3,358.81 | 2,971.69 | **13,065.48** |
| 310 | 6,421.65 | 5,491.94 | 5,347.04 | 5,232.92 | **22,493.55** |
| 312 | 1,833.28 | 1,701.63 | 1,684.36 | 1,849.85 | **7,069.12** |
| 313 | 1,779.03 | 1,511.48 | 1,370.26 | 1,443.20 | **6,103.97** |
| 314 | 1,773.82 | 1,363.27 | 1,243.90 | 1,266.17 | **5,647.16** |
| 315 | 6,285.97 | 5,512.26 | 5,614.56 | 5,352.87 | **22,765.67** |
| 316 | 2,493.71 | 2,488.83 | 2,445.10 | 2,059.36 | **9,487.00** |
| 317 | 6,509.94 | 5,682.63 | 5,515.16 | 5,321.03 | **23,028.76** |
| 318 | 434.20 | 423.33 | 545.24 | 455.28 | **1,858.05** |
| 319 | 2,900.10 | 2,588.79 | 2,631.51 | 2,347.23 | **10,467.62** |
| 398 | 2,214.52 | 2,207.89 | 1,898.48 | 1,952.62 | **8,273.51** |
| **GA - GEORGIA - Total** | **102,881.63** | **92,009.82** | **88,385.31** | **82,651.93** | **365,928.70** |

**STATE: GU - GUAM**

| REGISTRANT ZIP CODE 3 | 1ST QUARTER | 2ND QUARTER | 3RD QUARTER | 4TH QUARTER | TOTAL GRAMS |
|---|---|---|---|---|---|
| 969 | 930.91 | 1,022.54 | 961.07 | 684.38 | **3,598.89** |

| GU - GUAM - Total | 930.91 | 1,022.54 | 961.07 | 684.38 | 3,598.89 |
|---|---|---|---|---|---|

STATE: HI - HAWAII

| REGISTRANT ZIP CODE 3 | 1ST QUARTER | 2ND QUARTER | 3RD QUARTER | 4TH QUARTER | TOTAL GRAMS |
|---|---|---|---|---|---|
| 967 | 8,237.04 | 7,936.98 | 7,750.44 | 7,111.15 | 31,035.61 |
| 968 | 4,085.02 | 4,070.07 | 3,754.56 | 3,479.79 | 15,389.45 |
| HI - HAWAII - Total | 12,322.06 | 12,007.05 | 11,505.00 | 10,590.95 | 46,425.06 |

STATE: IA - IOWA

| REGISTRANT ZIP CODE 3 | 1ST QUARTER | 2ND QUARTER | 3RD QUARTER | 4TH QUARTER | TOTAL GRAMS |
|---|---|---|---|---|---|
| 500 | 1,155.74 | 969.87 | 1,285.41 | 1,042.05 | 4,453.07 |
| 501 | 1,015.10 | 887.68 | 1,322.04 | 847.65 | 4,072.48 |
| 502 | 899.96 | 888.35 | 963.36 | 840.50 | 3,592.16 |
| 503 | 1,913.10 | 1,775.97 | 1,929.89 | 1,464.18 | 7,083.13 |
| 504 | 472.40 | 412.18 | 313.83 | 364.67 | 1,563.09 |
| 505 | 767.08 | 503.99 | 759.48 | 645.90 | 2,676.45 |
| 506 | 805.39 | 726.47 | 960.29 | 765.21 | 3,257.36 |
| 507 | 527.82 | 464.49 | 604.27 | 416.79 | 2,013.36 |
| 508 | 209.96 | 145.87 | 201.48 | 214.38 | 771.69 |
| 510 | 473.91 | 437.03 | 445.87 | 372.94 | 1,729.74 |
| 511 | 708.87 | 608.69 | 565.59 | 631.99 | 2,515.14 |
| 512 | 118.24 | 129.29 | 121.56 | 54.15 | 423.23 |
| 513 | 362.64 | 290.25 | 398.88 | 332.59 | 1,384.36 |
| 514 | 332.79 | 243.09 | 211.25 | 294.48 | 1,081.60 |
| 515 | 818.46 | 808.42 | 802.34 | 723.60 | 3,152.82 |
| 516 | 232.78 | 193.85 | 337.07 | 240.88 | 1,004.57 |
| 520 | 1,201.64 | 959.33 | 955.19 | 948.28 | 4,064.45 |
| 521 | 300.76 | 280.68 | 184.50 | 233.17 | 999.11 |
| 522 | 1,101.00 | 1,073.03 | 1,086.58 | 841.18 | 4,101.79 |
| 523 | 675.07 | 519.09 | 567.85 | 554.07 | 2,316.08 |
| 524 | 718.83 | 678.12 | 834.12 | 687.89 | 2,918.96 |
| 525 | 925.37 | 861.37 | 1,225.68 | 777.19 | 3,789.61 |
| 526 | 785.02 | 792.57 | 895.24 | 692.42 | 3,165.24 |
| 527 | 1,371.81 | 1,119.04 | 1,563.82 | 1,201.72 | 5,256.39 |
| 528 | 885.16 | 856.20 | 836.41 | 877.76 | 3,455.53 |
| IA - IOWA - Total | 18,778.90 | 16,624.91 | 19,371.99 | 16,065.61 | 70,841.41 |

STATE: ID - IDAHO

| REGISTRANT ZIP CODE 3 | 1ST QUARTER | 2ND QUARTER | 3RD QUARTER | 4TH QUARTER | TOTAL GRAMS |
|---|---|---|---|---|---|
| 832 | 1,482.50 | 1,449.27 | 1,476.30 | 1,342.99 | 5,751.05 |
| 833 | 1,258.96 | 1,020.69 | 1,258.60 | 1,115.53 | 4,653.78 |
| 834 | 1,531.77 | 1,347.49 | 1,433.95 | 1,220.13 | 5,533.34 |
| 835 | 1,041.30 | 1,001.78 | 956.36 | 1,083.09 | 4,082.53 |

| | | | | | |
|---|---|---|---|---|---|
| 836 | 3,521.33 | 2,468.87 | 3,469.78 | 3,678.23 | **13,138.21** |
| 837 | 1,705.81 | 1,633.88 | 1,614.84 | 1,598.70 | **6,553.23** |
| 838 | 2,561.82 | 2,066.19 | 2,196.46 | 2,157.24 | **8,981.71** |
| **ID - IDAHO - Total** | **13,103.49** | **10,988.15** | **12,406.29** | **12,195.92** | **48,693.85** |

**STATE: IL - ILLINOIS**

| REGISTRANT ZIP CODE 3 | 1ST QUARTER | 2ND QUARTER | 3RD QUARTER | 4TH QUARTER | TOTAL GRAMS |
|---|---|---|---|---|---|
| 600 | 19,173.27 | 19,474.48 | 18,769.93 | 17,874.12 | **75,291.80** |
| 601 | 13,642.64 | 13,170.11 | 13,156.14 | 13,510.85 | **53,479.73** |
| 602 | 420.01 | 521.77 | 477.75 | 453.33 | **1,872.85** |
| 603 | 1,115.62 | 958.81 | 917.07 | 1,125.67 | **4,117.17** |
| 604 | 18,317.26 | 17,481.77 | 17,272.16 | 16,318.04 | **69,389.24** |
| 605 | 9,185.07 | 9,044.07 | 8,963.42 | 8,936.80 | **36,129.36** |
| 606 | 28,035.13 | 27,350.90 | 26,889.75 | 25,111.88 | **107,387.66** |
| 607 | 1,527.52 | 1,321.24 | 1,410.01 | 1,480.81 | **5,739.58** |
| 608 | 1,967.79 | 1,844.33 | 1,541.91 | 1,701.96 | **7,055.98** |
| 609 | 2,073.00 | 1,998.81 | 1,925.58 | 1,798.15 | **7,795.55** |
| 610 | 3,107.80 | 2,846.97 | 3,008.31 | 3,069.63 | **12,032.72** |
| 611 | 4,139.27 | 3,915.89 | 4,092.67 | 3,819.05 | **15,966.87** |
| 612 | 1,673.92 | 1,732.51 | 1,799.35 | 1,765.56 | **6,971.34** |
| 613 | 1,668.99 | 1,505.34 | 1,554.43 | 1,713.18 | **6,441.94** |
| 614 | 1,889.89 | 1,611.24 | 1,978.04 | 1,608.28 | **7,087.45** |
| 615 | 2,128.86 | 2,068.64 | 2,202.53 | 2,044.51 | **8,444.55** |
| 616 | 2,818.44 | 2,611.92 | 2,760.59 | 2,295.24 | **10,486.20** |
| 617 | 3,228.19 | 2,830.80 | 2,894.37 | 2,850.65 | **11,804.02** |
| 618 | 3,721.20 | 3,395.51 | 3,745.42 | 3,502.61 | **14,364.74** |
| 619 | 1,345.02 | 1,115.35 | 1,121.78 | 1,201.54 | **4,783.69** |
| 620 | 4,346.97 | 4,269.62 | 5,080.62 | 4,399.00 | **18,096.20** |
| 622 | 5,621.59 | 5,759.77 | 5,815.55 | 5,436.66 | **22,633.57** |
| 623 | 1,390.59 | 1,233.03 | 1,349.49 | 1,304.58 | **5,277.69** |
| 624 | 1,410.04 | 1,086.36 | 1,341.89 | 1,239.13 | **5,077.42** |
| 625 | 2,321.45 | 2,281.55 | 2,312.56 | 2,223.60 | **9,139.16** |
| 626 | 1,687.71 | 1,756.93 | 2,009.06 | 1,906.50 | **7,360.20** |
| 627 | 2,863.83 | 2,632.97 | 2,854.36 | 2,836.58 | **11,187.74** |
| 628 | 2,459.82 | 2,047.27 | 1,994.95 | 2,204.93 | **8,706.97** |
| 629 | 4,113.00 | 3,523.21 | 4,004.57 | 3,593.32 | **15,234.11** |
| **IL - ILLINOIS - Total** | **147,393.89** | **141,391.15** | **143,244.27** | **137,326.15** | **569,355.47** |

**STATE: IN - INDIANA**

| REGISTRANT ZIP CODE 3 | 1ST QUARTER | 2ND QUARTER | 3RD QUARTER | 4TH QUARTER | TOTAL GRAMS |
|---|---|---|---|---|---|
| 460 | 2,396.41 | 2,418.18 | 2,295.25 | 1,836.31 | **8,946.15** |
| 461 | 3,309.67 | 3,094.84 | 3,239.32 | 2,945.08 | **12,588.91** |

| | | | | | |
|---|---|---|---|---|---|
| 462 | 6,300.29 | 5,974.32 | 5,216.95 | 5,401.69 | **22,893.25** |
| 463 | 7,837.31 | 8,375.11 | 8,129.61 | 7,907.48 | **32,249.50** |
| 464 | 2,028.07 | 1,840.01 | 2,003.46 | 1,695.15 | **7,566.68** |
| 465 | 2,495.90 | 2,333.91 | 2,237.73 | 2,053.92 | **9,121.47** |
| 466 | 1,072.18 | 889.34 | 909.85 | 923.07 | **3,794.44** |
| 467 | 1,486.69 | 1,473.17 | 1,607.58 | 1,375.63 | **5,943.07** |
| 468 | 1,911.43 | 1,891.59 | 1,706.70 | 1,520.68 | **7,030.40** |
| 469 | 1,886.21 | 1,670.65 | 1,723.65 | 1,541.98 | **6,822.49** |
| 470 | 559.60 | 674.16 | 630.77 | 593.02 | **2,457.55** |
| 471 | 4,963.80 | 3,342.48 | 1,652.86 | 3,900.24 | **13,859.38** |
| 472 | 1,180.70 | 991.52 | 1,047.08 | 1,110.41 | **4,329.71** |
| 473 | 1,765.51 | 1,487.49 | 1,558.97 | 1,224.67 | **6,036.64** |
| 474 | 1,473.65 | 1,175.41 | 1,376.04 | 995.79 | **5,020.88** |
| 475 | 1,141.46 | 803.53 | 823.91 | 678.72 | **3,447.62** |
| 476 | 895.40 | 805.04 | 663.19 | 811.47 | **3,175.10** |
| 477 | 1,783.11 | 1,522.01 | 1,354.58 | 1,354.87 | **6,014.57** |
| 478 | 2,039.72 | 1,729.94 | 1,610.02 | 1,518.33 | **6,898.00** |
| 479 | 1,823.07 | 1,676.86 | 1,555.36 | 1,453.34 | **6,508.62** |
| **IN - INDIANA - Total** | **48,350.18** | **44,169.55** | **41,342.85** | **40,841.85** | **174,704.43** |

**STATE: KS - KANSAS**

| REGISTRANT ZIP CODE 3 | 1ST QUARTER | 2ND QUARTER | 3RD QUARTER | 4TH QUARTER | **TOTAL GRAMS** |
|---|---|---|---|---|---|
| 660 | 2,961.34 | 2,495.05 | 2,937.60 | 2,305.44 | **10,699.43** |
| 661 | 1,045.23 | 883.14 | 932.57 | 858.62 | **3,719.57** |
| 662 | 6,283.83 | 5,188.51 | 7,322.90 | 5,843.32 | **24,638.56** |
| 664 | 670.65 | 532.75 | 511.78 | 353.04 | **2,068.23** |
| 665 | 868.00 | 817.15 | 639.57 | 739.60 | **3,064.33** |
| 666 | 1,079.58 | 990.99 | 881.89 | 716.67 | **3,669.14** |
| 667 | 1,365.83 | 1,066.45 | 915.19 | 950.49 | **4,297.96** |
| 668 | 306.66 | 311.13 | 291.97 | 295.53 | **1,205.30** |
| 669 | 309.41 | 267.60 | 252.13 | 202.59 | **1,031.74** |
| 670 | 1,742.35 | 1,733.93 | 1,657.90 | 1,453.05 | **6,587.23** |
| 671 | 1,043.66 | 904.37 | 884.09 | 917.27 | **3,749.40** |
| 672 | 3,426.29 | 3,583.82 | 3,064.08 | 2,722.46 | **12,796.66** |
| 673 | 789.55 | 598.56 | 836.52 | 781.82 | **3,006.45** |
| 674 | 1,314.35 | 1,289.56 | 1,077.40 | 1,046.86 | **4,728.16** |
| 675 | 673.79 | 719.93 | 725.12 | 619.56 | **2,738.41** |
| 676 | 386.92 | 282.69 | 246.96 | 283.23 | **1,199.80** |
| 677 | 243.11 | 143.66 | 256.46 | 209.96 | **853.19** |
| 678 | 969.26 | 750.84 | 690.83 | 551.59 | **2,962.51** |
| 679 | 996.85 | 612.20 | 636.51 | 448.65 | **2,694.20** |

| KS - KANSAS - Total | 26,476.68 | 23,172.33 | 24,761.49 | 21,299.76 | 95,710.26 |
|---|---|---|---|---|---|

**STATE: KY - KENTUCKY**

| REGISTRANT ZIP CODE 3 | 1ST QUARTER | 2ND QUARTER | 3RD QUARTER | 4TH QUARTER | TOTAL GRAMS |
|---|---|---|---|---|---|
| 400 | 1,866.41 | 1,833.45 | 1,581.12 | 1,523.65 | **6,804.63** |
| 401 | 1,112.59 | 1,070.44 | 854.18 | 580.69 | **3,617.90** |
| 402 | 6,074.67 | 5,437.20 | 5,447.30 | 4,644.64 | **21,603.81** |
| 403 | 2,473.30 | 2,461.05 | 1,985.95 | 1,693.91 | **8,614.21** |
| 404 | 1,082.83 | 1,132.34 | 1,019.92 | 949.78 | **4,184.88** |
| 405 | 1,409.12 | 1,029.11 | 1,099.04 | 1,025.06 | **4,562.33** |
| 406 | 165.76 | 134.82 | 179.02 | 88.40 | **568.00** |
| 407 | 1,925.36 | 1,582.43 | 1,604.34 | 1,396.22 | **6,508.36** |
| 408 | 811.29 | 762.48 | 866.36 | 824.37 | **3,264.50** |
| 409 | 2,370.33 | 2,004.56 | 1,966.99 | 1,948.20 | **8,290.09** |
| 410 | 3,752.13 | 3,426.00 | 3,044.92 | 2,909.74 | **13,132.79** |
| 411 | 1,249.89 | 1,168.72 | 997.38 | 1,177.76 | **4,593.75** |
| 412 | 1,494.03 | 1,690.73 | 1,723.88 | 1,686.31 | **6,594.94** |
| 413 | 1,389.05 | 1,276.31 | 1,436.57 | 1,273.80 | **5,375.73** |
| 414 | 391.47 | 419.92 | 382.35 | 348.38 | **1,542.11** |
| 415 | 2,361.48 | 2,108.44 | 2,423.37 | 2,404.59 | **9,297.88** |
| 416 | 1,295.12 | 1,015.32 | 923.82 | 968.02 | **4,202.28** |
| 417 | 544.74 | 623.61 | 692.46 | 610.87 | **2,471.69** |
| 418 | 670.84 | 501.69 | 680.71 | 603.36 | **2,456.60** |
| 420 | 2,075.47 | 1,985.85 | 1,738.32 | 1,698.16 | **7,497.80** |
| 421 | 2,309.70 | 1,950.75 | 1,982.59 | 2,061.26 | **8,304.30** |
| 422 | 1,600.09 | 1,322.74 | 1,402.30 | 1,333.80 | **5,658.92** |
| 423 | 2,081.95 | 1,699.43 | 1,620.32 | 1,577.10 | **6,978.80** |
| 424 | 1,573.54 | 1,281.35 | 1,276.10 | 1,309.51 | **5,440.50** |
| 425 | 531.71 | 815.53 | 665.24 | 396.71 | **2,409.19** |
| 426 | 1,129.91 | 1,096.21 | 983.49 | 1,025.49 | **4,235.10** |
| 427 | 2,772.03 | 2,518.67 | 2,455.12 | 2,223.26 | **9,969.08** |
| **KY - KENTUCKY - Total** | **46,514.79** | **42,349.15** | **41,033.17** | **38,283.05** | **168,180.16** |

**STATE: LA - LOUISIANA**

| REGISTRANT ZIP CODE 3 | 1ST QUARTER | 2ND QUARTER | 3RD QUARTER | 4TH QUARTER | TOTAL GRAMS |
|---|---|---|---|---|---|
| 700 | 4,685.40 | 4,212.54 | 4,426.94 | 4,225.87 | **17,550.75** |
| 701 | 2,275.26 | 1,995.24 | 2,260.53 | 1,903.44 | **8,434.47** |
| 703 | 1,798.96 | 1,806.94 | 1,516.31 | 1,619.33 | **6,741.54** |
| 704 | 3,232.79 | 3,098.64 | 3,109.90 | 2,727.03 | **12,168.35** |
| 705 | 8,003.29 | 8,634.84 | 8,831.25 | 8,401.20 | **33,870.57** |
| 706 | 3,327.80 | 2,965.87 | 2,935.71 | 3,145.56 | **12,374.93** |
| 707 | 2,070.51 | 1,507.43 | 1,611.45 | 1,709.51 | **6,898.90** |

| | | | | | |
|---|---|---|---|---|---|
| 708 | 2,024.69 | 1,751.38 | 1,365.45 | 1,380.69 | **6,522.21** |
| 710 | 1,283.20 | 1,164.97 | 1,182.27 | 1,245.89 | **4,876.32** |
| 711 | 2,747.03 | 2,852.30 | 2,466.33 | 2,557.79 | **10,623.45** |
| 712 | 4,385.33 | 3,571.83 | 3,334.95 | 3,203.81 | **14,495.93** |
| 713 | 2,940.63 | 2,747.13 | 2,855.41 | 2,825.21 | **11,368.38** |
| 714 | 1,803.24 | 1,702.26 | 1,843.77 | 1,594.03 | **6,943.30** |
| **LA - LOUISIANA - Total** | **40,578.13** | **38,011.36** | **37,740.26** | **36,539.36** | **152,869.10** |

**STATE: MA - MASSACHUSETTS**

| REGISTRANT ZIP CODE 3 | 1ST QUARTER | 2ND QUARTER | 3RD QUARTER | 4TH QUARTER | TOTAL GRAMS |
|---|---|---|---|---|---|
| 010 | 2,658.59 | 2,586.53 | 2,513.43 | 2,513.97 | **10,272.52** |
| 011 | 1,011.67 | 1,036.72 | 946.87 | 1,009.67 | **4,004.93** |
| 012 | 475.75 | 507.30 | 480.48 | 537.47 | **2,000.99** |
| 013 | 425.55 | 521.48 | 418.81 | 402.17 | **1,768.02** |
| 014 | 1,491.99 | 1,853.11 | 1,494.55 | 1,569.28 | **6,408.92** |
| 015 | 2,515.82 | 2,890.01 | 2,154.17 | 2,752.57 | **10,312.57** |
| 016 | 2,007.13 | 1,773.36 | 1,826.76 | 1,790.74 | **7,397.99** |
| 017 | 2,067.74 | 2,144.00 | 1,931.90 | 2,037.65 | **8,181.29** |
| 018 | 7,678.26 | 7,476.92 | 6,791.33 | 7,264.20 | **29,210.71** |
| 019 | 2,822.76 | 2,479.65 | 2,716.42 | 2,689.93 | **10,708.76** |
| 020 | 2,019.98 | 2,174.08 | 1,998.84 | 2,148.29 | **8,341.19** |
| 021 | 6,699.87 | 7,100.02 | 5,852.69 | 6,140.70 | **25,793.27** |
| 022 | 121.06 | 237.43 | 184.22 | 210.85 | **753.56** |
| 023 | 2,553.67 | 2,576.72 | 2,210.12 | 2,553.65 | **9,894.16** |
| 024 | 1,290.51 | 1,570.38 | 1,449.45 | 1,437.58 | **5,747.93** |
| 025 | 824.15 | 678.07 | 654.22 | 790.63 | **2,947.07** |
| 026 | 847.80 | 1,217.39 | 1,138.14 | 982.98 | **4,186.31** |
| 027 | 3,667.73 | 3,330.00 | 3,041.19 | 2,999.02 | **13,037.94** |
| **MA - MASSACHUSETTS - Total** | **41,180.01** | **42,153.20** | **37,803.57** | **39,831.35** | **160,968.12** |

**STATE: MD - MARYLAND**

| REGISTRANT ZIP CODE 3 | 1ST QUARTER | 2ND QUARTER | 3RD QUARTER | 4TH QUARTER | TOTAL GRAMS |
|---|---|---|---|---|---|
| 206 | 3,966.48 | 3,651.31 | 3,629.25 | 3,651.17 | **14,898.21** |
| 207 | 11,954.27 | 13,227.07 | 11,333.66 | 11,209.26 | **47,724.27** |
| 208 | 5,219.05 | 5,728.47 | 5,492.48 | 4,855.52 | **21,295.52** |
| 209 | 2,383.98 | 2,059.75 | 1,736.22 | 2,018.70 | **8,198.65** |
| 210 | 5,397.47 | 5,088.66 | 4,764.92 | 5,054.26 | **20,305.31** |
| 211 | 2,537.39 | 2,896.26 | 2,640.40 | 2,670.13 | **10,744.18** |
| 212 | 9,953.23 | 9,276.49 | 8,069.07 | 8,314.21 | **35,613.00** |
| 214 | 1,095.66 | 1,043.03 | 1,110.89 | 1,027.46 | **4,277.04** |
| 215 | 999.89 | 754.55 | 770.19 | 731.73 | **3,256.36** |
| 216 | 1,491.02 | 1,310.99 | 1,244.81 | 1,427.83 | **5,474.66** |

| | | | | | |
|---|---|---|---|---|---|
| 217 | 4,307.99 | 4,417.91 | 3,637.37 | 3,908.69 | **16,271.96** |
| 218 | 1,243.55 | 1,307.38 | 1,105.44 | 1,027.03 | **4,683.40** |
| 219 | 921.21 | 967.53 | 715.29 | 750.11 | **3,354.14** |
| **MD - MARYLAND - Total** | **51,471.18** | **51,729.40** | **46,250.02** | **46,646.09** | **196,096.69** |

**STATE: ME - MAINE**

| REGISTRANT ZIP CODE 3 | 1ST QUARTER | 2ND QUARTER | 3RD QUARTER | 4TH QUARTER | TOTAL GRAMS |
|---|---|---|---|---|---|
| 039 | 40.33 | 117.14 | 80.85 | 112.72 | **351.03** |
| 040 | 1,988.72 | 1,591.52 | 1,784.57 | 1,726.05 | **7,090.86** |
| 041 | 536.11 | 393.92 | 467.41 | 410.89 | **1,808.33** |
| 042 | 1,558.44 | 1,510.84 | 1,471.24 | 1,350.48 | **5,891.01** |
| 043 | 758.35 | 755.59 | 832.28 | 739.71 | **3,085.93** |
| 044 | 1,483.20 | 1,142.31 | 1,136.51 | 1,060.93 | **4,822.95** |
| 045 | 247.15 | 289.15 | 212.90 | 174.02 | **923.22** |
| 046 | 664.20 | 618.44 | 713.10 | 746.81 | **2,742.54** |
| 047 | 1,007.22 | 898.84 | 933.00 | 800.23 | **3,639.29** |
| 048 | 220.64 | 181.89 | 252.39 | 185.03 | **839.96** |
| 049 | 1,180.99 | 1,022.30 | 1,109.33 | 924.51 | **4,237.13** |
| **ME - MAINE - Total** | **9,685.35** | **8,521.93** | **8,993.59** | **8,231.37** | **35,432.24** |

**STATE: MI - MICHIGAN**

| REGISTRANT ZIP CODE 3 | 1ST QUARTER | 2ND QUARTER | 3RD QUARTER | 4TH QUARTER | TOTAL GRAMS |
|---|---|---|---|---|---|
| 480 | 32,228.80 | 28,308.45 | 25,906.25 | 24,399.53 | **110,843.03** |
| 481 | 46,594.18 | 43,852.83 | 40,822.80 | 38,201.74 | **169,471.55** |
| 482 | 43,687.24 | 41,541.22 | 36,993.20 | 33,771.41 | **155,993.07** |
| 483 | 16,321.85 | 15,473.85 | 12,939.72 | 13,195.76 | **57,931.19** |
| 484 | 9,206.33 | 7,704.65 | 7,204.84 | 6,871.39 | **30,987.20** |
| 485 | 8,166.03 | 6,270.13 | 5,710.60 | 6,160.45 | **26,307.21** |
| 486 | 11,708.03 | 10,424.33 | 10,047.30 | 9,972.30 | **42,151.96** |
| 487 | 6,230.23 | 5,797.61 | 5,509.48 | 5,374.02 | **22,911.34** |
| 488 | 9,970.10 | 9,920.23 | 9,062.68 | 8,811.33 | **37,764.34** |
| 489 | 3,121.35 | 2,938.65 | 2,822.99 | 2,537.43 | **11,420.43** |
| 490 | 10,730.66 | 9,411.11 | 8,509.48 | 7,913.63 | **36,564.87** |
| 491 | 856.58 | 714.59 | 681.35 | 801.53 | **3,054.05** |
| 492 | 5,197.55 | 4,340.39 | 3,753.96 | 3,102.23 | **16,394.13** |
| 493 | 2,377.33 | 2,279.89 | 1,842.39 | 2,140.11 | **8,639.71** |
| 494 | 7,790.92 | 6,927.56 | 6,214.21 | 5,773.50 | **26,706.20** |
| 495 | 4,112.87 | 3,453.50 | 3,433.97 | 3,284.88 | **14,285.23** |
| 496 | 3,134.55 | 2,856.21 | 2,766.73 | 2,525.37 | **11,282.86** |
| 497 | 3,860.85 | 3,298.60 | 3,372.64 | 3,089.78 | **13,621.88** |
| 498 | 2,614.58 | 2,541.02 | 2,091.71 | 2,156.53 | **9,403.83** |
| 499 | 1,204.84 | 1,070.81 | 875.07 | 1,109.93 | **4,260.65** |

| MI - MICHIGAN - Total | 229,114.90 | 209,125.64 | 190,561.36 | 181,192.81 | 809,994.72 |
|---|---|---|---|---|---|

**STATE: MN - MINNESOTA**

| REGISTRANT ZIP CODE 3 | 1ST QUARTER | 2ND QUARTER | 3RD QUARTER | 4TH QUARTER | TOTAL GRAMS |
|---|---|---|---|---|---|
| 550 | 3,606.91 | 3,410.38 | 3,552.28 | 3,348.26 | **13,917.82** |
| 551 | 5,819.50 | 5,241.14 | 5,752.15 | 5,113.56 | **21,926.35** |
| 553 | 5,214.58 | 5,076.47 | 4,602.84 | 5,255.40 | **20,149.29** |
| 554 | 6,187.19 | 5,824.87 | 5,683.60 | 5,764.57 | **23,460.23** |
| 556 | 142.49 | 108.74 | 100.52 | 99.07 | **450.82** |
| 557 | 1,939.75 | 1,671.91 | 1,818.23 | 1,736.89 | **7,166.78** |
| 558 | 708.65 | 594.70 | 693.11 | 716.68 | **2,713.15** |
| 559 | 1,292.88 | 1,012.80 | 1,228.09 | 946.82 | **4,480.59** |
| 560 | 1,583.67 | 1,193.38 | 1,504.22 | 1,261.16 | **5,542.42** |
| 561 | 614.40 | 446.69 | 551.38 | 502.10 | **2,114.57** |
| 562 | 1,524.58 | 1,166.47 | 1,133.28 | 1,113.69 | **4,938.01** |
| 563 | 2,140.75 | 2,053.96 | 2,030.09 | 1,876.68 | **8,101.48** |
| 564 | 1,076.26 | 895.62 | 1,026.85 | 1,039.72 | **4,038.45** |
| 565 | 1,228.62 | 1,254.88 | 1,199.64 | 1,056.20 | **4,739.34** |
| 566 | 697.16 | 811.99 | 801.70 | 704.84 | **3,015.69** |
| 567 | 699.83 | 558.14 | 583.80 | 622.81 | **2,464.57** |
| **MN - MINNESOTA - Total** | **34,477.21** | **31,322.14** | **32,261.78** | **31,158.43** | **129,219.55** |

**STATE: MO - MISSOURI**

| REGISTRANT ZIP CODE 3 | 1ST QUARTER | 2ND QUARTER | 3RD QUARTER | 4TH QUARTER | TOTAL GRAMS |
|---|---|---|---|---|---|
| 630 | 7,973.94 | 6,538.03 | 6,458.24 | 6,096.55 | **27,066.76** |
| 631 | 15,692.43 | 16,444.24 | 19,274.13 | 16,642.86 | **68,053.66** |
| 633 | 3,626.25 | 3,050.70 | 3,195.46 | 3,323.79 | **13,196.20** |
| 634 | 618.07 | 570.92 | 526.36 | 482.31 | **2,197.66** |
| 635 | 773.54 | 729.52 | 828.79 | 691.76 | **3,023.60** |
| 636 | 1,819.99 | 1,821.34 | 1,728.29 | 1,670.24 | **7,039.86** |
| 637 | 1,122.41 | 723.17 | 1,019.39 | 937.07 | **3,802.04** |
| 638 | 2,291.35 | 2,276.90 | 2,197.02 | 2,213.51 | **8,978.78** |
| 639 | 1,324.03 | 1,226.61 | 1,145.01 | 1,003.39 | **4,699.04** |
| 640 | 5,256.27 | 4,994.14 | 5,639.41 | 4,803.71 | **20,693.52** |
| 641 | 4,722.97 | 4,843.80 | 4,840.03 | 4,488.38 | **18,895.19** |
| 644 | 645.90 | 582.17 | 696.13 | 629.13 | **2,553.32** |
| 645 | 625.64 | 607.22 | 605.13 | 619.67 | **2,457.66** |
| 646 | 629.87 | 600.89 | 686.51 | 571.46 | **2,488.73** |
| 647 | 1,447.17 | 1,409.11 | 1,367.77 | 1,405.05 | **5,629.10** |
| 648 | 1,587.85 | 1,351.84 | 1,498.81 | 1,383.50 | **5,822.00** |
| 650 | 1,222.82 | 1,040.76 | 989.83 | 1,095.35 | **4,348.76** |
| 651 | 657.69 | 625.64 | 691.11 | 531.98 | **2,506.43** |

| | | | | | |
|---|---|---|---|---|---|
| 652 | 1,912.13 | 1,624.65 | 1,815.51 | 1,537.82 | **6,890.11** |
| 653 | 1,052.36 | 788.43 | 1,165.63 | 844.06 | **3,850.49** |
| 654 | 627.66 | 773.07 | 615.88 | 641.29 | **2,657.89** |
| 655 | 1,090.10 | 1,104.04 | 1,088.01 | 1,066.71 | **4,348.86** |
| 656 | 1,367.48 | 1,088.28 | 1,315.10 | 1,207.82 | **4,978.68** |
| 657 | 2,446.10 | 2,057.02 | 2,356.68 | 2,518.58 | **9,378.38** |
| 658 | 2,030.45 | 1,763.08 | 1,902.87 | 1,827.04 | **7,523.44** |
| **MO - MISSOURI - Total** | **62,564.48** | **58,635.55** | **63,647.09** | **58,233.03** | **243,080.15** |

**STATE: MS - MISSISSIPPI**

| REGISTRANT ZIP CODE 3 | 1ST QUARTER | 2ND QUARTER | 3RD QUARTER | 4TH QUARTER | TOTAL GRAMS |
|---|---|---|---|---|---|
| 386 | 3,063.65 | 2,565.07 | 2,479.17 | 2,303.64 | **10,411.53** |
| 387 | 1,555.91 | 1,450.93 | 1,437.67 | 1,063.06 | **5,507.57** |
| 388 | 2,764.82 | 2,413.96 | 2,315.25 | 2,237.32 | **9,731.36** |
| 389 | 1,298.65 | 1,171.35 | 1,133.04 | 897.48 | **4,500.53** |
| 390 | 2,557.50 | 2,082.79 | 1,949.38 | 1,948.84 | **8,538.51** |
| 391 | 2,601.57 | 2,288.63 | 2,064.54 | 1,848.95 | **8,803.69** |
| 392 | 1,421.74 | 1,328.79 | 1,395.98 | 1,440.92 | **5,587.44** |
| 393 | 2,578.84 | 2,128.83 | 2,091.12 | 1,837.88 | **8,636.68** |
| 394 | 2,887.89 | 2,769.47 | 2,735.66 | 2,535.65 | **10,928.67** |
| 395 | 2,856.13 | 2,027.18 | 2,270.45 | 2,235.48 | **9,389.24** |
| 396 | 853.63 | 779.06 | 958.62 | 1,062.14 | **3,653.45** |
| 397 | 2,032.92 | 1,701.57 | 1,632.51 | 1,591.55 | **6,958.54** |
| **MS - MISSISSIPPI - Total** | **26,473.27** | **22,707.65** | **22,463.40** | **21,002.91** | **92,647.22** |

**STATE: MT - MONTANA**

| REGISTRANT ZIP CODE 3 | 1ST QUARTER | 2ND QUARTER | 3RD QUARTER | 4TH QUARTER | TOTAL GRAMS |
|---|---|---|---|---|---|
| 590 | 620.27 | 579.20 | 594.87 | 490.80 | **2,285.14** |
| 591 | 990.07 | 828.33 | 925.14 | 761.20 | **3,504.74** |
| 592 | 397.04 | 294.83 | 272.73 | 290.96 | **1,255.55** |
| 593 | 374.05 | 351.85 | 373.87 | 365.67 | **1,465.44** |
| 594 | 1,001.15 | 1,019.72 | 1,175.06 | 887.21 | **4,083.14** |
| 595 | 272.02 | 266.32 | 251.95 | 322.67 | **1,112.97** |
| 596 | 794.85 | 809.28 | 789.58 | 770.54 | **3,164.25** |
| 597 | 1,257.81 | 1,094.53 | 1,422.44 | 1,141.43 | **4,916.20** |
| 598 | 2,408.48 | 2,173.36 | 2,253.07 | 2,048.74 | **8,883.66** |
| 599 | 1,191.38 | 1,213.39 | 1,340.60 | 1,120.43 | **4,865.79** |
| **MT - MONTANA - Total** | **9,307.13** | **8,630.81** | **9,399.30** | **8,199.65** | **35,536.89** |

**STATE: NC - NORTH CAROLINA**

| REGISTRANT ZIP CODE 3 | 1ST QUARTER | 2ND QUARTER | 3RD QUARTER | 4TH QUARTER | TOTAL GRAMS |
|---|---|---|---|---|---|
| 270 | 1,146.52 | 1,083.31 | 996.38 | 845.91 | **4,072.12** |
| 271 | 1,191.37 | 920.24 | 951.57 | 896.89 | **3,960.07** |

| | | | | | |
|---|---|---|---|---|---|
| 272 | 2,211.62 | 1,946.74 | 1,882.09 | 2,530.10 | **8,570.56** |
| 273 | 1,556.21 | 1,567.23 | 1,126.66 | 1,321.80 | **5,571.90** |
| 274 | 1,878.71 | 1,800.46 | 1,264.96 | 1,472.80 | **6,416.94** |
| 275 | 4,793.13 | 4,159.72 | 3,831.97 | 4,198.32 | **16,983.15** |
| 276 | 1,494.38 | 1,438.15 | 1,150.15 | 1,897.63 | **5,980.30** |
| 277 | 1,172.65 | 1,182.06 | 942.25 | 1,147.45 | **4,444.41** |
| 278 | 2,867.88 | 2,614.43 | 2,511.20 | 2,664.51 | **10,658.01** |
| 279 | 1,305.57 | 1,144.24 | 1,246.92 | 1,028.97 | **4,725.69** |
| 280 | 2,741.02 | 2,388.13 | 2,128.37 | 2,382.61 | **9,640.12** |
| 281 | 1,980.66 | 2,027.22 | 1,726.77 | 1,683.40 | **7,418.04** |
| 282 | 2,388.13 | 2,075.93 | 1,547.17 | 1,934.92 | **7,946.15** |
| 283 | 6,587.46 | 5,478.27 | 5,642.31 | 5,847.48 | **23,555.52** |
| 284 | 2,146.89 | 2,187.54 | 2,042.73 | 1,923.97 | **8,301.13** |
| 285 | 3,321.90 | 2,413.83 | 2,662.66 | 2,779.25 | **11,177.65** |
| 286 | 3,229.40 | 2,782.76 | 2,484.48 | 2,556.88 | **11,053.53** |
| 287 | 2,149.04 | 1,788.52 | 1,663.22 | 1,736.83 | **7,337.60** |
| 288 | 793.08 | 592.65 | 564.28 | 599.64 | **2,549.64** |
| 289 | 861.82 | 712.17 | 613.84 | 710.89 | **2,898.72** |
| **NC - NORTH CAROLINA - Total** | **45,817.43** | **40,303.60** | **36,979.98** | **40,160.23** | **163,261.25** |

**STATE: ND - NORTH DAKOTA**

| REGISTRANT ZIP CODE 3 | 1ST QUARTER | 2ND QUARTER | 3RD QUARTER | 4TH QUARTER | TOTAL GRAMS |
|---|---|---|---|---|---|
| 580 | 446.99 | 325.61 | 328.01 | 372.03 | **1,472.63** |
| 581 | 851.38 | 618.08 | 598.61 | 595.56 | **2,663.63** |
| 582 | 695.77 | 755.18 | 551.62 | 624.33 | **2,626.90** |
| 583 | 556.95 | 402.24 | 327.09 | 354.72 | **1,641.00** |
| 584 | 199.60 | 179.86 | 203.11 | 220.79 | **803.36** |
| 585 | 963.37 | 1,052.14 | 938.50 | 905.06 | **3,859.08** |
| 586 | 191.26 | 240.23 | 258.11 | 216.10 | **905.71** |
| 587 | 499.11 | 596.52 | 387.68 | 369.27 | **1,852.57** |
| 588 | 670.71 | 649.39 | 768.52 | 861.94 | **2,950.56** |
| **ND - NORTH DAKOTA - Total** | **5,075.13** | **4,819.25** | **4,361.26** | **4,519.80** | **18,775.44** |

**STATE: NE - NEBRASKA**

| REGISTRANT ZIP CODE 3 | 1ST QUARTER | 2ND QUARTER | 3RD QUARTER | 4TH QUARTER | TOTAL GRAMS |
|---|---|---|---|---|---|
| 680 | 1,134.85 | 1,075.74 | 1,053.84 | 814.93 | **4,079.36** |
| 681 | 4,117.96 | 3,540.91 | 3,858.54 | 3,393.12 | **14,910.53** |
| 683 | 327.07 | 302.76 | 233.52 | 240.33 | **1,103.68** |
| 684 | 264.18 | 141.63 | 187.86 | 169.51 | **763.18** |
| 685 | 1,846.37 | 1,633.96 | 1,533.06 | 1,577.46 | **6,590.85** |
| 686 | 148.81 | 163.73 | 232.61 | 201.67 | **746.81** |
| 687 | 796.92 | 666.90 | 787.25 | 637.98 | **2,889.04** |

| | | | | | |
|---|---|---|---|---|---|
| 688 | 823.07 | 840.46 | 870.54 | 869.38 | **3,403.44** |
| 689 | 305.36 | 300.01 | 322.21 | 286.94 | **1,214.52** |
| 690 | 186.53 | 80.45 | 155.62 | 151.38 | **573.98** |
| 691 | 813.12 | 772.24 | 760.08 | 879.41 | **3,224.85** |
| 692 | 22.10 | 7.74 | 6.63 | 15.47 | **51.94** |
| 693 | 434.45 | 381.78 | 434.82 | 397.53 | **1,648.58** |
| **NE - NEBRASKA - Total** | **11,220.79** | **9,908.30** | **10,436.56** | **9,635.11** | **41,200.75** |

**STATE: NH - NEW HAMPSHIRE**

| REGISTRANT ZIP CODE 3 | 1ST QUARTER | 2ND QUARTER | 3RD QUARTER | 4TH QUARTER | TOTAL GRAMS |
|---|---|---|---|---|---|
| 030 | 1,753.51 | 1,661.63 | 1,752.97 | 1,780.29 | **6,948.40** |
| 031 | 1,153.30 | 1,297.39 | 1,357.29 | 1,162.78 | **4,970.76** |
| 032 | 860.67 | 773.64 | 817.49 | 652.66 | **3,104.46** |
| 033 | 502.29 | 580.02 | 407.65 | 515.11 | **2,005.07** |
| 034 | 372.21 | 389.71 | 459.51 | 313.45 | **1,534.86** |
| 035 | 332.58 | 213.06 | 258.45 | 241.34 | **1,045.43** |
| 036 | 24.31 | 22.10 | 22.10 | 24.31 | **92.82** |
| 037 | 480.50 | 418.89 | 388.04 | 411.89 | **1,699.32** |
| 038 | 1,480.24 | 1,286.23 | 1,073.16 | 1,136.32 | **4,975.94** |
| **NH - NEW HAMPSHIRE - Total** | **6,959.61** | **6,642.68** | **6,536.66** | **6,238.14** | **26,377.09** |

**STATE: NJ - NEW JERSEY**

| REGISTRANT ZIP CODE 3 | 1ST QUARTER | 2ND QUARTER | 3RD QUARTER | 4TH QUARTER | TOTAL GRAMS |
|---|---|---|---|---|---|
| 070 | 10,639.64 | 11,468.87 | 10,265.07 | 10,190.25 | **42,563.84** |
| 071 | 2,514.77 | 2,460.24 | 2,434.94 | 2,415.94 | **9,825.89** |
| 072 | 1,033.77 | 1,015.61 | 758.03 | 958.24 | **3,765.64** |
| 073 | 2,155.99 | 2,466.73 | 2,206.20 | 2,210.15 | **9,039.08** |
| 074 | 4,331.83 | 4,292.24 | 4,126.77 | 4,196.49 | **16,947.33** |
| 075 | 1,468.67 | 1,463.74 | 1,199.34 | 1,385.87 | **5,517.63** |
| 076 | 3,942.41 | 3,741.10 | 3,610.09 | 3,556.80 | **14,850.40** |
| 077 | 5,706.99 | 5,260.25 | 5,073.80 | 4,930.58 | **20,971.62** |
| 078 | 2,675.53 | 2,495.86 | 2,331.63 | 2,410.01 | **9,913.03** |
| 079 | 1,123.70 | 1,356.77 | 1,246.41 | 1,172.96 | **4,899.84** |
| 080 | 11,980.22 | 12,549.14 | 11,247.92 | 11,018.08 | **46,795.37** |
| 081 | 1,972.06 | 2,050.69 | 1,605.46 | 1,943.88 | **7,572.08** |
| 082 | 2,188.92 | 2,082.72 | 2,230.43 | 2,203.64 | **8,705.71** |
| 083 | 2,114.26 | 2,079.06 | 2,066.79 | 1,944.34 | **8,204.45** |
| 084 | 351.96 | 368.07 | 414.58 | 387.22 | **1,521.83** |
| 085 | 1,798.12 | 1,679.71 | 1,633.16 | 1,626.76 | **6,737.76** |
| 086 | 2,010.97 | 2,227.98 | 1,845.32 | 2,003.84 | **8,088.11** |
| 087 | 4,115.81 | 4,030.08 | 4,069.90 | 3,983.67 | **16,199.46** |
| 088 | 6,683.03 | 6,448.97 | 5,867.41 | 6,289.72 | **25,289.14** |

| | | | | | |
|---|---|---|---|---|---|
| 089 | 741.87 | 565.70 | 607.86 | 803.18 | **2,718.60** |
| **NJ - NEW JERSEY - Total** | **69,550.53** | **70,103.54** | **64,841.12** | **65,631.62** | **270,126.81** |

**STATE: NM - NEW MEXICO**

| REGISTRANT ZIP CODE 3 | 1ST QUARTER | 2ND QUARTER | 3RD QUARTER | 4TH QUARTER | TOTAL GRAMS |
|---|---|---|---|---|---|
| 870 | 1,771.86 | 1,535.09 | 1,664.73 | 1,499.19 | **6,470.86** |
| 871 | 5,540.00 | 4,586.53 | 4,909.55 | 4,345.16 | **19,381.24** |
| 873 | 1,694.87 | 1,220.37 | 1,571.59 | 1,334.28 | **5,821.10** |
| 874 | 1,203.58 | 520.40 | 645.34 | 623.61 | **2,992.93** |
| 875 | 1,165.92 | 942.93 | 970.60 | 723.45 | **3,802.90** |
| 877 | 243.84 | 244.02 | 295.59 | 297.43 | **1,080.88** |
| 878 | 114.93 | 53.04 | 47.61 | 86.19 | **301.77** |
| 879 | 95.03 | 68.51 | 164.28 | 169.06 | **496.88** |
| 880 | 2,277.50 | 1,945.65 | 2,100.48 | 2,329.49 | **8,653.12** |
| 881 | 902.93 | 711.65 | 890.76 | 912.19 | **3,417.53** |
| 882 | 2,697.64 | 2,335.49 | 2,411.62 | 2,353.24 | **9,797.99** |
| 883 | 942.96 | 674.26 | 962.12 | 758.23 | **3,337.58** |
| 884 | 64.09 | 163.69 | 139.24 | 85.09 | **452.11** |
| **NM - NEW MEXICO - Total** | **18,715.14** | **15,001.65** | **16,773.51** | **15,516.60** | **66,006.90** |

**STATE: NV - NEVADA**

| REGISTRANT ZIP CODE 3 | 1ST QUARTER | 2ND QUARTER | 3RD QUARTER | 4TH QUARTER | TOTAL GRAMS |
|---|---|---|---|---|---|
| 890 | 4,799.47 | 5,038.95 | 4,835.87 | 4,361.33 | **19,035.62** |
| 891 | 11,661.55 | 10,714.24 | 9,861.64 | 8,902.13 | **41,139.56** |
| 893 | 80.89 | 109.40 | 107.63 | 127.96 | **425.89** |
| 894 | 2,173.50 | 1,885.54 | 1,992.57 | 1,918.50 | **7,970.12** |
| 895 | 1,816.13 | 1,678.17 | 1,748.85 | 1,708.89 | **6,952.04** |
| 897 | 673.67 | 519.87 | 591.91 | 417.48 | **2,202.92** |
| 898 | 251.51 | 236.48 | 338.15 | 242.83 | **1,068.96** |
| **NV - NEVADA - Total** | **21,456.72** | **20,182.65** | **19,476.62** | **17,679.12** | **78,795.10** |

**STATE: NY - NEW YORK**

| REGISTRANT ZIP CODE 3 | 1ST QUARTER | 2ND QUARTER | 3RD QUARTER | 4TH QUARTER | TOTAL GRAMS |
|---|---|---|---|---|---|
| 063 | 0 | 4.42 | 2.21 | 0 | **6.63** |
| 100 | 12,172.44 | 11,818.15 | 11,337.69 | 10,580.43 | **45,908.72** |
| 101 | 684.33 | 604.33 | 591.16 | 431.92 | **2,311.73** |
| 102 | 22.10 | 8.28 | 32.05 | 19.79 | **82.22** |
| 103 | 2,649.07 | 2,854.59 | 2,599.22 | 2,442.60 | **10,545.48** |
| 104 | 9,130.76 | 9,403.97 | 7,964.54 | 8,231.65 | **34,730.91** |
| 105 | 2,990.04 | 3,442.35 | 3,033.41 | 2,862.12 | **12,327.92** |
| 106 | 604.18 | 518.72 | 522.59 | 517.53 | **2,163.01** |
| 107 | 1,249.97 | 1,289.77 | 1,311.55 | 1,274.87 | **5,126.16** |
| 108 | 341.72 | 451.59 | 388.32 | 274.40 | **1,456.03** |

| | | | | | |
|---|---|---|---|---|---|
| 109 | 3,958.69 | 3,808.49 | 3,604.48 | 3,732.64 | **15,104.30** |
| 110 | 1,414.16 | 1,460.27 | 1,169.76 | 1,177.05 | **5,221.23** |
| 111 | 1,187.00 | 1,277.06 | 1,309.19 | 1,130.74 | **4,903.99** |
| 112 | 15,716.06 | 16,083.73 | 14,498.74 | 14,549.98 | **60,848.52** |
| 113 | 5,703.69 | 6,020.88 | 5,523.21 | 5,320.97 | **22,568.75** |
| 114 | 3,100.20 | 3,566.35 | 2,753.03 | 2,617.30 | **12,036.89** |
| 115 | 3,810.43 | 3,847.09 | 3,875.85 | 3,153.61 | **14,686.98** |
| 116 | 826.86 | 878.70 | 754.93 | 744.06 | **3,204.55** |
| 117 | 7,506.41 | 7,366.91 | 6,796.54 | 6,686.98 | **28,356.83** |
| 118 | 478.25 | 512.90 | 523.96 | 467.02 | **1,982.14** |
| 119 | 1,120.36 | 1,213.15 | 1,167.13 | 1,066.86 | **4,567.50** |
| 120 | 1,680.57 | 1,420.76 | 1,436.50 | 1,587.43 | **6,125.26** |
| 121 | 831.35 | 576.46 | 635.39 | 596.81 | **2,640.01** |
| 122 | 541.84 | 609.93 | 618.95 | 632.64 | **2,403.35** |
| 123 | 815.24 | 559.88 | 586.95 | 704.37 | **2,666.45** |
| 124 | 943.78 | 1,048.20 | 1,166.92 | 912.94 | **4,071.84** |
| 125 | 2,358.10 | 2,655.31 | 2,253.88 | 2,534.21 | **9,801.49** |
| 126 | 370.37 | 427.00 | 398.46 | 372.95 | **1,568.78** |
| 127 | 1,308.18 | 1,314.81 | 1,327.15 | 1,289.67 | **5,239.82** |
| 128 | 1,667.17 | 1,400.04 | 1,434.49 | 1,335.86 | **5,837.56** |
| 129 | 1,418.29 | 1,256.69 | 1,251.33 | 1,354.64 | **5,280.95** |
| 130 | 3,072.21 | 2,904.44 | 2,839.97 | 2,954.77 | **11,771.39** |
| 131 | 1,077.85 | 978.30 | 933.82 | 1,001.68 | **3,991.66** |
| 132 | 1,775.49 | 1,494.72 | 1,470.30 | 1,534.66 | **6,275.17** |
| 133 | 905.62 | 838.06 | 664.55 | 679.30 | **3,087.53** |
| 134 | 953.98 | 828.49 | 946.37 | 773.97 | **3,502.80** |
| 135 | 450.30 | 480.23 | 351.77 | 456.10 | **1,738.40** |
| 136 | 3,572.39 | 3,950.75 | 3,766.16 | 3,875.92 | **15,165.22** |
| 137 | 863.44 | 822.08 | 682.62 | 634.20 | **3,002.34** |
| 138 | 1,123.60 | 1,001.32 | 932.27 | 985.95 | **4,043.15** |
| 139 | 837.03 | 874.97 | 870.46 | 708.95 | **3,291.41** |
| 140 | 4,395.56 | 4,464.38 | 4,487.63 | 4,495.55 | **17,843.12** |
| 141 | 2,205.63 | 2,002.85 | 2,103.39 | 1,978.91 | **8,290.78** |
| 142 | 9,096.16 | 9,164.91 | 9,140.17 | 8,838.32 | **36,239.55** |
| 143 | 596.90 | 784.55 | 623.22 | 532.26 | **2,536.93** |
| 144 | 2,414.58 | 2,344.31 | 2,227.46 | 1,969.72 | **8,956.06** |
| 145 | 1,859.10 | 1,832.19 | 1,693.09 | 1,638.33 | **7,022.70** |
| 146 | 3,783.83 | 3,479.84 | 3,410.06 | 3,445.75 | **14,119.47** |
| 147 | 1,117.08 | 1,090.65 | 989.16 | 998.37 | **4,195.26** |
| 148 | 1,874.17 | 1,754.93 | 1,955.93 | 1,804.81 | **7,389.84** |

| | | | | | |
|---|---|---|---|---|---|
| 149 | 388.77 | 399.11 | 279.94 | 289.89 | **1,357.70** |
| **NY - NEW YORK - Total** | **128,965.32** | **129,191.83** | **121,237.90** | **118,201.43** | **497,596.48** |

**STATE: OH - OHIO**

| REGISTRANT ZIP CODE 3 | 1ST QUARTER | 2ND QUARTER | 3RD QUARTER | 4TH QUARTER | **TOTAL GRAMS** |
|---|---|---|---|---|---|
| 430 | 4,029.00 | 4,387.06 | 3,764.69 | 3,071.88 | **15,252.64** |
| 431 | 2,955.66 | 2,672.19 | 2,949.12 | 2,470.08 | **11,047.04** |
| 432 | 11,847.56 | 7,992.16 | 4,475.47 | 4,082.71 | **28,397.90** |
| 433 | 1,008.72 | 865.63 | 798.14 | 738.65 | **3,411.13** |
| 434 | 1,191.82 | 1,079.02 | 1,184.98 | 921.42 | **4,377.23** |
| 435 | 2,927.20 | 2,660.15 | 2,365.85 | 2,110.96 | **10,064.16** |
| 436 | 5,769.57 | 5,336.43 | 5,302.62 | 4,339.81 | **20,748.43** |
| 437 | 1,520.95 | 1,253.25 | 1,468.19 | 1,152.31 | **5,394.69** |
| 438 | 306.10 | 298.55 | 279.94 | 267.79 | **1,152.37** |
| 439 | 2,002.44 | 2,015.60 | 1,849.73 | 1,570.45 | **7,438.22** |
| 440 | 4,643.98 | 4,151.29 | 4,512.00 | 4,194.17 | **17,501.44** |
| 441 | 8,624.38 | 8,725.08 | 8,111.20 | 7,163.25 | **32,623.91** |
| 442 | 2,192.99 | 2,250.37 | 2,172.38 | 1,959.40 | **8,575.15** |
| 443 | 1,252.66 | 1,142.78 | 1,073.12 | 868.73 | **4,337.29** |
| 444 | 2,677.51 | 3,476.43 | 2,926.71 | 2,055.19 | **11,135.82** |
| 445 | 1,544.48 | 1,869.62 | 1,763.47 | 1,245.77 | **6,423.33** |
| 446 | 2,149.61 | 1,837.21 | 1,967.24 | 1,791.98 | **7,746.04** |
| 447 | 1,908.92 | 1,555.37 | 1,517.08 | 1,622.54 | **6,603.90** |
| 448 | 1,637.84 | 1,545.92 | 1,778.74 | 1,621.27 | **6,583.76** |
| 449 | 829.99 | 945.55 | 846.47 | 772.98 | **3,394.98** |
| 450 | 3,837.57 | 3,699.07 | 3,039.30 | 3,151.63 | **13,727.57** |
| 451 | 1,939.64 | 1,390.86 | 1,709.19 | 1,417.74 | **6,457.43** |
| 452 | 5,955.39 | 5,070.05 | 5,137.62 | 4,280.52 | **20,443.58** |
| 453 | 3,386.08 | 3,048.05 | 3,148.14 | 2,587.96 | **12,170.24** |
| 454 | 3,881.36 | 3,600.79 | 3,358.71 | 3,196.98 | **14,037.84** |
| 455 | 657.28 | 625.75 | 571.39 | 619.54 | **2,473.96** |
| 456 | 3,038.85 | 2,702.11 | 2,673.38 | 2,571.44 | **10,985.77** |
| 457 | 1,365.18 | 1,333.47 | 1,240.59 | 1,161.64 | **5,100.89** |
| 458 | 2,839.03 | 2,369.64 | 2,481.20 | 2,259.24 | **9,949.11** |
| **OH - OHIO - Total** | **87,921.76** | **79,899.41** | **74,466.66** | **65,268.02** | **307,555.84** |

**STATE: OK - OKLAHOMA**

| REGISTRANT ZIP CODE 3 | 1ST QUARTER | 2ND QUARTER | 3RD QUARTER | 4TH QUARTER | **TOTAL GRAMS** |
|---|---|---|---|---|---|
| 730 | 13,124.28 | 13,747.19 | 13,504.01 | 12,249.36 | **52,624.84** |
| 731 | 13,274.64 | 12,167.17 | 12,027.83 | 10,698.52 | **48,168.15** |
| 734 | 3,875.59 | 3,760.12 | 3,593.17 | 3,325.46 | **14,554.33** |
| 735 | 9,604.81 | 9,123.74 | 8,465.57 | 7,843.38 | **35,037.50** |

| | | | | | |
|---|---|---|---|---|---|
| 736 | 2,170.50 | 2,319.77 | 2,073.33 | 1,921.68 | **8,485.28** |
| 737 | 1,969.00 | 1,744.98 | 1,816.14 | 1,696.25 | **7,226.37** |
| 738 | 626.19 | 553.61 | 466.51 | 355.74 | **2,002.04** |
| 739 | 495.24 | 495.24 | 367.24 | 434.47 | **1,792.20** |
| 740 | 10,357.73 | 9,674.38 | 9,727.65 | 9,883.33 | **39,643.10** |
| 741 | 3,767.35 | 3,492.79 | 3,990.31 | 3,864.00 | **15,114.46** |
| 743 | 3,569.61 | 3,617.93 | 3,760.49 | 3,380.70 | **14,328.72** |
| 744 | 6,490.61 | 6,078.68 | 6,177.42 | 5,701.60 | **24,448.31** |
| 745 | 2,083.02 | 1,897.46 | 1,961.46 | 1,944.89 | **7,886.83** |
| 746 | 1,772.50 | 1,538.23 | 1,594.59 | 1,549.28 | **6,454.60** |
| 747 | 6,401.94 | 6,673.40 | 6,371.72 | 5,897.65 | **25,344.71** |
| 748 | 8,599.29 | 8,235.73 | 8,921.13 | 8,179.92 | **33,936.07** |
| 749 | 3,261.00 | 2,465.37 | 2,638.86 | 2,683.06 | **11,048.29** |
| **OK - OKLAHOMA - Total** | **91,443.31** | **87,585.78** | **87,457.44** | **81,609.29** | **348,095.81** |

**STATE: OR - OREGON**

| REGISTRANT ZIP CODE 3 | 1ST QUARTER | 2ND QUARTER | 3RD QUARTER | 4TH QUARTER | **TOTAL GRAMS** |
|---|---|---|---|---|---|
| 970 | 6,383.42 | 5,694.98 | 5,707.54 | 4,850.18 | **22,636.11** |
| 971 | 2,247.17 | 2,154.47 | 2,167.65 | 2,005.36 | **8,574.65** |
| 972 | 7,467.57 | 6,978.60 | 6,871.89 | 6,279.17 | **27,597.24** |
| 973 | 4,611.93 | 4,156.71 | 4,251.83 | 4,182.54 | **17,203.01** |
| 974 | 4,816.88 | 4,345.72 | 4,504.27 | 3,930.36 | **17,597.23** |
| 975 | 3,628.32 | 3,332.62 | 3,085.29 | 3,082.31 | **13,128.54** |
| 976 | 476.49 | 436.43 | 433.96 | 447.18 | **1,794.06** |
| 977 | 1,728.97 | 1,490.38 | 1,635.67 | 1,529.37 | **6,384.39** |
| 978 | 1,652.97 | 1,434.15 | 1,424.11 | 1,332.59 | **5,843.83** |
| 979 | 282.63 | 194.49 | 274.94 | 274.72 | **1,026.77** |
| **OR - OREGON - Total** | **33,296.35** | **30,218.54** | **30,357.15** | **27,913.79** | **121,785.82** |

**STATE: PA - PENNSYLVANIA**

| REGISTRANT ZIP CODE 3 | 1ST QUARTER | 2ND QUARTER | 3RD QUARTER | 4TH QUARTER | **TOTAL GRAMS** |
|---|---|---|---|---|---|
| 150 | 3,377.25 | 3,002.01 | 2,874.45 | 2,971.97 | **12,225.67** |
| 151 | 2,753.58 | 2,706.86 | 2,287.69 | 2,488.77 | **10,236.90** |
| 152 | 4,685.46 | 4,732.43 | 4,527.68 | 4,432.10 | **18,377.66** |
| 153 | 1,295.78 | 1,378.43 | 1,296.08 | 1,175.50 | **5,145.78** |
| 154 | 1,044.23 | 968.50 | 958.56 | 990.40 | **3,961.69** |
| 155 | 3,119.60 | 2,355.90 | 2,724.17 | 2,477.76 | **10,677.43** |
| 156 | 1,843.68 | 1,769.85 | 1,698.83 | 1,840.90 | **7,153.25** |
| 157 | 1,466.41 | 1,168.48 | 1,283.81 | 1,409.09 | **5,327.79** |
| 158 | 629.50 | 600.04 | 802.27 | 504.08 | **2,535.90** |
| 159 | 1,342.02 | 1,189.27 | 1,244.87 | 1,324.37 | **5,100.53** |
| 160 | 1,387.98 | 1,281.98 | 1,554.35 | 1,380.35 | **5,604.66** |

| | | | | | |
|---|---|---|---|---|---|
| 161 | 1,570.51 | 1,396.81 | 1,496.35 | 1,443.96 | **5,907.62** |
| 162 | 1,116.52 | 1,171.33 | 1,202.28 | 1,070.15 | **4,560.29** |
| 163 | 1,197.80 | 1,217.52 | 1,221.52 | 973.29 | **4,610.14** |
| 164 | 688.21 | 614.11 | 621.57 | 470.17 | **2,394.06** |
| 165 | 1,674.76 | 1,404.31 | 1,180.18 | 1,319.82 | **5,579.08** |
| 166 | 1,574.88 | 1,342.48 | 1,372.00 | 1,238.12 | **5,527.48** |
| 167 | 492.85 | 508.32 | 507.22 | 480.70 | **1,989.09** |
| 168 | 586.21 | 617.38 | 709.48 | 578.21 | **2,491.28** |
| 169 | 526.71 | 416.77 | 383.05 | 399.99 | **1,726.52** |
| 170 | 4,351.21 | 4,156.78 | 3,978.77 | 3,841.39 | **16,328.15** |
| 171 | 1,109.67 | 1,254.58 | 858.34 | 1,071.16 | **4,293.74** |
| 172 | 1,366.55 | 1,180.78 | 1,095.16 | 1,157.22 | **4,799.71** |
| 173 | 2,312.40 | 2,153.42 | 2,013.65 | 2,115.09 | **8,594.57** |
| 174 | 1,331.19 | 1,192.04 | 1,273.63 | 1,091.25 | **4,888.11** |
| 175 | 1,291.05 | 1,255.32 | 1,286.89 | 1,064.33 | **4,897.60** |
| 176 | 1,285.44 | 1,382.99 | 1,072.96 | 1,008.02 | **4,749.41** |
| 177 | 1,009.85 | 962.72 | 980.22 | 971.21 | **3,924.00** |
| 178 | 1,448.83 | 1,563.08 | 1,539.10 | 1,552.89 | **6,103.91** |
| 179 | 1,375.55 | 1,405.77 | 1,229.54 | 1,345.85 | **5,356.71** |
| 180 | 2,863.94 | 2,603.22 | 2,528.29 | 2,408.00 | **10,403.45** |
| 181 | 1,253.91 | 1,532.53 | 1,088.90 | 1,243.87 | **5,119.21** |
| 182 | 1,119.97 | 978.52 | 858.43 | 981.56 | **3,938.48** |
| 183 | 1,726.91 | 1,620.55 | 1,727.46 | 1,594.19 | **6,669.11** |
| 184 | 1,690.40 | 1,614.36 | 1,651.65 | 1,598.90 | **6,555.31** |
| 185 | 1,433.92 | 1,572.41 | 1,379.39 | 1,368.44 | **5,754.16** |
| 186 | 1,872.32 | 2,121.05 | 1,924.64 | 1,879.79 | **7,797.80** |
| 187 | 2,869.83 | 3,981.93 | 3,546.02 | 3,278.15 | **13,675.94** |
| 188 | 840.28 | 899.58 | 1,132.15 | 1,151.47 | **4,023.48** |
| 189 | 3,403.42 | 2,860.08 | 2,888.08 | 3,173.63 | **12,325.21** |
| 190 | 12,758.96 | 13,112.03 | 11,732.96 | 11,441.58 | **49,045.53** |
| 191 | 22,016.80 | 22,948.73 | 20,580.07 | 20,064.76 | **85,610.35** |
| 193 | 3,148.78 | 2,745.68 | 2,527.59 | 2,633.30 | **11,055.35** |
| 194 | 7,080.81 | 6,108.81 | 5,827.78 | 5,211.46 | **24,228.85** |
| 195 | 1,448.39 | 1,477.20 | 1,215.97 | 1,370.76 | **5,512.32** |
| 196 | 2,135.85 | 1,999.58 | 1,831.50 | 1,919.32 | **7,886.26** |
| **PA - PENNSYLVANIA - Total** | **116,920.14** | **114,526.52** | **107,715.57** | **105,507.30** | **444,669.53** |

**STATE: PR - PUERTO RICO**

| REGISTRANT ZIP CODE 3 | 1ST QUARTER | 2ND QUARTER | 3RD QUARTER | 4TH QUARTER | **TOTAL GRAMS** |
|---|---|---|---|---|---|
| 006 | 4,543.97 | 4,725.19 | 4,853.38 | 4,087.58 | **18,210.12** |
| 007 | 4,015.75 | 4,528.49 | 4,227.92 | 3,634.51 | **16,406.68** |

| | | | | | |
|---|---|---|---|---|---|
| 009 | 2,928.38 | 3,152.71 | 3,253.27 | 2,948.27 | **12,282.63** |
| **PR - PUERTO RICO - Total** | **11,488.10** | **12,406.40** | **12,334.57** | **10,670.36** | **46,899.43** |

**STATE: RI - RHODE ISLAND**

| REGISTRANT ZIP CODE 3 | 1ST QUARTER | 2ND QUARTER | 3RD QUARTER | 4TH QUARTER | TOTAL GRAMS |
|---|---|---|---|---|---|
| 028 | 3,848.01 | 3,928.98 | 3,404.88 | 3,456.93 | **14,638.81** |
| 029 | 2,339.33 | 2,233.54 | 1,823.84 | 1,977.72 | **8,374.42** |
| **RI - RHODE ISLAND - Total** | **6,187.34** | **6,162.52** | **5,228.73** | **5,434.65** | **23,013.23** |

**STATE: SC - SOUTH CAROLINA**

| REGISTRANT ZIP CODE 3 | 1ST QUARTER | 2ND QUARTER | 3RD QUARTER | 4TH QUARTER | TOTAL GRAMS |
|---|---|---|---|---|---|
| 290 | 2,615.51 | 2,723.66 | 2,245.62 | 2,678.68 | **10,263.47** |
| 291 | 3,139.78 | 2,786.55 | 2,555.15 | 2,594.84 | **11,076.32** |
| 292 | 3,225.51 | 3,160.48 | 3,148.80 | 2,794.17 | **12,328.97** |
| 293 | 3,455.53 | 3,097.25 | 3,151.32 | 3,455.79 | **13,159.90** |
| 294 | 38,761.13 | 40,414.87 | 42,185.55 | 32,678.75 | **154,040.29** |
| 295 | 9,741.68 | 8,494.13 | 8,623.98 | 8,190.33 | **35,050.12** |
| 296 | 6,954.57 | 6,425.03 | 6,402.34 | 5,750.38 | **25,532.32** |
| 297 | 1,880.36 | 1,461.96 | 1,592.44 | 1,514.25 | **6,449.01** |
| 298 | 2,240.40 | 2,303.08 | 1,951.69 | 2,111.20 | **8,606.37** |
| 299 | 1,288.80 | 1,094.53 | 1,022.63 | 1,062.14 | **4,468.10** |
| **SC - SOUTH CAROLINA - Total** | **73,303.29** | **71,961.54** | **72,879.51** | **62,830.54** | **280,974.88** |

**STATE: SD - SOUTH DAKOTA**

| REGISTRANT ZIP CODE 3 | 1ST QUARTER | 2ND QUARTER | 3RD QUARTER | 4TH QUARTER | TOTAL GRAMS |
|---|---|---|---|---|---|
| 570 | 819.47 | 844.19 | 889.63 | 722.08 | **3,275.38** |
| 571 | 1,749.41 | 1,666.12 | 1,890.26 | 1,573.41 | **6,879.21** |
| 572 | 411.99 | 357.10 | 405.18 | 331.70 | **1,505.96** |
| 573 | 592.53 | 419.21 | 494.02 | 479.33 | **1,985.09** |
| 574 | 519.92 | 460.79 | 524.68 | 441.76 | **1,947.15** |
| 575 | 225.79 | 167.59 | 222.12 | 184.54 | **800.04** |
| 576 | 232.06 | 227.64 | 176.81 | 207.75 | **844.26** |
| 577 | 1,404.35 | 1,367.24 | 1,367.26 | 1,473.89 | **5,612.74** |
| **SD - SOUTH DAKOTA - Total** | **5,955.53** | **5,509.88** | **5,969.95** | **5,414.47** | **22,849.83** |

**STATE: TN - TENNESSEE**

| REGISTRANT ZIP CODE 3 | 1ST QUARTER | 2ND QUARTER | 3RD QUARTER | 4TH QUARTER | TOTAL GRAMS |
|---|---|---|---|---|---|
| 370 | 5,399.82 | 4,369.25 | 4,088.82 | 4,230.84 | **18,088.72** |
| 371 | 33,189.97 | 30,402.29 | 36,821.98 | 25,715.42 | **126,129.66** |
| 372 | 3,268.89 | 2,089.51 | 2,144.92 | 2,035.12 | **9,538.45** |
| 373 | 3,400.07 | 3,027.45 | 2,832.23 | 2,902.77 | **12,162.52** |
| 374 | 1,459.03 | 1,261.58 | 1,206.79 | 1,126.22 | **5,053.62** |
| 376 | 2,724.86 | 2,385.79 | 2,145.94 | 2,176.83 | **9,433.42** |
| 377 | 2,498.74 | 2,652.66 | 2,206.80 | 2,094.05 | **9,452.26** |

| | | | | | |
|---|---|---|---|---|---|
| 378 | 4,437.00 | 3,511.89 | 3,533.28 | 3,278.10 | **14,760.27** |
| 379 | 2,545.15 | 2,267.60 | 2,349.65 | 2,150.65 | **9,313.06** |
| 380 | 2,528.78 | 2,168.84 | 2,062.28 | 1,757.84 | **8,517.74** |
| 381 | 4,863.65 | 4,599.98 | 4,265.91 | 3,539.82 | **17,269.36** |
| 382 | 880.34 | 936.33 | 740.75 | 832.19 | **3,389.61** |
| 383 | 3,808.30 | 3,346.52 | 3,305.56 | 3,088.59 | **13,548.96** |
| 384 | 1,040.31 | 997.12 | 906.51 | 741.11 | **3,685.04** |
| 385 | 2,390.59 | 2,324.65 | 1,834.75 | 2,067.51 | **8,617.49** |
| **TN - TENNESSEE - Total** | **74,435.49** | **66,341.46** | **70,446.16** | **57,737.07** | **268,960.19** |

**STATE: TX - TEXAS**

| REGISTRANT ZIP CODE 3 | 1ST QUARTER | 2ND QUARTER | 3RD QUARTER | 4TH QUARTER | **TOTAL GRAMS** |
|---|---|---|---|---|---|
| 750 | 62,268.75 | 65,316.58 | 64,270.50 | 60,437.20 | **252,293.03** |
| 751 | 30,139.30 | 30,763.75 | 29,328.58 | 27,616.53 | **117,848.16** |
| 752 | 33,921.27 | 36,691.63 | 33,816.78 | 32,521.62 | **136,951.30** |
| 753 | 566.98 | 614.59 | 629.88 | 598.94 | **2,410.39** |
| 754 | 19,964.09 | 20,712.22 | 19,996.52 | 18,782.78 | **79,455.60** |
| 755 | 8,381.99 | 8,139.97 | 8,408.51 | 7,728.72 | **32,659.19** |
| 756 | 23,699.47 | 25,476.19 | 24,923.66 | 24,146.81 | **98,246.13** |
| 757 | 23,694.17 | 26,745.58 | 26,459.07 | 26,297.01 | **103,195.83** |
| 758 | 4,612.79 | 4,597.23 | 4,293.34 | 4,041.39 | **17,544.75** |
| 759 | 17,536.49 | 17,639.81 | 17,703.98 | 17,199.34 | **70,079.62** |
| 760 | 45,993.15 | 47,573.74 | 44,101.42 | 44,155.45 | **181,823.76** |
| 761 | 35,879.28 | 36,277.17 | 33,884.15 | 33,100.64 | **139,141.25** |
| 762 | 16,733.17 | 16,769.60 | 16,416.96 | 15,789.22 | **65,708.94** |
| 763 | 11,445.70 | 11,445.89 | 10,843.09 | 11,229.86 | **44,964.54** |
| 764 | 6,595.38 | 6,935.76 | 6,792.91 | 7,202.18 | **27,526.23** |
| 765 | 15,673.20 | 15,086.29 | 14,723.29 | 13,040.32 | **58,523.10** |
| 766 | 5,205.16 | 5,146.06 | 4,487.61 | 4,703.09 | **19,541.92** |
| 767 | 7,933.61 | 8,609.38 | 8,254.23 | 7,812.03 | **32,609.25** |
| 768 | 6,781.69 | 6,711.15 | 6,550.74 | 6,341.97 | **26,385.55** |
| 769 | 6,618.14 | 6,343.53 | 6,298.79 | 6,108.72 | **25,369.18** |
| 770 | 78,415.67 | 81,540.18 | 74,330.70 | 72,146.78 | **306,433.33** |
| 772 | 0 | 0 | 4.42 | 4.42 | **8.84** |
| 773 | 41,930.83 | 44,120.79 | 40,819.24 | 39,303.67 | **166,174.54** |
| 774 | 32,054.49 | 31,860.80 | 30,332.86 | 30,367.00 | **124,615.15** |
| 775 | 45,028.15 | 45,728.97 | 41,694.06 | 40,264.04 | **172,715.21** |
| 776 | 15,677.58 | 15,815.06 | 15,153.94 | 15,675.99 | **62,322.56** |
| 777 | 9,184.57 | 9,289.58 | 8,933.59 | 8,922.05 | **36,329.78** |
| 778 | 11,435.87 | 11,080.44 | 11,059.49 | 10,593.40 | **44,169.20** |
| 779 | 4,935.32 | 5,222.65 | 4,980.82 | 5,012.87 | **20,151.67** |

| | | | | | |
|---|---|---|---|---|---|
| 780 | 14,644.63 | 15,206.74 | 14,436.30 | 14,045.74 | **58,333.40** |
| 781 | 12,714.58 | 13,111.27 | 12,355.95 | 12,436.79 | **50,618.58** |
| 782 | 69,441.33 | 71,359.74 | 65,618.67 | 65,368.89 | **271,788.61** |
| 783 | 12,849.83 | 12,753.48 | 11,857.19 | 11,808.11 | **49,268.60** |
| 784 | 15,855.27 | 16,406.16 | 15,437.03 | 15,647.10 | **63,345.56** |
| 785 | 30,770.25 | 29,605.23 | 26,833.32 | 25,688.71 | **112,897.51** |
| 786 | 19,538.47 | 19,412.15 | 18,957.85 | 18,838.99 | **76,747.45** |
| 787 | 16,702.69 | 17,326.35 | 16,182.42 | 15,869.65 | **66,081.11** |
| 788 | 5,456.83 | 5,544.47 | 4,980.46 | 4,827.96 | **20,809.73** |
| 789 | 1,847.35 | 1,622.21 | 1,472.29 | 1,663.28 | **6,605.13** |
| 790 | 7,568.31 | 7,840.59 | 8,041.55 | 8,032.28 | **31,482.73** |
| 791 | 15,651.54 | 14,870.64 | 15,154.40 | 14,280.61 | **59,957.18** |
| 792 | 1,443.92 | 1,308.18 | 1,269.14 | 1,302.30 | **5,323.55** |
| 793 | 6,037.05 | 6,005.71 | 5,741.76 | 5,620.86 | **23,405.37** |
| 794 | 14,003.26 | 14,178.72 | 14,063.95 | 14,013.39 | **56,259.31** |
| 795 | 2,863.97 | 3,147.18 | 3,115.06 | 2,864.31 | **11,990.52** |
| 796 | 8,988.88 | 9,204.06 | 9,109.47 | 9,138.03 | **36,440.45** |
| 797 | 23,113.94 | 23,462.23 | 22,624.79 | 21,134.76 | **90,335.73** |
| 798 | 1,324.79 | 1,185.72 | 1,206.71 | 1,248.89 | **4,966.12** |
| 799 | 25,938.00 | 24,872.49 | 25,477.36 | 22,977.14 | **99,264.98** |
| **TX - TEXAS - Total** | **929,061.13** | **950,677.88** | **903,428.79** | **877,951.82** | **3,661,119.62** |

**STATE: UT - UTAH**

| REGISTRANT ZIP CODE 3 | 1ST QUARTER | 2ND QUARTER | 3RD QUARTER | 4TH QUARTER | TOTAL GRAMS |
|---|---|---|---|---|---|
| 840 | 7,339.37 | 6,088.65 | 5,991.64 | 5,864.27 | **25,283.92** |
| 841 | 3,640.41 | 2,927.03 | 2,942.99 | 2,861.59 | **12,372.02** |
| 843 | 833.16 | 665.97 | 696.52 | 672.35 | **2,868.00** |
| 844 | 1,019.18 | 838.77 | 756.23 | 780.78 | **3,394.97** |
| 845 | 512.74 | 475.17 | 383.67 | 422.31 | **1,793.90** |
| 846 | 1,945.27 | 1,591.16 | 1,833.53 | 1,806.32 | **7,176.28** |
| 847 | 1,589.41 | 1,351.46 | 1,517.88 | 1,489.96 | **5,948.72** |
| **UT - UTAH - Total** | **16,879.56** | **13,938.22** | **14,122.46** | **13,897.58** | **58,837.81** |

**STATE: VA - VIRGINIA**

| REGISTRANT ZIP CODE 3 | 1ST QUARTER | 2ND QUARTER | 3RD QUARTER | 4TH QUARTER | TOTAL GRAMS |
|---|---|---|---|---|---|
| 201 | 5,316.20 | 4,641.16 | 4,457.35 | 4,615.96 | **19,030.67** |
| 220 | 3,265.27 | 3,265.10 | 3,273.74 | 3,050.54 | **12,854.65** |
| 221 | 2,830.04 | 2,637.70 | 2,695.66 | 2,453.10 | **10,616.50** |
| 222 | 999.23 | 836.86 | 1,035.86 | 847.54 | **3,719.48** |
| 223 | 2,127.90 | 1,554.91 | 1,742.35 | 1,617.62 | **7,042.78** |
| 224 | 2,199.46 | 1,975.68 | 1,969.14 | 2,072.63 | **8,216.91** |
| 225 | 1,074.54 | 1,028.71 | 821.19 | 1,019.60 | **3,944.04** |

| | | | | | |
|---|---|---|---|---|---|
| 226 | 982.26 | 924.19 | 947.65 | 1,118.48 | **3,972.58** |
| 227 | 361.53 | 368.33 | 311.31 | 279.01 | **1,320.18** |
| 228 | 997.93 | 750.95 | 1,004.83 | 884.93 | **3,638.64** |
| 229 | 1,502.37 | 1,276.17 | 1,407.30 | 1,057.92 | **5,243.76** |
| 230 | 1,147.25 | 1,061.15 | 1,036.71 | 854.28 | **4,099.39** |
| 231 | 1,669.17 | 1,777.23 | 1,451.17 | 1,509.00 | **6,406.58** |
| 232 | 3,831.41 | 3,726.09 | 3,666.69 | 3,176.19 | **14,400.37** |
| 233 | 1,292.82 | 1,189.88 | 1,188.75 | 1,294.46 | **4,965.91** |
| 234 | 3,432.65 | 3,309.04 | 3,329.95 | 3,193.66 | **13,265.30** |
| 235 | 1,335.54 | 1,329.49 | 1,101.17 | 1,210.60 | **4,976.79** |
| 236 | 2,653.97 | 2,423.10 | 2,320.88 | 2,199.45 | **9,597.41** |
| 237 | 943.84 | 593.14 | 760.63 | 630.22 | **2,927.83** |
| 238 | 2,056.11 | 1,846.38 | 1,709.14 | 1,824.95 | **7,436.58** |
| 239 | 440.30 | 421.84 | 412.73 | 425.99 | **1,700.86** |
| 240 | 1,537.74 | 1,687.54 | 1,352.75 | 1,354.52 | **5,932.55** |
| 241 | 1,433.97 | 1,158.61 | 913.37 | 921.60 | **4,427.55** |
| 242 | 1,577.89 | 1,447.41 | 1,297.33 | 1,293.25 | **5,615.88** |
| 243 | 983.70 | 886.70 | 718.48 | 758.43 | **3,347.31** |
| 244 | 630.47 | 507.34 | 572.27 | 443.45 | **2,153.54** |
| 245 | 1,685.09 | 1,486.85 | 1,669.73 | 1,390.18 | **6,231.86** |
| 246 | 2,043.33 | 1,803.56 | 1,964.78 | 1,664.21 | **7,475.87** |
| **VA - VIRGINIA - Total** | **50,351.99** | **45,915.13** | **45,132.92** | **43,161.77** | **184,561.82** |

**STATE: VI - VIRGIN ISLANDS**

| REGISTRANT ZIP CODE 3 | 1ST QUARTER | 2ND QUARTER | 3RD QUARTER | 4TH QUARTER | **TOTAL GRAMS** |
|---|---|---|---|---|---|
| 008 | 634.30 | 834.39 | 457.64 | 852.07 | **2,778.39** |
| **VI - VIRGIN ISLANDS - Total** | **634.30** | **834.39** | **457.64** | **852.07** | **2,778.39** |

**STATE: VT - VERMONT**

| REGISTRANT ZIP CODE 3 | 1ST QUARTER | 2ND QUARTER | 3RD QUARTER | 4TH QUARTER | **TOTAL GRAMS** |
|---|---|---|---|---|---|
| 050 | 179.58 | 296.26 | 188.69 | 219.30 | **883.83** |
| 051 | 60.04 | 106.81 | 103.87 | 55.43 | **326.16** |
| 052 | 153.41 | 214.00 | 148.44 | 142.17 | **658.03** |
| 053 | 133.52 | 132.32 | 97.79 | 192.08 | **555.71** |
| 054 | 1,074.57 | 1,053.94 | 1,071.62 | 1,103.10 | **4,303.23** |
| 056 | 473.85 | 408.45 | 435.73 | 409.20 | **1,727.23** |
| 057 | 365.58 | 366.12 | 393.93 | 292.83 | **1,418.46** |
| 058 | 262.96 | 268.88 | 251.19 | 231.29 | **1,014.32** |
| **VT - VERMONT - Total** | **2,703.50** | **2,846.79** | **2,691.26** | **2,645.41** | **10,886.96** |

**STATE: WA - WASHINGTON**

| REGISTRANT ZIP CODE 3 | 1ST QUARTER | 2ND QUARTER | 3RD QUARTER | 4TH QUARTER | **TOTAL GRAMS** |
|---|---|---|---|---|---|
| 980 | 9,946.97 | 10,066.06 | 9,929.70 | 8,769.90 | **38,712.62** |

| | | | | | |
|---|---|---|---|---|---|
| 981 | 6,688.91 | 5,911.59 | 6,064.47 | 5,938.80 | **24,603.76** |
| 982 | 7,716.62 | 7,502.21 | 7,029.81 | 6,565.89 | **28,814.53** |
| 983 | 5,951.15 | 5,495.09 | 5,615.39 | 5,285.13 | **22,346.76** |
| 984 | 4,034.34 | 4,017.66 | 3,639.32 | 3,527.76 | **15,219.09** |
| 985 | 3,808.33 | 3,420.26 | 3,583.40 | 3,398.43 | **14,210.41** |
| 986 | 4,605.56 | 4,309.83 | 4,580.46 | 4,096.29 | **17,592.15** |
| 988 | 1,703.11 | 1,262.91 | 1,593.02 | 1,457.32 | **6,016.35** |
| 989 | 2,049.16 | 1,996.83 | 2,024.18 | 1,682.19 | **7,752.36** |
| 990 | 1,093.06 | 1,077.02 | 982.92 | 989.75 | **4,142.76** |
| 991 | 1,114.40 | 1,072.10 | 1,204.62 | 976.58 | **4,367.70** |
| 992 | 5,459.19 | 5,146.61 | 4,819.44 | 4,611.26 | **20,036.50** |
| 993 | 2,505.82 | 2,090.95 | 2,191.71 | 2,107.80 | **8,896.28** |
| 994 | 516.79 | 548.10 | 543.68 | 505.01 | **2,113.58** |
| **WA - WASHINGTON - Total** | **57,193.39** | **53,917.21** | **53,802.13** | **49,912.10** | **214,824.84** |

**STATE: WI - WISCONSIN**

| REGISTRANT ZIP CODE 3 | 1ST QUARTER | 2ND QUARTER | 3RD QUARTER | 4TH QUARTER | **TOTAL GRAMS** |
|---|---|---|---|---|---|
| 530 | 4,596.31 | 4,320.35 | 4,152.32 | 4,209.23 | **17,278.21** |
| 531 | 5,031.45 | 4,433.85 | 4,621.77 | 4,524.39 | **18,611.46** |
| 532 | 8,848.97 | 8,615.87 | 9,245.08 | 8,813.02 | **35,522.95** |
| 534 | 1,325.31 | 1,404.69 | 1,298.14 | 1,223.46 | **5,251.59** |
| 535 | 3,790.56 | 3,953.66 | 3,422.20 | 3,527.04 | **14,693.46** |
| 537 | 1,860.32 | 1,760.72 | 1,628.63 | 1,535.69 | **6,785.35** |
| 538 | 556.39 | 328.32 | 427.45 | 409.40 | **1,721.56** |
| 539 | 1,711.25 | 1,262.97 | 1,464.36 | 1,416.66 | **5,855.25** |
| 540 | 688.04 | 663.31 | 800.95 | 731.48 | **2,883.79** |
| 541 | 1,256.29 | 1,031.73 | 1,320.56 | 1,234.22 | **4,842.80** |
| 542 | 964.08 | 833.67 | 968.95 | 910.35 | **3,677.05** |
| 543 | 1,789.12 | 1,889.01 | 1,695.07 | 1,672.84 | **7,046.03** |
| 544 | 2,932.30 | 2,677.96 | 2,758.82 | 2,466.05 | **10,835.13** |
| 545 | 995.46 | 980.63 | 867.97 | 828.67 | **3,672.73** |
| 546 | 1,211.83 | 1,050.19 | 1,053.36 | 1,021.92 | **4,337.31** |
| 547 | 1,541.42 | 1,297.72 | 1,216.13 | 1,193.21 | **5,248.48** |
| 548 | 1,435.74 | 1,655.70 | 1,498.25 | 1,402.65 | **5,992.33** |
| 549 | 3,754.14 | 3,588.52 | 3,357.82 | 3,205.74 | **13,906.22** |
| **WI - WISCONSIN - Total** | **44,288.96** | **41,748.88** | **41,797.83** | **40,326.02** | **168,161.69** |

**STATE: WV - WEST VIRGINIA**

| REGISTRANT ZIP CODE 3 | 1ST QUARTER | 2ND QUARTER | 3RD QUARTER | 4TH QUARTER | **TOTAL GRAMS** |
|---|---|---|---|---|---|
| 247 | 1,109.45 | 1,214.13 | 1,039.11 | 854.57 | **4,217.25** |
| 248 | 3,121.69 | 4,203.79 | 3,660.02 | 3,788.11 | **14,773.61** |
| 249 | 464.93 | 326.76 | 430.63 | 357.52 | **1,579.83** |

| | | | | | |
|---|---|---|---|---|---|
| 250 | 1,333.37 | 1,280.91 | 1,421.77 | 1,476.42 | **5,512.47** |
| 251 | 882.88 | 982.49 | 1,191.24 | 1,316.11 | **4,372.72** |
| 252 | 1,356.76 | 1,077.63 | 1,222.19 | 1,248.71 | **4,905.28** |
| 253 | 2,086.15 | 2,047.32 | 2,351.59 | 2,071.39 | **8,556.44** |
| 254 | 1,306.66 | 1,054.27 | 1,221.25 | 1,044.17 | **4,626.35** |
| 255 | 2,173.00 | 2,285.92 | 2,287.45 | 2,845.97 | **9,592.35** |
| 256 | 5,645.70 | 6,623.67 | 6,610.77 | 6,922.03 | **25,802.18** |
| 257 | 1,183.34 | 1,199.70 | 1,163.42 | 1,135.72 | **4,682.18** |
| 258 | 2,075.04 | 2,348.95 | 2,172.23 | 2,142.48 | **8,738.70** |
| 259 | 731.53 | 745.35 | 683.47 | 727.49 | **2,887.83** |
| 260 | 1,220.07 | 1,142.77 | 1,195.61 | 1,252.70 | **4,811.16** |
| 261 | 1,531.31 | 1,197.20 | 1,422.81 | 1,254.49 | **5,405.81** |
| 262 | 2,036.41 | 1,763.47 | 1,950.95 | 1,723.87 | **7,474.69** |
| 263 | 1,244.50 | 1,227.06 | 1,193.44 | 1,064.98 | **4,729.99** |
| 264 | 392.47 | 349.38 | 354.16 | 461.91 | **1,557.93** |
| 265 | 2,973.46 | 2,975.49 | 2,699.54 | 2,949.90 | **11,598.40** |
| 266 | 1,160.30 | 972.44 | 1,109.47 | 912.77 | **4,154.99** |
| 267 | 503.72 | 536.65 | 547.16 | 620.36 | **2,207.89** |
| 268 | 279.04 | 254.53 | 272.02 | 311.62 | **1,117.22** |
| **WV - WEST VIRGINIA - Total** | **34,811.79** | **35,809.87** | **36,200.31** | **36,483.29** | **143,305.26** |

**STATE: WY - WYOMING**

| REGISTRANT ZIP CODE 3 | 1ST QUARTER | 2ND QUARTER | 3RD QUARTER | 4TH QUARTER | TOTAL GRAMS |
|---|---|---|---|---|---|
| 820 | 928.51 | 808.78 | 853.43 | 663.67 | **3,254.38** |
| 822 | 200.01 | 119.35 | 133.71 | 161.52 | **614.59** |
| 823 | 49.95 | 64.09 | 65.20 | 19.89 | **199.13** |
| 824 | 574.58 | 496.78 | 492.83 | 497.82 | **2,062.00** |
| 825 | 335.94 | 269.63 | 393.40 | 329.49 | **1,328.45** |
| 826 | 556.67 | 552.43 | 560.72 | 557.13 | **2,226.94** |
| 827 | 416.41 | 198.97 | 261.52 | 283.80 | **1,160.70** |
| 828 | 240.85 | 225.38 | 297.59 | 234.77 | **998.58** |
| 829 | 545.89 | 445.34 | 468.54 | 384.56 | **1,844.33** |
| 830 | 130.37 | 106.08 | 119.71 | 13.26 | **369.43** |
| 831 | 72.93 | 83.98 | 46.19 | 68.60 | **271.71** |
| **WY - WYOMING - Total** | **4,052.12** | **3,370.81** | **3,692.82** | **3,214.50** | **14,330.25** |

**DRUG: 9064 - BUPRENORPHINE**

**STATE: AK - ALASKA**

| REGISTRANT ZIP CODE 3 | 1ST QUARTER | 2ND QUARTER | 3RD QUARTER | 4TH QUARTER | TOTAL GRAMS |
|---|---|---|---|---|---|
| 995 | 1,143.68 | 1,166.34 | 1,117.42 | 1,069.35 | **4,496.79** |
| 996 | 726.47 | 740.25 | 736.86 | 749.49 | **2,953.06** |
| 997 | 222.24 | 232.80 | 227.25 | 248.65 | **930.95** |

| | | | | | |
|---|---|---|---|---|---|
| 998 | 301.39 | 329.81 | 363.65 | 379.86 | **1,374.70** |
| 999 | 190.06 | 201.08 | 198.54 | 219.21 | **808.89** |
| **AK - ALASKA - Total** | **2,583.83** | **2,670.28** | **2,643.71** | **2,666.55** | **10,564.38** |

**STATE: AL - ALABAMA**

| REGISTRANT ZIP CODE 3 | 1ST QUARTER | 2ND QUARTER | 3RD QUARTER | 4TH QUARTER | TOTAL GRAMS |
|---|---|---|---|---|---|
| 350 | 3,985.66 | 4,270.03 | 4,267.57 | 4,486.84 | **17,010.10** |
| 351 | 2,265.30 | 2,288.28 | 2,348.97 | 2,461.58 | **9,364.13** |
| 352 | 3,461.53 | 3,498.53 | 3,591.37 | 3,483.22 | **14,034.65** |
| 354 | 463.89 | 477.45 | 484.11 | 531.11 | **1,956.56** |
| 355 | 1,910.56 | 2,109.76 | 2,356.77 | 2,553.53 | **8,930.62** |
| 356 | 2,967.22 | 3,081.98 | 3,113.78 | 3,267.76 | **12,430.75** |
| 357 | 920.19 | 975.84 | 1,050.16 | 1,096.03 | **4,042.22** |
| 358 | 1,234.79 | 1,278.58 | 1,325.02 | 1,365.01 | **5,203.40** |
| 359 | 2,617.81 | 3,062.87 | 3,166.36 | 3,269.82 | **12,116.86** |
| 360 | 626.49 | 661.83 | 693.48 | 721.98 | **2,703.78** |
| 361 | 351.08 | 387.31 | 386.34 | 427.90 | **1,552.63** |
| 362 | 999.74 | 1,067.01 | 1,142.82 | 1,139.31 | **4,348.88** |
| 363 | 745.87 | 827.04 | 826.02 | 855.26 | **3,254.20** |
| 364 | 391.55 | 387.12 | 389.88 | 391.98 | **1,560.54** |
| 365 | 1,854.00 | 1,934.67 | 1,988.99 | 2,043.57 | **7,821.23** |
| 366 | 1,298.41 | 1,443.12 | 1,422.50 | 1,564.06 | **5,728.09** |
| 367 | 245.96 | 266.41 | 239.39 | 257.93 | **1,009.69** |
| 368 | 546.91 | 613.30 | 622.77 | 659.33 | **2,442.32** |
| 369 | 31.29 | 29.31 | 36.04 | 40.86 | **137.50** |
| **AL - ALABAMA - Total** | **26,918.25** | **28,660.45** | **29,452.35** | **30,617.09** | **115,648.15** |

**STATE: AR - ARKANSAS**

| REGISTRANT ZIP CODE 3 | 1ST QUARTER | 2ND QUARTER | 3RD QUARTER | 4TH QUARTER | TOTAL GRAMS |
|---|---|---|---|---|---|
| 716 | 121.01 | 141.69 | 128.77 | 129.14 | **520.61** |
| 717 | 175.80 | 193.35 | 207.44 | 204.05 | **780.64** |
| 718 | 213.63 | 226.65 | 253.25 | 243.75 | **937.28** |
| 719 | 870.94 | 930.73 | 910.38 | 936.91 | **3,648.96** |
| 720 | 683.91 | 713.88 | 711.27 | 695.02 | **2,804.08** |
| 721 | 1,069.05 | 1,123.70 | 1,153.36 | 1,155.48 | **4,501.58** |
| 722 | 696.71 | 764.04 | 820.24 | 899.74 | **3,180.72** |
| 723 | 146.71 | 163.91 | 144.81 | 149.79 | **605.23** |
| 724 | 730.95 | 862.54 | 879.11 | 963.81 | **3,436.40** |
| 725 | 217.85 | 263.62 | 274.90 | 305.42 | **1,061.79** |
| 726 | 268.16 | 325.86 | 357.64 | 390.85 | **1,342.52** |
| 727 | 837.53 | 938.33 | 1,008.71 | 1,018.40 | **3,802.97** |
| 728 | 136.97 | 176.35 | 168.11 | 182.07 | **663.50** |

| | | | | | |
|---|---|---|---|---|---|
| 729 | 396.40 | 424.38 | 468.37 | 498.83 | **1,787.98** |
| **AR - ARKANSAS - Total** | **6,565.61** | **7,249.03** | **7,486.34** | **7,773.26** | **29,074.25** |

**STATE: AS - AMERICAN SAMOA**

| REGISTRANT ZIP CODE 3 | 1ST QUARTER | 2ND QUARTER | 3RD QUARTER | 4TH QUARTER | TOTAL GRAMS |
|---|---|---|---|---|---|
| 967 | 0.02 | 0.00 | 0.02 | 0 | **0.04** |
| **AS - AMERICAN SAMOA - Tota** | **0.02** | **0.00** | **0.02** | **0.00** | **0.04** |

**STATE: AZ - ARIZONA**

| REGISTRANT ZIP CODE 3 | 1ST QUARTER | 2ND QUARTER | 3RD QUARTER | 4TH QUARTER | TOTAL GRAMS |
|---|---|---|---|---|---|
| 850 | 2,501.98 | 2,763.59 | 2,996.39 | 3,235.63 | **11,497.60** |
| 851 | 641.55 | 680.67 | 744.81 | 668.17 | **2,735.20** |
| 852 | 2,739.25 | 2,847.19 | 3,002.00 | 3,189.57 | **11,778.02** |
| 853 | 1,592.58 | 1,725.58 | 1,846.73 | 1,972.75 | **7,137.64** |
| 855 | 108.98 | 132.63 | 147.27 | 169.33 | **558.20** |
| 856 | 104.59 | 123.24 | 152.54 | 152.71 | **533.08** |
| 857 | 1,354.48 | 1,503.43 | 1,543.78 | 1,774.78 | **6,176.47** |
| 859 | 154.68 | 170.75 | 178.57 | 192.43 | **696.44** |
| 860 | 111.38 | 117.42 | 144.13 | 147.29 | **520.22** |
| 863 | 517.84 | 599.90 | 647.35 | 736.88 | **2,501.97** |
| 864 | 162.47 | 194.70 | 203.37 | 269.56 | **830.10** |
| 865 | 17.64 | 20.64 | 19.02 | 24.48 | **81.78** |
| **AZ - ARIZONA - Total** | **10,007.42** | **10,879.74** | **11,625.98** | **12,533.59** | **45,046.72** |

**STATE: CA - CALIFORNIA**

| REGISTRANT ZIP CODE 3 | 1ST QUARTER | 2ND QUARTER | 3RD QUARTER | 4TH QUARTER | TOTAL GRAMS |
|---|---|---|---|---|---|
| 900 | 998.17 | 994.76 | 1,100.75 | 1,141.25 | **4,234.93** |
| 902 | 836.30 | 841.03 | 859.06 | 956.93 | **3,493.32** |
| 903 | 31.00 | 31.90 | 47.18 | 35.78 | **145.86** |
| 904 | 222.05 | 240.93 | 233.91 | 216.45 | **913.34** |
| 905 | 190.73 | 193.15 | 197.55 | 191.54 | **772.98** |
| 906 | 294.77 | 281.98 | 314.30 | 277.33 | **1,168.38** |
| 907 | 643.03 | 660.95 | 702.73 | 645.52 | **2,652.22** |
| 908 | 328.72 | 343.95 | 334.71 | 345.88 | **1,353.26** |
| 910 | 115.69 | 119.88 | 118.55 | 122.75 | **476.88** |
| 911 | 178.43 | 191.70 | 168.10 | 181.03 | **719.27** |
| 912 | 287.37 | 269.94 | 299.26 | 284.36 | **1,140.93** |
| 913 | 1,186.72 | 1,275.76 | 1,352.25 | 1,330.66 | **5,145.39** |
| 914 | 527.75 | 541.86 | 559.09 | 603.96 | **2,232.65** |
| 915 | 214.47 | 216.51 | 220.97 | 207.72 | **859.68** |
| 916 | 131.43 | 128.88 | 138.84 | 159.38 | **558.53** |
| 917 | 871.79 | 858.73 | 919.88 | 896.17 | **3,546.57** |
| 918 | 23.08 | 5.98 | 6.11 | 8.36 | **43.53** |

| | | | | | |
|---|---|---|---|---|---|
| 919 | 243.83 | 246.60 | 260.29 | 293.85 | **1,044.57** |
| 920 | 1,441.73 | 1,483.74 | 1,632.20 | 1,627.79 | **6,185.47** |
| 921 | 1,195.82 | 1,228.16 | 1,273.62 | 1,338.97 | **5,036.56** |
| 922 | 388.70 | 416.42 | 417.17 | 521.02 | **1,743.31** |
| 923 | 451.17 | 498.92 | 526.90 | 598.68 | **2,075.66** |
| 924 | 55.08 | 65.95 | 60.74 | 66.45 | **248.23** |
| 925 | 1,401.21 | 1,452.01 | 1,622.48 | 1,636.94 | **6,112.65** |
| 926 | 1,791.38 | 1,869.50 | 1,892.81 | 1,976.94 | **7,530.64** |
| 927 | 335.87 | 344.12 | 333.22 | 352.54 | **1,365.75** |
| 928 | 822.93 | 866.70 | 880.57 | 865.49 | **3,435.69** |
| 930 | 668.32 | 658.11 | 704.80 | 678.25 | **2,709.49** |
| 931 | 453.73 | 476.23 | 500.97 | 560.83 | **1,991.76** |
| 932 | 398.37 | 412.13 | 431.07 | 466.73 | **1,708.30** |
| 933 | 282.53 | 264.46 | 285.68 | 275.07 | **1,107.73** |
| 934 | 874.69 | 961.72 | 920.04 | 828.95 | **3,585.40** |
| 935 | 482.96 | 523.19 | 507.78 | 523.88 | **2,037.81** |
| 936 | 180.26 | 176.65 | 191.96 | 200.20 | **749.07** |
| 937 | 299.63 | 322.71 | 333.79 | 357.59 | **1,313.72** |
| 939 | 686.97 | 713.97 | 742.98 | 784.63 | **2,928.56** |
| 940 | 677.37 | 726.59 | 774.24 | 827.16 | **3,005.36** |
| 941 | 1,205.09 | 1,363.64 | 1,306.13 | 1,465.62 | **5,340.48** |
| 943 | 115.04 | 119.87 | 122.34 | 150.23 | **507.48** |
| 944 | 145.74 | 153.13 | 184.93 | 192.66 | **676.45** |
| 945 | 2,766.32 | 2,903.85 | 3,157.50 | 3,353.07 | **12,180.75** |
| 946 | 403.86 | 462.36 | 533.23 | 587.77 | **1,987.23** |
| 947 | 115.99 | 137.52 | 129.92 | 133.39 | **516.82** |
| 948 | 125.92 | 137.70 | 143.44 | 176.92 | **583.98** |
| 949 | 901.07 | 919.80 | 955.99 | 980.00 | **3,756.86** |
| 950 | 1,080.50 | 1,129.00 | 1,188.56 | 1,299.94 | **4,698.00** |
| 951 | 666.10 | 685.01 | 738.62 | 813.17 | **2,902.89** |
| 952 | 360.56 | 364.97 | 355.38 | 375.96 | **1,456.87** |
| 953 | 1,074.31 | 1,132.12 | 1,195.87 | 1,299.49 | **4,701.78** |
| 954 | 2,272.76 | 2,333.19 | 2,494.17 | 2,559.53 | **9,659.65** |
| 955 | 1,370.63 | 1,484.58 | 1,576.59 | 1,642.22 | **6,074.02** |
| 956 | 2,273.00 | 2,377.76 | 2,571.36 | 2,651.32 | **9,873.44** |
| 957 | 301.73 | 308.39 | 313.01 | 326.66 | **1,249.79** |
| 958 | 2,154.44 | 2,174.50 | 2,295.19 | 2,468.78 | **9,092.91** |
| 959 | 1,411.80 | 1,490.64 | 1,585.44 | 1,799.17 | **6,287.05** |
| 960 | 1,023.18 | 1,090.88 | 1,080.34 | 1,174.66 | **4,369.06** |
| 961 | 151.22 | 167.88 | 171.53 | 220.34 | **710.97** |

| CA - CALIFORNIA - Total | 40,133.32 | 41,842.59 | 43,966.11 | 46,057.94 | 171,999.96 |
|---|---|---|---|---|---|

**STATE: CO - COLORADO**

| REGISTRANT ZIP CODE 3 | 1ST QUARTER | 2ND QUARTER | 3RD QUARTER | 4TH QUARTER | TOTAL GRAMS |
|---|---|---|---|---|---|
| 800 | 1,266.45 | 1,357.17 | 1,483.34 | 1,649.21 | **5,756.16** |
| 801 | 815.65 | 819.89 | 850.25 | 879.82 | **3,365.61** |
| 802 | 1,853.48 | 1,888.37 | 1,902.09 | 2,092.20 | **7,736.13** |
| 803 | 130.62 | 124.80 | 139.86 | 142.55 | **537.83** |
| 804 | 147.16 | 152.27 | 150.47 | 165.99 | **615.90** |
| 805 | 1,014.77 | 1,099.97 | 1,207.23 | 1,314.85 | **4,636.83** |
| 806 | 509.19 | 527.63 | 558.07 | 594.82 | **2,189.71** |
| 807 | 58.40 | 55.95 | 53.09 | 57.04 | **224.49** |
| 808 | 108.08 | 111.50 | 117.53 | 110.03 | **447.15** |
| 809 | 864.35 | 985.92 | 1,018.05 | 1,087.92 | **3,956.24** |
| 810 | 567.76 | 638.52 | 657.26 | 762.02 | **2,625.56** |
| 811 | 80.89 | 78.28 | 87.29 | 98.11 | **344.56** |
| 812 | 128.10 | 127.89 | 147.89 | 156.92 | **560.80** |
| 813 | 134.23 | 127.91 | 139.01 | 185.20 | **586.35** |
| 814 | 57.06 | 54.48 | 50.62 | 65.05 | **227.21** |
| 815 | 180.71 | 242.69 | 206.93 | 227.69 | **858.02** |
| 816 | 118.17 | 122.62 | 148.11 | 145.80 | **534.70** |
| **CO - COLORADO - Total** | **8,035.07** | **8,515.86** | **8,917.11** | **9,735.21** | **35,203.25** |

**STATE: CT - CONNECTICUT**

| REGISTRANT ZIP CODE 3 | 1ST QUARTER | 2ND QUARTER | 3RD QUARTER | 4TH QUARTER | TOTAL GRAMS |
|---|---|---|---|---|---|
| 060 | 3,261.83 | 3,289.13 | 3,305.60 | 3,450.14 | **13,306.70** |
| 061 | 1,498.61 | 1,549.74 | 1,583.20 | 1,677.51 | **6,309.05** |
| 062 | 788.04 | 832.54 | 835.15 | 887.13 | **3,342.85** |
| 063 | 1,468.05 | 1,599.35 | 1,624.98 | 1,840.89 | **6,533.28** |
| 064 | 3,691.24 | 3,837.94 | 3,703.45 | 4,025.59 | **15,258.21** |
| 065 | 2,146.69 | 2,316.21 | 2,191.33 | 2,239.29 | **8,893.52** |
| 066 | 284.39 | 315.81 | 324.29 | 378.77 | **1,303.26** |
| 067 | 1,496.79 | 1,592.47 | 1,597.59 | 1,770.14 | **6,456.99** |
| 068 | 694.01 | 747.03 | 709.80 | 766.23 | **2,917.07** |
| 069 | 180.27 | 198.24 | 195.20 | 193.53 | **767.24** |
| **CT - CONNECTICUT - Total** | **15,509.93** | **16,278.45** | **16,070.59** | **17,229.21** | **65,088.17** |

**STATE: DC - DISTRICT OF COLUMBIA**

| REGISTRANT ZIP CODE 3 | 1ST QUARTER | 2ND QUARTER | 3RD QUARTER | 4TH QUARTER | TOTAL GRAMS |
|---|---|---|---|---|---|
| 200 | 2,719.42 | 2,715.84 | 2,816.64 | 3,231.05 | **11,482.96** |
| 204 | 31.92 | 14.88 | 31.60 | 27.96 | **106.36** |
| **DC - DISTRICT OF COLUMBIA -** | **2,751.34** | **2,730.72** | **2,848.24** | **3,259.01** | **11,589.32** |

**STATE: DE - DELAWARE**

| REGISTRANT ZIP CODE 3 | 1ST QUARTER | 2ND QUARTER | 3RD QUARTER | 4TH QUARTER | TOTAL GRAMS |
|---|---|---|---|---|---|
| 197 | 1,073.25 | 1,165.75 | 1,109.59 | 1,169.58 | **4,518.17** |
| 198 | 987.88 | 1,054.75 | 1,024.89 | 1,056.55 | **4,124.06** |
| 199 | 1,690.21 | 1,842.89 | 1,832.83 | 1,897.44 | **7,263.37** |
| **DE - DELAWARE - Total** | **3,751.34** | **4,063.38** | **3,967.31** | **4,123.57** | **15,905.60** |

**STATE: FL - FLORIDA**

| REGISTRANT ZIP CODE 3 | 1ST QUARTER | 2ND QUARTER | 3RD QUARTER | 4TH QUARTER | TOTAL GRAMS |
|---|---|---|---|---|---|
| 320 | 3,083.10 | 3,176.99 | 3,236.22 | 3,245.76 | **12,742.08** |
| 321 | 2,323.34 | 2,490.04 | 2,637.47 | 2,867.37 | **10,318.24** |
| 322 | 3,764.56 | 4,110.90 | 4,260.25 | 4,454.43 | **16,590.15** |
| 323 | 704.91 | 712.08 | 651.88 | 723.59 | **2,792.46** |
| 324 | 900.91 | 1,011.24 | 1,026.82 | 996.66 | **3,935.63** |
| 325 | 2,135.14 | 2,403.81 | 2,475.08 | 2,494.39 | **9,508.43** |
| 326 | 2,606.10 | 2,834.44 | 3,085.25 | 3,078.10 | **11,603.88** |
| 327 | 2,638.74 | 2,802.32 | 2,729.55 | 2,785.45 | **10,956.05** |
| 328 | 1,133.08 | 1,212.37 | 1,279.23 | 1,331.69 | **4,956.38** |
| 329 | 2,908.70 | 3,079.80 | 3,150.77 | 3,313.62 | **12,452.89** |
| 330 | 1,590.00 | 1,602.53 | 1,682.70 | 1,678.78 | **6,554.01** |
| 331 | 835.69 | 881.40 | 859.22 | 902.00 | **3,478.31** |
| 333 | 1,246.12 | 1,249.67 | 1,391.97 | 1,368.51 | **5,256.27** |
| 334 | 3,807.47 | 3,939.72 | 4,072.17 | 4,207.22 | **16,026.58** |
| 335 | 1,477.09 | 1,497.48 | 1,485.55 | 1,556.93 | **6,017.05** |
| 336 | 1,462.87 | 1,445.53 | 1,455.84 | 1,546.93 | **5,911.18** |
| 337 | 2,332.21 | 2,497.49 | 2,551.32 | 2,654.33 | **10,035.35** |
| 338 | 965.79 | 1,020.30 | 1,052.46 | 1,098.64 | **4,137.19** |
| 339 | 2,653.07 | 2,601.90 | 2,601.22 | 2,742.62 | **10,598.81** |
| 341 | 971.47 | 1,034.43 | 1,086.98 | 1,191.27 | **4,284.14** |
| 342 | 3,534.46 | 3,687.54 | 3,615.47 | 3,851.66 | **14,689.13** |
| 344 | 1,384.29 | 1,510.79 | 1,510.38 | 1,574.32 | **5,979.78** |
| 346 | 1,988.83 | 2,130.79 | 2,145.41 | 2,279.69 | **8,544.72** |
| 347 | 1,085.53 | 1,111.71 | 1,150.50 | 1,254.75 | **4,602.50** |
| 349 | 1,468.92 | 1,544.17 | 1,548.01 | 1,585.78 | **6,146.88** |
| **FL - FLORIDA - Total** | **49,002.40** | **51,589.45** | **52,741.76** | **54,784.49** | **208,118.09** |

**STATE: GA - GEORGIA**

| REGISTRANT ZIP CODE 3 | 1ST QUARTER | 2ND QUARTER | 3RD QUARTER | 4TH QUARTER | TOTAL GRAMS |
|---|---|---|---|---|---|
| 300 | 2,455.21 | 2,504.83 | 2,481.27 | 2,611.36 | **10,052.67** |
| 301 | 2,242.72 | 2,258.58 | 2,335.48 | 2,436.06 | **9,272.84** |
| 302 | 1,438.70 | 1,521.14 | 1,526.06 | 1,587.75 | **6,073.65** |
| 303 | 629.83 | 701.09 | 687.51 | 759.70 | **2,778.13** |
| 304 | 746.52 | 736.60 | 766.86 | 749.97 | **2,999.95** |

| | | | | | |
|---|---|---|---|---|---|
| 305 | 930.19 | 999.20 | 1,006.52 | 1,085.47 | **4,021.38** |
| 306 | 837.25 | 933.20 | 953.31 | 985.92 | **3,709.68** |
| 307 | 565.53 | 666.46 | 865.47 | 1,143.79 | **3,241.25** |
| 308 | 107.22 | 117.55 | 120.19 | 130.30 | **475.26** |
| 309 | 350.14 | 345.47 | 383.00 | 397.66 | **1,476.27** |
| 310 | 1,112.49 | 1,147.32 | 1,055.15 | 1,119.93 | **4,434.89** |
| 312 | 388.89 | 430.43 | 468.97 | 468.16 | **1,756.45** |
| 313 | 715.64 | 796.18 | 813.19 | 764.95 | **3,089.96** |
| 314 | 837.01 | 844.92 | 826.56 | 861.85 | **3,370.35** |
| 315 | 1,797.97 | 2,005.00 | 2,065.33 | 2,139.88 | **8,008.18** |
| 316 | 156.44 | 188.20 | 196.46 | 192.47 | **733.56** |
| 317 | 449.39 | 533.35 | 509.48 | 546.56 | **2,038.78** |
| 318 | 18.67 | 19.87 | 14.60 | 11.99 | **65.14** |
| 319 | 288.58 | 276.78 | 269.59 | 295.48 | **1,130.44** |
| 398 | 80.67 | 95.35 | 111.55 | 133.50 | **421.07** |
| **GA - GEORGIA - Total** | **16,149.07** | **17,121.52** | **17,456.56** | **18,422.76** | **69,149.91** |

**STATE: GU - GUAM**

| REGISTRANT ZIP CODE 3 | 1ST QUARTER | 2ND QUARTER | 3RD QUARTER | 4TH QUARTER | TOTAL GRAMS |
|---|---|---|---|---|---|
| 969 | 0.62 | 0.25 | 1.44 | 0.00 | **2.31** |
| **GU - GUAM - Total** | **0.62** | **0.25** | **1.44** | **0.00** | **2.31** |

**STATE: HI - HAWAII**

| REGISTRANT ZIP CODE 3 | 1ST QUARTER | 2ND QUARTER | 3RD QUARTER | 4TH QUARTER | TOTAL GRAMS |
|---|---|---|---|---|---|
| 967 | 1,588.15 | 1,633.64 | 1,726.50 | 1,799.94 | **6,748.24** |
| 968 | 452.97 | 448.42 | 454.78 | 490.10 | **1,846.26** |
| **HI - HAWAII - Total** | **2,041.12** | **2,082.06** | **2,181.28** | **2,290.04** | **8,594.50** |

**STATE: IA - IOWA**

| REGISTRANT ZIP CODE 3 | 1ST QUARTER | 2ND QUARTER | 3RD QUARTER | 4TH QUARTER | TOTAL GRAMS |
|---|---|---|---|---|---|
| 500 | 90.85 | 91.21 | 96.60 | 102.13 | **380.79** |
| 501 | 59.66 | 72.40 | 87.93 | 113.88 | **333.86** |
| 502 | 77.93 | 79.12 | 92.80 | 90.26 | **340.11** |
| 503 | 416.79 | 368.32 | 373.60 | 364.23 | **1,522.94** |
| 504 | 169.93 | 184.06 | 194.23 | 211.46 | **759.68** |
| 505 | 63.19 | 61.55 | 78.33 | 70.43 | **273.50** |
| 506 | 16.43 | 33.23 | 41.35 | 50.33 | **141.34** |
| 507 | 61.24 | 61.80 | 49.22 | 58.03 | **230.28** |
| 508 | 4.46 | 9.03 | 7.04 | 6.52 | **27.05** |
| 510 | 100.16 | 95.83 | 105.64 | 107.06 | **408.69** |
| 511 | 140.70 | 161.52 | 169.56 | 182.58 | **654.36** |
| 512 | 43.02 | 39.81 | 43.43 | 30.08 | **156.34** |
| 513 | 20.30 | 22.67 | 33.73 | 37.96 | **114.66** |

| | | | | | |
|---|---|---|---|---|---|
| 514 | 11.42 | 18.15 | 18.96 | 15.50 | **64.03** |
| 515 | 63.60 | 72.49 | 74.86 | 74.73 | **285.69** |
| 516 | 5.33 | 5.53 | 5.64 | 5.89 | **22.39** |
| 520 | 61.61 | 59.96 | 64.40 | 61.01 | **246.99** |
| 521 | 4.11 | 8.40 | 3.63 | 7.58 | **23.72** |
| 522 | 122.03 | 152.04 | 154.41 | 178.50 | **606.97** |
| 523 | 13.69 | 20.02 | 19.63 | 22.92 | **76.26** |
| 524 | 54.14 | 72.41 | 89.69 | 93.34 | **309.57** |
| 525 | 44.21 | 61.70 | 63.07 | 69.16 | **238.15** |
| 526 | 47.45 | 50.75 | 51.56 | 66.86 | **216.62** |
| 527 | 135.32 | 135.25 | 120.29 | 118.29 | **509.14** |
| 528 | 80.12 | 73.31 | 78.41 | 85.30 | **317.13** |
| **IA - IOWA - Total** | **1,907.67** | **2,010.56** | **2,118.01** | **2,224.01** | **8,260.26** |

**STATE: ID - IDAHO**

| REGISTRANT ZIP CODE 3 | 1ST QUARTER | 2ND QUARTER | 3RD QUARTER | 4TH QUARTER | **TOTAL GRAMS** |
|---|---|---|---|---|---|
| 832 | 384.67 | 455.60 | 448.02 | 496.98 | **1,785.28** |
| 833 | 245.94 | 254.46 | 287.68 | 327.49 | **1,115.57** |
| 834 | 224.54 | 230.07 | 217.89 | 224.24 | **896.74** |
| 835 | 373.66 | 417.49 | 422.95 | 431.56 | **1,645.65** |
| 836 | 606.09 | 643.81 | 689.43 | 731.82 | **2,671.13** |
| 837 | 635.63 | 653.50 | 658.64 | 693.01 | **2,640.77** |
| 838 | 1,416.05 | 1,531.50 | 1,544.61 | 1,625.49 | **6,117.65** |
| **ID - IDAHO - Total** | **3,886.57** | **4,186.43** | **4,269.21** | **4,530.59** | **16,872.80** |

**STATE: IL - ILLINOIS**

| REGISTRANT ZIP CODE 3 | 1ST QUARTER | 2ND QUARTER | 3RD QUARTER | 4TH QUARTER | **TOTAL GRAMS** |
|---|---|---|---|---|---|
| 600 | 2,319.84 | 2,453.08 | 2,430.04 | 2,563.14 | **9,766.10** |
| 601 | 1,093.50 | 1,201.68 | 1,213.47 | 1,235.49 | **4,744.13** |
| 602 | 25.33 | 28.90 | 30.44 | 35.84 | **120.51** |
| 603 | 299.00 | 292.27 | 313.95 | 346.14 | **1,251.36** |
| 604 | 2,047.05 | 2,066.62 | 2,027.61 | 2,156.90 | **8,298.18** |
| 605 | 981.90 | 1,043.54 | 1,012.06 | 1,086.27 | **4,123.77** |
| 606 | 1,930.76 | 2,063.06 | 2,094.26 | 2,377.31 | **8,465.40** |
| 607 | 94.41 | 95.48 | 99.34 | 88.51 | **377.75** |
| 608 | 64.30 | 60.86 | 70.55 | 72.55 | **268.26** |
| 609 | 191.70 | 184.97 | 193.12 | 217.68 | **787.47** |
| 610 | 208.80 | 222.21 | 216.16 | 232.46 | **879.64** |
| 611 | 640.84 | 691.97 | 740.41 | 742.71 | **2,815.93** |
| 612 | 198.75 | 208.98 | 194.80 | 200.21 | **802.74** |
| 613 | 268.84 | 297.54 | 288.63 | 318.44 | **1,173.45** |
| 614 | 70.79 | 80.47 | 94.01 | 112.93 | **358.20** |

| | | | | | |
|---|---|---|---|---|---|
| 615 | 224.11 | 244.41 | 249.85 | 253.89 | **972.26** |
| 616 | 399.39 | 429.12 | 461.29 | 483.56 | **1,773.36** |
| 617 | 125.08 | 136.88 | 141.96 | 149.00 | **552.93** |
| 618 | 857.67 | 931.07 | 928.72 | 920.51 | **3,637.97** |
| 619 | 97.32 | 99.90 | 92.61 | 90.85 | **380.68** |
| 620 | 372.12 | 405.52 | 403.37 | 411.75 | **1,592.76** |
| 622 | 292.22 | 341.92 | 368.34 | 362.37 | **1,364.85** |
| 623 | 152.83 | 203.47 | 237.91 | 259.19 | **853.40** |
| 624 | 142.59 | 149.76 | 160.89 | 160.53 | **613.76** |
| 625 | 345.61 | 362.39 | 355.80 | 381.74 | **1,445.55** |
| 626 | 190.63 | 225.94 | 231.49 | 244.79 | **892.85** |
| 627 | 362.62 | 365.81 | 374.07 | 401.60 | **1,504.11** |
| 628 | 376.52 | 409.20 | 424.43 | 435.43 | **1,645.59** |
| 629 | 1,225.46 | 1,317.31 | 1,379.41 | 1,464.97 | **5,387.15** |
| **IL - ILLINOIS - Total** | **15,599.99** | **16,614.32** | **16,829.01** | **17,806.76** | **66,850.09** |

**STATE: IN - INDIANA**

| REGISTRANT ZIP CODE 3 | 1ST QUARTER | 2ND QUARTER | 3RD QUARTER | 4TH QUARTER | **TOTAL GRAMS** |
|---|---|---|---|---|---|
| 460 | 2,289.36 | 2,743.97 | 2,576.51 | 2,860.00 | **10,469.85** |
| 461 | 2,194.94 | 2,144.68 | 2,590.53 | 2,728.15 | **9,658.29** |
| 462 | 3,709.22 | 3,957.29 | 3,750.43 | 3,979.64 | **15,396.58** |
| 463 | 2,137.01 | 2,316.60 | 2,256.04 | 2,382.98 | **9,092.63** |
| 464 | 190.70 | 191.42 | 247.96 | 276.30 | **906.39** |
| 465 | 647.27 | 703.35 | 742.81 | 870.74 | **2,964.18** |
| 466 | 225.90 | 247.05 | 279.83 | 282.30 | **1,035.08** |
| 467 | 196.50 | 228.71 | 231.65 | 276.36 | **933.22** |
| 468 | 457.47 | 508.49 | 585.98 | 668.59 | **2,220.53** |
| 469 | 805.54 | 952.32 | 1,075.17 | 1,159.87 | **3,992.89** |
| 470 | 662.29 | 717.44 | 780.85 | 807.66 | **2,968.25** |
| 471 | 1,864.24 | 2,050.61 | 2,174.28 | 2,333.54 | **8,422.66** |
| 472 | 1,452.09 | 1,612.77 | 1,586.45 | 1,704.09 | **6,355.39** |
| 473 | 1,748.59 | 1,911.09 | 2,100.36 | 2,248.57 | **8,008.61** |
| 474 | 1,238.39 | 1,519.80 | 1,743.87 | 1,826.08 | **6,328.13** |
| 475 | 215.92 | 300.39 | 373.79 | 446.82 | **1,336.92** |
| 476 | 96.57 | 104.19 | 118.78 | 189.74 | **509.29** |
| 477 | 559.98 | 691.61 | 717.27 | 868.69 | **2,837.55** |
| 478 | 472.40 | 508.39 | 473.96 | 530.51 | **1,985.26** |
| 479 | 471.10 | 501.67 | 571.83 | 640.74 | **2,185.34** |
| **IN - INDIANA - Total** | **21,635.49** | **23,911.83** | **24,978.36** | **27,081.35** | **97,607.04** |

**STATE: KS - KANSAS**

| REGISTRANT ZIP CODE 3 | 1ST QUARTER | 2ND QUARTER | 3RD QUARTER | 4TH QUARTER | **TOTAL GRAMS** |
|---|---|---|---|---|---|

| | | | | | |
|---|---|---|---|---|---|
| 660 | 532.73 | 553.10 | 572.60 | 636.81 | **2,295.23** |
| 661 | 129.64 | 152.68 | 180.92 | 206.05 | **669.29** |
| 662 | 602.84 | 620.63 | 634.47 | 668.52 | **2,526.46** |
| 664 | 31.26 | 37.73 | 42.23 | 35.70 | **146.92** |
| 665 | 42.02 | 50.19 | 58.51 | 61.83 | **212.54** |
| 666 | 115.34 | 135.22 | 128.19 | 128.34 | **507.09** |
| 667 | 151.43 | 172.69 | 177.39 | 178.89 | **680.40** |
| 668 | 18.69 | 13.35 | 15.08 | 12.70 | **59.83** |
| 669 | 1.06 | 2.92 | 2.97 | 5.02 | **11.97** |
| 670 | 85.93 | 92.58 | 89.51 | 99.08 | **367.10** |
| 671 | 48.85 | 40.89 | 41.62 | 45.68 | **177.05** |
| 672 | 486.78 | 521.15 | 511.05 | 541.86 | **2,060.84** |
| 673 | 29.94 | 35.02 | 37.21 | 51.45 | **153.62** |
| 674 | 107.62 | 161.53 | 150.66 | 166.03 | **585.85** |
| 675 | 54.39 | 62.38 | 62.98 | 66.27 | **246.02** |
| 676 | 47.52 | 66.96 | 60.18 | 63.17 | **237.84** |
| 677 | 3.38 | 5.20 | 7.18 | 8.16 | **23.92** |
| 678 | 60.20 | 56.93 | 57.97 | 62.32 | **237.42** |
| 679 | 19.97 | 19.39 | 14.11 | 15.65 | **69.11** |
| **KS - KANSAS - Total** | **2,569.61** | **2,800.55** | **2,844.81** | **3,053.53** | **11,268.50** |

**STATE: KY - KENTUCKY**

| REGISTRANT ZIP CODE 3 | 1ST QUARTER | 2ND QUARTER | 3RD QUARTER | 4TH QUARTER | **TOTAL GRAMS** |
|---|---|---|---|---|---|
| 400 | 681.85 | 704.02 | 706.54 | 735.60 | **2,828.01** |
| 401 | 233.02 | 295.19 | 332.03 | 357.69 | **1,217.93** |
| 402 | 2,844.59 | 2,955.71 | 2,909.81 | 3,170.53 | **11,880.63** |
| 403 | 4,251.00 | 4,348.35 | 4,423.95 | 4,680.77 | **17,704.07** |
| 404 | 2,325.78 | 2,037.40 | 2,191.43 | 2,317.08 | **8,871.69** |
| 405 | 2,408.83 | 2,581.80 | 2,758.83 | 2,879.07 | **10,628.54** |
| 406 | 555.82 | 580.33 | 575.88 | 633.02 | **2,345.05** |
| 407 | 2,061.92 | 1,912.27 | 2,160.03 | 2,399.36 | **8,533.58** |
| 408 | 381.74 | 392.10 | 515.30 | 575.49 | **1,864.63** |
| 409 | 1,785.71 | 1,645.47 | 1,803.99 | 1,825.02 | **7,060.18** |
| 410 | 2,042.46 | 2,248.09 | 2,162.87 | 2,288.05 | **8,741.47** |
| 411 | 1,697.16 | 1,780.14 | 1,776.01 | 1,857.11 | **7,110.41** |
| 412 | 1,130.41 | 1,190.26 | 1,361.36 | 1,430.34 | **5,112.38** |
| 413 | 899.54 | 1,016.66 | 1,043.90 | 1,039.11 | **3,999.21** |
| 414 | 767.33 | 506.64 | 663.34 | 669.83 | **2,607.14** |
| 415 | 2,852.80 | 2,836.13 | 3,053.45 | 3,158.82 | **11,901.20** |
| 416 | 1,957.51 | 2,027.86 | 1,950.21 | 1,970.59 | **7,906.16** |
| 417 | 1,208.52 | 1,300.33 | 1,340.50 | 1,460.24 | **5,309.59** |

| | | | | | |
|---|---|---|---|---|---|
| 418 | 385.23 | 418.09 | 487.36 | 484.25 | **1,774.94** |
| 420 | 1,260.68 | 1,357.72 | 1,302.28 | 1,462.14 | **5,382.83** |
| 421 | 1,138.53 | 1,060.25 | 1,331.24 | 1,542.56 | **5,072.58** |
| 422 | 258.68 | 298.95 | 346.58 | 385.69 | **1,289.89** |
| 423 | 803.54 | 826.16 | 1,086.34 | 1,138.17 | **3,854.22** |
| 424 | 426.06 | 536.73 | 503.54 | 531.33 | **1,997.66** |
| 425 | 670.07 | 636.67 | 702.79 | 701.56 | **2,711.09** |
| 426 | 786.96 | 752.18 | 839.15 | 773.80 | **3,152.08** |
| 427 | 1,143.55 | 1,216.84 | 1,236.69 | 1,175.64 | **4,772.71** |
| **KY - KENTUCKY - Total** | **36,959.27** | **37,462.35** | **39,565.39** | **41,642.84** | **155,629.85** |

**STATE: LA - LOUISIANA**

| REGISTRANT ZIP CODE 3 | 1ST QUARTER | 2ND QUARTER | 3RD QUARTER | 4TH QUARTER | TOTAL GRAMS |
|---|---|---|---|---|---|
| 700 | 3,905.04 | 4,005.09 | 4,080.66 | 4,027.25 | **16,018.04** |
| 701 | 971.25 | 1,011.55 | 1,032.75 | 1,011.10 | **4,026.66** |
| 703 | 1,057.94 | 1,117.80 | 1,155.03 | 1,152.35 | **4,483.11** |
| 704 | 3,451.46 | 3,688.69 | 3,775.57 | 3,877.84 | **14,793.56** |
| 705 | 2,487.95 | 2,730.54 | 2,646.47 | 2,755.39 | **10,620.35** |
| 706 | 1,251.65 | 1,337.58 | 1,372.67 | 1,391.72 | **5,353.62** |
| 707 | 1,716.89 | 1,859.70 | 1,792.66 | 1,823.70 | **7,192.94** |
| 708 | 950.54 | 931.91 | 974.49 | 947.04 | **3,803.98** |
| 710 | 275.61 | 296.42 | 309.20 | 290.07 | **1,171.31** |
| 711 | 1,702.52 | 1,826.16 | 1,961.72 | 2,094.68 | **7,585.08** |
| 712 | 1,181.10 | 1,229.08 | 1,341.01 | 1,413.16 | **5,164.34** |
| 713 | 600.46 | 682.79 | 795.62 | 798.75 | **2,877.61** |
| 714 | 358.56 | 388.55 | 379.17 | 409.60 | **1,535.88** |
| **LA - LOUISIANA - Total** | **19,910.96** | **21,105.86** | **21,617.02** | **21,992.64** | **84,626.49** |

**STATE: MA - MASSACHUSETTS**

| REGISTRANT ZIP CODE 3 | 1ST QUARTER | 2ND QUARTER | 3RD QUARTER | 4TH QUARTER | TOTAL GRAMS |
|---|---|---|---|---|---|
| 010 | 3,588.74 | 3,641.86 | 3,613.30 | 3,713.77 | **14,557.67** |
| 011 | 1,325.40 | 1,346.09 | 1,359.70 | 1,355.21 | **5,386.39** |
| 012 | 1,345.23 | 1,389.58 | 1,407.26 | 1,525.50 | **5,667.57** |
| 013 | 958.71 | 1,023.16 | 1,019.22 | 1,055.34 | **4,056.42** |
| 014 | 1,135.97 | 1,189.10 | 1,167.74 | 1,275.02 | **4,767.83** |
| 015 | 1,450.17 | 1,601.60 | 1,514.25 | 1,672.42 | **6,238.44** |
| 016 | 2,110.08 | 2,192.21 | 2,097.44 | 2,224.38 | **8,624.11** |
| 017 | 1,415.73 | 1,464.54 | 1,453.81 | 1,503.36 | **5,837.44** |
| 018 | 3,415.46 | 3,609.51 | 3,494.61 | 3,816.88 | **14,336.45** |
| 019 | 2,908.09 | 3,038.33 | 3,001.68 | 3,064.18 | **12,012.28** |
| 020 | 1,638.26 | 1,706.04 | 1,669.35 | 1,783.44 | **6,797.09** |
| 021 | 8,909.13 | 9,360.12 | 9,315.75 | 9,946.34 | **37,531.34** |

| | | | | | |
|---|---|---|---|---|---|
| 022 | 91.79 | 117.52 | 114.11 | 131.08 | **454.51** |
| 023 | 3,061.33 | 3,156.48 | 3,199.20 | 3,316.48 | **12,733.48** |
| 024 | 978.92 | 1,061.43 | 1,022.00 | 1,063.08 | **4,125.43** |
| 025 | 1,046.63 | 1,042.06 | 1,013.52 | 1,050.20 | **4,152.41** |
| 026 | 1,388.95 | 1,397.91 | 1,392.05 | 1,529.33 | **5,708.24** |
| 027 | 5,424.34 | 5,608.17 | 5,523.89 | 5,868.00 | **22,424.40** |
| **MA - MASSACHUSETTS - Total** | **42,192.92** | **43,945.72** | **43,378.87** | **45,894.00** | **175,411.51** |

**STATE: MD - MARYLAND**

| REGISTRANT ZIP CODE 3 | 1ST QUARTER | 2ND QUARTER | 3RD QUARTER | 4TH QUARTER | TOTAL GRAMS |
|---|---|---|---|---|---|
| 206 | 1,750.67 | 1,890.31 | 1,990.22 | 2,138.51 | **7,769.71** |
| 207 | 1,085.34 | 1,106.01 | 1,192.87 | 1,241.01 | **4,625.23** |
| 208 | 931.84 | 997.63 | 977.19 | 1,042.64 | **3,949.30** |
| 209 | 443.00 | 441.11 | 411.04 | 388.92 | **1,684.07** |
| 210 | 3,478.29 | 3,703.15 | 3,724.23 | 4,016.44 | **14,922.12** |
| 211 | 1,515.18 | 1,574.62 | 1,509.44 | 1,606.37 | **6,205.61** |
| 212 | 11,191.52 | 11,697.17 | 11,837.29 | 12,292.56 | **47,018.54** |
| 214 | 569.87 | 580.51 | 603.63 | 742.57 | **2,496.58** |
| 215 | 1,266.76 | 1,354.66 | 1,479.12 | 1,560.39 | **5,660.93** |
| 216 | 761.02 | 794.34 | 841.73 | 873.04 | **3,270.13** |
| 217 | 2,177.15 | 2,306.95 | 2,449.43 | 2,582.58 | **9,516.10** |
| 218 | 832.43 | 862.52 | 865.99 | 932.12 | **3,493.06** |
| 219 | 1,319.09 | 1,320.20 | 1,303.66 | 1,429.17 | **5,372.12** |
| **MD - MARYLAND - Total** | **27,322.15** | **28,629.16** | **29,185.85** | **30,846.33** | **115,983.50** |

**STATE: ME - MAINE**

| REGISTRANT ZIP CODE 3 | 1ST QUARTER | 2ND QUARTER | 3RD QUARTER | 4TH QUARTER | TOTAL GRAMS |
|---|---|---|---|---|---|
| 039 | 65.60 | 71.95 | 79.86 | 82.64 | **300.05** |
| 040 | 2,437.98 | 2,517.17 | 2,591.13 | 2,671.27 | **10,217.56** |
| 041 | 988.86 | 1,036.95 | 1,125.02 | 1,157.32 | **4,308.15** |
| 042 | 1,520.94 | 1,570.47 | 1,626.81 | 1,688.32 | **6,406.54** |
| 043 | 838.40 | 853.45 | 906.42 | 977.97 | **3,576.23** |
| 044 | 1,111.19 | 1,160.61 | 1,247.18 | 1,272.51 | **4,791.48** |
| 045 | 365.94 | 385.09 | 392.03 | 424.65 | **1,567.71** |
| 046 | 699.26 | 729.06 | 804.14 | 807.77 | **3,040.23** |
| 047 | 430.46 | 461.59 | 417.57 | 436.01 | **1,745.64** |
| 048 | 787.14 | 818.54 | 907.74 | 959.37 | **3,472.79** |
| 049 | 1,203.25 | 1,307.35 | 1,429.70 | 1,512.01 | **5,452.32** |
| **ME - MAINE - Total** | **10,449.02** | **10,912.25** | **11,527.59** | **11,989.83** | **44,878.69** |

**STATE: MI - MICHIGAN**

| REGISTRANT ZIP CODE 3 | 1ST QUARTER | 2ND QUARTER | 3RD QUARTER | 4TH QUARTER | TOTAL GRAMS |
|---|---|---|---|---|---|
| 480 | 4,259.79 | 4,347.06 | 4,385.69 | 4,285.07 | **17,277.61** |

| | | | | | |
|---|---|---|---|---|---|
| 481 | 7,923.85 | 8,250.86 | 8,255.75 | 8,764.95 | **33,195.41** |
| 482 | 997.13 | 980.01 | 995.56 | 1,005.65 | **3,978.35** |
| 483 | 3,103.39 | 3,236.34 | 3,162.82 | 3,266.90 | **12,769.45** |
| 484 | 943.50 | 1,028.02 | 1,005.96 | 1,103.71 | **4,081.20** |
| 485 | 620.01 | 708.06 | 749.81 | 784.99 | **2,862.87** |
| 486 | 1,063.29 | 1,115.15 | 1,080.20 | 1,201.96 | **4,460.59** |
| 487 | 654.86 | 721.39 | 677.18 | 729.62 | **2,783.06** |
| 488 | 1,216.04 | 1,286.14 | 1,266.37 | 1,369.25 | **5,137.81** |
| 489 | 210.12 | 211.24 | 218.24 | 262.82 | **902.42** |
| 490 | 1,474.86 | 1,573.76 | 1,577.68 | 1,689.00 | **6,315.29** |
| 491 | 101.85 | 109.00 | 106.04 | 105.26 | **422.14** |
| 492 | 822.25 | 794.26 | 838.86 | 925.00 | **3,380.38** |
| 493 | 407.00 | 408.66 | 413.61 | 453.34 | **1,682.61** |
| 494 | 789.90 | 861.19 | 907.13 | 978.38 | **3,536.60** |
| 495 | 1,018.05 | 1,060.89 | 1,039.07 | 1,083.23 | **4,201.22** |
| 496 | 1,287.53 | 1,316.84 | 1,368.73 | 1,432.91 | **5,406.01** |
| 497 | 1,183.78 | 1,320.60 | 1,301.39 | 1,449.96 | **5,255.72** |
| 498 | 936.20 | 1,008.15 | 972.72 | 1,065.65 | **3,982.72** |
| 499 | 118.97 | 125.78 | 142.33 | 137.78 | **524.86** |
| **MI - MICHIGAN - Total** | **29,132.39** | **30,463.40** | **30,465.10** | **32,095.43** | **122,156.32** |

**STATE: MN - MINNESOTA**

| REGISTRANT ZIP CODE 3 | 1ST QUARTER | 2ND QUARTER | 3RD QUARTER | 4TH QUARTER | TOTAL GRAMS |
|---|---|---|---|---|---|
| 550 | 961.98 | 1,061.54 | 1,019.93 | 1,077.19 | **4,120.64** |
| 551 | 1,517.16 | 1,631.55 | 1,573.88 | 1,719.82 | **6,442.40** |
| 553 | 1,031.85 | 1,075.98 | 1,109.70 | 1,178.25 | **4,395.78** |
| 554 | 2,119.90 | 2,262.37 | 2,304.70 | 2,543.18 | **9,230.15** |
| 556 | 16.68 | 15.12 | 19.32 | 20.12 | **71.25** |
| 557 | 210.46 | 241.00 | 259.47 | 280.21 | **991.14** |
| 558 | 409.40 | 440.05 | 463.56 | 533.63 | **1,846.63** |
| 559 | 247.59 | 258.67 | 280.06 | 259.51 | **1,045.83** |
| 560 | 262.48 | 297.82 | 314.29 | 325.08 | **1,199.66** |
| 561 | 13.35 | 14.11 | 10.49 | 13.14 | **51.09** |
| 562 | 64.57 | 75.59 | 97.88 | 101.58 | **339.62** |
| 563 | 356.32 | 382.37 | 467.25 | 481.14 | **1,687.08** |
| 564 | 154.23 | 173.48 | 166.53 | 177.86 | **672.10** |
| 565 | 319.78 | 357.10 | 389.66 | 429.42 | **1,495.96** |
| 566 | 614.01 | 561.32 | 653.19 | 634.59 | **2,463.11** |
| 567 | 45.74 | 28.95 | 40.46 | 45.30 | **160.45** |
| **MN - MINNESOTA - Total** | **8,345.50** | **8,877.01** | **9,170.37** | **9,820.01** | **36,212.90** |

**STATE: MO - MISSOURI**

| REGISTRANT ZIP CODE 3 | 1ST QUARTER | 2ND QUARTER | 3RD QUARTER | 4TH QUARTER | **TOTAL GRAMS** |
|---|---|---|---|---|---|
| 630 | 1,041.26 | 1,025.24 | 1,027.61 | 1,073.70 | **4,167.82** |
| 631 | 2,023.83 | 2,295.97 | 2,639.42 | 2,887.31 | **9,846.54** |
| 633 | 661.47 | 761.14 | 741.81 | 829.11 | **2,993.53** |
| 634 | 84.59 | 111.57 | 159.58 | 170.21 | **525.96** |
| 635 | 25.68 | 30.91 | 38.33 | 53.52 | **148.44** |
| 636 | 675.62 | 701.82 | 659.89 | 713.62 | **2,750.95** |
| 637 | 190.20 | 192.86 | 265.67 | 300.93 | **949.66** |
| 638 | 230.55 | 246.44 | 258.76 | 274.42 | **1,010.16** |
| 639 | 259.40 | 240.69 | 337.54 | 376.80 | **1,214.43** |
| 640 | 768.54 | 817.07 | 827.80 | 841.51 | **3,254.93** |
| 641 | 720.56 | 781.59 | 791.80 | 840.59 | **3,134.54** |
| 644 | 61.70 | 70.15 | 65.33 | 64.65 | **261.83** |
| 645 | 258.21 | 285.00 | 342.06 | 431.04 | **1,316.31** |
| 646 | 28.24 | 31.11 | 38.87 | 44.29 | **142.51** |
| 647 | 110.38 | 116.15 | 149.21 | 170.21 | **545.95** |
| 648 | 294.77 | 338.96 | 383.57 | 482.31 | **1,499.60** |
| 650 | 168.88 | 212.16 | 254.89 | 269.21 | **905.13** |
| 651 | 137.58 | 139.30 | 156.63 | 164.09 | **597.61** |
| 652 | 584.62 | 651.28 | 689.07 | 797.73 | **2,722.70** |
| 653 | 176.66 | 207.38 | 261.07 | 294.06 | **939.17** |
| 654 | 165.33 | 203.48 | 299.63 | 390.35 | **1,058.79** |
| 655 | 232.74 | 288.07 | 330.93 | 394.52 | **1,246.25** |
| 656 | 232.49 | 239.43 | 285.33 | 341.03 | **1,098.28** |
| 657 | 488.30 | 583.04 | 704.26 | 733.64 | **2,509.24** |
| 658 | 1,818.32 | 2,005.34 | 2,262.45 | 2,713.35 | **8,799.45** |
| **MO - MISSOURI - Total** | **11,439.92** | **12,576.15** | **13,971.52** | **15,652.20** | **53,639.78** |

**STATE: MS - MISSISSIPPI**

| REGISTRANT ZIP CODE 3 | 1ST QUARTER | 2ND QUARTER | 3RD QUARTER | 4TH QUARTER | **TOTAL GRAMS** |
|---|---|---|---|---|---|
| 386 | 1,313.48 | 1,351.72 | 1,353.20 | 1,418.38 | **5,436.78** |
| 387 | 92.87 | 105.07 | 100.43 | 121.03 | **419.40** |
| 388 | 1,662.02 | 1,783.10 | 1,778.82 | 1,734.85 | **6,958.80** |
| 389 | 133.07 | 154.70 | 166.75 | 159.74 | **614.26** |
| 390 | 652.48 | 629.98 | 634.84 | 612.51 | **2,529.82** |
| 391 | 671.13 | 670.55 | 684.53 | 682.45 | **2,708.65** |
| 392 | 580.49 | 608.26 | 640.69 | 654.76 | **2,484.19** |
| 393 | 472.30 | 540.68 | 539.36 | 539.33 | **2,091.67** |
| 394 | 1,742.03 | 1,813.09 | 1,831.02 | 1,858.15 | **7,244.29** |
| 395 | 2,530.24 | 2,614.26 | 2,602.81 | 2,670.98 | **10,418.28** |
| 396 | 255.38 | 278.27 | 318.07 | 343.05 | **1,194.77** |

| | | | | | |
|---|---|---|---|---|---|
| 397 | 164.66 | 169.71 | 190.06 | 235.36 | **759.78** |
| **MS - MISSISSIPPI - Total** | **10,270.14** | **10,719.39** | **10,840.57** | **11,030.58** | **42,860.68** |

**STATE: MT - MONTANA**

| REGISTRANT ZIP CODE 3 | 1ST QUARTER | 2ND QUARTER | 3RD QUARTER | 4TH QUARTER | TOTAL GRAMS |
|---|---|---|---|---|---|
| 590 | 104.44 | 107.59 | 102.00 | 122.44 | **436.47** |
| 591 | 338.29 | 326.90 | 320.34 | 347.58 | **1,333.12** |
| 592 | 101.82 | 136.08 | 151.76 | 156.46 | **546.12** |
| 593 | 44.00 | 43.05 | 41.59 | 48.76 | **177.39** |
| 594 | 733.41 | 703.68 | 710.31 | 738.09 | **2,885.48** |
| 595 | 33.65 | 40.68 | 55.36 | 62.48 | **192.18** |
| 596 | 290.51 | 300.83 | 329.86 | 338.58 | **1,259.78** |
| 597 | 289.36 | 273.75 | 310.69 | 332.38 | **1,206.18** |
| 598 | 653.58 | 690.69 | 754.59 | 820.08 | **2,918.95** |
| 599 | 266.82 | 267.63 | 297.56 | 372.02 | **1,204.04** |
| **MT - MONTANA - Total** | **2,855.90** | **2,890.90** | **3,074.04** | **3,338.87** | **12,159.72** |

**STATE: NC - NORTH CAROLINA**

| REGISTRANT ZIP CODE 3 | 1ST QUARTER | 2ND QUARTER | 3RD QUARTER | 4TH QUARTER | TOTAL GRAMS |
|---|---|---|---|---|---|
| 270 | 659.11 | 674.24 | 756.53 | 872.03 | **2,961.92** |
| 271 | 707.78 | 717.11 | 739.22 | 815.77 | **2,979.89** |
| 272 | 1,477.24 | 1,631.91 | 1,700.54 | 1,899.93 | **6,709.62** |
| 273 | 1,084.92 | 1,157.89 | 1,214.39 | 1,299.82 | **4,757.02** |
| 274 | 737.83 | 798.21 | 722.96 | 834.16 | **3,093.16** |
| 275 | 2,283.15 | 2,459.71 | 2,535.92 | 2,745.11 | **10,023.89** |
| 276 | 1,158.14 | 1,205.48 | 1,156.12 | 1,329.42 | **4,849.16** |
| 277 | 1,527.62 | 1,605.49 | 1,670.01 | 1,826.83 | **6,629.95** |
| 278 | 1,268.43 | 2,099.66 | 1,465.83 | 1,554.38 | **6,388.30** |
| 279 | 459.68 | 482.68 | 525.91 | 517.45 | **1,985.72** |
| 280 | 3,072.84 | 3,246.95 | 3,207.46 | 3,549.50 | **13,076.75** |
| 281 | 2,202.56 | 2,371.93 | 2,500.85 | 2,944.46 | **10,019.80** |
| 282 | 1,981.65 | 1,964.95 | 1,939.20 | 2,189.82 | **8,075.62** |
| 283 | 3,127.25 | 4,161.13 | 3,441.04 | 3,472.99 | **14,202.42** |
| 284 | 3,425.65 | 3,593.96 | 3,594.37 | 4,409.54 | **15,023.52** |
| 285 | 1,245.39 | 1,458.46 | 1,542.57 | 1,655.40 | **5,901.81** |
| 286 | 4,881.59 | 5,208.04 | 5,876.44 | 6,295.94 | **22,262.01** |
| 287 | 2,547.29 | 2,668.41 | 2,783.76 | 3,155.24 | **11,154.71** |
| 288 | 2,045.87 | 2,141.26 | 2,177.53 | 2,589.75 | **8,954.42** |
| 289 | 209.10 | 240.08 | 246.43 | 279.58 | **975.19** |
| **NC - NORTH CAROLINA - Total** | **36,103.10** | **39,887.55** | **39,797.09** | **44,237.14** | **160,024.88** |

**STATE: ND - NORTH DAKOTA**

| REGISTRANT ZIP CODE 3 | 1ST QUARTER | 2ND QUARTER | 3RD QUARTER | 4TH QUARTER | TOTAL GRAMS |
|---|---|---|---|---|---|

| | | | | | |
|---|---|---|---|---|---|
| 580 | 8.83 | 14.19 | 19.06 | 18.58 | **60.67** |
| 581 | 276.28 | 273.60 | 291.18 | 289.49 | **1,130.55** |
| 582 | 69.10 | 89.30 | 108.55 | 130.59 | **397.54** |
| 583 | 73.53 | 101.08 | 105.41 | 123.09 | **403.12** |
| 584 | 12.28 | 20.98 | 17.55 | 19.47 | **70.27** |
| 585 | 415.28 | 427.36 | 448.84 | 419.25 | **1,710.73** |
| 586 | 55.04 | 61.00 | 74.38 | 77.25 | **267.67** |
| 587 | 33.94 | 47.00 | 61.99 | 72.70 | **215.63** |
| 588 | 82.37 | 100.80 | 103.60 | 110.34 | **397.10** |
| **ND - NORTH DAKOTA - Total** | **1,026.66** | **1,135.30** | **1,230.55** | **1,260.77** | **4,653.28** |

**STATE: NE - NEBRASKA**

| REGISTRANT ZIP CODE 3 | 1ST QUARTER | 2ND QUARTER | 3RD QUARTER | 4TH QUARTER | TOTAL GRAMS |
|---|---|---|---|---|---|
| 680 | 81.79 | 84.46 | 86.14 | 85.69 | **338.07** |
| 681 | 563.86 | 586.79 | 579.75 | 642.03 | **2,372.43** |
| 683 | 39.35 | 41.54 | 41.33 | 43.46 | **165.67** |
| 684 | 21.09 | 27.31 | 29.89 | 32.76 | **111.06** |
| 685 | 291.52 | 307.41 | 304.30 | 335.55 | **1,238.79** |
| 686 | 30.61 | 35.34 | 37.87 | 36.24 | **140.06** |
| 687 | 53.53 | 60.72 | 51.49 | 58.49 | **224.24** |
| 688 | 114.70 | 83.03 | 88.71 | 96.76 | **383.20** |
| 689 | 18.90 | 19.48 | 18.13 | 21.82 | **78.33** |
| 690 | 3.74 | 3.73 | 2.73 | 2.42 | **12.63** |
| 691 | 162.63 | 200.99 | 217.72 | 235.91 | **817.25** |
| 692 | 3.74 | 4.34 | 4.76 | 7.19 | **20.03** |
| 693 | 19.45 | 14.09 | 11.82 | 16.76 | **62.12** |
| **NE - NEBRASKA - Total** | **1,404.90** | **1,469.22** | **1,474.66** | **1,615.10** | **5,963.88** |

**STATE: NH - NEW HAMPSHIRE**

| REGISTRANT ZIP CODE 3 | 1ST QUARTER | 2ND QUARTER | 3RD QUARTER | 4TH QUARTER | TOTAL GRAMS |
|---|---|---|---|---|---|
| 030 | 1,296.16 | 1,414.11 | 1,331.67 | 1,488.24 | **5,530.18** |
| 031 | 1,240.96 | 1,372.39 | 1,361.04 | 1,423.60 | **5,397.99** |
| 032 | 1,288.02 | 1,367.90 | 1,426.61 | 1,513.20 | **5,595.74** |
| 033 | 852.84 | 923.20 | 864.87 | 960.86 | **3,601.78** |
| 034 | 338.90 | 350.93 | 324.80 | 370.93 | **1,385.56** |
| 035 | 322.37 | 366.22 | 385.81 | 408.55 | **1,482.95** |
| 036 | 11.67 | 11.61 | 12.94 | 13.57 | **49.79** |
| 037 | 1,153.15 | 1,192.81 | 1,041.46 | 1,119.48 | **4,506.90** |
| 038 | 1,715.99 | 1,826.86 | 1,804.82 | 1,946.95 | **7,294.62** |
| **NH - NEW HAMPSHIRE - Total** | **8,220.05** | **8,826.04** | **8,554.03** | **9,245.38** | **34,845.51** |

**STATE: NJ - NEW JERSEY**

| REGISTRANT ZIP CODE 3 | 1ST QUARTER | 2ND QUARTER | 3RD QUARTER | 4TH QUARTER | TOTAL GRAMS |
|---|---|---|---|---|---|

| | | | | | |
|---|---|---|---|---|---|
| 070 | 2,863.03 | 2,994.05 | 2,861.52 | 2,981.89 | **11,700.48** |
| 071 | 929.19 | 1,006.49 | 1,019.65 | 1,032.89 | **3,988.22** |
| 072 | 237.10 | 254.17 | 254.41 | 250.58 | **996.25** |
| 073 | 172.84 | 172.40 | 210.15 | 190.55 | **745.94** |
| 074 | 1,057.80 | 1,134.99 | 1,097.56 | 1,150.82 | **4,441.18** |
| 075 | 297.61 | 314.57 | 335.11 | 361.56 | **1,308.84** |
| 076 | 849.19 | 914.81 | 911.69 | 936.22 | **3,611.91** |
| 077 | 2,147.24 | 2,160.61 | 2,141.77 | 2,237.05 | **8,686.67** |
| 078 | 844.54 | 924.30 | 831.86 | 996.21 | **3,596.91** |
| 079 | 328.86 | 340.95 | 338.68 | 365.40 | **1,373.89** |
| 080 | 6,903.86 | 7,339.67 | 7,095.11 | 7,698.66 | **29,037.30** |
| 081 | 995.92 | 1,017.08 | 982.65 | 1,123.71 | **4,119.35** |
| 082 | 1,851.77 | 1,925.32 | 1,893.68 | 2,005.78 | **7,676.55** |
| 083 | 1,022.95 | 1,077.10 | 1,038.57 | 1,197.78 | **4,336.40** |
| 084 | 264.48 | 283.85 | 263.42 | 285.88 | **1,097.63** |
| 085 | 568.37 | 619.46 | 627.84 | 673.28 | **2,488.95** |
| 086 | 735.08 | 787.82 | 777.44 | 834.66 | **3,135.00** |
| 087 | 2,574.92 | 2,662.24 | 2,611.12 | 2,797.60 | **10,645.88** |
| 088 | 1,709.41 | 1,816.56 | 1,820.91 | 1,992.53 | **7,339.40** |
| 089 | 165.76 | 176.41 | 177.92 | 220.83 | **740.92** |
| **NJ - NEW JERSEY - Total** | **26,519.91** | **27,922.84** | **27,291.06** | **29,333.86** | **111,067.66** |

**STATE: NM - NEW MEXICO**

| REGISTRANT ZIP CODE 3 | 1ST QUARTER | 2ND QUARTER | 3RD QUARTER | 4TH QUARTER | **TOTAL GRAMS** |
|---|---|---|---|---|---|
| 870 | 428.38 | 474.84 | 493.23 | 502.93 | **1,899.37** |
| 871 | 3,781.72 | 4,027.27 | 4,145.25 | 4,202.21 | **16,156.46** |
| 873 | 130.68 | 146.26 | 138.69 | 145.29 | **560.93** |
| 874 | 315.14 | 323.06 | 353.53 | 319.53 | **1,311.26** |
| 875 | 1,998.02 | 2,156.06 | 2,242.90 | 2,300.93 | **8,697.91** |
| 877 | 201.57 | 210.39 | 230.81 | 236.59 | **879.37** |
| 878 | 73.69 | 104.48 | 138.49 | 144.92 | **461.58** |
| 879 | 11.76 | 12.66 | 19.92 | 16.50 | **60.84** |
| 880 | 361.73 | 398.25 | 441.28 | 466.24 | **1,667.51** |
| 881 | 33.16 | 31.87 | 41.00 | 32.36 | **138.39** |
| 882 | 397.35 | 430.94 | 443.12 | 480.45 | **1,751.86** |
| 883 | 137.59 | 144.21 | 164.45 | 135.20 | **581.46** |
| 884 | 4.02 | 6.98 | 8.58 | 9.02 | **28.60** |
| **NM - NEW MEXICO - Total** | **7,874.81** | **8,467.29** | **8,861.26** | **8,992.19** | **34,195.54** |

**STATE: NV - NEVADA**

| REGISTRANT ZIP CODE 3 | 1ST QUARTER | 2ND QUARTER | 3RD QUARTER | 4TH QUARTER | **TOTAL GRAMS** |
|---|---|---|---|---|---|
| 890 | 723.48 | 714.77 | 736.38 | 807.47 | **2,982.10** |

| | | | | | |
|---|---|---|---|---|---|
| 891 | 1,702.79 | 1,747.14 | 1,853.89 | 1,940.84 | **7,244.66** |
| 893 | 12.00 | 16.32 | 12.54 | 12.12 | **52.99** |
| 894 | 332.56 | 413.20 | 415.09 | 435.84 | **1,596.68** |
| 895 | 600.76 | 589.69 | 618.44 | 660.05 | **2,468.94** |
| 897 | 125.34 | 141.64 | 144.77 | 169.09 | **580.84** |
| 898 | 85.86 | 90.78 | 88.03 | 100.04 | **364.71** |
| **NV - NEVADA - Total** | **3,582.79** | **3,713.55** | **3,869.14** | **4,125.46** | **15,290.93** |

**STATE: NY - NEW YORK**

| REGISTRANT ZIP CODE 3 | 1ST QUARTER | 2ND QUARTER | 3RD QUARTER | 4TH QUARTER | **TOTAL GRAMS** |
|---|---|---|---|---|---|
| 100 | 2,650.76 | 2,797.01 | 2,651.53 | 2,903.32 | **11,002.62** |
| 101 | 66.79 | 83.77 | 93.87 | 105.73 | **350.17** |
| 102 | 1.39 | 2.24 | 0.98 | 3.95 | **8.56** |
| 103 | 1,838.50 | 1,878.79 | 1,913.82 | 1,940.77 | **7,571.88** |
| 104 | 2,467.85 | 2,518.21 | 2,580.42 | 2,777.05 | **10,343.53** |
| 105 | 1,077.49 | 1,069.98 | 1,137.71 | 1,181.78 | **4,466.96** |
| 106 | 232.47 | 224.69 | 225.89 | 207.88 | **890.93** |
| 107 | 360.74 | 389.72 | 401.68 | 416.92 | **1,569.05** |
| 108 | 101.12 | 103.10 | 94.21 | 95.30 | **393.72** |
| 109 | 1,474.58 | 1,565.67 | 1,567.65 | 1,643.01 | **6,250.91** |
| 110 | 580.58 | 591.58 | 596.27 | 572.45 | **2,340.89** |
| 111 | 348.17 | 398.38 | 416.88 | 435.40 | **1,598.83** |
| 112 | 3,010.34 | 3,314.23 | 3,357.94 | 3,496.53 | **13,179.04** |
| 113 | 1,050.75 | 1,063.83 | 1,050.51 | 1,134.40 | **4,299.49** |
| 114 | 476.65 | 470.33 | 453.65 | 493.99 | **1,894.61** |
| 115 | 1,446.02 | 1,514.40 | 1,570.18 | 1,676.58 | **6,207.19** |
| 116 | 116.90 | 125.78 | 134.20 | 133.19 | **510.07** |
| 117 | 5,951.47 | 6,178.61 | 6,037.25 | 6,455.19 | **24,622.51** |
| 118 | 497.67 | 537.41 | 499.65 | 533.51 | **2,068.24** |
| 119 | 1,138.03 | 1,213.28 | 1,225.25 | 1,259.93 | **4,836.50** |
| 120 | 1,911.10 | 2,019.97 | 2,070.07 | 2,073.77 | **8,074.92** |
| 121 | 920.15 | 1,006.79 | 1,048.80 | 1,088.46 | **4,064.20** |
| 122 | 778.67 | 880.19 | 932.05 | 985.72 | **3,576.63** |
| 123 | 709.26 | 762.94 | 784.01 | 842.81 | **3,099.03** |
| 124 | 896.66 | 983.90 | 955.95 | 1,056.16 | **3,892.67** |
| 125 | 1,591.55 | 1,627.39 | 1,680.32 | 1,778.66 | **6,677.93** |
| 126 | 323.69 | 337.43 | 319.89 | 377.55 | **1,358.56** |
| 127 | 887.30 | 895.52 | 866.09 | 968.39 | **3,617.30** |
| 128 | 1,522.15 | 1,581.51 | 1,616.49 | 1,693.01 | **6,413.16** |
| 129 | 1,381.63 | 1,405.52 | 1,507.47 | 1,582.48 | **5,877.11** |
| 130 | 1,116.05 | 1,189.99 | 1,220.52 | 1,400.92 | **4,927.47** |

| | | | | | |
|---|---|---|---|---|---|
| 131 | 398.14 | 425.02 | 417.60 | 462.62 | **1,703.38** |
| 132 | 1,166.12 | 1,261.94 | 1,259.11 | 1,440.87 | **5,128.03** |
| 133 | 361.59 | 360.07 | 376.87 | 379.30 | **1,477.83** |
| 134 | 763.28 | 826.15 | 897.62 | 960.87 | **3,447.92** |
| 135 | 411.12 | 411.70 | 374.93 | 392.62 | **1,590.38** |
| 136 | 1,026.38 | 1,209.39 | 1,218.92 | 1,348.12 | **4,802.81** |
| 137 | 371.61 | 390.96 | 456.12 | 461.04 | **1,679.73** |
| 138 | 822.32 | 898.82 | 926.95 | 985.38 | **3,633.47** |
| 139 | 492.79 | 573.11 | 570.49 | 675.07 | **2,311.46** |
| 140 | 2,612.32 | 2,784.42 | 2,670.02 | 2,843.23 | **10,910.00** |
| 141 | 1,008.47 | 1,135.44 | 1,129.44 | 1,198.31 | **4,471.66** |
| 142 | 3,639.54 | 3,845.78 | 3,839.34 | 4,212.57 | **15,537.23** |
| 143 | 907.03 | 964.28 | 940.57 | 983.88 | **3,795.76** |
| 144 | 954.71 | 1,000.54 | 986.90 | 1,046.71 | **3,988.85** |
| 145 | 721.32 | 742.91 | 758.79 | 813.17 | **3,036.19** |
| 146 | 2,357.06 | 2,488.73 | 2,632.79 | 2,766.07 | **10,244.65** |
| 147 | 895.17 | 944.46 | 902.54 | 934.97 | **3,677.14** |
| 148 | 1,285.15 | 1,404.62 | 1,368.60 | 1,441.75 | **5,500.13** |
| 149 | 286.81 | 351.20 | 343.90 | 374.43 | **1,356.34** |
| **NY - NEW YORK - Total** | **57,407.42** | **60,751.72** | **61,082.71** | **65,035.79** | **244,277.64** |

**STATE: OH - OHIO**

| REGISTRANT ZIP CODE 3 | 1ST QUARTER | 2ND QUARTER | 3RD QUARTER | 4TH QUARTER | **TOTAL GRAMS** |
|---|---|---|---|---|---|
| 430 | 3,208.93 | 3,330.53 | 3,384.53 | 3,495.53 | **13,419.51** |
| 431 | 2,542.38 | 2,661.12 | 2,583.68 | 2,649.38 | **10,436.56** |
| 432 | 5,304.15 | 5,600.83 | 5,819.28 | 6,340.33 | **23,064.59** |
| 433 | 1,157.82 | 1,196.46 | 1,282.87 | 1,381.03 | **5,018.18** |
| 434 | 339.38 | 374.71 | 341.96 | 370.99 | **1,427.05** |
| 435 | 646.06 | 649.77 | 628.81 | 690.66 | **2,615.30** |
| 436 | 2,118.98 | 2,280.22 | 2,122.37 | 2,291.92 | **8,813.49** |
| 437 | 1,366.03 | 1,423.48 | 1,449.75 | 1,563.54 | **5,802.80** |
| 438 | 64.44 | 63.78 | 75.28 | 80.74 | **284.25** |
| 439 | 1,924.58 | 2,032.12 | 1,911.17 | 1,978.12 | **7,845.99** |
| 440 | 2,280.69 | 2,379.24 | 2,216.48 | 2,368.85 | **9,245.26** |
| 441 | 3,200.89 | 3,369.22 | 3,353.67 | 3,563.94 | **13,487.72** |
| 442 | 1,614.40 | 1,704.65 | 1,692.94 | 1,777.16 | **6,789.15** |
| 443 | 835.94 | 879.88 | 849.84 | 945.32 | **3,510.98** |
| 444 | 1,410.03 | 1,516.04 | 1,465.90 | 1,587.57 | **5,979.54** |
| 445 | 1,550.45 | 1,669.64 | 1,686.54 | 1,731.57 | **6,638.21** |
| 446 | 781.15 | 806.24 | 829.49 | 873.70 | **3,290.58** |
| 447 | 823.25 | 817.17 | 861.99 | 890.21 | **3,392.62** |

| | | | | | |
|---|---|---|---|---|---|
| 448 | 1,149.03 | 1,258.79 | 1,166.67 | 1,287.97 | **4,862.46** |
| 449 | 942.83 | 978.87 | 984.77 | 971.94 | **3,878.40** |
| 450 | 2,876.37 | 3,217.55 | 3,282.42 | 3,405.74 | **12,782.08** |
| 451 | 2,269.16 | 2,282.94 | 2,510.07 | 2,710.11 | **9,772.28** |
| 452 | 5,000.42 | 4,566.12 | 4,719.05 | 4,946.45 | **19,232.04** |
| 453 | 1,091.24 | 1,205.31 | 1,238.42 | 1,307.41 | **4,842.38** |
| 454 | 2,224.11 | 2,278.30 | 2,098.22 | 2,296.00 | **8,896.62** |
| 455 | 772.68 | 770.16 | 752.65 | 731.31 | **3,026.81** |
| 456 | 4,888.97 | 5,052.96 | 5,015.07 | 5,219.46 | **20,176.46** |
| 457 | 1,059.92 | 1,136.32 | 1,069.32 | 1,195.27 | **4,460.82** |
| 458 | 489.96 | 550.67 | 611.34 | 729.71 | **2,381.68** |
| **OH - OHIO - Total** | **53,934.23** | **56,053.09** | **56,004.55** | **59,381.94** | **225,373.82** |

**STATE: OK - OKLAHOMA**

| REGISTRANT ZIP CODE 3 | 1ST QUARTER | 2ND QUARTER | 3RD QUARTER | 4TH QUARTER | TOTAL GRAMS |
|---|---|---|---|---|---|
| 730 | 1,837.03 | 1,962.91 | 2,059.95 | 2,417.19 | **8,277.08** |
| 731 | 2,818.65 | 3,043.86 | 3,085.64 | 3,262.82 | **12,210.98** |
| 734 | 123.62 | 149.59 | 154.25 | 182.47 | **609.93** |
| 735 | 574.85 | 603.13 | 646.58 | 676.74 | **2,501.30** |
| 736 | 141.05 | 148.27 | 157.83 | 153.76 | **600.90** |
| 737 | 215.72 | 223.08 | 239.37 | 241.19 | **919.36** |
| 738 | 65.38 | 71.70 | 83.00 | 75.25 | **295.32** |
| 739 | 14.42 | 14.55 | 18.13 | 15.28 | **62.38** |
| 740 | 2,328.52 | 2,539.24 | 2,769.73 | 3,031.13 | **10,668.61** |
| 741 | 1,625.64 | 1,734.65 | 1,882.33 | 1,997.78 | **7,240.40** |
| 743 | 233.70 | 240.72 | 281.17 | 328.81 | **1,084.40** |
| 744 | 898.84 | 1,009.31 | 1,053.38 | 1,159.14 | **4,120.67** |
| 745 | 237.37 | 273.00 | 335.75 | 332.91 | **1,179.03** |
| 746 | 201.49 | 197.86 | 205.90 | 229.83 | **835.09** |
| 747 | 125.52 | 158.12 | 173.85 | 242.90 | **700.39** |
| 748 | 672.99 | 734.44 | 761.24 | 851.76 | **3,020.43** |
| 749 | 258.28 | 254.36 | 302.59 | 328.72 | **1,143.96** |
| **OK - OKLAHOMA - Total** | **12,373.07** | **13,358.80** | **14,210.69** | **15,527.70** | **55,470.25** |

**STATE: OR - OREGON**

| REGISTRANT ZIP CODE 3 | 1ST QUARTER | 2ND QUARTER | 3RD QUARTER | 4TH QUARTER | TOTAL GRAMS |
|---|---|---|---|---|---|
| 970 | 1,865.61 | 2,010.05 | 1,992.27 | 2,256.59 | **8,124.51** |
| 971 | 769.18 | 855.81 | 910.18 | 1,084.25 | **3,619.41** |
| 972 | 3,054.90 | 3,111.23 | 3,255.16 | 3,530.60 | **12,951.90** |
| 973 | 1,543.33 | 1,712.90 | 1,761.51 | 1,880.92 | **6,898.67** |
| 974 | 1,687.04 | 1,828.69 | 1,923.28 | 2,118.87 | **7,557.88** |
| 975 | 1,069.09 | 1,110.02 | 1,199.20 | 1,304.96 | **4,683.27** |

| | | | | | |
|---|---|---|---|---|---|
| 976 | 44.93 | 53.40 | 61.22 | 84.86 | **244.41** |
| 977 | 408.71 | 453.78 | 463.64 | 502.79 | **1,828.93** |
| 978 | 653.61 | 707.97 | 766.62 | 898.72 | **3,026.93** |
| 979 | 46.58 | 55.90 | 68.37 | 67.88 | **238.73** |
| **OR - OREGON - Total** | **11,143.00** | **11,899.75** | **12,401.45** | **13,730.44** | **49,174.65** |

**STATE: PA - PENNSYLVANIA**

| REGISTRANT ZIP CODE 3 | 1ST QUARTER | 2ND QUARTER | 3RD QUARTER | 4TH QUARTER | **TOTAL GRAMS** |
|---|---|---|---|---|---|
| 150 | 3,488.37 | 3,760.84 | 3,770.91 | 4,042.23 | **15,062.35** |
| 151 | 2,728.19 | 3,029.40 | 3,016.20 | 2,968.77 | **11,742.57** |
| 152 | 6,181.73 | 6,291.22 | 5,692.04 | 6,119.86 | **24,284.86** |
| 153 | 1,977.62 | 1,849.33 | 1,984.74 | 2,125.40 | **7,937.08** |
| 154 | 1,374.97 | 1,584.48 | 1,829.68 | 1,986.47 | **6,775.60** |
| 155 | 476.90 | 481.62 | 553.45 | 675.73 | **2,187.70** |
| 156 | 3,330.61 | 3,579.06 | 3,558.55 | 3,682.73 | **14,150.95** |
| 157 | 713.70 | 758.45 | 775.04 | 845.39 | **3,092.59** |
| 158 | 695.63 | 721.25 | 708.85 | 746.97 | **2,872.70** |
| 159 | 1,287.55 | 1,423.35 | 1,402.28 | 1,516.93 | **5,630.11** |
| 160 | 1,490.02 | 1,537.51 | 1,577.95 | 1,672.68 | **6,278.16** |
| 161 | 1,278.74 | 1,382.97 | 1,437.06 | 1,505.25 | **5,604.02** |
| 162 | 844.85 | 896.90 | 911.35 | 909.24 | **3,562.34** |
| 163 | 736.56 | 879.82 | 888.96 | 989.72 | **3,495.05** |
| 164 | 284.09 | 289.83 | 286.83 | 270.71 | **1,131.45** |
| 165 | 1,838.68 | 1,973.44 | 1,908.30 | 1,969.70 | **7,690.12** |
| 166 | 1,940.23 | 2,089.89 | 2,111.08 | 2,212.02 | **8,353.21** |
| 167 | 144.95 | 155.16 | 151.30 | 231.88 | **683.29** |
| 168 | 694.56 | 795.52 | 801.99 | 852.63 | **3,144.70** |
| 169 | 149.12 | 148.82 | 172.51 | 204.53 | **674.98** |
| 170 | 1,832.29 | 1,934.94 | 1,953.51 | 2,143.92 | **7,864.66** |
| 171 | 470.48 | 557.52 | 540.98 | 641.18 | **2,210.16** |
| 172 | 841.10 | 926.94 | 943.04 | 1,034.89 | **3,745.97** |
| 173 | 1,030.80 | 1,101.26 | 1,137.70 | 1,170.01 | **4,439.76** |
| 174 | 533.92 | 598.77 | 572.98 | 638.05 | **2,343.71** |
| 175 | 327.75 | 379.13 | 387.28 | 427.50 | **1,521.66** |
| 176 | 492.72 | 547.71 | 534.73 | 548.37 | **2,123.54** |
| 177 | 1,616.49 | 1,834.57 | 1,818.60 | 2,023.20 | **7,292.87** |
| 178 | 926.52 | 1,009.21 | 990.36 | 1,099.96 | **4,026.06** |
| 179 | 512.48 | 541.74 | 528.99 | 667.18 | **2,250.39** |
| 180 | 857.75 | 980.23 | 1,088.71 | 983.06 | **3,909.75** |
| 181 | 493.02 | 558.89 | 550.42 | 636.46 | **2,238.79** |
| 182 | 444.25 | 465.29 | 429.06 | 467.90 | **1,806.50** |

| | | | | | |
|---|---|---|---|---|---|
| 183 | 476.34 | 533.24 | 499.90 | 575.72 | **2,085.20** |
| 184 | 1,010.33 | 1,163.16 | 1,255.78 | 1,332.15 | **4,761.43** |
| 185 | 2,196.31 | 2,184.37 | 2,205.88 | 2,304.68 | **8,891.24** |
| 186 | 1,011.20 | 1,063.39 | 971.01 | 1,082.25 | **4,127.84** |
| 187 | 2,062.66 | 2,156.39 | 2,043.86 | 2,264.29 | **8,527.19** |
| 188 | 368.50 | 484.32 | 484.77 | 519.65 | **1,857.23** |
| 189 | 1,123.94 | 1,190.04 | 1,182.28 | 1,251.00 | **4,747.26** |
| 190 | 5,370.76 | 5,641.32 | 5,520.78 | 5,964.03 | **22,496.90** |
| 191 | 8,069.36 | 9,214.39 | 9,240.37 | 10,344.06 | **36,868.18** |
| 193 | 979.94 | 989.46 | 984.33 | 1,152.26 | **4,105.99** |
| 194 | 1,801.73 | 1,795.44 | 1,761.51 | 1,900.49 | **7,259.16** |
| 195 | 295.10 | 314.00 | 317.04 | 356.77 | **1,282.91** |
| 196 | 664.17 | 699.97 | 688.17 | 759.64 | **2,811.95** |
| **PA - PENNSYLVANIA - Total** | **67,466.98** | **72,494.54** | **72,171.09** | **77,817.54** | **289,950.16** |

**STATE: PR - PUERTO RICO**

| REGISTRANT ZIP CODE 3 | 1ST QUARTER | 2ND QUARTER | 3RD QUARTER | 4TH QUARTER | TOTAL GRAMS |
|---|---|---|---|---|---|
| 006 | 504.01 | 483.11 | 472.26 | 495.10 | **1,954.47** |
| 007 | 1,184.84 | 1,150.89 | 1,192.57 | 1,201.68 | **4,729.98** |
| 009 | 445.14 | 457.02 | 457.86 | 456.66 | **1,816.68** |
| **PR - PUERTO RICO - Total** | **2,133.98** | **2,091.02** | **2,122.69** | **2,153.44** | **8,501.12** |

**STATE: RI - RHODE ISLAND**

| REGISTRANT ZIP CODE 3 | 1ST QUARTER | 2ND QUARTER | 3RD QUARTER | 4TH QUARTER | TOTAL GRAMS |
|---|---|---|---|---|---|
| 028 | 4,538.03 | 4,386.88 | 4,580.81 | 5,028.84 | **18,534.56** |
| 029 | 3,577.61 | 3,487.67 | 3,495.70 | 3,596.22 | **14,157.19** |
| **RI - RHODE ISLAND - Total** | **8,115.64** | **7,874.54** | **8,076.51** | **8,625.05** | **32,691.74** |

**STATE: SC - SOUTH CAROLINA**

| REGISTRANT ZIP CODE 3 | 1ST QUARTER | 2ND QUARTER | 3RD QUARTER | 4TH QUARTER | TOTAL GRAMS |
|---|---|---|---|---|---|
| 290 | 363.51 | 408.71 | 434.51 | 480.57 | **1,687.31** |
| 291 | 401.09 | 457.68 | 484.77 | 541.90 | **1,885.44** |
| 292 | 437.59 | 513.28 | 526.59 | 518.29 | **1,995.75** |
| 293 | 1,600.16 | 1,757.98 | 1,694.77 | 1,885.52 | **6,938.43** |
| 294 | 1,926.27 | 2,038.09 | 2,107.55 | 2,217.87 | **8,289.79** |
| 295 | 2,142.02 | 2,415.71 | 2,701.31 | 2,887.65 | **10,146.69** |
| 296 | 3,393.79 | 3,600.48 | 3,549.85 | 3,876.10 | **14,420.21** |
| 297 | 645.49 | 704.46 | 743.32 | 795.68 | **2,888.95** |
| 298 | 182.84 | 188.10 | 192.28 | 226.30 | **789.52** |
| 299 | 475.61 | 474.65 | 504.44 | 521.17 | **1,975.86** |
| **SC - SOUTH CAROLINA - Total** | **11,568.36** | **12,559.14** | **12,939.38** | **13,951.06** | **51,017.95** |

**STATE: SD - SOUTH DAKOTA**

| REGISTRANT ZIP CODE 3 | 1ST QUARTER | 2ND QUARTER | 3RD QUARTER | 4TH QUARTER | TOTAL GRAMS |
|---|---|---|---|---|---|

| | | | | | |
|---|---|---|---|---|---|
| 570 | 45.45 | 48.23 | 50.68 | 55.51 | **199.87** |
| 571 | 167.55 | 205.51 | 233.07 | 218.30 | **824.42** |
| 572 | 57.18 | 54.21 | 53.82 | 48.38 | **213.59** |
| 573 | 17.12 | 17.15 | 19.09 | 15.17 | **68.52** |
| 574 | 76.60 | 68.66 | 75.26 | 80.08 | **300.59** |
| 575 | 12.07 | 12.65 | 22.44 | 24.59 | **71.75** |
| 576 | 6.26 | 7.91 | 6.27 | 7.00 | **27.44** |
| 577 | 108.43 | 135.52 | 157.08 | 197.73 | **598.76** |
| **SD - SOUTH DAKOTA - Total** | **490.66** | **549.82** | **617.71** | **646.75** | **2,304.93** |

**STATE: TN - TENNESSEE**

| REGISTRANT ZIP CODE 3 | 1ST QUARTER | 2ND QUARTER | 3RD QUARTER | 4TH QUARTER | TOTAL GRAMS |
|---|---|---|---|---|---|
| 370 | 3,180.94 | 3,447.66 | 3,463.41 | 3,738.82 | **13,830.83** |
| 371 | 4,014.79 | 4,240.20 | 4,712.55 | 4,894.80 | **17,862.34** |
| 372 | 2,390.66 | 2,531.86 | 2,653.21 | 2,679.93 | **10,255.66** |
| 373 | 1,658.15 | 1,798.04 | 1,947.02 | 2,196.25 | **7,599.46** |
| 374 | 680.52 | 746.87 | 801.66 | 737.00 | **2,966.05** |
| 376 | 7,055.98 | 7,468.98 | 7,075.10 | 6,960.14 | **28,560.20** |
| 377 | 3,262.97 | 3,352.15 | 3,365.34 | 3,257.69 | **13,238.15** |
| 378 | 4,978.33 | 5,166.14 | 5,115.33 | 5,069.86 | **20,329.66** |
| 379 | 2,423.47 | 2,561.18 | 2,686.80 | 2,689.32 | **10,360.77** |
| 380 | 858.77 | 911.30 | 951.87 | 976.93 | **3,698.87** |
| 381 | 1,145.04 | 1,351.64 | 1,487.98 | 1,678.50 | **5,663.15** |
| 382 | 501.40 | 492.72 | 489.41 | 524.09 | **2,007.61** |
| 383 | 1,390.69 | 1,492.87 | 1,516.40 | 1,563.05 | **5,963.01** |
| 384 | 982.41 | 1,105.63 | 1,214.85 | 1,152.60 | **4,455.49** |
| 385 | 1,856.98 | 1,947.01 | 1,628.24 | 1,543.92 | **6,976.15** |
| **TN - TENNESSEE - Total** | **36,381.11** | **38,614.26** | **39,109.16** | **39,662.88** | **153,767.41** |

**STATE: TX - TEXAS**

| REGISTRANT ZIP CODE 3 | 1ST QUARTER | 2ND QUARTER | 3RD QUARTER | 4TH QUARTER | TOTAL GRAMS |
|---|---|---|---|---|---|
| 750 | 2,613.97 | 2,694.76 | 2,804.81 | 2,882.67 | **10,996.20** |
| 751 | 801.56 | 873.05 | 916.45 | 889.55 | **3,480.61** |
| 752 | 1,010.92 | 1,077.07 | 1,162.43 | 1,215.88 | **4,466.30** |
| 753 | 0.90 | 2.62 | 3.53 | 5.72 | **12.77** |
| 754 | 528.28 | 520.95 | 547.31 | 553.00 | **2,149.54** |
| 755 | 141.27 | 154.62 | 141.54 | 146.62 | **584.05** |
| 756 | 512.01 | 596.82 | 586.09 | 595.41 | **2,290.34** |
| 757 | 409.12 | 409.88 | 460.78 | 496.20 | **1,775.98** |
| 758 | 60.39 | 57.84 | 61.33 | 67.54 | **247.10** |
| 759 | 336.91 | 335.28 | 366.27 | 352.69 | **1,391.15** |
| 760 | 2,014.69 | 2,084.40 | 2,162.10 | 2,200.17 | **8,461.37** |

| | | | | | |
|---|---|---|---|---|---|
| 761 | 1,046.38 | 1,041.26 | 1,100.93 | 1,159.48 | **4,348.06** |
| 762 | 555.74 | 574.48 | 623.97 | 629.57 | **2,383.77** |
| 763 | 324.07 | 344.60 | 359.06 | 370.53 | **1,398.27** |
| 764 | 131.39 | 108.97 | 126.74 | 124.69 | **491.80** |
| 765 | 313.95 | 336.44 | 382.41 | 397.81 | **1,430.60** |
| 766 | 72.35 | 75.59 | 73.89 | 76.07 | **297.91** |
| 767 | 157.92 | 153.39 | 200.15 | 195.45 | **706.91** |
| 768 | 44.34 | 42.29 | 47.32 | 50.35 | **184.30** |
| 769 | 65.90 | 80.60 | 81.09 | 100.78 | **328.38** |
| 770 | 2,614.37 | 2,710.46 | 2,663.06 | 2,607.32 | **10,595.21** |
| 772 | 0.00 | 0 | 0 | 0.00 | **0.01** |
| 773 | 2,277.32 | 2,344.48 | 2,395.40 | 2,506.70 | **9,523.89** |
| 774 | 964.92 | 988.94 | 973.04 | 981.67 | **3,908.57** |
| 775 | 2,290.74 | 2,416.98 | 2,415.30 | 2,469.73 | **9,592.75** |
| 776 | 898.98 | 882.11 | 898.03 | 928.51 | **3,607.62** |
| 777 | 318.91 | 309.92 | 312.27 | 319.78 | **1,260.88** |
| 778 | 299.97 | 322.48 | 317.93 | 333.04 | **1,273.42** |
| 779 | 150.94 | 160.63 | 164.31 | 177.10 | **652.98** |
| 780 | 358.19 | 318.75 | 315.32 | 307.92 | **1,300.18** |
| 781 | 328.62 | 353.92 | 325.66 | 341.76 | **1,349.96** |
| 782 | 988.56 | 1,058.58 | 1,053.64 | 1,160.91 | **4,261.69** |
| 783 | 170.96 | 170.34 | 155.98 | 147.17 | **644.46** |
| 784 | 347.06 | 357.99 | 339.81 | 334.36 | **1,379.22** |
| 785 | 89.42 | 97.68 | 96.18 | 100.68 | **383.95** |
| 786 | 1,041.83 | 1,110.27 | 1,253.03 | 1,332.89 | **4,738.02** |
| 787 | 1,376.86 | 1,445.98 | 1,497.45 | 1,568.09 | **5,888.38** |
| 788 | 50.96 | 83.57 | 95.04 | 144.46 | **374.02** |
| 789 | 21.04 | 30.98 | 31.66 | 31.68 | **115.37** |
| 790 | 28.37 | 25.31 | 29.47 | 35.79 | **118.94** |
| 791 | 97.14 | 110.55 | 120.89 | 120.53 | **449.11** |
| 792 | 3.22 | 2.06 | 1.84 | 3.18 | **10.31** |
| 793 | 54.22 | 53.46 | 56.65 | 85.93 | **250.26** |
| 794 | 203.75 | 214.68 | 212.52 | 216.35 | **847.29** |
| 795 | 31.99 | 36.38 | 37.81 | 37.07 | **143.25** |
| 796 | 232.03 | 244.88 | 270.32 | 324.55 | **1,071.78** |
| 797 | 594.06 | 626.29 | 671.04 | 665.72 | **2,557.11** |
| 798 | 3.22 | 5.01 | 8.13 | 8.84 | **25.21** |
| 799 | 141.69 | 141.71 | 168.63 | 151.03 | **603.05** |
| **TX - TEXAS - Total** | **27,121.43** | **28,189.29** | **29,088.64** | **29,952.95** | **114,352.32** |

**STATE: UT - UTAH**

| REGISTRANT ZIP CODE 3 | 1ST QUARTER | 2ND QUARTER | 3RD QUARTER | 4TH QUARTER | TOTAL GRAMS |
|---|---|---|---|---|---|
| 840 | 4,309.06 | 4,437.39 | 4,667.55 | 4,907.13 | **18,321.13** |
| 841 | 2,424.30 | 2,540.81 | 2,620.44 | 2,725.50 | **10,311.05** |
| 843 | 354.22 | 393.73 | 401.16 | 446.87 | **1,595.98** |
| 844 | 836.70 | 881.04 | 922.19 | 942.91 | **3,582.84** |
| 845 | 269.29 | 252.57 | 265.10 | 295.68 | **1,082.64** |
| 846 | 1,131.55 | 1,196.76 | 1,240.78 | 1,294.49 | **4,863.58** |
| 847 | 421.08 | 451.19 | 445.60 | 500.04 | **1,817.91** |
| **UT - UTAH - Total** | **9,746.20** | **10,153.49** | **10,562.84** | **11,112.60** | **41,575.13** |

**STATE: VA - VIRGINIA**

| REGISTRANT ZIP CODE 3 | 1ST QUARTER | 2ND QUARTER | 3RD QUARTER | 4TH QUARTER | TOTAL GRAMS |
|---|---|---|---|---|---|
| 201 | 1,303.03 | 1,327.96 | 1,368.38 | 1,390.11 | **5,389.48** |
| 220 | 715.52 | 749.80 | 748.54 | 702.33 | **2,916.19** |
| 221 | 520.21 | 537.41 | 524.40 | 586.84 | **2,168.85** |
| 222 | 922.41 | 1,046.21 | 1,156.19 | 1,062.46 | **4,187.27** |
| 223 | 180.59 | 188.07 | 190.53 | 209.32 | **768.51** |
| 224 | 477.70 | 498.54 | 478.00 | 536.76 | **1,990.99** |
| 225 | 225.02 | 251.65 | 234.81 | 245.24 | **956.72** |
| 226 | 1,056.69 | 1,166.55 | 1,150.74 | 1,097.54 | **4,471.53** |
| 227 | 214.83 | 193.46 | 199.07 | 223.23 | **830.58** |
| 228 | 186.31 | 183.31 | 200.48 | 220.24 | **790.33** |
| 229 | 423.79 | 474.67 | 476.19 | 536.49 | **1,911.14** |
| 230 | 303.08 | 341.85 | 352.30 | 383.22 | **1,380.45** |
| 231 | 934.47 | 974.41 | 981.39 | 1,114.73 | **4,005.00** |
| 232 | 1,409.95 | 1,696.65 | 1,819.83 | 2,185.75 | **7,112.18** |
| 233 | 278.37 | 291.33 | 303.09 | 332.59 | **1,205.38** |
| 234 | 645.73 | 654.91 | 642.64 | 660.47 | **2,603.75** |
| 235 | 137.42 | 170.49 | 162.99 | 171.98 | **642.89** |
| 236 | 835.97 | 785.54 | 753.09 | 761.20 | **3,135.81** |
| 237 | 96.34 | 107.49 | 107.74 | 113.05 | **424.63** |
| 238 | 359.83 | 367.88 | 359.31 | 378.38 | **1,465.40** |
| 239 | 119.92 | 132.24 | 128.59 | 158.30 | **539.03** |
| 240 | 1,885.38 | 1,872.40 | 1,894.94 | 1,922.38 | **7,575.10** |
| 241 | 751.34 | 803.12 | 806.84 | 824.17 | **3,185.47** |
| 242 | 3,542.69 | 3,711.34 | 3,962.67 | 3,885.55 | **15,102.26** |
| 243 | 840.96 | 901.40 | 967.86 | 952.79 | **3,663.01** |
| 244 | 123.63 | 138.02 | 132.65 | 144.59 | **538.89** |
| 245 | 776.20 | 779.14 | 831.37 | 956.42 | **3,343.14** |
| 246 | 1,043.53 | 1,080.75 | 1,069.60 | 1,088.58 | **4,282.47** |
| **VA - VIRGINIA - Total** | **20,310.90** | **21,426.59** | **22,004.24** | **22,844.71** | **86,586.44** |

STATE: VI - VIRGIN ISLANDS

| REGISTRANT ZIP CODE 3 | 1ST QUARTER | 2ND QUARTER | 3RD QUARTER | 4TH QUARTER | TOTAL GRAMS |
|---|---|---|---|---|---|
| 008 | 12.94 | 10.37 | 14.07 | 11.75 | **49.12** |
| **VI - VIRGIN ISLANDS - Total** | **12.94** | **10.37** | **14.07** | **11.75** | **49.12** |

STATE: VT - VERMONT

| REGISTRANT ZIP CODE 3 | 1ST QUARTER | 2ND QUARTER | 3RD QUARTER | 4TH QUARTER | TOTAL GRAMS |
|---|---|---|---|---|---|
| 050 | 747.52 | 772.83 | 809.78 | 767.29 | **3,097.42** |
| 051 | 256.19 | 261.72 | 264.95 | 303.31 | **1,086.16** |
| 052 | 408.92 | 426.87 | 420.65 | 460.71 | **1,717.15** |
| 053 | 375.04 | 342.97 | 321.44 | 340.34 | **1,379.79** |
| 054 | 2,667.99 | 2,671.09 | 2,797.01 | 2,965.75 | **11,101.84** |
| 056 | 732.52 | 744.18 | 771.13 | 815.82 | **3,063.65** |
| 057 | 952.72 | 932.13 | 988.01 | 997.70 | **3,870.55** |
| 058 | 467.63 | 441.65 | 456.34 | 482.43 | **1,848.05** |
| **VT - VERMONT - Total** | **6,608.52** | **6,593.44** | **6,829.31** | **7,133.34** | **27,164.61** |

STATE: WA - WASHINGTON

| REGISTRANT ZIP CODE 3 | 1ST QUARTER | 2ND QUARTER | 3RD QUARTER | 4TH QUARTER | TOTAL GRAMS |
|---|---|---|---|---|---|
| 980 | 3,527.28 | 3,657.01 | 3,672.40 | 3,949.86 | **14,806.55** |
| 981 | 2,599.43 | 2,794.59 | 2,809.98 | 3,051.26 | **11,255.26** |
| 982 | 4,007.20 | 4,383.44 | 4,465.54 | 4,928.28 | **17,784.46** |
| 983 | 1,897.36 | 2,049.14 | 2,192.72 | 2,414.15 | **8,553.37** |
| 984 | 1,303.52 | 1,374.02 | 1,431.75 | 1,549.03 | **5,658.32** |
| 985 | 1,828.11 | 1,987.42 | 2,052.31 | 2,246.84 | **8,114.68** |
| 986 | 1,603.88 | 1,768.76 | 1,852.11 | 2,062.74 | **7,287.49** |
| 988 | 708.30 | 784.42 | 822.26 | 904.05 | **3,219.02** |
| 989 | 740.18 | 846.12 | 894.15 | 985.89 | **3,466.35** |
| 990 | 153.84 | 180.04 | 200.27 | 250.66 | **784.81** |
| 991 | 280.61 | 291.15 | 308.92 | 392.95 | **1,273.63** |
| 992 | 1,822.28 | 1,984.28 | 2,096.76 | 2,372.00 | **8,275.31** |
| 993 | 983.84 | 1,138.68 | 1,164.69 | 1,245.27 | **4,532.48** |
| 994 | 207.77 | 216.80 | 236.19 | 251.96 | **912.72** |
| **WA - WASHINGTON - Total** | **21,663.59** | **23,455.86** | **24,200.06** | **26,604.94** | **95,924.45** |

STATE: WI - WISCONSIN

| REGISTRANT ZIP CODE 3 | 1ST QUARTER | 2ND QUARTER | 3RD QUARTER | 4TH QUARTER | TOTAL GRAMS |
|---|---|---|---|---|---|
| 530 | 1,227.87 | 1,268.78 | 1,324.69 | 1,374.62 | **5,195.96** |
| 531 | 1,399.68 | 1,546.75 | 1,586.12 | 1,686.75 | **6,219.30** |
| 532 | 2,615.32 | 2,794.30 | 2,917.68 | 3,167.44 | **11,494.74** |
| 534 | 216.69 | 222.22 | 264.27 | 288.01 | **991.18** |
| 535 | 1,026.91 | 1,072.75 | 1,065.11 | 1,138.25 | **4,303.03** |
| 537 | 891.63 | 889.82 | 898.77 | 877.55 | **3,557.77** |

| | | | | | |
|---|---|---|---|---|---|
| 538 | 33.82 | 33.37 | 33.24 | 29.55 | **129.98** |
| 539 | 443.90 | 480.26 | 493.89 | 529.07 | **1,947.11** |
| 540 | 91.77 | 111.94 | 120.54 | 117.00 | **441.24** |
| 541 | 329.49 | 357.35 | 399.11 | 488.21 | **1,574.15** |
| 542 | 128.94 | 121.19 | 133.76 | 131.96 | **515.84** |
| 543 | 251.28 | 295.32 | 284.94 | 322.50 | **1,154.03** |
| 544 | 452.07 | 445.94 | 472.40 | 490.68 | **1,861.09** |
| 545 | 282.51 | 294.23 | 298.74 | 293.30 | **1,168.77** |
| 546 | 316.71 | 314.21 | 328.62 | 334.95 | **1,294.49** |
| 547 | 184.21 | 206.74 | 208.65 | 248.26 | **847.86** |
| 548 | 220.80 | 240.74 | 237.29 | 257.72 | **956.55** |
| 549 | 1,085.33 | 1,112.02 | 1,149.42 | 1,115.28 | **4,462.05** |
| **WI - WISCONSIN - Total** | **11,198.91** | **11,807.91** | **12,217.23** | **12,891.11** | **48,115.17** |

**STATE: WV - WEST VIRGINIA**

| REGISTRANT ZIP CODE 3 | 1ST QUARTER | 2ND QUARTER | 3RD QUARTER | 4TH QUARTER | **TOTAL GRAMS** |
|---|---|---|---|---|---|
| 247 | 279.28 | 286.46 | 281.36 | 323.64 | **1,170.74** |
| 248 | 278.86 | 265.30 | 270.29 | 271.39 | **1,085.84** |
| 249 | 99.76 | 95.41 | 93.17 | 100.71 | **389.04** |
| 250 | 730.23 | 719.97 | 731.81 | 715.77 | **2,897.78** |
| 251 | 236.82 | 303.80 | 309.70 | 343.59 | **1,193.91** |
| 252 | 256.88 | 196.67 | 205.82 | 258.83 | **918.21** |
| 253 | 1,367.42 | 1,388.88 | 1,412.77 | 1,385.52 | **5,554.59** |
| 254 | 1,190.58 | 1,218.33 | 1,241.53 | 1,358.35 | **5,008.79** |
| 255 | 1,359.04 | 1,390.06 | 1,382.94 | 1,425.02 | **5,557.06** |
| 256 | 805.35 | 626.96 | 653.54 | 632.62 | **2,718.48** |
| 257 | 1,035.87 | 1,186.43 | 1,309.57 | 1,396.97 | **4,928.85** |
| 258 | 2,351.28 | 2,435.05 | 2,460.08 | 2,485.35 | **9,731.76** |
| 259 | 313.88 | 322.69 | 334.83 | 318.94 | **1,290.33** |
| 260 | 1,713.18 | 1,676.38 | 1,843.19 | 1,989.00 | **7,221.75** |
| 261 | 579.11 | 585.14 | 626.40 | 647.66 | **2,438.31** |
| 262 | 513.00 | 441.50 | 354.88 | 353.80 | **1,663.18** |
| 263 | 894.74 | 874.62 | 850.24 | 903.23 | **3,522.83** |
| 264 | 140.24 | 129.48 | 128.15 | 130.41 | **528.27** |
| 265 | 1,243.93 | 1,238.82 | 1,284.30 | 1,305.00 | **5,072.05** |
| 266 | 100.54 | 105.64 | 118.69 | 168.39 | **493.26** |
| 267 | 236.55 | 255.07 | 314.61 | 319.56 | **1,125.79** |
| 268 | 9.34 | 9.75 | 9.05 | 6.02 | **34.17** |
| **WV - WEST VIRGINIA - Total** | **15,735.89** | **15,752.44** | **16,216.90** | **16,839.75** | **64,544.98** |

**STATE: WY - WYOMING**

| REGISTRANT ZIP CODE 3 | 1ST QUARTER | 2ND QUARTER | 3RD QUARTER | 4TH QUARTER | **TOTAL GRAMS** |
|---|---|---|---|---|---|

| | | | | | |
|---|---|---|---|---|---|
| 820 | 333.31 | 170.93 | 180.63 | 229.39 | **914.27** |
| 822 | 13.83 | 19.79 | 17.23 | 22.01 | **72.86** |
| 823 | 12.31 | 11.17 | 14.94 | 16.22 | **54.65** |
| 824 | 116.39 | 146.75 | 138.42 | 141.94 | **543.49** |
| 825 | 74.63 | 76.90 | 83.37 | 87.02 | **321.91** |
| 826 | 433.98 | 469.16 | 454.59 | 465.46 | **1,823.19** |
| 827 | 79.82 | 87.36 | 81.36 | 86.22 | **334.76** |
| 828 | 56.68 | 56.65 | 66.85 | 68.34 | **248.52** |
| 829 | 205.81 | 214.12 | 205.53 | 200.59 | **826.05** |
| 830 | 26.76 | 30.93 | 35.52 | 27.19 | **120.41** |
| 831 | 21.10 | 21.16 | 24.90 | 29.74 | **96.89** |
| **WY - WYOMING - Total** | **1,374.63** | **1,304.91** | **1,303.33** | **1,374.12** | **5,357.00** |

**DRUG: 9143 - OXYCODONE**

**STATE: AK - ALASKA**

| REGISTRANT ZIP CODE 3 | 1ST QUARTER | 2ND QUARTER | 3RD QUARTER | 4TH QUARTER | TOTAL GRAMS |
|---|---|---|---|---|---|
| 995 | 17,731.98 | 17,291.57 | 16,494.31 | 14,810.23 | **66,328.08** |
| 996 | 9,402.95 | 9,438.94 | 9,326.76 | 8,359.45 | **36,528.10** |
| 997 | 2,951.85 | 3,095.34 | 3,087.95 | 2,748.14 | **11,883.26** |
| 998 | 960.46 | 1,058.86 | 1,005.08 | 987.06 | **4,011.47** |
| 999 | 390.92 | 345.29 | 346.61 | 258.24 | **1,341.05** |
| **AK - ALASKA - Total** | **31,438.15** | **31,229.99** | **30,260.70** | **27,163.11** | **120,091.96** |

**STATE: AL - ALABAMA**

| REGISTRANT ZIP CODE 3 | 1ST QUARTER | 2ND QUARTER | 3RD QUARTER | 4TH QUARTER | TOTAL GRAMS |
|---|---|---|---|---|---|
| 350 | 24,916.15 | 23,722.92 | 22,773.36 | 22,046.61 | **93,459.04** |
| 351 | 14,828.92 | 14,450.77 | 13,965.97 | 13,714.06 | **56,959.72** |
| 352 | 20,580.40 | 19,532.33 | 19,131.67 | 18,825.08 | **78,069.48** |
| 354 | 7,934.30 | 7,761.05 | 7,179.33 | 7,375.86 | **30,250.55** |
| 355 | 11,695.35 | 11,223.39 | 10,821.40 | 10,355.04 | **44,095.17** |
| 356 | 18,134.23 | 17,072.31 | 16,800.96 | 16,187.18 | **68,194.68** |
| 357 | 9,773.97 | 9,400.31 | 8,774.43 | 8,811.62 | **36,760.33** |
| 358 | 12,736.54 | 11,841.25 | 11,420.02 | 10,083.14 | **46,080.96** |
| 359 | 18,912.07 | 18,804.42 | 17,583.77 | 17,730.28 | **73,030.55** |
| 360 | 7,445.77 | 6,787.96 | 6,536.43 | 6,566.66 | **27,336.83** |
| 361 | 5,020.83 | 4,803.05 | 4,348.78 | 4,551.98 | **18,724.64** |
| 362 | 9,290.63 | 9,084.90 | 8,629.85 | 8,497.91 | **35,503.28** |
| 363 | 8,896.36 | 8,659.80 | 8,492.09 | 8,210.60 | **34,258.85** |
| 364 | 5,012.38 | 4,593.56 | 4,061.47 | 4,186.25 | **17,853.66** |
| 365 | 15,288.78 | 14,436.00 | 13,800.92 | 13,553.88 | **57,079.58** |
| 366 | 11,897.96 | 11,370.59 | 10,845.18 | 10,644.94 | **44,758.68** |
| 367 | 1,647.17 | 1,669.44 | 1,591.04 | 1,526.34 | **6,433.99** |

| | | | | | |
|---|---|---|---|---|---|
| 368 | 5,973.38 | 5,952.79 | 5,758.46 | 6,108.33 | **23,792.96** |
| 369 | 644.15 | 576.50 | 547.35 | 558.71 | **2,326.72** |
| **AL - ALABAMA - Total** | **210,629.35** | **201,743.36** | **193,062.48** | **189,534.46** | **794,969.65** |

**STATE: AR - ARKANSAS**

| REGISTRANT ZIP CODE 3 | 1ST QUARTER | 2ND QUARTER | 3RD QUARTER | 4TH QUARTER | TOTAL GRAMS |
|---|---|---|---|---|---|
| 716 | 3,998.93 | 3,664.38 | 3,551.36 | 3,236.66 | **14,451.33** |
| 717 | 2,689.60 | 2,560.16 | 2,371.66 | 2,280.99 | **9,902.42** |
| 718 | 2,851.12 | 2,854.65 | 2,769.24 | 2,596.70 | **11,071.71** |
| 719 | 4,813.49 | 4,576.81 | 4,261.06 | 4,209.83 | **17,861.19** |
| 720 | 15,308.57 | 14,336.17 | 12,952.09 | 11,806.53 | **54,403.37** |
| 721 | 13,549.23 | 12,973.29 | 11,909.84 | 11,215.91 | **49,648.28** |
| 722 | 11,125.27 | 10,259.82 | 9,252.47 | 9,016.05 | **39,653.61** |
| 723 | 4,718.45 | 4,470.32 | 3,467.69 | 3,119.56 | **15,776.02** |
| 724 | 5,133.60 | 5,054.42 | 4,739.41 | 4,648.33 | **19,575.76** |
| 725 | 5,116.61 | 4,566.04 | 4,041.12 | 3,906.20 | **17,629.96** |
| 726 | 6,700.18 | 5,748.08 | 5,043.49 | 4,844.60 | **22,336.35** |
| 727 | 26,302.56 | 25,466.67 | 24,074.13 | 22,381.96 | **98,225.31** |
| 728 | 5,332.31 | 5,385.00 | 4,982.95 | 4,317.85 | **20,018.11** |
| 729 | 18,320.87 | 17,615.82 | 17,020.08 | 15,413.46 | **68,370.22** |
| **AR - ARKANSAS - Total** | **125,960.79** | **119,531.64** | **110,436.59** | **102,994.62** | **458,923.65** |

**STATE: AS - AMERICAN SAMOA**

| REGISTRANT ZIP CODE 3 | 1ST QUARTER | 2ND QUARTER | 3RD QUARTER | 4TH QUARTER | TOTAL GRAMS |
|---|---|---|---|---|---|
| 967 | 10.76 | 6.72 | 6.72 | 12.55 | **36.76** |
| **AS - AMERICAN SAMOA - Tota** | **10.76** | **6.72** | **6.72** | **12.55** | **36.76** |

**STATE: AZ - ARIZONA**

| REGISTRANT ZIP CODE 3 | 1ST QUARTER | 2ND QUARTER | 3RD QUARTER | 4TH QUARTER | TOTAL GRAMS |
|---|---|---|---|---|---|
| 850 | 77,662.97 | 73,470.57 | 70,864.29 | 69,461.82 | **291,459.65** |
| 851 | 19,348.62 | 17,730.32 | 17,064.86 | 17,083.17 | **71,226.96** |
| 852 | 89,316.22 | 82,326.72 | 80,947.38 | 73,567.70 | **326,158.02** |
| 853 | 68,505.95 | 63,791.44 | 60,200.74 | 60,184.77 | **252,682.91** |
| 855 | 6,650.23 | 6,209.12 | 6,104.72 | 5,987.84 | **24,951.91** |
| 856 | 12,583.68 | 11,899.90 | 11,784.64 | 11,042.49 | **47,310.71** |
| 857 | 58,723.72 | 55,111.97 | 53,619.40 | 52,417.78 | **219,872.87** |
| 859 | 6,306.40 | 6,058.46 | 5,984.00 | 5,751.35 | **24,100.21** |
| 860 | 5,488.97 | 5,155.40 | 4,694.34 | 4,385.02 | **19,723.73** |
| 863 | 12,294.39 | 12,071.29 | 11,809.75 | 11,258.56 | **47,433.99** |
| 864 | 16,179.39 | 14,024.56 | 13,032.67 | 13,346.81 | **56,583.42** |
| 865 | 241.16 | 222.33 | 214.71 | 175.27 | **853.47** |
| **AZ - ARIZONA - Total** | **373,301.70** | **348,072.07** | **336,321.51** | **324,662.57** | **1,382,357.85** |

**STATE: CA - CALIFORNIA**

| REGISTRANT ZIP CODE 3 | 1ST QUARTER | 2ND QUARTER | 3RD QUARTER | 4TH QUARTER | **TOTAL GRAMS** |
|---|---|---|---|---|---|
| 900 | 21,353.64 | 21,581.88 | 20,960.40 | 20,354.59 | **84,250.52** |
| 902 | 16,153.11 | 15,126.54 | 14,422.23 | 14,394.28 | **60,096.16** |
| 903 | 1,106.62 | 1,099.35 | 1,060.13 | 1,190.79 | **4,456.89** |
| 904 | 5,198.17 | 4,921.80 | 4,725.26 | 4,327.60 | **19,172.83** |
| 905 | 3,511.19 | 3,715.20 | 3,535.79 | 3,586.03 | **14,348.21** |
| 906 | 6,938.98 | 6,830.02 | 6,870.40 | 6,610.67 | **27,250.08** |
| 907 | 8,631.88 | 7,815.51 | 7,930.28 | 7,489.96 | **31,867.63** |
| 908 | 5,433.22 | 5,179.94 | 5,156.62 | 4,865.29 | **20,635.08** |
| 910 | 3,963.57 | 3,947.66 | 3,966.14 | 3,915.42 | **15,792.79** |
| 911 | 2,315.19 | 2,562.97 | 2,366.95 | 2,093.92 | **9,339.04** |
| 912 | 3,251.46 | 3,317.58 | 3,053.11 | 2,760.69 | **12,382.83** |
| 913 | 29,312.25 | 28,950.88 | 27,586.68 | 27,103.82 | **112,953.63** |
| 914 | 8,442.66 | 8,391.20 | 8,019.82 | 8,254.26 | **33,107.93** |
| 915 | 3,125.42 | 3,030.85 | 2,874.99 | 2,802.39 | **11,833.66** |
| 916 | 1,712.98 | 1,665.29 | 1,648.05 | 1,720.63 | **6,746.96** |
| 917 | 24,416.37 | 24,299.86 | 23,071.55 | 22,849.25 | **94,637.03** |
| 918 | 487.55 | 468.04 | 526.10 | 518.42 | **2,000.11** |
| 919 | 10,727.40 | 10,639.07 | 10,161.31 | 10,376.53 | **41,904.31** |
| 920 | 45,902.00 | 45,136.55 | 42,188.00 | 40,496.33 | **173,722.87** |
| 921 | 30,428.97 | 28,621.01 | 26,606.75 | 25,236.69 | **110,893.42** |
| 922 | 26,325.64 | 24,800.41 | 23,794.54 | 23,446.71 | **98,367.30** |
| 923 | 29,643.71 | 27,714.18 | 27,217.35 | 27,046.14 | **111,621.39** |
| 924 | 4,664.94 | 4,261.52 | 3,767.16 | 3,593.22 | **16,286.84** |
| 925 | 29,269.57 | 28,691.56 | 27,424.20 | 27,035.05 | **112,420.38** |
| 926 | 26,166.33 | 25,779.23 | 24,582.48 | 24,329.14 | **100,857.18** |
| 927 | 5,324.82 | 5,135.29 | 4,786.83 | 4,908.09 | **20,155.03** |
| 928 | 20,669.17 | 19,590.21 | 18,969.28 | 18,307.75 | **77,536.41** |
| 930 | 14,986.38 | 14,754.56 | 13,886.23 | 13,290.57 | **56,917.74** |
| 931 | 2,527.42 | 2,384.69 | 2,207.80 | 2,300.23 | **9,420.13** |
| 932 | 9,548.02 | 9,483.26 | 9,216.71 | 9,443.88 | **37,691.87** |
| 933 | 12,458.11 | 12,280.36 | 11,766.42 | 11,503.98 | **48,008.87** |
| 934 | 13,168.81 | 12,961.42 | 12,284.23 | 12,214.64 | **50,629.11** |
| 935 | 20,112.17 | 19,437.92 | 18,428.71 | 17,793.69 | **75,772.49** |
| 936 | 10,937.52 | 10,744.21 | 10,051.85 | 9,406.10 | **41,139.67** |
| 937 | 13,176.75 | 13,115.23 | 12,999.16 | 12,833.23 | **52,124.38** |
| 939 | 6,181.02 | 5,835.92 | 5,520.53 | 5,096.60 | **22,634.07** |
| 940 | 12,536.79 | 11,951.19 | 11,494.79 | 10,964.96 | **46,947.73** |
| 941 | 21,038.04 | 20,265.92 | 19,639.88 | 18,792.24 | **79,736.08** |
| 943 | 3,239.82 | 3,102.90 | 2,995.61 | 2,765.76 | **12,104.10** |

| | | | | | |
|---|---|---|---|---|---|
| 944 | 1,997.00 | 1,657.75 | 1,737.41 | 1,669.27 | **7,061.44** |
| 945 | 48,408.29 | 45,142.04 | 44,206.30 | 42,683.11 | **180,439.74** |
| 946 | 4,899.93 | 4,433.47 | 4,303.91 | 4,478.89 | **18,116.19** |
| 947 | 1,647.97 | 1,615.27 | 1,579.31 | 1,344.81 | **6,187.36** |
| 948 | 1,613.80 | 1,499.19 | 1,542.95 | 1,394.50 | **6,050.44** |
| 949 | 9,803.96 | 8,965.81 | 8,639.15 | 8,365.63 | **35,774.55** |
| 950 | 11,979.78 | 11,382.71 | 10,758.97 | 10,052.59 | **44,174.05** |
| 951 | 8,758.87 | 8,948.01 | 8,064.63 | 7,518.08 | **33,289.60** |
| 952 | 9,188.73 | 8,995.02 | 8,904.06 | 8,003.82 | **35,091.64** |
| 953 | 34,262.02 | 33,220.03 | 30,361.97 | 28,593.43 | **126,437.44** |
| 954 | 20,698.75 | 19,421.18 | 18,561.20 | 16,855.22 | **75,536.36** |
| 955 | 5,792.60 | 5,290.62 | 5,063.10 | 4,757.35 | **20,903.67** |
| 956 | 30,503.92 | 30,650.39 | 28,355.55 | 28,806.14 | **118,316.01** |
| 957 | 5,488.33 | 4,992.74 | 4,765.26 | 4,606.68 | **19,853.02** |
| 958 | 20,840.88 | 20,580.23 | 20,425.64 | 18,270.32 | **80,117.07** |
| 959 | 20,135.52 | 18,205.21 | 16,743.48 | 14,988.53 | **70,072.74** |
| 960 | 11,003.95 | 10,147.90 | 9,413.19 | 8,602.02 | **39,167.05** |
| 961 | 2,524.38 | 2,701.11 | 2,437.52 | 2,226.98 | **9,889.99** |
| **CA - CALIFORNIA - Total** | **763,936.38** | **737,439.87** | **703,627.93** | **679,236.91** | **2,884,241.08** |

**STATE: CO - COLORADO**

| REGISTRANT ZIP CODE 3 | 1ST QUARTER | 2ND QUARTER | 3RD QUARTER | 4TH QUARTER | TOTAL GRAMS |
|---|---|---|---|---|---|
| 800 | 32,329.03 | 31,185.36 | 33,451.30 | 33,806.36 | **130,772.05** |
| 801 | 24,191.48 | 24,379.91 | 23,977.83 | 24,420.25 | **96,969.48** |
| 802 | 39,612.67 | 37,732.92 | 34,870.12 | 32,871.01 | **145,086.72** |
| 803 | 3,396.69 | 3,172.86 | 2,884.02 | 3,022.99 | **12,476.57** |
| 804 | 6,052.96 | 5,198.41 | 5,371.27 | 5,346.72 | **21,969.36** |
| 805 | 17,353.24 | 16,629.97 | 16,289.19 | 15,907.15 | **66,179.55** |
| 806 | 8,994.47 | 8,908.46 | 8,465.94 | 8,146.75 | **34,515.63** |
| 807 | 1,997.07 | 1,848.10 | 1,717.21 | 1,795.86 | **7,358.23** |
| 808 | 4,680.64 | 4,366.25 | 4,434.93 | 4,085.92 | **17,567.74** |
| 809 | 25,593.02 | 23,821.48 | 22,301.18 | 22,450.58 | **94,166.25** |
| 810 | 18,319.59 | 17,263.30 | 16,874.78 | 16,171.40 | **68,629.07** |
| 811 | 2,127.08 | 2,019.47 | 1,953.61 | 1,787.65 | **7,887.81** |
| 812 | 3,203.45 | 2,907.81 | 2,918.02 | 2,935.93 | **11,965.21** |
| 813 | 1,431.06 | 1,400.77 | 1,376.83 | 1,255.14 | **5,463.80** |
| 814 | 2,171.60 | 2,062.60 | 1,985.79 | 1,917.84 | **8,137.83** |
| 815 | 4,645.76 | 4,494.01 | 4,014.99 | 4,054.66 | **17,209.42** |
| 816 | 3,551.63 | 3,276.00 | 3,276.82 | 3,258.45 | **13,362.90** |
| **CO - COLORADO - Total** | **199,651.44** | **190,667.68** | **186,163.83** | **183,234.65** | **759,717.61** |

**STATE: CT - CONNECTICUT**

| REGISTRANT ZIP CODE 3 | 1ST QUARTER | 2ND QUARTER | 3RD QUARTER | 4TH QUARTER | TOTAL GRAMS |
|---|---|---|---|---|---|
| 060 | 25,097.08 | 24,448.31 | 23,478.56 | 23,406.73 | **96,430.69** |
| 061 | 7,807.07 | 7,311.35 | 7,130.53 | 7,139.20 | **29,388.15** |
| 062 | 4,382.94 | 4,254.95 | 4,309.08 | 4,316.96 | **17,263.94** |
| 063 | 12,013.70 | 11,768.11 | 11,543.50 | 11,582.26 | **46,907.57** |
| 064 | 39,561.26 | 38,541.36 | 37,870.08 | 37,714.68 | **153,687.39** |
| 065 | 15,164.84 | 15,283.51 | 15,060.60 | 15,345.77 | **60,854.71** |
| 066 | 9,505.17 | 9,774.99 | 9,332.34 | 9,314.71 | **37,927.21** |
| 067 | 16,491.12 | 15,652.33 | 15,176.16 | 14,477.77 | **61,797.37** |
| 068 | 14,193.82 | 14,071.27 | 13,913.06 | 13,661.88 | **55,840.03** |
| 069 | 2,278.49 | 2,243.35 | 2,230.16 | 2,095.64 | **8,847.63** |
| **CT - CONNECTICUT - Total** | **146,495.48** | **143,349.53** | **140,044.07** | **139,055.60** | **568,944.68** |

**STATE: DC - DISTRICT OF COLUMBIA**

| REGISTRANT ZIP CODE 3 | 1ST QUARTER | 2ND QUARTER | 3RD QUARTER | 4TH QUARTER | TOTAL GRAMS |
|---|---|---|---|---|---|
| 200 | 11,922.01 | 11,791.10 | 11,212.82 | 11,185.23 | **46,111.16** |
| 203 | 36.31 | 22.86 | 22.41 | 8.07 | **89.65** |
| 204 | 1,021.47 | 777.80 | 822.81 | 806.67 | **3,428.75** |
| 205 | 0 | 1.79 | 0 | 0 | **1.79** |
| **DC - DISTRICT OF COLUMBIA -** | **12,979.79** | **12,593.56** | **12,058.04** | **11,999.97** | **49,631.36** |

**STATE: DE - DELAWARE**

| REGISTRANT ZIP CODE 3 | 1ST QUARTER | 2ND QUARTER | 3RD QUARTER | 4TH QUARTER | TOTAL GRAMS |
|---|---|---|---|---|---|
| 197 | 21,668.75 | 21,389.34 | 20,630.67 | 20,033.02 | **83,721.78** |
| 198 | 13,375.17 | 13,628.62 | 13,590.52 | 13,218.99 | **53,813.30** |
| 199 | 26,544.26 | 26,654.84 | 26,237.09 | 25,489.07 | **104,925.25** |
| **DE - DELAWARE - Total** | **61,588.18** | **61,672.80** | **60,458.28** | **58,741.07** | **242,460.33** |

**STATE: FL - FLORIDA**

| REGISTRANT ZIP CODE 3 | 1ST QUARTER | 2ND QUARTER | 3RD QUARTER | 4TH QUARTER | TOTAL GRAMS |
|---|---|---|---|---|---|
| 320 | 32,450.24 | 30,913.69 | 28,398.51 | 27,946.95 | **119,709.38** |
| 321 | 30,021.96 | 29,507.57 | 27,140.21 | 28,282.93 | **114,952.66** |
| 322 | 60,428.05 | 58,963.40 | 52,602.35 | 52,170.52 | **224,164.32** |
| 323 | 12,869.98 | 12,384.46 | 10,900.39 | 11,397.96 | **47,552.78** |
| 324 | 15,889.82 | 15,655.90 | 14,896.97 | 13,430.38 | **59,873.07** |
| 325 | 29,391.44 | 27,608.89 | 26,682.41 | 25,936.61 | **109,619.35** |
| 326 | 22,198.91 | 21,052.85 | 19,646.77 | 19,918.60 | **82,817.14** |
| 327 | 44,191.82 | 44,296.28 | 40,738.67 | 40,510.83 | **169,737.60** |
| 328 | 27,902.64 | 26,480.52 | 23,825.39 | 24,794.56 | **103,003.12** |
| 329 | 51,131.86 | 48,373.08 | 44,557.74 | 43,827.61 | **187,890.29** |
| 330 | 64,087.28 | 62,358.96 | 57,212.04 | 56,149.15 | **239,807.42** |
| 331 | 54,323.08 | 53,447.52 | 47,164.13 | 46,061.95 | **200,996.67** |
| 333 | 45,387.46 | 42,275.68 | 38,877.73 | 38,216.96 | **164,757.83** |

| | | | | | |
|---|---|---|---|---|---|
| 334 | 81,514.29 | 80,918.61 | 76,596.78 | 74,537.81 | **313,567.49** |
| 335 | 40,803.16 | 39,129.84 | 35,301.61 | 34,846.55 | **150,081.16** |
| 336 | 53,852.57 | 54,050.57 | 46,643.32 | 46,855.37 | **201,401.83** |
| 337 | 49,362.57 | 48,553.11 | 43,482.62 | 42,946.67 | **184,344.97** |
| 338 | 33,043.37 | 30,742.27 | 26,884.11 | 26,640.20 | **117,309.95** |
| 339 | 45,229.49 | 41,229.94 | 37,803.80 | 38,393.79 | **162,657.03** |
| 341 | 18,178.11 | 16,700.57 | 15,027.38 | 15,460.50 | **65,366.55** |
| 342 | 48,386.79 | 44,791.25 | 40,739.65 | 41,172.93 | **175,090.62** |
| 344 | 31,840.24 | 29,947.91 | 26,375.55 | 26,140.54 | **114,304.24** |
| 346 | 49,442.88 | 47,764.62 | 44,060.10 | 43,832.54 | **185,100.14** |
| 347 | 24,508.77 | 22,675.61 | 20,319.34 | 20,733.80 | **88,237.51** |
| 349 | 29,269.86 | 28,574.50 | 26,965.59 | 27,348.12 | **112,158.07** |
| **FL - FLORIDA - Total** | **995,706.65** | **958,397.59** | **872,843.15** | **867,553.82** | **3,694,501.21** |

**STATE: GA - GEORGIA**

| REGISTRANT ZIP CODE 3 | 1ST QUARTER | 2ND QUARTER | 3RD QUARTER | 4TH QUARTER | TOTAL GRAMS |
|---|---|---|---|---|---|
| 300 | 72,765.31 | 71,419.47 | 68,989.85 | 69,280.89 | **282,455.53** |
| 301 | 64,363.49 | 61,012.69 | 59,883.99 | 59,391.33 | **244,651.50** |
| 302 | 39,273.75 | 38,492.69 | 37,734.94 | 37,932.74 | **153,434.12** |
| 303 | 25,609.54 | 25,101.20 | 25,224.59 | 24,164.93 | **100,100.27** |
| 304 | 9,096.30 | 8,670.94 | 7,982.45 | 8,138.81 | **33,888.50** |
| 305 | 36,199.94 | 35,904.21 | 35,749.61 | 36,405.31 | **144,259.06** |
| 306 | 27,583.06 | 26,224.30 | 24,911.39 | 24,906.74 | **103,625.49** |
| 307 | 15,634.94 | 14,609.43 | 14,469.99 | 14,043.64 | **58,758.00** |
| 308 | 7,142.65 | 6,897.21 | 6,373.21 | 6,378.73 | **26,791.81** |
| 309 | 14,448.74 | 13,255.28 | 13,700.04 | 12,963.87 | **54,367.93** |
| 310 | 24,240.20 | 23,462.31 | 19,839.72 | 20,126.62 | **87,668.84** |
| 312 | 11,697.03 | 11,431.77 | 9,495.50 | 8,509.31 | **41,133.61** |
| 313 | 8,531.01 | 8,025.85 | 8,112.71 | 7,119.80 | **31,789.38** |
| 314 | 8,833.05 | 8,768.42 | 8,633.17 | 8,154.69 | **34,389.33** |
| 315 | 17,666.96 | 16,948.03 | 16,547.97 | 16,485.38 | **67,648.35** |
| 316 | 6,805.12 | 6,739.84 | 6,564.67 | 6,750.59 | **26,860.21** |
| 317 | 11,893.98 | 11,677.01 | 11,518.54 | 11,787.88 | **46,877.41** |
| 318 | 923.50 | 944.29 | 921.26 | 906.16 | **3,695.21** |
| 319 | 6,954.50 | 6,637.51 | 6,883.18 | 6,416.98 | **26,892.17** |
| 398 | 2,606.50 | 2,547.92 | 2,569.60 | 2,553.34 | **10,277.35** |
| **GA - GEORGIA - Total** | **412,269.59** | **398,770.38** | **386,106.36** | **382,417.75** | **1,579,564.08** |

**STATE: GU - GUAM**

| REGISTRANT ZIP CODE 3 | 1ST QUARTER | 2ND QUARTER | 3RD QUARTER | 4TH QUARTER | TOTAL GRAMS |
|---|---|---|---|---|---|
| 969 | 931.05 | 680.17 | 856.25 | 678.37 | **3,145.83** |
| **GU - GUAM - Total** | **931.05** | **680.17** | **856.25** | **678.37** | **3,145.83** |

**STATE: HI - HAWAII**

| REGISTRANT ZIP CODE 3 | 1ST QUARTER | 2ND QUARTER | 3RD QUARTER | 4TH QUARTER | TOTAL GRAMS |
|---|---|---|---|---|---|
| 967 | 27,192.66 | 26,667.74 | 24,659.52 | 23,987.83 | **102,507.75** |
| 968 | 11,812.55 | 11,750.55 | 11,385.10 | 10,487.33 | **45,435.53** |
| **HI - HAWAII - Total** | **39,005.21** | **38,418.29** | **36,044.62** | **34,475.17** | **147,943.28** |

**STATE: IA - IOWA**

| REGISTRANT ZIP CODE 3 | 1ST QUARTER | 2ND QUARTER | 3RD QUARTER | 4TH QUARTER | TOTAL GRAMS |
|---|---|---|---|---|---|
| 500 | 4,119.81 | 3,957.41 | 3,741.97 | 3,654.01 | **15,473.19** |
| 501 | 2,887.24 | 2,807.67 | 2,659.22 | 2,564.55 | **10,918.68** |
| 502 | 3,050.31 | 3,023.30 | 2,942.77 | 2,796.82 | **11,813.20** |
| 503 | 9,397.42 | 8,944.39 | 8,553.61 | 8,772.22 | **35,667.64** |
| 504 | 1,617.38 | 1,517.92 | 1,665.76 | 1,474.15 | **6,275.21** |
| 505 | 1,401.14 | 1,268.94 | 1,267.85 | 1,178.89 | **5,116.83** |
| 506 | 1,369.85 | 1,343.88 | 1,321.14 | 1,354.85 | **5,389.71** |
| 507 | 1,254.19 | 1,188.55 | 1,155.16 | 1,237.44 | **4,835.35** |
| 508 | 443.37 | 444.62 | 407.10 | 393.75 | **1,688.84** |
| 510 | 635.24 | 610.76 | 575.14 | 540.52 | **2,361.65** |
| 511 | 1,470.46 | 1,467.30 | 1,447.44 | 1,542.81 | **5,928.02** |
| 512 | 331.58 | 331.49 | 305.31 | 283.72 | **1,252.11** |
| 513 | 417.67 | 411.42 | 411.31 | 365.06 | **1,605.46** |
| 514 | 686.20 | 660.73 | 639.28 | 579.92 | **2,566.14** |
| 515 | 3,107.02 | 2,836.67 | 2,809.10 | 2,723.11 | **11,475.90** |
| 516 | 416.52 | 425.30 | 424.88 | 413.61 | **1,680.31** |
| 520 | 1,084.11 | 1,103.88 | 1,121.89 | 1,032.49 | **4,342.37** |
| 521 | 449.45 | 393.16 | 390.65 | 344.36 | **1,577.61** |
| 522 | 2,456.54 | 2,356.00 | 2,322.90 | 2,343.76 | **9,479.19** |
| 523 | 1,135.20 | 993.80 | 955.57 | 915.84 | **4,000.41** |
| 524 | 2,240.62 | 1,868.07 | 1,807.12 | 1,840.55 | **7,756.37** |
| 525 | 1,856.64 | 1,899.13 | 1,700.54 | 1,684.98 | **7,141.29** |
| 526 | 2,038.54 | 1,900.98 | 1,955.27 | 2,013.17 | **7,907.96** |
| 527 | 1,423.87 | 1,431.98 | 1,371.55 | 1,293.19 | **5,520.59** |
| 528 | 1,687.18 | 1,469.19 | 1,403.98 | 1,443.32 | **6,003.68** |
| **IA - IOWA - Total** | **46,977.54** | **44,656.56** | **43,356.50** | **42,787.09** | **177,777.69** |

**STATE: ID - IDAHO**

| REGISTRANT ZIP CODE 3 | 1ST QUARTER | 2ND QUARTER | 3RD QUARTER | 4TH QUARTER | TOTAL GRAMS |
|---|---|---|---|---|---|
| 832 | 6,036.34 | 5,928.86 | 6,039.26 | 6,107.07 | **24,111.53** |
| 833 | 6,444.85 | 6,164.08 | 6,007.90 | 5,893.61 | **24,510.44** |
| 834 | 5,448.18 | 5,186.19 | 5,060.75 | 5,210.22 | **20,905.35** |
| 835 | 1,757.45 | 1,755.52 | 1,554.51 | 1,580.77 | **6,648.25** |
| 836 | 17,055.63 | 15,631.17 | 15,310.02 | 15,627.05 | **63,623.87** |

| | | | | | |
|---|---|---|---|---|---|
| 837 | 12,327.25 | 11,689.63 | 11,470.03 | 10,503.85 | **45,990.76** |
| 838 | 7,092.23 | 6,882.51 | 6,680.21 | 6,480.39 | **27,135.33** |
| **ID - IDAHO - Total** | **56,161.93** | **53,237.95** | **52,122.67** | **51,402.97** | **212,925.53** |

**STATE: IL - ILLINOIS**

| REGISTRANT ZIP CODE 3 | 1ST QUARTER | 2ND QUARTER | 3RD QUARTER | 4TH QUARTER | **TOTAL GRAMS** |
|---|---|---|---|---|---|
| 600 | 22,308.20 | 22,540.37 | 21,753.61 | 21,516.56 | **88,118.75** |
| 601 | 14,367.74 | 13,318.35 | 13,069.62 | 13,353.19 | **54,108.89** |
| 602 | 406.28 | 410.75 | 383.84 | 343.03 | **1,543.89** |
| 603 | 561.42 | 581.22 | 530.57 | 535.02 | **2,208.24** |
| 604 | 19,212.71 | 18,363.25 | 17,310.26 | 17,527.50 | **72,413.73** |
| 605 | 9,812.34 | 8,772.28 | 8,729.03 | 8,856.43 | **36,170.09** |
| 606 | 11,916.23 | 11,263.95 | 10,545.38 | 10,782.43 | **44,507.99** |
| 607 | 1,372.18 | 1,398.87 | 1,283.72 | 1,390.87 | **5,445.65** |
| 608 | 541.51 | 654.52 | 585.77 | 584.45 | **2,366.24** |
| 609 | 2,837.05 | 2,784.76 | 2,440.53 | 2,520.26 | **10,582.60** |
| 610 | 2,832.27 | 2,758.16 | 2,733.00 | 2,593.76 | **10,917.18** |
| 611 | 3,126.17 | 2,974.04 | 3,137.80 | 3,060.40 | **12,298.41** |
| 612 | 1,439.96 | 1,243.16 | 1,246.84 | 1,242.41 | **5,172.37** |
| 613 | 1,675.08 | 1,724.80 | 1,594.97 | 1,606.79 | **6,601.62** |
| 614 | 1,158.66 | 1,039.28 | 1,005.61 | 938.64 | **4,142.20** |
| 615 | 2,037.97 | 1,921.61 | 1,784.90 | 1,828.85 | **7,573.34** |
| 616 | 2,568.97 | 2,465.81 | 2,654.71 | 2,450.27 | **10,139.76** |
| 617 | 2,043.34 | 1,833.11 | 1,836.63 | 1,824.28 | **7,537.36** |
| 618 | 3,623.37 | 3,520.54 | 3,501.18 | 3,412.97 | **14,058.06** |
| 619 | 860.62 | 712.60 | 755.63 | 712.00 | **3,040.85** |
| 620 | 7,951.70 | 7,396.46 | 7,067.22 | 6,703.87 | **29,119.25** |
| 622 | 9,079.48 | 8,545.85 | 7,675.53 | 7,530.01 | **32,830.87** |
| 623 | 1,785.82 | 1,751.50 | 1,689.01 | 1,736.60 | **6,962.93** |
| 624 | 2,025.45 | 1,823.13 | 1,952.15 | 1,834.01 | **7,634.74** |
| 625 | 1,797.90 | 1,704.40 | 1,650.40 | 1,584.12 | **6,736.82** |
| 626 | 1,793.33 | 1,767.72 | 1,565.00 | 1,464.96 | **6,591.01** |
| 627 | 1,552.34 | 1,397.08 | 1,318.63 | 1,231.59 | **5,499.64** |
| 628 | 4,333.70 | 4,290.34 | 3,977.07 | 3,943.55 | **16,544.66** |
| 629 | 7,633.41 | 6,892.46 | 6,958.63 | 6,651.03 | **28,135.53** |
| **IL - ILLINOIS - Total** | **142,655.20** | **135,850.40** | **130,737.24** | **129,759.85** | **539,002.68** |

**STATE: IN - INDIANA**

| REGISTRANT ZIP CODE 3 | 1ST QUARTER | 2ND QUARTER | 3RD QUARTER | 4TH QUARTER | **TOTAL GRAMS** |
|---|---|---|---|---|---|
| 460 | 11,991.33 | 11,103.41 | 10,850.68 | 10,991.55 | **44,936.97** |
| 461 | 15,742.94 | 15,181.10 | 18,112.13 | 18,838.02 | **67,874.19** |
| 462 | 38,302.20 | 36,828.83 | 32,070.61 | 32,464.02 | **139,665.66** |

| | | | | | |
|---|---|---|---|---|---|
| 463 | 16,434.76 | 14,761.67 | 14,722.69 | 14,623.67 | **60,542.79** |
| 464 | 1,733.35 | 1,726.80 | 1,708.56 | 1,798.81 | **6,967.52** |
| 465 | 16,409.43 | 15,202.02 | 14,398.62 | 13,709.23 | **59,719.30** |
| 466 | 6,174.72 | 5,794.56 | 5,036.78 | 4,869.89 | **21,875.94** |
| 467 | 7,890.09 | 7,329.38 | 7,195.02 | 7,420.45 | **29,834.94** |
| 468 | 10,494.79 | 10,162.99 | 10,112.72 | 10,769.27 | **41,539.77** |
| 469 | 9,043.55 | 8,282.94 | 8,004.41 | 7,880.04 | **33,210.93** |
| 470 | 6,689.74 | 6,345.04 | 6,519.51 | 6,303.48 | **25,857.77** |
| 471 | 15,490.28 | 14,397.82 | 13,325.30 | 13,146.51 | **56,359.90** |
| 472 | 6,473.12 | 6,128.64 | 5,849.66 | 5,721.17 | **24,172.59** |
| 473 | 11,370.55 | 11,066.88 | 11,100.38 | 11,242.52 | **44,780.34** |
| 474 | 7,400.02 | 7,070.39 | 6,819.29 | 6,401.10 | **27,690.80** |
| 475 | 4,971.36 | 4,699.75 | 4,225.30 | 4,272.13 | **18,168.53** |
| 476 | 3,851.90 | 3,833.49 | 3,471.44 | 3,427.29 | **14,584.13** |
| 477 | 10,653.68 | 10,245.04 | 9,397.93 | 9,165.56 | **39,462.20** |
| 478 | 2,783.73 | 2,635.81 | 2,661.94 | 2,541.40 | **10,622.88** |
| 479 | 8,429.73 | 7,681.56 | 6,731.76 | 5,960.89 | **28,803.94** |
| **IN - INDIANA - Total** | **212,331.25** | **200,478.12** | **192,314.74** | **191,547.00** | **796,671.10** |

**STATE: KS - KANSAS**

| REGISTRANT ZIP CODE 3 | 1ST QUARTER | 2ND QUARTER | 3RD QUARTER | 4TH QUARTER | **TOTAL GRAMS** |
|---|---|---|---|---|---|
| 660 | 15,084.01 | 14,598.89 | 13,493.38 | 14,097.08 | **57,273.36** |
| 661 | 7,575.55 | 6,506.58 | 6,129.04 | 6,570.27 | **26,781.44** |
| 662 | 13,509.23 | 13,289.37 | 13,017.47 | 13,356.98 | **53,173.05** |
| 664 | 2,166.96 | 1,947.94 | 2,015.42 | 1,781.08 | **7,911.41** |
| 665 | 2,860.69 | 2,705.74 | 2,647.10 | 2,508.14 | **10,721.68** |
| 666 | 5,788.68 | 5,720.74 | 5,667.81 | 5,621.60 | **22,798.82** |
| 667 | 4,885.64 | 4,652.20 | 4,418.62 | 4,427.59 | **18,384.05** |
| 668 | 1,533.42 | 1,440.30 | 1,269.65 | 1,282.03 | **5,525.40** |
| 669 | 688.78 | 696.45 | 600.74 | 649.29 | **2,635.26** |
| 670 | 8,094.72 | 8,033.45 | 7,470.68 | 7,671.11 | **31,269.96** |
| 671 | 3,600.62 | 3,322.79 | 3,152.21 | 3,223.25 | **13,298.86** |
| 672 | 23,488.96 | 23,046.98 | 22,265.76 | 22,276.90 | **91,078.60** |
| 673 | 2,244.64 | 2,128.45 | 2,098.29 | 1,828.30 | **8,299.68** |
| 674 | 4,881.92 | 4,898.45 | 4,683.83 | 4,558.33 | **19,022.53** |
| 675 | 4,811.59 | 4,905.79 | 4,882.16 | 4,709.15 | **19,308.69** |
| 676 | 2,236.89 | 2,117.79 | 2,056.23 | 2,059.70 | **8,470.60** |
| 677 | 1,243.79 | 1,352.68 | 1,250.42 | 1,234.00 | **5,080.90** |
| 678 | 4,367.30 | 4,153.64 | 3,956.12 | 3,895.48 | **16,372.54** |
| 679 | 1,023.80 | 992.93 | 1,067.80 | 925.17 | **4,009.70** |
| **KS - KANSAS - Total** | **110,087.19** | **106,511.16** | **102,142.72** | **102,675.44** | **421,416.52** |

**STATE: KY - KENTUCKY**

| REGISTRANT ZIP CODE 3 | 1ST QUARTER | 2ND QUARTER | 3RD QUARTER | 4TH QUARTER | TOTAL GRAMS |
|---|---|---|---|---|---|
| 400 | 8,023.85 | 7,660.33 | 7,247.72 | 7,391.74 | **30,323.64** |
| 401 | 4,364.17 | 4,207.56 | 4,172.13 | 3,829.58 | **16,573.44** |
| 402 | 33,611.28 | 31,978.70 | 30,549.64 | 31,194.86 | **127,334.49** |
| 403 | 13,523.73 | 13,011.54 | 12,234.73 | 12,383.38 | **51,153.38** |
| 404 | 7,094.17 | 6,981.77 | 6,539.01 | 6,453.22 | **27,068.16** |
| 405 | 10,185.92 | 9,723.89 | 9,574.25 | 9,496.73 | **38,980.79** |
| 406 | 1,379.77 | 1,299.42 | 1,273.28 | 1,285.32 | **5,237.79** |
| 407 | 6,211.53 | 5,989.92 | 5,544.31 | 5,686.12 | **23,431.88** |
| 408 | 1,148.61 | 1,112.43 | 1,113.05 | 1,083.55 | **4,457.65** |
| 409 | 3,700.83 | 3,571.07 | 3,470.32 | 3,302.36 | **14,044.58** |
| 410 | 27,702.63 | 27,046.07 | 26,215.36 | 26,066.81 | **107,030.86** |
| 411 | 4,673.04 | 4,347.07 | 4,078.71 | 4,052.30 | **17,151.11** |
| 412 | 1,922.98 | 1,935.35 | 2,038.66 | 1,959.54 | **7,856.54** |
| 413 | 1,590.52 | 1,517.26 | 1,388.38 | 1,394.99 | **5,891.15** |
| 414 | 819.20 | 731.34 | 736.06 | 741.76 | **3,028.36** |
| 415 | 3,673.60 | 3,501.64 | 3,516.85 | 3,502.29 | **14,194.38** |
| 416 | 3,038.78 | 3,172.18 | 3,063.07 | 3,126.84 | **12,400.88** |
| 417 | 4,191.57 | 4,263.69 | 4,116.35 | 4,020.08 | **16,591.69** |
| 418 | 1,719.80 | 1,631.22 | 1,701.43 | 1,615.51 | **6,667.97** |
| 420 | 11,938.02 | 11,694.41 | 11,084.18 | 10,831.32 | **45,547.93** |
| 421 | 9,766.79 | 9,686.06 | 9,104.12 | 8,947.14 | **37,504.11** |
| 422 | 5,752.53 | 5,499.32 | 5,399.53 | 5,048.71 | **21,700.09** |
| 423 | 6,561.88 | 6,244.91 | 6,130.81 | 5,621.35 | **24,558.94** |
| 424 | 7,882.47 | 7,642.34 | 7,277.40 | 6,961.82 | **29,764.04** |
| 425 | 2,892.62 | 2,728.54 | 2,601.29 | 2,617.37 | **10,839.82** |
| 426 | 3,460.63 | 3,239.85 | 2,914.27 | 3,065.40 | **12,680.15** |
| 427 | 7,148.75 | 6,932.86 | 6,740.13 | 6,363.44 | **27,185.18** |
| **KY - KENTUCKY - Total** | **193,979.66** | **187,350.73** | **179,825.04** | **178,043.55** | **739,198.98** |

**STATE: LA - LOUISIANA**

| REGISTRANT ZIP CODE 3 | 1ST QUARTER | 2ND QUARTER | 3RD QUARTER | 4TH QUARTER | TOTAL GRAMS |
|---|---|---|---|---|---|
| 700 | 29,615.27 | 25,682.51 | 24,313.18 | 23,547.47 | **103,158.42** |
| 701 | 12,503.91 | 12,274.11 | 12,313.31 | 12,092.14 | **49,183.47** |
| 703 | 7,835.13 | 7,550.34 | 7,253.09 | 7,130.76 | **29,769.32** |
| 704 | 24,853.75 | 23,657.22 | 22,975.66 | 22,252.72 | **93,739.35** |
| 705 | 19,641.07 | 19,338.02 | 19,316.63 | 18,102.81 | **76,398.53** |
| 706 | 6,027.13 | 5,801.35 | 5,810.85 | 5,592.60 | **23,231.94** |
| 707 | 19,936.29 | 18,758.92 | 18,665.21 | 18,709.39 | **76,069.81** |
| 708 | 13,483.77 | 12,950.65 | 12,605.55 | 12,437.92 | **51,477.89** |

| | | | | | |
|---|---|---|---|---|---|
| 710 | 3,940.85 | 3,864.56 | 3,863.27 | 3,847.00 | **15,515.68** |
| 711 | 14,031.39 | 13,328.37 | 13,445.10 | 13,412.60 | **54,217.47** |
| 712 | 6,195.32 | 5,942.72 | 6,091.14 | 5,926.94 | **24,156.12** |
| 713 | 10,510.93 | 10,044.15 | 9,628.29 | 9,738.91 | **39,922.28** |
| 714 | 5,029.04 | 5,056.01 | 4,834.38 | 4,945.37 | **19,864.80** |
| **LA - LOUISIANA - Total** | **173,603.85** | **164,248.94** | **161,115.66** | **157,736.64** | **656,705.08** |

**STATE: MA - MASSACHUSETTS**

| REGISTRANT ZIP CODE 3 | 1ST QUARTER | 2ND QUARTER | 3RD QUARTER | 4TH QUARTER | TOTAL GRAMS |
|---|---|---|---|---|---|
| 010 | 22,501.60 | 21,724.09 | 20,951.07 | 20,633.67 | **85,810.43** |
| 011 | 8,961.43 | 8,559.53 | 8,384.41 | 8,213.96 | **34,119.34** |
| 012 | 3,924.15 | 3,991.19 | 3,798.71 | 3,840.41 | **15,554.46** |
| 013 | 3,696.77 | 3,579.55 | 3,577.77 | 3,477.93 | **14,332.01** |
| 014 | 6,943.01 | 6,498.59 | 6,098.55 | 6,337.40 | **25,877.55** |
| 015 | 9,964.36 | 9,731.46 | 9,049.47 | 8,896.19 | **37,641.49** |
| 016 | 8,105.66 | 7,978.08 | 7,552.50 | 7,591.12 | **31,227.37** |
| 017 | 8,467.94 | 8,285.23 | 8,052.74 | 8,102.63 | **32,908.55** |
| 018 | 27,519.33 | 25,789.56 | 23,924.34 | 23,810.55 | **101,043.78** |
| 019 | 14,491.38 | 13,645.18 | 12,821.84 | 12,744.03 | **53,702.43** |
| 020 | 9,457.06 | 8,943.37 | 8,680.69 | 9,038.40 | **36,119.52** |
| 021 | 32,405.48 | 31,650.42 | 30,749.37 | 30,396.72 | **125,201.99** |
| 022 | 2,215.29 | 2,355.82 | 1,713.43 | 2,328.11 | **8,612.65** |
| 023 | 16,271.87 | 15,986.13 | 14,974.28 | 15,255.09 | **62,487.37** |
| 024 | 5,103.40 | 4,690.46 | 4,467.14 | 4,461.27 | **18,722.28** |
| 025 | 4,390.37 | 4,267.29 | 4,295.43 | 4,006.76 | **16,959.85** |
| 026 | 6,141.73 | 5,772.84 | 5,602.69 | 5,813.21 | **23,330.47** |
| 027 | 26,105.91 | 25,509.62 | 24,833.80 | 24,541.99 | **100,991.32** |
| **MA - MASSACHUSETTS - Total** | **216,666.74** | **208,958.42** | **199,528.24** | **199,489.45** | **824,642.85** |

**STATE: MD - MARYLAND**

| REGISTRANT ZIP CODE 3 | 1ST QUARTER | 2ND QUARTER | 3RD QUARTER | 4TH QUARTER | TOTAL GRAMS |
|---|---|---|---|---|---|
| 206 | 22,094.52 | 22,279.07 | 21,424.99 | 21,176.12 | **86,974.70** |
| 207 | 26,391.59 | 24,544.54 | 24,327.56 | 22,606.50 | **97,870.19** |
| 208 | 19,800.51 | 19,314.56 | 18,186.81 | 17,153.04 | **74,454.92** |
| 209 | 4,304.39 | 4,001.33 | 3,753.45 | 3,538.78 | **15,597.95** |
| 210 | 46,096.48 | 45,230.78 | 42,258.85 | 41,822.21 | **175,408.31** |
| 211 | 20,262.87 | 18,161.15 | 17,497.92 | 17,375.88 | **73,297.82** |
| 212 | 88,448.96 | 82,482.36 | 78,996.08 | 78,990.14 | **328,917.54** |
| 214 | 4,088.47 | 3,782.65 | 3,985.86 | 3,874.60 | **15,731.57** |
| 215 | 7,267.63 | 6,864.91 | 6,664.87 | 6,732.11 | **27,529.52** |
| 216 | 9,011.23 | 9,105.05 | 8,930.94 | 8,896.75 | **35,943.96** |
| 217 | 26,082.50 | 26,098.19 | 25,913.11 | 25,391.86 | **103,485.66** |

| | | | | | |
|---|---|---|---|---|---|
| 218 | 7,968.22 | 7,569.41 | 7,836.91 | 7,476.98 | **30,851.52** |
| 219 | 6,414.62 | 6,409.20 | 6,496.45 | 5,978.15 | **25,298.41** |
| **MD - MARYLAND - Total** | **288,231.97** | **275,843.20** | **266,273.79** | **261,013.12** | **1,091,362.08** |

**STATE: ME - MAINE**

| REGISTRANT ZIP CODE 3 | 1ST QUARTER | 2ND QUARTER | 3RD QUARTER | 4TH QUARTER | TOTAL GRAMS |
|---|---|---|---|---|---|
| 039 | 843.39 | 765.77 | 798.12 | 684.59 | **3,091.87** |
| 040 | 13,018.91 | 12,506.97 | 12,111.10 | 11,769.49 | **49,406.48** |
| 041 | 3,185.75 | 2,972.84 | 3,012.06 | 2,914.45 | **12,085.09** |
| 042 | 5,826.94 | 5,510.66 | 5,656.96 | 5,477.62 | **22,472.18** |
| 043 | 6,231.01 | 5,912.53 | 5,860.35 | 5,370.79 | **23,374.69** |
| 044 | 4,452.32 | 4,355.02 | 4,344.11 | 4,201.55 | **17,353.00** |
| 045 | 1,343.85 | 1,355.14 | 1,375.52 | 1,325.46 | **5,399.97** |
| 046 | 2,108.36 | 1,874.39 | 1,795.55 | 1,874.58 | **7,652.88** |
| 047 | 1,668.16 | 1,582.87 | 1,605.82 | 1,525.28 | **6,382.13** |
| 048 | 1,130.50 | 1,172.01 | 1,080.75 | 1,065.93 | **4,449.20** |
| 049 | 7,527.86 | 7,401.01 | 6,875.02 | 6,857.00 | **28,660.89** |
| **ME - MAINE - Total** | **47,337.06** | **45,409.21** | **44,515.35** | **43,066.75** | **180,328.38** |

**STATE: MI - MICHIGAN**

| REGISTRANT ZIP CODE 3 | 1ST QUARTER | 2ND QUARTER | 3RD QUARTER | 4TH QUARTER | TOTAL GRAMS |
|---|---|---|---|---|---|
| 480 | 44,044.27 | 41,608.50 | 40,221.77 | 38,001.29 | **163,875.83** |
| 481 | 48,223.28 | 43,869.20 | 41,631.79 | 39,351.12 | **173,075.40** |
| 482 | 37,038.47 | 33,126.90 | 29,911.17 | 29,090.32 | **129,166.86** |
| 483 | 31,394.04 | 28,254.70 | 26,511.22 | 25,980.31 | **112,140.28** |
| 484 | 7,021.74 | 6,809.61 | 6,469.59 | 6,380.20 | **26,681.15** |
| 485 | 3,586.29 | 3,480.25 | 3,192.17 | 3,036.65 | **13,295.36** |
| 486 | 7,605.78 | 7,429.81 | 7,066.79 | 6,929.29 | **29,031.67** |
| 487 | 3,742.59 | 3,527.06 | 3,446.60 | 3,270.11 | **13,986.37** |
| 488 | 9,967.83 | 9,156.94 | 8,664.72 | 8,636.56 | **36,426.05** |
| 489 | 4,572.53 | 4,245.86 | 4,053.00 | 3,900.15 | **16,771.54** |
| 490 | 15,769.23 | 14,701.80 | 13,872.30 | 13,866.85 | **58,210.18** |
| 491 | 1,946.75 | 1,782.13 | 1,683.66 | 1,821.94 | **7,234.48** |
| 492 | 8,744.36 | 8,136.57 | 7,268.11 | 7,029.80 | **31,178.85** |
| 493 | 2,795.29 | 2,564.75 | 2,612.71 | 2,509.58 | **10,482.33** |
| 494 | 6,369.39 | 5,850.14 | 5,718.43 | 5,537.86 | **23,475.81** |
| 495 | 6,866.28 | 6,185.83 | 5,815.07 | 5,820.37 | **24,687.54** |
| 496 | 7,897.57 | 7,436.19 | 7,287.37 | 7,058.63 | **29,679.77** |
| 497 | 6,962.21 | 6,845.27 | 6,369.88 | 5,927.01 | **26,104.37** |
| 498 | 4,051.31 | 3,747.58 | 3,594.94 | 3,390.18 | **14,784.01** |
| 499 | 1,328.21 | 1,354.54 | 1,284.06 | 1,076.54 | **5,043.35** |
| **MI - MICHIGAN - Total** | **259,927.45** | **240,113.62** | **226,675.36** | **218,614.77** | **945,331.20** |

**STATE: MN - MINNESOTA**

| REGISTRANT ZIP CODE 3 | 1ST QUARTER | 2ND QUARTER | 3RD QUARTER | 4TH QUARTER | TOTAL GRAMS |
|---|---|---|---|---|---|
| 550 | 14,794.78 | 14,392.55 | 13,846.19 | 13,765.61 | **56,799.13** |
| 551 | 22,575.77 | 22,097.92 | 22,027.66 | 20,410.38 | **87,111.73** |
| 553 | 20,513.47 | 19,461.00 | 18,590.00 | 18,305.48 | **76,869.94** |
| 554 | 34,292.99 | 33,098.69 | 31,723.86 | 31,533.98 | **130,649.52** |
| 556 | 209.42 | 192.52 | 189.25 | 208.53 | **799.72** |
| 557 | 4,296.54 | 4,103.24 | 3,982.77 | 4,005.27 | **16,387.82** |
| 558 | 2,315.95 | 2,334.21 | 2,223.88 | 2,166.93 | **9,040.96** |
| 559 | 6,017.93 | 5,513.34 | 5,180.75 | 5,047.08 | **21,759.09** |
| 560 | 4,213.46 | 3,843.77 | 3,720.13 | 3,638.17 | **15,415.53** |
| 561 | 1,059.19 | 1,049.56 | 1,016.29 | 955.06 | **4,080.10** |
| 562 | 1,969.61 | 1,884.38 | 1,888.60 | 1,752.87 | **7,495.46** |
| 563 | 6,524.00 | 6,304.54 | 5,801.06 | 5,763.19 | **24,392.79** |
| 564 | 2,993.20 | 2,932.41 | 2,820.40 | 2,733.25 | **11,479.26** |
| 565 | 2,662.09 | 2,524.47 | 2,340.25 | 2,110.22 | **9,637.04** |
| 566 | 1,151.45 | 1,133.83 | 1,097.46 | 1,041.05 | **4,423.78** |
| 567 | 1,419.88 | 1,310.37 | 1,272.58 | 1,183.56 | **5,186.38** |
| **MN - MINNESOTA - Total** | **127,009.73** | **122,176.79** | **117,721.14** | **114,620.63** | **481,528.29** |

**STATE: MO - MISSOURI**

| REGISTRANT ZIP CODE 3 | 1ST QUARTER | 2ND QUARTER | 3RD QUARTER | 4TH QUARTER | TOTAL GRAMS |
|---|---|---|---|---|---|
| 630 | 31,342.03 | 28,092.11 | 25,454.77 | 24,373.38 | **109,262.29** |
| 631 | 46,811.97 | 46,821.83 | 44,883.87 | 44,887.19 | **183,404.86** |
| 633 | 15,611.13 | 14,769.09 | 14,080.28 | 13,333.02 | **57,793.53** |
| 634 | 1,535.38 | 1,348.88 | 1,332.40 | 1,306.15 | **5,522.81** |
| 635 | 2,225.57 | 2,001.59 | 1,915.89 | 1,729.12 | **7,872.17** |
| 636 | 9,663.57 | 8,246.48 | 7,752.95 | 7,226.72 | **32,889.72** |
| 637 | 4,792.24 | 4,000.65 | 3,723.52 | 3,472.10 | **15,988.51** |
| 638 | 4,962.54 | 4,455.21 | 3,943.48 | 4,002.77 | **17,363.99** |
| 639 | 2,403.99 | 2,266.99 | 2,264.86 | 2,154.92 | **9,090.76** |
| 640 | 22,969.19 | 21,684.13 | 20,970.75 | 20,758.39 | **86,382.46** |
| 641 | 20,159.77 | 18,908.69 | 17,769.25 | 17,357.08 | **74,194.80** |
| 644 | 2,062.86 | 1,925.21 | 1,790.98 | 1,497.91 | **7,276.96** |
| 645 | 4,985.69 | 4,782.65 | 4,279.71 | 3,950.86 | **17,998.90** |
| 646 | 2,273.30 | 2,099.80 | 1,860.62 | 1,700.18 | **7,933.89** |
| 647 | 4,772.17 | 4,472.98 | 4,526.24 | 4,547.72 | **18,319.11** |
| 648 | 10,573.73 | 9,632.79 | 9,498.23 | 8,785.76 | **38,490.52** |
| 650 | 3,804.27 | 3,530.65 | 3,248.39 | 3,085.25 | **13,668.57** |
| 651 | 1,787.07 | 1,601.10 | 1,681.32 | 1,716.83 | **6,786.33** |
| 652 | 10,075.57 | 9,969.18 | 9,784.92 | 9,983.76 | **39,813.43** |

| | | | | | |
|---|---|---|---|---|---|
| 653 | 3,328.67 | 3,302.32 | 3,035.07 | 3,068.66 | **12,734.72** |
| 654 | 5,360.37 | 4,986.17 | 4,491.98 | 4,209.38 | **19,047.91** |
| 655 | 6,996.72 | 6,258.94 | 5,965.89 | 5,992.33 | **25,213.88** |
| 656 | 10,569.69 | 9,852.69 | 9,165.92 | 9,179.08 | **38,767.38** |
| 657 | 14,036.31 | 13,201.29 | 12,757.20 | 12,421.73 | **52,416.54** |
| 658 | 15,192.80 | 14,316.90 | 14,024.13 | 13,138.93 | **56,672.75** |
| **MO - MISSOURI - Total** | **258,296.60** | **242,528.32** | **230,202.63** | **223,879.23** | **954,906.78** |

**STATE: MS - MISSISSIPPI**

| REGISTRANT ZIP CODE 3 | 1ST QUARTER | 2ND QUARTER | 3RD QUARTER | 4TH QUARTER | **TOTAL GRAMS** |
|---|---|---|---|---|---|
| 386 | 13,315.82 | 12,921.26 | 13,019.98 | 12,655.48 | **51,912.53** |
| 387 | 1,540.55 | 1,526.34 | 1,530.57 | 1,539.71 | **6,137.16** |
| 388 | 8,665.68 | 8,388.70 | 8,494.66 | 7,908.57 | **33,457.61** |
| 389 | 1,562.73 | 1,531.70 | 1,466.02 | 1,475.06 | **6,035.51** |
| 390 | 5,169.41 | 5,101.85 | 4,885.86 | 4,681.99 | **19,839.12** |
| 391 | 4,472.01 | 4,431.01 | 4,268.93 | 4,278.84 | **17,450.80** |
| 392 | 5,722.10 | 5,970.43 | 5,900.84 | 5,571.13 | **23,164.50** |
| 393 | 5,743.87 | 5,395.06 | 5,264.02 | 5,016.54 | **21,419.49** |
| 394 | 19,325.49 | 18,204.52 | 17,920.84 | 17,571.13 | **73,021.98** |
| 395 | 20,877.15 | 19,959.32 | 18,736.36 | 18,260.34 | **77,833.18** |
| 396 | 4,189.73 | 4,123.92 | 4,022.12 | 3,881.27 | **16,217.05** |
| 397 | 3,021.45 | 2,798.57 | 3,056.17 | 2,809.99 | **11,686.18** |
| **MS - MISSISSIPPI - Total** | **93,605.99** | **90,352.67** | **88,566.38** | **85,650.06** | **358,175.10** |

**STATE: MT - MONTANA**

| REGISTRANT ZIP CODE 3 | 1ST QUARTER | 2ND QUARTER | 3RD QUARTER | 4TH QUARTER | **TOTAL GRAMS** |
|---|---|---|---|---|---|
| 590 | 2,365.28 | 2,079.34 | 1,976.10 | 1,902.91 | **8,323.62** |
| 591 | 8,057.77 | 7,350.58 | 7,085.84 | 7,005.36 | **29,499.55** |
| 592 | 1,135.22 | 1,138.98 | 1,011.48 | 1,009.66 | **4,295.33** |
| 593 | 1,449.42 | 1,280.47 | 1,259.39 | 1,242.63 | **5,231.91** |
| 594 | 3,620.18 | 3,248.65 | 3,132.02 | 3,196.04 | **13,196.90** |
| 595 | 639.50 | 569.48 | 474.79 | 474.62 | **2,158.39** |
| 596 | 2,649.02 | 2,646.55 | 2,496.01 | 2,170.86 | **9,962.43** |
| 597 | 3,969.71 | 3,644.18 | 3,467.50 | 3,362.20 | **14,443.59** |
| 598 | 5,407.01 | 4,988.73 | 4,927.88 | 5,023.29 | **20,346.91** |
| 599 | 4,365.81 | 4,432.65 | 4,360.40 | 4,237.20 | **17,396.05** |
| **MT - MONTANA - Total** | **33,658.91** | **31,379.60** | **30,191.39** | **29,624.77** | **124,854.68** |

**STATE: NC - NORTH CAROLINA**

| REGISTRANT ZIP CODE 3 | 1ST QUARTER | 2ND QUARTER | 3RD QUARTER | 4TH QUARTER | **TOTAL GRAMS** |
|---|---|---|---|---|---|
| 270 | 14,517.08 | 13,431.92 | 13,015.07 | 13,650.61 | **54,614.68** |
| 271 | 10,647.46 | 9,690.58 | 9,527.41 | 9,841.12 | **39,706.58** |
| 272 | 30,521.47 | 29,192.18 | 28,338.54 | 29,456.76 | **117,508.94** |

| | | | | | |
|---|---|---|---|---|---|
| 273 | 21,442.05 | 20,626.42 | 20,404.48 | 22,646.28 | **85,119.23** |
| 274 | 11,921.98 | 11,793.95 | 11,637.00 | 11,617.67 | **46,970.60** |
| 275 | 45,232.87 | 43,351.54 | 42,602.48 | 45,294.21 | **176,481.11** |
| 276 | 12,905.20 | 12,283.76 | 12,169.96 | 12,647.23 | **50,006.14** |
| 277 | 13,490.27 | 12,855.81 | 12,521.82 | 11,821.79 | **50,689.70** |
| 278 | 26,354.98 | 23,825.77 | 24,024.54 | 24,633.61 | **98,838.90** |
| 279 | 11,464.93 | 10,743.31 | 10,822.34 | 11,812.99 | **44,843.58** |
| 280 | 35,041.27 | 33,371.16 | 32,242.26 | 33,892.02 | **134,546.71** |
| 281 | 25,798.57 | 25,275.14 | 24,679.51 | 25,602.14 | **101,355.36** |
| 282 | 19,850.30 | 18,805.89 | 18,361.59 | 18,819.65 | **75,837.43** |
| 283 | 68,882.21 | 66,333.45 | 64,903.06 | 69,778.16 | **269,896.89** |
| 284 | 35,448.30 | 33,923.84 | 32,531.62 | 35,122.63 | **137,026.40** |
| 285 | 29,701.06 | 28,010.89 | 26,707.74 | 27,648.74 | **112,068.44** |
| 286 | 46,485.03 | 44,705.36 | 43,894.33 | 46,187.79 | **181,272.51** |
| 287 | 25,736.89 | 24,526.64 | 23,460.56 | 23,844.12 | **97,568.22** |
| 288 | 10,487.15 | 9,783.00 | 9,475.22 | 8,971.02 | **38,716.39** |
| 289 | 3,513.19 | 3,462.79 | 3,236.67 | 3,212.03 | **13,424.68** |
| **NC - NORTH CAROLINA - Total** | **499,442.28** | **475,993.43** | **464,556.21** | **486,500.56** | **1,926,492.48** |

**STATE: ND - NORTH DAKOTA**

| REGISTRANT ZIP CODE 3 | 1ST QUARTER | 2ND QUARTER | 3RD QUARTER | 4TH QUARTER | TOTAL GRAMS |
|---|---|---|---|---|---|
| 580 | 531.22 | 530.59 | 607.92 | 637.42 | **2,307.15** |
| 581 | 2,260.23 | 2,002.90 | 2,001.41 | 2,006.71 | **8,271.26** |
| 582 | 1,981.78 | 1,859.52 | 1,918.78 | 1,845.18 | **7,605.26** |
| 583 | 1,153.14 | 1,137.78 | 1,085.14 | 1,106.51 | **4,482.56** |
| 584 | 601.74 | 637.61 | 599.94 | 581.96 | **2,421.25** |
| 585 | 3,846.84 | 3,792.06 | 3,671.08 | 3,634.15 | **14,944.13** |
| 586 | 803.41 | 765.76 | 804.73 | 744.69 | **3,118.59** |
| 587 | 1,078.76 | 1,131.11 | 1,088.25 | 1,027.81 | **4,325.94** |
| 588 | 885.16 | 831.15 | 836.12 | 803.28 | **3,355.70** |
| **ND - NORTH DAKOTA - Total** | **13,142.28** | **12,688.48** | **12,613.37** | **12,387.71** | **50,831.84** |

**STATE: NE - NEBRASKA**

| REGISTRANT ZIP CODE 3 | 1ST QUARTER | 2ND QUARTER | 3RD QUARTER | 4TH QUARTER | TOTAL GRAMS |
|---|---|---|---|---|---|
| 680 | 3,950.13 | 3,967.23 | 3,870.92 | 3,928.28 | **15,716.56** |
| 681 | 16,629.72 | 16,103.94 | 15,499.01 | 15,375.74 | **63,608.40** |
| 683 | 1,009.39 | 1,026.72 | 955.22 | 995.64 | **3,986.97** |
| 684 | 891.98 | 919.45 | 851.19 | 1,030.70 | **3,693.32** |
| 685 | 4,853.06 | 4,519.99 | 4,262.40 | 4,310.20 | **17,945.66** |
| 686 | 599.46 | 581.60 | 552.87 | 459.96 | **2,193.89** |
| 687 | 1,246.70 | 1,213.92 | 1,127.25 | 1,137.65 | **4,725.52** |
| 688 | 2,813.68 | 2,775.37 | 2,722.55 | 2,635.45 | **10,947.05** |

| | | | | | |
|---|---|---|---|---|---|
| 689 | 1,161.85 | 1,220.98 | 1,186.19 | 1,106.08 | **4,675.10** |
| 690 | 567.13 | 547.85 | 550.36 | 553.77 | **2,219.11** |
| 691 | 1,669.18 | 1,691.36 | 1,611.62 | 1,586.45 | **6,558.61** |
| 692 | 97.90 | 94.80 | 70.87 | 100.86 | **364.43** |
| 693 | 1,884.93 | 1,811.59 | 1,693.27 | 1,636.50 | **7,026.30** |
| **NE - NEBRASKA - Total** | **37,375.11** | **36,474.80** | **34,953.72** | **34,857.29** | **143,660.91** |

**STATE: NH - NEW HAMPSHIRE**

| REGISTRANT ZIP CODE 3 | 1ST QUARTER | 2ND QUARTER | 3RD QUARTER | 4TH QUARTER | TOTAL GRAMS |
|---|---|---|---|---|---|
| 030 | 12,830.71 | 12,463.81 | 11,835.77 | 11,847.31 | **48,977.60** |
| 031 | 7,235.72 | 7,022.40 | 6,762.11 | 6,812.73 | **27,832.96** |
| 032 | 6,047.66 | 5,777.77 | 5,230.39 | 5,343.99 | **22,399.81** |
| 033 | 4,253.33 | 4,279.52 | 3,975.64 | 3,644.06 | **16,152.56** |
| 034 | 2,463.89 | 2,349.95 | 2,267.98 | 2,186.16 | **9,267.97** |
| 035 | 2,323.76 | 2,044.05 | 2,124.76 | 2,126.12 | **8,618.69** |
| 036 | 254.61 | 238.47 | 241.38 | 229.06 | **963.51** |
| 037 | 3,384.60 | 3,272.26 | 3,058.37 | 2,869.61 | **12,584.84** |
| 038 | 16,691.75 | 15,415.13 | 14,448.58 | 13,359.98 | **59,915.44** |
| **NH - NEW HAMPSHIRE - Total** | **55,486.03** | **52,863.35** | **49,944.98** | **48,419.02** | **206,713.38** |

**STATE: NJ - NEW JERSEY**

| REGISTRANT ZIP CODE 3 | 1ST QUARTER | 2ND QUARTER | 3RD QUARTER | 4TH QUARTER | TOTAL GRAMS |
|---|---|---|---|---|---|
| 070 | 61,555.63 | 60,159.87 | 58,094.91 | 56,617.77 | **236,428.18** |
| 071 | 14,734.48 | 14,298.73 | 13,463.74 | 13,660.80 | **56,157.75** |
| 072 | 5,330.55 | 5,158.27 | 5,219.90 | 5,112.48 | **20,821.20** |
| 073 | 9,045.56 | 8,975.58 | 8,509.00 | 8,371.10 | **34,901.24** |
| 074 | 18,899.71 | 18,579.88 | 17,467.45 | 17,075.23 | **72,022.27** |
| 075 | 7,377.62 | 6,946.62 | 6,907.09 | 6,435.28 | **27,666.60** |
| 076 | 21,915.87 | 21,478.70 | 21,188.92 | 21,689.73 | **86,273.23** |
| 077 | 29,511.10 | 28,930.95 | 27,389.29 | 27,552.86 | **113,384.20** |
| 078 | 11,974.63 | 11,865.43 | 11,743.34 | 11,600.77 | **47,184.17** |
| 079 | 6,322.72 | 6,015.62 | 5,687.70 | 5,877.19 | **23,903.23** |
| 080 | 95,462.88 | 92,023.81 | 88,502.61 | 86,144.85 | **362,134.15** |
| 081 | 13,377.36 | 13,181.11 | 12,898.86 | 12,998.56 | **52,455.89** |
| 082 | 20,554.65 | 18,667.77 | 18,236.44 | 17,693.87 | **75,152.72** |
| 083 | 13,713.88 | 13,177.18 | 12,596.64 | 12,190.45 | **51,678.14** |
| 084 | 3,012.87 | 3,001.06 | 3,372.68 | 3,422.69 | **12,809.31** |
| 085 | 10,396.17 | 9,763.19 | 9,198.06 | 8,997.66 | **38,355.07** |
| 086 | 12,650.37 | 12,507.25 | 12,144.80 | 12,422.40 | **49,724.83** |
| 087 | 23,608.27 | 21,780.14 | 21,184.11 | 20,731.57 | **87,304.10** |
| 088 | 42,702.85 | 40,904.24 | 39,587.57 | 38,503.75 | **161,698.41** |
| 089 | 3,481.82 | 3,462.87 | 3,477.67 | 3,358.85 | **13,781.21** |

| NJ - NEW JERSEY - Total | 425,629.00 | 410,878.28 | 396,870.77 | 390,457.85 | 1,623,835.91 |
|---|---|---|---|---|---|

STATE: NM - NEW MEXICO

| REGISTRANT ZIP CODE 3 | 1ST QUARTER | 2ND QUARTER | 3RD QUARTER | 4TH QUARTER | TOTAL GRAMS |
|---|---|---|---|---|---|
| 870 | 7,860.56 | 7,478.63 | 7,507.52 | 7,278.87 | **30,125.59** |
| 871 | 37,404.17 | 35,771.43 | 35,009.40 | 34,001.96 | **142,186.96** |
| 873 | 1,150.61 | 1,194.27 | 1,059.71 | 1,074.74 | **4,479.33** |
| 874 | 2,401.77 | 2,338.31 | 2,449.42 | 2,286.17 | **9,475.66** |
| 875 | 13,232.81 | 13,654.66 | 12,937.98 | 12,953.05 | **52,778.49** |
| 877 | 1,688.42 | 1,655.25 | 1,642.17 | 1,550.90 | **6,536.75** |
| 878 | 659.38 | 549.51 | 555.16 | 588.78 | **2,352.82** |
| 879 | 426.24 | 430.54 | 422.43 | 384.37 | **1,663.59** |
| 880 | 5,178.52 | 5,380.68 | 4,980.79 | 4,936.53 | **20,476.53** |
| 881 | 1,070.06 | 1,007.76 | 972.75 | 906.17 | **3,956.73** |
| 882 | 7,011.80 | 6,639.28 | 6,612.04 | 6,558.76 | **26,821.88** |
| 883 | 2,633.42 | 2,371.25 | 2,373.70 | 2,241.88 | **9,620.25** |
| 884 | 169.75 | 156.44 | 157.14 | 165.18 | **648.51** |
| NM - NEW MEXICO - Total | 80,887.52 | 78,628.00 | 76,680.20 | 74,927.37 | 311,123.09 |

STATE: NV - NEVADA

| REGISTRANT ZIP CODE 3 | 1ST QUARTER | 2ND QUARTER | 3RD QUARTER | 4TH QUARTER | TOTAL GRAMS |
|---|---|---|---|---|---|
| 890 | 37,030.34 | 35,249.18 | 33,507.71 | 33,968.78 | **139,756.01** |
| 891 | 87,848.87 | 81,677.70 | 81,628.23 | 79,841.56 | **330,996.36** |
| 893 | 486.12 | 502.17 | 451.39 | 412.39 | **1,852.07** |
| 894 | 12,332.12 | 12,035.78 | 11,462.08 | 11,249.07 | **47,079.05** |
| 895 | 14,448.01 | 13,566.55 | 13,029.06 | 12,481.58 | **53,525.19** |
| 897 | 4,659.39 | 4,365.91 | 4,086.83 | 4,043.37 | **17,155.51** |
| 898 | 1,145.57 | 1,137.09 | 1,156.83 | 1,140.31 | **4,579.81** |
| NV - NEVADA - Total | 157,950.42 | 148,534.39 | 145,322.13 | 143,137.06 | 594,943.99 |

STATE: NY - NEW YORK

| REGISTRANT ZIP CODE 3 | 1ST QUARTER | 2ND QUARTER | 3RD QUARTER | 4TH QUARTER | TOTAL GRAMS |
|---|---|---|---|---|---|
| 100 | 62,452.44 | 60,439.06 | 57,381.26 | 54,232.50 | **234,505.26** |
| 101 | 1,359.97 | 1,214.54 | 1,149.81 | 1,078.49 | **4,802.81** |
| 102 | 75.07 | 59.65 | 57.83 | 46.19 | **238.73** |
| 103 | 25,023.16 | 24,861.20 | 23,868.69 | 23,328.51 | **97,081.56** |
| 104 | 49,151.44 | 47,895.48 | 47,471.59 | 44,761.11 | **189,279.63** |
| 105 | 17,785.77 | 17,234.44 | 16,485.06 | 16,400.82 | **67,906.09** |
| 106 | 2,823.16 | 2,805.41 | 2,776.43 | 2,102.30 | **10,507.30** |
| 107 | 6,836.49 | 7,195.59 | 6,847.65 | 6,711.41 | **27,591.13** |
| 108 | 1,944.06 | 1,848.13 | 1,852.49 | 1,822.64 | **7,467.32** |
| 109 | 24,210.34 | 23,155.12 | 22,065.65 | 21,218.02 | **90,649.12** |
| 110 | 10,629.19 | 10,470.90 | 10,074.05 | 10,215.03 | **41,389.17** |

| | | | | | |
|---|---|---|---|---|---|
| 111 | 7,130.19 | 7,522.77 | 7,084.97 | 6,550.54 | **28,288.47** |
| 112 | 59,365.28 | 58,437.80 | 56,028.02 | 53,956.53 | **227,787.63** |
| 113 | 24,095.14 | 23,534.28 | 22,186.19 | 21,132.27 | **90,947.88** |
| 114 | 12,016.88 | 12,141.20 | 11,370.84 | 11,375.15 | **46,904.07** |
| 115 | 26,006.43 | 25,971.37 | 25,169.53 | 24,649.62 | **101,796.95** |
| 116 | 3,206.61 | 3,438.14 | 3,058.38 | 3,103.26 | **12,806.40** |
| 117 | 78,777.81 | 77,482.88 | 73,985.18 | 74,140.22 | **304,386.09** |
| 118 | 2,824.32 | 2,762.81 | 2,682.81 | 2,688.29 | **10,958.23** |
| 119 | 8,715.29 | 8,940.77 | 8,478.97 | 8,325.51 | **34,460.54** |
| 120 | 9,329.73 | 8,809.79 | 8,785.10 | 8,500.93 | **35,425.56** |
| 121 | 5,436.10 | 5,149.94 | 5,167.67 | 5,141.04 | **20,894.75** |
| 122 | 5,124.05 | 5,024.74 | 4,678.17 | 5,154.55 | **19,981.51** |
| 123 | 2,774.11 | 2,709.66 | 2,417.94 | 2,477.07 | **10,378.78** |
| 124 | 12,081.62 | 11,739.17 | 11,594.70 | 11,181.73 | **46,597.22** |
| 125 | 20,542.00 | 20,295.80 | 18,659.70 | 19,203.76 | **78,701.27** |
| 126 | 5,967.23 | 6,127.23 | 5,519.24 | 5,662.88 | **23,276.57** |
| 127 | 9,064.88 | 8,493.71 | 8,138.56 | 7,544.02 | **33,241.17** |
| 128 | 8,491.71 | 8,248.43 | 7,871.77 | 7,920.77 | **32,532.67** |
| 129 | 5,791.89 | 5,707.78 | 5,847.12 | 5,791.88 | **23,138.67** |
| 130 | 10,403.15 | 10,337.87 | 9,785.42 | 9,814.55 | **40,341.00** |
| 131 | 2,952.17 | 3,011.51 | 2,827.85 | 2,797.26 | **11,588.79** |
| 132 | 7,347.11 | 6,925.53 | 6,806.71 | 6,951.95 | **28,031.30** |
| 133 | 3,111.63 | 3,133.51 | 2,972.29 | 2,779.41 | **11,996.85** |
| 134 | 4,097.93 | 3,912.54 | 3,884.86 | 3,791.53 | **15,686.86** |
| 135 | 1,827.31 | 1,746.96 | 1,662.95 | 1,524.40 | **6,761.62** |
| 136 | 6,438.63 | 6,219.30 | 6,147.34 | 5,795.98 | **24,601.25** |
| 137 | 4,307.03 | 4,014.32 | 3,607.92 | 3,718.97 | **15,648.24** |
| 138 | 7,370.18 | 7,089.59 | 7,164.78 | 6,802.70 | **28,427.25** |
| 139 | 4,627.85 | 4,253.03 | 3,776.97 | 3,651.32 | **16,309.16** |
| 140 | 11,744.62 | 11,386.91 | 10,916.23 | 10,592.60 | **44,640.35** |
| 141 | 5,092.70 | 5,316.39 | 5,372.35 | 5,054.28 | **20,835.72** |
| 142 | 16,012.94 | 15,470.95 | 14,243.32 | 14,400.34 | **60,127.55** |
| 143 | 1,935.29 | 2,078.09 | 2,017.59 | 1,975.52 | **8,006.49** |
| 144 | 7,888.90 | 7,620.96 | 7,349.76 | 7,517.11 | **30,376.73** |
| 145 | 6,673.03 | 6,566.72 | 6,320.62 | 6,244.29 | **25,804.67** |
| 146 | 17,455.93 | 16,776.21 | 16,667.42 | 15,957.69 | **66,857.24** |
| 147 | 3,544.63 | 3,507.62 | 3,302.11 | 3,005.38 | **13,359.74** |
| 148 | 10,563.08 | 10,399.45 | 9,462.88 | 9,047.25 | **39,472.65** |
| 149 | 3,285.00 | 3,124.71 | 2,893.15 | 2,921.01 | **12,223.87** |
| **NY - NEW YORK - Total** | **645,711.48** | **632,609.94** | **605,937.90** | **590,760.57** | **2,475,019.89** |

**STATE: OH - OHIO**

| REGISTRANT ZIP CODE 3 | 1ST QUARTER | 2ND QUARTER | 3RD QUARTER | 4TH QUARTER | TOTAL GRAMS |
|---|---|---|---|---|---|
| 430 | 24,688.35 | 24,453.94 | 23,758.11 | 23,967.74 | **96,868.14** |
| 431 | 18,802.60 | 18,298.60 | 18,154.91 | 17,931.27 | **73,187.38** |
| 432 | 40,486.60 | 37,121.74 | 33,068.03 | 33,089.31 | **143,765.68** |
| 433 | 6,130.31 | 6,168.41 | 5,990.35 | 5,833.78 | **24,122.84** |
| 434 | 5,404.29 | 5,235.09 | 5,019.86 | 4,781.82 | **20,441.07** |
| 435 | 7,582.00 | 7,567.42 | 7,258.95 | 7,188.75 | **29,597.12** |
| 436 | 20,547.68 | 19,853.39 | 19,588.34 | 18,998.10 | **78,987.51** |
| 437 | 9,151.85 | 8,890.28 | 8,314.91 | 8,264.18 | **34,621.22** |
| 438 | 1,425.65 | 1,410.95 | 1,251.31 | 1,249.16 | **5,337.07** |
| 439 | 9,078.86 | 8,526.43 | 8,132.10 | 7,914.85 | **33,652.25** |
| 440 | 21,120.31 | 19,542.37 | 18,885.15 | 18,627.85 | **78,175.67** |
| 441 | 30,740.19 | 29,506.84 | 28,316.49 | 28,194.83 | **116,758.36** |
| 442 | 14,818.47 | 13,905.72 | 13,698.64 | 13,361.87 | **55,784.70** |
| 443 | 10,385.57 | 9,958.17 | 9,636.76 | 9,292.18 | **39,272.68** |
| 444 | 15,693.81 | 15,116.73 | 14,625.49 | 14,206.92 | **59,642.95** |
| 445 | 7,646.81 | 7,160.16 | 6,989.53 | 6,942.27 | **28,738.77** |
| 446 | 10,978.41 | 10,446.11 | 9,695.84 | 9,719.60 | **40,839.96** |
| 447 | 6,737.26 | 6,388.51 | 6,290.55 | 6,184.73 | **25,601.05** |
| 448 | 11,075.56 | 10,672.23 | 10,212.78 | 9,167.72 | **41,128.29** |
| 449 | 2,679.27 | 2,741.76 | 2,711.48 | 2,618.11 | **10,750.61** |
| 450 | 25,382.81 | 25,074.65 | 24,524.74 | 24,384.83 | **99,367.03** |
| 451 | 12,871.94 | 12,711.03 | 12,435.29 | 12,420.61 | **50,438.88** |
| 452 | 34,875.26 | 33,896.62 | 32,583.96 | 32,887.84 | **134,243.68** |
| 453 | 11,728.08 | 12,014.06 | 11,583.84 | 11,819.92 | **47,145.89** |
| 454 | 15,170.30 | 14,649.58 | 14,134.19 | 14,087.17 | **58,041.23** |
| 455 | 5,086.75 | 5,014.50 | 5,064.14 | 5,106.09 | **20,271.49** |
| 456 | 20,261.31 | 19,547.77 | 18,705.48 | 18,675.01 | **77,189.56** |
| 457 | 5,646.90 | 5,361.11 | 5,207.47 | 5,211.59 | **21,427.07** |
| 458 | 6,298.65 | 6,024.44 | 5,998.81 | 5,793.34 | **24,115.24** |
| **OH - OHIO - Total** | **412,495.88** | **397,258.59** | **381,837.49** | **377,921.46** | **1,569,513.43** |

**STATE: OK - OKLAHOMA**

| REGISTRANT ZIP CODE 3 | 1ST QUARTER | 2ND QUARTER | 3RD QUARTER | 4TH QUARTER | TOTAL GRAMS |
|---|---|---|---|---|---|
| 730 | 33,784.25 | 32,591.03 | 30,094.75 | 27,248.83 | **123,718.86** |
| 731 | 40,602.38 | 39,082.71 | 36,164.80 | 31,799.78 | **147,649.66** |
| 734 | 4,787.66 | 4,516.56 | 3,927.76 | 3,538.44 | **16,770.41** |
| 735 | 10,302.48 | 10,269.45 | 10,165.86 | 8,510.99 | **39,248.78** |
| 736 | 2,621.81 | 2,618.80 | 2,381.23 | 2,119.52 | **9,741.36** |
| 737 | 3,379.49 | 3,420.43 | 3,358.26 | 3,123.20 | **13,281.37** |

| | | | | | |
|---|---|---|---|---|---|
| 738 | 783.73 | 804.54 | 683.86 | 571.81 | **2,843.94** |
| 739 | 169.87 | 180.42 | 134.21 | 164.87 | **649.37** |
| 740 | 33,214.14 | 30,609.88 | 28,646.61 | 25,943.86 | **118,414.48** |
| 741 | 22,394.42 | 21,452.56 | 20,453.18 | 18,878.14 | **83,178.30** |
| 743 | 5,565.29 | 4,946.55 | 4,819.44 | 4,335.96 | **19,667.25** |
| 744 | 16,370.93 | 15,344.73 | 14,898.63 | 12,987.72 | **59,602.02** |
| 745 | 4,008.86 | 3,907.07 | 3,378.96 | 3,025.05 | **14,319.94** |
| 746 | 2,620.89 | 2,804.63 | 2,139.07 | 1,883.18 | **9,447.77** |
| 747 | 3,099.91 | 2,962.58 | 2,744.81 | 2,505.88 | **11,313.18** |
| 748 | 9,247.98 | 9,105.19 | 8,685.77 | 7,691.08 | **34,730.01** |
| 749 | 9,209.17 | 7,643.70 | 6,909.49 | 6,249.53 | **30,011.90** |
| **OK - OKLAHOMA - Total** | **202,163.26** | **192,260.83** | **179,586.69** | **160,577.83** | **734,588.62** |

**STATE: OR - OREGON**

| REGISTRANT ZIP CODE 3 | 1ST QUARTER | 2ND QUARTER | 3RD QUARTER | 4TH QUARTER | **TOTAL GRAMS** |
|---|---|---|---|---|---|
| 970 | 25,799.20 | 25,305.72 | 24,809.39 | 23,728.59 | **99,642.90** |
| 971 | 11,490.55 | 11,553.93 | 10,870.43 | 10,209.04 | **44,123.95** |
| 972 | 31,585.87 | 30,646.00 | 29,588.04 | 29,594.28 | **121,414.20** |
| 973 | 19,737.24 | 18,528.72 | 17,811.08 | 17,222.80 | **73,299.83** |
| 974 | 29,148.00 | 28,323.54 | 26,840.59 | 25,469.40 | **109,781.52** |
| 975 | 18,515.73 | 18,389.54 | 17,570.07 | 17,639.97 | **72,115.32** |
| 976 | 2,328.42 | 2,318.14 | 2,114.33 | 1,949.99 | **8,710.87** |
| 977 | 7,345.53 | 7,232.04 | 7,118.19 | 6,728.24 | **28,424.01** |
| 978 | 6,027.05 | 5,620.26 | 5,312.50 | 5,264.43 | **22,224.24** |
| 979 | 1,612.17 | 1,586.30 | 1,446.37 | 1,397.06 | **6,041.90** |
| **OR - OREGON - Total** | **153,589.77** | **149,504.19** | **143,480.99** | **139,203.81** | **585,778.76** |

**STATE: PA - PENNSYLVANIA**

| REGISTRANT ZIP CODE 3 | 1ST QUARTER | 2ND QUARTER | 3RD QUARTER | 4TH QUARTER | **TOTAL GRAMS** |
|---|---|---|---|---|---|
| 150 | 23,722.70 | 23,090.41 | 21,797.18 | 21,310.70 | **89,921.00** |
| 151 | 11,629.62 | 10,578.90 | 10,316.61 | 10,605.42 | **43,130.56** |
| 152 | 30,480.88 | 29,068.66 | 27,862.13 | 27,570.57 | **114,982.24** |
| 153 | 6,198.68 | 6,034.28 | 5,765.54 | 5,600.73 | **23,599.23** |
| 154 | 9,727.85 | 9,633.26 | 9,553.30 | 9,203.47 | **38,117.87** |
| 155 | 3,210.43 | 2,803.08 | 2,733.33 | 2,712.74 | **11,459.58** |
| 156 | 13,591.07 | 13,009.79 | 12,116.32 | 11,746.43 | **50,463.62** |
| 157 | 5,064.13 | 4,826.10 | 4,521.51 | 4,544.61 | **18,956.36** |
| 158 | 4,080.12 | 4,024.53 | 3,997.52 | 3,856.96 | **15,959.13** |
| 159 | 12,042.80 | 11,363.47 | 10,848.86 | 10,278.77 | **44,533.90** |
| 160 | 6,989.27 | 6,926.16 | 6,546.53 | 6,291.41 | **26,753.36** |
| 161 | 9,999.65 | 8,904.87 | 8,667.32 | 8,401.30 | **35,973.14** |
| 162 | 3,480.44 | 3,408.44 | 3,193.89 | 3,039.71 | **13,122.48** |

| | | | | | |
|---|---|---|---|---|---|
| 163 | 6,664.89 | 6,507.96 | 6,145.56 | 5,967.68 | **25,286.09** |
| 164 | 2,471.22 | 2,423.25 | 2,447.05 | 2,324.08 | **9,665.61** |
| 165 | 4,921.82 | 4,756.50 | 4,742.92 | 4,592.23 | **19,013.47** |
| 166 | 9,887.16 | 9,733.89 | 9,157.83 | 8,745.51 | **37,524.40** |
| 167 | 1,581.43 | 1,526.32 | 1,548.67 | 1,479.05 | **6,135.48** |
| 168 | 4,060.16 | 3,480.05 | 3,491.94 | 3,115.46 | **14,147.61** |
| 169 | 2,194.04 | 2,066.38 | 1,918.67 | 1,872.36 | **8,051.45** |
| 170 | 26,893.26 | 25,348.53 | 24,663.97 | 23,638.08 | **100,543.84** |
| 171 | 7,858.12 | 7,289.95 | 6,784.51 | 6,459.62 | **28,392.20** |
| 172 | 8,200.15 | 8,115.18 | 7,770.76 | 7,525.06 | **31,611.15** |
| 173 | 11,270.62 | 10,972.32 | 10,434.61 | 10,272.81 | **42,950.37** |
| 174 | 6,590.69 | 6,460.82 | 6,008.83 | 5,974.21 | **25,034.55** |
| 175 | 10,295.38 | 9,591.60 | 9,404.93 | 8,974.53 | **38,266.45** |
| 176 | 8,173.49 | 7,652.58 | 7,526.91 | 7,095.60 | **30,448.58** |
| 177 | 7,014.29 | 6,613.36 | 6,634.06 | 6,335.47 | **26,597.18** |
| 178 | 7,329.30 | 6,867.85 | 6,725.61 | 6,486.56 | **27,409.32** |
| 179 | 5,258.24 | 4,916.34 | 4,696.18 | 4,379.67 | **19,250.44** |
| 180 | 16,355.88 | 15,951.41 | 14,994.47 | 14,744.84 | **62,046.60** |
| 181 | 9,578.52 | 9,397.39 | 8,833.21 | 8,886.31 | **36,695.43** |
| 182 | 6,308.93 | 6,019.27 | 5,654.38 | 5,487.15 | **23,469.73** |
| 183 | 9,788.71 | 9,804.73 | 9,251.80 | 9,034.66 | **37,879.89** |
| 184 | 10,101.20 | 9,442.87 | 8,924.74 | 8,655.55 | **37,124.35** |
| 185 | 11,890.39 | 11,030.50 | 10,232.84 | 9,781.90 | **42,935.64** |
| 186 | 8,347.17 | 7,799.24 | 7,428.40 | 7,073.28 | **30,648.09** |
| 187 | 27,456.40 | 26,749.36 | 24,148.02 | 24,018.28 | **102,372.06** |
| 188 | 3,034.10 | 3,084.99 | 2,740.34 | 2,535.74 | **11,395.16** |
| 189 | 15,471.08 | 15,488.84 | 14,465.72 | 14,213.49 | **59,639.13** |
| 190 | 87,805.91 | 86,221.27 | 86,546.74 | 81,203.11 | **341,777.04** |
| 191 | 117,559.28 | 114,324.22 | 108,369.36 | 105,048.18 | **445,301.04** |
| 193 | 16,141.61 | 15,958.05 | 15,267.36 | 14,950.32 | **62,317.34** |
| 194 | 25,222.67 | 23,825.33 | 22,392.74 | 21,825.72 | **93,266.46** |
| 195 | 5,585.03 | 5,393.73 | 4,969.21 | 4,820.67 | **20,768.64** |
| 196 | 7,738.70 | 7,518.82 | 6,912.25 | 6,751.92 | **28,921.69** |
| **PA - PENNSYLVANIA - Total** | **649,267.48** | **626,004.85** | **599,154.66** | **579,431.92** | **2,453,858.91** |

**STATE: PR - PUERTO RICO**

| REGISTRANT ZIP CODE 3 | 1ST QUARTER | 2ND QUARTER | 3RD QUARTER | 4TH QUARTER | **TOTAL GRAMS** |
|---|---|---|---|---|---|
| 006 | 1,523.51 | 1,699.99 | 4,341.66 | 4,186.43 | **11,751.59** |
| 007 | 2,254.20 | 2,178.05 | 4,566.55 | 4,572.82 | **13,571.62** |
| 009 | 3,627.01 | 4,022.03 | 5,798.56 | 5,560.09 | **19,007.70** |
| **PR - PUERTO RICO - Total** | **7,404.73** | **7,900.07** | **14,706.77** | **14,319.35** | **44,330.91** |

**STATE: RI - RHODE ISLAND**

| REGISTRANT ZIP CODE 3 | 1ST QUARTER | 2ND QUARTER | 3RD QUARTER | 4TH QUARTER | TOTAL GRAMS |
|---|---|---|---|---|---|
| 028 | 21,673.83 | 20,218.35 | 19,454.50 | 19,547.41 | **80,894.10** |
| 029 | 14,193.43 | 13,529.59 | 12,621.21 | 12,683.56 | **53,027.80** |
| **RI - RHODE ISLAND - Total** | **35,867.26** | **33,747.94** | **32,075.72** | **32,230.97** | **133,921.89** |

**STATE: SC - SOUTH CAROLINA**

| REGISTRANT ZIP CODE 3 | 1ST QUARTER | 2ND QUARTER | 3RD QUARTER | 4TH QUARTER | TOTAL GRAMS |
|---|---|---|---|---|---|
| 290 | 11,440.22 | 11,072.67 | 10,939.33 | 10,777.02 | **44,229.24** |
| 291 | 14,009.46 | 13,922.53 | 13,063.63 | 13,151.19 | **54,146.80** |
| 292 | 9,252.13 | 9,015.38 | 9,013.15 | 8,808.83 | **36,089.48** |
| 293 | 23,725.59 | 21,428.91 | 20,153.90 | 19,125.44 | **84,433.83** |
| 294 | 36,711.88 | 34,272.01 | 33,423.32 | 32,368.77 | **136,775.97** |
| 295 | 47,616.54 | 44,743.13 | 42,806.05 | 43,481.62 | **178,647.33** |
| 296 | 39,021.21 | 36,458.43 | 35,429.64 | 34,227.37 | **145,136.65** |
| 297 | 9,607.17 | 8,949.68 | 8,418.13 | 8,736.22 | **35,711.20** |
| 298 | 8,843.48 | 8,351.87 | 8,051.72 | 7,625.17 | **32,872.24** |
| 299 | 6,583.61 | 6,557.74 | 6,650.28 | 6,479.97 | **26,271.60** |
| **SC - SOUTH CAROLINA - Total** | **206,811.28** | **194,772.35** | **187,949.15** | **184,781.57** | **774,314.35** |

**STATE: SD - SOUTH DAKOTA**

| REGISTRANT ZIP CODE 3 | 1ST QUARTER | 2ND QUARTER | 3RD QUARTER | 4TH QUARTER | TOTAL GRAMS |
|---|---|---|---|---|---|
| 570 | 2,577.84 | 2,400.97 | 2,420.19 | 2,365.46 | **9,764.46** |
| 571 | 4,801.78 | 4,752.14 | 4,266.07 | 4,172.98 | **17,992.97** |
| 572 | 658.03 | 701.66 | 680.40 | 636.43 | **2,676.51** |
| 573 | 885.83 | 812.34 | 797.03 | 780.76 | **3,275.96** |
| 574 | 878.12 | 873.29 | 817.38 | 775.80 | **3,344.59** |
| 575 | 655.39 | 624.41 | 616.12 | 532.75 | **2,428.66** |
| 576 | 177.33 | 132.86 | 135.82 | 132.28 | **578.29** |
| 577 | 4,582.00 | 4,189.99 | 4,007.81 | 3,800.26 | **16,580.07** |
| **SD - SOUTH DAKOTA - Total** | **15,216.32** | **14,487.66** | **13,740.82** | **13,196.71** | **56,641.51** |

**STATE: TN - TENNESSEE**

| REGISTRANT ZIP CODE 3 | 1ST QUARTER | 2ND QUARTER | 3RD QUARTER | 4TH QUARTER | TOTAL GRAMS |
|---|---|---|---|---|---|
| 370 | 56,545.11 | 54,315.16 | 51,427.23 | 48,757.14 | **211,044.63** |
| 371 | 43,686.24 | 42,368.14 | 41,225.90 | 40,244.76 | **167,525.04** |
| 372 | 27,807.50 | 27,428.23 | 25,974.25 | 26,315.67 | **107,525.66** |
| 373 | 40,621.03 | 38,628.14 | 36,949.93 | 36,936.88 | **153,135.98** |
| 374 | 15,142.97 | 14,509.15 | 13,141.92 | 13,186.01 | **55,980.04** |
| 376 | 34,084.11 | 33,234.77 | 31,791.06 | 30,630.22 | **129,740.17** |
| 377 | 33,037.44 | 32,398.45 | 30,874.31 | 29,920.40 | **126,230.60** |
| 378 | 44,679.20 | 43,376.02 | 40,148.89 | 39,056.26 | **167,260.38** |
| 379 | 22,816.34 | 22,468.44 | 21,397.08 | 20,838.08 | **87,519.95** |

| | | | | | |
|---|---|---|---|---|---|
| 380 | 14,327.47 | 13,846.13 | 12,890.19 | 12,574.77 | **53,638.56** |
| 381 | 26,212.99 | 26,146.93 | 28,584.04 | 24,682.05 | **105,626.01** |
| 382 | 7,252.93 | 6,779.23 | 6,321.60 | 5,853.48 | **26,207.23** |
| 383 | 14,370.49 | 13,872.71 | 12,813.53 | 12,077.43 | **53,134.15** |
| 384 | 12,974.14 | 13,069.77 | 12,632.69 | 12,711.46 | **51,388.06** |
| 385 | 27,433.21 | 26,967.71 | 24,467.19 | 24,050.79 | **102,918.90** |
| **TN - TENNESSEE - Total** | **420,991.18** | **409,408.99** | **390,639.81** | **377,835.40** | **1,598,875.37** |

**STATE: TX - TEXAS**

| REGISTRANT ZIP CODE 3 | 1ST QUARTER | 2ND QUARTER | 3RD QUARTER | 4TH QUARTER | TOTAL GRAMS |
|---|---|---|---|---|---|
| 750 | 33,365.25 | 31,958.30 | 31,100.89 | 30,798.82 | **127,223.26** |
| 751 | 7,815.18 | 7,565.07 | 7,720.93 | 7,502.90 | **30,604.09** |
| 752 | 9,958.25 | 9,510.31 | 8,802.27 | 8,806.43 | **37,077.27** |
| 753 | 185.40 | 213.23 | 154.52 | 210.76 | **763.90** |
| 754 | 4,765.86 | 4,496.82 | 4,270.45 | 4,285.95 | **17,819.08** |
| 755 | 1,459.63 | 1,532.33 | 1,283.70 | 1,151.73 | **5,427.38** |
| 756 | 2,692.04 | 2,797.66 | 2,436.94 | 2,492.72 | **10,419.37** |
| 757 | 4,488.87 | 4,231.86 | 4,395.93 | 4,196.34 | **17,313.00** |
| 758 | 838.32 | 792.66 | 769.76 | 722.61 | **3,123.35** |
| 759 | 1,640.32 | 1,524.45 | 1,601.03 | 1,626.40 | **6,392.20** |
| 760 | 24,107.86 | 22,534.57 | 21,361.60 | 21,163.23 | **89,167.26** |
| 761 | 17,506.12 | 16,641.04 | 15,520.46 | 15,496.70 | **65,164.33** |
| 762 | 7,931.28 | 7,542.92 | 7,256.46 | 7,227.25 | **29,957.91** |
| 763 | 2,388.69 | 2,538.36 | 2,564.37 | 2,279.81 | **9,771.23** |
| 764 | 1,294.72 | 1,255.29 | 1,243.80 | 1,235.93 | **5,029.74** |
| 765 | 5,838.69 | 5,446.36 | 5,262.79 | 4,467.31 | **21,015.16** |
| 766 | 1,488.93 | 1,367.60 | 1,298.45 | 1,300.76 | **5,455.74** |
| 767 | 2,252.01 | 2,294.09 | 2,139.15 | 2,259.70 | **8,944.95** |
| 768 | 875.70 | 891.36 | 890.77 | 837.12 | **3,494.95** |
| 769 | 870.81 | 858.29 | 860.38 | 790.22 | **3,379.70** |
| 770 | 46,528.58 | 50,396.22 | 46,338.29 | 48,181.48 | **191,444.57** |
| 773 | 12,591.86 | 12,446.37 | 12,514.74 | 12,653.33 | **50,206.30** |
| 774 | 14,655.29 | 14,823.63 | 15,290.06 | 17,801.81 | **62,570.78** |
| 775 | 12,145.65 | 12,931.38 | 13,351.58 | 14,383.86 | **52,812.47** |
| 776 | 2,507.08 | 2,232.95 | 2,381.25 | 2,306.71 | **9,427.99** |
| 777 | 983.37 | 836.39 | 1,064.23 | 1,121.81 | **4,005.80** |
| 778 | 1,195.87 | 1,211.44 | 1,161.97 | 1,151.16 | **4,720.44** |
| 779 | 3,097.54 | 3,074.43 | 3,302.80 | 3,145.97 | **12,620.73** |
| 780 | 3,266.38 | 3,121.63 | 3,069.11 | 2,752.91 | **12,210.03** |
| 781 | 3,711.70 | 3,742.29 | 3,543.18 | 3,163.71 | **14,160.87** |
| 782 | 21,444.39 | 20,386.68 | 18,676.44 | 17,362.05 | **77,869.55** |

| | | | | | |
|---|---|---|---|---|---|
| 783 | 1,112.82 | 1,039.57 | 1,090.45 | 1,111.10 | **4,353.94** |
| 784 | 1,678.71 | 1,581.83 | 1,523.45 | 1,591.72 | **6,375.71** |
| 785 | 2,882.42 | 2,789.01 | 2,680.02 | 2,735.74 | **11,087.19** |
| 786 | 16,572.71 | 16,112.75 | 14,357.97 | 13,364.24 | **60,407.68** |
| 787 | 13,477.69 | 13,430.46 | 11,784.96 | 11,392.41 | **50,085.51** |
| 788 | 541.39 | 494.45 | 517.99 | 411.30 | **1,965.13** |
| 789 | 653.28 | 628.59 | 625.95 | 605.85 | **2,513.67** |
| 790 | 1,203.16 | 1,197.82 | 1,081.48 | 1,004.54 | **4,487.01** |
| 791 | 2,734.36 | 2,893.19 | 2,574.50 | 2,789.73 | **10,991.77** |
| 792 | 354.99 | 305.21 | 266.62 | 290.82 | **1,217.65** |
| 793 | 621.90 | 713.75 | 641.74 | 639.76 | **2,617.15** |
| 794 | 2,547.36 | 2,389.88 | 2,265.42 | 2,225.97 | **9,428.64** |
| 795 | 618.72 | 488.91 | 583.79 | 575.24 | **2,266.65** |
| 796 | 1,440.94 | 1,382.91 | 1,320.70 | 1,190.55 | **5,335.10** |
| 797 | 3,235.86 | 3,085.52 | 2,996.74 | 2,844.55 | **12,162.67** |
| 798 | 144.36 | 127.11 | 141.00 | 129.81 | **542.29** |
| 799 | 4,205.07 | 4,006.90 | 4,269.26 | 3,444.15 | **15,925.38** |
| **TX - TEXAS - Total** | **307,917.35** | **303,863.86** | **290,350.34** | **289,225.01** | **1,191,356.55** |

**STATE: UT - UTAH**

| REGISTRANT ZIP CODE 3 | 1ST QUARTER | 2ND QUARTER | 3RD QUARTER | 4TH QUARTER | TOTAL GRAMS |
|---|---|---|---|---|---|
| 840 | 61,224.59 | 59,003.41 | 57,919.53 | 57,502.58 | **235,650.12** |
| 841 | 39,631.29 | 37,713.21 | 36,511.80 | 35,365.46 | **149,221.77** |
| 843 | 4,878.42 | 4,740.59 | 4,730.53 | 4,738.55 | **19,088.10** |
| 844 | 13,125.81 | 12,427.74 | 12,401.89 | 12,217.25 | **50,172.68** |
| 845 | 4,803.31 | 4,820.83 | 4,480.72 | 4,605.21 | **18,710.07** |
| 846 | 12,242.06 | 11,566.84 | 11,482.17 | 11,212.32 | **46,503.39** |
| 847 | 17,090.44 | 16,896.91 | 16,805.65 | 16,565.69 | **67,358.69** |
| **UT - UTAH - Total** | **152,995.93** | **147,169.53** | **144,332.30** | **142,207.06** | **586,704.81** |

**STATE: VA - VIRGINIA**

| REGISTRANT ZIP CODE 3 | 1ST QUARTER | 2ND QUARTER | 3RD QUARTER | 4TH QUARTER | TOTAL GRAMS |
|---|---|---|---|---|---|
| 201 | 17,248.15 | 17,738.14 | 16,825.83 | 17,087.91 | **68,900.03** |
| 220 | 10,023.86 | 9,595.57 | 9,488.60 | 8,992.98 | **38,101.00** |
| 221 | 9,343.45 | 9,150.28 | 9,246.16 | 9,156.36 | **36,896.24** |
| 222 | 2,857.56 | 2,927.39 | 2,809.37 | 2,726.07 | **11,320.38** |
| 223 | 5,120.26 | 5,154.36 | 4,772.89 | 4,744.95 | **19,792.46** |
| 224 | 8,593.37 | 8,007.64 | 7,789.40 | 7,562.07 | **31,952.48** |
| 225 | 3,384.06 | 3,106.79 | 3,237.26 | 3,242.63 | **12,970.74** |
| 226 | 8,287.14 | 7,983.41 | 7,690.13 | 7,250.38 | **31,211.06** |
| 227 | 1,693.85 | 1,702.40 | 1,666.58 | 1,761.82 | **6,824.65** |
| 228 | 4,797.05 | 4,877.63 | 4,889.04 | 4,779.30 | **19,343.02** |

| | | | | | |
|---|---|---|---|---|---|
| 229 | 7,981.94 | 7,441.66 | 7,427.43 | 7,185.61 | **30,036.64** |
| 230 | 5,402.89 | 5,279.29 | 5,286.94 | 5,196.64 | **21,165.76** |
| 231 | 11,246.44 | 10,566.97 | 9,653.25 | 9,802.41 | **41,269.07** |
| 232 | 21,870.63 | 20,237.99 | 19,972.18 | 19,520.36 | **81,601.16** |
| 233 | 8,425.37 | 8,461.08 | 8,040.68 | 7,777.28 | **32,704.41** |
| 234 | 17,477.28 | 17,111.75 | 15,841.75 | 15,382.40 | **65,813.18** |
| 235 | 6,665.22 | 6,062.30 | 5,831.27 | 5,751.47 | **24,310.27** |
| 236 | 10,458.07 | 9,979.34 | 9,886.19 | 9,239.94 | **39,563.54** |
| 237 | 5,340.53 | 4,678.98 | 4,757.25 | 4,424.27 | **19,201.03** |
| 238 | 14,892.35 | 14,443.32 | 14,287.26 | 15,315.50 | **58,938.43** |
| 239 | 2,462.24 | 2,446.57 | 2,322.29 | 2,343.99 | **9,575.09** |
| 240 | 15,732.51 | 15,303.55 | 14,970.90 | 15,151.19 | **61,158.14** |
| 241 | 14,329.94 | 13,570.39 | 13,178.18 | 13,488.95 | **54,567.47** |
| 242 | 19,962.60 | 19,253.76 | 18,534.21 | 18,912.38 | **76,662.94** |
| 243 | 7,921.17 | 7,693.45 | 7,607.83 | 8,097.19 | **31,319.63** |
| 244 | 4,143.90 | 4,024.55 | 3,905.27 | 3,862.25 | **15,935.97** |
| 245 | 11,558.39 | 11,147.87 | 11,010.65 | 10,851.43 | **44,568.34** |
| 246 | 5,304.21 | 4,877.53 | 4,618.02 | 4,689.58 | **19,489.35** |
| **VA - VIRGINIA - Total** | **262,524.42** | **252,823.97** | **245,546.79** | **244,297.29** | **1,005,192.47** |

**STATE: VI - VIRGIN ISLANDS**

| REGISTRANT ZIP CODE 3 | 1ST QUARTER | 2ND QUARTER | 3RD QUARTER | 4TH QUARTER | **TOTAL GRAMS** |
|---|---|---|---|---|---|
| 008 | 463.94 | 480.43 | 394.91 | 413.29 | **1,752.57** |
| **VI - VIRGIN ISLANDS - Total** | **463.94** | **480.43** | **394.91** | **413.29** | **1,752.57** |

**STATE: VT - VERMONT**

| REGISTRANT ZIP CODE 3 | 1ST QUARTER | 2ND QUARTER | 3RD QUARTER | 4TH QUARTER | **TOTAL GRAMS** |
|---|---|---|---|---|---|
| 050 | 1,824.90 | 1,941.17 | 1,737.97 | 1,628.37 | **7,132.42** |
| 051 | 1,622.42 | 1,479.92 | 1,501.34 | 1,460.16 | **6,063.84** |
| 052 | 1,165.15 | 1,160.78 | 1,209.62 | 1,094.30 | **4,629.85** |
| 053 | 1,467.67 | 1,559.32 | 1,551.50 | 1,423.89 | **6,002.38** |
| 054 | 9,252.65 | 8,721.03 | 8,300.89 | 8,184.29 | **34,458.84** |
| 056 | 1,916.40 | 1,866.31 | 1,804.41 | 1,721.56 | **7,308.68** |
| 057 | 1,807.96 | 1,675.85 | 1,533.78 | 1,602.55 | **6,620.15** |
| 058 | 1,784.37 | 1,619.75 | 1,693.14 | 1,704.84 | **6,802.09** |
| **VT - VERMONT - Total** | **20,841.53** | **20,024.13** | **19,332.64** | **18,819.96** | **79,018.26** |

**STATE: WA - WASHINGTON**

| REGISTRANT ZIP CODE 3 | 1ST QUARTER | 2ND QUARTER | 3RD QUARTER | 4TH QUARTER | **TOTAL GRAMS** |
|---|---|---|---|---|---|
| 980 | 43,903.02 | 43,249.32 | 41,153.86 | 41,090.48 | **169,396.68** |
| 981 | 28,299.31 | 26,618.30 | 26,144.51 | 26,037.17 | **107,099.29** |
| 982 | 34,718.88 | 33,395.09 | 33,290.16 | 31,918.25 | **133,322.38** |
| 983 | 30,509.47 | 29,908.79 | 28,476.16 | 27,988.08 | **116,882.49** |

| | | | | | |
|---|---|---|---|---|---|
| 984 | 21,847.83 | 20,838.68 | 19,879.95 | 19,300.27 | **81,866.74** |
| 985 | 17,029.56 | 16,967.22 | 16,484.26 | 16,050.83 | **66,531.87** |
| 986 | 25,897.21 | 25,182.83 | 25,098.42 | 24,083.53 | **100,261.99** |
| 988 | 8,082.40 | 8,050.45 | 7,553.16 | 7,513.00 | **31,199.01** |
| 989 | 9,656.94 | 9,708.96 | 9,337.24 | 9,001.06 | **37,704.20** |
| 990 | 2,817.79 | 2,676.19 | 2,690.37 | 2,750.30 | **10,934.65** |
| 991 | 4,523.91 | 4,350.18 | 4,301.32 | 4,119.03 | **17,294.44** |
| 992 | 16,184.73 | 16,291.36 | 14,872.59 | 14,571.37 | **61,920.05** |
| 993 | 13,686.14 | 12,985.49 | 12,156.01 | 12,152.65 | **50,980.30** |
| 994 | 1,059.94 | 1,186.32 | 1,003.84 | 1,098.41 | **4,348.52** |
| **WA - WASHINGTON - Total** | **258,217.13** | **251,409.19** | **242,441.87** | **237,674.43** | **989,742.63** |

**STATE: WI - WISCONSIN**

| REGISTRANT ZIP CODE 3 | 1ST QUARTER | 2ND QUARTER | 3RD QUARTER | 4TH QUARTER | TOTAL GRAMS |
|---|---|---|---|---|---|
| 530 | 19,322.95 | 18,062.30 | 17,151.97 | 16,721.60 | **71,258.82** |
| 531 | 23,321.77 | 21,506.57 | 19,554.20 | 18,461.18 | **82,843.72** |
| 532 | 55,875.88 | 50,681.22 | 46,883.57 | 45,020.66 | **198,461.33** |
| 534 | 4,592.85 | 4,208.28 | 4,253.03 | 4,170.42 | **17,224.58** |
| 535 | 14,775.15 | 14,030.78 | 13,491.09 | 12,912.00 | **55,209.02** |
| 537 | 11,128.28 | 10,335.65 | 10,069.81 | 9,725.74 | **41,259.48** |
| 538 | 983.21 | 885.79 | 917.63 | 992.00 | **3,778.63** |
| 539 | 5,737.57 | 5,580.19 | 5,483.87 | 5,170.20 | **21,971.84** |
| 540 | 2,763.24 | 2,864.80 | 2,582.59 | 2,679.70 | **10,890.33** |
| 541 | 3,136.02 | 2,979.30 | 2,913.41 | 2,810.42 | **11,839.16** |
| 542 | 1,832.27 | 1,769.62 | 1,834.01 | 1,623.85 | **7,059.75** |
| 543 | 3,530.80 | 3,406.39 | 3,155.52 | 3,294.18 | **13,386.90** |
| 544 | 6,504.04 | 6,124.10 | 5,956.36 | 5,682.19 | **24,266.69** |
| 545 | 2,386.22 | 2,387.35 | 2,104.86 | 1,945.11 | **8,823.53** |
| 546 | 5,812.57 | 5,593.57 | 5,355.91 | 5,326.27 | **22,088.32** |
| 547 | 3,657.99 | 3,464.88 | 3,353.04 | 3,197.26 | **13,673.17** |
| 548 | 3,999.33 | 3,927.25 | 4,076.95 | 3,865.96 | **15,869.49** |
| 549 | 10,315.70 | 9,778.29 | 9,494.85 | 9,218.47 | **38,807.31** |
| **WI - WISCONSIN - Total** | **179,675.85** | **167,586.32** | **158,632.67** | **152,817.21** | **658,712.06** |

**STATE: WV - WEST VIRGINIA**

| REGISTRANT ZIP CODE 3 | 1ST QUARTER | 2ND QUARTER | 3RD QUARTER | 4TH QUARTER | TOTAL GRAMS |
|---|---|---|---|---|---|
| 247 | 2,598.98 | 2,485.67 | 2,074.54 | 1,979.05 | **9,138.24** |
| 248 | 1,272.45 | 1,337.54 | 1,300.95 | 1,246.43 | **5,157.37** |
| 249 | 1,931.24 | 1,979.32 | 1,795.09 | 1,696.35 | **7,402.00** |
| 250 | 1,864.09 | 1,840.46 | 1,737.43 | 1,632.32 | **7,074.30** |
| 251 | 1,284.86 | 1,450.25 | 1,404.68 | 1,455.11 | **5,594.91** |
| 252 | 1,544.13 | 1,171.15 | 949.67 | 975.81 | **4,640.76** |

| | | | | | |
|---|---|---|---|---|---|
| 253 | 6,160.04 | 5,394.28 | 5,074.23 | 4,363.78 | **20,992.34** |
| 254 | 12,429.48 | 11,476.63 | 11,011.46 | 10,534.09 | **45,451.65** |
| 255 | 4,661.51 | 4,445.50 | 4,165.39 | 4,330.73 | **17,603.14** |
| 256 | 1,476.59 | 1,237.32 | 1,253.73 | 1,300.99 | **5,268.64** |
| 257 | 3,435.84 | 3,333.00 | 3,301.67 | 3,237.96 | **13,308.47** |
| 258 | 3,402.06 | 3,020.89 | 2,817.65 | 2,364.15 | **11,604.75** |
| 259 | 1,479.76 | 1,170.66 | 1,206.82 | 1,197.53 | **5,054.76** |
| 260 | 8,080.10 | 7,351.32 | 7,127.56 | 6,861.64 | **29,420.62** |
| 261 | 2,887.49 | 2,447.02 | 2,202.67 | 2,142.66 | **9,679.85** |
| 262 | 2,201.62 | 2,180.97 | 2,129.91 | 1,913.94 | **8,426.44** |
| 263 | 4,198.26 | 4,082.66 | 4,022.78 | 3,814.48 | **16,118.19** |
| 264 | 1,367.19 | 1,269.90 | 1,239.77 | 1,137.66 | **5,014.52** |
| 265 | 6,227.53 | 5,809.95 | 5,360.66 | 5,117.99 | **22,516.12** |
| 266 | 1,308.78 | 1,227.98 | 1,098.69 | 933.18 | **4,568.64** |
| 267 | 1,841.14 | 2,154.01 | 1,929.05 | 2,051.11 | **7,975.30** |
| 268 | 595.56 | 604.06 | 578.03 | 545.80 | **2,323.44** |
| **WV - WEST VIRGINIA - Total** | **72,248.71** | **67,470.52** | **63,782.46** | **60,832.77** | **264,334.46** |

**STATE: WY - WYOMING**

| REGISTRANT ZIP CODE 3 | 1ST QUARTER | 2ND QUARTER | 3RD QUARTER | 4TH QUARTER | TOTAL GRAMS |
|---|---|---|---|---|---|
| 820 | 4,547.70 | 4,458.90 | 4,239.01 | 3,928.65 | **17,174.25** |
| 822 | 580.75 | 546.46 | 507.78 | 685.73 | **2,320.73** |
| 823 | 332.56 | 357.83 | 357.42 | 372.69 | **1,420.50** |
| 824 | 2,817.29 | 2,657.31 | 2,602.77 | 2,654.85 | **10,732.22** |
| 825 | 2,172.65 | 1,736.53 | 1,772.31 | 1,832.52 | **7,513.99** |
| 826 | 4,361.07 | 4,231.51 | 4,065.71 | 4,228.93 | **16,887.22** |
| 827 | 1,119.12 | 1,060.07 | 993.11 | 981.64 | **4,153.94** |
| 828 | 1,356.35 | 1,308.24 | 1,163.56 | 1,139.08 | **4,967.23** |
| 829 | 5,733.66 | 5,688.87 | 5,434.99 | 5,519.40 | **22,376.93** |
| 830 | 214.76 | 170.34 | 208.80 | 198.09 | **791.99** |
| 831 | 724.87 | 646.47 | 617.46 | 577.66 | **2,566.46** |
| **WY - WYOMING - Total** | **23,960.77** | **22,862.52** | **21,962.93** | **22,119.24** | **90,905.46** |

**DRUG: 9150 - HYDROMORPHONE**

**STATE: AK - ALASKA**

| REGISTRANT ZIP CODE 3 | 1ST QUARTER | 2ND QUARTER | 3RD QUARTER | 4TH QUARTER | TOTAL GRAMS |
|---|---|---|---|---|---|
| 995 | 503.46 | 381.71 | 402.40 | 358.59 | **1,646.17** |
| 996 | 360.52 | 296.20 | 298.56 | 210.99 | **1,166.26** |
| 997 | 42.83 | 47.22 | 35.49 | 31.27 | **156.80** |
| 998 | 44.26 | 25.14 | 16.55 | 14.86 | **100.82** |
| 999 | 7.11 | 6.63 | 13.25 | 6.30 | **33.30** |
| **AK - ALASKA - Total** | **958.18** | **756.91** | **766.25** | **622.01** | **3,103.34** |

**STATE: AL - ALABAMA**

| REGISTRANT ZIP CODE 3 | 1ST QUARTER | 2ND QUARTER | 3RD QUARTER | 4TH QUARTER | TOTAL GRAMS |
|---|---|---|---|---|---|
| 350 | 201.26 | 154.00 | 157.71 | 130.01 | **642.98** |
| 351 | 125.40 | 118.78 | 106.19 | 95.55 | **445.93** |
| 352 | 324.30 | 260.59 | 298.57 | 307.47 | **1,190.93** |
| 354 | 101.50 | 124.82 | 107.98 | 110.52 | **444.81** |
| 355 | 57.80 | 44.69 | 41.41 | 35.12 | **179.01** |
| 356 | 148.40 | 168.83 | 149.01 | 146.86 | **613.10** |
| 357 | 107.12 | 93.87 | 91.26 | 84.80 | **377.06** |
| 358 | 219.61 | 222.90 | 243.40 | 183.33 | **869.25** |
| 359 | 119.56 | 125.86 | 123.00 | 97.67 | **466.08** |
| 360 | 131.63 | 145.41 | 120.98 | 109.67 | **507.70** |
| 361 | 115.64 | 127.57 | 113.15 | 131.71 | **488.07** |
| 362 | 81.26 | 73.22 | 65.41 | 56.35 | **276.25** |
| 363 | 190.67 | 172.77 | 219.34 | 158.18 | **740.96** |
| 364 | 84.65 | 77.78 | 90.22 | 90.66 | **343.32** |
| 365 | 367.50 | 509.20 | 275.97 | 246.29 | **1,398.97** |
| 366 | 278.97 | 267.66 | 268.75 | 239.42 | **1,054.81** |
| 367 | 18.22 | 22.33 | 16.54 | 16.86 | **73.95** |
| 368 | 98.69 | 105.16 | 102.12 | 99.07 | **405.03** |
| 369 | 5.14 | 6.38 | 6.36 | 5.41 | **23.29** |
| **AL - ALABAMA - Total** | **2,777.32** | **2,821.82** | **2,597.38** | **2,344.97** | **10,541.50** |

**STATE: AR - ARKANSAS**

| REGISTRANT ZIP CODE 3 | 1ST QUARTER | 2ND QUARTER | 3RD QUARTER | 4TH QUARTER | TOTAL GRAMS |
|---|---|---|---|---|---|
| 716 | 51.54 | 51.01 | 36.39 | 39.16 | **178.10** |
| 717 | 42.81 | 24.47 | 35.41 | 26.70 | **129.38** |
| 718 | 88.42 | 71.43 | 69.38 | 74.10 | **303.32** |
| 719 | 131.12 | 127.01 | 117.05 | 130.90 | **506.08** |
| 720 | 250.55 | 222.72 | 196.39 | 184.02 | **853.69** |
| 721 | 263.74 | 227.62 | 219.09 | 166.10 | **876.54** |
| 722 | 433.34 | 404.79 | 307.70 | 279.86 | **1,425.69** |
| 723 | 36.18 | 31.10 | 30.32 | 33.53 | **131.13** |
| 724 | 154.56 | 118.04 | 110.13 | 123.84 | **506.57** |
| 725 | 123.41 | 97.64 | 96.56 | 100.99 | **418.60** |
| 726 | 116.87 | 117.78 | 111.72 | 105.39 | **451.77** |
| 727 | 700.41 | 623.74 | 642.90 | 647.19 | **2,614.24** |
| 728 | 85.02 | 84.28 | 75.41 | 61.53 | **306.24** |
| 729 | 333.15 | 339.28 | 316.86 | 306.85 | **1,296.15** |
| **AR - ARKANSAS - Total** | **2,811.12** | **2,540.91** | **2,365.32** | **2,280.16** | **9,997.50** |

**STATE: AS - AMERICAN SAMOA**

| REGISTRANT ZIP CODE 3 | 1ST QUARTER | 2ND QUARTER | 3RD QUARTER | 4TH QUARTER | TOTAL GRAMS |
|---|---|---|---|---|---|
| 967 | 0.18 | 0.03 | 0.73 | 0.27 | **1.20** |
| **AS - AMERICAN SAMOA - Tota** | **0.18** | **0.03** | **0.73** | **0.27** | **1.20** |

**STATE: AZ - ARIZONA**

| REGISTRANT ZIP CODE 3 | 1ST QUARTER | 2ND QUARTER | 3RD QUARTER | 4TH QUARTER | TOTAL GRAMS |
|---|---|---|---|---|---|
| 850 | 1,031.77 | 982.06 | 953.66 | 968.43 | **3,935.92** |
| 851 | 172.72 | 189.04 | 153.39 | 168.35 | **683.50** |
| 852 | 1,777.03 | 1,471.75 | 1,485.34 | 1,286.75 | **6,020.86** |
| 853 | 1,101.39 | 943.38 | 936.43 | 881.15 | **3,862.35** |
| 855 | 81.19 | 71.77 | 69.79 | 75.56 | **298.30** |
| 856 | 190.95 | 198.95 | 169.25 | 153.00 | **712.15** |
| 857 | 951.37 | 903.00 | 891.22 | 855.42 | **3,601.01** |
| 859 | 95.69 | 108.30 | 85.42 | 76.85 | **366.26** |
| 860 | 104.91 | 105.41 | 84.97 | 79.70 | **374.99** |
| 863 | 272.69 | 271.21 | 253.87 | 232.87 | **1,030.64** |
| 864 | 367.69 | 342.00 | 341.28 | 277.27 | **1,328.24** |
| 865 | 1.51 | 1.25 | 1.15 | 0.89 | **4.80** |
| **AZ - ARIZONA - Total** | **6,148.90** | **5,588.13** | **5,425.76** | **5,056.22** | **22,219.02** |

**STATE: CA - CALIFORNIA**

| REGISTRANT ZIP CODE 3 | 1ST QUARTER | 2ND QUARTER | 3RD QUARTER | 4TH QUARTER | TOTAL GRAMS |
|---|---|---|---|---|---|
| 900 | 1,257.86 | 1,248.22 | 1,187.98 | 1,117.58 | **4,811.64** |
| 902 | 546.93 | 538.53 | 467.87 | 474.50 | **2,027.83** |
| 903 | 26.42 | 26.66 | 27.94 | 25.21 | **106.24** |
| 904 | 287.20 | 245.01 | 217.89 | 209.87 | **959.98** |
| 905 | 321.63 | 190.16 | 251.68 | 266.66 | **1,030.12** |
| 906 | 358.85 | 315.03 | 333.68 | 344.39 | **1,351.95** |
| 907 | 384.89 | 401.74 | 364.72 | 318.24 | **1,469.59** |
| 908 | 1,096.22 | 1,632.93 | 924.80 | 1,181.72 | **4,835.67** |
| 910 | 479.45 | 496.79 | 395.20 | 443.97 | **1,815.41** |
| 911 | 152.55 | 203.67 | 150.93 | 127.83 | **634.98** |
| 912 | 190.97 | 187.33 | 154.43 | 137.55 | **670.28** |
| 913 | 1,240.25 | 1,180.38 | 1,162.28 | 1,097.80 | **4,680.71** |
| 914 | 436.91 | 314.20 | 343.69 | 359.35 | **1,454.15** |
| 915 | 193.25 | 190.96 | 142.72 | 129.05 | **655.97** |
| 916 | 79.62 | 55.62 | 51.69 | 64.39 | **251.31** |
| 917 | 1,513.23 | 1,340.06 | 1,273.39 | 1,167.69 | **5,294.37** |
| 918 | 14.75 | 21.78 | 27.08 | 22.50 | **86.12** |
| 919 | 283.22 | 295.33 | 257.59 | 254.82 | **1,090.97** |
| 920 | 1,785.92 | 1,662.87 | 1,538.99 | 1,449.50 | **6,437.28** |
| 921 | 1,228.74 | 1,023.55 | 1,078.12 | 938.69 | **4,269.10** |

| | | | | | |
|---|---|---|---|---|---|
| 922 | 809.01 | 750.67 | 695.80 | 681.56 | **2,937.04** |
| 923 | 1,134.42 | 1,086.27 | 1,083.81 | 1,006.46 | **4,310.96** |
| 924 | 175.84 | 148.80 | 120.27 | 137.35 | **582.25** |
| 925 | 1,053.24 | 1,228.07 | 1,229.50 | 765.06 | **4,275.86** |
| 926 | 898.21 | 711.29 | 711.68 | 880.45 | **3,201.63** |
| 927 | 352.42 | 284.80 | 221.25 | 238.78 | **1,097.25** |
| 928 | 1,232.30 | 1,232.91 | 1,095.46 | 1,000.89 | **4,561.56** |
| 930 | 499.91 | 517.67 | 443.12 | 441.93 | **1,902.63** |
| 931 | 149.33 | 112.92 | 133.24 | 95.32 | **490.81** |
| 932 | 482.57 | 438.41 | 370.65 | 358.82 | **1,650.44** |
| 933 | 619.73 | 608.96 | 544.49 | 533.16 | **2,306.34** |
| 934 | 702.98 | 708.51 | 706.52 | 666.33 | **2,784.35** |
| 935 | 1,137.65 | 1,144.66 | 974.21 | 928.09 | **4,184.61** |
| 936 | 332.96 | 326.29 | 224.86 | 226.50 | **1,110.60** |
| 937 | 429.15 | 427.26 | 437.65 | 419.11 | **1,713.16** |
| 939 | 284.06 | 238.38 | 230.29 | 212.48 | **965.20** |
| 940 | 486.26 | 443.29 | 395.34 | 400.71 | **1,725.60** |
| 941 | 873.29 | 928.14 | 793.48 | 825.61 | **3,420.52** |
| 943 | 382.13 | 199.87 | 165.79 | 196.36 | **944.14** |
| 944 | 70.76 | 50.92 | 63.56 | 78.35 | **263.59** |
| 945 | 2,315.91 | 2,162.62 | 2,023.64 | 1,979.81 | **8,481.98** |
| 946 | 354.45 | 320.12 | 313.01 | 308.58 | **1,296.15** |
| 947 | 133.91 | 115.51 | 173.30 | 93.39 | **516.11** |
| 948 | 70.45 | 76.69 | 71.82 | 74.92 | **293.88** |
| 949 | 481.47 | 451.64 | 406.23 | 411.59 | **1,750.93** |
| 950 | 442.93 | 417.11 | 388.95 | 349.96 | **1,598.95** |
| 951 | 503.04 | 345.44 | 303.93 | 253.28 | **1,405.68** |
| 952 | 392.60 | 323.56 | 318.57 | 261.98 | **1,296.71** |
| 953 | 923.71 | 875.92 | 785.79 | 712.87 | **3,298.29** |
| 954 | 687.38 | 602.64 | 587.49 | 573.90 | **2,451.42** |
| 955 | 297.05 | 218.47 | 188.91 | 164.82 | **869.25** |
| 956 | 1,588.16 | 1,307.29 | 1,349.37 | 1,220.04 | **5,464.87** |
| 957 | 153.97 | 130.21 | 119.14 | 96.33 | **499.65** |
| 958 | 1,133.99 | 1,057.10 | 938.89 | 802.47 | **3,932.45** |
| 959 | 818.61 | 721.02 | 683.29 | 595.37 | **2,818.28** |
| 960 | 416.09 | 343.99 | 332.90 | 286.77 | **1,379.76** |
| 961 | 65.25 | 46.05 | 60.00 | 43.12 | **214.42** |
| **CA - CALIFORNIA - Total** | **34,764.05** | **32,674.26** | **30,038.86** | **28,453.83** | **125,931.00** |

**STATE: CO - COLORADO**

| REGISTRANT ZIP CODE 3 | 1ST QUARTER | 2ND QUARTER | 3RD QUARTER | 4TH QUARTER | **TOTAL GRAMS** |
|---|---|---|---|---|---|

| | | | | | |
|---|---|---|---|---|---|
| 800 | 1,273.30 | 949.36 | 1,089.89 | 995.36 | **4,307.92** |
| 801 | 1,298.11 | 1,418.69 | 1,154.31 | 1,345.80 | **5,216.92** |
| 802 | 1,079.22 | 916.33 | 906.30 | 850.60 | **3,752.45** |
| 803 | 85.35 | 92.42 | 65.07 | 73.62 | **316.45** |
| 804 | 225.57 | 182.99 | 200.66 | 186.36 | **795.58** |
| 805 | 596.12 | 480.67 | 454.09 | 472.11 | **2,002.99** |
| 806 | 308.33 | 271.78 | 279.33 | 280.32 | **1,139.75** |
| 807 | 63.19 | 59.55 | 74.37 | 72.13 | **269.24** |
| 808 | 116.16 | 114.07 | 117.01 | 95.05 | **442.29** |
| 809 | 703.89 | 657.09 | 631.10 | 591.21 | **2,583.29** |
| 810 | 309.40 | 300.26 | 277.06 | 258.85 | **1,145.57** |
| 811 | 28.91 | 26.95 | 26.73 | 17.86 | **100.45** |
| 812 | 95.90 | 93.19 | 75.15 | 83.50 | **347.74** |
| 813 | 60.94 | 47.50 | 35.01 | 39.61 | **183.06** |
| 814 | 58.35 | 45.86 | 46.26 | 44.70 | **195.18** |
| 815 | 126.99 | 95.80 | 109.59 | 101.11 | **433.49** |
| 816 | 77.81 | 81.93 | 95.59 | 90.37 | **345.71** |
| **CO - COLORADO - Total** | **6,507.54** | **5,834.45** | **5,637.52** | **5,598.57** | **23,578.07** |

**STATE: CT - CONNECTICUT**

| REGISTRANT ZIP CODE 3 | 1ST QUARTER | 2ND QUARTER | 3RD QUARTER | 4TH QUARTER | **TOTAL GRAMS** |
|---|---|---|---|---|---|
| 060 | 964.19 | 949.11 | 913.91 | 934.79 | **3,762.00** |
| 061 | 487.51 | 480.52 | 474.27 | 447.88 | **1,890.17** |
| 062 | 157.97 | 170.28 | 156.97 | 157.22 | **642.44** |
| 063 | 290.62 | 252.58 | 259.87 | 233.72 | **1,036.78** |
| 064 | 1,484.74 | 1,467.40 | 1,255.69 | 1,210.91 | **5,418.74** |
| 065 | 520.49 | 437.71 | 418.10 | 448.94 | **1,825.25** |
| 066 | 165.28 | 159.38 | 121.49 | 125.71 | **571.87** |
| 067 | 423.00 | 398.15 | 379.75 | 412.46 | **1,613.36** |
| 068 | 1,301.69 | 624.98 | 606.30 | 640.16 | **3,173.14** |
| 069 | 99.09 | 119.07 | 104.26 | 99.31 | **421.73** |
| **CT - CONNECTICUT - Total** | **5,894.59** | **5,059.17** | **4,690.61** | **4,711.11** | **20,355.47** |

**STATE: DC - DISTRICT OF COLUMBIA**

| REGISTRANT ZIP CODE 3 | 1ST QUARTER | 2ND QUARTER | 3RD QUARTER | 4TH QUARTER | **TOTAL GRAMS** |
|---|---|---|---|---|---|
| 200 | 448.36 | 366.88 | 440.42 | 400.39 | **1,656.05** |
| 203 | 0.71 | 0 | 0 | 0 | **0.71** |
| 204 | 10.64 | 7.71 | 11.88 | 6.64 | **36.87** |
| **DC - DISTRICT OF COLUMBIA -** | **459.71** | **374.60** | **452.30** | **407.02** | **1,693.63** |

**STATE: DE - DELAWARE**

| REGISTRANT ZIP CODE 3 | 1ST QUARTER | 2ND QUARTER | 3RD QUARTER | 4TH QUARTER | **TOTAL GRAMS** |
|---|---|---|---|---|---|
| 197 | 794.13 | 691.47 | 680.44 | 656.21 | **2,822.24** |

| | | | | | |
|---|---|---|---|---|---|
| 198 | 304.47 | 278.03 | 271.95 | 252.32 | **1,106.77** |
| 199 | 715.95 | 683.94 | 568.09 | 575.40 | **2,543.37** |
| **DE - DELAWARE - Total** | **1,814.54** | **1,653.44** | **1,520.48** | **1,483.92** | **6,472.38** |

**STATE: FL - FLORIDA**

| REGISTRANT ZIP CODE 3 | 1ST QUARTER | 2ND QUARTER | 3RD QUARTER | 4TH QUARTER | TOTAL GRAMS |
|---|---|---|---|---|---|
| 320 | 766.70 | 730.17 | 656.65 | 577.21 | **2,730.73** |
| 321 | 1,148.78 | 1,048.63 | 955.67 | 875.38 | **4,028.45** |
| 322 | 1,859.52 | 1,472.90 | 1,358.48 | 1,253.97 | **5,944.87** |
| 323 | 329.89 | 300.06 | 283.07 | 285.02 | **1,198.05** |
| 324 | 334.32 | 288.76 | 254.52 | 218.80 | **1,096.40** |
| 325 | 746.52 | 720.24 | 637.03 | 561.24 | **2,665.03** |
| 326 | 540.74 | 415.89 | 389.39 | 385.13 | **1,731.14** |
| 327 | 2,723.31 | 2,341.24 | 2,429.84 | 2,190.56 | **9,684.95** |
| 328 | 1,793.27 | 1,694.11 | 1,621.34 | 1,504.21 | **6,612.94** |
| 329 | 1,849.25 | 1,686.07 | 1,495.17 | 1,357.04 | **6,387.53** |
| 330 | 1,566.62 | 1,462.77 | 1,276.97 | 1,173.12 | **5,479.49** |
| 331 | 1,545.41 | 1,246.89 | 1,198.42 | 1,118.51 | **5,109.23** |
| 333 | 1,321.10 | 1,044.38 | 932.78 | 873.49 | **4,171.76** |
| 334 | 2,123.49 | 1,926.96 | 1,823.18 | 1,682.59 | **7,556.21** |
| 335 | 1,893.89 | 1,821.36 | 1,650.77 | 1,652.41 | **7,018.43** |
| 336 | 7,808.94 | 6,739.33 | 7,218.92 | 6,818.28 | **28,585.47** |
| 337 | 2,851.12 | 3,013.15 | 2,568.53 | 2,382.00 | **10,814.80** |
| 338 | 1,199.27 | 1,076.07 | 862.88 | 861.96 | **4,000.17** |
| 339 | 1,835.22 | 1,556.16 | 1,408.79 | 1,448.47 | **6,248.65** |
| 341 | 725.54 | 597.12 | 513.19 | 553.22 | **2,389.08** |
| 342 | 2,299.56 | 2,159.61 | 1,891.76 | 1,882.67 | **8,233.59** |
| 344 | 999.28 | 908.28 | 815.23 | 720.01 | **3,442.79** |
| 346 | 3,062.14 | 2,949.17 | 2,834.50 | 2,722.19 | **11,568.00** |
| 347 | 1,351.33 | 1,191.85 | 1,098.35 | 1,059.99 | **4,701.53** |
| 349 | 1,077.00 | 1,099.15 | 981.58 | 963.83 | **4,121.55** |
| **FL - FLORIDA - Total** | **43,752.23** | **39,490.32** | **37,156.97** | **35,121.31** | **155,520.83** |

**STATE: GA - GEORGIA**

| REGISTRANT ZIP CODE 3 | 1ST QUARTER | 2ND QUARTER | 3RD QUARTER | 4TH QUARTER | TOTAL GRAMS |
|---|---|---|---|---|---|
| 300 | 1,567.53 | 1,545.05 | 1,472.13 | 1,299.05 | **5,883.77** |
| 301 | 828.81 | 850.67 | 819.67 | 755.00 | **3,254.15** |
| 302 | 649.13 | 583.01 | 516.65 | 459.89 | **2,208.68** |
| 303 | 742.65 | 1,059.35 | 703.48 | 666.83 | **3,172.31** |
| 304 | 95.25 | 105.83 | 85.21 | 90.82 | **377.11** |
| 305 | 528.62 | 524.04 | 515.92 | 473.83 | **2,042.42** |
| 306 | 428.53 | 415.02 | 400.80 | 412.62 | **1,656.97** |

| | | | | | |
|---|---|---|---|---|---|
| 307 | 152.59 | 145.15 | 167.48 | 170.94 | **636.15** |
| 308 | 108.84 | 104.42 | 108.69 | 100.16 | **422.11** |
| 309 | 296.34 | 249.41 | 218.57 | 203.00 | **967.31** |
| 310 | 232.27 | 221.42 | 235.37 | 194.24 | **883.30** |
| 312 | 218.86 | 125.82 | 129.93 | 57.66 | **532.27** |
| 313 | 158.46 | 168.57 | 149.87 | 104.59 | **581.49** |
| 314 | 139.75 | 181.84 | 134.82 | 132.65 | **589.05** |
| 315 | 346.57 | 330.04 | 311.18 | 276.61 | **1,264.40** |
| 316 | 125.15 | 161.68 | 149.85 | 143.90 | **580.57** |
| 317 | 470.67 | 365.61 | 335.42 | 346.13 | **1,517.83** |
| 318 | 8.69 | 7.89 | 8.43 | 6.47 | **31.48** |
| 319 | 60.39 | 67.10 | 60.65 | 57.06 | **245.21** |
| 398 | 52.89 | 55.38 | 51.98 | 55.39 | **215.63** |
| **GA - GEORGIA - Total** | **7,211.99** | **7,267.29** | **6,576.09** | **6,006.85** | **27,062.23** |

**STATE: GU - GUAM**

| REGISTRANT ZIP CODE 3 | 1ST QUARTER | 2ND QUARTER | 3RD QUARTER | 4TH QUARTER | TOTAL GRAMS |
|---|---|---|---|---|---|
| 969 | 28.28 | 17.21 | 30.71 | 14.31 | **90.51** |
| **GU - GUAM - Total** | **28.28** | **17.21** | **30.71** | **14.31** | **90.51** |

**STATE: HI - HAWAII**

| REGISTRANT ZIP CODE 3 | 1ST QUARTER | 2ND QUARTER | 3RD QUARTER | 4TH QUARTER | TOTAL GRAMS |
|---|---|---|---|---|---|
| 967 | 311.82 | 285.73 | 282.03 | 254.00 | **1,133.58** |
| 968 | 234.88 | 242.57 | 210.25 | 240.17 | **927.87** |
| **HI - HAWAII - Total** | **546.70** | **528.30** | **492.28** | **494.18** | **2,061.45** |

**STATE: IA - IOWA**

| REGISTRANT ZIP CODE 3 | 1ST QUARTER | 2ND QUARTER | 3RD QUARTER | 4TH QUARTER | TOTAL GRAMS |
|---|---|---|---|---|---|
| 500 | 214.34 | 193.01 | 159.18 | 194.18 | **760.71** |
| 501 | 148.58 | 140.04 | 150.18 | 120.32 | **559.12** |
| 502 | 162.34 | 150.27 | 169.51 | 170.60 | **652.72** |
| 503 | 617.02 | 514.84 | 461.72 | 483.24 | **2,076.82** |
| 504 | 45.45 | 52.22 | 51.97 | 58.11 | **207.75** |
| 505 | 31.25 | 31.64 | 30.42 | 49.87 | **143.19** |
| 506 | 65.72 | 66.46 | 64.24 | 73.18 | **269.60** |
| 507 | 73.52 | 50.03 | 51.07 | 35.53 | **210.14** |
| 508 | 14.46 | 16.72 | 22.45 | 16.93 | **70.56** |
| 510 | 13.43 | 12.73 | 9.73 | 9.34 | **45.23** |
| 511 | 49.65 | 55.20 | 57.87 | 48.21 | **210.94** |
| 512 | 6.69 | 3.50 | 6.62 | 5.50 | **22.31** |
| 513 | 24.68 | 26.44 | 19.91 | 17.11 | **88.15** |
| 514 | 32.25 | 35.12 | 31.16 | 28.60 | **127.13** |
| 515 | 51.37 | 60.91 | 51.42 | 65.19 | **228.89** |

| | | | | | |
|---|---|---|---|---|---|
| 516 | 10.95 | 13.30 | 13.79 | 14.41 | **52.44** |
| 520 | 73.70 | 61.82 | 45.15 | 55.58 | **236.25** |
| 521 | 15.89 | 12.03 | 15.31 | 11.46 | **54.70** |
| 522 | 358.78 | 299.90 | 314.96 | 330.98 | **1,304.62** |
| 523 | 59.90 | 61.50 | 58.60 | 46.60 | **226.59** |
| 524 | 117.14 | 137.31 | 79.35 | 68.07 | **401.87** |
| 525 | 190.67 | 150.01 | 152.86 | 144.68 | **638.21** |
| 526 | 96.38 | 97.08 | 87.96 | 77.51 | **358.94** |
| 527 | 85.83 | 89.73 | 94.48 | 81.24 | **351.29** |
| 528 | 62.87 | 73.00 | 76.42 | 68.87 | **281.16** |
| **IA - IOWA - Total** | **2,622.86** | **2,404.82** | **2,276.33** | **2,275.32** | **9,579.33** |

**STATE: ID - IDAHO**

| REGISTRANT ZIP CODE 3 | 1ST QUARTER | 2ND QUARTER | 3RD QUARTER | 4TH QUARTER | TOTAL GRAMS |
|---|---|---|---|---|---|
| 832 | 152.89 | 144.57 | 130.32 | 111.75 | **539.52** |
| 833 | 154.81 | 145.19 | 180.48 | 147.06 | **627.53** |
| 834 | 92.60 | 111.72 | 82.81 | 71.35 | **358.49** |
| 835 | 98.22 | 83.82 | 95.65 | 94.72 | **372.41** |
| 836 | 618.99 | 545.91 | 507.28 | 454.53 | **2,126.72** |
| 837 | 507.49 | 562.77 | 463.73 | 538.47 | **2,072.46** |
| 838 | 238.65 | 278.53 | 205.60 | 197.93 | **920.70** |
| **ID - IDAHO - Total** | **1,863.65** | **1,872.51** | **1,665.87** | **1,615.81** | **7,017.83** |

**STATE: IL - ILLINOIS**

| REGISTRANT ZIP CODE 3 | 1ST QUARTER | 2ND QUARTER | 3RD QUARTER | 4TH QUARTER | TOTAL GRAMS |
|---|---|---|---|---|---|
| 600 | 1,652.66 | 1,580.20 | 1,439.48 | 1,383.90 | **6,056.24** |
| 601 | 1,070.31 | 988.13 | 884.25 | 1,035.31 | **3,977.99** |
| 602 | 40.77 | 74.68 | 56.16 | 40.43 | **212.05** |
| 603 | 99.61 | 86.99 | 102.81 | 75.22 | **364.64** |
| 604 | 1,104.91 | 1,179.77 | 1,113.46 | 1,025.36 | **4,423.50** |
| 605 | 697.81 | 679.24 | 628.11 | 557.36 | **2,562.52** |
| 606 | 1,452.28 | 1,372.99 | 1,276.78 | 1,279.85 | **5,381.90** |
| 607 | 56.67 | 49.76 | 56.00 | 67.49 | **229.92** |
| 608 | 72.06 | 73.87 | 68.44 | 74.81 | **289.18** |
| 609 | 109.75 | 125.24 | 106.88 | 109.90 | **451.77** |
| 610 | 57.81 | 59.53 | 64.66 | 64.82 | **246.82** |
| 611 | 278.99 | 139.98 | 181.15 | 190.24 | **790.37** |
| 612 | 73.12 | 63.79 | 67.98 | 68.09 | **272.98** |
| 613 | 67.88 | 62.99 | 58.50 | 49.50 | **238.88** |
| 614 | 66.85 | 39.88 | 39.59 | 33.92 | **180.24** |
| 615 | 82.61 | 65.07 | 58.84 | 61.90 | **268.42** |
| 616 | 157.65 | 150.84 | 167.13 | 128.98 | **604.60** |

| | | | | | |
|---|---|---|---|---|---|
| 617 | 185.73 | 171.15 | 161.27 | 152.96 | **671.11** |
| 618 | 225.68 | 211.77 | 230.32 | 202.95 | **870.72** |
| 619 | 47.25 | 44.62 | 41.86 | 41.63 | **175.35** |
| 620 | 226.00 | 152.62 | 113.57 | 109.04 | **601.23** |
| 622 | 250.48 | 181.42 | 201.57 | 158.67 | **792.14** |
| 623 | 90.30 | 77.97 | 57.82 | 51.05 | **277.14** |
| 624 | 94.30 | 71.81 | 89.58 | 75.36 | **331.04** |
| 625 | 203.62 | 178.42 | 162.91 | 167.88 | **712.83** |
| 626 | 76.71 | 57.39 | 63.25 | 58.80 | **256.15** |
| 627 | 146.96 | 139.44 | 125.15 | 156.80 | **568.36** |
| 628 | 101.65 | 66.67 | 76.48 | 80.05 | **324.85** |
| 629 | 103.97 | 99.22 | 96.86 | 94.80 | **394.86** |
| **IL - ILLINOIS - Total** | **8,894.40** | **8,245.46** | **7,790.86** | **7,597.06** | **32,527.78** |

**STATE: IN - INDIANA**

| REGISTRANT ZIP CODE 3 | 1ST QUARTER | 2ND QUARTER | 3RD QUARTER | 4TH QUARTER | **TOTAL GRAMS** |
|---|---|---|---|---|---|
| 460 | 401.93 | 311.26 | 289.39 | 279.02 | **1,281.60** |
| 461 | 264.84 | 244.74 | 267.79 | 293.89 | **1,071.25** |
| 462 | 1,314.35 | 1,255.05 | 1,074.00 | 949.71 | **4,593.11** |
| 463 | 430.27 | 388.21 | 404.43 | 337.53 | **1,560.44** |
| 464 | 40.48 | 41.33 | 33.81 | 36.22 | **151.83** |
| 465 | 341.21 | 339.38 | 302.19 | 237.09 | **1,219.88** |
| 466 | 102.42 | 101.51 | 60.70 | 83.99 | **348.62** |
| 467 | 240.66 | 253.88 | 220.31 | 203.63 | **918.48** |
| 468 | 406.48 | 348.61 | 286.25 | 312.07 | **1,353.41** |
| 469 | 129.56 | 123.03 | 97.50 | 96.49 | **446.58** |
| 470 | 27.67 | 45.18 | 29.60 | 28.58 | **131.04** |
| 471 | 238.67 | 149.38 | 136.45 | 127.63 | **652.13** |
| 472 | 174.09 | 154.12 | 126.49 | 130.50 | **585.19** |
| 473 | 234.83 | 253.74 | 265.26 | 221.91 | **975.73** |
| 474 | 183.60 | 127.50 | 137.76 | 123.91 | **572.77** |
| 475 | 255.77 | 238.78 | 164.15 | 196.01 | **854.70** |
| 476 | 87.20 | 89.54 | 95.54 | 100.31 | **372.59** |
| 477 | 250.36 | 232.76 | 256.72 | 269.55 | **1,009.39** |
| 478 | 106.87 | 95.39 | 78.08 | 75.85 | **356.19** |
| 479 | 200.26 | 193.99 | 168.79 | 144.57 | **707.60** |
| **IN - INDIANA - Total** | **5,431.52** | **4,987.37** | **4,495.20** | **4,248.44** | **19,162.53** |

**STATE: KS - KANSAS**

| REGISTRANT ZIP CODE 3 | 1ST QUARTER | 2ND QUARTER | 3RD QUARTER | 4TH QUARTER | **TOTAL GRAMS** |
|---|---|---|---|---|---|
| 660 | 356.83 | 368.16 | 324.17 | 306.60 | **1,355.77** |
| 661 | 154.50 | 103.39 | 160.02 | 119.69 | **537.60** |

| | | | | | |
|---|---|---|---|---|---|
| 662 | 603.19 | 650.32 | 699.01 | 565.15 | **2,517.67** |
| 664 | 86.76 | 81.03 | 87.02 | 81.22 | **336.03** |
| 665 | 106.30 | 103.32 | 101.72 | 104.88 | **416.22** |
| 666 | 75.18 | 93.56 | 88.09 | 90.30 | **347.13** |
| 667 | 96.62 | 86.08 | 96.24 | 92.88 | **371.82** |
| 668 | 25.11 | 28.77 | 21.54 | 23.57 | **98.99** |
| 669 | 15.98 | 22.66 | 12.43 | 11.26 | **62.33** |
| 670 | 139.26 | 126.32 | 120.68 | 127.35 | **513.61** |
| 671 | 112.75 | 103.08 | 73.72 | 90.21 | **379.76** |
| 672 | 607.70 | 475.92 | 546.92 | 534.98 | **2,165.52** |
| 673 | 31.43 | 29.44 | 38.82 | 26.69 | **126.38** |
| 674 | 120.94 | 138.65 | 119.32 | 132.30 | **511.20** |
| 675 | 233.17 | 217.08 | 241.08 | 241.85 | **933.18** |
| 676 | 82.04 | 73.21 | 63.47 | 75.60 | **294.32** |
| 677 | 17.52 | 20.22 | 16.75 | 18.60 | **73.09** |
| 678 | 65.81 | 56.96 | 55.55 | 48.81 | **227.14** |
| 679 | 26.17 | 22.57 | 24.84 | 42.58 | **116.16** |
| **KS - KANSAS - Total** | **2,957.25** | **2,800.73** | **2,891.40** | **2,734.53** | **11,383.91** |

**STATE: KY - KENTUCKY**

| REGISTRANT ZIP CODE 3 | 1ST QUARTER | 2ND QUARTER | 3RD QUARTER | 4TH QUARTER | **TOTAL GRAMS** |
|---|---|---|---|---|---|
| 400 | 85.66 | 67.73 | 64.82 | 66.65 | **284.87** |
| 401 | 40.06 | 43.98 | 38.76 | 32.73 | **155.53** |
| 402 | 799.99 | 841.40 | 601.10 | 627.89 | **2,870.39** |
| 403 | 62.12 | 64.23 | 57.61 | 51.17 | **235.14** |
| 404 | 22.75 | 21.97 | 21.38 | 8.68 | **74.78** |
| 405 | 315.91 | 226.41 | 246.05 | 307.44 | **1,095.82** |
| 406 | 43.06 | 18.26 | 14.89 | 14.76 | **90.97** |
| 407 | 18.68 | 12.51 | 14.58 | 11.41 | **57.18** |
| 408 | 7.88 | 5.50 | 3.42 | 6.19 | **22.99** |
| 409 | 13.47 | 7.77 | 17.36 | 3.64 | **42.24** |
| 410 | 142.77 | 107.53 | 111.77 | 134.08 | **496.15** |
| 411 | 35.75 | 34.70 | 28.26 | 33.25 | **131.95** |
| 412 | 4.73 | 5.73 | 3.34 | 3.47 | **17.28** |
| 413 | 6.12 | 5.85 | 1.71 | 5.38 | **19.06** |
| 414 | 0.18 | 0.05 | 0 | 0 | **0.23** |
| 415 | 8.86 | 10.01 | 21.47 | 18.19 | **58.53** |
| 416 | 5.44 | 5.18 | 2.00 | 1.11 | **13.74** |
| 417 | 39.39 | 38.11 | 47.46 | 40.74 | **165.69** |
| 418 | 1.06 | 2.56 | 3.42 | 3.13 | **10.16** |
| 420 | 192.42 | 219.04 | 187.39 | 159.03 | **757.88** |

| | | | | | |
|---|---|---|---|---|---|
| 421 | 176.50 | 157.80 | 189.51 | 351.31 | **875.13** |
| 422 | 145.62 | 140.13 | 140.13 | 87.29 | **513.17** |
| 423 | 120.67 | 114.01 | 117.52 | 114.75 | **466.96** |
| 424 | 161.97 | 179.64 | 138.01 | 137.08 | **616.70** |
| 425 | 15.14 | 11.80 | 28.04 | 20.57 | **75.54** |
| 426 | 4.92 | 8.42 | 4.79 | 4.17 | **22.30** |
| 427 | 74.00 | 74.94 | 71.77 | 70.64 | **291.36** |
| **KY - KENTUCKY - Total** | **2,545.12** | **2,425.27** | **2,176.57** | **2,314.77** | **9,461.73** |

**STATE: LA - LOUISIANA**

| REGISTRANT ZIP CODE 3 | 1ST QUARTER | 2ND QUARTER | 3RD QUARTER | 4TH QUARTER | **TOTAL GRAMS** |
|---|---|---|---|---|---|
| 700 | 202.17 | 181.17 | 177.86 | 162.01 | **723.22** |
| 701 | 208.53 | 155.07 | 223.54 | 151.33 | **738.47** |
| 703 | 110.73 | 99.03 | 90.61 | 97.66 | **398.03** |
| 704 | 373.44 | 325.68 | 338.75 | 297.38 | **1,335.25** |
| 705 | 689.93 | 853.52 | 744.04 | 662.69 | **2,950.18** |
| 706 | 148.75 | 151.91 | 133.05 | 115.56 | **549.27** |
| 707 | 391.89 | 390.10 | 298.88 | 292.22 | **1,373.09** |
| 708 | 231.87 | 190.68 | 201.19 | 166.10 | **789.85** |
| 710 | 34.84 | 37.40 | 34.06 | 38.44 | **144.75** |
| 711 | 261.02 | 149.50 | 165.23 | 118.94 | **694.69** |
| 712 | 94.18 | 92.29 | 109.48 | 86.67 | **382.61** |
| 713 | 139.42 | 147.81 | 132.11 | 142.86 | **562.21** |
| 714 | 54.26 | 56.37 | 51.77 | 68.07 | **230.46** |
| **LA - LOUISIANA - Total** | **2,941.03** | **2,830.54** | **2,700.56** | **2,399.95** | **10,872.08** |

**STATE: MA - MASSACHUSETTS**

| REGISTRANT ZIP CODE 3 | 1ST QUARTER | 2ND QUARTER | 3RD QUARTER | 4TH QUARTER | **TOTAL GRAMS** |
|---|---|---|---|---|---|
| 010 | 486.19 | 479.54 | 440.23 | 488.84 | **1,894.81** |
| 011 | 359.70 | 460.69 | 263.58 | 276.42 | **1,360.39** |
| 012 | 90.64 | 87.43 | 88.57 | 82.66 | **349.30** |
| 013 | 50.81 | 55.53 | 46.94 | 52.53 | **205.82** |
| 014 | 77.66 | 76.22 | 75.76 | 70.71 | **300.35** |
| 015 | 175.10 | 163.15 | 166.28 | 149.34 | **653.87** |
| 016 | 181.14 | 197.11 | 122.56 | 130.53 | **631.34** |
| 017 | 282.18 | 299.06 | 387.36 | 309.68 | **1,278.28** |
| 018 | 1,139.47 | 1,148.36 | 1,115.84 | 844.83 | **4,248.51** |
| 019 | 373.04 | 346.04 | 350.87 | 355.37 | **1,425.32** |
| 020 | 407.59 | 459.70 | 326.08 | 320.13 | **1,513.50** |
| 021 | 1,703.97 | 1,611.51 | 1,392.44 | 1,462.49 | **6,170.40** |
| 022 | 270.41 | 318.14 | 220.27 | 249.84 | **1,058.65** |
| 023 | 354.82 | 353.44 | 372.69 | 310.38 | **1,391.33** |

| | | | | | |
|---|---|---|---|---|---|
| 024 | 236.24 | 193.41 | 172.43 | 198.96 | **801.03** |
| 025 | 89.74 | 81.89 | 67.02 | 69.03 | **307.68** |
| 026 | 152.60 | 140.41 | 134.00 | 115.09 | **542.11** |
| 027 | 457.76 | 422.10 | 439.35 | 442.18 | **1,761.39** |
| **MA - MASSACHUSETTS - Total** | **6,889.07** | **6,893.72** | **6,182.28** | **5,929.01** | **25,894.08** |

**STATE: MD - MARYLAND**

| REGISTRANT ZIP CODE 3 | 1ST QUARTER | 2ND QUARTER | 3RD QUARTER | 4TH QUARTER | TOTAL GRAMS |
|---|---|---|---|---|---|
| 206 | 400.15 | 385.66 | 349.73 | 382.72 | **1,518.25** |
| 207 | 584.45 | 497.79 | 442.58 | 403.23 | **1,928.06** |
| 208 | 926.56 | 825.45 | 801.36 | 742.16 | **3,295.53** |
| 209 | 226.42 | 163.77 | 167.02 | 188.16 | **745.36** |
| 210 | 994.54 | 997.75 | 1,032.74 | 891.49 | **3,916.53** |
| 211 | 487.36 | 396.78 | 375.00 | 362.14 | **1,621.29** |
| 212 | 1,838.62 | 1,706.93 | 1,688.57 | 1,760.92 | **6,995.04** |
| 214 | 157.21 | 105.76 | 100.81 | 101.75 | **465.53** |
| 215 | 88.50 | 58.96 | 49.24 | 57.92 | **254.63** |
| 216 | 102.13 | 100.82 | 117.44 | 132.22 | **452.62** |
| 217 | 765.31 | 667.05 | 646.38 | 612.28 | **2,691.01** |
| 218 | 283.40 | 185.46 | 206.37 | 198.43 | **873.67** |
| 219 | 357.32 | 303.62 | 271.48 | 236.01 | **1,168.43** |
| **MD - MARYLAND - Total** | **7,211.98** | **6,395.81** | **6,248.74** | **6,069.43** | **25,925.96** |

**STATE: ME - MAINE**

| REGISTRANT ZIP CODE 3 | 1ST QUARTER | 2ND QUARTER | 3RD QUARTER | 4TH QUARTER | TOTAL GRAMS |
|---|---|---|---|---|---|
| 039 | 22.50 | 8.74 | 13.39 | 16.42 | **61.05** |
| 040 | 315.37 | 299.21 | 321.55 | 301.95 | **1,238.09** |
| 041 | 373.04 | 215.42 | 186.82 | 295.13 | **1,070.41** |
| 042 | 107.63 | 100.45 | 108.52 | 102.73 | **419.34** |
| 043 | 280.54 | 380.14 | 126.36 | 303.45 | **1,090.48** |
| 044 | 207.17 | 203.09 | 201.03 | 147.68 | **758.97** |
| 045 | 41.14 | 37.96 | 37.02 | 36.35 | **152.47** |
| 046 | 69.64 | 60.94 | 57.69 | 78.39 | **266.65** |
| 047 | 28.86 | 33.86 | 25.69 | 25.98 | **114.39** |
| 048 | 34.17 | 32.45 | 28.73 | 16.35 | **111.71** |
| 049 | 143.08 | 148.56 | 128.32 | 117.80 | **537.76** |
| **ME - MAINE - Total** | **1,623.13** | **1,520.84** | **1,235.11** | **1,442.22** | **5,821.30** |

**STATE: MI - MICHIGAN**

| REGISTRANT ZIP CODE 3 | 1ST QUARTER | 2ND QUARTER | 3RD QUARTER | 4TH QUARTER | TOTAL GRAMS |
|---|---|---|---|---|---|
| 480 | 1,220.93 | 932.23 | 893.62 | 834.95 | **3,881.73** |
| 481 | 954.39 | 806.98 | 669.16 | 695.14 | **3,125.67** |
| 482 | 832.53 | 790.64 | 719.00 | 734.85 | **3,077.03** |

| | | | | | |
|---|---|---|---|---|---|
| 483 | 850.66 | 1,267.30 | 942.56 | 972.66 | **4,033.18** |
| 484 | 297.27 | 252.85 | 236.49 | 248.66 | **1,035.27** |
| 485 | 170.46 | 213.72 | 171.71 | 133.64 | **689.53** |
| 486 | 384.86 | 456.14 | 324.93 | 276.76 | **1,442.70** |
| 487 | 107.56 | 118.25 | 86.00 | 89.14 | **400.94** |
| 488 | 293.51 | 276.98 | 266.50 | 279.11 | **1,116.10** |
| 489 | 196.33 | 195.79 | 146.34 | 138.50 | **676.97** |
| 490 | 464.53 | 421.21 | 434.51 | 403.49 | **1,723.73** |
| 491 | 40.69 | 50.78 | 38.23 | 34.31 | **164.02** |
| 492 | 163.24 | 157.72 | 133.15 | 109.98 | **564.09** |
| 493 | 108.95 | 101.87 | 99.60 | 88.70 | **399.12** |
| 494 | 250.02 | 241.98 | 199.62 | 233.69 | **925.31** |
| 495 | 433.77 | 385.63 | 364.92 | 393.42 | **1,577.74** |
| 496 | 374.63 | 371.15 | 354.71 | 331.17 | **1,431.67** |
| 497 | 204.53 | 217.97 | 182.00 | 141.35 | **745.84** |
| 498 | 259.81 | 210.39 | 166.71 | 94.68 | **731.59** |
| 499 | 45.85 | 34.90 | 40.85 | 42.07 | **163.66** |
| **MI - MICHIGAN - Total** | **7,654.53** | **7,504.48** | **6,470.61** | **6,276.27** | **27,905.89** |

**STATE: MN - MINNESOTA**

| REGISTRANT ZIP CODE 3 | 1ST QUARTER | 2ND QUARTER | 3RD QUARTER | 4TH QUARTER | TOTAL GRAMS |
|---|---|---|---|---|---|
| 550 | 470.51 | 469.66 | 504.75 | 489.97 | **1,934.89** |
| 551 | 863.73 | 908.10 | 852.99 | 887.75 | **3,512.57** |
| 553 | 585.77 | 546.80 | 533.90 | 514.52 | **2,180.99** |
| 554 | 1,892.28 | 1,643.74 | 1,754.78 | 1,701.54 | **6,992.35** |
| 556 | 6.43 | 3.92 | 4.68 | 3.76 | **18.79** |
| 557 | 158.67 | 141.05 | 148.31 | 140.46 | **588.49** |
| 558 | 162.04 | 175.39 | 137.36 | 110.47 | **585.26** |
| 559 | 503.17 | 459.76 | 499.31 | 469.62 | **1,931.86** |
| 560 | 122.83 | 123.46 | 122.88 | 128.06 | **497.23** |
| 561 | 13.22 | 13.34 | 19.06 | 12.39 | **58.01** |
| 562 | 47.73 | 40.88 | 47.04 | 35.16 | **170.82** |
| 563 | 246.26 | 209.68 | 122.17 | 148.48 | **726.58** |
| 564 | 80.93 | 89.37 | 85.38 | 86.53 | **342.21** |
| 565 | 142.31 | 153.40 | 90.51 | 73.31 | **459.53** |
| 566 | 39.55 | 36.28 | 42.27 | 36.97 | **155.07** |
| 567 | 13.44 | 21.99 | 19.11 | 25.82 | **80.35** |
| **MN - MINNESOTA - Total** | **5,348.87** | **5,036.82** | **4,984.49** | **4,864.83** | **20,235.00** |

**STATE: MO - MISSOURI**

| REGISTRANT ZIP CODE 3 | 1ST QUARTER | 2ND QUARTER | 3RD QUARTER | 4TH QUARTER | TOTAL GRAMS |
|---|---|---|---|---|---|
| 630 | 603.08 | 596.25 | 501.49 | 573.35 | **2,274.17** |

| | | | | | |
|---|---|---|---|---|---|
| 631 | 1,198.71 | 789.22 | 804.89 | 792.04 | **3,584.87** |
| 633 | 226.34 | 220.13 | 188.06 | 169.52 | **804.05** |
| 634 | 37.75 | 33.80 | 24.84 | 25.95 | **122.33** |
| 635 | 50.09 | 46.03 | 48.99 | 49.96 | **195.08** |
| 636 | 144.57 | 105.97 | 82.19 | 75.61 | **408.34** |
| 637 | 93.78 | 81.17 | 73.85 | 63.02 | **311.82** |
| 638 | 55.39 | 40.20 | 38.01 | 50.28 | **183.87** |
| 639 | 38.20 | 39.59 | 36.48 | 32.52 | **146.79** |
| 640 | 350.61 | 354.86 | 383.68 | 325.47 | **1,414.61** |
| 641 | 443.14 | 340.09 | 319.78 | 302.22 | **1,405.22** |
| 644 | 85.01 | 73.92 | 58.88 | 60.84 | **278.65** |
| 645 | 442.18 | 362.99 | 316.78 | 279.39 | **1,401.33** |
| 646 | 114.05 | 110.01 | 86.41 | 76.31 | **386.78** |
| 647 | 62.77 | 56.15 | 54.47 | 54.14 | **227.53** |
| 648 | 206.21 | 218.92 | 233.45 | 203.02 | **861.60** |
| 650 | 98.71 | 62.28 | 57.52 | 53.81 | **272.31** |
| 651 | 32.66 | 36.37 | 31.79 | 30.73 | **131.55** |
| 652 | 324.29 | 286.68 | 246.59 | 359.14 | **1,216.70** |
| 653 | 33.34 | 34.87 | 35.89 | 33.47 | **137.56** |
| 654 | 27.13 | 25.70 | 24.06 | 14.78 | **91.67** |
| 655 | 65.89 | 73.20 | 72.87 | 63.23 | **275.19** |
| 656 | 272.34 | 250.89 | 238.22 | 248.85 | **1,010.30** |
| 657 | 399.58 | 397.72 | 375.62 | 401.13 | **1,574.05** |
| 658 | 400.47 | 398.63 | 353.12 | 362.70 | **1,514.92** |
| **MO - MISSOURI - Total** | **5,806.27** | **5,035.62** | **4,687.94** | **4,701.49** | **20,231.31** |

**STATE: MS - MISSISSIPPI**

| REGISTRANT ZIP CODE 3 | 1ST QUARTER | 2ND QUARTER | 3RD QUARTER | 4TH QUARTER | TOTAL GRAMS |
|---|---|---|---|---|---|
| 386 | 162.95 | 165.20 | 104.31 | 138.70 | **571.15** |
| 387 | 6.44 | 3.91 | 7.34 | 8.53 | **26.22** |
| 388 | 127.69 | 110.20 | 76.99 | 98.92 | **413.80** |
| 389 | 22.65 | 20.54 | 21.14 | 28.84 | **93.18** |
| 390 | 84.32 | 86.75 | 82.92 | 94.92 | **348.90** |
| 391 | 3,043.29 | 5,025.51 | 4,183.05 | 3,281.35 | **15,533.21** |
| 392 | 303.54 | 185.14 | 231.23 | 151.99 | **871.90** |
| 393 | 68.22 | 59.56 | 54.66 | 50.61 | **233.05** |
| 394 | 258.55 | 230.55 | 184.04 | 244.67 | **917.81** |
| 395 | 363.43 | 301.13 | 395.98 | 301.09 | **1,361.64** |
| 396 | 30.87 | 32.25 | 30.81 | 23.62 | **117.56** |
| 397 | 25.44 | 30.24 | 16.83 | 25.61 | **98.11** |
| **MS - MISSISSIPPI - Total** | **4,497.39** | **6,250.99** | **5,389.31** | **4,448.85** | **20,586.53** |

**STATE: MT - MONTANA**

| REGISTRANT ZIP CODE 3 | 1ST QUARTER | 2ND QUARTER | 3RD QUARTER | 4TH QUARTER | TOTAL GRAMS |
|---|---|---|---|---|---|
| 590 | 96.72 | 80.21 | 79.50 | 75.91 | **332.35** |
| 591 | 472.63 | 446.18 | 450.02 | 435.77 | **1,804.60** |
| 592 | 38.37 | 34.70 | 34.11 | 39.91 | **147.10** |
| 593 | 60.73 | 52.35 | 52.25 | 54.84 | **220.16** |
| 594 | 97.97 | 79.17 | 83.87 | 75.73 | **336.74** |
| 595 | 9.26 | 16.46 | 13.38 | 11.61 | **50.72** |
| 596 | 118.24 | 71.86 | 63.26 | 53.62 | **306.98** |
| 597 | 316.15 | 152.14 | 177.33 | 174.94 | **820.57** |
| 598 | 278.31 | 297.49 | 277.53 | 250.04 | **1,103.36** |
| 599 | 133.19 | 113.12 | 127.23 | 117.03 | **490.57** |
| **MT - MONTANA - Total** | **1,621.56** | **1,343.70** | **1,358.48** | **1,289.40** | **5,613.14** |

**STATE: NC - NORTH CAROLINA**

| REGISTRANT ZIP CODE 3 | 1ST QUARTER | 2ND QUARTER | 3RD QUARTER | 4TH QUARTER | TOTAL GRAMS |
|---|---|---|---|---|---|
| 270 | 252.10 | 209.74 | 210.16 | 218.45 | **890.46** |
| 271 | 446.39 | 358.60 | 841.70 | 294.00 | **1,940.70** |
| 272 | 536.68 | 684.48 | 623.66 | 533.37 | **2,378.19** |
| 273 | 364.87 | 361.03 | 326.23 | 333.37 | **1,385.51** |
| 274 | 296.00 | 241.45 | 280.71 | 268.09 | **1,086.25** |
| 275 | 1,407.59 | 977.53 | 1,172.24 | 1,253.70 | **4,811.05** |
| 276 | 397.30 | 374.29 | 349.59 | 372.98 | **1,494.15** |
| 277 | 438.54 | 626.71 | 573.25 | 441.09 | **2,079.58** |
| 278 | 405.86 | 335.57 | 382.03 | 373.68 | **1,497.14** |
| 279 | 254.38 | 222.94 | 224.36 | 227.60 | **929.28** |
| 280 | 571.79 | 532.46 | 539.86 | 515.04 | **2,159.14** |
| 281 | 372.76 | 415.63 | 393.54 | 420.21 | **1,602.14** |
| 282 | 639.04 | 505.59 | 569.27 | 651.52 | **2,365.42** |
| 283 | 686.61 | 681.11 | 651.81 | 587.98 | **2,607.51** |
| 284 | 468.30 | 451.10 | 446.61 | 448.18 | **1,814.18** |
| 285 | 425.15 | 398.25 | 396.82 | 393.91 | **1,614.13** |
| 286 | 1,015.63 | 726.51 | 658.91 | 691.07 | **3,092.12** |
| 287 | 351.59 | 346.58 | 321.21 | 319.24 | **1,338.61** |
| 288 | 311.99 | 261.64 | 302.94 | 191.67 | **1,068.24** |
| 289 | 63.41 | 51.60 | 70.02 | 53.55 | **238.58** |
| **NC - NORTH CAROLINA - Total** | **9,705.96** | **8,762.82** | **9,334.90** | **8,588.70** | **36,392.38** |

**STATE: ND - NORTH DAKOTA**

| REGISTRANT ZIP CODE 3 | 1ST QUARTER | 2ND QUARTER | 3RD QUARTER | 4TH QUARTER | TOTAL GRAMS |
|---|---|---|---|---|---|
| 580 | 31.85 | 32.29 | 32.71 | 27.10 | **123.95** |
| 581 | 124.34 | 115.89 | 177.16 | 246.65 | **664.04** |

| | | | | | |
|---|---|---|---|---|---|
| 582 | 54.59 | 36.69 | 43.26 | 51.79 | **186.33** |
| 583 | 13.48 | 10.85 | 15.30 | 16.86 | **56.49** |
| 584 | 10.92 | 14.84 | 11.53 | 14.52 | **51.82** |
| 585 | 214.79 | 228.51 | 241.56 | 193.70 | **878.57** |
| 586 | 30.90 | 32.73 | 27.76 | 30.59 | **121.97** |
| 587 | 51.30 | 46.01 | 41.17 | 39.83 | **178.31** |
| 588 | 24.70 | 28.08 | 28.60 | 29.88 | **111.26** |
| **ND - NORTH DAKOTA - Total** | **556.87** | **545.90** | **619.05** | **650.92** | **2,372.74** |

**STATE: NE - NEBRASKA**

| REGISTRANT ZIP CODE 3 | 1ST QUARTER | 2ND QUARTER | 3RD QUARTER | 4TH QUARTER | TOTAL GRAMS |
|---|---|---|---|---|---|
| 680 | 157.83 | 162.28 | 167.01 | 169.80 | **656.93** |
| 681 | 490.37 | 467.82 | 498.82 | 440.29 | **1,897.30** |
| 683 | 27.89 | 16.53 | 13.83 | 14.74 | **72.98** |
| 684 | 23.49 | 24.17 | 22.95 | 25.12 | **95.73** |
| 685 | 107.48 | 110.74 | 134.78 | 125.56 | **478.55** |
| 686 | 23.60 | 25.99 | 20.33 | 26.16 | **96.09** |
| 687 | 44.22 | 42.86 | 29.06 | 27.78 | **143.92** |
| 688 | 97.37 | 96.68 | 89.54 | 103.35 | **386.94** |
| 689 | 27.05 | 45.64 | 23.06 | 24.15 | **119.90** |
| 690 | 12.67 | 14.88 | 11.04 | 14.48 | **53.06** |
| 691 | 32.14 | 38.20 | 33.83 | 45.84 | **150.02** |
| 692 | 3.37 | 2.93 | 3.92 | 5.59 | **15.81** |
| 693 | 36.43 | 34.26 | 38.91 | 36.00 | **145.60** |
| **NE - NEBRASKA - Total** | **1,083.90** | **1,082.98** | **1,087.07** | **1,058.86** | **4,312.82** |

**STATE: NH - NEW HAMPSHIRE**

| REGISTRANT ZIP CODE 3 | 1ST QUARTER | 2ND QUARTER | 3RD QUARTER | 4TH QUARTER | TOTAL GRAMS |
|---|---|---|---|---|---|
| 030 | 382.50 | 350.42 | 373.34 | 338.42 | **1,444.69** |
| 031 | 267.75 | 170.34 | 173.17 | 190.83 | **802.10** |
| 032 | 142.66 | 168.63 | 160.33 | 148.73 | **620.35** |
| 033 | 197.28 | 213.75 | 125.37 | 283.77 | **820.17** |
| 034 | 66.68 | 55.39 | 61.33 | 69.73 | **253.13** |
| 035 | 81.29 | 76.71 | 73.69 | 84.64 | **316.33** |
| 036 | 4.26 | 5.59 | 4.08 | 4.01 | **17.93** |
| 037 | 203.03 | 314.50 | 220.32 | 171.07 | **908.93** |
| 038 | 401.81 | 383.00 | 322.24 | 304.07 | **1,411.12** |
| **NH - NEW HAMPSHIRE - Total** | **1,747.28** | **1,738.34** | **1,513.86** | **1,595.28** | **6,594.75** |

**STATE: NJ - NEW JERSEY**

| REGISTRANT ZIP CODE 3 | 1ST QUARTER | 2ND QUARTER | 3RD QUARTER | 4TH QUARTER | TOTAL GRAMS |
|---|---|---|---|---|---|
| 070 | 1,268.54 | 1,188.68 | 1,026.36 | 1,041.23 | **4,524.81** |
| 071 | 201.07 | 194.22 | 203.12 | 217.06 | **815.48** |

| | | | | | |
|---|---|---|---|---|---|
| 072 | 93.94 | 81.75 | 96.18 | 91.36 | **363.24** |
| 073 | 51.06 | 50.93 | 56.53 | 44.38 | **202.90** |
| 074 | 658.75 | 732.42 | 814.21 | 817.63 | **3,023.00** |
| 075 | 178.11 | 77.40 | 209.82 | 136.57 | **601.90** |
| 076 | 794.11 | 756.37 | 734.20 | 699.02 | **2,983.70** |
| 077 | 636.95 | 573.69 | 594.05 | 589.32 | **2,394.01** |
| 078 | 316.58 | 296.25 | 309.30 | 304.51 | **1,226.65** |
| 079 | 413.05 | 310.58 | 349.92 | 359.86 | **1,433.41** |
| 080 | 917.26 | 893.16 | 880.09 | 852.82 | **3,543.33** |
| 081 | 195.79 | 169.79 | 141.29 | 146.04 | **652.92** |
| 082 | 137.77 | 126.37 | 145.35 | 105.08 | **514.57** |
| 083 | 110.01 | 108.68 | 100.29 | 101.83 | **420.80** |
| 084 | 14.97 | 22.67 | 25.69 | 23.37 | **86.70** |
| 085 | 172.28 | 178.22 | 163.52 | 173.01 | **687.03** |
| 086 | 107.99 | 110.39 | 112.97 | 112.09 | **443.45** |
| 087 | 377.63 | 324.18 | 344.16 | 322.54 | **1,368.51** |
| 088 | 1,026.67 | 835.28 | 842.39 | 872.20 | **3,576.54** |
| 089 | 150.72 | 127.00 | 143.65 | 127.90 | **549.27** |
| **NJ - NEW JERSEY - Total** | **7,823.25** | **7,158.03** | **7,293.09** | **7,137.84** | **29,412.21** |

**STATE: NM - NEW MEXICO**

| REGISTRANT ZIP CODE 3 | 1ST QUARTER | 2ND QUARTER | 3RD QUARTER | 4TH QUARTER | TOTAL GRAMS |
|---|---|---|---|---|---|
| 870 | 73.50 | 69.82 | 75.44 | 75.64 | **294.40** |
| 871 | 593.12 | 559.68 | 610.32 | 521.33 | **2,284.46** |
| 873 | 7.61 | 6.86 | 14.25 | 8.44 | **37.15** |
| 874 | 60.71 | 52.54 | 54.51 | 60.16 | **227.92** |
| 875 | 116.69 | 142.64 | 124.66 | 124.80 | **508.79** |
| 877 | 30.03 | 24.76 | 26.50 | 26.91 | **108.19** |
| 878 | 3.90 | 1.11 | 1.77 | 0.71 | **7.49** |
| 879 | 9.84 | 15.39 | 9.03 | 10.55 | **44.81** |
| 880 | 102.52 | 111.42 | 103.81 | 109.34 | **427.09** |
| 881 | 16.98 | 20.27 | 22.47 | 22.19 | **81.90** |
| 882 | 105.85 | 96.07 | 94.27 | 74.81 | **371.00** |
| 883 | 26.60 | 22.28 | 26.03 | 29.69 | **104.61** |
| 884 | 0.35 | 1.07 | 0.73 | 1.08 | **3.24** |
| **NM - NEW MEXICO - Total** | **1,147.70** | **1,123.91** | **1,163.79** | **1,065.66** | **4,501.06** |

**STATE: NV - NEVADA**

| REGISTRANT ZIP CODE 3 | 1ST QUARTER | 2ND QUARTER | 3RD QUARTER | 4TH QUARTER | TOTAL GRAMS |
|---|---|---|---|---|---|
| 890 | 537.88 | 556.88 | 561.08 | 508.21 | **2,164.05** |
| 891 | 1,433.39 | 1,178.73 | 1,288.24 | 1,292.13 | **5,192.48** |
| 893 | 1.06 | 0.99 | 0.75 | 1.20 | **4.01** |

| | | | | | |
|---|---|---|---|---|---|
| 894 | 156.86 | 159.12 | 159.45 | 159.41 | **634.85** |
| 895 | 252.62 | 177.87 | 195.79 | 189.78 | **816.05** |
| 897 | 88.97 | 99.07 | 83.40 | 72.80 | **344.25** |
| 898 | 30.80 | 27.03 | 22.95 | 18.49 | **99.27** |
| **NV - NEVADA - Total** | **2,501.58** | **2,199.70** | **2,311.65** | **2,242.03** | **9,254.96** |

**STATE: NY - NEW YORK**

| REGISTRANT ZIP CODE 3 | 1ST QUARTER | 2ND QUARTER | 3RD QUARTER | 4TH QUARTER | **TOTAL GRAMS** |
|---|---|---|---|---|---|
| 100 | 2,970.26 | 2,948.42 | 2,643.70 | 2,458.37 | **11,020.75** |
| 101 | 35.16 | 38.03 | 31.85 | 26.53 | **131.56** |
| 102 | 4.43 | 2.38 | 2.19 | 2.66 | **11.66** |
| 103 | 378.29 | 330.90 | 340.80 | 323.44 | **1,373.44** |
| 104 | 618.55 | 778.32 | 699.31 | 734.04 | **2,830.22** |
| 105 | 457.94 | 507.87 | 443.66 | 408.63 | **1,818.11** |
| 106 | 128.08 | 93.70 | 105.45 | 90.67 | **417.90** |
| 107 | 127.20 | 95.70 | 97.22 | 104.30 | **424.41** |
| 108 | 34.93 | 52.03 | 29.36 | 37.06 | **153.39** |
| 109 | 491.08 | 463.47 | 434.03 | 447.36 | **1,835.93** |
| 110 | 1,063.49 | 745.74 | 1,003.30 | 806.88 | **3,619.42** |
| 111 | 209.10 | 161.18 | 150.86 | 121.74 | **642.88** |
| 112 | 1,121.49 | 1,101.79 | 996.27 | 964.00 | **4,183.55** |
| 113 | 475.60 | 431.17 | 432.56 | 430.60 | **1,769.93** |
| 114 | 349.67 | 348.82 | 222.74 | 222.47 | **1,143.70** |
| 115 | 758.48 | 842.61 | 753.65 | 733.13 | **3,087.87** |
| 116 | 42.62 | 52.36 | 54.66 | 45.89 | **195.53** |
| 117 | 2,191.08 | 2,188.95 | 1,979.34 | 1,972.11 | **8,331.48** |
| 118 | 123.02 | 186.99 | 202.24 | 153.60 | **665.85** |
| 119 | 218.42 | 195.01 | 195.72 | 170.75 | **779.90** |
| 120 | 195.97 | 207.12 | 158.88 | 169.81 | **731.79** |
| 121 | 84.45 | 86.84 | 83.06 | 89.31 | **343.66** |
| 122 | 162.86 | 221.95 | 206.36 | 242.84 | **834.01** |
| 123 | 47.55 | 49.73 | 49.04 | 51.13 | **197.45** |
| 124 | 187.12 | 183.00 | 179.63 | 169.60 | **719.34** |
| 125 | 343.09 | 320.35 | 344.10 | 318.18 | **1,325.72** |
| 126 | 159.23 | 221.36 | 107.36 | 118.18 | **606.13** |
| 127 | 144.99 | 99.29 | 98.62 | 87.93 | **430.83** |
| 128 | 192.46 | 174.18 | 176.24 | 159.15 | **702.03** |
| 129 | 319.57 | 293.07 | 375.01 | 288.33 | **1,275.98** |
| 130 | 172.80 | 171.66 | 152.75 | 135.45 | **632.65** |
| 131 | 93.93 | 96.28 | 97.60 | 86.87 | **374.68** |
| 132 | 179.70 | 159.75 | 167.56 | 134.78 | **641.80** |

| | | | | | |
|---|---|---|---|---|---|
| 133 | 79.80 | 79.08 | 66.35 | 63.24 | **288.47** |
| 134 | 71.21 | 82.66 | 75.11 | 65.67 | **294.65** |
| 135 | 60.21 | 38.40 | 35.00 | 27.35 | **160.96** |
| 136 | 133.17 | 145.98 | 152.65 | 153.14 | **584.95** |
| 137 | 53.27 | 58.73 | 45.71 | 40.93 | **198.64** |
| 138 | 101.52 | 105.13 | 123.51 | 135.07 | **465.23** |
| 139 | 62.66 | 33.14 | 35.06 | 29.62 | **160.49** |
| 140 | 262.38 | 231.88 | 249.61 | 194.82 | **938.69** |
| 141 | 116.43 | 120.86 | 134.61 | 122.50 | **494.40** |
| 142 | 563.08 | 414.33 | 398.89 | 414.77 | **1,791.07** |
| 143 | 34.74 | 36.89 | 30.74 | 23.30 | **125.66** |
| 144 | 308.06 | 266.79 | 270.84 | 287.62 | **1,133.31** |
| 145 | 215.29 | 192.91 | 197.27 | 192.03 | **797.50** |
| 146 | 858.27 | 859.90 | 859.92 | 861.84 | **3,439.93** |
| 147 | 55.20 | 61.33 | 66.56 | 79.47 | **262.56** |
| 148 | 231.38 | 201.62 | 213.34 | 213.83 | **860.16** |
| 149 | 52.22 | 45.18 | 42.81 | 36.86 | **177.08** |
| **NY - NEW YORK - Total** | **17,341.51** | **16,824.83** | **16,013.10** | **15,247.83** | **65,427.26** |

**STATE: OH - OHIO**

| REGISTRANT ZIP CODE 3 | 1ST QUARTER | 2ND QUARTER | 3RD QUARTER | 4TH QUARTER | **TOTAL GRAMS** |
|---|---|---|---|---|---|
| 430 | 776.01 | 744.47 | 743.25 | 703.26 | **2,966.99** |
| 431 | 239.99 | 240.36 | 229.36 | 229.33 | **939.04** |
| 432 | 1,263.49 | 1,075.08 | 666.66 | 649.01 | **3,654.24** |
| 433 | 126.11 | 121.01 | 123.11 | 127.33 | **497.57** |
| 434 | 77.74 | 87.22 | 70.22 | 49.56 | **284.74** |
| 435 | 383.38 | 187.53 | 212.49 | 397.52 | **1,180.92** |
| 436 | 260.27 | 235.49 | 230.27 | 208.96 | **934.98** |
| 437 | 159.95 | 118.28 | 121.52 | 111.60 | **511.35** |
| 438 | 17.02 | 22.09 | 18.55 | 16.85 | **74.51** |
| 439 | 115.87 | 99.62 | 86.13 | 101.75 | **403.37** |
| 440 | 293.09 | 311.62 | 287.75 | 245.51 | **1,137.97** |
| 441 | 1,161.86 | 806.45 | 771.09 | 702.42 | **3,441.82** |
| 442 | 302.44 | 305.89 | 301.58 | 276.78 | **1,186.70** |
| 443 | 247.30 | 228.33 | 216.44 | 182.02 | **874.10** |
| 444 | 130.04 | 149.76 | 87.50 | 128.03 | **495.34** |
| 445 | 83.41 | 81.81 | 84.42 | 127.08 | **376.73** |
| 446 | 378.95 | 335.49 | 318.58 | 350.85 | **1,383.88** |
| 447 | 296.62 | 262.66 | 241.59 | 269.29 | **1,070.17** |
| 448 | 152.42 | 126.20 | 127.09 | 109.71 | **515.42** |
| 449 | 31.64 | 40.08 | 43.20 | 37.44 | **152.35** |

| | | | | | |
|---|---|---|---|---|---|
| 450 | 388.77 | 411.44 | 355.59 | 373.63 | **1,529.44** |
| 451 | 199.59 | 136.53 | 147.12 | 129.34 | **612.58** |
| 452 | 619.59 | 425.37 | 467.33 | 416.26 | **1,928.55** |
| 453 | 155.55 | 149.10 | 149.38 | 144.58 | **598.61** |
| 454 | 372.42 | 285.78 | 254.43 | 242.87 | **1,155.50** |
| 455 | 63.69 | 52.84 | 40.29 | 43.92 | **200.74** |
| 456 | 219.41 | 191.08 | 158.18 | 220.16 | **788.83** |
| 457 | 71.91 | 82.39 | 64.26 | 136.73 | **355.28** |
| 458 | 198.87 | 177.98 | 235.23 | 222.34 | **834.42** |
| **OH - OHIO - Total** | **8,787.42** | **7,491.97** | **6,852.60** | **6,954.14** | **30,086.13** |

**STATE: OK - OKLAHOMA**

| REGISTRANT ZIP CODE 3 | 1ST QUARTER | 2ND QUARTER | 3RD QUARTER | 4TH QUARTER | **TOTAL GRAMS** |
|---|---|---|---|---|---|
| 730 | 538.80 | 520.64 | 570.34 | 368.63 | **1,998.41** |
| 731 | 766.21 | 672.41 | 779.47 | 643.88 | **2,861.97** |
| 734 | 89.21 | 68.86 | 64.43 | 53.22 | **275.71** |
| 735 | 197.70 | 174.79 | 223.37 | 154.23 | **750.09** |
| 736 | 17.25 | 15.59 | 18.32 | 26.58 | **77.73** |
| 737 | 62.74 | 44.44 | 40.55 | 36.77 | **184.49** |
| 738 | 16.14 | 14.71 | 14.09 | 14.32 | **59.25** |
| 739 | 7.31 | 1.78 | 0.89 | 0.90 | **10.88** |
| 740 | 453.65 | 604.65 | 410.61 | 369.05 | **1,837.97** |
| 741 | 460.18 | 411.33 | 378.29 | 415.76 | **1,665.57** |
| 743 | 108.07 | 97.10 | 97.42 | 108.27 | **410.85** |
| 744 | 154.70 | 129.21 | 140.31 | 137.06 | **561.29** |
| 745 | 91.30 | 97.91 | 96.64 | 91.48 | **377.32** |
| 746 | 20.39 | 18.13 | 16.38 | 7.42 | **62.31** |
| 747 | 96.74 | 96.45 | 69.89 | 84.30 | **347.38** |
| 748 | 106.52 | 92.45 | 76.69 | 72.50 | **348.16** |
| 749 | 179.60 | 164.38 | 174.24 | 153.22 | **671.44** |
| **OK - OKLAHOMA - Total** | **3,366.51** | **3,224.82** | **3,171.93** | **2,737.57** | **12,500.83** |

**STATE: OR - OREGON**

| REGISTRANT ZIP CODE 3 | 1ST QUARTER | 2ND QUARTER | 3RD QUARTER | 4TH QUARTER | **TOTAL GRAMS** |
|---|---|---|---|---|---|
| 970 | 664.66 | 641.24 | 613.96 | 620.66 | **2,540.52** |
| 971 | 302.94 | 295.10 | 256.67 | 282.11 | **1,136.82** |
| 972 | 1,361.10 | 1,102.64 | 982.20 | 1,015.85 | **4,461.80** |
| 973 | 610.42 | 551.52 | 515.84 | 493.76 | **2,171.53** |
| 974 | 804.05 | 717.52 | 683.86 | 637.64 | **2,843.07** |
| 975 | 409.51 | 441.89 | 397.71 | 407.94 | **1,657.05** |
| 976 | 25.85 | 38.11 | 30.87 | 39.66 | **134.48** |
| 977 | 130.25 | 149.61 | 151.28 | 137.45 | **568.59** |

| | | | | | |
|---|---|---|---|---|---|
| 978 | 142.37 | 120.51 | 96.67 | 98.83 | **458.37** |
| 979 | 30.99 | 38.29 | 34.80 | 25.30 | **129.37** |
| **OR - OREGON - Total** | **4,482.13** | **4,096.43** | **3,763.84** | **3,759.19** | **16,101.59** |

**STATE: PA - PENNSYLVANIA**

| REGISTRANT ZIP CODE 3 | 1ST QUARTER | 2ND QUARTER | 3RD QUARTER | 4TH QUARTER | TOTAL GRAMS |
|---|---|---|---|---|---|
| 150 | 326.56 | 329.68 | 335.26 | 316.53 | **1,308.03** |
| 151 | 120.76 | 116.88 | 108.64 | 94.43 | **440.72** |
| 152 | 687.34 | 454.38 | 743.99 | 639.75 | **2,525.46** |
| 153 | 104.80 | 107.52 | 108.50 | 71.69 | **392.51** |
| 154 | 80.42 | 72.13 | 70.08 | 56.29 | **278.92** |
| 155 | 102.76 | 69.54 | 65.96 | 56.96 | **295.22** |
| 156 | 142.44 | 239.09 | 143.36 | 137.46 | **662.35** |
| 157 | 42.96 | 54.00 | 35.60 | 41.28 | **173.84** |
| 158 | 56.03 | 67.76 | 59.28 | 56.83 | **239.91** |
| 159 | 115.65 | 95.55 | 82.74 | 81.03 | **374.96** |
| 160 | 63.59 | 66.79 | 63.06 | 55.26 | **248.69** |
| 161 | 127.14 | 112.27 | 100.77 | 102.02 | **442.20** |
| 162 | 36.79 | 33.94 | 25.35 | 28.05 | **124.13** |
| 163 | 223.21 | 192.89 | 245.07 | 219.17 | **880.34** |
| 164 | 74.84 | 90.84 | 74.05 | 88.10 | **327.83** |
| 165 | 111.07 | 101.18 | 103.55 | 120.87 | **436.67** |
| 166 | 178.11 | 144.78 | 147.43 | 145.76 | **616.08** |
| 167 | 30.55 | 29.99 | 31.25 | 37.11 | **128.90** |
| 168 | 69.11 | 68.87 | 54.88 | 45.79 | **238.65** |
| 169 | 37.64 | 24.13 | 29.82 | 30.55 | **122.13** |
| 170 | 570.13 | 464.14 | 488.64 | 396.52 | **1,919.43** |
| 171 | 190.04 | 144.97 | 126.17 | 132.33 | **593.51** |
| 172 | 124.12 | 202.92 | 112.12 | 110.77 | **549.93** |
| 173 | 198.89 | 204.39 | 186.72 | 175.45 | **765.45** |
| 174 | 143.44 | 132.53 | 117.88 | 112.11 | **505.96** |
| 175 | 194.47 | 181.49 | 165.58 | 159.11 | **700.65** |
| 176 | 372.97 | 389.80 | 218.54 | 204.85 | **1,186.16** |
| 177 | 91.41 | 85.72 | 83.10 | 64.75 | **324.98** |
| 178 | 223.71 | 221.68 | 159.03 | 174.76 | **779.18** |
| 179 | 39.72 | 33.97 | 33.17 | 30.46 | **137.33** |
| 180 | 364.52 | 336.18 | 285.04 | 317.16 | **1,302.90** |
| 181 | 279.60 | 191.74 | 244.16 | 209.63 | **925.13** |
| 182 | 49.12 | 54.93 | 57.14 | 38.35 | **199.53** |
| 183 | 162.90 | 176.69 | 128.49 | 133.50 | **601.58** |
| 184 | 170.84 | 146.70 | 159.52 | 144.83 | **621.89** |

| | | | | | |
|---|---|---|---|---|---|
| 185 | 182.24 | 201.76 | 118.59 | 102.85 | **605.43** |
| 186 | 107.68 | 77.61 | 77.98 | 90.69 | **353.96** |
| 187 | 632.67 | 594.22 | 554.16 | 554.40 | **2,335.44** |
| 188 | 42.27 | 55.55 | 39.01 | 34.42 | **171.27** |
| 189 | 334.86 | 311.09 | 315.77 | 317.81 | **1,279.54** |
| 190 | 4,471.59 | 3,150.98 | 2,484.41 | 2,703.42 | **12,810.40** |
| 191 | 1,612.44 | 1,628.12 | 1,237.29 | 1,263.24 | **5,741.09** |
| 193 | 442.31 | 401.37 | 365.14 | 394.74 | **1,603.55** |
| 194 | 714.98 | 648.23 | 471.02 | 537.88 | **2,372.11** |
| 195 | 71.19 | 62.64 | 73.01 | 69.92 | **276.76** |
| 196 | 166.02 | 183.04 | 163.10 | 136.53 | **648.69** |
| **PA - PENNSYLVANIA - Total** | **14,685.90** | **12,754.69** | **11,093.41** | **11,035.38** | **49,569.38** |

**STATE: PR - PUERTO RICO**

| REGISTRANT ZIP CODE 3 | 1ST QUARTER | 2ND QUARTER | 3RD QUARTER | 4TH QUARTER | TOTAL GRAMS |
|---|---|---|---|---|---|
| 006 | 13.48 | 15.96 | 8.87 | 6.92 | **45.22** |
| 007 | 5.32 | 3.90 | 5.14 | 3.90 | **18.26** |
| 009 | 32.45 | 46.77 | 41.58 | 45.39 | **166.19** |
| **PR - PUERTO RICO - Total** | **51.25** | **66.63** | **55.59** | **56.21** | **229.67** |

**STATE: RI - RHODE ISLAND**

| REGISTRANT ZIP CODE 3 | 1ST QUARTER | 2ND QUARTER | 3RD QUARTER | 4TH QUARTER | TOTAL GRAMS |
|---|---|---|---|---|---|
| 028 | 467.72 | 456.21 | 390.08 | 365.62 | **1,679.63** |
| 029 | 351.10 | 284.95 | 229.75 | 203.35 | **1,069.14** |
| **RI - RHODE ISLAND - Total** | **818.83** | **741.16** | **619.82** | **568.97** | **2,748.77** |

**STATE: SC - SOUTH CAROLINA**

| REGISTRANT ZIP CODE 3 | 1ST QUARTER | 2ND QUARTER | 3RD QUARTER | 4TH QUARTER | TOTAL GRAMS |
|---|---|---|---|---|---|
| 290 | 397.42 | 416.42 | 412.27 | 352.37 | **1,578.47** |
| 291 | 597.76 | 579.79 | 637.11 | 579.61 | **2,394.27** |
| 292 | 512.07 | 562.89 | 538.42 | 435.68 | **2,049.07** |
| 293 | 249.77 | 210.89 | 216.98 | 179.74 | **857.38** |
| 294 | 914.05 | 890.67 | 883.35 | 869.57 | **3,557.64** |
| 295 | 922.66 | 885.07 | 831.89 | 757.01 | **3,396.63** |
| 296 | 787.43 | 655.66 | 642.51 | 575.07 | **2,660.67** |
| 297 | 182.19 | 165.42 | 160.66 | 161.48 | **669.76** |
| 298 | 221.77 | 205.57 | 231.50 | 222.27 | **881.11** |
| 299 | 297.54 | 407.64 | 388.51 | 282.94 | **1,376.64** |
| **SC - SOUTH CAROLINA - Total** | **5,082.66** | **4,980.02** | **4,943.20** | **4,415.74** | **19,421.63** |

**STATE: SD - SOUTH DAKOTA**

| REGISTRANT ZIP CODE 3 | 1ST QUARTER | 2ND QUARTER | 3RD QUARTER | 4TH QUARTER | TOTAL GRAMS |
|---|---|---|---|---|---|
| 570 | 66.36 | 61.26 | 61.23 | 78.12 | **266.98** |
| 571 | 284.35 | 237.33 | 278.40 | 244.88 | **1,044.96** |

| | | | | | |
|---|---|---|---|---|---|
| 572 | 20.03 | 26.23 | 23.79 | 20.86 | **90.91** |
| 573 | 14.21 | 14.10 | 13.42 | 16.58 | **58.31** |
| 574 | 13.32 | 16.13 | 14.83 | 7.75 | **52.03** |
| 575 | 18.76 | 20.40 | 17.99 | 19.05 | **76.20** |
| 576 | 7.87 | 9.22 | 3.69 | 31.08 | **51.86** |
| 577 | 100.90 | 89.01 | 73.69 | 73.46 | **337.06** |
| **SD - SOUTH DAKOTA - Total** | **525.80** | **473.70** | **487.04** | **491.78** | **1,978.32** |

**STATE: TN - TENNESSEE**

| REGISTRANT ZIP CODE 3 | 1ST QUARTER | 2ND QUARTER | 3RD QUARTER | 4TH QUARTER | TOTAL GRAMS |
|---|---|---|---|---|---|
| 370 | 570.93 | 407.47 | 443.47 | 409.04 | **1,830.91** |
| 371 | 394.77 | 332.05 | 425.11 | 404.80 | **1,556.73** |
| 372 | 759.79 | 559.02 | 480.89 | 514.72 | **2,314.42** |
| 373 | 467.79 | 432.54 | 442.61 | 487.15 | **1,830.10** |
| 374 | 323.50 | 268.09 | 220.31 | 215.15 | **1,027.06** |
| 376 | 310.40 | 311.17 | 280.09 | 271.09 | **1,172.74** |
| 377 | 102.13 | 102.22 | 125.80 | 85.36 | **415.51** |
| 378 | 197.02 | 221.40 | 192.56 | 174.36 | **785.35** |
| 379 | 357.97 | 201.20 | 325.40 | 640.15 | **1,524.72** |
| 380 | 224.50 | 180.57 | 221.70 | 206.37 | **833.14** |
| 381 | 3,559.89 | 1,172.79 | 1,521.81 | 1,215.20 | **7,469.68** |
| 382 | 53.44 | 45.12 | 37.23 | 31.99 | **167.78** |
| 383 | 88.09 | 63.15 | 83.32 | 77.53 | **312.10** |
| 384 | 56.95 | 76.46 | 49.72 | 54.09 | **237.22** |
| 385 | 198.47 | 132.45 | 99.94 | 83.42 | **514.28** |
| **TN - TENNESSEE - Total** | **7,665.65** | **4,505.69** | **4,949.97** | **4,870.43** | **21,991.74** |

**STATE: TX - TEXAS**

| REGISTRANT ZIP CODE 3 | 1ST QUARTER | 2ND QUARTER | 3RD QUARTER | 4TH QUARTER | TOTAL GRAMS |
|---|---|---|---|---|---|
| 750 | 2,055.81 | 2,009.12 | 1,895.56 | 1,811.80 | **7,772.29** |
| 751 | 438.55 | 414.57 | 382.99 | 384.30 | **1,620.41** |
| 752 | 1,330.75 | 859.34 | 850.79 | 1,299.74 | **4,340.62** |
| 753 | 177.04 | 93.22 | 132.69 | 66.03 | **468.98** |
| 754 | 426.89 | 413.89 | 375.54 | 369.49 | **1,585.82** |
| 755 | 107.53 | 100.68 | 68.81 | 66.79 | **343.82** |
| 756 | 1,035.36 | 125.26 | 1,028.53 | 487.14 | **2,676.29** |
| 757 | 337.15 | 360.73 | 275.61 | 354.48 | **1,327.98** |
| 758 | 46.71 | 35.78 | 25.80 | 33.98 | **142.27** |
| 759 | 159.02 | 156.99 | 152.21 | 137.92 | **606.14** |
| 760 | 951.81 | 856.14 | 772.41 | 772.96 | **3,353.33** |
| 761 | 1,021.87 | 1,064.13 | 970.53 | 1,082.29 | **4,138.82** |
| 762 | 666.68 | 701.41 | 517.27 | 773.52 | **2,658.88** |

| | | | | | |
|---|---|---|---|---|---|
| 763 | 241.43 | 232.96 | 197.95 | 240.53 | **912.87** |
| 764 | 92.05 | 102.23 | 90.80 | 91.69 | **376.77** |
| 765 | 403.30 | 368.86 | 321.40 | 331.90 | **1,425.46** |
| 766 | 52.47 | 38.19 | 39.94 | 45.53 | **176.12** |
| 767 | 69.29 | 74.83 | 69.05 | 69.77 | **282.94** |
| 768 | 130.64 | 38.88 | 41.09 | 51.93 | **262.54** |
| 769 | 39.59 | 40.66 | 34.54 | 49.08 | **163.87** |
| 770 | 2,364.35 | 2,038.17 | 1,852.94 | 1,867.41 | **8,122.87** |
| 773 | 968.39 | 864.34 | 835.41 | 765.11 | **3,433.25** |
| 774 | 639.79 | 574.98 | 594.56 | 564.55 | **2,373.89** |
| 775 | 975.19 | 1,095.68 | 1,095.42 | 968.03 | **4,134.32** |
| 776 | 448.21 | 406.32 | 388.22 | 407.11 | **1,649.87** |
| 777 | 249.79 | 210.01 | 246.36 | 286.64 | **992.80** |
| 778 | 148.80 | 115.30 | 96.06 | 110.18 | **470.34** |
| 779 | 370.73 | 406.99 | 346.78 | 311.52 | **1,436.02** |
| 780 | 192.55 | 197.14 | 208.82 | 201.40 | **799.90** |
| 781 | 307.99 | 276.20 | 291.16 | 253.11 | **1,128.46** |
| 782 | 1,563.31 | 1,653.78 | 1,420.36 | 1,309.67 | **5,947.13** |
| 783 | 119.72 | 104.03 | 88.62 | 90.02 | **402.38** |
| 784 | 256.40 | 223.25 | 234.19 | 190.41 | **904.24** |
| 785 | 314.20 | 294.38 | 267.93 | 280.94 | **1,157.44** |
| 786 | 790.16 | 749.16 | 648.13 | 627.98 | **2,815.43** |
| 787 | 823.19 | 831.53 | 807.11 | 823.62 | **3,285.45** |
| 788 | 35.93 | 26.94 | 32.06 | 31.92 | **126.85** |
| 789 | 31.40 | 22.88 | 29.40 | 26.23 | **109.92** |
| 790 | 75.69 | 57.93 | 48.65 | 51.70 | **233.97** |
| 791 | 175.14 | 168.23 | 141.63 | 138.63 | **623.63** |
| 792 | 10.59 | 7.09 | 6.61 | 6.03 | **30.32** |
| 793 | 75.28 | 69.58 | 56.29 | 70.25 | **271.40** |
| 794 | 384.34 | 315.84 | 279.39 | 313.96 | **1,293.54** |
| 795 | 39.85 | 42.67 | 37.28 | 33.06 | **152.85** |
| 796 | 104.08 | 63.29 | 66.13 | 74.29 | **307.79** |
| 797 | 202.84 | 181.67 | 173.13 | 155.79 | **713.43** |
| 798 | 12.94 | 15.52 | 8.16 | 11.57 | **48.19** |
| 799 | 406.62 | 338.00 | 401.40 | 317.32 | **1,463.33** |
| **TX - TEXAS - Total** | **21,871.45** | **19,438.77** | **18,945.69** | **18,809.33** | **79,065.23** |

**STATE: UT - UTAH**

| REGISTRANT ZIP CODE 3 | 1ST QUARTER | 2ND QUARTER | 3RD QUARTER | 4TH QUARTER | **TOTAL GRAMS** |
|---|---|---|---|---|---|
| 840 | 1,020.21 | 778.25 | 885.30 | 794.74 | **3,478.50** |
| 841 | 982.38 | 684.70 | 756.39 | 730.74 | **3,154.20** |

| | | | | | |
|---|---|---|---|---|---|
| 843 | 148.68 | 120.37 | 142.67 | 137.55 | **549.27** |
| 844 | 169.46 | 141.33 | 167.74 | 171.10 | **649.63** |
| 845 | 30.47 | 29.94 | 31.50 | 29.79 | **121.71** |
| 846 | 224.21 | 187.52 | 202.52 | 206.81 | **821.06** |
| 847 | 236.63 | 203.42 | 250.71 | 204.08 | **894.83** |
| **UT - UTAH - Total** | **2,812.04** | **2,145.52** | **2,436.83** | **2,274.81** | **9,669.19** |

**STATE: VA - VIRGINIA**

| REGISTRANT ZIP CODE 3 | 1ST QUARTER | 2ND QUARTER | 3RD QUARTER | 4TH QUARTER | TOTAL GRAMS |
|---|---|---|---|---|---|
| 201 | 958.88 | 853.08 | 756.41 | 826.20 | **3,394.57** |
| 220 | 427.75 | 479.82 | 419.66 | 388.88 | **1,716.11** |
| 221 | 330.33 | 293.08 | 288.88 | 264.54 | **1,176.83** |
| 222 | 124.95 | 134.59 | 133.07 | 142.77 | **535.38** |
| 223 | 182.00 | 183.86 | 180.96 | 149.53 | **696.34** |
| 224 | 300.94 | 278.03 | 286.46 | 283.18 | **1,148.61** |
| 225 | 122.63 | 121.95 | 103.40 | 80.27 | **428.24** |
| 226 | 374.66 | 349.67 | 387.07 | 312.88 | **1,424.28** |
| 227 | 44.54 | 48.57 | 44.13 | 51.76 | **189.00** |
| 228 | 247.65 | 249.28 | 247.57 | 244.07 | **988.56** |
| 229 | 438.83 | 410.66 | 427.06 | 361.33 | **1,637.88** |
| 230 | 265.62 | 219.30 | 209.99 | 186.71 | **881.62** |
| 231 | 508.94 | 445.93 | 428.52 | 441.59 | **1,824.99** |
| 232 | 1,223.17 | 1,120.99 | 900.94 | 833.02 | **4,078.11** |
| 233 | 367.90 | 309.77 | 334.59 | 350.98 | **1,363.24** |
| 234 | 843.42 | 765.22 | 722.28 | 725.97 | **3,056.89** |
| 235 | 412.63 | 364.19 | 413.46 | 366.72 | **1,557.00** |
| 236 | 494.06 | 391.53 | 419.23 | 369.74 | **1,674.56** |
| 237 | 237.65 | 205.76 | 161.79 | 155.18 | **760.38** |
| 238 | 221.52 | 233.06 | 208.32 | 215.70 | **878.60** |
| 239 | 51.26 | 55.43 | 55.45 | 50.36 | **212.49** |
| 240 | 526.00 | 519.65 | 449.36 | 468.93 | **1,963.93** |
| 241 | 348.14 | 328.70 | 290.01 | 270.66 | **1,237.51** |
| 242 | 173.35 | 155.07 | 197.31 | 200.09 | **725.82** |
| 243 | 110.67 | 93.17 | 93.45 | 98.13 | **395.42** |
| 244 | 130.49 | 135.22 | 124.86 | 123.61 | **514.19** |
| 245 | 282.43 | 299.68 | 261.17 | 240.47 | **1,083.73** |
| 246 | 44.46 | 58.09 | 60.06 | 55.80 | **218.41** |
| **VA - VIRGINIA - Total** | **9,794.87** | **9,103.35** | **8,605.45** | **8,259.04** | **35,762.70** |

**STATE: VI - VIRGIN ISLANDS**

| REGISTRANT ZIP CODE 3 | 1ST QUARTER | 2ND QUARTER | 3RD QUARTER | 4TH QUARTER | TOTAL GRAMS |
|---|---|---|---|---|---|
| 008 | 10.86 | 13.30 | 16.31 | 13.83 | **54.30** |

| VI - VIRGIN ISLANDS - Total | 10.86 | 13.30 | 16.31 | 13.83 | 54.30 |
|---|---|---|---|---|---|

STATE: VT - VERMONT

| REGISTRANT ZIP CODE 3 | 1ST QUARTER | 2ND QUARTER | 3RD QUARTER | 4TH QUARTER | TOTAL GRAMS |
|---|---|---|---|---|---|
| 050 | 53.97 | 51.32 | 49.10 | 41.19 | **195.58** |
| 051 | 48.90 | 44.32 | 38.04 | 34.68 | **165.94** |
| 052 | 34.07 | 33.86 | 24.38 | 22.07 | **114.37** |
| 053 | 32.12 | 33.35 | 41.44 | 26.70 | **133.62** |
| 054 | 573.89 | 523.24 | 489.14 | 538.31 | **2,124.57** |
| 056 | 115.35 | 126.85 | 122.89 | 114.00 | **479.08** |
| 057 | 172.87 | 235.93 | 108.84 | 100.91 | **618.55** |
| 058 | 44.06 | 38.57 | 42.01 | 42.13 | **166.76** |
| VT - VERMONT - Total | 1,075.22 | 1,087.44 | 915.83 | 919.98 | 3,998.48 |

STATE: WA - WASHINGTON

| REGISTRANT ZIP CODE 3 | 1ST QUARTER | 2ND QUARTER | 3RD QUARTER | 4TH QUARTER | TOTAL GRAMS |
|---|---|---|---|---|---|
| 980 | 1,605.71 | 1,586.58 | 1,422.75 | 1,428.00 | **6,043.05** |
| 981 | 1,807.34 | 1,317.16 | 1,252.07 | 1,191.64 | **5,568.22** |
| 982 | 1,239.83 | 1,138.90 | 1,147.12 | 1,114.32 | **4,640.17** |
| 983 | 749.25 | 740.04 | 710.35 | 737.64 | **2,937.28** |
| 984 | 451.17 | 513.33 | 361.89 | 438.97 | **1,765.37** |
| 985 | 646.15 | 620.21 | 628.92 | 609.83 | **2,505.11** |
| 986 | 1,056.55 | 862.01 | 942.26 | 1,063.50 | **3,924.31** |
| 988 | 164.02 | 178.75 | 165.90 | 163.24 | **671.91** |
| 989 | 327.60 | 253.92 | 282.71 | 261.74 | **1,125.98** |
| 990 | 83.47 | 77.28 | 95.82 | 94.23 | **350.80** |
| 991 | 116.91 | 125.56 | 119.45 | 118.41 | **480.34** |
| 992 | 900.45 | 1,044.45 | 734.31 | 701.06 | **3,380.28** |
| 993 | 492.12 | 423.09 | 370.33 | 366.56 | **1,652.11** |
| 994 | 74.92 | 61.65 | 60.34 | 44.65 | **241.55** |
| WA - WASHINGTON - Total | 9,715.50 | 8,942.94 | 8,294.23 | 8,333.80 | 35,286.47 |

STATE: WI - WISCONSIN

| REGISTRANT ZIP CODE 3 | 1ST QUARTER | 2ND QUARTER | 3RD QUARTER | 4TH QUARTER | TOTAL GRAMS |
|---|---|---|---|---|---|
| 530 | 374.26 | 292.26 | 236.08 | 232.99 | **1,135.59** |
| 531 | 403.95 | 369.24 | 311.59 | 227.65 | **1,312.43** |
| 532 | 790.21 | 740.87 | 592.65 | 612.18 | **2,735.91** |
| 534 | 60.92 | 44.58 | 67.69 | 62.15 | **235.35** |
| 535 | 174.61 | 228.45 | 188.78 | 204.23 | **796.06** |
| 537 | 286.95 | 215.91 | 190.66 | 210.16 | **903.68** |
| 538 | 21.63 | 30.40 | 23.76 | 33.05 | **108.84** |
| 539 | 63.31 | 76.59 | 69.87 | 61.20 | **270.97** |
| 540 | 85.03 | 83.51 | 89.01 | 102.72 | **360.27** |

| | | | | | |
|---|---|---|---|---|---|
| 541 | 53.60 | 53.31 | 44.87 | 40.66 | **192.44** |
| 542 | 61.03 | 86.28 | 20.87 | 18.08 | **186.25** |
| 543 | 231.14 | 114.62 | 106.34 | 123.87 | **575.97** |
| 544 | 163.01 | 135.16 | 164.81 | 141.04 | **604.02** |
| 545 | 44.71 | 72.73 | 58.24 | 61.63 | **237.30** |
| 546 | 171.56 | 147.83 | 132.31 | 99.62 | **551.32** |
| 547 | 113.36 | 115.17 | 98.54 | 101.68 | **428.74** |
| 548 | 172.65 | 138.32 | 136.69 | 121.60 | **569.25** |
| 549 | 281.44 | 320.25 | 268.31 | 176.88 | **1,046.87** |
| **WI - WISCONSIN - Total** | **3,553.35** | **3,265.46** | **2,801.07** | **2,631.38** | **12,251.25** |

**STATE: WV - WEST VIRGINIA**

| REGISTRANT ZIP CODE 3 | 1ST QUARTER | 2ND QUARTER | 3RD QUARTER | 4TH QUARTER | TOTAL GRAMS |
|---|---|---|---|---|---|
| 247 | 32.16 | 35.02 | 39.14 | 40.40 | **146.71** |
| 248 | 5.32 | 5.67 | 4.79 | 5.50 | **21.28** |
| 249 | 24.31 | 18.75 | 17.55 | 19.92 | **80.53** |
| 250 | 6.92 | 9.75 | 6.92 | 4.68 | **28.26** |
| 251 | 6.07 | 29.97 | 4.37 | 7.85 | **48.26** |
| 252 | 6.38 | 4.79 | 4.78 | 3.64 | **19.58** |
| 253 | 94.26 | 126.13 | 79.97 | 68.29 | **368.65** |
| 254 | 324.44 | 317.08 | 320.58 | 283.50 | **1,245.60** |
| 255 | 43.59 | 37.07 | 35.68 | 47.70 | **164.05** |
| 256 | 11.53 | 9.18 | 9.54 | 4.30 | **34.54** |
| 257 | 164.65 | 67.15 | 61.58 | 66.09 | **359.47** |
| 258 | 80.93 | 48.97 | 52.45 | 54.72 | **237.07** |
| 259 | 11.35 | 10.33 | 10.02 | 5.32 | **37.02** |
| 260 | 65.01 | 54.69 | 47.73 | 47.40 | **214.83** |
| 261 | 35.50 | 40.89 | 29.43 | 29.01 | **134.83** |
| 262 | 16.69 | 24.21 | 14.58 | 14.57 | **70.04** |
| 263 | 76.03 | 72.33 | 73.18 | 73.65 | **295.18** |
| 264 | 20.33 | 16.82 | 22.01 | 22.83 | **81.99** |
| 265 | 99.91 | 121.41 | 143.28 | 231.21 | **595.81** |
| 266 | 5.87 | 9.94 | 12.89 | 7.06 | **35.77** |
| 267 | 29.65 | 35.06 | 30.04 | 27.47 | **122.21** |
| 268 | 20.37 | 22.79 | 20.29 | 18.69 | **82.14** |
| **WV - WEST VIRGINIA - Total** | **1,181.25** | **1,118.00** | **1,040.79** | **1,083.81** | **4,423.85** |

**STATE: WY - WYOMING**

| REGISTRANT ZIP CODE 3 | 1ST QUARTER | 2ND QUARTER | 3RD QUARTER | 4TH QUARTER | TOTAL GRAMS |
|---|---|---|---|---|---|
| 820 | 126.29 | 134.07 | 109.46 | 102.89 | **472.70** |
| 822 | 4.71 | 9.39 | 7.39 | 10.77 | **32.26** |
| 823 | 6.67 | 9.53 | 7.05 | 4.15 | **27.40** |

| | | | | | |
|---|---|---|---|---|---|
| 824 | 110.36 | 107.79 | 85.79 | 95.44 | **399.38** |
| 825 | 37.22 | 32.19 | 26.28 | 31.61 | **127.30** |
| 826 | 87.99 | 80.48 | 68.90 | 60.17 | **297.54** |
| 827 | 39.09 | 37.27 | 42.93 | 21.49 | **140.78** |
| 828 | 29.80 | 37.21 | 37.70 | 42.60 | **147.30** |
| 829 | 92.84 | 92.71 | 77.27 | 91.86 | **354.68** |
| 830 | 6.98 | 5.07 | 8.17 | 9.84 | **30.05** |
| 831 | 20.08 | 18.52 | 13.56 | 23.18 | **75.34** |
| **WY - WYOMING - Total** | **562.03** | **564.22** | **484.48** | **494.00** | **2,104.73** |

**DRUG: 9193 - HYDROCODONE**

**STATE: AK - ALASKA**

| REGISTRANT ZIP CODE 3 | 1ST QUARTER | 2ND QUARTER | 3RD QUARTER | 4TH QUARTER | TOTAL GRAMS |
|---|---|---|---|---|---|
| 995 | 4,685.23 | 4,493.87 | 4,166.75 | 4,056.68 | **17,402.54** |
| 996 | 3,228.83 | 3,153.17 | 3,133.12 | 2,934.89 | **12,450.01** |
| 997 | 1,034.14 | 986.96 | 914.12 | 896.38 | **3,831.59** |
| 998 | 481.58 | 437.36 | 443.63 | 420.81 | **1,783.37** |
| 999 | 391.79 | 357.22 | 367.41 | 336.83 | **1,453.25** |
| **AK - ALASKA - Total** | **9,821.57** | **9,428.58** | **9,025.03** | **8,645.58** | **36,920.76** |

**STATE: AL - ALABAMA**

| REGISTRANT ZIP CODE 3 | 1ST QUARTER | 2ND QUARTER | 3RD QUARTER | 4TH QUARTER | TOTAL GRAMS |
|---|---|---|---|---|---|
| 350 | 25,179.18 | 24,443.33 | 23,566.32 | 23,206.14 | **96,394.96** |
| 351 | 13,391.26 | 13,053.32 | 12,720.57 | 12,847.03 | **52,012.18** |
| 352 | 22,108.33 | 21,010.41 | 20,495.30 | 20,320.65 | **83,934.69** |
| 354 | 10,133.85 | 9,732.04 | 9,322.54 | 9,044.39 | **38,232.82** |
| 355 | 10,580.61 | 10,272.78 | 10,260.94 | 10,253.64 | **41,367.97** |
| 356 | 21,501.72 | 20,924.83 | 20,592.54 | 21,126.87 | **84,145.96** |
| 357 | 8,956.43 | 8,305.27 | 7,869.69 | 8,063.63 | **33,195.02** |
| 358 | 9,588.77 | 9,294.93 | 8,640.26 | 8,555.92 | **36,079.87** |
| 359 | 18,763.83 | 19,018.86 | 18,614.18 | 18,434.24 | **74,831.11** |
| 360 | 10,222.05 | 9,524.61 | 9,118.26 | 8,832.51 | **37,697.43** |
| 361 | 8,019.43 | 7,462.92 | 7,212.11 | 6,681.28 | **29,375.74** |
| 362 | 8,689.52 | 8,689.76 | 8,528.89 | 8,554.19 | **34,462.36** |
| 363 | 8,606.62 | 8,467.70 | 8,314.55 | 8,054.07 | **33,442.94** |
| 364 | 5,441.44 | 4,999.58 | 4,748.20 | 5,008.08 | **20,197.30** |
| 365 | 17,534.26 | 16,373.58 | 15,865.09 | 15,687.97 | **65,460.90** |
| 366 | 13,937.75 | 12,635.34 | 12,282.63 | 12,041.03 | **50,896.75** |
| 367 | 2,970.03 | 2,808.66 | 2,791.69 | 2,799.22 | **11,369.60** |
| 368 | 6,938.04 | 6,905.35 | 6,834.21 | 6,880.79 | **27,558.39** |
| 369 | 786.94 | 852.06 | 828.45 | 866.23 | **3,333.68** |
| **AL - ALABAMA - Total** | **223,350.03** | **214,775.33** | **208,606.42** | **207,257.90** | **853,989.67** |

**STATE: AR - ARKANSAS**

| REGISTRANT ZIP CODE 3 | 1ST QUARTER | 2ND QUARTER | 3RD QUARTER | 4TH QUARTER | TOTAL GRAMS |
|---|---|---|---|---|---|
| 716 | 5,381.50 | 5,165.57 | 5,094.39 | 4,991.58 | **20,633.03** |
| 717 | 4,991.01 | 4,962.40 | 4,682.25 | 4,767.61 | **19,403.27** |
| 718 | 7,314.10 | 7,013.02 | 7,111.83 | 6,927.02 | **28,365.97** |
| 719 | 9,440.27 | 8,879.55 | 8,817.29 | 8,642.13 | **35,779.24** |
| 720 | 11,398.20 | 11,082.60 | 10,749.00 | 10,693.28 | **43,923.07** |
| 721 | 14,151.26 | 13,257.97 | 12,830.03 | 12,396.41 | **52,635.66** |
| 722 | 7,318.05 | 6,664.50 | 6,534.09 | 6,490.56 | **27,007.20** |
| 723 | 7,359.36 | 6,536.37 | 5,856.05 | 5,808.40 | **25,560.18** |
| 724 | 11,101.27 | 10,532.75 | 9,923.21 | 9,381.83 | **40,939.06** |
| 725 | 6,075.24 | 5,689.67 | 5,454.49 | 5,370.97 | **22,590.37** |
| 726 | 5,451.07 | 5,153.01 | 4,798.56 | 4,842.08 | **20,244.72** |
| 727 | 14,759.74 | 14,375.53 | 13,340.71 | 13,068.53 | **55,544.50** |
| 728 | 3,977.78 | 3,982.76 | 3,814.24 | 3,590.08 | **15,364.86** |
| 729 | 12,144.78 | 11,950.24 | 11,619.33 | 11,420.54 | **47,134.89** |
| **AR - ARKANSAS - Total** | **120,863.63** | **115,245.93** | **110,625.47** | **108,391.02** | **455,126.05** |

**STATE: AS - AMERICAN SAMOA**

| REGISTRANT ZIP CODE 3 | 1ST QUARTER | 2ND QUARTER | 3RD QUARTER | 4TH QUARTER | TOTAL GRAMS |
|---|---|---|---|---|---|
| 967 | 3.63 | 0 | 0 | 0 | **3.63** |
| **AS - AMERICAN SAMOA - Tota** | **3.63** | **0.00** | **0.00** | **0.00** | **3.63** |

**STATE: AZ - ARIZONA**

| REGISTRANT ZIP CODE 3 | 1ST QUARTER | 2ND QUARTER | 3RD QUARTER | 4TH QUARTER | TOTAL GRAMS |
|---|---|---|---|---|---|
| 850 | 13,445.84 | 12,453.57 | 11,780.68 | 11,754.99 | **49,435.09** |
| 851 | 5,277.26 | 4,599.31 | 4,349.60 | 4,700.31 | **18,926.48** |
| 852 | 23,301.13 | 21,636.06 | 22,476.53 | 19,233.67 | **86,647.39** |
| 853 | 16,364.24 | 14,632.97 | 13,727.34 | 14,799.18 | **59,523.72** |
| 855 | 1,836.74 | 1,790.77 | 1,710.07 | 1,696.69 | **7,034.26** |
| 856 | 3,481.23 | 3,361.17 | 3,178.96 | 3,194.50 | **13,215.85** |
| 857 | 12,042.65 | 11,235.08 | 10,699.15 | 11,004.14 | **44,981.02** |
| 859 | 2,078.02 | 2,224.91 | 2,182.80 | 2,114.69 | **8,600.41** |
| 860 | 1,798.99 | 1,759.48 | 1,733.23 | 1,626.00 | **6,917.70** |
| 863 | 6,177.20 | 6,220.34 | 5,933.01 | 5,870.85 | **24,201.39** |
| 864 | 7,632.80 | 7,184.82 | 6,872.28 | 7,171.52 | **28,861.42** |
| 865 | 84.46 | 93.20 | 96.38 | 90.97 | **365.01** |
| **AZ - ARIZONA - Total** | **93,520.55** | **87,191.66** | **84,740.02** | **83,257.51** | **348,709.74** |

**STATE: CA - CALIFORNIA**

| REGISTRANT ZIP CODE 3 | 1ST QUARTER | 2ND QUARTER | 3RD QUARTER | 4TH QUARTER | TOTAL GRAMS |
|---|---|---|---|---|---|
| 900 | 18,281.95 | 17,803.84 | 17,029.70 | 16,089.12 | **69,204.62** |
| 902 | 15,705.15 | 14,789.28 | 14,387.35 | 14,049.63 | **58,931.41** |

| | | | | | |
|---|---|---|---|---|---|
| 903 | 1,519.77 | 1,473.96 | 1,503.71 | 1,550.25 | **6,047.69** |
| 904 | 1,782.98 | 1,759.87 | 1,635.55 | 1,562.21 | **6,740.60** |
| 905 | 3,635.94 | 3,662.37 | 3,528.45 | 3,401.72 | **14,228.48** |
| 906 | 12,029.54 | 11,255.97 | 11,029.02 | 10,831.00 | **45,145.52** |
| 907 | 11,528.48 | 11,074.73 | 10,832.33 | 10,306.66 | **43,742.19** |
| 908 | 7,197.62 | 7,024.24 | 6,792.88 | 6,779.82 | **27,794.56** |
| 910 | 3,970.25 | 3,809.12 | 3,721.45 | 3,535.16 | **15,035.97** |
| 911 | 2,310.93 | 2,257.35 | 2,112.14 | 2,120.93 | **8,801.35** |
| 912 | 3,268.25 | 3,179.99 | 3,054.17 | 2,808.65 | **12,311.06** |
| 913 | 22,917.23 | 22,339.66 | 21,491.63 | 20,904.91 | **87,653.44** |
| 914 | 6,159.75 | 6,131.35 | 5,810.62 | 5,900.59 | **24,002.31** |
| 915 | 1,840.31 | 1,863.78 | 1,764.18 | 1,653.58 | **7,121.85** |
| 916 | 1,300.41 | 1,334.33 | 1,231.26 | 1,297.00 | **5,163.00** |
| 917 | 33,212.75 | 31,689.68 | 31,684.15 | 31,828.99 | **128,415.57** |
| 918 | 926.18 | 852.28 | 906.99 | 905.92 | **3,591.37** |
| 919 | 7,823.25 | 7,621.19 | 7,385.23 | 7,031.95 | **29,861.62** |
| 920 | 26,682.64 | 25,707.98 | 24,656.44 | 24,058.33 | **101,105.38** |
| 921 | 18,212.02 | 17,068.22 | 16,521.23 | 15,221.27 | **67,022.74** |
| 922 | 19,165.55 | 18,199.14 | 17,375.91 | 17,257.30 | **71,997.90** |
| 923 | 44,003.57 | 40,929.71 | 39,843.78 | 39,208.53 | **163,985.59** |
| 924 | 6,865.93 | 6,724.51 | 6,438.39 | 6,272.68 | **26,301.51** |
| 925 | 37,697.37 | 36,373.85 | 35,704.93 | 34,345.99 | **144,122.14** |
| 926 | 22,026.65 | 21,073.59 | 20,480.83 | 19,917.60 | **83,498.67** |
| 927 | 6,569.69 | 6,480.56 | 6,108.64 | 6,017.25 | **25,176.14** |
| 928 | 23,012.82 | 22,095.65 | 21,482.65 | 21,232.46 | **87,823.57** |
| 930 | 15,003.75 | 14,598.01 | 14,274.12 | 13,881.65 | **57,757.52** |
| 931 | 2,604.70 | 2,522.32 | 2,296.66 | 2,268.84 | **9,692.52** |
| 932 | 21,869.89 | 20,764.83 | 20,052.42 | 19,436.60 | **82,123.74** |
| 933 | 18,565.63 | 17,995.85 | 17,276.18 | 17,385.79 | **71,223.46** |
| 934 | 13,619.23 | 13,276.58 | 12,798.47 | 12,466.94 | **52,161.22** |
| 935 | 16,883.71 | 15,933.10 | 15,275.41 | 14,983.42 | **63,075.64** |
| 936 | 15,952.71 | 15,813.14 | 14,939.60 | 14,428.04 | **61,133.49** |
| 937 | 18,570.52 | 18,025.81 | 17,697.90 | 18,101.40 | **72,395.63** |
| 939 | 7,597.37 | 7,035.61 | 6,850.10 | 6,412.29 | **27,895.38** |
| 940 | 8,495.65 | 8,463.23 | 8,064.53 | 7,934.87 | **32,958.28** |
| 941 | 7,927.19 | 7,700.61 | 7,313.67 | 6,958.84 | **29,900.31** |
| 943 | 1,865.47 | 1,748.36 | 1,661.65 | 1,587.47 | **6,862.95** |
| 944 | 1,474.93 | 1,421.97 | 1,383.85 | 1,456.11 | **5,736.86** |
| 945 | 56,354.79 | 53,853.06 | 52,122.23 | 51,435.01 | **213,765.09** |
| 946 | 8,423.94 | 7,030.49 | 7,018.41 | 6,911.71 | **29,384.55** |

| | | | | | |
|---|---|---|---|---|---|
| 947 | 1,403.92 | 1,320.43 | 1,305.77 | 1,170.83 | **5,200.95** |
| 948 | 2,993.40 | 2,896.10 | 2,858.09 | 2,779.20 | **11,526.80** |
| 949 | 7,796.28 | 7,266.45 | 7,404.70 | 6,829.48 | **29,296.91** |
| 950 | 11,478.13 | 10,898.33 | 10,722.54 | 10,144.53 | **43,243.53** |
| 951 | 8,102.94 | 8,041.54 | 7,669.58 | 7,238.52 | **31,052.58** |
| 952 | 16,706.88 | 15,601.26 | 15,109.01 | 14,213.77 | **61,630.93** |
| 953 | 44,444.59 | 43,066.48 | 40,117.03 | 39,425.00 | **167,053.09** |
| 954 | 15,764.83 | 14,824.24 | 14,228.92 | 13,571.46 | **58,389.45** |
| 955 | 7,410.19 | 7,112.43 | 6,845.55 | 6,254.45 | **27,622.61** |
| 956 | 48,035.93 | 47,632.57 | 45,073.68 | 44,203.06 | **184,945.24** |
| 957 | 8,637.66 | 8,225.73 | 7,715.35 | 7,419.72 | **31,998.48** |
| 958 | 29,780.54 | 28,173.89 | 27,734.73 | 26,401.52 | **112,090.67** |
| 959 | 29,722.72 | 28,046.55 | 25,999.97 | 24,153.10 | **107,922.34** |
| 960 | 19,136.21 | 18,069.22 | 16,984.82 | 16,446.42 | **70,636.68** |
| 961 | 2,731.73 | 2,619.67 | 2,516.43 | 2,499.78 | **10,367.62** |
| **CA - CALIFORNIA - Total** | **829,002.43** | **794,554.06** | **765,824.97** | **744,489.33** | **3,133,870.80** |

**STATE: CO - COLORADO**

| REGISTRANT ZIP CODE 3 | 1ST QUARTER | 2ND QUARTER | 3RD QUARTER | 4TH QUARTER | **TOTAL GRAMS** |
|---|---|---|---|---|---|
| 800 | 8,222.23 | 7,622.88 | 8,314.32 | 8,334.59 | **32,494.03** |
| 801 | 6,716.33 | 6,510.61 | 6,289.50 | 6,308.03 | **25,824.47** |
| 802 | 9,205.31 | 8,946.21 | 7,428.62 | 7,348.51 | **32,928.65** |
| 803 | 825.56 | 810.83 | 777.71 | 819.59 | **3,233.69** |
| 804 | 2,105.90 | 1,718.64 | 1,790.96 | 1,696.80 | **7,312.30** |
| 805 | 6,110.83 | 5,685.64 | 5,506.69 | 5,531.25 | **22,834.41** |
| 806 | 2,438.37 | 2,303.23 | 2,185.62 | 2,217.08 | **9,144.30** |
| 807 | 804.56 | 744.41 | 751.64 | 730.20 | **3,030.80** |
| 808 | 1,530.52 | 1,481.18 | 1,413.92 | 1,382.82 | **5,808.43** |
| 809 | 7,121.38 | 6,661.35 | 6,282.55 | 6,269.69 | **26,334.97** |
| 810 | 5,128.35 | 5,012.78 | 4,853.62 | 4,633.21 | **19,627.95** |
| 811 | 676.72 | 671.54 | 643.56 | 586.53 | **2,578.35** |
| 812 | 1,308.13 | 1,260.18 | 1,228.30 | 1,183.15 | **4,979.76** |
| 813 | 984.58 | 952.21 | 921.64 | 911.01 | **3,769.43** |
| 814 | 1,037.89 | 969.83 | 882.72 | 892.56 | **3,783.00** |
| 815 | 2,383.89 | 2,242.79 | 2,175.66 | 2,116.80 | **8,919.14** |
| 816 | 1,383.51 | 1,246.68 | 1,208.47 | 1,207.76 | **5,046.41** |
| **CO - COLORADO - Total** | **57,984.06** | **54,840.98** | **52,655.47** | **52,169.59** | **217,650.10** |

**STATE: CT - CONNECTICUT**

| REGISTRANT ZIP CODE 3 | 1ST QUARTER | 2ND QUARTER | 3RD QUARTER | 4TH QUARTER | **TOTAL GRAMS** |
|---|---|---|---|---|---|
| 060 | 3,923.90 | 3,587.35 | 3,373.05 | 3,544.94 | **14,429.25** |
| 061 | 1,131.05 | 990.78 | 940.02 | 944.06 | **4,005.91** |

| | | | | | |
|---|---|---|---|---|---|
| 062 | 890.32 | 827.56 | 808.48 | 837.21 | **3,363.56** |
| 063 | 1,923.30 | 1,827.36 | 1,821.59 | 1,788.41 | **7,360.65** |
| 064 | 4,305.31 | 3,999.79 | 3,948.27 | 3,930.91 | **16,184.28** |
| 065 | 1,124.94 | 1,035.65 | 946.21 | 992.45 | **4,099.25** |
| 066 | 789.98 | 747.95 | 750.07 | 691.03 | **2,979.02** |
| 067 | 2,336.38 | 2,187.87 | 2,096.65 | 2,123.37 | **8,744.27** |
| 068 | 1,434.11 | 1,324.50 | 1,302.71 | 1,239.75 | **5,301.07** |
| 069 | 262.62 | 232.35 | 232.26 | 249.54 | **976.77** |
| **CT - CONNECTICUT - Total** | **18,121.90** | **16,761.16** | **16,219.30** | **16,341.67** | **67,444.03** |

**STATE: DC - DISTRICT OF COLUMBIA**

| REGISTRANT ZIP CODE 3 | 1ST QUARTER | 2ND QUARTER | 3RD QUARTER | 4TH QUARTER | TOTAL GRAMS |
|---|---|---|---|---|---|
| 200 | 1,231.72 | 1,061.68 | 1,003.86 | 986.43 | **4,283.69** |
| 203 | 15.14 | 5.75 | 6.96 | 6.05 | **33.90** |
| 204 | 110.79 | 43.59 | 67.20 | 61.15 | **282.72** |
| 205 | 0 | 0.30 | 0 | 1.36 | **1.66** |
| **DC - DISTRICT OF COLUMBIA -** | **1,357.64** | **1,111.33** | **1,078.02** | **1,054.99** | **4,601.98** |

**STATE: DE - DELAWARE**

| REGISTRANT ZIP CODE 3 | 1ST QUARTER | 2ND QUARTER | 3RD QUARTER | 4TH QUARTER | TOTAL GRAMS |
|---|---|---|---|---|---|
| 197 | 1,459.85 | 1,462.03 | 1,393.63 | 1,359.95 | **5,675.46** |
| 198 | 927.90 | 947.87 | 903.73 | 946.17 | **3,725.67** |
| 199 | 2,786.22 | 2,724.42 | 2,610.05 | 2,540.50 | **10,661.20** |
| **DE - DELAWARE - Total** | **5,173.97** | **5,134.33** | **4,907.42** | **4,846.62** | **20,062.33** |

**STATE: FL - FLORIDA**

| REGISTRANT ZIP CODE 3 | 1ST QUARTER | 2ND QUARTER | 3RD QUARTER | 4TH QUARTER | TOTAL GRAMS |
|---|---|---|---|---|---|
| 320 | 18,178.67 | 17,052.10 | 15,138.80 | 15,078.47 | **65,448.03** |
| 321 | 21,041.71 | 19,806.46 | 17,194.44 | 17,565.96 | **75,608.58** |
| 322 | 31,763.56 | 30,219.49 | 26,577.57 | 26,387.26 | **114,947.88** |
| 323 | 7,762.13 | 7,175.05 | 6,300.86 | 6,363.32 | **27,601.36** |
| 324 | 16,502.44 | 15,826.92 | 14,554.88 | 12,948.84 | **59,833.07** |
| 325 | 25,348.72 | 24,031.85 | 22,237.78 | 21,583.65 | **93,202.01** |
| 326 | 6,077.14 | 5,724.86 | 5,135.24 | 5,118.69 | **22,055.94** |
| 327 | 18,521.32 | 17,427.56 | 14,718.47 | 15,262.36 | **65,929.71** |
| 328 | 8,599.09 | 7,801.56 | 6,432.44 | 6,642.85 | **29,475.94** |
| 329 | 15,348.39 | 14,272.13 | 12,259.65 | 12,721.03 | **54,601.20** |
| 330 | 8,796.47 | 7,942.38 | 6,639.69 | 6,518.22 | **29,896.77** |
| 331 | 4,518.05 | 4,099.31 | 3,454.32 | 3,402.70 | **15,474.37** |
| 333 | 6,243.29 | 5,696.40 | 4,606.87 | 4,670.59 | **21,217.14** |
| 334 | 13,848.49 | 12,871.98 | 10,654.31 | 11,039.51 | **48,414.28** |
| 335 | 12,268.51 | 11,344.89 | 9,677.44 | 9,917.72 | **43,208.55** |
| 336 | 12,120.37 | 11,483.57 | 9,435.64 | 9,756.05 | **42,795.63** |

| | | | | | |
|---|---|---|---|---|---|
| 337 | 17,628.45 | 16,582.68 | 14,133.46 | 14,397.83 | **62,742.43** |
| 338 | 12,467.41 | 10,914.93 | 9,039.66 | 9,598.30 | **42,020.31** |
| 339 | 14,705.36 | 13,136.36 | 11,068.43 | 11,779.09 | **50,689.24** |
| 341 | 4,774.24 | 3,554.46 | 2,795.24 | 3,371.63 | **14,495.57** |
| 342 | 17,505.23 | 15,411.20 | 12,873.15 | 13,633.39 | **59,422.97** |
| 344 | 13,136.06 | 12,314.60 | 10,425.78 | 10,567.10 | **46,443.53** |
| 346 | 17,222.48 | 16,164.95 | 14,435.80 | 13,960.56 | **61,783.80** |
| 347 | 8,611.64 | 7,917.47 | 6,485.69 | 6,728.53 | **29,743.33** |
| 349 | 7,185.67 | 6,618.85 | 5,736.96 | 5,864.59 | **25,406.06** |
| **FL - FLORIDA - Total** | **340,174.90** | **315,391.99** | **272,012.56** | **274,878.25** | **1,202,457.69** |

**STATE: GA - GEORGIA**

| REGISTRANT ZIP CODE 3 | 1ST QUARTER | 2ND QUARTER | 3RD QUARTER | 4TH QUARTER | **TOTAL GRAMS** |
|---|---|---|---|---|---|
| 300 | 28,270.50 | 26,780.97 | 25,186.28 | 25,311.19 | **105,548.95** |
| 301 | 30,645.09 | 29,592.37 | 28,195.15 | 28,467.72 | **116,900.33** |
| 302 | 18,449.32 | 17,548.25 | 16,800.98 | 16,897.46 | **69,696.01** |
| 303 | 5,660.57 | 5,435.95 | 5,057.87 | 5,206.95 | **21,361.33** |
| 304 | 6,146.72 | 5,780.88 | 5,459.08 | 5,459.82 | **22,846.50** |
| 305 | 19,091.81 | 18,771.70 | 17,999.57 | 18,472.14 | **74,335.23** |
| 306 | 13,580.11 | 12,777.64 | 11,985.33 | 12,313.46 | **50,656.53** |
| 307 | 10,956.62 | 10,674.14 | 10,262.35 | 10,133.82 | **42,026.93** |
| 308 | 2,636.31 | 2,404.26 | 2,264.16 | 2,285.03 | **9,589.76** |
| 309 | 5,498.76 | 4,549.44 | 4,898.91 | 4,323.46 | **19,270.58** |
| 310 | 20,983.23 | 19,922.04 | 18,681.80 | 18,892.32 | **78,479.40** |
| 312 | 7,637.17 | 7,435.69 | 7,095.10 | 7,298.45 | **29,466.41** |
| 313 | 4,388.16 | 4,273.28 | 4,031.81 | 3,885.80 | **16,579.05** |
| 314 | 4,379.35 | 4,157.66 | 3,918.79 | 3,831.57 | **16,287.38** |
| 315 | 15,282.81 | 14,651.05 | 14,340.50 | 13,966.90 | **58,241.26** |
| 316 | 8,297.29 | 8,324.18 | 7,846.25 | 7,910.25 | **32,377.97** |
| 317 | 10,417.21 | 10,009.12 | 9,757.82 | 9,642.95 | **39,827.10** |
| 318 | 1,110.55 | 1,219.72 | 1,120.54 | 1,160.92 | **4,611.74** |
| 319 | 5,870.53 | 5,334.61 | 5,600.86 | 5,284.51 | **22,090.51** |
| 398 | 3,274.03 | 3,312.25 | 2,957.86 | 3,099.83 | **12,643.96** |
| **GA - GEORGIA - Total** | **222,576.16** | **212,955.19** | **203,461.01** | **203,844.55** | **842,836.92** |

**STATE: GU - GUAM**

| REGISTRANT ZIP CODE 3 | 1ST QUARTER | 2ND QUARTER | 3RD QUARTER | 4TH QUARTER | **TOTAL GRAMS** |
|---|---|---|---|---|---|
| 969 | 142.59 | 101.24 | 135.95 | 116.67 | **496.46** |
| **GU - GUAM - Total** | **142.59** | **101.24** | **135.95** | **116.67** | **496.46** |

**STATE: HI - HAWAII**

| REGISTRANT ZIP CODE 3 | 1ST QUARTER | 2ND QUARTER | 3RD QUARTER | 4TH QUARTER | **TOTAL GRAMS** |
|---|---|---|---|---|---|
| 967 | 12,175.50 | 11,526.88 | 10,755.66 | 10,550.65 | **45,008.69** |

| | | | | | |
|---|---|---|---|---|---|
| 968 | 4,128.86 | 4,043.67 | 3,650.38 | 3,579.63 | **15,402.53** |
| **HI - HAWAII - Total** | **16,304.36** | **15,570.55** | **14,406.04** | **14,130.27** | **60,411.22** |

**STATE: IA - IOWA**

| REGISTRANT ZIP CODE 3 | 1ST QUARTER | 2ND QUARTER | 3RD QUARTER | 4TH QUARTER | TOTAL GRAMS |
|---|---|---|---|---|---|
| 500 | 3,804.61 | 3,717.12 | 3,600.91 | 3,667.20 | **14,789.84** |
| 501 | 3,469.53 | 3,445.46 | 3,286.25 | 3,331.19 | **13,532.43** |
| 502 | 3,525.27 | 3,312.36 | 3,260.85 | 3,236.92 | **13,335.40** |
| 503 | 8,168.83 | 8,021.88 | 7,749.49 | 7,697.65 | **31,637.85** |
| 504 | 1,791.96 | 1,637.95 | 1,558.41 | 1,571.89 | **6,560.20** |
| 505 | 1,815.48 | 1,853.50 | 1,755.67 | 1,756.12 | **7,180.77** |
| 506 | 1,600.16 | 1,568.08 | 1,532.85 | 1,596.96 | **6,298.05** |
| 507 | 1,319.30 | 1,312.22 | 1,191.19 | 1,170.64 | **4,993.36** |
| 508 | 649.15 | 636.62 | 591.78 | 605.95 | **2,483.50** |
| 510 | 918.92 | 812.35 | 833.76 | 860.06 | **3,425.09** |
| 511 | 2,049.64 | 1,930.85 | 1,928.01 | 1,881.52 | **7,790.02** |
| 512 | 442.93 | 405.97 | 358.65 | 377.99 | **1,585.54** |
| 513 | 694.56 | 614.34 | 617.27 | 631.28 | **2,557.44** |
| 514 | 669.11 | 641.84 | 637.58 | 634.62 | **2,583.14** |
| 515 | 2,273.56 | 2,172.52 | 2,026.08 | 2,006.71 | **8,478.86** |
| 516 | 433.02 | 450.55 | 379.91 | 424.22 | **1,687.71** |
| 520 | 1,643.79 | 1,531.21 | 1,491.60 | 1,478.87 | **6,145.47** |
| 521 | 522.39 | 454.16 | 440.85 | 426.80 | **1,844.19** |
| 522 | 2,275.72 | 2,150.89 | 1,962.38 | 1,937.74 | **8,326.73** |
| 523 | 1,289.17 | 1,157.90 | 1,137.13 | 1,075.05 | **4,659.25** |
| 524 | 2,107.39 | 1,859.34 | 1,816.27 | 1,716.97 | **7,499.97** |
| 525 | 2,644.32 | 2,590.21 | 2,363.86 | 2,406.60 | **10,004.99** |
| 526 | 2,074.97 | 1,941.73 | 1,958.58 | 1,973.16 | **7,948.44** |
| 527 | 1,831.34 | 1,748.14 | 1,654.55 | 1,607.57 | **6,841.60** |
| 528 | 2,362.80 | 2,290.81 | 2,224.16 | 2,174.64 | **9,052.40** |
| **IA - IOWA - Total** | **50,377.92** | **48,258.00** | **46,358.03** | **46,248.30** | **191,242.24** |

**STATE: ID - IDAHO**

| REGISTRANT ZIP CODE 3 | 1ST QUARTER | 2ND QUARTER | 3RD QUARTER | 4TH QUARTER | TOTAL GRAMS |
|---|---|---|---|---|---|
| 832 | 5,265.29 | 5,308.49 | 5,191.53 | 5,232.65 | **20,997.96** |
| 833 | 6,318.72 | 6,159.73 | 6,007.24 | 5,824.53 | **24,310.22** |
| 834 | 4,399.87 | 4,335.65 | 4,269.16 | 4,285.16 | **17,289.84** |
| 835 | 3,463.53 | 3,451.09 | 3,142.84 | 3,236.68 | **13,294.15** |
| 836 | 14,809.13 | 14,375.12 | 13,921.75 | 13,577.91 | **56,683.91** |
| 837 | 9,224.97 | 8,842.55 | 8,712.55 | 8,387.97 | **35,168.03** |
| 838 | 8,717.15 | 8,492.02 | 8,132.74 | 7,979.08 | **33,320.99** |
| **ID - IDAHO - Total** | **52,198.65** | **50,964.65** | **49,377.82** | **48,523.97** | **201,065.09** |

**STATE: IL - ILLINOIS**

| REGISTRANT ZIP CODE 3 | 1ST QUARTER | 2ND QUARTER | 3RD QUARTER | 4TH QUARTER | **TOTAL GRAMS** |
|---|---|---|---|---|---|
| 600 | 32,082.91 | 30,891.27 | 29,510.98 | 29,919.87 | **122,405.04** |
| 601 | 27,558.63 | 26,825.87 | 25,644.23 | 26,048.77 | **106,077.51** |
| 602 | 876.39 | 816.84 | 800.23 | 858.88 | **3,352.34** |
| 603 | 1,224.70 | 1,255.02 | 1,206.90 | 1,140.96 | **4,827.58** |
| 604 | 35,402.69 | 34,175.34 | 32,591.10 | 32,763.36 | **134,932.50** |
| 605 | 16,859.96 | 16,378.27 | 15,619.77 | 15,783.20 | **64,641.20** |
| 606 | 29,110.73 | 27,645.45 | 26,362.17 | 26,479.75 | **109,598.10** |
| 607 | 2,919.92 | 2,822.03 | 2,718.31 | 2,745.78 | **11,206.04** |
| 608 | 2,245.50 | 2,202.64 | 2,088.84 | 2,048.33 | **8,585.31** |
| 609 | 5,412.10 | 5,273.45 | 4,923.16 | 5,022.88 | **20,631.59** |
| 610 | 5,727.25 | 5,642.55 | 5,630.09 | 5,594.57 | **22,594.47** |
| 611 | 8,958.47 | 8,894.22 | 8,563.35 | 8,552.86 | **34,968.90** |
| 612 | 2,997.04 | 2,758.03 | 2,657.01 | 2,531.37 | **10,943.44** |
| 613 | 3,809.50 | 3,833.59 | 3,508.18 | 3,678.06 | **14,829.33** |
| 614 | 3,781.27 | 3,580.50 | 3,399.72 | 3,320.45 | **14,081.94** |
| 615 | 4,671.82 | 4,656.97 | 4,389.25 | 4,489.04 | **18,207.08** |
| 616 | 7,163.53 | 6,711.99 | 6,717.84 | 6,756.61 | **27,349.97** |
| 617 | 4,287.35 | 4,048.48 | 4,023.72 | 4,040.28 | **16,399.83** |
| 618 | 7,922.04 | 7,313.28 | 7,192.68 | 7,031.43 | **29,459.44** |
| 619 | 2,341.20 | 2,103.56 | 2,015.76 | 1,940.88 | **8,401.40** |
| 620 | 10,834.29 | 10,150.77 | 9,821.90 | 9,568.64 | **40,375.60** |
| 622 | 9,886.47 | 9,671.50 | 9,018.14 | 9,038.43 | **37,614.53** |
| 623 | 3,450.51 | 3,344.67 | 3,143.33 | 3,209.06 | **13,147.55** |
| 624 | 4,010.75 | 3,807.92 | 3,644.50 | 3,579.52 | **15,042.70** |
| 625 | 4,912.38 | 4,730.64 | 4,309.67 | 4,574.14 | **18,526.83** |
| 626 | 4,344.61 | 4,067.70 | 3,708.95 | 3,650.42 | **15,771.69** |
| 627 | 5,365.22 | 5,275.20 | 4,858.45 | 4,681.47 | **20,180.35** |
| 628 | 7,095.04 | 6,705.93 | 6,637.17 | 6,579.57 | **27,017.71** |
| 629 | 15,548.91 | 14,785.20 | 14,381.26 | 14,182.27 | **58,897.64** |
| **IL - ILLINOIS - Total** | **270,801.21** | **260,368.88** | **249,086.69** | **249,810.82** | **1,030,067.59** |

**STATE: IN - INDIANA**

| REGISTRANT ZIP CODE 3 | 1ST QUARTER | 2ND QUARTER | 3RD QUARTER | 4TH QUARTER | **TOTAL GRAMS** |
|---|---|---|---|---|---|
| 460 | 12,919.05 | 12,039.67 | 11,937.28 | 12,012.44 | **48,908.44** |
| 461 | 16,821.18 | 16,043.70 | 17,423.74 | 17,555.89 | **67,844.50** |
| 462 | 29,171.09 | 28,371.35 | 25,835.67 | 26,040.76 | **109,418.87** |
| 463 | 25,806.55 | 24,334.22 | 23,533.49 | 24,000.81 | **97,675.07** |
| 464 | 3,099.80 | 2,965.98 | 3,034.30 | 2,897.24 | **11,997.33** |
| 465 | 9,201.04 | 8,631.26 | 8,353.07 | 8,459.56 | **34,644.93** |

| | | | | | |
|---|---|---|---|---|---|
| 466 | 3,333.46 | 3,088.76 | 2,952.36 | 2,795.44 | **12,170.03** |
| 467 | 5,742.17 | 5,604.74 | 5,380.33 | 5,317.96 | **22,045.20** |
| 468 | 7,362.85 | 7,003.86 | 6,934.34 | 7,111.98 | **28,413.04** |
| 469 | 11,948.98 | 11,508.53 | 10,937.05 | 10,860.26 | **45,254.82** |
| 470 | 3,013.34 | 2,991.75 | 2,997.04 | 2,906.71 | **11,908.84** |
| 471 | 15,002.12 | 14,052.90 | 13,380.29 | 13,272.64 | **55,707.95** |
| 472 | 6,634.16 | 6,358.32 | 5,980.06 | 6,058.62 | **25,031.16** |
| 473 | 12,381.45 | 11,951.19 | 11,513.23 | 11,862.18 | **47,708.04** |
| 474 | 8,163.66 | 7,812.91 | 7,251.82 | 7,268.06 | **30,496.45** |
| 475 | 6,425.54 | 6,251.49 | 5,783.92 | 5,810.14 | **24,271.09** |
| 476 | 4,547.05 | 4,386.91 | 4,266.33 | 4,186.42 | **17,386.71** |
| 477 | 11,187.92 | 10,771.68 | 10,339.72 | 10,124.36 | **42,423.68** |
| 478 | 4,537.15 | 4,270.30 | 4,070.07 | 4,144.74 | **17,022.26** |
| 479 | 8,862.76 | 8,355.49 | 8,131.68 | 7,842.43 | **33,192.35** |
| **IN - INDIANA - Total** | **206,161.32** | **196,794.99** | **190,035.80** | **190,528.67** | **783,520.77** |

**STATE: KS - KANSAS**

| REGISTRANT ZIP CODE 3 | 1ST QUARTER | 2ND QUARTER | 3RD QUARTER | 4TH QUARTER | TOTAL GRAMS |
|---|---|---|---|---|---|
| 660 | 9,540.51 | 9,110.07 | 8,782.10 | 8,964.41 | **36,397.09** |
| 661 | 2,993.11 | 2,818.46 | 2,539.02 | 2,609.37 | **10,959.95** |
| 662 | 7,087.61 | 7,012.23 | 6,725.75 | 6,826.11 | **27,651.70** |
| 664 | 1,279.10 | 1,136.49 | 1,107.67 | 1,020.68 | **4,543.94** |
| 665 | 1,715.46 | 1,740.91 | 1,603.07 | 1,629.72 | **6,689.16** |
| 666 | 4,019.45 | 3,893.01 | 3,781.40 | 3,667.72 | **15,361.58** |
| 667 | 4,431.73 | 4,242.77 | 4,109.35 | 4,219.52 | **17,003.38** |
| 668 | 1,204.76 | 1,115.28 | 1,052.20 | 1,094.40 | **4,466.64** |
| 669 | 605.43 | 592.78 | 547.07 | 573.04 | **2,318.31** |
| 670 | 5,872.22 | 5,668.87 | 5,455.58 | 5,422.15 | **22,418.82** |
| 671 | 3,133.65 | 2,831.47 | 2,723.61 | 2,688.20 | **11,376.94** |
| 672 | 16,584.28 | 16,156.38 | 15,622.07 | 15,435.72 | **63,798.44** |
| 673 | 2,340.05 | 2,276.77 | 2,111.24 | 1,974.83 | **8,702.89** |
| 674 | 4,088.87 | 3,977.25 | 3,880.18 | 3,862.90 | **15,809.20** |
| 675 | 3,283.69 | 3,115.23 | 2,988.26 | 2,915.63 | **12,302.81** |
| 676 | 1,260.31 | 1,170.94 | 1,097.69 | 1,078.72 | **4,607.65** |
| 677 | 606.00 | 594.98 | 526.08 | 536.27 | **2,263.32** |
| 678 | 2,129.79 | 2,143.13 | 2,053.83 | 2,025.46 | **8,352.21** |
| 679 | 910.15 | 808.24 | 843.26 | 812.78 | **3,374.44** |
| **KS - KANSAS - Total** | **73,086.15** | **70,405.26** | **67,549.43** | **67,357.62** | **278,398.45** |

**STATE: KY - KENTUCKY**

| REGISTRANT ZIP CODE 3 | 1ST QUARTER | 2ND QUARTER | 3RD QUARTER | 4TH QUARTER | TOTAL GRAMS |
|---|---|---|---|---|---|
| 400 | 7,662.13 | 7,104.34 | 6,758.21 | 6,823.18 | **28,347.85** |

| | | | | | |
|---|---|---|---|---|---|
| 401 | 4,457.31 | 4,302.62 | 4,217.27 | 3,838.94 | **16,816.13** |
| 402 | 33,952.83 | 31,759.34 | 29,708.75 | 30,142.69 | **125,563.60** |
| 403 | 10,658.49 | 10,246.92 | 9,828.86 | 9,953.13 | **40,687.40** |
| 404 | 6,768.91 | 6,560.38 | 6,349.18 | 6,435.28 | **26,113.75** |
| 405 | 6,938.15 | 6,846.90 | 6,768.29 | 6,526.61 | **27,079.95** |
| 406 | 1,848.24 | 1,763.72 | 1,693.27 | 1,642.54 | **6,947.77** |
| 407 | 6,004.59 | 5,800.69 | 5,762.44 | 5,814.09 | **23,381.80** |
| 408 | 1,612.10 | 1,607.84 | 1,543.31 | 1,629.93 | **6,393.18** |
| 409 | 7,589.90 | 7,131.45 | 7,365.72 | 7,376.42 | **29,463.49** |
| 410 | 9,499.45 | 9,053.13 | 8,771.47 | 8,976.65 | **36,300.71** |
| 411 | 3,483.96 | 3,410.78 | 3,197.54 | 3,219.45 | **13,311.73** |
| 412 | 2,932.94 | 2,730.19 | 2,787.94 | 2,910.18 | **11,361.26** |
| 413 | 2,849.52 | 2,751.79 | 2,775.21 | 2,815.31 | **11,191.84** |
| 414 | 1,145.32 | 1,251.99 | 1,239.05 | 1,370.29 | **5,006.65** |
| 415 | 6,158.82 | 5,748.06 | 5,579.62 | 5,580.22 | **23,066.72** |
| 416 | 4,153.65 | 4,143.07 | 4,210.33 | 4,420.22 | **16,927.27** |
| 417 | 4,548.97 | 4,550.17 | 4,474.84 | 4,388.99 | **17,962.97** |
| 418 | 1,959.12 | 1,941.90 | 1,948.26 | 1,880.64 | **7,729.91** |
| 420 | 13,104.12 | 12,759.64 | 12,266.58 | 12,006.94 | **50,137.29** |
| 421 | 11,840.65 | 11,932.76 | 11,554.26 | 11,612.68 | **46,940.35** |
| 422 | 6,116.72 | 5,953.57 | 5,936.17 | 5,845.92 | **23,852.38** |
| 423 | 8,989.63 | 8,733.95 | 8,339.97 | 8,578.90 | **34,642.45** |
| 424 | 9,029.88 | 8,781.85 | 8,605.89 | 8,332.17 | **34,749.79** |
| 425 | 2,679.68 | 2,697.30 | 2,599.86 | 2,691.03 | **10,667.87** |
| 426 | 3,639.98 | 3,314.89 | 3,369.65 | 3,606.51 | **13,931.03** |
| 427 | 8,465.90 | 8,184.21 | 8,129.63 | 8,185.87 | **32,965.60** |
| **KY - KENTUCKY - Total** | **188,090.96** | **181,063.43** | **175,781.57** | **176,604.80** | **721,540.77** |

**STATE: LA - LOUISIANA**

| REGISTRANT ZIP CODE 3 | 1ST QUARTER | 2ND QUARTER | 3RD QUARTER | 4TH QUARTER | **TOTAL GRAMS** |
|---|---|---|---|---|---|
| 700 | 15,202.70 | 13,544.13 | 13,030.68 | 12,770.16 | **54,547.67** |
| 701 | 6,465.37 | 5,991.67 | 5,772.40 | 5,513.78 | **23,743.22** |
| 703 | 9,869.58 | 9,082.27 | 8,708.46 | 8,345.57 | **36,005.88** |
| 704 | 15,093.68 | 14,288.55 | 14,065.26 | 14,026.07 | **57,473.56** |
| 705 | 24,620.00 | 24,675.27 | 23,631.08 | 24,168.11 | **97,094.46** |
| 706 | 6,971.10 | 6,914.29 | 6,859.67 | 6,870.05 | **27,615.12** |
| 707 | 12,842.38 | 12,134.67 | 12,162.58 | 12,246.04 | **49,385.67** |
| 708 | 10,866.48 | 10,035.54 | 9,888.33 | 9,279.78 | **40,070.14** |
| 710 | 4,399.49 | 4,127.68 | 4,110.41 | 4,261.56 | **16,899.15** |
| 711 | 12,922.41 | 12,452.92 | 12,189.63 | 12,128.61 | **49,693.57** |
| 712 | 10,780.79 | 10,014.69 | 9,909.36 | 10,143.45 | **40,848.29** |

| | | | | | |
|---|---|---|---|---|---|
| 713 | 12,263.06 | 11,630.28 | 11,232.16 | 11,345.47 | **46,470.97** |
| 714 | 4,905.70 | 4,876.34 | 4,648.84 | 4,571.87 | **19,002.74** |
| **LA - LOUISIANA - Total** | **147,202.74** | **139,768.31** | **136,208.86** | **135,670.52** | **558,850.42** |

**STATE: MA - MASSACHUSETTS**

| REGISTRANT ZIP CODE 3 | 1ST QUARTER | 2ND QUARTER | 3RD QUARTER | 4TH QUARTER | TOTAL GRAMS |
|---|---|---|---|---|---|
| 010 | 1,979.85 | 1,928.68 | 1,852.47 | 1,856.53 | **7,617.52** |
| 011 | 449.66 | 429.70 | 400.92 | 406.12 | **1,686.40** |
| 012 | 603.11 | 580.09 | 564.13 | 527.25 | **2,274.58** |
| 013 | 342.22 | 324.03 | 315.01 | 311.65 | **1,292.92** |
| 014 | 1,179.55 | 1,037.29 | 995.57 | 1,016.32 | **4,228.73** |
| 015 | 1,720.01 | 1,725.16 | 1,495.05 | 1,557.23 | **6,497.44** |
| 016 | 1,107.21 | 1,000.61 | 997.65 | 949.91 | **4,055.37** |
| 017 | 1,308.19 | 1,199.28 | 1,150.10 | 1,138.80 | **4,796.37** |
| 018 | 5,052.54 | 4,741.46 | 4,286.41 | 4,386.01 | **18,466.42** |
| 019 | 2,382.54 | 2,296.41 | 2,290.70 | 2,176.24 | **9,145.88** |
| 020 | 1,255.58 | 1,106.05 | 1,106.35 | 1,142.61 | **4,610.59** |
| 021 | 2,935.67 | 2,822.59 | 2,629.59 | 2,672.80 | **11,060.64** |
| 022 | 51.24 | 48.32 | 48.04 | 44.21 | **191.81** |
| 023 | 1,887.30 | 1,830.65 | 1,701.97 | 1,738.78 | **7,158.70** |
| 024 | 749.11 | 709.77 | 631.14 | 689.55 | **2,779.57** |
| 025 | 515.54 | 480.13 | 485.95 | 485.79 | **1,967.41** |
| 026 | 840.50 | 810.08 | 781.98 | 758.17 | **3,190.73** |
| 027 | 2,893.44 | 2,754.51 | 2,620.40 | 2,545.50 | **10,813.84** |
| **MA - MASSACHUSETTS - Total** | **27,253.25** | **25,824.79** | **24,353.43** | **24,403.47** | **101,834.93** |

**STATE: MD - MARYLAND**

| REGISTRANT ZIP CODE 3 | 1ST QUARTER | 2ND QUARTER | 3RD QUARTER | 4TH QUARTER | TOTAL GRAMS |
|---|---|---|---|---|---|
| 206 | 2,450.01 | 2,459.20 | 2,319.84 | 2,389.04 | **9,618.10** |
| 207 | 2,764.61 | 2,509.18 | 2,445.90 | 2,301.05 | **10,020.74** |
| 208 | 2,283.98 | 2,187.43 | 1,927.17 | 2,006.01 | **8,404.59** |
| 209 | 563.60 | 478.66 | 450.12 | 452.23 | **1,944.61** |
| 210 | 5,027.14 | 4,818.76 | 4,567.60 | 4,601.54 | **19,015.04** |
| 211 | 2,558.65 | 2,371.21 | 2,257.62 | 2,263.87 | **9,451.35** |
| 212 | 5,440.94 | 5,132.42 | 4,809.25 | 5,019.09 | **20,401.70** |
| 214 | 661.45 | 562.59 | 550.81 | 560.97 | **2,335.82** |
| 215 | 1,970.63 | 1,900.77 | 1,712.66 | 1,749.56 | **7,333.62** |
| 216 | 1,222.42 | 1,235.25 | 1,180.69 | 1,212.16 | **4,850.51** |
| 217 | 3,468.22 | 3,807.20 | 3,232.74 | 3,149.52 | **13,657.67** |
| 218 | 1,296.53 | 1,122.82 | 1,163.77 | 1,113.50 | **4,696.62** |
| 219 | 547.12 | 538.08 | 529.92 | 487.01 | **2,102.13** |
| **MD - MARYLAND - Total** | **30,255.29** | **29,123.57** | **27,148.09** | **27,305.54** | **113,832.50** |

**STATE: ME - MAINE**

| REGISTRANT ZIP CODE 3 | 1ST QUARTER | 2ND QUARTER | 3RD QUARTER | 4TH QUARTER | TOTAL GRAMS |
|---|---|---|---|---|---|
| 039 | 183.71 | 151.22 | 171.88 | 150.41 | **657.22** |
| 040 | 3,978.53 | 3,931.43 | 3,899.78 | 3,771.74 | **15,581.48** |
| 041 | 869.45 | 836.49 | 845.17 | 816.87 | **3,367.98** |
| 042 | 2,034.90 | 1,888.21 | 1,876.94 | 1,819.72 | **7,619.77** |
| 043 | 1,786.93 | 1,776.28 | 1,649.76 | 1,694.65 | **6,907.63** |
| 044 | 1,738.96 | 1,601.77 | 1,566.56 | 1,542.88 | **6,450.17** |
| 045 | 516.55 | 466.54 | 488.12 | 483.40 | **1,954.61** |
| 046 | 1,027.79 | 988.62 | 971.94 | 928.00 | **3,916.36** |
| 047 | 1,057.66 | 1,023.81 | 973.66 | 960.58 | **4,015.71** |
| 048 | 444.72 | 446.48 | 447.26 | 409.58 | **1,748.04** |
| 049 | 2,144.77 | 2,051.49 | 2,044.26 | 1,925.70 | **8,166.22** |
| **ME - MAINE - Total** | **15,783.98** | **15,162.34** | **14,935.32** | **14,503.53** | **60,385.18** |

**STATE: MI - MICHIGAN**

| REGISTRANT ZIP CODE 3 | 1ST QUARTER | 2ND QUARTER | 3RD QUARTER | 4TH QUARTER | TOTAL GRAMS |
|---|---|---|---|---|---|
| 480 | 44,949.18 | 42,447.20 | 39,135.22 | 38,828.36 | **165,359.96** |
| 481 | 57,765.79 | 53,050.87 | 49,445.43 | 49,374.06 | **209,636.14** |
| 482 | 24,921.25 | 22,724.02 | 20,610.86 | 20,385.49 | **88,641.62** |
| 483 | 29,140.85 | 26,668.45 | 24,571.81 | 24,954.30 | **105,335.41** |
| 484 | 16,058.08 | 15,015.81 | 14,035.49 | 13,784.60 | **58,893.98** |
| 485 | 10,139.99 | 9,137.52 | 8,637.37 | 8,491.01 | **36,405.90** |
| 486 | 17,749.34 | 16,488.72 | 15,514.92 | 15,013.53 | **64,766.51** |
| 487 | 9,239.42 | 8,327.89 | 7,732.56 | 7,747.99 | **33,047.87** |
| 488 | 17,513.39 | 16,094.17 | 15,233.90 | 15,115.62 | **63,957.09** |
| 489 | 7,350.14 | 6,659.56 | 6,220.65 | 6,048.15 | **26,278.50** |
| 490 | 23,486.77 | 21,346.31 | 19,884.23 | 19,798.13 | **84,515.43** |
| 491 | 2,330.25 | 2,083.55 | 2,129.63 | 2,023.49 | **8,566.92** |
| 492 | 10,537.33 | 9,414.46 | 8,557.45 | 8,381.88 | **36,891.12** |
| 493 | 7,483.81 | 6,905.64 | 6,747.64 | 6,616.04 | **27,753.13** |
| 494 | 18,766.09 | 17,738.90 | 16,577.60 | 16,204.26 | **69,286.85** |
| 495 | 13,754.98 | 12,543.49 | 11,913.38 | 11,863.22 | **50,075.08** |
| 496 | 9,296.56 | 8,705.33 | 8,366.09 | 8,041.66 | **34,409.64** |
| 497 | 7,731.08 | 7,284.68 | 6,887.92 | 6,455.49 | **28,359.17** |
| 498 | 6,848.14 | 6,275.91 | 6,097.23 | 5,802.29 | **25,023.57** |
| 499 | 2,162.10 | 1,914.70 | 1,832.89 | 1,740.07 | **7,649.77** |
| **MI - MICHIGAN - Total** | **337,224.55** | **310,827.19** | **290,132.27** | **286,669.65** | **1,224,853.66** |

**STATE: MN - MINNESOTA**

| REGISTRANT ZIP CODE 3 | 1ST QUARTER | 2ND QUARTER | 3RD QUARTER | 4TH QUARTER | TOTAL GRAMS |
|---|---|---|---|---|---|
| 550 | 3,987.02 | 3,830.91 | 3,793.02 | 3,651.37 | **15,262.33** |

| | | | | | |
|---|---|---|---|---|---|
| 551 | 4,354.69 | 4,140.58 | 3,911.36 | 3,942.73 | **16,349.36** |
| 553 | 5,416.38 | 5,180.37 | 5,081.72 | 4,971.05 | **20,649.52** |
| 554 | 6,799.26 | 6,508.68 | 6,173.94 | 6,248.11 | **25,729.99** |
| 556 | 230.26 | 191.87 | 212.44 | 179.97 | **814.53** |
| 557 | 2,897.71 | 2,844.59 | 2,752.54 | 2,657.47 | **11,152.31** |
| 558 | 2,080.44 | 2,075.55 | 1,959.08 | 1,914.24 | **8,029.31** |
| 559 | 1,712.92 | 1,599.70 | 1,563.30 | 1,409.25 | **6,285.17** |
| 560 | 1,683.70 | 1,520.74 | 1,501.16 | 1,429.89 | **6,135.48** |
| 561 | 676.32 | 668.56 | 643.13 | 626.94 | **2,614.94** |
| 562 | 1,286.68 | 1,229.12 | 1,149.43 | 1,160.81 | **4,826.04** |
| 563 | 3,198.30 | 3,045.62 | 2,990.29 | 2,913.78 | **12,147.98** |
| 564 | 1,521.56 | 1,515.54 | 1,452.76 | 1,385.26 | **5,875.12** |
| 565 | 1,707.96 | 1,628.09 | 1,622.27 | 1,503.47 | **6,461.79** |
| 566 | 1,061.07 | 1,032.38 | 1,034.03 | 946.79 | **4,074.27** |
| 567 | 768.50 | 692.98 | 656.14 | 693.97 | **2,811.60** |
| **MN - MINNESOTA - Total** | **39,382.77** | **37,705.28** | **36,496.62** | **35,635.09** | **149,219.75** |

**STATE: MO - MISSOURI**

| REGISTRANT ZIP CODE 3 | 1ST QUARTER | 2ND QUARTER | 3RD QUARTER | 4TH QUARTER | TOTAL GRAMS |
|---|---|---|---|---|---|
| 630 | 14,983.24 | 14,243.23 | 13,435.10 | 13,304.02 | **55,965.60** |
| 631 | 18,571.95 | 18,483.55 | 18,150.47 | 17,668.30 | **72,874.27** |
| 633 | 8,661.58 | 8,263.24 | 8,026.52 | 7,821.54 | **32,772.88** |
| 634 | 1,816.08 | 1,731.20 | 1,684.08 | 1,694.49 | **6,925.85** |
| 635 | 1,606.21 | 1,564.12 | 1,367.54 | 1,378.33 | **5,916.20** |
| 636 | 6,181.29 | 5,954.67 | 5,594.07 | 5,277.73 | **23,007.76** |
| 637 | 4,538.35 | 4,384.17 | 4,137.05 | 3,801.01 | **16,860.58** |
| 638 | 8,340.91 | 8,204.33 | 8,153.38 | 7,712.66 | **32,411.28** |
| 639 | 5,651.50 | 5,329.80 | 4,979.96 | 5,052.61 | **21,013.87** |
| 640 | 15,106.93 | 14,626.05 | 14,138.20 | 14,107.85 | **57,979.03** |
| 641 | 11,428.87 | 10,594.49 | 10,096.87 | 9,849.43 | **41,969.67** |
| 644 | 1,861.79 | 1,801.93 | 1,618.72 | 1,667.29 | **6,949.72** |
| 645 | 3,820.66 | 3,625.17 | 3,406.44 | 3,224.85 | **14,077.12** |
| 646 | 1,365.64 | 1,220.08 | 1,163.72 | 1,121.97 | **4,871.41** |
| 647 | 3,794.20 | 3,478.35 | 3,342.48 | 3,288.82 | **13,903.84** |
| 648 | 6,238.19 | 5,977.05 | 5,582.80 | 5,624.72 | **23,422.76** |
| 650 | 2,953.92 | 2,902.16 | 2,769.55 | 2,680.74 | **11,306.38** |
| 651 | 1,413.59 | 1,309.73 | 1,227.63 | 1,260.10 | **5,211.05** |
| 652 | 4,068.10 | 3,892.00 | 3,805.83 | 3,771.27 | **15,537.20** |
| 653 | 2,132.18 | 1,992.69 | 1,917.99 | 1,928.17 | **7,971.03** |
| 654 | 1,976.85 | 1,868.83 | 1,779.34 | 1,711.30 | **7,336.31** |
| 655 | 3,424.39 | 3,175.75 | 3,091.81 | 3,070.74 | **12,762.68** |

| | | | | | |
|---|---|---|---|---|---|
| 656 | 6,502.86 | 6,304.97 | 5,854.60 | 5,906.30 | **24,568.73** |
| 657 | 9,888.75 | 9,536.32 | 9,196.84 | 9,052.44 | **37,674.35** |
| 658 | 8,959.72 | 8,628.15 | 8,564.79 | 7,990.37 | **34,143.02** |
| **MO - MISSOURI - Total** | **155,287.75** | **149,092.02** | **143,085.79** | **139,967.05** | **587,432.60** |

**STATE: MS - MISSISSIPPI**

| REGISTRANT ZIP CODE 3 | 1ST QUARTER | 2ND QUARTER | 3RD QUARTER | 4TH QUARTER | TOTAL GRAMS |
|---|---|---|---|---|---|
| 386 | 9,192.02 | 8,389.49 | 7,924.36 | 7,772.59 | **33,278.45** |
| 387 | 2,556.01 | 2,376.15 | 2,404.62 | 2,344.70 | **9,681.47** |
| 388 | 10,975.42 | 10,680.27 | 10,173.16 | 10,145.32 | **41,974.17** |
| 389 | 3,026.61 | 2,946.44 | 2,681.66 | 2,653.36 | **11,308.07** |
| 390 | 6,380.19 | 6,025.50 | 5,793.00 | 5,460.63 | **23,659.32** |
| 391 | 6,626.55 | 6,118.16 | 6,164.09 | 6,102.99 | **25,011.78** |
| 392 | 6,683.20 | 6,022.88 | 5,842.16 | 5,453.10 | **24,001.34** |
| 393 | 9,491.90 | 8,824.74 | 8,083.56 | 7,666.87 | **34,067.07** |
| 394 | 18,148.08 | 16,626.91 | 16,122.23 | 15,408.95 | **66,306.16** |
| 395 | 16,837.82 | 15,939.43 | 14,625.31 | 13,762.47 | **61,165.03** |
| 396 | 4,639.47 | 4,177.37 | 4,023.05 | 4,017.74 | **16,857.63** |
| 397 | 4,476.49 | 4,119.90 | 4,187.77 | 3,966.39 | **16,750.56** |
| **MS - MISSISSIPPI - Total** | **99,033.75** | **92,247.24** | **88,024.95** | **84,755.11** | **364,061.06** |

**STATE: MT - MONTANA**

| REGISTRANT ZIP CODE 3 | 1ST QUARTER | 2ND QUARTER | 3RD QUARTER | 4TH QUARTER | TOTAL GRAMS |
|---|---|---|---|---|---|
| 590 | 1,175.82 | 1,070.40 | 1,098.58 | 1,058.33 | **4,403.12** |
| 591 | 3,300.25 | 3,061.41 | 3,017.34 | 2,948.62 | **12,327.62** |
| 592 | 488.16 | 472.12 | 440.32 | 444.97 | **1,845.56** |
| 593 | 690.57 | 649.68 | 638.25 | 635.31 | **2,613.80** |
| 594 | 2,766.69 | 2,632.57 | 2,622.12 | 2,598.05 | **10,619.42** |
| 595 | 598.21 | 547.98 | 567.51 | 515.27 | **2,228.96** |
| 596 | 2,681.27 | 2,591.97 | 2,551.35 | 2,193.15 | **10,017.73** |
| 597 | 2,652.65 | 2,588.88 | 2,556.17 | 2,575.46 | **10,373.17** |
| 598 | 5,746.76 | 5,469.74 | 5,324.64 | 5,293.58 | **21,834.74** |
| 599 | 3,644.91 | 3,518.50 | 3,540.78 | 3,428.28 | **14,132.46** |
| **MT - MONTANA - Total** | **23,745.27** | **22,603.24** | **22,357.05** | **21,691.01** | **90,396.57** |

**STATE: NC - NORTH CAROLINA**

| REGISTRANT ZIP CODE 3 | 1ST QUARTER | 2ND QUARTER | 3RD QUARTER | 4TH QUARTER | TOTAL GRAMS |
|---|---|---|---|---|---|
| 270 | 6,593.29 | 6,033.24 | 5,822.09 | 5,997.17 | **24,445.79** |
| 271 | 4,447.70 | 4,185.36 | 3,955.32 | 4,316.14 | **16,904.52** |
| 272 | 11,258.88 | 10,303.68 | 9,788.45 | 10,321.40 | **41,672.41** |
| 273 | 6,338.55 | 6,045.96 | 5,783.27 | 6,385.23 | **24,553.02** |
| 274 | 3,938.00 | 3,565.78 | 3,476.96 | 3,555.25 | **14,535.99** |
| 275 | 12,337.37 | 11,544.71 | 10,993.17 | 11,881.76 | **46,757.02** |

| | | | | | |
|---|---|---|---|---|---|
| 276 | 4,181.68 | 3,732.51 | 3,620.63 | 3,922.86 | **15,457.68** |
| 277 | 3,192.85 | 2,875.03 | 2,562.48 | 2,530.00 | **11,160.35** |
| 278 | 7,874.53 | 7,394.92 | 7,334.42 | 7,547.57 | **30,151.44** |
| 279 | 2,370.53 | 2,390.65 | 2,302.58 | 2,483.91 | **9,547.66** |
| 280 | 18,732.46 | 17,097.92 | 16,619.20 | 17,489.06 | **69,938.63** |
| 281 | 13,708.72 | 12,447.29 | 12,087.91 | 12,618.08 | **50,862.00** |
| 282 | 9,535.96 | 8,191.56 | 7,736.06 | 8,028.61 | **33,492.19** |
| 283 | 14,889.56 | 13,886.90 | 13,504.77 | 14,460.80 | **56,742.02** |
| 284 | 9,401.49 | 9,014.32 | 8,432.94 | 9,256.51 | **36,105.25** |
| 285 | 7,088.86 | 6,629.50 | 6,253.55 | 6,790.34 | **26,762.24** |
| 286 | 16,648.03 | 15,901.59 | 15,503.51 | 15,890.74 | **63,943.87** |
| 287 | 9,388.92 | 8,934.94 | 8,608.48 | 8,671.06 | **35,603.40** |
| 288 | 4,828.60 | 4,228.25 | 3,925.83 | 3,802.13 | **16,784.81** |
| 289 | 1,498.07 | 1,395.93 | 1,403.12 | 1,462.32 | **5,759.44** |
| **NC - NORTH CAROLINA - Total** | **168,254.03** | **155,800.03** | **149,714.73** | **157,410.94** | **631,179.73** |

**STATE: ND - NORTH DAKOTA**

| REGISTRANT ZIP CODE 3 | 1ST QUARTER | 2ND QUARTER | 3RD QUARTER | 4TH QUARTER | TOTAL GRAMS |
|---|---|---|---|---|---|
| 580 | 506.33 | 450.98 | 451.99 | 452.09 | **1,861.38** |
| 581 | 1,719.14 | 1,547.61 | 1,565.92 | 1,472.33 | **6,305.01** |
| 582 | 1,043.42 | 1,044.18 | 1,027.31 | 970.85 | **4,085.76** |
| 583 | 578.81 | 591.61 | 575.72 | 530.95 | **2,277.09** |
| 584 | 490.45 | 503.27 | 487.64 | 501.55 | **1,982.91** |
| 585 | 3,153.59 | 2,924.14 | 2,916.20 | 2,869.71 | **11,863.64** |
| 586 | 620.44 | 574.27 | 597.06 | 555.25 | **2,347.02** |
| 587 | 665.32 | 646.51 | 663.09 | 603.34 | **2,578.26** |
| 588 | 562.22 | 524.29 | 560.47 | 563.15 | **2,210.13** |
| **ND - NORTH DAKOTA - Total** | **9,339.72** | **8,806.86** | **8,845.41** | **8,519.22** | **35,511.20** |

**STATE: NE - NEBRASKA**

| REGISTRANT ZIP CODE 3 | 1ST QUARTER | 2ND QUARTER | 3RD QUARTER | 4TH QUARTER | TOTAL GRAMS |
|---|---|---|---|---|---|
| 680 | 3,121.28 | 3,047.63 | 2,984.55 | 2,919.14 | **12,072.59** |
| 681 | 9,673.14 | 9,393.83 | 8,825.45 | 8,813.39 | **36,705.81** |
| 683 | 1,221.04 | 1,213.86 | 1,106.83 | 1,135.36 | **4,677.09** |
| 684 | 857.33 | 850.81 | 831.08 | 813.46 | **3,352.69** |
| 685 | 4,420.66 | 4,413.31 | 4,202.53 | 4,200.99 | **17,237.49** |
| 686 | 688.20 | 709.46 | 695.48 | 699.59 | **2,792.73** |
| 687 | 1,668.72 | 1,664.70 | 1,584.49 | 1,567.76 | **6,485.66** |
| 688 | 2,605.42 | 2,546.34 | 2,432.89 | 2,369.53 | **9,954.18** |
| 689 | 1,054.76 | 994.85 | 1,055.58 | 1,030.81 | **4,135.99** |
| 690 | 473.75 | 382.15 | 398.36 | 428.37 | **1,682.63** |
| 691 | 1,427.98 | 1,335.14 | 1,214.60 | 1,255.86 | **5,233.58** |

| | | | | | |
|---|---|---|---|---|---|
| 692 | 124.41 | 135.65 | 125.62 | 101.78 | **487.46** |
| 693 | 1,033.95 | 1,047.95 | 929.38 | 947.36 | **3,958.65** |
| **NE - NEBRASKA - Total** | **28,370.64** | **27,735.66** | **26,386.85** | **26,283.41** | **108,776.55** |

**STATE: NH - NEW HAMPSHIRE**

| REGISTRANT ZIP CODE 3 | 1ST QUARTER | 2ND QUARTER | 3RD QUARTER | 4TH QUARTER | TOTAL GRAMS |
|---|---|---|---|---|---|
| 030 | 1,926.74 | 1,864.19 | 1,826.40 | 1,890.33 | **7,507.65** |
| 031 | 1,215.23 | 1,095.27 | 1,042.84 | 1,091.72 | **4,445.05** |
| 032 | 995.05 | 926.00 | 903.09 | 939.56 | **3,763.70** |
| 033 | 608.06 | 600.01 | 594.39 | 554.96 | **2,357.41** |
| 034 | 470.30 | 438.23 | 446.71 | 431.65 | **1,786.88** |
| 035 | 272.90 | 265.24 | 277.59 | 255.55 | **1,071.29** |
| 036 | 29.16 | 27.89 | 29.99 | 28.95 | **115.98** |
| 037 | 388.73 | 360.02 | 356.74 | 367.30 | **1,472.79** |
| 038 | 2,336.07 | 2,118.87 | 2,118.75 | 2,041.83 | **8,615.53** |
| **NH - NEW HAMPSHIRE - Total** | **8,242.23** | **7,695.72** | **7,596.49** | **7,601.84** | **31,136.29** |

**STATE: NJ - NEW JERSEY**

| REGISTRANT ZIP CODE 3 | 1ST QUARTER | 2ND QUARTER | 3RD QUARTER | 4TH QUARTER | TOTAL GRAMS |
|---|---|---|---|---|---|
| 070 | 3,541.21 | 3,320.98 | 2,920.79 | 3,197.17 | **12,980.16** |
| 071 | 298.69 | 272.47 | 255.38 | 248.54 | **1,075.08** |
| 072 | 194.42 | 178.16 | 175.75 | 173.19 | **721.53** |
| 073 | 273.45 | 313.66 | 293.43 | 269.34 | **1,149.89** |
| 074 | 1,844.67 | 1,740.13 | 1,718.91 | 1,792.43 | **7,096.14** |
| 075 | 250.21 | 209.77 | 179.40 | 233.17 | **872.55** |
| 076 | 1,214.16 | 1,026.14 | 1,092.83 | 1,139.67 | **4,472.80** |
| 077 | 2,641.27 | 2,465.04 | 2,334.25 | 2,456.15 | **9,896.70** |
| 078 | 1,625.34 | 1,523.70 | 1,481.20 | 1,552.39 | **6,182.62** |
| 079 | 633.73 | 528.34 | 545.78 | 555.64 | **2,263.49** |
| 080 | 7,250.72 | 6,939.11 | 6,508.05 | 6,532.02 | **27,229.89** |
| 081 | 534.25 | 528.69 | 481.53 | 481.01 | **2,025.49** |
| 082 | 1,895.16 | 1,727.51 | 1,795.05 | 1,589.26 | **7,006.98** |
| 083 | 1,538.46 | 1,514.72 | 1,459.44 | 1,441.25 | **5,953.87** |
| 084 | 274.02 | 237.07 | 234.51 | 244.81 | **990.42** |
| 085 | 852.92 | 730.48 | 702.06 | 701.51 | **2,986.96** |
| 086 | 914.00 | 878.15 | 800.62 | 845.99 | **3,438.76** |
| 087 | 2,139.52 | 1,949.41 | 1,916.11 | 1,856.49 | **7,861.53** |
| 088 | 2,443.45 | 2,283.74 | 2,093.00 | 2,202.29 | **9,022.47** |
| 089 | 149.30 | 167.63 | 122.08 | 119.89 | **558.90** |
| **NJ - NEW JERSEY - Total** | **30,508.96** | **28,534.89** | **27,110.18** | **27,632.20** | **113,786.23** |

**STATE: NM - NEW MEXICO**

| REGISTRANT ZIP CODE 3 | 1ST QUARTER | 2ND QUARTER | 3RD QUARTER | 4TH QUARTER | TOTAL GRAMS |
|---|---|---|---|---|---|

| | | | | | |
|---|---|---|---|---|---|
| 870 | 2,467.58 | 2,319.14 | 2,250.31 | 2,281.81 | **9,318.85** |
| 871 | 11,216.95 | 10,343.90 | 10,123.83 | 10,085.80 | **41,770.48** |
| 873 | 612.31 | 533.22 | 535.25 | 528.42 | **2,209.20** |
| 874 | 1,601.01 | 1,473.23 | 1,493.81 | 1,412.96 | **5,981.01** |
| 875 | 3,703.65 | 3,607.64 | 3,471.14 | 3,557.19 | **14,339.62** |
| 877 | 854.89 | 767.25 | 775.06 | 760.78 | **3,157.97** |
| 878 | 232.29 | 207.18 | 218.85 | 238.22 | **896.54** |
| 879 | 245.02 | 222.05 | 191.94 | 202.18 | **861.19** |
| 880 | 2,707.75 | 2,609.06 | 2,436.48 | 2,527.11 | **10,280.40** |
| 881 | 1,294.45 | 1,230.76 | 1,161.12 | 1,165.93 | **4,852.26** |
| 882 | 5,754.97 | 5,483.29 | 5,215.56 | 5,207.00 | **21,660.81** |
| 883 | 1,427.86 | 1,395.51 | 1,365.37 | 1,383.61 | **5,572.34** |
| 884 | 207.08 | 233.28 | 226.46 | 231.96 | **898.78** |
| **NM - NEW MEXICO - Total** | **32,325.82** | **30,425.51** | **29,465.17** | **29,582.97** | **121,799.47** |

**STATE: NV - NEVADA**

| REGISTRANT ZIP CODE 3 | 1ST QUARTER | 2ND QUARTER | 3RD QUARTER | 4TH QUARTER | TOTAL GRAMS |
|---|---|---|---|---|---|
| 890 | 20,355.19 | 19,600.05 | 19,031.05 | 18,956.74 | **77,943.03** |
| 891 | 40,514.97 | 38,103.36 | 37,358.69 | 36,113.26 | **152,090.28** |
| 893 | 458.29 | 468.93 | 413.32 | 424.93 | **1,765.47** |
| 894 | 7,266.91 | 7,185.63 | 6,714.94 | 6,803.04 | **27,970.51** |
| 895 | 7,345.27 | 7,191.74 | 7,084.80 | 6,875.61 | **28,497.42** |
| 897 | 2,376.07 | 2,414.81 | 2,332.12 | 2,249.26 | **9,372.26** |
| 898 | 858.27 | 876.14 | 862.22 | 827.55 | **3,424.18** |
| **NV - NEVADA - Total** | **79,174.96** | **75,840.66** | **73,797.14** | **72,250.39** | **301,063.15** |

**STATE: NY - NEW YORK**

| REGISTRANT ZIP CODE 3 | 1ST QUARTER | 2ND QUARTER | 3RD QUARTER | 4TH QUARTER | TOTAL GRAMS |
|---|---|---|---|---|---|
| 063 | 0.30 | 0 | 0.30 | 0.30 | **0.91** |
| 100 | 3,540.55 | 3,475.71 | 3,158.50 | 3,281.51 | **13,456.27** |
| 101 | 220.64 | 174.46 | 142.92 | 208.10 | **746.12** |
| 102 | 12.77 | 9.64 | 5.42 | 10.03 | **37.85** |
| 103 | 2,269.55 | 2,247.15 | 2,125.49 | 2,263.01 | **8,905.20** |
| 104 | 1,345.40 | 1,268.58 | 1,155.76 | 1,130.78 | **4,900.52** |
| 105 | 2,241.88 | 2,068.61 | 1,937.97 | 1,979.83 | **8,228.30** |
| 106 | 303.78 | 271.88 | 246.58 | 224.53 | **1,046.77** |
| 107 | 736.68 | 696.39 | 571.64 | 578.62 | **2,583.33** |
| 108 | 217.64 | 197.34 | 193.28 | 212.12 | **820.38** |
| 109 | 2,312.66 | 2,269.94 | 2,042.91 | 2,184.23 | **8,809.75** |
| 110 | 921.91 | 862.67 | 834.99 | 862.43 | **3,481.99** |
| 111 | 461.64 | 446.40 | 373.66 | 416.63 | **1,698.33** |
| 112 | 3,548.26 | 3,324.46 | 3,002.62 | 3,185.92 | **13,061.26** |

| | | | | | |
|---|---|---|---|---|---|
| 113 | 1,771.36 | 1,664.42 | 1,582.99 | 1,663.23 | **6,682.00** |
| 114 | 835.73 | 858.50 | 790.18 | 803.85 | **3,288.26** |
| 115 | 2,533.85 | 2,464.77 | 2,388.93 | 2,428.23 | **9,815.78** |
| 116 | 242.55 | 242.93 | 233.01 | 210.88 | **929.36** |
| 117 | 11,598.09 | 11,177.35 | 10,494.25 | 10,797.11 | **44,066.80** |
| 118 | 364.41 | 347.39 | 347.29 | 355.14 | **1,414.23** |
| 119 | 1,827.82 | 1,737.95 | 1,768.69 | 1,624.91 | **6,959.37** |
| 120 | 5,786.43 | 5,811.55 | 5,503.32 | 5,394.87 | **22,496.18** |
| 121 | 3,345.12 | 3,317.04 | 3,147.58 | 3,135.40 | **12,945.14** |
| 122 | 2,599.45 | 2,447.35 | 2,336.73 | 2,372.78 | **9,756.31** |
| 123 | 2,471.52 | 2,369.46 | 2,188.23 | 2,143.23 | **9,172.44** |
| 124 | 2,252.74 | 2,149.26 | 2,063.30 | 2,102.99 | **8,568.29** |
| 125 | 4,186.67 | 3,943.47 | 3,770.11 | 3,843.70 | **15,743.95** |
| 126 | 689.98 | 603.05 | 583.43 | 626.94 | **2,503.41** |
| 127 | 813.50 | 803.27 | 747.26 | 800.07 | **3,164.10** |
| 128 | 4,114.86 | 3,951.78 | 3,735.58 | 3,647.48 | **15,449.70** |
| 129 | 2,624.05 | 2,563.75 | 2,529.48 | 2,501.70 | **10,218.98** |
| 130 | 4,847.57 | 4,807.82 | 4,597.09 | 4,561.89 | **18,814.36** |
| 131 | 1,726.36 | 1,718.12 | 1,651.24 | 1,681.20 | **6,776.91** |
| 132 | 3,648.78 | 3,398.08 | 3,212.49 | 3,402.47 | **13,661.83** |
| 133 | 2,168.00 | 2,060.49 | 1,993.17 | 1,947.31 | **8,168.97** |
| 134 | 3,187.59 | 3,122.23 | 2,845.92 | 2,809.32 | **11,965.06** |
| 135 | 1,634.65 | 1,642.84 | 1,546.65 | 1,497.78 | **6,321.92** |
| 136 | 3,360.40 | 3,261.32 | 3,223.59 | 3,084.12 | **12,929.42** |
| 137 | 1,532.74 | 1,369.70 | 1,327.09 | 1,205.19 | **5,434.72** |
| 138 | 3,222.28 | 3,202.80 | 2,939.93 | 2,972.45 | **12,337.46** |
| 139 | 1,441.79 | 1,333.80 | 1,214.93 | 1,145.60 | **5,136.12** |
| 140 | 10,585.76 | 10,182.70 | 9,880.79 | 9,706.78 | **40,356.03** |
| 141 | 4,976.82 | 4,998.18 | 4,864.89 | 4,838.10 | **19,677.99** |
| 142 | 19,801.08 | 18,651.25 | 18,564.22 | 18,329.45 | **75,345.99** |
| 143 | 2,504.92 | 2,462.58 | 2,398.35 | 2,383.37 | **9,749.23** |
| 144 | 3,895.22 | 3,711.52 | 3,581.28 | 3,534.19 | **14,722.21** |
| 145 | 3,197.43 | 3,003.51 | 2,904.04 | 2,881.91 | **11,986.88** |
| 146 | 5,499.04 | 5,155.21 | 4,980.82 | 4,903.46 | **20,538.53** |
| 147 | 4,246.06 | 4,040.74 | 3,992.04 | 3,809.60 | **16,088.44** |
| 148 | 3,791.27 | 3,721.12 | 3,539.07 | 3,412.84 | **14,464.29** |
| 149 | 858.41 | 873.20 | 832.50 | 848.77 | **3,412.89** |
| **NY - NEW YORK - Total** | **152,317.93** | **146,483.73** | **140,092.49** | **139,946.35** | **578,840.51** |

**STATE: OH - OHIO**

| REGISTRANT ZIP CODE 3 | 1ST QUARTER | 2ND QUARTER | 3RD QUARTER | 4TH QUARTER | **TOTAL GRAMS** |
|---|---|---|---|---|---|

| | | | | | |
|---|---|---|---|---|---|
| 430 | 8,521.62 | 7,967.09 | 7,869.30 | 7,995.18 | **32,353.19** |
| 431 | 6,438.25 | 5,980.14 | 5,921.14 | 5,930.41 | **24,269.93** |
| 432 | 13,052.76 | 11,450.38 | 9,205.14 | 9,336.51 | **43,044.78** |
| 433 | 2,599.84 | 2,557.57 | 2,494.27 | 2,398.24 | **10,049.91** |
| 434 | 1,551.39 | 1,564.20 | 1,516.57 | 1,459.00 | **6,091.16** |
| 435 | 3,367.76 | 3,162.71 | 3,094.73 | 3,145.54 | **12,770.74** |
| 436 | 6,133.33 | 5,837.38 | 5,755.19 | 5,392.46 | **23,118.36** |
| 437 | 3,460.53 | 3,312.50 | 3,150.25 | 3,110.11 | **13,033.39** |
| 438 | 475.77 | 468.40 | 442.89 | 464.07 | **1,851.14** |
| 439 | 2,972.99 | 2,671.00 | 2,546.07 | 2,523.25 | **10,713.32** |
| 440 | 7,336.08 | 6,898.10 | 6,589.36 | 6,688.06 | **27,511.59** |
| 441 | 8,677.05 | 8,552.36 | 8,150.93 | 8,103.66 | **33,484.01** |
| 442 | 5,589.39 | 5,413.68 | 5,226.49 | 5,104.56 | **21,334.12** |
| 443 | 2,641.07 | 2,518.11 | 2,435.86 | 2,364.16 | **9,959.19** |
| 444 | 6,898.24 | 6,409.43 | 6,068.34 | 6,007.14 | **25,383.15** |
| 445 | 4,249.96 | 3,947.43 | 3,835.84 | 3,754.43 | **15,787.66** |
| 446 | 5,169.90 | 4,866.04 | 4,544.00 | 4,536.95 | **19,116.88** |
| 447 | 3,002.24 | 2,797.78 | 2,698.59 | 2,783.27 | **11,281.88** |
| 448 | 3,122.43 | 2,924.05 | 3,008.51 | 2,808.29 | **11,863.28** |
| 449 | 802.07 | 784.75 | 741.92 | 800.57 | **3,129.30** |
| 450 | 12,490.98 | 11,994.29 | 11,625.93 | 11,898.35 | **48,009.55** |
| 451 | 4,511.15 | 4,440.98 | 4,291.91 | 4,275.45 | **17,519.49** |
| 452 | 10,312.24 | 9,810.97 | 9,199.04 | 9,318.15 | **38,640.40** |
| 453 | 7,906.38 | 7,957.29 | 7,728.33 | 7,956.01 | **31,548.02** |
| 454 | 9,304.30 | 8,943.85 | 8,670.25 | 8,685.77 | **35,604.17** |
| 455 | 2,192.71 | 2,131.70 | 2,212.50 | 2,087.47 | **8,624.38** |
| 456 | 7,705.45 | 7,249.13 | 6,871.33 | 6,937.89 | **28,763.79** |
| 457 | 2,793.43 | 2,600.77 | 2,491.61 | 2,470.92 | **10,356.72** |
| 458 | 4,132.23 | 4,016.15 | 3,874.88 | 3,860.89 | **15,884.15** |
| **OH - OHIO - Total** | **157,411.53** | **149,228.23** | **142,261.15** | **142,196.74** | **591,097.65** |

**STATE: OK - OKLAHOMA**

| REGISTRANT ZIP CODE 3 | 1ST QUARTER | 2ND QUARTER | 3RD QUARTER | 4TH QUARTER | **TOTAL GRAMS** |
|---|---|---|---|---|---|
| 730 | 23,445.51 | 22,762.55 | 22,124.00 | 21,289.88 | **89,621.94** |
| 731 | 32,292.89 | 31,885.36 | 30,049.57 | 27,499.47 | **121,727.29** |
| 734 | 5,187.43 | 5,064.69 | 4,901.82 | 4,498.18 | **19,652.13** |
| 735 | 9,226.14 | 8,889.92 | 8,847.47 | 8,116.75 | **35,080.28** |
| 736 | 1,587.06 | 1,617.46 | 1,513.57 | 1,449.00 | **6,167.10** |
| 737 | 3,820.83 | 3,835.49 | 3,622.70 | 3,519.93 | **14,798.95** |
| 738 | 665.90 | 638.80 | 640.17 | 612.53 | **2,557.40** |
| 739 | 287.86 | 304.49 | 285.44 | 285.08 | **1,162.87** |

| | | | | | |
|---|---|---|---|---|---|
| 740 | 25,933.28 | 25,865.42 | 23,953.69 | 22,312.04 | **98,064.43** |
| 741 | 17,842.84 | 17,598.74 | 16,588.55 | 15,799.51 | **67,829.65** |
| 743 | 4,393.90 | 4,225.03 | 3,925.61 | 3,538.43 | **16,082.96** |
| 744 | 13,787.47 | 13,510.12 | 12,805.69 | 11,902.01 | **52,005.30** |
| 745 | 4,147.56 | 4,137.76 | 3,808.61 | 3,558.92 | **15,652.85** |
| 746 | 1,939.69 | 1,761.11 | 1,679.85 | 1,574.96 | **6,955.62** |
| 747 | 5,618.72 | 5,443.00 | 5,428.04 | 5,008.28 | **21,498.04** |
| 748 | 7,940.55 | 7,916.26 | 7,537.86 | 7,080.71 | **30,475.37** |
| 749 | 4,608.93 | 4,611.99 | 4,531.86 | 4,143.76 | **17,896.54** |
| **OK - OKLAHOMA - Total** | **162,726.57** | **160,068.20** | **152,244.49** | **142,189.44** | **617,228.70** |

**STATE: OR - OREGON**

| REGISTRANT ZIP CODE 3 | 1ST QUARTER | 2ND QUARTER | 3RD QUARTER | 4TH QUARTER | **TOTAL GRAMS** |
|---|---|---|---|---|---|
| 970 | 12,782.62 | 12,427.19 | 11,963.61 | 11,888.09 | **49,061.51** |
| 971 | 5,718.64 | 5,633.52 | 5,316.67 | 5,174.15 | **21,842.97** |
| 972 | 15,757.63 | 15,122.29 | 14,569.74 | 14,420.65 | **59,870.31** |
| 973 | 10,503.27 | 10,087.23 | 9,798.12 | 9,497.57 | **39,886.18** |
| 974 | 16,715.78 | 16,087.59 | 15,721.85 | 15,035.46 | **63,560.69** |
| 975 | 10,442.64 | 10,547.94 | 10,367.99 | 10,120.87 | **41,479.43** |
| 976 | 2,388.94 | 2,422.23 | 2,261.49 | 2,067.78 | **9,140.43** |
| 977 | 4,791.73 | 4,689.87 | 4,598.53 | 4,482.81 | **18,562.94** |
| 978 | 3,829.04 | 3,700.51 | 3,506.81 | 3,409.80 | **14,446.16** |
| 979 | 1,671.53 | 1,613.24 | 1,486.74 | 1,567.83 | **6,339.34** |
| **OR - OREGON - Total** | **84,601.82** | **82,331.60** | **79,591.55** | **77,665.00** | **324,189.97** |

**STATE: PA - PENNSYLVANIA**

| REGISTRANT ZIP CODE 3 | 1ST QUARTER | 2ND QUARTER | 3RD QUARTER | 4TH QUARTER | **TOTAL GRAMS** |
|---|---|---|---|---|---|
| 150 | 7,525.69 | 7,106.04 | 6,545.62 | 6,692.14 | **27,869.49** |
| 151 | 5,915.52 | 5,178.59 | 4,902.33 | 4,784.10 | **20,780.55** |
| 152 | 10,313.12 | 9,743.96 | 9,095.79 | 9,252.61 | **38,405.49** |
| 153 | 3,510.97 | 2,788.15 | 3,349.86 | 2,740.15 | **12,389.13** |
| 154 | 4,345.79 | 4,122.16 | 3,947.38 | 3,907.62 | **16,322.94** |
| 155 | 1,164.46 | 1,101.98 | 1,096.81 | 1,078.66 | **4,441.92** |
| 156 | 4,766.62 | 4,400.75 | 4,370.22 | 4,232.09 | **17,769.69** |
| 157 | 2,145.33 | 2,049.75 | 1,948.74 | 1,967.13 | **8,110.96** |
| 158 | 1,876.58 | 1,763.66 | 1,770.84 | 1,747.91 | **7,158.98** |
| 159 | 2,182.64 | 2,040.50 | 1,916.96 | 1,935.11 | **8,075.21** |
| 160 | 2,615.31 | 2,479.38 | 2,408.27 | 2,332.10 | **9,835.06** |
| 161 | 3,591.24 | 3,325.08 | 3,250.43 | 3,135.92 | **13,302.67** |
| 162 | 1,511.84 | 1,431.26 | 1,400.80 | 1,257.55 | **5,601.44** |
| 163 | 2,996.50 | 2,874.36 | 2,723.75 | 2,646.86 | **11,241.46** |
| 164 | 1,996.48 | 1,866.73 | 1,829.37 | 1,802.70 | **7,495.27** |

| | | | | | |
|---|---|---|---|---|---|
| 165 | 6,790.17 | 6,328.37 | 6,226.99 | 5,969.32 | **25,314.85** |
| 166 | 4,655.15 | 4,348.25 | 4,217.58 | 3,960.01 | **17,180.99** |
| 167 | 1,272.60 | 1,176.21 | 1,123.64 | 1,088.54 | **4,661.00** |
| 168 | 1,309.93 | 1,216.62 | 1,169.60 | 1,091.27 | **4,787.43** |
| 169 | 781.25 | 718.56 | 712.79 | 706.80 | **2,919.41** |
| 170 | 7,097.38 | 6,553.97 | 6,422.50 | 6,362.28 | **26,436.13** |
| 171 | 1,656.25 | 1,560.54 | 1,438.28 | 1,389.89 | **6,044.96** |
| 172 | 1,326.34 | 1,225.57 | 1,145.69 | 1,141.89 | **4,839.50** |
| 173 | 2,247.36 | 2,170.93 | 2,090.82 | 2,019.10 | **8,528.20** |
| 174 | 1,214.53 | 1,139.26 | 1,107.00 | 1,102.42 | **4,563.21** |
| 175 | 2,440.92 | 2,318.77 | 2,181.37 | 2,166.82 | **9,107.89** |
| 176 | 1,792.36 | 1,707.16 | 1,657.72 | 1,573.14 | **6,730.38** |
| 177 | 2,656.92 | 2,478.68 | 2,363.48 | 2,354.35 | **9,853.44** |
| 178 | 1,534.38 | 1,415.97 | 1,356.74 | 1,350.30 | **5,657.39** |
| 179 | 1,515.72 | 1,381.27 | 1,129.60 | 1,088.58 | **5,115.18** |
| 180 | 2,755.13 | 2,737.54 | 2,586.53 | 2,627.38 | **10,706.58** |
| 181 | 1,141.16 | 1,064.75 | 1,010.19 | 1,027.49 | **4,243.59** |
| 182 | 1,582.13 | 1,567.62 | 1,453.04 | 1,396.13 | **5,998.92** |
| 183 | 1,391.95 | 1,413.61 | 1,273.49 | 1,289.16 | **5,368.21** |
| 184 | 2,613.06 | 2,467.27 | 2,462.19 | 2,362.32 | **9,904.85** |
| 185 | 3,285.88 | 3,110.07 | 2,910.05 | 2,834.55 | **12,140.56** |
| 186 | 2,541.80 | 2,339.54 | 2,321.10 | 2,148.38 | **9,350.82** |
| 187 | 6,806.91 | 7,321.56 | 6,217.50 | 6,707.52 | **27,053.49** |
| 188 | 910.97 | 879.02 | 829.57 | 834.85 | **3,454.42** |
| 189 | 2,219.20 | 2,256.35 | 2,103.61 | 2,147.21 | **8,726.36** |
| 190 | 8,691.38 | 9,038.26 | 10,617.32 | 6,649.07 | **34,996.03** |
| 191 | 5,601.89 | 5,512.83 | 5,115.44 | 4,958.57 | **21,188.73** |
| 193 | 2,264.65 | 2,188.06 | 2,058.32 | 2,051.57 | **8,562.61** |
| 194 | 3,516.37 | 3,354.09 | 3,109.01 | 3,029.58 | **13,009.05** |
| 195 | 1,516.00 | 1,459.31 | 1,324.86 | 1,318.79 | **5,618.96** |
| 196 | 1,771.30 | 1,700.35 | 1,643.12 | 1,593.25 | **6,708.03** |
| **PA - PENNSYLVANIA - Total** | **143,359.13** | **136,422.73** | **131,936.33** | **125,853.23** | **537,571.41** |

**STATE: PR - PUERTO RICO**

| REGISTRANT ZIP CODE 3 | 1ST QUARTER | 2ND QUARTER | 3RD QUARTER | 4TH QUARTER | **TOTAL GRAMS** |
|---|---|---|---|---|---|
| 006 | 134.16 | 125.34 | 265.80 | 206.89 | **732.18** |
| 007 | 128.55 | 108.40 | 153.97 | 138.50 | **529.42** |
| 009 | 110.77 | 111.07 | 187.30 | 169.91 | **579.04** |
| **PR - PUERTO RICO - Total** | **373.47** | **344.81** | **607.07** | **515.30** | **1,840.64** |

**STATE: RI - RHODE ISLAND**

| REGISTRANT ZIP CODE 3 | 1ST QUARTER | 2ND QUARTER | 3RD QUARTER | 4TH QUARTER | **TOTAL GRAMS** |
|---|---|---|---|---|---|

| | | | | | |
|---|---|---|---|---|---|
| 028 | 5,459.77 | 5,147.28 | 5,019.09 | 4,971.96 | **20,598.11** |
| 029 | 3,683.38 | 3,529.49 | 3,338.58 | 3,199.83 | **13,751.29** |
| **RI - RHODE ISLAND - Total** | **9,143.16** | **8,676.78** | **8,357.67** | **8,171.79** | **34,349.40** |

**STATE: SC - SOUTH CAROLINA**

| REGISTRANT ZIP CODE 3 | 1ST QUARTER | 2ND QUARTER | 3RD QUARTER | 4TH QUARTER | TOTAL GRAMS |
|---|---|---|---|---|---|
| 290 | 6,777.78 | 6,162.40 | 6,129.81 | 6,214.94 | **25,284.94** |
| 291 | 7,582.15 | 7,198.39 | 6,769.32 | 6,842.14 | **28,392.00** |
| 292 | 5,273.66 | 4,631.20 | 4,800.19 | 4,538.01 | **19,243.07** |
| 293 | 19,271.76 | 18,470.54 | 17,000.62 | 16,731.84 | **71,474.76** |
| 294 | 17,616.11 | 16,484.41 | 15,596.83 | 15,639.81 | **65,337.16** |
| 295 | 20,795.70 | 19,476.66 | 18,870.30 | 18,948.19 | **78,090.84** |
| 296 | 33,601.44 | 31,784.32 | 29,719.73 | 29,912.58 | **125,018.06** |
| 297 | 5,973.42 | 5,241.39 | 4,923.58 | 5,133.72 | **21,272.12** |
| 298 | 3,087.67 | 2,955.35 | 2,947.00 | 2,739.13 | **11,729.16** |
| 299 | 2,908.48 | 2,572.75 | 2,572.24 | 2,471.67 | **10,525.14** |
| **SC - SOUTH CAROLINA - Total** | **122,888.18** | **114,977.41** | **109,329.62** | **109,172.04** | **456,367.25** |

**STATE: SD - SOUTH DAKOTA**

| REGISTRANT ZIP CODE 3 | 1ST QUARTER | 2ND QUARTER | 3RD QUARTER | 4TH QUARTER | TOTAL GRAMS |
|---|---|---|---|---|---|
| 570 | 2,110.78 | 2,089.31 | 2,045.30 | 2,060.07 | **8,305.46** |
| 571 | 3,977.07 | 3,822.49 | 3,678.11 | 3,689.35 | **15,167.02** |
| 572 | 874.39 | 791.59 | 794.91 | 787.99 | **3,248.89** |
| 573 | 1,057.38 | 1,032.30 | 937.67 | 1,002.15 | **4,029.49** |
| 574 | 732.90 | 705.65 | 710.85 | 653.20 | **2,802.61** |
| 575 | 910.45 | 906.65 | 854.11 | 842.35 | **3,513.55** |
| 576 | 213.90 | 209.18 | 201.45 | 198.92 | **823.45** |
| 577 | 2,658.71 | 2,489.52 | 2,447.85 | 2,345.37 | **9,941.46** |
| **SD - SOUTH DAKOTA - Total** | **12,535.57** | **12,046.69** | **11,670.25** | **11,579.41** | **47,831.94** |

**STATE: TN - TENNESSEE**

| REGISTRANT ZIP CODE 3 | 1ST QUARTER | 2ND QUARTER | 3RD QUARTER | 4TH QUARTER | TOTAL GRAMS |
|---|---|---|---|---|---|
| 370 | 26,913.83 | 25,157.56 | 23,490.50 | 23,047.85 | **98,609.75** |
| 371 | 26,892.38 | 25,516.36 | 24,347.69 | 23,994.69 | **100,751.12** |
| 372 | 12,229.17 | 11,249.77 | 10,213.31 | 10,313.64 | **44,005.89** |
| 373 | 24,246.57 | 23,444.57 | 22,309.50 | 22,226.75 | **92,227.39** |
| 374 | 8,678.83 | 8,223.06 | 7,738.71 | 7,555.00 | **32,195.61** |
| 376 | 15,459.25 | 14,759.50 | 13,873.67 | 13,650.02 | **57,742.44** |
| 377 | 12,372.88 | 11,995.12 | 10,992.32 | 11,108.41 | **46,468.73** |
| 378 | 19,663.64 | 19,267.14 | 17,998.07 | 17,641.09 | **74,569.95** |
| 379 | 9,505.25 | 9,266.18 | 8,695.59 | 8,782.60 | **36,249.62** |
| 380 | 12,482.88 | 11,944.97 | 10,679.92 | 10,710.75 | **45,818.53** |
| 381 | 15,360.33 | 14,323.75 | 15,982.25 | 11,389.26 | **57,055.60** |

| | | | | | |
|---|---|---|---|---|---|
| 382 | 7,205.37 | 6,947.14 | 6,260.98 | 5,984.11 | **26,397.61** |
| 383 | 15,047.30 | 14,565.81 | 13,375.68 | 12,882.42 | **55,871.20** |
| 384 | 8,015.69 | 7,873.11 | 7,377.27 | 7,477.97 | **30,744.04** |
| 385 | 7,356.78 | 7,177.90 | 6,530.13 | 6,569.45 | **27,634.26** |
| **TN - TENNESSEE - Total** | **221,430.15** | **211,711.96** | **199,865.59** | **193,334.02** | **826,341.72** |

**STATE: TX - TEXAS**

| REGISTRANT ZIP CODE 3 | 1ST QUARTER | 2ND QUARTER | 3RD QUARTER | 4TH QUARTER | TOTAL GRAMS |
|---|---|---|---|---|---|
| 750 | 47,474.07 | 45,645.12 | 44,320.21 | 44,843.08 | **182,282.47** |
| 751 | 20,945.04 | 20,408.93 | 20,169.83 | 20,316.63 | **81,840.44** |
| 752 | 25,663.90 | 24,280.04 | 23,052.71 | 23,150.83 | **96,147.48** |
| 753 | 283.55 | 285.45 | 273.14 | 297.48 | **1,139.62** |
| 754 | 13,479.64 | 12,906.84 | 12,618.63 | 12,471.01 | **51,476.11** |
| 755 | 4,624.92 | 4,493.68 | 4,489.09 | 4,375.46 | **17,983.15** |
| 756 | 9,879.46 | 9,757.97 | 9,594.65 | 9,831.52 | **39,063.61** |
| 757 | 14,064.01 | 13,914.98 | 13,316.80 | 13,613.61 | **54,909.40** |
| 758 | 2,723.86 | 2,584.34 | 2,511.23 | 2,583.95 | **10,403.39** |
| 759 | 8,180.67 | 7,883.72 | 7,595.62 | 7,382.29 | **31,042.29** |
| 760 | 36,520.33 | 36,187.92 | 35,036.01 | 35,872.96 | **143,617.21** |
| 761 | 31,233.43 | 30,834.12 | 29,440.48 | 29,162.09 | **120,670.11** |
| 762 | 13,545.35 | 13,214.95 | 12,818.36 | 12,676.07 | **52,254.73** |
| 763 | 6,460.12 | 6,434.50 | 6,450.53 | 6,021.04 | **25,366.19** |
| 764 | 4,218.50 | 4,393.95 | 4,003.39 | 4,346.19 | **16,962.04** |
| 765 | 12,129.68 | 11,629.44 | 11,381.25 | 10,372.55 | **45,512.93** |
| 766 | 3,605.06 | 3,558.56 | 3,448.49 | 3,274.20 | **13,886.31** |
| 767 | 9,520.72 | 9,314.38 | 8,890.78 | 8,727.27 | **36,453.16** |
| 768 | 2,911.81 | 2,955.63 | 2,884.48 | 2,703.42 | **11,455.34** |
| 769 | 3,796.48 | 3,580.13 | 3,588.96 | 3,478.20 | **14,443.76** |
| 770 | 50,661.89 | 48,932.98 | 47,508.62 | 46,887.85 | **193,991.34** |
| 772 | 0.61 | 1.21 | 0 | 0.30 | **2.12** |
| 773 | 28,508.76 | 27,792.90 | 26,977.72 | 28,060.74 | **111,340.13** |
| 774 | 17,891.38 | 17,270.52 | 17,543.69 | 17,627.34 | **70,332.93** |
| 775 | 32,078.38 | 30,874.85 | 30,189.86 | 29,538.76 | **122,681.85** |
| 776 | 8,463.36 | 8,216.31 | 8,183.12 | 8,046.56 | **32,909.34** |
| 777 | 3,923.14 | 3,899.13 | 3,934.37 | 4,073.59 | **15,830.22** |
| 778 | 5,677.76 | 5,449.34 | 5,107.29 | 4,954.31 | **21,188.69** |
| 779 | 6,798.16 | 6,779.02 | 6,564.37 | 6,232.47 | **26,374.02** |
| 780 | 8,648.07 | 8,032.91 | 8,072.33 | 7,964.26 | **32,717.57** |
| 781 | 9,988.24 | 10,047.17 | 9,518.67 | 9,474.72 | **39,028.80** |
| 782 | 41,719.29 | 41,486.39 | 40,333.81 | 39,662.95 | **163,202.43** |
| 783 | 5,981.82 | 5,756.90 | 5,650.58 | 5,560.28 | **22,949.57** |

| | | | | | |
|---|---|---|---|---|---|
| 784 | 11,496.79 | 10,462.41 | 10,091.13 | 9,746.39 | **41,796.73** |
| 785 | 7,831.05 | 7,166.64 | 7,251.39 | 7,040.80 | **29,289.88** |
| 786 | 20,191.86 | 19,965.83 | 19,130.51 | 19,181.67 | **78,469.87** |
| 787 | 17,992.00 | 17,319.84 | 16,259.33 | 15,909.51 | **67,480.68** |
| 788 | 2,672.75 | 1,949.55 | 1,898.48 | 1,749.29 | **8,270.07** |
| 789 | 1,468.00 | 1,486.49 | 1,482.67 | 1,360.24 | **5,797.40** |
| 790 | 3,857.66 | 3,797.42 | 3,556.31 | 3,623.20 | **14,834.60** |
| 791 | 7,567.43 | 8,066.35 | 7,408.62 | 7,512.89 | **30,555.29** |
| 792 | 920.38 | 849.60 | 767.39 | 844.00 | **3,381.37** |
| 793 | 3,232.07 | 3,155.48 | 3,013.73 | 2,976.61 | **12,377.89** |
| 794 | 10,573.90 | 10,421.24 | 9,976.04 | 9,855.94 | **40,827.11** |
| 795 | 2,544.96 | 2,572.88 | 2,412.71 | 2,479.36 | **10,009.90** |
| 796 | 5,461.12 | 5,177.26 | 4,868.14 | 4,895.19 | **20,401.72** |
| 797 | 10,780.34 | 10,094.48 | 9,762.14 | 9,455.27 | **40,092.23** |
| 798 | 313.54 | 321.72 | 327.88 | 314.02 | **1,277.16** |
| 799 | 7,196.23 | 6,947.66 | 6,793.79 | 6,088.15 | **27,025.83** |
| **TX - TEXAS - Total** | **605,701.53** | **588,559.11** | **570,469.31** | **566,616.51** | **2,331,346.46** |

**STATE: UT - UTAH**

| REGISTRANT ZIP CODE 3 | 1ST QUARTER | 2ND QUARTER | 3RD QUARTER | 4TH QUARTER | **TOTAL GRAMS** |
|---|---|---|---|---|---|
| 840 | 20,285.17 | 19,090.90 | 18,620.71 | 19,029.40 | **77,026.19** |
| 841 | 13,182.89 | 12,595.49 | 12,085.28 | 11,992.30 | **49,855.96** |
| 843 | 2,512.80 | 2,328.69 | 2,454.15 | 2,329.81 | **9,625.45** |
| 844 | 4,031.05 | 3,787.98 | 3,617.62 | 3,695.16 | **15,131.81** |
| 845 | 1,553.35 | 1,695.93 | 1,471.87 | 1,675.43 | **6,396.57** |
| 846 | 4,857.38 | 4,660.10 | 4,518.08 | 4,623.53 | **18,659.08** |
| 847 | 6,703.14 | 6,574.30 | 6,342.35 | 6,433.97 | **26,053.76** |
| **UT - UTAH - Total** | **53,125.77** | **50,733.38** | **49,110.06** | **49,779.60** | **202,748.82** |

**STATE: VA - VIRGINIA**

| REGISTRANT ZIP CODE 3 | 1ST QUARTER | 2ND QUARTER | 3RD QUARTER | 4TH QUARTER | **TOTAL GRAMS** |
|---|---|---|---|---|---|
| 201 | 3,730.57 | 3,455.17 | 3,316.27 | 3,475.42 | **13,977.44** |
| 220 | 1,688.46 | 1,486.82 | 1,504.02 | 1,396.21 | **6,075.52** |
| 221 | 1,398.28 | 1,411.53 | 1,343.32 | 1,395.21 | **5,548.34** |
| 222 | 463.85 | 416.00 | 408.70 | 398.78 | **1,687.33** |
| 223 | 839.42 | 690.84 | 636.48 | 727.69 | **2,894.44** |
| 224 | 2,328.94 | 2,106.55 | 2,014.65 | 2,061.03 | **8,511.17** |
| 225 | 1,061.72 | 956.72 | 937.31 | 923.88 | **3,879.63** |
| 226 | 2,265.42 | 2,108.56 | 1,978.56 | 2,069.31 | **8,421.85** |
| 227 | 467.80 | 459.75 | 405.63 | 373.58 | **1,706.77** |
| 228 | 1,888.29 | 1,757.53 | 1,727.00 | 1,726.03 | **7,098.84** |
| 229 | 2,143.42 | 2,002.40 | 1,927.76 | 1,832.62 | **7,906.20** |

| | | | | | |
|---|---|---|---|---|---|
| 230 | 1,905.49 | 1,786.70 | 1,803.27 | 1,770.88 | **7,266.33** |
| 231 | 3,379.27 | 3,181.55 | 3,046.16 | 3,119.29 | **12,726.27** |
| 232 | 5,351.87 | 4,771.88 | 4,507.14 | 4,521.91 | **19,152.80** |
| 233 | 2,454.74 | 2,271.30 | 2,149.58 | 2,191.34 | **9,066.96** |
| 234 | 5,460.76 | 5,190.03 | 4,776.20 | 4,695.96 | **20,122.95** |
| 235 | 1,941.22 | 1,709.15 | 1,618.78 | 1,627.95 | **6,897.11** |
| 236 | 3,320.39 | 3,045.55 | 2,896.43 | 2,828.91 | **12,091.27** |
| 237 | 1,829.56 | 1,665.72 | 1,564.91 | 1,398.39 | **6,458.57** |
| 238 | 2,697.39 | 2,525.69 | 2,472.81 | 2,438.72 | **10,134.62** |
| 239 | 1,089.29 | 975.07 | 930.57 | 1,012.58 | **4,007.51** |
| 240 | 7,805.80 | 7,711.76 | 7,377.46 | 7,453.40 | **30,348.41** |
| 241 | 7,671.42 | 7,213.68 | 6,856.02 | 6,956.96 | **28,698.09** |
| 242 | 11,544.02 | 10,776.48 | 10,504.30 | 10,730.98 | **43,555.78** |
| 243 | 4,104.39 | 4,023.78 | 3,826.13 | 4,185.52 | **16,139.83** |
| 244 | 2,412.71 | 2,216.16 | 2,190.20 | 2,157.42 | **8,976.49** |
| 245 | 6,170.80 | 5,870.09 | 5,572.90 | 5,701.35 | **23,315.14** |
| 246 | 3,629.01 | 3,487.26 | 3,352.96 | 3,401.51 | **13,870.75** |
| **VA - VIRGINIA - Total** | **91,044.29** | **85,273.72** | **81,645.52** | **82,572.85** | **340,536.39** |

**STATE: VI - VIRGIN ISLANDS**

| REGISTRANT ZIP CODE 3 | 1ST QUARTER | 2ND QUARTER | 3RD QUARTER | 4TH QUARTER | TOTAL GRAMS |
|---|---|---|---|---|---|
| 008 | 285.51 | 221.22 | 224.79 | 237.30 | **968.81** |
| **VI - VIRGIN ISLANDS - Total** | **285.51** | **221.22** | **224.79** | **237.30** | **968.81** |

**STATE: VT - VERMONT**

| REGISTRANT ZIP CODE 3 | 1ST QUARTER | 2ND QUARTER | 3RD QUARTER | 4TH QUARTER | TOTAL GRAMS |
|---|---|---|---|---|---|
| 050 | 283.11 | 315.78 | 319.82 | 264.75 | **1,183.46** |
| 051 | 177.85 | 165.85 | 149.96 | 151.13 | **644.80** |
| 052 | 232.05 | 219.02 | 219.79 | 198.75 | **869.60** |
| 053 | 285.70 | 236.86 | 240.37 | 244.14 | **1,007.06** |
| 054 | 1,715.77 | 1,675.89 | 1,621.54 | 1,590.78 | **6,603.99** |
| 056 | 691.69 | 682.20 | 669.72 | 674.18 | **2,717.78** |
| 057 | 360.80 | 320.63 | 324.83 | 322.29 | **1,328.56** |
| 058 | 517.14 | 520.24 | 483.40 | 537.50 | **2,058.29** |
| **VT - VERMONT - Total** | **4,264.10** | **4,136.47** | **4,029.43** | **3,983.52** | **16,413.53** |

**STATE: WA - WASHINGTON**

| REGISTRANT ZIP CODE 3 | 1ST QUARTER | 2ND QUARTER | 3RD QUARTER | 4TH QUARTER | TOTAL GRAMS |
|---|---|---|---|---|---|
| 980 | 15,204.69 | 14,549.79 | 14,093.99 | 13,905.68 | **57,754.15** |
| 981 | 7,147.93 | 7,032.39 | 6,489.34 | 6,720.64 | **27,390.31** |
| 982 | 13,380.68 | 12,903.69 | 12,614.07 | 12,075.76 | **50,974.19** |
| 983 | 11,158.25 | 11,187.14 | 10,706.61 | 10,818.46 | **43,870.45** |
| 984 | 6,110.41 | 5,957.77 | 5,593.37 | 5,426.97 | **23,088.51** |

| | | | | | |
|---|---|---|---|---|---|
| 985 | 7,724.74 | 7,460.42 | 7,235.71 | 7,093.23 | **29,514.09** |
| 986 | 11,390.58 | 10,991.73 | 10,909.25 | 10,702.26 | **43,993.82** |
| 988 | 4,158.66 | 3,976.61 | 3,864.75 | 3,764.54 | **15,764.57** |
| 989 | 4,947.05 | 4,858.94 | 4,696.26 | 4,536.22 | **19,038.48** |
| 990 | 3,252.96 | 3,155.09 | 3,154.60 | 3,206.92 | **12,769.56** |
| 991 | 4,060.28 | 3,913.36 | 3,747.90 | 3,730.78 | **15,452.30** |
| 992 | 17,136.28 | 16,995.27 | 16,035.39 | 15,958.16 | **66,125.09** |
| 993 | 9,449.14 | 9,179.76 | 8,662.18 | 8,514.10 | **35,805.18** |
| 994 | 1,965.33 | 1,894.59 | 1,740.08 | 1,826.91 | **7,426.91** |
| **WA - WASHINGTON - Total** | **117,086.98** | **114,056.54** | **109,543.49** | **108,280.60** | **448,967.62** |

**STATE: WI - WISCONSIN**

| REGISTRANT ZIP CODE 3 | 1ST QUARTER | 2ND QUARTER | 3RD QUARTER | 4TH QUARTER | TOTAL GRAMS |
|---|---|---|---|---|---|
| 530 | 6,561.20 | 6,557.67 | 6,456.35 | 6,522.91 | **26,098.12** |
| 531 | 8,915.70 | 8,897.25 | 8,792.40 | 8,676.58 | **35,281.93** |
| 532 | 9,506.47 | 9,601.47 | 9,176.29 | 8,945.41 | **37,229.63** |
| 534 | 2,133.15 | 2,162.36 | 2,129.45 | 2,015.87 | **8,440.82** |
| 535 | 6,714.62 | 6,660.14 | 6,557.25 | 6,530.42 | **26,462.42** |
| 537 | 3,154.55 | 3,175.60 | 3,002.42 | 2,931.32 | **12,263.89** |
| 538 | 708.16 | 663.49 | 637.41 | 666.81 | **2,675.87** |
| 539 | 3,296.03 | 3,228.16 | 3,194.52 | 3,114.80 | **12,833.51** |
| 540 | 779.81 | 730.25 | 726.05 | 768.91 | **3,005.02** |
| 541 | 2,224.03 | 2,285.52 | 2,189.99 | 2,080.55 | **8,780.09** |
| 542 | 1,267.89 | 1,250.13 | 1,259.16 | 1,226.16 | **5,003.34** |
| 543 | 2,363.29 | 2,315.71 | 2,273.22 | 2,256.71 | **9,208.94** |
| 544 | 4,483.82 | 4,296.94 | 4,237.80 | 4,139.44 | **17,158.00** |
| 545 | 1,244.89 | 1,277.35 | 1,283.96 | 1,169.34 | **4,975.55** |
| 546 | 3,956.72 | 3,812.71 | 3,649.49 | 3,661.51 | **15,080.44** |
| 547 | 2,624.89 | 2,415.35 | 2,351.13 | 2,404.02 | **9,795.39** |
| 548 | 2,781.29 | 2,761.90 | 2,663.24 | 2,680.25 | **10,886.68** |
| 549 | 6,113.31 | 5,944.56 | 6,012.10 | 5,958.45 | **24,028.42** |
| **WI - WISCONSIN - Total** | **68,829.82** | **68,036.57** | **66,592.22** | **65,749.46** | **269,208.06** |

**STATE: WV - WEST VIRGINIA**

| REGISTRANT ZIP CODE 3 | 1ST QUARTER | 2ND QUARTER | 3RD QUARTER | 4TH QUARTER | TOTAL GRAMS |
|---|---|---|---|---|---|
| 247 | 1,862.66 | 1,695.26 | 1,514.43 | 1,223.58 | **6,295.93** |
| 248 | 989.31 | 659.98 | 640.12 | 670.01 | **2,959.42** |
| 249 | 1,009.37 | 1,019.56 | 971.64 | 894.87 | **3,895.45** |
| 250 | 2,275.13 | 2,123.34 | 2,037.10 | 1,872.45 | **8,308.02** |
| 251 | 1,733.02 | 1,703.46 | 1,599.10 | 1,601.13 | **6,636.70** |
| 252 | 1,250.90 | 989.19 | 901.06 | 816.29 | **3,957.44** |
| 253 | 4,912.75 | 4,423.27 | 4,075.46 | 3,564.40 | **16,975.88** |

| | | | | | |
|---|---|---|---|---|---|
| 254 | 2,074.84 | 2,018.66 | 1,970.16 | 1,879.19 | **7,942.84** |
| 255 | 4,694.52 | 4,450.24 | 4,232.93 | 4,277.94 | **17,655.63** |
| 256 | 2,380.12 | 1,941.86 | 1,740.15 | 1,953.30 | **8,015.42** |
| 257 | 3,872.27 | 3,389.95 | 3,377.44 | 3,343.38 | **13,983.03** |
| 258 | 3,442.95 | 3,156.37 | 2,857.09 | 2,662.24 | **12,118.65** |
| 259 | 1,111.41 | 1,040.62 | 959.13 | 922.59 | **4,033.76** |
| 260 | 3,039.88 | 2,910.45 | 2,862.12 | 2,772.20 | **11,584.64** |
| 261 | 3,100.21 | 3,014.80 | 2,657.64 | 2,440.19 | **11,212.84** |
| 262 | 2,009.92 | 1,935.41 | 1,832.64 | 1,735.92 | **7,513.89** |
| 263 | 4,329.29 | 4,192.57 | 4,240.69 | 3,867.87 | **16,630.43** |
| 264 | 1,128.51 | 1,088.30 | 1,002.97 | 1,048.46 | **4,268.25** |
| 265 | 3,514.52 | 3,334.37 | 3,153.51 | 3,239.11 | **13,241.51** |
| 266 | 1,085.70 | 1,063.61 | 989.20 | 941.57 | **4,080.08** |
| 267 | 685.11 | 685.68 | 612.01 | 650.61 | **2,633.40** |
| 268 | 326.41 | 302.75 | 290.65 | 276.58 | **1,196.40** |
| **WV - WEST VIRGINIA - Total** | **50,828.80** | **47,139.71** | **44,517.22** | **42,653.89** | **185,139.63** |

**STATE: WY - WYOMING**

| REGISTRANT ZIP CODE 3 | 1ST QUARTER | 2ND QUARTER | 3RD QUARTER | 4TH QUARTER | TOTAL GRAMS |
|---|---|---|---|---|---|
| 820 | 1,877.06 | 1,912.64 | 1,794.65 | 1,762.35 | **7,346.70** |
| 822 | 428.80 | 404.72 | 410.97 | 382.87 | **1,627.35** |
| 823 | 198.63 | 199.89 | 179.67 | 182.99 | **761.17** |
| 824 | 1,076.33 | 1,035.40 | 1,073.41 | 1,041.16 | **4,226.30** |
| 825 | 698.61 | 656.03 | 605.96 | 706.14 | **2,666.75** |
| 826 | 1,913.43 | 1,925.16 | 1,890.89 | 1,847.22 | **7,576.70** |
| 827 | 668.19 | 651.68 | 641.50 | 660.16 | **2,621.53** |
| 828 | 1,053.55 | 987.18 | 1,004.09 | 939.96 | **3,984.78** |
| 829 | 1,579.78 | 1,500.65 | 1,363.17 | 1,445.02 | **5,888.63** |
| 830 | 171.89 | 167.71 | 205.06 | 158.11 | **702.77** |
| 831 | 373.87 | 416.27 | 351.44 | 322.10 | **1,463.67** |
| **WY - WYOMING - Total** | **10,040.15** | **9,857.33** | **9,520.81** | **9,448.06** | **38,866.35** |

**DRUG: 9230 - MEPERIDINE (PETHIDINE)**

**STATE: AK - ALASKA**

| REGISTRANT ZIP CODE 3 | 1ST QUARTER | 2ND QUARTER | 3RD QUARTER | 4TH QUARTER | TOTAL GRAMS |
|---|---|---|---|---|---|
| 995 | 465.05 | 247.29 | 202.62 | 162.86 | **1,077.83** |
| 996 | 140.53 | 119.83 | 82.25 | 80.07 | **422.68** |
| 997 | 53.82 | 70.59 | 97.50 | 40.31 | **262.21** |
| 998 | 5.45 | 8.17 | 5.45 | 5.45 | **24.51** |
| 999 | 2.29 | 1.09 | 0 | 5.45 | **8.82** |
| **AK - ALASKA - Total** | **667.13** | **446.97** | **387.82** | **294.13** | **1,796.05** |

**STATE: AL - ALABAMA**

| REGISTRANT ZIP CODE 3 | 1ST QUARTER | 2ND QUARTER | 3RD QUARTER | 4TH QUARTER | TOTAL GRAMS |
|---|---|---|---|---|---|
| 350 | 735.33 | 515.25 | 413.96 | 444.47 | **2,109.01** |
| 351 | 418.32 | 287.60 | 248.38 | 210.79 | **1,165.09** |
| 352 | 730.43 | 612.45 | 602.97 | 400.35 | **2,346.19** |
| 354 | 604.39 | 634.67 | 424.86 | 296.85 | **1,960.77** |
| 355 | 302.54 | 178.66 | 153.06 | 131.27 | **765.53** |
| 356 | 450.72 | 379.21 | 372.57 | 344.79 | **1,547.28** |
| 357 | 307.75 | 167.22 | 163.41 | 164.50 | **802.87** |
| 358 | 364.94 | 358.40 | 379.10 | 298.49 | **1,400.94** |
| 359 | 306.22 | 199.88 | 309.38 | 237.48 | **1,052.97** |
| 360 | 394.68 | 286.22 | 219.51 | 186.83 | **1,087.24** |
| 361 | 255.57 | 260.36 | 204.80 | 187.37 | **908.10** |
| 362 | 353.50 | 227.13 | 218.96 | 160.14 | **959.74** |
| 363 | 625.26 | 515.47 | 363.85 | 308.29 | **1,812.87** |
| 364 | 258.18 | 285.96 | 217.88 | 146.52 | **908.54** |
| 365 | 572.47 | 590.99 | 431.94 | 321.37 | **1,916.76** |
| 366 | 735.83 | 957.69 | 767.14 | 306.66 | **2,767.32** |
| 367 | 123.64 | 115.47 | 101.31 | 107.85 | **448.28** |
| 368 | 359.49 | 354.05 | 264.17 | 254.91 | **1,232.63** |
| 369 | 3.27 | 17.97 | 3.27 | 8.72 | **33.23** |
| **AL - ALABAMA - Total** | **7,902.52** | **6,944.66** | **5,860.51** | **4,517.64** | **25,225.33** |

**STATE: AR - ARKANSAS**

| REGISTRANT ZIP CODE 3 | 1ST QUARTER | 2ND QUARTER | 3RD QUARTER | 4TH QUARTER | TOTAL GRAMS |
|---|---|---|---|---|---|
| 716 | 514.95 | 412.00 | 460.59 | 326.81 | **1,714.35** |
| 717 | 244.02 | 157.57 | 148.70 | 129.09 | **679.38** |
| 718 | 51.75 | 53.92 | 64.82 | 59.37 | **229.86** |
| 719 | 340.97 | 272.34 | 261.45 | 153.06 | **1,027.83** |
| 720 | 993.29 | 754.22 | 799.60 | 800.69 | **3,347.80** |
| 721 | 1,039.53 | 1,337.97 | 974.99 | 857.34 | **4,209.82** |
| 722 | 821.58 | 773.39 | 744.59 | 563.21 | **2,902.77** |
| 723 | 184.65 | 239.66 | 173.21 | 90.42 | **687.94** |
| 724 | 616.59 | 462.44 | 456.45 | 302.85 | **1,838.32** |
| 725 | 209.05 | 165.59 | 175.93 | 102.40 | **652.97** |
| 726 | 223.87 | 144.34 | 151.97 | 115.47 | **635.65** |
| 727 | 1,003.86 | 898.19 | 714.63 | 593.71 | **3,210.39** |
| 728 | 199.36 | 136.17 | 154.69 | 49.57 | **539.79** |
| 729 | 308.84 | 221.14 | 266.35 | 171.58 | **967.91** |
| **AR - ARKANSAS - Total** | **6,752.29** | **6,028.95** | **5,547.97** | **4,315.56** | **22,644.77** |

**STATE: AZ - ARIZONA**

| REGISTRANT ZIP CODE 3 | 1ST QUARTER | 2ND QUARTER | 3RD QUARTER | 4TH QUARTER | TOTAL GRAMS |
|---|---|---|---|---|---|

| | | | | | |
|---|---|---|---|---|---|
| 850 | 1,096.76 | 888.15 | 908.43 | 931.42 | **3,824.75** |
| 851 | 173.76 | 165.04 | 117.65 | 104.58 | **561.03** |
| 852 | 1,824.90 | 1,910.28 | 1,544.49 | 1,075.23 | **6,354.91** |
| 853 | 554.49 | 615.82 | 519.13 | 463.07 | **2,152.52** |
| 855 | 139.44 | 160.68 | 116.56 | 84.97 | **501.66** |
| 856 | 112.75 | 174.30 | 191.19 | 90.96 | **569.20** |
| 857 | 936.86 | 694.48 | 752.21 | 615.50 | **2,999.05** |
| 859 | 29.96 | 27.23 | 5.45 | 19.61 | **82.25** |
| 860 | 50.66 | 51.79 | 65.95 | 38.80 | **207.20** |
| 863 | 110.57 | 103.49 | 67.63 | 45.25 | **326.94** |
| 864 | 355.35 | 167.22 | 145.98 | 104.04 | **772.58** |
| 865 | 2.72 | 2.18 | 4.90 | 1.09 | **10.89** |
| **AZ - ARIZONA - Total** | **5,388.22** | **4,960.67** | **4,439.57** | **3,574.52** | **18,362.98** |

**STATE: CA - CALIFORNIA**

| REGISTRANT ZIP CODE 3 | 1ST QUARTER | 2ND QUARTER | 3RD QUARTER | 4TH QUARTER | **TOTAL GRAMS** |
|---|---|---|---|---|---|
| 900 | 806.68 | 719.53 | 607.87 | 483.14 | **2,617.22** |
| 902 | 529.98 | 608.96 | 435.21 | 438.47 | **2,012.62** |
| 903 | 2.18 | 11.44 | 2.18 | 4.36 | **20.15** |
| 904 | 164.50 | 75.17 | 90.42 | 139.44 | **469.52** |
| 905 | 239.12 | 386.73 | 278.88 | 181.93 | **1,086.65** |
| 906 | 153.06 | 117.00 | 71.35 | 103.49 | **444.90** |
| 907 | 215.15 | 108.94 | 59.92 | 43.58 | **427.58** |
| 908 | 197.94 | 88.78 | 49.02 | 63.73 | **399.47** |
| 910 | 93.14 | 84.97 | 41.40 | 54.47 | **273.98** |
| 911 | 131.27 | 67.00 | 115.47 | 127.46 | **441.20** |
| 912 | 215.15 | 138.90 | 209.16 | 129.64 | **692.84** |
| 913 | 721.06 | 833.37 | 553.95 | 583.91 | **2,692.28** |
| 914 | 248.81 | 180.84 | 96.95 | 150.33 | **676.94** |
| 915 | 89.33 | 140.53 | 131.81 | 153.60 | **515.27** |
| 916 | 2.18 | 19.61 | 8.72 | 4.36 | **34.86** |
| 917 | 1,236.44 | 918.89 | 963.01 | 803.96 | **3,922.29** |
| 918 | 96.95 | 45.75 | 176.48 | 19.61 | **338.80** |
| 919 | 131.27 | 135.08 | 121.47 | 104.04 | **491.85** |
| 920 | 1,607.05 | 1,066.17 | 621.49 | 651.45 | **3,946.15** |
| 921 | 976.08 | 792.41 | 407.97 | 485.86 | **2,662.32** |
| 922 | 275.07 | 244.56 | 354.05 | 188.46 | **1,062.14** |
| 923 | 674.32 | 831.19 | 574.65 | 437.38 | **2,517.55** |
| 924 | 26.15 | 45.21 | 4.36 | 14.16 | **89.87** |
| 925 | 867.14 | 943.94 | 820.30 | 386.18 | **3,017.57** |
| 926 | 675.41 | 587.17 | 413.42 | 312.65 | **1,988.65** |

| | | | | | |
|---|---|---|---|---|---|
| 927 | 782.72 | 209.16 | 92.05 | 163.41 | **1,247.33** |
| 928 | 629.11 | 550.13 | 404.16 | 283.24 | **1,866.64** |
| 930 | 149.79 | 179.75 | 149.24 | 82.79 | **561.57** |
| 931 | 174.84 | 217.33 | 154.69 | 176.48 | **723.35** |
| 932 | 266.90 | 490.76 | 242.93 | 276.16 | **1,276.75** |
| 933 | 682.49 | 794.70 | 677.05 | 572.47 | **2,726.71** |
| 934 | 284.87 | 279.42 | 162.32 | 188.46 | **915.08** |
| 935 | 232.04 | 77.35 | 117.00 | 83.88 | **510.26** |
| 936 | 281.60 | 266.35 | 118.74 | 94.78 | **761.47** |
| 937 | 474.64 | 401.11 | 216.24 | 153.60 | **1,245.59** |
| 939 | 204.48 | 173.76 | 189.55 | 195.00 | **762.78** |
| 940 | 308.84 | 308.84 | 219.51 | 210.25 | **1,047.43** |
| 941 | 647.09 | 257.09 | 480.41 | 153.06 | **1,537.65** |
| 943 | 364.40 | 285.96 | 133.99 | 65.91 | **850.26** |
| 944 | 0.54 | 21.79 | 13.07 | 7.63 | **43.03** |
| 945 | 835.55 | 1,013.12 | 765.29 | 630.20 | **3,244.16** |
| 946 | 8.72 | 31.59 | 7.08 | 14.71 | **62.09** |
| 947 | 8.72 | 0 | 0 | 0 | **8.72** |
| 948 | 0 | 10.89 | 2.72 | 0 | **13.62** |
| 949 | 95.32 | 26.69 | 46.30 | 27.23 | **195.54** |
| 950 | 162.32 | 194.45 | 127.46 | 84.43 | **568.65** |
| 951 | 220.05 | 194.45 | 63.73 | 75.17 | **553.40** |
| 952 | 66.45 | 97.50 | 67.00 | 53.38 | **284.33** |
| 953 | 800.36 | 1,060.51 | 509.28 | 431.94 | **2,802.09** |
| 954 | 84.97 | 94.23 | 89.33 | 74.62 | **343.15** |
| 955 | 47.39 | 39.22 | 27.23 | 37.04 | **150.88** |
| 956 | 550.13 | 508.19 | 468.98 | 386.18 | **1,913.49** |
| 957 | 18.52 | 51.20 | 14.16 | 26.69 | **110.57** |
| 958 | 798.08 | 375.29 | 178.66 | 275.61 | **1,627.64** |
| 959 | 251.10 | 218.42 | 233.67 | 179.75 | **882.94** |
| 960 | 131.27 | 43.03 | 31.05 | 45.75 | **251.10** |
| 961 | 28.32 | 11.98 | 35.40 | 33.77 | **109.48** |
| **CA - CALIFORNIA - Total** | **19,967.05** | **17,676.42** | **13,247.78** | **11,149.21** | **62,040.45** |

**STATE: CO - COLORADO**

| REGISTRANT ZIP CODE 3 | 1ST QUARTER | 2ND QUARTER | 3RD QUARTER | 4TH QUARTER | **TOTAL GRAMS** |
|---|---|---|---|---|---|
| 800 | 114.38 | 175.39 | 117.11 | 58.28 | **465.16** |
| 801 | 353.07 | 354.16 | 241.30 | 230.40 | **1,178.92** |
| 802 | 192.17 | 166.13 | 104.04 | 117.11 | **579.44** |
| 803 | 49.02 | 48.48 | 21.79 | 58.83 | **178.11** |
| 804 | 51.20 | 65.36 | 46.84 | 40.31 | **203.71** |

| | | | | | |
|---|---|---|---|---|---|
| 805 | 90.96 | 129.09 | 105.67 | 60.46 | **386.18** |
| 806 | 68.41 | 44.12 | 76.26 | 53.38 | **242.17** |
| 807 | 18.52 | 23.97 | 15.25 | 5.99 | **63.73** |
| 808 | 69.50 | 40.85 | 70.26 | 47.93 | **228.55** |
| 809 | 217.88 | 164.50 | 168.31 | 170.49 | **721.17** |
| 810 | 102.40 | 76.80 | 80.61 | 91.51 | **351.32** |
| 811 | 17.43 | 8.72 | 13.62 | 7.08 | **46.84** |
| 812 | 49.02 | 21.24 | 44.12 | 22.88 | **137.26** |
| 813 | 10.35 | 11.44 | 15.25 | 11.44 | **48.48** |
| 814 | 86.06 | 90.42 | 125.28 | 55.56 | **357.32** |
| 815 | 51.64 | 65.36 | 75.17 | 33.23 | **225.39** |
| 816 | 61.55 | 85.52 | 52.83 | 76.26 | **276.16** |
| **CO - COLORADO - Total** | **1,603.56** | **1,571.53** | **1,373.70** | **1,141.12** | **5,689.91** |

**STATE: CT - CONNECTICUT**

| REGISTRANT ZIP CODE 3 | 1ST QUARTER | 2ND QUARTER | 3RD QUARTER | 4TH QUARTER | TOTAL GRAMS |
|---|---|---|---|---|---|
| 060 | 99.68 | 70.26 | 42.49 | 68.63 | **281.06** |
| 061 | 74.62 | 65.36 | 65.36 | 85.52 | **290.86** |
| 062 | 23.97 | 9.26 | 8.72 | 6.54 | **48.48** |
| 063 | 175.93 | 82.79 | 208.62 | 166.67 | **634.02** |
| 064 | 276.16 | 244.56 | 168.85 | 185.19 | **874.77** |
| 065 | 75.38 | 45.75 | 61.01 | 70.26 | **252.41** |
| 066 | 64.27 | 38.13 | 77.35 | 31.59 | **211.34** |
| 067 | 129.09 | 129.64 | 113.30 | 100.22 | **472.24** |
| 068 | 177.57 | 193.36 | 119.83 | 119.29 | **610.05** |
| 069 | 4.90 | 5.99 | 6.54 | 2.18 | **19.61** |
| **CT - CONNECTICUT - Total** | **1,101.58** | **885.12** | **872.04** | **836.10** | **3,694.83** |

**STATE: DC - DISTRICT OF COLUMBIA**

| REGISTRANT ZIP CODE 3 | 1ST QUARTER | 2ND QUARTER | 3RD QUARTER | 4TH QUARTER | TOTAL GRAMS |
|---|---|---|---|---|---|
| 200 | 144.34 | 162.32 | 130.18 | 196.63 | **633.47** |
| 204 | 0 | 2.18 | 0 | 0 | **2.18** |
| **DC - DISTRICT OF COLUMBIA -** | **144.34** | **164.50** | **130.18** | **196.63** | **635.65** |

**STATE: DE - DELAWARE**

| REGISTRANT ZIP CODE 3 | 1ST QUARTER | 2ND QUARTER | 3RD QUARTER | 4TH QUARTER | TOTAL GRAMS |
|---|---|---|---|---|---|
| 197 | 118.63 | 26.15 | 37.58 | 58.83 | **241.19** |
| 198 | 41.40 | 8.72 | 3.27 | 17.43 | **70.81** |
| 199 | 63.73 | 71.90 | 56.65 | 56.10 | **248.38** |
| **DE - DELAWARE - Total** | **223.76** | **106.76** | **97.50** | **132.36** | **560.37** |

**STATE: FL - FLORIDA**

| REGISTRANT ZIP CODE 3 | 1ST QUARTER | 2ND QUARTER | 3RD QUARTER | 4TH QUARTER | TOTAL GRAMS |
|---|---|---|---|---|---|
| 320 | 290.32 | 246.74 | 260.91 | 189.01 | **986.97** |

| | | | | | |
|---|---|---|---|---|---|
| 321 | 217.88 | 258.73 | 168.85 | 194.45 | **839.91** |
| 322 | 461.35 | 425.95 | 302.30 | 284.87 | **1,474.47** |
| 323 | 528.02 | 349.69 | 232.58 | 218.42 | **1,328.71** |
| 324 | 121.47 | 161.23 | 113.30 | 87.69 | **483.68** |
| 325 | 597.52 | 448.28 | 368.75 | 285.96 | **1,700.51** |
| 326 | 297.94 | 92.60 | 99.13 | 113.30 | **602.97** |
| 327 | 291.95 | 177.57 | 176.48 | 149.24 | **795.24** |
| 328 | 162.32 | 125.28 | 86.61 | 78.44 | **452.64** |
| 329 | 413.96 | 357.32 | 274.52 | 221.69 | **1,267.49** |
| 330 | 872.59 | 611.68 | 424.86 | 307.20 | **2,216.33** |
| 331 | 854.07 | 633.47 | 588.81 | 431.39 | **2,507.74** |
| 333 | 651.77 | 715.72 | 405.79 | 446.64 | **2,219.93** |
| 334 | 689.57 | 643.82 | 483.68 | 378.56 | **2,195.64** |
| 335 | 380.19 | 370.93 | 268.10 | 198.27 | **1,217.49** |
| 336 | 420.39 | 574.21 | 433.57 | 317.55 | **1,745.72** |
| 337 | 136.17 | 181.93 | 136.72 | 121.47 | **576.28** |
| 338 | 457.86 | 411.24 | 368.75 | 297.94 | **1,535.80** |
| 339 | 487.06 | 373.66 | 362.22 | 359.49 | **1,582.43** |
| 341 | 165.59 | 95.87 | 52.29 | 102.95 | **416.69** |
| 342 | 206.98 | 197.72 | 162.32 | 107.30 | **674.32** |
| 344 | 334.98 | 257.09 | 246.74 | 147.61 | **986.43** |
| 346 | 143.25 | 229.86 | 156.87 | 143.25 | **673.23** |
| 347 | 204.26 | 130.18 | 87.15 | 73.53 | **495.12** |
| 349 | 248.38 | 258.51 | 232.58 | 217.88 | **957.34** |
| **FL - FLORIDA - Total** | **9,635.85** | **8,329.25** | **6,493.87** | **5,474.11** | **29,933.08** |

**STATE: GA - GEORGIA**

| REGISTRANT ZIP CODE 3 | 1ST QUARTER | 2ND QUARTER | 3RD QUARTER | 4TH QUARTER | TOTAL GRAMS |
|---|---|---|---|---|---|
| 300 | 1,382.20 | 999.50 | 852.44 | 695.02 | **3,929.16** |
| 301 | 928.91 | 965.73 | 656.35 | 601.34 | **3,152.32** |
| 302 | 612.77 | 663.97 | 410.69 | 416.14 | **2,103.58** |
| 303 | 532.27 | 508.74 | 411.78 | 243.48 | **1,696.27** |
| 304 | 428.67 | 240.75 | 257.09 | 171.58 | **1,098.09** |
| 305 | 567.02 | 568.11 | 503.84 | 269.62 | **1,908.59** |
| 306 | 323.00 | 376.38 | 295.77 | 221.69 | **1,216.83** |
| 307 | 618.77 | 556.13 | 513.10 | 429.21 | **2,117.20** |
| 308 | 184.65 | 190.10 | 161.23 | 133.99 | **669.97** |
| 309 | 480.20 | 355.68 | 341.52 | 300.12 | **1,477.52** |
| 310 | 748.23 | 815.81 | 492.62 | 492.94 | **2,549.60** |
| 312 | 252.19 | 187.92 | 183.56 | 130.18 | **753.85** |
| 313 | 264.17 | 212.97 | 189.55 | 143.25 | **809.95** |

| | | | | | |
|---|---|---|---|---|---|
| 314 | 279.97 | 283.24 | 268.53 | 142.71 | **974.45** |
| 315 | 461.35 | 642.19 | 441.20 | 346.42 | **1,891.16** |
| 316 | 784.79 | 771.02 | 638.90 | 554.49 | **2,749.19** |
| 317 | 892.63 | 678.59 | 508.19 | 465.16 | **2,544.58** |
| 318 | 29.96 | 44.66 | 35.40 | 27.23 | **137.26** |
| 319 | 307.73 | 243.19 | 376.16 | 188.46 | **1,115.54** |
| 398 | 191.14 | 162.30 | 92.05 | 119.29 | **564.78** |
| **GA - GEORGIA - Total** | **10,270.61** | **9,466.97** | **7,629.96** | **6,092.33** | **33,459.87** |

**STATE: GU - GUAM**

| REGISTRANT ZIP CODE 3 | 1ST QUARTER | 2ND QUARTER | 3RD QUARTER | 4TH QUARTER | TOTAL GRAMS |
|---|---|---|---|---|---|
| 969 | 14.16 | 35.95 | 40.85 | 29.96 | **120.92** |
| **GU - GUAM - Total** | **14.16** | **35.95** | **40.85** | **29.96** | **120.92** |

**STATE: HI - HAWAII**

| REGISTRANT ZIP CODE 3 | 1ST QUARTER | 2ND QUARTER | 3RD QUARTER | 4TH QUARTER | TOTAL GRAMS |
|---|---|---|---|---|---|
| 967 | 120.38 | 246.20 | 228.22 | 131.27 | **726.07** |
| 968 | 126.80 | 156.11 | 124.19 | 101.31 | **508.41** |
| **HI - HAWAII - Total** | **247.18** | **402.31** | **352.41** | **232.58** | **1,234.48** |

**STATE: IA - IOWA**

| REGISTRANT ZIP CODE 3 | 1ST QUARTER | 2ND QUARTER | 3RD QUARTER | 4TH QUARTER | TOTAL GRAMS |
|---|---|---|---|---|---|
| 500 | 42.49 | 241.49 | 35.95 | 29.96 | **349.89** |
| 501 | 32.68 | 38.13 | 50.11 | 23.97 | **144.89** |
| 502 | 115.47 | 113.84 | 102.95 | 98.04 | **430.30** |
| 503 | 393.81 | 476.60 | 359.49 | 437.93 | **1,667.83** |
| 504 | 1.09 | 5.99 | 7.63 | 9.26 | **23.97** |
| 505 | 76.26 | 27.23 | 81.16 | 61.55 | **246.20** |
| 506 | 45.75 | 36.49 | 38.13 | 10.89 | **131.27** |
| 507 | 370.93 | 106.76 | 103.49 | 198.81 | **779.99** |
| 508 | 21.24 | 4.90 | 2.18 | 17.97 | **46.30** |
| 510 | 12.53 | 31.59 | 10.35 | 31.59 | **86.06** |
| 511 | 39.22 | 96.95 | 30.50 | 46.84 | **213.52** |
| 512 | 2.18 | 5.45 | 0 | 2.18 | **9.80** |
| 513 | 50.11 | 25.60 | 1.09 | 2.18 | **78.98** |
| 514 | 8.17 | 5.45 | 3.27 | 8.72 | **25.60** |
| 515 | 17.95 | 41.40 | 13.62 | 25.60 | **98.57** |
| 516 | 0 | 1.09 | 4.36 | 2.18 | **7.63** |
| 520 | 14.16 | 15.25 | 14.16 | 14.16 | **57.74** |
| 521 | 1.09 | 1.09 | 2.18 | 0 | **4.36** |
| 522 | 101.31 | 114.93 | 39.76 | 52.83 | **308.84** |
| 523 | 63.18 | 43.58 | 59.92 | 95.87 | **262.54** |
| 524 | 240.21 | 177.57 | 234.76 | 183.56 | **836.10** |

| | | | | | |
|---|---|---|---|---|---|
| 525 | 128.00 | 76.80 | 72.99 | 108.94 | **386.73** |
| 526 | 26.15 | 117.65 | 8.72 | 46.84 | **199.36** |
| 527 | 37.58 | 30.50 | 8.72 | 11.44 | **88.24** |
| 528 | 19.61 | 53.92 | 9.80 | 19.61 | **102.95** |
| **IA - IOWA - Total** | **1,861.18** | **1,890.26** | **1,295.27** | **1,540.92** | **6,587.62** |

**STATE: ID - IDAHO**

| REGISTRANT ZIP CODE 3 | 1ST QUARTER | 2ND QUARTER | 3RD QUARTER | 4TH QUARTER | TOTAL GRAMS |
|---|---|---|---|---|---|
| 832 | 101.86 | 108.94 | 61.01 | 89.33 | **361.13** |
| 833 | 77.89 | 84.43 | 25.06 | 56.10 | **243.48** |
| 834 | 123.10 | 120.38 | 118.20 | 125.28 | **486.95** |
| 835 | 45.54 | 27.78 | 30.50 | 5.99 | **109.81** |
| 836 | 228.77 | 290.32 | 207.53 | 251.65 | **978.26** |
| 837 | 102.95 | 76.26 | 77.35 | 117.11 | **373.66** |
| 838 | 61.77 | 69.72 | 53.92 | 49.02 | **234.43** |
| **ID - IDAHO - Total** | **741.86** | **777.81** | **573.56** | **694.48** | **2,787.71** |

**STATE: IL - ILLINOIS**

| REGISTRANT ZIP CODE 3 | 1ST QUARTER | 2ND QUARTER | 3RD QUARTER | 4TH QUARTER | TOTAL GRAMS |
|---|---|---|---|---|---|
| 600 | 273.43 | 263.63 | 228.77 | 181.93 | **947.76** |
| 601 | 449.37 | 447.19 | 265.70 | 179.75 | **1,342.00** |
| 602 | 1.09 | 1.09 | 0 | 0.54 | **2.72** |
| 603 | 0 | 3.27 | 1.09 | 2.18 | **6.54** |
| 604 | 245.11 | 266.90 | 230.40 | 179.75 | **922.16** |
| 605 | 100.33 | 156.87 | 96.95 | 116.56 | **470.72** |
| 606 | 186.28 | 132.36 | 176.48 | 100.77 | **595.89** |
| 607 | 6.54 | 8.72 | 8.72 | 8.72 | **32.68** |
| 608 | 52.29 | 43.58 | 61.01 | 47.93 | **204.80** |
| 609 | 4.90 | 8.72 | 0 | 4.36 | **17.97** |
| 610 | 132.58 | 107.15 | 89.33 | 70.81 | **399.87** |
| 611 | 59.92 | 45.75 | 38.13 | 42.49 | **186.28** |
| 612 | 7.08 | 6.54 | 9.26 | 7.08 | **29.96** |
| 613 | 17.43 | 53.38 | 15.80 | 52.29 | **138.90** |
| 614 | 18.52 | 28.87 | 13.07 | 26.15 | **86.61** |
| 615 | 43.58 | 33.77 | 55.56 | 4.90 | **137.81** |
| 616 | 89.33 | 126.37 | 87.15 | 37.04 | **339.89** |
| 617 | 44.01 | 60.18 | 42.49 | 14.16 | **160.84** |
| 618 | 205.35 | 126.37 | 85.52 | 86.06 | **503.29** |
| 619 | 32.68 | 45.75 | 51.20 | 35.95 | **165.59** |
| 620 | 26.15 | 10.89 | 16.34 | 5.45 | **58.83** |
| 622 | 200.45 | 317.55 | 218.42 | 129.64 | **866.05** |
| 623 | 70.26 | 102.95 | 37.04 | 39.76 | **250.01** |

| | | | | | |
|---|---|---|---|---|---|
| 624 | 9.80 | 16.89 | 34.86 | 2.72 | **64.27** |
| 625 | 29.41 | 1.09 | 2.18 | 7.63 | **40.31** |
| 626 | 59.92 | 43.58 | 48.48 | 33.77 | **185.74** |
| 627 | 68.74 | 107.85 | 62.64 | 65.36 | **304.59** |
| 628 | 102.40 | 50.37 | 74.08 | 43.58 | **270.43** |
| 629 | 281.06 | 201.53 | 163.41 | 234.22 | **880.22** |
| **IL - ILLINOIS - Total** | **2,818.00** | **2,819.13** | **2,214.05** | **1,761.52** | **9,612.69** |

**STATE: IN - INDIANA**

| REGISTRANT ZIP CODE 3 | 1ST QUARTER | 2ND QUARTER | 3RD QUARTER | 4TH QUARTER | TOTAL GRAMS |
|---|---|---|---|---|---|
| 460 | 278.88 | 162.32 | 292.50 | 194.45 | **928.15** |
| 461 | 423.22 | 223.32 | 318.10 | 224.96 | **1,189.60** |
| 462 | 870.08 | 936.32 | 794.15 | 523.99 | **3,124.55** |
| 463 | 233.13 | 187.92 | 147.61 | 99.13 | **667.79** |
| 464 | 49.57 | 49.02 | 51.20 | 24.51 | **174.30** |
| 465 | 71.90 | 64.27 | 59.37 | 41.40 | **236.94** |
| 466 | 28.32 | 17.43 | 31.05 | 18.52 | **95.32** |
| 467 | 56.21 | 34.32 | 19.39 | 20.70 | **130.62** |
| 468 | 187.37 | 170.49 | 143.80 | 122.01 | **623.67** |
| 469 | 110.57 | 116.02 | 46.30 | 32.14 | **305.03** |
| 470 | 48.46 | 128.81 | 57.74 | 42.49 | **277.49** |
| 471 | 128.70 | 235.31 | 138.90 | 80.07 | **582.97** |
| 472 | 125.82 | 120.92 | 139.44 | 72.99 | **459.17** |
| 473 | 111.66 | 81.70 | 76.26 | 82.25 | **351.87** |
| 474 | 25.60 | 34.86 | 33.77 | 11.44 | **105.67** |
| 475 | 47.93 | 59.63 | 51.20 | 38.67 | **197.44** |
| 476 | 46.84 | 147.61 | 73.53 | 30.50 | **298.49** |
| 477 | 138.79 | 145.98 | 94.23 | 81.70 | **460.70** |
| 478 | 91.51 | 90.42 | 73.53 | 55.56 | **311.02** |
| 479 | 72.99 | 53.38 | 55.01 | 42.49 | **223.87** |
| **IN - INDIANA - Total** | **3,147.55** | **3,060.03** | **2,697.07** | **1,839.95** | **10,744.61** |

**STATE: KS - KANSAS**

| REGISTRANT ZIP CODE 3 | 1ST QUARTER | 2ND QUARTER | 3RD QUARTER | 4TH QUARTER | TOTAL GRAMS |
|---|---|---|---|---|---|
| 660 | 143.80 | 265.81 | 195.00 | 113.84 | **718.44** |
| 661 | 35.95 | 26.69 | 24.51 | 19.61 | **106.76** |
| 662 | 213.52 | 144.34 | 128.55 | 110.57 | **596.98** |
| 664 | 49.81 | 29.59 | 22.31 | 11.98 | **113.69** |
| 665 | 16.34 | 49.02 | 38.13 | 19.61 | **123.10** |
| 666 | 40.85 | 76.26 | 21.79 | 16.34 | **155.24** |
| 667 | 62.64 | 77.61 | 95.32 | 82.25 | **317.81** |
| 668 | 21.79 | 22.88 | 15.80 | 13.62 | **74.08** |

| | | | | | |
|---|---|---|---|---|---|
| 669 | 10.35 | 14.71 | 13.62 | 11.44 | **50.11** |
| 670 | 111.12 | 97.50 | 84.43 | 55.01 | **348.06** |
| 671 | 72.99 | 29.41 | 45.21 | 33.77 | **181.38** |
| 672 | 300.67 | 240.21 | 278.88 | 181.38 | **1,001.14** |
| 673 | 57.74 | 41.40 | 28.32 | 11.98 | **139.44** |
| 674 | 72.99 | 78.44 | 44.66 | 77.35 | **273.43** |
| 675 | 109.48 | 112.21 | 88.24 | 89.87 | **399.80** |
| 676 | 101.31 | 53.38 | 77.35 | 62.09 | **294.13** |
| 677 | 10.35 | 30.50 | 27.78 | 7.63 | **76.26** |
| 678 | 85.52 | 71.90 | 82.79 | 27.23 | **267.44** |
| 679 | 27.23 | 50.11 | 54.47 | 53.92 | **185.74** |
| **KS - KANSAS - Total** | **1,544.43** | **1,511.94** | **1,367.14** | **999.50** | **5,423.02** |

**STATE: KY - KENTUCKY**

| REGISTRANT ZIP CODE 3 | 1ST QUARTER | 2ND QUARTER | 3RD QUARTER | 4TH QUARTER | **TOTAL GRAMS** |
|---|---|---|---|---|---|
| 400 | 53.90 | 32.14 | 21.79 | 15.25 | **123.08** |
| 401 | 61.55 | 34.32 | 9.80 | 7.08 | **112.75** |
| 402 | 502.75 | 271.25 | 319.73 | 201.53 | **1,295.27** |
| 403 | 112.75 | 81.70 | 71.90 | 65.91 | **332.26** |
| 404 | 43.03 | 51.20 | 47.39 | 48.48 | **190.10** |
| 405 | 137.81 | 64.82 | 31.59 | 81.70 | **315.92** |
| 406 | 17.43 | 4.36 | 0.54 | 6.54 | **28.87** |
| 407 | 41.40 | 38.67 | 28.32 | 28.32 | **136.72** |
| 408 | 35.40 | 138.35 | 70.16 | 38.67 | **282.58** |
| 409 | 47.93 | 47.39 | 29.41 | 9.26 | **133.99** |
| 410 | 238.25 | 57.85 | 64.82 | 60.46 | **421.37** |
| 411 | 45.75 | 54.47 | 52.29 | 37.04 | **189.55** |
| 412 | 0 | 0 | 0.54 | 0.54 | **1.09** |
| 413 | 12.53 | 15.80 | 2.72 | 9.26 | **40.31** |
| 415 | 38.13 | 26.15 | 31.05 | 19.06 | **114.38** |
| 416 | 30.18 | 46.84 | 27.23 | 16.34 | **120.59** |
| 417 | 19.06 | 178.66 | 0 | 7.63 | **205.35** |
| 418 | 61.01 | 69.72 | 29.96 | 44.66 | **205.35** |
| 420 | 448.36 | 310.73 | 286.51 | 177.02 | **1,222.63** |
| 421 | 294.78 | 264.72 | 234.22 | 190.10 | **983.81** |
| 422 | 160.14 | 171.58 | 104.04 | 73.53 | **509.28** |
| 423 | 118.20 | 81.14 | 61.01 | 96.95 | **357.29** |
| 424 | 132.90 | 139.44 | 118.20 | 99.68 | **490.22** |
| 425 | 92.60 | 70.81 | 42.49 | 53.38 | **259.27** |
| 426 | 1.09 | 17.43 | 31.59 | 23.42 | **73.53** |
| 427 | 90.96 | 143.25 | 93.14 | 75.71 | **403.07** |

| KY - KENTUCKY - Total | 2,837.89 | 2,412.77 | 1,810.43 | 1,487.54 | 8,548.63 |
|---|---|---|---|---|---|

STATE: LA - LOUISIANA

| REGISTRANT ZIP CODE 3 | 1ST QUARTER | 2ND QUARTER | 3RD QUARTER | 4TH QUARTER | TOTAL GRAMS |
|---|---|---|---|---|---|
| 700 | 813.44 | 577.37 | 515.27 | 498.93 | **2,405.01** |
| 701 | 407.64 | 165.59 | 109.48 | 108.94 | **791.65** |
| 703 | 363.85 | 361.89 | 276.16 | 192.27 | **1,194.17** |
| 704 | 607.76 | 602.90 | 473.33 | 413.96 | **2,097.96** |
| 705 | 1,270.67 | 1,307.75 | 946.67 | 727.16 | **4,252.24** |
| 706 | 484.97 | 494.29 | 407.97 | 287.60 | **1,674.83** |
| 707 | 344.79 | 398.17 | 344.24 | 284.33 | **1,371.52** |
| 708 | 697.90 | 525.08 | 467.34 | 308.29 | **1,998.61** |
| 710 | 226.59 | 295.22 | 206.98 | 206.44 | **935.23** |
| 711 | 669.42 | 772.37 | 555.58 | 370.39 | **2,367.76** |
| 712 | 546.04 | 514.73 | 483.14 | 259.27 | **1,803.18** |
| 713 | 275.61 | 279.42 | 231.49 | 165.59 | **952.11** |
| 714 | 257.64 | 194.45 | 130.73 | 145.98 | **728.79** |
| LA - LOUISIANA - Total | 6,966.31 | 6,489.23 | 5,148.39 | 3,969.14 | 22,573.07 |

STATE: MA - MASSACHUSETTS

| REGISTRANT ZIP CODE 3 | 1ST QUARTER | 2ND QUARTER | 3RD QUARTER | 4TH QUARTER | TOTAL GRAMS |
|---|---|---|---|---|---|
| 010 | 256.87 | 240.75 | 220.05 | 236.39 | **954.07** |
| 011 | 83.88 | 30.50 | 6.54 | 23.42 | **144.34** |
| 012 | 0 | 34.86 | 0 | 4.90 | **39.76** |
| 013 | 27.78 | 17.43 | 9.80 | 4.36 | **59.37** |
| 014 | 98.04 | 70.81 | 71.90 | 98.04 | **338.80** |
| 015 | 40.31 | 38.13 | 37.04 | 39.76 | **155.24** |
| 016 | 82.68 | 27.23 | 20.70 | 26.69 | **157.31** |
| 017 | 116.24 | 113.84 | 111.66 | 145.43 | **487.17** |
| 018 | 90.42 | 129.64 | 129.09 | 108.39 | **457.54** |
| 019 | 184.87 | 154.69 | 176.48 | 103.49 | **619.53** |
| 020 | 118.74 | 71.90 | 123.10 | 71.35 | **385.09** |
| 021 | 451.26 | 306.88 | 207.53 | 200.99 | **1,166.66** |
| 022 | 27.02 | 43.03 | 16.34 | 14.16 | **100.55** |
| 023 | 151.75 | 175.39 | 174.30 | 173.21 | **674.65** |
| 024 | 101.75 | 101.31 | 75.71 | 71.35 | **350.13** |
| 025 | 96.41 | 67.54 | 66.45 | 70.81 | **301.21** |
| 026 | 37.04 | 27.23 | 11.44 | 19.61 | **95.32** |
| 027 | 132.47 | 142.16 | 115.47 | 90.96 | **481.07** |
| MA - MASSACHUSETTS - Total | 2,097.53 | 1,793.33 | 1,573.60 | 1,503.34 | 6,967.80 |

STATE: MD - MARYLAND

| REGISTRANT ZIP CODE 3 | 1ST QUARTER | 2ND QUARTER | 3RD QUARTER | 4TH QUARTER | TOTAL GRAMS |
|---|---|---|---|---|---|

| | | | | | |
|---|---|---|---|---|---|
| 206 | 97.50 | 77.89 | 107.30 | 100.77 | **383.46** |
| 207 | 209.70 | 224.41 | 171.03 | 139.98 | **745.13** |
| 208 | 315.27 | 419.95 | 181.93 | 250.01 | **1,167.16** |
| 209 | 39.76 | 7.08 | 5.99 | 39.76 | **92.60** |
| 210 | 329.97 | 388.91 | 382.92 | 349.14 | **1,450.94** |
| 211 | 134.54 | 137.81 | 146.52 | 125.28 | **544.14** |
| 212 | 546.87 | 479.87 | 408.52 | 306.11 | **1,741.37** |
| 214 | 159.05 | 131.81 | 44.66 | 95.87 | **431.39** |
| 215 | 159.05 | 78.98 | 86.06 | 38.67 | **362.76** |
| 216 | 70.26 | 63.73 | 53.92 | 56.65 | **244.56** |
| 217 | 55.01 | 87.15 | 52.29 | 34.32 | **228.77** |
| 218 | 98.04 | 69.72 | 80.61 | 49.02 | **297.40** |
| 219 | 2.18 | 0.54 | 4.90 | 1.09 | **8.72** |
| **MD - MARYLAND - Total** | **2,217.20** | **2,167.86** | **1,726.66** | **1,586.67** | **7,698.40** |

**STATE: ME - MAINE**

| REGISTRANT ZIP CODE 3 | 1ST QUARTER | 2ND QUARTER | 3RD QUARTER | 4TH QUARTER | TOTAL GRAMS |
|---|---|---|---|---|---|
| 039 | 7.41 | 13.51 | 10.89 | 9.80 | **41.61** |
| 040 | 99.57 | 60.46 | 103.49 | 87.69 | **351.21** |
| 041 | 5.45 | 62.64 | 22.88 | 39.22 | **130.18** |
| 042 | 15.25 | 13.62 | 20.70 | 10.35 | **59.92** |
| 043 | 0.87 | 0 | 9.80 | 0 | **10.68** |
| 044 | 85.73 | 27.23 | 21.79 | 19.61 | **154.36** |
| 046 | 27.23 | 52.29 | 29.41 | 46.30 | **155.24** |
| 047 | 76.26 | 87.15 | 79.52 | 86.61 | **329.54** |
| 048 | 4.36 | 4.90 | 4.90 | 0 | **14.16** |
| 049 | 31.05 | 13.07 | 13.62 | 8.72 | **66.45** |
| **ME - MAINE - Total** | **353.18** | **334.87** | **317.01** | **308.29** | **1,313.35** |

**STATE: MI - MICHIGAN**

| REGISTRANT ZIP CODE 3 | 1ST QUARTER | 2ND QUARTER | 3RD QUARTER | 4TH QUARTER | TOTAL GRAMS |
|---|---|---|---|---|---|
| 480 | 349.69 | 286.51 | 236.39 | 243.48 | **1,116.06** |
| 481 | 370.93 | 344.24 | 206.65 | 186.28 | **1,108.11** |
| 482 | 32.68 | 71.90 | 39.22 | 44.66 | **188.46** |
| 483 | 100.11 | 82.79 | 58.83 | 73.53 | **315.27** |
| 484 | 77.35 | 133.45 | 124.19 | 131.81 | **466.80** |
| 485 | 43.58 | 32.68 | 42.49 | 52.29 | **171.03** |
| 486 | 330.06 | 391.09 | 184.65 | 219.51 | **1,125.30** |
| 487 | 218.96 | 9.80 | 13.62 | 20.15 | **262.54** |
| 488 | 221.80 | 265.81 | 174.30 | 208.07 | **869.97** |
| 489 | 76.80 | 43.58 | 60.46 | 126.37 | **307.20** |
| 490 | 170.49 | 230.40 | 136.72 | 148.16 | **685.76** |

| | | | | | |
|---|---|---|---|---|---|
| 491 | 4.36 | 0 | 0 | 1.09 | **5.45** |
| 492 | 139.55 | 96.95 | 92.60 | 131.81 | **460.91** |
| 493 | 130.73 | 93.69 | 43.58 | 97.50 | **365.49** |
| 494 | 69.07 | 53.92 | 49.02 | 69.72 | **241.73** |
| 495 | 123.64 | 70.81 | 72.44 | 23.97 | **290.86** |
| 496 | 71.90 | 57.74 | 54.47 | 47.39 | **231.49** |
| 497 | 181.93 | 173.21 | 155.78 | 206.44 | **717.35** |
| 498 | 139.22 | 73.53 | 57.41 | 21.79 | **291.95** |
| 499 | 37.04 | 32.68 | 35.95 | 33.23 | **138.90** |
| **MI - MICHIGAN - Total** | **2,889.87** | **2,544.78** | **1,838.76** | **2,087.24** | **9,360.65** |

**STATE: MN - MINNESOTA**

| REGISTRANT ZIP CODE 3 | 1ST QUARTER | 2ND QUARTER | 3RD QUARTER | 4TH QUARTER | TOTAL GRAMS |
|---|---|---|---|---|---|
| 550 | 83.34 | 78.44 | 59.92 | 31.05 | **252.74** |
| 551 | 20.70 | 15.80 | 42.49 | 26.69 | **105.67** |
| 553 | 68.09 | 65.91 | 46.84 | 58.83 | **239.66** |
| 554 | 79.96 | 155.78 | 43.03 | 84.43 | **363.20** |
| 556 | 4.36 | 4.36 | 4.36 | 4.36 | **17.43** |
| 557 | 40.42 | 56.65 | 35.40 | 41.40 | **173.86** |
| 558 | 67.00 | 40.31 | 38.67 | 17.97 | **163.95** |
| 559 | 56.10 | 67.54 | 22.33 | 28.32 | **174.30** |
| 560 | 26.69 | 27.78 | 13.07 | 8.17 | **75.71** |
| 561 | 1.09 | 1.09 | 4.90 | 0 | **7.08** |
| 562 | 11.98 | 33.77 | 23.42 | 23.97 | **93.14** |
| 563 | 49.57 | 59.92 | 31.59 | 65.36 | **206.44** |
| 564 | 40.57 | 33.44 | 5.45 | 4.90 | **84.36** |
| 565 | 11.98 | 8.72 | 24.51 | 13.07 | **58.28** |
| 566 | 7.08 | 1.09 | 9.26 | 20.15 | **37.58** |
| 567 | 17.43 | 31.05 | 18.52 | 31.05 | **98.04** |
| **MN - MINNESOTA - Total** | **586.35** | **681.62** | **423.77** | **459.72** | **2,151.45** |

**STATE: MO - MISSOURI**

| REGISTRANT ZIP CODE 3 | 1ST QUARTER | 2ND QUARTER | 3RD QUARTER | 4TH QUARTER | TOTAL GRAMS |
|---|---|---|---|---|---|
| 630 | 288.14 | 205.89 | 114.38 | 165.59 | **774.00** |
| 631 | 541.42 | 553.95 | 364.94 | 450.46 | **1,910.76** |
| 633 | 64.27 | 77.35 | 62.64 | 25.60 | **229.86** |
| 634 | 40.85 | 21.79 | 28.32 | 18.52 | **109.48** |
| 635 | 46.84 | 44.12 | 37.58 | 42.49 | **171.03** |
| 636 | 53.92 | 48.48 | 34.86 | 61.55 | **198.81** |
| 637 | 35.95 | 35.95 | 40.85 | 15.80 | **128.55** |
| 638 | 84.43 | 76.26 | 39.22 | 39.76 | **239.66** |
| 639 | 41.94 | 59.92 | 57.19 | 67.54 | **226.59** |

| | | | | | |
|---|---|---|---|---|---|
| 640 | 312.11 | 193.91 | 251.65 | 239.12 | **996.78** |
| 641 | 203.71 | 326.27 | 204.26 | 166.67 | **900.91** |
| 644 | 44.12 | 19.06 | 31.59 | 15.25 | **110.03** |
| 645 | 53.92 | 29.96 | 11.98 | 34.86 | **130.73** |
| 646 | 7.63 | 10.89 | 28.32 | 7.63 | **54.47** |
| 647 | 53.38 | 40.31 | 33.23 | 49.02 | **175.93** |
| 648 | 141.62 | 109.48 | 98.59 | 85.52 | **435.21** |
| 650 | 32.14 | 65.36 | 32.14 | 36.49 | **166.13** |
| 651 | 77.35 | 89.87 | 55.01 | 73.53 | **295.77** |
| 652 | 366.03 | 285.42 | 241.84 | 205.35 | **1,098.63** |
| 653 | 104.80 | 79.74 | 83.88 | 83.88 | **352.30** |
| 654 | 52.29 | 70.81 | 66.45 | 56.65 | **246.20** |
| 655 | 190.64 | 157.96 | 102.40 | 117.65 | **568.65** |
| 656 | 92.60 | 43.58 | 70.81 | 68.63 | **275.61** |
| 657 | 245.65 | 202.62 | 205.89 | 178.11 | **832.28** |
| 658 | 113.30 | 245.65 | 183.56 | 143.80 | **686.31** |
| **MO - MISSOURI - Total** | **3,289.04** | **3,094.59** | **2,481.60** | **2,449.46** | **11,314.68** |

**STATE: MS - MISSISSIPPI**

| REGISTRANT ZIP CODE 3 | 1ST QUARTER | 2ND QUARTER | 3RD QUARTER | 4TH QUARTER | **TOTAL GRAMS** |
|---|---|---|---|---|---|
| 386 | 480.96 | 419.41 | 436.29 | 299.58 | **1,636.24** |
| 387 | 256.46 | 339.25 | 231.71 | 173.76 | **1,001.18** |
| 388 | 314.72 | 245.35 | 203.71 | 183.56 | **947.34** |
| 389 | 186.09 | 189.55 | 117.00 | 165.04 | **657.68** |
| 390 | 236.07 | 216.79 | 155.78 | 159.59 | **768.23** |
| 391 | 273.43 | 286.51 | 259.82 | 105.67 | **925.42** |
| 392 | 333.35 | 293.04 | 314.83 | 261.99 | **1,203.21** |
| 393 | 394.35 | 509.81 | 317.55 | 251.65 | **1,473.36** |
| 394 | 323.83 | 487.91 | 379.65 | 340.97 | **1,532.36** |
| 395 | 826.75 | 525.51 | 567.02 | 535.43 | **2,454.71** |
| 396 | 101.31 | 75.71 | 78.44 | 45.21 | **300.67** |
| 397 | 200.21 | 126.48 | 144.30 | 91.51 | **562.49** |
| **MS - MISSISSIPPI - Total** | **3,927.52** | **3,715.31** | **3,206.10** | **2,613.96** | **13,462.89** |

**STATE: MT - MONTANA**

| REGISTRANT ZIP CODE 3 | 1ST QUARTER | 2ND QUARTER | 3RD QUARTER | 4TH QUARTER | **TOTAL GRAMS** |
|---|---|---|---|---|---|
| 590 | 20.26 | 9.80 | 22.88 | 6.54 | **59.48** |
| 591 | 113.62 | 79.52 | 41.40 | 52.29 | **286.83** |
| 592 | 27.78 | 14.71 | 21.79 | 14.16 | **78.44** |
| 593 | 4.36 | 34.86 | 16.34 | 17.43 | **72.99** |
| 594 | 168.85 | 89.33 | 95.87 | 62.09 | **416.14** |
| 595 | 8.72 | 7.63 | 13.07 | 19.61 | **49.02** |

| | | | | | |
|---|---|---|---|---|---|
| 596 | 80.61 | 89.33 | 81.70 | 26.15 | **277.79** |
| 597 | 41.40 | 45.21 | 45.75 | 25.06 | **157.41** |
| 598 | 81.16 | 100.22 | 72.44 | 61.01 | **314.83** |
| 599 | 97.50 | 106.21 | 76.80 | 40.85 | **321.37** |
| **MT - MONTANA - Total** | **644.26** | **576.82** | **488.04** | **325.18** | **2,034.30** |

**STATE: NC - NORTH CAROLINA**

| REGISTRANT ZIP CODE 3 | 1ST QUARTER | 2ND QUARTER | 3RD QUARTER | 4TH QUARTER | TOTAL GRAMS |
|---|---|---|---|---|---|
| 270 | 123.10 | 124.73 | 113.30 | 116.56 | **477.69** |
| 271 | 166.67 | 142.71 | 83.88 | 70.26 | **463.53** |
| 272 | 334.44 | 263.08 | 247.83 | 226.05 | **1,071.40** |
| 273 | 191.73 | 135.08 | 100.77 | 128.55 | **556.13** |
| 274 | 130.73 | 123.64 | 80.61 | 82.79 | **417.78** |
| 275 | 419.95 | 451.55 | 478.78 | 289.23 | **1,639.51** |
| 276 | 147.61 | 183.02 | 216.24 | 169.94 | **716.81** |
| 277 | 21.79 | 25.06 | 40.31 | 16.34 | **103.49** |
| 278 | 291.52 | 366.03 | 324.63 | 287.60 | **1,269.78** |
| 279 | 237.48 | 228.22 | 203.71 | 200.45 | **869.87** |
| 280 | 438.58 | 388.36 | 325.18 | 311.02 | **1,463.14** |
| 281 | 478.24 | 398.17 | 388.36 | 413.42 | **1,678.18** |
| 282 | 395.12 | 408.52 | 163.95 | 183.56 | **1,151.14** |
| 283 | 337.60 | 290.32 | 301.21 | 245.65 | **1,174.78** |
| 284 | 202.51 | 228.22 | 148.70 | 223.87 | **803.31** |
| 285 | 274.52 | 232.04 | 188.46 | 189.01 | **884.03** |
| 286 | 551.44 | 563.21 | 428.67 | 345.88 | **1,889.19** |
| 287 | 279.97 | 214.61 | 265.26 | 234.22 | **994.05** |
| 288 | 13.07 | 27.23 | 43.03 | 9.80 | **93.14** |
| 289 | 9.80 | 16.34 | 4.36 | 8.72 | **39.22** |
| **NC - NORTH CAROLINA - Total** | **5,045.88** | **4,810.14** | **4,147.25** | **3,752.90** | **17,756.16** |

**STATE: ND - NORTH DAKOTA**

| REGISTRANT ZIP CODE 3 | 1ST QUARTER | 2ND QUARTER | 3RD QUARTER | 4TH QUARTER | TOTAL GRAMS |
|---|---|---|---|---|---|
| 580 | 4.36 | 5.45 | 20.15 | 26.15 | **56.10** |
| 581 | 35.40 | 31.05 | 20.70 | 7.63 | **94.78** |
| 582 | 14.71 | 8.72 | 27.78 | 9.80 | **61.01** |
| 583 | 28.32 | 25.60 | 33.77 | 19.06 | **106.76** |
| 584 | 5.66 | 11.44 | 5.99 | 15.25 | **38.35** |
| 585 | 40.52 | 50.11 | 72.44 | 68.63 | **231.71** |
| 586 | 13.73 | 9.80 | 5.45 | 4.36 | **33.33** |
| 587 | 35.95 | 8.72 | 29.41 | 34.32 | **108.39** |
| 588 | 1.31 | 9.80 | 4.36 | 7.08 | **22.55** |
| **ND - NORTH DAKOTA - Total** | **179.96** | **160.68** | **220.05** | **192.27** | **752.98** |

**STATE: NE - NEBRASKA**

| REGISTRANT ZIP CODE 3 | 1ST QUARTER | 2ND QUARTER | 3RD QUARTER | 4TH QUARTER | TOTAL GRAMS |
|---|---|---|---|---|---|
| 680 | 43.03 | 64.27 | 57.19 | 44.12 | **208.62** |
| 681 | 244.02 | 107.85 | 102.95 | 140.53 | **595.34** |
| 683 | 2.72 | 5.45 | 3.27 | 7.08 | **18.52** |
| 684 | 7.63 | 2.18 | 3.27 | 26.15 | **39.22** |
| 685 | 83.75 | 56.10 | 72.99 | 27.78 | **240.62** |
| 686 | 7.63 | 11.44 | 11.98 | 8.72 | **39.76** |
| 687 | 17.97 | 50.66 | 19.61 | 26.69 | **114.93** |
| 688 | 60.46 | 52.83 | 52.29 | 29.41 | **195.00** |
| 689 | 47.28 | 76.26 | 52.83 | 59.92 | **236.29** |
| 690 | 11.33 | 11.44 | 9.80 | 4.36 | **36.93** |
| 691 | 27.23 | 14.71 | 35.40 | 26.69 | **104.04** |
| 692 | 14.16 | 10.89 | 8.17 | 10.89 | **44.12** |
| 693 | 29.41 | 10.89 | 9.26 | 13.62 | **63.18** |
| **NE - NEBRASKA - Total** | **596.63** | **474.97** | **439.02** | **425.95** | **1,936.56** |

**STATE: NH - NEW HAMPSHIRE**

| REGISTRANT ZIP CODE 3 | 1ST QUARTER | 2ND QUARTER | 3RD QUARTER | 4TH QUARTER | TOTAL GRAMS |
|---|---|---|---|---|---|
| 030 | 46.84 | 91.51 | 28.87 | 58.83 | **226.05** |
| 031 | 59.70 | 49.24 | 63.18 | 52.29 | **224.41** |
| 032 | 108.83 | 88.24 | 67.54 | 73.53 | **338.14** |
| 033 | 217.88 | 187.37 | 164.50 | 180.84 | **750.58** |
| 034 | 28.32 | 31.59 | 25.06 | 15.25 | **100.22** |
| 035 | 5.45 | 12.53 | 1.09 | 4.36 | **23.42** |
| 037 | 26.15 | 17.43 | 29.41 | 21.79 | **94.78** |
| 038 | 151.42 | 95.87 | 94.23 | 89.87 | **431.39** |
| **NH - NEW HAMPSHIRE - Total** | **644.58** | **573.77** | **473.88** | **496.76** | **2,188.99** |

**STATE: NJ - NEW JERSEY**

| REGISTRANT ZIP CODE 3 | 1ST QUARTER | 2ND QUARTER | 3RD QUARTER | 4TH QUARTER | TOTAL GRAMS |
|---|---|---|---|---|---|
| 070 | 258.51 | 204.26 | 248.38 | 186.83 | **897.97** |
| 071 | 8.72 | 21.24 | 5.99 | 9.26 | **45.21** |
| 072 | 3.27 | 1.09 | 1.09 | 43.58 | **49.02** |
| 073 | 38.67 | 49.57 | 31.59 | 0 | **119.83** |
| 074 | 55.23 | 62.64 | 33.77 | 28.32 | **179.96** |
| 075 | 80.29 | 26.15 | 14.16 | 23.42 | **144.02** |
| 076 | 106.21 | 139.55 | 135.08 | 177.57 | **558.41** |
| 077 | 197.61 | 214.06 | 230.40 | 124.19 | **766.27** |
| 078 | 77.89 | 89.33 | 81.16 | 51.75 | **300.12** |
| 079 | 76.26 | 47.93 | 60.46 | 47.39 | **232.04** |
| 080 | 281.93 | 211.34 | 147.07 | 110.57 | **750.91** |

| | | | | | |
|---|---|---|---|---|---|
| 081 | 10.89 | 4.90 | 17.43 | 4.90 | **38.13** |
| 082 | 23.97 | 74.08 | 33.77 | 47.39 | **179.20** |
| 083 | 13.07 | 13.62 | 7.08 | 5.99 | **39.76** |
| 084 | 0 | 0 | 4.36 | 1.63 | **5.99** |
| 085 | 52.29 | 37.04 | 51.20 | 26.15 | **166.67** |
| 086 | 30.50 | 24.51 | 29.41 | 13.07 | **97.50** |
| 087 | 51.64 | 62.09 | 86.61 | 28.32 | **228.66** |
| 088 | 414.40 | 328.45 | 262.54 | 358.40 | **1,363.79** |
| 089 | 14.16 | 43.03 | 13.62 | 35.95 | **106.76** |
| **NJ - NEW JERSEY - Total** | **1,795.51** | **1,654.87** | **1,495.17** | **1,324.68** | **6,270.22** |

**STATE: NM - NEW MEXICO**

| REGISTRANT ZIP CODE 3 | 1ST QUARTER | 2ND QUARTER | 3RD QUARTER | 4TH QUARTER | **TOTAL GRAMS** |
|---|---|---|---|---|---|
| 870 | 117.65 | 137.26 | 63.18 | 68.63 | **386.73** |
| 871 | 686.85 | 570.29 | 514.73 | 401.98 | **2,173.85** |
| 873 | 0.54 | 1.09 | 9.80 | 4.36 | **15.80** |
| 874 | 13.62 | 37.04 | 26.15 | 13.07 | **89.87** |
| 875 | 171.03 | 166.13 | 161.23 | 68.63 | **567.02** |
| 877 | 33.23 | 20.15 | 32.68 | 19.61 | **105.67** |
| 878 | 8.72 | 0 | 17.43 | 0 | **26.15** |
| 879 | 0 | 6.54 | 21.79 | 8.72 | **37.04** |
| 880 | 355.90 | 276.70 | 164.50 | 110.57 | **907.67** |
| 881 | 73.53 | 85.52 | 95.32 | 31.05 | **285.42** |
| 882 | 143.80 | 152.51 | 76.80 | 60.46 | **433.57** |
| 883 | 2.72 | 30.50 | 14.16 | 13.62 | **61.01** |
| 884 | 0 | 1.09 | 0 | 4.36 | **5.45** |
| **NM - NEW MEXICO - Total** | **1,607.59** | **1,484.82** | **1,197.77** | **805.05** | **5,095.22** |

**STATE: NV - NEVADA**

| REGISTRANT ZIP CODE 3 | 1ST QUARTER | 2ND QUARTER | 3RD QUARTER | 4TH QUARTER | **TOTAL GRAMS** |
|---|---|---|---|---|---|
| 890 | 317.55 | 225.50 | 168.85 | 200.45 | **912.35** |
| 891 | 1,050.27 | 876.40 | 715.72 | 602.97 | **3,245.36** |
| 893 | 6.54 | 11.98 | 2.18 | 2.18 | **22.88** |
| 894 | 88.24 | 38.13 | 28.32 | 28.32 | **183.02** |
| 895 | 181.93 | 131.27 | 90.96 | 97.50 | **501.66** |
| 897 | 45.75 | 77.35 | 31.59 | 16.34 | **171.03** |
| 898 | 14.71 | 31.59 | 17.97 | 12.53 | **76.80** |
| **NV - NEVADA - Total** | **1,704.98** | **1,392.22** | **1,055.60** | **960.28** | **5,113.09** |

**STATE: NY - NEW YORK**

| REGISTRANT ZIP CODE 3 | 1ST QUARTER | 2ND QUARTER | 3RD QUARTER | 4TH QUARTER | **TOTAL GRAMS** |
|---|---|---|---|---|---|
| 100 | 677.92 | 615.50 | 464.62 | 272.89 | **2,030.92** |
| 101 | 38.13 | 55.56 | 32.68 | 74.08 | **200.45** |

| | | | | | |
|---|---|---|---|---|---|
| 103 | 120.48 | 177.57 | 40.31 | 114.38 | **452.74** |
| 104 | 108.94 | 116.02 | 168.85 | 69.72 | **463.53** |
| 105 | 131.27 | 178.66 | 159.05 | 100.77 | **569.74** |
| 106 | 18.52 | 5.45 | 14.71 | 4.36 | **43.03** |
| 107 | 32.14 | 71.90 | 50.11 | 46.84 | **200.99** |
| 108 | 5.66 | 28.87 | 11.44 | 9.80 | **55.78** |
| 109 | 43.03 | 56.10 | 81.70 | 117.65 | **298.49** |
| 110 | 64.82 | 138.35 | 87.15 | 67.54 | **357.86** |
| 111 | 0 | 2.18 | 3.81 | 4.90 | **10.89** |
| 112 | 294.68 | 105.67 | 135.63 | 116.02 | **651.99** |
| 113 | 72.99 | 93.69 | 93.14 | 114.93 | **374.75** |
| 114 | 51.20 | 38.13 | 27.23 | 38.67 | **155.24** |
| 115 | 140.75 | 145.43 | 210.79 | 239.66 | **736.64** |
| 117 | 147.61 | 193.36 | 161.23 | 106.76 | **608.96** |
| 118 | 69.72 | 44.12 | 78.44 | 61.55 | **253.82** |
| 119 | 3.81 | 19.61 | 10.89 | 5.99 | **40.31** |
| 120 | 79.31 | 75.17 | 52.29 | 91.51 | **298.27** |
| 121 | 48.48 | 91.51 | 105.12 | 23.97 | **269.08** |
| 122 | 57.74 | 22.33 | 27.23 | 16.89 | **124.19** |
| 123 | 89.87 | 136.72 | 87.15 | 79.52 | **393.26** |
| 124 | 78.44 | 56.65 | 66.45 | 87.15 | **288.68** |
| 125 | 51.20 | 51.20 | 20.70 | 77.89 | **200.99** |
| 126 | 50.66 | 45.21 | 103.49 | 9.80 | **209.16** |
| 127 | 16.89 | 18.52 | 30.50 | 13.62 | **79.52** |
| 128 | 119.50 | 57.74 | 130.18 | 117.11 | **424.53** |
| 129 | 39.65 | 96.95 | 23.97 | 35.40 | **195.98** |
| 130 | 219.51 | 271.78 | 221.14 | 175.39 | **887.82** |
| 131 | 40.31 | 44.66 | 53.92 | 9.80 | **148.70** |
| 132 | 55.01 | 59.37 | 67.00 | 61.55 | **242.93** |
| 133 | 59.89 | 43.84 | 32.38 | 53.38 | **189.49** |
| 134 | 32.35 | 23.09 | 18.52 | 44.12 | **118.09** |
| 135 | 9.26 | 28.32 | 14.71 | 13.62 | **65.91** |
| 136 | 9.80 | 10.35 | 8.17 | 14.16 | **42.49** |
| 137 | 17.43 | 2.18 | 4.36 | 3.27 | **27.23** |
| 138 | 67.54 | 74.62 | 26.15 | 23.97 | **192.27** |
| 139 | 13.07 | 3.27 | 3.27 | 19.61 | **39.22** |
| 140 | 52.29 | 62.09 | 49.02 | 35.95 | **199.36** |
| 141 | 71.90 | 78.44 | 34.86 | 39.22 | **224.41** |
| 142 | 977.13 | 349.69 | 81.70 | 147.07 | **1,555.58** |
| 143 | 61.01 | 86.06 | 86.06 | 54.47 | **287.60** |

| | | | | | |
|---|---|---|---|---|---|
| 144 | 16.34 | 20.70 | 22.88 | 8.17 | **68.09** |
| 145 | 49.57 | 76.80 | 24.51 | 23.97 | **174.84** |
| 146 | 137.26 | 105.12 | 144.89 | 68.09 | **455.36** |
| 147 | 45.75 | 61.01 | 40.31 | 22.88 | **169.94** |
| 148 | 196.09 | 138.35 | 99.13 | 110.57 | **544.14** |
| 149 | 43.03 | 25.60 | 20.70 | 10.89 | **100.22** |
| **NY - NEW YORK - Total** | **4,827.94** | **4,303.49** | **3,532.54** | **3,059.51** | **15,723.47** |

**STATE: OH - OHIO**

| REGISTRANT ZIP CODE 3 | 1ST QUARTER | 2ND QUARTER | 3RD QUARTER | 4TH QUARTER | TOTAL GRAMS |
|---|---|---|---|---|---|
| 430 | 203.71 | 139.44 | 101.86 | 101.31 | **546.32** |
| 431 | 267.44 | 115.47 | 138.35 | 185.19 | **706.46** |
| 432 | 674.65 | 324.63 | 154.69 | 190.64 | **1,344.62** |
| 433 | 50.11 | 10.89 | 4.36 | 10.89 | **76.26** |
| 434 | 132.90 | 56.65 | 58.83 | 71.90 | **320.28** |
| 435 | 49.02 | 80.61 | 36.49 | 54.47 | **220.60** |
| 436 | 91.51 | 46.84 | 57.85 | 48.48 | **244.67** |
| 437 | 250.01 | 393.26 | 102.40 | 137.81 | **883.48** |
| 439 | 99.68 | 91.51 | 62.09 | 28.87 | **282.15** |
| 440 | 205.46 | 233.13 | 286.51 | 197.72 | **922.81** |
| 441 | 927.82 | 774.00 | 574.10 | 729.34 | **3,005.26** |
| 442 | 103.27 | 57.74 | 47.93 | 57.74 | **266.68** |
| 443 | 73.53 | 51.20 | 34.86 | 23.42 | **183.02** |
| 444 | 44.66 | 80.61 | 40.31 | 45.21 | **210.79** |
| 445 | 49.78 | 55.01 | 62.09 | 46.30 | **213.19** |
| 446 | 89.05 | 81.70 | 56.65 | 64.27 | **291.67** |
| 447 | 43.47 | 38.13 | 52.29 | 33.77 | **167.65** |
| 448 | 221.40 | 140.53 | 163.12 | 131.27 | **656.33** |
| 449 | 87.15 | 64.82 | 14.71 | 20.70 | **187.37** |
| 450 | 254.37 | 150.33 | 124.19 | 112.21 | **641.10** |
| 451 | 93.14 | 66.45 | 128.00 | 57.19 | **344.79** |
| 452 | 890.24 | 683.02 | 544.83 | 636.74 | **2,754.83** |
| 453 | 109.70 | 153.06 | 122.01 | 106.76 | **491.53** |
| 454 | 293.91 | 170.16 | 201.32 | 178.44 | **843.83** |
| 455 | 17.43 | 13.07 | 21.79 | 68.63 | **120.92** |
| 456 | 43.58 | 42.49 | 14.71 | 10.35 | **111.12** |
| 457 | 11.98 | 40.85 | 11.44 | 9.80 | **74.08** |
| 458 | 139.44 | 149.79 | 115.47 | 78.44 | **483.14** |
| **OH - OHIO - Total** | **5,518.43** | **4,305.41** | **3,333.24** | **3,437.85** | **16,594.92** |

**STATE: OK - OKLAHOMA**

| REGISTRANT ZIP CODE 3 | 1ST QUARTER | 2ND QUARTER | 3RD QUARTER | 4TH QUARTER | TOTAL GRAMS |
|---|---|---|---|---|---|

| | | | | | |
|---|---|---|---|---|---|
| 730 | 1,060.51 | 892.74 | 872.59 | 646.54 | **3,472.38** |
| 731 | 1,779.49 | 1,606.68 | 1,581.23 | 1,130.23 | **6,097.62** |
| 734 | 41.29 | 33.23 | 38.13 | 40.85 | **153.49** |
| 735 | 614.08 | 491.31 | 471.37 | 339.89 | **1,916.65** |
| 736 | 58.83 | 49.02 | 63.18 | 33.77 | **204.80** |
| 737 | 86.87 | 87.96 | 92.60 | 77.89 | **345.31** |
| 738 | 17.97 | 4.90 | 8.17 | 15.25 | **46.30** |
| 739 | 32.68 | 20.70 | 20.15 | 13.07 | **86.61** |
| 740 | 1,070.31 | 792.52 | 673.23 | 483.68 | **3,019.75** |
| 741 | 671.06 | 614.41 | 516.91 | 399.80 | **2,202.17** |
| 743 | 123.10 | 129.64 | 114.93 | 94.78 | **462.44** |
| 744 | 464.62 | 302.85 | 266.35 | 136.17 | **1,169.99** |
| 745 | 65.91 | 81.70 | 76.80 | 30.50 | **254.91** |
| 746 | 29.41 | 42.49 | 17.43 | 16.89 | **106.21** |
| 747 | 153.60 | 118.20 | 145.98 | 96.95 | **514.73** |
| 748 | 330.08 | 189.01 | 177.02 | 192.27 | **888.39** |
| 749 | 152.47 | 90.94 | 41.94 | 46.84 | **332.19** |
| **OK - OKLAHOMA - Total** | **6,752.27** | **5,548.27** | **5,178.02** | **3,795.38** | **21,273.95** |

**STATE: OR - OREGON**

| REGISTRANT ZIP CODE 3 | 1ST QUARTER | 2ND QUARTER | 3RD QUARTER | 4TH QUARTER | **TOTAL GRAMS** |
|---|---|---|---|---|---|
| 970 | 128.66 | 148.70 | 132.36 | 40.31 | **450.02** |
| 971 | 148.92 | 138.35 | 102.40 | 94.23 | **483.90** |
| 972 | 265.26 | 339.34 | 318.10 | 214.06 | **1,136.76** |
| 973 | 193.91 | 149.79 | 173.21 | 104.04 | **620.94** |
| 974 | 110.24 | 206.11 | 169.40 | 177.02 | **662.78** |
| 975 | 211.99 | 142.71 | 153.60 | 107.85 | **616.15** |
| 976 | 29.20 | 20.70 | 9.26 | 14.71 | **73.86** |
| 977 | 39.22 | 60.46 | 39.76 | 53.38 | **192.82** |
| 978 | 69.72 | 79.52 | 60.46 | 65.91 | **275.61** |
| 979 | 2.72 | 13.62 | 20.70 | 20.70 | **57.74** |
| **OR - OREGON - Total** | **1,199.84** | **1,299.30** | **1,179.25** | **892.20** | **4,570.58** |

**STATE: PA - PENNSYLVANIA**

| REGISTRANT ZIP CODE 3 | 1ST QUARTER | 2ND QUARTER | 3RD QUARTER | 4TH QUARTER | **TOTAL GRAMS** |
|---|---|---|---|---|---|
| 150 | 91.51 | 67.54 | 55.56 | 76.80 | **291.41** |
| 151 | 54.36 | 111.12 | 39.22 | 96.41 | **301.10** |
| 152 | 186.94 | 218.96 | 196.09 | 103.49 | **705.48** |
| 153 | 38.67 | 52.29 | 35.40 | 0.54 | **126.91** |
| 154 | 15.47 | 31.59 | 9.26 | 13.07 | **69.39** |
| 155 | 14.16 | 2.18 | 5.45 | 7.63 | **29.41** |
| 156 | 54.90 | 50.11 | 32.68 | 29.96 | **167.65** |

| | | | | | |
|---|---|---|---|---|---|
| 157 | 11.44 | 22.33 | 7.63 | 21.24 | **62.64** |
| 158 | 76.04 | 83.88 | 76.26 | 67.54 | **303.72** |
| 159 | 65.36 | 50.11 | 60.46 | 46.84 | **222.78** |
| 160 | 26.15 | 17.43 | 27.23 | 33.77 | **104.58** |
| 161 | 39.76 | 25.06 | 26.69 | 14.16 | **105.67** |
| 162 | 5.99 | 9.26 | 0 | 0 | **15.25** |
| 163 | 35.40 | 19.61 | 41.94 | 23.42 | **120.38** |
| 164 | 57.19 | 31.05 | 30.50 | 35.40 | **154.15** |
| 165 | 43.03 | 56.65 | 50.66 | 40.31 | **190.64** |
| 166 | 67.43 | 50.11 | 40.31 | 18.52 | **176.37** |
| 167 | 1.09 | 1.09 | 5.45 | 0 | **7.63** |
| 168 | 34.86 | 28.87 | 44.66 | 9.80 | **118.20** |
| 169 | 0 | 20.70 | 2.18 | 1.09 | **23.97** |
| 170 | 128.00 | 175.39 | 145.43 | 118.74 | **567.56** |
| 171 | 42.49 | 52.83 | 36.49 | 34.86 | **166.67** |
| 172 | 4.79 | 12.64 | 1.09 | 6.54 | **25.06** |
| 173 | 198.81 | 160.14 | 121.47 | 165.04 | **645.45** |
| 174 | 133.99 | 137.26 | 71.90 | 9.80 | **352.96** |
| 175 | 33.77 | 41.94 | 37.04 | 40.85 | **153.60** |
| 176 | 37.04 | 54.47 | 32.68 | 17.43 | **141.62** |
| 177 | 43.58 | 29.96 | 21.79 | 38.13 | **133.45** |
| 178 | 34.86 | 41.40 | 29.41 | 34.32 | **139.98** |
| 179 | 8.72 | 23.97 | 0 | 4.90 | **37.58** |
| 180 | 90.96 | 59.92 | 43.03 | 76.26 | **270.17** |
| 181 | 108.39 | 89.33 | 98.59 | 72.44 | **368.75** |
| 182 | 34.86 | 37.58 | 33.77 | 36.49 | **142.71** |
| 183 | 10.89 | 0.54 | 10.89 | 0 | **22.33** |
| 184 | 32.68 | 9.80 | 17.43 | 14.16 | **74.08** |
| 185 | 87.15 | 37.04 | 39.22 | 27.78 | **191.19** |
| 186 | 34.86 | 13.62 | 9.80 | 17.43 | **75.71** |
| 187 | 314.07 | 286.51 | 180.29 | 159.05 | **939.91** |
| 188 | 168.31 | 64.51 | 106.98 | 108.39 | **448.19** |
| 189 | 26.15 | 38.67 | 11.98 | 13.07 | **89.87** |
| 190 | 464.18 | 461.90 | 334.98 | 324.63 | **1,585.69** |
| 191 | 289.12 | 238.03 | 165.59 | 153.60 | **846.34** |
| 193 | 120.38 | 90.96 | 128.00 | 81.70 | **421.04** |
| 194 | 90.42 | 62.64 | 94.23 | 98.04 | **345.33** |
| 195 | 30.50 | 26.15 | 8.72 | 21.79 | **87.15** |
| 196 | 44.12 | 2.18 | 57.74 | 52.29 | **156.33** |
| **PA - PENNSYLVANIA - Total** | **3,532.84** | **3,199.30** | **2,626.16** | **2,367.76** | **11,726.05** |

**STATE: PR - PUERTO RICO**

| REGISTRANT ZIP CODE 3 | 1ST QUARTER | 2ND QUARTER | 3RD QUARTER | 4TH QUARTER | TOTAL GRAMS |
|---|---|---|---|---|---|
| 006 | 774.00 | 513.64 | 1,347.01 | 1,229.36 | **3,864.01** |
| 007 | 473.33 | 491.31 | 1,228.27 | 962.46 | **3,155.37** |
| 009 | 1,198.86 | 1,219.01 | 1,660.75 | 1,384.60 | **5,463.22** |
| **PR - PUERTO RICO - Total** | **2,446.19** | **2,223.96** | **4,236.03** | **3,576.42** | **12,482.60** |

**STATE: RI - RHODE ISLAND**

| REGISTRANT ZIP CODE 3 | 1ST QUARTER | 2ND QUARTER | 3RD QUARTER | 4TH QUARTER | TOTAL GRAMS |
|---|---|---|---|---|---|
| 028 | 80.07 | 62.64 | 52.83 | 15.25 | **210.79** |
| 029 | 96.95 | 163.41 | 117.65 | 69.18 | **447.19** |
| **RI - RHODE ISLAND - Total** | **177.02** | **226.05** | **170.49** | **84.43** | **657.98** |

**STATE: SC - SOUTH CAROLINA**

| REGISTRANT ZIP CODE 3 | 1ST QUARTER | 2ND QUARTER | 3RD QUARTER | 4TH QUARTER | TOTAL GRAMS |
|---|---|---|---|---|---|
| 290 | 595.89 | 489.13 | 472.24 | 313.74 | **1,871.00** |
| 291 | 585.43 | 437.93 | 320.28 | 409.61 | **1,753.24** |
| 292 | 739.69 | 604.06 | 546.87 | 406.88 | **2,297.49** |
| 293 | 275.07 | 267.44 | 293.59 | 196.09 | **1,032.18** |
| 294 | 851.35 | 830.65 | 652.54 | 745.68 | **3,080.21** |
| 295 | 537.06 | 532.16 | 460.26 | 384.55 | **1,914.03** |
| 296 | 883.16 | 782.50 | 784.35 | 592.08 | **3,042.08** |
| 297 | 196.63 | 150.33 | 139.44 | 77.35 | **563.75** |
| 298 | 346.42 | 311.56 | 362.76 | 218.96 | **1,239.71** |
| 299 | 123.64 | 136.17 | 139.44 | 72.99 | **472.24** |
| **SC - SOUTH CAROLINA - Total** | **5,134.33** | **4,541.93** | **4,171.76** | **3,417.91** | **17,265.94** |

**STATE: SD - SOUTH DAKOTA**

| REGISTRANT ZIP CODE 3 | 1ST QUARTER | 2ND QUARTER | 3RD QUARTER | 4TH QUARTER | TOTAL GRAMS |
|---|---|---|---|---|---|
| 570 | 138.35 | 113.84 | 131.27 | 100.22 | **483.68** |
| 571 | 166.67 | 233.67 | 118.74 | 62.09 | **581.18** |
| 572 | 37.58 | 32.68 | 46.84 | 43.03 | **160.14** |
| 573 | 53.38 | 51.20 | 26.15 | 14.71 | **145.43** |
| 574 | 27.23 | 14.71 | 15.80 | 25.06 | **82.79** |
| 575 | 0.54 | 0 | 0 | 8.17 | **8.72** |
| 576 | 0 | 2.18 | 0 | 1.09 | **3.27** |
| 577 | 108.07 | 146.52 | 87.69 | 75.71 | **417.99** |
| **SD - SOUTH DAKOTA - Total** | **531.83** | **594.80** | **426.49** | **330.08** | **1,883.20** |

**STATE: TN - TENNESSEE**

| REGISTRANT ZIP CODE 3 | 1ST QUARTER | 2ND QUARTER | 3RD QUARTER | 4TH QUARTER | TOTAL GRAMS |
|---|---|---|---|---|---|
| 370 | 1,118.79 | 931.96 | 817.03 | 477.69 | **3,345.47** |
| 371 | 1,004.40 | 861.15 | 821.39 | 450.46 | **3,137.40** |
| 372 | 747.86 | 493.49 | 305.03 | 291.95 | **1,838.32** |

| | | | | | |
|---|---|---|---|---|---|
| 373 | 640.49 | 638.09 | 447.19 | 415.60 | **2,141.36** |
| 374 | 403.07 | 307.20 | 179.20 | 220.60 | **1,110.07** |
| 376 | 213.52 | 311.56 | 181.93 | 173.76 | **880.76** |
| 377 | 294.68 | 211.34 | 164.50 | 114.93 | **785.44** |
| 378 | 318.10 | 275.61 | 164.50 | 169.94 | **928.15** |
| 379 | 297.38 | 302.30 | 195.54 | 172.12 | **967.34** |
| 380 | 356.23 | 284.33 | 215.41 | 166.67 | **1,022.64** |
| 381 | 775.64 | 661.25 | 612.77 | 450.46 | **2,500.12** |
| 382 | 157.13 | 107.56 | 108.39 | 82.25 | **455.34** |
| 383 | 455.36 | 391.63 | 333.89 | 264.72 | **1,445.60** |
| 384 | 257.09 | 227.68 | 209.70 | 142.71 | **837.18** |
| 385 | 283.78 | 144.34 | 133.99 | 106.21 | **668.33** |
| **TN - TENNESSEE - Total** | **7,323.50** | **6,149.50** | **4,890.47** | **3,700.06** | **22,063.53** |

**STATE: TX - TEXAS**

| REGISTRANT ZIP CODE 3 | 1ST QUARTER | 2ND QUARTER | 3RD QUARTER | 4TH QUARTER | TOTAL GRAMS |
|---|---|---|---|---|---|
| 750 | 786.42 | 782.43 | 588.96 | 563.21 | **2,721.02** |
| 751 | 241.84 | 168.85 | 100.77 | 92.60 | **604.06** |
| 752 | 589.90 | 628.13 | 252.74 | 208.07 | **1,678.84** |
| 753 | 28.87 | 12.64 | 5.45 | 5.45 | **52.40** |
| 754 | 185.74 | 118.20 | 142.71 | 83.34 | **529.98** |
| 755 | 67.00 | 98.04 | 75.17 | 67.54 | **307.75** |
| 756 | 210.40 | 364.96 | 167.11 | 121.70 | **864.18** |
| 757 | 200.25 | 268.10 | 197.98 | 144.34 | **810.67** |
| 758 | 10.02 | 18.85 | 11.44 | 14.16 | **54.47** |
| 759 | 196.35 | 115.47 | 96.41 | 100.22 | **508.45** |
| 760 | 406.03 | 512.55 | 503.84 | 301.21 | **1,723.63** |
| 761 | 369.84 | 366.68 | 220.05 | 135.63 | **1,092.21** |
| 762 | 241.30 | 200.90 | 132.36 | 131.81 | **706.37** |
| 763 | 61.01 | 271.80 | 97.50 | 56.10 | **486.41** |
| 764 | 74.08 | 71.83 | 52.83 | 23.97 | **222.71** |
| 765 | 124.73 | 137.81 | 98.04 | 54.47 | **415.05** |
| 766 | 25.06 | 14.71 | 21.79 | 33.77 | **95.32** |
| 767 | 70.81 | 72.44 | 70.81 | 89.87 | **303.94** |
| 768 | 68.63 | 43.58 | 59.37 | 29.41 | **200.99** |
| 769 | 91.51 | 92.05 | 104.04 | 88.78 | **376.38** |
| 770 | 2,474.52 | 1,658.03 | 1,079.03 | 1,933.64 | **7,145.21** |
| 773 | 585.28 | 398.71 | 340.97 | 324.51 | **1,649.48** |
| 774 | 334.76 | 212.54 | 181.93 | 159.05 | **888.28** |
| 775 | 497.84 | 439.56 | 379.10 | 265.26 | **1,581.77** |
| 776 | 139.98 | 81.16 | 86.71 | 63.18 | **371.04** |

| | | | | | |
|---|---|---|---|---|---|
| 777 | 104.04 | 50.66 | 61.01 | 80.07 | **295.77** |
| 778 | 190.10 | 116.56 | 156.87 | 118.20 | **581.73** |
| 779 | 49.57 | 26.15 | 30.50 | 66.45 | **172.67** |
| 780 | 260.91 | 230.40 | 160.68 | 174.30 | **826.29** |
| 781 | 52.29 | 65.91 | 43.58 | 38.13 | **199.90** |
| 782 | 950.20 | 916.71 | 677.00 | 510.92 | **3,054.83** |
| 783 | 32.14 | 6.54 | 31.59 | 15.25 | **85.52** |
| 784 | 283.24 | 399.26 | 122.55 | 99.13 | **904.18** |
| 785 | 1,355.18 | 871.22 | 811.58 | 757.66 | **3,795.64** |
| 786 | 219.62 | 168.85 | 222.78 | 106.76 | **718.01** |
| 787 | 351.43 | 244.02 | 184.10 | 149.24 | **928.80** |
| 788 | 89.87 | 269.08 | 77.35 | 84.97 | **521.27** |
| 789 | 15.58 | 29.96 | 11.98 | 32.68 | **90.20** |
| 790 | 13.07 | 29.96 | 34.86 | 15.80 | **93.69** |
| 791 | 83.34 | 55.56 | 58.83 | 53.92 | **251.65** |
| 792 | 5.45 | 11.44 | 3.81 | 4.90 | **25.60** |
| 793 | 111.66 | 99.68 | 86.61 | 112.75 | **410.69** |
| 794 | 298.49 | 174.84 | 147.61 | 120.38 | **741.32** |
| 795 | 32.14 | 10.89 | 21.24 | 61.01 | **125.28** |
| 796 | 188.46 | 75.17 | 52.83 | 78.98 | **395.44** |
| 797 | 549.05 | 370.93 | 342.06 | 287.05 | **1,549.09** |
| 798 | 0 | 13.07 | 0 | 13.07 | **26.15** |
| 799 | 218.96 | 227.68 | 126.37 | 107.85 | **680.86** |
| **TX - TEXAS - Total** | **13,536.92** | **11,614.55** | **8,532.90** | **8,180.78** | **41,865.15** |

**STATE: UT - UTAH**

| REGISTRANT ZIP CODE 3 | 1ST QUARTER | 2ND QUARTER | 3RD QUARTER | 4TH QUARTER | **TOTAL GRAMS** |
|---|---|---|---|---|---|
| 840 | 691.73 | 729.86 | 477.15 | 497.84 | **2,396.58** |
| 841 | 398.71 | 392.18 | 324.09 | 236.39 | **1,351.37** |
| 843 | 46.84 | 47.39 | 38.13 | 38.67 | **171.03** |
| 844 | 111.66 | 171.03 | 87.15 | 181.38 | **551.22** |
| 845 | 101.31 | 135.08 | 74.08 | 94.78 | **405.25** |
| 846 | 173.21 | 147.07 | 175.93 | 125.28 | **621.49** |
| 847 | 186.83 | 248.38 | 203.17 | 197.72 | **836.10** |
| **UT - UTAH - Total** | **1,710.30** | **1,870.98** | **1,379.69** | **1,372.07** | **6,333.04** |

**STATE: VA - VIRGINIA**

| REGISTRANT ZIP CODE 3 | 1ST QUARTER | 2ND QUARTER | 3RD QUARTER | 4TH QUARTER | **TOTAL GRAMS** |
|---|---|---|---|---|---|
| 201 | 368.75 | 287.60 | 219.51 | 182.47 | **1,058.33** |
| 220 | 268.64 | 238.57 | 181.93 | 177.57 | **866.71** |
| 221 | 98.59 | 166.13 | 112.21 | 161.77 | **538.70** |
| 222 | 37.04 | 19.61 | 14.16 | 6.54 | **77.35** |

| | | | | | |
|---|---|---|---|---|---|
| 223 | 185.19 | 101.31 | 197.72 | 172.67 | **656.89** |
| 224 | 145.43 | 163.41 | 137.81 | 127.46 | **574.10** |
| 225 | 40.31 | 9.80 | 43.58 | 44.66 | **138.35** |
| 226 | 144.34 | 162.32 | 149.24 | 150.88 | **606.78** |
| 227 | 28.32 | 17.43 | 19.61 | 9.80 | **75.17** |
| 228 | 104.58 | 55.56 | 52.29 | 83.34 | **295.77** |
| 229 | 293.04 | 313.74 | 248.92 | 183.56 | **1,039.26** |
| 230 | 240.75 | 141.62 | 82.79 | 108.94 | **574.10** |
| 231 | 464.07 | 355.14 | 368.21 | 281.06 | **1,468.48** |
| 232 | 805.59 | 679.23 | 616.04 | 508.19 | **2,609.05** |
| 233 | 198.81 | 209.16 | 209.16 | 169.94 | **787.07** |
| 234 | 477.47 | 482.05 | 449.37 | 346.42 | **1,755.31** |
| 235 | 308.08 | 179.20 | 144.89 | 102.40 | **734.57** |
| 236 | 289.34 | 299.36 | 184.98 | 130.73 | **904.40** |
| 237 | 153.82 | 96.52 | 43.03 | 89.33 | **382.70** |
| 238 | 233.67 | 166.67 | 186.83 | 209.70 | **796.88** |
| 239 | 69.18 | 99.13 | 46.84 | 86.06 | **301.21** |
| 240 | 150.66 | 88.24 | 103.49 | 82.79 | **425.18** |
| 241 | 118.74 | 177.02 | 147.07 | 107.85 | **550.68** |
| 242 | 261.99 | 147.07 | 137.81 | 144.89 | **691.75** |
| 243 | 42.49 | 37.04 | 60.46 | 39.76 | **179.75** |
| 244 | 104.58 | 66.45 | 59.92 | 56.65 | **287.60** |
| 245 | 78.44 | 148.16 | 81.70 | 107.30 | **415.60** |
| 246 | 4.90 | 1.09 | 0 | 1.09 | **7.08** |
| **VA - VIRGINIA - Total** | **5,716.82** | **4,908.61** | **4,299.55** | **3,873.82** | **18,798.80** |

**STATE: VI - VIRGIN ISLANDS**

| REGISTRANT ZIP CODE 3 | 1ST QUARTER | 2ND QUARTER | 3RD QUARTER | 4TH QUARTER | **TOTAL GRAMS** |
|---|---|---|---|---|---|
| 008 | 120.92 | 155.78 | 153.06 | 30.50 | **460.26** |
| **VI - VIRGIN ISLANDS - Total** | **120.92** | **155.78** | **153.06** | **30.50** | **460.26** |

**STATE: VT - VERMONT**

| REGISTRANT ZIP CODE 3 | 1ST QUARTER | 2ND QUARTER | 3RD QUARTER | 4TH QUARTER | **TOTAL GRAMS** |
|---|---|---|---|---|---|
| 050 | 6.75 | 0 | 4.36 | 1.09 | **12.20** |
| 051 | 1.09 | 0 | 1.09 | 0 | **2.18** |
| 053 | 0 | 0 | 4.36 | 0 | **4.36** |
| 054 | 229.31 | 254.37 | 128.55 | 182.47 | **794.70** |
| 056 | 30.50 | 11.44 | 0 | 2.18 | **44.12** |
| 057 | 82.25 | 33.77 | 38.67 | 36.49 | **191.19** |
| 058 | 4.36 | 2.18 | 1.09 | 0 | **7.63** |
| **VT - VERMONT - Total** | **354.26** | **301.76** | **178.11** | **222.23** | **1,056.37** |

**STATE: WA - WASHINGTON**

| REGISTRANT ZIP CODE 3 | 1ST QUARTER | 2ND QUARTER | 3RD QUARTER | 4TH QUARTER | TOTAL GRAMS |
|---|---|---|---|---|---|
| 980 | 544.69 | 574.10 | 452.09 | 452.09 | **2,022.97** |
| 981 | 424.09 | 347.51 | 224.41 | 210.25 | **1,206.26** |
| 982 | 484.77 | 701.56 | 495.67 | 391.09 | **2,073.08** |
| 983 | 290.86 | 323.54 | 324.09 | 253.82 | **1,192.32** |
| 984 | 146.96 | 144.34 | 115.47 | 107.85 | **514.62** |
| 985 | 334.44 | 174.84 | 171.58 | 218.96 | **899.82** |
| 986 | 154.36 | 185.19 | 144.34 | 160.14 | **644.04** |
| 988 | 167.11 | 181.93 | 78.44 | 112.21 | **539.68** |
| 989 | 154.15 | 123.10 | 82.25 | 102.40 | **461.90** |
| 990 | 9.80 | 17.43 | 13.07 | 30.50 | **70.81** |
| 991 | 88.57 | 116.02 | 115.47 | 67.00 | **387.05** |
| 992 | 217.88 | 196.09 | 139.98 | 133.65 | **687.60** |
| 993 | 198.05 | 186.83 | 89.33 | 121.47 | **595.67** |
| 994 | 43.03 | 17.43 | 32.68 | 17.43 | **110.57** |
| **WA - WASHINGTON - Total** | **3,258.76** | **3,289.91** | **2,478.87** | **2,378.85** | **11,406.39** |

**STATE: WI - WISCONSIN**

| REGISTRANT ZIP CODE 3 | 1ST QUARTER | 2ND QUARTER | 3RD QUARTER | 4TH QUARTER | TOTAL GRAMS |
|---|---|---|---|---|---|
| 530 | 168.31 | 131.27 | 129.09 | 77.89 | **506.56** |
| 531 | 235.31 | 167.76 | 68.09 | 96.95 | **568.11** |
| 532 | 119.29 | 195.54 | 63.73 | 58.39 | **436.95** |
| 534 | 11.98 | 14.71 | 13.07 | 13.07 | **52.83** |
| 535 | 84.43 | 141.07 | 81.16 | 51.20 | **357.86** |
| 537 | 69.18 | 97.39 | 61.01 | 47.93 | **275.50** |
| 538 | 5.45 | 0 | 4.36 | 2.18 | **11.98** |
| 539 | 30.50 | 12.53 | 12.53 | 3.27 | **58.83** |
| 540 | 2.18 | 2.18 | 2.18 | 9.80 | **16.34** |
| 541 | 3.53 | 13.07 | 9.80 | 23.97 | **50.37** |
| 542 | 49.02 | 33.77 | 46.30 | 42.49 | **171.58** |
| 543 | 153.60 | 157.96 | 118.20 | 152.51 | **582.27** |
| 544 | 138.35 | 126.37 | 77.89 | 74.08 | **416.69** |
| 545 | 4.36 | 7.08 | 0 | 1.63 | **13.07** |
| 546 | 27.23 | 21.79 | 28.87 | 10.89 | **88.78** |
| 547 | 29.96 | 21.24 | 41.40 | 17.43 | **110.03** |
| 548 | 14.92 | 10.35 | 7.63 | 11.44 | **44.34** |
| 549 | 97.50 | 75.17 | 45.75 | 53.38 | **271.80** |
| **WI - WISCONSIN - Total** | **1,245.09** | **1,229.25** | **811.04** | **748.51** | **4,033.89** |

**STATE: WV - WEST VIRGINIA**

| REGISTRANT ZIP CODE 3 | 1ST QUARTER | 2ND QUARTER | 3RD QUARTER | 4TH QUARTER | TOTAL GRAMS |
|---|---|---|---|---|---|
| 247 | 52.29 | 32.68 | 3.81 | 0 | **88.78** |

| | | | | | |
|---|---|---|---|---|---|
| 248 | 10.89 | 18.52 | 13.07 | 10.89 | **53.38** |
| 249 | 47.93 | 43.03 | 21.24 | 17.97 | **130.18** |
| 250 | 1.09 | 4.36 | 0 | 0 | **5.45** |
| 251 | 6.54 | 17.43 | 1.09 | 0.54 | **25.60** |
| 252 | 0 | 0 | 4.36 | 0 | **4.36** |
| 253 | 115.47 | 41.94 | 49.02 | 22.88 | **229.31** |
| 254 | 52.83 | 51.75 | 26.69 | 21.79 | **153.06** |
| 255 | 61.01 | 30.50 | 9.80 | 13.07 | **114.38** |
| 256 | 0 | 17.43 | 0 | 8.72 | **26.15** |
| 257 | 55.56 | 48.48 | 65.36 | 35.40 | **204.80** |
| 258 | 22.88 | 6.54 | 15.25 | 8.72 | **53.38** |
| 259 | 5.99 | 11.98 | 6.54 | 1.09 | **25.60** |
| 260 | 65.36 | 67.54 | 30.50 | 70.26 | **233.67** |
| 261 | 78.44 | 85.52 | 65.36 | 40.31 | **269.62** |
| 262 | 0.54 | 4.36 | 4.36 | 5.45 | **14.71** |
| 263 | 73.42 | 25.06 | 23.97 | 32.68 | **155.13** |
| 264 | 3.81 | 3.81 | 3.81 | 1.09 | **12.53** |
| 265 | 46.84 | 44.12 | 30.50 | 15.25 | **136.72** |
| 266 | 0 | 4.36 | 4.36 | 0 | **8.72** |
| 267 | 8.72 | 0 | 5.45 | 8.72 | **22.88** |
| 268 | 5.45 | 7.63 | 0 | 1.09 | **14.16** |
| **WV - WEST VIRGINIA - Total** | **715.07** | **567.02** | **384.55** | **315.92** | **1,982.55** |

**STATE: WY - WYOMING**

| REGISTRANT ZIP CODE 3 | 1ST QUARTER | 2ND QUARTER | 3RD QUARTER | 4TH QUARTER | TOTAL GRAMS |
|---|---|---|---|---|---|
| 820 | 58.39 | 28.32 | 39.22 | 33.77 | **159.70** |
| 822 | 52.29 | 57.19 | 40.31 | 65.36 | **215.15** |
| 823 | 47.93 | 26.15 | 43.58 | 17.43 | **135.08** |
| 824 | 33.23 | 21.79 | 28.32 | 16.34 | **99.68** |
| 825 | 37.04 | 39.22 | 32.14 | 32.14 | **140.53** |
| 826 | 62.64 | 34.32 | 34.32 | 57.74 | **189.01** |
| 827 | 11.55 | 20.15 | 13.07 | 13.62 | **58.39** |
| 828 | 11.44 | 41.40 | 30.50 | 20.15 | **103.49** |
| 829 | 10.35 | 20.15 | 35.95 | 35.95 | **102.40** |
| 830 | 3.27 | 1.09 | 1.09 | 2.18 | **7.63** |
| 831 | 5.45 | 16.89 | 12.53 | 7.63 | **42.49** |
| **WY - WYOMING - Total** | **333.57** | **306.66** | **311.02** | **302.30** | **1,253.54** |

**DRUG: 9250B - METHADONE**

**STATE: AK - ALASKA**

| REGISTRANT ZIP CODE 3 | 1ST QUARTER | 2ND QUARTER | 3RD QUARTER | 4TH QUARTER | TOTAL GRAMS |
|---|---|---|---|---|---|
| 995 | 5,098.71 | 4,891.56 | 5,683.73 | 3,971.76 | **19,645.76** |

| | | | | | |
|---|---|---|---|---|---|
| 996 | 2,197.90 | 2,130.10 | 2,394.13 | 2,959.03 | **9,681.16** |
| 997 | 732.78 | 747.90 | 727.59 | 713.19 | **2,921.47** |
| 998 | 160.99 | 146.23 | 120.74 | 136.84 | **564.81** |
| 999 | 48.30 | 38.46 | 38.91 | 44.72 | **170.38** |
| **AK - ALASKA - Total** | **8,238.68** | **7,954.26** | **8,965.11** | **7,825.55** | **32,983.59** |

**STATE: AL - ALABAMA**

| REGISTRANT ZIP CODE 3 | 1ST QUARTER | 2ND QUARTER | 3RD QUARTER | 4TH QUARTER | TOTAL GRAMS |
|---|---|---|---|---|---|
| 350 | 10,657.22 | 12,842.87 | 10,773.32 | 12,154.63 | **46,428.04** |
| 351 | 1,184.63 | 1,130.97 | 1,033.93 | 923.92 | **4,273.44** |
| 352 | 11,437.86 | 11,631.67 | 9,364.55 | 14,333.92 | **46,768.00** |
| 354 | 4,157.98 | 3,949.58 | 4,121.75 | 3,654.97 | **15,884.28** |
| 355 | 6,116.35 | 5,092.27 | 5,472.83 | 5,858.77 | **22,540.22** |
| 356 | 2,007.93 | 2,249.95 | 2,906.35 | 2,203.35 | **9,367.59** |
| 357 | 889.93 | 906.03 | 887.24 | 533.51 | **3,216.71** |
| 358 | 8,515.22 | 11,651.80 | 11,594.73 | 12,738.04 | **44,499.80** |
| 359 | 2,483.30 | 3,115.20 | 3,430.47 | 2,554.41 | **11,583.37** |
| 360 | 467.32 | 446.75 | 409.19 | 387.72 | **1,710.99** |
| 361 | 1,325.50 | 1,313.07 | 2,320.88 | 1,278.01 | **6,237.46** |
| 362 | 2,848.40 | 2,816.56 | 3,945.20 | 2,319.00 | **11,929.15** |
| 363 | 2,185.73 | 1,694.44 | 2,212.75 | 1,824.13 | **7,917.05** |
| 364 | 299.00 | 272.70 | 263.85 | 258.48 | **1,094.03** |
| 365 | 3,278.87 | 3,338.80 | 2,558.88 | 3,069.22 | **12,245.77** |
| 366 | 9,140.32 | 6,025.13 | 11,902.23 | 10,321.38 | **37,389.05** |
| 367 | 224.05 | 168.59 | 183.80 | 195.87 | **772.31** |
| 368 | 688.51 | 615.08 | 617.58 | 608.91 | **2,530.08** |
| 369 | 6.26 | 9.84 | 8.94 | 12.97 | **38.01** |
| **AL - ALABAMA - Total** | **67,914.39** | **69,271.28** | **74,008.47** | **75,231.20** | **286,425.34** |

**STATE: AR - ARKANSAS**

| REGISTRANT ZIP CODE 3 | 1ST QUARTER | 2ND QUARTER | 3RD QUARTER | 4TH QUARTER | TOTAL GRAMS |
|---|---|---|---|---|---|
| 716 | 180.67 | 150.71 | 150.71 | 160.54 | **642.63** |
| 717 | 333.61 | 314.83 | 285.76 | 238.36 | **1,172.56** |
| 718 | 1,624.68 | 1,615.29 | 1,598.29 | 1,226.67 | **6,064.93** |
| 719 | 348.82 | 290.68 | 295.78 | 291.57 | **1,226.85** |
| 720 | 628.76 | 527.07 | 1,412.17 | 469.56 | **3,037.56** |
| 721 | 2,567.20 | 1,991.65 | 1,793.45 | 1,641.40 | **7,993.70** |
| 722 | 1,270.67 | 1,226.76 | 714.63 | 1,117.91 | **4,329.97** |
| 723 | 184.25 | 183.80 | 144.00 | 143.10 | **655.15** |
| 724 | 2,424.27 | 3,225.65 | 2,288.77 | 2,269.99 | **10,208.68** |
| 725 | 265.64 | 223.60 | 216.44 | 210.18 | **915.87** |
| 726 | 415.00 | 331.82 | 255.35 | 225.84 | **1,228.01** |

| | | | | | |
|---|---|---|---|---|---|
| 727 | 3,486.01 | 2,430.62 | 3,471.17 | 2,905.10 | **12,292.90** |
| 728 | 168.86 | 189.97 | 180.22 | 110.46 | **649.51** |
| 729 | 947.62 | 875.62 | 809.61 | 636.37 | **3,269.21** |
| **AR - ARKANSAS - Total** | **14,846.06** | **13,578.07** | **13,616.35** | **11,647.06** | **53,687.52** |

**STATE: AZ - ARIZONA**

| REGISTRANT ZIP CODE 3 | 1ST QUARTER | 2ND QUARTER | 3RD QUARTER | 4TH QUARTER | **TOTAL GRAMS** |
|---|---|---|---|---|---|
| 850 | 32,446.77 | 33,402.08 | 33,049.87 | 32,040.18 | **130,938.91** |
| 851 | 482.35 | 709.71 | 949.94 | 950.48 | **3,092.48** |
| 852 | 19,277.99 | 18,360.33 | 18,178.77 | 15,900.02 | **71,717.11** |
| 853 | 5,991.41 | 5,625.15 | 9,838.67 | 8,354.14 | **29,809.37** |
| 855 | 442.28 | 635.47 | 343.90 | 686.00 | **2,107.65** |
| 856 | 464.01 | 398.28 | 349.26 | 271.54 | **1,483.09** |
| 857 | 25,926.51 | 26,373.44 | 25,784.93 | 21,769.16 | **99,854.04** |
| 859 | 355.17 | 301.23 | 291.57 | 264.30 | **1,212.27** |
| 860 | 165.04 | 905.13 | 911.93 | 885.01 | **2,867.12** |
| 863 | 1,298.04 | 1,226.67 | 1,196.71 | 1,145.73 | **4,867.15** |
| 864 | 1,339.81 | 1,263.61 | 1,220.86 | 1,192.68 | **5,016.96** |
| 865 | 9.39 | 2.24 | 9.39 | 4.02 | **25.04** |
| **AZ - ARIZONA - Total** | **88,198.78** | **89,203.34** | **92,125.79** | **83,463.26** | **352,991.18** |

**STATE: CA - CALIFORNIA**

| REGISTRANT ZIP CODE 3 | 1ST QUARTER | 2ND QUARTER | 3RD QUARTER | 4TH QUARTER | **TOTAL GRAMS** |
|---|---|---|---|---|---|
| 900 | 17,028.48 | 14,617.27 | 15,194.34 | 10,572.08 | **57,412.17** |
| 902 | 3,059.74 | 2,139.14 | 3,566.51 | 2,504.77 | **11,270.16** |
| 903 | 707.02 | 1,579.06 | 1,246.79 | 344.34 | **3,877.22** |
| 904 | 230.58 | 230.31 | 221.10 | 191.22 | **873.20** |
| 905 | 297.03 | 313.49 | 341.39 | 345.24 | **1,297.15** |
| 906 | 5,445.11 | 5,040.39 | 6,324.39 | 4,224.25 | **21,034.14** |
| 907 | 3,811.58 | 4,506.61 | 3,308.12 | 2,881.76 | **14,508.06** |
| 908 | 1,603.93 | 1,192.24 | 1,965.27 | 1,355.73 | **6,117.16** |
| 910 | 431.73 | 389.33 | 329.23 | 345.24 | **1,495.53** |
| 911 | 636.19 | 626.97 | 1,049.49 | 586.10 | **2,898.75** |
| 912 | 1,385.43 | 1,434.89 | 980.98 | 1,228.14 | **5,029.43** |
| 913 | 5,356.38 | 5,020.36 | 5,819.15 | 4,346.61 | **20,542.49** |
| 914 | 3,073.61 | 2,738.12 | 3,025.84 | 2,068.30 | **10,905.87** |
| 915 | 202.22 | 207.77 | 184.34 | 161.98 | **756.30** |
| 916 | 761.49 | 793.24 | 665.34 | 911.84 | **3,131.92** |
| 917 | 9,736.89 | 7,565.91 | 7,607.32 | 6,407.03 | **31,317.15** |
| 918 | 38.91 | 27.28 | 32.20 | 37.56 | **135.95** |
| 919 | 2,817.27 | 2,729.89 | 2,796.52 | 2,200.67 | **10,544.35** |
| 920 | 11,952.05 | 12,243.80 | 11,983.71 | 8,905.54 | **45,085.09** |

| | | | | | |
|---|---|---|---|---|---|
| 921 | 10,449.54 | 10,645.77 | 10,084.54 | 7,610.36 | **38,790.22** |
| 922 | 7,571.36 | 3,246.85 | 5,972.71 | 6,237.10 | **23,028.03** |
| 923 | 7,937.25 | 7,784.95 | 8,137.70 | 5,477.84 | **29,337.74** |
| 924 | 194.08 | 186.12 | 145.79 | 141.94 | **667.94** |
| 925 | 7,271.83 | 7,650.16 | 8,177.32 | 5,577.30 | **28,676.61** |
| 926 | 2,465.06 | 1,972.60 | 1,922.78 | 2,044.42 | **8,404.86** |
| 927 | 3,196.05 | 2,907.52 | 3,334.95 | 2,333.94 | **11,772.45** |
| 928 | 5,246.76 | 4,155.76 | 5,187.26 | 3,827.21 | **18,416.99** |
| 930 | 10,389.26 | 7,541.13 | 10,137.67 | 6,290.05 | **34,358.11** |
| 931 | 2,739.64 | 1,830.84 | 2,671.66 | 1,843.54 | **9,085.67** |
| 932 | 7,276.21 | 7,824.84 | 8,048.53 | 5,934.79 | **29,084.37** |
| 933 | 9,257.93 | 10,488.45 | 9,051.60 | 6,902.53 | **35,700.51** |
| 934 | 6,448.53 | 5,823.71 | 5,783.46 | 4,658.57 | **22,714.27** |
| 935 | 3,307.94 | 2,533.57 | 3,588.96 | 2,860.74 | **12,291.20** |
| 936 | 2,061.81 | 2,105.69 | 1,731.84 | 1,821.17 | **7,720.52** |
| 937 | 14,545.63 | 17,080.89 | 15,522.67 | 12,377.69 | **59,526.88** |
| 939 | 2,244.68 | 2,101.57 | 2,492.69 | 1,597.22 | **8,436.16** |
| 940 | 4,411.63 | 3,806.03 | 4,425.67 | 3,493.26 | **16,136.59** |
| 941 | 20,618.60 | 16,744.87 | 19,002.60 | 17,376.49 | **73,742.56** |
| 943 | 504.44 | 432.98 | 470.72 | 419.38 | **1,827.53** |
| 944 | 290.50 | 249.81 | 242.02 | 259.11 | **1,041.44** |
| 945 | 22,219.40 | 18,518.64 | 20,873.06 | 16,231.84 | **77,842.94** |
| 946 | 9,109.82 | 8,748.22 | 8,103.98 | 7,145.81 | **33,107.83** |
| 947 | 1,044.12 | 1,381.58 | 1,010.49 | 982.50 | **4,418.69** |
| 948 | 2,736.60 | 2,670.50 | 2,645.99 | 2,609.32 | **10,662.41** |
| 949 | 3,074.69 | 2,940.33 | 2,996.51 | 2,801.89 | **11,813.41** |
| 950 | 7,455.09 | 6,431.00 | 6,690.83 | 8,352.44 | **28,929.37** |
| 951 | 2,673.63 | 2,665.22 | 2,612.81 | 2,538.58 | **10,490.24** |
| 952 | 14,706.17 | 13,413.23 | 14,989.34 | 10,064.06 | **53,172.80** |
| 953 | 16,694.24 | 15,592.43 | 15,015.37 | 14,003.71 | **61,305.75** |
| 954 | 6,491.82 | 5,921.91 | 5,831.31 | 6,063.58 | **24,308.63** |
| 955 | 1,127.93 | 1,112.01 | 1,049.40 | 893.15 | **4,182.48** |
| 956 | 12,665.42 | 12,124.49 | 13,650.24 | 9,450.14 | **47,890.29** |
| 957 | 601.93 | 510.26 | 487.54 | 429.04 | **2,028.77** |
| 958 | 19,476.81 | 17,089.03 | 19,258.13 | 12,818.45 | **68,642.43** |
| 959 | 11,294.66 | 9,836.16 | 10,054.04 | 8,438.49 | **39,623.35** |
| 960 | 3,069.31 | 3,254.63 | 2,793.75 | 2,360.41 | **11,478.10** |
| 961 | 276.37 | 260.36 | 217.79 | 211.53 | **966.04** |
| **CA - CALIFORNIA - Total** | **331,722.38** | **302,980.16** | **321,055.74** | **254,097.99** | **1,209,856.28** |

**STATE: CO - COLORADO**

| REGISTRANT ZIP CODE 3 | 1ST QUARTER | 2ND QUARTER | 3RD QUARTER | 4TH QUARTER | TOTAL GRAMS |
|---|---|---|---|---|---|
| 800 | 9,220.74 | 8,487.74 | 10,289.59 | 8,043.74 | **36,041.82** |
| 801 | 2,645.81 | 2,683.38 | 3,380.03 | 2,538.04 | **11,247.26** |
| 802 | 18,599.23 | 19,443.27 | 17,741.77 | 15,418.65 | **71,202.92** |
| 803 | 394.07 | 693.07 | 822.85 | 594.78 | **2,504.77** |
| 804 | 248.64 | 218.23 | 214.92 | 194.98 | **876.78** |
| 805 | 3,928.92 | 3,294.34 | 3,039.89 | 2,594.03 | **12,857.18** |
| 806 | 520.63 | 681.89 | 831.70 | 856.92 | **2,891.15** |
| 807 | 46.06 | 54.56 | 28.62 | 41.59 | **170.83** |
| 808 | 220.47 | 184.43 | 184.25 | 146.23 | **735.38** |
| 809 | 4,346.07 | 4,246.07 | 4,196.26 | 4,415.65 | **17,204.05** |
| 810 | 2,530.53 | 2,590.81 | 3,066.72 | 3,573.22 | **11,761.27** |
| 811 | 578.68 | 304.10 | 584.49 | 313.04 | **1,780.30** |
| 812 | 161.89 | 154.28 | 149.81 | 161.89 | **627.87** |
| 813 | 730.72 | 506.95 | 504.89 | 715.07 | **2,457.63** |
| 814 | 152.05 | 69.58 | 56.35 | 54.56 | **332.54** |
| 815 | 1,981.72 | 1,083.21 | 1,941.83 | 1,937.27 | **6,944.03** |
| 816 | 105.09 | 78.71 | 112.87 | 101.69 | **398.37** |
| **CO - COLORADO - Total** | **46,411.33** | **44,774.62** | **47,146.83** | **41,701.36** | **180,034.13** |

**STATE: CT - CONNECTICUT**

| REGISTRANT ZIP CODE 3 | 1ST QUARTER | 2ND QUARTER | 3RD QUARTER | 4TH QUARTER | TOTAL GRAMS |
|---|---|---|---|---|---|
| 060 | 13,925.81 | 17,130.17 | 11,158.00 | 10,732.80 | **52,946.78** |
| 061 | 17,616.55 | 14,009.34 | 11,115.33 | 9,747.98 | **52,489.21** |
| 062 | 5,130.73 | 4,818.13 | 4,953.19 | 4,547.13 | **19,449.18** |
| 063 | 6,870.24 | 6,859.51 | 6,981.24 | 7,398.30 | **28,109.29** |
| 064 | 8,271.95 | 8,186.62 | 14,932.72 | 8,139.13 | **39,530.42** |
| 065 | 19,877.86 | 33,391.44 | 32,625.57 | 19,589.15 | **105,484.02** |
| 066 | 19,975.53 | 8,811.63 | 19,914.26 | 9,172.61 | **57,874.03** |
| 067 | 7,333.63 | 7,726.01 | 7,761.25 | 5,828.36 | **28,649.24** |
| 068 | 6,615.52 | 5,101.57 | 4,758.48 | 6,492.36 | **22,967.92** |
| 069 | 1,933.87 | 3,705.95 | 1,912.14 | 1,892.10 | **9,444.06** |
| **CT - CONNECTICUT - Total** | **107,551.69** | **109,740.38** | **116,112.17** | **83,539.91** | **416,944.15** |

**STATE: DC - DISTRICT OF COLUMBIA**

| REGISTRANT ZIP CODE 3 | 1ST QUARTER | 2ND QUARTER | 3RD QUARTER | 4TH QUARTER | TOTAL GRAMS |
|---|---|---|---|---|---|
| 200 | 8,511.56 | 8,093.12 | 9,777.34 | 6,411.02 | **32,793.04** |
| 204 | 821.06 | 584.04 | 609.98 | 603.72 | **2,618.80** |
| **DC - DISTRICT OF COLUMBIA -** | **9,332.62** | **8,677.16** | **10,387.32** | **7,014.74** | **35,411.84** |

**STATE: DE - DELAWARE**

| REGISTRANT ZIP CODE 3 | 1ST QUARTER | 2ND QUARTER | 3RD QUARTER | 4TH QUARTER | TOTAL GRAMS |
|---|---|---|---|---|---|
| 197 | 13,051.62 | 14,494.20 | 10,722.34 | 11,531.14 | **49,799.30** |

| | | | | | |
|---|---|---|---|---|---|
| 198 | 6,929.45 | 6,893.59 | 5,864.40 | 5,490.45 | **25,177.90** |
| 199 | 11,408.61 | 13,448.20 | 12,819.79 | 12,260.79 | **49,937.39** |
| **DE - DELAWARE - Total** | **31,389.68** | **34,835.99** | **29,406.53** | **29,282.39** | **124,914.59** |

**STATE: FL - FLORIDA**

| REGISTRANT ZIP CODE 3 | 1ST QUARTER | 2ND QUARTER | 3RD QUARTER | 4TH QUARTER | **TOTAL GRAMS** |
|---|---|---|---|---|---|
| 320 | 7,459.03 | 5,709.49 | 7,034.72 | 6,343.80 | **26,547.04** |
| 321 | 8,483.83 | 10,253.67 | 8,149.77 | 7,480.31 | **34,367.59** |
| 322 | 21,460.20 | 20,422.51 | 19,792.79 | 19,250.80 | **80,926.29** |
| 323 | 3,468.84 | 3,120.92 | 2,468.45 | 4,163.61 | **13,221.83** |
| 324 | 4,460.55 | 2,548.59 | 4,376.84 | 6,089.97 | **17,475.95** |
| 325 | 11,761.63 | 12,916.03 | 14,203.61 | 11,110.33 | **49,991.59** |
| 326 | 4,189.64 | 2,856.98 | 3,821.59 | 4,326.93 | **15,195.14** |
| 327 | 2,303.62 | 2,218.11 | 1,978.06 | 1,978.41 | **8,478.20** |
| 328 | 9,803.72 | 12,936.77 | 6,982.45 | 13,923.46 | **43,646.41** |
| 329 | 6,868.99 | 3,596.38 | 5,238.95 | 5,996.59 | **21,700.92** |
| 330 | 5,624.97 | 3,755.76 | 5,292.88 | 5,926.74 | **20,600.36** |
| 331 | 2,462.37 | 1,451.16 | 3,198.46 | 2,422.12 | **9,534.13** |
| 333 | 3,202.22 | 2,689.01 | 2,169.28 | 3,088.36 | **11,148.87** |
| 334 | 8,413.98 | 7,569.31 | 9,425.99 | 8,786.05 | **34,195.33** |
| 335 | 2,162.30 | 1,987.98 | 1,742.74 | 1,728.16 | **7,621.18** |
| 336 | 10,970.44 | 13,925.54 | 10,853.19 | 11,631.30 | **47,380.47** |
| 337 | 20,265.85 | 12,153.38 | 20,468.97 | 18,536.62 | **71,424.82** |
| 338 | 3,019.05 | 3,378.69 | 2,222.49 | 2,868.07 | **11,488.30** |
| 339 | 13,764.91 | 6,843.95 | 13,892.27 | 14,223.91 | **48,725.03** |
| 341 | 3,252.40 | 3,350.06 | 3,538.87 | 3,243.09 | **13,384.43** |
| 342 | 13,443.19 | 8,588.65 | 13,696.57 | 12,728.92 | **48,457.34** |
| 344 | 10,983.32 | 8,267.30 | 7,469.04 | 9,721.14 | **36,440.81** |
| 346 | 15,267.41 | 10,924.47 | 16,372.35 | 15,853.69 | **58,417.91** |
| 347 | 4,344.37 | 2,571.85 | 3,906.29 | 3,929.01 | **14,751.52** |
| 349 | 2,582.58 | 2,643.13 | 2,366.85 | 2,316.05 | **9,908.61** |
| **FL - FLORIDA - Total** | **200,019.40** | **166,679.70** | **190,663.49** | **197,667.47** | **755,030.06** |

**STATE: GA - GEORGIA**

| REGISTRANT ZIP CODE 3 | 1ST QUARTER | 2ND QUARTER | 3RD QUARTER | 4TH QUARTER | **TOTAL GRAMS** |
|---|---|---|---|---|---|
| 300 | 15,151.67 | 13,483.53 | 15,241.29 | 13,076.66 | **56,953.16** |
| 301 | 14,776.20 | 11,905.09 | 13,421.90 | 12,017.87 | **52,121.07** |
| 302 | 8,192.61 | 7,892.54 | 7,938.60 | 7,702.93 | **31,726.69** |
| 303 | 4,841.03 | 4,325.32 | 4,368.52 | 4,472.27 | **18,007.13** |
| 304 | 2,245.93 | 1,605.72 | 1,375.05 | 1,273.18 | **6,499.87** |
| 305 | 14,778.44 | 12,276.62 | 13,741.20 | 11,549.21 | **52,345.48** |
| 306 | 5,899.64 | 4,891.56 | 4,759.37 | 4,859.72 | **20,410.30** |

| | | | | | |
|---|---|---|---|---|---|
| 307 | 27,441.98 | 23,340.71 | 25,954.15 | 26,591.67 | **103,328.51** |
| 308 | 1,069.70 | 411.87 | 1,022.75 | 1,605.00 | **4,109.32** |
| 309 | 2,684.15 | 1,983.69 | 3,176.73 | 3,258.75 | **11,103.32** |
| 310 | 5,589.55 | 4,422.27 | 5,290.91 | 4,205.29 | **19,508.03** |
| 312 | 2,322.49 | 2,846.25 | 2,143.43 | 2,636.69 | **9,948.86** |
| 313 | 375.92 | 346.67 | 337.28 | 307.05 | **1,366.91** |
| 314 | 4,185.79 | 4,447.05 | 3,949.22 | 3,560.61 | **16,142.67** |
| 315 | 2,546.36 | 2,140.12 | 2,754.57 | 2,917.98 | **10,359.03** |
| 316 | 2,092.09 | 928.83 | 2,087.08 | 2,097.90 | **7,205.91** |
| 317 | 3,224.31 | 3,013.86 | 2,955.19 | 2,359.07 | **11,552.43** |
| 318 | 118.33 | 106.79 | 100.17 | 99.73 | **425.02** |
| 319 | 3,509.45 | 2,813.51 | 3,140.06 | 2,269.81 | **11,732.83** |
| 398 | 1,004.41 | 1,018.72 | 1,046.90 | 1,318.35 | **4,388.37** |
| **GA - GEORGIA - Total** | **122,050.06** | **104,200.73** | **114,804.38** | **108,179.74** | **449,234.91** |

**STATE: GU - GUAM**

| REGISTRANT ZIP CODE 3 | 1ST QUARTER | 2ND QUARTER | 3RD QUARTER | 4TH QUARTER | TOTAL GRAMS |
|---|---|---|---|---|---|
| 969 | 42.48 | 43.83 | 33.54 | 43.38 | **163.23** |
| **GU - GUAM - Total** | **42.48** | **43.83** | **33.54** | **43.38** | **163.23** |

**STATE: HI - HAWAII**

| REGISTRANT ZIP CODE 3 | 1ST QUARTER | 2ND QUARTER | 3RD QUARTER | 4TH QUARTER | TOTAL GRAMS |
|---|---|---|---|---|---|
| 967 | 4,094.29 | 3,562.13 | 4,784.06 | 4,498.03 | **16,938.50** |
| 968 | 4,219.24 | 1,412.35 | 3,442.37 | 1,882.53 | **10,956.49** |
| **HI - HAWAII - Total** | **8,313.54** | **4,974.47** | **8,226.42** | **6,380.56** | **27,894.99** |

**STATE: IA - IOWA**

| REGISTRANT ZIP CODE 3 | 1ST QUARTER | 2ND QUARTER | 3RD QUARTER | 4TH QUARTER | TOTAL GRAMS |
|---|---|---|---|---|---|
| 500 | 405.16 | 366.26 | 371.09 | 461.33 | **1,603.84** |
| 501 | 158.58 | 158.76 | 158.76 | 167.25 | **643.34** |
| 502 | 212.87 | 212.60 | 214.66 | 198.56 | **838.68** |
| 503 | 4,536.31 | 4,915.89 | 4,852.30 | 3,577.60 | **17,882.10** |
| 504 | 166.81 | 148.47 | 151.15 | 141.05 | **607.48** |
| 505 | 194.71 | 194.08 | 182.90 | 186.93 | **758.63** |
| 506 | 768.56 | 766.77 | 426.63 | 436.47 | **2,398.42** |
| 507 | 269.30 | 243.28 | 507.57 | 333.79 | **1,353.94** |
| 508 | 68.42 | 66.63 | 57.69 | 60.37 | **253.12** |
| 510 | 134.16 | 148.65 | 141.67 | 141.32 | **565.80** |
| 511 | 350.07 | 791.81 | 329.59 | 321.36 | **1,792.82** |
| 512 | 10.29 | 11.18 | 18.34 | 16.99 | **56.79** |
| 513 | 32.65 | 36.67 | 27.28 | 23.70 | **120.30** |
| 514 | 33.36 | 39.80 | 29.07 | 113.59 | **215.82** |
| 515 | 232.10 | 222.71 | 194.08 | 178.88 | **827.77** |

| | | | | | |
|---|---|---|---|---|---|
| 516 | 37.56 | 39.80 | 37.12 | 24.15 | **138.63** |
| 520 | 98.03 | 103.57 | 223.24 | 222.97 | **647.81** |
| 521 | 10.73 | 11.63 | 12.97 | 19.68 | **55.01** |
| 522 | 216.36 | 213.40 | 189.61 | 148.92 | **768.29** |
| 523 | 1,153.95 | 1,126.59 | 1,145.10 | 795.03 | **4,220.67** |
| 524 | 1,209.85 | 917.56 | 1,462.08 | 1,066.12 | **4,655.62** |
| 525 | 144.89 | 299.18 | 121.64 | 303.65 | **869.36** |
| 526 | 158.04 | 242.38 | 152.76 | 329.32 | **882.50** |
| 527 | 122.98 | 118.87 | 115.38 | 96.60 | **453.82** |
| 528 | 1,737.37 | 1,719.04 | 1,377.82 | 1,718.14 | **6,552.37** |
| **IA - IOWA - Total** | **12,463.11** | **13,115.57** | **12,500.49** | **11,083.76** | **49,162.93** |

**STATE: ID - IDAHO**

| REGISTRANT ZIP CODE 3 | 1ST QUARTER | 2ND QUARTER | 3RD QUARTER | 4TH QUARTER | TOTAL GRAMS |
|---|---|---|---|---|---|
| 832 | 721.78 | 721.78 | 677.06 | 692.98 | **2,813.60** |
| 833 | 390.67 | 376.54 | 303.20 | 270.11 | **1,340.53** |
| 834 | 406.68 | 393.36 | 402.48 | 369.21 | **1,571.73** |
| 835 | 352.30 | 333.16 | 313.93 | 326.55 | **1,325.95** |
| 836 | 3,648.26 | 2,858.59 | 2,935.78 | 2,848.57 | **12,291.20** |
| 837 | 5,213.01 | 4,173.54 | 4,317.54 | 4,431.93 | **18,136.02** |
| 838 | 1,066.57 | 1,000.65 | 944.49 | 973.55 | **3,985.27** |
| **ID - IDAHO - Total** | **11,799.28** | **9,857.63** | **9,894.48** | **9,912.90** | **41,464.29** |

**STATE: IL - ILLINOIS**

| REGISTRANT ZIP CODE 3 | 1ST QUARTER | 2ND QUARTER | 3RD QUARTER | 4TH QUARTER | TOTAL GRAMS |
|---|---|---|---|---|---|
| 600 | 8,838.37 | 8,666.83 | 7,929.12 | 8,074.29 | **33,508.61** |
| 601 | 7,134.18 | 6,775.71 | 7,888.52 | 5,394.93 | **27,193.34** |
| 602 | 3,780.18 | 3,932.23 | 4,766.08 | 2,837.22 | **15,315.71** |
| 603 | 46.96 | 54.83 | 40.70 | 40.25 | **182.73** |
| 604 | 6,605.86 | 6,915.32 | 7,409.12 | 6,011.98 | **26,942.28** |
| 605 | 6,448.36 | 4,762.50 | 6,244.07 | 4,703.11 | **22,158.04** |
| 606 | 58,165.52 | 53,493.89 | 58,478.73 | 51,126.32 | **221,264.45** |
| 607 | 1,147.52 | 60.46 | 55.99 | 36.85 | **1,300.82** |
| 608 | 4,011.83 | 3,981.42 | 3,528.41 | 3,459.99 | **14,981.65** |
| 609 | 1,126.77 | 1,125.16 | 994.48 | 1,134.19 | **4,380.59** |
| 610 | 1,118.09 | 414.11 | 1,131.68 | 769.18 | **3,433.06** |
| 611 | 2,254.34 | 2,932.56 | 2,944.63 | 2,813.87 | **10,945.40** |
| 612 | 1,113.08 | 897.62 | 1,059.42 | 687.79 | **3,757.91** |
| 613 | 532.62 | 969.80 | 578.23 | 619.82 | **2,700.46** |
| 614 | 136.84 | 112.25 | 127.45 | 114.93 | **491.47** |
| 615 | 175.75 | 153.39 | 134.16 | 135.95 | **599.25** |
| 616 | 8,316.85 | 6,192.68 | 8,523.47 | 7,190.59 | **30,223.59** |

| | | | | | |
|---|---|---|---|---|---|
| 617 | 1,766.17 | 461.69 | 1,684.78 | 1,398.57 | **5,311.22** |
| 618 | 2,022.60 | 855.76 | 2,506.91 | 2,409.96 | **7,795.23** |
| 619 | 66.63 | 55.01 | 59.12 | 40.07 | **220.83** |
| 620 | 2,749.03 | 3,252.66 | 3,549.07 | 4,140.89 | **13,691.65** |
| 622 | 883.22 | 1,421.47 | 853.17 | 815.60 | **3,973.46** |
| 623 | 362.14 | 346.85 | 324.40 | 301.32 | **1,334.71** |
| 624 | 145.34 | 142.66 | 156.25 | 156.97 | **601.22** |
| 625 | 1,948.45 | 1,512.70 | 2,132.52 | 1,482.02 | **7,075.69** |
| 626 | 173.96 | 135.05 | 134.61 | 130.14 | **573.76** |
| 627 | 3,418.84 | 2,693.49 | 3,350.87 | 3,703.26 | **13,166.46** |
| 628 | 195.43 | 187.38 | 202.58 | 186.57 | **771.96** |
| 629 | 475.37 | 473.41 | 466.25 | 398.81 | **1,813.84** |
| **IL - ILLINOIS - Total** | **125,160.28** | **112,978.85** | **127,254.80** | **110,315.45** | **475,709.38** |

**STATE: IN - INDIANA**

| REGISTRANT ZIP CODE 3 | 1ST QUARTER | 2ND QUARTER | 3RD QUARTER | 4TH QUARTER | TOTAL GRAMS |
|---|---|---|---|---|---|
| 460 | 612.66 | 541.11 | 466.25 | 468.04 | **2,088.07** |
| 461 | 1,179.27 | 1,078.20 | 1,845.77 | 2,205.32 | **6,308.56** |
| 462 | 18,727.13 | 16,119.68 | 17,729.78 | 11,658.76 | **64,235.35** |
| 463 | 2,941.95 | 3,111.26 | 2,832.74 | 2,306.30 | **11,192.25** |
| 464 | 5,159.35 | 4,277.92 | 6,377.97 | 6,369.92 | **22,185.14** |
| 465 | 1,838.89 | 1,665.82 | 1,392.94 | 1,282.21 | **6,179.86** |
| 466 | 3,702.55 | 4,112.54 | 3,621.43 | 3,162.60 | **14,599.11** |
| 467 | 894.04 | 869.45 | 833.58 | 834.83 | **3,431.90** |
| 468 | 6,401.40 | 6,739.21 | 5,187.25 | 6,505.51 | **24,833.37** |
| 469 | 7,363.60 | 7,226.04 | 7,269.15 | 7,735.40 | **29,594.18** |
| 470 | 6,768.01 | 4,146.53 | 5,434.55 | 4,147.33 | **20,496.43** |
| 471 | 12,683.67 | 12,808.61 | 10,978.40 | 8,301.82 | **44,772.50** |
| 472 | 437.81 | 331.38 | 347.03 | 321.98 | **1,438.20** |
| 473 | 5,806.98 | 6,218.14 | 7,050.02 | 3,801.20 | **22,876.34** |
| 474 | 735.20 | 965.50 | 1,126.94 | 1,104.14 | **3,931.78** |
| 475 | 299.18 | 253.12 | 227.09 | 224.85 | **1,004.23** |
| 476 | 242.38 | 245.07 | 238.36 | 224.05 | **949.85** |
| 477 | 4,115.13 | 3,510.07 | 3,895.29 | 2,277.59 | **13,798.09** |
| 478 | 139.53 | 533.06 | 469.56 | 1,078.20 | **2,220.35** |
| 479 | 1,017.29 | 1,219.34 | 812.47 | 1,139.47 | **4,188.56** |
| **IN - INDIANA - Total** | **81,066.00** | **75,972.04** | **78,136.57** | **65,149.51** | **300,324.12** |

**STATE: KS - KANSAS**

| REGISTRANT ZIP CODE 3 | 1ST QUARTER | 2ND QUARTER | 3RD QUARTER | 4TH QUARTER | TOTAL GRAMS |
|---|---|---|---|---|---|
| 660 | 1,760.27 | 1,722.79 | 1,603.66 | 1,657.05 | **6,743.78** |
| 661 | 3,545.85 | 4,144.47 | 3,438.07 | 3,704.07 | **14,832.46** |

| | | | | | |
|---|---|---|---|---|---|
| 662 | 1,823.06 | 1,713.76 | 1,439.45 | 1,685.41 | **6,661.67** |
| 664 | 103.75 | 92.12 | 76.92 | 88.10 | **360.89** |
| 665 | 129.60 | 140.51 | 121.19 | 130.94 | **522.24** |
| 666 | 2,096.12 | 1,118.09 | 1,368.97 | 1,888.35 | **6,471.52** |
| 667 | 364.02 | 333.16 | 309.46 | 332.72 | **1,339.36** |
| 668 | 92.12 | 96.60 | 83.63 | 79.60 | **351.95** |
| 669 | 88.99 | 81.39 | 75.13 | 74.68 | **320.20** |
| 670 | 548.09 | 525.91 | 512.04 | 501.76 | **2,087.80** |
| 671 | 276.64 | 243.46 | 208.84 | 213.76 | **942.70** |
| 672 | 8,895.79 | 11,585.97 | 9,377.78 | 9,741.62 | **39,601.17** |
| 673 | 115.82 | 114.04 | 133.27 | 113.14 | **476.27** |
| 674 | 250.07 | 238.80 | 215.55 | 188.72 | **893.15** |
| 675 | 188.27 | 178.88 | 179.33 | 183.80 | **730.28** |
| 676 | 78.71 | 67.97 | 61.71 | 79.60 | **288.00** |
| 677 | 32.20 | 37.56 | 27.28 | 35.33 | **132.37** |
| 678 | 156.97 | 154.73 | 175.75 | 146.06 | **633.50** |
| 679 | 29.96 | 31.75 | 24.15 | 25.94 | **111.80** |
| **KS - KANSAS - Total** | **20,576.30** | **22,621.97** | **19,432.18** | **20,870.64** | **83,501.09** |

**STATE: KY - KENTUCKY**

| REGISTRANT ZIP CODE 3 | 1ST QUARTER | 2ND QUARTER | 3RD QUARTER | 4TH QUARTER | **TOTAL GRAMS** |
|---|---|---|---|---|---|
| 400 | 548.80 | 379.67 | 517.86 | 498.18 | **1,944.52** |
| 401 | 175.75 | 177.09 | 173.07 | 168.15 | **694.05** |
| 402 | 11,740.43 | 9,212.86 | 9,639.22 | 8,380.71 | **38,973.21** |
| 403 | 1,642.12 | 2,461.66 | 1,810.71 | 2,177.33 | **8,091.82** |
| 404 | 937.42 | 1,109.50 | 1,525.67 | 1,161.38 | **4,733.97** |
| 405 | 4,204.29 | 3,895.42 | 4,188.75 | 3,840.82 | **16,129.28** |
| 406 | 1,566.99 | 1,719.04 | 1,877.70 | 1,605.00 | **6,768.73** |
| 407 | 2,186.36 | 2,161.32 | 1,741.40 | 2,142.54 | **8,231.61** |
| 408 | 80.05 | 79.15 | 56.79 | 61.71 | **277.71** |
| 409 | 165.02 | 147.58 | 148.47 | 153.84 | **614.90** |
| 410 | 11,288.31 | 10,561.25 | 10,061.82 | 9,590.20 | **41,501.59** |
| 411 | 2,285.46 | 2,275.35 | 4,730.75 | 2,611.20 | **11,902.76** |
| 412 | 1,242.77 | 862.65 | 1,237.85 | 1,232.48 | **4,575.75** |
| 413 | 195.43 | 158.31 | 172.62 | 151.60 | **677.96** |
| 414 | 86.76 | 74.68 | 55.01 | 76.47 | **292.92** |
| 415 | 1,204.31 | 1,217.28 | 1,580.40 | 840.74 | **4,842.73** |
| 416 | 164.12 | 151.42 | 139.97 | 147.13 | **602.65** |
| 417 | 2,577.66 | 2,537.41 | 2,545.91 | 2,007.48 | **9,668.46** |
| 418 | 37.12 | 36.22 | 23.25 | 33.09 | **129.69** |
| 420 | 1,694.26 | 2,369.80 | 1,410.74 | 2,400.57 | **7,875.37** |

| | | | | | |
|---|---|---|---|---|---|
| 421 | 2,473.02 | 2,602.70 | 2,178.94 | 2,111.68 | **9,366.34** |
| 422 | 1,718.14 | 1,710.09 | 1,692.65 | 1,703.38 | **6,824.27** |
| 423 | 770.70 | 760.24 | 1,296.88 | 737.43 | **3,565.26** |
| 424 | 356.69 | 367.87 | 321.36 | 283.52 | **1,329.44** |
| 425 | 119.85 | 431.01 | 690.92 | 900.21 | **2,142.00** |
| 426 | 87.20 | 90.60 | 72.89 | 63.95 | **314.65** |
| 427 | 1,686.39 | 2,398.78 | 1,309.40 | 1,650.62 | **7,045.19** |
| **KY - KENTUCKY - Total** | **51,235.42** | **49,948.97** | **51,201.01** | **46,731.41** | **199,116.82** |

**STATE: LA - LOUISIANA**

| REGISTRANT ZIP CODE 3 | 1ST QUARTER | 2ND QUARTER | 3RD QUARTER | 4TH QUARTER | TOTAL GRAMS |
|---|---|---|---|---|---|
| 700 | 6,570.80 | 7,973.58 | 5,999.55 | 6,930.35 | **27,474.27** |
| 701 | 5,769.33 | 3,121.37 | 5,231.08 | 5,318.37 | **19,440.14** |
| 703 | 158.93 | 170.38 | 131.92 | 165.46 | **626.71** |
| 704 | 3,940.10 | 3,903.70 | 3,810.77 | 3,842.07 | **15,496.64** |
| 705 | 3,465.17 | 3,398.27 | 4,254.48 | 2,351.20 | **13,469.13** |
| 706 | 2,042.63 | 2,126.44 | 1,856.33 | 1,924.75 | **7,950.14** |
| 707 | 380.57 | 348.64 | 355.97 | 372.70 | **1,457.87** |
| 708 | 3,676.25 | 3,867.92 | 3,842.70 | 2,607.44 | **13,994.32** |
| 710 | 396.22 | 391.75 | 370.28 | 353.65 | **1,511.89** |
| 711 | 4,474.33 | 3,725.18 | 3,852.68 | 3,737.20 | **15,789.38** |
| 712 | 988.76 | 1,611.08 | 930.62 | 913.99 | **4,444.45** |
| 713 | 3,873.65 | 3,819.98 | 3,781.08 | 3,772.58 | **15,247.28** |
| 714 | 377.88 | 358.21 | 329.68 | 326.90 | **1,392.67** |
| **LA - LOUISIANA - Total** | **36,114.62** | **34,816.49** | **34,747.14** | **32,616.66** | **138,294.90** |

**STATE: MA - MASSACHUSETTS**

| REGISTRANT ZIP CODE 3 | 1ST QUARTER | 2ND QUARTER | 3RD QUARTER | 4TH QUARTER | TOTAL GRAMS |
|---|---|---|---|---|---|
| 010 | 11,239.03 | 11,124.64 | 11,786.22 | 9,847.52 | **43,997.41** |
| 011 | 7,643.63 | 6,089.25 | 7,874.30 | 6,420.99 | **28,028.17** |
| 012 | 5,385.09 | 3,465.89 | 5,487.32 | 3,497.46 | **17,835.77** |
| 013 | 1,436.05 | 1,116.30 | 1,401.08 | 1,243.13 | **5,196.55** |
| 014 | 6,409.27 | 5,234.30 | 6,478.14 | 4,830.65 | **22,952.36** |
| 015 | 4,443.11 | 4,237.22 | 4,285.96 | 3,213.76 | **16,180.05** |
| 016 | 9,106.53 | 8,707.94 | 9,066.01 | 7,394.91 | **34,275.40** |
| 017 | 3,815.06 | 3,721.24 | 3,666.59 | 2,915.12 | **14,118.01** |
| 018 | 18,906.72 | 14,903.66 | 16,880.28 | 11,396.53 | **62,087.19** |
| 019 | 13,175.23 | 12,415.97 | 12,955.83 | 11,367.38 | **49,914.41** |
| 020 | 654.88 | 602.56 | 480.83 | 628.49 | **2,366.76** |
| 021 | 24,928.99 | 20,611.68 | 24,915.57 | 19,408.32 | **89,864.56** |
| 022 | 278.25 | 297.03 | 289.34 | 347.30 | **1,211.91** |
| 023 | 9,502.02 | 8,445.37 | 9,453.90 | 6,759.25 | **34,160.53** |

| | | | | | |
|---|---|---|---|---|---|
| 024 | 1,268.44 | 954.86 | 1,260.48 | 1,222.82 | **4,706.60** |
| 025 | 3,563.02 | 2,451.28 | 3,565.08 | 3,469.56 | **13,048.94** |
| 026 | 2,858.06 | 2,452.36 | 3,194.26 | 1,785.22 | **10,289.89** |
| 027 | 19,037.57 | 15,971.66 | 19,244.00 | 14,307.45 | **68,560.68** |
| **MA - MASSACHUSETTS - Total** | **143,650.94** | **122,803.21** | **142,285.19** | **110,055.87** | **518,795.21** |

**STATE: MD - MARYLAND**

| REGISTRANT ZIP CODE 3 | 1ST QUARTER | 2ND QUARTER | 3RD QUARTER | 4TH QUARTER | TOTAL GRAMS |
|---|---|---|---|---|---|
| 206 | 4,768.85 | 3,769.09 | 5,096.47 | 3,827.09 | **17,461.51** |
| 207 | 2,487.59 | 3,318.49 | 2,229.47 | 2,807.07 | **10,842.63** |
| 208 | 2,706.54 | 2,781.41 | 3,170.55 | 2,541.80 | **11,200.30** |
| 209 | 834.92 | 1,406.00 | 934.65 | 775.44 | **3,951.01** |
| 210 | 35,257.16 | 26,778.11 | 31,467.41 | 33,954.73 | **127,457.41** |
| 211 | 4,858.92 | 4,868.49 | 5,722.19 | 4,581.47 | **20,031.07** |
| 212 | 101,461.27 | 94,715.58 | 99,609.16 | 96,376.53 | **392,162.54** |
| 214 | 2,344.85 | 1,788.00 | 2,337.78 | 2,316.50 | **8,787.12** |
| 215 | 3,473.67 | 3,455.34 | 3,486.64 | 2,826.39 | **13,242.04** |
| 216 | 3,641.28 | 2,513.26 | 3,691.55 | 2,490.46 | **12,336.55** |
| 217 | 13,183.63 | 13,235.60 | 13,824.03 | 11,400.83 | **51,644.09** |
| 218 | 4,750.52 | 4,811.60 | 5,080.64 | 4,061.65 | **18,704.41** |
| 219 | 7,893.53 | 6,509.89 | 7,209.67 | 6,703.08 | **28,316.17** |
| **MD - MARYLAND - Total** | **187,662.74** | **169,950.86** | **183,860.20** | **174,663.05** | **716,136.84** |

**STATE: ME - MAINE**

| REGISTRANT ZIP CODE 3 | 1ST QUARTER | 2ND QUARTER | 3RD QUARTER | 4TH QUARTER | TOTAL GRAMS |
|---|---|---|---|---|---|
| 039 | 78.26 | 69.32 | 78.71 | 66.19 | **292.47** |
| 040 | 4,782.00 | 3,631.35 | 3,664.80 | 3,623.21 | **15,701.37** |
| 041 | 5,006.67 | 4,178.64 | 5,975.13 | 2,974.68 | **18,135.12** |
| 042 | 1,986.46 | 1,371.03 | 1,945.59 | 1,374.07 | **6,677.14** |
| 043 | 598.26 | 605.51 | 541.74 | 528.68 | **2,274.19** |
| 044 | 10,878.23 | 7,824.93 | 9,479.30 | 8,103.80 | **36,286.26** |
| 045 | 91.68 | 83.63 | 88.55 | 75.13 | **338.98** |
| 046 | 1,393.65 | 1,397.95 | 1,389.90 | 1,079.54 | **5,261.04** |
| 047 | 75.13 | 64.84 | 85.86 | 64.84 | **290.68** |
| 048 | 1,593.82 | 1,519.59 | 1,499.19 | 1,975.19 | **6,587.79** |
| 049 | 4,326.39 | 2,597.34 | 5,081.80 | 2,467.65 | **14,473.18** |
| **ME - MAINE - Total** | **30,810.56** | **23,344.11** | **29,830.57** | **22,332.99** | **106,318.22** |

**STATE: MI - MICHIGAN**

| REGISTRANT ZIP CODE 3 | 1ST QUARTER | 2ND QUARTER | 3RD QUARTER | 4TH QUARTER | TOTAL GRAMS |
|---|---|---|---|---|---|
| 480 | 16,152.15 | 15,202.07 | 16,132.34 | 15,494.05 | **62,980.62** |
| 481 | 11,080.18 | 13,340.42 | 10,089.64 | 10,444.18 | **44,954.42** |
| 482 | 15,121.96 | 15,534.19 | 14,943.81 | 14,225.07 | **59,825.04** |

| | | | | | |
|---|---|---|---|---|---|
| 483 | 10,969.10 | 12,088.98 | 12,762.01 | 11,650.19 | **47,470.28** |
| 484 | 1,054.68 | 903.97 | 902.00 | 877.41 | **3,738.06** |
| 485 | 9,550.85 | 11,755.99 | 8,696.70 | 9,495.84 | **39,499.39** |
| 486 | 4,841.83 | 4,705.89 | 5,410.85 | 3,804.42 | **18,762.99** |
| 487 | 544.42 | 502.65 | 461.06 | 459.72 | **1,967.86** |
| 488 | 5,816.19 | 5,210.15 | 5,094.86 | 5,358.71 | **21,479.91** |
| 489 | 7,211.75 | 6,602.46 | 7,888.25 | 5,821.92 | **27,524.38** |
| 490 | 10,596.94 | 10,150.37 | 10,094.29 | 8,252.00 | **39,093.60** |
| 491 | 458.83 | 402.48 | 398.90 | 382.53 | **1,642.74** |
| 492 | 5,275.62 | 4,930.83 | 4,546.24 | 3,077.18 | **17,829.86** |
| 493 | 966.76 | 953.97 | 856.84 | 773.48 | **3,551.04** |
| 494 | 9,912.10 | 9,899.40 | 9,000.44 | 8,722.73 | **37,534.66** |
| 495 | 8,460.04 | 9,103.38 | 9,084.78 | 6,712.56 | **33,360.76** |
| 496 | 2,517.91 | 2,296.46 | 2,209.79 | 2,082.52 | **9,106.69** |
| 497 | 5,065.52 | 3,243.99 | 4,922.33 | 3,037.83 | **16,269.67** |
| 498 | 838.23 | 748.17 | 628.32 | 490.58 | **2,705.29** |
| 499 | 151.51 | 127.00 | 128.35 | 70.21 | **477.07** |
| **MI - MICHIGAN - Total** | **126,586.59** | **127,702.82** | **124,251.80** | **111,233.13** | **489,774.33** |

**STATE: MN - MINNESOTA**

| REGISTRANT ZIP CODE 3 | 1ST QUARTER | 2ND QUARTER | 3RD QUARTER | 4TH QUARTER | **TOTAL GRAMS** |
|---|---|---|---|---|---|
| 550 | 911.48 | 900.12 | 817.84 | 767.22 | **3,396.66** |
| 551 | 12,991.52 | 11,864.84 | 12,929.63 | 12,382.88 | **50,168.86** |
| 553 | 2,373.92 | 3,084.88 | 3,074.95 | 2,265.52 | **10,799.25** |
| 554 | 26,522.63 | 29,354.57 | 27,492.25 | 22,942.08 | **106,311.51** |
| 556 | 13.42 | 20.12 | 22.81 | 15.20 | **71.55** |
| 557 | 331.91 | 304.99 | 288.00 | 247.30 | **1,172.20** |
| 558 | 4,076.85 | 4,006.11 | 2,786.95 | 4,030.35 | **14,900.26** |
| 559 | 905.94 | 2,170.17 | 1,516.54 | 875.53 | **5,468.18** |
| 560 | 236.39 | 211.79 | 223.06 | 195.07 | **866.32** |
| 561 | 37.92 | 34.70 | 47.22 | 33.54 | **153.39** |
| 562 | 99.46 | 97.49 | 94.90 | 98.83 | **390.67** |
| 563 | 1,944.87 | 2,108.73 | 2,798.31 | 1,478.35 | **8,330.26** |
| 564 | 3,878.03 | 3,852.54 | 3,873.83 | 3,846.64 | **15,451.03** |
| 565 | 205.26 | 188.09 | 176.02 | 135.77 | **705.14** |
| 566 | 214.12 | 108.67 | 97.49 | 86.04 | **506.32** |
| 567 | 104.64 | 112.69 | 98.83 | 84.79 | **400.96** |
| **MN - MINNESOTA - Total** | **54,848.36** | **58,420.51** | **56,338.61** | **49,485.09** | **219,092.58** |

**STATE: MO - MISSOURI**

| REGISTRANT ZIP CODE 3 | 1ST QUARTER | 2ND QUARTER | 3RD QUARTER | 4TH QUARTER | **TOTAL GRAMS** |
|---|---|---|---|---|---|
| 630 | 5,301.29 | 4,213.52 | 4,962.94 | 3,195.51 | **17,673.25** |

| | | | | | |
|---|---|---|---|---|---|
| 631 | 9,563.78 | 12,392.00 | 10,251.47 | 9,014.99 | **41,222.24** |
| 633 | 963.18 | 1,494.54 | 1,933.78 | 1,637.74 | **6,029.24** |
| 634 | 204.82 | 192.30 | 216.00 | 205.71 | **818.82** |
| 635 | 50.53 | 46.96 | 33.54 | 23.70 | **154.73** |
| 636 | 288.71 | 269.57 | 244.17 | 250.88 | **1,053.33** |
| 637 | 1,538.99 | 2,010.16 | 1,038.76 | 1,143.49 | **5,731.40** |
| 638 | 127.36 | 124.32 | 115.38 | 122.53 | **489.59** |
| 639 | 1,249.48 | 977.58 | 737.43 | 754.87 | **3,719.36** |
| 640 | 966.67 | 954.86 | 908.89 | 863.90 | **3,694.32** |
| 641 | 3,785.82 | 5,047.99 | 4,291.05 | 4,394.72 | **17,519.58** |
| 644 | 101.07 | 81.39 | 57.69 | 81.39 | **321.54** |
| 645 | 2,236.00 | 2,166.24 | 2,155.50 | 1,426.12 | **7,983.86** |
| 646 | 132.37 | 115.38 | 84.52 | 72.45 | **404.72** |
| 647 | 161.53 | 142.03 | 131.48 | 128.79 | **563.83** |
| 648 | 1,637.47 | 1,939.95 | 1,410.74 | 2,252.10 | **7,240.26** |
| 650 | 157.41 | 133.27 | 119.85 | 119.85 | **530.38** |
| 651 | 110.91 | 99.10 | 87.20 | 95.07 | **392.28** |
| 652 | 1,697.12 | 1,369.77 | 1,326.40 | 1,193.93 | **5,587.23** |
| 653 | 89.89 | 78.71 | 86.31 | 92.12 | **347.03** |
| 654 | 129.69 | 116.27 | 114.93 | 88.99 | **449.88** |
| 655 | 200.79 | 188.27 | 161.89 | 164.12 | **715.07** |
| 656 | 440.31 | 388.08 | 353.02 | 330.21 | **1,511.63** |
| 657 | 1,587.56 | 1,379.43 | 1,324.34 | 1,468.87 | **5,760.20** |
| 658 | 4,361.36 | 4,571.28 | 3,691.28 | 4,530.40 | **17,154.32** |
| **MO - MISSOURI - Total** | **37,084.11** | **40,492.97** | **35,838.54** | **33,652.49** | **147,068.11** |

**STATE: MS - MISSISSIPPI**

| REGISTRANT ZIP CODE 3 | 1ST QUARTER | 2ND QUARTER | 3RD QUARTER | 4TH QUARTER | TOTAL GRAMS |
|---|---|---|---|---|---|
| 386 | 986.08 | 1,258.87 | 1,323.26 | 1,489.18 | **5,057.38** |
| 387 | 286.66 | 254.90 | 202.58 | 162.33 | **906.47** |
| 388 | 121.46 | 111.80 | 95.79 | 91.41 | **420.46** |
| 389 | 96.60 | 88.99 | 95.25 | 59.92 | **340.77** |
| 390 | 196.86 | 181.56 | 180.67 | 180.67 | **739.76** |
| 391 | 184.25 | 131.83 | 162.78 | 114.93 | **593.79** |
| 392 | 1,253.50 | 1,774.49 | 1,430.59 | 1,599.28 | **6,057.86** |
| 393 | 766.05 | 1,219.96 | 629.66 | 931.07 | **3,546.74** |
| 394 | 392.64 | 351.50 | 333.16 | 250.43 | **1,327.74** |
| 395 | 2,464.43 | 2,646.08 | 1,905.07 | 2,441.71 | **9,457.30** |
| 396 | 165.02 | 146.23 | 117.61 | 133.71 | **562.58** |
| 397 | 86.13 | 80.23 | 84.61 | 81.75 | **332.72** |
| **MS - MISSISSIPPI - Total** | **6,999.66** | **8,246.46** | **6,561.05** | **7,536.39** | **29,343.56** |

**STATE: MT - MONTANA**

| REGISTRANT ZIP CODE 3 | 1ST QUARTER | 2ND QUARTER | 3RD QUARTER | 4TH QUARTER | TOTAL GRAMS |
|---|---|---|---|---|---|
| 590 | 53.22 | 52.32 | 55.01 | 58.58 | **219.13** |
| 591 | 1,463.95 | 1,477.10 | 2,133.68 | 1,507.51 | **6,582.25** |
| 592 | 22.81 | 19.68 | 21.91 | 26.83 | **91.23** |
| 593 | 44.72 | 33.54 | 53.22 | 38.91 | **170.38** |
| 594 | 158.13 | 135.05 | 109.21 | 114.21 | **516.61** |
| 595 | 32.20 | 27.73 | 25.94 | 9.84 | **95.70** |
| 596 | 253.12 | 201.24 | 195.43 | 124.77 | **774.55** |
| 597 | 972.66 | 1,230.96 | 938.40 | 1,205.20 | **4,347.23** |
| 598 | 2,215.61 | 2,434.47 | 2,597.07 | 2,591.79 | **9,838.94** |
| 599 | 2,349.68 | 2,364.88 | 2,352.09 | 2,317.03 | **9,383.69** |
| **MT - MONTANA - Total** | **7,566.09** | **7,976.97** | **8,481.95** | **7,994.68** | **32,019.70** |

**STATE: NC - NORTH CAROLINA**

| REGISTRANT ZIP CODE 3 | 1ST QUARTER | 2ND QUARTER | 3RD QUARTER | 4TH QUARTER | TOTAL GRAMS |
|---|---|---|---|---|---|
| 270 | 538.97 | 444.07 | 438.26 | 425.29 | **1,846.58** |
| 271 | 3,947.88 | 5,407.10 | 4,931.72 | 3,318.05 | **17,604.74** |
| 272 | 6,781.61 | 6,192.02 | 6,445.93 | 6,043.73 | **25,463.29** |
| 273 | 5,168.74 | 4,068.63 | 4,595.87 | 4,404.92 | **18,238.16** |
| 274 | 9,287.00 | 3,551.22 | 6,622.14 | 6,535.02 | **25,995.38** |
| 275 | 7,627.32 | 6,184.24 | 8,277.95 | 7,119.87 | **29,209.38** |
| 276 | 6,010.37 | 5,420.51 | 6,185.58 | 5,602.16 | **23,218.62** |
| 277 | 7,066.21 | 6,240.85 | 7,365.83 | 6,348.36 | **27,021.26** |
| 278 | 5,343.32 | 4,964.01 | 5,812.88 | 4,336.59 | **20,456.81** |
| 279 | 886.80 | 1,340.97 | 817.03 | 1,126.94 | **4,171.75** |
| 280 | 9,069.84 | 8,925.40 | 9,498.98 | 8,532.75 | **36,026.97** |
| 281 | 6,349.26 | 5,430.35 | 6,768.37 | 6,553.72 | **25,101.69** |
| 282 | 7,237.66 | 7,176.49 | 7,336.05 | 6,386.55 | **28,136.75** |
| 283 | 9,924.80 | 10,050.46 | 11,839.35 | 8,698.13 | **40,512.74** |
| 284 | 6,223.48 | 6,245.42 | 5,232.71 | 6,599.69 | **24,301.29** |
| 285 | 5,175.36 | 3,915.24 | 4,940.58 | 4,345.00 | **18,376.16** |
| 286 | 14,105.76 | 14,429.53 | 11,553.68 | 12,143.54 | **52,232.51** |
| 287 | 9,127.44 | 7,621.36 | 7,669.57 | 7,723.77 | **32,142.14** |
| 288 | 14,889.61 | 11,421.04 | 15,280.47 | 11,718.88 | **53,310.00** |
| 289 | 1,043.32 | 1,432.38 | 1,048.95 | 960.59 | **4,485.24** |
| **NC - NORTH CAROLINA - Total** | **135,804.74** | **120,461.28** | **132,661.89** | **118,923.54** | **507,851.46** |

**STATE: ND - NORTH DAKOTA**

| REGISTRANT ZIP CODE 3 | 1ST QUARTER | 2ND QUARTER | 3RD QUARTER | 4TH QUARTER | TOTAL GRAMS |
|---|---|---|---|---|---|
| 580 | 75.58 | 75.58 | 80.94 | 59.30 | **291.40** |
| 581 | 1,192.50 | 1,704.73 | 2,217.31 | 1,675.75 | **6,790.28** |

| | | | | | |
|---|---|---|---|---|---|
| 582 | 207.86 | 188.72 | 165.91 | 202.13 | **764.62** |
| 583 | 29.52 | 25.04 | 26.83 | 26.83 | **108.22** |
| 584 | 30.86 | 31.75 | 25.76 | 23.97 | **112.34** |
| 585 | 792.26 | 808.98 | 1,341.78 | 1,064.34 | **4,007.36** |
| 586 | 27.28 | 43.38 | 56.79 | 41.59 | **169.04** |
| 587 | 765.16 | 758.54 | 756.13 | 1,119.70 | **3,399.52** |
| 588 | 55.01 | 55.01 | 63.06 | 58.14 | **231.20** |
| **ND - NORTH DAKOTA - Total** | **3,176.01** | **3,691.73** | **4,734.51** | **4,271.74** | **15,873.99** |

**STATE: NE - NEBRASKA**

| REGISTRANT ZIP CODE 3 | 1ST QUARTER | 2ND QUARTER | 3RD QUARTER | 4TH QUARTER | TOTAL GRAMS |
|---|---|---|---|---|---|
| 680 | 305.35 | 301.41 | 286.30 | 246.23 | **1,139.29** |
| 681 | 4,778.42 | 3,836.26 | 5,298.87 | 4,153.95 | **18,067.51** |
| 683 | 40.96 | 52.77 | 48.74 | 45.17 | **187.65** |
| 684 | 31.75 | 42.04 | 32.65 | 36.22 | **142.66** |
| 685 | 1,278.56 | 1,601.33 | 1,615.29 | 1,724.73 | **6,219.91** |
| 686 | 59.92 | 63.06 | 55.45 | 55.45 | **233.89** |
| 687 | 165.91 | 144.89 | 148.02 | 126.11 | **584.94** |
| 688 | 214.21 | 177.54 | 186.48 | 149.81 | **728.04** |
| 689 | 40.16 | 47.22 | 44.27 | 47.22 | **178.88** |
| 690 | 17.89 | 18.78 | 15.20 | 13.42 | **65.29** |
| 691 | 99.99 | 98.21 | 97.85 | 113.23 | **409.28** |
| 692 | 0.89 | 2.24 | 4.02 | 1.79 | **8.94** |
| 693 | 113.86 | 98.56 | 99.90 | 75.31 | **387.63** |
| **NE - NEBRASKA - Total** | **7,147.88** | **6,484.31** | **7,933.06** | **6,788.64** | **28,353.89** |

**STATE: NH - NEW HAMPSHIRE**

| REGISTRANT ZIP CODE 3 | 1ST QUARTER | 2ND QUARTER | 3RD QUARTER | 4TH QUARTER | TOTAL GRAMS |
|---|---|---|---|---|---|
| 030 | 3,409.99 | 3,300.60 | 3,769.81 | 2,611.29 | **13,091.69** |
| 031 | 9,971.76 | 7,402.95 | 10,703.46 | 9,111.34 | **37,189.51** |
| 032 | 537.53 | 1,221.75 | 484.05 | 1,188.03 | **3,431.37** |
| 033 | 5,947.67 | 6,021.46 | 3,114.21 | 5,764.94 | **20,848.29** |
| 034 | 1,971.26 | 3,829.82 | 1,967.23 | 1,955.16 | **9,723.47** |
| 035 | 196.86 | 184.69 | 183.80 | 178.43 | **743.78** |
| 036 | 28.17 | 29.96 | 33.54 | 25.04 | **116.72** |
| 037 | 3,217.69 | 2,630.25 | 2,876.66 | 1,888.88 | **10,613.49** |
| 038 | 6,813.36 | 6,574.20 | 6,341.83 | 4,382.11 | **24,111.50** |
| **NH - NEW HAMPSHIRE - Total** | **32,094.29** | **31,195.69** | **29,474.59** | **27,105.24** | **119,869.81** |

**STATE: NJ - NEW JERSEY**

| REGISTRANT ZIP CODE 3 | 1ST QUARTER | 2ND QUARTER | 3RD QUARTER | 4TH QUARTER | TOTAL GRAMS |
|---|---|---|---|---|---|
| 070 | 7,705.26 | 9,324.03 | 8,224.99 | 8,155.59 | **33,409.86** |
| 071 | 9,071.54 | 13,429.15 | 10,614.11 | 13,489.52 | **46,604.32** |

| | | | | | |
|---|---|---|---|---|---|
| 072 | 1,578.17 | 1,573.70 | 1,584.43 | 965.06 | **5,701.35** |
| 073 | 5,560.48 | 5,695.45 | 5,707.61 | 3,104.82 | **20,068.37** |
| 074 | 702.19 | 602.83 | 570.63 | 540.22 | **2,415.86** |
| 075 | 3,181.83 | 3,444.78 | 4,371.38 | 2,930.32 | **13,928.31** |
| 076 | 2,508.79 | 2,081.72 | 2,430.80 | 1,652.67 | **8,673.98** |
| 077 | 9,515.52 | 10,573.60 | 8,637.76 | 12,874.08 | **41,600.96** |
| 078 | 3,007.51 | 2,984.61 | 2,967.44 | 2,973.61 | **11,933.17** |
| 079 | 221.99 | 237.46 | 197.93 | 238.00 | **895.38** |
| 080 | 3,599.16 | 2,657.98 | 3,485.12 | 3,400.87 | **13,143.12** |
| 081 | 24,879.44 | 20,136.52 | 25,393.45 | 20,095.56 | **90,504.96** |
| 082 | 7,381.75 | 9,355.60 | 7,937.62 | 9,258.83 | **33,933.80** |
| 083 | 4,920.54 | 6,245.60 | 5,745.18 | 7,500.89 | **24,412.20** |
| 084 | 124.77 | 141.32 | 146.68 | 154.28 | **567.05** |
| 085 | 382.27 | 396.04 | 399.53 | 365.99 | **1,543.82** |
| 086 | 5,006.76 | 5,263.99 | 6,507.65 | 5,034.85 | **21,813.25** |
| 087 | 10,974.74 | 11,627.65 | 11,243.06 | 12,312.31 | **46,157.75** |
| 088 | 11,130.18 | 8,016.78 | 11,243.77 | 9,141.75 | **39,532.48** |
| 089 | 3,882.86 | 1,869.12 | 3,978.74 | 1,944.52 | **11,675.23** |
| **NJ - NEW JERSEY - Total** | **115,335.74** | **115,657.90** | **121,387.88** | **116,133.73** | **468,515.25** |

**STATE: NM - NEW MEXICO**

| REGISTRANT ZIP CODE 3 | 1ST QUARTER | 2ND QUARTER | 3RD QUARTER | 4TH QUARTER | **TOTAL GRAMS** |
|---|---|---|---|---|---|
| 870 | 1,810.27 | 1,245.00 | 1,253.50 | 1,372.46 | **5,681.23** |
| 871 | 29,759.82 | 30,418.81 | 33,869.50 | 28,930.80 | **122,978.93** |
| 873 | 36.67 | 39.35 | 42.04 | 44.72 | **162.78** |
| 874 | 643.97 | 1,370.85 | 1,250.55 | 452.57 | **3,717.93** |
| 875 | 5,682.57 | 5,279.91 | 5,477.75 | 8,133.67 | **24,573.91** |
| 877 | 66.19 | 45.61 | 51.88 | 34.88 | **198.56** |
| 878 | 13.42 | 14.31 | 12.52 | 16.99 | **57.24** |
| 879 | 8.50 | 6.71 | 7.16 | 3.58 | **25.94** |
| 880 | 2,199.87 | 2,345.83 | 2,332.42 | 1,616.63 | **8,494.74** |
| 881 | 25.94 | 25.49 | 26.38 | 25.76 | **103.57** |
| 882 | 1,359.31 | 1,012.19 | 1,347.41 | 1,195.37 | **4,914.28** |
| 883 | 156.07 | 141.32 | 147.13 | 115.82 | **560.34** |
| 884 | 4.02 | 3.58 | 3.58 | 5.81 | **16.99** |
| **NM - NEW MEXICO - Total** | **41,766.60** | **41,948.97** | **45,821.81** | **41,949.06** | **171,486.44** |

**STATE: NV - NEVADA**

| REGISTRANT ZIP CODE 3 | 1ST QUARTER | 2ND QUARTER | 3RD QUARTER | 4TH QUARTER | **TOTAL GRAMS** |
|---|---|---|---|---|---|
| 890 | 9,152.84 | 8,308.26 | 9,752.54 | 9,371.17 | **36,584.81** |
| 891 | 17,218.36 | 15,173.23 | 14,964.83 | 14,351.63 | **61,708.05** |
| 893 | 35.78 | 33.09 | 33.09 | 26.83 | **128.79** |

| | | | | | |
|---|---|---|---|---|---|
| 894 | 4,835.13 | 3,885.63 | 4,715.28 | 3,697.00 | **17,133.04** |
| 895 | 3,107.59 | 4,542.75 | 2,875.94 | 4,397.32 | **14,923.60** |
| 897 | 1,498.03 | 1,437.66 | 1,405.10 | 1,509.75 | **5,850.54** |
| 898 | 59.92 | 57.24 | 64.40 | 56.79 | **238.36** |
| **NV - NEVADA - Total** | **35,907.66** | **33,437.86** | **33,811.18** | **33,410.49** | **136,567.19** |

**STATE: NY - NEW YORK**

| REGISTRANT ZIP CODE 3 | 1ST QUARTER | 2ND QUARTER | 3RD QUARTER | 4TH QUARTER | TOTAL GRAMS |
|---|---|---|---|---|---|
| 100 | 85,397.53 | 77,501.96 | 81,097.52 | 59,732.60 | **303,729.61** |
| 101 | 1,337.75 | 1,356.09 | 1,353.23 | 1,337.31 | **5,384.38** |
| 102 | 15.20 | 16.10 | 13.42 | 15.20 | **59.92** |
| 103 | 11,251.64 | 9,838.13 | 9,874.89 | 8,313.90 | **39,278.56** |
| 104 | 57,101.18 | 53,588.69 | 71,522.57 | 39,530.33 | **221,742.78** |
| 105 | 4,649.27 | 4,959.63 | 6,134.42 | 4,761.07 | **20,504.39** |
| 106 | 120.12 | 120.83 | 113.14 | 111.35 | **465.45** |
| 107 | 4,760.89 | 6,565.70 | 4,824.84 | 3,893.59 | **20,045.02** |
| 108 | 1,190.00 | 2,003.01 | 1,285.25 | 1,385.43 | **5,863.69** |
| 109 | 2,927.64 | 3,729.74 | 3,154.82 | 3,396.48 | **13,208.68** |
| 110 | 3,111.51 | 3,221.43 | 3,303.47 | 3,338.35 | **12,974.76** |
| 111 | 144.89 | 174.41 | 171.72 | 131.48 | **622.50** |
| 112 | 31,146.98 | 28,685.38 | 32,557.75 | 26,220.31 | **118,610.42** |
| 113 | 6,171.63 | 6,635.20 | 7,191.15 | 8,302.62 | **28,300.60** |
| 114 | 6,817.56 | 5,705.91 | 6,143.19 | 4,988.34 | **23,655.00** |
| 115 | 2,750.46 | 6,266.35 | 2,743.39 | 4,724.31 | **16,484.51** |
| 116 | 128.79 | 110.64 | 113.59 | 110.01 | **463.03** |
| 117 | 9,800.84 | 11,005.95 | 10,222.54 | 8,187.07 | **39,216.40** |
| 118 | 130.14 | 117.17 | 117.61 | 114.30 | **479.22** |
| 119 | 2,540.10 | 2,513.71 | 2,527.04 | 1,817.87 | **9,398.71** |
| 120 | 1,524.59 | 1,054.32 | 1,451.61 | 1,385.43 | **5,415.95** |
| 121 | 710.78 | 673.93 | 937.60 | 629.21 | **2,951.52** |
| 122 | 6,964.96 | 6,074.85 | 7,235.79 | 5,201.11 | **25,476.72** |
| 123 | 417.68 | 464.19 | 438.70 | 540.31 | **1,860.89** |
| 124 | 3,059.12 | 3,192.02 | 2,587.95 | 3,441.29 | **12,280.38** |
| 125 | 3,350.42 | 3,592.00 | 6,293.89 | 4,427.28 | **17,663.60** |
| 126 | 3,415.98 | 3,053.03 | 2,851.35 | 3,135.14 | **12,455.50** |
| 127 | 338.98 | 327.80 | 295.78 | 257.50 | **1,220.05** |
| 128 | 771.15 | 683.77 | 665.43 | 676.97 | **2,797.33** |
| 129 | 1,208.87 | 571.43 | 1,206.10 | 1,025.88 | **4,012.28** |
| 130 | 1,779.41 | 1,122.47 | 2,193.87 | 1,399.47 | **6,495.22** |
| 131 | 854.15 | 882.86 | 847.71 | 823.74 | **3,408.47** |
| 132 | 7,988.24 | 6,829.01 | 7,884.49 | 6,803.61 | **29,505.36** |

| | | | | | |
|---|---|---|---|---|---|
| 133 | 195.87 | 183.80 | 211.70 | 188.09 | **779.47** |
| 134 | 1,612.42 | 1,592.93 | 1,609.65 | 1,594.36 | **6,409.36** |
| 135 | 221.01 | 479.13 | 721.15 | 442.73 | **1,864.02** |
| 136 | 767.22 | 801.92 | 1,026.77 | 628.76 | **3,224.67** |
| 137 | 216.89 | 175.75 | 165.46 | 180.22 | **738.33** |
| 138 | 491.92 | 447.65 | 461.06 | 478.06 | **1,878.69** |
| 139 | 1,289.72 | 1,606.34 | 1,621.99 | 1,589.35 | **6,107.41** |
| 140 | 1,187.85 | 1,162.63 | 1,231.95 | 1,253.32 | **4,835.75** |
| 141 | 475.64 | 616.69 | 702.19 | 540.40 | **2,334.92** |
| 142 | 15,914.71 | 13,966.41 | 17,101.38 | 11,346.69 | **58,329.19** |
| 143 | 804.16 | 850.93 | 1,469.59 | 830.36 | **3,955.04** |
| 144 | 630.02 | 606.40 | 577.16 | 635.74 | **2,449.31** |
| 145 | 358.21 | 334.15 | 350.34 | 334.86 | **1,377.55** |
| 146 | 4,950.33 | 4,720.91 | 6,467.76 | 5,392.82 | **21,531.82** |
| 147 | 492.01 | 432.08 | 370.19 | 315.45 | **1,609.74** |
| 148 | 687.08 | 622.41 | 588.25 | 575.10 | **2,472.84** |
| 149 | 223.15 | 200.79 | 134.61 | 132.37 | **690.92** |
| **NY - NEW YORK - Total** | **294,396.67** | **281,438.64** | **314,167.06** | **236,617.55** | **1,126,619.92** |

**STATE: OH - OHIO**

| REGISTRANT ZIP CODE 3 | 1ST QUARTER | 2ND QUARTER | 3RD QUARTER | 4TH QUARTER | **TOTAL GRAMS** |
|---|---|---|---|---|---|
| 430 | 1,397.77 | 1,351.26 | 1,372.55 | 1,223.72 | **5,345.29** |
| 431 | 760.24 | 687.61 | 643.79 | 673.39 | **2,765.04** |
| 432 | 14,841.76 | 14,452.63 | 13,653.41 | 13,382.48 | **56,330.28** |
| 433 | 601.04 | 587.35 | 555.60 | 511.33 | **2,255.32** |
| 434 | 254.90 | 312.06 | 269.93 | 264.38 | **1,101.27** |
| 435 | 274.04 | 220.47 | 224.23 | 203.48 | **922.22** |
| 436 | 4,900.95 | 3,904.06 | 4,796.49 | 5,231.88 | **18,833.38** |
| 437 | 299.45 | 497.11 | 451.49 | 669.10 | **1,917.15** |
| 438 | 102.41 | 96.51 | 74.68 | 87.20 | **360.80** |
| 439 | 399.17 | 368.14 | 364.65 | 398.28 | **1,530.23** |
| 440 | 1,326.31 | 1,276.67 | 1,213.25 | 1,225.69 | **5,041.91** |
| 441 | 9,054.28 | 8,109.44 | 8,420.69 | 8,671.74 | **34,256.15** |
| 442 | 733.50 | 708.45 | 1,324.79 | 1,889.69 | **4,656.43** |
| 443 | 4,118.62 | 4,094.92 | 3,703.80 | 3,579.12 | **15,496.46** |
| 444 | 1,506.62 | 1,676.73 | 1,486.22 | 1,415.57 | **6,085.14** |
| 445 | 3,969.88 | 4,062.19 | 5,075.00 | 6,075.93 | **19,183.00** |
| 446 | 571.34 | 504.71 | 502.12 | 498.90 | **2,077.07** |
| 447 | 1,703.74 | 1,691.76 | 1,498.57 | 1,692.56 | **6,586.63** |
| 448 | 642.27 | 583.95 | 582.25 | 566.78 | **2,375.26** |
| 449 | 132.46 | 119.94 | 108.67 | 104.20 | **465.27** |

| | | | | | |
|---|---|---|---|---|---|
| 450 | 3,361.14 | 3,519.82 | 3,570.27 | 3,589.05 | **14,040.28** |
| 451 | 732.07 | 744.14 | 692.27 | 639.05 | **2,807.52** |
| 452 | 12,586.45 | 14,025.98 | 13,891.00 | 13,550.90 | **54,054.33** |
| 453 | 758.81 | 769.36 | 743.07 | 740.92 | **3,012.16** |
| 454 | 4,249.47 | 5,597.42 | 5,217.93 | 6,004.74 | **21,069.57** |
| 455 | 150.71 | 158.31 | 140.15 | 151.51 | **600.68** |
| 456 | 950.75 | 1,276.76 | 1,774.94 | 2,158.37 | **6,160.81** |
| 457 | 348.37 | 238.80 | 300.25 | 338.08 | **1,225.51** |
| 458 | 779.11 | 1,189.64 | 620.80 | 791.90 | **3,381.46** |
| **OH - OHIO - Total** | **71,507.63** | **72,826.19** | **73,272.85** | **76,329.93** | **293,936.59** |

**STATE: OK - OKLAHOMA**

| REGISTRANT ZIP CODE 3 | 1ST QUARTER | 2ND QUARTER | 3RD QUARTER | 4TH QUARTER | TOTAL GRAMS |
|---|---|---|---|---|---|
| 730 | 974.18 | 813.64 | 726.97 | 593.88 | **3,108.67** |
| 731 | 11,245.02 | 9,588.42 | 10,800.95 | 10,584.15 | **42,218.54** |
| 734 | 2,033.60 | 1,127.39 | 1,834.06 | 1,680.31 | **6,675.35** |
| 735 | 1,083.57 | 1,101.01 | 1,035.72 | 1,747.21 | **4,967.50** |
| 736 | 62.61 | 65.29 | 50.98 | 37.12 | **216.00** |
| 737 | 136.49 | 136.40 | 118.06 | 96.86 | **487.81** |
| 738 | 60.82 | 60.82 | 61.71 | 60.37 | **243.72** |
| 739 | 21.91 | 22.81 | 20.12 | 16.10 | **80.94** |
| 740 | 2,543.85 | 2,514.16 | 2,320.34 | 2,025.82 | **9,404.17** |
| 741 | 11,554.66 | 8,811.27 | 12,379.75 | 9,887.68 | **42,633.36** |
| 743 | 1,235.61 | 142.21 | 1,201.18 | 621.61 | **3,200.61** |
| 744 | 566.87 | 510.70 | 435.57 | 317.33 | **1,830.48** |
| 745 | 430.21 | 418.58 | 508.91 | 546.48 | **1,904.18** |
| 746 | 1,071.94 | 1,033.93 | 1,102.35 | 1,110.40 | **4,318.61** |
| 747 | 165.46 | 189.52 | 151.06 | 131.83 | **637.89** |
| 748 | 231.20 | 242.38 | 211.97 | 173.07 | **858.62** |
| 749 | 3,668.83 | 3,094.62 | 3,633.50 | 4,293.57 | **14,690.52** |
| **OK - OKLAHOMA - Total** | **37,086.83** | **29,873.14** | **36,593.21** | **33,923.79** | **137,476.97** |

**STATE: OR - OREGON**

| REGISTRANT ZIP CODE 3 | 1ST QUARTER | 2ND QUARTER | 3RD QUARTER | 4TH QUARTER | TOTAL GRAMS |
|---|---|---|---|---|---|
| 970 | 2,059.53 | 1,959.09 | 1,940.67 | 1,874.57 | **7,833.87** |
| 971 | 1,201.54 | 1,128.73 | 1,123.37 | 1,033.48 | **4,487.12** |
| 972 | 35,717.00 | 26,034.16 | 32,155.82 | 26,009.51 | **119,916.49** |
| 973 | 9,241.39 | 8,846.42 | 8,997.75 | 7,615.73 | **34,701.29** |
| 974 | 6,471.79 | 6,965.32 | 6,579.12 | 6,834.47 | **26,850.69** |
| 975 | 7,016.03 | 6,166.62 | 7,163.34 | 5,357.37 | **25,703.36** |
| 976 | 253.12 | 240.15 | 228.97 | 203.48 | **925.70** |
| 977 | 1,195.10 | 1,592.03 | 1,185.44 | 1,235.88 | **5,208.45** |

| | | | | | |
|---|---|---|---|---|---|
| 978 | 391.30 | 373.86 | 339.87 | 289.43 | **1,394.46** |
| 979 | 137.29 | 121.64 | 127.90 | 105.99 | **492.81** |
| **OR - OREGON - Total** | **63,684.08** | **53,428.02** | **59,842.24** | **50,559.90** | **227,514.24** |

**STATE: PA - PENNSYLVANIA**

| REGISTRANT ZIP CODE 3 | 1ST QUARTER | 2ND QUARTER | 3RD QUARTER | 4TH QUARTER | TOTAL GRAMS |
|---|---|---|---|---|---|
| 150 | 3,294.34 | 4,835.93 | 2,754.30 | 3,710.42 | **14,595.00** |
| 151 | 474.39 | 441.65 | 391.93 | 394.79 | **1,702.76** |
| 152 | 21,109.54 | 21,852.52 | 20,927.44 | 20,246.62 | **84,136.12** |
| 153 | 5,207.11 | 3,187.19 | 4,914.82 | 3,373.77 | **16,682.89** |
| 154 | 3,032.91 | 3,395.14 | 2,768.17 | 1,988.25 | **11,184.47** |
| 155 | 54.56 | 72.00 | 67.35 | 75.58 | **269.48** |
| 156 | 7,646.49 | 8,396.18 | 8,012.04 | 7,280.86 | **31,335.57** |
| 157 | 997.26 | 538.88 | 1,032.14 | 567.77 | **3,136.03** |
| 158 | 133.44 | 145.61 | 133.53 | 125.13 | **537.71** |
| 159 | 3,092.48 | 3,796.64 | 3,085.68 | 3,065.65 | **13,040.44** |
| 160 | 4,153.95 | 3,671.15 | 4,399.20 | 2,132.79 | **14,357.09** |
| 161 | 6,170.91 | 4,820.55 | 5,210.60 | 5,320.34 | **21,522.39** |
| 162 | 232.90 | 231.65 | 198.56 | 182.46 | **845.57** |
| 163 | 1,576.83 | 2,142.71 | 1,541.77 | 2,264.89 | **7,526.20** |
| 164 | 97.94 | 93.91 | 91.68 | 89.44 | **372.96** |
| 165 | 3,665.97 | 3,712.03 | 3,723.39 | 2,407.10 | **13,508.48** |
| 166 | 6,227.71 | 4,535.77 | 5,990.33 | 4,983.87 | **21,737.68** |
| 167 | 163.23 | 173.96 | 148.47 | 156.07 | **641.73** |
| 168 | 3,015.47 | 2,569.70 | 3,008.76 | 2,028.95 | **10,622.88** |
| 169 | 118.51 | 126.74 | 96.06 | 75.58 | **416.88** |
| 170 | 6,609.79 | 6,540.52 | 5,874.33 | 5,701.71 | **24,726.36** |
| 171 | 3,844.58 | 1,977.70 | 4,666.08 | 1,990.49 | **12,478.85** |
| 172 | 459.27 | 451.22 | 431.82 | 409.28 | **1,751.59** |
| 173 | 1,894.34 | 1,850.33 | 1,780.84 | 2,342.88 | **7,868.39** |
| 174 | 2,666.21 | 3,337.10 | 4,020.60 | 2,547.70 | **12,571.60** |
| 175 | 349.71 | 363.39 | 364.02 | 309.46 | **1,386.59** |
| 176 | 5,581.41 | 4,688.53 | 5,570.32 | 4,871.89 | **20,712.16** |
| 177 | 3,710.69 | 5,103.89 | 4,678.70 | 3,106.16 | **16,599.44** |
| 178 | 1,106.64 | 1,057.00 | 1,151.36 | 998.24 | **4,313.24** |
| 179 | 142.75 | 143.55 | 127.90 | 116.72 | **530.92** |
| 180 | 5,245.92 | 3,995.20 | 4,158.96 | 3,995.28 | **17,395.36** |
| 181 | 1,883.16 | 1,827.80 | 1,679.95 | 1,108.88 | **6,499.78** |
| 182 | 176.20 | 160.10 | 148.29 | 144.89 | **629.48** |
| 183 | 3,321.80 | 3,304.54 | 3,269.48 | 2,567.11 | **12,462.93** |
| 184 | 249.54 | 255.26 | 243.72 | 209.74 | **958.26** |

| | | | | | |
|---|---|---|---|---|---|
| 185 | 2,584.37 | 3,049.28 | 2,459.33 | 2,502.08 | **10,595.06** |
| 186 | 407.40 | 410.98 | 380.57 | 361.78 | **1,560.73** |
| 187 | 5,802.96 | 4,769.30 | 5,695.99 | 4,447.23 | **20,715.47** |
| 188 | 161.44 | 182.90 | 118.96 | 126.56 | **589.86** |
| 189 | 6,222.52 | 5,676.40 | 6,280.57 | 5,035.83 | **23,215.31** |
| 190 | 13,243.74 | 14,301.99 | 14,547.33 | 9,893.50 | **51,986.55** |
| 191 | 40,701.05 | 31,284.54 | 38,375.77 | 34,929.74 | **145,291.09** |
| 193 | 3,867.83 | 4,592.48 | 3,877.13 | 3,790.65 | **16,128.09** |
| 194 | 6,067.52 | 5,629.17 | 5,554.31 | 4,897.56 | **22,148.56** |
| 195 | 135.05 | 152.94 | 128.79 | 140.06 | **556.85** |
| 196 | 2,592.90 | 3,478.16 | 2,959.21 | 3,213.65 | **12,243.92** |
| **PA - PENNSYLVANIA - Total** | **189,494.72** | **177,324.20** | **187,040.52** | **160,229.35** | **714,088.79** |

**STATE: PR - PUERTO RICO**

| REGISTRANT ZIP CODE 3 | 1ST QUARTER | 2ND QUARTER | 3RD QUARTER | 4TH QUARTER | TOTAL GRAMS |
|---|---|---|---|---|---|
| 006 | 1,079.54 | 1,395.26 | 1,618.42 | 1,616.63 | **5,709.85** |
| 007 | 3,886.62 | 5,380.89 | 5,377.58 | 4,906.68 | **19,551.76** |
| 009 | 15,444.05 | 14,784.88 | 14,751.79 | 16,592.82 | **61,573.54** |
| **PR - PUERTO RICO - Total** | **20,410.21** | **21,561.03** | **21,747.78** | **23,116.13** | **86,835.15** |

**STATE: RI - RHODE ISLAND**

| REGISTRANT ZIP CODE 3 | 1ST QUARTER | 2ND QUARTER | 3RD QUARTER | 4TH QUARTER | TOTAL GRAMS |
|---|---|---|---|---|---|
| 028 | 20,999.44 | 19,169.32 | 18,644.84 | 19,189.89 | **78,003.49** |
| 029 | 23,272.38 | 28,473.31 | 18,480.00 | 20,921.80 | **91,147.50** |
| **RI - RHODE ISLAND - Total** | **44,271.82** | **47,642.63** | **37,124.84** | **40,111.69** | **169,150.99** |

**STATE: SC - SOUTH CAROLINA**

| REGISTRANT ZIP CODE 3 | 1ST QUARTER | 2ND QUARTER | 3RD QUARTER | 4TH QUARTER | TOTAL GRAMS |
|---|---|---|---|---|---|
| 290 | 937.33 | 851.47 | 760.24 | 784.66 | **3,333.70** |
| 291 | 3,325.38 | 3,544.51 | 2,142.98 | 2,122.41 | **11,135.28** |
| 292 | 2,149.24 | 2,231.89 | 2,563.53 | 1,738.71 | **8,683.37** |
| 293 | 5,435.27 | 5,809.40 | 5,409.33 | 4,654.01 | **21,308.01** |
| 294 | 8,224.63 | 6,063.76 | 9,316.16 | 6,560.69 | **30,165.25** |
| 295 | 15,212.67 | 9,933.83 | 13,576.90 | 10,298.66 | **49,022.06** |
| 296 | 11,897.40 | 11,835.95 | 12,612.29 | 9,175.56 | **45,521.20** |
| 297 | 4,402.86 | 6,286.56 | 4,992.63 | 5,121.78 | **20,803.83** |
| 298 | 1,521.37 | 1,552.68 | 1,499.01 | 988.76 | **5,561.83** |
| 299 | 892.61 | 832.24 | 1,008.88 | 793.78 | **3,527.51** |
| **SC - SOUTH CAROLINA - Total** | **53,998.77** | **48,942.28** | **53,881.97** | **42,239.02** | **199,062.05** |

**STATE: SD - SOUTH DAKOTA**

| REGISTRANT ZIP CODE 3 | 1ST QUARTER | 2ND QUARTER | 3RD QUARTER | 4TH QUARTER | TOTAL GRAMS |
|---|---|---|---|---|---|
| 570 | 333.88 | 305.80 | 308.75 | 295.51 | **1,243.93** |
| 571 | 1,195.46 | 1,216.21 | 1,307.17 | 784.84 | **4,503.66** |

| | | | | | |
|---|---|---|---|---|---|
| 572 | 22.00 | 25.31 | 19.68 | 29.60 | **96.60** |
| 573 | 78.71 | 81.39 | 87.20 | 85.42 | **332.72** |
| 574 | 34.88 | 36.67 | 24.60 | 14.31 | **110.46** |
| 575 | 30.95 | 27.73 | 30.41 | 32.65 | **121.73** |
| 576 | 11.63 | 16.10 | 9.84 | 6.26 | **43.83** |
| 577 | 311.07 | 285.05 | 269.12 | 272.70 | **1,137.95** |
| **SD - SOUTH DAKOTA - Total** | **2,018.57** | **1,994.24** | **2,056.76** | **1,521.28** | **7,590.86** |

**STATE: TN - TENNESSEE**

| REGISTRANT ZIP CODE 3 | 1ST QUARTER | 2ND QUARTER | 3RD QUARTER | 4TH QUARTER | TOTAL GRAMS |
|---|---|---|---|---|---|
| 370 | 1,385.34 | 1,305.56 | 1,161.38 | 1,002.53 | **4,854.80** |
| 371 | 1,131.24 | 1,061.92 | 1,021.40 | 957.46 | **4,172.02** |
| 372 | 5,568.02 | 4,798.19 | 6,511.11 | 5,898.80 | **22,776.12** |
| 373 | 848.88 | 758.00 | 667.49 | 655.60 | **2,929.96** |
| 374 | 7,220.22 | 7,532.91 | 7,608.93 | 6,983.83 | **29,345.89** |
| 376 | 1,239.19 | 1,349.20 | 1,621.90 | 1,690.06 | **5,900.36** |
| 377 | 149.81 | 143.55 | 122.98 | 117.61 | **533.96** |
| 378 | 206.61 | 182.99 | 221.63 | 190.06 | **801.29** |
| 379 | 11,077.23 | 11,070.35 | 11,120.88 | 9,530.46 | **42,798.92** |
| 380 | 3,166.80 | 3,160.18 | 3,495.49 | 2,539.65 | **12,362.13** |
| 381 | 9,286.64 | 8,686.59 | 8,717.27 | 8,931.75 | **35,622.25** |
| 382 | 3,181.38 | 2,267.30 | 3,327.62 | 2,591.08 | **11,367.38** |
| 383 | 3,896.63 | 2,620.59 | 4,020.60 | 2,986.31 | **13,524.13** |
| 384 | 1,465.83 | 1,035.00 | 1,406.98 | 1,370.58 | **5,278.39** |
| 385 | 180.22 | 201.78 | 158.76 | 151.60 | **692.36** |
| **TN - TENNESSEE - Total** | **50,004.04** | **46,174.12** | **51,184.43** | **45,597.37** | **192,959.96** |

**STATE: TX - TEXAS**

| REGISTRANT ZIP CODE 3 | 1ST QUARTER | 2ND QUARTER | 3RD QUARTER | 4TH QUARTER | TOTAL GRAMS |
|---|---|---|---|---|---|
| 750 | 7,335.78 | 7,278.81 | 6,913.09 | 6,051.69 | **27,579.36** |
| 751 | 1,045.20 | 963.72 | 1,165.40 | 970.69 | **4,145.01** |
| 752 | 8,884.27 | 6,803.87 | 9,188.08 | 7,453.15 | **32,329.37** |
| 753 | 24.24 | 19.14 | 14.04 | 21.91 | **79.33** |
| 754 | 942.52 | 826.69 | 1,078.47 | 769.45 | **3,617.13** |
| 755 | 284.42 | 232.10 | 215.10 | 194.08 | **925.70** |
| 756 | 3,501.40 | 3,143.19 | 3,197.03 | 3,321.80 | **13,163.42** |
| 757 | 4,695.78 | 5,086.90 | 4,603.75 | 4,314.41 | **18,700.83** |
| 758 | 250.43 | 219.58 | 213.76 | 162.33 | **846.10** |
| 759 | 1,323.44 | 1,303.59 | 1,357.88 | 1,577.45 | **5,562.36** |
| 760 | 2,990.87 | 2,181.71 | 2,730.33 | 2,733.47 | **10,636.38** |
| 761 | 11,037.16 | 10,777.88 | 10,130.69 | 10,531.11 | **42,476.84** |
| 762 | 2,257.56 | 1,767.51 | 2,870.04 | 2,122.23 | **9,017.34** |

| | | | | | |
|---|---|---|---|---|---|
| 763 | 678.40 | 858.00 | 847.62 | 787.07 | **3,171.10** |
| 764 | 214.92 | 196.77 | 173.07 | 174.41 | **759.17** |
| 765 | 860.32 | 629.57 | 791.54 | 337.81 | **2,619.25** |
| 766 | 199.00 | 192.12 | 160.10 | 173.96 | **725.18** |
| 767 | 2,182.34 | 1,591.58 | 2,145.13 | 901.02 | **6,820.07** |
| 768 | 272.97 | 236.57 | 230.13 | 182.90 | **922.57** |
| 769 | 912.82 | 776.61 | 751.65 | 735.38 | **3,176.46** |
| 770 | 13,894.22 | 12,176.36 | 13,167.16 | 11,551.07 | **50,788.81** |
| 772 | 0 | 429.31 | 429.31 | 429.31 | **1,287.94** |
| 773 | 3,891.44 | 3,908.44 | 3,927.67 | 3,746.33 | **15,473.88** |
| 774 | 853.97 | 875.80 | 810.24 | 777.50 | **3,317.51** |
| 775 | 4,800.96 | 4,602.49 | 5,283.31 | 3,908.08 | **18,594.84** |
| 776 | 1,001.10 | 957.01 | 1,620.47 | 992.87 | **4,571.46** |
| 777 | 1,026.15 | 995.02 | 1,650.26 | 1,690.86 | **5,362.29** |
| 778 | 218.50 | 311.70 | 323.59 | 356.06 | **1,209.85** |
| 779 | 1,149.30 | 1,161.29 | 1,105.93 | 657.47 | **4,073.99** |
| 780 | 1,882.09 | 1,434.80 | 1,780.57 | 1,767.60 | **6,865.06** |
| 781 | 2,007.21 | 1,551.43 | 1,728.61 | 1,305.47 | **6,592.71** |
| 782 | 29,435.15 | 25,817.93 | 31,151.68 | 28,702.28 | **115,107.04** |
| 783 | 275.74 | 292.92 | 217.34 | 217.79 | **1,003.79** |
| 784 | 6,731.70 | 6,203.11 | 5,790.26 | 6,863.98 | **25,589.05** |
| 785 | 1,337.84 | 1,867.06 | 1,673.51 | 921.68 | **5,800.09** |
| 786 | 2,255.86 | 1,506.97 | 1,642.39 | 1,489.80 | **6,895.02** |
| 787 | 7,089.37 | 6,650.76 | 6,869.35 | 6,324.66 | **26,934.14** |
| 788 | 67.53 | 75.22 | 68.24 | 80.94 | **291.93** |
| 789 | 38.91 | 50.09 | 51.88 | 49.64 | **190.51** |
| 790 | 241.94 | 242.65 | 225.66 | 212.87 | **923.11** |
| 791 | 1,209.05 | 1,025.52 | 1,212.45 | 801.20 | **4,248.22** |
| 792 | 49.19 | 58.14 | 50.09 | 65.74 | **223.15** |
| 793 | 108.67 | 124.14 | 116.27 | 100.17 | **449.26** |
| 794 | 1,472.36 | 768.92 | 1,914.46 | 1,797.92 | **5,953.66** |
| 795 | 179.77 | 156.16 | 164.12 | 122.09 | **622.14** |
| 796 | 430.65 | 341.39 | 381.91 | 374.40 | **1,528.35** |
| 797 | 3,052.05 | 1,493.83 | 1,984.58 | 1,400.27 | **7,930.73** |
| 798 | 13.42 | 8.94 | 8.94 | 8.94 | **40.25** |
| 799 | 2,640.00 | 2,069.46 | 2,784.80 | 2,008.02 | **9,502.28** |
| **TX - TEXAS - Total** | **137,248.00** | **122,242.73** | **136,911.97** | **122,241.36** | **518,644.06** |

**STATE: UT - UTAH**

| REGISTRANT ZIP CODE 3 | 1ST QUARTER | 2ND QUARTER | 3RD QUARTER | 4TH QUARTER | **TOTAL GRAMS** |
|---|---|---|---|---|---|
| 840 | 7,645.24 | 6,280.83 | 6,545.40 | 6,235.22 | **26,706.69** |

| | | | | | |
|---|---|---|---|---|---|
| 841 | 13,777.52 | 10,791.65 | 12,833.66 | 7,721.36 | **45,124.18** |
| 843 | 295.60 | 282.00 | 248.20 | 210.18 | **1,035.98** |
| 844 | 2,753.05 | 2,574.44 | 2,527.04 | 1,660.01 | **9,514.54** |
| 845 | 181.21 | 193.64 | 257.32 | 525.46 | **1,157.62** |
| 846 | 1,264.41 | 681.26 | 1,753.74 | 670.17 | **4,369.59** |
| 847 | 2,221.60 | 2,544.03 | 2,173.57 | 2,675.60 | **9,614.80** |
| **UT - UTAH - Total** | **28,138.63** | **23,347.86** | **26,338.92** | **19,698.00** | **97,523.41** |

**STATE: VA - VIRGINIA**

| REGISTRANT ZIP CODE 3 | 1ST QUARTER | 2ND QUARTER | 3RD QUARTER | 4TH QUARTER | TOTAL GRAMS |
|---|---|---|---|---|---|
| 201 | 3,662.75 | 2,496.72 | 3,769.18 | 1,964.91 | **11,893.55** |
| 220 | 1,549.46 | 1,847.11 | 1,393.92 | 1,432.83 | **6,223.32** |
| 221 | 414.46 | 408.65 | 346.04 | 361.61 | **1,530.77** |
| 222 | 83.63 | 101.78 | 98.21 | 85.33 | **368.94** |
| 223 | 668.12 | 687.79 | 251.68 | 688.51 | **2,296.10** |
| 224 | 4,351.79 | 4,000.29 | 3,221.63 | 3,586.99 | **15,160.71** |
| 225 | 137.11 | 136.84 | 120.39 | 118.96 | **513.30** |
| 226 | 4,900.24 | 5,786.95 | 5,530.52 | 4,584.87 | **20,802.58** |
| 227 | 152.32 | 138.18 | 159.92 | 131.92 | **582.34** |
| 228 | 3,985.71 | 2,200.94 | 3,931.78 | 2,148.38 | **12,266.81** |
| 229 | 3,951.91 | 4,589.17 | 4,501.07 | 3,934.64 | **16,976.79** |
| 230 | 3,029.60 | 3,357.49 | 3,710.24 | 3,241.84 | **13,339.17** |
| 231 | 1,349.02 | 1,178.82 | 1,450.90 | 1,310.74 | **5,289.48** |
| 232 | 13,581.37 | 14,432.58 | 14,751.34 | 14,783.27 | **57,548.56** |
| 233 | 3,435.66 | 3,861.30 | 3,825.97 | 3,096.41 | **14,219.35** |
| 234 | 5,756.18 | 6,706.30 | 6,095.87 | 5,696.08 | **24,254.43** |
| 235 | 1,418.79 | 1,370.04 | 1,899.08 | 1,404.21 | **6,092.12** |
| 236 | 5,339.93 | 5,263.01 | 4,978.50 | 5,240.83 | **20,822.26** |
| 237 | 1,366.11 | 801.29 | 746.20 | 735.64 | **3,649.24** |
| 238 | 2,109.00 | 1,833.07 | 2,081.81 | 1,421.02 | **7,444.90** |
| 239 | 162.42 | 137.74 | 142.93 | 122.98 | **566.07** |
| 240 | 4,970.40 | 3,762.11 | 4,569.85 | 4,059.32 | **17,361.69** |
| 241 | 898.07 | 811.22 | 778.40 | 794.50 | **3,282.18** |
| 242 | 615.35 | 538.52 | 562.13 | 550.50 | **2,266.50** |
| 243 | 6,322.78 | 5,863.51 | 4,817.95 | 5,096.56 | **22,100.80** |
| 244 | 493.80 | 561.41 | 888.23 | 722.23 | **2,665.67** |
| 245 | 4,068.00 | 3,112.06 | 3,716.59 | 3,829.82 | **14,726.47** |
| 246 | 4,213.07 | 3,225.12 | 4,238.38 | 2,403.25 | **14,079.82** |
| **VA - VIRGINIA - Total** | **82,987.04** | **79,210.03** | **82,578.70** | **73,548.15** | **318,323.92** |

**STATE: VI - VIRGIN ISLANDS**

| REGISTRANT ZIP CODE 3 | 1ST QUARTER | 2ND QUARTER | 3RD QUARTER | 4TH QUARTER | TOTAL GRAMS |
|---|---|---|---|---|---|

| | | | | | |
|---|---|---|---|---|---|
| 008 | 1.79 | 3.58 | 9.84 | 549.16 | **564.37** |
| **VI - VIRGIN ISLANDS - Total** | **1.79** | **3.58** | **9.84** | **549.16** | **564.37** |

**STATE: VT - VERMONT**

| REGISTRANT ZIP CODE 3 | 1ST QUARTER | 2ND QUARTER | 3RD QUARTER | 4TH QUARTER | TOTAL GRAMS |
|---|---|---|---|---|---|
| 050 | 426.36 | 438.70 | 426.27 | 326.99 | **1,618.33** |
| 051 | 242.83 | 207.50 | 223.15 | 167.25 | **840.74** |
| 052 | 147.13 | 151.60 | 148.38 | 141.49 | **588.60** |
| 053 | 2,041.47 | 1,813.40 | 2,400.57 | 1,465.47 | **7,720.91** |
| 054 | 7,531.03 | 5,754.84 | 7,662.06 | 7,295.89 | **28,243.81** |
| 056 | 2,672.47 | 1,780.30 | 2,433.22 | 1,952.92 | **8,838.91** |
| 057 | 2,489.03 | 2,272.85 | 2,233.76 | 2,218.56 | **9,214.20** |
| 058 | 4,534.25 | 3,176.55 | 4,102.34 | 3,023.43 | **14,836.58** |
| **VT - VERMONT - Total** | **20,084.56** | **15,595.74** | **19,629.75** | **16,592.01** | **71,902.07** |

**STATE: WA - WASHINGTON**

| REGISTRANT ZIP CODE 3 | 1ST QUARTER | 2ND QUARTER | 3RD QUARTER | 4TH QUARTER | TOTAL GRAMS |
|---|---|---|---|---|---|
| 980 | 16,519.75 | 13,546.58 | 16,695.85 | 15,512.12 | **62,274.30** |
| 981 | 21,197.91 | 17,368.35 | 21,450.66 | 20,081.96 | **80,098.89** |
| 982 | 13,637.27 | 14,161.12 | 14,837.65 | 11,509.32 | **54,145.37** |
| 983 | 3,222.88 | 2,994.00 | 2,759.76 | 2,712.98 | **11,689.63** |
| 984 | 13,015.76 | 11,590.71 | 12,362.84 | 11,058.54 | **48,027.85** |
| 985 | 7,186.15 | 3,456.05 | 8,615.04 | 9,109.46 | **28,366.70** |
| 986 | 7,565.64 | 6,455.33 | 7,983.68 | 6,701.11 | **28,705.77** |
| 988 | 655.60 | 588.43 | 550.86 | 517.05 | **2,311.93** |
| 989 | 1,735.58 | 2,057.03 | 2,046.92 | 1,288.38 | **7,127.92** |
| 990 | 284.60 | 249.09 | 249.98 | 242.38 | **1,026.06** |
| 991 | 513.39 | 459.36 | 466.25 | 402.03 | **1,841.03** |
| 992 | 11,747.23 | 9,892.96 | 12,024.67 | 8,515.94 | **42,180.80** |
| 993 | 833.13 | 731.98 | 643.07 | 606.31 | **2,814.50** |
| 994 | 262.95 | 229.68 | 183.71 | 201.78 | **878.12** |
| **WA - WASHINGTON - Total** | **98,377.83** | **83,780.68** | **100,870.97** | **88,459.38** | **371,488.86** |

**STATE: WI - WISCONSIN**

| REGISTRANT ZIP CODE 3 | 1ST QUARTER | 2ND QUARTER | 3RD QUARTER | 4TH QUARTER | TOTAL GRAMS |
|---|---|---|---|---|---|
| 530 | 3,380.21 | 2,339.75 | 2,845.00 | 2,054.88 | **10,619.84** |
| 531 | 4,578.79 | 3,956.02 | 4,441.32 | 3,187.82 | **16,163.95** |
| 532 | 14,409.05 | 14,252.98 | 16,922.14 | 8,353.70 | **53,937.87** |
| 534 | 3,347.38 | 2,523.46 | 3,311.96 | 2,142.54 | **11,325.34** |
| 535 | 3,670.98 | 2,972.09 | 3,509.36 | 2,298.07 | **12,450.50** |
| 537 | 10,301.07 | 9,587.25 | 10,241.07 | 8,528.04 | **38,657.43** |
| 538 | 105.99 | 98.83 | 102.41 | 82.28 | **389.51** |
| 539 | 443.18 | 393.09 | 372.07 | 350.60 | **1,558.94** |

| | | | | | |
|---|---|---|---|---|---|
| 540 | 229.68 | 198.11 | 222.53 | 196.32 | **846.64** |
| 541 | 213.31 | 165.02 | 179.33 | 159.56 | **717.22** |
| 542 | 84.97 | 72.45 | 79.15 | 59.48 | **296.05** |
| 543 | 4,448.92 | 3,759.79 | 4,403.67 | 2,175.99 | **14,788.37** |
| 544 | 2,001.49 | 1,923.14 | 2,352.54 | 1,479.25 | **7,756.42** |
| 545 | 48.74 | 54.11 | 46.96 | 49.64 | **199.45** |
| 546 | 3,313.22 | 4,306.18 | 3,355.52 | 3,280.39 | **14,255.30** |
| 547 | 1,985.12 | 2,376.87 | 2,647.69 | 2,232.51 | **9,242.19** |
| 548 | 300.25 | 317.96 | 307.85 | 280.39 | **1,206.46** |
| 549 | 8,653.23 | 9,129.50 | 7,800.60 | 6,971.31 | **32,554.64** |
| **WI - WISCONSIN - Total** | **61,515.58** | **58,426.59** | **63,141.16** | **43,882.77** | **226,966.10** |

**STATE: WV - WEST VIRGINIA**

| REGISTRANT ZIP CODE 3 | 1ST QUARTER | 2ND QUARTER | 3RD QUARTER | 4TH QUARTER | TOTAL GRAMS |
|---|---|---|---|---|---|
| 247 | 29.07 | 37.56 | 33.09 | 44.27 | **144.00** |
| 248 | 17.89 | 19.68 | 16.55 | 16.10 | **70.21** |
| 249 | 32.20 | 28.17 | 29.07 | 23.25 | **112.69** |
| 250 | 28.62 | 29.07 | 30.41 | 26.65 | **114.75** |
| 251 | 33.99 | 31.75 | 27.28 | 31.75 | **124.77** |
| 252 | 42.93 | 20.12 | 21.47 | 20.57 | **105.09** |
| 253 | 2,252.01 | 1,236.60 | 2,111.23 | 1,871.35 | **7,471.19** |
| 254 | 3,481.00 | 3,338.80 | 3,224.31 | 3,121.46 | **13,165.57** |
| 255 | 92.30 | 93.46 | 102.86 | 67.08 | **355.70** |
| 256 | 2,692.14 | 1,938.16 | 2,701.54 | 1,798.19 | **9,130.04** |
| 257 | 5,301.11 | 5,318.46 | 5,516.93 | 3,414.46 | **19,550.96** |
| 258 | 3,148.74 | 2,712.72 | 3,502.02 | 2,314.71 | **11,678.18** |
| 259 | 38.46 | 31.30 | 41.14 | 47.40 | **158.31** |
| 260 | 1,260.21 | 1,236.51 | 1,434.98 | 889.84 | **4,821.53** |
| 261 | 1,090.72 | 939.12 | 1,228.91 | 650.68 | **3,909.42** |
| 262 | 107.33 | 110.01 | 80.05 | 79.15 | **376.54** |
| 263 | 1,041.98 | 1,462.34 | 1,228.37 | 1,009.96 | **4,742.65** |
| 264 | 62.61 | 55.01 | 55.27 | 52.32 | **225.21** |
| 265 | 1,812.59 | 1,392.67 | 1,837.10 | 1,361.01 | **6,403.37** |
| 266 | 15.20 | 16.55 | 18.78 | 17.89 | **68.42** |
| 267 | 83.63 | 76.47 | 66.63 | 70.21 | **296.94** |
| 268 | 46.33 | 37.12 | 28.62 | 31.30 | **143.37** |
| **WV - WEST VIRGINIA - Total** | **22,711.05** | **20,161.65** | **23,336.60** | **16,959.61** | **83,168.91** |

**STATE: WY - WYOMING**

| REGISTRANT ZIP CODE 3 | 1ST QUARTER | 2ND QUARTER | 3RD QUARTER | 4TH QUARTER | TOTAL GRAMS |
|---|---|---|---|---|---|
| 820 | 172.62 | 200.61 | 138.18 | 151.15 | **662.57** |
| 822 | 30.23 | 37.56 | 25.94 | 27.28 | **121.01** |

| | | | | | |
|---|---|---|---|---|---|
| 823 | 17.89 | 13.42 | 9.39 | 17.44 | **58.14** |
| 824 | 70.21 | 47.40 | 47.40 | 46.06 | **211.08** |
| 825 | 130.58 | 88.55 | 91.68 | 75.58 | **386.38** |
| 826 | 218.68 | 201.24 | 207.95 | 215.55 | **843.42** |
| 827 | 42.93 | 39.35 | 34.88 | 31.75 | **148.92** |
| 828 | 65.29 | 53.66 | 56.35 | 61.27 | **236.57** |
| 829 | 247.75 | 189.61 | 210.63 | 216.00 | **863.99** |
| 830 | 29.07 | 40.70 | 29.52 | 29.52 | **128.79** |
| 831 | 23.25 | 19.68 | 25.94 | 27.73 | **96.60** |
| **WY - WYOMING - Total** | **1,048.51** | **931.79** | **877.85** | **899.32** | **3,757.46** |

**DRUG: 9300 - MORPHINE**

**STATE: AK - ALASKA**

| REGISTRANT ZIP CODE 3 | 1ST QUARTER | 2ND QUARTER | 3RD QUARTER | 4TH QUARTER | **TOTAL GRAMS** |
|---|---|---|---|---|---|
| 995 | 5,079.76 | 5,071.05 | 4,859.17 | 4,367.73 | **19,377.71** |
| 996 | 3,891.86 | 3,700.74 | 3,681.76 | 3,130.08 | **14,404.44** |
| 997 | 1,235.70 | 1,107.36 | 1,238.71 | 1,026.64 | **4,608.40** |
| 998 | 416.58 | 382.18 | 391.58 | 434.17 | **1,624.51** |
| 999 | 175.56 | 223.07 | 167.94 | 158.15 | **724.71** |
| **AK - ALASKA - Total** | **10,799.45** | **10,484.39** | **10,339.16** | **9,116.76** | **40,739.77** |

**STATE: AL - ALABAMA**

| REGISTRANT ZIP CODE 3 | 1ST QUARTER | 2ND QUARTER | 3RD QUARTER | 4TH QUARTER | **TOTAL GRAMS** |
|---|---|---|---|---|---|
| 350 | 7,364.11 | 7,050.50 | 6,898.20 | 6,702.44 | **28,015.25** |
| 351 | 3,949.06 | 3,905.48 | 3,656.05 | 3,459.88 | **14,970.47** |
| 352 | 6,104.91 | 5,792.83 | 5,494.67 | 5,607.54 | **22,999.95** |
| 354 | 4,092.91 | 3,571.68 | 3,677.79 | 3,550.74 | **14,893.11** |
| 355 | 2,088.64 | 2,154.52 | 2,020.64 | 1,920.68 | **8,184.48** |
| 356 | 4,230.36 | 4,248.64 | 4,102.53 | 4,095.09 | **16,676.62** |
| 357 | 2,288.95 | 2,207.52 | 2,320.02 | 2,181.20 | **8,997.70** |
| 358 | 3,836.36 | 3,518.64 | 3,399.66 | 3,294.09 | **14,048.75** |
| 359 | 5,473.78 | 5,348.69 | 5,235.26 | 4,870.72 | **20,928.44** |
| 360 | 3,428.21 | 3,077.90 | 2,673.15 | 2,858.48 | **12,037.73** |
| 361 | 2,547.69 | 2,450.68 | 2,318.79 | 2,504.89 | **9,822.05** |
| 362 | 2,251.04 | 2,098.83 | 1,922.27 | 1,971.67 | **8,243.80** |
| 363 | 1,998.43 | 1,888.58 | 1,829.22 | 1,811.60 | **7,527.84** |
| 364 | 1,260.03 | 976.22 | 930.73 | 956.90 | **4,123.89** |
| 365 | 4,418.08 | 4,137.85 | 4,040.33 | 4,152.88 | **16,749.13** |
| 366 | 4,112.19 | 4,042.75 | 3,607.56 | 3,756.51 | **15,519.02** |
| 367 | 588.05 | 513.38 | 565.27 | 587.30 | **2,254.00** |
| 368 | 3,528.27 | 3,391.80 | 3,383.53 | 3,449.13 | **13,752.74** |
| 369 | 188.45 | 136.18 | 113.21 | 133.77 | **571.61** |

| | | | | | |
|---|---|---|---|---|---|
| **AL - ALABAMA - Total** | **63,749.51** | **60,512.68** | **58,188.90** | **57,865.49** | **240,316.58** |

**STATE: AR - ARKANSAS**

| REGISTRANT ZIP CODE 3 | 1ST QUARTER | 2ND QUARTER | 3RD QUARTER | 4TH QUARTER | TOTAL GRAMS |
|---|---|---|---|---|---|
| 716 | 1,176.83 | 1,186.71 | 1,015.22 | 1,010.10 | **4,388.86** |
| 717 | 1,220.78 | 1,371.91 | 1,164.77 | 1,092.09 | **4,849.55** |
| 718 | 919.69 | 794.56 | 787.23 | 732.82 | **3,234.30** |
| 719 | 2,487.88 | 2,245.03 | 2,222.00 | 2,156.47 | **9,111.38** |
| 720 | 4,435.47 | 4,336.62 | 3,884.36 | 3,619.90 | **16,276.35** |
| 721 | 4,900.53 | 4,741.96 | 4,233.75 | 4,074.54 | **17,950.77** |
| 722 | 2,804.70 | 2,781.86 | 2,187.94 | 2,278.42 | **10,052.92** |
| 723 | 1,132.62 | 1,022.33 | 929.84 | 960.01 | **4,044.80** |
| 724 | 2,892.25 | 2,495.17 | 2,384.80 | 2,474.46 | **10,246.68** |
| 725 | 1,595.68 | 1,377.61 | 1,318.06 | 1,237.47 | **5,528.81** |
| 726 | 2,610.17 | 2,105.60 | 1,976.38 | 1,975.60 | **8,667.75** |
| 727 | 7,441.59 | 6,706.68 | 6,489.23 | 5,870.76 | **26,508.26** |
| 728 | 1,453.16 | 1,470.16 | 1,353.37 | 1,332.52 | **5,609.22** |
| 729 | 3,831.08 | 3,455.53 | 3,193.90 | 2,866.12 | **13,346.64** |
| **AR - ARKANSAS - Total** | **38,902.44** | **36,091.73** | **33,140.85** | **31,681.30** | **139,816.32** |

**STATE: AS - AMERICAN SAMOA**

| REGISTRANT ZIP CODE 3 | 1ST QUARTER | 2ND QUARTER | 3RD QUARTER | 4TH QUARTER | TOTAL GRAMS |
|---|---|---|---|---|---|
| 967 | 6.77 | 25.12 | 12.18 | 4.51 | **48.58** |
| **AS - AMERICAN SAMOA - Tota** | **6.77** | **25.12** | **12.18** | **4.51** | **48.58** |

**STATE: AZ - ARIZONA**

| REGISTRANT ZIP CODE 3 | 1ST QUARTER | 2ND QUARTER | 3RD QUARTER | 4TH QUARTER | TOTAL GRAMS |
|---|---|---|---|---|---|
| 850 | 22,052.89 | 20,125.97 | 18,673.63 | 18,146.75 | **78,999.24** |
| 851 | 5,498.82 | 4,861.45 | 4,481.87 | 4,396.16 | **19,238.29** |
| 852 | 30,427.31 | 27,387.48 | 26,838.44 | 22,418.06 | **107,071.31** |
| 853 | 19,826.86 | 17,788.81 | 16,105.89 | 16,545.68 | **70,267.24** |
| 855 | 3,011.93 | 2,753.63 | 2,705.32 | 2,453.20 | **10,924.08** |
| 856 | 5,560.06 | 5,472.12 | 5,099.09 | 4,672.12 | **20,803.40** |
| 857 | 30,978.76 | 28,642.72 | 26,887.67 | 25,275.45 | **111,784.60** |
| 859 | 3,200.55 | 2,761.40 | 2,759.21 | 2,493.37 | **11,214.53** |
| 860 | 2,594.74 | 2,266.26 | 2,232.36 | 1,883.88 | **8,977.23** |
| 863 | 6,648.76 | 6,661.55 | 5,876.54 | 5,545.75 | **24,732.60** |
| 864 | 6,776.97 | 5,759.08 | 5,115.12 | 5,045.56 | **22,696.73** |
| 865 | 164.11 | 166.65 | 135.08 | 127.09 | **592.93** |
| **AZ - ARIZONA - Total** | **136,741.78** | **124,647.12** | **116,910.22** | **109,003.06** | **487,302.18** |

**STATE: CA - CALIFORNIA**

| REGISTRANT ZIP CODE 3 | 1ST QUARTER | 2ND QUARTER | 3RD QUARTER | 4TH QUARTER | TOTAL GRAMS |
|---|---|---|---|---|---|
| 900 | 5,966.28 | 5,901.25 | 5,983.42 | 6,285.96 | **24,136.91** |

| | | | | | |
|---|---|---|---|---|---|
| 902 | 4,763.52 | 4,407.27 | 4,079.15 | 3,957.72 | **17,207.66** |
| 903 | 245.05 | 187.17 | 204.02 | 229.31 | **865.55** |
| 904 | 1,309.03 | 1,098.17 | 1,074.38 | 1,023.07 | **4,504.64** |
| 905 | 2,117.46 | 2,184.57 | 1,832.23 | 1,920.27 | **8,054.53** |
| 906 | 6,856.20 | 6,734.67 | 6,288.01 | 5,569.37 | **25,448.25** |
| 907 | 4,229.22 | 4,173.71 | 3,660.05 | 3,598.10 | **15,661.08** |
| 908 | 4,429.18 | 4,158.32 | 4,102.90 | 4,362.58 | **17,052.98** |
| 910 | 2,469.78 | 2,158.77 | 2,208.41 | 1,987.56 | **8,824.53** |
| 911 | 1,438.90 | 1,297.64 | 1,265.56 | 1,248.88 | **5,250.98** |
| 912 | 3,269.39 | 2,939.82 | 2,914.87 | 2,640.55 | **11,764.63** |
| 913 | 11,083.63 | 10,563.41 | 10,492.46 | 9,855.89 | **41,995.40** |
| 914 | 4,177.54 | 3,859.55 | 3,769.80 | 3,779.85 | **15,586.75** |
| 915 | 888.68 | 929.47 | 892.29 | 715.03 | **3,425.47** |
| 916 | 349.47 | 387.52 | 434.58 | 391.79 | **1,563.36** |
| 917 | 14,378.29 | 13,840.06 | 12,851.40 | 13,061.00 | **54,130.75** |
| 918 | 218.80 | 181.94 | 157.04 | 126.75 | **684.53** |
| 919 | 4,004.22 | 3,824.20 | 3,594.70 | 3,319.53 | **14,742.65** |
| 920 | 20,135.24 | 18,934.70 | 16,892.86 | 16,091.60 | **72,054.41** |
| 921 | 13,590.40 | 12,342.07 | 11,478.38 | 11,066.75 | **48,477.60** |
| 922 | 9,179.86 | 8,254.64 | 8,560.35 | 8,000.92 | **33,995.76** |
| 923 | 17,509.40 | 15,947.86 | 15,608.01 | 14,912.50 | **63,977.77** |
| 924 | 2,009.79 | 1,978.94 | 1,506.30 | 1,729.36 | **7,224.40** |
| 925 | 12,632.26 | 12,355.72 | 11,841.44 | 11,887.94 | **48,717.37** |
| 926 | 11,222.53 | 10,670.26 | 9,914.03 | 9,978.72 | **41,785.54** |
| 927 | 3,028.70 | 2,817.43 | 2,632.68 | 2,670.96 | **11,149.78** |
| 928 | 10,811.78 | 10,258.28 | 10,175.17 | 9,616.37 | **40,861.60** |
| 930 | 6,357.76 | 5,974.64 | 6,082.02 | 5,466.47 | **23,880.89** |
| 931 | 1,056.76 | 728.28 | 747.98 | 764.81 | **3,297.84** |
| 932 | 6,380.65 | 6,272.64 | 6,102.84 | 5,667.84 | **24,423.97** |
| 933 | 7,699.65 | 7,423.13 | 7,198.23 | 6,682.05 | **29,003.07** |
| 934 | 7,146.34 | 7,007.65 | 6,246.62 | 6,505.49 | **26,906.11** |
| 935 | 9,249.91 | 8,942.96 | 8,201.86 | 7,482.04 | **33,876.78** |
| 936 | 5,989.50 | 5,934.19 | 5,288.18 | 5,203.61 | **22,415.47** |
| 937 | 8,345.03 | 7,771.14 | 7,721.96 | 7,517.98 | **31,356.12** |
| 939 | 3,131.99 | 2,769.39 | 2,699.79 | 2,460.63 | **11,061.80** |
| 940 | 6,226.90 | 6,078.17 | 5,771.45 | 6,696.88 | **24,773.41** |
| 941 | 11,320.06 | 11,246.79 | 10,225.22 | 10,686.41 | **43,478.46** |
| 943 | 1,663.34 | 1,467.72 | 1,342.52 | 1,328.38 | **5,801.96** |
| 944 | 1,496.43 | 1,523.21 | 1,214.39 | 1,007.40 | **5,241.42** |
| 945 | 32,522.84 | 28,609.39 | 29,252.11 | 27,857.96 | **118,242.30** |

| | | | | | |
|---|---|---|---|---|---|
| 946 | 3,583.00 | 3,003.51 | 2,789.39 | 2,965.47 | **12,341.38** |
| 947 | 721.22 | 532.39 | 488.21 | 667.51 | **2,409.32** |
| 948 | 1,078.47 | 964.42 | 1,018.47 | 1,029.36 | **4,090.71** |
| 949 | 5,639.70 | 5,014.03 | 4,679.94 | 4,069.60 | **19,403.26** |
| 950 | 5,373.48 | 4,851.92 | 4,656.81 | 4,623.82 | **19,506.03** |
| 951 | 3,150.79 | 3,198.45 | 2,725.28 | 2,926.55 | **12,001.07** |
| 952 | 7,032.70 | 6,822.00 | 6,185.16 | 6,109.54 | **26,149.40** |
| 953 | 18,557.75 | 18,668.24 | 16,926.37 | 16,236.49 | **70,388.84** |
| 954 | 10,602.57 | 9,821.68 | 9,857.97 | 8,598.01 | **38,880.23** |
| 955 | 4,342.85 | 4,093.89 | 4,222.26 | 3,728.09 | **16,387.09** |
| 956 | 22,139.07 | 20,798.78 | 19,798.86 | 20,167.15 | **82,903.85** |
| 957 | 2,562.70 | 2,446.35 | 2,355.98 | 2,365.31 | **9,730.34** |
| 958 | 14,422.37 | 13,035.77 | 12,802.28 | 11,571.45 | **51,831.87** |
| 959 | 10,930.72 | 9,591.85 | 9,443.66 | 8,843.96 | **38,810.18** |
| 960 | 7,602.74 | 6,856.22 | 6,281.22 | 5,947.96 | **26,688.14** |
| 961 | 1,459.64 | 1,448.76 | 1,368.10 | 1,320.31 | **5,596.80** |
| **CA - CALIFORNIA - Total** | **400,101.55** | **375,284.98** | **358,109.62** | **346,526.82** | **1,480,022.97** |

**STATE: CO - COLORADO**

| REGISTRANT ZIP CODE 3 | 1ST QUARTER | 2ND QUARTER | 3RD QUARTER | 4TH QUARTER | **TOTAL GRAMS** |
|---|---|---|---|---|---|
| 800 | 11,877.90 | 10,618.19 | 12,794.11 | 12,863.88 | **48,154.08** |
| 801 | 9,062.17 | 8,703.57 | 8,602.30 | 8,243.72 | **34,611.76** |
| 802 | 13,779.66 | 12,991.16 | 10,701.98 | 9,032.78 | **46,505.57** |
| 803 | 1,001.93 | 853.36 | 813.69 | 848.33 | **3,517.31** |
| 804 | 3,005.61 | 2,549.57 | 2,656.95 | 2,539.45 | **10,751.58** |
| 805 | 6,674.91 | 6,239.58 | 5,901.46 | 5,319.85 | **24,135.79** |
| 806 | 2,808.76 | 2,876.26 | 2,545.20 | 2,461.57 | **10,691.79** |
| 807 | 1,113.68 | 1,003.29 | 911.46 | 943.37 | **3,971.80** |
| 808 | 1,294.24 | 1,311.60 | 1,353.08 | 1,157.92 | **5,116.85** |
| 809 | 8,239.41 | 7,425.48 | 6,692.28 | 6,529.82 | **28,886.99** |
| 810 | 4,250.99 | 3,827.04 | 3,973.70 | 3,755.12 | **15,806.85** |
| 811 | 585.86 | 558.16 | 566.57 | 473.25 | **2,183.83** |
| 812 | 1,238.81 | 1,127.33 | 1,030.28 | 1,000.56 | **4,396.98** |
| 813 | 531.60 | 521.54 | 483.36 | 483.72 | **2,020.22** |
| 814 | 703.96 | 620.70 | 632.51 | 466.90 | **2,424.08** |
| 815 | 2,148.74 | 1,981.41 | 1,820.40 | 1,795.91 | **7,746.46** |
| 816 | 1,148.12 | 1,045.65 | 1,052.47 | 1,034.49 | **4,280.73** |
| **CO - COLORADO - Total** | **69,466.35** | **64,253.88** | **62,531.80** | **58,950.62** | **255,202.66** |

**STATE: CT - CONNECTICUT**

| REGISTRANT ZIP CODE 3 | 1ST QUARTER | 2ND QUARTER | 3RD QUARTER | 4TH QUARTER | **TOTAL GRAMS** |
|---|---|---|---|---|---|
| 060 | 5,538.72 | 5,520.16 | 5,091.02 | 5,239.78 | **21,389.68** |

| | | | | | |
|---|---|---|---|---|---|
| 061 | 1,666.04 | 1,770.44 | 1,588.69 | 1,528.99 | **6,554.16** |
| 062 | 1,407.53 | 1,371.52 | 1,452.65 | 1,429.71 | **5,661.40** |
| 063 | 1,924.70 | 1,892.78 | 1,718.27 | 1,751.52 | **7,287.26** |
| 064 | 9,655.75 | 9,166.34 | 9,243.27 | 9,366.10 | **37,431.47** |
| 065 | 3,877.42 | 3,748.33 | 3,264.21 | 3,150.28 | **14,040.23** |
| 066 | 1,367.36 | 1,283.57 | 1,147.08 | 1,146.76 | **4,944.78** |
| 067 | 3,058.45 | 2,960.35 | 2,764.50 | 2,510.37 | **11,293.67** |
| 068 | 4,111.23 | 3,351.68 | 3,213.62 | 2,861.52 | **13,538.06** |
| 069 | 241.65 | 294.07 | 248.54 | 205.90 | **990.16** |
| **CT - CONNECTICUT - Total** | **32,848.85** | **31,359.24** | **29,731.85** | **29,190.94** | **123,130.87** |

**STATE: DC - DISTRICT OF COLUMBIA**

| REGISTRANT ZIP CODE 3 | 1ST QUARTER | 2ND QUARTER | 3RD QUARTER | 4TH QUARTER | TOTAL GRAMS |
|---|---|---|---|---|---|
| 200 | 2,495.26 | 2,274.50 | 2,202.83 | 1,918.98 | **8,891.58** |
| 203 | 18.05 | 0 | 20.30 | 12.48 | **50.84** |
| 204 | 1,174.62 | 454.77 | 766.06 | 537.08 | **2,932.54** |
| 205 | 0 | 0.38 | 0 | 0.38 | **0.75** |
| **DC - DISTRICT OF COLUMBIA -** | **3,687.93** | **2,729.65** | **2,989.20** | **2,468.92** | **11,875.70** |

**STATE: DE - DELAWARE**

| REGISTRANT ZIP CODE 3 | 1ST QUARTER | 2ND QUARTER | 3RD QUARTER | 4TH QUARTER | TOTAL GRAMS |
|---|---|---|---|---|---|
| 197 | 7,930.05 | 7,376.98 | 6,820.55 | 6,275.44 | **28,403.03** |
| 198 | 4,524.30 | 4,030.98 | 3,835.72 | 3,712.26 | **16,103.26** |
| 199 | 7,571.22 | 7,278.16 | 6,868.71 | 6,603.68 | **28,321.76** |
| **DE - DELAWARE - Total** | **20,025.57** | **18,686.12** | **17,524.99** | **16,591.38** | **72,828.05** |

**STATE: FL - FLORIDA**

| REGISTRANT ZIP CODE 3 | 1ST QUARTER | 2ND QUARTER | 3RD QUARTER | 4TH QUARTER | TOTAL GRAMS |
|---|---|---|---|---|---|
| 320 | 11,994.79 | 11,304.11 | 9,971.16 | 9,868.91 | **43,138.97** |
| 321 | 14,539.81 | 13,467.90 | 12,368.21 | 12,777.10 | **53,153.03** |
| 322 | 20,245.34 | 19,538.56 | 18,057.95 | 16,410.71 | **74,252.55** |
| 323 | 4,662.70 | 4,768.66 | 4,316.76 | 4,523.30 | **18,271.42** |
| 324 | 4,141.15 | 4,220.88 | 4,368.09 | 3,973.79 | **16,703.92** |
| 325 | 7,927.34 | 7,570.93 | 6,814.80 | 6,382.19 | **28,695.26** |
| 326 | 8,605.18 | 7,634.59 | 7,421.55 | 6,936.19 | **30,597.51** |
| 327 | 19,411.68 | 18,654.18 | 17,246.30 | 17,636.29 | **72,948.45** |
| 328 | 10,471.82 | 9,911.06 | 9,074.29 | 8,725.54 | **38,182.72** |
| 329 | 17,385.49 | 16,621.35 | 15,603.49 | 14,545.96 | **64,156.29** |
| 330 | 17,984.43 | 18,330.15 | 17,856.97 | 16,930.30 | **71,101.85** |
| 331 | 7,109.50 | 7,058.29 | 6,497.32 | 6,324.30 | **26,989.41** |
| 333 | 17,628.43 | 15,818.33 | 14,238.65 | 13,388.52 | **61,073.92** |
| 334 | 26,752.00 | 25,886.79 | 24,942.65 | 24,053.65 | **101,635.08** |
| 335 | 15,951.71 | 15,560.99 | 13,659.71 | 13,108.40 | **58,280.80** |

| | | | | | |
|---|---|---|---|---|---|
| 336 | 25,050.03 | 24,276.35 | 21,508.50 | 20,616.29 | **91,451.16** |
| 337 | 23,996.52 | 23,613.16 | 21,507.25 | 20,785.50 | **89,902.42** |
| 338 | 8,005.74 | 7,519.82 | 6,300.44 | 6,491.54 | **28,317.54** |
| 339 | 20,954.16 | 19,358.13 | 17,290.05 | 16,363.22 | **73,965.56** |
| 341 | 6,401.14 | 6,008.64 | 5,235.87 | 5,068.71 | **22,714.36** |
| 342 | 20,382.77 | 19,048.42 | 16,747.18 | 16,882.49 | **73,060.86** |
| 344 | 12,932.85 | 12,126.94 | 10,459.66 | 10,144.80 | **45,664.26** |
| 346 | 25,260.27 | 24,137.87 | 22,147.61 | 22,428.64 | **93,974.39** |
| 347 | 9,589.50 | 8,951.92 | 8,265.28 | 8,547.67 | **35,354.38** |
| 349 | 13,643.55 | 13,463.77 | 12,561.72 | 13,069.33 | **52,738.37** |
| **FL - FLORIDA - Total** | **371,027.93** | **354,851.78** | **324,461.45** | **315,983.33** | **1,366,324.49** |

**STATE: GA - GEORGIA**

| REGISTRANT ZIP CODE 3 | 1ST QUARTER | 2ND QUARTER | 3RD QUARTER | 4TH QUARTER | TOTAL GRAMS |
|---|---|---|---|---|---|
| 300 | 21,275.41 | 20,619.93 | 20,040.73 | 18,850.39 | **80,786.46** |
| 301 | 15,327.96 | 14,607.94 | 14,113.72 | 13,745.19 | **57,794.80** |
| 302 | 10,273.00 | 9,834.22 | 9,129.45 | 9,086.72 | **38,323.39** |
| 303 | 5,943.53 | 5,543.07 | 5,530.87 | 4,946.56 | **21,964.04** |
| 304 | 2,174.37 | 2,027.15 | 2,169.86 | 2,101.97 | **8,473.35** |
| 305 | 8,276.19 | 8,395.90 | 7,808.25 | 7,959.53 | **32,439.87** |
| 306 | 4,304.16 | 3,858.66 | 3,739.71 | 3,640.93 | **15,543.45** |
| 307 | 3,096.22 | 2,869.54 | 2,828.08 | 2,772.71 | **11,566.55** |
| 308 | 1,811.51 | 1,860.59 | 1,788.68 | 1,755.75 | **7,216.54** |
| 309 | 4,344.94 | 3,964.18 | 3,855.71 | 3,463.78 | **15,628.60** |
| 310 | 5,961.82 | 5,656.46 | 5,295.03 | 5,199.59 | **22,112.90** |
| 312 | 2,506.33 | 2,461.06 | 2,846.53 | 2,200.06 | **10,013.97** |
| 313 | 2,482.51 | 2,181.14 | 2,442.94 | 1,957.94 | **9,064.53** |
| 314 | 2,421.69 | 2,335.12 | 2,420.54 | 2,382.84 | **9,560.20** |
| 315 | 4,044.00 | 3,718.11 | 3,716.32 | 3,304.18 | **14,782.61** |
| 316 | 2,900.14 | 2,900.75 | 2,938.60 | 2,883.51 | **11,623.01** |
| 317 | 2,971.75 | 3,131.92 | 2,897.26 | 2,892.37 | **11,893.30** |
| 318 | 454.86 | 409.82 | 391.19 | 462.12 | **1,717.99** |
| 319 | 3,385.89 | 3,389.26 | 3,125.25 | 2,898.93 | **12,799.34** |
| 398 | 849.00 | 869.24 | 794.00 | 763.45 | **3,275.69** |
| **GA - GEORGIA - Total** | **104,805.28** | **100,634.05** | **97,872.74** | **93,268.52** | **396,580.59** |

**STATE: GU - GUAM**

| REGISTRANT ZIP CODE 3 | 1ST QUARTER | 2ND QUARTER | 3RD QUARTER | 4TH QUARTER | TOTAL GRAMS |
|---|---|---|---|---|---|
| 969 | 1,005.28 | 826.94 | 769.30 | 765.17 | **3,366.70** |
| **GU - GUAM - Total** | **1,005.28** | **826.94** | **769.30** | **765.17** | **3,366.70** |

**STATE: HI - HAWAII**

| REGISTRANT ZIP CODE 3 | 1ST QUARTER | 2ND QUARTER | 3RD QUARTER | 4TH QUARTER | TOTAL GRAMS |
|---|---|---|---|---|---|

| | | | | | |
|---|---|---|---|---|---|
| 967 | 11,693.82 | 12,115.84 | 10,771.07 | 9,904.86 | **44,485.59** |
| 968 | 6,816.77 | 6,666.33 | 6,228.34 | 5,945.65 | **25,657.08** |
| **HI - HAWAII - Total** | **18,510.58** | **18,782.17** | **16,999.40** | **15,850.51** | **70,142.67** |

**STATE: IA - IOWA**

| REGISTRANT ZIP CODE 3 | 1ST QUARTER | 2ND QUARTER | 3RD QUARTER | 4TH QUARTER | TOTAL GRAMS |
|---|---|---|---|---|---|
| 500 | 2,213.15 | 2,165.22 | 1,945.75 | 1,973.96 | **8,298.08** |
| 501 | 2,353.03 | 2,392.79 | 2,308.00 | 2,125.97 | **9,179.79** |
| 502 | 1,866.36 | 1,767.58 | 1,754.57 | 1,686.07 | **7,074.57** |
| 503 | 4,763.91 | 4,372.68 | 4,051.90 | 4,298.51 | **17,487.00** |
| 504 | 1,139.36 | 1,049.08 | 1,005.54 | 935.59 | **4,129.58** |
| 505 | 1,265.25 | 1,242.85 | 1,301.95 | 1,262.46 | **5,072.51** |
| 506 | 1,030.48 | 982.02 | 1,029.76 | 951.86 | **3,994.12** |
| 507 | 672.78 | 701.74 | 669.74 | 632.75 | **2,677.02** |
| 508 | 396.33 | 417.89 | 422.82 | 431.64 | **1,668.69** |
| 510 | 521.32 | 447.80 | 395.98 | 407.49 | **1,772.60** |
| 511 | 1,004.97 | 885.88 | 876.20 | 928.39 | **3,695.44** |
| 512 | 245.27 | 234.57 | 308.23 | 271.31 | **1,059.38** |
| 513 | 346.00 | 367.83 | 394.53 | 286.34 | **1,394.69** |
| 514 | 607.02 | 622.70 | 476.41 | 489.00 | **2,195.13** |
| 515 | 1,178.32 | 1,101.13 | 1,045.80 | 998.96 | **4,324.21** |
| 516 | 185.44 | 145.51 | 128.89 | 102.87 | **562.71** |
| 520 | 1,285.45 | 1,240.41 | 1,264.84 | 1,196.80 | **4,987.49** |
| 521 | 278.21 | 216.32 | 234.91 | 251.94 | **981.37** |
| 522 | 2,113.17 | 1,664.39 | 1,972.52 | 1,727.87 | **7,477.95** |
| 523 | 733.20 | 748.62 | 725.58 | 685.24 | **2,892.65** |
| 524 | 1,319.90 | 1,341.56 | 1,237.38 | 1,294.79 | **5,193.63** |
| 525 | 1,577.08 | 1,567.21 | 1,543.21 | 1,373.49 | **6,061.00** |
| 526 | 1,144.32 | 1,058.57 | 1,060.37 | 1,053.26 | **4,316.52** |
| 527 | 986.54 | 1,022.67 | 1,008.31 | 1,023.60 | **4,041.12** |
| 528 | 1,351.72 | 1,176.44 | 1,050.36 | 1,365.88 | **4,944.40** |
| **IA - IOWA - Total** | **30,578.58** | **28,933.47** | **28,213.55** | **27,756.04** | **115,481.64** |

**STATE: ID - IDAHO**

| REGISTRANT ZIP CODE 3 | 1ST QUARTER | 2ND QUARTER | 3RD QUARTER | 4TH QUARTER | TOTAL GRAMS |
|---|---|---|---|---|---|
| 832 | 2,525.72 | 2,574.08 | 2,338.43 | 2,345.24 | **9,783.47** |
| 833 | 2,750.10 | 2,360.12 | 2,138.45 | 2,083.17 | **9,331.84** |
| 834 | 1,788.93 | 1,748.84 | 1,537.21 | 1,526.79 | **6,601.76** |
| 835 | 1,630.74 | 1,486.62 | 1,321.83 | 1,328.90 | **5,768.08** |
| 836 | 7,343.62 | 7,046.12 | 6,916.30 | 6,287.36 | **27,593.40** |
| 837 | 5,490.22 | 5,701.78 | 5,857.10 | 5,073.74 | **22,122.85** |
| 838 | 4,120.24 | 4,014.10 | 3,866.51 | 3,802.21 | **15,803.07** |

| ID - IDAHO - Total | 25,649.58 | 24,931.66 | 23,975.83 | 22,447.40 | 97,004.47 |
|---|---|---|---|---|---|

**STATE: IL - ILLINOIS**

| REGISTRANT ZIP CODE 3 | 1ST QUARTER | 2ND QUARTER | 3RD QUARTER | 4TH QUARTER | TOTAL GRAMS |
|---|---|---|---|---|---|
| 600 | 18,788.57 | 17,858.79 | 18,025.30 | 16,811.51 | **71,484.17** |
| 601 | 8,232.80 | 7,684.22 | 7,554.77 | 7,405.74 | **30,877.53** |
| 602 | 327.55 | 279.85 | 317.91 | 255.48 | **1,180.79** |
| 603 | 506.64 | 484.31 | 433.49 | 429.67 | **1,854.11** |
| 604 | 10,083.61 | 10,005.98 | 9,405.62 | 9,357.58 | **38,852.79** |
| 605 | 5,589.97 | 5,064.45 | 4,811.48 | 4,817.62 | **20,283.52** |
| 606 | 10,185.85 | 9,725.26 | 9,068.97 | 9,475.42 | **38,455.49** |
| 607 | 671.80 | 730.40 | 691.57 | 775.21 | **2,868.98** |
| 608 | 491.83 | 398.01 | 415.45 | 476.04 | **1,781.33** |
| 609 | 1,539.14 | 1,484.53 | 1,603.27 | 1,506.67 | **6,133.62** |
| 610 | 2,745.28 | 2,744.12 | 2,752.26 | 2,660.76 | **10,902.42** |
| 611 | 3,474.69 | 3,081.82 | 3,089.22 | 2,933.13 | **12,578.87** |
| 612 | 1,299.56 | 1,282.88 | 1,231.84 | 1,387.45 | **5,201.72** |
| 613 | 1,100.85 | 1,112.38 | 989.92 | 1,003.87 | **4,207.02** |
| 614 | 841.63 | 814.25 | 786.75 | 773.42 | **3,216.04** |
| 615 | 1,820.17 | 1,706.04 | 1,593.37 | 1,782.68 | **6,902.26** |
| 616 | 3,109.16 | 3,112.13 | 3,077.96 | 3,580.59 | **12,879.84** |
| 617 | 1,474.57 | 1,388.20 | 1,304.85 | 1,378.86 | **5,546.48** |
| 618 | 2,483.49 | 2,485.15 | 2,346.42 | 2,491.43 | **9,806.50** |
| 619 | 795.49 | 741.74 | 712.25 | 738.34 | **2,987.83** |
| 620 | 3,321.27 | 2,955.24 | 2,896.45 | 2,842.42 | **12,015.38** |
| 622 | 3,630.57 | 3,434.24 | 2,880.15 | 2,882.78 | **12,827.74** |
| 623 | 1,131.79 | 1,154.79 | 1,046.35 | 1,005.13 | **4,338.06** |
| 624 | 1,085.45 | 1,056.43 | 1,189.78 | 1,026.79 | **4,358.46** |
| 625 | 1,914.18 | 1,818.43 | 1,768.96 | 1,868.73 | **7,370.30** |
| 626 | 1,354.44 | 1,457.35 | 1,245.96 | 1,382.07 | **5,439.81** |
| 627 | 1,538.31 | 1,513.61 | 1,586.45 | 1,613.60 | **6,251.97** |
| 628 | 1,788.80 | 1,977.47 | 1,744.16 | 1,746.73 | **7,257.16** |
| 629 | 3,844.96 | 3,797.28 | 3,452.15 | 3,491.07 | **14,585.46** |
| **IL - ILLINOIS - Total** | **95,172.42** | **91,349.37** | **88,023.10** | **87,900.76** | **362,445.64** |

**STATE: IN - INDIANA**

| REGISTRANT ZIP CODE 3 | 1ST QUARTER | 2ND QUARTER | 3RD QUARTER | 4TH QUARTER | TOTAL GRAMS |
|---|---|---|---|---|---|
| 460 | 4,543.62 | 4,140.93 | 4,299.06 | 4,181.72 | **17,165.33** |
| 461 | 6,179.61 | 6,125.22 | 6,983.10 | 7,111.01 | **26,398.93** |
| 462 | 14,901.50 | 14,807.36 | 11,928.73 | 11,815.53 | **53,453.12** |
| 463 | 6,298.92 | 5,953.06 | 5,585.93 | 5,335.35 | **23,173.27** |
| 464 | 546.94 | 457.06 | 423.14 | 343.90 | **1,771.05** |

| | | | | | |
|---|---|---|---|---|---|
| 465 | 5,736.87 | 5,205.76 | 4,897.19 | 4,864.29 | **20,704.11** |
| 466 | 1,771.16 | 1,565.86 | 1,373.73 | 1,369.14 | **6,079.89** |
| 467 | 2,838.42 | 3,018.90 | 2,592.31 | 2,826.96 | **11,276.59** |
| 468 | 3,697.77 | 3,662.41 | 3,780.38 | 3,766.37 | **14,906.93** |
| 469 | 4,247.71 | 3,850.55 | 3,614.85 | 3,478.33 | **15,191.44** |
| 470 | 1,486.18 | 1,332.81 | 1,278.07 | 1,252.45 | **5,349.51** |
| 471 | 4,118.47 | 3,546.72 | 3,490.15 | 3,395.62 | **14,550.96** |
| 472 | 2,318.78 | 2,107.75 | 1,986.14 | 2,009.38 | **8,422.05** |
| 473 | 4,888.00 | 4,832.05 | 4,664.79 | 4,641.28 | **19,026.11** |
| 474 | 1,930.46 | 1,739.30 | 1,667.60 | 1,597.53 | **6,934.89** |
| 475 | 1,920.42 | 1,734.92 | 1,571.94 | 1,385.45 | **6,612.73** |
| 476 | 1,276.96 | 1,111.92 | 1,104.09 | 1,006.47 | **4,499.44** |
| 477 | 3,633.36 | 3,448.52 | 3,152.70 | 2,852.88 | **13,087.46** |
| 478 | 1,767.75 | 1,348.65 | 1,519.57 | 1,313.50 | **5,949.48** |
| 479 | 3,235.20 | 3,253.78 | 2,474.32 | 2,166.19 | **11,129.49** |
| **IN - INDIANA - Total** | **77,338.11** | **73,243.52** | **68,387.78** | **66,713.35** | **285,682.76** |

**STATE: KS - KANSAS**

| REGISTRANT ZIP CODE 3 | 1ST QUARTER | 2ND QUARTER | 3RD QUARTER | 4TH QUARTER | **TOTAL GRAMS** |
|---|---|---|---|---|---|
| 660 | 6,028.43 | 5,602.52 | 5,239.76 | 5,559.33 | **22,430.04** |
| 661 | 1,558.11 | 1,153.59 | 1,016.09 | 1,029.97 | **4,757.76** |
| 662 | 6,176.01 | 5,935.84 | 5,795.81 | 5,644.06 | **23,551.72** |
| 664 | 618.88 | 627.54 | 660.89 | 610.40 | **2,517.72** |
| 665 | 1,107.90 | 1,028.73 | 901.57 | 946.26 | **3,984.47** |
| 666 | 2,645.51 | 2,591.86 | 2,665.06 | 2,629.54 | **10,531.97** |
| 667 | 2,476.69 | 2,479.65 | 2,266.74 | 2,223.88 | **9,446.96** |
| 668 | 738.78 | 670.67 | 657.71 | 659.88 | **2,727.05** |
| 669 | 671.08 | 678.41 | 666.77 | 609.59 | **2,625.85** |
| 670 | 2,301.87 | 2,271.60 | 2,083.61 | 2,109.95 | **8,767.03** |
| 671 | 1,373.99 | 1,225.11 | 1,125.20 | 1,043.08 | **4,767.37** |
| 672 | 7,388.21 | 7,215.23 | 6,675.48 | 6,255.47 | **27,534.39** |
| 673 | 1,364.16 | 1,213.14 | 1,097.53 | 1,031.96 | **4,706.80** |
| 674 | 2,075.68 | 2,042.65 | 1,957.23 | 1,909.30 | **7,984.86** |
| 675 | 1,199.15 | 1,252.94 | 1,235.03 | 1,134.81 | **4,821.93** |
| 676 | 970.04 | 991.27 | 883.00 | 894.82 | **3,739.12** |
| 677 | 543.83 | 675.40 | 488.80 | 484.14 | **2,192.17** |
| 678 | 893.16 | 889.37 | 871.64 | 801.35 | **3,455.53** |
| 679 | 342.80 | 368.87 | 371.91 | 318.84 | **1,402.42** |
| **KS - KANSAS - Total** | **40,474.30** | **38,914.39** | **36,659.84** | **35,896.64** | **151,945.16** |

**STATE: KY - KENTUCKY**

| REGISTRANT ZIP CODE 3 | 1ST QUARTER | 2ND QUARTER | 3RD QUARTER | 4TH QUARTER | **TOTAL GRAMS** |
|---|---|---|---|---|---|

| | | | | | |
|---|---|---|---|---|---|
| 400 | 1,231.92 | 1,182.49 | 1,135.15 | 1,150.35 | **4,699.92** |
| 401 | 1,161.71 | 960.42 | 987.48 | 943.80 | **4,053.40** |
| 402 | 7,066.03 | 6,732.58 | 6,072.32 | 6,388.98 | **26,259.91** |
| 403 | 2,555.90 | 2,473.06 | 2,210.98 | 2,286.97 | **9,526.91** |
| 404 | 1,296.50 | 1,226.04 | 1,181.34 | 1,139.93 | **4,843.81** |
| 405 | 3,460.82 | 3,136.01 | 2,848.59 | 3,198.33 | **12,643.74** |
| 406 | 275.88 | 259.25 | 217.29 | 226.00 | **978.42** |
| 407 | 1,155.25 | 1,004.99 | 1,065.57 | 995.58 | **4,221.39** |
| 408 | 271.80 | 274.30 | 254.58 | 230.98 | **1,031.66** |
| 409 | 524.30 | 518.37 | 441.85 | 470.22 | **1,954.73** |
| 410 | 5,601.92 | 5,305.71 | 4,901.78 | 4,889.44 | **20,698.85** |
| 411 | 1,120.88 | 1,117.66 | 937.30 | 1,022.86 | **4,198.69** |
| 412 | 333.33 | 309.66 | 250.02 | 267.59 | **1,160.59** |
| 413 | 349.00 | 335.13 | 263.35 | 292.90 | **1,240.37** |
| 414 | 162.63 | 163.33 | 162.35 | 153.37 | **641.69** |
| 415 | 422.08 | 427.14 | 422.34 | 396.94 | **1,668.51** |
| 416 | 287.41 | 291.54 | 288.37 | 312.63 | **1,179.95** |
| 417 | 962.52 | 1,106.49 | 1,028.77 | 1,083.80 | **4,181.57** |
| 418 | 152.54 | 131.27 | 135.95 | 130.55 | **550.31** |
| 420 | 2,691.11 | 2,629.55 | 2,624.60 | 2,521.19 | **10,466.45** |
| 421 | 2,649.58 | 2,541.82 | 2,262.03 | 2,201.26 | **9,654.70** |
| 422 | 1,519.28 | 1,343.28 | 1,357.47 | 1,090.53 | **5,310.57** |
| 423 | 1,791.02 | 1,718.05 | 1,546.10 | 1,581.71 | **6,636.88** |
| 424 | 1,904.13 | 1,739.37 | 1,592.83 | 1,576.29 | **6,812.62** |
| 425 | 618.54 | 622.34 | 671.71 | 581.65 | **2,494.23** |
| 426 | 419.16 | 455.91 | 435.95 | 367.28 | **1,678.31** |
| 427 | 1,845.62 | 1,710.37 | 1,697.34 | 1,628.70 | **6,882.03** |
| **KY - KENTUCKY - Total** | **41,830.87** | **39,716.11** | **36,993.42** | **37,129.80** | **155,670.20** |

**STATE: LA - LOUISIANA**

| REGISTRANT ZIP CODE 3 | 1ST QUARTER | 2ND QUARTER | 3RD QUARTER | 4TH QUARTER | **TOTAL GRAMS** |
|---|---|---|---|---|---|
| 700 | 5,931.58 | 5,093.71 | 4,950.72 | 4,691.06 | **20,667.06** |
| 701 | 3,388.01 | 3,020.69 | 3,226.99 | 2,580.53 | **12,216.22** |
| 703 | 1,764.00 | 1,656.45 | 1,441.85 | 1,349.21 | **6,211.51** |
| 704 | 5,731.14 | 5,416.63 | 4,928.21 | 4,912.91 | **20,988.89** |
| 705 | 5,224.26 | 5,372.05 | 5,523.20 | 4,929.93 | **21,049.45** |
| 706 | 1,677.70 | 1,778.23 | 1,679.81 | 1,579.15 | **6,714.89** |
| 707 | 5,649.02 | 5,415.84 | 5,333.19 | 4,897.29 | **21,295.34** |
| 708 | 4,270.92 | 4,275.24 | 3,687.07 | 3,853.38 | **16,086.60** |
| 710 | 1,686.29 | 1,642.71 | 1,582.34 | 1,469.93 | **6,381.27** |
| 711 | 5,679.45 | 5,286.61 | 5,735.02 | 5,852.11 | **22,553.19** |

| | | | | | |
|---|---|---|---|---|---|
| 712 | 2,029.57 | 1,958.67 | 1,903.11 | 1,958.32 | **7,849.66** |
| 713 | 3,911.78 | 4,138.83 | 3,815.86 | 3,659.04 | **15,525.50** |
| 714 | 1,814.50 | 1,853.20 | 1,824.31 | 1,727.79 | **7,219.80** |
| **LA - LOUISIANA - Total** | **48,758.22** | **46,908.85** | **45,631.68** | **43,460.65** | **184,759.40** |

**STATE: MA - MASSACHUSETTS**

| REGISTRANT ZIP CODE 3 | 1ST QUARTER | 2ND QUARTER | 3RD QUARTER | 4TH QUARTER | TOTAL GRAMS |
|---|---|---|---|---|---|
| 010 | 6,676.75 | 6,544.78 | 6,266.12 | 6,361.54 | **25,849.20** |
| 011 | 2,928.45 | 3,014.81 | 2,559.75 | 2,573.43 | **11,076.43** |
| 012 | 1,156.89 | 1,209.59 | 1,112.86 | 1,087.48 | **4,566.83** |
| 013 | 1,307.86 | 1,153.44 | 1,147.82 | 1,173.68 | **4,782.80** |
| 014 | 2,211.58 | 2,154.91 | 1,962.83 | 1,868.93 | **8,198.24** |
| 015 | 3,046.34 | 2,803.89 | 2,806.88 | 2,581.71 | **11,238.83** |
| 016 | 1,894.56 | 1,911.72 | 1,904.42 | 1,654.84 | **7,365.54** |
| 017 | 2,902.45 | 3,049.74 | 3,285.35 | 3,148.28 | **12,385.82** |
| 018 | 8,829.01 | 8,399.16 | 7,431.07 | 7,244.97 | **31,904.20** |
| 019 | 3,802.71 | 3,436.21 | 3,344.14 | 3,326.92 | **13,909.98** |
| 020 | 2,069.27 | 1,867.73 | 1,832.54 | 1,953.50 | **7,723.04** |
| 021 | 7,211.99 | 6,956.31 | 6,032.31 | 6,128.62 | **26,329.23** |
| 022 | 839.18 | 825.57 | 639.11 | 839.55 | **3,143.40** |
| 023 | 4,497.08 | 4,480.28 | 4,099.05 | 5,169.28 | **18,245.69** |
| 024 | 1,298.20 | 1,238.80 | 1,186.07 | 1,148.09 | **4,871.17** |
| 025 | 1,042.01 | 1,000.55 | 971.23 | 972.57 | **3,986.36** |
| 026 | 1,488.85 | 1,416.27 | 1,393.51 | 1,211.55 | **5,510.18** |
| 027 | 7,655.33 | 7,615.69 | 7,482.33 | 7,147.30 | **29,900.64** |
| **MA - MASSACHUSETTS - Total** | **60,858.52** | **59,079.46** | **55,457.37** | **55,592.24** | **230,987.59** |

**STATE: MD - MARYLAND**

| REGISTRANT ZIP CODE 3 | 1ST QUARTER | 2ND QUARTER | 3RD QUARTER | 4TH QUARTER | TOTAL GRAMS |
|---|---|---|---|---|---|
| 206 | 4,366.81 | 4,302.09 | 3,985.85 | 3,864.78 | **16,519.52** |
| 207 | 5,737.12 | 4,901.66 | 4,767.20 | 3,979.93 | **19,385.90** |
| 208 | 5,490.21 | 6,049.09 | 4,543.02 | 4,607.61 | **20,689.93** |
| 209 | 912.82 | 768.02 | 755.27 | 734.15 | **3,170.25** |
| 210 | 11,179.51 | 10,557.03 | 9,567.70 | 9,459.66 | **40,763.89** |
| 211 | 5,077.55 | 4,541.44 | 4,203.84 | 3,934.47 | **17,757.29** |
| 212 | 16,598.21 | 15,331.77 | 14,174.19 | 13,952.96 | **60,057.14** |
| 214 | 955.55 | 850.11 | 848.13 | 920.45 | **3,574.24** |
| 215 | 1,755.12 | 1,517.50 | 1,381.13 | 1,310.14 | **5,963.88** |
| 216 | 2,464.96 | 2,259.28 | 2,115.39 | 2,041.13 | **8,880.76** |
| 217 | 7,059.78 | 6,836.64 | 6,407.39 | 6,154.36 | **26,458.18** |
| 218 | 2,103.51 | 1,781.59 | 2,018.55 | 1,705.05 | **7,608.69** |
| 219 | 3,676.87 | 3,427.19 | 2,989.85 | 2,641.78 | **12,735.69** |

| MD - MARYLAND - Total | 67,378.01 | 63,123.40 | 57,757.50 | 55,306.46 | 243,565.36 |
|---|---|---|---|---|---|

**STATE: ME - MAINE**

| REGISTRANT ZIP CODE 3 | 1ST QUARTER | 2ND QUARTER | 3RD QUARTER | 4TH QUARTER | TOTAL GRAMS |
|---|---|---|---|---|---|
| 039 | 357.67 | 253.97 | 251.97 | 253.52 | **1,117.12** |
| 040 | 3,556.59 | 3,339.64 | 3,285.36 | 3,310.31 | **13,491.90** |
| 041 | 981.25 | 1,100.71 | 851.98 | 823.13 | **3,757.07** |
| 042 | 1,527.65 | 1,434.18 | 1,575.76 | 1,666.69 | **6,204.28** |
| 043 | 2,514.05 | 2,588.29 | 2,405.11 | 2,159.06 | **9,666.51** |
| 044 | 1,319.54 | 1,105.39 | 1,134.42 | 1,040.45 | **4,599.81** |
| 045 | 366.23 | 392.42 | 398.75 | 356.60 | **1,514.00** |
| 046 | 589.06 | 521.38 | 558.37 | 657.50 | **2,326.30** |
| 047 | 594.58 | 629.35 | 509.11 | 514.21 | **2,247.25** |
| 048 | 581.73 | 597.96 | 471.89 | 403.72 | **2,055.30** |
| 049 | 2,344.64 | 2,405.19 | 2,331.73 | 2,391.19 | **9,472.75** |
| ME - MAINE - Total | 14,733.01 | 14,368.48 | 13,774.44 | 13,576.37 | 56,452.30 |

**STATE: MI - MICHIGAN**

| REGISTRANT ZIP CODE 3 | 1ST QUARTER | 2ND QUARTER | 3RD QUARTER | 4TH QUARTER | TOTAL GRAMS |
|---|---|---|---|---|---|
| 480 | 18,554.16 | 17,835.08 | 16,176.43 | 15,259.21 | **67,824.89** |
| 481 | 19,619.29 | 18,104.82 | 17,250.16 | 16,562.96 | **71,537.23** |
| 482 | 10,519.17 | 10,639.73 | 9,189.25 | 9,596.47 | **39,944.63** |
| 483 | 13,275.14 | 12,556.65 | 11,497.98 | 10,894.87 | **48,224.65** |
| 484 | 6,270.94 | 6,106.26 | 5,819.88 | 5,898.33 | **24,095.41** |
| 485 | 3,706.41 | 3,476.27 | 3,186.11 | 2,979.14 | **13,347.93** |
| 486 | 11,012.29 | 10,546.76 | 10,026.93 | 9,438.87 | **41,024.84** |
| 487 | 5,179.35 | 4,847.97 | 4,629.31 | 4,545.79 | **19,202.43** |
| 488 | 8,398.86 | 7,861.86 | 7,608.72 | 7,487.87 | **31,357.30** |
| 489 | 3,655.57 | 3,830.12 | 3,719.15 | 3,091.70 | **14,296.54** |
| 490 | 14,105.42 | 13,309.92 | 12,496.61 | 11,774.11 | **51,686.05** |
| 491 | 1,685.65 | 1,548.76 | 1,531.25 | 1,598.26 | **6,363.92** |
| 492 | 5,836.64 | 5,138.98 | 4,423.27 | 4,246.85 | **19,645.74** |
| 493 | 3,299.52 | 3,207.04 | 2,970.17 | 2,908.93 | **12,385.65** |
| 494 | 9,008.21 | 8,944.54 | 8,098.39 | 8,437.14 | **34,488.28** |
| 495 | 7,406.22 | 6,828.33 | 6,902.87 | 6,517.59 | **27,655.02** |
| 496 | 8,090.26 | 7,647.47 | 7,264.97 | 6,627.47 | **29,630.17** |
| 497 | 7,214.37 | 6,871.09 | 6,522.55 | 5,870.05 | **26,478.06** |
| 498 | 4,030.66 | 3,271.30 | 3,298.94 | 2,995.92 | **13,596.82** |
| 499 | 1,295.90 | 1,174.14 | 1,142.25 | 1,129.93 | **4,742.21** |
| MI - MICHIGAN - Total | 162,164.04 | 153,747.08 | 143,755.19 | 137,861.48 | 597,527.79 |

**STATE: MN - MINNESOTA**

| REGISTRANT ZIP CODE 3 | 1ST QUARTER | 2ND QUARTER | 3RD QUARTER | 4TH QUARTER | TOTAL GRAMS |
|---|---|---|---|---|---|

| | | | | | |
|---|---|---|---|---|---|
| 550 | 4,448.72 | 4,323.24 | 4,189.38 | 4,092.24 | **17,053.58** |
| 551 | 5,549.83 | 5,469.21 | 5,611.89 | 4,933.02 | **21,563.94** |
| 553 | 5,540.66 | 5,364.16 | 5,111.12 | 4,802.11 | **20,818.05** |
| 554 | 10,197.22 | 9,115.43 | 8,673.70 | 9,189.65 | **37,176.00** |
| 556 | 158.48 | 158.12 | 183.10 | 155.39 | **655.09** |
| 557 | 1,587.21 | 1,338.37 | 1,393.41 | 1,326.72 | **5,645.71** |
| 558 | 1,391.50 | 1,382.24 | 1,233.25 | 1,091.32 | **5,098.31** |
| 559 | 1,857.49 | 1,779.07 | 1,701.23 | 1,645.80 | **6,983.59** |
| 560 | 1,800.48 | 1,463.87 | 1,386.77 | 1,287.97 | **5,939.10** |
| 561 | 640.67 | 736.42 | 805.91 | 739.64 | **2,922.63** |
| 562 | 1,023.65 | 900.39 | 924.93 | 919.30 | **3,768.27** |
| 563 | 3,021.75 | 3,191.18 | 2,973.19 | 2,870.55 | **12,056.66** |
| 564 | 1,384.55 | 1,392.42 | 1,312.60 | 1,257.83 | **5,347.39** |
| 565 | 1,359.36 | 1,265.70 | 1,240.46 | 1,169.11 | **5,034.62** |
| 566 | 666.68 | 716.37 | 671.36 | 572.77 | **2,627.18** |
| 567 | 677.46 | 562.88 | 587.46 | 441.02 | **2,268.83** |
| **MN - MINNESOTA - Total** | **41,305.71** | **39,159.05** | **37,999.77** | **36,494.43** | **154,958.95** |

**STATE: MO - MISSOURI**

| REGISTRANT ZIP CODE 3 | 1ST QUARTER | 2ND QUARTER | 3RD QUARTER | 4TH QUARTER | **TOTAL GRAMS** |
|---|---|---|---|---|---|
| 630 | 9,606.70 | 8,795.28 | 7,934.93 | 7,541.10 | **33,878.02** |
| 631 | 14,113.28 | 13,052.25 | 13,396.57 | 13,032.34 | **53,594.45** |
| 633 | 4,816.63 | 4,476.35 | 4,157.91 | 4,307.92 | **17,758.80** |
| 634 | 577.39 | 548.07 | 486.72 | 594.37 | **2,206.56** |
| 635 | 1,092.81 | 1,077.78 | 1,119.54 | 1,015.69 | **4,305.82** |
| 636 | 2,827.24 | 2,274.61 | 2,193.21 | 1,999.73 | **9,294.78** |
| 637 | 1,317.26 | 1,172.46 | 1,080.70 | 1,121.70 | **4,692.12** |
| 638 | 1,240.17 | 1,077.59 | 1,064.19 | 1,094.00 | **4,475.95** |
| 639 | 1,339.33 | 1,159.91 | 1,151.24 | 1,072.20 | **4,722.69** |
| 640 | 6,899.20 | 6,562.12 | 6,301.59 | 6,530.96 | **26,293.87** |
| 641 | 6,240.08 | 5,760.53 | 5,400.72 | 4,669.40 | **22,070.73** |
| 644 | 833.00 | 700.93 | 669.15 | 645.93 | **2,849.02** |
| 645 | 1,310.62 | 1,262.18 | 1,068.08 | 972.07 | **4,612.95** |
| 646 | 771.23 | 757.27 | 647.16 | 492.07 | **2,667.73** |
| 647 | 2,066.23 | 1,770.22 | 1,872.46 | 1,722.10 | **7,431.01** |
| 648 | 3,470.86 | 3,440.69 | 3,408.86 | 3,529.69 | **13,850.10** |
| 650 | 1,599.86 | 1,525.85 | 1,315.57 | 1,342.23 | **5,783.50** |
| 651 | 1,037.92 | 929.53 | 812.90 | 936.50 | **3,716.85** |
| 652 | 4,135.24 | 4,041.89 | 3,787.12 | 3,513.28 | **15,477.53** |
| 653 | 897.68 | 853.06 | 732.02 | 732.49 | **3,215.24** |
| 654 | 1,177.85 | 1,036.66 | 1,013.71 | 946.01 | **4,174.23** |

| | | | | | |
|---|---|---|---|---|---|
| 655 | 1,878.33 | 1,741.18 | 1,525.83 | 1,615.36 | **6,760.70** |
| 656 | 4,090.88 | 3,676.23 | 3,902.49 | 3,726.35 | **15,395.96** |
| 657 | 6,939.13 | 6,432.58 | 6,508.87 | 6,312.72 | **26,193.31** |
| 658 | 6,890.25 | 6,382.16 | 6,222.38 | 5,986.13 | **25,480.92** |
| **MO - MISSOURI - Total** | **87,169.18** | **80,507.38** | **77,773.94** | **75,452.35** | **320,902.84** |

**STATE: MS - MISSISSIPPI**

| REGISTRANT ZIP CODE 3 | 1ST QUARTER | 2ND QUARTER | 3RD QUARTER | 4TH QUARTER | TOTAL GRAMS |
|---|---|---|---|---|---|
| 386 | 3,427.31 | 3,469.17 | 3,418.02 | 3,167.57 | **13,482.07** |
| 387 | 307.46 | 327.99 | 324.99 | 340.31 | **1,300.75** |
| 388 | 3,952.14 | 3,691.26 | 3,470.83 | 3,316.81 | **14,431.03** |
| 389 | 772.81 | 814.59 | 680.87 | 589.24 | **2,857.52** |
| 390 | 2,695.66 | 2,591.01 | 2,350.20 | 2,194.12 | **9,830.99** |
| 391 | 4,888.78 | 5,338.29 | 5,365.61 | 4,798.11 | **20,390.79** |
| 392 | 3,090.49 | 3,079.30 | 3,020.50 | 2,618.04 | **11,808.33** |
| 393 | 2,893.97 | 2,569.51 | 2,350.46 | 2,168.36 | **9,982.29** |
| 394 | 4,794.16 | 4,608.72 | 4,458.28 | 4,566.40 | **18,427.57** |
| 395 | 5,017.67 | 4,825.08 | 4,371.35 | 3,767.77 | **17,981.88** |
| 396 | 1,782.34 | 1,535.38 | 1,549.83 | 1,431.70 | **6,299.26** |
| 397 | 1,326.73 | 1,274.03 | 1,179.52 | 1,176.64 | **4,956.92** |
| **MS - MISSISSIPPI - Total** | **34,949.52** | **34,124.33** | **32,540.47** | **30,135.07** | **131,749.38** |

**STATE: MT - MONTANA**

| REGISTRANT ZIP CODE 3 | 1ST QUARTER | 2ND QUARTER | 3RD QUARTER | 4TH QUARTER | TOTAL GRAMS |
|---|---|---|---|---|---|
| 590 | 1,082.87 | 1,046.85 | 1,051.35 | 780.65 | **3,961.72** |
| 591 | 3,353.59 | 2,935.06 | 2,752.98 | 2,575.41 | **11,617.04** |
| 592 | 557.52 | 557.98 | 543.94 | 485.15 | **2,144.60** |
| 593 | 553.20 | 557.23 | 534.11 | 465.13 | **2,109.67** |
| 594 | 1,726.02 | 1,652.48 | 1,533.31 | 1,574.75 | **6,486.55** |
| 595 | 240.46 | 247.00 | 243.62 | 204.58 | **935.66** |
| 596 | 1,819.75 | 1,809.35 | 1,892.78 | 1,163.19 | **6,685.06** |
| 597 | 1,717.45 | 1,636.77 | 1,719.77 | 1,723.11 | **6,797.10** |
| 598 | 2,621.04 | 2,503.49 | 2,505.84 | 2,277.33 | **9,907.70** |
| 599 | 2,296.51 | 2,302.22 | 2,111.90 | 1,876.87 | **8,587.51** |
| **MT - MONTANA - Total** | **15,968.41** | **15,248.43** | **14,889.61** | **13,126.17** | **59,232.61** |

**STATE: NC - NORTH CAROLINA**

| REGISTRANT ZIP CODE 3 | 1ST QUARTER | 2ND QUARTER | 3RD QUARTER | 4TH QUARTER | TOTAL GRAMS |
|---|---|---|---|---|---|
| 270 | 3,446.73 | 3,257.98 | 3,120.57 | 3,168.76 | **12,994.04** |
| 271 | 3,256.59 | 2,924.83 | 2,730.92 | 2,816.48 | **11,728.83** |
| 272 | 6,416.86 | 5,961.45 | 6,148.96 | 5,871.04 | **24,398.32** |
| 273 | 4,041.00 | 4,229.99 | 3,979.66 | 4,565.64 | **16,816.29** |
| 274 | 2,940.18 | 2,595.07 | 2,706.78 | 2,591.50 | **10,833.53** |

| | | | | | |
|---|---|---|---|---|---|
| 275 | 10,753.70 | 10,517.92 | 10,052.23 | 11,172.32 | **42,496.17** |
| 276 | 3,420.73 | 3,381.36 | 3,294.97 | 3,274.46 | **13,371.52** |
| 277 | 5,234.87 | 4,677.04 | 4,161.41 | 4,324.80 | **18,398.13** |
| 278 | 6,162.56 | 6,092.64 | 6,108.44 | 6,271.92 | **24,635.56** |
| 279 | 2,107.23 | 2,190.73 | 2,275.51 | 2,301.35 | **8,874.82** |
| 280 | 8,872.87 | 8,207.31 | 7,736.19 | 7,841.00 | **32,657.37** |
| 281 | 7,062.25 | 6,832.87 | 6,455.52 | 6,598.69 | **26,949.35** |
| 282 | 4,989.48 | 4,752.92 | 5,297.73 | 4,537.76 | **19,577.90** |
| 283 | 8,245.73 | 7,705.17 | 7,348.83 | 7,567.07 | **30,866.80** |
| 284 | 6,249.16 | 5,871.21 | 5,998.51 | 6,360.29 | **24,479.17** |
| 285 | 5,966.54 | 5,594.61 | 5,610.13 | 6,203.03 | **23,374.31** |
| 286 | 9,890.92 | 10,130.03 | 9,616.52 | 9,937.81 | **39,575.30** |
| 287 | 6,898.39 | 6,510.15 | 6,223.22 | 6,265.03 | **25,896.79** |
| 288 | 3,279.83 | 2,894.96 | 2,953.05 | 2,994.85 | **12,122.69** |
| 289 | 735.23 | 795.60 | 815.57 | 822.73 | **3,169.13** |
| **NC - NORTH CAROLINA - Total** | **109,970.87** | **105,123.85** | **102,634.74** | **105,486.56** | **423,216.02** |

**STATE: ND - NORTH DAKOTA**

| REGISTRANT ZIP CODE 3 | 1ST QUARTER | 2ND QUARTER | 3RD QUARTER | 4TH QUARTER | **TOTAL GRAMS** |
|---|---|---|---|---|---|
| 580 | 328.53 | 264.32 | 306.85 | 286.04 | **1,185.74** |
| 581 | 1,220.74 | 995.00 | 1,064.39 | 1,211.66 | **4,491.78** |
| 582 | 861.96 | 801.76 | 827.84 | 865.67 | **3,357.23** |
| 583 | 327.34 | 297.00 | 344.49 | 277.10 | **1,245.92** |
| 584 | 252.07 | 274.21 | 303.16 | 264.68 | **1,094.12** |
| 585 | 1,859.16 | 1,818.70 | 1,667.37 | 1,516.96 | **6,862.20** |
| 586 | 244.89 | 249.96 | 221.16 | 208.31 | **924.32** |
| 587 | 346.27 | 355.13 | 370.84 | 358.98 | **1,431.22** |
| 588 | 230.97 | 191.86 | 244.89 | 267.56 | **935.28** |
| **ND - NORTH DAKOTA - Total** | **5,671.92** | **5,247.94** | **5,351.00** | **5,256.95** | **21,527.80** |

**STATE: NE - NEBRASKA**

| REGISTRANT ZIP CODE 3 | 1ST QUARTER | 2ND QUARTER | 3RD QUARTER | 4TH QUARTER | **TOTAL GRAMS** |
|---|---|---|---|---|---|
| 680 | 2,155.53 | 2,154.06 | 2,121.08 | 2,195.85 | **8,626.52** |
| 681 | 5,893.72 | 5,953.89 | 5,716.79 | 5,516.71 | **23,081.11** |
| 683 | 584.11 | 637.12 | 541.40 | 508.78 | **2,271.41** |
| 684 | 506.04 | 585.30 | 490.62 | 565.07 | **2,147.03** |
| 685 | 2,460.59 | 2,492.57 | 2,299.47 | 2,141.23 | **9,393.87** |
| 686 | 420.54 | 402.62 | 367.79 | 362.65 | **1,553.61** |
| 687 | 930.82 | 918.60 | 841.27 | 861.88 | **3,552.57** |
| 688 | 1,755.02 | 1,667.11 | 1,581.14 | 1,487.04 | **6,490.30** |
| 689 | 588.88 | 628.94 | 576.13 | 571.22 | **2,365.16** |
| 690 | 310.74 | 299.74 | 339.94 | 305.66 | **1,256.08** |

| | | | | | |
|---|---|---|---|---|---|
| 691 | 701.77 | 740.71 | 662.41 | 598.52 | **2,703.41** |
| 692 | 54.74 | 60.19 | 44.59 | 40.12 | **199.65** |
| 693 | 969.83 | 909.40 | 850.03 | 814.02 | **3,543.28** |
| **NE - NEBRASKA - Total** | **17,332.34** | **17,450.26** | **16,432.67** | **15,968.73** | **67,184.01** |

**STATE: NH - NEW HAMPSHIRE**

| REGISTRANT ZIP CODE 3 | 1ST QUARTER | 2ND QUARTER | 3RD QUARTER | 4TH QUARTER | TOTAL GRAMS |
|---|---|---|---|---|---|
| 030 | 4,185.20 | 4,296.68 | 3,990.04 | 4,142.38 | **16,614.30** |
| 031 | 2,535.56 | 2,613.73 | 2,334.55 | 2,396.04 | **9,879.87** |
| 032 | 1,847.16 | 1,749.86 | 1,656.49 | 1,590.99 | **6,844.49** |
| 033 | 1,896.59 | 1,825.07 | 1,787.55 | 1,844.80 | **7,354.00** |
| 034 | 780.92 | 769.11 | 721.42 | 716.72 | **2,988.17** |
| 035 | 650.77 | 625.03 | 559.20 | 522.46 | **2,357.46** |
| 036 | 142.33 | 191.59 | 155.74 | 158.75 | **648.40** |
| 037 | 1,056.42 | 1,171.15 | 912.39 | 825.95 | **3,965.91** |
| 038 | 3,910.43 | 3,851.02 | 3,516.95 | 3,448.49 | **14,726.89** |
| **NH - NEW HAMPSHIRE - Total** | **17,005.38** | **17,093.23** | **15,634.32** | **15,646.56** | **65,379.49** |

**STATE: NJ - NEW JERSEY**

| REGISTRANT ZIP CODE 3 | 1ST QUARTER | 2ND QUARTER | 3RD QUARTER | 4TH QUARTER | TOTAL GRAMS |
|---|---|---|---|---|---|
| 070 | 10,075.08 | 9,641.50 | 8,867.20 | 8,892.59 | **37,476.37** |
| 071 | 1,360.98 | 1,508.97 | 1,283.44 | 1,185.24 | **5,338.64** |
| 072 | 529.14 | 524.33 | 510.14 | 595.98 | **2,159.60** |
| 073 | 750.19 | 739.75 | 623.42 | 694.37 | **2,807.72** |
| 074 | 4,788.60 | 4,245.08 | 4,117.57 | 4,107.59 | **17,258.84** |
| 075 | 681.40 | 734.18 | 712.29 | 748.53 | **2,876.40** |
| 076 | 5,229.64 | 5,169.08 | 5,134.38 | 5,413.46 | **20,946.56** |
| 077 | 5,503.84 | 5,399.60 | 4,785.78 | 4,861.40 | **20,550.63** |
| 078 | 2,709.91 | 2,399.12 | 2,376.98 | 2,054.55 | **9,540.56** |
| 079 | 1,884.85 | 2,063.63 | 1,823.90 | 2,043.58 | **7,815.95** |
| 080 | 16,609.39 | 16,057.69 | 15,211.00 | 14,340.72 | **62,218.80** |
| 081 | 1,468.91 | 1,362.37 | 1,364.43 | 1,284.53 | **5,480.23** |
| 082 | 2,928.02 | 2,793.36 | 2,745.94 | 2,423.64 | **10,890.97** |
| 083 | 1,976.19 | 1,947.67 | 1,767.68 | 1,674.66 | **7,366.20** |
| 084 | 453.73 | 383.23 | 341.55 | 403.27 | **1,581.78** |
| 085 | 1,827.44 | 1,765.30 | 1,698.92 | 1,486.44 | **6,778.10** |
| 086 | 2,104.26 | 1,818.77 | 1,818.59 | 2,044.68 | **7,786.29** |
| 087 | 4,789.34 | 4,795.17 | 4,530.83 | 4,475.17 | **18,590.51** |
| 088 | 8,835.97 | 8,432.13 | 8,074.31 | 8,087.16 | **33,429.58** |
| 089 | 660.71 | 566.83 | 599.25 | 511.55 | **2,338.34** |
| **NJ - NEW JERSEY - Total** | **75,167.59** | **72,347.77** | **68,387.60** | **67,329.10** | **283,232.06** |

**STATE: NM - NEW MEXICO**

| REGISTRANT ZIP CODE 3 | 1ST QUARTER | 2ND QUARTER | 3RD QUARTER | 4TH QUARTER | TOTAL GRAMS |
|---|---|---|---|---|---|
| 870 | 1,785.07 | 1,574.62 | 1,603.77 | 1,634.14 | **6,597.60** |
| 871 | 10,943.00 | 10,131.85 | 9,689.59 | 9,879.45 | **40,643.88** |
| 873 | 773.57 | 791.44 | 815.13 | 742.98 | **3,123.13** |
| 874 | 1,071.72 | 841.04 | 935.47 | 872.43 | **3,720.66** |
| 875 | 2,490.95 | 2,370.75 | 2,083.76 | 2,232.01 | **9,177.47** |
| 877 | 340.70 | 298.02 | 335.24 | 306.25 | **1,280.20** |
| 878 | 95.14 | 109.94 | 90.60 | 90.42 | **386.11** |
| 879 | 208.30 | 200.67 | 173.15 | 188.60 | **770.72** |
| 880 | 1,897.82 | 1,843.54 | 1,812.17 | 1,855.31 | **7,408.84** |
| 881 | 483.96 | 444.82 | 460.24 | 413.12 | **1,802.15** |
| 882 | 2,237.74 | 2,217.91 | 2,036.43 | 2,021.68 | **8,513.76** |
| 883 | 632.63 | 587.55 | 544.30 | 573.49 | **2,337.98** |
| 884 | 66.85 | 52.31 | 42.64 | 44.57 | **206.37** |
| **NM - NEW MEXICO - Total** | **23,027.43** | **21,464.48** | **20,622.48** | **20,854.45** | **85,968.84** |

**STATE: NV - NEVADA**

| REGISTRANT ZIP CODE 3 | 1ST QUARTER | 2ND QUARTER | 3RD QUARTER | 4TH QUARTER | TOTAL GRAMS |
|---|---|---|---|---|---|
| 890 | 13,269.88 | 11,969.46 | 11,248.89 | 10,698.54 | **47,186.76** |
| 891 | 27,046.54 | 25,032.08 | 25,034.29 | 22,280.29 | **99,393.20** |
| 893 | 82.57 | 73.38 | 48.66 | 69.30 | **273.91** |
| 894 | 3,858.84 | 3,645.09 | 3,477.59 | 3,278.35 | **14,259.87** |
| 895 | 4,605.82 | 4,425.03 | 4,327.48 | 4,332.87 | **17,691.19** |
| 897 | 1,451.98 | 1,442.89 | 1,503.72 | 1,592.30 | **5,990.89** |
| 898 | 407.22 | 316.75 | 297.02 | 283.67 | **1,304.66** |
| **NV - NEVADA - Total** | **50,722.85** | **46,904.66** | **45,937.64** | **42,535.32** | **186,100.47** |

**STATE: NY - NEW YORK**

| REGISTRANT ZIP CODE 3 | 1ST QUARTER | 2ND QUARTER | 3RD QUARTER | 4TH QUARTER | TOTAL GRAMS |
|---|---|---|---|---|---|
| 100 | 12,272.44 | 11,276.83 | 10,461.21 | 10,377.81 | **44,388.30** |
| 101 | 235.01 | 177.36 | 199.16 | 159.83 | **771.37** |
| 102 | 15.79 | 4.51 | 6.77 | 6.77 | **33.84** |
| 103 | 5,085.75 | 5,310.09 | 5,439.58 | 4,571.67 | **20,407.10** |
| 104 | 5,761.14 | 5,659.14 | 5,643.87 | 4,713.93 | **21,778.09** |
| 105 | 2,352.17 | 2,168.19 | 2,396.77 | 1,775.75 | **8,692.89** |
| 106 | 401.48 | 381.30 | 467.08 | 392.86 | **1,642.73** |
| 107 | 713.28 | 825.86 | 736.33 | 685.42 | **2,960.88** |
| 108 | 234.31 | 241.01 | 214.79 | 268.31 | **958.43** |
| 109 | 2,741.78 | 2,850.60 | 2,710.13 | 2,427.40 | **10,729.91** |
| 110 | 3,354.53 | 3,059.72 | 3,443.51 | 3,430.62 | **13,288.38** |
| 111 | 643.91 | 700.53 | 533.78 | 503.59 | **2,381.80** |
| 112 | 7,786.53 | 7,472.77 | 6,805.09 | 6,995.92 | **29,060.32** |

| | | | | | |
|---|---|---|---|---|---|
| 113 | 2,841.52 | 2,706.43 | 2,671.65 | 2,642.11 | **10,861.71** |
| 114 | 1,391.86 | 1,277.90 | 1,244.17 | 1,150.40 | **5,064.34** |
| 115 | 4,448.55 | 4,404.49 | 4,006.34 | 3,972.35 | **16,831.74** |
| 116 | 462.52 | 473.93 | 435.81 | 332.18 | **1,704.44** |
| 117 | 15,972.60 | 16,074.73 | 14,967.74 | 14,840.79 | **61,855.86** |
| 118 | 390.87 | 517.34 | 374.19 | 460.96 | **1,743.37** |
| 119 | 1,813.70 | 1,813.78 | 1,728.14 | 1,804.06 | **7,159.69** |
| 120 | 3,440.29 | 3,440.80 | 3,401.63 | 3,166.18 | **13,448.90** |
| 121 | 1,596.33 | 1,370.83 | 1,544.15 | 1,460.07 | **5,971.39** |
| 122 | 2,466.03 | 2,539.99 | 2,222.59 | 2,340.95 | **9,569.56** |
| 123 | 873.64 | 754.09 | 735.94 | 672.50 | **3,036.17** |
| 124 | 2,122.63 | 1,991.56 | 2,272.38 | 2,090.63 | **8,477.20** |
| 125 | 3,164.10 | 3,150.08 | 2,849.26 | 2,710.74 | **11,874.18** |
| 126 | 860.32 | 929.80 | 784.22 | 832.81 | **3,407.16** |
| 127 | 1,288.76 | 1,173.74 | 1,010.63 | 982.43 | **4,455.56** |
| 128 | 2,800.31 | 2,534.25 | 2,712.07 | 2,488.89 | **10,535.52** |
| 129 | 2,144.30 | 1,972.95 | 2,054.86 | 1,955.72 | **8,127.82** |
| 130 | 4,150.83 | 3,958.30 | 3,869.60 | 3,672.75 | **15,651.48** |
| 131 | 1,406.52 | 1,439.05 | 1,439.25 | 1,374.96 | **5,659.77** |
| 132 | 3,594.78 | 3,597.54 | 3,235.62 | 3,041.97 | **13,469.91** |
| 133 | 1,308.15 | 1,191.11 | 1,183.93 | 1,060.76 | **4,743.96** |
| 134 | 1,718.49 | 1,565.56 | 1,679.95 | 1,584.07 | **6,548.06** |
| 135 | 706.89 | 685.69 | 554.39 | 505.77 | **2,452.74** |
| 136 | 2,079.90 | 2,075.40 | 2,057.98 | 2,069.86 | **8,283.14** |
| 137 | 869.77 | 712.29 | 730.18 | 679.50 | **2,991.74** |
| 138 | 3,143.28 | 2,616.00 | 2,706.55 | 2,865.49 | **11,331.32** |
| 139 | 757.68 | 686.85 | 624.52 | 550.00 | **2,619.04** |
| 140 | 4,497.58 | 4,530.08 | 4,132.83 | 4,008.48 | **17,168.97** |
| 141 | 1,908.48 | 1,899.02 | 1,989.26 | 1,862.64 | **7,659.40** |
| 142 | 6,174.69 | 5,807.64 | 5,321.30 | 5,429.25 | **22,732.89** |
| 143 | 824.49 | 763.42 | 667.03 | 643.34 | **2,898.29** |
| 144 | 2,788.05 | 2,884.53 | 2,632.17 | 2,570.95 | **10,875.70** |
| 145 | 2,214.32 | 2,165.26 | 2,068.32 | 2,115.24 | **8,563.14** |
| 146 | 5,587.04 | 5,437.23 | 5,153.73 | 5,107.87 | **21,285.87** |
| 147 | 1,999.42 | 1,945.88 | 1,894.35 | 1,804.58 | **7,644.23** |
| 148 | 3,877.31 | 3,617.23 | 3,343.13 | 3,466.94 | **14,304.62** |
| 149 | 860.61 | 750.95 | 685.45 | 755.72 | **3,052.73** |
| **NY - NEW YORK - Total** | **140,144.76** | **135,583.64** | **130,043.41** | **125,383.84** | **531,155.65** |

**STATE: OH - OHIO**

| REGISTRANT ZIP CODE 3 | 1ST QUARTER | 2ND QUARTER | 3RD QUARTER | 4TH QUARTER | **TOTAL GRAMS** |
|---|---|---|---|---|---|

| | | | | | |
|---|---|---|---|---|---|
| 430 | 6,409.30 | 5,851.53 | 5,799.42 | 6,027.91 | **24,088.15** |
| 431 | 3,724.82 | 3,425.58 | 3,073.92 | 3,277.70 | **13,502.01** |
| 432 | 10,225.85 | 9,198.67 | 7,309.13 | 7,001.68 | **33,735.33** |
| 433 | 1,361.13 | 1,241.51 | 1,198.58 | 1,234.72 | **5,035.93** |
| 434 | 1,186.46 | 1,213.16 | 1,002.56 | 1,105.96 | **4,508.15** |
| 435 | 3,176.46 | 2,852.63 | 2,646.94 | 2,940.39 | **11,616.42** |
| 436 | 4,986.13 | 4,560.83 | 4,400.59 | 3,948.89 | **17,896.44** |
| 437 | 1,798.97 | 1,913.88 | 1,808.27 | 1,656.92 | **7,178.04** |
| 438 | 312.73 | 287.54 | 296.89 | 310.69 | **1,207.85** |
| 439 | 2,151.10 | 2,016.92 | 2,054.61 | 2,055.16 | **8,277.78** |
| 440 | 5,046.11 | 4,859.48 | 4,678.17 | 4,786.43 | **19,370.19** |
| 441 | 7,568.79 | 7,355.50 | 7,173.37 | 7,122.27 | **29,219.94** |
| 442 | 4,806.05 | 4,495.93 | 4,129.29 | 4,076.49 | **17,507.76** |
| 443 | 2,221.64 | 2,287.15 | 2,078.79 | 1,990.58 | **8,578.16** |
| 444 | 3,550.20 | 3,276.91 | 3,085.39 | 3,064.16 | **12,976.66** |
| 445 | 2,252.63 | 2,234.91 | 2,142.45 | 2,357.82 | **8,987.80** |
| 446 | 3,114.58 | 2,871.64 | 2,832.79 | 2,949.17 | **11,768.18** |
| 447 | 2,498.70 | 2,276.92 | 2,314.05 | 2,339.64 | **9,429.31** |
| 448 | 2,672.76 | 2,531.77 | 2,490.08 | 2,246.42 | **9,941.03** |
| 449 | 640.05 | 653.67 | 602.67 | 725.80 | **2,622.19** |
| 450 | 7,099.78 | 6,780.24 | 6,373.65 | 6,469.25 | **26,722.92** |
| 451 | 2,747.47 | 2,665.36 | 2,541.41 | 2,629.42 | **10,583.66** |
| 452 | 8,249.03 | 7,293.51 | 6,953.75 | 7,086.22 | **29,582.51** |
| 453 | 4,306.10 | 4,296.17 | 4,226.49 | 4,602.92 | **17,431.68** |
| 454 | 5,328.51 | 4,620.14 | 4,286.08 | 4,030.14 | **18,264.87** |
| 455 | 829.76 | 821.14 | 803.87 | 678.02 | **3,132.79** |
| 456 | 4,971.58 | 4,953.80 | 4,569.37 | 4,382.09 | **18,876.84** |
| 457 | 1,087.67 | 1,283.07 | 1,141.34 | 1,364.03 | **4,876.11** |
| 458 | 2,102.03 | 2,261.83 | 2,058.19 | 2,164.35 | **8,586.40** |
| **OH - OHIO - Total** | **106,426.38** | **100,381.39** | **94,072.11** | **94,625.23** | **395,505.12** |

**STATE: OK - OKLAHOMA**

| REGISTRANT ZIP CODE 3 | 1ST QUARTER | 2ND QUARTER | 3RD QUARTER | 4TH QUARTER | **TOTAL GRAMS** |
|---|---|---|---|---|---|
| 730 | 13,727.85 | 12,519.56 | 11,686.40 | 9,645.90 | **47,579.70** |
| 731 | 18,222.66 | 16,620.24 | 15,997.05 | 14,357.05 | **65,197.00** |
| 734 | 2,151.08 | 2,011.68 | 1,731.35 | 1,515.60 | **7,409.72** |
| 735 | 4,535.31 | 4,387.69 | 4,209.92 | 3,540.94 | **16,673.86** |
| 736 | 856.49 | 789.11 | 765.84 | 640.00 | **3,051.44** |
| 737 | 1,240.56 | 1,118.21 | 1,094.06 | 938.07 | **4,390.90** |
| 738 | 258.73 | 237.47 | 201.04 | 184.73 | **881.97** |
| 739 | 61.42 | 72.30 | 68.92 | 41.94 | **244.58** |

| | | | | | |
|---|---|---|---|---|---|
| 740 | 15,977.29 | 13,368.84 | 11,941.27 | 10,076.96 | **51,364.36** |
| 741 | 8,386.05 | 8,000.10 | 7,056.96 | 6,239.08 | **29,682.19** |
| 743 | 2,192.44 | 1,839.62 | 1,669.86 | 1,415.25 | **7,117.17** |
| 744 | 5,378.25 | 4,455.91 | 4,176.97 | 3,329.13 | **17,340.26** |
| 745 | 1,587.15 | 1,579.18 | 1,320.85 | 1,202.81 | **5,689.99** |
| 746 | 919.62 | 803.18 | 669.73 | 744.83 | **3,137.37** |
| 747 | 994.10 | 947.50 | 908.48 | 789.34 | **3,639.43** |
| 748 | 3,055.74 | 2,945.55 | 2,690.84 | 2,229.10 | **10,921.23** |
| 749 | 1,631.05 | 1,292.91 | 1,292.22 | 1,006.66 | **5,222.86** |
| **OK - OKLAHOMA - Total** | **81,175.79** | **72,989.07** | **67,481.79** | **57,897.39** | **279,544.04** |

**STATE: OR - OREGON**

| REGISTRANT ZIP CODE 3 | 1ST QUARTER | 2ND QUARTER | 3RD QUARTER | 4TH QUARTER | TOTAL GRAMS |
|---|---|---|---|---|---|
| 970 | 10,107.50 | 9,612.71 | 9,258.78 | 8,865.43 | **37,844.41** |
| 971 | 5,413.72 | 5,138.83 | 4,993.01 | 4,877.47 | **20,423.04** |
| 972 | 15,282.34 | 14,115.40 | 13,763.56 | 13,493.46 | **56,654.76** |
| 973 | 7,008.18 | 6,627.21 | 6,723.97 | 6,305.25 | **26,664.61** |
| 974 | 11,799.90 | 11,306.56 | 10,879.74 | 9,735.53 | **43,721.73** |
| 975 | 6,083.45 | 6,283.36 | 6,032.03 | 5,996.22 | **24,395.06** |
| 976 | 1,211.04 | 1,263.60 | 1,167.56 | 1,178.45 | **4,820.64** |
| 977 | 2,847.40 | 3,422.16 | 3,307.15 | 3,177.33 | **12,754.05** |
| 978 | 2,188.63 | 2,059.39 | 1,953.62 | 1,905.08 | **8,106.71** |
| 979 | 460.69 | 360.17 | 335.14 | 311.27 | **1,467.27** |
| **OR - OREGON - Total** | **62,402.85** | **60,189.39** | **58,414.56** | **55,845.49** | **236,852.29** |

**STATE: PA - PENNSYLVANIA**

| REGISTRANT ZIP CODE 3 | 1ST QUARTER | 2ND QUARTER | 3RD QUARTER | 4TH QUARTER | TOTAL GRAMS |
|---|---|---|---|---|---|
| 150 | 10,482.04 | 9,827.26 | 9,161.65 | 8,465.90 | **37,936.86** |
| 151 | 3,123.51 | 2,939.83 | 2,792.36 | 2,907.71 | **11,763.40** |
| 152 | 8,973.84 | 8,503.11 | 8,193.50 | 7,740.60 | **33,411.05** |
| 153 | 2,339.40 | 2,032.79 | 2,032.56 | 1,798.55 | **8,203.30** |
| 154 | 2,087.94 | 2,074.05 | 2,044.94 | 2,125.01 | **8,331.94** |
| 155 | 1,068.07 | 935.86 | 957.35 | 875.34 | **3,836.63** |
| 156 | 5,007.01 | 4,845.11 | 4,573.36 | 4,535.98 | **18,961.45** |
| 157 | 2,764.99 | 2,464.54 | 2,225.27 | 2,269.19 | **9,723.98** |
| 158 | 1,778.22 | 1,807.38 | 1,693.54 | 1,569.45 | **6,848.59** |
| 159 | 2,417.97 | 2,188.63 | 2,132.79 | 2,027.36 | **8,766.74** |
| 160 | 3,182.55 | 2,905.24 | 2,676.33 | 2,674.79 | **11,438.92** |
| 161 | 3,992.45 | 3,668.73 | 3,542.02 | 3,274.94 | **14,478.14** |
| 162 | 1,380.61 | 1,384.97 | 1,242.97 | 1,247.04 | **5,255.59** |
| 163 | 4,349.33 | 3,708.93 | 3,736.33 | 3,799.60 | **15,594.20** |
| 164 | 1,467.90 | 1,438.06 | 1,336.28 | 1,375.33 | **5,617.57** |

| | | | | | |
|---|---|---|---|---|---|
| 165 | 3,603.91 | 3,743.02 | 3,643.46 | 3,255.67 | **14,246.06** |
| 166 | 4,070.49 | 3,698.53 | 3,831.14 | 3,765.88 | **15,366.04** |
| 167 | 1,748.18 | 1,706.29 | 1,431.30 | 1,300.71 | **6,186.49** |
| 168 | 1,124.96 | 1,111.91 | 1,100.94 | 959.49 | **4,297.30** |
| 169 | 675.41 | 607.71 | 623.49 | 584.34 | **2,490.94** |
| 170 | 10,957.36 | 10,001.33 | 9,529.46 | 9,294.14 | **39,782.28** |
| 171 | 2,474.92 | 2,063.47 | 1,932.13 | 2,114.96 | **8,585.48** |
| 172 | 2,542.96 | 2,497.03 | 2,346.66 | 2,361.51 | **9,748.17** |
| 173 | 3,839.40 | 3,951.38 | 3,667.73 | 3,440.36 | **14,898.86** |
| 174 | 2,590.53 | 2,289.80 | 2,149.49 | 1,989.98 | **9,019.80** |
| 175 | 3,444.34 | 3,191.66 | 3,070.51 | 3,097.46 | **12,803.96** |
| 176 | 3,153.90 | 2,969.80 | 2,942.13 | 2,807.59 | **11,873.42** |
| 177 | 1,914.88 | 1,929.06 | 1,723.52 | 1,715.93 | **7,283.39** |
| 178 | 2,654.94 | 2,539.87 | 2,581.02 | 2,338.30 | **10,114.12** |
| 179 | 1,369.50 | 1,277.40 | 1,192.58 | 1,109.71 | **4,949.19** |
| 180 | 3,910.21 | 4,013.31 | 3,550.35 | 3,770.11 | **15,243.98** |
| 181 | 3,407.02 | 3,207.69 | 3,296.14 | 3,112.84 | **13,023.70** |
| 182 | 1,826.73 | 1,535.80 | 1,642.06 | 1,495.51 | **6,500.10** |
| 183 | 2,389.86 | 2,514.42 | 2,248.99 | 2,095.92 | **9,249.19** |
| 184 | 2,587.04 | 2,475.58 | 2,419.11 | 2,216.53 | **9,698.26** |
| 185 | 3,176.76 | 2,989.13 | 2,765.52 | 2,462.33 | **11,393.74** |
| 186 | 2,176.83 | 2,040.91 | 2,049.94 | 2,025.50 | **8,293.18** |
| 187 | 7,887.62 | 7,681.21 | 6,975.11 | 6,718.74 | **29,262.69** |
| 188 | 860.10 | 1,041.17 | 841.41 | 925.59 | **3,668.27** |
| 189 | 3,906.94 | 4,115.63 | 3,888.78 | 3,575.13 | **15,486.48** |
| 190 | 45,538.10 | 33,700.69 | 54,308.33 | 22,274.96 | **155,822.08** |
| 191 | 13,832.03 | 12,980.48 | 11,200.94 | 11,404.60 | **49,418.05** |
| 193 | 5,295.98 | 4,750.35 | 4,386.54 | 4,593.56 | **19,026.43** |
| 194 | 8,279.56 | 7,568.89 | 6,886.26 | 6,366.32 | **29,101.03** |
| 195 | 1,629.17 | 1,576.79 | 1,682.41 | 1,570.83 | **6,459.21** |
| 196 | 2,047.16 | 2,067.96 | 1,930.70 | 1,921.74 | **7,967.56** |
| **PA - PENNSYLVANIA - Total** | **209,332.61** | **188,562.77** | **200,179.40** | **163,353.02** | **761,427.79** |

**STATE: PR - PUERTO RICO**

| REGISTRANT ZIP CODE 3 | 1ST QUARTER | 2ND QUARTER | 3RD QUARTER | 4TH QUARTER | **TOTAL GRAMS** |
|---|---|---|---|---|---|
| 006 | 308.28 | 267.57 | 350.78 | 374.32 | **1,300.95** |
| 007 | 364.99 | 300.80 | 477.04 | 593.18 | **1,736.02** |
| 009 | 758.02 | 776.97 | 1,177.14 | 1,085.95 | **3,798.08** |
| **PR - PUERTO RICO - Total** | **1,431.29** | **1,345.34** | **2,004.96** | **2,053.46** | **6,835.05** |

**STATE: RI - RHODE ISLAND**

| REGISTRANT ZIP CODE 3 | 1ST QUARTER | 2ND QUARTER | 3RD QUARTER | 4TH QUARTER | **TOTAL GRAMS** |
|---|---|---|---|---|---|

| | | | | | |
|---|---|---|---|---|---|
| 028 | 6,508.98 | 5,929.53 | 5,515.80 | 5,666.58 | **23,620.89** |
| 029 | 3,853.02 | 3,562.40 | 3,292.58 | 3,610.87 | **14,318.86** |
| **RI - RHODE ISLAND - Total** | **10,361.99** | **9,491.94** | **8,808.37** | **9,277.45** | **37,939.75** |

**STATE: SC - SOUTH CAROLINA**

| REGISTRANT ZIP CODE 3 | 1ST QUARTER | 2ND QUARTER | 3RD QUARTER | 4TH QUARTER | TOTAL GRAMS |
|---|---|---|---|---|---|
| 290 | 5,030.14 | 5,154.29 | 4,998.91 | 4,960.18 | **20,143.52** |
| 291 | 4,779.98 | 4,715.35 | 4,415.99 | 4,388.33 | **18,299.65** |
| 292 | 4,665.30 | 4,418.78 | 4,389.11 | 3,883.85 | **17,357.04** |
| 293 | 5,599.60 | 5,534.89 | 5,102.09 | 5,119.39 | **21,355.97** |
| 294 | 11,897.10 | 11,043.90 | 11,069.87 | 11,321.20 | **45,332.07** |
| 295 | 9,209.66 | 8,603.49 | 8,277.60 | 8,474.89 | **34,565.64** |
| 296 | 12,390.17 | 12,125.71 | 11,623.49 | 11,358.69 | **47,498.06** |
| 297 | 2,490.57 | 2,452.64 | 2,157.09 | 2,225.74 | **9,326.05** |
| 298 | 2,133.30 | 2,066.88 | 2,134.66 | 2,163.85 | **8,498.68** |
| 299 | 2,556.08 | 2,248.06 | 2,374.80 | 2,436.48 | **9,615.42** |
| **SC - SOUTH CAROLINA - Total** | **60,751.90** | **58,363.99** | **56,543.60** | **56,332.60** | **231,992.09** |

**STATE: SD - SOUTH DAKOTA**

| REGISTRANT ZIP CODE 3 | 1ST QUARTER | 2ND QUARTER | 3RD QUARTER | 4TH QUARTER | TOTAL GRAMS |
|---|---|---|---|---|---|
| 570 | 1,215.64 | 1,326.93 | 1,269.24 | 1,313.49 | **5,125.31** |
| 571 | 2,522.77 | 2,422.08 | 2,379.04 | 2,205.28 | **9,529.17** |
| 572 | 369.06 | 316.42 | 352.48 | 317.23 | **1,355.18** |
| 573 | 450.59 | 417.36 | 360.97 | 379.46 | **1,608.38** |
| 574 | 412.48 | 406.33 | 394.83 | 388.25 | **1,601.88** |
| 575 | 419.48 | 367.84 | 393.00 | 364.21 | **1,544.53** |
| 576 | 141.03 | 36.60 | 50.80 | 43.25 | **271.68** |
| 577 | 2,153.47 | 1,942.54 | 1,955.50 | 1,593.29 | **7,644.79** |
| **SD - SOUTH DAKOTA - Total** | **7,684.52** | **7,236.09** | **7,155.86** | **6,604.44** | **28,680.91** |

**STATE: TN - TENNESSEE**

| REGISTRANT ZIP CODE 3 | 1ST QUARTER | 2ND QUARTER | 3RD QUARTER | 4TH QUARTER | TOTAL GRAMS |
|---|---|---|---|---|---|
| 370 | 14,867.01 | 13,428.11 | 12,760.12 | 11,619.62 | **52,674.86** |
| 371 | 14,497.49 | 13,295.38 | 12,571.84 | 12,144.64 | **52,509.36** |
| 372 | 8,436.46 | 7,616.08 | 7,190.52 | 6,844.38 | **30,087.44** |
| 373 | 11,025.94 | 10,011.75 | 9,107.35 | 8,719.38 | **38,864.42** |
| 374 | 3,621.64 | 3,239.62 | 2,855.97 | 2,559.56 | **12,276.79** |
| 376 | 8,591.92 | 8,163.54 | 7,644.37 | 7,740.91 | **32,140.74** |
| 377 | 7,074.84 | 7,013.49 | 6,489.96 | 6,361.46 | **26,939.75** |
| 378 | 10,634.81 | 9,839.21 | 9,670.20 | 9,759.20 | **39,903.42** |
| 379 | 7,203.25 | 6,781.20 | 6,329.94 | 6,882.85 | **27,197.25** |
| 380 | 4,316.24 | 4,119.28 | 3,629.34 | 3,429.09 | **15,493.96** |
| 381 | 33,964.70 | 29,511.80 | 39,562.27 | 12,900.94 | **115,939.71** |

| | | | | | |
|---|---|---|---|---|---|
| 382 | 2,569.76 | 2,270.62 | 1,892.30 | 1,878.73 | **8,611.41** |
| 383 | 6,372.44 | 5,916.01 | 5,531.67 | 4,717.55 | **22,537.67** |
| 384 | 3,734.52 | 3,684.49 | 3,535.37 | 3,359.32 | **14,313.71** |
| 385 | 7,294.11 | 7,304.68 | 6,645.78 | 6,132.50 | **27,377.07** |
| **TN - TENNESSEE - Total** | **144,205.13** | **132,195.26** | **135,417.02** | **105,050.14** | **516,867.55** |

**STATE: TX - TEXAS**

| REGISTRANT ZIP CODE 3 | 1ST QUARTER | 2ND QUARTER | 3RD QUARTER | 4TH QUARTER | TOTAL GRAMS |
|---|---|---|---|---|---|
| 750 | 15,624.09 | 15,335.26 | 14,044.36 | 13,262.18 | **58,265.89** |
| 751 | 5,054.72 | 5,081.67 | 4,794.75 | 4,441.38 | **19,372.52** |
| 752 | 7,690.40 | 7,325.85 | 6,610.90 | 6,293.82 | **27,920.97** |
| 753 | 209.18 | 350.24 | 366.12 | 209.85 | **1,135.39** |
| 754 | 3,951.05 | 3,894.92 | 3,435.72 | 3,325.57 | **14,607.26** |
| 755 | 1,185.11 | 1,188.28 | 1,112.59 | 1,131.57 | **4,617.55** |
| 756 | 3,107.31 | 3,276.66 | 2,815.21 | 3,579.24 | **12,778.43** |
| 757 | 3,583.36 | 3,451.57 | 3,438.42 | 3,408.45 | **13,881.80** |
| 758 | 777.98 | 664.83 | 675.55 | 645.44 | **2,763.80** |
| 759 | 2,397.42 | 2,244.30 | 2,239.61 | 2,054.99 | **8,936.31** |
| 760 | 11,350.31 | 10,785.74 | 10,034.96 | 9,575.79 | **41,746.80** |
| 761 | 12,351.35 | 11,276.64 | 11,495.95 | 11,047.29 | **46,171.24** |
| 762 | 4,050.56 | 3,742.82 | 3,566.75 | 3,324.63 | **14,684.76** |
| 763 | 2,494.06 | 2,401.19 | 2,369.58 | 2,195.71 | **9,460.55** |
| 764 | 1,066.39 | 1,068.06 | 931.04 | 882.93 | **3,948.43** |
| 765 | 3,633.70 | 3,495.26 | 3,356.32 | 2,964.43 | **13,449.71** |
| 766 | 1,113.74 | 1,041.15 | 1,039.62 | 1,015.00 | **4,209.51** |
| 767 | 2,831.06 | 2,618.45 | 2,289.79 | 2,521.77 | **10,261.08** |
| 768 | 1,008.99 | 875.24 | 817.52 | 840.06 | **3,541.80** |
| 769 | 1,282.68 | 1,109.00 | 1,005.17 | 913.00 | **4,309.85** |
| 770 | 12,100.78 | 11,781.59 | 11,266.86 | 11,966.30 | **47,115.52** |
| 773 | 8,112.87 | 7,819.16 | 7,169.70 | 7,129.39 | **30,231.12** |
| 774 | 5,584.56 | 5,776.87 | 5,556.92 | 5,193.94 | **22,112.29** |
| 775 | 10,809.36 | 10,006.41 | 9,294.34 | 8,755.01 | **38,865.13** |
| 776 | 2,811.32 | 2,112.20 | 2,879.92 | 2,106.91 | **9,910.34** |
| 777 | 1,496.26 | 1,328.79 | 1,792.97 | 1,718.63 | **6,336.65** |
| 778 | 2,038.85 | 1,847.38 | 1,794.15 | 1,851.39 | **7,531.76** |
| 779 | 2,264.70 | 2,200.37 | 2,349.76 | 2,181.98 | **8,996.81** |
| 780 | 2,471.73 | 2,379.26 | 2,385.84 | 2,258.02 | **9,494.85** |
| 781 | 2,606.50 | 2,440.41 | 2,523.41 | 2,304.62 | **9,874.93** |
| 782 | 14,709.72 | 14,193.46 | 12,879.60 | 12,311.27 | **54,094.05** |
| 783 | 1,416.23 | 1,261.15 | 1,284.42 | 1,318.35 | **5,280.15** |
| 784 | 2,728.13 | 2,703.75 | 2,502.12 | 2,384.38 | **10,318.38** |

| | | | | | |
|---|---|---|---|---|---|
| 785 | 2,813.37 | 2,463.50 | 2,563.05 | 2,404.30 | **10,244.22** |
| 786 | 7,970.08 | 7,770.94 | 6,943.17 | 6,289.36 | **28,973.55** |
| 787 | 7,313.04 | 6,483.85 | 6,067.34 | 5,909.58 | **25,773.80** |
| 788 | 569.96 | 490.93 | 365.74 | 389.04 | **1,815.68** |
| 789 | 413.78 | 410.75 | 445.25 | 359.88 | **1,629.65** |
| 790 | 1,416.45 | 1,321.33 | 1,266.73 | 1,164.32 | **5,168.84** |
| 791 | 3,246.78 | 3,340.85 | 3,268.50 | 3,379.54 | **13,235.66** |
| 792 | 279.16 | 271.71 | 256.27 | 236.71 | **1,043.85** |
| 793 | 684.99 | 690.37 | 696.26 | 602.58 | **2,674.20** |
| 794 | 3,042.25 | 2,958.46 | 2,806.19 | 2,915.61 | **11,722.51** |
| 795 | 530.55 | 440.27 | 443.69 | 467.62 | **1,882.12** |
| 796 | 1,557.54 | 1,361.61 | 1,335.49 | 1,264.92 | **5,519.56** |
| 797 | 2,579.36 | 2,497.48 | 2,147.89 | 2,342.43 | **9,567.16** |
| 798 | 112.28 | 90.20 | 89.53 | 111.97 | **403.98** |
| 799 | 3,377.37 | 2,948.98 | 2,813.46 | 2,702.13 | **11,841.94** |
| **TX - TEXAS - Total** | **189,821.39** | **180,619.18** | **171,628.50** | **165,653.28** | **707,722.35** |

**STATE: UT - UTAH**

| REGISTRANT ZIP CODE 3 | 1ST QUARTER | 2ND QUARTER | 3RD QUARTER | 4TH QUARTER | TOTAL GRAMS |
|---|---|---|---|---|---|
| 840 | 17,999.08 | 16,741.17 | 15,817.88 | 15,560.53 | **66,118.66** |
| 841 | 12,545.47 | 11,532.11 | 11,618.48 | 10,674.41 | **46,370.46** |
| 843 | 1,755.82 | 1,603.71 | 1,639.80 | 1,612.27 | **6,611.60** |
| 844 | 4,021.99 | 3,826.17 | 3,724.98 | 3,661.46 | **15,234.60** |
| 845 | 1,201.37 | 1,137.57 | 1,155.06 | 1,172.25 | **4,666.25** |
| 846 | 2,983.35 | 2,818.14 | 2,878.67 | 2,896.62 | **11,576.78** |
| 847 | 5,268.20 | 5,163.05 | 5,006.67 | 4,694.34 | **20,132.27** |
| **UT - UTAH - Total** | **45,775.28** | **42,821.91** | **41,841.55** | **40,271.89** | **170,710.63** |

**STATE: VA - VIRGINIA**

| REGISTRANT ZIP CODE 3 | 1ST QUARTER | 2ND QUARTER | 3RD QUARTER | 4TH QUARTER | TOTAL GRAMS |
|---|---|---|---|---|---|
| 201 | 5,483.58 | 5,906.66 | 5,518.76 | 5,989.11 | **22,898.12** |
| 220 | 2,328.98 | 2,288.10 | 2,351.29 | 2,026.49 | **8,994.86** |
| 221 | 2,456.21 | 2,550.49 | 2,420.00 | 2,699.56 | **10,126.27** |
| 222 | 576.48 | 598.68 | 519.15 | 464.94 | **2,159.26** |
| 223 | 1,275.86 | 1,345.29 | 1,006.10 | 1,128.54 | **4,755.80** |
| 224 | 2,431.98 | 2,444.39 | 2,301.43 | 2,223.71 | **9,401.51** |
| 225 | 852.38 | 886.09 | 687.84 | 837.74 | **3,264.05** |
| 226 | 2,785.75 | 2,728.77 | 2,592.39 | 2,490.44 | **10,597.36** |
| 227 | 466.64 | 518.52 | 470.19 | 488.00 | **1,943.36** |
| 228 | 1,801.83 | 1,671.98 | 1,900.41 | 2,169.03 | **7,543.26** |
| 229 | 2,170.01 | 2,324.79 | 2,222.90 | 2,056.28 | **8,773.98** |
| 230 | 1,864.07 | 1,704.29 | 1,859.78 | 1,976.64 | **7,404.78** |

| | | | | | |
|---|---|---|---|---|---|
| 231 | 2,895.84 | 2,798.62 | 2,760.37 | 2,598.58 | **11,053.40** |
| 232 | 7,142.37 | 6,175.11 | 5,654.04 | 5,354.71 | **24,326.23** |
| 233 | 2,313.71 | 2,326.42 | 2,306.90 | 2,309.30 | **9,256.34** |
| 234 | 5,047.07 | 5,005.22 | 4,567.76 | 4,518.49 | **19,138.53** |
| 235 | 1,892.92 | 1,684.70 | 1,652.60 | 1,685.69 | **6,915.91** |
| 236 | 3,305.37 | 2,867.35 | 2,980.58 | 3,042.85 | **12,196.15** |
| 237 | 1,789.55 | 1,592.04 | 1,265.94 | 1,230.70 | **5,878.23** |
| 238 | 2,800.37 | 2,946.34 | 2,766.77 | 2,523.00 | **11,036.47** |
| 239 | 654.44 | 695.49 | 663.17 | 644.27 | **2,657.37** |
| 240 | 3,703.82 | 3,456.47 | 3,447.88 | 3,521.73 | **14,129.91** |
| 241 | 4,081.07 | 3,858.26 | 3,410.68 | 3,642.21 | **14,992.22** |
| 242 | 3,732.47 | 3,666.60 | 3,431.27 | 3,286.59 | **14,116.93** |
| 243 | 2,115.62 | 1,934.35 | 1,944.33 | 2,134.84 | **8,129.14** |
| 244 | 1,229.81 | 1,154.21 | 1,114.41 | 971.06 | **4,469.49** |
| 245 | 2,646.53 | 2,894.63 | 2,773.50 | 2,799.59 | **11,114.25** |
| 246 | 814.26 | 804.88 | 861.96 | 679.84 | **3,160.94** |
| **VA - VIRGINIA - Total** | **70,659.02** | **68,828.75** | **65,452.40** | **65,493.94** | **270,434.11** |

**STATE: VI - VIRGIN ISLANDS**

| REGISTRANT ZIP CODE 3 | 1ST QUARTER | 2ND QUARTER | 3RD QUARTER | 4TH QUARTER | **TOTAL GRAMS** |
|---|---|---|---|---|---|
| 008 | 96.93 | 133.71 | 134.38 | 106.74 | **471.76** |
| **VI - VIRGIN ISLANDS - Total** | **96.93** | **133.71** | **134.38** | **106.74** | **471.76** |

**STATE: VT - VERMONT**

| REGISTRANT ZIP CODE 3 | 1ST QUARTER | 2ND QUARTER | 3RD QUARTER | 4TH QUARTER | **TOTAL GRAMS** |
|---|---|---|---|---|---|
| 050 | 1,012.96 | 952.61 | 881.83 | 1,001.13 | **3,848.53** |
| 051 | 511.31 | 418.28 | 537.03 | 428.87 | **1,895.48** |
| 052 | 424.57 | 384.24 | 446.01 | 436.09 | **1,690.91** |
| 053 | 617.70 | 638.18 | 542.96 | 550.75 | **2,349.59** |
| 054 | 3,899.80 | 3,961.33 | 3,696.45 | 3,489.03 | **15,046.61** |
| 056 | 830.46 | 684.71 | 627.68 | 669.27 | **2,812.11** |
| 057 | 1,504.25 | 1,560.66 | 1,489.35 | 1,472.60 | **6,026.86** |
| 058 | 783.31 | 802.65 | 691.26 | 788.79 | **3,066.02** |
| **VT - VERMONT - Total** | **9,584.36** | **9,402.67** | **8,912.58** | **8,836.52** | **36,736.12** |

**STATE: WA - WASHINGTON**

| REGISTRANT ZIP CODE 3 | 1ST QUARTER | 2ND QUARTER | 3RD QUARTER | 4TH QUARTER | **TOTAL GRAMS** |
|---|---|---|---|---|---|
| 980 | 13,840.49 | 13,289.94 | 12,338.49 | 12,078.90 | **51,547.82** |
| 981 | 10,103.52 | 10,043.34 | 8,983.72 | 8,546.86 | **37,677.44** |
| 982 | 10,763.67 | 10,855.08 | 10,923.24 | 9,774.35 | **42,316.35** |
| 983 | 10,992.83 | 10,779.16 | 10,953.75 | 10,599.39 | **43,325.13** |
| 984 | 6,500.27 | 6,077.83 | 5,573.85 | 5,574.17 | **23,726.13** |
| 985 | 5,921.12 | 5,760.76 | 5,635.57 | 5,616.89 | **22,934.34** |

| | | | | | |
|---|---|---|---|---|---|
| 986 | 12,690.12 | 11,801.77 | 12,153.47 | 11,151.69 | **47,797.06** |
| 988 | 2,821.10 | 2,832.25 | 2,673.38 | 2,622.41 | **10,949.14** |
| 989 | 3,229.23 | 3,064.61 | 2,744.50 | 2,692.32 | **11,730.66** |
| 990 | 1,609.75 | 1,546.63 | 1,451.17 | 1,525.19 | **6,132.74** |
| 991 | 2,167.90 | 2,195.03 | 1,902.15 | 1,854.17 | **8,119.26** |
| 992 | 9,558.57 | 9,675.20 | 8,553.62 | 8,458.52 | **36,245.91** |
| 993 | 5,298.69 | 5,062.15 | 4,534.85 | 4,170.71 | **19,066.40** |
| 994 | 924.00 | 1,117.88 | 1,022.52 | 945.62 | **4,010.01** |
| **WA - WASHINGTON - Total** | **96,421.27** | **94,101.65** | **89,444.28** | **85,611.21** | **365,578.40** |

**STATE: WI - WISCONSIN**

| REGISTRANT ZIP CODE 3 | 1ST QUARTER | 2ND QUARTER | 3RD QUARTER | 4TH QUARTER | TOTAL GRAMS |
|---|---|---|---|---|---|
| 530 | 7,608.91 | 7,522.66 | 7,007.30 | 7,248.94 | **29,387.81** |
| 531 | 7,592.73 | 7,509.60 | 6,486.11 | 6,501.57 | **28,090.02** |
| 532 | 11,990.73 | 10,838.44 | 10,158.08 | 9,788.04 | **42,775.29** |
| 534 | 1,441.84 | 1,406.68 | 1,341.45 | 1,293.03 | **5,483.00** |
| 535 | 7,438.31 | 7,033.25 | 6,726.26 | 6,286.55 | **27,484.37** |
| 537 | 5,987.26 | 5,857.90 | 5,228.00 | 5,196.25 | **22,269.40** |
| 538 | 633.30 | 627.73 | 578.93 | 657.50 | **2,497.45** |
| 539 | 3,279.74 | 3,199.88 | 3,064.71 | 3,262.93 | **12,807.25** |
| 540 | 938.32 | 949.31 | 894.36 | 850.90 | **3,632.90** |
| 541 | 1,202.16 | 1,179.98 | 1,175.22 | 1,154.95 | **4,712.30** |
| 542 | 1,051.76 | 1,109.37 | 1,093.19 | 1,022.88 | **4,277.19** |
| 543 | 2,192.40 | 1,850.72 | 1,719.26 | 1,888.30 | **7,650.68** |
| 544 | 3,060.10 | 2,930.05 | 2,897.27 | 2,886.07 | **11,773.48** |
| 545 | 856.17 | 864.17 | 845.60 | 838.19 | **3,404.14** |
| 546 | 3,206.19 | 3,159.65 | 2,922.21 | 2,848.76 | **12,136.81** |
| 547 | 2,473.39 | 2,373.82 | 2,325.07 | 2,330.25 | **9,502.54** |
| 548 | 1,944.13 | 1,907.62 | 1,961.77 | 1,900.56 | **7,714.08** |
| 549 | 5,416.58 | 5,477.30 | 4,920.51 | 5,141.69 | **20,956.07** |
| **WI - WISCONSIN - Total** | **68,314.02** | **65,798.11** | **61,345.30** | **61,097.36** | **256,554.79** |

**STATE: WV - WEST VIRGINIA**

| REGISTRANT ZIP CODE 3 | 1ST QUARTER | 2ND QUARTER | 3RD QUARTER | 4TH QUARTER | TOTAL GRAMS |
|---|---|---|---|---|---|
| 247 | 568.89 | 620.92 | 575.20 | 591.44 | **2,356.45** |
| 248 | 189.79 | 208.36 | 145.10 | 223.68 | **766.93** |
| 249 | 399.73 | 364.25 | 403.18 | 269.49 | **1,436.64** |
| 250 | 590.02 | 538.96 | 407.13 | 511.74 | **2,047.85** |
| 251 | 232.55 | 352.68 | 288.76 | 266.28 | **1,140.28** |
| 252 | 485.07 | 469.60 | 347.13 | 408.10 | **1,709.91** |
| 253 | 1,280.45 | 1,080.51 | 1,115.34 | 1,034.44 | **4,510.74** |
| 254 | 3,247.63 | 3,102.35 | 3,424.42 | 2,575.85 | **12,350.24** |

| | | | | | |
|---|---|---|---|---|---|
| 255 | 909.12 | 857.11 | 818.02 | 807.40 | **3,391.66** |
| 256 | 322.52 | 312.31 | 377.43 | 278.34 | **1,290.61** |
| 257 | 1,235.85 | 1,077.68 | 960.15 | 1,083.05 | **4,356.74** |
| 258 | 977.78 | 979.96 | 972.45 | 896.23 | **3,826.42** |
| 259 | 270.49 | 195.97 | 185.50 | 218.42 | **870.37** |
| 260 | 1,922.00 | 1,591.62 | 1,590.00 | 1,703.21 | **6,806.83** |
| 261 | 832.07 | 1,063.06 | 871.68 | 955.99 | **3,722.80** |
| 262 | 731.76 | 561.59 | 591.21 | 604.13 | **2,488.68** |
| 263 | 1,583.21 | 1,500.82 | 1,359.16 | 1,396.82 | **5,840.01** |
| 264 | 354.61 | 360.24 | 379.57 | 362.95 | **1,457.38** |
| 265 | 1,661.62 | 1,468.24 | 1,491.53 | 1,362.31 | **5,983.70** |
| 266 | 431.36 | 459.74 | 375.69 | 326.32 | **1,593.11** |
| 267 | 546.02 | 577.08 | 524.49 | 504.22 | **2,151.80** |
| 268 | 242.35 | 269.89 | 339.61 | 303.73 | **1,155.58** |
| **WV - WEST VIRGINIA - Total** | **19,014.89** | **18,012.96** | **17,542.76** | **16,684.12** | **71,254.72** |

**STATE: WY - WYOMING**

| REGISTRANT ZIP CODE 3 | 1ST QUARTER | 2ND QUARTER | 3RD QUARTER | 4TH QUARTER | TOTAL GRAMS |
|---|---|---|---|---|---|
| 820 | 1,547.70 | 1,326.28 | 1,378.57 | 1,081.79 | **5,334.35** |
| 822 | 342.71 | 246.66 | 236.22 | 235.23 | **1,060.82** |
| 823 | 128.53 | 95.67 | 114.95 | 106.69 | **445.84** |
| 824 | 841.50 | 776.88 | 766.03 | 697.06 | **3,081.47** |
| 825 | 625.48 | 646.77 | 620.99 | 519.44 | **2,412.68** |
| 826 | 1,181.16 | 1,191.95 | 1,246.55 | 1,020.83 | **4,640.49** |
| 827 | 352.44 | 401.41 | 322.33 | 346.97 | **1,423.15** |
| 828 | 985.76 | 880.87 | 902.28 | 975.67 | **3,744.58** |
| 829 | 2,018.60 | 1,931.78 | 1,557.52 | 1,574.74 | **7,082.64** |
| 830 | 76.72 | 81.33 | 74.85 | 86.01 | **318.91** |
| 831 | 247.26 | 271.77 | 209.19 | 214.09 | **942.30** |
| **WY - WYOMING - Total** | **8,347.86** | **7,851.37** | **7,429.48** | **6,858.52** | **30,487.23** |

**DRUG: 9652 - OXYMORPHONE**

**STATE: AK - ALASKA**

| REGISTRANT ZIP CODE 3 | 1ST QUARTER | 2ND QUARTER | 3RD QUARTER | 4TH QUARTER | TOTAL GRAMS |
|---|---|---|---|---|---|
| 995 | 76.01 | 59.24 | 54.86 | 36.04 | **226.15** |
| 996 | 79.66 | 85.91 | 48.62 | 53.53 | **267.72** |
| 997 | 10.17 | 8.92 | 18.73 | 4.28 | **42.11** |
| 998 | 0 | 0 | 0.54 | 0 | **0.54** |
| 999 | 2.14 | 4.28 | 0 | 0 | **6.42** |
| **AK - ALASKA - Total** | **167.98** | **158.35** | **122.75** | **93.85** | **542.93** |

**STATE: AL - ALABAMA**

| REGISTRANT ZIP CODE 3 | 1ST QUARTER | 2ND QUARTER | 3RD QUARTER | 4TH QUARTER | TOTAL GRAMS |
|---|---|---|---|---|---|

| | | | | | |
|---|---|---|---|---|---|
| 350 | 215.31 | 143.63 | 123.47 | 112.72 | **595.12** |
| 351 | 129.18 | 111.56 | 93.27 | 102.46 | **436.46** |
| 352 | 86.44 | 74.27 | 77.70 | 84.44 | **322.85** |
| 354 | 67.93 | 73.96 | 93.72 | 82.03 | **317.63** |
| 355 | 88.27 | 73.33 | 81.72 | 62.63 | **305.95** |
| 356 | 146.62 | 172.18 | 134.53 | 129.53 | **582.85** |
| 357 | 143.54 | 121.68 | 93.05 | 90.19 | **448.46** |
| 358 | 197.51 | 173.60 | 148.40 | 150.23 | **669.74** |
| 359 | 402.02 | 391.85 | 353.27 | 370.58 | **1,517.73** |
| 360 | 33.45 | 32.25 | 27.03 | 33.23 | **125.96** |
| 361 | 24.98 | 28.64 | 13.20 | 26.58 | **93.40** |
| 362 | 116.42 | 107.50 | 106.78 | 112.05 | **442.75** |
| 363 | 61.38 | 58.97 | 61.29 | 61.29 | **242.92** |
| 364 | 21.05 | 18.47 | 18.20 | 13.02 | **70.74** |
| 365 | 222.04 | 214.01 | 184.31 | 157.37 | **777.73** |
| 366 | 226.24 | 178.60 | 149.96 | 164.32 | **719.12** |
| 367 | 12.49 | 15.57 | 7.36 | 6.82 | **42.24** |
| 368 | 174.58 | 167.63 | 169.50 | 188.68 | **700.39** |
| 369 | 14.90 | 8.21 | 6.07 | 9.99 | **39.16** |
| **AL - ALABAMA - Total** | **2,384.36** | **2,165.88** | **1,942.82** | **1,958.16** | **8,451.22** |

**STATE: AR - ARKANSAS**

| REGISTRANT ZIP CODE 3 | 1ST QUARTER | 2ND QUARTER | 3RD QUARTER | 4TH QUARTER | **TOTAL GRAMS** |
|---|---|---|---|---|---|
| 716 | 23.64 | 14.27 | 10.17 | 8.92 | **57.01** |
| 717 | 23.37 | 16.91 | 11.33 | 11.86 | **63.47** |
| 718 | 2.14 | 0 | 0 | 0 | **2.14** |
| 719 | 40.90 | 23.60 | 11.33 | 11.24 | **87.07** |
| 720 | 128.55 | 102.73 | 97.55 | 81.14 | **409.96** |
| 721 | 101.92 | 83.81 | 60.08 | 28.19 | **274.01** |
| 722 | 66.55 | 52.99 | 30.78 | 31.58 | **181.90** |
| 723 | 9.63 | 9.99 | 4.73 | 7.32 | **31.67** |
| 724 | 30.06 | 30.15 | 13.11 | 10.79 | **84.13** |
| 725 | 21.05 | 20.25 | 12.49 | 18.11 | **71.90** |
| 726 | 9.19 | 6.78 | 13.02 | 3.03 | **32.03** |
| 727 | 176.41 | 124.76 | 133.37 | 118.47 | **553.01** |
| 728 | 15.97 | 5.89 | 0 | 0 | **21.86** |
| 729 | 60.75 | 39.25 | 33.36 | 37.83 | **171.19** |
| **AR - ARKANSAS - Total** | **710.16** | **531.38** | **431.33** | **368.48** | **2,041.35** |

**STATE: AZ - ARIZONA**

| REGISTRANT ZIP CODE 3 | 1ST QUARTER | 2ND QUARTER | 3RD QUARTER | 4TH QUARTER | **TOTAL GRAMS** |
|---|---|---|---|---|---|
| 850 | 547.30 | 425.84 | 425.80 | 383.38 | **1,782.33** |

| | | | | | |
|---|---|---|---|---|---|
| 851 | 89.66 | 50.40 | 43.80 | 44.20 | **228.07** |
| 852 | 823.99 | 786.25 | 800.08 | 563.63 | **2,973.95** |
| 853 | 545.30 | 455.42 | 388.02 | 394.75 | **1,783.49** |
| 855 | 57.36 | 43.18 | 50.05 | 37.38 | **187.97** |
| 856 | 64.90 | 53.70 | 44.96 | 44.25 | **207.81** |
| 857 | 373.06 | 240.29 | 219.95 | 200.77 | **1,034.06** |
| 859 | 9.05 | 12.76 | 15.70 | 12.58 | **50.09** |
| 860 | 32.29 | 29.97 | 25.60 | 20.16 | **108.03** |
| 863 | 95.54 | 48.35 | 57.09 | 42.55 | **243.54** |
| 864 | 36.67 | 38.00 | 22.84 | 20.96 | **118.47** |
| **AZ - ARIZONA - Total** | **2,675.12** | **2,184.17** | **2,093.89** | **1,764.62** | **8,717.81** |

**STATE: CA - CALIFORNIA**

| REGISTRANT ZIP CODE 3 | 1ST QUARTER | 2ND QUARTER | 3RD QUARTER | 4TH QUARTER | **TOTAL GRAMS** |
|---|---|---|---|---|---|
| 900 | 388.96 | 280.57 | 309.29 | 312.77 | **1,291.58** |
| 902 | 216.16 | 203.84 | 180.11 | 175.30 | **775.41** |
| 903 | 30.33 | 9.81 | 33.01 | 24.98 | **98.13** |
| 904 | 63.61 | 37.47 | 43.00 | 58.34 | **202.42** |
| 905 | 7.94 | 8.65 | 19.98 | 17.40 | **53.97** |
| 906 | 42.82 | 31.22 | 28.81 | 32.92 | **135.78** |
| 907 | 91.53 | 66.10 | 29.80 | 27.66 | **215.09** |
| 908 | 63.07 | 87.07 | 39.70 | 56.29 | **246.13** |
| 910 | 5.71 | 11.78 | 13.74 | 16.59 | **47.82** |
| 911 | 96.88 | 94.92 | 81.54 | 76.72 | **350.06** |
| 912 | 35.86 | 32.74 | 31.13 | 21.14 | **120.88** |
| 913 | 264.51 | 237.12 | 186.63 | 184.75 | **873.01** |
| 914 | 163.08 | 178.60 | 184.40 | 172.53 | **698.60** |
| 915 | 10.97 | 7.85 | 8.21 | 4.46 | **31.49** |
| 916 | 13.78 | 12.13 | 4.46 | 17.49 | **47.86** |
| 917 | 173.60 | 165.48 | 153.26 | 156.83 | **649.18** |
| 918 | 0 | 0 | 0 | 0.45 | **0.45** |
| 919 | 89.39 | 85.73 | 71.86 | 75.47 | **322.45** |
| 920 | 650.39 | 695.61 | 519.43 | 511.62 | **2,377.05** |
| 921 | 267.72 | 163.79 | 135.42 | 134.80 | **701.73** |
| 922 | 164.50 | 139.97 | 152.46 | 125.34 | **582.27** |
| 923 | 111.47 | 99.29 | 85.11 | 57.72 | **353.58** |
| 924 | 36.58 | 26.67 | 23.19 | 28.64 | **115.08** |
| 925 | 77.88 | 50.05 | 51.21 | 76.19 | **255.32** |
| 926 | 392.35 | 335.16 | 373.30 | 346.36 | **1,447.16** |
| 927 | 37.74 | 28.55 | 27.88 | 32.16 | **126.32** |
| 928 | 270.48 | 214.55 | 216.33 | 191.98 | **893.35** |

| | | | | | |
|---|---|---|---|---|---|
| 930 | 174.32 | 138.54 | 153.89 | 79.49 | **546.23** |
| 931 | 2.68 | 1.78 | 0.89 | 0.89 | **6.24** |
| 932 | 67.89 | 55.49 | 66.42 | 87.25 | **277.04** |
| 933 | 9.10 | 11.15 | 14.27 | 3.12 | **37.65** |
| 934 | 156.92 | 136.05 | 127.12 | 147.82 | **567.91** |
| 935 | 101.39 | 67.62 | 64.94 | 62.54 | **296.49** |
| 936 | 267.59 | 225.61 | 172.58 | 159.11 | **824.88** |
| 937 | 291.72 | 292.88 | 331.50 | 271.64 | **1,187.74** |
| 939 | 15.17 | 24.35 | 21.14 | 5.00 | **65.66** |
| 940 | 215.44 | 190.11 | 217.05 | 219.64 | **842.23** |
| 941 | 89.66 | 48.71 | 52.54 | 48.53 | **239.44** |
| 943 | 10.53 | 8.92 | 5.80 | 11.86 | **37.11** |
| 944 | 63.07 | 30.96 | 28.37 | 33.90 | **156.30** |
| 945 | 549.31 | 429.14 | 425.93 | 354.12 | **1,758.51** |
| 946 | 1.34 | 2.23 | 0.71 | 1.74 | **6.02** |
| 947 | 1.61 | 0 | 0.80 | 0.27 | **2.68** |
| 948 | 2.05 | 0 | 0 | 0.45 | **2.50** |
| 949 | 76.41 | 65.66 | 52.90 | 64.94 | **259.91** |
| 950 | 111.78 | 92.42 | 88.41 | 86.09 | **378.70** |
| 951 | 40.14 | 29.75 | 21.10 | 22.12 | **113.12** |
| 952 | 57.54 | 38.81 | 45.05 | 27.74 | **169.14** |
| 953 | 120.52 | 88.94 | 92.96 | 108.03 | **410.46** |
| 954 | 105.45 | 75.03 | 88.59 | 55.31 | **324.37** |
| 955 | 30.78 | 34.61 | 29.35 | 36.67 | **131.41** |
| 956 | 143.45 | 123.91 | 122.40 | 85.20 | **474.95** |
| 957 | 41.93 | 29.44 | 15.25 | 14.10 | **100.72** |
| 958 | 119.99 | 95.95 | 63.34 | 51.03 | **330.30** |
| 959 | 117.85 | 110.98 | 78.33 | 78.06 | **385.21** |
| 960 | 24.09 | 24.98 | 20.25 | 21.37 | **90.68** |
| 961 | 11.95 | 1.61 | 3.75 | 3.48 | **20.79** |
| **CA - CALIFORNIA - Total** | **6,788.93** | **5,780.36** | **5,408.89** | **5,078.37** | **23,056.55** |

**STATE: CO - COLORADO**

| REGISTRANT ZIP CODE 3 | 1ST QUARTER | 2ND QUARTER | 3RD QUARTER | 4TH QUARTER | TOTAL GRAMS |
|---|---|---|---|---|---|
| 800 | 243.10 | 218.30 | 207.23 | 243.77 | **912.40** |
| 801 | 335.34 | 294.57 | 281.55 | 276.28 | **1,187.74** |
| 802 | 333.56 | 302.02 | 266.29 | 231.01 | **1,132.88** |
| 803 | 46.21 | 46.75 | 40.50 | 40.23 | **173.69** |
| 804 | 22.03 | 18.38 | 32.29 | 19.63 | **92.33** |
| 805 | 195.68 | 190.42 | 148.76 | 121.59 | **656.45** |
| 806 | 86.80 | 78.77 | 71.32 | 69.32 | **306.21** |

| | | | | | |
|---|---|---|---|---|---|
| 807 | 14.94 | 11.02 | 14.50 | 12.98 | **53.44** |
| 808 | 40.50 | 32.74 | 39.74 | 35.86 | **148.85** |
| 809 | 176.99 | 161.02 | 119.63 | 130.43 | **588.07** |
| 810 | 51.65 | 39.61 | 49.51 | 42.24 | **183.01** |
| 811 | 3.57 | 2.68 | 3.57 | 1.78 | **11.60** |
| 812 | 13.56 | 11.78 | 11.24 | 9.99 | **46.57** |
| 813 | 5.35 | 2.23 | 1.78 | 1.78 | **11.15** |
| 814 | 2.68 | 2.68 | 2.68 | 3.12 | **11.15** |
| 815 | 23.37 | 7.05 | 7.32 | 5.35 | **43.09** |
| 816 | 8.39 | 8.47 | 14.72 | 7.67 | **39.25** |
| **CO - COLORADO - Total** | **1,603.73** | **1,428.48** | **1,312.64** | **1,253.04** | **5,597.88** |

**STATE: CT - CONNECTICUT**

| REGISTRANT ZIP CODE 3 | 1ST QUARTER | 2ND QUARTER | 3RD QUARTER | 4TH QUARTER | TOTAL GRAMS |
|---|---|---|---|---|---|
| 060 | 189.30 | 176.50 | 191.44 | 176.90 | **734.15** |
| 061 | 38.09 | 25.87 | 21.59 | 48.98 | **134.53** |
| 062 | 46.30 | 26.58 | 39.88 | 24.62 | **137.38** |
| 063 | 67.71 | 53.97 | 63.96 | 59.59 | **245.24** |
| 064 | 494.31 | 546.46 | 479.59 | 486.86 | **2,007.23** |
| 065 | 199.65 | 223.07 | 178.46 | 180.16 | **781.35** |
| 066 | 125.88 | 146.57 | 116.15 | 94.29 | **482.89** |
| 067 | 90.64 | 79.53 | 61.60 | 58.79 | **290.56** |
| 068 | 166.02 | 139.75 | 92.60 | 95.01 | **493.38** |
| 069 | 17.13 | 16.24 | 15.34 | 18.38 | **67.09** |
| **CT - CONNECTICUT - Total** | **1,435.03** | **1,434.54** | **1,260.63** | **1,243.59** | **5,373.79** |

**STATE: DC - DISTRICT OF COLUMBIA**

| REGISTRANT ZIP CODE 3 | 1ST QUARTER | 2ND QUARTER | 3RD QUARTER | 4TH QUARTER | TOTAL GRAMS |
|---|---|---|---|---|---|
| 200 | 98.22 | 61.02 | 50.58 | 52.05 | **261.88** |
| **DC - DISTRICT OF COLUMBIA -** | **98.22** | **61.02** | **50.58** | **52.05** | **261.88** |

**STATE: DE - DELAWARE**

| REGISTRANT ZIP CODE 3 | 1ST QUARTER | 2ND QUARTER | 3RD QUARTER | 4TH QUARTER | TOTAL GRAMS |
|---|---|---|---|---|---|
| 197 | 595.52 | 548.02 | 463.85 | 481.02 | **2,088.41** |
| 198 | 336.37 | 364.33 | 418.71 | 425.80 | **1,545.21** |
| 199 | 539.14 | 574.47 | 581.74 | 540.79 | **2,236.14** |
| **DE - DELAWARE - Total** | **1,471.03** | **1,486.82** | **1,464.29** | **1,447.61** | **5,869.75** |

**STATE: FL - FLORIDA**

| REGISTRANT ZIP CODE 3 | 1ST QUARTER | 2ND QUARTER | 3RD QUARTER | 4TH QUARTER | TOTAL GRAMS |
|---|---|---|---|---|---|
| 320 | 466.35 | 463.36 | 426.07 | 385.70 | **1,741.47** |
| 321 | 202.46 | 188.41 | 170.26 | 150.81 | **711.94** |
| 322 | 833.31 | 811.81 | 702.71 | 658.64 | **3,006.47** |
| 323 | 283.02 | 302.73 | 256.88 | 276.95 | **1,119.59** |

| | | | | | |
|---|---|---|---|---|---|
| 324 | 256.84 | 220.21 | 226.91 | 215.44 | **919.40** |
| 325 | 104.33 | 88.18 | 63.29 | 52.63 | **308.44** |
| 326 | 301.98 | 249.56 | 242.74 | 237.83 | **1,032.12** |
| 327 | 520.41 | 502.21 | 474.42 | 462.87 | **1,959.90** |
| 328 | 340.69 | 294.75 | 242.25 | 236.32 | **1,114.01** |
| 329 | 857.04 | 812.79 | 690.31 | 682.41 | **3,042.55** |
| 330 | 513.40 | 555.56 | 580.27 | 475.76 | **2,124.98** |
| 331 | 159.06 | 98.67 | 107.05 | 111.96 | **476.74** |
| 333 | 541.59 | 303.67 | 281.77 | 284.85 | **1,411.88** |
| 334 | 1,171.15 | 1,168.03 | 1,043.09 | 928.59 | **4,310.85** |
| 335 | 331.06 | 355.55 | 285.65 | 290.29 | **1,262.54** |
| 336 | 440.79 | 459.70 | 391.72 | 420.89 | **1,713.10** |
| 337 | 475.58 | 481.11 | 465.14 | 483.79 | **1,905.61** |
| 338 | 179.13 | 182.61 | 145.68 | 168.07 | **675.50** |
| 339 | 402.02 | 338.33 | 299.52 | 213.12 | **1,253.00** |
| 341 | 118.29 | 95.54 | 63.87 | 71.19 | **348.90** |
| 342 | 730.14 | 654.18 | 668.27 | 642.22 | **2,694.81** |
| 344 | 174.14 | 161.74 | 134.80 | 139.88 | **610.55** |
| 346 | 1,119.59 | 1,013.34 | 897.27 | 895.85 | **3,926.04** |
| 347 | 306.75 | 242.34 | 242.65 | 225.88 | **1,017.62** |
| 349 | 591.69 | 596.32 | 534.86 | 567.20 | **2,290.07** |
| **FL - FLORIDA - Total** | **11,420.80** | **10,640.70** | **9,637.45** | **9,279.13** | **40,978.08** |

**STATE: GA - GEORGIA**

| REGISTRANT ZIP CODE 3 | 1ST QUARTER | 2ND QUARTER | 3RD QUARTER | 4TH QUARTER | **TOTAL GRAMS** |
|---|---|---|---|---|---|
| 300 | 617.78 | 593.74 | 646.73 | 595.34 | **2,453.59** |
| 301 | 582.14 | 552.57 | 491.50 | 513.58 | **2,139.79** |
| 302 | 258.98 | 226.86 | 225.17 | 178.38 | **889.38** |
| 303 | 174.05 | 149.34 | 133.82 | 148.80 | **606.00** |
| 304 | 74.49 | 62.71 | 60.93 | 48.53 | **246.67** |
| 305 | 245.24 | 224.63 | 207.06 | 186.18 | **863.11** |
| 306 | 150.05 | 123.20 | 113.52 | 85.95 | **472.72** |
| 307 | 114.46 | 87.29 | 113.16 | 90.33 | **405.24** |
| 308 | 36.49 | 29.89 | 31.22 | 35.33 | **132.92** |
| 309 | 32.38 | 27.30 | 25.42 | 28.73 | **113.83** |
| 310 | 147.46 | 127.08 | 91.71 | 94.47 | **460.73** |
| 312 | 33.54 | 51.65 | 39.16 | 45.32 | **169.68** |
| 313 | 105.04 | 111.29 | 118.29 | 73.96 | **408.58** |
| 314 | 42.37 | 30.42 | 26.50 | 38.00 | **137.29** |
| 315 | 99.92 | 76.01 | 102.23 | 79.04 | **357.20** |
| 316 | 171.10 | 137.25 | 112.94 | 117.22 | **538.52** |

| | | | | | |
|---|---|---|---|---|---|
| 317 | 120.70 | 82.47 | 83.32 | 77.03 | **363.53** |
| 318 | 1.78 | 2.68 | 3.57 | 5.35 | **13.38** |
| 319 | 167.80 | 152.64 | 123.29 | 122.57 | **566.31** |
| 398 | 24.27 | 17.75 | 15.52 | 13.65 | **71.19** |
| **GA - GEORGIA - Total** | **3,200.05** | **2,866.76** | **2,765.06** | **2,577.77** | **11,409.65** |

**STATE: HI - HAWAII**

| REGISTRANT ZIP CODE 3 | 1ST QUARTER | 2ND QUARTER | 3RD QUARTER | 4TH QUARTER | TOTAL GRAMS |
|---|---|---|---|---|---|
| 967 | 158.44 | 111.69 | 115.62 | 114.10 | **499.84** |
| 968 | 97.15 | 69.58 | 90.19 | 103.93 | **360.85** |
| **HI - HAWAII - Total** | **255.59** | **181.27** | **205.81** | **218.03** | **860.70** |

**STATE: IA - IOWA**

| REGISTRANT ZIP CODE 3 | 1ST QUARTER | 2ND QUARTER | 3RD QUARTER | 4TH QUARTER | TOTAL GRAMS |
|---|---|---|---|---|---|
| 500 | 42.06 | 61.47 | 44.92 | 36.58 | **185.02** |
| 501 | 41.57 | 17.40 | 11.15 | 12.22 | **82.34** |
| 502 | 87.07 | 52.90 | 40.50 | 49.42 | **229.89** |
| 503 | 170.61 | 158.44 | 122.57 | 124.27 | **575.90** |
| 504 | 36.04 | 30.42 | 30.02 | 25.20 | **121.68** |
| 505 | 15.97 | 6.69 | 5.62 | 13.38 | **41.66** |
| 508 | 3.12 | 2.01 | 2.68 | 2.23 | **10.04** |
| 510 | 12.67 | 10.62 | 9.28 | 8.47 | **41.04** |
| 511 | 13.38 | 24.27 | 19.45 | 16.33 | **73.42** |
| 513 | 1.61 | 1.07 | 1.61 | 1.61 | **5.89** |
| 514 | 0 | 4.91 | 6.78 | 10.17 | **21.86** |
| 515 | 44.16 | 42.95 | 35.77 | 28.73 | **151.61** |
| 520 | 2.41 | 2.41 | 2.41 | 2.94 | **10.17** |
| 522 | 1.34 | 2.68 | 0.80 | 0.45 | **5.26** |
| 523 | 3.21 | 3.66 | 2.14 | 3.66 | **12.67** |
| 524 | 16.06 | 12.49 | 11.60 | 9.81 | **49.96** |
| 525 | 16.59 | 8.03 | 5.89 | 2.14 | **32.65** |
| 526 | 0 | 6.69 | 8.56 | 1.61 | **16.86** |
| 527 | 0.80 | 6.51 | 4.82 | 3.57 | **15.70** |
| 528 | 17.66 | 8.83 | 10.71 | 4.91 | **42.11** |
| **IA - IOWA - Total** | **526.34** | **464.43** | **377.27** | **357.69** | **1,725.72** |

**STATE: ID - IDAHO**

| REGISTRANT ZIP CODE 3 | 1ST QUARTER | 2ND QUARTER | 3RD QUARTER | 4TH QUARTER | TOTAL GRAMS |
|---|---|---|---|---|---|
| 832 | 37.96 | 43.85 | 50.31 | 38.81 | **170.93** |
| 833 | 78.01 | 52.95 | 44.03 | 32.65 | **207.64** |
| 834 | 37.91 | 28.90 | 29.97 | 32.65 | **129.44** |
| 835 | 24.80 | 28.55 | 32.47 | 26.67 | **112.49** |
| 836 | 315.94 | 316.78 | 274.72 | 307.86 | **1,215.31** |

| | | | | | |
|---|---|---|---|---|---|
| 837 | 251.22 | 224.81 | 187.97 | 183.42 | **847.41** |
| 838 | 51.74 | 54.95 | 49.51 | 47.10 | **203.31** |
| **ID - IDAHO - Total** | **797.58** | **750.79** | **668.99** | **669.16** | **2,886.52** |

**STATE: IL - ILLINOIS**

| REGISTRANT ZIP CODE 3 | 1ST QUARTER | 2ND QUARTER | 3RD QUARTER | 4TH QUARTER | TOTAL GRAMS |
|---|---|---|---|---|---|
| 600 | 652.35 | 546.23 | 558.99 | 610.02 | **2,367.59** |
| 601 | 244.52 | 192.65 | 180.29 | 236.90 | **854.36** |
| 602 | 36.00 | 49.47 | 52.05 | 31.18 | **168.70** |
| 603 | 24.80 | 30.33 | 18.02 | 30.24 | **103.39** |
| 604 | 306.61 | 304.70 | 246.89 | 298.59 | **1,156.79** |
| 605 | 190.82 | 169.99 | 173.78 | 177.80 | **712.39** |
| 606 | 188.95 | 168.79 | 160.71 | 136.05 | **654.49** |
| 607 | 16.50 | 12.58 | 10.35 | 5.98 | **45.41** |
| 608 | 0.54 | 0.54 | 0 | 0 | **1.07** |
| 609 | 11.60 | 7.58 | 8.92 | 7.14 | **35.24** |
| 610 | 19.89 | 23.37 | 14.54 | 25.96 | **83.77** |
| 611 | 21.41 | 7.23 | 13.02 | 8.56 | **50.23** |
| 612 | 5.89 | 3.35 | 5.58 | 2.45 | **17.26** |
| 613 | 34.44 | 33.45 | 49.60 | 35.51 | **153.00** |
| 614 | 10.71 | 0 | 10.71 | 3.57 | **24.98** |
| 615 | 6.24 | 16.77 | 8.74 | 11.60 | **43.36** |
| 616 | 12.67 | 10.62 | 8.03 | 10.79 | **42.11** |
| 617 | 13.83 | 9.28 | 16.06 | 11.15 | **50.31** |
| 618 | 12.67 | 28.64 | 17.04 | 27.83 | **86.18** |
| 619 | 6.02 | 6.16 | 7.76 | 5.53 | **25.47** |
| 620 | 33.63 | 28.90 | 22.48 | 17.84 | **102.86** |
| 622 | 53.88 | 47.64 | 32.65 | 27.30 | **161.47** |
| 623 | 20.07 | 13.83 | 13.65 | 7.76 | **55.31** |
| 624 | 21.32 | 24.18 | 20.16 | 12.85 | **78.50** |
| 625 | 3.30 | 8.65 | 4.55 | 0.71 | **17.22** |
| 626 | 21.95 | 16.41 | 19.27 | 22.30 | **79.93** |
| 627 | 42.20 | 32.83 | 31.58 | 22.21 | **128.82** |
| 628 | 16.06 | 2.14 | 6.47 | 0 | **24.67** |
| 629 | 59.37 | 35.42 | 33.90 | 27.03 | **155.72** |
| **IL - ILLINOIS - Total** | **2,088.23** | **1,831.70** | **1,745.80** | **1,814.84** | **7,480.57** |

**STATE: IN - INDIANA**

| REGISTRANT ZIP CODE 3 | 1ST QUARTER | 2ND QUARTER | 3RD QUARTER | 4TH QUARTER | TOTAL GRAMS |
|---|---|---|---|---|---|
| 460 | 126.81 | 105.45 | 95.50 | 130.56 | **458.32** |
| 461 | 242.21 | 208.31 | 311.21 | 281.59 | **1,043.31** |
| 462 | 504.84 | 445.87 | 354.21 | 275.97 | **1,580.89** |

| | | | | | |
|---|---|---|---|---|---|
| 463 | 234.00 | 199.38 | 163.34 | 163.34 | **760.07** |
| 464 | 1.52 | 7.14 | 4.82 | 2.14 | **15.61** |
| 465 | 185.38 | 187.79 | 158.66 | 145.23 | **677.06** |
| 466 | 35.15 | 21.86 | 31.05 | 21.95 | **110.00** |
| 467 | 47.82 | 43.27 | 47.46 | 43.71 | **182.26** |
| 468 | 61.02 | 47.82 | 54.02 | 37.11 | **199.96** |
| 469 | 58.25 | 35.24 | 46.84 | 47.50 | **187.83** |
| 470 | 67.62 | 68.78 | 68.51 | 66.64 | **271.56** |
| 471 | 159.33 | 115.35 | 80.38 | 74.04 | **429.10** |
| 472 | 67.71 | 43.00 | 29.62 | 59.50 | **199.83** |
| 473 | 92.06 | 94.29 | 81.76 | 95.72 | **363.84** |
| 474 | 69.05 | 78.86 | 107.41 | 92.06 | **347.38** |
| 475 | 66.91 | 56.92 | 61.11 | 48.80 | **233.73** |
| 476 | 68.69 | 53.62 | 73.15 | 59.59 | **255.05** |
| 477 | 181.90 | 138.63 | 127.39 | 106.96 | **554.89** |
| 478 | 177.53 | 194.30 | 157.54 | 147.55 | **676.93** |
| 479 | 55.85 | 72.62 | 52.63 | 42.82 | **223.92** |
| **IN - INDIANA - Total** | **2,503.63** | **2,218.47** | **2,106.60** | **1,942.82** | **8,771.53** |

**STATE: KS - KANSAS**

| REGISTRANT ZIP CODE 3 | 1ST QUARTER | 2ND QUARTER | 3RD QUARTER | 4TH QUARTER | **TOTAL GRAMS** |
|---|---|---|---|---|---|
| 660 | 240.60 | 212.94 | 231.95 | 237.48 | **922.97** |
| 661 | 92.78 | 83.05 | 40.95 | 76.90 | **293.68** |
| 662 | 145.90 | 133.82 | 146.80 | 122.57 | **549.09** |
| 664 | 17.31 | 23.02 | 30.33 | 27.74 | **98.40** |
| 665 | 13.38 | 11.78 | 7.85 | 13.83 | **46.84** |
| 666 | 37.65 | 38.09 | 24.18 | 30.60 | **130.51** |
| 667 | 30.87 | 22.66 | 27.74 | 22.48 | **103.75** |
| 668 | 8.03 | 4.46 | 4.46 | 5.35 | **22.30** |
| 669 | 0.80 | 1.07 | 0.80 | 5.35 | **8.03** |
| 670 | 55.76 | 71.46 | 55.49 | 47.10 | **229.80** |
| 671 | 24.80 | 14.01 | 13.38 | 12.49 | **64.68** |
| 672 | 102.32 | 103.39 | 78.77 | 78.50 | **363.00** |
| 673 | 10.53 | 2.68 | 4.01 | 0 | **17.22** |
| 674 | 22.93 | 27.57 | 24.18 | 26.41 | **101.07** |
| 675 | 41.13 | 35.51 | 21.68 | 21.59 | **119.90** |
| 676 | 22.30 | 19.80 | 13.92 | 14.27 | **70.30** |
| 677 | 0.89 | 0.45 | 0 | 0 | **1.34** |
| 678 | 17.31 | 13.92 | 17.93 | 14.99 | **64.14** |
| 679 | 3.21 | 3.21 | 1.07 | 3.21 | **10.71** |
| **KS - KANSAS - Total** | **888.49** | **822.87** | **745.48** | **760.87** | **3,217.72** |

**STATE: KY - KENTUCKY**

| REGISTRANT ZIP CODE 3 | 1ST QUARTER | 2ND QUARTER | 3RD QUARTER | 4TH QUARTER | TOTAL GRAMS |
|---|---|---|---|---|---|
| 400 | 38.85 | 33.72 | 19.54 | 19.36 | **111.47** |
| 401 | 56.02 | 24.35 | 11.42 | 10.35 | **102.15** |
| 402 | 231.72 | 187.52 | 177.62 | 172.04 | **768.90** |
| 403 | 210.58 | 200.45 | 165.57 | 143.94 | **720.55** |
| 404 | 101.70 | 104.69 | 74.31 | 64.90 | **345.60** |
| 405 | 141.13 | 128.19 | 116.06 | 100.18 | **485.57** |
| 406 | 6.38 | 8.83 | 6.07 | 6.42 | **27.70** |
| 407 | 262.81 | 238.50 | 253.18 | 266.02 | **1,020.52** |
| 408 | 12.04 | 8.92 | 7.58 | 5.00 | **33.54** |
| 409 | 106.83 | 140.77 | 138.36 | 122.04 | **508.01** |
| 410 | 153.93 | 131.05 | 111.96 | 125.34 | **522.28** |
| 411 | 67.22 | 69.23 | 56.38 | 61.02 | **253.85** |
| 412 | 7.49 | 6.69 | 14.59 | 8.30 | **37.07** |
| 413 | 5.62 | 4.55 | 4.01 | 4.01 | **18.20** |
| 414 | 4.55 | 4.95 | 6.02 | 4.15 | **19.67** |
| 415 | 85.37 | 81.63 | 73.87 | 69.72 | **310.58** |
| 416 | 73.60 | 64.45 | 72.75 | 72.30 | **283.11** |
| 417 | 17.80 | 11.29 | 6.20 | 8.03 | **43.31** |
| 418 | 0.80 | 1.07 | 2.01 | 0.80 | **4.68** |
| 420 | 53.53 | 44.87 | 39.52 | 28.28 | **166.20** |
| 421 | 109.95 | 121.77 | 100.14 | 91.89 | **423.75** |
| 422 | 115.26 | 73.46 | 65.84 | 68.16 | **322.72** |
| 423 | 19.89 | 12.31 | 12.04 | 9.46 | **53.70** |
| 424 | 46.21 | 39.70 | 36.75 | 21.95 | **144.61** |
| 425 | 47.91 | 50.54 | 49.07 | 56.20 | **203.71** |
| 426 | 18.60 | 11.55 | 9.95 | 9.63 | **49.73** |
| 427 | 124.98 | 102.01 | 98.67 | 82.21 | **407.87** |
| **KY - KENTUCKY - Total** | **2,120.79** | **1,907.09** | **1,729.47** | **1,631.70** | **7,389.04** |

**STATE: LA - LOUISIANA**

| REGISTRANT ZIP CODE 3 | 1ST QUARTER | 2ND QUARTER | 3RD QUARTER | 4TH QUARTER | TOTAL GRAMS |
|---|---|---|---|---|---|
| 700 | 502.03 | 460.23 | 444.44 | 438.73 | **1,845.44** |
| 701 | 117.44 | 166.64 | 190.60 | 216.78 | **691.47** |
| 703 | 159.60 | 148.71 | 185.02 | 127.17 | **620.50** |
| 704 | 623.80 | 621.21 | 565.01 | 570.32 | **2,380.35** |
| 705 | 362.01 | 324.90 | 297.34 | 229.89 | **1,214.15** |
| 706 | 26.58 | 38.18 | 35.55 | 32.47 | **132.79** |
| 707 | 1,190.86 | 1,117.58 | 1,136.80 | 1,088.45 | **4,533.70** |
| 708 | 791.52 | 712.97 | 613.14 | 580.49 | **2,698.11** |

| | | | | | |
|---|---|---|---|---|---|
| 710 | 73.06 | 40.23 | 34.08 | 39.61 | **186.98** |
| 711 | 322.18 | 279.49 | 226.91 | 212.23 | **1,040.81** |
| 712 | 19.80 | 17.84 | 12.31 | 16.95 | **66.91** |
| 713 | 39.79 | 38.63 | 33.10 | 30.78 | **142.29** |
| 714 | 13.56 | 17.13 | 13.02 | 8.83 | **52.54** |
| **LA - LOUISIANA - Total** | **4,242.25** | **3,983.76** | **3,787.32** | **3,592.71** | **15,606.04** |

**STATE: MA - MASSACHUSETTS**

| REGISTRANT ZIP CODE 3 | 1ST QUARTER | 2ND QUARTER | 3RD QUARTER | 4TH QUARTER | TOTAL GRAMS |
|---|---|---|---|---|---|
| 010 | 49.51 | 45.23 | 52.90 | 40.59 | **188.23** |
| 011 | 3.57 | 5.26 | 5.89 | 5.26 | **19.98** |
| 012 | 17.13 | 19.27 | 12.85 | 12.85 | **62.09** |
| 013 | 25.60 | 23.28 | 18.73 | 22.21 | **89.83** |
| 014 | 5.62 | 5.89 | 3.75 | 5.62 | **20.88** |
| 015 | 79.49 | 71.19 | 55.13 | 50.31 | **256.12** |
| 016 | 11.78 | 6.24 | 4.46 | 7.14 | **29.62** |
| 017 | 11.24 | 7.14 | 8.92 | 16.06 | **43.36** |
| 018 | 222.67 | 236.58 | 220.93 | 165.98 | **846.16** |
| 019 | 76.81 | 57.05 | 66.95 | 53.75 | **254.56** |
| 020 | 65.93 | 51.21 | 45.50 | 49.51 | **212.14** |
| 021 | 65.84 | 73.73 | 79.44 | 89.66 | **308.67** |
| 022 | 37.47 | 26.67 | 28.55 | 33.90 | **126.59** |
| 023 | 27.03 | 36.22 | 32.65 | 29.35 | **125.25** |
| 024 | 28.73 | 24.00 | 19.36 | 23.02 | **95.10** |
| 025 | 23.55 | 14.72 | 25.87 | 14.27 | **78.42** |
| 026 | 31.67 | 26.94 | 12.94 | 29.44 | **100.99** |
| 027 | 344.08 | 304.74 | 296.53 | 319.73 | **1,265.09** |
| **MA - MASSACHUSETTS - Total** | **1,127.70** | **1,035.37** | **991.35** | **968.64** | **4,123.06** |

**STATE: MD - MARYLAND**

| REGISTRANT ZIP CODE 3 | 1ST QUARTER | 2ND QUARTER | 3RD QUARTER | 4TH QUARTER | TOTAL GRAMS |
|---|---|---|---|---|---|
| 206 | 872.92 | 940.41 | 889.47 | 909.23 | **3,612.02** |
| 207 | 553.59 | 473.30 | 433.51 | 407.98 | **1,868.39** |
| 208 | 432.01 | 379.45 | 358.85 | 287.12 | **1,457.43** |
| 209 | 40.77 | 60.93 | 68.25 | 68.51 | **238.46** |
| 210 | 1,083.41 | 1,067.53 | 1,012.89 | 1,037.24 | **4,201.08** |
| 211 | 559.61 | 459.03 | 413.80 | 389.98 | **1,822.43** |
| 212 | 678.49 | 561.49 | 551.76 | 548.95 | **2,340.69** |
| 214 | 122.22 | 125.38 | 106.61 | 101.21 | **455.42** |
| 215 | 240.42 | 182.17 | 172.89 | 167.89 | **763.37** |
| 216 | 186.05 | 217.54 | 183.06 | 195.19 | **781.84** |
| 217 | 1,329.54 | 1,145.55 | 1,152.10 | 1,173.74 | **4,800.93** |

| | | | | | |
|---|---|---|---|---|---|
| 218 | 62.63 | 89.03 | 68.78 | 62.54 | **282.97** |
| 219 | 195.24 | 199.83 | 217.58 | 205.36 | **818.01** |
| **MD - MARYLAND - Total** | **6,356.89** | **5,901.64** | **5,629.54** | **5,554.96** | **23,443.03** |

**STATE: ME - MAINE**

| REGISTRANT ZIP CODE 3 | 1ST QUARTER | 2ND QUARTER | 3RD QUARTER | 4TH QUARTER | TOTAL GRAMS |
|---|---|---|---|---|---|
| 040 | 120.12 | 97.60 | 91.26 | 78.73 | **387.71** |
| 041 | 12.85 | 6.07 | 7.36 | 4.46 | **30.73** |
| 042 | 113.52 | 94.12 | 91.93 | 79.98 | **379.54** |
| 043 | 179.40 | 115.13 | 98.67 | 112.72 | **505.91** |
| 044 | 6.69 | 5.08 | 1.61 | 2.41 | **15.79** |
| 045 | 31.85 | 28.77 | 26.23 | 24.80 | **111.65** |
| 046 | 3.30 | 3.30 | 2.41 | 2.94 | **11.95** |
| 047 | 2.14 | 0 | 0 | 0 | **2.14** |
| 048 | 9.37 | 3.84 | 3.75 | 4.55 | **21.50** |
| 049 | 111.51 | 81.81 | 76.01 | 51.92 | **321.25** |
| **ME - MAINE - Total** | **590.75** | **435.70** | **399.21** | **362.50** | **1,788.17** |

**STATE: MI - MICHIGAN**

| REGISTRANT ZIP CODE 3 | 1ST QUARTER | 2ND QUARTER | 3RD QUARTER | 4TH QUARTER | TOTAL GRAMS |
|---|---|---|---|---|---|
| 480 | 3,182.97 | 2,859.85 | 2,665.77 | 2,111.20 | **10,819.79** |
| 481 | 2,740.44 | 2,374.06 | 2,114.68 | 1,856.42 | **9,085.59** |
| 482 | 5,967.93 | 4,739.10 | 3,977.78 | 3,402.92 | **18,087.73** |
| 483 | 1,148.40 | 1,149.38 | 946.12 | 841.25 | **4,085.15** |
| 484 | 115.71 | 80.56 | 72.62 | 84.30 | **353.18** |
| 485 | 102.23 | 88.14 | 77.43 | 86.80 | **354.61** |
| 486 | 89.34 | 77.52 | 86.04 | 65.57 | **318.48** |
| 487 | 29.53 | 28.90 | 28.73 | 21.77 | **108.93** |
| 488 | 125.79 | 110.98 | 106.96 | 101.74 | **445.47** |
| 489 | 74.49 | 66.91 | 58.57 | 54.24 | **254.20** |
| 490 | 224.41 | 192.69 | 200.90 | 186.14 | **804.14** |
| 491 | 32.03 | 52.54 | 31.40 | 31.22 | **147.20** |
| 492 | 101.97 | 81.00 | 82.34 | 88.14 | **353.45** |
| 493 | 40.32 | 42.46 | 30.51 | 32.65 | **145.95** |
| 494 | 76.19 | 75.47 | 69.49 | 74.58 | **295.73** |
| 495 | 54.02 | 48.44 | 48.98 | 49.15 | **200.59** |
| 496 | 102.41 | 118.92 | 87.07 | 106.43 | **414.83** |
| 497 | 131.54 | 82.92 | 73.64 | 74.36 | **362.46** |
| 498 | 3.57 | 5.26 | 7.40 | 2.14 | **18.38** |
| 499 | 0.89 | 0 | 0 | 0 | **0.89** |
| **MI - MICHIGAN - Total** | **14,344.17** | **12,275.12** | **10,766.44** | **9,271.02** | **46,656.74** |

**STATE: MN - MINNESOTA**

| REGISTRANT ZIP CODE 3 | 1ST QUARTER | 2ND QUARTER | 3RD QUARTER | 4TH QUARTER | TOTAL GRAMS |
|---|---|---|---|---|---|
| 550 | 37.74 | 34.08 | 35.24 | 38.27 | **145.32** |
| 551 | 52.63 | 63.79 | 43.09 | 51.74 | **211.25** |
| 553 | 50.67 | 40.32 | 50.58 | 41.39 | **182.97** |
| 554 | 23.55 | 16.15 | 17.31 | 19.54 | **76.54** |
| 557 | 10.88 | 12.94 | 14.10 | 12.67 | **50.58** |
| 558 | 4.28 | 6.42 | 5.35 | 6.42 | **22.48** |
| 559 | 5.62 | 0 | 4.01 | 0 | **9.63** |
| 560 | 4.91 | 4.91 | 2.68 | 7.58 | **20.07** |
| 562 | 0 | 1.78 | 2.23 | 1.78 | **5.80** |
| 563 | 21.41 | 25.87 | 24.53 | 28.55 | **100.36** |
| 565 | 3.57 | 1.78 | 0 | 1.34 | **6.69** |
| 567 | 13.02 | 6.42 | 8.21 | 6.07 | **33.72** |
| **MN - MINNESOTA - Total** | **228.29** | **214.46** | **207.32** | **215.35** | **865.43** |

**STATE: MO - MISSOURI**

| REGISTRANT ZIP CODE 3 | 1ST QUARTER | 2ND QUARTER | 3RD QUARTER | 4TH QUARTER | TOTAL GRAMS |
|---|---|---|---|---|---|
| 630 | 264.06 | 185.38 | 169.10 | 134.44 | **752.98** |
| 631 | 513.31 | 513.76 | 391.01 | 623.40 | **2,041.48** |
| 633 | 155.58 | 131.67 | 135.69 | 133.64 | **556.58** |
| 634 | 54.42 | 42.73 | 32.96 | 31.05 | **161.16** |
| 635 | 3.08 | 3.48 | 4.82 | 1.78 | **13.16** |
| 636 | 49.96 | 33.19 | 39.92 | 49.87 | **172.93** |
| 637 | 11.69 | 3.84 | 4.64 | 6.16 | **26.32** |
| 638 | 5.17 | 2.32 | 2.32 | 2.68 | **12.49** |
| 639 | 3.57 | 2.14 | 0.54 | 0 | **6.24** |
| 640 | 133.28 | 128.60 | 121.55 | 105.36 | **488.78** |
| 641 | 167.45 | 135.91 | 151.12 | 124.36 | **578.84** |
| 644 | 3.75 | 1.07 | 5.35 | 4.82 | **14.99** |
| 645 | 64.77 | 35.15 | 43.18 | 19.89 | **162.99** |
| 646 | 19.72 | 20.96 | 22.39 | 23.02 | **86.09** |
| 647 | 42.20 | 40.23 | 43.09 | 36.67 | **162.18** |
| 648 | 35.51 | 21.50 | 13.65 | 4.64 | **75.29** |
| 650 | 46.26 | 40.50 | 30.82 | 34.97 | **152.55** |
| 651 | 50.85 | 51.03 | 31.98 | 34.52 | **168.38** |
| 652 | 43.18 | 39.92 | 32.74 | 28.81 | **144.65** |
| 653 | 12.85 | 2.23 | 2.68 | 1.34 | **19.09** |
| 654 | 10.30 | 12.04 | 16.73 | 12.31 | **51.38** |
| 655 | 22.57 | 18.20 | 37.83 | 27.12 | **105.71** |
| 656 | 42.91 | 27.48 | 19.27 | 38.49 | **128.15** |
| 657 | 145.72 | 114.72 | 111.16 | 82.16 | **453.77** |

| | | | | | |
|---|---|---|---|---|---|
| 658 | 118.74 | 92.06 | 96.26 | 68.96 | **376.02** |
| **MO - MISSOURI - Total** | **2,020.87** | **1,700.12** | **1,560.77** | **1,630.45** | **6,912.21** |

**STATE: MS - MISSISSIPPI**

| REGISTRANT ZIP CODE 3 | 1ST QUARTER | 2ND QUARTER | 3RD QUARTER | 4TH QUARTER | TOTAL GRAMS |
|---|---|---|---|---|---|
| 386 | 79.62 | 60.62 | 57.23 | 38.14 | **235.60** |
| 387 | 14.63 | 4.28 | 0.54 | 0 | **19.45** |
| 388 | 230.65 | 204.51 | 197.73 | 187.39 | **820.29** |
| 389 | 21.86 | 12.31 | 9.72 | 10.62 | **54.51** |
| 390 | 70.92 | 49.15 | 23.73 | 37.29 | **181.10** |
| 391 | 43.36 | 21.50 | 24.62 | 35.24 | **124.72** |
| 392 | 63.74 | 50.23 | 37.07 | 34.61 | **185.65** |
| 393 | 43.13 | 18.20 | 18.78 | 26.50 | **106.61** |
| 394 | 134.22 | 68.96 | 51.47 | 103.57 | **358.22** |
| 395 | 964.45 | 884.96 | 814.44 | 789.37 | **3,453.23** |
| 396 | 53.26 | 34.52 | 34.35 | 32.92 | **155.05** |
| 397 | 44.61 | 42.29 | 39.97 | 14.63 | **141.49** |
| **MS - MISSISSIPPI - Total** | **1,764.44** | **1,451.54** | **1,309.65** | **1,310.27** | **5,835.90** |

**STATE: MT - MONTANA**

| REGISTRANT ZIP CODE 3 | 1ST QUARTER | 2ND QUARTER | 3RD QUARTER | 4TH QUARTER | TOTAL GRAMS |
|---|---|---|---|---|---|
| 590 | 0 | 0 | 0 | 1.34 | **1.34** |
| 591 | 27.92 | 24.89 | 31.67 | 21.86 | **106.34** |
| 592 | 0 | 0 | 1.61 | 0 | **1.61** |
| 593 | 0.45 | 2.23 | 0 | 0 | **2.68** |
| 594 | 15.52 | 22.66 | 14.18 | 11.51 | **63.87** |
| 596 | 3.21 | 4.82 | 10.79 | 3.30 | **22.12** |
| 597 | 6.96 | 2.59 | 6.42 | 2.85 | **18.82** |
| 598 | 11.42 | 9.81 | 10.04 | 4.10 | **35.37** |
| 599 | 67.80 | 58.97 | 63.70 | 75.56 | **266.02** |
| **MT - MONTANA - Total** | **133.28** | **125.96** | **138.41** | **120.52** | **518.18** |

**STATE: NC - NORTH CAROLINA**

| REGISTRANT ZIP CODE 3 | 1ST QUARTER | 2ND QUARTER | 3RD QUARTER | 4TH QUARTER | TOTAL GRAMS |
|---|---|---|---|---|---|
| 270 | 295.64 | 218.21 | 223.47 | 194.75 | **932.07** |
| 271 | 285.65 | 217.32 | 198.94 | 154.51 | **856.42** |
| 272 | 626.43 | 506.26 | 479.15 | 490.88 | **2,102.72** |
| 273 | 355.77 | 251.13 | 231.14 | 185.47 | **1,023.51** |
| 274 | 248.32 | 247.33 | 237.97 | 264.86 | **998.48** |
| 275 | 865.34 | 719.70 | 690.35 | 751.10 | **3,026.49** |
| 276 | 254.25 | 262.63 | 236.14 | 228.20 | **981.22** |
| 277 | 217.36 | 153.08 | 162.45 | 195.28 | **728.17** |
| 278 | 1,481.47 | 1,422.54 | 1,266.16 | 1,409.87 | **5,580.04** |

| | | | | | |
|---|---|---|---|---|---|
| 279 | 208.84 | 156.88 | 149.34 | 152.28 | **667.34** |
| 280 | 1,380.93 | 1,376.24 | 1,320.13 | 1,354.21 | **5,431.51** |
| 281 | 662.12 | 694.86 | 656.01 | 691.29 | **2,704.27** |
| 282 | 488.78 | 471.92 | 428.52 | 407.24 | **1,796.47** |
| 283 | 778.85 | 645.17 | 648.38 | 535.13 | **2,607.52** |
| 284 | 504.30 | 455.15 | 405.68 | 357.91 | **1,723.05** |
| 285 | 751.42 | 650.92 | 576.70 | 631.07 | **2,610.11** |
| 286 | 1,283.38 | 1,167.89 | 1,115.48 | 1,134.80 | **4,701.55** |
| 287 | 196.13 | 189.71 | 210.22 | 182.61 | **778.67** |
| 288 | 23.69 | 20.07 | 17.13 | 20.30 | **81.18** |
| 289 | 135.60 | 129.71 | 137.12 | 162.27 | **564.70** |
| **NC - NORTH CAROLINA - Total** | **11,044.24** | **9,956.72** | **9,390.47** | **9,504.03** | **39,895.46** |

**STATE: ND - NORTH DAKOTA**

| REGISTRANT ZIP CODE 3 | 1ST QUARTER | 2ND QUARTER | 3RD QUARTER | 4TH QUARTER | **TOTAL GRAMS** |
|---|---|---|---|---|---|
| 580 | 0.89 | 0 | 0 | 0 | **0.89** |
| 581 | 6.60 | 3.57 | 2.85 | 3.21 | **16.24** |
| 582 | 0.45 | 0.45 | 1.34 | 1.43 | **3.66** |
| 583 | 1.34 | 4.01 | 1.34 | 3.12 | **9.81** |
| 584 | 5.35 | 2.68 | 0.89 | 0 | **8.92** |
| 585 | 36.40 | 43.18 | 37.02 | 26.50 | **143.09** |
| 586 | 3.21 | 3.57 | 5.35 | 6.69 | **18.82** |
| 587 | 15.39 | 16.95 | 16.06 | 19.63 | **68.02** |
| 588 | 12.31 | 6.42 | 6.42 | 6.42 | **31.58** |
| **ND - NORTH DAKOTA - Total** | **81.94** | **80.82** | **71.28** | **67.00** | **301.04** |

**STATE: NE - NEBRASKA**

| REGISTRANT ZIP CODE 3 | 1ST QUARTER | 2ND QUARTER | 3RD QUARTER | 4TH QUARTER | **TOTAL GRAMS** |
|---|---|---|---|---|---|
| 680 | 17.84 | 17.93 | 21.77 | 20.96 | **78.50** |
| 681 | 316.61 | 281.23 | 254.78 | 227.22 | **1,079.84** |
| 683 | 7.49 | 1.07 | 0 | 0 | **8.56** |
| 684 | 17.75 | 15.79 | 18.56 | 17.98 | **70.07** |
| 685 | 153.04 | 102.73 | 109.15 | 89.75 | **454.66** |
| 686 | 36.22 | 34.79 | 39.70 | 39.83 | **150.54** |
| 687 | 38.00 | 44.87 | 41.66 | 38.63 | **163.17** |
| 688 | 5.62 | 9.28 | 8.43 | 6.16 | **29.48** |
| 689 | 3.30 | 3.21 | 2.41 | 2.41 | **11.33** |
| 691 | 5.00 | 2.68 | 1.78 | 2.14 | **11.60** |
| 693 | 16.95 | 12.76 | 18.51 | 8.97 | **57.18** |
| **NE - NEBRASKA - Total** | **617.82** | **526.34** | **516.75** | **454.03** | **2,114.95** |

**STATE: NH - NEW HAMPSHIRE**

| REGISTRANT ZIP CODE 3 | 1ST QUARTER | 2ND QUARTER | 3RD QUARTER | 4TH QUARTER | **TOTAL GRAMS** |
|---|---|---|---|---|---|

| | | | | | |
|---|---|---|---|---|---|
| 030 | 90.64 | 125.61 | 117.13 | 84.48 | **417.86** |
| 031 | 43.89 | 62.89 | 43.53 | 40.28 | **190.60** |
| 032 | 53.39 | 52.14 | 54.33 | 37.91 | **197.78** |
| 033 | 79.04 | 82.88 | 70.39 | 49.42 | **281.73** |
| 034 | 15.97 | 5.17 | 9.10 | 4.28 | **34.52** |
| 035 | 15.25 | 6.78 | 9.99 | 7.49 | **39.52** |
| 037 | 5.35 | 5.53 | 1.34 | 0 | **12.22** |
| 038 | 166.73 | 133.10 | 104.15 | 99.11 | **503.10** |
| **NH - NEW HAMPSHIRE - Total** | **470.27** | **474.11** | **409.96** | **322.98** | **1,677.33** |

**STATE: NJ - NEW JERSEY**

| REGISTRANT ZIP CODE 3 | 1ST QUARTER | 2ND QUARTER | 3RD QUARTER | 4TH QUARTER | **TOTAL GRAMS** |
|---|---|---|---|---|---|
| 070 | 644.99 | 548.02 | 463.54 | 498.24 | **2,154.78** |
| 071 | 48.89 | 59.95 | 51.21 | 40.68 | **200.72** |
| 072 | 27.21 | 26.50 | 25.16 | 18.56 | **97.42** |
| 073 | 14.27 | 6.78 | 6.60 | 8.39 | **36.04** |
| 074 | 206.61 | 188.59 | 185.56 | 162.85 | **743.61** |
| 075 | 54.60 | 52.37 | 59.77 | 77.70 | **244.44** |
| 076 | 255.94 | 215.17 | 209.82 | 230.25 | **911.19** |
| 077 | 348.63 | 326.15 | 321.96 | 298.14 | **1,294.88** |
| 078 | 74.58 | 93.05 | 82.70 | 63.34 | **313.66** |
| 079 | 92.69 | 87.25 | 76.10 | 106.12 | **362.15** |
| 080 | 785.54 | 752.17 | 671.80 | 690.35 | **2,899.86** |
| 081 | 65.03 | 36.13 | 36.04 | 44.78 | **181.99** |
| 082 | 82.52 | 62.89 | 57.99 | 59.95 | **263.35** |
| 083 | 51.47 | 46.03 | 43.36 | 37.47 | **178.33** |
| 084 | 5.89 | 1.78 | 3.57 | 2.68 | **13.92** |
| 085 | 143.54 | 117.31 | 74.13 | 76.90 | **411.88** |
| 086 | 45.23 | 43.71 | 38.27 | 49.24 | **176.46** |
| 087 | 256.88 | 246.67 | 210.18 | 170.12 | **883.85** |
| 088 | 503.50 | 437.40 | 379.10 | 356.93 | **1,676.92** |
| 089 | 29.89 | 21.95 | 27.12 | 30.69 | **109.64** |
| **NJ - NEW JERSEY - Total** | **3,737.90** | **3,369.86** | **3,023.95** | **3,023.37** | **13,155.09** |

**STATE: NM - NEW MEXICO**

| REGISTRANT ZIP CODE 3 | 1ST QUARTER | 2ND QUARTER | 3RD QUARTER | 4TH QUARTER | **TOTAL GRAMS** |
|---|---|---|---|---|---|
| 870 | 68.29 | 67.84 | 51.25 | 56.83 | **244.21** |
| 871 | 305.86 | 253.09 | 182.17 | 183.77 | **924.88** |
| 873 | 22.61 | 17.80 | 11.78 | 9.10 | **61.29** |
| 874 | 8.12 | 5.22 | 4.01 | 2.94 | **20.30** |
| 875 | 50.05 | 52.90 | 60.31 | 40.14 | **203.40** |
| 877 | 8.30 | 5.17 | 4.10 | 5.62 | **23.19** |

| | | | | | |
|---|---|---|---|---|---|
| 879 | 0.89 | 2.68 | 2.68 | 3.57 | **9.81** |
| 880 | 27.48 | 20.61 | 30.15 | 12.67 | **90.90** |
| 882 | 47.64 | 35.95 | 27.12 | 20.96 | **131.67** |
| 883 | 17.04 | 15.08 | 15.34 | 13.92 | **61.38** |
| **NM - NEW MEXICO - Total** | **556.27** | **476.34** | **388.91** | **349.52** | **1,771.04** |

**STATE: NV - NEVADA**

| REGISTRANT ZIP CODE 3 | 1ST QUARTER | 2ND QUARTER | 3RD QUARTER | 4TH QUARTER | TOTAL GRAMS |
|---|---|---|---|---|---|
| 890 | 616.17 | 437.31 | 477.72 | 425.40 | **1,956.60** |
| 891 | 1,147.15 | 1,007.58 | 889.74 | 776.08 | **3,820.55** |
| 894 | 85.11 | 74.67 | 70.21 | 59.68 | **289.66** |
| 895 | 87.47 | 72.26 | 67.00 | 57.90 | **284.62** |
| 897 | 34.79 | 36.58 | 37.65 | 25.25 | **134.26** |
| **NV - NEVADA - Total** | **1,970.69** | **1,628.39** | **1,542.31** | **1,344.31** | **6,485.70** |

**STATE: NY - NEW YORK**

| REGISTRANT ZIP CODE 3 | 1ST QUARTER | 2ND QUARTER | 3RD QUARTER | 4TH QUARTER | TOTAL GRAMS |
|---|---|---|---|---|---|
| 100 | 1,222.62 | 1,036.26 | 834.56 | 755.52 | **3,848.97** |
| 101 | 30.51 | 25.16 | 14.27 | 19.98 | **89.92** |
| 103 | 562.38 | 668.27 | 546.19 | 577.46 | **2,354.30** |
| 104 | 352.29 | 277.53 | 322.32 | 324.64 | **1,276.77** |
| 105 | 280.65 | 245.95 | 263.08 | 219.55 | **1,009.23** |
| 106 | 39.61 | 34.08 | 31.94 | 33.54 | **139.17** |
| 107 | 64.86 | 68.07 | 61.02 | 50.00 | **243.94** |
| 108 | 25.42 | 12.27 | 29.44 | 30.33 | **97.46** |
| 109 | 502.43 | 522.10 | 490.92 | 547.04 | **2,062.49** |
| 110 | 118.83 | 131.50 | 117.85 | 113.65 | **481.82** |
| 111 | 80.87 | 97.60 | 158.97 | 85.46 | **422.90** |
| 112 | 1,231.41 | 1,219.05 | 1,208.35 | 1,019.76 | **4,678.57** |
| 113 | 269.24 | 241.40 | 232.57 | 213.66 | **956.87** |
| 114 | 127.30 | 107.50 | 109.01 | 257.73 | **601.54** |
| 115 | 348.14 | 342.12 | 340.25 | 347.65 | **1,378.16** |
| 116 | 61.02 | 79.40 | 79.75 | 72.44 | **292.61** |
| 117 | 1,089.74 | 979.39 | 859.45 | 922.92 | **3,851.51** |
| 118 | 24.00 | 11.15 | 8.03 | 9.63 | **52.81** |
| 119 | 73.15 | 81.72 | 79.22 | 66.73 | **300.82** |
| 120 | 118.20 | 101.79 | 104.91 | 87.60 | **412.51** |
| 121 | 44.34 | 26.58 | 40.86 | 19.80 | **131.58** |
| 122 | 25.78 | 18.02 | 26.32 | 24.35 | **94.47** |
| 123 | 16.41 | 20.61 | 19.63 | 15.25 | **71.90** |
| 124 | 227.57 | 222.53 | 185.20 | 189.62 | **824.92** |
| 125 | 360.41 | 357.46 | 306.35 | 303.58 | **1,327.80** |

| | | | | | |
|---|---|---|---|---|---|
| 126 | 134.89 | 150.05 | 116.51 | 180.92 | **582.36** |
| 127 | 141.00 | 103.53 | 107.05 | 85.46 | **437.04** |
| 128 | 122.31 | 169.23 | 143.90 | 171.86 | **607.30** |
| 129 | 293.59 | 251.75 | 268.61 | 257.19 | **1,071.14** |
| 130 | 321.56 | 305.32 | 293.59 | 270.57 | **1,191.04** |
| 131 | 70.74 | 73.51 | 65.93 | 72.30 | **282.48** |
| 132 | 77.12 | 71.46 | 64.10 | 46.97 | **259.65** |
| 133 | 76.63 | 49.15 | 47.99 | 48.71 | **222.49** |
| 134 | 43.09 | 42.55 | 41.30 | 24.40 | **151.34** |
| 135 | 35.24 | 9.72 | 17.93 | 12.22 | **75.11** |
| 136 | 147.82 | 145.50 | 118.92 | 124.27 | **536.51** |
| 137 | 114.81 | 116.33 | 77.61 | 80.82 | **389.58** |
| 138 | 172.93 | 149.03 | 118.02 | 126.41 | **566.39** |
| 139 | 169.14 | 174.05 | 160.76 | 165.04 | **668.99** |
| 140 | 572.68 | 469.16 | 456.84 | 456.18 | **1,954.86** |
| 141 | 236.94 | 242.74 | 272.09 | 236.23 | **988.00** |
| 142 | 590.21 | 497.48 | 450.06 | 528.44 | **2,066.19** |
| 143 | 214.33 | 200.41 | 205.81 | 224.23 | **844.77** |
| 144 | 86.62 | 76.63 | 83.05 | 82.52 | **328.83** |
| 145 | 67.71 | 43.49 | 32.83 | 59.10 | **203.13** |
| 146 | 79.75 | 73.78 | 86.18 | 72.08 | **311.79** |
| 147 | 65.12 | 70.39 | 63.96 | 68.16 | **267.63** |
| 148 | 237.03 | 208.39 | 163.48 | 193.18 | **802.09** |
| 149 | 26.05 | 18.56 | 21.32 | 13.11 | **79.04** |
| **NY - NEW YORK - Total** | **11,394.53** | **10,639.72** | **9,948.30** | **9,908.29** | **41,890.83** |

**STATE: OH - OHIO**

| REGISTRANT ZIP CODE 3 | 1ST QUARTER | 2ND QUARTER | 3RD QUARTER | 4TH QUARTER | TOTAL GRAMS |
|---|---|---|---|---|---|
| 430 | 261.16 | 220.17 | 173.38 | 187.34 | **842.05** |
| 431 | 104.02 | 101.92 | 113.83 | 94.29 | **414.07** |
| 432 | 344.58 | 318.70 | 101.57 | 122.01 | **886.85** |
| 433 | 45.68 | 46.84 | 36.80 | 41.26 | **170.57** |
| 434 | 190.64 | 157.54 | 148.45 | 84.57 | **581.20** |
| 435 | 111.65 | 110.93 | 79.40 | 84.57 | **386.55** |
| 436 | 316.70 | 255.72 | 205.63 | 176.90 | **954.95** |
| 437 | 52.37 | 22.48 | 25.69 | 23.19 | **123.73** |
| 438 | 30.33 | 23.37 | 29.26 | 19.09 | **102.06** |
| 439 | 197.87 | 191.36 | 185.82 | 168.03 | **743.07** |
| 440 | 164.99 | 137.03 | 143.00 | 107.05 | **552.08** |
| 441 | 123.51 | 94.12 | 83.14 | 70.30 | **371.07** |
| 442 | 168.38 | 142.33 | 137.03 | 134.22 | **581.96** |

| | | | | | |
|---|---|---|---|---|---|
| 443 | 54.42 | 53.17 | 44.03 | 37.20 | **188.81** |
| 444 | 166.20 | 133.77 | 114.59 | 115.84 | **530.40** |
| 445 | 107.59 | 109.19 | 91.22 | 73.87 | **381.86** |
| 446 | 196.26 | 213.66 | 150.10 | 150.76 | **710.78** |
| 447 | 123.20 | 105.54 | 112.49 | 101.79 | **443.02** |
| 448 | 372.63 | 360.41 | 286.19 | 166.38 | **1,185.60** |
| 449 | 16.95 | 15.08 | 27.39 | 24.18 | **83.59** |
| 450 | 425.93 | 370.27 | 344.22 | 349.61 | **1,490.03** |
| 451 | 193.18 | 171.06 | 149.83 | 155.23 | **669.30** |
| 452 | 210.31 | 189.04 | 192.02 | 158.21 | **749.59** |
| 453 | 169.10 | 168.83 | 156.52 | 150.90 | **645.35** |
| 454 | 153.84 | 152.10 | 144.88 | 138.99 | **589.81** |
| 455 | 22.03 | 36.04 | 27.12 | 17.08 | **102.28** |
| 456 | 814.53 | 805.61 | 825.37 | 807.08 | **3,252.60** |
| 457 | 46.43 | 35.51 | 42.91 | 32.38 | **157.23** |
| 458 | 20.07 | 30.96 | 21.59 | 23.11 | **95.72** |
| **OH - OHIO - Total** | **5,204.56** | **4,772.73** | **4,193.45** | **3,815.44** | **17,986.18** |

**STATE: OK - OKLAHOMA**

| REGISTRANT ZIP CODE 3 | 1ST QUARTER | 2ND QUARTER | 3RD QUARTER | 4TH QUARTER | TOTAL GRAMS |
|---|---|---|---|---|---|
| 730 | 933.23 | 817.83 | 614.03 | 378.79 | **2,743.88** |
| 731 | 624.92 | 595.92 | 417.15 | 250.99 | **1,888.98** |
| 734 | 46.03 | 35.68 | 28.81 | 36.04 | **146.57** |
| 735 | 85.20 | 69.94 | 69.67 | 49.15 | **273.96** |
| 736 | 31.85 | 15.43 | 14.54 | 8.47 | **70.30** |
| 737 | 16.24 | 18.29 | 20.34 | 11.42 | **66.28** |
| 738 | 7.05 | 5.80 | 3.03 | 3.21 | **19.09** |
| 740 | 323.74 | 249.48 | 177.08 | 164.68 | **914.98** |
| 741 | 214.10 | 179.54 | 130.34 | 96.12 | **620.10** |
| 743 | 60.13 | 61.38 | 51.74 | 59.50 | **232.75** |
| 744 | 89.30 | 82.88 | 67.18 | 45.14 | **284.49** |
| 745 | 148.62 | 139.26 | 97.42 | 108.75 | **494.04** |
| 746 | 24.44 | 23.64 | 10.26 | 13.29 | **71.64** |
| 747 | 7.14 | 7.32 | 9.37 | 6.07 | **29.89** |
| 748 | 163.52 | 118.74 | 92.42 | 45.23 | **419.91** |
| 749 | 44.96 | 34.17 | 24.35 | 25.51 | **129.00** |
| **OK - OKLAHOMA - Total** | **2,820.46** | **2,455.28** | **1,827.73** | **1,302.38** | **8,405.86** |

**STATE: OR - OREGON**

| REGISTRANT ZIP CODE 3 | 1ST QUARTER | 2ND QUARTER | 3RD QUARTER | 4TH QUARTER | TOTAL GRAMS |
|---|---|---|---|---|---|
| 970 | 191.18 | 161.92 | 166.91 | 168.74 | **688.75** |
| 971 | 39.43 | 31.22 | 25.42 | 19.63 | **115.71** |

| | | | | | |
|---|---|---|---|---|---|
| 972 | 160.09 | 174.54 | 161.20 | 141.93 | **637.76** |
| 973 | 96.12 | 107.59 | 91.08 | 84.08 | **378.87** |
| 974 | 339.00 | 282.04 | 263.04 | 256.03 | **1,140.10** |
| 975 | 55.00 | 54.06 | 52.72 | 40.59 | **202.37** |
| 976 | 9.81 | 5.80 | 7.23 | 3.03 | **25.87** |
| 977 | 76.99 | 66.28 | 49.42 | 73.87 | **266.56** |
| 978 | 57.90 | 57.54 | 48.44 | 46.75 | **210.62** |
| 979 | 16.15 | 8.03 | 4.91 | 1.78 | **30.87** |
| **OR - OREGON - Total** | **1,041.66** | **949.02** | **870.38** | **836.43** | **3,697.49** |

**STATE: PA - PENNSYLVANIA**

| REGISTRANT ZIP CODE 3 | 1ST QUARTER | 2ND QUARTER | 3RD QUARTER | 4TH QUARTER | **TOTAL GRAMS** |
|---|---|---|---|---|---|
| 150 | 918.55 | 773.58 | 706.78 | 687.81 | **3,086.72** |
| 151 | 550.69 | 537.18 | 456.67 | 451.63 | **1,996.16** |
| 152 | 1,412.60 | 1,319.73 | 1,166.69 | 1,108.92 | **5,007.94** |
| 153 | 373.39 | 344.40 | 370.53 | 300.50 | **1,388.82** |
| 154 | 858.42 | 865.83 | 821.00 | 801.06 | **3,346.31** |
| 155 | 256.43 | 167.05 | 143.14 | 88.23 | **654.85** |
| 156 | 937.95 | 873.63 | 862.62 | 895.09 | **3,569.29** |
| 157 | 306.70 | 187.61 | 195.55 | 162.45 | **852.31** |
| 158 | 316.70 | 316.25 | 225.61 | 216.11 | **1,074.67** |
| 159 | 748.65 | 595.03 | 526.16 | 499.31 | **2,369.15** |
| 160 | 405.68 | 374.24 | 349.84 | 338.64 | **1,468.40** |
| 161 | 1,051.43 | 854.27 | 699.58 | 638.57 | **3,243.85** |
| 162 | 333.42 | 258.26 | 224.36 | 215.98 | **1,032.03** |
| 163 | 452.52 | 433.78 | 367.81 | 382.26 | **1,636.38** |
| 164 | 97.37 | 76.19 | 82.65 | 70.48 | **326.69** |
| 165 | 157.68 | 141.58 | 135.06 | 104.29 | **538.61** |
| 166 | 432.67 | 351.18 | 286.59 | 278.25 | **1,348.68** |
| 167 | 166.29 | 146.93 | 121.59 | 79.93 | **514.74** |
| 168 | 55.22 | 29.62 | 29.57 | 22.70 | **137.12** |
| 169 | 16.59 | 19.27 | 6.69 | 8.03 | **50.58** |
| 170 | 535.48 | 457.60 | 367.05 | 345.51 | **1,705.65** |
| 171 | 172.53 | 89.66 | 90.10 | 100.90 | **453.19** |
| 172 | 284.13 | 299.83 | 250.59 | 259.82 | **1,094.38** |
| 173 | 345.91 | 276.55 | 176.10 | 206.61 | **1,005.17** |
| 174 | 209.20 | 191.13 | 145.28 | 123.56 | **669.16** |
| 175 | 173.51 | 131.58 | 129.76 | 129.27 | **564.12** |
| 176 | 87.07 | 97.55 | 102.41 | 82.61 | **369.64** |
| 177 | 42.20 | 20.96 | 38.36 | 25.78 | **127.30** |
| 178 | 19.63 | 18.91 | 12.58 | 13.65 | **64.77** |

| | | | | | |
|---|---|---|---|---|---|
| 179 | 17.40 | 19.36 | 9.46 | 14.10 | **60.31** |
| 180 | 85.20 | 81.63 | 54.91 | 69.00 | **290.74** |
| 181 | 61.11 | 53.57 | 54.46 | 41.75 | **210.89** |
| 182 | 28.64 | 11.60 | 49.07 | 34.70 | **124.00** |
| 183 | 29.80 | 26.50 | 21.68 | 11.78 | **89.75** |
| 184 | 46.66 | 58.16 | 50.40 | 54.51 | **209.73** |
| 185 | 43.53 | 53.70 | 46.39 | 23.46 | **167.09** |
| 186 | 67.53 | 48.98 | 37.74 | 47.10 | **201.35** |
| 187 | 561.67 | 572.15 | 512.42 | 526.87 | **2,173.11** |
| 188 | 27.66 | 20.52 | 24.98 | 21.41 | **94.56** |
| 189 | 107.01 | 142.25 | 135.06 | 133.59 | **517.91** |
| 190 | 876.58 | 724.30 | 702.71 | 689.46 | **2,993.04** |
| 191 | 259.87 | 249.52 | 194.03 | 191.44 | **894.87** |
| 193 | 209.38 | 203.49 | 214.06 | 205.90 | **832.82** |
| 194 | 174.00 | 153.53 | 149.07 | 120.52 | **597.13** |
| 195 | 40.90 | 23.73 | 36.40 | 31.13 | **132.16** |
| 196 | 84.21 | 82.88 | 83.05 | 72.97 | **323.12** |
| **PA - PENNSYLVANIA - Total** | **14,439.75** | **12,775.23** | **11,466.62** | **10,927.65** | **49,609.24** |

**STATE: RI - RHODE ISLAND**

| REGISTRANT ZIP CODE 3 | 1ST QUARTER | 2ND QUARTER | 3RD QUARTER | 4TH QUARTER | **TOTAL GRAMS** |
|---|---|---|---|---|---|
| 028 | 116.87 | 86.62 | 101.43 | 83.59 | **388.51** |
| 029 | 114.19 | 81.09 | 57.81 | 39.70 | **292.79** |
| **RI - RHODE ISLAND - Total** | **231.05** | **167.71** | **159.24** | **123.29** | **681.30** |

**STATE: SC - SOUTH CAROLINA**

| REGISTRANT ZIP CODE 3 | 1ST QUARTER | 2ND QUARTER | 3RD QUARTER | 4TH QUARTER | **TOTAL GRAMS** |
|---|---|---|---|---|---|
| 290 | 381.11 | 395.56 | 353.18 | 375.48 | **1,505.33** |
| 291 | 363.62 | 378.25 | 362.91 | 320.35 | **1,425.13** |
| 292 | 719.84 | 643.29 | 635.09 | 603.19 | **2,601.41** |
| 293 | 56.38 | 44.43 | 35.15 | 38.45 | **174.41** |
| 294 | 655.29 | 581.29 | 606.32 | 604.89 | **2,447.79** |
| 295 | 447.03 | 371.38 | 301.53 | 323.30 | **1,443.24** |
| 296 | 389.31 | 339.58 | 360.19 | 319.15 | **1,408.22** |
| 297 | 147.95 | 193.45 | 165.75 | 157.28 | **664.44** |
| 298 | 67.18 | 54.73 | 58.43 | 47.15 | **227.49** |
| 299 | 149.16 | 130.92 | 138.99 | 118.38 | **537.45** |
| **SC - SOUTH CAROLINA - Total** | **3,376.87** | **3,132.88** | **3,017.53** | **2,907.62** | **12,434.89** |

**STATE: SD - SOUTH DAKOTA**

| REGISTRANT ZIP CODE 3 | 1ST QUARTER | 2ND QUARTER | 3RD QUARTER | 4TH QUARTER | **TOTAL GRAMS** |
|---|---|---|---|---|---|
| 570 | 42.02 | 29.08 | 29.26 | 35.42 | **135.78** |
| 571 | 85.11 | 80.02 | 62.22 | 80.11 | **307.46** |

| | | | | | |
|---|---|---|---|---|---|
| 572 | 3.57 | 0.89 | 0.89 | 0 | **5.35** |
| 573 | 0 | 1.34 | 1.34 | 0.45 | **3.12** |
| 574 | 7.85 | 8.74 | 7.85 | 7.32 | **31.76** |
| 575 | 6.60 | 4.46 | 9.46 | 9.99 | **30.51** |
| 576 | 2.68 | 5.35 | 2.68 | 4.46 | **15.17** |
| 577 | 187.07 | 158.26 | 124.98 | 154.60 | **624.92** |
| **SD - SOUTH DAKOTA - Total** | **334.89** | **288.15** | **238.68** | **292.34** | **1,154.07** |

**STATE: TN - TENNESSEE**

| REGISTRANT ZIP CODE 3 | 1ST QUARTER | 2ND QUARTER | 3RD QUARTER | 4TH QUARTER | TOTAL GRAMS |
|---|---|---|---|---|---|
| 370 | 1,949.37 | 1,841.61 | 1,685.27 | 1,530.89 | **7,007.13** |
| 371 | 1,040.28 | 979.88 | 978.01 | 923.64 | **3,921.81** |
| 372 | 762.43 | 729.16 | 671.35 | 697.27 | **2,860.21** |
| 373 | 797.27 | 678.26 | 592.13 | 656.81 | **2,724.47** |
| 374 | 458.36 | 423.61 | 367.81 | 407.96 | **1,657.74** |
| 376 | 662.25 | 570.81 | 581.02 | 535.53 | **2,349.61** |
| 377 | 3,235.96 | 2,672.69 | 2,327.31 | 2,328.65 | **10,564.61** |
| 378 | 2,977.34 | 2,760.92 | 2,435.97 | 2,338.19 | **10,512.42** |
| 379 | 1,006.47 | 969.09 | 956.42 | 891.07 | **3,823.05** |
| 380 | 79.04 | 71.55 | 51.30 | 53.17 | **255.05** |
| 381 | 119.81 | 67.53 | 74.80 | 61.47 | **323.61** |
| 382 | 102.06 | 108.21 | 82.07 | 86.36 | **378.70** |
| 383 | 189.88 | 237.39 | 179.76 | 138.36 | **745.39** |
| 384 | 146.93 | 121.41 | 137.56 | 133.37 | **539.27** |
| 385 | 640.53 | 573.09 | 511.98 | 476.16 | **2,201.75** |
| **TN - TENNESSEE - Total** | **14,167.98** | **12,805.20** | **11,632.76** | **11,258.88** | **49,864.82** |

**STATE: TX - TEXAS**

| REGISTRANT ZIP CODE 3 | 1ST QUARTER | 2ND QUARTER | 3RD QUARTER | 4TH QUARTER | TOTAL GRAMS |
|---|---|---|---|---|---|
| 750 | 786.92 | 670.99 | 599.27 | 515.54 | **2,572.73** |
| 751 | 113.16 | 103.93 | 57.09 | 100.58 | **374.77** |
| 752 | 139.76 | 106.34 | 78.61 | 88.41 | **413.12** |
| 754 | 56.20 | 43.04 | 27.48 | 35.51 | **162.23** |
| 755 | 1.78 | 0 | 0 | 0.54 | **2.32** |
| 756 | 9.10 | 0.89 | 0 | 1.78 | **11.78** |
| 757 | 25.34 | 30.24 | 21.68 | 31.40 | **108.66** |
| 758 | 16.59 | 13.02 | 18.56 | 14.54 | **62.71** |
| 759 | 36.84 | 24.09 | 33.36 | 26.85 | **121.15** |
| 760 | 346.54 | 306.57 | 252.33 | 245.73 | **1,151.17** |
| 761 | 246.71 | 164.95 | 124.36 | 111.29 | **647.31** |
| 762 | 89.30 | 107.59 | 106.25 | 88.32 | **391.45** |
| 763 | 136.80 | 124.09 | 132.79 | 110.89 | **504.57** |

| | | | | | |
|---|---|---|---|---|---|
| 764 | 15.70 | 21.77 | 17.75 | 14.81 | **70.03** |
| 765 | 50.23 | 51.12 | 24.80 | 24.18 | **150.32** |
| 766 | 0 | 0 | 0 | 5.35 | **5.35** |
| 767 | 22.48 | 10.35 | 10.97 | 9.77 | **53.57** |
| 768 | 8.30 | 14.90 | 16.77 | 6.24 | **46.21** |
| 769 | 30.96 | 21.59 | 16.55 | 10.84 | **79.93** |
| 770 | 300.46 | 305.46 | 272.48 | 244.17 | **1,122.56** |
| 773 | 197.15 | 183.33 | 135.87 | 152.64 | **668.99** |
| 774 | 258.89 | 233.28 | 234.62 | 210.62 | **937.42** |
| 775 | 354.61 | 332.93 | 297.69 | 291.67 | **1,276.91** |
| 776 | 119.63 | 125.03 | 115.79 | 90.19 | **450.64** |
| 777 | 87.11 | 39.39 | 40.90 | 33.19 | **200.59** |
| 778 | 26.36 | 28.01 | 18.11 | 16.06 | **88.54** |
| 779 | 31.31 | 35.55 | 26.81 | 26.67 | **120.34** |
| 780 | 66.46 | 53.79 | 66.02 | 63.25 | **249.52** |
| 781 | 77.79 | 81.00 | 66.82 | 73.51 | **299.12** |
| 782 | 640.84 | 522.06 | 572.82 | 459.07 | **2,194.79** |
| 783 | 6.78 | 8.92 | 3.57 | 10.26 | **29.53** |
| 784 | 75.20 | 36.93 | 52.72 | 40.59 | **205.45** |
| 785 | 0 | 1.78 | 0.89 | 4.28 | **6.96** |
| 786 | 480.57 | 427.81 | 319.46 | 227.49 | **1,455.33** |
| 787 | 394.26 | 313.17 | 265.36 | 186.81 | **1,159.60** |
| 788 | 23.73 | 7.58 | 6.78 | 7.14 | **45.23** |
| 789 | 15.34 | 16.95 | 8.92 | 10.71 | **51.92** |
| 790 | 9.72 | 9.86 | 8.79 | 13.11 | **41.48** |
| 791 | 75.38 | 76.14 | 62.94 | 67.44 | **281.90** |
| 793 | 4.01 | 0 | 0 | 0 | **4.01** |
| 794 | 29.48 | 17.93 | 5.80 | 6.69 | **59.90** |
| 795 | 0.80 | 0.89 | 1.43 | 1.69 | **4.82** |
| 796 | 19.80 | 7.67 | 12.76 | 12.40 | **52.63** |
| 797 | 20.70 | 13.92 | 4.64 | 4.46 | **43.71** |
| 799 | 86.80 | 71.37 | 60.57 | 37.56 | **256.30** |
| **TX - TEXAS - Total** | **5,535.93** | **4,766.22** | **4,201.18** | **3,734.24** | **18,237.57** |

**STATE: UT - UTAH**

| REGISTRANT ZIP CODE 3 | 1ST QUARTER | 2ND QUARTER | 3RD QUARTER | 4TH QUARTER | **TOTAL GRAMS** |
|---|---|---|---|---|---|
| 840 | 1,571.05 | 1,324.68 | 1,301.66 | 1,252.37 | **5,449.77** |
| 841 | 413.00 | 427.90 | 343.19 | 360.72 | **1,544.80** |
| 843 | 67.18 | 76.10 | 85.91 | 76.01 | **305.19** |
| 844 | 366.79 | 362.15 | 309.47 | 329.45 | **1,367.86** |
| 845 | 67.98 | 59.77 | 61.11 | 43.36 | **232.21** |

| | | | | | |
|---|---|---|---|---|---|
| 846 | 262.95 | 230.12 | 218.70 | 225.79 | **937.55** |
| 847 | 234.76 | 196.35 | 182.43 | 229.27 | **842.81** |
| **UT - UTAH - Total** | **2,983.69** | **2,677.06** | **2,502.47** | **2,516.97** | **10,680.19** |

**STATE: VA - VIRGINIA**

| REGISTRANT ZIP CODE 3 | 1ST QUARTER | 2ND QUARTER | 3RD QUARTER | 4TH QUARTER | TOTAL GRAMS |
|---|---|---|---|---|---|
| 201 | 643.74 | 717.83 | 636.07 | 709.93 | **2,707.57** |
| 220 | 414.29 | 361.35 | 302.96 | 322.49 | **1,401.09** |
| 221 | 501.14 | 412.19 | 450.33 | 374.95 | **1,738.61** |
| 222 | 126.77 | 136.31 | 152.50 | 129.76 | **545.34** |
| 223 | 135.78 | 100.67 | 81.89 | 101.43 | **419.78** |
| 224 | 740.98 | 805.57 | 676.17 | 660.73 | **2,883.45** |
| 225 | 117.49 | 102.86 | 91.80 | 68.25 | **380.39** |
| 226 | 351.89 | 282.53 | 269.73 | 224.50 | **1,128.64** |
| 227 | 34.79 | 57.09 | 51.65 | 40.77 | **184.31** |
| 228 | 24.27 | 25.34 | 33.63 | 25.78 | **109.01** |
| 229 | 94.74 | 81.81 | 76.54 | 90.01 | **343.10** |
| 230 | 165.57 | 78.95 | 98.40 | 72.88 | **415.81** |
| 231 | 171.82 | 132.43 | 139.08 | 161.60 | **604.93** |
| 232 | 190.73 | 159.51 | 158.66 | 140.86 | **649.76** |
| 233 | 206.70 | 235.29 | 229.09 | 228.82 | **899.91** |
| 234 | 470.40 | 435.70 | 350.46 | 305.68 | **1,562.25** |
| 235 | 166.91 | 177.22 | 132.48 | 136.67 | **613.27** |
| 236 | 140.91 | 121.82 | 128.86 | 115.30 | **506.89** |
| 237 | 96.79 | 96.44 | 69.54 | 100.45 | **363.22** |
| 238 | 251.13 | 176.81 | 179.31 | 172.00 | **779.25** |
| 239 | 54.24 | 38.45 | 32.47 | 24.00 | **149.16** |
| 240 | 227.40 | 179.94 | 185.02 | 148.80 | **741.16** |
| 241 | 147.20 | 114.86 | 123.91 | 113.74 | **499.71** |
| 242 | 289.17 | 267.32 | 248.67 | 274.59 | **1,079.75** |
| 243 | 84.57 | 83.50 | 101.70 | 80.47 | **350.24** |
| 244 | 27.57 | 18.20 | 20.61 | 22.75 | **89.12** |
| 245 | 203.13 | 206.25 | 146.04 | 129.31 | **684.73** |
| 246 | 68.78 | 79.75 | 78.42 | 51.65 | **278.60** |
| **VA - VIRGINIA - Total** | **6,148.89** | **5,685.98** | **5,245.99** | **5,028.19** | **22,109.05** |

**STATE: VT - VERMONT**

| REGISTRANT ZIP CODE 3 | 1ST QUARTER | 2ND QUARTER | 3RD QUARTER | 4TH QUARTER | TOTAL GRAMS |
|---|---|---|---|---|---|
| 050 | 2.68 | 3.57 | 1.78 | 4.46 | **12.49** |
| 052 | 32.12 | 17.84 | 18.73 | 23.19 | **91.89** |
| 054 | 107.68 | 79.40 | 72.71 | 82.61 | **342.39** |
| 056 | 14.99 | 12.85 | 12.85 | 14.99 | **55.67** |

| | | | | | |
|---|---|---|---|---|---|
| 057 | 7.49 | 12.67 | 11.06 | 10.97 | **42.20** |
| 058 | 4.46 | 1.34 | 3.12 | 3.66 | **12.58** |
| **VT - VERMONT - Total** | **169.41** | **127.66** | **120.26** | **139.88** | **557.21** |

**STATE: WA - WASHINGTON**

| REGISTRANT ZIP CODE 3 | 1ST QUARTER | 2ND QUARTER | 3RD QUARTER | 4TH QUARTER | TOTAL GRAMS |
|---|---|---|---|---|---|
| 980 | 395.29 | 328.11 | 326.06 | 283.06 | **1,332.53** |
| 981 | 102.50 | 77.61 | 87.07 | 104.46 | **371.65** |
| 982 | 255.90 | 219.90 | 221.20 | 252.11 | **949.11** |
| 983 | 96.26 | 104.42 | 86.40 | 94.12 | **381.19** |
| 984 | 76.27 | 56.69 | 58.16 | 62.98 | **254.11** |
| 985 | 148.67 | 120.75 | 118.96 | 74.67 | **463.04** |
| 986 | 238.82 | 210.62 | 186.98 | 222.94 | **859.36** |
| 988 | 23.28 | 23.91 | 19.49 | 18.07 | **84.75** |
| 989 | 44.87 | 39.97 | 41.57 | 52.54 | **178.96** |
| 990 | 14.72 | 7.23 | 7.94 | 4.64 | **34.52** |
| 992 | 136.98 | 106.74 | 115.66 | 107.99 | **467.37** |
| 993 | 90.01 | 79.75 | 66.37 | 68.87 | **305.01** |
| 994 | 13.74 | 12.13 | 9.72 | 7.49 | **43.09** |
| **WA - WASHINGTON - Total** | **1,637.32** | **1,387.84** | **1,345.60** | **1,353.94** | **5,724.69** |

**STATE: WI - WISCONSIN**

| REGISTRANT ZIP CODE 3 | 1ST QUARTER | 2ND QUARTER | 3RD QUARTER | 4TH QUARTER | TOTAL GRAMS |
|---|---|---|---|---|---|
| 530 | 115.26 | 107.77 | 108.84 | 113.34 | **445.20** |
| 531 | 250.81 | 178.06 | 152.46 | 139.08 | **720.42** |
| 532 | 129.22 | 102.90 | 89.25 | 79.40 | **400.78** |
| 534 | 77.52 | 62.63 | 62.36 | 58.61 | **261.12** |
| 535 | 43.98 | 44.52 | 25.87 | 33.63 | **148.00** |
| 537 | 16.50 | 5.89 | 6.69 | 5.89 | **34.97** |
| 538 | 2.14 | 1.74 | 0.40 | 0 | **4.28** |
| 539 | 20.43 | 25.42 | 14.63 | 15.34 | **75.83** |
| 540 | 16.06 | 11.06 | 9.63 | 6.96 | **43.71** |
| 541 | 23.91 | 19.45 | 20.65 | 20.61 | **84.62** |
| 542 | 15.61 | 16.86 | 14.81 | 11.95 | **59.24** |
| 543 | 32.20 | 23.69 | 21.81 | 22.79 | **100.50** |
| 544 | 90.73 | 74.80 | 76.94 | 64.41 | **306.88** |
| 545 | 11.24 | 14.27 | 10.35 | 7.94 | **43.80** |
| 546 | 7.14 | 4.01 | 0 | 0 | **11.15** |
| 547 | 0.27 | 0 | 0 | 0 | **0.27** |
| 548 | 18.82 | 22.03 | 17.04 | 18.82 | **76.72** |
| 549 | 159.33 | 113.21 | 141.09 | 109.19 | **522.82** |
| **WI - WISCONSIN - Total** | **1,031.18** | **828.31** | **772.83** | **707.97** | **3,340.29** |

**STATE: WV - WEST VIRGINIA**

| REGISTRANT ZIP CODE 3 | 1ST QUARTER | 2ND QUARTER | 3RD QUARTER | 4TH QUARTER | TOTAL GRAMS |
|---|---|---|---|---|---|
| 247 | 74.27 | 54.37 | 35.46 | 33.41 | **197.51** |
| 248 | 79.62 | 70.48 | 61.55 | 64.86 | **276.51** |
| 249 | 38.90 | 39.25 | 44.16 | 29.44 | **151.75** |
| 250 | 15.08 | 14.72 | 10.44 | 9.10 | **49.33** |
| 251 | 19.89 | 15.43 | 15.08 | 9.81 | **60.22** |
| 252 | 4.28 | 0.89 | 0 | 1.78 | **6.96** |
| 253 | 102.41 | 110.35 | 99.11 | 70.12 | **382.00** |
| 254 | 623.27 | 611.89 | 488.34 | 469.20 | **2,192.69** |
| 255 | 123.96 | 86.94 | 101.57 | 124.31 | **436.77** |
| 256 | 35.86 | 32.74 | 18.11 | 14.81 | **101.52** |
| 257 | 46.52 | 50.23 | 36.13 | 52.37 | **185.24** |
| 258 | 8.21 | 10.35 | 11.51 | 10.71 | **40.77** |
| 259 | 14.90 | 13.02 | 14.10 | 15.70 | **57.72** |
| 260 | 144.07 | 127.04 | 115.04 | 139.57 | **525.71** |
| 261 | 18.20 | 17.13 | 20.74 | 21.41 | **77.48** |
| 262 | 18.02 | 4.28 | 2.68 | 4.19 | **29.17** |
| 263 | 22.30 | 22.75 | 8.92 | 7.05 | **61.02** |
| 264 | 5.35 | 3.57 | 3.75 | 5.17 | **17.84** |
| 265 | 31.49 | 26.50 | 28.01 | 29.17 | **115.17** |
| 266 | 2.41 | 5.35 | 0.98 | 1.96 | **10.71** |
| 267 | 48.89 | 59.24 | 25.87 | 49.51 | **183.50** |
| 268 | 2.23 | 0 | 0 | 0 | **2.23** |
| **WV - WEST VIRGINIA - Total** | **1,480.13** | **1,376.51** | **1,141.53** | **1,163.66** | **5,161.82** |

**STATE: WY - WYOMING**

| REGISTRANT ZIP CODE 3 | 1ST QUARTER | 2ND QUARTER | 3RD QUARTER | 4TH QUARTER | TOTAL GRAMS |
|---|---|---|---|---|---|
| 820 | 32.29 | 15.25 | 18.64 | 18.29 | **84.48** |
| 822 | 0 | 0 | 0.45 | 0 | **0.45** |
| 823 | 3.57 | 1.34 | 0 | 0 | **4.91** |
| 824 | 5.71 | 10.53 | 6.07 | 3.21 | **25.51** |
| 825 | 50.98 | 21.77 | 23.69 | 24.27 | **120.70** |
| 826 | 36.49 | 25.83 | 24.80 | 28.06 | **115.17** |
| 827 | 21.23 | 17.40 | 13.65 | 13.74 | **66.02** |
| 828 | 13.92 | 7.32 | 6.42 | 8.56 | **36.22** |
| 829 | 11.78 | 12.31 | 8.03 | 9.63 | **41.75** |
| 830 | 0 | 3.57 | 10.71 | 4.46 | **18.73** |
| 831 | 18.82 | 9.90 | 3.21 | 2.41 | **34.35** |
| **WY - WYOMING - Total** | **194.79** | **125.21** | **115.66** | **112.63** | **548.28** |

**DRUG: 9780 - TAPENTADOL**

**STATE: AK - ALASKA**

| REGISTRANT ZIP CODE 3 | 1ST QUARTER | 2ND QUARTER | 3RD QUARTER | 4TH QUARTER | TOTAL GRAMS |
|---|---|---|---|---|---|
| 995 | 1,148.00 | 1,249.50 | 1,205.00 | 946.50 | **4,549.00** |
| 996 | 858.00 | 755.00 | 804.00 | 583.50 | **3,000.50** |
| 997 | 95.00 | 138.00 | 99.00 | 82.00 | **414.00** |
| 998 | 3.00 | 5.00 | 0 | 25.00 | **33.00** |
| 999 | 0 | 0 | 5.00 | 9.00 | **14.00** |
| **AK - ALASKA - Total** | **2,104.00** | **2,147.50** | **2,113.00** | **1,646.00** | **8,010.50** |

**STATE: AL - ALABAMA**

| REGISTRANT ZIP CODE 3 | 1ST QUARTER | 2ND QUARTER | 3RD QUARTER | 4TH QUARTER | TOTAL GRAMS |
|---|---|---|---|---|---|
| 350 | 6,487.00 | 6,998.00 | 6,104.00 | 5,023.50 | **24,612.50** |
| 351 | 3,494.00 | 3,811.00 | 3,131.00 | 2,715.50 | **13,151.50** |
| 352 | 3,444.50 | 3,488.00 | 3,522.00 | 3,096.00 | **13,550.50** |
| 354 | 2,851.50 | 2,764.00 | 2,620.50 | 2,635.00 | **10,871.00** |
| 355 | 1,943.50 | 1,921.50 | 1,734.00 | 1,671.00 | **7,270.00** |
| 356 | 3,424.00 | 3,386.00 | 2,948.00 | 2,566.00 | **12,324.00** |
| 357 | 1,457.00 | 1,670.00 | 1,396.50 | 1,174.50 | **5,698.00** |
| 358 | 2,220.50 | 2,121.50 | 1,871.00 | 1,989.00 | **8,202.00** |
| 359 | 1,915.50 | 1,836.50 | 1,584.50 | 1,800.00 | **7,136.50** |
| 360 | 3,426.00 | 3,220.50 | 3,221.50 | 2,946.50 | **12,814.50** |
| 361 | 2,221.00 | 2,085.00 | 2,165.50 | 1,998.50 | **8,470.00** |
| 362 | 1,331.50 | 1,087.00 | 1,088.50 | 1,279.50 | **4,786.50** |
| 363 | 1,091.00 | 1,117.00 | 1,049.50 | 889.50 | **4,147.00** |
| 364 | 271.00 | 309.00 | 304.00 | 291.00 | **1,175.00** |
| 365 | 2,759.50 | 2,963.00 | 2,710.00 | 2,511.50 | **10,944.00** |
| 366 | 2,392.50 | 2,142.00 | 2,066.00 | 1,757.00 | **8,357.50** |
| 367 | 390.50 | 383.00 | 484.50 | 600.50 | **1,858.50** |
| 368 | 2,006.00 | 1,816.00 | 2,149.00 | 1,986.50 | **7,957.50** |
| 369 | 219.00 | 253.00 | 287.50 | 218.00 | **977.50** |
| **AL - ALABAMA - Total** | **43,345.50** | **43,372.00** | **40,437.50** | **37,149.00** | **164,304.00** |

**STATE: AR - ARKANSAS**

| REGISTRANT ZIP CODE 3 | 1ST QUARTER | 2ND QUARTER | 3RD QUARTER | 4TH QUARTER | TOTAL GRAMS |
|---|---|---|---|---|---|
| 716 | 432.00 | 432.00 | 347.00 | 271.50 | **1,482.50** |
| 717 | 699.00 | 789.50 | 650.00 | 656.00 | **2,794.50** |
| 718 | 170.00 | 80.00 | 127.00 | 166.50 | **543.50** |
| 719 | 886.50 | 778.00 | 633.00 | 540.50 | **2,838.00** |
| 720 | 2,919.00 | 2,479.00 | 2,480.00 | 2,010.50 | **9,888.50** |
| 721 | 2,071.50 | 2,085.50 | 1,826.00 | 1,517.00 | **7,500.00** |
| 722 | 1,150.50 | 1,159.50 | 991.00 | 997.50 | **4,298.50** |
| 723 | 205.00 | 219.00 | 173.00 | 167.00 | **764.00** |

| | | | | | |
|---|---|---|---|---|---|
| 724 | 1,269.50 | 1,016.00 | 951.00 | 809.50 | **4,046.00** |
| 725 | 767.00 | 667.50 | 491.50 | 376.00 | **2,302.00** |
| 726 | 855.50 | 749.00 | 706.50 | 564.50 | **2,875.50** |
| 727 | 2,668.50 | 2,763.50 | 2,499.50 | 2,339.00 | **10,270.50** |
| 728 | 368.00 | 372.50 | 424.50 | 479.50 | **1,644.50** |
| 729 | 1,119.50 | 782.00 | 803.00 | 804.50 | **3,509.00** |
| **AR - ARKANSAS - Total** | **15,581.50** | **14,373.00** | **13,103.00** | **11,699.50** | **54,757.00** |

**STATE: AZ - ARIZONA**

| REGISTRANT ZIP CODE 3 | 1ST QUARTER | 2ND QUARTER | 3RD QUARTER | 4TH QUARTER | TOTAL GRAMS |
|---|---|---|---|---|---|
| 850 | 7,083.50 | 6,717.00 | 5,377.50 | 4,723.00 | **23,901.00** |
| 851 | 881.50 | 717.00 | 764.50 | 728.50 | **3,091.50** |
| 852 | 12,336.50 | 12,096.50 | 12,014.00 | 10,026.50 | **46,473.50** |
| 853 | 5,280.50 | 5,101.50 | 4,523.00 | 4,655.00 | **19,560.00** |
| 855 | 270.00 | 292.50 | 211.50 | 363.50 | **1,137.50** |
| 856 | 873.00 | 989.00 | 867.50 | 973.50 | **3,703.00** |
| 857 | 3,754.00 | 3,946.00 | 3,928.50 | 3,639.00 | **15,267.50** |
| 859 | 46.00 | 73.00 | 161.50 | 221.00 | **501.50** |
| 860 | 253.50 | 250.00 | 130.50 | 144.50 | **778.50** |
| 863 | 550.00 | 534.50 | 401.50 | 457.50 | **1,943.50** |
| 864 | 325.00 | 234.00 | 184.00 | 164.00 | **907.00** |
| **AZ - ARIZONA - Total** | **31,653.50** | **30,951.00** | **28,564.00** | **26,096.00** | **117,264.50** |

**STATE: CA - CALIFORNIA**

| REGISTRANT ZIP CODE 3 | 1ST QUARTER | 2ND QUARTER | 3RD QUARTER | 4TH QUARTER | TOTAL GRAMS |
|---|---|---|---|---|---|
| 900 | 1,857.00 | 1,571.50 | 1,666.00 | 1,867.50 | **6,962.00** |
| 902 | 2,289.00 | 2,037.50 | 1,937.00 | 1,800.50 | **8,064.00** |
| 903 | 65.50 | 80.00 | 32.50 | 45.00 | **223.00** |
| 904 | 1,398.50 | 1,346.00 | 1,220.50 | 1,248.00 | **5,213.00** |
| 905 | 308.50 | 472.50 | 395.00 | 409.00 | **1,585.00** |
| 906 | 780.00 | 818.00 | 648.50 | 725.50 | **2,972.00** |
| 907 | 1,207.50 | 1,237.00 | 1,136.50 | 974.50 | **4,555.50** |
| 908 | 598.00 | 684.50 | 570.50 | 602.00 | **2,455.00** |
| 910 | 521.00 | 387.00 | 292.50 | 356.50 | **1,557.00** |
| 911 | 507.00 | 555.50 | 523.50 | 545.50 | **2,131.50** |
| 912 | 178.00 | 282.50 | 176.50 | 195.50 | **832.50** |
| 913 | 4,356.00 | 4,170.50 | 4,178.00 | 3,958.50 | **16,663.00** |
| 914 | 1,086.00 | 1,055.00 | 1,017.50 | 920.00 | **4,078.50** |
| 915 | 137.00 | 147.00 | 153.00 | 167.50 | **604.50** |
| 916 | 98.00 | 58.00 | 103.00 | 137.00 | **396.00** |
| 917 | 5,751.00 | 5,029.50 | 4,693.50 | 4,838.50 | **20,312.50** |
| 918 | 15.00 | 53.00 | 65.00 | 35.00 | **168.00** |

| | | | | | |
|---|---|---|---|---|---|
| 919 | 1,351.50 | 1,412.50 | 1,354.50 | 1,186.00 | **5,304.50** |
| 920 | 7,205.00 | 7,402.50 | 6,756.00 | 6,728.00 | **28,091.50** |
| 921 | 2,850.50 | 2,449.50 | 2,526.50 | 2,282.00 | **10,108.50** |
| 922 | 1,695.00 | 1,750.00 | 1,680.50 | 1,538.50 | **6,664.00** |
| 923 | 3,031.50 | 2,709.00 | 2,641.00 | 2,691.50 | **11,073.00** |
| 924 | 830.50 | 671.00 | 399.00 | 266.00 | **2,166.50** |
| 925 | 5,114.50 | 4,934.50 | 5,085.50 | 4,980.00 | **20,114.50** |
| 926 | 5,119.40 | 5,391.43 | 5,222.95 | 5,146.45 | **20,880.23** |
| 927 | 925.00 | 995.00 | 918.00 | 959.00 | **3,797.00** |
| 928 | 3,352.50 | 3,358.50 | 2,923.50 | 3,084.00 | **12,718.50** |
| 930 | 2,521.50 | 2,594.00 | 2,288.50 | 2,228.00 | **9,632.00** |
| 931 | 371.50 | 380.50 | 329.00 | 305.50 | **1,386.50** |
| 932 | 2,930.50 | 2,472.50 | 2,344.00 | 2,270.00 | **10,017.00** |
| 933 | 1,310.00 | 1,434.50 | 1,290.00 | 1,208.50 | **5,243.00** |
| 934 | 1,757.50 | 2,070.00 | 1,559.50 | 1,601.00 | **6,988.00** |
| 935 | 1,410.00 | 1,330.00 | 1,236.00 | 1,185.50 | **5,161.50** |
| 936 | 2,133.50 | 2,314.00 | 1,782.50 | 1,910.00 | **8,140.00** |
| 937 | 2,693.00 | 2,677.00 | 3,074.00 | 2,795.50 | **11,239.50** |
| 939 | 1,334.50 | 751.50 | 955.50 | 698.00 | **3,739.50** |
| 940 | 1,137.00 | 1,215.50 | 1,170.00 | 1,174.00 | **4,696.50** |
| 941 | 495.50 | 571.50 | 629.00 | 690.50 | **2,386.50** |
| 943 | 0 | 78.00 | 67.00 | 11.00 | **156.00** |
| 944 | 176.00 | 205.00 | 237.00 | 209.50 | **827.50** |
| 945 | 3,490.00 | 3,184.50 | 3,404.00 | 3,235.00 | **13,313.50** |
| 946 | 249.50 | 289.50 | 292.00 | 203.50 | **1,034.50** |
| 947 | 70.00 | 93.00 | 54.00 | 61.00 | **278.00** |
| 948 | 24.00 | 21.00 | 13.50 | 7.50 | **66.00** |
| 949 | 206.50 | 261.50 | 306.00 | 206.50 | **980.50** |
| 950 | 731.50 | 645.00 | 704.00 | 508.00 | **2,588.50** |
| 951 | 372.00 | 390.00 | 270.00 | 345.00 | **1,377.00** |
| 952 | 301.50 | 347.00 | 389.50 | 440.50 | **1,478.50** |
| 953 | 1,907.00 | 1,468.00 | 1,532.50 | 1,547.50 | **6,455.00** |
| 954 | 458.00 | 360.50 | 359.00 | 534.00 | **1,711.50** |
| 955 | 45.00 | 34.00 | 9.00 | 3.00 | **91.00** |
| 956 | 2,027.00 | 1,828.50 | 2,137.50 | 1,977.00 | **7,970.00** |
| 957 | 673.50 | 561.00 | 441.50 | 436.00 | **2,112.00** |
| 958 | 1,474.50 | 1,671.00 | 1,471.00 | 1,486.50 | **6,103.00** |
| 959 | 1,768.50 | 1,425.00 | 1,408.50 | 1,281.50 | **5,883.50** |
| 960 | 984.00 | 782.00 | 701.00 | 482.00 | **2,949.00** |
| 961 | 5.00 | 0 | 12.00 | 6.00 | **23.00** |

| CA - CALIFORNIA - Total | 85,686.40 | 82,514.43 | 78,783.45 | 76,734.95 | 323,719.23 |
|---|---|---|---|---|---|

**STATE: CO - COLORADO**

| REGISTRANT ZIP CODE 3 | 1ST QUARTER | 2ND QUARTER | 3RD QUARTER | 4TH QUARTER | TOTAL GRAMS |
|---|---|---|---|---|---|
| 800 | 3,241.50 | 3,331.00 | 3,127.00 | 3,245.50 | **12,945.00** |
| 801 | 5,723.00 | 5,262.00 | 5,431.50 | 5,956.00 | **22,372.50** |
| 802 | 3,705.50 | 2,907.50 | 2,916.00 | 2,564.50 | **12,093.50** |
| 803 | 705.50 | 686.50 | 593.00 | 671.50 | **2,656.50** |
| 804 | 731.00 | 606.00 | 593.50 | 596.50 | **2,527.00** |
| 805 | 2,859.00 | 3,153.00 | 3,231.00 | 3,322.00 | **12,565.00** |
| 806 | 1,530.50 | 1,352.00 | 1,388.00 | 1,415.50 | **5,686.00** |
| 807 | 266.00 | 217.00 | 203.00 | 238.50 | **924.50** |
| 808 | 511.50 | 399.00 | 399.00 | 437.00 | **1,746.50** |
| 809 | 3,895.00 | 3,946.00 | 3,786.00 | 3,393.00 | **15,020.00** |
| 810 | 2,041.00 | 1,806.00 | 1,743.00 | 1,424.00 | **7,014.00** |
| 811 | 81.50 | 123.00 | 76.00 | 115.50 | **396.00** |
| 812 | 473.00 | 384.00 | 277.00 | 470.00 | **1,604.00** |
| 813 | 260.50 | 188.00 | 202.50 | 205.50 | **856.50** |
| 814 | 53.00 | 79.00 | 72.00 | 83.00 | **287.00** |
| 815 | 536.50 | 569.00 | 390.00 | 357.50 | **1,853.00** |
| 816 | 133.00 | 135.50 | 152.00 | 267.00 | **687.50** |
| **CO - COLORADO - Total** | **26,747.00** | **25,144.50** | **24,580.50** | **24,762.50** | **101,234.50** |

**STATE: CT - CONNECTICUT**

| REGISTRANT ZIP CODE 3 | 1ST QUARTER | 2ND QUARTER | 3RD QUARTER | 4TH QUARTER | TOTAL GRAMS |
|---|---|---|---|---|---|
| 060 | 1,357.50 | 1,387.00 | 1,228.50 | 1,306.50 | **5,279.50** |
| 061 | 332.50 | 440.00 | 362.00 | 378.50 | **1,513.00** |
| 062 | 551.00 | 505.50 | 554.00 | 482.00 | **2,092.50** |
| 063 | 1,005.50 | 1,113.50 | 1,068.00 | 1,398.00 | **4,585.00** |
| 064 | 2,228.00 | 2,314.00 | 2,243.50 | 2,185.00 | **8,970.50** |
| 065 | 530.00 | 590.50 | 632.00 | 647.50 | **2,400.00** |
| 066 | 225.00 | 313.50 | 384.50 | 338.50 | **1,261.50** |
| 067 | 836.50 | 974.00 | 696.50 | 669.00 | **3,176.00** |
| 068 | 1,008.50 | 1,164.00 | 1,053.50 | 1,136.50 | **4,362.50** |
| 069 | 166.50 | 92.50 | 106.00 | 132.00 | **497.00** |
| **CT - CONNECTICUT - Total** | **8,241.00** | **8,894.50** | **8,328.50** | **8,673.50** | **34,137.50** |

**STATE: DC - DISTRICT OF COLUMBIA**

| REGISTRANT ZIP CODE 3 | 1ST QUARTER | 2ND QUARTER | 3RD QUARTER | 4TH QUARTER | TOTAL GRAMS |
|---|---|---|---|---|---|
| 200 | 864.00 | 860.00 | 807.00 | 765.00 | **3,296.00** |
| 204 | 20.00 | 20.00 | 20.00 | 10.00 | **70.00** |
| **DC - DISTRICT OF COLUMBIA -** | **884.00** | **880.00** | **827.00** | **775.00** | **3,366.00** |

**STATE: DE - DELAWARE**

| REGISTRANT ZIP CODE 3 | 1ST QUARTER | 2ND QUARTER | 3RD QUARTER | 4TH QUARTER | TOTAL GRAMS |
|---|---|---|---|---|---|
| 197 | 1,943.50 | 2,107.50 | 2,229.00 | 2,179.50 | **8,459.50** |
| 198 | 1,403.50 | 1,354.00 | 1,228.00 | 1,090.00 | **5,075.50** |
| 199 | 2,184.50 | 2,173.50 | 2,156.00 | 1,855.50 | **8,369.50** |
| **DE - DELAWARE - Total** | **5,531.50** | **5,635.00** | **5,613.00** | **5,125.00** | **21,904.50** |

**STATE: FL - FLORIDA**

| REGISTRANT ZIP CODE 3 | 1ST QUARTER | 2ND QUARTER | 3RD QUARTER | 4TH QUARTER | TOTAL GRAMS |
|---|---|---|---|---|---|
| 320 | 2,520.50 | 2,434.00 | 2,004.00 | 2,257.50 | **9,216.00** |
| 321 | 1,991.50 | 1,660.00 | 1,470.50 | 1,712.00 | **6,834.00** |
| 322 | 4,285.50 | 4,360.00 | 3,803.50 | 3,277.00 | **15,726.00** |
| 323 | 2,115.50 | 2,598.50 | 2,287.00 | 2,218.00 | **9,219.00** |
| 324 | 1,278.50 | 1,363.50 | 1,232.00 | 1,082.50 | **4,956.50** |
| 325 | 4,178.00 | 3,402.00 | 3,086.50 | 2,709.50 | **13,376.00** |
| 326 | 1,468.50 | 1,493.50 | 1,417.50 | 1,448.50 | **5,828.00** |
| 327 | 4,788.00 | 4,796.00 | 3,937.00 | 4,199.50 | **17,720.50** |
| 328 | 2,040.00 | 2,314.50 | 2,014.00 | 2,022.50 | **8,391.00** |
| 329 | 3,356.00 | 2,977.50 | 2,967.50 | 2,675.50 | **11,976.50** |
| 330 | 2,310.00 | 2,369.50 | 2,239.50 | 2,381.50 | **9,300.50** |
| 331 | 364.50 | 447.00 | 453.50 | 467.00 | **1,732.00** |
| 333 | 1,748.50 | 1,844.50 | 1,684.50 | 1,495.50 | **6,773.00** |
| 334 | 6,446.50 | 6,301.00 | 5,887.00 | 5,870.00 | **24,504.50** |
| 335 | 5,200.00 | 4,911.00 | 4,202.50 | 4,398.50 | **18,712.00** |
| 336 | 3,129.00 | 3,319.50 | 3,076.50 | 2,704.50 | **12,229.50** |
| 337 | 2,739.00 | 3,113.50 | 2,780.50 | 2,717.50 | **11,350.50** |
| 338 | 2,619.00 | 2,349.00 | 2,105.50 | 2,170.00 | **9,243.50** |
| 339 | 8,097.50 | 8,074.00 | 7,126.00 | 6,499.50 | **29,797.00** |
| 341 | 3,512.50 | 3,422.00 | 2,633.50 | 2,950.50 | **12,518.50** |
| 342 | 4,556.00 | 4,525.50 | 3,610.00 | 3,853.00 | **16,544.50** |
| 344 | 1,859.00 | 1,909.00 | 1,503.50 | 1,684.00 | **6,955.50** |
| 346 | 3,305.00 | 2,899.50 | 2,644.50 | 2,696.00 | **11,545.00** |
| 347 | 2,305.00 | 2,485.00 | 2,031.00 | 2,087.50 | **8,908.50** |
| 349 | 1,963.50 | 2,062.00 | 2,085.50 | 1,800.50 | **7,911.50** |
| **FL - FLORIDA - Total** | **78,177.00** | **77,431.50** | **68,283.00** | **67,378.00** | **291,269.50** |

**STATE: GA - GEORGIA**

| REGISTRANT ZIP CODE 3 | 1ST QUARTER | 2ND QUARTER | 3RD QUARTER | 4TH QUARTER | TOTAL GRAMS |
|---|---|---|---|---|---|
| 300 | 9,939.00 | 9,732.00 | 9,458.00 | 9,057.00 | **38,186.00** |
| 301 | 7,715.50 | 7,952.00 | 7,320.50 | 7,142.00 | **30,130.00** |
| 302 | 4,220.50 | 4,270.00 | 4,471.50 | 4,594.50 | **17,556.50** |
| 303 | 2,862.00 | 2,601.00 | 2,526.50 | 2,802.50 | **10,792.00** |
| 304 | 3,185.50 | 2,659.50 | 2,685.50 | 2,676.00 | **11,206.50** |

| | | | | | |
|---|---|---|---|---|---|
| 305 | 2,862.00 | 2,950.50 | 2,814.00 | 2,943.00 | **11,569.50** |
| 306 | 1,510.00 | 1,696.00 | 1,530.00 | 1,523.50 | **6,259.50** |
| 307 | 1,265.50 | 1,080.50 | 1,000.50 | 1,006.50 | **4,353.00** |
| 308 | 492.50 | 553.00 | 492.00 | 543.50 | **2,081.00** |
| 309 | 833.50 | 876.50 | 791.50 | 801.50 | **3,303.00** |
| 310 | 2,851.00 | 2,774.00 | 2,621.00 | 2,694.00 | **10,940.00** |
| 312 | 682.00 | 716.00 | 675.00 | 677.00 | **2,750.00** |
| 313 | 3,011.50 | 3,629.00 | 3,483.50 | 3,211.50 | **13,335.50** |
| 314 | 1,722.00 | 1,834.50 | 1,715.50 | 1,622.00 | **6,894.00** |
| 315 | 5,270.50 | 5,744.00 | 5,511.50 | 5,243.00 | **21,769.00** |
| 316 | 1,253.50 | 1,079.00 | 1,007.50 | 1,191.50 | **4,531.50** |
| 317 | 1,210.00 | 1,059.00 | 1,098.00 | 1,235.50 | **4,602.50** |
| 318 | 147.00 | 177.50 | 173.50 | 177.50 | **675.50** |
| 319 | 797.00 | 817.50 | 727.50 | 692.00 | **3,034.00** |
| 398 | 276.50 | 276.50 | 277.50 | 240.00 | **1,070.50** |
| **GA - GEORGIA - Total** | **52,107.00** | **52,478.00** | **50,380.50** | **50,074.00** | **205,039.50** |

**STATE: HI - HAWAII**

| REGISTRANT ZIP CODE 3 | 1ST QUARTER | 2ND QUARTER | 3RD QUARTER | 4TH QUARTER | **TOTAL GRAMS** |
|---|---|---|---|---|---|
| 967 | 1,261.00 | 1,002.00 | 875.50 | 883.00 | **4,021.50** |
| 968 | 1,240.00 | 1,029.00 | 921.00 | 870.50 | **4,060.50** |
| **HI - HAWAII - Total** | **2,501.00** | **2,031.00** | **1,796.50** | **1,753.50** | **8,082.00** |

**STATE: IA - IOWA**

| REGISTRANT ZIP CODE 3 | 1ST QUARTER | 2ND QUARTER | 3RD QUARTER | 4TH QUARTER | **TOTAL GRAMS** |
|---|---|---|---|---|---|
| 500 | 1,564.50 | 1,245.00 | 1,195.00 | 1,225.50 | **5,230.00** |
| 501 | 777.00 | 739.00 | 763.00 | 831.00 | **3,110.00** |
| 502 | 1,287.00 | 1,119.50 | 1,196.50 | 1,238.00 | **4,841.00** |
| 503 | 2,006.50 | 1,767.50 | 1,654.00 | 1,728.50 | **7,156.50** |
| 504 | 69.00 | 107.00 | 87.00 | 68.00 | **331.00** |
| 505 | 195.50 | 202.00 | 220.00 | 184.00 | **801.50** |
| 506 | 372.00 | 409.00 | 327.00 | 374.00 | **1,482.00** |
| 507 | 273.00 | 236.00 | 243.50 | 288.50 | **1,041.00** |
| 508 | 175.00 | 142.00 | 138.00 | 123.00 | **578.00** |
| 510 | 197.00 | 158.00 | 221.00 | 245.00 | **821.00** |
| 511 | 305.50 | 312.00 | 339.50 | 385.50 | **1,342.50** |
| 512 | 127.50 | 115.00 | 85.00 | 127.50 | **455.00** |
| 513 | 156.00 | 120.50 | 258.50 | 209.50 | **744.50** |
| 514 | 20.00 | 15.00 | 0 | 30.00 | **65.00** |
| 515 | 334.50 | 451.00 | 486.00 | 398.00 | **1,669.50** |
| 516 | 133.00 | 285.50 | 211.00 | 248.00 | **877.50** |
| 520 | 264.00 | 374.00 | 315.00 | 226.00 | **1,179.00** |

| | | | | | |
|---|---|---|---|---|---|
| 521 | 80.00 | 45.00 | 52.50 | 42.50 | **220.00** |
| 522 | 212.00 | 150.00 | 65.00 | 55.00 | **482.00** |
| 523 | 277.50 | 182.50 | 136.50 | 100.00 | **696.50** |
| 524 | 376.50 | 273.50 | 365.00 | 269.00 | **1,284.00** |
| 525 | 352.00 | 273.50 | 182.00 | 206.00 | **1,013.50** |
| 526 | 140.50 | 77.50 | 105.00 | 66.00 | **389.00** |
| 527 | 84.00 | 136.00 | 124.50 | 164.00 | **508.50** |
| 528 | 221.00 | 230.00 | 194.00 | 142.00 | **787.00** |
| **IA - IOWA - Total** | **10,000.50** | **9,166.00** | **8,964.50** | **8,974.50** | **37,105.50** |

**STATE: ID - IDAHO**

| REGISTRANT ZIP CODE 3 | 1ST QUARTER | 2ND QUARTER | 3RD QUARTER | 4TH QUARTER | **TOTAL GRAMS** |
|---|---|---|---|---|---|
| 832 | 1,341.00 | 1,350.50 | 1,415.50 | 1,111.00 | **5,218.00** |
| 833 | 389.00 | 426.00 | 406.00 | 449.50 | **1,670.50** |
| 834 | 686.00 | 772.00 | 854.00 | 850.00 | **3,162.00** |
| 835 | 183.00 | 195.00 | 115.00 | 103.00 | **596.00** |
| 836 | 1,378.50 | 1,509.50 | 1,387.00 | 1,263.00 | **5,538.00** |
| 837 | 994.50 | 1,041.50 | 789.00 | 629.00 | **3,454.00** |
| 838 | 508.00 | 545.00 | 631.00 | 504.00 | **2,188.00** |
| **ID - IDAHO - Total** | **5,480.00** | **5,839.50** | **5,597.50** | **4,909.50** | **21,826.50** |

**STATE: IL - ILLINOIS**

| REGISTRANT ZIP CODE 3 | 1ST QUARTER | 2ND QUARTER | 3RD QUARTER | 4TH QUARTER | **TOTAL GRAMS** |
|---|---|---|---|---|---|
| 600 | 4,875.00 | 4,931.50 | 4,680.00 | 4,731.50 | **19,218.00** |
| 601 | 2,569.00 | 2,755.00 | 2,334.00 | 2,584.50 | **10,242.50** |
| 602 | 253.50 | 223.50 | 196.50 | 169.50 | **843.00** |
| 603 | 355.50 | 377.50 | 419.00 | 376.00 | **1,528.00** |
| 604 | 3,254.00 | 3,351.00 | 3,337.50 | 3,067.50 | **13,010.00** |
| 605 | 3,173.50 | 2,899.00 | 3,046.00 | 3,181.50 | **12,300.00** |
| 606 | 1,938.00 | 1,863.00 | 1,596.00 | 1,556.00 | **6,953.00** |
| 607 | 330.00 | 404.50 | 252.50 | 229.50 | **1,216.50** |
| 608 | 313.50 | 180.50 | 204.00 | 264.50 | **962.50** |
| 609 | 597.50 | 584.00 | 705.50 | 621.00 | **2,508.00** |
| 610 | 700.50 | 806.50 | 823.50 | 748.50 | **3,079.00** |
| 611 | 1,116.00 | 925.00 | 848.50 | 829.50 | **3,719.00** |
| 612 | 150.50 | 151.50 | 125.00 | 153.00 | **580.00** |
| 613 | 57.00 | 128.00 | 208.00 | 135.00 | **528.00** |
| 614 | 80.00 | 88.00 | 80.00 | 15.50 | **263.50** |
| 615 | 453.00 | 423.00 | 393.00 | 470.00 | **1,739.00** |
| 616 | 306.50 | 232.00 | 301.00 | 236.00 | **1,075.50** |
| 617 | 755.00 | 602.50 | 704.00 | 703.00 | **2,764.50** |
| 618 | 1,880.50 | 1,663.00 | 1,580.50 | 1,638.50 | **6,762.50** |

| | | | | | |
|---|---|---|---|---|---|
| 619 | 215.00 | 253.00 | 190.00 | 251.50 | **909.50** |
| 620 | 1,000.00 | 942.00 | 828.00 | 986.00 | **3,756.00** |
| 622 | 2,997.50 | 2,887.50 | 2,597.50 | 2,739.00 | **11,221.50** |
| 623 | 279.00 | 344.00 | 272.00 | 264.00 | **1,159.00** |
| 624 | 105.00 | 165.00 | 126.00 | 136.50 | **532.50** |
| 625 | 769.00 | 770.50 | 660.50 | 577.50 | **2,777.50** |
| 626 | 113.00 | 132.00 | 106.00 | 109.00 | **460.00** |
| 627 | 131.50 | 188.00 | 152.00 | 187.00 | **658.50** |
| 628 | 352.50 | 433.00 | 374.50 | 455.00 | **1,615.00** |
| 629 | 360.00 | 469.00 | 390.50 | 375.50 | **1,595.00** |
| **IL - ILLINOIS - Total** | **29,481.00** | **29,173.00** | **27,531.50** | **27,791.50** | **113,977.00** |

**STATE: IN - INDIANA**

| REGISTRANT ZIP CODE 3 | 1ST QUARTER | 2ND QUARTER | 3RD QUARTER | 4TH QUARTER | TOTAL GRAMS |
|---|---|---|---|---|---|
| 460 | 1,397.50 | 1,374.50 | 1,099.00 | 1,141.50 | **5,012.50** |
| 461 | 2,189.00 | 1,935.50 | 2,293.00 | 2,285.50 | **8,703.00** |
| 462 | 1,885.00 | 1,963.00 | 1,638.50 | 1,671.50 | **7,158.00** |
| 463 | 3,830.00 | 3,523.50 | 3,399.50 | 3,452.50 | **14,205.50** |
| 464 | 412.00 | 331.00 | 338.50 | 335.50 | **1,417.00** |
| 465 | 2,371.00 | 2,318.50 | 2,053.00 | 2,076.00 | **8,818.50** |
| 466 | 629.50 | 712.00 | 694.00 | 578.50 | **2,614.00** |
| 467 | 2,602.00 | 2,524.00 | 2,282.50 | 2,489.00 | **9,897.50** |
| 468 | 3,681.00 | 3,730.50 | 3,444.00 | 3,416.00 | **14,271.50** |
| 469 | 1,021.00 | 1,297.00 | 1,050.50 | 1,124.00 | **4,492.50** |
| 470 | 295.50 | 336.50 | 269.00 | 265.00 | **1,166.00** |
| 471 | 930.50 | 930.50 | 794.00 | 669.50 | **3,324.50** |
| 472 | 616.00 | 583.00 | 554.00 | 618.50 | **2,371.50** |
| 473 | 534.00 | 602.00 | 671.00 | 663.00 | **2,470.00** |
| 474 | 688.50 | 642.50 | 656.00 | 619.00 | **2,606.00** |
| 475 | 654.50 | 600.50 | 757.50 | 736.00 | **2,748.50** |
| 476 | 508.00 | 611.50 | 548.00 | 660.50 | **2,328.00** |
| 477 | 1,216.50 | 1,341.00 | 1,331.00 | 1,162.50 | **5,051.00** |
| 478 | 4,919.00 | 4,841.00 | 4,327.00 | 4,311.50 | **18,398.50** |
| 479 | 977.00 | 828.50 | 771.50 | 686.50 | **3,263.50** |
| **IN - INDIANA - Total** | **31,357.50** | **31,026.50** | **28,971.50** | **28,962.00** | **120,317.50** |

**STATE: KS - KANSAS**

| REGISTRANT ZIP CODE 3 | 1ST QUARTER | 2ND QUARTER | 3RD QUARTER | 4TH QUARTER | TOTAL GRAMS |
|---|---|---|---|---|---|
| 660 | 1,838.50 | 1,795.50 | 1,648.50 | 1,514.00 | **6,796.50** |
| 661 | 213.50 | 161.00 | 188.00 | 183.50 | **746.00** |
| 662 | 1,486.50 | 1,313.50 | 1,378.50 | 1,204.50 | **5,383.00** |
| 664 | 196.00 | 185.00 | 138.00 | 165.00 | **684.00** |

| | | | | | |
|---|---|---|---|---|---|
| 665 | 259.00 | 270.00 | 327.50 | 216.50 | **1,073.00** |
| 666 | 882.00 | 762.00 | 735.00 | 770.00 | **3,149.00** |
| 667 | 354.00 | 247.00 | 195.50 | 210.00 | **1,006.50** |
| 668 | 102.50 | 90.00 | 104.50 | 128.00 | **425.00** |
| 669 | 0 | 0 | 7.50 | 11.00 | **18.50** |
| 670 | 94.00 | 96.00 | 161.50 | 116.00 | **467.50** |
| 671 | 156.00 | 118.00 | 114.00 | 146.00 | **534.00** |
| 672 | 616.50 | 608.00 | 663.00 | 575.00 | **2,462.50** |
| 673 | 310.00 | 205.00 | 187.00 | 136.00 | **838.00** |
| 674 | 299.50 | 201.00 | 207.00 | 164.00 | **871.50** |
| 675 | 121.00 | 169.00 | 138.00 | 150.00 | **578.00** |
| 676 | 191.50 | 90.00 | 74.00 | 94.00 | **449.50** |
| 677 | 69.00 | 106.00 | 68.00 | 69.00 | **312.00** |
| 678 | 65.00 | 30.00 | 42.00 | 24.00 | **161.00** |
| 679 | 5.00 | 9.00 | 0 | 0 | **14.00** |
| **KS - KANSAS - Total** | **7,259.50** | **6,456.00** | **6,377.50** | **5,876.50** | **25,969.50** |

**STATE: KY - KENTUCKY**

| REGISTRANT ZIP CODE 3 | 1ST QUARTER | 2ND QUARTER | 3RD QUARTER | 4TH QUARTER | TOTAL GRAMS |
|---|---|---|---|---|---|
| 400 | 530.50 | 530.00 | 584.50 | 492.50 | **2,137.50** |
| 401 | 244.00 | 344.50 | 243.50 | 278.00 | **1,110.00** |
| 402 | 2,283.00 | 2,116.50 | 1,735.50 | 1,748.50 | **7,883.50** |
| 403 | 599.00 | 656.50 | 519.00 | 498.50 | **2,273.00** |
| 404 | 430.00 | 328.00 | 311.00 | 318.50 | **1,387.50** |
| 405 | 605.50 | 727.00 | 494.50 | 591.00 | **2,418.00** |
| 406 | 52.00 | 28.00 | 65.00 | 30.00 | **175.00** |
| 407 | 85.50 | 175.50 | 186.00 | 127.50 | **574.50** |
| 408 | 6.00 | 0 | 3.00 | 46.00 | **55.00** |
| 409 | 15.00 | 20.00 | 22.50 | 35.00 | **92.50** |
| 410 | 2,118.50 | 2,071.00 | 1,788.50 | 1,958.00 | **7,936.00** |
| 411 | 194.00 | 194.50 | 143.00 | 171.00 | **702.50** |
| 412 | 45.00 | 35.50 | 30.50 | 59.50 | **170.50** |
| 413 | 7.50 | 0 | 0 | 0 | **7.50** |
| 414 | 0 | 0 | 0 | 10.00 | **10.00** |
| 415 | 152.50 | 128.00 | 116.00 | 124.00 | **520.50** |
| 416 | 223.00 | 131.00 | 118.50 | 88.50 | **561.00** |
| 417 | 43.50 | 88.50 | 47.50 | 54.00 | **233.50** |
| 418 | 70.00 | 43.00 | 39.00 | 35.00 | **187.00** |
| 420 | 1,482.50 | 1,684.50 | 1,682.00 | 1,511.50 | **6,360.50** |
| 421 | 403.00 | 368.00 | 285.50 | 515.50 | **1,572.00** |
| 422 | 859.00 | 465.00 | 1,005.00 | 581.00 | **2,910.00** |

| | | | | | |
|---|---|---|---|---|---|
| 423 | 1,116.50 | 1,069.00 | 1,110.50 | 1,140.00 | **4,436.00** |
| 424 | 556.50 | 429.00 | 433.00 | 417.00 | **1,835.50** |
| 425 | 35.00 | 50.00 | 30.00 | 35.00 | **150.00** |
| 426 | 21.00 | 25.00 | 15.00 | 25.00 | **86.00** |
| 427 | 592.50 | 541.00 | 406.50 | 460.50 | **2,000.50** |
| **KY - KENTUCKY - Total** | **12,770.50** | **12,249.00** | **11,415.00** | **11,351.00** | **47,785.50** |

**STATE: LA - LOUISIANA**

| REGISTRANT ZIP CODE 3 | 1ST QUARTER | 2ND QUARTER | 3RD QUARTER | 4TH QUARTER | TOTAL GRAMS |
|---|---|---|---|---|---|
| 700 | 1,768.50 | 1,635.50 | 1,682.50 | 1,635.00 | **6,721.50** |
| 701 | 160.00 | 177.00 | 126.50 | 165.00 | **628.50** |
| 703 | 501.00 | 492.50 | 594.50 | 403.00 | **1,991.00** |
| 704 | 2,480.00 | 2,537.00 | 2,560.00 | 2,889.00 | **10,466.00** |
| 705 | 1,305.00 | 1,536.50 | 1,262.00 | 1,279.50 | **5,383.00** |
| 706 | 431.00 | 414.00 | 397.00 | 392.50 | **1,634.50** |
| 707 | 2,551.50 | 2,529.00 | 2,549.50 | 2,468.00 | **10,098.00** |
| 708 | 1,214.50 | 1,182.50 | 1,056.00 | 949.00 | **4,402.00** |
| 710 | 237.50 | 285.50 | 315.50 | 221.50 | **1,060.00** |
| 711 | 1,115.00 | 1,220.50 | 1,214.50 | 1,260.50 | **4,810.50** |
| 712 | 387.50 | 404.00 | 411.00 | 386.50 | **1,589.00** |
| 713 | 769.50 | 711.50 | 576.50 | 677.50 | **2,735.00** |
| 714 | 309.50 | 453.00 | 357.50 | 403.50 | **1,523.50** |
| **LA - LOUISIANA - Total** | **13,230.50** | **13,578.50** | **13,103.00** | **13,130.50** | **53,042.50** |

**STATE: MA - MASSACHUSETTS**

| REGISTRANT ZIP CODE 3 | 1ST QUARTER | 2ND QUARTER | 3RD QUARTER | 4TH QUARTER | TOTAL GRAMS |
|---|---|---|---|---|---|
| 010 | 611.50 | 829.50 | 858.00 | 862.50 | **3,161.50** |
| 011 | 230.50 | 319.50 | 236.00 | 269.00 | **1,055.00** |
| 012 | 58.00 | 85.00 | 70.00 | 70.00 | **283.00** |
| 013 | 68.00 | 91.00 | 80.50 | 139.50 | **379.00** |
| 014 | 434.50 | 270.00 | 239.00 | 277.50 | **1,221.00** |
| 015 | 519.50 | 482.00 | 453.00 | 512.00 | **1,966.50** |
| 016 | 207.50 | 260.50 | 187.00 | 164.00 | **819.00** |
| 017 | 499.50 | 551.50 | 596.50 | 667.50 | **2,315.00** |
| 018 | 8,339.50 | 8,288.50 | 8,395.50 | 8,075.50 | **33,099.00** |
| 019 | 1,124.50 | 1,201.00 | 1,202.50 | 1,497.50 | **5,025.50** |
| 020 | 368.00 | 451.00 | 337.50 | 308.00 | **1,464.50** |
| 021 | 742.00 | 749.50 | 697.50 | 738.00 | **2,927.00** |
| 022 | 80.50 | 74.50 | 133.50 | 72.50 | **361.00** |
| 023 | 531.00 | 684.00 | 647.00 | 748.00 | **2,610.00** |
| 024 | 137.50 | 284.00 | 260.00 | 281.00 | **962.50** |
| 025 | 202.50 | 166.00 | 80.50 | 81.00 | **530.00** |

| | | | | | |
|---|---|---|---|---|---|
| 026 | 222.50 | 253.50 | 291.00 | 312.50 | **1,079.50** |
| 027 | 1,486.00 | 1,989.00 | 1,800.00 | 1,523.50 | **6,798.50** |
| **MA - MASSACHUSETTS - Total** | **15,863.00** | **17,030.00** | **16,565.00** | **16,599.50** | **66,057.50** |

**STATE: MD - MARYLAND**

| REGISTRANT ZIP CODE 3 | 1ST QUARTER | 2ND QUARTER | 3RD QUARTER | 4TH QUARTER | TOTAL GRAMS |
|---|---|---|---|---|---|
| 206 | 2,140.00 | 2,089.50 | 2,186.50 | 2,179.50 | **8,595.50** |
| 207 | 1,919.50 | 2,315.00 | 2,094.00 | 1,992.50 | **8,321.00** |
| 208 | 2,398.50 | 3,171.50 | 2,738.00 | 2,408.00 | **10,716.00** |
| 209 | 304.00 | 320.50 | 278.50 | 340.00 | **1,243.00** |
| 210 | 5,648.50 | 5,694.50 | 5,221.00 | 5,169.00 | **21,733.00** |
| 211 | 2,324.50 | 2,529.50 | 2,478.50 | 2,462.50 | **9,795.00** |
| 212 | 2,840.50 | 2,990.00 | 2,994.50 | 2,930.00 | **11,755.00** |
| 214 | 837.50 | 791.00 | 924.00 | 800.50 | **3,353.00** |
| 215 | 511.00 | 266.00 | 252.00 | 291.00 | **1,320.00** |
| 216 | 1,335.50 | 1,421.50 | 1,384.50 | 1,386.00 | **5,527.50** |
| 217 | 4,045.50 | 3,910.00 | 3,915.50 | 3,860.00 | **15,731.00** |
| 218 | 636.00 | 817.50 | 820.50 | 778.50 | **3,052.50** |
| 219 | 394.00 | 422.00 | 343.00 | 341.00 | **1,500.00** |
| **MD - MARYLAND - Total** | **25,335.00** | **26,738.50** | **25,630.50** | **24,938.50** | **102,642.50** |

**STATE: ME - MAINE**

| REGISTRANT ZIP CODE 3 | 1ST QUARTER | 2ND QUARTER | 3RD QUARTER | 4TH QUARTER | TOTAL GRAMS |
|---|---|---|---|---|---|
| 039 | 69.00 | 54.00 | 73.50 | 68.00 | **264.50** |
| 040 | 245.00 | 326.00 | 325.50 | 204.50 | **1,101.00** |
| 041 | 26.00 | 63.00 | 25.00 | 45.00 | **159.00** |
| 042 | 290.50 | 239.00 | 314.00 | 271.50 | **1,115.00** |
| 043 | 31.00 | 29.50 | 28.50 | 55.00 | **144.00** |
| 044 | 58.00 | 27.00 | 57.00 | 71.00 | **213.00** |
| 045 | 53.00 | 80.00 | 64.00 | 86.50 | **283.50** |
| 046 | 9.00 | 30.00 | 42.50 | 32.50 | **114.00** |
| 047 | 117.00 | 199.00 | 170.00 | 157.00 | **643.00** |
| 048 | 18.00 | 27.00 | 27.00 | 27.00 | **99.00** |
| 049 | 119.00 | 173.00 | 162.00 | 106.00 | **560.00** |
| **ME - MAINE - Total** | **1,035.50** | **1,247.50** | **1,289.00** | **1,124.00** | **4,696.00** |

**STATE: MI - MICHIGAN**

| REGISTRANT ZIP CODE 3 | 1ST QUARTER | 2ND QUARTER | 3RD QUARTER | 4TH QUARTER | TOTAL GRAMS |
|---|---|---|---|---|---|
| 480 | 2,430.50 | 2,163.00 | 1,889.00 | 1,764.50 | **8,247.00** |
| 481 | 2,820.00 | 2,821.50 | 2,508.50 | 2,679.50 | **10,829.50** |
| 482 | 228.50 | 271.50 | 195.00 | 235.50 | **930.50** |
| 483 | 2,228.50 | 1,924.00 | 1,633.50 | 1,637.50 | **7,423.50** |
| 484 | 326.00 | 337.00 | 452.00 | 477.00 | **1,592.00** |

| | | | | | |
|---|---|---|---|---|---|
| 485 | 542.00 | 493.00 | 480.50 | 537.00 | **2,052.50** |
| 486 | 609.50 | 640.00 | 566.50 | 449.50 | **2,265.50** |
| 487 | 457.00 | 286.00 | 415.00 | 328.00 | **1,486.00** |
| 488 | 1,500.50 | 1,510.00 | 1,514.50 | 1,363.50 | **5,888.50** |
| 489 | 389.00 | 315.50 | 270.50 | 293.50 | **1,268.50** |
| 490 | 1,855.00 | 1,931.00 | 1,986.50 | 1,682.00 | **7,454.50** |
| 491 | 368.50 | 393.00 | 382.50 | 413.00 | **1,557.00** |
| 492 | 458.50 | 360.00 | 409.50 | 434.50 | **1,662.50** |
| 493 | 1,057.50 | 867.00 | 787.00 | 784.00 | **3,495.50** |
| 494 | 610.00 | 619.00 | 676.00 | 598.50 | **2,503.50** |
| 495 | 1,054.50 | 764.00 | 789.50 | 752.00 | **3,360.00** |
| 496 | 561.00 | 606.50 | 574.50 | 729.00 | **2,471.00** |
| 497 | 361.00 | 549.00 | 506.00 | 599.00 | **2,015.00** |
| 498 | 262.00 | 175.00 | 155.00 | 125.00 | **717.00** |
| 499 | 55.00 | 70.00 | 137.00 | 49.00 | **311.00** |
| **MI - MICHIGAN - Total** | **18,174.50** | **17,096.00** | **16,328.50** | **15,931.50** | **67,530.50** |

**STATE: MN - MINNESOTA**

| REGISTRANT ZIP CODE 3 | 1ST QUARTER | 2ND QUARTER | 3RD QUARTER | 4TH QUARTER | **TOTAL GRAMS** |
|---|---|---|---|---|---|
| 550 | 644.50 | 563.00 | 476.50 | 434.50 | **2,118.50** |
| 551 | 539.00 | 634.00 | 638.50 | 757.50 | **2,569.00** |
| 553 | 555.00 | 399.50 | 435.50 | 325.50 | **1,715.50** |
| 554 | 846.00 | 709.50 | 596.00 | 641.50 | **2,793.00** |
| 556 | 5.00 | 0 | 10.00 | 20.00 | **35.00** |
| 557 | 18.00 | 46.00 | 28.00 | 10.00 | **102.00** |
| 558 | 61.00 | 83.00 | 141.00 | 97.00 | **382.00** |
| 559 | 120.00 | 135.00 | 137.50 | 144.00 | **536.50** |
| 560 | 234.00 | 190.00 | 263.00 | 216.00 | **903.00** |
| 561 | 288.00 | 178.00 | 292.50 | 260.00 | **1,018.50** |
| 562 | 76.50 | 62.00 | 69.50 | 73.00 | **281.00** |
| 563 | 155.00 | 198.00 | 144.50 | 250.00 | **747.50** |
| 564 | 74.00 | 40.00 | 42.00 | 36.00 | **192.00** |
| 565 | 147.00 | 148.00 | 133.00 | 125.00 | **553.00** |
| 566 | 34.50 | 85.50 | 76.00 | 85.50 | **281.50** |
| 567 | 165.00 | 173.00 | 205.50 | 179.00 | **722.50** |
| **MN - MINNESOTA - Total** | **3,962.50** | **3,644.50** | **3,689.00** | **3,654.50** | **14,950.50** |

**STATE: MO - MISSOURI**

| REGISTRANT ZIP CODE 3 | 1ST QUARTER | 2ND QUARTER | 3RD QUARTER | 4TH QUARTER | **TOTAL GRAMS** |
|---|---|---|---|---|---|
| 630 | 2,657.50 | 2,323.00 | 2,217.00 | 1,974.00 | **9,171.50** |
| 631 | 4,915.00 | 4,718.50 | 4,812.00 | 5,580.50 | **20,026.00** |
| 633 | 2,345.50 | 2,549.50 | 2,342.00 | 2,245.00 | **9,482.00** |

| | | | | | |
|---|---|---|---|---|---|
| 634 | 264.00 | 318.00 | 250.00 | 270.00 | **1,102.00** |
| 635 | 223.00 | 199.00 | 239.00 | 138.50 | **799.50** |
| 636 | 341.50 | 330.00 | 205.00 | 297.00 | **1,173.50** |
| 637 | 1,024.50 | 905.00 | 608.50 | 615.50 | **3,153.50** |
| 638 | 600.50 | 689.00 | 391.00 | 429.50 | **2,110.00** |
| 639 | 234.00 | 314.00 | 248.50 | 257.50 | **1,054.00** |
| 640 | 1,674.50 | 1,672.00 | 1,632.50 | 1,507.50 | **6,486.50** |
| 641 | 1,229.00 | 1,046.00 | 894.50 | 902.50 | **4,072.00** |
| 644 | 69.00 | 74.00 | 85.50 | 97.00 | **325.50** |
| 645 | 539.50 | 451.50 | 425.50 | 231.00 | **1,647.50** |
| 646 | 58.50 | 44.00 | 47.00 | 55.00 | **204.50** |
| 647 | 261.50 | 378.50 | 328.00 | 333.50 | **1,301.50** |
| 648 | 734.50 | 574.50 | 727.50 | 684.00 | **2,720.50** |
| 650 | 267.00 | 512.50 | 433.50 | 323.50 | **1,536.50** |
| 651 | 495.50 | 473.00 | 511.50 | 481.00 | **1,961.00** |
| 652 | 534.50 | 455.50 | 411.00 | 393.00 | **1,794.00** |
| 653 | 173.50 | 336.50 | 217.00 | 296.50 | **1,023.50** |
| 654 | 154.00 | 219.00 | 209.50 | 235.00 | **817.50** |
| 655 | 378.00 | 391.50 | 379.00 | 358.00 | **1,506.50** |
| 656 | 542.50 | 666.00 | 521.00 | 423.50 | **2,153.00** |
| 657 | 480.00 | 636.50 | 704.00 | 708.00 | **2,528.50** |
| 658 | 731.50 | 926.50 | 755.00 | 686.50 | **3,099.50** |
| **MO - MISSOURI - Total** | **20,928.50** | **21,203.50** | **19,595.00** | **19,523.00** | **81,250.00** |

**STATE: MS - MISSISSIPPI**

| REGISTRANT ZIP CODE 3 | 1ST QUARTER | 2ND QUARTER | 3RD QUARTER | 4TH QUARTER | **TOTAL GRAMS** |
|---|---|---|---|---|---|
| 386 | 844.50 | 707.00 | 759.00 | 720.00 | **3,030.50** |
| 387 | 206.00 | 219.00 | 232.00 | 260.00 | **917.00** |
| 388 | 1,077.50 | 1,036.50 | 1,113.00 | 1,032.00 | **4,259.00** |
| 389 | 196.00 | 178.50 | 117.50 | 111.00 | **603.00** |
| 390 | 1,016.00 | 941.50 | 934.00 | 894.50 | **3,786.00** |
| 391 | 829.00 | 749.50 | 704.50 | 589.50 | **2,872.50** |
| 392 | 945.00 | 915.00 | 835.00 | 790.00 | **3,485.00** |
| 393 | 590.50 | 627.00 | 558.00 | 594.00 | **2,369.50** |
| 394 | 1,297.50 | 1,083.00 | 1,038.00 | 1,041.50 | **4,460.00** |
| 395 | 2,150.00 | 1,786.00 | 1,753.00 | 1,376.00 | **7,065.00** |
| 396 | 194.50 | 229.00 | 301.00 | 250.00 | **974.50** |
| 397 | 531.50 | 478.00 | 590.00 | 564.50 | **2,164.00** |
| **MS - MISSISSIPPI - Total** | **9,878.00** | **8,950.00** | **8,935.00** | **8,223.00** | **35,986.00** |

**STATE: MT - MONTANA**

| REGISTRANT ZIP CODE 3 | 1ST QUARTER | 2ND QUARTER | 3RD QUARTER | 4TH QUARTER | **TOTAL GRAMS** |
|---|---|---|---|---|---|

| | | | | | |
|---|---|---|---|---|---|
| 590 | 161.00 | 166.00 | 155.00 | 186.00 | **668.00** |
| 591 | 346.50 | 267.50 | 310.00 | 328.50 | **1,252.50** |
| 593 | 95.00 | 92.00 | 68.00 | 60.00 | **315.00** |
| 594 | 219.00 | 179.00 | 178.00 | 146.00 | **722.00** |
| 595 | 32.00 | 56.00 | 75.50 | 83.00 | **246.50** |
| 596 | 68.00 | 95.00 | 88.00 | 95.00 | **346.00** |
| 597 | 291.50 | 421.50 | 370.00 | 349.50 | **1,432.50** |
| 598 | 200.50 | 179.00 | 190.00 | 259.50 | **829.00** |
| 599 | 77.00 | 92.00 | 90.00 | 57.00 | **316.00** |
| **MT - MONTANA - Total** | **1,490.50** | **1,548.00** | **1,524.50** | **1,564.50** | **6,127.50** |

**STATE: NC - NORTH CAROLINA**

| REGISTRANT ZIP CODE 3 | 1ST QUARTER | 2ND QUARTER | 3RD QUARTER | 4TH QUARTER | **TOTAL GRAMS** |
|---|---|---|---|---|---|
| 270 | 3,586.00 | 4,136.00 | 3,769.50 | 3,737.00 | **15,228.50** |
| 271 | 4,988.50 | 4,537.50 | 4,299.00 | 4,442.50 | **18,267.50** |
| 272 | 6,880.00 | 6,472.50 | 6,212.00 | 6,544.00 | **26,108.50** |
| 273 | 3,924.00 | 4,190.50 | 3,858.00 | 4,250.50 | **16,223.00** |
| 274 | 2,829.50 | 2,719.00 | 2,948.50 | 2,813.50 | **11,310.50** |
| 275 | 8,688.50 | 9,622.50 | 9,704.50 | 10,624.50 | **38,640.00** |
| 276 | 3,919.00 | 3,731.50 | 3,885.00 | 3,830.50 | **15,366.00** |
| 277 | 1,887.00 | 2,189.50 | 1,765.00 | 2,093.00 | **7,934.50** |
| 278 | 3,329.50 | 3,466.00 | 3,937.00 | 3,983.50 | **14,716.00** |
| 279 | 1,425.00 | 1,737.00 | 1,598.00 | 1,535.00 | **6,295.00** |
| 280 | 5,161.50 | 5,309.50 | 5,179.50 | 5,576.00 | **21,226.50** |
| 281 | 3,996.00 | 4,306.50 | 4,446.50 | 4,428.00 | **17,177.00** |
| 282 | 3,465.00 | 3,741.00 | 2,922.00 | 2,992.00 | **13,120.00** |
| 283 | 4,775.50 | 5,102.50 | 4,998.50 | 5,811.50 | **20,688.00** |
| 284 | 4,859.00 | 5,041.50 | 4,815.50 | 5,010.00 | **19,726.00** |
| 285 | 6,873.50 | 6,584.00 | 7,038.50 | 6,579.50 | **27,075.50** |
| 286 | 6,677.00 | 6,608.00 | 6,325.50 | 6,581.00 | **26,191.50** |
| 287 | 2,612.50 | 2,896.00 | 2,564.00 | 3,138.00 | **11,210.50** |
| 288 | 975.00 | 1,177.00 | 1,430.50 | 1,076.50 | **4,659.00** |
| 289 | 214.50 | 198.00 | 162.00 | 95.50 | **670.00** |
| **NC - NORTH CAROLINA - Total** | **81,066.50** | **83,766.00** | **81,859.00** | **85,142.00** | **331,833.50** |

**STATE: ND - NORTH DAKOTA**

| REGISTRANT ZIP CODE 3 | 1ST QUARTER | 2ND QUARTER | 3RD QUARTER | 4TH QUARTER | **TOTAL GRAMS** |
|---|---|---|---|---|---|
| 580 | 94.00 | 115.50 | 136.00 | 132.50 | **478.00** |
| 581 | 22.50 | 21.00 | 42.50 | 58.00 | **144.00** |
| 582 | 256.50 | 282.00 | 280.00 | 274.50 | **1,093.00** |
| 583 | 0 | 15.00 | 10.00 | 44.50 | **69.50** |
| 584 | 41.50 | 41.00 | 32.00 | 24.00 | **138.50** |

| | | | | | |
|---|---|---|---|---|---|
| 585 | 894.50 | 975.00 | 950.00 | 844.00 | **3,663.50** |
| 586 | 175.00 | 238.50 | 135.00 | 179.50 | **728.00** |
| 587 | 241.50 | 150.00 | 187.00 | 195.00 | **773.50** |
| 588 | 33.00 | 45.00 | 50.00 | 10.00 | **138.00** |
| **ND - NORTH DAKOTA - Total** | **1,758.50** | **1,883.00** | **1,822.50** | **1,762.00** | **7,226.00** |

**STATE: NE - NEBRASKA**

| REGISTRANT ZIP CODE 3 | 1ST QUARTER | 2ND QUARTER | 3RD QUARTER | 4TH QUARTER | TOTAL GRAMS |
|---|---|---|---|---|---|
| 680 | 711.50 | 799.00 | 1,032.50 | 1,011.50 | **3,554.50** |
| 681 | 4,192.00 | 4,043.50 | 4,446.50 | 4,758.50 | **17,440.50** |
| 683 | 184.00 | 153.00 | 184.00 | 176.00 | **697.00** |
| 684 | 190.00 | 214.00 | 228.50 | 221.50 | **854.00** |
| 685 | 1,493.00 | 1,307.00 | 1,147.00 | 1,394.50 | **5,341.50** |
| 686 | 359.00 | 423.00 | 541.00 | 395.50 | **1,718.50** |
| 687 | 288.50 | 342.00 | 293.00 | 322.50 | **1,246.00** |
| 688 | 973.00 | 1,229.50 | 950.00 | 959.00 | **4,111.50** |
| 689 | 137.50 | 249.00 | 181.00 | 171.00 | **738.50** |
| 690 | 52.00 | 42.00 | 70.00 | 78.00 | **242.00** |
| 691 | 225.00 | 242.00 | 255.50 | 297.00 | **1,019.50** |
| 692 | 10.00 | 0 | 0 | 5.00 | **15.00** |
| 693 | 31.00 | 9.00 | 19.00 | 17.00 | **76.00** |
| **NE - NEBRASKA - Total** | **8,846.50** | **9,053.00** | **9,348.00** | **9,807.00** | **37,054.50** |

**STATE: NH - NEW HAMPSHIRE**

| REGISTRANT ZIP CODE 3 | 1ST QUARTER | 2ND QUARTER | 3RD QUARTER | 4TH QUARTER | TOTAL GRAMS |
|---|---|---|---|---|---|
| 030 | 1,562.00 | 1,729.00 | 1,463.50 | 1,614.00 | **6,368.50** |
| 031 | 500.00 | 567.00 | 730.50 | 617.00 | **2,414.50** |
| 032 | 765.50 | 839.50 | 936.00 | 874.50 | **3,415.50** |
| 033 | 612.00 | 498.00 | 519.50 | 552.00 | **2,181.50** |
| 034 | 52.50 | 57.50 | 111.50 | 141.50 | **363.00** |
| 035 | 337.50 | 342.00 | 377.00 | 296.00 | **1,352.50** |
| 036 | 6.00 | 0 | 0 | 5.00 | **11.00** |
| 037 | 105.00 | 71.00 | 116.50 | 94.50 | **387.00** |
| 038 | 1,836.50 | 1,914.00 | 2,134.00 | 1,913.50 | **7,798.00** |
| **NH - NEW HAMPSHIRE - Total** | **5,777.00** | **6,018.00** | **6,388.50** | **6,108.00** | **24,291.50** |

**STATE: NJ - NEW JERSEY**

| REGISTRANT ZIP CODE 3 | 1ST QUARTER | 2ND QUARTER | 3RD QUARTER | 4TH QUARTER | TOTAL GRAMS |
|---|---|---|---|---|---|
| 070 | 5,441.50 | 5,136.00 | 4,155.00 | 4,546.00 | **19,278.50** |
| 071 | 467.50 | 439.50 | 414.50 | 361.00 | **1,682.50** |
| 072 | 341.00 | 233.00 | 176.50 | 253.50 | **1,004.00** |
| 073 | 215.00 | 258.50 | 352.00 | 272.50 | **1,098.00** |
| 074 | 1,823.50 | 1,627.50 | 1,712.50 | 1,872.50 | **7,036.00** |

| | | | | | |
|---|---|---|---|---|---|
| 075 | 152.00 | 308.00 | 240.50 | 337.00 | **1,037.50** |
| 076 | 1,997.50 | 2,146.50 | 2,145.00 | 1,890.50 | **8,179.50** |
| 077 | 3,540.50 | 3,366.50 | 3,248.50 | 3,220.00 | **13,375.50** |
| 078 | 1,585.00 | 1,664.50 | 1,593.00 | 1,509.50 | **6,352.00** |
| 079 | 885.00 | 803.00 | 759.00 | 799.50 | **3,246.50** |
| 080 | 5,820.50 | 6,338.00 | 5,868.50 | 6,145.50 | **24,172.50** |
| 081 | 395.50 | 279.50 | 276.00 | 342.00 | **1,293.00** |
| 082 | 1,528.00 | 1,652.50 | 1,521.00 | 1,192.00 | **5,893.50** |
| 083 | 1,420.00 | 1,306.50 | 1,424.00 | 1,146.50 | **5,297.00** |
| 084 | 0 | 20.00 | 16.00 | 6.00 | **42.00** |
| 085 | 1,631.50 | 1,474.50 | 1,271.50 | 1,161.50 | **5,539.00** |
| 086 | 1,165.00 | 1,098.00 | 836.00 | 963.50 | **4,062.50** |
| 087 | 2,471.00 | 2,527.50 | 2,140.00 | 2,312.00 | **9,450.50** |
| 088 | 3,911.50 | 4,004.50 | 3,794.50 | 3,396.00 | **15,106.50** |
| 089 | 378.00 | 389.00 | 383.00 | 444.50 | **1,594.50** |
| **NJ - NEW JERSEY - Total** | **35,169.50** | **35,073.00** | **32,327.00** | **32,171.50** | **134,741.00** |

**STATE: NM - NEW MEXICO**

| REGISTRANT ZIP CODE 3 | 1ST QUARTER | 2ND QUARTER | 3RD QUARTER | 4TH QUARTER | **TOTAL GRAMS** |
|---|---|---|---|---|---|
| 870 | 199.00 | 187.00 | 244.50 | 162.00 | **792.50** |
| 871 | 1,235.00 | 1,074.50 | 1,070.00 | 1,077.50 | **4,457.00** |
| 873 | 12.00 | 42.00 | 12.00 | 12.00 | **78.00** |
| 874 | 161.00 | 163.00 | 87.00 | 112.00 | **523.00** |
| 875 | 187.00 | 112.00 | 205.50 | 182.50 | **687.00** |
| 877 | 31.00 | 34.00 | 40.00 | 28.00 | **133.00** |
| 878 | 42.00 | 26.00 | 40.00 | 60.00 | **168.00** |
| 879 | 22.50 | 22.50 | 35.00 | 15.00 | **95.00** |
| 880 | 443.00 | 511.00 | 465.00 | 468.50 | **1,887.50** |
| 881 | 82.00 | 109.00 | 46.00 | 56.00 | **293.00** |
| 882 | 323.50 | 194.50 | 169.50 | 137.00 | **824.50** |
| 883 | 15.00 | 15.00 | 45.00 | 55.00 | **130.00** |
| 884 | 45.00 | 25.00 | 38.00 | 28.00 | **136.00** |
| **NM - NEW MEXICO - Total** | **2,798.00** | **2,515.50** | **2,497.50** | **2,393.50** | **10,204.50** |

**STATE: NV - NEVADA**

| REGISTRANT ZIP CODE 3 | 1ST QUARTER | 2ND QUARTER | 3RD QUARTER | 4TH QUARTER | **TOTAL GRAMS** |
|---|---|---|---|---|---|
| 890 | 2,337.50 | 2,152.00 | 2,162.50 | 2,315.50 | **8,967.50** |
| 891 | 6,208.50 | 5,480.00 | 4,825.00 | 4,563.50 | **21,077.00** |
| 893 | 18.00 | 9.00 | 0 | 0 | **27.00** |
| 894 | 454.00 | 455.50 | 438.00 | 521.00 | **1,868.50** |
| 895 | 543.00 | 436.50 | 402.50 | 567.00 | **1,949.00** |
| 897 | 181.00 | 167.00 | 138.00 | 78.50 | **564.50** |

| | | | | | |
|---|---|---|---|---|---|
| 898 | 90.50 | 77.00 | 92.00 | 54.00 | **313.50** |
| **NV - NEVADA - Total** | **9,832.50** | **8,777.00** | **8,058.00** | **8,099.50** | **34,767.00** |

**STATE: NY - NEW YORK**

| REGISTRANT ZIP CODE 3 | 1ST QUARTER | 2ND QUARTER | 3RD QUARTER | 4TH QUARTER | TOTAL GRAMS |
|---|---|---|---|---|---|
| 100 | 2,827.00 | 3,015.00 | 2,843.50 | 2,922.50 | **11,608.00** |
| 101 | 8.00 | 47.50 | 31.00 | 31.00 | **117.50** |
| 103 | 1,827.50 | 1,980.00 | 1,713.50 | 2,031.00 | **7,552.00** |
| 104 | 1,093.50 | 1,124.50 | 1,260.00 | 1,236.00 | **4,714.00** |
| 105 | 1,301.50 | 1,133.00 | 993.50 | 1,026.50 | **4,454.50** |
| 106 | 120.50 | 177.50 | 117.50 | 81.50 | **497.00** |
| 107 | 466.00 | 544.00 | 428.50 | 391.00 | **1,829.50** |
| 108 | 95.00 | 215.00 | 155.50 | 138.50 | **604.00** |
| 109 | 1,600.00 | 1,505.50 | 1,268.00 | 1,229.50 | **5,603.00** |
| 110 | 1,739.50 | 1,475.00 | 1,656.50 | 1,600.00 | **6,471.00** |
| 111 | 1,087.00 | 1,098.50 | 975.50 | 1,107.50 | **4,268.50** |
| 112 | 3,105.00 | 3,309.50 | 3,030.50 | 2,981.50 | **12,426.50** |
| 113 | 2,342.50 | 2,278.00 | 2,090.00 | 2,096.00 | **8,806.50** |
| 114 | 732.50 | 770.50 | 661.00 | 802.50 | **2,966.50** |
| 115 | 3,417.50 | 3,961.50 | 3,727.50 | 3,451.00 | **14,557.50** |
| 116 | 56.50 | 83.50 | 66.00 | 82.00 | **288.00** |
| 117 | 10,901.00 | 12,388.00 | 11,258.00 | 11,915.00 | **46,462.00** |
| 118 | 480.50 | 715.50 | 647.50 | 683.50 | **2,527.00** |
| 119 | 1,211.00 | 1,390.00 | 1,221.50 | 1,400.50 | **5,223.00** |
| 120 | 519.00 | 556.50 | 527.00 | 443.50 | **2,046.00** |
| 121 | 280.50 | 278.00 | 303.50 | 276.00 | **1,138.00** |
| 122 | 1,009.50 | 1,027.00 | 1,078.50 | 868.50 | **3,983.50** |
| 123 | 152.00 | 258.50 | 190.00 | 244.00 | **844.50** |
| 124 | 582.00 | 546.00 | 524.50 | 472.00 | **2,124.50** |
| 125 | 1,577.00 | 1,442.00 | 1,340.50 | 1,460.00 | **5,819.50** |
| 126 | 118.00 | 118.00 | 192.00 | 195.50 | **623.50** |
| 127 | 291.00 | 411.00 | 394.50 | 412.50 | **1,509.00** |
| 128 | 1,191.50 | 1,136.50 | 1,151.00 | 1,196.50 | **4,675.50** |
| 129 | 730.00 | 558.50 | 579.50 | 795.00 | **2,663.00** |
| 130 | 4,301.50 | 4,570.00 | 4,267.00 | 4,281.50 | **17,420.00** |
| 131 | 1,047.00 | 937.50 | 894.50 | 913.00 | **3,792.00** |
| 132 | 1,730.00 | 1,396.00 | 1,500.00 | 1,570.00 | **6,196.00** |
| 133 | 1,327.00 | 1,393.00 | 1,423.50 | 1,280.00 | **5,423.50** |
| 134 | 1,478.50 | 1,357.50 | 1,381.50 | 1,586.50 | **5,804.00** |
| 135 | 704.50 | 599.50 | 632.50 | 561.00 | **2,497.50** |
| 136 | 3,110.00 | 2,410.00 | 2,397.50 | 2,446.00 | **10,363.50** |

| | | | | | |
|---|---|---|---|---|---|
| 137 | 657.50 | 770.50 | 773.00 | 685.00 | **2,886.00** |
| 138 | 1,266.50 | 1,315.00 | 1,307.00 | 1,287.00 | **5,175.50** |
| 139 | 944.50 | 1,036.00 | 1,041.50 | 995.00 | **4,017.00** |
| 140 | 5,087.50 | 4,749.50 | 4,423.50 | 4,015.50 | **18,276.00** |
| 141 | 2,173.00 | 1,759.50 | 1,604.00 | 1,661.00 | **7,197.50** |
| 142 | 5,552.50 | 5,268.50 | 4,833.50 | 4,648.50 | **20,303.00** |
| 143 | 811.00 | 723.50 | 720.50 | 554.00 | **2,809.00** |
| 144 | 1,638.50 | 1,557.50 | 1,494.00 | 1,439.00 | **6,129.00** |
| 145 | 1,450.50 | 1,514.00 | 1,503.00 | 1,559.00 | **6,026.50** |
| 146 | 1,994.00 | 1,800.50 | 1,912.50 | 1,734.50 | **7,441.50** |
| 147 | 463.00 | 620.50 | 565.00 | 523.50 | **2,172.00** |
| 148 | 1,650.50 | 1,732.50 | 1,638.00 | 1,588.50 | **6,609.50** |
| 149 | 195.00 | 168.00 | 159.00 | 146.50 | **668.50** |
| **NY - NEW YORK - Total** | **78,445.00** | **79,223.00** | **74,897.00** | **75,045.50** | **307,610.50** |

**STATE: OH - OHIO**

| REGISTRANT ZIP CODE 3 | 1ST QUARTER | 2ND QUARTER | 3RD QUARTER | 4TH QUARTER | **TOTAL GRAMS** |
|---|---|---|---|---|---|
| 430 | 2,558.00 | 2,652.50 | 2,585.00 | 2,794.00 | **10,589.50** |
| 431 | 1,934.00 | 1,774.50 | 1,958.50 | 2,016.00 | **7,683.00** |
| 432 | 5,687.45 | 4,277.45 | 3,188.83 | 2,742.33 | **15,896.05** |
| 433 | 496.00 | 582.00 | 502.00 | 475.50 | **2,055.50** |
| 434 | 895.00 | 856.50 | 791.50 | 739.50 | **3,282.50** |
| 435 | 2,775.50 | 2,733.00 | 2,497.50 | 2,364.50 | **10,370.50** |
| 436 | 4,653.50 | 4,097.00 | 3,617.00 | 3,277.00 | **15,644.50** |
| 437 | 259.00 | 271.00 | 211.00 | 221.00 | **962.00** |
| 438 | 139.00 | 114.50 | 201.50 | 149.50 | **604.50** |
| 439 | 988.50 | 888.50 | 847.50 | 706.00 | **3,430.50** |
| 440 | 4,691.50 | 4,677.00 | 4,120.00 | 4,071.00 | **17,559.50** |
| 441 | 2,830.50 | 2,929.00 | 2,462.50 | 2,480.00 | **10,702.00** |
| 442 | 2,300.00 | 2,290.00 | 2,323.50 | 2,284.00 | **9,197.50** |
| 443 | 592.50 | 628.50 | 748.50 | 709.00 | **2,678.50** |
| 444 | 1,035.00 | 1,204.00 | 1,095.50 | 972.50 | **4,307.00** |
| 445 | 791.00 | 803.50 | 701.00 | 675.50 | **2,971.00** |
| 446 | 2,581.50 | 2,506.00 | 2,417.50 | 2,499.00 | **10,004.00** |
| 447 | 997.50 | 1,097.50 | 1,300.50 | 1,179.50 | **4,575.00** |
| 448 | 572.50 | 573.00 | 627.00 | 540.00 | **2,312.50** |
| 449 | 164.00 | 224.50 | 243.50 | 178.50 | **810.50** |
| 450 | 2,766.00 | 2,865.50 | 2,858.50 | 2,723.00 | **11,213.00** |
| 451 | 1,192.00 | 1,157.00 | 1,081.00 | 1,184.50 | **4,614.50** |
| 452 | 2,457.00 | 2,472.00 | 2,347.50 | 2,623.50 | **9,900.00** |
| 453 | 1,374.50 | 1,647.50 | 1,449.00 | 1,406.00 | **5,877.00** |

| | | | | | |
|---|---|---|---|---|---|
| 454 | 1,381.50 | 1,764.00 | 1,657.50 | 1,652.50 | **6,455.50** |
| 455 | 338.00 | 256.00 | 457.50 | 471.00 | **1,522.50** |
| 456 | 1,196.00 | 1,270.00 | 1,204.00 | 1,101.50 | **4,771.50** |
| 457 | 220.50 | 180.00 | 194.00 | 138.50 | **733.00** |
| 458 | 790.00 | 815.00 | 751.00 | 647.50 | **3,003.50** |
| **OH - OHIO - Total** | **48,657.45** | **47,606.95** | **44,439.83** | **43,022.33** | **183,726.55** |

**STATE: OK - OKLAHOMA**

| REGISTRANT ZIP CODE 3 | 1ST QUARTER | 2ND QUARTER | 3RD QUARTER | 4TH QUARTER | **TOTAL GRAMS** |
|---|---|---|---|---|---|
| 730 | 4,718.00 | 4,522.00 | 3,842.00 | 3,056.50 | **16,138.50** |
| 731 | 2,718.00 | 3,021.00 | 2,378.00 | 1,811.50 | **9,928.50** |
| 734 | 985.00 | 888.50 | 736.50 | 492.00 | **3,102.00** |
| 735 | 900.00 | 712.00 | 642.50 | 500.50 | **2,755.00** |
| 736 | 194.00 | 167.00 | 104.00 | 144.00 | **609.00** |
| 737 | 168.50 | 158.50 | 140.00 | 143.50 | **610.50** |
| 738 | 46.00 | 55.00 | 55.00 | 26.00 | **182.00** |
| 739 | 20.00 | 15.00 | 32.50 | 6.00 | **73.50** |
| 740 | 4,364.50 | 3,373.50 | 2,984.00 | 2,208.50 | **12,930.50** |
| 741 | 3,055.50 | 2,328.50 | 1,877.50 | 1,050.00 | **8,311.50** |
| 743 | 380.50 | 287.00 | 370.00 | 204.00 | **1,241.50** |
| 744 | 708.50 | 749.50 | 618.00 | 462.50 | **2,538.50** |
| 745 | 250.50 | 240.00 | 241.50 | 195.00 | **927.00** |
| 746 | 123.00 | 187.50 | 91.00 | 43.00 | **444.50** |
| 747 | 30.00 | 65.50 | 77.00 | 46.00 | **218.50** |
| 748 | 910.50 | 776.50 | 654.00 | 330.00 | **2,671.00** |
| 749 | 99.00 | 71.00 | 48.00 | 46.00 | **264.00** |
| **OK - OKLAHOMA - Total** | **19,671.50** | **17,618.00** | **14,891.50** | **10,765.00** | **62,946.00** |

**STATE: OR - OREGON**

| REGISTRANT ZIP CODE 3 | 1ST QUARTER | 2ND QUARTER | 3RD QUARTER | 4TH QUARTER | **TOTAL GRAMS** |
|---|---|---|---|---|---|
| 970 | 1,554.50 | 1,448.50 | 1,419.00 | 1,357.50 | **5,779.50** |
| 971 | 278.00 | 245.00 | 277.00 | 188.00 | **988.00** |
| 972 | 770.00 | 677.00 | 684.50 | 597.50 | **2,729.00** |
| 973 | 1,161.50 | 1,207.50 | 1,148.50 | 871.50 | **4,389.00** |
| 974 | 934.50 | 935.50 | 882.50 | 840.00 | **3,592.50** |
| 975 | 1,281.00 | 1,127.00 | 1,101.50 | 1,058.50 | **4,568.00** |
| 976 | 191.00 | 210.50 | 110.00 | 97.00 | **608.50** |
| 977 | 363.00 | 463.00 | 432.00 | 318.00 | **1,576.00** |
| 978 | 181.50 | 146.00 | 77.00 | 67.00 | **471.50** |
| 979 | 12.00 | 48.50 | 23.00 | 9.00 | **92.50** |
| **OR - OREGON - Total** | **6,727.00** | **6,508.50** | **6,155.00** | **5,404.00** | **24,794.50** |

**STATE: PA - PENNSYLVANIA**

| REGISTRANT ZIP CODE 3 | 1ST QUARTER | 2ND QUARTER | 3RD QUARTER | 4TH QUARTER | **TOTAL GRAMS** |
|---|---|---|---|---|---|
| 150 | 842.00 | 1,173.50 | 1,225.50 | 1,176.00 | **4,417.00** |
| 151 | 692.00 | 771.50 | 538.50 | 623.00 | **2,625.00** |
| 152 | 508.50 | 637.00 | 534.00 | 645.00 | **2,324.50** |
| 153 | 638.00 | 553.50 | 379.00 | 401.00 | **1,971.50** |
| 154 | 123.00 | 114.00 | 188.00 | 115.00 | **540.00** |
| 155 | 515.50 | 367.00 | 287.50 | 226.50 | **1,396.50** |
| 156 | 356.50 | 382.00 | 412.00 | 323.50 | **1,474.00** |
| 157 | 178.50 | 209.00 | 112.50 | 130.00 | **630.00** |
| 158 | 199.00 | 218.00 | 233.00 | 144.00 | **794.00** |
| 159 | 779.50 | 515.50 | 461.00 | 424.00 | **2,180.00** |
| 160 | 276.50 | 268.00 | 312.50 | 256.50 | **1,113.50** |
| 161 | 659.50 | 482.50 | 567.50 | 621.50 | **2,331.00** |
| 162 | 302.00 | 301.50 | 307.50 | 356.50 | **1,267.50** |
| 163 | 416.00 | 492.00 | 509.00 | 512.50 | **1,929.50** |
| 164 | 240.00 | 165.00 | 183.50 | 234.00 | **822.50** |
| 165 | 636.50 | 634.00 | 653.50 | 537.00 | **2,461.00** |
| 166 | 705.50 | 696.00 | 650.00 | 670.00 | **2,721.50** |
| 167 | 85.00 | 92.50 | 70.00 | 45.00 | **292.50** |
| 168 | 787.00 | 715.50 | 631.00 | 660.50 | **2,794.00** |
| 169 | 84.00 | 43.00 | 60.00 | 50.00 | **237.00** |
| 170 | 3,140.50 | 2,680.00 | 2,092.00 | 2,115.00 | **10,027.50** |
| 171 | 823.50 | 736.00 | 545.50 | 645.00 | **2,750.00** |
| 172 | 880.50 | 1,046.00 | 720.00 | 972.00 | **3,618.50** |
| 173 | 2,604.00 | 2,344.00 | 2,014.50 | 2,143.50 | **9,106.00** |
| 174 | 1,634.00 | 1,197.00 | 760.00 | 667.50 | **4,258.50** |
| 175 | 337.50 | 383.50 | 392.50 | 352.50 | **1,466.00** |
| 176 | 383.00 | 373.00 | 452.00 | 317.50 | **1,525.50** |
| 177 | 275.00 | 104.00 | 145.00 | 115.00 | **639.00** |
| 178 | 266.50 | 240.00 | 234.00 | 212.50 | **953.00** |
| 179 | 363.50 | 408.50 | 372.00 | 318.00 | **1,462.00** |
| 180 | 5,155.00 | 5,010.00 | 5,003.50 | 4,538.50 | **19,707.00** |
| 181 | 2,310.00 | 2,204.00 | 2,289.50 | 2,066.50 | **8,870.00** |
| 182 | 442.00 | 382.00 | 591.00 | 514.00 | **1,929.00** |
| 183 | 2,558.00 | 2,812.50 | 2,430.00 | 2,490.50 | **10,291.00** |
| 184 | 1,027.00 | 1,197.00 | 1,012.50 | 1,052.00 | **4,288.50** |
| 185 | 539.00 | 531.50 | 532.00 | 644.50 | **2,247.00** |
| 186 | 218.00 | 336.00 | 412.50 | 335.50 | **1,302.00** |
| 187 | 5,018.00 | 5,629.50 | 5,821.00 | 5,811.50 | **22,280.00** |
| 188 | 101.00 | 112.00 | 95.00 | 93.50 | **401.50** |

| | | | | | |
|---|---|---|---|---|---|
| 189 | 1,786.50 | 1,619.50 | 1,580.50 | 1,495.00 | **6,481.50** |
| 190 | 6,555.00 | 6,917.00 | 6,664.00 | 6,811.00 | **26,947.00** |
| 191 | 1,810.00 | 2,055.50 | 1,641.50 | 1,820.00 | **7,327.00** |
| 193 | 1,736.00 | 1,587.50 | 1,559.50 | 1,760.50 | **6,643.50** |
| 194 | 1,938.50 | 1,976.50 | 2,014.50 | 2,020.50 | **7,950.00** |
| 195 | 1,036.00 | 1,009.50 | 914.00 | 967.00 | **3,926.50** |
| 196 | 910.00 | 1,069.50 | 937.00 | 964.00 | **3,880.50** |
| **PA - PENNSYLVANIA - Total** | **52,872.50** | **52,792.50** | **49,541.00** | **49,394.50** | **204,600.50** |

**STATE: PR - PUERTO RICO**

| REGISTRANT ZIP CODE 3 | 1ST QUARTER | 2ND QUARTER | 3RD QUARTER | 4TH QUARTER | TOTAL GRAMS |
|---|---|---|---|---|---|
| 006 | 38.00 | 30.00 | 30.00 | 35.00 | **133.00** |
| 007 | 90.50 | 70.00 | 20.00 | 25.00 | **205.50** |
| 009 | 358.50 | 474.50 | 392.00 | 392.50 | **1,617.50** |
| **PR - PUERTO RICO - Total** | **487.00** | **574.50** | **442.00** | **452.50** | **1,956.00** |

**STATE: RI - RHODE ISLAND**

| REGISTRANT ZIP CODE 3 | 1ST QUARTER | 2ND QUARTER | 3RD QUARTER | 4TH QUARTER | TOTAL GRAMS |
|---|---|---|---|---|---|
| 028 | 1,120.50 | 1,579.00 | 1,345.00 | 1,292.50 | **5,337.00** |
| 029 | 607.50 | 584.50 | 470.50 | 569.00 | **2,231.50** |
| **RI - RHODE ISLAND - Total** | **1,728.00** | **2,163.50** | **1,815.50** | **1,861.50** | **7,568.50** |

**STATE: SC - SOUTH CAROLINA**

| REGISTRANT ZIP CODE 3 | 1ST QUARTER | 2ND QUARTER | 3RD QUARTER | 4TH QUARTER | TOTAL GRAMS |
|---|---|---|---|---|---|
| 290 | 2,325.50 | 2,413.50 | 2,711.00 | 2,696.00 | **10,146.00** |
| 291 | 2,419.00 | 2,449.00 | 2,519.50 | 2,199.50 | **9,587.00** |
| 292 | 2,857.00 | 3,199.00 | 3,051.00 | 3,002.00 | **12,109.00** |
| 293 | 3,252.50 | 3,275.50 | 2,930.50 | 2,804.50 | **12,263.00** |
| 294 | 3,822.50 | 3,597.00 | 3,461.50 | 3,271.50 | **14,152.50** |
| 295 | 4,176.50 | 4,057.00 | 3,983.00 | 4,073.50 | **16,290.00** |
| 296 | 11,426.00 | 12,894.50 | 11,484.00 | 10,746.50 | **46,551.00** |
| 297 | 2,088.00 | 1,508.50 | 1,451.00 | 1,565.00 | **6,612.50** |
| 298 | 1,647.00 | 1,834.50 | 1,793.50 | 1,654.50 | **6,929.50** |
| 299 | 1,670.00 | 1,697.50 | 1,615.50 | 1,607.50 | **6,590.50** |
| **SC - SOUTH CAROLINA - Total** | **35,684.00** | **36,926.00** | **35,000.50** | **33,620.50** | **141,231.00** |

**STATE: SD - SOUTH DAKOTA**

| REGISTRANT ZIP CODE 3 | 1ST QUARTER | 2ND QUARTER | 3RD QUARTER | 4TH QUARTER | TOTAL GRAMS |
|---|---|---|---|---|---|
| 570 | 665.50 | 664.50 | 640.50 | 663.50 | **2,634.00** |
| 571 | 1,210.00 | 1,161.50 | 930.50 | 1,024.00 | **4,326.00** |
| 572 | 285.00 | 199.00 | 244.50 | 211.50 | **940.00** |
| 573 | 293.00 | 302.50 | 247.00 | 261.00 | **1,103.50** |
| 574 | 602.50 | 539.00 | 456.50 | 534.00 | **2,132.00** |
| 575 | 60.00 | 70.00 | 110.00 | 107.50 | **347.50** |

| | | | | | |
|---|---|---|---|---|---|
| 576 | 171.50 | 129.00 | 125.00 | 67.00 | **492.50** |
| 577 | 840.00 | 725.00 | 718.00 | 609.50 | **2,892.50** |
| **SD - SOUTH DAKOTA - Total** | **4,127.50** | **3,790.50** | **3,472.00** | **3,478.00** | **14,868.00** |

**STATE: TN - TENNESSEE**

| REGISTRANT ZIP CODE 3 | 1ST QUARTER | 2ND QUARTER | 3RD QUARTER | 4TH QUARTER | TOTAL GRAMS |
|---|---|---|---|---|---|
| 370 | 4,698.00 | 4,990.00 | 4,186.50 | 4,412.50 | **18,287.00** |
| 371 | 4,248.00 | 4,829.00 | 4,391.50 | 4,542.00 | **18,010.50** |
| 372 | 1,770.00 | 2,023.50 | 1,968.00 | 1,635.00 | **7,396.50** |
| 373 | 3,189.00 | 2,979.00 | 2,731.50 | 2,561.50 | **11,461.00** |
| 374 | 1,772.50 | 1,721.50 | 1,363.00 | 1,423.50 | **6,280.50** |
| 376 | 1,710.00 | 2,040.50 | 2,418.50 | 2,378.50 | **8,547.50** |
| 377 | 634.50 | 671.00 | 738.00 | 638.50 | **2,682.00** |
| 378 | 1,411.50 | 1,355.00 | 1,194.00 | 1,192.50 | **5,153.00** |
| 379 | 953.00 | 830.50 | 838.50 | 762.00 | **3,384.00** |
| 380 | 1,165.50 | 1,185.00 | 943.50 | 729.50 | **4,023.50** |
| 381 | 1,256.00 | 1,368.00 | 968.50 | 981.50 | **4,574.00** |
| 382 | 737.50 | 742.50 | 576.00 | 525.00 | **2,581.00** |
| 383 | 1,269.00 | 1,330.00 | 1,429.00 | 1,182.00 | **5,210.00** |
| 384 | 865.50 | 682.50 | 662.00 | 699.50 | **2,909.50** |
| 385 | 779.00 | 798.50 | 715.00 | 678.00 | **2,970.50** |
| **TN - TENNESSEE - Total** | **26,459.00** | **27,546.50** | **25,123.50** | **24,341.50** | **103,470.50** |

**STATE: TX - TEXAS**

| REGISTRANT ZIP CODE 3 | 1ST QUARTER | 2ND QUARTER | 3RD QUARTER | 4TH QUARTER | TOTAL GRAMS |
|---|---|---|---|---|---|
| 750 | 10,695.00 | 10,768.00 | 10,421.50 | 10,133.00 | **42,017.50** |
| 751 | 3,211.50 | 2,589.50 | 2,892.50 | 2,891.50 | **11,585.00** |
| 752 | 3,212.00 | 2,841.00 | 2,472.00 | 2,884.00 | **11,409.00** |
| 753 | 3.00 | 3.00 | 0 | 0 | **6.00** |
| 754 | 839.00 | 952.50 | 838.50 | 795.00 | **3,425.00** |
| 755 | 64.00 | 99.00 | 94.00 | 72.00 | **329.00** |
| 756 | 533.50 | 483.00 | 369.50 | 441.00 | **1,827.00** |
| 757 | 1,071.50 | 1,135.50 | 911.50 | 880.00 | **3,998.50** |
| 758 | 370.50 | 395.00 | 278.00 | 299.50 | **1,343.00** |
| 759 | 294.00 | 279.50 | 264.50 | 209.50 | **1,047.50** |
| 760 | 5,892.50 | 5,299.50 | 4,960.00 | 4,821.00 | **20,973.00** |
| 761 | 3,722.00 | 3,227.50 | 3,093.50 | 3,217.00 | **13,260.00** |
| 762 | 2,083.00 | 1,978.00 | 1,828.50 | 1,791.00 | **7,680.50** |
| 763 | 892.50 | 983.00 | 1,125.00 | 954.50 | **3,955.00** |
| 764 | 191.00 | 182.00 | 214.00 | 271.00 | **858.00** |
| 765 | 906.00 | 1,018.00 | 1,018.00 | 941.50 | **3,883.50** |
| 766 | 139.00 | 126.00 | 114.00 | 184.00 | **563.00** |

| | | | | | |
|---|---|---|---|---|---|
| 767 | 226.50 | 343.50 | 247.50 | 342.00 | **1,159.50** |
| 768 | 27.00 | 19.00 | 25.00 | 23.00 | **94.00** |
| 769 | 306.00 | 223.00 | 266.00 | 345.00 | **1,140.00** |
| 770 | 3,956.00 | 4,075.00 | 4,023.50 | 3,373.00 | **15,427.50** |
| 773 | 3,946.00 | 3,504.00 | 3,883.50 | 3,415.00 | **14,748.50** |
| 774 | 4,014.50 | 4,037.95 | 4,039.00 | 4,010.00 | **16,101.45** |
| 775 | 4,128.00 | 3,809.00 | 3,512.00 | 3,830.00 | **15,279.00** |
| 776 | 918.00 | 648.50 | 706.50 | 513.00 | **2,786.00** |
| 777 | 300.50 | 218.00 | 227.50 | 193.50 | **939.50** |
| 778 | 534.50 | 534.50 | 527.50 | 272.50 | **1,869.00** |
| 779 | 112.50 | 98.00 | 176.00 | 207.00 | **593.50** |
| 780 | 1,459.00 | 1,338.50 | 1,315.50 | 1,171.50 | **5,284.50** |
| 781 | 1,918.00 | 1,887.50 | 1,441.50 | 1,510.00 | **6,757.00** |
| 782 | 10,204.50 | 9,668.00 | 9,395.00 | 7,770.50 | **37,038.00** |
| 783 | 409.00 | 463.00 | 349.00 | 476.00 | **1,697.00** |
| 784 | 1,089.00 | 1,058.00 | 1,006.00 | 1,013.00 | **4,166.00** |
| 785 | 404.00 | 326.00 | 400.50 | 392.00 | **1,522.50** |
| 786 | 3,282.00 | 2,943.50 | 3,014.50 | 2,896.00 | **12,136.00** |
| 787 | 4,257.50 | 4,096.50 | 3,554.50 | 3,324.00 | **15,232.50** |
| 788 | 149.00 | 235.50 | 253.00 | 163.00 | **800.50** |
| 789 | 186.00 | 217.00 | 174.00 | 183.00 | **760.00** |
| 790 | 299.00 | 370.50 | 287.50 | 354.00 | **1,311.00** |
| 791 | 1,027.00 | 964.00 | 840.50 | 805.50 | **3,637.00** |
| 792 | 0 | 0 | 23.00 | 5.00 | **28.00** |
| 793 | 111.00 | 83.00 | 68.00 | 92.00 | **354.00** |
| 794 | 1,039.00 | 898.00 | 794.00 | 908.00 | **3,639.00** |
| 795 | 16.00 | 0 | 6.00 | 16.00 | **38.00** |
| 796 | 60.00 | 72.00 | 66.00 | 39.50 | **237.50** |
| 797 | 260.50 | 204.50 | 203.00 | 162.50 | **830.50** |
| 799 | 2,347.50 | 1,975.50 | 1,791.00 | 1,897.00 | **8,011.00** |
| **TX - TEXAS - Total** | **81,107.50** | **76,671.45** | **73,511.50** | **70,488.00** | **301,778.45** |

**STATE: UT - UTAH**

| REGISTRANT ZIP CODE 3 | 1ST QUARTER | 2ND QUARTER | 3RD QUARTER | 4TH QUARTER | TOTAL GRAMS |
|---|---|---|---|---|---|
| 840 | 4,791.00 | 4,532.00 | 4,597.50 | 4,654.00 | **18,574.50** |
| 841 | 1,425.00 | 1,558.00 | 1,410.50 | 1,359.00 | **5,752.50** |
| 843 | 513.50 | 490.00 | 427.50 | 357.00 | **1,788.00** |
| 844 | 887.50 | 794.50 | 838.00 | 754.50 | **3,274.50** |
| 845 | 654.00 | 669.50 | 625.00 | 844.00 | **2,792.50** |
| 846 | 965.50 | 1,005.50 | 754.00 | 898.50 | **3,623.50** |
| 847 | 1,833.00 | 1,446.50 | 1,539.00 | 1,604.00 | **6,422.50** |

| UT - UTAH - Total | 11,069.50 | 10,496.00 | 10,191.50 | 10,471.00 | 42,228.00 |
|---|---|---|---|---|---|

STATE: VA - VIRGINIA

| REGISTRANT ZIP CODE 3 | 1ST QUARTER | 2ND QUARTER | 3RD QUARTER | 4TH QUARTER | TOTAL GRAMS |
|---|---|---|---|---|---|
| 201 | 3,748.50 | 3,713.50 | 3,454.50 | 3,548.00 | **14,464.50** |
| 220 | 1,647.00 | 1,434.00 | 1,444.50 | 1,477.00 | **6,002.50** |
| 221 | 1,633.50 | 1,651.50 | 1,467.50 | 1,426.50 | **6,179.00** |
| 222 | 1,295.00 | 1,080.50 | 985.00 | 960.00 | **4,320.50** |
| 223 | 985.50 | 1,003.50 | 844.50 | 864.50 | **3,698.00** |
| 224 | 1,327.00 | 1,246.00 | 1,118.50 | 1,091.00 | **4,782.50** |
| 225 | 465.50 | 471.50 | 455.50 | 426.00 | **1,818.50** |
| 226 | 977.00 | 1,116.50 | 944.50 | 892.00 | **3,930.00** |
| 227 | 140.00 | 140.50 | 141.00 | 215.00 | **636.50** |
| 228 | 522.50 | 731.00 | 925.50 | 922.50 | **3,101.50** |
| 229 | 407.50 | 472.50 | 345.00 | 403.00 | **1,628.00** |
| 230 | 463.00 | 300.00 | 224.50 | 249.00 | **1,236.50** |
| 231 | 1,303.50 | 1,111.00 | 1,038.50 | 955.00 | **4,408.00** |
| 232 | 1,541.00 | 1,506.50 | 1,441.50 | 1,248.00 | **5,737.00** |
| 233 | 2,079.00 | 1,646.50 | 1,667.00 | 1,507.50 | **6,900.00** |
| 234 | 5,532.50 | 4,923.00 | 4,551.50 | 4,048.00 | **19,055.00** |
| 235 | 1,050.50 | 1,015.50 | 874.00 | 713.00 | **3,653.00** |
| 236 | 1,649.00 | 1,466.00 | 1,102.00 | 1,004.50 | **5,221.50** |
| 237 | 580.50 | 908.00 | 516.00 | 492.00 | **2,496.50** |
| 238 | 897.50 | 1,006.00 | 952.00 | 1,039.50 | **3,895.00** |
| 239 | 162.00 | 251.50 | 236.50 | 261.00 | **911.00** |
| 240 | 917.00 | 1,173.50 | 1,006.50 | 1,214.00 | **4,311.00** |
| 241 | 954.00 | 1,109.50 | 1,029.50 | 1,138.00 | **4,231.00** |
| 242 | 729.50 | 830.50 | 930.00 | 907.00 | **3,397.00** |
| 243 | 1,030.00 | 727.00 | 723.50 | 944.00 | **3,424.50** |
| 244 | 363.00 | 331.50 | 384.00 | 349.50 | **1,428.00** |
| 245 | 1,132.00 | 1,226.00 | 1,122.50 | 1,143.50 | **4,624.00** |
| 246 | 153.00 | 176.00 | 118.00 | 179.50 | **626.50** |
| **VA - VIRGINIA - Total** | **33,686.00** | **32,769.00** | **30,043.50** | **29,618.50** | **126,117.00** |

STATE: VI - VIRGIN ISLANDS

| REGISTRANT ZIP CODE 3 | 1ST QUARTER | 2ND QUARTER | 3RD QUARTER | 4TH QUARTER | TOTAL GRAMS |
|---|---|---|---|---|---|
| 008 | 0 | 0 | 5.00 | 0 | **5.00** |
| **VI - VIRGIN ISLANDS - Total** | **0.00** | **0.00** | **5.00** | **0.00** | **5.00** |

STATE: VT - VERMONT

| REGISTRANT ZIP CODE 3 | 1ST QUARTER | 2ND QUARTER | 3RD QUARTER | 4TH QUARTER | TOTAL GRAMS |
|---|---|---|---|---|---|
| 050 | 18.00 | 59.00 | 90.00 | 23.00 | **190.00** |
| 051 | 0 | 6.00 | 11.00 | 0 | **17.00** |

| | | | | | |
|---|---|---|---|---|---|
| 052 | 141.00 | 174.50 | 184.00 | 164.00 | **663.50** |
| 054 | 258.00 | 350.50 | 277.50 | 250.50 | **1,136.50** |
| 056 | 106.00 | 80.00 | 104.00 | 89.00 | **379.00** |
| 057 | 343.00 | 234.00 | 335.50 | 312.00 | **1,224.50** |
| 058 | 50.00 | 63.00 | 98.00 | 67.00 | **278.00** |
| **VT - VERMONT - Total** | **916.00** | **967.00** | **1,100.00** | **905.50** | **3,888.50** |

**STATE: WA - WASHINGTON**

| REGISTRANT ZIP CODE 3 | 1ST QUARTER | 2ND QUARTER | 3RD QUARTER | 4TH QUARTER | TOTAL GRAMS |
|---|---|---|---|---|---|
| 980 | 2,394.50 | 2,252.00 | 2,285.00 | 2,128.00 | **9,059.50** |
| 981 | 852.00 | 942.50 | 929.50 | 1,057.00 | **3,781.00** |
| 982 | 926.00 | 848.00 | 970.00 | 780.50 | **3,524.50** |
| 983 | 2,264.50 | 2,183.50 | 2,085.00 | 1,927.50 | **8,460.50** |
| 984 | 666.50 | 764.00 | 651.50 | 697.50 | **2,779.50** |
| 985 | 799.00 | 798.50 | 570.00 | 696.50 | **2,864.00** |
| 986 | 1,092.50 | 1,140.50 | 1,024.50 | 1,444.00 | **4,701.50** |
| 988 | 497.50 | 460.50 | 491.00 | 393.00 | **1,842.00** |
| 989 | 440.50 | 335.50 | 269.00 | 347.50 | **1,392.50** |
| 990 | 304.00 | 403.00 | 415.00 | 573.50 | **1,695.50** |
| 991 | 202.00 | 199.00 | 156.00 | 102.00 | **659.00** |
| 992 | 2,865.00 | 2,973.50 | 3,061.00 | 3,118.00 | **12,017.50** |
| 993 | 1,063.50 | 1,141.00 | 936.50 | 801.50 | **3,942.50** |
| 994 | 59.50 | 41.00 | 28.00 | 60.50 | **189.00** |
| **WA - WASHINGTON - Total** | **14,427.00** | **14,482.50** | **13,872.00** | **14,127.00** | **56,908.50** |

**STATE: WI - WISCONSIN**

| REGISTRANT ZIP CODE 3 | 1ST QUARTER | 2ND QUARTER | 3RD QUARTER | 4TH QUARTER | TOTAL GRAMS |
|---|---|---|---|---|---|
| 530 | 2,219.50 | 2,091.50 | 2,096.50 | 2,252.50 | **8,660.00** |
| 531 | 1,949.00 | 1,547.00 | 1,597.00 | 1,543.50 | **6,636.50** |
| 532 | 1,966.00 | 1,955.00 | 1,548.50 | 1,634.50 | **7,104.00** |
| 534 | 554.50 | 606.50 | 569.00 | 520.00 | **2,250.00** |
| 535 | 805.00 | 690.50 | 653.50 | 655.00 | **2,804.00** |
| 537 | 175.00 | 133.50 | 102.15 | 142.50 | **553.15** |
| 538 | 44.00 | 38.50 | 29.00 | 9.00 | **120.50** |
| 539 | 179.00 | 148.50 | 265.00 | 214.00 | **806.50** |
| 540 | 517.00 | 444.50 | 467.00 | 255.00 | **1,683.50** |
| 541 | 331.00 | 301.50 | 319.50 | 338.50 | **1,290.50** |
| 542 | 174.50 | 243.50 | 214.00 | 240.00 | **872.00** |
| 543 | 376.00 | 441.00 | 514.00 | 477.50 | **1,808.50** |
| 544 | 737.50 | 613.00 | 676.00 | 646.50 | **2,673.00** |
| 545 | 279.00 | 234.00 | 127.00 | 134.00 | **774.00** |
| 546 | 63.00 | 49.00 | 42.00 | 25.00 | **179.00** |

| | | | | | |
|---|---|---|---|---|---|
| 547 | 205.00 | 332.50 | 209.50 | 207.50 | **954.50** |
| 548 | 155.00 | 132.00 | 118.00 | 69.00 | **474.00** |
| 549 | 1,176.50 | 1,215.00 | 1,227.50 | 1,103.00 | **4,722.00** |
| **WI - WISCONSIN - Total** | **11,906.50** | **11,217.00** | **10,775.15** | **10,467.00** | **44,365.65** |

**STATE: WV - WEST VIRGINIA**

| REGISTRANT ZIP CODE 3 | 1ST QUARTER | 2ND QUARTER | 3RD QUARTER | 4TH QUARTER | TOTAL GRAMS |
|---|---|---|---|---|---|
| 247 | 202.00 | 110.00 | 112.00 | 93.00 | **517.00** |
| 249 | 46.00 | 86.00 | 54.00 | 60.00 | **246.00** |
| 250 | 82.50 | 127.50 | 36.50 | 71.00 | **317.50** |
| 251 | 65.50 | 50.00 | 45.00 | 125.00 | **285.50** |
| 252 | 15.00 | 22.50 | 42.00 | 55.50 | **135.00** |
| 253 | 211.00 | 232.50 | 180.00 | 107.00 | **730.50** |
| 254 | 1,222.00 | 1,095.00 | 959.00 | 798.50 | **4,074.50** |
| 255 | 362.00 | 491.00 | 424.50 | 450.00 | **1,727.50** |
| 256 | 30.00 | 29.00 | 18.00 | 28.00 | **105.00** |
| 257 | 487.00 | 483.50 | 404.00 | 290.00 | **1,664.50** |
| 258 | 88.50 | 94.50 | 72.50 | 18.00 | **273.50** |
| 259 | 22.00 | 17.50 | 10.00 | 5.00 | **54.50** |
| 260 | 899.50 | 744.50 | 774.00 | 804.00 | **3,222.00** |
| 261 | 402.00 | 373.50 | 244.00 | 207.00 | **1,226.50** |
| 262 | 49.00 | 15.00 | 30.00 | 15.00 | **109.00** |
| 263 | 113.00 | 55.50 | 15.00 | 9.00 | **192.50** |
| 265 | 243.00 | 252.00 | 182.00 | 172.00 | **849.00** |
| 266 | 21.00 | 35.50 | 44.50 | 40.50 | **141.50** |
| 267 | 60.00 | 72.50 | 82.50 | 67.00 | **282.00** |
| 268 | 77.00 | 32.00 | 54.00 | 57.00 | **220.00** |
| **WV - WEST VIRGINIA - Total** | **4,698.00** | **4,419.50** | **3,783.50** | **3,472.50** | **16,373.50** |

**STATE: WY - WYOMING**

| REGISTRANT ZIP CODE 3 | 1ST QUARTER | 2ND QUARTER | 3RD QUARTER | 4TH QUARTER | TOTAL GRAMS |
|---|---|---|---|---|---|
| 820 | 937.50 | 809.00 | 910.50 | 896.00 | **3,553.00** |
| 822 | 44.50 | 39.00 | 64.00 | 32.00 | **179.50** |
| 823 | 15.00 | 35.00 | 37.50 | 0 | **87.50** |
| 824 | 59.00 | 68.00 | 78.00 | 43.00 | **248.00** |
| 825 | 0 | 0 | 5.00 | 20.00 | **25.00** |
| 826 | 263.00 | 175.00 | 195.00 | 113.00 | **746.00** |
| 827 | 251.50 | 221.00 | 207.00 | 182.50 | **862.00** |
| 828 | 88.00 | 65.00 | 40.00 | 93.00 | **286.00** |
| 829 | 111.00 | 95.00 | 81.00 | 102.50 | **389.50** |
| 830 | 6.00 | 0 | 0 | 0 | **6.00** |
| 831 | 5.00 | 3.00 | 6.00 | 0 | **14.00** |

| WY - WYOMING - Total | 1,780.50 | 1,510.00 | 1,624.00 | 1,482.00 | 6,396.50 |
|---|---|---|---|---|---|

**DRUG: 9801 - FENTANYL BASE**

**STATE: AK - ALASKA**

| REGISTRANT ZIP CODE 3 | 1ST QUARTER | 2ND QUARTER | 3RD QUARTER | 4TH QUARTER | TOTAL GRAMS |
|---|---|---|---|---|---|
| 995 | 92.61 | 73.72 | 73.15 | 64.53 | **304.01** |
| 996 | 87.38 | 78.33 | 71.53 | 52.17 | **289.41** |
| 997 | 24.30 | 22.74 | 19.71 | 17.61 | **84.36** |
| 998 | 11.86 | 7.65 | 7.74 | 7.39 | **34.64** |
| 999 | 8.42 | 7.02 | 4.59 | 4.71 | **24.74** |
| **AK - ALASKA - Total** | **224.57** | **189.47** | **176.71** | **146.41** | **737.16** |

**STATE: AL - ALABAMA**

| REGISTRANT ZIP CODE 3 | 1ST QUARTER | 2ND QUARTER | 3RD QUARTER | 4TH QUARTER | TOTAL GRAMS |
|---|---|---|---|---|---|
| 350 | 142.17 | 130.11 | 117.87 | 102.89 | **493.04** |
| 351 | 81.61 | 77.13 | 68.42 | 60.90 | **288.06** |
| 352 | 134.90 | 120.22 | 110.32 | 103.03 | **468.47** |
| 354 | 77.59 | 74.17 | 69.19 | 65.39 | **286.35** |
| 355 | 52.55 | 48.28 | 48.00 | 39.38 | **188.21** |
| 356 | 80.91 | 65.20 | 61.89 | 64.28 | **272.28** |
| 357 | 32.31 | 26.12 | 21.16 | 18.97 | **98.56** |
| 358 | 60.71 | 57.09 | 50.10 | 43.38 | **211.28** |
| 359 | 107.78 | 100.62 | 90.63 | 80.44 | **379.48** |
| 360 | 70.65 | 59.87 | 53.36 | 49.04 | **232.93** |
| 361 | 71.29 | 64.71 | 63.95 | 58.13 | **258.09** |
| 362 | 40.76 | 39.76 | 37.33 | 34.04 | **151.90** |
| 363 | 76.60 | 73.95 | 73.29 | 65.45 | **289.30** |
| 364 | 31.05 | 27.06 | 25.57 | 23.44 | **107.12** |
| 365 | 83.37 | 69.32 | 68.48 | 69.33 | **290.49** |
| 366 | 68.15 | 62.86 | 54.55 | 50.48 | **236.04** |
| 367 | 22.79 | 20.23 | 20.07 | 23.08 | **86.16** |
| 368 | 57.71 | 55.28 | 52.85 | 48.38 | **214.21** |
| 369 | 5.80 | 5.83 | 5.35 | 4.18 | **21.16** |
| **AL - ALABAMA - Total** | **1,298.70** | **1,177.82** | **1,092.40** | **1,004.20** | **4,573.12** |

**STATE: AR - ARKANSAS**

| REGISTRANT ZIP CODE 3 | 1ST QUARTER | 2ND QUARTER | 3RD QUARTER | 4TH QUARTER | TOTAL GRAMS |
|---|---|---|---|---|---|
| 716 | 36.72 | 28.61 | 26.07 | 22.93 | **114.33** |
| 717 | 27.60 | 24.89 | 23.39 | 22.86 | **98.74** |
| 718 | 21.51 | 20.00 | 19.70 | 17.01 | **78.21** |
| 719 | 64.73 | 54.28 | 54.87 | 50.17 | **224.05** |
| 720 | 86.18 | 74.06 | 63.06 | 56.27 | **279.57** |
| 721 | 114.22 | 93.88 | 81.03 | 75.95 | **365.08** |

| | | | | | |
|---|---|---|---|---|---|
| 722 | 80.42 | 69.88 | 60.43 | 58.34 | **269.07** |
| 723 | 21.45 | 18.64 | 16.03 | 14.86 | **70.99** |
| 724 | 52.73 | 49.53 | 44.30 | 42.68 | **189.24** |
| 725 | 42.07 | 37.32 | 34.49 | 33.78 | **147.66** |
| 726 | 35.81 | 30.90 | 30.94 | 26.43 | **124.09** |
| 727 | 126.86 | 104.20 | 96.46 | 92.61 | **420.13** |
| 728 | 29.11 | 23.46 | 20.89 | 19.41 | **92.87** |
| 729 | 43.70 | 41.02 | 41.98 | 40.47 | **167.16** |
| **AR - ARKANSAS - Total** | **783.11** | **670.68** | **613.65** | **573.76** | **2,641.19** |

**STATE: AS - AMERICAN SAMOA**

| REGISTRANT ZIP CODE 3 | 1ST QUARTER | 2ND QUARTER | 3RD QUARTER | 4TH QUARTER | TOTAL GRAMS |
|---|---|---|---|---|---|
| 967 | 0.01 | 0.05 | 0.26 | 0.05 | **0.37** |
| **AS - AMERICAN SAMOA - Tota** | **0.01** | **0.05** | **0.26** | **0.05** | **0.37** |

**STATE: AZ - ARIZONA**

| REGISTRANT ZIP CODE 3 | 1ST QUARTER | 2ND QUARTER | 3RD QUARTER | 4TH QUARTER | TOTAL GRAMS |
|---|---|---|---|---|---|
| 850 | 241.13 | 223.95 | 192.21 | 184.40 | **841.70** |
| 851 | 55.52 | 42.87 | 32.82 | 36.09 | **167.30** |
| 852 | 500.19 | 413.11 | 419.79 | 309.69 | **1,642.77** |
| 853 | 244.96 | 207.15 | 180.22 | 180.25 | **812.58** |
| 855 | 21.17 | 18.40 | 15.12 | 13.71 | **68.39** |
| 856 | 52.65 | 45.90 | 42.45 | 41.27 | **182.27** |
| 857 | 228.10 | 205.58 | 186.39 | 191.08 | **811.15** |
| 859 | 13.08 | 12.59 | 13.96 | 13.45 | **53.07** |
| 860 | 25.09 | 20.92 | 17.76 | 15.39 | **79.16** |
| 863 | 51.36 | 49.83 | 48.75 | 41.02 | **190.95** |
| 864 | 59.79 | 42.78 | 35.61 | 33.62 | **171.79** |
| 865 | 0.78 | 1.51 | 0.62 | 1.55 | **4.47** |
| **AZ - ARIZONA - Total** | **1,493.82** | **1,284.59** | **1,185.69** | **1,061.51** | **5,025.61** |

**STATE: CA - CALIFORNIA**

| REGISTRANT ZIP CODE 3 | 1ST QUARTER | 2ND QUARTER | 3RD QUARTER | 4TH QUARTER | TOTAL GRAMS |
|---|---|---|---|---|---|
| 900 | 154.28 | 144.35 | 124.45 | 117.39 | **540.47** |
| 902 | 84.73 | 77.16 | 70.20 | 68.41 | **300.50** |
| 903 | 4.12 | 5.29 | 4.10 | 4.59 | **18.10** |
| 904 | 60.19 | 55.85 | 51.74 | 55.63 | **223.40** |
| 905 | 32.97 | 38.06 | 31.08 | 32.89 | **135.00** |
| 906 | 74.57 | 70.14 | 69.18 | 63.39 | **277.28** |
| 907 | 69.21 | 61.86 | 57.24 | 55.08 | **243.40** |
| 908 | 152.82 | 218.71 | 116.99 | 177.88 | **666.39** |
| 910 | 46.80 | 48.67 | 43.41 | 41.12 | **180.00** |
| 911 | 20.69 | 21.39 | 17.10 | 15.91 | **75.09** |

| | | | | | |
|---|---|---|---|---|---|
| 912 | 35.01 | 38.08 | 33.89 | 33.13 | **140.12** |
| 913 | 188.75 | 176.87 | 162.39 | 158.99 | **687.00** |
| 914 | 69.75 | 67.29 | 65.34 | 63.67 | **266.05** |
| 915 | 11.12 | 13.48 | 8.97 | 9.77 | **43.34** |
| 916 | 7.43 | 6.16 | 6.19 | 4.91 | **24.68** |
| 917 | 265.76 | 262.56 | 242.35 | 237.47 | **1,008.14** |
| 918 | 6.56 | 7.06 | 6.78 | 4.45 | **24.85** |
| 919 | 43.80 | 41.55 | 33.95 | 29.32 | **148.61** |
| 920 | 298.09 | 276.46 | 237.69 | 227.27 | **1,039.52** |
| 921 | 328.04 | 249.60 | 261.24 | 225.15 | **1,064.03** |
| 922 | 196.24 | 176.05 | 160.11 | 140.15 | **672.55** |
| 923 | 239.19 | 202.41 | 184.75 | 170.04 | **796.39** |
| 924 | 41.46 | 33.95 | 26.14 | 25.71 | **127.27** |
| 925 | 236.74 | 224.46 | 212.49 | 202.53 | **876.21** |
| 926 | 239.23 | 230.53 | 208.02 | 190.11 | **867.90** |
| 927 | 42.71 | 49.29 | 39.67 | 42.10 | **173.78** |
| 928 | 189.80 | 175.62 | 166.09 | 165.67 | **697.18** |
| 930 | 103.94 | 100.11 | 88.59 | 77.91 | **370.54** |
| 931 | 21.21 | 20.52 | 17.87 | 18.58 | **78.18** |
| 932 | 102.52 | 89.66 | 74.34 | 74.71 | **341.23** |
| 933 | 125.31 | 108.78 | 96.20 | 86.92 | **417.20** |
| 934 | 106.17 | 99.31 | 89.57 | 92.17 | **387.21** |
| 935 | 82.92 | 72.61 | 69.82 | 61.38 | **286.73** |
| 936 | 61.60 | 55.20 | 45.42 | 46.96 | **209.19** |
| 937 | 95.92 | 99.47 | 85.77 | 79.84 | **361.00** |
| 939 | 29.76 | 28.71 | 24.45 | 21.00 | **103.93** |
| 940 | 110.99 | 100.07 | 92.44 | 83.18 | **386.69** |
| 941 | 124.35 | 99.46 | 93.92 | 91.50 | **409.22** |
| 943 | 33.03 | 31.65 | 26.07 | 28.24 | **118.98** |
| 944 | 14.78 | 13.74 | 12.88 | 11.74 | **53.14** |
| 945 | 404.44 | 336.43 | 323.91 | 306.91 | **1,371.69** |
| 946 | 42.27 | 35.74 | 38.16 | 33.41 | **149.58** |
| 947 | 13.90 | 12.78 | 10.22 | 10.06 | **46.95** |
| 948 | 10.85 | 11.71 | 11.50 | 10.80 | **44.86** |
| 949 | 51.85 | 43.80 | 42.63 | 36.29 | **174.56** |
| 950 | 101.30 | 89.73 | 84.23 | 82.85 | **358.10** |
| 951 | 57.92 | 63.23 | 52.77 | 45.59 | **219.52** |
| 952 | 95.88 | 82.99 | 86.95 | 74.95 | **340.77** |
| 953 | 233.12 | 215.23 | 196.71 | 189.21 | **834.26** |
| 954 | 95.85 | 96.27 | 81.53 | 74.99 | **348.64** |

| | | | | | |
|---|---|---|---|---|---|
| 955 | 41.06 | 36.50 | 33.85 | 34.23 | **145.64** |
| 956 | 288.60 | 256.60 | 229.88 | 232.20 | **1,007.28** |
| 957 | 46.13 | 35.79 | 34.97 | 30.51 | **147.41** |
| 958 | 183.89 | 161.44 | 147.55 | 137.29 | **630.16** |
| 959 | 152.99 | 137.82 | 121.28 | 116.33 | **528.43** |
| 960 | 106.64 | 88.55 | 73.00 | 56.71 | **324.91** |
| 961 | 16.35 | 13.69 | 12.19 | 10.39 | **52.62** |
| **CA - CALIFORNIA - Total** | **6,095.60** | **5,610.45** | **5,040.20** | **4,819.59** | **21,565.83** |

**STATE: CO - COLORADO**

| REGISTRANT ZIP CODE 3 | 1ST QUARTER | 2ND QUARTER | 3RD QUARTER | 4TH QUARTER | **TOTAL GRAMS** |
|---|---|---|---|---|---|
| 800 | 175.51 | 158.86 | 167.25 | 154.07 | **655.69** |
| 801 | 146.49 | 131.22 | 124.05 | 115.70 | **517.46** |
| 802 | 191.85 | 154.84 | 139.50 | 128.42 | **614.61** |
| 803 | 28.18 | 26.50 | 23.09 | 23.09 | **100.85** |
| 804 | 39.37 | 37.39 | 39.21 | 34.87 | **150.85** |
| 805 | 123.23 | 108.48 | 111.08 | 93.30 | **436.09** |
| 806 | 47.41 | 43.25 | 37.80 | 31.96 | **160.41** |
| 807 | 21.19 | 20.57 | 18.42 | 16.15 | **76.33** |
| 808 | 27.24 | 22.62 | 20.05 | 19.05 | **88.97** |
| 809 | 128.24 | 121.07 | 112.88 | 101.60 | **463.78** |
| 810 | 63.13 | 50.58 | 47.54 | 43.90 | **205.15** |
| 811 | 10.61 | 12.02 | 10.99 | 10.98 | **44.61** |
| 812 | 20.18 | 18.76 | 17.26 | 16.64 | **72.85** |
| 813 | 13.81 | 11.51 | 11.38 | 10.20 | **46.90** |
| 814 | 12.63 | 9.82 | 9.44 | 8.46 | **40.36** |
| 815 | 47.78 | 41.00 | 31.70 | 27.29 | **147.77** |
| 816 | 18.03 | 16.41 | 14.74 | 14.12 | **63.30** |
| **CO - COLORADO - Total** | **1,114.87** | **984.90** | **936.41** | **849.80** | **3,885.98** |

**STATE: CT - CONNECTICUT**

| REGISTRANT ZIP CODE 3 | 1ST QUARTER | 2ND QUARTER | 3RD QUARTER | 4TH QUARTER | **TOTAL GRAMS** |
|---|---|---|---|---|---|
| 060 | 155.06 | 137.40 | 130.58 | 120.52 | **543.57** |
| 061 | 42.66 | 33.92 | 35.38 | 33.30 | **145.26** |
| 062 | 37.13 | 35.76 | 30.52 | 29.37 | **132.77** |
| 063 | 62.25 | 54.30 | 52.82 | 56.75 | **226.11** |
| 064 | 216.66 | 191.03 | 206.26 | 189.26 | **803.21** |
| 065 | 77.60 | 73.29 | 62.65 | 68.14 | **281.69** |
| 066 | 33.48 | 29.27 | 26.89 | 25.93 | **115.57** |
| 067 | 77.89 | 70.30 | 67.69 | 65.23 | **281.11** |
| 068 | 153.79 | 73.85 | 68.07 | 62.83 | **358.54** |
| 069 | 11.68 | 10.16 | 10.91 | 9.52 | **42.28** |

| | | | | | |
|---|---|---|---|---|---|
| **CT - CONNECTICUT - Total** | **868.21** | **709.28** | **691.77** | **660.86** | **2,930.12** |

**STATE: DC - DISTRICT OF COLUMBIA**

| REGISTRANT ZIP CODE 3 | 1ST QUARTER | 2ND QUARTER | 3RD QUARTER | 4TH QUARTER | TOTAL GRAMS |
|---|---|---|---|---|---|
| 200 | 73.21 | 69.29 | 65.86 | 63.22 | **271.58** |
| 203 | 0.04 | 0 | 0 | 0 | **0.04** |
| 204 | 5.42 | 4.45 | 2.51 | 3.33 | **15.71** |
| **DC - DISTRICT OF COLUMBIA -** | **78.67** | **73.74** | **68.38** | **66.54** | **287.34** |

**STATE: DE - DELAWARE**

| REGISTRANT ZIP CODE 3 | 1ST QUARTER | 2ND QUARTER | 3RD QUARTER | 4TH QUARTER | TOTAL GRAMS |
|---|---|---|---|---|---|
| 197 | 125.03 | 107.72 | 93.20 | 83.33 | **409.28** |
| 198 | 63.40 | 52.76 | 48.78 | 43.59 | **208.53** |
| 199 | 125.97 | 99.91 | 88.70 | 75.63 | **390.21** |
| **DE - DELAWARE - Total** | **314.40** | **260.38** | **230.68** | **202.56** | **1,008.02** |

**STATE: FL - FLORIDA**

| REGISTRANT ZIP CODE 3 | 1ST QUARTER | 2ND QUARTER | 3RD QUARTER | 4TH QUARTER | TOTAL GRAMS |
|---|---|---|---|---|---|
| 320 | 191.05 | 173.51 | 150.88 | 135.46 | **650.90** |
| 321 | 219.23 | 191.76 | 168.04 | 164.38 | **743.41** |
| 322 | 368.04 | 327.59 | 273.71 | 242.97 | **1,212.32** |
| 323 | 77.79 | 79.63 | 70.94 | 69.54 | **297.90** |
| 324 | 115.69 | 105.72 | 97.99 | 86.79 | **406.19** |
| 325 | 135.02 | 118.89 | 106.65 | 97.35 | **457.91** |
| 326 | 121.51 | 106.17 | 99.16 | 92.96 | **419.80** |
| 327 | 297.46 | 269.02 | 234.67 | 223.47 | **1,024.62** |
| 328 | 146.98 | 148.18 | 121.88 | 126.13 | **543.17** |
| 329 | 309.56 | 278.37 | 239.78 | 228.00 | **1,055.73** |
| 330 | 243.52 | 218.90 | 193.19 | 191.32 | **846.93** |
| 331 | 192.19 | 182.52 | 159.88 | 144.16 | **678.75** |
| 333 | 217.83 | 191.85 | 171.83 | 169.51 | **751.01** |
| 334 | 450.62 | 398.90 | 350.14 | 345.80 | **1,545.47** |
| 335 | 190.26 | 168.99 | 141.68 | 133.53 | **634.46** |
| 336 | 431.34 | 442.88 | 278.82 | 308.71 | **1,461.77** |
| 337 | 271.04 | 249.24 | 194.66 | 181.74 | **896.68** |
| 338 | 161.52 | 119.80 | 101.69 | 100.40 | **483.41** |
| 339 | 256.09 | 225.85 | 195.75 | 180.25 | **857.94** |
| 341 | 95.00 | 73.14 | 66.35 | 65.66 | **300.14** |
| 342 | 296.87 | 247.51 | 209.22 | 205.60 | **959.20** |
| 344 | 154.70 | 128.52 | 98.95 | 85.40 | **467.58** |
| 346 | 273.48 | 246.48 | 201.55 | 197.09 | **918.59** |
| 347 | 125.61 | 107.54 | 90.38 | 92.98 | **416.51** |
| 349 | 174.65 | 149.47 | 128.45 | 127.19 | **579.76** |

| FL - FLORIDA - Total | 5,517.05 | 4,950.45 | 4,146.25 | 3,996.39 | 18,610.14 |
|---|---|---|---|---|---|

STATE: GA - GEORGIA

| REGISTRANT ZIP CODE 3 | 1ST QUARTER | 2ND QUARTER | 3RD QUARTER | 4TH QUARTER | TOTAL GRAMS |
|---|---|---|---|---|---|
| 300 | 493.12 | 442.75 | 416.94 | 400.58 | **1,753.40** |
| 301 | 321.11 | 288.00 | 263.69 | 258.04 | **1,130.83** |
| 302 | 208.79 | 189.57 | 180.07 | 175.95 | **754.39** |
| 303 | 159.12 | 150.38 | 152.08 | 143.28 | **604.85** |
| 304 | 67.94 | 57.62 | 49.30 | 52.42 | **227.28** |
| 305 | 187.90 | 177.72 | 170.56 | 166.48 | **702.67** |
| 306 | 103.73 | 91.53 | 83.64 | 79.68 | **358.58** |
| 307 | 53.39 | 45.71 | 40.39 | 39.33 | **178.82** |
| 308 | 51.28 | 48.95 | 45.69 | 42.49 | **188.42** |
| 309 | 94.28 | 79.19 | 80.35 | 69.67 | **323.49** |
| 310 | 170.35 | 159.66 | 142.07 | 136.84 | **608.91** |
| 312 | 64.03 | 61.81 | 54.22 | 47.17 | **227.22** |
| 313 | 46.57 | 39.63 | 39.01 | 32.60 | **157.80** |
| 314 | 57.82 | 57.54 | 53.07 | 44.72 | **213.15** |
| 315 | 110.93 | 100.47 | 88.02 | 80.73 | **380.15** |
| 316 | 65.77 | 60.06 | 57.92 | 57.30 | **241.05** |
| 317 | 116.95 | 111.27 | 101.36 | 96.64 | **426.22** |
| 318 | 7.41 | 6.31 | 6.47 | 5.42 | **25.60** |
| 319 | 57.16 | 44.95 | 41.06 | 35.29 | **178.45** |
| 398 | 29.70 | 26.18 | 26.78 | 25.99 | **108.65** |
| **GA - GEORGIA - Total** | **2,467.35** | **2,239.30** | **2,092.68** | **1,990.60** | **8,789.93** |

STATE: GU - GUAM

| REGISTRANT ZIP CODE 3 | 1ST QUARTER | 2ND QUARTER | 3RD QUARTER | 4TH QUARTER | TOTAL GRAMS |
|---|---|---|---|---|---|
| 969 | 8.96 | 7.20 | 9.21 | 7.39 | **32.77** |
| **GU - GUAM - Total** | **8.96** | **7.20** | **9.21** | **7.39** | **32.77** |

STATE: HI - HAWAII

| REGISTRANT ZIP CODE 3 | 1ST QUARTER | 2ND QUARTER | 3RD QUARTER | 4TH QUARTER | TOTAL GRAMS |
|---|---|---|---|---|---|
| 967 | 102.14 | 92.21 | 86.75 | 82.79 | **363.88** |
| 968 | 67.84 | 67.03 | 61.02 | 60.53 | **256.42** |
| **HI - HAWAII - Total** | **169.98** | **159.24** | **147.76** | **143.32** | **620.30** |

STATE: IA - IOWA

| REGISTRANT ZIP CODE 3 | 1ST QUARTER | 2ND QUARTER | 3RD QUARTER | 4TH QUARTER | TOTAL GRAMS |
|---|---|---|---|---|---|
| 500 | 63.48 | 58.53 | 55.68 | 57.41 | **235.11** |
| 501 | 68.79 | 62.03 | 60.30 | 54.63 | **245.75** |
| 502 | 63.75 | 59.07 | 57.70 | 54.41 | **234.93** |
| 503 | 90.51 | 78.07 | 74.54 | 72.47 | **315.59** |
| 504 | 34.38 | 30.84 | 33.41 | 27.92 | **126.55** |

| | | | | | |
|---|---|---|---|---|---|
| 505 | 40.28 | 38.28 | 37.45 | 36.50 | **152.51** |
| 506 | 42.37 | 41.17 | 42.54 | 42.39 | **168.47** |
| 507 | 18.76 | 16.69 | 14.63 | 13.44 | **63.52** |
| 508 | 8.85 | 9.88 | 9.34 | 7.78 | **35.85** |
| 510 | 11.17 | 10.43 | 9.40 | 9.08 | **40.08** |
| 511 | 24.16 | 22.11 | 19.19 | 21.03 | **86.49** |
| 512 | 10.04 | 8.02 | 8.44 | 7.98 | **34.48** |
| 513 | 11.81 | 10.28 | 10.28 | 9.93 | **42.30** |
| 514 | 22.09 | 19.56 | 20.01 | 19.30 | **80.96** |
| 515 | 29.04 | 27.41 | 25.20 | 22.37 | **104.01** |
| 516 | 6.71 | 7.50 | 6.77 | 7.08 | **28.06** |
| 520 | 35.99 | 36.52 | 33.00 | 32.97 | **138.48** |
| 521 | 13.76 | 13.44 | 13.75 | 14.51 | **55.45** |
| 522 | 28.70 | 31.07 | 31.55 | 30.29 | **121.61** |
| 523 | 23.27 | 21.53 | 21.63 | 18.50 | **84.94** |
| 524 | 29.14 | 24.44 | 24.92 | 24.17 | **102.67** |
| 525 | 40.24 | 35.40 | 32.38 | 31.51 | **139.53** |
| 526 | 26.77 | 25.57 | 23.85 | 21.84 | **98.03** |
| 527 | 20.87 | 19.23 | 17.81 | 17.70 | **75.61** |
| 528 | 30.63 | 29.50 | 27.22 | 25.51 | **112.87** |
| **IA - IOWA - Total** | **795.57** | **736.56** | **711.00** | **680.72** | **2,923.85** |

**STATE: ID - IDAHO**

| REGISTRANT ZIP CODE 3 | 1ST QUARTER | 2ND QUARTER | 3RD QUARTER | 4TH QUARTER | TOTAL GRAMS |
|---|---|---|---|---|---|
| 832 | 58.47 | 51.09 | 51.46 | 51.89 | **212.90** |
| 833 | 56.16 | 46.57 | 44.54 | 39.24 | **186.51** |
| 834 | 51.78 | 52.30 | 53.41 | 52.59 | **210.08** |
| 835 | 40.73 | 39.33 | 36.62 | 36.21 | **152.89** |
| 836 | 159.33 | 135.82 | 121.00 | 117.57 | **533.72** |
| 837 | 116.66 | 103.22 | 102.40 | 93.17 | **415.44** |
| 838 | 77.55 | 68.59 | 66.34 | 59.29 | **271.77** |
| **ID - IDAHO - Total** | **560.67** | **496.92** | **475.76** | **449.96** | **1,983.31** |

**STATE: IL - ILLINOIS**

| REGISTRANT ZIP CODE 3 | 1ST QUARTER | 2ND QUARTER | 3RD QUARTER | 4TH QUARTER | TOTAL GRAMS |
|---|---|---|---|---|---|
| 600 | 394.14 | 350.05 | 335.39 | 306.34 | **1,385.92** |
| 601 | 243.52 | 207.90 | 202.04 | 191.81 | **845.26** |
| 602 | 12.03 | 12.11 | 8.71 | 9.48 | **42.33** |
| 603 | 13.24 | 13.45 | 9.88 | 10.54 | **47.11** |
| 604 | 309.26 | 273.44 | 255.86 | 247.00 | **1,085.56** |
| 605 | 188.66 | 156.50 | 151.00 | 138.49 | **634.65** |
| 606 | 259.22 | 237.73 | 219.49 | 210.90 | **927.33** |

| | | | | | |
|---|---|---|---|---|---|
| 607 | 26.45 | 23.22 | 20.96 | 20.90 | **91.52** |
| 608 | 18.76 | 13.89 | 13.81 | 11.01 | **57.48** |
| 609 | 41.42 | 36.35 | 31.02 | 30.33 | **139.13** |
| 610 | 93.79 | 83.88 | 87.49 | 84.41 | **349.57** |
| 611 | 106.40 | 82.23 | 81.72 | 81.65 | **352.00** |
| 612 | 28.99 | 23.15 | 23.24 | 23.61 | **99.00** |
| 613 | 33.54 | 31.12 | 24.41 | 24.36 | **113.43** |
| 614 | 39.97 | 39.22 | 32.65 | 32.24 | **144.08** |
| 615 | 61.34 | 57.89 | 49.99 | 47.20 | **216.43** |
| 616 | 115.69 | 94.29 | 92.42 | 89.23 | **391.63** |
| 617 | 70.50 | 60.26 | 56.00 | 56.03 | **242.79** |
| 618 | 70.10 | 56.16 | 52.09 | 51.46 | **229.81** |
| 619 | 28.68 | 24.50 | 21.93 | 21.65 | **96.76** |
| 620 | 96.49 | 81.42 | 75.26 | 74.20 | **327.37** |
| 622 | 93.43 | 75.12 | 68.74 | 64.59 | **301.88** |
| 623 | 40.83 | 36.93 | 32.04 | 33.60 | **143.40** |
| 624 | 42.04 | 33.11 | 29.26 | 28.31 | **132.72** |
| 625 | 77.37 | 63.41 | 63.32 | 63.22 | **267.32** |
| 626 | 50.08 | 43.55 | 40.63 | 40.45 | **174.71** |
| 627 | 69.31 | 65.74 | 53.49 | 52.00 | **240.54** |
| 628 | 48.06 | 43.84 | 39.68 | 36.94 | **168.52** |
| 629 | 78.42 | 66.27 | 64.08 | 58.12 | **266.89** |
| **IL - ILLINOIS - Total** | **2,751.72** | **2,386.75** | **2,236.62** | **2,140.06** | **9,515.16** |

**STATE: IN - INDIANA**

| REGISTRANT ZIP CODE 3 | 1ST QUARTER | 2ND QUARTER | 3RD QUARTER | 4TH QUARTER | TOTAL GRAMS |
|---|---|---|---|---|---|
| 460 | 107.65 | 100.94 | 92.39 | 88.58 | **389.57** |
| 461 | 119.36 | 112.22 | 119.05 | 114.32 | **464.95** |
| 462 | 313.76 | 283.90 | 262.82 | 228.10 | **1,088.57** |
| 463 | 168.93 | 150.27 | 144.49 | 144.04 | **607.73** |
| 464 | 14.61 | 13.09 | 10.38 | 9.94 | **48.02** |
| 465 | 125.78 | 110.15 | 98.80 | 94.15 | **428.88** |
| 466 | 39.88 | 31.81 | 30.70 | 27.82 | **130.21** |
| 467 | 57.85 | 51.90 | 48.21 | 51.38 | **209.34** |
| 468 | 99.20 | 86.04 | 83.00 | 81.09 | **349.33** |
| 469 | 87.34 | 67.64 | 60.80 | 61.80 | **277.58** |
| 470 | 41.70 | 41.40 | 37.71 | 35.30 | **156.11** |
| 471 | 81.77 | 69.64 | 65.87 | 64.00 | **281.27** |
| 472 | 48.96 | 39.31 | 34.86 | 31.47 | **154.60** |
| 473 | 92.64 | 82.14 | 78.64 | 78.59 | **332.00** |
| 474 | 56.83 | 53.06 | 50.98 | 50.06 | **210.93** |

| | | | | | |
|---|---|---|---|---|---|
| 475 | 48.35 | 42.06 | 37.80 | 35.16 | **163.37** |
| 476 | 18.03 | 17.77 | 15.18 | 15.40 | **66.38** |
| 477 | 47.98 | 43.35 | 41.32 | 43.57 | **176.22** |
| 478 | 33.85 | 27.57 | 28.06 | 23.67 | **113.15** |
| 479 | 58.67 | 53.88 | 48.27 | 34.97 | **195.79** |
| **IN - INDIANA - Total** | **1,663.14** | **1,478.14** | **1,389.31** | **1,313.43** | **5,844.02** |

**STATE: KS - KANSAS**

| REGISTRANT ZIP CODE 3 | 1ST QUARTER | 2ND QUARTER | 3RD QUARTER | 4TH QUARTER | **TOTAL GRAMS** |
|---|---|---|---|---|---|
| 660 | 134.32 | 124.59 | 121.28 | 112.67 | **492.86** |
| 661 | 51.77 | 36.73 | 34.26 | 31.11 | **153.87** |
| 662 | 139.86 | 131.61 | 119.83 | 128.39 | **519.69** |
| 664 | 24.74 | 19.13 | 22.78 | 19.21 | **85.85** |
| 665 | 39.90 | 39.54 | 33.36 | 35.89 | **148.69** |
| 666 | 56.08 | 54.56 | 50.85 | 47.28 | **208.77** |
| 667 | 57.91 | 55.61 | 50.64 | 52.01 | **216.17** |
| 668 | 10.11 | 8.12 | 7.63 | 6.45 | **32.30** |
| 669 | 16.89 | 18.07 | 15.61 | 15.58 | **66.15** |
| 670 | 52.53 | 48.98 | 43.29 | 43.38 | **188.19** |
| 671 | 29.12 | 26.50 | 24.64 | 22.96 | **103.23** |
| 672 | 119.00 | 106.00 | 98.39 | 91.86 | **415.25** |
| 673 | 22.08 | 17.89 | 16.02 | 15.05 | **71.04** |
| 674 | 65.64 | 57.60 | 57.46 | 57.82 | **238.52** |
| 675 | 29.23 | 26.45 | 28.16 | 30.41 | **114.26** |
| 676 | 31.72 | 29.62 | 27.14 | 28.12 | **116.61** |
| 677 | 17.45 | 18.12 | 14.24 | 16.13 | **65.93** |
| 678 | 45.52 | 41.99 | 40.19 | 39.77 | **167.47** |
| 679 | 16.14 | 12.60 | 12.08 | 11.32 | **52.14** |
| **KS - KANSAS - Total** | **960.00** | **873.71** | **817.86** | **805.40** | **3,456.97** |

**STATE: KY - KENTUCKY**

| REGISTRANT ZIP CODE 3 | 1ST QUARTER | 2ND QUARTER | 3RD QUARTER | 4TH QUARTER | **TOTAL GRAMS** |
|---|---|---|---|---|---|
| 400 | 38.20 | 33.19 | 34.57 | 33.72 | **139.68** |
| 401 | 17.96 | 17.51 | 18.06 | 16.13 | **69.66** |
| 402 | 179.86 | 159.74 | 170.79 | 143.80 | **654.20** |
| 403 | 72.56 | 66.05 | 59.91 | 52.59 | **251.11** |
| 404 | 37.08 | 34.07 | 31.07 | 27.78 | **129.99** |
| 405 | 78.76 | 72.25 | 56.92 | 61.15 | **269.07** |
| 406 | 6.73 | 7.67 | 6.11 | 5.99 | **26.50** |
| 407 | 32.39 | 28.93 | 29.17 | 25.87 | **116.36** |
| 408 | 6.47 | 7.17 | 5.20 | 5.75 | **24.59** |
| 409 | 12.58 | 10.67 | 10.44 | 8.50 | **42.20** |

| | | | | | |
|---|---|---|---|---|---|
| 410 | 128.79 | 116.49 | 104.94 | 92.12 | **442.33** |
| 411 | 28.86 | 26.85 | 23.01 | 23.00 | **101.72** |
| 412 | 11.83 | 10.88 | 9.05 | 9.29 | **41.05** |
| 413 | 7.71 | 7.56 | 7.28 | 6.19 | **28.74** |
| 414 | 2.29 | 2.02 | 2.23 | 2.09 | **8.62** |
| 415 | 13.03 | 11.41 | 9.60 | 9.52 | **43.56** |
| 416 | 10.84 | 11.41 | 10.32 | 10.11 | **42.67** |
| 417 | 25.89 | 32.95 | 26.94 | 24.91 | **110.69** |
| 418 | 3.00 | 2.89 | 2.24 | 2.29 | **10.42** |
| 420 | 53.80 | 45.37 | 44.07 | 37.58 | **180.82** |
| 421 | 59.80 | 54.78 | 50.31 | 42.53 | **207.42** |
| 422 | 25.86 | 20.29 | 22.38 | 16.19 | **84.72** |
| 423 | 33.18 | 31.90 | 29.21 | 25.03 | **119.32** |
| 424 | 37.86 | 35.96 | 29.82 | 28.23 | **131.87** |
| 425 | 19.97 | 18.17 | 18.26 | 15.24 | **71.63** |
| 426 | 14.94 | 13.00 | 9.56 | 11.97 | **49.47** |
| 427 | 44.36 | 41.47 | 36.52 | 36.92 | **159.27** |
| **KY - KENTUCKY - Total** | **1,004.59** | **920.63** | **857.97** | **774.49** | **3,557.68** |

**STATE: LA - LOUISIANA**

| REGISTRANT ZIP CODE 3 | 1ST QUARTER | 2ND QUARTER | 3RD QUARTER | 4TH QUARTER | **TOTAL GRAMS** |
|---|---|---|---|---|---|
| 700 | 87.90 | 74.79 | 71.22 | 72.68 | **306.58** |
| 701 | 58.57 | 56.00 | 51.25 | 44.99 | **210.81** |
| 703 | 66.63 | 56.05 | 45.47 | 44.97 | **213.11** |
| 704 | 99.98 | 86.76 | 81.33 | 78.31 | **346.38** |
| 705 | 163.23 | 167.34 | 154.93 | 139.03 | **624.53** |
| 706 | 42.01 | 34.74 | 31.19 | 33.81 | **141.76** |
| 707 | 92.47 | 90.29 | 87.44 | 81.31 | **351.50** |
| 708 | 93.30 | 75.46 | 67.15 | 64.09 | **300.00** |
| 710 | 31.81 | 27.21 | 29.37 | 24.32 | **112.71** |
| 711 | 75.47 | 69.11 | 67.07 | 66.09 | **277.74** |
| 712 | 93.53 | 83.19 | 82.43 | 82.85 | **341.99** |
| 713 | 75.49 | 65.81 | 59.97 | 57.65 | **258.92** |
| 714 | 34.19 | 28.99 | 26.74 | 27.57 | **117.49** |
| **LA - LOUISIANA - Total** | **1,014.57** | **915.75** | **855.53** | **817.67** | **3,603.52** |

**STATE: MA - MASSACHUSETTS**

| REGISTRANT ZIP CODE 3 | 1ST QUARTER | 2ND QUARTER | 3RD QUARTER | 4TH QUARTER | **TOTAL GRAMS** |
|---|---|---|---|---|---|
| 010 | 132.69 | 116.81 | 107.35 | 105.95 | **462.78** |
| 011 | 38.55 | 30.47 | 30.19 | 28.97 | **128.18** |
| 012 | 23.15 | 22.98 | 22.02 | 21.43 | **89.59** |
| 013 | 16.51 | 13.39 | 11.63 | 12.06 | **53.59** |

| | | | | | |
|---|---|---|---|---|---|
| 014 | 34.41 | 30.17 | 27.95 | 27.97 | **120.50** |
| 015 | 51.12 | 50.82 | 43.65 | 45.68 | **191.27** |
| 016 | 42.85 | 37.42 | 32.51 | 36.96 | **149.74** |
| 017 | 53.21 | 51.47 | 48.73 | 52.79 | **206.20** |
| 018 | 163.66 | 159.88 | 141.42 | 135.02 | **599.99** |
| 019 | 81.68 | 71.60 | 71.81 | 65.89 | **290.97** |
| 020 | 47.60 | 54.65 | 39.20 | 40.46 | **181.90** |
| 021 | 162.62 | 152.02 | 137.81 | 133.33 | **585.79** |
| 022 | 19.60 | 22.32 | 19.18 | 20.73 | **81.83** |
| 023 | 81.64 | 78.04 | 71.18 | 71.22 | **302.07** |
| 024 | 34.66 | 33.35 | 32.09 | 31.44 | **131.54** |
| 025 | 20.91 | 23.37 | 21.14 | 19.36 | **84.78** |
| 026 | 22.89 | 18.20 | 19.74 | 19.20 | **80.03** |
| 027 | 146.60 | 136.69 | 126.05 | 125.06 | **534.40** |
| **MA - MASSACHUSETTS - Total** | **1,174.34** | **1,103.65** | **1,003.65** | **993.50** | **4,275.14** |

**STATE: MD - MARYLAND**

| REGISTRANT ZIP CODE 3 | 1ST QUARTER | 2ND QUARTER | 3RD QUARTER | 4TH QUARTER | **TOTAL GRAMS** |
|---|---|---|---|---|---|
| 206 | 74.43 | 71.43 | 60.53 | 59.49 | **265.89** |
| 207 | 91.10 | 93.29 | 85.11 | 67.26 | **336.75** |
| 208 | 135.35 | 122.85 | 107.42 | 93.15 | **458.77** |
| 209 | 33.14 | 31.06 | 26.33 | 24.91 | **115.44** |
| 210 | 175.92 | 153.34 | 140.64 | 137.11 | **607.01** |
| 211 | 89.01 | 70.92 | 70.42 | 66.86 | **297.21** |
| 212 | 256.60 | 234.22 | 210.08 | 206.96 | **907.86** |
| 214 | 22.66 | 21.95 | 18.12 | 19.03 | **81.77** |
| 215 | 35.46 | 25.90 | 24.13 | 19.65 | **105.14** |
| 216 | 37.89 | 31.45 | 27.01 | 26.39 | **122.74** |
| 217 | 153.21 | 133.65 | 128.08 | 122.68 | **537.61** |
| 218 | 42.25 | 34.39 | 35.13 | 31.39 | **143.15** |
| 219 | 30.17 | 23.37 | 21.78 | 16.07 | **91.39** |
| **MD - MARYLAND - Total** | **1,177.18** | **1,047.83** | **954.78** | **890.95** | **4,070.73** |

**STATE: ME - MAINE**

| REGISTRANT ZIP CODE 3 | 1ST QUARTER | 2ND QUARTER | 3RD QUARTER | 4TH QUARTER | **TOTAL GRAMS** |
|---|---|---|---|---|---|
| 039 | 2.58 | 2.02 | 3.05 | 3.43 | **11.08** |
| 040 | 73.52 | 66.63 | 67.61 | 68.62 | **276.38** |
| 041 | 36.64 | 29.68 | 30.92 | 31.08 | **128.32** |
| 042 | 31.12 | 31.01 | 29.10 | 31.53 | **122.76** |
| 043 | 42.42 | 33.58 | 35.21 | 30.14 | **141.35** |
| 044 | 35.61 | 31.44 | 28.49 | 27.67 | **123.21** |
| 045 | 6.54 | 5.98 | 6.43 | 6.14 | **25.08** |

| | | | | | |
|---|---|---|---|---|---|
| 046 | 12.28 | 11.26 | 11.90 | 11.37 | **46.80** |
| 047 | 18.31 | 16.77 | 16.11 | 14.47 | **65.65** |
| 048 | 8.02 | 8.86 | 7.61 | 4.81 | **29.30** |
| 049 | 36.63 | 36.37 | 30.01 | 28.79 | **131.80** |
| **ME - MAINE - Total** | **303.68** | **273.58** | **266.44** | **258.04** | **1,101.74** |

**STATE: MI - MICHIGAN**

| REGISTRANT ZIP CODE 3 | 1ST QUARTER | 2ND QUARTER | 3RD QUARTER | 4TH QUARTER | TOTAL GRAMS |
|---|---|---|---|---|---|
| 480 | 292.01 | 255.91 | 243.99 | 221.66 | **1,013.58** |
| 481 | 381.94 | 343.78 | 301.85 | 278.19 | **1,305.76** |
| 482 | 76.00 | 74.92 | 65.13 | 63.76 | **279.81** |
| 483 | 241.53 | 228.40 | 206.06 | 190.00 | **866.00** |
| 484 | 108.20 | 95.01 | 90.16 | 81.00 | **374.38** |
| 485 | 61.42 | 55.18 | 49.09 | 46.72 | **212.41** |
| 486 | 178.00 | 166.50 | 154.10 | 144.72 | **643.32** |
| 487 | 80.97 | 69.77 | 66.70 | 66.88 | **284.33** |
| 488 | 138.07 | 123.19 | 109.81 | 106.28 | **477.35** |
| 489 | 54.05 | 45.05 | 44.38 | 38.50 | **181.98** |
| 490 | 282.40 | 264.65 | 246.76 | 228.10 | **1,021.91** |
| 491 | 30.07 | 26.65 | 25.11 | 25.07 | **106.90** |
| 492 | 89.51 | 77.94 | 69.04 | 66.25 | **302.73** |
| 493 | 53.09 | 45.03 | 41.81 | 39.72 | **179.65** |
| 494 | 154.52 | 137.13 | 130.86 | 120.18 | **542.69** |
| 495 | 171.90 | 143.18 | 126.51 | 119.00 | **560.60** |
| 496 | 125.22 | 111.16 | 102.99 | 93.37 | **432.75** |
| 497 | 88.43 | 71.16 | 70.89 | 63.07 | **293.54** |
| 498 | 70.13 | 59.79 | 59.64 | 56.02 | **245.58** |
| 499 | 22.81 | 19.83 | 19.38 | 18.67 | **80.70** |
| **MI - MICHIGAN - Total** | **2,700.25** | **2,414.26** | **2,224.25** | **2,067.19** | **9,405.95** |

**STATE: MN - MINNESOTA**

| REGISTRANT ZIP CODE 3 | 1ST QUARTER | 2ND QUARTER | 3RD QUARTER | 4TH QUARTER | TOTAL GRAMS |
|---|---|---|---|---|---|
| 550 | 97.86 | 78.33 | 74.36 | 79.25 | **329.80** |
| 551 | 127.19 | 118.59 | 107.81 | 103.87 | **457.46** |
| 553 | 118.49 | 102.86 | 98.04 | 95.31 | **414.71** |
| 554 | 201.50 | 152.91 | 198.13 | 159.48 | **712.01** |
| 556 | 2.53 | 2.57 | 2.30 | 1.97 | **9.36** |
| 557 | 34.66 | 31.59 | 29.05 | 27.04 | **122.35** |
| 558 | 25.85 | 22.00 | 20.61 | 18.84 | **87.29** |
| 559 | 55.51 | 47.09 | 47.70 | 44.80 | **195.09** |
| 560 | 40.13 | 34.33 | 31.08 | 32.18 | **137.73** |
| 561 | 19.64 | 17.49 | 17.57 | 15.49 | **70.19** |

| | | | | | |
|---|---|---|---|---|---|
| 562 | 31.22 | 26.93 | 24.60 | 24.09 | **106.84** |
| 563 | 64.90 | 60.58 | 47.24 | 47.92 | **220.64** |
| 564 | 28.39 | 27.46 | 24.83 | 22.45 | **103.13** |
| 565 | 32.71 | 28.86 | 25.37 | 23.61 | **110.55** |
| 566 | 14.74 | 13.36 | 12.96 | 10.18 | **51.25** |
| 567 | 27.55 | 23.06 | 22.15 | 21.81 | **94.58** |
| **MN - MINNESOTA - Total** | **922.88** | **788.02** | **783.81** | **728.27** | **3,222.99** |

**STATE: MO - MISSOURI**

| REGISTRANT ZIP CODE 3 | 1ST QUARTER | 2ND QUARTER | 3RD QUARTER | 4TH QUARTER | TOTAL GRAMS |
|---|---|---|---|---|---|
| 630 | 189.22 | 163.87 | 145.72 | 132.20 | **631.01** |
| 631 | 300.28 | 280.50 | 294.90 | 283.12 | **1,158.80** |
| 633 | 105.25 | 86.08 | 77.53 | 69.83 | **338.69** |
| 634 | 26.35 | 25.00 | 23.34 | 24.95 | **99.65** |
| 635 | 28.38 | 26.70 | 23.85 | 22.95 | **101.88** |
| 636 | 49.58 | 42.04 | 39.41 | 35.71 | **166.73** |
| 637 | 32.31 | 26.64 | 24.21 | 22.45 | **105.60** |
| 638 | 36.45 | 29.58 | 28.76 | 30.07 | **124.85** |
| 639 | 15.22 | 12.99 | 11.86 | 11.51 | **51.58** |
| 640 | 184.41 | 171.46 | 158.45 | 155.31 | **669.63** |
| 641 | 148.93 | 140.81 | 125.43 | 122.14 | **537.32** |
| 644 | 27.91 | 24.83 | 24.43 | 21.16 | **98.34** |
| 645 | 40.78 | 35.25 | 29.04 | 26.81 | **131.88** |
| 646 | 28.29 | 23.47 | 18.32 | 16.07 | **86.15** |
| 647 | 49.11 | 40.58 | 43.37 | 38.93 | **171.99** |
| 648 | 71.01 | 67.07 | 62.97 | 63.06 | **264.11** |
| 650 | 43.23 | 39.47 | 37.00 | 31.26 | **150.96** |
| 651 | 24.74 | 22.81 | 22.65 | 21.55 | **91.74** |
| 652 | 79.80 | 75.50 | 65.14 | 65.13 | **285.57** |
| 653 | 29.62 | 27.21 | 25.12 | 24.28 | **106.22** |
| 654 | 32.18 | 30.56 | 23.69 | 22.63 | **109.06** |
| 655 | 36.50 | 30.42 | 28.10 | 27.36 | **122.38** |
| 656 | 76.46 | 68.79 | 64.12 | 67.57 | **276.94** |
| 657 | 115.56 | 106.06 | 105.98 | 100.58 | **428.17** |
| 658 | 138.35 | 121.14 | 108.90 | 97.99 | **466.38** |
| **MO - MISSOURI - Total** | **1,909.91** | **1,718.82** | **1,612.29** | **1,534.63** | **6,775.65** |

**STATE: MS - MISSISSIPPI**

| REGISTRANT ZIP CODE 3 | 1ST QUARTER | 2ND QUARTER | 3RD QUARTER | 4TH QUARTER | TOTAL GRAMS |
|---|---|---|---|---|---|
| 386 | 98.04 | 84.98 | 83.01 | 75.78 | **341.80** |
| 387 | 22.21 | 19.40 | 16.50 | 17.40 | **75.50** |
| 388 | 95.93 | 86.99 | 79.13 | 75.15 | **337.19** |

| | | | | | |
|---|---|---|---|---|---|
| 389 | 24.14 | 21.70 | 19.64 | 20.39 | **85.86** |
| 390 | 65.26 | 54.02 | 55.65 | 48.71 | **223.65** |
| 391 | 289.50 | 272.24 | 403.91 | 260.05 | **1,225.71** |
| 392 | 85.98 | 79.48 | 78.45 | 70.31 | **314.22** |
| 393 | 75.17 | 60.10 | 54.56 | 51.02 | **240.85** |
| 394 | 132.50 | 115.27 | 102.82 | 97.85 | **448.44** |
| 395 | 91.42 | 78.42 | 74.93 | 69.42 | **314.19** |
| 396 | 48.04 | 38.36 | 34.74 | 35.21 | **156.35** |
| 397 | 32.32 | 29.93 | 26.37 | 27.34 | **115.96** |
| **MS - MISSISSIPPI - Total** | **1,060.52** | **940.87** | **1,029.70** | **848.63** | **3,879.72** |

**STATE: MT - MONTANA**

| REGISTRANT ZIP CODE 3 | 1ST QUARTER | 2ND QUARTER | 3RD QUARTER | 4TH QUARTER | TOTAL GRAMS |
|---|---|---|---|---|---|
| 590 | 15.83 | 13.10 | 11.79 | 12.87 | **53.59** |
| 591 | 59.78 | 52.56 | 50.90 | 49.51 | **212.75** |
| 592 | 11.71 | 10.85 | 11.19 | 9.64 | **43.40** |
| 593 | 12.70 | 12.00 | 10.89 | 9.86 | **45.45** |
| 594 | 38.84 | 37.74 | 34.89 | 34.37 | **145.83** |
| 595 | 9.68 | 10.55 | 9.75 | 7.67 | **37.64** |
| 596 | 24.71 | 25.16 | 23.04 | 22.78 | **95.69** |
| 597 | 32.07 | 28.33 | 25.63 | 26.42 | **112.45** |
| 598 | 50.70 | 54.55 | 47.82 | 42.25 | **195.32** |
| 599 | 35.91 | 28.98 | 27.44 | 26.18 | **118.50** |
| **MT - MONTANA - Total** | **291.92** | **273.83** | **253.34** | **241.54** | **1,060.63** |

**STATE: NC - NORTH CAROLINA**

| REGISTRANT ZIP CODE 3 | 1ST QUARTER | 2ND QUARTER | 3RD QUARTER | 4TH QUARTER | TOTAL GRAMS |
|---|---|---|---|---|---|
| 270 | 62.05 | 53.36 | 44.45 | 43.31 | **203.18** |
| 271 | 82.22 | 83.48 | 69.56 | 56.11 | **291.37** |
| 272 | 153.06 | 127.49 | 125.09 | 121.97 | **527.62** |
| 273 | 86.10 | 76.21 | 71.36 | 74.30 | **307.97** |
| 274 | 49.25 | 45.57 | 40.32 | 41.81 | **176.95** |
| 275 | 237.21 | 219.06 | 207.33 | 198.62 | **862.21** |
| 276 | 101.14 | 106.67 | 100.36 | 100.46 | **408.63** |
| 277 | 81.40 | 60.32 | 63.04 | 55.75 | **260.51** |
| 278 | 164.61 | 150.20 | 147.16 | 143.07 | **605.04** |
| 279 | 56.75 | 49.64 | 49.81 | 51.69 | **207.88** |
| 280 | 181.92 | 166.04 | 160.45 | 159.64 | **668.06** |
| 281 | 144.33 | 124.57 | 117.25 | 117.21 | **503.37** |
| 282 | 125.33 | 106.40 | 98.41 | 89.47 | **419.61** |
| 283 | 176.65 | 156.15 | 156.42 | 146.85 | **636.07** |
| 284 | 154.41 | 134.30 | 127.79 | 122.51 | **539.01** |

| | | | | | |
|---|---|---|---|---|---|
| 285 | 167.15 | 139.15 | 137.72 | 130.41 | **574.44** |
| 286 | 225.50 | 212.72 | 195.75 | 193.16 | **827.13** |
| 287 | 118.47 | 110.82 | 102.76 | 97.48 | **429.53** |
| 288 | 57.18 | 51.83 | 45.27 | 43.48 | **197.76** |
| 289 | 16.79 | 13.07 | 13.89 | 11.90 | **55.65** |
| **NC - NORTH CAROLINA - Total** | **2,441.52** | **2,187.04** | **2,074.21** | **1,999.21** | **8,701.98** |

**STATE: ND - NORTH DAKOTA**

| REGISTRANT ZIP CODE 3 | 1ST QUARTER | 2ND QUARTER | 3RD QUARTER | 4TH QUARTER | TOTAL GRAMS |
|---|---|---|---|---|---|
| 580 | 11.49 | 8.98 | 8.20 | 7.64 | **36.32** |
| 581 | 34.37 | 38.13 | 27.35 | 27.46 | **127.31** |
| 582 | 34.04 | 30.24 | 30.94 | 27.98 | **123.21** |
| 583 | 16.82 | 17.23 | 16.80 | 14.77 | **65.62** |
| 584 | 13.72 | 13.98 | 12.39 | 10.18 | **50.28** |
| 585 | 51.08 | 50.01 | 48.81 | 45.32 | **195.22** |
| 586 | 14.70 | 11.20 | 12.02 | 11.09 | **49.01** |
| 587 | 15.39 | 14.51 | 13.41 | 11.65 | **54.96** |
| 588 | 9.31 | 8.94 | 10.19 | 9.39 | **37.82** |
| **ND - NORTH DAKOTA - Total** | **200.92** | **193.22** | **180.11** | **165.49** | **739.73** |

**STATE: NE - NEBRASKA**

| REGISTRANT ZIP CODE 3 | 1ST QUARTER | 2ND QUARTER | 3RD QUARTER | 4TH QUARTER | TOTAL GRAMS |
|---|---|---|---|---|---|
| 680 | 61.22 | 49.34 | 52.04 | 53.37 | **215.97** |
| 681 | 121.75 | 122.35 | 98.58 | 99.93 | **442.61** |
| 683 | 22.65 | 21.29 | 18.82 | 19.49 | **82.24** |
| 684 | 16.67 | 14.45 | 16.51 | 13.83 | **61.46** |
| 685 | 49.60 | 48.28 | 47.26 | 43.47 | **188.61** |
| 686 | 12.88 | 13.40 | 11.63 | 11.75 | **49.66** |
| 687 | 32.41 | 30.44 | 29.70 | 27.31 | **119.86** |
| 688 | 57.25 | 51.24 | 50.23 | 51.16 | **209.88** |
| 689 | 22.21 | 20.96 | 22.24 | 21.38 | **86.80** |
| 690 | 17.75 | 16.80 | 15.64 | 17.87 | **68.07** |
| 691 | 32.65 | 30.83 | 29.32 | 25.79 | **118.59** |
| 692 | 1.30 | 1.25 | 1.28 | 1.42 | **5.25** |
| 693 | 31.97 | 31.65 | 27.91 | 27.41 | **118.94** |
| **NE - NEBRASKA - Total** | **480.32** | **452.28** | **421.16** | **414.18** | **1,767.93** |

**STATE: NH - NEW HAMPSHIRE**

| REGISTRANT ZIP CODE 3 | 1ST QUARTER | 2ND QUARTER | 3RD QUARTER | 4TH QUARTER | TOTAL GRAMS |
|---|---|---|---|---|---|
| 030 | 88.71 | 83.16 | 74.32 | 72.31 | **318.50** |
| 031 | 42.62 | 37.18 | 35.73 | 32.78 | **148.32** |
| 032 | 21.37 | 21.31 | 18.56 | 18.21 | **79.45** |
| 033 | 40.10 | 38.68 | 36.41 | 33.49 | **148.68** |

| | | | | | |
|---|---|---|---|---|---|
| 034 | 9.77 | 9.25 | 8.01 | 7.45 | **34.48** |
| 035 | 14.04 | 12.73 | 10.78 | 10.51 | **48.06** |
| 036 | 1.56 | 0.69 | 0.63 | 0.45 | **3.34** |
| 037 | 20.00 | 23.46 | 20.35 | 17.19 | **80.99** |
| 038 | 68.69 | 65.63 | 57.22 | 50.30 | **241.84** |
| **NH - NEW HAMPSHIRE - Total** | **306.85** | **292.11** | **262.01** | **242.68** | **1,103.65** |

**STATE: NJ - NEW JERSEY**

| REGISTRANT ZIP CODE 3 | 1ST QUARTER | 2ND QUARTER | 3RD QUARTER | 4TH QUARTER | **TOTAL GRAMS** |
|---|---|---|---|---|---|
| 070 | 335.33 | 315.42 | 273.57 | 270.22 | **1,194.54** |
| 071 | 29.30 | 32.04 | 30.06 | 31.00 | **122.39** |
| 072 | 12.17 | 13.15 | 12.00 | 10.97 | **48.29** |
| 073 | 12.75 | 13.68 | 11.33 | 11.62 | **49.38** |
| 074 | 152.78 | 122.95 | 118.36 | 125.39 | **519.48** |
| 075 | 25.71 | 16.28 | 15.48 | 17.28 | **74.74** |
| 076 | 180.85 | 175.36 | 178.97 | 173.96 | **709.14** |
| 077 | 160.77 | 151.63 | 132.67 | 130.71 | **575.79** |
| 078 | 60.27 | 55.74 | 55.25 | 52.14 | **223.41** |
| 079 | 62.34 | 58.31 | 49.53 | 52.66 | **222.84** |
| 080 | 315.09 | 273.69 | 250.56 | 248.62 | **1,087.96** |
| 081 | 22.46 | 21.43 | 22.03 | 17.56 | **83.47** |
| 082 | 50.11 | 44.88 | 42.89 | 38.81 | **176.69** |
| 083 | 29.59 | 28.36 | 22.46 | 20.96 | **101.38** |
| 084 | 8.30 | 8.45 | 8.55 | 8.33 | **33.63** |
| 085 | 50.64 | 45.84 | 43.89 | 45.29 | **185.66** |
| 086 | 60.08 | 51.31 | 47.28 | 45.36 | **204.03** |
| 087 | 129.71 | 103.41 | 105.82 | 93.51 | **432.45** |
| 088 | 614.84 | 226.43 | 214.16 | 209.45 | **1,264.88** |
| 089 | 20.61 | 23.80 | 22.57 | 23.63 | **90.60** |
| **NJ - NEW JERSEY - Total** | **2,333.70** | **1,782.18** | **1,657.43** | **1,627.45** | **7,400.76** |

**STATE: NM - NEW MEXICO**

| REGISTRANT ZIP CODE 3 | 1ST QUARTER | 2ND QUARTER | 3RD QUARTER | 4TH QUARTER | **TOTAL GRAMS** |
|---|---|---|---|---|---|
| 870 | 23.16 | 16.51 | 17.67 | 16.33 | **73.66** |
| 871 | 170.70 | 141.43 | 135.25 | 135.48 | **582.87** |
| 873 | 7.42 | 6.53 | 6.04 | 5.65 | **25.64** |
| 874 | 25.54 | 18.70 | 19.64 | 19.40 | **83.28** |
| 875 | 50.95 | 43.73 | 41.36 | 37.77 | **173.81** |
| 877 | 5.72 | 5.23 | 4.86 | 4.41 | **20.22** |
| 878 | 1.89 | 1.44 | 1.26 | 1.09 | **5.69** |
| 879 | 4.35 | 3.54 | 3.02 | 2.56 | **13.47** |
| 880 | 27.91 | 24.91 | 22.50 | 22.03 | **97.35** |

| | | | | | |
|---|---|---|---|---|---|
| 881 | 13.07 | 10.59 | 7.70 | 8.55 | **39.91** |
| 882 | 46.01 | 43.09 | 40.40 | 37.24 | **166.74** |
| 883 | 15.58 | 14.17 | 13.71 | 11.57 | **55.04** |
| 884 | 3.71 | 3.36 | 2.77 | 2.85 | **12.69** |
| **NM - NEW MEXICO - Total** | **396.02** | **333.23** | **316.18** | **304.93** | **1,350.36** |

**STATE: NV - NEVADA**

| REGISTRANT ZIP CODE 3 | 1ST QUARTER | 2ND QUARTER | 3RD QUARTER | 4TH QUARTER | TOTAL GRAMS |
|---|---|---|---|---|---|
| 890 | 122.09 | 110.08 | 97.41 | 85.13 | **414.71** |
| 891 | 249.11 | 215.16 | 204.17 | 189.40 | **857.84** |
| 893 | 1.49 | 2.44 | 1.90 | 1.50 | **7.34** |
| 894 | 36.08 | 32.52 | 26.59 | 24.49 | **119.68** |
| 895 | 51.32 | 45.70 | 42.28 | 39.70 | **179.00** |
| 897 | 14.37 | 12.44 | 11.37 | 11.09 | **49.28** |
| 898 | 7.42 | 8.18 | 7.90 | 9.30 | **32.79** |
| **NV - NEVADA - Total** | **481.88** | **426.52** | **391.63** | **360.60** | **1,660.62** |

**STATE: NY - NEW YORK**

| REGISTRANT ZIP CODE 3 | 1ST QUARTER | 2ND QUARTER | 3RD QUARTER | 4TH QUARTER | TOTAL GRAMS |
|---|---|---|---|---|---|
| 100 | 353.57 | 339.86 | 279.76 | 292.55 | **1,265.74** |
| 101 | 6.47 | 5.71 | 5.63 | 3.91 | **21.72** |
| 102 | 0.49 | 0.32 | 0.11 | 0.20 | **1.11** |
| 103 | 130.45 | 117.52 | 110.98 | 114.84 | **473.79** |
| 104 | 136.82 | 125.04 | 104.75 | 106.18 | **472.78** |
| 105 | 99.17 | 101.21 | 88.17 | 91.33 | **379.88** |
| 106 | 19.78 | 15.72 | 12.82 | 13.54 | **61.87** |
| 107 | 30.73 | 26.78 | 24.67 | 24.15 | **106.33** |
| 108 | 9.33 | 6.70 | 5.60 | 4.74 | **26.37** |
| 109 | 102.73 | 94.34 | 89.37 | 91.90 | **378.34** |
| 110 | 71.02 | 66.50 | 62.78 | 65.64 | **265.93** |
| 111 | 15.12 | 17.72 | 13.06 | 13.36 | **59.26** |
| 112 | 222.30 | 198.93 | 163.73 | 162.71 | **747.68** |
| 113 | 83.92 | 88.85 | 77.26 | 81.93 | **331.96** |
| 114 | 49.71 | 59.05 | 54.44 | 38.10 | **201.31** |
| 115 | 148.15 | 121.90 | 123.62 | 121.12 | **514.79** |
| 116 | 11.11 | 10.75 | 9.36 | 9.16 | **40.39** |
| 117 | 401.73 | 387.66 | 337.08 | 339.45 | **1,465.92** |
| 118 | 17.03 | 12.44 | 12.17 | 14.07 | **55.70** |
| 119 | 54.70 | 50.59 | 50.97 | 42.01 | **198.27** |
| 120 | 94.61 | 83.14 | 81.08 | 78.97 | **337.79** |
| 121 | 38.88 | 36.32 | 35.92 | 31.69 | **142.80** |
| 122 | 67.23 | 56.60 | 52.11 | 56.44 | **232.38** |

| | | | | | |
|---|---|---|---|---|---|
| 123 | 28.42 | 25.18 | 25.00 | 23.41 | **102.01** |
| 124 | 45.83 | 42.39 | 44.26 | 42.24 | **174.72** |
| 125 | 83.34 | 79.85 | 76.53 | 69.99 | **309.71** |
| 126 | 29.01 | 26.40 | 26.78 | 25.25 | **107.43** |
| 127 | 29.22 | 23.36 | 18.37 | 17.89 | **88.85** |
| 128 | 85.78 | 78.53 | 74.51 | 76.03 | **314.85** |
| 129 | 46.89 | 41.99 | 36.13 | 33.70 | **158.70** |
| 130 | 86.04 | 78.13 | 73.05 | 69.29 | **306.51** |
| 131 | 26.45 | 26.26 | 22.49 | 18.46 | **93.66** |
| 132 | 101.25 | 81.13 | 72.73 | 72.44 | **327.55** |
| 133 | 35.83 | 29.27 | 30.87 | 26.78 | **122.75** |
| 134 | 52.67 | 47.18 | 44.06 | 41.25 | **185.16** |
| 135 | 23.86 | 19.48 | 17.57 | 14.06 | **74.96** |
| 136 | 58.09 | 52.51 | 51.88 | 50.03 | **212.52** |
| 137 | 27.75 | 25.10 | 21.67 | 24.52 | **99.05** |
| 138 | 73.92 | 64.77 | 63.49 | 60.13 | **262.31** |
| 139 | 32.24 | 28.87 | 23.40 | 20.20 | **104.72** |
| 140 | 93.49 | 86.43 | 81.62 | 74.80 | **336.34** |
| 141 | 49.38 | 45.92 | 43.81 | 37.29 | **176.41** |
| 142 | 150.18 | 125.85 | 113.32 | 104.01 | **493.36** |
| 143 | 17.52 | 14.28 | 13.67 | 10.29 | **55.76** |
| 144 | 67.55 | 62.67 | 61.22 | 57.40 | **248.84** |
| 145 | 62.87 | 58.09 | 53.25 | 50.47 | **224.69** |
| 146 | 132.57 | 121.71 | 119.84 | 111.55 | **485.67** |
| 147 | 48.98 | 42.95 | 37.74 | 37.72 | **167.38** |
| 148 | 88.43 | 78.10 | 69.78 | 64.27 | **300.58** |
| 149 | 24.74 | 22.96 | 19.42 | 19.01 | **86.14** |
| **NY - NEW YORK - Total** | **3,767.36** | **3,452.98** | **3,131.91** | **3,050.47** | **13,402.72** |

**STATE: OH - OHIO**

| REGISTRANT ZIP CODE 3 | 1ST QUARTER | 2ND QUARTER | 3RD QUARTER | 4TH QUARTER | **TOTAL GRAMS** |
|---|---|---|---|---|---|
| 430 | 119.69 | 106.62 | 103.99 | 97.14 | **427.44** |
| 431 | 73.90 | 60.32 | 58.20 | 52.85 | **245.26** |
| 432 | 242.24 | 194.34 | 121.55 | 118.12 | **676.25** |
| 433 | 29.20 | 28.95 | 27.29 | 25.33 | **110.77** |
| 434 | 29.59 | 22.14 | 20.61 | 16.03 | **88.37** |
| 435 | 73.40 | 66.13 | 56.89 | 58.54 | **254.96** |
| 436 | 135.98 | 128.21 | 108.39 | 97.00 | **469.57** |
| 437 | 32.65 | 30.56 | 28.92 | 28.47 | **120.62** |
| 438 | 3.57 | 2.81 | 2.72 | 2.79 | **11.88** |
| 439 | 32.72 | 28.46 | 28.56 | 25.65 | **115.39** |

| | | | | | |
|---|---|---|---|---|---|
| 440 | 101.44 | 91.98 | 81.06 | 78.28 | **352.76** |
| 441 | 157.32 | 166.79 | 148.15 | 131.54 | **603.80** |
| 442 | 67.48 | 58.34 | 50.21 | 47.20 | **223.23** |
| 443 | 35.06 | 30.25 | 24.95 | 24.71 | **114.98** |
| 444 | 45.82 | 42.04 | 35.96 | 31.24 | **155.06** |
| 445 | 50.60 | 49.89 | 38.95 | 36.66 | **176.10** |
| 446 | 59.61 | 51.90 | 47.91 | 51.21 | **210.62** |
| 447 | 46.05 | 42.84 | 42.18 | 40.76 | **171.83** |
| 448 | 44.89 | 42.61 | 41.22 | 33.85 | **162.57** |
| 449 | 15.26 | 14.32 | 15.02 | 14.48 | **59.08** |
| 450 | 126.21 | 118.13 | 109.55 | 103.17 | **457.06** |
| 451 | 41.61 | 37.56 | 29.68 | 30.52 | **139.36** |
| 452 | 159.90 | 151.01 | 127.19 | 129.01 | **567.11** |
| 453 | 92.31 | 90.41 | 92.22 | 89.75 | **364.70** |
| 454 | 90.62 | 83.83 | 74.01 | 71.60 | **320.06** |
| 455 | 18.61 | 16.82 | 14.42 | 13.98 | **63.83** |
| 456 | 75.15 | 70.89 | 65.61 | 60.73 | **272.39** |
| 457 | 23.89 | 19.29 | 19.57 | 18.16 | **80.91** |
| 458 | 46.08 | 45.70 | 39.42 | 35.79 | **166.99** |
| **OH - OHIO - Total** | **2,070.84** | **1,893.13** | **1,654.43** | **1,564.55** | **7,182.96** |

**STATE: OK - OKLAHOMA**

| REGISTRANT ZIP CODE 3 | 1ST QUARTER | 2ND QUARTER | 3RD QUARTER | 4TH QUARTER | **TOTAL GRAMS** |
|---|---|---|---|---|---|
| 730 | 216.90 | 197.87 | 174.29 | 146.46 | **735.52** |
| 731 | 261.36 | 214.96 | 197.00 | 173.36 | **846.67** |
| 734 | 39.76 | 33.05 | 30.37 | 25.81 | **128.99** |
| 735 | 101.67 | 92.04 | 87.56 | 77.20 | **358.46** |
| 736 | 29.13 | 26.74 | 25.60 | 20.69 | **102.16** |
| 737 | 38.12 | 35.45 | 33.75 | 27.27 | **134.58** |
| 738 | 9.39 | 8.48 | 8.25 | 7.20 | **33.33** |
| 739 | 2.64 | 2.89 | 2.84 | 3.33 | **11.69** |
| 740 | 243.62 | 195.84 | 163.46 | 121.87 | **724.79** |
| 741 | 179.71 | 152.93 | 134.22 | 106.38 | **573.24** |
| 743 | 47.15 | 39.25 | 37.06 | 28.70 | **152.16** |
| 744 | 84.16 | 71.13 | 64.11 | 51.17 | **270.57** |
| 745 | 41.35 | 38.40 | 31.12 | 26.16 | **137.03** |
| 746 | 20.68 | 19.56 | 16.40 | 17.16 | **73.80** |
| 747 | 28.18 | 26.61 | 26.50 | 22.48 | **103.77** |
| 748 | 57.49 | 54.40 | 45.30 | 41.38 | **198.56** |
| 749 | 21.54 | 15.08 | 15.14 | 11.80 | **63.55** |
| **OK - OKLAHOMA - Total** | **1,422.86** | **1,224.66** | **1,092.96** | **908.41** | **4,648.89** |

**STATE: OR - OREGON**

| REGISTRANT ZIP CODE 3 | 1ST QUARTER | 2ND QUARTER | 3RD QUARTER | 4TH QUARTER | TOTAL GRAMS |
|---|---|---|---|---|---|
| 970 | 120.04 | 115.97 | 106.90 | 104.83 | **447.74** |
| 971 | 71.25 | 67.39 | 61.70 | 57.42 | **257.75** |
| 972 | 170.49 | 174.64 | 151.55 | 144.74 | **641.43** |
| 973 | 110.46 | 93.04 | 88.56 | 82.88 | **374.94** |
| 974 | 144.28 | 128.67 | 115.35 | 102.95 | **491.25** |
| 975 | 80.93 | 71.69 | 63.13 | 60.48 | **276.23** |
| 976 | 11.41 | 9.74 | 9.68 | 9.15 | **39.98** |
| 977 | 49.19 | 46.77 | 44.31 | 42.46 | **182.73** |
| 978 | 38.10 | 36.91 | 35.15 | 32.60 | **142.75** |
| 979 | 12.65 | 11.60 | 12.68 | 10.45 | **47.38** |
| **OR - OREGON - Total** | **808.80** | **756.41** | **689.02** | **647.95** | **2,902.19** |

**STATE: PA - PENNSYLVANIA**

| REGISTRANT ZIP CODE 3 | 1ST QUARTER | 2ND QUARTER | 3RD QUARTER | 4TH QUARTER | TOTAL GRAMS |
|---|---|---|---|---|---|
| 150 | 192.93 | 165.59 | 149.18 | 144.46 | **652.15** |
| 151 | 105.25 | 94.90 | 86.96 | 81.25 | **368.37** |
| 152 | 202.91 | 193.20 | 171.81 | 169.59 | **737.51** |
| 153 | 39.83 | 37.08 | 35.44 | 32.17 | **144.53** |
| 154 | 42.99 | 40.56 | 35.49 | 32.78 | **151.81** |
| 155 | 34.89 | 29.21 | 26.69 | 20.75 | **111.55** |
| 156 | 99.96 | 91.06 | 83.53 | 81.00 | **355.55** |
| 157 | 66.87 | 65.65 | 59.69 | 51.37 | **243.58** |
| 158 | 52.12 | 50.60 | 48.66 | 44.75 | **196.14** |
| 159 | 67.42 | 55.32 | 53.85 | 44.76 | **221.36** |
| 160 | 76.89 | 71.14 | 72.13 | 66.87 | **287.03** |
| 161 | 70.52 | 63.91 | 57.04 | 55.53 | **247.00** |
| 162 | 34.23 | 32.57 | 28.86 | 24.92 | **120.58** |
| 163 | 85.04 | 77.97 | 66.91 | 66.89 | **296.82** |
| 164 | 23.57 | 24.04 | 20.30 | 18.87 | **86.78** |
| 165 | 62.51 | 59.14 | 53.36 | 53.24 | **228.26** |
| 166 | 97.01 | 82.12 | 78.33 | 69.73 | **327.19** |
| 167 | 16.98 | 14.25 | 13.13 | 11.98 | **56.34** |
| 168 | 39.82 | 30.25 | 31.07 | 24.62 | **125.76** |
| 169 | 18.66 | 18.23 | 13.54 | 14.86 | **65.29** |
| 170 | 187.58 | 173.47 | 163.62 | 149.13 | **673.81** |
| 171 | 43.60 | 41.99 | 30.77 | 29.42 | **145.78** |
| 172 | 45.91 | 47.34 | 45.24 | 41.41 | **179.91** |
| 173 | 88.75 | 79.48 | 69.94 | 69.69 | **307.87** |
| 174 | 57.12 | 56.43 | 47.19 | 40.70 | **201.44** |

| | | | | | |
|---|---|---|---|---|---|
| 175 | 51.23 | 50.68 | 43.62 | 42.78 | **188.32** |
| 176 | 50.26 | 49.52 | 45.24 | 43.51 | **188.53** |
| 177 | 72.85 | 64.72 | 59.21 | 56.47 | **253.25** |
| 178 | 92.40 | 79.60 | 78.52 | 75.81 | **326.34** |
| 179 | 34.67 | 32.00 | 31.88 | 29.48 | **128.02** |
| 180 | 129.63 | 114.57 | 103.90 | 108.98 | **457.09** |
| 181 | 81.80 | 78.96 | 73.18 | 69.99 | **303.93** |
| 182 | 49.32 | 45.42 | 46.75 | 44.08 | **185.57** |
| 183 | 52.68 | 46.39 | 43.90 | 41.02 | **183.99** |
| 184 | 61.69 | 56.01 | 55.48 | 45.29 | **218.46** |
| 185 | 74.62 | 64.58 | 59.70 | 58.85 | **257.75** |
| 186 | 63.17 | 62.69 | 52.20 | 50.87 | **228.93** |
| 187 | 259.31 | 237.80 | 212.88 | 229.89 | **939.88** |
| 188 | 30.85 | 23.40 | 23.05 | 23.41 | **100.72** |
| 189 | 99.34 | 90.75 | 86.77 | 82.19 | **359.05** |
| 190 | 625.60 | 496.98 | 602.38 | 393.12 | **2,118.08** |
| 191 | 294.62 | 283.02 | 244.78 | 240.87 | **1,063.29** |
| 193 | 94.81 | 85.57 | 79.85 | 78.62 | **338.84** |
| 194 | 153.90 | 144.59 | 132.83 | 133.88 | **565.20** |
| 195 | 43.62 | 38.27 | 37.18 | 34.28 | **153.36** |
| 196 | 44.69 | 41.26 | 39.93 | 39.23 | **165.12** |
| **PA - PENNSYLVANIA - Total** | **4,314.42** | **3,882.29** | **3,696.00** | **3,363.39** | **15,256.11** |

**STATE: PR - PUERTO RICO**

| REGISTRANT ZIP CODE 3 | 1ST QUARTER | 2ND QUARTER | 3RD QUARTER | 4TH QUARTER | **TOTAL GRAMS** |
|---|---|---|---|---|---|
| 006 | 13.11 | 14.38 | 41.65 | 43.06 | **112.20** |
| 007 | 29.66 | 27.91 | 58.82 | 59.54 | **175.93** |
| 009 | 35.05 | 39.11 | 61.16 | 65.36 | **200.68** |
| **PR - PUERTO RICO - Total** | **77.83** | **81.40** | **161.63** | **167.96** | **488.81** |

**STATE: RI - RHODE ISLAND**

| REGISTRANT ZIP CODE 3 | 1ST QUARTER | 2ND QUARTER | 3RD QUARTER | 4TH QUARTER | **TOTAL GRAMS** |
|---|---|---|---|---|---|
| 028 | 99.63 | 82.39 | 87.49 | 82.67 | **352.18** |
| 029 | 63.12 | 56.97 | 52.30 | 50.39 | **222.78** |
| **RI - RHODE ISLAND - Total** | **162.75** | **139.36** | **139.79** | **133.06** | **574.96** |

**STATE: SC - SOUTH CAROLINA**

| REGISTRANT ZIP CODE 3 | 1ST QUARTER | 2ND QUARTER | 3RD QUARTER | 4TH QUARTER | **TOTAL GRAMS** |
|---|---|---|---|---|---|
| 290 | 72.63 | 67.53 | 68.37 | 62.31 | **270.85** |
| 291 | 93.08 | 76.36 | 72.67 | 72.99 | **315.10** |
| 292 | 79.37 | 71.69 | 72.93 | 62.11 | **286.10** |
| 293 | 101.15 | 86.59 | 81.64 | 78.26 | **347.64** |
| 294 | 186.69 | 162.31 | 159.88 | 150.01 | **658.89** |

| | | | | | |
|---|---|---|---|---|---|
| 295 | 210.05 | 178.91 | 172.47 | 168.07 | **729.50** |
| 296 | 240.69 | 207.02 | 198.44 | 174.07 | **820.23** |
| 297 | 52.62 | 42.93 | 39.47 | 39.49 | **174.51** |
| 298 | 65.12 | 62.25 | 57.23 | 57.14 | **241.74** |
| 299 | 70.42 | 49.80 | 55.11 | 56.44 | **231.77** |
| **SC - SOUTH CAROLINA - Total** | **1,171.83** | **1,005.39** | **978.22** | **920.89** | **4,076.33** |

**STATE: SD - SOUTH DAKOTA**

| REGISTRANT ZIP CODE 3 | 1ST QUARTER | 2ND QUARTER | 3RD QUARTER | 4TH QUARTER | TOTAL GRAMS |
|---|---|---|---|---|---|
| 570 | 43.40 | 41.86 | 41.81 | 39.09 | **166.16** |
| 571 | 100.61 | 95.74 | 83.01 | 79.64 | **359.01** |
| 572 | 17.77 | 16.19 | 16.37 | 13.02 | **63.34** |
| 573 | 23.16 | 20.05 | 18.99 | 17.74 | **79.94** |
| 574 | 17.63 | 15.83 | 15.86 | 13.81 | **63.14** |
| 575 | 13.65 | 13.43 | 12.19 | 13.81 | **53.08** |
| 576 | 5.14 | 2.94 | 2.51 | 2.56 | **13.14** |
| 577 | 36.95 | 27.34 | 25.42 | 20.46 | **110.17** |
| **SD - SOUTH DAKOTA - Total** | **258.33** | **233.37** | **216.15** | **200.14** | **907.99** |

**STATE: TN - TENNESSEE**

| REGISTRANT ZIP CODE 3 | 1ST QUARTER | 2ND QUARTER | 3RD QUARTER | 4TH QUARTER | TOTAL GRAMS |
|---|---|---|---|---|---|
| 370 | 183.33 | 162.13 | 145.01 | 128.83 | **619.30** |
| 371 | 152.66 | 136.48 | 123.29 | 115.66 | **528.09** |
| 372 | 150.12 | 135.96 | 106.12 | 100.90 | **493.10** |
| 373 | 119.03 | 105.05 | 94.07 | 87.40 | **405.54** |
| 374 | 58.85 | 41.70 | 36.33 | 33.45 | **170.33** |
| 376 | 116.82 | 111.14 | 100.41 | 90.39 | **418.76** |
| 377 | 56.56 | 50.80 | 48.04 | 46.39 | **201.79** |
| 378 | 111.06 | 99.93 | 90.42 | 92.97 | **394.38** |
| 379 | 88.09 | 83.24 | 72.90 | 77.52 | **321.75** |
| 380 | 82.08 | 71.36 | 62.26 | 62.31 | **278.01** |
| 381 | 435.66 | 310.40 | 421.15 | 231.90 | **1,399.12** |
| 382 | 27.40 | 23.63 | 21.39 | 18.24 | **90.66** |
| 383 | 68.62 | 62.22 | 53.25 | 51.62 | **235.71** |
| 384 | 37.66 | 41.07 | 33.17 | 32.43 | **144.33** |
| 385 | 80.05 | 74.12 | 63.34 | 57.95 | **275.46** |
| **TN - TENNESSEE - Total** | **1,767.99** | **1,509.25** | **1,471.14** | **1,227.94** | **5,976.32** |

**STATE: TX - TEXAS**

| REGISTRANT ZIP CODE 3 | 1ST QUARTER | 2ND QUARTER | 3RD QUARTER | 4TH QUARTER | TOTAL GRAMS |
|---|---|---|---|---|---|
| 750 | 454.76 | 402.01 | 375.79 | 369.46 | **1,602.01** |
| 751 | 131.72 | 117.74 | 113.95 | 106.86 | **470.27** |
| 752 | 191.71 | 181.11 | 153.85 | 141.69 | **668.36** |

| | | | | | |
|---|---|---|---|---|---|
| 753 | 12.76 | 11.77 | 8.97 | 11.48 | **44.98** |
| 754 | 131.48 | 120.19 | 107.36 | 99.66 | **458.69** |
| 755 | 22.28 | 22.46 | 24.71 | 19.39 | **88.84** |
| 756 | 68.09 | 115.26 | 61.12 | 73.23 | **317.69** |
| 757 | 125.82 | 123.18 | 109.73 | 110.73 | **469.45** |
| 758 | 20.35 | 18.27 | 14.83 | 16.53 | **69.98** |
| 759 | 50.96 | 45.01 | 40.95 | 40.88 | **177.80** |
| 760 | 311.90 | 265.84 | 237.10 | 225.98 | **1,040.83** |
| 761 | 284.57 | 244.38 | 238.25 | 216.06 | **983.26** |
| 762 | 116.99 | 111.49 | 105.79 | 99.39 | **433.67** |
| 763 | 77.69 | 66.47 | 63.40 | 60.88 | **268.45** |
| 764 | 48.34 | 43.68 | 38.73 | 36.02 | **166.77** |
| 765 | 74.33 | 64.31 | 62.24 | 54.78 | **255.66** |
| 766 | 25.86 | 22.41 | 21.45 | 17.92 | **87.64** |
| 767 | 45.10 | 41.50 | 30.99 | 29.40 | **147.00** |
| 768 | 36.55 | 32.18 | 31.29 | 30.30 | **130.32** |
| 769 | 31.33 | 20.67 | 18.50 | 17.28 | **87.78** |
| 770 | 332.39 | 299.70 | 271.32 | 261.91 | **1,165.32** |
| 772 | 0.03 | 0 | 0.01 | 0 | **0.04** |
| 773 | 233.68 | 203.16 | 189.55 | 179.24 | **805.64** |
| 774 | 170.17 | 138.74 | 131.94 | 126.50 | **567.35** |
| 775 | 203.45 | 185.29 | 172.43 | 168.06 | **729.23** |
| 776 | 52.60 | 47.11 | 46.16 | 45.23 | **191.10** |
| 777 | 28.01 | 21.73 | 24.10 | 23.34 | **97.19** |
| 778 | 56.64 | 51.61 | 48.48 | 45.00 | **201.73** |
| 779 | 51.89 | 48.71 | 49.12 | 42.38 | **192.10** |
| 780 | 71.91 | 67.82 | 64.10 | 58.39 | **262.21** |
| 781 | 77.40 | 69.42 | 64.97 | 56.50 | **268.28** |
| 782 | 381.57 | 375.39 | 340.70 | 287.23 | **1,384.89** |
| 783 | 40.05 | 37.29 | 31.20 | 29.76 | **138.30** |
| 784 | 84.36 | 78.10 | 70.50 | 66.15 | **299.10** |
| 785 | 113.53 | 111.08 | 103.08 | 96.13 | **423.82** |
| 786 | 207.87 | 171.78 | 145.37 | 125.47 | **650.49** |
| 787 | 184.44 | 168.69 | 149.18 | 137.12 | **639.44** |
| 788 | 17.32 | 15.92 | 13.11 | 9.42 | **55.77** |
| 789 | 11.73 | 10.06 | 9.45 | 6.33 | **37.57** |
| 790 | 55.27 | 48.91 | 42.84 | 40.57 | **187.59** |
| 791 | 100.47 | 93.19 | 83.63 | 77.41 | **354.70** |
| 792 | 9.68 | 9.41 | 8.42 | 8.16 | **35.67** |
| 793 | 29.17 | 26.06 | 23.94 | 23.89 | **103.06** |

| | | | | | |
|---|---|---|---|---|---|
| 794 | 99.28 | 83.30 | 81.20 | 76.27 | **340.04** |
| 795 | 30.08 | 27.64 | 24.80 | 22.19 | **104.71** |
| 796 | 59.91 | 52.95 | 50.28 | 47.45 | **210.59** |
| 797 | 51.95 | 43.83 | 37.42 | 35.27 | **168.47** |
| 798 | 4.86 | 4.47 | 3.98 | 4.51 | **17.82** |
| 799 | 114.16 | 100.32 | 91.45 | 80.56 | **386.50** |
| **TX - TEXAS - Total** | **5,136.47** | **4,661.60** | **4,231.75** | **3,958.35** | **17,988.16** |

**STATE: UT - UTAH**

| REGISTRANT ZIP CODE 3 | 1ST QUARTER | 2ND QUARTER | 3RD QUARTER | 4TH QUARTER | TOTAL GRAMS |
|---|---|---|---|---|---|
| 840 | 261.58 | 240.59 | 227.64 | 221.07 | **950.88** |
| 841 | 186.76 | 157.56 | 145.64 | 141.79 | **631.76** |
| 843 | 50.32 | 38.64 | 36.13 | 32.51 | **157.59** |
| 844 | 57.82 | 51.99 | 50.44 | 47.48 | **207.73** |
| 845 | 25.21 | 23.53 | 23.69 | 21.79 | **94.21** |
| 846 | 65.24 | 64.72 | 61.35 | 62.70 | **254.01** |
| 847 | 107.97 | 98.60 | 93.68 | 92.55 | **392.80** |
| **UT - UTAH - Total** | **754.91** | **675.62** | **638.57** | **619.90** | **2,688.99** |

**STATE: VA - VIRGINIA**

| REGISTRANT ZIP CODE 3 | 1ST QUARTER | 2ND QUARTER | 3RD QUARTER | 4TH QUARTER | TOTAL GRAMS |
|---|---|---|---|---|---|
| 201 | 107.92 | 93.36 | 87.19 | 87.80 | **376.27** |
| 220 | 51.86 | 42.51 | 40.32 | 41.23 | **175.93** |
| 221 | 32.40 | 29.61 | 29.05 | 28.53 | **119.60** |
| 222 | 12.72 | 12.32 | 11.66 | 8.80 | **45.51** |
| 223 | 23.84 | 21.29 | 20.73 | 16.96 | **82.82** |
| 224 | 38.25 | 34.16 | 28.99 | 29.90 | **131.30** |
| 225 | 16.27 | 13.23 | 14.24 | 11.30 | **55.05** |
| 226 | 49.51 | 44.57 | 39.96 | 38.03 | **172.06** |
| 227 | 8.43 | 9.25 | 7.81 | 7.47 | **32.96** |
| 228 | 44.83 | 40.19 | 38.35 | 40.63 | **164.00** |
| 229 | 64.77 | 63.74 | 56.00 | 48.87 | **233.37** |
| 230 | 48.38 | 45.00 | 41.08 | 43.28 | **177.73** |
| 231 | 70.54 | 61.27 | 53.98 | 54.79 | **240.58** |
| 232 | 140.77 | 124.33 | 115.86 | 106.54 | **487.50** |
| 233 | 61.79 | 58.55 | 55.29 | 52.60 | **228.23** |
| 234 | 123.60 | 116.10 | 107.79 | 99.07 | **446.56** |
| 235 | 38.96 | 43.90 | 37.69 | 37.50 | **158.05** |
| 236 | 70.70 | 64.52 | 59.37 | 55.32 | **249.90** |
| 237 | 35.98 | 33.53 | 25.29 | 26.10 | **120.89** |
| 238 | 49.55 | 41.26 | 39.73 | 35.49 | **166.02** |
| 239 | 12.04 | 12.20 | 10.99 | 12.45 | **47.69** |

| | | | | | |
|---|---|---|---|---|---|
| 240 | 86.68 | 73.18 | 74.19 | 74.60 | **308.64** |
| 241 | 63.45 | 58.99 | 52.86 | 48.87 | **224.16** |
| 242 | 70.61 | 62.98 | 61.01 | 57.42 | **252.01** |
| 243 | 33.94 | 33.92 | 26.75 | 27.24 | **121.84** |
| 244 | 43.17 | 39.68 | 39.10 | 35.27 | **157.23** |
| 245 | 71.04 | 67.44 | 64.13 | 57.71 | **260.32** |
| 246 | 20.40 | 18.58 | 17.10 | 16.54 | **72.62** |
| **VA - VIRGINIA - Total** | **1,492.39** | **1,359.62** | **1,256.52** | **1,200.31** | **5,308.84** |

**STATE: VI - VIRGIN ISLANDS**

| REGISTRANT ZIP CODE 3 | 1ST QUARTER | 2ND QUARTER | 3RD QUARTER | 4TH QUARTER | TOTAL GRAMS |
|---|---|---|---|---|---|
| 008 | 3.93 | 4.58 | 5.41 | 4.01 | **17.93** |
| **VI - VIRGIN ISLANDS - Total** | **3.93** | **4.58** | **5.41** | **4.01** | **17.93** |

**STATE: VT - VERMONT**

| REGISTRANT ZIP CODE 3 | 1ST QUARTER | 2ND QUARTER | 3RD QUARTER | 4TH QUARTER | TOTAL GRAMS |
|---|---|---|---|---|---|
| 050 | 7.91 | 7.63 | 7.89 | 5.84 | **29.26** |
| 051 | 6.10 | 4.72 | 3.51 | 3.48 | **17.82** |
| 052 | 11.14 | 10.36 | 9.71 | 9.72 | **40.94** |
| 053 | 4.60 | 4.68 | 4.38 | 4.10 | **17.76** |
| 054 | 70.77 | 66.08 | 56.73 | 59.59 | **253.17** |
| 056 | 22.22 | 19.80 | 18.50 | 18.76 | **79.27** |
| 057 | 27.95 | 27.15 | 23.04 | 21.04 | **99.18** |
| 058 | 16.69 | 14.66 | 15.92 | 12.89 | **60.16** |
| **VT - VERMONT - Total** | **167.38** | **155.09** | **139.68** | **135.42** | **597.57** |

**STATE: WA - WASHINGTON**

| REGISTRANT ZIP CODE 3 | 1ST QUARTER | 2ND QUARTER | 3RD QUARTER | 4TH QUARTER | TOTAL GRAMS |
|---|---|---|---|---|---|
| 980 | 165.48 | 160.34 | 151.95 | 143.93 | **621.70** |
| 981 | 157.36 | 151.64 | 131.45 | 130.56 | **571.01** |
| 982 | 133.60 | 136.66 | 121.95 | 122.31 | **514.52** |
| 983 | 136.20 | 125.31 | 118.14 | 106.41 | **486.06** |
| 984 | 82.91 | 74.15 | 63.48 | 56.66 | **277.19** |
| 985 | 69.54 | 66.93 | 61.84 | 63.63 | **261.95** |
| 986 | 150.92 | 144.07 | 147.94 | 142.40 | **585.34** |
| 988 | 34.88 | 33.64 | 30.17 | 28.70 | **127.39** |
| 989 | 60.72 | 60.63 | 51.74 | 42.80 | **215.89** |
| 990 | 21.63 | 22.83 | 22.60 | 16.21 | **83.27** |
| 991 | 32.46 | 30.98 | 28.94 | 24.80 | **117.18** |
| 992 | 155.87 | 139.47 | 131.54 | 117.45 | **544.33** |
| 993 | 104.28 | 96.13 | 90.78 | 87.53 | **378.71** |
| 994 | 20.85 | 19.32 | 16.26 | 17.76 | **74.18** |
| **WA - WASHINGTON - Total** | **1,326.71** | **1,262.09** | **1,168.78** | **1,101.16** | **4,858.73** |

**STATE: WI - WISCONSIN**

| REGISTRANT ZIP CODE 3 | 1ST QUARTER | 2ND QUARTER | 3RD QUARTER | 4TH QUARTER | TOTAL GRAMS |
|---|---|---|---|---|---|
| 530 | 132.67 | 112.96 | 103.07 | 100.31 | **449.01** |
| 531 | 135.54 | 127.69 | 103.41 | 89.94 | **456.59** |
| 532 | 206.78 | 180.03 | 157.88 | 132.97 | **677.66** |
| 534 | 24.10 | 20.74 | 18.52 | 15.64 | **78.99** |
| 535 | 116.34 | 108.86 | 98.84 | 92.17 | **416.22** |
| 537 | 84.01 | 73.13 | 70.33 | 63.18 | **290.65** |
| 538 | 18.72 | 16.29 | 19.17 | 17.38 | **71.56** |
| 539 | 53.90 | 49.29 | 46.05 | 44.27 | **193.51** |
| 540 | 20.29 | 17.49 | 16.55 | 17.76 | **72.09** |
| 541 | 35.73 | 32.44 | 31.87 | 27.12 | **127.16** |
| 542 | 20.71 | 21.47 | 21.44 | 16.64 | **80.27** |
| 543 | 51.13 | 40.15 | 37.90 | 35.92 | **165.11** |
| 544 | 60.25 | 56.00 | 56.03 | 54.36 | **226.63** |
| 545 | 15.08 | 12.57 | 12.38 | 12.16 | **52.19** |
| 546 | 83.31 | 74.46 | 66.03 | 63.26 | **287.07** |
| 547 | 66.35 | 54.48 | 52.82 | 56.27 | **229.92** |
| 548 | 35.63 | 33.06 | 31.66 | 28.31 | **128.65** |
| 549 | 155.33 | 137.29 | 132.46 | 129.37 | **554.45** |
| **WI - WISCONSIN - Total** | **1,315.89** | **1,168.39** | **1,076.41** | **997.05** | **4,557.74** |

**STATE: WV - WEST VIRGINIA**

| REGISTRANT ZIP CODE 3 | 1ST QUARTER | 2ND QUARTER | 3RD QUARTER | 4TH QUARTER | TOTAL GRAMS |
|---|---|---|---|---|---|
| 247 | 14.06 | 13.36 | 12.17 | 11.87 | **51.46** |
| 248 | 5.67 | 4.58 | 5.24 | 3.93 | **19.43** |
| 249 | 6.13 | 6.45 | 5.78 | 3.96 | **22.31** |
| 250 | 15.12 | 13.37 | 12.70 | 11.54 | **52.73** |
| 251 | 6.22 | 11.54 | 9.14 | 8.27 | **35.16** |
| 252 | 12.47 | 9.73 | 7.55 | 7.37 | **37.12** |
| 253 | 60.72 | 48.96 | 40.96 | 41.47 | **192.11** |
| 254 | 60.29 | 55.49 | 47.79 | 49.63 | **213.20** |
| 255 | 24.79 | 22.54 | 19.88 | 19.20 | **86.41** |
| 256 | 7.56 | 8.61 | 7.22 | 7.66 | **31.06** |
| 257 | 34.78 | 38.91 | 33.28 | 39.94 | **146.90** |
| 258 | 18.28 | 16.46 | 15.17 | 16.69 | **66.61** |
| 259 | 9.86 | 8.03 | 6.73 | 7.01 | **31.64** |
| 260 | 35.78 | 31.16 | 29.92 | 28.04 | **124.89** |
| 261 | 26.91 | 19.12 | 16.65 | 16.06 | **78.73** |
| 262 | 19.34 | 17.68 | 17.17 | 14.23 | **68.42** |
| 263 | 27.05 | 24.60 | 24.24 | 24.26 | **100.15** |

| | | | | | |
|---|---|---|---|---|---|
| 264 | 8.96 | 10.36 | 10.45 | 8.69 | **38.46** |
| 265 | 53.67 | 51.01 | 44.83 | 47.33 | **196.85** |
| 266 | 7.80 | 5.98 | 5.32 | 5.38 | **24.47** |
| 267 | 16.94 | 16.97 | 15.54 | 14.20 | **63.66** |
| 268 | 8.00 | 8.61 | 7.25 | 5.04 | **28.91** |
| **WV - WEST VIRGINIA - Total** | **480.43** | **443.49** | **394.99** | **391.76** | **1,710.68** |

**STATE: WY - WYOMING**

| REGISTRANT ZIP CODE 3 | 1ST QUARTER | 2ND QUARTER | 3RD QUARTER | 4TH QUARTER | **TOTAL GRAMS** |
|---|---|---|---|---|---|
| 820 | 27.25 | 22.73 | 20.83 | 20.09 | **90.89** |
| 822 | 6.08 | 6.38 | 5.15 | 5.49 | **23.11** |
| 823 | 1.50 | 1.24 | 1.22 | 0.78 | **4.75** |
| 824 | 18.99 | 16.35 | 18.63 | 14.81 | **68.78** |
| 825 | 7.68 | 5.44 | 5.49 | 5.34 | **23.94** |
| 826 | 23.16 | 22.43 | 21.34 | 19.80 | **86.73** |
| 827 | 5.48 | 5.68 | 6.16 | 4.11 | **21.44** |
| 828 | 8.20 | 7.05 | 6.82 | 5.85 | **27.92** |
| 829 | 31.07 | 24.87 | 22.89 | 24.68 | **103.51** |
| 830 | 5.62 | 4.21 | 4.27 | 3.84 | **17.94** |
| 831 | 6.03 | 4.52 | 4.94 | 6.21 | **21.71** |
| **WY - WYOMING - Total** | **141.06** | **120.91** | **117.74** | **111.00** | **490.71** |

**DRUG: 1100 - AMPHETAMINE**

| STATE | 1ST QUARTER | 2ND QUARTER | 3RD QUARTER | 4TH QUARTER | TOTAL GRAMS |
|---|---|---|---|---|---|
| ALABAMA | 135,476.45 | 134,539.67 | 137,334.74 | 137,252.15 | 544,603.01 |
| ALASKA | 8,200.25 | 8,513.41 | 8,518.59 | 8,668.44 | 33,900.69 |
| AMERICAN SAMOA | 0 | 2.50 | 0 | 0 | 2.50 |
| ARIZONA | 87,488.48 | 94,365.76 | 102,044.82 | 92,476.65 | 376,375.70 |
| ARKANSAS | 52,324.62 | 51,480.47 | 52,927.92 | 52,539.62 | 209,272.63 |
| CALIFORNIA | 300,208.74 | 309,327.49 | 305,214.17 | 313,452.33 | 1,228,202.73 |
| COLORADO | 83,248.50 | 84,854.33 | 87,509.13 | 89,339.44 | 344,951.40 |
| CONNECTICUT | 55,629.20 | 55,915.61 | 57,420.84 | 58,251.72 | 227,217.37 |
| DELAWARE | 21,085.06 | 21,261.45 | 21,178.96 | 21,541.78 | 85,067.25 |
| DISTRICT OF COLUMBIA | 12,090.86 | 12,441.63 | 12,465.45 | 12,530.71 | 49,528.65 |
| FLORIDA | 319,019.15 | 323,318.57 | 320,630.26 | 323,934.22 | 1,286,902.21 |
| GEORGIA | 196,409.28 | 192,243.75 | 198,303.85 | 199,123.65 | 786,080.52 |
| GUAM | 371.55 | 474.05 | 488.00 | 425.77 | 1,759.37 |
| HAWAII | 10,119.13 | 10,318.54 | 10,676.67 | 10,119.02 | 41,233.36 |
| IDAHO | 27,251.22 | 27,959.13 | 28,207.65 | 29,402.52 | 112,820.52 |
| ILLINOIS | 196,812.32 | 197,343.37 | 201,042.12 | 204,928.58 | 800,126.39 |
| INDIANA | 134,905.57 | 134,666.73 | 137,057.34 | 138,147.04 | 544,776.67 |
| IOWA | 51,563.94 | 51,941.54 | 53,160.32 | 54,137.53 | 210,803.33 |
| KANSAS | 62,589.40 | 63,212.10 | 64,115.24 | 64,634.36 | 254,551.10 |
| KENTUCKY | 72,196.77 | 70,509.45 | 71,489.75 | 72,311.88 | 286,507.85 |
| LOUISIANA | 162,113.52 | 161,690.52 | 168,800.94 | 169,361.04 | 661,966.01 |
| MAINE | 23,492.06 | 23,529.79 | 24,881.99 | 24,503.48 | 96,407.31 |
| MARYLAND | 94,580.51 | 97,001.98 | 95,864.34 | 98,045.05 | 385,491.88 |
| MASSACHUSETTS | 180,782.88 | 181,178.42 | 183,546.84 | 186,618.99 | 732,127.12 |
| MICHIGAN | 244,857.64 | 243,689.95 | 243,316.89 | 246,963.00 | 978,827.49 |
| MINNESOTA | 110,209.54 | 110,828.85 | 110,480.36 | 115,207.26 | 446,726.01 |
| MISSISSIPPI | 61,959.09 | 60,749.14 | 62,922.53 | 61,716.51 | 247,347.27 |
| MISSOURI | 136,734.01 | 137,180.84 | 141,555.73 | 154,782.23 | 570,252.80 |
| MONTANA | 14,318.39 | 14,715.49 | 14,864.09 | 15,189.87 | 59,087.84 |
| NEBRASKA | 26,760.12 | 27,240.52 | 27,621.89 | 28,395.64 | 110,018.17 |
| NEVADA | 26,446.23 | 26,727.17 | 27,429.70 | 27,450.72 | 108,053.81 |
| NEW HAMPSHIRE | 39,302.49 | 40,236.40 | 41,310.22 | 40,604.80 | 161,453.90 |
| NEW JERSEY | 106,182.82 | 109,314.85 | 110,044.20 | 112,697.38 | 438,239.25 |
| NEW MEXICO | 14,967.28 | 15,203.27 | 15,463.42 | 15,562.75 | 61,196.72 |
| NEW YORK | 242,980.79 | 250,546.17 | 249,153.25 | 256,466.39 | 999,146.60 |
| NORTH CAROLINA | 212,289.15 | 208,548.54 | 212,799.85 | 220,673.55 | 854,311.09 |
| NORTH DAKOTA | 9,199.58 | 9,183.41 | 9,486.72 | 9,637.35 | 37,507.06 |
| OHIO | 213,799.88 | 209,380.92 | 201,041.51 | 207,477.66 | 831,699.97 |
| OKLAHOMA | 54,762.92 | 54,197.33 | 54,995.55 | 55,124.71 | 219,080.50 |
| OREGON | 60,579.93 | 63,167.43 | 62,182.52 | 63,991.50 | 249,921.37 |
| PENNSYLVANIA | 213,400.03 | 215,641.98 | 214,275.61 | 215,447.84 | 858,765.46 |

| | | | | | |
|---|---|---|---|---|---|
| PUERTO RICO | 9,570.15 | 9,204.07 | 9,292.09 | 9,789.29 | 37,855.60 |
| RHODE ISLAND | 38,092.55 | 38,387.90 | 39,379.68 | 38,655.81 | 154,515.93 |
| SOUTH CAROLINA | 139,289.57 | 135,695.31 | 141,608.66 | 141,163.81 | 557,757.34 |
| SOUTH DAKOTA | 15,798.14 | 14,841.50 | 15,203.28 | 15,328.63 | 61,171.55 |
| TENNESSEE | 133,694.24 | 133,302.95 | 137,110.71 | 135,800.52 | 539,908.42 |
| TEXAS | 432,137.12 | 436,425.19 | 453,570.03 | 459,055.68 | 1,781,188.02 |
| UTAH | 86,977.90 | 87,846.30 | 90,042.90 | 93,821.93 | 358,689.03 |
| VERMONT | 10,075.97 | 10,030.16 | 10,158.72 | 10,272.84 | 40,537.69 |
| VIRGIN ISLANDS | 308.38 | 333.01 | 348.04 | 335.67 | 1,325.10 |
| VIRGINIA | 182,308.82 | 181,772.40 | 185,252.82 | 184,566.91 | 733,900.96 |
| WASHINGTON | 112,692.56 | 116,489.87 | 114,444.53 | 118,917.24 | 462,544.20 |
| WEST VIRGINIA | 23,523.40 | 23,603.65 | 23,593.42 | 23,708.16 | 94,428.63 |
| WISCONSIN | 126,465.87 | 125,370.76 | 125,884.09 | 126,925.65 | 504,646.38 |
| WYOMING | 6,870.46 | 6,776.74 | 6,930.96 | 7,044.55 | 27,622.71 |
| **1100 - AMPHETAMINE - Total** | **5,394,514.46** | **5,424,722.33** | **5,494,643.85** | **5,574,521.82** | **21,888,402.46** |

**DRUG: 1105 - METHAMPHETAMINE**

| STATE | 1ST QUARTER | 2ND QUARTER | 3RD QUARTER | 4TH QUARTER | TOTAL GRAMS |
|---|---|---|---|---|---|
| ALABAMA | 30.14 | 33.35 | 34.55 | 37.77 | 135.81 |
| ALASKA | 2.01 | 2.01 | 3.62 | 0 | 7.63 |
| ARIZONA | 36.97 | 40.18 | 63.89 | 38.97 | 180.01 |
| ARKANSAS | 8.84 | 7.23 | 2.41 | 9.64 | 28.13 |
| CALIFORNIA | 272.41 | 274.01 | 273.20 | 260.34 | 1,079.95 |
| COLORADO | 56.66 | 59.07 | 63.49 | 56.25 | 235.47 |
| CONNECTICUT | 24.51 | 24.91 | 20.49 | 26.12 | 96.03 |
| DELAWARE | 3.21 | 5.63 | 4.42 | 4.42 | 17.68 |
| DISTRICT OF COLUMBIA | 5.22 | 11.65 | 7.23 | 5.62 | 29.73 |
| FLORIDA | 49.82 | 47.01 | 52.64 | 47.01 | 196.48 |
| GEORGIA | 24.11 | 16.88 | 22.50 | 25.31 | 88.80 |
| HAWAII | 5.22 | 4.82 | 4.82 | 7.23 | 22.10 |
| IDAHO | 15.67 | 12.05 | 14.06 | 13.66 | 55.44 |
| ILLINOIS | 55.45 | 51.84 | 51.83 | 48.62 | 207.74 |
| INDIANA | 25.72 | 27.73 | 24.51 | 24.51 | 102.47 |
| IOWA | 17.28 | 17.68 | 21.30 | 14.46 | 70.72 |
| KANSAS | 14.47 | 15.67 | 15.67 | 15.67 | 61.48 |
| KENTUCKY | 10.85 | 11.65 | 14.06 | 10.45 | 47.01 |
| LOUISIANA | 15.67 | 15.27 | 15.67 | 13.66 | 60.27 |
| MAINE | 10.45 | 10.45 | 20.09 | 12.05 | 53.04 |
| MARYLAND | 36.97 | 33.35 | 38.57 | 45.40 | 154.29 |
| MASSACHUSETTS | 43.00 | 37.77 | 31.74 | 26.92 | 139.43 |
| MICHIGAN | 30.54 | 25.31 | 24.51 | 20.89 | 101.26 |

| | | | | | |
|---|---|---|---|---|---|
| MINNESOTA | 77.14 | 71.12 | 73.93 | 78.75 | 300.95 |
| MISSISSIPPI | 6.43 | 6.03 | 8.44 | 10.04 | 30.94 |
| MISSOURI | 27.73 | 44.60 | 41.78 | 58.25 | 172.36 |
| MONTANA | 0 | 0.40 | 0 | 0 | 0.40 |
| NEBRASKA | 5.22 | 6.03 | 7.63 | 1.21 | 20.09 |
| NEVADA | 19.29 | 25.72 | 22.90 | 14.06 | 81.98 |
| NEW HAMPSHIRE | 5.63 | 5.22 | 4.82 | 4.02 | 19.69 |
| NEW JERSEY | 43.40 | 32.55 | 33.75 | 29.73 | 139.43 |
| NEW MEXICO | 4.82 | 3.21 | 6.03 | 8.84 | 22.90 |
| NEW YORK | 97.64 | 90.41 | 89.60 | 87.99 | 365.64 |
| NORTH CAROLINA | 50.23 | 51.83 | 50.23 | 38.97 | 191.26 |
| NORTH DAKOTA | 3.62 | 7.23 | 4.02 | 4.42 | 19.28 |
| OHIO | 20.09 | 71.53 | 33.75 | 32.15 | 157.52 |
| OKLAHOMA | 16.07 | 12.46 | 15.27 | 10.85 | 54.65 |
| OREGON | 39.78 | 27.32 | 34.95 | 40.17 | 142.23 |
| PENNSYLVANIA | 122.55 | 124.16 | 108.09 | 126.56 | 481.36 |
| RHODE ISLAND | 3.62 | 4.42 | 2.01 | 4.82 | 14.87 |
| SOUTH CAROLINA | 16.07 | 19.29 | 14.06 | 23.31 | 72.73 |
| SOUTH DAKOTA | 10.85 | 6.83 | 5.63 | 8.03 | 31.34 |
| TENNESSEE | 14.47 | 13.26 | 18.08 | 13.26 | 59.07 |
| TEXAS | 106.88 | 108.49 | 103.66 | 105.27 | 424.31 |
| UTAH | 28.93 | 23.71 | 21.70 | 20.09 | 94.43 |
| VERMONT | 2.01 | 1.21 | 2.81 | 2.41 | 8.44 |
| VIRGINIA | 34.96 | 28.93 | 32.95 | 29.33 | 126.17 |
| WASHINGTON | 92.02 | 93.63 | 90.00 | 77.94 | 353.59 |
| WEST VIRGINIA | 4.82 | 3.62 | 3.21 | 6.83 | 18.48 |
| WISCONSIN | 32.14 | 35.76 | 35.36 | 34.95 | 138.21 |
| WYOMING | 4.82 | 3.62 | 5.22 | 4.82 | 18.48 |
| **1105 - METHAMPHETAMINE -** | **1,686.41** | **1,708.10** | **1,695.18** | **1,642.07** | **6,731.76** |

**DRUG: 1205 - LISDEXAMFETAMINE**

| STATE | 1ST QUARTER | 2ND QUARTER | 3RD QUARTER | 4TH QUARTER | TOTAL GRAMS |
|---|---|---|---|---|---|
| ALABAMA | 72,123.93 | 69,118.13 | 70,050.78 | 72,556.75 | 283,849.59 |
| ALASKA | 2,562.72 | 2,630.36 | 2,740.19 | 2,695.10 | 10,628.37 |
| ARIZONA | 38,400.29 | 41,600.65 | 48,559.82 | 40,465.84 | 169,026.61 |
| ARKANSAS | 30,656.30 | 28,890.06 | 28,698.50 | 29,708.18 | 117,953.04 |
| CALIFORNIA | 112,339.92 | 112,485.61 | 110,815.58 | 116,614.45 | 452,255.56 |
| COLORADO | 28,951.83 | 28,504.38 | 28,739.66 | 30,308.05 | 116,503.91 |
| CONNECTICUT | 36,169.40 | 36,193.68 | 35,273.35 | 37,166.63 | 144,803.07 |
| DELAWARE | 14,356.54 | 13,805.61 | 13,252.94 | 14,334.57 | 55,749.65 |
| DISTRICT OF COLUMBIA | 5,532.42 | 5,735.33 | 5,554.38 | 5,786.20 | 22,608.33 |

| | | | | | |
|---|---|---|---|---|---|
| FLORIDA | 138,767.77 | 133,537.99 | 128,723.45 | 140,070.45 | 541,099.66 |
| GEORGIA | 100,588.82 | 95,591.18 | 96,744.25 | 99,001.36 | 391,925.61 |
| GUAM | 9.25 | 18.50 | 8.67 | 7.52 | 43.94 |
| HAWAII | 1,452.19 | 1,582.26 | 1,573.01 | 1,507.68 | 6,115.14 |
| IDAHO | 9,651.96 | 9,595.88 | 9,329.38 | 10,012.11 | 38,589.33 |
| ILLINOIS | 71,443.91 | 71,857.83 | 71,109.77 | 74,655.83 | 289,067.34 |
| INDIANA | 85,840.33 | 81,939.32 | 81,501.12 | 84,682.40 | 333,963.17 |
| IOWA | 34,637.44 | 34,215.43 | 33,062.70 | 34,930.54 | 136,846.10 |
| KANSAS | 31,682.77 | 31,863.72 | 31,166.53 | 33,139.58 | 127,852.60 |
| KENTUCKY | 42,106.49 | 40,397.62 | 40,230.79 | 41,066.46 | 163,801.36 |
| LOUISIANA | 90,976.44 | 87,437.36 | 87,415.08 | 90,414.84 | 356,243.72 |
| MAINE | 16,908.85 | 16,845.26 | 16,438.27 | 16,795.54 | 66,987.92 |
| MARYLAND | 56,495.81 | 57,306.97 | 54,140.71 | 58,491.00 | 226,434.49 |
| MASSACHUSETTS | 36,835.38 | 37,767.27 | 36,205.25 | 39,647.25 | 150,455.15 |
| MICHIGAN | 61,743.97 | 60,558.49 | 58,509.50 | 63,991.62 | 244,803.58 |
| MINNESOTA | 30,854.35 | 30,869.38 | 30,182.02 | 32,355.68 | 124,261.44 |
| MISSISSIPPI | 39,409.66 | 36,095.99 | 36,796.64 | 37,321.56 | 149,623.84 |
| MISSOURI | 65,805.12 | 63,269.40 | 62,838.31 | 76,370.48 | 268,283.32 |
| MONTANA | 6,157.34 | 5,952.70 | 6,096.06 | 6,433.10 | 24,639.20 |
| NEBRASKA | 17,319.15 | 16,736.10 | 17,063.94 | 18,331.38 | 69,450.56 |
| NEVADA | 7,995.86 | 7,875.63 | 7,723.47 | 7,886.44 | 31,481.41 |
| NEW HAMPSHIRE | 13,841.45 | 14,374.46 | 13,759.36 | 14,562.92 | 56,538.18 |
| NEW JERSEY | 48,479.93 | 49,337.40 | 46,962.10 | 49,997.07 | 194,776.49 |
| NEW MEXICO | 4,145.56 | 3,690.59 | 3,864.60 | 3,977.91 | 15,678.65 |
| NEW YORK | 86,529.15 | 87,894.32 | 83,500.76 | 90,616.00 | 348,540.24 |
| NORTH CAROLINA | 107,240.44 | 104,919.37 | 99,882.38 | 111,037.40 | 423,079.59 |
| NORTH DAKOTA | 6,038.83 | 5,649.19 | 5,725.50 | 6,108.78 | 23,522.31 |
| OHIO | 109,273.88 | 100,450.21 | 92,434.78 | 98,111.14 | 400,270.01 |
| OKLAHOMA | 38,073.52 | 36,397.88 | 36,120.95 | 38,071.56 | 148,663.91 |
| OREGON | 15,509.27 | 15,870.09 | 15,413.88 | 15,945.15 | 62,738.39 |
| PENNSYLVANIA | 88,934.33 | 89,087.52 | 86,022.44 | 91,627.69 | 355,671.98 |
| PUERTO RICO | 2,270.20 | 2,042.43 | 1,992.71 | 2,172.50 | 8,477.84 |
| RHODE ISLAND | 7,395.06 | 7,403.15 | 7,167.28 | 7,492.75 | 29,458.24 |
| SOUTH CAROLINA | 61,834.65 | 60,357.57 | 58,239.07 | 62,860.86 | 243,292.14 |
| SOUTH DAKOTA | 10,579.23 | 10,189.59 | 9,770.47 | 10,443.95 | 40,983.24 |
| TENNESSEE | 68,950.48 | 66,341.86 | 65,924.39 | 68,314.63 | 269,531.35 |
| TEXAS | 272,010.78 | 262,408.59 | 257,910.38 | 272,618.33 | 1,064,948.09 |
| UTAH | 22,578.56 | 22,089.45 | 21,898.79 | 23,710.32 | 90,277.12 |
| VERMONT | 7,231.45 | 7,107.16 | 6,995.59 | 7,368.46 | 28,702.67 |
| VIRGIN ISLANDS | 70.53 | 58.97 | 80.36 | 67.64 | 277.49 |

| | | | | | |
|---|---|---|---|---|---|
| VIRGINIA | 67,860.17 | 66,442.17 | 64,874.90 | 69,065.29 | 268,242.53 |
| WASHINGTON | 35,036.73 | 35,734.76 | 34,641.37 | 37,186.34 | 142,599.20 |
| WEST VIRGINIA | 19,119.50 | 18,848.37 | 17,940.18 | 19,243.21 | 75,151.27 |
| WISCONSIN | 72,286.20 | 72,258.45 | 71,146.19 | 76,018.99 | 291,709.84 |
| WYOMING | 2,750.60 | 2,647.70 | 2,486.41 | 2,633.82 | 10,518.53 |
| **1205 - LISDEXAMFETAMINE - T** | **2,455,816.67** | **2,401,879.32** | **2,355,298.97** | **2,496,001.33** | **9,708,996.30** |

**DRUG: 1724 - METHYLPHENIDATE (DL;D;L;ISOMERS)**

| STATE | 1ST QUARTER | 2ND QUARTER | 3RD QUARTER | 4TH QUARTER | TOTAL GRAMS |
|---|---|---|---|---|---|
| ALABAMA | 73,954.47 | 67,629.75 | 67,718.28 | 71,802.60 | 281,105.10 |
| ALASKA | 8,236.69 | 8,011.60 | 7,459.48 | 8,286.37 | 31,994.14 |
| AMERICAN SAMOA | 12.97 | 7.78 | 6.92 | 16.43 | 44.10 |
| ARIZONA | 77,304.46 | 82,216.33 | 94,718.85 | 76,779.75 | 331,019.40 |
| ARKANSAS | 39,785.47 | 37,383.60 | 36,914.01 | 38,137.82 | 152,220.89 |
| CALIFORNIA | 268,922.66 | 263,072.88 | 245,924.94 | 264,375.09 | 1,042,295.57 |
| COLORADO | 62,927.66 | 60,547.88 | 58,822.20 | 62,726.88 | 245,024.63 |
| CONNECTICUT | 58,262.20 | 54,993.48 | 50,379.41 | 54,485.01 | 218,120.11 |
| DELAWARE | 18,553.13 | 17,466.85 | 16,661.60 | 17,816.85 | 70,498.43 |
| DISTRICT OF COLUMBIA | 8,490.85 | 7,763.08 | 7,340.67 | 8,046.73 | 31,641.33 |
| FLORIDA | 209,254.86 | 197,980.76 | 184,626.22 | 193,019.84 | 784,881.68 |
| GEORGIA | 167,815.59 | 150,931.34 | 152,737.31 | 159,353.22 | 630,837.46 |
| GUAM | 404.01 | 477.07 | 675.57 | 428.08 | 1,984.74 |
| HAWAII | 11,227.21 | 10,823.48 | 11,014.27 | 10,683.77 | 43,748.73 |
| IDAHO | 28,691.29 | 27,986.90 | 26,288.20 | 27,501.42 | 110,467.80 |
| ILLINOIS | 173,329.01 | 164,316.41 | 157,083.11 | 168,351.34 | 663,079.86 |
| INDIANA | 120,590.42 | 113,821.67 | 109,218.78 | 114,266.85 | 457,897.73 |
| IOWA | 81,724.04 | 80,095.38 | 75,296.80 | 79,857.27 | 316,973.49 |
| KANSAS | 49,155.41 | 48,312.66 | 46,037.33 | 49,753.01 | 193,258.41 |
| KENTUCKY | 76,769.40 | 70,662.09 | 70,582.99 | 72,879.85 | 290,894.33 |
| LOUISIANA | 86,052.29 | 77,650.93 | 77,108.30 | 85,027.56 | 325,839.09 |
| MAINE | 36,018.64 | 35,549.13 | 33,511.13 | 33,426.83 | 138,505.73 |
| MARYLAND | 101,577.01 | 99,931.99 | 89,718.61 | 97,356.66 | 388,584.27 |
| MASSACHUSETTS | 146,426.72 | 143,662.81 | 129,424.98 | 142,116.33 | 561,630.85 |
| MICHIGAN | 188,466.09 | 179,211.28 | 166,223.22 | 178,149.49 | 712,050.08 |
| MINNESOTA | 103,660.03 | 98,883.75 | 94,073.07 | 102,086.78 | 398,703.63 |
| MISSISSIPPI | 42,505.17 | 39,683.90 | 40,091.23 | 42,642.84 | 164,923.13 |
| MISSOURI | 123,539.76 | 121,203.09 | 121,659.82 | 148,942.62 | 515,345.28 |
| MONTANA | 17,912.93 | 16,945.32 | 15,852.89 | 16,972.77 | 67,683.91 |
| NEBRASKA | 31,178.63 | 29,875.99 | 29,465.08 | 31,109.35 | 121,629.05 |
| NEVADA | 14,970.31 | 14,279.54 | 14,408.94 | 14,557.37 | 58,216.16 |
| NEW HAMPSHIRE | 33,912.80 | 34,077.66 | 31,849.73 | 33,123.67 | 132,963.86 |

| | | | | | |
|---|---|---|---|---|---|
| NEW JERSEY | 92,399.11 | 91,074.92 | 81,589.86 | 90,439.94 | 355,503.83 |
| NEW MEXICO | 19,346.00 | 17,761.48 | 17,259.56 | 18,074.75 | 72,441.79 |
| NEW YORK | 231,770.97 | 231,423.99 | 207,622.54 | 226,850.77 | 897,668.27 |
| NORTH CAROLINA | 193,329.00 | 181,622.18 | 163,836.01 | 182,231.37 | 721,018.56 |
| NORTH DAKOTA | 15,661.85 | 14,816.96 | 14,297.70 | 15,483.09 | 60,259.60 |
| OHIO | 223,286.55 | 206,612.44 | 183,164.36 | 199,316.48 | 812,379.83 |
| OKLAHOMA | 50,133.42 | 45,323.93 | 44,368.37 | 46,713.07 | 186,538.79 |
| OREGON | 61,940.45 | 62,899.40 | 58,891.81 | 62,070.04 | 245,801.71 |
| PENNSYLVANIA | 225,860.97 | 219,876.07 | 205,477.41 | 219,873.88 | 871,088.33 |
| PUERTO RICO | 15,201.86 | 14,619.82 | 13,373.30 | 14,194.99 | 57,389.98 |
| RHODE ISLAND | 19,259.60 | 18,695.05 | 16,735.99 | 17,847.06 | 72,537.70 |
| SOUTH CAROLINA | 89,615.93 | 84,394.62 | 79,721.60 | 86,314.11 | 340,046.25 |
| SOUTH DAKOTA | 22,043.10 | 19,277.23 | 19,977.33 | 20,400.21 | 81,697.86 |
| TENNESSEE | 83,708.72 | 78,659.73 | 77,707.66 | 81,088.40 | 321,164.51 |
| TEXAS | 335,585.23 | 321,501.38 | 295,348.98 | 331,196.25 | 1,283,631.84 |
| UTAH | 47,173.78 | 45,120.91 | 43,905.93 | 47,503.23 | 183,703.86 |
| VERMONT | 19,113.46 | 18,889.50 | 17,964.74 | 18,957.69 | 74,925.40 |
| VIRGIN ISLANDS | 135.03 | 159.83 | 132.68 | 158.62 | 586.16 |
| VIRGINIA | 135,949.94 | 131,084.77 | 123,784.72 | 129,736.33 | 520,555.76 |
| WASHINGTON | 114,614.62 | 114,297.71 | 105,303.23 | 113,412.05 | 447,627.62 |
| WEST VIRGINIA | 32,337.10 | 30,539.97 | 28,472.70 | 29,392.91 | 120,742.68 |
| WISCONSIN | 109,059.24 | 100,475.74 | 96,366.22 | 103,108.32 | 409,009.53 |
| WYOMING | 7,817.34 | 7,546.33 | 7,028.17 | 7,895.84 | 30,287.69 |
| **1724 - METHYLPHENIDATE (DI** | **4,606,975.47** | **4,412,130.36** | **4,165,224.79** | **4,466,339.90** | **17,650,670.52** |

**DRUG: 9050 - CODEINE**

| STATE | 1ST QUARTER | 2ND QUARTER | 3RD QUARTER | 4TH QUARTER | TOTAL GRAMS |
|---|---|---|---|---|---|
| ALABAMA | 54,727.52 | 48,860.49 | 49,149.79 | 48,867.23 | 201,605.04 |
| ALASKA | 6,246.24 | 6,092.57 | 6,095.09 | 5,135.85 | 23,569.75 |
| AMERICAN SAMOA | 190.07 | 353.62 | 181.23 | 335.94 | 1,060.85 |
| ARIZONA | 67,250.60 | 57,385.77 | 57,667.06 | 54,979.04 | 237,282.47 |
| ARKANSAS | 67,887.27 | 60,785.00 | 58,982.89 | 54,092.64 | 241,747.80 |
| CALIFORNIA | 430,883.25 | 403,723.32 | 399,591.36 | 370,959.56 | 1,605,157.49 |
| COLORADO | 24,603.70 | 21,420.62 | 20,764.32 | 17,943.44 | 84,732.08 |
| CONNECTICUT | 20,352.91 | 20,383.90 | 19,023.12 | 18,720.29 | 78,480.23 |
| DELAWARE | 8,417.06 | 8,170.89 | 7,638.85 | 7,722.29 | 31,949.10 |
| DISTRICT OF COLUMBIA | 11,055.64 | 10,557.62 | 10,837.50 | 9,011.16 | 41,461.92 |
| FLORIDA | 170,848.17 | 152,311.43 | 140,653.09 | 136,684.76 | 600,497.44 |
| GEORGIA | 102,881.63 | 92,009.82 | 88,385.31 | 82,651.93 | 365,928.70 |
| GUAM | 930.91 | 1,022.54 | 961.07 | 684.38 | 3,598.89 |
| HAWAII | 12,322.06 | 12,007.05 | 11,505.00 | 10,590.95 | 46,425.06 |

| | | | | | |
|---|---|---|---|---|---|
| IDAHO | 13,103.49 | 10,988.15 | 12,406.29 | 12,195.92 | 48,693.85 |
| ILLINOIS | 147,393.89 | 141,391.15 | 143,244.27 | 137,326.15 | 569,355.47 |
| INDIANA | 48,350.18 | 44,169.55 | 41,342.85 | 40,841.85 | 174,704.43 |
| IOWA | 18,778.90 | 16,624.91 | 19,371.99 | 16,065.61 | 70,841.41 |
| KANSAS | 26,476.68 | 23,172.33 | 24,761.49 | 21,299.76 | 95,710.26 |
| KENTUCKY | 46,514.79 | 42,349.15 | 41,033.17 | 38,283.05 | 168,180.16 |
| LOUISIANA | 40,578.13 | 38,011.36 | 37,740.26 | 36,539.36 | 152,869.10 |
| MAINE | 9,685.35 | 8,521.93 | 8,993.59 | 8,231.37 | 35,432.24 |
| MARYLAND | 51,471.18 | 51,729.40 | 46,250.02 | 46,646.09 | 196,096.69 |
| MASSACHUSETTS | 41,180.01 | 42,153.20 | 37,803.57 | 39,831.35 | 160,968.12 |
| MICHIGAN | 229,114.90 | 209,125.64 | 190,561.36 | 181,192.81 | 809,994.72 |
| MINNESOTA | 34,477.21 | 31,322.14 | 32,261.78 | 31,158.43 | 129,219.55 |
| MISSISSIPPI | 26,473.27 | 22,707.65 | 22,463.40 | 21,002.91 | 92,647.22 |
| MISSOURI | 62,564.48 | 58,635.55 | 63,647.09 | 58,233.03 | 243,080.15 |
| MONTANA | 9,307.13 | 8,630.81 | 9,399.30 | 8,199.65 | 35,536.89 |
| NEBRASKA | 11,220.79 | 9,908.30 | 10,436.56 | 9,635.11 | 41,200.75 |
| NEVADA | 21,456.72 | 20,182.65 | 19,476.62 | 17,679.12 | 78,795.10 |
| NEW HAMPSHIRE | 6,959.61 | 6,642.68 | 6,536.66 | 6,238.14 | 26,377.09 |
| NEW JERSEY | 69,550.53 | 70,103.54 | 64,841.12 | 65,631.62 | 270,126.81 |
| NEW MEXICO | 18,715.14 | 15,001.65 | 16,773.51 | 15,516.60 | 66,006.90 |
| NEW YORK | 128,965.32 | 129,191.83 | 121,237.90 | 118,201.43 | 497,596.48 |
| NORTH CAROLINA | 45,817.43 | 40,303.60 | 36,979.98 | 40,160.23 | 163,261.25 |
| NORTH DAKOTA | 5,075.13 | 4,819.25 | 4,361.26 | 4,519.80 | 18,775.44 |
| OHIO | 87,921.76 | 79,899.41 | 74,466.66 | 65,268.02 | 307,555.84 |
| OKLAHOMA | 91,443.31 | 87,585.78 | 87,457.44 | 81,609.29 | 348,095.81 |
| OREGON | 33,296.35 | 30,218.54 | 30,357.15 | 27,913.79 | 121,785.82 |
| PENNSYLVANIA | 116,920.14 | 114,526.52 | 107,715.57 | 105,507.30 | 444,669.53 |
| PUERTO RICO | 11,488.10 | 12,406.40 | 12,334.57 | 10,670.36 | 46,899.43 |
| RHODE ISLAND | 6,187.34 | 6,162.52 | 5,228.73 | 5,434.65 | 23,013.23 |
| SOUTH CAROLINA | 73,303.29 | 71,961.54 | 72,879.51 | 62,830.54 | 280,974.88 |
| SOUTH DAKOTA | 5,955.53 | 5,509.88 | 5,969.95 | 5,414.47 | 22,849.83 |
| TENNESSEE | 74,435.49 | 66,341.46 | 70,446.16 | 57,737.07 | 268,960.19 |
| TEXAS | 929,061.13 | 950,677.88 | 903,428.79 | 877,951.82 | 3,661,119.62 |
| UTAH | 16,879.56 | 13,938.22 | 14,122.46 | 13,897.58 | 58,837.81 |
| VERMONT | 2,703.50 | 2,846.79 | 2,691.26 | 2,645.41 | 10,886.96 |
| VIRGIN ISLANDS | 634.30 | 834.39 | 457.64 | 852.07 | 2,778.39 |
| VIRGINIA | 50,351.99 | 45,915.13 | 45,132.92 | 43,161.77 | 184,561.82 |
| WASHINGTON | 57,193.39 | 53,917.21 | 53,802.13 | 49,912.10 | 214,824.84 |
| WEST VIRGINIA | 34,811.79 | 35,809.87 | 36,200.31 | 36,483.29 | 143,305.26 |
| WISCONSIN | 44,288.96 | 41,748.88 | 41,797.83 | 40,326.02 | 168,161.69 |

| | | | | | |
|---|---|---|---|---|---|
| WYOMING | 4,052.12 | 3,370.81 | 3,692.82 | 3,214.50 | 14,330.25 |
| **9050 - CODEINE - Total** | **3,732,755.34** | **3,564,442.25** | **3,451,541.66** | **3,283,838.88** | **14,032,578.12** |

**DRUG: 9064 - BUPRENORPHINE**

| STATE | 1ST QUARTER | 2ND QUARTER | 3RD QUARTER | 4TH QUARTER | TOTAL GRAMS |
|---|---|---|---|---|---|
| ALABAMA | 26,918.25 | 28,660.45 | 29,452.35 | 30,617.09 | 115,648.15 |
| ALASKA | 2,583.83 | 2,670.28 | 2,643.71 | 2,666.55 | 10,564.38 |
| AMERICAN SAMOA | 0.02 | 0.00 | 0.02 | 0 | 0.04 |
| ARIZONA | 10,007.42 | 10,879.74 | 11,625.98 | 12,533.59 | 45,046.72 |
| ARKANSAS | 6,565.61 | 7,249.03 | 7,486.34 | 7,773.26 | 29,074.25 |
| CALIFORNIA | 40,133.32 | 41,842.59 | 43,966.11 | 46,057.94 | 171,999.96 |
| COLORADO | 8,035.07 | 8,515.86 | 8,917.11 | 9,735.21 | 35,203.25 |
| CONNECTICUT | 15,509.93 | 16,278.45 | 16,070.59 | 17,229.21 | 65,088.17 |
| DELAWARE | 3,751.34 | 4,063.38 | 3,967.31 | 4,123.57 | 15,905.60 |
| DISTRICT OF COLUMBIA | 2,751.34 | 2,730.72 | 2,848.24 | 3,259.01 | 11,589.32 |
| FLORIDA | 49,002.40 | 51,589.45 | 52,741.76 | 54,784.49 | 208,118.09 |
| GEORGIA | 16,149.07 | 17,121.52 | 17,456.56 | 18,422.76 | 69,149.91 |
| GUAM | 0.62 | 0.25 | 1.44 | 0.00 | 2.31 |
| HAWAII | 2,041.12 | 2,082.06 | 2,181.28 | 2,290.04 | 8,594.50 |
| IDAHO | 3,886.57 | 4,186.43 | 4,269.21 | 4,530.59 | 16,872.80 |
| ILLINOIS | 15,599.99 | 16,614.32 | 16,829.01 | 17,806.76 | 66,850.09 |
| INDIANA | 21,635.49 | 23,911.83 | 24,978.36 | 27,081.35 | 97,607.04 |
| IOWA | 1,907.67 | 2,010.56 | 2,118.01 | 2,224.01 | 8,260.26 |
| KANSAS | 2,569.61 | 2,800.55 | 2,844.81 | 3,053.53 | 11,268.50 |
| KENTUCKY | 36,959.27 | 37,462.35 | 39,565.39 | 41,642.84 | 155,629.85 |
| LOUISIANA | 19,910.96 | 21,105.86 | 21,617.02 | 21,992.64 | 84,626.49 |
| MAINE | 10,449.02 | 10,912.25 | 11,527.59 | 11,989.83 | 44,878.69 |
| MARYLAND | 27,322.15 | 28,629.16 | 29,185.85 | 30,846.33 | 115,983.50 |
| MASSACHUSETTS | 42,192.92 | 43,945.72 | 43,378.87 | 45,894.00 | 175,411.51 |
| MICHIGAN | 29,132.39 | 30,463.40 | 30,465.10 | 32,095.43 | 122,156.32 |
| MINNESOTA | 8,345.50 | 8,877.01 | 9,170.37 | 9,820.01 | 36,212.90 |
| MISSISSIPPI | 10,270.14 | 10,719.39 | 10,840.57 | 11,030.58 | 42,860.68 |
| MISSOURI | 11,439.92 | 12,576.15 | 13,971.52 | 15,652.20 | 53,639.78 |
| MONTANA | 2,855.90 | 2,890.90 | 3,074.04 | 3,338.87 | 12,159.72 |
| NEBRASKA | 1,404.90 | 1,469.22 | 1,474.66 | 1,615.10 | 5,963.88 |
| NEVADA | 3,582.79 | 3,713.55 | 3,869.14 | 4,125.46 | 15,290.93 |
| NEW HAMPSHIRE | 8,220.05 | 8,826.04 | 8,554.03 | 9,245.38 | 34,845.51 |
| NEW JERSEY | 26,519.91 | 27,922.84 | 27,291.06 | 29,333.86 | 111,067.66 |
| NEW MEXICO | 7,874.81 | 8,467.29 | 8,861.26 | 8,992.19 | 34,195.54 |
| NEW YORK | 57,407.42 | 60,751.72 | 61,082.71 | 65,035.79 | 244,277.64 |
| NORTH CAROLINA | 36,103.10 | 39,887.55 | 39,797.09 | 44,237.14 | 160,024.88 |

| | | | | | |
|---|---|---|---|---|---|
| NORTH DAKOTA | 1,026.66 | 1,135.30 | 1,230.55 | 1,260.77 | 4,653.28 |
| OHIO | 53,934.23 | 56,053.09 | 56,004.55 | 59,381.94 | 225,373.82 |
| OKLAHOMA | 12,373.07 | 13,358.80 | 14,210.69 | 15,527.70 | 55,470.25 |
| OREGON | 11,143.00 | 11,899.75 | 12,401.45 | 13,730.44 | 49,174.65 |
| PENNSYLVANIA | 67,466.98 | 72,494.54 | 72,171.09 | 77,817.54 | 289,950.16 |
| PUERTO RICO | 2,133.98 | 2,091.02 | 2,122.69 | 2,153.44 | 8,501.12 |
| RHODE ISLAND | 8,115.64 | 7,874.54 | 8,076.51 | 8,625.05 | 32,691.74 |
| SOUTH CAROLINA | 11,568.36 | 12,559.14 | 12,939.38 | 13,951.06 | 51,017.95 |
| SOUTH DAKOTA | 490.66 | 549.82 | 617.71 | 646.75 | 2,304.93 |
| TENNESSEE | 36,381.11 | 38,614.26 | 39,109.16 | 39,662.88 | 153,767.41 |
| TEXAS | 27,121.43 | 28,189.29 | 29,088.64 | 29,952.95 | 114,352.32 |
| UTAH | 9,746.20 | 10,153.49 | 10,562.84 | 11,112.60 | 41,575.13 |
| VERMONT | 6,608.52 | 6,593.44 | 6,829.31 | 7,133.34 | 27,164.61 |
| VIRGIN ISLANDS | 12.94 | 10.37 | 14.07 | 11.75 | 49.12 |
| VIRGINIA | 20,310.90 | 21,426.59 | 22,004.24 | 22,844.71 | 86,586.44 |
| WASHINGTON | 21,663.59 | 23,455.86 | 24,200.06 | 26,604.94 | 95,924.45 |
| WEST VIRGINIA | 15,735.89 | 15,752.44 | 16,216.90 | 16,839.75 | 64,544.98 |
| WISCONSIN | 11,198.91 | 11,807.91 | 12,217.23 | 12,891.11 | 48,115.17 |
| WYOMING | 1,374.63 | 1,304.91 | 1,303.33 | 1,374.12 | 5,357.00 |
| **9064 - BUPRENORPHINE - Tota** | **887,446.54** | **937,152.47** | **955,444.87** | **1,012,599.44** | **3,792,643.31** |

**DRUG: 9143 - OXYCODONE**

| STATE | 1ST QUARTER | 2ND QUARTER | 3RD QUARTER | 4TH QUARTER | TOTAL GRAMS |
|---|---|---|---|---|---|
| ALABAMA | 210,629.35 | 201,743.36 | 193,062.48 | 189,534.46 | 794,969.65 |
| ALASKA | 31,438.15 | 31,229.99 | 30,260.70 | 27,163.11 | 120,091.96 |
| AMERICAN SAMOA | 10.76 | 6.72 | 6.72 | 12.55 | 36.76 |
| ARIZONA | 373,301.70 | 348,072.07 | 336,321.51 | 324,662.57 | 1,382,357.85 |
| ARKANSAS | 125,960.79 | 119,531.64 | 110,436.59 | 102,994.62 | 458,923.65 |
| CALIFORNIA | 763,936.38 | 737,439.87 | 703,627.93 | 679,236.91 | 2,884,241.08 |
| COLORADO | 199,651.44 | 190,667.68 | 186,163.83 | 183,234.65 | 759,717.61 |
| CONNECTICUT | 146,495.48 | 143,349.53 | 140,044.07 | 139,055.60 | 568,944.68 |
| DELAWARE | 61,588.18 | 61,672.80 | 60,458.28 | 58,741.07 | 242,460.33 |
| DISTRICT OF COLUMBIA | 12,979.79 | 12,593.56 | 12,058.04 | 11,999.97 | 49,631.36 |
| FLORIDA | 995,706.65 | 958,397.59 | 872,843.15 | 867,553.82 | 3,694,501.21 |
| GEORGIA | 412,269.59 | 398,770.38 | 386,106.36 | 382,417.75 | 1,579,564.08 |
| GUAM | 931.05 | 680.17 | 856.25 | 678.37 | 3,145.83 |
| HAWAII | 39,005.21 | 38,418.29 | 36,044.62 | 34,475.17 | 147,943.28 |
| IDAHO | 56,161.93 | 53,237.95 | 52,122.67 | 51,402.97 | 212,925.53 |
| ILLINOIS | 142,655.20 | 135,850.40 | 130,737.24 | 129,759.85 | 539,002.68 |
| INDIANA | 212,331.25 | 200,478.12 | 192,314.74 | 191,547.00 | 796,671.10 |
| IOWA | 46,977.54 | 44,656.56 | 43,356.50 | 42,787.09 | 177,777.69 |

| | | | | | |
|---|---|---|---|---|---|
| KANSAS | 110,087.19 | 106,511.16 | 102,142.72 | 102,675.44 | 421,416.52 |
| KENTUCKY | 193,979.66 | 187,350.73 | 179,825.04 | 178,043.55 | 739,198.98 |
| LOUISIANA | 173,603.85 | 164,248.94 | 161,115.66 | 157,736.64 | 656,705.08 |
| MAINE | 47,337.06 | 45,409.21 | 44,515.35 | 43,066.75 | 180,328.38 |
| MARYLAND | 288,231.97 | 275,843.20 | 266,273.79 | 261,013.12 | 1,091,362.08 |
| MASSACHUSETTS | 216,666.74 | 208,958.42 | 199,528.24 | 199,489.45 | 824,642.85 |
| MICHIGAN | 259,927.45 | 240,113.62 | 226,675.36 | 218,614.77 | 945,331.20 |
| MINNESOTA | 127,009.73 | 122,176.79 | 117,721.14 | 114,620.63 | 481,528.29 |
| MISSISSIPPI | 93,605.99 | 90,352.67 | 88,566.38 | 85,650.06 | 358,175.10 |
| MISSOURI | 258,296.60 | 242,528.32 | 230,202.63 | 223,879.23 | 954,906.78 |
| MONTANA | 33,658.91 | 31,379.60 | 30,191.39 | 29,624.77 | 124,854.68 |
| NEBRASKA | 37,375.11 | 36,474.80 | 34,953.72 | 34,857.29 | 143,660.91 |
| NEVADA | 157,950.42 | 148,534.39 | 145,322.13 | 143,137.06 | 594,943.99 |
| NEW HAMPSHIRE | 55,486.03 | 52,863.35 | 49,944.98 | 48,419.02 | 206,713.38 |
| NEW JERSEY | 425,629.00 | 410,878.28 | 396,870.77 | 390,457.85 | 1,623,835.91 |
| NEW MEXICO | 80,887.52 | 78,628.00 | 76,680.20 | 74,927.37 | 311,123.09 |
| NEW YORK | 645,711.48 | 632,609.94 | 605,937.90 | 590,760.57 | 2,475,019.89 |
| NORTH CAROLINA | 499,442.28 | 475,993.43 | 464,556.21 | 486,500.56 | 1,926,492.48 |
| NORTH DAKOTA | 13,142.28 | 12,688.48 | 12,613.37 | 12,387.71 | 50,831.84 |
| OHIO | 412,495.88 | 397,258.59 | 381,837.49 | 377,921.46 | 1,569,513.43 |
| OKLAHOMA | 202,163.26 | 192,260.83 | 179,586.69 | 160,577.83 | 734,588.62 |
| OREGON | 153,589.77 | 149,504.19 | 143,480.99 | 139,203.81 | 585,778.76 |
| PENNSYLVANIA | 649,267.48 | 626,004.85 | 599,154.66 | 579,431.92 | 2,453,858.91 |
| PUERTO RICO | 7,404.73 | 7,900.07 | 14,706.77 | 14,319.35 | 44,330.91 |
| RHODE ISLAND | 35,867.26 | 33,747.94 | 32,075.72 | 32,230.97 | 133,921.89 |
| SOUTH CAROLINA | 206,811.28 | 194,772.35 | 187,949.15 | 184,781.57 | 774,314.35 |
| SOUTH DAKOTA | 15,216.32 | 14,487.66 | 13,740.82 | 13,196.71 | 56,641.51 |
| TENNESSEE | 420,991.18 | 409,408.99 | 390,639.81 | 377,835.40 | 1,598,875.37 |
| TEXAS | 307,917.35 | 303,863.86 | 290,350.34 | 289,225.01 | 1,191,356.55 |
| UTAH | 152,995.93 | 147,169.53 | 144,332.30 | 142,207.06 | 586,704.81 |
| VERMONT | 20,841.53 | 20,024.13 | 19,332.64 | 18,819.96 | 79,018.26 |
| VIRGIN ISLANDS | 463.94 | 480.43 | 394.91 | 413.29 | 1,752.57 |
| VIRGINIA | 262,524.42 | 252,823.97 | 245,546.79 | 244,297.29 | 1,005,192.47 |
| WASHINGTON | 258,217.13 | 251,409.19 | 242,441.87 | 237,674.43 | 989,742.63 |
| WEST VIRGINIA | 72,248.71 | 67,470.52 | 63,782.46 | 60,832.77 | 264,334.46 |
| WISCONSIN | 179,675.85 | 167,586.32 | 158,632.67 | 152,817.21 | 658,712.06 |
| WYOMING | 23,960.77 | 22,862.52 | 21,962.93 | 22,119.24 | 90,905.46 |
| **9143 - OXYCODONE - Total** | **10,932,712.46** | **10,499,375.98** | **10,050,407.68** | **9,861,024.63** | **41,343,520.76** |

**DRUG: 9150 - HYDROMORPHONE**

| STATE | 1ST QUARTER | 2ND QUARTER | 3RD QUARTER | 4TH QUARTER | TOTAL GRAMS |
|---|---|---|---|---|---|

| | | | | | |
|---|---|---|---|---|---|
| ALABAMA | 2,777.32 | 2,821.82 | 2,597.38 | 2,344.97 | 10,541.50 |
| ALASKA | 958.18 | 756.91 | 766.25 | 622.01 | 3,103.34 |
| AMERICAN SAMOA | 0.18 | 0.03 | 0.73 | 0.27 | 1.20 |
| ARIZONA | 6,148.90 | 5,588.13 | 5,425.76 | 5,056.22 | 22,219.02 |
| ARKANSAS | 2,811.12 | 2,540.91 | 2,365.32 | 2,280.16 | 9,997.50 |
| CALIFORNIA | 34,764.05 | 32,674.26 | 30,038.86 | 28,453.83 | 125,931.00 |
| COLORADO | 6,507.54 | 5,834.45 | 5,637.52 | 5,598.57 | 23,578.07 |
| CONNECTICUT | 5,894.59 | 5,059.17 | 4,690.61 | 4,711.11 | 20,355.47 |
| DELAWARE | 1,814.54 | 1,653.44 | 1,520.48 | 1,483.92 | 6,472.38 |
| DISTRICT OF COLUMBIA | 459.71 | 374.60 | 452.30 | 407.02 | 1,693.63 |
| FLORIDA | 43,752.23 | 39,490.32 | 37,156.97 | 35,121.31 | 155,520.83 |
| GEORGIA | 7,211.99 | 7,267.29 | 6,576.09 | 6,006.85 | 27,062.23 |
| GUAM | 28.28 | 17.21 | 30.71 | 14.31 | 90.51 |
| HAWAII | 546.70 | 528.30 | 492.28 | 494.18 | 2,061.45 |
| IDAHO | 1,863.65 | 1,872.51 | 1,665.87 | 1,615.81 | 7,017.83 |
| ILLINOIS | 8,894.40 | 8,245.46 | 7,790.86 | 7,597.06 | 32,527.78 |
| INDIANA | 5,431.52 | 4,987.37 | 4,495.20 | 4,248.44 | 19,162.53 |
| IOWA | 2,622.86 | 2,404.82 | 2,276.33 | 2,275.32 | 9,579.33 |
| KANSAS | 2,957.25 | 2,800.73 | 2,891.40 | 2,734.53 | 11,383.91 |
| KENTUCKY | 2,545.12 | 2,425.27 | 2,176.57 | 2,314.77 | 9,461.73 |
| LOUISIANA | 2,941.03 | 2,830.54 | 2,700.56 | 2,399.95 | 10,872.08 |
| MAINE | 1,623.13 | 1,520.84 | 1,235.11 | 1,442.22 | 5,821.30 |
| MARYLAND | 7,211.98 | 6,395.81 | 6,248.74 | 6,069.43 | 25,925.96 |
| MASSACHUSETTS | 6,889.07 | 6,893.72 | 6,182.28 | 5,929.01 | 25,894.08 |
| MICHIGAN | 7,654.53 | 7,504.48 | 6,470.61 | 6,276.27 | 27,905.89 |
| MINNESOTA | 5,348.87 | 5,036.82 | 4,984.49 | 4,864.83 | 20,235.00 |
| MISSISSIPPI | 4,497.39 | 6,250.99 | 5,389.31 | 4,448.85 | 20,586.53 |
| MISSOURI | 5,806.27 | 5,035.62 | 4,687.94 | 4,701.49 | 20,231.31 |
| MONTANA | 1,621.56 | 1,343.70 | 1,358.48 | 1,289.40 | 5,613.14 |
| NEBRASKA | 1,083.90 | 1,082.98 | 1,087.07 | 1,058.86 | 4,312.82 |
| NEVADA | 2,501.58 | 2,199.70 | 2,311.65 | 2,242.03 | 9,254.96 |
| NEW HAMPSHIRE | 1,747.28 | 1,738.34 | 1,513.86 | 1,595.28 | 6,594.75 |
| NEW JERSEY | 7,823.25 | 7,158.03 | 7,293.09 | 7,137.84 | 29,412.21 |
| NEW MEXICO | 1,147.70 | 1,123.91 | 1,163.79 | 1,065.66 | 4,501.06 |
| NEW YORK | 17,341.51 | 16,824.83 | 16,013.10 | 15,247.83 | 65,427.26 |
| NORTH CAROLINA | 9,705.96 | 8,762.82 | 9,334.90 | 8,588.70 | 36,392.38 |
| NORTH DAKOTA | 556.87 | 545.90 | 619.05 | 650.92 | 2,372.74 |
| OHIO | 8,787.42 | 7,491.97 | 6,852.60 | 6,954.14 | 30,086.13 |
| OKLAHOMA | 3,366.51 | 3,224.82 | 3,171.93 | 2,737.57 | 12,500.83 |
| OREGON | 4,482.13 | 4,096.43 | 3,763.84 | 3,759.19 | 16,101.59 |

| | | | | | |
|---|---|---|---|---|---|
| PENNSYLVANIA | 14,685.90 | 12,754.69 | 11,093.41 | 11,035.38 | 49,569.38 |
| PUERTO RICO | 51.25 | 66.63 | 55.59 | 56.21 | 229.67 |
| RHODE ISLAND | 818.83 | 741.16 | 619.82 | 568.97 | 2,748.77 |
| SOUTH CAROLINA | 5,082.66 | 4,980.02 | 4,943.20 | 4,415.74 | 19,421.63 |
| SOUTH DAKOTA | 525.80 | 473.70 | 487.04 | 491.78 | 1,978.32 |
| TENNESSEE | 7,665.65 | 4,505.69 | 4,949.97 | 4,870.43 | 21,991.74 |
| TEXAS | 21,871.45 | 19,438.77 | 18,945.69 | 18,809.33 | 79,065.23 |
| UTAH | 2,812.04 | 2,145.52 | 2,436.83 | 2,274.81 | 9,669.19 |
| VERMONT | 1,075.22 | 1,087.44 | 915.83 | 919.98 | 3,998.48 |
| VIRGIN ISLANDS | 10.86 | 13.30 | 16.31 | 13.83 | 54.30 |
| VIRGINIA | 9,794.87 | 9,103.35 | 8,605.45 | 8,259.04 | 35,762.70 |
| WASHINGTON | 9,715.50 | 8,942.94 | 8,294.23 | 8,333.80 | 35,286.47 |
| WEST VIRGINIA | 1,181.25 | 1,118.00 | 1,040.79 | 1,083.81 | 4,423.85 |
| WISCONSIN | 3,553.35 | 3,265.46 | 2,801.07 | 2,631.38 | 12,251.25 |
| WYOMING | 562.03 | 564.22 | 484.48 | 494.00 | 2,104.73 |
| **9150 - HYDROMORPHONE - To** | **319,534.72** | **293,606.14** | **277,119.57** | **266,098.58** | **1,156,359.01** |

**DRUG: 9193 - HYDROCODONE**

| STATE | 1ST QUARTER | 2ND QUARTER | 3RD QUARTER | 4TH QUARTER | TOTAL GRAMS |
|---|---|---|---|---|---|
| ALABAMA | 223,350.03 | 214,775.33 | 208,606.42 | 207,257.90 | 853,989.67 |
| ALASKA | 9,821.57 | 9,428.58 | 9,025.03 | 8,645.58 | 36,920.76 |
| AMERICAN SAMOA | 3.63 | 0 | 0 | 0 | 3.63 |
| ARIZONA | 93,520.55 | 87,191.66 | 84,740.02 | 83,257.51 | 348,709.74 |
| ARKANSAS | 120,863.63 | 115,245.93 | 110,625.47 | 108,391.02 | 455,126.05 |
| CALIFORNIA | 829,002.43 | 794,554.06 | 765,824.97 | 744,489.33 | 3,133,870.80 |
| COLORADO | 57,984.06 | 54,840.98 | 52,655.47 | 52,169.59 | 217,650.10 |
| CONNECTICUT | 18,121.90 | 16,761.16 | 16,219.30 | 16,341.67 | 67,444.03 |
| DELAWARE | 5,173.97 | 5,134.33 | 4,907.42 | 4,846.62 | 20,062.33 |
| DISTRICT OF COLUMBIA | 1,357.64 | 1,111.33 | 1,078.02 | 1,054.99 | 4,601.98 |
| FLORIDA | 340,174.90 | 315,391.99 | 272,012.56 | 274,878.25 | 1,202,457.69 |
| GEORGIA | 222,576.16 | 212,955.19 | 203,461.01 | 203,844.55 | 842,836.92 |
| GUAM | 142.59 | 101.24 | 135.95 | 116.67 | 496.46 |
| HAWAII | 16,304.36 | 15,570.55 | 14,406.04 | 14,130.27 | 60,411.22 |
| IDAHO | 52,198.65 | 50,964.65 | 49,377.82 | 48,523.97 | 201,065.09 |
| ILLINOIS | 270,801.21 | 260,368.88 | 249,086.69 | 249,810.82 | 1,030,067.59 |
| INDIANA | 206,161.32 | 196,794.99 | 190,035.80 | 190,528.67 | 783,520.77 |
| IOWA | 50,377.92 | 48,258.00 | 46,358.03 | 46,248.30 | 191,242.24 |
| KANSAS | 73,086.15 | 70,405.26 | 67,549.43 | 67,357.62 | 278,398.45 |
| KENTUCKY | 188,090.96 | 181,063.43 | 175,781.57 | 176,604.80 | 721,540.77 |
| LOUISIANA | 147,202.74 | 139,768.31 | 136,208.86 | 135,670.52 | 558,850.42 |
| MAINE | 15,783.98 | 15,162.34 | 14,935.32 | 14,503.53 | 60,385.18 |

| | | | | | |
|---|---|---|---|---|---|
| MARYLAND | 30,255.29 | 29,123.57 | 27,148.09 | 27,305.54 | 113,832.50 |
| MASSACHUSETTS | 27,253.25 | 25,824.79 | 24,353.43 | 24,403.47 | 101,834.93 |
| MICHIGAN | 337,224.55 | 310,827.19 | 290,132.27 | 286,669.65 | 1,224,853.66 |
| MINNESOTA | 39,382.77 | 37,705.28 | 36,496.62 | 35,635.09 | 149,219.75 |
| MISSISSIPPI | 99,033.75 | 92,247.24 | 88,024.95 | 84,755.11 | 364,061.06 |
| MISSOURI | 155,287.75 | 149,092.02 | 143,085.79 | 139,967.05 | 587,432.60 |
| MONTANA | 23,745.27 | 22,603.24 | 22,357.05 | 21,691.01 | 90,396.57 |
| NEBRASKA | 28,370.64 | 27,735.66 | 26,386.85 | 26,283.41 | 108,776.55 |
| NEVADA | 79,174.96 | 75,840.66 | 73,797.14 | 72,250.39 | 301,063.15 |
| NEW HAMPSHIRE | 8,242.23 | 7,695.72 | 7,596.49 | 7,601.84 | 31,136.29 |
| NEW JERSEY | 30,508.96 | 28,534.89 | 27,110.18 | 27,632.20 | 113,786.23 |
| NEW MEXICO | 32,325.82 | 30,425.51 | 29,465.17 | 29,582.97 | 121,799.47 |
| NEW YORK | 152,317.93 | 146,483.73 | 140,092.49 | 139,946.35 | 578,840.51 |
| NORTH CAROLINA | 168,254.03 | 155,800.03 | 149,714.73 | 157,410.94 | 631,179.73 |
| NORTH DAKOTA | 9,339.72 | 8,806.86 | 8,845.41 | 8,519.22 | 35,511.20 |
| OHIO | 157,411.53 | 149,228.23 | 142,261.15 | 142,196.74 | 591,097.65 |
| OKLAHOMA | 162,726.57 | 160,068.20 | 152,244.49 | 142,189.44 | 617,228.70 |
| OREGON | 84,601.82 | 82,331.60 | 79,591.55 | 77,665.00 | 324,189.97 |
| PENNSYLVANIA | 143,359.13 | 136,422.73 | 131,936.33 | 125,853.23 | 537,571.41 |
| PUERTO RICO | 373.47 | 344.81 | 607.07 | 515.30 | 1,840.64 |
| RHODE ISLAND | 9,143.16 | 8,676.78 | 8,357.67 | 8,171.79 | 34,349.40 |
| SOUTH CAROLINA | 122,888.18 | 114,977.41 | 109,329.62 | 109,172.04 | 456,367.25 |
| SOUTH DAKOTA | 12,535.57 | 12,046.69 | 11,670.25 | 11,579.41 | 47,831.94 |
| TENNESSEE | 221,430.15 | 211,711.96 | 199,865.59 | 193,334.02 | 826,341.72 |
| TEXAS | 605,701.53 | 588,559.11 | 570,469.31 | 566,616.51 | 2,331,346.46 |
| UTAH | 53,125.77 | 50,733.38 | 49,110.06 | 49,779.60 | 202,748.82 |
| VERMONT | 4,264.10 | 4,136.47 | 4,029.43 | 3,983.52 | 16,413.53 |
| VIRGIN ISLANDS | 285.51 | 221.22 | 224.79 | 237.30 | 968.81 |
| VIRGINIA | 91,044.29 | 85,273.72 | 81,645.52 | 82,572.85 | 340,536.39 |
| WASHINGTON | 117,086.98 | 114,056.54 | 109,543.49 | 108,280.60 | 448,967.62 |
| WEST VIRGINIA | 50,828.80 | 47,139.71 | 44,517.22 | 42,653.89 | 185,139.63 |
| WISCONSIN | 68,829.82 | 68,036.57 | 66,592.22 | 65,749.46 | 269,208.06 |
| WYOMING | 10,040.15 | 9,857.33 | 9,520.81 | 9,448.06 | 38,866.35 |
| **9193 - HYDROCODONE - Total** | **6,078,493.82** | **5,798,417.00** | **5,539,154.44** | **5,478,325.20** | **22,894,390.46** |

**DRUG: 9230 - MEPERIDINE (PETHIDINE)**

| STATE | 1ST QUARTER | 2ND QUARTER | 3RD QUARTER | 4TH QUARTER | TOTAL GRAMS |
|---|---|---|---|---|---|
| ALABAMA | 7,902.52 | 6,944.66 | 5,860.51 | 4,517.64 | 25,225.33 |
| ALASKA | 667.13 | 446.97 | 387.82 | 294.13 | 1,796.05 |
| ARIZONA | 5,388.22 | 4,960.67 | 4,439.57 | 3,574.52 | 18,362.98 |
| ARKANSAS | 6,752.29 | 6,028.95 | 5,547.97 | 4,315.56 | 22,644.77 |

| | | | | | |
|---|---|---|---|---|---|
| CALIFORNIA | 19,967.05 | 17,676.42 | 13,247.78 | 11,149.21 | 62,040.45 |
| COLORADO | 1,603.56 | 1,571.53 | 1,373.70 | 1,141.12 | 5,689.91 |
| CONNECTICUT | 1,101.58 | 885.12 | 872.04 | 836.10 | 3,694.83 |
| DELAWARE | 223.76 | 106.76 | 97.50 | 132.36 | 560.37 |
| DISTRICT OF COLUMBIA | 144.34 | 164.50 | 130.18 | 196.63 | 635.65 |
| FLORIDA | 9,635.85 | 8,329.25 | 6,493.87 | 5,474.11 | 29,933.08 |
| GEORGIA | 10,270.61 | 9,466.97 | 7,629.96 | 6,092.33 | 33,459.87 |
| GUAM | 14.16 | 35.95 | 40.85 | 29.96 | 120.92 |
| HAWAII | 247.18 | 402.31 | 352.41 | 232.58 | 1,234.48 |
| IDAHO | 741.86 | 777.81 | 573.56 | 694.48 | 2,787.71 |
| ILLINOIS | 2,818.00 | 2,819.13 | 2,214.05 | 1,761.52 | 9,612.69 |
| INDIANA | 3,147.55 | 3,060.03 | 2,697.07 | 1,839.95 | 10,744.61 |
| IOWA | 1,861.18 | 1,890.26 | 1,295.27 | 1,540.92 | 6,587.62 |
| KANSAS | 1,544.43 | 1,511.94 | 1,367.14 | 999.50 | 5,423.02 |
| KENTUCKY | 2,837.89 | 2,412.77 | 1,810.43 | 1,487.54 | 8,548.63 |
| LOUISIANA | 6,966.31 | 6,489.23 | 5,148.39 | 3,969.14 | 22,573.07 |
| MAINE | 353.18 | 334.87 | 317.01 | 308.29 | 1,313.35 |
| MARYLAND | 2,217.20 | 2,167.86 | 1,726.66 | 1,586.67 | 7,698.40 |
| MASSACHUSETTS | 2,097.53 | 1,793.33 | 1,573.60 | 1,503.34 | 6,967.80 |
| MICHIGAN | 2,889.87 | 2,544.78 | 1,838.76 | 2,087.24 | 9,360.65 |
| MINNESOTA | 586.35 | 681.62 | 423.77 | 459.72 | 2,151.45 |
| MISSISSIPPI | 3,927.52 | 3,715.31 | 3,206.10 | 2,613.96 | 13,462.89 |
| MISSOURI | 3,289.04 | 3,094.59 | 2,481.60 | 2,449.46 | 11,314.68 |
| MONTANA | 644.26 | 576.82 | 488.04 | 325.18 | 2,034.30 |
| NEBRASKA | 596.63 | 474.97 | 439.02 | 425.95 | 1,936.56 |
| NEVADA | 1,704.98 | 1,392.22 | 1,055.60 | 960.28 | 5,113.09 |
| NEW HAMPSHIRE | 644.58 | 573.77 | 473.88 | 496.76 | 2,188.99 |
| NEW JERSEY | 1,795.51 | 1,654.87 | 1,495.17 | 1,324.68 | 6,270.22 |
| NEW MEXICO | 1,607.59 | 1,484.82 | 1,197.77 | 805.05 | 5,095.22 |
| NEW YORK | 4,827.94 | 4,303.49 | 3,532.54 | 3,059.51 | 15,723.47 |
| NORTH CAROLINA | 5,045.88 | 4,810.14 | 4,147.25 | 3,752.90 | 17,756.16 |
| NORTH DAKOTA | 179.96 | 160.68 | 220.05 | 192.27 | 752.98 |
| OHIO | 5,518.43 | 4,305.41 | 3,333.24 | 3,437.85 | 16,594.92 |
| OKLAHOMA | 6,752.27 | 5,548.27 | 5,178.02 | 3,795.38 | 21,273.95 |
| OREGON | 1,199.84 | 1,299.30 | 1,179.25 | 892.20 | 4,570.58 |
| PENNSYLVANIA | 3,532.84 | 3,199.30 | 2,626.16 | 2,367.76 | 11,726.05 |
| PUERTO RICO | 2,446.19 | 2,223.96 | 4,236.03 | 3,576.42 | 12,482.60 |
| RHODE ISLAND | 177.02 | 226.05 | 170.49 | 84.43 | 657.98 |
| SOUTH CAROLINA | 5,134.33 | 4,541.93 | 4,171.76 | 3,417.91 | 17,265.94 |
| SOUTH DAKOTA | 531.83 | 594.80 | 426.49 | 330.08 | 1,883.20 |

| | | | | | |
|---|---|---|---|---|---|
| TENNESSEE | 7,323.50 | 6,149.50 | 4,890.47 | 3,700.06 | 22,063.53 |
| TEXAS | 13,536.92 | 11,614.55 | 8,532.90 | 8,180.78 | 41,865.15 |
| UTAH | 1,710.30 | 1,870.98 | 1,379.69 | 1,372.07 | 6,333.04 |
| VERMONT | 354.26 | 301.76 | 178.11 | 222.23 | 1,056.37 |
| VIRGIN ISLANDS | 120.92 | 155.78 | 153.06 | 30.50 | 460.26 |
| VIRGINIA | 5,716.82 | 4,908.61 | 4,299.55 | 3,873.82 | 18,798.80 |
| WASHINGTON | 3,258.76 | 3,289.91 | 2,478.87 | 2,378.85 | 11,406.39 |
| WEST VIRGINIA | 715.07 | 567.02 | 384.55 | 315.92 | 1,982.55 |
| WISCONSIN | 1,245.09 | 1,229.25 | 811.04 | 748.51 | 4,033.89 |
| WYOMING | 333.57 | 306.66 | 311.02 | 302.30 | 1,253.54 |
| **9230 - MEPERIDINE (PETHIDIN** | **175,851.45** | **158,078.40** | **130,937.57** | **111,657.62** | **576,525.04** |

**DRUG: 9250B - METHADONE**

| STATE | 1ST QUARTER | 2ND QUARTER | 3RD QUARTER | 4TH QUARTER | TOTAL GRAMS |
|---|---|---|---|---|---|
| ALABAMA | 67,914.39 | 69,271.28 | 74,008.47 | 75,231.20 | 286,425.34 |
| ALASKA | 8,238.68 | 7,954.26 | 8,965.11 | 7,825.55 | 32,983.59 |
| ARIZONA | 88,198.78 | 89,203.34 | 92,125.79 | 83,463.26 | 352,991.18 |
| ARKANSAS | 14,846.06 | 13,578.07 | 13,616.35 | 11,647.06 | 53,687.52 |
| CALIFORNIA | 331,722.38 | 302,980.16 | 321,055.74 | 254,097.99 | 1,209,856.28 |
| COLORADO | 46,411.33 | 44,774.62 | 47,146.83 | 41,701.36 | 180,034.13 |
| CONNECTICUT | 107,551.69 | 109,740.38 | 116,112.17 | 83,539.91 | 416,944.15 |
| DELAWARE | 31,389.68 | 34,835.99 | 29,406.53 | 29,282.39 | 124,914.59 |
| DISTRICT OF COLUMBIA | 9,332.62 | 8,677.16 | 10,387.32 | 7,014.74 | 35,411.84 |
| FLORIDA | 200,019.40 | 166,679.70 | 190,663.49 | 197,667.47 | 755,030.06 |
| GEORGIA | 122,050.06 | 104,200.73 | 114,804.38 | 108,179.74 | 449,234.91 |
| GUAM | 42.48 | 43.83 | 33.54 | 43.38 | 163.23 |
| HAWAII | 8,313.54 | 4,974.47 | 8,226.42 | 6,380.56 | 27,894.99 |
| IDAHO | 11,799.28 | 9,857.63 | 9,894.48 | 9,912.90 | 41,464.29 |
| ILLINOIS | 125,160.28 | 112,978.85 | 127,254.80 | 110,315.45 | 475,709.38 |
| INDIANA | 81,066.00 | 75,972.04 | 78,136.57 | 65,149.51 | 300,324.12 |
| IOWA | 12,463.11 | 13,115.57 | 12,500.49 | 11,083.76 | 49,162.93 |
| KANSAS | 20,576.30 | 22,621.97 | 19,432.18 | 20,870.64 | 83,501.09 |
| KENTUCKY | 51,235.42 | 49,948.97 | 51,201.01 | 46,731.41 | 199,116.82 |
| LOUISIANA | 36,114.62 | 34,816.49 | 34,747.14 | 32,616.66 | 138,294.90 |
| MAINE | 30,810.56 | 23,344.11 | 29,830.57 | 22,332.99 | 106,318.22 |
| MARYLAND | 187,662.74 | 169,950.86 | 183,860.20 | 174,663.05 | 716,136.84 |
| MASSACHUSETTS | 143,650.94 | 122,803.21 | 142,285.19 | 110,055.87 | 518,795.21 |
| MICHIGAN | 126,586.59 | 127,702.82 | 124,251.80 | 111,233.13 | 489,774.33 |
| MINNESOTA | 54,848.36 | 58,420.51 | 56,338.61 | 49,485.09 | 219,092.58 |
| MISSISSIPPI | 6,999.66 | 8,246.46 | 6,561.05 | 7,536.39 | 29,343.56 |
| MISSOURI | 37,084.11 | 40,492.97 | 35,838.54 | 33,652.49 | 147,068.11 |

| | | | | | |
|---|---|---|---|---|---|
| MONTANA | 7,566.09 | 7,976.97 | 8,481.95 | 7,994.68 | 32,019.70 |
| NEBRASKA | 7,147.88 | 6,484.31 | 7,933.06 | 6,788.64 | 28,353.89 |
| NEVADA | 35,907.66 | 33,437.86 | 33,811.18 | 33,410.49 | 136,567.19 |
| NEW HAMPSHIRE | 32,094.29 | 31,195.69 | 29,474.59 | 27,105.24 | 119,869.81 |
| NEW JERSEY | 115,335.74 | 115,657.90 | 121,387.88 | 116,133.73 | 468,515.25 |
| NEW MEXICO | 41,766.60 | 41,948.97 | 45,821.81 | 41,949.06 | 171,486.44 |
| NEW YORK | 294,396.67 | 281,438.64 | 314,167.06 | 236,617.55 | 1,126,619.92 |
| NORTH CAROLINA | 135,804.74 | 120,461.28 | 132,661.89 | 118,923.54 | 507,851.46 |
| NORTH DAKOTA | 3,176.01 | 3,691.73 | 4,734.51 | 4,271.74 | 15,873.99 |
| OHIO | 71,507.63 | 72,826.19 | 73,272.85 | 76,329.93 | 293,936.59 |
| OKLAHOMA | 37,086.83 | 29,873.14 | 36,593.21 | 33,923.79 | 137,476.97 |
| OREGON | 63,684.08 | 53,428.02 | 59,842.24 | 50,559.90 | 227,514.24 |
| PENNSYLVANIA | 189,494.72 | 177,324.20 | 187,040.52 | 160,229.35 | 714,088.79 |
| PUERTO RICO | 20,410.21 | 21,561.03 | 21,747.78 | 23,116.13 | 86,835.15 |
| RHODE ISLAND | 44,271.82 | 47,642.63 | 37,124.84 | 40,111.69 | 169,150.99 |
| SOUTH CAROLINA | 53,998.77 | 48,942.28 | 53,881.97 | 42,239.02 | 199,062.05 |
| SOUTH DAKOTA | 2,018.57 | 1,994.24 | 2,056.76 | 1,521.28 | 7,590.86 |
| TENNESSEE | 50,004.04 | 46,174.12 | 51,184.43 | 45,597.37 | 192,959.96 |
| TEXAS | 137,248.00 | 122,242.73 | 136,911.97 | 122,241.36 | 518,644.06 |
| UTAH | 28,138.63 | 23,347.86 | 26,338.92 | 19,698.00 | 97,523.41 |
| VERMONT | 20,084.56 | 15,595.74 | 19,629.75 | 16,592.01 | 71,902.07 |
| VIRGIN ISLANDS | 1.79 | 3.58 | 9.84 | 549.16 | 564.37 |
| VIRGINIA | 82,987.04 | 79,210.03 | 82,578.70 | 73,548.15 | 318,323.92 |
| WASHINGTON | 98,377.83 | 83,780.68 | 100,870.97 | 88,459.38 | 371,488.86 |
| WEST VIRGINIA | 22,711.05 | 20,161.65 | 23,336.60 | 16,959.61 | 83,168.91 |
| WISCONSIN | 61,515.58 | 58,426.59 | 63,141.16 | 43,882.77 | 226,966.10 |
| WYOMING | 1,048.51 | 931.79 | 877.85 | 899.32 | 3,757.46 |
| **9250B - METHADONE - Total** | **3,619,874.42** | **3,372,946.30** | **3,613,599.05** | **3,161,366.86** | **13,767,786.63** |

**DRUG: 9300 - MORPHINE**

| STATE | 1ST QUARTER | 2ND QUARTER | 3RD QUARTER | 4TH QUARTER | TOTAL GRAMS |
|---|---|---|---|---|---|
| ALABAMA | 63,749.51 | 60,512.68 | 58,188.90 | 57,865.49 | 240,316.58 |
| ALASKA | 10,799.45 | 10,484.39 | 10,339.16 | 9,116.76 | 40,739.77 |
| AMERICAN SAMOA | 6.77 | 25.12 | 12.18 | 4.51 | 48.58 |
| ARIZONA | 136,741.78 | 124,647.12 | 116,910.22 | 109,003.06 | 487,302.18 |
| ARKANSAS | 38,902.44 | 36,091.73 | 33,140.85 | 31,681.30 | 139,816.32 |
| CALIFORNIA | 400,101.55 | 375,284.98 | 358,109.62 | 346,526.82 | 1,480,022.97 |
| COLORADO | 69,466.35 | 64,253.88 | 62,531.80 | 58,950.62 | 255,202.66 |
| CONNECTICUT | 32,848.85 | 31,359.24 | 29,731.85 | 29,190.94 | 123,130.87 |
| DELAWARE | 20,025.57 | 18,686.12 | 17,524.99 | 16,591.38 | 72,828.05 |
| DISTRICT OF COLUMBIA | 3,687.93 | 2,729.65 | 2,989.20 | 2,468.92 | 11,875.70 |

| | | | | | |
|---|---|---|---|---|---|
| FLORIDA | 371,027.93 | 354,851.78 | 324,461.45 | 315,983.33 | 1,366,324.49 |
| GEORGIA | 104,805.28 | 100,634.05 | 97,872.74 | 93,268.52 | 396,580.59 |
| GUAM | 1,005.28 | 826.94 | 769.30 | 765.17 | 3,366.70 |
| HAWAII | 18,510.58 | 18,782.17 | 16,999.40 | 15,850.51 | 70,142.67 |
| IDAHO | 25,649.58 | 24,931.66 | 23,975.83 | 22,447.40 | 97,004.47 |
| ILLINOIS | 95,172.42 | 91,349.37 | 88,023.10 | 87,900.76 | 362,445.64 |
| INDIANA | 77,338.11 | 73,243.52 | 68,387.78 | 66,713.35 | 285,682.76 |
| IOWA | 30,578.58 | 28,933.47 | 28,213.55 | 27,756.04 | 115,481.64 |
| KANSAS | 40,474.30 | 38,914.39 | 36,659.84 | 35,896.64 | 151,945.16 |
| KENTUCKY | 41,830.87 | 39,716.11 | 36,993.42 | 37,129.80 | 155,670.20 |
| LOUISIANA | 48,758.22 | 46,908.85 | 45,631.68 | 43,460.65 | 184,759.40 |
| MAINE | 14,733.01 | 14,368.48 | 13,774.44 | 13,576.37 | 56,452.30 |
| MARYLAND | 67,378.01 | 63,123.40 | 57,757.50 | 55,306.46 | 243,565.36 |
| MASSACHUSETTS | 60,858.52 | 59,079.46 | 55,457.37 | 55,592.24 | 230,987.59 |
| MICHIGAN | 162,164.04 | 153,747.08 | 143,755.19 | 137,861.48 | 597,527.79 |
| MINNESOTA | 41,305.71 | 39,159.05 | 37,999.77 | 36,494.43 | 154,958.95 |
| MISSISSIPPI | 34,949.52 | 34,124.33 | 32,540.47 | 30,135.07 | 131,749.38 |
| MISSOURI | 87,169.18 | 80,507.38 | 77,773.94 | 75,452.35 | 320,902.84 |
| MONTANA | 15,968.41 | 15,248.43 | 14,889.61 | 13,126.17 | 59,232.61 |
| NEBRASKA | 17,332.34 | 17,450.26 | 16,432.67 | 15,968.73 | 67,184.01 |
| NEVADA | 50,722.85 | 46,904.66 | 45,937.64 | 42,535.32 | 186,100.47 |
| NEW HAMPSHIRE | 17,005.38 | 17,093.23 | 15,634.32 | 15,646.56 | 65,379.49 |
| NEW JERSEY | 75,167.59 | 72,347.77 | 68,387.60 | 67,329.10 | 283,232.06 |
| NEW MEXICO | 23,027.43 | 21,464.48 | 20,622.48 | 20,854.45 | 85,968.84 |
| NEW YORK | 140,144.76 | 135,583.64 | 130,043.41 | 125,383.84 | 531,155.65 |
| NORTH CAROLINA | 109,970.87 | 105,123.85 | 102,634.74 | 105,486.56 | 423,216.02 |
| NORTH DAKOTA | 5,671.92 | 5,247.94 | 5,351.00 | 5,256.95 | 21,527.80 |
| OHIO | 106,426.38 | 100,381.39 | 94,072.11 | 94,625.23 | 395,505.12 |
| OKLAHOMA | 81,175.79 | 72,989.07 | 67,481.79 | 57,897.39 | 279,544.04 |
| OREGON | 62,402.85 | 60,189.39 | 58,414.56 | 55,845.49 | 236,852.29 |
| PENNSYLVANIA | 209,332.61 | 188,562.77 | 200,179.40 | 163,353.02 | 761,427.79 |
| PUERTO RICO | 1,431.29 | 1,345.34 | 2,004.96 | 2,053.46 | 6,835.05 |
| RHODE ISLAND | 10,361.99 | 9,491.94 | 8,808.37 | 9,277.45 | 37,939.75 |
| SOUTH CAROLINA | 60,751.90 | 58,363.99 | 56,543.60 | 56,332.60 | 231,992.09 |
| SOUTH DAKOTA | 7,684.52 | 7,236.09 | 7,155.86 | 6,604.44 | 28,680.91 |
| TENNESSEE | 144,205.13 | 132,195.26 | 135,417.02 | 105,050.14 | 516,867.55 |
| TEXAS | 189,821.39 | 180,619.18 | 171,628.50 | 165,653.28 | 707,722.35 |
| UTAH | 45,775.28 | 42,821.91 | 41,841.55 | 40,271.89 | 170,710.63 |
| VERMONT | 9,584.36 | 9,402.67 | 8,912.58 | 8,836.52 | 36,736.12 |
| VIRGIN ISLANDS | 96.93 | 133.71 | 134.38 | 106.74 | 471.76 |

| | | | | | |
|---|---|---|---|---|---|
| VIRGINIA | 70,659.02 | 68,828.75 | 65,452.40 | 65,493.94 | 270,434.11 |
| WASHINGTON | 96,421.27 | 94,101.65 | 89,444.28 | 85,611.21 | 365,578.40 |
| WEST VIRGINIA | 19,014.89 | 18,012.96 | 17,542.76 | 16,684.12 | 71,254.72 |
| WISCONSIN | 68,314.02 | 65,798.11 | 61,345.30 | 61,097.36 | 256,554.79 |
| WYOMING | 8,347.86 | 7,851.37 | 7,429.48 | 6,858.52 | 30,487.23 |
| **9300 - MORPHINE - Total** | **3,746,858.35** | **3,542,066.20** | **3,390,267.87** | **3,226,230.83** | **13,905,423.24** |

**DRUG: 9652 - OXYMORPHONE**

| STATE | 1ST QUARTER | 2ND QUARTER | 3RD QUARTER | 4TH QUARTER | TOTAL GRAMS |
|---|---|---|---|---|---|
| ALABAMA | 2,384.36 | 2,165.88 | 1,942.82 | 1,958.16 | 8,451.22 |
| ALASKA | 167.98 | 158.35 | 122.75 | 93.85 | 542.93 |
| ARIZONA | 2,675.12 | 2,184.17 | 2,093.89 | 1,764.62 | 8,717.81 |
| ARKANSAS | 710.16 | 531.38 | 431.33 | 368.48 | 2,041.35 |
| CALIFORNIA | 6,788.93 | 5,780.36 | 5,408.89 | 5,078.37 | 23,056.55 |
| COLORADO | 1,603.73 | 1,428.48 | 1,312.64 | 1,253.04 | 5,597.88 |
| CONNECTICUT | 1,435.03 | 1,434.54 | 1,260.63 | 1,243.59 | 5,373.79 |
| DELAWARE | 1,471.03 | 1,486.82 | 1,464.29 | 1,447.61 | 5,869.75 |
| DISTRICT OF COLUMBIA | 98.22 | 61.02 | 50.58 | 52.05 | 261.88 |
| FLORIDA | 11,420.80 | 10,640.70 | 9,637.45 | 9,279.13 | 40,978.08 |
| GEORGIA | 3,200.05 | 2,866.76 | 2,765.06 | 2,577.77 | 11,409.65 |
| HAWAII | 255.59 | 181.27 | 205.81 | 218.03 | 860.70 |
| IDAHO | 797.58 | 750.79 | 668.99 | 669.16 | 2,886.52 |
| ILLINOIS | 2,088.23 | 1,831.70 | 1,745.80 | 1,814.84 | 7,480.57 |
| INDIANA | 2,503.63 | 2,218.47 | 2,106.60 | 1,942.82 | 8,771.53 |
| IOWA | 526.34 | 464.43 | 377.27 | 357.69 | 1,725.72 |
| KANSAS | 888.49 | 822.87 | 745.48 | 760.87 | 3,217.72 |
| KENTUCKY | 2,120.79 | 1,907.09 | 1,729.47 | 1,631.70 | 7,389.04 |
| LOUISIANA | 4,242.25 | 3,983.76 | 3,787.32 | 3,592.71 | 15,606.04 |
| MAINE | 590.75 | 435.70 | 399.21 | 362.50 | 1,788.17 |
| MARYLAND | 6,356.89 | 5,901.64 | 5,629.54 | 5,554.96 | 23,443.03 |
| MASSACHUSETTS | 1,127.70 | 1,035.37 | 991.35 | 968.64 | 4,123.06 |
| MICHIGAN | 14,344.17 | 12,275.12 | 10,766.44 | 9,271.02 | 46,656.74 |
| MINNESOTA | 228.29 | 214.46 | 207.32 | 215.35 | 865.43 |
| MISSISSIPPI | 1,764.44 | 1,451.54 | 1,309.65 | 1,310.27 | 5,835.90 |
| MISSOURI | 2,020.87 | 1,700.12 | 1,560.77 | 1,630.45 | 6,912.21 |
| MONTANA | 133.28 | 125.96 | 138.41 | 120.52 | 518.18 |
| NEBRASKA | 617.82 | 526.34 | 516.75 | 454.03 | 2,114.95 |
| NEVADA | 1,970.69 | 1,628.39 | 1,542.31 | 1,344.31 | 6,485.70 |
| NEW HAMPSHIRE | 470.27 | 474.11 | 409.96 | 322.98 | 1,677.33 |
| NEW JERSEY | 3,737.90 | 3,369.86 | 3,023.95 | 3,023.37 | 13,155.09 |
| NEW MEXICO | 556.27 | 476.34 | 388.91 | 349.52 | 1,771.04 |

| | | | | | |
|---|---|---|---|---|---|
| NEW YORK | 11,394.53 | 10,639.72 | 9,948.30 | 9,908.29 | 41,890.83 |
| NORTH CAROLINA | 11,044.24 | 9,956.72 | 9,390.47 | 9,504.03 | 39,895.46 |
| NORTH DAKOTA | 81.94 | 80.82 | 71.28 | 67.00 | 301.04 |
| OHIO | 5,204.56 | 4,772.73 | 4,193.45 | 3,815.44 | 17,986.18 |
| OKLAHOMA | 2,820.46 | 2,455.28 | 1,827.73 | 1,302.38 | 8,405.86 |
| OREGON | 1,041.66 | 949.02 | 870.38 | 836.43 | 3,697.49 |
| PENNSYLVANIA | 14,439.75 | 12,775.23 | 11,466.62 | 10,927.65 | 49,609.24 |
| RHODE ISLAND | 231.05 | 167.71 | 159.24 | 123.29 | 681.30 |
| SOUTH CAROLINA | 3,376.87 | 3,132.88 | 3,017.53 | 2,907.62 | 12,434.89 |
| SOUTH DAKOTA | 334.89 | 288.15 | 238.68 | 292.34 | 1,154.07 |
| TENNESSEE | 14,167.98 | 12,805.20 | 11,632.76 | 11,258.88 | 49,864.82 |
| TEXAS | 5,535.93 | 4,766.22 | 4,201.18 | 3,734.24 | 18,237.57 |
| UTAH | 2,983.69 | 2,677.06 | 2,502.47 | 2,516.97 | 10,680.19 |
| VERMONT | 169.41 | 127.66 | 120.26 | 139.88 | 557.21 |
| VIRGINIA | 6,148.89 | 5,685.98 | 5,245.99 | 5,028.19 | 22,109.05 |
| WASHINGTON | 1,637.32 | 1,387.84 | 1,345.60 | 1,353.94 | 5,724.69 |
| WEST VIRGINIA | 1,480.13 | 1,376.51 | 1,141.53 | 1,163.66 | 5,161.82 |
| WISCONSIN | 1,031.18 | 828.31 | 772.83 | 707.97 | 3,340.29 |
| WYOMING | 194.79 | 125.21 | 115.66 | 112.63 | 548.28 |
| **9652 - OXYMORPHONE - Total** | **162,616.91** | **145,512.07** | **133,003.60** | **126,733.24** | **567,865.82** |

**DRUG: 9780 - TAPENTADOL**

| STATE | 1ST QUARTER | 2ND QUARTER | 3RD QUARTER | 4TH QUARTER | TOTAL GRAMS |
|---|---|---|---|---|---|
| ALABAMA | 43,345.50 | 43,372.00 | 40,437.50 | 37,149.00 | 164,304.00 |
| ALASKA | 2,104.00 | 2,147.50 | 2,113.00 | 1,646.00 | 8,010.50 |
| ARIZONA | 31,653.50 | 30,951.00 | 28,564.00 | 26,096.00 | 117,264.50 |
| ARKANSAS | 15,581.50 | 14,373.00 | 13,103.00 | 11,699.50 | 54,757.00 |
| CALIFORNIA | 85,686.40 | 82,514.43 | 78,783.45 | 76,734.95 | 323,719.23 |
| COLORADO | 26,747.00 | 25,144.50 | 24,580.50 | 24,762.50 | 101,234.50 |
| CONNECTICUT | 8,241.00 | 8,894.50 | 8,328.50 | 8,673.50 | 34,137.50 |
| DELAWARE | 5,531.50 | 5,635.00 | 5,613.00 | 5,125.00 | 21,904.50 |
| DISTRICT OF COLUMBIA | 884.00 | 880.00 | 827.00 | 775.00 | 3,366.00 |
| FLORIDA | 78,177.00 | 77,431.50 | 68,283.00 | 67,378.00 | 291,269.50 |
| GEORGIA | 52,107.00 | 52,478.00 | 50,380.50 | 50,074.00 | 205,039.50 |
| HAWAII | 2,501.00 | 2,031.00 | 1,796.50 | 1,753.50 | 8,082.00 |
| IDAHO | 5,480.00 | 5,839.50 | 5,597.50 | 4,909.50 | 21,826.50 |
| ILLINOIS | 29,481.00 | 29,173.00 | 27,531.50 | 27,791.50 | 113,977.00 |
| INDIANA | 31,357.50 | 31,026.50 | 28,971.50 | 28,962.00 | 120,317.50 |
| IOWA | 10,000.50 | 9,166.00 | 8,964.50 | 8,974.50 | 37,105.50 |
| KANSAS | 7,259.50 | 6,456.00 | 6,377.50 | 5,876.50 | 25,969.50 |
| KENTUCKY | 12,770.50 | 12,249.00 | 11,415.00 | 11,351.00 | 47,785.50 |

| | | | | | |
|---|---|---|---|---|---|
| LOUISIANA | 13,230.50 | 13,578.50 | 13,103.00 | 13,130.50 | 53,042.50 |
| MAINE | 1,035.50 | 1,247.50 | 1,289.00 | 1,124.00 | 4,696.00 |
| MARYLAND | 25,335.00 | 26,738.50 | 25,630.50 | 24,938.50 | 102,642.50 |
| MASSACHUSETTS | 15,863.00 | 17,030.00 | 16,565.00 | 16,599.50 | 66,057.50 |
| MICHIGAN | 18,174.50 | 17,096.00 | 16,328.50 | 15,931.50 | 67,530.50 |
| MINNESOTA | 3,962.50 | 3,644.50 | 3,689.00 | 3,654.50 | 14,950.50 |
| MISSISSIPPI | 9,878.00 | 8,950.00 | 8,935.00 | 8,223.00 | 35,986.00 |
| MISSOURI | 20,928.50 | 21,203.50 | 19,595.00 | 19,523.00 | 81,250.00 |
| MONTANA | 1,490.50 | 1,548.00 | 1,524.50 | 1,564.50 | 6,127.50 |
| NEBRASKA | 8,846.50 | 9,053.00 | 9,348.00 | 9,807.00 | 37,054.50 |
| NEVADA | 9,832.50 | 8,777.00 | 8,058.00 | 8,099.50 | 34,767.00 |
| NEW HAMPSHIRE | 5,777.00 | 6,018.00 | 6,388.50 | 6,108.00 | 24,291.50 |
| NEW JERSEY | 35,169.50 | 35,073.00 | 32,327.00 | 32,171.50 | 134,741.00 |
| NEW MEXICO | 2,798.00 | 2,515.50 | 2,497.50 | 2,393.50 | 10,204.50 |
| NEW YORK | 78,445.00 | 79,223.00 | 74,897.00 | 75,045.50 | 307,610.50 |
| NORTH CAROLINA | 81,066.50 | 83,766.00 | 81,859.00 | 85,142.00 | 331,833.50 |
| NORTH DAKOTA | 1,758.50 | 1,883.00 | 1,822.50 | 1,762.00 | 7,226.00 |
| OHIO | 48,657.45 | 47,606.95 | 44,439.83 | 43,022.33 | 183,726.55 |
| OKLAHOMA | 19,671.50 | 17,618.00 | 14,891.50 | 10,765.00 | 62,946.00 |
| OREGON | 6,727.00 | 6,508.50 | 6,155.00 | 5,404.00 | 24,794.50 |
| PENNSYLVANIA | 52,872.50 | 52,792.50 | 49,541.00 | 49,394.50 | 204,600.50 |
| PUERTO RICO | 487.00 | 574.50 | 442.00 | 452.50 | 1,956.00 |
| RHODE ISLAND | 1,728.00 | 2,163.50 | 1,815.50 | 1,861.50 | 7,568.50 |
| SOUTH CAROLINA | 35,684.00 | 36,926.00 | 35,000.50 | 33,620.50 | 141,231.00 |
| SOUTH DAKOTA | 4,127.50 | 3,790.50 | 3,472.00 | 3,478.00 | 14,868.00 |
| TENNESSEE | 26,459.00 | 27,546.50 | 25,123.50 | 24,341.50 | 103,470.50 |
| TEXAS | 81,107.50 | 76,671.45 | 73,511.50 | 70,488.00 | 301,778.45 |
| UTAH | 11,069.50 | 10,496.00 | 10,191.50 | 10,471.00 | 42,228.00 |
| VERMONT | 916.00 | 967.00 | 1,100.00 | 905.50 | 3,888.50 |
| VIRGIN ISLANDS | 0 | 0 | 5.00 | 0 | 5.00 |
| VIRGINIA | 33,686.00 | 32,769.00 | 30,043.50 | 29,618.50 | 126,117.00 |
| WASHINGTON | 14,427.00 | 14,482.50 | 13,872.00 | 14,127.00 | 56,908.50 |
| WEST VIRGINIA | 4,698.00 | 4,419.50 | 3,783.50 | 3,472.50 | 16,373.50 |
| WISCONSIN | 11,906.50 | 11,217.00 | 10,775.15 | 10,467.00 | 44,365.65 |
| WYOMING | 1,780.50 | 1,510.00 | 1,624.00 | 1,482.00 | 6,396.50 |
| **9780 - TAPENTADOL - Total** | **1,138,504.85** | **1,125,166.83** | **1,061,311.43** | **1,038,321.28** | **4,363,304.38** |

**DRUG: 9801 - FENTANYL BASE**

| STATE | 1ST QUARTER | 2ND QUARTER | 3RD QUARTER | 4TH QUARTER | TOTAL GRAMS |
|---|---|---|---|---|---|
| ALABAMA | 1,298.70 | 1,177.82 | 1,092.40 | 1,004.20 | 4,573.12 |
| ALASKA | 224.57 | 189.47 | 176.71 | 146.41 | 737.16 |

| | | | | | |
|---|---|---|---|---|---|
| AMERICAN SAMOA | 0.01 | 0.05 | 0.26 | 0.05 | 0.37 |
| ARIZONA | 1,493.82 | 1,284.59 | 1,185.69 | 1,061.51 | 5,025.61 |
| ARKANSAS | 783.11 | 670.68 | 613.65 | 573.76 | 2,641.19 |
| CALIFORNIA | 6,095.60 | 5,610.45 | 5,040.20 | 4,819.59 | 21,565.83 |
| COLORADO | 1,114.87 | 984.90 | 936.41 | 849.80 | 3,885.98 |
| CONNECTICUT | 868.21 | 709.28 | 691.77 | 660.86 | 2,930.12 |
| DELAWARE | 314.40 | 260.38 | 230.68 | 202.56 | 1,008.02 |
| DISTRICT OF COLUMBIA | 78.67 | 73.74 | 68.38 | 66.54 | 287.34 |
| FLORIDA | 5,517.05 | 4,950.45 | 4,146.25 | 3,996.39 | 18,610.14 |
| GEORGIA | 2,467.35 | 2,239.30 | 2,092.68 | 1,990.60 | 8,789.93 |
| GUAM | 8.96 | 7.20 | 9.21 | 7.39 | 32.77 |
| HAWAII | 169.98 | 159.24 | 147.76 | 143.32 | 620.30 |
| IDAHO | 560.67 | 496.92 | 475.76 | 449.96 | 1,983.31 |
| ILLINOIS | 2,751.72 | 2,386.75 | 2,236.62 | 2,140.06 | 9,515.16 |
| INDIANA | 1,663.14 | 1,478.14 | 1,389.31 | 1,313.43 | 5,844.02 |
| IOWA | 795.57 | 736.56 | 711.00 | 680.72 | 2,923.85 |
| KANSAS | 960.00 | 873.71 | 817.86 | 805.40 | 3,456.97 |
| KENTUCKY | 1,004.59 | 920.63 | 857.97 | 774.49 | 3,557.68 |
| LOUISIANA | 1,014.57 | 915.75 | 855.53 | 817.67 | 3,603.52 |
| MAINE | 303.68 | 273.58 | 266.44 | 258.04 | 1,101.74 |
| MARYLAND | 1,177.18 | 1,047.83 | 954.78 | 890.95 | 4,070.73 |
| MASSACHUSETTS | 1,174.34 | 1,103.65 | 1,003.65 | 993.50 | 4,275.14 |
| MICHIGAN | 2,700.25 | 2,414.26 | 2,224.25 | 2,067.19 | 9,405.95 |
| MINNESOTA | 922.88 | 788.02 | 783.81 | 728.27 | 3,222.99 |
| MISSISSIPPI | 1,060.52 | 940.87 | 1,029.70 | 848.63 | 3,879.72 |
| MISSOURI | 1,909.91 | 1,718.82 | 1,612.29 | 1,534.63 | 6,775.65 |
| MONTANA | 291.92 | 273.83 | 253.34 | 241.54 | 1,060.63 |
| NEBRASKA | 480.32 | 452.28 | 421.16 | 414.18 | 1,767.93 |
| NEVADA | 481.88 | 426.52 | 391.63 | 360.60 | 1,660.62 |
| NEW HAMPSHIRE | 306.85 | 292.11 | 262.01 | 242.68 | 1,103.65 |
| NEW JERSEY | 2,333.70 | 1,782.18 | 1,657.43 | 1,627.45 | 7,400.76 |
| NEW MEXICO | 396.02 | 333.23 | 316.18 | 304.93 | 1,350.36 |
| NEW YORK | 3,767.36 | 3,452.98 | 3,131.91 | 3,050.47 | 13,402.72 |
| NORTH CAROLINA | 2,441.52 | 2,187.04 | 2,074.21 | 1,999.21 | 8,701.98 |
| NORTH DAKOTA | 200.92 | 193.22 | 180.11 | 165.49 | 739.73 |
| OHIO | 2,070.84 | 1,893.13 | 1,654.43 | 1,564.55 | 7,182.96 |
| OKLAHOMA | 1,422.86 | 1,224.66 | 1,092.96 | 908.41 | 4,648.89 |
| OREGON | 808.80 | 756.41 | 689.02 | 647.95 | 2,902.19 |
| PENNSYLVANIA | 4,314.42 | 3,882.29 | 3,696.00 | 3,363.39 | 15,256.11 |
| PUERTO RICO | 77.83 | 81.40 | 161.63 | 167.96 | 488.81 |

| | | | | | |
|---|---|---|---|---|---|
| RHODE ISLAND | 162.75 | 139.36 | 139.79 | 133.06 | 574.96 |
| SOUTH CAROLINA | 1,171.83 | 1,005.39 | 978.22 | 920.89 | 4,076.33 |
| SOUTH DAKOTA | 258.33 | 233.37 | 216.15 | 200.14 | 907.99 |
| TENNESSEE | 1,767.99 | 1,509.25 | 1,471.14 | 1,227.94 | 5,976.32 |
| TEXAS | 5,136.47 | 4,661.60 | 4,231.75 | 3,958.35 | 17,988.16 |
| UTAH | 754.91 | 675.62 | 638.57 | 619.90 | 2,688.99 |
| VERMONT | 167.38 | 155.09 | 139.68 | 135.42 | 597.57 |
| VIRGIN ISLANDS | 3.93 | 4.58 | 5.41 | 4.01 | 17.93 |
| VIRGINIA | 1,492.39 | 1,359.62 | 1,256.52 | 1,200.31 | 5,308.84 |
| WASHINGTON | 1,326.71 | 1,262.09 | 1,168.78 | 1,101.16 | 4,858.73 |
| WEST VIRGINIA | 480.43 | 443.49 | 394.99 | 391.76 | 1,710.68 |
| WISCONSIN | 1,315.89 | 1,168.39 | 1,076.41 | 997.05 | 4,557.74 |
| WYOMING | 141.06 | 120.91 | 117.74 | 111.00 | 490.71 |
| **9801 - FENTANYL BASE - Total** | **72,009.63** | **64,383.09** | **59,468.16** | **55,885.75** | **251,746.63** |

**DRUG: 1100 - AMPHETAMINE**

| STATE NAME | Q1 CALC GRAMS | Q2 CALC GRAMS | Q3 CALC GRAMS | Q4 CALC GRAMS | TOTAL GRAMS |
|---|---|---|---|---|---|
| ALABAMA | 2,834.39 | 2,814.79 | 2,873.27 | 2,871.54 | 11,394.00 |
| ALASKA | 1,154.59 | 1,198.68 | 1,199.41 | 1,220.51 | 4,773.19 |
| AMERICAN SAMOA | 0.00 | 3.81 | 0.00 | 0.00 | 3.81 |
| ARIZONA | 1,368.71 | 1,476.31 | 1,596.44 | 1,446.75 | 5,888.21 |
| ARKANSAS | 1,794.45 | 1,765.50 | 1,815.14 | 1,801.82 | 7,176.90 |
| CALIFORNIA | 805.84 | 830.32 | 819.28 | 841.39 | 3,296.84 |
| COLORADO | 1,655.30 | 1,687.23 | 1,740.02 | 1,776.42 | 6,858.98 |
| CONNECTICUT | 1,556.45 | 1,564.47 | 1,606.58 | 1,629.83 | 6,357.34 |
| DELAWARE | 2,348.17 | 2,367.82 | 2,358.63 | 2,399.04 | 9,473.66 |
| DISTRICT OF COLUMBIA | 2,009.37 | 2,067.67 | 2,071.63 | 2,082.47 | 8,231.14 |
| FLORIDA | 1,696.79 | 1,719.66 | 1,705.36 | 1,722.93 | 6,844.75 |
| GEORGIA | 2,027.42 | 1,984.42 | 2,046.98 | 2,055.44 | 8,114.25 |
| GUAM | 208.23 | 265.68 | 273.50 | 238.62 | 986.03 |
| HAWAII | 743.89 | 758.55 | 784.88 | 743.88 | 3,031.19 |
| IDAHO | 1,738.42 | 1,783.58 | 1,799.44 | 1,875.66 | 7,197.10 |
| ILLINOIS | 1,533.93 | 1,538.06 | 1,566.89 | 1,597.18 | 6,236.06 |
| INDIANA | 2,080.66 | 2,076.97 | 2,113.84 | 2,130.65 | 8,402.12 |
| IOWA | 1,692.64 | 1,705.04 | 1,745.05 | 1,777.12 | 6,919.85 |
| KANSAS | 2,193.72 | 2,215.54 | 2,247.20 | 2,265.39 | 8,921.86 |
| KENTUCKY | 1,663.76 | 1,624.88 | 1,647.47 | 1,666.42 | 6,602.53 |
| LOUISIANA | 3,576.00 | 3,566.67 | 3,723.52 | 3,735.87 | 14,602.07 |
| MAINE | 1,768.50 | 1,771.34 | 1,873.13 | 1,844.64 | 7,257.61 |
| MARYLAND | 1,638.17 | 1,680.11 | 1,660.40 | 1,698.18 | 6,676.86 |
| MASSACHUSETTS | 2,761.04 | 2,767.08 | 2,803.26 | 2,850.18 | 11,181.56 |
| MICHIGAN | 2,477.40 | 2,465.59 | 2,461.81 | 2,498.70 | 9,903.51 |
| MINNESOTA | 2,077.89 | 2,089.56 | 2,082.99 | 2,172.11 | 8,422.56 |
| MISSISSIPPI | 2,088.06 | 2,047.29 | 2,120.53 | 2,079.89 | 8,335.78 |
| MISSOURI | 2,283.11 | 2,290.57 | 2,363.62 | 2,584.47 | 9,521.79 |
| MONTANA | 1,447.16 | 1,487.29 | 1,502.31 | 1,535.24 | 5,972.00 |
| NEBRASKA | 1,465.23 | 1,491.54 | 1,512.42 | 1,554.78 | 6,023.97 |
| NEVADA | 979.29 | 989.69 | 1,015.71 | 1,016.49 | 4,001.18 |
| NEW HAMPSHIRE | 2,985.45 | 3,056.39 | 3,137.95 | 3,084.37 | 12,264.15 |
| NEW JERSEY | 1,207.74 | 1,243.36 | 1,251.66 | 1,281.83 | 4,984.58 |
| NEW MEXICO | 726.86 | 738.32 | 750.95 | 755.77 | 2,971.90 |
| NEW YORK | 1,253.89 | 1,292.93 | 1,285.75 | 1,323.49 | 5,156.06 |
| NORTH CAROLINA | 2,226.31 | 2,187.08 | 2,231.66 | 2,314.24 | 8,959.28 |
| NORTH DAKOTA | 1,367.78 | 1,365.38 | 1,410.47 | 1,432.87 | 5,576.50 |
| OHIO | 1,853.25 | 1,814.94 | 1,742.66 | 1,798.44 | 7,209.29 |
| OKLAHOMA | 1,459.82 | 1,444.74 | 1,466.02 | 1,469.46 | 5,840.04 |
| OREGON | 1,581.28 | 1,648.82 | 1,623.11 | 1,670.33 | 6,523.53 |
| PENNSYLVANIA | 1,680.00 | 1,697.65 | 1,686.89 | 1,696.12 | 6,760.67 |

| | | | | | |
|---|---|---|---|---|---|
| PUERTO RICO | 256.86 | 247.04 | 249.40 | 262.74 | 1,016.04 |
| RHODE ISLAND | 3,619.01 | 3,647.07 | 3,741.30 | 3,672.53 | 14,679.91 |
| SOUTH CAROLINA | 3,011.43 | 2,933.72 | 3,061.57 | 3,051.95 | 12,058.67 |
| SOUTH DAKOTA | 1,940.37 | 1,822.88 | 1,867.31 | 1,882.71 | 7,513.27 |
| TENNESSEE | 2,106.71 | 2,100.55 | 2,160.55 | 2,139.90 | 8,507.71 |
| TEXAS | 1,718.54 | 1,735.60 | 1,803.78 | 1,825.59 | 7,083.51 |
| UTAH | 3,146.94 | 3,178.36 | 3,257.84 | 3,394.57 | 12,977.71 |
| VERMONT | 1,610.25 | 1,602.93 | 1,623.47 | 1,641.71 | 6,478.35 |
| VIRGIN ISLANDS | 280.79 | 303.22 | 316.91 | 305.64 | 1,206.55 |
| VIRGINIA | 2,278.57 | 2,271.86 | 2,315.36 | 2,306.79 | 9,172.59 |
| WASHINGTON | 1,675.84 | 1,732.31 | 1,701.89 | 1,768.41 | 6,878.45 |
| WEST VIRGINIA | 1,269.48 | 1,273.81 | 1,273.26 | 1,279.45 | 5,096.00 |
| WISCONSIN | 2,223.78 | 2,204.52 | 2,213.55 | 2,231.86 | 8,873.71 |
| WYOMING | 1,218.98 | 1,202.35 | 1,229.71 | 1,249.86 | 4,900.89 |
| **1100 - AMPHETAMINE - Total** | **1,724.45** | **1,734.11** | **1,756.46** | **1,781.99** | **6,997.01** |

**DRUG: 1105 - METHAMPHETAMINE**

| STATE NAME | Q1 CALC GRAMS | Q2 CALC GRAMS | Q3 CALC GRAMS | Q4 CALC GRAMS | TOTAL GRAMS |
|---|---|---|---|---|---|
| ALABAMA | 0.63 | 0.70 | 0.72 | 0.79 | 2.84 |
| ALASKA | 0.28 | 0.28 | 0.51 | 0.00 | 1.07 |
| ARIZONA | 0.58 | 0.63 | 1.00 | 0.61 | 2.82 |
| ARKANSAS | 0.30 | 0.25 | 0.08 | 0.33 | 0.96 |
| CALIFORNIA | 0.73 | 0.74 | 0.73 | 0.70 | 2.90 |
| COLORADO | 1.13 | 1.17 | 1.26 | 1.12 | 4.68 |
| CONNECTICUT | 0.69 | 0.70 | 0.57 | 0.73 | 2.69 |
| DELAWARE | 0.36 | 0.63 | 0.49 | 0.49 | 1.97 |
| DISTRICT OF COLUMBIA | 0.87 | 1.94 | 1.20 | 0.93 | 4.94 |
| FLORIDA | 0.27 | 0.25 | 0.28 | 0.25 | 1.05 |
| GEORGIA | 0.25 | 0.17 | 0.23 | 0.26 | 0.92 |
| HAWAII | 0.38 | 0.35 | 0.35 | 0.53 | 1.62 |
| IDAHO | 1.00 | 0.77 | 0.90 | 0.87 | 3.54 |
| ILLINOIS | 0.43 | 0.40 | 0.40 | 0.38 | 1.62 |
| INDIANA | 0.40 | 0.43 | 0.38 | 0.38 | 1.58 |
| IOWA | 0.57 | 0.58 | 0.70 | 0.47 | 2.32 |
| KANSAS | 0.51 | 0.55 | 0.55 | 0.55 | 2.15 |
| KENTUCKY | 0.25 | 0.27 | 0.32 | 0.24 | 1.08 |
| LOUISIANA | 0.35 | 0.34 | 0.35 | 0.30 | 1.33 |
| MAINE | 0.79 | 0.79 | 1.51 | 0.91 | 3.99 |
| MARYLAND | 0.64 | 0.58 | 0.67 | 0.79 | 2.67 |
| MASSACHUSETTS | 0.66 | 0.58 | 0.48 | 0.41 | 2.13 |
| MICHIGAN | 0.31 | 0.26 | 0.25 | 0.21 | 1.02 |

| | | | | | |
|---|---|---|---|---|---|
| MINNESOTA | 1.45 | 1.34 | 1.39 | 1.48 | 5.67 |
| MISSISSIPPI | 0.22 | 0.20 | 0.28 | 0.34 | 1.04 |
| MISSOURI | 0.46 | 0.74 | 0.70 | 0.97 | 2.88 |
| MONTANA | 0.00 | 0.04 | 0.00 | 0.00 | 0.04 |
| NEBRASKA | 0.29 | 0.33 | 0.42 | 0.07 | 1.10 |
| NEVADA | 0.71 | 0.95 | 0.85 | 0.52 | 3.04 |
| NEW HAMPSHIRE | 0.43 | 0.40 | 0.37 | 0.31 | 1.50 |
| NEW JERSEY | 0.49 | 0.37 | 0.38 | 0.34 | 1.59 |
| NEW MEXICO | 0.23 | 0.16 | 0.29 | 0.43 | 1.11 |
| NEW YORK | 0.50 | 0.47 | 0.46 | 0.45 | 1.89 |
| NORTH CAROLINA | 0.53 | 0.54 | 0.53 | 0.41 | 2.01 |
| NORTH DAKOTA | 0.54 | 1.08 | 0.60 | 0.66 | 2.87 |
| OHIO | 0.17 | 0.62 | 0.29 | 0.28 | 1.37 |
| OKLAHOMA | 0.43 | 0.33 | 0.41 | 0.29 | 1.46 |
| OREGON | 1.04 | 0.71 | 0.91 | 1.05 | 3.71 |
| PENNSYLVANIA | 0.96 | 0.98 | 0.85 | 1.00 | 3.79 |
| RHODE ISLAND | 0.34 | 0.42 | 0.19 | 0.46 | 1.41 |
| SOUTH CAROLINA | 0.35 | 0.42 | 0.30 | 0.50 | 1.57 |
| SOUTH DAKOTA | 1.33 | 0.84 | 0.69 | 0.99 | 3.85 |
| TENNESSEE | 0.23 | 0.21 | 0.28 | 0.21 | 0.93 |
| TEXAS | 0.43 | 0.43 | 0.41 | 0.42 | 1.69 |
| UTAH | 1.05 | 0.86 | 0.78 | 0.73 | 3.42 |
| VERMONT | 0.32 | 0.19 | 0.45 | 0.39 | 1.35 |
| VIRGINIA | 0.44 | 0.36 | 0.41 | 0.37 | 1.58 |
| WASHINGTON | 1.37 | 1.39 | 1.34 | 1.16 | 5.26 |
| WEST VIRGINIA | 0.26 | 0.20 | 0.17 | 0.37 | 1.00 |
| WISCONSIN | 0.57 | 0.63 | 0.62 | 0.61 | 2.43 |
| WYOMING | 0.86 | 0.64 | 0.93 | 0.86 | 3.28 |
| **1105 - METHAMPHETAMINE -** | **0.54** | **0.55** | **0.54** | **0.52** | **2.15** |

**DRUG: 1205 - LISDEXAMFETAMINE**

| STATE NAME | Q1 CALC GRAMS | Q2 CALC GRAMS | Q3 CALC GRAMS | Q4 CALC GRAMS | TOTAL GRAMS |
|---|---|---|---|---|---|
| ALABAMA | 1,508.95 | 1,446.07 | 1,465.58 | 1,518.01 | 5,938.60 |
| ALASKA | 360.83 | 370.35 | 385.82 | 379.47 | 1,496.47 |
| ARIZONA | 600.75 | 650.82 | 759.69 | 633.07 | 2,644.34 |
| ARKANSAS | 1,051.34 | 990.77 | 984.20 | 1,018.83 | 4,045.14 |
| CALIFORNIA | 301.55 | 301.94 | 297.46 | 313.03 | 1,213.98 |
| COLORADO | 575.68 | 566.78 | 571.46 | 602.64 | 2,316.55 |
| CONNECTICUT | 1,011.99 | 1,012.67 | 986.92 | 1,039.89 | 4,051.46 |
| DELAWARE | 1,598.84 | 1,537.49 | 1,475.94 | 1,596.39 | 6,208.66 |
| DISTRICT OF COLUMBIA | 919.43 | 953.15 | 923.08 | 961.61 | 3,757.27 |

| | | | | | |
|---|---|---|---|---|---|
| FLORIDA | 738.08 | 710.26 | 684.65 | 745.00 | 2,877.99 |
| GEORGIA | 1,038.32 | 986.73 | 998.63 | 1,021.93 | 4,045.62 |
| GUAM | 5.18 | 10.37 | 4.86 | 4.21 | 24.62 |
| HAWAII | 106.75 | 116.32 | 115.64 | 110.83 | 449.54 |
| IDAHO | 615.72 | 612.15 | 595.14 | 638.70 | 2,461.71 |
| ILLINOIS | 556.82 | 560.05 | 554.22 | 581.86 | 2,252.95 |
| INDIANA | 1,323.92 | 1,263.75 | 1,257.00 | 1,306.06 | 5,150.73 |
| IOWA | 1,137.01 | 1,123.16 | 1,085.32 | 1,146.63 | 4,492.13 |
| KANSAS | 1,110.46 | 1,116.80 | 1,092.37 | 1,161.52 | 4,481.15 |
| KENTUCKY | 970.34 | 930.96 | 927.11 | 946.37 | 3,774.78 |
| LOUISIANA | 2,006.82 | 1,928.75 | 1,928.26 | 1,994.43 | 7,858.25 |
| MAINE | 1,272.91 | 1,268.12 | 1,237.49 | 1,264.38 | 5,042.90 |
| MARYLAND | 978.53 | 992.58 | 937.74 | 1,013.09 | 3,921.93 |
| MASSACHUSETTS | 562.58 | 576.81 | 552.95 | 605.52 | 2,297.86 |
| MICHIGAN | 624.71 | 612.71 | 591.98 | 647.45 | 2,476.86 |
| MINNESOTA | 581.73 | 582.01 | 569.05 | 610.03 | 2,342.82 |
| MISSISSIPPI | 1,328.13 | 1,216.46 | 1,240.07 | 1,257.76 | 5,042.43 |
| MISSOURI | 1,098.78 | 1,056.44 | 1,049.24 | 1,275.19 | 4,479.66 |
| MONTANA | 622.32 | 601.64 | 616.13 | 650.19 | 2,490.28 |
| NEBRASKA | 948.30 | 916.37 | 934.32 | 1,003.72 | 3,802.72 |
| NEVADA | 296.08 | 291.63 | 286.00 | 292.03 | 1,165.74 |
| NEW HAMPSHIRE | 1,051.41 | 1,091.89 | 1,045.17 | 1,106.21 | 4,294.68 |
| NEW JERSEY | 551.42 | 561.17 | 534.15 | 568.67 | 2,215.41 |
| NEW MEXICO | 201.32 | 179.23 | 187.68 | 193.18 | 761.40 |
| NEW YORK | 446.53 | 453.58 | 430.90 | 467.62 | 1,798.63 |
| NORTH CAROLINA | 1,124.65 | 1,100.30 | 1,047.48 | 1,164.47 | 4,436.90 |
| NORTH DAKOTA | 897.85 | 839.92 | 851.26 | 908.25 | 3,497.27 |
| OHIO | 947.20 | 870.72 | 801.24 | 850.44 | 3,469.60 |
| OKLAHOMA | 1,014.93 | 970.26 | 962.88 | 1,014.88 | 3,962.94 |
| OREGON | 404.83 | 414.25 | 402.34 | 416.21 | 1,637.62 |
| PENNSYLVANIA | 700.14 | 701.35 | 677.22 | 721.34 | 2,800.04 |
| PUERTO RICO | 60.93 | 54.82 | 53.48 | 58.31 | 227.54 |
| RHODE ISLAND | 702.57 | 703.34 | 680.93 | 711.86 | 2,798.70 |
| SOUTH CAROLINA | 1,336.86 | 1,304.93 | 1,259.12 | 1,359.05 | 5,259.96 |
| SOUTH DAKOTA | 1,299.37 | 1,251.52 | 1,200.04 | 1,282.76 | 5,033.68 |
| TENNESSEE | 1,086.50 | 1,045.39 | 1,038.82 | 1,076.48 | 4,247.19 |
| TEXAS | 1,081.74 | 1,043.56 | 1,025.67 | 1,084.16 | 4,235.13 |
| UTAH | 816.91 | 799.22 | 792.32 | 857.86 | 3,266.31 |
| VERMONT | 1,155.66 | 1,135.80 | 1,117.97 | 1,177.56 | 4,586.99 |
| VIRGIN ISLANDS | 64.22 | 53.69 | 73.17 | 61.59 | 252.66 |

| | | | | | |
|---|---|---|---|---|---|
| VIRGINIA | 848.14 | 830.42 | 810.83 | 863.21 | 3,352.60 |
| WASHINGTON | 521.03 | 531.41 | 515.15 | 552.99 | 2,120.58 |
| WEST VIRGINIA | 1,031.82 | 1,017.18 | 968.17 | 1,038.49 | 4,055.67 |
| WISCONSIN | 1,271.08 | 1,270.59 | 1,251.04 | 1,336.72 | 5,129.43 |
| WYOMING | 488.02 | 469.76 | 441.15 | 467.30 | 1,866.23 |
| **1205 - LISDEXAMFETAMINE - T** | **785.04** | **767.80** | **752.91** | **797.89** | **3,103.65** |

**DRUG: 1724 - METHYLPHENIDATE (DL;D;L;ISOMERS)**

| STATE NAME | Q1 CALC GRAMS | Q2 CALC GRAMS | Q3 CALC GRAMS | Q4 CALC GRAMS | TOTAL GRAMS |
|---|---|---|---|---|---|
| ALABAMA | 1,547.25 | 1,414.93 | 1,416.78 | 1,502.23 | 5,881.18 |
| ALASKA | 1,159.72 | 1,128.03 | 1,050.29 | 1,166.72 | 4,504.75 |
| AMERICAN SAMOA | 19.76 | 11.86 | 10.54 | 25.03 | 67.20 |
| ARIZONA | 1,209.39 | 1,286.23 | 1,481.83 | 1,201.18 | 5,178.64 |
| ARKANSAS | 1,364.42 | 1,282.05 | 1,265.95 | 1,307.92 | 5,220.34 |
| CALIFORNIA | 721.86 | 706.16 | 660.13 | 709.66 | 2,797.81 |
| COLORADO | 1,251.25 | 1,203.93 | 1,169.61 | 1,247.25 | 4,872.04 |
| CONNECTICUT | 1,630.12 | 1,538.67 | 1,409.57 | 1,524.44 | 6,102.80 |
| DELAWARE | 2,066.20 | 1,945.23 | 1,855.55 | 1,984.21 | 7,851.18 |
| DISTRICT OF COLUMBIA | 1,411.09 | 1,290.14 | 1,219.94 | 1,337.28 | 5,258.45 |
| FLORIDA | 1,112.98 | 1,053.02 | 981.99 | 1,026.63 | 4,174.61 |
| GEORGIA | 1,732.26 | 1,557.98 | 1,576.62 | 1,644.91 | 6,511.77 |
| GUAM | 226.43 | 267.37 | 378.62 | 239.92 | 1,112.34 |
| HAWAII | 825.35 | 795.67 | 809.69 | 785.40 | 3,216.11 |
| IDAHO | 1,830.29 | 1,785.35 | 1,676.99 | 1,754.39 | 7,047.02 |
| ILLINOIS | 1,350.90 | 1,280.66 | 1,224.28 | 1,312.10 | 5,167.94 |
| INDIANA | 1,859.87 | 1,755.48 | 1,684.49 | 1,762.34 | 7,062.18 |
| IOWA | 2,682.68 | 2,629.22 | 2,471.70 | 2,621.40 | 10,405.01 |
| KANSAS | 1,722.87 | 1,693.33 | 1,613.58 | 1,743.81 | 6,773.59 |
| KENTUCKY | 1,769.14 | 1,628.40 | 1,626.57 | 1,679.50 | 6,703.61 |
| LOUISIANA | 1,898.20 | 1,712.87 | 1,700.90 | 1,875.59 | 7,187.57 |
| MAINE | 2,711.51 | 2,676.16 | 2,522.74 | 2,516.40 | 10,426.81 |
| MARYLAND | 1,759.35 | 1,730.86 | 1,553.96 | 1,686.25 | 6,730.42 |
| MASSACHUSETTS | 2,236.33 | 2,194.12 | 1,976.67 | 2,170.50 | 8,577.62 |
| MICHIGAN | 1,906.85 | 1,813.21 | 1,681.80 | 1,802.47 | 7,204.33 |
| MINNESOTA | 1,954.40 | 1,864.35 | 1,773.65 | 1,924.74 | 7,517.14 |
| MISSISSIPPI | 1,432.45 | 1,337.38 | 1,351.10 | 1,437.09 | 5,558.03 |
| MISSOURI | 2,062.80 | 2,023.79 | 2,031.41 | 2,486.97 | 8,604.97 |
| MONTANA | 1,810.46 | 1,712.66 | 1,602.25 | 1,715.44 | 6,840.80 |
| NEBRASKA | 1,707.16 | 1,635.84 | 1,613.34 | 1,703.37 | 6,659.71 |
| NEVADA | 554.34 | 528.76 | 533.56 | 539.05 | 2,155.71 |
| NEW HAMPSHIRE | 2,576.04 | 2,588.56 | 2,419.33 | 2,516.10 | 10,100.03 |

| | | | | | |
|---|---|---|---|---|---|
| NEW JERSEY | 1,050.96 | 1,035.90 | 928.01 | 1,028.67 | 4,043.54 |
| NEW MEXICO | 939.50 | 862.55 | 838.18 | 877.77 | 3,517.99 |
| NEW YORK | 1,196.05 | 1,194.26 | 1,071.43 | 1,170.66 | 4,632.38 |
| NORTH CAROLINA | 2,027.47 | 1,904.70 | 1,718.17 | 1,911.09 | 7,561.43 |
| NORTH DAKOTA | 2,328.58 | 2,202.97 | 2,125.76 | 2,302.01 | 8,959.32 |
| OHIO | 1,935.48 | 1,790.94 | 1,587.69 | 1,727.70 | 7,041.82 |
| OKLAHOMA | 1,336.41 | 1,208.20 | 1,182.73 | 1,245.23 | 4,972.58 |
| OREGON | 1,616.79 | 1,641.82 | 1,537.21 | 1,620.17 | 6,416.00 |
| PENNSYLVANIA | 1,778.10 | 1,730.98 | 1,617.63 | 1,730.97 | 6,857.68 |
| PUERTO RICO | 408.02 | 392.40 | 358.94 | 380.99 | 1,540.34 |
| RHODE ISLAND | 1,829.77 | 1,776.14 | 1,590.02 | 1,695.57 | 6,891.50 |
| SOUTH CAROLINA | 1,937.49 | 1,824.60 | 1,723.57 | 1,866.10 | 7,351.77 |
| SOUTH DAKOTA | 2,707.40 | 2,367.69 | 2,453.67 | 2,505.61 | 10,034.37 |
| TENNESSEE | 1,319.06 | 1,239.50 | 1,224.49 | 1,277.77 | 5,060.81 |
| TEXAS | 1,334.57 | 1,278.56 | 1,174.56 | 1,317.12 | 5,104.80 |
| UTAH | 1,706.79 | 1,632.52 | 1,588.56 | 1,718.71 | 6,646.58 |
| VERMONT | 3,054.53 | 3,018.74 | 2,870.95 | 3,029.64 | 11,973.87 |
| VIRGIN ISLANDS | 122.95 | 145.53 | 120.81 | 144.43 | 533.72 |
| VIRGINIA | 1,699.16 | 1,638.35 | 1,547.11 | 1,621.50 | 6,506.11 |
| WASHINGTON | 1,704.42 | 1,699.71 | 1,565.95 | 1,686.54 | 6,656.63 |
| WEST VIRGINIA | 1,745.13 | 1,648.14 | 1,536.58 | 1,586.24 | 6,516.09 |
| WISCONSIN | 1,917.70 | 1,766.77 | 1,694.50 | 1,813.06 | 7,192.03 |
| WYOMING | 1,386.97 | 1,338.89 | 1,246.96 | 1,400.90 | 5,373.72 |
| **1724 - METHYLPHENIDATE (DI** | **1,472.70** | **1,410.41** | **1,331.49** | **1,427.74** | **5,642.34** |

**DRUG: 9050 - CODEINE**

| STATE NAME | Q1 CALC GRAMS | Q2 CALC GRAMS | Q3 CALC GRAMS | Q4 CALC GRAMS | TOTAL GRAMS |
|---|---|---|---|---|---|
| ALABAMA | 1,144.99 | 1,022.24 | 1,028.30 | 1,022.38 | 4,217.91 |
| ALASKA | 879.47 | 857.83 | 858.18 | 723.12 | 3,318.60 |
| AMERICAN SAMOA | 289.62 | 538.82 | 276.14 | 511.88 | 1,616.46 |
| ARIZONA | 1,052.10 | 897.77 | 902.17 | 860.12 | 3,712.17 |
| ARKANSAS | 2,328.16 | 2,084.59 | 2,022.79 | 1,855.08 | 8,290.62 |
| CALIFORNIA | 1,156.61 | 1,083.71 | 1,072.61 | 995.76 | 4,308.69 |
| COLORADO | 489.22 | 425.93 | 412.88 | 356.79 | 1,684.80 |
| CONNECTICUT | 569.46 | 570.32 | 532.25 | 523.78 | 2,195.81 |
| DELAWARE | 937.38 | 909.97 | 850.71 | 860.01 | 3,558.07 |
| DISTRICT OF COLUMBIA | 1,837.33 | 1,754.57 | 1,801.08 | 1,497.56 | 6,890.53 |
| FLORIDA | 908.70 | 810.11 | 748.10 | 727.00 | 3,193.91 |
| GEORGIA | 1,061.99 | 949.76 | 912.35 | 853.17 | 3,777.27 |
| GUAM | 521.72 | 573.07 | 538.63 | 383.56 | 2,016.98 |
| HAWAII | 905.83 | 882.68 | 845.77 | 778.57 | 3,412.85 |

| | | | | | |
|---|---|---|---|---|---|
| IDAHO | 835.90 | 700.96 | 791.43 | 778.01 | 3,106.30 |
| ILLINOIS | 1,148.77 | 1,101.98 | 1,116.42 | 1,070.30 | 4,437.47 |
| INDIANA | 745.71 | 681.23 | 637.63 | 629.91 | 2,694.48 |
| IOWA | 616.44 | 545.73 | 635.91 | 527.37 | 2,325.45 |
| KANSAS | 927.99 | 812.18 | 867.87 | 746.54 | 3,354.58 |
| KENTUCKY | 1,071.93 | 975.93 | 945.60 | 882.23 | 3,875.68 |
| LOUISIANA | 895.10 | 838.48 | 832.50 | 806.01 | 3,372.08 |
| MAINE | 729.12 | 641.54 | 677.04 | 619.66 | 2,667.37 |
| MARYLAND | 891.50 | 895.97 | 801.07 | 807.93 | 3,396.47 |
| MASSACHUSETTS | 628.93 | 643.79 | 577.36 | 608.33 | 2,458.42 |
| MICHIGAN | 2,318.12 | 2,115.88 | 1,928.05 | 1,833.26 | 8,195.31 |
| MINNESOTA | 650.03 | 590.55 | 608.26 | 587.46 | 2,436.30 |
| MISSISSIPPI | 892.17 | 765.26 | 757.03 | 707.81 | 3,122.28 |
| MISSOURI | 1,044.67 | 979.07 | 1,062.75 | 972.34 | 4,058.83 |
| MONTANA | 940.67 | 872.31 | 949.99 | 828.74 | 3,591.71 |
| NEBRASKA | 614.39 | 542.52 | 571.45 | 527.56 | 2,255.92 |
| NEVADA | 794.53 | 747.35 | 721.21 | 654.65 | 2,917.74 |
| NEW HAMPSHIRE | 528.66 | 504.58 | 496.53 | 473.85 | 2,003.62 |
| NEW JERSEY | 791.08 | 797.37 | 737.51 | 746.50 | 3,072.45 |
| NEW MEXICO | 908.86 | 728.53 | 814.57 | 753.53 | 3,205.50 |
| NEW YORK | 665.52 | 666.69 | 625.64 | 609.97 | 2,567.83 |
| NORTH CAROLINA | 480.49 | 422.67 | 387.81 | 421.17 | 1,712.14 |
| NORTH DAKOTA | 754.56 | 716.52 | 648.43 | 672.00 | 2,791.51 |
| OHIO | 762.12 | 692.58 | 645.49 | 565.75 | 2,665.94 |
| OKLAHOMA | 2,437.61 | 2,334.78 | 2,331.36 | 2,175.46 | 9,279.21 |
| OREGON | 869.11 | 788.77 | 792.39 | 728.62 | 3,178.90 |
| PENNSYLVANIA | 920.46 | 901.61 | 848.00 | 830.61 | 3,500.68 |
| PUERTO RICO | 308.34 | 332.99 | 331.06 | 286.39 | 1,258.78 |
| RHODE ISLAND | 587.83 | 585.48 | 496.76 | 516.32 | 2,186.39 |
| SOUTH CAROLINA | 1,584.81 | 1,555.80 | 1,575.65 | 1,358.39 | 6,074.65 |
| SOUTH DAKOTA | 731.48 | 676.74 | 733.25 | 665.02 | 2,806.48 |
| TENNESSEE | 1,172.93 | 1,045.39 | 1,110.07 | 909.80 | 4,238.19 |
| TEXAS | 3,694.73 | 3,780.70 | 3,592.80 | 3,491.48 | 14,559.71 |
| UTAH | 610.72 | 504.30 | 510.96 | 502.83 | 2,128.81 |
| VERMONT | 432.05 | 454.95 | 430.09 | 422.76 | 1,739.85 |
| VIRGIN ISLANDS | 577.55 | 759.74 | 416.70 | 775.84 | 2,529.83 |
| VIRGINIA | 629.32 | 573.87 | 564.09 | 539.45 | 2,306.73 |
| WASHINGTON | 850.52 | 801.80 | 800.09 | 742.24 | 3,194.64 |
| WEST VIRGINIA | 1,878.68 | 1,932.54 | 1,953.61 | 1,968.88 | 7,733.71 |
| WISCONSIN | 778.78 | 734.11 | 734.97 | 709.09 | 2,956.96 |

| | | | | | |
|---|---|---|---|---|---|
| WYOMING | 718.94 | 598.06 | 655.19 | 570.33 | 2,542.51 |
| **9050 - CODEINE - Total** | **1,193.24** | **1,139.44** | **1,103.35** | **1,049.74** | **4,485.76** |

**DRUG: 9064 - BUPRENORPHINE**

| STATE NAME | Q1 CALC GRAMS | Q2 CALC GRAMS | Q3 CALC GRAMS | Q4 CALC GRAMS | TOTAL GRAMS |
|---|---|---|---|---|---|
| ALABAMA | 563.17 | 599.62 | 616.19 | 640.56 | 2,419.55 |
| ALASKA | 363.80 | 375.97 | 372.23 | 375.45 | 1,487.46 |
| AMERICAN SAMOA | 0.03 | 0.00 | 0.03 | 0.00 | 0.06 |
| ARIZONA | 156.56 | 170.21 | 181.88 | 196.08 | 704.73 |
| ARKANSAS | 225.16 | 248.60 | 256.74 | 266.58 | 997.09 |
| CALIFORNIA | 107.73 | 112.32 | 118.02 | 123.63 | 461.70 |
| COLORADO | 159.77 | 169.33 | 177.31 | 193.57 | 699.98 |
| CONNECTICUT | 433.95 | 455.46 | 449.64 | 482.06 | 1,821.11 |
| DELAWARE | 417.77 | 452.53 | 441.83 | 459.23 | 1,771.36 |
| DISTRICT OF COLUMBIA | 457.24 | 453.82 | 473.35 | 541.61 | 1,926.02 |
| FLORIDA | 260.63 | 274.39 | 280.52 | 291.39 | 1,106.93 |
| GEORGIA | 166.70 | 176.74 | 180.19 | 190.17 | 713.79 |
| GUAM | 0.35 | 0.14 | 0.81 | 0.00 | 1.29 |
| HAWAII | 150.05 | 153.06 | 160.35 | 168.35 | 631.81 |
| IDAHO | 247.93 | 267.06 | 272.34 | 289.02 | 1,076.36 |
| ILLINOIS | 121.58 | 129.49 | 131.16 | 138.78 | 521.02 |
| INDIANA | 333.69 | 368.79 | 385.24 | 417.68 | 1,505.40 |
| IOWA | 62.62 | 66.00 | 69.53 | 73.01 | 271.15 |
| KANSAS | 90.06 | 98.16 | 99.71 | 107.02 | 394.95 |
| KENTUCKY | 851.72 | 863.31 | 911.78 | 959.65 | 3,586.46 |
| LOUISIANA | 439.21 | 465.57 | 476.84 | 485.13 | 1,866.74 |
| MAINE | 786.61 | 821.48 | 867.81 | 902.60 | 3,378.50 |
| MARYLAND | 473.23 | 495.87 | 505.51 | 534.27 | 2,008.88 |
| MASSACHUSETTS | 644.40 | 671.17 | 662.51 | 700.93 | 2,679.01 |
| MICHIGAN | 294.75 | 308.22 | 308.24 | 324.73 | 1,235.94 |
| MINNESOTA | 157.35 | 167.37 | 172.90 | 185.15 | 682.76 |
| MISSISSIPPI | 346.11 | 361.25 | 365.33 | 371.74 | 1,444.44 |
| MISSOURI | 191.02 | 209.99 | 233.29 | 261.35 | 895.65 |
| MONTANA | 288.65 | 292.18 | 310.69 | 337.46 | 1,228.98 |
| NEBRASKA | 76.92 | 80.45 | 80.74 | 88.43 | 326.55 |
| NEVADA | 132.67 | 137.51 | 143.27 | 152.76 | 566.22 |
| NEW HAMPSHIRE | 624.40 | 670.43 | 649.77 | 702.29 | 2,646.89 |
| NEW JERSEY | 301.64 | 317.60 | 310.41 | 333.65 | 1,263.30 |
| NEW MEXICO | 382.42 | 411.20 | 430.33 | 436.69 | 1,660.64 |
| NEW YORK | 296.25 | 313.51 | 315.22 | 335.61 | 1,260.59 |
| NORTH CAROLINA | 378.62 | 418.31 | 417.36 | 463.92 | 1,678.20 |

| | | | | | |
|---|---|---|---|---|---|
| NORTH DAKOTA | 152.64 | 168.79 | 182.96 | 187.45 | 691.84 |
| OHIO | 467.51 | 485.88 | 485.46 | 514.73 | 1,953.57 |
| OKLAHOMA | 329.83 | 356.11 | 378.82 | 413.92 | 1,478.67 |
| OREGON | 290.86 | 310.61 | 323.71 | 358.40 | 1,283.57 |
| PENNSYLVANIA | 531.14 | 570.72 | 568.17 | 612.62 | 2,282.64 |
| PUERTO RICO | 57.28 | 56.12 | 56.97 | 57.80 | 228.17 |
| RHODE ISLAND | 771.03 | 748.13 | 767.32 | 819.43 | 3,105.91 |
| SOUTH CAROLINA | 250.11 | 271.53 | 279.75 | 301.62 | 1,103.00 |
| SOUTH DAKOTA | 60.26 | 67.53 | 75.87 | 79.44 | 283.10 |
| TENNESSEE | 573.28 | 608.47 | 616.27 | 625.00 | 2,423.02 |
| TEXAS | 107.86 | 112.10 | 115.68 | 119.12 | 454.76 |
| UTAH | 352.63 | 367.36 | 382.17 | 402.06 | 1,504.23 |
| VERMONT | 1,056.11 | 1,053.70 | 1,091.40 | 1,139.98 | 4,341.19 |
| VIRGIN ISLANDS | 11.79 | 9.44 | 12.81 | 10.70 | 44.73 |
| VIRGINIA | 253.85 | 267.80 | 275.02 | 285.52 | 1,082.19 |
| WASHINGTON | 322.16 | 348.81 | 359.88 | 395.64 | 1,426.48 |
| WEST VIRGINIA | 849.21 | 850.11 | 875.17 | 908.79 | 3,483.28 |
| WISCONSIN | 196.92 | 207.63 | 214.83 | 226.68 | 846.06 |
| WYOMING | 243.89 | 231.52 | 231.24 | 243.80 | 950.45 |
| **9064 - BUPRENORPHINE - Tota** | **283.69** | **299.58** | **305.42** | **323.69** | **1,212.38** |

**DRUG: 9143 - OXYCODONE**

| STATE NAME | Q1 CALC GRAMS | Q2 CALC GRAMS | Q3 CALC GRAMS | Q4 CALC GRAMS | TOTAL GRAMS |
|---|---|---|---|---|---|
| ALABAMA | 4,406.72 | 4,220.81 | 4,039.19 | 3,965.38 | 16,632.08 |
| ALASKA | 4,426.47 | 4,397.16 | 4,260.68 | 3,824.55 | 16,908.86 |
| AMERICAN SAMOA | 16.39 | 10.25 | 10.25 | 19.12 | 56.01 |
| ARIZONA | 5,840.12 | 5,445.42 | 5,261.59 | 5,079.19 | 21,626.32 |
| ARKANSAS | 4,319.76 | 4,099.28 | 3,787.37 | 3,532.15 | 15,738.56 |
| CALIFORNIA | 2,050.62 | 1,979.49 | 1,888.73 | 1,823.26 | 7,742.11 |
| COLORADO | 3,969.85 | 3,791.22 | 3,701.66 | 3,643.42 | 15,106.14 |
| CONNECTICUT | 4,098.81 | 4,010.79 | 3,918.31 | 3,890.65 | 15,918.56 |
| DELAWARE | 6,858.88 | 6,868.30 | 6,733.04 | 6,541.80 | 27,002.02 |
| DISTRICT OF COLUMBIA | 2,157.10 | 2,092.92 | 2,003.92 | 1,994.27 | 8,248.21 |
| FLORIDA | 5,295.94 | 5,097.50 | 4,642.46 | 4,614.33 | 19,650.23 |
| GEORGIA | 4,255.62 | 4,116.27 | 3,985.55 | 3,947.48 | 16,304.92 |
| GUAM | 521.80 | 381.20 | 479.88 | 380.19 | 1,763.06 |
| HAWAII | 2,867.40 | 2,824.25 | 2,649.75 | 2,534.38 | 10,875.78 |
| IDAHO | 3,582.71 | 3,396.18 | 3,325.04 | 3,279.13 | 13,583.06 |
| ILLINOIS | 1,111.83 | 1,058.80 | 1,018.95 | 1,011.33 | 4,200.91 |
| INDIANA | 3,274.80 | 3,091.98 | 2,966.08 | 2,954.24 | 12,287.10 |
| IOWA | 1,542.09 | 1,465.90 | 1,423.23 | 1,404.53 | 5,835.75 |

| | | | | | |
|---|---|---|---|---|---|
| KANSAS | 3,858.49 | 3,733.15 | 3,580.04 | 3,598.71 | 14,770.39 |
| KENTUCKY | 4,470.23 | 4,317.47 | 4,144.04 | 4,102.98 | 17,034.72 |
| LOUISIANA | 3,829.46 | 3,623.11 | 3,553.99 | 3,479.45 | 14,486.02 |
| MAINE | 3,563.57 | 3,418.44 | 3,351.15 | 3,242.10 | 13,575.25 |
| MARYLAND | 4,992.28 | 4,777.70 | 4,611.96 | 4,520.84 | 18,902.78 |
| MASSACHUSETTS | 3,309.09 | 3,191.36 | 3,047.34 | 3,046.74 | 12,594.53 |
| MICHIGAN | 2,629.88 | 2,429.40 | 2,293.44 | 2,211.89 | 9,564.61 |
| MINNESOTA | 2,394.64 | 2,303.52 | 2,219.51 | 2,161.05 | 9,078.72 |
| MISSISSIPPI | 3,154.59 | 3,044.95 | 2,984.75 | 2,886.47 | 12,070.75 |
| MISSOURI | 4,312.90 | 4,049.61 | 3,843.80 | 3,738.22 | 15,944.54 |
| MONTANA | 3,401.90 | 3,171.53 | 3,051.44 | 2,994.17 | 12,619.04 |
| NEBRASKA | 2,046.45 | 1,997.15 | 1,913.87 | 1,908.59 | 7,866.05 |
| NEVADA | 5,848.82 | 5,500.15 | 5,381.20 | 5,300.29 | 22,030.47 |
| NEW HAMPSHIRE | 4,214.76 | 4,015.54 | 3,793.86 | 3,677.94 | 15,702.10 |
| NEW JERSEY | 4,841.15 | 4,673.38 | 4,514.05 | 4,441.11 | 18,469.69 |
| NEW MEXICO | 3,928.14 | 3,818.42 | 3,723.82 | 3,638.70 | 15,109.08 |
| NEW YORK | 3,332.17 | 3,264.56 | 3,126.92 | 3,048.60 | 12,772.25 |
| NORTH CAROLINA | 5,237.72 | 4,991.81 | 4,871.87 | 5,102.00 | 20,203.41 |
| NORTH DAKOTA | 1,953.98 | 1,886.51 | 1,875.34 | 1,841.79 | 7,557.62 |
| OHIO | 3,575.57 | 3,443.49 | 3,309.82 | 3,275.88 | 13,604.76 |
| OKLAHOMA | 5,389.08 | 5,125.11 | 4,787.25 | 4,280.53 | 19,581.98 |
| OREGON | 4,009.05 | 3,902.41 | 3,745.19 | 3,633.55 | 15,290.20 |
| PENNSYLVANIA | 5,111.38 | 4,928.25 | 4,716.87 | 4,561.60 | 19,318.10 |
| PUERTO RICO | 198.74 | 212.04 | 394.73 | 384.33 | 1,189.84 |
| RHODE ISLAND | 3,407.60 | 3,206.25 | 3,047.38 | 3,062.13 | 12,723.36 |
| SOUTH CAROLINA | 4,471.24 | 4,210.96 | 4,063.45 | 3,994.96 | 16,740.61 |
| SOUTH DAKOTA | 1,868.91 | 1,779.42 | 1,687.69 | 1,620.86 | 6,956.88 |
| TENNESSEE | 6,633.85 | 6,451.34 | 6,155.58 | 5,953.82 | 25,194.59 |
| TEXAS | 1,224.54 | 1,208.42 | 1,154.68 | 1,150.20 | 4,737.84 |
| UTAH | 5,535.54 | 5,324.73 | 5,222.08 | 5,145.19 | 21,227.54 |
| VERMONT | 3,330.70 | 3,200.07 | 3,089.56 | 3,007.63 | 12,627.95 |
| VIRGIN ISLANDS | 422.43 | 437.45 | 359.58 | 376.31 | 1,595.78 |
| VIRGINIA | 3,281.14 | 3,159.90 | 3,068.94 | 3,053.33 | 12,563.30 |
| WASHINGTON | 3,839.92 | 3,738.68 | 3,605.33 | 3,534.43 | 14,718.37 |
| WEST VIRGINIA | 3,899.03 | 3,641.16 | 3,442.13 | 3,282.94 | 14,265.26 |
| WISCONSIN | 3,159.42 | 2,946.84 | 2,789.40 | 2,687.14 | 11,582.80 |
| WYOMING | 4,251.18 | 4,056.33 | 3,896.72 | 3,924.45 | 16,128.68 |
| **9143 - OXYCODONE - Total** | **3,494.83** | **3,356.31** | **3,212.79** | **3,152.25** | **13,216.17** |

**DRUG: 9150 - HYDROMORPHONE**

| STATE NAME | Q1 CALC GRAMS | Q2 CALC GRAMS | Q3 CALC GRAMS | Q4 CALC GRAMS | TOTAL GRAMS |
|---|---|---|---|---|---|

| | | | | | |
|---|---|---|---|---|---|
| ALABAMA | 58.11 | 59.04 | 54.34 | 49.06 | 220.55 |
| ALASKA | 134.91 | 106.57 | 107.89 | 87.58 | 436.95 |
| AMERICAN SAMOA | 0.27 | 0.05 | 1.11 | 0.41 | 1.83 |
| ARIZONA | 96.20 | 87.42 | 84.88 | 79.10 | 347.61 |
| ARKANSAS | 96.41 | 87.14 | 81.12 | 78.20 | 342.86 |
| CALIFORNIA | 93.32 | 87.71 | 80.63 | 76.38 | 338.03 |
| COLORADO | 129.40 | 116.01 | 112.10 | 111.32 | 468.82 |
| CONNECTICUT | 164.93 | 141.55 | 131.24 | 131.81 | 569.53 |
| DELAWARE | 202.08 | 184.14 | 169.33 | 165.26 | 720.81 |
| DISTRICT OF COLUMBIA | 76.40 | 62.25 | 75.17 | 67.64 | 281.46 |
| FLORIDA | 232.71 | 210.04 | 197.63 | 186.80 | 827.18 |
| GEORGIA | 74.45 | 75.02 | 67.88 | 62.01 | 279.35 |
| GUAM | 15.85 | 9.65 | 17.21 | 8.02 | 50.73 |
| HAWAII | 40.19 | 38.84 | 36.19 | 36.33 | 151.54 |
| IDAHO | 118.89 | 119.45 | 106.27 | 103.08 | 447.69 |
| ILLINOIS | 69.32 | 64.26 | 60.72 | 59.21 | 253.52 |
| INDIANA | 83.77 | 76.92 | 69.33 | 65.52 | 295.54 |
| IOWA | 86.10 | 78.94 | 74.72 | 74.69 | 314.45 |
| KANSAS | 103.65 | 98.16 | 101.34 | 95.84 | 399.00 |
| KENTUCKY | 58.65 | 55.89 | 50.16 | 53.34 | 218.04 |
| LOUISIANA | 64.88 | 62.44 | 59.57 | 52.94 | 239.82 |
| MAINE | 122.19 | 114.49 | 92.98 | 108.57 | 438.23 |
| MARYLAND | 124.91 | 110.78 | 108.23 | 105.12 | 449.05 |
| MASSACHUSETTS | 105.21 | 105.29 | 94.42 | 90.55 | 395.47 |
| MICHIGAN | 77.45 | 75.93 | 65.47 | 63.50 | 282.34 |
| MINNESOTA | 100.85 | 94.96 | 93.98 | 91.72 | 381.51 |
| MISSISSIPPI | 151.57 | 210.66 | 181.62 | 149.93 | 693.78 |
| MISSOURI | 96.95 | 84.08 | 78.28 | 78.50 | 337.81 |
| MONTANA | 163.89 | 135.81 | 137.30 | 130.32 | 567.32 |
| NEBRASKA | 59.35 | 59.30 | 59.52 | 57.98 | 236.15 |
| NEVADA | 92.63 | 81.45 | 85.60 | 83.02 | 342.71 |
| NEW HAMPSHIRE | 132.72 | 132.05 | 114.99 | 121.18 | 500.94 |
| NEW JERSEY | 88.98 | 81.42 | 82.95 | 81.19 | 334.54 |
| NEW MEXICO | 55.74 | 54.58 | 56.52 | 51.75 | 218.59 |
| NEW YORK | 89.49 | 86.82 | 82.64 | 78.69 | 337.64 |
| NORTH CAROLINA | 101.79 | 91.90 | 97.90 | 90.07 | 381.65 |
| NORTH DAKOTA | 82.79 | 81.16 | 92.04 | 96.78 | 352.78 |
| OHIO | 76.17 | 64.94 | 59.40 | 60.28 | 260.79 |
| OKLAHOMA | 89.74 | 85.96 | 84.55 | 72.98 | 333.24 |
| OREGON | 116.99 | 106.93 | 98.24 | 98.12 | 420.29 |

| | | | | | |
|---|---|---|---|---|---|
| PENNSYLVANIA | 115.62 | 100.41 | 87.33 | 86.88 | 390.24 |
| PUERTO RICO | 1.38 | 1.79 | 1.49 | 1.51 | 6.16 |
| RHODE ISLAND | 77.79 | 70.41 | 58.89 | 54.06 | 261.15 |
| SOUTH CAROLINA | 109.89 | 107.67 | 106.87 | 95.47 | 419.89 |
| SOUTH DAKOTA | 64.58 | 58.18 | 59.82 | 60.40 | 242.98 |
| TENNESSEE | 120.79 | 71.00 | 78.00 | 76.75 | 346.54 |
| TEXAS | 86.98 | 77.30 | 75.34 | 74.80 | 314.43 |
| UTAH | 101.74 | 77.63 | 88.17 | 82.30 | 349.84 |
| VERMONT | 171.83 | 173.78 | 146.36 | 147.02 | 639.00 |
| VIRGIN ISLANDS | 9.89 | 12.11 | 14.85 | 12.59 | 49.45 |
| VIRGINIA | 122.42 | 113.78 | 107.55 | 103.22 | 446.98 |
| WASHINGTON | 144.48 | 132.99 | 123.34 | 123.93 | 524.74 |
| WEST VIRGINIA | 63.75 | 60.33 | 56.17 | 58.49 | 238.74 |
| WISCONSIN | 62.48 | 57.42 | 49.25 | 46.27 | 215.43 |
| WYOMING | 99.72 | 100.10 | 85.96 | 87.65 | 373.43 |
| **9150 - HYDROMORPHONE - To** | **102.14** | **93.86** | **88.59** | **85.06** | **369.65** |

**DRUG: 9193 - HYDROCODONE**

| STATE NAME | Q1 CALC GRAMS | Q2 CALC GRAMS | Q3 CALC GRAMS | Q4 CALC GRAMS | TOTAL GRAMS |
|---|---|---|---|---|---|
| ALABAMA | 4,672.85 | 4,493.46 | 4,364.39 | 4,336.18 | 17,866.88 |
| ALASKA | 1,382.87 | 1,327.54 | 1,270.72 | 1,217.29 | 5,198.42 |
| AMERICAN SAMOA | 5.53 | 0.00 | 0.00 | 0.00 | 5.53 |
| ARIZONA | 1,463.08 | 1,364.07 | 1,325.72 | 1,302.52 | 5,455.39 |
| ARKANSAS | 4,144.96 | 3,952.30 | 3,793.85 | 3,717.22 | 15,608.33 |
| CALIFORNIA | 2,225.27 | 2,132.80 | 2,055.69 | 1,998.42 | 8,412.18 |
| COLORADO | 1,152.95 | 1,090.45 | 1,047.00 | 1,037.33 | 4,327.73 |
| CONNECTICUT | 507.03 | 468.96 | 453.80 | 457.23 | 1,887.02 |
| DELAWARE | 576.21 | 571.79 | 546.52 | 539.75 | 2,234.28 |
| DISTRICT OF COLUMBIA | 225.63 | 184.69 | 179.16 | 175.33 | 764.80 |
| FLORIDA | 1,809.31 | 1,677.50 | 1,446.77 | 1,462.02 | 6,395.61 |
| GEORGIA | 2,297.52 | 2,198.21 | 2,100.21 | 2,104.17 | 8,700.11 |
| GUAM | 79.92 | 56.74 | 76.19 | 65.39 | 278.24 |
| HAWAII | 1,198.58 | 1,144.64 | 1,059.03 | 1,038.76 | 4,441.02 |
| IDAHO | 3,329.88 | 3,251.16 | 3,149.94 | 3,095.47 | 12,826.45 |
| ILLINOIS | 2,110.58 | 2,029.28 | 1,941.34 | 1,946.99 | 8,028.19 |
| INDIANA | 3,179.64 | 3,035.18 | 2,930.93 | 2,938.53 | 12,084.28 |
| IOWA | 1,653.71 | 1,584.12 | 1,521.75 | 1,518.15 | 6,277.74 |
| KANSAS | 2,561.62 | 2,467.66 | 2,367.57 | 2,360.84 | 9,757.69 |
| KENTUCKY | 4,334.53 | 4,172.58 | 4,050.86 | 4,069.83 | 16,627.79 |
| LOUISIANA | 3,247.09 | 3,083.10 | 3,004.58 | 2,992.71 | 12,327.48 |
| MAINE | 1,188.23 | 1,141.43 | 1,124.34 | 1,091.84 | 4,545.84 |

| | | | | | |
|---|---|---|---|---|---|
| MARYLAND | 524.03 | 504.43 | 470.21 | 472.94 | 1,971.62 |
| MASSACHUSETTS | 416.23 | 394.41 | 371.94 | 372.71 | 1,555.29 |
| MICHIGAN | 3,411.95 | 3,144.87 | 2,935.48 | 2,900.45 | 12,392.74 |
| MINNESOTA | 742.52 | 710.89 | 688.11 | 671.86 | 2,813.38 |
| MISSISSIPPI | 3,337.51 | 3,108.80 | 2,966.50 | 2,856.31 | 12,269.11 |
| MISSOURI | 2,592.91 | 2,489.46 | 2,389.17 | 2,337.10 | 9,808.65 |
| MONTANA | 2,399.93 | 2,284.51 | 2,259.62 | 2,192.31 | 9,136.37 |
| NEBRASKA | 1,553.41 | 1,518.65 | 1,444.79 | 1,439.13 | 5,955.98 |
| NEVADA | 2,931.81 | 2,808.34 | 2,732.67 | 2,675.39 | 11,148.21 |
| NEW HAMPSHIRE | 626.09 | 584.57 | 577.04 | 577.44 | 2,365.13 |
| NEW JERSEY | 347.01 | 324.56 | 308.35 | 314.29 | 1,294.22 |
| NEW MEXICO | 1,569.84 | 1,477.56 | 1,430.92 | 1,436.64 | 5,914.95 |
| NEW YORK | 786.03 | 755.92 | 722.94 | 722.19 | 2,987.09 |
| NORTH CAROLINA | 1,764.50 | 1,633.90 | 1,570.08 | 1,650.79 | 6,619.27 |
| NORTH DAKOTA | 1,388.62 | 1,309.39 | 1,315.12 | 1,266.63 | 5,279.76 |
| OHIO | 1,364.46 | 1,293.53 | 1,233.14 | 1,232.58 | 5,123.72 |
| OKLAHOMA | 4,337.81 | 4,266.95 | 4,058.39 | 3,790.35 | 16,453.50 |
| OREGON | 2,208.31 | 2,149.05 | 2,077.53 | 2,027.24 | 8,462.12 |
| PENNSYLVANIA | 1,128.60 | 1,073.99 | 1,038.67 | 990.78 | 4,232.05 |
| PUERTO RICO | 10.02 | 9.25 | 16.29 | 13.83 | 49.40 |
| RHODE ISLAND | 868.65 | 824.34 | 794.03 | 776.37 | 3,263.39 |
| SOUTH CAROLINA | 2,656.83 | 2,485.80 | 2,363.70 | 2,360.29 | 9,866.62 |
| SOUTH DAKOTA | 1,539.66 | 1,479.61 | 1,433.38 | 1,422.22 | 5,874.86 |
| TENNESSEE | 3,489.23 | 3,336.09 | 3,149.42 | 3,046.50 | 13,021.24 |
| TEXAS | 2,408.78 | 2,340.61 | 2,268.67 | 2,253.35 | 9,271.40 |
| UTAH | 1,922.14 | 1,835.58 | 1,776.85 | 1,801.07 | 7,335.65 |
| VERMONT | 681.45 | 661.05 | 643.95 | 636.61 | 2,623.06 |
| VIRGIN ISLANDS | 259.97 | 201.43 | 204.68 | 216.07 | 882.14 |
| VIRGINIA | 1,137.91 | 1,065.79 | 1,020.44 | 1,032.03 | 4,256.16 |
| WASHINGTON | 1,741.19 | 1,696.12 | 1,629.01 | 1,610.23 | 6,676.56 |
| WEST VIRGINIA | 2,743.06 | 2,543.98 | 2,402.45 | 2,301.89 | 9,991.38 |
| WISCONSIN | 1,210.30 | 1,196.36 | 1,170.96 | 1,156.14 | 4,733.76 |
| WYOMING | 1,781.35 | 1,748.91 | 1,689.21 | 1,676.30 | 6,895.77 |
| **9193 - HYDROCODONE - Total** | **1,943.10** | **1,853.56** | **1,770.69** | **1,751.24** | **7,318.59** |

**DRUG: 9230 - MEPERIDINE (PETHIDINE)**

| STATE NAME | Q1 CALC GRAMS | Q2 CALC GRAMS | Q3 CALC GRAMS | Q4 CALC GRAMS | TOTAL GRAMS |
|---|---|---|---|---|---|
| ALABAMA | 165.33 | 145.29 | 122.61 | 94.52 | 527.76 |
| ALASKA | 93.93 | 62.93 | 54.60 | 41.41 | 252.88 |
| ARIZONA | 84.30 | 77.61 | 69.45 | 55.92 | 287.28 |
| ARKANSAS | 231.57 | 206.76 | 190.26 | 148.00 | 776.59 |

| | | | | | |
|---|---|---|---|---|---|
| CALIFORNIA | 53.60 | 47.45 | 35.56 | 29.93 | 166.53 |
| COLORADO | 31.89 | 31.25 | 27.31 | 22.69 | 113.14 |
| CONNECTICUT | 30.82 | 24.76 | 24.40 | 23.39 | 103.38 |
| DELAWARE | 24.92 | 11.89 | 10.86 | 14.74 | 62.41 |
| DISTRICT OF COLUMBIA | 23.99 | 27.34 | 21.63 | 32.68 | 105.64 |
| FLORIDA | 51.25 | 44.30 | 34.54 | 29.12 | 159.21 |
| GEORGIA | 106.02 | 97.72 | 78.76 | 62.89 | 345.39 |
| GUAM | 7.94 | 20.15 | 22.90 | 16.79 | 67.77 |
| HAWAII | 18.17 | 29.57 | 25.91 | 17.10 | 90.75 |
| IDAHO | 47.33 | 49.62 | 36.59 | 44.30 | 177.84 |
| ILLINOIS | 21.96 | 21.97 | 17.26 | 13.73 | 74.92 |
| INDIANA | 48.54 | 47.20 | 41.60 | 28.38 | 165.71 |
| IOWA | 61.10 | 62.05 | 42.52 | 50.58 | 216.25 |
| KANSAS | 54.13 | 52.99 | 47.92 | 35.03 | 190.07 |
| KENTUCKY | 65.40 | 55.60 | 41.72 | 34.28 | 197.00 |
| LOUISIANA | 153.67 | 143.14 | 113.57 | 87.55 | 497.93 |
| MAINE | 26.59 | 25.21 | 23.86 | 23.21 | 98.87 |
| MARYLAND | 38.40 | 37.55 | 29.91 | 27.48 | 133.34 |
| MASSACHUSETTS | 32.03 | 27.39 | 24.03 | 22.96 | 106.42 |
| MICHIGAN | 29.24 | 25.75 | 18.60 | 21.12 | 94.71 |
| MINNESOTA | 11.05 | 12.85 | 7.99 | 8.67 | 40.56 |
| MISSISSIPPI | 132.36 | 125.21 | 108.05 | 88.09 | 453.71 |
| MISSOURI | 54.92 | 51.67 | 41.44 | 40.90 | 188.93 |
| MONTANA | 65.11 | 58.30 | 49.33 | 32.87 | 205.61 |
| NEBRASKA | 32.67 | 26.01 | 24.04 | 23.32 | 106.03 |
| NEVADA | 63.13 | 51.55 | 39.09 | 35.56 | 189.34 |
| NEW HAMPSHIRE | 48.96 | 43.58 | 36.00 | 37.73 | 166.28 |
| NEW JERSEY | 20.42 | 18.82 | 17.01 | 15.07 | 71.32 |
| NEW MEXICO | 78.07 | 72.11 | 58.17 | 39.10 | 247.44 |
| NEW YORK | 24.91 | 22.21 | 18.23 | 15.79 | 81.14 |
| NORTH CAROLINA | 52.92 | 50.44 | 43.49 | 39.36 | 186.21 |
| NORTH DAKOTA | 26.76 | 23.89 | 32.72 | 28.59 | 111.95 |
| OHIO | 47.83 | 37.32 | 28.89 | 29.80 | 143.85 |
| OKLAHOMA | 180.00 | 147.90 | 138.03 | 101.17 | 567.10 |
| OREGON | 31.32 | 33.91 | 30.78 | 23.29 | 119.30 |
| PENNSYLVANIA | 27.81 | 25.19 | 20.67 | 18.64 | 92.31 |
| PUERTO RICO | 65.66 | 59.69 | 113.69 | 95.99 | 335.03 |
| RHODE ISLAND | 16.82 | 21.48 | 16.20 | 8.02 | 62.51 |
| SOUTH CAROLINA | 111.00 | 98.20 | 90.19 | 73.90 | 373.29 |
| SOUTH DAKOTA | 65.32 | 73.05 | 52.38 | 40.54 | 231.30 |

| | | | | | |
|---|---|---|---|---|---|
| TENNESSEE | 115.40 | 96.90 | 77.06 | 58.30 | 347.67 |
| TEXAS | 53.83 | 46.19 | 33.93 | 32.53 | 166.49 |
| UTAH | 61.88 | 67.69 | 49.92 | 49.64 | 229.14 |
| VERMONT | 56.62 | 48.22 | 28.46 | 35.52 | 168.82 |
| VIRGIN ISLANDS | 110.10 | 141.84 | 139.36 | 27.77 | 419.09 |
| VIRGINIA | 71.45 | 61.35 | 53.74 | 48.42 | 234.95 |
| WASHINGTON | 48.46 | 48.92 | 36.86 | 35.38 | 169.62 |
| WEST VIRGINIA | 38.59 | 30.60 | 20.75 | 17.05 | 106.99 |
| WISCONSIN | 21.89 | 21.62 | 14.26 | 13.16 | 70.93 |
| WYOMING | 59.18 | 54.41 | 55.18 | 53.64 | 222.41 |
| **9230 - MEPERIDINE (PETHIDIN** | **56.21** | **50.53** | **41.86** | **35.69** | **184.30** |

**DRUG: 9250B - METHADONE**

| STATE NAME | Q1 CALC GRAMS | Q2 CALC GRAMS | Q3 CALC GRAMS | Q4 CALC GRAMS | TOTAL GRAMS |
|---|---|---|---|---|---|
| ALABAMA | 1,420.88 | 1,449.27 | 1,548.38 | 1,573.96 | 5,992.49 |
| ALASKA | 1,160.00 | 1,119.95 | 1,262.28 | 1,101.83 | 4,644.07 |
| ARIZONA | 1,379.83 | 1,395.54 | 1,441.26 | 1,305.74 | 5,522.38 |
| ARKANSAS | 509.14 | 465.65 | 466.97 | 399.43 | 1,841.19 |
| CALIFORNIA | 890.44 | 813.28 | 861.80 | 682.07 | 3,247.59 |
| COLORADO | 922.84 | 890.29 | 937.46 | 829.19 | 3,579.78 |
| CONNECTICUT | 3,009.20 | 3,070.44 | 3,248.71 | 2,337.37 | 11,665.72 |
| DELAWARE | 3,495.77 | 3,879.57 | 3,274.91 | 3,261.08 | 13,911.33 |
| DISTRICT OF COLUMBIA | 1,550.98 | 1,442.05 | 1,726.26 | 1,165.78 | 5,885.07 |
| FLORIDA | 1,063.86 | 886.53 | 1,014.10 | 1,051.35 | 4,015.84 |
| GEORGIA | 1,259.85 | 1,075.60 | 1,185.06 | 1,116.68 | 4,637.19 |
| GUAM | 23.81 | 24.56 | 18.80 | 24.31 | 91.48 |
| HAWAII | 611.15 | 365.69 | 604.75 | 469.06 | 2,050.65 |
| IDAHO | 752.71 | 628.84 | 631.19 | 632.37 | 2,645.11 |
| ILLINOIS | 975.48 | 880.54 | 991.80 | 859.78 | 3,707.61 |
| INDIANA | 1,250.28 | 1,171.72 | 1,205.10 | 1,004.80 | 4,631.91 |
| IOWA | 409.12 | 430.53 | 410.34 | 363.84 | 1,613.83 |
| KANSAS | 721.19 | 792.89 | 681.09 | 731.50 | 2,926.66 |
| KENTUCKY | 1,180.71 | 1,151.07 | 1,179.92 | 1,076.92 | 4,588.61 |
| LOUISIANA | 796.64 | 768.00 | 766.47 | 719.48 | 3,050.60 |
| MAINE | 2,319.44 | 1,757.36 | 2,245.67 | 1,681.24 | 8,003.71 |
| MARYLAND | 3,250.39 | 2,943.61 | 3,184.52 | 3,025.23 | 12,403.75 |
| MASSACHUSETTS | 2,193.94 | 1,875.54 | 2,173.08 | 1,680.85 | 7,923.41 |
| MICHIGAN | 1,280.77 | 1,292.06 | 1,257.15 | 1,125.43 | 4,955.40 |
| MINNESOTA | 1,034.11 | 1,101.46 | 1,062.21 | 932.99 | 4,130.76 |
| MISSISSIPPI | 235.89 | 277.91 | 221.11 | 253.98 | 988.90 |
| MISSOURI | 619.21 | 676.13 | 598.41 | 561.91 | 2,455.67 |

| | | | | | |
|---|---|---|---|---|---|
| MONTANA | 764.70 | 806.23 | 857.27 | 808.02 | 3,236.23 |
| NEBRASKA | 391.38 | 355.04 | 434.37 | 371.71 | 1,552.50 |
| NEVADA | 1,329.64 | 1,238.19 | 1,252.01 | 1,237.17 | 5,057.01 |
| NEW HAMPSHIRE | 2,437.91 | 2,369.65 | 2,238.91 | 2,058.93 | 9,105.40 |
| NEW JERSEY | 1,311.84 | 1,315.51 | 1,380.68 | 1,320.92 | 5,328.95 |
| NEW MEXICO | 2,028.31 | 2,037.17 | 2,225.25 | 2,037.17 | 8,327.90 |
| NEW YORK | 1,519.22 | 1,452.35 | 1,621.25 | 1,221.06 | 5,813.88 |
| NORTH CAROLINA | 1,424.20 | 1,263.30 | 1,391.24 | 1,247.17 | 5,325.91 |
| NORTH DAKOTA | 472.21 | 548.88 | 703.92 | 635.12 | 2,360.13 |
| OHIO | 619.84 | 631.27 | 635.14 | 661.64 | 2,547.88 |
| OKLAHOMA | 988.63 | 796.33 | 975.47 | 904.31 | 3,664.73 |
| OREGON | 1,662.30 | 1,394.60 | 1,562.02 | 1,319.73 | 5,938.65 |
| PENNSYLVANIA | 1,491.80 | 1,395.99 | 1,472.48 | 1,261.41 | 5,621.69 |
| PUERTO RICO | 547.81 | 578.70 | 583.71 | 620.44 | 2,330.65 |
| RHODE ISLAND | 4,206.08 | 4,526.33 | 3,527.08 | 3,810.84 | 16,070.33 |
| SOUTH CAROLINA | 1,167.45 | 1,058.13 | 1,164.92 | 913.20 | 4,303.71 |
| SOUTH DAKOTA | 247.93 | 244.94 | 252.62 | 186.85 | 932.33 |
| TENNESSEE | 787.95 | 727.60 | 806.55 | 718.51 | 3,040.60 |
| TEXAS | 545.81 | 486.14 | 544.48 | 486.13 | 2,062.57 |
| UTAH | 1,018.08 | 844.75 | 952.97 | 712.69 | 3,528.49 |
| VERMONT | 3,209.72 | 2,492.36 | 3,137.04 | 2,651.58 | 11,490.71 |
| VIRGIN ISLANDS | 1.63 | 3.26 | 8.96 | 500.03 | 513.88 |
| VIRGINIA | 1,037.21 | 990.00 | 1,032.10 | 919.23 | 3,978.54 |
| WASHINGTON | 1,462.97 | 1,245.89 | 1,500.04 | 1,315.47 | 5,524.38 |
| WEST VIRGINIA | 1,225.64 | 1,088.06 | 1,259.40 | 915.25 | 4,488.35 |
| WISCONSIN | 1,081.69 | 1,027.37 | 1,110.27 | 771.63 | 3,990.97 |
| WYOMING | 186.03 | 165.32 | 155.75 | 159.56 | 666.66 |
| **9250B - METHADONE - Total** | **1,157.16** | **1,078.22** | **1,155.15** | **1,010.59** | **4,401.11** |

**DRUG: 9300 - MORPHINE**

| STATE NAME | Q1 CALC GRAMS | Q2 CALC GRAMS | Q3 CALC GRAMS | Q4 CALC GRAMS | TOTAL GRAMS |
|---|---|---|---|---|---|
| ALABAMA | 1,333.75 | 1,266.03 | 1,217.41 | 1,210.64 | 5,027.82 |
| ALASKA | 1,520.56 | 1,476.19 | 1,455.75 | 1,283.63 | 5,736.13 |
| AMERICAN SAMOA | 10.31 | 38.27 | 18.56 | 6.88 | 74.02 |
| ARIZONA | 2,139.26 | 1,950.04 | 1,829.00 | 1,705.30 | 7,623.61 |
| ARKANSAS | 1,334.14 | 1,237.75 | 1,136.55 | 1,086.49 | 4,794.93 |
| CALIFORNIA | 1,073.98 | 1,007.37 | 961.27 | 930.17 | 3,972.79 |
| COLORADO | 1,381.26 | 1,277.62 | 1,243.38 | 1,172.17 | 5,074.42 |
| CONNECTICUT | 919.08 | 877.40 | 831.87 | 816.74 | 3,445.09 |
| DELAWARE | 2,230.18 | 2,081.01 | 1,951.70 | 1,847.73 | 8,110.62 |
| DISTRICT OF COLUMBIA | 612.90 | 453.64 | 496.77 | 410.31 | 1,973.62 |

| | | | | | |
|---|---|---|---|---|---|
| FLORIDA | 1,973.42 | 1,887.38 | 1,725.74 | 1,680.65 | 7,267.18 |
| GEORGIA | 1,081.84 | 1,038.79 | 1,010.28 | 962.76 | 4,093.67 |
| GUAM | 563.40 | 463.46 | 431.15 | 428.84 | 1,886.84 |
| HAWAII | 1,360.77 | 1,380.74 | 1,249.68 | 1,165.22 | 5,156.41 |
| IDAHO | 1,636.25 | 1,590.45 | 1,529.48 | 1,431.98 | 6,188.16 |
| ILLINOIS | 741.76 | 711.96 | 686.04 | 685.09 | 2,824.85 |
| INDIANA | 1,192.79 | 1,129.64 | 1,054.75 | 1,028.92 | 4,406.10 |
| IOWA | 1,003.78 | 949.77 | 926.14 | 911.12 | 3,790.81 |
| KANSAS | 1,418.60 | 1,363.93 | 1,284.90 | 1,258.15 | 5,325.58 |
| KENTUCKY | 963.99 | 915.25 | 852.51 | 855.65 | 3,587.39 |
| LOUISIANA | 1,075.54 | 1,034.75 | 1,006.57 | 958.68 | 4,075.54 |
| MAINE | 1,109.11 | 1,081.67 | 1,036.95 | 1,022.04 | 4,249.77 |
| MARYLAND | 1,167.01 | 1,093.32 | 1,000.38 | 957.93 | 4,218.64 |
| MASSACHUSETTS | 929.47 | 902.30 | 846.98 | 849.04 | 3,527.81 |
| MICHIGAN | 1,640.73 | 1,555.57 | 1,454.48 | 1,394.85 | 6,045.62 |
| MINNESOTA | 778.78 | 738.30 | 716.45 | 688.06 | 2,921.59 |
| MISSISSIPPI | 1,177.82 | 1,150.01 | 1,096.64 | 1,015.57 | 4,440.05 |
| MISSOURI | 1,455.51 | 1,344.27 | 1,298.63 | 1,259.86 | 5,358.27 |
| MONTANA | 1,613.92 | 1,541.16 | 1,504.89 | 1,326.66 | 5,986.63 |
| NEBRASKA | 949.02 | 955.48 | 899.76 | 874.36 | 3,678.61 |
| NEVADA | 1,878.24 | 1,736.86 | 1,701.05 | 1,575.06 | 6,891.20 |
| NEW HAMPSHIRE | 1,291.74 | 1,298.41 | 1,187.59 | 1,188.52 | 4,966.27 |
| NEW JERSEY | 854.96 | 822.89 | 777.85 | 765.81 | 3,221.51 |
| NEW MEXICO | 1,118.28 | 1,042.38 | 1,001.49 | 1,012.76 | 4,174.91 |
| NEW YORK | 723.21 | 699.67 | 671.08 | 647.04 | 2,741.01 |
| NORTH CAROLINA | 1,153.28 | 1,102.45 | 1,076.35 | 1,106.25 | 4,438.33 |
| NORTH DAKOTA | 843.29 | 780.26 | 795.58 | 781.60 | 3,200.73 |
| OHIO | 922.52 | 870.12 | 815.43 | 820.22 | 3,428.29 |
| OKLAHOMA | 2,163.91 | 1,945.67 | 1,798.87 | 1,543.37 | 7,451.82 |
| OREGON | 1,628.86 | 1,571.08 | 1,524.76 | 1,457.70 | 6,182.40 |
| PENNSYLVANIA | 1,647.98 | 1,484.47 | 1,575.92 | 1,286.00 | 5,994.37 |
| PUERTO RICO | 38.42 | 36.11 | 53.81 | 55.11 | 183.45 |
| RHODE ISLAND | 984.45 | 901.79 | 836.85 | 881.41 | 3,604.50 |
| SOUTH CAROLINA | 1,313.45 | 1,261.82 | 1,222.47 | 1,217.91 | 5,015.65 |
| SOUTH DAKOTA | 943.84 | 888.76 | 878.90 | 811.18 | 3,522.67 |
| TENNESSEE | 2,272.34 | 2,083.09 | 2,133.86 | 1,655.35 | 8,144.64 |
| TEXAS | 754.89 | 718.29 | 682.54 | 658.78 | 2,814.50 |
| UTAH | 1,656.19 | 1,549.34 | 1,513.87 | 1,457.08 | 6,176.47 |
| VERMONT | 1,531.68 | 1,502.65 | 1,424.32 | 1,412.17 | 5,870.82 |
| VIRGIN ISLANDS | 88.26 | 121.74 | 122.36 | 97.19 | 429.56 |

| | | | | | |
|---|---|---|---|---|---|
| VIRGINIA | 883.12 | 860.25 | 818.05 | 818.57 | 3,379.99 |
| WASHINGTON | 1,433.87 | 1,399.38 | 1,330.12 | 1,273.12 | 5,436.48 |
| WEST VIRGINIA | 1,026.17 | 972.10 | 946.72 | 900.39 | 3,845.38 |
| WISCONSIN | 1,201.23 | 1,156.99 | 1,078.70 | 1,074.34 | 4,511.26 |
| WYOMING | 1,481.10 | 1,393.01 | 1,318.16 | 1,216.86 | 5,409.12 |
| **9300 - MORPHINE - Total** | **1,197.75** | **1,132.28** | **1,083.76** | **1,031.32** | **4,445.11** |

**DRUG: 9652 - OXYMORPHONE**

| STATE NAME | Q1 CALC GRAMS | Q2 CALC GRAMS | Q3 CALC GRAMS | Q4 CALC GRAMS | TOTAL GRAMS |
|---|---|---|---|---|---|
| ALABAMA | 49.88 | 45.31 | 40.65 | 40.97 | 176.81 |
| ALASKA | 23.65 | 22.30 | 17.28 | 13.21 | 76.44 |
| ARIZONA | 41.85 | 34.17 | 32.76 | 27.61 | 136.39 |
| ARKANSAS | 24.35 | 18.22 | 14.79 | 12.64 | 70.01 |
| CALIFORNIA | 18.22 | 15.52 | 14.52 | 13.63 | 61.89 |
| COLORADO | 31.89 | 28.40 | 26.10 | 24.92 | 111.31 |
| CONNECTICUT | 40.15 | 40.14 | 35.27 | 34.79 | 150.35 |
| DELAWARE | 163.82 | 165.58 | 163.07 | 161.22 | 653.70 |
| DISTRICT OF COLUMBIA | 16.32 | 10.14 | 8.41 | 8.65 | 43.52 |
| FLORIDA | 60.74 | 56.60 | 51.26 | 49.35 | 217.95 |
| GEORGIA | 33.03 | 29.59 | 28.54 | 26.61 | 117.78 |
| HAWAII | 18.79 | 13.33 | 15.13 | 16.03 | 63.27 |
| IDAHO | 50.88 | 47.89 | 42.68 | 42.69 | 184.14 |
| ILLINOIS | 16.28 | 14.28 | 13.61 | 14.14 | 58.30 |
| INDIANA | 38.61 | 34.22 | 32.49 | 29.96 | 135.28 |
| IOWA | 17.28 | 15.25 | 12.38 | 11.74 | 56.65 |
| KANSAS | 31.14 | 28.84 | 26.13 | 26.67 | 112.78 |
| KENTUCKY | 48.87 | 43.95 | 39.86 | 37.60 | 170.28 |
| LOUISIANA | 93.58 | 87.88 | 83.54 | 79.25 | 344.25 |
| MAINE | 44.47 | 32.80 | 30.05 | 27.29 | 134.61 |
| MARYLAND | 110.10 | 102.22 | 97.51 | 96.21 | 406.04 |
| MASSACHUSETTS | 17.22 | 15.81 | 15.14 | 14.79 | 62.97 |
| MICHIGAN | 145.13 | 124.20 | 108.93 | 93.80 | 472.06 |
| MINNESOTA | 4.30 | 4.04 | 3.91 | 4.06 | 16.32 |
| MISSISSIPPI | 59.46 | 48.92 | 44.14 | 44.16 | 196.67 |
| MISSOURI | 33.74 | 28.39 | 26.06 | 27.22 | 115.42 |
| MONTANA | 13.47 | 12.73 | 13.99 | 12.18 | 52.37 |
| NEBRASKA | 33.83 | 28.82 | 28.29 | 24.86 | 115.80 |
| NEVADA | 72.97 | 60.30 | 57.11 | 49.78 | 240.16 |
| NEW HAMPSHIRE | 35.72 | 36.01 | 31.14 | 24.53 | 127.41 |
| NEW JERSEY | 42.52 | 38.33 | 34.39 | 34.39 | 149.63 |
| NEW MEXICO | 27.01 | 23.13 | 18.89 | 16.97 | 86.01 |

| | | | | | |
|---|---|---|---|---|---|
| NEW YORK | 58.80 | 54.91 | 51.34 | 51.13 | 216.18 |
| NORTH CAROLINA | 115.82 | 104.42 | 98.48 | 99.67 | 418.39 |
| NORTH DAKOTA | 12.18 | 12.02 | 10.60 | 9.96 | 44.76 |
| OHIO | 45.11 | 41.37 | 36.35 | 33.07 | 155.91 |
| OKLAHOMA | 75.19 | 65.45 | 48.72 | 34.72 | 224.08 |
| OREGON | 27.19 | 24.77 | 22.72 | 21.83 | 96.51 |
| PENNSYLVANIA | 113.68 | 100.57 | 90.27 | 86.03 | 390.55 |
| RHODE ISLAND | 21.95 | 15.93 | 15.13 | 11.71 | 64.73 |
| SOUTH CAROLINA | 73.01 | 67.73 | 65.24 | 62.86 | 268.84 |
| SOUTH DAKOTA | 41.13 | 35.39 | 29.32 | 35.91 | 141.75 |
| TENNESSEE | 223.25 | 201.78 | 183.31 | 177.41 | 785.75 |
| TEXAS | 22.02 | 18.95 | 16.71 | 14.85 | 72.53 |
| UTAH | 107.95 | 96.86 | 90.54 | 91.07 | 386.42 |
| VERMONT | 27.07 | 20.40 | 19.22 | 22.35 | 89.05 |
| VIRGINIA | 76.85 | 71.07 | 65.57 | 62.84 | 276.33 |
| WASHINGTON | 24.35 | 20.64 | 20.01 | 20.13 | 85.13 |
| WEST VIRGINIA | 79.88 | 74.29 | 61.60 | 62.80 | 278.57 |
| WISCONSIN | 18.13 | 14.57 | 13.59 | 12.45 | 58.74 |
| WYOMING | 34.56 | 22.21 | 20.52 | 19.98 | 97.28 |
| **9652 - OXYMORPHONE - Total** | **51.98** | **46.52** | **42.52** | **40.51** | **181.53** |

**DRUG: 9780 - TAPENTADOL**

| STATE NAME | Q1 CALC GRAMS | Q2 CALC GRAMS | Q3 CALC GRAMS | Q4 CALC GRAMS | TOTAL GRAMS |
|---|---|---|---|---|---|
| ALABAMA | 906.86 | 907.41 | 846.02 | 777.22 | 3,437.51 |
| ALASKA | 296.24 | 302.37 | 297.51 | 231.76 | 1,127.87 |
| ARIZONA | 495.20 | 484.21 | 446.87 | 408.26 | 1,834.55 |
| ARKANSAS | 534.36 | 492.92 | 449.36 | 401.23 | 1,877.86 |
| CALIFORNIA | 230.01 | 221.49 | 211.48 | 205.98 | 868.95 |
| COLORADO | 531.83 | 499.97 | 488.76 | 492.37 | 2,012.94 |
| CONNECTICUT | 230.58 | 248.86 | 233.02 | 242.68 | 955.14 |
| DELAWARE | 616.03 | 627.55 | 625.10 | 570.75 | 2,439.43 |
| DISTRICT OF COLUMBIA | 146.91 | 146.25 | 137.44 | 128.80 | 559.39 |
| FLORIDA | 415.81 | 411.84 | 363.18 | 358.37 | 1,549.20 |
| GEORGIA | 537.87 | 541.70 | 520.05 | 516.88 | 2,116.50 |
| HAWAII | 183.86 | 149.31 | 132.07 | 128.91 | 594.13 |
| IDAHO | 349.58 | 372.52 | 357.08 | 313.19 | 1,392.37 |
| ILLINOIS | 229.77 | 227.37 | 214.58 | 216.60 | 888.32 |
| INDIANA | 483.63 | 478.52 | 446.83 | 446.68 | 1,855.66 |
| IOWA | 328.28 | 300.88 | 294.27 | 294.60 | 1,218.03 |
| KANSAS | 254.44 | 226.28 | 223.53 | 205.97 | 910.21 |
| KENTUCKY | 294.29 | 282.28 | 263.06 | 261.58 | 1,101.21 |

| | | | | | |
|---|---|---|---|---|---|
| LOUISIANA | 291.85 | 299.52 | 289.03 | 289.64 | 1,170.05 |
| MAINE | 77.95 | 93.91 | 97.04 | 84.62 | 353.52 |
| MARYLAND | 438.81 | 463.12 | 443.93 | 431.94 | 1,777.81 |
| MASSACHUSETTS | 242.27 | 260.09 | 252.99 | 253.52 | 1,008.88 |
| MICHIGAN | 183.88 | 172.97 | 165.21 | 161.19 | 683.26 |
| MINNESOTA | 74.71 | 68.71 | 69.55 | 68.90 | 281.88 |
| MISSISSIPPI | 332.90 | 301.62 | 301.12 | 277.12 | 1,212.75 |
| MISSOURI | 349.45 | 354.05 | 327.19 | 325.98 | 1,356.67 |
| MONTANA | 150.64 | 156.46 | 154.08 | 158.12 | 619.31 |
| NEBRASKA | 484.38 | 495.69 | 511.84 | 536.98 | 2,028.89 |
| NEVADA | 364.09 | 325.01 | 298.38 | 299.92 | 1,287.40 |
| NEW HAMPSHIRE | 438.83 | 457.13 | 485.28 | 463.97 | 1,845.20 |
| NEW JERSEY | 400.02 | 398.92 | 367.69 | 365.92 | 1,532.56 |
| NEW MEXICO | 135.88 | 122.16 | 121.29 | 116.24 | 495.56 |
| NEW YORK | 404.81 | 408.83 | 386.50 | 387.27 | 1,587.41 |
| NORTH CAROLINA | 850.16 | 878.47 | 858.47 | 892.90 | 3,479.99 |
| NORTH DAKOTA | 261.45 | 279.96 | 270.97 | 261.97 | 1,074.35 |
| OHIO | 421.77 | 412.66 | 385.21 | 372.92 | 1,592.57 |
| OKLAHOMA | 524.38 | 469.64 | 396.96 | 286.96 | 1,677.96 |
| OREGON | 175.59 | 169.89 | 160.66 | 141.06 | 647.19 |
| PENNSYLVANIA | 416.24 | 415.61 | 390.01 | 388.86 | 1,610.73 |
| PUERTO RICO | 13.07 | 15.42 | 11.86 | 12.15 | 52.50 |
| RHODE ISLAND | 164.17 | 205.55 | 172.48 | 176.85 | 719.05 |
| SOUTH CAROLINA | 771.49 | 798.34 | 756.71 | 726.87 | 3,053.40 |
| SOUTH DAKOTA | 506.95 | 465.56 | 426.44 | 427.18 | 1,826.13 |
| TENNESSEE | 416.93 | 434.07 | 395.89 | 383.57 | 1,630.46 |
| TEXAS | 322.55 | 304.91 | 292.34 | 280.32 | 1,200.13 |
| UTAH | 400.51 | 379.76 | 368.74 | 378.85 | 1,527.85 |
| VERMONT | 146.39 | 154.54 | 175.79 | 144.71 | 621.42 |
| VIRGIN ISLANDS | 0.00 | 0.00 | 4.55 | 0.00 | 4.55 |
| VIRGINIA | 421.02 | 409.56 | 375.50 | 370.18 | 1,576.26 |
| WASHINGTON | 214.54 | 215.37 | 206.29 | 210.08 | 846.28 |
| WEST VIRGINIA | 253.54 | 238.51 | 204.18 | 187.40 | 883.62 |
| WISCONSIN | 209.36 | 197.24 | 189.47 | 184.05 | 780.13 |
| WYOMING | 315.90 | 267.91 | 288.13 | 262.94 | 1,134.88 |
| **9780 - TAPENTADOL - Total** | **363.94** | **359.68** | **339.27** | **331.92** | **1,394.81** |

**DRUG: 9801 - FENTANYL BASE**

| STATE NAME | Q1 CALC GRAMS | Q2 CALC GRAMS | Q3 CALC GRAMS | Q4 CALC GRAMS | TOTAL GRAMS |
|---|---|---|---|---|---|
| ALABAMA | 27.17 | 24.64 | 22.85 | 21.01 | 95.68 |
| ALASKA | 31.62 | 26.68 | 24.88 | 20.61 | 103.79 |

| | | | | | |
|---|---|---|---|---|---|
| AMERICAN SAMOA | 0.01 | 0.08 | 0.40 | 0.08 | 0.56 |
| ARIZONA | 23.37 | 20.10 | 18.55 | 16.61 | 78.62 |
| ARKANSAS | 26.86 | 23.00 | 21.04 | 19.68 | 90.58 |
| CALIFORNIA | 16.36 | 15.06 | 13.53 | 12.94 | 57.89 |
| COLORADO | 22.17 | 19.58 | 18.62 | 16.90 | 77.27 |
| CONNECTICUT | 24.29 | 19.85 | 19.36 | 18.49 | 81.98 |
| DELAWARE | 35.01 | 29.00 | 25.69 | 22.56 | 112.26 |
| DISTRICT OF COLUMBIA | 13.07 | 12.26 | 11.36 | 11.06 | 47.75 |
| FLORIDA | 29.34 | 26.33 | 22.05 | 21.26 | 98.98 |
| GEORGIA | 25.47 | 23.12 | 21.60 | 20.55 | 90.73 |
| GUAM | 5.02 | 4.04 | 5.16 | 4.14 | 18.37 |
| HAWAII | 12.50 | 11.71 | 10.86 | 10.54 | 45.60 |
| IDAHO | 35.77 | 31.70 | 30.35 | 28.70 | 126.52 |
| ILLINOIS | 21.45 | 18.60 | 17.43 | 16.68 | 74.16 |
| INDIANA | 25.65 | 22.80 | 21.43 | 20.26 | 90.13 |
| IOWA | 26.12 | 24.18 | 23.34 | 22.35 | 95.98 |
| KANSAS | 33.65 | 30.62 | 28.67 | 28.23 | 121.16 |
| KENTUCKY | 23.15 | 21.22 | 19.77 | 17.85 | 81.99 |
| LOUISIANA | 22.38 | 20.20 | 18.87 | 18.04 | 79.49 |
| MAINE | 22.86 | 20.60 | 20.06 | 19.43 | 82.94 |
| MARYLAND | 20.39 | 18.15 | 16.54 | 15.43 | 70.51 |
| MASSACHUSETTS | 17.94 | 16.86 | 15.33 | 15.17 | 65.29 |
| MICHIGAN | 27.32 | 24.43 | 22.50 | 20.92 | 95.17 |
| MINNESOTA | 17.40 | 14.86 | 14.78 | 13.73 | 60.77 |
| MISSISSIPPI | 35.74 | 31.71 | 34.70 | 28.60 | 130.75 |
| MISSOURI | 31.89 | 28.70 | 26.92 | 25.62 | 113.14 |
| MONTANA | 29.50 | 27.68 | 25.60 | 24.41 | 107.20 |
| NEBRASKA | 26.30 | 24.76 | 23.06 | 22.68 | 96.80 |
| NEVADA | 17.84 | 15.79 | 14.50 | 13.35 | 61.49 |
| NEW HAMPSHIRE | 23.31 | 22.19 | 19.90 | 18.43 | 83.83 |
| NEW JERSEY | 26.54 | 20.27 | 18.85 | 18.51 | 84.18 |
| NEW MEXICO | 19.23 | 16.18 | 15.35 | 14.81 | 65.58 |
| NEW YORK | 19.44 | 17.82 | 16.16 | 15.74 | 69.16 |
| NORTH CAROLINA | 25.60 | 22.94 | 21.75 | 20.97 | 91.26 |
| NORTH DAKOTA | 29.87 | 28.73 | 26.78 | 24.60 | 109.98 |
| OHIO | 17.95 | 16.41 | 14.34 | 13.56 | 62.26 |
| OKLAHOMA | 37.93 | 32.65 | 29.14 | 24.22 | 123.93 |
| OREGON | 21.11 | 19.74 | 17.99 | 16.91 | 75.75 |
| PENNSYLVANIA | 33.97 | 30.56 | 29.10 | 26.48 | 120.10 |
| PUERTO RICO | 2.09 | 2.18 | 4.34 | 4.51 | 13.12 |

| | | | | | |
|---|---|---|---|---|---|
| RHODE ISLAND | 15.46 | 13.24 | 13.28 | 12.64 | 54.62 |
| SOUTH CAROLINA | 25.33 | 21.74 | 21.15 | 19.91 | 88.13 |
| SOUTH DAKOTA | 31.73 | 28.66 | 26.55 | 24.58 | 111.52 |
| TENNESSEE | 27.86 | 23.78 | 23.18 | 19.35 | 94.17 |
| TEXAS | 20.43 | 18.54 | 16.83 | 15.74 | 71.54 |
| UTAH | 27.31 | 24.44 | 23.10 | 22.43 | 97.29 |
| VERMONT | 26.75 | 24.78 | 22.32 | 21.64 | 95.50 |
| VIRGIN ISLANDS | 3.58 | 4.17 | 4.92 | 3.66 | 16.33 |
| VIRGINIA | 18.65 | 16.99 | 15.70 | 15.00 | 66.35 |
| WASHINGTON | 19.73 | 18.77 | 17.38 | 16.38 | 72.25 |
| WEST VIRGINIA | 25.93 | 23.93 | 21.32 | 21.14 | 92.32 |
| WISCONSIN | 23.14 | 20.54 | 18.93 | 17.53 | 80.14 |
| WYOMING | 25.03 | 21.45 | 20.89 | 19.69 | 87.06 |
| **9801 - FENTANYL BASE - Total** | **23.02** | **20.58** | **19.01** | **17.86** | **80.48** |

**DRUG: 1100 - AMPHETAMINE**

| RANK | STATE NAME | STATE POPULATION | TOTAL GRAMS | TOTAL GRAMS/100K POPULATION |
|---|---|---|---|---|
| 1 | RHODE ISLAND | 1,052,567 | 154,515.93 | 14,679.91 |
| 2 | LOUISIANA | 4,533,372 | 661,966.01 | 14,602.07 |
| 3 | UTAH | 2,763,885 | 358,689.03 | 12,977.71 |
| 4 | NEW HAMPSHIRE | 1,316,470 | 161,453.90 | 12,264.15 |
| 5 | SOUTH CAROLINA | 4,625,364 | 557,757.34 | 12,058.67 |
| 6 | ALABAMA | 4,779,736 | 544,603.01 | 11,394.00 |
| 7 | MASSACHUSETTS | 6,547,629 | 732,127.12 | 11,181.56 |
| 8 | MICHIGAN | 9,883,640 | 978,827.49 | 9,903.51 |
| 9 | MISSOURI | 5,988,927 | 570,252.80 | 9,521.79 |
| 10 | DELAWARE | 897,934 | 85,067.25 | 9,473.66 |
| 11 | VIRGINIA | 8,001,024 | 733,900.96 | 9,172.59 |
| 12 | NORTH CAROLINA | 9,535,483 | 854,311.09 | 8,959.28 |
| 13 | KANSAS | 2,853,118 | 254,551.10 | 8,921.86 |
| 14 | WISCONSIN | 5,686,986 | 504,646.38 | 8,873.71 |
| 15 | TENNESSEE | 6,346,105 | 539,908.42 | 8,507.71 |
| 16 | MINNESOTA | 5,303,925 | 446,726.01 | 8,422.56 |
| 17 | INDIANA | 6,483,802 | 544,776.67 | 8,402.12 |
| 18 | MISSISSIPPI | 2,967,297 | 247,347.27 | 8,335.78 |
| 19 | DISTRICT OF COLUMBIA | 601,723 | 49,528.65 | 8,231.14 |
| 20 | GEORGIA | 9,687,653 | 786,080.52 | 8,114.25 |
| 21 | SOUTH DAKOTA | 814,180 | 61,171.55 | 7,513.27 |
| 22 | MAINE | 1,328,361 | 96,407.31 | 7,257.61 |
| 23 | OHIO | 11,536,504 | 831,699.97 | 7,209.29 |
| 24 | IDAHO | 1,567,582 | 112,820.52 | 7,197.10 |
| 25 | ARKANSAS | 2,915,918 | 209,272.63 | 7,176.90 |
| 26 | TEXAS | 25,145,561 | 1,781,188.02 | 7,083.51 |
| 27 | IOWA | 3,046,355 | 210,803.33 | 6,919.85 |
| 28 | WASHINGTON | 6,724,540 | 462,544.20 | 6,878.45 |
| 29 | COLORADO | 5,029,196 | 344,951.40 | 6,858.98 |
| 30 | FLORIDA | 18,801,310 | 1,286,902.21 | 6,844.75 |
| 31 | PENNSYLVANIA | 12,702,379 | 858,765.46 | 6,760.67 |
| 32 | MARYLAND | 5,773,552 | 385,491.88 | 6,676.86 |
| 33 | KENTUCKY | 4,339,367 | 286,507.85 | 6,602.53 |
| 34 | OREGON | 3,831,074 | 249,921.37 | 6,523.53 |
| 35 | VERMONT | 625,741 | 40,537.69 | 6,478.35 |
| 36 | CONNECTICUT | 3,574,097 | 227,217.37 | 6,357.34 |
| 37 | ILLINOIS | 12,830,632 | 800,126.39 | 6,236.06 |
| 38 | NEBRASKA | 1,826,341 | 110,018.17 | 6,023.97 |
| 39 | MONTANA | 989,415 | 59,087.84 | 5,972.00 |
| 40 | ARIZONA | 6,392,017 | 376,375.70 | 5,888.21 |
| 41 | OKLAHOMA | 3,751,351 | 219,080.50 | 5,840.04 |
| 42 | NORTH DAKOTA | 672,591 | 37,507.06 | 5,576.50 |

| | | | | |
|---|---|---|---|---|
| 43 | NEW YORK | 19,378,102 | 999,146.60 | 5,156.06 |
| 44 | WEST VIRGINIA | 1,852,994 | 94,428.63 | 5,096.00 |
| 45 | NEW JERSEY | 8,791,894 | 438,239.25 | 4,984.58 |
| 46 | WYOMING | 563,626 | 27,622.71 | 4,900.89 |
| 47 | ALASKA | 710,231 | 33,900.69 | 4,773.19 |
| 48 | NEVADA | 2,700,551 | 108,053.81 | 4,001.18 |
| 49 | CALIFORNIA | 37,253,956 | 1,228,202.73 | 3,296.84 |
| 50 | HAWAII | 1,360,301 | 41,233.36 | 3,031.19 |
| 51 | NEW MEXICO | 2,059,179 | 61,196.72 | 2,971.90 |
| 52 | VIRGIN ISLANDS | 109,825 | 1,325.10 | 1,206.55 |
| 53 | PUERTO RICO | 3,725,789 | 37,855.60 | 1,016.04 |
| 54 | GUAM | 178,430 | 1,759.37 | 986.03 |
| 55 | AMERICAN SAMOA | 65,628 | 2.50 | 3.81 |
| **1100 - AMPHETAMINE - Total** | | **312,825,210** | 21,888,402.46 | **6,997.01** |

**DRUG: 1105 - METHAMPHETAMINE**

| RANK | STATE NAME | STATE POPULATION | TOTAL GRAMS | TOTAL GRAMS/100K POPULATION |
|---|---|---|---|---|
| 1 | MINNESOTA | 5,303,925 | 300.95 | 5.67 |
| 2 | WASHINGTON | 6,724,540 | 353.59 | 5.26 |
| 3 | DISTRICT OF COLUMBIA | 601,723 | 29.73 | 4.94 |
| 4 | COLORADO | 5,029,196 | 235.47 | 4.68 |
| 5 | MAINE | 1,328,361 | 53.04 | 3.99 |
| 6 | SOUTH DAKOTA | 814,180 | 31.34 | 3.85 |
| 7 | PENNSYLVANIA | 12,702,379 | 481.36 | 3.79 |
| 8 | OREGON | 3,831,074 | 142.23 | 3.71 |
| 9 | IDAHO | 1,567,582 | 55.44 | 3.54 |
| 10 | UTAH | 2,763,885 | 94.43 | 3.42 |
| 11 | WYOMING | 563,626 | 18.48 | 3.28 |
| 12 | NEVADA | 2,700,551 | 81.98 | 3.04 |
| 13 | CALIFORNIA | 37,253,956 | 1,079.95 | 2.90 |
| 14 | MISSOURI | 5,988,927 | 172.36 | 2.88 |
| 15 | NORTH DAKOTA | 672,591 | 19.28 | 2.87 |
| 16 | ALABAMA | 4,779,736 | 135.81 | 2.84 |
| 17 | ARIZONA | 6,392,017 | 180.01 | 2.82 |
| 18 | CONNECTICUT | 3,574,097 | 96.03 | 2.69 |
| 19 | MARYLAND | 5,773,552 | 154.29 | 2.67 |
| 20 | WISCONSIN | 5,686,986 | 138.21 | 2.43 |
| 21 | IOWA | 3,046,355 | 70.72 | 2.32 |
| 22 | KANSAS | 2,853,118 | 61.48 | 2.15 |
| 23 | MASSACHUSETTS | 6,547,629 | 139.43 | 2.13 |
| 24 | NORTH CAROLINA | 9,535,483 | 191.26 | 2.01 |
| 25 | DELAWARE | 897,934 | 17.68 | 1.97 |

| | | | | |
|---|---|---|---|---|
| 26 | NEW YORK | 19,378,102 | 365.64 | 1.89 |
| 27 | TEXAS | 25,145,561 | 424.31 | 1.69 |
| 28 | HAWAII | 1,360,301 | 22.10 | 1.62 |
| 29 | ILLINOIS | 12,830,632 | 207.74 | 1.62 |
| 30 | NEW JERSEY | 8,791,894 | 139.43 | 1.59 |
| 31 | INDIANA | 6,483,802 | 102.47 | 1.58 |
| 32 | VIRGINIA | 8,001,024 | 126.17 | 1.58 |
| 33 | SOUTH CAROLINA | 4,625,364 | 72.73 | 1.57 |
| 34 | NEW HAMPSHIRE | 1,316,470 | 19.69 | 1.50 |
| 35 | OKLAHOMA | 3,751,351 | 54.65 | 1.46 |
| 36 | RHODE ISLAND | 1,052,567 | 14.87 | 1.41 |
| 37 | OHIO | 11,536,504 | 157.52 | 1.37 |
| 38 | VERMONT | 625,741 | 8.44 | 1.35 |
| 39 | LOUISIANA | 4,533,372 | 60.27 | 1.33 |
| 40 | NEW MEXICO | 2,059,179 | 22.90 | 1.11 |
| 41 | NEBRASKA | 1,826,341 | 20.09 | 1.10 |
| 42 | KENTUCKY | 4,339,367 | 47.01 | 1.08 |
| 43 | ALASKA | 710,231 | 7.63 | 1.07 |
| 44 | FLORIDA | 18,801,310 | 196.48 | 1.05 |
| 45 | MISSISSIPPI | 2,967,297 | 30.94 | 1.04 |
| 46 | MICHIGAN | 9,883,640 | 101.26 | 1.02 |
| 47 | WEST VIRGINIA | 1,852,994 | 18.48 | 1.00 |
| 48 | ARKANSAS | 2,915,918 | 28.13 | 0.96 |
| 49 | TENNESSEE | 6,346,105 | 59.07 | 0.93 |
| 50 | GEORGIA | 9,687,653 | 88.80 | 0.92 |
| 51 | MONTANA | 989,415 | 0.40 | 0.04 |
| **1105 - METHAMPHETAMINE - Total** | | **312,825,210** | 6,731.76 | **2.15** |

**DRUG: 1205 - LISDEXAMFETAMINE**

| RANK | STATE NAME | STATE POPULATION | TOTAL GRAMS | TOTAL GRAMS/100K POPULATION |
|---|---|---|---|---|
| 1 | LOUISIANA | 4,533,372 | 356,243.72 | 7,858.25 |
| 2 | DELAWARE | 897,934 | 55,749.65 | 6,208.66 |
| 3 | ALABAMA | 4,779,736 | 283,849.59 | 5,938.60 |
| 4 | SOUTH CAROLINA | 4,625,364 | 243,292.14 | 5,259.96 |
| 5 | INDIANA | 6,483,802 | 333,963.17 | 5,150.73 |
| 6 | WISCONSIN | 5,686,986 | 291,709.84 | 5,129.43 |
| 7 | MAINE | 1,328,361 | 66,987.92 | 5,042.90 |
| 8 | MISSISSIPPI | 2,967,297 | 149,623.84 | 5,042.43 |
| 9 | SOUTH DAKOTA | 814,180 | 40,983.24 | 5,033.68 |
| 10 | VERMONT | 625,741 | 28,702.67 | 4,586.99 |
| 11 | IOWA | 3,046,355 | 136,846.10 | 4,492.13 |
| 12 | KANSAS | 2,853,118 | 127,852.60 | 4,481.15 |

| | | | | |
|---|---|---|---|---|
| 13 | MISSOURI | 5,988,927 | 268,283.32 | 4,479.66 |
| 14 | NORTH CAROLINA | 9,535,483 | 423,079.59 | 4,436.90 |
| 15 | NEW HAMPSHIRE | 1,316,470 | 56,538.18 | 4,294.68 |
| 16 | TENNESSEE | 6,346,105 | 269,531.35 | 4,247.19 |
| 17 | TEXAS | 25,145,561 | 1,064,948.09 | 4,235.13 |
| 18 | WEST VIRGINIA | 1,852,994 | 75,151.27 | 4,055.67 |
| 19 | CONNECTICUT | 3,574,097 | 144,803.07 | 4,051.46 |
| 20 | GEORGIA | 9,687,653 | 391,925.61 | 4,045.62 |
| 21 | ARKANSAS | 2,915,918 | 117,953.04 | 4,045.14 |
| 22 | OKLAHOMA | 3,751,351 | 148,663.91 | 3,962.94 |
| 23 | MARYLAND | 5,773,552 | 226,434.49 | 3,921.93 |
| 24 | NEBRASKA | 1,826,341 | 69,450.56 | 3,802.72 |
| 25 | KENTUCKY | 4,339,367 | 163,801.36 | 3,774.78 |
| 26 | DISTRICT OF COLUMBIA | 601,723 | 22,608.33 | 3,757.27 |
| 27 | NORTH DAKOTA | 672,591 | 23,522.31 | 3,497.27 |
| 28 | OHIO | 11,536,504 | 400,270.01 | 3,469.60 |
| 29 | VIRGINIA | 8,001,024 | 268,242.53 | 3,352.60 |
| 30 | UTAH | 2,763,885 | 90,277.12 | 3,266.31 |
| 31 | FLORIDA | 18,801,310 | 541,099.66 | 2,877.99 |
| 32 | PENNSYLVANIA | 12,702,379 | 355,671.98 | 2,800.04 |
| 33 | RHODE ISLAND | 1,052,567 | 29,458.24 | 2,798.70 |
| 34 | ARIZONA | 6,392,017 | 169,026.61 | 2,644.34 |
| 35 | MONTANA | 989,415 | 24,639.20 | 2,490.28 |
| 36 | MICHIGAN | 9,883,640 | 244,803.58 | 2,476.86 |
| 37 | IDAHO | 1,567,582 | 38,589.33 | 2,461.71 |
| 38 | MINNESOTA | 5,303,925 | 124,261.44 | 2,342.82 |
| 39 | COLORADO | 5,029,196 | 116,503.91 | 2,316.55 |
| 40 | MASSACHUSETTS | 6,547,629 | 150,455.15 | 2,297.86 |
| 41 | ILLINOIS | 12,830,632 | 289,067.34 | 2,252.95 |
| 42 | NEW JERSEY | 8,791,894 | 194,776.49 | 2,215.41 |
| 43 | WASHINGTON | 6,724,540 | 142,599.20 | 2,120.58 |
| 44 | WYOMING | 563,626 | 10,518.53 | 1,866.23 |
| 45 | NEW YORK | 19,378,102 | 348,540.24 | 1,798.63 |
| 46 | OREGON | 3,831,074 | 62,738.39 | 1,637.62 |
| 47 | ALASKA | 710,231 | 10,628.37 | 1,496.47 |
| 48 | CALIFORNIA | 37,253,956 | 452,255.56 | 1,213.98 |
| 49 | NEVADA | 2,700,551 | 31,481.41 | 1,165.74 |
| 50 | NEW MEXICO | 2,059,179 | 15,678.65 | 761.40 |
| 51 | HAWAII | 1,360,301 | 6,115.14 | 449.54 |
| 52 | VIRGIN ISLANDS | 109,825 | 277.49 | 252.66 |
| 53 | PUERTO RICO | 3,725,789 | 8,477.84 | 227.54 |

| | | | | |
|---|---|---|---|---|
| 54 | GUAM | 178,430 | 43.94 | 24.62 |
| **1205 - LISDEXAMFETAMINE - Total** | | **312,825,210** | 9,708,996.30 | **3,103.65** |

**DRUG: 1724 - METHYLPHENIDATE (DL;D;L;ISOMERS)**

| RANK | STATE NAME | STATE POPULATION | TOTAL GRAMS | TOTAL GRAMS/100K POPULATION |
|---|---|---|---|---|
| 1 | VERMONT | 625,741 | 74,925.40 | 11,973.87 |
| 2 | MAINE | 1,328,361 | 138,505.73 | 10,426.81 |
| 3 | IOWA | 3,046,355 | 316,973.49 | 10,405.01 |
| 4 | NEW HAMPSHIRE | 1,316,470 | 132,963.86 | 10,100.03 |
| 5 | SOUTH DAKOTA | 814,180 | 81,697.86 | 10,034.37 |
| 6 | NORTH DAKOTA | 672,591 | 60,259.60 | 8,959.32 |
| 7 | MISSOURI | 5,988,927 | 515,345.28 | 8,604.97 |
| 8 | MASSACHUSETTS | 6,547,629 | 561,630.85 | 8,577.62 |
| 9 | DELAWARE | 897,934 | 70,498.43 | 7,851.18 |
| 10 | NORTH CAROLINA | 9,535,483 | 721,018.56 | 7,561.43 |
| 11 | MINNESOTA | 5,303,925 | 398,703.63 | 7,517.14 |
| 12 | SOUTH CAROLINA | 4,625,364 | 340,046.25 | 7,351.77 |
| 13 | MICHIGAN | 9,883,640 | 712,050.08 | 7,204.33 |
| 14 | WISCONSIN | 5,686,986 | 409,009.53 | 7,192.03 |
| 15 | LOUISIANA | 4,533,372 | 325,839.09 | 7,187.57 |
| 16 | INDIANA | 6,483,802 | 457,897.73 | 7,062.18 |
| 17 | IDAHO | 1,567,582 | 110,467.80 | 7,047.02 |
| 18 | OHIO | 11,536,504 | 812,379.83 | 7,041.82 |
| 19 | RHODE ISLAND | 1,052,567 | 72,537.70 | 6,891.50 |
| 20 | PENNSYLVANIA | 12,702,379 | 871,088.33 | 6,857.68 |
| 21 | MONTANA | 989,415 | 67,683.91 | 6,840.80 |
| 22 | KANSAS | 2,853,118 | 193,258.41 | 6,773.59 |
| 23 | MARYLAND | 5,773,552 | 388,584.27 | 6,730.42 |
| 24 | KENTUCKY | 4,339,367 | 290,894.33 | 6,703.61 |
| 25 | NEBRASKA | 1,826,341 | 121,629.05 | 6,659.71 |
| 26 | WASHINGTON | 6,724,540 | 447,627.62 | 6,656.63 |
| 27 | UTAH | 2,763,885 | 183,703.86 | 6,646.58 |
| 28 | WEST VIRGINIA | 1,852,994 | 120,742.68 | 6,516.09 |
| 29 | GEORGIA | 9,687,653 | 630,837.46 | 6,511.77 |
| 30 | VIRGINIA | 8,001,024 | 520,555.76 | 6,506.11 |
| 31 | OREGON | 3,831,074 | 245,801.71 | 6,416.00 |
| 32 | CONNECTICUT | 3,574,097 | 218,120.11 | 6,102.80 |
| 33 | ALABAMA | 4,779,736 | 281,105.10 | 5,881.18 |
| 34 | MISSISSIPPI | 2,967,297 | 164,923.13 | 5,558.03 |
| 35 | WYOMING | 563,626 | 30,287.69 | 5,373.72 |
| 36 | DISTRICT OF COLUMBIA | 601,723 | 31,641.33 | 5,258.45 |
| 37 | ARKANSAS | 2,915,918 | 152,220.89 | 5,220.34 |

| | | | | |
|---|---|---|---|---|
| 38 | ARIZONA | 6,392,017 | 331,019.40 | 5,178.64 |
| 39 | ILLINOIS | 12,830,632 | 663,079.86 | 5,167.94 |
| 40 | TEXAS | 25,145,561 | 1,283,631.84 | 5,104.80 |
| 41 | TENNESSEE | 6,346,105 | 321,164.51 | 5,060.81 |
| 42 | OKLAHOMA | 3,751,351 | 186,538.79 | 4,972.58 |
| 43 | COLORADO | 5,029,196 | 245,024.63 | 4,872.04 |
| 44 | NEW YORK | 19,378,102 | 897,668.27 | 4,632.38 |
| 45 | ALASKA | 710,231 | 31,994.14 | 4,504.75 |
| 46 | FLORIDA | 18,801,310 | 784,881.68 | 4,174.61 |
| 47 | NEW JERSEY | 8,791,894 | 355,503.83 | 4,043.54 |
| 48 | NEW MEXICO | 2,059,179 | 72,441.79 | 3,517.99 |
| 49 | HAWAII | 1,360,301 | 43,748.73 | 3,216.11 |
| 50 | CALIFORNIA | 37,253,956 | 1,042,295.57 | 2,797.81 |
| 51 | NEVADA | 2,700,551 | 58,216.16 | 2,155.71 |
| 52 | PUERTO RICO | 3,725,789 | 57,389.98 | 1,540.34 |
| 53 | GUAM | 178,430 | 1,984.74 | 1,112.34 |
| 54 | VIRGIN ISLANDS | 109,825 | 586.16 | 533.72 |
| 55 | AMERICAN SAMOA | 65,628 | 44.10 | 67.20 |
| **1724 - METHYLPHENIDATE (DL;D;L;ISOMERS) - Total** | | **312,825,210** | 17,650,670.52 | **5,642.34** |

**DRUG: 9050 - CODEINE**

| RANK | STATE NAME | STATE POPULATION | TOTAL GRAMS | TOTAL GRAMS/100K POPULATION |
|---|---|---|---|---|
| 1 | TEXAS | 25,145,561 | 3,661,119.62 | 14,559.71 |
| 2 | OKLAHOMA | 3,751,351 | 348,095.81 | 9,279.21 |
| 3 | ARKANSAS | 2,915,918 | 241,747.80 | 8,290.62 |
| 4 | MICHIGAN | 9,883,640 | 809,994.72 | 8,195.31 |
| 5 | WEST VIRGINIA | 1,852,994 | 143,305.26 | 7,733.71 |
| 6 | DISTRICT OF COLUMBIA | 601,723 | 41,461.92 | 6,890.53 |
| 7 | SOUTH CAROLINA | 4,625,364 | 280,974.88 | 6,074.65 |
| 8 | ILLINOIS | 12,830,632 | 569,355.47 | 4,437.47 |
| 9 | CALIFORNIA | 37,253,956 | 1,605,157.49 | 4,308.69 |
| 10 | TENNESSEE | 6,346,105 | 268,960.19 | 4,238.19 |
| 11 | ALABAMA | 4,779,736 | 201,605.04 | 4,217.91 |
| 12 | MISSOURI | 5,988,927 | 243,080.15 | 4,058.83 |
| 13 | KENTUCKY | 4,339,367 | 168,180.16 | 3,875.68 |
| 14 | GEORGIA | 9,687,653 | 365,928.70 | 3,777.27 |
| 15 | ARIZONA | 6,392,017 | 237,282.47 | 3,712.17 |
| 16 | MONTANA | 989,415 | 35,536.89 | 3,591.71 |
| 17 | DELAWARE | 897,934 | 31,949.10 | 3,558.07 |
| 18 | PENNSYLVANIA | 12,702,379 | 444,669.53 | 3,500.68 |
| 19 | HAWAII | 1,360,301 | 46,425.06 | 3,412.85 |
| 20 | MARYLAND | 5,773,552 | 196,096.69 | 3,396.47 |

| | | | | |
|---|---|---|---|---|
| 21 | LOUISIANA | 4,533,372 | 152,869.10 | 3,372.08 |
| 22 | KANSAS | 2,853,118 | 95,710.26 | 3,354.58 |
| 23 | ALASKA | 710,231 | 23,569.75 | 3,318.60 |
| 24 | NEW MEXICO | 2,059,179 | 66,006.90 | 3,205.50 |
| 25 | WASHINGTON | 6,724,540 | 214,824.84 | 3,194.64 |
| 26 | FLORIDA | 18,801,310 | 600,497.44 | 3,193.91 |
| 27 | OREGON | 3,831,074 | 121,785.82 | 3,178.90 |
| 28 | MISSISSIPPI | 2,967,297 | 92,647.22 | 3,122.28 |
| 29 | IDAHO | 1,567,582 | 48,693.85 | 3,106.30 |
| 30 | NEW JERSEY | 8,791,894 | 270,126.81 | 3,072.45 |
| 31 | WISCONSIN | 5,686,986 | 168,161.69 | 2,956.96 |
| 32 | NEVADA | 2,700,551 | 78,795.10 | 2,917.74 |
| 33 | SOUTH DAKOTA | 814,180 | 22,849.83 | 2,806.48 |
| 34 | NORTH DAKOTA | 672,591 | 18,775.44 | 2,791.51 |
| 35 | INDIANA | 6,483,802 | 174,704.43 | 2,694.48 |
| 36 | MAINE | 1,328,361 | 35,432.24 | 2,667.37 |
| 37 | OHIO | 11,536,504 | 307,555.84 | 2,665.94 |
| 38 | NEW YORK | 19,378,102 | 497,596.48 | 2,567.83 |
| 39 | WYOMING | 563,626 | 14,330.25 | 2,542.51 |
| 40 | VIRGIN ISLANDS | 109,825 | 2,778.39 | 2,529.83 |
| 41 | MASSACHUSETTS | 6,547,629 | 160,968.12 | 2,458.42 |
| 42 | MINNESOTA | 5,303,925 | 129,219.55 | 2,436.30 |
| 43 | IOWA | 3,046,355 | 70,841.41 | 2,325.45 |
| 44 | VIRGINIA | 8,001,024 | 184,561.82 | 2,306.73 |
| 45 | NEBRASKA | 1,826,341 | 41,200.75 | 2,255.92 |
| 46 | CONNECTICUT | 3,574,097 | 78,480.23 | 2,195.81 |
| 47 | RHODE ISLAND | 1,052,567 | 23,013.23 | 2,186.39 |
| 48 | UTAH | 2,763,885 | 58,837.81 | 2,128.81 |
| 49 | GUAM | 178,430 | 3,598.89 | 2,016.98 |
| 50 | NEW HAMPSHIRE | 1,316,470 | 26,377.09 | 2,003.62 |
| 51 | VERMONT | 625,741 | 10,886.96 | 1,739.85 |
| 52 | NORTH CAROLINA | 9,535,483 | 163,261.25 | 1,712.14 |
| 53 | COLORADO | 5,029,196 | 84,732.08 | 1,684.80 |
| 54 | AMERICAN SAMOA | 65,628 | 1,060.85 | 1,616.46 |
| 55 | PUERTO RICO | 3,725,789 | 46,899.43 | 1,258.78 |
| **9050 - CODEINE - Total** | | **312,825,210** | 14,032,578.12 | **4,485.76** |

**DRUG: 9064 - BUPRENORPHINE**

| RANK | STATE NAME | STATE POPULATION | TOTAL GRAMS | TOTAL GRAMS/100K POPULATION |
|---|---|---|---|---|
| 1 | VERMONT | 625,741 | 27,164.61 | 4,341.19 |
| 2 | KENTUCKY | 4,339,367 | 155,629.85 | 3,586.46 |
| 3 | WEST VIRGINIA | 1,852,994 | 64,544.98 | 3,483.28 |

| | | | | | |
|---|---|---|---|---|---|
| 4 | MAINE | | 1,328,361 | 44,878.69 | 3,378.50 |
| 5 | RHODE ISLAND | | 1,052,567 | 32,691.74 | 3,105.91 |
| 6 | MASSACHUSETTS | | 6,547,629 | 175,411.51 | 2,679.01 |
| 7 | NEW HAMPSHIRE | | 1,316,470 | 34,845.51 | 2,646.89 |
| 8 | TENNESSEE | | 6,346,105 | 153,767.41 | 2,423.02 |
| 9 | ALABAMA | | 4,779,736 | 115,648.15 | 2,419.55 |
| 10 | PENNSYLVANIA | | 12,702,379 | 289,950.16 | 2,282.64 |
| 11 | MARYLAND | | 5,773,552 | 115,983.50 | 2,008.88 |
| 12 | OHIO | | 11,536,504 | 225,373.82 | 1,953.57 |
| 13 | DISTRICT OF COLUMBIA | | 601,723 | 11,589.32 | 1,926.02 |
| 14 | LOUISIANA | | 4,533,372 | 84,626.49 | 1,866.74 |
| 15 | CONNECTICUT | | 3,574,097 | 65,088.17 | 1,821.11 |
| 16 | DELAWARE | | 897,934 | 15,905.60 | 1,771.36 |
| 17 | NORTH CAROLINA | | 9,535,483 | 160,024.88 | 1,678.20 |
| 18 | NEW MEXICO | | 2,059,179 | 34,195.54 | 1,660.64 |
| 19 | INDIANA | | 6,483,802 | 97,607.04 | 1,505.40 |
| 20 | UTAH | | 2,763,885 | 41,575.13 | 1,504.23 |
| 21 | ALASKA | | 710,231 | 10,564.38 | 1,487.46 |
| 22 | OKLAHOMA | | 3,751,351 | 55,470.25 | 1,478.67 |
| 23 | MISSISSIPPI | | 2,967,297 | 42,860.68 | 1,444.44 |
| 24 | WASHINGTON | | 6,724,540 | 95,924.45 | 1,426.48 |
| 25 | OREGON | | 3,831,074 | 49,174.65 | 1,283.57 |
| 26 | NEW JERSEY | | 8,791,894 | 111,067.66 | 1,263.30 |
| 27 | NEW YORK | | 19,378,102 | 244,277.64 | 1,260.59 |
| 28 | MICHIGAN | | 9,883,640 | 122,156.32 | 1,235.94 |
| 29 | MONTANA | | 989,415 | 12,159.72 | 1,228.98 |
| 30 | FLORIDA | | 18,801,310 | 208,118.09 | 1,106.93 |
| 31 | SOUTH CAROLINA | | 4,625,364 | 51,017.95 | 1,103.00 |
| 32 | VIRGINIA | | 8,001,024 | 86,586.44 | 1,082.19 |
| 33 | IDAHO | | 1,567,582 | 16,872.80 | 1,076.36 |
| 34 | ARKANSAS | | 2,915,918 | 29,074.25 | 997.09 |
| 35 | WYOMING | | 563,626 | 5,357.00 | 950.45 |
| 36 | MISSOURI | | 5,988,927 | 53,639.78 | 895.65 |
| 37 | WISCONSIN | | 5,686,986 | 48,115.17 | 846.06 |
| 38 | GEORGIA | | 9,687,653 | 69,149.91 | 713.79 |
| 39 | ARIZONA | | 6,392,017 | 45,046.72 | 704.73 |
| 40 | COLORADO | | 5,029,196 | 35,203.25 | 699.98 |
| 41 | NORTH DAKOTA | | 672,591 | 4,653.28 | 691.84 |
| 42 | MINNESOTA | | 5,303,925 | 36,212.90 | 682.76 |
| 43 | HAWAII | | 1,360,301 | 8,594.50 | 631.81 |
| 44 | NEVADA | | 2,700,551 | 15,290.93 | 566.22 |

| | | | | |
|---|---|---|---|---|
| 45 | ILLINOIS | 12,830,632 | 66,850.09 | 521.02 |
| 46 | CALIFORNIA | 37,253,956 | 171,999.96 | 461.70 |
| 47 | TEXAS | 25,145,561 | 114,352.32 | 454.76 |
| 48 | KANSAS | 2,853,118 | 11,268.50 | 394.95 |
| 49 | NEBRASKA | 1,826,341 | 5,963.88 | 326.55 |
| 50 | SOUTH DAKOTA | 814,180 | 2,304.93 | 283.10 |
| 51 | IOWA | 3,046,355 | 8,260.26 | 271.15 |
| 52 | PUERTO RICO | 3,725,789 | 8,501.12 | 228.17 |
| 53 | VIRGIN ISLANDS | 109,825 | 49.12 | 44.73 |
| 54 | GUAM | 178,430 | 2.31 | 1.29 |
| 55 | AMERICAN SAMOA | 65,628 | 0.04 | 0.06 |
| **9064 - BUPRENORPHINE - Total** | | **312,825,210** | 3,792,643.31 | **1,212.38** |

**DRUG: 9143 - OXYCODONE**

| RANK | STATE NAME | STATE POPULATION | TOTAL GRAMS | TOTAL GRAMS/100K POPULATION |
|---|---|---|---|---|
| 1 | DELAWARE | 897,934 | 242,460.33 | 27,002.02 |
| 2 | TENNESSEE | 6,346,105 | 1,598,875.37 | 25,194.59 |
| 3 | NEVADA | 2,700,551 | 594,943.99 | 22,030.47 |
| 4 | ARIZONA | 6,392,017 | 1,382,357.85 | 21,626.32 |
| 5 | UTAH | 2,763,885 | 586,704.81 | 21,227.54 |
| 6 | NORTH CAROLINA | 9,535,483 | 1,926,492.48 | 20,203.41 |
| 7 | FLORIDA | 18,801,310 | 3,694,501.21 | 19,650.23 |
| 8 | OKLAHOMA | 3,751,351 | 734,588.62 | 19,581.98 |
| 9 | PENNSYLVANIA | 12,702,379 | 2,453,858.91 | 19,318.10 |
| 10 | MARYLAND | 5,773,552 | 1,091,362.08 | 18,902.78 |
| 11 | NEW JERSEY | 8,791,894 | 1,623,835.91 | 18,469.69 |
| 12 | KENTUCKY | 4,339,367 | 739,198.98 | 17,034.72 |
| 13 | ALASKA | 710,231 | 120,091.96 | 16,908.86 |
| 14 | SOUTH CAROLINA | 4,625,364 | 774,314.35 | 16,740.61 |
| 15 | ALABAMA | 4,779,736 | 794,969.65 | 16,632.08 |
| 16 | GEORGIA | 9,687,653 | 1,579,564.08 | 16,304.92 |
| 17 | WYOMING | 563,626 | 90,905.46 | 16,128.68 |
| 18 | MISSOURI | 5,988,927 | 954,906.78 | 15,944.54 |
| 19 | CONNECTICUT | 3,574,097 | 568,944.68 | 15,918.56 |
| 20 | ARKANSAS | 2,915,918 | 458,923.65 | 15,738.56 |
| 21 | NEW HAMPSHIRE | 1,316,470 | 206,713.38 | 15,702.10 |
| 22 | OREGON | 3,831,074 | 585,778.76 | 15,290.20 |
| 23 | NEW MEXICO | 2,059,179 | 311,123.09 | 15,109.08 |
| 24 | COLORADO | 5,029,196 | 759,717.61 | 15,106.14 |
| 25 | KANSAS | 2,853,118 | 421,416.52 | 14,770.39 |
| 26 | WASHINGTON | 6,724,540 | 989,742.63 | 14,718.37 |
| 27 | LOUISIANA | 4,533,372 | 656,705.08 | 14,486.02 |

| | | | | |
|---|---|---|---|---|
| 28 | WEST VIRGINIA | 1,852,994 | 264,334.46 | 14,265.26 |
| 29 | OHIO | 11,536,504 | 1,569,513.43 | 13,604.76 |
| 30 | IDAHO | 1,567,582 | 212,925.53 | 13,583.06 |
| 31 | MAINE | 1,328,361 | 180,328.38 | 13,575.25 |
| 32 | NEW YORK | 19,378,102 | 2,475,019.89 | 12,772.25 |
| 33 | RHODE ISLAND | 1,052,567 | 133,921.89 | 12,723.36 |
| 34 | VERMONT | 625,741 | 79,018.26 | 12,627.95 |
| 35 | MONTANA | 989,415 | 124,854.68 | 12,619.04 |
| 36 | MASSACHUSETTS | 6,547,629 | 824,642.85 | 12,594.53 |
| 37 | VIRGINIA | 8,001,024 | 1,005,192.47 | 12,563.30 |
| 38 | INDIANA | 6,483,802 | 796,671.10 | 12,287.10 |
| 39 | MISSISSIPPI | 2,967,297 | 358,175.10 | 12,070.75 |
| 40 | WISCONSIN | 5,686,986 | 658,712.06 | 11,582.80 |
| 41 | HAWAII | 1,360,301 | 147,943.28 | 10,875.78 |
| 42 | MICHIGAN | 9,883,640 | 945,331.20 | 9,564.61 |
| 43 | MINNESOTA | 5,303,925 | 481,528.29 | 9,078.72 |
| 44 | DISTRICT OF COLUMBIA | 601,723 | 49,631.36 | 8,248.21 |
| 45 | NEBRASKA | 1,826,341 | 143,660.91 | 7,866.05 |
| 46 | CALIFORNIA | 37,253,956 | 2,884,241.08 | 7,742.11 |
| 47 | NORTH DAKOTA | 672,591 | 50,831.84 | 7,557.62 |
| 48 | SOUTH DAKOTA | 814,180 | 56,641.51 | 6,956.88 |
| 49 | IOWA | 3,046,355 | 177,777.69 | 5,835.75 |
| 50 | TEXAS | 25,145,561 | 1,191,356.55 | 4,737.84 |
| 51 | ILLINOIS | 12,830,632 | 539,002.68 | 4,200.91 |
| 52 | GUAM | 178,430 | 3,145.83 | 1,763.06 |
| 53 | VIRGIN ISLANDS | 109,825 | 1,752.57 | 1,595.78 |
| 54 | PUERTO RICO | 3,725,789 | 44,330.91 | 1,189.84 |
| 55 | AMERICAN SAMOA | 65,628 | 36.76 | 56.01 |
| **9143 - OXYCODONE - Total** | | **312,825,210** | 41,343,520.76 | **13,216.17** |

**DRUG: 9150 - HYDROMORPHONE**

| RANK | STATE NAME | STATE POPULATION | TOTAL GRAMS | TOTAL GRAMS/100K POPULATION |
|---|---|---|---|---|
| 1 | FLORIDA | 18,801,310 | 155,520.83 | 827.18 |
| 2 | DELAWARE | 897,934 | 6,472.38 | 720.81 |
| 3 | MISSISSIPPI | 2,967,297 | 20,586.53 | 693.78 |
| 4 | VERMONT | 625,741 | 3,998.48 | 639.00 |
| 5 | CONNECTICUT | 3,574,097 | 20,355.47 | 569.53 |
| 6 | MONTANA | 989,415 | 5,613.14 | 567.32 |
| 7 | WASHINGTON | 6,724,540 | 35,286.47 | 524.74 |
| 8 | NEW HAMPSHIRE | 1,316,470 | 6,594.75 | 500.94 |
| 9 | COLORADO | 5,029,196 | 23,578.07 | 468.82 |
| 10 | MARYLAND | 5,773,552 | 25,925.96 | 449.05 |

| | | | | | |
|---|---|---|---|---|---|
| 11 | IDAHO | | 1,567,582 | 7,017.83 | 447.69 |
| 12 | VIRGINIA | | 8,001,024 | 35,762.70 | 446.98 |
| 13 | MAINE | | 1,328,361 | 5,821.30 | 438.23 |
| 14 | ALASKA | | 710,231 | 3,103.34 | 436.95 |
| 15 | OREGON | | 3,831,074 | 16,101.59 | 420.29 |
| 16 | SOUTH CAROLINA | | 4,625,364 | 19,421.63 | 419.89 |
| 17 | KANSAS | | 2,853,118 | 11,383.91 | 399.00 |
| 18 | MASSACHUSETTS | | 6,547,629 | 25,894.08 | 395.47 |
| 19 | PENNSYLVANIA | | 12,702,379 | 49,569.38 | 390.24 |
| 20 | NORTH CAROLINA | | 9,535,483 | 36,392.38 | 381.65 |
| 21 | MINNESOTA | | 5,303,925 | 20,235.00 | 381.51 |
| 22 | WYOMING | | 563,626 | 2,104.73 | 373.43 |
| 23 | NORTH DAKOTA | | 672,591 | 2,372.74 | 352.78 |
| 24 | UTAH | | 2,763,885 | 9,669.19 | 349.84 |
| 25 | ARIZONA | | 6,392,017 | 22,219.02 | 347.61 |
| 26 | TENNESSEE | | 6,346,105 | 21,991.74 | 346.54 |
| 27 | ARKANSAS | | 2,915,918 | 9,997.50 | 342.86 |
| 28 | NEVADA | | 2,700,551 | 9,254.96 | 342.71 |
| 29 | CALIFORNIA | | 37,253,956 | 125,931.00 | 338.03 |
| 30 | MISSOURI | | 5,988,927 | 20,231.31 | 337.81 |
| 31 | NEW YORK | | 19,378,102 | 65,427.26 | 337.64 |
| 32 | NEW JERSEY | | 8,791,894 | 29,412.21 | 334.54 |
| 33 | OKLAHOMA | | 3,751,351 | 12,500.83 | 333.24 |
| 34 | IOWA | | 3,046,355 | 9,579.33 | 314.45 |
| 35 | TEXAS | | 25,145,561 | 79,065.23 | 314.43 |
| 36 | INDIANA | | 6,483,802 | 19,162.53 | 295.54 |
| 37 | MICHIGAN | | 9,883,640 | 27,905.89 | 282.34 |
| 38 | DISTRICT OF COLUMBIA | | 601,723 | 1,693.63 | 281.46 |
| 39 | GEORGIA | | 9,687,653 | 27,062.23 | 279.35 |
| 40 | RHODE ISLAND | | 1,052,567 | 2,748.77 | 261.15 |
| 41 | OHIO | | 11,536,504 | 30,086.13 | 260.79 |
| 42 | ILLINOIS | | 12,830,632 | 32,527.78 | 253.52 |
| 43 | SOUTH DAKOTA | | 814,180 | 1,978.32 | 242.98 |
| 44 | LOUISIANA | | 4,533,372 | 10,872.08 | 239.82 |
| 45 | WEST VIRGINIA | | 1,852,994 | 4,423.85 | 238.74 |
| 46 | NEBRASKA | | 1,826,341 | 4,312.82 | 236.15 |
| 47 | ALABAMA | | 4,779,736 | 10,541.50 | 220.55 |
| 48 | NEW MEXICO | | 2,059,179 | 4,501.06 | 218.59 |
| 49 | KENTUCKY | | 4,339,367 | 9,461.73 | 218.04 |
| 50 | WISCONSIN | | 5,686,986 | 12,251.25 | 215.43 |
| 51 | HAWAII | | 1,360,301 | 2,061.45 | 151.54 |

| | | | | |
|---|---|---|---|---|
| 52 | GUAM | 178,430 | 90.51 | 50.73 |
| 53 | VIRGIN ISLANDS | 109,825 | 54.30 | 49.45 |
| 54 | PUERTO RICO | 3,725,789 | 229.67 | 6.16 |
| 55 | AMERICAN SAMOA | 65,628 | 1.20 | 1.83 |
| **9150 - HYDROMORPHONE - Total** | | **312,825,210** | 1,156,359.01 | **369.65** |

**DRUG: 9193 - HYDROCODONE**

| RANK | STATE NAME | STATE POPULATION | TOTAL GRAMS | TOTAL GRAMS/100K POPULATION |
|---|---|---|---|---|
| 1 | ALABAMA | 4,779,736 | 853,989.67 | 17,866.88 |
| 2 | KENTUCKY | 4,339,367 | 721,540.77 | 16,627.79 |
| 3 | OKLAHOMA | 3,751,351 | 617,228.70 | 16,453.50 |
| 4 | ARKANSAS | 2,915,918 | 455,126.05 | 15,608.33 |
| 5 | TENNESSEE | 6,346,105 | 826,341.72 | 13,021.24 |
| 6 | IDAHO | 1,567,582 | 201,065.09 | 12,826.45 |
| 7 | MICHIGAN | 9,883,640 | 1,224,853.66 | 12,392.74 |
| 8 | LOUISIANA | 4,533,372 | 558,850.42 | 12,327.48 |
| 9 | MISSISSIPPI | 2,967,297 | 364,061.06 | 12,269.11 |
| 10 | INDIANA | 6,483,802 | 783,520.77 | 12,084.28 |
| 11 | NEVADA | 2,700,551 | 301,063.15 | 11,148.21 |
| 12 | WEST VIRGINIA | 1,852,994 | 185,139.63 | 9,991.38 |
| 13 | SOUTH CAROLINA | 4,625,364 | 456,367.25 | 9,866.62 |
| 14 | MISSOURI | 5,988,927 | 587,432.60 | 9,808.65 |
| 15 | KANSAS | 2,853,118 | 278,398.45 | 9,757.69 |
| 16 | TEXAS | 25,145,561 | 2,331,346.46 | 9,271.40 |
| 17 | MONTANA | 989,415 | 90,396.57 | 9,136.37 |
| 18 | GEORGIA | 9,687,653 | 842,836.92 | 8,700.11 |
| 19 | OREGON | 3,831,074 | 324,189.97 | 8,462.12 |
| 20 | CALIFORNIA | 37,253,956 | 3,133,870.80 | 8,412.18 |
| 21 | ILLINOIS | 12,830,632 | 1,030,067.59 | 8,028.19 |
| 22 | UTAH | 2,763,885 | 202,748.82 | 7,335.65 |
| 23 | WYOMING | 563,626 | 38,866.35 | 6,895.77 |
| 24 | WASHINGTON | 6,724,540 | 448,967.62 | 6,676.56 |
| 25 | NORTH CAROLINA | 9,535,483 | 631,179.73 | 6,619.27 |
| 26 | FLORIDA | 18,801,310 | 1,202,457.69 | 6,395.61 |
| 27 | IOWA | 3,046,355 | 191,242.24 | 6,277.74 |
| 28 | NEBRASKA | 1,826,341 | 108,776.55 | 5,955.98 |
| 29 | NEW MEXICO | 2,059,179 | 121,799.47 | 5,914.95 |
| 30 | SOUTH DAKOTA | 814,180 | 47,831.94 | 5,874.86 |
| 31 | ARIZONA | 6,392,017 | 348,709.74 | 5,455.39 |
| 32 | NORTH DAKOTA | 672,591 | 35,511.20 | 5,279.76 |
| 33 | ALASKA | 710,231 | 36,920.76 | 5,198.42 |
| 34 | OHIO | 11,536,504 | 591,097.65 | 5,123.72 |

| | | | | |
|---|---|---|---|---|
| 35 | WISCONSIN | 5,686,986 | 269,208.06 | 4,733.76 |
| 36 | MAINE | 1,328,361 | 60,385.18 | 4,545.84 |
| 37 | HAWAII | 1,360,301 | 60,411.22 | 4,441.02 |
| 38 | COLORADO | 5,029,196 | 217,650.10 | 4,327.73 |
| 39 | VIRGINIA | 8,001,024 | 340,536.39 | 4,256.16 |
| 40 | PENNSYLVANIA | 12,702,379 | 537,571.41 | 4,232.05 |
| 41 | RHODE ISLAND | 1,052,567 | 34,349.40 | 3,263.39 |
| 42 | NEW YORK | 19,378,102 | 578,840.51 | 2,987.09 |
| 43 | MINNESOTA | 5,303,925 | 149,219.75 | 2,813.38 |
| 44 | VERMONT | 625,741 | 16,413.53 | 2,623.06 |
| 45 | NEW HAMPSHIRE | 1,316,470 | 31,136.29 | 2,365.13 |
| 46 | DELAWARE | 897,934 | 20,062.33 | 2,234.28 |
| 47 | MARYLAND | 5,773,552 | 113,832.50 | 1,971.62 |
| 48 | CONNECTICUT | 3,574,097 | 67,444.03 | 1,887.02 |
| 49 | MASSACHUSETTS | 6,547,629 | 101,834.93 | 1,555.29 |
| 50 | NEW JERSEY | 8,791,894 | 113,786.23 | 1,294.22 |
| 51 | VIRGIN ISLANDS | 109,825 | 968.81 | 882.14 |
| 52 | DISTRICT OF COLUMBIA | 601,723 | 4,601.98 | 764.80 |
| 53 | GUAM | 178,430 | 496.46 | 278.24 |
| 54 | PUERTO RICO | 3,725,789 | 1,840.64 | 49.40 |
| 55 | AMERICAN SAMOA | 65,628 | 3.63 | 5.53 |
| **9193 - HYDROCODONE - Total** | | **312,825,210** | 22,894,390.46 | **7,318.59** |

**DRUG: 9230 - MEPERIDINE (PETHIDINE)**

| RANK | STATE NAME | STATE POPULATION | TOTAL GRAMS | TOTAL GRAMS/100K POPULATION |
|---|---|---|---|---|
| 1 | ARKANSAS | 2,915,918 | 22,644.77 | 776.59 |
| 2 | OKLAHOMA | 3,751,351 | 21,273.95 | 567.10 |
| 3 | ALABAMA | 4,779,736 | 25,225.33 | 527.76 |
| 4 | LOUISIANA | 4,533,372 | 22,573.07 | 497.93 |
| 5 | MISSISSIPPI | 2,967,297 | 13,462.89 | 453.71 |
| 6 | VIRGIN ISLANDS | 109,825 | 460.26 | 419.09 |
| 7 | SOUTH CAROLINA | 4,625,364 | 17,265.94 | 373.29 |
| 8 | TENNESSEE | 6,346,105 | 22,063.53 | 347.67 |
| 9 | GEORGIA | 9,687,653 | 33,459.87 | 345.39 |
| 10 | PUERTO RICO | 3,725,789 | 12,482.60 | 335.03 |
| 11 | ARIZONA | 6,392,017 | 18,362.98 | 287.28 |
| 12 | ALASKA | 710,231 | 1,796.05 | 252.88 |
| 13 | NEW MEXICO | 2,059,179 | 5,095.22 | 247.44 |
| 14 | VIRGINIA | 8,001,024 | 18,798.80 | 234.95 |
| 15 | SOUTH DAKOTA | 814,180 | 1,883.20 | 231.30 |
| 16 | UTAH | 2,763,885 | 6,333.04 | 229.14 |
| 17 | WYOMING | 563,626 | 1,253.54 | 222.41 |

| | | | | |
|---|---|---|---|---|
| 18 | IOWA | 3,046,355 | 6,587.62 | 216.25 |
| 19 | MONTANA | 989,415 | 2,034.30 | 205.61 |
| 20 | KENTUCKY | 4,339,367 | 8,548.63 | 197.00 |
| 21 | KANSAS | 2,853,118 | 5,423.02 | 190.07 |
| 22 | NEVADA | 2,700,551 | 5,113.09 | 189.34 |
| 23 | MISSOURI | 5,988,927 | 11,314.68 | 188.93 |
| 24 | NORTH CAROLINA | 9,535,483 | 17,756.16 | 186.21 |
| 25 | IDAHO | 1,567,582 | 2,787.71 | 177.84 |
| 26 | WASHINGTON | 6,724,540 | 11,406.39 | 169.62 |
| 27 | VERMONT | 625,741 | 1,056.37 | 168.82 |
| 28 | CALIFORNIA | 37,253,956 | 62,040.45 | 166.53 |
| 29 | TEXAS | 25,145,561 | 41,865.15 | 166.49 |
| 30 | NEW HAMPSHIRE | 1,316,470 | 2,188.99 | 166.28 |
| 31 | INDIANA | 6,483,802 | 10,744.61 | 165.71 |
| 32 | FLORIDA | 18,801,310 | 29,933.08 | 159.21 |
| 33 | OHIO | 11,536,504 | 16,594.92 | 143.85 |
| 34 | MARYLAND | 5,773,552 | 7,698.40 | 133.34 |
| 35 | OREGON | 3,831,074 | 4,570.58 | 119.30 |
| 36 | COLORADO | 5,029,196 | 5,689.91 | 113.14 |
| 37 | NORTH DAKOTA | 672,591 | 752.98 | 111.95 |
| 38 | WEST VIRGINIA | 1,852,994 | 1,982.55 | 106.99 |
| 39 | MASSACHUSETTS | 6,547,629 | 6,967.80 | 106.42 |
| 40 | NEBRASKA | 1,826,341 | 1,936.56 | 106.03 |
| 41 | DISTRICT OF COLUMBIA | 601,723 | 635.65 | 105.64 |
| 42 | CONNECTICUT | 3,574,097 | 3,694.83 | 103.38 |
| 43 | MAINE | 1,328,361 | 1,313.35 | 98.87 |
| 44 | MICHIGAN | 9,883,640 | 9,360.65 | 94.71 |
| 45 | PENNSYLVANIA | 12,702,379 | 11,726.05 | 92.31 |
| 46 | HAWAII | 1,360,301 | 1,234.48 | 90.75 |
| 47 | NEW YORK | 19,378,102 | 15,723.47 | 81.14 |
| 48 | ILLINOIS | 12,830,632 | 9,612.69 | 74.92 |
| 49 | NEW JERSEY | 8,791,894 | 6,270.22 | 71.32 |
| 50 | WISCONSIN | 5,686,986 | 4,033.89 | 70.93 |
| 51 | GUAM | 178,430 | 120.92 | 67.77 |
| 52 | RHODE ISLAND | 1,052,567 | 657.98 | 62.51 |
| 53 | DELAWARE | 897,934 | 560.37 | 62.41 |
| 54 | MINNESOTA | 5,303,925 | 2,151.45 | 40.56 |
| **9230 - MEPERIDINE (PETHIDINE) - Total** | | **312,825,210** | **576,525.04** | **184.30** |

**DRUG: 9250B - METHADONE**

| RANK | STATE NAME | STATE POPULATION | TOTAL GRAMS | TOTAL GRAMS/100K POPULATION |
|---|---|---|---|---|
| 1 | RHODE ISLAND | 1,052,567 | 169,150.99 | 16,070.33 |

| | | | | | |
|---|---|---|---|---|---|
| 2 | DELAWARE | | 897,934 | 124,914.59 | 13,911.33 |
| 3 | MARYLAND | | 5,773,552 | 716,136.84 | 12,403.75 |
| 4 | CONNECTICUT | | 3,574,097 | 416,944.15 | 11,665.72 |
| 5 | VERMONT | | 625,741 | 71,902.07 | 11,490.71 |
| 6 | NEW HAMPSHIRE | | 1,316,470 | 119,869.81 | 9,105.40 |
| 7 | NEW MEXICO | | 2,059,179 | 171,486.44 | 8,327.90 |
| 8 | MAINE | | 1,328,361 | 106,318.22 | 8,003.71 |
| 9 | MASSACHUSETTS | | 6,547,629 | 518,795.21 | 7,923.41 |
| 10 | ALABAMA | | 4,779,736 | 286,425.34 | 5,992.49 |
| 11 | OREGON | | 3,831,074 | 227,514.24 | 5,938.65 |
| 12 | DISTRICT OF COLUMBIA | | 601,723 | 35,411.84 | 5,885.07 |
| 13 | NEW YORK | | 19,378,102 | 1,126,619.92 | 5,813.88 |
| 14 | PENNSYLVANIA | | 12,702,379 | 714,088.79 | 5,621.69 |
| 15 | WASHINGTON | | 6,724,540 | 371,488.86 | 5,524.38 |
| 16 | ARIZONA | | 6,392,017 | 352,991.18 | 5,522.38 |
| 17 | NEW JERSEY | | 8,791,894 | 468,515.25 | 5,328.95 |
| 18 | NORTH CAROLINA | | 9,535,483 | 507,851.46 | 5,325.91 |
| 19 | NEVADA | | 2,700,551 | 136,567.19 | 5,057.01 |
| 20 | MICHIGAN | | 9,883,640 | 489,774.33 | 4,955.40 |
| 21 | ALASKA | | 710,231 | 32,983.59 | 4,644.07 |
| 22 | GEORGIA | | 9,687,653 | 449,234.91 | 4,637.19 |
| 23 | INDIANA | | 6,483,802 | 300,324.12 | 4,631.91 |
| 24 | KENTUCKY | | 4,339,367 | 199,116.82 | 4,588.61 |
| 25 | WEST VIRGINIA | | 1,852,994 | 83,168.91 | 4,488.35 |
| 26 | SOUTH CAROLINA | | 4,625,364 | 199,062.05 | 4,303.71 |
| 27 | MINNESOTA | | 5,303,925 | 219,092.58 | 4,130.76 |
| 28 | FLORIDA | | 18,801,310 | 755,030.06 | 4,015.84 |
| 29 | WISCONSIN | | 5,686,986 | 226,966.10 | 3,990.97 |
| 30 | VIRGINIA | | 8,001,024 | 318,323.92 | 3,978.54 |
| 31 | ILLINOIS | | 12,830,632 | 475,709.38 | 3,707.61 |
| 32 | OKLAHOMA | | 3,751,351 | 137,476.97 | 3,664.73 |
| 33 | COLORADO | | 5,029,196 | 180,034.13 | 3,579.78 |
| 34 | UTAH | | 2,763,885 | 97,523.41 | 3,528.49 |
| 35 | CALIFORNIA | | 37,253,956 | 1,209,856.28 | 3,247.59 |
| 36 | MONTANA | | 989,415 | 32,019.70 | 3,236.23 |
| 37 | LOUISIANA | | 4,533,372 | 138,294.90 | 3,050.60 |
| 38 | TENNESSEE | | 6,346,105 | 192,959.96 | 3,040.60 |
| 39 | KANSAS | | 2,853,118 | 83,501.09 | 2,926.66 |
| 40 | IDAHO | | 1,567,582 | 41,464.29 | 2,645.11 |
| 41 | OHIO | | 11,536,504 | 293,936.59 | 2,547.88 |
| 42 | MISSOURI | | 5,988,927 | 147,068.11 | 2,455.67 |

| | | | | |
|---|---|---|---|---|
| 43 | NORTH DAKOTA | 672,591 | 15,873.99 | 2,360.13 |
| 44 | PUERTO RICO | 3,725,789 | 86,835.15 | 2,330.65 |
| 45 | TEXAS | 25,145,561 | 518,644.06 | 2,062.57 |
| 46 | HAWAII | 1,360,301 | 27,894.99 | 2,050.65 |
| 47 | ARKANSAS | 2,915,918 | 53,687.52 | 1,841.19 |
| 48 | IOWA | 3,046,355 | 49,162.93 | 1,613.83 |
| 49 | NEBRASKA | 1,826,341 | 28,353.89 | 1,552.50 |
| 50 | MISSISSIPPI | 2,967,297 | 29,343.56 | 988.90 |
| 51 | SOUTH DAKOTA | 814,180 | 7,590.86 | 932.33 |
| 52 | WYOMING | 563,626 | 3,757.46 | 666.66 |
| 53 | VIRGIN ISLANDS | 109,825 | 564.37 | 513.88 |
| 54 | GUAM | 178,430 | 163.23 | 91.48 |
| **9250B - METHADONE - Total** | | **312,825,210** | 13,767,786.63 | **4,401.11** |

**DRUG: 9300 - MORPHINE**

| RANK | STATE NAME | STATE POPULATION | TOTAL GRAMS | TOTAL GRAMS/100K POPULATION |
|---|---|---|---|---|
| 1 | TENNESSEE | 6,346,105 | 516,867.55 | 8,144.64 |
| 2 | DELAWARE | 897,934 | 72,828.05 | 8,110.62 |
| 3 | ARIZONA | 6,392,017 | 487,302.18 | 7,623.61 |
| 4 | OKLAHOMA | 3,751,351 | 279,544.04 | 7,451.82 |
| 5 | FLORIDA | 18,801,310 | 1,366,324.49 | 7,267.18 |
| 6 | NEVADA | 2,700,551 | 186,100.47 | 6,891.20 |
| 7 | IDAHO | 1,567,582 | 97,004.47 | 6,188.16 |
| 8 | OREGON | 3,831,074 | 236,852.29 | 6,182.40 |
| 9 | UTAH | 2,763,885 | 170,710.63 | 6,176.47 |
| 10 | MICHIGAN | 9,883,640 | 597,527.79 | 6,045.62 |
| 11 | PENNSYLVANIA | 12,702,379 | 761,427.79 | 5,994.37 |
| 12 | MONTANA | 989,415 | 59,232.61 | 5,986.63 |
| 13 | VERMONT | 625,741 | 36,736.12 | 5,870.82 |
| 14 | ALASKA | 710,231 | 40,739.77 | 5,736.13 |
| 15 | WASHINGTON | 6,724,540 | 365,578.40 | 5,436.48 |
| 16 | WYOMING | 563,626 | 30,487.23 | 5,409.12 |
| 17 | MISSOURI | 5,988,927 | 320,902.84 | 5,358.27 |
| 18 | KANSAS | 2,853,118 | 151,945.16 | 5,325.58 |
| 19 | HAWAII | 1,360,301 | 70,142.67 | 5,156.41 |
| 20 | COLORADO | 5,029,196 | 255,202.66 | 5,074.42 |
| 21 | ALABAMA | 4,779,736 | 240,316.58 | 5,027.82 |
| 22 | SOUTH CAROLINA | 4,625,364 | 231,992.09 | 5,015.65 |
| 23 | NEW HAMPSHIRE | 1,316,470 | 65,379.49 | 4,966.27 |
| 24 | ARKANSAS | 2,915,918 | 139,816.32 | 4,794.93 |
| 25 | WISCONSIN | 5,686,986 | 256,554.79 | 4,511.26 |
| 26 | MISSISSIPPI | 2,967,297 | 131,749.38 | 4,440.05 |

| | | | | |
|---|---|---|---|---|
| 27 | NORTH CAROLINA | 9,535,483 | 423,216.02 | 4,438.33 |
| 28 | INDIANA | 6,483,802 | 285,682.76 | 4,406.10 |
| 29 | MAINE | 1,328,361 | 56,452.30 | 4,249.77 |
| 30 | MARYLAND | 5,773,552 | 243,565.36 | 4,218.64 |
| 31 | NEW MEXICO | 2,059,179 | 85,968.84 | 4,174.91 |
| 32 | GEORGIA | 9,687,653 | 396,580.59 | 4,093.67 |
| 33 | LOUISIANA | 4,533,372 | 184,759.40 | 4,075.54 |
| 34 | CALIFORNIA | 37,253,956 | 1,480,022.97 | 3,972.79 |
| 35 | WEST VIRGINIA | 1,852,994 | 71,254.72 | 3,845.38 |
| 36 | IOWA | 3,046,355 | 115,481.64 | 3,790.81 |
| 37 | NEBRASKA | 1,826,341 | 67,184.01 | 3,678.61 |
| 38 | RHODE ISLAND | 1,052,567 | 37,939.75 | 3,604.50 |
| 39 | KENTUCKY | 4,339,367 | 155,670.20 | 3,587.39 |
| 40 | MASSACHUSETTS | 6,547,629 | 230,987.59 | 3,527.81 |
| 41 | SOUTH DAKOTA | 814,180 | 28,680.91 | 3,522.67 |
| 42 | CONNECTICUT | 3,574,097 | 123,130.87 | 3,445.09 |
| 43 | OHIO | 11,536,504 | 395,505.12 | 3,428.29 |
| 44 | VIRGINIA | 8,001,024 | 270,434.11 | 3,379.99 |
| 45 | NEW JERSEY | 8,791,894 | 283,232.06 | 3,221.51 |
| 46 | NORTH DAKOTA | 672,591 | 21,527.80 | 3,200.73 |
| 47 | MINNESOTA | 5,303,925 | 154,958.95 | 2,921.59 |
| 48 | ILLINOIS | 12,830,632 | 362,445.64 | 2,824.85 |
| 49 | TEXAS | 25,145,561 | 707,722.35 | 2,814.50 |
| 50 | NEW YORK | 19,378,102 | 531,155.65 | 2,741.01 |
| 51 | DISTRICT OF COLUMBIA | 601,723 | 11,875.70 | 1,973.62 |
| 52 | GUAM | 178,430 | 3,366.70 | 1,886.84 |
| 53 | VIRGIN ISLANDS | 109,825 | 471.76 | 429.56 |
| 54 | PUERTO RICO | 3,725,789 | 6,835.05 | 183.45 |
| 55 | AMERICAN SAMOA | 65,628 | 48.58 | 74.02 |
| **9300 - MORPHINE - Total** | | **312,825,210** | 13,905,423.24 | **4,445.11** |

**DRUG: 9652 - OXYMORPHONE**

| RANK | STATE NAME | STATE POPULATION | TOTAL GRAMS | TOTAL GRAMS/100K POPULATION |
|---|---|---|---|---|
| 1 | TENNESSEE | 6,346,105 | 49,864.82 | 785.75 |
| 2 | DELAWARE | 897,934 | 5,869.75 | 653.70 |
| 3 | MICHIGAN | 9,883,640 | 46,656.74 | 472.06 |
| 4 | NORTH CAROLINA | 9,535,483 | 39,895.46 | 418.39 |
| 5 | MARYLAND | 5,773,552 | 23,443.03 | 406.04 |
| 6 | PENNSYLVANIA | 12,702,379 | 49,609.24 | 390.55 |
| 7 | UTAH | 2,763,885 | 10,680.19 | 386.42 |
| 8 | LOUISIANA | 4,533,372 | 15,606.04 | 344.25 |
| 9 | WEST VIRGINIA | 1,852,994 | 5,161.82 | 278.57 |

| | | | | | |
|---|---|---|---|---|---|
| 10 | VIRGINIA | | 8,001,024 | 22,109.05 | 276.33 |
| 11 | SOUTH CAROLINA | | 4,625,364 | 12,434.89 | 268.84 |
| 12 | NEVADA | | 2,700,551 | 6,485.70 | 240.16 |
| 13 | OKLAHOMA | | 3,751,351 | 8,405.86 | 224.08 |
| 14 | FLORIDA | | 18,801,310 | 40,978.08 | 217.95 |
| 15 | NEW YORK | | 19,378,102 | 41,890.83 | 216.18 |
| 16 | MISSISSIPPI | | 2,967,297 | 5,835.90 | 196.67 |
| 17 | IDAHO | | 1,567,582 | 2,886.52 | 184.14 |
| 18 | ALABAMA | | 4,779,736 | 8,451.22 | 176.81 |
| 19 | KENTUCKY | | 4,339,367 | 7,389.04 | 170.28 |
| 20 | OHIO | | 11,536,504 | 17,986.18 | 155.91 |
| 21 | CONNECTICUT | | 3,574,097 | 5,373.79 | 150.35 |
| 22 | NEW JERSEY | | 8,791,894 | 13,155.09 | 149.63 |
| 23 | SOUTH DAKOTA | | 814,180 | 1,154.07 | 141.75 |
| 24 | ARIZONA | | 6,392,017 | 8,717.81 | 136.39 |
| 25 | INDIANA | | 6,483,802 | 8,771.53 | 135.28 |
| 26 | MAINE | | 1,328,361 | 1,788.17 | 134.61 |
| 27 | NEW HAMPSHIRE | | 1,316,470 | 1,677.33 | 127.41 |
| 28 | GEORGIA | | 9,687,653 | 11,409.65 | 117.78 |
| 29 | NEBRASKA | | 1,826,341 | 2,114.95 | 115.80 |
| 30 | MISSOURI | | 5,988,927 | 6,912.21 | 115.42 |
| 31 | KANSAS | | 2,853,118 | 3,217.72 | 112.78 |
| 32 | COLORADO | | 5,029,196 | 5,597.88 | 111.31 |
| 33 | WYOMING | | 563,626 | 548.28 | 97.28 |
| 34 | OREGON | | 3,831,074 | 3,697.49 | 96.51 |
| 35 | VERMONT | | 625,741 | 557.21 | 89.05 |
| 36 | NEW MEXICO | | 2,059,179 | 1,771.04 | 86.01 |
| 37 | WASHINGTON | | 6,724,540 | 5,724.69 | 85.13 |
| 38 | ALASKA | | 710,231 | 542.93 | 76.44 |
| 39 | TEXAS | | 25,145,561 | 18,237.57 | 72.53 |
| 40 | ARKANSAS | | 2,915,918 | 2,041.35 | 70.01 |
| 41 | RHODE ISLAND | | 1,052,567 | 681.30 | 64.73 |
| 42 | HAWAII | | 1,360,301 | 860.70 | 63.27 |
| 43 | MASSACHUSETTS | | 6,547,629 | 4,123.06 | 62.97 |
| 44 | CALIFORNIA | | 37,253,956 | 23,056.55 | 61.89 |
| 45 | WISCONSIN | | 5,686,986 | 3,340.29 | 58.74 |
| 46 | ILLINOIS | | 12,830,632 | 7,480.57 | 58.30 |
| 47 | IOWA | | 3,046,355 | 1,725.72 | 56.65 |
| 48 | MONTANA | | 989,415 | 518.18 | 52.37 |
| 49 | NORTH DAKOTA | | 672,591 | 301.04 | 44.76 |
| 50 | DISTRICT OF COLUMBIA | | 601,723 | 261.88 | 43.52 |

| | | | | |
|---|---|---|---|---|
| 51 | MINNESOTA | 5,303,925 | 865.43 | 16.32 |
| **9652 - OXYMORPHONE - Total** | | **312,825,210** | **567,865.82** | **181.53** |

**DRUG: 9780 - TAPENTADOL**

| RANK | STATE NAME | STATE POPULATION | TOTAL GRAMS | TOTAL GRAMS/100K POPULATION |
|---|---|---|---|---|
| 1 | NORTH CAROLINA | 9,535,483 | 331,833.50 | 3,479.99 |
| 2 | ALABAMA | 4,779,736 | 164,304.00 | 3,437.51 |
| 3 | SOUTH CAROLINA | 4,625,364 | 141,231.00 | 3,053.40 |
| 4 | DELAWARE | 897,934 | 21,904.50 | 2,439.43 |
| 5 | GEORGIA | 9,687,653 | 205,039.50 | 2,116.50 |
| 6 | NEBRASKA | 1,826,341 | 37,054.50 | 2,028.89 |
| 7 | COLORADO | 5,029,196 | 101,234.50 | 2,012.94 |
| 8 | ARKANSAS | 2,915,918 | 54,757.00 | 1,877.86 |
| 9 | INDIANA | 6,483,802 | 120,317.50 | 1,855.66 |
| 10 | NEW HAMPSHIRE | 1,316,470 | 24,291.50 | 1,845.20 |
| 11 | ARIZONA | 6,392,017 | 117,264.50 | 1,834.55 |
| 12 | SOUTH DAKOTA | 814,180 | 14,868.00 | 1,826.13 |
| 13 | MARYLAND | 5,773,552 | 102,642.50 | 1,777.81 |
| 14 | OKLAHOMA | 3,751,351 | 62,946.00 | 1,677.96 |
| 15 | TENNESSEE | 6,346,105 | 103,470.50 | 1,630.46 |
| 16 | PENNSYLVANIA | 12,702,379 | 204,600.50 | 1,610.73 |
| 17 | OHIO | 11,536,504 | 183,726.55 | 1,592.57 |
| 18 | NEW YORK | 19,378,102 | 307,610.50 | 1,587.41 |
| 19 | VIRGINIA | 8,001,024 | 126,117.00 | 1,576.26 |
| 20 | FLORIDA | 18,801,310 | 291,269.50 | 1,549.20 |
| 21 | NEW JERSEY | 8,791,894 | 134,741.00 | 1,532.56 |
| 22 | UTAH | 2,763,885 | 42,228.00 | 1,527.85 |
| 23 | IDAHO | 1,567,582 | 21,826.50 | 1,392.37 |
| 24 | MISSOURI | 5,988,927 | 81,250.00 | 1,356.67 |
| 25 | NEVADA | 2,700,551 | 34,767.00 | 1,287.40 |
| 26 | IOWA | 3,046,355 | 37,105.50 | 1,218.03 |
| 27 | MISSISSIPPI | 2,967,297 | 35,986.00 | 1,212.75 |
| 28 | TEXAS | 25,145,561 | 301,778.45 | 1,200.13 |
| 29 | LOUISIANA | 4,533,372 | 53,042.50 | 1,170.05 |
| 30 | WYOMING | 563,626 | 6,396.50 | 1,134.88 |
| 31 | ALASKA | 710,231 | 8,010.50 | 1,127.87 |
| 32 | KENTUCKY | 4,339,367 | 47,785.50 | 1,101.21 |
| 33 | NORTH DAKOTA | 672,591 | 7,226.00 | 1,074.35 |
| 34 | MASSACHUSETTS | 6,547,629 | 66,057.50 | 1,008.88 |
| 35 | CONNECTICUT | 3,574,097 | 34,137.50 | 955.14 |
| 36 | KANSAS | 2,853,118 | 25,969.50 | 910.21 |
| 37 | ILLINOIS | 12,830,632 | 113,977.00 | 888.32 |

| | | | | |
|---|---|---|---|---|
| 38 | WEST VIRGINIA | 1,852,994 | 16,373.50 | 883.62 |
| 39 | CALIFORNIA | 37,253,956 | 323,719.23 | 868.95 |
| 40 | WASHINGTON | 6,724,540 | 56,908.50 | 846.28 |
| 41 | WISCONSIN | 5,686,986 | 44,365.65 | 780.13 |
| 42 | RHODE ISLAND | 1,052,567 | 7,568.50 | 719.05 |
| 43 | MICHIGAN | 9,883,640 | 67,530.50 | 683.26 |
| 44 | OREGON | 3,831,074 | 24,794.50 | 647.19 |
| 45 | VERMONT | 625,741 | 3,888.50 | 621.42 |
| 46 | MONTANA | 989,415 | 6,127.50 | 619.31 |
| 47 | HAWAII | 1,360,301 | 8,082.00 | 594.13 |
| 48 | DISTRICT OF COLUMBIA | 601,723 | 3,366.00 | 559.39 |
| 49 | NEW MEXICO | 2,059,179 | 10,204.50 | 495.56 |
| 50 | MAINE | 1,328,361 | 4,696.00 | 353.52 |
| 51 | MINNESOTA | 5,303,925 | 14,950.50 | 281.88 |
| 52 | PUERTO RICO | 3,725,789 | 1,956.00 | 52.50 |
| 53 | VIRGIN ISLANDS | 109,825 | 5.00 | 4.55 |
| **9780 - TAPENTADOL - Total** | | **312,825,210** | 4,363,304.38 | **1,394.81** |

**DRUG: 9801 - FENTANYL BASE**

| RANK | STATE NAME | STATE POPULATION | TOTAL GRAMS | TOTAL GRAMS/100K POPULATION |
|---|---|---|---|---|
| 1 | MISSISSIPPI | 2,967,297 | 3,879.72 | 130.75 |
| 2 | IDAHO | 1,567,582 | 1,983.31 | 126.52 |
| 3 | OKLAHOMA | 3,751,351 | 4,648.89 | 123.93 |
| 4 | KANSAS | 2,853,118 | 3,456.97 | 121.16 |
| 5 | PENNSYLVANIA | 12,702,379 | 15,256.11 | 120.10 |
| 6 | MISSOURI | 5,988,927 | 6,775.65 | 113.14 |
| 7 | DELAWARE | 897,934 | 1,008.02 | 112.26 |
| 8 | SOUTH DAKOTA | 814,180 | 907.99 | 111.52 |
| 9 | NORTH DAKOTA | 672,591 | 739.73 | 109.98 |
| 10 | MONTANA | 989,415 | 1,060.63 | 107.20 |
| 11 | ALASKA | 710,231 | 737.16 | 103.79 |
| 12 | FLORIDA | 18,801,310 | 18,610.14 | 98.98 |
| 13 | UTAH | 2,763,885 | 2,688.99 | 97.29 |
| 14 | NEBRASKA | 1,826,341 | 1,767.93 | 96.80 |
| 15 | IOWA | 3,046,355 | 2,923.85 | 95.98 |
| 16 | ALABAMA | 4,779,736 | 4,573.12 | 95.68 |
| 17 | VERMONT | 625,741 | 597.57 | 95.50 |
| 18 | MICHIGAN | 9,883,640 | 9,405.95 | 95.17 |
| 19 | TENNESSEE | 6,346,105 | 5,976.32 | 94.17 |
| 20 | WEST VIRGINIA | 1,852,994 | 1,710.68 | 92.32 |
| 21 | NORTH CAROLINA | 9,535,483 | 8,701.98 | 91.26 |
| 22 | GEORGIA | 9,687,653 | 8,789.93 | 90.73 |

| | | | | | |
|---|---|---|---|---|---|
| 23 | ARKANSAS | | 2,915,918 | 2,641.19 | 90.58 |
| 24 | INDIANA | | 6,483,802 | 5,844.02 | 90.13 |
| 25 | SOUTH CAROLINA | | 4,625,364 | 4,076.33 | 88.13 |
| 26 | WYOMING | | 563,626 | 490.71 | 87.06 |
| 27 | NEW JERSEY | | 8,791,894 | 7,400.76 | 84.18 |
| 28 | NEW HAMPSHIRE | | 1,316,470 | 1,103.65 | 83.83 |
| 29 | MAINE | | 1,328,361 | 1,101.74 | 82.94 |
| 30 | KENTUCKY | | 4,339,367 | 3,557.68 | 81.99 |
| 31 | CONNECTICUT | | 3,574,097 | 2,930.12 | 81.98 |
| 32 | WISCONSIN | | 5,686,986 | 4,557.74 | 80.14 |
| 33 | LOUISIANA | | 4,533,372 | 3,603.52 | 79.49 |
| 34 | ARIZONA | | 6,392,017 | 5,025.61 | 78.62 |
| 35 | COLORADO | | 5,029,196 | 3,885.98 | 77.27 |
| 36 | OREGON | | 3,831,074 | 2,902.19 | 75.75 |
| 37 | ILLINOIS | | 12,830,632 | 9,515.16 | 74.16 |
| 38 | WASHINGTON | | 6,724,540 | 4,858.73 | 72.25 |
| 39 | TEXAS | | 25,145,561 | 17,988.16 | 71.54 |
| 40 | MARYLAND | | 5,773,552 | 4,070.73 | 70.51 |
| 41 | NEW YORK | | 19,378,102 | 13,402.72 | 69.16 |
| 42 | VIRGINIA | | 8,001,024 | 5,308.84 | 66.35 |
| 43 | NEW MEXICO | | 2,059,179 | 1,350.36 | 65.58 |
| 44 | MASSACHUSETTS | | 6,547,629 | 4,275.14 | 65.29 |
| 45 | OHIO | | 11,536,504 | 7,182.96 | 62.26 |
| 46 | NEVADA | | 2,700,551 | 1,660.62 | 61.49 |
| 47 | MINNESOTA | | 5,303,925 | 3,222.99 | 60.77 |
| 48 | CALIFORNIA | | 37,253,956 | 21,565.83 | 57.89 |
| 49 | RHODE ISLAND | | 1,052,567 | 574.96 | 54.62 |
| 50 | DISTRICT OF COLUMBIA | | 601,723 | 287.34 | 47.75 |
| 51 | HAWAII | | 1,360,301 | 620.30 | 45.60 |
| 52 | GUAM | | 178,430 | 32.77 | 18.37 |
| 53 | VIRGIN ISLANDS | | 109,825 | 17.93 | 16.33 |
| 54 | PUERTO RICO | | 3,725,789 | 488.81 | 13.12 |
| 55 | AMERICAN SAMOA | | 65,628 | 0.37 | 0.56 |
| **9801 - FENTANYL BASE - Total** | | | **312,825,210** | 251,746.63 | **80.48** |

**STATE: AK - ALASKA**

**BUSINESS ACTIVITY: A - PHARMACIES**

| DRUG NAME | DRUG CODE | NUMBER OF REGISTRANT SOLD TO | TOTAL GRAMS | AVG GRAMS |
|---|---|---|---|---|
| AMPHETAMINE | 1100 | 108 | 27,406.71 | 253.77 |
| METHAMPHETAMINE | 1105 | 4 | 7.63 | 1.91 |
| LISDEXAMFETAMINE | 1205 | 100 | 9,553.10 | 95.53 |
| METHYLPHENIDATE (DL;D;L;ISOMERS) | 1724 | 108 | 24,299.27 | 224.99 |
| CODEINE | 9050 | 110 | 17,228.42 | 156.62 |
| BUPRENORPHINE | 9064 | 105 | 9,581.95 | 91.26 |
| OXYCODONE | 9143 | 113 | 108,084.87 | 956.50 |
| HYDROMORPHONE | 9150 | 97 | 2,526.35 | 26.04 |
| HYDROCODONE | 9193 | 111 | 32,667.02 | 294.30 |
| MEPERIDINE (PETHIDINE) | 9230 | 49 | 1,453.99 | 29.67 |
| METHADONE | 9250B | 94 | 12,887.68 | 137.10 |
| MORPHINE | 9300 | 110 | 34,063.89 | 309.67 |
| OXYMORPHONE | 9652 | 22 | 541.15 | 24.60 |
| TAPENTADOL | 9780 | 55 | 7,538.50 | 137.06 |
| FENTANYL BASE | 9801 | 105 | 601.99 | 5.73 |

**BUSINESS ACTIVITY: B - HOSPITALS**

| DRUG NAME | DRUG CODE | NUMBER OF REGISTRANT SOLD TO | TOTAL GRAMS | AVG GRAMS |
|---|---|---|---|---|
| AMPHETAMINE | 1100 | 26 | 6,493.98 | 249.77 |
| LISDEXAMFETAMINE | 1205 | 17 | 1,075.27 | 63.25 |
| METHYLPHENIDATE (DL;D;L;ISOMERS) | 1724 | 31 | 7,694.88 | 248.22 |
| CODEINE | 9050 | 40 | 6,256.74 | 156.42 |
| BUPRENORPHINE | 9064 | 32 | 936.83 | 29.28 |
| OXYCODONE | 9143 | 47 | 11,948.96 | 254.23 |
| HYDROMORPHONE | 9150 | 40 | 545.68 | 13.64 |
| HYDROCODONE | 9193 | 47 | 4,108.74 | 87.42 |
| MEPERIDINE (PETHIDINE) | 9230 | 26 | 285.85 | 10.99 |
| METHADONE | 9250B | 29 | 1,062.73 | 36.65 |
| MORPHINE | 9300 | 49 | 6,627.46 | 135.25 |
| OXYMORPHONE | 9652 | 1 | 1.78 | 1.78 |
| TAPENTADOL | 9780 | 5 | 472.00 | 94.40 |
| FENTANYL BASE | 9801 | 51 | 126.46 | 2.48 |

**BUSINESS ACTIVITY: C - PRACTITIONERS**

| DRUG NAME | DRUG CODE | NUMBER OF REGISTRANT SOLD TO | TOTAL GRAMS | AVG GRAMS |
|---|---|---|---|---|
| CODEINE | 9050 | 9 | 71.32 | 7.92 |
| BUPRENORPHINE | 9064 | 51 | 4.73 | 0.09 |
| OXYCODONE | 9143 | 7 | 56.79 | 8.11 |
| HYDROMORPHONE | 9150 | 44 | 31.31 | 0.71 |
| HYDROCODONE | 9193 | 40 | 132.70 | 3.32 |
| MEPERIDINE (PETHIDINE) | 9230 | 15 | 56.21 | 3.75 |

| | | | | |
|---|---|---|---|---|
| METHADONE | 9250B | 1 | 0.36 | 0.36 |
| MORPHINE | 9300 | 44 | 47.40 | 1.08 |
| FENTANYL BASE | 9801 | 72 | 8.68 | 0.12 |

**BUSINESS ACTIVITY: M - MID-LEVEL PRACTITIONERS**

| DRUG NAME | DRUG CODE | NUMBER OF REGISTRANT SOLD TO | TOTAL GRAMS | AVG GRAMS |
|---|---|---|---|---|
| CODEINE | 9050 | 3 | 13.26 | 4.42 |
| OXYCODONE | 9143 | 1 | 1.34 | 1.34 |
| HYDROCODONE | 9193 | 6 | 12.30 | 2.05 |
| MORPHINE | 9300 | 4 | 1.02 | 0.25 |
| FENTANYL BASE | 9801 | 5 | 0.02 | 0.00 |

**BUSINESS ACTIVITY: NTP - NARCOTIC TREATMENT PROGRAMS**

| DRUG NAME | DRUG CODE | NUMBER OF REGISTRANT SOLD TO | TOTAL GRAMS | AVG GRAMS |
|---|---|---|---|---|
| BUPRENORPHINE | 9064 | 2 | 40.86 | 20.43 |
| METHADONE | 9250B | 4 | 19,032.83 | 4,758.21 |

**STATE: AL - ALABAMA**

**BUSINESS ACTIVITY: A - PHARMACIES**

| DRUG NAME | DRUG CODE | NUMBER OF REGISTRANT SOLD TO | TOTAL GRAMS | AVG GRAMS |
|---|---|---|---|---|
| AMPHETAMINE | 1100 | 1,345 | 535,561.66 | 398.19 |
| METHAMPHETAMINE | 1105 | 33 | 135.81 | 4.12 |
| LISDEXAMFETAMINE | 1205 | 1,317 | 280,699.35 | 213.14 |
| METHYLPHENIDATE (DL;D;L;ISOMERS) | 1724 | 1,332 | 273,932.72 | 205.66 |
| CODEINE | 9050 | 1,326 | 195,634.76 | 147.54 |
| BUPRENORPHINE | 9064 | 1,273 | 113,904.52 | 89.48 |
| OXYCODONE | 9143 | 1,352 | 762,943.93 | 564.31 |
| HYDROMORPHONE | 9150 | 1,041 | 8,368.91 | 8.04 |
| HYDROCODONE | 9193 | 1,368 | 815,499.10 | 596.13 |
| MEPERIDINE (PETHIDINE) | 9230 | 767 | 13,421.54 | 17.50 |
| METHADONE | 9250B | 1,105 | 56,126.37 | 50.79 |
| MORPHINE | 9300 | 1,314 | 219,148.35 | 166.78 |
| OXYMORPHONE | 9652 | 551 | 8,425.80 | 15.29 |
| TAPENTADOL | 9780 | 1,030 | 163,382.50 | 158.62 |
| FENTANYL BASE | 9801 | 1,279 | 4,143.90 | 3.24 |

**BUSINESS ACTIVITY: B - HOSPITALS**

| DRUG NAME | DRUG CODE | NUMBER OF REGISTRANT SOLD TO | TOTAL GRAMS | AVG GRAMS |
|---|---|---|---|---|
| AMPHETAMINE | 1100 | 72 | 9,039.10 | 125.54 |
| LISDEXAMFETAMINE | 1205 | 29 | 3,126.36 | 107.81 |
| METHYLPHENIDATE (DL;D;L;ISOMERS) | 1724 | 85 | 7,151.88 | 84.14 |
| CODEINE | 9050 | 119 | 4,432.93 | 37.25 |
| BUPRENORPHINE | 9064 | 104 | 1,328.62 | 12.78 |
| OXYCODONE | 9143 | 148 | 31,567.10 | 213.29 |

| | | | | |
|---|---|---|---|---|
| HYDROMORPHONE | 9150 | 137 | 2,070.27 | 15.11 |
| HYDROCODONE | 9193 | 150 | 37,594.01 | 250.63 |
| MEPERIDINE (PETHIDINE) | 9230 | 121 | 10,171.58 | 84.06 |
| METHADONE | 9250B | 114 | 2,855.82 | 25.05 |
| MORPHINE | 9300 | 148 | 20,769.91 | 140.34 |
| OXYMORPHONE | 9652 | 6 | 25.42 | 4.24 |
| TAPENTADOL | 9780 | 30 | 921.50 | 30.72 |
| FENTANYL BASE | 9801 | 133 | 399.07 | 3.00 |

**BUSINESS ACTIVITY: C - PRACTITIONERS**

| DRUG NAME | DRUG CODE | NUMBER OF REGISTRANT SOLD TO | TOTAL GRAMS | AVG GRAMS |
|---|---|---|---|---|
| AMPHETAMINE | 1100 | 1 | 2.25 | 2.25 |
| LISDEXAMFETAMINE | 1205 | 1 | 23.88 | 23.88 |
| METHYLPHENIDATE (DL;D;L;ISOMERS) | 1724 | 1 | 20.49 | 20.49 |
| CODEINE | 9050 | 90 | 1,535.19 | 17.06 |
| BUPRENORPHINE | 9064 | 303 | 49.09 | 0.16 |
| OXYCODONE | 9143 | 34 | 458.63 | 13.49 |
| HYDROMORPHONE | 9150 | 87 | 101.61 | 1.17 |
| HYDROCODONE | 9193 | 136 | 896.27 | 6.59 |
| MEPERIDINE (PETHIDINE) | 9230 | 107 | 1,632.21 | 15.25 |
| METHADONE | 9250B | 1 | 0.36 | 0.36 |
| MORPHINE | 9300 | 110 | 398.31 | 3.62 |
| FENTANYL BASE | 9801 | 252 | 29.95 | 0.12 |

**BUSINESS ACTIVITY: D - TEACHING INSTITUTIONS**

| DRUG NAME | DRUG CODE | NUMBER OF REGISTRANT SOLD TO | TOTAL GRAMS | AVG GRAMS |
|---|---|---|---|---|
| BUPRENORPHINE | 9064 | 1 | 0.02 | 0.02 |
| HYDROMORPHONE | 9150 | 1 | 0.71 | 0.71 |
| HYDROCODONE | 9193 | 1 | 0.29 | 0.29 |
| FENTANYL BASE | 9801 | 1 | 0.20 | 0.20 |

**BUSINESS ACTIVITY: M - MID-LEVEL PRACTITIONERS**

| DRUG NAME | DRUG CODE | NUMBER OF REGISTRANT SOLD TO | TOTAL GRAMS | AVG GRAMS |
|---|---|---|---|---|
| CODEINE | 9050 | 2 | 2.17 | 1.08 |
| BUPRENORPHINE | 9064 | 1 | 2.64 | 2.64 |

**BUSINESS ACTIVITY: NTP - NARCOTIC TREATMENT PROGRAMS**

| DRUG NAME | DRUG CODE | NUMBER OF REGISTRANT SOLD TO | TOTAL GRAMS | AVG GRAMS |
|---|---|---|---|---|
| BUPRENORPHINE | 9064 | 10 | 363.25 | 36.32 |
| METHADONE | 9250B | 26 | 227,442.79 | 8,747.80 |

**STATE: AR - ARKANSAS**

**BUSINESS ACTIVITY: A - PHARMACIES**

| DRUG NAME | DRUG CODE | NUMBER OF REGISTRANT SOLD TO | TOTAL GRAMS | AVG GRAMS |
|---|---|---|---|---|
| AMPHETAMINE | 1100 | 749 | 205,172.85 | 273.93 |

| | | | | |
|---|---|---|---|---|
| METHAMPHETAMINE | 1105 | 13 | 28.13 | 2.16 |
| LISDEXAMFETAMINE | 1205 | 743 | 116,847.31 | 157.26 |
| METHYLPHENIDATE (DL;D;L;ISOMERS) | 1724 | 752 | 148,140.10 | 196.99 |
| CODEINE | 9050 | 756 | 237,197.64 | 313.75 |
| BUPRENORPHINE | 9064 | 703 | 27,928.74 | 39.73 |
| OXYCODONE | 9143 | 761 | 439,716.52 | 577.81 |
| HYDROMORPHONE | 9150 | 709 | 8,329.52 | 11.75 |
| HYDROCODONE | 9193 | 763 | 436,803.17 | 572.48 |
| MEPERIDINE (PETHIDINE) | 9230 | 607 | 16,907.75 | 27.85 |
| METHADONE | 9250B | 667 | 22,498.01 | 33.73 |
| MORPHINE | 9300 | 758 | 128,188.55 | 169.11 |
| OXYMORPHONE | 9652 | 245 | 2,011.91 | 8.21 |
| TAPENTADOL | 9780 | 542 | 53,559.50 | 98.82 |
| FENTANYL BASE | 9801 | 736 | 2,312.70 | 3.14 |

**BUSINESS ACTIVITY: B - HOSPITALS**

| DRUG NAME | DRUG CODE | NUMBER OF REGISTRANT SOLD TO | TOTAL GRAMS | AVG GRAMS |
|---|---|---|---|---|
| AMPHETAMINE | 1100 | 47 | 4,096.50 | 87.16 |
| LISDEXAMFETAMINE | 1205 | 25 | 1,058.50 | 42.34 |
| METHYLPHENIDATE (DL;D;L;ISOMERS) | 1724 | 74 | 4,032.02 | 54.49 |
| CODEINE | 9050 | 89 | 4,039.28 | 45.39 |
| BUPRENORPHINE | 9064 | 47 | 1,122.05 | 23.87 |
| OXYCODONE | 9143 | 124 | 19,178.89 | 154.67 |
| HYDROMORPHONE | 9150 | 108 | 1,644.70 | 15.23 |
| HYDROCODONE | 9193 | 124 | 18,138.55 | 146.28 |
| MEPERIDINE (PETHIDINE) | 9230 | 99 | 5,184.92 | 52.37 |
| METHADONE | 9250B | 89 | 1,935.21 | 21.74 |
| MORPHINE | 9300 | 124 | 11,560.21 | 93.23 |
| OXYMORPHONE | 9652 | 5 | 29.44 | 5.89 |
| TAPENTADOL | 9780 | 21 | 1,197.50 | 57.02 |
| FENTANYL BASE | 9801 | 123 | 315.60 | 2.57 |

**BUSINESS ACTIVITY: C - PRACTITIONERS**

| DRUG NAME | DRUG CODE | NUMBER OF REGISTRANT SOLD TO | TOTAL GRAMS | AVG GRAMS |
|---|---|---|---|---|
| AMPHETAMINE | 1100 | 1 | 3.27 | 3.27 |
| LISDEXAMFETAMINE | 1205 | 1 | 47.23 | 47.23 |
| METHYLPHENIDATE (DL;D;L;ISOMERS) | 1724 | 1 | 48.77 | 48.77 |
| CODEINE | 9050 | 25 | 510.01 | 20.40 |
| BUPRENORPHINE | 9064 | 163 | 11.22 | 0.07 |
| OXYCODONE | 9143 | 8 | 28.24 | 3.53 |
| HYDROMORPHONE | 9150 | 55 | 23.28 | 0.42 |
| HYDROCODONE | 9193 | 83 | 184.34 | 2.22 |

| | | | | |
|---|---|---|---|---|
| MEPERIDINE (PETHIDINE) | 9230 | 35 | 552.10 | 15.77 |
| METHADONE | 9250B | 1 | 3.85 | 3.85 |
| MORPHINE | 9300 | 76 | 67.55 | 0.89 |
| FENTANYL BASE | 9801 | 152 | 12.88 | 0.08 |

**BUSINESS ACTIVITY: D - TEACHING INSTITUTIONS**

| DRUG NAME | DRUG CODE | NUMBER OF REGISTRANT SOLD TO | TOTAL GRAMS | AVG GRAMS |
|---|---|---|---|---|
| BUPRENORPHINE | 9064 | 1 | 0.00 | 0.00 |

**BUSINESS ACTIVITY: M - MID-LEVEL PRACTITIONERS**

| DRUG NAME | DRUG CODE | NUMBER OF REGISTRANT SOLD TO | TOTAL GRAMS | AVG GRAMS |
|---|---|---|---|---|
| CODEINE | 9050 | 1 | 0.87 | 0.87 |

**BUSINESS ACTIVITY: NTP - NARCOTIC TREATMENT PROGRAMS**

| DRUG NAME | DRUG CODE | NUMBER OF REGISTRANT SOLD TO | TOTAL GRAMS | AVG GRAMS |
|---|---|---|---|---|
| BUPRENORPHINE | 9064 | 1 | 12.24 | 12.24 |
| METHADONE | 9250B | 6 | 29,250.46 | 4,875.08 |

**STATE: AS - AMERICAN SAMOA**

**BUSINESS ACTIVITY: B - HOSPITALS**

| DRUG NAME | DRUG CODE | NUMBER OF REGISTRANT SOLD TO | TOTAL GRAMS | AVG GRAMS |
|---|---|---|---|---|
| AMPHETAMINE | 1100 | 1 | 2.50 | 2.50 |
| METHYLPHENIDATE (DL;D;L;ISOMERS) | 1724 | 1 | 44.10 | 44.10 |
| CODEINE | 9050 | 1 | 1,060.85 | 1,060.85 |
| OXYCODONE | 9143 | 1 | 36.76 | 36.76 |
| HYDROMORPHONE | 9150 | 1 | 1.20 | 1.20 |
| HYDROCODONE | 9193 | 1 | 3.63 | 3.63 |
| MORPHINE | 9300 | 1 | 48.58 | 48.58 |
| FENTANYL BASE | 9801 | 1 | 0.37 | 0.37 |

**BUSINESS ACTIVITY: C - PRACTITIONERS**

| DRUG NAME | DRUG CODE | NUMBER OF REGISTRANT SOLD TO | TOTAL GRAMS | AVG GRAMS |
|---|---|---|---|---|
| BUPRENORPHINE | 9064 | 1 | 0.04 | 0.04 |

**STATE: AZ - ARIZONA**

**BUSINESS ACTIVITY: A - PHARMACIES**

| DRUG NAME | DRUG CODE | NUMBER OF REGISTRANT SOLD TO | TOTAL GRAMS | AVG GRAMS |
|---|---|---|---|---|
| AMPHETAMINE | 1100 | 1,090 | 368,078.69 | 337.69 |
| METHAMPHETAMINE | 1105 | 32 | 180.01 | 5.63 |
| LISDEXAMFETAMINE | 1205 | 1,025 | 167,313.70 | 163.23 |
| METHYLPHENIDATE (DL;D;L;ISOMERS) | 1724 | 1,094 | 316,426.47 | 289.24 |
| CODEINE | 9050 | 1,104 | 219,297.31 | 198.64 |
| BUPRENORPHINE | 9064 | 1,067 | 40,934.18 | 38.36 |
| OXYCODONE | 9143 | 1,114 | 1,325,818.08 | 1,190.14 |
| HYDROMORPHONE | 9150 | 1,012 | 19,539.73 | 19.31 |
| HYDROCODONE | 9193 | 1,114 | 331,368.84 | 297.46 |

| | | | | |
|---|---|---|---|---|
| MEPERIDINE (PETHIDINE) | 9230 | 487 | 10,518.46 | 21.60 |
| METHADONE | 9250B | 909 | 36,901.78 | 40.60 |
| MORPHINE | 9300 | 1,102 | 446,500.14 | 405.17 |
| OXYMORPHONE | 9652 | 536 | 8,696.15 | 16.22 |
| TAPENTADOL | 9780 | 768 | 116,667.00 | 151.91 |
| FENTANYL BASE | 9801 | 1,019 | 4,479.19 | 4.40 |

**BUSINESS ACTIVITY: B - HOSPITALS**

| DRUG NAME | DRUG CODE | NUMBER OF REGISTRANT SOLD TO | TOTAL GRAMS | AVG GRAMS |
|---|---|---|---|---|
| AMPHETAMINE | 1100 | 95 | 8,297.02 | 87.34 |
| LISDEXAMFETAMINE | 1205 | 43 | 1,712.91 | 39.84 |
| METHYLPHENIDATE (DL;D;L;ISOMERS) | 1724 | 111 | 14,592.93 | 131.47 |
| CODEINE | 9050 | 126 | 15,054.29 | 119.48 |
| BUPRENORPHINE | 9064 | 76 | 3,143.45 | 41.36 |
| OXYCODONE | 9143 | 157 | 56,284.54 | 358.50 |
| HYDROMORPHONE | 9150 | 123 | 2,410.56 | 19.60 |
| HYDROCODONE | 9193 | 150 | 16,314.99 | 108.77 |
| MEPERIDINE (PETHIDINE) | 9230 | 87 | 4,336.80 | 49.85 |
| METHADONE | 9250B | 110 | 3,862.76 | 35.12 |
| MORPHINE | 9300 | 153 | 40,407.75 | 264.10 |
| OXYMORPHONE | 9652 | 8 | 21.41 | 2.68 |
| TAPENTADOL | 9780 | 10 | 597.50 | 59.75 |
| FENTANYL BASE | 9801 | 130 | 491.53 | 3.78 |

**BUSINESS ACTIVITY: C - PRACTITIONERS**

| DRUG NAME | DRUG CODE | NUMBER OF REGISTRANT SOLD TO | TOTAL GRAMS | AVG GRAMS |
|---|---|---|---|---|
| CODEINE | 9050 | 105 | 2,734.13 | 26.04 |
| BUPRENORPHINE | 9064 | 660 | 84.32 | 0.13 |
| OXYCODONE | 9143 | 109 | 254.79 | 2.34 |
| HYDROMORPHONE | 9150 | 360 | 268.71 | 0.75 |
| HYDROCODONE | 9193 | 326 | 1,025.04 | 3.14 |
| MEPERIDINE (PETHIDINE) | 9230 | 262 | 3,507.72 | 13.39 |
| METHADONE | 9250B | 22 | 135.14 | 6.14 |
| MORPHINE | 9300 | 334 | 394.09 | 1.18 |
| OXYMORPHONE | 9652 | 1 | 0.25 | 0.25 |
| FENTANYL BASE | 9801 | 740 | 54.89 | 0.07 |

**BUSINESS ACTIVITY: M - MID-LEVEL PRACTITIONERS**

| DRUG NAME | DRUG CODE | NUMBER OF REGISTRANT SOLD TO | TOTAL GRAMS | AVG GRAMS |
|---|---|---|---|---|
| CODEINE | 9050 | 25 | 196.74 | 7.87 |
| BUPRENORPHINE | 9064 | 3 | 8.16 | 2.72 |
| OXYCODONE | 9143 | 1 | 0.45 | 0.45 |
| HYDROMORPHONE | 9150 | 1 | 0.02 | 0.02 |

| | | | | |
|---|---|---|---|---|
| HYDROCODONE | 9193 | 1 | 0.88 | 0.88 |
| MORPHINE | 9300 | 1 | 0.19 | 0.19 |
| FENTANYL BASE | 9801 | 2 | 0.01 | 0.00 |

**BUSINESS ACTIVITY: NTP - NARCOTIC TREATMENT PROGRAMS**

| DRUG NAME | DRUG CODE | NUMBER OF REGISTRANT SOLD TO | TOTAL GRAMS | AVG GRAMS |
|---|---|---|---|---|
| BUPRENORPHINE | 9064 | 17 | 876.61 | 51.57 |
| METHADONE | 9250B | 48 | 312,091.49 | 6,501.91 |

**STATE: CA - CALIFORNIA**

**BUSINESS ACTIVITY: A - PHARMACIES**

| DRUG NAME | DRUG CODE | NUMBER OF REGISTRANT SOLD TO | TOTAL GRAMS | AVG GRAMS |
|---|---|---|---|---|
| AMPHETAMINE | 1100 | 5,469 | 1,182,729.85 | 216.26 |
| METHAMPHETAMINE | 1105 | 269 | 1,059.86 | 3.94 |
| LISDEXAMFETAMINE | 1205 | 4,817 | 444,384.31 | 92.25 |
| METHYLPHENIDATE (DL;D;L;ISOMERS) | 1724 | 5,360 | 998,655.09 | 186.32 |
| CODEINE | 9050 | 6,121 | 1,516,865.84 | 247.81 |
| BUPRENORPHINE | 9064 | 5,174 | 163,041.89 | 31.51 |
| OXYCODONE | 9143 | 5,782 | 2,737,503.11 | 473.45 |
| HYDROMORPHONE | 9150 | 5,014 | 103,481.09 | 20.64 |
| HYDROCODONE | 9193 | 6,055 | 2,965,542.79 | 489.77 |
| MEPERIDINE (PETHIDINE) | 9230 | 761 | 21,674.21 | 28.48 |
| METHADONE | 9250B | 4,550 | 269,052.25 | 59.13 |
| MORPHINE | 9300 | 5,415 | 1,336,574.16 | 246.83 |
| OXYMORPHONE | 9652 | 1,207 | 22,898.29 | 18.97 |
| TAPENTADOL | 9780 | 2,902 | 321,217.50 | 110.69 |
| FENTANYL BASE | 9801 | 4,842 | 18,484.00 | 3.82 |

**BUSINESS ACTIVITY: B - HOSPITALS**

| DRUG NAME | DRUG CODE | NUMBER OF REGISTRANT SOLD TO | TOTAL GRAMS | AVG GRAMS |
|---|---|---|---|---|
| AMPHETAMINE | 1100 | 254 | 44,561.59 | 175.44 |
| METHAMPHETAMINE | 1105 | 2 | 20.09 | 10.04 |
| LISDEXAMFETAMINE | 1205 | 93 | 7,772.55 | 83.58 |
| METHYLPHENIDATE (DL;D;L;ISOMERS) | 1724 | 402 | 43,561.96 | 108.36 |
| CODEINE | 9050 | 567 | 66,045.99 | 116.48 |
| BUPRENORPHINE | 9064 | 372 | 6,471.77 | 17.40 |
| OXYCODONE | 9143 | 649 | 146,121.10 | 225.15 |
| HYDROMORPHONE | 9150 | 664 | 21,144.53 | 31.84 |
| HYDROCODONE | 9193 | 735 | 157,416.36 | 214.17 |
| MEPERIDINE (PETHIDINE) | 9230 | 597 | 30,231.14 | 50.64 |
| METHADONE | 9250B | 527 | 38,549.59 | 73.15 |
| MORPHINE | 9300 | 764 | 142,105.66 | 186.00 |
| OXYMORPHONE | 9652 | 29 | 158.26 | 5.46 |

| | | | | |
|---|---|---|---|---|
| TAPENTADOL | 9780 | 66 | 2,501.73 | 37.90 |
| FENTANYL BASE | 9801 | 839 | 2,536.90 | 3.02 |

**BUSINESS ACTIVITY: C - PRACTITIONERS**

| DRUG NAME | DRUG CODE | NUMBER OF REGISTRANT SOLD TO | TOTAL GRAMS | AVG GRAMS |
|---|---|---|---|---|
| AMPHETAMINE | 1100 | 16 | 911.30 | 56.96 |
| LISDEXAMFETAMINE | 1205 | 7 | 98.70 | 14.10 |
| METHYLPHENIDATE (DL;D;L;ISOMERS) | 1724 | 3 | 78.52 | 26.17 |
| CODEINE | 9050 | 698 | 21,685.14 | 31.07 |
| BUPRENORPHINE | 9064 | 3,047 | 1,108.07 | 0.36 |
| OXYCODONE | 9143 | 223 | 616.87 | 2.77 |
| HYDROMORPHONE | 9150 | 1,509 | 1,304.96 | 0.86 |
| HYDROCODONE | 9193 | 2,084 | 10,911.64 | 5.24 |
| MEPERIDINE (PETHIDINE) | 9230 | 787 | 10,135.11 | 12.88 |
| METHADONE | 9250B | 152 | 304.81 | 2.01 |
| MORPHINE | 9300 | 979 | 1,343.15 | 1.37 |
| FENTANYL BASE | 9801 | 2,567 | 544.94 | 0.21 |

**BUSINESS ACTIVITY: D - TEACHING INSTITUTIONS**

| DRUG NAME | DRUG CODE | NUMBER OF REGISTRANT SOLD TO | TOTAL GRAMS | AVG GRAMS |
|---|---|---|---|---|
| BUPRENORPHINE | 9064 | 6 | 0.29 | 0.05 |
| HYDROMORPHONE | 9150 | 3 | 0.43 | 0.14 |

**BUSINESS ACTIVITY: M - MID-LEVEL PRACTITIONERS**

| DRUG NAME | DRUG CODE | NUMBER OF REGISTRANT SOLD TO | TOTAL GRAMS | AVG GRAMS |
|---|---|---|---|---|
| CODEINE | 9050 | 49 | 560.52 | 11.44 |
| BUPRENORPHINE | 9064 | 4 | 2.51 | 0.63 |

**BUSINESS ACTIVITY: NTP - NARCOTIC TREATMENT PROGRAMS**

| DRUG NAME | DRUG CODE | NUMBER OF REGISTRANT SOLD TO | TOTAL GRAMS | AVG GRAMS |
|---|---|---|---|---|
| BUPRENORPHINE | 9064 | 92 | 1,375.44 | 14.95 |
| METHADONE | 9250B | 151 | 901,949.63 | 5,973.18 |

**STATE: CO - COLORADO**

**BUSINESS ACTIVITY: A - PHARMACIES**

| DRUG NAME | DRUG CODE | NUMBER OF REGISTRANT SOLD TO | TOTAL GRAMS | AVG GRAMS |
|---|---|---|---|---|
| AMPHETAMINE | 1100 | 795 | 297,114.98 | 373.73 |
| METHAMPHETAMINE | 1105 | 81 | 214.98 | 2.65 |
| LISDEXAMFETAMINE | 1205 | 760 | 109,131.98 | 143.59 |
| METHYLPHENIDATE (DL;D;L;ISOMERS) | 1724 | 792 | 202,215.90 | 255.32 |
| CODEINE | 9050 | 788 | 70,978.48 | 90.07 |
| BUPRENORPHINE | 9064 | 777 | 30,271.97 | 38.96 |
| OXYCODONE | 9143 | 802 | 650,037.29 | 810.52 |
| HYDROMORPHONE | 9150 | 785 | 17,980.93 | 22.91 |
| HYDROCODONE | 9193 | 805 | 185,358.58 | 230.26 |

| | | | | |
|---|---|---|---|---|
| MEPERIDINE (PETHIDINE) | 9230 | 209 | 4,238.76 | 20.28 |
| METHADONE | 9250B | 663 | 24,732.57 | 37.30 |
| MORPHINE | 9300 | 799 | 204,367.81 | 255.78 |
| OXYMORPHONE | 9652 | 381 | 5,363.26 | 14.08 |
| TAPENTADOL | 9780 | 626 | 96,773.00 | 154.59 |
| FENTANYL BASE | 9801 | 760 | 3,069.42 | 4.04 |

**BUSINESS ACTIVITY: B - HOSPITALS**

| DRUG NAME | DRUG CODE | NUMBER OF REGISTRANT SOLD TO | TOTAL GRAMS | AVG GRAMS |
|---|---|---|---|---|
| AMPHETAMINE | 1100 | 122 | 47,835.16 | 392.09 |
| METHAMPHETAMINE | 1105 | 6 | 20.49 | 3.42 |
| LISDEXAMFETAMINE | 1205 | 74 | 7,371.93 | 99.62 |
| METHYLPHENIDATE (DL;D;L;ISOMERS) | 1724 | 134 | 42,777.08 | 319.23 |
| CODEINE | 9050 | 153 | 12,893.25 | 84.27 |
| BUPRENORPHINE | 9064 | 117 | 4,116.08 | 35.18 |
| OXYCODONE | 9143 | 211 | 109,030.95 | 516.73 |
| HYDROMORPHONE | 9150 | 201 | 5,232.09 | 26.03 |
| HYDROCODONE | 9193 | 224 | 31,360.24 | 140.00 |
| MEPERIDINE (PETHIDINE) | 9230 | 94 | 1,181.65 | 12.57 |
| METHADONE | 9250B | 135 | 7,027.03 | 52.05 |
| MORPHINE | 9300 | 211 | 50,388.25 | 238.81 |
| OXYMORPHONE | 9652 | 31 | 234.62 | 7.57 |
| TAPENTADOL | 9780 | 60 | 4,461.50 | 74.36 |
| FENTANYL BASE | 9801 | 212 | 744.03 | 3.51 |

**BUSINESS ACTIVITY: C - PRACTITIONERS**

| DRUG NAME | DRUG CODE | NUMBER OF REGISTRANT SOLD TO | TOTAL GRAMS | AVG GRAMS |
|---|---|---|---|---|
| AMPHETAMINE | 1100 | 1 | 1.25 | 1.25 |
| METHYLPHENIDATE (DL;D;L;ISOMERS) | 1724 | 1 | 31.65 | 31.65 |
| CODEINE | 9050 | 58 | 849.51 | 14.65 |
| BUPRENORPHINE | 9064 | 797 | 188.56 | 0.24 |
| OXYCODONE | 9143 | 116 | 647.57 | 5.58 |
| HYDROMORPHONE | 9150 | 493 | 364.72 | 0.74 |
| HYDROCODONE | 9193 | 339 | 930.68 | 2.75 |
| MEPERIDINE (PETHIDINE) | 9230 | 92 | 269.51 | 2.93 |
| METHADONE | 9250B | 36 | 54.56 | 1.52 |
| MORPHINE | 9300 | 315 | 433.41 | 1.38 |
| FENTANYL BASE | 9801 | 591 | 70.89 | 0.12 |

**BUSINESS ACTIVITY: M - MID-LEVEL PRACTITIONERS**

| DRUG NAME | DRUG CODE | NUMBER OF REGISTRANT SOLD TO | TOTAL GRAMS | AVG GRAMS |
|---|---|---|---|---|
| CODEINE | 9050 | 13 | 10.83 | 0.83 |
| BUPRENORPHINE | 9064 | 2 | 1.66 | 0.83 |

| | | | | |
|---|---|---|---|---|
| OXYCODONE | 9143 | 1 | 1.79 | 1.79 |
| HYDROMORPHONE | 9150 | 6 | 0.34 | 0.06 |
| HYDROCODONE | 9193 | 1 | 0.61 | 0.61 |
| MORPHINE | 9300 | 27 | 13.19 | 0.49 |
| FENTANYL BASE | 9801 | 49 | 1.64 | 0.03 |

**BUSINESS ACTIVITY: NTP - NARCOTIC TREATMENT PROGRAMS**

| DRUG NAME | DRUG CODE | NUMBER OF REGISTRANT SOLD TO | TOTAL GRAMS | AVG GRAMS |
|---|---|---|---|---|
| BUPRENORPHINE | 9064 | 17 | 624.99 | 36.76 |
| METHADONE | 9250B | 24 | 148,219.97 | 6,175.83 |

**STATE: CT - CONNECTICUT**

**BUSINESS ACTIVITY: A - PHARMACIES**

| DRUG NAME | DRUG CODE | NUMBER OF REGISTRANT SOLD TO | TOTAL GRAMS | AVG GRAMS |
|---|---|---|---|---|
| AMPHETAMINE | 1100 | 700 | 222,506.94 | 317.87 |
| METHAMPHETAMINE | 1105 | 29 | 83.58 | 2.88 |
| LISDEXAMFETAMINE | 1205 | 689 | 143,049.69 | 207.62 |
| METHYLPHENIDATE (DL;D;L;ISOMERS) | 1724 | 701 | 213,683.13 | 304.83 |
| CODEINE | 9050 | 696 | 76,056.84 | 109.28 |
| BUPRENORPHINE | 9064 | 694 | 62,488.09 | 90.04 |
| OXYCODONE | 9143 | 705 | 550,665.71 | 781.09 |
| HYDROMORPHONE | 9150 | 681 | 17,245.39 | 25.32 |
| HYDROCODONE | 9193 | 701 | 65,368.17 | 93.25 |
| MEPERIDINE (PETHIDINE) | 9230 | 111 | 3,160.82 | 28.48 |
| METHADONE | 9250B | 528 | 18,637.24 | 35.30 |
| MORPHINE | 9300 | 675 | 112,246.76 | 166.29 |
| OXYMORPHONE | 9652 | 286 | 5,355.37 | 18.73 |
| TAPENTADOL | 9780 | 408 | 33,659.00 | 82.50 |
| FENTANYL BASE | 9801 | 643 | 2,626.65 | 4.08 |

**BUSINESS ACTIVITY: B - HOSPITALS**

| DRUG NAME | DRUG CODE | NUMBER OF REGISTRANT SOLD TO | TOTAL GRAMS | AVG GRAMS |
|---|---|---|---|---|
| AMPHETAMINE | 1100 | 39 | 4,707.26 | 120.70 |
| METHAMPHETAMINE | 1105 | 1 | 12.45 | 12.45 |
| LISDEXAMFETAMINE | 1205 | 21 | 1,753.38 | 83.49 |
| METHYLPHENIDATE (DL;D;L;ISOMERS) | 1724 | 53 | 4,436.98 | 83.72 |
| CODEINE | 9050 | 60 | 2,217.40 | 36.96 |
| BUPRENORPHINE | 9064 | 62 | 2,495.09 | 40.24 |
| OXYCODONE | 9143 | 74 | 17,567.10 | 237.39 |
| HYDROMORPHONE | 9150 | 63 | 2,974.48 | 47.21 |
| HYDROCODONE | 9193 | 56 | 1,706.00 | 30.46 |
| MEPERIDINE (PETHIDINE) | 9230 | 41 | 348.82 | 8.51 |
| METHADONE | 9250B | 51 | 3,901.91 | 76.51 |

| | | | | |
|---|---|---|---|---|
| MORPHINE | 9300 | 64 | 10,626.32 | 166.04 |
| OXYMORPHONE | 9652 | 5 | 18.42 | 3.68 |
| TAPENTADOL | 9780 | 6 | 478.50 | 79.75 |
| FENTANYL BASE | 9801 | 67 | 271.62 | 4.05 |

**BUSINESS ACTIVITY: C - PRACTITIONERS**

| DRUG NAME | DRUG CODE | NUMBER OF REGISTRANT SOLD TO | TOTAL GRAMS | AVG GRAMS |
|---|---|---|---|---|
| AMPHETAMINE | 1100 | 1 | 3.17 | 3.17 |
| CODEINE | 9050 | 45 | 203.82 | 4.53 |
| BUPRENORPHINE | 9064 | 344 | 25.76 | 0.07 |
| OXYCODONE | 9143 | 52 | 711.87 | 13.69 |
| HYDROMORPHONE | 9150 | 168 | 135.60 | 0.81 |
| HYDROCODONE | 9193 | 101 | 369.87 | 3.66 |
| MEPERIDINE (PETHIDINE) | 9230 | 25 | 185.19 | 7.41 |
| METHADONE | 9250B | 19 | 87.29 | 4.59 |
| MORPHINE | 9300 | 118 | 257.79 | 2.18 |
| FENTANYL BASE | 9801 | 235 | 31.78 | 0.14 |

**BUSINESS ACTIVITY: M - MID-LEVEL PRACTITIONERS**

| DRUG NAME | DRUG CODE | NUMBER OF REGISTRANT SOLD TO | TOTAL GRAMS | AVG GRAMS |
|---|---|---|---|---|
| CODEINE | 9050 | 4 | 2.17 | 0.54 |
| FENTANYL BASE | 9801 | 1 | 0.08 | 0.08 |

**BUSINESS ACTIVITY: NTP - NARCOTIC TREATMENT PROGRAMS**

| DRUG NAME | DRUG CODE | NUMBER OF REGISTRANT SOLD TO | TOTAL GRAMS | AVG GRAMS |
|---|---|---|---|---|
| BUPRENORPHINE | 9064 | 4 | 79.22 | 19.81 |
| METHADONE | 9250B | 42 | 394,317.71 | 9,388.52 |

**STATE: DC - DISTRICT OF COLUMBIA**

**BUSINESS ACTIVITY: A - PHARMACIES**

| DRUG NAME | DRUG CODE | NUMBER OF REGISTRANT SOLD TO | TOTAL GRAMS | AVG GRAMS |
|---|---|---|---|---|
| AMPHETAMINE | 1100 | 148 | 46,912.83 | 316.98 |
| METHAMPHETAMINE | 1105 | 8 | 29.73 | 3.72 |
| LISDEXAMFETAMINE | 1205 | 138 | 22,359.75 | 162.03 |
| METHYLPHENIDATE (DL;D;L;ISOMERS) | 1724 | 147 | 28,800.05 | 195.92 |
| CODEINE | 9050 | 157 | 39,752.03 | 253.20 |
| BUPRENORPHINE | 9064 | 134 | 11,335.96 | 84.60 |
| OXYCODONE | 9143 | 153 | 41,509.65 | 271.30 |
| HYDROMORPHONE | 9150 | 114 | 859.76 | 7.54 |
| HYDROCODONE | 9193 | 149 | 4,178.26 | 28.04 |
| MEPERIDINE (PETHIDINE) | 9230 | 15 | 295.77 | 19.72 |
| METHADONE | 9250B | 47 | 1,015.14 | 21.60 |
| MORPHINE | 9300 | 111 | 7,383.42 | 66.52 |
| OXYMORPHONE | 9652 | 25 | 258.31 | 10.33 |

| | | | | |
|---|---|---|---|---|
| TAPENTADOL | 9780 | 48 | 3,055.00 | 63.65 |
| FENTANYL BASE | 9801 | 96 | 151.49 | 1.58 |

**BUSINESS ACTIVITY: B - HOSPITALS**

| DRUG NAME | DRUG CODE | NUMBER OF REGISTRANT SOLD TO | TOTAL GRAMS | AVG GRAMS |
|---|---|---|---|---|
| AMPHETAMINE | 1100 | 9 | 2,613.31 | 290.37 |
| LISDEXAMFETAMINE | 1205 | 5 | 248.58 | 49.72 |
| METHYLPHENIDATE (DL;D;L;ISOMERS) | 1724 | 15 | 2,829.78 | 188.65 |
| CODEINE | 9050 | 15 | 1,675.62 | 111.71 |
| BUPRENORPHINE | 9064 | 22 | 248.75 | 11.31 |
| OXYCODONE | 9143 | 22 | 8,110.06 | 368.64 |
| HYDROMORPHONE | 9150 | 25 | 831.54 | 33.26 |
| HYDROCODONE | 9193 | 18 | 406.61 | 22.59 |
| MEPERIDINE (PETHIDINE) | 9230 | 11 | 319.19 | 29.02 |
| METHADONE | 9250B | 14 | 785.15 | 56.08 |
| MORPHINE | 9300 | 22 | 4,489.82 | 204.08 |
| OXYMORPHONE | 9652 | 1 | 3.57 | 3.57 |
| TAPENTADOL | 9780 | 3 | 311.00 | 103.67 |
| FENTANYL BASE | 9801 | 22 | 135.33 | 6.15 |

**BUSINESS ACTIVITY: C - PRACTITIONERS**

| DRUG NAME | DRUG CODE | NUMBER OF REGISTRANT SOLD TO | TOTAL GRAMS | AVG GRAMS |
|---|---|---|---|---|
| AMPHETAMINE | 1100 | 1 | 2.50 | 2.50 |
| METHYLPHENIDATE (DL;D;L;ISOMERS) | 1724 | 1 | 11.50 | 11.50 |
| CODEINE | 9050 | 8 | 32.06 | 4.01 |
| BUPRENORPHINE | 9064 | 16 | 1.60 | 0.10 |
| OXYCODONE | 9143 | 6 | 11.65 | 1.94 |
| HYDROMORPHONE | 9150 | 3 | 2.32 | 0.77 |
| HYDROCODONE | 9193 | 7 | 17.10 | 2.44 |
| MEPERIDINE (PETHIDINE) | 9230 | 3 | 20.70 | 6.90 |
| MORPHINE | 9300 | 6 | 2.47 | 0.41 |
| FENTANYL BASE | 9801 | 11 | 0.49 | 0.04 |

**BUSINESS ACTIVITY: D - TEACHING INSTITUTIONS**

| DRUG NAME | DRUG CODE | NUMBER OF REGISTRANT SOLD TO | TOTAL GRAMS | AVG GRAMS |
|---|---|---|---|---|
| BUPRENORPHINE | 9064 | 1 | 0.00 | 0.00 |

**BUSINESS ACTIVITY: M - MID-LEVEL PRACTITIONERS**

| DRUG NAME | DRUG CODE | NUMBER OF REGISTRANT SOLD TO | TOTAL GRAMS | AVG GRAMS |
|---|---|---|---|---|
| CODEINE | 9050 | 1 | 2.21 | 2.21 |
| FENTANYL BASE | 9801 | 1 | 0.03 | 0.03 |

**BUSINESS ACTIVITY: NTP - NARCOTIC TREATMENT PROGRAMS**

| DRUG NAME | DRUG CODE | NUMBER OF REGISTRANT SOLD TO | TOTAL GRAMS | AVG GRAMS |
|---|---|---|---|---|
| BUPRENORPHINE | 9064 | 1 | 3.00 | 3.00 |

| | | | | |
|---|---|---|---|---|
| METHADONE | 9250B | 5 | 33,611.55 | 6,722.31 |

**STATE: DE - DELAWARE**

**BUSINESS ACTIVITY: A - PHARMACIES**

| DRUG NAME | DRUG CODE | NUMBER OF REGISTRANT SOLD TO | TOTAL GRAMS | AVG GRAMS |
|---|---|---|---|---|
| AMPHETAMINE | 1100 | 207 | 82,997.66 | 400.95 |
| METHAMPHETAMINE | 1105 | 4 | 17.68 | 4.42 |
| LISDEXAMFETAMINE | 1205 | 202 | 55,192.36 | 273.23 |
| METHYLPHENIDATE (DL;D;L;ISOMERS) | 1724 | 205 | 68,692.65 | 335.09 |
| CODEINE | 9050 | 203 | 29,994.02 | 147.75 |
| BUPRENORPHINE | 9064 | 206 | 12,562.51 | 60.98 |
| OXYCODONE | 9143 | 206 | 233,433.22 | 1,133.17 |
| HYDROMORPHONE | 9150 | 198 | 5,959.87 | 30.10 |
| HYDROCODONE | 9193 | 205 | 19,137.47 | 93.35 |
| MEPERIDINE (PETHIDINE) | 9230 | 18 | 265.26 | 14.74 |
| METHADONE | 9250B | 160 | 8,741.42 | 54.63 |
| MORPHINE | 9300 | 203 | 67,839.35 | 334.18 |
| OXYMORPHONE | 9652 | 121 | 5,858.87 | 48.42 |
| TAPENTADOL | 9780 | 156 | 21,632.00 | 138.67 |
| FENTANYL BASE | 9801 | 198 | 921.39 | 4.65 |

**BUSINESS ACTIVITY: B - HOSPITALS**

| DRUG NAME | DRUG CODE | NUMBER OF REGISTRANT SOLD TO | TOTAL GRAMS | AVG GRAMS |
|---|---|---|---|---|
| AMPHETAMINE | 1100 | 16 | 2,069.59 | 129.35 |
| LISDEXAMFETAMINE | 1205 | 8 | 557.29 | 69.66 |
| METHYLPHENIDATE (DL;D;L;ISOMERS) | 1724 | 17 | 1,805.77 | 106.22 |
| CODEINE | 9050 | 18 | 1,197.37 | 66.52 |
| BUPRENORPHINE | 9064 | 18 | 883.89 | 49.10 |
| OXYCODONE | 9143 | 26 | 9,023.21 | 347.05 |
| HYDROMORPHONE | 9150 | 25 | 478.61 | 19.14 |
| HYDROCODONE | 9193 | 24 | 864.80 | 36.03 |
| MEPERIDINE (PETHIDINE) | 9230 | 17 | 294.02 | 17.30 |
| METHADONE | 9250B | 18 | 1,014.34 | 56.35 |
| MORPHINE | 9300 | 23 | 4,932.84 | 214.47 |
| OXYMORPHONE | 9652 | 5 | 10.88 | 2.18 |
| TAPENTADOL | 9780 | 5 | 272.50 | 54.50 |
| FENTANYL BASE | 9801 | 35 | 82.06 | 2.34 |

**BUSINESS ACTIVITY: C - PRACTITIONERS**

| DRUG NAME | DRUG CODE | NUMBER OF REGISTRANT SOLD TO | TOTAL GRAMS | AVG GRAMS |
|---|---|---|---|---|
| CODEINE | 9050 | 13 | 757.06 | 58.24 |
| BUPRENORPHINE | 9064 | 68 | 18.71 | 0.28 |
| OXYCODONE | 9143 | 5 | 3.90 | 0.78 |

| | | | | |
|---|---|---|---|---|
| HYDROMORPHONE | 9150 | 36 | 33.90 | 0.94 |
| HYDROCODONE | 9193 | 23 | 60.06 | 2.61 |
| MEPERIDINE (PETHIDINE) | 9230 | 1 | 1.09 | 1.09 |
| METHADONE | 9250B | 6 | 4.83 | 0.80 |
| MORPHINE | 9300 | 11 | 55.68 | 5.06 |
| FENTANYL BASE | 9801 | 54 | 4.57 | 0.08 |

**BUSINESS ACTIVITY: M - MID-LEVEL PRACTITIONERS**

| DRUG NAME | DRUG CODE | NUMBER OF REGISTRANT SOLD TO | TOTAL GRAMS | AVG GRAMS |
|---|---|---|---|---|
| CODEINE | 9050 | 1 | 0.65 | 0.65 |
| MORPHINE | 9300 | 1 | 0.19 | 0.19 |

**BUSINESS ACTIVITY: NTP - NARCOTIC TREATMENT PROGRAMS**

| DRUG NAME | DRUG CODE | NUMBER OF REGISTRANT SOLD TO | TOTAL GRAMS | AVG GRAMS |
|---|---|---|---|---|
| BUPRENORPHINE | 9064 | 17 | 2,440.49 | 143.56 |
| METHADONE | 9250B | 15 | 115,154.00 | 7,676.93 |

**STATE: FL - FLORIDA**

**BUSINESS ACTIVITY: A - PHARMACIES**

| DRUG NAME | DRUG CODE | NUMBER OF REGISTRANT SOLD TO | TOTAL GRAMS | AVG GRAMS |
|---|---|---|---|---|
| AMPHETAMINE | 1100 | 4,331 | 1,255,941.60 | 289.99 |
| METHAMPHETAMINE | 1105 | 76 | 190.86 | 2.51 |
| LISDEXAMFETAMINE | 1205 | 3,970 | 533,514.62 | 134.39 |
| METHYLPHENIDATE (DL;D;L;ISOMERS) | 1724 | 4,166 | 755,015.58 | 181.23 |
| CODEINE | 9050 | 4,370 | 581,073.01 | 132.97 |
| BUPRENORPHINE | 9064 | 3,993 | 198,252.91 | 49.65 |
| OXYCODONE | 9143 | 4,514 | 3,571,232.50 | 791.15 |
| HYDROMORPHONE | 9150 | 3,973 | 143,347.92 | 36.08 |
| HYDROCODONE | 9193 | 4,448 | 1,149,565.30 | 258.45 |
| MEPERIDINE (PETHIDINE) | 9230 | 1,112 | 18,260.64 | 16.42 |
| METHADONE | 9250B | 3,325 | 180,193.34 | 54.19 |
| MORPHINE | 9300 | 4,215 | 1,279,349.30 | 303.52 |
| OXYMORPHONE | 9652 | 1,697 | 40,885.84 | 24.09 |
| TAPENTADOL | 9780 | 2,569 | 286,054.00 | 111.35 |
| FENTANYL BASE | 9801 | 3,959 | 16,658.78 | 4.21 |

**BUSINESS ACTIVITY: B - HOSPITALS**

| DRUG NAME | DRUG CODE | NUMBER OF REGISTRANT SOLD TO | TOTAL GRAMS | AVG GRAMS |
|---|---|---|---|---|
| AMPHETAMINE | 1100 | 191 | 30,846.02 | 161.50 |
| METHAMPHETAMINE | 1105 | 1 | 5.63 | 5.63 |
| LISDEXAMFETAMINE | 1205 | 87 | 7,463.85 | 85.79 |
| METHYLPHENIDATE (DL;D;L;ISOMERS) | 1724 | 271 | 28,904.44 | 106.66 |
| CODEINE | 9050 | 395 | 17,882.59 | 45.27 |
| BUPRENORPHINE | 9064 | 295 | 7,323.99 | 24.83 |

| | | | | |
|---|---|---|---|---|
| OXYCODONE | 9143 | 580 | 122,879.66 | 211.86 |
| HYDROMORPHONE | 9150 | 591 | 11,752.01 | 19.88 |
| HYDROCODONE | 9193 | 567 | 49,567.19 | 87.42 |
| MEPERIDINE (PETHIDINE) | 9230 | 464 | 8,840.17 | 19.05 |
| METHADONE | 9250B | 321 | 8,942.51 | 27.86 |
| MORPHINE | 9300 | 631 | 86,258.78 | 136.70 |
| OXYMORPHONE | 9652 | 22 | 92.24 | 4.19 |
| TAPENTADOL | 9780 | 83 | 5,215.50 | 62.84 |
| FENTANYL BASE | 9801 | 792 | 1,873.63 | 2.37 |

**BUSINESS ACTIVITY: C - PRACTITIONERS**

| DRUG NAME | DRUG CODE | NUMBER OF REGISTRANT SOLD TO | TOTAL GRAMS | AVG GRAMS |
|---|---|---|---|---|
| AMPHETAMINE | 1100 | 19 | 114.59 | 6.03 |
| LISDEXAMFETAMINE | 1205 | 8 | 121.20 | 15.15 |
| METHYLPHENIDATE (DL;D;L;ISOMERS) | 1724 | 6 | 961.66 | 160.28 |
| CODEINE | 9050 | 158 | 1,541.84 | 9.76 |
| BUPRENORPHINE | 9064 | 2,114 | 496.24 | 0.23 |
| OXYCODONE | 9143 | 74 | 389.05 | 5.26 |
| HYDROMORPHONE | 9150 | 631 | 420.90 | 0.67 |
| HYDROCODONE | 9193 | 876 | 3,325.21 | 3.80 |
| MEPERIDINE (PETHIDINE) | 9230 | 254 | 2,832.27 | 11.15 |
| METHADONE | 9250B | 31 | 61.89 | 2.00 |
| MORPHINE | 9300 | 413 | 716.41 | 1.73 |
| FENTANYL BASE | 9801 | 1,245 | 77.73 | 0.06 |

**BUSINESS ACTIVITY: D - TEACHING INSTITUTIONS**

| DRUG NAME | DRUG CODE | NUMBER OF REGISTRANT SOLD TO | TOTAL GRAMS | AVG GRAMS |
|---|---|---|---|---|
| BUPRENORPHINE | 9064 | 1 | 0.00 | 0.00 |

**BUSINESS ACTIVITY: M - MID-LEVEL PRACTITIONERS**

| DRUG NAME | DRUG CODE | NUMBER OF REGISTRANT SOLD TO | TOTAL GRAMS | AVG GRAMS |
|---|---|---|---|---|
| BUPRENORPHINE | 9064 | 2 | 1.29 | 0.65 |

**BUSINESS ACTIVITY: NTP - NARCOTIC TREATMENT PROGRAMS**

| DRUG NAME | DRUG CODE | NUMBER OF REGISTRANT SOLD TO | TOTAL GRAMS | AVG GRAMS |
|---|---|---|---|---|
| BUPRENORPHINE | 9064 | 54 | 2,043.64 | 37.85 |
| METHADONE | 9250B | 58 | 565,832.32 | 9,755.73 |

**STATE: GA - GEORGIA**

**BUSINESS ACTIVITY: A - PHARMACIES**

| DRUG NAME | DRUG CODE | NUMBER OF REGISTRANT SOLD TO | TOTAL GRAMS | AVG GRAMS |
|---|---|---|---|---|
| AMPHETAMINE | 1100 | 2,413 | 768,553.75 | 318.51 |
| METHAMPHETAMINE | 1105 | 31 | 77.95 | 2.51 |
| LISDEXAMFETAMINE | 1205 | 2,312 | 385,377.07 | 166.69 |
| METHYLPHENIDATE (DL;D;L;ISOMERS) | 1724 | 2,397 | 610,937.76 | 254.88 |

| | | | | |
|---|---|---|---|---|
| CODEINE | 9050 | 2,372 | 351,636.29 | 148.24 |
| BUPRENORPHINE | 9064 | 2,285 | 65,140.88 | 28.51 |
| OXYCODONE | 9143 | 2,433 | 1,518,244.84 | 624.02 |
| HYDROMORPHONE | 9150 | 2,069 | 21,876.46 | 10.57 |
| HYDROCODONE | 9193 | 2,444 | 801,140.24 | 327.80 |
| MEPERIDINE (PETHIDINE) | 9230 | 1,215 | 24,322.85 | 20.02 |
| METHADONE | 9250B | 1,935 | 79,762.59 | 41.22 |
| MORPHINE | 9300 | 2,333 | 362,140.56 | 155.23 |
| OXYMORPHONE | 9652 | 817 | 11,392.61 | 13.94 |
| TAPENTADOL | 9780 | 1,663 | 201,658.00 | 121.26 |
| FENTANYL BASE | 9801 | 2,277 | 7,638.53 | 3.35 |

**BUSINESS ACTIVITY: B - HOSPITALS**

| DRUG NAME | DRUG CODE | NUMBER OF REGISTRANT SOLD TO | TOTAL GRAMS | AVG GRAMS |
|---|---|---|---|---|
| AMPHETAMINE | 1100 | 97 | 17,494.47 | 180.36 |
| METHAMPHETAMINE | 1105 | 2 | 10.85 | 5.42 |
| LISDEXAMFETAMINE | 1205 | 55 | 6,545.83 | 119.02 |
| METHYLPHENIDATE (DL;D;L;ISOMERS) | 1724 | 143 | 19,898.93 | 139.15 |
| CODEINE | 9050 | 193 | 11,307.08 | 58.59 |
| BUPRENORPHINE | 9064 | 114 | 2,066.80 | 18.13 |
| OXYCODONE | 9143 | 227 | 58,890.71 | 259.43 |
| HYDROMORPHONE | 9150 | 196 | 4,965.95 | 25.34 |
| HYDROCODONE | 9193 | 221 | 38,260.00 | 173.12 |
| MEPERIDINE (PETHIDINE) | 9230 | 162 | 7,226.26 | 44.61 |
| METHADONE | 9250B | 163 | 3,558.07 | 21.83 |
| MORPHINE | 9300 | 220 | 33,598.04 | 152.72 |
| OXYMORPHONE | 9652 | 7 | 17.04 | 2.43 |
| TAPENTADOL | 9780 | 55 | 3,381.50 | 61.48 |
| FENTANYL BASE | 9801 | 207 | 1,085.56 | 5.24 |

**BUSINESS ACTIVITY: C - PRACTITIONERS**

| DRUG NAME | DRUG CODE | NUMBER OF REGISTRANT SOLD TO | TOTAL GRAMS | AVG GRAMS |
|---|---|---|---|---|
| AMPHETAMINE | 1100 | 7 | 32.30 | 4.61 |
| LISDEXAMFETAMINE | 1205 | 1 | 2.72 | 2.72 |
| METHYLPHENIDATE (DL;D;L;ISOMERS) | 1724 | 2 | 0.78 | 0.39 |
| CODEINE | 9050 | 216 | 2,968.22 | 13.74 |
| BUPRENORPHINE | 9064 | 835 | 326.78 | 0.39 |
| OXYCODONE | 9143 | 110 | 2,428.52 | 22.08 |
| HYDROMORPHONE | 9150 | 343 | 219.82 | 0.64 |
| HYDROCODONE | 9193 | 449 | 3,436.67 | 7.65 |
| MEPERIDINE (PETHIDINE) | 9230 | 211 | 1,910.76 | 9.06 |
| METHADONE | 9250B | 12 | 47.85 | 3.99 |

| | | | | |
|---|---|---|---|---|
| MORPHINE | 9300 | 314 | 841.98 | 2.68 |
| FENTANYL BASE | 9801 | 835 | 65.85 | 0.08 |

**BUSINESS ACTIVITY: M - MID-LEVEL PRACTITIONERS**

| DRUG NAME | DRUG CODE | NUMBER OF REGISTRANT SOLD TO | TOTAL GRAMS | AVG GRAMS |
|---|---|---|---|---|
| CODEINE | 9050 | 23 | 17.11 | 0.74 |

**BUSINESS ACTIVITY: NTP - NARCOTIC TREATMENT PROGRAMS**

| DRUG NAME | DRUG CODE | NUMBER OF REGISTRANT SOLD TO | TOTAL GRAMS | AVG GRAMS |
|---|---|---|---|---|
| BUPRENORPHINE | 9064 | 33 | 1,615.45 | 48.95 |
| METHADONE | 9250B | 74 | 365,866.39 | 4,944.14 |

**STATE: GU - GUAM**

**BUSINESS ACTIVITY: A - PHARMACIES**

| DRUG NAME | DRUG CODE | NUMBER OF REGISTRANT SOLD TO | TOTAL GRAMS | AVG GRAMS |
|---|---|---|---|---|
| AMPHETAMINE | 1100 | 11 | 1,070.01 | 97.27 |
| LISDEXAMFETAMINE | 1205 | 3 | 35.84 | 11.95 |
| METHYLPHENIDATE (DL;D;L;ISOMERS) | 1724 | 10 | 677.29 | 67.73 |
| CODEINE | 9050 | 18 | 3,015.68 | 167.54 |
| BUPRENORPHINE | 9064 | 3 | 2.16 | 0.72 |
| OXYCODONE | 9143 | 12 | 2,335.35 | 194.61 |
| HYDROMORPHONE | 9150 | 4 | 66.32 | 16.58 |
| HYDROCODONE | 9193 | 11 | 298.10 | 27.10 |
| METHADONE | 9250B | 2 | 127.00 | 63.50 |
| MORPHINE | 9300 | 10 | 2,822.41 | 282.24 |
| FENTANYL BASE | 9801 | 7 | 15.54 | 2.22 |

**BUSINESS ACTIVITY: B - HOSPITALS**

| DRUG NAME | DRUG CODE | NUMBER OF REGISTRANT SOLD TO | TOTAL GRAMS | AVG GRAMS |
|---|---|---|---|---|
| AMPHETAMINE | 1100 | 4 | 689.37 | 172.34 |
| LISDEXAMFETAMINE | 1205 | 1 | 8.09 | 8.09 |
| METHYLPHENIDATE (DL;D;L;ISOMERS) | 1724 | 4 | 1,307.45 | 326.86 |
| CODEINE | 9050 | 6 | 578.79 | 96.46 |
| BUPRENORPHINE | 9064 | 2 | 0.02 | 0.01 |
| OXYCODONE | 9143 | 6 | 810.48 | 135.08 |
| HYDROMORPHONE | 9150 | 8 | 23.49 | 2.94 |
| HYDROCODONE | 9193 | 6 | 187.46 | 31.24 |
| MEPERIDINE (PETHIDINE) | 9230 | 3 | 19.61 | 6.54 |
| METHADONE | 9250B | 3 | 36.22 | 12.07 |
| MORPHINE | 9300 | 7 | 542.36 | 77.48 |
| FENTANYL BASE | 9801 | 7 | 16.78 | 2.40 |

**BUSINESS ACTIVITY: C - PRACTITIONERS**

| DRUG NAME | DRUG CODE | NUMBER OF REGISTRANT SOLD TO | TOTAL GRAMS | AVG GRAMS |
|---|---|---|---|---|
| CODEINE | 9050 | 1 | 4.42 | 4.42 |

| | | | | |
|---|---|---|---|---|
| BUPRENORPHINE | 9064 | 2 | 0.12 | 0.06 |
| HYDROMORPHONE | 9150 | 3 | 0.71 | 0.24 |
| HYDROCODONE | 9193 | 2 | 10.90 | 5.45 |
| MEPERIDINE (PETHIDINE) | 9230 | 4 | 101.31 | 25.33 |
| MORPHINE | 9300 | 4 | 1.93 | 0.48 |
| FENTANYL BASE | 9801 | 5 | 0.45 | 0.09 |

**STATE: HI - HAWAII**

**BUSINESS ACTIVITY: A - PHARMACIES**

| DRUG NAME | DRUG CODE | NUMBER OF REGISTRANT SOLD TO | TOTAL GRAMS | AVG GRAMS |
|---|---|---|---|---|
| AMPHETAMINE | 1100 | 219 | 32,601.50 | 148.87 |
| METHAMPHETAMINE | 1105 | 4 | 22.10 | 5.52 |
| LISDEXAMFETAMINE | 1205 | 148 | 4,643.88 | 31.38 |
| METHYLPHENIDATE (DL;D;L;ISOMERS) | 1724 | 212 | 32,605.68 | 153.80 |
| CODEINE | 9050 | 227 | 42,160.52 | 185.73 |
| BUPRENORPHINE | 9064 | 200 | 8,265.51 | 41.33 |
| OXYCODONE | 9143 | 225 | 130,238.78 | 578.84 |
| HYDROMORPHONE | 9150 | 143 | 1,585.92 | 11.09 |
| HYDROCODONE | 9193 | 227 | 54,078.77 | 238.23 |
| MEPERIDINE (PETHIDINE) | 9230 | 24 | 816.16 | 34.01 |
| METHADONE | 9250B | 155 | 10,352.50 | 66.79 |
| MORPHINE | 9300 | 206 | 62,223.21 | 302.05 |
| OXYMORPHONE | 9652 | 41 | 608.95 | 14.85 |
| TAPENTADOL | 9780 | 65 | 7,545.00 | 116.08 |
| FENTANYL BASE | 9801 | 180 | 479.51 | 2.66 |

**BUSINESS ACTIVITY: B - HOSPITALS**

| DRUG NAME | DRUG CODE | NUMBER OF REGISTRANT SOLD TO | TOTAL GRAMS | AVG GRAMS |
|---|---|---|---|---|
| AMPHETAMINE | 1100 | 17 | 8,631.86 | 507.76 |
| LISDEXAMFETAMINE | 1205 | 12 | 1,471.26 | 122.61 |
| METHYLPHENIDATE (DL;D;L;ISOMERS) | 1724 | 23 | 11,143.05 | 484.48 |
| CODEINE | 9050 | 38 | 3,613.89 | 95.10 |
| BUPRENORPHINE | 9064 | 23 | 324.33 | 14.10 |
| OXYCODONE | 9143 | 39 | 16,044.10 | 411.39 |
| HYDROMORPHONE | 9150 | 36 | 450.48 | 12.51 |
| HYDROCODONE | 9193 | 37 | 4,670.95 | 126.24 |
| MEPERIDINE (PETHIDINE) | 9230 | 26 | 267.44 | 10.29 |
| METHADONE | 9250B | 27 | 903.25 | 33.45 |
| MORPHINE | 9300 | 41 | 7,697.49 | 187.74 |
| OXYMORPHONE | 9652 | 3 | 249.34 | 83.11 |
| TAPENTADOL | 9780 | 8 | 537.00 | 67.13 |
| FENTANYL BASE | 9801 | 44 | 137.09 | 3.12 |

**BUSINESS ACTIVITY: C - PRACTITIONERS**

| DRUG NAME | DRUG CODE | NUMBER OF REGISTRANT SOLD TO | TOTAL GRAMS | AVG GRAMS |
|---|---|---|---|---|
| CODEINE | 9050 | 15 | 650.65 | 43.38 |
| BUPRENORPHINE | 9064 | 82 | 4.65 | 0.06 |
| OXYCODONE | 9143 | 18 | 1,660.41 | 92.24 |
| HYDROMORPHONE | 9150 | 38 | 25.05 | 0.66 |
| HYDROCODONE | 9193 | 51 | 1,661.50 | 32.58 |
| MEPERIDINE (PETHIDINE) | 9230 | 18 | 150.88 | 8.38 |
| METHADONE | 9250B | 2 | 3.40 | 1.70 |
| MORPHINE | 9300 | 20 | 221.02 | 11.05 |
| OXYMORPHONE | 9652 | 1 | 2.41 | 2.41 |
| FENTANYL BASE | 9801 | 53 | 3.68 | 0.07 |

**BUSINESS ACTIVITY: M - MID-LEVEL PRACTITIONERS**

| DRUG NAME | DRUG CODE | NUMBER OF REGISTRANT SOLD TO | TOTAL GRAMS | AVG GRAMS |
|---|---|---|---|---|
| MORPHINE | 9300 | 1 | 0.94 | 0.94 |
| FENTANYL BASE | 9801 | 1 | 0.02 | 0.02 |

**BUSINESS ACTIVITY: NTP - NARCOTIC TREATMENT PROGRAMS**

| DRUG NAME | DRUG CODE | NUMBER OF REGISTRANT SOLD TO | TOTAL GRAMS | AVG GRAMS |
|---|---|---|---|---|
| METHADONE | 9250B | 4 | 16,635.84 | 4,158.96 |

**STATE: IA - IOWA**

**BUSINESS ACTIVITY: A - PHARMACIES**

| DRUG NAME | DRUG CODE | NUMBER OF REGISTRANT SOLD TO | TOTAL GRAMS | AVG GRAMS |
|---|---|---|---|---|
| AMPHETAMINE | 1100 | 693 | 205,396.16 | 296.39 |
| METHAMPHETAMINE | 1105 | 18 | 70.32 | 3.91 |
| LISDEXAMFETAMINE | 1205 | 669 | 135,066.71 | 201.89 |
| METHYLPHENIDATE (DL;D;L;ISOMERS) | 1724 | 697 | 309,846.68 | 444.54 |
| CODEINE | 9050 | 689 | 68,251.26 | 99.06 |
| BUPRENORPHINE | 9064 | 566 | 7,048.17 | 12.45 |
| OXYCODONE | 9143 | 699 | 165,463.13 | 236.71 |
| HYDROMORPHONE | 9150 | 636 | 7,551.21 | 11.87 |
| HYDROCODONE | 9193 | 708 | 178,882.83 | 252.66 |
| MEPERIDINE (PETHIDINE) | 9230 | 105 | 2,277.34 | 21.69 |
| METHADONE | 9250B | 512 | 12,999.48 | 25.39 |
| MORPHINE | 9300 | 692 | 102,100.29 | 147.54 |
| OXYMORPHONE | 9652 | 136 | 1,713.41 | 12.60 |
| TAPENTADOL | 9780 | 387 | 36,323.00 | 93.86 |
| FENTANYL BASE | 9801 | 674 | 2,608.86 | 3.87 |

**BUSINESS ACTIVITY: B - HOSPITALS**

| DRUG NAME | DRUG CODE | NUMBER OF REGISTRANT SOLD TO | TOTAL GRAMS | AVG GRAMS |
|---|---|---|---|---|
| AMPHETAMINE | 1100 | 47 | 5,407.17 | 115.05 |

| | | | | |
|---|---|---|---|---|
| METHAMPHETAMINE | 1105 | 1 | 0.40 | 0.40 |
| LISDEXAMFETAMINE | 1205 | 33 | 1,779.39 | 53.92 |
| METHYLPHENIDATE (DL;D;L;ISOMERS) | 1724 | 79 | 7,126.80 | 90.21 |
| CODEINE | 9050 | 109 | 2,342.47 | 21.49 |
| BUPRENORPHINE | 9064 | 66 | 454.96 | 6.89 |
| OXYCODONE | 9143 | 143 | 12,214.68 | 85.42 |
| HYDROMORPHONE | 9150 | 141 | 1,985.76 | 14.08 |
| HYDROCODONE | 9193 | 146 | 11,987.39 | 82.11 |
| MEPERIDINE (PETHIDINE) | 9230 | 105 | 3,259.04 | 31.04 |
| METHADONE | 9250B | 73 | 1,042.15 | 14.28 |
| MORPHINE | 9300 | 144 | 13,314.78 | 92.46 |
| OXYMORPHONE | 9652 | 5 | 12.31 | 2.46 |
| TAPENTADOL | 9780 | 34 | 782.50 | 23.01 |
| FENTANYL BASE | 9801 | 150 | 279.76 | 1.87 |

**BUSINESS ACTIVITY: C - PRACTITIONERS**

| DRUG NAME | DRUG CODE | NUMBER OF REGISTRANT SOLD TO | TOTAL GRAMS | AVG GRAMS |
|---|---|---|---|---|
| CODEINE | 9050 | 14 | 247.67 | 17.69 |
| BUPRENORPHINE | 9064 | 273 | 17.32 | 0.06 |
| OXYCODONE | 9143 | 7 | 99.87 | 14.27 |
| HYDROMORPHONE | 9150 | 67 | 42.37 | 0.63 |
| HYDROCODONE | 9193 | 106 | 372.03 | 3.51 |
| MEPERIDINE (PETHIDINE) | 9230 | 20 | 1,051.25 | 52.56 |
| MORPHINE | 9300 | 72 | 66.57 | 0.92 |
| FENTANYL BASE | 9801 | 109 | 35.23 | 0.32 |

**BUSINESS ACTIVITY: D - TEACHING INSTITUTIONS**

| DRUG NAME | DRUG CODE | NUMBER OF REGISTRANT SOLD TO | TOTAL GRAMS | AVG GRAMS |
|---|---|---|---|---|
| BUPRENORPHINE | 9064 | 1 | 0.00 | 0.00 |

**BUSINESS ACTIVITY: M - MID-LEVEL PRACTITIONERS**

| DRUG NAME | DRUG CODE | NUMBER OF REGISTRANT SOLD TO | TOTAL GRAMS | AVG GRAMS |
|---|---|---|---|---|
| BUPRENORPHINE | 9064 | 1 | 0.00 | 0.00 |
| FENTANYL BASE | 9801 | 1 | 0.01 | 0.01 |

**BUSINESS ACTIVITY: NTP - NARCOTIC TREATMENT PROGRAMS**

| DRUG NAME | DRUG CODE | NUMBER OF REGISTRANT SOLD TO | TOTAL GRAMS | AVG GRAMS |
|---|---|---|---|---|
| BUPRENORPHINE | 9064 | 10 | 739.80 | 73.98 |
| METHADONE | 9250B | 14 | 35,121.30 | 2,508.66 |

**STATE: ID - IDAHO**

**BUSINESS ACTIVITY: A - PHARMACIES**

| DRUG NAME | DRUG CODE | NUMBER OF REGISTRANT SOLD TO | TOTAL GRAMS | AVG GRAMS |
|---|---|---|---|---|
| AMPHETAMINE | 1100 | 316 | 111,181.69 | 351.84 |
| METHAMPHETAMINE | 1105 | 15 | 55.44 | 3.70 |

| | | | | |
|---|---|---|---|---|
| LISDEXAMFETAMINE | 1205 | 302 | 38,453.48 | 127.33 |
| METHYLPHENIDATE (DL;D;L;ISOMERS) | 1724 | 314 | 108,724.50 | 346.26 |
| CODEINE | 9050 | 315 | 44,193.05 | 140.30 |
| BUPRENORPHINE | 9064 | 307 | 16,217.93 | 52.83 |
| OXYCODONE | 9143 | 316 | 201,970.59 | 639.15 |
| HYDROMORPHONE | 9150 | 301 | 6,076.56 | 20.19 |
| HYDROCODONE | 9193 | 319 | 191,811.91 | 601.29 |
| MEPERIDINE (PETHIDINE) | 9230 | 101 | 2,125.81 | 21.05 |
| METHADONE | 9250B | 293 | 19,855.95 | 67.77 |
| MORPHINE | 9300 | 321 | 88,357.06 | 275.26 |
| OXYMORPHONE | 9652 | 147 | 2,868.41 | 19.51 |
| TAPENTADOL | 9780 | 204 | 21,668.50 | 106.22 |
| FENTANYL BASE | 9801 | 311 | 1,791.41 | 5.76 |

**BUSINESS ACTIVITY: B - HOSPITALS**

| DRUG NAME | DRUG CODE | NUMBER OF REGISTRANT SOLD TO | TOTAL GRAMS | AVG GRAMS |
|---|---|---|---|---|
| AMPHETAMINE | 1100 | 23 | 1,632.29 | 70.97 |
| LISDEXAMFETAMINE | 1205 | 10 | 135.85 | 13.59 |
| METHYLPHENIDATE (DL;D;L;ISOMERS) | 1724 | 36 | 1,743.31 | 48.43 |
| CODEINE | 9050 | 46 | 1,794.34 | 39.01 |
| BUPRENORPHINE | 9064 | 33 | 467.11 | 14.15 |
| OXYCODONE | 9143 | 59 | 10,875.96 | 184.34 |
| HYDROMORPHONE | 9150 | 53 | 856.39 | 16.16 |
| HYDROCODONE | 9193 | 61 | 8,939.75 | 146.55 |
| MEPERIDINE (PETHIDINE) | 9230 | 40 | 550.79 | 13.77 |
| METHADONE | 9250B | 39 | 1,105.48 | 28.35 |
| MORPHINE | 9300 | 59 | 8,509.67 | 144.23 |
| OXYMORPHONE | 9652 | 4 | 18.11 | 4.53 |
| TAPENTADOL | 9780 | 8 | 158.00 | 19.75 |
| FENTANYL BASE | 9801 | 57 | 172.47 | 3.03 |

**BUSINESS ACTIVITY: C - PRACTITIONERS**

| DRUG NAME | DRUG CODE | NUMBER OF REGISTRANT SOLD TO | TOTAL GRAMS | AVG GRAMS |
|---|---|---|---|---|
| AMPHETAMINE | 1100 | 2 | 6.53 | 3.27 |
| CODEINE | 9050 | 20 | 2,637.16 | 131.86 |
| BUPRENORPHINE | 9064 | 136 | 14.13 | 0.10 |
| OXYCODONE | 9143 | 15 | 78.98 | 5.27 |
| HYDROMORPHONE | 9150 | 79 | 84.78 | 1.07 |
| HYDROCODONE | 9193 | 93 | 311.76 | 3.35 |
| MEPERIDINE (PETHIDINE) | 9230 | 19 | 106.76 | 5.62 |
| METHADONE | 9250B | 5 | 3.22 | 0.64 |
| MORPHINE | 9300 | 71 | 137.69 | 1.94 |

| | | | | |
|---|---|---|---|---|
| FENTANYL BASE | 9801 | 128 | 19.28 | 0.15 |

**BUSINESS ACTIVITY: D - TEACHING INSTITUTIONS**

| DRUG NAME | DRUG CODE | NUMBER OF REGISTRANT SOLD TO | TOTAL GRAMS | AVG GRAMS |
|---|---|---|---|---|
| BUPRENORPHINE | 9064 | 1 | 0.00 | 0.00 |

**BUSINESS ACTIVITY: M - MID-LEVEL PRACTITIONERS**

| DRUG NAME | DRUG CODE | NUMBER OF REGISTRANT SOLD TO | TOTAL GRAMS | AVG GRAMS |
|---|---|---|---|---|
| CODEINE | 9050 | 3 | 69.30 | 23.10 |
| HYDROMORPHONE | 9150 | 1 | 0.10 | 0.10 |
| HYDROCODONE | 9193 | 2 | 1.66 | 0.83 |
| MEPERIDINE (PETHIDINE) | 9230 | 3 | 4.36 | 1.45 |
| MORPHINE | 9300 | 2 | 0.05 | 0.03 |
| FENTANYL BASE | 9801 | 3 | 0.16 | 0.05 |

**BUSINESS ACTIVITY: NTP - NARCOTIC TREATMENT PROGRAMS**

| DRUG NAME | DRUG CODE | NUMBER OF REGISTRANT SOLD TO | TOTAL GRAMS | AVG GRAMS |
|---|---|---|---|---|
| BUPRENORPHINE | 9064 | 4 | 173.63 | 43.41 |
| METHADONE | 9250B | 4 | 20,499.65 | 5,124.91 |

**STATE: IL - ILLINOIS**

**BUSINESS ACTIVITY: A - PHARMACIES**

| DRUG NAME | DRUG CODE | NUMBER OF REGISTRANT SOLD TO | TOTAL GRAMS | AVG GRAMS |
|---|---|---|---|---|
| AMPHETAMINE | 1100 | 2,124 | 790,152.37 | 372.01 |
| METHAMPHETAMINE | 1105 | 59 | 204.12 | 3.46 |
| LISDEXAMFETAMINE | 1205 | 1,911 | 286,934.15 | 150.15 |
| METHYLPHENIDATE (DL;D;L;ISOMERS) | 1724 | 2,087 | 652,101.61 | 312.46 |
| CODEINE | 9050 | 2,175 | 554,385.10 | 254.89 |
| BUPRENORPHINE | 9064 | 1,906 | 62,310.89 | 32.69 |
| OXYCODONE | 9143 | 2,076 | 513,545.03 | 247.37 |
| HYDROMORPHONE | 9150 | 1,764 | 26,065.60 | 14.78 |
| HYDROCODONE | 9193 | 2,188 | 974,995.92 | 445.61 |
| MEPERIDINE (PETHIDINE) | 9230 | 175 | 4,897.29 | 27.98 |
| METHADONE | 9250B | 1,425 | 39,182.05 | 27.50 |
| MORPHINE | 9300 | 1,968 | 326,726.89 | 166.02 |
| OXYMORPHONE | 9652 | 599 | 7,283.37 | 12.16 |
| TAPENTADOL | 9780 | 1,142 | 112,005.00 | 98.08 |
| FENTANYL BASE | 9801 | 1,942 | 8,404.94 | 4.33 |

**BUSINESS ACTIVITY: B - HOSPITALS**

| DRUG NAME | DRUG CODE | NUMBER OF REGISTRANT SOLD TO | TOTAL GRAMS | AVG GRAMS |
|---|---|---|---|---|
| AMPHETAMINE | 1100 | 135 | 9,957.16 | 73.76 |
| METHAMPHETAMINE | 1105 | 2 | 3.62 | 1.81 |
| LISDEXAMFETAMINE | 1205 | 53 | 2,133.19 | 40.25 |
| METHYLPHENIDATE (DL;D;L;ISOMERS) | 1724 | 190 | 10,927.14 | 57.51 |

| | | | | |
|---|---|---|---|---|
| CODEINE | 9050 | 239 | 11,092.73 | 46.41 |
| BUPRENORPHINE | 9064 | 162 | 2,227.74 | 13.75 |
| OXYCODONE | 9143 | 234 | 25,312.40 | 108.17 |
| HYDROMORPHONE | 9150 | 235 | 6,114.84 | 26.02 |
| HYDROCODONE | 9193 | 256 | 52,016.97 | 203.19 |
| MEPERIDINE (PETHIDINE) | 9230 | 169 | 3,522.54 | 20.84 |
| METHADONE | 9250B | 189 | 5,752.54 | 30.44 |
| MORPHINE | 9300 | 242 | 35,354.87 | 146.09 |
| OXYMORPHONE | 9652 | 15 | 197.20 | 13.15 |
| TAPENTADOL | 9780 | 43 | 1,972.00 | 45.86 |
| FENTANYL BASE | 9801 | 237 | 1,044.76 | 4.41 |

**BUSINESS ACTIVITY: C - PRACTITIONERS**

| DRUG NAME | DRUG CODE | NUMBER OF REGISTRANT SOLD TO | TOTAL GRAMS | AVG GRAMS |
|---|---|---|---|---|
| AMPHETAMINE | 1100 | 3 | 16.86 | 5.62 |
| METHYLPHENIDATE (DL;D;L;ISOMERS) | 1724 | 1 | 51.11 | 51.11 |
| CODEINE | 9050 | 214 | 3,857.88 | 18.03 |
| BUPRENORPHINE | 9064 | 911 | 72.88 | 0.08 |
| OXYCODONE | 9143 | 47 | 145.25 | 3.09 |
| HYDROMORPHONE | 9150 | 391 | 343.22 | 0.88 |
| HYDROCODONE | 9193 | 540 | 3,044.81 | 5.64 |
| MEPERIDINE (PETHIDINE) | 9230 | 108 | 1,192.87 | 11.05 |
| METHADONE | 9250B | 25 | 48.48 | 1.94 |
| MORPHINE | 9300 | 267 | 326.99 | 1.22 |
| FENTANYL BASE | 9801 | 630 | 63.38 | 0.10 |

**BUSINESS ACTIVITY: D - TEACHING INSTITUTIONS**

| DRUG NAME | DRUG CODE | NUMBER OF REGISTRANT SOLD TO | TOTAL GRAMS | AVG GRAMS |
|---|---|---|---|---|
| CODEINE | 9050 | 1 | 11.96 | 11.96 |
| BUPRENORPHINE | 9064 | 2 | 0.87 | 0.44 |
| HYDROMORPHONE | 9150 | 1 | 4.13 | 4.13 |
| HYDROCODONE | 9193 | 1 | 9.89 | 9.89 |
| METHADONE | 9250B | 1 | 23.08 | 23.08 |
| MORPHINE | 9300 | 1 | 36.89 | 36.89 |
| FENTANYL BASE | 9801 | 1 | 2.07 | 2.07 |

**BUSINESS ACTIVITY: M - MID-LEVEL PRACTITIONERS**

| DRUG NAME | DRUG CODE | NUMBER OF REGISTRANT SOLD TO | TOTAL GRAMS | AVG GRAMS |
|---|---|---|---|---|
| CODEINE | 9050 | 6 | 7.80 | 1.30 |

**BUSINESS ACTIVITY: NTP - NARCOTIC TREATMENT PROGRAMS**

| DRUG NAME | DRUG CODE | NUMBER OF REGISTRANT SOLD TO | TOTAL GRAMS | AVG GRAMS |
|---|---|---|---|---|
| BUPRENORPHINE | 9064 | 13 | 2,237.71 | 172.13 |
| METHADONE | 9250B | 83 | 430,703.24 | 5,189.20 |

**STATE: IN - INDIANA**

**BUSINESS ACTIVITY: A - PHARMACIES**

| DRUG NAME | DRUG CODE | NUMBER OF REGISTRANT SOLD TO | TOTAL GRAMS | AVG GRAMS |
|---|---|---|---|---|
| AMPHETAMINE | 1100 | 1,133 | 535,506.80 | 472.65 |
| METHAMPHETAMINE | 1105 | 25 | 102.47 | 4.10 |
| LISDEXAMFETAMINE | 1205 | 1,125 | 329,076.49 | 292.51 |
| METHYLPHENIDATE (DL;D;L;ISOMERS) | 1724 | 1,135 | 449,639.07 | 396.16 |
| CODEINE | 9050 | 1,116 | 169,795.59 | 152.15 |
| BUPRENORPHINE | 9064 | 1,108 | 93,581.04 | 84.46 |
| OXYCODONE | 9143 | 1,143 | 763,241.37 | 667.75 |
| HYDROMORPHONE | 9150 | 1,002 | 15,157.50 | 15.13 |
| HYDROCODONE | 9193 | 1,156 | 745,666.23 | 645.04 |
| MEPERIDINE (PETHIDINE) | 9230 | 261 | 3,689.28 | 14.14 |
| METHADONE | 9250B | 967 | 51,047.43 | 52.79 |
| MORPHINE | 9300 | 1,114 | 261,368.41 | 234.62 |
| OXYMORPHONE | 9652 | 487 | 8,724.16 | 17.91 |
| TAPENTADOL | 9780 | 785 | 119,129.50 | 151.76 |
| FENTANYL BASE | 9801 | 1,083 | 5,105.52 | 4.71 |

**BUSINESS ACTIVITY: B - HOSPITALS**

| DRUG NAME | DRUG CODE | NUMBER OF REGISTRANT SOLD TO | TOTAL GRAMS | AVG GRAMS |
|---|---|---|---|---|
| AMPHETAMINE | 1100 | 128 | 9,269.25 | 72.42 |
| LISDEXAMFETAMINE | 1205 | 76 | 4,886.68 | 64.30 |
| METHYLPHENIDATE (DL;D;L;ISOMERS) | 1724 | 150 | 8,258.66 | 55.06 |
| CODEINE | 9050 | 159 | 4,423.59 | 27.82 |
| BUPRENORPHINE | 9064 | 138 | 2,517.86 | 18.25 |
| OXYCODONE | 9143 | 249 | 33,389.89 | 134.10 |
| HYDROMORPHONE | 9150 | 243 | 3,883.05 | 15.98 |
| HYDROCODONE | 9193 | 263 | 37,192.71 | 141.42 |
| MEPERIDINE (PETHIDINE) | 9230 | 199 | 6,901.74 | 34.68 |
| METHADONE | 9250B | 159 | 3,323.67 | 20.90 |
| MORPHINE | 9300 | 263 | 24,167.21 | 91.89 |
| OXYMORPHONE | 9652 | 12 | 47.37 | 3.95 |
| TAPENTADOL | 9780 | 28 | 1,188.00 | 42.43 |
| FENTANYL BASE | 9801 | 290 | 722.70 | 2.49 |

**BUSINESS ACTIVITY: C - PRACTITIONERS**

| DRUG NAME | DRUG CODE | NUMBER OF REGISTRANT SOLD TO | TOTAL GRAMS | AVG GRAMS |
|---|---|---|---|---|
| AMPHETAMINE | 1100 | 1 | 0.63 | 0.63 |
| CODEINE | 9050 | 46 | 473.94 | 10.30 |
| BUPRENORPHINE | 9064 | 498 | 90.27 | 0.18 |
| OXYCODONE | 9143 | 14 | 39.85 | 2.85 |

| | | | | |
|---|---|---|---|---|
| HYDROMORPHONE | 9150 | 185 | 121.99 | 0.66 |
| HYDROCODONE | 9193 | 259 | 661.83 | 2.56 |
| MEPERIDINE (PETHIDINE) | 9230 | 51 | 153.60 | 3.01 |
| METHADONE | 9250B | 4 | 1.97 | 0.49 |
| MORPHINE | 9300 | 147 | 147.14 | 1.00 |
| FENTANYL BASE | 9801 | 268 | 15.80 | 0.06 |

**BUSINESS ACTIVITY: M - MID-LEVEL PRACTITIONERS**

| DRUG NAME | DRUG CODE | NUMBER OF REGISTRANT SOLD TO | TOTAL GRAMS | AVG GRAMS |
|---|---|---|---|---|
| CODEINE | 9050 | 4 | 11.32 | 2.83 |

**BUSINESS ACTIVITY: NTP - NARCOTIC TREATMENT PROGRAMS**

| DRUG NAME | DRUG CODE | NUMBER OF REGISTRANT SOLD TO | TOTAL GRAMS | AVG GRAMS |
|---|---|---|---|---|
| BUPRENORPHINE | 9064 | 9 | 1,417.87 | 157.54 |
| METHADONE | 9250B | 19 | 245,951.06 | 12,944.79 |

**STATE: KS - KANSAS**

**BUSINESS ACTIVITY: A - PHARMACIES**

| DRUG NAME | DRUG CODE | NUMBER OF REGISTRANT SOLD TO | TOTAL GRAMS | AVG GRAMS |
|---|---|---|---|---|
| AMPHETAMINE | 1100 | 627 | 245,451.29 | 391.47 |
| METHAMPHETAMINE | 1105 | 13 | 57.86 | 4.45 |
| LISDEXAMFETAMINE | 1205 | 613 | 125,699.75 | 205.06 |
| METHYLPHENIDATE (DL;D;L;ISOMERS) | 1724 | 624 | 184,884.10 | 296.29 |
| CODEINE | 9050 | 608 | 90,384.89 | 148.66 |
| BUPRENORPHINE | 9064 | 545 | 10,232.46 | 18.78 |
| OXYCODONE | 9143 | 630 | 393,201.39 | 624.13 |
| HYDROMORPHONE | 9150 | 553 | 9,733.70 | 17.60 |
| HYDROCODONE | 9193 | 634 | 262,648.46 | 414.27 |
| MEPERIDINE (PETHIDINE) | 9230 | 197 | 3,611.28 | 18.33 |
| METHADONE | 9250B | 496 | 22,110.02 | 44.58 |
| MORPHINE | 9300 | 613 | 138,103.39 | 225.29 |
| OXYMORPHONE | 9652 | 201 | 3,024.53 | 15.05 |
| TAPENTADOL | 9780 | 301 | 25,053.00 | 83.23 |
| FENTANYL BASE | 9801 | 604 | 3,071.41 | 5.09 |

**BUSINESS ACTIVITY: B - HOSPITALS**

| DRUG NAME | DRUG CODE | NUMBER OF REGISTRANT SOLD TO | TOTAL GRAMS | AVG GRAMS |
|---|---|---|---|---|
| AMPHETAMINE | 1100 | 50 | 9,096.04 | 181.92 |
| METHAMPHETAMINE | 1105 | 1 | 3.62 | 3.62 |
| LISDEXAMFETAMINE | 1205 | 34 | 2,152.84 | 63.32 |
| METHYLPHENIDATE (DL;D;L;ISOMERS) | 1724 | 84 | 8,342.66 | 99.32 |
| CODEINE | 9050 | 148 | 4,360.88 | 29.47 |
| BUPRENORPHINE | 9064 | 47 | 715.49 | 15.22 |
| OXYCODONE | 9143 | 194 | 28,062.01 | 144.65 |

| | | | | |
|---|---|---|---|---|
| HYDROMORPHONE | 9150 | 172 | 1,581.41 | 9.19 |
| HYDROCODONE | 9193 | 201 | 15,572.20 | 77.47 |
| MEPERIDINE (PETHIDINE) | 9230 | 132 | 1,709.34 | 12.95 |
| METHADONE | 9250B | 90 | 1,533.80 | 17.04 |
| MORPHINE | 9300 | 208 | 13,774.61 | 66.22 |
| OXYMORPHONE | 9652 | 9 | 193.18 | 21.46 |
| TAPENTADOL | 9780 | 33 | 916.50 | 27.77 |
| FENTANYL BASE | 9801 | 206 | 372.38 | 1.81 |

**BUSINESS ACTIVITY: C - PRACTITIONERS**

| DRUG NAME | DRUG CODE | NUMBER OF REGISTRANT SOLD TO | TOTAL GRAMS | AVG GRAMS |
|---|---|---|---|---|
| AMPHETAMINE | 1100 | 1 | 3.77 | 3.77 |
| METHYLPHENIDATE (DL;D;L;ISOMERS) | 1724 | 1 | 31.65 | 31.65 |
| CODEINE | 9050 | 29 | 964.49 | 33.26 |
| BUPRENORPHINE | 9064 | 280 | 65.25 | 0.23 |
| OXYCODONE | 9143 | 7 | 153.11 | 21.87 |
| HYDROMORPHONE | 9150 | 97 | 68.53 | 0.71 |
| HYDROCODONE | 9193 | 100 | 177.80 | 1.78 |
| MEPERIDINE (PETHIDINE) | 9230 | 20 | 102.40 | 5.12 |
| METHADONE | 9250B | 1 | 1.79 | 1.79 |
| MORPHINE | 9300 | 52 | 61.68 | 1.19 |
| FENTANYL BASE | 9801 | 109 | 12.70 | 0.12 |

**BUSINESS ACTIVITY: D - TEACHING INSTITUTIONS**

| DRUG NAME | DRUG CODE | NUMBER OF REGISTRANT SOLD TO | TOTAL GRAMS | AVG GRAMS |
|---|---|---|---|---|
| BUPRENORPHINE | 9064 | 1 | 0.00 | 0.00 |

**BUSINESS ACTIVITY: M - MID-LEVEL PRACTITIONERS**

| DRUG NAME | DRUG CODE | NUMBER OF REGISTRANT SOLD TO | TOTAL GRAMS | AVG GRAMS |
|---|---|---|---|---|
| HYDROMORPHONE | 9150 | 8 | 0.27 | 0.03 |
| MORPHINE | 9300 | 24 | 5.49 | 0.23 |
| FENTANYL BASE | 9801 | 36 | 0.49 | 0.01 |

**BUSINESS ACTIVITY: NTP - NARCOTIC TREATMENT PROGRAMS**

| DRUG NAME | DRUG CODE | NUMBER OF REGISTRANT SOLD TO | TOTAL GRAMS | AVG GRAMS |
|---|---|---|---|---|
| BUPRENORPHINE | 9064 | 6 | 255.30 | 42.55 |
| METHADONE | 9250B | 10 | 59,855.48 | 5,985.55 |

**STATE: KY - KENTUCKY**

**BUSINESS ACTIVITY: A - PHARMACIES**

| DRUG NAME | DRUG CODE | NUMBER OF REGISTRANT SOLD TO | TOTAL GRAMS | AVG GRAMS |
|---|---|---|---|---|
| AMPHETAMINE | 1100 | 1,217 | 278,149.28 | 228.55 |
| METHAMPHETAMINE | 1105 | 21 | 47.01 | 2.24 |
| LISDEXAMFETAMINE | 1205 | 1,139 | 161,825.22 | 142.08 |
| METHYLPHENIDATE (DL;D;L;ISOMERS) | 1724 | 1,212 | 282,140.45 | 232.79 |

| | | | | |
|---|---|---|---|---|
| CODEINE | 9050 | 1,163 | 164,055.91 | 141.06 |
| BUPRENORPHINE | 9064 | 1,128 | 153,392.31 | 135.99 |
| OXYCODONE | 9143 | 1,228 | 707,528.00 | 576.16 |
| HYDROMORPHONE | 9150 | 789 | 6,981.12 | 8.85 |
| HYDROCODONE | 9193 | 1,239 | 695,694.45 | 561.50 |
| MEPERIDINE (PETHIDINE) | 9230 | 232 | 3,838.19 | 16.54 |
| METHADONE | 9250B | 947 | 39,086.53 | 41.27 |
| MORPHINE | 9300 | 1,176 | 137,635.02 | 117.04 |
| OXYMORPHONE | 9652 | 477 | 7,356.21 | 15.42 |
| TAPENTADOL | 9780 | 528 | 45,918.00 | 86.97 |
| FENTANYL BASE | 9801 | 1,108 | 3,054.26 | 2.76 |

**BUSINESS ACTIVITY: B - HOSPITALS**

| DRUG NAME | DRUG CODE | NUMBER OF REGISTRANT SOLD TO | TOTAL GRAMS | AVG GRAMS |
|---|---|---|---|---|
| AMPHETAMINE | 1100 | 49 | 8,356.59 | 170.54 |
| LISDEXAMFETAMINE | 1205 | 26 | 1,976.14 | 76.01 |
| METHYLPHENIDATE (DL;D;L;ISOMERS) | 1724 | 83 | 8,753.88 | 105.47 |
| CODEINE | 9050 | 122 | 3,630.04 | 29.75 |
| BUPRENORPHINE | 9064 | 89 | 1,678.82 | 18.86 |
| OXYCODONE | 9143 | 145 | 31,652.74 | 218.29 |
| HYDROMORPHONE | 9150 | 131 | 2,374.20 | 18.12 |
| HYDROCODONE | 9193 | 143 | 25,293.38 | 176.88 |
| MEPERIDINE (PETHIDINE) | 9230 | 113 | 4,275.78 | 37.84 |
| METHADONE | 9250B | 101 | 2,607.84 | 25.82 |
| MORPHINE | 9300 | 144 | 17,674.55 | 122.74 |
| OXYMORPHONE | 9652 | 13 | 32.83 | 2.53 |
| TAPENTADOL | 9780 | 20 | 1,867.50 | 93.38 |
| FENTANYL BASE | 9801 | 142 | 477.79 | 3.36 |

**BUSINESS ACTIVITY: C - PRACTITIONERS**

| DRUG NAME | DRUG CODE | NUMBER OF REGISTRANT SOLD TO | TOTAL GRAMS | AVG GRAMS |
|---|---|---|---|---|
| AMPHETAMINE | 1100 | 1 | 1.97 | 1.97 |
| CODEINE | 9050 | 15 | 494.21 | 32.95 |
| BUPRENORPHINE | 9064 | 302 | 81.44 | 0.27 |
| OXYCODONE | 9143 | 19 | 18.24 | 0.96 |
| HYDROMORPHONE | 9150 | 83 | 106.41 | 1.28 |
| HYDROCODONE | 9193 | 130 | 552.94 | 4.25 |
| MEPERIDINE (PETHIDINE) | 9230 | 47 | 434.66 | 9.25 |
| MORPHINE | 9300 | 113 | 360.63 | 3.19 |
| FENTANYL BASE | 9801 | 280 | 25.63 | 0.09 |

**BUSINESS ACTIVITY: NTP - NARCOTIC TREATMENT PROGRAMS**

| DRUG NAME | DRUG CODE | NUMBER OF REGISTRANT SOLD TO | TOTAL GRAMS | AVG GRAMS |
|---|---|---|---|---|

| | | | | |
|---|---|---|---|---|
| BUPRENORPHINE | 9064 | 18 | 477.28 | 26.52 |
| METHADONE | 9250B | 28 | 157,422.45 | 5,622.23 |

**STATE: LA - LOUISIANA**

**BUSINESS ACTIVITY: A - PHARMACIES**

| DRUG NAME | DRUG CODE | NUMBER OF REGISTRANT SOLD TO | TOTAL GRAMS | AVG GRAMS |
|---|---|---|---|---|
| AMPHETAMINE | 1100 | 1,230 | 654,482.80 | 532.10 |
| METHAMPHETAMINE | 1105 | 28 | 60.27 | 2.15 |
| LISDEXAMFETAMINE | 1205 | 1,208 | 354,195.51 | 293.21 |
| METHYLPHENIDATE (DL;D;L;ISOMERS) | 1724 | 1,224 | 321,243.86 | 262.45 |
| CODEINE | 9050 | 1,228 | 149,078.98 | 121.40 |
| BUPRENORPHINE | 9064 | 1,099 | 83,362.67 | 75.85 |
| OXYCODONE | 9143 | 1,241 | 631,264.48 | 508.67 |
| HYDROMORPHONE | 9150 | 934 | 8,707.01 | 9.32 |
| HYDROCODONE | 9193 | 1,253 | 527,188.94 | 420.74 |
| MEPERIDINE (PETHIDINE) | 9230 | 802 | 14,779.38 | 18.43 |
| METHADONE | 9250B | 710 | 26,374.60 | 37.15 |
| MORPHINE | 9300 | 1,120 | 168,893.40 | 150.80 |
| OXYMORPHONE | 9652 | 454 | 15,555.28 | 34.26 |
| TAPENTADOL | 9780 | 583 | 52,487.50 | 90.03 |
| FENTANYL BASE | 9801 | 1,082 | 3,227.06 | 2.98 |

**BUSINESS ACTIVITY: B - HOSPITALS**

| DRUG NAME | DRUG CODE | NUMBER OF REGISTRANT SOLD TO | TOTAL GRAMS | AVG GRAMS |
|---|---|---|---|---|
| AMPHETAMINE | 1100 | 56 | 7,483.22 | 133.63 |
| LISDEXAMFETAMINE | 1205 | 28 | 2,043.58 | 72.99 |
| METHYLPHENIDATE (DL;D;L;ISOMERS) | 1724 | 103 | 4,595.23 | 44.61 |
| CODEINE | 9050 | 139 | 2,848.03 | 20.49 |
| BUPRENORPHINE | 9064 | 98 | 810.64 | 8.27 |
| OXYCODONE | 9143 | 190 | 25,417.47 | 133.78 |
| HYDROMORPHONE | 9150 | 186 | 2,119.97 | 11.40 |
| HYDROCODONE | 9193 | 202 | 31,202.96 | 154.47 |
| MEPERIDINE (PETHIDINE) | 9230 | 157 | 6,920.02 | 44.08 |
| METHADONE | 9250B | 128 | 2,237.34 | 17.48 |
| MORPHINE | 9300 | 196 | 15,767.25 | 80.45 |
| OXYMORPHONE | 9652 | 15 | 50.76 | 3.38 |
| TAPENTADOL | 9780 | 14 | 555.00 | 39.64 |
| FENTANYL BASE | 9801 | 191 | 357.69 | 1.87 |

**BUSINESS ACTIVITY: C - PRACTITIONERS**

| DRUG NAME | DRUG CODE | NUMBER OF REGISTRANT SOLD TO | TOTAL GRAMS | AVG GRAMS |
|---|---|---|---|---|
| LISDEXAMFETAMINE | 1205 | 1 | 4.62 | 4.62 |
| CODEINE | 9050 | 37 | 942.10 | 25.46 |

| | | | | |
|---|---|---|---|---|
| BUPRENORPHINE | 9064 | 330 | 15.81 | 0.05 |
| OXYCODONE | 9143 | 14 | 23.13 | 1.65 |
| HYDROMORPHONE | 9150 | 88 | 45.10 | 0.51 |
| HYDROCODONE | 9193 | 213 | 458.52 | 2.15 |
| MEPERIDINE (PETHIDINE) | 9230 | 106 | 873.68 | 8.24 |
| METHADONE | 9250B | 3 | 6.71 | 2.24 |
| MORPHINE | 9300 | 128 | 98.75 | 0.77 |
| FENTANYL BASE | 9801 | 266 | 18.74 | 0.07 |

**BUSINESS ACTIVITY: D - TEACHING INSTITUTIONS**

| DRUG NAME | DRUG CODE | NUMBER OF REGISTRANT SOLD TO | TOTAL GRAMS | AVG GRAMS |
|---|---|---|---|---|
| FENTANYL BASE | 9801 | 1 | 0.03 | 0.03 |

**BUSINESS ACTIVITY: NTP - NARCOTIC TREATMENT PROGRAMS**

| DRUG NAME | DRUG CODE | NUMBER OF REGISTRANT SOLD TO | TOTAL GRAMS | AVG GRAMS |
|---|---|---|---|---|
| BUPRENORPHINE | 9064 | 9 | 437.37 | 48.60 |
| METHADONE | 9250B | 10 | 109,676.25 | 10,967.62 |

**STATE: MA - MASSACHUSETTS**

**BUSINESS ACTIVITY: A - PHARMACIES**

| DRUG NAME | DRUG CODE | NUMBER OF REGISTRANT SOLD TO | TOTAL GRAMS | AVG GRAMS |
|---|---|---|---|---|
| AMPHETAMINE | 1100 | 1,265 | 704,635.48 | 557.02 |
| METHAMPHETAMINE | 1105 | 40 | 134.61 | 3.37 |
| LISDEXAMFETAMINE | 1205 | 1,199 | 146,648.36 | 122.31 |
| METHYLPHENIDATE (DL;D;L;ISOMERS) | 1724 | 1,255 | 535,484.66 | 426.68 |
| CODEINE | 9050 | 1,199 | 152,677.79 | 127.34 |
| BUPRENORPHINE | 9064 | 1,250 | 164,947.20 | 131.96 |
| OXYCODONE | 9143 | 1,268 | 762,766.91 | 601.55 |
| HYDROMORPHONE | 9150 | 1,168 | 18,259.23 | 15.63 |
| HYDROCODONE | 9193 | 1,254 | 97,843.02 | 78.02 |
| MEPERIDINE (PETHIDINE) | 9230 | 166 | 5,105.57 | 30.76 |
| METHADONE | 9250B | 1,040 | 56,697.00 | 54.52 |
| MORPHINE | 9300 | 1,188 | 204,749.94 | 172.35 |
| OXYMORPHONE | 9652 | 184 | 3,718.59 | 20.21 |
| TAPENTADOL | 9780 | 471 | 63,273.50 | 134.34 |
| FENTANYL BASE | 9801 | 1,111 | 3,603.34 | 3.24 |

**BUSINESS ACTIVITY: B - HOSPITALS**

| DRUG NAME | DRUG CODE | NUMBER OF REGISTRANT SOLD TO | TOTAL GRAMS | AVG GRAMS |
|---|---|---|---|---|
| AMPHETAMINE | 1100 | 144 | 27,491.64 | 190.91 |
| METHAMPHETAMINE | 1105 | 1 | 0.80 | 0.80 |
| LISDEXAMFETAMINE | 1205 | 72 | 3,806.79 | 52.87 |
| METHYLPHENIDATE (DL;D;L;ISOMERS) | 1724 | 157 | 26,146.19 | 166.54 |
| CODEINE | 9050 | 160 | 7,145.63 | 44.66 |

| | | | | |
|---|---|---|---|---|
| BUPRENORPHINE | 9064 | 159 | 9,725.63 | 61.17 |
| OXYCODONE | 9143 | 196 | 61,845.73 | 315.54 |
| HYDROMORPHONE | 9150 | 171 | 7,467.80 | 43.67 |
| HYDROCODONE | 9193 | 173 | 3,558.25 | 20.57 |
| MEPERIDINE (PETHIDINE) | 9230 | 107 | 1,536.50 | 14.36 |
| METHADONE | 9250B | 149 | 11,464.43 | 76.94 |
| MORPHINE | 9300 | 187 | 26,066.45 | 139.39 |
| OXYMORPHONE | 9652 | 16 | 404.48 | 25.28 |
| TAPENTADOL | 9780 | 17 | 2,784.00 | 163.76 |
| FENTANYL BASE | 9801 | 183 | 644.44 | 3.52 |

**BUSINESS ACTIVITY: C - PRACTITIONERS**

| DRUG NAME | DRUG CODE | NUMBER OF REGISTRANT SOLD TO | TOTAL GRAMS | AVG GRAMS |
|---|---|---|---|---|
| METHAMPHETAMINE | 1105 | 1 | 4.02 | 4.02 |
| CODEINE | 9050 | 26 | 1,144.27 | 44.01 |
| BUPRENORPHINE | 9064 | 578 | 58.43 | 0.10 |
| OXYCODONE | 9143 | 21 | 30.21 | 1.44 |
| HYDROMORPHONE | 9150 | 237 | 167.06 | 0.70 |
| HYDROCODONE | 9193 | 131 | 433.66 | 3.31 |
| MEPERIDINE (PETHIDINE) | 9230 | 26 | 325.72 | 12.53 |
| METHADONE | 9250B | 29 | 59.21 | 2.04 |
| MORPHINE | 9300 | 125 | 171.19 | 1.37 |
| FENTANYL BASE | 9801 | 301 | 27.35 | 0.09 |

**BUSINESS ACTIVITY: D - TEACHING INSTITUTIONS**

| DRUG NAME | DRUG CODE | NUMBER OF REGISTRANT SOLD TO | TOTAL GRAMS | AVG GRAMS |
|---|---|---|---|---|
| BUPRENORPHINE | 9064 | 1 | 0.01 | 0.01 |

**BUSINESS ACTIVITY: M - MID-LEVEL PRACTITIONERS**

| DRUG NAME | DRUG CODE | NUMBER OF REGISTRANT SOLD TO | TOTAL GRAMS | AVG GRAMS |
|---|---|---|---|---|
| CODEINE | 9050 | 1 | 0.43 | 0.43 |

**BUSINESS ACTIVITY: NTP - NARCOTIC TREATMENT PROGRAMS**

| DRUG NAME | DRUG CODE | NUMBER OF REGISTRANT SOLD TO | TOTAL GRAMS | AVG GRAMS |
|---|---|---|---|---|
| BUPRENORPHINE | 9064 | 37 | 680.24 | 18.38 |
| METHADONE | 9250B | 78 | 450,574.57 | 5,776.60 |

**STATE: MD - MARYLAND**

**BUSINESS ACTIVITY: A - PHARMACIES**

| DRUG NAME | DRUG CODE | NUMBER OF REGISTRANT SOLD TO | TOTAL GRAMS | AVG GRAMS |
|---|---|---|---|---|
| AMPHETAMINE | 1100 | 1,334 | 375,249.98 | 281.30 |
| METHAMPHETAMINE | 1105 | 39 | 154.29 | 3.96 |
| LISDEXAMFETAMINE | 1205 | 1,298 | 224,318.99 | 172.82 |
| METHYLPHENIDATE (DL;D;L;ISOMERS) | 1724 | 1,322 | 374,557.60 | 283.33 |
| CODEINE | 9050 | 1,341 | 185,094.50 | 138.03 |

| | | | | |
|---|---|---|---|---|
| BUPRENORPHINE | 9064 | 1,288 | 106,477.03 | 82.67 |
| OXYCODONE | 9143 | 1,365 | 1,044,758.61 | 765.39 |
| HYDROMORPHONE | 9150 | 1,215 | 21,280.10 | 17.51 |
| HYDROCODONE | 9193 | 1,331 | 108,934.26 | 81.84 |
| MEPERIDINE (PETHIDINE) | 9230 | 283 | 5,606.47 | 19.81 |
| METHADONE | 9250B | 994 | 41,629.49 | 41.88 |
| MORPHINE | 9300 | 1,258 | 224,612.70 | 178.55 |
| OXYMORPHONE | 9652 | 757 | 23,188.71 | 30.63 |
| TAPENTADOL | 9780 | 816 | 100,455.00 | 123.11 |
| FENTANYL BASE | 9801 | 1,169 | 3,558.97 | 3.04 |

**BUSINESS ACTIVITY: B - HOSPITALS**

| DRUG NAME | DRUG CODE | NUMBER OF REGISTRANT SOLD TO | TOTAL GRAMS | AVG GRAMS |
|---|---|---|---|---|
| AMPHETAMINE | 1100 | 53 | 10,237.12 | 193.15 |
| LISDEXAMFETAMINE | 1205 | 25 | 2,115.50 | 84.62 |
| METHYLPHENIDATE (DL;D;L;ISOMERS) | 1724 | 67 | 14,026.67 | 209.35 |
| CODEINE | 9050 | 84 | 4,346.01 | 51.74 |
| BUPRENORPHINE | 9064 | 72 | 2,051.90 | 28.50 |
| OXYCODONE | 9143 | 133 | 43,986.07 | 330.72 |
| HYDROMORPHONE | 9150 | 130 | 4,400.75 | 33.85 |
| HYDROCODONE | 9193 | 116 | 3,569.96 | 30.78 |
| MEPERIDINE (PETHIDINE) | 9230 | 86 | 1,530.57 | 17.80 |
| METHADONE | 9250B | 64 | 5,765.45 | 90.09 |
| MORPHINE | 9300 | 120 | 17,968.53 | 149.74 |
| OXYMORPHONE | 9652 | 6 | 109.37 | 18.23 |
| TAPENTADOL | 9780 | 16 | 1,914.50 | 119.66 |
| FENTANYL BASE | 9801 | 203 | 460.12 | 2.27 |

**BUSINESS ACTIVITY: C - PRACTITIONERS**

| DRUG NAME | DRUG CODE | NUMBER OF REGISTRANT SOLD TO | TOTAL GRAMS | AVG GRAMS |
|---|---|---|---|---|
| AMPHETAMINE | 1100 | 2 | 4.78 | 2.39 |
| CODEINE | 9050 | 178 | 6,649.25 | 37.36 |
| BUPRENORPHINE | 9064 | 593 | 252.90 | 0.43 |
| OXYCODONE | 9143 | 62 | 2,616.50 | 42.20 |
| HYDROMORPHONE | 9150 | 253 | 245.12 | 0.97 |
| HYDROCODONE | 9193 | 249 | 1,328.28 | 5.33 |
| MEPERIDINE (PETHIDINE) | 9230 | 62 | 557.00 | 8.98 |
| METHADONE | 9250B | 18 | 88.46 | 4.91 |
| MORPHINE | 9300 | 165 | 984.06 | 5.96 |
| OXYMORPHONE | 9652 | 12 | 144.95 | 12.08 |
| TAPENTADOL | 9780 | 7 | 273.00 | 39.00 |
| FENTANYL BASE | 9801 | 367 | 51.62 | 0.14 |

**BUSINESS ACTIVITY: D - TEACHING INSTITUTIONS**

| DRUG NAME | DRUG CODE | NUMBER OF REGISTRANT SOLD TO | TOTAL GRAMS | AVG GRAMS |
|---|---|---|---|---|
| MEPERIDINE (PETHIDINE) | 9230 | 1 | 4.36 | 4.36 |
| MORPHINE | 9300 | 1 | 0.08 | 0.08 |
| FENTANYL BASE | 9801 | 1 | 0.02 | 0.02 |

**BUSINESS ACTIVITY: M - MID-LEVEL PRACTITIONERS**

| DRUG NAME | DRUG CODE | NUMBER OF REGISTRANT SOLD TO | TOTAL GRAMS | AVG GRAMS |
|---|---|---|---|---|
| CODEINE | 9050 | 6 | 6.93 | 1.16 |
| BUPRENORPHINE | 9064 | 3 | 2.58 | 0.86 |
| OXYCODONE | 9143 | 1 | 0.90 | 0.90 |

**BUSINESS ACTIVITY: NTP - NARCOTIC TREATMENT PROGRAMS**

| DRUG NAME | DRUG CODE | NUMBER OF REGISTRANT SOLD TO | TOTAL GRAMS | AVG GRAMS |
|---|---|---|---|---|
| BUPRENORPHINE | 9064 | 52 | 7,199.09 | 138.44 |
| METHADONE | 9250B | 84 | 668,653.44 | 7,960.16 |

**STATE: ME - MAINE**

**BUSINESS ACTIVITY: A - PHARMACIES**

| DRUG NAME | DRUG CODE | NUMBER OF REGISTRANT SOLD TO | TOTAL GRAMS | AVG GRAMS |
|---|---|---|---|---|
| AMPHETAMINE | 1100 | 379 | 94,855.24 | 250.28 |
| METHAMPHETAMINE | 1105 | 10 | 53.04 | 5.30 |
| LISDEXAMFETAMINE | 1205 | 375 | 66,592.50 | 177.58 |
| METHYLPHENIDATE (DL;D;L;ISOMERS) | 1724 | 381 | 136,275.89 | 357.68 |
| CODEINE | 9050 | 341 | 34,290.99 | 100.56 |
| BUPRENORPHINE | 9064 | 384 | 43,235.25 | 112.59 |
| OXYCODONE | 9143 | 382 | 171,544.25 | 449.07 |
| HYDROMORPHONE | 9150 | 354 | 4,816.31 | 13.61 |
| HYDROCODONE | 9193 | 383 | 57,052.90 | 148.96 |
| MEPERIDINE (PETHIDINE) | 9230 | 53 | 1,137.20 | 21.46 |
| METHADONE | 9250B | 338 | 14,636.68 | 43.30 |
| MORPHINE | 9300 | 368 | 51,741.73 | 140.60 |
| OXYMORPHONE | 9652 | 133 | 1,787.90 | 13.44 |
| TAPENTADOL | 9780 | 102 | 4,651.00 | 45.60 |
| FENTANYL BASE | 9801 | 359 | 970.29 | 2.70 |

**BUSINESS ACTIVITY: B - HOSPITALS**

| DRUG NAME | DRUG CODE | NUMBER OF REGISTRANT SOLD TO | TOTAL GRAMS | AVG GRAMS |
|---|---|---|---|---|
| AMPHETAMINE | 1100 | 24 | 1,552.07 | 64.67 |
| LISDEXAMFETAMINE | 1205 | 13 | 395.42 | 30.42 |
| METHYLPHENIDATE (DL;D;L;ISOMERS) | 1724 | 38 | 2,229.84 | 58.68 |
| CODEINE | 9050 | 38 | 851.57 | 22.41 |
| BUPRENORPHINE | 9064 | 41 | 1,116.27 | 27.23 |
| OXYCODONE | 9143 | 48 | 8,763.51 | 182.57 |

| | | | | |
|---|---|---|---|---|
| HYDROMORPHONE | 9150 | 41 | 958.91 | 23.39 |
| HYDROCODONE | 9193 | 45 | 3,238.50 | 71.97 |
| MEPERIDINE (PETHIDINE) | 9230 | 33 | 130.94 | 3.97 |
| METHADONE | 9250B | 37 | 1,115.23 | 30.14 |
| MORPHINE | 9300 | 43 | 4,645.62 | 108.04 |
| OXYMORPHONE | 9652 | 1 | 0.27 | 0.27 |
| TAPENTADOL | 9780 | 1 | 45.00 | 45.00 |
| FENTANYL BASE | 9801 | 43 | 125.36 | 2.92 |

**BUSINESS ACTIVITY: C - PRACTITIONERS**

| DRUG NAME | DRUG CODE | NUMBER OF REGISTRANT SOLD TO | TOTAL GRAMS | AVG GRAMS |
|---|---|---|---|---|
| CODEINE | 9050 | 6 | 277.24 | 46.21 |
| BUPRENORPHINE | 9064 | 161 | 10.49 | 0.07 |
| OXYCODONE | 9143 | 3 | 20.62 | 6.87 |
| HYDROMORPHONE | 9150 | 76 | 46.09 | 0.61 |
| HYDROCODONE | 9193 | 52 | 93.78 | 1.80 |
| MEPERIDINE (PETHIDINE) | 9230 | 4 | 45.21 | 11.30 |
| METHADONE | 9250B | 1 | 0.27 | 0.27 |
| MORPHINE | 9300 | 38 | 64.94 | 1.71 |
| FENTANYL BASE | 9801 | 48 | 6.10 | 0.13 |

**BUSINESS ACTIVITY: D - TEACHING INSTITUTIONS**

| DRUG NAME | DRUG CODE | NUMBER OF REGISTRANT SOLD TO | TOTAL GRAMS | AVG GRAMS |
|---|---|---|---|---|
| BUPRENORPHINE | 9064 | 1 | 0.01 | 0.01 |

**BUSINESS ACTIVITY: M - MID-LEVEL PRACTITIONERS**

| DRUG NAME | DRUG CODE | NUMBER OF REGISTRANT SOLD TO | TOTAL GRAMS | AVG GRAMS |
|---|---|---|---|---|
| CODEINE | 9050 | 3 | 12.44 | 4.15 |

**BUSINESS ACTIVITY: NTP - NARCOTIC TREATMENT PROGRAMS**

| DRUG NAME | DRUG CODE | NUMBER OF REGISTRANT SOLD TO | TOTAL GRAMS | AVG GRAMS |
|---|---|---|---|---|
| BUPRENORPHINE | 9064 | 5 | 516.66 | 103.33 |
| METHADONE | 9250B | 10 | 90,566.05 | 9,056.60 |

**STATE: MI - MICHIGAN**

**BUSINESS ACTIVITY: A - PHARMACIES**

| DRUG NAME | DRUG CODE | NUMBER OF REGISTRANT SOLD TO | TOTAL GRAMS | AVG GRAMS |
|---|---|---|---|---|
| AMPHETAMINE | 1100 | 2,376 | 959,614.12 | 403.88 |
| METHAMPHETAMINE | 1105 | 29 | 100.45 | 3.46 |
| LISDEXAMFETAMINE | 1205 | 2,152 | 240,905.83 | 111.95 |
| METHYLPHENIDATE (DL;D;L;ISOMERS) | 1724 | 2,298 | 694,758.73 | 302.33 |
| CODEINE | 9050 | 2,420 | 788,648.21 | 325.89 |
| BUPRENORPHINE | 9064 | 2,192 | 118,762.03 | 54.18 |
| OXYCODONE | 9143 | 2,418 | 894,155.00 | 369.79 |
| HYDROMORPHONE | 9150 | 1,837 | 22,766.01 | 12.39 |

| | | | | |
|---|---|---|---|---|
| HYDROCODONE | 9193 | 2,456 | 1,166,285.44 | 474.87 |
| MEPERIDINE (PETHIDINE) | 9230 | 217 | 5,897.77 | 27.18 |
| METHADONE | 9250B | 1,819 | 103,856.48 | 57.10 |
| MORPHINE | 9300 | 2,259 | 550,960.91 | 243.90 |
| OXYMORPHONE | 9652 | 831 | 46,343.44 | 55.77 |
| TAPENTADOL | 9780 | 841 | 66,759.50 | 79.38 |
| FENTANYL BASE | 9801 | 2,106 | 8,375.77 | 3.98 |

**BUSINESS ACTIVITY: B - HOSPITALS**

| DRUG NAME | DRUG CODE | NUMBER OF REGISTRANT SOLD TO | TOTAL GRAMS | AVG GRAMS |
|---|---|---|---|---|
| AMPHETAMINE | 1100 | 112 | 19,196.96 | 171.40 |
| METHAMPHETAMINE | 1105 | 1 | 0.80 | 0.80 |
| LISDEXAMFETAMINE | 1205 | 61 | 3,891.19 | 63.79 |
| METHYLPHENIDATE (DL;D;L;ISOMERS) | 1724 | 154 | 17,291.35 | 112.28 |
| CODEINE | 9050 | 188 | 17,946.68 | 95.46 |
| BUPRENORPHINE | 9064 | 125 | 2,802.41 | 22.42 |
| OXYCODONE | 9143 | 192 | 50,964.54 | 265.44 |
| HYDROMORPHONE | 9150 | 184 | 4,884.48 | 26.55 |
| HYDROCODONE | 9193 | 203 | 57,384.53 | 282.68 |
| MEPERIDINE (PETHIDINE) | 9230 | 119 | 2,670.91 | 22.44 |
| METHADONE | 9250B | 162 | 7,195.09 | 44.41 |
| MORPHINE | 9300 | 197 | 45,992.82 | 233.47 |
| OXYMORPHONE | 9652 | 17 | 313.31 | 18.43 |
| TAPENTADOL | 9780 | 15 | 771.00 | 51.40 |
| FENTANYL BASE | 9801 | 190 | 962.07 | 5.06 |

**BUSINESS ACTIVITY: C - PRACTITIONERS**

| DRUG NAME | DRUG CODE | NUMBER OF REGISTRANT SOLD TO | TOTAL GRAMS | AVG GRAMS |
|---|---|---|---|---|
| AMPHETAMINE | 1100 | 5 | 16.41 | 3.28 |
| LISDEXAMFETAMINE | 1205 | 2 | 6.56 | 3.28 |
| CODEINE | 9050 | 136 | 2,450.04 | 18.02 |
| BUPRENORPHINE | 9064 | 773 | 374.46 | 0.48 |
| OXYCODONE | 9143 | 40 | 211.66 | 5.29 |
| HYDROMORPHONE | 9150 | 367 | 247.86 | 0.68 |
| HYDROCODONE | 9193 | 283 | 1,159.51 | 4.10 |
| MEPERIDINE (PETHIDINE) | 9230 | 109 | 790.89 | 7.26 |
| METHADONE | 9250B | 8 | 30.32 | 3.79 |
| MORPHINE | 9300 | 292 | 569.21 | 1.95 |
| FENTANYL BASE | 9801 | 537 | 66.59 | 0.12 |

**BUSINESS ACTIVITY: D - TEACHING INSTITUTIONS**

| DRUG NAME | DRUG CODE | NUMBER OF REGISTRANT SOLD TO | TOTAL GRAMS | AVG GRAMS |
|---|---|---|---|---|
| CODEINE | 9050 | 1 | 611.21 | 611.21 |

| | | | | |
|---|---|---|---|---|
| BUPRENORPHINE | 9064 | 1 | 1.56 | 1.56 |
| HYDROMORPHONE | 9150 | 1 | 7.54 | 7.54 |
| HYDROCODONE | 9193 | 1 | 24.18 | 24.18 |
| MEPERIDINE (PETHIDINE) | 9230 | 1 | 1.09 | 1.09 |
| METHADONE | 9250B | 1 | 25.40 | 25.40 |
| MORPHINE | 9300 | 1 | 4.85 | 4.85 |
| FENTANYL BASE | 9801 | 2 | 1.51 | 0.76 |

**BUSINESS ACTIVITY: M - MID-LEVEL PRACTITIONERS**

| DRUG NAME | DRUG CODE | NUMBER OF REGISTRANT SOLD TO | TOTAL GRAMS | AVG GRAMS |
|---|---|---|---|---|
| CODEINE | 9050 | 2 | 338.58 | 169.29 |
| BUPRENORPHINE | 9064 | 2 | 0.64 | 0.32 |

**BUSINESS ACTIVITY: NTP - NARCOTIC TREATMENT PROGRAMS**

| DRUG NAME | DRUG CODE | NUMBER OF REGISTRANT SOLD TO | TOTAL GRAMS | AVG GRAMS |
|---|---|---|---|---|
| BUPRENORPHINE | 9064 | 11 | 215.22 | 19.57 |
| METHADONE | 9250B | 42 | 378,667.05 | 9,015.88 |

**STATE: MN - MINNESOTA**

**BUSINESS ACTIVITY: A - PHARMACIES**

| DRUG NAME | DRUG CODE | NUMBER OF REGISTRANT SOLD TO | TOTAL GRAMS | AVG GRAMS |
|---|---|---|---|---|
| AMPHETAMINE | 1100 | 1,027 | 439,492.69 | 427.94 |
| METHAMPHETAMINE | 1105 | 49 | 233.04 | 4.76 |
| LISDEXAMFETAMINE | 1205 | 1,005 | 122,699.99 | 122.09 |
| METHYLPHENIDATE (DL;D;L;ISOMERS) | 1724 | 1,028 | 391,173.73 | 380.52 |
| CODEINE | 9050 | 1,023 | 122,931.81 | 120.17 |
| BUPRENORPHINE | 9064 | 926 | 32,584.60 | 35.19 |
| OXYCODONE | 9143 | 1,040 | 450,637.78 | 433.31 |
| HYDROMORPHONE | 9150 | 979 | 15,891.57 | 16.23 |
| HYDROCODONE | 9193 | 1,043 | 138,540.20 | 132.83 |
| MEPERIDINE (PETHIDINE) | 9230 | 89 | 1,094.39 | 12.30 |
| METHADONE | 9250B | 837 | 25,493.53 | 30.46 |
| MORPHINE | 9300 | 1,016 | 138,289.02 | 136.11 |
| OXYMORPHONE | 9652 | 108 | 840.80 | 7.79 |
| TAPENTADOL | 9780 | 269 | 14,798.00 | 55.01 |
| FENTANYL BASE | 9801 | 963 | 2,766.26 | 2.87 |

**BUSINESS ACTIVITY: B - HOSPITALS**

| DRUG NAME | DRUG CODE | NUMBER OF REGISTRANT SOLD TO | TOTAL GRAMS | AVG GRAMS |
|---|---|---|---|---|
| AMPHETAMINE | 1100 | 75 | 7,218.12 | 96.24 |
| METHAMPHETAMINE | 1105 | 3 | 67.91 | 22.64 |
| LISDEXAMFETAMINE | 1205 | 46 | 1,561.45 | 33.94 |
| METHYLPHENIDATE (DL;D;L;ISOMERS) | 1724 | 106 | 7,510.23 | 70.85 |
| CODEINE | 9050 | 123 | 4,553.83 | 37.02 |

| | | | | |
|---|---|---|---|---|
| BUPRENORPHINE | 9064 | 82 | 1,347.39 | 16.43 |
| OXYCODONE | 9143 | 152 | 30,603.81 | 201.34 |
| HYDROMORPHONE | 9150 | 154 | 4,199.95 | 27.27 |
| HYDROCODONE | 9193 | 150 | 10,130.24 | 67.53 |
| MEPERIDINE (PETHIDINE) | 9230 | 96 | 758.68 | 7.90 |
| METHADONE | 9250B | 98 | 2,860.65 | 29.19 |
| MORPHINE | 9300 | 156 | 16,474.29 | 105.60 |
| OXYMORPHONE | 9652 | 6 | 24.62 | 4.10 |
| TAPENTADOL | 9780 | 13 | 152.50 | 11.73 |
| FENTANYL BASE | 9801 | 150 | 420.84 | 2.81 |

**BUSINESS ACTIVITY: C - PRACTITIONERS**

| DRUG NAME | DRUG CODE | NUMBER OF REGISTRANT SOLD TO | TOTAL GRAMS | AVG GRAMS |
|---|---|---|---|---|
| AMPHETAMINE | 1100 | 1 | 15.21 | 15.21 |
| METHYLPHENIDATE (DL;D;L;ISOMERS) | 1724 | 2 | 19.66 | 9.83 |
| CODEINE | 9050 | 46 | 1,723.08 | 37.46 |
| BUPRENORPHINE | 9064 | 562 | 40.71 | 0.07 |
| OXYCODONE | 9143 | 56 | 286.70 | 5.12 |
| HYDROMORPHONE | 9150 | 297 | 143.47 | 0.48 |
| HYDROCODONE | 9193 | 264 | 549.32 | 2.08 |
| MEPERIDINE (PETHIDINE) | 9230 | 45 | 298.38 | 6.63 |
| METHADONE | 9250B | 4 | 6.53 | 1.63 |
| MORPHINE | 9300 | 219 | 195.64 | 0.89 |
| FENTANYL BASE | 9801 | 374 | 35.88 | 0.10 |

**BUSINESS ACTIVITY: D - TEACHING INSTITUTIONS**

| DRUG NAME | DRUG CODE | NUMBER OF REGISTRANT SOLD TO | TOTAL GRAMS | AVG GRAMS |
|---|---|---|---|---|
| BUPRENORPHINE | 9064 | 1 | 0.00 | 0.00 |

**BUSINESS ACTIVITY: M - MID-LEVEL PRACTITIONERS**

| DRUG NAME | DRUG CODE | NUMBER OF REGISTRANT SOLD TO | TOTAL GRAMS | AVG GRAMS |
|---|---|---|---|---|
| CODEINE | 9050 | 3 | 10.83 | 3.61 |
| FENTANYL BASE | 9801 | 1 | 0.01 | 0.01 |

**BUSINESS ACTIVITY: NTP - NARCOTIC TREATMENT PROGRAMS**

| DRUG NAME | DRUG CODE | NUMBER OF REGISTRANT SOLD TO | TOTAL GRAMS | AVG GRAMS |
|---|---|---|---|---|
| BUPRENORPHINE | 9064 | 14 | 2,240.19 | 160.01 |
| METHADONE | 9250B | 15 | 190,731.87 | 12,715.46 |

**STATE: MO - MISSOURI**

**BUSINESS ACTIVITY: A - PHARMACIES**

| DRUG NAME | DRUG CODE | NUMBER OF REGISTRANT SOLD TO | TOTAL GRAMS | AVG GRAMS |
|---|---|---|---|---|
| AMPHETAMINE | 1100 | 1,319 | 560,171.60 | 424.69 |
| METHAMPHETAMINE | 1105 | 30 | 172.36 | 5.75 |
| LISDEXAMFETAMINE | 1205 | 1,277 | 265,255.40 | 207.72 |

| | | | | |
|---|---|---|---|---|
| METHYLPHENIDATE (DL;D;L;ISOMERS) | 1724 | 1,323 | 506,083.09 | 382.53 |
| CODEINE | 9050 | 1,312 | 233,745.86 | 178.16 |
| BUPRENORPHINE | 9064 | 1,259 | 51,105.45 | 40.59 |
| OXYCODONE | 9143 | 1,334 | 908,874.82 | 681.32 |
| HYDROMORPHONE | 9150 | 1,148 | 17,222.63 | 15.00 |
| HYDROCODONE | 9193 | 1,334 | 562,527.62 | 421.68 |
| MEPERIDINE (PETHIDINE) | 9230 | 422 | 8,324.46 | 19.73 |
| METHADONE | 9250B | 1,105 | 39,680.77 | 35.91 |
| MORPHINE | 9300 | 1,303 | 293,622.40 | 225.34 |
| OXYMORPHONE | 9652 | 457 | 6,881.44 | 15.06 |
| TAPENTADOL | 9780 | 793 | 79,994.50 | 100.88 |
| FENTANYL BASE | 9801 | 1,266 | 6,131.77 | 4.84 |

**BUSINESS ACTIVITY: B - HOSPITALS**

| DRUG NAME | DRUG CODE | NUMBER OF REGISTRANT SOLD TO | TOTAL GRAMS | AVG GRAMS |
|---|---|---|---|---|
| AMPHETAMINE | 1100 | 100 | 10,079.30 | 100.79 |
| LISDEXAMFETAMINE | 1205 | 46 | 3,027.91 | 65.82 |
| METHYLPHENIDATE (DL;D;L;ISOMERS) | 1724 | 125 | 9,262.19 | 74.10 |
| CODEINE | 9050 | 168 | 5,915.60 | 35.21 |
| BUPRENORPHINE | 9064 | 98 | 1,291.89 | 13.18 |
| OXYCODONE | 9143 | 230 | 45,906.22 | 199.59 |
| HYDROMORPHONE | 9150 | 187 | 2,882.75 | 15.42 |
| HYDROCODONE | 9193 | 241 | 23,325.19 | 96.79 |
| MEPERIDINE (PETHIDINE) | 9230 | 159 | 2,689.56 | 16.92 |
| METHADONE | 9250B | 119 | 3,453.41 | 29.02 |
| MORPHINE | 9300 | 231 | 27,120.34 | 117.40 |
| OXYMORPHONE | 9652 | 14 | 30.78 | 2.20 |
| TAPENTADOL | 9780 | 29 | 1,255.50 | 43.29 |
| FENTANYL BASE | 9801 | 261 | 626.80 | 2.40 |

**BUSINESS ACTIVITY: C - PRACTITIONERS**

| DRUG NAME | DRUG CODE | NUMBER OF REGISTRANT SOLD TO | TOTAL GRAMS | AVG GRAMS |
|---|---|---|---|---|
| AMPHETAMINE | 1100 | 1 | 1.90 | 1.90 |
| CODEINE | 9050 | 52 | 3,226.41 | 62.05 |
| BUPRENORPHINE | 9064 | 453 | 54.65 | 0.12 |
| OXYCODONE | 9143 | 10 | 125.73 | 12.57 |
| HYDROMORPHONE | 9150 | 133 | 120.98 | 0.91 |
| HYDROCODONE | 9193 | 220 | 1,565.55 | 7.12 |
| MEPERIDINE (PETHIDINE) | 9230 | 17 | 300.67 | 17.69 |
| METHADONE | 9250B | 7 | 13.15 | 1.88 |
| MORPHINE | 9300 | 123 | 123.97 | 1.01 |
| FENTANYL BASE | 9801 | 153 | 14.51 | 0.09 |

**BUSINESS ACTIVITY: D - TEACHING INSTITUTIONS**

| DRUG NAME | DRUG CODE | NUMBER OF REGISTRANT SOLD TO | TOTAL GRAMS | AVG GRAMS |
|---|---|---|---|---|
| CODEINE | 9050 | 1 | 192.28 | 192.28 |
| BUPRENORPHINE | 9064 | 2 | 1.22 | 0.61 |
| HYDROMORPHONE | 9150 | 2 | 4.79 | 2.39 |
| HYDROCODONE | 9193 | 2 | 14.24 | 7.12 |
| MORPHINE | 9300 | 2 | 14.39 | 7.20 |
| FENTANYL BASE | 9801 | 3 | 1.89 | 0.63 |

**BUSINESS ACTIVITY: M - MID-LEVEL PRACTITIONERS**

| DRUG NAME | DRUG CODE | NUMBER OF REGISTRANT SOLD TO | TOTAL GRAMS | AVG GRAMS |
|---|---|---|---|---|
| HYDROMORPHONE | 9150 | 5 | 0.16 | 0.03 |
| MORPHINE | 9300 | 44 | 21.73 | 0.49 |
| FENTANYL BASE | 9801 | 63 | 0.69 | 0.01 |

**BUSINESS ACTIVITY: NTP - NARCOTIC TREATMENT PROGRAMS**

| DRUG NAME | DRUG CODE | NUMBER OF REGISTRANT SOLD TO | TOTAL GRAMS | AVG GRAMS |
|---|---|---|---|---|
| BUPRENORPHINE | 9064 | 16 | 1,186.57 | 74.16 |
| METHADONE | 9250B | 16 | 103,920.78 | 6,495.05 |

**STATE: MS - MISSISSIPPI**

**BUSINESS ACTIVITY: A - PHARMACIES**

| DRUG NAME | DRUG CODE | NUMBER OF REGISTRANT SOLD TO | TOTAL GRAMS | AVG GRAMS |
|---|---|---|---|---|
| AMPHETAMINE | 1100 | 781 | 240,506.87 | 307.95 |
| METHAMPHETAMINE | 1105 | 6 | 30.94 | 5.16 |
| LISDEXAMFETAMINE | 1205 | 758 | 145,611.83 | 192.10 |
| METHYLPHENIDATE (DL;D;L;ISOMERS) | 1724 | 770 | 159,790.15 | 207.52 |
| CODEINE | 9050 | 759 | 87,988.78 | 115.93 |
| BUPRENORPHINE | 9064 | 710 | 41,557.25 | 58.53 |
| OXYCODONE | 9143 | 792 | 344,134.51 | 434.51 |
| HYDROMORPHONE | 9150 | 573 | 18,997.44 | 33.15 |
| HYDROCODONE | 9193 | 800 | 346,071.35 | 432.59 |
| MEPERIDINE (PETHIDINE) | 9230 | 450 | 8,023.68 | 17.83 |
| METHADONE | 9250B | 429 | 8,946.50 | 20.85 |
| MORPHINE | 9300 | 748 | 121,860.94 | 162.92 |
| OXYMORPHONE | 9652 | 222 | 5,785.40 | 26.06 |
| TAPENTADOL | 9780 | 435 | 34,749.00 | 79.88 |
| FENTANYL BASE | 9801 | 751 | 3,555.25 | 4.73 |

**BUSINESS ACTIVITY: B - HOSPITALS**

| DRUG NAME | DRUG CODE | NUMBER OF REGISTRANT SOLD TO | TOTAL GRAMS | AVG GRAMS |
|---|---|---|---|---|
| AMPHETAMINE | 1100 | 54 | 6,840.40 | 126.67 |
| LISDEXAMFETAMINE | 1205 | 35 | 4,012.01 | 114.63 |
| METHYLPHENIDATE (DL;D;L;ISOMERS) | 1724 | 64 | 5,132.98 | 80.20 |

| | | | | |
|---|---|---|---|---|
| CODEINE | 9050 | 111 | 3,981.98 | 35.87 |
| BUPRENORPHINE | 9064 | 56 | 1,120.14 | 20.00 |
| OXYCODONE | 9143 | 135 | 14,017.54 | 103.83 |
| HYDROMORPHONE | 9150 | 123 | 1,569.01 | 12.76 |
| HYDROCODONE | 9193 | 150 | 17,902.26 | 119.35 |
| MEPERIDINE (PETHIDINE) | 9230 | 116 | 5,017.07 | 43.25 |
| METHADONE | 9250B | 59 | 473.41 | 8.02 |
| MORPHINE | 9300 | 142 | 9,834.76 | 69.26 |
| OXYMORPHONE | 9652 | 7 | 50.49 | 7.21 |
| TAPENTADOL | 9780 | 18 | 1,237.00 | 68.72 |
| FENTANYL BASE | 9801 | 137 | 316.88 | 2.31 |

**BUSINESS ACTIVITY: C - PRACTITIONERS**

| DRUG NAME | DRUG CODE | NUMBER OF REGISTRANT SOLD TO | TOTAL GRAMS | AVG GRAMS |
|---|---|---|---|---|
| CODEINE | 9050 | 12 | 674.08 | 56.17 |
| BUPRENORPHINE | 9064 | 118 | 6.32 | 0.05 |
| OXYCODONE | 9143 | 12 | 23.04 | 1.92 |
| HYDROMORPHONE | 9150 | 36 | 20.09 | 0.56 |
| HYDROCODONE | 9193 | 40 | 87.45 | 2.19 |
| MEPERIDINE (PETHIDINE) | 9230 | 39 | 422.13 | 10.82 |
| METHADONE | 9250B | 2 | 0.89 | 0.45 |
| MORPHINE | 9300 | 51 | 53.69 | 1.05 |
| FENTANYL BASE | 9801 | 107 | 7.60 | 0.07 |

**BUSINESS ACTIVITY: D - TEACHING INSTITUTIONS**

| DRUG NAME | DRUG CODE | NUMBER OF REGISTRANT SOLD TO | TOTAL GRAMS | AVG GRAMS |
|---|---|---|---|---|
| BUPRENORPHINE | 9064 | 1 | 0.01 | 0.01 |

**BUSINESS ACTIVITY: M - MID-LEVEL PRACTITIONERS**

| DRUG NAME | DRUG CODE | NUMBER OF REGISTRANT SOLD TO | TOTAL GRAMS | AVG GRAMS |
|---|---|---|---|---|
| CODEINE | 9050 | 4 | 2.38 | 0.60 |

**BUSINESS ACTIVITY: NTP - NARCOTIC TREATMENT PROGRAMS**

| DRUG NAME | DRUG CODE | NUMBER OF REGISTRANT SOLD TO | TOTAL GRAMS | AVG GRAMS |
|---|---|---|---|---|
| BUPRENORPHINE | 9064 | 4 | 176.95 | 44.24 |
| METHADONE | 9250B | 5 | 19,922.76 | 3,984.55 |

**STATE: MT - MONTANA**

**BUSINESS ACTIVITY: A - PHARMACIES**

| DRUG NAME | DRUG CODE | NUMBER OF REGISTRANT SOLD TO | TOTAL GRAMS | AVG GRAMS |
|---|---|---|---|---|
| AMPHETAMINE | 1100 | 239 | 55,076.55 | 230.45 |
| METHAMPHETAMINE | 1105 | 1 | 0.40 | 0.40 |
| LISDEXAMFETAMINE | 1205 | 235 | 23,192.22 | 98.69 |
| METHYLPHENIDATE (DL;D;L;ISOMERS) | 1724 | 239 | 62,227.15 | 260.36 |
| CODEINE | 9050 | 235 | 31,526.36 | 134.15 |

| | | | | |
|---|---|---|---|---|
| BUPRENORPHINE | 9064 | 227 | 10,345.94 | 45.58 |
| OXYCODONE | 9143 | 241 | 111,951.20 | 464.53 |
| HYDROMORPHONE | 9150 | 228 | 4,775.36 | 20.94 |
| HYDROCODONE | 9193 | 241 | 77,078.54 | 319.83 |
| MEPERIDINE (PETHIDINE) | 9230 | 88 | 1,286.01 | 14.61 |
| METHADONE | 9250B | 194 | 5,919.68 | 30.51 |
| MORPHINE | 9300 | 241 | 48,924.03 | 203.00 |
| OXYMORPHONE | 9652 | 57 | 516.12 | 9.05 |
| TAPENTADOL | 9780 | 97 | 5,934.50 | 61.18 |
| FENTANYL BASE | 9801 | 237 | 851.71 | 3.59 |

**BUSINESS ACTIVITY: B - HOSPITALS**

| DRUG NAME | DRUG CODE | NUMBER OF REGISTRANT SOLD TO | TOTAL GRAMS | AVG GRAMS |
|---|---|---|---|---|
| AMPHETAMINE | 1100 | 37 | 4,011.29 | 108.41 |
| LISDEXAMFETAMINE | 1205 | 25 | 1,446.98 | 57.88 |
| METHYLPHENIDATE (DL;D;L;ISOMERS) | 1724 | 45 | 5,456.76 | 121.26 |
| CODEINE | 9050 | 58 | 3,789.20 | 65.33 |
| BUPRENORPHINE | 9064 | 29 | 877.63 | 30.26 |
| OXYCODONE | 9143 | 81 | 12,897.20 | 159.22 |
| HYDROMORPHONE | 9150 | 69 | 791.43 | 11.47 |
| HYDROCODONE | 9193 | 84 | 13,143.14 | 156.47 |
| MEPERIDINE (PETHIDINE) | 9230 | 50 | 712.89 | 14.26 |
| METHADONE | 9250B | 32 | 796.55 | 24.89 |
| MORPHINE | 9300 | 86 | 10,248.61 | 119.17 |
| OXYMORPHONE | 9652 | 2 | 2.05 | 1.03 |
| TAPENTADOL | 9780 | 8 | 193.00 | 24.13 |
| FENTANYL BASE | 9801 | 84 | 201.56 | 2.40 |

**BUSINESS ACTIVITY: C - PRACTITIONERS**

| DRUG NAME | DRUG CODE | NUMBER OF REGISTRANT SOLD TO | TOTAL GRAMS | AVG GRAMS |
|---|---|---|---|---|
| CODEINE | 9050 | 11 | 221.33 | 20.12 |
| BUPRENORPHINE | 9064 | 138 | 6.94 | 0.05 |
| OXYCODONE | 9143 | 2 | 6.28 | 3.14 |
| HYDROMORPHONE | 9150 | 63 | 46.35 | 0.74 |
| HYDROCODONE | 9193 | 64 | 174.89 | 2.73 |
| MEPERIDINE (PETHIDINE) | 9230 | 7 | 35.40 | 5.06 |
| METHADONE | 9250B | 2 | 9.84 | 4.92 |
| MORPHINE | 9300 | 52 | 58.74 | 1.13 |
| FENTANYL BASE | 9801 | 86 | 7.34 | 0.09 |

**BUSINESS ACTIVITY: M - MID-LEVEL PRACTITIONERS**

| DRUG NAME | DRUG CODE | NUMBER OF REGISTRANT SOLD TO | TOTAL GRAMS | AVG GRAMS |
|---|---|---|---|---|
| MORPHINE | 9300 | 4 | 1.24 | 0.31 |

| | | | | |
|---|---|---|---|---|
| FENTANYL BASE | 9801 | 3 | 0.02 | 0.01 |

**BUSINESS ACTIVITY: NTP - NARCOTIC TREATMENT PROGRAMS**

| DRUG NAME | DRUG CODE | NUMBER OF REGISTRANT SOLD TO | TOTAL GRAMS | AVG GRAMS |
|---|---|---|---|---|
| BUPRENORPHINE | 9064 | 4 | 929.21 | 232.30 |
| METHADONE | 9250B | 4 | 25,293.63 | 6,323.41 |

**STATE: NC - NORTH CAROLINA**

**BUSINESS ACTIVITY: A - PHARMACIES**

| DRUG NAME | DRUG CODE | NUMBER OF REGISTRANT SOLD TO | TOTAL GRAMS | AVG GRAMS |
|---|---|---|---|---|
| AMPHETAMINE | 1100 | 2,361 | 824,015.81 | 349.01 |
| METHAMPHETAMINE | 1105 | 69 | 191.26 | 2.77 |
| LISDEXAMFETAMINE | 1205 | 2,318 | 412,935.67 | 178.14 |
| METHYLPHENIDATE (DL;D;L;ISOMERS) | 1724 | 2,350 | 690,010.25 | 293.62 |
| CODEINE | 9050 | 2,170 | 155,969.26 | 71.88 |
| BUPRENORPHINE | 9064 | 2,264 | 142,020.19 | 62.73 |
| OXYCODONE | 9143 | 2,375 | 1,832,036.43 | 771.38 |
| HYDROMORPHONE | 9150 | 2,141 | 29,333.91 | 13.70 |
| HYDROCODONE | 9193 | 2,382 | 592,805.94 | 248.87 |
| MEPERIDINE (PETHIDINE) | 9230 | 877 | 15,385.24 | 17.54 |
| METHADONE | 9250B | 2,037 | 81,656.48 | 40.09 |
| MORPHINE | 9300 | 2,297 | 378,607.69 | 164.83 |
| OXYMORPHONE | 9652 | 1,276 | 39,398.70 | 30.88 |
| TAPENTADOL | 9780 | 1,839 | 319,518.50 | 173.75 |
| FENTANYL BASE | 9801 | 2,204 | 7,615.58 | 3.46 |

**BUSINESS ACTIVITY: B - HOSPITALS**

| DRUG NAME | DRUG CODE | NUMBER OF REGISTRANT SOLD TO | TOTAL GRAMS | AVG GRAMS |
|---|---|---|---|---|
| AMPHETAMINE | 1100 | 146 | 30,294.33 | 207.50 |
| LISDEXAMFETAMINE | 1205 | 84 | 10,143.92 | 120.76 |
| METHYLPHENIDATE (DL;D;L;ISOMERS) | 1724 | 175 | 30,997.41 | 177.13 |
| CODEINE | 9050 | 172 | 6,875.73 | 39.98 |
| BUPRENORPHINE | 9064 | 285 | 3,819.13 | 13.40 |
| OXYCODONE | 9143 | 261 | 94,423.77 | 361.78 |
| HYDROMORPHONE | 9150 | 318 | 6,924.78 | 21.78 |
| HYDROCODONE | 9193 | 307 | 37,759.82 | 123.00 |
| MEPERIDINE (PETHIDINE) | 9230 | 160 | 2,081.69 | 13.01 |
| METHADONE | 9250B | 177 | 7,394.68 | 41.78 |
| MORPHINE | 9300 | 315 | 44,310.55 | 140.67 |
| OXYMORPHONE | 9652 | 48 | 496.76 | 10.35 |
| TAPENTADOL | 9780 | 83 | 12,315.00 | 148.37 |
| FENTANYL BASE | 9801 | 340 | 1,051.64 | 3.09 |

**BUSINESS ACTIVITY: C - PRACTITIONERS**

| DRUG NAME | DRUG CODE | NUMBER OF REGISTRANT SOLD TO | TOTAL GRAMS | AVG GRAMS |
|---|---|---|---|---|
| AMPHETAMINE | 1100 | 1 | 0.95 | 0.95 |
| METHYLPHENIDATE (DL;D;L;ISOMERS) | 1724 | 1 | 10.90 | 10.90 |
| CODEINE | 9050 | 69 | 369.26 | 5.35 |
| BUPRENORPHINE | 9064 | 1,003 | 189.96 | 0.19 |
| OXYCODONE | 9143 | 13 | 32.27 | 2.48 |
| HYDROMORPHONE | 9150 | 312 | 132.56 | 0.42 |
| HYDROCODONE | 9193 | 254 | 613.67 | 2.42 |
| MEPERIDINE (PETHIDINE) | 9230 | 66 | 289.23 | 4.38 |
| METHADONE | 9250B | 6 | 9.12 | 1.52 |
| MORPHINE | 9300 | 179 | 186.39 | 1.04 |
| FENTANYL BASE | 9801 | 458 | 30.44 | 0.07 |

**BUSINESS ACTIVITY: D - TEACHING INSTITUTIONS**

| DRUG NAME | DRUG CODE | NUMBER OF REGISTRANT SOLD TO | TOTAL GRAMS | AVG GRAMS |
|---|---|---|---|---|
| BUPRENORPHINE | 9064 | 3 | 0.02 | 0.01 |
| HYDROMORPHONE | 9150 | 3 | 0.28 | 0.09 |
| HYDROCODONE | 9193 | 1 | 0.30 | 0.30 |
| MORPHINE | 9300 | 2 | 0.26 | 0.13 |
| FENTANYL BASE | 9801 | 2 | 0.04 | 0.02 |

**BUSINESS ACTIVITY: M - MID-LEVEL PRACTITIONERS**

| DRUG NAME | DRUG CODE | NUMBER OF REGISTRANT SOLD TO | TOTAL GRAMS | AVG GRAMS |
|---|---|---|---|---|
| CODEINE | 9050 | 39 | 47.00 | 1.21 |
| BUPRENORPHINE | 9064 | 5 | 0.18 | 0.04 |
| HYDROMORPHONE | 9150 | 9 | 0.84 | 0.09 |
| MORPHINE | 9300 | 79 | 111.12 | 1.41 |
| FENTANYL BASE | 9801 | 84 | 4.28 | 0.05 |

**BUSINESS ACTIVITY: NTP - NARCOTIC TREATMENT PROGRAMS**

| DRUG NAME | DRUG CODE | NUMBER OF REGISTRANT SOLD TO | TOTAL GRAMS | AVG GRAMS |
|---|---|---|---|---|
| BUPRENORPHINE | 9064 | 55 | 13,995.40 | 254.46 |
| METHADONE | 9250B | 71 | 418,791.17 | 5,898.47 |

**STATE: ND - NORTH DAKOTA**

**BUSINESS ACTIVITY: A - PHARMACIES**

| DRUG NAME | DRUG CODE | NUMBER OF REGISTRANT SOLD TO | TOTAL GRAMS | AVG GRAMS |
|---|---|---|---|---|
| AMPHETAMINE | 1100 | 171 | 34,654.61 | 202.66 |
| METHAMPHETAMINE | 1105 | 4 | 14.06 | 3.51 |
| LISDEXAMFETAMINE | 1205 | 166 | 22,062.03 | 132.90 |
| METHYLPHENIDATE (DL;D;L;ISOMERS) | 1724 | 170 | 54,587.50 | 321.10 |
| CODEINE | 9050 | 165 | 15,906.08 | 96.40 |
| BUPRENORPHINE | 9064 | 136 | 4,306.25 | 31.66 |
| OXYCODONE | 9143 | 170 | 42,595.65 | 250.56 |

| | | | | |
|---|---|---|---|---|
| HYDROMORPHONE | 9150 | 153 | 1,712.72 | 11.19 |
| HYDROCODONE | 9193 | 175 | 28,462.56 | 162.64 |
| MEPERIDINE (PETHIDINE) | 9230 | 39 | 524.75 | 13.46 |
| METHADONE | 9250B | 126 | 3,098.56 | 24.59 |
| MORPHINE | 9300 | 167 | 16,685.46 | 99.91 |
| OXYMORPHONE | 9652 | 25 | 292.12 | 11.68 |
| TAPENTADOL | 9780 | 70 | 6,260.50 | 89.44 |
| FENTANYL BASE | 9801 | 163 | 585.56 | 3.59 |

**BUSINESS ACTIVITY: B - HOSPITALS**

| DRUG NAME | DRUG CODE | NUMBER OF REGISTRANT SOLD TO | TOTAL GRAMS | AVG GRAMS |
|---|---|---|---|---|
| AMPHETAMINE | 1100 | 23 | 2,850.98 | 123.96 |
| METHAMPHETAMINE | 1105 | 1 | 5.22 | 5.22 |
| LISDEXAMFETAMINE | 1205 | 17 | 1,460.28 | 85.90 |
| METHYLPHENIDATE (DL;D;L;ISOMERS) | 1724 | 27 | 5,671.24 | 210.05 |
| CODEINE | 9050 | 40 | 2,811.49 | 70.29 |
| BUPRENORPHINE | 9064 | 18 | 310.57 | 17.25 |
| OXYCODONE | 9143 | 53 | 8,218.31 | 155.06 |
| HYDROMORPHONE | 9150 | 52 | 650.85 | 12.52 |
| HYDROCODONE | 9193 | 54 | 6,891.02 | 127.61 |
| MEPERIDINE (PETHIDINE) | 9230 | 27 | 187.37 | 6.94 |
| METHADONE | 9250B | 24 | 396.49 | 16.52 |
| MORPHINE | 9300 | 56 | 4,804.29 | 85.79 |
| OXYMORPHONE | 9652 | 1 | 8.92 | 8.92 |
| TAPENTADOL | 9780 | 7 | 965.50 | 137.93 |
| FENTANYL BASE | 9801 | 52 | 148.82 | 2.86 |

**BUSINESS ACTIVITY: C - PRACTITIONERS**

| DRUG NAME | DRUG CODE | NUMBER OF REGISTRANT SOLD TO | TOTAL GRAMS | AVG GRAMS |
|---|---|---|---|---|
| CODEINE | 9050 | 9 | 57.88 | 6.43 |
| BUPRENORPHINE | 9064 | 65 | 35.73 | 0.55 |
| OXYCODONE | 9143 | 6 | 17.89 | 2.98 |
| HYDROMORPHONE | 9150 | 25 | 9.16 | 0.37 |
| HYDROCODONE | 9193 | 27 | 157.62 | 5.84 |
| MEPERIDINE (PETHIDINE) | 9230 | 12 | 40.85 | 3.40 |
| METHADONE | 9250B | 1 | 0.45 | 0.45 |
| MORPHINE | 9300 | 34 | 38.05 | 1.12 |
| FENTANYL BASE | 9801 | 45 | 5.35 | 0.12 |

**BUSINESS ACTIVITY: M - MID-LEVEL PRACTITIONERS**

| DRUG NAME | DRUG CODE | NUMBER OF REGISTRANT SOLD TO | TOTAL GRAMS | AVG GRAMS |
|---|---|---|---|---|
| AMPHETAMINE | 1100 | 1 | 1.47 | 1.47 |
| METHYLPHENIDATE (DL;D;L;ISOMERS) | 1724 | 1 | 0.86 | 0.86 |

| | | | | |
|---|---|---|---|---|
| BUPRENORPHINE | 9064 | 1 | 0.72 | 0.72 |

**BUSINESS ACTIVITY: NTP - NARCOTIC TREATMENT PROGRAMS**

| DRUG NAME | DRUG CODE | NUMBER OF REGISTRANT SOLD TO | TOTAL GRAMS | AVG GRAMS |
|---|---|---|---|---|
| METHADONE | 9250B | 3 | 12,378.50 | 4,126.17 |

**STATE: NE - NEBRASKA**

**BUSINESS ACTIVITY: A - PHARMACIES**

| DRUG NAME | DRUG CODE | NUMBER OF REGISTRANT SOLD TO | TOTAL GRAMS | AVG GRAMS |
|---|---|---|---|---|
| AMPHETAMINE | 1100 | 425 | 106,247.19 | 249.99 |
| METHAMPHETAMINE | 1105 | 8 | 20.09 | 2.51 |
| LISDEXAMFETAMINE | 1205 | 418 | 68,763.26 | 164.51 |
| METHYLPHENIDATE (DL;D;L;ISOMERS) | 1724 | 433 | 117,609.12 | 271.61 |
| CODEINE | 9050 | 420 | 37,762.18 | 89.91 |
| BUPRENORPHINE | 9064 | 369 | 5,611.34 | 15.21 |
| OXYCODONE | 9143 | 438 | 134,598.30 | 307.30 |
| HYDROMORPHONE | 9150 | 374 | 3,515.69 | 9.40 |
| HYDROCODONE | 9193 | 442 | 102,181.78 | 231.18 |
| MEPERIDINE (PETHIDINE) | 9230 | 80 | 1,003.10 | 12.54 |
| METHADONE | 9250B | 334 | 8,764.94 | 26.24 |
| MORPHINE | 9300 | 436 | 59,148.84 | 135.66 |
| OXYMORPHONE | 9652 | 130 | 2,063.65 | 15.87 |
| TAPENTADOL | 9780 | 301 | 35,915.50 | 119.32 |
| FENTANYL BASE | 9801 | 411 | 1,564.25 | 3.81 |

**BUSINESS ACTIVITY: B - HOSPITALS**

| DRUG NAME | DRUG CODE | NUMBER OF REGISTRANT SOLD TO | TOTAL GRAMS | AVG GRAMS |
|---|---|---|---|---|
| AMPHETAMINE | 1100 | 41 | 3,768.94 | 91.93 |
| LISDEXAMFETAMINE | 1205 | 21 | 666.55 | 31.74 |
| METHYLPHENIDATE (DL;D;L;ISOMERS) | 1724 | 59 | 4,019.93 | 68.13 |
| CODEINE | 9050 | 73 | 2,105.38 | 28.84 |
| BUPRENORPHINE | 9064 | 38 | 330.86 | 8.71 |
| OXYCODONE | 9143 | 124 | 8,966.55 | 72.31 |
| HYDROMORPHONE | 9150 | 109 | 757.11 | 6.95 |
| HYDROCODONE | 9193 | 129 | 6,418.01 | 49.75 |
| MEPERIDINE (PETHIDINE) | 9230 | 98 | 812.65 | 8.29 |
| METHADONE | 9250B | 46 | 959.05 | 20.85 |
| MORPHINE | 9300 | 124 | 7,986.33 | 64.41 |
| OXYMORPHONE | 9652 | 9 | 48.08 | 5.34 |
| TAPENTADOL | 9780 | 40 | 1,079.00 | 26.98 |
| FENTANYL BASE | 9801 | 132 | 191.42 | 1.45 |

**BUSINESS ACTIVITY: C - PRACTITIONERS**

| DRUG NAME | DRUG CODE | NUMBER OF REGISTRANT SOLD TO | TOTAL GRAMS | AVG GRAMS |
|---|---|---|---|---|

| | | | | |
|---|---|---|---|---|
| AMPHETAMINE | 1100 | 1 | 2.04 | 2.04 |
| LISDEXAMFETAMINE | 1205 | 1 | 20.75 | 20.75 |
| CODEINE | 9050 | 29 | 1,326.69 | 45.75 |
| BUPRENORPHINE | 9064 | 169 | 18.30 | 0.11 |
| OXYCODONE | 9143 | 7 | 96.06 | 13.72 |
| HYDROMORPHONE | 9150 | 58 | 39.22 | 0.68 |
| HYDROCODONE | 9193 | 66 | 176.77 | 2.68 |
| MEPERIDINE (PETHIDINE) | 9230 | 19 | 120.81 | 6.36 |
| METHADONE | 9250B | 3 | 8.50 | 2.83 |
| MORPHINE | 9300 | 43 | 42.67 | 0.99 |
| OXYMORPHONE | 9652 | 1 | 3.21 | 3.21 |
| TAPENTADOL | 9780 | 1 | 60.00 | 60.00 |
| FENTANYL BASE | 9801 | 95 | 11.74 | 0.12 |

**BUSINESS ACTIVITY: D - TEACHING INSTITUTIONS**

| DRUG NAME | DRUG CODE | NUMBER OF REGISTRANT SOLD TO | TOTAL GRAMS | AVG GRAMS |
|---|---|---|---|---|
| HYDROMORPHONE | 9150 | 1 | 0.44 | 0.44 |

**BUSINESS ACTIVITY: M - MID-LEVEL PRACTITIONERS**

| DRUG NAME | DRUG CODE | NUMBER OF REGISTRANT SOLD TO | TOTAL GRAMS | AVG GRAMS |
|---|---|---|---|---|
| CODEINE | 9050 | 5 | 6.50 | 1.30 |
| HYDROMORPHONE | 9150 | 8 | 0.36 | 0.05 |
| MORPHINE | 9300 | 22 | 6.17 | 0.28 |
| FENTANYL BASE | 9801 | 29 | 0.52 | 0.02 |

**BUSINESS ACTIVITY: NTP - NARCOTIC TREATMENT PROGRAMS**

| DRUG NAME | DRUG CODE | NUMBER OF REGISTRANT SOLD TO | TOTAL GRAMS | AVG GRAMS |
|---|---|---|---|---|
| BUPRENORPHINE | 9064 | 1 | 3.38 | 3.38 |
| METHADONE | 9250B | 3 | 18,621.41 | 6,207.14 |

**STATE: NH - NEW HAMPSHIRE**

**BUSINESS ACTIVITY: A - PHARMACIES**

| DRUG NAME | DRUG CODE | NUMBER OF REGISTRANT SOLD TO | TOTAL GRAMS | AVG GRAMS |
|---|---|---|---|---|
| AMPHETAMINE | 1100 | 274 | 158,538.42 | 578.61 |
| METHAMPHETAMINE | 1105 | 7 | 19.69 | 2.81 |
| LISDEXAMFETAMINE | 1205 | 272 | 56,084.95 | 206.19 |
| METHYLPHENIDATE (DL;D;L;ISOMERS) | 1724 | 274 | 130,759.86 | 477.23 |
| CODEINE | 9050 | 265 | 25,908.75 | 97.77 |
| BUPRENORPHINE | 9064 | 271 | 33,650.54 | 124.17 |
| OXYCODONE | 9143 | 273 | 197,100.97 | 721.98 |
| HYDROMORPHONE | 9150 | 270 | 5,628.07 | 20.84 |
| HYDROCODONE | 9193 | 273 | 29,904.39 | 109.54 |
| MEPERIDINE (PETHIDINE) | 9230 | 52 | 1,823.61 | 35.07 |
| METHADONE | 9250B | 257 | 17,270.15 | 67.20 |

| | | | | |
|---|---|---|---|---|
| MORPHINE | 9300 | 276 | 60,853.75 | 220.48 |
| OXYMORPHONE | 9652 | 110 | 1,677.33 | 15.25 |
| TAPENTADOL | 9780 | 203 | 23,990.00 | 118.18 |
| FENTANYL BASE | 9801 | 258 | 965.01 | 3.74 |

**BUSINESS ACTIVITY: B - HOSPITALS**

| DRUG NAME | DRUG CODE | NUMBER OF REGISTRANT SOLD TO | TOTAL GRAMS | AVG GRAMS |
|---|---|---|---|---|
| AMPHETAMINE | 1100 | 22 | 2,915.49 | 132.52 |
| LISDEXAMFETAMINE | 1205 | 11 | 453.23 | 41.20 |
| METHYLPHENIDATE (DL;D;L;ISOMERS) | 1724 | 33 | 2,204.01 | 66.79 |
| CODEINE | 9050 | 34 | 459.14 | 13.50 |
| BUPRENORPHINE | 9064 | 29 | 636.27 | 21.94 |
| OXYCODONE | 9143 | 45 | 9,600.30 | 213.34 |
| HYDROMORPHONE | 9150 | 40 | 905.07 | 22.63 |
| HYDROCODONE | 9193 | 41 | 1,224.50 | 29.87 |
| MEPERIDINE (PETHIDINE) | 9230 | 25 | 293.48 | 11.74 |
| METHADONE | 9250B | 34 | 1,284.54 | 37.78 |
| MORPHINE | 9300 | 43 | 4,478.49 | 104.15 |
| TAPENTADOL | 9780 | 2 | 301.50 | 150.75 |
| FENTANYL BASE | 9801 | 49 | 127.74 | 2.61 |

**BUSINESS ACTIVITY: C - PRACTITIONERS**

| DRUG NAME | DRUG CODE | NUMBER OF REGISTRANT SOLD TO | TOTAL GRAMS | AVG GRAMS |
|---|---|---|---|---|
| CODEINE | 9050 | 1 | 4.78 | 4.78 |
| BUPRENORPHINE | 9064 | 197 | 11.22 | 0.06 |
| OXYCODONE | 9143 | 9 | 12.10 | 1.34 |
| HYDROMORPHONE | 9150 | 88 | 61.61 | 0.70 |
| HYDROCODONE | 9193 | 10 | 7.39 | 0.74 |
| MEPERIDINE (PETHIDINE) | 9230 | 8 | 71.90 | 8.99 |
| METHADONE | 9250B | 6 | 5.55 | 0.92 |
| MORPHINE | 9300 | 51 | 47.25 | 0.93 |
| FENTANYL BASE | 9801 | 100 | 10.89 | 0.11 |

**BUSINESS ACTIVITY: M - MID-LEVEL PRACTITIONERS**

| DRUG NAME | DRUG CODE | NUMBER OF REGISTRANT SOLD TO | TOTAL GRAMS | AVG GRAMS |
|---|---|---|---|---|
| CODEINE | 9050 | 1 | 4.42 | 4.42 |
| FENTANYL BASE | 9801 | 1 | 0.01 | 0.01 |

**BUSINESS ACTIVITY: NTP - NARCOTIC TREATMENT PROGRAMS**

| DRUG NAME | DRUG CODE | NUMBER OF REGISTRANT SOLD TO | TOTAL GRAMS | AVG GRAMS |
|---|---|---|---|---|
| BUPRENORPHINE | 9064 | 5 | 547.48 | 109.50 |
| METHADONE | 9250B | 9 | 101,309.58 | 11,256.62 |

**STATE: NJ - NEW JERSEY**

**BUSINESS ACTIVITY: A - PHARMACIES**

| DRUG NAME | DRUG CODE | NUMBER OF REGISTRANT SOLD TO | TOTAL GRAMS | AVG GRAMS |
|---|---|---|---|---|
| AMPHETAMINE | 1100 | 2,085 | 434,940.52 | 208.60 |
| METHAMPHETAMINE | 1105 | 52 | 139.43 | 2.68 |
| LISDEXAMFETAMINE | 1205 | 1,889 | 193,393.53 | 102.38 |
| METHYLPHENIDATE (DL;D;L;ISOMERS) | 1724 | 2,044 | 351,924.03 | 172.17 |
| CODEINE | 9050 | 2,100 | 260,507.88 | 124.05 |
| BUPRENORPHINE | 9064 | 1,936 | 107,789.89 | 55.68 |
| OXYCODONE | 9143 | 2,165 | 1,590,234.19 | 734.52 |
| HYDROMORPHONE | 9150 | 1,770 | 24,740.05 | 13.98 |
| HYDROCODONE | 9193 | 2,088 | 111,658.42 | 53.48 |
| MEPERIDINE (PETHIDINE) | 9230 | 180 | 4,752.07 | 26.40 |
| METHADONE | 9250B | 1,199 | 35,353.04 | 29.49 |
| MORPHINE | 9300 | 1,872 | 268,314.43 | 143.33 |
| OXYMORPHONE | 9652 | 670 | 13,135.99 | 19.61 |
| TAPENTADOL | 9780 | 1,212 | 133,297.50 | 109.98 |
| FENTANYL BASE | 9801 | 1,790 | 6,346.22 | 3.55 |

**BUSINESS ACTIVITY: B - HOSPITALS**

| DRUG NAME | DRUG CODE | NUMBER OF REGISTRANT SOLD TO | TOTAL GRAMS | AVG GRAMS |
|---|---|---|---|---|
| AMPHETAMINE | 1100 | 53 | 3,287.83 | 62.03 |
| LISDEXAMFETAMINE | 1205 | 23 | 1,382.96 | 60.13 |
| METHYLPHENIDATE (DL;D;L;ISOMERS) | 1724 | 102 | 3,579.80 | 35.10 |
| CODEINE | 9050 | 106 | 4,094.83 | 38.63 |
| BUPRENORPHINE | 9064 | 88 | 943.38 | 10.72 |
| OXYCODONE | 9143 | 124 | 33,433.54 | 269.63 |
| HYDROMORPHONE | 9150 | 111 | 4,517.07 | 40.69 |
| HYDROCODONE | 9193 | 92 | 1,223.64 | 13.30 |
| MEPERIDINE (PETHIDINE) | 9230 | 76 | 1,192.97 | 15.70 |
| METHADONE | 9250B | 103 | 3,938.49 | 38.24 |
| MORPHINE | 9300 | 120 | 14,745.60 | 122.88 |
| OXYMORPHONE | 9652 | 4 | 19.09 | 4.77 |
| TAPENTADOL | 9780 | 30 | 1,443.50 | 48.12 |
| FENTANYL BASE | 9801 | 110 | 992.08 | 9.02 |

**BUSINESS ACTIVITY: C - PRACTITIONERS**

| DRUG NAME | DRUG CODE | NUMBER OF REGISTRANT SOLD TO | TOTAL GRAMS | AVG GRAMS |
|---|---|---|---|---|
| AMPHETAMINE | 1100 | 2 | 10.90 | 5.45 |
| CODEINE | 9050 | 240 | 4,959.15 | 20.66 |
| BUPRENORPHINE | 9064 | 606 | 727.95 | 1.20 |
| OXYCODONE | 9143 | 114 | 168.18 | 1.48 |
| HYDROMORPHONE | 9150 | 284 | 155.08 | 0.55 |
| HYDROCODONE | 9193 | 268 | 904.17 | 3.37 |

| | | | | |
|---|---|---|---|---|
| MEPERIDINE (PETHIDINE) | 9230 | 91 | 325.18 | 3.57 |
| METHADONE | 9250B | 22 | 62.88 | 2.86 |
| MORPHINE | 9300 | 178 | 172.03 | 0.97 |
| FENTANYL BASE | 9801 | 516 | 62.46 | 0.12 |

**BUSINESS ACTIVITY: D - TEACHING INSTITUTIONS**

| DRUG NAME | DRUG CODE | NUMBER OF REGISTRANT SOLD TO | TOTAL GRAMS | AVG GRAMS |
|---|---|---|---|---|
| BUPRENORPHINE | 9064 | 3 | 0.01 | 0.00 |

**BUSINESS ACTIVITY: M - MID-LEVEL PRACTITIONERS**

| DRUG NAME | DRUG CODE | NUMBER OF REGISTRANT SOLD TO | TOTAL GRAMS | AVG GRAMS |
|---|---|---|---|---|
| CODEINE | 9050 | 10 | 564.96 | 56.50 |
| BUPRENORPHINE | 9064 | 2 | 0.85 | 0.42 |

**BUSINESS ACTIVITY: NTP - NARCOTIC TREATMENT PROGRAMS**

| DRUG NAME | DRUG CODE | NUMBER OF REGISTRANT SOLD TO | TOTAL GRAMS | AVG GRAMS |
|---|---|---|---|---|
| BUPRENORPHINE | 9064 | 24 | 1,605.58 | 66.90 |
| METHADONE | 9250B | 33 | 429,160.85 | 13,004.87 |

**STATE: NM - NEW MEXICO**

**BUSINESS ACTIVITY: A - PHARMACIES**

| DRUG NAME | DRUG CODE | NUMBER OF REGISTRANT SOLD TO | TOTAL GRAMS | AVG GRAMS |
|---|---|---|---|---|
| AMPHETAMINE | 1100 | 305 | 58,563.20 | 192.01 |
| METHAMPHETAMINE | 1105 | 13 | 22.90 | 1.76 |
| LISDEXAMFETAMINE | 1205 | 272 | 15,249.12 | 56.06 |
| METHYLPHENIDATE (DL;D;L;ISOMERS) | 1724 | 311 | 66,225.81 | 212.94 |
| CODEINE | 9050 | 315 | 57,099.08 | 181.27 |
| BUPRENORPHINE | 9064 | 311 | 31,976.84 | 102.82 |
| OXYCODONE | 9143 | 314 | 288,341.87 | 918.29 |
| HYDROMORPHONE | 9150 | 266 | 3,731.75 | 14.03 |
| HYDROCODONE | 9193 | 315 | 111,824.69 | 355.00 |
| MEPERIDINE (PETHIDINE) | 9230 | 162 | 4,085.70 | 25.22 |
| METHADONE | 9250B | 254 | 7,075.42 | 27.86 |
| MORPHINE | 9300 | 312 | 72,835.08 | 233.45 |
| OXYMORPHONE | 9652 | 130 | 1,766.13 | 13.59 |
| TAPENTADOL | 9780 | 141 | 10,168.50 | 72.12 |
| FENTANYL BASE | 9801 | 297 | 1,079.99 | 3.64 |

**BUSINESS ACTIVITY: B - HOSPITALS**

| DRUG NAME | DRUG CODE | NUMBER OF REGISTRANT SOLD TO | TOTAL GRAMS | AVG GRAMS |
|---|---|---|---|---|
| AMPHETAMINE | 1100 | 41 | 2,629.01 | 64.12 |
| LISDEXAMFETAMINE | 1205 | 12 | 429.53 | 35.79 |
| METHYLPHENIDATE (DL;D;L;ISOMERS) | 1724 | 58 | 6,215.98 | 107.17 |
| CODEINE | 9050 | 63 | 8,541.68 | 135.58 |
| BUPRENORPHINE | 9064 | 51 | 1,695.79 | 33.25 |

| | | | | |
|---|---|---|---|---|
| OXYCODONE | 9143 | 86 | 22,760.60 | 264.66 |
| HYDROMORPHONE | 9150 | 63 | 711.80 | 11.30 |
| HYDROCODONE | 9193 | 91 | 9,886.96 | 108.65 |
| MEPERIDINE (PETHIDINE) | 9230 | 45 | 969.22 | 21.54 |
| METHADONE | 9250B | 44 | 2,032.70 | 46.20 |
| MORPHINE | 9300 | 89 | 13,069.11 | 146.84 |
| OXYMORPHONE | 9652 | 3 | 4.91 | 1.64 |
| TAPENTADOL | 9780 | 1 | 36.00 | 36.00 |
| FENTANYL BASE | 9801 | 91 | 253.51 | 2.79 |

**BUSINESS ACTIVITY: C - PRACTITIONERS**

| DRUG NAME | DRUG CODE | NUMBER OF REGISTRANT SOLD TO | TOTAL GRAMS | AVG GRAMS |
|---|---|---|---|---|
| CODEINE | 9050 | 12 | 347.93 | 28.99 |
| BUPRENORPHINE | 9064 | 134 | 10.71 | 0.08 |
| OXYCODONE | 9143 | 3 | 20.62 | 6.87 |
| HYDROMORPHONE | 9150 | 58 | 55.21 | 0.95 |
| HYDROCODONE | 9193 | 45 | 82.93 | 1.84 |
| MEPERIDINE (PETHIDINE) | 9230 | 7 | 40.31 | 5.76 |
| MORPHINE | 9300 | 37 | 43.57 | 1.18 |
| FENTANYL BASE | 9801 | 104 | 15.73 | 0.15 |

**BUSINESS ACTIVITY: D - TEACHING INSTITUTIONS**

| DRUG NAME | DRUG CODE | NUMBER OF REGISTRANT SOLD TO | TOTAL GRAMS | AVG GRAMS |
|---|---|---|---|---|
| BUPRENORPHINE | 9064 | 1 | 0.02 | 0.02 |
| MORPHINE | 9300 | 1 | 0.04 | 0.04 |

**BUSINESS ACTIVITY: M - MID-LEVEL PRACTITIONERS**

| DRUG NAME | DRUG CODE | NUMBER OF REGISTRANT SOLD TO | TOTAL GRAMS | AVG GRAMS |
|---|---|---|---|---|
| AMPHETAMINE | 1100 | 1 | 4.50 | 4.50 |
| CODEINE | 9050 | 7 | 18.21 | 2.60 |
| BUPRENORPHINE | 9064 | 6 | 2.35 | 0.39 |
| HYDROMORPHONE | 9150 | 5 | 2.31 | 0.46 |
| HYDROCODONE | 9193 | 2 | 4.90 | 2.45 |
| MORPHINE | 9300 | 33 | 21.05 | 0.64 |
| FENTANYL BASE | 9801 | 47 | 1.13 | 0.02 |

**BUSINESS ACTIVITY: NTP - NARCOTIC TREATMENT PROGRAMS**

| DRUG NAME | DRUG CODE | NUMBER OF REGISTRANT SOLD TO | TOTAL GRAMS | AVG GRAMS |
|---|---|---|---|---|
| BUPRENORPHINE | 9064 | 4 | 509.84 | 127.46 |
| METHADONE | 9250B | 16 | 162,378.32 | 10,148.65 |

**STATE: NV - NEVADA**

**BUSINESS ACTIVITY: A - PHARMACIES**

| DRUG NAME | DRUG CODE | NUMBER OF REGISTRANT SOLD TO | TOTAL GRAMS | AVG GRAMS |
|---|---|---|---|---|
| AMPHETAMINE | 1100 | 458 | 104,503.92 | 228.17 |

| | | | | |
|---|---|---|---|---|
| METHAMPHETAMINE | 1105 | 20 | 81.98 | 4.10 |
| LISDEXAMFETAMINE | 1205 | 420 | 30,982.11 | 73.77 |
| METHYLPHENIDATE (DL;D;L;ISOMERS) | 1724 | 440 | 53,267.37 | 121.06 |
| CODEINE | 9050 | 462 | 76,087.90 | 164.69 |
| BUPRENORPHINE | 9064 | 432 | 14,500.54 | 33.57 |
| OXYCODONE | 9143 | 465 | 561,088.85 | 1,206.64 |
| HYDROMORPHONE | 9150 | 423 | 7,428.95 | 17.56 |
| HYDROCODONE | 9193 | 465 | 279,813.77 | 601.75 |
| MEPERIDINE (PETHIDINE) | 9230 | 138 | 2,140.62 | 15.51 |
| METHADONE | 9250B | 431 | 32,117.37 | 74.52 |
| MORPHINE | 9300 | 462 | 166,347.02 | 360.06 |
| OXYMORPHONE | 9652 | 278 | 6,466.16 | 23.26 |
| TAPENTADOL | 9780 | 313 | 34,130.00 | 109.04 |
| FENTANYL BASE | 9801 | 427 | 1,339.69 | 3.14 |

**BUSINESS ACTIVITY: B - HOSPITALS**

| DRUG NAME | DRUG CODE | NUMBER OF REGISTRANT SOLD TO | TOTAL GRAMS | AVG GRAMS |
|---|---|---|---|---|
| AMPHETAMINE | 1100 | 36 | 3,547.31 | 98.54 |
| LISDEXAMFETAMINE | 1205 | 11 | 493.70 | 44.88 |
| METHYLPHENIDATE (DL;D;L;ISOMERS) | 1724 | 52 | 4,536.36 | 87.24 |
| CODEINE | 9050 | 47 | 2,541.57 | 54.08 |
| BUPRENORPHINE | 9064 | 36 | 509.73 | 14.16 |
| OXYCODONE | 9143 | 92 | 33,509.54 | 364.23 |
| HYDROMORPHONE | 9150 | 79 | 1,634.40 | 20.69 |
| HYDROCODONE | 9193 | 97 | 20,707.75 | 213.48 |
| MEPERIDINE (PETHIDINE) | 9230 | 75 | 2,013.71 | 26.85 |
| METHADONE | 9250B | 54 | 1,996.48 | 36.97 |
| MORPHINE | 9300 | 102 | 19,478.17 | 190.96 |
| OXYMORPHONE | 9652 | 6 | 19.54 | 3.26 |
| TAPENTADOL | 9780 | 13 | 617.00 | 47.46 |
| FENTANYL BASE | 9801 | 120 | 293.21 | 2.44 |

**BUSINESS ACTIVITY: C - PRACTITIONERS**

| DRUG NAME | DRUG CODE | NUMBER OF REGISTRANT SOLD TO | TOTAL GRAMS | AVG GRAMS |
|---|---|---|---|---|
| AMPHETAMINE | 1100 | 1 | 2.58 | 2.58 |
| LISDEXAMFETAMINE | 1205 | 2 | 5.60 | 2.80 |
| METHYLPHENIDATE (DL;D;L;ISOMERS) | 1724 | 1 | 412.42 | 412.42 |
| CODEINE | 9050 | 25 | 136.09 | 5.44 |
| BUPRENORPHINE | 9064 | 241 | 65.42 | 0.27 |
| OXYCODONE | 9143 | 20 | 216.06 | 10.80 |
| HYDROMORPHONE | 9150 | 124 | 187.35 | 1.51 |
| HYDROCODONE | 9193 | 112 | 502.28 | 4.48 |

| | | | | |
|---|---|---|---|---|
| MEPERIDINE (PETHIDINE) | 9230 | 55 | 958.76 | 17.43 |
| METHADONE | 9250B | 18 | 52.14 | 2.90 |
| MORPHINE | 9300 | 78 | 227.54 | 2.92 |
| TAPENTADOL | 9780 | 1 | 10.00 | 10.00 |
| FENTANYL BASE | 9801 | 178 | 26.87 | 0.15 |

**BUSINESS ACTIVITY: M - MID-LEVEL PRACTITIONERS**

| DRUG NAME | DRUG CODE | NUMBER OF REGISTRANT SOLD TO | TOTAL GRAMS | AVG GRAMS |
|---|---|---|---|---|
| CODEINE | 9050 | 6 | 29.54 | 4.92 |
| BUPRENORPHINE | 9064 | 2 | 1.74 | 0.87 |
| OXYCODONE | 9143 | 2 | 129.54 | 64.77 |
| HYDROMORPHONE | 9150 | 2 | 4.26 | 2.13 |
| HYDROCODONE | 9193 | 2 | 39.35 | 19.68 |
| METHADONE | 9250B | 2 | 42.48 | 21.24 |
| MORPHINE | 9300 | 3 | 47.75 | 15.92 |
| TAPENTADOL | 9780 | 1 | 10.00 | 10.00 |
| FENTANYL BASE | 9801 | 3 | 0.85 | 0.28 |

**BUSINESS ACTIVITY: NTP - NARCOTIC TREATMENT PROGRAMS**

| DRUG NAME | DRUG CODE | NUMBER OF REGISTRANT SOLD TO | TOTAL GRAMS | AVG GRAMS |
|---|---|---|---|---|
| BUPRENORPHINE | 9064 | 7 | 213.50 | 30.50 |
| METHADONE | 9250B | 14 | 102,358.71 | 7,311.34 |

**STATE: NY - NEW YORK**

**BUSINESS ACTIVITY: A - PHARMACIES**

| DRUG NAME | DRUG CODE | NUMBER OF REGISTRANT SOLD TO | TOTAL GRAMS | AVG GRAMS |
|---|---|---|---|---|
| AMPHETAMINE | 1100 | 4,823 | 990,069.62 | 205.28 |
| METHAMPHETAMINE | 1105 | 98 | 365.64 | 3.73 |
| LISDEXAMFETAMINE | 1205 | 3,936 | 347,036.32 | 88.17 |
| METHYLPHENIDATE (DL;D;L;ISOMERS) | 1724 | 4,714 | 886,922.74 | 188.15 |
| CODEINE | 9050 | 5,009 | 482,419.58 | 96.31 |
| BUPRENORPHINE | 9064 | 4,225 | 236,237.31 | 55.91 |
| OXYCODONE | 9143 | 5,182 | 2,392,089.63 | 461.62 |
| HYDROMORPHONE | 9150 | 3,918 | 50,877.48 | 12.99 |
| HYDROCODONE | 9193 | 4,858 | 555,535.29 | 114.35 |
| MEPERIDINE (PETHIDINE) | 9230 | 325 | 9,899.04 | 30.46 |
| METHADONE | 9250B | 3,192 | 117,122.31 | 36.69 |
| MORPHINE | 9300 | 4,178 | 476,949.74 | 114.16 |
| OXYMORPHONE | 9652 | 1,601 | 41,726.15 | 26.06 |
| TAPENTADOL | 9780 | 2,514 | 301,767.50 | 120.03 |
| FENTANYL BASE | 9801 | 4,043 | 11,290.18 | 2.79 |

**BUSINESS ACTIVITY: B - HOSPITALS**

| DRUG NAME | DRUG CODE | NUMBER OF REGISTRANT SOLD TO | TOTAL GRAMS | AVG GRAMS |
|---|---|---|---|---|

| | | | | |
|---|---|---|---|---|
| AMPHETAMINE | 1100 | 181 | 9,067.74 | 50.10 |
| LISDEXAMFETAMINE | 1205 | 84 | 1,502.44 | 17.89 |
| METHYLPHENIDATE (DL;D;L;ISOMERS) | 1724 | 245 | 10,738.53 | 43.83 |
| CODEINE | 9050 | 264 | 10,833.37 | 41.04 |
| BUPRENORPHINE | 9064 | 229 | 5,065.52 | 22.12 |
| OXYCODONE | 9143 | 328 | 82,238.16 | 250.73 |
| HYDROMORPHONE | 9150 | 275 | 14,157.63 | 51.48 |
| HYDROCODONE | 9193 | 243 | 21,838.70 | 89.87 |
| MEPERIDINE (PETHIDINE) | 9230 | 177 | 4,317.61 | 24.39 |
| METHADONE | 9250B | 246 | 35,454.69 | 144.12 |
| MORPHINE | 9300 | 319 | 53,542.16 | 167.84 |
| OXYMORPHONE | 9652 | 30 | 164.68 | 5.49 |
| TAPENTADOL | 9780 | 81 | 5,843.00 | 72.14 |
| FENTANYL BASE | 9801 | 283 | 1,971.46 | 6.97 |

**BUSINESS ACTIVITY: C - PRACTITIONERS**

| DRUG NAME | DRUG CODE | NUMBER OF REGISTRANT SOLD TO | TOTAL GRAMS | AVG GRAMS |
|---|---|---|---|---|
| AMPHETAMINE | 1100 | 4 | 8.51 | 2.13 |
| LISDEXAMFETAMINE | 1205 | 1 | 1.47 | 1.47 |
| METHYLPHENIDATE (DL;D;L;ISOMERS) | 1724 | 1 | 7.00 | 7.00 |
| CODEINE | 9050 | 136 | 4,307.93 | 31.68 |
| BUPRENORPHINE | 9064 | 1,136 | 184.75 | 0.16 |
| OXYCODONE | 9143 | 163 | 691.65 | 4.24 |
| HYDROMORPHONE | 9150 | 579 | 392.11 | 0.68 |
| HYDROCODONE | 9193 | 407 | 1,462.89 | 3.59 |
| MEPERIDINE (PETHIDINE) | 9230 | 172 | 1,497.56 | 8.71 |
| METHADONE | 9250B | 39 | 93.73 | 2.40 |
| MORPHINE | 9300 | 422 | 663.72 | 1.57 |
| FENTANYL BASE | 9801 | 1,112 | 140.79 | 0.13 |

**BUSINESS ACTIVITY: D - TEACHING INSTITUTIONS**

| DRUG NAME | DRUG CODE | NUMBER OF REGISTRANT SOLD TO | TOTAL GRAMS | AVG GRAMS |
|---|---|---|---|---|
| AMPHETAMINE | 1100 | 1 | 0.73 | 0.73 |
| BUPRENORPHINE | 9064 | 7 | 0.14 | 0.02 |
| FENTANYL BASE | 9801 | 1 | 0.01 | 0.01 |

**BUSINESS ACTIVITY: M - MID-LEVEL PRACTITIONERS**

| DRUG NAME | DRUG CODE | NUMBER OF REGISTRANT SOLD TO | TOTAL GRAMS | AVG GRAMS |
|---|---|---|---|---|
| CODEINE | 9050 | 4 | 35.60 | 8.90 |
| BUPRENORPHINE | 9064 | 4 | 3.71 | 0.93 |
| OXYCODONE | 9143 | 1 | 0.45 | 0.45 |
| HYDROMORPHONE | 9150 | 1 | 0.04 | 0.04 |
| HYDROCODONE | 9193 | 1 | 3.63 | 3.63 |

| | | | | |
|---|---|---|---|---|
| MEPERIDINE (PETHIDINE) | 9230 | 1 | 9.26 | 9.26 |
| METHADONE | 9250B | 1 | 4.47 | 4.47 |
| MORPHINE | 9300 | 1 | 0.04 | 0.04 |
| FENTANYL BASE | 9801 | 1 | 0.28 | 0.28 |

**BUSINESS ACTIVITY: NTP - NARCOTIC TREATMENT PROGRAMS**

| DRUG NAME | DRUG CODE | NUMBER OF REGISTRANT SOLD TO | TOTAL GRAMS | AVG GRAMS |
|---|---|---|---|---|
| BUPRENORPHINE | 9064 | 42 | 2,786.21 | 66.34 |
| METHADONE | 9250B | 119 | 973,944.72 | 8,184.41 |

**STATE: OH - OHIO**

**BUSINESS ACTIVITY: A - PHARMACIES**

| DRUG NAME | DRUG CODE | NUMBER OF REGISTRANT SOLD TO | TOTAL GRAMS | AVG GRAMS |
|---|---|---|---|---|
| AMPHETAMINE | 1100 | 2,223 | 816,549.89 | 367.32 |
| METHAMPHETAMINE | 1105 | 44 | 155.10 | 3.53 |
| LISDEXAMFETAMINE | 1205 | 2,163 | 396,473.12 | 183.30 |
| METHYLPHENIDATE (DL;D;L;ISOMERS) | 1724 | 2,220 | 797,052.31 | 359.03 |
| CODEINE | 9050 | 2,170 | 299,656.61 | 138.09 |
| BUPRENORPHINE | 9064 | 2,100 | 207,498.26 | 98.81 |
| OXYCODONE | 9143 | 2,230 | 1,486,158.65 | 666.44 |
| HYDROMORPHONE | 9150 | 1,825 | 22,448.82 | 12.30 |
| HYDROCODONE | 9193 | 2,235 | 568,817.03 | 254.50 |
| MEPERIDINE (PETHIDINE) | 9230 | 348 | 6,958.71 | 20.00 |
| METHADONE | 9250B | 1,872 | 68,886.78 | 36.80 |
| MORPHINE | 9300 | 2,143 | 356,441.77 | 166.33 |
| OXYMORPHONE | 9652 | 911 | 17,796.19 | 19.53 |
| TAPENTADOL | 9780 | 1,492 | 180,234.50 | 120.80 |
| FENTANYL BASE | 9801 | 2,037 | 6,092.28 | 2.99 |

**BUSINESS ACTIVITY: B - HOSPITALS**

| DRUG NAME | DRUG CODE | NUMBER OF REGISTRANT SOLD TO | TOTAL GRAMS | AVG GRAMS |
|---|---|---|---|---|
| AMPHETAMINE | 1100 | 163 | 15,124.92 | 92.79 |
| METHAMPHETAMINE | 1105 | 1 | 2.41 | 2.41 |
| LISDEXAMFETAMINE | 1205 | 68 | 3,779.04 | 55.57 |
| METHYLPHENIDATE (DL;D;L;ISOMERS) | 1724 | 216 | 15,225.04 | 70.49 |
| CODEINE | 9050 | 266 | 7,263.10 | 27.30 |
| BUPRENORPHINE | 9064 | 259 | 7,406.94 | 28.60 |
| OXYCODONE | 9143 | 393 | 83,279.89 | 211.91 |
| HYDROMORPHONE | 9150 | 368 | 7,485.37 | 20.34 |
| HYDROCODONE | 9193 | 394 | 21,465.76 | 54.48 |
| MEPERIDINE (PETHIDINE) | 9230 | 283 | 9,118.00 | 32.22 |
| METHADONE | 9250B | 253 | 7,529.68 | 29.76 |
| MORPHINE | 9300 | 409 | 38,704.98 | 94.63 |

| | | | | |
|---|---|---|---|---|
| OXYMORPHONE | 9652 | 36 | 189.99 | 5.28 |
| TAPENTADOL | 9780 | 63 | 3,492.05 | 55.43 |
| FENTANYL BASE | 9801 | 471 | 1,059.48 | 2.25 |

**BUSINESS ACTIVITY: C - PRACTITIONERS**

| DRUG NAME | DRUG CODE | NUMBER OF REGISTRANT SOLD TO | TOTAL GRAMS | AVG GRAMS |
|---|---|---|---|---|
| AMPHETAMINE | 1100 | 5 | 25.15 | 5.03 |
| LISDEXAMFETAMINE | 1205 | 2 | 17.85 | 8.92 |
| METHYLPHENIDATE (DL;D;L;ISOMERS) | 1724 | 2 | 102.48 | 51.24 |
| CODEINE | 9050 | 15 | 636.14 | 42.41 |
| BUPRENORPHINE | 9064 | 935 | 114.58 | 0.12 |
| OXYCODONE | 9143 | 26 | 74.88 | 2.88 |
| HYDROMORPHONE | 9150 | 298 | 151.66 | 0.51 |
| HYDROCODONE | 9193 | 279 | 814.86 | 2.92 |
| MEPERIDINE (PETHIDINE) | 9230 | 61 | 518.22 | 8.50 |
| METHADONE | 9250B | 9 | 7.42 | 0.82 |
| MORPHINE | 9300 | 177 | 348.52 | 1.97 |
| FENTANYL BASE | 9801 | 400 | 29.96 | 0.07 |

**BUSINESS ACTIVITY: D - TEACHING INSTITUTIONS**

| DRUG NAME | DRUG CODE | NUMBER OF REGISTRANT SOLD TO | TOTAL GRAMS | AVG GRAMS |
|---|---|---|---|---|
| BUPRENORPHINE | 9064 | 4 | 0.14 | 0.04 |
| HYDROMORPHONE | 9150 | 2 | 0.07 | 0.04 |
| FENTANYL BASE | 9801 | 1 | 0.11 | 0.11 |

**BUSINESS ACTIVITY: M - MID-LEVEL PRACTITIONERS**

| DRUG NAME | DRUG CODE | NUMBER OF REGISTRANT SOLD TO | TOTAL GRAMS | AVG GRAMS |
|---|---|---|---|---|
| BUPRENORPHINE | 9064 | 4 | 5.41 | 1.35 |
| HYDROMORPHONE | 9150 | 3 | 0.21 | 0.07 |
| MORPHINE | 9300 | 46 | 9.85 | 0.21 |
| FENTANYL BASE | 9801 | 125 | 1.14 | 0.01 |

**BUSINESS ACTIVITY: NTP - NARCOTIC TREATMENT PROGRAMS**

| DRUG NAME | DRUG CODE | NUMBER OF REGISTRANT SOLD TO | TOTAL GRAMS | AVG GRAMS |
|---|---|---|---|---|
| BUPRENORPHINE | 9064 | 29 | 10,348.48 | 356.84 |
| METHADONE | 9250B | 30 | 217,512.71 | 7,250.42 |

**STATE: OK - OKLAHOMA**

**BUSINESS ACTIVITY: A - PHARMACIES**

| DRUG NAME | DRUG CODE | NUMBER OF REGISTRANT SOLD TO | TOTAL GRAMS | AVG GRAMS |
|---|---|---|---|---|
| AMPHETAMINE | 1100 | 837 | 211,979.35 | 253.26 |
| METHAMPHETAMINE | 1105 | 20 | 53.44 | 2.67 |
| LISDEXAMFETAMINE | 1205 | 811 | 144,201.84 | 177.81 |
| METHYLPHENIDATE (DL;D;L;ISOMERS) | 1724 | 833 | 177,797.35 | 213.44 |
| CODEINE | 9050 | 866 | 317,177.90 | 366.26 |

| | | | | |
|---|---|---|---|---|
| BUPRENORPHINE | 9064 | 823 | 53,264.84 | 64.72 |
| OXYCODONE | 9143 | 856 | 705,257.64 | 823.90 |
| HYDROMORPHONE | 9150 | 732 | 10,588.52 | 14.47 |
| HYDROCODONE | 9193 | 867 | 581,863.49 | 671.12 |
| MEPERIDINE (PETHIDINE) | 9230 | 561 | 16,574.19 | 29.54 |
| METHADONE | 9250B | 681 | 22,776.25 | 33.45 |
| MORPHINE | 9300 | 838 | 256,268.85 | 305.81 |
| OXYMORPHONE | 9652 | 374 | 8,381.15 | 22.41 |
| TAPENTADOL | 9780 | 546 | 62,260.50 | 114.03 |
| FENTANYL BASE | 9801 | 823 | 4,199.90 | 5.10 |

**BUSINESS ACTIVITY: B - HOSPITALS**

| DRUG NAME | DRUG CODE | NUMBER OF REGISTRANT SOLD TO | TOTAL GRAMS | AVG GRAMS |
|---|---|---|---|---|
| AMPHETAMINE | 1100 | 67 | 7,100.17 | 105.97 |
| METHAMPHETAMINE | 1105 | 1 | 1.21 | 1.21 |
| LISDEXAMFETAMINE | 1205 | 44 | 4,371.48 | 99.35 |
| METHYLPHENIDATE (DL;D;L;ISOMERS) | 1724 | 95 | 8,741.43 | 92.02 |
| CODEINE | 9050 | 153 | 28,661.77 | 187.33 |
| BUPRENORPHINE | 9064 | 82 | 1,357.80 | 16.56 |
| OXYCODONE | 9143 | 197 | 28,953.64 | 146.97 |
| HYDROMORPHONE | 9150 | 170 | 1,836.76 | 10.80 |
| HYDROCODONE | 9193 | 203 | 34,678.94 | 170.83 |
| MEPERIDINE (PETHIDINE) | 9230 | 148 | 4,447.57 | 30.05 |
| METHADONE | 9250B | 90 | 1,455.46 | 16.17 |
| MORPHINE | 9300 | 202 | 22,585.22 | 111.81 |
| OXYMORPHONE | 9652 | 11 | 24.71 | 2.25 |
| TAPENTADOL | 9780 | 24 | 685.50 | 28.56 |
| FENTANYL BASE | 9801 | 199 | 434.51 | 2.18 |

**BUSINESS ACTIVITY: C - PRACTITIONERS**

| DRUG NAME | DRUG CODE | NUMBER OF REGISTRANT SOLD TO | TOTAL GRAMS | AVG GRAMS |
|---|---|---|---|---|
| AMPHETAMINE | 1100 | 1 | 0.99 | 0.99 |
| LISDEXAMFETAMINE | 1205 | 4 | 90.59 | 22.65 |
| CODEINE | 9050 | 60 | 2,248.78 | 37.48 |
| BUPRENORPHINE | 9064 | 279 | 368.68 | 1.32 |
| OXYCODONE | 9143 | 11 | 377.34 | 34.30 |
| HYDROMORPHONE | 9150 | 58 | 72.77 | 1.25 |
| HYDROCODONE | 9193 | 68 | 678.33 | 9.98 |
| MEPERIDINE (PETHIDINE) | 9230 | 37 | 252.19 | 6.82 |
| METHADONE | 9250B | 6 | 25.22 | 4.20 |
| MORPHINE | 9300 | 76 | 600.78 | 7.91 |
| FENTANYL BASE | 9801 | 146 | 13.21 | 0.09 |

**BUSINESS ACTIVITY: D - TEACHING INSTITUTIONS**

| DRUG NAME | DRUG CODE | NUMBER OF REGISTRANT SOLD TO | TOTAL GRAMS | AVG GRAMS |
|---|---|---|---|---|
| BUPRENORPHINE | 9064 | 2 | 0.56 | 0.28 |
| HYDROMORPHONE | 9150 | 1 | 2.13 | 2.13 |
| HYDROCODONE | 9193 | 1 | 7.94 | 7.94 |
| METHADONE | 9250B | 1 | 44.45 | 44.45 |
| MORPHINE | 9300 | 1 | 79.71 | 79.71 |
| FENTANYL BASE | 9801 | 1 | 0.57 | 0.57 |

**BUSINESS ACTIVITY: M - MID-LEVEL PRACTITIONERS**

| DRUG NAME | DRUG CODE | NUMBER OF REGISTRANT SOLD TO | TOTAL GRAMS | AVG GRAMS |
|---|---|---|---|---|
| CODEINE | 9050 | 9 | 7.36 | 0.82 |
| HYDROMORPHONE | 9150 | 11 | 0.65 | 0.06 |
| MORPHINE | 9300 | 31 | 9.47 | 0.31 |
| FENTANYL BASE | 9801 | 54 | 0.71 | 0.01 |

**BUSINESS ACTIVITY: NTP - NARCOTIC TREATMENT PROGRAMS**

| DRUG NAME | DRUG CODE | NUMBER OF REGISTRANT SOLD TO | TOTAL GRAMS | AVG GRAMS |
|---|---|---|---|---|
| BUPRENORPHINE | 9064 | 12 | 478.38 | 39.87 |
| METHADONE | 9250B | 19 | 113,175.59 | 5,956.61 |

**STATE: OR - OREGON**

**BUSINESS ACTIVITY: A - PHARMACIES**

| DRUG NAME | DRUG CODE | NUMBER OF REGISTRANT SOLD TO | TOTAL GRAMS | AVG GRAMS |
|---|---|---|---|---|
| AMPHETAMINE | 1100 | 661 | 246,538.80 | 372.98 |
| METHAMPHETAMINE | 1105 | 29 | 142.23 | 4.90 |
| LISDEXAMFETAMINE | 1205 | 622 | 62,263.68 | 100.10 |
| METHYLPHENIDATE (DL;D;L;ISOMERS) | 1724 | 659 | 241,072.10 | 365.82 |
| CODEINE | 9050 | 656 | 113,489.59 | 173.00 |
| BUPRENORPHINE | 9064 | 658 | 42,320.04 | 64.32 |
| OXYCODONE | 9143 | 673 | 553,179.00 | 821.96 |
| HYDROMORPHONE | 9150 | 653 | 13,332.12 | 20.42 |
| HYDROCODONE | 9193 | 674 | 303,934.27 | 450.94 |
| MEPERIDINE (PETHIDINE) | 9230 | 149 | 3,353.75 | 22.51 |
| METHADONE | 9250B | 622 | 46,495.74 | 74.75 |
| MORPHINE | 9300 | 672 | 217,003.99 | 322.92 |
| OXYMORPHONE | 9652 | 253 | 3,695.52 | 14.61 |
| TAPENTADOL | 9780 | 303 | 24,593.00 | 81.17 |
| FENTANYL BASE | 9801 | 620 | 2,563.77 | 4.14 |

**BUSINESS ACTIVITY: B - HOSPITALS**

| DRUG NAME | DRUG CODE | NUMBER OF REGISTRANT SOLD TO | TOTAL GRAMS | AVG GRAMS |
|---|---|---|---|---|
| AMPHETAMINE | 1100 | 49 | 3,371.34 | 68.80 |
| LISDEXAMFETAMINE | 1205 | 17 | 471.73 | 27.75 |

| | | | | |
|---|---|---|---|---|
| METHYLPHENIDATE (DL;D;L;ISOMERS) | 1724 | 57 | 4,663.19 | 81.81 |
| CODEINE | 9050 | 83 | 3,439.39 | 41.44 |
| BUPRENORPHINE | 9064 | 66 | 1,257.55 | 19.05 |
| OXYCODONE | 9143 | 93 | 32,431.13 | 348.72 |
| HYDROMORPHONE | 9150 | 78 | 2,577.87 | 33.05 |
| HYDROCODONE | 9193 | 99 | 19,460.31 | 196.57 |
| MEPERIDINE (PETHIDINE) | 9230 | 59 | 779.45 | 13.21 |
| METHADONE | 9250B | 66 | 3,190.22 | 48.34 |
| MORPHINE | 9300 | 93 | 19,639.33 | 211.18 |
| OXYMORPHONE | 9652 | 3 | 1.96 | 0.65 |
| TAPENTADOL | 9780 | 5 | 201.50 | 40.30 |
| FENTANYL BASE | 9801 | 80 | 302.93 | 3.79 |

**BUSINESS ACTIVITY: C - PRACTITIONERS**

| DRUG NAME | DRUG CODE | NUMBER OF REGISTRANT SOLD TO | TOTAL GRAMS | AVG GRAMS |
|---|---|---|---|---|
| AMPHETAMINE | 1100 | 1 | 11.23 | 11.23 |
| LISDEXAMFETAMINE | 1205 | 1 | 2.98 | 2.98 |
| METHYLPHENIDATE (DL;D;L;ISOMERS) | 1724 | 2 | 66.42 | 33.21 |
| CODEINE | 9050 | 63 | 4,822.57 | 76.55 |
| BUPRENORPHINE | 9064 | 581 | 155.27 | 0.27 |
| OXYCODONE | 9143 | 59 | 168.63 | 2.86 |
| HYDROMORPHONE | 9150 | 352 | 191.52 | 0.54 |
| HYDROCODONE | 9193 | 290 | 776.72 | 2.68 |
| MEPERIDINE (PETHIDINE) | 9230 | 74 | 433.03 | 5.85 |
| METHADONE | 9250B | 19 | 35.87 | 1.89 |
| MORPHINE | 9300 | 216 | 208.03 | 0.96 |
| FENTANYL BASE | 9801 | 381 | 35.46 | 0.09 |

**BUSINESS ACTIVITY: D - TEACHING INSTITUTIONS**

| DRUG NAME | DRUG CODE | NUMBER OF REGISTRANT SOLD TO | TOTAL GRAMS | AVG GRAMS |
|---|---|---|---|---|
| BUPRENORPHINE | 9064 | 1 | 0.01 | 0.01 |
| FENTANYL BASE | 9801 | 1 | 0.02 | 0.02 |

**BUSINESS ACTIVITY: M - MID-LEVEL PRACTITIONERS**

| DRUG NAME | DRUG CODE | NUMBER OF REGISTRANT SOLD TO | TOTAL GRAMS | AVG GRAMS |
|---|---|---|---|---|
| CODEINE | 9050 | 4 | 34.27 | 8.57 |
| BUPRENORPHINE | 9064 | 6 | 9.88 | 1.65 |
| HYDROMORPHONE | 9150 | 2 | 0.09 | 0.04 |
| HYDROCODONE | 9193 | 8 | 18.68 | 2.33 |
| MEPERIDINE (PETHIDINE) | 9230 | 1 | 4.36 | 4.36 |
| MORPHINE | 9300 | 5 | 0.94 | 0.19 |
| FENTANYL BASE | 9801 | 1 | 0.01 | 0.01 |

**BUSINESS ACTIVITY: NTP - NARCOTIC TREATMENT PROGRAMS**

| DRUG NAME | DRUG CODE | NUMBER OF REGISTRANT SOLD TO | TOTAL GRAMS | AVG GRAMS |
|---|---|---|---|---|
| BUPRENORPHINE | 9064 | 17 | 5,431.90 | 319.52 |
| METHADONE | 9250B | 19 | 177,792.41 | 9,357.50 |

**STATE: PA - PENNSYLVANIA**

**BUSINESS ACTIVITY: A - PHARMACIES**

| DRUG NAME | DRUG CODE | NUMBER OF REGISTRANT SOLD TO | TOTAL GRAMS | AVG GRAMS |
|---|---|---|---|---|
| AMPHETAMINE | 1100 | 2,915 | 846,387.57 | 290.36 |
| METHAMPHETAMINE | 1105 | 80 | 477.34 | 5.97 |
| LISDEXAMFETAMINE | 1205 | 2,708 | 352,202.22 | 130.06 |
| METHYLPHENIDATE (DL;D;L;ISOMERS) | 1724 | 2,879 | 858,323.24 | 298.13 |
| CODEINE | 9050 | 2,923 | 418,698.30 | 143.24 |
| BUPRENORPHINE | 9064 | 2,758 | 279,903.83 | 101.49 |
| OXYCODONE | 9143 | 2,980 | 2,346,458.00 | 787.40 |
| HYDROMORPHONE | 9150 | 2,459 | 39,924.92 | 16.24 |
| HYDROCODONE | 9193 | 2,959 | 513,515.89 | 173.54 |
| MEPERIDINE (PETHIDINE) | 9230 | 361 | 9,475.82 | 26.25 |
| METHADONE | 9250B | 2,168 | 93,327.51 | 43.05 |
| MORPHINE | 9300 | 2,865 | 703,926.41 | 245.70 |
| OXYMORPHONE | 9652 | 1,428 | 49,103.96 | 34.39 |
| TAPENTADOL | 9780 | 1,502 | 203,133.50 | 135.24 |
| FENTANYL BASE | 9801 | 2,759 | 13,859.21 | 5.02 |

**BUSINESS ACTIVITY: B - HOSPITALS**

| DRUG NAME | DRUG CODE | NUMBER OF REGISTRANT SOLD TO | TOTAL GRAMS | AVG GRAMS |
|---|---|---|---|---|
| AMPHETAMINE | 1100 | 172 | 12,369.64 | 71.92 |
| LISDEXAMFETAMINE | 1205 | 64 | 3,459.93 | 54.06 |
| METHYLPHENIDATE (DL;D;L;ISOMERS) | 1724 | 236 | 12,745.42 | 54.01 |
| CODEINE | 9050 | 256 | 12,932.42 | 50.52 |
| BUPRENORPHINE | 9064 | 215 | 8,338.32 | 38.78 |
| OXYCODONE | 9143 | 325 | 107,212.92 | 329.89 |
| HYDROMORPHONE | 9150 | 300 | 9,219.66 | 30.73 |
| HYDROCODONE | 9193 | 288 | 22,895.04 | 79.50 |
| MEPERIDINE (PETHIDINE) | 9230 | 191 | 1,913.29 | 10.02 |
| METHADONE | 9250B | 245 | 12,737.07 | 51.99 |
| MORPHINE | 9300 | 331 | 57,164.89 | 172.70 |
| OXYMORPHONE | 9652 | 47 | 505.29 | 10.75 |
| TAPENTADOL | 9780 | 52 | 1,467.00 | 28.21 |
| FENTANYL BASE | 9801 | 357 | 1,329.88 | 3.73 |

**BUSINESS ACTIVITY: C - PRACTITIONERS**

| DRUG NAME | DRUG CODE | NUMBER OF REGISTRANT SOLD TO | TOTAL GRAMS | AVG GRAMS |
|---|---|---|---|---|
| AMPHETAMINE | 1100 | 1 | 8.25 | 8.25 |

| | | | | |
|---|---|---|---|---|
| METHAMPHETAMINE | 1105 | 1 | 4.02 | 4.02 |
| LISDEXAMFETAMINE | 1205 | 1 | 9.83 | 9.83 |
| METHYLPHENIDATE (DL;D;L;ISOMERS) | 1724 | 1 | 19.67 | 19.67 |
| CODEINE | 9050 | 217 | 12,163.25 | 56.05 |
| BUPRENORPHINE | 9064 | 1,034 | 865.15 | 0.84 |
| OXYCODONE | 9143 | 58 | 187.54 | 3.23 |
| HYDROMORPHONE | 9150 | 392 | 421.50 | 1.08 |
| HYDROCODONE | 9193 | 373 | 1,106.09 | 2.97 |
| MEPERIDINE (PETHIDINE) | 9230 | 77 | 336.94 | 4.38 |
| METHADONE | 9250B | 41 | 151.24 | 3.69 |
| MORPHINE | 9300 | 241 | 298.89 | 1.24 |
| FENTANYL BASE | 9801 | 588 | 64.20 | 0.11 |

**BUSINESS ACTIVITY: D - TEACHING INSTITUTIONS**

| DRUG NAME | DRUG CODE | NUMBER OF REGISTRANT SOLD TO | TOTAL GRAMS | AVG GRAMS |
|---|---|---|---|---|
| CODEINE | 9050 | 1 | 851.62 | 851.62 |
| BUPRENORPHINE | 9064 | 3 | 1.42 | 0.47 |
| OXYCODONE | 9143 | 1 | 0.45 | 0.45 |
| HYDROMORPHONE | 9150 | 1 | 3.31 | 3.31 |
| HYDROCODONE | 9193 | 1 | 54.40 | 54.40 |
| METHADONE | 9250B | 2 | 33.81 | 16.90 |
| MORPHINE | 9300 | 2 | 37.60 | 18.80 |
| FENTANYL BASE | 9801 | 3 | 2.81 | 0.94 |

**BUSINESS ACTIVITY: M - MID-LEVEL PRACTITIONERS**

| DRUG NAME | DRUG CODE | NUMBER OF REGISTRANT SOLD TO | TOTAL GRAMS | AVG GRAMS |
|---|---|---|---|---|
| CODEINE | 9050 | 19 | 23.94 | 1.26 |

**BUSINESS ACTIVITY: NTP - NARCOTIC TREATMENT PROGRAMS**

| DRUG NAME | DRUG CODE | NUMBER OF REGISTRANT SOLD TO | TOTAL GRAMS | AVG GRAMS |
|---|---|---|---|---|
| BUPRENORPHINE | 9064 | 17 | 841.44 | 49.50 |
| METHADONE | 9250B | 88 | 607,839.16 | 6,907.26 |

**STATE: PR - PUERTO RICO**

**BUSINESS ACTIVITY: A - PHARMACIES**

| DRUG NAME | DRUG CODE | NUMBER OF REGISTRANT SOLD TO | TOTAL GRAMS | AVG GRAMS |
|---|---|---|---|---|
| AMPHETAMINE | 1100 | 643 | 37,734.02 | 58.68 |
| LISDEXAMFETAMINE | 1205 | 270 | 8,477.84 | 31.40 |
| METHYLPHENIDATE (DL;D;L;ISOMERS) | 1724 | 683 | 57,151.20 | 83.68 |
| CODEINE | 9050 | 1,002 | 43,411.89 | 43.33 |
| BUPRENORPHINE | 9064 | 267 | 7,885.10 | 29.53 |
| OXYCODONE | 9143 | 785 | 39,157.21 | 49.88 |
| HYDROMORPHONE | 9150 | 27 | 138.13 | 5.12 |
| HYDROCODONE | 9193 | 462 | 1,695.86 | 3.67 |

| | | | | |
|---|---|---|---|---|
| MEPERIDINE (PETHIDINE) | 9230 | 644 | 4,500.75 | 6.99 |
| METHADONE | 9250B | 9 | 193.37 | 21.49 |
| MORPHINE | 9300 | 135 | 3,933.12 | 29.13 |
| TAPENTADOL | 9780 | 42 | 1,403.00 | 33.40 |
| FENTANYL BASE | 9801 | 674 | 370.67 | 0.55 |

**BUSINESS ACTIVITY: B - HOSPITALS**

| DRUG NAME | DRUG CODE | NUMBER OF REGISTRANT SOLD TO | TOTAL GRAMS | AVG GRAMS |
|---|---|---|---|---|
| AMPHETAMINE | 1100 | 7 | 121.57 | 17.37 |
| METHYLPHENIDATE (DL;D;L;ISOMERS) | 1724 | 19 | 238.78 | 12.57 |
| CODEINE | 9050 | 75 | 3,487.54 | 46.50 |
| BUPRENORPHINE | 9064 | 22 | 614.11 | 27.91 |
| OXYCODONE | 9143 | 65 | 5,173.70 | 79.60 |
| HYDROMORPHONE | 9150 | 4 | 91.54 | 22.89 |
| HYDROCODONE | 9193 | 25 | 144.79 | 5.79 |
| MEPERIDINE (PETHIDINE) | 9230 | 90 | 7,932.28 | 88.14 |
| METHADONE | 9250B | 4 | 99.64 | 24.91 |
| MORPHINE | 9300 | 78 | 2,901.18 | 37.19 |
| TAPENTADOL | 9780 | 4 | 553.00 | 138.25 |
| FENTANYL BASE | 9801 | 82 | 117.24 | 1.43 |

**BUSINESS ACTIVITY: C - PRACTITIONERS**

| DRUG NAME | DRUG CODE | NUMBER OF REGISTRANT SOLD TO | TOTAL GRAMS | AVG GRAMS |
|---|---|---|---|---|
| BUPRENORPHINE | 9064 | 20 | 1.91 | 0.10 |
| MEPERIDINE (PETHIDINE) | 9230 | 4 | 49.57 | 12.39 |
| MORPHINE | 9300 | 1 | 0.75 | 0.75 |
| FENTANYL BASE | 9801 | 10 | 0.90 | 0.09 |

**BUSINESS ACTIVITY: NTP - NARCOTIC TREATMENT PROGRAMS**

| DRUG NAME | DRUG CODE | NUMBER OF REGISTRANT SOLD TO | TOTAL GRAMS | AVG GRAMS |
|---|---|---|---|---|
| METHADONE | 9250B | 5 | 86,542.14 | 17,308.43 |

**STATE: RI - RHODE ISLAND**

**BUSINESS ACTIVITY: A - PHARMACIES**

| DRUG NAME | DRUG CODE | NUMBER OF REGISTRANT SOLD TO | TOTAL GRAMS | AVG GRAMS |
|---|---|---|---|---|
| AMPHETAMINE | 1100 | 235 | 151,923.64 | 646.48 |
| METHAMPHETAMINE | 1105 | 7 | 14.87 | 2.12 |
| LISDEXAMFETAMINE | 1205 | 219 | 29,018.31 | 132.50 |
| METHYLPHENIDATE (DL;D;L;ISOMERS) | 1724 | 227 | 69,766.29 | 307.34 |
| CODEINE | 9050 | 209 | 22,399.55 | 107.17 |
| BUPRENORPHINE | 9064 | 233 | 31,350.23 | 134.55 |
| OXYCODONE | 9143 | 234 | 125,578.21 | 536.66 |
| HYDROMORPHONE | 9150 | 180 | 2,138.57 | 11.88 |
| HYDROCODONE | 9193 | 234 | 32,479.06 | 138.80 |

| | | | | |
|---|---|---|---|---|
| MEPERIDINE (PETHIDINE) | 9230 | 18 | 424.86 | 23.60 |
| METHADONE | 9250B | 165 | 5,418.10 | 32.84 |
| MORPHINE | 9300 | 223 | 33,476.48 | 150.12 |
| OXYMORPHONE | 9652 | 58 | 680.40 | 11.73 |
| TAPENTADOL | 9780 | 96 | 7,568.50 | 78.84 |
| FENTANYL BASE | 9801 | 200 | 477.05 | 2.39 |

**BUSINESS ACTIVITY: B - HOSPITALS**

| DRUG NAME | DRUG CODE | NUMBER OF REGISTRANT SOLD TO | TOTAL GRAMS | AVG GRAMS |
|---|---|---|---|---|
| AMPHETAMINE | 1100 | 16 | 2,592.30 | 162.02 |
| LISDEXAMFETAMINE | 1205 | 7 | 439.93 | 62.85 |
| METHYLPHENIDATE (DL;D;L;ISOMERS) | 1724 | 20 | 2,771.40 | 138.57 |
| CODEINE | 9050 | 16 | 613.68 | 38.36 |
| BUPRENORPHINE | 9064 | 15 | 690.12 | 46.01 |
| OXYCODONE | 9143 | 22 | 8,336.33 | 378.92 |
| HYDROMORPHONE | 9150 | 17 | 577.00 | 33.94 |
| HYDROCODONE | 9193 | 21 | 1,778.79 | 84.70 |
| MEPERIDINE (PETHIDINE) | 9230 | 12 | 160.14 | 13.34 |
| METHADONE | 9250B | 14 | 1,453.31 | 103.81 |
| MORPHINE | 9300 | 19 | 4,447.76 | 234.09 |
| OXYMORPHONE | 9652 | 1 | 0.89 | 0.89 |
| FENTANYL BASE | 9801 | 16 | 90.64 | 5.66 |

**BUSINESS ACTIVITY: C - PRACTITIONERS**

| DRUG NAME | DRUG CODE | NUMBER OF REGISTRANT SOLD TO | TOTAL GRAMS | AVG GRAMS |
|---|---|---|---|---|
| BUPRENORPHINE | 9064 | 81 | 7.99 | 0.10 |
| OXYCODONE | 9143 | 3 | 7.35 | 2.45 |
| HYDROMORPHONE | 9150 | 37 | 33.20 | 0.90 |
| HYDROCODONE | 9193 | 27 | 91.55 | 3.39 |
| MEPERIDINE (PETHIDINE) | 9230 | 8 | 72.99 | 9.12 |
| METHADONE | 9250B | 2 | 11.63 | 5.81 |
| MORPHINE | 9300 | 12 | 15.51 | 1.29 |
| FENTANYL BASE | 9801 | 37 | 7.28 | 0.20 |

**BUSINESS ACTIVITY: NTP - NARCOTIC TREATMENT PROGRAMS**

| DRUG NAME | DRUG CODE | NUMBER OF REGISTRANT SOLD TO | TOTAL GRAMS | AVG GRAMS |
|---|---|---|---|---|
| BUPRENORPHINE | 9064 | 15 | 643.40 | 42.89 |
| METHADONE | 9250B | 21 | 162,267.95 | 7,727.05 |

**STATE: SC - SOUTH CAROLINA**

**BUSINESS ACTIVITY: A - PHARMACIES**

| DRUG NAME | DRUG CODE | NUMBER OF REGISTRANT SOLD TO | TOTAL GRAMS | AVG GRAMS |
|---|---|---|---|---|
| AMPHETAMINE | 1100 | 1,193 | 548,608.65 | 459.86 |
| METHAMPHETAMINE | 1105 | 23 | 59.07 | 2.57 |

| | | | | |
|---|---|---|---|---|
| LISDEXAMFETAMINE | 1205 | 1,163 | 241,631.35 | 207.77 |
| METHYLPHENIDATE (DL;D;L;ISOMERS) | 1724 | 1,187 | 331,277.49 | 279.09 |
| CODEINE | 9050 | 1,144 | 276,075.42 | 241.32 |
| BUPRENORPHINE | 9064 | 1,107 | 48,453.32 | 43.77 |
| OXYCODONE | 9143 | 1,196 | 747,107.15 | 624.67 |
| HYDROMORPHONE | 9150 | 1,063 | 16,444.97 | 15.47 |
| HYDROCODONE | 9193 | 1,208 | 443,187.66 | 366.88 |
| MEPERIDINE (PETHIDINE) | 9230 | 638 | 13,826.46 | 21.67 |
| METHADONE | 9250B | 901 | 31,248.99 | 34.68 |
| MORPHINE | 9300 | 1,155 | 212,957.89 | 184.38 |
| OXYMORPHONE | 9652 | 599 | 12,424.63 | 20.74 |
| TAPENTADOL | 9780 | 894 | 140,171.00 | 156.79 |
| FENTANYL BASE | 9801 | 1,127 | 3,674.56 | 3.26 |

**BUSINESS ACTIVITY: B - HOSPITALS**

| DRUG NAME | DRUG CODE | NUMBER OF REGISTRANT SOLD TO | TOTAL GRAMS | AVG GRAMS |
|---|---|---|---|---|
| AMPHETAMINE | 1100 | 59 | 9,124.96 | 154.66 |
| METHAMPHETAMINE | 1105 | 1 | 13.66 | 13.66 |
| LISDEXAMFETAMINE | 1205 | 24 | 1,601.34 | 66.72 |
| METHYLPHENIDATE (DL;D;L;ISOMERS) | 1724 | 78 | 8,768.76 | 112.42 |
| CODEINE | 9050 | 85 | 4,596.76 | 54.08 |
| BUPRENORPHINE | 9064 | 66 | 1,137.26 | 17.23 |
| OXYCODONE | 9143 | 137 | 27,182.95 | 198.42 |
| HYDROMORPHONE | 9150 | 137 | 2,855.93 | 20.85 |
| HYDROCODONE | 9193 | 140 | 12,998.16 | 92.84 |
| MEPERIDINE (PETHIDINE) | 9230 | 101 | 2,880.63 | 28.52 |
| METHADONE | 9250B | 79 | 1,884.41 | 23.85 |
| MORPHINE | 9300 | 144 | 18,920.34 | 131.39 |
| OXYMORPHONE | 9652 | 5 | 10.26 | 2.05 |
| TAPENTADOL | 9780 | 31 | 1,060.00 | 34.19 |
| FENTANYL BASE | 9801 | 171 | 389.57 | 2.28 |

**BUSINESS ACTIVITY: C - PRACTITIONERS**

| DRUG NAME | DRUG CODE | NUMBER OF REGISTRANT SOLD TO | TOTAL GRAMS | AVG GRAMS |
|---|---|---|---|---|
| AMPHETAMINE | 1100 | 3 | 23.73 | 7.91 |
| LISDEXAMFETAMINE | 1205 | 2 | 59.46 | 29.73 |
| CODEINE | 9050 | 16 | 302.27 | 18.89 |
| BUPRENORPHINE | 9064 | 427 | 70.17 | 0.16 |
| OXYCODONE | 9143 | 8 | 24.25 | 3.03 |
| HYDROMORPHONE | 9150 | 132 | 120.55 | 0.91 |
| HYDROCODONE | 9193 | 74 | 181.43 | 2.45 |
| MEPERIDINE (PETHIDINE) | 9230 | 65 | 558.85 | 8.60 |

| | | | | |
|---|---|---|---|---|
| METHADONE | 9250B | 4 | 1.79 | 0.45 |
| MORPHINE | 9300 | 79 | 91.73 | 1.16 |
| FENTANYL BASE | 9801 | 237 | 11.46 | 0.05 |

**BUSINESS ACTIVITY: D - TEACHING INSTITUTIONS**

| DRUG NAME | DRUG CODE | NUMBER OF REGISTRANT SOLD TO | TOTAL GRAMS | AVG GRAMS |
|---|---|---|---|---|
| BUPRENORPHINE | 9064 | 1 | 0.04 | 0.04 |
| HYDROMORPHONE | 9150 | 1 | 0.18 | 0.18 |

**BUSINESS ACTIVITY: M - MID-LEVEL PRACTITIONERS**

| DRUG NAME | DRUG CODE | NUMBER OF REGISTRANT SOLD TO | TOTAL GRAMS | AVG GRAMS |
|---|---|---|---|---|
| CODEINE | 9050 | 1 | 0.43 | 0.43 |
| MORPHINE | 9300 | 52 | 22.13 | 0.43 |
| FENTANYL BASE | 9801 | 62 | 0.75 | 0.01 |

**BUSINESS ACTIVITY: NTP - NARCOTIC TREATMENT PROGRAMS**

| DRUG NAME | DRUG CODE | NUMBER OF REGISTRANT SOLD TO | TOTAL GRAMS | AVG GRAMS |
|---|---|---|---|---|
| BUPRENORPHINE | 9064 | 11 | 1,357.15 | 123.38 |
| METHADONE | 9250B | 21 | 165,926.85 | 7,901.28 |

**STATE: SD - SOUTH DAKOTA**

**BUSINESS ACTIVITY: A - PHARMACIES**

| DRUG NAME | DRUG CODE | NUMBER OF REGISTRANT SOLD TO | TOTAL GRAMS | AVG GRAMS |
|---|---|---|---|---|
| AMPHETAMINE | 1100 | 190 | 58,518.84 | 307.99 |
| METHAMPHETAMINE | 1105 | 5 | 30.94 | 6.19 |
| LISDEXAMFETAMINE | 1205 | 186 | 40,406.30 | 217.24 |
| METHYLPHENIDATE (DL;D;L;ISOMERS) | 1724 | 190 | 78,199.50 | 411.58 |
| CODEINE | 9050 | 191 | 20,546.13 | 107.57 |
| BUPRENORPHINE | 9064 | 140 | 2,015.75 | 14.40 |
| OXYCODONE | 9143 | 190 | 50,827.46 | 267.51 |
| HYDROMORPHONE | 9150 | 162 | 1,372.98 | 8.48 |
| HYDROCODONE | 9193 | 193 | 39,730.53 | 205.86 |
| MEPERIDINE (PETHIDINE) | 9230 | 43 | 1,072.49 | 24.94 |
| METHADONE | 9250B | 144 | 3,955.31 | 27.47 |
| MORPHINE | 9300 | 187 | 22,506.22 | 120.35 |
| OXYMORPHONE | 9652 | 50 | 1,136.13 | 22.72 |
| TAPENTADOL | 9780 | 131 | 14,280.00 | 109.01 |
| FENTANYL BASE | 9801 | 188 | 773.81 | 4.12 |

**BUSINESS ACTIVITY: B - HOSPITALS**

| DRUG NAME | DRUG CODE | NUMBER OF REGISTRANT SOLD TO | TOTAL GRAMS | AVG GRAMS |
|---|---|---|---|---|
| AMPHETAMINE | 1100 | 26 | 2,652.71 | 102.03 |
| METHAMPHETAMINE | 1105 | 1 | 0.40 | 0.40 |
| LISDEXAMFETAMINE | 1205 | 17 | 576.94 | 33.94 |
| METHYLPHENIDATE (DL;D;L;ISOMERS) | 1724 | 35 | 3,498.36 | 99.95 |

| | | | | |
|---|---|---|---|---|
| CODEINE | 9050 | 49 | 2,248.27 | 45.88 |
| BUPRENORPHINE | 9064 | 22 | 286.88 | 13.04 |
| OXYCODONE | 9143 | 69 | 5,811.36 | 84.22 |
| HYDROMORPHONE | 9150 | 61 | 548.88 | 9.00 |
| HYDROCODONE | 9193 | 72 | 8,032.20 | 111.56 |
| MEPERIDINE (PETHIDINE) | 9230 | 36 | 517.13 | 14.36 |
| METHADONE | 9250B | 27 | 337.01 | 12.48 |
| MORPHINE | 9300 | 74 | 6,121.85 | 82.73 |
| OXYMORPHONE | 9652 | 7 | 17.93 | 2.56 |
| TAPENTADOL | 9780 | 16 | 588.00 | 36.75 |
| FENTANYL BASE | 9801 | 72 | 129.45 | 1.80 |

**BUSINESS ACTIVITY: C - PRACTITIONERS**

| DRUG NAME | DRUG CODE | NUMBER OF REGISTRANT SOLD TO | TOTAL GRAMS | AVG GRAMS |
|---|---|---|---|---|
| CODEINE | 9050 | 3 | 55.43 | 18.48 |
| BUPRENORPHINE | 9064 | 65 | 2.30 | 0.04 |
| OXYCODONE | 9143 | 3 | 2.69 | 0.90 |
| HYDROMORPHONE | 9150 | 24 | 56.10 | 2.34 |
| HYDROCODONE | 9193 | 27 | 69.20 | 2.56 |
| MEPERIDINE (PETHIDINE) | 9230 | 9 | 293.59 | 32.62 |
| MORPHINE | 9300 | 19 | 51.34 | 2.70 |
| FENTANYL BASE | 9801 | 45 | 4.58 | 0.10 |

**BUSINESS ACTIVITY: D - TEACHING INSTITUTIONS**

| DRUG NAME | DRUG CODE | NUMBER OF REGISTRANT SOLD TO | TOTAL GRAMS | AVG GRAMS |
|---|---|---|---|---|
| BUPRENORPHINE | 9064 | 1 | 0.00 | 0.00 |

**BUSINESS ACTIVITY: M - MID-LEVEL PRACTITIONERS**

| DRUG NAME | DRUG CODE | NUMBER OF REGISTRANT SOLD TO | TOTAL GRAMS | AVG GRAMS |
|---|---|---|---|---|
| HYDROMORPHONE | 9150 | 4 | 0.35 | 0.09 |
| MORPHINE | 9300 | 4 | 1.50 | 0.38 |
| FENTANYL BASE | 9801 | 6 | 0.15 | 0.03 |

**BUSINESS ACTIVITY: NTP - NARCOTIC TREATMENT PROGRAMS**

| DRUG NAME | DRUG CODE | NUMBER OF REGISTRANT SOLD TO | TOTAL GRAMS | AVG GRAMS |
|---|---|---|---|---|
| METHADONE | 9250B | 1 | 3,298.55 | 3,298.55 |

**STATE: TN - TENNESSEE**

**BUSINESS ACTIVITY: A - PHARMACIES**

| DRUG NAME | DRUG CODE | NUMBER OF REGISTRANT SOLD TO | TOTAL GRAMS | AVG GRAMS |
|---|---|---|---|---|
| AMPHETAMINE | 1100 | 1,529 | 532,953.09 | 348.56 |
| METHAMPHETAMINE | 1105 | 18 | 59.07 | 3.28 |
| LISDEXAMFETAMINE | 1205 | 1,486 | 266,131.23 | 179.09 |
| METHYLPHENIDATE (DL;D;L;ISOMERS) | 1724 | 1,515 | 316,032.55 | 208.60 |
| CODEINE | 9050 | 1,481 | 261,657.33 | 176.68 |

| | | | | |
|---|---|---|---|---|
| BUPRENORPHINE | 9064 | 1,398 | 150,298.47 | 107.51 |
| OXYCODONE | 9143 | 1,541 | 1,531,351.74 | 993.74 |
| HYDROMORPHONE | 9150 | 1,293 | 17,742.17 | 13.72 |
| HYDROCODONE | 9193 | 1,550 | 785,658.83 | 506.88 |
| MEPERIDINE (PETHIDINE) | 9230 | 883 | 17,462.14 | 19.78 |
| METHADONE | 9250B | 994 | 32,986.01 | 33.19 |
| MORPHINE | 9300 | 1,521 | 486,216.20 | 319.67 |
| OXYMORPHONE | 9652 | 892 | 49,790.20 | 55.82 |
| TAPENTADOL | 9780 | 1,077 | 101,743.00 | 94.47 |
| FENTANYL BASE | 9801 | 1,447 | 5,277.34 | 3.65 |

**BUSINESS ACTIVITY: B - HOSPITALS**

| DRUG NAME | DRUG CODE | NUMBER OF REGISTRANT SOLD TO | TOTAL GRAMS | AVG GRAMS |
|---|---|---|---|---|
| AMPHETAMINE | 1100 | 82 | 5,885.02 | 71.77 |
| LISDEXAMFETAMINE | 1205 | 39 | 2,567.92 | 65.84 |
| METHYLPHENIDATE (DL;D;L;ISOMERS) | 1724 | 116 | 4,819.11 | 41.54 |
| CODEINE | 9050 | 133 | 4,060.05 | 30.53 |
| BUPRENORPHINE | 9064 | 130 | 2,467.06 | 18.98 |
| OXYCODONE | 9143 | 183 | 66,993.77 | 366.09 |
| HYDROMORPHONE | 9150 | 170 | 4,060.64 | 23.89 |
| HYDROCODONE | 9193 | 197 | 39,592.99 | 200.98 |
| MEPERIDINE (PETHIDINE) | 9230 | 136 | 3,671.61 | 27.00 |
| METHADONE | 9250B | 136 | 1,980.34 | 14.56 |
| MORPHINE | 9300 | 189 | 30,439.08 | 161.05 |
| OXYMORPHONE | 9652 | 23 | 74.62 | 3.24 |
| TAPENTADOL | 9780 | 36 | 1,727.50 | 47.99 |
| FENTANYL BASE | 9801 | 177 | 656.38 | 3.71 |

**BUSINESS ACTIVITY: C - PRACTITIONERS**

| DRUG NAME | DRUG CODE | NUMBER OF REGISTRANT SOLD TO | TOTAL GRAMS | AVG GRAMS |
|---|---|---|---|---|
| AMPHETAMINE | 1100 | 11 | 1,068.75 | 97.16 |
| LISDEXAMFETAMINE | 1205 | 7 | 832.20 | 118.89 |
| METHYLPHENIDATE (DL;D;L;ISOMERS) | 1724 | 3 | 312.86 | 104.29 |
| CODEINE | 9050 | 56 | 3,191.58 | 56.99 |
| BUPRENORPHINE | 9064 | 585 | 63.27 | 0.11 |
| OXYCODONE | 9143 | 85 | 529.87 | 6.23 |
| HYDROMORPHONE | 9150 | 245 | 188.54 | 0.77 |
| HYDROCODONE | 9193 | 283 | 1,088.69 | 3.85 |
| MEPERIDINE (PETHIDINE) | 9230 | 131 | 914.53 | 6.98 |
| METHADONE | 9250B | 6 | 12.16 | 2.03 |
| MORPHINE | 9300 | 172 | 185.39 | 1.08 |
| FENTANYL BASE | 9801 | 429 | 41.05 | 0.10 |

**BUSINESS ACTIVITY: D - TEACHING INSTITUTIONS**

| DRUG NAME | DRUG CODE | NUMBER OF REGISTRANT SOLD TO | TOTAL GRAMS | AVG GRAMS |
|---|---|---|---|---|
| BUPRENORPHINE | 9064 | 1 | 0.02 | 0.02 |
| FENTANYL BASE | 9801 | 1 | 0.01 | 0.01 |

**BUSINESS ACTIVITY: M - MID-LEVEL PRACTITIONERS**

| DRUG NAME | DRUG CODE | NUMBER OF REGISTRANT SOLD TO | TOTAL GRAMS | AVG GRAMS |
|---|---|---|---|---|
| AMPHETAMINE | 1100 | 1 | 1.56 | 1.56 |
| CODEINE | 9050 | 3 | 51.22 | 17.07 |
| HYDROMORPHONE | 9150 | 3 | 0.40 | 0.13 |
| HYDROCODONE | 9193 | 1 | 1.21 | 1.21 |
| MEPERIDINE (PETHIDINE) | 9230 | 7 | 15.25 | 2.18 |
| MORPHINE | 9300 | 65 | 26.88 | 0.41 |
| FENTANYL BASE | 9801 | 85 | 1.55 | 0.02 |

**BUSINESS ACTIVITY: NTP - NARCOTIC TREATMENT PROGRAMS**

| DRUG NAME | DRUG CODE | NUMBER OF REGISTRANT SOLD TO | TOTAL GRAMS | AVG GRAMS |
|---|---|---|---|---|
| BUPRENORPHINE | 9064 | 12 | 938.58 | 78.22 |
| METHADONE | 9250B | 13 | 157,981.45 | 12,152.42 |

**STATE: TX - TEXAS**

**BUSINESS ACTIVITY: A - PHARMACIES**

| DRUG NAME | DRUG CODE | NUMBER OF REGISTRANT SOLD TO | TOTAL GRAMS | AVG GRAMS |
|---|---|---|---|---|
| AMPHETAMINE | 1100 | 4,607 | 1,743,754.07 | 378.50 |
| METHAMPHETAMINE | 1105 | 149 | 424.31 | 2.85 |
| LISDEXAMFETAMINE | 1205 | 4,404 | 1,055,317.50 | 239.63 |
| METHYLPHENIDATE (DL;D;L;ISOMERS) | 1724 | 4,546 | 1,235,718.65 | 271.83 |
| CODEINE | 9050 | 4,960 | 3,550,465.50 | 715.82 |
| BUPRENORPHINE | 9064 | 4,181 | 108,166.38 | 25.87 |
| OXYCODONE | 9143 | 4,534 | 1,136,473.23 | 250.66 |
| HYDROMORPHONE | 9150 | 3,894 | 65,454.01 | 16.81 |
| HYDROCODONE | 9193 | 4,943 | 2,202,422.66 | 445.56 |
| MEPERIDINE (PETHIDINE) | 9230 | 904 | 15,463.42 | 17.11 |
| METHADONE | 9250B | 3,376 | 112,154.00 | 33.22 |
| MORPHINE | 9300 | 4,317 | 621,261.52 | 143.91 |
| OXYMORPHONE | 9652 | 1,197 | 17,889.68 | 14.95 |
| TAPENTADOL | 9780 | 2,609 | 296,937.00 | 113.81 |
| FENTANYL BASE | 9801 | 4,270 | 15,267.69 | 3.58 |

**BUSINESS ACTIVITY: B - HOSPITALS**

| DRUG NAME | DRUG CODE | NUMBER OF REGISTRANT SOLD TO | TOTAL GRAMS | AVG GRAMS |
|---|---|---|---|---|
| AMPHETAMINE | 1100 | 290 | 37,402.57 | 128.97 |
| LISDEXAMFETAMINE | 1205 | 129 | 9,553.10 | 74.06 |
| METHYLPHENIDATE (DL;D;L;ISOMERS) | 1724 | 430 | 47,403.08 | 110.24 |

| | | | | |
|---|---|---|---|---|
| CODEINE | 9050 | 919 | 106,420.75 | 115.80 |
| BUPRENORPHINE | 9064 | 325 | 4,357.23 | 13.41 |
| OXYCODONE | 9143 | 758 | 54,821.55 | 72.32 |
| HYDROMORPHONE | 9150 | 1,091 | 13,123.44 | 12.03 |
| HYDROCODONE | 9193 | 1,215 | 126,889.29 | 104.44 |
| MEPERIDINE (PETHIDINE) | 9230 | 773 | 23,416.62 | 30.29 |
| METHADONE | 9250B | 500 | 10,120.42 | 20.24 |
| MORPHINE | 9300 | 1,298 | 85,808.09 | 66.11 |
| OXYMORPHONE | 9652 | 35 | 339.86 | 9.71 |
| TAPENTADOL | 9780 | 93 | 4,826.45 | 51.90 |
| FENTANYL BASE | 9801 | 1,325 | 2,600.86 | 1.96 |

**BUSINESS ACTIVITY: C - PRACTITIONERS**

| DRUG NAME | DRUG CODE | NUMBER OF REGISTRANT SOLD TO | TOTAL GRAMS | AVG GRAMS |
|---|---|---|---|---|
| AMPHETAMINE | 1100 | 8 | 31.39 | 3.92 |
| LISDEXAMFETAMINE | 1205 | 6 | 77.48 | 12.91 |
| METHYLPHENIDATE (DL;D;L;ISOMERS) | 1724 | 5 | 510.12 | 102.02 |
| CODEINE | 9050 | 393 | 4,159.69 | 10.58 |
| BUPRENORPHINE | 9064 | 2,060 | 614.22 | 0.30 |
| OXYCODONE | 9143 | 17 | 61.77 | 3.63 |
| HYDROMORPHONE | 9150 | 613 | 481.42 | 0.79 |
| HYDROCODONE | 9193 | 856 | 2,030.34 | 2.37 |
| MEPERIDINE (PETHIDINE) | 9230 | 372 | 2,969.32 | 7.98 |
| METHADONE | 9250B | 31 | 63.86 | 2.06 |
| MORPHINE | 9300 | 463 | 515.96 | 1.11 |
| OXYMORPHONE | 9652 | 1 | 8.03 | 8.03 |
| TAPENTADOL | 9780 | 1 | 15.00 | 15.00 |
| FENTANYL BASE | 9801 | 1,323 | 83.76 | 0.06 |

**BUSINESS ACTIVITY: D - TEACHING INSTITUTIONS**

| DRUG NAME | DRUG CODE | NUMBER OF REGISTRANT SOLD TO | TOTAL GRAMS | AVG GRAMS |
|---|---|---|---|---|
| BUPRENORPHINE | 9064 | 3 | 0.94 | 0.31 |
| HYDROMORPHONE | 9150 | 1 | 5.04 | 5.04 |
| HYDROCODONE | 9193 | 1 | 2.96 | 2.96 |
| MEPERIDINE (PETHIDINE) | 9230 | 1 | 2.18 | 2.18 |
| METHADONE | 9250B | 1 | 6.98 | 6.98 |
| MORPHINE | 9300 | 2 | 7.51 | 3.75 |
| FENTANYL BASE | 9801 | 3 | 2.74 | 0.91 |

**BUSINESS ACTIVITY: M - MID-LEVEL PRACTITIONERS**

| DRUG NAME | DRUG CODE | NUMBER OF REGISTRANT SOLD TO | TOTAL GRAMS | AVG GRAMS |
|---|---|---|---|---|
| CODEINE | 9050 | 71 | 73.68 | 1.04 |
| BUPRENORPHINE | 9064 | 1 | 0.00 | 0.00 |

| | | | | |
|---|---|---|---|---|
| HYDROMORPHONE | 9150 | 15 | 1.32 | 0.09 |
| HYDROCODONE | 9193 | 1 | 1.21 | 1.21 |
| MEPERIDINE (PETHIDINE) | 9230 | 4 | 13.62 | 3.40 |
| MORPHINE | 9300 | 321 | 129.27 | 0.40 |
| FENTANYL BASE | 9801 | 589 | 33.11 | 0.06 |

**BUSINESS ACTIVITY: NTP - NARCOTIC TREATMENT PROGRAMS**

| DRUG NAME | DRUG CODE | NUMBER OF REGISTRANT SOLD TO | TOTAL GRAMS | AVG GRAMS |
|---|---|---|---|---|
| BUPRENORPHINE | 9064 | 36 | 1,213.55 | 33.71 |
| METHADONE | 9250B | 93 | 396,298.80 | 4,261.28 |

**STATE: UT - UTAH**

**BUSINESS ACTIVITY: A - PHARMACIES**

| DRUG NAME | DRUG CODE | NUMBER OF REGISTRANT SOLD TO | TOTAL GRAMS | AVG GRAMS |
|---|---|---|---|---|
| AMPHETAMINE | 1100 | 544 | 354,420.63 | 651.51 |
| METHAMPHETAMINE | 1105 | 30 | 94.43 | 3.15 |
| LISDEXAMFETAMINE | 1205 | 521 | 89,374.26 | 171.54 |
| METHYLPHENIDATE (DL;D;L;ISOMERS) | 1724 | 543 | 179,388.78 | 330.37 |
| CODEINE | 9050 | 526 | 56,900.99 | 108.18 |
| BUPRENORPHINE | 9064 | 533 | 40,424.24 | 75.84 |
| OXYCODONE | 9143 | 547 | 563,666.03 | 1,030.47 |
| HYDROMORPHONE | 9150 | 508 | 8,102.85 | 15.95 |
| HYDROCODONE | 9193 | 548 | 192,510.03 | 351.30 |
| MEPERIDINE (PETHIDINE) | 9230 | 231 | 4,816.13 | 20.85 |
| METHADONE | 9250B | 492 | 31,285.66 | 63.59 |
| MORPHINE | 9300 | 533 | 157,907.27 | 296.26 |
| OXYMORPHONE | 9652 | 367 | 10,630.77 | 28.97 |
| TAPENTADOL | 9780 | 375 | 39,586.50 | 105.56 |
| FENTANYL BASE | 9801 | 523 | 2,475.05 | 4.73 |

**BUSINESS ACTIVITY: B - HOSPITALS**

| DRUG NAME | DRUG CODE | NUMBER OF REGISTRANT SOLD TO | TOTAL GRAMS | AVG GRAMS |
|---|---|---|---|---|
| AMPHETAMINE | 1100 | 31 | 4,262.30 | 137.49 |
| LISDEXAMFETAMINE | 1205 | 15 | 862.53 | 57.50 |
| METHYLPHENIDATE (DL;D;L;ISOMERS) | 1724 | 44 | 4,310.92 | 97.98 |
| CODEINE | 9050 | 36 | 1,430.03 | 39.72 |
| BUPRENORPHINE | 9064 | 44 | 841.21 | 19.12 |
| OXYCODONE | 9143 | 90 | 23,027.35 | 255.86 |
| HYDROMORPHONE | 9150 | 82 | 1,492.85 | 18.21 |
| HYDROCODONE | 9193 | 116 | 9,901.99 | 85.36 |
| MEPERIDINE (PETHIDINE) | 9230 | 73 | 1,340.43 | 18.36 |
| METHADONE | 9250B | 55 | 1,171.93 | 21.31 |
| MORPHINE | 9300 | 102 | 12,682.43 | 124.34 |

| | | | | |
|---|---|---|---|---|
| OXYMORPHONE | 9652 | 9 | 49.42 | 5.49 |
| TAPENTADOL | 9780 | 19 | 2,641.50 | 139.03 |
| FENTANYL BASE | 9801 | 104 | 201.37 | 1.94 |

**BUSINESS ACTIVITY: C - PRACTITIONERS**

| DRUG NAME | DRUG CODE | NUMBER OF REGISTRANT SOLD TO | TOTAL GRAMS | AVG GRAMS |
|---|---|---|---|---|
| AMPHETAMINE | 1100 | 2 | 6.10 | 3.05 |
| LISDEXAMFETAMINE | 1205 | 1 | 40.34 | 40.34 |
| METHYLPHENIDATE (DL;D;L;ISOMERS) | 1724 | 1 | 4.15 | 4.15 |
| CODEINE | 9050 | 11 | 506.79 | 46.07 |
| BUPRENORPHINE | 9064 | 190 | 18.48 | 0.10 |
| OXYCODONE | 9143 | 9 | 11.43 | 1.27 |
| HYDROMORPHONE | 9150 | 82 | 73.45 | 0.90 |
| HYDROCODONE | 9193 | 78 | 336.80 | 4.32 |
| MEPERIDINE (PETHIDINE) | 9230 | 15 | 124.19 | 8.28 |
| METHADONE | 9250B | 6 | 14.31 | 2.39 |
| MORPHINE | 9300 | 98 | 112.21 | 1.15 |
| FENTANYL BASE | 9801 | 190 | 12.00 | 0.06 |

**BUSINESS ACTIVITY: D - TEACHING INSTITUTIONS**

| DRUG NAME | DRUG CODE | NUMBER OF REGISTRANT SOLD TO | TOTAL GRAMS | AVG GRAMS |
|---|---|---|---|---|
| BUPRENORPHINE | 9064 | 1 | 0.01 | 0.01 |

**BUSINESS ACTIVITY: M - MID-LEVEL PRACTITIONERS**

| DRUG NAME | DRUG CODE | NUMBER OF REGISTRANT SOLD TO | TOTAL GRAMS | AVG GRAMS |
|---|---|---|---|---|
| BUPRENORPHINE | 9064 | 1 | 0.42 | 0.42 |
| HYDROMORPHONE | 9150 | 1 | 0.04 | 0.04 |
| MEPERIDINE (PETHIDINE) | 9230 | 3 | 52.29 | 17.43 |
| MORPHINE | 9300 | 3 | 8.72 | 2.91 |
| FENTANYL BASE | 9801 | 6 | 0.57 | 0.10 |

**BUSINESS ACTIVITY: NTP - NARCOTIC TREATMENT PROGRAMS**

| DRUG NAME | DRUG CODE | NUMBER OF REGISTRANT SOLD TO | TOTAL GRAMS | AVG GRAMS |
|---|---|---|---|---|
| BUPRENORPHINE | 9064 | 12 | 290.78 | 24.23 |
| METHADONE | 9250B | 16 | 65,051.50 | 4,065.72 |

**STATE: VA - VIRGINIA**

**BUSINESS ACTIVITY: A - PHARMACIES**

| DRUG NAME | DRUG CODE | NUMBER OF REGISTRANT SOLD TO | TOTAL GRAMS | AVG GRAMS |
|---|---|---|---|---|
| AMPHETAMINE | 1100 | 1,596 | 703,182.32 | 440.59 |
| METHAMPHETAMINE | 1105 | 42 | 126.17 | 3.00 |
| LISDEXAMFETAMINE | 1205 | 1,542 | 260,281.43 | 168.79 |
| METHYLPHENIDATE (DL;D;L;ISOMERS) | 1724 | 1,583 | 485,850.87 | 306.92 |
| CODEINE | 9050 | 1,562 | 173,802.06 | 111.27 |
| BUPRENORPHINE | 9064 | 1,491 | 80,679.63 | 54.11 |

| | | | | |
|---|---|---|---|---|
| OXYCODONE | 9143 | 1,598 | 945,287.75 | 591.54 |
| HYDROMORPHONE | 9150 | 1,505 | 29,288.35 | 19.46 |
| HYDROCODONE | 9193 | 1,607 | 323,450.80 | 201.28 |
| MEPERIDINE (PETHIDINE) | 9230 | 639 | 14,739.24 | 23.07 |
| METHADONE | 9250B | 1,242 | 49,869.51 | 40.15 |
| MORPHINE | 9300 | 1,523 | 244,402.01 | 160.47 |
| OXYMORPHONE | 9652 | 844 | 22,047.58 | 26.12 |
| TAPENTADOL | 9780 | 1,050 | 123,618.00 | 117.73 |
| FENTANYL BASE | 9801 | 1,462 | 4,583.83 | 3.14 |

**BUSINESS ACTIVITY: B - HOSPITALS**

| DRUG NAME | DRUG CODE | NUMBER OF REGISTRANT SOLD TO | TOTAL GRAMS | AVG GRAMS |
|---|---|---|---|---|
| AMPHETAMINE | 1100 | 91 | 30,622.67 | 336.51 |
| LISDEXAMFETAMINE | 1205 | 45 | 7,923.44 | 176.08 |
| METHYLPHENIDATE (DL;D;L;ISOMERS) | 1724 | 122 | 34,665.63 | 284.14 |
| CODEINE | 9050 | 126 | 6,052.29 | 48.03 |
| BUPRENORPHINE | 9064 | 115 | 2,811.63 | 24.45 |
| OXYCODONE | 9143 | 176 | 59,683.18 | 339.11 |
| HYDROMORPHONE | 9150 | 174 | 6,267.76 | 36.02 |
| HYDROCODONE | 9193 | 175 | 16,067.06 | 91.81 |
| MEPERIDINE (PETHIDINE) | 9230 | 125 | 3,421.40 | 27.37 |
| METHADONE | 9250B | 123 | 3,401.70 | 27.66 |
| MORPHINE | 9300 | 183 | 25,743.03 | 140.67 |
| OXYMORPHONE | 9652 | 14 | 45.94 | 3.28 |
| TAPENTADOL | 9780 | 29 | 2,310.00 | 79.66 |
| FENTANYL BASE | 9801 | 187 | 684.97 | 3.66 |

**BUSINESS ACTIVITY: C - PRACTITIONERS**

| DRUG NAME | DRUG CODE | NUMBER OF REGISTRANT SOLD TO | TOTAL GRAMS | AVG GRAMS |
|---|---|---|---|---|
| AMPHETAMINE | 1100 | 3 | 95.97 | 31.99 |
| LISDEXAMFETAMINE | 1205 | 1 | 37.66 | 37.66 |
| METHYLPHENIDATE (DL;D;L;ISOMERS) | 1724 | 2 | 39.26 | 19.63 |
| CODEINE | 9050 | 146 | 4,206.21 | 28.81 |
| BUPRENORPHINE | 9064 | 883 | 77.30 | 0.09 |
| OXYCODONE | 9143 | 28 | 221.53 | 7.91 |
| HYDROMORPHONE | 9150 | 345 | 205.21 | 0.59 |
| HYDROCODONE | 9193 | 221 | 1,016.43 | 4.60 |
| MEPERIDINE (PETHIDINE) | 9230 | 112 | 638.16 | 5.70 |
| METHADONE | 9250B | 27 | 58.63 | 2.17 |
| MORPHINE | 9300 | 211 | 254.95 | 1.21 |
| OXYMORPHONE | 9652 | 1 | 15.52 | 15.52 |
| TAPENTADOL | 9780 | 2 | 189.00 | 94.50 |

| | | | | |
|---|---|---|---|---|
| FENTANYL BASE | 9801 | 465 | 38.70 | 0.08 |

**BUSINESS ACTIVITY: D - TEACHING INSTITUTIONS**

| DRUG NAME | DRUG CODE | NUMBER OF REGISTRANT SOLD TO | TOTAL GRAMS | AVG GRAMS |
|---|---|---|---|---|
| CODEINE | 9050 | 1 | 477.76 | 477.76 |
| BUPRENORPHINE | 9064 | 1 | 0.57 | 0.57 |
| HYDROMORPHONE | 9150 | 1 | 1.38 | 1.38 |
| HYDROCODONE | 9193 | 1 | 2.10 | 2.10 |
| METHADONE | 9250B | 1 | 2.15 | 2.15 |
| MORPHINE | 9300 | 1 | 31.17 | 31.17 |
| FENTANYL BASE | 9801 | 1 | 1.16 | 1.16 |

**BUSINESS ACTIVITY: M - MID-LEVEL PRACTITIONERS**

| DRUG NAME | DRUG CODE | NUMBER OF REGISTRANT SOLD TO | TOTAL GRAMS | AVG GRAMS |
|---|---|---|---|---|
| CODEINE | 9050 | 21 | 23.49 | 1.12 |
| BUPRENORPHINE | 9064 | 1 | 0.02 | 0.02 |
| MORPHINE | 9300 | 8 | 2.96 | 0.37 |
| FENTANYL BASE | 9801 | 14 | 0.19 | 0.01 |

**BUSINESS ACTIVITY: NTP - NARCOTIC TREATMENT PROGRAMS**

| DRUG NAME | DRUG CODE | NUMBER OF REGISTRANT SOLD TO | TOTAL GRAMS | AVG GRAMS |
|---|---|---|---|---|
| BUPRENORPHINE | 9064 | 28 | 3,017.29 | 107.76 |
| METHADONE | 9250B | 44 | 264,991.94 | 6,022.54 |

**STATE: VI - VIRGIN ISLANDS**

**BUSINESS ACTIVITY: A - PHARMACIES**

| DRUG NAME | DRUG CODE | NUMBER OF REGISTRANT SOLD TO | TOTAL GRAMS | AVG GRAMS |
|---|---|---|---|---|
| AMPHETAMINE | 1100 | 19 | 1,325.10 | 69.74 |
| LISDEXAMFETAMINE | 1205 | 14 | 277.49 | 19.82 |
| METHYLPHENIDATE (DL;D;L;ISOMERS) | 1724 | 16 | 585.73 | 36.61 |
| CODEINE | 9050 | 20 | 2,705.46 | 135.27 |
| BUPRENORPHINE | 9064 | 11 | 48.92 | 4.45 |
| OXYCODONE | 9143 | 20 | 1,735.18 | 86.76 |
| HYDROMORPHONE | 9150 | 13 | 46.50 | 3.58 |
| HYDROCODONE | 9193 | 20 | 946.58 | 47.33 |
| MEPERIDINE (PETHIDINE) | 9230 | 9 | 209.16 | 23.24 |
| METHADONE | 9250B | 3 | 27.73 | 9.24 |
| MORPHINE | 9300 | 12 | 461.06 | 38.42 |
| TAPENTADOL | 9780 | 1 | 5.00 | 5.00 |
| FENTANYL BASE | 9801 | 13 | 15.49 | 1.19 |

**BUSINESS ACTIVITY: B - HOSPITALS**

| DRUG NAME | DRUG CODE | NUMBER OF REGISTRANT SOLD TO | TOTAL GRAMS | AVG GRAMS |
|---|---|---|---|---|
| METHYLPHENIDATE (DL;D;L;ISOMERS) | 1724 | 1 | 0.43 | 0.43 |
| CODEINE | 9050 | 2 | 72.93 | 36.47 |

| | | | | |
|---|---|---|---|---|
| OXYCODONE | 9143 | 2 | 17.39 | 8.70 |
| HYDROMORPHONE | 9150 | 2 | 7.80 | 3.90 |
| HYDROCODONE | 9193 | 2 | 22.24 | 11.12 |
| MEPERIDINE (PETHIDINE) | 9230 | 2 | 251.10 | 125.55 |
| MORPHINE | 9300 | 2 | 10.68 | 5.34 |
| FENTANYL BASE | 9801 | 3 | 2.38 | 0.79 |

**BUSINESS ACTIVITY: C - PRACTITIONERS**

| DRUG NAME | DRUG CODE | NUMBER OF REGISTRANT SOLD TO | TOTAL GRAMS | AVG GRAMS |
|---|---|---|---|---|
| BUPRENORPHINE | 9064 | 5 | 0.21 | 0.04 |
| MORPHINE | 9300 | 1 | 0.02 | 0.02 |
| FENTANYL BASE | 9801 | 2 | 0.07 | 0.03 |

**BUSINESS ACTIVITY: NTP - NARCOTIC TREATMENT PROGRAMS**

| DRUG NAME | DRUG CODE | NUMBER OF REGISTRANT SOLD TO | TOTAL GRAMS | AVG GRAMS |
|---|---|---|---|---|
| METHADONE | 9250B | 1 | 536.64 | 536.64 |

**STATE: VT - VERMONT**

**BUSINESS ACTIVITY: A - PHARMACIES**

| DRUG NAME | DRUG CODE | NUMBER OF REGISTRANT SOLD TO | TOTAL GRAMS | AVG GRAMS |
|---|---|---|---|---|
| AMPHETAMINE | 1100 | 172 | 39,381.80 | 228.96 |
| METHAMPHETAMINE | 1105 | 4 | 8.44 | 2.11 |
| LISDEXAMFETAMINE | 1205 | 172 | 28,343.09 | 164.79 |
| METHYLPHENIDATE (DL;D;L;ISOMERS) | 1724 | 172 | 72,744.85 | 422.94 |
| CODEINE | 9050 | 155 | 10,522.47 | 67.89 |
| BUPRENORPHINE | 9064 | 171 | 22,196.91 | 129.81 |
| OXYCODONE | 9143 | 172 | 73,951.45 | 429.95 |
| HYDROMORPHONE | 9150 | 169 | 3,404.85 | 20.15 |
| HYDROCODONE | 9193 | 173 | 15,583.34 | 90.08 |
| MEPERIDINE (PETHIDINE) | 9230 | 22 | 374.75 | 17.03 |
| METHADONE | 9250B | 163 | 11,809.21 | 72.45 |
| MORPHINE | 9300 | 170 | 32,839.68 | 193.17 |
| OXYMORPHONE | 9652 | 25 | 548.64 | 21.95 |
| TAPENTADOL | 9780 | 44 | 3,808.50 | 86.56 |
| FENTANYL BASE | 9801 | 161 | 547.79 | 3.40 |

**BUSINESS ACTIVITY: B - HOSPITALS**

| DRUG NAME | DRUG CODE | NUMBER OF REGISTRANT SOLD TO | TOTAL GRAMS | AVG GRAMS |
|---|---|---|---|---|
| AMPHETAMINE | 1100 | 12 | 1,155.90 | 96.32 |
| LISDEXAMFETAMINE | 1205 | 6 | 359.58 | 59.93 |
| METHYLPHENIDATE (DL;D;L;ISOMERS) | 1724 | 16 | 2,180.55 | 136.28 |
| CODEINE | 9050 | 13 | 351.23 | 27.02 |
| BUPRENORPHINE | 9064 | 17 | 1,182.14 | 69.54 |
| OXYCODONE | 9143 | 18 | 5,066.81 | 281.49 |

| | | | | |
|---|---|---|---|---|
| HYDROMORPHONE | 9150 | 16 | 564.03 | 35.25 |
| HYDROCODONE | 9193 | 17 | 808.55 | 47.56 |
| MEPERIDINE (PETHIDINE) | 9230 | 12 | 681.62 | 56.80 |
| METHADONE | 9250B | 16 | 643.70 | 40.23 |
| MORPHINE | 9300 | 17 | 3,875.88 | 227.99 |
| OXYMORPHONE | 9652 | 1 | 8.56 | 8.56 |
| TAPENTADOL | 9780 | 1 | 80.00 | 80.00 |
| FENTANYL BASE | 9801 | 17 | 47.28 | 2.78 |

**BUSINESS ACTIVITY: C - PRACTITIONERS**

| DRUG NAME | DRUG CODE | NUMBER OF REGISTRANT SOLD TO | TOTAL GRAMS | AVG GRAMS |
|---|---|---|---|---|
| CODEINE | 9050 | 1 | 4.42 | 4.42 |
| BUPRENORPHINE | 9064 | 98 | 6.59 | 0.07 |
| HYDROMORPHONE | 9150 | 52 | 29.59 | 0.57 |
| HYDROCODONE | 9193 | 14 | 21.65 | 1.55 |
| METHADONE | 9250B | 1 | 0.18 | 0.18 |
| MORPHINE | 9300 | 23 | 20.56 | 0.89 |
| FENTANYL BASE | 9801 | 37 | 2.50 | 0.07 |

**BUSINESS ACTIVITY: D - TEACHING INSTITUTIONS**

| DRUG NAME | DRUG CODE | NUMBER OF REGISTRANT SOLD TO | TOTAL GRAMS | AVG GRAMS |
|---|---|---|---|---|
| BUPRENORPHINE | 9064 | 1 | 0.00 | 0.00 |

**BUSINESS ACTIVITY: M - MID-LEVEL PRACTITIONERS**

| DRUG NAME | DRUG CODE | NUMBER OF REGISTRANT SOLD TO | TOTAL GRAMS | AVG GRAMS |
|---|---|---|---|---|
| CODEINE | 9050 | 3 | 8.84 | 2.95 |

**BUSINESS ACTIVITY: NTP - NARCOTIC TREATMENT PROGRAMS**

| DRUG NAME | DRUG CODE | NUMBER OF REGISTRANT SOLD TO | TOTAL GRAMS | AVG GRAMS |
|---|---|---|---|---|
| BUPRENORPHINE | 9064 | 10 | 3,778.98 | 377.90 |
| METHADONE | 9250B | 10 | 59,448.98 | 5,944.90 |

**STATE: WA - WASHINGTON**

**BUSINESS ACTIVITY: A - PHARMACIES**

| DRUG NAME | DRUG CODE | NUMBER OF REGISTRANT SOLD TO | TOTAL GRAMS | AVG GRAMS |
|---|---|---|---|---|
| AMPHETAMINE | 1100 | 1,204 | 447,715.75 | 371.86 |
| METHAMPHETAMINE | 1105 | 96 | 353.19 | 3.68 |
| LISDEXAMFETAMINE | 1205 | 1,133 | 139,132.48 | 122.80 |
| METHYLPHENIDATE (DL;D;L;ISOMERS) | 1724 | 1,204 | 424,904.58 | 352.91 |
| CODEINE | 9050 | 1,219 | 200,782.80 | 164.71 |
| BUPRENORPHINE | 9064 | 1,200 | 90,235.71 | 75.20 |
| OXYCODONE | 9143 | 1,226 | 921,189.05 | 751.38 |
| HYDROMORPHONE | 9150 | 1,166 | 29,106.65 | 24.96 |
| HYDROCODONE | 9193 | 1,240 | 420,969.87 | 339.49 |
| MEPERIDINE (PETHIDINE) | 9230 | 396 | 8,187.62 | 20.68 |

| | | | | |
|---|---|---|---|---|
| METHADONE | 9250B | 1,143 | 81,137.46 | 70.99 |
| MORPHINE | 9300 | 1,213 | 330,565.58 | 272.52 |
| OXYMORPHONE | 9652 | 434 | 5,678.04 | 13.08 |
| TAPENTADOL | 9780 | 639 | 55,160.50 | 86.32 |
| FENTANYL BASE | 9801 | 1,113 | 4,194.22 | 3.77 |

**BUSINESS ACTIVITY: B - HOSPITALS**

| DRUG NAME | DRUG CODE | NUMBER OF REGISTRANT SOLD TO | TOTAL GRAMS | AVG GRAMS |
|---|---|---|---|---|
| AMPHETAMINE | 1100 | 95 | 14,825.57 | 156.06 |
| METHAMPHETAMINE | 1105 | 1 | 0.40 | 0.40 |
| LISDEXAMFETAMINE | 1205 | 37 | 3,441.75 | 93.02 |
| METHYLPHENIDATE (DL;D;L;ISOMERS) | 1724 | 111 | 22,717.59 | 204.66 |
| CODEINE | 9050 | 156 | 7,135.27 | 45.74 |
| BUPRENORPHINE | 9064 | 122 | 3,506.49 | 28.74 |
| OXYCODONE | 9143 | 225 | 68,489.55 | 304.40 |
| HYDROMORPHONE | 9150 | 192 | 5,925.98 | 30.86 |
| HYDROCODONE | 9193 | 213 | 26,871.11 | 126.16 |
| MEPERIDINE (PETHIDINE) | 9230 | 158 | 2,293.35 | 14.51 |
| METHADONE | 9250B | 124 | 8,267.12 | 66.67 |
| MORPHINE | 9300 | 224 | 34,485.77 | 153.95 |
| OXYMORPHONE | 9652 | 9 | 46.66 | 5.18 |
| TAPENTADOL | 9780 | 23 | 1,748.00 | 76.00 |
| FENTANYL BASE | 9801 | 230 | 622.23 | 2.71 |

**BUSINESS ACTIVITY: C - PRACTITIONERS**

| DRUG NAME | DRUG CODE | NUMBER OF REGISTRANT SOLD TO | TOTAL GRAMS | AVG GRAMS |
|---|---|---|---|---|
| AMPHETAMINE | 1100 | 2 | 2.89 | 1.44 |
| LISDEXAMFETAMINE | 1205 | 1 | 24.97 | 24.97 |
| METHYLPHENIDATE (DL;D;L;ISOMERS) | 1724 | 1 | 5.45 | 5.45 |
| CODEINE | 9050 | 96 | 6,889.64 | 71.77 |
| BUPRENORPHINE | 9064 | 857 | 197.65 | 0.23 |
| OXYCODONE | 9143 | 39 | 55.96 | 1.43 |
| HYDROMORPHONE | 9150 | 446 | 253.62 | 0.57 |
| HYDROCODONE | 9193 | 398 | 1,126.46 | 2.83 |
| MEPERIDINE (PETHIDINE) | 9230 | 92 | 912.35 | 9.92 |
| METHADONE | 9250B | 43 | 147.93 | 3.44 |
| MORPHINE | 9300 | 340 | 524.41 | 1.54 |
| FENTANYL BASE | 9801 | 644 | 41.84 | 0.06 |

**BUSINESS ACTIVITY: D - TEACHING INSTITUTIONS**

| DRUG NAME | DRUG CODE | NUMBER OF REGISTRANT SOLD TO | TOTAL GRAMS | AVG GRAMS |
|---|---|---|---|---|
| BUPRENORPHINE | 9064 | 1 | 0.01 | 0.01 |
| HYDROMORPHONE | 9150 | 1 | 0.04 | 0.04 |

**BUSINESS ACTIVITY: M - MID-LEVEL PRACTITIONERS**

| DRUG NAME | DRUG CODE | NUMBER OF REGISTRANT SOLD TO | TOTAL GRAMS | AVG GRAMS |
|---|---|---|---|---|
| CODEINE | 9050 | 5 | 17.13 | 3.43 |
| BUPRENORPHINE | 9064 | 11 | 90.77 | 8.25 |
| OXYCODONE | 9143 | 3 | 8.07 | 2.69 |
| HYDROMORPHONE | 9150 | 2 | 0.18 | 0.09 |
| HYDROCODONE | 9193 | 1 | 0.18 | 0.18 |
| MEPERIDINE (PETHIDINE) | 9230 | 4 | 13.07 | 3.27 |
| MORPHINE | 9300 | 3 | 2.63 | 0.88 |
| FENTANYL BASE | 9801 | 11 | 0.43 | 0.04 |

**BUSINESS ACTIVITY: NTP - NARCOTIC TREATMENT PROGRAMS**

| DRUG NAME | DRUG CODE | NUMBER OF REGISTRANT SOLD TO | TOTAL GRAMS | AVG GRAMS |
|---|---|---|---|---|
| BUPRENORPHINE | 9064 | 15 | 1,893.82 | 126.25 |
| METHADONE | 9250B | 26 | 281,936.35 | 10,843.71 |

**STATE: WI - WISCONSIN**

**BUSINESS ACTIVITY: A - PHARMACIES**

| DRUG NAME | DRUG CODE | NUMBER OF REGISTRANT SOLD TO | TOTAL GRAMS | AVG GRAMS |
|---|---|---|---|---|
| AMPHETAMINE | 1100 | 1,095 | 492,962.17 | 450.19 |
| METHAMPHETAMINE | 1105 | 36 | 136.21 | 3.78 |
| LISDEXAMFETAMINE | 1205 | 1,087 | 289,089.89 | 265.95 |
| METHYLPHENIDATE (DL;D;L;ISOMERS) | 1724 | 1,094 | 400,189.86 | 365.80 |
| CODEINE | 9050 | 1,091 | 163,076.46 | 149.47 |
| BUPRENORPHINE | 9064 | 1,032 | 43,120.24 | 41.78 |
| OXYCODONE | 9143 | 1,103 | 617,883.03 | 560.18 |
| HYDROMORPHONE | 9150 | 915 | 8,248.67 | 9.01 |
| HYDROCODONE | 9193 | 1,109 | 255,190.32 | 230.11 |
| MEPERIDINE (PETHIDINE) | 9230 | 94 | 1,313.50 | 13.97 |
| METHADONE | 9250B | 846 | 24,799.57 | 29.31 |
| MORPHINE | 9300 | 1,089 | 230,297.62 | 211.48 |
| OXYMORPHONE | 9652 | 321 | 3,327.89 | 10.37 |
| TAPENTADOL | 9780 | 575 | 43,930.00 | 76.40 |
| FENTANYL BASE | 9801 | 1,052 | 3,972.17 | 3.78 |

**BUSINESS ACTIVITY: B - HOSPITALS**

| DRUG NAME | DRUG CODE | NUMBER OF REGISTRANT SOLD TO | TOTAL GRAMS | AVG GRAMS |
|---|---|---|---|---|
| AMPHETAMINE | 1100 | 104 | 11,684.21 | 112.35 |
| METHAMPHETAMINE | 1105 | 1 | 2.01 | 2.01 |
| LISDEXAMFETAMINE | 1205 | 67 | 2,619.95 | 39.10 |
| METHYLPHENIDATE (DL;D;L;ISOMERS) | 1724 | 128 | 8,812.31 | 68.85 |
| CODEINE | 9050 | 156 | 4,582.04 | 29.37 |
| BUPRENORPHINE | 9064 | 102 | 2,759.77 | 27.06 |

| | | | | |
|---|---|---|---|---|
| OXYCODONE | 9143 | 173 | 40,326.45 | 233.10 |
| HYDROMORPHONE | 9150 | 161 | 3,765.96 | 23.39 |
| HYDROCODONE | 9193 | 170 | 13,398.95 | 78.82 |
| MEPERIDINE (PETHIDINE) | 9230 | 109 | 2,268.41 | 20.81 |
| METHADONE | 9250B | 121 | 2,319.84 | 19.17 |
| MORPHINE | 9300 | 167 | 25,926.13 | 155.25 |
| OXYMORPHONE | 9652 | 6 | 7.58 | 1.26 |
| TAPENTADOL | 9780 | 25 | 435.65 | 17.43 |
| FENTANYL BASE | 9801 | 164 | 540.33 | 3.29 |

**BUSINESS ACTIVITY: C - PRACTITIONERS**

| DRUG NAME | DRUG CODE | NUMBER OF REGISTRANT SOLD TO | TOTAL GRAMS | AVG GRAMS |
|---|---|---|---|---|
| METHYLPHENIDATE (DL;D;L;ISOMERS) | 1724 | 2 | 7.35 | 3.67 |
| CODEINE | 9050 | 55 | 495.08 | 9.00 |
| BUPRENORPHINE | 9064 | 562 | 201.15 | 0.36 |
| OXYCODONE | 9143 | 58 | 502.13 | 8.66 |
| HYDROMORPHONE | 9150 | 322 | 236.58 | 0.73 |
| HYDROCODONE | 9193 | 236 | 618.79 | 2.62 |
| MEPERIDINE (PETHIDINE) | 9230 | 60 | 451.98 | 7.53 |
| METHADONE | 9250B | 20 | 46.69 | 2.33 |
| MORPHINE | 9300 | 220 | 330.86 | 1.50 |
| OXYMORPHONE | 9652 | 1 | 4.82 | 4.82 |
| FENTANYL BASE | 9801 | 334 | 45.22 | 0.14 |

**BUSINESS ACTIVITY: M - MID-LEVEL PRACTITIONERS**

| DRUG NAME | DRUG CODE | NUMBER OF REGISTRANT SOLD TO | TOTAL GRAMS | AVG GRAMS |
|---|---|---|---|---|
| CODEINE | 9050 | 2 | 8.10 | 4.05 |
| OXYCODONE | 9143 | 1 | 0.45 | 0.45 |
| HYDROMORPHONE | 9150 | 1 | 0.04 | 0.04 |
| MORPHINE | 9300 | 1 | 0.19 | 0.19 |
| FENTANYL BASE | 9801 | 1 | 0.01 | 0.01 |

**BUSINESS ACTIVITY: NTP - NARCOTIC TREATMENT PROGRAMS**

| DRUG NAME | DRUG CODE | NUMBER OF REGISTRANT SOLD TO | TOTAL GRAMS | AVG GRAMS |
|---|---|---|---|---|
| BUPRENORPHINE | 9064 | 12 | 2,034.01 | 169.50 |
| METHADONE | 9250B | 20 | 199,800.02 | 9,990.00 |

**STATE: WV - WEST VIRGINIA**

**BUSINESS ACTIVITY: A - PHARMACIES**

| DRUG NAME | DRUG CODE | NUMBER OF REGISTRANT SOLD TO | TOTAL GRAMS | AVG GRAMS |
|---|---|---|---|---|
| AMPHETAMINE | 1100 | 647 | 92,489.99 | 142.95 |
| METHAMPHETAMINE | 1105 | 3 | 18.48 | 6.16 |
| LISDEXAMFETAMINE | 1205 | 636 | 74,392.22 | 116.97 |
| METHYLPHENIDATE (DL;D;L;ISOMERS) | 1724 | 653 | 118,494.31 | 181.46 |

| | | | | |
|---|---|---|---|---|
| CODEINE | 9050 | 632 | 140,835.68 | 222.84 |
| BUPRENORPHINE | 9064 | 581 | 60,503.12 | 104.14 |
| OXYCODONE | 9143 | 660 | 249,779.20 | 378.45 |
| HYDROMORPHONE | 9150 | 424 | 3,471.25 | 8.19 |
| HYDROCODONE | 9193 | 659 | 170,304.66 | 258.43 |
| MEPERIDINE (PETHIDINE) | 9230 | 68 | 976.52 | 14.36 |
| METHADONE | 9250B | 375 | 10,577.98 | 28.21 |
| MORPHINE | 9300 | 622 | 61,625.64 | 99.08 |
| OXYMORPHONE | 9652 | 200 | 5,138.76 | 25.69 |
| TAPENTADOL | 9780 | 208 | 16,212.00 | 77.94 |
| FENTANYL BASE | 9801 | 586 | 1,485.68 | 2.54 |

**BUSINESS ACTIVITY: B - HOSPITALS**

| DRUG NAME | DRUG CODE | NUMBER OF REGISTRANT SOLD TO | TOTAL GRAMS | AVG GRAMS |
|---|---|---|---|---|
| AMPHETAMINE | 1100 | 34 | 1,938.64 | 57.02 |
| LISDEXAMFETAMINE | 1205 | 21 | 759.05 | 36.15 |
| METHYLPHENIDATE (DL;D;L;ISOMERS) | 1724 | 51 | 2,248.38 | 44.09 |
| CODEINE | 9050 | 71 | 2,354.74 | 33.17 |
| BUPRENORPHINE | 9064 | 51 | 3,189.82 | 62.55 |
| OXYCODONE | 9143 | 76 | 14,554.81 | 191.51 |
| HYDROMORPHONE | 9150 | 69 | 940.33 | 13.63 |
| HYDROCODONE | 9193 | 78 | 14,685.73 | 188.28 |
| MEPERIDINE (PETHIDINE) | 9230 | 48 | 803.96 | 16.75 |
| METHADONE | 9250B | 47 | 788.50 | 16.78 |
| MORPHINE | 9300 | 74 | 9,593.38 | 129.64 |
| OXYMORPHONE | 9652 | 6 | 23.06 | 3.84 |
| TAPENTADOL | 9780 | 9 | 161.50 | 17.94 |
| FENTANYL BASE | 9801 | 73 | 220.92 | 3.03 |

**BUSINESS ACTIVITY: C - PRACTITIONERS**

| DRUG NAME | DRUG CODE | NUMBER OF REGISTRANT SOLD TO | TOTAL GRAMS | AVG GRAMS |
|---|---|---|---|---|
| CODEINE | 9050 | 18 | 111.38 | 6.19 |
| BUPRENORPHINE | 9064 | 111 | 8.91 | 0.08 |
| OXYCODONE | 9143 | 1 | 0.45 | 0.45 |
| HYDROMORPHONE | 9150 | 25 | 12.27 | 0.49 |
| HYDROCODONE | 9193 | 43 | 149.24 | 3.47 |
| MEPERIDINE (PETHIDINE) | 9230 | 7 | 202.08 | 28.87 |
| MORPHINE | 9300 | 30 | 22.55 | 0.75 |
| FENTANYL BASE | 9801 | 49 | 3.54 | 0.07 |

**BUSINESS ACTIVITY: D - TEACHING INSTITUTIONS**

| DRUG NAME | DRUG CODE | NUMBER OF REGISTRANT SOLD TO | TOTAL GRAMS | AVG GRAMS |
|---|---|---|---|---|
| BUPRENORPHINE | 9064 | 1 | 0.00 | 0.00 |

**BUSINESS ACTIVITY: M - MID-LEVEL PRACTITIONERS**

| DRUG NAME | DRUG CODE | NUMBER OF REGISTRANT SOLD TO | TOTAL GRAMS | AVG GRAMS |
|---|---|---|---|---|
| CODEINE | 9050 | 4 | 3.47 | 0.87 |
| MORPHINE | 9300 | 35 | 13.15 | 0.38 |
| FENTANYL BASE | 9801 | 50 | 0.55 | 0.01 |

**BUSINESS ACTIVITY: NTP - NARCOTIC TREATMENT PROGRAMS**

| DRUG NAME | DRUG CODE | NUMBER OF REGISTRANT SOLD TO | TOTAL GRAMS | AVG GRAMS |
|---|---|---|---|---|
| BUPRENORPHINE | 9064 | 7 | 843.12 | 120.45 |
| METHADONE | 9250B | 9 | 71,802.43 | 7,978.05 |

**STATE: WY - WYOMING**

**BUSINESS ACTIVITY: A - PHARMACIES**

| DRUG NAME | DRUG CODE | NUMBER OF REGISTRANT SOLD TO | TOTAL GRAMS | AVG GRAMS |
|---|---|---|---|---|
| AMPHETAMINE | 1100 | 116 | 25,878.61 | 223.09 |
| METHAMPHETAMINE | 1105 | 5 | 18.48 | 3.70 |
| LISDEXAMFETAMINE | 1205 | 114 | 10,095.94 | 88.56 |
| METHYLPHENIDATE (DL;D;L;ISOMERS) | 1724 | 116 | 28,334.55 | 244.26 |
| CODEINE | 9050 | 117 | 12,909.45 | 110.34 |
| BUPRENORPHINE | 9064 | 117 | 5,026.19 | 42.96 |
| OXYCODONE | 9143 | 118 | 84,962.70 | 720.02 |
| HYDROMORPHONE | 9150 | 114 | 1,863.76 | 16.35 |
| HYDROCODONE | 9193 | 118 | 34,325.53 | 290.89 |
| MEPERIDINE (PETHIDINE) | 9230 | 45 | 992.97 | 22.07 |
| METHADONE | 9250B | 99 | 3,374.84 | 34.09 |
| MORPHINE | 9300 | 118 | 24,680.88 | 209.16 |
| OXYMORPHONE | 9652 | 41 | 548.28 | 13.37 |
| TAPENTADOL | 9780 | 54 | 6,245.00 | 115.65 |
| FENTANYL BASE | 9801 | 116 | 434.29 | 3.74 |

**BUSINESS ACTIVITY: B - HOSPITALS**

| DRUG NAME | DRUG CODE | NUMBER OF REGISTRANT SOLD TO | TOTAL GRAMS | AVG GRAMS |
|---|---|---|---|---|
| AMPHETAMINE | 1100 | 17 | 1,744.11 | 102.59 |
| LISDEXAMFETAMINE | 1205 | 10 | 422.59 | 42.26 |
| METHYLPHENIDATE (DL;D;L;ISOMERS) | 1724 | 20 | 1,953.15 | 97.66 |
| CODEINE | 9050 | 29 | 1,228.37 | 42.36 |
| BUPRENORPHINE | 9064 | 19 | 326.85 | 17.20 |
| OXYCODONE | 9143 | 45 | 5,928.23 | 131.74 |
| HYDROMORPHONE | 9150 | 42 | 231.11 | 5.50 |
| HYDROCODONE | 9193 | 46 | 4,402.13 | 95.70 |
| MEPERIDINE (PETHIDINE) | 9230 | 32 | 249.68 | 7.80 |
| METHADONE | 9250B | 15 | 382.62 | 25.51 |
| MORPHINE | 9300 | 47 | 5,788.77 | 123.17 |

| | | | | |
|---|---|---|---|---|
| TAPENTADOL | 9780 | 5 | 151.50 | 30.30 |
| FENTANYL BASE | 9801 | 46 | 54.45 | 1.18 |

**BUSINESS ACTIVITY: C - PRACTITIONERS**

| DRUG NAME | DRUG CODE | NUMBER OF REGISTRANT SOLD TO | TOTAL GRAMS | AVG GRAMS |
|---|---|---|---|---|
| CODEINE | 9050 | 8 | 183.44 | 22.93 |
| BUPRENORPHINE | 9064 | 62 | 3.96 | 0.06 |
| OXYCODONE | 9143 | 1 | 14.52 | 14.52 |
| HYDROMORPHONE | 9150 | 19 | 9.86 | 0.52 |
| HYDROCODONE | 9193 | 27 | 138.51 | 5.13 |
| MEPERIDINE (PETHIDINE) | 9230 | 3 | 10.89 | 3.63 |
| MORPHINE | 9300 | 32 | 17.58 | 0.55 |
| FENTANYL BASE | 9801 | 38 | 1.97 | 0.05 |

**BUSINESS ACTIVITY: M - MID-LEVEL PRACTITIONERS**

| DRUG NAME | DRUG CODE | NUMBER OF REGISTRANT SOLD TO | TOTAL GRAMS | AVG GRAMS |
|---|---|---|---|---|
| CODEINE | 9050 | 2 | 9.00 | 4.50 |
| HYDROCODONE | 9193 | 1 | 0.18 | 0.18 |

**ENTIRE UNITED STATES**

**BUSINESS ACTIVITY: A - PHARMACIES**

| DRUG NAME | DRUG CODE | NUMBER OF REGISTRANT SOLD TO | TOTAL GRAMS | AVG GRAMS |
|---|---|---|---|---|
| AMPHETAMINE | 1100 | 64,973 | 21,340,409.48 | 328.45 |
| METHAMPHETAMINE | 1105 | 1,829 | 6,551.75 | 3.58 |
| LISDEXAMFETAMINE | 1205 | 60,432 | 9,572,196.56 | 158.40 |
| METHYLPHENIDATE (DL;D;L;ISOMERS) | 1724 | 64,241 | 17,067,203.87 | 265.67 |
| CODEINE | 9050 | 66,002 | 13,454,714.20 | 203.85 |
| BUPRENORPHINE | 9064 | 60,254 | 3,588,355.59 | 59.55 |
| OXYCODONE | 9143 | 66,390 | 39,354,889.48 | 592.78 |
| HYDROMORPHONE | 9150 | 55,738 | 955,536.23 | 17.14 |
| HYDROCODONE | 9193 | 66,537 | 21,691,001.12 | 326.00 |
| MEPERIDINE (PETHIDINE) | 9230 | 16,922 | 347,366.89 | 20.53 |
| METHADONE | 9250B | 49,371 | 2,161,326.37 | 43.78 |
| MORPHINE | 9300 | 61,972 | 12,617,308.23 | 203.60 |
| OXYMORPHONE | 9652 | 23,393 | 563,228.34 | 24.08 |
| TAPENTADOL | 9780 | 36,817 | 4,281,878.00 | 116.30 |
| FENTANYL BASE | 9801 | 59,631 | 219,711.21 | 3.68 |

**BUSINESS ACTIVITY: B - HOSPITALS**

| DRUG NAME | DRUG CODE | NUMBER OF REGISTRANT SOLD TO | TOTAL GRAMS | AVG GRAMS |
|---|---|---|---|---|
| AMPHETAMINE | 1100 | 3,935 | 545,546.62 | 138.64 |
| METHAMPHETAMINE | 1105 | 28 | 171.97 | 6.14 |
| LISDEXAMFETAMINE | 1205 | 1,926 | 135,273.65 | 70.24 |
| METHYLPHENIDATE (DL;D;L;ISOMERS) | 1724 | 5,391 | 580,711.91 | 107.72 |
| CODEINE | 9050 | 7,135 | 462,472.47 | 64.82 |
| BUPRENORPHINE | 9064 | 5,025 | 115,509.99 | 22.99 |
| OXYCODONE | 9143 | 8,852 | 1,973,845.19 | 222.98 |
| HYDROMORPHONE | 9150 | 8,619 | 192,427.89 | 22.33 |
| HYDROCODONE | 9193 | 9,319 | 1,155,101.35 | 123.95 |
| MEPERIDINE (PETHIDINE) | 9230 | 6,500 | 188,909.17 | 29.06 |
| METHADONE | 9250B | 5,743 | 236,381.71 | 41.16 |
| MORPHINE | 9300 | 9,697 | 1,274,221.30 | 131.40 |
| OXYMORPHONE | 9652 | 579 | 4,458.29 | 7.70 |
| TAPENTADOL | 9780 | 1,341 | 80,869.38 | 60.31 |
| FENTANYL BASE | 9801 | 10,138 | 29,916.36 | 2.95 |

**BUSINESS ACTIVITY: C - PRACTITIONERS**

| DRUG NAME | DRUG CODE | NUMBER OF REGISTRANT SOLD TO | TOTAL GRAMS | AVG GRAMS |
|---|---|---|---|---|
| AMPHETAMINE | 1100 | 110 | 2,438.09 | 22.16 |
| METHAMPHETAMINE | 1105 | 2 | 8.03 | 4.02 |
| LISDEXAMFETAMINE | 1205 | 51 | 1,526.09 | 29.92 |

| | | | | |
|---|---|---|---|---|
| METHYLPHENIDATE (DL;D;L;ISOMERS) | 1724 | 41 | 2,753.88 | 67.17 |
| CODEINE | 9050 | 3,959 | 111,012.92 | 28.04 |
| BUPRENORPHINE | 9064 | 26,879 | 7,503.76 | 0.28 |
| OXYCODONE | 9143 | 1,831 | 14,642.65 | 8.00 |
| HYDROMORPHONE | 9150 | 11,182 | 8,352.43 | 0.75 |
| HYDROCODONE | 9193 | 11,737 | 48,086.88 | 4.10 |
| MEPERIDINE (PETHIDINE) | 9230 | 4,051 | 40,129.14 | 9.91 |
| METHADONE | 9250B | 700 | 1,784.46 | 2.55 |
| MORPHINE | 9300 | 8,086 | 13,223.36 | 1.64 |
| OXYMORPHONE | 9652 | 18 | 179.19 | 9.95 |
| TAPENTADOL | 9780 | 12 | 547.00 | 45.58 |
| FENTANYL BASE | 9801 | 18,521 | 2,056.49 | 0.11 |

**BUSINESS ACTIVITY: D - TEACHING INSTITUTIONS**

| DRUG NAME | DRUG CODE | NUMBER OF REGISTRANT SOLD TO | TOTAL GRAMS | AVG GRAMS |
|---|---|---|---|---|
| AMPHETAMINE | 1100 | 1 | 0.73 | 0.73 |
| CODEINE | 9050 | 5 | 2,144.83 | 428.97 |
| BUPRENORPHINE | 9064 | 57 | 7.90 | 0.14 |
| OXYCODONE | 9143 | 1 | 0.45 | 0.45 |
| HYDROMORPHONE | 9150 | 20 | 30.45 | 1.52 |
| HYDROCODONE | 9193 | 10 | 116.31 | 11.63 |
| MEPERIDINE (PETHIDINE) | 9230 | 3 | 7.63 | 2.54 |
| METHADONE | 9250B | 7 | 135.86 | 19.41 |
| MORPHINE | 9300 | 14 | 212.50 | 15.18 |
| FENTANYL BASE | 9801 | 23 | 13.17 | 0.57 |

**BUSINESS ACTIVITY: M - MID-LEVEL PRACTITIONERS**

| DRUG NAME | DRUG CODE | NUMBER OF REGISTRANT SOLD TO | TOTAL GRAMS | AVG GRAMS |
|---|---|---|---|---|
| AMPHETAMINE | 1100 | 3 | 7.54 | 2.51 |
| METHYLPHENIDATE (DL;D;L;ISOMERS) | 1724 | 1 | 0.86 | 0.86 |
| CODEINE | 9050 | 370 | 2,233.71 | 6.04 |
| BUPRENORPHINE | 9064 | 62 | 135.53 | 2.19 |
| OXYCODONE | 9143 | 11 | 142.99 | 13.00 |
| HYDROMORPHONE | 9150 | 88 | 11.99 | 0.14 |
| HYDROCODONE | 9193 | 27 | 84.79 | 3.14 |
| MEPERIDINE (PETHIDINE) | 9230 | 23 | 112.21 | 4.88 |
| METHADONE | 9250B | 3 | 46.96 | 15.65 |
| MORPHINE | 9300 | 820 | 457.84 | 0.56 |
| TAPENTADOL | 9780 | 1 | 10.00 | 10.00 |
| FENTANYL BASE | 9801 | 1,335 | 49.39 | 0.04 |

**BUSINESS ACTIVITY: NTP - NARCOTIC TREATMENT PROGRAMS**

| DRUG NAME | DRUG CODE | NUMBER OF REGISTRANT SOLD TO | TOTAL GRAMS | AVG GRAMS |
|---|---|---|---|---|
| BUPRENORPHINE | 9064 | 831 | 81,130.54 | 97.63 |
| METHADONE | 9250B | 1,583 | 11,368,111.28 | 7,181.37 |

Apr 22, 2020 1:26:28 PM
