## Supplementary material for "Methadone Distribution Trends from 2017-2019 in the United States": Raw data from 2019

### DEPARTMENT OF JUSTICE DRUG ENFORCEMENT ADMINISTRATION

#### DEA DIVERSION CONTROL DIVISION

#### ARCOS DRUG RETAIL SUMMARY REPORTS

REPORT 01 by GRAMS - RETAIL DRUG DISTRIBUTION BY ZIP CODE WITHIN STATE

REPORT 02 by GRAMS - RETAIL DRUG DISTRIBUTION BY STATE WITHIN DRUG CODE

REPORT 03 by GRAMS - QUARTERLY DRUG DISTRIBUTION BY STATE PER 100K POPULATION

REPORT 04 by GRAMS - CUMULATIVE DISTRIBUTION BY STATE PER 100K POPULATION

REPORT 05 by GRAMS - STATISTICAL SUMMARY FOR RETAIL DRUG PURCHASES

REPORT 07 by GRAMS - UNITED STATES SUMMARY FOR RETAIL DRUG PURCHASES

DATE RANGE: 01/01/2019 TO 12/31/2019

**DISCLAIMER**: Automated Reports and Consolidated Ordering System (ARCOS) is a data collection system in which manufacturers and distributors report their controlled substances transactions to the Drug Enforcement Administration (DEA). ARCOS provides an acquisition/distribution transactional records of applicable activities to the DEA involving certain controlled substances, in accordance with Title 21, United States Code, Section 827(d)(1), and Title 21, Code of Federal Regulations, Section 1304.33. This information is collected and compiled by DEA in accordance with law for determining quota, distribution trends, internal audits, and other analyses.

Recipients should be aware, prior to use, analysis, studies, or publication of ARCOS data, the information contained in ARCOS generated reports are complete and accurate as to what was provided by DEA registrants to ARCOS in accordance with reporting requirements. This information is susceptible to future updates and corrections, without notice, as new information is obtained. DEA assumes no liability for analysis, conclusions or policy decisions of third parties based on internal interpretation of provided data.

**DRUG: 1100 - AMPHETAMINE**

**STATE: AK - ALASKA**

| REGISTRANT ZIP CODE 3 | 1ST QUARTER | 2ND QUARTER | 3RD QUARTER | 4TH QUARTER | TOTAL GRAMS |
|---|---|---|---|---|---|
| 995 | 5,035.10 | 5,108.64 | 5,028.09 | 5,154.09 | **20,325.92** |
| 996 | 1,619.57 | 1,720.28 | 1,806.04 | 1,770.28 | **6,916.17** |
| 997 | 1,080.50 | 1,165.21 | 1,212.56 | 1,126.45 | **4,584.72** |
| 998 | 502.84 | 493.86 | 514.38 | 519.54 | **2,030.62** |
| 999 | 170.95 | 187.50 | 194.46 | 180.25 | **733.16** |
| **AK ALASKA - Total** | **8,408.96** | **8,675.49** | **8,755.54** | **8,750.60** | **34,590.59** |

**STATE: AL - ALABAMA**

| REGISTRANT ZIP CODE 3 | 1ST QUARTER | 2ND QUARTER | 3RD QUARTER | 4TH QUARTER | TOTAL GRAMS |
|---|---|---|---|---|---|
| 350 | 18,174.04 | 18,874.20 | 18,735.77 | 18,736.29 | **74,520.29** |
| 351 | 9,165.47 | 9,494.72 | 9,501.77 | 9,517.19 | **37,679.15** |
| 352 | 20,084.37 | 20,963.11 | 20,648.79 | 20,646.00 | **82,342.27** |
| 354 | 8,289.56 | 8,193.40 | 8,379.94 | 8,178.62 | **33,041.51** |
| 355 | 3,346.17 | 3,306.59 | 3,561.83 | 3,465.88 | **13,680.48** |
| 356 | 10,428.23 | 10,479.43 | 10,806.92 | 10,918.70 | **42,633.28** |
| 357 | 4,885.07 | 5,068.13 | 5,190.22 | 5,213.96 | **20,357.38** |
| 358 | 6,089.46 | 6,152.91 | 6,149.64 | 6,137.00 | **24,529.02** |
| 359 | 5,504.65 | 5,409.97 | 5,668.54 | 5,470.11 | **22,053.26** |
| 360 | 6,469.98 | 6,591.79 | 6,477.49 | 6,718.09 | **26,257.35** |
| 361 | 4,966.18 | 5,189.99 | 5,159.08 | 5,232.07 | **20,547.32** |
| 362 | 2,506.98 | 2,581.24 | 2,549.56 | 2,559.57 | **10,197.35** |
| 363 | 5,649.52 | 5,317.64 | 5,170.88 | 4,878.53 | **21,016.58** |
| 364 | 1,387.46 | 1,321.01 | 1,311.15 | 1,422.49 | **5,442.11** |
| 365 | 12,080.39 | 12,750.07 | 12,484.69 | 12,261.03 | **49,576.19** |
| 366 | 8,255.31 | 8,160.31 | 8,052.50 | 8,222.61 | **32,690.73** |
| 367 | 1,583.86 | 1,466.78 | 1,430.55 | 1,339.47 | **5,820.65** |
| 368 | 6,524.99 | 6,648.92 | 6,226.89 | 6,680.50 | **26,081.30** |
| 369 | 165.85 | 145.37 | 133.01 | 152.17 | **596.40** |
| **AL ALABAMA - Total** | **135,557.55** | **138,115.57** | **137,639.22** | **137,750.28** | **549,062.62** |

**STATE: AR - ARKANSAS**

| REGISTRANT ZIP CODE 3 | 1ST QUARTER | 2ND QUARTER | 3RD QUARTER | 4TH QUARTER | TOTAL GRAMS |
|---|---|---|---|---|---|
| 716 | 3,051.71 | 2,803.83 | 2,777.95 | 2,881.37 | **11,514.86** |
| 717 | 963.73 | 980.67 | 977.15 | 1,041.70 | **3,963.26** |
| 718 | 920.05 | 976.76 | 935.95 | 964.77 | **3,797.53** |
| 719 | 3,578.89 | 3,421.07 | 3,578.95 | 3,652.54 | **14,231.45** |
| 720 | 8,652.92 | 8,446.64 | 8,770.94 | 8,903.41 | **34,773.91** |
| 721 | 6,657.23 | 6,399.96 | 6,392.27 | 6,607.32 | **26,056.77** |
| 722 | 7,770.39 | 7,630.26 | 7,802.78 | 7,534.07 | **30,737.50** |
| 723 | 1,653.92 | 1,643.49 | 1,633.31 | 1,664.58 | **6,595.31** |
| 724 | 2,953.06 | 2,770.28 | 2,903.39 | 3,124.78 | **11,751.51** |
| 725 | 750.38 | 760.39 | 740.61 | 837.97 | **3,089.35** |

| | | | | | |
|---|---|---|---|---|---|
| 726 | 1,569.81 | 1,675.33 | 1,575.85 | 1,731.09 | **6,552.08** |
| 727 | 13,227.31 | 13,823.11 | 13,870.20 | 14,354.55 | **55,275.17** |
| 728 | 1,080.97 | 1,074.22 | 1,035.99 | 1,133.81 | **4,325.00** |
| 729 | 1,974.10 | 2,020.60 | 2,056.82 | 2,112.50 | **8,164.01** |
| **AR ARKANSAS - Total** | **54,804.47** | **54,426.63** | **55,052.16** | **56,544.45** | **220,827.70** |

**STATE: AS - AMERICAN SAMOA**

| REGISTRANT ZIP CODE 3 | 1ST QUARTER | 2ND QUARTER | 3RD QUARTER | 4TH QUARTER | TOTAL GRAMS |
|---|---|---|---|---|---|
| 967 | 3.75 | 0 | 0 | 0 | **3.75** |
| **AS - AMERICAN SAMOA - Tota** | **3.75** | **0.00** | **0.00** | **0.00** | **3.75** |

**STATE: AZ - ARIZONA**

| REGISTRANT ZIP CODE 3 | 1ST QUARTER | 2ND QUARTER | 3RD QUARTER | 4TH QUARTER | TOTAL GRAMS |
|---|---|---|---|---|---|
| 850 | 19,866.06 | 20,000.58 | 20,468.01 | 21,648.96 | **81,983.61** |
| 851 | 3,848.52 | 3,944.09 | 4,055.26 | 4,370.74 | **16,218.61** |
| 852 | 38,852.85 | 40,700.39 | 40,457.67 | 42,773.98 | **162,784.89** |
| 853 | 11,959.11 | 12,015.82 | 12,252.17 | 13,030.09 | **49,257.20** |
| 855 | 439.42 | 484.30 | 558.51 | 558.63 | **2,040.86** |
| 856 | 1,828.46 | 1,747.67 | 1,799.92 | 1,864.85 | **7,240.90** |
| 857 | 11,228.40 | 11,431.27 | 11,373.04 | 11,761.53 | **45,794.24** |
| 859 | 493.23 | 569.57 | 587.53 | 549.48 | **2,199.81** |
| 860 | 1,402.29 | 1,347.44 | 1,364.01 | 1,412.61 | **5,526.35** |
| 863 | 1,734.13 | 1,834.44 | 1,840.60 | 2,074.90 | **7,484.07** |
| 864 | 1,020.89 | 960.92 | 1,029.89 | 1,056.83 | **4,068.52** |
| 865 | 71.90 | 67.51 | 92.69 | 87.52 | **319.61** |
| **AZ ARIZONA - Total** | **92,745.26** | **95,104.00** | **95,879.30** | **101,190.12** | **384,918.68** |

**STATE: CA - CALIFORNIA**

| REGISTRANT ZIP CODE 3 | 1ST QUARTER | 2ND QUARTER | 3RD QUARTER | 4TH QUARTER | TOTAL GRAMS |
|---|---|---|---|---|---|
| 900 | 22,849.41 | 23,401.28 | 23,923.83 | 24,628.73 | **94,803.24** |
| 902 | 13,113.72 | 13,594.50 | 13,629.39 | 13,686.05 | **54,023.65** |
| 903 | 229.69 | 250.72 | 241.82 | 331.05 | **1,053.27** |
| 904 | 4,291.44 | 4,226.42 | 4,364.15 | 4,525.09 | **17,407.10** |
| 905 | 1,908.12 | 1,911.08 | 1,968.41 | 2,076.75 | **7,864.36** |
| 906 | 2,447.02 | 2,548.48 | 2,661.09 | 2,675.86 | **10,332.45** |
| 907 | 3,483.45 | 3,715.11 | 3,524.16 | 3,633.81 | **14,356.52** |
| 908 | 3,334.31 | 3,349.52 | 3,331.41 | 3,647.83 | **13,663.06** |
| 910 | 1,630.09 | 1,555.99 | 1,559.40 | 1,426.18 | **6,171.67** |
| 911 | 2,823.24 | 2,762.64 | 2,770.24 | 2,898.44 | **11,254.56** |
| 912 | 1,708.06 | 1,692.41 | 1,829.57 | 1,811.78 | **7,041.83** |
| 913 | 15,645.88 | 15,350.81 | 15,866.09 | 15,683.29 | **62,546.08** |
| 914 | 4,933.22 | 5,207.37 | 5,209.81 | 5,248.04 | **20,598.43** |
| 915 | 1,775.06 | 1,876.29 | 1,839.56 | 1,847.76 | **7,338.67** |

| | | | | | |
|---|---|---|---|---|---|
| 916 | 1,737.40 | 1,950.87 | 1,941.72 | 1,929.86 | **7,559.85** |
| 917 | 7,273.18 | 7,291.33 | 7,285.78 | 7,468.43 | **29,318.71** |
| 918 | 395.06 | 449.61 | 489.63 | 512.58 | **1,846.88** |
| 919 | 2,856.71 | 2,770.68 | 2,764.56 | 2,949.55 | **11,341.50** |
| 920 | 26,980.67 | 28,137.62 | 28,067.89 | 28,471.25 | **111,657.43** |
| 921 | 18,666.99 | 18,776.20 | 19,121.52 | 19,131.71 | **75,696.42** |
| 922 | 3,570.12 | 3,570.18 | 3,545.11 | 3,604.73 | **14,290.14** |
| 923 | 4,357.05 | 4,274.67 | 4,351.47 | 4,489.69 | **17,472.88** |
| 924 | 309.39 | 284.18 | 303.42 | 319.64 | **1,216.63** |
| 925 | 6,519.54 | 6,411.61 | 6,510.58 | 6,701.32 | **26,143.04** |
| 926 | 21,190.69 | 21,487.89 | 21,894.80 | 21,725.84 | **86,299.22** |
| 927 | 4,824.82 | 4,921.07 | 4,835.61 | 4,820.48 | **19,401.98** |
| 928 | 8,300.33 | 8,726.61 | 8,790.37 | 8,786.07 | **34,603.37** |
| 930 | 5,842.96 | 5,834.94 | 6,007.23 | 6,456.28 | **24,141.40** |
| 931 | 3,458.15 | 3,440.84 | 3,398.48 | 3,341.73 | **13,639.21** |
| 932 | 1,871.62 | 1,887.83 | 1,927.42 | 2,019.65 | **7,706.53** |
| 933 | 2,059.02 | 1,989.78 | 1,967.11 | 1,974.17 | **7,990.08** |
| 934 | 3,181.36 | 3,275.05 | 3,155.29 | 3,413.58 | **13,025.27** |
| 935 | 2,240.18 | 2,237.81 | 2,275.87 | 2,252.61 | **9,006.47** |
| 936 | 2,177.77 | 2,214.86 | 2,187.94 | 2,233.75 | **8,814.32** |
| 937 | 3,800.16 | 3,725.60 | 3,786.20 | 3,852.59 | **15,164.56** |
| 939 | 1,706.48 | 1,888.30 | 1,840.32 | 1,847.23 | **7,282.34** |
| 940 | 6,077.08 | 6,383.99 | 6,508.43 | 6,608.07 | **25,577.57** |
| 941 | 12,508.26 | 13,215.67 | 13,441.72 | 13,147.78 | **52,313.43** |
| 943 | 981.35 | 1,152.33 | 1,101.49 | 1,046.23 | **4,281.40** |
| 944 | 1,333.71 | 1,405.41 | 1,375.57 | 1,363.66 | **5,478.35** |
| 945 | 16,375.74 | 16,579.87 | 17,305.20 | 17,209.60 | **67,470.40** |
| 946 | 3,565.91 | 4,015.60 | 3,786.33 | 4,086.20 | **15,454.04** |
| 947 | 1,379.98 | 1,500.91 | 1,443.46 | 1,501.69 | **5,826.04** |
| 948 | 729.52 | 715.81 | 747.07 | 752.85 | **2,945.25** |
| 949 | 4,931.59 | 5,044.45 | 5,247.57 | 5,174.87 | **20,398.48** |
| 950 | 7,144.17 | 7,215.99 | 7,387.99 | 7,141.31 | **28,889.46** |
| 951 | 4,172.14 | 4,336.32 | 4,305.14 | 4,307.55 | **17,121.14** |
| 952 | 1,984.33 | 1,961.69 | 1,966.85 | 2,103.56 | **8,016.44** |
| 953 | 4,918.39 | 4,961.76 | 5,004.28 | 5,062.17 | **19,946.60** |
| 954 | 3,949.06 | 3,878.72 | 4,095.64 | 3,843.09 | **15,766.51** |
| 955 | 895.58 | 946.94 | 976.01 | 902.92 | **3,721.45** |
| 956 | 10,344.13 | 10,754.03 | 11,058.75 | 11,374.26 | **43,531.17** |
| 957 | 2,458.98 | 2,503.98 | 2,592.66 | 2,552.52 | **10,108.15** |
| 958 | 5,047.95 | 5,160.32 | 5,360.68 | 5,341.75 | **20,910.69** |

| | | | | | |
|---|---|---|---|---|---|
| 959 | 3,933.21 | 4,021.47 | 3,899.64 | 4,002.17 | **15,856.49** |
| 960 | 1,821.70 | 1,935.97 | 2,016.82 | 2,046.18 | **7,820.66** |
| 961 | 614.62 | 672.85 | 739.00 | 724.72 | **2,751.19** |
| **CA - CALIFORNIA - Total** | **308,659.71** | **315,354.24** | **319,527.54** | **322,716.52** | **1,266,258.02** |

**STATE: CO - COLORADO**

| REGISTRANT ZIP CODE 3 | 1ST QUARTER | 2ND QUARTER | 3RD QUARTER | 4TH QUARTER | TOTAL GRAMS |
|---|---|---|---|---|---|
| 800 | 14,402.48 | 15,134.28 | 15,766.41 | 16,435.05 | **61,738.21** |
| 801 | 16,071.22 | 16,854.61 | 17,515.48 | 17,904.29 | **68,345.60** |
| 802 | 22,000.68 | 23,518.32 | 23,424.53 | 24,767.48 | **93,711.00** |
| 803 | 3,814.05 | 3,959.04 | 3,753.79 | 4,081.14 | **15,608.01** |
| 804 | 3,420.81 | 3,498.95 | 3,512.19 | 3,702.69 | **14,134.64** |
| 805 | 8,861.82 | 9,241.83 | 9,513.88 | 9,935.38 | **37,552.91** |
| 806 | 2,692.62 | 2,703.32 | 2,734.73 | 2,894.45 | **11,025.12** |
| 807 | 352.67 | 393.82 | 373.88 | 372.73 | **1,493.11** |
| 808 | 1,341.50 | 1,321.06 | 1,411.18 | 1,503.95 | **5,577.69** |
| 809 | 8,160.98 | 8,326.14 | 8,708.82 | 8,968.65 | **34,164.58** |
| 810 | 1,557.80 | 1,602.45 | 1,600.30 | 1,805.56 | **6,566.11** |
| 811 | 314.89 | 306.00 | 352.22 | 358.96 | **1,332.07** |
| 812 | 586.97 | 601.73 | 634.37 | 727.31 | **2,550.38** |
| 813 | 757.18 | 788.38 | 849.30 | 814.08 | **3,208.95** |
| 814 | 686.88 | 644.16 | 720.36 | 702.18 | **2,753.58** |
| 815 | 1,925.51 | 1,941.61 | 1,953.48 | 2,020.70 | **7,841.29** |
| 816 | 2,620.60 | 2,689.89 | 2,796.24 | 2,992.15 | **11,098.89** |
| **CO - COLORADO - Total** | **89,568.67** | **93,525.58** | **95,621.16** | **99,986.74** | **378,702.16** |

**STATE: CT - CONNECTICUT**

| REGISTRANT ZIP CODE 3 | 1ST QUARTER | 2ND QUARTER | 3RD QUARTER | 4TH QUARTER | TOTAL GRAMS |
|---|---|---|---|---|---|
| 060 | 9,455.58 | 10,166.96 | 9,903.79 | 10,618.98 | **40,145.31** |
| 061 | 3,470.50 | 3,830.64 | 3,482.18 | 3,838.28 | **14,621.61** |
| 062 | 2,642.75 | 2,786.07 | 2,698.75 | 2,840.97 | **10,968.54** |
| 063 | 5,951.08 | 6,196.44 | 6,167.65 | 6,436.95 | **24,752.12** |
| 064 | 13,266.65 | 13,985.88 | 13,986.56 | 14,771.22 | **56,010.30** |
| 065 | 4,360.44 | 4,582.97 | 4,469.47 | 4,902.73 | **18,315.61** |
| 066 | 1,766.30 | 1,863.04 | 1,961.41 | 2,085.56 | **7,676.31** |
| 067 | 3,757.48 | 3,930.46 | 3,924.35 | 4,231.80 | **15,844.10** |
| 068 | 9,967.40 | 10,679.22 | 10,870.81 | 11,384.28 | **42,901.71** |
| 069 | 2,056.47 | 2,136.30 | 2,123.30 | 2,377.98 | **8,694.04** |
| **CT CONNECTICUT Total** | **56,694.65** | **60,157.98** | **59,588.28** | **63,488.75** | **239,929.66** |

**STATE: DC - DISTRICT OF COLUMBIA**

| REGISTRANT ZIP CODE 3 | 1ST QUARTER | 2ND QUARTER | 3RD QUARTER | 4TH QUARTER | TOTAL GRAMS |
|---|---|---|---|---|---|
| 200 | 11,950.70 | 12,471.80 | 12,327.00 | 12,525.69 | **49,275.19** |

| | | | | | |
|---|---|---|---|---|---|
| 203 | 154.53 | 93.53 | 130.88 | 72.16 | **451.10** |
| 204 | 359.96 | 487.17 | 753.92 | 374.26 | **1,975.32** |
| 205 | 0 | 0 | 2.50 | 0 | **2.50** |
| **DC - DISTRICT OF COLUMBIA** | **12,465.18** | **13,052.50** | **13,214.31** | **12,972.12** | **51,704.11** |

**STATE: DE - DELAWARE**

| REGISTRANT ZIP CODE 3 | 1ST QUARTER | 2ND QUARTER | 3RD QUARTER | 4TH QUARTER | TOTAL GRAMS |
|---|---|---|---|---|---|
| 197 | 8,848.09 | 9,027.77 | 9,153.23 | 9,107.43 | **36,136.52** |
| 198 | 6,527.85 | 6,565.96 | 6,599.51 | 6,764.97 | **26,458.28** |
| 199 | 5,815.34 | 6,108.80 | 6,201.29 | 6,357.86 | **24,483.28** |
| **DE DELAWARE Total** | **21,191.28** | **21,702.52** | **21,954.03** | **22,230.26** | **87,078.09** |

**STATE: FL - FLORIDA**

| REGISTRANT ZIP CODE 3 | 1ST QUARTER | 2ND QUARTER | 3RD QUARTER | 4TH QUARTER | TOTAL GRAMS |
|---|---|---|---|---|---|
| 320 | 10,697.84 | 10,772.32 | 10,860.08 | 11,347.51 | **43,677.75** |
| 321 | 8,598.84 | 8,888.58 | 8,917.92 | 9,586.61 | **35,991.95** |
| 322 | 23,355.75 | 25,004.79 | 23,797.92 | 25,169.90 | **97,328.36** |
| 323 | 9,686.34 | 10,068.02 | 9,348.39 | 10,347.89 | **39,450.64** |
| 324 | 7,569.04 | 7,962.74 | 7,852.57 | 8,117.75 | **31,502.10** |
| 325 | 19,637.05 | 20,698.80 | 19,757.87 | 20,612.94 | **80,706.67** |
| 326 | 6,554.18 | 6,347.50 | 6,438.46 | 7,035.56 | **26,375.70** |
| 327 | 19,289.14 | 19,871.60 | 18,696.45 | 20,843.23 | **78,700.43** |
| 328 | 13,908.88 | 14,902.20 | 14,211.99 | 15,400.52 | **58,423.60** |
| 329 | 14,334.48 | 14,250.40 | 14,595.23 | 15,038.01 | **58,218.11** |
| 330 | 16,023.02 | 16,699.06 | 15,546.38 | 17,367.56 | **65,636.02** |
| 331 | 19,237.29 | 19,926.27 | 17,461.52 | 21,116.68 | **77,741.76** |
| 333 | 13,577.66 | 14,292.86 | 12,895.71 | 15,024.05 | **55,790.29** |
| 334 | 27,000.47 | 27,809.42 | 26,284.00 | 29,264.66 | **110,358.56** |
| 335 | 12,363.67 | 12,639.50 | 12,408.63 | 13,521.09 | **50,932.89** |
| 336 | 16,309.27 | 17,138.65 | 16,075.79 | 17,829.84 | **67,353.55** |
| 337 | 16,126.77 | 16,974.15 | 16,429.76 | 17,790.35 | **67,321.02** |
| 338 | 7,972.97 | 7,956.69 | 7,885.69 | 8,813.09 | **32,628.44** |
| 339 | 14,690.75 | 14,517.07 | 14,064.59 | 15,322.29 | **58,594.70** |
| 341 | 8,265.66 | 8,576.51 | 7,779.29 | 8,804.14 | **33,425.60** |
| 342 | 14,726.55 | 15,553.12 | 14,421.19 | 16,555.22 | **61,256.07** |
| 344 | 4,428.73 | 4,492.46 | 4,437.21 | 5,151.03 | **18,509.43** |
| 346 | 10,581.03 | 10,797.41 | 10,493.78 | 11,834.72 | **43,706.94** |
| 347 | 9,542.49 | 9,536.55 | 9,110.13 | 10,016.28 | **38,205.44** |
| 349 | 7,747.32 | 7,872.04 | 7,289.03 | 8,236.78 | **31,145.17** |
| **FL FLORIDA - Total** | **332,225.20** | **343,548.71** | **327,059.59** | **360,147.70** | **1,362,981.20** |

**STATE: GA - GEORGIA**

| REGISTRANT ZIP CODE 3 | 1ST QUARTER | 2ND QUARTER | 3RD QUARTER | 4TH QUARTER | TOTAL GRAMS |
|---|---|---|---|---|---|

| | | | | | |
|---|---|---|---|---|---|
| 300 | 41,047.83 | 42,294.74 | 41,838.49 | 44,065.44 | **169,246.50** |
| 301 | 23,866.38 | 24,644.28 | 24,962.29 | 25,898.43 | **99,371.38** |
| 302 | 14,907.54 | 15,612.11 | 15,566.46 | 15,653.79 | **61,739.90** |
| 303 | 24,266.64 | 26,469.34 | 26,043.76 | 26,562.61 | **103,342.35** |
| 304 | 7,041.53 | 6,729.31 | 7,400.89 | 6,821.86 | **27,993.59** |
| 305 | 12,743.92 | 13,245.59 | 13,459.62 | 13,735.47 | **53,184.61** |
| 306 | 8,914.53 | 8,737.88 | 8,984.43 | 9,321.73 | **35,958.56** |
| 307 | 6,589.49 | 6,606.73 | 6,662.94 | 6,888.55 | **26,747.71** |
| 308 | 3,826.11 | 4,034.94 | 4,089.87 | 4,354.66 | **16,305.58** |
| 309 | 5,922.27 | 5,597.19 | 5,813.10 | 5,474.62 | **22,807.18** |
| 310 | 7,679.39 | 7,505.81 | 7,815.51 | 8,028.80 | **31,029.51** |
| 312 | 2,732.96 | 2,770.84 | 2,735.25 | 2,943.30 | **11,182.35** |
| 313 | 6,072.08 | 6,113.79 | 6,336.85 | 6,214.81 | **24,737.52** |
| 314 | 6,719.51 | 6,862.10 | 6,899.67 | 7,123.91 | **27,605.18** |
| 315 | 8,706.08 | 8,582.15 | 9,064.88 | 8,854.16 | **35,207.27** |
| 316 | 3,476.16 | 3,435.81 | 3,547.14 | 3,685.42 | **14,144.53** |
| 317 | 7,296.09 | 7,209.46 | 7,466.72 | 7,580.80 | **29,553.07** |
| 318 | 544.08 | 579.66 | 503.82 | 530.23 | **2,157.78** |
| 319 | 6,406.51 | 6,842.66 | 6,685.67 | 6,191.73 | **26,126.56** |
| 398 | 1,557.62 | 1,522.44 | 1,579.31 | 1,573.85 | **6,233.22** |
| **GA GEORGIA Total** | **200,316.71** | **205,396.82** | **207,456.66** | **211,504.15** | **824,674.34** |

**STATE: GU - GUAM**

| REGISTRANT ZIP CODE 3 | 1ST QUARTER | 2ND QUARTER | 3RD QUARTER | 4TH QUARTER | TOTAL GRAMS |
|---|---|---|---|---|---|
| 969 | 422.85 | 432.20 | 452.03 | 408.34 | **1,715.41** |
| **GU - GUAM - Total** | **422.85** | **432.20** | **452.03** | **408.34** | **1,715.41** |

**STATE: HI - HAWAII**

| REGISTRANT ZIP CODE 3 | 1ST QUARTER | 2ND QUARTER | 3RD QUARTER | 4TH QUARTER | TOTAL GRAMS |
|---|---|---|---|---|---|
| 967 | 5,289.65 | 5,386.31 | 5,487.67 | 5,642.02 | **21,805.65** |
| 968 | 5,276.74 | 5,317.19 | 5,391.47 | 5,272.43 | **21,257.83** |
| **HI - HAWAII - Total** | **10,566.38** | **10,703.49** | **10,879.15** | **10,914.45** | **43,063.47** |

**STATE: IA - IOWA**

| REGISTRANT ZIP CODE 3 | 1ST QUARTER | 2ND QUARTER | 3RD QUARTER | 4TH QUARTER | TOTAL GRAMS |
|---|---|---|---|---|---|
| 500 | 4,430.10 | 4,421.31 | 4,424.51 | 4,678.56 | **17,954.48** |
| 501 | 1,963.84 | 1,918.51 | 2,026.82 | 2,055.17 | **7,964.34** |
| 502 | 3,279.97 | 3,197.95 | 3,393.73 | 3,431.62 | **13,303.26** |
| 503 | 5,975.01 | 6,125.81 | 6,025.11 | 6,276.13 | **24,402.06** |
| 504 | 935.52 | 939.06 | 927.04 | 955.21 | **3,756.83** |
| 505 | 1,281.06 | 1,305.60 | 1,349.35 | 1,397.69 | **5,333.69** |
| 506 | 1,718.47 | 1,645.28 | 1,716.20 | 1,793.25 | **6,873.21** |
| 507 | 1,340.30 | 1,355.45 | 1,225.87 | 1,278.59 | **5,200.21** |

| | | | | | |
|---|---|---|---|---|---|
| 508 | 180.01 | 156.09 | 187.29 | 192.80 | **716.19** |
| 510 | 1,058.09 | 1,056.83 | 1,107.85 | 1,056.74 | **4,279.52** |
| 511 | 2,983.69 | 2,979.29 | 3,057.27 | 3,033.78 | **12,054.03** |
| 512 | 590.39 | 503.05 | 547.93 | 544.49 | **2,185.85** |
| 513 | 721.06 | 746.55 | 777.56 | 745.19 | **2,990.36** |
| 514 | 371.62 | 409.00 | 435.39 | 430.28 | **1,646.29** |
| 515 | 2,277.87 | 2,332.08 | 2,326.24 | 2,365.42 | **9,301.60** |
| 516 | 180.29 | 161.74 | 184.90 | 193.83 | **720.76** |
| 520 | 3,443.89 | 3,507.59 | 3,519.65 | 3,648.64 | **14,119.77** |
| 521 | 521.91 | 503.22 | 540.27 | 552.27 | **2,117.67** |
| 522 | 5,391.11 | 5,585.71 | 5,620.10 | 5,797.07 | **22,393.98** |
| 523 | 2,652.04 | 2,621.39 | 2,651.50 | 2,676.80 | **10,601.73** |
| 524 | 4,213.96 | 4,333.82 | 4,503.58 | 4,425.36 | **17,476.72** |
| 525 | 972.88 | 936.42 | 968.74 | 972.56 | **3,850.59** |
| 526 | 1,583.97 | 1,607.81 | 1,580.84 | 1,662.87 | **6,435.50** |
| 527 | 3,161.93 | 3,199.74 | 3,185.81 | 3,291.39 | **12,838.88** |
| 528 | 2,674.66 | 2,677.26 | 2,610.31 | 2,719.07 | **10,681.29** |
| **IA IOWA Total** | **53,903.62** | **54,226.56** | **54,893.88** | **56,174.78** | **219,198.84** |

**STATE: ID - IDAHO**

| REGISTRANT ZIP CODE 3 | 1ST QUARTER | 2ND QUARTER | 3RD QUARTER | 4TH QUARTER | **TOTAL GRAMS** |
|---|---|---|---|---|---|
| 832 | 2,569.48 | 2,478.05 | 2,765.49 | 2,598.12 | **10,411.14** |
| 833 | 2,166.28 | 2,186.45 | 2,203.67 | 2,264.02 | **8,820.42** |
| 834 | 5,576.62 | 5,581.31 | 5,590.02 | 5,852.24 | **22,600.20** |
| 835 | 491.64 | 480.08 | 490.03 | 458.21 | **1,919.95** |
| 836 | 8,398.51 | 8,620.26 | 8,942.19 | 8,963.80 | **34,924.76** |
| 837 | 7,942.10 | 8,329.16 | 8,424.32 | 8,481.59 | **33,177.17** |
| 838 | 2,408.18 | 2,468.85 | 2,542.72 | 2,664.86 | **10,084.61** |
| **ID IDAHO Total** | **29,552.81** | **30,144.15** | **30,958.45** | **31,282.85** | **121,938.25** |

**STATE: IL - ILLINOIS**

| REGISTRANT ZIP CODE 3 | 1ST QUARTER | 2ND QUARTER | 3RD QUARTER | 4TH QUARTER | **TOTAL GRAMS** |
|---|---|---|---|---|---|
| 600 | 34,212.83 | 35,956.85 | 37,025.34 | 36,906.17 | **144,101.20** |
| 601 | 24,603.11 | 25,423.79 | 26,669.59 | 26,449.81 | **103,146.30** |
| 602 | 1,599.69 | 1,648.62 | 1,620.43 | 1,713.49 | **6,582.23** |
| 603 | 1,372.10 | 1,446.58 | 1,513.39 | 1,482.70 | **5,814.77** |
| 604 | 19,738.28 | 20,623.31 | 21,486.39 | 21,296.98 | **83,144.97** |
| 605 | 18,388.75 | 19,422.43 | 20,171.27 | 20,366.95 | **78,349.41** |
| 606 | 39,828.30 | 42,194.61 | 42,743.32 | 44,048.26 | **168,814.50** |
| 607 | 2,337.22 | 2,536.16 | 2,623.07 | 2,817.02 | **10,313.48** |
| 608 | 766.16 | 743.95 | 878.23 | 786.90 | **3,175.23** |
| 609 | 2,120.12 | 2,106.94 | 2,295.25 | 2,170.40 | **8,692.70** |

| | | | | | |
|---|---|---|---|---|---|
| 610 | 3,432.04 | 3,650.74 | 3,768.99 | 3,864.56 | **14,716.33** |
| 611 | 4,202.21 | 4,296.18 | 4,556.01 | 4,137.57 | **17,191.97** |
| 612 | 2,999.04 | 3,180.66 | 3,266.59 | 3,268.78 | **12,715.06** |
| 613 | 2,702.57 | 2,801.61 | 2,842.63 | 2,905.91 | **11,252.71** |
| 614 | 1,992.14 | 1,943.56 | 2,057.36 | 2,152.75 | **8,145.80** |
| 615 | 3,352.15 | 3,473.09 | 3,446.52 | 3,508.44 | **13,780.19** |
| 616 | 4,172.49 | 4,448.66 | 4,385.77 | 4,613.81 | **17,620.73** |
| 617 | 3,838.11 | 4,006.19 | 4,104.46 | 4,232.35 | **16,181.10** |
| 618 | 3,974.72 | 4,013.24 | 4,168.02 | 4,199.43 | **16,355.41** |
| 619 | 1,184.39 | 1,090.53 | 1,088.27 | 1,137.98 | **4,501.16** |
| 620 | 3,862.50 | 3,922.17 | 3,737.13 | 3,821.08 | **15,342.89** |
| 622 | 5,939.51 | 5,872.02 | 5,955.83 | 6,109.55 | **23,876.91** |
| 623 | 2,027.23 | 1,990.24 | 1,944.06 | 2,050.55 | **8,012.09** |
| 624 | 1,236.95 | 1,192.06 | 1,267.15 | 1,302.43 | **4,998.59** |
| 625 | 1,605.30 | 1,612.75 | 1,649.37 | 1,651.26 | **6,518.68** |
| 626 | 1,617.46 | 1,598.73 | 1,631.83 | 1,677.99 | **6,526.01** |
| 627 | 2,473.52 | 2,523.03 | 2,494.50 | 2,510.27 | **10,001.33** |
| 628 | 2,476.26 | 2,406.04 | 2,388.26 | 2,568.72 | **9,839.28** |
| 629 | 3,787.56 | 3,775.94 | 3,760.46 | 4,052.38 | **15,376.33** |
| **IL ILLINOIS Total** | **201,842.70** | **209,900.70** | **215,539.49** | **217,804.48** | **845,087.37** |

**STATE: IN - INDIANA**

| REGISTRANT ZIP CODE 3 | 1ST QUARTER | 2ND QUARTER | 3RD QUARTER | 4TH QUARTER | **TOTAL GRAMS** |
|---|---|---|---|---|---|
| 460 | 16,554.85 | 16,797.05 | 18,526.61 | 17,346.32 | **69,224.83** |
| 461 | 9,928.27 | 10,430.24 | 11,137.25 | 10,574.82 | **42,070.58** |
| 462 | 17,377.74 | 17,970.64 | 18,951.75 | 17,956.48 | **72,256.61** |
| 463 | 12,808.17 | 13,531.41 | 14,465.12 | 14,129.98 | **54,934.68** |
| 464 | 1,396.67 | 1,469.46 | 1,546.84 | 1,491.44 | **5,904.41** |
| 465 | 17,652.50 | 17,983.52 | 19,189.70 | 17,931.29 | **72,757.00** |
| 466 | 4,991.68 | 5,074.82 | 5,232.87 | 4,900.32 | **20,199.70** |
| 467 | 3,747.89 | 3,907.81 | 4,131.86 | 4,076.14 | **15,863.71** |
| 468 | 6,401.24 | 6,630.68 | 7,192.27 | 6,586.42 | **26,810.62** |
| 469 | 5,516.35 | 5,805.40 | 6,173.71 | 5,828.16 | **23,323.63** |
| 470 | 1,876.15 | 1,933.92 | 1,954.83 | 1,966.68 | **7,731.59** |
| 471 | 6,221.01 | 6,283.59 | 6,660.60 | 6,445.44 | **25,610.63** |
| 472 | 2,622.72 | 2,726.67 | 2,886.84 | 2,759.47 | **10,995.69** |
| 473 | 5,366.37 | 5,233.03 | 5,789.21 | 5,503.58 | **21,892.18** |
| 474 | 4,054.54 | 4,301.13 | 4,401.13 | 4,539.35 | **17,296.15** |
| 475 | 2,436.23 | 2,449.10 | 2,564.97 | 2,651.95 | **10,102.26** |
| 476 | 2,844.42 | 3,010.70 | 3,086.22 | 3,113.22 | **12,054.56** |
| 477 | 6,318.66 | 6,568.05 | 6,832.08 | 6,378.26 | **26,097.05** |

| | | | | | |
|---|---|---|---|---|---|
| 478 | 2,266.14 | 2,361.33 | 2,459.42 | 2,493.09 | **9,579.99** |
| 479 | 7,323.60 | 7,344.40 | 7,683.99 | 7,440.62 | **29,792.61** |
| **IN INDIANA Total** | **137,705.23** | **141,812.97** | **150,867.28** | **144,113.02** | **574,498.49** |

**STATE: KS - KANSAS**

| REGISTRANT ZIP CODE 3 | 1ST QUARTER | 2ND QUARTER | 3RD QUARTER | 4TH QUARTER | TOTAL GRAMS |
|---|---|---|---|---|---|
| 660 | 15,540.13 | 15,483.48 | 15,947.03 | 16,291.62 | **63,262.25** |
| 661 | 2,098.08 | 2,112.11 | 2,338.45 | 2,321.70 | **8,870.35** |
| 662 | 19,883.31 | 20,482.77 | 20,888.45 | 21,083.66 | **82,338.19** |
| 664 | 960.84 | 929.42 | 891.59 | 977.71 | **3,759.55** |
| 665 | 1,649.43 | 1,633.53 | 1,633.49 | 1,902.25 | **6,818.69** |
| 666 | 4,993.93 | 5,363.30 | 5,528.67 | 5,580.30 | **21,466.21** |
| 667 | 1,820.77 | 1,809.87 | 1,852.85 | 2,025.95 | **7,509.45** |
| 668 | 571.74 | 637.98 | 618.53 | 665.85 | **2,494.10** |
| 669 | 108.77 | 126.69 | 116.77 | 100.24 | **452.47** |
| 670 | 2,751.63 | 2,813.50 | 2,631.26 | 2,985.29 | **11,181.68** |
| 671 | 1,016.87 | 1,020.96 | 1,051.95 | 1,046.20 | **4,135.99** |
| 672 | 8,001.17 | 8,374.68 | 8,190.50 | 8,384.01 | **32,950.37** |
| 673 | 811.93 | 870.06 | 972.63 | 886.44 | **3,541.05** |
| 674 | 1,346.43 | 1,374.31 | 1,376.06 | 1,399.39 | **5,496.20** |
| 675 | 1,277.38 | 1,237.16 | 1,344.27 | 1,362.26 | **5,221.06** |
| 676 | 769.39 | 768.35 | 793.59 | 818.68 | **3,150.00** |
| 677 | 436.48 | 393.76 | 415.80 | 410.04 | **1,656.08** |
| 678 | 694.03 | 693.85 | 694.15 | 743.12 | **2,825.16** |
| 679 | 187.59 | 219.59 | 266.52 | 262.91 | **936.61** |
| **KS - KANSAS - Total** | **64,919.88** | **66,345.39** | **67,552.57** | **69,247.62** | **268,065.45** |

**STATE: KY - KENTUCKY**

| REGISTRANT ZIP CODE 3 | 1ST QUARTER | 2ND QUARTER | 3RD QUARTER | 4TH QUARTER | TOTAL GRAMS |
|---|---|---|---|---|---|
| 400 | 4,168.70 | 4,209.78 | 4,304.10 | 4,482.42 | **17,165.01** |
| 401 | 1,670.47 | 1,548.43 | 1,973.79 | 1,614.90 | **6,807.58** |
| 402 | 17,214.34 | 17,634.85 | 17,715.80 | 18,222.31 | **70,787.31** |
| 403 | 4,520.90 | 4,623.14 | 4,859.59 | 4,992.76 | **18,996.38** |
| 404 | 1,879.05 | 1,839.46 | 1,996.25 | 2,053.89 | **7,768.65** |
| 405 | 7,867.85 | 7,802.47 | 8,179.12 | 8,484.19 | **32,333.63** |
| 406 | 786.86 | 776.53 | 817.30 | 877.09 | **3,257.78** |
| 407 | 645.95 | 590.96 | 664.09 | 753.99 | **2,654.98** |
| 408 | 176.58 | 129.47 | 173.69 | 166.96 | **646.70** |
| 409 | 187.04 | 205.34 | 215.53 | 215.72 | **823.63** |
| 410 | 10,379.87 | 10,424.88 | 10,795.72 | 10,972.89 | **42,573.35** |
| 411 | 1,520.10 | 1,460.29 | 1,446.43 | 1,492.19 | **5,919.00** |
| 412 | 382.95 | 346.74 | 403.69 | 415.35 | **1,548.74** |

| | | | | | |
|---|---|---|---|---|---|
| 413 | 184.18 | 145.37 | 188.03 | 162.79 | **680.37** |
| 414 | 125.10 | 139.78 | 127.37 | 129.72 | **521.97** |
| 415 | 607.98 | 555.36 | 593.70 | 607.04 | **2,364.07** |
| 416 | 352.56 | 370.55 | 405.15 | 364.68 | **1,492.94** |
| 417 | 355.95 | 348.94 | 387.38 | 359.00 | **1,451.28** |
| 418 | 112.79 | 94.57 | 80.58 | 85.18 | **373.12** |
| 420 | 4,893.36 | 4,523.96 | 4,733.82 | 4,704.71 | **18,855.85** |
| 421 | 3,193.34 | 3,127.49 | 3,107.93 | 3,183.91 | **12,612.67** |
| 422 | 2,131.56 | 1,842.60 | 2,306.87 | 1,862.26 | **8,143.29** |
| 423 | 3,713.24 | 3,396.19 | 3,638.06 | 3,380.76 | **14,128.24** |
| 424 | 3,568.84 | 3,647.39 | 3,697.92 | 3,876.66 | **14,790.80** |
| 425 | 408.43 | 375.78 | 389.05 | 392.38 | **1,565.64** |
| 426 | 239.39 | 200.59 | 261.62 | 238.02 | **939.62** |
| 427 | 2,603.55 | 2,590.54 | 2,693.93 | 2,930.77 | **10,818.79** |
| **KY - KENTUCKY - Total** | **73,890.93** | **72,951.47** | **76,156.50** | **77,022.51** | **300,021.42** |

**STATE: LA - LOUISIANA**

| REGISTRANT ZIP CODE 3 | 1ST QUARTER | 2ND QUARTER | 3RD QUARTER | 4TH QUARTER | **TOTAL GRAMS** |
|---|---|---|---|---|---|
| 700 | 20,504.91 | 20,399.21 | 19,720.26 | 20,475.07 | **81,099.45** |
| 701 | 11,771.07 | 11,614.45 | 11,499.35 | 12,459.21 | **47,344.08** |
| 703 | 6,504.09 | 6,275.67 | 6,403.93 | 6,603.78 | **25,787.46** |
| 704 | 21,263.73 | 21,298.62 | 21,687.81 | 21,589.37 | **85,839.53** |
| 705 | 35,232.77 | 35,565.02 | 35,556.65 | 37,467.15 | **143,821.59** |
| 706 | 13,002.88 | 13,253.74 | 13,239.51 | 14,104.48 | **53,600.61** |
| 707 | 21,463.23 | 21,181.26 | 21,648.10 | 22,087.47 | **86,380.07** |
| 708 | 17,255.99 | 16,758.76 | 16,299.24 | 17,747.12 | **68,061.11** |
| 710 | 2,033.98 | 2,223.09 | 2,124.96 | 2,135.85 | **8,517.88** |
| 711 | 8,228.14 | 8,065.38 | 8,188.70 | 8,448.34 | **32,930.56** |
| 712 | 7,399.61 | 7,479.35 | 7,417.83 | 7,839.93 | **30,136.71** |
| 713 | 6,095.35 | 5,854.69 | 6,011.83 | 6,106.38 | **24,068.24** |
| 714 | 2,403.22 | 2,338.42 | 2,342.17 | 2,380.09 | **9,463.89** |
| **LA - LOUISIANA - Total** | **173,158.96** | **172,307.65** | **172,140.33** | **179,444.24** | **697,051.17** |

**STATE: MA - MASSACHUSETTS**

| REGISTRANT ZIP CODE 3 | 1ST QUARTER | 2ND QUARTER | 3RD QUARTER | 4TH QUARTER | **TOTAL GRAMS** |
|---|---|---|---|---|---|
| 010 | 8,860.53 | 9,363.88 | 8,906.18 | 9,892.11 | **37,022.70** |
| 011 | 2,393.96 | 2,571.19 | 2,417.81 | 2,622.96 | **10,005.93** |
| 012 | 2,401.41 | 2,519.43 | 2,476.42 | 2,738.73 | **10,136.00** |
| 013 | 2,714.14 | 2,691.39 | 2,663.29 | 2,816.29 | **10,885.11** |
| 014 | 5,462.38 | 5,710.57 | 5,630.00 | 5,718.82 | **22,521.77** |
| 015 | 7,626.59 | 8,052.48 | 7,977.93 | 8,474.52 | **32,131.51** |
| 016 | 5,579.50 | 5,840.14 | 5,747.62 | 5,933.67 | **23,100.93** |

| | | | | | |
|---|---|---|---|---|---|
| 017 | 11,275.74 | 11,922.38 | 11,748.04 | 12,136.88 | **47,083.03** |
| 018 | 19,654.21 | 21,828.52 | 20,762.97 | 22,205.00 | **84,450.70** |
| 019 | 16,910.71 | 18,870.95 | 18,337.27 | 18,845.44 | **72,964.38** |
| 020 | 14,277.45 | 14,828.80 | 14,835.23 | 15,153.49 | **59,094.97** |
| 021 | 34,576.41 | 37,289.27 | 35,784.78 | 37,670.94 | **145,321.40** |
| 022 | 1,761.71 | 1,790.64 | 1,807.30 | 1,792.51 | **7,152.16** |
| 023 | 13,467.83 | 14,655.51 | 14,375.82 | 14,643.84 | **57,143.00** |
| 024 | 10,996.10 | 11,562.42 | 11,390.32 | 11,804.13 | **45,752.97** |
| 025 | 4,088.28 | 4,305.03 | 4,639.98 | 4,467.90 | **17,501.19** |
| 026 | 5,143.09 | 5,761.82 | 6,093.50 | 5,797.44 | **22,795.85** |
| 027 | 13,155.45 | 14,030.47 | 13,297.59 | 14,063.02 | **54,546.53** |
| **MA - MASSACHUSETTS - Total** | **180,345.49** | **193,594.89** | **188,892.05** | **196,777.68** | **759,610.12** |

**STATE: MD - MARYLAND**

| REGISTRANT ZIP CODE 3 | 1ST QUARTER | 2ND QUARTER | 3RD QUARTER | 4TH QUARTER | **TOTAL GRAMS** |
|---|---|---|---|---|---|
| 206 | 4,933.10 | 5,132.02 | 4,922.73 | 5,268.50 | **20,256.34** |
| 207 | 6,158.42 | 6,299.66 | 6,474.49 | 6,485.51 | **25,418.08** |
| 208 | 14,918.67 | 15,000.14 | 15,505.71 | 15,485.24 | **60,909.75** |
| 209 | 2,645.14 | 2,853.42 | 2,917.68 | 2,943.32 | **11,359.56** |
| 210 | 18,761.10 | 19,612.64 | 19,118.28 | 20,005.33 | **77,497.36** |
| 211 | 8,472.29 | 8,633.40 | 8,849.44 | 9,241.95 | **35,197.08** |
| 212 | 18,017.97 | 18,610.82 | 18,482.89 | 19,503.75 | **74,615.43** |
| 214 | 3,089.07 | 2,977.43 | 3,424.20 | 3,160.43 | **12,651.13** |
| 215 | 1,902.41 | 1,932.59 | 1,957.06 | 2,014.34 | **7,806.40** |
| 216 | 3,239.44 | 3,430.47 | 3,466.55 | 3,450.80 | **13,587.26** |
| 217 | 11,433.81 | 11,854.31 | 11,938.13 | 12,234.29 | **47,460.54** |
| 218 | 2,157.07 | 2,337.45 | 2,335.30 | 2,352.41 | **9,182.23** |
| 219 | 2,793.92 | 3,008.73 | 2,540.23 | 2,890.47 | **11,233.34** |
| **MD MARYLAND Total** | **98,522.40** | **101,683.07** | **101,932.68** | **105,036.34** | **407,174.51** |

**STATE: ME - MAINE**

| REGISTRANT ZIP CODE 3 | 1ST QUARTER | 2ND QUARTER | 3RD QUARTER | 4TH QUARTER | **TOTAL GRAMS** |
|---|---|---|---|---|---|
| 039 | 1,035.81 | 1,082.18 | 1,107.15 | 1,056.65 | **4,281.79** |
| 040 | 8,786.06 | 9,359.30 | 9,573.18 | 9,621.87 | **37,340.42** |
| 041 | 4,101.75 | 4,452.34 | 4,319.91 | 4,254.54 | **17,128.55** |
| 042 | 2,209.11 | 2,424.49 | 2,515.38 | 2,557.01 | **9,705.99** |
| 043 | 1,713.55 | 1,763.32 | 1,900.90 | 1,797.73 | **7,175.50** |
| 044 | 2,211.62 | 2,255.87 | 2,305.05 | 2,297.11 | **9,069.64** |
| 045 | 888.14 | 880.04 | 952.56 | 905.78 | **3,626.52** |
| 046 | 691.31 | 834.11 | 862.42 | 885.14 | **3,272.98** |
| 047 | 483.75 | 515.24 | 581.39 | 593.93 | **2,174.30** |
| 048 | 649.69 | 697.86 | 795.18 | 677.34 | **2,820.07** |

| | | | | | |
|---|---|---|---|---|---|
| 049 | 2,024.76 | 2,095.27 | 2,144.46 | 2,147.99 | **8,412.49** |
| **ME MAINE Total** | **24,795.54** | **26,360.02** | **27,057.58** | **26,795.10** | **105,008.24** |

**STATE: MI - MICHIGAN**

| REGISTRANT ZIP CODE 3 | 1ST QUARTER | 2ND QUARTER | 3RD QUARTER | 4TH QUARTER | TOTAL GRAMS |
|---|---|---|---|---|---|
| 480 | 39,828.65 | 40,290.93 | 42,842.28 | 40,832.22 | **163,794.08** |
| 481 | 42,254.52 | 42,989.83 | 45,143.15 | 43,164.33 | **173,551.83** |
| 482 | 6,081.02 | 6,500.70 | 6,756.53 | 6,670.04 | **26,008.29** |
| 483 | 33,689.84 | 34,779.87 | 37,147.80 | 34,965.93 | **140,583.44** |
| 484 | 9,061.96 | 8,750.72 | 9,514.79 | 9,173.47 | **36,500.94** |
| 485 | 2,752.87 | 2,828.18 | 3,060.94 | 2,725.80 | **11,367.79** |
| 486 | 5,165.93 | 5,102.95 | 5,532.60 | 5,196.03 | **20,997.51** |
| 487 | 2,923.32 | 2,881.07 | 3,125.93 | 2,984.47 | **11,914.79** |
| 488 | 15,282.22 | 15,172.57 | 17,160.14 | 14,729.43 | **62,344.36** |
| 489 | 4,468.35 | 4,326.04 | 4,725.35 | 4,473.77 | **17,993.51** |
| 490 | 18,033.04 | 17,847.64 | 19,183.12 | 17,466.48 | **72,530.27** |
| 491 | 2,311.40 | 2,247.62 | 2,567.11 | 2,262.73 | **9,388.85** |
| 492 | 4,348.64 | 4,210.74 | 4,701.73 | 4,232.07 | **17,493.18** |
| 493 | 7,338.37 | 7,355.63 | 8,587.95 | 6,915.14 | **30,197.10** |
| 494 | 19,287.13 | 19,575.84 | 21,049.23 | 18,446.44 | **78,358.63** |
| 495 | 19,540.37 | 19,959.20 | 21,437.89 | 18,931.17 | **79,868.62** |
| 496 | 5,688.31 | 5,835.39 | 6,573.13 | 5,810.20 | **23,907.04** |
| 497 | 3,367.58 | 3,539.40 | 4,095.28 | 3,418.16 | **14,420.42** |
| 498 | 3,436.77 | 3,283.87 | 3,765.94 | 3,317.97 | **13,804.54** |
| 499 | 1,448.32 | 1,496.92 | 1,504.43 | 1,617.28 | **6,066.95** |
| **MI - MICHIGAN Total** | **246,308.62** | **248,975.11** | **268,475.31** | **247,333.12** | **1,011,092.15** |

**STATE: MN - MINNESOTA**

| REGISTRANT ZIP CODE 3 | 1ST QUARTER | 2ND QUARTER | 3RD QUARTER | 4TH QUARTER | TOTAL GRAMS |
|---|---|---|---|---|---|
| 550 | 13,051.16 | 13,403.21 | 13,300.73 | 13,922.45 | **53,677.55** |
| 551 | 18,233.60 | 18,537.33 | 18,596.43 | 19,371.16 | **74,738.52** |
| 553 | 23,412.19 | 24,059.51 | 24,255.15 | 25,503.29 | **97,230.14** |
| 554 | 27,383.51 | 28,249.85 | 28,144.21 | 29,195.68 | **112,973.24** |
| 556 | 101.19 | 119.12 | 131.38 | 135.42 | **487.10** |
| 557 | 3,416.59 | 3,413.59 | 3,648.40 | 3,687.11 | **14,165.70** |
| 558 | 2,177.22 | 2,127.27 | 2,121.22 | 2,143.98 | **8,569.69** |
| 559 | 5,116.29 | 5,546.61 | 5,386.22 | 5,781.99 | **21,831.11** |
| 560 | 5,241.94 | 5,363.45 | 5,530.96 | 5,655.81 | **21,792.15** |
| 561 | 908.25 | 908.56 | 919.08 | 895.30 | **3,631.20** |
| 562 | 1,675.28 | 1,658.47 | 1,736.47 | 1,751.24 | **6,821.46** |
| 563 | 4,437.88 | 4,556.91 | 4,556.13 | 4,688.99 | **18,239.91** |
| 564 | 2,420.49 | 2,526.88 | 2,598.28 | 2,450.10 | **9,995.75** |

| | | | | | |
|---|---|---|---|---|---|
| 565 | 2,895.43 | 2,999.33 | 3,028.77 | 3,002.21 | **11,925.73** |
| 566 | 1,383.98 | 1,391.29 | 1,445.19 | 1,516.11 | **5,736.56** |
| 567 | 821.94 | 871.72 | 921.64 | 967.03 | **3,582.33** |
| **MN MINNESOTA Total** | **112,676.93** | **115,733.08** | **116,320.27** | **120,667.86** | **465,398.14** |

**STATE: MO - MISSOURI**

| REGISTRANT ZIP CODE 3 | 1ST QUARTER | 2ND QUARTER | 3RD QUARTER | 4TH QUARTER | TOTAL GRAMS |
|---|---|---|---|---|---|
| 630 | 20,112.58 | 20,617.91 | 19,940.41 | 20,376.98 | **81,047.88** |
| 631 | 53,312.32 | 53,379.03 | 50,473.37 | 51,733.17 | **208,897.88** |
| 633 | 15,262.45 | 15,569.82 | 15,866.16 | 16,039.61 | **62,738.03** |
| 634 | 1,155.21 | 1,136.95 | 1,288.00 | 1,226.90 | **4,807.05** |
| 635 | 1,323.80 | 1,415.27 | 1,330.13 | 1,379.89 | **5,449.09** |
| 636 | 2,049.69 | 1,938.89 | 2,069.01 | 2,001.85 | **8,059.44** |
| 637 | 2,178.81 | 2,077.89 | 2,194.17 | 2,267.08 | **8,717.95** |
| 638 | 1,615.01 | 1,565.66 | 1,644.55 | 1,719.30 | **6,544.52** |
| 639 | 642.64 | 537.12 | 561.60 | 634.45 | **2,375.81** |
| 640 | 15,170.83 | 15,871.94 | 15,967.85 | 16,445.70 | **63,456.32** |
| 641 | 17,009.56 | 15,997.28 | 15,622.51 | 15,777.16 | **64,406.52** |
| 644 | 706.03 | 708.70 | 668.18 | 749.32 | **2,832.23** |
| 645 | 1,158.39 | 1,125.48 | 1,181.16 | 1,274.14 | **4,739.17** |
| 646 | 418.51 | 388.97 | 396.08 | 394.32 | **1,597.87** |
| 647 | 1,116.38 | 1,033.62 | 1,136.75 | 1,188.07 | **4,474.83** |
| 648 | 1,714.14 | 1,784.77 | 1,750.22 | 1,831.00 | **7,080.13** |
| 650 | 1,179.44 | 1,208.15 | 1,223.80 | 1,175.31 | **4,786.69** |
| 651 | 1,205.67 | 1,142.87 | 1,175.43 | 1,215.33 | **4,739.29** |
| 652 | 5,563.23 | 5,633.53 | 5,659.22 | 5,777.51 | **22,633.49** |
| 653 | 1,049.07 | 1,090.29 | 1,070.03 | 1,078.33 | **4,287.71** |
| 654 | 738.06 | 758.72 | 959.66 | 908.12 | **3,364.57** |
| 655 | 622.96 | 599.44 | 599.16 | 639.22 | **2,460.78** |
| 656 | 1,364.61 | 1,434.07 | 1,376.78 | 1,473.95 | **5,649.41** |
| 657 | 2,059.76 | 2,078.76 | 2,143.04 | 2,183.27 | **8,464.84** |
| 658 | 3,672.76 | 3,769.61 | 3,943.51 | 3,916.15 | **15,302.03** |
| **MO MISSOURI - Total** | **152,401.88** | **152,864.74** | **150,240.80** | **153,406.12** | **608,913.54** |

**STATE: MS - MISSISSIPPI**

| REGISTRANT ZIP CODE 3 | 1ST QUARTER | 2ND QUARTER | 3RD QUARTER | 4TH QUARTER | TOTAL GRAMS |
|---|---|---|---|---|---|
| 386 | 9,905.88 | 9,811.54 | 9,769.86 | 10,313.52 | **39,800.79** |
| 387 | 1,691.57 | 1,468.25 | 1,520.31 | 1,564.22 | **6,244.35** |
| 388 | 5,208.89 | 5,071.26 | 5,190.60 | 5,510.54 | **20,981.29** |
| 389 | 1,437.46 | 1,483.53 | 1,534.15 | 1,591.65 | **6,046.79** |
| 390 | 5,163.17 | 4,962.12 | 5,281.34 | 5,288.86 | **20,695.49** |
| 391 | 6,363.45 | 6,498.79 | 6,516.10 | 6,705.44 | **26,083.78** |

| | | | | | |
|---|---|---|---|---|---|
| 392 | 5,410.20 | 5,444.89 | 5,057.70 | 5,184.44 | **21,097.23** |
| 393 | 3,807.19 | 3,782.16 | 3,974.42 | 4,001.56 | **15,565.33** |
| 394 | 9,050.37 | 8,621.23 | 8,995.11 | 9,167.25 | **35,833.96** |
| 395 | 9,384.30 | 8,869.49 | 9,245.17 | 9,266.95 | **36,765.90** |
| 396 | 2,662.40 | 2,458.22 | 2,756.04 | 2,607.07 | **10,483.72** |
| 397 | 3,546.16 | 3,293.31 | 3,239.52 | 3,575.97 | **13,654.95** |
| **MS MISSISSIPPI - Total** | **63,631.03** | **61,764.81** | **63,080.31** | **64,777.45** | **253,253.60** |

**STATE: MT - MONTANA**

| REGISTRANT ZIP CODE 3 | 1ST QUARTER | 2ND QUARTER | 3RD QUARTER | 4TH QUARTER | TOTAL GRAMS |
|---|---|---|---|---|---|
| 590 | 582.25 | 712.86 | 684.52 | 717.73 | **2,697.37** |
| 591 | 2,680.60 | 2,675.29 | 2,666.20 | 2,769.96 | **10,792.04** |
| 592 | 362.83 | 319.49 | 379.56 | 393.28 | **1,455.16** |
| 593 | 405.45 | 377.20 | 386.12 | 413.20 | **1,581.97** |
| 594 | 1,131.45 | 1,211.27 | 1,188.36 | 1,296.39 | **4,827.48** |
| 595 | 260.06 | 215.32 | 234.88 | 236.45 | **946.71** |
| 596 | 1,121.47 | 1,271.65 | 1,307.56 | 1,271.93 | **4,972.61** |
| 597 | 3,867.67 | 3,811.16 | 4,073.43 | 3,931.80 | **15,684.07** |
| 598 | 3,841.25 | 3,775.77 | 3,955.27 | 3,918.66 | **15,490.95** |
| 599 | 1,354.79 | 1,405.08 | 1,551.70 | 1,459.99 | **5,771.57** |
| **MT - MONTANA Total** | **15,607.82** | **15,775.09** | **16,427.60** | **16,409.41** | **64,219.93** |

**STATE: NC - NORTH CAROLINA**

| REGISTRANT ZIP CODE 3 | 1ST QUARTER | 2ND QUARTER | 3RD QUARTER | 4TH QUARTER | TOTAL GRAMS |
|---|---|---|---|---|---|
| 270 | 4,394.84 | 4,439.94 | 4,620.89 | 4,485.90 | **17,941.57** |
| 271 | 6,039.61 | 6,301.45 | 6,144.61 | 6,033.39 | **24,519.06** |
| 272 | 13,317.89 | 13,358.09 | 13,166.37 | 12,797.87 | **52,640.22** |
| 273 | 6,467.96 | 6,764.56 | 6,731.43 | 6,642.47 | **26,606.41** |
| 274 | 8,560.08 | 8,611.42 | 8,796.31 | 8,721.19 | **34,689.00** |
| 275 | 27,043.57 | 27,816.30 | 28,139.77 | 27,648.25 | **110,647.89** |
| 276 | 16,981.99 | 16,916.87 | 17,332.95 | 17,040.44 | **68,272.25** |
| 277 | 6,229.53 | 6,405.27 | 6,387.90 | 6,803.36 | **25,826.06** |
| 278 | 10,102.58 | 10,333.17 | 10,341.68 | 10,341.95 | **41,119.38** |
| 279 | 3,911.08 | 4,266.93 | 4,027.54 | 3,617.88 | **15,823.43** |
| 280 | 20,039.21 | 20,761.08 | 20,640.85 | 20,422.95 | **81,864.09** |
| 281 | 13,142.66 | 13,691.21 | 14,021.81 | 14,010.41 | **54,866.08** |
| 282 | 20,801.09 | 22,039.44 | 22,082.75 | 22,498.15 | **87,421.44** |
| 283 | 17,242.21 | 17,291.29 | 17,241.17 | 16,505.31 | **68,279.98** |
| 284 | 19,555.33 | 20,710.03 | 20,929.14 | 20,436.29 | **81,630.79** |
| 285 | 12,700.49 | 13,013.98 | 13,314.34 | 11,921.60 | **50,950.42** |
| 286 | 9,012.58 | 9,182.38 | 9,118.53 | 9,347.17 | **36,660.66** |
| 287 | 7,851.89 | 8,174.80 | 8,450.04 | 8,180.56 | **32,657.29** |

| | | | | | |
|---|---|---|---|---|---|
| 288 | 4,597.79 | 4,714.84 | 4,961.89 | 4,969.01 | **19,243.53** |
| 289 | 406.60 | 459.28 | 455.61 | 461.23 | **1,782.73** |
| **NC - NORTH CAROLINA Total** | **228,398.99** | **235,252.31** | **236,905.59** | **232,885.37** | **933,442.27** |

**STATE: ND - NORTH DAKOTA**

| REGISTRANT ZIP CODE 3 | 1ST QUARTER | 2ND QUARTER | 3RD QUARTER | 4TH QUARTER | TOTAL GRAMS |
|---|---|---|---|---|---|
| 580 | 809.55 | 819.41 | 857.88 | 945.53 | **3,432.37** |
| 581 | 2,411.70 | 2,329.89 | 2,443.43 | 2,546.96 | **9,731.97** |
| 582 | 1,548.23 | 1,424.59 | 1,411.84 | 1,536.73 | **5,921.39** |
| 583 | 486.51 | 410.37 | 419.45 | 411.11 | **1,727.43** |
| 584 | 282.49 | 286.26 | 281.91 | 300.99 | **1,151.66** |
| 585 | 1,632.90 | 1,558.32 | 1,623.05 | 1,609.72 | **6,423.98** |
| 586 | 433.30 | 438.95 | 481.26 | 486.21 | **1,839.72** |
| 587 | 1,456.83 | 1,485.21 | 1,609.78 | 1,490.35 | **6,042.16** |
| 588 | 824.71 | 849.25 | 914.80 | 891.06 | **3,479.82** |
| **ND NORTH DAKOTA Total** | **9,886.22** | **9,602.26** | **10,043.39** | **10,218.65** | **39,750.51** |

**STATE: NE - NEBRASKA**

| REGISTRANT ZIP CODE 3 | 1ST QUARTER | 2ND QUARTER | 3RD QUARTER | 4TH QUARTER | TOTAL GRAMS |
|---|---|---|---|---|---|
| 680 | 2,553.63 | 2,581.96 | 2,683.58 | 2,872.08 | **10,691.26** |
| 681 | 13,635.36 | 14,194.87 | 14,499.80 | 14,735.74 | **57,065.78** |
| 683 | 572.38 | 564.98 | 639.34 | 608.63 | **2,385.32** |
| 684 | 485.69 | 459.90 | 494.01 | 577.81 | **2,017.41** |
| 685 | 5,884.20 | 5,898.66 | 6,002.69 | 6,222.19 | **24,007.75** |
| 686 | 515.20 | 517.69 | 515.76 | 505.73 | **2,054.39** |
| 687 | 1,109.79 | 1,126.26 | 1,113.92 | 1,179.82 | **4,529.79** |
| 688 | 1,768.35 | 1,776.40 | 1,892.58 | 1,887.37 | **7,324.70** |
| 689 | 542.83 | 510.09 | 538.10 | 530.93 | **2,121.95** |
| 690 | 161.06 | 162.64 | 182.56 | 202.58 | **708.85** |
| 691 | 752.91 | 718.11 | 730.63 | 689.58 | **2,891.24** |
| 692 | 14.41 | 19.08 | 17.52 | 12.83 | **63.83** |
| 693 | 552.44 | 600.70 | 625.01 | 612.27 | **2,390.43** |
| **NE NEBRASKA Total** | **28,548.26** | **29,131.35** | **29,935.52** | **30,637.56** | **118,252.69** |

**STATE: NH - NEW HAMPSHIRE**

| REGISTRANT ZIP CODE 3 | 1ST QUARTER | 2ND QUARTER | 3RD QUARTER | 4TH QUARTER | TOTAL GRAMS |
|---|---|---|---|---|---|
| 030 | 12,606.75 | 13,170.73 | 13,263.47 | 13,513.16 | **52,554.11** |
| 031 | 6,609.14 | 6,825.05 | 6,879.85 | 7,225.74 | **27,539.79** |
| 032 | 3,047.03 | 3,233.77 | 3,342.05 | 3,452.88 | **13,075.73** |
| 033 | 2,256.74 | 2,344.32 | 2,449.22 | 2,364.99 | **9,415.27** |
| 034 | 1,581.76 | 1,732.38 | 1,692.21 | 1,796.44 | **6,802.79** |
| 035 | 655.26 | 752.20 | 735.20 | 730.11 | **2,872.77** |
| 036 | 118.95 | 140.75 | 136.87 | 130.69 | **527.26** |

| | | | | | |
|---|---|---|---|---|---|
| 037 | 1,789.23 | 1,903.07 | 1,923.04 | 1,940.70 | **7,556.04** |
| 038 | 12,481.14 | 13,146.55 | 13,319.12 | 13,337.56 | **52,284.37** |
| **NH - NEW HAMPSHIRE Total** | **41,145.99** | **43,248.82** | **43,741.04** | **44,492.27** | **172,628.12** |

**STATE: NJ - NEW JERSEY**

| REGISTRANT ZIP CODE 3 | 1ST QUARTER | 2ND QUARTER | 3RD QUARTER | 4TH QUARTER | TOTAL GRAMS |
|---|---|---|---|---|---|
| 070 | 17,885.15 | 18,343.61 | 18,683.54 | 19,813.09 | **74,725.38** |
| 071 | 1,455.04 | 1,307.07 | 1,334.25 | 1,591.71 | **5,688.08** |
| 072 | 690.12 | 669.09 | 695.42 | 750.41 | **2,805.04** |
| 073 | 1,962.83 | 1,943.76 | 2,005.41 | 2,210.88 | **8,122.88** |
| 074 | 6,948.12 | 6,822.78 | 6,728.21 | 7,541.77 | **28,040.88** |
| 075 | 805.44 | 807.33 | 851.51 | 981.15 | **3,445.42** |
| 076 | 6,348.16 | 6,182.94 | 6,649.61 | 6,951.35 | **26,132.06** |
| 077 | 9,534.06 | 9,422.91 | 9,966.24 | 10,156.54 | **39,079.75** |
| 078 | 5,805.49 | 5,719.01 | 6,008.06 | 6,439.61 | **23,972.17** |
| 079 | 4,889.00 | 4,924.32 | 5,225.31 | 5,093.12 | **20,131.76** |
| 080 | 24,261.74 | 24,757.25 | 25,203.55 | 25,822.95 | **100,045.49** |
| 081 | 1,879.80 | 2,010.72 | 2,068.15 | 2,089.33 | **8,048.00** |
| 082 | 3,985.76 | 4,273.31 | 4,645.49 | 4,350.19 | **17,254.75** |
| 083 | 2,850.32 | 2,744.15 | 2,817.25 | 2,870.65 | **11,282.38** |
| 084 | 381.97 | 439.40 | 532.95 | 500.04 | **1,854.36** |
| 085 | 3,704.74 | 3,559.16 | 3,669.17 | 3,904.30 | **14,837.37** |
| 086 | 2,825.95 | 2,858.46 | 2,895.74 | 2,924.06 | **11,504.20** |
| 087 | 6,350.51 | 6,575.86 | 6,686.68 | 7,097.16 | **26,710.22** |
| 088 | 9,826.86 | 9,774.73 | 10,161.36 | 10,152.53 | **39,915.48** |
| 089 | 1,064.44 | 1,039.28 | 1,061.38 | 1,071.62 | **4,236.72** |
| **NJ NEW JERSEY - Total** | **113,455.51** | **114,175.15** | **117,889.28** | **122,312.45** | **467,832.40** |

**STATE: NM - NEW MEXICO**

| REGISTRANT ZIP CODE 3 | 1ST QUARTER | 2ND QUARTER | 3RD QUARTER | 4TH QUARTER | TOTAL GRAMS |
|---|---|---|---|---|---|
| 870 | 686.96 | 684.95 | 635.70 | 642.61 | **2,650.23** |
| 871 | 6,421.87 | 6,429.96 | 6,620.51 | 6,857.50 | **26,329.84** |
| 873 | 104.56 | 132.41 | 128.61 | 129.03 | **494.61** |
| 874 | 670.46 | 700.15 | 709.10 | 691.67 | **2,771.38** |
| 875 | 1,961.35 | 1,988.08 | 2,093.44 | 2,100.62 | **8,143.48** |
| 877 | 179.98 | 187.04 | 153.60 | 158.86 | **679.48** |
| 878 | 45.57 | 63.22 | 50.14 | 51.80 | **210.73** |
| 879 | 67.10 | 72.45 | 50.46 | 63.96 | **253.97** |
| 880 | 2,284.13 | 2,114.91 | 2,112.85 | 2,258.17 | **8,770.06** |
| 881 | 357.59 | 380.00 | 373.51 | 390.31 | **1,501.41** |
| 882 | 2,201.80 | 2,153.74 | 2,049.23 | 2,174.77 | **8,579.54** |
| 883 | 943.27 | 952.26 | 920.23 | 971.90 | **3,787.66** |

| | | | | | |
|---|---|---|---|---|---|
| 884 | 53.69 | 54.61 | 40.45 | 56.86 | **205.61** |
| **NM NEW MEXICO Total** | **15,978.33** | **15,913.78** | **15,937.83** | **16,548.06** | **64,378.00** |

**STATE: NV - NEVADA**

| REGISTRANT ZIP CODE 3 | 1ST QUARTER | 2ND QUARTER | 3RD QUARTER | 4TH QUARTER | TOTAL GRAMS |
|---|---|---|---|---|---|
| 890 | 6,537.18 | 6,466.01 | 6,591.17 | 7,158.32 | **26,752.67** |
| 891 | 15,613.34 | 15,567.53 | 15,842.01 | 16,585.31 | **63,608.19** |
| 893 | 29.64 | 39.67 | 42.63 | 52.38 | **164.31** |
| 894 | 1,683.45 | 1,752.38 | 1,778.30 | 1,903.17 | **7,117.30** |
| 895 | 2,974.60 | 3,084.95 | 3,136.38 | 3,280.09 | **12,476.02** |
| 897 | 575.39 | 508.66 | 518.75 | 578.49 | **2,181.28** |
| 898 | 552.13 | 501.94 | 477.15 | 534.89 | **2,066.11** |
| **NV NEVADA Total** | **27,965.72** | **27,921.14** | **28,386.39** | **30,092.64** | **114,365.89** |

**STATE: NY - NEW YORK**

| REGISTRANT ZIP CODE 3 | 1ST QUARTER | 2ND QUARTER | 3RD QUARTER | 4TH QUARTER | TOTAL GRAMS |
|---|---|---|---|---|---|
| 100 | 52,836.66 | 53,909.62 | 53,234.95 | 57,744.71 | **217,725.94** |
| 101 | 2,906.43 | 2,995.62 | 2,697.69 | 2,739.68 | **11,339.42** |
| 102 | 416.59 | 453.06 | 420.18 | 487.83 | **1,777.66** |
| 103 | 3,799.19 | 3,869.71 | 3,990.59 | 4,238.72 | **15,898.21** |
| 104 | 4,113.15 | 3,828.61 | 4,174.65 | 4,366.39 | **16,482.81** |
| 105 | 10,320.35 | 9,658.81 | 10,365.11 | 10,708.81 | **41,053.08** |
| 106 | 1,180.33 | 1,179.44 | 1,304.12 | 1,274.42 | **4,938.31** |
| 107 | 1,890.28 | 1,908.74 | 1,948.12 | 2,143.54 | **7,890.68** |
| 108 | 724.14 | 800.78 | 807.11 | 803.76 | **3,135.78** |
| 109 | 7,060.43 | 6,855.44 | 7,388.97 | 7,409.97 | **28,714.80** |
| 110 | 2,895.65 | 2,738.62 | 2,852.48 | 3,071.28 | **11,558.03** |
| 111 | 3,162.94 | 2,923.16 | 2,948.65 | 3,216.31 | **12,251.06** |
| 112 | 17,077.98 | 16,637.37 | 16,810.27 | 18,327.27 | **68,852.90** |
| 113 | 6,050.67 | 5,642.94 | 6,180.85 | 6,438.35 | **24,312.81** |
| 114 | 1,357.99 | 1,455.44 | 1,450.66 | 1,442.68 | **5,706.77** |
| 115 | 8,014.65 | 7,840.83 | 8,251.67 | 8,151.58 | **32,258.74** |
| 116 | 552.20 | 563.94 | 642.58 | 629.95 | **2,388.67** |
| 117 | 22,933.38 | 22,718.21 | 24,414.28 | 23,934.02 | **93,999.88** |
| 118 | 1,137.67 | 1,170.83 | 1,155.43 | 1,123.44 | **4,587.36** |
| 119 | 3,400.05 | 3,357.56 | 4,093.79 | 3,652.10 | **14,503.50** |
| 120 | 5,886.53 | 6,297.48 | 6,279.74 | 6,350.61 | **24,814.36** |
| 121 | 4,269.23 | 4,317.71 | 4,547.87 | 4,560.26 | **17,695.07** |
| 122 | 3,183.92 | 3,381.44 | 3,254.36 | 3,407.93 | **13,227.64** |
| 123 | 2,459.68 | 2,710.87 | 2,745.51 | 2,924.38 | **10,840.45** |
| 124 | 2,155.90 | 2,326.37 | 2,405.03 | 2,375.29 | **9,262.59** |
| 125 | 6,067.50 | 5,899.74 | 6,489.75 | 6,495.90 | **24,952.88** |

| | | | | | |
|---|---|---|---|---|---|
| 126 | 1,352.14 | 1,425.36 | 1,448.09 | 1,624.88 | **5,850.47** |
| 127 | 1,373.63 | 1,340.21 | 1,456.02 | 1,413.98 | **5,583.84** |
| 128 | 3,747.98 | 3,988.43 | 4,052.93 | 4,031.09 | **15,820.43** |
| 129 | 2,758.04 | 2,705.90 | 2,698.97 | 2,826.79 | **10,989.70** |
| 130 | 6,593.53 | 6,727.58 | 6,889.08 | 7,011.59 | **27,221.78** |
| 131 | 2,017.16 | 2,218.76 | 1,978.40 | 2,206.15 | **8,420.47** |
| 132 | 3,652.56 | 3,683.14 | 3,573.75 | 4,077.00 | **14,986.46** |
| 133 | 1,177.65 | 1,227.48 | 1,179.89 | 1,245.77 | **4,830.79** |
| 134 | 2,202.97 | 2,251.57 | 2,241.37 | 2,386.41 | **9,082.33** |
| 135 | 1,010.67 | 1,019.51 | 935.32 | 1,051.80 | **4,017.29** |
| 136 | 3,041.50 | 3,204.75 | 3,241.51 | 3,358.91 | **12,846.68** |
| 137 | 1,281.21 | 1,309.11 | 1,273.85 | 1,303.87 | **5,168.05** |
| 138 | 1,644.41 | 1,678.72 | 1,728.28 | 1,839.27 | **6,890.68** |
| 139 | 1,033.17 | 1,015.80 | 1,020.49 | 1,097.01 | **4,166.47** |
| 140 | 6,818.25 | 7,042.19 | 7,062.04 | 7,588.57 | **28,511.06** |
| 141 | 2,935.71 | 2,896.20 | 3,009.47 | 3,308.97 | **12,150.36** |
| 142 | 10,038.03 | 10,225.04 | 10,306.83 | 11,083.83 | **41,653.73** |
| 143 | 1,299.45 | 1,309.22 | 1,349.12 | 1,419.78 | **5,377.56** |
| 144 | 6,046.90 | 6,214.24 | 6,367.87 | 6,673.60 | **25,302.61** |
| 145 | 5,631.06 | 5,672.19 | 5,468.42 | 5,725.75 | **22,497.42** |
| 146 | 10,674.54 | 10,972.94 | 11,003.34 | 11,786.43 | **44,437.25** |
| 147 | 1,619.28 | 1,659.22 | 1,675.94 | 1,765.37 | **6,719.81** |
| 148 | 3,809.38 | 3,942.11 | 3,778.37 | 4,006.29 | **15,536.15** |
| 149 | 939.89 | 960.20 | 957.35 | 923.21 | **3,780.65** |
| **NY - NEW YORK - Total** | **258,552.62** | **260,132.23** | **265,551.13** | **277,775.49** | **1,062,011.46** |

**STATE: OH - OHIO**

| REGISTRANT ZIP CODE 3 | 1ST QUARTER | 2ND QUARTER | 3RD QUARTER | 4TH QUARTER | TOTAL GRAMS |
|---|---|---|---|---|---|
| 430 | 17,005.37 | 17,484.60 | 18,692.83 | 18,458.94 | **71,641.74** |
| 431 | 6,528.91 | 6,700.62 | 7,106.53 | 7,120.79 | **27,456.85** |
| 432 | 14,900.84 | 15,095.46 | 15,620.93 | 16,046.09 | **61,663.32** |
| 433 | 2,421.37 | 2,380.99 | 2,500.67 | 2,411.17 | **9,714.20** |
| 434 | 2,298.72 | 2,337.81 | 2,584.93 | 2,498.05 | **9,719.50** |
| 435 | 5,871.44 | 6,094.92 | 6,290.79 | 6,385.13 | **24,642.28** |
| 436 | 9,251.93 | 9,395.60 | 10,040.22 | 9,849.25 | **38,536.99** |
| 437 | 1,727.34 | 1,722.10 | 1,713.28 | 1,847.50 | **7,010.22** |
| 438 | 250.74 | 275.70 | 250.96 | 291.28 | **1,068.68** |
| 439 | 2,302.38 | 2,279.60 | 2,257.63 | 2,371.66 | **9,211.28** |
| 440 | 16,237.36 | 16,702.98 | 17,299.12 | 17,755.62 | **67,995.08** |
| 441 | 22,021.81 | 22,856.31 | 23,738.73 | 23,976.06 | **92,592.90** |
| 442 | 8,769.25 | 8,809.44 | 9,169.85 | 9,214.72 | **35,963.26** |

| | | | | | |
|---|---|---|---|---|---|
| 443 | 4,152.60 | 4,069.79 | 4,263.25 | 4,342.78 | **16,828.42** |
| 444 | 4,863.76 | 4,955.15 | 5,157.87 | 5,277.13 | **20,253.91** |
| 445 | 5,145.19 | 5,144.07 | 5,391.74 | 5,566.31 | **21,247.31** |
| 446 | 6,006.95 | 5,813.91 | 6,008.65 | 6,202.26 | **24,031.78** |
| 447 | 5,365.21 | 5,220.68 | 5,340.67 | 5,470.72 | **21,397.29** |
| 448 | 3,582.45 | 3,551.80 | 3,799.03 | 3,904.05 | **14,837.32** |
| 449 | 965.51 | 992.39 | 1,065.05 | 1,007.90 | **4,030.85** |
| 450 | 15,722.42 | 15,684.76 | 16,540.14 | 16,885.52 | **64,832.83** |
| 451 | 5,820.66 | 5,596.79 | 6,126.50 | 5,675.50 | **23,219.46** |
| 452 | 22,295.02 | 22,654.02 | 23,361.18 | 23,579.25 | **91,889.47** |
| 453 | 5,329.27 | 5,499.18 | 5,809.63 | 5,792.51 | **22,430.58** |
| 454 | 6,686.10 | 6,691.32 | 7,139.08 | 6,986.56 | **27,503.05** |
| 455 | 1,461.10 | 1,450.63 | 1,502.46 | 1,496.36 | **5,910.55** |
| 456 | 3,915.44 | 4,021.68 | 4,068.11 | 4,222.85 | **16,228.09** |
| 457 | 2,767.98 | 2,658.77 | 2,633.64 | 2,822.79 | **10,883.18** |
| 458 | 3,783.14 | 3,848.25 | 4,103.16 | 4,145.74 | **15,880.30** |
| **OH - OHIO Total** | **207,450.24** | **209,989.32** | **219,576.63** | **221,604.47** | **858,620.65** |

**STATE: OK - OKLAHOMA**

| REGISTRANT ZIP CODE 3 | 1ST QUARTER | 2ND QUARTER | 3RD QUARTER | 4TH QUARTER | TOTAL GRAMS |
|---|---|---|---|---|---|
| 730 | 11,478.88 | 11,486.78 | 11,944.56 | 12,256.43 | **47,166.65** |
| 731 | 9,622.61 | 10,339.63 | 10,375.51 | 10,553.24 | **40,890.99** |
| 734 | 1,011.01 | 1,009.31 | 1,003.25 | 1,091.11 | **4,114.68** |
| 735 | 2,450.10 | 2,328.23 | 2,750.11 | 2,521.33 | **10,049.77** |
| 736 | 318.40 | 302.80 | 314.52 | 295.54 | **1,231.27** |
| 737 | 1,095.41 | 1,179.74 | 1,109.92 | 1,173.14 | **4,558.20** |
| 738 | 160.95 | 170.40 | 187.82 | 208.22 | **727.39** |
| 739 | 80.42 | 63.65 | 69.44 | 81.42 | **294.92** |
| 740 | 12,475.25 | 12,550.70 | 12,576.50 | 12,752.27 | **50,354.73** |
| 741 | 9,763.90 | 9,645.55 | 9,573.20 | 9,481.40 | **38,464.06** |
| 743 | 1,112.21 | 992.65 | 1,112.55 | 1,099.59 | **4,317.01** |
| 744 | 2,558.84 | 2,516.83 | 2,604.29 | 2,620.74 | **10,300.71** |
| 745 | 501.60 | 471.80 | 533.78 | 512.84 | **2,020.02** |
| 746 | 256.67 | 269.49 | 318.29 | 324.44 | **1,168.89** |
| 747 | 667.89 | 758.16 | 783.24 | 777.97 | **2,987.25** |
| 748 | 1,420.91 | 1,482.76 | 1,459.52 | 1,538.32 | **5,901.51** |
| 749 | 453.73 | 383.48 | 408.89 | 423.94 | **1,670.04** |
| **OK - OKLAHOMA Total** | **55,428.77** | **55,951.98** | **57,125.39** | **57,711.93** | **226,218.08** |

**STATE: OR - OREGON**

| REGISTRANT ZIP CODE 3 | 1ST QUARTER | 2ND QUARTER | 3RD QUARTER | 4TH QUARTER | TOTAL GRAMS |
|---|---|---|---|---|---|
| 970 | 13,618.64 | 14,321.88 | 14,175.06 | 14,747.51 | **56,863.09** |

| | | | | | |
|---|---|---|---|---|---|
| 971 | 5,264.30 | 5,544.08 | 5,560.26 | 5,879.90 | **22,248.54** |
| 972 | 20,649.32 | 21,598.02 | 22,139.22 | 23,293.02 | **87,679.59** |
| 973 | 8,787.16 | 8,865.25 | 8,848.40 | 9,385.73 | **35,886.54** |
| 974 | 7,601.99 | 7,783.73 | 7,660.06 | 8,210.36 | **31,256.13** |
| 975 | 2,525.56 | 2,692.61 | 2,826.15 | 3,000.02 | **11,044.34** |
| 976 | 484.82 | 468.54 | 494.20 | 512.39 | **1,959.95** |
| 977 | 4,176.04 | 4,418.98 | 4,537.05 | 4,594.94 | **17,727.00** |
| 978 | 1,018.83 | 1,045.12 | 1,134.21 | 1,134.11 | **4,332.28** |
| 979 | 420.87 | 428.57 | 413.00 | 392.47 | **1,654.92** |
| **OR OREGON Total** | **64,547.54** | **67,166.78** | **67,787.61** | **71,150.45** | **270,652.38** |

**STATE: PA - PENNSYLVANIA**

| REGISTRANT ZIP CODE 3 | 1ST QUARTER | 2ND QUARTER | 3RD QUARTER | 4TH QUARTER | **TOTAL GRAMS** |
|---|---|---|---|---|---|
| 150 | 6,690.38 | 6,721.02 | 6,882.82 | 6,959.80 | **27,254.02** |
| 151 | 5,678.73 | 5,471.27 | 5,978.80 | 6,027.64 | **23,156.44** |
| 152 | 11,751.59 | 11,912.23 | 12,088.95 | 12,533.08 | **48,285.84** |
| 153 | 2,628.09 | 2,567.30 | 2,668.82 | 2,637.85 | **10,502.06** |
| 154 | 1,924.83 | 1,920.24 | 1,870.38 | 1,951.05 | **7,666.50** |
| 155 | 1,043.58 | 1,023.40 | 1,127.19 | 1,082.85 | **4,277.01** |
| 156 | 4,378.20 | 4,250.62 | 4,449.80 | 4,418.16 | **17,496.77** |
| 157 | 1,343.55 | 1,209.07 | 1,333.66 | 1,545.06 | **5,431.33** |
| 158 | 1,288.97 | 1,282.67 | 1,398.36 | 1,400.23 | **5,370.23** |
| 159 | 1,437.65 | 1,400.86 | 1,475.81 | 1,394.94 | **5,709.26** |
| 160 | 3,439.32 | 3,365.38 | 3,549.84 | 3,463.60 | **13,818.14** |
| 161 | 2,593.02 | 2,622.62 | 2,770.12 | 2,737.33 | **10,723.09** |
| 162 | 973.06 | 965.27 | 1,000.61 | 958.08 | **3,897.02** |
| 163 | 1,832.91 | 1,836.82 | 1,906.87 | 1,805.22 | **7,381.81** |
| 164 | 1,000.33 | 1,010.91 | 1,059.54 | 1,150.53 | **4,221.31** |
| 165 | 4,414.54 | 4,401.49 | 4,708.87 | 4,657.85 | **18,182.77** |
| 166 | 2,789.82 | 2,691.98 | 2,739.76 | 2,780.78 | **11,002.33** |
| 167 | 617.15 | 619.55 | 665.13 | 627.71 | **2,529.54** |
| 168 | 2,952.07 | 2,621.63 | 2,752.46 | 3,144.70 | **11,470.85** |
| 169 | 513.20 | 504.32 | 541.08 | 545.12 | **2,103.71** |
| 170 | 11,934.40 | 12,098.76 | 12,369.18 | 12,608.50 | **49,010.84** |
| 171 | 3,066.45 | 3,007.87 | 3,119.13 | 3,076.23 | **12,269.68** |
| 172 | 3,497.53 | 3,565.00 | 3,598.67 | 3,764.91 | **14,426.11** |
| 173 | 5,675.61 | 6,027.66 | 6,026.34 | 6,344.02 | **24,073.64** |
| 174 | 4,160.67 | 4,166.13 | 4,082.88 | 4,171.87 | **16,581.54** |
| 175 | 3,957.67 | 3,685.09 | 3,969.09 | 3,716.01 | **15,327.85** |
| 176 | 4,227.95 | 4,365.46 | 4,380.50 | 4,420.15 | **17,394.07** |
| 177 | 2,387.23 | 2,334.03 | 2,498.52 | 2,559.92 | **9,779.70** |

| | | | | | |
|---|---|---|---|---|---|
| 178 | 2,702.11 | 2,635.77 | 2,681.01 | 2,804.81 | **10,823.69** |
| 179 | 830.36 | 863.56 | 774.41 | 867.78 | **3,336.12** |
| 180 | 5,956.39 | 5,859.67 | 6,142.25 | 6,180.48 | **24,138.80** |
| 181 | 2,825.17 | 2,835.96 | 2,913.38 | 3,112.51 | **11,687.02** |
| 182 | 914.34 | 906.14 | 891.08 | 1,033.21 | **3,744.76** |
| 183 | 1,545.03 | 1,544.44 | 1,589.50 | 1,669.36 | **6,348.33** |
| 184 | 2,383.76 | 2,347.18 | 2,401.15 | 2,522.56 | **9,654.66** |
| 185 | 3,716.36 | 3,693.33 | 3,654.62 | 3,735.51 | **14,799.82** |
| 186 | 1,851.45 | 1,903.37 | 2,021.05 | 1,913.94 | **7,689.81** |
| 187 | 18,679.26 | 20,028.45 | 18,982.27 | 23,883.65 | **81,573.64** |
| 188 | 593.04 | 607.85 | 650.50 | 707.64 | **2,559.03** |
| 189 | 7,693.23 | 7,693.62 | 7,920.18 | 7,844.79 | **31,151.81** |
| 190 | 23,308.23 | 22,876.56 | 23,591.71 | 23,597.09 | **93,373.59** |
| 191 | 20,586.52 | 20,783.10 | 21,058.12 | 21,744.06 | **84,171.80** |
| 193 | 10,270.36 | 10,411.52 | 10,794.64 | 11,094.39 | **42,570.91** |
| 194 | 11,508.35 | 11,648.30 | 11,867.95 | 12,068.24 | **47,092.84** |
| 195 | 1,927.31 | 2,040.60 | 2,041.36 | 2,123.58 | **8,132.85** |
| 196 | 3,305.89 | 3,206.58 | 3,409.92 | 3,106.03 | **13,028.43** |
| **PA - PENNSYLVANIA - Total** | **218,795.65** | **219,534.66** | **224,398.25** | **232,492.82** | **895,221.38** |

**STATE: PR - PUERTO RICO**

| REGISTRANT ZIP CODE 3 | 1ST QUARTER | 2ND QUARTER | 3RD QUARTER | 4TH QUARTER | **TOTAL GRAMS** |
|---|---|---|---|---|---|
| 006 | 2,023.44 | 2,133.85 | 1,590.98 | 2,338.56 | **8,086.84** |
| 007 | 2,352.30 | 2,269.97 | 1,571.58 | 2,294.63 | **8,488.48** |
| 009 | 5,005.39 | 5,001.48 | 4,030.40 | 5,295.02 | **19,332.29** |
| **PR - PUERTO RICO - Total** | **9,381.13** | **9,405.29** | **7,192.96** | **9,928.22** | **35,907.60** |

**STATE: RI - RHODE ISLAND**

| REGISTRANT ZIP CODE 3 | 1ST QUARTER | 2ND QUARTER | 3RD QUARTER | 4TH QUARTER | **TOTAL GRAMS** |
|---|---|---|---|---|---|
| 028 | 23,953.25 | 25,567.85 | 25,795.17 | 26,247.12 | **101,563.39** |
| 029 | 13,248.41 | 14,705.57 | 13,748.83 | 14,719.26 | **56,422.06** |
| **RI RHODE ISLAND Total** | **37,201.65** | **40,273.42** | **39,544.00** | **40,966.39** | **157,985.45** |

**STATE: SC - SOUTH CAROLINA**

| REGISTRANT ZIP CODE 3 | 1ST QUARTER | 2ND QUARTER | 3RD QUARTER | 4TH QUARTER | **TOTAL GRAMS** |
|---|---|---|---|---|---|
| 290 | 12,304.31 | 12,937.95 | 12,933.66 | 13,374.47 | **51,550.39** |
| 291 | 9,838.15 | 9,973.84 | 10,118.75 | 9,875.96 | **39,806.70** |
| 292 | 10,416.46 | 10,702.07 | 10,514.43 | 10,764.88 | **42,397.85** |
| 293 | 8,624.67 | 9,018.17 | 8,857.91 | 9,249.76 | **35,750.51** |
| 294 | 34,518.44 | 36,980.28 | 36,171.29 | 36,604.05 | **144,274.06** |
| 295 | 18,943.41 | 19,493.68 | 19,796.91 | 19,051.01 | **77,285.00** |
| 296 | 29,173.37 | 31,238.73 | 30,518.17 | 31,481.95 | **122,412.23** |
| 297 | 8,705.75 | 8,630.22 | 8,957.87 | 9,448.99 | **35,742.83** |

| | | | | | |
|---|---|---|---|---|---|
| 298 | 4,517.78 | 4,560.76 | 4,406.38 | 4,638.52 | **18,123.43** |
| 299 | 4,245.96 | 4,479.99 | 4,381.51 | 4,506.96 | **17,614.42** |
| **SC - SOUTH CAROLINA Total** | **141,288.30** | **148,015.69** | **146,656.89** | **148,996.55** | **584,957.42** |

**STATE: SD - SOUTH DAKOTA**

| REGISTRANT ZIP CODE 3 | 1ST QUARTER | 2ND QUARTER | 3RD QUARTER | 4TH QUARTER | **TOTAL GRAMS** |
|---|---|---|---|---|---|
| 570 | 2,361.06 | 2,188.96 | 2,284.17 | 2,387.22 | **9,221.41** |
| 571 | 7,107.49 | 7,930.98 | 7,205.59 | 7,713.82 | **29,957.89** |
| 572 | 697.72 | 671.27 | 706.55 | 707.67 | **2,783.21** |
| 573 | 601.00 | 607.30 | 645.46 | 627.80 | **2,481.54** |
| 574 | 718.64 | 701.90 | 748.75 | 757.95 | **2,927.24** |
| 575 | 303.57 | 254.08 | 258.99 | 275.93 | **1,092.58** |
| 576 | 52.58 | 40.09 | 50.13 | 48.68 | **191.49** |
| 577 | 2,674.55 | 2,738.87 | 2,841.57 | 2,816.56 | **11,071.54** |
| **SD SOUTH DAKOTA - Total** | **14,516.62** | **15,133.45** | **14,741.20** | **15,335.62** | **59,726.90** |

**STATE: TN - TENNESSEE**

| REGISTRANT ZIP CODE 3 | 1ST QUARTER | 2ND QUARTER | 3RD QUARTER | 4TH QUARTER | **TOTAL GRAMS** |
|---|---|---|---|---|---|
| 370 | 21,282.65 | 21,752.03 | 22,550.17 | 22,420.82 | **88,005.67** |
| 371 | 13,635.12 | 13,974.33 | 14,020.83 | 14,444.26 | **56,074.54** |
| 372 | 15,272.70 | 15,655.71 | 15,685.94 | 16,161.03 | **62,775.39** |
| 373 | 12,145.43 | 13,029.18 | 12,828.34 | 13,383.29 | **51,386.24** |
| 374 | 8,856.40 | 9,663.85 | 9,194.65 | 9,222.71 | **36,937.61** |
| 376 | 4,104.31 | 4,406.47 | 4,347.55 | 4,500.34 | **17,358.68** |
| 377 | 4,514.94 | 4,805.89 | 4,871.64 | 4,935.21 | **19,127.67** |
| 378 | 7,171.25 | 7,515.67 | 7,477.40 | 7,607.62 | **29,771.94** |
| 379 | 11,833.71 | 12,320.05 | 12,308.12 | 12,799.65 | **49,261.54** |
| 380 | 10,867.49 | 10,703.52 | 10,911.53 | 10,945.80 | **43,428.34** |
| 381 | 12,797.69 | 13,112.95 | 13,439.46 | 13,227.96 | **52,578.05** |
| 382 | 3,315.24 | 3,056.99 | 3,186.48 | 3,131.18 | **12,689.90** |
| 383 | 4,927.38 | 4,616.13 | 4,640.93 | 4,909.03 | **19,093.48** |
| 384 | 3,371.21 | 3,392.47 | 3,497.00 | 3,567.92 | **13,828.60** |
| 385 | 2,258.38 | 2,163.59 | 2,285.59 | 2,258.16 | **8,965.72** |
| **TN TENNESSEE Total** | **136,353.89** | **140,168.83** | **141,245.65** | **143,514.98** | **561,283.36** |

**STATE: TX - TEXAS**

| REGISTRANT ZIP CODE 3 | 1ST QUARTER | 2ND QUARTER | 3RD QUARTER | 4TH QUARTER | **TOTAL GRAMS** |
|---|---|---|---|---|---|
| 750 | 57,517.36 | 59,323.28 | 60,226.08 | 62,953.44 | **240,020.17** |
| 751 | 10,358.18 | 10,747.21 | 10,869.04 | 11,402.31 | **43,376.74** |
| 752 | 29,194.82 | 30,445.87 | 30,595.92 | 32,545.37 | **122,781.97** |
| 753 | 68.17 | 53.01 | 56.36 | 73.51 | **251.05** |
| 754 | 4,820.50 | 4,848.45 | 4,953.98 | 5,198.69 | **19,821.63** |
| 755 | 1,276.30 | 1,295.82 | 1,368.60 | 1,453.33 | **5,394.06** |

| | | | | | |
|---|---|---|---|---|---|
| 756 | 4,372.44 | 4,168.73 | 4,276.80 | 4,667.77 | **17,485.73** |
| 757 | 5,661.92 | 5,565.56 | 5,934.92 | 6,016.14 | **23,178.53** |
| 758 | 838.20 | 813.48 | 852.17 | 820.53 | **3,324.37** |
| 759 | 2,623.47 | 2,648.29 | 2,871.28 | 3,003.56 | **11,146.60** |
| 760 | 26,272.54 | 26,813.97 | 26,694.97 | 28,210.71 | **107,992.19** |
| 761 | 16,246.37 | 16,562.37 | 16,559.94 | 17,598.83 | **66,967.50** |
| 762 | 13,840.87 | 13,786.47 | 14,296.41 | 14,945.80 | **56,869.55** |
| 763 | 3,621.27 | 3,770.65 | 3,638.13 | 3,938.90 | **14,968.94** |
| 764 | 1,668.23 | 1,422.41 | 1,435.08 | 1,577.71 | **6,103.43** |
| 765 | 6,244.11 | 6,325.81 | 6,450.26 | 6,384.04 | **25,404.22** |
| 766 | 1,219.37 | 1,163.19 | 1,215.50 | 1,220.76 | **4,818.82** |
| 767 | 3,936.26 | 3,819.29 | 4,003.93 | 4,093.66 | **15,853.14** |
| 768 | 853.29 | 849.49 | 840.68 | 900.38 | **3,443.84** |
| 769 | 1,908.37 | 1,997.22 | 2,052.29 | 2,036.98 | **7,994.87** |
| 770 | 43,310.36 | 43,550.32 | 42,939.89 | 46,924.95 | **176,725.52** |
| 772 | 28.78 | 19.08 | 20.34 | 27.59 | **95.79** |
| 773 | 32,113.76 | 32,640.60 | 33,265.73 | 34,817.75 | **132,837.84** |
| 774 | 23,699.68 | 23,655.39 | 24,919.94 | 25,890.04 | **98,165.05** |
| 775 | 25,468.91 | 26,810.85 | 26,388.30 | 28,038.78 | **106,706.83** |
| 776 | 5,302.30 | 5,438.31 | 5,389.46 | 5,837.45 | **21,967.52** |
| 777 | 2,659.46 | 2,906.47 | 2,774.40 | 2,885.42 | **11,225.75** |
| 778 | 6,326.47 | 6,231.48 | 6,587.08 | 6,781.71 | **25,926.75** |
| 779 | 2,375.14 | 2,463.53 | 2,467.54 | 2,651.39 | **9,957.60** |
| 780 | 5,555.70 | 5,707.25 | 6,079.85 | 6,144.98 | **23,487.77** |
| 781 | 7,006.19 | 7,175.55 | 7,315.82 | 7,663.29 | **29,160.84** |
| 782 | 30,120.75 | 29,593.49 | 30,742.90 | 31,534.90 | **121,992.05** |
| 783 | 2,232.85 | 2,115.78 | 2,226.85 | 2,388.59 | **8,964.07** |
| 784 | 5,112.94 | 5,265.37 | 5,215.11 | 5,453.50 | **21,046.92** |
| 785 | 3,759.15 | 3,592.38 | 3,745.68 | 3,909.46 | **15,006.67** |
| 786 | 20,187.23 | 20,463.92 | 21,300.77 | 21,664.57 | **83,616.49** |
| 787 | 32,582.89 | 34,213.91 | 34,599.71 | 35,295.42 | **136,691.92** |
| 788 | 743.20 | 689.46 | 693.51 | 728.62 | **2,854.78** |
| 789 | 833.29 | 832.82 | 896.66 | 896.46 | **3,459.22** |
| 790 | 939.79 | 905.66 | 918.34 | 1,029.03 | **3,792.82** |
| 791 | 2,208.32 | 2,360.27 | 2,349.34 | 2,353.23 | **9,271.16** |
| 792 | 82.12 | 105.86 | 108.89 | 97.54 | **394.41** |
| 793 | 479.43 | 553.23 | 463.70 | 558.76 | **2,055.11** |
| 794 | 4,965.15 | 4,847.59 | 4,813.93 | 5,053.07 | **19,679.75** |
| 795 | 542.90 | 533.54 | 535.60 | 570.92 | **2,182.95** |
| 796 | 2,513.77 | 2,547.06 | 2,418.02 | 2,662.55 | **10,141.41** |

| | | | | | |
|---|---|---|---|---|---|
| 797 | 6,916.40 | 7,188.20 | 7,160.80 | 7,192.60 | **28,458.00** |
| 798 | 219.00 | 246.36 | 268.62 | 234.30 | **968.29** |
| 799 | 3,657.02 | 3,733.25 | 3,843.71 | 3,586.48 | **14,820.47** |
| **TX - TEXAS Total** | **464,484.94** | **472,807.55** | **479,642.83** | **501,915.77** | **1,918,851.09** |

**STATE: UT - UTAH**

| REGISTRANT ZIP CODE 3 | 1ST QUARTER | 2ND QUARTER | 3RD QUARTER | 4TH QUARTER | TOTAL GRAMS |
|---|---|---|---|---|---|
| 840 | 46,675.10 | 48,890.29 | 50,219.05 | 52,094.24 | **197,878.67** |
| 841 | 17,912.77 | 18,407.95 | 18,830.80 | 20,064.22 | **75,215.73** |
| 843 | 4,897.65 | 4,788.67 | 4,934.31 | 5,158.91 | **19,779.54** |
| 844 | 4,672.17 | 4,775.69 | 5,041.66 | 5,218.26 | **19,707.78** |
| 845 | 667.75 | 727.06 | 793.93 | 736.81 | **2,925.55** |
| 846 | 9,810.36 | 9,688.73 | 10,117.26 | 10,550.39 | **40,166.74** |
| 847 | 7,259.35 | 7,229.01 | 7,239.74 | 7,325.54 | **29,053.63** |
| **UT - UTAH - Total** | **91,895.15** | **94,507.39** | **97,176.75** | **101,148.36** | **384,727.64** |

**STATE: VA - VIRGINIA**

| REGISTRANT ZIP CODE 3 | 1ST QUARTER | 2ND QUARTER | 3RD QUARTER | 4TH QUARTER | TOTAL GRAMS |
|---|---|---|---|---|---|
| 201 | 19,261.39 | 19,826.51 | 20,233.46 | 20,512.74 | **79,834.10** |
| 220 | 9,812.62 | 10,068.39 | 11,031.53 | 10,218.43 | **41,130.96** |
| 221 | 7,145.86 | 7,611.23 | 7,876.68 | 7,691.04 | **30,324.82** |
| 222 | 4,779.26 | 5,300.72 | 5,179.99 | 5,283.35 | **20,543.33** |
| 223 | 4,405.48 | 4,508.32 | 4,765.74 | 4,848.91 | **18,528.45** |
| 224 | 4,754.82 | 5,057.53 | 5,010.04 | 5,286.08 | **20,108.47** |
| 225 | 2,267.30 | 2,263.14 | 2,252.37 | 2,396.41 | **9,179.21** |
| 226 | 3,476.03 | 3,731.78 | 3,659.87 | 3,784.63 | **14,652.30** |
| 227 | 1,148.44 | 1,194.36 | 1,119.42 | 1,136.66 | **4,598.89** |
| 228 | 2,950.32 | 3,106.56 | 3,021.55 | 3,218.17 | **12,296.60** |
| 229 | 6,319.64 | 6,757.49 | 6,813.23 | 6,907.39 | **26,797.75** |
| 230 | 4,990.84 | 5,411.46 | 5,088.25 | 5,358.92 | **20,849.47** |
| 231 | 12,177.07 | 13,397.76 | 13,293.15 | 13,957.27 | **52,825.24** |
| 232 | 16,576.73 | 17,525.61 | 17,138.53 | 17,699.94 | **68,940.81** |
| 233 | 9,625.67 | 10,326.47 | 9,881.37 | 10,450.41 | **40,283.92** |
| 234 | 24,444.97 | 26,777.33 | 26,063.71 | 25,996.45 | **103,282.46** |
| 235 | 5,758.82 | 6,212.29 | 6,279.62 | 6,091.51 | **24,342.24** |
| 236 | 10,008.76 | 10,197.27 | 10,238.79 | 10,347.27 | **40,792.09** |
| 237 | 4,205.65 | 4,083.93 | 4,389.99 | 3,904.73 | **16,584.30** |
| 238 | 7,257.73 | 7,446.60 | 7,096.99 | 6,898.65 | **28,699.97** |
| 239 | 1,071.47 | 925.42 | 1,026.60 | 939.88 | **3,963.36** |
| 240 | 5,637.55 | 5,857.53 | 5,735.12 | 5,701.96 | **22,932.17** |
| 241 | 2,735.44 | 2,869.05 | 2,848.11 | 3,013.47 | **11,466.06** |
| 242 | 1,329.07 | 1,369.74 | 1,317.80 | 1,309.95 | **5,326.56** |

| | | | | | |
|---|---|---|---|---|---|
| 243 | 982.57 | 1,027.78 | 1,049.43 | 969.92 | **4,029.70** |
| 244 | 3,455.83 | 3,799.87 | 3,688.58 | 3,773.15 | **14,717.43** |
| 245 | 5,676.85 | 5,636.53 | 5,574.88 | 5,326.57 | **22,214.85** |
| 246 | 448.65 | 441.37 | 477.56 | 439.67 | **1,807.26** |
| **VA VIRGINIA Total** | **182,704.81** | **192,732.07** | **192,152.37** | **193,463.51** | **761,052.77** |

**STATE: VI - VIRGIN ISLANDS**

| REGISTRANT ZIP CODE 3 | 1ST QUARTER | 2ND QUARTER | 3RD QUARTER | 4TH QUARTER | TOTAL GRAMS |
|---|---|---|---|---|---|
| 008 | 335.93 | 426.39 | 317.39 | 380.70 | **1,460.40** |
| **VI - VIRGIN ISLANDS Total** | **335.93** | **426.39** | **317.39** | **380.70** | **1,460.40** |

**STATE: VT - VERMONT**

| REGISTRANT ZIP CODE 3 | 1ST QUARTER | 2ND QUARTER | 3RD QUARTER | 4TH QUARTER | TOTAL GRAMS |
|---|---|---|---|---|---|
| 050 | 644.51 | 623.46 | 619.52 | 622.54 | **2,510.03** |
| 051 | 534.91 | 506.24 | 528.34 | 493.84 | **2,063.34** |
| 052 | 738.02 | 813.40 | 857.45 | 859.38 | **3,268.26** |
| 053 | 1,560.75 | 1,579.42 | 1,663.34 | 1,559.80 | **6,363.31** |
| 054 | 3,463.58 | 3,672.19 | 3,658.35 | 3,842.89 | **14,637.01** |
| 056 | 1,578.94 | 1,742.80 | 1,696.86 | 1,657.23 | **6,675.84** |
| 057 | 844.89 | 924.19 | 905.98 | 907.12 | **3,582.19** |
| 058 | 728.50 | 784.31 | 840.27 | 825.45 | **3,178.54** |
| **VT VERMONT Total** | **10,094.12** | **10,646.01** | **10,770.11** | **10,768.26** | **42,278.50** |

**STATE: WA - WASHINGTON**

| REGISTRANT ZIP CODE 3 | 1ST QUARTER | 2ND QUARTER | 3RD QUARTER | 4TH QUARTER | TOTAL GRAMS |
|---|---|---|---|---|---|
| 980 | 28,944.64 | 30,556.17 | 30,412.11 | 31,358.27 | **121,271.18** |
| 981 | 19,116.32 | 20,330.02 | 20,539.42 | 20,923.73 | **80,909.50** |
| 982 | 17,192.06 | 17,932.04 | 17,637.09 | 18,318.62 | **71,079.81** |
| 983 | 11,388.11 | 11,814.68 | 11,634.13 | 12,191.62 | **47,028.54** |
| 984 | 7,087.44 | 7,660.27 | 7,435.68 | 7,712.26 | **29,895.65** |
| 985 | 5,985.83 | 6,148.23 | 6,053.03 | 6,650.40 | **24,837.50** |
| 986 | 8,621.44 | 8,954.34 | 8,845.11 | 9,337.58 | **35,758.46** |
| 988 | 2,109.22 | 2,050.40 | 2,125.46 | 2,159.67 | **8,444.75** |
| 989 | 2,621.81 | 2,614.84 | 2,621.16 | 2,762.34 | **10,620.14** |
| 990 | 1,132.89 | 1,121.10 | 1,150.61 | 1,200.31 | **4,604.91** |
| 991 | 1,246.32 | 1,038.52 | 1,156.93 | 1,197.83 | **4,639.60** |
| 992 | 6,404.53 | 6,668.51 | 6,891.56 | 7,166.31 | **27,130.91** |
| 993 | 5,532.99 | 6,021.02 | 5,693.68 | 6,023.59 | **23,271.29** |
| 994 | 230.72 | 237.89 | 242.55 | 237.26 | **948.42** |
| **WA WASHINGTON Total** | **117,614.32** | **123,148.03** | **122,438.52** | **127,239.79** | **490,440.66** |

**STATE: WI - WISCONSIN**

| REGISTRANT ZIP CODE 3 | 1ST QUARTER | 2ND QUARTER | 3RD QUARTER | 4TH QUARTER | TOTAL GRAMS |
|---|---|---|---|---|---|
| 530 | 14,366.94 | 14,889.19 | 14,974.11 | 14,973.93 | **59,204.16** |

| | | | | | |
|---|---|---|---|---|---|
| 531 | 15,117.22 | 15,201.56 | 15,458.77 | 15,440.61 | **61,218.15** |
| 532 | 13,460.15 | 13,718.51 | 13,488.04 | 13,579.14 | **54,245.83** |
| 534 | 4,144.38 | 4,344.58 | 4,116.46 | 4,140.83 | **16,746.25** |
| 535 | 10,277.88 | 10,367.35 | 10,315.30 | 10,476.86 | **41,437.38** |
| 537 | 7,978.24 | 8,183.70 | 8,194.06 | 8,374.01 | **32,730.00** |
| 538 | 1,119.96 | 1,105.91 | 1,040.65 | 1,166.68 | **4,433.20** |
| 539 | 3,397.27 | 3,647.51 | 3,612.50 | 3,585.63 | **14,242.91** |
| 540 | 2,317.99 | 2,439.00 | 2,469.75 | 2,537.87 | **9,764.61** |
| 541 | 6,443.66 | 6,580.97 | 6,991.26 | 6,768.82 | **26,784.72** |
| 542 | 2,530.63 | 2,579.21 | 2,598.75 | 2,560.01 | **10,268.61** |
| 543 | 8,330.32 | 8,270.54 | 8,289.21 | 8,245.12 | **33,135.18** |
| 544 | 8,064.65 | 8,286.83 | 8,391.70 | 8,335.05 | **33,078.23** |
| 545 | 2,194.48 | 2,166.75 | 2,281.74 | 2,114.92 | **8,757.89** |
| 546 | 4,069.16 | 4,153.65 | 4,213.06 | 4,065.48 | **16,501.35** |
| 547 | 4,165.80 | 4,203.35 | 4,175.73 | 4,382.10 | **16,926.98** |
| 548 | 2,316.07 | 2,255.87 | 2,367.30 | 2,429.20 | **9,368.45** |
| 549 | 14,749.64 | 14,928.99 | 15,307.86 | 14,947.99 | **59,934.48** |
| **WI - WISCONSIN Total** | **125,044.43** | **127,323.46** | **128,286.26** | **128,124.25** | **508,778.40** |

**STATE: WV - WEST VIRGINIA**

| REGISTRANT ZIP CODE 3 | 1ST QUARTER | 2ND QUARTER | 3RD QUARTER | 4TH QUARTER | TOTAL GRAMS |
|---|---|---|---|---|---|
| 247 | 305.76 | 307.35 | 335.96 | 345.53 | **1,294.60** |
| 248 | 240.07 | 227.97 | 202.19 | 188.31 | **858.54** |
| 249 | 374.54 | 406.00 | 456.94 | 474.42 | **1,711.91** |
| 250 | 904.60 | 934.56 | 953.75 | 947.85 | **3,740.76** |
| 251 | 744.98 | 787.48 | 779.35 | 709.41 | **3,021.23** |
| 252 | 396.29 | 390.84 | 360.84 | 401.43 | **1,549.39** |
| 253 | 3,421.05 | 3,190.05 | 3,263.46 | 3,314.14 | **13,188.70** |
| 254 | 2,732.96 | 2,786.69 | 2,991.99 | 2,870.47 | **11,382.11** |
| 255 | 2,457.00 | 2,480.14 | 2,413.77 | 2,441.87 | **9,792.77** |
| 256 | 326.05 | 301.38 | 286.42 | 325.83 | **1,239.68** |
| 257 | 1,855.38 | 1,867.64 | 2,006.54 | 2,006.55 | **7,736.11** |
| 258 | 1,134.35 | 1,078.28 | 1,106.04 | 1,204.88 | **4,523.55** |
| 259 | 234.97 | 253.39 | 243.98 | 201.88 | **934.21** |
| 260 | 1,534.52 | 1,421.59 | 1,553.65 | 1,458.94 | **5,968.71** |
| 261 | 2,302.67 | 2,091.24 | 1,925.35 | 2,041.44 | **8,360.70** |
| 262 | 376.12 | 310.80 | 332.53 | 334.78 | **1,354.23** |
| 263 | 1,052.91 | 1,042.39 | 1,081.44 | 1,179.80 | **4,356.55** |
| 264 | 217.09 | 207.06 | 208.95 | 242.66 | **875.76** |
| 265 | 2,874.24 | 2,770.25 | 2,843.29 | 2,876.45 | **11,364.22** |
| 266 | 138.27 | 132.50 | 156.72 | 152.20 | **579.69** |

| | | | | | |
|---|---|---|---|---|---|
| 267 | 343.87 | 365.32 | 345.51 | 367.40 | **1,422.11** |
| 268 | 265.08 | 225.50 | 241.17 | 257.51 | **989.25** |
| **WV WEST VIRGINIA Total** | **24,232.75** | **23,578.42** | **24,089.83** | **24,343.76** | **96,244.76** |

**STATE: WY - WYOMING**

| REGISTRANT ZIP CODE 3 | 1ST QUARTER | 2ND QUARTER | 3RD QUARTER | 4TH QUARTER | TOTAL GRAMS |
|---|---|---|---|---|---|
| 820 | 1,481.27 | 1,509.59 | 1,507.39 | 1,555.97 | **6,054.23** |
| 822 | 86.80 | 102.25 | 69.80 | 86.73 | **345.58** |
| 823 | 126.53 | 119.89 | 137.52 | 159.12 | **543.07** |
| 824 | 862.76 | 881.94 | 966.76 | 886.78 | **3,598.23** |
| 825 | 323.08 | 331.75 | 313.78 | 336.11 | **1,304.72** |
| 826 | 1,083.70 | 1,136.72 | 1,105.84 | 1,162.91 | **4,489.18** |
| 827 | 562.39 | 601.19 | 599.05 | 626.07 | **2,388.69** |
| 828 | 582.98 | 518.17 | 488.94 | 452.88 | **2,042.96** |
| 829 | 1,119.51 | 1,202.60 | 1,363.43 | 1,351.33 | **5,036.87** |
| 830 | 632.01 | 594.11 | 695.91 | 657.59 | **2,579.62** |
| 831 | 125.44 | 172.76 | 200.87 | 158.65 | **657.72** |
| **WY - WYOMING Total** | **6,986.48** | **7,170.96** | **7,449.29** | **7,434.14** | **29,040.88** |

**DRUG: 1105 - METHAMPHETAMINE**

**STATE: AK - ALASKA**

| REGISTRANT ZIP CODE 3 | 1ST QUARTER | 2ND QUARTER | 3RD QUARTER | 4TH QUARTER | TOTAL GRAMS |
|---|---|---|---|---|---|
| 996 | 0.40 | 0 | 0 | 0 | **0.40** |
| 997 | 0 | 0 | 2.01 | 3.21 | **5.22** |
| **AK ALASKA - Total** | **0.40** | **0.00** | **2.01** | **3.21** | **5.62** |

**STATE: AL - ALABAMA**

| REGISTRANT ZIP CODE 3 | 1ST QUARTER | 2ND QUARTER | 3RD QUARTER | 4TH QUARTER | TOTAL GRAMS |
|---|---|---|---|---|---|
| 350 | 3.62 | 4.02 | 3.62 | 3.21 | **14.46** |
| 351 | 2.41 | 1.21 | 4.02 | 0 | **7.63** |
| 352 | 3.62 | 3.62 | 5.62 | 8.44 | **21.29** |
| 354 | 1.21 | 1.61 | 1.21 | 0.80 | **4.82** |
| 355 | 3.62 | 2.41 | 3.21 | 3.21 | **12.45** |
| 356 | 6.43 | 6.83 | 8.03 | 7.23 | **28.52** |
| 357 | 0 | 0.40 | 0.40 | 0.80 | **1.61** |
| 359 | 0 | 2.41 | 0.80 | 0.40 | **3.62** |
| 362 | 2.81 | 4.82 | 0.80 | 4.82 | **13.26** |
| 363 | 0.80 | 0 | 0 | 0.40 | **1.21** |
| 365 | 1.61 | 1.21 | 2.81 | 0 | **5.63** |
| 366 | 2.41 | 4.02 | 0.40 | 1.21 | **8.04** |
| **AL ALABAMA - Total** | **28.53** | **32.54** | **30.93** | **30.53** | **122.54** |

**STATE: AR - ARKANSAS**

| REGISTRANT ZIP CODE 3 | 1ST QUARTER | 2ND QUARTER | 3RD QUARTER | 4TH QUARTER | TOTAL GRAMS |
|---|---|---|---|---|---|

| | | | | | |
|---|---|---|---|---|---|
| 717 | 0.80 | 0 | 0 | 0 | **0.80** |
| 720 | 2.01 | 0.80 | 0 | 0 | **2.81** |
| 722 | 0.80 | 0.80 | 1.21 | 0.80 | **3.62** |
| 727 | 0 | 0 | 0.40 | 0 | **0.40** |
| 729 | 0 | 1.21 | 0 | 0 | **1.21** |
| **AR ARKANSAS - Total** | **3.62** | **2.81** | **1.61** | **0.80** | **8.84** |

**STATE: AZ - ARIZONA**

| REGISTRANT ZIP CODE 3 | 1ST QUARTER | 2ND QUARTER | 3RD QUARTER | 4TH QUARTER | **TOTAL GRAMS** |
|---|---|---|---|---|---|
| 850 | 5.63 | 2.81 | 5.22 | 6.03 | **19.69** |
| 851 | 0 | 0.80 | 2.81 | 2.81 | **6.43** |
| 852 | 14.06 | 10.05 | 10.85 | 8.44 | **43.39** |
| 853 | 4.02 | 2.01 | 0.40 | 0 | **6.43** |
| 855 | 1.21 | 0 | 0 | 0 | **1.21** |
| 856 | 0 | 0 | 0 | 0.40 | **0.40** |
| 857 | 0.80 | 0.40 | 0.40 | 0 | **1.61** |
| 863 | 0.40 | 0 | 0.40 | 0 | **0.80** |
| 864 | 0 | 0 | 1.61 | 0.80 | **2.41** |
| **AZ ARIZONA - Total** | **26.12** | **16.07** | **21.70** | **18.48** | **82.37** |

**STATE: CA - CALIFORNIA**

| REGISTRANT ZIP CODE 3 | 1ST QUARTER | 2ND QUARTER | 3RD QUARTER | 4TH QUARTER | **TOTAL GRAMS** |
|---|---|---|---|---|---|
| 900 | 24.91 | 9.24 | 10.85 | 10.44 | **55.44** |
| 902 | 2.81 | 10.04 | 6.03 | 7.23 | **26.11** |
| 904 | 0.80 | 2.01 | 1.21 | 3.21 | **7.23** |
| 906 | 0 | 0.40 | 0 | 0 | **0.40** |
| 907 | 0 | 0 | 0 | 0.40 | **0.40** |
| 908 | 0.80 | 2.81 | 1.61 | 3.62 | **8.84** |
| 911 | 1.21 | 0.80 | 1.21 | 3.62 | **6.83** |
| 912 | 4.42 | 5.22 | 6.43 | 5.62 | **21.69** |
| 913 | 21.70 | 27.72 | 15.27 | 15.27 | **79.95** |
| 914 | 4.42 | 4.82 | 5.22 | 2.41 | **16.87** |
| 915 | 0 | 3.62 | 5.63 | 7.23 | **16.48** |
| 917 | 18.88 | 7.63 | 8.04 | 8.03 | **42.58** |
| 919 | 2.81 | 4.02 | 2.81 | 2.81 | **12.45** |
| 920 | 35.35 | 24.50 | 21.69 | 31.33 | **112.88** |
| 921 | 9.64 | 10.85 | 11.25 | 11.25 | **42.99** |
| 922 | 1.21 | 2.01 | 6.03 | 6.43 | **15.67** |
| 923 | 4.02 | 4.42 | 2.41 | 6.83 | **17.68** |
| 925 | 7.23 | 23.30 | 22.50 | 25.71 | **78.74** |
| 926 | 16.07 | 14.46 | 19.28 | 13.66 | **63.48** |
| 927 | 2.81 | 6.43 | 3.21 | 4.02 | **16.47** |

| | | | | | |
|---|---|---|---|---|---|
| 928 | 10.85 | 13.66 | 12.86 | 16.07 | **53.43** |
| 930 | 0 | 0 | 0.80 | 0 | **0.80** |
| 931 | 2.01 | 0.40 | 0 | 0.40 | **2.81** |
| 934 | 0.40 | 0 | 0.80 | 0.40 | **1.61** |
| 935 | 3.21 | 0 | 2.01 | 2.41 | **7.63** |
| 937 | 2.01 | 1.61 | 0.40 | 2.01 | **6.03** |
| 939 | 0.80 | 0.80 | 0 | 0.80 | **2.41** |
| 940 | 7.23 | 6.83 | 3.21 | 4.42 | **21.70** |
| 941 | 5.22 | 7.23 | 10.44 | 13.26 | **36.16** |
| 943 | 0 | 0.40 | 0.40 | 0 | **0.80** |
| 945 | 4.42 | 9.24 | 8.84 | 9.24 | **31.74** |
| 946 | 2.81 | 2.81 | 2.01 | 1.21 | **8.84** |
| 947 | 0 | 0.80 | 0.40 | 0 | **1.21** |
| 949 | 2.81 | 2.01 | 1.21 | 1.61 | **7.63** |
| 950 | 8.44 | 5.62 | 5.22 | 8.44 | **27.72** |
| 951 | 15.67 | 3.62 | 5.62 | 4.82 | **29.73** |
| 953 | 2.01 | 4.02 | 3.62 | 2.41 | **12.05** |
| 955 | 0.80 | 1.21 | 0.40 | 1.21 | **3.62** |
| 956 | 7.23 | 7.23 | 8.03 | 6.43 | **28.93** |
| 957 | 3.21 | 2.81 | 5.22 | 2.81 | **14.06** |
| 958 | 0.80 | 0 | 1.61 | 0 | **2.41** |
| 959 | 1.61 | 1.61 | 0.80 | 0.80 | **4.82** |
| 960 | 0 | 0.80 | 1.21 | 3.62 | **5.62** |
| 961 | 0 | 0 | 0.80 | 0.80 | **1.61** |
| **CA - CALIFORNIA - Total** | **240.65** | **237.04** | **226.58** | **252.29** | **956.56** |

**STATE: CO - COLORADO**

| REGISTRANT ZIP CODE 3 | 1ST QUARTER | 2ND QUARTER | 3RD QUARTER | 4TH QUARTER | TOTAL GRAMS |
|---|---|---|---|---|---|
| 800 | 7.63 | 9.64 | 9.64 | 4.82 | **31.74** |
| 801 | 15.27 | 11.65 | 14.46 | 8.03 | **49.41** |
| 802 | 16.88 | 12.46 | 15.67 | 15.67 | **60.67** |
| 803 | 2.81 | 2.81 | 0.40 | 6.03 | **12.05** |
| 804 | 0.80 | 2.41 | 1.61 | 2.01 | **6.83** |
| 805 | 0 | 0.80 | 0.80 | 1.21 | **2.81** |
| 806 | 1.21 | 0 | 0 | 0 | **1.21** |
| 808 | 0 | 1.61 | 3.21 | 2.41 | **7.23** |
| 809 | 3.21 | 1.21 | 3.62 | 3.21 | **11.25** |
| 810 | 2.01 | 4.02 | 2.01 | 0.40 | **8.44** |
| 816 | 0 | 0 | 0.40 | 0 | **0.40** |
| **CO - COLORADO - Total** | **49.82** | **46.61** | **51.82** | **43.79** | **192.04** |

**STATE: CT - CONNECTICUT**

| REGISTRANT ZIP CODE 3 | 1ST QUARTER | 2ND QUARTER | 3RD QUARTER | 4TH QUARTER | TOTAL GRAMS |
|---|---|---|---|---|---|
| 060 | 7.64 | 8.44 | 9.64 | 7.63 | **33.35** |
| 061 | 1.61 | 1.21 | 0 | 0 | **2.81** |
| 064 | 1.61 | 0.80 | 1.21 | 2.01 | **5.62** |
| 065 | 3.62 | 3.21 | 2.41 | 2.41 | **11.65** |
| 066 | 0.80 | 0.40 | 0 | 0 | **1.21** |
| 067 | 1.61 | 0.40 | 1.21 | 0.80 | **4.02** |
| 068 | 6.03 | 8.04 | 5.22 | 6.43 | **25.71** |
| 069 | 2.01 | 0.40 | 0.40 | 1.61 | **4.42** |
| **CT CONNECTICUT Total** | **24.91** | **22.90** | **20.09** | **20.89** | **88.79** |

**STATE: DC - DISTRICT OF COLUMBIA**

| REGISTRANT ZIP CODE 3 | 1ST QUARTER | 2ND QUARTER | 3RD QUARTER | 4TH QUARTER | TOTAL GRAMS |
|---|---|---|---|---|---|
| 200 | 4.82 | 4.42 | 4.82 | 6.03 | **20.09** |
| **DC - DISTRICT OF COLUMBIA** | **4.82** | **4.42** | **4.82** | **6.03** | **20.09** |

**STATE: DE - DELAWARE**

| REGISTRANT ZIP CODE 3 | 1ST QUARTER | 2ND QUARTER | 3RD QUARTER | 4TH QUARTER | TOTAL GRAMS |
|---|---|---|---|---|---|
| 197 | 0 | 0.40 | 0.40 | 0.80 | **1.61** |
| 198 | 3.62 | 0.80 | 3.21 | 2.41 | **10.04** |
| 199 | 0.80 | 1.61 | 1.61 | 0.80 | **4.82** |
| **DE DELAWARE Total** | **4.42** | **2.81** | **5.22** | **4.02** | **16.47** |

**STATE: FL - FLORIDA**

| REGISTRANT ZIP CODE 3 | 1ST QUARTER | 2ND QUARTER | 3RD QUARTER | 4TH QUARTER | TOTAL GRAMS |
|---|---|---|---|---|---|
| 320 | 2.01 | 2.41 | 0.80 | 1.21 | **6.43** |
| 321 | 0.80 | 0 | 1.61 | 0.40 | **2.81** |
| 322 | 4.82 | 2.01 | 2.81 | 3.21 | **12.86** |
| 323 | 0 | 0 | 0 | 1.21 | **1.21** |
| 324 | 1.61 | 0.80 | 1.61 | 1.61 | **5.63** |
| 325 | 4.02 | 7.23 | 0.80 | 2.01 | **14.06** |
| 326 | 4.02 | 1.21 | 2.41 | 2.81 | **10.45** |
| 327 | 8.84 | 4.82 | 8.84 | 11.25 | **33.74** |
| 328 | 2.81 | 2.01 | 2.81 | 4.82 | **12.45** |
| 329 | 5.63 | 5.62 | 6.03 | 2.81 | **20.09** |
| 330 | 1.61 | 3.21 | 3.62 | 6.03 | **14.47** |
| 331 | 2.41 | 3.21 | 3.21 | 2.41 | **11.25** |
| 333 | 0.80 | 0 | 1.21 | 0.40 | **2.41** |
| 334 | 2.01 | 2.41 | 3.62 | 0.80 | **8.84** |
| 335 | 0 | 0.80 | 0.80 | 1.61 | **3.21** |
| 336 | 2.01 | 1.21 | 3.21 | 1.21 | **7.63** |
| 337 | 1.21 | 0.40 | 0.80 | 0.40 | **2.81** |
| 338 | 1.21 | 1.21 | 0 | 0 | **2.41** |

| | | | | | |
|---|---|---|---|---|---|
| 339 | 4.02 | 3.62 | 4.82 | 3.62 | **16.07** |
| 341 | 0.80 | 2.01 | 1.61 | 0 | **4.42** |
| 342 | 2.41 | 1.61 | 2.41 | 4.82 | **11.25** |
| 346 | 3.21 | 0.80 | 1.21 | 1.61 | **6.83** |
| 347 | 0 | 0 | 0 | 0.40 | **0.40** |
| **FL FLORIDA - Total** | **56.25** | **46.61** | **54.24** | **54.64** | **211.74** |

**STATE: GA - GEORGIA**

| REGISTRANT ZIP CODE 3 | 1ST QUARTER | 2ND QUARTER | 3RD QUARTER | 4TH QUARTER | **TOTAL GRAMS** |
|---|---|---|---|---|---|
| 300 | 4.42 | 6.83 | 7.63 | 5.63 | **24.51** |
| 301 | 5.62 | 3.62 | 3.62 | 8.03 | **20.89** |
| 302 | 4.82 | 3.21 | 2.41 | 2.41 | **12.85** |
| 303 | 3.62 | 6.43 | 2.41 | 6.83 | **19.29** |
| 304 | 1.61 | 1.21 | 2.01 | 1.21 | **6.03** |
| 306 | 0.40 | 0 | 0 | 0 | **0.40** |
| 308 | 0 | 0 | 0 | 0.40 | **0.40** |
| 310 | 0.80 | 0 | 0 | 0.80 | **1.61** |
| 312 | 1.61 | 0 | 0 | 0 | **1.61** |
| 314 | 0 | 1.21 | 0 | 0 | **1.21** |
| 315 | 1.61 | 2.01 | 1.61 | 1.61 | **6.83** |
| 317 | 0 | 0 | 0 | 0.80 | **0.80** |
| **GA GEORGIA Total** | **24.51** | **24.51** | **19.69** | **27.72** | **96.42** |

**STATE: HI - HAWAII**

| REGISTRANT ZIP CODE 3 | 1ST QUARTER | 2ND QUARTER | 3RD QUARTER | 4TH QUARTER | **TOTAL GRAMS** |
|---|---|---|---|---|---|
| 967 | 2.41 | 1.61 | 0.40 | 1.21 | **5.62** |
| 968 | 3.21 | 2.81 | 3.21 | 3.62 | **12.86** |
| **HI - HAWAII - Total** | **5.63** | **4.42** | **3.62** | **4.82** | **18.48** |

**STATE: IA - IOWA**

| REGISTRANT ZIP CODE 3 | 1ST QUARTER | 2ND QUARTER | 3RD QUARTER | 4TH QUARTER | **TOTAL GRAMS** |
|---|---|---|---|---|---|
| 500 | 1.61 | 0 | 0 | 1.21 | **2.81** |
| 502 | 5.22 | 7.23 | 6.03 | 9.24 | **27.72** |
| 503 | 0.80 | 0.80 | 1.61 | 1.21 | **4.42** |
| 504 | 3.21 | 2.41 | 1.61 | 2.01 | **9.24** |
| 505 | 0.80 | 1.21 | 0.40 | 0.40 | **2.81** |
| 513 | 0 | 0 | 0.40 | 0 | **0.40** |
| 516 | 0 | 0 | 0 | 0.80 | **0.80** |
| 528 | 2.01 | 1.61 | 2.41 | 2.01 | **8.03** |
| **IA IOWA Total** | **13.66** | **13.26** | **12.45** | **16.87** | **56.25** |

**STATE: ID - IDAHO**

| REGISTRANT ZIP CODE 3 | 1ST QUARTER | 2ND QUARTER | 3RD QUARTER | 4TH QUARTER | **TOTAL GRAMS** |
|---|---|---|---|---|---|
| 832 | 0 | 0 | 0 | 3.62 | **3.62** |

| | | | | | |
|---|---|---|---|---|---|
| 833 | 0.40 | 0 | 0 | 0 | **0.40** |
| 834 | 1.61 | 1.61 | 0 | 0.40 | **3.62** |
| 836 | 4.82 | 6.03 | 7.23 | 6.43 | **24.51** |
| 837 | 0.40 | 0.80 | 0.80 | 0.80 | **2.81** |
| 838 | 4.02 | 3.21 | 5.62 | 4.82 | **17.67** |
| **ID IDAHO Total** | **11.25** | **11.65** | **13.66** | **16.07** | **52.62** |

**STATE: IL - ILLINOIS**

| REGISTRANT ZIP CODE 3 | 1ST QUARTER | 2ND QUARTER | 3RD QUARTER | 4TH QUARTER | TOTAL GRAMS |
|---|---|---|---|---|---|
| 600 | 17.68 | 16.88 | 17.68 | 13.66 | **65.89** |
| 601 | 9.24 | 8.44 | 6.03 | 6.43 | **30.13** |
| 602 | 1.61 | 3.21 | 2.01 | 2.01 | **8.84** |
| 604 | 4.42 | 2.41 | 2.81 | 2.01 | **11.65** |
| 605 | 12.86 | 11.25 | 10.85 | 12.05 | **47.00** |
| 606 | 12.86 | 12.45 | 8.84 | 11.65 | **45.80** |
| 607 | 2.41 | 0 | 2.41 | 3.21 | **8.03** |
| 611 | 0 | 0.40 | 1.61 | 1.61 | **3.62** |
| 615 | 0.80 | 0.80 | 0.40 | 0 | **2.01** |
| 623 | 0 | 0 | 0.40 | 0.80 | **1.21** |
| 627 | 0.80 | 0.80 | 0.80 | 2.01 | **4.42** |
| **IL ILLINOIS Total** | **62.67** | **56.65** | **53.83** | **55.44** | **228.59** |

**STATE: IN - INDIANA**

| REGISTRANT ZIP CODE 3 | 1ST QUARTER | 2ND QUARTER | 3RD QUARTER | 4TH QUARTER | TOTAL GRAMS |
|---|---|---|---|---|---|
| 460 | 2.41 | 6.03 | 4.02 | 13.66 | **26.11** |
| 462 | 0.40 | 0.80 | 0 | 1.61 | **2.81** |
| 463 | 1.61 | 2.01 | 1.21 | 4.02 | **8.84** |
| 465 | 0.80 | 0.40 | 1.21 | 2.81 | **5.22** |
| 469 | 1.21 | 0.80 | 1.61 | 2.81 | **6.43** |
| 471 | 0.40 | 0.40 | 0 | 0 | **0.80** |
| 473 | 0.40 | 2.01 | 0.80 | 0 | **3.21** |
| 474 | 1.21 | 4.02 | 0.40 | 3.62 | **9.24** |
| 477 | 1.21 | 1.61 | 1.61 | 1.61 | **6.03** |
| 478 | 0 | 0 | 0.80 | 0 | **0.80** |
| 479 | 0.40 | 0.80 | 0 | 0 | **1.21** |
| **IN INDIANA Total** | **10.05** | **18.88** | **11.65** | **30.13** | **70.71** |

**STATE: KS - KANSAS**

| REGISTRANT ZIP CODE 3 | 1ST QUARTER | 2ND QUARTER | 3RD QUARTER | 4TH QUARTER | TOTAL GRAMS |
|---|---|---|---|---|---|
| 660 | 3.62 | 4.42 | 3.62 | 4.82 | **16.47** |
| 662 | 1.21 | 0.80 | 0.80 | 0.40 | **3.21** |
| 664 | 0 | 0 | 0.40 | 0 | **0.40** |
| 666 | 0.40 | 0.40 | 0 | 0.80 | **1.61** |

| | | | | | |
|---|---|---|---|---|---|
| 670 | 1.21 | 1.61 | 2.01 | 1.61 | **6.43** |
| 671 | 0 | 0.40 | 1.21 | 0.80 | **2.41** |
| 672 | 6.83 | 5.62 | 4.82 | 5.22 | **22.50** |
| **KS - KANSAS - Total** | **13.26** | **13.26** | **12.86** | **13.66** | **53.03** |

**STATE: KY - KENTUCKY**

| REGISTRANT ZIP CODE 3 | 1ST QUARTER | 2ND QUARTER | 3RD QUARTER | 4TH QUARTER | TOTAL GRAMS |
|---|---|---|---|---|---|
| 402 | 0.40 | 1.61 | 1.61 | 0.80 | **4.42** |
| 403 | 2.01 | 0.80 | 0.40 | 0 | **3.21** |
| 405 | 6.83 | 6.03 | 6.03 | 3.21 | **22.10** |
| 408 | 1.61 | 2.41 | 3.21 | 0 | **7.23** |
| 410 | 0.40 | 0 | 0 | 0.40 | **0.80** |
| 417 | 0 | 0 | 0.40 | 0.40 | **0.80** |
| 425 | 1.21 | 0 | 0 | 0 | **1.21** |
| **KY - KENTUCKY - Total** | **12.46** | **10.85** | **11.65** | **4.82** | **39.77** |

**STATE: LA - LOUISIANA**

| REGISTRANT ZIP CODE 3 | 1ST QUARTER | 2ND QUARTER | 3RD QUARTER | 4TH QUARTER | TOTAL GRAMS |
|---|---|---|---|---|---|
| 703 | 2.01 | 0.40 | 1.21 | 1.21 | **4.82** |
| 705 | 0.80 | 0 | 0 | 0 | **0.80** |
| 706 | 1.61 | 2.41 | 1.21 | 1.21 | **6.43** |
| 707 | 0 | 0.40 | 0 | 0 | **0.40** |
| 708 | 7.23 | 6.83 | 4.02 | 5.62 | **23.70** |
| 711 | 7.63 | 2.81 | 2.81 | 2.81 | **16.07** |
| 713 | 0.80 | 0 | 0 | 0 | **0.80** |
| 714 | 0.40 | 0 | 0 | 0 | **0.40** |
| **LA - LOUISIANA - Total** | **20.49** | **12.86** | **9.24** | **10.85** | **53.44** |

**STATE: MA - MASSACHUSETTS**

| REGISTRANT ZIP CODE 3 | 1ST QUARTER | 2ND QUARTER | 3RD QUARTER | 4TH QUARTER | TOTAL GRAMS |
|---|---|---|---|---|---|
| 010 | 2.81 | 1.61 | 1.21 | 1.61 | **7.23** |
| 013 | 2.01 | 0.80 | 0 | 0 | **2.81** |
| 014 | 2.41 | 0.80 | 2.01 | 2.81 | **8.04** |
| 017 | 2.01 | 2.41 | 2.41 | 3.21 | **10.04** |
| 018 | 3.62 | 2.41 | 5.22 | 4.02 | **15.27** |
| 019 | 4.42 | 5.63 | 3.21 | 6.83 | **20.09** |
| 021 | 2.81 | 3.62 | 3.21 | 4.82 | **14.46** |
| 023 | 0.80 | 0.40 | 0.80 | 0 | **2.01** |
| 024 | 2.81 | 4.82 | 4.02 | 1.61 | **13.26** |
| 027 | 1.21 | 1.61 | 0.40 | 1.61 | **4.82** |
| **MA - MASSACHUSETTS - Total** | **24.91** | **24.11** | **22.50** | **26.52** | **98.04** |

**STATE: MD - MARYLAND**

| REGISTRANT ZIP CODE 3 | 1ST QUARTER | 2ND QUARTER | 3RD QUARTER | 4TH QUARTER | TOTAL GRAMS |
|---|---|---|---|---|---|

| | | | | | |
|---|---|---|---|---|---|
| 206 | 2.01 | 0.40 | 0.40 | 0.80 | **3.62** |
| 207 | 0.80 | 1.61 | 0.80 | 0.80 | **4.02** |
| 208 | 8.03 | 8.44 | 10.45 | 6.03 | **32.94** |
| 209 | 0 | 0 | 2.41 | 2.41 | **4.82** |
| 210 | 5.22 | 5.22 | 6.83 | 3.21 | **20.49** |
| 211 | 10.45 | 7.63 | 12.46 | 10.45 | **40.98** |
| 212 | 1.61 | 3.21 | 6.43 | 5.22 | **16.47** |
| 214 | 2.81 | 0.40 | 0.40 | 1.21 | **4.82** |
| 217 | 1.21 | 0.80 | 1.21 | 0.80 | **4.02** |
| 218 | 0 | 0.40 | 0 | 0 | **0.40** |
| **MD MARYLAND Total** | **32.14** | **28.12** | **41.38** | **30.93** | **132.58** |

**STATE: ME - MAINE**

| REGISTRANT ZIP CODE 3 | 1ST QUARTER | 2ND QUARTER | 3RD QUARTER | 4TH QUARTER | TOTAL GRAMS |
|---|---|---|---|---|---|
| 040 | 1.61 | 2.01 | 1.61 | 2.01 | **7.23** |
| 042 | 7.23 | 6.43 | 2.81 | 6.03 | **22.50** |
| 043 | 0 | 0 | 0.40 | 0 | **0.40** |
| 045 | 4.02 | 5.22 | 4.02 | 2.01 | **15.27** |
| 048 | 0.40 | 0 | 0 | 0 | **0.40** |
| **ME MAINE Total** | **13.26** | **13.66** | **8.84** | **10.04** | **45.80** |

**STATE: MI - MICHIGAN**

| REGISTRANT ZIP CODE 3 | 1ST QUARTER | 2ND QUARTER | 3RD QUARTER | 4TH QUARTER | TOTAL GRAMS |
|---|---|---|---|---|---|
| 480 | 5.22 | 2.01 | 2.81 | 0 | **10.04** |
| 481 | 2.01 | 2.41 | 1.61 | 1.61 | **7.63** |
| 482 | 0 | 0.40 | 0.80 | 0 | **1.21** |
| 483 | 1.61 | 1.21 | 2.01 | 1.21 | **6.03** |
| 484 | 0 | 2.41 | 1.61 | 1.61 | **5.62** |
| 485 | 0.40 | 0 | 0 | 0 | **0.40** |
| 486 | 4.02 | 6.03 | 8.04 | 4.02 | **22.10** |
| 488 | 0.80 | 1.21 | 0 | 0 | **2.01** |
| 490 | 6.83 | 6.43 | 5.62 | 7.23 | **26.11** |
| 494 | 0.40 | 0.40 | 0.80 | 0.80 | **2.41** |
| 495 | 0.40 | 0.40 | 0 | 0.40 | **1.21** |
| 496 | 0 | 0.40 | 0 | 0 | **0.40** |
| 498 | 0.40 | 0 | 0 | 0 | **0.40** |
| **MI - MICHIGAN Total** | **22.10** | **23.30** | **23.30** | **16.87** | **85.58** |

**STATE: MN - MINNESOTA**

| REGISTRANT ZIP CODE 3 | 1ST QUARTER | 2ND QUARTER | 3RD QUARTER | 4TH QUARTER | TOTAL GRAMS |
|---|---|---|---|---|---|
| 550 | 1.61 | 1.21 | 1.21 | 2.01 | **6.03** |
| 551 | 8.84 | 8.44 | 9.64 | 7.23 | **34.15** |
| 553 | 4.02 | 5.22 | 5.63 | 8.04 | **22.90** |

| | | | | | |
|---|---|---|---|---|---|
| 554 | 18.08 | 24.11 | 25.31 | 31.74 | **99.24** |
| 557 | 3.21 | 3.62 | 5.22 | 5.22 | **17.28** |
| 559 | 4.42 | 6.43 | 5.62 | 5.22 | **21.69** |
| 560 | 2.41 | 1.61 | 2.41 | 1.61 | **8.03** |
| 563 | 1.61 | 0.80 | 0 | 0 | **2.41** |
| 565 | 2.01 | 0.80 | 0 | 0 | **2.81** |
| **MN MINNESOTA Total** | **46.21** | **52.23** | **55.04** | **61.07** | **214.55** |

**STATE: MO - MISSOURI**

| REGISTRANT ZIP CODE 3 | 1ST QUARTER | 2ND QUARTER | 3RD QUARTER | 4TH QUARTER | **TOTAL GRAMS** |
|---|---|---|---|---|---|
| 630 | 0.40 | 0.80 | 0 | 0 | **1.21** |
| 631 | 49.81 | 39.37 | 41.38 | 34.95 | **165.51** |
| 633 | 0.40 | 0 | 0 | 1.21 | **1.61** |
| 636 | 0 | 0.40 | 0.40 | 0 | **0.80** |
| 637 | 0 | 0 | 1.61 | 0 | **1.61** |
| 639 | 0.40 | 2.01 | 0.80 | 2.01 | **5.22** |
| 640 | 3.21 | 0 | 3.62 | 2.01 | **8.84** |
| 641 | 5.63 | 1.61 | 1.61 | 0.40 | **9.24** |
| 645 | 1.21 | 1.21 | 1.61 | 1.61 | **5.63** |
| 646 | 0 | 0 | 0.40 | 0 | **0.40** |
| 648 | 0 | 0 | 0 | 0.80 | **0.80** |
| 655 | 0.80 | 1.61 | 0.80 | 0 | **3.21** |
| **MO MISSOURI - Total** | **61.87** | **47.00** | **52.23** | **42.99** | **204.08** |

**STATE: MS - MISSISSIPPI**

| REGISTRANT ZIP CODE 3 | 1ST QUARTER | 2ND QUARTER | 3RD QUARTER | 4TH QUARTER | **TOTAL GRAMS** |
|---|---|---|---|---|---|
| 390 | 1.61 | 0.40 | 1.21 | 0.80 | **4.02** |
| 391 | 0 | 0 | 6.03 | 5.62 | **11.65** |
| 392 | 4.82 | 6.83 | 0 | 0 | **11.65** |
| 396 | 0.40 | 0 | 0 | 0 | **0.40** |
| 397 | 1.21 | 0 | 0.40 | 0 | **1.61** |
| **MS MISSISSIPPI - Total** | **8.03** | **7.23** | **7.63** | **6.43** | **29.33** |

**STATE: NC - NORTH CAROLINA**

| REGISTRANT ZIP CODE 3 | 1ST QUARTER | 2ND QUARTER | 3RD QUARTER | 4TH QUARTER | **TOTAL GRAMS** |
|---|---|---|---|---|---|
| 270 | 0 | 0.40 | 0 | 0 | **0.40** |
| 272 | 2.41 | 3.62 | 0.80 | 0 | **6.83** |
| 274 | 2.01 | 1.21 | 1.61 | 1.61 | **6.43** |
| 275 | 23.71 | 22.90 | 12.45 | 17.28 | **76.34** |
| 276 | 4.42 | 6.43 | 2.01 | 4.42 | **17.28** |
| 277 | 0 | 0 | 0.40 | 3.21 | **3.62** |
| 278 | 0.80 | 1.21 | 3.62 | 2.01 | **7.63** |
| 280 | 0.80 | 0.80 | 0 | 0.80 | **2.41** |

| | | | | | |
|---|---|---|---|---|---|
| 281 | 3.62 | 1.61 | 5.62 | 9.24 | **20.09** |
| 282 | 2.01 | 3.62 | 2.01 | 4.42 | **12.05** |
| 283 | 0 | 0 | 1.61 | 0.80 | **2.41** |
| 284 | 1.21 | 0.40 | 0 | 0 | **1.61** |
| 285 | 0 | 0 | 1.21 | 4.82 | **6.03** |
| 287 | 3.21 | 2.01 | 0.40 | 0 | **5.62** |
| 288 | 1.21 | 2.81 | 0.80 | 1.61 | **6.43** |
| **NC - NORTH CAROLINA Total** | **45.41** | **47.01** | **32.54** | **50.22** | **175.18** |

**STATE: ND - NORTH DAKOTA**

| REGISTRANT ZIP CODE 3 | 1ST QUARTER | 2ND QUARTER | 3RD QUARTER | 4TH QUARTER | TOTAL GRAMS |
|---|---|---|---|---|---|
| 581 | 2.01 | 2.81 | 11.25 | 3.62 | **19.69** |
| 584 | 3.21 | 4.02 | 0 | 0 | **7.23** |
| 585 | 0 | 0 | 2.01 | 1.61 | **3.62** |
| 587 | 2.01 | 0 | 0 | 0 | **2.01** |
| **ND NORTH DAKOTA Total** | **7.23** | **6.83** | **13.26** | **5.22** | **32.54** |

**STATE: NE - NEBRASKA**

| REGISTRANT ZIP CODE 3 | 1ST QUARTER | 2ND QUARTER | 3RD QUARTER | 4TH QUARTER | TOTAL GRAMS |
|---|---|---|---|---|---|
| 680 | 0.80 | 0 | 0 | 0 | **0.80** |
| 681 | 1.61 | 2.81 | 2.01 | 1.21 | **7.63** |
| 684 | 2.41 | 1.61 | 1.61 | 2.01 | **7.63** |
| 685 | 0.40 | 0.80 | 1.61 | 0.80 | **3.62** |
| 687 | 0 | 0 | 0 | 1.61 | **1.61** |
| 688 | 0 | 0 | 0.80 | 0 | **0.80** |
| 691 | 0 | 0.80 | 0 | 0 | **0.80** |
| **NE NEBRASKA Total** | **5.22** | **6.03** | **6.03** | **5.62** | **22.90** |

**STATE: NH - NEW HAMPSHIRE**

| REGISTRANT ZIP CODE 3 | 1ST QUARTER | 2ND QUARTER | 3RD QUARTER | 4TH QUARTER | TOTAL GRAMS |
|---|---|---|---|---|---|
| 031 | 1.21 | 2.81 | 3.21 | 2.81 | **10.04** |
| 033 | 0.80 | 1.61 | 0 | 0.40 | **2.81** |
| 037 | 0.80 | 4.42 | 3.21 | 2.81 | **11.25** |
| **NH - NEW HAMPSHIRE Total** | **2.81** | **8.84** | **6.43** | **6.03** | **24.10** |

**STATE: NJ - NEW JERSEY**

| REGISTRANT ZIP CODE 3 | 1ST QUARTER | 2ND QUARTER | 3RD QUARTER | 4TH QUARTER | TOTAL GRAMS |
|---|---|---|---|---|---|
| 070 | 3.62 | 3.21 | 3.62 | 5.22 | **15.67** |
| 074 | 0.40 | 0 | 3.21 | 2.41 | **6.03** |
| 076 | 5.63 | 8.04 | 7.63 | 14.46 | **35.76** |
| 077 | 0 | 0 | 1.61 | 2.41 | **4.02** |
| 078 | 6.43 | 4.42 | 1.61 | 2.81 | **15.27** |
| 079 | 1.21 | 1.21 | 2.01 | 2.41 | **6.83** |
| 080 | 3.21 | 4.02 | 4.42 | 5.22 | **16.87** |

| | | | | | |
|---|---|---|---|---|---|
| 081 | 1.61 | 0.80 | 0.40 | 1.21 | **4.02** |
| 082 | 0.40 | 1.21 | 0 | 0.80 | **2.41** |
| 083 | 0 | 0 | 0.40 | 0.80 | **1.21** |
| 085 | 1.61 | 1.61 | 1.61 | 0.40 | **5.22** |
| 086 | 0.80 | 0 | 0.40 | 0 | **1.21** |
| 087 | 2.41 | 3.21 | 4.42 | 5.62 | **15.67** |
| 088 | 6.43 | 2.81 | 0.80 | 0.40 | **10.45** |
| **NJ NEW JERSEY - Total** | **33.75** | **30.54** | **32.14** | **44.19** | **140.63** |

**STATE: NM - NEW MEXICO**

| REGISTRANT ZIP CODE 3 | 1ST QUARTER | 2ND QUARTER | 3RD QUARTER | 4TH QUARTER | TOTAL GRAMS |
|---|---|---|---|---|---|
| 870 | 0 | 0 | 0 | 1.61 | **1.61** |
| 871 | 1.61 | 1.61 | 3.21 | 2.01 | **8.44** |
| 880 | 3.62 | 1.61 | 2.01 | 3.62 | **10.85** |
| 882 | 0 | 0 | 0.40 | 0.40 | **0.80** |
| 883 | 0.40 | 1.61 | 2.01 | 2.81 | **6.83** |
| **NM NEW MEXICO Total** | **5.63** | **4.82** | **7.63** | **10.45** | **28.52** |

**STATE: NV - NEVADA**

| REGISTRANT ZIP CODE 3 | 1ST QUARTER | 2ND QUARTER | 3RD QUARTER | 4TH QUARTER | TOTAL GRAMS |
|---|---|---|---|---|---|
| 891 | 10.45 | 6.43 | 8.84 | 8.04 | **33.75** |
| 895 | 0.80 | 1.21 | 2.01 | 2.01 | **6.03** |
| **NV NEVADA Total** | **11.25** | **7.63** | **10.85** | **10.04** | **39.78** |

**STATE: NY - NEW YORK**

| REGISTRANT ZIP CODE 3 | 1ST QUARTER | 2ND QUARTER | 3RD QUARTER | 4TH QUARTER | TOTAL GRAMS |
|---|---|---|---|---|---|
| 100 | 15.27 | 20.89 | 21.69 | 20.89 | **78.74** |
| 101 | 0.40 | 0 | 0.40 | 0.80 | **1.61** |
| 104 | 0.80 | 4.02 | 2.01 | 0.80 | **7.63** |
| 105 | 5.63 | 1.21 | 2.41 | 0.40 | **9.64** |
| 106 | 8.04 | 5.22 | 7.63 | 4.02 | **24.91** |
| 107 | 0 | 0 | 1.21 | 0 | **1.21** |
| 108 | 0 | 0 | 0 | 1.21 | **1.21** |
| 109 | 1.61 | 2.41 | 2.41 | 1.21 | **7.63** |
| 110 | 2.41 | 2.41 | 1.61 | 2.81 | **9.24** |
| 111 | 0 | 1.21 | 0 | 1.61 | **2.81** |
| 112 | 3.21 | 1.21 | 2.81 | 5.22 | **12.45** |
| 113 | 2.41 | 2.01 | 2.81 | 3.21 | **10.44** |
| 114 | 2.01 | 1.61 | 3.21 | 0 | **6.83** |
| 115 | 4.02 | 2.41 | 7.23 | 3.21 | **16.87** |
| 116 | 0.80 | 1.21 | 0.40 | 0 | **2.41** |
| 117 | 13.66 | 10.45 | 19.28 | 13.26 | **56.65** |
| 119 | 0.40 | 0 | 0 | 0 | **0.40** |

| | | | | | |
|---|---|---|---|---|---|
| 120 | 1.61 | 2.81 | 2.41 | 3.21 | **10.05** |
| 122 | 1.21 | 0 | 0 | 0.40 | **1.61** |
| 123 | 3.21 | 3.21 | 4.42 | 3.21 | **14.06** |
| 124 | 0.80 | 0 | 0 | 0 | **0.80** |
| 126 | 0 | 0 | 1.21 | 0 | **1.21** |
| 127 | 0.40 | 0.40 | 1.61 | 1.21 | **3.62** |
| 128 | 0 | 0 | 0 | 1.61 | **1.61** |
| 130 | 0.80 | 0 | 0 | 0 | **0.80** |
| 137 | 0.40 | 0 | 0 | 0 | **0.40** |
| 139 | 0.80 | 0.80 | 0.40 | 1.61 | **3.62** |
| 140 | 4.82 | 6.03 | 3.62 | 4.02 | **18.48** |
| 141 | 1.21 | 1.61 | 0 | 0.40 | **3.21** |
| 142 | 0 | 0.40 | 0 | 0 | **0.40** |
| 146 | 1.61 | 2.81 | 0.80 | 2.01 | **7.23** |
| 148 | 2.01 | 3.21 | 2.41 | 2.81 | **10.44** |
| 149 | 2.81 | 0 | 1.61 | 1.21 | **5.63** |
| **NY - NEW YORK - Total** | **82.37** | **77.54** | **93.61** | **80.35** | **333.87** |

**STATE: OH - OHIO**

| REGISTRANT ZIP CODE 3 | 1ST QUARTER | 2ND QUARTER | 3RD QUARTER | 4TH QUARTER | **TOTAL GRAMS** |
|---|---|---|---|---|---|
| 430 | 4.42 | 3.62 | 6.43 | 4.02 | **18.48** |
| 431 | 1.21 | 2.01 | 1.21 | 0.80 | **5.22** |
| 432 | 4.42 | 2.81 | 3.62 | 6.83 | **17.68** |
| 433 | 0 | 0.40 | 0 | 0 | **0.40** |
| 434 | 0.80 | 0.80 | 1.21 | 0.40 | **3.21** |
| 435 | 0 | 0 | 0.80 | 0 | **0.80** |
| 440 | 4.02 | 4.82 | 3.21 | 3.62 | **15.67** |
| 441 | 4.02 | 2.41 | 2.01 | 0 | **8.44** |
| 445 | 3.21 | 2.81 | 2.01 | 4.42 | **12.45** |
| 448 | 1.21 | 1.61 | 2.01 | 0 | **4.82** |
| 450 | 8.04 | 3.21 | 3.21 | 3.21 | **17.68** |
| 451 | 0.80 | 0.80 | 0 | 0 | **1.61** |
| 452 | 4.82 | 1.61 | 2.01 | 1.21 | **9.64** |
| 453 | 1.21 | 1.61 | 2.01 | 1.61 | **6.43** |
| 454 | 6.03 | 3.21 | 4.02 | 4.82 | **18.08** |
| 456 | 0.80 | 0 | 0 | 0 | **0.80** |
| **OH - OHIO Total** | **45.00** | **31.74** | **33.75** | **30.94** | **141.43** |

**STATE: OK - OKLAHOMA**

| REGISTRANT ZIP CODE 3 | 1ST QUARTER | 2ND QUARTER | 3RD QUARTER | 4TH QUARTER | **TOTAL GRAMS** |
|---|---|---|---|---|---|
| 730 | 0.40 | 0 | 0 | 0.40 | **0.80** |
| 731 | 2.01 | 2.41 | 1.61 | 0.40 | **6.43** |

| | | | | | |
|---|---|---|---|---|---|
| 734 | 0 | 0.40 | 0 | 0 | **0.40** |
| 740 | 4.42 | 5.63 | 5.62 | 4.42 | **20.09** |
| 743 | 0 | 1.61 | 0.80 | 1.61 | **4.02** |
| 745 | 0 | 0 | 0 | 0.40 | **0.40** |
| 747 | 1.21 | 0 | 0 | 0.80 | **2.01** |
| 748 | 0.40 | 0.80 | 0.40 | 0.80 | **2.41** |
| **OK - OKLAHOMA Total** | **8.44** | **10.85** | **8.44** | **8.84** | **36.56** |

**STATE: OR - OREGON**

| REGISTRANT ZIP CODE 3 | 1ST QUARTER | 2ND QUARTER | 3RD QUARTER | 4TH QUARTER | TOTAL GRAMS |
|---|---|---|---|---|---|
| 970 | 14.46 | 9.24 | 10.44 | 13.26 | **47.40** |
| 971 | 0 | 0 | 0.40 | 0.80 | **1.21** |
| 972 | 7.63 | 12.45 | 11.65 | 16.47 | **48.21** |
| 973 | 0.40 | 1.61 | 1.61 | 1.21 | **4.82** |
| 974 | 0.40 | 0.40 | 0.40 | 4.42 | **5.62** |
| 975 | 0.40 | 3.62 | 3.62 | 2.41 | **10.04** |
| 977 | 1.21 | 0.80 | 0 | 0.40 | **2.41** |
| 978 | 14.06 | 6.83 | 8.84 | 12.05 | **41.78** |
| **OR OREGON Total** | **38.57** | **34.95** | **36.96** | **51.02** | **161.50** |

**STATE: PA - PENNSYLVANIA**

| REGISTRANT ZIP CODE 3 | 1ST QUARTER | 2ND QUARTER | 3RD QUARTER | 4TH QUARTER | TOTAL GRAMS |
|---|---|---|---|---|---|
| 150 | 4.02 | 4.82 | 5.62 | 4.82 | **19.28** |
| 151 | 7.63 | 6.43 | 6.43 | 7.63 | **28.12** |
| 152 | 7.63 | 9.64 | 10.04 | 5.63 | **32.95** |
| 156 | 0.80 | 0.40 | 1.61 | 0.80 | **3.62** |
| 161 | 0 | 0.80 | 1.21 | 0 | **2.01** |
| 168 | 0.40 | 4.42 | 4.42 | 3.62 | **12.86** |
| 170 | 1.21 | 1.21 | 0 | 0.80 | **3.21** |
| 171 | 2.41 | 0.80 | 2.81 | 2.41 | **8.44** |
| 172 | 0.80 | 0.40 | 0 | 0 | **1.21** |
| 173 | 0.40 | 0.80 | 0 | 0.80 | **2.01** |
| 174 | 1.61 | 0.80 | 0.80 | 2.01 | **5.22** |
| 175 | 0 | 0 | 0.80 | 0 | **0.80** |
| 178 | 3.62 | 2.81 | 0 | 0 | **6.43** |
| 179 | 1.61 | 2.41 | 3.21 | 2.41 | **9.64** |
| 180 | 1.61 | 0 | 0 | 0 | **1.61** |
| 181 | 1.21 | 1.21 | 0 | 2.41 | **4.82** |
| 183 | 2.81 | 0 | 0 | 2.81 | **5.62** |
| 184 | 4.42 | 2.01 | 1.61 | 2.81 | **10.85** |
| 185 | 0 | 0 | 0 | 0.80 | **0.80** |
| 186 | 1.61 | 2.01 | 1.61 | 2.81 | **8.03** |

| | | | | | |
|---|---|---|---|---|---|
| 187 | 22.50 | 45.01 | 39.38 | 49.02 | **155.91** |
| 189 | 5.63 | 6.03 | 3.21 | 4.02 | **18.88** |
| 190 | 3.62 | 6.43 | 7.23 | 8.44 | **25.71** |
| 191 | 2.41 | 1.61 | 0.80 | 0.40 | **5.22** |
| 193 | 2.41 | 2.41 | 2.41 | 2.41 | **9.64** |
| 194 | 2.81 | 1.21 | 0.80 | 1.61 | **6.43** |
| 195 | 1.21 | 0.80 | 0 | 0.40 | **2.41** |
| 196 | 0.80 | 0 | 0.80 | 0.80 | **2.41** |
| **PA - PENNSYLVANIA - Total** | **85.18** | **104.47** | **94.82** | **109.69** | **394.15** |

**STATE: RI - RHODE ISLAND**

| REGISTRANT ZIP CODE 3 | 1ST QUARTER | 2ND QUARTER | 3RD QUARTER | 4TH QUARTER | TOTAL GRAMS |
|---|---|---|---|---|---|
| 028 | 0.80 | 1.21 | 2.41 | 2.81 | **7.23** |
| 029 | 0 | 0 | 0.80 | 0 | **0.80** |
| **RI RHODE ISLAND Total** | **0.80** | **1.21** | **3.21** | **2.81** | **8.04** |

**STATE: SC - SOUTH CAROLINA**

| REGISTRANT ZIP CODE 3 | 1ST QUARTER | 2ND QUARTER | 3RD QUARTER | 4TH QUARTER | TOTAL GRAMS |
|---|---|---|---|---|---|
| 290 | 0.80 | 1.21 | 0.40 | 0 | **2.41** |
| 291 | 0.40 | 0 | 0 | 0 | **0.40** |
| 292 | 2.41 | 6.43 | 3.62 | 4.42 | **16.88** |
| 293 | 1.21 | 0.80 | 2.01 | 0 | **4.02** |
| 294 | 3.62 | 0.40 | 2.41 | 2.81 | **9.24** |
| 295 | 1.61 | 1.21 | 1.61 | 3.62 | **8.04** |
| 296 | 2.41 | 2.01 | 1.61 | 2.41 | **8.44** |
| 297 | 0 | 0 | 0 | 0.40 | **0.40** |
| **SC - SOUTH CAROLINA Total** | **12.46** | **12.05** | **11.65** | **13.66** | **49.82** |

**STATE: SD - SOUTH DAKOTA**

| REGISTRANT ZIP CODE 3 | 1ST QUARTER | 2ND QUARTER | 3RD QUARTER | 4TH QUARTER | TOTAL GRAMS |
|---|---|---|---|---|---|
| 571 | 5.22 | 4.82 | 6.83 | 6.43 | **23.30** |
| **SD SOUTH DAKOTA - Total** | **5.22** | **4.82** | **6.83** | **6.43** | **23.30** |

**STATE: TN - TENNESSEE**

| REGISTRANT ZIP CODE 3 | 1ST QUARTER | 2ND QUARTER | 3RD QUARTER | 4TH QUARTER | TOTAL GRAMS |
|---|---|---|---|---|---|
| 370 | 0.80 | 0.40 | 0 | 0 | **1.21** |
| 372 | 1.61 | 1.61 | 2.01 | 3.62 | **8.84** |
| 373 | 0 | 0 | 0 | 0.40 | **0.40** |
| 374 | 0 | 1.21 | 0 | 0 | **1.21** |
| 377 | 0 | 1.21 | 0 | 0 | **1.21** |
| 378 | 0.80 | 0 | 3.21 | 1.21 | **5.22** |
| 379 | 0 | 1.21 | 2.81 | 2.01 | **6.03** |
| 380 | 0.80 | 1.61 | 2.01 | 2.41 | **6.83** |
| 381 | 6.03 | 5.63 | 6.83 | 4.42 | **22.90** |

| | | | | | |
|---|---|---|---|---|---|
| 383 | 0 | 1.61 | 0 | 0 | **1.61** |
| **TN TENNESSEE Total** | **10.05** | **14.46** | **16.87** | **14.06** | **55.45** |

**STATE: TX - TEXAS**

| REGISTRANT ZIP CODE 3 | 1ST QUARTER | 2ND QUARTER | 3RD QUARTER | 4TH QUARTER | TOTAL GRAMS |
|---|---|---|---|---|---|
| 750 | 17.68 | 20.49 | 20.49 | 24.91 | **83.57** |
| 751 | 0.80 | 0 | 0 | 0 | **0.80** |
| 752 | 11.25 | 12.46 | 13.26 | 14.86 | **51.83** |
| 754 | 0.80 | 0.80 | 1.21 | 0.80 | **3.62** |
| 756 | 0.40 | 0 | 2.41 | 1.21 | **4.02** |
| 757 | 2.41 | 2.81 | 2.01 | 2.81 | **10.04** |
| 760 | 2.81 | 1.21 | 1.21 | 4.42 | **9.64** |
| 761 | 1.21 | 1.21 | 2.41 | 0.40 | **5.22** |
| 762 | 0.80 | 0 | 2.01 | 1.61 | **4.42** |
| 764 | 0.40 | 1.61 | 0.40 | 0.80 | **3.21** |
| 767 | 0 | 0.40 | 0.40 | 0.80 | **1.61** |
| 769 | 0.40 | 0 | 0 | 0 | **0.40** |
| 770 | 9.24 | 11.65 | 7.23 | 10.04 | **38.16** |
| 773 | 1.61 | 2.01 | 0.80 | 3.62 | **8.04** |
| 774 | 4.42 | 3.62 | 2.81 | 2.41 | **13.26** |
| 775 | 1.61 | 2.01 | 2.01 | 1.61 | **7.23** |
| 780 | 0.40 | 0.40 | 0 | 0 | **0.80** |
| 781 | 0.40 | 0 | 0 | 1.21 | **1.61** |
| 782 | 3.62 | 2.01 | 3.62 | 2.81 | **12.05** |
| 784 | 2.41 | 1.21 | 2.81 | 0.80 | **7.23** |
| 785 | 3.62 | 2.01 | 1.61 | 2.01 | **9.24** |
| 786 | 2.81 | 4.42 | 1.61 | 4.42 | **13.26** |
| 787 | 8.04 | 12.05 | 8.44 | 8.44 | **36.96** |
| 788 | 4.02 | 1.61 | 3.62 | 3.62 | **12.85** |
| 790 | 2.01 | 1.61 | 1.21 | 1.61 | **6.43** |
| 797 | 0.80 | 1.61 | 0.80 | 0 | **3.21** |
| 799 | 0 | 0 | 0.40 | 0 | **0.40** |
| **TX - TEXAS Total** | **83.97** | **87.19** | **82.76** | **95.21** | **349.13** |

**STATE: UT - UTAH**

| REGISTRANT ZIP CODE 3 | 1ST QUARTER | 2ND QUARTER | 3RD QUARTER | 4TH QUARTER | TOTAL GRAMS |
|---|---|---|---|---|---|
| 840 | 8.44 | 5.22 | 14.06 | 11.25 | **38.97** |
| 841 | 2.41 | 3.21 | 2.01 | 4.42 | **12.05** |
| 844 | 0 | 1.21 | 2.81 | 1.21 | **5.22** |
| 846 | 7.23 | 7.23 | 6.43 | 6.83 | **27.72** |
| 847 | 8.03 | 4.82 | 5.62 | 1.61 | **20.09** |
| **UT - UTAH - Total** | **26.11** | **21.70** | **30.93** | **25.31** | **104.05** |

**STATE: VA - VIRGINIA**

| REGISTRANT ZIP CODE 3 | 1ST QUARTER | 2ND QUARTER | 3RD QUARTER | 4TH QUARTER | TOTAL GRAMS |
|---|---|---|---|---|---|
| 201 | 5.22 | 6.03 | 6.03 | 1.21 | **18.48** |
| 220 | 0 | 0 | 0 | 1.21 | **1.21** |
| 221 | 2.81 | 3.21 | 3.62 | 4.02 | **13.66** |
| 223 | 2.01 | 2.01 | 3.21 | 1.61 | **8.84** |
| 228 | 0.40 | 0.40 | 0.40 | 0 | **1.21** |
| 230 | 0 | 2.41 | 0 | 0 | **2.41** |
| 231 | 4.42 | 4.02 | 4.02 | 5.63 | **18.08** |
| 232 | 4.42 | 4.02 | 2.81 | 5.63 | **16.88** |
| 233 | 0 | 0.80 | 0.40 | 1.61 | **2.81** |
| 234 | 2.41 | 0.80 | 0.80 | 1.21 | **5.22** |
| 235 | 1.21 | 2.01 | 1.61 | 2.01 | **6.83** |
| 237 | 1.61 | 0.80 | 0.80 | 1.21 | **4.42** |
| 238 | 2.81 | 3.21 | 1.21 | 2.41 | **9.64** |
| 240 | 3.21 | 0.40 | 2.41 | 0.40 | **6.43** |
| 241 | 0.80 | 1.61 | 1.61 | 1.21 | **5.22** |
| 242 | 0.80 | 0.40 | 0.80 | 0 | **2.01** |
| 245 | 0.80 | 0.80 | 0.80 | 0.40 | **2.81** |
| **VA VIRGINIA Total** | **32.95** | **32.95** | **30.54** | **29.73** | **126.16** |

**STATE: VT - VERMONT**

| REGISTRANT ZIP CODE 3 | 1ST QUARTER | 2ND QUARTER | 3RD QUARTER | 4TH QUARTER | TOTAL GRAMS |
|---|---|---|---|---|---|
| 050 | 0 | 0 | 0 | 1.21 | **1.21** |
| 054 | 0.80 | 0.80 | 0 | 0.80 | **2.41** |
| 056 | 0 | 0 | 2.41 | 0.40 | **2.81** |
| 058 | 0.80 | 0 | 0 | 0 | **0.80** |
| **VT VERMONT Total** | **1.61** | **0.80** | **2.41** | **2.41** | **7.23** |

**STATE: WA - WASHINGTON**

| REGISTRANT ZIP CODE 3 | 1ST QUARTER | 2ND QUARTER | 3RD QUARTER | 4TH QUARTER | TOTAL GRAMS |
|---|---|---|---|---|---|
| 980 | 27.32 | 22.50 | 26.11 | 21.69 | **97.62** |
| 981 | 10.85 | 7.23 | 16.07 | 21.29 | **55.44** |
| 982 | 10.44 | 10.45 | 10.85 | 10.04 | **41.78** |
| 983 | 11.25 | 12.45 | 12.45 | 12.85 | **49.01** |
| 984 | 0.80 | 2.01 | 3.21 | 7.63 | **13.66** |
| 985 | 3.21 | 2.01 | 4.42 | 4.42 | **14.06** |
| 986 | 14.86 | 7.63 | 10.85 | 9.64 | **42.98** |
| 988 | 0 | 0.80 | 1.61 | 1.21 | **3.62** |
| 990 | 0.40 | 0 | 0 | 0.80 | **1.21** |
| 992 | 4.82 | 2.41 | 4.42 | 3.21 | **14.86** |
| 993 | 0 | 0.80 | 0 | 0.40 | **1.21** |

| | | | | | |
|---|---|---|---|---|---|
| 994 | 0 | 0 | 0 | 3.21 | **3.21** |
| **WA WASHINGTON Total** | **83.96** | **68.30** | **89.98** | **96.41** | **338.65** |

**STATE: WI - WISCONSIN**

| REGISTRANT ZIP CODE 3 | 1ST QUARTER | 2ND QUARTER | 3RD QUARTER | 4TH QUARTER | TOTAL GRAMS |
|---|---|---|---|---|---|
| 531 | 1.21 | 1.21 | 2.41 | 4.82 | **9.64** |
| 532 | 5.22 | 4.42 | 1.21 | 4.42 | **15.27** |
| 535 | 2.01 | 0 | 0.40 | 1.21 | **3.62** |
| 537 | 5.22 | 7.63 | 9.64 | 6.43 | **28.92** |
| 539 | 0.40 | 0.40 | 1.61 | 0.80 | **3.21** |
| 540 | 0 | 1.21 | 0.80 | 0 | **2.01** |
| 542 | 6.03 | 5.22 | 5.22 | 5.63 | **22.10** |
| 544 | 2.41 | 2.81 | 1.21 | 0 | **6.43** |
| 549 | 4.42 | 2.81 | 3.62 | 3.62 | **14.46** |
| **WI - WISCONSIN Total** | **26.92** | **25.71** | **26.11** | **26.92** | **105.66** |

**STATE: WV - WEST VIRGINIA**

| REGISTRANT ZIP CODE 3 | 1ST QUARTER | 2ND QUARTER | 3RD QUARTER | 4TH QUARTER | TOTAL GRAMS |
|---|---|---|---|---|---|
| 258 | 1.21 | 1.21 | 2.01 | 1.61 | **6.03** |
| 260 | 2.41 | 0 | 0 | 0 | **2.41** |
| 265 | 1.21 | 2.01 | 1.21 | 1.21 | **5.63** |
| **WV WEST VIRGINIA Total** | **4.82** | **3.21** | **3.21** | **2.81** | **14.06** |

**STATE: WY - WYOMING**

| REGISTRANT ZIP CODE 3 | 1ST QUARTER | 2ND QUARTER | 3RD QUARTER | 4TH QUARTER | TOTAL GRAMS |
|---|---|---|---|---|---|
| 820 | 2.41 | 2.81 | 1.61 | 3.62 | **10.44** |
| 824 | 0 | 0 | 0 | 0.80 | **0.80** |
| 826 | 1.21 | 1.21 | 2.01 | 1.21 | **5.62** |
| 827 | 0.80 | 0.80 | 0.80 | 0.40 | **2.81** |
| **WY - WYOMING Total** | **4.42** | **4.82** | **4.42** | **6.03** | **19.68** |

**DRUG: 1205 - LISDEXAMFETAMINE**

**STATE: AK - ALASKA**

| REGISTRANT ZIP CODE 3 | 1ST QUARTER | 2ND QUARTER | 3RD QUARTER | 4TH QUARTER | TOTAL GRAMS |
|---|---|---|---|---|---|
| 995 | 1,342.93 | 1,432.53 | 1,407.10 | 1,585.73 | **5,768.28** |
| 996 | 628.39 | 819.75 | 814.54 | 857.90 | **3,120.58** |
| 997 | 328.36 | 329.52 | 350.33 | 432.42 | **1,440.63** |
| 998 | 197.71 | 211.58 | 225.46 | 207.54 | **842.29** |
| 999 | 124.87 | 131.23 | 128.92 | 134.70 | **519.71** |
| **AK ALASKA - Total** | **2,622.26** | **2,924.61** | **2,926.34** | **3,218.28** | **11,691.49** |

**STATE: AL - ALABAMA**

| REGISTRANT ZIP CODE 3 | 1ST QUARTER | 2ND QUARTER | 3RD QUARTER | 4TH QUARTER | TOTAL GRAMS |
|---|---|---|---|---|---|
| 350 | 6,974.20 | 6,778.80 | 6,653.93 | 7,207.75 | **27,614.68** |
| 351 | 4,736.37 | 4,384.89 | 4,686.08 | 4,704.58 | **18,511.92** |

| | | | | | |
|---|---|---|---|---|---|
| 352 | 9,863.54 | 9,768.16 | 9,857.18 | 10,203.47 | **39,692.35** |
| 354 | 4,643.88 | 4,162.32 | 4,513.80 | 4,602.83 | **17,922.83** |
| 355 | 1,379.92 | 1,304.19 | 1,333.68 | 1,358.54 | **5,376.33** |
| 356 | 4,070.40 | 4,012.59 | 4,180.82 | 4,288.35 | **16,552.16** |
| 357 | 2,501.44 | 2,398.54 | 2,635.56 | 2,638.45 | **10,173.98** |
| 358 | 2,847.14 | 2,939.64 | 3,071.45 | 3,146.02 | **12,004.25** |
| 359 | 2,476.00 | 2,542.48 | 2,566.19 | 2,611.28 | **10,195.95** |
| 360 | 3,457.04 | 3,390.56 | 3,286.50 | 3,406.17 | **13,540.26** |
| 361 | 2,855.81 | 2,495.66 | 2,583.53 | 2,515.89 | **10,450.89** |
| 362 | 1,769.56 | 1,811.19 | 1,838.94 | 1,893.86 | **7,313.54** |
| 363 | 4,273.89 | 4,118.96 | 4,007.97 | 4,042.08 | **16,442.90** |
| 364 | 1,123.25 | 842.29 | 875.24 | 1,037.11 | **3,877.89** |
| 365 | 8,021.14 | 7,893.38 | 7,869.68 | 8,121.73 | **31,905.92** |
| 366 | 6,193.76 | 5,871.76 | 5,839.97 | 6,245.21 | **24,150.71** |
| 367 | 668.86 | 551.51 | 609.32 | 598.33 | **2,428.02** |
| 368 | 4,690.70 | 4,308.58 | 4,216.66 | 4,645.03 | **17,860.98** |
| 369 | 137.59 | 133.54 | 118.51 | 157.24 | **546.88** |
| **AL ALABAMA - Total** | **72,684.51** | **69,709.03** | **70,744.99** | **73,423.90** | **286,562.44** |

**STATE: AR - ARKANSAS**

| REGISTRANT ZIP CODE 3 | 1ST QUARTER | 2ND QUARTER | 3RD QUARTER | 4TH QUARTER | **TOTAL GRAMS** |
|---|---|---|---|---|---|
| 716 | 1,322.69 | 1,156.20 | 1,166.61 | 1,265.46 | **4,910.96** |
| 717 | 649.21 | 593.13 | 622.04 | 736.50 | **2,600.87** |
| 718 | 793.15 | 704.13 | 657.30 | 689.10 | **2,843.67** |
| 719 | 1,822.75 | 1,763.21 | 1,725.05 | 1,802.52 | **7,113.52** |
| 720 | 4,376.22 | 4,303.95 | 4,301.06 | 4,465.82 | **17,447.06** |
| 721 | 3,196.31 | 2,951.20 | 2,950.04 | 3,120.58 | **12,218.14** |
| 722 | 3,649.60 | 3,577.86 | 3,587.11 | 3,514.27 | **14,328.84** |
| 723 | 1,085.09 | 955.02 | 926.69 | 1,115.73 | **4,082.54** |
| 724 | 2,552.89 | 2,349.98 | 2,420.50 | 2,602.03 | **9,925.40** |
| 725 | 605.85 | 600.07 | 597.18 | 612.21 | **2,415.30** |
| 726 | 737.08 | 782.17 | 713.95 | 842.87 | **3,076.07** |
| 727 | 5,369.39 | 5,268.23 | 5,383.27 | 5,724.92 | **21,745.81** |
| 728 | 1,079.31 | 986.82 | 1,022.66 | 1,128.45 | **4,217.24** |
| 729 | 2,489.30 | 2,305.46 | 2,316.45 | 2,394.49 | **9,505.70** |
| **AR ARKANSAS - Total** | **29,728.85** | **28,297.42** | **28,389.91** | **30,014.95** | **116,431.13** |

**STATE: AZ - ARIZONA**

| REGISTRANT ZIP CODE 3 | 1ST QUARTER | 2ND QUARTER | 3RD QUARTER | 4TH QUARTER | **TOTAL GRAMS** |
|---|---|---|---|---|---|
| 850 | 7,166.71 | 6,999.63 | 7,039.52 | 7,352.28 | **28,558.14** |
| 851 | 1,600.76 | 1,627.35 | 1,658.57 | 1,836.62 | **6,723.30** |
| 852 | 18,177.78 | 18,456.42 | 18,920.06 | 20,088.40 | **75,642.65** |

| | | | | | |
|---|---|---|---|---|---|
| 853 | 4,596.47 | 4,743.89 | 4,745.62 | 5,127.17 | **19,213.15** |
| 855 | 152.04 | 151.46 | 141.63 | 167.07 | **612.21** |
| 856 | 606.43 | 674.64 | 614.52 | 596.60 | **2,492.19** |
| 857 | 4,827.71 | 4,853.73 | 4,878.59 | 5,230.07 | **19,790.10** |
| 859 | 207.54 | 235.29 | 219.68 | 223.72 | **886.23** |
| 860 | 420.28 | 391.37 | 365.36 | 488.49 | **1,665.51** |
| 863 | 797.20 | 814.54 | 875.82 | 918.60 | **3,406.17** |
| 864 | 336.45 | 335.30 | 376.92 | 374.03 | **1,422.70** |
| 865 | 2.31 | 3.47 | 2.31 | 0 | **8.09** |
| **AZ ARIZONA - Total** | **38,891.68** | **39,287.10** | **39,838.61** | **42,403.06** | **160,420.44** |

**STATE: CA - CALIFORNIA**

| REGISTRANT ZIP CODE 3 | 1ST QUARTER | 2ND QUARTER | 3RD QUARTER | 4TH QUARTER | TOTAL GRAMS |
|---|---|---|---|---|---|
| 900 | 6,462.00 | 6,861.47 | 6,785.74 | 7,160.35 | **27,269.56** |
| 902 | 4,994.78 | 5,156.65 | 4,987.27 | 5,167.06 | **20,305.76** |
| 903 | 105.21 | 85.56 | 105.21 | 88.45 | **384.44** |
| 904 | 1,227.31 | 1,272.40 | 1,270.09 | 1,385.13 | **5,154.92** |
| 905 | 708.75 | 856.74 | 829.57 | 921.49 | **3,316.56** |
| 906 | 1,083.36 | 1,034.80 | 985.66 | 1,152.15 | **4,255.97** |
| 907 | 1,492.08 | 1,529.07 | 1,551.62 | 1,577.06 | **6,149.83** |
| 908 | 849.81 | 866.57 | 880.45 | 993.18 | **3,590.00** |
| 910 | 758.47 | 740.55 | 802.98 | 778.70 | **3,080.69** |
| 911 | 1,516.36 | 1,442.36 | 1,325.01 | 1,472.42 | **5,756.14** |
| 912 | 499.48 | 515.67 | 541.68 | 507.57 | **2,064.40** |
| 913 | 5,959.63 | 6,023.22 | 5,970.62 | 5,982.76 | **23,936.23** |
| 914 | 1,373.57 | 1,392.06 | 1,353.91 | 1,305.93 | **5,425.47** |
| 915 | 543.41 | 498.32 | 483.29 | 590.24 | **2,115.27** |
| 916 | 545.15 | 664.82 | 616.25 | 685.63 | **2,511.84** |
| 917 | 2,934.44 | 3,031.56 | 2,760.43 | 3,092.26 | **11,818.68** |
| 918 | 104.64 | 120.24 | 133.54 | 125.45 | **483.87** |
| 919 | 864.26 | 805.29 | 896.06 | 857.90 | **3,423.51** |
| 920 | 13,770.34 | 13,962.85 | 14,051.30 | 13,847.23 | **55,631.72** |
| 921 | 5,040.45 | 5,004.03 | 5,060.69 | 5,103.47 | **20,208.64** |
| 922 | 1,326.74 | 1,209.39 | 1,157.93 | 1,233.09 | **4,927.15** |
| 923 | 1,906.57 | 1,850.50 | 2,018.73 | 1,947.04 | **7,722.84** |
| 924 | 139.32 | 151.46 | 121.40 | 143.95 | **556.13** |
| 925 | 2,117.00 | 2,029.71 | 2,001.38 | 2,091.57 | **8,239.66** |
| 926 | 7,709.54 | 7,921.13 | 7,824.58 | 8,058.14 | **31,513.39** |
| 927 | 1,378.77 | 1,296.10 | 1,283.96 | 1,256.79 | **5,215.62** |
| 928 | 3,191.11 | 3,349.51 | 3,130.99 | 3,356.45 | **13,028.06** |
| 930 | 3,021.15 | 3,065.66 | 3,009.59 | 3,176.08 | **12,272.48** |

| | | | | | |
|---|---|---|---|---|---|
| 931 | 1,045.78 | 1,092.61 | 925.54 | 1,021.50 | **4,085.43** |
| 932 | 1,024.97 | 997.22 | 972.36 | 1,081.63 | **4,076.18** |
| 933 | 887.38 | 859.06 | 882.76 | 905.30 | **3,534.50** |
| 934 | 2,012.37 | 2,051.68 | 1,861.48 | 2,123.36 | **8,048.89** |
| 935 | 1,188.57 | 1,090.87 | 1,177.59 | 1,155.62 | **4,612.66** |
| 936 | 1,372.41 | 1,277.02 | 1,205.92 | 1,298.99 | **5,154.34** |
| 937 | 1,885.18 | 1,837.20 | 1,833.16 | 1,792.69 | **7,348.23** |
| 939 | 789.11 | 785.06 | 811.07 | 803.56 | **3,188.80** |
| 940 | 2,387.55 | 2,530.92 | 2,529.19 | 2,595.67 | **10,043.33** |
| 941 | 4,184.29 | 4,404.54 | 4,490.10 | 4,591.85 | **17,670.78** |
| 943 | 430.11 | 498.32 | 487.34 | 465.37 | **1,881.14** |
| 944 | 506.42 | 516.82 | 533.59 | 541.68 | **2,098.50** |
| 945 | 5,106.36 | 5,144.51 | 5,400.03 | 5,517.39 | **21,168.29** |
| 946 | 1,199.56 | 1,249.27 | 1,263.73 | 1,312.87 | **5,025.42** |
| 947 | 583.30 | 567.69 | 597.76 | 563.07 | **2,311.82** |
| 948 | 93.07 | 119.09 | 119.67 | 127.76 | **459.59** |
| 949 | 1,470.69 | 1,529.07 | 1,523.29 | 1,627.35 | **6,150.41** |
| 950 | 2,325.12 | 2,410.68 | 2,477.16 | 2,374.83 | **9,587.79** |
| 951 | 1,279.34 | 1,342.35 | 1,325.01 | 1,426.17 | **5,372.86** |
| 952 | 909.35 | 816.86 | 727.83 | 802.40 | **3,256.44** |
| 953 | 2,322.81 | 2,330.90 | 2,145.91 | 2,174.81 | **8,974.42** |
| 954 | 683.31 | 653.25 | 729.56 | 738.23 | **2,804.36** |
| 955 | 164.76 | 195.40 | 166.49 | 189.62 | **716.27** |
| 956 | 3,335.06 | 3,646.65 | 3,491.72 | 3,520.63 | **13,994.07** |
| 957 | 1,068.33 | 1,075.84 | 1,015.72 | 1,141.17 | **4,301.06** |
| 958 | 1,469.53 | 1,546.42 | 1,578.21 | 1,634.87 | **6,229.03** |
| 959 | 1,219.21 | 1,376.46 | 1,329.05 | 1,250.43 | **5,175.15** |
| 960 | 743.44 | 838.25 | 765.98 | 774.65 | **3,122.32** |
| 961 | 149.15 | 176.90 | 158.40 | 206.96 | **691.41** |
| **CA - CALIFORNIA - Total** | **113,460.22** | **115,698.62** | **114,465.53** | **117,817.94** | **461,442.31** |

**STATE: CO - COLORADO**

| REGISTRANT ZIP CODE 3 | 1ST QUARTER | 2ND QUARTER | 3RD QUARTER | 4TH QUARTER | **TOTAL GRAMS** |
|---|---|---|---|---|---|
| 800 | 4,284.30 | 4,357.72 | 4,517.85 | 4,657.75 | **17,817.62** |
| 801 | 5,380.95 | 5,217.93 | 5,582.13 | 5,556.12 | **21,737.14** |
| 802 | 6,019.18 | 6,382.22 | 6,186.83 | 6,587.45 | **25,175.68** |
| 803 | 1,717.54 | 1,785.17 | 1,741.24 | 1,895.59 | **7,139.54** |
| 804 | 1,008.21 | 922.07 | 982.77 | 982.77 | **3,895.82** |
| 805 | 3,241.98 | 3,316.56 | 3,517.16 | 3,591.74 | **13,667.44** |
| 806 | 1,087.41 | 1,151.00 | 1,146.95 | 1,268.35 | **4,653.71** |
| 807 | 175.74 | 160.13 | 152.04 | 149.73 | **637.64** |

| | | | | | |
|---|---|---|---|---|---|
| 808 | 476.35 | 511.04 | 491.39 | 512.20 | **1,990.98** |
| 809 | 3,213.66 | 3,075.49 | 3,248.92 | 3,389.40 | **12,927.47** |
| 810 | 693.72 | 630.71 | 609.32 | 672.33 | **2,606.07** |
| 811 | 111.57 | 101.17 | 101.17 | 97.70 | **411.61** |
| 812 | 118.51 | 147.42 | 161.29 | 168.23 | **595.44** |
| 813 | 195.40 | 179.21 | 228.93 | 222.57 | **826.10** |
| 814 | 160.13 | 157.82 | 182.68 | 159.56 | **660.19** |
| 815 | 449.18 | 515.09 | 554.98 | 586.77 | **2,106.02** |
| 816 | 765.98 | 809.34 | 823.21 | 816.86 | **3,215.39** |
| **CO - COLORADO - Total** | **29,099.82** | **29,420.09** | **30,228.85** | **31,315.10** | **120,063.85** |

**STATE: CT - CONNECTICUT**

| REGISTRANT ZIP CODE 3 | 1ST QUARTER | 2ND QUARTER | 3RD QUARTER | 4TH QUARTER | TOTAL GRAMS |
|---|---|---|---|---|---|
| 060 | 5,945.18 | 6,320.37 | 6,061.38 | 6,362.57 | **24,689.49** |
| 061 | 2,283.50 | 2,208.34 | 2,095.61 | 2,216.44 | **8,803.88** |
| 062 | 1,834.89 | 1,829.69 | 1,639.49 | 1,960.92 | **7,264.98** |
| 063 | 3,687.70 | 3,916.63 | 3,801.01 | 3,991.20 | **15,396.54** |
| 064 | 7,200.81 | 7,301.98 | 7,088.08 | 7,585.83 | **29,176.71** |
| 065 | 1,818.12 | 1,884.03 | 1,762.63 | 2,191.58 | **7,656.36** |
| 066 | 1,481.67 | 1,432.53 | 1,383.97 | 1,422.70 | **5,720.88** |
| 067 | 2,906.69 | 2,950.04 | 2,788.18 | 3,043.12 | **11,688.03** |
| 068 | 7,427.43 | 7,492.18 | 7,483.50 | 7,805.51 | **30,208.62** |
| 069 | 1,201.29 | 1,194.35 | 1,138.28 | 1,123.25 | **4,657.17** |
| **CT CONNECTICUT Total** | **35,787.28** | **36,530.14** | **35,242.13** | **37,703.10** | **145,262.66** |

**STATE: DC - DISTRICT OF COLUMBIA**

| REGISTRANT ZIP CODE 3 | 1ST QUARTER | 2ND QUARTER | 3RD QUARTER | 4TH QUARTER | TOTAL GRAMS |
|---|---|---|---|---|---|
| 200 | 5,844.01 | 6,087.97 | 6,226.72 | 6,437.14 | **24,595.84** |
| 203 | 0 | 21.39 | 40.47 | 22.55 | **84.40** |
| 204 | 32.37 | 30.64 | 161.29 | 17.34 | **241.65** |
| **DC - DISTRICT OF COLUMBIA** | **5,876.39** | **6,140.00** | **6,428.47** | **6,477.03** | **24,921.89** |

**STATE: DE - DELAWARE**

| REGISTRANT ZIP CODE 3 | 1ST QUARTER | 2ND QUARTER | 3RD QUARTER | 4TH QUARTER | TOTAL GRAMS |
|---|---|---|---|---|---|
| 197 | 5,119.08 | 5,073.98 | 4,897.66 | 5,221.40 | **20,312.12** |
| 198 | 3,248.34 | 3,057.57 | 3,189.96 | 3,314.83 | **12,810.70** |
| 199 | 5,346.85 | 5,256.66 | 5,286.15 | 5,445.70 | **21,335.36** |
| **DE DELAWARE Total** | **13,714.27** | **13,388.22** | **13,373.77** | **13,981.93** | **54,458.18** |

**STATE: FL - FLORIDA**

| REGISTRANT ZIP CODE 3 | 1ST QUARTER | 2ND QUARTER | 3RD QUARTER | 4TH QUARTER | TOTAL GRAMS |
|---|---|---|---|---|---|
| 320 | 4,668.74 | 4,741.58 | 4,695.33 | 5,039.88 | **19,145.52** |
| 321 | 3,007.85 | 2,954.09 | 2,757.54 | 2,833.27 | **11,552.75** |
| 322 | 8,250.64 | 8,000.90 | 7,892.22 | 8,751.86 | **32,895.62** |

| | | | | | |
|---|---|---|---|---|---|
| 323 | 5,332.97 | 5,200.01 | 5,148.56 | 5,563.06 | **21,244.60** |
| 324 | 2,797.43 | 2,744.82 | 2,744.24 | 2,733.83 | **11,020.32** |
| 325 | 8,070.28 | 8,130.40 | 7,776.02 | 8,035.59 | **32,012.29** |
| 326 | 3,167.41 | 2,981.84 | 2,922.30 | 3,174.35 | **12,245.89** |
| 327 | 8,987.72 | 8,817.18 | 8,833.95 | 9,135.14 | **35,773.98** |
| 328 | 7,477.46 | 7,220.18 | 6,904.94 | 7,352.85 | **28,955.44** |
| 329 | 6,306.49 | 6,248.10 | 6,118.03 | 6,397.83 | **25,070.46** |
| 330 | 6,681.10 | 6,299.56 | 5,779.84 | 6,489.75 | **25,250.25** |
| 331 | 8,324.06 | 8,221.74 | 7,719.37 | 8,579.58 | **32,844.75** |
| 333 | 3,537.97 | 3,528.14 | 3,388.82 | 3,639.14 | **14,094.08** |
| 334 | 9,326.49 | 9,036.86 | 8,751.86 | 9,632.30 | **36,747.50** |
| 335 | 6,918.12 | 6,804.24 | 6,636.59 | 6,890.95 | **27,249.90** |
| 336 | 6,683.41 | 6,337.13 | 6,124.39 | 6,298.98 | **25,443.92** |
| 337 | 4,872.98 | 4,867.02 | 4,730.01 | 5,058.95 | **19,528.97** |
| 338 | 5,600.05 | 5,234.12 | 4,932.93 | 5,497.15 | **21,264.25** |
| 339 | 5,175.73 | 4,852.57 | 4,851.99 | 5,432.41 | **20,312.70** |
| 341 | 2,362.69 | 2,121.63 | 2,138.97 | 2,471.96 | **9,095.25** |
| 342 | 5,616.24 | 5,711.05 | 5,480.39 | 6,070.05 | **22,877.73** |
| 344 | 2,447.68 | 2,314.13 | 2,277.14 | 2,337.84 | **9,376.78** |
| 346 | 4,314.36 | 4,408.01 | 4,131.10 | 4,390.67 | **17,244.14** |
| 347 | 5,306.38 | 4,908.07 | 4,777.42 | 5,112.72 | **20,104.58** |
| 349 | 2,874.89 | 2,986.46 | 2,824.02 | 3,134.46 | **11,819.83** |
| **FL FLORIDA - Total** | **138,109.16** | **134,669.84** | **130,337.96** | **140,054.55** | **543,171.52** |

**STATE: GA - GEORGIA**

| REGISTRANT ZIP CODE 3 | 1ST QUARTER | 2ND QUARTER | 3RD QUARTER | 4TH QUARTER | TOTAL GRAMS |
|---|---|---|---|---|---|
| 300 | 22,883.51 | 22,122.15 | 22,681.18 | 23,213.61 | **90,900.44** |
| 301 | 11,456.21 | 11,181.03 | 11,808.27 | 12,081.13 | **46,526.64** |
| 302 | 7,510.10 | 6,916.39 | 7,426.85 | 7,696.82 | **29,550.16** |
| 303 | 12,264.39 | 12,160.33 | 12,148.19 | 12,936.72 | **49,509.64** |
| 304 | 2,601.45 | 2,617.06 | 2,538.44 | 2,828.07 | **10,585.01** |
| 305 | 6,218.04 | 5,877.54 | 6,066.58 | 6,333.66 | **24,495.83** |
| 306 | 5,149.71 | 4,745.04 | 4,918.47 | 5,350.32 | **20,163.55** |
| 307 | 2,374.26 | 2,186.37 | 2,295.64 | 2,408.94 | **9,265.21** |
| 308 | 1,998.49 | 1,817.55 | 1,918.14 | 2,075.96 | **7,810.13** |
| 309 | 2,796.85 | 2,551.73 | 2,740.19 | 2,706.09 | **10,794.86** |
| 310 | 4,767.01 | 4,830.60 | 5,028.31 | 5,019.64 | **19,645.57** |
| 312 | 2,086.94 | 2,170.19 | 1,990.40 | 2,148.80 | **8,396.32** |
| 313 | 2,476.00 | 2,326.27 | 2,422.82 | 2,570.23 | **9,795.33** |
| 314 | 3,109.60 | 2,939.06 | 2,885.88 | 3,096.88 | **12,031.42** |
| 315 | 3,016.53 | 2,819.39 | 3,035.03 | 3,109.02 | **11,979.97** |

| | | | | | |
|---|---|---|---|---|---|
| 316 | 1,412.88 | 1,278.76 | 1,304.77 | 1,379.35 | **5,375.75** |
| 317 | 3,085.32 | 2,622.26 | 2,822.28 | 2,857.55 | **11,387.41** |
| 318 | 251.47 | 213.32 | 175.74 | 219.68 | **860.21** |
| 319 | 2,032.02 | 2,181.75 | 2,066.71 | 1,978.84 | **8,259.31** |
| 398 | 383.86 | 367.09 | 379.23 | 428.95 | **1,559.14** |
| **GA GEORGIA Total** | **97,874.64** | **93,923.91** | **96,653.12** | **100,440.25** | **388,891.92** |

**STATE: GU - GUAM**

| REGISTRANT ZIP CODE 3 | 1ST QUARTER | 2ND QUARTER | 3RD QUARTER | 4TH QUARTER | TOTAL GRAMS |
|---|---|---|---|---|---|
| 969 | 11.56 | 25.44 | 0.58 | 19.08 | **56.65** |
| **GU - GUAM - Total** | **11.56** | **25.44** | **0.58** | **19.08** | **56.65** |

**STATE: HI - HAWAII**

| REGISTRANT ZIP CODE 3 | 1ST QUARTER | 2ND QUARTER | 3RD QUARTER | 4TH QUARTER | TOTAL GRAMS |
|---|---|---|---|---|---|
| 967 | 792.00 | 881.60 | 878.13 | 998.96 | **3,550.69** |
| 968 | 687.36 | 751.53 | 840.56 | 923.80 | **3,203.25** |
| **HI - HAWAII - Total** | **1,479.36** | **1,633.13** | **1,718.69** | **1,922.76** | **6,753.94** |

**STATE: IA - IOWA**

| REGISTRANT ZIP CODE 3 | 1ST QUARTER | 2ND QUARTER | 3RD QUARTER | 4TH QUARTER | TOTAL GRAMS |
|---|---|---|---|---|---|
| 500 | 2,379.46 | 2,280.60 | 2,375.41 | 2,559.25 | **9,594.73** |
| 501 | 1,379.35 | 1,375.88 | 1,416.35 | 1,436.00 | **5,607.57** |
| 502 | 1,594.98 | 1,571.85 | 1,523.87 | 1,626.77 | **6,317.48** |
| 503 | 2,639.03 | 2,676.60 | 2,752.91 | 3,009.01 | **11,077.55** |
| 504 | 893.74 | 848.07 | 892.59 | 888.54 | **3,522.94** |
| 505 | 1,510.00 | 1,529.65 | 1,421.55 | 1,573.59 | **6,034.79** |
| 506 | 1,346.97 | 1,326.74 | 1,370.10 | 1,415.19 | **5,459.00** |
| 507 | 1,150.42 | 1,178.75 | 1,151.58 | 1,123.25 | **4,603.99** |
| 508 | 226.04 | 205.23 | 203.49 | 216.79 | **851.54** |
| 510 | 1,057.92 | 1,011.10 | 1,009.94 | 1,041.74 | **4,120.70** |
| 511 | 2,530.34 | 2,603.76 | 2,565.61 | 2,749.44 | **10,449.16** |
| 512 | 450.34 | 404.67 | 437.62 | 423.75 | **1,716.38** |
| 513 | 580.41 | 580.99 | 525.49 | 549.20 | **2,236.09** |
| 514 | 312.17 | 295.41 | 300.61 | 299.46 | **1,207.65** |
| 515 | 1,743.55 | 1,851.65 | 1,797.31 | 1,933.74 | **7,326.26** |
| 516 | 187.88 | 212.74 | 149.73 | 186.73 | **737.08** |
| 520 | 1,688.63 | 1,714.07 | 1,710.02 | 1,774.19 | **6,886.91** |
| 521 | 165.91 | 189.62 | 203.49 | 234.71 | **793.73** |
| 522 | 2,350.55 | 2,154.58 | 2,229.73 | 2,396.22 | **9,131.09** |
| 523 | 1,737.77 | 1,744.71 | 1,762.63 | 1,806.56 | **7,051.66** |
| 524 | 2,370.79 | 2,379.46 | 2,385.24 | 2,415.30 | **9,550.79** |
| 525 | 970.05 | 997.80 | 988.55 | 1,011.68 | **3,968.08** |
| 526 | 1,053.88 | 1,091.45 | 1,026.13 | 1,046.94 | **4,218.40** |

| | | | | | |
|---|---|---|---|---|---|
| 527 | 1,604.81 | 1,742.39 | 1,616.95 | 1,699.61 | **6,663.76** |
| 528 | 1,412.30 | 1,316.33 | 1,377.61 | 1,482.25 | **5,588.49** |
| **IA IOWA Total** | **33,337.29** | **33,284.11** | **33,194.50** | **34,899.90** | **134,715.80** |

**STATE: ID - IDAHO**

| REGISTRANT ZIP CODE 3 | 1ST QUARTER | 2ND QUARTER | 3RD QUARTER | 4TH QUARTER | TOTAL GRAMS |
|---|---|---|---|---|---|
| 832 | 670.60 | 665.97 | 637.07 | 728.98 | **2,702.62** |
| 833 | 765.40 | 816.86 | 820.90 | 843.45 | **3,246.61** |
| 834 | 1,737.77 | 1,837.20 | 1,719.27 | 1,769.56 | **7,063.80** |
| 835 | 139.32 | 167.65 | 146.84 | 178.63 | **632.44** |
| 836 | 3,217.13 | 3,350.09 | 3,360.50 | 3,527.57 | **13,455.28** |
| 837 | 2,159.78 | 2,291.01 | 2,197.36 | 2,373.10 | **9,021.25** |
| 838 | 1,026.71 | 1,056.19 | 1,090.87 | 1,167.18 | **4,340.95** |
| **ID IDAHO Total** | **9,716.70** | **10,184.97** | **9,972.80** | **10,588.48** | **40,462.95** |

**STATE: IL - ILLINOIS**

| REGISTRANT ZIP CODE 3 | 1ST QUARTER | 2ND QUARTER | 3RD QUARTER | 4TH QUARTER | TOTAL GRAMS |
|---|---|---|---|---|---|
| 600 | 15,440.47 | 15,258.95 | 15,505.22 | 15,959.03 | **62,163.67** |
| 601 | 9,646.75 | 10,110.97 | 10,147.97 | 10,399.44 | **40,305.13** |
| 602 | 730.14 | 763.09 | 769.45 | 801.82 | **3,064.51** |
| 603 | 618.57 | 711.64 | 644.00 | 735.92 | **2,710.13** |
| 604 | 7,366.15 | 7,492.18 | 7,565.02 | 7,737.87 | **30,161.21** |
| 605 | 8,463.96 | 8,670.34 | 8,848.40 | 9,216.07 | **35,198.77** |
| 606 | 11,586.86 | 12,107.15 | 12,385.79 | 12,963.31 | **49,043.11** |
| 607 | 668.28 | 617.41 | 650.94 | 697.77 | **2,634.40** |
| 608 | 151.46 | 148.57 | 157.24 | 177.48 | **634.75** |
| 609 | 1,092.61 | 1,053.88 | 1,096.08 | 1,116.89 | **4,359.45** |
| 610 | 892.01 | 901.84 | 938.83 | 1,027.28 | **3,759.96** |
| 611 | 998.96 | 1,056.19 | 1,045.20 | 1,026.71 | **4,127.06** |
| 612 | 601.80 | 664.82 | 645.16 | 660.77 | **2,572.55** |
| 613 | 661.92 | 624.93 | 618.57 | 705.86 | **2,611.28** |
| 614 | 714.53 | 710.48 | 686.78 | 765.98 | **2,877.78** |
| 615 | 631.29 | 645.16 | 619.72 | 677.53 | **2,573.70** |
| 616 | 1,164.29 | 1,173.54 | 1,213.43 | 1,330.79 | **4,882.05** |
| 617 | 1,486.87 | 1,347.55 | 1,396.69 | 1,602.49 | **5,833.61** |
| 618 | 1,408.83 | 1,325.01 | 1,390.91 | 1,514.62 | **5,639.37** |
| 619 | 256.10 | 238.18 | 219.68 | 279.22 | **993.18** |
| 620 | 1,080.47 | 1,090.30 | 1,049.25 | 1,163.14 | **4,383.15** |
| 622 | 1,852.23 | 1,937.21 | 1,960.34 | 1,942.42 | **7,692.20** |
| 623 | 835.35 | 842.29 | 869.46 | 909.35 | **3,456.46** |
| 624 | 266.50 | 220.26 | 267.08 | 260.15 | **1,013.99** |
| 625 | 468.26 | 441.09 | 484.45 | 459.59 | **1,853.39** |

| | | | | | |
|---|---|---|---|---|---|
| 626 | 411.61 | 366.52 | 369.41 | 428.37 | **1,575.90** |
| 627 | 628.39 | 749.22 | 671.17 | 700.08 | **2,748.87** |
| 628 | 441.09 | 438.78 | 491.96 | 468.26 | **1,840.09** |
| 629 | 1,087.41 | 1,083.36 | 1,143.48 | 1,218.06 | **4,532.30** |
| **IL ILLINOIS Total** | **71,653.18** | **72,790.88** | **73,851.70** | **76,946.27** | **295,242.03** |

**STATE: IN - INDIANA**

| REGISTRANT ZIP CODE 3 | 1ST QUARTER | 2ND QUARTER | 3RD QUARTER | 4TH QUARTER | TOTAL GRAMS |
|---|---|---|---|---|---|
| 460 | 10,182.65 | 10,010.96 | 10,340.47 | 10,656.12 | **41,190.20** |
| 461 | 6,830.83 | 6,547.56 | 6,650.46 | 6,865.52 | **26,894.37** |
| 462 | 8,579.00 | 8,517.15 | 8,762.26 | 8,720.06 | **34,578.47** |
| 463 | 8,272.61 | 8,173.18 | 8,213.64 | 8,558.77 | **33,218.20** |
| 464 | 665.39 | 655.57 | 628.39 | 691.99 | **2,641.34** |
| 465 | 7,381.76 | 7,470.79 | 7,438.99 | 7,597.39 | **29,888.93** |
| 466 | 2,351.13 | 2,428.60 | 2,258.06 | 2,434.38 | **9,472.17** |
| 467 | 2,544.22 | 2,419.35 | 2,528.61 | 2,632.09 | **10,124.27** |
| 468 | 3,498.66 | 3,613.13 | 3,628.73 | 3,880.21 | **14,620.73** |
| 469 | 6,130.17 | 5,853.84 | 5,997.21 | 5,685.04 | **23,666.26** |
| 470 | 698.34 | 655.57 | 653.83 | 685.05 | **2,692.79** |
| 471 | 3,677.87 | 3,581.91 | 3,640.87 | 3,803.32 | **14,703.97** |
| 472 | 1,538.32 | 1,521.56 | 1,554.51 | 1,502.48 | **6,116.88** |
| 473 | 3,178.39 | 2,958.72 | 3,298.06 | 3,357.03 | **12,792.20** |
| 474 | 2,758.69 | 2,815.93 | 2,525.72 | 2,886.45 | **10,986.79** |
| 475 | 1,590.35 | 1,504.79 | 1,583.42 | 1,737.77 | **6,416.33** |
| 476 | 2,248.23 | 2,310.67 | 2,284.65 | 2,539.59 | **9,383.14** |
| 477 | 4,682.03 | 4,595.90 | 4,916.74 | 4,876.85 | **19,071.52** |
| 478 | 1,172.96 | 1,144.64 | 1,066.02 | 1,231.35 | **4,614.97** |
| 479 | 3,284.76 | 3,117.69 | 3,224.64 | 3,371.48 | **12,998.58** |
| **IN INDIANA Total** | **81,266.41** | **79,897.47** | **81,195.30** | **83,712.93** | **326,072.10** |

**STATE: KS - KANSAS**

| REGISTRANT ZIP CODE 3 | 1ST QUARTER | 2ND QUARTER | 3RD QUARTER | 4TH QUARTER | TOTAL GRAMS |
|---|---|---|---|---|---|
| 660 | 7,150.52 | 7,234.34 | 7,367.88 | 7,654.62 | **29,407.37** |
| 661 | 907.04 | 922.07 | 949.24 | 949.82 | **3,728.17** |
| 662 | 7,362.68 | 7,820.54 | 7,559.81 | 7,858.69 | **30,601.72** |
| 664 | 468.26 | 478.67 | 527.23 | 506.42 | **1,980.57** |
| 665 | 906.46 | 955.60 | 819.17 | 945.77 | **3,627.00** |
| 666 | 3,598.09 | 3,857.66 | 4,024.73 | 4,135.15 | **15,615.64** |
| 667 | 1,113.42 | 1,151.00 | 1,200.14 | 1,204.18 | **4,668.74** |
| 668 | 546.88 | 523.18 | 562.49 | 626.66 | **2,259.21** |
| 669 | 100.59 | 102.32 | 104.64 | 143.37 | **450.92** |
| 670 | 1,635.44 | 1,693.83 | 1,678.22 | 1,697.88 | **6,705.38** |

| | | | | | |
|---|---|---|---|---|---|
| 671 | 545.73 | 581.57 | 611.63 | 581.57 | **2,320.49** |
| 672 | 3,679.61 | 4,020.11 | 4,085.43 | 4,150.76 | **15,935.90** |
| 673 | 550.93 | 554.40 | 582.15 | 582.72 | **2,270.20** |
| 674 | 964.27 | 985.08 | 872.93 | 1,013.41 | **3,835.69** |
| 675 | 751.53 | 771.76 | 805.87 | 839.40 | **3,168.57** |
| 676 | 479.24 | 471.73 | 499.48 | 512.20 | **1,962.65** |
| 677 | 107.53 | 95.39 | 115.04 | 116.20 | **434.15** |
| 678 | 302.35 | 352.06 | 324.89 | 354.95 | **1,334.25** |
| 679 | 131.81 | 133.54 | 136.43 | 171.70 | **573.48** |
| **KS - KANSAS - Total** | **31,302.38** | **32,704.85** | **32,827.41** | **34,045.47** | **130,880.11** |

**STATE: KY - KENTUCKY**

| REGISTRANT ZIP CODE 3 | 1ST QUARTER | 2ND QUARTER | 3RD QUARTER | 4TH QUARTER | **TOTAL GRAMS** |
|---|---|---|---|---|---|
| 400 | 3,124.63 | 2,959.87 | 2,806.10 | 2,703.20 | **11,593.80** |
| 401 | 883.91 | 788.53 | 624.35 | 690.25 | **2,987.04** |
| 402 | 9,678.55 | 9,341.52 | 8,607.91 | 8,744.34 | **36,372.32** |
| 403 | 2,554.05 | 2,570.81 | 2,581.79 | 2,551.16 | **10,257.81** |
| 404 | 984.50 | 1,030.17 | 981.61 | 1,077.00 | **4,073.29** |
| 405 | 3,882.52 | 3,906.22 | 3,986.00 | 4,569.30 | **16,344.04** |
| 406 | 446.29 | 417.97 | 443.98 | 480.98 | **1,789.22** |
| 407 | 345.13 | 338.19 | 367.67 | 397.73 | **1,448.72** |
| 408 | 99.43 | 79.78 | 100.59 | 104.64 | **384.44** |
| 409 | 127.76 | 129.49 | 108.68 | 107.53 | **473.46** |
| 410 | 4,233.43 | 4,127.06 | 3,994.09 | 4,484.90 | **16,839.47** |
| 411 | 694.88 | 612.21 | 631.86 | 677.53 | **2,616.48** |
| 412 | 203.49 | 199.44 | 234.71 | 198.29 | **835.93** |
| 413 | 65.90 | 75.73 | 65.33 | 57.23 | **264.19** |
| 414 | 87.29 | 91.92 | 84.98 | 95.96 | **360.16** |
| 415 | 449.18 | 410.45 | 431.84 | 416.81 | **1,708.29** |
| 416 | 216.79 | 194.82 | 190.77 | 235.86 | **838.25** |
| 417 | 265.35 | 223.15 | 248.00 | 286.16 | **1,022.66** |
| 418 | 46.25 | 49.72 | 46.25 | 50.29 | **192.51** |
| 420 | 3,924.72 | 3,633.36 | 3,892.35 | 3,717.76 | **15,168.19** |
| 421 | 1,601.34 | 1,521.56 | 1,659.73 | 1,781.70 | **6,564.33** |
| 422 | 771.76 | 750.37 | 939.41 | 660.19 | **3,121.74** |
| 423 | 1,941.26 | 1,835.47 | 1,963.23 | 1,998.49 | **7,738.45** |
| 424 | 2,319.92 | 2,280.03 | 2,276.56 | 2,251.70 | **9,128.20** |
| 425 | 319.69 | 319.69 | 307.55 | 308.13 | **1,255.06** |
| 426 | 117.35 | 124.29 | 96.54 | 111.57 | **449.76** |
| 427 | 1,414.03 | 1,330.79 | 1,240.02 | 1,271.24 | **5,256.09** |
| **KY - KENTUCKY - Total** | **40,799.41** | **39,342.60** | **38,911.91** | **40,029.96** | **159,083.87** |

**STATE: LA - LOUISIANA**

| REGISTRANT ZIP CODE 3 | 1ST QUARTER | 2ND QUARTER | 3RD QUARTER | 4TH QUARTER | TOTAL GRAMS |
|---|---|---|---|---|---|
| 700 | 8,069.12 | 7,284.64 | 7,613.00 | 8,325.22 | **31,291.97** |
| 701 | 4,645.61 | 4,397.61 | 4,492.42 | 4,924.83 | **18,460.47** |
| 703 | 5,336.44 | 4,840.43 | 5,191.34 | 5,487.33 | **20,855.54** |
| 704 | 8,818.92 | 8,313.08 | 8,846.09 | 9,258.27 | **35,236.35** |
| 705 | 16,959.14 | 15,689.63 | 16,091.41 | 17,038.92 | **65,779.11** |
| 706 | 8,737.98 | 8,365.11 | 8,310.77 | 8,984.25 | **34,398.11** |
| 707 | 11,707.68 | 11,250.40 | 11,657.96 | 11,725.02 | **46,341.07** |
| 708 | 8,553.57 | 7,286.37 | 7,763.30 | 7,890.49 | **31,493.73** |
| 710 | 1,952.24 | 1,983.46 | 2,021.04 | 2,090.41 | **8,047.15** |
| 711 | 5,959.05 | 5,601.21 | 5,914.54 | 6,229.03 | **23,703.83** |
| 712 | 4,121.85 | 3,922.41 | 4,073.29 | 4,418.42 | **16,535.97** |
| 713 | 3,358.18 | 3,162.79 | 3,226.95 | 3,451.26 | **13,199.18** |
| 714 | 1,829.69 | 1,633.13 | 1,692.10 | 1,834.89 | **6,989.81** |
| **LA - LOUISIANA - Total** | **90,049.48** | **83,730.27** | **86,894.21** | **91,658.33** | **352,332.29** |

**STATE: MA - MASSACHUSETTS**

| REGISTRANT ZIP CODE 3 | 1ST QUARTER | 2ND QUARTER | 3RD QUARTER | 4TH QUARTER | TOTAL GRAMS |
|---|---|---|---|---|---|
| 010 | 2,041.27 | 1,959.76 | 1,940.68 | 2,149.95 | **8,091.67** |
| 011 | 822.06 | 776.97 | 751.53 | 919.18 | **3,269.73** |
| 012 | 594.29 | 572.90 | 537.63 | 622.61 | **2,327.43** |
| 013 | 446.87 | 455.54 | 447.45 | 518.56 | **1,868.42** |
| 014 | 1,494.39 | 1,536.01 | 1,551.04 | 1,630.24 | **6,211.68** |
| 015 | 1,647.59 | 1,578.21 | 1,606.54 | 1,756.27 | **6,588.61** |
| 016 | 1,026.71 | 998.96 | 953.29 | 1,096.08 | **4,075.03** |
| 017 | 2,645.96 | 2,710.71 | 2,600.87 | 2,856.39 | **10,813.94** |
| 018 | 4,405.12 | 4,616.13 | 4,625.38 | 4,698.22 | **18,344.85** |
| 019 | 3,696.95 | 4,037.45 | 3,986.57 | 4,150.18 | **15,871.15** |
| 020 | 3,236.78 | 3,366.85 | 3,388.82 | 3,420.62 | **13,413.08** |
| 021 | 6,337.71 | 6,747.01 | 6,564.90 | 7,327.42 | **26,977.04** |
| 022 | 506.99 | 431.84 | 482.71 | 509.31 | **1,930.85** |
| 023 | 2,594.51 | 2,607.23 | 2,503.17 | 2,880.09 | **10,585.01** |
| 024 | 2,822.28 | 3,011.90 | 3,056.41 | 3,200.94 | **12,091.54** |
| 025 | 753.26 | 727.25 | 944.62 | 841.14 | **3,266.27** |
| 026 | 888.54 | 986.24 | 1,203.03 | 1,056.77 | **4,134.57** |
| 027 | 2,467.33 | 2,639.03 | 2,615.90 | 2,837.31 | **10,559.57** |
| **MA - MASSACHUSETTS - Total** | **38,428.62** | **39,759.98** | **39,760.56** | **42,471.27** | **160,420.43** |

**STATE: MD - MARYLAND**

| REGISTRANT ZIP CODE 3 | 1ST QUARTER | 2ND QUARTER | 3RD QUARTER | 4TH QUARTER | TOTAL GRAMS |
|---|---|---|---|---|---|
| 206 | 3,188.22 | 3,232.74 | 3,050.06 | 3,213.08 | **12,684.09** |

| | | | | | |
|---|---|---|---|---|---|
| 207 | 3,956.52 | 3,966.92 | 3,943.80 | 4,003.92 | **15,871.16** |
| 208 | 7,690.46 | 7,829.79 | 7,604.33 | 8,078.37 | **31,202.95** |
| 209 | 1,487.45 | 1,542.37 | 1,470.69 | 1,414.61 | **5,915.12** |
| 210 | 10,553.22 | 10,724.91 | 10,490.78 | 11,303.01 | **43,071.92** |
| 211 | 5,087.86 | 5,298.86 | 5,017.33 | 5,563.63 | **20,967.69** |
| 212 | 9,779.78 | 9,992.46 | 9,453.67 | 10,589.64 | **39,815.54** |
| 214 | 1,527.92 | 1,507.11 | 1,643.54 | 1,651.05 | **6,329.62** |
| 215 | 1,863.22 | 1,814.66 | 1,828.53 | 1,791.53 | **7,297.93** |
| 216 | 2,254.59 | 2,188.11 | 2,108.91 | 2,289.28 | **8,840.88** |
| 217 | 6,003.57 | 6,109.36 | 6,204.17 | 6,449.86 | **24,766.96** |
| 218 | 2,305.46 | 2,316.45 | 2,243.03 | 2,265.00 | **9,129.93** |
| 219 | 1,209.96 | 1,259.68 | 1,247.54 | 1,335.41 | **5,052.59** |
| **MD MARYLAND Total** | **56,908.22** | **57,783.41** | **56,306.36** | **59,948.39** | **230,946.38** |

**STATE: ME - MAINE**

| REGISTRANT ZIP CODE 3 | 1ST QUARTER | 2ND QUARTER | 3RD QUARTER | 4TH QUARTER | TOTAL GRAMS |
|---|---|---|---|---|---|
| 039 | 603.54 | 587.35 | 590.82 | 567.69 | **2,349.40** |
| 040 | 5,413.91 | 5,655.55 | 5,747.47 | 5,656.13 | **22,473.06** |
| 041 | 2,067.86 | 2,059.77 | 2,010.63 | 2,075.38 | **8,213.64** |
| 042 | 1,855.70 | 2,000.23 | 1,855.12 | 2,077.69 | **7,788.74** |
| 043 | 843.45 | 858.48 | 894.32 | 907.62 | **3,503.86** |
| 044 | 1,863.22 | 1,815.81 | 1,769.56 | 2,053.41 | **7,502.00** |
| 045 | 391.95 | 356.11 | 378.08 | 374.61 | **1,500.75** |
| 046 | 728.98 | 767.72 | 785.64 | 739.97 | **3,022.31** |
| 047 | 785.06 | 838.25 | 797.20 | 749.22 | **3,169.72** |
| 048 | 316.80 | 371.72 | 361.89 | 364.78 | **1,415.19** |
| 049 | 1,613.48 | 1,710.60 | 1,805.41 | 1,788.06 | **6,917.54** |
| **ME MAINE Total** | **16,483.94** | **17,021.58** | **16,996.14** | **17,354.56** | **67,856.22** |

**STATE: MI - MICHIGAN**

| REGISTRANT ZIP CODE 3 | 1ST QUARTER | 2ND QUARTER | 3RD QUARTER | 4TH QUARTER | TOTAL GRAMS |
|---|---|---|---|---|---|
| 480 | 7,896.85 | 8,007.26 | 7,941.36 | 8,428.70 | **32,274.17** |
| 481 | 7,545.94 | 7,841.35 | 7,545.36 | 8,289.95 | **31,222.60** |
| 482 | 1,504.79 | 1,671.29 | 1,648.16 | 1,675.91 | **6,500.16** |
| 483 | 7,178.27 | 7,273.65 | 7,304.29 | 7,696.82 | **29,453.04** |
| 484 | 1,900.21 | 1,875.36 | 1,918.14 | 2,070.75 | **7,764.46** |
| 485 | 779.28 | 830.15 | 762.51 | 754.42 | **3,126.36** |
| 486 | 2,530.34 | 2,514.16 | 2,395.07 | 2,488.14 | **9,927.71** |
| 487 | 998.96 | 1,030.17 | 987.97 | 1,049.25 | **4,066.36** |
| 488 | 4,795.34 | 4,757.18 | 4,660.06 | 5,205.21 | **19,417.80** |
| 489 | 1,241.18 | 1,181.06 | 1,182.21 | 1,352.18 | **4,956.63** |
| 490 | 5,640.52 | 5,872.92 | 5,603.52 | 6,269.49 | **23,386.46** |

| | | | | | |
|---|---|---|---|---|---|
| 491 | 872.35 | 814.54 | 780.44 | 803.56 | **3,270.89** |
| 492 | 1,549.31 | 1,518.09 | 1,427.91 | 1,563.18 | **6,058.49** |
| 493 | 1,895.01 | 1,955.71 | 1,867.84 | 1,977.68 | **7,696.25** |
| 494 | 5,287.88 | 5,471.14 | 5,327.19 | 5,796.61 | **21,882.82** |
| 495 | 4,227.07 | 4,425.36 | 4,282.56 | 4,490.10 | **17,425.09** |
| 496 | 2,462.13 | 2,606.07 | 2,586.42 | 2,645.39 | **10,300.01** |
| 497 | 1,339.46 | 1,500.75 | 1,455.66 | 1,527.92 | **5,823.78** |
| 498 | 1,182.79 | 1,277.60 | 1,302.46 | 1,273.55 | **5,036.41** |
| 499 | 351.48 | 363.05 | 367.09 | 433.00 | **1,514.62** |
| **MI - MICHIGAN Total** | **61,179.17** | **62,786.86** | **61,346.24** | **65,791.83** | **251,104.09** |

**STATE: MN - MINNESOTA**

| REGISTRANT ZIP CODE 3 | 1ST QUARTER | 2ND QUARTER | 3RD QUARTER | 4TH QUARTER | TOTAL GRAMS |
|---|---|---|---|---|---|
| 550 | 2,939.06 | 3,050.06 | 2,928.65 | 3,272.05 | **12,189.82** |
| 551 | 4,253.66 | 4,314.36 | 4,369.86 | 4,725.39 | **17,663.27** |
| 553 | 5,892.57 | 5,920.90 | 5,953.27 | 6,488.02 | **24,254.76** |
| 554 | 6,204.17 | 6,460.27 | 6,491.48 | 7,227.98 | **26,383.91** |
| 556 | 21.97 | 13.87 | 27.17 | 14.45 | **77.47** |
| 557 | 1,067.75 | 1,085.09 | 1,093.19 | 1,300.15 | **4,546.18** |
| 558 | 492.54 | 469.42 | 482.14 | 521.45 | **1,965.54** |
| 559 | 1,196.09 | 1,329.63 | 1,369.52 | 1,471.84 | **5,367.08** |
| 560 | 2,789.91 | 2,870.27 | 2,986.46 | 3,192.85 | **11,839.49** |
| 561 | 642.85 | 620.88 | 648.63 | 687.36 | **2,599.72** |
| 562 | 670.02 | 684.47 | 722.05 | 799.51 | **2,876.05** |
| 563 | 1,401.31 | 1,482.83 | 1,451.61 | 1,602.49 | **5,938.24** |
| 564 | 856.74 | 791.42 | 828.42 | 877.56 | **3,354.14** |
| 565 | 1,363.74 | 1,403.63 | 1,522.14 | 1,548.15 | **5,837.65** |
| 566 | 850.96 | 746.33 | 724.36 | 927.27 | **3,248.92** |
| 567 | 452.65 | 513.93 | 487.92 | 542.84 | **1,997.34** |
| **MN MINNESOTA Total** | **31,096.00** | **31,757.35** | **32,086.86** | **35,199.35** | **130,139.56** |

**STATE: MO - MISSOURI**

| REGISTRANT ZIP CODE 3 | 1ST QUARTER | 2ND QUARTER | 3RD QUARTER | 4TH QUARTER | TOTAL GRAMS |
|---|---|---|---|---|---|
| 630 | 7,493.33 | 7,419.34 | 7,315.28 | 7,723.42 | **29,951.36** |
| 631 | 29,842.68 | 27,680.01 | 25,567.63 | 26,439.98 | **109,530.29** |
| 633 | 7,136.07 | 7,268.45 | 7,414.13 | 7,650.00 | **29,468.65** |
| 634 | 504.68 | 497.17 | 605.27 | 588.51 | **2,195.62** |
| 635 | 505.84 | 472.89 | 511.62 | 565.38 | **2,055.72** |
| 636 | 783.90 | 774.65 | 827.84 | 815.70 | **3,202.10** |
| 637 | 546.30 | 491.39 | 553.82 | 561.34 | **2,152.84** |
| 638 | 1,113.42 | 994.33 | 1,059.08 | 1,108.80 | **4,275.63** |
| 639 | 412.76 | 378.08 | 365.36 | 392.53 | **1,548.73** |

| | | | | | |
|---|---|---|---|---|---|
| 640 | 5,651.51 | 5,924.37 | 5,634.16 | 5,942.29 | **23,152.33** |
| 641 | 7,330.31 | 6,098.96 | 5,691.39 | 5,895.46 | **25,016.12** |
| 644 | 368.83 | 388.48 | 386.17 | 408.14 | **1,551.62** |
| 645 | 1,030.75 | 1,041.16 | 952.71 | 1,047.52 | **4,072.14** |
| 646 | 182.10 | 190.77 | 182.10 | 180.95 | **735.92** |
| 647 | 713.38 | 705.86 | 746.33 | 757.89 | **2,923.45** |
| 648 | 1,037.11 | 957.33 | 1,016.30 | 1,044.63 | **4,055.37** |
| 650 | 499.48 | 512.20 | 513.35 | 465.95 | **1,990.98** |
| 651 | 707.02 | 678.11 | 705.86 | 765.98 | **2,856.97** |
| 652 | 2,165.56 | 2,165.56 | 2,203.14 | 2,289.85 | **8,824.12** |
| 653 | 441.67 | 443.40 | 383.86 | 382.12 | **1,651.05** |
| 654 | 441.67 | 374.03 | 398.89 | 449.18 | **1,663.77** |
| 655 | 547.46 | 544.57 | 540.52 | 526.65 | **2,159.20** |
| 656 | 716.27 | 711.06 | 719.16 | 680.42 | **2,826.91** |
| 657 | 1,142.33 | 1,186.84 | 1,185.11 | 1,200.14 | **4,714.41** |
| 658 | 1,817.55 | 1,755.11 | 1,697.88 | 1,641.80 | **6,912.34** |
| **MO MISSOURI - Total** | **73,131.96** | **69,654.11** | **67,176.95** | **69,524.62** | **279,487.65** |

**STATE: MS - MISSISSIPPI**

| REGISTRANT ZIP CODE 3 | 1ST QUARTER | 2ND QUARTER | 3RD QUARTER | 4TH QUARTER | TOTAL GRAMS |
|---|---|---|---|---|---|
| 386 | 6,469.52 | 6,221.51 | 6,196.08 | 6,856.84 | **25,743.95** |
| 387 | 598.91 | 461.32 | 510.46 | 475.78 | **2,046.47** |
| 388 | 2,803.21 | 2,688.17 | 2,721.12 | 2,954.67 | **11,167.16** |
| 389 | 738.81 | 775.81 | 755.58 | 757.89 | **3,028.09** |
| 390 | 2,758.69 | 2,412.99 | 2,587.00 | 2,713.60 | **10,472.28** |
| 391 | 3,045.43 | 2,899.17 | 3,011.32 | 3,050.06 | **12,005.98** |
| 392 | 2,838.47 | 2,731.52 | 2,726.90 | 2,890.50 | **11,187.39** |
| 393 | 2,286.39 | 2,191.58 | 2,370.21 | 2,532.08 | **9,380.25** |
| 394 | 6,400.72 | 5,951.54 | 6,166.01 | 6,446.97 | **24,965.25** |
| 395 | 5,801.81 | 5,264.18 | 5,387.31 | 5,681.57 | **22,134.87** |
| 396 | 1,736.03 | 1,546.42 | 1,596.13 | 1,722.74 | **6,601.32** |
| 397 | 1,947.62 | 1,681.11 | 1,693.25 | 1,776.50 | **7,098.49** |
| **MS MISSISSIPPI - Total** | **37,425.62** | **34,825.32** | **35,721.38** | **37,859.19** | **145,831.51** |

**STATE: MT - MONTANA**

| REGISTRANT ZIP CODE 3 | 1ST QUARTER | 2ND QUARTER | 3RD QUARTER | 4TH QUARTER | TOTAL GRAMS |
|---|---|---|---|---|---|
| 590 | 246.85 | 256.68 | 209.85 | 267.08 | **980.46** |
| 591 | 1,252.16 | 1,196.09 | 1,200.71 | 1,331.94 | **4,980.91** |
| 592 | 145.68 | 128.92 | 138.17 | 153.20 | **565.96** |
| 593 | 263.61 | 248.00 | 225.46 | 253.79 | **990.86** |
| 594 | 696.03 | 733.61 | 741.12 | 775.81 | **2,946.58** |
| 595 | 174.59 | 175.16 | 206.96 | 230.66 | **787.37** |

| | | | | | |
|---|---|---|---|---|---|
| 596 | 465.95 | 475.20 | 472.89 | 463.06 | **1,877.09** |
| 597 | 1,429.06 | 1,303.62 | 1,419.24 | 1,287.43 | **5,439.34** |
| 598 | 1,383.97 | 1,477.05 | 1,423.28 | 1,545.84 | **5,830.14** |
| 599 | 477.51 | 557.87 | 509.88 | 515.09 | **2,060.35** |
| **MT - MONTANA Total** | **6,535.42** | **6,552.19** | **6,547.56** | **6,823.89** | **26,459.06** |

**STATE: NC - NORTH CAROLINA**

| REGISTRANT ZIP CODE 3 | 1ST QUARTER | 2ND QUARTER | 3RD QUARTER | 4TH QUARTER | **TOTAL GRAMS** |
|---|---|---|---|---|---|
| 270 | 2,470.22 | 2,546.53 | 2,606.65 | 2,799.16 | **10,422.56** |
| 271 | 3,645.50 | 3,555.32 | 3,662.26 | 3,705.04 | **14,568.12** |
| 272 | 7,182.89 | 6,807.71 | 6,608.84 | 6,531.95 | **27,131.39** |
| 273 | 3,782.51 | 3,795.23 | 3,883.10 | 3,807.94 | **15,268.78** |
| 274 | 4,227.07 | 4,053.06 | 4,251.35 | 4,149.02 | **16,680.50** |
| 275 | 13,619.46 | 13,152.93 | 13,122.87 | 13,524.65 | **53,419.91** |
| 276 | 6,256.20 | 6,012.82 | 6,320.95 | 6,598.43 | **25,188.40** |
| 277 | 2,445.94 | 2,478.31 | 2,542.48 | 2,666.78 | **10,133.51** |
| 278 | 5,686.19 | 5,280.37 | 5,291.93 | 5,395.99 | **21,654.47** |
| 279 | 1,530.23 | 1,504.22 | 1,401.31 | 1,249.27 | **5,685.04** |
| 280 | 9,919.04 | 9,895.92 | 9,805.15 | 10,206.36 | **39,826.47** |
| 281 | 7,530.33 | 7,212.95 | 7,178.27 | 7,837.88 | **29,759.43** |
| 282 | 8,241.97 | 8,320.59 | 8,298.05 | 8,896.38 | **33,756.99** |
| 283 | 9,506.28 | 8,605.60 | 8,433.90 | 8,392.86 | **34,938.63** |
| 284 | 6,808.86 | 6,590.34 | 6,623.29 | 6,684.57 | **26,707.06** |
| 285 | 5,810.48 | 5,802.97 | 5,887.95 | 4,946.22 | **22,447.62** |
| 286 | 9,350.77 | 9,554.84 | 9,521.31 | 9,731.74 | **38,158.65** |
| 287 | 4,069.25 | 3,966.92 | 4,088.32 | 4,216.08 | **16,340.57** |
| 288 | 1,798.47 | 1,841.25 | 1,874.20 | 1,979.99 | **7,493.91** |
| 289 | 253.79 | 283.27 | 280.38 | 295.99 | **1,113.42** |
| **NC - NORTH CAROLINA Total** | **114,135.44** | **111,261.13** | **111,682.56** | **113,616.31** | **450,695.43** |

**STATE: ND - NORTH DAKOTA**

| REGISTRANT ZIP CODE 3 | 1ST QUARTER | 2ND QUARTER | 3RD QUARTER | 4TH QUARTER | **TOTAL GRAMS** |
|---|---|---|---|---|---|
| 580 | 568.85 | 604.69 | 545.73 | 598.91 | **2,318.18** |
| 581 | 1,769.56 | 1,775.92 | 1,786.91 | 1,963.81 | **7,296.20** |
| 582 | 1,276.44 | 1,166.61 | 1,202.45 | 1,272.40 | **4,917.90** |
| 583 | 305.24 | 282.11 | 268.24 | 328.36 | **1,183.95** |
| 584 | 202.34 | 231.82 | 209.27 | 234.71 | **878.13** |
| 585 | 618.57 | 528.96 | 620.30 | 612.79 | **2,380.62** |
| 586 | 160.71 | 168.23 | 167.07 | 160.71 | **656.72** |
| 587 | 966.01 | 923.23 | 979.30 | 981.61 | **3,850.15** |
| 588 | 282.11 | 287.89 | 284.43 | 298.30 | **1,152.73** |
| **ND NORTH DAKOTA Total** | **6,149.83** | **5,969.46** | **6,063.69** | **6,451.60** | **24,634.58** |

**STATE: NE - NEBRASKA**

| REGISTRANT ZIP CODE 3 | 1ST QUARTER | 2ND QUARTER | 3RD QUARTER | 4TH QUARTER | TOTAL GRAMS |
|---|---|---|---|---|---|
| 680 | 1,632.55 | 1,644.69 | 1,526.76 | 1,743.55 | **6,547.56** |
| 681 | 6,791.52 | 7,017.56 | 7,033.16 | 7,724.57 | **28,566.81** |
| 683 | 448.61 | 464.21 | 457.86 | 506.99 | **1,877.67** |
| 684 | 353.80 | 335.88 | 385.01 | 382.70 | **1,457.39** |
| 685 | 4,146.18 | 4,270.84 | 4,273.68 | 4,469.96 | **17,160.65** |
| 686 | 450.92 | 469.42 | 387.33 | 419.70 | **1,727.36** |
| 687 | 553.82 | 567.12 | 578.68 | 613.36 | **2,312.98** |
| 688 | 1,328.47 | 1,292.05 | 1,329.05 | 1,475.89 | **5,425.47** |
| 689 | 441.09 | 459.01 | 433.00 | 500.63 | **1,833.73** |
| 690 | 156.67 | 160.13 | 168.81 | 186.15 | **671.75** |
| 691 | 338.77 | 361.89 | 387.33 | 426.64 | **1,514.62** |
| 692 | 10.41 | 14.45 | 9.25 | 13.30 | **47.40** |
| 693 | 480.40 | 462.48 | 450.92 | 472.31 | **1,866.11** |
| **NE NEBRASKA Total** | **17,133.19** | **17,519.74** | **17,420.83** | **18,935.75** | **71,009.51** |

**STATE: NH - NEW HAMPSHIRE**

| REGISTRANT ZIP CODE 3 | 1ST QUARTER | 2ND QUARTER | 3RD QUARTER | 4TH QUARTER | TOTAL GRAMS |
|---|---|---|---|---|---|
| 030 | 4,099.31 | 4,135.73 | 3,994.67 | 4,222.44 | **16,452.15** |
| 031 | 2,358.65 | 2,330.32 | 2,340.73 | 2,323.96 | **9,353.66** |
| 032 | 1,114.58 | 1,127.30 | 1,181.06 | 1,190.31 | **4,613.24** |
| 033 | 924.38 | 890.27 | 869.46 | 949.82 | **3,633.94** |
| 034 | 395.42 | 408.14 | 425.48 | 454.96 | **1,684.01** |
| 035 | 302.92 | 298.30 | 277.49 | 303.50 | **1,182.21** |
| 036 | 35.26 | 34.11 | 38.15 | 37.00 | **144.53** |
| 037 | 835.93 | 834.78 | 839.40 | 970.63 | **3,480.74** |
| 038 | 4,288.92 | 4,347.31 | 4,420.15 | 4,590.69 | **17,647.08** |
| **NH - NEW HAMPSHIRE Total** | **14,355.38** | **14,406.25** | **14,386.60** | **15,043.32** | **58,191.55** |

**STATE: NJ - NEW JERSEY**

| REGISTRANT ZIP CODE 3 | 1ST QUARTER | 2ND QUARTER | 3RD QUARTER | 4TH QUARTER | TOTAL GRAMS |
|---|---|---|---|---|---|
| 070 | 8,752.43 | 8,548.94 | 8,462.81 | 8,822.38 | **34,586.57** |
| 071 | 553.82 | 443.98 | 453.81 | 552.66 | **2,004.27** |
| 072 | 384.44 | 294.83 | 279.22 | 338.77 | **1,297.26** |
| 073 | 667.71 | 660.77 | 646.32 | 680.42 | **2,655.21** |
| 074 | 3,458.19 | 3,597.52 | 3,463.40 | 3,758.23 | **14,277.34** |
| 075 | 341.66 | 306.39 | 310.44 | 326.63 | **1,285.12** |
| 076 | 3,235.05 | 3,258.75 | 3,396.92 | 3,412.52 | **13,303.24** |
| 077 | 3,220.68 | 3,187.64 | 3,195.74 | 3,246.61 | **12,850.67** |
| 078 | 2,668.51 | 2,892.81 | 2,667.93 | 2,929.23 | **11,158.49** |
| 079 | 3,170.30 | 3,182.44 | 3,201.52 | 3,174.93 | **12,729.18** |

| | | | | | |
|---|---|---|---|---|---|
| 080 | 8,525.24 | 8,485.35 | 8,390.54 | 8,980.21 | **34,381.34** |
| 081 | 566.54 | 559.02 | 544.57 | 575.79 | **2,245.92** |
| 082 | 1,142.33 | 1,171.23 | 1,249.27 | 1,092.61 | **4,655.44** |
| 083 | 755.58 | 827.26 | 709.91 | 767.72 | **3,060.46** |
| 084 | 125.45 | 117.93 | 141.63 | 83.82 | **468.84** |
| 085 | 2,154.58 | 2,144.17 | 2,064.97 | 2,223.95 | **8,587.68** |
| 086 | 1,519.25 | 1,474.16 | 1,296.10 | 1,366.63 | **5,656.13** |
| 087 | 2,169.61 | 2,172.50 | 2,170.19 | 2,312.98 | **8,825.27** |
| 088 | 4,002.19 | 4,054.22 | 4,055.37 | 4,363.50 | **16,475.27** |
| 089 | 448.03 | 427.22 | 410.45 | 443.40 | **1,729.10** |
| **NJ NEW JERSEY - Total** | **47,861.56** | **47,807.14** | **47,111.10** | **49,452.99** | **192,232.79** |

**STATE: NM - NEW MEXICO**

| REGISTRANT ZIP CODE 3 | 1ST QUARTER | 2ND QUARTER | 3RD QUARTER | 4TH QUARTER | TOTAL GRAMS |
|---|---|---|---|---|---|
| 870 | 204.07 | 232.97 | 197.71 | 220.83 | **855.59** |
| 871 | 1,637.76 | 1,623.30 | 1,647.01 | 1,603.65 | **6,511.72** |
| 873 | 37.58 | 19.66 | 14.45 | 27.75 | **99.43** |
| 874 | 154.93 | 127.18 | 145.68 | 157.82 | **585.62** |
| 875 | 390.80 | 437.04 | 405.83 | 360.16 | **1,593.82** |
| 877 | 75.15 | 58.97 | 50.29 | 78.62 | **263.04** |
| 878 | 38.73 | 33.53 | 50.29 | 47.40 | **169.96** |
| 879 | 24.86 | 15.61 | 18.50 | 24.28 | **83.25** |
| 880 | 416.23 | 404.67 | 424.90 | 524.91 | **1,770.72** |
| 881 | 127.76 | 138.74 | 69.37 | 108.10 | **443.98** |
| 882 | 554.98 | 516.24 | 469.42 | 514.51 | **2,055.15** |
| 883 | 166.49 | 168.81 | 154.93 | 187.30 | **677.53** |
| 884 | 20.81 | 24.28 | 16.76 | 20.81 | **82.67** |
| **NM NEW MEXICO Total** | **3,850.15** | **3,801.01** | **3,665.15** | **3,876.16** | **15,192.47** |

**STATE: NV - NEVADA**

| REGISTRANT ZIP CODE 3 | 1ST QUARTER | 2ND QUARTER | 3RD QUARTER | 4TH QUARTER | TOTAL GRAMS |
|---|---|---|---|---|---|
| 890 | 2,026.82 | 1,921.60 | 1,912.35 | 1,968.43 | **7,829.21** |
| 891 | 4,081.96 | 3,952.47 | 3,759.96 | 4,206.83 | **16,001.23** |
| 893 | 11.56 | 12.72 | 8.67 | 6.94 | **39.89** |
| 894 | 767.14 | 729.56 | 686.20 | 744.59 | **2,927.50** |
| 895 | 1,019.77 | 1,022.08 | 976.41 | 1,087.41 | **4,105.67** |
| 897 | 207.54 | 223.15 | 195.98 | 198.87 | **825.53** |
| 898 | 69.95 | 97.70 | 81.51 | 120.82 | **369.98** |
| **NV NEVADA Total** | **8,184.74** | **7,959.28** | **7,621.09** | **8,333.89** | **32,099.00** |

**STATE: NY - NEW YORK**

| REGISTRANT ZIP CODE 3 | 1ST QUARTER | 2ND QUARTER | 3RD QUARTER | 4TH QUARTER | TOTAL GRAMS |
|---|---|---|---|---|---|
| 100 | 17,853.46 | 18,737.38 | 19,212.00 | 20,316.75 | **76,119.58** |

| | | | | | |
|---|---|---|---|---|---|
| 101 | 905.30 | 955.02 | 960.22 | 853.85 | **3,674.40** |
| 102 | 143.37 | 150.31 | 139.32 | 137.01 | **570.01** |
| 103 | 1,087.41 | 980.46 | 1,004.16 | 1,138.86 | **4,210.88** |
| 104 | 1,109.37 | 1,046.94 | 1,040.58 | 1,089.14 | **4,286.03** |
| 105 | 5,074.56 | 5,133.53 | 5,259.55 | 5,469.40 | **20,937.05** |
| 106 | 572.90 | 556.71 | 561.91 | 588.51 | **2,280.03** |
| 107 | 783.90 | 645.74 | 635.33 | 844.03 | **2,909.00** |
| 108 | 391.37 | 395.42 | 364.78 | 437.62 | **1,589.20** |
| 109 | 2,628.62 | 2,676.60 | 2,623.42 | 2,840.21 | **10,768.85** |
| 110 | 1,437.16 | 1,418.08 | 1,349.29 | 1,573.59 | **5,778.11** |
| 111 | 763.09 | 767.14 | 892.59 | 912.24 | **3,335.06** |
| 112 | 5,882.17 | 5,735.33 | 5,691.39 | 6,242.32 | **23,551.22** |
| 113 | 1,838.36 | 1,622.73 | 1,788.06 | 1,748.17 | **6,997.32** |
| 114 | 553.82 | 460.17 | 459.01 | 538.21 | **2,011.21** |
| 115 | 4,380.84 | 4,227.07 | 4,166.37 | 4,524.79 | **17,299.06** |
| 116 | 325.47 | 313.91 | 327.20 | 352.06 | **1,318.65** |
| 117 | 8,855.91 | 8,674.97 | 9,052.47 | 9,540.38 | **36,123.73** |
| 118 | 537.05 | 559.02 | 527.23 | 549.77 | **2,173.08** |
| 119 | 886.81 | 1,005.89 | 1,238.87 | 1,067.17 | **4,198.74** |
| 120 | 1,821.02 | 1,914.09 | 1,898.48 | 1,852.23 | **7,485.82** |
| 121 | 1,177.01 | 1,097.23 | 1,131.34 | 1,153.31 | **4,558.90** |
| 122 | 748.06 | 720.31 | 718.58 | 815.12 | **3,002.07** |
| 123 | 783.90 | 856.17 | 701.24 | 818.01 | **3,159.32** |
| 124 | 485.03 | 452.65 | 474.62 | 457.28 | **1,869.58** |
| 125 | 2,307.20 | 2,171.34 | 2,389.87 | 2,593.93 | **9,462.34** |
| 126 | 535.90 | 522.60 | 505.26 | 565.38 | **2,129.14** |
| 127 | 363.62 | 343.97 | 390.80 | 359.58 | **1,457.97** |
| 128 | 1,445.83 | 1,488.03 | 1,443.52 | 1,540.64 | **5,918.01** |
| 129 | 432.42 | 445.14 | 501.21 | 506.99 | **1,885.76** |
| 130 | 1,732.57 | 1,800.20 | 1,879.40 | 1,728.52 | **7,140.69** |
| 131 | 744.01 | 744.59 | 756.15 | 734.19 | **2,978.95** |
| 132 | 967.16 | 820.32 | 790.26 | 933.05 | **3,510.80** |
| 133 | 481.56 | 486.76 | 527.23 | 478.09 | **1,973.63** |
| 134 | 653.25 | 669.44 | 667.71 | 744.59 | **2,734.99** |
| 135 | 203.49 | 215.05 | 184.99 | 220.83 | **824.37** |
| 136 | 1,258.52 | 1,229.04 | 1,295.52 | 1,264.88 | **5,047.97** |
| 137 | 582.15 | 538.79 | 502.95 | 506.42 | **2,130.30** |
| 138 | 727.25 | 838.25 | 761.94 | 787.37 | **3,114.80** |
| 139 | 537.63 | 563.07 | 519.13 | 586.77 | **2,206.61** |
| 140 | 1,531.39 | 1,616.95 | 1,625.04 | 1,713.49 | **6,486.86** |

| | | | | | |
|---|---|---|---|---|---|
| 141 | 609.32 | 591.97 | 630.13 | 663.08 | **2,494.50** |
| 142 | 2,201.98 | 2,152.27 | 2,279.45 | 2,373.10 | **9,006.80** |
| 143 | 164.18 | 160.13 | 158.40 | 168.23 | **650.94** |
| 144 | 2,252.28 | 2,278.87 | 2,292.74 | 2,577.75 | **9,401.64** |
| 145 | 1,845.87 | 1,734.88 | 1,779.39 | 1,866.68 | **7,226.83** |
| 146 | 2,774.88 | 2,948.31 | 2,758.12 | 3,175.50 | **11,656.81** |
| 147 | 585.62 | 588.51 | 570.58 | 574.63 | **2,319.34** |
| 148 | 1,176.43 | 1,237.71 | 1,187.42 | 1,311.13 | **4,912.69** |
| 149 | 306.97 | 365.94 | 285.00 | 291.94 | **1,249.85** |
| **NY - NEW YORK - Total** | **87,447.45** | **87,654.99** | **88,900.22** | **94,126.82** | **358,129.48** |

**STATE: OH - OHIO**

| REGISTRANT ZIP CODE 3 | 1ST QUARTER | 2ND QUARTER | 3RD QUARTER | 4TH QUARTER | **TOTAL GRAMS** |
|---|---|---|---|---|---|
| 430 | 9,353.08 | 9,203.93 | 9,276.77 | 9,625.37 | **37,459.15** |
| 431 | 3,705.04 | 3,666.89 | 3,649.55 | 3,817.19 | **14,838.67** |
| 432 | 6,645.84 | 6,490.91 | 6,511.14 | 6,821.00 | **26,468.89** |
| 433 | 855.01 | 808.18 | 848.07 | 931.90 | **3,443.16** |
| 434 | 954.44 | 909.35 | 994.91 | 1,015.72 | **3,874.43** |
| 435 | 2,085.21 | 2,123.36 | 2,006.59 | 2,194.47 | **8,409.62** |
| 436 | 2,697.41 | 2,653.48 | 2,583.53 | 2,652.90 | **10,587.32** |
| 437 | 978.15 | 1,022.66 | 1,051.56 | 1,008.21 | **4,060.57** |
| 438 | 185.57 | 169.38 | 172.27 | 191.35 | **718.58** |
| 439 | 1,156.20 | 1,193.78 | 1,149.26 | 1,245.23 | **4,744.47** |
| 440 | 9,112.01 | 9,029.92 | 8,958.24 | 9,517.84 | **36,618.01** |
| 441 | 10,310.41 | 10,310.41 | 10,114.44 | 10,372.85 | **41,108.11** |
| 442 | 4,698.22 | 4,561.79 | 4,815.57 | 4,984.96 | **19,060.54** |
| 443 | 1,896.75 | 1,838.36 | 1,837.78 | 1,953.98 | **7,526.86** |
| 444 | 2,132.61 | 2,113.53 | 2,071.91 | 2,188.11 | **8,506.16** |
| 445 | 1,919.87 | 1,918.14 | 1,907.73 | 2,038.96 | **7,784.69** |
| 446 | 3,033.29 | 3,118.85 | 2,971.43 | 3,191.11 | **12,314.69** |
| 447 | 2,548.26 | 2,460.97 | 2,590.47 | 2,516.47 | **10,116.17** |
| 448 | 1,760.89 | 1,824.48 | 1,790.38 | 1,840.67 | **7,216.42** |
| 449 | 741.70 | 682.74 | 620.88 | 769.45 | **2,814.77** |
| 450 | 6,354.01 | 6,253.31 | 6,159.66 | 6,692.66 | **25,459.64** |
| 451 | 2,239.56 | 2,134.35 | 2,297.37 | 2,240.72 | **8,911.99** |
| 452 | 7,312.39 | 7,401.99 | 7,451.13 | 7,906.67 | **30,072.18** |
| 453 | 3,294.59 | 3,107.29 | 3,296.33 | 3,435.07 | **13,133.28** |
| 454 | 3,621.80 | 3,544.33 | 3,446.05 | 3,636.83 | **14,249.01** |
| 455 | 672.91 | 634.18 | 561.34 | 675.22 | **2,543.64** |
| 456 | 1,763.78 | 1,677.07 | 1,612.90 | 1,757.42 | **6,811.17** |
| 457 | 1,412.30 | 1,193.20 | 1,225.57 | 1,307.66 | **5,138.73** |

| | | | | | |
|---|---|---|---|---|---|
| 458 | 1,116.89 | 1,112.84 | 1,061.97 | 1,148.68 | **4,440.39** |
| **OH - OHIO Total** | **94,558.20** | **93,159.66** | **93,034.79** | **97,678.67** | **378,431.31** |

**STATE: OK - OKLAHOMA**

| REGISTRANT ZIP CODE 3 | 1ST QUARTER | 2ND QUARTER | 3RD QUARTER | 4TH QUARTER | TOTAL GRAMS |
|---|---|---|---|---|---|
| 730 | 8,922.40 | 8,853.02 | 8,844.93 | 9,483.15 | **36,103.50** |
| 731 | 6,822.97 | 7,033.16 | 6,990.39 | 7,356.32 | **28,202.84** |
| 734 | 731.87 | 616.25 | 660.19 | 719.16 | **2,727.48** |
| 735 | 1,342.35 | 1,191.46 | 1,322.69 | 1,392.64 | **5,249.15** |
| 736 | 372.30 | 298.88 | 281.53 | 361.31 | **1,314.02** |
| 737 | 901.26 | 879.29 | 875.82 | 943.46 | **3,599.83** |
| 738 | 99.43 | 98.86 | 114.46 | 105.79 | **418.54** |
| 739 | 59.54 | 67.06 | 78.04 | 89.03 | **293.67** |
| 740 | 7,035.48 | 6,796.14 | 7,133.18 | 7,523.97 | **28,488.77** |
| 741 | 4,199.90 | 4,227.07 | 4,352.51 | 4,340.95 | **17,120.43** |
| 743 | 1,115.73 | 1,084.52 | 1,056.19 | 1,186.84 | **4,443.28** |
| 744 | 1,897.32 | 1,798.47 | 1,763.21 | 1,828.53 | **7,287.53** |
| 745 | 665.39 | 594.29 | 686.20 | 750.95 | **2,696.84** |
| 746 | 434.15 | 422.59 | 371.14 | 403.51 | **1,631.40** |
| 747 | 659.61 | 530.70 | 612.79 | 667.13 | **2,470.22** |
| 748 | 1,705.40 | 1,677.65 | 1,529.07 | 1,770.72 | **6,682.84** |
| 749 | 611.63 | 528.38 | 513.93 | 523.76 | **2,177.70** |
| **OK - OKLAHOMA Total** | **37,576.73** | **36,697.79** | **37,186.28** | **39,447.23** | **150,908.03** |

**STATE: OR - OREGON**

| REGISTRANT ZIP CODE 3 | 1ST QUARTER | 2ND QUARTER | 3RD QUARTER | 4TH QUARTER | TOTAL GRAMS |
|---|---|---|---|---|---|
| 970 | 3,541.44 | 3,738.57 | 3,762.27 | 3,905.07 | **14,947.35** |
| 971 | 1,085.09 | 1,153.31 | 1,087.41 | 1,167.76 | **4,493.57** |
| 972 | 4,612.66 | 4,971.66 | 4,841.01 | 5,097.69 | **19,523.02** |
| 973 | 1,625.04 | 1,708.86 | 1,714.07 | 1,882.29 | **6,930.26** |
| 974 | 1,796.16 | 1,825.64 | 1,803.67 | 2,043.58 | **7,469.05** |
| 975 | 602.38 | 651.52 | 687.36 | 777.54 | **2,718.80** |
| 976 | 95.39 | 88.45 | 91.34 | 92.50 | **367.67** |
| 977 | 1,311.71 | 1,295.52 | 1,235.40 | 1,368.94 | **5,211.57** |
| 978 | 450.34 | 419.12 | 470.00 | 468.84 | **1,808.30** |
| 979 | 100.59 | 85.56 | 102.32 | 130.07 | **418.54** |
| **OR OREGON Total** | **15,220.79** | **15,938.22** | **15,794.85** | **16,934.28** | **63,888.14** |

**STATE: PA - PENNSYLVANIA**

| REGISTRANT ZIP CODE 3 | 1ST QUARTER | 2ND QUARTER | 3RD QUARTER | 4TH QUARTER | TOTAL GRAMS |
|---|---|---|---|---|---|
| 150 | 2,843.10 | 3,093.99 | 2,750.60 | 2,992.25 | **11,679.93** |
| 151 | 2,198.51 | 2,204.30 | 2,246.50 | 2,386.97 | **9,036.28** |
| 152 | 4,879.74 | 4,961.25 | 4,913.27 | 5,304.07 | **20,058.34** |

| | | | | | |
|---|---|---|---|---|---|
| 153 | 1,155.62 | 1,119.78 | 1,167.76 | 1,248.70 | **4,691.86** |
| 154 | 849.81 | 862.53 | 755.58 | 833.04 | **3,300.95** |
| 155 | 297.72 | 331.83 | 376.34 | 360.16 | **1,366.05** |
| 156 | 1,940.10 | 1,971.32 | 1,906.57 | 2,087.52 | **7,905.52** |
| 157 | 1,321.54 | 1,227.88 | 1,161.98 | 1,256.79 | **4,968.19** |
| 158 | 1,075.27 | 1,086.83 | 1,129.61 | 1,124.40 | **4,416.11** |
| 159 | 775.81 | 802.98 | 798.93 | 830.15 | **3,207.88** |
| 160 | 1,549.31 | 1,523.87 | 1,577.63 | 1,598.45 | **6,249.26** |
| 161 | 1,719.85 | 1,838.94 | 1,764.36 | 1,906.57 | **7,229.72** |
| 162 | 675.80 | 725.52 | 660.19 | 730.14 | **2,791.64** |
| 163 | 1,117.47 | 1,090.30 | 1,107.06 | 1,134.23 | **4,449.06** |
| 164 | 734.77 | 675.22 | 645.16 | 668.28 | **2,723.43** |
| 165 | 2,894.55 | 2,869.69 | 2,861.02 | 2,860.44 | **11,485.69** |
| 166 | 1,480.51 | 1,576.48 | 1,437.16 | 1,502.48 | **5,996.63** |
| 167 | 512.20 | 567.12 | 593.13 | 571.74 | **2,244.18** |
| 168 | 1,508.26 | 1,330.21 | 1,311.71 | 1,460.28 | **5,610.46** |
| 169 | 375.77 | 396.58 | 344.55 | 415.08 | **1,531.97** |
| 170 | 4,728.28 | 4,696.48 | 4,612.08 | 4,873.38 | **18,910.23** |
| 171 | 1,236.56 | 1,233.09 | 1,165.45 | 1,226.73 | **4,861.82** |
| 172 | 1,368.36 | 1,310.55 | 1,399.58 | 1,404.78 | **5,483.28** |
| 173 | 2,228.00 | 2,204.87 | 2,138.39 | 2,344.20 | **8,915.46** |
| 174 | 1,176.43 | 1,218.63 | 1,212.85 | 1,312.87 | **4,920.79** |
| 175 | 1,423.86 | 1,464.91 | 1,327.32 | 1,439.47 | **5,655.55** |
| 176 | 1,655.68 | 1,504.22 | 1,582.84 | 1,637.76 | **6,380.49** |
| 177 | 799.51 | 748.64 | 785.64 | 881.60 | **3,215.39** |
| 178 | 839.40 | 790.26 | 832.46 | 798.93 | **3,261.06** |
| 179 | 241.07 | 309.28 | 289.05 | 305.81 | **1,145.22** |
| 180 | 2,228.58 | 2,149.38 | 2,071.33 | 2,380.04 | **8,829.32** |
| 181 | 922.07 | 856.17 | 892.01 | 986.24 | **3,656.48** |
| 182 | 261.88 | 265.93 | 250.32 | 237.02 | **1,015.14** |
| 183 | 583.30 | 466.53 | 556.71 | 554.40 | **2,160.94** |
| 184 | 578.10 | 587.35 | 597.76 | 617.99 | **2,381.19** |
| 185 | 483.87 | 470.00 | 450.34 | 517.98 | **1,922.18** |
| 186 | 396.00 | 438.78 | 414.50 | 434.15 | **1,683.43** |
| 187 | 10,848.62 | 12,301.39 | 14,360.58 | 15,027.71 | **52,538.31** |
| 188 | 186.15 | 188.46 | 200.02 | 243.38 | **818.01** |
| 189 | 2,309.51 | 2,400.27 | 2,266.73 | 2,444.78 | **9,421.30** |
| 190 | 8,104.96 | 8,086.46 | 7,769.66 | 8,264.52 | **32,225.61** |
| 191 | 6,676.48 | 6,955.12 | 6,771.29 | 7,595.08 | **27,997.96** |
| 193 | 4,635.78 | 4,630.58 | 4,458.89 | 4,816.73 | **18,541.98** |

| | | | | | |
|---|---|---|---|---|---|
| 194 | 4,014.90 | 3,855.35 | 3,954.78 | 4,313.78 | **16,138.82** |
| 195 | 941.15 | 914.55 | 885.65 | 944.04 | **3,685.39** |
| 196 | 1,848.19 | 1,738.35 | 1,715.22 | 1,764.36 | **7,066.12** |
| **PA - PENNSYLVANIA - Total** | **90,622.38** | **92,042.19** | **92,470.56** | **98,639.47** | **373,774.60** |

**STATE: PR - PUERTO RICO**

| REGISTRANT ZIP CODE 3 | 1ST QUARTER | 2ND QUARTER | 3RD QUARTER | 4TH QUARTER | TOTAL GRAMS |
|---|---|---|---|---|---|
| 006 | 424.33 | 382.12 | 347.44 | 392.53 | **1,546.42** |
| 007 | 319.69 | 234.13 | 286.74 | 280.96 | **1,121.51** |
| 009 | 1,588.62 | 1,438.89 | 1,356.80 | 1,662.62 | **6,046.93** |
| **PR - PUERTO RICO - Total** | **2,332.63** | **2,055.15** | **1,990.98** | **2,336.10** | **8,714.86** |

**STATE: RI - RHODE ISLAND**

| REGISTRANT ZIP CODE 3 | 1ST QUARTER | 2ND QUARTER | 3RD QUARTER | 4TH QUARTER | TOTAL GRAMS |
|---|---|---|---|---|---|
| 028 | 5,105.20 | 5,060.69 | 5,295.40 | 5,560.74 | **21,022.03** |
| 029 | 2,389.29 | 2,415.30 | 2,207.19 | 2,663.88 | **9,675.66** |
| **RI RHODE ISLAND Total** | **7,494.49** | **7,475.99** | **7,502.58** | **8,224.63** | **30,697.69** |

**STATE: SC - SOUTH CAROLINA**

| REGISTRANT ZIP CODE 3 | 1ST QUARTER | 2ND QUARTER | 3RD QUARTER | 4TH QUARTER | TOTAL GRAMS |
|---|---|---|---|---|---|
| 290 | 5,274.01 | 5,189.60 | 5,090.75 | 5,518.54 | **21,072.90** |
| 291 | 3,497.51 | 3,494.04 | 3,225.22 | 3,569.77 | **13,786.53** |
| 292 | 4,898.82 | 4,505.71 | 4,618.44 | 4,998.25 | **19,021.22** |
| 293 | 4,913.27 | 4,947.96 | 4,579.13 | 5,238.74 | **19,679.10** |
| 294 | 13,872.69 | 13,545.78 | 13,016.85 | 13,796.49 | **54,231.81** |
| 295 | 7,856.96 | 7,500.27 | 7,370.20 | 7,934.42 | **30,661.85** |
| 296 | 14,957.18 | 14,631.13 | 14,540.37 | 15,615.06 | **59,743.74** |
| 297 | 3,540.86 | 3,510.22 | 3,458.19 | 3,768.06 | **14,277.34** |
| 298 | 2,266.15 | 2,269.62 | 2,282.34 | 2,319.92 | **9,138.03** |
| 299 | 1,911.78 | 1,878.83 | 1,800.20 | 1,765.52 | **7,356.32** |
| **SC - SOUTH CAROLINA Total** | **62,989.23** | **61,473.16** | **59,981.69** | **64,524.76** | **248,968.84** |

**STATE: SD - SOUTH DAKOTA**

| REGISTRANT ZIP CODE 3 | 1ST QUARTER | 2ND QUARTER | 3RD QUARTER | 4TH QUARTER | TOTAL GRAMS |
|---|---|---|---|---|---|
| 570 | 1,686.32 | 1,605.96 | 1,607.70 | 1,786.91 | **6,686.88** |
| 571 | 4,787.82 | 4,724.23 | 4,887.84 | 4,807.48 | **19,207.37** |
| 572 | 432.42 | 439.93 | 500.63 | 481.56 | **1,854.54** |
| 573 | 937.10 | 896.06 | 908.77 | 957.91 | **3,699.84** |
| 574 | 517.98 | 482.14 | 486.76 | 572.32 | **2,059.19** |
| 575 | 275.75 | 291.94 | 294.25 | 335.30 | **1,197.25** |
| 576 | 26.59 | 20.81 | 30.06 | 17.92 | **95.39** |
| 577 | 971.79 | 1,027.28 | 1,020.35 | 1,066.02 | **4,085.43** |
| **SD SOUTH DAKOTA - Total** | **9,635.77** | **9,488.36** | **9,736.36** | **10,025.41** | **38,885.90** |

**STATE: TN - TENNESSEE**

| REGISTRANT ZIP CODE 3 | 1ST QUARTER | 2ND QUARTER | 3RD QUARTER | 4TH QUARTER | TOTAL GRAMS |
|---|---|---|---|---|---|
| 370 | 10,813.94 | 10,368.80 | 10,639.35 | 11,230.17 | **43,052.26** |
| 371 | 6,919.28 | 6,892.11 | 6,912.34 | 7,223.94 | **27,947.67** |
| 372 | 7,369.62 | 7,174.22 | 7,312.39 | 7,542.47 | **29,398.70** |
| 373 | 5,232.38 | 4,977.44 | 5,201.74 | 5,454.37 | **20,865.94** |
| 374 | 3,102.66 | 2,945.42 | 3,056.41 | 3,104.98 | **12,209.47** |
| 376 | 2,252.28 | 2,212.39 | 2,221.64 | 2,394.49 | **9,080.79** |
| 377 | 2,813.61 | 2,651.17 | 2,762.74 | 2,784.13 | **11,011.65** |
| 378 | 4,058.26 | 3,875.00 | 4,102.20 | 4,470.45 | **16,505.91** |
| 379 | 5,626.65 | 5,679.25 | 5,760.77 | 6,004.72 | **23,071.39** |
| 380 | 5,223.13 | 5,031.20 | 5,288.46 | 5,060.11 | **20,602.91** |
| 381 | 7,245.80 | 7,060.18 | 7,043.90 | 7,118.23 | **28,468.12** |
| 382 | 921.49 | 691.99 | 760.20 | 842.87 | **3,216.55** |
| 383 | 2,207.19 | 2,037.22 | 2,079.43 | 2,237.25 | **8,561.08** |
| 384 | 1,723.89 | 1,615.79 | 1,719.27 | 1,792.11 | **6,851.06** |
| 385 | 1,535.43 | 1,496.12 | 1,562.03 | 1,647.01 | **6,240.59** |
| **TN TENNESSEE Total** | **67,045.62** | **64,708.31** | **66,422.87** | **68,907.29** | **267,084.09** |

**STATE: TX - TEXAS**

| REGISTRANT ZIP CODE 3 | 1ST QUARTER | 2ND QUARTER | 3RD QUARTER | 4TH QUARTER | TOTAL GRAMS |
|---|---|---|---|---|---|
| 750 | 30,240.41 | 30,166.41 | 29,965.24 | 31,331.86 | **121,703.92** |
| 751 | 7,574.84 | 7,117.57 | 7,268.45 | 7,638.44 | **29,599.30** |
| 752 | 14,954.29 | 14,969.90 | 14,942.15 | 15,405.79 | **60,272.13** |
| 753 | 48.56 | 43.36 | 47.40 | 43.94 | **183.26** |
| 754 | 2,769.68 | 2,621.11 | 2,600.29 | 2,831.53 | **10,822.61** |
| 755 | 961.96 | 888.54 | 856.74 | 993.75 | **3,701.00** |
| 756 | 3,517.74 | 3,247.77 | 3,128.68 | 3,443.74 | **13,337.92** |
| 757 | 3,640.87 | 3,458.19 | 3,459.93 | 3,461.66 | **14,020.66** |
| 758 | 792.00 | 705.28 | 733.03 | 671.75 | **2,902.06** |
| 759 | 2,115.27 | 1,898.48 | 1,804.83 | 1,914.67 | **7,733.24** |
| 760 | 13,122.87 | 12,721.09 | 12,415.85 | 13,331.56 | **51,591.38** |
| 761 | 7,092.13 | 7,060.34 | 6,822.16 | 7,322.79 | **28,297.42** |
| 762 | 6,036.52 | 5,896.04 | 5,897.20 | 6,455.64 | **24,285.40** |
| 763 | 1,785.17 | 1,617.52 | 1,617.52 | 1,696.72 | **6,716.94** |
| 764 | 632.44 | 578.68 | 589.66 | 685.05 | **2,485.83** |
| 765 | 2,781.82 | 3,051.21 | 2,969.12 | 2,606.07 | **11,408.23** |
| 766 | 734.19 | 628.39 | 702.39 | 727.25 | **2,792.22** |
| 767 | 2,804.94 | 2,725.16 | 2,624.57 | 3,061.62 | **11,216.30** |
| 768 | 372.30 | 368.25 | 372.30 | 389.64 | **1,502.48** |
| 769 | 1,375.30 | 1,434.84 | 1,246.96 | 1,440.63 | **5,497.73** |
| 770 | 25,958.02 | 25,386.68 | 24,612.36 | 26,567.74 | **102,524.81** |

| | | | | | |
|---|---|---|---|---|---|
| 773 | 19,255.46 | 18,874.97 | 18,709.63 | 19,807.44 | **76,647.49** |
| 774 | 15,622.00 | 15,194.20 | 15,065.29 | 15,983.31 | **61,864.79** |
| 775 | 12,096.16 | 11,794.40 | 11,316.89 | 12,094.43 | **47,301.88** |
| 776 | 3,555.32 | 3,413.68 | 3,135.61 | 3,414.26 | **13,518.87** |
| 777 | 1,624.46 | 1,655.10 | 1,497.28 | 1,703.08 | **6,479.92** |
| 778 | 5,135.84 | 4,743.89 | 4,747.94 | 5,168.21 | **19,795.88** |
| 779 | 1,830.26 | 1,649.90 | 1,525.61 | 1,754.53 | **6,760.30** |
| 780 | 4,872.23 | 4,693.02 | 4,772.79 | 5,084.39 | **19,422.43** |
| 781 | 3,949.00 | 3,788.87 | 3,626.42 | 4,032.25 | **15,396.54** |
| 782 | 16,335.37 | 16,135.93 | 15,781.55 | 16,873.00 | **65,125.86** |
| 783 | 2,068.44 | 1,783.44 | 1,696.15 | 1,921.60 | **7,469.63** |
| 784 | 3,799.85 | 3,396.34 | 3,283.61 | 3,427.55 | **13,907.35** |
| 785 | 7,283.48 | 6,495.53 | 6,503.05 | 6,946.45 | **27,228.51** |
| 786 | 9,258.27 | 9,182.54 | 8,956.50 | 9,792.44 | **37,189.75** |
| 787 | 14,080.20 | 14,394.11 | 14,358.85 | 15,002.27 | **57,835.44** |
| 788 | 727.25 | 596.60 | 652.67 | 665.39 | **2,641.92** |
| 789 | 417.97 | 375.19 | 373.45 | 402.94 | **1,569.54** |
| 790 | 524.34 | 523.76 | 551.51 | 648.63 | **2,248.23** |
| 791 | 1,274.13 | 1,405.36 | 1,434.27 | 1,393.80 | **5,507.56** |
| 792 | 77.47 | 80.36 | 76.89 | 87.87 | **322.58** |
| 793 | 321.42 | 387.91 | 330.67 | 322.00 | **1,362.00** |
| 794 | 3,248.92 | 3,191.11 | 3,128.68 | 3,234.47 | **12,803.18** |
| 795 | 402.36 | 375.19 | 378.66 | 409.87 | **1,566.07** |
| 796 | 1,383.97 | 1,234.24 | 1,220.37 | 1,320.96 | **5,159.54** |
| 797 | 3,003.81 | 2,715.91 | 2,764.47 | 3,019.99 | **11,504.19** |
| 798 | 84.40 | 63.59 | 61.28 | 53.76 | **263.04** |
| 799 | 2,864.49 | 2,673.71 | 2,490.45 | 2,633.82 | **10,662.48** |
| **TX - TEXAS Total** | **264,408.18** | **257,403.65** | **253,117.37** | **269,220.59** | **1,044,149.80** |

**STATE: UT - UTAH**

| REGISTRANT ZIP CODE 3 | 1ST QUARTER | 2ND QUARTER | 3RD QUARTER | 4TH QUARTER | **TOTAL GRAMS** |
|---|---|---|---|---|---|
| 840 | 13,326.95 | 13,571.48 | 13,965.74 | 14,776.81 | **55,640.98** |
| 841 | 4,008.55 | 4,147.29 | 4,105.67 | 4,272.16 | **16,533.66** |
| 843 | 701.81 | 774.65 | 794.31 | 892.01 | **3,162.79** |
| 844 | 1,151.00 | 1,130.55 | 1,170.46 | 1,222.71 | **4,674.72** |
| 845 | 124.29 | 150.88 | 138.17 | 129.49 | **542.84** |
| 846 | 2,048.79 | 2,060.35 | 2,051.68 | 2,141.28 | **8,302.09** |
| 847 | 954.44 | 932.48 | 896.06 | 981.04 | **3,764.01** |
| **UT - UTAH - Total** | **22,315.83** | **22,767.67** | **23,122.08** | **24,415.50** | **92,621.09** |

**STATE: VA - VIRGINIA**

| REGISTRANT ZIP CODE 3 | 1ST QUARTER | 2ND QUARTER | 3RD QUARTER | 4TH QUARTER | **TOTAL GRAMS** |
|---|---|---|---|---|---|

| | | | | | |
|---|---|---|---|---|---|
| 201 | 7,266.72 | 7,456.33 | 7,431.48 | 8,003.79 | **30,158.32** |
| 220 | 3,514.27 | 3,763.43 | 4,029.36 | 3,803.32 | **15,110.38** |
| 221 | 2,845.41 | 2,987.62 | 2,978.37 | 3,107.87 | **11,919.27** |
| 222 | 2,108.33 | 2,503.17 | 2,408.94 | 2,456.93 | **9,477.37** |
| 223 | 1,916.98 | 1,979.99 | 1,814.66 | 2,056.30 | **7,767.93** |
| 224 | 2,191.00 | 2,121.05 | 2,230.31 | 2,394.49 | **8,936.85** |
| 225 | 1,115.73 | 1,034.22 | 1,147.53 | 1,098.39 | **4,395.87** |
| 226 | 1,514.62 | 1,530.81 | 1,541.79 | 1,631.98 | **6,219.20** |
| 227 | 548.04 | 573.48 | 524.34 | 557.87 | **2,203.72** |
| 228 | 1,020.92 | 1,041.74 | 975.83 | 1,183.37 | **4,221.86** |
| 229 | 2,356.34 | 2,406.63 | 2,368.48 | 2,724.01 | **9,855.45** |
| 230 | 1,655.10 | 1,760.89 | 1,647.01 | 1,804.83 | **6,867.83** |
| 231 | 4,557.74 | 4,582.02 | 4,686.66 | 4,871.07 | **18,697.49** |
| 232 | 5,237.59 | 5,537.62 | 5,256.66 | 5,741.11 | **21,772.98** |
| 233 | 3,115.38 | 3,100.93 | 2,892.23 | 3,038.49 | **12,147.04** |
| 234 | 6,947.61 | 7,021.60 | 6,500.73 | 6,907.14 | **27,377.08** |
| 235 | 1,789.80 | 1,781.13 | 1,734.88 | 1,738.92 | **7,044.73** |
| 236 | 3,234.47 | 3,281.30 | 3,243.14 | 3,300.95 | **13,059.86** |
| 237 | 1,185.68 | 1,229.04 | 1,307.08 | 1,209.39 | **4,931.19** |
| 238 | 2,056.30 | 2,230.31 | 1,967.27 | 2,241.87 | **8,495.76** |
| 239 | 548.62 | 473.46 | 509.31 | 551.51 | **2,082.89** |
| 240 | 3,195.74 | 3,054.68 | 3,007.85 | 3,155.85 | **12,414.12** |
| 241 | 1,749.91 | 1,727.94 | 1,686.32 | 1,919.87 | **7,084.04** |
| 242 | 903.57 | 921.49 | 816.86 | 785.64 | **3,427.55** |
| 243 | 912.24 | 916.29 | 915.71 | 957.33 | **3,701.57** |
| 244 | 1,334.25 | 1,299.57 | 1,218.63 | 1,462.59 | **5,315.05** |
| 245 | 3,132.72 | 2,910.73 | 3,144.86 | 3,037.34 | **12,225.66** |
| 246 | 552.09 | 490.23 | 461.32 | 535.90 | **2,039.54** |
| **VA VIRGINIA Total** | **68,507.16** | **69,717.70** | **68,447.62** | **72,278.11** | **278,950.59** |

**STATE: VI - VIRGIN ISLANDS**

| REGISTRANT ZIP CODE 3 | 1ST QUARTER | 2ND QUARTER | 3RD QUARTER | 4TH QUARTER | **TOTAL GRAMS** |
|---|---|---|---|---|---|
| 008 | 83.25 | 66.48 | 67.64 | 106.95 | **324.31** |
| **VI - VIRGIN ISLANDS Total** | **83.25** | **66.48** | **67.64** | **106.95** | **324.31** |

**STATE: VT - VERMONT**

| REGISTRANT ZIP CODE 3 | 1ST QUARTER | 2ND QUARTER | 3RD QUARTER | 4TH QUARTER | **TOTAL GRAMS** |
|---|---|---|---|---|---|
| 050 | 319.69 | 328.94 | 327.78 | 300.61 | **1,277.02** |
| 051 | 293.10 | 246.27 | 375.19 | 242.22 | **1,156.78** |
| 052 | 603.54 | 659.03 | 657.30 | 723.78 | **2,643.65** |
| 053 | 646.32 | 607.01 | 708.75 | 616.83 | **2,578.90** |
| 054 | 2,852.35 | 2,822.86 | 2,895.70 | 2,955.25 | **11,526.16** |

| | | | | | |
|---|---|---|---|---|---|
| 056 | 737.66 | 710.48 | 735.34 | 675.80 | **2,859.28** |
| 057 | 911.66 | 927.27 | 853.85 | 911.66 | **3,604.45** |
| 058 | 845.18 | 901.84 | 981.04 | 1,013.99 | **3,742.04** |
| **VT VERMONT Total** | **7,209.49** | **7,203.70** | **7,534.96** | **7,440.15** | **29,388.29** |

**STATE: WA - WASHINGTON**

| REGISTRANT ZIP CODE 3 | 1ST QUARTER | 2ND QUARTER | 3RD QUARTER | 4TH QUARTER | TOTAL GRAMS |
|---|---|---|---|---|---|
| 980 | 9,689.07 | 10,301.16 | 10,245.67 | 10,769.42 | **41,005.33** |
| 981 | 5,998.37 | 6,316.32 | 6,412.86 | 6,755.68 | **25,483.23** |
| 982 | 3,609.66 | 3,646.65 | 3,796.96 | 3,930.50 | **14,983.77** |
| 983 | 3,455.30 | 3,745.51 | 3,864.02 | 3,810.26 | **14,875.09** |
| 984 | 1,962.07 | 2,171.92 | 2,073.64 | 2,131.45 | **8,339.09** |
| 985 | 1,963.81 | 2,103.13 | 2,067.86 | 2,280.60 | **8,415.40** |
| 986 | 2,517.63 | 2,736.73 | 2,653.48 | 2,905.53 | **10,813.36** |
| 988 | 326.63 | 387.33 | 433.00 | 379.81 | **1,526.76** |
| 989 | 763.67 | 779.86 | 700.66 | 882.76 | **3,126.94** |
| 990 | 522.02 | 469.42 | 459.59 | 502.37 | **1,953.40** |
| 991 | 419.12 | 350.91 | 369.98 | 454.39 | **1,594.40** |
| 992 | 2,483.52 | 2,526.30 | 2,510.11 | 2,624.57 | **10,144.50** |
| 993 | 1,760.89 | 1,922.76 | 1,847.03 | 2,074.22 | **7,604.91** |
| 994 | 90.76 | 105.21 | 82.67 | 140.48 | **419.12** |
| **WA WASHINGTON Total** | **35,562.52** | **37,563.20** | **37,517.53** | **39,642.05** | **150,285.30** |

**STATE: WI - WISCONSIN**

| REGISTRANT ZIP CODE 3 | 1ST QUARTER | 2ND QUARTER | 3RD QUARTER | 4TH QUARTER | TOTAL GRAMS |
|---|---|---|---|---|---|
| 530 | 8,154.68 | 8,274.35 | 8,151.21 | 9,043.80 | **33,624.03** |
| 531 | 9,251.91 | 9,517.26 | 9,371.00 | 9,757.75 | **37,897.92** |
| 532 | 10,082.06 | 9,836.95 | 9,643.29 | 10,150.86 | **39,713.16** |
| 534 | 2,691.63 | 2,767.94 | 2,608.39 | 2,743.08 | **10,811.05** |
| 535 | 5,613.93 | 5,921.48 | 5,694.86 | 6,092.02 | **23,322.29** |
| 537 | 3,445.48 | 3,657.64 | 3,433.34 | 3,805.05 | **14,341.50** |
| 538 | 777.54 | 861.95 | 871.20 | 910.51 | **3,421.20** |
| 539 | 2,258.06 | 2,438.43 | 2,348.24 | 2,421.08 | **9,465.81** |
| 540 | 775.23 | 856.74 | 865.42 | 933.63 | **3,431.02** |
| 541 | 3,881.94 | 4,032.25 | 4,096.99 | 4,341.53 | **16,352.71** |
| 542 | 1,793.27 | 1,787.49 | 1,789.80 | 1,878.25 | **7,248.80** |
| 543 | 5,670.58 | 5,784.47 | 5,914.54 | 5,867.72 | **23,237.31** |
| 544 | 5,302.91 | 5,608.73 | 5,405.24 | 5,672.32 | **21,989.19** |
| 545 | 1,188.00 | 1,226.15 | 1,355.07 | 1,297.83 | **5,067.05** |
| 546 | 2,148.80 | 2,189.84 | 2,129.14 | 2,130.30 | **8,598.08** |
| 547 | 1,457.97 | 1,534.86 | 1,505.95 | 1,677.07 | **6,175.84** |
| 548 | 1,656.83 | 1,675.91 | 1,692.10 | 1,858.01 | **6,882.86** |

| | | | | | |
|---|---|---|---|---|---|
| 549 | 7,869.10 | 8,162.77 | 8,328.69 | 8,506.16 | **32,866.72** |
| **WI - WISCONSIN Total** | **74,019.92** | **76,135.19** | **75,204.45** | **79,086.97** | **304,446.54** |

**STATE: WV - WEST VIRGINIA**

| REGISTRANT ZIP CODE 3 | 1ST QUARTER | 2ND QUARTER | 3RD QUARTER | 4TH QUARTER | TOTAL GRAMS |
|---|---|---|---|---|---|
| 247 | 367.09 | 379.81 | 364.20 | 456.12 | **1,567.23** |
| 248 | 297.14 | 279.22 | 257.83 | 238.18 | **1,072.38** |
| 249 | 373.45 | 357.27 | 311.02 | 319.11 | **1,360.85** |
| 250 | 738.23 | 719.73 | 705.86 | 759.62 | **2,923.45** |
| 251 | 609.90 | 638.22 | 547.46 | 639.96 | **2,435.54** |
| 252 | 352.64 | 347.44 | 372.30 | 397.73 | **1,470.11** |
| 253 | 1,755.69 | 1,698.46 | 1,643.54 | 1,700.77 | **6,798.46** |
| 254 | 1,259.10 | 1,305.35 | 1,271.24 | 1,355.07 | **5,190.76** |
| 255 | 1,747.60 | 1,607.12 | 1,617.52 | 1,732.57 | **6,704.80** |
| 256 | 297.72 | 286.74 | 300.61 | 252.05 | **1,137.12** |
| 257 | 1,497.86 | 1,530.23 | 1,642.38 | 1,642.96 | **6,313.43** |
| 258 | 1,112.26 | 985.66 | 1,001.85 | 1,102.44 | **4,202.21** |
| 259 | 287.32 | 287.32 | 269.97 | 280.96 | **1,125.56** |
| 260 | 1,025.55 | 1,083.36 | 920.34 | 1,115.73 | **4,144.98** |
| 261 | 1,827.37 | 1,687.47 | 1,520.98 | 1,607.70 | **6,643.53** |
| 262 | 616.83 | 624.35 | 576.37 | 633.60 | **2,451.14** |
| 263 | 1,159.67 | 1,171.23 | 1,100.70 | 1,175.86 | **4,607.46** |
| 264 | 334.14 | 309.86 | 334.72 | 352.64 | **1,331.36** |
| 265 | 1,993.87 | 1,991.55 | 2,074.22 | 2,332.63 | **8,392.28** |
| 266 | 198.87 | 212.16 | 200.60 | 191.93 | **803.56** |
| 267 | 694.88 | 652.10 | 615.68 | 671.17 | **2,633.82** |
| 268 | 388.48 | 372.30 | 377.50 | 396.00 | **1,534.28** |
| **WV WEST VIRGINIA Total** | **18,935.67** | **18,526.95** | **18,026.89** | **19,354.79** | **74,844.29** |

**STATE: WY - WYOMING**

| REGISTRANT ZIP CODE 3 | 1ST QUARTER | 2ND QUARTER | 3RD QUARTER | 4TH QUARTER | TOTAL GRAMS |
|---|---|---|---|---|---|
| 820 | 550.93 | 626.66 | 644.58 | 653.25 | **2,475.42** |
| 822 | 88.45 | 65.90 | 70.53 | 66.48 | **291.36** |
| 823 | 23.12 | 22.55 | 20.23 | 21.39 | **87.29** |
| 824 | 556.13 | 494.85 | 583.30 | 522.60 | **2,156.89** |
| 825 | 78.62 | 79.20 | 83.25 | 86.14 | **327.20** |
| 826 | 322.00 | 328.36 | 339.34 | 384.44 | **1,374.14** |
| 827 | 303.50 | 319.11 | 339.92 | 326.63 | **1,289.16** |
| 828 | 171.12 | 162.45 | 165.91 | 180.95 | **680.42** |
| 829 | 378.66 | 366.52 | 387.33 | 371.14 | **1,503.64** |
| 830 | 137.59 | 106.37 | 119.67 | 106.95 | **470.57** |
| 831 | 28.91 | 57.23 | 54.34 | 53.19 | **193.66** |

| | | | | | |
|---|---|---|---|---|---|
| WY - WYOMING Total | 2,639.03 | 2,629.20 | 2,808.41 | 2,773.15 | 10,849.78 |

**DRUG: 1724 - METHYLPHENIDATE (DL;D;L;ISOMERS)**

**STATE: AK - ALASKA**

| REGISTRANT ZIP CODE 3 | 1ST QUARTER | 2ND QUARTER | 3RD QUARTER | 4TH QUARTER | TOTAL GRAMS |
|---|---|---|---|---|---|
| 995 | 4,126.30 | 4,202.95 | 4,220.23 | 4,562.62 | **17,112.10** |
| 996 | 1,478.54 | 1,585.62 | 1,403.23 | 1,680.10 | **6,147.48** |
| 997 | 1,359.37 | 1,213.51 | 1,394.52 | 1,350.05 | **5,317.45** |
| 998 | 528.62 | 530.37 | 597.44 | 618.35 | **2,274.77** |
| 999 | 341.53 | 425.12 | 411.99 | 416.50 | **1,595.15** |
| AK ALASKA - Total | 7,834.35 | 7,957.58 | 8,027.40 | 8,627.62 | 32,446.95 |

**STATE: AL - ALABAMA**

| REGISTRANT ZIP CODE 3 | 1ST QUARTER | 2ND QUARTER | 3RD QUARTER | 4TH QUARTER | TOTAL GRAMS |
|---|---|---|---|---|---|
| 350 | 8,975.50 | 8,121.79 | 8,679.42 | 9,258.08 | **35,034.79** |
| 351 | 4,867.18 | 4,590.15 | 4,574.75 | 5,069.50 | **19,101.58** |
| 352 | 8,698.80 | 8,354.20 | 8,039.55 | 9,032.70 | **34,125.24** |
| 354 | 2,689.73 | 2,334.48 | 2,614.60 | 2,700.27 | **10,339.08** |
| 355 | 2,121.03 | 1,886.68 | 2,211.18 | 2,366.31 | **8,585.19** |
| 356 | 4,263.83 | 3,961.22 | 4,049.58 | 4,222.06 | **16,496.70** |
| 357 | 2,847.07 | 2,781.37 | 2,731.63 | 3,072.53 | **11,432.60** |
| 358 | 2,930.05 | 2,990.24 | 3,240.38 | 2,981.23 | **12,141.90** |
| 359 | 3,889.33 | 3,739.52 | 3,867.57 | 4,145.54 | **15,641.96** |
| 360 | 3,674.66 | 3,420.72 | 3,671.04 | 3,799.10 | **14,565.51** |
| 361 | 3,008.94 | 2,727.57 | 2,831.54 | 2,857.68 | **11,425.73** |
| 362 | 2,095.93 | 2,009.66 | 2,102.06 | 2,329.21 | **8,536.86** |
| 363 | 5,598.23 | 5,020.75 | 5,570.23 | 5,480.97 | **21,670.18** |
| 364 | 1,517.64 | 1,396.58 | 1,429.14 | 1,587.94 | **5,931.29** |
| 365 | 7,040.79 | 6,446.49 | 6,837.90 | 7,202.79 | **27,527.97** |
| 366 | 5,042.06 | 4,465.93 | 4,609.72 | 5,217.65 | **19,335.36** |
| 367 | 696.53 | 554.55 | 598.28 | 612.94 | **2,462.30** |
| 368 | 4,094.09 | 3,650.37 | 3,919.04 | 4,154.26 | **15,817.76** |
| 369 | 157.56 | 176.95 | 122.22 | 177.48 | **634.22** |
| AL ALABAMA - Total | 74,208.96 | 68,629.22 | 71,699.82 | 76,268.23 | 290,806.23 |

**STATE: AR - ARKANSAS**

| REGISTRANT ZIP CODE 3 | 1ST QUARTER | 2ND QUARTER | 3RD QUARTER | 4TH QUARTER | TOTAL GRAMS |
|---|---|---|---|---|---|
| 716 | 1,599.12 | 1,486.28 | 1,378.32 | 1,487.63 | **5,951.35** |
| 717 | 856.35 | 784.80 | 771.25 | 870.88 | **3,283.27** |
| 718 | 1,055.37 | 902.59 | 836.26 | 947.84 | **3,742.06** |
| 719 | 2,321.03 | 2,143.51 | 2,042.69 | 2,326.46 | **8,833.69** |
| 720 | 6,378.15 | 6,120.11 | 6,360.85 | 6,654.60 | **25,513.71** |
| 721 | 4,770.00 | 4,264.84 | 4,248.34 | 4,627.49 | **17,910.67** |

| | | | | | |
|---|---|---|---|---|---|
| 722 | 4,319.56 | 3,825.73 | 4,083.65 | 4,014.65 | **16,243.58** |
| 723 | 1,473.79 | 1,225.50 | 1,244.40 | 1,344.70 | **5,288.40** |
| 724 | 3,443.85 | 2,954.47 | 3,174.53 | 3,281.57 | **12,854.42** |
| 725 | 1,159.70 | 1,007.25 | 1,075.38 | 1,146.75 | **4,389.08** |
| 726 | 1,219.23 | 1,109.01 | 1,134.68 | 1,180.83 | **4,643.76** |
| 727 | 7,140.96 | 7,016.76 | 7,228.58 | 7,489.36 | **28,875.66** |
| 728 | 1,437.59 | 1,238.87 | 1,279.26 | 1,334.58 | **5,290.31** |
| 729 | 2,966.47 | 2,619.77 | 2,704.40 | 2,842.01 | **11,132.64** |
| **AR ARKANSAS - Total** | **40,141.16** | **36,699.49** | **37,562.60** | **39,549.34** | **153,952.59** |

**STATE: AS - AMERICAN SAMOA**

| REGISTRANT ZIP CODE 3 | 1ST QUARTER | 2ND QUARTER | 3RD QUARTER | 4TH QUARTER | TOTAL GRAMS |
|---|---|---|---|---|---|
| 967 | 8.65 | 14.70 | 2.59 | 9.08 | **35.02** |
| **AS - AMERICAN SAMOA - Tota** | **8.65** | **14.70** | **2.59** | **9.08** | **35.02** |

**STATE: AZ - ARIZONA**

| REGISTRANT ZIP CODE 3 | 1ST QUARTER | 2ND QUARTER | 3RD QUARTER | 4TH QUARTER | TOTAL GRAMS |
|---|---|---|---|---|---|
| 850 | 12,254.16 | 12,561.93 | 11,551.30 | 12,507.59 | **48,874.99** |
| 851 | 4,333.61 | 4,338.70 | 4,155.09 | 4,484.46 | **17,311.86** |
| 852 | 27,016.95 | 28,075.29 | 26,838.40 | 29,240.69 | **111,171.33** |
| 853 | 12,107.06 | 11,944.99 | 11,282.10 | 12,263.07 | **47,597.22** |
| 855 | 777.01 | 698.84 | 786.93 | 666.15 | **2,928.93** |
| 856 | 2,625.30 | 2,552.16 | 2,543.85 | 2,612.51 | **10,333.82** |
| 857 | 11,510.77 | 11,741.61 | 10,910.56 | 11,803.41 | **45,966.35** |
| 859 | 637.83 | 782.96 | 713.09 | 645.77 | **2,779.65** |
| 860 | 1,393.15 | 1,409.90 | 1,338.11 | 1,469.05 | **5,610.21** |
| 863 | 1,866.21 | 2,195.62 | 1,940.14 | 2,172.74 | **8,174.71** |
| 864 | 1,003.86 | 1,128.65 | 1,078.99 | 978.34 | **4,189.84** |
| 865 | 452.81 | 261.88 | 344.02 | 358.45 | **1,417.17** |
| **AZ ARIZONA - Total** | **75,978.71** | **77,692.53** | **73,482.60** | **79,202.24** | **306,356.07** |

**STATE: CA - CALIFORNIA**

| REGISTRANT ZIP CODE 3 | 1ST QUARTER | 2ND QUARTER | 3RD QUARTER | 4TH QUARTER | TOTAL GRAMS |
|---|---|---|---|---|---|
| 900 | 9,660.38 | 10,080.97 | 9,374.46 | 10,514.36 | **39,630.16** |
| 902 | 7,334.07 | 7,389.58 | 6,853.19 | 7,763.44 | **29,340.28** |
| 903 | 427.48 | 400.54 | 336.82 | 377.27 | **1,542.11** |
| 904 | 1,412.31 | 1,472.60 | 1,440.46 | 1,620.03 | **5,945.40** |
| 905 | 1,290.26 | 1,280.90 | 1,166.26 | 1,261.56 | **4,998.98** |
| 906 | 3,444.94 | 3,243.27 | 3,216.78 | 3,345.60 | **13,250.60** |
| 907 | 3,316.91 | 3,244.84 | 3,144.37 | 3,522.49 | **13,228.61** |
| 908 | 2,156.59 | 2,189.60 | 2,019.35 | 2,386.42 | **8,751.96** |
| 910 | 1,275.86 | 1,332.37 | 1,272.84 | 1,331.85 | **5,212.93** |
| 911 | 2,307.66 | 2,474.52 | 2,344.98 | 2,539.54 | **9,666.71** |

| | | | | | |
|---|---|---|---|---|---|
| 912 | 940.23 | 990.85 | 971.52 | 1,067.39 | **3,969.99** |
| 913 | 11,324.72 | 11,163.52 | 10,686.96 | 11,867.06 | **45,042.25** |
| 914 | 2,678.62 | 2,781.63 | 2,713.90 | 2,980.94 | **11,155.09** |
| 915 | 712.43 | 720.94 | 691.83 | 792.87 | **2,918.08** |
| 916 | 840.98 | 847.06 | 799.31 | 1,034.42 | **3,521.77** |
| 917 | 8,072.63 | 8,165.38 | 7,986.03 | 8,721.81 | **32,945.85** |
| 918 | 312.39 | 246.64 | 328.30 | 302.03 | **1,189.37** |
| 919 | 2,892.96 | 2,909.19 | 2,663.15 | 3,185.37 | **11,650.68** |
| 920 | 26,333.20 | 26,329.35 | 25,677.52 | 26,597.26 | **104,937.33** |
| 921 | 11,504.59 | 12,052.69 | 10,834.81 | 11,808.42 | **46,200.52** |
| 922 | 4,102.30 | 4,166.14 | 3,537.16 | 4,256.88 | **16,062.48** |
| 923 | 6,931.29 | 7,034.23 | 6,827.01 | 7,612.41 | **28,404.94** |
| 924 | 770.62 | 675.83 | 646.25 | 774.65 | **2,867.35** |
| 925 | 8,736.38 | 8,900.65 | 8,184.12 | 9,405.31 | **35,226.46** |
| 926 | 11,751.70 | 12,072.88 | 11,484.92 | 12,195.35 | **47,504.85** |
| 927 | 2,812.28 | 2,673.22 | 2,728.07 | 2,851.81 | **11,065.39** |
| 928 | 7,681.48 | 7,784.21 | 7,575.26 | 8,124.35 | **31,165.30** |
| 930 | 5,403.44 | 5,642.67 | 5,116.14 | 5,823.19 | **21,985.45** |
| 931 | 1,676.77 | 1,583.17 | 1,601.00 | 1,567.69 | **6,428.63** |
| 932 | 3,347.89 | 3,218.29 | 3,045.39 | 3,254.74 | **12,866.31** |
| 933 | 2,893.72 | 2,739.32 | 2,622.59 | 2,831.87 | **11,087.50** |
| 934 | 3,294.70 | 3,183.92 | 2,773.76 | 3,374.45 | **12,626.82** |
| 935 | 3,096.84 | 3,510.81 | 2,994.88 | 3,219.78 | **12,822.31** |
| 936 | 3,515.64 | 3,732.41 | 3,268.66 | 3,766.20 | **14,282.91** |
| 937 | 4,838.83 | 4,971.94 | 4,780.30 | 5,039.03 | **19,630.11** |
| 939 | 1,989.88 | 2,054.29 | 1,958.37 | 2,069.55 | **8,072.09** |
| 940 | 5,500.68 | 5,199.92 | 5,014.95 | 5,742.88 | **21,458.43** |
| 941 | 5,874.14 | 6,209.74 | 6,004.32 | 6,249.06 | **24,337.26** |
| 943 | 1,045.29 | 1,253.84 | 1,114.17 | 1,199.89 | **4,613.18** |
| 944 | 969.94 | 1,101.57 | 1,017.64 | 1,191.42 | **4,280.56** |
| 945 | 17,153.09 | 16,602.45 | 16,337.92 | 17,365.26 | **67,458.71** |
| 946 | 2,760.24 | 3,073.63 | 3,047.89 | 3,139.88 | **12,021.64** |
| 947 | 848.60 | 847.60 | 863.59 | 854.54 | **3,414.33** |
| 948 | 597.99 | 584.57 | 671.17 | 713.64 | **2,567.38** |
| 949 | 3,745.32 | 3,810.73 | 3,684.59 | 3,985.60 | **15,226.24** |
| 950 | 6,728.78 | 6,128.69 | 6,633.68 | 6,318.39 | **25,809.53** |
| 951 | 3,920.86 | 3,902.34 | 3,640.16 | 3,867.98 | **15,331.35** |
| 952 | 2,644.47 | 2,644.86 | 2,464.57 | 2,812.48 | **10,566.38** |
| 953 | 5,497.01 | 5,354.67 | 5,320.45 | 5,403.91 | **21,576.04** |
| 954 | 3,284.09 | 3,300.75 | 3,307.44 | 3,561.13 | **13,453.41** |

| | | | | | |
|---|---|---|---|---|---|
| 955 | 1,036.23 | 1,052.89 | 1,060.10 | 1,089.39 | **4,238.60** |
| 956 | 13,410.37 | 12,467.60 | 13,045.95 | 13,761.82 | **52,685.74** |
| 957 | 3,340.36 | 3,272.91 | 3,129.32 | 3,468.92 | **13,211.50** |
| 958 | 5,492.83 | 5,547.42 | 5,386.19 | 5,768.50 | **22,194.95** |
| 959 | 3,641.20 | 3,409.56 | 3,097.05 | 3,349.98 | **13,497.79** |
| 960 | 2,702.17 | 2,654.75 | 2,681.18 | 2,759.35 | **10,797.45** |
| 961 | 414.84 | 436.36 | 380.44 | 451.45 | **1,683.10** |
| **CA - CALIFORNIA - Total** | **261,691.50** | **262,117.30** | **251,569.47** | **272,242.86** | **1,047,621.13** |

**STATE: CO - COLORADO**

| REGISTRANT ZIP CODE 3 | 1ST QUARTER | 2ND QUARTER | 3RD QUARTER | 4TH QUARTER | TOTAL GRAMS |
|---|---|---|---|---|---|
| 800 | 10,588.55 | 10,450.34 | 10,345.56 | 11,589.50 | **42,973.95** |
| 801 | 11,185.32 | 11,546.58 | 11,317.48 | 12,680.16 | **46,729.54** |
| 802 | 10,551.66 | 10,795.42 | 10,391.94 | 11,410.17 | **43,149.19** |
| 803 | 2,095.48 | 2,308.84 | 2,125.88 | 2,367.48 | **8,897.68** |
| 804 | 1,832.53 | 1,585.56 | 1,892.19 | 1,970.23 | **7,280.51** |
| 805 | 7,007.61 | 7,131.45 | 6,951.85 | 7,820.27 | **28,911.18** |
| 806 | 2,311.42 | 2,266.88 | 2,306.92 | 2,620.06 | **9,505.28** |
| 807 | 435.06 | 469.41 | 446.12 | 514.24 | **1,864.83** |
| 808 | 1,571.17 | 1,286.30 | 1,744.83 | 1,239.37 | **5,841.68** |
| 809 | 7,437.57 | 7,039.64 | 7,438.06 | 8,102.83 | **30,018.11** |
| 810 | 1,885.53 | 1,895.64 | 1,662.19 | 2,093.57 | **7,536.92** |
| 811 | 292.81 | 280.31 | 314.87 | 305.22 | **1,193.20** |
| 812 | 565.90 | 561.94 | 579.62 | 613.04 | **2,320.50** |
| 813 | 506.39 | 507.48 | 561.61 | 593.26 | **2,168.73** |
| 814 | 676.34 | 629.10 | 620.56 | 713.43 | **2,639.42** |
| 815 | 1,972.63 | 1,843.62 | 1,749.67 | 1,979.83 | **7,545.74** |
| 816 | 1,302.56 | 1,182.65 | 1,216.52 | 1,207.01 | **4,908.74** |
| **CO - COLORADO - Total** | **62,218.53** | **61,781.15** | **61,665.86** | **67,819.66** | **253,485.20** |

**STATE: CT - CONNECTICUT**

| REGISTRANT ZIP CODE 3 | 1ST QUARTER | 2ND QUARTER | 3RD QUARTER | 4TH QUARTER | TOTAL GRAMS |
|---|---|---|---|---|---|
| 060 | 11,775.12 | 11,689.62 | 11,015.39 | 12,052.55 | **46,532.68** |
| 061 | 4,295.37 | 4,253.70 | 3,838.46 | 4,254.73 | **16,642.27** |
| 062 | 2,748.43 | 2,525.43 | 2,511.24 | 2,836.34 | **10,621.43** |
| 063 | 4,652.24 | 4,885.59 | 4,283.08 | 4,922.12 | **18,743.02** |
| 064 | 11,905.89 | 12,093.72 | 11,262.38 | 12,465.63 | **47,727.62** |
| 065 | 3,584.22 | 3,500.75 | 3,132.47 | 3,590.70 | **13,808.14** |
| 066 | 1,603.45 | 1,611.30 | 1,374.54 | 1,634.11 | **6,223.40** |
| 067 | 4,534.75 | 4,620.20 | 4,258.78 | 4,695.24 | **18,108.97** |
| 068 | 8,498.78 | 8,304.86 | 8,061.05 | 8,728.39 | **33,593.09** |
| 069 | 1,236.50 | 1,095.36 | 1,134.36 | 1,222.62 | **4,688.85** |

| CT CONNECTICUT Total | 54,834.76 | 54,580.53 | 50,871.75 | 56,402.44 | 216,689.47 |
|---|---|---|---|---|---|

**STATE: DC - DISTRICT OF COLUMBIA**

| REGISTRANT ZIP CODE 3 | 1ST QUARTER | 2ND QUARTER | 3RD QUARTER | 4TH QUARTER | TOTAL GRAMS |
|---|---|---|---|---|---|
| 200 | 7,638.30 | 7,661.52 | 7,170.42 | 8,106.32 | **30,576.57** |
| 203 | 320.46 | 188.79 | 300.47 | 69.60 | **879.32** |
| 204 | 379.26 | 425.52 | 775.82 | 274.82 | **1,855.41** |
| 205 | 0 | 0 | 6.83 | 0.43 | **7.26** |
| **DC - DISTRICT OF COLUMBIA** | **8,338.02** | **8,275.83** | **8,253.54** | **8,451.18** | **33,318.56** |

**STATE: DE - DELAWARE**

| REGISTRANT ZIP CODE 3 | 1ST QUARTER | 2ND QUARTER | 3RD QUARTER | 4TH QUARTER | TOTAL GRAMS |
|---|---|---|---|---|---|
| 197 | 6,751.31 | 6,678.37 | 6,222.34 | 7,016.83 | **26,668.86** |
| 198 | 4,868.49 | 4,921.08 | 4,493.53 | 4,982.37 | **19,265.47** |
| 199 | 6,435.91 | 5,734.10 | 5,786.56 | 6,280.50 | **24,237.08** |
| **DE DELAWARE Total** | **18,055.71** | **17,333.55** | **16,502.44** | **18,279.70** | **70,171.41** |

**STATE: FL - FLORIDA**

| REGISTRANT ZIP CODE 3 | 1ST QUARTER | 2ND QUARTER | 3RD QUARTER | 4TH QUARTER | TOTAL GRAMS |
|---|---|---|---|---|---|
| 320 | 8,001.31 | 7,813.41 | 7,635.58 | 8,495.39 | **31,945.68** |
| 321 | 4,763.48 | 4,791.51 | 4,526.78 | 4,832.18 | **18,913.96** |
| 322 | 11,736.40 | 11,392.50 | 11,741.86 | 12,194.93 | **47,065.70** |
| 323 | 4,785.93 | 4,660.69 | 4,524.53 | 5,116.13 | **19,087.28** |
| 324 | 5,132.18 | 4,818.07 | 4,895.04 | 5,413.93 | **20,259.22** |
| 325 | 12,838.75 | 11,668.95 | 11,112.34 | 12,657.58 | **48,277.62** |
| 326 | 6,020.56 | 5,858.19 | 5,534.82 | 6,311.28 | **23,724.85** |
| 327 | 11,404.37 | 11,447.83 | 10,763.83 | 12,485.61 | **46,101.64** |
| 328 | 7,678.23 | 7,488.97 | 6,973.75 | 7,889.38 | **30,030.32** |
| 329 | 9,195.92 | 8,971.52 | 8,699.56 | 9,286.52 | **36,153.52** |
| 330 | 9,190.99 | 9,034.32 | 7,919.00 | 9,437.67 | **35,581.98** |
| 331 | 10,728.22 | 10,394.02 | 9,296.46 | 10,918.28 | **41,336.98** |
| 333 | 4,398.07 | 4,577.83 | 4,021.92 | 4,680.96 | **17,678.79** |
| 334 | 12,811.00 | 12,310.69 | 11,689.16 | 13,198.11 | **50,008.96** |
| 335 | 9,673.82 | 9,594.28 | 9,251.07 | 10,927.62 | **39,446.80** |
| 336 | 8,835.53 | 8,668.36 | 8,552.61 | 9,382.26 | **35,438.76** |
| 337 | 6,777.91 | 6,738.11 | 6,439.04 | 6,864.41 | **26,819.47** |
| 338 | 8,080.47 | 7,383.81 | 7,150.55 | 8,320.82 | **30,935.66** |
| 339 | 8,543.39 | 8,157.44 | 7,959.92 | 8,980.82 | **33,641.57** |
| 341 | 3,203.86 | 3,051.61 | 2,723.82 | 3,356.87 | **12,336.17** |
| 342 | 9,449.49 | 9,466.41 | 8,871.56 | 9,752.49 | **37,539.96** |
| 344 | 5,189.84 | 5,064.47 | 4,720.57 | 5,152.12 | **20,126.99** |
| 346 | 7,562.04 | 7,684.26 | 6,976.80 | 8,176.62 | **30,399.73** |
| 347 | 6,764.11 | 6,690.43 | 6,346.77 | 7,175.21 | **26,976.52** |

| | | | | | |
|---|---|---|---|---|---|
| 349 | 4,565.58 | 4,536.62 | 4,113.81 | 4,495.41 | **17,711.42** |
| **FL FLORIDA - Total** | **197,331.46** | **192,264.30** | **182,441.17** | **205,502.62** | **777,539.54** |
| **STATE: GA - GEORGIA** | | | | | |
| REGISTRANT ZIP CODE 3 | 1ST QUARTER | 2ND QUARTER | 3RD QUARTER | 4TH QUARTER | **TOTAL GRAMS** |
| 300 | 31,404.11 | 28,978.18 | 29,537.40 | 32,873.26 | **122,792.96** |
| 301 | 19,735.94 | 17,785.77 | 18,851.78 | 20,446.09 | **76,819.57** |
| 302 | 14,280.01 | 12,622.77 | 13,295.66 | 13,918.13 | **54,116.56** |
| 303 | 12,756.36 | 11,915.46 | 11,961.29 | 13,576.37 | **50,209.48** |
| 304 | 3,831.33 | 3,531.80 | 3,739.15 | 4,132.76 | **15,235.04** |
| 305 | 9,846.50 | 8,765.09 | 9,119.47 | 9,848.34 | **37,579.41** |
| 306 | 6,366.78 | 5,511.79 | 5,815.01 | 6,239.57 | **23,933.15** |
| 307 | 6,642.77 | 6,141.93 | 6,083.88 | 6,801.77 | **25,670.35** |
| 308 | 3,207.59 | 2,973.51 | 2,991.86 | 3,626.31 | **12,799.26** |
| 309 | 4,864.47 | 4,074.69 | 4,906.57 | 3,985.78 | **17,831.51** |
| 310 | 11,833.17 | 10,303.81 | 10,733.24 | 11,586.40 | **44,456.61** |
| 312 | 3,649.59 | 3,421.33 | 3,650.54 | 3,768.05 | **14,489.50** |
| 313 | 5,762.17 | 4,484.44 | 4,766.33 | 5,470.73 | **20,483.67** |
| 314 | 4,183.26 | 3,652.93 | 3,676.69 | 4,058.18 | **15,571.07** |
| 315 | 8,603.09 | 7,416.52 | 7,891.80 | 8,457.52 | **32,368.93** |
| 316 | 3,885.01 | 3,499.29 | 3,602.17 | 3,866.92 | **14,853.39** |
| 317 | 8,589.56 | 7,394.80 | 7,701.58 | 7,949.02 | **31,634.96** |
| 318 | 554.01 | 488.80 | 430.41 | 453.31 | **1,926.53** |
| 319 | 5,442.88 | 4,986.93 | 4,878.05 | 4,817.01 | **20,124.86** |
| 398 | 1,574.51 | 1,247.11 | 1,460.76 | 1,379.28 | **5,661.66** |
| **GA GEORGIA Total** | **167,013.09** | **149,196.94** | **155,093.66** | **167,254.80** | **638,558.48** |
| **STATE: GU - GUAM** | | | | | |
| REGISTRANT ZIP CODE 3 | 1ST QUARTER | 2ND QUARTER | 3RD QUARTER | 4TH QUARTER | **TOTAL GRAMS** |
| 969 | 343.40 | 453.68 | 405.68 | 319.84 | **1,522.60** |
| **GU - GUAM - Total** | **343.40** | **453.68** | **405.68** | **319.84** | **1,522.60** |
| **STATE: HI - HAWAII** | | | | | |
| REGISTRANT ZIP CODE 3 | 1ST QUARTER | 2ND QUARTER | 3RD QUARTER | 4TH QUARTER | **TOTAL GRAMS** |
| 967 | 5,357.88 | 5,513.85 | 5,211.58 | 5,678.16 | **21,761.48** |
| 968 | 5,179.06 | 5,253.49 | 5,320.60 | 5,249.67 | **21,002.82** |
| **HI - HAWAII - Total** | **10,536.94** | **10,767.34** | **10,532.17** | **10,927.84** | **42,764.29** |
| **STATE: IA - IOWA** | | | | | |
| REGISTRANT ZIP CODE 3 | 1ST QUARTER | 2ND QUARTER | 3RD QUARTER | 4TH QUARTER | **TOTAL GRAMS** |
| 500 | 6,670.39 | 6,432.65 | 6,420.89 | 7,214.05 | **26,737.98** |
| 501 | 3,668.17 | 3,664.40 | 3,590.89 | 3,838.22 | **14,761.68** |
| 502 | 4,604.54 | 4,486.42 | 4,394.34 | 4,690.99 | **18,176.29** |
| 503 | 7,449.44 | 7,449.37 | 7,200.90 | 7,792.64 | **29,892.36** |

| | | | | | |
|---|---|---|---|---|---|
| 504 | 1,990.62 | 1,955.15 | 2,025.97 | 2,053.37 | **8,025.11** |
| 505 | 2,139.48 | 2,080.50 | 1,999.67 | 2,225.91 | **8,445.56** |
| 506 | 3,167.62 | 3,104.02 | 3,035.78 | 3,521.43 | **12,828.85** |
| 507 | 1,777.24 | 1,757.37 | 1,627.09 | 1,866.67 | **7,028.37** |
| 508 | 532.41 | 622.05 | 505.47 | 669.25 | **2,329.18** |
| 510 | 2,117.72 | 1,981.65 | 1,814.64 | 2,103.44 | **8,017.45** |
| 511 | 3,891.31 | 3,978.26 | 3,943.56 | 4,143.32 | **15,956.45** |
| 512 | 1,061.12 | 960.87 | 945.65 | 1,013.96 | **3,981.60** |
| 513 | 931.10 | 922.24 | 904.55 | 931.47 | **3,689.36** |
| 514 | 964.42 | 933.65 | 822.37 | 876.95 | **3,597.39** |
| 515 | 3,182.51 | 3,063.45 | 2,893.73 | 3,302.88 | **12,442.57** |
| 516 | 459.82 | 379.29 | 386.07 | 375.33 | **1,600.51** |
| 520 | 3,648.47 | 3,682.30 | 3,559.47 | 3,760.71 | **14,650.95** |
| 521 | 1,164.47 | 1,141.19 | 1,102.04 | 1,234.56 | **4,642.26** |
| 522 | 6,806.04 | 6,891.44 | 6,783.93 | 7,337.33 | **27,818.74** |
| 523 | 3,843.94 | 3,766.85 | 3,895.14 | 4,091.56 | **15,597.49** |
| 524 | 5,991.73 | 5,843.09 | 5,859.15 | 6,319.31 | **24,013.28** |
| 525 | 2,065.53 | 2,090.70 | 2,105.20 | 2,360.21 | **8,621.64** |
| 526 | 2,894.01 | 2,868.17 | 2,816.05 | 3,225.15 | **11,803.38** |
| 527 | 4,432.45 | 4,472.69 | 4,089.08 | 4,563.89 | **17,558.10** |
| 528 | 3,407.93 | 3,276.18 | 3,189.23 | 3,639.40 | **13,512.75** |
| **IA IOWA Total** | **78,862.48** | **77,803.95** | **75,910.86** | **83,152.00** | **315,729.29** |

**STATE: ID - IDAHO**

| REGISTRANT ZIP CODE 3 | 1ST QUARTER | 2ND QUARTER | 3RD QUARTER | 4TH QUARTER | TOTAL GRAMS |
|---|---|---|---|---|---|
| 832 | 2,737.18 | 2,687.31 | 2,500.54 | 3,025.94 | **10,950.97** |
| 833 | 2,803.04 | 2,594.35 | 2,587.52 | 2,716.38 | **10,701.30** |
| 834 | 3,456.08 | 3,568.55 | 3,436.56 | 3,973.03 | **14,434.22** |
| 835 | 744.18 | 786.75 | 769.72 | 863.17 | **3,163.82** |
| 836 | 9,174.40 | 8,868.74 | 8,492.81 | 9,260.02 | **35,795.96** |
| 837 | 6,648.48 | 6,565.32 | 6,237.25 | 6,663.22 | **26,114.27** |
| 838 | 2,838.83 | 2,956.72 | 2,744.89 | 3,272.26 | **11,812.70** |
| **ID IDAHO Total** | **28,402.19** | **28,027.75** | **26,769.29** | **29,774.01** | **112,973.23** |

**STATE: IL - ILLINOIS**

| REGISTRANT ZIP CODE 3 | 1ST QUARTER | 2ND QUARTER | 3RD QUARTER | 4TH QUARTER | TOTAL GRAMS |
|---|---|---|---|---|---|
| 600 | 25,451.95 | 25,346.90 | 25,155.47 | 27,227.88 | **103,182.19** |
| 601 | 18,464.97 | 17,763.64 | 17,819.13 | 18,784.06 | **72,831.81** |
| 602 | 1,431.32 | 1,374.54 | 1,442.53 | 1,566.82 | **5,815.20** |
| 603 | 1,328.13 | 1,360.81 | 1,326.25 | 1,432.25 | **5,447.45** |
| 604 | 13,201.04 | 12,674.16 | 12,229.85 | 13,316.50 | **51,421.55** |
| 605 | 14,856.88 | 14,243.41 | 14,308.05 | 15,825.66 | **59,234.00** |

| | | | | | |
|---|---|---|---|---|---|
| 606 | 16,491.34 | 16,662.55 | 15,529.85 | 17,252.20 | **65,935.94** |
| 607 | 1,306.50 | 1,204.10 | 1,256.99 | 1,298.56 | **5,066.16** |
| 608 | 705.99 | 601.14 | 584.34 | 618.86 | **2,510.33** |
| 609 | 2,289.13 | 2,145.23 | 2,204.88 | 2,135.03 | **8,774.27** |
| 610 | 3,737.93 | 3,574.47 | 3,375.03 | 3,800.39 | **14,487.83** |
| 611 | 3,872.26 | 3,826.36 | 3,784.85 | 3,728.43 | **15,211.89** |
| 612 | 2,953.44 | 2,869.76 | 2,891.91 | 2,981.84 | **11,696.95** |
| 613 | 2,177.74 | 2,096.85 | 2,035.47 | 2,132.75 | **8,442.81** |
| 614 | 2,088.70 | 2,207.96 | 2,065.05 | 2,230.30 | **8,592.02** |
| 615 | 4,183.98 | 3,781.92 | 3,797.24 | 3,838.76 | **15,601.90** |
| 616 | 5,021.11 | 4,688.26 | 4,413.51 | 4,843.13 | **18,966.00** |
| 617 | 5,675.90 | 5,480.74 | 5,598.06 | 5,896.31 | **22,651.01** |
| 618 | 6,150.71 | 5,759.89 | 5,697.06 | 5,882.42 | **23,490.08** |
| 619 | 2,254.80 | 2,140.13 | 1,996.00 | 2,137.08 | **8,528.01** |
| 620 | 5,846.97 | 5,282.25 | 5,166.28 | 5,511.80 | **21,807.30** |
| 622 | 7,835.60 | 7,253.24 | 7,217.99 | 7,542.90 | **29,849.73** |
| 623 | 2,194.94 | 2,127.03 | 2,091.95 | 2,115.29 | **8,529.21** |
| 624 | 2,229.17 | 1,908.89 | 2,043.32 | 2,076.12 | **8,257.50** |
| 625 | 2,724.21 | 2,521.95 | 2,497.98 | 2,588.72 | **10,332.87** |
| 626 | 3,905.80 | 3,624.25 | 3,511.07 | 3,908.30 | **14,949.41** |
| 627 | 4,713.46 | 4,289.06 | 4,040.10 | 4,349.24 | **17,391.86** |
| 628 | 3,550.37 | 3,378.01 | 3,326.24 | 3,566.82 | **13,821.44** |
| 629 | 4,249.80 | 3,834.49 | 3,846.59 | 3,993.85 | **15,924.73** |
| **IL ILLINOIS Total** | **170,894.16** | **164,021.96** | **161,253.04** | **172,582.29** | **668,751.45** |

**STATE: IN - INDIANA**

| REGISTRANT ZIP CODE 3 | 1ST QUARTER | 2ND QUARTER | 3RD QUARTER | 4TH QUARTER | **TOTAL GRAMS** |
|---|---|---|---|---|---|
| 460 | 13,870.19 | 13,304.89 | 14,040.99 | 14,486.70 | **55,702.77** |
| 461 | 11,082.03 | 10,325.99 | 10,972.72 | 11,396.29 | **43,777.03** |
| 462 | 15,426.30 | 14,434.70 | 14,558.39 | 15,255.80 | **59,675.19** |
| 463 | 8,990.16 | 9,000.35 | 8,738.43 | 9,474.40 | **36,203.34** |
| 464 | 938.38 | 936.36 | 777.85 | 920.73 | **3,573.31** |
| 465 | 8,186.05 | 8,094.16 | 8,230.26 | 8,375.83 | **32,886.30** |
| 466 | 2,645.24 | 2,689.80 | 2,808.19 | 2,545.46 | **10,688.69** |
| 467 | 4,488.03 | 4,091.48 | 4,268.69 | 4,769.21 | **17,617.42** |
| 468 | 6,637.18 | 6,217.53 | 6,585.39 | 6,723.14 | **26,163.24** |
| 469 | 4,955.75 | 4,648.96 | 4,967.73 | 5,079.10 | **19,651.54** |
| 470 | 1,532.22 | 1,383.52 | 1,519.77 | 1,565.89 | **6,001.41** |
| 471 | 4,681.94 | 4,383.07 | 4,449.46 | 4,819.51 | **18,333.97** |
| 472 | 3,681.84 | 3,631.08 | 3,582.34 | 3,830.83 | **14,726.09** |
| 473 | 5,077.16 | 4,716.32 | 4,878.19 | 5,141.45 | **19,813.12** |

| | | | | | |
|---|---|---|---|---|---|
| 474 | 3,681.77 | 3,605.93 | 3,708.32 | 4,098.19 | **15,094.21** |
| 475 | 2,864.28 | 2,724.61 | 2,698.89 | 3,028.08 | **11,315.86** |
| 476 | 2,919.93 | 2,663.48 | 2,613.29 | 2,747.35 | **10,944.04** |
| 477 | 5,533.10 | 5,124.23 | 5,374.54 | 5,853.71 | **21,885.59** |
| 478 | 3,283.54 | 2,829.73 | 3,158.70 | 3,117.04 | **12,389.01** |
| 479 | 5,719.00 | 5,374.93 | 5,437.13 | 5,739.42 | **22,270.48** |
| **IN INDIANA Total** | **116,194.08** | **110,181.12** | **113,369.27** | **118,968.14** | **458,712.61** |

**STATE: KS - KANSAS**

| REGISTRANT ZIP CODE 3 | 1ST QUARTER | 2ND QUARTER | 3RD QUARTER | 4TH QUARTER | TOTAL GRAMS |
|---|---|---|---|---|---|
| 660 | 9,957.90 | 9,832.49 | 9,845.03 | 10,640.30 | **40,275.72** |
| 661 | 1,327.26 | 1,215.02 | 1,172.83 | 1,234.20 | **4,949.32** |
| 662 | 8,544.85 | 8,665.93 | 8,326.61 | 11,484.17 | **37,021.56** |
| 664 | 1,538.47 | 1,597.08 | 1,334.96 | 1,495.97 | **5,966.48** |
| 665 | 1,741.33 | 1,675.74 | 1,668.32 | 1,933.73 | **7,019.12** |
| 666 | 5,067.05 | 5,109.75 | 5,202.61 | 5,538.75 | **20,918.16** |
| 667 | 1,936.03 | 1,904.75 | 1,910.47 | 1,957.60 | **7,708.85** |
| 668 | 782.25 | 732.40 | 770.25 | 908.02 | **3,192.91** |
| 669 | 269.19 | 259.61 | 243.69 | 303.45 | **1,075.94** |
| 670 | 2,780.89 | 2,683.64 | 2,545.29 | 3,020.31 | **11,030.14** |
| 671 | 1,242.06 | 1,215.32 | 1,213.44 | 1,331.50 | **5,002.32** |
| 672 | 6,307.36 | 5,919.20 | 6,238.92 | 6,857.23 | **25,322.71** |
| 673 | 1,465.95 | 1,337.89 | 1,418.92 | 1,426.60 | **5,649.36** |
| 674 | 1,974.37 | 1,979.88 | 1,892.97 | 2,123.47 | **7,970.71** |
| 675 | 1,774.67 | 1,550.06 | 1,581.05 | 1,874.83 | **6,780.61** |
| 676 | 1,030.81 | 895.71 | 931.92 | 962.79 | **3,821.24** |
| 677 | 324.54 | 365.33 | 361.85 | 373.86 | **1,425.59** |
| 678 | 815.12 | 817.48 | 806.68 | 865.97 | **3,305.26** |
| 679 | 169.50 | 127.20 | 142.42 | 150.14 | **589.27** |
| **KS - KANSAS - Total** | **49,049.62** | **47,884.49** | **47,608.23** | **54,482.90** | **199,025.25** |

**STATE: KY - KENTUCKY**

| REGISTRANT ZIP CODE 3 | 1ST QUARTER | 2ND QUARTER | 3RD QUARTER | 4TH QUARTER | TOTAL GRAMS |
|---|---|---|---|---|---|
| 400 | 4,108.17 | 3,807.81 | 3,991.89 | 4,166.42 | **16,074.29** |
| 401 | 2,070.26 | 1,588.25 | 2,187.89 | 1,926.61 | **7,773.00** |
| 402 | 13,475.12 | 12,418.71 | 12,733.31 | 14,197.09 | **52,824.22** |
| 403 | 5,973.02 | 5,840.32 | 5,772.93 | 6,503.80 | **24,090.07** |
| 404 | 2,241.18 | 2,017.02 | 2,282.46 | 2,367.70 | **8,908.36** |
| 405 | 5,574.51 | 5,105.79 | 5,325.47 | 6,035.13 | **22,040.90** |
| 406 | 1,068.27 | 969.87 | 992.65 | 1,128.85 | **4,159.64** |
| 407 | 1,674.61 | 1,573.50 | 1,660.90 | 1,731.01 | **6,640.02** |
| 408 | 295.74 | 283.92 | 265.64 | 295.26 | **1,140.56** |

| | | | | | |
|---|---|---|---|---|---|
| 409 | 699.55 | 650.58 | 690.93 | 723.51 | **2,764.56** |
| 410 | 10,576.63 | 9,541.34 | 9,736.55 | 10,693.19 | **40,547.72** |
| 411 | 2,181.19 | 2,140.98 | 1,974.79 | 2,467.87 | **8,764.83** |
| 412 | 616.97 | 693.79 | 666.66 | 744.71 | **2,722.12** |
| 413 | 416.69 | 350.22 | 397.54 | 410.02 | **1,574.46** |
| 414 | 239.06 | 230.66 | 212.76 | 258.48 | **940.96** |
| 415 | 1,124.56 | 1,099.66 | 1,038.92 | 1,182.55 | **4,445.69** |
| 416 | 685.56 | 659.02 | 550.05 | 640.42 | **2,535.06** |
| 417 | 962.29 | 811.97 | 770.62 | 831.01 | **3,375.89** |
| 418 | 389.39 | 274.80 | 336.12 | 283.69 | **1,284.00** |
| 420 | 4,518.78 | 4,120.93 | 4,238.33 | 4,370.58 | **17,248.61** |
| 421 | 3,093.52 | 2,931.06 | 3,080.35 | 3,133.75 | **12,238.69** |
| 422 | 2,388.08 | 2,453.79 | 2,739.68 | 2,244.53 | **9,826.08** |
| 423 | 2,846.17 | 2,674.65 | 2,557.82 | 2,837.33 | **10,915.97** |
| 424 | 3,438.06 | 3,050.61 | 3,150.78 | 3,274.62 | **12,914.07** |
| 425 | 1,027.32 | 916.44 | 1,003.06 | 1,017.49 | **3,964.31** |
| 426 | 788.09 | 712.12 | 712.93 | 752.12 | **2,965.27** |
| 427 | 2,786.26 | 2,715.52 | 2,668.01 | 2,955.18 | **11,124.96** |
| **KY - KENTUCKY - Total** | **75,259.04** | **69,633.34** | **71,739.04** | **77,172.91** | **293,804.33** |

**STATE: LA - LOUISIANA**

| REGISTRANT ZIP CODE 3 | 1ST QUARTER | 2ND QUARTER | 3RD QUARTER | 4TH QUARTER | TOTAL GRAMS |
|---|---|---|---|---|---|
| 700 | 9,519.08 | 8,458.02 | 8,938.87 | 9,571.06 | **36,487.02** |
| 701 | 4,613.30 | 4,402.42 | 4,037.08 | 4,894.21 | **17,947.01** |
| 703 | 4,656.41 | 3,886.73 | 4,064.97 | 4,742.63 | **17,350.73** |
| 704 | 9,859.33 | 8,642.92 | 8,759.63 | 9,813.38 | **37,075.26** |
| 705 | 13,574.21 | 11,291.98 | 12,211.64 | 13,736.38 | **50,814.20** |
| 706 | 8,008.22 | 6,885.92 | 6,965.77 | 7,360.60 | **29,220.51** |
| 707 | 10,399.11 | 9,558.01 | 9,789.54 | 10,555.49 | **40,302.15** |
| 708 | 7,264.18 | 5,986.82 | 6,477.54 | 7,011.47 | **26,740.01** |
| 710 | 1,787.03 | 1,663.96 | 1,577.36 | 1,707.36 | **6,735.71** |
| 711 | 4,610.27 | 4,157.11 | 4,129.82 | 4,804.02 | **17,701.21** |
| 712 | 5,012.62 | 4,130.08 | 4,342.97 | 4,672.37 | **18,158.05** |
| 713 | 3,829.91 | 3,149.19 | 3,414.26 | 3,822.38 | **14,215.73** |
| 714 | 2,675.33 | 2,176.18 | 2,513.53 | 2,582.87 | **9,947.92** |
| **LA - LOUISIANA - Total** | **85,808.99** | **74,389.33** | **77,222.99** | **85,274.21** | **322,695.52** |

**STATE: MA - MASSACHUSETTS**

| REGISTRANT ZIP CODE 3 | 1ST QUARTER | 2ND QUARTER | 3RD QUARTER | 4TH QUARTER | TOTAL GRAMS |
|---|---|---|---|---|---|
| 010 | 9,934.11 | 10,213.86 | 9,111.70 | 10,294.05 | **39,553.72** |
| 011 | 3,914.64 | 4,028.71 | 3,733.10 | 4,152.58 | **15,829.03** |
| 012 | 4,100.86 | 4,167.66 | 3,765.99 | 4,085.41 | **16,119.92** |

| | | | | | |
|---|---|---|---|---|---|
| 013 | 2,087.08 | 2,104.21 | 1,944.58 | 2,122.06 | **8,257.93** |
| 014 | 5,947.00 | 5,748.76 | 5,588.69 | 5,713.52 | **22,997.97** |
| 015 | 8,116.33 | 8,214.70 | 7,242.32 | 8,522.35 | **32,095.69** |
| 016 | 3,893.36 | 4,062.21 | 3,704.20 | 4,188.20 | **15,847.98** |
| 017 | 10,531.41 | 10,496.02 | 9,695.66 | 10,712.76 | **41,435.85** |
| 018 | 15,542.46 | 15,105.29 | 13,705.01 | 16,179.51 | **60,532.28** |
| 019 | 10,902.53 | 11,291.40 | 10,204.44 | 11,520.93 | **43,919.29** |
| 020 | 9,911.42 | 10,063.05 | 9,533.67 | 10,440.14 | **39,948.29** |
| 021 | 21,167.31 | 21,333.43 | 18,941.45 | 21,974.91 | **83,417.10** |
| 022 | 1,290.05 | 1,073.73 | 1,160.80 | 1,138.17 | **4,662.76** |
| 023 | 8,953.58 | 9,122.74 | 8,420.74 | 9,563.59 | **36,060.66** |
| 024 | 11,795.66 | 11,921.46 | 11,011.91 | 12,063.53 | **46,792.56** |
| 025 | 2,846.99 | 2,814.34 | 2,774.46 | 2,781.59 | **11,217.38** |
| 026 | 3,746.95 | 3,588.35 | 3,918.08 | 3,963.00 | **15,216.38** |
| 027 | 9,201.58 | 9,276.28 | 8,358.71 | 9,403.51 | **36,240.09** |
| **MA - MASSACHUSETTS - Total** | **143,883.34** | **144,626.21** | **132,815.51** | **148,819.79** | **570,144.86** |

**STATE: MD - MARYLAND**

| REGISTRANT ZIP CODE 3 | 1ST QUARTER | 2ND QUARTER | 3RD QUARTER | 4TH QUARTER | **TOTAL GRAMS** |
|---|---|---|---|---|---|
| 206 | 3,837.16 | 3,853.04 | 3,848.00 | 4,073.17 | **15,611.37** |
| 207 | 6,730.70 | 6,351.06 | 6,157.47 | 5,920.97 | **25,160.20** |
| 208 | 15,933.95 | 16,406.97 | 14,697.18 | 16,283.80 | **63,321.91** |
| 209 | 2,761.26 | 2,915.86 | 2,788.25 | 2,869.60 | **11,334.96** |
| 210 | 17,150.56 | 17,421.39 | 16,049.02 | 18,134.15 | **68,755.11** |
| 211 | 8,422.79 | 8,589.16 | 7,701.53 | 8,871.95 | **33,585.43** |
| 212 | 19,828.65 | 18,923.49 | 17,588.75 | 20,249.24 | **76,590.12** |
| 214 | 1,946.44 | 1,924.40 | 1,792.93 | 2,031.19 | **7,694.97** |
| 215 | 3,488.79 | 3,393.26 | 3,472.48 | 3,606.34 | **13,960.88** |
| 216 | 3,769.26 | 3,687.03 | 3,276.09 | 4,166.22 | **14,898.60** |
| 217 | 11,700.84 | 11,478.14 | 10,823.78 | 12,261.08 | **46,263.84** |
| 218 | 3,309.11 | 3,332.22 | 3,003.38 | 3,473.62 | **13,118.33** |
| 219 | 1,821.81 | 1,782.60 | 1,826.21 | 1,809.79 | **7,240.42** |
| **MD MARYLAND Total** | **100,701.31** | **100,058.63** | **93,025.07** | **103,751.13** | **397,536.14** |

**STATE: ME - MAINE**

| REGISTRANT ZIP CODE 3 | 1ST QUARTER | 2ND QUARTER | 3RD QUARTER | 4TH QUARTER | **TOTAL GRAMS** |
|---|---|---|---|---|---|
| 039 | 770.07 | 724.55 | 726.29 | 705.43 | **2,926.33** |
| 040 | 7,764.45 | 7,873.35 | 7,775.92 | 7,960.04 | **31,373.76** |
| 041 | 2,702.42 | 2,949.56 | 2,890.15 | 2,943.03 | **11,485.15** |
| 042 | 3,183.42 | 3,282.87 | 3,009.14 | 3,332.89 | **12,808.31** |
| 043 | 2,741.22 | 2,912.42 | 2,963.23 | 2,871.14 | **11,488.01** |
| 044 | 6,253.10 | 5,923.85 | 6,278.97 | 6,322.01 | **24,777.93** |

| | | | | | |
|---|---|---|---|---|---|
| 045 | 1,366.66 | 1,350.19 | 1,393.54 | 1,448.95 | **5,559.33** |
| 046 | 1,861.02 | 1,891.97 | 2,040.91 | 1,977.96 | **7,771.87** |
| 047 | 1,731.65 | 1,679.15 | 1,923.03 | 1,983.57 | **7,317.39** |
| 048 | 1,334.54 | 1,376.04 | 1,393.26 | 1,336.43 | **5,440.26** |
| 049 | 5,079.28 | 4,831.38 | 5,017.16 | 4,605.96 | **19,533.78** |
| **ME MAINE Total** | **34,787.81** | **34,795.32** | **35,411.59** | **35,487.40** | **140,482.12** |

**STATE: MI - MICHIGAN**

| REGISTRANT ZIP CODE 3 | 1ST QUARTER | 2ND QUARTER | 3RD QUARTER | 4TH QUARTER | TOTAL GRAMS |
|---|---|---|---|---|---|
| 480 | 21,802.26 | 21,269.58 | 20,129.80 | 21,725.06 | **84,926.70** |
| 481 | 25,469.96 | 25,317.41 | 24,644.96 | 26,254.16 | **101,686.48** |
| 482 | 4,224.46 | 4,298.39 | 3,733.49 | 3,892.35 | **16,148.69** |
| 483 | 18,368.56 | 18,309.86 | 17,926.05 | 18,866.13 | **73,470.60** |
| 484 | 6,045.25 | 5,866.98 | 5,840.65 | 5,971.74 | **23,724.62** |
| 485 | 2,041.23 | 2,016.27 | 1,863.56 | 1,831.04 | **7,752.10** |
| 486 | 6,785.47 | 6,562.25 | 6,682.70 | 6,351.49 | **26,381.90** |
| 487 | 3,859.01 | 3,600.02 | 3,545.31 | 3,597.35 | **14,601.69** |
| 488 | 12,056.59 | 11,855.49 | 12,924.98 | 11,745.80 | **48,582.85** |
| 489 | 2,876.18 | 2,725.96 | 2,774.22 | 2,738.31 | **11,114.67** |
| 490 | 13,165.39 | 12,902.29 | 13,546.07 | 12,656.41 | **52,270.16** |
| 491 | 1,023.15 | 884.06 | 1,035.30 | 993.82 | **3,936.33** |
| 492 | 5,550.55 | 5,045.61 | 5,181.33 | 4,925.50 | **20,703.00** |
| 493 | 5,140.61 | 5,101.31 | 5,593.22 | 4,988.32 | **20,823.45** |
| 494 | 15,794.20 | 15,515.29 | 16,367.28 | 14,936.78 | **62,613.54** |
| 495 | 12,255.84 | 11,695.65 | 12,610.99 | 11,591.90 | **48,154.37** |
| 496 | 6,233.03 | 6,139.75 | 6,418.15 | 6,152.42 | **24,943.35** |
| 497 | 4,860.16 | 4,703.40 | 4,875.13 | 4,762.11 | **19,200.81** |
| 498 | 6,270.63 | 6,230.11 | 6,403.58 | 6,404.36 | **25,308.68** |
| 499 | 1,354.98 | 1,223.86 | 1,190.08 | 1,471.21 | **5,240.14** |
| **MI - MICHIGAN Total** | **175,177.52** | **171,263.53** | **173,286.83** | **171,856.27** | **691,584.16** |

**STATE: MN - MINNESOTA**

| REGISTRANT ZIP CODE 3 | 1ST QUARTER | 2ND QUARTER | 3RD QUARTER | 4TH QUARTER | TOTAL GRAMS |
|---|---|---|---|---|---|
| 550 | 11,318.55 | 11,362.99 | 10,446.49 | 11,730.48 | **44,858.51** |
| 551 | 14,564.74 | 14,945.72 | 13,892.04 | 15,031.70 | **58,434.20** |
| 553 | 21,088.41 | 20,910.13 | 19,472.45 | 21,777.09 | **83,248.08** |
| 554 | 19,800.53 | 19,969.78 | 18,530.36 | 20,456.35 | **78,757.02** |
| 556 | 154.18 | 162.23 | 130.77 | 137.72 | **584.91** |
| 557 | 2,301.17 | 2,283.50 | 2,071.58 | 2,298.26 | **8,954.50** |
| 558 | 2,682.02 | 2,655.57 | 2,453.72 | 2,680.55 | **10,471.85** |
| 559 | 7,935.23 | 8,471.61 | 7,842.25 | 8,632.75 | **32,881.84** |
| 560 | 4,428.20 | 4,445.20 | 4,297.65 | 4,575.02 | **17,746.07** |

| | | | | | |
|---|---|---|---|---|---|
| 561 | 1,408.80 | 1,368.46 | 1,382.15 | 1,425.77 | **5,585.18** |
| 562 | 2,396.85 | 2,186.63 | 2,051.34 | 2,389.78 | **9,024.61** |
| 563 | 6,001.74 | 5,833.96 | 5,548.49 | 6,191.25 | **23,575.44** |
| 564 | 2,052.79 | 2,056.74 | 2,016.90 | 2,217.18 | **8,343.60** |
| 565 | 3,683.23 | 3,704.59 | 3,532.84 | 3,915.03 | **14,835.69** |
| 566 | 1,063.82 | 1,031.02 | 958.43 | 1,106.74 | **4,160.01** |
| 567 | 943.11 | 1,014.29 | 956.78 | 1,035.45 | **3,949.64** |
| **MN MINNESOTA Total** | **101,823.36** | **102,402.43** | **95,584.23** | **105,601.12** | **405,411.14** |

**STATE: MO - MISSOURI**

| REGISTRANT ZIP CODE 3 | 1ST QUARTER | 2ND QUARTER | 3RD QUARTER | 4TH QUARTER | TOTAL GRAMS |
|---|---|---|---|---|---|
| 630 | 16,716.53 | 16,079.95 | 15,262.14 | 17,133.18 | **65,191.81** |
| 631 | 56,081.98 | 57,209.11 | 52,810.57 | 58,724.23 | **224,825.89** |
| 633 | 12,232.01 | 11,590.40 | 12,033.82 | 13,028.09 | **48,884.33** |
| 634 | 1,428.85 | 1,221.03 | 1,335.56 | 1,324.96 | **5,310.41** |
| 635 | 797.60 | 825.82 | 797.44 | 857.59 | **3,278.45** |
| 636 | 2,299.78 | 2,117.00 | 1,978.38 | 2,475.11 | **8,870.27** |
| 637 | 2,084.46 | 1,923.06 | 1,880.51 | 2,183.69 | **8,071.72** |
| 638 | 2,309.81 | 1,945.98 | 2,090.20 | 2,354.54 | **8,700.54** |
| 639 | 848.27 | 723.82 | 787.06 | 937.07 | **3,296.22** |
| 640 | 9,688.69 | 9,975.26 | 9,436.40 | 10,311.76 | **39,412.11** |
| 641 | 9,771.99 | 7,965.78 | 7,119.93 | 7,797.36 | **32,655.05** |
| 644 | 1,083.44 | 1,039.75 | 1,059.34 | 1,159.68 | **4,342.21** |
| 645 | 2,096.19 | 1,942.03 | 1,867.55 | 2,014.70 | **7,920.47** |
| 646 | 555.18 | 498.81 | 483.74 | 505.01 | **2,042.74** |
| 647 | 990.97 | 995.57 | 1,070.90 | 1,114.95 | **4,172.39** |
| 648 | 3,109.08 | 2,833.06 | 2,811.80 | 3,359.38 | **12,113.33** |
| 650 | 1,410.15 | 1,358.07 | 1,341.89 | 1,476.57 | **5,586.68** |
| 651 | 1,768.96 | 1,592.50 | 1,589.64 | 1,906.34 | **6,857.44** |
| 652 | 3,966.53 | 3,945.96 | 3,870.35 | 4,370.65 | **16,153.49** |
| 653 | 1,164.25 | 1,160.96 | 1,028.43 | 1,302.48 | **4,656.12** |
| 654 | 1,227.20 | 1,297.08 | 1,491.13 | 1,349.53 | **5,364.94** |
| 655 | 1,323.09 | 1,229.35 | 1,238.73 | 1,439.67 | **5,230.84** |
| 656 | 1,668.09 | 1,595.14 | 1,727.26 | 1,759.33 | **6,749.82** |
| 657 | 3,276.87 | 3,171.49 | 3,259.82 | 3,538.07 | **13,246.26** |
| 658 | 4,386.77 | 4,044.97 | 4,108.37 | 4,235.25 | **16,775.35** |
| **MO MISSOURI - Total** | **142,286.74** | **138,281.95** | **132,480.98** | **146,659.18** | **559,708.85** |

**STATE: MS - MISSISSIPPI**

| REGISTRANT ZIP CODE 3 | 1ST QUARTER | 2ND QUARTER | 3RD QUARTER | 4TH QUARTER | TOTAL GRAMS |
|---|---|---|---|---|---|
| 386 | 6,503.65 | 5,646.64 | 5,966.23 | 6,831.40 | **24,947.92** |
| 387 | 620.57 | 576.35 | 611.18 | 654.58 | **2,462.68** |

| | | | | | |
|---|---|---|---|---|---|
| 388 | 3,407.17 | 3,071.36 | 3,170.86 | 3,909.93 | **13,559.31** |
| 389 | 1,200.13 | 1,080.73 | 1,131.45 | 1,204.66 | **4,616.97** |
| 390 | 3,451.92 | 3,080.88 | 3,205.06 | 3,495.98 | **13,233.85** |
| 391 | 3,643.46 | 3,066.85 | 3,372.52 | 3,680.76 | **13,763.59** |
| 392 | 3,274.77 | 2,889.61 | 2,963.27 | 3,567.14 | **12,694.80** |
| 393 | 2,505.90 | 2,257.04 | 2,379.74 | 2,644.84 | **9,787.52** |
| 394 | 6,904.21 | 6,326.57 | 6,676.11 | 7,464.15 | **27,371.04** |
| 395 | 9,146.23 | 7,874.47 | 8,104.68 | 9,475.79 | **34,601.16** |
| 396 | 1,812.53 | 1,672.06 | 1,801.14 | 1,930.38 | **7,216.11** |
| 397 | 1,637.34 | 1,497.54 | 1,542.94 | 1,701.52 | **6,379.33** |
| **MS MISSISSIPPI - Total** | **44,107.88** | **39,040.08** | **40,925.18** | **46,561.13** | **170,634.28** |

**STATE: MT - MONTANA**

| REGISTRANT ZIP CODE 3 | 1ST QUARTER | 2ND QUARTER | 3RD QUARTER | 4TH QUARTER | TOTAL GRAMS |
|---|---|---|---|---|---|
| 590 | 771.12 | 836.00 | 804.60 | 876.05 | **3,287.76** |
| 591 | 4,338.20 | 4,311.45 | 3,751.97 | 4,467.48 | **16,869.10** |
| 592 | 507.01 | 547.29 | 584.77 | 565.86 | **2,204.93** |
| 593 | 740.38 | 741.88 | 685.23 | 676.83 | **2,844.32** |
| 594 | 2,231.52 | 2,389.29 | 2,394.82 | 2,261.51 | **9,277.15** |
| 595 | 312.39 | 314.80 | 303.17 | 343.86 | **1,274.22** |
| 596 | 1,251.46 | 1,323.40 | 1,304.12 | 1,449.26 | **5,328.23** |
| 597 | 2,737.97 | 2,455.69 | 2,445.71 | 2,645.50 | **10,284.87** |
| 598 | 2,665.44 | 2,727.26 | 2,639.37 | 2,812.66 | **10,844.74** |
| 599 | 1,423.16 | 1,495.68 | 1,462.79 | 1,502.04 | **5,883.68** |
| **MT - MONTANA Total** | **16,978.65** | **17,142.74** | **16,376.55** | **17,601.06** | **68,098.99** |

**STATE: NC - NORTH CAROLINA**

| REGISTRANT ZIP CODE 3 | 1ST QUARTER | 2ND QUARTER | 3RD QUARTER | 4TH QUARTER | TOTAL GRAMS |
|---|---|---|---|---|---|
| 270 | 4,992.95 | 4,676.27 | 4,428.17 | 5,097.16 | **19,194.55** |
| 271 | 5,929.70 | 5,519.53 | 4,989.98 | 5,639.39 | **22,078.61** |
| 272 | 12,199.40 | 11,387.67 | 11,158.70 | 11,624.86 | **46,370.63** |
| 273 | 7,020.06 | 6,576.83 | 6,300.63 | 6,841.20 | **26,738.72** |
| 274 | 6,143.08 | 5,333.57 | 5,235.82 | 5,694.91 | **22,407.38** |
| 275 | 26,380.88 | 25,299.60 | 23,638.37 | 25,663.56 | **100,982.41** |
| 276 | 10,278.20 | 9,503.57 | 9,270.52 | 10,479.89 | **39,532.18** |
| 277 | 5,783.49 | 5,772.66 | 5,369.67 | 5,896.84 | **22,822.66** |
| 278 | 9,376.83 | 8,842.16 | 8,319.06 | 9,384.02 | **35,922.07** |
| 279 | 2,903.26 | 2,773.07 | 2,546.59 | 2,406.61 | **10,629.52** |
| 280 | 17,658.19 | 16,522.12 | 16,116.86 | 17,785.18 | **68,082.36** |
| 281 | 13,549.55 | 12,616.27 | 12,111.21 | 13,635.91 | **51,912.93** |
| 282 | 13,411.30 | 13,176.38 | 12,521.99 | 13,920.84 | **53,030.52** |
| 283 | 17,360.65 | 16,775.20 | 14,850.30 | 15,999.89 | **64,986.04** |

| | | | | | |
|---|---|---|---|---|---|
| 284 | 8,899.74 | 8,424.32 | 8,176.30 | 8,839.54 | **34,339.89** |
| 285 | 11,643.80 | 11,078.56 | 11,341.07 | 10,427.24 | **44,490.66** |
| 286 | 11,038.50 | 10,365.07 | 10,087.61 | 11,107.29 | **42,598.48** |
| 287 | 8,818.77 | 8,909.82 | 8,599.22 | 9,138.24 | **35,466.05** |
| 288 | 3,657.63 | 3,790.67 | 3,728.96 | 3,952.08 | **15,129.34** |
| 289 | 527.85 | 599.59 | 530.04 | 493.83 | **2,151.31** |
| **NC - NORTH CAROLINA Total** | **197,573.85** | **187,942.92** | **179,321.06** | **194,028.48** | **758,866.32** |

**STATE: ND - NORTH DAKOTA**

| REGISTRANT ZIP CODE 3 | 1ST QUARTER | 2ND QUARTER | 3RD QUARTER | 4TH QUARTER | TOTAL GRAMS |
|---|---|---|---|---|---|
| 580 | 1,602.72 | 1,674.77 | 1,598.28 | 1,787.10 | **6,662.87** |
| 581 | 3,674.36 | 3,384.12 | 3,613.13 | 3,730.09 | **14,401.69** |
| 582 | 2,272.53 | 2,210.70 | 2,012.42 | 2,472.06 | **8,967.72** |
| 583 | 1,032.01 | 971.12 | 908.88 | 909.16 | **3,821.17** |
| 584 | 655.52 | 578.51 | 625.04 | 619.51 | **2,478.58** |
| 585 | 2,775.69 | 2,784.27 | 2,618.42 | 3,072.91 | **11,251.29** |
| 586 | 832.96 | 847.38 | 789.22 | 884.51 | **3,354.07** |
| 587 | 1,781.12 | 1,685.54 | 1,719.62 | 1,550.00 | **6,736.29** |
| 588 | 696.47 | 701.70 | 660.67 | 708.12 | **2,766.95** |
| **ND NORTH DAKOTA Total** | **15,323.39** | **14,838.10** | **14,545.69** | **15,733.46** | **60,440.63** |

**STATE: NE - NEBRASKA**

| REGISTRANT ZIP CODE 3 | 1ST QUARTER | 2ND QUARTER | 3RD QUARTER | 4TH QUARTER | TOTAL GRAMS |
|---|---|---|---|---|---|
| 680 | 3,233.96 | 3,145.54 | 3,162.21 | 3,541.29 | **13,083.00** |
| 681 | 12,446.53 | 12,509.98 | 12,330.67 | 13,376.18 | **50,663.36** |
| 683 | 850.96 | 789.27 | 765.47 | 859.09 | **3,264.79** |
| 684 | 743.26 | 721.67 | 709.88 | 831.49 | **3,006.29** |
| 685 | 5,901.85 | 5,658.26 | 5,669.55 | 6,117.29 | **23,346.96** |
| 686 | 752.41 | 749.74 | 749.37 | 810.67 | **3,062.19** |
| 687 | 1,574.10 | 1,464.09 | 1,509.09 | 1,544.98 | **6,092.26** |
| 688 | 2,427.92 | 2,166.66 | 2,238.06 | 2,344.56 | **9,177.20** |
| 689 | 1,081.90 | 878.41 | 978.11 | 1,006.91 | **3,945.33** |
| 690 | 298.38 | 247.23 | 235.77 | 273.86 | **1,055.24** |
| 691 | 1,032.68 | 967.79 | 995.89 | 1,084.06 | **4,080.42** |
| 692 | 32.43 | 25.25 | 21.71 | 17.81 | **97.19** |
| 693 | 923.40 | 792.74 | 817.72 | 785.95 | **3,319.81** |
| **NE NEBRASKA Total** | **31,299.78** | **30,116.62** | **30,183.50** | **32,594.14** | **124,194.04** |

**STATE: NH - NEW HAMPSHIRE**

| REGISTRANT ZIP CODE 3 | 1ST QUARTER | 2ND QUARTER | 3RD QUARTER | 4TH QUARTER | TOTAL GRAMS |
|---|---|---|---|---|---|
| 030 | 8,621.10 | 8,740.44 | 8,331.17 | 8,796.32 | **34,489.02** |
| 031 | 4,968.98 | 4,913.35 | 4,794.51 | 5,124.58 | **19,801.42** |
| 032 | 3,136.12 | 3,198.79 | 3,186.18 | 3,325.82 | **12,846.92** |

| | | | | | |
|---|---|---|---|---|---|
| 033 | 2,358.97 | 2,493.32 | 2,289.17 | 2,470.65 | **9,612.10** |
| 034 | 2,013.21 | 1,903.65 | 1,874.13 | 2,101.63 | **7,892.61** |
| 035 | 1,894.11 | 1,721.26 | 1,833.03 | 1,739.84 | **7,188.23** |
| 036 | 89.58 | 104.80 | 111.81 | 84.48 | **390.67** |
| 037 | 3,327.42 | 3,371.34 | 3,153.50 | 3,502.04 | **13,354.30** |
| 038 | 7,691.53 | 7,789.69 | 7,556.69 | 8,068.80 | **31,106.71** |
| **NH - NEW HAMPSHIRE Total** | **34,101.02** | **34,236.64** | **33,130.18** | **35,214.16** | **136,682.00** |

**STATE: NJ - NEW JERSEY**

| REGISTRANT ZIP CODE 3 | 1ST QUARTER | 2ND QUARTER | 3RD QUARTER | 4TH QUARTER | TOTAL GRAMS |
|---|---|---|---|---|---|
| 070 | 16,441.88 | 15,138.65 | 14,240.01 | 16,367.36 | **62,187.90** |
| 071 | 1,849.06 | 1,727.29 | 1,639.30 | 1,818.90 | **7,034.55** |
| 072 | 943.15 | 847.41 | 771.32 | 895.19 | **3,457.07** |
| 073 | 1,109.27 | 1,043.36 | 936.28 | 1,120.37 | **4,209.28** |
| 074 | 5,119.57 | 4,920.44 | 4,566.92 | 5,484.27 | **20,091.20** |
| 075 | 827.81 | 904.86 | 714.30 | 855.58 | **3,302.54** |
| 076 | 4,761.47 | 4,712.20 | 4,580.91 | 4,927.37 | **18,981.94** |
| 077 | 6,421.36 | 6,356.04 | 5,818.56 | 6,424.40 | **25,020.35** |
| 078 | 4,684.96 | 4,359.82 | 4,075.76 | 4,635.68 | **17,756.22** |
| 079 | 4,849.79 | 4,851.94 | 4,506.00 | 5,300.19 | **19,507.93** |
| 080 | 17,493.70 | 17,121.50 | 15,862.13 | 18,296.19 | **68,773.52** |
| 081 | 1,555.09 | 1,577.21 | 1,431.60 | 1,757.84 | **6,321.74** |
| 082 | 2,584.35 | 2,735.37 | 2,647.67 | 2,548.89 | **10,516.28** |
| 083 | 2,438.08 | 2,336.82 | 2,093.79 | 2,529.92 | **9,398.61** |
| 084 | 246.49 | 239.55 | 327.82 | 285.52 | **1,099.38** |
| 085 | 3,941.69 | 3,717.37 | 3,649.52 | 3,926.48 | **15,235.06** |
| 086 | 2,996.98 | 3,031.05 | 2,422.03 | 3,115.67 | **11,565.73** |
| 087 | 5,164.99 | 4,785.66 | 4,663.73 | 5,097.39 | **19,711.78** |
| 088 | 8,424.87 | 8,365.06 | 7,561.83 | 8,480.01 | **32,831.77** |
| 089 | 902.81 | 724.96 | 741.32 | 829.14 | **3,198.24** |
| **NJ NEW JERSEY - Total** | **92,757.36** | **89,496.56** | **83,250.81** | **94,696.34** | **360,201.08** |

**STATE: NM - NEW MEXICO**

| REGISTRANT ZIP CODE 3 | 1ST QUARTER | 2ND QUARTER | 3RD QUARTER | 4TH QUARTER | TOTAL GRAMS |
|---|---|---|---|---|---|
| 870 | 1,303.77 | 1,253.15 | 1,255.87 | 1,299.75 | **5,112.54** |
| 871 | 7,843.68 | 8,196.77 | 7,508.54 | 8,253.27 | **31,802.25** |
| 873 | 499.89 | 520.06 | 434.87 | 477.63 | **1,932.45** |
| 874 | 1,393.90 | 1,378.23 | 1,255.28 | 1,350.58 | **5,377.99** |
| 875 | 1,827.99 | 1,889.90 | 1,891.51 | 2,021.77 | **7,631.17** |
| 877 | 247.17 | 256.03 | 265.07 | 215.94 | **984.21** |
| 878 | 121.15 | 111.38 | 93.09 | 121.41 | **447.03** |
| 879 | 92.40 | 62.00 | 68.28 | 72.55 | **295.24** |

| | | | | | |
|---|---|---|---|---|---|
| 880 | 1,722.58 | 1,644.52 | 1,553.51 | 1,826.85 | **6,747.46** |
| 881 | 520.98 | 332.49 | 379.96 | 406.36 | **1,639.79** |
| 882 | 1,909.38 | 1,695.77 | 1,565.77 | 1,801.86 | **6,972.79** |
| 883 | 567.92 | 561.19 | 634.36 | 637.20 | **2,400.66** |
| 884 | 105.03 | 82.48 | 84.76 | 85.95 | **358.21** |
| **NM NEW MEXICO Total** | **18,155.83** | **17,983.96** | **16,990.89** | **18,571.10** | **71,701.78** |

**STATE: NV - NEVADA**

| REGISTRANT ZIP CODE 3 | 1ST QUARTER | 2ND QUARTER | 3RD QUARTER | 4TH QUARTER | TOTAL GRAMS |
|---|---|---|---|---|---|
| 890 | 3,431.27 | 3,551.17 | 3,274.87 | 3,600.65 | **13,857.97** |
| 891 | 6,530.62 | 6,407.76 | 6,149.10 | 6,739.36 | **25,826.84** |
| 893 | 29.47 | 21.36 | 25.51 | 25.34 | **101.68** |
| 894 | 1,767.99 | 1,762.05 | 1,729.64 | 1,925.14 | **7,184.82** |
| 895 | 2,215.55 | 2,277.30 | 2,249.66 | 2,402.36 | **9,144.87** |
| 897 | 436.05 | 450.73 | 459.50 | 520.38 | **1,866.65** |
| 898 | 493.26 | 397.65 | 372.36 | 391.50 | **1,654.78** |
| **NV NEVADA Total** | **14,904.21** | **14,868.03** | **14,260.64** | **15,604.73** | **59,637.61** |

**STATE: NY - NEW YORK**

| REGISTRANT ZIP CODE 3 | 1ST QUARTER | 2ND QUARTER | 3RD QUARTER | 4TH QUARTER | TOTAL GRAMS |
|---|---|---|---|---|---|
| 100 | 22,493.68 | 22,364.78 | 20,910.35 | 24,500.48 | **90,269.29** |
| 101 | 1,176.15 | 1,237.86 | 1,025.19 | 1,148.58 | **4,587.78** |
| 102 | 172.03 | 131.57 | 156.36 | 170.83 | **630.78** |
| 103 | 2,752.45 | 2,624.96 | 2,442.88 | 2,608.44 | **10,428.74** |
| 104 | 7,293.52 | 6,699.51 | 6,333.67 | 7,024.03 | **27,350.73** |
| 105 | 9,961.28 | 10,035.60 | 9,121.49 | 10,801.50 | **39,919.87** |
| 106 | 1,125.53 | 1,041.12 | 994.16 | 1,078.79 | **4,239.59** |
| 107 | 1,751.69 | 1,670.30 | 1,629.78 | 1,915.65 | **6,967.43** |
| 108 | 1,002.06 | 909.83 | 783.01 | 1,031.11 | **3,726.01** |
| 109 | 6,768.34 | 6,505.14 | 6,249.53 | 6,617.16 | **26,140.17** |
| 110 | 3,361.07 | 3,113.17 | 2,742.19 | 3,434.53 | **12,650.95** |
| 111 | 1,322.95 | 1,240.79 | 1,017.23 | 1,280.82 | **4,861.79** |
| 112 | 12,089.66 | 11,535.58 | 10,614.35 | 12,139.03 | **46,378.63** |
| 113 | 4,706.55 | 4,596.46 | 4,140.29 | 4,946.14 | **18,389.45** |
| 114 | 2,225.94 | 2,164.13 | 1,872.98 | 2,035.67 | **8,298.73** |
| 115 | 8,377.82 | 8,078.02 | 7,160.64 | 8,615.18 | **32,231.66** |
| 116 | 984.84 | 937.05 | 805.27 | 952.49 | **3,679.65** |
| 117 | 19,638.13 | 18,897.63 | 18,387.95 | 20,493.19 | **77,416.90** |
| 118 | 1,324.14 | 1,439.92 | 1,149.92 | 1,309.27 | **5,223.25** |
| 119 | 2,726.23 | 2,606.15 | 2,828.88 | 2,749.25 | **10,910.51** |
| 120 | 7,531.74 | 7,542.95 | 7,118.74 | 7,552.00 | **29,745.44** |
| 121 | 4,979.30 | 4,868.41 | 4,530.11 | 4,820.43 | **19,198.26** |

| | | | | | |
|---|---|---|---|---|---|
| 122 | 2,813.88 | 2,802.96 | 2,711.29 | 2,780.91 | **11,109.04** |
| 123 | 3,130.09 | 3,185.69 | 2,996.74 | 3,167.51 | **12,480.03** |
| 124 | 1,867.57 | 1,625.48 | 1,747.40 | 1,756.30 | **6,996.76** |
| 125 | 5,244.35 | 4,779.59 | 4,727.63 | 5,366.08 | **20,117.65** |
| 126 | 1,044.77 | 1,002.60 | 948.33 | 1,084.07 | **4,079.77** |
| 127 | 927.56 | 955.38 | 1,038.59 | 1,034.54 | **3,956.07** |
| 128 | 5,093.43 | 5,096.81 | 4,721.89 | 4,867.81 | **19,779.93** |
| 129 | 4,180.16 | 4,208.22 | 4,100.90 | 4,254.79 | **16,744.07** |
| 130 | 5,883.37 | 6,325.14 | 6,006.43 | 6,203.00 | **24,417.94** |
| 131 | 2,225.01 | 2,219.73 | 2,134.32 | 2,346.24 | **8,925.31** |
| 132 | 3,337.12 | 3,330.85 | 3,041.31 | 3,463.63 | **13,172.91** |
| 133 | 1,912.47 | 1,991.03 | 1,821.37 | 1,994.51 | **7,719.39** |
| 134 | 2,824.60 | 2,812.56 | 2,547.47 | 2,819.37 | **11,004.00** |
| 135 | 1,033.73 | 1,048.79 | 1,010.60 | 1,166.53 | **4,259.65** |
| 136 | 5,289.52 | 5,496.85 | 5,514.75 | 5,519.48 | **21,820.60** |
| 137 | 1,392.93 | 1,391.97 | 1,244.45 | 1,276.70 | **5,306.05** |
| 138 | 2,667.10 | 2,569.68 | 2,368.53 | 2,627.79 | **10,233.10** |
| 139 | 1,092.39 | 1,097.32 | 1,024.65 | 1,220.01 | **4,434.38** |
| 140 | 5,653.52 | 5,542.69 | 5,128.93 | 5,844.42 | **22,169.56** |
| 141 | 2,788.80 | 2,605.90 | 2,492.06 | 2,668.23 | **10,554.99** |
| 142 | 7,204.88 | 7,339.67 | 6,956.87 | 7,826.75 | **29,328.16** |
| 143 | 683.02 | 673.59 | 622.88 | 725.90 | **2,705.39** |
| 144 | 7,641.33 | 7,427.94 | 7,092.87 | 8,054.92 | **30,217.06** |
| 145 | 7,884.19 | 7,940.01 | 7,597.08 | 7,748.92 | **31,170.21** |
| 146 | 12,568.69 | 12,477.36 | 11,703.84 | 12,926.87 | **49,676.76** |
| 147 | 2,101.68 | 2,190.09 | 2,091.24 | 2,283.41 | **8,666.43** |
| 148 | 5,210.40 | 5,315.60 | 4,696.83 | 5,364.23 | **20,587.05** |
| 149 | 1,114.35 | 1,274.66 | 1,097.26 | 1,260.59 | **4,746.86** |
| **NY - NEW YORK - Total** | **228,576.00** | **224,969.15** | **211,201.46** | **234,878.10** | **899,624.71** |

**STATE: OH - OHIO**

| REGISTRANT ZIP CODE 3 | 1ST QUARTER | 2ND QUARTER | 3RD QUARTER | 4TH QUARTER | TOTAL GRAMS |
|---|---|---|---|---|---|
| 430 | 16,793.46 | 15,753.31 | 15,897.80 | 17,275.87 | **65,720.44** |
| 431 | 8,130.26 | 7,676.34 | 7,775.79 | 8,454.40 | **32,036.80** |
| 432 | 10,893.59 | 10,219.89 | 10,390.98 | 10,875.87 | **42,380.32** |
| 433 | 2,820.99 | 2,591.56 | 2,561.59 | 2,760.50 | **10,734.64** |
| 434 | 2,813.26 | 2,686.02 | 2,820.96 | 2,755.87 | **11,076.10** |
| 435 | 6,201.71 | 6,243.22 | 6,163.45 | 6,420.57 | **25,028.95** |
| 436 | 6,655.00 | 7,010.86 | 6,907.17 | 7,131.45 | **27,704.48** |
| 437 | 2,459.87 | 2,203.92 | 2,137.89 | 2,394.51 | **9,196.19** |
| 438 | 334.17 | 395.54 | 397.38 | 411.70 | **1,538.80** |

| | | | | | |
|---|---|---|---|---|---|
| 439 | 2,017.62 | 1,860.32 | 1,826.43 | 2,066.63 | **7,771.01** |
| 440 | 13,975.67 | 13,451.43 | 13,039.62 | 14,564.62 | **55,031.33** |
| 441 | 16,083.29 | 15,160.49 | 15,016.29 | 16,763.06 | **63,023.13** |
| 442 | 8,420.02 | 7,865.48 | 7,578.39 | 8,656.64 | **32,520.53** |
| 443 | 3,836.52 | 3,704.21 | 3,443.60 | 4,134.70 | **15,119.03** |
| 444 | 3,594.74 | 3,302.32 | 3,270.36 | 3,469.38 | **13,636.80** |
| 445 | 2,709.14 | 2,606.13 | 2,515.21 | 2,932.90 | **10,763.38** |
| 446 | 5,610.22 | 5,662.39 | 5,498.83 | 6,216.84 | **22,988.28** |
| 447 | 4,575.42 | 4,207.46 | 4,054.52 | 4,677.59 | **17,515.00** |
| 448 | 4,428.74 | 4,190.41 | 4,152.84 | 4,472.62 | **17,244.61** |
| 449 | 1,577.06 | 1,487.75 | 1,436.73 | 1,720.22 | **6,221.76** |
| 450 | 16,002.98 | 14,644.87 | 15,156.13 | 16,477.28 | **62,281.25** |
| 451 | 6,635.49 | 5,958.72 | 6,064.32 | 6,362.51 | **25,021.04** |
| 452 | 20,270.31 | 18,096.06 | 18,258.77 | 20,600.38 | **77,225.53** |
| 453 | 7,935.34 | 7,622.95 | 7,696.81 | 8,550.78 | **31,805.89** |
| 454 | 9,357.11 | 8,988.82 | 9,163.28 | 9,800.91 | **37,310.12** |
| 455 | 2,410.32 | 2,095.23 | 2,095.10 | 2,266.99 | **8,867.64** |
| 456 | 6,752.30 | 6,321.78 | 6,565.23 | 6,885.00 | **26,524.31** |
| 457 | 2,413.54 | 2,131.56 | 2,081.43 | 2,384.72 | **9,011.25** |
| 458 | 5,140.56 | 4,859.65 | 4,908.13 | 5,244.73 | **20,153.07** |
| **OH - OHIO Total** | **200,848.71** | **188,998.68** | **188,875.03** | **206,729.24** | **785,451.66** |

**STATE: OK - OKLAHOMA**

| REGISTRANT ZIP CODE 3 | 1ST QUARTER | 2ND QUARTER | 3RD QUARTER | 4TH QUARTER | **TOTAL GRAMS** |
|---|---|---|---|---|---|
| 730 | 9,550.04 | 9,229.59 | 9,520.49 | 10,332.61 | **38,632.73** |
| 731 | 7,091.95 | 6,828.66 | 6,742.35 | 7,487.14 | **28,150.10** |
| 734 | 698.11 | 612.06 | 647.09 | 751.94 | **2,709.20** |
| 735 | 2,949.36 | 2,535.96 | 2,876.56 | 2,614.61 | **10,976.49** |
| 736 | 751.46 | 823.86 | 734.20 | 854.22 | **3,163.73** |
| 737 | 1,232.14 | 1,045.84 | 1,003.62 | 1,211.52 | **4,493.11** |
| 738 | 197.90 | 152.83 | 192.89 | 165.19 | **708.81** |
| 739 | 45.83 | 34.85 | 41.85 | 46.18 | **168.71** |
| 740 | 10,120.94 | 9,368.70 | 9,554.08 | 10,236.09 | **39,279.81** |
| 741 | 5,431.29 | 5,224.95 | 4,928.66 | 5,559.45 | **21,144.34** |
| 743 | 1,683.53 | 1,372.51 | 1,546.24 | 1,680.02 | **6,282.31** |
| 744 | 3,030.92 | 2,681.93 | 2,684.03 | 3,119.56 | **11,516.44** |
| 745 | 766.21 | 626.30 | 685.56 | 734.39 | **2,812.46** |
| 746 | 708.64 | 709.90 | 656.24 | 739.42 | **2,814.20** |
| 747 | 822.11 | 741.55 | 779.57 | 847.93 | **3,191.16** |
| 748 | 1,693.23 | 1,602.08 | 1,604.15 | 1,732.36 | **6,631.82** |
| 749 | 694.81 | 709.47 | 702.52 | 635.13 | **2,741.93** |

| OK - OKLAHOMA Total | 47,468.47 | 44,301.03 | 44,900.09 | 48,747.77 | 185,417.36 |
|---|---|---|---|---|---|

STATE: OR - OREGON

| REGISTRANT ZIP CODE 3 | 1ST QUARTER | 2ND QUARTER | 3RD QUARTER | 4TH QUARTER | TOTAL GRAMS |
|---|---|---|---|---|---|
| 970 | 12,265.64 | 12,802.65 | 11,972.28 | 13,446.86 | **50,487.42** |
| 971 | 5,576.66 | 5,438.42 | 5,173.26 | 5,719.58 | **21,907.93** |
| 972 | 16,627.29 | 16,929.93 | 16,221.55 | 18,084.26 | **67,863.03** |
| 973 | 11,091.76 | 11,173.18 | 10,208.78 | 11,401.09 | **43,874.81** |
| 974 | 7,861.17 | 7,823.35 | 7,330.85 | 8,211.58 | **31,226.95** |
| 975 | 3,575.31 | 3,392.65 | 3,224.32 | 3,608.70 | **13,800.99** |
| 976 | 653.43 | 632.82 | 587.04 | 672.47 | **2,545.75** |
| 977 | 3,238.45 | 3,267.12 | 3,144.71 | 3,607.71 | **13,258.00** |
| 978 | 1,836.77 | 1,642.24 | 1,608.19 | 1,786.18 | **6,873.38** |
| 979 | 578.80 | 563.29 | 535.00 | 587.69 | **2,264.79** |
| OR OREGON Total | 63,305.28 | 63,665.64 | 60,005.98 | 67,126.13 | 254,103.03 |

STATE: PA - PENNSYLVANIA

| REGISTRANT ZIP CODE 3 | 1ST QUARTER | 2ND QUARTER | 3RD QUARTER | 4TH QUARTER | TOTAL GRAMS |
|---|---|---|---|---|---|
| 150 | 7,612.50 | 7,485.42 | 6,958.93 | 7,592.41 | **29,649.26** |
| 151 | 6,535.84 | 5,942.02 | 5,931.05 | 6,450.34 | **24,859.26** |
| 152 | 11,804.56 | 11,905.19 | 11,548.20 | 12,474.19 | **47,732.14** |
| 153 | 3,522.20 | 3,313.76 | 3,143.97 | 3,474.38 | **13,454.31** |
| 154 | 1,632.74 | 1,607.49 | 1,607.42 | 1,599.92 | **6,447.56** |
| 155 | 1,068.06 | 1,101.73 | 1,034.95 | 1,141.22 | **4,345.96** |
| 156 | 5,194.78 | 4,629.14 | 4,693.69 | 4,899.80 | **19,417.41** |
| 157 | 2,019.41 | 2,029.85 | 2,031.78 | 2,089.81 | **8,170.85** |
| 158 | 2,145.80 | 2,140.14 | 2,106.05 | 2,191.30 | **8,583.29** |
| 159 | 2,104.08 | 2,079.82 | 2,218.40 | 2,027.39 | **8,429.69** |
| 160 | 3,720.29 | 3,688.65 | 3,717.80 | 4,050.69 | **15,177.43** |
| 161 | 3,945.98 | 3,648.08 | 3,643.78 | 3,960.97 | **15,198.81** |
| 162 | 1,491.80 | 1,503.24 | 1,337.47 | 1,409.87 | **5,742.39** |
| 163 | 2,086.46 | 2,105.94 | 2,168.67 | 2,130.76 | **8,491.83** |
| 164 | 1,181.20 | 1,053.64 | 1,110.97 | 1,247.68 | **4,593.49** |
| 165 | 4,032.76 | 4,016.03 | 3,948.59 | 4,168.94 | **16,166.31** |
| 166 | 3,309.14 | 3,162.72 | 3,023.63 | 3,354.64 | **12,850.13** |
| 167 | 870.69 | 867.35 | 783.48 | 826.16 | **3,347.68** |
| 168 | 3,372.27 | 3,111.12 | 2,958.06 | 3,312.78 | **12,754.23** |
| 169 | 789.56 | 798.11 | 773.58 | 869.14 | **3,230.38** |
| 170 | 14,982.87 | 14,189.72 | 14,029.86 | 15,038.28 | **58,240.73** |
| 171 | 2,997.46 | 2,772.38 | 2,703.58 | 2,963.76 | **11,437.18** |
| 172 | 3,760.49 | 3,622.07 | 3,344.05 | 3,954.48 | **14,681.09** |
| 173 | 6,466.21 | 6,127.13 | 5,802.94 | 6,304.44 | **24,700.73** |

| | | | | | |
|---|---|---|---|---|---|
| 174 | 4,371.04 | 4,242.11 | 3,965.01 | 4,336.60 | **16,914.75** |
| 175 | 4,592.09 | 4,472.48 | 4,204.22 | 4,638.20 | **17,906.98** |
| 176 | 4,538.83 | 4,271.55 | 4,254.75 | 4,557.68 | **17,622.81** |
| 177 | 3,005.51 | 2,740.17 | 2,649.47 | 2,923.93 | **11,319.07** |
| 178 | 3,446.64 | 3,248.60 | 3,114.17 | 3,419.43 | **13,228.84** |
| 179 | 1,305.96 | 1,265.30 | 1,134.67 | 1,273.76 | **4,979.69** |
| 180 | 5,700.51 | 5,783.00 | 5,437.61 | 6,091.91 | **23,013.03** |
| 181 | 2,808.57 | 2,861.84 | 2,654.54 | 3,168.60 | **11,493.55** |
| 182 | 1,290.46 | 1,291.83 | 1,128.20 | 1,287.67 | **4,998.16** |
| 183 | 1,570.55 | 1,599.07 | 1,460.85 | 1,604.36 | **6,234.84** |
| 184 | 2,295.55 | 2,109.55 | 2,149.78 | 2,324.10 | **8,878.99** |
| 185 | 2,590.21 | 2,540.78 | 2,420.99 | 2,621.21 | **10,173.19** |
| 186 | 2,111.56 | 2,070.19 | 1,827.66 | 2,123.81 | **8,133.22** |
| 187 | 22,582.04 | 25,675.46 | 26,941.17 | 28,283.22 | **103,481.88** |
| 188 | 1,065.71 | 999.81 | 956.69 | 1,077.96 | **4,100.18** |
| 189 | 5,874.96 | 5,955.45 | 5,771.90 | 6,205.77 | **23,808.08** |
| 190 | 18,318.09 | 18,033.69 | 17,104.86 | 18,886.96 | **72,343.60** |
| 191 | 13,403.26 | 13,165.80 | 11,801.19 | 13,692.28 | **52,062.52** |
| 193 | 8,324.16 | 8,284.51 | 7,657.49 | 8,771.36 | **33,037.51** |
| 194 | 8,426.72 | 8,460.77 | 7,892.06 | 8,897.92 | **33,677.47** |
| 195 | 2,554.85 | 2,444.47 | 2,371.01 | 2,608.03 | **9,978.37** |
| 196 | 3,396.02 | 3,277.09 | 3,314.37 | 3,455.17 | **13,442.65** |
| **PA - PENNSYLVANIA - Total** | **220,220.47** | **217,694.25** | **210,833.55** | **229,783.27** | **878,531.54** |

**STATE: PR - PUERTO RICO**

| REGISTRANT ZIP CODE 3 | 1ST QUARTER | 2ND QUARTER | 3RD QUARTER | 4TH QUARTER | TOTAL GRAMS |
|---|---|---|---|---|---|
| 006 | 3,763.58 | 3,578.51 | 2,512.63 | 3,740.91 | **13,595.63** |
| 007 | 4,704.45 | 3,849.55 | 2,656.19 | 4,307.43 | **15,517.62** |
| 009 | 7,046.68 | 5,805.04 | 4,754.50 | 6,667.18 | **24,273.40** |
| **PR - PUERTO RICO - Total** | **15,514.70** | **13,233.10** | **9,923.32** | **14,715.52** | **53,386.65** |

**STATE: RI - RHODE ISLAND**

| REGISTRANT ZIP CODE 3 | 1ST QUARTER | 2ND QUARTER | 3RD QUARTER | 4TH QUARTER | TOTAL GRAMS |
|---|---|---|---|---|---|
| 028 | 12,820.39 | 12,558.11 | 11,674.96 | 13,054.61 | **50,108.06** |
| 029 | 6,180.64 | 6,005.53 | 5,242.78 | 6,373.20 | **23,802.15** |
| **RI RHODE ISLAND Total** | **19,001.03** | **18,563.64** | **16,917.74** | **19,427.80** | **73,910.21** |

**STATE: SC - SOUTH CAROLINA**

| REGISTRANT ZIP CODE 3 | 1ST QUARTER | 2ND QUARTER | 3RD QUARTER | 4TH QUARTER | TOTAL GRAMS |
|---|---|---|---|---|---|
| 290 | 7,220.75 | 6,781.93 | 6,398.25 | 7,324.06 | **27,724.98** |
| 291 | 5,797.05 | 5,137.77 | 5,110.41 | 5,562.38 | **21,607.62** |
| 292 | 6,515.01 | 6,020.86 | 5,894.81 | 6,505.07 | **24,935.75** |
| 293 | 7,942.12 | 7,227.08 | 6,922.72 | 8,147.19 | **30,239.11** |

| | | | | | |
|---|---|---|---|---|---|
| 294 | 18,737.84 | 18,158.58 | 16,474.45 | 18,073.05 | **71,443.92** |
| 295 | 11,037.20 | 10,569.22 | 10,183.07 | 11,203.77 | **42,993.26** |
| 296 | 20,238.55 | 18,760.12 | 18,199.39 | 20,652.33 | **77,850.39** |
| 297 | 7,741.55 | 7,022.70 | 6,762.83 | 7,912.78 | **29,439.86** |
| 298 | 3,230.54 | 2,886.81 | 3,016.87 | 3,372.04 | **12,506.26** |
| 299 | 2,913.82 | 2,992.69 | 2,642.34 | 3,097.00 | **11,645.85** |
| **SC - SOUTH CAROLINA Total** | **91,374.42** | **85,557.76** | **81,605.14** | **91,849.67** | **350,387.00** |

**STATE: SD - SOUTH DAKOTA**

| REGISTRANT ZIP CODE 3 | 1ST QUARTER | 2ND QUARTER | 3RD QUARTER | 4TH QUARTER | TOTAL GRAMS |
|---|---|---|---|---|---|
| 570 | 2,740.65 | 2,584.90 | 2,629.02 | 2,745.44 | **10,700.01** |
| 571 | 9,098.68 | 9,077.60 | 8,882.78 | 9,595.46 | **36,654.52** |
| 572 | 1,031.77 | 1,097.32 | 1,023.45 | 1,103.53 | **4,256.07** |
| 573 | 1,658.34 | 1,528.27 | 1,586.41 | 1,772.06 | **6,545.07** |
| 574 | 1,203.58 | 1,135.77 | 1,136.24 | 1,311.65 | **4,787.23** |
| 575 | 979.73 | 829.70 | 854.84 | 946.75 | **3,611.01** |
| 576 | 197.53 | 173.19 | 197.34 | 202.52 | **770.58** |
| 577 | 2,891.92 | 2,749.70 | 2,802.64 | 3,020.13 | **11,464.39** |
| **SD SOUTH DAKOTA - Total** | **19,802.18** | **19,176.45** | **19,112.72** | **20,697.54** | **78,788.90** |

**STATE: TN - TENNESSEE**

| REGISTRANT ZIP CODE 3 | 1ST QUARTER | 2ND QUARTER | 3RD QUARTER | 4TH QUARTER | TOTAL GRAMS |
|---|---|---|---|---|---|
| 370 | 12,493.80 | 11,890.48 | 12,371.17 | 13,289.75 | **50,045.19** |
| 371 | 8,528.69 | 7,961.13 | 8,184.75 | 8,729.17 | **33,403.74** |
| 372 | 5,727.42 | 5,312.53 | 5,305.10 | 5,829.22 | **22,174.26** |
| 373 | 8,460.22 | 7,817.29 | 7,957.53 | 8,706.53 | **32,941.57** |
| 374 | 3,447.95 | 3,269.37 | 3,479.38 | 3,826.65 | **14,023.35** |
| 376 | 4,733.78 | 4,351.46 | 4,536.76 | 4,711.06 | **18,333.06** |
| 377 | 4,672.00 | 4,405.06 | 4,389.95 | 4,900.73 | **18,367.74** |
| 378 | 6,731.57 | 6,683.85 | 6,625.62 | 7,180.14 | **27,221.18** |
| 379 | 6,192.62 | 5,659.98 | 6,224.19 | 6,564.67 | **24,641.46** |
| 380 | 5,857.07 | 5,713.44 | 5,754.06 | 6,308.25 | **23,632.82** |
| 381 | 7,402.83 | 7,312.70 | 6,997.33 | 7,513.27 | **29,226.13** |
| 382 | 1,572.37 | 1,273.15 | 1,355.41 | 1,390.69 | **5,591.62** |
| 383 | 3,891.12 | 3,322.96 | 3,380.34 | 3,712.40 | **14,306.82** |
| 384 | 1,871.89 | 1,647.22 | 1,714.64 | 1,709.93 | **6,943.69** |
| 385 | 2,132.30 | 1,803.52 | 2,095.52 | 2,123.60 | **8,154.95** |
| **TN TENNESSEE Total** | **83,715.64** | **78,424.14** | **80,371.73** | **86,496.07** | **329,007.58** |

**STATE: TX - TEXAS**

| REGISTRANT ZIP CODE 3 | 1ST QUARTER | 2ND QUARTER | 3RD QUARTER | 4TH QUARTER | TOTAL GRAMS |
|---|---|---|---|---|---|
| 750 | 32,137.56 | 29,964.31 | 29,623.29 | 33,796.26 | **125,521.43** |
| 751 | 8,750.92 | 7,739.27 | 7,874.58 | 8,649.86 | **33,014.63** |

| | | | | | |
|---|---|---|---|---|---|
| 752 | 11,681.18 | 11,271.62 | 10,730.39 | 12,312.39 | **45,995.57** |
| 753 | 48.94 | 44.40 | 53.09 | 47.52 | **193.95** |
| 754 | 5,085.84 | 4,644.79 | 4,623.58 | 5,449.79 | **19,804.01** |
| 755 | 1,346.44 | 1,328.68 | 1,267.93 | 1,587.58 | **5,530.63** |
| 756 | 4,346.85 | 3,939.11 | 3,789.40 | 4,462.79 | **16,538.15** |
| 757 | 5,563.91 | 4,948.63 | 5,036.03 | 5,505.38 | **21,053.96** |
| 758 | 1,171.17 | 1,018.43 | 1,125.42 | 1,237.76 | **4,552.78** |
| 759 | 4,385.97 | 3,935.83 | 3,819.02 | 4,588.71 | **16,729.53** |
| 760 | 17,913.85 | 16,551.38 | 15,789.32 | 18,239.28 | **68,493.82** |
| 761 | 10,525.09 | 10,065.30 | 9,380.20 | 11,049.91 | **41,020.50** |
| 762 | 8,281.96 | 7,622.22 | 7,239.54 | 8,731.06 | **31,874.78** |
| 763 | 3,545.29 | 3,327.68 | 3,186.60 | 3,544.54 | **13,604.11** |
| 764 | 1,263.14 | 1,243.92 | 1,261.12 | 1,345.93 | **5,114.10** |
| 765 | 8,516.04 | 9,860.62 | 8,657.19 | 8,975.07 | **36,008.91** |
| 766 | 1,440.86 | 1,229.38 | 1,273.22 | 1,357.44 | **5,300.90** |
| 767 | 4,178.62 | 3,691.75 | 3,837.55 | 4,252.90 | **15,960.82** |
| 768 | 1,189.18 | 1,119.60 | 1,089.06 | 1,171.31 | **4,569.15** |
| 769 | 2,376.04 | 2,199.74 | 2,112.66 | 2,422.76 | **9,111.20** |
| 770 | 23,894.72 | 22,469.39 | 21,525.01 | 24,905.52 | **92,794.64** |
| 772 | 6.92 | 9.86 | 1.56 | 5.19 | **23.52** |
| 773 | 17,934.94 | 15,942.83 | 16,318.94 | 18,848.30 | **69,045.02** |
| 774 | 17,732.89 | 16,392.64 | 15,731.74 | 18,281.98 | **68,139.25** |
| 775 | 16,492.44 | 15,060.13 | 14,785.11 | 17,462.01 | **63,799.70** |
| 776 | 3,414.24 | 3,056.69 | 2,904.44 | 3,416.44 | **12,791.81** |
| 777 | 1,584.97 | 1,612.25 | 1,407.39 | 1,632.76 | **6,237.38** |
| 778 | 4,921.91 | 4,488.81 | 4,705.54 | 5,392.24 | **19,508.50** |
| 779 | 2,353.83 | 2,051.15 | 2,123.90 | 2,634.98 | **9,163.86** |
| 780 | 5,986.08 | 5,567.43 | 5,674.01 | 6,292.00 | **23,519.53** |
| 781 | 6,417.61 | 5,983.63 | 5,657.29 | 6,705.55 | **24,764.08** |
| 782 | 25,891.86 | 24,230.37 | 24,024.54 | 26,974.09 | **101,120.86** |
| 783 | 3,295.06 | 2,904.77 | 2,785.99 | 3,509.40 | **12,495.21** |
| 784 | 5,389.62 | 5,069.29 | 5,241.70 | 5,949.65 | **21,650.26** |
| 785 | 10,922.59 | 9,310.89 | 9,743.37 | 10,751.98 | **40,728.83** |
| 786 | 13,746.49 | 12,661.08 | 12,589.17 | 14,129.71 | **53,126.45** |
| 787 | 13,305.48 | 12,795.40 | 12,828.36 | 14,324.96 | **53,254.19** |
| 788 | 1,422.12 | 1,124.27 | 1,202.63 | 1,208.33 | **4,957.35** |
| 789 | 772.03 | 679.99 | 683.41 | 781.61 | **2,917.05** |
| 790 | 1,297.12 | 1,075.59 | 1,248.69 | 1,283.78 | **4,905.18** |
| 791 | 2,520.92 | 2,146.42 | 2,212.68 | 2,299.42 | **9,179.45** |
| 792 | 285.66 | 268.76 | 239.19 | 315.29 | **1,108.91** |

| | | | | | |
|---|---|---|---|---|---|
| 793 | 1,167.94 | 1,089.14 | 852.64 | 971.21 | **4,080.93** |
| 794 | 5,070.81 | 4,371.02 | 4,174.66 | 4,941.05 | **18,557.54** |
| 795 | 1,087.54 | 1,050.69 | 1,051.93 | 1,090.34 | **4,280.51** |
| 796 | 3,523.00 | 3,214.42 | 2,951.68 | 3,433.30 | **13,122.40** |
| 797 | 4,409.95 | 4,033.05 | 3,789.13 | 4,311.98 | **16,544.11** |
| 798 | 193.05 | 180.34 | 166.03 | 146.83 | **686.24** |
| 799 | 8,394.13 | 7,580.26 | 7,536.85 | 7,722.85 | **31,234.09** |
| **TX - TEXAS Total** | **337,184.79** | **312,167.22** | **305,926.76** | **348,451.01** | **1,303,729.79** |

**STATE: UT - UTAH**

| REGISTRANT ZIP CODE 3 | 1ST QUARTER | 2ND QUARTER | 3RD QUARTER | 4TH QUARTER | TOTAL GRAMS |
|---|---|---|---|---|---|
| 840 | 24,389.99 | 24,346.73 | 24,042.48 | 27,815.43 | **100,594.63** |
| 841 | 8,895.11 | 9,033.72 | 8,742.07 | 9,746.88 | **36,417.78** |
| 843 | 2,193.33 | 2,158.53 | 2,109.81 | 2,409.20 | **8,870.87** |
| 844 | 3,026.91 | 2,911.46 | 2,816.61 | 3,220.55 | **11,975.54** |
| 845 | 705.01 | 627.96 | 622.65 | 641.89 | **2,597.51** |
| 846 | 4,843.04 | 4,844.93 | 4,924.88 | 5,290.24 | **19,903.09** |
| 847 | 3,010.12 | 3,041.36 | 3,094.12 | 3,254.09 | **12,399.69** |
| **UT - UTAH - Total** | **47,063.52** | **46,964.69** | **46,352.63** | **52,378.27** | **192,759.11** |

**STATE: VA - VIRGINIA**

| REGISTRANT ZIP CODE 3 | 1ST QUARTER | 2ND QUARTER | 3RD QUARTER | 4TH QUARTER | TOTAL GRAMS |
|---|---|---|---|---|---|
| 201 | 12,309.89 | 11,979.45 | 11,809.57 | 13,274.02 | **49,372.93** |
| 220 | 8,849.67 | 8,975.85 | 8,932.62 | 9,235.18 | **35,993.32** |
| 221 | 5,552.73 | 5,792.51 | 5,973.64 | 5,899.69 | **23,218.57** |
| 222 | 2,775.98 | 3,130.54 | 2,818.54 | 3,196.62 | **11,921.68** |
| 223 | 3,357.43 | 3,504.74 | 3,255.36 | 3,943.50 | **14,061.04** |
| 224 | 4,780.78 | 4,622.64 | 4,471.26 | 4,797.87 | **18,672.56** |
| 225 | 2,522.39 | 2,321.40 | 2,349.81 | 2,623.22 | **9,816.82** |
| 226 | 3,107.86 | 2,856.71 | 3,006.33 | 3,190.97 | **12,161.86** |
| 227 | 1,025.73 | 839.69 | 804.31 | 850.39 | **3,520.13** |
| 228 | 1,901.34 | 1,847.24 | 1,917.16 | 2,024.93 | **7,690.67** |
| 229 | 6,206.10 | 5,956.45 | 5,681.40 | 6,520.05 | **24,364.01** |
| 230 | 3,561.19 | 3,680.19 | 3,255.38 | 3,545.91 | **14,042.67** |
| 231 | 8,364.47 | 8,283.11 | 7,768.61 | 9,349.62 | **33,765.81** |
| 232 | 9,439.00 | 9,025.78 | 8,596.51 | 9,553.41 | **36,614.69** |
| 233 | 6,125.03 | 6,143.14 | 5,178.78 | 6,027.60 | **23,474.56** |
| 234 | 12,220.28 | 12,298.49 | 11,306.16 | 12,168.90 | **47,993.83** |
| 235 | 2,924.34 | 3,133.16 | 2,952.40 | 2,802.66 | **11,812.55** |
| 236 | 7,127.08 | 6,997.97 | 6,338.92 | 6,899.87 | **27,363.85** |
| 237 | 2,871.08 | 2,415.53 | 2,658.57 | 2,540.52 | **10,485.71** |
| 238 | 5,048.02 | 4,601.16 | 4,324.20 | 4,450.81 | **18,424.19** |

| | | | | | |
|---|---|---|---|---|---|
| 239 | 1,321.46 | 1,128.89 | 1,109.07 | 1,226.45 | **4,785.87** |
| 240 | 5,827.26 | 5,509.76 | 5,564.38 | 5,713.69 | **22,615.09** |
| 241 | 3,526.91 | 3,319.63 | 3,288.69 | 3,706.07 | **13,841.31** |
| 242 | 2,368.47 | 2,148.46 | 2,107.17 | 2,324.26 | **8,948.36** |
| 243 | 2,099.18 | 1,839.90 | 1,837.66 | 1,939.20 | **7,715.94** |
| 244 | 2,598.78 | 2,429.47 | 2,460.76 | 2,680.21 | **10,169.23** |
| 245 | 6,480.53 | 5,802.07 | 5,944.69 | 5,951.31 | **24,178.59** |
| 246 | 985.78 | 990.87 | 964.26 | 934.38 | **3,875.28** |
| **VA VIRGINIA Total** | **135,278.76** | **131,574.81** | **126,676.22** | **137,371.32** | **530,901.12** |

**STATE: VI - VIRGIN ISLANDS**

| REGISTRANT ZIP CODE 3 | 1ST QUARTER | 2ND QUARTER | 3RD QUARTER | 4TH QUARTER | TOTAL GRAMS |
|---|---|---|---|---|---|
| 008 | 95.20 | 155.20 | 84.26 | 135.00 | **469.65** |
| **VI - VIRGIN ISLANDS Total** | **95.20** | **155.20** | **84.26** | **135.00** | **469.65** |

**STATE: VT - VERMONT**

| REGISTRANT ZIP CODE 3 | 1ST QUARTER | 2ND QUARTER | 3RD QUARTER | 4TH QUARTER | TOTAL GRAMS |
|---|---|---|---|---|---|
| 050 | 1,399.21 | 1,568.64 | 1,358.82 | 1,486.38 | **5,813.04** |
| 051 | 750.48 | 749.67 | 831.45 | 776.25 | **3,107.84** |
| 052 | 1,250.24 | 1,212.61 | 1,200.81 | 1,209.59 | **4,873.25** |
| 053 | 1,148.73 | 1,162.61 | 1,147.87 | 1,164.95 | **4,624.16** |
| 054 | 6,303.86 | 6,538.06 | 6,510.19 | 6,563.99 | **25,916.11** |
| 056 | 3,036.45 | 3,154.16 | 2,973.23 | 2,890.98 | **12,054.81** |
| 057 | 2,418.87 | 2,397.04 | 2,401.40 | 2,348.36 | **9,565.67** |
| 058 | 2,101.32 | 2,005.53 | 2,152.50 | 2,231.49 | **8,490.84** |
| **VT VERMONT Total** | **18,409.16** | **18,788.31** | **18,576.26** | **18,672.00** | **74,445.73** |

**STATE: WA - WASHINGTON**

| REGISTRANT ZIP CODE 3 | 1ST QUARTER | 2ND QUARTER | 3RD QUARTER | 4TH QUARTER | TOTAL GRAMS |
|---|---|---|---|---|---|
| 980 | 23,638.47 | 24,865.99 | 23,615.86 | 26,151.48 | **98,271.81** |
| 981 | 13,588.23 | 14,655.99 | 14,049.08 | 15,328.85 | **57,622.14** |
| 982 | 15,533.54 | 16,296.13 | 14,898.37 | 16,390.12 | **63,118.16** |
| 983 | 12,852.02 | 13,073.79 | 11,752.91 | 13,456.26 | **51,134.98** |
| 984 | 7,265.73 | 8,041.14 | 7,036.08 | 7,704.59 | **30,047.55** |
| 985 | 7,629.49 | 7,770.66 | 6,987.13 | 7,873.79 | **30,261.06** |
| 986 | 9,079.38 | 9,268.36 | 8,771.56 | 9,403.60 | **36,522.91** |
| 988 | 2,462.28 | 2,454.66 | 2,431.95 | 2,494.51 | **9,843.39** |
| 989 | 3,441.00 | 3,490.12 | 3,152.41 | 3,641.68 | **13,725.22** |
| 990 | 1,855.25 | 1,780.22 | 1,695.87 | 1,779.98 | **7,111.31** |
| 991 | 1,357.49 | 1,233.26 | 1,107.20 | 1,314.18 | **5,012.12** |
| 992 | 8,304.77 | 8,539.74 | 7,923.04 | 8,798.15 | **33,565.69** |
| 993 | 4,988.31 | 5,271.99 | 4,834.42 | 5,378.02 | **20,472.74** |
| 994 | 596.46 | 601.09 | 568.86 | 661.39 | **2,427.79** |

| WA WASHINGTON Total | 112,592.41 | 117,343.15 | 108,824.74 | 120,376.59 | 459,136.89 |
|---|---|---|---|---|---|

**STATE: WI - WISCONSIN**

| REGISTRANT ZIP CODE 3 | 1ST QUARTER | 2ND QUARTER | 3RD QUARTER | 4TH QUARTER | TOTAL GRAMS |
|---|---|---|---|---|---|
| 530 | 12,559.18 | 12,571.00 | 12,118.94 | 13,393.54 | **50,642.67** |
| 531 | 11,632.50 | 11,233.57 | 10,662.87 | 11,708.37 | **45,237.31** |
| 532 | 11,436.19 | 11,325.60 | 10,427.41 | 11,805.96 | **44,995.17** |
| 534 | 2,311.45 | 2,277.91 | 2,256.60 | 2,341.89 | **9,187.85** |
| 535 | 10,318.46 | 10,241.77 | 9,821.40 | 10,592.23 | **40,973.85** |
| 537 | 6,134.17 | 6,302.34 | 5,965.44 | 6,553.02 | **24,954.97** |
| 538 | 903.67 | 885.46 | 866.58 | 1,019.44 | **3,675.15** |
| 539 | 3,406.86 | 3,497.69 | 3,407.11 | 3,587.52 | **13,899.17** |
| 540 | 2,308.98 | 2,331.71 | 2,185.22 | 2,420.04 | **9,245.95** |
| 541 | 3,989.94 | 3,789.96 | 3,846.67 | 4,316.51 | **15,943.09** |
| 542 | 2,027.97 | 2,010.12 | 1,984.16 | 2,259.44 | **8,281.69** |
| 543 | 3,885.66 | 3,752.10 | 3,688.10 | 3,921.14 | **15,247.00** |
| 544 | 7,350.35 | 7,345.85 | 6,784.03 | 7,592.52 | **29,072.75** |
| 545 | 1,379.28 | 1,344.21 | 1,433.63 | 1,377.15 | **5,534.26** |
| 546 | 5,112.05 | 5,097.63 | 4,895.60 | 5,038.05 | **20,143.34** |
| 547 | 5,332.96 | 5,355.74 | 5,342.72 | 5,730.98 | **21,762.39** |
| 548 | 2,738.35 | 2,724.32 | 2,812.00 | 3,069.02 | **11,343.69** |
| 549 | 10,458.77 | 9,843.15 | 10,330.74 | 10,399.48 | **41,032.15** |
| WI - WISCONSIN Total | 103,286.79 | 101,930.14 | 98,829.22 | 107,126.30 | 411,172.45 |

**STATE: WV - WEST VIRGINIA**

| REGISTRANT ZIP CODE 3 | 1ST QUARTER | 2ND QUARTER | 3RD QUARTER | 4TH QUARTER | TOTAL GRAMS |
|---|---|---|---|---|---|
| 247 | 981.90 | 860.59 | 875.93 | 899.49 | **3,617.92** |
| 248 | 491.39 | 358.10 | 281.01 | 297.86 | **1,428.36** |
| 249 | 658.95 | 583.55 | 655.95 | 636.64 | **2,535.09** |
| 250 | 1,125.13 | 959.47 | 1,094.88 | 1,255.74 | **4,435.22** |
| 251 | 979.62 | 968.91 | 966.37 | 1,096.59 | **4,011.50** |
| 252 | 742.90 | 658.30 | 658.71 | 671.37 | **2,731.28** |
| 253 | 2,787.85 | 2,595.48 | 2,487.07 | 2,807.24 | **10,677.64** |
| 254 | 2,710.61 | 2,536.54 | 2,577.39 | 2,724.50 | **10,549.04** |
| 255 | 2,983.63 | 2,908.54 | 2,699.13 | 3,052.44 | **11,643.75** |
| 256 | 934.49 | 785.59 | 805.52 | 723.47 | **3,249.08** |
| 257 | 2,161.02 | 2,174.05 | 2,308.50 | 2,277.23 | **8,920.80** |
| 258 | 1,837.06 | 1,550.01 | 1,530.08 | 1,714.48 | **6,631.63** |
| 259 | 605.07 | 629.63 | 653.84 | 619.60 | **2,508.14** |
| 260 | 1,888.88 | 1,836.06 | 1,699.52 | 1,929.71 | **7,354.16** |
| 261 | 1,846.61 | 1,784.79 | 1,710.79 | 1,881.62 | **7,223.81** |
| 262 | 841.77 | 799.06 | 811.47 | 923.11 | **3,375.41** |

| | | | | | |
|---|---|---|---|---|---|
| 263 | 1,354.98 | 1,270.02 | 1,266.56 | 1,538.22 | **5,429.78** |
| 264 | 441.96 | 392.43 | 380.04 | 456.68 | **1,671.11** |
| 265 | 2,936.13 | 2,855.30 | 2,726.20 | 3,174.80 | **11,692.43** |
| 266 | 429.33 | 401.49 | 379.75 | 438.06 | **1,648.63** |
| 267 | 849.97 | 759.59 | 769.20 | 753.26 | **3,132.01** |
| 268 | 523.78 | 525.88 | 504.94 | 584.35 | **2,138.95** |
| **WV WEST VIRGINIA Total** | **30,113.03** | **28,193.39** | **27,842.85** | **30,456.46** | **116,605.74** |

**STATE: WY - WYOMING**

| REGISTRANT ZIP CODE 3 | 1ST QUARTER | 2ND QUARTER | 3RD QUARTER | 4TH QUARTER | TOTAL GRAMS |
|---|---|---|---|---|---|
| 820 | 2,180.88 | 2,124.43 | 2,028.63 | 2,041.54 | **8,375.48** |
| 822 | 259.48 | 206.74 | 197.84 | 175.76 | **839.83** |
| 823 | 103.33 | 106.45 | 91.23 | 125.04 | **426.05** |
| 824 | 723.97 | 677.04 | 767.36 | 760.54 | **2,928.90** |
| 825 | 415.80 | 467.12 | 482.52 | 466.56 | **1,832.00** |
| 826 | 1,556.54 | 1,534.34 | 1,230.82 | 1,525.07 | **5,846.77** |
| 827 | 654.92 | 686.70 | 698.69 | 671.83 | **2,712.13** |
| 828 | 581.13 | 512.14 | 470.41 | 525.95 | **2,089.63** |
| 829 | 759.80 | 801.53 | 763.01 | 912.19 | **3,236.52** |
| 830 | 394.75 | 393.72 | 357.91 | 417.15 | **1,563.53** |
| 831 | 160.29 | 199.24 | 210.39 | 207.24 | **777.16** |
| **WY - WYOMING Total** | **7,790.88** | **7,709.43** | **7,298.82** | **7,828.86** | **30,627.99** |

**DRUG: 9050 - CODEINE**

**STATE: AK - ALASKA**

| REGISTRANT ZIP CODE 3 | 1ST QUARTER | 2ND QUARTER | 3RD QUARTER | 4TH QUARTER | TOTAL GRAMS |
|---|---|---|---|---|---|
| 995 | 1,932.72 | 1,767.80 | 2,022.57 | 1,759.05 | **7,482.14** |
| 996 | 1,111.03 | 891.71 | 1,121.21 | 881.78 | **4,005.74** |
| 997 | 1,575.05 | 1,618.14 | 1,676.35 | 1,690.53 | **6,560.07** |
| 998 | 225.24 | 315.85 | 271.28 | 247.53 | **1,059.90** |
| 999 | 237.95 | 232.22 | 203.68 | 240.16 | **914.01** |
| **AK ALASKA - Total** | **5,081.99** | **4,825.72** | **5,295.10** | **4,819.05** | **20,021.86** |

**STATE: AL - ALABAMA**

| REGISTRANT ZIP CODE 3 | 1ST QUARTER | 2ND QUARTER | 3RD QUARTER | 4TH QUARTER | TOTAL GRAMS |
|---|---|---|---|---|---|
| 350 | 5,173.56 | 5,190.75 | 5,195.69 | 4,773.92 | **20,333.92** |
| 351 | 3,261.15 | 3,473.34 | 3,292.93 | 3,450.89 | **13,478.32** |
| 352 | 4,518.85 | 4,322.45 | 4,065.43 | 4,416.73 | **17,323.45** |
| 354 | 1,468.62 | 1,853.88 | 1,595.76 | 1,453.49 | **6,371.75** |
| 355 | 1,894.42 | 1,896.63 | 1,880.89 | 1,719.05 | **7,390.99** |
| 356 | 5,308.77 | 5,029.99 | 4,802.53 | 5,139.41 | **20,280.71** |
| 357 | 596.08 | 671.23 | 436.77 | 795.00 | **2,499.08** |
| 358 | 626.68 | 639.47 | 637.00 | 842.56 | **2,745.71** |

| | | | | | |
|---|---|---|---|---|---|
| 359 | 1,782.74 | 2,078.02 | 1,917.81 | 1,945.48 | **7,724.05** |
| 360 | 3,589.44 | 3,466.39 | 2,790.39 | 2,753.15 | **12,599.36** |
| 361 | 1,974.32 | 1,962.27 | 1,804.72 | 1,841.58 | **7,582.89** |
| 362 | 1,143.07 | 1,049.13 | 1,085.82 | 1,211.50 | **4,489.51** |
| 363 | 1,704.14 | 1,590.39 | 1,488.80 | 1,546.83 | **6,330.16** |
| 364 | 1,587.79 | 1,582.61 | 1,349.99 | 1,192.90 | **5,713.29** |
| 365 | 3,180.10 | 3,268.16 | 3,269.92 | 3,098.09 | **12,816.27** |
| 366 | 3,550.59 | 3,355.79 | 2,946.25 | 3,235.23 | **13,087.87** |
| 367 | 2,047.12 | 1,965.14 | 1,911.74 | 1,910.63 | **7,834.63** |
| 368 | 1,708.79 | 1,926.98 | 1,778.90 | 1,770.10 | **7,184.77** |
| 369 | 428.76 | 380.14 | 486.22 | 419.92 | **1,715.04** |
| **AL ALABAMA - Total** | **45,544.99** | **45,702.76** | **42,737.56** | **43,516.47** | **177,501.78** |

**STATE: AR - ARKANSAS**

| REGISTRANT ZIP CODE 3 | 1ST QUARTER | 2ND QUARTER | 3RD QUARTER | 4TH QUARTER | TOTAL GRAMS |
|---|---|---|---|---|---|
| 716 | 2,919.50 | 2,742.51 | 2,773.45 | 2,760.36 | **11,195.82** |
| 717 | 2,614.31 | 2,512.51 | 2,621.02 | 2,466.82 | **10,214.66** |
| 718 | 4,811.38 | 4,774.92 | 4,865.62 | 4,609.45 | **19,061.37** |
| 719 | 4,170.08 | 3,659.55 | 3,694.35 | 3,341.55 | **14,865.53** |
| 720 | 4,343.89 | 4,170.56 | 4,211.01 | 4,147.61 | **16,873.07** |
| 721 | 5,199.45 | 4,626.74 | 4,931.00 | 4,902.40 | **19,659.59** |
| 722 | 2,164.09 | 2,018.37 | 2,195.70 | 1,969.80 | **8,347.95** |
| 723 | 5,161.49 | 4,945.83 | 5,026.48 | 5,008.60 | **20,142.40** |
| 724 | 6,964.04 | 6,578.71 | 6,164.57 | 5,992.59 | **25,699.92** |
| 725 | 3,530.08 | 3,367.26 | 3,144.57 | 3,230.86 | **13,272.77** |
| 726 | 1,839.52 | 1,936.02 | 1,924.58 | 1,862.00 | **7,562.12** |
| 727 | 3,318.91 | 3,054.03 | 2,969.05 | 3,025.42 | **12,367.41** |
| 728 | 2,296.84 | 2,299.61 | 2,129.61 | 2,055.58 | **8,781.64** |
| 729 | 3,794.55 | 3,373.16 | 3,517.73 | 2,972.39 | **13,657.82** |
| **AR ARKANSAS - Total** | **53,128.12** | **50,059.78** | **50,168.75** | **48,345.43** | **201,702.07** |

**STATE: AS - AMERICAN SAMOA**

| REGISTRANT ZIP CODE 3 | 1ST QUARTER | 2ND QUARTER | 3RD QUARTER | 4TH QUARTER | TOTAL GRAMS |
|---|---|---|---|---|---|
| 967 | 274.05 | 293.94 | 243.11 | 388.98 | **1,200.08** |
| **AS - AMERICAN SAMOA - Tota** | **274.05** | **293.94** | **243.11** | **388.98** | **1,200.08** |

**STATE: AZ - ARIZONA**

| REGISTRANT ZIP CODE 3 | 1ST QUARTER | 2ND QUARTER | 3RD QUARTER | 4TH QUARTER | TOTAL GRAMS |
|---|---|---|---|---|---|
| 850 | 12,216.00 | 13,202.06 | 12,239.96 | 12,634.71 | **50,292.73** |
| 851 | 2,197.43 | 1,972.36 | 1,837.46 | 2,034.62 | **8,041.87** |
| 852 | 14,060.91 | 12,396.64 | 10,152.96 | 10,271.50 | **46,882.01** |
| 853 | 7,784.14 | 7,604.49 | 6,967.23 | 7,081.89 | **29,437.76** |
| 855 | 784.57 | 786.24 | 697.27 | 780.67 | **3,048.76** |

| | | | | | |
|---|---|---|---|---|---|
| 856 | 1,672.58 | 1,548.18 | 1,611.92 | 1,718.27 | **6,550.95** |
| 857 | 6,450.24 | 6,341.15 | 6,077.66 | 6,218.62 | **25,087.67** |
| 859 | 508.84 | 613.99 | 492.39 | 396.12 | **2,011.34** |
| 860 | 605.36 | 855.19 | 636.48 | 680.37 | **2,777.41** |
| 863 | 1,687.27 | 1,497.28 | 1,548.52 | 1,399.27 | **6,132.35** |
| 864 | 2,066.67 | 1,965.52 | 1,941.63 | 1,888.29 | **7,862.10** |
| 865 | 726.73 | 587.50 | 583.08 | 558.77 | **2,456.07** |
| **AZ ARIZONA - Total** | **50,760.76** | **49,370.61** | **44,786.56** | **45,663.09** | **190,581.01** |

**STATE: CA - CALIFORNIA**

| REGISTRANT ZIP CODE 3 | 1ST QUARTER | 2ND QUARTER | 3RD QUARTER | 4TH QUARTER | **TOTAL GRAMS** |
|---|---|---|---|---|---|
| 900 | 19,362.12 | 18,761.32 | 18,237.10 | 18,393.53 | **74,754.07** |
| 902 | 18,709.41 | 17,022.71 | 17,745.33 | 17,088.35 | **70,565.81** |
| 903 | 2,237.28 | 2,453.87 | 2,215.63 | 2,427.79 | **9,334.58** |
| 904 | 1,078.94 | 890.32 | 977.33 | 850.43 | **3,797.03** |
| 905 | 1,617.80 | 1,521.71 | 1,740.65 | 1,638.75 | **6,518.92** |
| 906 | 5,472.58 | 5,077.31 | 4,959.78 | 5,191.23 | **20,700.89** |
| 907 | 6,932.26 | 6,595.59 | 6,509.22 | 6,996.73 | **27,033.81** |
| 908 | 5,451.53 | 5,089.48 | 4,913.21 | 4,689.88 | **20,144.09** |
| 910 | 2,580.49 | 2,422.62 | 2,396.29 | 2,361.49 | **9,760.90** |
| 911 | 1,579.16 | 1,588.08 | 1,496.50 | 1,465.94 | **6,129.68** |
| 912 | 2,113.70 | 1,975.79 | 2,080.25 | 1,749.70 | **7,919.44** |
| 913 | 11,123.01 | 10,563.08 | 10,875.67 | 10,546.13 | **43,107.90** |
| 914 | 3,448.64 | 3,134.31 | 3,085.22 | 3,097.91 | **12,766.08** |
| 915 | 1,156.54 | 1,169.17 | 1,307.68 | 1,277.98 | **4,911.37** |
| 916 | 1,151.31 | 1,122.43 | 1,112.19 | 1,153.05 | **4,538.97** |
| 917 | 16,997.03 | 16,057.97 | 15,254.33 | 14,827.73 | **63,137.06** |
| 918 | 608.05 | 571.77 | 562.47 | 704.10 | **2,446.39** |
| 919 | 3,247.92 | 3,164.44 | 3,170.94 | 2,745.24 | **12,328.54** |
| 920 | 11,562.35 | 10,896.77 | 12,137.16 | 11,719.08 | **46,315.36** |
| 921 | 8,381.42 | 7,195.66 | 7,090.46 | 7,174.58 | **29,842.13** |
| 922 | 8,244.15 | 7,996.77 | 7,226.22 | 7,434.06 | **30,901.20** |
| 923 | 14,560.06 | 14,037.52 | 13,869.26 | 13,473.72 | **55,940.56** |
| 924 | 2,214.52 | 2,137.17 | 2,119.57 | 2,132.75 | **8,604.01** |
| 925 | 13,684.46 | 13,633.96 | 12,996.07 | 13,115.39 | **53,429.88** |
| 926 | 9,240.43 | 8,542.16 | 8,614.82 | 8,526.48 | **34,923.88** |
| 927 | 3,113.56 | 2,883.61 | 2,689.35 | 2,794.63 | **11,481.15** |
| 928 | 9,924.70 | 9,607.96 | 8,932.25 | 9,325.38 | **37,790.29** |
| 930 | 5,356.68 | 5,879.34 | 5,160.51 | 5,581.59 | **21,978.12** |
| 931 | 1,462.12 | 1,255.94 | 1,248.68 | 1,141.95 | **5,108.69** |
| 932 | 7,753.85 | 7,525.00 | 7,411.58 | 7,405.90 | **30,096.33** |

| | | | | | |
|---|---|---|---|---|---|
| 933 | 5,876.13 | 5,670.45 | 5,342.54 | 4,840.82 | **21,729.94** |
| 934 | 4,541.57 | 3,778.30 | 3,893.36 | 4,122.08 | **16,335.31** |
| 935 | 7,638.65 | 7,795.41 | 7,258.10 | 6,953.73 | **29,645.89** |
| 936 | 6,175.72 | 5,743.48 | 5,791.77 | 5,590.07 | **23,301.04** |
| 937 | 8,395.55 | 7,732.35 | 7,667.15 | 7,662.00 | **31,457.05** |
| 939 | 2,945.23 | 3,083.77 | 3,110.67 | 3,344.43 | **12,484.10** |
| 940 | 4,527.03 | 4,309.01 | 4,234.34 | 4,209.74 | **17,280.12** |
| 941 | 7,307.42 | 6,690.86 | 6,610.10 | 6,321.83 | **26,930.22** |
| 943 | 482.56 | 546.43 | 446.59 | 500.06 | **1,975.64** |
| 944 | 770.01 | 776.33 | 720.15 | 707.41 | **2,973.91** |
| 945 | 23,629.56 | 21,528.09 | 22,526.97 | 22,063.50 | **89,748.11** |
| 946 | 3,227.76 | 2,835.14 | 2,892.75 | 3,085.70 | **12,041.35** |
| 947 | 702.70 | 775.90 | 805.82 | 832.27 | **3,116.69** |
| 948 | 1,127.88 | 1,105.22 | 936.73 | 910.37 | **4,080.19** |
| 949 | 2,766.64 | 2,772.19 | 2,523.87 | 2,673.21 | **10,735.92** |
| 950 | 4,547.56 | 4,153.48 | 4,575.46 | 4,370.32 | **17,646.82** |
| 951 | 4,725.78 | 4,844.66 | 4,485.07 | 4,683.24 | **18,738.75** |
| 952 | 7,588.70 | 7,297.05 | 6,803.55 | 6,969.04 | **28,658.33** |
| 953 | 13,760.03 | 12,844.65 | 12,810.02 | 12,535.92 | **51,950.61** |
| 954 | 4,440.92 | 3,738.52 | 3,928.46 | 3,505.68 | **15,613.58** |
| 955 | 2,339.55 | 2,142.63 | 2,195.54 | 2,182.40 | **8,860.12** |
| 956 | 11,795.24 | 10,919.34 | 10,481.92 | 10,429.32 | **43,625.82** |
| 957 | 1,949.06 | 1,932.62 | 1,784.02 | 1,968.17 | **7,633.87** |
| 958 | 6,530.99 | 5,466.90 | 5,561.01 | 5,292.97 | **22,851.88** |
| 959 | 5,310.78 | 5,351.58 | 5,715.63 | 5,617.10 | **21,995.09** |
| 960 | 3,248.08 | 2,859.52 | 3,165.72 | 3,270.40 | **12,543.73** |
| 961 | 916.27 | 1,216.66 | 941.87 | 802.27 | **3,877.06** |
| **CA - CALIFORNIA - Total** | **347,633.47** | **328,684.36** | **325,344.88** | **322,475.55** | **1,324,138.27** |

**STATE: CO - COLORADO**

| REGISTRANT ZIP CODE 3 | 1ST QUARTER | 2ND QUARTER | 3RD QUARTER | 4TH QUARTER | **TOTAL GRAMS** |
|---|---|---|---|---|---|
| 800 | 2,818.30 | 2,700.65 | 2,927.88 | 2,723.52 | **11,170.35** |
| 801 | 2,090.67 | 2,122.66 | 1,910.16 | 2,214.56 | **8,338.05** |
| 802 | 2,582.10 | 2,970.22 | 2,608.35 | 2,940.41 | **11,101.09** |
| 803 | 301.90 | 206.78 | 305.70 | 237.37 | **1,051.74** |
| 804 | 385.16 | 301.74 | 379.25 | 399.86 | **1,466.01** |
| 805 | 1,650.41 | 1,744.88 | 1,597.17 | 1,742.51 | **6,734.97** |
| 806 | 599.67 | 667.19 | 700.53 | 569.39 | **2,536.77** |
| 807 | 358.04 | 331.52 | 346.99 | 300.57 | **1,337.11** |
| 808 | 499.56 | 529.10 | 478.85 | 536.59 | **2,044.10** |
| 809 | 1,929.90 | 1,779.86 | 1,877.46 | 1,779.67 | **7,366.89** |

| | | | | | |
|---|---|---|---|---|---|
| 810 | 1,557.19 | 1,613.17 | 1,348.71 | 1,421.78 | **5,940.84** |
| 811 | 238.69 | 242.19 | 342.10 | 251.95 | **1,074.93** |
| 812 | 465.86 | 434.09 | 398.99 | 452.06 | **1,751.00** |
| 813 | 224.96 | 122.93 | 234.27 | 178.28 | **760.44** |
| 814 | 267.08 | 281.97 | 324.98 | 324.94 | **1,198.97** |
| 815 | 615.23 | 579.76 | 507.71 | 572.91 | **2,275.61** |
| 816 | 315.94 | 277.74 | 260.87 | 267.06 | **1,121.61** |
| **CO - COLORADO - Total** | **16,900.66** | **16,906.46** | **16,549.95** | **16,913.43** | **67,270.50** |

**STATE: CT - CONNECTICUT**

| REGISTRANT ZIP CODE 3 | 1ST QUARTER | 2ND QUARTER | 3RD QUARTER | 4TH QUARTER | **TOTAL GRAMS** |
|---|---|---|---|---|---|
| 060 | 3,007.05 | 3,745.57 | 3,529.15 | 3,113.29 | **13,395.06** |
| 061 | 1,140.91 | 1,338.93 | 1,241.00 | 1,147.67 | **4,868.50** |
| 062 | 554.45 | 621.39 | 506.26 | 504.72 | **2,186.82** |
| 063 | 1,350.80 | 1,339.47 | 1,251.16 | 1,278.66 | **5,220.09** |
| 064 | 3,624.84 | 4,330.90 | 3,913.15 | 3,937.94 | **15,806.84** |
| 065 | 1,110.50 | 1,227.73 | 1,057.99 | 983.49 | **4,379.71** |
| 066 | 1,045.93 | 1,063.84 | 990.23 | 1,027.60 | **4,127.61** |
| 067 | 1,813.74 | 2,099.81 | 1,619.82 | 1,961.86 | **7,495.23** |
| 068 | 1,917.92 | 2,277.32 | 1,787.03 | 1,859.70 | **7,841.97** |
| 069 | 442.37 | 530.59 | 367.33 | 519.91 | **1,860.20** |
| **CT CONNECTICUT Total** | **16,008.51** | **18,575.55** | **16,263.14** | **16,334.85** | **67,182.04** |

**STATE: DC - DISTRICT OF COLUMBIA**

| REGISTRANT ZIP CODE 3 | 1ST QUARTER | 2ND QUARTER | 3RD QUARTER | 4TH QUARTER | **TOTAL GRAMS** |
|---|---|---|---|---|---|
| 200 | 8,224.52 | 8,524.22 | 7,829.93 | 7,908.74 | **32,487.41** |
| 203 | 24.31 | 18.59 | 19.89 | 0 | **62.79** |
| 204 | 82.68 | 281.59 | 69.06 | 103.50 | **536.83** |
| 205 | 0 | 0 | 4.42 | 0 | **4.42** |
| **DC - DISTRICT OF COLUMBIA** | **8,331.51** | **8,824.41** | **7,923.30** | **8,012.24** | **33,091.45** |

**STATE: DE - DELAWARE**

| REGISTRANT ZIP CODE 3 | 1ST QUARTER | 2ND QUARTER | 3RD QUARTER | 4TH QUARTER | **TOTAL GRAMS** |
|---|---|---|---|---|---|
| 197 | 2,562.68 | 2,478.47 | 2,641.13 | 2,270.23 | **9,952.50** |
| 198 | 1,838.77 | 1,879.76 | 1,756.46 | 1,693.15 | **7,168.14** |
| 199 | 2,480.71 | 2,608.21 | 2,471.33 | 2,506.75 | **10,067.00** |
| **DE DELAWARE Total** | **6,882.16** | **6,966.43** | **6,868.92** | **6,470.13** | **27,187.64** |

**STATE: FL - FLORIDA**

| REGISTRANT ZIP CODE 3 | 1ST QUARTER | 2ND QUARTER | 3RD QUARTER | 4TH QUARTER | **TOTAL GRAMS** |
|---|---|---|---|---|---|
| 320 | 4,294.30 | 4,155.24 | 4,230.09 | 4,629.83 | **17,309.46** |
| 321 | 4,565.52 | 4,514.41 | 3,915.83 | 4,399.61 | **17,395.36** |
| 322 | 4,741.75 | 4,730.67 | 4,589.04 | 4,508.62 | **18,570.07** |
| 323 | 2,729.36 | 2,739.67 | 2,536.12 | 2,723.15 | **10,728.29** |

| | | | | | |
|---|---|---|---|---|---|
| 324 | 3,867.80 | 4,165.84 | 3,127.40 | 3,263.44 | **14,424.48** |
| 325 | 3,229.32 | 3,660.84 | 3,606.07 | 3,251.83 | **13,748.05** |
| 326 | 2,097.43 | 2,488.42 | 2,398.57 | 2,298.29 | **9,282.70** |
| 327 | 5,903.11 | 5,747.40 | 5,769.23 | 5,739.23 | **23,158.96** |
| 328 | 4,202.15 | 4,125.67 | 4,329.88 | 4,016.35 | **16,674.05** |
| 329 | 3,912.37 | 4,056.37 | 3,732.66 | 3,946.58 | **15,647.97** |
| 330 | 9,270.08 | 9,052.66 | 9,195.73 | 8,688.26 | **36,206.73** |
| 331 | 10,501.13 | 10,323.40 | 10,869.37 | 10,523.52 | **42,217.42** |
| 333 | 5,056.26 | 4,795.95 | 5,023.74 | 4,718.57 | **19,594.52** |
| 334 | 8,761.01 | 8,835.05 | 8,899.70 | 8,824.50 | **35,320.26** |
| 335 | 6,282.74 | 6,003.03 | 6,017.33 | 6,031.58 | **24,334.68** |
| 336 | 6,745.76 | 6,396.27 | 6,708.72 | 6,435.63 | **26,286.38** |
| 337 | 7,766.80 | 6,763.67 | 7,001.55 | 6,979.91 | **28,511.92** |
| 338 | 6,688.00 | 6,458.17 | 6,346.79 | 6,050.42 | **25,543.39** |
| 339 | 6,537.37 | 5,655.59 | 5,352.81 | 5,469.90 | **23,015.66** |
| 341 | 1,835.70 | 1,453.18 | 1,287.05 | 1,643.96 | **6,219.89** |
| 342 | 4,211.63 | 4,162.31 | 4,203.79 | 4,265.15 | **16,842.88** |
| 344 | 5,433.41 | 5,621.05 | 5,254.70 | 5,932.25 | **22,241.41** |
| 346 | 6,031.30 | 5,932.92 | 5,608.84 | 6,362.62 | **23,935.69** |
| 347 | 3,536.68 | 3,578.50 | 3,268.45 | 3,554.87 | **13,938.49** |
| 349 | 2,652.32 | 2,561.39 | 2,479.00 | 2,784.50 | **10,477.21** |
| **FL FLORIDA - Total** | **130,853.28** | **127,977.69** | **125,752.44** | **127,042.54** | **511,625.95** |

**STATE: GA - GEORGIA**

| REGISTRANT ZIP CODE 3 | 1ST QUARTER | 2ND QUARTER | 3RD QUARTER | 4TH QUARTER | **TOTAL GRAMS** |
|---|---|---|---|---|---|
| 300 | 10,835.59 | 11,884.97 | 10,856.38 | 11,126.84 | **44,703.78** |
| 301 | 6,787.36 | 7,315.66 | 7,541.19 | 7,235.38 | **28,879.59** |
| 302 | 8,974.20 | 8,957.19 | 9,123.42 | 9,023.35 | **36,078.17** |
| 303 | 5,064.63 | 5,141.54 | 4,723.32 | 4,707.77 | **19,637.26** |
| 304 | 2,979.99 | 3,018.54 | 2,923.08 | 2,864.93 | **11,786.54** |
| 305 | 5,645.95 | 5,322.23 | 5,329.98 | 5,469.89 | **21,768.05** |
| 306 | 5,080.33 | 5,038.79 | 4,989.78 | 5,033.80 | **20,142.70** |
| 307 | 2,187.92 | 2,253.76 | 1,948.20 | 1,787.97 | **8,177.85** |
| 308 | 2,430.84 | 2,577.54 | 2,330.54 | 2,485.80 | **9,824.72** |
| 309 | 2,827.85 | 2,649.60 | 2,639.16 | 2,467.06 | **10,583.67** |
| 310 | 5,088.25 | 5,635.76 | 5,199.86 | 5,414.24 | **21,338.12** |
| 312 | 1,587.21 | 1,797.63 | 1,653.65 | 1,524.05 | **6,562.54** |
| 313 | 1,107.26 | 1,318.34 | 1,301.22 | 1,208.04 | **4,934.86** |
| 314 | 1,023.20 | 1,255.23 | 1,221.08 | 996.90 | **4,496.41** |
| 315 | 4,549.78 | 5,008.77 | 5,208.20 | 4,801.09 | **19,567.84** |
| 316 | 2,196.30 | 2,186.00 | 1,936.39 | 1,854.20 | **8,172.89** |

| | | | | | |
|---|---|---|---|---|---|
| 317 | 5,282.19 | 5,340.24 | 5,305.19 | 5,234.82 | **21,162.44** |
| 318 | 467.72 | 465.23 | 304.99 | 397.82 | **1,635.76** |
| 319 | 2,349.22 | 2,353.85 | 2,321.21 | 2,014.51 | **9,038.79** |
| 398 | 1,720.56 | 1,898.48 | 1,875.27 | 1,821.12 | **7,315.43** |
| **GA GEORGIA Total** | **78,186.36** | **81,419.32** | **78,732.14** | **77,469.58** | **315,807.40** |

**STATE: GU - GUAM**

| REGISTRANT ZIP CODE 3 | 1ST QUARTER | 2ND QUARTER | 3RD QUARTER | 4TH QUARTER | TOTAL GRAMS |
|---|---|---|---|---|---|
| 969 | 616.62 | 799.46 | 846.55 | 596.73 | **2,859.35** |
| **GU - GUAM - Total** | **616.62** | **799.46** | **846.55** | **596.73** | **2,859.35** |

**STATE: HI - HAWAII**

| REGISTRANT ZIP CODE 3 | 1ST QUARTER | 2ND QUARTER | 3RD QUARTER | 4TH QUARTER | TOTAL GRAMS |
|---|---|---|---|---|---|
| 967 | 7,069.59 | 6,655.93 | 6,521.91 | 6,532.41 | **26,779.84** |
| 968 | 3,579.46 | 3,066.96 | 3,201.59 | 3,478.31 | **13,326.32** |
| **HI - HAWAII - Total** | **10,649.05** | **9,722.89** | **9,723.51** | **10,010.72** | **40,106.16** |

**STATE: IA - IOWA**

| REGISTRANT ZIP CODE 3 | 1ST QUARTER | 2ND QUARTER | 3RD QUARTER | 4TH QUARTER | TOTAL GRAMS |
|---|---|---|---|---|---|
| 500 | 857.51 | 903.04 | 749.75 | 827.75 | **3,338.06** |
| 501 | 939.19 | 788.18 | 789.11 | 699.22 | **3,215.70** |
| 502 | 749.58 | 740.55 | 847.03 | 672.68 | **3,009.83** |
| 503 | 1,336.18 | 1,490.75 | 1,510.98 | 1,575.54 | **5,913.46** |
| 504 | 261.16 | 229.85 | 263.55 | 302.97 | **1,057.52** |
| 505 | 503.08 | 534.29 | 503.48 | 548.04 | **2,088.89** |
| 506 | 549.30 | 635.77 | 553.99 | 459.70 | **2,198.76** |
| 507 | 434.56 | 511.44 | 529.10 | 318.44 | **1,793.54** |
| 508 | 170.36 | 161.34 | 159.13 | 151.39 | **642.22** |
| 510 | 349.55 | 350.65 | 355.07 | 347.79 | **1,403.07** |
| 511 | 455.92 | 522.30 | 551.75 | 619.35 | **2,149.31** |
| 512 | 145.87 | 100.82 | 72.93 | 98.61 | **418.24** |
| 513 | 266.50 | 203.13 | 221.56 | 203.88 | **895.07** |
| 514 | 183.80 | 204.06 | 297.90 | 222.48 | **908.24** |
| 515 | 604.27 | 795.53 | 583.19 | 654.01 | **2,637.00** |
| 516 | 219.86 | 240.88 | 180.47 | 218.78 | **859.99** |
| 520 | 887.15 | 824.80 | 723.05 | 646.34 | **3,081.34** |
| 521 | 205.54 | 190.07 | 228.75 | 234.27 | **858.62** |
| 522 | 722.08 | 767.82 | 819.31 | 665.41 | **2,974.62** |
| 523 | 430.88 | 407.30 | 528.20 | 396.15 | **1,762.53** |
| 524 | 645.25 | 641.84 | 522.49 | 599.85 | **2,409.43** |
| 525 | 751.77 | 727.06 | 624.84 | 702.40 | **2,806.07** |
| 526 | 607.22 | 685.92 | 749.03 | 587.58 | **2,629.75** |
| 527 | 1,149.69 | 1,132.48 | 1,027.14 | 951.63 | **4,260.94** |

| | | | | | |
|---|---|---|---|---|---|
| 528 | 751.45 | 511.31 | 653.82 | 697.53 | **2,614.11** |
| **IA IOWA Total** | **14,177.70** | **14,301.18** | **14,045.63** | **13,401.78** | **55,926.30** |

**STATE: ID - IDAHO**

| REGISTRANT ZIP CODE 3 | 1ST QUARTER | 2ND QUARTER | 3RD QUARTER | 4TH QUARTER | TOTAL GRAMS |
|---|---|---|---|---|---|
| 832 | 1,356.44 | 1,428.90 | 1,193.45 | 1,282.53 | **5,261.33** |
| 833 | 879.59 | 909.81 | 955.86 | 1,068.57 | **3,813.83** |
| 834 | 1,010.91 | 1,135.92 | 1,148.67 | 1,070.99 | **4,366.49** |
| 835 | 854.23 | 967.01 | 946.16 | 995.25 | **3,762.65** |
| 836 | 3,223.22 | 3,448.76 | 3,432.90 | 3,508.16 | **13,613.04** |
| 837 | 1,386.96 | 1,435.61 | 1,517.23 | 1,472.28 | **5,812.08** |
| 838 | 2,133.53 | 1,903.46 | 2,024.55 | 2,211.73 | **8,273.26** |
| **ID IDAHO Total** | **10,844.89** | **11,229.46** | **11,218.81** | **11,609.51** | **44,902.67** |

**STATE: IL - ILLINOIS**

| REGISTRANT ZIP CODE 3 | 1ST QUARTER | 2ND QUARTER | 3RD QUARTER | 4TH QUARTER | TOTAL GRAMS |
|---|---|---|---|---|---|
| 600 | 17,009.23 | 17,574.09 | 17,324.58 | 18,385.41 | **70,293.31** |
| 601 | 11,421.30 | 11,724.21 | 11,542.58 | 11,305.00 | **45,993.09** |
| 602 | 458.60 | 491.75 | 420.56 | 506.30 | **1,877.19** |
| 603 | 971.14 | 848.71 | 835.96 | 937.73 | **3,593.55** |
| 604 | 15,347.26 | 15,571.49 | 15,336.02 | 15,161.34 | **61,416.12** |
| 605 | 8,541.03 | 7,942.70 | 8,356.51 | 8,045.98 | **32,886.22** |
| 606 | 22,159.74 | 22,438.21 | 22,329.57 | 20,960.01 | **87,887.53** |
| 607 | 1,367.30 | 1,207.25 | 1,578.57 | 1,320.82 | **5,473.94** |
| 608 | 1,431.40 | 1,589.21 | 1,487.49 | 1,380.32 | **5,888.42** |
| 609 | 1,538.94 | 1,660.23 | 1,633.63 | 1,618.43 | **6,451.23** |
| 610 | 2,626.18 | 2,603.68 | 2,720.62 | 2,516.00 | **10,466.49** |
| 611 | 3,698.18 | 3,684.35 | 3,741.98 | 3,649.79 | **14,774.29** |
| 612 | 1,354.40 | 1,584.96 | 1,415.73 | 1,653.31 | **6,008.40** |
| 613 | 1,457.35 | 1,240.95 | 1,510.03 | 1,187.36 | **5,395.69** |
| 614 | 1,542.65 | 1,529.45 | 1,515.25 | 1,519.70 | **6,107.05** |
| 615 | 1,708.39 | 1,796.06 | 1,796.00 | 1,764.91 | **7,065.37** |
| 616 | 2,273.56 | 2,348.53 | 2,045.08 | 2,395.20 | **9,062.36** |
| 617 | 2,338.51 | 2,516.23 | 2,440.13 | 2,170.58 | **9,465.45** |
| 618 | 3,307.22 | 3,329.85 | 3,566.66 | 3,165.78 | **13,369.51** |
| 619 | 1,004.47 | 878.13 | 927.67 | 1,044.44 | **3,854.70** |
| 620 | 4,383.27 | 4,432.89 | 4,180.00 | 4,082.32 | **17,078.47** |
| 622 | 5,371.58 | 5,343.87 | 5,246.61 | 5,267.87 | **21,229.92** |
| 623 | 1,261.30 | 1,021.60 | 1,094.71 | 1,156.06 | **4,533.68** |
| 624 | 1,551.41 | 1,137.20 | 1,297.42 | 1,350.37 | **5,336.40** |
| 625 | 1,890.20 | 1,894.94 | 1,879.11 | 1,726.74 | **7,390.98** |
| 626 | 1,798.94 | 1,777.14 | 1,761.42 | 1,617.56 | **6,955.07** |

| | | | | | |
|---|---|---|---|---|---|
| 627 | 2,495.14 | 2,423.16 | 2,222.71 | 2,529.60 | **9,670.61** |
| 628 | 1,781.33 | 1,778.74 | 1,852.71 | 1,763.27 | **7,176.05** |
| 629 | 3,315.03 | 3,464.57 | 3,254.57 | 3,163.02 | **13,197.19** |
| **IL ILLINOIS Total** | **125,405.08** | **125,834.10** | **125,313.89** | **123,345.23** | **499,898.30** |

**STATE: IN - INDIANA**

| REGISTRANT ZIP CODE 3 | 1ST QUARTER | 2ND QUARTER | 3RD QUARTER | 4TH QUARTER | TOTAL GRAMS |
|---|---|---|---|---|---|
| 460 | 2,032.82 | 1,686.68 | 2,060.81 | 1,955.73 | **7,736.04** |
| 461 | 2,607.61 | 2,523.87 | 2,611.81 | 2,328.02 | **10,071.31** |
| 462 | 4,870.87 | 4,184.93 | 4,145.87 | 4,077.38 | **17,279.04** |
| 463 | 7,574.60 | 7,374.40 | 7,133.15 | 7,106.63 | **29,188.78** |
| 464 | 1,853.35 | 1,682.07 | 1,599.01 | 1,618.90 | **6,753.32** |
| 465 | 1,865.59 | 1,694.21 | 1,925.59 | 1,829.54 | **7,314.93** |
| 466 | 680.04 | 624.16 | 626.75 | 443.49 | **2,374.43** |
| 467 | 1,230.24 | 1,020.85 | 1,318.12 | 1,076.20 | **4,645.41** |
| 468 | 1,362.94 | 1,324.01 | 1,387.89 | 1,237.53 | **5,312.36** |
| 469 | 1,335.26 | 1,421.07 | 1,467.75 | 1,354.58 | **5,578.67** |
| 470 | 610.04 | 508.69 | 471.19 | 589.30 | **2,179.23** |
| 471 | 4,253.67 | 5,156.71 | 2,766.66 | 1,655.63 | **13,832.66** |
| 472 | 939.35 | 834.77 | 1,039.71 | 1,045.04 | **3,858.87** |
| 473 | 1,359.79 | 1,307.27 | 1,502.45 | 1,423.67 | **5,593.18** |
| 474 | 1,066.00 | 1,029.44 | 986.03 | 1,026.40 | **4,107.86** |
| 475 | 734.30 | 890.29 | 579.65 | 702.25 | **2,906.50** |
| 476 | 503.97 | 647.74 | 606.84 | 624.35 | **2,382.91** |
| 477 | 1,263.15 | 1,378.71 | 1,455.33 | 1,365.09 | **5,462.29** |
| 478 | 1,716.12 | 1,707.35 | 1,588.34 | 1,531.38 | **6,543.18** |
| 479 | 1,118.13 | 1,204.63 | 1,279.98 | 1,171.96 | **4,774.70** |
| **IN INDIANA Total** | **38,977.83** | **38,201.85** | **36,552.92** | **34,163.06** | **147,895.67** |

**STATE: KS - KANSAS**

| REGISTRANT ZIP CODE 3 | 1ST QUARTER | 2ND QUARTER | 3RD QUARTER | 4TH QUARTER | TOTAL GRAMS |
|---|---|---|---|---|---|
| 660 | 2,481.73 | 2,227.50 | 2,329.56 | 2,198.24 | **9,237.03** |
| 661 | 941.10 | 725.45 | 784.10 | 817.92 | **3,268.57** |
| 662 | 4,460.20 | 4,755.28 | 5,869.99 | 6,487.53 | **21,573.00** |
| 664 | 391.53 | 353.42 | 351.32 | 367.32 | **1,463.58** |
| 665 | 565.73 | 737.76 | 664.71 | 677.99 | **2,646.19** |
| 666 | 742.47 | 800.75 | 911.94 | 784.35 | **3,239.51** |
| 667 | 780.71 | 716.98 | 793.22 | 696.55 | **2,987.46** |
| 668 | 382.08 | 269.56 | 219.54 | 278.33 | **1,149.51** |
| 669 | 248.73 | 292.10 | 270.00 | 349.38 | **1,160.20** |
| 670 | 1,295.09 | 1,458.24 | 1,401.39 | 1,109.72 | **5,264.44** |
| 671 | 634.83 | 749.48 | 673.05 | 772.61 | **2,829.98** |

| | | | | | |
|---|---|---|---|---|---|
| 672 | 2,594.75 | 2,431.87 | 2,490.85 | 2,491.05 | **10,008.52** |
| 673 | 698.94 | 616.06 | 699.60 | 709.99 | **2,724.59** |
| 674 | 943.96 | 986.69 | 1,033.45 | 826.66 | **3,790.76** |
| 675 | 462.46 | 554.36 | 534.29 | 498.00 | **2,049.10** |
| 676 | 279.93 | 285.26 | 251.74 | 222.42 | **1,039.35** |
| 677 | 156.92 | 156.92 | 203.55 | 132.38 | **649.77** |
| 678 | 518.35 | 563.38 | 498.55 | 588.80 | **2,169.07** |
| 679 | 413.29 | 471.95 | 543.68 | 434.38 | **1,863.30** |
| **KS - KANSAS - Total** | **18,992.79** | **19,153.00** | **20,524.53** | **20,443.61** | **79,113.93** |

**STATE: KY - KENTUCKY**

| REGISTRANT ZIP CODE 3 | 1ST QUARTER | 2ND QUARTER | 3RD QUARTER | 4TH QUARTER | **TOTAL GRAMS** |
|---|---|---|---|---|---|
| 400 | 1,452.99 | 1,414.75 | 1,337.24 | 1,347.52 | **5,552.49** |
| 401 | 695.18 | 615.67 | 614.54 | 502.42 | **2,427.81** |
| 402 | 4,566.09 | 4,029.95 | 3,806.28 | 4,076.33 | **16,478.65** |
| 403 | 1,534.08 | 1,892.84 | 1,721.34 | 1,649.18 | **6,797.44** |
| 404 | 968.23 | 1,076.87 | 970.96 | 938.41 | **3,954.47** |
| 405 | 953.07 | 951.08 | 924.84 | 763.35 | **3,592.33** |
| 406 | 215.67 | 160.33 | 85.56 | 67.88 | **529.43** |
| 407 | 1,295.48 | 1,378.45 | 1,282.40 | 1,204.04 | **5,160.38** |
| 408 | 690.93 | 861.94 | 877.41 | 908.35 | **3,338.63** |
| 409 | 1,700.67 | 1,624.42 | 1,828.33 | 1,867.72 | **7,021.14** |
| 410 | 2,830.26 | 2,728.07 | 2,729.15 | 2,379.99 | **10,667.48** |
| 411 | 888.12 | 860.72 | 897.92 | 939.03 | **3,585.80** |
| 412 | 1,715.04 | 1,403.41 | 1,442.75 | 1,421.09 | **5,982.30** |
| 413 | 1,281.64 | 1,287.16 | 1,191.09 | 1,183.03 | **4,942.91** |
| 414 | 379.92 | 323.34 | 371.96 | 336.60 | **1,411.81** |
| 415 | 2,045.45 | 2,329.63 | 1,973.33 | 2,267.56 | **8,615.96** |
| 416 | 1,091.97 | 771.32 | 722.70 | 755.85 | **3,341.85** |
| 417 | 638.89 | 570.21 | 745.91 | 590.10 | **2,545.10** |
| 418 | 689.55 | 642.03 | 561.37 | 560.72 | **2,453.67** |
| 420 | 1,764.76 | 1,654.17 | 1,633.72 | 1,738.05 | **6,790.70** |
| 421 | 2,008.08 | 1,995.32 | 1,946.32 | 2,130.62 | **8,080.34** |
| 422 | 1,325.77 | 1,154.78 | 1,185.52 | 1,251.10 | **4,917.17** |
| 423 | 1,602.55 | 1,509.05 | 1,409.76 | 1,568.00 | **6,089.36** |
| 424 | 1,201.31 | 1,237.20 | 975.43 | 1,064.74 | **4,478.67** |
| 425 | 536.82 | 552.53 | 489.54 | 619.93 | **2,198.81** |
| 426 | 1,002.28 | 1,158.09 | 860.83 | 965.81 | **3,987.02** |
| 427 | 2,069.70 | 2,279.27 | 2,231.37 | 2,199.58 | **8,779.92** |
| **KY - KENTUCKY - Total** | **37,144.49** | **36,462.58** | **34,817.59** | **35,296.99** | **143,721.65** |

**STATE: LA - LOUISIANA**

| REGISTRANT ZIP CODE 3 | 1ST QUARTER | 2ND QUARTER | 3RD QUARTER | 4TH QUARTER | TOTAL GRAMS |
|---|---|---|---|---|---|
| 700 | 3,646.69 | 3,554.61 | 3,400.48 | 3,443.68 | **14,045.45** |
| 701 | 1,970.42 | 1,722.75 | 1,625.12 | 1,750.17 | **7,068.46** |
| 703 | 1,577.09 | 1,685.45 | 1,607.04 | 1,500.21 | **6,369.80** |
| 704 | 2,853.76 | 2,801.44 | 2,759.86 | 2,633.01 | **11,048.08** |
| 705 | 7,679.32 | 8,334.26 | 8,344.12 | 7,861.10 | **32,218.81** |
| 706 | 2,895.35 | 2,811.87 | 2,764.21 | 2,534.61 | **11,006.05** |
| 707 | 1,472.86 | 1,417.35 | 1,626.34 | 1,118.89 | **5,635.44** |
| 708 | 1,522.66 | 1,284.28 | 1,361.46 | 1,400.38 | **5,568.79** |
| 710 | 1,175.14 | 1,205.74 | 1,096.20 | 1,240.96 | **4,718.04** |
| 711 | 2,424.60 | 2,095.49 | 2,416.18 | 2,309.80 | **9,246.07** |
| 712 | 3,217.33 | 3,370.85 | 3,056.23 | 2,887.14 | **12,531.55** |
| 713 | 2,506.18 | 2,411.45 | 2,527.95 | 2,275.19 | **9,720.76** |
| 714 | 1,616.97 | 1,633.26 | 1,654.44 | 1,668.09 | **6,572.77** |
| **LA - LOUISIANA - Total** | **34,558.38** | **34,328.82** | **34,239.63** | **32,623.23** | **135,750.06** |

**STATE: MA - MASSACHUSETTS**

| REGISTRANT ZIP CODE 3 | 1ST QUARTER | 2ND QUARTER | 3RD QUARTER | 4TH QUARTER | TOTAL GRAMS |
|---|---|---|---|---|---|
| 010 | 2,191.28 | 2,321.44 | 2,430.18 | 2,431.97 | **9,374.88** |
| 011 | 747.13 | 965.48 | 847.71 | 827.75 | **3,388.07** |
| 012 | 493.74 | 537.56 | 389.68 | 592.57 | **2,013.55** |
| 013 | 445.15 | 392.24 | 383.95 | 401.37 | **1,622.71** |
| 014 | 1,338.71 | 1,421.19 | 1,226.06 | 1,320.10 | **5,306.06** |
| 015 | 1,997.40 | 2,389.35 | 2,063.07 | 2,056.59 | **8,506.41** |
| 016 | 1,555.39 | 1,641.00 | 1,582.81 | 1,614.27 | **6,393.48** |
| 017 | 1,976.14 | 2,363.83 | 1,813.95 | 2,063.12 | **8,217.04** |
| 018 | 6,605.66 | 7,046.87 | 6,688.81 | 6,592.30 | **26,933.63** |
| 019 | 2,036.65 | 2,788.41 | 2,180.07 | 2,064.88 | **9,070.01** |
| 020 | 1,475.70 | 1,851.50 | 1,715.30 | 2,052.35 | **7,094.85** |
| 021 | 5,044.88 | 5,876.09 | 5,183.53 | 5,218.15 | **21,322.65** |
| 022 | 176.86 | 226.50 | 186.53 | 127.88 | **717.77** |
| 023 | 1,925.01 | 2,230.06 | 2,070.38 | 2,073.17 | **8,298.63** |
| 024 | 1,077.54 | 1,693.78 | 1,232.41 | 1,353.50 | **5,357.23** |
| 025 | 601.97 | 693.22 | 597.11 | 602.41 | **2,494.70** |
| 026 | 762.80 | 1,205.17 | 967.95 | 843.66 | **3,779.58** |
| 027 | 2,792.27 | 3,503.27 | 2,543.38 | 2,978.92 | **11,817.84** |
| **MA - MASSACHUSETTS - Total** | **33,244.30** | **39,146.96** | **34,102.87** | **35,214.96** | **141,709.08** |

**STATE: MD - MARYLAND**

| REGISTRANT ZIP CODE 3 | 1ST QUARTER | 2ND QUARTER | 3RD QUARTER | 4TH QUARTER | TOTAL GRAMS |
|---|---|---|---|---|---|
| 206 | 3,352.41 | 3,536.09 | 3,645.23 | 3,475.86 | **14,009.59** |
| 207 | 10,007.79 | 10,759.32 | 10,676.93 | 10,412.45 | **41,856.49** |

| | | | | | |
|---|---|---|---|---|---|
| 208 | 4,820.91 | 5,000.81 | 4,414.00 | 4,595.10 | **18,830.81** |
| 209 | 1,621.84 | 1,930.81 | 1,856.04 | 1,581.99 | **6,990.67** |
| 210 | 4,558.12 | 4,816.19 | 5,121.62 | 5,290.17 | **19,786.11** |
| 211 | 2,546.47 | 2,468.34 | 2,499.80 | 2,499.03 | **10,013.64** |
| 212 | 7,553.01 | 7,516.20 | 7,597.23 | 7,448.68 | **30,115.11** |
| 214 | 921.89 | 1,296.88 | 870.41 | 872.56 | **3,961.73** |
| 215 | 774.09 | 673.49 | 812.58 | 665.59 | **2,925.75** |
| 216 | 1,201.12 | 1,124.79 | 1,256.98 | 1,160.91 | **4,743.80** |
| 217 | 3,722.59 | 3,606.08 | 3,788.89 | 3,708.47 | **14,826.04** |
| 218 | 1,055.08 | 1,092.99 | 1,181.47 | 1,130.07 | **4,459.60** |
| 219 | 805.93 | 669.64 | 691.36 | 646.26 | **2,813.19** |
| **MD MARYLAND Total** | **42,941.25** | **44,491.61** | **44,412.52** | **43,487.14** | **175,332.52** |

**STATE: ME - MAINE**

| REGISTRANT ZIP CODE 3 | 1ST QUARTER | 2ND QUARTER | 3RD QUARTER | 4TH QUARTER | TOTAL GRAMS |
|---|---|---|---|---|---|
| 039 | 90.61 | 58.57 | 138.13 | 130.58 | **417.89** |
| 040 | 1,432.03 | 1,517.08 | 1,670.65 | 1,518.37 | **6,138.13** |
| 041 | 316.75 | 408.49 | 472.86 | 314.94 | **1,513.04** |
| 042 | 1,123.34 | 1,197.28 | 1,233.21 | 1,319.84 | **4,873.67** |
| 043 | 692.22 | 697.34 | 733.81 | 644.69 | **2,768.07** |
| 044 | 1,032.29 | 1,044.99 | 920.68 | 875.37 | **3,873.32** |
| 045 | 180.67 | 186.38 | 225.69 | 191.64 | **784.38** |
| 046 | 652.60 | 722.04 | 686.12 | 715.88 | **2,776.64** |
| 047 | 823.06 | 779.96 | 623.17 | 779.96 | **3,006.14** |
| 048 | 189.16 | 200.66 | 203.24 | 177.20 | **770.26** |
| 049 | 881.99 | 839.44 | 970.37 | 896.52 | **3,588.31** |
| **ME MAINE Total** | **7,414.71** | **7,652.23** | **7,877.92** | **7,564.99** | **30,509.85** |

**STATE: MI - MICHIGAN**

| REGISTRANT ZIP CODE 3 | 1ST QUARTER | 2ND QUARTER | 3RD QUARTER | 4TH QUARTER | TOTAL GRAMS |
|---|---|---|---|---|---|
| 480 | 23,103.01 | 23,361.53 | 23,914.22 | 22,353.19 | **92,731.95** |
| 481 | 36,463.79 | 35,192.59 | 37,817.47 | 34,464.85 | **143,938.71** |
| 482 | 33,069.91 | 33,013.68 | 30,799.17 | 32,447.00 | **129,329.77** |
| 483 | 12,335.81 | 12,000.19 | 12,668.59 | 11,580.68 | **48,585.27** |
| 484 | 5,858.93 | 6,016.84 | 6,200.94 | 6,001.59 | **24,078.31** |
| 485 | 5,543.84 | 5,831.99 | 5,990.19 | 5,815.68 | **23,181.70** |
| 486 | 8,696.06 | 9,385.40 | 9,164.45 | 8,431.55 | **35,677.46** |
| 487 | 4,611.74 | 4,855.86 | 5,229.91 | 4,572.79 | **19,270.30** |
| 488 | 7,582.83 | 8,008.44 | 9,248.57 | 7,531.30 | **32,371.15** |
| 489 | 2,460.99 | 2,277.38 | 2,574.38 | 1,958.88 | **9,271.63** |
| 490 | 7,159.92 | 7,366.19 | 8,477.25 | 6,934.10 | **29,937.46** |
| 491 | 537.60 | 628.96 | 702.99 | 499.48 | **2,369.03** |

| | | | | | |
|---|---|---|---|---|---|
| 492 | 3,114.20 | 3,397.07 | 3,088.94 | 3,118.04 | **12,718.25** |
| 493 | 1,637.87 | 1,826.92 | 2,071.59 | 1,475.59 | **7,011.97** |
| 494 | 5,125.85 | 5,014.41 | 5,783.45 | 4,835.86 | **20,759.56** |
| 495 | 3,099.60 | 2,946.10 | 3,346.30 | 2,657.00 | **12,049.01** |
| 496 | 2,471.33 | 2,339.67 | 2,403.54 | 2,140.73 | **9,355.27** |
| 497 | 2,884.96 | 2,744.96 | 3,072.22 | 2,519.22 | **11,221.36** |
| 498 | 1,660.49 | 1,875.23 | 2,053.42 | 1,943.36 | **7,532.50** |
| 499 | 871.51 | 971.07 | 1,054.97 | 952.53 | **3,850.08** |
| **MI - MICHIGAN Total** | **168,290.24** | **169,054.48** | **175,662.59** | **162,233.42** | **675,240.73** |

**STATE: MN - MINNESOTA**

| REGISTRANT ZIP CODE 3 | 1ST QUARTER | 2ND QUARTER | 3RD QUARTER | 4TH QUARTER | TOTAL GRAMS |
|---|---|---|---|---|---|
| 550 | 2,755.18 | 2,946.02 | 2,656.71 | 2,437.18 | **10,795.09** |
| 551 | 4,234.86 | 4,490.70 | 4,234.19 | 4,163.68 | **17,123.42** |
| 553 | 4,210.79 | 3,982.03 | 4,084.13 | 4,078.02 | **16,354.97** |
| 554 | 5,088.25 | 4,717.27 | 4,667.54 | 4,681.73 | **19,154.79** |
| 556 | 104.21 | 100.00 | 110.84 | 74.39 | **389.45** |
| 557 | 1,623.06 | 1,300.71 | 1,476.45 | 1,602.86 | **6,003.08** |
| 558 | 612.16 | 630.78 | 567.79 | 648.46 | **2,459.18** |
| 559 | 759.56 | 990.52 | 839.87 | 829.84 | **3,419.79** |
| 560 | 1,143.79 | 1,103.10 | 1,163.37 | 1,028.86 | **4,439.12** |
| 561 | 356.48 | 378.24 | 462.13 | 436.98 | **1,633.83** |
| 562 | 898.34 | 817.71 | 803.19 | 855.02 | **3,374.27** |
| 563 | 1,642.71 | 1,739.46 | 1,647.35 | 1,491.26 | **6,520.78** |
| 564 | 774.65 | 861.26 | 865.16 | 942.12 | **3,443.20** |
| 565 | 946.90 | 896.73 | 1,042.34 | 1,042.93 | **3,928.90** |
| 566 | 718.65 | 715.96 | 642.85 | 635.37 | **2,712.83** |
| 567 | 575.29 | 458.37 | 478.74 | 536.64 | **2,049.04** |
| **MN MINNESOTA Total** | **26,444.88** | **26,128.86** | **25,742.67** | **25,485.33** | **103,801.74** |

**STATE: MO - MISSOURI**

| REGISTRANT ZIP CODE 3 | 1ST QUARTER | 2ND QUARTER | 3RD QUARTER | 4TH QUARTER | TOTAL GRAMS |
|---|---|---|---|---|---|
| 630 | 5,524.21 | 6,080.78 | 5,466.34 | 5,978.49 | **23,049.82** |
| 631 | 16,421.29 | 17,479.68 | 19,133.29 | 20,134.43 | **73,168.69** |
| 633 | 2,827.24 | 2,927.60 | 2,890.69 | 2,792.24 | **11,437.77** |
| 634 | 484.01 | 448.33 | 525.07 | 373.86 | **1,831.27** |
| 635 | 632.27 | 521.58 | 589.12 | 548.10 | **2,291.08** |
| 636 | 1,537.67 | 1,597.12 | 1,549.90 | 1,653.78 | **6,338.48** |
| 637 | 699.17 | 888.70 | 770.85 | 915.30 | **3,274.01** |
| 638 | 1,936.41 | 2,132.75 | 2,046.64 | 2,001.43 | **8,117.23** |
| 639 | 980.18 | 922.72 | 971.43 | 850.89 | **3,725.21** |
| 640 | 4,700.01 | 4,507.08 | 4,285.36 | 4,433.85 | **17,926.31** |

| | | | | | |
|---|---|---|---|---|---|
| 641 | 4,196.61 | 3,938.73 | 3,172.92 | 3,349.33 | **14,657.59** |
| 644 | 587.17 | 501.98 | 616.12 | 564.63 | **2,269.91** |
| 645 | 524.43 | 555.18 | 519.44 | 523.72 | **2,122.76** |
| 646 | 415.50 | 490.64 | 540.22 | 405.55 | **1,851.92** |
| 647 | 1,439.95 | 1,217.31 | 1,271.90 | 1,039.40 | **4,968.55** |
| 648 | 1,218.04 | 1,231.03 | 1,247.78 | 1,287.76 | **4,984.61** |
| 650 | 952.94 | 814.50 | 948.50 | 847.94 | **3,563.88** |
| 651 | 561.29 | 471.60 | 568.18 | 534.47 | **2,135.53** |
| 652 | 1,543.74 | 1,490.70 | 1,482.45 | 1,532.52 | **6,049.42** |
| 653 | 901.19 | 957.69 | 847.94 | 736.13 | **3,442.95** |
| 654 | 605.01 | 550.30 | 646.01 | 497.27 | **2,298.60** |
| 655 | 1,032.94 | 1,026.81 | 1,147.22 | 1,002.51 | **4,209.49** |
| 656 | 1,277.62 | 1,148.81 | 1,159.49 | 1,232.98 | **4,818.89** |
| 657 | 2,780.10 | 2,757.01 | 2,322.56 | 2,210.22 | **10,069.89** |
| 658 | 1,723.11 | 1,887.49 | 2,066.03 | 1,945.77 | **7,622.40** |
| **MO MISSOURI - Total** | **55,502.10** | **56,546.13** | **56,785.45** | **57,392.58** | **226,226.25** |

**STATE: MS - MISSISSIPPI**

| REGISTRANT ZIP CODE 3 | 1ST QUARTER | 2ND QUARTER | 3RD QUARTER | 4TH QUARTER | TOTAL GRAMS |
|---|---|---|---|---|---|
| 386 | 2,237.26 | 2,363.42 | 1,899.07 | 1,814.95 | **8,314.70** |
| 387 | 1,288.49 | 1,225.68 | 1,102.99 | 996.76 | **4,613.92** |
| 388 | 2,087.71 | 1,954.73 | 1,932.49 | 1,928.88 | **7,903.82** |
| 389 | 974.65 | 875.20 | 1,019.96 | 872.99 | **3,742.80** |
| 390 | 1,711.08 | 1,602.22 | 1,458.64 | 1,443.07 | **6,215.01** |
| 391 | 1,767.48 | 1,924.37 | 1,797.10 | 1,569.81 | **7,058.75** |
| 392 | 1,092.65 | 1,163.58 | 1,054.42 | 1,054.71 | **4,365.36** |
| 393 | 1,889.08 | 1,689.99 | 1,687.73 | 1,535.90 | **6,802.70** |
| 394 | 2,405.11 | 2,369.11 | 2,156.12 | 2,014.52 | **8,944.87** |
| 395 | 2,209.43 | 1,914.00 | 1,865.31 | 1,746.86 | **7,735.60** |
| 396 | 995.83 | 866.54 | 878.96 | 953.66 | **3,694.99** |
| 397 | 1,710.06 | 1,560.64 | 1,393.64 | 1,487.57 | **6,151.91** |
| **MS MISSISSIPPI - Total** | **20,368.82** | **19,509.49** | **18,246.43** | **17,419.68** | **75,544.42** |

**STATE: MT - MONTANA**

| REGISTRANT ZIP CODE 3 | 1ST QUARTER | 2ND QUARTER | 3RD QUARTER | 4TH QUARTER | TOTAL GRAMS |
|---|---|---|---|---|---|
| 590 | 544.23 | 499.20 | 534.27 | 431.52 | **2,009.22** |
| 591 | 793.91 | 814.76 | 851.97 | 708.68 | **3,169.32** |
| 592 | 309.21 | 225.77 | 352.84 | 232.58 | **1,120.40** |
| 593 | 301.85 | 305.70 | 332.23 | 287.29 | **1,227.06** |
| 594 | 965.78 | 922.86 | 882.72 | 816.05 | **3,587.40** |
| 595 | 322.67 | 282.89 | 280.68 | 245.32 | **1,131.57** |
| 596 | 664.69 | 606.61 | 682.47 | 705.21 | **2,658.97** |

| | | | | | |
|---|---|---|---|---|---|
| 597 | 1,067.30 | 1,081.95 | 1,089.40 | 1,059.33 | **4,297.97** |
| 598 | 1,726.43 | 1,752.09 | 2,085.01 | 1,807.24 | **7,370.77** |
| 599 | 933.12 | 1,027.66 | 892.01 | 847.80 | **3,700.59** |
| **MT - MONTANA Total** | **7,629.17** | **7,519.49** | **7,983.60** | **7,141.01** | **30,273.27** |

**STATE: NC - NORTH CAROLINA**

| REGISTRANT ZIP CODE 3 | 1ST QUARTER | 2ND QUARTER | 3RD QUARTER | 4TH QUARTER | TOTAL GRAMS |
|---|---|---|---|---|---|
| 270 | 1,045.65 | 922.52 | 987.67 | 867.74 | **3,823.58** |
| 271 | 828.01 | 895.52 | 996.07 | 725.77 | **3,445.37** |
| 272 | 2,073.75 | 2,273.85 | 1,785.90 | 1,989.74 | **8,123.24** |
| 273 | 1,136.15 | 1,531.95 | 1,423.47 | 1,258.47 | **5,350.04** |
| 274 | 1,329.03 | 1,456.68 | 1,164.38 | 1,283.25 | **5,233.34** |
| 275 | 3,486.57 | 3,942.99 | 3,886.92 | 3,665.26 | **14,981.74** |
| 276 | 1,221.83 | 1,550.78 | 1,174.00 | 1,307.47 | **5,254.07** |
| 277 | 1,075.52 | 1,087.50 | 1,120.29 | 939.41 | **4,222.72** |
| 278 | 2,411.32 | 2,371.31 | 2,324.39 | 1,999.94 | **9,106.96** |
| 279 | 1,173.68 | 1,135.93 | 1,096.89 | 934.41 | **4,340.91** |
| 280 | 2,085.51 | 2,142.88 | 2,486.05 | 2,011.49 | **8,725.93** |
| 281 | 1,635.81 | 1,754.52 | 1,768.80 | 2,059.06 | **7,218.19** |
| 282 | 1,711.54 | 1,881.73 | 1,635.13 | 1,873.51 | **7,101.91** |
| 283 | 5,884.62 | 5,650.03 | 5,643.16 | 5,123.75 | **22,301.56** |
| 284 | 1,842.37 | 2,054.54 | 2,106.05 | 1,827.92 | **7,830.88** |
| 285 | 2,556.01 | 2,515.39 | 2,565.48 | 2,099.22 | **9,736.10** |
| 286 | 2,556.95 | 2,721.29 | 2,560.66 | 2,318.41 | **10,157.31** |
| 287 | 1,518.46 | 1,679.58 | 1,668.28 | 1,898.40 | **6,764.71** |
| 288 | 548.43 | 630.11 | 678.49 | 605.26 | **2,462.29** |
| 289 | 635.88 | 744.70 | 523.51 | 571.12 | **2,475.21** |
| **NC - NORTH CAROLINA Total** | **36,757.08** | **38,943.79** | **37,595.58** | **35,359.60** | **148,656.06** |

**STATE: ND - NORTH DAKOTA**

| REGISTRANT ZIP CODE 3 | 1ST QUARTER | 2ND QUARTER | 3RD QUARTER | 4TH QUARTER | TOTAL GRAMS |
|---|---|---|---|---|---|
| 580 | 259.49 | 302.21 | 345.69 | 184.73 | **1,092.12** |
| 581 | 618.95 | 649.10 | 544.84 | 504.05 | **2,316.94** |
| 582 | 532.21 | 713.72 | 459.69 | 515.84 | **2,221.45** |
| 583 | 329.30 | 330.41 | 243.11 | 192.28 | **1,095.10** |
| 584 | 212.35 | 203.11 | 225.43 | 182.07 | **822.96** |
| 585 | 809.65 | 709.76 | 761.58 | 715.46 | **2,996.45** |
| 586 | 190.61 | 244.67 | 188.40 | 216.59 | **840.28** |
| 587 | 392.84 | 354.81 | 318.44 | 451.41 | **1,517.49** |
| 588 | 702.58 | 698.38 | 804.43 | 775.54 | **2,980.93** |
| **ND NORTH DAKOTA Total** | **4,048.00** | **4,206.17** | **3,891.61** | **3,737.96** | **15,883.73** |

**STATE: NE - NEBRASKA**

| REGISTRANT ZIP CODE 3 | 1ST QUARTER | 2ND QUARTER | 3RD QUARTER | 4TH QUARTER | TOTAL GRAMS |
|---|---|---|---|---|---|
| 680 | 943.13 | 727.42 | 982.70 | 867.03 | **3,520.29** |
| 681 | 2,821.27 | 3,054.20 | 2,994.81 | 2,666.69 | **11,536.97** |
| 683 | 222.62 | 200.18 | 242.83 | 262.61 | **928.25** |
| 684 | 214.56 | 152.68 | 224.40 | 168.33 | **759.97** |
| 685 | 1,381.09 | 1,442.56 | 1,345.20 | 1,154.32 | **5,323.17** |
| 686 | 111.61 | 113.44 | 154.59 | 93.01 | **472.65** |
| 687 | 548.29 | 516.24 | 444.01 | 542.80 | **2,051.34** |
| 688 | 822.50 | 847.90 | 834.42 | 728.23 | **3,233.04** |
| 689 | 217.97 | 298.55 | 207.75 | 251.95 | **976.21** |
| 690 | 133.52 | 193.01 | 107.36 | 105.15 | **539.03** |
| 691 | 751.80 | 722.88 | 781.67 | 667.81 | **2,924.17** |
| 692 | 8.84 | 11.05 | 11.05 | 11.05 | **41.99** |
| 693 | 341.27 | 343.48 | 381.97 | 387.50 | **1,454.21** |
| **NE NEBRASKA Total** | **8,518.47** | **8,623.59** | **8,712.76** | **7,906.48** | **33,761.30** |

**STATE: NH - NEW HAMPSHIRE**

| REGISTRANT ZIP CODE 3 | 1ST QUARTER | 2ND QUARTER | 3RD QUARTER | 4TH QUARTER | TOTAL GRAMS |
|---|---|---|---|---|---|
| 030 | 1,669.36 | 1,700.82 | 1,426.93 | 1,621.72 | **6,418.83** |
| 031 | 1,173.22 | 1,251.56 | 1,040.20 | 1,129.83 | **4,594.80** |
| 032 | 725.14 | 865.93 | 615.36 | 686.45 | **2,892.88** |
| 033 | 502.86 | 436.11 | 362.80 | 396.69 | **1,698.45** |
| 034 | 319.86 | 328.37 | 299.82 | 327.07 | **1,275.12** |
| 035 | 211.42 | 197.97 | 311.88 | 262.44 | **983.72** |
| 036 | 22.10 | 30.94 | 49.73 | 24.31 | **127.08** |
| 037 | 422.21 | 322.48 | 437.69 | 323.49 | **1,505.87** |
| 038 | 1,073.16 | 1,212.77 | 906.20 | 1,187.50 | **4,379.63** |
| **NH - NEW HAMPSHIRE Total** | **6,119.31** | **6,346.95** | **5,450.60** | **5,959.51** | **23,876.38** |

**STATE: NJ - NEW JERSEY**

| REGISTRANT ZIP CODE 3 | 1ST QUARTER | 2ND QUARTER | 3RD QUARTER | 4TH QUARTER | TOTAL GRAMS |
|---|---|---|---|---|---|
| 070 | 9,648.25 | 9,587.02 | 9,822.32 | 9,530.42 | **38,588.01** |
| 071 | 2,307.49 | 1,989.50 | 2,366.33 | 2,039.50 | **8,702.82** |
| 072 | 795.34 | 1,044.79 | 796.90 | 957.42 | **3,594.45** |
| 073 | 2,146.40 | 1,930.71 | 1,789.05 | 1,817.39 | **7,683.55** |
| 074 | 3,709.30 | 3,657.64 | 3,588.41 | 3,269.39 | **14,224.73** |
| 075 | 1,247.38 | 1,275.66 | 1,141.04 | 1,165.91 | **4,829.99** |
| 076 | 3,054.15 | 3,508.98 | 3,028.89 | 2,995.21 | **12,587.22** |
| 077 | 4,829.85 | 4,753.33 | 4,992.01 | 4,627.81 | **19,202.99** |
| 078 | 2,265.21 | 2,251.27 | 2,354.75 | 2,368.23 | **9,239.47** |
| 079 | 1,053.05 | 1,033.64 | 1,130.52 | 1,009.34 | **4,226.56** |
| 080 | 10,221.88 | 9,905.22 | 10,290.82 | 9,979.27 | **40,397.20** |

| | | | | | |
|---|---|---|---|---|---|
| 081 | 1,500.66 | 1,352.57 | 1,556.14 | 1,581.04 | **5,990.41** |
| 082 | 1,962.22 | 2,057.27 | 1,887.02 | 1,877.83 | **7,784.35** |
| 083 | 1,830.05 | 1,855.16 | 1,536.41 | 1,799.35 | **7,020.97** |
| 084 | 280.77 | 387.13 | 292.93 | 427.65 | **1,388.49** |
| 085 | 1,428.84 | 1,447.94 | 1,515.57 | 1,617.96 | **6,010.31** |
| 086 | 1,756.24 | 1,600.02 | 1,559.04 | 1,652.44 | **6,567.73** |
| 087 | 3,687.99 | 3,515.24 | 3,682.11 | 3,812.77 | **14,698.11** |
| 088 | 5,610.76 | 5,536.18 | 5,405.31 | 5,884.77 | **22,437.03** |
| 089 | 513.02 | 547.18 | 453.26 | 436.61 | **1,950.07** |
| **NJ NEW JERSEY - Total** | **59,848.83** | **59,236.44** | **59,188.83** | **58,850.32** | **237,124.43** |

**STATE: NM - NEW MEXICO**

| REGISTRANT ZIP CODE 3 | 1ST QUARTER | 2ND QUARTER | 3RD QUARTER | 4TH QUARTER | **TOTAL GRAMS** |
|---|---|---|---|---|---|
| 870 | 1,317.11 | 1,340.47 | 1,078.07 | 1,357.94 | **5,093.58** |
| 871 | 4,540.19 | 4,100.15 | 3,894.72 | 3,866.56 | **16,401.62** |
| 873 | 1,436.23 | 1,308.69 | 969.17 | 1,235.19 | **4,949.28** |
| 874 | 570.75 | 523.33 | 619.93 | 523.96 | **2,237.97** |
| 875 | 880.63 | 684.82 | 745.13 | 726.21 | **3,036.80** |
| 877 | 351.05 | 386.76 | 294.41 | 294.97 | **1,327.19** |
| 878 | 106.08 | 33.15 | 36.91 | 59.67 | **235.82** |
| 879 | 130.40 | 155.25 | 76.43 | 99.45 | **461.53** |
| 880 | 2,139.97 | 2,130.64 | 2,441.36 | 2,391.70 | **9,103.67** |
| 881 | 739.37 | 910.56 | 794.53 | 873.65 | **3,318.11** |
| 882 | 1,936.68 | 2,057.79 | 2,041.21 | 2,076.79 | **8,112.47** |
| 883 | 685.66 | 862.11 | 730.39 | 743.13 | **3,021.29** |
| 884 | 119.35 | 123.77 | 71.83 | 137.03 | **451.97** |
| **NM NEW MEXICO Total** | **14,953.48** | **14,617.48** | **13,794.09** | **14,386.26** | **57,751.31** |

**STATE: NV - NEVADA**

| REGISTRANT ZIP CODE 3 | 1ST QUARTER | 2ND QUARTER | 3RD QUARTER | 4TH QUARTER | **TOTAL GRAMS** |
|---|---|---|---|---|---|
| 890 | 4,157.30 | 4,502.59 | 3,907.29 | 4,591.77 | **17,158.96** |
| 891 | 8,593.01 | 8,575.06 | 9,005.40 | 8,445.12 | **34,618.58** |
| 893 | 55.25 | 46.41 | 69.84 | 27.18 | **198.69** |
| 894 | 1,553.03 | 1,386.23 | 1,375.29 | 1,305.03 | **5,619.58** |
| 895 | 1,418.29 | 1,367.47 | 1,557.44 | 1,668.82 | **6,012.03** |
| 897 | 428.73 | 459.49 | 381.78 | 511.24 | **1,781.24** |
| 898 | 222.30 | 193.01 | 193.75 | 192.83 | **801.88** |
| **NV NEVADA Total** | **16,427.91** | **16,530.27** | **16,490.78** | **16,741.99** | **66,190.95** |

**STATE: NY - NEW YORK**

| REGISTRANT ZIP CODE 3 | 1ST QUARTER | 2ND QUARTER | 3RD QUARTER | 4TH QUARTER | **TOTAL GRAMS** |
|---|---|---|---|---|---|
| 100 | 9,918.34 | 8,744.60 | 10,011.46 | 9,079.02 | **37,753.43** |
| 101 | 593.95 | 540.04 | 393.82 | 405.74 | **1,933.54** |

| | | | | | |
|---|---|---|---|---|---|
| 102 | 46.59 | 35.26 | 14.35 | 13.26 | **109.47** |
| 103 | 2,372.88 | 2,402.25 | 2,263.29 | 2,391.30 | **9,429.72** |
| 104 | 6,862.10 | 6,018.62 | 5,805.84 | 5,994.13 | **24,680.69** |
| 105 | 2,689.05 | 2,552.68 | 2,535.68 | 2,661.29 | **10,438.70** |
| 106 | 481.80 | 490.82 | 475.35 | 589.08 | **2,037.06** |
| 107 | 1,078.10 | 1,222.97 | 1,161.59 | 979.79 | **4,442.45** |
| 108 | 336.20 | 350.18 | 372.01 | 360.31 | **1,418.71** |
| 109 | 3,366.14 | 3,215.92 | 3,113.63 | 3,569.74 | **13,265.43** |
| 110 | 1,072.60 | 802.03 | 993.61 | 784.13 | **3,652.37** |
| 111 | 1,109.37 | 1,175.58 | 885.51 | 1,075.38 | **4,245.84** |
| 112 | 12,474.38 | 11,161.42 | 12,605.08 | 11,553.38 | **47,794.26** |
| 113 | 5,126.02 | 4,248.77 | 4,470.43 | 4,613.24 | **18,458.46** |
| 114 | 2,408.74 | 2,000.88 | 2,096.10 | 2,089.82 | **8,595.53** |
| 115 | 3,232.94 | 2,800.24 | 2,946.40 | 3,062.07 | **12,041.65** |
| 116 | 746.46 | 600.50 | 706.50 | 582.54 | **2,636.00** |
| 117 | 6,277.64 | 6,425.81 | 6,373.08 | 6,916.06 | **25,992.60** |
| 118 | 274.91 | 431.92 | 368.79 | 495.83 | **1,571.45** |
| 119 | 815.22 | 746.78 | 907.64 | 1,004.59 | **3,474.22** |
| 120 | 1,346.56 | 1,490.53 | 1,403.00 | 1,355.22 | **5,595.31** |
| 121 | 595.61 | 744.43 | 788.16 | 574.25 | **2,702.45** |
| 122 | 438.73 | 499.45 | 502.59 | 559.99 | **2,000.76** |
| 123 | 550.68 | 606.95 | 416.32 | 542.20 | **2,116.15** |
| 124 | 939.28 | 696.25 | 856.22 | 868.28 | **3,360.03** |
| 125 | 2,204.25 | 2,249.52 | 2,266.52 | 2,421.91 | **9,142.20** |
| 126 | 300.85 | 342.01 | 335.19 | 345.87 | **1,323.92** |
| 127 | 1,289.59 | 1,229.00 | 1,259.76 | 1,268.78 | **5,047.13** |
| 128 | 1,223.24 | 1,265.49 | 1,425.00 | 1,228.39 | **5,142.12** |
| 129 | 1,027.01 | 1,166.77 | 1,096.79 | 1,136.91 | **4,427.49** |
| 130 | 2,588.13 | 2,617.31 | 2,791.13 | 2,509.75 | **10,506.33** |
| 131 | 698.46 | 839.15 | 800.77 | 909.43 | **3,247.81** |
| 132 | 1,279.78 | 1,214.75 | 1,041.23 | 1,235.82 | **4,771.59** |
| 133 | 693.67 | 710.51 | 667.15 | 793.01 | **2,864.35** |
| 134 | 816.53 | 810.63 | 697.99 | 824.97 | **3,150.11** |
| 135 | 407.39 | 321.93 | 360.42 | 404.25 | **1,493.99** |
| 136 | 3,397.60 | 3,592.82 | 3,757.98 | 3,725.97 | **14,474.37** |
| 137 | 641.48 | 672.68 | 558.69 | 578.67 | **2,451.51** |
| 138 | 940.66 | 1,010.82 | 828.50 | 986.60 | **3,766.58** |
| 139 | 680.43 | 660.25 | 656.93 | 625.82 | **2,623.43** |
| 140 | 4,030.72 | 4,434.14 | 4,226.42 | 4,421.08 | **17,112.36** |
| 141 | 1,734.83 | 1,942.81 | 1,847.02 | 2,041.43 | **7,566.09** |

| | | | | | |
|---|---|---|---|---|---|
| 142 | 7,918.27 | 8,148.31 | 7,637.50 | 8,244.81 | **31,948.90** |
| 143 | 593.22 | 520.27 | 477.93 | 602.80 | **2,194.22** |
| 144 | 2,056.49 | 1,815.18 | 1,956.16 | 1,968.15 | **7,795.98** |
| 145 | 1,583.85 | 1,798.31 | 1,717.81 | 1,423.72 | **6,523.69** |
| 146 | 3,109.30 | 3,234.54 | 3,093.40 | 2,993.75 | **12,430.99** |
| 147 | 881.26 | 1,020.12 | 985.77 | 989.54 | **3,876.69** |
| 148 | 1,414.97 | 1,464.13 | 1,599.59 | 1,415.34 | **5,894.03** |
| 149 | 332.06 | 186.75 | 307.20 | 170.36 | **996.38** |
| **NY - NEW YORK - Total** | **106,998.32** | **103,273.09** | **104,859.32** | **105,387.82** | **420,518.55** |

**STATE: OH - OHIO**

| REGISTRANT ZIP CODE 3 | 1ST QUARTER | 2ND QUARTER | 3RD QUARTER | 4TH QUARTER | **TOTAL GRAMS** |
|---|---|---|---|---|---|
| 430 | 2,890.71 | 3,165.24 | 3,045.52 | 3,144.73 | **12,246.19** |
| 431 | 2,044.60 | 2,085.57 | 2,162.36 | 2,392.97 | **8,685.50** |
| 432 | 4,203.76 | 3,660.50 | 4,007.19 | 3,527.40 | **15,398.85** |
| 433 | 647.96 | 666.03 | 535.48 | 600.37 | **2,449.85** |
| 434 | 924.44 | 814.60 | 1,038.16 | 826.93 | **3,604.13** |
| 435 | 1,872.02 | 1,883.10 | 1,750.74 | 1,765.66 | **7,271.52** |
| 436 | 3,951.86 | 4,114.47 | 3,950.79 | 3,742.19 | **15,759.31** |
| 437 | 1,113.46 | 1,062.54 | 1,130.57 | 1,187.76 | **4,494.34** |
| 438 | 272.39 | 233.62 | 205.72 | 200.74 | **912.47** |
| 439 | 1,695.58 | 1,677.64 | 1,578.92 | 1,703.05 | **6,655.19** |
| 440 | 3,958.91 | 3,542.89 | 3,496.76 | 3,844.99 | **14,843.54** |
| 441 | 7,287.93 | 7,447.85 | 7,358.90 | 7,447.70 | **29,542.38** |
| 442 | 1,698.32 | 1,936.78 | 1,780.91 | 1,635.01 | **7,051.02** |
| 443 | 823.39 | 981.51 | 799.98 | 860.93 | **3,465.81** |
| 444 | 1,784.21 | 1,770.02 | 1,844.30 | 1,806.99 | **7,205.53** |
| 445 | 1,041.57 | 1,273.28 | 1,170.81 | 1,071.18 | **4,556.84** |
| 446 | 1,708.55 | 1,481.68 | 1,629.91 | 1,584.81 | **6,404.95** |
| 447 | 1,412.02 | 1,337.24 | 1,326.40 | 1,689.86 | **5,765.52** |
| 448 | 1,461.23 | 1,514.34 | 1,399.53 | 1,495.29 | **5,870.38** |
| 449 | 645.53 | 709.16 | 567.25 | 653.27 | **2,575.21** |
| 450 | 2,850.85 | 2,593.38 | 2,901.50 | 2,910.41 | **11,256.14** |
| 451 | 1,236.52 | 1,177.59 | 1,201.44 | 1,233.95 | **4,849.49** |
| 452 | 4,059.48 | 3,793.29 | 4,241.64 | 3,653.44 | **15,747.85** |
| 453 | 2,615.15 | 2,444.41 | 2,457.12 | 2,608.70 | **10,125.38** |
| 454 | 3,178.31 | 3,202.22 | 2,865.45 | 3,125.92 | **12,371.90** |
| 455 | 513.08 | 453.51 | 472.28 | 551.59 | **1,990.46** |
| 456 | 2,379.62 | 2,448.14 | 2,153.32 | 2,291.47 | **9,272.54** |
| 457 | 1,027.40 | 926.84 | 1,181.29 | 1,065.98 | **4,201.51** |
| 458 | 2,005.05 | 1,937.37 | 2,086.20 | 1,849.26 | **7,877.88** |

| OH - OHIO Total | 61,303.89 | 60,334.80 | 60,340.45 | 60,472.55 | 242,451.69 |
|---|---|---|---|---|---|

STATE: OK - OKLAHOMA

| REGISTRANT ZIP CODE 3 | 1ST QUARTER | 2ND QUARTER | 3RD QUARTER | 4TH QUARTER | TOTAL GRAMS |
|---|---|---|---|---|---|
| 730 | 10,587.41 | 10,207.77 | 9,954.33 | 9,355.52 | **40,105.03** |
| 731 | 8,398.76 | 8,337.10 | 7,735.40 | 7,993.96 | **32,465.22** |
| 734 | 2,996.90 | 3,344.06 | 3,246.82 | 3,192.78 | **12,780.55** |
| 735 | 6,618.42 | 6,740.22 | 6,154.05 | 6,392.64 | **25,905.33** |
| 736 | 1,650.94 | 1,432.14 | 1,620.00 | 1,460.88 | **6,163.97** |
| 737 | 1,664.57 | 1,638.05 | 1,770.58 | 1,656.65 | **6,729.85** |
| 738 | 531.14 | 425.07 | 457.22 | 422.22 | **1,835.64** |
| 739 | 398.00 | 347.17 | 434.47 | 335.94 | **1,515.57** |
| 740 | 8,458.62 | 9,347.93 | 9,241.13 | 9,439.83 | **36,487.50** |
| 741 | 4,087.66 | 4,418.64 | 4,568.76 | 4,415.40 | **17,490.46** |
| 743 | 3,237.71 | 3,244.85 | 3,364.53 | 3,377.76 | **13,224.85** |
| 744 | 5,211.78 | 5,801.42 | 5,211.20 | 5,184.74 | **21,409.13** |
| 745 | 1,804.55 | 1,772.50 | 1,918.37 | 1,907.32 | **7,402.73** |
| 746 | 1,606.74 | 1,710.62 | 1,523.82 | 1,668.63 | **6,509.81** |
| 747 | 5,100.38 | 5,101.48 | 4,823.54 | 5,028.26 | **20,053.67** |
| 748 | 7,040.89 | 7,291.93 | 7,166.69 | 7,070.12 | **28,569.63** |
| 749 | 2,362.60 | 2,541.62 | 2,315.17 | 2,226.96 | **9,446.34** |
| **OK - OKLAHOMA Total** | **71,757.05** | **73,702.57** | **71,506.08** | **71,129.59** | **288,095.29** |

STATE: OR - OREGON

| REGISTRANT ZIP CODE 3 | 1ST QUARTER | 2ND QUARTER | 3RD QUARTER | 4TH QUARTER | TOTAL GRAMS |
|---|---|---|---|---|---|
| 970 | 4,671.91 | 4,787.24 | 4,856.99 | 4,218.28 | **18,534.41** |
| 971 | 1,986.50 | 1,936.25 | 1,797.47 | 1,687.55 | **7,407.77** |
| 972 | 6,079.08 | 6,538.30 | 5,962.20 | 5,735.94 | **24,315.52** |
| 973 | 3,598.23 | 3,683.09 | 3,820.47 | 3,330.57 | **14,432.37** |
| 974 | 4,034.98 | 3,963.84 | 3,987.04 | 3,720.75 | **15,706.62** |
| 975 | 2,523.07 | 2,607.73 | 2,488.75 | 2,638.43 | **10,257.98** |
| 976 | 377.93 | 396.40 | 448.76 | 425.97 | **1,649.05** |
| 977 | 1,374.13 | 1,310.71 | 1,362.51 | 1,267.01 | **5,314.36** |
| 978 | 1,314.92 | 1,192.44 | 1,092.52 | 1,208.60 | **4,808.48** |
| 979 | 238.30 | 249.08 | 286.39 | 332.18 | **1,105.95** |
| **OR OREGON Total** | **26,199.06** | **26,665.08** | **26,103.10** | **24,565.28** | **103,532.51** |

STATE: PA - PENNSYLVANIA

| REGISTRANT ZIP CODE 3 | 1ST QUARTER | 2ND QUARTER | 3RD QUARTER | 4TH QUARTER | TOTAL GRAMS |
|---|---|---|---|---|---|
| 150 | 2,368.18 | 2,626.56 | 2,211.56 | 2,660.17 | **9,866.48** |
| 151 | 2,299.20 | 2,241.49 | 2,213.50 | 2,562.20 | **9,316.39** |
| 152 | 4,334.85 | 3,831.19 | 3,691.57 | 4,288.57 | **16,146.18** |
| 153 | 1,011.21 | 1,333.70 | 894.73 | 1,153.39 | **4,393.03** |

| | | | | | |
|---|---|---|---|---|---|
| 154 | 695.52 | 776.86 | 741.15 | 832.27 | **3,045.80** |
| 155 | 2,302.09 | 2,084.70 | 2,376.15 | 2,204.54 | **8,967.48** |
| 156 | 1,411.45 | 1,591.13 | 1,581.61 | 1,504.91 | **6,089.10** |
| 157 | 1,261.40 | 860.60 | 1,371.05 | 1,155.20 | **4,648.26** |
| 158 | 683.47 | 566.70 | 609.31 | 641.66 | **2,501.13** |
| 159 | 1,003.84 | 1,024.46 | 1,094.26 | 1,033.76 | **4,156.32** |
| 160 | 1,192.92 | 1,348.69 | 1,302.22 | 1,278.41 | **5,122.24** |
| 161 | 1,231.70 | 1,332.39 | 1,156.40 | 1,159.21 | **4,879.70** |
| 162 | 748.51 | 493.52 | 589.68 | 513.33 | **2,345.04** |
| 163 | 1,043.20 | 1,021.70 | 1,028.96 | 976.20 | **4,070.06** |
| 164 | 440.06 | 509.31 | 515.81 | 462.20 | **1,927.38** |
| 165 | 1,179.14 | 1,120.67 | 1,214.70 | 1,096.73 | **4,611.24** |
| 166 | 1,222.39 | 1,072.15 | 997.56 | 966.75 | **4,258.85** |
| 167 | 353.62 | 423.23 | 429.86 | 328.20 | **1,534.91** |
| 168 | 539.55 | 442.65 | 634.99 | 444.14 | **2,061.33** |
| 169 | 378.47 | 292.83 | 320.45 | 352.32 | **1,344.07** |
| 170 | 3,030.88 | 3,093.79 | 3,110.19 | 3,306.23 | **12,541.09** |
| 171 | 762.25 | 693.83 | 818.88 | 750.60 | **3,025.57** |
| 172 | 970.85 | 900.32 | 807.13 | 893.69 | **3,571.98** |
| 173 | 2,096.06 | 1,788.04 | 1,907.97 | 1,744.33 | **7,536.40** |
| 174 | 1,178.69 | 907.06 | 796.53 | 919.18 | **3,801.46** |
| 175 | 1,147.76 | 1,100.33 | 1,017.04 | 1,306.89 | **4,572.01** |
| 176 | 949.08 | 1,002.83 | 715.02 | 1,027.75 | **3,694.67** |
| 177 | 707.66 | 894.33 | 836.51 | 668.61 | **3,107.11** |
| 178 | 1,099.42 | 1,249.62 | 1,100.20 | 957.68 | **4,406.91** |
| 179 | 1,086.73 | 1,046.69 | 1,096.85 | 1,016.97 | **4,247.24** |
| 180 | 2,262.27 | 2,342.44 | 2,021.58 | 2,443.03 | **9,069.32** |
| 181 | 1,119.38 | 1,281.08 | 1,216.79 | 921.12 | **4,538.37** |
| 182 | 727.83 | 799.03 | 676.10 | 647.00 | **2,849.95** |
| 183 | 1,669.28 | 1,516.55 | 1,382.60 | 1,579.73 | **6,148.16** |
| 184 | 1,453.47 | 1,541.30 | 1,500.06 | 1,407.47 | **5,902.30** |
| 185 | 1,329.03 | 1,155.18 | 1,257.64 | 1,131.49 | **4,873.34** |
| 186 | 1,580.74 | 1,490.79 | 1,582.02 | 1,571.82 | **6,225.37** |
| 187 | 3,030.43 | 3,283.70 | 2,839.60 | 3,728.24 | **12,881.97** |
| 188 | 918.25 | 941.62 | 1,030.39 | 909.19 | **3,799.45** |
| 189 | 2,982.48 | 3,422.24 | 3,517.10 | 3,809.41 | **13,731.24** |
| 190 | 10,444.77 | 10,250.90 | 10,497.60 | 10,706.95 | **41,900.21** |
| 191 | 17,801.68 | 15,963.18 | 15,993.67 | 15,274.44 | **65,032.97** |
| 193 | 2,319.59 | 2,482.87 | 2,513.77 | 2,544.71 | **9,860.94** |
| 194 | 4,873.57 | 4,042.70 | 4,151.33 | 3,834.08 | **16,901.68** |

| | | | | | |
|---|---|---|---|---|---|
| 195 | 1,143.86 | 1,153.27 | 1,187.50 | 1,101.61 | **4,586.24** |
| 196 | 1,626.47 | 1,706.69 | 1,623.74 | 1,640.06 | **6,596.96** |
| **PA - PENNSYLVANIA - Total** | **94,013.24** | **91,044.89** | **90,173.31** | **91,456.44** | **366,687.88** |

**STATE: PR - PUERTO RICO**

| REGISTRANT ZIP CODE 3 | 1ST QUARTER | 2ND QUARTER | 3RD QUARTER | 4TH QUARTER | TOTAL GRAMS |
|---|---|---|---|---|---|
| 006 | 4,056.64 | 3,745.01 | 3,172.60 | 3,727.33 | **14,701.59** |
| 007 | 3,405.76 | 3,290.84 | 2,854.34 | 3,428.97 | **12,979.92** |
| 009 | 2,505.15 | 2,533.88 | 2,205.92 | 2,299.61 | **9,544.56** |
| **PR - PUERTO RICO - Total** | **9,967.55** | **9,569.73** | **8,232.87** | **9,455.91** | **37,226.06** |

**STATE: RI - RHODE ISLAND**

| REGISTRANT ZIP CODE 3 | 1ST QUARTER | 2ND QUARTER | 3RD QUARTER | 4TH QUARTER | TOTAL GRAMS |
|---|---|---|---|---|---|
| 028 | 2,932.43 | 3,201.09 | 2,876.90 | 3,010.37 | **12,020.78** |
| 029 | 1,544.82 | 1,601.56 | 1,573.76 | 1,590.79 | **6,310.94** |
| **RI RHODE ISLAND Total** | **4,477.25** | **4,802.65** | **4,450.66** | **4,601.16** | **18,331.72** |

**STATE: SC - SOUTH CAROLINA**

| REGISTRANT ZIP CODE 3 | 1ST QUARTER | 2ND QUARTER | 3RD QUARTER | 4TH QUARTER | TOTAL GRAMS |
|---|---|---|---|---|---|
| 290 | 2,301.96 | 2,286.30 | 2,164.05 | 2,197.53 | **8,949.84** |
| 291 | 2,403.22 | 2,655.80 | 2,429.44 | 2,231.90 | **9,720.36** |
| 292 | 2,833.60 | 2,671.57 | 3,028.33 | 2,473.82 | **11,007.32** |
| 293 | 3,131.66 | 2,952.99 | 2,966.50 | 2,915.99 | **11,967.14** |
| 294 | 30,540.30 | 31,391.54 | 30,740.26 | 28,372.64 | **121,044.75** |
| 295 | 7,977.37 | 8,292.06 | 7,656.28 | 7,739.51 | **31,665.23** |
| 296 | 5,400.82 | 5,527.39 | 5,680.94 | 4,934.75 | **21,543.90** |
| 297 | 1,287.18 | 1,418.67 | 1,359.16 | 1,419.52 | **5,484.53** |
| 298 | 1,658.63 | 1,856.12 | 1,914.75 | 1,767.08 | **7,196.58** |
| 299 | 1,144.01 | 969.58 | 909.32 | 1,231.03 | **4,253.94** |
| **SC - SOUTH CAROLINA Total** | **58,678.76** | **60,022.03** | **58,849.03** | **55,283.77** | **232,833.58** |

**STATE: SD - SOUTH DAKOTA**

| REGISTRANT ZIP CODE 3 | 1ST QUARTER | 2ND QUARTER | 3RD QUARTER | 4TH QUARTER | TOTAL GRAMS |
|---|---|---|---|---|---|
| 570 | 845.77 | 778.23 | 743.29 | 750.99 | **3,118.28** |
| 571 | 1,242.94 | 1,246.54 | 1,279.88 | 1,316.27 | **5,085.62** |
| 572 | 249.74 | 288.42 | 279.58 | 330.63 | **1,148.37** |
| 573 | 466.19 | 433.78 | 517.71 | 381.97 | **1,799.65** |
| 574 | 480.68 | 440.54 | 504.90 | 553.98 | **1,980.11** |
| 575 | 197.80 | 172.39 | 190.07 | 174.60 | **734.86** |
| 576 | 194.49 | 170.18 | 179.02 | 154.71 | **698.39** |
| 577 | 1,110.11 | 1,162.53 | 1,316.55 | 1,184.75 | **4,773.93** |
| **SD SOUTH DAKOTA - Total** | **4,787.72** | **4,692.60** | **5,010.99** | **4,847.90** | **19,339.21** |

**STATE: TN - TENNESSEE**

| REGISTRANT ZIP CODE 3 | 1ST QUARTER | 2ND QUARTER | 3RD QUARTER | 4TH QUARTER | TOTAL GRAMS |
|---|---|---|---|---|---|

| | | | | | |
|---|---|---|---|---|---|
| 370 | 3,528.33 | 3,889.12 | 3,896.83 | 3,583.50 | **14,897.78** |
| 371 | 29,419.60 | 21,655.45 | 28,075.46 | 21,961.61 | **101,112.11** |
| 372 | 1,796.88 | 1,748.72 | 1,848.59 | 1,581.13 | **6,975.33** |
| 373 | 2,806.33 | 2,635.09 | 2,573.33 | 2,914.20 | **10,928.95** |
| 374 | 1,046.75 | 1,153.39 | 1,228.91 | 1,333.87 | **4,762.92** |
| 376 | 1,827.72 | 2,093.30 | 1,898.74 | 1,676.02 | **7,495.78** |
| 377 | 1,800.32 | 2,265.21 | 1,940.79 | 2,067.88 | **8,074.21** |
| 378 | 2,748.60 | 3,003.77 | 3,018.43 | 3,108.21 | **11,879.00** |
| 379 | 1,792.65 | 2,031.36 | 1,793.37 | 1,734.36 | **7,351.73** |
| 380 | 1,475.12 | 1,438.60 | 1,377.40 | 1,333.06 | **5,624.18** |
| 381 | 3,254.43 | 3,410.84 | 3,152.22 | 4,581.55 | **14,399.03** |
| 382 | 676.47 | 654.19 | 557.72 | 629.88 | **2,518.26** |
| 383 | 2,914.63 | 3,194.70 | 2,845.45 | 3,017.66 | **11,972.45** |
| 384 | 893.99 | 951.91 | 773.94 | 699.68 | **3,319.51** |
| 385 | 1,886.68 | 1,962.21 | 1,757.64 | 1,631.82 | **7,238.36** |
| **TN TENNESSEE Total** | **57,868.48** | **52,087.86** | **56,738.81** | **51,854.44** | **218,549.59** |

**STATE: TX - TEXAS**

| REGISTRANT ZIP CODE 3 | 1ST QUARTER | 2ND QUARTER | 3RD QUARTER | 4TH QUARTER | **TOTAL GRAMS** |
|---|---|---|---|---|---|
| 750 | 55,536.08 | 58,801.47 | 57,111.49 | 56,619.27 | **228,068.32** |
| 751 | 27,271.85 | 27,584.94 | 27,869.34 | 26,775.31 | **109,501.43** |
| 752 | 30,271.39 | 30,905.34 | 30,345.01 | 28,391.92 | **119,913.66** |
| 753 | 603.36 | 560.90 | 570.21 | 594.52 | **2,328.98** |
| 754 | 17,787.25 | 17,992.45 | 17,494.91 | 17,514.54 | **70,789.14** |
| 755 | 7,417.35 | 7,523.39 | 7,743.74 | 6,708.30 | **29,392.79** |
| 756 | 22,932.43 | 23,130.53 | 24,179.01 | 23,071.42 | **93,313.39** |
| 757 | 25,546.99 | 25,297.68 | 26,525.48 | 24,840.86 | **102,211.00** |
| 758 | 4,456.89 | 4,109.98 | 4,101.14 | 3,933.09 | **16,601.09** |
| 759 | 16,922.77 | 18,716.26 | 18,502.41 | 17,825.47 | **71,966.90** |
| 760 | 39,314.18 | 41,372.10 | 41,553.98 | 40,233.78 | **162,474.04** |
| 761 | 30,288.10 | 31,341.23 | 30,960.14 | 29,818.74 | **122,408.22** |
| 762 | 14,556.97 | 14,912.80 | 15,506.34 | 14,391.00 | **59,367.11** |
| 763 | 10,372.17 | 10,714.37 | 10,943.15 | 9,650.13 | **41,679.82** |
| 764 | 6,314.26 | 7,102.16 | 6,245.08 | 6,189.39 | **25,850.88** |
| 765 | 12,223.58 | 11,937.28 | 11,519.29 | 11,277.71 | **46,957.87** |
| 766 | 4,291.56 | 4,814.88 | 4,723.06 | 4,711.62 | **18,541.12** |
| 767 | 7,629.75 | 7,414.89 | 7,342.36 | 7,130.88 | **29,517.87** |
| 768 | 5,885.50 | 5,820.30 | 5,746.26 | 5,231.59 | **22,683.65** |
| 769 | 5,083.41 | 5,041.24 | 4,609.83 | 4,130.20 | **18,864.67** |
| 770 | 66,189.74 | 69,900.75 | 67,228.19 | 66,593.88 | **269,912.57** |
| 772 | 2.21 | 4.42 | 0 | 4.42 | **11.05** |

| | | | | | |
|---|---|---|---|---|---|
| 773 | 37,009.88 | 38,292.75 | 37,609.66 | 37,476.20 | **150,388.49** |
| 774 | 27,912.85 | 28,922.42 | 28,821.86 | 28,887.53 | **114,544.66** |
| 775 | 36,672.29 | 37,295.23 | 36,862.05 | 35,424.60 | **146,254.17** |
| 776 | 14,682.53 | 14,903.88 | 13,883.29 | 13,499.37 | **56,969.06** |
| 777 | 8,720.57 | 9,231.35 | 8,854.32 | 8,660.09 | **35,466.34** |
| 778 | 9,921.48 | 10,318.60 | 9,878.33 | 10,485.94 | **40,604.35** |
| 779 | 5,060.66 | 5,266.85 | 5,307.53 | 4,826.04 | **20,461.08** |
| 780 | 12,605.31 | 14,067.69 | 13,669.06 | 13,049.41 | **53,391.47** |
| 781 | 11,687.52 | 12,020.51 | 12,426.59 | 12,002.49 | **48,137.11** |
| 782 | 61,595.78 | 61,195.25 | 60,632.08 | 57,367.55 | **240,790.66** |
| 783 | 12,394.61 | 11,300.79 | 11,733.42 | 11,686.79 | **47,115.60** |
| 784 | 15,516.42 | 15,149.12 | 15,521.97 | 15,099.10 | **61,286.61** |
| 785 | 24,728.87 | 25,702.15 | 25,026.19 | 25,328.36 | **100,785.57** |
| 786 | 17,484.47 | 18,778.04 | 19,240.67 | 18,766.29 | **74,269.47** |
| 787 | 14,444.16 | 15,327.90 | 16,039.44 | 14,911.49 | **60,722.98** |
| 788 | 4,194.77 | 4,169.35 | 3,855.70 | 3,748.69 | **15,968.52** |
| 789 | 1,655.36 | 1,654.44 | 1,726.71 | 1,560.69 | **6,597.21** |
| 790 | 7,594.81 | 8,273.02 | 7,849.74 | 7,774.39 | **31,491.97** |
| 791 | 13,423.56 | 14,156.83 | 14,052.79 | 13,214.30 | **54,847.48** |
| 792 | 1,294.92 | 1,322.00 | 1,101.72 | 1,000.06 | **4,718.71** |
| 793 | 6,380.56 | 6,046.02 | 5,020.91 | 5,256.68 | **22,704.16** |
| 794 | 12,662.58 | 14,181.44 | 13,208.98 | 13,244.77 | **53,297.77** |
| 795 | 2,865.94 | 3,066.51 | 3,037.78 | 2,873.70 | **11,843.94** |
| 796 | 9,175.74 | 9,142.91 | 9,392.18 | 9,135.90 | **36,846.73** |
| 797 | 19,851.78 | 21,062.09 | 20,268.92 | 18,837.75 | **80,020.53** |
| 798 | 1,164.72 | 1,218.87 | 1,112.79 | 1,222.19 | **4,718.56** |
| 799 | 21,597.18 | 22,336.70 | 22,993.81 | 19,859.40 | **86,787.09** |
| **TX - TEXAS Total** | **823,197.09** | **849,402.09** | **839,948.89** | **810,837.80** | **3,323,385.87** |

**STATE: UT - UTAH**

| REGISTRANT ZIP CODE 3 | 1ST QUARTER | 2ND QUARTER | 3RD QUARTER | 4TH QUARTER | TOTAL GRAMS |
|---|---|---|---|---|---|
| 840 | 5,418.39 | 5,864.98 | 5,547.79 | 5,610.12 | **22,441.28** |
| 841 | 2,701.11 | 2,517.33 | 2,647.72 | 2,358.06 | **10,224.22** |
| 843 | 557.84 | 666.89 | 573.33 | 541.46 | **2,339.52** |
| 844 | 694.18 | 687.12 | 585.64 | 674.44 | **2,641.38** |
| 845 | 346.99 | 382.53 | 331.52 | 358.04 | **1,419.07** |
| 846 | 1,692.52 | 1,736.03 | 1,694.14 | 1,638.79 | **6,761.48** |
| 847 | 1,205.93 | 1,317.78 | 1,126.45 | 1,244.66 | **4,894.83** |
| **UT - UTAH - Total** | **12,616.96** | **13,172.67** | **12,506.58** | **12,425.58** | **50,721.79** |

**STATE: VA - VIRGINIA**

| REGISTRANT ZIP CODE 3 | 1ST QUARTER | 2ND QUARTER | 3RD QUARTER | 4TH QUARTER | TOTAL GRAMS |
|---|---|---|---|---|---|

| | | | | | |
|---|---|---|---|---|---|
| 201 | 4,475.57 | 4,567.95 | 4,130.13 | 4,748.51 | **17,922.16** |
| 220 | 2,722.93 | 2,708.15 | 2,821.76 | 2,470.25 | **10,723.09** |
| 221 | 2,277.98 | 2,595.72 | 2,397.47 | 2,310.18 | **9,581.35** |
| 222 | 752.13 | 952.55 | 963.19 | 837.68 | **3,505.54** |
| 223 | 1,479.70 | 1,517.15 | 1,415.28 | 1,526.31 | **5,938.45** |
| 224 | 1,749.36 | 1,929.85 | 1,916.46 | 2,060.07 | **7,655.74** |
| 225 | 866.35 | 941.83 | 843.60 | 793.97 | **3,445.75** |
| 226 | 987.40 | 1,092.96 | 896.53 | 1,014.68 | **3,991.58** |
| 227 | 258.65 | 340.79 | 302.67 | 320.91 | **1,223.01** |
| 228 | 879.40 | 822.76 | 798.55 | 675.71 | **3,176.41** |
| 229 | 1,052.19 | 1,145.46 | 1,159.03 | 1,069.81 | **4,426.49** |
| 230 | 913.85 | 1,065.13 | 858.16 | 836.06 | **3,673.19** |
| 231 | 1,336.03 | 1,624.98 | 1,454.85 | 1,466.75 | **5,882.62** |
| 232 | 3,372.33 | 3,529.03 | 3,069.55 | 3,279.66 | **13,250.56** |
| 233 | 1,100.22 | 1,077.59 | 1,075.59 | 1,130.52 | **4,383.92** |
| 234 | 2,634.51 | 2,855.60 | 3,007.52 | 3,035.24 | **11,532.87** |
| 235 | 988.00 | 1,168.87 | 1,115.58 | 961.86 | **4,234.31** |
| 236 | 1,880.15 | 2,122.83 | 1,932.33 | 2,073.92 | **8,009.23** |
| 237 | 711.15 | 549.02 | 513.02 | 521.68 | **2,294.86** |
| 238 | 1,589.86 | 1,819.72 | 1,597.71 | 1,738.90 | **6,746.19** |
| 239 | 403.93 | 333.35 | 357.74 | 442.02 | **1,537.04** |
| 240 | 920.12 | 1,338.27 | 1,229.83 | 1,184.79 | **4,673.01** |
| 241 | 643.45 | 1,017.69 | 917.95 | 896.30 | **3,475.38** |
| 242 | 1,545.59 | 1,205.85 | 1,174.64 | 1,114.98 | **5,041.06** |
| 243 | 820.49 | 712.77 | 760.94 | 640.37 | **2,934.57** |
| 244 | 435.03 | 526.45 | 551.20 | 525.72 | **2,038.40** |
| 245 | 1,382.04 | 1,557.84 | 1,386.69 | 1,251.61 | **5,578.17** |
| 246 | 1,219.07 | 1,187.73 | 1,241.01 | 972.29 | **4,620.10** |
| **VA VIRGINIA Total** | **39,397.47** | **42,307.87** | **39,888.97** | **39,900.74** | **161,495.05** |

**STATE: VI - VIRGIN ISLANDS**

| REGISTRANT ZIP CODE 3 | 1ST QUARTER | 2ND QUARTER | 3RD QUARTER | 4TH QUARTER | **TOTAL GRAMS** |
|---|---|---|---|---|---|
| 008 | 580.22 | 563.65 | 707.38 | 532.63 | **2,383.89** |
| **VI - VIRGIN ISLANDS Total** | **580.22** | **563.65** | **707.38** | **532.63** | **2,383.89** |

**STATE: VT - VERMONT**

| REGISTRANT ZIP CODE 3 | 1ST QUARTER | 2ND QUARTER | 3RD QUARTER | 4TH QUARTER | **TOTAL GRAMS** |
|---|---|---|---|---|---|
| 050 | 157.05 | 190.94 | 169.84 | 188.06 | **705.89** |
| 051 | 97.79 | 67.11 | 70.72 | 48.99 | **284.61** |
| 052 | 162.07 | 141.98 | 141.25 | 224.86 | **670.16** |
| 053 | 125.42 | 86.92 | 122.83 | 105.06 | **440.23** |
| 054 | 904.86 | 1,047.59 | 815.23 | 854.96 | **3,622.64** |

| | | | | | |
|---|---|---|---|---|---|
| 056 | 436.85 | 360.22 | 363.73 | 364.85 | **1,525.65** |
| 057 | 332.15 | 239.42 | 347.53 | 288.22 | **1,207.33** |
| 058 | 289.94 | 217.48 | 226.87 | 206.79 | **941.08** |
| **VT VERMONT Total** | **2,506.12** | **2,351.66** | **2,258.02** | **2,281.78** | **9,397.58** |

**STATE: WA - WASHINGTON**

| REGISTRANT ZIP CODE 3 | 1ST QUARTER | 2ND QUARTER | 3RD QUARTER | 4TH QUARTER | TOTAL GRAMS |
|---|---|---|---|---|---|
| 980 | 8,158.54 | 7,891.14 | 7,595.18 | 7,677.08 | **31,321.95** |
| 981 | 4,944.82 | 4,944.15 | 4,926.24 | 4,668.22 | **19,483.43** |
| 982 | 5,885.14 | 6,327.67 | 5,941.75 | 5,791.10 | **23,945.66** |
| 983 | 5,002.38 | 5,062.06 | 4,791.02 | 4,661.30 | **19,516.75** |
| 984 | 3,170.85 | 3,078.12 | 3,004.56 | 3,097.55 | **12,351.07** |
| 985 | 3,094.67 | 3,089.36 | 3,101.17 | 2,678.14 | **11,963.34** |
| 986 | 3,415.21 | 3,616.06 | 3,477.77 | 3,740.43 | **14,249.47** |
| 988 | 1,208.12 | 1,336.66 | 1,246.72 | 1,190.23 | **4,981.73** |
| 989 | 1,580.82 | 1,646.52 | 1,610.22 | 1,449.02 | **6,286.58** |
| 990 | 898.12 | 898.76 | 868.00 | 1,022.89 | **3,687.76** |
| 991 | 1,043.71 | 813.60 | 949.47 | 954.53 | **3,761.32** |
| 992 | 4,473.09 | 4,548.22 | 4,551.76 | 4,161.33 | **17,734.41** |
| 993 | 1,876.83 | 1,929.07 | 2,050.72 | 1,842.22 | **7,698.84** |
| 994 | 534.10 | 561.55 | 484.01 | 518.45 | **2,098.11** |
| **WA WASHINGTON Total** | **45,286.41** | **45,742.94** | **44,598.58** | **43,452.50** | **179,080.43** |

**STATE: WI - WISCONSIN**

| REGISTRANT ZIP CODE 3 | 1ST QUARTER | 2ND QUARTER | 3RD QUARTER | 4TH QUARTER | TOTAL GRAMS |
|---|---|---|---|---|---|
| 530 | 3,872.81 | 3,829.74 | 3,934.74 | 3,653.07 | **15,290.36** |
| 531 | 4,140.05 | 4,197.41 | 4,584.90 | 4,093.42 | **17,015.79** |
| 532 | 8,455.69 | 8,240.30 | 7,746.46 | 7,972.73 | **32,415.18** |
| 534 | 1,103.39 | 1,244.92 | 1,165.26 | 1,068.57 | **4,582.14** |
| 535 | 3,204.69 | 3,121.46 | 3,354.30 | 3,049.50 | **12,729.95** |
| 537 | 1,267.23 | 1,278.75 | 1,374.30 | 1,361.69 | **5,281.97** |
| 538 | 282.16 | 301.00 | 360.75 | 264.58 | **1,208.49** |
| 539 | 1,163.03 | 1,241.17 | 1,201.54 | 1,129.48 | **4,735.22** |
| 540 | 735.95 | 604.24 | 692.85 | 631.32 | **2,664.36** |
| 541 | 1,050.25 | 959.16 | 913.59 | 1,021.40 | **3,944.41** |
| 542 | 741.00 | 773.00 | 773.93 | 848.82 | **3,136.75** |
| 543 | 1,317.95 | 1,463.61 | 1,476.02 | 1,386.59 | **5,644.18** |
| 544 | 2,037.18 | 2,166.17 | 2,298.16 | 2,067.41 | **8,568.92** |
| 545 | 815.83 | 730.24 | 864.70 | 800.95 | **3,211.72** |
| 546 | 872.48 | 893.66 | 1,010.88 | 839.02 | **3,616.04** |
| 547 | 1,407.12 | 1,021.38 | 1,109.11 | 1,062.41 | **4,600.02** |
| 548 | 1,359.12 | 1,341.73 | 1,295.65 | 1,437.18 | **5,433.67** |

| | | | | | |
|---|---|---|---|---|---|
| 549 | 2,926.64 | 2,809.16 | 2,870.39 | 2,781.18 | **11,387.37** |
| **WI - WISCONSIN Total** | **36,752.57** | **36,217.11** | **37,027.55** | **35,469.33** | **145,466.57** |

**STATE: WV - WEST VIRGINIA**

| REGISTRANT ZIP CODE 3 | 1ST QUARTER | 2ND QUARTER | 3RD QUARTER | 4TH QUARTER | TOTAL GRAMS |
|---|---|---|---|---|---|
| 247 | 1,021.32 | 908.47 | 1,078.46 | 1,041.69 | **4,049.94** |
| 248 | 4,468.00 | 4,170.46 | 3,456.60 | 2,417.03 | **14,512.09** |
| 249 | 316.07 | 397.30 | 306.10 | 380.50 | **1,399.97** |
| 250 | 1,421.35 | 1,308.51 | 1,538.23 | 1,437.67 | **5,705.76** |
| 251 | 1,507.86 | 1,771.97 | 1,989.08 | 2,115.96 | **7,384.87** |
| 252 | 1,131.49 | 1,359.30 | 1,325.60 | 1,132.12 | **4,948.52** |
| 253 | 2,220.30 | 2,154.84 | 2,436.74 | 2,255.79 | **9,067.66** |
| 254 | 1,076.92 | 1,076.51 | 1,140.87 | 1,382.42 | **4,676.71** |
| 255 | 2,668.63 | 2,795.09 | 2,678.14 | 2,753.78 | **10,895.65** |
| 256 | 6,849.10 | 6,871.20 | 6,729.75 | 5,774.99 | **26,225.05** |
| 257 | 1,052.06 | 912.58 | 1,132.66 | 1,070.59 | **4,167.89** |
| 258 | 1,934.62 | 1,966.78 | 1,966.69 | 1,928.48 | **7,796.57** |
| 259 | 707.71 | 798.57 | 850.33 | 859.53 | **3,216.15** |
| 260 | 1,229.09 | 1,233.39 | 1,363.69 | 1,211.64 | **5,037.82** |
| 261 | 1,296.29 | 1,199.60 | 2,194.43 | 1,441.15 | **6,131.48** |
| 262 | 1,497.71 | 1,456.76 | 1,479.29 | 1,417.78 | **5,851.53** |
| 263 | 1,077.32 | 1,228.53 | 1,163.54 | 1,122.45 | **4,591.84** |
| 264 | 362.46 | 360.25 | 388.98 | 346.99 | **1,458.67** |
| 265 | 2,691.38 | 2,798.36 | 2,766.84 | 2,711.94 | **10,968.52** |
| 266 | 806.69 | 919.40 | 782.73 | 1,045.38 | **3,554.19** |
| 267 | 416.23 | 446.79 | 508.67 | 380.49 | **1,752.18** |
| 268 | 245.32 | 243.11 | 223.22 | 194.49 | **906.14** |
| **WV WEST VIRGINIA Total** | **35,997.92** | **36,377.77** | **37,500.65** | **34,422.85** | **144,299.19** |

**STATE: WY - WYOMING**

| REGISTRANT ZIP CODE 3 | 1ST QUARTER | 2ND QUARTER | 3RD QUARTER | 4TH QUARTER | TOTAL GRAMS |
|---|---|---|---|---|---|
| 820 | 660.35 | 738.82 | 763.01 | 596.09 | **2,758.27** |
| 822 | 68.51 | 144.85 | 101.66 | 106.08 | **421.12** |
| 823 | 35.36 | 44.20 | 22.10 | 26.52 | **128.19** |
| 824 | 489.98 | 634.25 | 626.89 | 502.76 | **2,253.88** |
| 825 | 336.12 | 277.55 | 384.56 | 238.69 | **1,236.92** |
| 826 | 448.28 | 464.58 | 475.35 | 541.79 | **1,930.00** |
| 827 | 268.15 | 237.39 | 219.53 | 239.24 | **964.31** |
| 828 | 279.88 | 203.28 | 150.17 | 254.63 | **887.96** |
| 829 | 380.32 | 484.01 | 426.55 | 413.29 | **1,704.17** |
| 830 | 57.46 | 108.29 | 133.15 | 86.19 | **385.10** |
| 831 | 44.20 | 39.96 | 30.94 | 37.13 | **152.24** |

| WY - WYOMING Total | 3,068.62 | 3,377.20 | 3,333.91 | 3,042.42 | 12,822.15 |
|---|---|---|---|---|---|

**DRUG: 9064 - BUPRENORPHINE**

**STATE: AK - ALASKA**

| REGISTRANT ZIP CODE 3 | 1ST QUARTER | 2ND QUARTER | 3RD QUARTER | 4TH QUARTER | TOTAL GRAMS |
|---|---|---|---|---|---|
| 995 | 1,046.17 | 1,056.94 | 1,073.22 | 1,062.21 | **4,238.53** |
| 996 | 733.77 | 670.36 | 672.35 | 708.44 | **2,784.92** |
| 997 | 231.72 | 181.41 | 191.03 | 174.80 | **778.96** |
| 998 | 399.77 | 382.30 | 376.44 | 368.97 | **1,527.48** |
| 999 | 245.28 | 302.90 | 303.61 | 305.64 | **1,157.43** |
| AK ALASKA - Total | 2,656.71 | 2,593.91 | 2,616.65 | 2,620.06 | 10,487.33 |

**STATE: AL - ALABAMA**

| REGISTRANT ZIP CODE 3 | 1ST QUARTER | 2ND QUARTER | 3RD QUARTER | 4TH QUARTER | TOTAL GRAMS |
|---|---|---|---|---|---|
| 350 | 4,306.86 | 4,416.78 | 4,622.68 | 4,496.14 | **17,842.46** |
| 351 | 2,432.61 | 2,752.26 | 2,800.89 | 2,747.01 | **10,732.77** |
| 352 | 3,872.37 | 4,053.01 | 3,930.90 | 3,722.13 | **15,578.41** |
| 354 | 552.50 | 733.97 | 711.40 | 713.54 | **2,711.41** |
| 355 | 2,427.31 | 2,475.28 | 2,464.92 | 2,528.81 | **9,896.32** |
| 356 | 3,516.65 | 3,490.46 | 3,668.02 | 3,884.19 | **14,559.33** |
| 357 | 1,106.05 | 1,127.47 | 1,197.22 | 1,231.82 | **4,662.56** |
| 358 | 1,494.11 | 1,411.14 | 1,406.19 | 1,424.96 | **5,736.40** |
| 359 | 3,239.44 | 3,476.05 | 3,482.08 | 3,661.90 | **13,859.47** |
| 360 | 783.86 | 866.32 | 917.60 | 833.48 | **3,401.26** |
| 361 | 424.46 | 496.80 | 504.90 | 522.89 | **1,949.05** |
| 362 | 1,154.48 | 1,270.94 | 1,281.88 | 1,374.83 | **5,082.13** |
| 363 | 896.22 | 942.69 | 978.36 | 1,017.76 | **3,835.04** |
| 364 | 432.43 | 455.12 | 428.27 | 460.04 | **1,775.86** |
| 365 | 2,077.29 | 2,252.40 | 2,250.25 | 2,292.04 | **8,871.98** |
| 366 | 1,617.44 | 1,786.29 | 1,844.10 | 1,932.16 | **7,179.99** |
| 367 | 242.14 | 235.19 | 247.57 | 261.36 | **986.26** |
| 368 | 627.26 | 686.33 | 669.53 | 655.74 | **2,638.86** |
| 369 | 38.13 | 37.50 | 31.57 | 33.85 | **141.05** |
| AL ALABAMA - Total | 31,241.60 | 32,965.99 | 33,438.34 | 33,794.65 | 131,440.58 |

**STATE: AR - ARKANSAS**

| REGISTRANT ZIP CODE 3 | 1ST QUARTER | 2ND QUARTER | 3RD QUARTER | 4TH QUARTER | TOTAL GRAMS |
|---|---|---|---|---|---|
| 716 | 131.89 | 174.32 | 173.92 | 197.83 | **677.96** |
| 717 | 219.31 | 242.73 | 241.70 | 279.99 | **983.74** |
| 718 | 273.86 | 277.37 | 237.59 | 263.16 | **1,051.97** |
| 719 | 953.75 | 969.48 | 1,036.07 | 1,073.18 | **4,032.47** |
| 720 | 736.29 | 797.00 | 795.86 | 833.46 | **3,162.61** |
| 721 | 1,128.26 | 1,218.26 | 1,207.22 | 1,278.13 | **4,831.87** |

| | | | | | |
|---|---|---|---|---|---|
| 722 | 976.47 | 1,051.99 | 1,107.47 | 1,043.07 | **4,178.99** |
| 723 | 146.95 | 160.37 | 151.38 | 165.88 | **624.58** |
| 724 | 1,047.31 | 1,082.86 | 1,101.70 | 1,134.64 | **4,366.51** |
| 725 | 307.05 | 301.76 | 395.90 | 399.26 | **1,403.97** |
| 726 | 416.71 | 439.70 | 499.11 | 523.01 | **1,878.54** |
| 727 | 1,049.70 | 1,112.05 | 1,206.76 | 1,271.17 | **4,639.68** |
| 728 | 207.19 | 223.77 | 223.93 | 245.28 | **900.18** |
| 729 | 548.93 | 637.12 | 668.35 | 701.69 | **2,556.08** |
| **AR ARKANSAS - Total** | **8,143.68** | **8,688.79** | **9,046.96** | **9,409.73** | **35,289.16** |

**STATE: AZ - ARIZONA**

| REGISTRANT ZIP CODE 3 | 1ST QUARTER | 2ND QUARTER | 3RD QUARTER | 4TH QUARTER | **TOTAL GRAMS** |
|---|---|---|---|---|---|
| 850 | 3,430.70 | 3,634.07 | 3,876.18 | 4,054.31 | **14,995.26** |
| 851 | 703.92 | 717.15 | 769.80 | 790.26 | **2,981.13** |
| 852 | 3,277.51 | 3,320.34 | 3,348.13 | 3,507.06 | **13,453.04** |
| 853 | 2,028.05 | 2,084.20 | 2,147.54 | 2,336.08 | **8,595.86** |
| 855 | 164.74 | 174.49 | 182.54 | 195.72 | **717.49** |
| 856 | 170.13 | 172.74 | 226.06 | 216.18 | **785.11** |
| 857 | 1,921.22 | 2,006.93 | 2,187.97 | 2,210.79 | **8,326.90** |
| 859 | 205.66 | 216.29 | 224.37 | 217.65 | **863.98** |
| 860 | 167.00 | 185.36 | 194.29 | 209.18 | **755.83** |
| 863 | 774.10 | 793.41 | 915.86 | 925.49 | **3,408.86** |
| 864 | 296.39 | 343.72 | 366.44 | 355.39 | **1,361.94** |
| 865 | 24.84 | 32.76 | 35.94 | 43.38 | **136.92** |
| **AZ ARIZONA - Total** | **13,164.25** | **13,681.46** | **14,475.12** | **15,061.49** | **56,382.32** |

**STATE: CA - CALIFORNIA**

| REGISTRANT ZIP CODE 3 | 1ST QUARTER | 2ND QUARTER | 3RD QUARTER | 4TH QUARTER | **TOTAL GRAMS** |
|---|---|---|---|---|---|
| 900 | 1,132.07 | 1,165.55 | 1,262.47 | 1,252.61 | **4,812.71** |
| 902 | 925.89 | 1,016.51 | 1,034.25 | 1,030.74 | **4,007.38** |
| 903 | 23.17 | 32.13 | 41.63 | 34.17 | **131.09** |
| 904 | 199.87 | 230.69 | 229.49 | 257.84 | **917.89** |
| 905 | 205.27 | 209.99 | 224.13 | 218.76 | **858.14** |
| 906 | 291.34 | 287.98 | 324.61 | 323.39 | **1,227.32** |
| 907 | 655.18 | 732.79 | 881.31 | 912.76 | **3,182.04** |
| 908 | 349.58 | 308.42 | 293.54 | 333.06 | **1,284.60** |
| 910 | 127.08 | 130.59 | 123.70 | 125.19 | **506.56** |
| 911 | 172.35 | 179.37 | 203.52 | 178.73 | **733.98** |
| 912 | 295.53 | 331.80 | 285.26 | 291.04 | **1,203.62** |
| 913 | 1,366.79 | 1,402.76 | 1,554.56 | 1,575.65 | **5,899.76** |
| 914 | 657.38 | 616.51 | 680.30 | 680.98 | **2,635.17** |
| 915 | 181.75 | 172.08 | 163.95 | 170.59 | **688.37** |

| | | | | | |
|---|---|---|---|---|---|
| 916 | 179.58 | 184.34 | 197.34 | 191.37 | **752.64** |
| 917 | 913.47 | 981.80 | 1,059.65 | 1,032.18 | **3,987.09** |
| 918 | 14.05 | 15.07 | 17.41 | 19.93 | **66.46** |
| 919 | 317.09 | 351.62 | 377.32 | 371.35 | **1,417.37** |
| 920 | 1,677.08 | 1,665.69 | 1,772.47 | 1,860.59 | **6,975.83** |
| 921 | 1,406.99 | 1,471.13 | 1,624.68 | 1,700.57 | **6,203.37** |
| 922 | 573.51 | 614.66 | 671.00 | 767.93 | **2,627.11** |
| 923 | 606.52 | 700.40 | 715.20 | 867.45 | **2,889.57** |
| 924 | 77.33 | 89.67 | 83.16 | 105.78 | **355.93** |
| 925 | 1,674.03 | 1,754.86 | 1,867.72 | 1,960.30 | **7,256.91** |
| 926 | 1,930.84 | 2,007.86 | 2,144.02 | 2,084.34 | **8,167.05** |
| 927 | 397.40 | 405.80 | 474.21 | 825.51 | **2,102.92** |
| 928 | 903.67 | 869.48 | 988.29 | 1,029.75 | **3,791.18** |
| 930 | 711.28 | 769.13 | 851.58 | 823.43 | **3,155.42** |
| 931 | 519.94 | 492.90 | 547.40 | 573.05 | **2,133.30** |
| 932 | 499.19 | 534.24 | 596.17 | 666.65 | **2,296.27** |
| 933 | 319.98 | 415.60 | 438.74 | 467.63 | **1,641.95** |
| 934 | 931.36 | 942.98 | 1,007.62 | 1,058.52 | **3,940.49** |
| 935 | 526.60 | 565.47 | 598.54 | 599.70 | **2,290.30** |
| 936 | 190.91 | 193.55 | 217.38 | 216.24 | **818.07** |
| 937 | 371.00 | 416.58 | 476.86 | 461.35 | **1,725.79** |
| 939 | 855.48 | 829.84 | 936.80 | 989.95 | **3,612.07** |
| 940 | 822.63 | 854.70 | 878.00 | 854.67 | **3,410.01** |
| 941 | 1,495.96 | 1,658.18 | 1,528.76 | 1,644.74 | **6,327.64** |
| 943 | 141.79 | 163.26 | 172.74 | 184.50 | **662.29** |
| 944 | 212.45 | 207.03 | 203.55 | 232.26 | **855.29** |
| 945 | 3,539.49 | 3,615.02 | 3,915.03 | 4,118.00 | **15,187.54** |
| 946 | 619.23 | 658.07 | 664.04 | 655.99 | **2,597.33** |
| 947 | 140.19 | 133.01 | 152.99 | 154.19 | **580.37** |
| 948 | 175.07 | 196.84 | 209.50 | 210.66 | **792.07** |
| 949 | 1,045.12 | 1,021.89 | 1,090.58 | 1,141.72 | **4,299.31** |
| 950 | 1,328.50 | 1,333.56 | 1,409.60 | 1,479.51 | **5,551.16** |
| 951 | 803.91 | 825.23 | 863.36 | 919.50 | **3,411.99** |
| 952 | 379.78 | 409.20 | 445.67 | 497.39 | **1,732.03** |
| 953 | 1,348.35 | 1,449.89 | 1,568.64 | 1,530.90 | **5,897.77** |
| 954 | 2,619.80 | 2,716.62 | 2,848.65 | 2,854.72 | **11,039.79** |
| 955 | 1,777.53 | 1,807.58 | 1,800.52 | 1,907.08 | **7,292.71** |
| 956 | 2,866.67 | 2,976.00 | 3,244.29 | 3,230.39 | **12,317.35** |
| 957 | 328.88 | 353.37 | 370.23 | 372.33 | **1,424.82** |
| 958 | 2,528.95 | 2,738.35 | 2,817.12 | 2,913.74 | **10,998.17** |

| | | | | | |
|---|---|---|---|---|---|
| 959 | 1,867.11 | 1,980.55 | 2,123.78 | 2,320.93 | **8,292.37** |
| 960 | 1,281.40 | 1,356.08 | 1,500.28 | 1,702.34 | **5,840.10** |
| 961 | 279.10 | 301.22 | 315.46 | 369.95 | **1,265.73** |
| **CA - CALIFORNIA - Total** | **47,782.46** | **49,845.50** | **53,089.05** | **55,354.57** | **206,071.57** |

**STATE: CO - COLORADO**

| REGISTRANT ZIP CODE 3 | 1ST QUARTER | 2ND QUARTER | 3RD QUARTER | 4TH QUARTER | TOTAL GRAMS |
|---|---|---|---|---|---|
| 800 | 1,649.28 | 1,703.84 | 1,637.27 | 1,828.71 | **6,819.11** |
| 801 | 933.61 | 949.97 | 973.98 | 997.43 | **3,855.00** |
| 802 | 2,132.30 | 2,237.54 | 2,286.51 | 2,513.15 | **9,169.50** |
| 803 | 139.26 | 147.60 | 161.37 | 177.79 | **626.02** |
| 804 | 157.80 | 145.35 | 175.03 | 192.67 | **670.85** |
| 805 | 1,380.82 | 1,449.92 | 1,535.73 | 1,694.69 | **6,061.16** |
| 806 | 613.14 | 604.96 | 569.49 | 613.30 | **2,400.89** |
| 807 | 54.27 | 65.99 | 57.56 | 68.89 | **246.71** |
| 808 | 104.77 | 100.76 | 120.74 | 129.40 | **455.67** |
| 809 | 1,102.38 | 1,206.33 | 1,219.86 | 1,262.57 | **4,791.14** |
| 810 | 827.43 | 979.51 | 991.13 | 1,079.93 | **3,878.01** |
| 811 | 111.76 | 128.14 | 139.81 | 151.92 | **531.63** |
| 812 | 140.47 | 150.95 | 150.32 | 147.82 | **589.56** |
| 813 | 157.33 | 145.37 | 161.25 | 153.29 | **617.23** |
| 814 | 54.21 | 61.42 | 74.09 | 82.44 | **272.16** |
| 815 | 241.15 | 235.51 | 281.58 | 294.06 | **1,052.30** |
| 816 | 153.92 | 147.81 | 151.34 | 169.65 | **622.73** |
| **CO - COLORADO - Total** | **9,953.90** | **10,460.96** | **10,687.08** | **11,557.72** | **42,659.66** |

**STATE: CT - CONNECTICUT**

| REGISTRANT ZIP CODE 3 | 1ST QUARTER | 2ND QUARTER | 3RD QUARTER | 4TH QUARTER | TOTAL GRAMS |
|---|---|---|---|---|---|
| 060 | 3,499.65 | 3,420.51 | 3,716.71 | 3,582.01 | **14,218.88** |
| 061 | 1,823.99 | 1,674.60 | 1,821.22 | 1,899.15 | **7,218.96** |
| 062 | 934.00 | 941.78 | 978.02 | 973.40 | **3,827.20** |
| 063 | 2,034.17 | 2,015.60 | 2,209.37 | 2,211.86 | **8,471.01** |
| 064 | 4,182.45 | 4,158.46 | 4,355.95 | 4,351.78 | **17,048.64** |
| 065 | 2,244.55 | 2,286.51 | 2,396.29 | 2,412.10 | **9,339.45** |
| 066 | 378.73 | 383.72 | 406.36 | 425.63 | **1,594.44** |
| 067 | 1,839.14 | 1,740.84 | 1,898.22 | 1,968.99 | **7,447.20** |
| 068 | 812.69 | 794.35 | 865.18 | 874.83 | **3,347.05** |
| 069 | 165.58 | 131.41 | 149.82 | 157.21 | **604.01** |
| **CT CONNECTICUT Total** | **17,914.95** | **17,547.79** | **18,797.14** | **18,856.96** | **73,116.83** |

**STATE: DC - DISTRICT OF COLUMBIA**

| REGISTRANT ZIP CODE 3 | 1ST QUARTER | 2ND QUARTER | 3RD QUARTER | 4TH QUARTER | TOTAL GRAMS |
|---|---|---|---|---|---|
| 200 | 3,342.20 | 3,544.64 | 3,568.90 | 3,355.45 | **13,811.18** |

| | | | | | |
|---|---|---|---|---|---|
| 204 | 33.48 | 38.72 | 39.00 | 18.46 | **129.66** |
| **DC - DISTRICT OF COLUMBIA** | **3,375.68** | **3,583.36** | **3,607.90** | **3,373.91** | **13,940.85** |

**STATE: DE - DELAWARE**

| REGISTRANT ZIP CODE 3 | 1ST QUARTER | 2ND QUARTER | 3RD QUARTER | 4TH QUARTER | TOTAL GRAMS |
|---|---|---|---|---|---|
| 197 | 1,219.10 | 1,086.76 | 1,209.67 | 1,338.74 | **4,854.26** |
| 198 | 1,116.67 | 1,069.70 | 1,136.14 | 1,095.08 | **4,417.60** |
| 199 | 1,988.59 | 2,010.33 | 2,041.32 | 2,064.30 | **8,104.53** |
| **DE DELAWARE Total** | **4,324.36** | **4,166.79** | **4,387.13** | **4,498.12** | **17,376.39** |

**STATE: FL - FLORIDA**

| REGISTRANT ZIP CODE 3 | 1ST QUARTER | 2ND QUARTER | 3RD QUARTER | 4TH QUARTER | TOTAL GRAMS |
|---|---|---|---|---|---|
| 320 | 3,440.63 | 3,537.58 | 3,741.18 | 3,777.58 | **14,496.97** |
| 321 | 2,892.17 | 3,033.77 | 3,306.40 | 3,151.37 | **12,383.72** |
| 322 | 4,399.20 | 4,597.76 | 5,013.69 | 4,949.99 | **18,960.63** |
| 323 | 774.02 | 926.45 | 969.87 | 1,049.04 | **3,719.38** |
| 324 | 1,106.72 | 1,106.28 | 1,132.37 | 1,136.05 | **4,481.43** |
| 325 | 2,529.72 | 2,738.30 | 2,898.19 | 2,870.36 | **11,036.56** |
| 326 | 3,135.10 | 3,107.19 | 3,281.25 | 3,215.32 | **12,738.86** |
| 327 | 2,720.21 | 2,743.88 | 2,824.70 | 2,815.15 | **11,103.94** |
| 328 | 1,474.89 | 1,549.70 | 1,756.90 | 1,754.46 | **6,535.95** |
| 329 | 3,386.99 | 3,631.93 | 3,615.43 | 3,636.37 | **14,270.72** |
| 330 | 1,718.67 | 1,634.44 | 1,779.44 | 1,709.79 | **6,842.34** |
| 331 | 940.24 | 954.40 | 1,061.24 | 962.17 | **3,918.06** |
| 333 | 1,256.59 | 1,392.83 | 1,480.72 | 1,420.78 | **5,550.92** |
| 334 | 4,271.54 | 4,526.02 | 4,486.98 | 4,456.51 | **17,741.05** |
| 335 | 1,597.22 | 1,700.33 | 1,764.50 | 1,714.38 | **6,776.43** |
| 336 | 1,661.65 | 1,519.80 | 1,730.00 | 1,657.39 | **6,568.84** |
| 337 | 2,711.86 | 2,738.14 | 2,880.06 | 2,753.62 | **11,083.68** |
| 338 | 1,202.40 | 1,252.99 | 1,315.10 | 1,346.00 | **5,116.49** |
| 339 | 2,892.08 | 2,873.65 | 2,870.56 | 2,880.10 | **11,516.39** |
| 341 | 1,224.57 | 1,312.27 | 1,350.98 | 1,403.77 | **5,291.59** |
| 342 | 3,745.60 | 3,966.55 | 3,965.49 | 4,029.23 | **15,706.87** |
| 344 | 1,545.39 | 1,628.69 | 1,651.42 | 1,684.36 | **6,509.87** |
| 346 | 2,304.60 | 2,353.87 | 2,517.67 | 2,630.86 | **9,807.00** |
| 347 | 1,253.04 | 1,390.57 | 1,447.88 | 1,590.33 | **5,681.83** |
| 349 | 1,664.33 | 1,744.13 | 1,838.31 | 1,834.93 | **7,081.70** |
| **FL FLORIDA - Total** | **55,849.42** | **57,961.54** | **60,680.34** | **60,429.93** | **234,921.23** |

**STATE: GA - GEORGIA**

| REGISTRANT ZIP CODE 3 | 1ST QUARTER | 2ND QUARTER | 3RD QUARTER | 4TH QUARTER | TOTAL GRAMS |
|---|---|---|---|---|---|
| 300 | 2,632.80 | 2,760.31 | 2,883.09 | 2,890.14 | **11,166.34** |
| 301 | 2,464.77 | 2,563.53 | 2,699.14 | 2,847.71 | **10,575.15** |

| | | | | | |
|---|---|---|---|---|---|
| 302 | 1,639.22 | 1,698.25 | 1,875.49 | 1,815.65 | **7,028.61** |
| 303 | 775.94 | 842.24 | 992.25 | 940.70 | **3,551.13** |
| 304 | 707.43 | 775.53 | 782.59 | 827.77 | **3,093.31** |
| 305 | 1,096.74 | 1,222.63 | 1,287.97 | 1,264.08 | **4,871.44** |
| 306 | 1,102.97 | 1,086.29 | 1,120.87 | 1,169.49 | **4,479.62** |
| 307 | 1,128.63 | 1,021.41 | 619.17 | 680.62 | **3,449.84** |
| 308 | 130.62 | 137.37 | 136.89 | 154.59 | **559.47** |
| 309 | 405.19 | 430.68 | 443.77 | 382.73 | **1,662.37** |
| 310 | 1,190.46 | 1,181.53 | 1,226.26 | 1,287.26 | **4,885.51** |
| 312 | 516.16 | 483.48 | 502.72 | 494.38 | **1,996.75** |
| 313 | 843.43 | 784.61 | 868.57 | 831.63 | **3,328.23** |
| 314 | 911.62 | 901.53 | 898.78 | 839.32 | **3,551.26** |
| 315 | 2,337.31 | 2,308.35 | 2,461.02 | 2,533.65 | **9,640.33** |
| 316 | 225.77 | 223.68 | 239.68 | 244.99 | **934.13** |
| 317 | 591.37 | 612.72 | 623.93 | 620.53 | **2,448.55** |
| 318 | 9.91 | 14.35 | 23.81 | 22.04 | **70.11** |
| 319 | 288.40 | 290.99 | 311.47 | 292.86 | **1,183.72** |
| 398 | 143.55 | 151.72 | 178.65 | 190.61 | **664.52** |
| **GA GEORGIA Total** | **19,142.31** | **19,491.18** | **20,176.14** | **20,330.75** | **79,140.38** |

**STATE: GU - GUAM**

| REGISTRANT ZIP CODE 3 | 1ST QUARTER | 2ND QUARTER | 3RD QUARTER | 4TH QUARTER | TOTAL GRAMS |
|---|---|---|---|---|---|
| 969 | 2.66 | 4.34 | 12.48 | 0.83 | **20.31** |
| **GU - GUAM - Total** | **2.66** | **4.34** | **12.48** | **0.83** | **20.31** |

**STATE: HI - HAWAII**

| REGISTRANT ZIP CODE 3 | 1ST QUARTER | 2ND QUARTER | 3RD QUARTER | 4TH QUARTER | TOTAL GRAMS |
|---|---|---|---|---|---|
| 967 | 1,676.69 | 1,704.77 | 1,780.24 | 1,745.72 | **6,907.42** |
| 968 | 500.63 | 514.99 | 484.75 | 472.06 | **1,972.42** |
| **HI - HAWAII - Total** | **2,177.32** | **2,219.76** | **2,264.99** | **2,217.78** | **8,879.84** |

**STATE: IA - IOWA**

| REGISTRANT ZIP CODE 3 | 1ST QUARTER | 2ND QUARTER | 3RD QUARTER | 4TH QUARTER | TOTAL GRAMS |
|---|---|---|---|---|---|
| 500 | 104.91 | 104.52 | 123.15 | 103.00 | **435.57** |
| 501 | 134.10 | 145.70 | 144.13 | 129.31 | **553.25** |
| 502 | 95.15 | 92.03 | 111.42 | 106.07 | **404.67** |
| 503 | 389.77 | 440.60 | 481.50 | 416.06 | **1,727.94** |
| 504 | 203.81 | 226.60 | 219.77 | 208.42 | **858.61** |
| 505 | 75.44 | 78.22 | 79.37 | 65.01 | **298.05** |
| 506 | 71.20 | 80.87 | 107.98 | 102.34 | **362.39** |
| 507 | 57.52 | 54.03 | 64.60 | 65.66 | **241.80** |
| 508 | 4.70 | 7.13 | 8.92 | 3.72 | **24.46** |
| 510 | 106.97 | 110.91 | 100.33 | 100.41 | **418.62** |

| | | | | | |
|---|---|---|---|---|---|
| 511 | 197.50 | 197.81 | 206.63 | 216.42 | **818.36** |
| 512 | 38.93 | 48.33 | 46.41 | 45.68 | **179.35** |
| 513 | 39.42 | 38.98 | 37.70 | 35.73 | **151.83** |
| 514 | 10.91 | 17.06 | 10.77 | 7.46 | **46.20** |
| 515 | 92.42 | 97.40 | 102.52 | 99.91 | **392.25** |
| 516 | 8.30 | 6.19 | 5.90 | 8.84 | **29.24** |
| 520 | 68.34 | 64.03 | 54.74 | 55.36 | **242.47** |
| 521 | 7.05 | 10.01 | 10.07 | 12.39 | **39.52** |
| 522 | 204.73 | 206.95 | 264.82 | 252.01 | **928.52** |
| 523 | 25.58 | 27.70 | 37.31 | 31.15 | **121.74** |
| 524 | 88.44 | 110.84 | 115.75 | 104.24 | **419.27** |
| 525 | 81.97 | 79.04 | 84.85 | 86.28 | **332.15** |
| 526 | 52.85 | 62.78 | 67.09 | 61.80 | **244.52** |
| 527 | 120.65 | 118.77 | 133.98 | 168.13 | **541.52** |
| 528 | 87.20 | 88.94 | 98.71 | 93.39 | **368.24** |
| **IA IOWA Total** | **2,367.86** | **2,515.46** | **2,718.40** | **2,578.79** | **10,180.51** |

**STATE: ID - IDAHO**

| REGISTRANT ZIP CODE 3 | 1ST QUARTER | 2ND QUARTER | 3RD QUARTER | 4TH QUARTER | TOTAL GRAMS |
|---|---|---|---|---|---|
| 832 | 488.62 | 564.53 | 552.58 | 605.51 | **2,211.25** |
| 833 | 354.81 | 380.87 | 409.52 | 413.47 | **1,558.68** |
| 834 | 225.82 | 237.23 | 253.94 | 247.92 | **964.90** |
| 835 | 449.15 | 489.55 | 492.78 | 493.48 | **1,924.96** |
| 836 | 769.07 | 825.91 | 803.96 | 847.47 | **3,246.41** |
| 837 | 718.76 | 778.44 | 797.69 | 808.44 | **3,103.34** |
| 838 | 1,613.47 | 1,763.87 | 1,710.87 | 1,824.67 | **6,912.88** |
| **ID IDAHO Total** | **4,619.70** | **5,040.41** | **5,021.35** | **5,240.96** | **19,922.41** |

**STATE: IL - ILLINOIS**

| REGISTRANT ZIP CODE 3 | 1ST QUARTER | 2ND QUARTER | 3RD QUARTER | 4TH QUARTER | TOTAL GRAMS |
|---|---|---|---|---|---|
| 600 | 2,631.59 | 2,725.86 | 2,871.98 | 2,934.20 | **11,163.63** |
| 601 | 1,242.61 | 1,351.28 | 1,411.19 | 1,414.13 | **5,419.21** |
| 602 | 37.23 | 42.21 | 48.38 | 40.74 | **168.56** |
| 603 | 373.94 | 462.15 | 335.56 | 285.99 | **1,457.64** |
| 604 | 2,208.18 | 2,325.97 | 2,339.83 | 2,367.40 | **9,241.38** |
| 605 | 1,063.93 | 1,091.14 | 1,076.05 | 1,110.38 | **4,341.50** |
| 606 | 2,543.47 | 2,760.04 | 2,782.56 | 2,926.29 | **11,012.37** |
| 607 | 105.23 | 98.06 | 122.07 | 119.83 | **445.19** |
| 608 | 72.07 | 93.79 | 101.15 | 92.73 | **359.74** |
| 609 | 229.67 | 223.11 | 240.80 | 267.21 | **960.80** |
| 610 | 230.14 | 244.60 | 280.42 | 289.72 | **1,044.88** |
| 611 | 791.41 | 852.46 | 840.95 | 825.37 | **3,310.19** |

| | | | | | |
|---|---|---|---|---|---|
| 612 | 211.43 | 196.71 | 196.81 | 204.06 | **809.01** |
| 613 | 338.96 | 382.06 | 393.42 | 381.02 | **1,495.47** |
| 614 | 121.27 | 122.96 | 124.55 | 124.49 | **493.27** |
| 615 | 273.48 | 287.95 | 297.58 | 319.80 | **1,178.81** |
| 616 | 500.74 | 531.93 | 620.09 | 564.93 | **2,217.70** |
| 617 | 158.28 | 185.87 | 236.38 | 240.11 | **820.64** |
| 618 | 915.15 | 823.45 | 664.44 | 620.81 | **3,023.86** |
| 619 | 98.06 | 99.46 | 96.49 | 106.29 | **400.31** |
| 620 | 443.16 | 496.69 | 510.86 | 553.62 | **2,004.32** |
| 622 | 389.86 | 426.30 | 432.85 | 428.64 | **1,677.64** |
| 623 | 281.30 | 300.56 | 337.82 | 314.35 | **1,234.03** |
| 624 | 172.26 | 181.34 | 195.27 | 199.42 | **748.28** |
| 625 | 388.30 | 392.37 | 389.97 | 413.43 | **1,584.08** |
| 626 | 259.86 | 264.82 | 264.40 | 328.34 | **1,117.42** |
| 627 | 441.07 | 488.07 | 500.34 | 542.38 | **1,971.86** |
| 628 | 477.70 | 516.53 | 539.69 | 521.36 | **2,055.27** |
| 629 | 1,517.56 | 1,619.04 | 1,724.97 | 1,802.53 | **6,664.10** |
| **IL ILLINOIS Total** | **18,517.89** | **19,586.80** | **19,976.88** | **20,339.60** | **78,421.17** |

**STATE: IN - INDIANA**

| REGISTRANT ZIP CODE 3 | 1ST QUARTER | 2ND QUARTER | 3RD QUARTER | 4TH QUARTER | **TOTAL GRAMS** |
|---|---|---|---|---|---|
| 460 | 2,884.27 | 2,697.20 | 3,072.70 | 3,183.62 | **11,837.78** |
| 461 | 2,595.14 | 2,699.39 | 2,873.68 | 2,904.87 | **11,073.08** |
| 462 | 4,148.70 | 4,247.35 | 4,515.20 | 4,636.31 | **17,547.55** |
| 463 | 2,407.76 | 2,627.05 | 2,711.69 | 2,715.17 | **10,461.68** |
| 464 | 278.39 | 303.63 | 321.66 | 351.59 | **1,255.28** |
| 465 | 907.59 | 1,036.20 | 1,070.24 | 1,088.89 | **4,102.93** |
| 466 | 267.61 | 306.38 | 330.92 | 369.51 | **1,274.42** |
| 467 | 309.05 | 311.57 | 365.30 | 420.65 | **1,406.57** |
| 468 | 781.88 | 839.05 | 846.86 | 883.55 | **3,351.34** |
| 469 | 1,250.76 | 1,432.40 | 1,567.60 | 1,682.85 | **5,933.61** |
| 470 | 830.66 | 822.58 | 807.49 | 832.98 | **3,293.72** |
| 471 | 2,514.31 | 2,792.63 | 2,875.07 | 2,884.15 | **11,066.17** |
| 472 | 1,709.61 | 1,806.59 | 1,849.68 | 1,941.57 | **7,307.44** |
| 473 | 2,436.17 | 2,034.96 | 2,631.78 | 2,892.70 | **9,995.60** |
| 474 | 1,850.85 | 2,125.66 | 2,189.10 | 2,260.54 | **8,426.15** |
| 475 | 479.57 | 576.61 | 639.41 | 709.96 | **2,405.54** |
| 476 | 180.00 | 176.57 | 198.49 | 223.47 | **778.53** |
| 477 | 910.00 | 986.75 | 1,114.93 | 1,236.95 | **4,248.63** |
| 478 | 527.67 | 583.22 | 600.93 | 611.21 | **2,323.03** |
| 479 | 641.87 | 729.27 | 784.94 | 814.77 | **2,970.84** |

| IN INDIANA Total | 27,911.85 | 29,135.06 | 31,367.65 | 32,645.30 | 121,059.87 |
|---|---|---|---|---|---|

STATE: KS - KANSAS

| REGISTRANT ZIP CODE 3 | 1ST QUARTER | 2ND QUARTER | 3RD QUARTER | 4TH QUARTER | TOTAL GRAMS |
|---|---|---|---|---|---|
| 660 | 623.32 | 659.03 | 669.08 | 659.79 | **2,611.22** |
| 661 | 181.19 | 189.07 | 196.73 | 202.08 | **769.08** |
| 662 | 667.93 | 663.18 | 774.51 | 806.72 | **2,912.34** |
| 664 | 35.20 | 31.66 | 40.39 | 36.41 | **143.67** |
| 665 | 60.39 | 66.42 | 58.50 | 57.08 | **242.38** |
| 666 | 124.13 | 157.26 | 123.47 | 158.81 | **563.67** |
| 667 | 190.02 | 214.40 | 209.32 | 205.68 | **819.42** |
| 668 | 11.54 | 14.78 | 15.03 | 10.48 | **51.84** |
| 669 | 5.14 | 6.37 | 10.52 | 7.08 | **29.10** |
| 670 | 96.23 | 86.77 | 94.80 | 102.15 | **379.95** |
| 671 | 49.61 | 44.05 | 42.07 | 42.73 | **178.45** |
| 672 | 553.07 | 561.03 | 560.00 | 551.15 | **2,225.26** |
| 673 | 55.00 | 58.62 | 67.79 | 74.91 | **256.31** |
| 674 | 152.65 | 163.05 | 164.40 | 175.04 | **655.14** |
| 675 | 62.86 | 56.21 | 57.40 | 64.66 | **241.13** |
| 676 | 69.21 | 67.72 | 69.94 | 73.25 | **280.12** |
| 677 | 9.84 | 10.84 | 11.54 | 12.79 | **45.01** |
| 678 | 64.86 | 68.06 | 65.76 | 65.64 | **264.32** |
| 679 | 13.78 | 18.18 | 11.10 | 12.30 | **55.36** |
| **KS - KANSAS - Total** | **3,025.98** | **3,136.68** | **3,242.35** | **3,318.77** | **12,723.79** |

STATE: KY - KENTUCKY

| REGISTRANT ZIP CODE 3 | 1ST QUARTER | 2ND QUARTER | 3RD QUARTER | 4TH QUARTER | TOTAL GRAMS |
|---|---|---|---|---|---|
| 400 | 777.89 | 796.73 | 849.82 | 805.53 | **3,229.97** |
| 401 | 304.74 | 338.47 | 379.26 | 388.77 | **1,411.24** |
| 402 | 3,355.03 | 3,631.96 | 3,704.73 | 3,907.08 | **14,598.80** |
| 403 | 4,693.04 | 4,658.05 | 4,820.65 | 5,024.06 | **19,195.80** |
| 404 | 2,349.31 | 2,451.42 | 2,428.46 | 2,467.98 | **9,697.17** |
| 405 | 2,813.05 | 2,744.80 | 3,002.56 | 3,172.79 | **11,733.20** |
| 406 | 606.03 | 660.88 | 672.60 | 637.47 | **2,576.98** |
| 407 | 2,369.36 | 2,519.92 | 2,668.49 | 2,733.49 | **10,291.26** |
| 408 | 522.89 | 564.46 | 545.26 | 598.70 | **2,231.32** |
| 409 | 1,952.20 | 1,986.89 | 2,204.08 | 2,258.29 | **8,401.46** |
| 410 | 2,387.08 | 2,451.19 | 2,613.71 | 2,853.57 | **10,305.55** |
| 411 | 1,915.27 | 2,019.01 | 2,026.53 | 2,053.13 | **8,013.95** |
| 412 | 1,467.34 | 1,590.99 | 1,598.00 | 1,640.29 | **6,296.62** |
| 413 | 1,032.32 | 1,118.43 | 1,159.95 | 1,174.87 | **4,485.57** |
| 414 | 665.88 | 669.03 | 707.06 | 716.55 | **2,758.52** |

| | | | | | |
|---|---|---|---|---|---|
| 415 | 3,172.22 | 3,141.45 | 3,175.08 | 3,207.36 | **12,696.12** |
| 416 | 2,024.43 | 1,987.28 | 1,902.98 | 1,894.61 | **7,809.31** |
| 417 | 1,537.97 | 1,575.88 | 1,573.81 | 1,549.03 | **6,236.70** |
| 418 | 547.34 | 612.82 | 655.07 | 729.85 | **2,545.08** |
| 420 | 1,527.44 | 1,665.62 | 1,825.94 | 1,881.24 | **6,900.24** |
| 421 | 1,469.25 | 1,613.16 | 1,633.57 | 1,745.90 | **6,461.88** |
| 422 | 439.86 | 514.43 | 498.11 | 480.31 | **1,932.71** |
| 423 | 1,183.00 | 1,253.76 | 1,367.02 | 1,575.59 | **5,379.38** |
| 424 | 560.59 | 629.32 | 682.20 | 662.43 | **2,534.54** |
| 425 | 674.06 | 881.67 | 952.48 | 1,043.78 | **3,551.99** |
| 426 | 767.12 | 741.57 | 669.94 | 588.75 | **2,767.38** |
| 427 | 1,211.83 | 1,363.74 | 1,492.50 | 1,539.23 | **5,607.29** |
| **KY - KENTUCKY - Total** | **42,326.54** | **44,182.94** | **45,809.89** | **47,330.66** | **179,650.03** |

**STATE: LA - LOUISIANA**

| REGISTRANT ZIP CODE 3 | 1ST QUARTER | 2ND QUARTER | 3RD QUARTER | 4TH QUARTER | **TOTAL GRAMS** |
|---|---|---|---|---|---|
| 700 | 4,056.42 | 4,099.20 | 4,073.78 | 4,115.13 | **16,344.51** |
| 701 | 1,165.69 | 1,344.51 | 1,460.71 | 1,554.74 | **5,525.66** |
| 703 | 1,211.38 | 1,260.31 | 1,239.11 | 1,264.66 | **4,975.46** |
| 704 | 4,026.19 | 4,236.70 | 4,283.47 | 4,283.96 | **16,830.32** |
| 705 | 2,801.49 | 2,942.55 | 2,918.56 | 2,887.34 | **11,549.94** |
| 706 | 1,413.12 | 1,514.75 | 1,492.84 | 1,484.19 | **5,904.90** |
| 707 | 1,863.69 | 1,957.68 | 1,985.96 | 1,967.55 | **7,774.88** |
| 708 | 969.68 | 1,019.14 | 1,090.52 | 1,081.96 | **4,161.31** |
| 710 | 223.89 | 261.50 | 297.77 | 324.21 | **1,107.36** |
| 711 | 2,059.59 | 2,135.72 | 2,177.12 | 2,203.35 | **8,575.77** |
| 712 | 1,353.38 | 1,437.20 | 1,477.81 | 1,548.88 | **5,817.27** |
| 713 | 862.58 | 928.42 | 1,025.96 | 1,159.16 | **3,976.12** |
| 714 | 479.81 | 589.85 | 588.30 | 593.82 | **2,251.79** |
| **LA - LOUISIANA - Total** | **22,486.90** | **23,727.54** | **24,111.89** | **24,468.96** | **94,795.28** |

**STATE: MA - MASSACHUSETTS**

| REGISTRANT ZIP CODE 3 | 1ST QUARTER | 2ND QUARTER | 3RD QUARTER | 4TH QUARTER | **TOTAL GRAMS** |
|---|---|---|---|---|---|
| 010 | 3,702.20 | 3,627.06 | 3,702.92 | 3,780.06 | **14,812.24** |
| 011 | 1,427.01 | 1,442.08 | 1,495.40 | 1,547.69 | **5,912.17** |
| 012 | 1,503.14 | 1,500.61 | 1,566.88 | 1,593.78 | **6,164.41** |
| 013 | 1,110.07 | 1,048.29 | 1,120.74 | 1,174.57 | **4,453.67** |
| 014 | 1,313.95 | 1,264.22 | 1,396.92 | 1,392.62 | **5,367.71** |
| 015 | 1,771.62 | 1,661.24 | 1,760.91 | 1,764.66 | **6,958.43** |
| 016 | 2,267.92 | 2,295.12 | 2,425.18 | 2,501.01 | **9,489.23** |
| 017 | 1,542.88 | 1,501.84 | 1,554.76 | 1,565.98 | **6,165.45** |
| 018 | 3,970.06 | 4,074.90 | 4,328.36 | 4,722.47 | **17,095.79** |

| | | | | | |
|---|---|---|---|---|---|
| 019 | 3,174.65 | 3,101.26 | 3,307.45 | 3,377.99 | **12,961.35** |
| 020 | 1,864.67 | 1,826.04 | 1,964.45 | 1,966.44 | **7,621.60** |
| 021 | 10,021.84 | 10,014.50 | 10,265.58 | 10,589.62 | **40,891.54** |
| 022 | 158.29 | 139.01 | 151.00 | 157.84 | **606.14** |
| 023 | 3,355.71 | 3,345.06 | 3,528.02 | 3,699.77 | **13,928.56** |
| 024 | 1,100.51 | 1,035.83 | 1,079.06 | 1,090.04 | **4,305.45** |
| 025 | 1,083.09 | 1,095.91 | 1,138.13 | 1,127.80 | **4,444.93** |
| 026 | 1,616.61 | 1,611.72 | 1,661.96 | 1,705.21 | **6,595.50** |
| 027 | 6,018.12 | 5,806.15 | 6,121.94 | 6,470.46 | **24,416.66** |
| **MA - MASSACHUSETTS - Total** | **47,002.35** | **46,390.84** | **48,569.65** | **50,227.99** | **192,190.83** |

**STATE: MD - MARYLAND**

| REGISTRANT ZIP CODE 3 | 1ST QUARTER | 2ND QUARTER | 3RD QUARTER | 4TH QUARTER | TOTAL GRAMS |
|---|---|---|---|---|---|
| 206 | 2,313.09 | 2,402.10 | 2,596.95 | 2,718.79 | **10,030.93** |
| 207 | 1,415.54 | 1,283.31 | 1,478.50 | 1,418.15 | **5,595.50** |
| 208 | 1,122.66 | 1,128.93 | 1,168.92 | 1,177.95 | **4,598.45** |
| 209 | 401.35 | 393.19 | 411.05 | 386.58 | **1,592.17** |
| 210 | 4,270.12 | 4,451.80 | 4,688.70 | 4,803.23 | **18,213.84** |
| 211 | 1,644.19 | 1,671.81 | 1,740.93 | 1,758.36 | **6,815.29** |
| 212 | 12,438.70 | 12,525.68 | 13,003.96 | 13,473.09 | **51,441.43** |
| 214 | 983.84 | 992.68 | 1,084.83 | 1,095.23 | **4,156.58** |
| 215 | 1,679.18 | 1,743.40 | 1,789.45 | 1,898.01 | **7,110.04** |
| 216 | 872.07 | 915.44 | 976.50 | 1,030.65 | **3,794.66** |
| 217 | 2,660.44 | 2,731.49 | 2,890.90 | 2,951.47 | **11,234.31** |
| 218 | 914.63 | 943.21 | 993.89 | 993.25 | **3,844.98** |
| 219 | 1,345.47 | 1,267.49 | 1,451.41 | 1,444.25 | **5,508.62** |
| **MD MARYLAND Total** | **32,061.26** | **32,450.53** | **34,275.99** | **35,149.01** | **133,936.79** |

**STATE: ME - MAINE**

| REGISTRANT ZIP CODE 3 | 1ST QUARTER | 2ND QUARTER | 3RD QUARTER | 4TH QUARTER | TOTAL GRAMS |
|---|---|---|---|---|---|
| 039 | 85.66 | 79.74 | 111.08 | 116.56 | **393.03** |
| 040 | 2,874.95 | 3,027.08 | 3,205.58 | 3,218.59 | **12,326.20** |
| 041 | 1,172.55 | 1,216.58 | 1,294.96 | 1,320.17 | **5,004.26** |
| 042 | 1,787.91 | 1,823.96 | 1,922.88 | 1,943.65 | **7,478.40** |
| 043 | 1,057.28 | 1,161.48 | 1,192.92 | 1,256.44 | **4,668.12** |
| 044 | 1,346.79 | 1,455.31 | 1,563.58 | 1,559.01 | **5,924.68** |
| 045 | 448.38 | 485.44 | 524.45 | 525.53 | **1,983.80** |
| 046 | 826.53 | 911.80 | 953.09 | 995.87 | **3,687.29** |
| 047 | 397.58 | 416.76 | 451.60 | 477.73 | **1,743.67** |
| 048 | 908.80 | 989.63 | 1,021.81 | 1,149.21 | **4,069.45** |
| 049 | 1,632.21 | 1,797.97 | 1,817.16 | 1,836.63 | **7,083.97** |
| **ME MAINE Total** | **12,538.63** | **13,365.76** | **14,059.11** | **14,399.38** | **54,362.88** |

**STATE: MI - MICHIGAN**

| REGISTRANT ZIP CODE 3 | 1ST QUARTER | 2ND QUARTER | 3RD QUARTER | 4TH QUARTER | TOTAL GRAMS |
|---|---|---|---|---|---|
| 480 | 4,465.79 | 4,458.92 | 4,601.09 | 4,763.61 | **18,289.41** |
| 481 | 8,767.32 | 8,712.34 | 8,979.24 | 9,218.94 | **35,677.84** |
| 482 | 996.11 | 1,000.80 | 1,044.47 | 1,066.81 | **4,108.19** |
| 483 | 3,319.81 | 3,346.68 | 3,317.39 | 3,367.45 | **13,351.33** |
| 484 | 1,240.49 | 1,167.51 | 1,175.29 | 1,281.83 | **4,865.12** |
| 485 | 829.25 | 889.90 | 881.43 | 972.77 | **3,573.35** |
| 486 | 1,277.42 | 1,181.53 | 1,234.64 | 1,324.13 | **5,017.71** |
| 487 | 772.33 | 738.06 | 788.95 | 805.55 | **3,104.89** |
| 488 | 1,470.41 | 1,465.41 | 1,567.20 | 1,609.38 | **6,112.40** |
| 489 | 257.87 | 238.30 | 256.07 | 270.52 | **1,022.76** |
| 490 | 1,719.57 | 1,729.44 | 1,791.95 | 1,811.36 | **7,052.32** |
| 491 | 127.82 | 118.82 | 122.36 | 121.58 | **490.58** |
| 492 | 942.30 | 968.04 | 963.98 | 1,003.20 | **3,877.52** |
| 493 | 476.63 | 476.44 | 480.68 | 510.81 | **1,944.57** |
| 494 | 983.68 | 1,012.46 | 1,043.88 | 1,099.35 | **4,139.37** |
| 495 | 1,111.00 | 1,111.34 | 1,216.53 | 1,273.10 | **4,711.97** |
| 496 | 1,468.28 | 1,485.84 | 1,554.10 | 1,606.99 | **6,115.21** |
| 497 | 1,413.94 | 1,504.43 | 1,589.94 | 1,726.69 | **6,235.00** |
| 498 | 1,075.73 | 978.48 | 1,095.21 | 1,064.09 | **4,213.51** |
| 499 | 137.55 | 169.56 | 230.58 | 254.45 | **792.13** |
| **MI - MICHIGAN Total** | **32,853.28** | **32,754.31** | **33,934.97** | **35,152.61** | **134,695.17** |

**STATE: MN - MINNESOTA**

| REGISTRANT ZIP CODE 3 | 1ST QUARTER | 2ND QUARTER | 3RD QUARTER | 4TH QUARTER | TOTAL GRAMS |
|---|---|---|---|---|---|
| 550 | 1,116.28 | 1,171.02 | 1,176.26 | 1,215.23 | **4,678.79** |
| 551 | 1,717.67 | 1,835.11 | 1,849.51 | 1,951.42 | **7,353.70** |
| 553 | 1,255.96 | 1,294.89 | 1,347.62 | 1,344.61 | **5,243.08** |
| 554 | 2,624.41 | 2,688.02 | 2,817.67 | 3,043.99 | **11,174.09** |
| 556 | 18.86 | 28.46 | 30.26 | 26.00 | **103.58** |
| 557 | 330.09 | 372.47 | 391.15 | 437.27 | **1,530.99** |
| 558 | 536.34 | 563.97 | 541.26 | 570.77 | **2,212.34** |
| 559 | 275.77 | 295.79 | 265.57 | 283.31 | **1,120.45** |
| 560 | 338.34 | 325.66 | 318.72 | 341.54 | **1,324.26** |
| 561 | 9.59 | 15.51 | 14.98 | 11.37 | **51.45** |
| 562 | 115.51 | 126.16 | 144.55 | 147.50 | **533.71** |
| 563 | 500.36 | 511.11 | 524.92 | 571.12 | **2,107.51** |
| 564 | 184.34 | 202.12 | 224.45 | 258.30 | **869.21** |
| 565 | 465.57 | 470.02 | 464.66 | 493.62 | **1,893.88** |
| 566 | 713.34 | 805.57 | 814.56 | 795.29 | **3,128.77** |

| | | | | | |
|---|---|---|---|---|---|
| 567 | 52.31 | 64.33 | 98.60 | 110.20 | **325.45** |
| **MN MINNESOTA Total** | **10,254.75** | **10,770.22** | **11,024.75** | **11,601.55** | **43,651.27** |

**STATE: MO - MISSOURI**

| REGISTRANT ZIP CODE 3 | 1ST QUARTER | 2ND QUARTER | 3RD QUARTER | 4TH QUARTER | TOTAL GRAMS |
|---|---|---|---|---|---|
| 630 | 1,037.94 | 1,162.32 | 1,196.17 | 1,105.31 | **4,501.74** |
| 631 | 2,950.34 | 3,117.90 | 3,373.32 | 3,436.21 | **12,877.78** |
| 633 | 828.27 | 887.90 | 825.93 | 828.97 | **3,371.08** |
| 634 | 178.50 | 211.66 | 213.50 | 216.79 | **820.45** |
| 635 | 58.02 | 74.98 | 88.64 | 95.97 | **317.62** |
| 636 | 742.19 | 781.15 | 851.67 | 878.60 | **3,253.61** |
| 637 | 347.84 | 344.61 | 382.28 | 378.50 | **1,453.23** |
| 638 | 279.81 | 303.85 | 325.72 | 309.02 | **1,218.40** |
| 639 | 443.90 | 468.32 | 499.13 | 472.57 | **1,883.91** |
| 640 | 902.60 | 957.14 | 1,027.18 | 1,008.78 | **3,895.70** |
| 641 | 872.00 | 890.98 | 1,003.81 | 951.11 | **3,717.89** |
| 644 | 68.07 | 60.88 | 63.10 | 57.61 | **249.66** |
| 645 | 511.90 | 566.65 | 641.63 | 687.40 | **2,407.59** |
| 646 | 52.40 | 49.93 | 64.87 | 54.02 | **221.22** |
| 647 | 182.08 | 177.98 | 208.39 | 203.66 | **772.12** |
| 648 | 437.75 | 586.84 | 627.80 | 631.77 | **2,284.15** |
| 650 | 272.81 | 294.42 | 285.69 | 296.69 | **1,149.61** |
| 651 | 193.84 | 198.44 | 206.97 | 211.27 | **810.52** |
| 652 | 835.75 | 938.44 | 990.96 | 1,062.44 | **3,827.59** |
| 653 | 267.85 | 325.94 | 328.83 | 337.04 | **1,259.66** |
| 654 | 417.77 | 483.46 | 519.35 | 507.49 | **1,928.08** |
| 655 | 406.47 | 453.02 | 472.18 | 508.77 | **1,840.43** |
| 656 | 338.71 | 380.72 | 438.27 | 446.66 | **1,604.36** |
| 657 | 723.29 | 850.33 | 869.21 | 913.22 | **3,356.06** |
| 658 | 2,700.74 | 2,905.97 | 3,144.07 | 2,962.04 | **11,712.82** |
| **MO MISSOURI - Total** | **16,050.86** | **17,473.83** | **18,648.66** | **18,561.92** | **70,735.27** |

**STATE: MS - MISSISSIPPI**

| REGISTRANT ZIP CODE 3 | 1ST QUARTER | 2ND QUARTER | 3RD QUARTER | 4TH QUARTER | TOTAL GRAMS |
|---|---|---|---|---|---|
| 386 | 1,424.58 | 1,472.05 | 1,565.99 | 1,490.79 | **5,953.40** |
| 387 | 117.70 | 118.85 | 118.50 | 127.87 | **482.93** |
| 388 | 1,867.88 | 1,744.05 | 1,867.36 | 1,901.14 | **7,380.43** |
| 389 | 175.53 | 178.60 | 203.43 | 191.14 | **748.70** |
| 390 | 637.37 | 652.25 | 719.61 | 720.92 | **2,730.15** |
| 391 | 677.90 | 684.53 | 732.63 | 714.20 | **2,809.25** |
| 392 | 685.10 | 684.00 | 688.27 | 695.77 | **2,753.14** |
| 393 | 547.61 | 540.26 | 582.16 | 672.19 | **2,342.23** |

| | | | | | |
|---|---|---|---|---|---|
| 394 | 1,896.53 | 2,028.20 | 2,037.24 | 2,107.36 | **8,069.32** |
| 395 | 2,625.84 | 2,857.95 | 2,728.59 | 2,800.98 | **11,013.36** |
| 396 | 330.65 | 341.03 | 349.87 | 372.08 | **1,393.64** |
| 397 | 260.32 | 308.37 | 340.22 | 368.43 | **1,277.34** |
| **MS MISSISSIPPI - Total** | **11,247.01** | **11,610.13** | **11,933.86** | **12,162.88** | **46,953.89** |

**STATE: MT - MONTANA**

| REGISTRANT ZIP CODE 3 | 1ST QUARTER | 2ND QUARTER | 3RD QUARTER | 4TH QUARTER | TOTAL GRAMS |
|---|---|---|---|---|---|
| 590 | 120.24 | 116.00 | 137.94 | 147.53 | **521.72** |
| 591 | 397.80 | 428.89 | 491.92 | 505.31 | **1,823.93** |
| 592 | 141.76 | 141.49 | 165.41 | 171.46 | **620.12** |
| 593 | 67.07 | 75.71 | 76.33 | 75.69 | **294.80** |
| 594 | 770.19 | 824.42 | 889.28 | 933.91 | **3,417.80** |
| 595 | 66.48 | 88.72 | 116.15 | 122.85 | **394.20** |
| 596 | 368.13 | 396.80 | 425.02 | 435.86 | **1,625.82** |
| 597 | 331.01 | 345.64 | 354.75 | 380.41 | **1,411.81** |
| 598 | 839.90 | 932.30 | 898.10 | 962.29 | **3,632.59** |
| 599 | 331.46 | 417.59 | 432.51 | 387.35 | **1,568.91** |
| **MT - MONTANA Total** | **3,434.05** | **3,767.57** | **3,987.40** | **4,122.69** | **15,311.71** |

**STATE: NC - NORTH CAROLINA**

| REGISTRANT ZIP CODE 3 | 1ST QUARTER | 2ND QUARTER | 3RD QUARTER | 4TH QUARTER | TOTAL GRAMS |
|---|---|---|---|---|---|
| 270 | 1,061.94 | 1,220.79 | 1,334.84 | 1,398.61 | **5,016.18** |
| 271 | 941.62 | 956.69 | 1,059.82 | 1,085.13 | **4,043.26** |
| 272 | 2,288.92 | 2,373.85 | 2,736.43 | 2,701.81 | **10,101.01** |
| 273 | 1,676.46 | 1,714.04 | 1,895.19 | 1,894.78 | **7,180.47** |
| 274 | 856.02 | 877.34 | 912.68 | 935.35 | **3,581.38** |
| 275 | 3,469.67 | 3,534.00 | 3,799.70 | 3,505.35 | **14,308.73** |
| 276 | 1,504.39 | 1,645.01 | 1,730.55 | 1,575.97 | **6,455.91** |
| 277 | 2,139.30 | 2,245.58 | 2,372.26 | 2,263.81 | **9,020.96** |
| 278 | 1,872.04 | 2,066.73 | 2,194.22 | 2,128.57 | **8,261.56** |
| 279 | 666.54 | 671.33 | 720.42 | 680.77 | **2,739.06** |
| 280 | 4,468.16 | 4,895.48 | 5,290.88 | 4,689.94 | **19,344.46** |
| 281 | 3,057.10 | 2,651.41 | 2,962.00 | 2,914.81 | **11,585.32** |
| 282 | 2,251.08 | 1,992.49 | 2,470.92 | 2,473.25 | **9,187.74** |
| 283 | 4,625.14 | 4,799.97 | 5,219.52 | 5,066.34 | **19,710.97** |
| 284 | 5,243.48 | 5,230.62 | 5,336.19 | 4,806.68 | **20,616.96** |
| 285 | 2,028.31 | 2,044.30 | 2,294.05 | 2,115.43 | **8,482.10** |
| 286 | 7,322.35 | 7,723.97 | 7,838.78 | 7,088.52 | **29,973.62** |
| 287 | 3,456.99 | 3,791.81 | 4,061.02 | 3,822.50 | **15,132.32** |
| 288 | 2,255.53 | 2,071.61 | 2,051.12 | 2,156.32 | **8,534.58** |
| 289 | 255.48 | 265.34 | 273.03 | 255.08 | **1,048.93** |

| NC - NORTH CAROLINA Total | 51,440.53 | 52,772.36 | 56,553.60 | 53,559.03 | 214,325.52 |
|---|---|---|---|---|---|

**STATE: ND - NORTH DAKOTA**

| REGISTRANT ZIP CODE 3 | 1ST QUARTER | 2ND QUARTER | 3RD QUARTER | 4TH QUARTER | TOTAL GRAMS |
|---|---|---|---|---|---|
| 580 | 18.38 | 21.26 | 29.09 | 27.94 | **96.67** |
| 581 | 281.88 | 347.50 | 368.72 | 375.09 | **1,373.20** |
| 582 | 160.23 | 187.29 | 208.12 | 193.37 | **749.01** |
| 583 | 125.89 | 151.32 | 155.88 | 177.00 | **610.09** |
| 584 | 20.05 | 21.63 | 36.30 | 28.82 | **106.80** |
| 585 | 407.73 | 392.84 | 416.59 | 427.71 | **1,644.87** |
| 586 | 75.05 | 89.54 | 92.44 | 94.67 | **351.69** |
| 587 | 91.31 | 103.17 | 119.19 | 147.35 | **461.02** |
| 588 | 131.31 | 139.38 | 171.19 | 164.10 | **605.98** |
| **ND NORTH DAKOTA Total** | **1,311.83** | **1,453.93** | **1,597.52** | **1,636.04** | **5,999.33** |

**STATE: NE - NEBRASKA**

| REGISTRANT ZIP CODE 3 | 1ST QUARTER | 2ND QUARTER | 3RD QUARTER | 4TH QUARTER | TOTAL GRAMS |
|---|---|---|---|---|---|
| 680 | 82.43 | 83.08 | 103.05 | 99.45 | **368.02** |
| 681 | 665.42 | 647.93 | 730.95 | 735.77 | **2,780.07** |
| 683 | 49.18 | 50.33 | 49.50 | 56.67 | **205.68** |
| 684 | 24.47 | 30.08 | 33.81 | 34.73 | **123.09** |
| 685 | 327.33 | 325.24 | 365.13 | 394.42 | **1,412.13** |
| 686 | 40.06 | 48.30 | 55.91 | 48.17 | **192.44** |
| 687 | 58.78 | 71.22 | 79.50 | 76.17 | **285.66** |
| 688 | 116.93 | 127.34 | 167.64 | 151.64 | **563.55** |
| 689 | 23.56 | 27.19 | 29.07 | 27.78 | **107.60** |
| 690 | 1.70 | 1.02 | 0.50 | 1.31 | **4.52** |
| 691 | 288.89 | 260.93 | 304.88 | 357.70 | **1,212.39** |
| 692 | 4.14 | 2.01 | 1.92 | 2.17 | **10.24** |
| 693 | 23.60 | 15.76 | 32.37 | 52.34 | **124.07** |
| **NE NEBRASKA Total** | **1,706.50** | **1,690.42** | **1,954.24** | **2,038.30** | **7,389.45** |

**STATE: NH - NEW HAMPSHIRE**

| REGISTRANT ZIP CODE 3 | 1ST QUARTER | 2ND QUARTER | 3RD QUARTER | 4TH QUARTER | TOTAL GRAMS |
|---|---|---|---|---|---|
| 030 | 1,568.33 | 1,637.44 | 1,709.99 | 1,800.02 | **6,715.78** |
| 031 | 1,470.98 | 1,528.01 | 1,615.04 | 1,702.72 | **6,316.74** |
| 032 | 1,498.72 | 1,474.69 | 1,539.09 | 1,566.67 | **6,079.16** |
| 033 | 957.40 | 1,055.86 | 1,075.97 | 1,049.26 | **4,138.49** |
| 034 | 411.75 | 421.82 | 467.43 | 532.89 | **1,833.88** |
| 035 | 471.18 | 507.46 | 571.95 | 505.58 | **2,056.17** |
| 036 | 13.80 | 11.41 | 13.28 | 16.42 | **54.91** |
| 037 | 1,139.98 | 1,170.25 | 1,168.87 | 1,206.21 | **4,685.31** |
| 038 | 2,041.30 | 2,107.40 | 2,203.03 | 2,214.77 | **8,566.50** |

| NH - NEW HAMPSHIRE Total | 9,573.43 | 9,914.33 | 10,364.65 | 10,594.53 | 40,446.94 |
|---|---|---|---|---|---|

**STATE: NJ - NEW JERSEY**

| REGISTRANT ZIP CODE 3 | 1ST QUARTER | 2ND QUARTER | 3RD QUARTER | 4TH QUARTER | TOTAL GRAMS |
|---|---|---|---|---|---|
| 070 | 3,008.42 | 2,872.20 | 3,075.10 | 3,167.05 | **12,122.78** |
| 071 | 1,020.14 | 1,137.98 | 1,253.47 | 1,306.76 | **4,718.35** |
| 072 | 255.26 | 241.73 | 249.13 | 259.09 | **1,005.22** |
| 073 | 200.79 | 203.18 | 215.22 | 203.33 | **822.52** |
| 074 | 1,170.68 | 1,159.86 | 1,275.61 | 1,271.52 | **4,877.68** |
| 075 | 390.11 | 390.47 | 473.03 | 468.40 | **1,722.01** |
| 076 | 924.47 | 924.12 | 1,071.92 | 1,050.20 | **3,970.72** |
| 077 | 2,250.79 | 2,324.08 | 2,536.67 | 2,533.22 | **9,644.76** |
| 078 | 948.87 | 1,019.85 | 1,036.71 | 1,113.47 | **4,118.90** |
| 079 | 385.22 | 382.22 | 398.23 | 390.13 | **1,555.81** |
| 080 | 7,620.26 | 7,442.17 | 7,901.84 | 8,204.25 | **31,168.53** |
| 081 | 1,202.56 | 1,252.89 | 1,320.52 | 1,450.53 | **5,226.50** |
| 082 | 1,961.34 | 2,130.95 | 2,256.87 | 2,217.39 | **8,566.54** |
| 083 | 1,212.03 | 1,255.63 | 1,295.74 | 1,422.80 | **5,186.19** |
| 084 | 283.19 | 283.04 | 309.44 | 375.41 | **1,251.07** |
| 085 | 701.88 | 724.91 | 781.45 | 767.73 | **2,975.98** |
| 086 | 851.55 | 848.40 | 958.62 | 999.61 | **3,658.17** |
| 087 | 2,771.99 | 2,826.08 | 2,942.40 | 2,989.85 | **11,530.32** |
| 088 | 1,851.11 | 1,881.66 | 2,000.97 | 2,056.83 | **7,790.58** |
| 089 | 215.09 | 241.34 | 261.83 | 274.18 | **992.45** |
| NJ NEW JERSEY - Total | 29,225.78 | 29,542.76 | 31,614.77 | 32,521.75 | 122,905.06 |

**STATE: NM - NEW MEXICO**

| REGISTRANT ZIP CODE 3 | 1ST QUARTER | 2ND QUARTER | 3RD QUARTER | 4TH QUARTER | TOTAL GRAMS |
|---|---|---|---|---|---|
| 870 | 529.30 | 525.39 | 530.80 | 540.28 | **2,125.78** |
| 871 | 4,395.69 | 4,417.52 | 4,406.86 | 4,614.55 | **17,834.61** |
| 873 | 147.34 | 164.41 | 150.63 | 178.02 | **640.40** |
| 874 | 311.77 | 320.11 | 316.57 | 331.74 | **1,280.18** |
| 875 | 2,187.27 | 2,282.35 | 2,180.87 | 2,217.21 | **8,867.70** |
| 877 | 231.73 | 252.11 | 257.17 | 240.64 | **981.64** |
| 878 | 182.28 | 168.52 | 194.66 | 218.30 | **763.77** |
| 879 | 20.65 | 15.55 | 21.29 | 34.34 | **91.83** |
| 880 | 453.12 | 485.21 | 496.69 | 508.96 | **1,943.99** |
| 881 | 31.43 | 42.03 | 41.04 | 46.78 | **161.29** |
| 882 | 495.53 | 544.47 | 512.24 | 493.42 | **2,045.66** |
| 883 | 152.45 | 158.77 | 169.25 | 173.17 | **653.63** |
| 884 | 12.97 | 14.34 | 18.65 | 23.79 | **69.75** |
| NM NEW MEXICO Total | 9,151.52 | 9,390.80 | 9,296.73 | 9,621.18 | 37,460.23 |

**STATE: NV - NEVADA**

| REGISTRANT ZIP CODE 3 | 1ST QUARTER | 2ND QUARTER | 3RD QUARTER | 4TH QUARTER | TOTAL GRAMS |
|---|---|---|---|---|---|
| 890 | 890.73 | 859.27 | 910.66 | 932.62 | **3,593.29** |
| 891 | 2,005.76 | 2,117.29 | 2,132.42 | 2,225.84 | **8,481.31** |
| 893 | 11.51 | 16.82 | 18.62 | 15.06 | **61.99** |
| 894 | 470.80 | 443.92 | 480.60 | 512.41 | **1,907.72** |
| 895 | 668.58 | 626.84 | 669.43 | 665.23 | **2,630.09** |
| 897 | 172.85 | 191.84 | 203.45 | 228.50 | **796.65** |
| 898 | 109.21 | 91.40 | 89.76 | 95.83 | **386.20** |
| **NV NEVADA Total** | **4,329.44** | **4,347.38** | **4,504.95** | **4,675.48** | **17,857.25** |

**STATE: NY - NEW YORK**

| REGISTRANT ZIP CODE 3 | 1ST QUARTER | 2ND QUARTER | 3RD QUARTER | 4TH QUARTER | TOTAL GRAMS |
|---|---|---|---|---|---|
| 100 | 3,184.00 | 2,951.78 | 3,445.94 | 3,513.24 | **13,094.95** |
| 101 | 100.29 | 83.23 | 104.70 | 85.18 | **373.39** |
| 102 | 2.92 | 3.79 | 1.97 | 1.55 | **10.22** |
| 103 | 1,985.04 | 1,816.84 | 2,022.19 | 2,099.77 | **7,923.84** |
| 104 | 2,736.87 | 2,151.87 | 2,618.65 | 2,714.07 | **10,221.45** |
| 105 | 1,273.36 | 1,271.64 | 1,438.79 | 1,424.07 | **5,407.86** |
| 106 | 230.55 | 194.96 | 234.86 | 241.77 | **902.14** |
| 107 | 436.82 | 415.76 | 442.16 | 457.71 | **1,752.46** |
| 108 | 117.91 | 116.35 | 114.54 | 98.63 | **447.42** |
| 109 | 1,699.90 | 1,534.77 | 1,775.09 | 1,810.95 | **6,820.71** |
| 110 | 579.96 | 559.92 | 614.31 | 574.75 | **2,328.94** |
| 111 | 414.72 | 415.88 | 430.76 | 391.49 | **1,652.84** |
| 112 | 3,567.92 | 3,301.17 | 3,889.32 | 4,099.02 | **14,857.43** |
| 113 | 1,103.10 | 1,045.14 | 1,153.57 | 1,203.05 | **4,504.86** |
| 114 | 499.46 | 479.74 | 525.80 | 523.20 | **2,028.20** |
| 115 | 1,737.87 | 1,702.56 | 1,907.78 | 1,906.73 | **7,254.94** |
| 116 | 136.60 | 143.34 | 113.67 | 126.67 | **520.28** |
| 117 | 6,456.23 | 6,187.03 | 6,873.76 | 7,030.44 | **26,547.47** |
| 118 | 562.24 | 465.84 | 528.56 | 572.21 | **2,128.85** |
| 119 | 1,354.54 | 1,258.38 | 1,397.18 | 1,504.02 | **5,514.12** |
| 120 | 2,204.44 | 2,258.08 | 2,259.70 | 2,347.94 | **9,070.16** |
| 121 | 1,174.42 | 1,224.02 | 1,319.38 | 1,262.87 | **4,980.69** |
| 122 | 1,094.06 | 1,035.95 | 1,190.18 | 1,250.08 | **4,570.27** |
| 123 | 879.19 | 931.26 | 961.46 | 1,024.36 | **3,796.27** |
| 124 | 1,110.97 | 1,157.90 | 1,190.55 | 1,224.60 | **4,684.02** |
| 125 | 1,865.06 | 1,763.37 | 2,043.71 | 2,146.04 | **7,818.17** |
| 126 | 427.06 | 426.45 | 550.85 | 622.23 | **2,026.59** |
| 127 | 1,014.78 | 1,036.51 | 1,110.61 | 1,118.68 | **4,280.57** |

| | | | | | |
|---|---|---|---|---|---|
| 128 | 1,772.76 | 1,795.98 | 1,738.87 | 1,804.91 | **7,112.52** |
| 129 | 1,743.59 | 1,778.36 | 1,763.56 | 1,841.61 | **7,127.13** |
| 130 | 1,547.95 | 1,501.71 | 1,645.08 | 1,678.87 | **6,373.61** |
| 131 | 552.25 | 477.37 | 512.37 | 526.73 | **2,068.71** |
| 132 | 1,606.59 | 1,516.15 | 1,576.44 | 1,677.14 | **6,376.32** |
| 133 | 448.90 | 521.45 | 532.39 | 549.62 | **2,052.37** |
| 134 | 1,005.59 | 1,048.90 | 1,078.04 | 1,171.12 | **4,303.66** |
| 135 | 397.95 | 387.18 | 406.89 | 480.89 | **1,672.90** |
| 136 | 1,540.99 | 1,615.06 | 1,725.38 | 1,876.13 | **6,757.55** |
| 137 | 540.65 | 518.45 | 571.74 | 583.40 | **2,214.23** |
| 138 | 1,038.83 | 990.49 | 990.40 | 1,040.50 | **4,060.21** |
| 139 | 708.57 | 815.78 | 793.19 | 908.89 | **3,226.43** |
| 140 | 2,995.46 | 3,057.49 | 2,920.22 | 3,297.37 | **12,270.54** |
| 141 | 1,242.64 | 1,261.44 | 1,245.25 | 1,469.95 | **5,219.27** |
| 142 | 4,443.33 | 4,648.58 | 4,581.70 | 5,121.86 | **18,795.48** |
| 143 | 1,040.99 | 1,111.12 | 1,078.01 | 1,178.35 | **4,408.47** |
| 144 | 1,157.67 | 1,206.03 | 1,250.28 | 1,366.45 | **4,980.43** |
| 145 | 904.62 | 868.22 | 962.46 | 1,056.95 | **3,792.24** |
| 146 | 2,996.17 | 3,087.49 | 3,211.80 | 3,310.44 | **12,605.90** |
| 147 | 990.29 | 1,049.41 | 994.85 | 1,110.26 | **4,144.81** |
| 148 | 1,570.51 | 1,563.40 | 1,633.57 | 1,699.69 | **6,467.18** |
| 149 | 410.41 | 403.52 | 415.24 | 501.06 | **1,730.24** |
| **NY - NEW YORK - Total** | **68,607.00** | **67,157.11** | **71,887.74** | **75,627.52** | **283,279.37** |

**STATE: OH - OHIO**

| REGISTRANT ZIP CODE 3 | 1ST QUARTER | 2ND QUARTER | 3RD QUARTER | 4TH QUARTER | **TOTAL GRAMS** |
|---|---|---|---|---|---|
| 430 | 3,469.76 | 3,417.19 | 3,319.25 | 3,240.04 | **13,446.24** |
| 431 | 2,720.62 | 2,646.94 | 2,778.15 | 2,747.82 | **10,893.52** |
| 432 | 6,348.30 | 6,204.69 | 6,288.16 | 6,328.62 | **25,169.77** |
| 433 | 1,399.51 | 1,372.99 | 1,330.77 | 1,466.95 | **5,570.22** |
| 434 | 376.27 | 366.63 | 345.32 | 377.32 | **1,465.53** |
| 435 | 687.73 | 610.79 | 619.74 | 655.63 | **2,573.89** |
| 436 | 2,330.84 | 2,254.45 | 2,223.98 | 2,392.76 | **9,202.02** |
| 437 | 1,592.74 | 1,672.98 | 1,692.87 | 1,745.81 | **6,704.40** |
| 438 | 81.25 | 89.76 | 94.89 | 111.17 | **377.07** |
| 439 | 1,930.48 | 1,912.14 | 1,951.75 | 1,919.98 | **7,714.35** |
| 440 | 2,382.93 | 2,456.83 | 2,544.81 | 2,528.79 | **9,913.36** |
| 441 | 3,703.48 | 3,843.47 | 3,825.72 | 4,001.31 | **15,373.98** |
| 442 | 1,816.24 | 1,924.41 | 1,941.06 | 2,057.50 | **7,739.20** |
| 443 | 963.64 | 1,027.08 | 1,020.70 | 1,192.99 | **4,204.41** |
| 444 | 1,562.64 | 1,564.01 | 1,650.87 | 1,701.54 | **6,479.07** |

| | | | | | |
|---|---|---|---|---|---|
| 445 | 1,703.87 | 1,801.37 | 1,933.25 | 1,939.16 | **7,377.64** |
| 446 | 888.87 | 880.06 | 922.73 | 936.04 | **3,627.70** |
| 447 | 932.92 | 947.05 | 942.66 | 1,082.02 | **3,904.65** |
| 448 | 1,282.31 | 1,324.98 | 1,363.48 | 1,397.52 | **5,368.30** |
| 449 | 1,012.41 | 1,030.26 | 964.99 | 985.73 | **3,993.39** |
| 450 | 3,518.86 | 3,636.76 | 3,641.13 | 3,904.82 | **14,701.57** |
| 451 | 2,750.22 | 2,858.12 | 3,100.04 | 2,921.90 | **11,630.28** |
| 452 | 4,672.99 | 4,533.48 | 4,498.05 | 4,534.44 | **18,238.96** |
| 453 | 1,294.52 | 1,344.42 | 1,356.31 | 1,480.84 | **5,476.10** |
| 454 | 2,324.24 | 2,403.28 | 2,350.58 | 2,520.02 | **9,598.14** |
| 455 | 727.13 | 739.43 | 802.68 | 886.41 | **3,155.65** |
| 456 | 5,220.73 | 5,544.72 | 5,568.66 | 5,552.04 | **21,886.14** |
| 457 | 1,209.12 | 1,244.99 | 1,353.13 | 1,346.42 | **5,153.66** |
| 458 | 837.84 | 896.39 | 897.59 | 970.74 | **3,602.56** |
| **OH - OHIO Total** | **59,742.45** | **60,549.65** | **61,323.32** | **62,926.34** | **244,541.76** |

**STATE: OK - OKLAHOMA**

| REGISTRANT ZIP CODE 3 | 1ST QUARTER | 2ND QUARTER | 3RD QUARTER | 4TH QUARTER | TOTAL GRAMS |
|---|---|---|---|---|---|
| 730 | 2,583.39 | 2,590.23 | 2,850.76 | 2,898.00 | **10,922.38** |
| 731 | 3,340.11 | 3,406.23 | 3,520.95 | 3,469.21 | **13,736.50** |
| 734 | 225.83 | 215.37 | 217.78 | 218.35 | **877.32** |
| 735 | 667.24 | 722.34 | 748.01 | 759.39 | **2,896.99** |
| 736 | 133.05 | 129.58 | 153.18 | 153.43 | **569.25** |
| 737 | 257.48 | 273.20 | 294.53 | 290.36 | **1,115.57** |
| 738 | 83.79 | 96.96 | 88.77 | 96.84 | **366.36** |
| 739 | 25.79 | 18.63 | 18.03 | 21.23 | **83.68** |
| 740 | 3,072.54 | 3,245.50 | 3,357.16 | 3,547.44 | **13,222.64** |
| 741 | 2,110.09 | 2,180.10 | 2,480.53 | 2,527.77 | **9,298.48** |
| 743 | 377.45 | 392.62 | 419.02 | 481.03 | **1,670.13** |
| 744 | 1,239.14 | 1,301.94 | 1,450.34 | 1,621.26 | **5,612.68** |
| 745 | 373.92 | 367.92 | 393.36 | 403.01 | **1,538.20** |
| 746 | 223.62 | 250.92 | 235.95 | 239.25 | **949.75** |
| 747 | 250.27 | 276.79 | 283.87 | 312.86 | **1,123.80** |
| 748 | 888.31 | 900.22 | 929.91 | 955.59 | **3,674.02** |
| 749 | 353.85 | 399.91 | 459.81 | 572.60 | **1,786.18** |
| **OK - OKLAHOMA Total** | **16,205.87** | **16,768.47** | **17,901.95** | **18,567.63** | **69,443.93** |

**STATE: OR - OREGON**

| REGISTRANT ZIP CODE 3 | 1ST QUARTER | 2ND QUARTER | 3RD QUARTER | 4TH QUARTER | TOTAL GRAMS |
|---|---|---|---|---|---|
| 970 | 2,160.25 | 2,333.16 | 2,391.44 | 2,472.05 | **9,356.91** |
| 971 | 1,096.93 | 1,230.29 | 1,258.30 | 1,337.59 | **4,923.12** |
| 972 | 3,558.59 | 3,875.43 | 4,024.06 | 4,246.41 | **15,704.49** |

| | | | | | |
|---|---|---|---|---|---|
| 973 | 1,954.91 | 2,123.97 | 2,091.69 | 2,297.44 | **8,468.01** |
| 974 | 2,065.50 | 2,204.08 | 2,281.51 | 2,472.73 | **9,023.82** |
| 975 | 1,360.50 | 1,524.29 | 1,531.53 | 1,560.51 | **5,976.83** |
| 976 | 90.83 | 113.50 | 114.11 | 140.56 | **459.00** |
| 977 | 508.64 | 557.74 | 563.32 | 630.77 | **2,260.47** |
| 978 | 946.92 | 1,033.11 | 1,029.03 | 1,080.47 | **4,089.54** |
| 979 | 84.38 | 92.29 | 108.71 | 100.58 | **385.96** |
| **OR OREGON Total** | **13,827.45** | **15,087.87** | **15,393.72** | **16,339.10** | **60,648.14** |

**STATE: PA - PENNSYLVANIA**

| REGISTRANT ZIP CODE 3 | 1ST QUARTER | 2ND QUARTER | 3RD QUARTER | 4TH QUARTER | TOTAL GRAMS |
|---|---|---|---|---|---|
| 150 | 4,200.95 | 4,263.90 | 4,351.48 | 4,532.27 | **17,348.60** |
| 151 | 3,236.40 | 2,901.12 | 3,311.72 | 3,417.41 | **12,866.65** |
| 152 | 6,447.38 | 6,436.30 | 6,477.29 | 6,561.61 | **25,922.58** |
| 153 | 2,097.97 | 2,098.64 | 2,178.15 | 2,084.27 | **8,459.03** |
| 154 | 2,251.65 | 2,287.07 | 2,402.83 | 2,433.01 | **9,374.56** |
| 155 | 983.59 | 1,171.22 | 1,371.71 | 1,249.83 | **4,776.35** |
| 156 | 3,887.88 | 3,400.13 | 3,421.90 | 3,567.52 | **14,277.44** |
| 157 | 883.80 | 953.87 | 1,032.16 | 1,071.20 | **3,941.04** |
| 158 | 683.26 | 719.33 | 728.97 | 767.59 | **2,899.16** |
| 159 | 1,512.86 | 1,493.37 | 1,477.45 | 1,456.17 | **5,939.84** |
| 160 | 1,671.24 | 1,752.28 | 1,828.84 | 1,877.53 | **7,129.89** |
| 161 | 1,595.90 | 1,591.84 | 1,675.53 | 1,689.34 | **6,552.62** |
| 162 | 906.54 | 917.69 | 981.09 | 1,019.73 | **3,825.06** |
| 163 | 1,016.84 | 1,011.84 | 1,105.52 | 1,080.60 | **4,214.81** |
| 164 | 305.85 | 328.52 | 333.69 | 332.61 | **1,300.68** |
| 165 | 1,938.78 | 1,993.89 | 1,992.01 | 2,011.51 | **7,936.20** |
| 166 | 2,198.22 | 2,328.52 | 2,342.47 | 2,479.43 | **9,348.64** |
| 167 | 269.56 | 322.65 | 347.67 | 359.89 | **1,299.77** |
| 168 | 953.27 | 1,010.37 | 1,131.27 | 1,233.13 | **4,328.03** |
| 169 | 240.22 | 254.54 | 308.69 | 350.56 | **1,154.00** |
| 170 | 2,260.19 | 2,350.22 | 2,323.23 | 2,464.88 | **9,398.52** |
| 171 | 652.96 | 696.92 | 747.92 | 779.36 | **2,877.17** |
| 172 | 1,045.82 | 1,107.07 | 1,124.53 | 1,162.30 | **4,439.72** |
| 173 | 1,242.73 | 1,263.72 | 1,303.00 | 1,386.15 | **5,195.60** |
| 174 | 676.86 | 616.87 | 695.48 | 780.74 | **2,769.96** |
| 175 | 451.81 | 490.80 | 499.42 | 515.46 | **1,957.49** |
| 176 | 577.08 | 605.55 | 644.04 | 678.16 | **2,504.84** |
| 177 | 2,037.64 | 2,178.23 | 2,209.30 | 2,193.62 | **8,618.80** |
| 178 | 1,073.63 | 1,183.76 | 1,217.81 | 1,333.60 | **4,808.80** |
| 179 | 606.15 | 605.66 | 586.48 | 685.50 | **2,483.80** |

| | | | | | |
|---|---|---|---|---|---|
| 180 | 977.69 | 982.10 | 990.53 | 1,057.99 | **4,008.31** |
| 181 | 655.75 | 651.82 | 780.84 | 819.25 | **2,907.67** |
| 182 | 485.26 | 515.18 | 501.15 | 506.00 | **2,007.59** |
| 183 | 573.68 | 551.15 | 560.29 | 616.01 | **2,301.13** |
| 184 | 1,192.35 | 1,175.52 | 1,331.63 | 1,528.63 | **5,228.13** |
| 185 | 2,541.16 | 2,676.90 | 2,605.72 | 2,474.00 | **10,297.78** |
| 186 | 1,076.20 | 1,110.37 | 1,138.22 | 1,306.69 | **4,631.47** |
| 187 | 2,140.86 | 2,398.42 | 2,227.07 | 2,465.97 | **9,232.31** |
| 188 | 512.28 | 535.08 | 536.90 | 573.23 | **2,157.50** |
| 189 | 1,240.82 | 1,315.99 | 1,409.77 | 1,362.04 | **5,328.63** |
| 190 | 5,878.98 | 5,968.26 | 6,177.74 | 6,274.01 | **24,298.98** |
| 191 | 10,360.83 | 10,553.75 | 11,156.16 | 12,185.40 | **44,256.14** |
| 193 | 1,139.30 | 1,125.73 | 1,201.46 | 1,190.01 | **4,656.49** |
| 194 | 1,911.30 | 1,860.68 | 1,911.35 | 1,986.38 | **7,669.72** |
| 195 | 349.85 | 375.30 | 390.17 | 420.51 | **1,535.84** |
| 196 | 733.84 | 747.11 | 731.16 | 853.03 | **3,065.13** |
| **PA - PENNSYLVANIA - Total** | **79,677.19** | **80,879.27** | **83,801.84** | **87,174.14** | **331,532.43** |

**STATE: PR - PUERTO RICO**

| REGISTRANT ZIP CODE 3 | 1ST QUARTER | 2ND QUARTER | 3RD QUARTER | 4TH QUARTER | TOTAL GRAMS |
|---|---|---|---|---|---|
| 006 | 494.88 | 479.87 | 569.43 | 508.38 | **2,052.56** |
| 007 | 1,209.66 | 1,285.44 | 1,051.35 | 1,288.98 | **4,835.43** |
| 009 | 444.69 | 475.29 | 463.51 | 488.74 | **1,872.23** |
| **PR - PUERTO RICO - Total** | **2,149.24** | **2,240.60** | **2,084.30** | **2,286.09** | **8,760.22** |

**STATE: RI - RHODE ISLAND**

| REGISTRANT ZIP CODE 3 | 1ST QUARTER | 2ND QUARTER | 3RD QUARTER | 4TH QUARTER | TOTAL GRAMS |
|---|---|---|---|---|---|
| 028 | 5,030.96 | 4,915.35 | 5,291.85 | 5,291.37 | **20,529.54** |
| 029 | 3,676.95 | 3,527.27 | 3,783.52 | 3,828.02 | **14,815.75** |
| **RI RHODE ISLAND Total** | **8,707.91** | **8,442.62** | **9,075.37** | **9,119.39** | **35,345.29** |

**STATE: SC - SOUTH CAROLINA**

| REGISTRANT ZIP CODE 3 | 1ST QUARTER | 2ND QUARTER | 3RD QUARTER | 4TH QUARTER | TOTAL GRAMS |
|---|---|---|---|---|---|
| 290 | 589.71 | 616.76 | 628.86 | 627.40 | **2,462.73** |
| 291 | 543.16 | 618.18 | 633.76 | 656.29 | **2,451.39** |
| 292 | 496.87 | 550.68 | 560.48 | 602.99 | **2,211.02** |
| 293 | 1,972.39 | 2,132.07 | 2,342.18 | 2,413.46 | **8,860.10** |
| 294 | 2,318.97 | 2,342.28 | 2,425.87 | 2,525.73 | **9,612.85** |
| 295 | 3,437.22 | 3,722.04 | 4,013.37 | 3,914.70 | **15,087.33** |
| 296 | 4,034.60 | 4,231.82 | 4,411.38 | 4,322.37 | **17,000.16** |
| 297 | 881.29 | 965.35 | 1,021.29 | 1,017.99 | **3,885.92** |
| 298 | 230.29 | 227.52 | 246.14 | 314.45 | **1,018.39** |
| 299 | 517.10 | 582.75 | 536.74 | 491.02 | **2,127.62** |

| SC - SOUTH CAROLINA Total | 15,021.60 | 15,989.45 | 16,820.07 | 16,886.39 | 64,717.50 |
|---|---|---|---|---|---|

**STATE: SD - SOUTH DAKOTA**

| REGISTRANT ZIP CODE 3 | 1ST QUARTER | 2ND QUARTER | 3RD QUARTER | 4TH QUARTER | TOTAL GRAMS |
|---|---|---|---|---|---|
| 570 | 61.25 | 72.51 | 83.92 | 73.00 | **290.68** |
| 571 | 297.96 | 291.18 | 343.12 | 349.83 | **1,282.09** |
| 572 | 47.81 | 50.48 | 61.44 | 57.03 | **216.76** |
| 573 | 15.47 | 19.86 | 26.38 | 38.11 | **99.82** |
| 574 | 77.54 | 80.73 | 99.32 | 96.34 | **353.94** |
| 575 | 28.90 | 31.47 | 34.74 | 47.91 | **143.02** |
| 576 | 6.48 | 6.85 | 9.08 | 11.30 | **33.71** |
| 577 | 245.33 | 318.60 | 392.30 | 452.43 | **1,408.66** |
| SD SOUTH DAKOTA - Total | 780.73 | 871.68 | 1,050.30 | 1,125.97 | 3,828.67 |

**STATE: TN - TENNESSEE**

| REGISTRANT ZIP CODE 3 | 1ST QUARTER | 2ND QUARTER | 3RD QUARTER | 4TH QUARTER | TOTAL GRAMS |
|---|---|---|---|---|---|
| 370 | 3,856.70 | 4,313.07 | 4,615.65 | 4,770.98 | **17,556.39** |
| 371 | 4,421.15 | 5,138.01 | 5,241.62 | 5,371.14 | **20,171.92** |
| 372 | 2,420.15 | 2,508.97 | 2,717.66 | 2,859.12 | **10,505.89** |
| 373 | 2,208.06 | 2,577.50 | 2,603.04 | 2,645.18 | **10,033.78** |
| 374 | 772.58 | 832.42 | 792.14 | 816.69 | **3,213.83** |
| 376 | 6,570.35 | 7,791.86 | 7,425.74 | 7,392.99 | **29,180.94** |
| 377 | 3,255.20 | 3,498.61 | 3,460.17 | 3,387.98 | **13,601.97** |
| 378 | 5,106.02 | 5,388.96 | 5,380.31 | 5,469.64 | **21,344.93** |
| 379 | 2,782.27 | 2,952.62 | 2,930.17 | 2,951.27 | **11,616.33** |
| 380 | 993.81 | 1,041.83 | 1,102.27 | 1,108.78 | **4,246.69** |
| 381 | 1,974.10 | 2,124.12 | 2,266.07 | 2,301.88 | **8,666.17** |
| 382 | 542.51 | 536.07 | 567.39 | 627.03 | **2,273.00** |
| 383 | 1,597.48 | 1,695.46 | 1,872.62 | 1,873.81 | **7,039.38** |
| 384 | 1,220.10 | 1,279.72 | 1,250.13 | 1,343.96 | **5,093.91** |
| 385 | 1,420.93 | 1,435.79 | 1,459.64 | 1,488.64 | **5,805.00** |
| TN TENNESSEE Total | 39,141.41 | 43,115.01 | 43,684.62 | 44,409.10 | 170,350.15 |

**STATE: TX - TEXAS**

| REGISTRANT ZIP CODE 3 | 1ST QUARTER | 2ND QUARTER | 3RD QUARTER | 4TH QUARTER | TOTAL GRAMS |
|---|---|---|---|---|---|
| 750 | 2,861.87 | 2,853.61 | 3,084.62 | 3,076.53 | **11,876.63** |
| 751 | 900.63 | 935.84 | 971.95 | 929.64 | **3,738.06** |
| 752 | 1,330.98 | 1,405.89 | 1,466.19 | 1,412.96 | **5,616.02** |
| 753 | 2.02 | 3.17 | 2.56 | 3.99 | **11.75** |
| 754 | 517.66 | 546.69 | 592.00 | 629.43 | **2,285.77** |
| 755 | 145.72 | 156.11 | 163.35 | 148.35 | **613.53** |
| 756 | 593.35 | 638.06 | 644.38 | 604.13 | **2,479.92** |
| 757 | 572.74 | 616.13 | 675.96 | 662.49 | **2,527.31** |

| | | | | | |
|---|---|---|---|---|---|
| 758 | 67.34 | 69.96 | 74.22 | 90.64 | **302.16** |
| 759 | 344.17 | 373.02 | 370.09 | 346.43 | **1,433.72** |
| 760 | 2,254.63 | 2,316.98 | 2,355.83 | 2,315.65 | **9,243.08** |
| 761 | 1,190.33 | 1,253.88 | 1,262.75 | 1,288.27 | **4,995.23** |
| 762 | 611.79 | 635.31 | 641.22 | 643.98 | **2,532.30** |
| 763 | 365.65 | 399.56 | 377.87 | 385.49 | **1,528.58** |
| 764 | 134.00 | 130.74 | 147.72 | 148.14 | **560.60** |
| 765 | 424.33 | 396.84 | 425.51 | 401.29 | **1,647.97** |
| 766 | 89.37 | 79.24 | 83.41 | 98.54 | **350.56** |
| 767 | 223.08 | 219.50 | 267.09 | 288.69 | **998.36** |
| 768 | 54.37 | 59.58 | 47.42 | 52.47 | **213.85** |
| 769 | 112.49 | 132.74 | 123.56 | 128.77 | **497.57** |
| 770 | 2,827.80 | 2,864.28 | 3,043.18 | 3,001.48 | **11,736.75** |
| 772 | 0.00 | 0.00 | 0 | 0 | **0.00** |
| 773 | 2,389.72 | 2,430.66 | 2,490.82 | 2,479.71 | **9,790.91** |
| 774 | 962.02 | 982.14 | 1,045.41 | 955.45 | **3,945.02** |
| 775 | 2,378.37 | 2,461.67 | 2,556.52 | 2,505.40 | **9,901.95** |
| 776 | 920.90 | 986.29 | 937.99 | 996.29 | **3,841.47** |
| 777 | 310.84 | 383.94 | 342.29 | 356.15 | **1,393.22** |
| 778 | 316.52 | 326.21 | 323.63 | 333.93 | **1,300.28** |
| 779 | 193.06 | 191.27 | 216.13 | 224.70 | **825.15** |
| 780 | 337.70 | 349.53 | 369.64 | 357.88 | **1,414.76** |
| 781 | 394.42 | 370.84 | 400.56 | 395.45 | **1,561.27** |
| 782 | 1,196.77 | 1,277.35 | 1,326.25 | 1,200.44 | **5,000.81** |
| 783 | 164.56 | 164.05 | 185.00 | 167.80 | **681.42** |
| 784 | 377.35 | 358.73 | 383.68 | 380.63 | **1,500.39** |
| 785 | 121.05 | 111.12 | 173.22 | 138.62 | **544.00** |
| 786 | 1,398.07 | 1,188.89 | 1,497.77 | 1,437.25 | **5,521.98** |
| 787 | 1,621.09 | 1,600.24 | 1,733.85 | 1,676.25 | **6,631.43** |
| 788 | 158.82 | 91.33 | 93.35 | 68.96 | **412.45** |
| 789 | 31.18 | 36.56 | 32.71 | 30.91 | **131.37** |
| 790 | 36.59 | 44.30 | 70.48 | 63.17 | **214.54** |
| 791 | 158.41 | 177.95 | 191.29 | 212.21 | **739.85** |
| 792 | 2.96 | 1.63 | 2.36 | 5.73 | **12.68** |
| 793 | 68.57 | 71.90 | 54.85 | 52.35 | **247.66** |
| 794 | 213.47 | 226.16 | 229.01 | 238.08 | **906.71** |
| 795 | 37.95 | 42.03 | 43.05 | 45.80 | **168.82** |
| 796 | 316.46 | 340.55 | 340.40 | 358.14 | **1,355.55** |
| 797 | 715.09 | 754.14 | 724.18 | 718.42 | **2,911.82** |
| 798 | 6.67 | 6.86 | 7.15 | 10.46 | **31.15** |

| | | | | | |
|---|---|---|---|---|---|
| 799 | 166.89 | 187.88 | 227.78 | 184.57 | **767.13** |
| **TX - TEXAS Total** | **30,619.84** | **31,251.33** | **32,820.21** | **32,252.14** | **126,943.52** |

**STATE: UT - UTAH**

| REGISTRANT ZIP CODE 3 | 1ST QUARTER | 2ND QUARTER | 3RD QUARTER | 4TH QUARTER | TOTAL GRAMS |
|---|---|---|---|---|---|
| 840 | 4,951.38 | 4,886.11 | 4,880.44 | 5,132.00 | **19,849.92** |
| 841 | 2,618.87 | 2,646.93 | 2,707.25 | 3,023.41 | **10,996.46** |
| 843 | 414.17 | 405.58 | 402.52 | 418.89 | **1,641.16** |
| 844 | 963.88 | 966.95 | 992.55 | 973.34 | **3,896.72** |
| 845 | 309.15 | 320.55 | 336.31 | 344.12 | **1,310.13** |
| 846 | 1,278.80 | 1,232.69 | 1,364.08 | 1,404.20 | **5,279.78** |
| 847 | 556.97 | 563.57 | 597.97 | 651.23 | **2,369.73** |
| **UT - UTAH - Total** | **11,093.22** | **11,022.38** | **11,281.12** | **11,947.19** | **45,343.90** |

**STATE: VA - VIRGINIA**

| REGISTRANT ZIP CODE 3 | 1ST QUARTER | 2ND QUARTER | 3RD QUARTER | 4TH QUARTER | TOTAL GRAMS |
|---|---|---|---|---|---|
| 201 | 1,728.59 | 1,626.61 | 1,780.01 | 1,694.81 | **6,830.02** |
| 220 | 715.78 | 718.56 | 759.76 | 715.16 | **2,909.26** |
| 221 | 678.50 | 683.66 | 721.38 | 656.74 | **2,740.28** |
| 222 | 161.96 | 155.71 | 233.29 | 197.37 | **748.33** |
| 223 | 198.81 | 251.62 | 238.70 | 219.33 | **908.46** |
| 224 | 555.99 | 588.15 | 634.05 | 663.63 | **2,441.82** |
| 225 | 259.29 | 290.11 | 294.80 | 285.82 | **1,130.02** |
| 226 | 1,146.99 | 1,200.16 | 1,302.56 | 1,242.29 | **4,892.00** |
| 227 | 242.75 | 211.09 | 247.09 | 247.75 | **948.68** |
| 228 | 227.09 | 233.24 | 258.45 | 266.33 | **985.11** |
| 229 | 550.02 | 607.67 | 666.57 | 707.78 | **2,532.04** |
| 230 | 392.64 | 397.06 | 426.87 | 458.22 | **1,674.78** |
| 231 | 1,145.79 | 1,258.01 | 1,225.74 | 1,385.94 | **5,015.48** |
| 232 | 2,240.77 | 2,421.87 | 2,495.13 | 2,665.83 | **9,823.61** |
| 233 | 398.40 | 402.07 | 447.08 | 423.84 | **1,671.39** |
| 234 | 698.54 | 679.18 | 775.26 | 757.54 | **2,910.51** |
| 235 | 175.69 | 211.81 | 243.53 | 287.38 | **918.41** |
| 236 | 700.23 | 732.51 | 795.87 | 811.25 | **3,039.86** |
| 237 | 114.05 | 116.55 | 135.26 | 157.56 | **523.41** |
| 238 | 492.60 | 458.98 | 543.74 | 536.25 | **2,031.57** |
| 239 | 147.75 | 139.24 | 150.29 | 154.41 | **591.69** |
| 240 | 2,238.67 | 2,350.45 | 2,521.50 | 2,417.17 | **9,527.79** |
| 241 | 906.27 | 947.59 | 1,056.68 | 1,066.02 | **3,976.55** |
| 242 | 4,180.32 | 4,685.36 | 4,572.79 | 4,572.65 | **18,011.12** |
| 243 | 1,514.00 | 1,692.48 | 1,869.50 | 1,694.64 | **6,770.62** |
| 244 | 171.75 | 190.57 | 209.76 | 230.51 | **802.59** |

| | | | | | |
|---|---|---|---|---|---|
| 245 | 1,084.60 | 1,162.24 | 1,273.47 | 1,218.90 | **4,739.21** |
| 246 | 1,213.20 | 1,244.41 | 1,321.52 | 1,248.13 | **5,027.26** |
| **VA VIRGINIA Total** | **24,281.03** | **25,656.95** | **27,200.65** | **26,983.25** | **104,121.87** |

**STATE: VI - VIRGIN ISLANDS**

| REGISTRANT ZIP CODE 3 | 1ST QUARTER | 2ND QUARTER | 3RD QUARTER | 4TH QUARTER | TOTAL GRAMS |
|---|---|---|---|---|---|
| 008 | 17.74 | 19.02 | 23.63 | 23.79 | **84.19** |
| **VI - VIRGIN ISLANDS Total** | **17.74** | **19.02** | **23.63** | **23.79** | **84.19** |

**STATE: VT - VERMONT**

| REGISTRANT ZIP CODE 3 | 1ST QUARTER | 2ND QUARTER | 3RD QUARTER | 4TH QUARTER | TOTAL GRAMS |
|---|---|---|---|---|---|
| 050 | 798.52 | 804.40 | 747.08 | 757.31 | **3,107.31** |
| 051 | 338.94 | 382.57 | 414.09 | 412.94 | **1,548.54** |
| 052 | 525.48 | 582.31 | 608.53 | 626.83 | **2,343.14** |
| 053 | 329.21 | 322.59 | 318.17 | 337.26 | **1,307.23** |
| 054 | 3,091.83 | 3,069.49 | 3,241.15 | 3,352.28 | **12,754.74** |
| 056 | 867.21 | 770.71 | 813.21 | 863.65 | **3,314.77** |
| 057 | 1,046.71 | 1,053.50 | 1,030.75 | 1,088.52 | **4,219.49** |
| 058 | 480.27 | 507.19 | 461.54 | 537.78 | **1,986.78** |
| **VT VERMONT Total** | **7,478.16** | **7,492.76** | **7,634.51** | **7,976.57** | **30,581.99** |

**STATE: WA - WASHINGTON**

| REGISTRANT ZIP CODE 3 | 1ST QUARTER | 2ND QUARTER | 3RD QUARTER | 4TH QUARTER | TOTAL GRAMS |
|---|---|---|---|---|---|
| 980 | 4,028.89 | 4,181.81 | 4,231.35 | 4,499.59 | **16,941.63** |
| 981 | 3,201.31 | 3,307.81 | 3,548.19 | 3,777.98 | **13,835.29** |
| 982 | 5,073.24 | 5,264.18 | 5,590.24 | 6,048.31 | **21,975.98** |
| 983 | 2,518.08 | 2,646.23 | 2,814.97 | 3,106.57 | **11,085.84** |
| 984 | 1,635.92 | 1,713.75 | 1,769.75 | 1,853.73 | **6,973.14** |
| 985 | 2,419.78 | 2,604.37 | 2,819.78 | 2,966.00 | **10,809.94** |
| 986 | 2,271.36 | 2,486.97 | 2,674.36 | 2,937.53 | **10,370.22** |
| 988 | 979.77 | 1,027.72 | 1,097.50 | 1,210.15 | **4,315.14** |
| 989 | 1,087.71 | 1,166.74 | 1,212.19 | 1,282.08 | **4,748.71** |
| 990 | 287.17 | 298.53 | 305.53 | 330.42 | **1,221.65** |
| 991 | 464.21 | 512.81 | 535.82 | 603.77 | **2,116.62** |
| 992 | 2,343.02 | 2,489.13 | 2,597.09 | 2,748.71 | **10,177.96** |
| 993 | 1,335.19 | 1,340.32 | 1,485.52 | 1,659.71 | **5,820.74** |
| 994 | 275.63 | 283.63 | 324.69 | 344.72 | **1,228.67** |
| **WA WASHINGTON Total** | **27,921.27** | **29,324.00** | **31,006.99** | **33,369.27** | **121,621.53** |

**STATE: WI - WISCONSIN**

| REGISTRANT ZIP CODE 3 | 1ST QUARTER | 2ND QUARTER | 3RD QUARTER | 4TH QUARTER | TOTAL GRAMS |
|---|---|---|---|---|---|
| 530 | 1,396.41 | 1,419.02 | 1,479.43 | 1,457.40 | **5,752.25** |
| 531 | 1,770.14 | 1,841.16 | 1,946.26 | 2,091.82 | **7,649.38** |
| 532 | 3,253.26 | 3,282.63 | 3,414.17 | 3,508.53 | **13,458.59** |

| | | | | | |
|---|---|---|---|---|---|
| 534 | 325.79 | 356.26 | 389.31 | 415.85 | **1,487.21** |
| 535 | 1,194.80 | 1,245.01 | 1,269.02 | 1,336.84 | **5,045.68** |
| 537 | 905.92 | 968.22 | 954.89 | 975.14 | **3,804.17** |
| 538 | 33.16 | 29.24 | 27.38 | 35.85 | **125.62** |
| 539 | 577.25 | 603.23 | 641.29 | 665.11 | **2,486.88** |
| 540 | 111.45 | 119.03 | 120.94 | 128.12 | **479.54** |
| 541 | 486.80 | 548.48 | 537.22 | 562.84 | **2,135.33** |
| 542 | 131.59 | 131.60 | 150.86 | 147.97 | **562.03** |
| 543 | 350.64 | 361.79 | 411.82 | 420.33 | **1,544.58** |
| 544 | 553.07 | 480.22 | 583.24 | 556.12 | **2,172.65** |
| 545 | 294.41 | 304.29 | 293.38 | 296.15 | **1,188.22** |
| 546 | 321.64 | 341.51 | 347.38 | 360.91 | **1,371.44** |
| 547 | 282.14 | 349.14 | 402.64 | 387.68 | **1,421.60** |
| 548 | 293.09 | 310.11 | 365.46 | 397.43 | **1,366.10** |
| 549 | 1,152.40 | 1,187.38 | 1,248.99 | 1,224.76 | **4,813.52** |
| **WI - WISCONSIN Total** | **13,433.94** | **13,878.31** | **14,583.69** | **14,968.85** | **56,864.79** |

**STATE: WV - WEST VIRGINIA**

| REGISTRANT ZIP CODE 3 | 1ST QUARTER | 2ND QUARTER | 3RD QUARTER | 4TH QUARTER | TOTAL GRAMS |
|---|---|---|---|---|---|
| 247 | 355.24 | 424.56 | 586.62 | 677.69 | **2,044.11** |
| 248 | 535.76 | 578.33 | 552.04 | 527.25 | **2,193.38** |
| 249 | 107.70 | 104.80 | 120.92 | 98.54 | **431.96** |
| 250 | 719.82 | 559.81 | 630.59 | 582.99 | **2,493.21** |
| 251 | 413.34 | 407.08 | 465.89 | 450.84 | **1,737.14** |
| 252 | 351.70 | 308.67 | 281.36 | 291.96 | **1,233.69** |
| 253 | 1,381.85 | 1,448.49 | 1,519.98 | 1,485.79 | **5,836.11** |
| 254 | 1,442.42 | 1,435.62 | 1,567.73 | 1,643.19 | **6,088.95** |
| 255 | 1,481.85 | 1,404.87 | 1,530.32 | 1,530.16 | **5,947.20** |
| 256 | 1,327.23 | 1,423.98 | 1,514.35 | 1,388.88 | **5,654.44** |
| 257 | 1,638.36 | 2,392.97 | 2,540.60 | 2,813.57 | **9,385.49** |
| 258 | 2,403.35 | 2,479.58 | 2,430.45 | 2,456.29 | **9,769.68** |
| 259 | 224.43 | 297.45 | 283.14 | 271.66 | **1,076.68** |
| 260 | 1,891.39 | 1,989.73 | 1,855.27 | 1,904.38 | **7,640.76** |
| 261 | 666.77 | 632.75 | 713.27 | 763.82 | **2,776.60** |
| 262 | 353.79 | 346.95 | 293.08 | 250.17 | **1,243.99** |
| 263 | 900.13 | 943.41 | 953.15 | 932.64 | **3,729.33** |
| 264 | 117.69 | 153.53 | 140.80 | 142.05 | **554.07** |
| 265 | 1,243.97 | 1,254.06 | 1,294.78 | 1,365.41 | **5,158.22** |
| 266 | 153.96 | 177.49 | 180.62 | 175.51 | **687.57** |
| 267 | 362.57 | 333.91 | 309.38 | 346.73 | **1,352.60** |
| 268 | 11.30 | 12.22 | 15.43 | 18.45 | **57.40** |

| WV WEST VIRGINIA Total | 18,084.61 | 19,110.25 | 19,779.77 | 20,117.97 | 77,092.60 |
|---|---|---|---|---|---|

STATE: WY - WYOMING

| REGISTRANT ZIP CODE 3 | 1ST QUARTER | 2ND QUARTER | 3RD QUARTER | 4TH QUARTER | TOTAL GRAMS |
|---|---|---|---|---|---|
| 820 | 237.70 | 249.90 | 247.69 | 268.94 | **1,004.23** |
| 822 | 20.26 | 22.22 | 19.65 | 22.95 | **85.07** |
| 823 | 13.20 | 13.35 | 18.28 | 21.12 | **65.94** |
| 824 | 145.21 | 161.52 | 152.60 | 174.30 | **633.62** |
| 825 | 96.56 | 103.30 | 121.09 | 132.86 | **453.81** |
| 826 | 468.09 | 482.09 | 482.26 | 490.89 | **1,923.34** |
| 827 | 96.33 | 93.62 | 98.58 | 103.79 | **392.31** |
| 828 | 92.55 | 87.89 | 92.72 | 96.64 | **369.80** |
| 829 | 204.85 | 217.64 | 202.99 | 211.02 | **836.50** |
| 830 | 28.79 | 25.49 | 27.91 | 29.26 | **111.45** |
| 831 | 25.53 | 22.55 | 24.18 | 20.18 | **92.43** |
| WY - WYOMING Total | 1,429.05 | 1,479.57 | 1,487.94 | 1,571.94 | 5,968.49 |

DRUG: 9143 - OXYCODONE

STATE: AK - ALASKA

| REGISTRANT ZIP CODE 3 | 1ST QUARTER | 2ND QUARTER | 3RD QUARTER | 4TH QUARTER | TOTAL GRAMS |
|---|---|---|---|---|---|
| 995 | 12,440.30 | 11,888.58 | 11,758.51 | 10,984.34 | **47,071.73** |
| 996 | 7,663.58 | 7,430.31 | 7,441.14 | 6,594.33 | **29,129.36** |
| 997 | 2,150.45 | 2,210.07 | 2,322.15 | 2,207.82 | **8,890.50** |
| 998 | 886.47 | 852.68 | 838.64 | 763.80 | **3,341.60** |
| 999 | 238.94 | 279.26 | 299.61 | 255.34 | **1,073.15** |
| AK ALASKA - Total | 23,379.75 | 22,660.91 | 22,660.05 | 20,805.64 | 89,506.34 |

STATE: AL - ALABAMA

| REGISTRANT ZIP CODE 3 | 1ST QUARTER | 2ND QUARTER | 3RD QUARTER | 4TH QUARTER | TOTAL GRAMS |
|---|---|---|---|---|---|
| 350 | 20,553.66 | 20,589.89 | 19,660.48 | 20,024.85 | **80,828.88** |
| 351 | 12,669.31 | 12,669.04 | 12,217.49 | 12,263.02 | **49,818.86** |
| 352 | 17,418.70 | 17,663.23 | 16,865.58 | 17,089.45 | **69,036.96** |
| 354 | 6,595.21 | 6,657.44 | 6,675.14 | 6,377.72 | **26,305.49** |
| 355 | 9,447.75 | 9,328.49 | 9,573.20 | 9,594.62 | **37,944.06** |
| 356 | 14,580.08 | 14,570.39 | 14,598.94 | 14,327.11 | **58,076.51** |
| 357 | 8,333.46 | 7,728.67 | 8,132.85 | 8,089.34 | **32,284.32** |
| 358 | 9,742.27 | 9,595.57 | 9,538.79 | 9,180.52 | **38,057.16** |
| 359 | 16,540.74 | 16,816.67 | 16,485.90 | 16,567.60 | **66,410.91** |
| 360 | 6,019.32 | 5,951.00 | 5,868.69 | 5,960.86 | **23,799.86** |
| 361 | 4,197.36 | 4,109.42 | 4,123.09 | 4,252.48 | **16,682.36** |
| 362 | 7,624.17 | 7,041.84 | 6,599.77 | 6,589.33 | **27,855.11** |
| 363 | 7,764.75 | 7,753.75 | 7,959.62 | 7,931.68 | **31,409.80** |
| 364 | 4,170.01 | 3,730.88 | 3,865.52 | 3,861.18 | **15,627.59** |

| | | | | | |
|---|---|---|---|---|---|
| 365 | 11,847.79 | 11,696.46 | 11,600.89 | 11,524.40 | **46,669.54** |
| 366 | 9,982.93 | 9,355.95 | 9,350.32 | 9,416.77 | **38,105.97** |
| 367 | 1,447.49 | 1,323.95 | 1,308.77 | 1,282.12 | **5,362.33** |
| 368 | 5,622.46 | 5,680.01 | 5,646.60 | 5,898.38 | **22,847.46** |
| 369 | 464.56 | 486.30 | 461.06 | 465.08 | **1,876.99** |
| **AL ALABAMA - Total** | **175,022.03** | **172,748.95** | **170,532.67** | **170,696.51** | **689,000.17** |

**STATE: AR - ARKANSAS**

| REGISTRANT ZIP CODE 3 | 1ST QUARTER | 2ND QUARTER | 3RD QUARTER | 4TH QUARTER | TOTAL GRAMS |
|---|---|---|---|---|---|
| 716 | 3,099.02 | 3,196.04 | 2,895.10 | 2,901.88 | **12,092.05** |
| 717 | 2,103.41 | 2,121.55 | 1,957.65 | 1,921.55 | **8,104.16** |
| 718 | 2,447.23 | 2,117.67 | 1,797.13 | 1,608.72 | **7,970.75** |
| 719 | 4,044.48 | 3,983.88 | 3,914.30 | 3,947.01 | **15,889.67** |
| 720 | 11,157.51 | 10,758.03 | 10,452.48 | 9,890.39 | **42,258.41** |
| 721 | 10,313.40 | 9,897.89 | 9,434.59 | 9,398.13 | **39,044.01** |
| 722 | 8,107.00 | 7,903.59 | 7,955.84 | 7,630.51 | **31,596.94** |
| 723 | 2,989.02 | 3,128.26 | 3,362.50 | 3,587.50 | **13,067.28** |
| 724 | 4,351.23 | 4,187.37 | 4,338.66 | 4,125.56 | **17,002.82** |
| 725 | 3,750.33 | 3,471.23 | 3,459.19 | 3,356.94 | **14,037.69** |
| 726 | 4,415.75 | 4,325.05 | 4,368.57 | 4,383.49 | **17,492.87** |
| 727 | 20,309.81 | 19,396.19 | 18,780.52 | 17,077.33 | **75,563.86** |
| 728 | 3,937.40 | 3,495.42 | 3,278.02 | 3,212.39 | **13,923.23** |
| 729 | 13,276.57 | 11,544.61 | 11,050.61 | 10,644.40 | **46,516.18** |
| **AR ARKANSAS - Total** | **94,302.17** | **89,526.78** | **87,045.18** | **83,685.79** | **354,559.92** |

**STATE: AS - AMERICAN SAMOA**

| REGISTRANT ZIP CODE 3 | 1ST QUARTER | 2ND QUARTER | 3RD QUARTER | 4TH QUARTER | TOTAL GRAMS |
|---|---|---|---|---|---|
| 967 | 10.76 | 15.69 | 6.72 | 21.96 | **55.13** |
| **AS - AMERICAN SAMOA - Tota** | **10.76** | **15.69** | **6.72** | **21.96** | **55.13** |

**STATE: AZ - ARIZONA**

| REGISTRANT ZIP CODE 3 | 1ST QUARTER | 2ND QUARTER | 3RD QUARTER | 4TH QUARTER | TOTAL GRAMS |
|---|---|---|---|---|---|
| 850 | 64,222.44 | 62,571.08 | 61,505.03 | 61,236.13 | **249,534.68** |
| 851 | 15,730.88 | 14,919.84 | 14,832.97 | 15,297.36 | **60,781.05** |
| 852 | 67,436.64 | 65,394.07 | 63,895.00 | 64,543.47 | **261,269.18** |
| 853 | 56,011.19 | 54,041.60 | 52,232.02 | 53,635.75 | **215,920.56** |
| 855 | 5,403.77 | 5,286.45 | 5,520.77 | 5,370.25 | **21,581.23** |
| 856 | 10,221.18 | 9,557.93 | 9,625.54 | 9,096.49 | **38,501.15** |
| 857 | 49,389.79 | 47,573.55 | 47,287.94 | 45,337.43 | **189,588.72** |
| 859 | 5,580.53 | 5,226.07 | 5,754.78 | 5,123.87 | **21,685.26** |
| 860 | 3,492.46 | 3,516.77 | 3,502.68 | 3,285.45 | **13,797.35** |
| 863 | 10,574.28 | 10,687.39 | 10,383.85 | 10,552.88 | **42,198.41** |
| 864 | 12,598.34 | 12,399.79 | 12,110.43 | 11,820.41 | **48,928.97** |

| | | | | | |
|---|---|---|---|---|---|
| 865 | 184.68 | 171.68 | 157.78 | 186.47 | **700.61** |
| **AZ ARIZONA - Total** | **300,846.19** | **291,346.21** | **286,808.81** | **285,485.97** | **1,164,487.18** |

**STATE: CA - CALIFORNIA**

| REGISTRANT ZIP CODE 3 | 1ST QUARTER | 2ND QUARTER | 3RD QUARTER | 4TH QUARTER | TOTAL GRAMS |
|---|---|---|---|---|---|
| 900 | 18,519.75 | 18,604.41 | 17,937.69 | 17,843.94 | **72,905.79** |
| 902 | 13,241.74 | 13,410.80 | 12,782.50 | 13,097.84 | **52,532.88** |
| 903 | 1,165.83 | 1,034.47 | 1,120.95 | 1,188.76 | **4,510.02** |
| 904 | 3,947.30 | 3,766.77 | 3,716.79 | 3,907.05 | **15,337.91** |
| 905 | 3,243.22 | 3,294.56 | 3,167.49 | 3,251.32 | **12,956.58** |
| 906 | 6,546.72 | 6,228.08 | 6,214.27 | 6,244.65 | **25,233.73** |
| 907 | 6,651.97 | 6,653.58 | 6,106.93 | 6,201.45 | **25,613.93** |
| 908 | 4,443.95 | 4,366.23 | 4,010.58 | 4,032.59 | **16,853.36** |
| 910 | 3,398.43 | 3,702.30 | 3,309.50 | 3,201.71 | **13,611.95** |
| 911 | 2,097.72 | 1,957.21 | 2,113.10 | 2,107.50 | **8,275.52** |
| 912 | 2,651.14 | 2,742.19 | 2,344.14 | 2,294.39 | **10,031.87** |
| 913 | 24,734.70 | 23,769.52 | 22,789.97 | 23,020.01 | **94,314.21** |
| 914 | 7,481.97 | 6,981.96 | 6,786.32 | 6,218.00 | **27,468.26** |
| 915 | 2,592.39 | 2,569.37 | 2,570.76 | 2,572.50 | **10,305.01** |
| 916 | 1,620.81 | 1,517.12 | 1,662.00 | 1,615.66 | **6,415.59** |
| 917 | 20,112.83 | 21,094.64 | 20,673.28 | 20,933.91 | **82,814.67** |
| 918 | 522.10 | 485.61 | 584.38 | 587.12 | **2,179.21** |
| 919 | 9,097.76 | 9,035.80 | 8,835.86 | 8,701.73 | **35,671.14** |
| 920 | 36,513.40 | 37,251.99 | 35,964.40 | 34,284.01 | **144,013.80** |
| 921 | 23,122.03 | 22,583.58 | 21,929.51 | 20,818.56 | **88,453.69** |
| 922 | 21,730.99 | 21,455.64 | 19,939.83 | 19,461.50 | **82,587.96** |
| 923 | 24,772.95 | 24,428.73 | 24,264.58 | 24,193.74 | **97,660.01** |
| 924 | 3,335.14 | 3,243.60 | 3,041.71 | 3,000.65 | **12,621.10** |
| 925 | 25,282.59 | 25,008.98 | 24,545.09 | 24,863.35 | **99,700.02** |
| 926 | 21,800.90 | 20,901.00 | 20,295.77 | 19,769.62 | **82,767.30** |
| 927 | 4,801.62 | 4,872.93 | 4,784.49 | 4,936.54 | **19,395.58** |
| 928 | 16,326.87 | 15,712.27 | 15,867.06 | 15,676.29 | **63,582.49** |
| 930 | 12,042.80 | 11,884.22 | 11,640.35 | 11,237.76 | **46,805.12** |
| 931 | 1,991.40 | 1,801.60 | 1,762.40 | 1,748.12 | **7,303.53** |
| 932 | 8,481.60 | 8,535.98 | 8,557.52 | 8,176.56 | **33,751.67** |
| 933 | 10,829.00 | 10,986.91 | 10,697.93 | 10,530.49 | **43,044.32** |
| 934 | 10,967.37 | 10,998.00 | 11,123.02 | 10,795.43 | **43,883.83** |
| 935 | 16,154.97 | 16,700.51 | 15,774.03 | 15,615.57 | **64,245.09** |
| 936 | 8,305.03 | 8,300.60 | 8,041.85 | 7,442.74 | **32,090.21** |
| 937 | 11,846.34 | 11,732.37 | 11,535.02 | 10,919.47 | **46,033.20** |
| 939 | 4,260.48 | 4,013.51 | 3,712.03 | 3,589.63 | **15,575.64** |

| | | | | | |
|---|---|---|---|---|---|
| 940 | 10,229.78 | 10,036.84 | 9,929.15 | 9,699.99 | **39,895.77** |
| 941 | 16,825.41 | 16,612.66 | 15,641.25 | 15,448.84 | **64,528.15** |
| 943 | 2,599.47 | 2,781.75 | 2,560.02 | 2,617.69 | **10,558.93** |
| 944 | 1,542.63 | 1,462.83 | 1,368.80 | 1,354.19 | **5,728.44** |
| 945 | 37,589.96 | 35,414.82 | 35,116.09 | 33,462.11 | **141,582.98** |
| 946 | 3,660.05 | 3,813.99 | 3,690.99 | 3,709.62 | **14,874.65** |
| 947 | 914.38 | 813.05 | 874.98 | 846.02 | **3,448.43** |
| 948 | 1,278.57 | 1,268.30 | 1,244.04 | 1,201.71 | **4,992.63** |
| 949 | 7,757.95 | 7,895.86 | 7,241.81 | 7,269.88 | **30,165.48** |
| 950 | 9,307.32 | 9,701.97 | 9,173.57 | 9,124.93 | **37,307.79** |
| 951 | 6,748.69 | 6,890.30 | 6,337.06 | 6,412.96 | **26,389.00** |
| 952 | 7,438.62 | 6,676.38 | 5,973.39 | 6,179.79 | **26,268.18** |
| 953 | 25,622.97 | 25,817.19 | 24,170.19 | 23,489.51 | **99,099.85** |
| 954 | 15,439.51 | 15,472.69 | 14,975.05 | 14,805.43 | **60,692.68** |
| 955 | 4,235.89 | 4,228.67 | 4,126.70 | 3,856.06 | **16,447.32** |
| 956 | 26,164.37 | 26,115.32 | 25,048.61 | 24,260.10 | **101,588.40** |
| 957 | 4,159.41 | 4,427.63 | 4,235.04 | 4,007.58 | **16,829.65** |
| 958 | 15,895.11 | 15,752.61 | 15,178.71 | 14,808.99 | **61,635.41** |
| 959 | 12,950.05 | 12,456.07 | 11,362.21 | 11,122.24 | **47,890.58** |
| 960 | 7,686.14 | 7,325.87 | 7,376.79 | 6,768.03 | **29,156.83** |
| 961 | 2,013.27 | 1,963.92 | 1,815.73 | 1,922.28 | **7,715.20** |
| **CA - CALIFORNIA - Total** | **614,695.39** | **608,555.76** | **587,643.27** | **576,448.13** | **2,387,342.55** |

**STATE: CO - COLORADO**

| REGISTRANT ZIP CODE 3 | 1ST QUARTER | 2ND QUARTER | 3RD QUARTER | 4TH QUARTER | **TOTAL GRAMS** |
|---|---|---|---|---|---|
| 800 | 30,399.68 | 30,122.23 | 29,677.76 | 29,305.38 | **119,505.05** |
| 801 | 21,675.74 | 21,735.36 | 22,219.28 | 22,212.15 | **87,842.54** |
| 802 | 30,160.45 | 30,613.46 | 30,274.13 | 29,447.64 | **120,495.69** |
| 803 | 2,740.82 | 2,565.83 | 2,517.97 | 2,487.30 | **10,311.92** |
| 804 | 4,762.00 | 4,515.36 | 4,685.73 | 4,699.15 | **18,662.24** |
| 805 | 14,425.58 | 14,160.04 | 13,696.60 | 13,428.56 | **55,710.78** |
| 806 | 7,447.86 | 7,042.11 | 6,800.05 | 6,779.18 | **28,069.19** |
| 807 | 1,755.69 | 1,829.19 | 1,836.81 | 1,752.85 | **7,174.53** |
| 808 | 4,064.44 | 4,053.90 | 3,954.09 | 3,926.68 | **15,999.11** |
| 809 | 20,588.66 | 20,774.24 | 21,223.20 | 21,179.69 | **83,765.78** |
| 810 | 15,748.89 | 15,366.29 | 15,244.18 | 15,196.61 | **61,555.97** |
| 811 | 1,713.55 | 1,643.16 | 1,669.19 | 1,669.95 | **6,695.85** |
| 812 | 2,615.65 | 2,701.72 | 2,641.32 | 2,585.29 | **10,543.98** |
| 813 | 1,104.53 | 1,180.51 | 1,215.34 | 984.33 | **4,484.71** |
| 814 | 1,738.69 | 1,700.65 | 1,738.32 | 1,639.78 | **6,817.44** |
| 815 | 3,702.08 | 3,661.00 | 3,686.02 | 3,442.77 | **14,491.87** |

| | | | | | |
|---|---|---|---|---|---|
| 816 | 2,979.49 | 2,963.65 | 2,853.82 | 2,769.06 | **11,566.03** |
| **CO - COLORADO - Total** | **167,623.81** | **166,628.71** | **165,933.81** | **163,506.36** | **663,692.69** |

**STATE: CT - CONNECTICUT**

| REGISTRANT ZIP CODE 3 | 1ST QUARTER | 2ND QUARTER | 3RD QUARTER | 4TH QUARTER | TOTAL GRAMS |
|---|---|---|---|---|---|
| 060 | 21,214.37 | 21,217.74 | 21,037.44 | 20,082.23 | **83,551.78** |
| 061 | 6,552.90 | 6,688.85 | 5,987.73 | 6,481.11 | **25,710.60** |
| 062 | 3,921.07 | 3,760.95 | 3,702.09 | 3,638.31 | **15,022.43** |
| 063 | 10,613.93 | 10,453.39 | 9,841.70 | 9,843.57 | **40,752.59** |
| 064 | 34,770.21 | 33,998.09 | 34,392.60 | 34,181.19 | **137,342.10** |
| 065 | 14,130.70 | 13,938.49 | 13,983.02 | 14,102.09 | **56,154.29** |
| 066 | 8,761.92 | 8,348.70 | 8,445.65 | 8,231.86 | **33,788.14** |
| 067 | 13,964.74 | 13,487.86 | 13,352.44 | 13,246.71 | **54,051.76** |
| 068 | 12,384.21 | 11,907.70 | 12,471.94 | 12,119.26 | **48,883.11** |
| 069 | 2,216.52 | 2,106.57 | 2,211.43 | 2,469.46 | **9,003.97** |
| **CT CONNECTICUT Total** | **128,530.58** | **125,908.35** | **125,426.03** | **124,395.80** | **504,260.77** |

**STATE: DC - DISTRICT OF COLUMBIA**

| REGISTRANT ZIP CODE 3 | 1ST QUARTER | 2ND QUARTER | 3RD QUARTER | 4TH QUARTER | TOTAL GRAMS |
|---|---|---|---|---|---|
| 200 | 10,852.09 | 10,260.50 | 10,050.68 | 9,953.40 | **41,116.68** |
| 203 | 15.24 | 17.03 | 27.79 | 6.28 | **66.34** |
| 204 | 812.59 | 759.87 | 1,039.76 | 406.74 | **3,018.96** |
| 205 | 1.34 | 0 | 0.90 | 0 | **2.24** |
| **DC - DISTRICT OF COLUMBIA** | **11,681.27** | **11,037.41** | **11,119.13** | **10,366.42** | **44,204.22** |

**STATE: DE - DELAWARE**

| REGISTRANT ZIP CODE 3 | 1ST QUARTER | 2ND QUARTER | 3RD QUARTER | 4TH QUARTER | TOTAL GRAMS |
|---|---|---|---|---|---|
| 197 | 18,419.42 | 17,908.05 | 17,306.42 | 15,878.23 | **69,512.12** |
| 198 | 11,942.31 | 11,427.63 | 11,556.37 | 10,885.16 | **45,811.47** |
| 199 | 23,166.82 | 22,994.39 | 22,686.33 | 21,476.07 | **90,323.60** |
| **DE DELAWARE Total** | **53,528.55** | **52,330.07** | **51,549.11** | **48,239.45** | **205,647.19** |

**STATE: FL - FLORIDA**

| REGISTRANT ZIP CODE 3 | 1ST QUARTER | 2ND QUARTER | 3RD QUARTER | 4TH QUARTER | TOTAL GRAMS |
|---|---|---|---|---|---|
| 320 | 26,653.39 | 26,773.55 | 26,932.47 | 27,319.54 | **107,678.94** |
| 321 | 26,735.06 | 26,804.23 | 26,999.97 | 27,365.60 | **107,904.85** |
| 322 | 50,412.65 | 50,528.53 | 51,165.61 | 50,532.82 | **202,639.61** |
| 323 | 10,931.56 | 11,163.85 | 10,875.21 | 11,270.01 | **44,240.63** |
| 324 | 14,283.70 | 14,210.63 | 14,349.48 | 13,997.63 | **56,841.45** |
| 325 | 25,742.07 | 25,519.68 | 24,897.86 | 24,721.83 | **100,881.44** |
| 326 | 19,186.37 | 19,115.27 | 18,744.07 | 19,093.02 | **76,138.73** |
| 327 | 38,486.66 | 37,250.72 | 39,028.94 | 38,571.87 | **153,338.19** |
| 328 | 24,403.90 | 25,870.54 | 25,359.29 | 25,670.43 | **101,304.17** |
| 329 | 41,938.78 | 41,360.38 | 41,735.84 | 41,377.90 | **166,412.91** |

| | | | | | |
|---|---|---|---|---|---|
| 330 | 55,206.15 | 52,719.38 | 54,147.44 | 54,402.30 | **216,475.26** |
| 331 | 44,191.96 | 44,060.36 | 43,245.19 | 44,875.29 | **176,372.80** |
| 333 | 36,194.42 | 36,570.61 | 35,656.25 | 36,176.51 | **144,597.79** |
| 334 | 73,132.63 | 75,441.85 | 75,337.70 | 77,368.18 | **301,280.35** |
| 335 | 33,881.66 | 33,692.26 | 33,518.53 | 34,147.99 | **135,240.44** |
| 336 | 44,530.69 | 43,625.90 | 46,416.18 | 48,947.13 | **183,519.90** |
| 337 | 41,225.40 | 40,789.12 | 41,859.24 | 42,678.23 | **166,551.99** |
| 338 | 26,591.67 | 25,158.48 | 25,717.77 | 26,024.71 | **103,492.63** |
| 339 | 37,965.62 | 37,124.11 | 37,031.87 | 37,279.29 | **149,400.89** |
| 341 | 15,757.27 | 14,790.25 | 14,534.89 | 15,074.37 | **60,156.79** |
| 342 | 39,938.14 | 38,791.83 | 37,302.40 | 38,566.94 | **154,599.31** |
| 344 | 25,255.83 | 24,310.86 | 24,403.19 | 25,509.07 | **99,478.95** |
| 346 | 43,319.96 | 44,053.42 | 44,673.49 | 45,762.72 | **177,809.58** |
| 347 | 20,700.68 | 20,961.21 | 21,635.21 | 21,726.91 | **85,024.00** |
| 349 | 27,308.52 | 27,219.69 | 27,125.14 | 27,026.30 | **108,679.65** |
| **FL FLORIDA - Total** | **843,974.72** | **837,906.70** | **842,693.23** | **855,486.60** | **3,380,061.24** |

**STATE: GA - GEORGIA**

| REGISTRANT ZIP CODE 3 | 1ST QUARTER | 2ND QUARTER | 3RD QUARTER | 4TH QUARTER | TOTAL GRAMS |
|---|---|---|---|---|---|
| 300 | 66,723.69 | 66,037.73 | 65,850.82 | 81,226.10 | **279,838.34** |
| 301 | 56,240.13 | 55,832.05 | 55,833.50 | 54,898.01 | **222,803.68** |
| 302 | 36,931.96 | 37,067.10 | 36,738.38 | 35,134.55 | **145,871.99** |
| 303 | 23,106.25 | 22,639.15 | 22,943.06 | 22,495.39 | **91,183.85** |
| 304 | 7,161.07 | 7,455.53 | 7,172.27 | 7,285.24 | **29,074.11** |
| 305 | 33,005.82 | 32,604.68 | 33,224.77 | 33,154.33 | **131,989.61** |
| 306 | 24,370.33 | 24,321.03 | 24,136.65 | 24,082.99 | **96,911.00** |
| 307 | 13,247.55 | 12,688.14 | 12,294.85 | 11,988.22 | **50,218.75** |
| 308 | 6,056.62 | 6,199.99 | 6,132.58 | 6,259.66 | **24,648.85** |
| 309 | 12,409.84 | 12,318.03 | 12,801.52 | 11,724.94 | **49,254.34** |
| 310 | 18,824.59 | 18,986.10 | 19,068.05 | 18,106.88 | **74,985.62** |
| 312 | 8,216.76 | 7,903.09 | 7,951.41 | 7,981.59 | **32,052.85** |
| 313 | 6,554.02 | 6,336.94 | 6,111.77 | 5,929.45 | **24,932.18** |
| 314 | 7,570.50 | 7,611.19 | 7,386.03 | 7,200.53 | **29,768.25** |
| 315 | 15,399.43 | 14,795.57 | 14,305.45 | 13,771.74 | **58,272.19** |
| 316 | 6,564.61 | 6,731.63 | 6,789.00 | 6,877.65 | **26,962.89** |
| 317 | 11,371.46 | 11,486.11 | 11,442.94 | 11,644.33 | **45,944.85** |
| 318 | 935.35 | 897.61 | 890.59 | 884.40 | **3,607.94** |
| 319 | 6,433.98 | 6,321.32 | 6,842.52 | 6,096.16 | **25,693.98** |
| 398 | 2,381.62 | 2,491.93 | 2,623.14 | 2,440.76 | **9,937.45** |
| **GA GEORGIA Total** | **363,505.56** | **360,724.91** | **360,539.31** | **369,182.95** | **1,453,952.73** |

**STATE: GU - GUAM**

| REGISTRANT ZIP CODE 3 | 1ST QUARTER | 2ND QUARTER | 3RD QUARTER | 4TH QUARTER | TOTAL GRAMS |
|---|---|---|---|---|---|
| 969 | 732.22 | 762.11 | 724.64 | 537.54 | **2,756.52** |
| **GU - GUAM - Total** | **732.22** | **762.11** | **724.64** | **537.54** | **2,756.52** |

**STATE: HI - HAWAII**

| REGISTRANT ZIP CODE 3 | 1ST QUARTER | 2ND QUARTER | 3RD QUARTER | 4TH QUARTER | TOTAL GRAMS |
|---|---|---|---|---|---|
| 967 | 21,846.65 | 22,323.27 | 21,543.90 | 20,018.81 | **85,732.62** |
| 968 | 9,378.40 | 9,762.73 | 9,662.62 | 9,115.02 | **37,918.77** |
| **HI - HAWAII - Total** | **31,225.04** | **32,086.00** | **31,206.51** | **29,133.83** | **123,651.39** |

**STATE: IA - IOWA**

| REGISTRANT ZIP CODE 3 | 1ST QUARTER | 2ND QUARTER | 3RD QUARTER | 4TH QUARTER | TOTAL GRAMS |
|---|---|---|---|---|---|
| 500 | 3,331.76 | 3,279.03 | 3,156.38 | 3,132.00 | **12,899.18** |
| 501 | 2,578.50 | 2,355.93 | 2,436.20 | 2,242.93 | **9,613.56** |
| 502 | 2,528.11 | 2,553.19 | 2,657.06 | 2,442.13 | **10,180.48** |
| 503 | 7,858.27 | 7,976.36 | 7,503.03 | 7,130.09 | **30,467.76** |
| 504 | 1,319.86 | 1,445.75 | 1,349.43 | 1,391.67 | **5,506.72** |
| 505 | 1,092.57 | 1,165.17 | 1,113.02 | 1,029.38 | **4,400.14** |
| 506 | 1,224.50 | 1,209.67 | 1,290.52 | 1,265.21 | **4,989.89** |
| 507 | 1,105.03 | 1,208.95 | 1,110.21 | 1,121.78 | **4,545.98** |
| 508 | 352.72 | 308.00 | 343.93 | 332.87 | **1,337.52** |
| 510 | 548.15 | 534.97 | 537.69 | 555.35 | **2,176.17** |
| 511 | 1,385.98 | 1,346.50 | 1,481.63 | 1,343.64 | **5,557.76** |
| 512 | 272.24 | 263.98 | 263.27 | 257.58 | **1,057.07** |
| 513 | 400.26 | 451.65 | 423.99 | 392.35 | **1,668.25** |
| 514 | 568.55 | 555.22 | 498.89 | 522.27 | **2,144.93** |
| 515 | 2,303.93 | 2,247.07 | 2,149.99 | 2,081.15 | **8,782.14** |
| 516 | 355.33 | 367.14 | 382.05 | 349.08 | **1,453.60** |
| 520 | 984.72 | 1,062.41 | 1,025.55 | 1,153.80 | **4,226.48** |
| 521 | 344.67 | 338.55 | 386.30 | 342.54 | **1,412.06** |
| 522 | 2,139.89 | 2,160.63 | 2,083.38 | 2,374.71 | **8,758.61** |
| 523 | 878.76 | 854.25 | 824.15 | 792.25 | **3,349.40** |
| 524 | 1,682.99 | 1,726.15 | 1,680.28 | 1,643.63 | **6,733.04** |
| 525 | 1,513.43 | 1,519.89 | 1,439.44 | 1,506.34 | **5,979.10** |
| 526 | 1,726.07 | 1,614.15 | 1,623.19 | 1,661.17 | **6,624.58** |
| 527 | 1,191.66 | 1,159.06 | 1,039.57 | 1,079.38 | **4,469.68** |
| 528 | 1,179.69 | 1,135.69 | 1,072.33 | 1,113.20 | **4,500.91** |
| **IA IOWA Total** | **38,867.65** | **38,839.37** | **37,871.48** | **37,256.51** | **152,835.01** |

**STATE: ID - IDAHO**

| REGISTRANT ZIP CODE 3 | 1ST QUARTER | 2ND QUARTER | 3RD QUARTER | 4TH QUARTER | TOTAL GRAMS |
|---|---|---|---|---|---|
| 832 | 5,804.54 | 5,889.43 | 5,901.58 | 5,908.14 | **23,503.68** |
| 833 | 5,707.89 | 5,690.33 | 5,898.98 | 5,721.54 | **23,018.74** |

| | | | | | |
|---|---|---|---|---|---|
| 834 | 4,465.50 | 4,894.04 | 4,790.83 | 4,909.05 | **19,059.43** |
| 835 | 1,560.90 | 1,563.98 | 1,598.42 | 1,569.78 | **6,293.08** |
| 836 | 14,096.27 | 14,201.95 | 13,733.63 | 13,281.66 | **55,313.52** |
| 837 | 9,504.25 | 10,004.01 | 9,472.68 | 9,181.61 | **38,162.54** |
| 838 | 5,970.97 | 5,943.37 | 6,030.71 | 5,934.96 | **23,880.01** |
| **ID IDAHO Total** | **47,110.32** | **48,187.10** | **47,426.83** | **46,506.74** | **189,231.00** |

**STATE: IL - ILLINOIS**

| REGISTRANT ZIP CODE 3 | 1ST QUARTER | 2ND QUARTER | 3RD QUARTER | 4TH QUARTER | **TOTAL GRAMS** |
|---|---|---|---|---|---|
| 600 | 18,937.11 | 18,750.04 | 18,668.18 | 18,114.02 | **74,469.34** |
| 601 | 12,155.21 | 11,750.13 | 11,747.55 | 11,312.88 | **46,965.76** |
| 602 | 329.75 | 302.77 | 307.26 | 343.14 | **1,282.92** |
| 603 | 547.76 | 493.95 | 446.85 | 483.53 | **1,972.09** |
| 604 | 15,223.22 | 15,059.27 | 14,906.53 | 14,565.43 | **59,754.44** |
| 605 | 7,608.07 | 7,061.82 | 7,209.31 | 7,253.09 | **29,132.29** |
| 606 | 9,839.75 | 10,143.68 | 9,752.77 | 9,611.67 | **39,347.88** |
| 607 | 1,331.48 | 1,289.23 | 1,397.64 | 1,301.89 | **5,320.23** |
| 608 | 572.89 | 532.76 | 537.51 | 534.53 | **2,177.69** |
| 609 | 2,498.73 | 2,215.37 | 2,166.32 | 2,141.52 | **9,021.95** |
| 610 | 2,234.74 | 2,296.71 | 2,226.19 | 2,182.05 | **8,939.70** |
| 611 | 2,692.11 | 2,543.21 | 2,737.03 | 2,547.65 | **10,520.01** |
| 612 | 995.56 | 1,012.15 | 1,074.26 | 1,017.89 | **4,099.86** |
| 613 | 1,459.60 | 1,417.95 | 1,393.61 | 1,451.08 | **5,722.24** |
| 614 | 882.60 | 802.60 | 861.01 | 771.69 | **3,317.90** |
| 615 | 1,561.57 | 1,712.76 | 1,602.18 | 1,585.78 | **6,462.29** |
| 616 | 2,257.39 | 2,233.93 | 2,372.83 | 2,429.72 | **9,293.87** |
| 617 | 1,668.38 | 1,645.13 | 1,570.94 | 1,570.25 | **6,454.69** |
| 618 | 3,016.63 | 3,152.93 | 3,387.98 | 3,224.37 | **12,781.91** |
| 619 | 641.59 | 696.53 | 648.04 | 605.33 | **2,591.49** |
| 620 | 6,161.92 | 5,881.81 | 5,753.22 | 5,641.73 | **23,438.68** |
| 622 | 6,825.23 | 6,800.04 | 6,911.39 | 6,875.74 | **27,412.40** |
| 623 | 1,521.53 | 1,591.55 | 1,622.87 | 1,507.39 | **6,243.33** |
| 624 | 1,743.53 | 1,709.34 | 1,695.16 | 1,594.45 | **6,742.47** |
| 625 | 1,388.10 | 1,423.37 | 1,465.95 | 1,396.16 | **5,673.58** |
| 626 | 1,268.82 | 1,219.94 | 1,261.03 | 1,195.67 | **4,945.45** |
| 627 | 1,246.03 | 1,133.64 | 1,091.08 | 1,201.55 | **4,672.30** |
| 628 | 3,747.23 | 3,611.61 | 3,647.40 | 3,570.30 | **14,576.54** |
| 629 | 6,092.96 | 5,971.84 | 5,899.31 | 5,802.55 | **23,766.66** |
| **IL ILLINOIS Total** | **116,449.46** | **114,456.05** | **114,361.40** | **111,833.05** | **457,099.97** |

**STATE: IN - INDIANA**

| REGISTRANT ZIP CODE 3 | 1ST QUARTER | 2ND QUARTER | 3RD QUARTER | 4TH QUARTER | **TOTAL GRAMS** |
|---|---|---|---|---|---|

| | | | | | |
|---|---|---|---|---|---|
| 460 | 10,391.53 | 10,370.82 | 10,544.60 | 10,658.44 | **41,965.39** |
| 461 | 17,723.23 | 17,371.49 | 16,460.69 | 15,805.57 | **67,360.98** |
| 462 | 29,829.84 | 28,709.57 | 29,719.54 | 29,234.57 | **117,493.52** |
| 463 | 13,106.96 | 12,962.11 | 12,869.60 | 13,040.36 | **51,979.04** |
| 464 | 1,716.94 | 1,663.05 | 1,681.93 | 1,741.58 | **6,803.50** |
| 465 | 12,403.94 | 12,537.70 | 12,448.58 | 11,758.91 | **49,149.13** |
| 466 | 4,542.26 | 4,447.11 | 4,498.90 | 4,311.43 | **17,799.70** |
| 467 | 6,751.41 | 6,876.48 | 7,188.03 | 7,302.61 | **28,118.54** |
| 468 | 9,799.55 | 10,023.37 | 10,648.20 | 10,358.86 | **40,829.98** |
| 469 | 7,172.00 | 7,304.40 | 7,577.69 | 7,323.25 | **29,377.34** |
| 470 | 5,918.70 | 6,049.15 | 5,947.48 | 5,296.42 | **23,211.75** |
| 471 | 12,451.34 | 12,387.48 | 11,988.03 | 11,822.71 | **48,649.55** |
| 472 | 5,318.95 | 5,235.48 | 5,232.19 | 5,056.23 | **20,842.84** |
| 473 | 10,629.03 | 10,290.53 | 10,499.67 | 10,271.82 | **41,691.06** |
| 474 | 5,700.49 | 5,911.25 | 6,219.68 | 5,852.72 | **23,684.14** |
| 475 | 3,823.82 | 3,989.86 | 4,015.02 | 4,094.41 | **15,923.11** |
| 476 | 3,099.14 | 3,021.98 | 3,102.81 | 2,892.20 | **12,116.13** |
| 477 | 8,855.01 | 8,538.16 | 8,345.12 | 8,058.25 | **33,796.56** |
| 478 | 2,374.30 | 2,435.30 | 2,546.07 | 2,486.95 | **9,842.61** |
| 479 | 5,437.83 | 5,078.80 | 4,920.33 | 4,662.86 | **20,099.82** |
| **IN INDIANA Total** | **177,046.26** | **175,204.12** | **176,454.16** | **172,030.16** | **700,734.70** |

**STATE: KS - KANSAS**

| REGISTRANT ZIP CODE 3 | 1ST QUARTER | 2ND QUARTER | 3RD QUARTER | 4TH QUARTER | **TOTAL GRAMS** |
|---|---|---|---|---|---|
| 660 | 12,734.94 | 12,525.94 | 12,834.68 | 12,242.27 | **50,337.82** |
| 661 | 5,488.41 | 5,348.22 | 5,360.82 | 5,137.01 | **21,334.46** |
| 662 | 12,346.96 | 12,114.99 | 11,590.98 | 11,753.06 | **47,805.99** |
| 664 | 1,839.37 | 1,786.00 | 1,724.76 | 1,777.08 | **7,127.20** |
| 665 | 2,441.33 | 2,235.43 | 2,301.27 | 2,143.44 | **9,121.47** |
| 666 | 4,923.52 | 5,080.26 | 4,935.64 | 5,089.71 | **20,029.13** |
| 667 | 4,031.13 | 3,731.86 | 3,661.04 | 3,527.14 | **14,951.17** |
| 668 | 1,193.60 | 1,080.49 | 1,012.60 | 987.66 | **4,274.36** |
| 669 | 596.24 | 540.63 | 568.20 | 576.95 | **2,282.02** |
| 670 | 7,006.26 | 6,874.10 | 6,811.66 | 6,659.29 | **27,351.32** |
| 671 | 2,916.98 | 2,885.05 | 2,868.10 | 2,958.00 | **11,628.13** |
| 672 | 19,581.41 | 19,673.39 | 19,301.48 | 19,041.62 | **77,597.90** |
| 673 | 1,813.30 | 1,732.73 | 1,737.13 | 1,624.31 | **6,907.47** |
| 674 | 4,307.66 | 4,439.17 | 4,214.56 | 4,036.96 | **16,998.35** |
| 675 | 4,388.28 | 4,447.42 | 4,344.81 | 4,487.86 | **17,668.37** |
| 676 | 1,794.83 | 1,927.27 | 1,747.19 | 1,711.56 | **7,180.85** |
| 677 | 1,121.84 | 1,072.46 | 1,074.48 | 1,070.60 | **4,339.38** |

| | | | | | |
|---|---|---|---|---|---|
| 678 | 3,558.98 | 3,569.93 | 3,339.30 | 3,256.00 | **13,724.21** |
| 679 | 851.19 | 926.28 | 908.23 | 880.25 | **3,565.95** |
| **KS - KANSAS - Total** | **92,936.22** | **91,991.63** | **90,336.94** | **88,960.78** | **364,225.57** |

**STATE: KY - KENTUCKY**

| REGISTRANT ZIP CODE 3 | 1ST QUARTER | 2ND QUARTER | 3RD QUARTER | 4TH QUARTER | TOTAL GRAMS |
|---|---|---|---|---|---|
| 400 | 6,961.37 | 6,993.75 | 6,937.77 | 7,090.24 | **27,983.12** |
| 401 | 3,929.84 | 3,679.82 | 3,867.94 | 3,761.78 | **15,239.38** |
| 402 | 29,140.05 | 27,946.72 | 28,132.31 | 27,881.56 | **113,100.64** |
| 403 | 12,004.46 | 11,834.94 | 11,714.66 | 11,672.83 | **47,226.90** |
| 404 | 6,422.69 | 6,364.16 | 6,335.96 | 6,395.16 | **25,517.98** |
| 405 | 8,870.45 | 8,585.53 | 8,820.91 | 8,858.48 | **35,135.36** |
| 406 | 1,146.24 | 1,141.13 | 1,326.13 | 1,312.14 | **4,925.64** |
| 407 | 5,088.87 | 5,107.32 | 5,382.17 | 5,312.77 | **20,891.12** |
| 408 | 1,109.19 | 1,093.67 | 1,168.41 | 1,162.37 | **4,533.64** |
| 409 | 3,348.56 | 3,378.55 | 3,352.90 | 3,398.89 | **13,478.90** |
| 410 | 23,054.48 | 22,496.64 | 22,667.13 | 22,496.59 | **90,714.84** |
| 411 | 3,924.09 | 3,862.29 | 3,570.79 | 3,626.42 | **14,983.59** |
| 412 | 1,919.54 | 1,778.33 | 1,675.64 | 1,689.42 | **7,062.92** |
| 413 | 1,280.79 | 1,364.76 | 1,374.94 | 1,296.94 | **5,317.43** |
| 414 | 725.36 | 759.16 | 793.68 | 797.58 | **3,075.77** |
| 415 | 3,249.81 | 3,430.23 | 3,274.91 | 3,216.37 | **13,171.32** |
| 416 | 3,224.57 | 3,329.45 | 3,417.67 | 3,374.94 | **13,346.63** |
| 417 | 3,845.10 | 3,567.77 | 3,702.00 | 3,785.51 | **14,900.38** |
| 418 | 1,736.15 | 1,619.67 | 1,711.02 | 1,580.13 | **6,646.97** |
| 420 | 10,555.59 | 9,994.92 | 10,122.43 | 9,997.82 | **40,670.76** |
| 421 | 8,627.67 | 8,632.18 | 8,817.59 | 8,494.18 | **34,571.61** |
| 422 | 4,587.82 | 4,619.96 | 4,660.92 | 4,082.65 | **17,951.34** |
| 423 | 5,346.24 | 5,293.85 | 5,308.23 | 5,161.61 | **21,109.92** |
| 424 | 6,215.34 | 6,348.08 | 6,285.38 | 6,080.47 | **24,929.28** |
| 425 | 2,564.99 | 2,651.82 | 2,381.51 | 2,357.40 | **9,955.72** |
| 426 | 2,898.08 | 2,856.69 | 2,665.64 | 2,625.58 | **11,045.99** |
| 427 | 6,452.18 | 6,152.54 | 6,068.23 | 6,190.07 | **24,863.02** |
| **KY - KENTUCKY - Total** | **168,229.50** | **164,883.93** | **165,536.85** | **163,699.89** | **662,350.17** |

**STATE: LA - LOUISIANA**

| REGISTRANT ZIP CODE 3 | 1ST QUARTER | 2ND QUARTER | 3RD QUARTER | 4TH QUARTER | TOTAL GRAMS |
|---|---|---|---|---|---|
| 700 | 21,853.74 | 21,387.74 | 20,923.54 | 19,944.51 | **84,109.53** |
| 701 | 12,483.55 | 11,923.36 | 12,204.13 | 12,345.93 | **48,956.97** |
| 703 | 6,517.62 | 6,773.17 | 6,817.59 | 6,569.51 | **26,677.90** |
| 704 | 20,505.73 | 20,112.64 | 20,081.62 | 19,983.94 | **80,683.93** |
| 705 | 16,903.77 | 16,513.90 | 16,642.76 | 16,346.17 | **66,406.61** |

| | | | | | |
|---|---|---|---|---|---|
| 706 | 5,108.20 | 5,155.13 | 4,670.29 | 4,639.63 | **19,573.24** |
| 707 | 17,696.95 | 17,350.13 | 17,676.45 | 16,204.99 | **68,928.52** |
| 708 | 11,756.30 | 11,106.05 | 11,170.19 | 11,811.40 | **45,843.95** |
| 710 | 3,665.90 | 3,983.18 | 3,918.58 | 3,740.67 | **15,308.34** |
| 711 | 12,869.92 | 12,612.99 | 12,684.18 | 12,442.97 | **50,610.06** |
| 712 | 5,784.18 | 5,793.01 | 5,898.21 | 5,786.02 | **23,261.42** |
| 713 | 8,809.86 | 8,251.92 | 8,458.20 | 8,176.33 | **33,696.32** |
| 714 | 4,705.66 | 5,160.64 | 4,750.95 | 4,534.31 | **19,151.56** |
| **LA - LOUISIANA - Total** | **148,661.39** | **146,123.87** | **145,896.69** | **142,526.39** | **583,208.34** |

**STATE: MA - MASSACHUSETTS**

| REGISTRANT ZIP CODE 3 | 1ST QUARTER | 2ND QUARTER | 3RD QUARTER | 4TH QUARTER | **TOTAL GRAMS** |
|---|---|---|---|---|---|
| 010 | 19,578.78 | 18,833.98 | 18,073.32 | 18,074.18 | **74,560.26** |
| 011 | 7,934.97 | 7,433.90 | 7,252.92 | 7,480.95 | **30,102.74** |
| 012 | 3,676.81 | 3,569.61 | 3,590.14 | 3,452.09 | **14,288.65** |
| 013 | 3,514.16 | 3,381.28 | 3,241.58 | 3,207.07 | **13,344.08** |
| 014 | 5,925.12 | 5,618.86 | 5,927.16 | 5,680.58 | **23,151.73** |
| 015 | 8,710.32 | 8,387.72 | 8,322.00 | 8,404.96 | **33,825.00** |
| 016 | 7,088.09 | 7,330.12 | 6,895.60 | 7,185.48 | **28,499.29** |
| 017 | 7,773.87 | 7,573.26 | 7,483.58 | 7,664.63 | **30,495.34** |
| 018 | 21,944.36 | 21,575.49 | 21,742.91 | 20,379.88 | **85,642.64** |
| 019 | 11,662.14 | 11,696.93 | 11,473.16 | 10,911.17 | **45,743.39** |
| 020 | 8,073.76 | 8,195.85 | 8,103.24 | 7,918.02 | **32,290.87** |
| 021 | 28,239.90 | 27,536.13 | 26,768.82 | 26,456.24 | **109,001.10** |
| 022 | 2,299.13 | 2,315.88 | 2,561.86 | 2,095.82 | **9,272.70** |
| 023 | 14,218.43 | 13,854.73 | 13,724.24 | 12,982.51 | **54,779.91** |
| 024 | 4,179.68 | 4,053.74 | 4,160.23 | 4,287.01 | **16,680.66** |
| 025 | 3,839.29 | 3,868.49 | 3,943.54 | 3,874.48 | **15,525.80** |
| 026 | 5,341.61 | 5,219.32 | 5,431.16 | 5,377.55 | **21,369.64** |
| 027 | 23,297.37 | 23,297.50 | 23,563.26 | 24,088.71 | **94,246.84** |
| **MA - MASSACHUSETTS - Total** | **187,297.80** | **183,742.77** | **182,258.72** | **179,521.34** | **732,820.63** |

**STATE: MD - MARYLAND**

| REGISTRANT ZIP CODE 3 | 1ST QUARTER | 2ND QUARTER | 3RD QUARTER | 4TH QUARTER | **TOTAL GRAMS** |
|---|---|---|---|---|---|
| 206 | 19,693.87 | 19,355.66 | 18,505.78 | 18,300.55 | **75,855.86** |
| 207 | 21,893.63 | 21,536.29 | 21,784.34 | 21,272.00 | **86,486.26** |
| 208 | 16,112.67 | 16,602.24 | 15,903.64 | 15,940.28 | **64,558.83** |
| 209 | 3,195.68 | 3,341.62 | 3,405.95 | 3,136.62 | **13,079.87** |
| 210 | 39,354.70 | 39,820.09 | 39,564.52 | 39,839.22 | **158,578.53** |
| 211 | 15,869.78 | 15,760.91 | 15,689.59 | 15,658.33 | **62,978.61** |
| 212 | 72,751.15 | 71,339.35 | 70,946.39 | 71,370.64 | **286,407.52** |
| 214 | 3,394.96 | 3,376.06 | 3,625.41 | 3,556.31 | **13,952.74** |

| | | | | | |
|---|---|---|---|---|---|
| 215 | 6,394.66 | 6,582.96 | 6,578.66 | 6,936.78 | **26,493.07** |
| 216 | 8,216.66 | 8,257.52 | 8,051.69 | 8,213.84 | **32,739.71** |
| 217 | 23,952.01 | 23,488.25 | 23,342.21 | 22,874.89 | **93,657.36** |
| 218 | 7,243.33 | 7,195.76 | 7,007.73 | 6,920.09 | **28,366.91** |
| 219 | 5,600.62 | 5,448.89 | 5,194.96 | 5,574.04 | **21,818.50** |
| **MD MARYLAND Total** | **243,673.71** | **242,105.60** | **239,600.87** | **239,593.58** | **964,973.75** |

**STATE: ME - MAINE**

| REGISTRANT ZIP CODE 3 | 1ST QUARTER | 2ND QUARTER | 3RD QUARTER | 4TH QUARTER | TOTAL GRAMS |
|---|---|---|---|---|---|
| 039 | 670.29 | 727.61 | 677.75 | 609.72 | **2,685.37** |
| 040 | 11,212.05 | 11,120.54 | 11,128.59 | 10,301.23 | **43,762.41** |
| 041 | 2,795.73 | 2,788.90 | 2,839.75 | 2,760.38 | **11,184.76** |
| 042 | 5,089.62 | 5,157.83 | 5,055.92 | 5,001.83 | **20,305.20** |
| 043 | 5,303.97 | 5,229.54 | 5,300.06 | 4,813.19 | **20,646.77** |
| 044 | 3,959.89 | 3,711.57 | 4,106.51 | 3,924.90 | **15,702.86** |
| 045 | 1,284.78 | 1,303.53 | 1,354.56 | 1,412.09 | **5,354.96** |
| 046 | 1,593.55 | 1,876.87 | 1,780.11 | 1,610.41 | **6,860.94** |
| 047 | 1,369.61 | 1,407.01 | 1,521.88 | 1,278.58 | **5,577.09** |
| 048 | 972.16 | 1,015.46 | 988.33 | 970.46 | **3,946.41** |
| 049 | 6,545.77 | 6,428.48 | 6,757.86 | 6,046.37 | **25,778.47** |
| **ME MAINE Total** | **40,797.41** | **40,767.34** | **41,511.33** | **38,729.16** | **161,805.24** |

**STATE: MI - MICHIGAN**

| REGISTRANT ZIP CODE 3 | 1ST QUARTER | 2ND QUARTER | 3RD QUARTER | 4TH QUARTER | TOTAL GRAMS |
|---|---|---|---|---|---|
| 480 | 35,285.32 | 35,716.06 | 35,951.54 | 35,088.44 | **142,041.35** |
| 481 | 35,592.61 | 34,952.14 | 35,025.77 | 34,198.56 | **139,769.07** |
| 482 | 25,626.38 | 25,064.67 | 24,741.42 | 23,887.01 | **99,319.47** |
| 483 | 24,160.64 | 23,939.94 | 24,200.14 | 23,528.37 | **95,829.09** |
| 484 | 5,737.31 | 5,672.75 | 5,753.94 | 5,741.12 | **22,905.13** |
| 485 | 2,449.73 | 2,352.01 | 2,436.84 | 2,301.77 | **9,540.35** |
| 486 | 6,694.32 | 6,676.36 | 6,725.66 | 6,410.47 | **26,506.83** |
| 487 | 2,993.16 | 2,986.75 | 3,147.77 | 2,981.21 | **12,108.89** |
| 488 | 7,650.10 | 7,632.57 | 8,036.78 | 7,253.21 | **30,572.66** |
| 489 | 3,388.55 | 3,463.87 | 3,378.50 | 3,343.07 | **13,573.99** |
| 490 | 12,608.73 | 12,276.97 | 13,051.79 | 12,092.71 | **50,030.20** |
| 491 | 1,597.15 | 1,622.95 | 1,716.64 | 1,713.76 | **6,650.50** |
| 492 | 6,365.79 | 6,238.15 | 6,191.60 | 5,770.33 | **24,565.87** |
| 493 | 2,262.21 | 2,254.92 | 2,514.28 | 2,199.89 | **9,231.29** |
| 494 | 5,143.56 | 5,271.89 | 5,407.45 | 5,003.90 | **20,826.80** |
| 495 | 5,406.74 | 5,649.37 | 5,363.07 | 5,153.00 | **21,572.19** |
| 496 | 6,582.10 | 6,686.09 | 6,905.64 | 6,336.79 | **26,510.63** |
| 497 | 5,044.32 | 5,115.15 | 5,515.73 | 4,940.21 | **20,615.40** |

| | | | | | |
|---|---|---|---|---|---|
| 498 | 2,929.00 | 3,048.90 | 3,097.51 | 2,816.96 | **11,892.36** |
| 499 | 997.96 | 972.83 | 973.42 | 1,006.45 | **3,950.65** |
| **MI - MICHIGAN Total** | **198,515.69** | **197,594.33** | **200,135.47** | **191,767.23** | **788,012.72** |

**STATE: MN - MINNESOTA**

| REGISTRANT ZIP CODE 3 | 1ST QUARTER | 2ND QUARTER | 3RD QUARTER | 4TH QUARTER | TOTAL GRAMS |
|---|---|---|---|---|---|
| 550 | 12,459.51 | 12,572.70 | 12,186.70 | 12,291.13 | **49,510.04** |
| 551 | 18,386.41 | 18,770.59 | 17,411.62 | 16,982.58 | **71,551.20** |
| 553 | 15,867.62 | 15,739.69 | 15,262.84 | 15,218.35 | **62,088.50** |
| 554 | 28,178.20 | 27,554.64 | 26,705.38 | 26,342.65 | **108,780.87** |
| 556 | 158.46 | 197.63 | 210.23 | 176.92 | **743.24** |
| 557 | 3,739.97 | 3,777.04 | 3,616.44 | 3,402.40 | **14,535.86** |
| 558 | 1,904.37 | 2,035.25 | 1,850.76 | 1,863.63 | **7,654.00** |
| 559 | 4,501.49 | 4,585.07 | 3,917.33 | 4,123.65 | **17,127.54** |
| 560 | 3,194.03 | 3,244.77 | 3,216.52 | 3,172.04 | **12,827.36** |
| 561 | 802.47 | 877.88 | 843.21 | 846.65 | **3,370.21** |
| 562 | 1,624.12 | 1,599.43 | 1,514.41 | 1,491.79 | **6,229.76** |
| 563 | 5,246.44 | 5,336.13 | 5,301.23 | 5,151.43 | **21,035.23** |
| 564 | 2,329.56 | 2,476.93 | 2,409.91 | 2,232.87 | **9,449.27** |
| 565 | 2,009.75 | 1,913.18 | 1,758.46 | 1,751.49 | **7,432.88** |
| 566 | 968.80 | 984.83 | 1,043.48 | 1,029.03 | **4,026.14** |
| 567 | 1,150.54 | 1,181.38 | 1,117.73 | 1,152.62 | **4,602.28** |
| **MN MINNESOTA Total** | **102,521.74** | **102,847.14** | **98,366.26** | **97,229.24** | **400,964.38** |

**STATE: MO - MISSOURI**

| REGISTRANT ZIP CODE 3 | 1ST QUARTER | 2ND QUARTER | 3RD QUARTER | 4TH QUARTER | TOTAL GRAMS |
|---|---|---|---|---|---|
| 630 | 21,612.52 | 21,831.99 | 21,384.89 | 20,565.57 | **85,394.97** |
| 631 | 38,100.64 | 37,381.56 | 36,079.35 | 36,040.95 | **147,602.50** |
| 633 | 11,888.02 | 11,962.44 | 11,602.05 | 11,406.32 | **46,858.83** |
| 634 | 1,265.60 | 1,125.58 | 1,214.73 | 1,216.53 | **4,822.43** |
| 635 | 1,717.00 | 1,607.22 | 1,528.38 | 1,421.45 | **6,274.06** |
| 636 | 6,457.72 | 6,122.99 | 5,909.20 | 5,784.16 | **24,274.07** |
| 637 | 2,964.33 | 2,812.59 | 2,843.92 | 2,669.30 | **11,290.15** |
| 638 | 3,431.96 | 3,301.03 | 3,217.37 | 3,100.37 | **13,050.73** |
| 639 | 1,944.19 | 1,878.76 | 1,600.14 | 1,675.38 | **7,098.47** |
| 640 | 18,591.03 | 18,479.14 | 18,170.07 | 17,943.28 | **73,183.52** |
| 641 | 15,990.61 | 14,388.89 | 13,849.42 | 13,443.36 | **57,672.27** |
| 644 | 1,476.15 | 1,467.40 | 1,366.68 | 1,409.65 | **5,719.88** |
| 645 | 3,747.32 | 3,641.79 | 3,925.55 | 3,775.03 | **15,089.69** |
| 646 | 1,615.95 | 1,663.59 | 1,687.37 | 1,603.45 | **6,570.36** |
| 647 | 4,203.91 | 3,992.73 | 3,910.46 | 3,633.32 | **15,740.43** |
| 648 | 8,061.98 | 5,499.36 | 5,357.26 | 5,200.21 | **24,118.81** |

| | | | | | |
|---|---|---|---|---|---|
| 650 | 2,899.46 | 2,984.79 | 2,872.48 | 2,804.28 | **11,561.01** |
| 651 | 1,589.10 | 1,430.35 | 1,441.79 | 1,365.23 | **5,826.47** |
| 652 | 9,178.63 | 9,371.55 | 9,258.82 | 9,279.39 | **37,088.39** |
| 653 | 2,984.54 | 3,098.16 | 2,828.54 | 2,741.50 | **11,652.74** |
| 654 | 3,957.19 | 4,055.68 | 4,135.78 | 4,010.97 | **16,159.63** |
| 655 | 5,618.00 | 5,821.38 | 5,582.39 | 5,624.01 | **22,645.78** |
| 656 | 8,528.46 | 8,334.24 | 8,182.00 | 7,872.85 | **32,917.55** |
| 657 | 11,613.08 | 11,337.51 | 11,343.16 | 11,136.35 | **45,430.09** |
| 658 | 12,368.23 | 11,725.62 | 11,407.19 | 10,725.79 | **46,226.83** |
| **MO MISSOURI - Total** | **201,805.63** | **195,316.33** | **190,698.98** | **186,448.70** | **774,269.64** |

**STATE: MS - MISSISSIPPI**

| REGISTRANT ZIP CODE 3 | 1ST QUARTER | 2ND QUARTER | 3RD QUARTER | 4TH QUARTER | TOTAL GRAMS |
|---|---|---|---|---|---|
| 386 | 11,939.04 | 11,596.00 | 11,624.83 | 12,066.57 | **47,226.43** |
| 387 | 1,588.30 | 1,465.27 | 1,550.43 | 1,412.56 | **6,016.56** |
| 388 | 7,727.27 | 7,474.73 | 7,330.52 | 7,513.28 | **30,045.79** |
| 389 | 1,409.08 | 1,351.10 | 1,357.02 | 1,383.77 | **5,500.97** |
| 390 | 4,356.06 | 4,135.23 | 4,060.89 | 3,993.07 | **16,545.25** |
| 391 | 4,238.11 | 3,935.45 | 4,092.04 | 3,898.55 | **16,164.14** |
| 392 | 5,082.10 | 4,798.10 | 4,836.10 | 4,658.29 | **19,374.58** |
| 393 | 4,872.75 | 4,964.03 | 4,944.72 | 4,713.76 | **19,495.26** |
| 394 | 16,178.75 | 16,171.63 | 16,988.06 | 16,867.11 | **66,205.54** |
| 395 | 16,859.86 | 15,949.80 | 16,503.00 | 16,307.22 | **65,619.88** |
| 396 | 3,691.86 | 3,511.22 | 3,632.03 | 3,559.99 | **14,395.09** |
| 397 | 2,407.52 | 2,284.43 | 2,288.17 | 2,328.08 | **9,308.20** |
| **MS MISSISSIPPI - Total** | **80,350.69** | **77,636.97** | **79,207.80** | **78,702.24** | **315,897.71** |

**STATE: MT - MONTANA**

| REGISTRANT ZIP CODE 3 | 1ST QUARTER | 2ND QUARTER | 3RD QUARTER | 4TH QUARTER | TOTAL GRAMS |
|---|---|---|---|---|---|
| 590 | 1,709.82 | 1,707.93 | 1,848.27 | 1,746.32 | **7,012.35** |
| 591 | 6,788.74 | 6,611.51 | 6,061.96 | 6,074.68 | **25,536.89** |
| 592 | 972.21 | 965.19 | 992.89 | 884.98 | **3,815.26** |
| 593 | 1,128.36 | 1,120.63 | 1,036.22 | 1,016.07 | **4,301.29** |
| 594 | 2,825.17 | 3,079.77 | 3,140.74 | 2,698.43 | **11,744.12** |
| 595 | 485.99 | 426.65 | 452.43 | 451.84 | **1,816.91** |
| 596 | 1,929.56 | 2,013.52 | 2,105.31 | 1,958.57 | **8,006.96** |
| 597 | 3,232.05 | 3,162.34 | 3,260.83 | 3,115.34 | **12,770.55** |
| 598 | 4,733.60 | 4,826.50 | 4,631.78 | 4,564.39 | **18,756.27** |
| 599 | 4,114.17 | 4,141.64 | 4,115.40 | 4,089.61 | **16,460.83** |
| **MT - MONTANA Total** | **27,919.67** | **28,055.69** | **27,645.84** | **26,600.23** | **110,221.43** |

**STATE: NC - NORTH CAROLINA**

| REGISTRANT ZIP CODE 3 | 1ST QUARTER | 2ND QUARTER | 3RD QUARTER | 4TH QUARTER | TOTAL GRAMS |
|---|---|---|---|---|---|

| | | | | | |
|---|---|---|---|---|---|
| 270 | 13,477.80 | 13,951.83 | 13,839.42 | 13,128.31 | **54,397.36** |
| 271 | 9,383.79 | 9,684.73 | 9,866.67 | 9,477.51 | **38,412.70** |
| 272 | 30,697.17 | 29,226.12 | 30,391.16 | 26,418.96 | **116,733.41** |
| 273 | 22,519.40 | 21,764.10 | 21,782.58 | 18,650.13 | **84,716.21** |
| 274 | 11,651.89 | 10,895.14 | 11,206.27 | 10,677.45 | **44,430.75** |
| 275 | 46,435.23 | 48,023.37 | 48,209.04 | 39,868.84 | **182,536.48** |
| 276 | 12,727.56 | 12,186.97 | 12,258.82 | 11,559.01 | **48,732.35** |
| 277 | 12,346.80 | 11,946.70 | 12,034.63 | 11,235.04 | **47,563.17** |
| 278 | 25,101.52 | 24,717.23 | 24,752.48 | 22,398.35 | **96,969.58** |
| 279 | 12,225.01 | 12,031.29 | 11,332.40 | 9,656.96 | **45,245.65** |
| 280 | 34,149.73 | 33,669.33 | 33,571.89 | 30,674.25 | **132,065.20** |
| 281 | 25,094.85 | 24,611.25 | 25,032.75 | 23,319.28 | **98,058.14** |
| 282 | 17,297.28 | 17,880.89 | 18,039.39 | 18,437.46 | **71,655.03** |
| 283 | 73,543.29 | 72,274.06 | 72,537.89 | 60,583.04 | **278,938.28** |
| 284 | 35,423.22 | 34,366.60 | 34,295.36 | 29,048.60 | **133,133.78** |
| 285 | 28,318.45 | 27,678.31 | 27,611.33 | 25,542.45 | **109,150.54** |
| 286 | 46,815.57 | 45,131.55 | 44,503.99 | 40,728.32 | **177,179.43** |
| 287 | 22,042.77 | 21,839.01 | 22,160.48 | 20,302.93 | **86,345.18** |
| 288 | 8,213.84 | 7,714.89 | 7,914.14 | 7,594.67 | **31,437.53** |
| 289 | 3,063.21 | 2,913.53 | 3,033.62 | 3,188.90 | **12,199.27** |
| **NC - NORTH CAROLINA Total** | **490,528.39** | **482,506.89** | **484,374.30** | **432,490.46** | **1,889,900.04** |

**STATE: ND - NORTH DAKOTA**

| REGISTRANT ZIP CODE 3 | 1ST QUARTER | 2ND QUARTER | 3RD QUARTER | 4TH QUARTER | **TOTAL GRAMS** |
|---|---|---|---|---|---|
| 580 | 540.01 | 519.61 | 489.27 | 492.85 | **2,041.74** |
| 581 | 1,878.71 | 1,883.71 | 1,847.48 | 2,012.10 | **7,622.00** |
| 582 | 1,685.83 | 1,645.36 | 1,683.04 | 1,630.33 | **6,644.56** |
| 583 | 1,040.78 | 1,105.61 | 997.48 | 868.08 | **4,011.95** |
| 584 | 577.40 | 545.25 | 562.33 | 507.01 | **2,191.99** |
| 585 | 3,346.01 | 2,924.33 | 2,926.63 | 2,809.25 | **12,006.21** |
| 586 | 726.08 | 725.36 | 774.63 | 777.71 | **3,003.78** |
| 587 | 972.88 | 965.58 | 978.10 | 963.31 | **3,879.87** |
| 588 | 805.40 | 800.54 | 786.26 | 825.07 | **3,217.27** |
| **ND NORTH DAKOTA Total** | **11,573.09** | **11,115.35** | **11,045.22** | **10,885.71** | **44,619.37** |

**STATE: NE - NEBRASKA**

| REGISTRANT ZIP CODE 3 | 1ST QUARTER | 2ND QUARTER | 3RD QUARTER | 4TH QUARTER | **TOTAL GRAMS** |
|---|---|---|---|---|---|
| 680 | 3,418.46 | 3,277.67 | 3,356.22 | 3,508.47 | **13,560.82** |
| 681 | 13,645.86 | 13,290.33 | 12,702.08 | 12,298.79 | **51,937.07** |
| 683 | 912.88 | 922.46 | 1,039.07 | 1,010.36 | **3,884.77** |
| 684 | 787.50 | 767.17 | 768.85 | 817.06 | **3,140.58** |
| 685 | 3,823.15 | 3,546.64 | 3,580.18 | 3,477.55 | **14,427.52** |

| | | | | | |
|---|---|---|---|---|---|
| 686 | 484.45 | 470.14 | 412.25 | 463.84 | **1,830.69** |
| 687 | 1,140.79 | 1,006.26 | 1,009.24 | 1,023.69 | **4,179.98** |
| 688 | 2,500.72 | 2,378.52 | 2,490.60 | 2,450.70 | **9,820.55** |
| 689 | 1,037.85 | 1,128.18 | 1,008.30 | 1,035.34 | **4,209.68** |
| 690 | 488.85 | 487.48 | 513.04 | 462.38 | **1,951.76** |
| 691 | 1,438.77 | 1,449.27 | 1,428.79 | 1,361.31 | **5,678.14** |
| 692 | 56.93 | 72.54 | 79.74 | 82.70 | **291.91** |
| 693 | 1,515.73 | 1,454.06 | 1,502.94 | 1,357.36 | **5,830.09** |
| **NE NEBRASKA Total** | **31,251.94** | **30,250.71** | **29,891.34** | **29,349.56** | **120,743.56** |

**STATE: NH - NEW HAMPSHIRE**

| REGISTRANT ZIP CODE 3 | 1ST QUARTER | 2ND QUARTER | 3RD QUARTER | 4TH QUARTER | TOTAL GRAMS |
|---|---|---|---|---|---|
| 030 | 11,008.84 | 10,763.69 | 10,729.10 | 10,577.80 | **43,079.42** |
| 031 | 6,140.48 | 5,974.61 | 6,288.21 | 5,921.74 | **24,325.04** |
| 032 | 5,234.56 | 5,254.16 | 5,057.33 | 4,849.57 | **20,395.62** |
| 033 | 3,675.88 | 3,415.08 | 3,445.93 | 3,325.27 | **13,862.17** |
| 034 | 2,130.52 | 2,069.41 | 2,107.72 | 2,180.67 | **8,488.32** |
| 035 | 1,907.92 | 1,955.96 | 2,033.65 | 1,824.87 | **7,722.40** |
| 036 | 229.06 | 209.56 | 167.65 | 140.30 | **746.56** |
| 037 | 2,833.42 | 2,933.35 | 2,954.53 | 2,838.63 | **11,559.93** |
| 038 | 12,482.76 | 12,098.31 | 12,215.34 | 11,785.85 | **48,582.26** |
| **NH - NEW HAMPSHIRE Total** | **45,643.45** | **44,674.12** | **44,999.46** | **43,444.69** | **178,761.72** |

**STATE: NJ - NEW JERSEY**

| REGISTRANT ZIP CODE 3 | 1ST QUARTER | 2ND QUARTER | 3RD QUARTER | 4TH QUARTER | TOTAL GRAMS |
|---|---|---|---|---|---|
| 070 | 51,944.05 | 48,623.63 | 51,631.66 | 51,336.27 | **203,535.62** |
| 071 | 12,454.23 | 11,056.50 | 12,293.35 | 12,082.37 | **47,886.45** |
| 072 | 4,508.09 | 4,204.12 | 4,926.05 | 5,100.04 | **18,738.31** |
| 073 | 7,771.19 | 7,535.67 | 8,316.03 | 7,919.18 | **31,542.08** |
| 074 | 16,220.96 | 15,089.13 | 15,153.52 | 15,190.43 | **61,654.03** |
| 075 | 6,328.88 | 5,505.62 | 5,950.59 | 6,339.25 | **24,124.34** |
| 076 | 20,081.73 | 20,167.88 | 18,973.17 | 19,222.20 | **78,444.98** |
| 077 | 24,818.90 | 23,903.68 | 24,361.03 | 23,946.72 | **97,030.32** |
| 078 | 10,782.45 | 10,546.70 | 10,624.69 | 10,169.79 | **42,123.63** |
| 079 | 5,317.81 | 5,313.53 | 5,357.95 | 5,177.34 | **21,166.63** |
| 080 | 80,862.04 | 79,130.10 | 79,623.19 | 78,742.31 | **318,357.64** |
| 081 | 12,426.67 | 12,376.73 | 12,548.06 | 13,046.17 | **50,397.63** |
| 082 | 16,425.78 | 16,234.02 | 16,827.61 | 16,340.24 | **65,827.64** |
| 083 | 11,688.06 | 11,443.35 | 11,320.72 | 11,101.75 | **45,553.87** |
| 084 | 3,173.94 | 3,233.88 | 3,403.35 | 2,794.98 | **12,606.16** |
| 085 | 8,728.05 | 8,385.55 | 8,803.34 | 8,992.95 | **34,909.89** |
| 086 | 11,867.95 | 11,280.37 | 11,838.93 | 11,760.26 | **46,747.51** |

| | | | | | |
|---|---|---|---|---|---|
| 087 | 19,281.24 | 18,929.26 | 19,163.00 | 19,211.68 | **76,585.19** |
| 088 | 33,942.19 | 32,664.46 | 33,619.31 | 33,195.07 | **133,421.03** |
| 089 | 2,987.49 | 2,845.20 | 2,915.69 | 2,891.91 | **11,640.29** |
| **NJ NEW JERSEY - Total** | **361,611.68** | **348,469.38** | **357,651.24** | **354,560.93** | **1,422,293.24** |

**STATE: NM - NEW MEXICO**

| REGISTRANT ZIP CODE 3 | 1ST QUARTER | 2ND QUARTER | 3RD QUARTER | 4TH QUARTER | TOTAL GRAMS |
|---|---|---|---|---|---|
| 870 | 6,625.79 | 6,274.35 | 6,006.95 | 5,967.20 | **24,874.29** |
| 871 | 31,804.02 | 31,066.61 | 30,100.65 | 29,805.44 | **122,776.71** |
| 873 | 1,035.41 | 1,021.34 | 967.10 | 948.23 | **3,972.08** |
| 874 | 2,129.53 | 2,046.58 | 2,017.65 | 1,896.56 | **8,090.32** |
| 875 | 12,507.52 | 12,188.45 | 11,810.18 | 11,772.04 | **48,278.19** |
| 877 | 1,605.90 | 1,537.45 | 1,560.34 | 1,454.81 | **6,158.51** |
| 878 | 569.99 | 546.19 | 539.02 | 563.68 | **2,218.89** |
| 879 | 330.51 | 351.23 | 297.86 | 327.22 | **1,306.82** |
| 880 | 4,796.60 | 4,269.39 | 4,119.20 | 4,021.64 | **17,206.83** |
| 881 | 928.47 | 892.26 | 737.06 | 762.61 | **3,320.41** |
| 882 | 6,123.69 | 6,075.13 | 5,790.23 | 5,701.07 | **23,690.13** |
| 883 | 2,164.46 | 2,011.83 | 2,007.98 | 2,030.70 | **8,214.97** |
| 884 | 149.27 | 171.68 | 134.25 | 171.90 | **627.10** |
| **NM NEW MEXICO Total** | **70,771.16** | **68,452.51** | **66,088.46** | **65,423.12** | **270,735.26** |

**STATE: NV - NEVADA**

| REGISTRANT ZIP CODE 3 | 1ST QUARTER | 2ND QUARTER | 3RD QUARTER | 4TH QUARTER | TOTAL GRAMS |
|---|---|---|---|---|---|
| 890 | 32,289.35 | 32,053.30 | 31,621.14 | 32,584.52 | **128,548.31** |
| 891 | 76,418.89 | 78,065.48 | 75,469.52 | 76,774.03 | **306,727.91** |
| 893 | 421.36 | 366.35 | 358.38 | 330.81 | **1,476.89** |
| 894 | 10,620.95 | 9,987.56 | 9,965.30 | 10,111.90 | **40,685.71** |
| 895 | 11,209.25 | 10,672.50 | 10,421.15 | 10,206.44 | **42,509.34** |
| 897 | 3,601.34 | 3,419.28 | 3,454.38 | 3,415.28 | **13,890.28** |
| 898 | 1,074.21 | 1,063.26 | 1,097.77 | 1,006.06 | **4,241.30** |
| **NV NEVADA Total** | **135,635.35** | **135,627.73** | **132,387.63** | **134,429.04** | **538,079.75** |

**STATE: NY - NEW YORK**

| REGISTRANT ZIP CODE 3 | 1ST QUARTER | 2ND QUARTER | 3RD QUARTER | 4TH QUARTER | TOTAL GRAMS |
|---|---|---|---|---|---|
| 100 | 50,663.78 | 42,062.43 | 45,120.85 | 44,873.52 | **182,720.58** |
| 101 | 1,009.20 | 920.77 | 1,014.00 | 1,154.40 | **4,098.38** |
| 102 | 57.39 | 50.67 | 49.55 | 47.98 | **205.59** |
| 103 | 21,406.43 | 19,926.81 | 20,809.39 | 20,411.54 | **82,554.17** |
| 104 | 43,667.58 | 34,016.88 | 38,191.51 | 36,069.29 | **151,945.27** |
| 105 | 15,250.76 | 13,282.21 | 14,203.51 | 14,053.05 | **56,789.53** |
| 106 | 1,925.42 | 2,052.47 | 1,975.11 | 2,109.39 | **8,062.39** |
| 107 | 6,254.47 | 5,443.75 | 5,860.95 | 6,086.79 | **23,645.96** |

| | | | | | |
|---|---|---|---|---|---|
| 108 | 1,805.30 | 1,590.07 | 1,574.45 | 1,699.12 | **6,668.94** |
| 109 | 19,743.73 | 18,044.58 | 18,741.41 | 17,528.65 | **74,058.37** |
| 110 | 9,139.06 | 8,269.53 | 8,568.98 | 8,606.92 | **34,584.49** |
| 111 | 6,189.11 | 5,541.91 | 5,715.03 | 5,171.43 | **22,617.48** |
| 112 | 51,878.93 | 42,738.79 | 44,646.54 | 44,278.18 | **183,542.44** |
| 113 | 20,432.11 | 18,422.05 | 21,108.87 | 20,450.90 | **80,413.93** |
| 114 | 10,667.50 | 9,719.61 | 10,243.61 | 10,099.49 | **40,730.21** |
| 115 | 22,636.18 | 20,142.17 | 19,947.50 | 20,086.03 | **82,811.88** |
| 116 | 2,861.19 | 2,409.50 | 2,865.59 | 2,737.74 | **10,874.01** |
| 117 | 68,352.72 | 64,874.04 | 66,437.20 | 66,516.88 | **266,180.84** |
| 118 | 2,315.18 | 2,385.12 | 2,239.52 | 2,377.79 | **9,317.60** |
| 119 | 7,725.32 | 7,512.86 | 7,982.05 | 7,779.00 | **30,999.23** |
| 120 | 8,116.77 | 8,173.46 | 8,123.69 | 8,032.10 | **32,446.02** |
| 121 | 4,964.32 | 4,945.03 | 4,894.03 | 4,693.86 | **19,497.24** |
| 122 | 4,480.72 | 4,426.25 | 4,432.99 | 4,201.65 | **17,541.61** |
| 123 | 2,418.14 | 2,441.37 | 2,313.03 | 2,706.82 | **9,879.35** |
| 124 | 10,339.63 | 9,908.49 | 10,111.51 | 9,738.56 | **40,098.19** |
| 125 | 17,866.69 | 15,665.03 | 16,400.15 | 15,444.40 | **65,376.27** |
| 126 | 5,154.11 | 4,242.47 | 4,762.20 | 4,762.11 | **18,920.89** |
| 127 | 7,464.26 | 6,676.46 | 6,365.70 | 5,851.36 | **26,357.78** |
| 128 | 7,546.01 | 7,529.74 | 7,154.01 | 7,294.15 | **29,523.91** |
| 129 | 4,936.45 | 5,118.04 | 5,236.84 | 5,208.40 | **20,499.73** |
| 130 | 9,034.85 | 8,699.16 | 8,815.05 | 8,472.63 | **35,021.70** |
| 131 | 2,559.89 | 2,584.01 | 2,480.01 | 2,451.26 | **10,075.17** |
| 132 | 5,790.96 | 5,912.44 | 5,778.30 | 5,877.70 | **23,359.40** |
| 133 | 2,576.00 | 2,654.25 | 2,703.52 | 2,507.80 | **10,441.56** |
| 134 | 3,358.27 | 3,625.16 | 3,391.43 | 3,234.72 | **13,609.56** |
| 135 | 1,524.85 | 1,452.79 | 1,400.64 | 1,392.61 | **5,770.89** |
| 136 | 5,169.99 | 5,303.75 | 5,253.68 | 5,201.03 | **20,928.46** |
| 137 | 3,418.56 | 3,228.11 | 2,954.49 | 3,110.56 | **12,711.72** |
| 138 | 6,672.12 | 6,505.55 | 5,894.08 | 5,669.50 | **24,741.24** |
| 139 | 3,498.39 | 3,270.01 | 3,050.08 | 2,788.12 | **12,606.60** |
| 140 | 10,182.20 | 10,197.26 | 9,546.89 | 9,633.87 | **39,560.23** |
| 141 | 4,383.48 | 4,293.42 | 4,306.59 | 4,207.88 | **17,191.37** |
| 142 | 13,487.16 | 12,881.76 | 12,948.16 | 12,707.26 | **52,024.34** |
| 143 | 1,932.00 | 1,850.38 | 1,852.58 | 1,743.86 | **7,378.82** |
| 144 | 6,663.47 | 6,731.19 | 6,782.06 | 6,706.57 | **26,883.29** |
| 145 | 5,731.89 | 5,543.44 | 5,273.28 | 5,252.53 | **21,801.14** |
| 146 | 15,315.65 | 15,147.41 | 14,818.45 | 15,403.25 | **60,684.75** |
| 147 | 2,875.85 | 2,886.31 | 2,783.74 | 2,806.49 | **11,352.39** |

| | | | | | |
|---|---|---|---|---|---|
| 148 | 8,461.17 | 8,378.50 | 7,926.94 | 7,696.25 | **32,462.87** |
| 149 | 2,643.74 | 2,501.10 | 2,405.72 | 2,307.96 | **9,858.52** |
| **NY - NEW YORK - Total** | **552,548.94** | **502,178.55** | **517,455.44** | **509,243.36** | **2,081,426.29** |

**STATE: OH - OHIO**

| REGISTRANT ZIP CODE 3 | 1ST QUARTER | 2ND QUARTER | 3RD QUARTER | 4TH QUARTER | TOTAL GRAMS |
|---|---|---|---|---|---|
| 430 | 20,968.28 | 21,099.00 | 20,987.02 | 20,941.59 | **83,995.89** |
| 431 | 16,106.57 | 16,131.13 | 15,953.54 | 15,871.19 | **64,062.43** |
| 432 | 30,122.41 | 28,469.21 | 28,493.40 | 28,784.98 | **115,870.00** |
| 433 | 5,378.73 | 5,462.98 | 5,475.01 | 5,451.57 | **21,768.29** |
| 434 | 4,217.28 | 4,315.10 | 3,964.82 | 2,957.24 | **15,454.44** |
| 435 | 6,542.91 | 6,494.94 | 6,303.70 | 5,960.67 | **25,302.23** |
| 436 | 16,826.67 | 17,192.88 | 17,183.33 | 16,315.73 | **67,518.62** |
| 437 | 7,435.32 | 7,462.74 | 7,284.86 | 7,268.24 | **29,451.16** |
| 438 | 1,144.53 | 1,218.05 | 1,137.02 | 1,164.61 | **4,664.21** |
| 439 | 7,192.35 | 6,794.36 | 6,642.46 | 5,264.92 | **25,894.10** |
| 440 | 16,487.69 | 16,172.82 | 16,080.95 | 15,426.17 | **64,167.61** |
| 441 | 25,684.14 | 24,797.23 | 24,788.41 | 24,044.74 | **99,314.52** |
| 442 | 12,009.39 | 11,775.27 | 11,331.51 | 11,366.80 | **46,482.97** |
| 443 | 8,705.31 | 8,292.51 | 8,215.17 | 8,110.30 | **33,323.29** |
| 444 | 13,265.68 | 12,928.43 | 12,982.28 | 11,655.80 | **50,832.19** |
| 445 | 6,503.41 | 6,644.05 | 6,691.54 | 6,565.11 | **26,404.11** |
| 446 | 8,792.49 | 8,687.17 | 8,493.45 | 8,358.18 | **34,331.28** |
| 447 | 5,646.85 | 5,760.16 | 5,658.64 | 5,714.82 | **22,780.46** |
| 448 | 8,539.30 | 8,257.16 | 7,591.65 | 6,250.32 | **30,638.42** |
| 449 | 2,330.37 | 2,270.98 | 2,163.35 | 2,103.84 | **8,868.54** |
| 450 | 22,643.74 | 22,260.26 | 21,923.15 | 21,387.17 | **88,214.33** |
| 451 | 11,121.82 | 10,929.10 | 11,054.26 | 10,660.95 | **43,766.14** |
| 452 | 29,796.70 | 29,314.86 | 28,605.04 | 27,308.47 | **115,025.08** |
| 453 | 10,677.02 | 10,614.17 | 10,609.19 | 10,502.23 | **42,402.60** |
| 454 | 13,053.80 | 12,592.02 | 12,721.52 | 12,232.44 | **50,599.78** |
| 455 | 4,589.95 | 4,762.10 | 4,640.87 | 4,504.93 | **18,497.84** |
| 456 | 17,441.03 | 17,308.42 | 17,322.49 | 16,612.18 | **68,684.12** |
| 457 | 4,859.83 | 4,624.68 | 4,694.19 | 4,653.39 | **18,832.08** |
| 458 | 5,333.22 | 5,339.37 | 5,368.28 | 5,261.29 | **21,302.16** |
| **OH - OHIO Total** | **343,416.78** | **337,971.12** | **334,361.12** | **322,699.86** | **1,338,448.88** |

**STATE: OK - OKLAHOMA**

| REGISTRANT ZIP CODE 3 | 1ST QUARTER | 2ND QUARTER | 3RD QUARTER | 4TH QUARTER | TOTAL GRAMS |
|---|---|---|---|---|---|
| 730 | 23,883.18 | 24,201.76 | 24,201.33 | 23,768.19 | **96,054.46** |
| 731 | 28,239.09 | 27,502.07 | 28,249.40 | 27,856.26 | **111,846.81** |
| 734 | 3,216.87 | 2,943.97 | 3,027.29 | 3,038.33 | **12,226.46** |

| | | | | | |
|---|---|---|---|---|---|
| 735 | 7,752.91 | 7,764.05 | 7,905.23 | 7,756.21 | **31,178.40** |
| 736 | 2,030.25 | 1,995.53 | 2,030.65 | 2,086.90 | **8,143.34** |
| 737 | 2,933.19 | 2,921.40 | 2,962.03 | 2,917.66 | **11,734.28** |
| 738 | 550.49 | 582.28 | 620.18 | 634.71 | **2,387.66** |
| 739 | 148.58 | 155.27 | 167.65 | 158.46 | **629.96** |
| 740 | 23,575.11 | 23,342.73 | 22,885.86 | 22,305.02 | **92,108.71** |
| 741 | 16,011.04 | 15,572.40 | 15,721.40 | 14,932.07 | **62,236.92** |
| 743 | 3,690.78 | 3,473.57 | 3,538.43 | 3,534.84 | **14,237.62** |
| 744 | 11,514.52 | 11,668.83 | 11,960.20 | 11,740.02 | **46,883.57** |
| 745 | 2,692.86 | 2,603.94 | 2,771.59 | 2,585.70 | **10,654.09** |
| 746 | 1,737.05 | 1,674.49 | 1,617.46 | 1,759.65 | **6,788.65** |
| 747 | 2,300.46 | 2,123.25 | 1,889.35 | 1,966.23 | **8,279.30** |
| 748 | 7,234.86 | 7,102.80 | 7,052.14 | 7,054.05 | **28,443.83** |
| 749 | 5,861.26 | 5,305.65 | 5,037.75 | 4,717.23 | **20,921.90** |
| **OK - OKLAHOMA Total** | **143,372.51** | **140,933.98** | **141,637.92** | **138,811.54** | **564,755.96** |

**STATE: OR - OREGON**

| REGISTRANT ZIP CODE 3 | 1ST QUARTER | 2ND QUARTER | 3RD QUARTER | 4TH QUARTER | **TOTAL GRAMS** |
|---|---|---|---|---|---|
| 970 | 21,856.83 | 21,472.31 | 20,942.85 | 21,068.71 | **85,340.71** |
| 971 | 9,439.01 | 9,420.65 | 9,375.73 | 9,260.17 | **37,495.55** |
| 972 | 26,951.37 | 27,669.71 | 26,780.15 | 26,622.93 | **108,024.16** |
| 973 | 15,630.56 | 15,616.17 | 15,527.00 | 15,425.74 | **62,199.47** |
| 974 | 22,904.51 | 22,481.78 | 22,170.99 | 21,523.45 | **89,080.74** |
| 975 | 16,726.15 | 17,109.73 | 17,099.04 | 16,771.63 | **67,706.55** |
| 976 | 1,871.03 | 1,923.68 | 1,936.37 | 1,991.23 | **7,722.30** |
| 977 | 6,115.17 | 6,446.57 | 6,357.59 | 6,281.33 | **25,200.66** |
| 978 | 4,818.61 | 4,539.76 | 4,230.43 | 4,079.02 | **17,667.82** |
| 979 | 1,308.23 | 1,285.87 | 1,270.51 | 1,117.17 | **4,981.78** |
| **OR OREGON Total** | **127,621.47** | **127,966.22** | **125,690.66** | **124,141.38** | **505,419.73** |

**STATE: PA - PENNSYLVANIA**

| REGISTRANT ZIP CODE 3 | 1ST QUARTER | 2ND QUARTER | 3RD QUARTER | 4TH QUARTER | **TOTAL GRAMS** |
|---|---|---|---|---|---|
| 150 | 19,636.62 | 19,272.33 | 18,754.55 | 18,237.39 | **75,900.90** |
| 151 | 9,465.42 | 8,983.58 | 9,056.51 | 9,208.82 | **36,714.34** |
| 152 | 24,166.39 | 24,190.21 | 24,281.81 | 23,118.78 | **95,757.19** |
| 153 | 5,127.19 | 5,072.73 | 5,333.48 | 5,396.31 | **20,929.70** |
| 154 | 8,381.09 | 8,356.81 | 7,721.02 | 7,479.25 | **31,938.17** |
| 155 | 2,475.51 | 2,432.11 | 2,413.19 | 2,270.03 | **9,590.84** |
| 156 | 10,726.02 | 10,241.14 | 9,876.10 | 9,477.26 | **40,320.52** |
| 157 | 4,106.26 | 3,786.53 | 3,790.36 | 3,815.68 | **15,498.82** |
| 158 | 3,499.95 | 3,542.17 | 3,608.03 | 3,356.92 | **14,007.06** |
| 159 | 8,683.19 | 7,504.22 | 7,291.70 | 6,806.31 | **30,285.42** |

| | | | | | |
|---|---|---|---|---|---|
| 160 | 5,856.48 | 6,105.14 | 6,063.26 | 5,990.29 | **24,015.17** |
| 161 | 7,845.39 | 7,514.12 | 7,681.76 | 7,260.78 | **30,302.04** |
| 162 | 2,732.35 | 2,478.66 | 2,333.38 | 2,073.39 | **9,617.78** |
| 163 | 5,523.96 | 5,464.85 | 5,432.63 | 5,277.81 | **21,699.25** |
| 164 | 2,138.91 | 2,073.58 | 2,108.13 | 2,072.97 | **8,393.58** |
| 165 | 4,232.94 | 4,160.74 | 4,327.15 | 4,093.02 | **16,813.85** |
| 166 | 8,111.78 | 7,837.47 | 7,462.43 | 6,900.76 | **30,312.43** |
| 167 | 1,416.34 | 1,282.74 | 1,350.97 | 1,202.80 | **5,252.85** |
| 168 | 2,920.38 | 2,845.20 | 2,808.16 | 2,748.20 | **11,321.94** |
| 169 | 1,635.91 | 1,614.71 | 1,682.55 | 1,469.16 | **6,402.32** |
| 170 | 21,658.50 | 21,108.91 | 20,189.78 | 19,438.67 | **82,395.87** |
| 171 | 5,818.55 | 5,527.80 | 5,324.43 | 5,123.67 | **21,794.45** |
| 172 | 6,859.56 | 6,450.58 | 6,151.81 | 5,992.30 | **25,454.25** |
| 173 | 9,371.45 | 9,359.67 | 9,096.14 | 8,881.66 | **36,708.92** |
| 174 | 5,298.13 | 5,031.10 | 5,199.57 | 5,139.15 | **20,667.95** |
| 175 | 8,479.18 | 8,277.03 | 8,169.52 | 7,811.52 | **32,737.25** |
| 176 | 6,882.02 | 6,504.76 | 6,733.55 | 6,750.98 | **26,871.31** |
| 177 | 5,748.90 | 5,709.48 | 5,693.97 | 5,662.98 | **22,815.33** |
| 178 | 5,914.53 | 5,836.95 | 5,526.77 | 5,349.02 | **22,627.27** |
| 179 | 3,903.68 | 3,676.34 | 3,580.22 | 3,511.63 | **14,671.86** |
| 180 | 13,585.64 | 13,261.51 | 12,796.79 | 12,481.88 | **52,125.82** |
| 181 | 8,410.62 | 8,200.83 | 8,484.60 | 8,150.33 | **33,246.38** |
| 182 | 4,744.12 | 4,875.18 | 4,253.53 | 4,394.13 | **18,266.96** |
| 183 | 8,305.09 | 7,532.26 | 7,186.78 | 6,539.08 | **29,563.22** |
| 184 | 8,049.89 | 7,495.63 | 7,376.58 | 7,467.30 | **30,389.40** |
| 185 | 8,780.53 | 8,152.56 | 8,035.93 | 7,566.73 | **32,535.74** |
| 186 | 6,312.56 | 6,388.60 | 6,186.57 | 5,970.31 | **24,858.04** |
| 187 | 20,875.96 | 23,273.95 | 21,549.16 | 22,058.16 | **87,757.23** |
| 188 | 2,438.49 | 2,361.10 | 2,206.81 | 2,376.64 | **9,383.04** |
| 189 | 12,600.83 | 11,788.19 | 11,965.69 | 11,713.76 | **48,068.47** |
| 190 | 77,674.44 | 77,013.10 | 74,433.26 | 80,237.70 | **309,358.50** |
| 191 | 94,729.71 | 88,831.54 | 88,958.95 | 87,504.15 | **360,024.35** |
| 193 | 13,500.85 | 13,297.18 | 13,283.70 | 12,675.74 | **52,757.48** |
| 194 | 19,681.33 | 18,942.25 | 18,888.94 | 18,394.28 | **75,906.80** |
| 195 | 4,370.28 | 4,184.56 | 4,272.43 | 3,971.10 | **16,798.38** |
| 196 | 6,082.05 | 5,987.26 | 5,914.55 | 5,283.13 | **23,267.00** |
| **PA - PENNSYLVANIA - Total** | **528,758.98** | **513,827.35** | **504,837.17** | **498,701.92** | **2,046,125.42** |

**STATE: PR - PUERTO RICO**

| REGISTRANT ZIP CODE 3 | 1ST QUARTER | 2ND QUARTER | 3RD QUARTER | 4TH QUARTER | **TOTAL GRAMS** |
|---|---|---|---|---|---|
| 006 | 3,967.42 | 3,599.90 | 3,275.63 | 3,720.88 | **14,563.82** |

| | | | | | |
|---|---|---|---|---|---|
| 007 | 3,957.29 | 3,421.04 | 3,169.76 | 3,639.39 | **14,187.47** |
| 009 | 4,796.72 | 4,563.14 | 4,370.17 | 4,543.06 | **18,273.09** |
| **PR - PUERTO RICO - Total** | **12,721.42** | **11,584.08** | **10,815.56** | **11,903.32** | **47,024.38** |

**STATE: RI - RHODE ISLAND**

| REGISTRANT ZIP CODE 3 | 1ST QUARTER | 2ND QUARTER | 3RD QUARTER | 4TH QUARTER | TOTAL GRAMS |
|---|---|---|---|---|---|
| 028 | 18,196.51 | 17,920.07 | 17,958.72 | 18,135.23 | **72,210.53** |
| 029 | 11,564.69 | 11,346.56 | 11,545.29 | 11,607.10 | **46,063.64** |
| **RI RHODE ISLAND Total** | **29,761.19** | **29,266.63** | **29,504.01** | **29,742.33** | **118,274.17** |

**STATE: SC - SOUTH CAROLINA**

| REGISTRANT ZIP CODE 3 | 1ST QUARTER | 2ND QUARTER | 3RD QUARTER | 4TH QUARTER | TOTAL GRAMS |
|---|---|---|---|---|---|
| 290 | 10,571.36 | 10,581.34 | 10,461.45 | 10,595.01 | **42,209.16** |
| 291 | 12,571.73 | 12,086.00 | 12,344.93 | 12,105.91 | **49,108.58** |
| 292 | 8,635.86 | 8,664.25 | 8,286.43 | 8,181.73 | **33,768.26** |
| 293 | 17,749.03 | 16,748.33 | 15,600.94 | 15,213.47 | **65,311.76** |
| 294 | 30,712.46 | 30,601.51 | 29,161.45 | 27,987.30 | **118,462.72** |
| 295 | 42,991.62 | 41,413.77 | 41,009.86 | 38,434.65 | **163,849.90** |
| 296 | 31,328.43 | 29,711.60 | 29,069.25 | 29,133.16 | **119,242.44** |
| 297 | 8,007.87 | 7,880.23 | 7,968.87 | 8,011.30 | **31,868.27** |
| 298 | 7,382.59 | 7,367.83 | 7,568.30 | 7,504.40 | **29,823.12** |
| 299 | 6,538.58 | 6,623.35 | 6,419.36 | 6,359.85 | **25,941.15** |
| **SC - SOUTH CAROLINA Total** | **176,489.52** | **171,678.21** | **167,890.84** | **163,526.79** | **679,585.37** |

**STATE: SD - SOUTH DAKOTA**

| REGISTRANT ZIP CODE 3 | 1ST QUARTER | 2ND QUARTER | 3RD QUARTER | 4TH QUARTER | TOTAL GRAMS |
|---|---|---|---|---|---|
| 570 | 2,140.68 | 2,159.20 | 2,076.50 | 2,132.73 | **8,509.10** |
| 571 | 7,124.14 | 4,262.38 | 5,049.55 | 3,425.57 | **19,861.63** |
| 572 | 533.29 | 522.24 | 593.32 | 517.72 | **2,166.57** |
| 573 | 663.87 | 702.20 | 639.65 | 607.56 | **2,613.28** |
| 574 | 695.64 | 699.46 | 671.08 | 674.53 | **2,740.71** |
| 575 | 450.94 | 532.35 | 518.04 | 504.90 | **2,006.23** |
| 576 | 126.14 | 145.46 | 128.96 | 116.37 | **516.92** |
| 577 | 3,458.48 | 3,236.69 | 3,458.93 | 3,119.85 | **13,273.95** |
| **SD SOUTH DAKOTA - Total** | **15,193.18** | **12,259.99** | **13,136.02** | **11,099.21** | **51,688.40** |

**STATE: TN - TENNESSEE**

| REGISTRANT ZIP CODE 3 | 1ST QUARTER | 2ND QUARTER | 3RD QUARTER | 4TH QUARTER | TOTAL GRAMS |
|---|---|---|---|---|---|
| 370 | 45,165.47 | 44,481.16 | 43,481.82 | 42,388.40 | **175,516.86** |
| 371 | 38,002.62 | 36,788.39 | 35,744.43 | 35,374.11 | **145,909.54** |
| 372 | 24,397.15 | 24,155.26 | 24,009.82 | 24,717.44 | **97,279.67** |
| 373 | 34,471.20 | 33,852.16 | 33,251.23 | 33,138.24 | **134,712.83** |
| 374 | 12,251.42 | 12,096.08 | 12,078.25 | 12,129.42 | **48,555.18** |
| 376 | 28,879.57 | 28,964.54 | 28,692.64 | 28,525.78 | **115,062.53** |

| | | | | | |
|---|---|---|---|---|---|
| 377 | 26,046.46 | 25,674.37 | 24,682.17 | 23,790.18 | **100,193.17** |
| 378 | 35,657.07 | 34,822.79 | 34,783.61 | 34,182.75 | **139,446.22** |
| 379 | 19,286.81 | 19,389.85 | 18,868.19 | 18,825.97 | **76,370.82** |
| 380 | 12,047.65 | 11,981.07 | 12,013.71 | 11,861.06 | **47,903.48** |
| 381 | 26,079.68 | 27,264.72 | 25,611.96 | 32,669.39 | **111,625.75** |
| 382 | 5,537.66 | 5,215.13 | 5,112.76 | 5,140.53 | **21,006.07** |
| 383 | 11,255.77 | 11,054.67 | 10,778.65 | 11,213.66 | **44,302.74** |
| 384 | 12,090.37 | 11,584.13 | 11,010.03 | 10,718.68 | **45,403.21** |
| 385 | 22,066.71 | 20,078.19 | 19,590.81 | 18,778.35 | **80,514.06** |
| **TN TENNESSEE Total** | **353,235.60** | **347,402.51** | **339,710.06** | **343,453.96** | **1,383,802.14** |

**STATE: TX - TEXAS**

| REGISTRANT ZIP CODE 3 | 1ST QUARTER | 2ND QUARTER | 3RD QUARTER | 4TH QUARTER | **TOTAL GRAMS** |
|---|---|---|---|---|---|
| 750 | 28,295.59 | 27,690.63 | 27,401.58 | 26,289.31 | **109,677.11** |
| 751 | 7,098.98 | 6,972.38 | 7,079.29 | 6,604.35 | **27,754.99** |
| 752 | 8,093.19 | 7,746.79 | 7,647.59 | 7,397.96 | **30,885.54** |
| 753 | 212.34 | 193.95 | 230.26 | 213.47 | **850.02** |
| 754 | 4,112.01 | 3,920.90 | 4,104.94 | 3,711.48 | **15,849.33** |
| 755 | 1,103.63 | 1,018.24 | 898.17 | 959.07 | **3,979.11** |
| 756 | 2,198.08 | 2,313.69 | 2,237.36 | 2,161.15 | **8,910.27** |
| 757 | 3,907.97 | 4,087.12 | 4,189.31 | 3,878.00 | **16,062.41** |
| 758 | 666.99 | 661.38 | 653.07 | 530.10 | **2,511.55** |
| 759 | 1,430.74 | 1,348.35 | 1,405.92 | 1,331.01 | **5,516.02** |
| 760 | 17,977.88 | 16,718.16 | 16,275.20 | 16,479.25 | **67,450.49** |
| 761 | 13,193.15 | 12,417.00 | 12,565.73 | 11,821.82 | **49,997.71** |
| 762 | 6,443.06 | 6,347.19 | 6,236.49 | 6,007.81 | **25,034.55** |
| 763 | 1,963.58 | 1,797.81 | 1,625.32 | 1,610.92 | **6,997.62** |
| 764 | 1,159.55 | 1,129.53 | 1,061.67 | 1,032.65 | **4,383.40** |
| 765 | 4,241.05 | 4,279.75 | 3,913.16 | 3,929.42 | **16,363.38** |
| 766 | 1,149.25 | 1,130.15 | 1,024.05 | 868.85 | **4,172.30** |
| 767 | 2,013.01 | 1,964.99 | 1,983.80 | 1,856.26 | **7,818.05** |
| 768 | 713.68 | 700.56 | 620.40 | 680.84 | **2,715.48** |
| 769 | 718.84 | 740.52 | 692.67 | 652.36 | **2,804.39** |
| 770 | 43,214.41 | 45,296.65 | 48,791.97 | 35,968.58 | **173,271.60** |
| 773 | 12,595.58 | 12,680.98 | 11,983.77 | 10,433.77 | **47,694.11** |
| 774 | 16,437.96 | 14,443.22 | 17,536.86 | 15,582.34 | **64,000.37** |
| 775 | 13,286.31 | 14,234.46 | 14,564.96 | 12,796.05 | **54,881.78** |
| 776 | 2,223.14 | 2,187.12 | 2,372.16 | 2,187.82 | **8,970.25** |
| 777 | 1,043.60 | 1,116.44 | 1,186.44 | 1,131.25 | **4,477.73** |
| 778 | 1,179.65 | 1,153.71 | 1,105.05 | 1,095.30 | **4,533.72** |
| 779 | 2,641.28 | 2,500.57 | 2,442.25 | 2,444.38 | **10,028.49** |

| | | | | | |
|---|---|---|---|---|---|
| 780 | 2,659.31 | 2,371.76 | 2,375.41 | 2,194.11 | **9,600.58** |
| 781 | 3,109.63 | 2,916.90 | 2,953.56 | 2,978.00 | **11,958.09** |
| 782 | 15,635.13 | 14,901.35 | 14,838.50 | 13,972.84 | **59,347.82** |
| 783 | 1,013.29 | 1,096.17 | 1,110.18 | 1,082.85 | **4,302.50** |
| 784 | 1,493.87 | 1,461.51 | 1,396.74 | 1,484.07 | **5,836.19** |
| 785 | 2,614.68 | 2,198.29 | 2,367.97 | 2,203.69 | **9,384.63** |
| 786 | 12,086.02 | 11,486.76 | 11,597.51 | 11,890.09 | **47,060.38** |
| 787 | 9,614.11 | 9,506.79 | 9,223.17 | 9,415.41 | **37,759.48** |
| 788 | 404.95 | 395.85 | 345.21 | 331.25 | **1,477.26** |
| 789 | 579.21 | 627.22 | 657.25 | 674.93 | **2,538.61** |
| 790 | 986.30 | 924.35 | 961.48 | 972.44 | **3,844.57** |
| 791 | 2,429.39 | 2,600.19 | 2,381.03 | 2,333.28 | **9,743.90** |
| 792 | 208.57 | 230.88 | 198.20 | 214.62 | **852.26** |
| 793 | 549.90 | 584.94 | 422.69 | 406.29 | **1,963.82** |
| 794 | 2,105.91 | 2,213.75 | 2,224.59 | 2,189.87 | **8,734.13** |
| 795 | 476.53 | 520.31 | 475.97 | 427.56 | **1,900.37** |
| 796 | 1,158.47 | 1,052.53 | 1,155.08 | 943.76 | **4,309.83** |
| 797 | 1,923.80 | 1,446.53 | 1,613.51 | 1,408.33 | **6,392.16** |
| 798 | 90.16 | 122.09 | 99.32 | 103.48 | **415.04** |
| 799 | 3,602.66 | 3,274.25 | 3,733.07 | 2,990.52 | **13,600.49** |
| **TX - TEXAS Total** | **262,056.40** | **256,724.63** | **261,959.88** | **237,872.95** | **1,018,613.87** |

**STATE: UT - UTAH**

| REGISTRANT ZIP CODE 3 | 1ST QUARTER | 2ND QUARTER | 3RD QUARTER | 4TH QUARTER | TOTAL GRAMS |
|---|---|---|---|---|---|
| 840 | 53,510.01 | 53,432.15 | 53,733.98 | 54,143.31 | **214,819.45** |
| 841 | 33,216.35 | 32,738.62 | 32,393.57 | 32,524.73 | **130,873.28** |
| 843 | 4,206.97 | 4,480.96 | 4,422.54 | 4,505.08 | **17,615.55** |
| 844 | 11,374.28 | 11,548.75 | 11,705.43 | 11,795.59 | **46,424.05** |
| 845 | 4,169.25 | 4,223.78 | 4,189.41 | 4,071.22 | **16,653.66** |
| 846 | 10,474.43 | 10,982.99 | 11,089.14 | 11,151.32 | **43,697.88** |
| 847 | 15,855.90 | 15,402.06 | 15,501.31 | 15,453.99 | **62,213.26** |
| **UT - UTAH - Total** | **132,807.19** | **132,809.31** | **133,035.40** | **133,645.24** | **532,297.13** |

**STATE: VA - VIRGINIA**

| REGISTRANT ZIP CODE 3 | 1ST QUARTER | 2ND QUARTER | 3RD QUARTER | 4TH QUARTER | TOTAL GRAMS |
|---|---|---|---|---|---|
| 201 | 16,202.14 | 15,656.94 | 15,461.70 | 15,203.12 | **62,523.90** |
| 220 | 7,989.58 | 7,517.43 | 8,402.98 | 8,727.80 | **32,637.79** |
| 221 | 8,483.50 | 9,425.01 | 9,419.76 | 7,607.92 | **34,936.20** |
| 222 | 2,368.73 | 2,388.72 | 2,426.66 | 2,533.23 | **9,717.35** |
| 223 | 4,324.26 | 4,492.65 | 4,308.72 | 4,360.97 | **17,486.61** |
| 224 | 7,282.04 | 7,186.98 | 7,366.26 | 7,346.04 | **29,181.32** |
| 225 | 3,010.37 | 3,032.32 | 2,936.12 | 2,945.16 | **11,923.97** |

| | | | | | |
|---|---|---|---|---|---|
| 226 | 6,432.15 | 6,360.52 | 6,401.72 | 6,210.94 | **25,405.33** |
| 227 | 1,618.09 | 1,659.96 | 1,627.86 | 1,695.44 | **6,601.36** |
| 228 | 4,543.71 | 4,514.30 | 4,445.44 | 4,392.27 | **17,895.72** |
| 229 | 6,597.32 | 6,798.35 | 6,530.88 | 6,829.53 | **26,756.08** |
| 230 | 4,798.96 | 4,739.55 | 4,805.72 | 4,795.89 | **19,140.13** |
| 231 | 9,003.31 | 9,460.43 | 9,603.62 | 9,690.31 | **37,757.66** |
| 232 | 18,278.16 | 18,600.65 | 17,977.64 | 18,183.83 | **73,040.28** |
| 233 | 7,388.32 | 7,503.09 | 7,681.41 | 7,678.78 | **30,251.61** |
| 234 | 14,720.85 | 14,200.01 | 14,390.50 | 13,671.05 | **56,982.41** |
| 235 | 5,492.34 | 5,277.96 | 5,110.22 | 4,941.00 | **20,821.52** |
| 236 | 8,605.15 | 8,913.32 | 8,818.09 | 8,826.08 | **35,162.65** |
| 237 | 4,152.90 | 4,023.69 | 3,910.16 | 3,813.06 | **15,899.81** |
| 238 | 14,539.54 | 14,891.15 | 14,674.39 | 13,177.66 | **57,282.73** |
| 239 | 2,327.05 | 2,371.06 | 2,126.95 | 2,166.72 | **8,991.78** |
| 240 | 14,648.17 | 14,319.66 | 14,307.48 | 13,090.78 | **56,366.08** |
| 241 | 12,903.72 | 12,826.87 | 12,319.42 | 12,313.87 | **50,363.88** |
| 242 | 19,301.48 | 19,074.22 | 18,535.85 | 17,812.01 | **74,723.56** |
| 243 | 8,605.27 | 8,559.59 | 8,616.10 | 7,780.49 | **33,561.46** |
| 244 | 3,787.89 | 3,977.46 | 3,991.90 | 4,012.87 | **15,770.12** |
| 245 | 11,109.30 | 10,983.24 | 10,646.80 | 10,030.76 | **42,770.11** |
| 246 | 5,102.73 | 4,983.76 | 4,807.14 | 4,256.74 | **19,150.37** |
| **VA VIRGINIA Total** | **233,617.07** | **233,738.89** | **231,651.51** | **224,094.34** | **923,101.80** |

**STATE: VI - VIRGIN ISLANDS**

| REGISTRANT ZIP CODE 3 | 1ST QUARTER | 2ND QUARTER | 3RD QUARTER | 4TH QUARTER | TOTAL GRAMS |
|---|---|---|---|---|---|
| 008 | 440.18 | 457.44 | 420.68 | 381.01 | **1,699.32** |
| **VI - VIRGIN ISLANDS Total** | **440.18** | **457.44** | **420.68** | **381.01** | **1,699.32** |

**STATE: VT - VERMONT**

| REGISTRANT ZIP CODE 3 | 1ST QUARTER | 2ND QUARTER | 3RD QUARTER | 4TH QUARTER | TOTAL GRAMS |
|---|---|---|---|---|---|
| 050 | 1,549.29 | 1,832.70 | 1,594.32 | 1,469.95 | **6,446.25** |
| 051 | 1,289.57 | 1,166.56 | 1,271.31 | 1,181.85 | **4,909.28** |
| 052 | 1,049.16 | 1,114.42 | 1,066.59 | 1,054.07 | **4,284.24** |
| 053 | 1,224.44 | 1,098.94 | 1,174.97 | 1,114.00 | **4,612.35** |
| 054 | 6,904.79 | 6,838.15 | 6,793.84 | 6,341.09 | **26,877.87** |
| 056 | 1,586.43 | 1,641.50 | 1,657.63 | 1,624.69 | **6,510.25** |
| 057 | 1,408.68 | 1,662.12 | 1,613.36 | 1,710.79 | **6,394.93** |
| 058 | 1,577.94 | 1,569.60 | 1,626.30 | 1,674.52 | **6,448.36** |
| **VT VERMONT Total** | **16,590.29** | **16,923.99** | **16,798.31** | **16,170.95** | **66,483.54** |

**STATE: WA - WASHINGTON**

| REGISTRANT ZIP CODE 3 | 1ST QUARTER | 2ND QUARTER | 3RD QUARTER | 4TH QUARTER | TOTAL GRAMS |
|---|---|---|---|---|---|
| 980 | 37,410.54 | 37,094.98 | 36,750.72 | 36,818.10 | **148,074.34** |

| | | | | | |
|---|---|---|---|---|---|
| 981 | 23,468.76 | 23,477.79 | 22,735.22 | 21,908.81 | **91,590.58** |
| 982 | 29,221.59 | 29,280.11 | 29,669.98 | 28,661.64 | **116,833.32** |
| 983 | 26,408.88 | 26,394.26 | 26,398.58 | 26,561.00 | **105,762.71** |
| 984 | 18,079.66 | 18,127.75 | 18,096.21 | 17,986.83 | **72,290.45** |
| 985 | 15,200.65 | 14,809.07 | 14,966.87 | 14,924.43 | **59,901.03** |
| 986 | 22,103.95 | 22,200.40 | 21,640.53 | 21,717.44 | **87,662.31** |
| 988 | 7,030.27 | 6,676.53 | 6,608.19 | 6,524.57 | **26,839.56** |
| 989 | 8,234.86 | 8,518.35 | 8,481.43 | 8,329.42 | **33,564.06** |
| 990 | 2,540.45 | 2,473.06 | 2,573.46 | 2,568.82 | **10,155.79** |
| 991 | 4,005.89 | 3,793.30 | 4,005.63 | 3,809.10 | **15,613.93** |
| 992 | 13,005.20 | 13,449.82 | 13,156.13 | 13,267.86 | **52,879.01** |
| 993 | 10,780.51 | 10,870.24 | 10,387.19 | 10,532.39 | **42,570.33** |
| 994 | 915.74 | 971.17 | 841.29 | 834.38 | **3,562.58** |
| **WA WASHINGTON Total** | **218,406.95** | **218,136.83** | **216,311.43** | **214,444.80** | **867,300.00** |

**STATE: WI - WISCONSIN**

| REGISTRANT ZIP CODE 3 | 1ST QUARTER | 2ND QUARTER | 3RD QUARTER | 4TH QUARTER | **TOTAL GRAMS** |
|---|---|---|---|---|---|
| 530 | 14,323.48 | 14,013.90 | 13,497.55 | 13,280.39 | **55,115.32** |
| 531 | 16,653.17 | 15,960.26 | 15,237.12 | 14,885.72 | **62,736.26** |
| 532 | 38,776.24 | 36,828.82 | 35,029.63 | 34,678.64 | **145,313.34** |
| 534 | 3,796.17 | 3,579.26 | 3,453.94 | 3,320.22 | **14,149.59** |
| 535 | 12,104.49 | 11,939.66 | 11,528.41 | 11,016.33 | **46,588.89** |
| 537 | 8,796.26 | 8,684.56 | 8,112.28 | 8,171.31 | **33,764.40** |
| 538 | 885.09 | 833.29 | 822.52 | 860.01 | **3,400.90** |
| 539 | 4,753.07 | 4,858.53 | 4,833.34 | 4,536.44 | **18,981.38** |
| 540 | 2,269.97 | 2,323.56 | 2,306.83 | 2,331.62 | **9,231.98** |
| 541 | 2,326.09 | 2,310.87 | 2,385.51 | 2,263.67 | **9,286.14** |
| 542 | 1,386.26 | 1,347.19 | 1,397.29 | 1,353.33 | **5,484.07** |
| 543 | 3,010.46 | 2,818.20 | 2,619.35 | 2,722.60 | **11,170.61** |
| 544 | 5,171.80 | 4,830.78 | 4,821.48 | 4,628.01 | **19,452.07** |
| 545 | 1,711.74 | 1,765.08 | 1,874.48 | 1,647.27 | **6,998.57** |
| 546 | 4,725.31 | 5,015.55 | 4,259.45 | 4,196.05 | **18,196.36** |
| 547 | 3,163.97 | 3,174.41 | 3,066.79 | 3,012.65 | **12,417.82** |
| 548 | 3,546.21 | 3,601.72 | 3,586.40 | 3,675.70 | **14,410.03** |
| 549 | 8,156.60 | 8,265.76 | 7,908.17 | 7,791.03 | **32,121.55** |
| **WI - WISCONSIN Total** | **135,556.37** | **132,151.39** | **126,740.56** | **124,370.98** | **518,819.30** |

**STATE: WV - WEST VIRGINIA**

| REGISTRANT ZIP CODE 3 | 1ST QUARTER | 2ND QUARTER | 3RD QUARTER | 4TH QUARTER | **TOTAL GRAMS** |
|---|---|---|---|---|---|
| 247 | 1,821.19 | 1,823.80 | 1,852.81 | 1,887.03 | **7,384.84** |
| 248 | 1,453.67 | 1,325.06 | 1,022.89 | 772.96 | **4,574.59** |
| 249 | 1,580.53 | 1,494.21 | 1,438.30 | 1,510.71 | **6,023.74** |

| | | | | | |
|---|---|---|---|---|---|
| 250 | 1,588.82 | 1,457.40 | 1,463.88 | 1,442.44 | **5,952.54** |
| 251 | 1,219.27 | 1,261.21 | 1,233.28 | 1,306.60 | **5,020.36** |
| 252 | 901.23 | 802.82 | 676.06 | 634.28 | **3,014.39** |
| 253 | 3,831.81 | 3,584.31 | 3,556.88 | 3,626.87 | **14,599.87** |
| 254 | 9,736.29 | 9,369.88 | 9,175.82 | 8,724.92 | **37,006.90** |
| 255 | 4,148.85 | 4,008.56 | 3,989.53 | 3,506.04 | **15,652.98** |
| 256 | 1,375.01 | 1,209.13 | 1,208.08 | 1,001.55 | **4,793.77** |
| 257 | 3,002.61 | 3,069.47 | 3,064.42 | 3,069.64 | **12,206.13** |
| 258 | 2,500.79 | 2,082.16 | 2,325.77 | 2,343.78 | **9,252.50** |
| 259 | 1,049.86 | 1,151.16 | 1,168.44 | 1,082.36 | **4,451.81** |
| 260 | 6,468.43 | 6,015.23 | 6,197.82 | 5,395.85 | **24,077.33** |
| 261 | 2,072.95 | 2,006.57 | 1,871.98 | 1,774.72 | **7,726.23** |
| 262 | 1,730.00 | 1,791.04 | 1,691.87 | 1,727.24 | **6,940.16** |
| 263 | 3,676.05 | 3,460.26 | 3,368.25 | 3,211.65 | **13,716.21** |
| 264 | 1,019.05 | 1,027.04 | 913.27 | 939.31 | **3,898.67** |
| 265 | 4,741.14 | 4,268.65 | 4,257.71 | 4,360.26 | **17,627.76** |
| 266 | 883.24 | 902.10 | 815.05 | 818.44 | **3,418.84** |
| 267 | 1,908.20 | 1,985.89 | 2,018.00 | 1,780.82 | **7,692.90** |
| 268 | 545.63 | 501.27 | 546.76 | 509.58 | **2,103.25** |
| **WV WEST VIRGINIA Total** | **57,254.62** | **54,597.23** | **53,856.88** | **51,427.06** | **217,135.79** |

**STATE: WY - WYOMING**

| REGISTRANT ZIP CODE 3 | 1ST QUARTER | 2ND QUARTER | 3RD QUARTER | 4TH QUARTER | TOTAL GRAMS |
|---|---|---|---|---|---|
| 820 | 3,738.61 | 3,480.85 | 3,373.67 | 3,105.39 | **13,698.53** |
| 822 | 471.05 | 516.12 | 524.59 | 513.47 | **2,025.23** |
| 823 | 341.10 | 389.72 | 387.95 | 448.05 | **1,566.82** |
| 824 | 2,340.75 | 2,306.38 | 2,465.73 | 2,357.47 | **9,470.33** |
| 825 | 1,628.80 | 1,682.67 | 1,580.31 | 1,412.72 | **6,304.50** |
| 826 | 4,326.07 | 4,069.83 | 4,170.68 | 3,927.08 | **16,493.66** |
| 827 | 952.61 | 956.43 | 1,013.88 | 945.28 | **3,868.20** |
| 828 | 1,067.11 | 1,015.13 | 988.06 | 973.38 | **4,043.67** |
| 829 | 4,894.06 | 4,987.92 | 4,996.11 | 5,040.94 | **19,919.03** |
| 830 | 223.50 | 203.63 | 193.62 | 158.70 | **779.45** |
| 831 | 500.52 | 588.82 | 556.73 | 480.17 | **2,126.23** |
| **WY - WYOMING Total** | **20,484.18** | **20,197.49** | **20,251.33** | **19,362.65** | **80,295.64** |

**DRUG: 9150 - HYDROMORPHONE**

**STATE: AK - ALASKA**

| REGISTRANT ZIP CODE 3 | 1ST QUARTER | 2ND QUARTER | 3RD QUARTER | 4TH QUARTER | TOTAL GRAMS |
|---|---|---|---|---|---|
| 995 | 308.97 | 246.48 | 235.70 | 228.64 | **1,019.78** |
| 996 | 207.18 | 196.09 | 198.37 | 153.24 | **754.88** |
| 997 | 18.81 | 27.88 | 31.43 | 19.86 | **97.99** |

| | | | | | |
|---|---|---|---|---|---|
| 998 | 13.55 | 20.16 | 17.81 | 16.30 | **67.82** |
| 999 | 18.14 | 5.97 | 3.68 | 7.44 | **35.23** |
| **AK ALASKA - Total** | **566.64** | **496.58** | **487.00** | **425.49** | **1,975.71** |

**STATE: AL - ALABAMA**

| REGISTRANT ZIP CODE 3 | 1ST QUARTER | 2ND QUARTER | 3RD QUARTER | 4TH QUARTER | TOTAL GRAMS |
|---|---|---|---|---|---|
| 350 | 138.53 | 130.91 | 129.85 | 134.39 | **533.69** |
| 351 | 91.35 | 88.02 | 84.75 | 95.14 | **359.26** |
| 352 | 300.39 | 290.53 | 310.09 | 298.18 | **1,199.19** |
| 354 | 111.79 | 113.49 | 104.85 | 100.17 | **430.31** |
| 355 | 36.34 | 35.61 | 39.31 | 41.69 | **152.94** |
| 356 | 131.68 | 124.63 | 124.19 | 120.04 | **500.54** |
| 357 | 83.79 | 72.25 | 85.58 | 83.52 | **325.14** |
| 358 | 178.80 | 153.91 | 173.81 | 122.22 | **628.73** |
| 359 | 94.91 | 104.61 | 118.06 | 112.37 | **429.96** |
| 360 | 84.34 | 71.93 | 72.47 | 70.77 | **299.51** |
| 361 | 91.26 | 113.82 | 90.04 | 80.04 | **375.15** |
| 362 | 48.04 | 54.93 | 55.92 | 53.84 | **212.74** |
| 363 | 193.20 | 151.83 | 151.63 | 155.03 | **651.69** |
| 364 | 80.47 | 78.55 | 59.44 | 58.69 | **277.15** |
| 365 | 269.48 | 332.86 | 243.67 | 148.43 | **994.45** |
| 366 | 215.21 | 290.22 | 182.31 | 178.22 | **865.96** |
| 367 | 19.13 | 11.67 | 14.64 | 9.52 | **54.96** |
| 368 | 86.65 | 77.58 | 74.35 | 67.92 | **306.50** |
| 369 | 6.83 | 7.05 | 4.92 | 4.88 | **23.67** |
| **AL ALABAMA - Total** | **2,262.18** | **2,304.41** | **2,119.89** | **1,935.05** | **8,621.53** |

**STATE: AR - ARKANSAS**

| REGISTRANT ZIP CODE 3 | 1ST QUARTER | 2ND QUARTER | 3RD QUARTER | 4TH QUARTER | TOTAL GRAMS |
|---|---|---|---|---|---|
| 716 | 34.82 | 35.49 | 31.67 | 31.97 | **133.95** |
| 717 | 31.31 | 40.43 | 34.22 | 35.77 | **141.74** |
| 718 | 65.65 | 46.48 | 53.18 | 47.07 | **212.39** |
| 719 | 141.44 | 117.81 | 129.25 | 109.44 | **497.94** |
| 720 | 151.37 | 131.20 | 147.12 | 141.31 | **571.01** |
| 721 | 167.50 | 162.71 | 148.14 | 152.26 | **630.61** |
| 722 | 258.48 | 290.28 | 355.86 | 249.03 | **1,153.65** |
| 723 | 30.12 | 27.70 | 41.93 | 32.97 | **132.72** |
| 724 | 103.96 | 125.66 | 125.24 | 118.38 | **473.24** |
| 725 | 95.76 | 90.34 | 102.73 | 102.10 | **390.93** |
| 726 | 89.41 | 85.15 | 77.20 | 77.81 | **329.56** |
| 727 | 575.03 | 574.52 | 596.79 | 440.26 | **2,186.61** |
| 728 | 65.11 | 47.13 | 38.22 | 46.69 | **197.14** |

| | | | | | |
|---|---|---|---|---|---|
| 729 | 287.38 | 246.25 | 204.30 | 212.63 | **950.56** |
| **AR ARKANSAS - Total** | **2,097.34** | **2,021.15** | **2,085.85** | **1,797.70** | **8,002.04** |

**STATE: AS - AMERICAN SAMOA**

| REGISTRANT ZIP CODE 3 | 1ST QUARTER | 2ND QUARTER | 3RD QUARTER | 4TH QUARTER | TOTAL GRAMS |
|---|---|---|---|---|---|
| 967 | 2.84 | 2.13 | 1.46 | 1.33 | **7.76** |
| **AS - AMERICAN SAMOA - Tota** | **2.84** | **2.13** | **1.46** | **1.33** | **7.76** |

**STATE: AZ - ARIZONA**

| REGISTRANT ZIP CODE 3 | 1ST QUARTER | 2ND QUARTER | 3RD QUARTER | 4TH QUARTER | TOTAL GRAMS |
|---|---|---|---|---|---|
| 850 | 885.90 | 779.48 | 788.17 | 814.29 | **3,267.83** |
| 851 | 152.27 | 132.14 | 132.18 | 138.47 | **555.05** |
| 852 | 1,327.09 | 1,163.55 | 1,240.39 | 1,244.41 | **4,975.43** |
| 853 | 827.12 | 727.89 | 725.30 | 768.56 | **3,048.87** |
| 855 | 58.50 | 57.33 | 47.66 | 52.73 | **216.23** |
| 856 | 156.27 | 135.08 | 125.96 | 134.70 | **552.02** |
| 857 | 812.52 | 788.12 | 789.15 | 856.59 | **3,246.39** |
| 859 | 82.92 | 75.37 | 80.64 | 73.28 | **312.22** |
| 860 | 61.08 | 72.17 | 68.40 | 70.67 | **272.32** |
| 863 | 247.02 | 221.41 | 228.34 | 233.35 | **930.12** |
| 864 | 270.15 | 252.37 | 259.85 | 256.41 | **1,038.79** |
| 865 | 1.11 | 0.66 | 0.66 | 0.37 | **2.81** |
| **AZ ARIZONA - Total** | **4,881.95** | **4,405.59** | **4,486.70** | **4,643.83** | **18,418.08** |

**STATE: CA - CALIFORNIA**

| REGISTRANT ZIP CODE 3 | 1ST QUARTER | 2ND QUARTER | 3RD QUARTER | 4TH QUARTER | TOTAL GRAMS |
|---|---|---|---|---|---|
| 900 | 1,032.29 | 1,031.01 | 1,066.46 | 1,129.44 | **4,259.21** |
| 902 | 390.06 | 425.68 | 430.05 | 482.27 | **1,728.06** |
| 903 | 20.52 | 28.04 | 25.78 | 22.87 | **97.22** |
| 904 | 173.85 | 199.55 | 212.14 | 217.97 | **803.51** |
| 905 | 223.02 | 208.30 | 313.58 | 357.37 | **1,102.26** |
| 906 | 270.47 | 243.33 | 250.29 | 276.87 | **1,040.95** |
| 907 | 327.04 | 271.08 | 296.92 | 300.79 | **1,195.83** |
| 908 | 1,186.27 | 984.21 | 971.91 | 970.06 | **4,112.45** |
| 910 | 329.56 | 283.65 | 289.06 | 276.20 | **1,178.47** |
| 911 | 112.84 | 122.67 | 132.15 | 123.15 | **490.81** |
| 912 | 116.16 | 100.96 | 97.02 | 115.36 | **429.50** |
| 913 | 989.21 | 895.14 | 904.95 | 893.72 | **3,683.02** |
| 914 | 316.82 | 286.21 | 292.28 | 243.77 | **1,139.08** |
| 915 | 117.85 | 147.01 | 138.36 | 149.43 | **552.65** |
| 916 | 59.48 | 56.88 | 48.41 | 47.11 | **211.88** |
| 917 | 1,073.59 | 1,140.23 | 1,194.93 | 1,156.92 | **4,565.67** |
| 918 | 30.52 | 48.37 | 27.66 | 21.10 | **127.65** |

| | | | | | |
|---|---|---|---|---|---|
| 919 | 245.01 | 292.16 | 229.58 | 239.29 | **1,006.03** |
| 920 | 1,291.51 | 1,317.21 | 1,218.81 | 1,168.81 | **4,996.34** |
| 921 | 874.06 | 924.06 | 820.92 | 868.31 | **3,487.34** |
| 922 | 558.28 | 552.87 | 524.12 | 508.93 | **2,144.20** |
| 923 | 889.82 | 987.10 | 873.13 | 863.85 | **3,613.91** |
| 924 | 118.69 | 115.69 | 101.06 | 89.09 | **424.53** |
| 925 | 834.63 | 1,097.33 | 868.52 | 861.76 | **3,662.24** |
| 926 | 906.90 | 560.73 | 583.51 | 620.37 | **2,671.51** |
| 927 | 206.66 | 196.89 | 231.68 | 201.58 | **836.81** |
| 928 | 872.57 | 896.28 | 953.22 | 868.42 | **3,590.49** |
| 930 | 385.26 | 389.86 | 353.11 | 324.87 | **1,453.10** |
| 931 | 91.93 | 82.58 | 97.02 | 76.35 | **347.88** |
| 932 | 274.84 | 311.88 | 327.31 | 299.23 | **1,213.26** |
| 933 | 419.46 | 502.06 | 511.26 | 432.85 | **1,865.64** |
| 934 | 583.82 | 558.35 | 548.49 | 569.83 | **2,260.48** |
| 935 | 889.64 | 844.36 | 814.53 | 782.47 | **3,331.00** |
| 936 | 186.90 | 195.34 | 190.21 | 183.52 | **755.98** |
| 937 | 384.67 | 424.51 | 382.50 | 364.73 | **1,556.40** |
| 939 | 100.73 | 129.44 | 115.38 | 114.91 | **460.46** |
| 940 | 356.33 | 357.44 | 400.12 | 390.97 | **1,504.86** |
| 941 | 748.09 | 661.39 | 604.73 | 629.98 | **2,644.20** |
| 943 | 186.87 | 228.72 | 245.85 | 254.49 | **915.93** |
| 944 | 42.80 | 45.05 | 40.49 | 42.84 | **171.18** |
| 945 | 1,741.99 | 1,828.39 | 1,692.58 | 1,633.81 | **6,896.77** |
| 946 | 263.07 | 289.86 | 203.21 | 197.04 | **953.18** |
| 947 | 73.75 | 68.82 | 90.20 | 81.99 | **314.77** |
| 948 | 58.42 | 66.19 | 61.59 | 55.06 | **241.26** |
| 949 | 332.18 | 316.83 | 352.20 | 342.89 | **1,344.10** |
| 950 | 365.05 | 322.53 | 335.97 | 337.36 | **1,360.92** |
| 951 | 243.34 | 242.12 | 243.86 | 225.92 | **955.24** |
| 952 | 242.05 | 227.12 | 199.78 | 209.12 | **878.07** |
| 953 | 711.25 | 706.44 | 669.68 | 607.56 | **2,694.93** |
| 954 | 500.36 | 514.86 | 513.76 | 567.51 | **2,096.49** |
| 955 | 200.01 | 162.97 | 164.06 | 152.37 | **679.40** |
| 956 | 1,119.02 | 1,061.73 | 1,095.69 | 1,022.32 | **4,298.76** |
| 957 | 91.98 | 90.84 | 107.49 | 95.03 | **385.34** |
| 958 | 793.66 | 666.10 | 817.31 | 699.68 | **2,976.75** |
| 959 | 531.44 | 526.45 | 478.98 | 474.89 | **2,011.76** |
| 960 | 249.33 | 228.75 | 232.06 | 230.32 | **940.46** |
| 961 | 38.79 | 39.18 | 38.86 | 37.85 | **154.69** |

| CA - CALIFORNIA - Total | 25,774.72 | 25,502.79 | 25,024.81 | 24,512.55 | 100,814.87 |
|---|---|---|---|---|---|

STATE: CO - COLORADO

| REGISTRANT ZIP CODE 3 | 1ST QUARTER | 2ND QUARTER | 3RD QUARTER | 4TH QUARTER | TOTAL GRAMS |
|---|---|---|---|---|---|
| 800 | 942.03 | 878.08 | 882.12 | 826.01 | **3,528.24** |
| 801 | 1,153.99 | 1,290.08 | 1,234.45 | 1,239.39 | **4,917.90** |
| 802 | 805.57 | 737.10 | 765.44 | 803.07 | **3,111.18** |
| 803 | 64.53 | 59.65 | 73.14 | 68.96 | **266.27** |
| 804 | 156.79 | 170.25 | 182.15 | 166.75 | **675.94** |
| 805 | 386.74 | 380.38 | 367.94 | 367.51 | **1,502.58** |
| 806 | 227.32 | 223.73 | 198.70 | 208.80 | **858.55** |
| 807 | 67.86 | 66.88 | 64.22 | 66.10 | **265.06** |
| 808 | 96.20 | 94.75 | 87.17 | 99.46 | **377.58** |
| 809 | 522.33 | 537.56 | 549.77 | 495.78 | **2,105.43** |
| 810 | 242.35 | 218.03 | 266.06 | 281.60 | **1,008.04** |
| 811 | 12.79 | 13.58 | 18.19 | 17.81 | **62.39** |
| 812 | 82.87 | 65.34 | 74.03 | 77.37 | **299.61** |
| 813 | 36.15 | 39.01 | 50.28 | 50.59 | **176.03** |
| 814 | 41.75 | 45.34 | 44.17 | 38.59 | **169.85** |
| 815 | 100.59 | 83.18 | 108.57 | 183.36 | **475.70** |
| 816 | 70.46 | 73.33 | 84.81 | 79.24 | **307.85** |
| **CO - COLORADO - Total** | **5,010.30** | **4,976.28** | **5,051.22** | **5,070.39** | **20,108.19** |

STATE: CT - CONNECTICUT

| REGISTRANT ZIP CODE 3 | 1ST QUARTER | 2ND QUARTER | 3RD QUARTER | 4TH QUARTER | TOTAL GRAMS |
|---|---|---|---|---|---|
| 060 | 830.16 | 880.87 | 860.31 | 829.66 | **3,401.01** |
| 061 | 423.18 | 417.58 | 434.85 | 485.15 | **1,760.76** |
| 062 | 135.89 | 129.10 | 120.69 | 120.72 | **506.40** |
| 063 | 219.81 | 205.94 | 258.15 | 221.86 | **905.77** |
| 064 | 1,235.45 | 1,216.90 | 1,266.08 | 1,483.65 | **5,202.07** |
| 065 | 423.17 | 418.33 | 395.27 | 457.52 | **1,694.28** |
| 066 | 127.47 | 116.80 | 129.26 | 144.12 | **517.65** |
| 067 | 371.96 | 371.73 | 323.07 | 362.32 | **1,429.08** |
| 068 | 561.94 | 503.05 | 537.71 | 547.90 | **2,150.61** |
| 069 | 119.47 | 86.98 | 100.73 | 105.42 | **412.60** |
| **CT CONNECTICUT Total** | **4,448.49** | **4,347.28** | **4,426.13** | **4,758.33** | **17,980.22** |

STATE: DC - DISTRICT OF COLUMBIA

| REGISTRANT ZIP CODE 3 | 1ST QUARTER | 2ND QUARTER | 3RD QUARTER | 4TH QUARTER | TOTAL GRAMS |
|---|---|---|---|---|---|
| 200 | 390.33 | 373.33 | 359.38 | 334.77 | **1,457.81** |
| 203 | 0 | 0 | 0.89 | 0 | **0.89** |
| 204 | 10.98 | 9.95 | 15.02 | 9.66 | **45.61** |
| **DC - DISTRICT OF COLUMBIA** | **401.31** | **383.28** | **375.29** | **344.43** | **1,504.31** |

**STATE: DE - DELAWARE**

| REGISTRANT ZIP CODE 3 | 1ST QUARTER | 2ND QUARTER | 3RD QUARTER | 4TH QUARTER | TOTAL GRAMS |
|---|---|---|---|---|---|
| 197 | 534.73 | 473.42 | 481.67 | 443.60 | **1,933.42** |
| 198 | 233.62 | 236.20 | 221.54 | 236.59 | **927.95** |
| 199 | 487.11 | 493.02 | 467.14 | 478.87 | **1,926.13** |
| **DE DELAWARE Total** | **1,255.46** | **1,202.64** | **1,170.35** | **1,159.06** | **4,787.51** |

**STATE: FL - FLORIDA**

| REGISTRANT ZIP CODE 3 | 1ST QUARTER | 2ND QUARTER | 3RD QUARTER | 4TH QUARTER | TOTAL GRAMS |
|---|---|---|---|---|---|
| 320 | 535.55 | 544.58 | 498.72 | 490.19 | **2,069.05** |
| 321 | 728.94 | 790.04 | 801.75 | 788.14 | **3,108.86** |
| 322 | 1,264.14 | 1,276.91 | 1,173.16 | 1,288.14 | **5,002.36** |
| 323 | 282.85 | 275.09 | 266.40 | 281.51 | **1,105.85** |
| 324 | 203.81 | 192.12 | 179.21 | 187.24 | **762.38** |
| 325 | 606.51 | 595.85 | 558.51 | 561.94 | **2,322.81** |
| 326 | 340.74 | 385.54 | 344.69 | 340.07 | **1,411.05** |
| 327 | 2,190.21 | 1,800.16 | 2,222.40 | 1,832.50 | **8,045.26** |
| 328 | 1,594.17 | 1,570.19 | 1,471.35 | 1,424.44 | **6,060.15** |
| 329 | 1,444.27 | 1,295.46 | 1,225.34 | 1,177.49 | **5,142.55** |
| 330 | 1,089.07 | 1,016.69 | 1,023.47 | 978.68 | **4,107.91** |
| 331 | 1,048.37 | 1,051.75 | 961.09 | 890.48 | **3,951.69** |
| 333 | 801.21 | 814.66 | 775.70 | 718.83 | **3,110.40** |
| 334 | 1,490.24 | 1,431.72 | 1,494.35 | 1,435.59 | **5,851.89** |
| 335 | 1,510.56 | 1,470.82 | 1,477.08 | 1,470.23 | **5,928.69** |
| 336 | 7,452.18 | 6,240.82 | 7,174.60 | 6,433.19 | **27,300.78** |
| 337 | 2,257.88 | 2,192.44 | 2,236.19 | 2,017.81 | **8,704.33** |
| 338 | 805.28 | 747.75 | 747.11 | 737.51 | **3,037.64** |
| 339 | 1,367.44 | 1,461.42 | 1,317.52 | 1,371.37 | **5,517.75** |
| 341 | 525.40 | 425.25 | 442.41 | 444.70 | **1,837.76** |
| 342 | 1,769.35 | 1,741.40 | 1,655.67 | 1,690.06 | **6,856.48** |
| 344 | 696.05 | 635.50 | 603.16 | 614.02 | **2,548.74** |
| 346 | 2,516.38 | 2,430.28 | 2,805.10 | 2,778.79 | **10,530.55** |
| 347 | 953.25 | 978.55 | 979.32 | 842.94 | **3,754.06** |
| 349 | 954.35 | 963.92 | 977.67 | 957.85 | **3,853.80** |
| **FL FLORIDA - Total** | **34,428.22** | **32,328.92** | **33,411.95** | **31,753.69** | **131,922.78** |

**STATE: GA - GEORGIA**

| REGISTRANT ZIP CODE 3 | 1ST QUARTER | 2ND QUARTER | 3RD QUARTER | 4TH QUARTER | TOTAL GRAMS |
|---|---|---|---|---|---|
| 300 | 1,356.88 | 1,298.05 | 1,286.13 | 1,410.62 | **5,351.69** |
| 301 | 703.15 | 704.96 | 737.28 | 759.11 | **2,904.51** |
| 302 | 441.29 | 430.19 | 429.69 | 404.21 | **1,705.38** |
| 303 | 833.80 | 764.59 | 601.11 | 619.62 | **2,819.12** |

| | | | | | |
|---|---|---|---|---|---|
| 304 | 82.99 | 81.94 | 88.23 | 88.87 | **342.03** |
| 305 | 403.30 | 372.41 | 405.34 | 422.30 | **1,603.36** |
| 306 | 400.25 | 395.84 | 379.71 | 396.29 | **1,572.09** |
| 307 | 176.25 | 156.84 | 148.23 | 171.97 | **653.29** |
| 308 | 107.23 | 114.07 | 115.05 | 101.64 | **438.00** |
| 309 | 244.11 | 229.13 | 219.99 | 225.70 | **918.93** |
| 310 | 202.65 | 167.98 | 164.21 | 146.50 | **681.34** |
| 312 | 73.03 | 71.86 | 62.47 | 49.39 | **256.75** |
| 313 | 71.74 | 72.92 | 74.31 | 86.66 | **305.64** |
| 314 | 109.52 | 119.74 | 126.82 | 117.95 | **474.03** |
| 315 | 250.45 | 218.29 | 212.33 | 224.92 | **905.99** |
| 316 | 120.93 | 124.92 | 117.74 | 104.15 | **467.73** |
| 317 | 352.16 | 392.54 | 371.82 | 334.69 | **1,451.22** |
| 318 | 5.76 | 8.95 | 7.23 | 7.40 | **29.35** |
| 319 | 54.58 | 56.37 | 57.43 | 56.38 | **224.76** |
| 398 | 38.64 | 41.12 | 43.25 | 45.95 | **168.96** |
| **GA GEORGIA Total** | **6,028.73** | **5,822.72** | **5,648.38** | **5,774.32** | **23,274.15** |

**STATE: GU - GUAM**

| REGISTRANT ZIP CODE 3 | 1ST QUARTER | 2ND QUARTER | 3RD QUARTER | 4TH QUARTER | **TOTAL GRAMS** |
|---|---|---|---|---|---|
| 969 | 26.17 | 21.07 | 12.50 | 12.86 | **72.61** |
| **GU - GUAM - Total** | **26.17** | **21.07** | **12.50** | **12.86** | **72.61** |

**STATE: HI - HAWAII**

| REGISTRANT ZIP CODE 3 | 1ST QUARTER | 2ND QUARTER | 3RD QUARTER | 4TH QUARTER | **TOTAL GRAMS** |
|---|---|---|---|---|---|
| 967 | 252.65 | 278.18 | 274.32 | 244.17 | **1,049.31** |
| 968 | 228.87 | 227.33 | 229.33 | 271.22 | **956.75** |
| **HI - HAWAII - Total** | **481.52** | **505.51** | **503.65** | **515.39** | **2,006.06** |

**STATE: IA - IOWA**

| REGISTRANT ZIP CODE 3 | 1ST QUARTER | 2ND QUARTER | 3RD QUARTER | 4TH QUARTER | **TOTAL GRAMS** |
|---|---|---|---|---|---|
| 500 | 185.02 | 182.40 | 186.54 | 176.59 | **730.54** |
| 501 | 105.97 | 101.77 | 95.26 | 93.32 | **396.32** |
| 502 | 165.79 | 150.36 | 162.93 | 147.74 | **626.83** |
| 503 | 483.80 | 432.52 | 386.99 | 421.92 | **1,725.23** |
| 504 | 43.95 | 45.50 | 53.53 | 59.70 | **202.69** |
| 505 | 30.95 | 31.77 | 33.61 | 39.50 | **135.84** |
| 506 | 55.17 | 59.67 | 60.61 | 51.57 | **227.02** |
| 507 | 32.05 | 48.39 | 43.16 | 33.76 | **157.36** |
| 508 | 13.78 | 11.31 | 14.64 | 13.44 | **53.16** |
| 510 | 8.43 | 9.15 | 12.90 | 18.57 | **49.05** |
| 511 | 48.21 | 52.88 | 50.65 | 30.61 | **182.35** |
| 512 | 4.16 | 4.42 | 6.34 | 4.66 | **19.58** |

| | | | | | |
|---|---|---|---|---|---|
| 513 | 13.73 | 15.47 | 13.29 | 13.63 | **56.12** |
| 514 | 24.78 | 25.96 | 27.73 | 28.05 | **106.51** |
| 515 | 45.61 | 50.35 | 45.55 | 45.47 | **186.98** |
| 516 | 12.24 | 10.64 | 10.64 | 10.55 | **44.06** |
| 520 | 45.90 | 40.01 | 37.72 | 38.43 | **162.06** |
| 521 | 9.39 | 8.64 | 9.50 | 13.30 | **40.82** |
| 522 | 253.51 | 261.57 | 242.13 | 259.07 | **1,016.28** |
| 523 | 51.51 | 41.97 | 35.66 | 35.70 | **164.84** |
| 524 | 78.37 | 81.14 | 66.60 | 78.19 | **304.29** |
| 525 | 150.76 | 128.26 | 114.89 | 118.57 | **512.47** |
| 526 | 78.51 | 78.85 | 67.07 | 61.86 | **286.28** |
| 527 | 58.41 | 57.50 | 61.77 | 51.52 | **229.21** |
| 528 | 57.60 | 35.18 | 53.10 | 51.62 | **197.50** |
| **IA IOWA Total** | **2,057.58** | **1,965.70** | **1,892.79** | **1,897.33** | **7,813.40** |

**STATE: ID - IDAHO**

| REGISTRANT ZIP CODE 3 | 1ST QUARTER | 2ND QUARTER | 3RD QUARTER | 4TH QUARTER | TOTAL GRAMS |
|---|---|---|---|---|---|
| 832 | 92.13 | 94.80 | 95.71 | 79.04 | **361.68** |
| 833 | 117.96 | 117.89 | 118.95 | 102.10 | **456.90** |
| 834 | 58.99 | 70.80 | 72.00 | 53.87 | **255.67** |
| 835 | 89.54 | 106.53 | 80.41 | 96.71 | **373.19** |
| 836 | 403.85 | 423.42 | 439.13 | 380.08 | **1,646.48** |
| 837 | 390.61 | 356.23 | 371.30 | 366.49 | **1,484.62** |
| 838 | 187.06 | 181.01 | 167.21 | 176.42 | **711.71** |
| **ID IDAHO Total** | **1,340.15** | **1,350.69** | **1,344.70** | **1,254.71** | **5,290.25** |

**STATE: IL - ILLINOIS**

| REGISTRANT ZIP CODE 3 | 1ST QUARTER | 2ND QUARTER | 3RD QUARTER | 4TH QUARTER | TOTAL GRAMS |
|---|---|---|---|---|---|
| 600 | 1,269.63 | 1,464.56 | 1,395.19 | 1,364.82 | **5,494.20** |
| 601 | 812.11 | 793.20 | 946.47 | 797.10 | **3,348.89** |
| 602 | 45.84 | 47.05 | 46.20 | 54.37 | **193.46** |
| 603 | 77.11 | 87.62 | 81.11 | 50.31 | **296.15** |
| 604 | 978.98 | 913.25 | 923.85 | 900.68 | **3,716.76** |
| 605 | 482.45 | 466.77 | 503.12 | 502.38 | **1,954.72** |
| 606 | 1,121.21 | 1,272.53 | 1,204.48 | 1,164.58 | **4,762.80** |
| 607 | 63.57 | 50.41 | 60.91 | 44.92 | **219.81** |
| 608 | 66.85 | 55.84 | 69.51 | 60.24 | **252.45** |
| 609 | 113.96 | 111.71 | 123.97 | 111.16 | **460.79** |
| 610 | 64.73 | 60.81 | 57.57 | 57.14 | **240.24** |
| 611 | 144.58 | 127.36 | 168.21 | 115.27 | **555.43** |
| 612 | 47.20 | 58.33 | 43.61 | 45.63 | **194.76** |
| 613 | 51.98 | 45.45 | 43.53 | 53.17 | **194.13** |

| | | | | | |
|---|---|---|---|---|---|
| 614 | 38.90 | 31.99 | 32.17 | 24.67 | **127.73** |
| 615 | 59.99 | 59.74 | 55.75 | 45.28 | **220.75** |
| 616 | 132.42 | 128.63 | 140.85 | 135.25 | **537.15** |
| 617 | 146.13 | 142.66 | 146.72 | 140.51 | **576.02** |
| 618 | 162.49 | 187.01 | 186.96 | 178.06 | **714.52** |
| 619 | 32.30 | 36.98 | 35.47 | 37.06 | **141.82** |
| 620 | 108.84 | 101.62 | 96.20 | 89.61 | **396.26** |
| 622 | 163.92 | 157.98 | 156.89 | 175.44 | **654.22** |
| 623 | 49.43 | 38.99 | 35.28 | 32.98 | **156.69** |
| 624 | 87.78 | 70.59 | 84.30 | 79.15 | **321.82** |
| 625 | 152.52 | 140.23 | 140.78 | 145.28 | **578.81** |
| 626 | 51.15 | 42.46 | 46.33 | 48.14 | **188.07** |
| 627 | 106.27 | 134.40 | 106.90 | 116.73 | **464.30** |
| 628 | 69.50 | 58.56 | 56.60 | 61.39 | **246.05** |
| 629 | 86.28 | 72.25 | 67.46 | 67.96 | **293.96** |
| **IL ILLINOIS Total** | **6,788.14** | **6,958.96** | **7,056.39** | **6,699.29** | **27,502.77** |

**STATE: IN - INDIANA**

| REGISTRANT ZIP CODE 3 | 1ST QUARTER | 2ND QUARTER | 3RD QUARTER | 4TH QUARTER | TOTAL GRAMS |
|---|---|---|---|---|---|
| 460 | 346.67 | 329.41 | 249.98 | 302.82 | **1,228.89** |
| 461 | 249.08 | 233.09 | 251.52 | 285.53 | **1,019.22** |
| 462 | 926.49 | 836.62 | 753.77 | 737.41 | **3,254.29** |
| 463 | 286.74 | 308.77 | 309.10 | 312.16 | **1,216.76** |
| 464 | 34.34 | 30.06 | 31.16 | 28.11 | **123.66** |
| 465 | 239.12 | 285.15 | 313.73 | 245.58 | **1,083.58** |
| 466 | 69.17 | 71.13 | 66.36 | 53.37 | **260.04** |
| 467 | 212.16 | 202.38 | 233.46 | 203.50 | **851.50** |
| 468 | 316.38 | 344.35 | 361.02 | 303.35 | **1,325.10** |
| 469 | 118.03 | 105.54 | 104.41 | 99.43 | **427.42** |
| 470 | 24.68 | 27.87 | 19.84 | 17.40 | **89.80** |
| 471 | 101.94 | 115.87 | 132.85 | 135.44 | **486.10** |
| 472 | 116.40 | 122.97 | 109.78 | 109.09 | **458.24** |
| 473 | 197.65 | 210.17 | 199.88 | 215.54 | **823.25** |
| 474 | 129.88 | 116.19 | 117.84 | 121.58 | **485.50** |
| 475 | 164.84 | 176.92 | 185.66 | 174.76 | **702.18** |
| 476 | 75.11 | 95.49 | 74.24 | 82.94 | **327.78** |
| 477 | 441.39 | 655.88 | 423.21 | 390.95 | **1,911.43** |
| 478 | 68.09 | 77.48 | 75.78 | 68.35 | **289.70** |
| 479 | 142.78 | 117.47 | 104.04 | 101.97 | **466.26** |
| **IN INDIANA Total** | **4,260.94** | **4,462.79** | **4,117.66** | **3,989.31** | **16,830.70** |

**STATE: KS - KANSAS**

| REGISTRANT ZIP CODE 3 | 1ST QUARTER | 2ND QUARTER | 3RD QUARTER | 4TH QUARTER | TOTAL GRAMS |
|---|---|---|---|---|---|
| 660 | 289.11 | 302.62 | 300.42 | 315.09 | **1,207.24** |
| 661 | 120.86 | 115.41 | 125.19 | 112.46 | **473.92** |
| 662 | 588.23 | 517.05 | 578.74 | 622.01 | **2,306.03** |
| 664 | 72.73 | 71.95 | 83.09 | 74.35 | **302.12** |
| 665 | 86.33 | 92.17 | 86.45 | 63.91 | **328.86** |
| 666 | 93.98 | 87.33 | 85.28 | 81.39 | **347.97** |
| 667 | 70.46 | 66.72 | 64.17 | 61.55 | **262.90** |
| 668 | 21.12 | 28.34 | 13.29 | 13.74 | **76.49** |
| 669 | 8.18 | 9.46 | 8.31 | 6.14 | **32.10** |
| 670 | 94.88 | 104.70 | 103.34 | 103.92 | **406.84** |
| 671 | 65.27 | 78.61 | 70.08 | 61.13 | **275.10** |
| 672 | 423.76 | 440.31 | 416.78 | 431.56 | **1,712.40** |
| 673 | 27.40 | 28.81 | 29.21 | 29.52 | **114.95** |
| 674 | 122.91 | 124.09 | 112.73 | 124.30 | **484.02** |
| 675 | 233.14 | 225.14 | 220.02 | 209.96 | **888.26** |
| 676 | 61.23 | 72.91 | 65.13 | 53.40 | **252.67** |
| 677 | 18.30 | 15.95 | 10.04 | 11.98 | **56.27** |
| 678 | 56.01 | 44.61 | 49.14 | 49.82 | **199.59** |
| 679 | 21.08 | 19.54 | 19.59 | 14.81 | **75.01** |
| **KS - KANSAS - Total** | **2,474.96** | **2,445.73** | **2,441.01** | **2,441.07** | **9,802.77** |

**STATE: KY - KENTUCKY**

| REGISTRANT ZIP CODE 3 | 1ST QUARTER | 2ND QUARTER | 3RD QUARTER | 4TH QUARTER | TOTAL GRAMS |
|---|---|---|---|---|---|
| 400 | 67.73 | 83.85 | 81.88 | 86.81 | **320.27** |
| 401 | 37.81 | 32.23 | 39.88 | 29.89 | **139.81** |
| 402 | 597.81 | 539.82 | 786.06 | 527.97 | **2,451.66** |
| 403 | 47.91 | 59.49 | 53.49 | 63.78 | **224.67** |
| 404 | 11.85 | 12.61 | 13.47 | 14.92 | **52.85** |
| 405 | 233.89 | 208.13 | 291.97 | 211.46 | **945.44** |
| 406 | 11.00 | 14.52 | 16.28 | 11.41 | **53.22** |
| 407 | 10.47 | 12.80 | 17.46 | 16.84 | **57.56** |
| 408 | 4.57 | 2.74 | 8.02 | 4.12 | **19.45** |
| 409 | 8.01 | 6.26 | 8.75 | 5.50 | **28.51** |
| 410 | 113.13 | 87.26 | 91.02 | 99.19 | **390.60** |
| 411 | 37.65 | 32.13 | 55.14 | 70.29 | **195.20** |
| 412 | 3.75 | 3.53 | 5.15 | 2.41 | **14.84** |
| 413 | 2.31 | 3.75 | 4.02 | 3.06 | **13.14** |
| 414 | 0.30 | 1.26 | 0.62 | 0.20 | **2.38** |
| 415 | 20.88 | 24.26 | 21.63 | 22.65 | **89.43** |
| 416 | 2.49 | 2.50 | 4.27 | 2.28 | **11.54** |

| | | | | | |
|---|---|---|---|---|---|
| 417 | 64.33 | 59.72 | 48.57 | 33.35 | **205.98** |
| 418 | 3.08 | 4.62 | 3.34 | 3.12 | **14.17** |
| 420 | 135.65 | 157.22 | 146.90 | 149.29 | **589.06** |
| 421 | 372.13 | 323.48 | 192.41 | 196.32 | **1,084.35** |
| 422 | 108.87 | 117.83 | 123.60 | 132.55 | **482.85** |
| 423 | 106.95 | 103.36 | 103.99 | 90.18 | **404.47** |
| 424 | 127.25 | 155.29 | 154.93 | 170.28 | **607.75** |
| 425 | 12.44 | 11.43 | 17.37 | 9.06 | **50.30** |
| 426 | 3.94 | 4.79 | 2.94 | 7.99 | **19.66** |
| 427 | 68.05 | 72.10 | 61.07 | 71.16 | **272.38** |
| **KY - KENTUCKY - Total** | **2,214.27** | **2,136.96** | **2,354.21** | **2,036.10** | **8,741.55** |

**STATE: LA - LOUISIANA**

| REGISTRANT ZIP CODE 3 | 1ST QUARTER | 2ND QUARTER | 3RD QUARTER | 4TH QUARTER | TOTAL GRAMS |
|---|---|---|---|---|---|
| 700 | 166.38 | 155.96 | 153.57 | 159.08 | **635.00** |
| 701 | 160.04 | 154.06 | 155.13 | 162.06 | **631.29** |
| 703 | 98.04 | 77.08 | 82.67 | 72.57 | **330.36** |
| 704 | 295.15 | 277.15 | 286.85 | 309.35 | **1,168.50** |
| 705 | 676.37 | 808.36 | 722.21 | 439.17 | **2,646.11** |
| 706 | 105.74 | 115.88 | 121.61 | 123.96 | **467.19** |
| 707 | 350.98 | 256.72 | 346.04 | 150.85 | **1,104.59** |
| 708 | 189.33 | 175.17 | 201.08 | 169.78 | **735.37** |
| 710 | 28.13 | 30.16 | 36.68 | 27.32 | **122.28** |
| 711 | 137.99 | 116.55 | 104.72 | 106.72 | **465.99** |
| 712 | 64.08 | 79.08 | 81.85 | 88.92 | **313.93** |
| 713 | 118.14 | 110.97 | 103.60 | 103.61 | **436.33** |
| 714 | 54.39 | 47.21 | 52.28 | 54.50 | **208.38** |
| **LA - LOUISIANA - Total** | **2,444.77** | **2,404.35** | **2,448.31** | **1,967.88** | **9,265.32** |

**STATE: MA - MASSACHUSETTS**

| REGISTRANT ZIP CODE 3 | 1ST QUARTER | 2ND QUARTER | 3RD QUARTER | 4TH QUARTER | TOTAL GRAMS |
|---|---|---|---|---|---|
| 010 | 419.85 | 419.87 | 408.63 | 403.99 | **1,652.35** |
| 011 | 222.56 | 296.19 | 273.69 | 267.60 | **1,060.04** |
| 012 | 95.63 | 80.96 | 91.13 | 87.52 | **355.24** |
| 013 | 61.40 | 60.88 | 56.12 | 68.76 | **247.17** |
| 014 | 58.11 | 72.29 | 77.47 | 58.38 | **266.25** |
| 015 | 159.09 | 149.08 | 139.15 | 137.20 | **584.52** |
| 016 | 155.44 | 124.42 | 115.68 | 128.86 | **524.40** |
| 017 | 314.20 | 323.81 | 342.60 | 340.81 | **1,321.42** |
| 018 | 772.66 | 946.32 | 786.38 | 1,405.65 | **3,911.01** |
| 019 | 356.83 | 319.30 | 308.30 | 279.33 | **1,263.75** |
| 020 | 359.57 | 274.48 | 320.77 | 378.24 | **1,333.06** |

| | | | | | |
|---|---|---|---|---|---|
| 021 | 1,375.42 | 1,313.10 | 1,375.30 | 1,384.59 | **5,448.41** |
| 022 | 260.93 | 287.69 | 223.99 | 225.45 | **998.05** |
| 023 | 285.33 | 337.95 | 270.85 | 275.99 | **1,170.12** |
| 024 | 174.87 | 186.03 | 166.25 | 191.86 | **719.02** |
| 025 | 83.70 | 70.97 | 62.09 | 78.07 | **294.84** |
| 026 | 100.07 | 106.12 | 111.71 | 134.68 | **452.59** |
| 027 | 394.55 | 356.06 | 406.39 | 439.40 | **1,596.41** |
| **MA - MASSACHUSETTS - Total** | **5,650.22** | **5,725.52** | **5,536.52** | **6,286.38** | **23,198.64** |

**STATE: MD - MARYLAND**

| REGISTRANT ZIP CODE 3 | 1ST QUARTER | 2ND QUARTER | 3RD QUARTER | 4TH QUARTER | **TOTAL GRAMS** |
|---|---|---|---|---|---|
| 206 | 319.16 | 337.57 | 318.48 | 306.00 | **1,281.21** |
| 207 | 423.79 | 396.60 | 383.69 | 398.31 | **1,602.39** |
| 208 | 711.34 | 673.44 | 663.66 | 688.11 | **2,736.55** |
| 209 | 161.93 | 179.80 | 174.79 | 171.88 | **688.40** |
| 210 | 733.36 | 823.61 | 760.51 | 841.37 | **3,158.84** |
| 211 | 344.10 | 371.40 | 361.65 | 325.96 | **1,403.12** |
| 212 | 1,503.88 | 1,548.12 | 1,537.41 | 1,517.11 | **6,106.51** |
| 214 | 92.70 | 85.21 | 87.00 | 79.17 | **344.08** |
| 215 | 53.57 | 59.67 | 63.23 | 57.07 | **233.55** |
| 216 | 110.51 | 109.45 | 111.90 | 107.27 | **439.12** |
| 217 | 565.97 | 542.25 | 566.63 | 550.25 | **2,225.09** |
| 218 | 218.96 | 211.28 | 286.69 | 236.06 | **952.98** |
| 219 | 182.86 | 182.41 | 161.46 | 159.30 | **686.03** |
| **MD MARYLAND Total** | **5,422.12** | **5,520.79** | **5,477.11** | **5,437.84** | **21,857.86** |

**STATE: ME - MAINE**

| REGISTRANT ZIP CODE 3 | 1ST QUARTER | 2ND QUARTER | 3RD QUARTER | 4TH QUARTER | **TOTAL GRAMS** |
|---|---|---|---|---|---|
| 039 | 14.93 | 15.90 | 21.46 | 12.08 | **64.37** |
| 040 | 315.16 | 276.02 | 303.46 | 312.15 | **1,206.78** |
| 041 | 201.97 | 299.73 | 293.38 | 281.92 | **1,077.00** |
| 042 | 85.09 | 92.39 | 104.26 | 94.00 | **375.74** |
| 043 | 290.44 | 110.19 | 293.07 | 204.11 | **897.81** |
| 044 | 132.05 | 144.32 | 137.64 | 144.49 | **558.50** |
| 045 | 42.20 | 37.95 | 36.71 | 44.06 | **160.92** |
| 046 | 70.06 | 80.09 | 89.92 | 82.72 | **322.79** |
| 047 | 16.02 | 20.90 | 23.50 | 24.64 | **85.05** |
| 048 | 13.62 | 18.68 | 17.88 | 24.66 | **74.84** |
| 049 | 115.37 | 127.34 | 124.40 | 110.14 | **477.24** |
| **ME MAINE Total** | **1,296.89** | **1,223.50** | **1,445.67** | **1,334.98** | **5,301.05** |

**STATE: MI - MICHIGAN**

| REGISTRANT ZIP CODE 3 | 1ST QUARTER | 2ND QUARTER | 3RD QUARTER | 4TH QUARTER | **TOTAL GRAMS** |
|---|---|---|---|---|---|

| | | | | | |
|---|---|---|---|---|---|
| 480 | 774.77 | 815.22 | 808.59 | 830.28 | **3,228.87** |
| 481 | 604.78 | 660.00 | 598.46 | 660.90 | **2,524.14** |
| 482 | 621.28 | 558.65 | 530.52 | 535.15 | **2,245.60** |
| 483 | 856.96 | 659.39 | 896.60 | 1,150.13 | **3,563.08** |
| 484 | 203.39 | 214.55 | 268.15 | 192.03 | **878.12** |
| 485 | 140.45 | 111.20 | 109.95 | 135.78 | **497.39** |
| 486 | 314.78 | 301.81 | 308.36 | 324.70 | **1,249.65** |
| 487 | 89.36 | 96.11 | 103.08 | 102.00 | **390.55** |
| 488 | 206.93 | 257.01 | 247.91 | 248.18 | **960.04** |
| 489 | 128.29 | 109.04 | 104.84 | 104.87 | **447.04** |
| 490 | 380.10 | 357.71 | 377.59 | 365.34 | **1,480.74** |
| 491 | 27.78 | 38.63 | 26.95 | 16.98 | **110.34** |
| 492 | 96.31 | 103.67 | 101.33 | 91.02 | **392.32** |
| 493 | 95.31 | 92.83 | 85.87 | 83.04 | **357.06** |
| 494 | 239.27 | 209.77 | 203.17 | 201.56 | **853.77** |
| 495 | 355.49 | 410.01 | 306.71 | 289.45 | **1,361.66** |
| 496 | 312.15 | 296.19 | 314.97 | 272.56 | **1,195.88** |
| 497 | 146.96 | 177.25 | 155.13 | 133.84 | **613.17** |
| 498 | 94.43 | 98.74 | 128.20 | 128.09 | **449.46** |
| 499 | 32.23 | 32.66 | 24.16 | 28.73 | **117.78** |
| **MI - MICHIGAN Total** | **5,721.03** | **5,600.44** | **5,700.56** | **5,894.62** | **22,916.66** |

**STATE: MN - MINNESOTA**

| REGISTRANT ZIP CODE 3 | 1ST QUARTER | 2ND QUARTER | 3RD QUARTER | 4TH QUARTER | TOTAL GRAMS |
|---|---|---|---|---|---|
| 550 | 415.11 | 409.11 | 407.11 | 419.98 | **1,651.30** |
| 551 | 815.11 | 870.64 | 930.31 | 860.38 | **3,476.43** |
| 553 | 467.03 | 461.90 | 422.92 | 427.83 | **1,779.69** |
| 554 | 1,454.37 | 1,523.93 | 1,561.52 | 1,499.50 | **6,039.32** |
| 556 | 5.57 | 4.22 | 5.80 | 5.54 | **21.13** |
| 557 | 136.70 | 140.50 | 119.85 | 116.48 | **513.52** |
| 558 | 111.62 | 105.13 | 106.06 | 120.54 | **443.33** |
| 559 | 388.67 | 442.25 | 495.89 | 487.53 | **1,814.34** |
| 560 | 128.26 | 129.75 | 124.36 | 131.43 | **513.79** |
| 561 | 14.19 | 16.44 | 17.77 | 12.98 | **61.38** |
| 562 | 33.60 | 32.39 | 39.82 | 43.28 | **149.09** |
| 563 | 132.04 | 113.71 | 112.97 | 100.74 | **459.46** |
| 564 | 74.07 | 69.02 | 69.05 | 60.40 | **272.53** |
| 565 | 70.43 | 75.97 | 72.46 | 63.67 | **282.53** |
| 566 | 43.75 | 38.26 | 38.52 | 40.45 | **160.98** |
| 567 | 24.04 | 22.60 | 21.08 | 15.49 | **83.21** |
| **MN MINNESOTA Total** | **4,314.55** | **4,455.81** | **4,545.47** | **4,406.20** | **17,722.03** |

**STATE: MO - MISSOURI**

| REGISTRANT ZIP CODE 3 | 1ST QUARTER | 2ND QUARTER | 3RD QUARTER | 4TH QUARTER | TOTAL GRAMS |
|---|---|---|---|---|---|
| 630 | 372.32 | 339.99 | 317.68 | 371.17 | **1,401.16** |
| 631 | 807.43 | 771.14 | 817.27 | 792.35 | **3,188.20** |
| 633 | 148.17 | 159.97 | 155.23 | 159.86 | **623.24** |
| 634 | 21.76 | 22.92 | 20.29 | 18.72 | **83.69** |
| 635 | 37.42 | 44.25 | 39.43 | 33.87 | **154.98** |
| 636 | 89.29 | 87.03 | 88.66 | 93.79 | **358.77** |
| 637 | 54.06 | 62.40 | 65.96 | 52.90 | **235.32** |
| 638 | 46.59 | 57.18 | 44.62 | 39.99 | **188.37** |
| 639 | 30.43 | 20.57 | 24.04 | 21.22 | **96.26** |
| 640 | 306.94 | 257.80 | 280.80 | 275.69 | **1,121.22** |
| 641 | 300.55 | 305.58 | 292.07 | 301.75 | **1,199.95** |
| 644 | 37.19 | 23.95 | 28.74 | 29.05 | **118.94** |
| 645 | 152.80 | 101.39 | 112.90 | 111.83 | **478.91** |
| 646 | 51.21 | 28.61 | 30.87 | 26.71 | **137.41** |
| 647 | 44.75 | 49.22 | 45.84 | 38.10 | **177.90** |
| 648 | 166.76 | 117.14 | 96.37 | 105.90 | **486.17** |
| 650 | 54.12 | 43.64 | 48.81 | 41.85 | **188.41** |
| 651 | 27.15 | 24.59 | 33.48 | 32.86 | **118.09** |
| 652 | 299.73 | 367.07 | 497.85 | 388.74 | **1,553.39** |
| 653 | 37.72 | 41.54 | 32.63 | 35.32 | **147.21** |
| 654 | 27.23 | 21.34 | 20.00 | 19.57 | **88.15** |
| 655 | 57.62 | 53.11 | 62.09 | 53.95 | **226.77** |
| 656 | 212.92 | 210.92 | 198.05 | 192.50 | **814.40** |
| 657 | 340.85 | 281.62 | 251.56 | 256.90 | **1,130.93** |
| 658 | 306.11 | 322.65 | 283.22 | 241.32 | **1,153.29** |
| **MO MISSOURI - Total** | **4,031.12** | **3,815.62** | **3,888.47** | **3,735.92** | **15,471.13** |

**STATE: MS - MISSISSIPPI**

| REGISTRANT ZIP CODE 3 | 1ST QUARTER | 2ND QUARTER | 3RD QUARTER | 4TH QUARTER | TOTAL GRAMS |
|---|---|---|---|---|---|
| 386 | 109.19 | 108.68 | 95.94 | 88.55 | **402.36** |
| 387 | 4.05 | 6.29 | 5.67 | 6.41 | **22.41** |
| 388 | 94.78 | 91.44 | 79.36 | 62.45 | **328.03** |
| 389 | 23.98 | 18.88 | 14.63 | 20.37 | **77.86** |
| 390 | 68.09 | 58.64 | 56.67 | 60.44 | **243.85** |
| 391 | 4,076.38 | 4,066.44 | 4,081.75 | 5,479.22 | **17,703.80** |
| 392 | 165.34 | 152.08 | 182.50 | 276.37 | **776.29** |
| 393 | 54.16 | 49.50 | 44.61 | 46.24 | **194.52** |
| 394 | 212.05 | 184.45 | 213.82 | 231.59 | **841.91** |
| 395 | 247.61 | 253.66 | 244.93 | 224.88 | **971.08** |

| | | | | | |
|---|---|---|---|---|---|
| 396 | 24.49 | 16.22 | 35.45 | 70.92 | **147.08** |
| 397 | 24.33 | 26.35 | 22.26 | 24.54 | **97.47** |
| **MS MISSISSIPPI - Total** | **5,104.46** | **5,032.64** | **5,077.58** | **6,591.99** | **21,806.67** |

**STATE: MT - MONTANA**

| REGISTRANT ZIP CODE 3 | 1ST QUARTER | 2ND QUARTER | 3RD QUARTER | 4TH QUARTER | TOTAL GRAMS |
|---|---|---|---|---|---|
| 590 | 72.52 | 81.76 | 80.50 | 93.08 | **327.85** |
| 591 | 439.41 | 455.17 | 402.79 | 412.80 | **1,710.17** |
| 592 | 42.75 | 41.32 | 33.03 | 36.71 | **153.81** |
| 593 | 62.87 | 68.45 | 53.94 | 50.42 | **235.67** |
| 594 | 77.16 | 99.64 | 78.48 | 82.25 | **337.54** |
| 595 | 11.99 | 11.91 | 13.82 | 15.60 | **53.32** |
| 596 | 57.57 | 65.75 | 57.12 | 65.05 | **245.49** |
| 597 | 154.92 | 147.34 | 148.36 | 144.78 | **595.41** |
| 598 | 271.68 | 281.13 | 238.20 | 254.98 | **1,046.00** |
| 599 | 131.80 | 122.50 | 131.35 | 154.00 | **539.65** |
| **MT - MONTANA Total** | **1,322.67** | **1,374.96** | **1,237.59** | **1,309.68** | **5,244.91** |

**STATE: NC - NORTH CAROLINA**

| REGISTRANT ZIP CODE 3 | 1ST QUARTER | 2ND QUARTER | 3RD QUARTER | 4TH QUARTER | TOTAL GRAMS |
|---|---|---|---|---|---|
| 270 | 203.76 | 203.80 | 175.69 | 189.61 | **772.87** |
| 271 | 300.43 | 294.52 | 346.08 | 281.16 | **1,222.20** |
| 272 | 523.24 | 564.92 | 517.75 | 541.73 | **2,147.64** |
| 273 | 330.98 | 332.20 | 321.62 | 287.52 | **1,272.31** |
| 274 | 262.01 | 225.16 | 277.84 | 248.13 | **1,013.14** |
| 275 | 1,058.05 | 1,090.51 | 1,098.64 | 1,048.05 | **4,295.25** |
| 276 | 373.01 | 382.73 | 415.55 | 347.47 | **1,518.75** |
| 277 | 631.05 | 372.99 | 385.95 | 547.50 | **1,937.49** |
| 278 | 395.79 | 347.10 | 380.34 | 278.59 | **1,401.81** |
| 279 | 232.88 | 224.83 | 227.05 | 145.39 | **830.14** |
| 280 | 542.89 | 532.43 | 551.83 | 532.43 | **2,159.59** |
| 281 | 409.37 | 375.14 | 389.71 | 390.90 | **1,565.12** |
| 282 | 423.03 | 422.22 | 486.98 | 501.86 | **1,834.08** |
| 283 | 619.37 | 605.17 | 591.30 | 551.40 | **2,367.25** |
| 284 | 425.77 | 496.43 | 473.89 | 451.35 | **1,847.44** |
| 285 | 370.84 | 378.13 | 423.90 | 390.50 | **1,563.37** |
| 286 | 721.23 | 699.26 | 709.10 | 748.81 | **2,878.40** |
| 287 | 297.52 | 333.80 | 339.31 | 321.48 | **1,292.11** |
| 288 | 213.58 | 213.54 | 215.64 | 196.23 | **838.99** |
| 289 | 57.44 | 43.35 | 36.08 | 37.89 | **174.76** |
| **NC - NORTH CAROLINA Total** | **8,392.23** | **8,138.22** | **8,364.25** | **8,038.01** | **32,932.71** |

**STATE: ND - NORTH DAKOTA**

| REGISTRANT ZIP CODE 3 | 1ST QUARTER | 2ND QUARTER | 3RD QUARTER | 4TH QUARTER | TOTAL GRAMS |
|---|---|---|---|---|---|
| 580 | 29.21 | 33.01 | 32.50 | 32.97 | **127.69** |
| 581 | 164.52 | 117.22 | 140.22 | 133.31 | **555.28** |
| 582 | 63.38 | 70.45 | 45.38 | 38.62 | **217.84** |
| 583 | 17.17 | 21.94 | 16.03 | 14.11 | **69.26** |
| 584 | 13.66 | 14.35 | 12.52 | 17.02 | **57.54** |
| 585 | 186.27 | 129.23 | 149.17 | 146.14 | **610.80** |
| 586 | 29.56 | 26.48 | 23.90 | 23.27 | **103.20** |
| 587 | 38.11 | 34.19 | 40.90 | 40.22 | **153.43** |
| 588 | 33.65 | 29.35 | 29.53 | 25.89 | **118.42** |
| **ND NORTH DAKOTA Total** | **575.55** | **476.22** | **490.15** | **471.55** | **2,013.46** |

**STATE: NE - NEBRASKA**

| REGISTRANT ZIP CODE 3 | 1ST QUARTER | 2ND QUARTER | 3RD QUARTER | 4TH QUARTER | TOTAL GRAMS |
|---|---|---|---|---|---|
| 680 | 137.53 | 129.11 | 137.63 | 141.75 | **546.03** |
| 681 | 392.69 | 387.53 | 341.76 | 390.10 | **1,512.08** |
| 683 | 12.46 | 14.36 | 9.56 | 8.31 | **44.69** |
| 684 | 17.35 | 17.08 | 16.62 | 15.23 | **66.27** |
| 685 | 94.47 | 71.48 | 88.32 | 67.71 | **321.98** |
| 686 | 19.84 | 19.39 | 23.81 | 10.91 | **73.95** |
| 687 | 23.59 | 18.32 | 17.31 | 22.79 | **82.01** |
| 688 | 82.48 | 92.47 | 79.28 | 87.32 | **341.54** |
| 689 | 22.92 | 19.87 | 22.34 | 19.27 | **84.40** |
| 690 | 12.65 | 9.77 | 12.44 | 11.07 | **45.94** |
| 691 | 30.74 | 34.90 | 23.34 | 26.74 | **115.73** |
| 692 | 2.66 | 5.00 | 3.59 | 3.77 | **15.02** |
| 693 | 38.95 | 33.41 | 41.14 | 39.22 | **152.72** |
| **NE NEBRASKA Total** | **888.34** | **852.69** | **817.15** | **844.17** | **3,402.35** |

**STATE: NH - NEW HAMPSHIRE**

| REGISTRANT ZIP CODE 3 | 1ST QUARTER | 2ND QUARTER | 3RD QUARTER | 4TH QUARTER | TOTAL GRAMS |
|---|---|---|---|---|---|
| 030 | 303.56 | 319.12 | 337.82 | 332.94 | **1,293.44** |
| 031 | 184.35 | 154.56 | 217.40 | 207.89 | **764.20** |
| 032 | 140.10 | 131.63 | 133.55 | 149.64 | **554.92** |
| 033 | 306.10 | 278.12 | 240.72 | 119.12 | **944.07** |
| 034 | 54.36 | 66.32 | 57.93 | 62.34 | **240.94** |
| 035 | 65.36 | 66.36 | 84.14 | 84.09 | **299.96** |
| 036 | 6.38 | 6.70 | 19.04 | 20.53 | **52.66** |
| 037 | 153.05 | 196.56 | 198.06 | 215.20 | **762.87** |
| 038 | 299.59 | 273.23 | 278.91 | 294.88 | **1,146.61** |
| **NH - NEW HAMPSHIRE Total** | **1,512.86** | **1,492.60** | **1,567.57** | **1,486.64** | **6,059.66** |

**STATE: NJ - NEW JERSEY**

| REGISTRANT ZIP CODE 3 | 1ST QUARTER | 2ND QUARTER | 3RD QUARTER | 4TH QUARTER | TOTAL GRAMS |
|---|---|---|---|---|---|
| 070 | 959.03 | 983.35 | 1,029.37 | 1,002.96 | **3,974.71** |
| 071 | 194.50 | 190.93 | 164.13 | 179.86 | **729.41** |
| 072 | 89.89 | 71.78 | 89.31 | 105.12 | **356.11** |
| 073 | 41.72 | 34.46 | 37.81 | 46.51 | **160.50** |
| 074 | 763.97 | 676.42 | 884.53 | 594.43 | **2,919.34** |
| 075 | 191.22 | 108.69 | 158.68 | 177.44 | **636.03** |
| 076 | 711.80 | 730.13 | 752.51 | 707.02 | **2,901.46** |
| 077 | 497.48 | 448.45 | 557.30 | 476.56 | **1,979.79** |
| 078 | 332.42 | 273.96 | 260.68 | 268.62 | **1,135.68** |
| 079 | 339.81 | 330.96 | 331.80 | 309.97 | **1,312.55** |
| 080 | 771.60 | 785.63 | 804.16 | 819.15 | **3,180.55** |
| 081 | 135.14 | 219.95 | 116.91 | 120.16 | **592.16** |
| 082 | 114.09 | 123.32 | 124.38 | 107.03 | **468.82** |
| 083 | 93.56 | 114.03 | 92.15 | 104.90 | **404.66** |
| 084 | 15.49 | 27.05 | 21.57 | 20.51 | **84.62** |
| 085 | 172.80 | 137.93 | 138.49 | 175.22 | **624.44** |
| 086 | 111.01 | 113.80 | 101.95 | 106.62 | **433.38** |
| 087 | 269.75 | 297.60 | 320.46 | 307.50 | **1,195.30** |
| 088 | 770.75 | 746.32 | 756.97 | 735.14 | **3,009.18** |
| 089 | 130.30 | 108.97 | 109.97 | 92.80 | **442.06** |
| **NJ NEW JERSEY - Total** | **6,706.35** | **6,523.72** | **6,853.15** | **6,457.52** | **26,540.73** |

**STATE: NM - NEW MEXICO**

| REGISTRANT ZIP CODE 3 | 1ST QUARTER | 2ND QUARTER | 3RD QUARTER | 4TH QUARTER | TOTAL GRAMS |
|---|---|---|---|---|---|
| 870 | 56.21 | 61.33 | 59.30 | 59.72 | **236.56** |
| 871 | 525.39 | 411.46 | 433.49 | 485.42 | **1,855.75** |
| 873 | 15.74 | 14.38 | 14.84 | 12.50 | **57.46** |
| 874 | 48.14 | 63.04 | 47.12 | 45.21 | **203.51** |
| 875 | 118.97 | 108.24 | 119.83 | 117.64 | **464.68** |
| 877 | 23.21 | 31.46 | 31.42 | 30.85 | **116.95** |
| 878 | 0.89 | 1.42 | 1.60 | 1.60 | **5.51** |
| 879 | 9.40 | 7.62 | 3.26 | 2.91 | **23.19** |
| 880 | 98.26 | 83.72 | 98.84 | 88.43 | **369.24** |
| 881 | 26.06 | 19.25 | 17.46 | 21.08 | **83.85** |
| 882 | 81.76 | 74.08 | 76.52 | 75.29 | **307.64** |
| 883 | 23.08 | 24.60 | 26.85 | 24.16 | **98.68** |
| 884 | 1.21 | 1.67 | 2.53 | 1.69 | **7.09** |
| **NM NEW MEXICO Total** | **1,028.30** | **902.27** | **933.04** | **966.50** | **3,830.12** |

**STATE: NV - NEVADA**

| REGISTRANT ZIP CODE 3 | 1ST QUARTER | 2ND QUARTER | 3RD QUARTER | 4TH QUARTER | TOTAL GRAMS |
|---|---|---|---|---|---|

| | | | | | |
|---|---|---|---|---|---|
| 890 | 506.23 | 457.62 | 461.74 | 478.34 | **1,903.93** |
| 891 | 1,249.12 | 1,272.96 | 1,230.27 | 959.90 | **4,712.25** |
| 893 | 0.57 | 1.61 | 0.92 | 1.77 | **4.88** |
| 894 | 161.20 | 149.95 | 149.50 | 153.31 | **613.96** |
| 895 | 165.94 | 139.55 | 152.90 | 155.99 | **614.37** |
| 897 | 75.58 | 67.20 | 73.76 | 67.65 | **284.18** |
| 898 | 11.15 | 14.60 | 10.58 | 13.25 | **49.57** |
| **NV NEVADA Total** | **2,169.78** | **2,103.50** | **2,079.67** | **1,830.20** | **8,183.15** |

**STATE: NY - NEW YORK**

| REGISTRANT ZIP CODE 3 | 1ST QUARTER | 2ND QUARTER | 3RD QUARTER | 4TH QUARTER | TOTAL GRAMS |
|---|---|---|---|---|---|
| 100 | 2,187.85 | 1,773.91 | 2,004.16 | 2,140.42 | **8,106.34** |
| 101 | 38.21 | 26.22 | 31.08 | 31.78 | **127.29** |
| 102 | 3.79 | 1.42 | 4.11 | 2.13 | **11.45** |
| 103 | 300.12 | 262.81 | 367.64 | 349.54 | **1,280.11** |
| 104 | 728.23 | 846.49 | 722.76 | 522.51 | **2,819.99** |
| 105 | 387.93 | 339.99 | 381.27 | 366.39 | **1,475.58** |
| 106 | 83.86 | 85.92 | 75.48 | 97.43 | **342.69** |
| 107 | 94.17 | 91.13 | 87.49 | 95.90 | **368.70** |
| 108 | 51.72 | 35.02 | 43.45 | 46.66 | **176.84** |
| 109 | 522.94 | 355.56 | 396.53 | 413.90 | **1,688.93** |
| 110 | 780.27 | 858.52 | 928.87 | 903.22 | **3,470.89** |
| 111 | 110.44 | 117.53 | 95.13 | 108.31 | **431.42** |
| 112 | 880.20 | 761.53 | 839.11 | 922.75 | **3,403.60** |
| 113 | 430.12 | 349.42 | 443.39 | 401.78 | **1,624.71** |
| 114 | 220.11 | 157.13 | 191.75 | 187.11 | **756.10** |
| 115 | 718.01 | 648.62 | 641.40 | 673.87 | **2,681.90** |
| 116 | 32.80 | 32.04 | 37.59 | 34.05 | **136.48** |
| 117 | 1,792.86 | 1,718.64 | 1,817.83 | 1,786.68 | **7,116.01** |
| 118 | 191.86 | 107.78 | 108.32 | 102.65 | **510.61** |
| 119 | 162.00 | 136.09 | 164.82 | 125.70 | **588.60** |
| 120 | 163.27 | 156.48 | 159.56 | 189.69 | **669.00** |
| 121 | 78.33 | 86.67 | 77.47 | 67.34 | **309.82** |
| 122 | 168.70 | 239.04 | 136.99 | 149.29 | **694.02** |
| 123 | 41.29 | 43.70 | 53.26 | 49.92 | **188.17** |
| 124 | 161.97 | 150.43 | 147.83 | 148.18 | **608.40** |
| 125 | 279.88 | 280.56 | 320.38 | 283.39 | **1,164.21** |
| 126 | 122.40 | 103.29 | 126.52 | 116.07 | **468.28** |
| 127 | 74.23 | 86.63 | 86.13 | 84.75 | **331.74** |
| 128 | 160.62 | 187.63 | 168.99 | 151.61 | **668.85** |
| 129 | 272.93 | 255.93 | 242.88 | 262.29 | **1,034.02** |

| | | | | | |
|---|---|---|---|---|---|
| 130 | 141.70 | 137.82 | 146.86 | 136.98 | **563.36** |
| 131 | 87.73 | 103.65 | 107.10 | 83.30 | **381.78** |
| 132 | 127.35 | 100.28 | 99.66 | 111.55 | **438.84** |
| 133 | 52.77 | 68.29 | 82.74 | 80.85 | **284.65** |
| 134 | 69.56 | 71.48 | 66.49 | 65.91 | **273.44** |
| 135 | 24.37 | 24.15 | 32.91 | 27.90 | **109.32** |
| 136 | 136.26 | 114.15 | 131.26 | 130.22 | **511.88** |
| 137 | 36.34 | 41.36 | 42.80 | 48.72 | **169.21** |
| 138 | 127.74 | 105.77 | 97.55 | 89.68 | **420.74** |
| 139 | 28.81 | 27.44 | 24.57 | 43.77 | **124.59** |
| 140 | 196.14 | 188.18 | 175.10 | 191.03 | **750.44** |
| 141 | 107.23 | 98.40 | 105.13 | 99.79 | **410.55** |
| 142 | 427.27 | 498.46 | 498.71 | 431.47 | **1,855.91** |
| 143 | 24.41 | 30.72 | 34.66 | 31.32 | **121.11** |
| 144 | 250.33 | 251.54 | 271.28 | 267.12 | **1,040.27** |
| 145 | 188.97 | 189.87 | 187.46 | 181.43 | **747.73** |
| 146 | 803.55 | 865.57 | 851.30 | 893.28 | **3,413.71** |
| 147 | 75.06 | 75.11 | 63.96 | 56.34 | **270.46** |
| 148 | 210.45 | 206.67 | 205.96 | 202.93 | **826.01** |
| 149 | 36.21 | 34.94 | 40.26 | 44.22 | **155.64** |
| **NY - NEW YORK - Total** | **14,393.39** | **13,529.97** | **14,167.94** | **14,033.12** | **56,124.42** |

**STATE: OH - OHIO**

| REGISTRANT ZIP CODE 3 | 1ST QUARTER | 2ND QUARTER | 3RD QUARTER | 4TH QUARTER | TOTAL GRAMS |
|---|---|---|---|---|---|
| 430 | 488.13 | 653.59 | 556.59 | 680.70 | **2,379.02** |
| 431 | 210.19 | 209.19 | 242.47 | 240.53 | **902.38** |
| 432 | 633.17 | 653.00 | 715.27 | 711.37 | **2,712.81** |
| 433 | 112.51 | 109.55 | 109.86 | 123.68 | **455.59** |
| 434 | 46.14 | 53.29 | 50.36 | 42.08 | **191.88** |
| 435 | 123.54 | 296.66 | 443.25 | 132.39 | **995.84** |
| 436 | 158.50 | 185.96 | 162.80 | 150.65 | **657.91** |
| 437 | 137.96 | 108.07 | 103.16 | 104.24 | **453.43** |
| 438 | 15.69 | 15.07 | 19.06 | 22.08 | **71.90** |
| 439 | 95.93 | 71.93 | 80.08 | 66.58 | **314.52** |
| 440 | 244.76 | 223.20 | 219.33 | 225.03 | **912.33** |
| 441 | 609.77 | 633.94 | 720.29 | 637.85 | **2,601.84** |
| 442 | 250.17 | 231.98 | 240.45 | 243.77 | **966.36** |
| 443 | 223.15 | 229.59 | 200.29 | 239.91 | **892.94** |
| 444 | 157.53 | 108.64 | 89.22 | 82.57 | **437.96** |
| 445 | 74.17 | 49.77 | 46.73 | 47.97 | **218.64** |
| 446 | 287.48 | 312.45 | 303.94 | 309.67 | **1,213.54** |

| | | | | | |
|---|---|---|---|---|---|
| 447 | 235.91 | 213.81 | 229.99 | 235.52 | **915.24** |
| 448 | 102.31 | 110.00 | 110.04 | 100.44 | **422.79** |
| 449 | 48.67 | 21.21 | 28.38 | 25.78 | **124.05** |
| 450 | 383.54 | 287.09 | 403.70 | 312.43 | **1,386.75** |
| 451 | 121.96 | 106.97 | 83.32 | 95.91 | **408.16** |
| 452 | 381.75 | 391.31 | 439.25 | 354.67 | **1,566.98** |
| 453 | 129.76 | 143.78 | 142.60 | 135.86 | **552.00** |
| 454 | 241.32 | 214.91 | 267.07 | 237.79 | **961.09** |
| 455 | 41.26 | 51.69 | 42.68 | 50.26 | **185.89** |
| 456 | 109.78 | 129.86 | 142.86 | 149.39 | **531.89** |
| 457 | 69.95 | 74.08 | 67.11 | 64.60 | **275.74** |
| 458 | 208.58 | 178.53 | 173.90 | 172.96 | **733.98** |
| **OH - OHIO Total** | **5,943.59** | **6,069.12** | **6,434.05** | **5,996.69** | **24,443.46** |

**STATE: OK - OKLAHOMA**

| REGISTRANT ZIP CODE 3 | 1ST QUARTER | 2ND QUARTER | 3RD QUARTER | 4TH QUARTER | TOTAL GRAMS |
|---|---|---|---|---|---|
| 730 | 356.43 | 402.98 | 357.00 | 452.35 | **1,568.76** |
| 731 | 603.64 | 613.17 | 597.36 | 655.13 | **2,469.29** |
| 734 | 61.79 | 56.60 | 46.32 | 40.79 | **205.51** |
| 735 | 140.62 | 149.57 | 146.94 | 149.68 | **586.81** |
| 736 | 15.92 | 21.49 | 14.05 | 20.40 | **71.86** |
| 737 | 44.09 | 41.36 | 37.05 | 40.25 | **162.76** |
| 738 | 13.56 | 11.52 | 25.68 | 15.21 | **65.96** |
| 739 | 0.37 | 0.40 | 0.68 | 0.05 | **1.50** |
| 740 | 329.23 | 357.13 | 352.83 | 340.42 | **1,379.60** |
| 741 | 334.73 | 333.08 | 288.40 | 321.74 | **1,277.95** |
| 743 | 91.24 | 67.99 | 69.60 | 64.49 | **293.33** |
| 744 | 126.94 | 129.71 | 140.22 | 140.75 | **537.62** |
| 745 | 86.61 | 80.60 | 72.43 | 68.06 | **307.70** |
| 746 | 10.91 | 6.98 | 11.38 | 9.46 | **38.72** |
| 747 | 69.99 | 59.72 | 61.84 | 61.56 | **253.11** |
| 748 | 75.60 | 67.72 | 65.08 | 69.26 | **277.67** |
| 749 | 150.75 | 136.36 | 140.94 | 152.73 | **580.78** |
| **OK - OKLAHOMA Total** | **2,512.41** | **2,536.40** | **2,427.80** | **2,602.31** | **10,078.91** |

**STATE: OR - OREGON**

| REGISTRANT ZIP CODE 3 | 1ST QUARTER | 2ND QUARTER | 3RD QUARTER | 4TH QUARTER | TOTAL GRAMS |
|---|---|---|---|---|---|
| 970 | 533.01 | 526.02 | 534.98 | 522.88 | **2,116.89** |
| 971 | 253.63 | 248.99 | 247.81 | 257.28 | **1,007.71** |
| 972 | 942.80 | 990.99 | 1,139.70 | 1,099.04 | **4,172.54** |
| 973 | 506.82 | 463.37 | 440.29 | 426.07 | **1,836.56** |
| 974 | 535.81 | 536.02 | 571.62 | 533.52 | **2,176.99** |

| | | | | | |
|---|---|---|---|---|---|
| 975 | 408.75 | 439.83 | 412.83 | 410.59 | **1,672.00** |
| 976 | 30.58 | 30.65 | 33.20 | 38.45 | **132.88** |
| 977 | 134.93 | 120.17 | 116.65 | 134.78 | **506.52** |
| 978 | 94.03 | 85.16 | 89.62 | 86.55 | **355.35** |
| 979 | 27.37 | 21.35 | 18.47 | 16.66 | **83.84** |
| **OR OREGON Total** | **3,467.73** | **3,462.56** | **3,605.16** | **3,525.82** | **14,061.27** |

**STATE: PA - PENNSYLVANIA**

| REGISTRANT ZIP CODE 3 | 1ST QUARTER | 2ND QUARTER | 3RD QUARTER | 4TH QUARTER | TOTAL GRAMS |
|---|---|---|---|---|---|
| 150 | 268.00 | 260.66 | 302.71 | 314.75 | **1,146.12** |
| 151 | 87.67 | 92.13 | 86.17 | 87.58 | **353.55** |
| 152 | 574.91 | 515.56 | 611.31 | 558.86 | **2,260.64** |
| 153 | 55.26 | 36.07 | 42.03 | 42.82 | **176.18** |
| 154 | 59.33 | 60.25 | 49.20 | 49.15 | **217.93** |
| 155 | 43.47 | 42.73 | 42.94 | 33.40 | **162.54** |
| 156 | 94.98 | 110.77 | 108.38 | 121.88 | **436.00** |
| 157 | 33.25 | 27.62 | 39.43 | 44.00 | **144.29** |
| 158 | 49.88 | 68.49 | 66.63 | 64.19 | **249.18** |
| 159 | 72.30 | 61.72 | 78.90 | 53.99 | **266.92** |
| 160 | 56.08 | 57.02 | 74.55 | 76.83 | **264.49** |
| 161 | 89.03 | 86.62 | 79.08 | 61.56 | **316.29** |
| 162 | 21.84 | 24.88 | 20.71 | 25.28 | **92.72** |
| 163 | 178.89 | 174.43 | 182.69 | 153.80 | **689.80** |
| 164 | 93.49 | 102.44 | 105.35 | 104.71 | **405.99** |
| 165 | 92.54 | 71.57 | 90.50 | 84.69 | **339.30** |
| 166 | 128.31 | 131.09 | 110.58 | 107.41 | **477.40** |
| 167 | 33.88 | 26.99 | 22.21 | 24.94 | **108.02** |
| 168 | 42.70 | 49.83 | 44.15 | 45.26 | **181.94** |
| 169 | 30.84 | 29.77 | 33.31 | 33.91 | **127.84** |
| 170 | 401.09 | 355.36 | 392.63 | 366.91 | **1,516.00** |
| 171 | 125.84 | 113.42 | 127.81 | 87.70 | **454.78** |
| 172 | 112.05 | 105.48 | 100.91 | 99.81 | **418.26** |
| 173 | 180.54 | 179.25 | 186.42 | 180.75 | **726.97** |
| 174 | 95.38 | 131.31 | 95.76 | 100.36 | **422.82** |
| 175 | 125.51 | 145.59 | 140.91 | 153.96 | **565.97** |
| 176 | 155.52 | 229.96 | 212.95 | 189.05 | **787.48** |
| 177 | 67.75 | 69.58 | 83.81 | 76.00 | **297.14** |
| 178 | 155.66 | 182.57 | 183.29 | 219.79 | **741.32** |
| 179 | 31.62 | 31.02 | 31.59 | 69.69 | **163.93** |
| 180 | 328.04 | 283.45 | 336.28 | 264.46 | **1,212.23** |
| 181 | 190.77 | 284.07 | 153.90 | 123.88 | **752.62** |

| | | | | | |
|---|---|---|---|---|---|
| 182 | 33.16 | 38.92 | 45.06 | 31.08 | **148.23** |
| 183 | 110.50 | 119.22 | 118.04 | 114.84 | **462.60** |
| 184 | 137.44 | 155.89 | 138.22 | 129.91 | **561.47** |
| 185 | 105.68 | 106.51 | 112.91 | 118.07 | **443.17** |
| 186 | 83.19 | 63.24 | 71.92 | 65.34 | **283.69** |
| 187 | 478.46 | 483.20 | 461.10 | 506.97 | **1,929.72** |
| 188 | 19.81 | 31.53 | 21.71 | 47.61 | **120.66** |
| 189 | 296.41 | 255.96 | 277.77 | 272.73 | **1,102.87** |
| 190 | 3,247.60 | 2,558.60 | 2,395.71 | 3,593.88 | **11,795.78** |
| 191 | 1,116.65 | 1,181.61 | 996.14 | 1,059.33 | **4,353.74** |
| 193 | 348.39 | 322.05 | 309.77 | 298.55 | **1,278.76** |
| 194 | 402.68 | 465.76 | 467.67 | 512.14 | **1,848.26** |
| 195 | 82.45 | 81.09 | 79.00 | 79.51 | **322.05** |
| 196 | 97.06 | 140.07 | 113.04 | 108.75 | **458.91** |
| **PA - PENNSYLVANIA - Total** | **10,635.89** | **10,145.37** | **9,845.17** | **10,960.10** | **41,586.54** |

**STATE: PR - PUERTO RICO**

| REGISTRANT ZIP CODE 3 | 1ST QUARTER | 2ND QUARTER | 3RD QUARTER | 4TH QUARTER | TOTAL GRAMS |
|---|---|---|---|---|---|
| 006 | 19.51 | 15.25 | 18.09 | 12.24 | **65.08** |
| 007 | 5.32 | 4.96 | 3.37 | 4.61 | **18.26** |
| 009 | 29.72 | 33.69 | 28.89 | 31.37 | **123.66** |
| **PR - PUERTO RICO - Total** | **54.54** | **53.91** | **50.34** | **48.21** | **207.00** |

**STATE: RI - RHODE ISLAND**

| REGISTRANT ZIP CODE 3 | 1ST QUARTER | 2ND QUARTER | 3RD QUARTER | 4TH QUARTER | TOTAL GRAMS |
|---|---|---|---|---|---|
| 028 | 355.28 | 364.25 | 352.91 | 677.24 | **1,749.68** |
| 029 | 250.93 | 248.34 | 242.11 | 264.92 | **1,006.30** |
| **RI RHODE ISLAND Total** | **606.21** | **612.59** | **595.02** | **942.16** | **2,755.98** |

**STATE: SC - SOUTH CAROLINA**

| REGISTRANT ZIP CODE 3 | 1ST QUARTER | 2ND QUARTER | 3RD QUARTER | 4TH QUARTER | TOTAL GRAMS |
|---|---|---|---|---|---|
| 290 | 331.23 | 348.77 | 312.77 | 303.41 | **1,296.19** |
| 291 | 535.72 | 569.42 | 488.10 | 498.98 | **2,092.22** |
| 292 | 402.92 | 432.94 | 419.89 | 383.17 | **1,638.92** |
| 293 | 165.22 | 161.84 | 144.96 | 154.08 | **626.10** |
| 294 | 684.56 | 831.68 | 773.77 | 735.68 | **3,025.69** |
| 295 | 761.04 | 733.45 | 711.16 | 634.07 | **2,839.71** |
| 296 | 585.87 | 551.13 | 518.63 | 632.22 | **2,287.85** |
| 297 | 158.16 | 115.49 | 127.39 | 128.30 | **529.34** |
| 298 | 227.46 | 217.23 | 229.02 | 216.40 | **890.10** |
| 299 | 345.84 | 347.04 | 356.70 | 242.93 | **1,292.50** |
| **SC - SOUTH CAROLINA Total** | **4,198.02** | **4,309.00** | **4,082.37** | **3,929.24** | **16,518.63** |

**STATE: SD - SOUTH DAKOTA**

| REGISTRANT ZIP CODE 3 | 1ST QUARTER | 2ND QUARTER | 3RD QUARTER | 4TH QUARTER | TOTAL GRAMS |
|---|---|---|---|---|---|
| 570 | 54.42 | 61.15 | 61.58 | 70.11 | **247.24** |
| 571 | 186.80 | 206.46 | 220.95 | 223.13 | **837.34** |
| 572 | 17.31 | 22.72 | 17.68 | 19.55 | **77.26** |
| 573 | 15.51 | 17.78 | 21.16 | 20.34 | **74.80** |
| 574 | 8.81 | 10.58 | 12.79 | 13.59 | **45.77** |
| 575 | 14.85 | 10.85 | 13.55 | 8.96 | **48.22** |
| 576 | 5.44 | 3.80 | 5.01 | 5.04 | **19.30** |
| 577 | 68.96 | 72.06 | 71.60 | 60.33 | **272.96** |
| **SD SOUTH DAKOTA - Total** | **372.11** | **405.41** | **424.32** | **421.05** | **1,622.88** |

**STATE: TN - TENNESSEE**

| REGISTRANT ZIP CODE 3 | 1ST QUARTER | 2ND QUARTER | 3RD QUARTER | 4TH QUARTER | TOTAL GRAMS |
|---|---|---|---|---|---|
| 370 | 298.25 | 385.47 | 337.64 | 252.29 | **1,273.65** |
| 371 | 256.40 | 282.32 | 274.74 | 253.04 | **1,066.50** |
| 372 | 430.94 | 515.61 | 391.12 | 551.15 | **1,888.82** |
| 373 | 377.01 | 459.48 | 458.58 | 340.59 | **1,635.67** |
| 374 | 195.08 | 179.24 | 174.57 | 194.33 | **743.21** |
| 376 | 219.38 | 217.30 | 229.41 | 195.57 | **861.65** |
| 377 | 73.49 | 88.79 | 89.00 | 107.12 | **358.40** |
| 378 | 148.15 | 139.65 | 157.18 | 163.39 | **608.37** |
| 379 | 616.21 | 437.21 | 496.42 | 580.22 | **2,130.06** |
| 380 | 182.44 | 143.79 | 133.72 | 187.35 | **647.30** |
| 381 | 2,120.01 | 1,715.15 | 1,682.76 | 2,920.47 | **8,438.40** |
| 382 | 30.79 | 26.15 | 36.42 | 27.09 | **120.44** |
| 383 | 72.40 | 74.68 | 68.87 | 74.81 | **290.76** |
| 384 | 41.96 | 57.74 | 66.34 | 49.95 | **215.99** |
| 385 | 98.26 | 77.62 | 72.60 | 79.90 | **328.37** |
| **TN TENNESSEE Total** | **5,160.77** | **4,800.19** | **4,669.36** | **5,977.27** | **20,607.60** |

**STATE: TX - TEXAS**

| REGISTRANT ZIP CODE 3 | 1ST QUARTER | 2ND QUARTER | 3RD QUARTER | 4TH QUARTER | TOTAL GRAMS |
|---|---|---|---|---|---|
| 750 | 1,663.35 | 1,671.13 | 1,579.82 | 1,513.31 | **6,427.61** |
| 751 | 354.59 | 390.81 | 355.54 | 340.62 | **1,441.56** |
| 752 | 779.66 | 1,656.84 | 1,628.99 | 1,923.29 | **5,988.78** |
| 753 | 82.38 | 85.51 | 98.11 | 101.26 | **367.27** |
| 754 | 336.51 | 347.84 | 317.38 | 318.93 | **1,320.65** |
| 755 | 75.21 | 55.40 | 87.57 | 62.26 | **280.44** |
| 756 | 671.81 | 1,026.83 | 161.05 | 335.32 | **2,195.00** |
| 757 | 233.58 | 319.30 | 228.41 | 307.83 | **1,089.12** |
| 758 | 27.27 | 21.29 | 30.63 | 31.08 | **110.27** |
| 759 | 119.66 | 140.85 | 119.25 | 106.67 | **486.44** |

| | | | | | |
|---|---|---|---|---|---|
| 760 | 696.58 | 647.40 | 641.56 | 630.01 | **2,615.55** |
| 761 | 704.47 | 940.38 | 785.69 | 631.64 | **3,062.19** |
| 762 | 448.07 | 789.55 | 761.30 | 584.20 | **2,583.13** |
| 763 | 174.25 | 162.69 | 139.50 | 161.23 | **637.67** |
| 764 | 73.17 | 74.25 | 76.91 | 67.22 | **291.54** |
| 765 | 350.74 | 315.01 | 299.00 | 291.08 | **1,255.83** |
| 766 | 36.93 | 41.04 | 45.44 | 39.25 | **162.67** |
| 767 | 61.94 | 58.92 | 77.51 | 83.42 | **281.79** |
| 768 | 45.58 | 47.48 | 233.36 | 221.27 | **547.69** |
| 769 | 42.50 | 33.27 | 34.16 | 33.66 | **143.58** |
| 770 | 1,751.58 | 1,726.73 | 1,704.59 | 1,919.60 | **7,102.51** |
| 773 | 766.05 | 696.68 | 683.73 | 667.83 | **2,814.29** |
| 774 | 489.32 | 545.06 | 546.68 | 673.78 | **2,254.84** |
| 775 | 742.53 | 703.94 | 679.05 | 644.77 | **2,770.28** |
| 776 | 374.93 | 328.47 | 311.28 | 318.71 | **1,333.39** |
| 777 | 248.41 | 296.64 | 273.43 | 300.87 | **1,119.35** |
| 778 | 82.25 | 64.80 | 77.49 | 77.96 | **302.51** |
| 779 | 277.78 | 296.12 | 260.75 | 263.61 | **1,098.26** |
| 780 | 169.87 | 147.98 | 156.27 | 160.13 | **634.25** |
| 781 | 240.87 | 224.43 | 227.57 | 229.04 | **921.91** |
| 782 | 1,249.89 | 1,173.40 | 1,204.05 | 1,111.83 | **4,739.18** |
| 783 | 99.29 | 94.99 | 113.95 | 89.16 | **397.39** |
| 784 | 198.48 | 180.39 | 171.12 | 183.29 | **733.27** |
| 785 | 263.78 | 242.87 | 238.98 | 257.29 | **1,002.92** |
| 786 | 552.46 | 538.11 | 548.60 | 511.93 | **2,151.10** |
| 787 | 451.74 | 671.17 | 662.41 | 485.30 | **2,270.62** |
| 788 | 37.00 | 32.80 | 31.70 | 25.01 | **126.50** |
| 789 | 28.56 | 24.19 | 21.07 | 20.12 | **93.95** |
| 790 | 39.86 | 38.09 | 38.63 | 37.78 | **154.36** |
| 791 | 110.38 | 110.22 | 118.67 | 110.06 | **449.32** |
| 792 | 2.85 | 4.96 | 7.86 | 10.11 | **25.79** |
| 793 | 60.14 | 62.01 | 57.96 | 65.85 | **245.96** |
| 794 | 287.14 | 291.88 | 358.87 | 358.31 | **1,296.20** |
| 795 | 30.45 | 29.81 | 28.01 | 28.32 | **116.60** |
| 796 | 65.33 | 61.42 | 63.14 | 64.14 | **254.03** |
| 797 | 134.25 | 105.55 | 116.27 | 128.51 | **484.58** |
| 798 | 10.31 | 6.34 | 6.54 | 8.55 | **31.73** |
| 799 | 294.61 | 278.62 | 312.85 | 296.70 | **1,182.77** |
| **TX - TEXAS Total** | **16,038.37** | **17,803.44** | **16,722.72** | **16,832.13** | **67,396.65** |

**STATE: UT - UTAH**

| REGISTRANT ZIP CODE 3 | 1ST QUARTER | 2ND QUARTER | 3RD QUARTER | 4TH QUARTER | **TOTAL GRAMS** |
|---|---|---|---|---|---|
| 840 | 813.87 | 746.96 | 1,075.54 | 792.56 | **3,428.93** |
| 841 | 677.98 | 726.36 | 619.56 | 816.17 | **2,840.07** |
| 843 | 140.26 | 127.22 | 116.49 | 132.83 | **516.80** |
| 844 | 168.33 | 161.53 | 166.23 | 186.76 | **682.86** |
| 845 | 20.25 | 20.78 | 24.93 | 19.31 | **85.28** |
| 846 | 209.26 | 213.48 | 205.72 | 196.43 | **824.89** |
| 847 | 217.56 | 254.52 | 228.37 | 230.73 | **931.19** |
| **UT - UTAH - Total** | **2,247.51** | **2,250.85** | **2,436.85** | **2,374.80** | **9,310.02** |

**STATE: VA - VIRGINIA**

| REGISTRANT ZIP CODE 3 | 1ST QUARTER | 2ND QUARTER | 3RD QUARTER | 4TH QUARTER | **TOTAL GRAMS** |
|---|---|---|---|---|---|
| 201 | 729.06 | 723.86 | 775.93 | 642.34 | **2,871.19** |
| 220 | 345.33 | 287.18 | 343.09 | 318.71 | **1,294.32** |
| 221 | 256.67 | 245.06 | 230.08 | 235.95 | **967.76** |
| 222 | 139.16 | 152.93 | 130.11 | 140.97 | **563.17** |
| 223 | 142.79 | 111.34 | 147.56 | 145.29 | **546.98** |
| 224 | 270.92 | 230.32 | 239.09 | 263.48 | **1,003.81** |
| 225 | 80.04 | 81.04 | 82.01 | 94.82 | **337.91** |
| 226 | 310.22 | 308.28 | 314.20 | 289.02 | **1,221.72** |
| 227 | 46.13 | 51.67 | 56.17 | 56.20 | **210.16** |
| 228 | 207.20 | 231.14 | 219.76 | 218.11 | **876.21** |
| 229 | 401.42 | 319.74 | 384.03 | 421.65 | **1,526.84** |
| 230 | 184.30 | 176.01 | 171.91 | 173.85 | **706.06** |
| 231 | 409.09 | 404.53 | 388.57 | 382.95 | **1,585.14** |
| 232 | 840.15 | 725.13 | 761.63 | 739.06 | **3,065.97** |
| 233 | 342.44 | 418.90 | 341.36 | 394.38 | **1,497.08** |
| 234 | 653.06 | 648.45 | 635.37 | 640.14 | **2,577.01** |
| 235 | 328.89 | 385.38 | 359.87 | 338.73 | **1,412.87** |
| 236 | 367.38 | 325.33 | 333.30 | 350.40 | **1,376.41** |
| 237 | 146.90 | 152.18 | 147.65 | 138.69 | **585.42** |
| 238 | 285.71 | 204.57 | 194.09 | 184.68 | **869.06** |
| 239 | 60.77 | 57.10 | 53.36 | 40.92 | **212.15** |
| 240 | 416.88 | 457.99 | 495.32 | 393.37 | **1,763.56** |
| 241 | 258.19 | 276.16 | 256.82 | 251.68 | **1,042.85** |
| 242 | 143.34 | 174.15 | 178.95 | 186.62 | **683.07** |
| 243 | 115.55 | 109.59 | 94.55 | 90.55 | **410.24** |
| 244 | 125.92 | 159.28 | 134.89 | 134.13 | **554.21** |
| 245 | 250.75 | 234.30 | 237.48 | 237.04 | **959.56** |
| 246 | 58.83 | 50.72 | 42.80 | 35.37 | **187.72** |
| **VA VIRGINIA Total** | **7,917.08** | **7,702.34** | **7,749.94** | **7,539.10** | **30,908.45** |

**STATE: VI - VIRGIN ISLANDS**

| REGISTRANT ZIP CODE 3 | 1ST QUARTER | 2ND QUARTER | 3RD QUARTER | 4TH QUARTER | TOTAL GRAMS |
|---|---|---|---|---|---|
| 008 | 12.57 | 20.44 | 7.91 | 19.95 | **60.87** |
| **VI - VIRGIN ISLANDS Total** | **12.57** | **20.44** | **7.91** | **19.95** | **60.87** |

**STATE: VT - VERMONT**

| REGISTRANT ZIP CODE 3 | 1ST QUARTER | 2ND QUARTER | 3RD QUARTER | 4TH QUARTER | TOTAL GRAMS |
|---|---|---|---|---|---|
| 050 | 36.64 | 45.66 | 28.52 | 34.17 | **145.00** |
| 051 | 32.51 | 32.41 | 32.54 | 33.17 | **130.63** |
| 052 | 15.79 | 20.35 | 21.41 | 20.10 | **77.64** |
| 053 | 27.10 | 21.43 | 23.16 | 19.58 | **91.27** |
| 054 | 474.96 | 468.44 | 547.22 | 479.71 | **1,970.32** |
| 056 | 109.83 | 140.41 | 104.21 | 100.85 | **455.30** |
| 057 | 127.68 | 176.44 | 180.41 | 125.84 | **610.38** |
| 058 | 40.45 | 47.82 | 59.09 | 52.21 | **199.58** |
| **VT VERMONT Total** | **864.97** | **952.96** | **996.56** | **865.62** | **3,680.11** |

**STATE: WA - WASHINGTON**

| REGISTRANT ZIP CODE 3 | 1ST QUARTER | 2ND QUARTER | 3RD QUARTER | 4TH QUARTER | TOTAL GRAMS |
|---|---|---|---|---|---|
| 980 | 1,254.83 | 1,304.97 | 1,267.18 | 1,254.48 | **5,081.46** |
| 981 | 1,234.42 | 1,301.59 | 1,066.37 | 1,240.41 | **4,842.79** |
| 982 | 957.14 | 1,056.09 | 1,049.34 | 975.65 | **4,038.22** |
| 983 | 647.36 | 712.11 | 759.48 | 762.99 | **2,881.95** |
| 984 | 358.83 | 394.41 | 385.70 | 414.63 | **1,553.57** |
| 985 | 567.89 | 577.28 | 564.37 | 581.19 | **2,290.74** |
| 986 | 765.06 | 773.22 | 740.83 | 790.24 | **3,069.35** |
| 988 | 142.88 | 171.73 | 187.05 | 168.13 | **669.79** |
| 989 | 248.21 | 250.56 | 238.77 | 218.46 | **956.01** |
| 990 | 81.14 | 82.69 | 92.86 | 87.14 | **343.82** |
| 991 | 91.49 | 99.10 | 98.42 | 95.68 | **384.69** |
| 992 | 604.98 | 657.18 | 655.32 | 662.54 | **2,580.01** |
| 993 | 351.85 | 341.14 | 320.81 | 304.29 | **1,318.08** |
| 994 | 52.60 | 55.47 | 39.00 | 141.62 | **288.68** |
| **WA WASHINGTON Total** | **7,358.68** | **7,777.55** | **7,465.50** | **7,697.44** | **30,299.17** |

**STATE: WI - WISCONSIN**

| REGISTRANT ZIP CODE 3 | 1ST QUARTER | 2ND QUARTER | 3RD QUARTER | 4TH QUARTER | TOTAL GRAMS |
|---|---|---|---|---|---|
| 530 | 212.79 | 182.43 | 204.57 | 257.96 | **857.77** |
| 531 | 332.03 | 283.88 | 227.59 | 242.69 | **1,086.19** |
| 532 | 590.34 | 512.97 | 564.97 | 412.23 | **2,080.51** |
| 534 | 51.71 | 52.63 | 41.11 | 35.64 | **181.09** |
| 535 | 208.51 | 203.16 | 219.66 | 186.92 | **818.24** |
| 537 | 205.07 | 206.18 | 192.83 | 232.21 | **836.29** |

| | | | | | |
|---|---|---|---|---|---|
| 538 | 27.95 | 31.87 | 21.25 | 21.09 | **102.16** |
| 539 | 43.15 | 48.95 | 65.18 | 59.69 | **216.97** |
| 540 | 80.41 | 73.63 | 78.04 | 83.44 | **315.52** |
| 541 | 38.46 | 35.35 | 43.02 | 38.04 | **154.86** |
| 542 | 15.91 | 21.12 | 24.27 | 16.38 | **77.68** |
| 543 | 93.24 | 94.46 | 148.99 | 121.67 | **458.36** |
| 544 | 303.84 | 262.01 | 114.97 | 97.92 | **778.74** |
| 545 | 51.45 | 62.67 | 58.25 | 61.78 | **234.16** |
| 546 | 106.13 | 107.64 | 109.43 | 126.55 | **449.74** |
| 547 | 92.87 | 129.67 | 163.16 | 86.61 | **472.30** |
| 548 | 121.33 | 105.95 | 102.53 | 110.08 | **439.89** |
| 549 | 160.06 | 229.64 | 232.00 | 200.64 | **822.34** |
| **WI - WISCONSIN Total** | **2,735.25** | **2,644.19** | **2,611.82** | **2,391.55** | **10,382.81** |

**STATE: WV - WEST VIRGINIA**

| REGISTRANT ZIP CODE 3 | 1ST QUARTER | 2ND QUARTER | 3RD QUARTER | 4TH QUARTER | TOTAL GRAMS |
|---|---|---|---|---|---|
| 247 | 51.41 | 41.33 | 43.07 | 42.10 | **177.92** |
| 248 | 6.38 | 4.88 | 7.45 | 5.14 | **23.85** |
| 249 | 18.92 | 13.78 | 15.63 | 12.90 | **61.24** |
| 250 | 5.64 | 5.25 | 4.50 | 4.33 | **19.72** |
| 251 | 6.91 | 7.70 | 10.77 | 11.46 | **36.83** |
| 252 | 2.99 | 0.95 | 1.76 | 0.92 | **6.63** |
| 253 | 87.77 | 111.62 | 50.18 | 61.64 | **311.21** |
| 254 | 273.35 | 254.93 | 262.64 | 245.40 | **1,036.32** |
| 255 | 52.02 | 32.03 | 27.36 | 26.94 | **138.35** |
| 256 | 4.26 | 5.36 | 4.95 | 3.95 | **18.52** |
| 257 | 68.71 | 84.56 | 79.19 | 85.96 | **318.42** |
| 258 | 87.06 | 52.89 | 67.43 | 107.62 | **315.00** |
| 259 | 4.96 | 6.52 | 7.45 | 7.36 | **26.29** |
| 260 | 45.13 | 46.18 | 63.46 | 47.86 | **202.63** |
| 261 | 25.77 | 31.24 | 26.58 | 23.56 | **107.14** |
| 262 | 10.29 | 20.21 | 13.39 | 9.80 | **53.69** |
| 263 | 53.51 | 54.63 | 50.51 | 41.95 | **200.60** |
| 264 | 26.64 | 22.80 | 13.39 | 11.72 | **74.55** |
| 265 | 91.06 | 155.55 | 157.20 | 121.62 | **525.43** |
| 266 | 2.84 | 1.17 | 2.77 | 1.76 | **8.54** |
| 267 | 23.92 | 26.27 | 26.99 | 31.23 | **108.41** |
| 268 | 18.12 | 17.63 | 24.85 | 24.48 | **85.08** |
| **WV WEST VIRGINIA Total** | **967.65** | **997.47** | **961.53** | **929.70** | **3,856.36** |

**STATE: WY - WYOMING**

| REGISTRANT ZIP CODE 3 | 1ST QUARTER | 2ND QUARTER | 3RD QUARTER | 4TH QUARTER | TOTAL GRAMS |
|---|---|---|---|---|---|

| | | | | | |
|---|---|---|---|---|---|
| 820 | 87.56 | 95.24 | 130.33 | 91.59 | **404.72** |
| 822 | 7.63 | 4.37 | 9.16 | 4.62 | **25.78** |
| 823 | 3.81 | 6.56 | 4.61 | 4.26 | **19.24** |
| 824 | 100.25 | 86.85 | 95.69 | 98.22 | **381.01** |
| 825 | 28.01 | 25.29 | 18.74 | 12.11 | **84.15** |
| 826 | 69.23 | 65.02 | 77.58 | 63.74 | **275.57** |
| 827 | 20.39 | 27.83 | 43.70 | 46.16 | **138.07** |
| 828 | 45.04 | 44.46 | 42.34 | 44.53 | **176.37** |
| 829 | 65.54 | 67.62 | 114.89 | 212.16 | **460.21** |
| 830 | 7.88 | 7.93 | 9.06 | 7.37 | **32.24** |
| 831 | 24.38 | 22.87 | 27.40 | 24.56 | **99.21** |
| **WY - WYOMING Total** | **459.72** | **454.05** | **573.50** | **609.31** | **2,096.58** |

**DRUG: 9193 - HYDROCODONE**

**STATE: AK - ALASKA**

| REGISTRANT ZIP CODE 3 | 1ST QUARTER | 2ND QUARTER | 3RD QUARTER | 4TH QUARTER | TOTAL GRAMS |
|---|---|---|---|---|---|
| 995 | 3,799.54 | 3,792.68 | 3,673.07 | 3,688.56 | **14,953.85** |
| 996 | 2,922.80 | 2,958.83 | 2,944.98 | 2,776.56 | **11,603.17** |
| 997 | 806.05 | 787.36 | 865.79 | 766.34 | **3,225.54** |
| 998 | 405.76 | 391.28 | 380.27 | 363.89 | **1,541.20** |
| 999 | 347.01 | 307.54 | 332.35 | 341.18 | **1,328.08** |
| **AK ALASKA - Total** | **8,281.17** | **8,237.70** | **8,196.46** | **7,936.52** | **32,651.84** |

**STATE: AL - ALABAMA**

| REGISTRANT ZIP CODE 3 | 1ST QUARTER | 2ND QUARTER | 3RD QUARTER | 4TH QUARTER | TOTAL GRAMS |
|---|---|---|---|---|---|
| 350 | 21,659.54 | 21,477.22 | 20,702.26 | 21,027.66 | **84,866.68** |
| 351 | 11,920.16 | 11,813.42 | 11,756.53 | 11,576.19 | **47,066.30** |
| 352 | 18,572.93 | 17,841.10 | 17,574.42 | 17,343.51 | **71,331.95** |
| 354 | 8,764.73 | 8,512.60 | 8,642.08 | 8,097.99 | **34,017.41** |
| 355 | 9,182.33 | 9,334.42 | 9,155.71 | 8,903.37 | **36,575.82** |
| 356 | 19,342.88 | 19,497.51 | 19,003.04 | 18,857.69 | **76,701.13** |
| 357 | 7,419.52 | 7,300.43 | 7,360.26 | 7,026.94 | **29,107.15** |
| 358 | 7,993.48 | 7,574.88 | 7,812.95 | 7,120.02 | **30,501.34** |
| 359 | 17,008.12 | 17,449.29 | 16,703.14 | 16,970.81 | **68,131.36** |
| 360 | 8,428.58 | 8,239.72 | 7,886.76 | 7,906.96 | **32,462.02** |
| 361 | 6,422.11 | 6,096.47 | 6,242.01 | 6,138.40 | **24,898.99** |
| 362 | 7,704.34 | 7,589.57 | 8,048.27 | 7,738.67 | **31,080.85** |
| 363 | 7,999.25 | 7,675.58 | 7,781.09 | 7,603.17 | **31,059.08** |
| 364 | 4,799.08 | 4,476.00 | 4,821.64 | 4,448.23 | **18,544.95** |
| 365 | 15,175.00 | 14,839.94 | 14,850.33 | 14,515.40 | **59,380.66** |
| 366 | 11,835.57 | 11,237.08 | 11,323.23 | 11,246.93 | **45,642.82** |
| 367 | 2,686.17 | 2,364.61 | 2,200.08 | 2,259.91 | **9,510.78** |

| | | | | | |
|---|---|---|---|---|---|
| 368 | 6,464.29 | 6,522.39 | 6,307.45 | 6,218.80 | **25,512.94** |
| 369 | 728.00 | 703.33 | 731.04 | 745.94 | **2,908.31** |
| **AL ALABAMA - Total** | **194,106.07** | **190,545.56** | **188,902.31** | **185,746.59** | **759,300.53** |

**STATE: AR - ARKANSAS**

| REGISTRANT ZIP CODE 3 | 1ST QUARTER | 2ND QUARTER | 3RD QUARTER | 4TH QUARTER | TOTAL GRAMS |
|---|---|---|---|---|---|
| 716 | 4,882.78 | 4,823.23 | 4,654.45 | 4,544.57 | **18,905.03** |
| 717 | 4,283.62 | 4,300.32 | 4,310.43 | 4,360.85 | **17,255.21** |
| 718 | 6,455.45 | 6,479.19 | 6,102.64 | 5,974.11 | **25,011.39** |
| 719 | 8,432.47 | 7,942.63 | 8,298.46 | 8,141.59 | **32,815.15** |
| 720 | 10,313.83 | 10,074.17 | 10,099.15 | 9,793.13 | **40,280.28** |
| 721 | 11,833.99 | 11,761.74 | 11,789.06 | 11,873.81 | **47,258.61** |
| 722 | 6,201.24 | 6,237.44 | 6,046.30 | 5,943.78 | **24,428.75** |
| 723 | 5,684.15 | 5,676.52 | 5,756.66 | 5,790.94 | **22,908.28** |
| 724 | 9,352.67 | 9,182.79 | 8,975.45 | 8,844.93 | **36,355.85** |
| 725 | 4,941.38 | 5,093.15 | 5,038.92 | 4,827.54 | **19,900.98** |
| 726 | 4,405.65 | 4,451.00 | 4,383.44 | 4,503.73 | **17,743.82** |
| 727 | 12,211.19 | 11,793.72 | 11,774.48 | 11,492.94 | **47,272.33** |
| 728 | 3,674.77 | 3,559.12 | 3,538.65 | 3,334.77 | **14,107.30** |
| 729 | 11,090.00 | 10,933.95 | 10,979.06 | 10,742.55 | **43,745.56** |
| **AR ARKANSAS - Total** | **103,763.17** | **102,308.99** | **101,747.15** | **100,169.23** | **407,988.54** |

**STATE: AS - AMERICAN SAMOA**

| REGISTRANT ZIP CODE 3 | 1ST QUARTER | 2ND QUARTER | 3RD QUARTER | 4TH QUARTER | TOTAL GRAMS |
|---|---|---|---|---|---|
| 967 | 0 | 0.61 | 0 | 3.03 | **3.63** |
| **AS - AMERICAN SAMOA - Tota** | **0.00** | **0.61** | **0.00** | **3.03** | **3.63** |

**STATE: AZ - ARIZONA**

| REGISTRANT ZIP CODE 3 | 1ST QUARTER | 2ND QUARTER | 3RD QUARTER | 4TH QUARTER | TOTAL GRAMS |
|---|---|---|---|---|---|
| 850 | 10,835.69 | 10,647.25 | 10,551.46 | 10,671.77 | **42,706.18** |
| 851 | 4,560.32 | 4,177.50 | 4,103.64 | 4,350.35 | **17,191.81** |
| 852 | 18,105.58 | 17,688.74 | 16,771.69 | 17,348.06 | **69,914.07** |
| 853 | 13,896.87 | 13,304.88 | 12,831.70 | 13,496.31 | **53,529.76** |
| 855 | 1,569.12 | 1,571.36 | 1,594.69 | 1,605.73 | **6,340.90** |
| 856 | 3,131.61 | 2,962.17 | 2,939.48 | 2,860.90 | **11,894.15** |
| 857 | 10,117.68 | 10,106.57 | 9,727.62 | 9,939.08 | **39,890.95** |
| 859 | 1,926.57 | 2,009.09 | 2,240.87 | 1,936.15 | **8,112.68** |
| 860 | 1,447.03 | 1,437.90 | 1,343.88 | 1,313.52 | **5,542.34** |
| 863 | 5,380.13 | 5,553.46 | 5,467.57 | 5,548.44 | **21,949.60** |
| 864 | 6,938.64 | 6,664.03 | 6,634.03 | 6,725.20 | **26,961.90** |
| 865 | 65.21 | 69.46 | 84.27 | 87.16 | **306.10** |
| **AZ ARIZONA - Total** | **77,974.44** | **76,192.40** | **74,290.92** | **75,882.68** | **304,340.44** |

**STATE: CA - CALIFORNIA**

| REGISTRANT ZIP CODE 3 | 1ST QUARTER | 2ND QUARTER | 3RD QUARTER | 4TH QUARTER | TOTAL GRAMS |
|---|---|---|---|---|---|
| 900 | 14,679.26 | 14,315.62 | 14,074.37 | 13,805.72 | **56,874.98** |
| 902 | 12,868.94 | 12,646.39 | 12,392.46 | 12,497.56 | **50,405.36** |
| 903 | 1,363.40 | 1,295.52 | 1,238.63 | 1,225.49 | **5,123.05** |
| 904 | 1,390.02 | 1,286.60 | 1,216.07 | 1,258.06 | **5,150.75** |
| 905 | 3,271.77 | 3,070.92 | 3,029.03 | 2,981.74 | **12,353.47** |
| 906 | 9,874.93 | 9,776.68 | 9,786.90 | 9,758.85 | **39,197.35** |
| 907 | 9,553.82 | 9,410.44 | 9,028.85 | 8,996.68 | **36,989.79** |
| 908 | 6,168.74 | 5,993.98 | 5,825.70 | 5,750.94 | **23,739.35** |
| 910 | 3,309.71 | 3,267.69 | 3,242.70 | 3,093.39 | **12,913.49** |
| 911 | 1,979.77 | 1,776.52 | 1,861.48 | 1,825.10 | **7,442.87** |
| 912 | 2,413.89 | 2,256.83 | 2,274.78 | 2,338.17 | **9,283.67** |
| 913 | 19,328.45 | 18,610.86 | 18,553.97 | 17,960.57 | **74,453.84** |
| 914 | 5,392.00 | 4,981.58 | 5,043.48 | 4,708.60 | **20,125.66** |
| 915 | 1,539.58 | 1,587.31 | 1,629.42 | 1,561.79 | **6,318.11** |
| 916 | 1,129.64 | 1,099.11 | 1,150.16 | 1,176.18 | **4,555.09** |
| 917 | 29,584.45 | 28,491.71 | 26,315.85 | 26,167.69 | **110,559.70** |
| 918 | 776.57 | 769.53 | 806.46 | 753.15 | **3,105.71** |
| 919 | 6,564.49 | 6,393.25 | 6,215.78 | 6,267.61 | **25,441.13** |
| 920 | 21,575.84 | 21,725.41 | 21,327.91 | 20,608.66 | **85,237.82** |
| 921 | 14,475.97 | 13,769.39 | 13,425.01 | 12,938.48 | **54,608.85** |
| 922 | 16,200.56 | 15,858.70 | 15,493.20 | 15,734.80 | **63,287.25** |
| 923 | 36,410.29 | 35,863.01 | 35,586.85 | 35,328.63 | **143,188.79** |
| 924 | 5,806.70 | 5,455.74 | 5,679.08 | 5,579.63 | **22,521.15** |
| 925 | 32,075.44 | 32,244.58 | 31,509.45 | 31,269.78 | **127,099.25** |
| 926 | 18,019.00 | 17,868.72 | 17,760.06 | 17,418.64 | **71,066.42** |
| 927 | 5,630.77 | 5,599.57 | 5,464.68 | 5,475.30 | **22,170.32** |
| 928 | 19,217.45 | 19,183.16 | 19,132.11 | 18,764.58 | **76,297.30** |
| 930 | 12,855.96 | 12,938.26 | 12,673.64 | 12,404.76 | **50,872.62** |
| 931 | 2,125.62 | 2,061.35 | 2,032.61 | 1,932.56 | **8,152.15** |
| 932 | 18,340.83 | 18,339.08 | 18,110.13 | 18,158.71 | **72,948.74** |
| 933 | 16,249.33 | 16,471.34 | 16,218.46 | 15,648.29 | **64,587.41** |
| 934 | 11,417.17 | 11,571.94 | 11,244.16 | 11,286.14 | **45,519.41** |
| 935 | 14,116.65 | 13,962.10 | 13,660.46 | 13,643.94 | **55,383.16** |
| 936 | 13,151.19 | 13,239.83 | 12,850.36 | 12,770.59 | **52,011.97** |
| 937 | 16,533.32 | 16,656.51 | 16,391.14 | 15,838.65 | **65,419.63** |
| 939 | 5,833.44 | 5,629.61 | 5,410.59 | 5,333.85 | **22,207.49** |
| 940 | 7,159.13 | 6,999.82 | 7,029.59 | 6,593.74 | **27,782.28** |
| 941 | 6,376.68 | 6,141.85 | 5,857.26 | 5,769.88 | **24,145.67** |
| 943 | 1,335.31 | 1,584.15 | 1,445.42 | 1,335.19 | **5,700.07** |

| | | | | | |
|---|---|---|---|---|---|
| 944 | 1,295.40 | 1,264.00 | 1,202.60 | 1,149.15 | **4,911.15** |
| 945 | 46,887.00 | 45,830.58 | 45,350.78 | 44,302.01 | **182,370.37** |
| 946 | 6,010.56 | 5,970.87 | 5,630.06 | 5,330.60 | **22,942.09** |
| 947 | 906.69 | 892.10 | 862.70 | 815.19 | **3,476.68** |
| 948 | 2,373.99 | 2,599.28 | 2,445.98 | 2,191.19 | **9,610.44** |
| 949 | 6,551.93 | 6,345.62 | 6,358.45 | 6,200.15 | **25,456.16** |
| 950 | 9,332.82 | 9,131.10 | 9,010.98 | 8,945.91 | **36,420.82** |
| 951 | 6,511.29 | 6,295.15 | 6,363.84 | 5,973.79 | **25,144.07** |
| 952 | 13,325.10 | 12,708.04 | 11,726.23 | 12,218.97 | **49,978.34** |
| 953 | 36,254.78 | 36,834.83 | 35,408.54 | 35,531.92 | **144,030.07** |
| 954 | 12,951.41 | 12,639.54 | 12,355.47 | 12,189.43 | **50,135.85** |
| 955 | 5,868.06 | 5,686.69 | 5,392.46 | 5,210.31 | **22,157.52** |
| 956 | 41,005.76 | 41,379.50 | 40,011.14 | 39,660.48 | **162,056.88** |
| 957 | 6,933.32 | 6,974.61 | 6,944.55 | 6,819.27 | **27,671.75** |
| 958 | 23,968.35 | 23,764.69 | 23,433.96 | 22,284.90 | **93,451.91** |
| 959 | 21,846.85 | 21,660.18 | 19,920.61 | 19,253.99 | **82,681.62** |
| 960 | 14,732.24 | 14,373.84 | 14,004.20 | 13,474.64 | **56,584.93** |
| 961 | 2,300.12 | 2,102.85 | 2,058.41 | 2,050.46 | **8,511.85** |
| **CA - CALIFORNIA - Total** | **685,151.80** | **676,648.13** | **660,469.26** | **649,564.43** | **2,671,833.62** |

**STATE: CO - COLORADO**

| REGISTRANT ZIP CODE 3 | 1ST QUARTER | 2ND QUARTER | 3RD QUARTER | 4TH QUARTER | **TOTAL GRAMS** |
|---|---|---|---|---|---|
| 800 | 7,578.88 | 7,376.04 | 7,247.89 | 7,164.82 | **29,367.64** |
| 801 | 5,885.65 | 5,935.75 | 5,902.71 | 5,858.09 | **23,582.20** |
| 802 | 6,741.61 | 6,603.86 | 6,488.07 | 6,351.37 | **26,184.91** |
| 803 | 742.35 | 683.22 | 645.08 | 691.05 | **2,761.70** |
| 804 | 1,537.46 | 1,395.31 | 1,440.00 | 1,390.70 | **5,763.47** |
| 805 | 5,099.40 | 4,996.45 | 5,077.43 | 4,974.59 | **20,147.86** |
| 806 | 2,047.53 | 2,004.46 | 1,984.79 | 1,901.29 | **7,938.07** |
| 807 | 714.52 | 690.54 | 694.22 | 663.11 | **2,762.39** |
| 808 | 1,280.67 | 1,243.89 | 1,200.27 | 1,179.86 | **4,904.68** |
| 809 | 5,909.72 | 5,871.93 | 5,826.00 | 5,781.40 | **23,389.04** |
| 810 | 4,436.63 | 4,370.86 | 4,407.73 | 4,231.47 | **17,446.69** |
| 811 | 584.32 | 557.97 | 615.89 | 557.16 | **2,315.35** |
| 812 | 1,126.37 | 1,151.59 | 1,126.92 | 1,100.45 | **4,505.33** |
| 813 | 844.67 | 848.16 | 896.15 | 778.99 | **3,367.97** |
| 814 | 865.11 | 776.00 | 814.18 | 751.41 | **3,206.70** |
| 815 | 2,004.96 | 1,944.92 | 1,887.09 | 1,871.45 | **7,708.42** |
| 816 | 1,097.48 | 1,050.73 | 1,034.00 | 1,012.88 | **4,195.09** |
| **CO - COLORADO - Total** | **48,497.32** | **47,501.70** | **47,288.42** | **46,260.08** | **189,547.52** |

**STATE: CT - CONNECTICUT**

| REGISTRANT ZIP CODE 3 | 1ST QUARTER | 2ND QUARTER | 3RD QUARTER | 4TH QUARTER | TOTAL GRAMS |
|---|---|---|---|---|---|
| 060 | 3,223.51 | 3,157.95 | 3,027.89 | 3,088.95 | **12,498.31** |
| 061 | 866.82 | 875.44 | 721.43 | 775.23 | **3,238.93** |
| 062 | 810.82 | 752.46 | 748.31 | 753.93 | **3,065.52** |
| 063 | 1,613.67 | 1,601.89 | 1,555.48 | 1,584.37 | **6,355.41** |
| 064 | 3,747.72 | 3,629.08 | 3,589.43 | 3,547.95 | **14,514.18** |
| 065 | 954.35 | 875.66 | 844.03 | 937.90 | **3,611.94** |
| 066 | 672.33 | 649.26 | 631.68 | 621.37 | **2,574.64** |
| 067 | 1,954.59 | 1,889.68 | 1,918.12 | 1,868.00 | **7,630.39** |
| 068 | 1,157.18 | 1,044.52 | 1,113.67 | 1,144.44 | **4,459.81** |
| 069 | 242.64 | 206.78 | 230.00 | 207.65 | **887.08** |
| **CT CONNECTICUT Total** | **15,243.64** | **14,682.74** | **14,380.04** | **14,529.80** | **58,836.21** |

**STATE: DC - DISTRICT OF COLUMBIA**

| REGISTRANT ZIP CODE 3 | 1ST QUARTER | 2ND QUARTER | 3RD QUARTER | 4TH QUARTER | TOTAL GRAMS |
|---|---|---|---|---|---|
| 200 | 972.72 | 910.17 | 853.69 | 853.93 | **3,590.52** |
| 203 | 4.54 | 1.51 | 14.23 | 1.21 | **21.49** |
| 204 | 77.49 | 58.12 | 83.55 | 49.95 | **269.10** |
| 205 | 3.33 | 0 | 0.61 | 0 | **3.94** |
| **DC - DISTRICT OF COLUMBIA** | **1,058.09** | **969.80** | **952.07** | **905.09** | **3,885.04** |

**STATE: DE - DELAWARE**

| REGISTRANT ZIP CODE 3 | 1ST QUARTER | 2ND QUARTER | 3RD QUARTER | 4TH QUARTER | TOTAL GRAMS |
|---|---|---|---|---|---|
| 197 | 1,245.28 | 1,270.05 | 1,181.78 | 1,157.63 | **4,854.74** |
| 198 | 798.20 | 774.23 | 811.25 | 703.38 | **3,087.06** |
| 199 | 2,413.62 | 2,667.46 | 2,437.74 | 2,318.30 | **9,837.12** |
| **DE DELAWARE Total** | **4,457.11** | **4,711.74** | **4,430.76** | **4,179.32** | **17,778.93** |

**STATE: FL - FLORIDA**

| REGISTRANT ZIP CODE 3 | 1ST QUARTER | 2ND QUARTER | 3RD QUARTER | 4TH QUARTER | TOTAL GRAMS |
|---|---|---|---|---|---|
| 320 | 14,462.01 | 14,359.78 | 14,241.55 | 14,144.71 | **57,208.05** |
| 321 | 16,815.58 | 16,793.51 | 16,378.48 | 16,691.36 | **66,678.94** |
| 322 | 25,469.03 | 25,705.93 | 25,240.67 | 25,008.76 | **101,424.38** |
| 323 | 6,419.90 | 6,373.82 | 6,451.18 | 6,526.12 | **25,771.02** |
| 324 | 13,307.22 | 12,891.45 | 12,733.22 | 12,610.35 | **51,542.24** |
| 325 | 21,859.96 | 20,888.71 | 20,733.00 | 20,528.67 | **84,010.35** |
| 326 | 5,045.90 | 4,740.83 | 4,924.83 | 4,993.70 | **19,705.26** |
| 327 | 14,606.36 | 13,994.22 | 14,177.66 | 13,964.93 | **56,743.17** |
| 328 | 6,166.74 | 6,187.86 | 5,954.96 | 5,991.38 | **24,300.94** |
| 329 | 12,063.19 | 11,885.99 | 11,856.43 | 11,568.61 | **47,374.22** |
| 330 | 6,739.62 | 6,052.67 | 6,113.05 | 6,126.73 | **25,032.08** |
| 331 | 3,330.61 | 3,236.47 | 3,061.36 | 3,104.74 | **12,733.18** |
| 333 | 4,774.81 | 4,518.00 | 4,370.92 | 4,396.53 | **18,060.25** |

| | | | | | |
|---|---|---|---|---|---|
| 334 | 11,499.96 | 10,380.28 | 10,371.15 | 10,498.78 | **42,750.16** |
| 335 | 10,025.93 | 9,466.10 | 9,558.43 | 9,866.67 | **38,917.12** |
| 336 | 9,232.69 | 9,034.04 | 8,844.34 | 8,832.17 | **35,943.24** |
| 337 | 13,732.42 | 13,321.82 | 13,110.66 | 12,777.74 | **52,942.64** |
| 338 | 9,719.18 | 8,908.94 | 8,953.44 | 8,902.99 | **36,484.55** |
| 339 | 12,127.20 | 11,235.89 | 11,089.13 | 11,556.76 | **46,008.99** |
| 341 | 3,713.16 | 3,159.79 | 2,895.36 | 3,283.50 | **13,051.81** |
| 342 | 13,917.97 | 12,927.65 | 12,614.58 | 13,213.49 | **52,673.68** |
| 344 | 10,468.31 | 10,133.87 | 9,834.57 | 10,286.47 | **40,723.22** |
| 346 | 13,947.59 | 13,497.70 | 13,158.43 | 13,554.53 | **54,158.25** |
| 347 | 6,710.37 | 6,391.97 | 6,480.98 | 6,517.42 | **26,100.75** |
| 349 | 5,973.24 | 5,848.73 | 5,597.84 | 5,623.94 | **23,043.75** |
| **FL FLORIDA - Total** | **272,128.93** | **261,936.03** | **258,746.21** | **260,571.08** | **1,053,382.25** |

**STATE: GA - GEORGIA**

| REGISTRANT ZIP CODE 3 | 1ST QUARTER | 2ND QUARTER | 3RD QUARTER | 4TH QUARTER | TOTAL GRAMS |
|---|---|---|---|---|---|
| 300 | 23,763.11 | 23,363.34 | 22,971.82 | 24,592.78 | **94,691.05** |
| 301 | 27,198.06 | 26,315.26 | 26,602.88 | 26,210.73 | **106,326.92** |
| 302 | 16,041.14 | 15,545.62 | 15,784.86 | 15,327.51 | **62,699.12** |
| 303 | 4,844.32 | 4,596.94 | 4,597.18 | 4,518.06 | **18,556.51** |
| 304 | 5,136.41 | 5,192.55 | 5,012.95 | 4,934.18 | **20,276.09** |
| 305 | 17,259.35 | 17,033.21 | 16,847.98 | 16,759.47 | **67,900.01** |
| 306 | 11,458.24 | 11,270.41 | 10,887.01 | 10,924.27 | **44,539.93** |
| 307 | 9,667.37 | 9,430.34 | 9,288.07 | 9,139.55 | **37,525.33** |
| 308 | 2,139.78 | 2,199.13 | 2,180.56 | 2,221.24 | **8,740.70** |
| 309 | 4,219.78 | 4,353.19 | 4,048.25 | 3,700.93 | **16,322.15** |
| 310 | 17,967.25 | 17,330.66 | 17,221.75 | 17,441.60 | **69,961.25** |
| 312 | 6,790.23 | 6,794.14 | 6,739.31 | 6,799.41 | **27,123.08** |
| 313 | 3,803.22 | 3,613.12 | 3,534.71 | 3,634.63 | **14,585.68** |
| 314 | 3,807.51 | 3,550.37 | 3,490.57 | 3,403.12 | **14,251.57** |
| 315 | 13,510.98 | 13,758.44 | 13,180.18 | 12,530.31 | **52,979.90** |
| 316 | 8,009.52 | 7,692.75 | 7,937.07 | 7,911.59 | **31,550.93** |
| 317 | 9,553.46 | 9,258.92 | 9,470.39 | 9,263.38 | **37,546.15** |
| 318 | 1,115.79 | 1,066.33 | 1,090.69 | 1,008.83 | **4,281.65** |
| 319 | 5,149.91 | 5,137.27 | 4,981.54 | 4,735.40 | **20,004.12** |
| 398 | 2,940.13 | 3,014.81 | 2,949.31 | 2,887.02 | **11,791.27** |
| **GA GEORGIA Total** | **194,375.53** | **190,516.79** | **188,817.10** | **187,944.01** | **761,653.42** |

**STATE: GU - GUAM**

| REGISTRANT ZIP CODE 3 | 1ST QUARTER | 2ND QUARTER | 3RD QUARTER | 4TH QUARTER | TOTAL GRAMS |
|---|---|---|---|---|---|
| 969 | 126.82 | 100.90 | 120.90 | 98.30 | **446.92** |
| **GU - GUAM - Total** | **126.82** | **100.90** | **120.90** | **98.30** | **446.92** |

**STATE: HI - HAWAII**

| REGISTRANT ZIP CODE 3 | 1ST QUARTER | 2ND QUARTER | 3RD QUARTER | 4TH QUARTER | TOTAL GRAMS |
|---|---|---|---|---|---|
| 967 | 9,824.28 | 9,693.30 | 9,881.73 | 9,709.26 | **39,108.57** |
| 968 | 3,458.87 | 3,234.99 | 3,229.65 | 3,097.09 | **13,020.61** |
| **HI - HAWAII - Total** | **13,283.15** | **12,928.29** | **13,111.38** | **12,806.35** | **52,129.17** |

**STATE: IA - IOWA**

| REGISTRANT ZIP CODE 3 | 1ST QUARTER | 2ND QUARTER | 3RD QUARTER | 4TH QUARTER | TOTAL GRAMS |
|---|---|---|---|---|---|
| 500 | 3,192.92 | 3,255.70 | 3,209.43 | 3,214.22 | **12,872.27** |
| 501 | 3,020.16 | 2,940.16 | 2,827.87 | 2,847.54 | **11,635.72** |
| 502 | 3,009.14 | 2,948.58 | 2,905.21 | 2,843.89 | **11,706.82** |
| 503 | 7,109.78 | 6,991.00 | 6,822.65 | 6,642.25 | **27,565.67** |
| 504 | 1,330.90 | 1,401.35 | 1,387.86 | 1,354.16 | **5,474.26** |
| 505 | 1,679.75 | 1,630.63 | 1,621.17 | 1,641.57 | **6,573.11** |
| 506 | 1,336.80 | 1,289.85 | 1,321.14 | 1,321.79 | **5,269.57** |
| 507 | 1,117.25 | 1,138.38 | 1,175.75 | 1,199.07 | **4,630.45** |
| 508 | 500.36 | 517.61 | 525.05 | 485.55 | **2,028.56** |
| 510 | 776.99 | 761.29 | 760.55 | 770.29 | **3,069.12** |
| 511 | 1,752.01 | 1,798.16 | 1,781.77 | 1,653.81 | **6,985.75** |
| 512 | 388.22 | 331.55 | 334.21 | 322.89 | **1,376.87** |
| 513 | 513.52 | 490.47 | 499.98 | 532.65 | **2,036.63** |
| 514 | 510.76 | 487.88 | 519.31 | 492.91 | **2,010.85** |
| 515 | 1,828.77 | 1,738.39 | 1,678.25 | 1,586.53 | **6,831.94** |
| 516 | 367.10 | 345.51 | 292.31 | 323.79 | **1,328.70** |
| 520 | 1,393.30 | 1,313.89 | 1,286.38 | 1,285.20 | **5,278.76** |
| 521 | 369.97 | 390.74 | 369.52 | 389.82 | **1,520.04** |
| 522 | 1,763.52 | 1,858.66 | 1,664.66 | 1,666.22 | **6,953.06** |
| 523 | 1,040.86 | 984.95 | 1,016.85 | 899.26 | **3,941.91** |
| 524 | 1,570.23 | 1,515.76 | 1,590.88 | 1,476.97 | **6,153.84** |
| 525 | 2,098.66 | 2,145.69 | 2,078.07 | 2,042.09 | **8,364.51** |
| 526 | 1,755.59 | 1,615.08 | 1,614.00 | 1,625.61 | **6,610.28** |
| 527 | 1,431.64 | 1,416.90 | 1,413.38 | 1,383.68 | **5,645.60** |
| 528 | 1,891.43 | 1,851.38 | 1,818.10 | 1,791.44 | **7,352.36** |
| **IA IOWA Total** | **41,749.61** | **41,159.53** | **40,514.33** | **39,793.18** | **163,216.64** |

**STATE: ID - IDAHO**

| REGISTRANT ZIP CODE 3 | 1ST QUARTER | 2ND QUARTER | 3RD QUARTER | 4TH QUARTER | TOTAL GRAMS |
|---|---|---|---|---|---|
| 832 | 4,848.37 | 4,996.04 | 4,904.12 | 4,950.54 | **19,699.08** |
| 833 | 5,521.89 | 5,578.23 | 5,617.79 | 5,321.26 | **22,039.17** |
| 834 | 3,864.73 | 4,026.85 | 3,995.47 | 3,902.11 | **15,789.16** |
| 835 | 2,867.78 | 3,039.40 | 2,943.53 | 2,960.59 | **11,811.30** |
| 836 | 12,813.14 | 12,610.20 | 12,507.23 | 12,180.24 | **50,110.82** |

| | | | | | |
|---|---|---|---|---|---|
| 837 | 7,757.79 | 7,602.85 | 7,594.12 | 7,368.45 | **30,323.21** |
| 838 | 7,608.56 | 7,557.68 | 7,923.49 | 7,672.31 | **30,762.04** |
| **ID IDAHO Total** | **45,282.27** | **45,411.26** | **45,485.77** | **44,355.48** | **180,534.78** |

**STATE: IL - ILLINOIS**

| REGISTRANT ZIP CODE 3 | 1ST QUARTER | 2ND QUARTER | 3RD QUARTER | 4TH QUARTER | TOTAL GRAMS |
|---|---|---|---|---|---|
| 600 | 27,654.89 | 27,292.37 | 27,065.02 | 26,425.18 | **108,437.46** |
| 601 | 24,158.53 | 23,990.39 | 24,029.98 | 23,547.94 | **95,726.85** |
| 602 | 814.89 | 739.83 | 738.66 | 733.03 | **3,026.41** |
| 603 | 981.16 | 986.48 | 940.76 | 942.92 | **3,851.33** |
| 604 | 29,712.87 | 30,129.04 | 29,957.42 | 29,373.62 | **119,172.95** |
| 605 | 14,239.05 | 13,990.73 | 14,068.85 | 13,737.93 | **56,036.56** |
| 606 | 23,830.56 | 23,521.81 | 23,408.89 | 22,756.66 | **93,517.93** |
| 607 | 2,483.99 | 2,421.89 | 2,489.36 | 2,370.41 | **9,765.65** |
| 608 | 1,877.68 | 1,859.52 | 1,861.23 | 1,764.57 | **7,362.99** |
| 609 | 4,504.00 | 4,354.15 | 4,375.36 | 4,255.40 | **17,488.91** |
| 610 | 5,025.13 | 5,142.07 | 5,098.91 | 4,970.96 | **20,237.08** |
| 611 | 8,221.94 | 8,011.31 | 7,991.75 | 7,574.31 | **31,799.30** |
| 612 | 2,294.32 | 2,254.10 | 2,288.34 | 2,162.31 | **8,999.07** |
| 613 | 3,373.73 | 3,472.45 | 3,410.09 | 3,255.03 | **13,511.30** |
| 614 | 3,030.45 | 3,042.70 | 2,878.95 | 2,836.42 | **11,788.53** |
| 615 | 4,145.73 | 4,103.39 | 4,160.66 | 3,939.49 | **16,349.27** |
| 616 | 6,321.08 | 6,347.64 | 6,333.43 | 6,347.14 | **25,349.29** |
| 617 | 3,730.34 | 3,676.80 | 3,648.08 | 3,587.58 | **14,642.81** |
| 618 | 6,619.47 | 6,477.79 | 6,668.64 | 6,250.57 | **26,016.47** |
| 619 | 1,891.21 | 1,785.12 | 1,790.37 | 1,733.95 | **7,200.65** |
| 620 | 8,912.75 | 8,866.61 | 8,739.43 | 8,458.89 | **34,977.69** |
| 622 | 8,613.64 | 8,430.09 | 8,298.46 | 8,253.58 | **33,595.77** |
| 623 | 2,995.13 | 2,916.19 | 2,756.80 | 2,817.82 | **11,485.94** |
| 624 | 3,321.46 | 3,287.24 | 3,297.01 | 3,269.72 | **13,175.42** |
| 625 | 3,949.40 | 3,989.63 | 3,932.57 | 3,895.82 | **15,767.42** |
| 626 | 3,418.23 | 3,301.63 | 3,370.96 | 3,339.95 | **13,430.77** |
| 627 | 4,387.63 | 4,275.52 | 4,254.15 | 4,146.06 | **17,063.36** |
| 628 | 6,137.16 | 6,178.84 | 5,958.61 | 5,853.78 | **24,128.39** |
| 629 | 13,135.36 | 13,039.85 | 12,478.57 | 12,572.21 | **51,226.00** |
| **IL ILLINOIS Total** | **229,781.79** | **227,885.18** | **226,291.32** | **221,173.29** | **905,131.57** |

**STATE: IN - INDIANA**

| REGISTRANT ZIP CODE 3 | 1ST QUARTER | 2ND QUARTER | 3RD QUARTER | 4TH QUARTER | TOTAL GRAMS |
|---|---|---|---|---|---|
| 460 | 11,272.36 | 11,372.61 | 11,328.10 | 11,151.49 | **45,124.56** |
| 461 | 16,224.97 | 16,341.65 | 16,079.13 | 15,617.54 | **64,263.29** |
| 462 | 23,996.02 | 23,481.97 | 23,151.41 | 22,458.91 | **93,088.32** |

| | | | | | |
|---|---|---|---|---|---|
| 463 | 21,753.28 | 21,964.40 | 21,536.48 | 20,851.14 | **86,105.29** |
| 464 | 2,753.34 | 2,699.06 | 2,773.68 | 2,670.79 | **10,896.86** |
| 465 | 7,557.21 | 7,366.95 | 7,420.26 | 7,062.47 | **29,406.89** |
| 466 | 2,681.93 | 2,472.92 | 2,473.61 | 2,311.15 | **9,939.61** |
| 467 | 4,882.97 | 4,874.32 | 4,972.58 | 4,914.54 | **19,644.41** |
| 468 | 6,371.52 | 6,432.88 | 6,565.44 | 5,882.04 | **25,251.88** |
| 469 | 10,290.91 | 10,268.00 | 10,482.72 | 9,699.08 | **40,740.71** |
| 470 | 2,772.63 | 2,705.28 | 2,764.01 | 2,602.59 | **10,844.50** |
| 471 | 12,992.43 | 12,935.65 | 12,948.57 | 12,415.12 | **51,291.77** |
| 472 | 5,465.94 | 5,414.18 | 5,406.27 | 5,267.48 | **21,553.87** |
| 473 | 11,092.62 | 10,729.00 | 11,092.80 | 10,497.37 | **43,411.78** |
| 474 | 6,874.63 | 6,772.28 | 6,618.83 | 6,532.66 | **26,798.41** |
| 475 | 5,471.19 | 5,373.18 | 5,519.53 | 5,566.22 | **21,930.13** |
| 476 | 4,033.47 | 3,941.66 | 3,662.67 | 3,709.80 | **15,347.60** |
| 477 | 9,788.03 | 9,630.02 | 9,512.82 | 9,058.12 | **37,988.99** |
| 478 | 3,742.89 | 3,826.01 | 3,855.71 | 3,727.05 | **15,151.65** |
| 479 | 7,172.57 | 7,094.21 | 7,042.70 | 6,709.71 | **28,019.19** |
| **IN INDIANA Total** | **177,190.90** | **175,696.23** | **175,207.32** | **168,705.29** | **696,799.74** |

**STATE: KS - KANSAS**

| REGISTRANT ZIP CODE 3 | 1ST QUARTER | 2ND QUARTER | 3RD QUARTER | 4TH QUARTER | **TOTAL GRAMS** |
|---|---|---|---|---|---|
| 660 | 8,234.26 | 8,085.67 | 8,127.59 | 8,044.16 | **32,491.67** |
| 661 | 2,394.96 | 2,327.07 | 2,219.80 | 2,219.59 | **9,161.43** |
| 662 | 6,442.88 | 6,495.76 | 6,377.54 | 6,248.84 | **25,565.02** |
| 664 | 1,054.88 | 1,037.71 | 1,017.90 | 987.54 | **4,098.03** |
| 665 | 1,531.40 | 1,518.71 | 1,433.08 | 1,499.51 | **5,982.70** |
| 666 | 3,397.64 | 3,498.16 | 3,224.55 | 3,406.88 | **13,527.25** |
| 667 | 3,865.01 | 3,743.56 | 3,859.92 | 3,679.74 | **15,148.23** |
| 668 | 964.09 | 949.96 | 933.77 | 883.40 | **3,731.21** |
| 669 | 509.50 | 518.09 | 480.94 | 464.08 | **1,972.61** |
| 670 | 5,027.35 | 4,865.60 | 4,773.72 | 4,861.87 | **19,528.54** |
| 671 | 2,497.59 | 2,559.13 | 2,516.83 | 2,547.60 | **10,121.15** |
| 672 | 13,997.45 | 13,797.05 | 13,665.26 | 13,583.10 | **55,042.86** |
| 673 | 1,944.75 | 1,960.22 | 1,963.40 | 1,907.58 | **7,775.95** |
| 674 | 3,707.91 | 3,627.92 | 3,532.76 | 3,482.60 | **14,351.20** |
| 675 | 2,853.76 | 2,771.06 | 2,765.04 | 2,795.27 | **11,185.13** |
| 676 | 1,017.31 | 1,019.37 | 1,033.55 | 1,002.92 | **4,073.16** |
| 677 | 524.49 | 497.21 | 497.90 | 514.02 | **2,033.61** |
| 678 | 1,896.90 | 1,985.01 | 1,779.01 | 1,846.30 | **7,507.23** |
| 679 | 762.07 | 748.57 | 715.22 | 750.78 | **2,976.64** |
| **KS - KANSAS - Total** | **62,624.21** | **62,005.83** | **60,917.79** | **60,725.79** | **246,273.62** |

**STATE: KY - KENTUCKY**

| REGISTRANT ZIP CODE 3 | 1ST QUARTER | 2ND QUARTER | 3RD QUARTER | 4TH QUARTER | TOTAL GRAMS |
|---|---|---|---|---|---|
| 400 | 6,619.25 | 6,594.66 | 6,579.25 | 6,558.27 | **26,351.43** |
| 401 | 4,051.16 | 3,971.37 | 4,206.25 | 3,772.02 | **16,000.80** |
| 402 | 29,257.45 | 28,606.25 | 28,488.08 | 28,283.40 | **114,635.18** |
| 403 | 9,380.70 | 9,304.47 | 9,517.21 | 9,397.80 | **37,600.18** |
| 404 | 6,185.17 | 6,112.07 | 6,130.80 | 6,104.29 | **24,532.33** |
| 405 | 6,152.63 | 6,177.66 | 6,132.57 | 6,005.75 | **24,468.61** |
| 406 | 1,621.82 | 1,535.66 | 1,572.27 | 1,529.38 | **6,259.13** |
| 407 | 5,382.94 | 5,412.45 | 5,541.99 | 5,330.59 | **21,667.96** |
| 408 | 1,511.71 | 1,510.52 | 1,469.88 | 1,441.50 | **5,933.62** |
| 409 | 7,409.39 | 6,914.27 | 6,863.06 | 6,594.53 | **27,781.25** |
| 410 | 8,203.30 | 8,270.09 | 8,248.57 | 8,136.79 | **32,858.76** |
| 411 | 3,234.14 | 3,005.75 | 2,872.14 | 2,938.16 | **12,050.19** |
| 412 | 2,948.44 | 2,558.29 | 2,605.30 | 2,538.63 | **10,650.66** |
| 413 | 2,673.07 | 2,715.44 | 2,774.09 | 2,780.36 | **10,942.95** |
| 414 | 1,276.55 | 1,220.30 | 1,219.11 | 1,112.03 | **4,827.99** |
| 415 | 5,151.62 | 4,706.33 | 4,794.68 | 4,883.77 | **19,536.40** |
| 416 | 4,134.66 | 3,883.22 | 3,778.57 | 3,695.68 | **15,492.12** |
| 417 | 4,349.47 | 4,052.39 | 4,357.56 | 4,247.43 | **17,006.85** |
| 418 | 1,879.62 | 1,715.76 | 1,760.68 | 1,802.15 | **7,158.20** |
| 420 | 11,802.38 | 11,351.09 | 11,503.47 | 11,103.95 | **45,760.90** |
| 421 | 10,862.50 | 10,694.53 | 10,985.04 | 10,843.72 | **43,385.78** |
| 422 | 5,467.12 | 5,516.79 | 5,598.67 | 5,367.23 | **21,949.81** |
| 423 | 8,309.83 | 8,181.85 | 7,868.69 | 7,658.54 | **32,018.91** |
| 424 | 7,711.23 | 7,889.28 | 7,515.60 | 7,548.42 | **30,664.52** |
| 425 | 2,736.42 | 2,661.39 | 2,661.62 | 2,569.55 | **10,628.98** |
| 426 | 3,312.52 | 3,439.54 | 3,295.02 | 3,080.85 | **13,127.93** |
| 427 | 7,875.62 | 7,652.89 | 7,415.69 | 7,483.59 | **30,427.79** |
| **KY - KENTUCKY - Total** | **169,500.70** | **165,654.30** | **165,755.86** | **162,808.39** | **663,719.26** |

**STATE: LA - LOUISIANA**

| REGISTRANT ZIP CODE 3 | 1ST QUARTER | 2ND QUARTER | 3RD QUARTER | 4TH QUARTER | TOTAL GRAMS |
|---|---|---|---|---|---|
| 700 | 11,935.98 | 11,637.43 | 11,365.47 | 10,976.13 | **45,915.02** |
| 701 | 5,780.48 | 5,691.17 | 5,315.71 | 5,242.45 | **22,029.81** |
| 703 | 7,885.84 | 7,741.06 | 7,206.90 | 6,638.81 | **29,472.62** |
| 704 | 13,379.90 | 13,049.42 | 12,981.66 | 12,755.73 | **52,166.71** |
| 705 | 22,041.90 | 21,436.47 | 20,796.84 | 19,842.89 | **84,118.10** |
| 706 | 6,164.15 | 6,211.21 | 5,907.95 | 5,823.47 | **24,106.79** |
| 707 | 11,189.30 | 10,961.17 | 11,015.66 | 10,385.59 | **43,551.72** |
| 708 | 8,534.70 | 8,042.80 | 7,825.93 | 7,989.83 | **32,393.26** |

| | | | | | |
|---|---|---|---|---|---|
| 710 | 3,726.51 | 3,983.31 | 3,912.96 | 3,787.58 | **15,410.36** |
| 711 | 11,381.48 | 11,221.95 | 11,006.67 | 10,844.06 | **44,454.15** |
| 712 | 9,704.16 | 9,598.37 | 9,485.05 | 9,497.88 | **38,285.47** |
| 713 | 10,444.41 | 9,971.08 | 9,929.96 | 9,271.42 | **39,616.86** |
| 714 | 4,535.46 | 4,634.82 | 4,573.76 | 4,480.81 | **18,224.85** |
| **LA - LOUISIANA - Total** | **126,704.27** | **124,180.25** | **121,324.53** | **117,536.67** | **489,745.71** |

**STATE: MA - MASSACHUSETTS**

| REGISTRANT ZIP CODE 3 | 1ST QUARTER | 2ND QUARTER | 3RD QUARTER | 4TH QUARTER | TOTAL GRAMS |
|---|---|---|---|---|---|
| 010 | 1,693.72 | 1,623.31 | 1,587.22 | 1,567.22 | **6,471.46** |
| 011 | 389.11 | 357.47 | 340.06 | 359.88 | **1,446.51** |
| 012 | 524.33 | 507.06 | 497.53 | 502.10 | **2,031.03** |
| 013 | 271.90 | 309.37 | 302.36 | 281.96 | **1,165.58** |
| 014 | 919.81 | 965.64 | 958.32 | 940.01 | **3,783.79** |
| 015 | 1,437.68 | 1,396.06 | 1,352.29 | 1,402.09 | **5,588.12** |
| 016 | 888.28 | 919.85 | 870.68 | 817.54 | **3,496.35** |
| 017 | 1,041.97 | 982.66 | 1,040.11 | 1,027.74 | **4,092.48** |
| 018 | 4,053.01 | 4,046.75 | 3,922.49 | 3,862.59 | **15,884.84** |
| 019 | 2,081.47 | 2,000.65 | 2,018.81 | 1,954.09 | **8,055.02** |
| 020 | 1,069.75 | 1,008.10 | 1,029.53 | 1,023.62 | **4,131.00** |
| 021 | 2,500.80 | 2,413.72 | 2,359.14 | 2,380.02 | **9,653.69** |
| 022 | 60.91 | 50.91 | 44.78 | 54.86 | **211.46** |
| 023 | 1,564.50 | 1,518.10 | 1,503.64 | 1,512.69 | **6,098.93** |
| 024 | 645.54 | 647.70 | 609.45 | 629.14 | **2,531.83** |
| 025 | 466.36 | 428.89 | 430.18 | 445.74 | **1,771.16** |
| 026 | 710.38 | 732.31 | 737.36 | 684.25 | **2,864.30** |
| 027 | 2,521.16 | 2,453.98 | 2,461.92 | 2,453.02 | **9,890.07** |
| **MA - MASSACHUSETTS - Total** | **22,840.66** | **22,362.53** | **22,065.87** | **21,898.59** | **89,167.65** |

**STATE: MD - MARYLAND**

| REGISTRANT ZIP CODE 3 | 1ST QUARTER | 2ND QUARTER | 3RD QUARTER | 4TH QUARTER | TOTAL GRAMS |
|---|---|---|---|---|---|
| 206 | 2,258.00 | 2,158.00 | 2,204.26 | 2,108.35 | **8,728.62** |
| 207 | 2,322.95 | 2,275.42 | 2,196.55 | 2,157.41 | **8,952.32** |
| 208 | 1,895.12 | 1,810.45 | 1,645.82 | 1,725.12 | **7,076.51** |
| 209 | 411.14 | 442.68 | 371.68 | 378.97 | **1,604.47** |
| 210 | 4,366.86 | 4,257.24 | 4,351.29 | 4,353.07 | **17,328.46** |
| 211 | 2,194.70 | 2,113.17 | 2,001.41 | 1,999.29 | **8,308.57** |
| 212 | 4,615.90 | 4,482.57 | 4,468.02 | 4,228.22 | **17,794.71** |
| 214 | 522.77 | 517.10 | 548.41 | 502.41 | **2,090.69** |
| 215 | 1,574.14 | 1,650.71 | 1,614.79 | 1,580.34 | **6,419.98** |
| 216 | 1,071.14 | 1,123.42 | 1,121.76 | 1,130.05 | **4,446.36** |
| 217 | 3,015.25 | 2,884.37 | 2,911.80 | 2,922.69 | **11,734.11** |

| | | | | | |
|---|---|---|---|---|---|
| 218 | 1,072.15 | 1,043.12 | 1,119.76 | 1,023.89 | **4,258.92** |
| 219 | 455.98 | 444.33 | 443.34 | 443.16 | **1,786.81** |
| **MD MARYLAND Total** | **25,776.10** | **25,202.59** | **24,998.90** | **24,552.96** | **100,530.54** |

**STATE: ME - MAINE**

| REGISTRANT ZIP CODE 3 | 1ST QUARTER | 2ND QUARTER | 3RD QUARTER | 4TH QUARTER | TOTAL GRAMS |
|---|---|---|---|---|---|
| 039 | 140.75 | 138.01 | 165.71 | 129.70 | **574.17** |
| 040 | 3,711.64 | 3,539.90 | 3,608.56 | 3,241.32 | **14,101.43** |
| 041 | 723.08 | 723.24 | 686.45 | 705.31 | **2,838.09** |
| 042 | 1,635.40 | 1,662.03 | 1,678.80 | 1,614.99 | **6,591.23** |
| 043 | 1,591.09 | 1,587.22 | 1,577.65 | 1,491.90 | **6,247.86** |
| 044 | 1,415.25 | 1,417.43 | 1,459.12 | 1,372.36 | **5,664.17** |
| 045 | 488.20 | 502.39 | 495.51 | 486.97 | **1,973.07** |
| 046 | 812.61 | 842.67 | 840.20 | 824.30 | **3,319.78** |
| 047 | 932.79 | 886.02 | 983.29 | 863.12 | **3,665.22** |
| 048 | 392.11 | 373.93 | 408.02 | 372.07 | **1,546.13** |
| 049 | 1,859.06 | 1,862.73 | 1,852.72 | 1,711.16 | **7,285.66** |
| **ME MAINE Total** | **13,701.99** | **13,535.58** | **13,756.03** | **12,813.19** | **53,806.80** |

**STATE: MI - MICHIGAN**

| REGISTRANT ZIP CODE 3 | 1ST QUARTER | 2ND QUARTER | 3RD QUARTER | 4TH QUARTER | TOTAL GRAMS |
|---|---|---|---|---|---|
| 480 | 34,946.11 | 35,028.83 | 35,462.87 | 33,590.14 | **139,027.95** |
| 481 | 44,720.35 | 44,707.15 | 44,338.94 | 41,935.56 | **175,702.01** |
| 482 | 17,407.68 | 16,435.39 | 16,871.92 | 15,964.77 | **66,679.76** |
| 483 | 22,481.30 | 22,600.75 | 23,240.95 | 21,705.27 | **90,028.27** |
| 484 | 12,934.45 | 12,520.40 | 12,670.32 | 11,935.91 | **50,061.08** |
| 485 | 7,730.60 | 7,633.65 | 7,506.99 | 7,062.01 | **29,933.26** |
| 486 | 14,079.49 | 14,161.47 | 14,098.63 | 13,183.54 | **55,523.14** |
| 487 | 6,954.73 | 6,946.31 | 6,898.50 | 6,608.65 | **27,408.19** |
| 488 | 13,886.07 | 13,675.88 | 14,193.47 | 11,913.84 | **53,669.26** |
| 489 | 5,582.36 | 5,336.45 | 5,298.46 | 4,848.42 | **21,065.69** |
| 490 | 18,127.99 | 18,173.33 | 18,706.08 | 16,902.54 | **71,909.94** |
| 491 | 1,843.08 | 1,814.36 | 1,911.27 | 1,638.65 | **7,207.35** |
| 492 | 7,705.42 | 7,536.50 | 7,712.26 | 6,791.26 | **29,745.44** |
| 493 | 6,083.51 | 6,161.92 | 6,225.98 | 5,270.87 | **23,742.28** |
| 494 | 14,936.43 | 14,453.94 | 14,676.37 | 12,938.70 | **57,005.44** |
| 495 | 10,696.33 | 10,591.55 | 10,284.21 | 9,173.36 | **40,745.45** |
| 496 | 7,228.27 | 7,293.63 | 7,727.48 | 6,688.15 | **28,937.54** |
| 497 | 5,776.23 | 5,792.08 | 6,299.50 | 5,342.83 | **23,210.64** |
| 498 | 5,408.14 | 5,162.81 | 5,607.05 | 4,992.20 | **21,170.20** |
| 499 | 1,551.77 | 1,592.85 | 1,620.00 | 1,535.09 | **6,299.71** |
| **MI - MICHIGAN Total** | **260,080.32** | **257,619.24** | **261,351.26** | **240,021.77** | **1,019,072.59** |

**STATE: MN - MINNESOTA**

| REGISTRANT ZIP CODE 3 | 1ST QUARTER | 2ND QUARTER | 3RD QUARTER | 4TH QUARTER | TOTAL GRAMS |
|---|---|---|---|---|---|
| 550 | 3,383.75 | 3,345.10 | 3,276.72 | 3,200.96 | **13,206.53** |
| 551 | 3,537.62 | 3,616.35 | 3,407.19 | 3,399.72 | **13,960.88** |
| 553 | 4,581.62 | 4,514.42 | 4,475.78 | 4,430.90 | **18,002.72** |
| 554 | 5,632.53 | 5,594.50 | 5,345.96 | 5,290.63 | **21,863.61** |
| 556 | 179.99 | 181.29 | 177.90 | 168.93 | **708.10** |
| 557 | 2,515.86 | 2,497.63 | 2,470.24 | 2,402.61 | **9,886.34** |
| 558 | 1,778.41 | 1,736.86 | 1,592.99 | 1,548.62 | **6,656.89** |
| 559 | 1,291.68 | 1,297.49 | 1,236.96 | 1,242.72 | **5,068.85** |
| 560 | 1,305.70 | 1,330.89 | 1,295.36 | 1,262.24 | **5,194.19** |
| 561 | 511.70 | 532.85 | 524.41 | 507.32 | **2,076.29** |
| 562 | 1,041.34 | 1,035.49 | 1,010.28 | 1,000.18 | **4,087.30** |
| 563 | 2,592.57 | 2,585.39 | 2,438.86 | 2,399.11 | **10,015.93** |
| 564 | 1,217.91 | 1,228.13 | 1,223.00 | 1,176.34 | **4,845.38** |
| 565 | 1,430.91 | 1,363.95 | 1,342.87 | 1,295.65 | **5,433.38** |
| 566 | 882.70 | 920.67 | 920.42 | 877.53 | **3,601.31** |
| 567 | 617.36 | 603.92 | 592.32 | 563.31 | **2,376.91** |
| **MN MINNESOTA Total** | **32,501.64** | **32,384.94** | **31,331.28** | **30,766.76** | **126,984.62** |

**STATE: MO - MISSOURI**

| REGISTRANT ZIP CODE 3 | 1ST QUARTER | 2ND QUARTER | 3RD QUARTER | 4TH QUARTER | TOTAL GRAMS |
|---|---|---|---|---|---|
| 630 | 12,217.90 | 12,318.53 | 12,165.56 | 12,136.96 | **48,838.95** |
| 631 | 17,775.35 | 17,733.79 | 17,168.74 | 18,128.72 | **70,806.60** |
| 633 | 7,370.98 | 7,357.88 | 7,223.85 | 7,045.63 | **28,998.35** |
| 634 | 1,567.28 | 1,584.11 | 1,605.74 | 1,560.65 | **6,317.79** |
| 635 | 1,346.58 | 1,265.37 | 1,226.18 | 1,182.57 | **5,020.69** |
| 636 | 5,004.48 | 4,749.88 | 4,702.44 | 4,585.13 | **19,041.93** |
| 637 | 3,387.23 | 3,280.97 | 3,299.35 | 3,116.12 | **13,083.67** |
| 638 | 7,154.01 | 6,747.70 | 6,593.11 | 6,369.49 | **26,864.31** |
| 639 | 4,537.39 | 4,348.43 | 4,298.33 | 4,174.88 | **17,359.03** |
| 640 | 13,206.29 | 13,209.14 | 12,860.98 | 12,745.92 | **52,022.33** |
| 641 | 9,152.33 | 8,763.07 | 8,311.65 | 8,290.43 | **34,517.48** |
| 644 | 1,562.85 | 1,496.14 | 1,522.34 | 1,516.56 | **6,097.90** |
| 645 | 3,056.97 | 3,015.21 | 2,984.52 | 2,918.55 | **11,975.25** |
| 646 | 1,023.69 | 970.25 | 1,049.03 | 970.77 | **4,013.73** |
| 647 | 2,994.20 | 3,037.76 | 3,109.63 | 3,018.58 | **12,160.17** |
| 648 | 5,319.71 | 4,964.96 | 5,052.79 | 4,884.94 | **20,222.40** |
| 650 | 2,437.53 | 2,448.79 | 2,520.33 | 2,415.67 | **9,822.33** |
| 651 | 1,107.88 | 1,101.65 | 1,043.14 | 1,081.59 | **4,334.26** |
| 652 | 3,459.79 | 3,509.95 | 3,361.26 | 3,374.00 | **13,705.00** |

| | | | | | |
|---|---|---|---|---|---|
| 653 | 1,804.17 | 1,848.15 | 1,812.15 | 1,739.03 | **7,203.49** |
| 654 | 1,702.26 | 1,759.03 | 1,839.66 | 1,769.48 | **7,070.42** |
| 655 | 2,886.24 | 2,906.36 | 3,030.93 | 2,899.57 | **11,723.10** |
| 656 | 5,552.62 | 5,399.19 | 5,457.88 | 5,453.75 | **21,863.44** |
| 657 | 8,485.09 | 8,408.42 | 8,335.05 | 8,225.23 | **33,453.79** |
| 658 | 7,544.76 | 7,479.39 | 7,205.25 | 7,156.26 | **29,385.65** |
| **MO MISSOURI - Total** | **131,657.60** | **129,704.10** | **127,779.91** | **126,760.46** | **515,902.06** |

**STATE: MS - MISSISSIPPI**

| REGISTRANT ZIP CODE 3 | 1ST QUARTER | 2ND QUARTER | 3RD QUARTER | 4TH QUARTER | TOTAL GRAMS |
|---|---|---|---|---|---|
| 386 | 7,661.97 | 7,403.73 | 7,200.07 | 7,424.93 | **29,690.70** |
| 387 | 2,210.77 | 2,198.28 | 2,222.79 | 2,110.23 | **8,742.07** |
| 388 | 9,532.06 | 9,467.04 | 9,191.78 | 9,434.39 | **37,625.27** |
| 389 | 2,370.07 | 2,442.77 | 2,316.68 | 2,203.74 | **9,333.26** |
| 390 | 5,059.98 | 4,847.73 | 4,745.46 | 4,779.29 | **19,432.45** |
| 391 | 5,873.18 | 5,386.90 | 5,729.11 | 5,695.12 | **22,684.32** |
| 392 | 4,976.76 | 4,900.52 | 4,579.60 | 4,533.18 | **18,990.06** |
| 393 | 7,369.27 | 7,236.81 | 6,888.11 | 6,991.62 | **28,485.81** |
| 394 | 14,456.38 | 14,236.99 | 14,167.99 | 14,166.71 | **57,028.07** |
| 395 | 12,344.45 | 11,836.73 | 11,635.44 | 11,233.00 | **47,049.63** |
| 396 | 3,706.60 | 3,664.96 | 3,790.26 | 3,612.42 | **14,774.23** |
| 397 | 3,666.37 | 3,452.22 | 3,332.25 | 3,167.68 | **13,618.51** |
| **MS MISSISSIPPI - Total** | **79,227.87** | **77,074.67** | **75,799.54** | **75,352.30** | **307,454.38** |

**STATE: MT - MONTANA**

| REGISTRANT ZIP CODE 3 | 1ST QUARTER | 2ND QUARTER | 3RD QUARTER | 4TH QUARTER | TOTAL GRAMS |
|---|---|---|---|---|---|
| 590 | 935.40 | 956.06 | 998.78 | 930.73 | **3,820.97** |
| 591 | 2,823.20 | 2,847.23 | 2,754.85 | 2,716.91 | **11,142.19** |
| 592 | 387.99 | 406.23 | 414.17 | 432.25 | **1,640.64** |
| 593 | 639.58 | 602.64 | 570.40 | 580.21 | **2,392.84** |
| 594 | 2,424.77 | 2,512.20 | 2,527.01 | 2,390.32 | **9,854.29** |
| 595 | 473.73 | 484.72 | 444.73 | 459.92 | **1,863.10** |
| 596 | 2,056.43 | 1,964.65 | 1,952.56 | 1,878.25 | **7,851.89** |
| 597 | 2,305.51 | 2,381.32 | 2,377.91 | 2,090.57 | **9,155.31** |
| 598 | 5,139.28 | 4,911.42 | 4,819.90 | 4,784.65 | **19,655.25** |
| 599 | 3,237.59 | 3,313.68 | 3,229.01 | 3,213.72 | **12,994.00** |
| **MT - MONTANA Total** | **20,423.48** | **20,380.14** | **20,089.33** | **19,477.54** | **80,370.49** |

**STATE: NC - NORTH CAROLINA**

| REGISTRANT ZIP CODE 3 | 1ST QUARTER | 2ND QUARTER | 3RD QUARTER | 4TH QUARTER | TOTAL GRAMS |
|---|---|---|---|---|---|
| 270 | 6,115.14 | 5,711.58 | 5,780.73 | 5,544.63 | **23,152.08** |
| 271 | 4,008.36 | 3,927.12 | 3,861.67 | 3,675.99 | **15,473.13** |
| 272 | 10,652.45 | 9,786.09 | 9,550.10 | 9,293.24 | **39,281.87** |

| | | | | | |
|---|---|---|---|---|---|
| 273 | 6,474.96 | 6,141.98 | 5,987.89 | 5,632.09 | **24,236.92** |
| 274 | 3,637.58 | 3,390.76 | 3,406.79 | 3,342.40 | **13,777.54** |
| 275 | 12,269.29 | 11,522.01 | 11,100.16 | 10,586.73 | **45,478.20** |
| 276 | 3,832.67 | 3,302.07 | 3,275.76 | 3,392.19 | **13,802.69** |
| 277 | 2,606.10 | 2,461.72 | 2,472.45 | 2,235.61 | **9,775.88** |
| 278 | 7,744.30 | 7,221.04 | 7,233.44 | 6,789.33 | **28,988.11** |
| 279 | 2,502.33 | 2,476.54 | 2,437.15 | 2,073.81 | **9,489.84** |
| 280 | 17,204.08 | 16,276.13 | 16,155.49 | 15,585.25 | **65,220.95** |
| 281 | 12,383.72 | 11,616.01 | 11,613.24 | 11,385.47 | **46,998.44** |
| 282 | 7,755.68 | 7,390.32 | 7,428.95 | 7,768.80 | **30,343.76** |
| 283 | 15,152.92 | 14,055.78 | 14,264.43 | 12,556.76 | **56,029.88** |
| 284 | 9,075.92 | 8,803.86 | 8,667.11 | 7,959.88 | **34,506.78** |
| 285 | 7,059.07 | 6,654.77 | 6,524.51 | 6,191.01 | **26,429.36** |
| 286 | 16,247.21 | 15,227.05 | 14,794.53 | 14,277.42 | **60,546.21** |
| 287 | 8,393.23 | 8,187.10 | 8,266.68 | 7,744.75 | **32,591.76** |
| 288 | 3,755.46 | 3,432.35 | 3,330.94 | 3,335.23 | **13,853.98** |
| 289 | 1,385.93 | 1,428.36 | 1,413.66 | 1,486.74 | **5,714.68** |
| **NC - NORTH CAROLINA Total** | **158,256.39** | **149,012.63** | **147,565.69** | **140,857.33** | **595,692.04** |

**STATE: ND - NORTH DAKOTA**

| REGISTRANT ZIP CODE 3 | 1ST QUARTER | 2ND QUARTER | 3RD QUARTER | 4TH QUARTER | TOTAL GRAMS |
|---|---|---|---|---|---|
| 580 | 412.82 | 401.39 | 407.94 | 432.10 | **1,654.25** |
| 581 | 1,374.35 | 1,254.91 | 1,333.62 | 1,297.91 | **5,260.78** |
| 582 | 951.08 | 887.21 | 854.94 | 809.85 | **3,503.08** |
| 583 | 483.17 | 493.97 | 522.03 | 480.72 | **1,979.89** |
| 584 | 446.13 | 471.75 | 423.13 | 393.00 | **1,734.01** |
| 585 | 2,641.19 | 2,437.30 | 2,421.61 | 2,428.03 | **9,928.14** |
| 586 | 581.31 | 530.22 | 546.50 | 540.21 | **2,198.24** |
| 587 | 629.84 | 559.30 | 587.07 | 534.72 | **2,310.93** |
| 588 | 530.55 | 518.79 | 483.05 | 474.18 | **2,006.57** |
| **ND NORTH DAKOTA Total** | **8,050.44** | **7,554.84** | **7,579.90** | **7,390.71** | **30,575.89** |

**STATE: NE - NEBRASKA**

| REGISTRANT ZIP CODE 3 | 1ST QUARTER | 2ND QUARTER | 3RD QUARTER | 4TH QUARTER | TOTAL GRAMS |
|---|---|---|---|---|---|
| 680 | 2,828.16 | 2,764.48 | 2,624.98 | 2,726.77 | **10,944.39** |
| 681 | 7,945.79 | 7,768.10 | 7,517.42 | 7,430.01 | **30,661.32** |
| 683 | 1,014.68 | 980.11 | 1,040.01 | 1,031.00 | **4,065.79** |
| 684 | 801.75 | 754.68 | 757.55 | 747.50 | **3,061.49** |
| 685 | 3,848.12 | 3,727.77 | 3,591.14 | 3,535.49 | **14,702.52** |
| 686 | 599.65 | 615.98 | 585.18 | 603.34 | **2,404.16** |
| 687 | 1,411.13 | 1,391.40 | 1,365.17 | 1,342.41 | **5,510.11** |
| 688 | 2,165.17 | 2,225.32 | 2,164.81 | 2,235.26 | **8,790.57** |

| | | | | | |
|---|---|---|---|---|---|
| 689 | 956.75 | 930.08 | 927.23 | 893.07 | **3,707.13** |
| 690 | 368.15 | 373.25 | 337.94 | 350.44 | **1,429.78** |
| 691 | 1,102.40 | 1,099.60 | 1,091.60 | 1,058.16 | **4,351.76** |
| 692 | 60.68 | 70.13 | 64.36 | 64.20 | **259.37** |
| 693 | 867.65 | 900.76 | 895.31 | 847.51 | **3,511.23** |
| **NE NEBRASKA Total** | **23,970.08** | **23,601.67** | **22,962.70** | **22,865.16** | **93,399.61** |

**STATE: NH - NEW HAMPSHIRE**

| REGISTRANT ZIP CODE 3 | 1ST QUARTER | 2ND QUARTER | 3RD QUARTER | 4TH QUARTER | **TOTAL GRAMS** |
|---|---|---|---|---|---|
| 030 | 1,827.27 | 1,768.24 | 1,678.82 | 1,682.99 | **6,957.32** |
| 031 | 1,015.71 | 972.87 | 1,010.27 | 964.21 | **3,963.06** |
| 032 | 929.35 | 820.08 | 871.52 | 882.42 | **3,503.37** |
| 033 | 595.70 | 567.25 | 531.71 | 544.45 | **2,239.11** |
| 034 | 428.77 | 416.86 | 429.39 | 417.44 | **1,692.46** |
| 035 | 270.14 | 229.62 | 292.44 | 248.28 | **1,040.48** |
| 036 | 27.73 | 24.97 | 23.38 | 26.09 | **102.16** |
| 037 | 351.90 | 322.80 | 324.97 | 319.71 | **1,319.39** |
| 038 | 1,939.26 | 1,917.56 | 1,857.68 | 1,856.96 | **7,571.46** |
| **NH - NEW HAMPSHIRE Total** | **7,385.84** | **7,040.25** | **7,020.17** | **6,942.55** | **28,388.81** |

**STATE: NJ - NEW JERSEY**

| REGISTRANT ZIP CODE 3 | 1ST QUARTER | 2ND QUARTER | 3RD QUARTER | 4TH QUARTER | **TOTAL GRAMS** |
|---|---|---|---|---|---|
| 070 | 3,010.04 | 2,690.05 | 2,785.34 | 2,765.70 | **11,251.13** |
| 071 | 250.66 | 239.39 | 220.97 | 212.67 | **923.69** |
| 072 | 154.20 | 162.69 | 154.68 | 134.38 | **605.96** |
| 073 | 277.90 | 242.42 | 234.68 | 229.89 | **984.88** |
| 074 | 1,717.95 | 1,519.84 | 1,561.04 | 1,546.91 | **6,345.74** |
| 075 | 170.17 | 167.63 | 182.06 | 170.80 | **690.66** |
| 076 | 1,028.72 | 1,026.17 | 991.52 | 989.41 | **4,035.82** |
| 077 | 2,373.78 | 2,081.94 | 2,088.89 | 2,126.55 | **8,671.16** |
| 078 | 1,361.18 | 1,348.12 | 1,419.31 | 1,369.37 | **5,497.98** |
| 079 | 568.96 | 466.11 | 507.20 | 461.70 | **2,003.96** |
| 080 | 6,219.84 | 6,184.74 | 6,084.38 | 6,307.50 | **24,796.46** |
| 081 | 476.12 | 466.60 | 444.79 | 437.99 | **1,825.50** |
| 082 | 1,530.38 | 1,569.11 | 1,641.42 | 1,575.79 | **6,316.69** |
| 083 | 1,354.27 | 1,404.47 | 1,322.65 | 1,345.27 | **5,426.67** |
| 084 | 249.27 | 238.84 | 265.88 | 252.53 | **1,006.53** |
| 085 | 675.21 | 609.14 | 603.06 | 611.74 | **2,499.16** |
| 086 | 807.62 | 739.34 | 777.14 | 736.52 | **3,060.61** |
| 087 | 1,774.65 | 1,701.66 | 1,781.76 | 1,781.58 | **7,039.65** |
| 088 | 2,082.96 | 1,920.62 | 1,944.42 | 1,991.53 | **7,939.53** |
| 089 | 129.18 | 110.45 | 103.54 | 128.13 | **471.31** |

| NJ NEW JERSEY - Total | 26,213.06 | 24,889.34 | 25,114.73 | 25,175.95 | 101,393.08 |
|---|---|---|---|---|---|

**STATE: NM - NEW MEXICO**

| REGISTRANT ZIP CODE 3 | 1ST QUARTER | 2ND QUARTER | 3RD QUARTER | 4TH QUARTER | TOTAL GRAMS |
|---|---|---|---|---|---|
| 870 | 2,105.23 | 1,965.77 | 1,995.11 | 1,910.96 | **7,977.07** |
| 871 | 9,310.14 | 9,197.81 | 8,715.10 | 8,842.77 | **36,065.81** |
| 873 | 504.66 | 486.67 | 455.79 | 460.63 | **1,907.75** |
| 874 | 1,370.83 | 1,265.68 | 1,207.07 | 1,155.69 | **4,999.28** |
| 875 | 3,251.22 | 3,340.92 | 3,117.86 | 3,197.20 | **12,907.20** |
| 877 | 712.48 | 757.89 | 714.92 | 710.56 | **2,895.86** |
| 878 | 227.11 | 205.23 | 213.65 | 206.65 | **852.64** |
| 879 | 193.44 | 192.34 | 168.50 | 168.92 | **723.19** |
| 880 | 2,470.92 | 2,361.10 | 2,170.87 | 2,179.29 | **9,182.19** |
| 881 | 1,153.57 | 1,065.20 | 1,077.06 | 1,080.91 | **4,376.73** |
| 882 | 4,935.48 | 4,930.43 | 4,869.56 | 4,782.39 | **19,517.85** |
| 883 | 1,270.04 | 1,289.81 | 1,259.41 | 1,270.20 | **5,089.45** |
| 884 | 254.61 | 223.31 | 211.97 | 228.40 | **918.30** |
| NM NEW MEXICO Total | 27,759.73 | 27,282.16 | 26,176.86 | 26,194.58 | 107,413.33 |

**STATE: NV - NEVADA**

| REGISTRANT ZIP CODE 3 | 1ST QUARTER | 2ND QUARTER | 3RD QUARTER | 4TH QUARTER | TOTAL GRAMS |
|---|---|---|---|---|---|
| 890 | 18,154.32 | 18,437.90 | 17,945.11 | 18,526.35 | **73,063.68** |
| 891 | 34,547.00 | 34,033.42 | 33,026.53 | 33,461.56 | **135,068.51** |
| 893 | 433.70 | 420.70 | 386.23 | 356.58 | **1,597.20** |
| 894 | 6,496.41 | 5,995.02 | 5,940.56 | 6,082.99 | **24,514.97** |
| 895 | 6,421.36 | 6,212.13 | 6,110.18 | 5,905.95 | **24,649.62** |
| 897 | 2,104.52 | 2,026.66 | 1,990.71 | 1,997.07 | **8,118.95** |
| 898 | 831.95 | 782.44 | 776.62 | 746.99 | **3,137.99** |
| NV NEVADA Total | 68,989.24 | 67,908.27 | 66,175.94 | 67,077.48 | 270,150.93 |

**STATE: NY - NEW YORK**

| REGISTRANT ZIP CODE 3 | 1ST QUARTER | 2ND QUARTER | 3RD QUARTER | 4TH QUARTER | TOTAL GRAMS |
|---|---|---|---|---|---|
| 063 | 0 | 0 | 0.30 | 0 | **0.30** |
| 100 | 3,041.04 | 2,661.68 | 2,613.77 | 2,732.66 | **11,049.14** |
| 101 | 144.82 | 135.25 | 173.08 | 114.31 | **567.46** |
| 102 | 8.11 | 9.03 | 8.64 | 8.72 | **34.50** |
| 103 | 2,023.98 | 2,057.50 | 1,931.32 | 1,945.70 | **7,958.51** |
| 104 | 1,128.20 | 863.35 | 975.68 | 981.48 | **3,948.71** |
| 105 | 1,880.63 | 1,609.19 | 1,681.05 | 1,780.38 | **6,951.25** |
| 106 | 221.50 | 191.93 | 204.42 | 202.38 | **820.22** |
| 107 | 536.71 | 501.83 | 507.46 | 531.22 | **2,077.22** |
| 108 | 178.66 | 191.03 | 160.75 | 190.32 | **720.78** |
| 109 | 1,922.89 | 1,751.67 | 1,823.81 | 1,890.80 | **7,389.17** |

| | | | | | |
|---|---|---|---|---|---|
| 110 | 760.76 | 729.43 | 636.81 | 795.43 | **2,922.44** |
| 111 | 377.15 | 386.30 | 386.29 | 344.22 | **1,493.95** |
| 112 | 2,937.61 | 2,418.60 | 2,589.45 | 2,552.96 | **10,498.62** |
| 113 | 1,545.33 | 1,426.41 | 1,384.73 | 1,394.00 | **5,750.48** |
| 114 | 732.36 | 686.27 | 635.38 | 651.93 | **2,705.93** |
| 115 | 2,193.67 | 2,080.84 | 2,070.31 | 2,095.49 | **8,440.31** |
| 116 | 209.26 | 174.34 | 195.07 | 197.26 | **775.93** |
| 117 | 9,906.44 | 9,283.88 | 9,327.24 | 9,371.05 | **37,888.60** |
| 118 | 326.76 | 291.20 | 292.50 | 307.65 | **1,218.11** |
| 119 | 1,544.06 | 1,441.98 | 1,628.28 | 1,472.54 | **6,086.86** |
| 120 | 5,044.85 | 4,914.38 | 4,847.78 | 4,621.21 | **19,428.22** |
| 121 | 3,055.96 | 2,944.17 | 2,804.68 | 2,822.93 | **11,627.75** |
| 122 | 2,154.13 | 2,112.18 | 2,027.62 | 1,987.19 | **8,281.12** |
| 123 | 2,157.10 | 2,079.08 | 1,919.94 | 2,092.79 | **8,248.92** |
| 124 | 1,995.42 | 1,825.61 | 1,957.11 | 1,808.15 | **7,586.28** |
| 125 | 3,445.32 | 3,280.22 | 3,272.52 | 3,282.94 | **13,281.00** |
| 126 | 547.43 | 480.96 | 492.09 | 539.16 | **2,059.64** |
| 127 | 693.79 | 679.73 | 671.76 | 647.73 | **2,693.00** |
| 128 | 3,406.13 | 3,212.82 | 3,283.59 | 3,195.89 | **13,098.44** |
| 129 | 2,309.27 | 2,324.16 | 2,333.26 | 2,233.14 | **9,199.82** |
| 130 | 4,382.55 | 4,275.60 | 4,261.40 | 4,099.62 | **17,019.16** |
| 131 | 1,529.37 | 1,534.99 | 1,512.29 | 1,489.51 | **6,066.16** |
| 132 | 2,936.14 | 2,846.56 | 2,831.35 | 2,793.21 | **11,407.26** |
| 133 | 1,907.71 | 1,774.49 | 1,807.76 | 1,754.53 | **7,244.50** |
| 134 | 2,800.16 | 2,671.85 | 2,628.72 | 2,492.45 | **10,593.18** |
| 135 | 1,386.49 | 1,339.68 | 1,336.57 | 1,329.20 | **5,391.93** |
| 136 | 2,838.15 | 2,887.10 | 2,884.75 | 2,836.36 | **11,446.36** |
| 137 | 1,202.77 | 1,165.43 | 1,125.06 | 1,057.83 | **4,551.08** |
| 138 | 2,689.78 | 2,871.64 | 2,494.59 | 2,436.42 | **10,492.43** |
| 139 | 1,121.23 | 1,109.51 | 1,052.24 | 1,026.12 | **4,309.11** |
| 140 | 9,264.42 | 9,244.83 | 9,018.61 | 9,081.06 | **36,608.92** |
| 141 | 4,487.32 | 4,386.39 | 4,353.72 | 4,334.45 | **17,561.88** |
| 142 | 17,651.59 | 16,914.47 | 17,191.44 | 16,955.78 | **68,713.28** |
| 143 | 2,299.24 | 2,171.99 | 2,201.97 | 2,130.92 | **8,804.11** |
| 144 | 3,181.07 | 3,146.81 | 3,363.05 | 3,005.34 | **12,696.27** |
| 145 | 2,612.37 | 2,447.23 | 2,438.86 | 2,467.39 | **9,965.84** |
| 146 | 4,690.39 | 4,550.60 | 4,378.14 | 4,350.66 | **17,969.79** |
| 147 | 3,636.30 | 3,668.00 | 3,732.71 | 3,596.08 | **14,633.10** |
| 148 | 3,209.03 | 3,142.50 | 3,059.95 | 2,972.23 | **12,383.71** |
| 149 | 830.27 | 784.11 | 747.06 | 725.30 | **3,086.72** |

| NY - NEW YORK - Total | 131,085.68 | 125,678.80 | 125,256.94 | 123,726.07 | 505,747.50 |
|---|---|---|---|---|---|

STATE: OH - OHIO

| REGISTRANT ZIP CODE 3 | 1ST QUARTER | 2ND QUARTER | 3RD QUARTER | 4TH QUARTER | TOTAL GRAMS |
|---|---|---|---|---|---|
| 430 | 7,166.55 | 7,161.69 | 7,115.73 | 7,136.98 | **28,580.95** |
| 431 | 5,603.70 | 5,545.47 | 5,742.61 | 5,584.11 | **22,475.90** |
| 432 | 8,703.94 | 8,505.22 | 8,522.23 | 8,582.10 | **34,313.49** |
| 433 | 2,338.65 | 2,228.29 | 2,252.38 | 2,194.82 | **9,014.14** |
| 434 | 1,289.05 | 1,296.25 | 1,325.46 | 1,151.42 | **5,062.18** |
| 435 | 2,873.55 | 2,909.35 | 2,872.30 | 2,744.54 | **11,399.75** |
| 436 | 4,981.41 | 5,046.83 | 4,989.73 | 4,718.14 | **19,736.12** |
| 437 | 2,833.88 | 2,856.50 | 2,777.33 | 2,790.29 | **11,258.00** |
| 438 | 406.23 | 430.20 | 400.54 | 418.74 | **1,655.71** |
| 439 | 2,382.38 | 2,177.68 | 2,193.16 | 2,028.41 | **8,781.62** |
| 440 | 6,026.96 | 6,037.41 | 5,919.99 | 5,747.76 | **23,732.12** |
| 441 | 7,630.58 | 7,734.50 | 7,495.22 | 7,387.17 | **30,247.47** |
| 442 | 4,695.01 | 4,648.85 | 4,473.95 | 4,472.76 | **18,290.57** |
| 443 | 2,216.18 | 2,162.10 | 2,106.18 | 2,027.61 | **8,512.06** |
| 444 | 5,663.24 | 5,457.16 | 5,511.82 | 5,292.01 | **21,924.23** |
| 445 | 3,600.85 | 3,557.24 | 3,517.08 | 3,435.53 | **14,110.69** |
| 446 | 4,225.91 | 4,183.14 | 4,012.54 | 4,069.29 | **16,490.87** |
| 447 | 2,555.95 | 2,528.52 | 2,436.83 | 2,516.71 | **10,038.01** |
| 448 | 2,685.82 | 2,579.25 | 2,574.57 | 2,496.23 | **10,335.86** |
| 449 | 771.39 | 744.76 | 751.77 | 733.15 | **3,001.07** |
| 450 | 10,811.28 | 11,029.73 | 11,067.20 | 10,813.39 | **43,721.59** |
| 451 | 4,019.18 | 3,882.34 | 3,996.52 | 3,844.46 | **15,742.50** |
| 452 | 8,429.80 | 8,283.72 | 8,235.74 | 7,835.32 | **32,784.57** |
| 453 | 7,345.59 | 7,402.25 | 7,377.51 | 7,200.11 | **29,325.46** |
| 454 | 8,076.18 | 7,989.83 | 7,759.95 | 7,312.08 | **31,138.04** |
| 455 | 2,039.79 | 2,095.12 | 2,002.91 | 2,011.30 | **8,149.13** |
| 456 | 6,392.79 | 6,366.26 | 6,327.84 | 6,148.53 | **25,235.42** |
| 457 | 2,273.18 | 2,328.06 | 2,288.97 | 2,198.30 | **9,088.50** |
| 458 | 3,607.16 | 3,543.91 | 3,585.22 | 3,484.54 | **14,220.82** |
| OH - OHIO Total | 131,646.17 | 130,711.60 | 129,633.27 | 126,375.80 | 518,366.85 |

STATE: OK - OKLAHOMA

| REGISTRANT ZIP CODE 3 | 1ST QUARTER | 2ND QUARTER | 3RD QUARTER | 4TH QUARTER | TOTAL GRAMS |
|---|---|---|---|---|---|
| 730 | 19,384.82 | 19,262.72 | 19,083.59 | 19,108.53 | **76,839.67** |
| 731 | 26,494.88 | 26,399.35 | 26,418.34 | 26,416.06 | **105,728.64** |
| 734 | 3,956.94 | 4,013.82 | 3,938.93 | 3,905.04 | **15,814.74** |
| 735 | 7,817.88 | 7,572.11 | 7,856.99 | 7,279.15 | **30,526.13** |
| 736 | 1,447.61 | 1,454.60 | 1,436.87 | 1,452.82 | **5,791.90** |

| | | | | | |
|---|---|---|---|---|---|
| 737 | 3,200.34 | 3,187.15 | 3,215.63 | 3,137.55 | **12,740.67** |
| 738 | 607.63 | 603.25 | 656.48 | 627.66 | **2,495.02** |
| 739 | 294.96 | 275.29 | 259.29 | 288.24 | **1,117.77** |
| 740 | 20,710.14 | 20,224.14 | 20,299.98 | 19,823.11 | **81,057.37** |
| 741 | 14,173.82 | 14,245.49 | 13,855.68 | 13,762.14 | **56,037.14** |
| 743 | 3,374.39 | 3,259.78 | 3,346.82 | 3,259.61 | **13,240.60** |
| 744 | 10,980.18 | 10,981.71 | 11,167.76 | 10,925.79 | **44,055.44** |
| 745 | 3,355.69 | 3,332.21 | 3,340.93 | 3,231.79 | **13,260.61** |
| 746 | 1,545.03 | 1,494.66 | 1,479.62 | 1,477.07 | **5,996.39** |
| 747 | 4,555.51 | 4,385.83 | 4,520.71 | 4,290.71 | **17,752.76** |
| 748 | 6,594.84 | 6,659.22 | 6,405.58 | 6,312.51 | **25,972.16** |
| 749 | 4,031.00 | 3,958.83 | 4,017.63 | 3,763.30 | **15,770.75** |
| **OK - OKLAHOMA Total** | **132,525.66** | **131,310.16** | **131,300.83** | **129,061.11** | **524,197.75** |

**STATE: OR - OREGON**

| REGISTRANT ZIP CODE 3 | 1ST QUARTER | 2ND QUARTER | 3RD QUARTER | 4TH QUARTER | TOTAL GRAMS |
|---|---|---|---|---|---|
| 970 | 11,089.26 | 11,053.93 | 10,876.62 | 10,959.69 | **43,979.50** |
| 971 | 4,838.43 | 4,758.31 | 4,771.64 | 4,639.90 | **19,008.28** |
| 972 | 13,448.43 | 13,592.98 | 13,169.53 | 13,118.21 | **53,329.15** |
| 973 | 8,900.84 | 8,738.16 | 8,914.93 | 8,683.72 | **35,237.65** |
| 974 | 14,038.39 | 13,800.03 | 13,377.76 | 13,234.16 | **54,450.34** |
| 975 | 9,931.91 | 9,787.38 | 9,855.85 | 9,838.63 | **39,413.76** |
| 976 | 2,050.17 | 2,026.36 | 2,004.71 | 2,051.78 | **8,133.02** |
| 977 | 4,343.80 | 4,279.47 | 4,258.65 | 4,193.04 | **17,074.96** |
| 978 | 3,235.92 | 3,182.38 | 3,092.20 | 2,918.83 | **12,429.33** |
| 979 | 1,469.21 | 1,471.67 | 1,496.55 | 1,439.55 | **5,876.98** |
| **OR OREGON Total** | **73,346.36** | **72,690.66** | **71,818.44** | **71,077.51** | **288,932.98** |

**STATE: PA - PENNSYLVANIA**

| REGISTRANT ZIP CODE 3 | 1ST QUARTER | 2ND QUARTER | 3RD QUARTER | 4TH QUARTER | TOTAL GRAMS |
|---|---|---|---|---|---|
| 150 | 5,922.61 | 5,995.59 | 5,430.50 | 5,437.01 | **22,785.71** |
| 151 | 4,395.78 | 4,549.97 | 4,119.83 | 4,182.63 | **17,248.20** |
| 152 | 8,195.29 | 8,091.81 | 8,015.79 | 7,501.31 | **31,804.20** |
| 153 | 2,489.95 | 3,087.93 | 2,460.33 | 3,178.37 | **11,216.58** |
| 154 | 3,675.19 | 3,593.17 | 3,539.11 | 3,417.00 | **14,224.48** |
| 155 | 1,005.28 | 962.28 | 1,010.59 | 927.39 | **3,905.53** |
| 156 | 3,806.23 | 3,727.46 | 3,563.18 | 3,525.22 | **14,622.08** |
| 157 | 1,811.13 | 1,678.92 | 1,605.01 | 1,580.85 | **6,675.91** |
| 158 | 1,585.29 | 1,526.02 | 1,535.15 | 1,610.72 | **6,257.17** |
| 159 | 1,742.58 | 1,662.92 | 1,608.13 | 1,644.37 | **6,658.00** |
| 160 | 2,163.58 | 2,185.46 | 2,198.03 | 2,174.67 | **8,721.74** |
| 161 | 2,990.24 | 2,836.90 | 2,799.47 | 2,815.15 | **11,441.75** |

| | | | | | |
|---|---|---|---|---|---|
| 162 | 1,145.86 | 1,099.68 | 1,091.00 | 960.45 | **4,296.99** |
| 163 | 2,454.64 | 2,354.55 | 2,419.63 | 2,294.06 | **9,522.90** |
| 164 | 1,657.26 | 1,634.46 | 1,665.11 | 1,680.96 | **6,637.79** |
| 165 | 5,616.37 | 5,461.43 | 5,605.87 | 5,322.28 | **22,005.96** |
| 166 | 3,736.16 | 3,627.78 | 3,481.46 | 3,360.58 | **14,205.97** |
| 167 | 1,013.01 | 952.75 | 967.00 | 938.59 | **3,871.35** |
| 168 | 994.68 | 978.96 | 944.96 | 994.77 | **3,913.37** |
| 169 | 670.76 | 641.02 | 624.03 | 642.54 | **2,578.35** |
| 170 | 6,000.24 | 5,574.67 | 5,521.52 | 5,422.71 | **22,519.14** |
| 171 | 1,328.88 | 1,248.59 | 1,249.93 | 1,188.28 | **5,015.67** |
| 172 | 1,050.99 | 989.85 | 991.39 | 970.36 | **4,002.59** |
| 173 | 1,886.34 | 1,818.70 | 1,859.66 | 1,778.27 | **7,342.97** |
| 174 | 981.70 | 964.73 | 989.80 | 958.99 | **3,895.23** |
| 175 | 2,052.29 | 1,912.44 | 1,838.96 | 1,841.83 | **7,645.53** |
| 176 | 1,523.09 | 1,456.04 | 1,401.19 | 1,421.52 | **5,801.85** |
| 177 | 2,169.97 | 2,052.18 | 2,103.13 | 2,063.92 | **8,389.19** |
| 178 | 1,245.09 | 1,142.94 | 1,102.47 | 1,094.16 | **4,584.66** |
| 179 | 959.86 | 874.26 | 821.79 | 833.29 | **3,489.19** |
| 180 | 2,404.01 | 2,331.17 | 2,196.45 | 2,202.24 | **9,133.86** |
| 181 | 1,014.46 | 909.59 | 874.13 | 931.15 | **3,729.34** |
| 182 | 1,246.60 | 1,162.44 | 1,102.29 | 1,137.92 | **4,649.26** |
| 183 | 1,179.31 | 1,069.66 | 1,088.21 | 1,002.74 | **4,339.92** |
| 184 | 2,134.78 | 2,021.78 | 2,018.07 | 1,939.16 | **8,113.79** |
| 185 | 2,597.37 | 2,371.75 | 2,327.03 | 2,351.86 | **9,648.02** |
| 186 | 1,989.83 | 1,878.73 | 1,849.87 | 1,840.59 | **7,559.02** |
| 187 | 5,576.51 | 5,101.36 | 5,953.58 | 5,616.74 | **22,248.19** |
| 188 | 770.94 | 756.49 | 725.79 | 753.48 | **3,006.69** |
| 189 | 2,018.39 | 1,907.97 | 1,940.89 | 1,957.67 | **7,824.91** |
| 190 | 7,411.42 | 9,425.04 | 6,251.29 | 10,977.95 | **34,065.69** |
| 191 | 4,486.33 | 4,131.88 | 4,056.19 | 4,054.87 | **16,729.27** |
| 193 | 1,908.65 | 1,743.50 | 1,857.62 | 1,804.68 | **7,314.44** |
| 194 | 2,722.61 | 2,738.30 | 2,739.83 | 2,543.67 | **10,744.41** |
| 195 | 1,199.71 | 1,197.57 | 1,176.19 | 1,137.82 | **4,711.28** |
| 196 | 1,529.84 | 1,465.28 | 1,494.86 | 1,401.49 | **5,891.47** |
| **PA - PENNSYLVANIA - Total** | **116,461.12** | **114,895.96** | **110,216.30** | **113,416.28** | **454,989.65** |

**STATE: PR - PUERTO RICO**

| REGISTRANT ZIP CODE 3 | 1ST QUARTER | 2ND QUARTER | 3RD QUARTER | 4TH QUARTER | TOTAL GRAMS |
|---|---|---|---|---|---|
| 006 | 244.13 | 194.55 | 133.61 | 177.97 | **750.25** |
| 007 | 144.88 | 109.99 | 94.74 | 108.00 | **457.61** |
| 009 | 178.71 | 101.95 | 114.37 | 119.60 | **514.63** |

| PR - PUERTO RICO - Total | 567.72 | 406.49 | 342.72 | 405.56 | 1,722.49 |
|---|---|---|---|---|---|

STATE: RI - RHODE ISLAND

| REGISTRANT ZIP CODE 3 | 1ST QUARTER | 2ND QUARTER | 3RD QUARTER | 4TH QUARTER | TOTAL GRAMS |
|---|---|---|---|---|---|
| 028 | 4,635.55 | 4,608.61 | 4,584.81 | 4,595.11 | **18,424.08** |
| 029 | 3,045.61 | 2,986.74 | 2,974.19 | 2,950.43 | **11,956.97** |
| RI RHODE ISLAND Total | 7,681.15 | 7,595.34 | 7,559.00 | 7,545.55 | 30,381.05 |

STATE: SC - SOUTH CAROLINA

| REGISTRANT ZIP CODE 3 | 1ST QUARTER | 2ND QUARTER | 3RD QUARTER | 4TH QUARTER | TOTAL GRAMS |
|---|---|---|---|---|---|
| 290 | 6,230.59 | 5,930.71 | 5,821.64 | 5,853.14 | **23,836.08** |
| 291 | 6,658.04 | 6,349.35 | 6,421.40 | 6,147.88 | **25,576.66** |
| 292 | 4,636.63 | 4,522.54 | 4,456.86 | 4,313.85 | **17,929.89** |
| 293 | 15,481.65 | 14,822.37 | 13,980.29 | 13,622.82 | **57,907.14** |
| 294 | 15,084.45 | 15,465.28 | 14,027.58 | 13,826.07 | **58,403.39** |
| 295 | 18,021.94 | 17,482.86 | 17,129.58 | 16,150.41 | **68,784.79** |
| 296 | 27,712.50 | 26,514.27 | 26,146.65 | 25,801.91 | **106,175.34** |
| 297 | 4,887.08 | 4,655.66 | 4,651.83 | 4,753.79 | **18,948.37** |
| 298 | 2,779.89 | 2,746.72 | 2,677.93 | 2,691.94 | **10,896.48** |
| 299 | 2,543.09 | 2,425.34 | 2,324.13 | 2,329.18 | **9,621.74** |
| SC - SOUTH CAROLINA Total | 104,035.86 | 100,915.12 | 97,637.89 | 95,491.00 | 398,079.87 |

STATE: SD - SOUTH DAKOTA

| REGISTRANT ZIP CODE 3 | 1ST QUARTER | 2ND QUARTER | 3RD QUARTER | 4TH QUARTER | TOTAL GRAMS |
|---|---|---|---|---|---|
| 570 | 1,844.90 | 1,782.31 | 1,724.15 | 1,702.87 | **7,054.24** |
| 571 | 3,245.28 | 3,204.72 | 3,284.64 | 3,163.36 | **12,898.00** |
| 572 | 678.56 | 669.23 | 695.98 | 635.63 | **2,679.40** |
| 573 | 861.72 | 842.33 | 870.43 | 849.75 | **3,424.24** |
| 574 | 597.39 | 589.74 | 574.18 | 578.31 | **2,339.62** |
| 575 | 787.00 | 750.14 | 728.30 | 704.76 | **2,970.21** |
| 576 | 168.90 | 158.83 | 164.00 | 148.59 | **640.33** |
| 577 | 2,191.73 | 2,117.48 | 2,262.42 | 2,135.20 | **8,706.83** |
| SD SOUTH DAKOTA - Total | 10,375.49 | 10,114.80 | 10,304.10 | 9,918.48 | 40,712.87 |

STATE: TN - TENNESSEE

| REGISTRANT ZIP CODE 3 | 1ST QUARTER | 2ND QUARTER | 3RD QUARTER | 4TH QUARTER | TOTAL GRAMS |
|---|---|---|---|---|---|
| 370 | 21,921.01 | 21,654.12 | 21,400.73 | 21,146.79 | **86,122.65** |
| 371 | 22,928.92 | 22,078.56 | 22,308.56 | 21,987.04 | **89,303.08** |
| 372 | 9,385.11 | 9,384.91 | 9,156.30 | 9,021.67 | **36,947.99** |
| 373 | 20,976.65 | 20,981.52 | 20,868.28 | 20,555.36 | **83,381.80** |
| 374 | 7,132.32 | 7,159.12 | 7,117.08 | 7,138.85 | **28,547.37** |
| 376 | 12,709.11 | 12,809.17 | 12,723.92 | 12,605.28 | **50,847.46** |
| 377 | 10,622.46 | 10,414.97 | 10,326.95 | 10,439.50 | **41,803.88** |
| 378 | 16,575.17 | 16,146.79 | 16,440.60 | 16,423.79 | **65,586.35** |

| | | | | | |
|---|---|---|---|---|---|
| 379 | 8,126.56 | 8,212.87 | 8,223.64 | 8,093.01 | **32,656.08** |
| 380 | 10,009.57 | 9,649.25 | 9,510.39 | 9,439.24 | **38,608.45** |
| 381 | 14,203.36 | 14,416.59 | 11,870.93 | 20,908.77 | **61,399.65** |
| 382 | 5,699.43 | 5,497.00 | 5,526.06 | 5,248.02 | **21,970.51** |
| 383 | 12,574.15 | 12,163.35 | 11,498.15 | 11,628.81 | **47,864.46** |
| 384 | 6,836.12 | 6,922.28 | 6,743.79 | 6,427.82 | **26,930.01** |
| 385 | 5,950.69 | 5,855.90 | 5,603.38 | 5,660.57 | **23,070.54** |
| **TN TENNESSEE Total** | **185,650.62** | **183,346.41** | **179,318.75** | **186,724.50** | **735,040.29** |

**STATE: TX - TEXAS**

| REGISTRANT ZIP CODE 3 | 1ST QUARTER | 2ND QUARTER | 3RD QUARTER | 4TH QUARTER | TOTAL GRAMS |
|---|---|---|---|---|---|
| 750 | 41,551.83 | 41,017.66 | 40,476.97 | 40,400.34 | **163,446.79** |
| 751 | 18,856.00 | 18,591.08 | 18,545.37 | 18,629.36 | **74,621.80** |
| 752 | 21,270.09 | 20,934.08 | 20,345.76 | 19,873.15 | **82,423.09** |
| 753 | 254.75 | 250.80 | 239.48 | 216.78 | **961.81** |
| 754 | 11,688.70 | 11,448.91 | 11,900.88 | 11,541.47 | **46,579.96** |
| 755 | 4,163.43 | 3,829.17 | 3,442.10 | 3,555.21 | **14,989.91** |
| 756 | 9,334.15 | 9,101.51 | 8,741.11 | 8,781.87 | **35,958.64** |
| 757 | 12,913.27 | 12,651.95 | 12,564.81 | 12,256.96 | **50,387.00** |
| 758 | 2,442.88 | 2,446.72 | 2,354.17 | 2,280.00 | **9,523.77** |
| 759 | 6,966.61 | 6,431.92 | 6,217.75 | 5,877.85 | **25,494.12** |
| 760 | 32,429.75 | 32,343.33 | 31,854.63 | 31,746.85 | **128,374.56** |
| 761 | 27,101.36 | 26,684.77 | 26,488.12 | 25,219.54 | **105,493.79** |
| 762 | 12,151.28 | 11,643.62 | 11,881.59 | 11,253.03 | **46,929.51** |
| 763 | 5,245.83 | 4,913.90 | 4,785.86 | 4,775.66 | **19,721.25** |
| 764 | 3,646.42 | 3,767.40 | 3,593.89 | 3,415.90 | **14,423.60** |
| 765 | 9,771.16 | 9,794.52 | 9,247.27 | 9,362.83 | **38,175.77** |
| 766 | 2,998.21 | 3,126.66 | 2,869.04 | 2,952.09 | **11,946.00** |
| 767 | 8,248.98 | 8,029.65 | 7,829.43 | 7,741.64 | **31,849.69** |
| 768 | 2,417.11 | 2,560.67 | 2,597.71 | 2,537.60 | **10,113.09** |
| 769 | 3,102.19 | 2,911.84 | 3,008.00 | 2,601.36 | **11,623.40** |
| 770 | 44,230.66 | 42,240.83 | 40,496.39 | 38,880.38 | **165,848.26** |
| 772 | 0 | 0 | 2.12 | 0.91 | **3.03** |
| 773 | 24,931.81 | 24,603.33 | 24,551.92 | 23,585.68 | **97,672.73** |
| 774 | 16,848.05 | 16,449.37 | 16,768.72 | 16,544.47 | **66,610.62** |
| 775 | 27,708.26 | 27,012.16 | 25,747.42 | 24,877.29 | **105,345.13** |
| 776 | 7,708.30 | 7,211.65 | 7,296.72 | 7,384.93 | **29,601.59** |
| 777 | 3,773.93 | 4,286.85 | 4,082.50 | 4,132.76 | **16,276.05** |
| 778 | 4,685.07 | 4,502.54 | 4,560.48 | 4,518.40 | **18,266.48** |
| 779 | 5,849.30 | 5,800.87 | 5,825.32 | 5,700.47 | **23,175.95** |
| 780 | 7,754.42 | 7,344.24 | 7,505.53 | 7,354.83 | **29,959.03** |

| | | | | | |
|---|---|---|---|---|---|
| 781 | 9,041.04 | 8,589.30 | 8,621.40 | 8,751.38 | **35,003.12** |
| 782 | 36,677.56 | 35,594.53 | 35,557.66 | 35,248.54 | **143,078.29** |
| 783 | 5,264.71 | 5,107.01 | 5,138.57 | 4,964.47 | **20,474.76** |
| 784 | 9,042.15 | 8,676.41 | 8,381.30 | 8,151.29 | **34,251.16** |
| 785 | 6,772.02 | 6,121.75 | 6,380.07 | 6,029.06 | **25,302.89** |
| 786 | 18,025.40 | 17,120.40 | 17,483.20 | 17,294.24 | **69,923.25** |
| 787 | 14,338.93 | 14,476.24 | 13,955.03 | 13,753.87 | **56,524.07** |
| 788 | 1,689.89 | 1,497.08 | 1,483.81 | 1,363.64 | **6,034.42** |
| 789 | 1,284.92 | 1,292.75 | 1,294.22 | 1,299.34 | **5,171.23** |
| 790 | 3,514.37 | 3,326.55 | 3,232.36 | 3,265.88 | **13,339.16** |
| 791 | 6,773.51 | 7,056.47 | 6,347.11 | 6,008.81 | **26,185.90** |
| 792 | 769.43 | 732.85 | 769.14 | 756.75 | **3,028.17** |
| 793 | 2,522.08 | 2,523.95 | 2,420.64 | 2,462.67 | **9,929.34** |
| 794 | 9,574.16 | 9,069.75 | 9,036.10 | 8,636.14 | **36,316.15** |
| 795 | 2,283.84 | 2,333.06 | 2,264.61 | 2,114.33 | **8,995.84** |
| 796 | 4,578.73 | 4,373.09 | 4,495.07 | 4,275.74 | **17,722.63** |
| 797 | 7,234.52 | 5,660.66 | 5,638.75 | 5,655.97 | **24,189.91** |
| 798 | 275.04 | 249.43 | 277.97 | 241.68 | **1,044.11** |
| 799 | 5,675.95 | 5,418.66 | 5,640.57 | 5,194.07 | **21,929.25** |
| **TX - TEXAS Total** | **525,382.04** | **511,151.94** | **504,238.60** | **493,467.49** | **2,034,240.07** |

**STATE: UT - UTAH**

| REGISTRANT ZIP CODE 3 | 1ST QUARTER | 2ND QUARTER | 3RD QUARTER | 4TH QUARTER | **TOTAL GRAMS** |
|---|---|---|---|---|---|
| 840 | 17,563.97 | 17,547.79 | 17,546.73 | 17,527.22 | **70,185.71** |
| 841 | 11,024.34 | 10,989.73 | 10,550.71 | 11,004.51 | **43,569.30** |
| 843 | 2,173.51 | 2,178.04 | 2,121.57 | 2,286.40 | **8,759.53** |
| 844 | 3,281.13 | 3,367.31 | 3,285.17 | 3,290.03 | **13,223.64** |
| 845 | 1,523.67 | 1,574.69 | 1,499.46 | 1,486.13 | **6,083.95** |
| 846 | 4,304.70 | 4,391.14 | 4,244.70 | 4,209.01 | **17,149.55** |
| 847 | 6,061.47 | 6,060.63 | 6,062.95 | 6,135.84 | **24,320.90** |
| **UT - UTAH - Total** | **45,932.81** | **46,109.33** | **45,311.30** | **45,939.13** | **183,292.58** |

**STATE: VA - VIRGINIA**

| REGISTRANT ZIP CODE 3 | 1ST QUARTER | 2ND QUARTER | 3RD QUARTER | 4TH QUARTER | **TOTAL GRAMS** |
|---|---|---|---|---|---|
| 201 | 3,265.89 | 3,198.83 | 3,113.41 | 3,146.92 | **12,725.05** |
| 220 | 1,345.05 | 1,207.50 | 1,255.83 | 1,175.02 | **4,983.40** |
| 221 | 1,265.97 | 1,301.55 | 1,307.32 | 1,289.39 | **5,164.23** |
| 222 | 370.91 | 378.37 | 334.14 | 324.05 | **1,407.46** |
| 223 | 657.96 | 648.32 | 635.50 | 645.19 | **2,586.98** |
| 224 | 2,064.07 | 2,000.24 | 1,940.43 | 2,048.80 | **8,053.53** |
| 225 | 889.89 | 870.44 | 797.05 | 833.95 | **3,391.33** |
| 226 | 1,943.78 | 1,939.96 | 1,873.46 | 1,803.57 | **7,560.77** |

| | | | | | |
|---|---|---|---|---|---|
| 227 | 371.15 | 366.52 | 365.83 | 345.11 | **1,448.61** |
| 228 | 1,665.63 | 1,666.10 | 1,643.70 | 1,678.48 | **6,653.91** |
| 229 | 1,780.87 | 1,739.79 | 1,715.33 | 1,772.64 | **7,008.62** |
| 230 | 1,757.46 | 1,714.22 | 1,636.99 | 1,643.50 | **6,752.17** |
| 231 | 2,992.92 | 2,858.49 | 2,722.08 | 2,929.00 | **11,502.49** |
| 232 | 4,403.64 | 4,154.19 | 4,054.28 | 3,832.46 | **16,444.57** |
| 233 | 2,139.87 | 2,094.06 | 2,055.36 | 2,070.42 | **8,359.70** |
| 234 | 4,681.37 | 4,396.49 | 4,276.54 | 4,379.11 | **17,733.50** |
| 235 | 1,574.59 | 1,468.56 | 1,437.46 | 1,464.49 | **5,945.10** |
| 236 | 2,818.38 | 2,646.58 | 2,520.20 | 2,482.12 | **10,467.28** |
| 237 | 1,363.60 | 1,300.88 | 1,291.92 | 1,239.71 | **5,196.11** |
| 238 | 2,427.89 | 2,369.64 | 2,237.27 | 2,156.54 | **9,191.34** |
| 239 | 973.79 | 919.22 | 879.04 | 844.50 | **3,616.56** |
| 240 | 7,085.81 | 7,017.96 | 6,876.11 | 6,490.75 | **27,470.63** |
| 241 | 6,595.92 | 6,440.46 | 6,592.08 | 6,059.40 | **25,687.86** |
| 242 | 10,605.49 | 10,459.07 | 9,768.70 | 9,173.31 | **40,006.58** |
| 243 | 4,200.45 | 4,091.01 | 4,041.05 | 3,736.89 | **16,069.40** |
| 244 | 2,100.28 | 2,124.58 | 2,034.52 | 2,048.84 | **8,308.22** |
| 245 | 5,754.75 | 5,736.92 | 5,484.30 | 5,279.85 | **22,255.81** |
| 246 | 3,142.18 | 3,188.24 | 3,148.97 | 2,978.11 | **12,457.50** |
| **VA VIRGINIA Total** | **80,239.56** | **78,298.17** | **76,038.87** | **73,872.10** | **308,448.70** |

**STATE: VI - VIRGIN ISLANDS**

| REGISTRANT ZIP CODE 3 | 1ST QUARTER | 2ND QUARTER | 3RD QUARTER | 4TH QUARTER | **TOTAL GRAMS** |
|---|---|---|---|---|---|
| 008 | 256.51 | 186.01 | 181.09 | 200.01 | **823.62** |
| **VI - VIRGIN ISLANDS Total** | **256.51** | **186.01** | **181.09** | **200.01** | **823.62** |

**STATE: VT - VERMONT**

| REGISTRANT ZIP CODE 3 | 1ST QUARTER | 2ND QUARTER | 3RD QUARTER | 4TH QUARTER | **TOTAL GRAMS** |
|---|---|---|---|---|---|
| 050 | 296.44 | 301.80 | 263.48 | 273.39 | **1,135.11** |
| 051 | 151.74 | 132.44 | 151.66 | 136.31 | **572.14** |
| 052 | 192.52 | 209.23 | 236.85 | 221.70 | **860.29** |
| 053 | 207.31 | 204.34 | 191.61 | 192.46 | **795.71** |
| 054 | 1,398.10 | 1,409.21 | 1,395.92 | 1,328.09 | **5,531.32** |
| 056 | 646.54 | 633.37 | 641.72 | 610.02 | **2,531.64** |
| 057 | 293.72 | 312.50 | 318.71 | 320.23 | **1,245.15** |
| 058 | 473.80 | 483.85 | 483.90 | 477.06 | **1,918.62** |
| **VT VERMONT Total** | **3,660.15** | **3,686.74** | **3,683.83** | **3,559.26** | **14,589.98** |

**STATE: WA - WASHINGTON**

| REGISTRANT ZIP CODE 3 | 1ST QUARTER | 2ND QUARTER | 3RD QUARTER | 4TH QUARTER | **TOTAL GRAMS** |
|---|---|---|---|---|---|
| 980 | 12,738.03 | 12,330.58 | 12,249.74 | 12,005.25 | **49,323.59** |
| 981 | 5,787.62 | 5,870.65 | 5,574.94 | 5,503.75 | **22,736.96** |

| | | | | | |
|---|---|---|---|---|---|
| 982 | 11,353.45 | 11,269.32 | 11,085.60 | 10,905.93 | **44,614.29** |
| 983 | 9,992.38 | 9,896.95 | 9,885.00 | 9,794.22 | **39,568.54** |
| 984 | 5,144.13 | 4,845.18 | 4,843.02 | 4,859.84 | **19,692.17** |
| 985 | 6,676.19 | 6,637.86 | 6,638.98 | 6,484.04 | **26,437.07** |
| 986 | 9,938.19 | 9,982.12 | 9,633.20 | 9,742.21 | **39,295.71** |
| 988 | 3,600.65 | 3,435.61 | 3,418.30 | 3,324.70 | **13,779.27** |
| 989 | 4,280.06 | 4,211.86 | 4,102.68 | 4,031.53 | **16,626.13** |
| 990 | 2,983.54 | 2,960.10 | 2,929.68 | 2,895.07 | **11,768.39** |
| 991 | 3,661.21 | 3,534.81 | 3,490.77 | 3,604.03 | **14,290.82** |
| 992 | 15,042.26 | 14,932.93 | 14,774.86 | 14,372.42 | **59,122.47** |
| 993 | 7,791.62 | 7,651.71 | 7,673.77 | 7,587.65 | **30,704.76** |
| 994 | 1,691.09 | 1,670.13 | 1,650.21 | 1,620.76 | **6,632.19** |
| **WA WASHINGTON Total** | **100,680.42** | **99,229.81** | **97,950.74** | **96,731.38** | **394,592.36** |

**STATE: WI - WISCONSIN**

| REGISTRANT ZIP CODE 3 | 1ST QUARTER | 2ND QUARTER | 3RD QUARTER | 4TH QUARTER | TOTAL GRAMS |
|---|---|---|---|---|---|
| 530 | 5,945.70 | 5,951.40 | 6,003.40 | 5,951.49 | **23,852.00** |
| 531 | 8,064.82 | 8,162.88 | 8,072.25 | 7,959.71 | **32,259.66** |
| 532 | 8,330.92 | 8,215.93 | 7,983.76 | 7,694.18 | **32,224.79** |
| 534 | 1,929.78 | 1,882.64 | 1,714.37 | 1,754.27 | **7,281.07** |
| 535 | 6,035.79 | 5,992.27 | 5,851.03 | 5,716.03 | **23,595.12** |
| 537 | 2,746.43 | 2,739.14 | 2,587.79 | 2,616.58 | **10,689.93** |
| 538 | 625.97 | 628.52 | 621.90 | 641.58 | **2,517.97** |
| 539 | 2,901.61 | 2,900.73 | 2,930.96 | 2,753.14 | **11,486.44** |
| 540 | 676.64 | 689.25 | 662.74 | 672.96 | **2,701.60** |
| 541 | 1,812.16 | 1,961.77 | 1,908.78 | 1,862.54 | **7,545.25** |
| 542 | 1,096.54 | 1,156.91 | 1,178.45 | 1,094.39 | **4,526.29** |
| 543 | 2,157.28 | 2,082.50 | 1,941.47 | 1,927.42 | **8,108.68** |
| 544 | 3,882.99 | 3,791.48 | 3,752.65 | 3,644.67 | **15,071.78** |
| 545 | 1,099.49 | 1,109.30 | 1,182.54 | 1,100.08 | **4,491.41** |
| 546 | 3,095.16 | 3,224.97 | 3,116.38 | 2,958.46 | **12,394.98** |
| 547 | 2,153.62 | 2,172.71 | 2,213.77 | 2,169.18 | **8,709.28** |
| 548 | 2,413.99 | 2,457.23 | 2,520.66 | 2,447.63 | **9,839.51** |
| 549 | 5,269.06 | 5,403.90 | 5,242.29 | 5,107.83 | **21,023.08** |
| **WI - WISCONSIN Total** | **60,237.95** | **60,523.54** | **59,485.20** | **58,072.15** | **238,318.84** |

**STATE: WV - WEST VIRGINIA**

| REGISTRANT ZIP CODE 3 | 1ST QUARTER | 2ND QUARTER | 3RD QUARTER | 4TH QUARTER | TOTAL GRAMS |
|---|---|---|---|---|---|
| 247 | 1,129.07 | 1,178.08 | 1,109.80 | 1,202.28 | **4,619.22** |
| 248 | 603.43 | 626.67 | 521.96 | 408.32 | **2,160.39** |
| 249 | 872.70 | 855.78 | 828.49 | 828.02 | **3,384.99** |
| 250 | 1,851.40 | 1,782.56 | 1,816.83 | 1,702.34 | **7,153.14** |

| | | | | | |
|---|---|---|---|---|---|
| 251 | 1,534.77 | 1,563.01 | 1,479.99 | 1,505.70 | **6,083.47** |
| 252 | 813.10 | 748.20 | 736.50 | 669.37 | **2,967.16** |
| 253 | 3,440.34 | 3,268.07 | 3,351.06 | 3,204.83 | **13,264.30** |
| 254 | 1,734.95 | 1,683.33 | 1,694.27 | 1,665.39 | **6,777.94** |
| 255 | 4,108.72 | 4,027.41 | 3,860.21 | 3,485.63 | **15,481.97** |
| 256 | 1,963.79 | 1,865.28 | 1,697.39 | 1,528.06 | **7,054.51** |
| 257 | 3,070.15 | 2,904.09 | 3,147.76 | 3,071.95 | **12,193.96** |
| 258 | 2,680.71 | 2,516.33 | 2,565.17 | 2,407.65 | **10,169.86** |
| 259 | 841.32 | 831.35 | 761.81 | 775.42 | **3,209.89** |
| 260 | 2,555.64 | 2,527.60 | 2,535.86 | 2,356.23 | **9,975.33** |
| 261 | 2,383.77 | 2,269.76 | 2,112.69 | 1,698.93 | **8,465.15** |
| 262 | 1,617.74 | 1,573.72 | 1,472.15 | 1,469.17 | **6,132.77** |
| 263 | 3,727.74 | 3,572.45 | 3,553.93 | 3,443.00 | **14,297.12** |
| 264 | 996.42 | 905.89 | 928.68 | 873.02 | **3,704.01** |
| 265 | 2,841.21 | 2,605.35 | 2,635.55 | 2,566.14 | **10,648.24** |
| 266 | 904.36 | 863.42 | 862.02 | 797.91 | **3,427.70** |
| 267 | 632.90 | 604.89 | 648.68 | 615.92 | **2,502.40** |
| 268 | 246.49 | 234.46 | 238.89 | 244.28 | **964.12** |
| **WV WEST VIRGINIA Total** | **40,550.69** | **39,007.70** | **38,559.69** | **36,519.54** | **154,637.62** |

**STATE: WY - WYOMING**

| REGISTRANT ZIP CODE 3 | 1ST QUARTER | 2ND QUARTER | 3RD QUARTER | 4TH QUARTER | TOTAL GRAMS |
|---|---|---|---|---|---|
| 820 | 1,693.49 | 1,626.93 | 1,655.16 | 1,534.87 | **6,510.45** |
| 822 | 327.24 | 362.04 | 353.77 | 360.50 | **1,403.55** |
| 823 | 152.68 | 151.92 | 155.67 | 163.81 | **624.07** |
| 824 | 921.59 | 926.87 | 962.28 | 971.09 | **3,781.83** |
| 825 | 637.29 | 626.99 | 608.69 | 572.13 | **2,445.11** |
| 826 | 1,717.59 | 1,758.13 | 1,708.96 | 1,590.95 | **6,775.63** |
| 827 | 590.54 | 620.54 | 611.17 | 625.67 | **2,447.92** |
| 828 | 836.35 | 752.89 | 838.25 | 750.89 | **3,178.39** |
| 829 | 1,275.21 | 1,281.10 | 1,349.87 | 1,311.79 | **5,217.97** |
| 830 | 159.12 | 151.25 | 164.48 | 131.11 | **605.96** |
| 831 | 284.58 | 341.23 | 402.78 | 324.92 | **1,353.51** |
| **WY - WYOMING Total** | **8,595.68** | **8,599.90** | **8,811.08** | **8,337.73** | **34,344.39** |

**DRUG: 9230 - MEPERIDINE (PETHIDINE)**

**STATE: AK - ALASKA**

| REGISTRANT ZIP CODE 3 | 1ST QUARTER | 2ND QUARTER | 3RD QUARTER | 4TH QUARTER | TOTAL GRAMS |
|---|---|---|---|---|---|
| 995 | 195.00 | 141.07 | 34.86 | 76.26 | **447.19** |
| 996 | 116.02 | 75.71 | 28.87 | 14.16 | **234.76** |
| 997 | 39.76 | 14.71 | 9.26 | 2.72 | **66.45** |
| 998 | 5.99 | 5.45 | 0 | 0.54 | **11.98** |

| | | | | | |
|---|---|---|---|---|---|
| 999 | 0 | 0.54 | 1.09 | 0 | **1.63** |
| **AK ALASKA - Total** | **356.77** | **237.48** | **74.08** | **93.69** | **762.02** |

**STATE: AL - ALABAMA**

| REGISTRANT ZIP CODE 3 | 1ST QUARTER | 2ND QUARTER | 3RD QUARTER | 4TH QUARTER | TOTAL GRAMS |
|---|---|---|---|---|---|
| 350 | 414.51 | 240.21 | 203.71 | 166.67 | **1,025.10** |
| 351 | 223.32 | 124.73 | 26.69 | 15.25 | **390.00** |
| 352 | 412.87 | 303.94 | 272.45 | 173.21 | **1,162.47** |
| 354 | 349.14 | 238.57 | 198.27 | 164.50 | **950.48** |
| 355 | 175.93 | 170.49 | 78.44 | 71.90 | **496.76** |
| 356 | 335.53 | 195.54 | 181.93 | 107.30 | **820.30** |
| 357 | 175.39 | 104.04 | 38.67 | 38.13 | **356.23** |
| 358 | 294.13 | 151.42 | 250.01 | 96.95 | **792.52** |
| 359 | 198.81 | 136.72 | 118.96 | 117.11 | **571.60** |
| 360 | 236.39 | 130.18 | 76.80 | 45.75 | **489.13** |
| 361 | 150.33 | 120.38 | 52.83 | 64.82 | **388.36** |
| 362 | 132.90 | 149.24 | 108.94 | 51.20 | **442.29** |
| 363 | 376.38 | 277.79 | 248.38 | 123.64 | **1,026.19** |
| 364 | 143.25 | 156.33 | 110.57 | 86.28 | **496.43** |
| 365 | 302.85 | 297.94 | 196.09 | 174.30 | **971.18** |
| 366 | 380.19 | 390.54 | 226.05 | 181.82 | **1,178.59** |
| 367 | 112.75 | 79.52 | 77.35 | 45.75 | **315.37** |
| 368 | 179.75 | 190.64 | 77.35 | 89.33 | **537.06** |
| 369 | 3.27 | 0 | 0 | 0 | **3.27** |
| **AL ALABAMA - Total** | **4,597.71** | **3,458.22** | **2,543.47** | **1,813.92** | **12,413.32** |

**STATE: AR - ARKANSAS**

| REGISTRANT ZIP CODE 3 | 1ST QUARTER | 2ND QUARTER | 3RD QUARTER | 4TH QUARTER | TOTAL GRAMS |
|---|---|---|---|---|---|
| 716 | 399.26 | 283.78 | 182.47 | 114.38 | **979.89** |
| 717 | 164.50 | 91.51 | 78.44 | 24.51 | **358.95** |
| 718 | 38.13 | 38.13 | 12.53 | 19.61 | **108.39** |
| 719 | 178.11 | 124.19 | 59.37 | 37.04 | **398.71** |
| 720 | 750.03 | 561.03 | 278.34 | 143.25 | **1,732.65** |
| 721 | 952.11 | 604.60 | 430.30 | 290.86 | **2,277.88** |
| 722 | 498.93 | 321.37 | 219.29 | 272.34 | **1,311.93** |
| 723 | 130.18 | 58.28 | 49.57 | 60.46 | **298.49** |
| 724 | 387.82 | 198.81 | 139.98 | 83.88 | **810.50** |
| 725 | 81.70 | 58.28 | 61.01 | 30.50 | **231.49** |
| 726 | 173.21 | 77.89 | 87.69 | 20.15 | **358.95** |
| 727 | 492.94 | 350.23 | 388.91 | 267.44 | **1,499.52** |
| 728 | 101.31 | 47.93 | 45.75 | 27.78 | **222.78** |
| 729 | 210.79 | 123.10 | 139.44 | 47.93 | **521.27** |

| AR ARKANSAS - Total | 4,559.03 | 2,939.13 | 2,173.09 | 1,440.15 | 11,111.41 |
|---|---|---|---|---|---|

**STATE: AZ - ARIZONA**

| REGISTRANT ZIP CODE 3 | 1ST QUARTER | 2ND QUARTER | 3RD QUARTER | 4TH QUARTER | TOTAL GRAMS |
|---|---|---|---|---|---|
| 850 | 619.61 | 648.72 | 669.86 | 414.55 | **2,352.74** |
| 851 | 88.78 | 62.09 | 44.12 | 49.57 | **244.56** |
| 852 | 1,299.86 | 821.15 | 915.38 | 736.24 | **3,772.64** |
| 853 | 385.18 | 512.14 | 383.66 | 265.81 | **1,546.78** |
| 855 | 62.64 | 86.61 | 68.09 | 28.87 | **246.20** |
| 856 | 164.50 | 134.54 | 56.10 | 104.58 | **459.72** |
| 857 | 451.39 | 340.82 | 296.14 | 178.44 | **1,266.79** |
| 859 | 5.45 | 3.81 | 0.54 | 0 | **9.80** |
| 860 | 29.41 | 50.11 | 23.97 | 9.26 | **112.75** |
| 863 | 50.66 | 21.87 | 28.37 | 19.06 | **119.96** |
| 864 | 93.69 | 68.09 | 79.52 | 88.78 | **330.08** |
| 865 | 2.18 | 0 | 0 | 1.09 | **3.27** |
| **AZ ARIZONA - Total** | **3,253.35** | **2,749.95** | **2,565.74** | **1,896.25** | **10,465.30** |

**STATE: CA - CALIFORNIA**

| REGISTRANT ZIP CODE 3 | 1ST QUARTER | 2ND QUARTER | 3RD QUARTER | 4TH QUARTER | TOTAL GRAMS |
|---|---|---|---|---|---|
| 900 | 476.60 | 430.85 | 480.41 | 221.14 | **1,609.01** |
| 902 | 338.25 | 296.31 | 360.58 | 252.74 | **1,247.88** |
| 903 | 12.53 | 1.09 | 7.63 | 19.61 | **40.85** |
| 904 | 67.00 | 60.46 | 51.75 | 59.92 | **239.12** |
| 905 | 282.15 | 204.80 | 316.68 | 206.44 | **1,010.07** |
| 906 | 86.61 | 65.91 | 45.21 | 45.75 | **243.48** |
| 907 | 37.04 | 30.50 | 31.59 | 21.79 | **120.92** |
| 908 | 44.66 | 67.54 | 31.05 | 47.39 | **190.64** |
| 910 | 39.76 | 40.85 | 21.79 | 32.68 | **135.08** |
| 911 | 90.96 | 32.14 | 48.48 | 16.34 | **187.92** |
| 912 | 156.33 | 101.86 | 250.56 | 96.95 | **605.69** |
| 913 | 516.91 | 448.28 | 406.34 | 316.25 | **1,687.77** |
| 914 | 67.54 | 54.47 | 279.86 | 161.34 | **563.21** |
| 915 | 144.34 | 104.58 | 211.34 | 77.35 | **537.61** |
| 916 | 0 | 0.54 | 2.18 | 1.09 | **3.81** |
| 917 | 887.84 | 568.11 | 691.75 | 415.60 | **2,563.30** |
| 918 | 34.86 | 21.79 | 32.68 | 47.93 | **137.26** |
| 919 | 99.13 | 62.09 | 65.36 | 46.84 | **273.43** |
| 920 | 620.94 | 422.13 | 673.89 | 342.61 | **2,059.57** |
| 921 | 442.83 | 557.22 | 393.26 | 468.43 | **1,861.74** |
| 922 | 141.07 | 209.70 | 150.55 | 108.39 | **609.72** |
| 923 | 314.28 | 526.17 | 384.55 | 235.31 | **1,460.31** |

| | | | | | |
|---|---|---|---|---|---|
| 924 | 33.77 | 10.35 | 19.61 | 7.08 | **70.81** |
| 925 | 473.33 | 390.00 | 957.89 | 453.18 | **2,274.40** |
| 926 | 534.34 | 253.82 | 322.89 | 262.54 | **1,373.59** |
| 927 | 139.44 | 89.33 | 137.81 | 131.27 | **497.84** |
| 928 | 321.37 | 309.38 | 361.13 | 138.35 | **1,130.23** |
| 930 | 96.95 | 76.80 | 119.72 | 57.08 | **350.56** |
| 931 | 200.45 | 145.43 | 212.43 | 197.72 | **756.03** |
| 932 | 195.54 | 153.60 | 159.27 | 146.52 | **654.93** |
| 933 | 628.02 | 497.84 | 416.14 | 471.15 | **2,013.17** |
| 934 | 197.72 | 101.86 | 113.30 | 50.11 | **462.98** |
| 935 | 49.57 | 41.94 | 32.68 | 28.87 | **153.06** |
| 936 | 125.82 | 112.21 | 137.26 | 99.68 | **474.97** |
| 937 | 175.93 | 163.95 | 180.29 | 148.16 | **668.33** |
| 939 | 73.53 | 45.21 | 33.23 | 26.69 | **178.66** |
| 940 | 180.84 | 119.83 | 304.92 | 215.70 | **821.28** |
| 941 | 510.37 | 141.62 | 205.89 | 181.93 | **1,039.81** |
| 943 | 115.47 | 66.45 | 84.43 | 57.19 | **323.54** |
| 944 | 2.72 | 10.89 | 5.99 | 4.90 | **24.51** |
| 945 | 485.32 | 429.76 | 262.54 | 117.22 | **1,294.83** |
| 946 | 20.70 | 3.81 | 18.52 | 5.99 | **49.02** |
| 947 | 35.40 | 7.08 | 1.63 | 3.27 | **47.39** |
| 948 | 1.09 | 0 | 1.63 | 0 | **2.72** |
| 949 | 17.43 | 9.26 | 15.14 | 15.25 | **57.08** |
| 950 | 88.24 | 81.70 | 53.82 | 71.68 | **295.44** |
| 951 | 78.44 | 82.25 | 33.23 | 62.42 | **256.33** |
| 952 | 39.76 | 56.65 | 43.58 | 44.66 | **184.65** |
| 953 | 431.94 | 400.35 | 502.20 | 385.09 | **1,719.58** |
| 954 | 64.27 | 80.07 | 21.90 | 23.97 | **190.20** |
| 955 | 33.77 | 56.65 | 37.04 | 2.18 | **129.64** |
| 956 | 233.13 | 257.09 | 286.51 | 138.35 | **915.08** |
| 957 | 5.45 | 4.90 | 15.25 | 0 | **25.60** |
| 958 | 105.67 | 119.83 | 155.24 | 151.42 | **532.16** |
| 959 | 140.53 | 122.01 | 98.04 | 44.12 | **404.70** |
| 960 | 46.84 | 34.86 | 51.75 | 37.58 | **171.03** |
| 961 | 11.44 | 33.23 | 20.15 | 8.72 | **73.53** |
| **CA - CALIFORNIA - Total** | **10,796.25** | **8,817.40** | **10,360.50** | **7,031.92** | **37,006.07** |

**STATE: CO - COLORADO**

| REGISTRANT ZIP CODE 3 | 1ST QUARTER | 2ND QUARTER | 3RD QUARTER | 4TH QUARTER | **TOTAL GRAMS** |
|---|---|---|---|---|---|
| 800 | 81.70 | 93.69 | 34.86 | 60.46 | **270.71** |
| 801 | 244.02 | 167.22 | 20.15 | 41.40 | **472.79** |

| | | | | | |
|---|---|---|---|---|---|
| 802 | 103.49 | 70.81 | 56.10 | 21.79 | **252.19** |
| 803 | 2.18 | 16.34 | 0 | 1.63 | **20.15** |
| 804 | 44.66 | 26.69 | 45.21 | 24.51 | **141.07** |
| 805 | 67.54 | 27.78 | 37.04 | 5.45 | **137.81** |
| 806 | 43.58 | 28.87 | 14.16 | 0.54 | **87.15** |
| 807 | 2.18 | 2.72 | 1.09 | 0.54 | **6.54** |
| 808 | 49.57 | 42.49 | 9.26 | 1.09 | **102.40** |
| 809 | 152.51 | 134.54 | 66.67 | 20.70 | **374.42** |
| 810 | 46.30 | 28.87 | 8.72 | 19.06 | **102.95** |
| 811 | 17.97 | 17.97 | 9.26 | 9.80 | **55.01** |
| 812 | 27.23 | 50.11 | 11.44 | 0 | **88.78** |
| 813 | 15.25 | 15.25 | 1.09 | 0.54 | **32.14** |
| 814 | 74.08 | 47.93 | 33.77 | 39.22 | **195.00** |
| 815 | 5.99 | 5.45 | 9.80 | 9.80 | **31.05** |
| 816 | 83.34 | 28.32 | 8.17 | 9.80 | **129.64** |
| **CO - COLORADO - Total** | **1,061.60** | **805.05** | **366.79** | **266.35** | **2,499.79** |

**STATE: CT - CONNECTICUT**

| REGISTRANT ZIP CODE 3 | 1ST QUARTER | 2ND QUARTER | 3RD QUARTER | 4TH QUARTER | **TOTAL GRAMS** |
|---|---|---|---|---|---|
| 060 | 13.62 | 18.52 | 8.17 | 1.09 | **41.40** |
| 061 | 74.08 | 21.24 | 10.35 | 16.34 | **122.01** |
| 062 | 5.45 | 4.36 | 0 | 0.54 | **10.35** |
| 063 | 106.76 | 97.50 | 11.11 | 10.89 | **226.26** |
| 064 | 210.79 | 128.00 | 7.08 | 7.63 | **353.50** |
| 065 | 27.78 | 41.40 | 10.35 | 10.89 | **90.42** |
| 066 | 56.65 | 29.41 | 16.89 | 6.54 | **109.48** |
| 067 | 83.34 | 61.01 | 1.09 | 0 | **145.43** |
| 068 | 63.73 | 51.20 | 9.26 | 3.27 | **127.46** |
| 069 | 2.18 | 2.18 | 2.18 | 1.09 | **7.63** |
| **CT CONNECTICUT Total** | **644.37** | **454.81** | **76.47** | **58.28** | **1,233.94** |

**STATE: DC - DISTRICT OF COLUMBIA**

| REGISTRANT ZIP CODE 3 | 1ST QUARTER | 2ND QUARTER | 3RD QUARTER | 4TH QUARTER | **TOTAL GRAMS** |
|---|---|---|---|---|---|
| 200 | 121.47 | 95.87 | 149.79 | 69.72 | **436.84** |
| 204 | 0 | 0 | 4.36 | 0 | **4.36** |
| **DC - DISTRICT OF COLUMBIA** | **121.47** | **95.87** | **154.15** | **69.72** | **441.20** |

**STATE: DE - DELAWARE**

| REGISTRANT ZIP CODE 3 | 1ST QUARTER | 2ND QUARTER | 3RD QUARTER | 4TH QUARTER | **TOTAL GRAMS** |
|---|---|---|---|---|---|
| 197 | 26.15 | 37.04 | 15.25 | 4.36 | **82.79** |
| 198 | 3.27 | 2.18 | 5.45 | 0 | **10.89** |
| 199 | 62.64 | 94.23 | 7.84 | 26.69 | **191.40** |
| **DE DELAWARE Total** | **92.05** | **133.45** | **28.54** | **31.05** | **285.09** |

**STATE: FL - FLORIDA**

| REGISTRANT ZIP CODE 3 | 1ST QUARTER | 2ND QUARTER | 3RD QUARTER | 4TH QUARTER | TOTAL GRAMS |
|---|---|---|---|---|---|
| 320 | 153.06 | 69.72 | 81.16 | 24.51 | **328.45** |
| 321 | 159.05 | 87.69 | 100.77 | 20.04 | **367.56** |
| 322 | 147.07 | 131.71 | 80.61 | 43.03 | **402.42** |
| 323 | 210.25 | 111.66 | 189.01 | 27.78 | **538.70** |
| 324 | 62.64 | 80.61 | 62.09 | 60.46 | **265.81** |
| 325 | 378.01 | 221.69 | 117.65 | 109.48 | **826.84** |
| 326 | 114.38 | 69.72 | 53.92 | 40.31 | **278.34** |
| 327 | 143.80 | 56.10 | 43.03 | 29.96 | **272.89** |
| 328 | 102.40 | 68.09 | 37.58 | 18.52 | **226.59** |
| 329 | 203.17 | 145.98 | 70.70 | 95.32 | **515.17** |
| 330 | 368.21 | 173.21 | 224.41 | 184.10 | **949.94** |
| 331 | 369.30 | 485.32 | 412.87 | 293.26 | **1,560.75** |
| 333 | 379.10 | 419.41 | 315.92 | 206.98 | **1,321.41** |
| 334 | 356.77 | 251.10 | 178.66 | 107.52 | **894.05** |
| 335 | 208.07 | 107.30 | 63.18 | 42.70 | **421.26** |
| 336 | 163.41 | 280.51 | 148.16 | 138.35 | **730.43** |
| 337 | 106.21 | 115.47 | 96.41 | 57.19 | **375.29** |
| 338 | 336.62 | 333.89 | 295.77 | 228.77 | **1,195.04** |
| 339 | 294.68 | 192.27 | 139.00 | 196.74 | **822.70** |
| 341 | 70.81 | 65.36 | 18.52 | 11.44 | **166.13** |
| 342 | 140.53 | 117.65 | 31.59 | 59.92 | **349.69** |
| 344 | 67.54 | 59.92 | 26.15 | 9.80 | **163.41** |
| 346 | 116.02 | 72.44 | 43.03 | 70.26 | **301.76** |
| 347 | 83.88 | 59.92 | 28.32 | 11.44 | **183.56** |
| 349 | 150.88 | 158.50 | 90.64 | 69.39 | **469.41** |
| **FL FLORIDA - Total** | **4,885.85** | **3,935.26** | **2,949.16** | **2,157.29** | **13,927.55** |

**STATE: GA - GEORGIA**

| REGISTRANT ZIP CODE 3 | 1ST QUARTER | 2ND QUARTER | 3RD QUARTER | 4TH QUARTER | TOTAL GRAMS |
|---|---|---|---|---|---|
| 300 | 712.45 | 567.02 | 579.77 | 236.83 | **2,096.07** |
| 301 | 541.96 | 416.14 | 279.97 | 58.83 | **1,296.90** |
| 302 | 443.92 | 261.99 | 123.54 | 83.34 | **912.79** |
| 303 | 296.31 | 209.16 | 233.13 | 96.95 | **835.55** |
| 304 | 145.43 | 89.33 | 45.75 | 35.40 | **315.92** |
| 305 | 346.42 | 192.27 | 99.13 | 82.79 | **720.62** |
| 306 | 208.62 | 116.56 | 152.51 | 73.53 | **551.22** |
| 307 | 527.26 | 238.57 | 177.57 | 167.65 | **1,111.05** |
| 308 | 106.76 | 67.54 | 26.69 | 9.26 | **210.25** |
| 309 | 289.77 | 127.46 | 121.47 | 73.53 | **612.23** |

| | | | | | |
|---|---|---|---|---|---|
| 310 | 340.43 | 362.22 | 287.60 | 165.59 | **1,155.83** |
| 312 | 106.76 | 110.57 | 74.62 | 39.76 | **331.71** |
| 313 | 116.56 | 132.36 | 22.88 | 10.24 | **282.04** |
| 314 | 204.26 | 168.85 | 80.61 | 112.31 | **566.04** |
| 315 | 373.66 | 197.18 | 123.64 | 100.22 | **794.70** |
| 316 | 554.49 | 351.32 | 237.48 | 142.16 | **1,285.46** |
| 317 | 462.98 | 251.65 | 163.41 | 124.19 | **1,002.23** |
| 318 | 19.06 | 14.71 | 9.80 | 1.63 | **45.21** |
| 319 | 199.36 | 129.64 | 53.92 | 87.15 | **470.07** |
| 398 | 62.64 | 40.31 | 40.31 | 30.50 | **173.76** |
| **GA GEORGIA Total** | **6,059.10** | **4,044.85** | **2,933.80** | **1,731.89** | **14,769.64** |

**STATE: GU - GUAM**

| REGISTRANT ZIP CODE 3 | 1ST QUARTER | 2ND QUARTER | 3RD QUARTER | 4TH QUARTER | TOTAL GRAMS |
|---|---|---|---|---|---|
| 969 | 14.16 | 16.34 | 34.86 | 19.61 | **84.97** |
| **GU - GUAM - Total** | **14.16** | **16.34** | **34.86** | **19.61** | **84.97** |

**STATE: HI - HAWAII**

| REGISTRANT ZIP CODE 3 | 1ST QUARTER | 2ND QUARTER | 3RD QUARTER | 4TH QUARTER | TOTAL GRAMS |
|---|---|---|---|---|---|
| 967 | 136.72 | 105.12 | 103.49 | 44.12 | **389.45** |
| 968 | 93.69 | 61.01 | 80.07 | 28.32 | **263.08** |
| **HI - HAWAII - Total** | **230.40** | **166.13** | **183.56** | **72.44** | **652.54** |

**STATE: IA - IOWA**

| REGISTRANT ZIP CODE 3 | 1ST QUARTER | 2ND QUARTER | 3RD QUARTER | 4TH QUARTER | TOTAL GRAMS |
|---|---|---|---|---|---|
| 500 | 39.22 | 34.86 | 18.52 | 41.94 | **134.54** |
| 501 | 10.89 | 11.44 | 2.72 | 1.63 | **26.69** |
| 502 | 100.22 | 99.68 | 35.95 | 31.05 | **266.90** |
| 503 | 538.70 | 246.74 | 287.05 | 369.84 | **1,442.33** |
| 504 | 0 | 2.18 | 5.45 | 5.99 | **13.62** |
| 505 | 35.40 | 77.89 | 93.69 | 54.47 | **261.45** |
| 506 | 17.97 | 19.61 | 2.18 | 7.08 | **46.84** |
| 507 | 225.50 | 116.02 | 313.74 | 179.20 | **834.46** |
| 508 | 1.09 | 0 | 1.09 | 2.72 | **4.90** |
| 510 | 9.80 | 1.09 | 5.99 | 1.09 | **17.97** |
| 511 | 54.47 | 5.45 | 19.61 | 15.80 | **95.32** |
| 512 | 2.18 | 1.63 | 7.63 | 0 | **11.44** |
| 513 | 52.29 | 74.08 | 38.13 | 21.79 | **186.28** |
| 514 | 1.09 | 4.36 | 2.18 | 2.72 | **10.35** |
| 515 | 14.71 | 8.17 | 29.41 | 3.81 | **56.10** |
| 516 | 5.45 | 0 | 2.18 | 4.36 | **11.98** |
| 520 | 5.99 | 4.90 | 5.45 | 3.81 | **20.15** |
| 521 | 0 | 0 | 2.18 | 0 | **2.18** |

| | | | | | |
|---|---|---|---|---|---|
| 522 | 56.10 | 81.70 | 52.29 | 39.76 | **229.86** |
| 523 | 36.49 | 26.69 | 10.89 | 0 | **74.08** |
| 524 | 139.44 | 170.49 | 164.50 | 122.01 | **596.43** |
| 525 | 80.61 | 61.01 | 15.25 | 5.45 | **162.32** |
| 526 | 28.87 | 4.90 | 2.18 | 10.02 | **45.97** |
| 527 | 26.69 | 37.04 | 10.35 | 14.16 | **88.24** |
| 528 | 19.61 | 7.63 | 26.15 | 27.23 | **80.61** |
| **IA IOWA Total** | **1,502.79** | **1,097.55** | **1,154.74** | **965.95** | **4,721.02** |

**STATE: ID - IDAHO**

| REGISTRANT ZIP CODE 3 | 1ST QUARTER | 2ND QUARTER | 3RD QUARTER | 4TH QUARTER | TOTAL GRAMS |
|---|---|---|---|---|---|
| 832 | 22.88 | 39.76 | 26.69 | 23.64 | **112.97** |
| 833 | 39.76 | 40.85 | 17.97 | 22.01 | **120.59** |
| 834 | 150.33 | 73.53 | 76.80 | 56.43 | **357.10** |
| 835 | 11.44 | 4.36 | 49.02 | 3.27 | **68.09** |
| 836 | 233.67 | 150.33 | 16.34 | 11.44 | **411.78** |
| 837 | 107.30 | 58.83 | 47.61 | 28.87 | **242.60** |
| 838 | 33.77 | 51.20 | 21.24 | 9.80 | **116.02** |
| **ID IDAHO Total** | **599.16** | **418.86** | **255.68** | **155.45** | **1,429.15** |

**STATE: IL - ILLINOIS**

| REGISTRANT ZIP CODE 3 | 1ST QUARTER | 2ND QUARTER | 3RD QUARTER | 4TH QUARTER | TOTAL GRAMS |
|---|---|---|---|---|---|
| 600 | 210.79 | 106.76 | 112.21 | 70.48 | **500.24** |
| 601 | 218.42 | 253.82 | 79.52 | 61.01 | **612.77** |
| 602 | 4.36 | 2.18 | 0 | 2.18 | **8.72** |
| 604 | 123.64 | 120.92 | 55.01 | 51.75 | **351.32** |
| 605 | 66.45 | 88.24 | 38.67 | 68.63 | **261.99** |
| 606 | 62.09 | 145.98 | 88.78 | 29.96 | **326.81** |
| 607 | 15.25 | 0 | 0 | 0.54 | **15.80** |
| 608 | 117.65 | 43.58 | 34.86 | 0 | **196.09** |
| 609 | 0.54 | 0.54 | 1.09 | 0 | **2.18** |
| 610 | 92.60 | 71.90 | 113.30 | 59.92 | **337.71** |
| 611 | 41.94 | 24.51 | 17.97 | 7.63 | **92.05** |
| 612 | 12.53 | 4.36 | 16.34 | 8.72 | **41.94** |
| 613 | 52.29 | 6.54 | 19.06 | 11.44 | **89.33** |
| 614 | 29.41 | 17.97 | 25.06 | 6.54 | **78.98** |
| 615 | 1.63 | 8.72 | 2.18 | 2.18 | **14.71** |
| 616 | 38.67 | 27.78 | 30.50 | 22.33 | **119.29** |
| 617 | 27.78 | 32.68 | 5.99 | 3.81 | **70.26** |
| 618 | 49.02 | 47.39 | 137.81 | 9.80 | **244.02** |
| 619 | 20.15 | 31.05 | 37.04 | 30.50 | **118.74** |
| 620 | 20.70 | 0.54 | 3.27 | 5.45 | **29.96** |

| | | | | | |
|---|---|---|---|---|---|
| 622 | 105.12 | 98.59 | 49.57 | 25.06 | **278.34** |
| 623 | 41.40 | 37.58 | 29.41 | 7.63 | **116.02** |
| 624 | 10.35 | 2.72 | 10.89 | 2.18 | **26.15** |
| 625 | 2.72 | 0.54 | 16.34 | 7.08 | **26.69** |
| 626 | 35.95 | 35.40 | 27.23 | 8.72 | **107.30** |
| 627 | 57.19 | 16.89 | 26.15 | 19.61 | **119.83** |
| 628 | 46.84 | 28.87 | 60.46 | 10.35 | **146.52** |
| 629 | 119.29 | 158.50 | 123.64 | 106.76 | **508.19** |
| **IL ILLINOIS Total** | **1,624.80** | **1,414.55** | **1,162.36** | **640.23** | **4,841.95** |

**STATE: IN - INDIANA**

| REGISTRANT ZIP CODE 3 | 1ST QUARTER | 2ND QUARTER | 3RD QUARTER | 4TH QUARTER | **TOTAL GRAMS** |
|---|---|---|---|---|---|
| 460 | 174.30 | 147.61 | 137.26 | 159.27 | **618.44** |
| 461 | 292.50 | 183.02 | 149.79 | 139.98 | **765.29** |
| 462 | 592.08 | 357.32 | 494.03 | 376.92 | **1,820.35** |
| 463 | 77.89 | 66.45 | 59.37 | 29.96 | **233.67** |
| 464 | 14.16 | 25.06 | 28.32 | 20.15 | **87.69** |
| 465 | 56.10 | 25.06 | 50.11 | 17.43 | **148.70** |
| 466 | 20.70 | 18.52 | 27.23 | 72.44 | **138.90** |
| 467 | 17.97 | 8.72 | 14.16 | 9.26 | **50.11** |
| 468 | 81.16 | 77.89 | 55.01 | 56.10 | **270.17** |
| 469 | 41.40 | 15.80 | 19.61 | 3.81 | **80.61** |
| 470 | 95.87 | 7.63 | 36.49 | 28.87 | **168.85** |
| 471 | 113.84 | 39.22 | 39.76 | 44.12 | **236.94** |
| 472 | 56.65 | 77.35 | 37.58 | 61.55 | **233.13** |
| 473 | 62.64 | 75.71 | 76.26 | 38.67 | **253.28** |
| 474 | 20.70 | 8.17 | 6.54 | 7.63 | **43.03** |
| 475 | 31.59 | 56.10 | 29.96 | 18.52 | **136.17** |
| 476 | 31.05 | 18.52 | 40.31 | 9.26 | **99.13** |
| 477 | 25.60 | 113.84 | 46.84 | 20.70 | **206.98** |
| 478 | 101.31 | 31.59 | 39.22 | 2.72 | **174.84** |
| 479 | 65.91 | 34.32 | 22.88 | 20.70 | **143.80** |
| **IN INDIANA Total** | **1,973.40** | **1,387.86** | **1,410.74** | **1,138.07** | **5,910.08** |

**STATE: KS - KANSAS**

| REGISTRANT ZIP CODE 3 | 1ST QUARTER | 2ND QUARTER | 3RD QUARTER | 4TH QUARTER | **TOTAL GRAMS** |
|---|---|---|---|---|---|
| 660 | 102.95 | 168.31 | 125.82 | 37.04 | **434.12** |
| 661 | 11.98 | 1.09 | 6.54 | 6.54 | **26.15** |
| 662 | 108.94 | 74.08 | 40.85 | 28.32 | **252.19** |
| 664 | 3.27 | 6.54 | 0.54 | 7.63 | **17.97** |
| 665 | 11.98 | 19.61 | 4.36 | 1.09 | **37.04** |
| 666 | 11.44 | 30.50 | 28.32 | 13.07 | **83.34** |

| | | | | | |
|---|---|---|---|---|---|
| 667 | 106.76 | 50.66 | 28.32 | 11.98 | **197.72** |
| 668 | 4.36 | 6.54 | 3.27 | 21.79 | **35.95** |
| 669 | 18.52 | 2.72 | 10.35 | 4.36 | **35.95** |
| 670 | 68.09 | 21.79 | 5.99 | 16.89 | **112.75** |
| 671 | 41.40 | 23.97 | 12.53 | 4.36 | **82.25** |
| 672 | 222.23 | 77.35 | 155.78 | 111.66 | **567.02** |
| 673 | 11.44 | 19.61 | 4.36 | 19.61 | **55.01** |
| 674 | 56.10 | 53.92 | 9.26 | 17.43 | **136.72** |
| 675 | 159.05 | 71.90 | 18.52 | 11.98 | **261.45** |
| 676 | 58.28 | 31.59 | 7.08 | 5.45 | **102.40** |
| 677 | 10.89 | 3.81 | 4.36 | 3.27 | **22.33** |
| 678 | 51.20 | 32.68 | 33.77 | 37.58 | **155.24** |
| 679 | 69.72 | 37.04 | 52.83 | 21.24 | **180.84** |
| **KS - KANSAS - Total** | **1,128.59** | **733.69** | **552.86** | **381.28** | **2,796.43** |

**STATE: KY - KENTUCKY**

| REGISTRANT ZIP CODE 3 | 1ST QUARTER | 2ND QUARTER | 3RD QUARTER | 4TH QUARTER | **TOTAL GRAMS** |
|---|---|---|---|---|---|
| 400 | 38.67 | 26.69 | 6.54 | 9.80 | **81.70** |
| 401 | 7.08 | 1.09 | 4.36 | 5.99 | **18.52** |
| 402 | 180.29 | 31.05 | 102.40 | 66.13 | **379.87** |
| 403 | 20.70 | 21.79 | 23.42 | 16.89 | **82.79** |
| 404 | 17.43 | 18.52 | 27.45 | 14.16 | **77.56** |
| 405 | 38.13 | 61.01 | 48.48 | 45.75 | **193.36** |
| 406 | 2.72 | 2.72 | 0 | 1.09 | **6.54** |
| 407 | 25.60 | 16.34 | 19.61 | 23.97 | **85.52** |
| 408 | 15.25 | 16.34 | 34.32 | 29.41 | **95.32** |
| 409 | 10.89 | 13.07 | 20.15 | 20.70 | **64.82** |
| 410 | 82.25 | 52.29 | 29.09 | 16.89 | **180.51** |
| 411 | 0 | 1.09 | 9.80 | 0.54 | **11.44** |
| 412 | 0 | 0 | 0.54 | 0 | **0.54** |
| 413 | 4.90 | 7.08 | 3.27 | 2.72 | **17.97** |
| 415 | 9.26 | 7.08 | 34.86 | 17.97 | **69.18** |
| 416 | 25.60 | 5.99 | 27.23 | 10.35 | **69.18** |
| 417 | 9.26 | 5.99 | 6.54 | 9.26 | **31.05** |
| 418 | 36.49 | 43.03 | 56.65 | 33.23 | **169.40** |
| 420 | 202.62 | 177.02 | 181.93 | 51.20 | **612.77** |
| 421 | 178.11 | 166.67 | 100.22 | 63.73 | **508.74** |
| 422 | 81.16 | 68.09 | 39.22 | 11.98 | **200.45** |
| 423 | 61.55 | 22.88 | 27.78 | 38.67 | **150.88** |
| 424 | 52.83 | 35.40 | 15.80 | 11.98 | **116.02** |
| 425 | 47.39 | 33.77 | 31.59 | 50.66 | **163.41** |

| | | | | | |
|---|---|---|---|---|---|
| 426 | 7.63 | 20.70 | 25.06 | 10.35 | **63.73** |
| 427 | 62.09 | 76.26 | 71.35 | 33.77 | **243.48** |
| **KY - KENTUCKY - Total** | **1,217.92** | **931.96** | **947.65** | **597.20** | **3,694.72** |

**STATE: LA - LOUISIANA**

| REGISTRANT ZIP CODE 3 | 1ST QUARTER | 2ND QUARTER | 3RD QUARTER | 4TH QUARTER | TOTAL GRAMS |
|---|---|---|---|---|---|
| 700 | 432.48 | 322.46 | 191.40 | 114.38 | **1,060.72** |
| 701 | 78.44 | 50.11 | 30.50 | 28.87 | **187.92** |
| 703 | 181.93 | 160.14 | 84.43 | 70.59 | **497.08** |
| 704 | 342.06 | 237.48 | 187.92 | 209.70 | **977.17** |
| 705 | 647.09 | 496.21 | 380.74 | 319.19 | **1,843.22** |
| 706 | 320.82 | 171.58 | 111.66 | 136.17 | **740.23** |
| 707 | 354.05 | 239.66 | 184.65 | 59.92 | **838.27** |
| 708 | 241.84 | 225.50 | 128.00 | 126.91 | **722.26** |
| 710 | 167.76 | 115.47 | 88.24 | 7.63 | **379.10** |
| 711 | 364.40 | 253.28 | 227.68 | 99.68 | **945.03** |
| 712 | 337.71 | 204.80 | 263.85 | 131.81 | **938.17** |
| 713 | 179.75 | 124.73 | 81.16 | 133.99 | **519.63** |
| 714 | 94.78 | 104.58 | 75.71 | 64.82 | **339.89** |
| **LA - LOUISIANA - Total** | **3,743.09** | **2,706.01** | **2,035.93** | **1,503.66** | **9,988.70** |

**STATE: MA - MASSACHUSETTS**

| REGISTRANT ZIP CODE 3 | 1ST QUARTER | 2ND QUARTER | 3RD QUARTER | 4TH QUARTER | TOTAL GRAMS |
|---|---|---|---|---|---|
| 010 | 196.63 | 93.69 | 55.56 | 44.12 | **390.00** |
| 011 | 2.72 | 6.54 | 11.44 | 0 | **20.70** |
| 012 | 5.45 | 0.54 | 0 | 0 | **5.99** |
| 013 | 4.36 | 4.36 | 0.54 | 0 | **9.26** |
| 014 | 62.09 | 62.64 | 2.72 | 0 | **127.46** |
| 015 | 27.23 | 28.32 | 32.68 | 6.54 | **94.78** |
| 016 | 31.59 | 6.54 | 4.90 | 4.36 | **47.39** |
| 017 | 106.76 | 67.54 | 15.80 | 7.30 | **197.39** |
| 018 | 86.06 | 40.31 | 38.35 | 20.70 | **185.41** |
| 019 | 108.39 | 100.77 | 92.60 | 80.61 | **382.37** |
| 020 | 71.90 | 53.92 | 1.09 | 2.61 | **129.53** |
| 021 | 117.65 | 116.56 | 86.06 | 81.70 | **401.98** |
| 022 | 28.32 | 20.15 | 16.34 | 1.63 | **66.45** |
| 023 | 148.70 | 92.05 | 33.77 | 35.95 | **310.47** |
| 024 | 106.21 | 37.58 | 43.58 | 58.83 | **246.20** |
| 025 | 57.19 | 47.93 | 3.27 | 0 | **108.39** |
| 026 | 17.43 | 8.72 | 1.09 | 0 | **27.23** |
| 027 | 108.39 | 89.87 | 15.69 | 4.90 | **218.86** |
| **MA - MASSACHUSETTS - Total** | **1,287.10** | **878.04** | **455.47** | **349.25** | **2,969.85** |

**STATE: MD - MARYLAND**

| REGISTRANT ZIP CODE 3 | 1ST QUARTER | 2ND QUARTER | 3RD QUARTER | 4TH QUARTER | TOTAL GRAMS |
|---|---|---|---|---|---|
| 206 | 114.38 | 96.41 | 62.64 | 61.55 | **334.98** |
| 207 | 129.09 | 71.90 | 86.61 | 76.26 | **363.85** |
| 208 | 186.83 | 218.96 | 82.25 | 43.03 | **531.07** |
| 209 | 13.62 | 10.35 | 11.98 | 34.32 | **70.26** |
| 210 | 306.66 | 330.08 | 219.51 | 89.33 | **945.58** |
| 211 | 107.30 | 54.47 | 13.62 | 0.54 | **175.93** |
| 212 | 265.81 | 296.85 | 184.65 | 97.50 | **844.81** |
| 214 | 64.27 | 44.12 | 96.41 | 17.97 | **222.78** |
| 215 | 45.75 | 8.72 | 97.50 | 4.36 | **156.33** |
| 216 | 33.77 | 37.04 | 50.66 | 18.52 | **139.98** |
| 217 | 19.61 | 26.15 | 9.26 | 8.17 | **63.18** |
| 218 | 87.15 | 57.74 | 22.88 | 6.54 | **174.30** |
| 219 | 0 | 0.54 | 0 | 0 | **0.54** |
| **MD MARYLAND Total** | **1,374.25** | **1,253.33** | **937.95** | **458.08** | **4,023.61** |

**STATE: ME - MAINE**

| REGISTRANT ZIP CODE 3 | 1ST QUARTER | 2ND QUARTER | 3RD QUARTER | 4TH QUARTER | TOTAL GRAMS |
|---|---|---|---|---|---|
| 039 | 4.36 | 4.36 | 4.36 | 3.27 | **16.34** |
| 040 | 79.52 | 39.22 | 25.82 | 10.35 | **154.91** |
| 041 | 21.24 | 1.63 | 7.63 | 22.88 | **53.38** |
| 042 | 19.06 | 1.63 | 3.27 | 1.09 | **25.06** |
| 043 | 10.35 | 2.72 | 1.09 | 0 | **14.16** |
| 044 | 2.72 | 14.71 | 14.71 | 5.45 | **37.58** |
| 045 | 4.36 | 1.09 | 0 | 0 | **5.45** |
| 046 | 17.43 | 15.80 | 25.06 | 14.16 | **72.44** |
| 047 | 17.43 | 4.90 | 3.27 | 1.63 | **27.23** |
| 048 | 0.54 | 0 | 0 | 0 | **0.54** |
| 049 | 7.08 | 2.18 | 8.17 | 4.90 | **22.33** |
| **ME MAINE Total** | **184.10** | **88.24** | **93.36** | **63.73** | **429.43** |

**STATE: MI - MICHIGAN**

| REGISTRANT ZIP CODE 3 | 1ST QUARTER | 2ND QUARTER | 3RD QUARTER | 4TH QUARTER | TOTAL GRAMS |
|---|---|---|---|---|---|
| 480 | 145.43 | 159.59 | 66.34 | 52.29 | **423.66** |
| 481 | 185.74 | 153.06 | 166.35 | 35.95 | **541.09** |
| 482 | 34.86 | 41.94 | 25.06 | 23.42 | **125.28** |
| 483 | 37.58 | 36.49 | 46.52 | 17.97 | **138.57** |
| 484 | 149.79 | 101.31 | 25.06 | 9.26 | **285.42** |
| 485 | 1.09 | 58.83 | 5.45 | 1.63 | **67.00** |
| 486 | 180.84 | 135.08 | 116.02 | 135.63 | **567.56** |
| 487 | 19.61 | 2.18 | 9.80 | 28.32 | **59.92** |

| | | | | | |
|---|---|---|---|---|---|
| 488 | 200.99 | 99.13 | 30.50 | 15.25 | **345.88** |
| 489 | 83.88 | 51.75 | 41.94 | 16.34 | **193.91** |
| 490 | 160.68 | 133.99 | 38.67 | 21.79 | **355.14** |
| 491 | 0.54 | 0 | 1.63 | 0 | **2.18** |
| 492 | 52.29 | 16.34 | 10.02 | 13.07 | **91.73** |
| 493 | 22.33 | 39.22 | 44.12 | 8.72 | **114.38** |
| 494 | 54.47 | 37.58 | 58.83 | 16.89 | **167.76** |
| 495 | 101.31 | 37.04 | 81.16 | 20.15 | **239.66** |
| 496 | 47.39 | 56.10 | 39.22 | 17.43 | **160.14** |
| 497 | 183.56 | 175.93 | 101.31 | 59.37 | **520.18** |
| 498 | 59.37 | 33.23 | 50.11 | 62.09 | **204.80** |
| 499 | 13.07 | 16.34 | 0.54 | 1.09 | **31.05** |
| **MI - MICHIGAN Total** | **1,734.83** | **1,385.14** | **958.65** | **556.67** | **4,635.29** |

**STATE: MN - MINNESOTA**

| REGISTRANT ZIP CODE 3 | 1ST QUARTER | 2ND QUARTER | 3RD QUARTER | 4TH QUARTER | TOTAL GRAMS |
|---|---|---|---|---|---|
| 550 | 46.84 | 38.67 | 8.72 | 4.90 | **99.13** |
| 551 | 24.51 | 23.97 | 13.07 | 15.80 | **77.35** |
| 553 | 54.47 | 35.95 | 47.93 | 31.05 | **169.40** |
| 554 | 40.31 | 34.86 | 47.39 | 25.06 | **147.61** |
| 556 | 4.90 | 0 | 0 | 0 | **4.90** |
| 557 | 35.40 | 42.49 | 34.86 | 18.52 | **131.27** |
| 558 | 67.00 | 25.06 | 10.35 | 4.36 | **106.76** |
| 559 | 18.52 | 44.66 | 17.97 | 15.25 | **96.41** |
| 560 | 22.33 | 13.07 | 5.99 | 7.63 | **49.02** |
| 561 | 1.09 | 2.18 | 1.63 | 0 | **4.90** |
| 562 | 9.80 | 9.26 | 9.26 | 2.18 | **30.50** |
| 563 | 22.88 | 33.23 | 57.74 | 17.43 | **131.27** |
| 564 | 4.90 | 4.90 | 4.90 | 2.72 | **17.43** |
| 565 | 11.98 | 11.44 | 3.81 | 4.36 | **31.59** |
| 566 | 1.63 | 4.36 | 0.54 | 1.09 | **7.63** |
| 567 | 23.97 | 16.34 | 0 | 14.16 | **54.47** |
| **MN MINNESOTA Total** | **390.54** | **340.43** | **264.17** | **164.50** | **1,159.64** |

**STATE: MO - MISSOURI**

| REGISTRANT ZIP CODE 3 | 1ST QUARTER | 2ND QUARTER | 3RD QUARTER | 4TH QUARTER | TOTAL GRAMS |
|---|---|---|---|---|---|
| 630 | 141.07 | 126.37 | 88.78 | 69.18 | **425.40** |
| 631 | 392.72 | 229.31 | 151.42 | 86.61 | **860.06** |
| 633 | 36.49 | 25.06 | 49.57 | 14.71 | **125.82** |
| 634 | 7.08 | 1.09 | 9.80 | 7.08 | **25.06** |
| 635 | 14.71 | 24.51 | 8.72 | 25.06 | **72.99** |
| 636 | 49.02 | 26.15 | 7.63 | 5.45 | **88.24** |

| | | | | | |
|---|---|---|---|---|---|
| 637 | 16.34 | 13.07 | 20.70 | 10.35 | **60.46** |
| 638 | 42.49 | 29.41 | 57.19 | 8.72 | **137.81** |
| 639 | 40.31 | 8.72 | 12.53 | 0.54 | **62.09** |
| 640 | 64.27 | 28.87 | 96.41 | 25.60 | **215.15** |
| 641 | 214.06 | 133.45 | 69.18 | 46.84 | **463.53** |
| 644 | 13.62 | 3.81 | 21.79 | 14.16 | **53.38** |
| 645 | 11.98 | 80.07 | 9.80 | 11.98 | **113.84** |
| 646 | 32.68 | 17.43 | 4.36 | 3.27 | **57.74** |
| 647 | 15.25 | 4.36 | 9.80 | 2.18 | **31.59** |
| 648 | 76.80 | 53.92 | 48.48 | 24.51 | **203.71** |
| 650 | 61.01 | 4.90 | 1.63 | 0.54 | **68.09** |
| 651 | 33.77 | 82.79 | 46.84 | 33.23 | **196.63** |
| 652 | 209.16 | 130.18 | 261.45 | 29.41 | **630.20** |
| 653 | 63.18 | 68.63 | 27.23 | 52.29 | **211.34** |
| 654 | 18.52 | 63.18 | 26.15 | 0 | **107.85** |
| 655 | 99.13 | 97.50 | 89.33 | 37.58 | **323.54** |
| 656 | 45.75 | 14.16 | 9.80 | 0 | **69.72** |
| 657 | 165.59 | 99.13 | 104.58 | 14.16 | **383.46** |
| 658 | 148.16 | 153.60 | 78.98 | 43.03 | **423.77** |
| **MO MISSOURI - Total** | **2,013.17** | **1,519.68** | **1,312.15** | **566.48** | **5,411.47** |

**STATE: MS - MISSISSIPPI**

| REGISTRANT ZIP CODE 3 | 1ST QUARTER | 2ND QUARTER | 3RD QUARTER | 4TH QUARTER | **TOTAL GRAMS** |
|---|---|---|---|---|---|
| 386 | 306.66 | 165.80 | 236.07 | 134.54 | **843.07** |
| 387 | 241.30 | 149.24 | 138.35 | 166.67 | **695.57** |
| 388 | 161.77 | 128.55 | 101.31 | 120.38 | **512.01** |
| 389 | 117.11 | 85.52 | 77.35 | 44.12 | **324.09** |
| 390 | 184.65 | 124.73 | 66.45 | 43.58 | **419.41** |
| 391 | 146.52 | 85.52 | 79.52 | 102.95 | **414.51** |
| 392 | 245.11 | 169.40 | 161.77 | 171.58 | **747.86** |
| 393 | 221.69 | 233.13 | 170.49 | 116.02 | **741.32** |
| 394 | 298.49 | 182.47 | 141.29 | 139.77 | **762.02** |
| 395 | 468.43 | 314.83 | 273.98 | 177.57 | **1,234.81** |
| 396 | 68.63 | 53.38 | 47.93 | 32.68 | **202.62** |
| 397 | 70.81 | 76.26 | 77.89 | 25.60 | **250.56** |
| **MS MISSISSIPPI - Total** | **2,531.16** | **1,768.82** | **1,572.40** | **1,275.44** | **7,147.83** |

**STATE: MT - MONTANA**

| REGISTRANT ZIP CODE 3 | 1ST QUARTER | 2ND QUARTER | 3RD QUARTER | 4TH QUARTER | **TOTAL GRAMS** |
|---|---|---|---|---|---|
| 590 | 5.45 | 9.80 | 2.18 | 0 | **17.43** |
| 591 | 29.96 | 27.23 | 14.71 | 49.57 | **121.47** |
| 592 | 20.70 | 4.36 | 1.09 | 1.09 | **27.23** |

| | | | | | |
|---|---|---|---|---|---|
| 593 | 44.12 | 0.54 | 0 | 2.18 | **46.84** |
| 594 | 53.38 | 61.01 | 65.91 | 30.50 | **210.79** |
| 595 | 4.36 | 0 | 15.25 | 4.36 | **23.97** |
| 596 | 10.89 | 7.63 | 8.72 | 6.54 | **33.77** |
| 597 | 24.51 | 16.89 | 27.78 | 9.80 | **78.98** |
| 598 | 34.86 | 40.31 | 13.62 | 3.81 | **92.60** |
| 599 | 65.91 | 45.21 | 23.42 | 3.81 | **138.35** |
| **MT - MONTANA Total** | **294.13** | **212.97** | **172.67** | **111.66** | **791.43** |

**STATE: NC - NORTH CAROLINA**

| REGISTRANT ZIP CODE 3 | 1ST QUARTER | 2ND QUARTER | 3RD QUARTER | 4TH QUARTER | TOTAL GRAMS |
|---|---|---|---|---|---|
| 270 | 51.75 | 34.86 | 1.09 | 17.43 | **105.12** |
| 271 | 74.08 | 36.49 | 23.42 | 123.10 | **257.09** |
| 272 | 172.12 | 75.71 | 43.03 | 35.95 | **326.81** |
| 273 | 142.71 | 82.25 | 70.26 | 31.59 | **326.81** |
| 274 | 126.37 | 80.07 | 22.33 | 7.63 | **236.39** |
| 275 | 355.68 | 215.15 | 106.21 | 50.66 | **727.70** |
| 276 | 199.36 | 108.94 | 71.35 | 38.13 | **417.78** |
| 277 | 2.72 | 71.90 | 11.98 | 11.44 | **98.04** |
| 278 | 179.20 | 165.59 | 92.60 | 21.24 | **458.63** |
| 279 | 154.69 | 152.51 | 77.35 | 14.71 | **399.26** |
| 280 | 227.68 | 110.57 | 93.69 | 60.46 | **492.40** |
| 281 | 468.43 | 192.82 | 57.74 | 52.83 | **771.82** |
| 282 | 131.27 | 113.30 | 45.21 | 16.34 | **306.11** |
| 283 | 223.87 | 112.75 | 116.02 | 135.63 | **588.26** |
| 284 | 191.19 | 63.73 | 46.30 | 13.07 | **314.28** |
| 285 | 118.20 | 130.18 | 50.11 | 30.50 | **328.99** |
| 286 | 360.58 | 229.31 | 26.15 | 59.81 | **675.85** |
| 287 | 162.32 | 86.61 | 61.55 | 61.01 | **371.48** |
| 288 | 27.23 | 7.08 | 35.95 | 32.68 | **102.95** |
| 289 | 8.72 | 4.36 | 1.09 | 0 | **14.16** |
| **NC - NORTH CAROLINA Total** | **3,378.15** | **2,074.17** | **1,053.43** | **814.20** | **7,319.95** |

**STATE: ND - NORTH DAKOTA**

| REGISTRANT ZIP CODE 3 | 1ST QUARTER | 2ND QUARTER | 3RD QUARTER | 4TH QUARTER | TOTAL GRAMS |
|---|---|---|---|---|---|
| 580 | 26.15 | 4.36 | 17.97 | 4.36 | **52.83** |
| 581 | 4.36 | 33.23 | 8.17 | 3.27 | **49.02** |
| 582 | 0 | 1.09 | 3.81 | 6.54 | **11.44** |
| 583 | 22.88 | 26.15 | 14.16 | 1.63 | **64.82** |
| 584 | 4.90 | 4.90 | 2.18 | 4.36 | **16.34** |
| 585 | 57.74 | 39.22 | 19.61 | 11.44 | **128.00** |
| 586 | 8.72 | 4.36 | 3.27 | 0 | **16.34** |

| | | | | | |
|---|---|---|---|---|---|
| 587 | 26.15 | 21.24 | 10.57 | 8.72 | **66.67** |
| 588 | 3.27 | 1.63 | 0 | 2.18 | **7.08** |
| **ND NORTH DAKOTA Total** | **154.15** | **136.17** | **79.74** | **42.49** | **412.55** |

**STATE: NE - NEBRASKA**

| REGISTRANT ZIP CODE 3 | 1ST QUARTER | 2ND QUARTER | 3RD QUARTER | 4TH QUARTER | TOTAL GRAMS |
|---|---|---|---|---|---|
| 680 | 40.85 | 15.80 | 4.90 | 10.89 | **72.44** |
| 681 | 178.66 | 47.39 | 58.28 | 39.76 | **324.09** |
| 683 | 5.99 | 4.36 | 4.36 | 4.36 | **19.06** |
| 684 | 6.54 | 2.72 | 9.26 | 4.36 | **22.88** |
| 685 | 27.78 | 10.35 | 53.92 | 14.71 | **106.76** |
| 686 | 7.63 | 6.54 | 19.61 | 3.27 | **37.04** |
| 687 | 13.62 | 9.80 | 7.63 | 10.35 | **41.40** |
| 688 | 55.56 | 38.13 | 33.23 | 16.34 | **143.25** |
| 689 | 33.77 | 29.96 | 22.88 | 17.43 | **104.04** |
| 690 | 22.88 | 5.99 | 6.54 | 4.36 | **39.76** |
| 691 | 20.15 | 17.97 | 16.89 | 5.99 | **61.01** |
| 692 | 10.89 | 4.90 | 11.98 | 2.18 | **29.96** |
| 693 | 14.71 | 13.07 | 13.62 | 14.16 | **55.56** |
| **NE NEBRASKA Total** | **439.02** | **206.98** | **263.08** | **148.16** | **1,057.24** |

**STATE: NH - NEW HAMPSHIRE**

| REGISTRANT ZIP CODE 3 | 1ST QUARTER | 2ND QUARTER | 3RD QUARTER | 4TH QUARTER | TOTAL GRAMS |
|---|---|---|---|---|---|
| 030 | 44.66 | 10.24 | 10.68 | 4.90 | **70.48** |
| 031 | 56.65 | 32.68 | 2.53 | 2.18 | **94.03** |
| 032 | 28.32 | 15.80 | 21.79 | 8.72 | **74.62** |
| 033 | 149.24 | 120.92 | 38.13 | 14.25 | **322.54** |
| 034 | 68.09 | 13.07 | 45.75 | 2.18 | **129.09** |
| 035 | 0.54 | 2.72 | 1.09 | 4.36 | **8.72** |
| 037 | 22.88 | 2.18 | 4.36 | 11.98 | **41.40** |
| 038 | 45.75 | 46.30 | 7.63 | 5.45 | **105.12** |
| **NH - NEW HAMPSHIRE Total** | **416.14** | **243.91** | **131.95** | **54.01** | **846.01** |

**STATE: NJ - NEW JERSEY**

| REGISTRANT ZIP CODE 3 | 1ST QUARTER | 2ND QUARTER | 3RD QUARTER | 4TH QUARTER | TOTAL GRAMS |
|---|---|---|---|---|---|
| 070 | 184.10 | 101.86 | 52.83 | 67.21 | **406.01** |
| 071 | 2.72 | 10.89 | 4.36 | 8.17 | **26.15** |
| 072 | 0 | 15.80 | 16.34 | 22.88 | **55.01** |
| 073 | 0.54 | 0 | 0 | 0 | **0.54** |
| 074 | 29.96 | 47.39 | 52.83 | 31.59 | **161.77** |
| 075 | 11.98 | 14.71 | 5.45 | 13.62 | **45.75** |
| 076 | 99.68 | 116.56 | 39.76 | 28.87 | **284.87** |
| 077 | 47.39 | 27.78 | 21.79 | 23.42 | **120.38** |

| | | | | | |
|---|---|---|---|---|---|
| 078 | 18.52 | 10.35 | 6.54 | 4.58 | **39.98** |
| 079 | 45.21 | 14.71 | 19.50 | 21.24 | **100.66** |
| 080 | 150.88 | 143.80 | 95.87 | 32.68 | **423.22** |
| 081 | 9.26 | 0.54 | 2.72 | 0.54 | **13.07** |
| 082 | 25.06 | 23.97 | 5.99 | 5.45 | **60.46** |
| 083 | 9.80 | 0.54 | 1.09 | 1.09 | **12.53** |
| 084 | 0.54 | 1.09 | 0.54 | 3.81 | **5.99** |
| 085 | 65.36 | 17.43 | 13.62 | 9.26 | **105.67** |
| 086 | 35.40 | 13.07 | 3.27 | 2.72 | **54.47** |
| 087 | 52.83 | 45.75 | 105.67 | 108.94 | **313.20** |
| 088 | 432.48 | 243.48 | 106.76 | 29.96 | **812.67** |
| 089 | 20.15 | 10.89 | 0.54 | 0.54 | **32.14** |
| **NJ NEW JERSEY - Total** | **1,241.89** | **860.61** | **555.47** | **416.58** | **3,074.54** |

**STATE: NM - NEW MEXICO**

| REGISTRANT ZIP CODE 3 | 1ST QUARTER | 2ND QUARTER | 3RD QUARTER | 4TH QUARTER | **TOTAL GRAMS** |
|---|---|---|---|---|---|
| 870 | 47.93 | 26.15 | 17.43 | 2.18 | **93.69** |
| 871 | 307.20 | 228.22 | 192.27 | 136.72 | **864.42** |
| 874 | 15.80 | 5.45 | 0.54 | 2.18 | **23.97** |
| 875 | 46.84 | 35.40 | 5.45 | 23.42 | **111.12** |
| 877 | 22.33 | 10.89 | 11.98 | 0 | **45.21** |
| 878 | 5.45 | 4.36 | 0 | 0 | **9.80** |
| 880 | 55.01 | 63.73 | 53.38 | 53.38 | **225.50** |
| 881 | 5.45 | 16.34 | 14.71 | 0 | **36.49** |
| 882 | 80.07 | 82.25 | 41.40 | 25.60 | **229.31** |
| 883 | 4.36 | 4.36 | 19.06 | 0.54 | **28.32** |
| 884 | 0 | 0 | 1.09 | 0 | **1.09** |
| **NM NEW MEXICO Total** | **590.44** | **477.15** | **357.32** | **244.02** | **1,668.92** |

**STATE: NV - NEVADA**

| REGISTRANT ZIP CODE 3 | 1ST QUARTER | 2ND QUARTER | 3RD QUARTER | 4TH QUARTER | **TOTAL GRAMS** |
|---|---|---|---|---|---|
| 890 | 157.96 | 105.67 | 208.62 | 84.43 | **556.67** |
| 891 | 461.35 | 467.89 | 505.47 | 297.40 | **1,732.11** |
| 893 | 13.07 | 7.63 | 3.27 | 0 | **23.97** |
| 894 | 36.49 | 26.69 | 4.90 | 11.44 | **79.52** |
| 895 | 133.99 | 80.07 | 87.69 | 60.46 | **362.22** |
| 897 | 34.32 | 3.81 | 21.79 | 1.09 | **61.01** |
| 898 | 6.54 | 22.33 | 14.71 | 5.45 | **49.02** |
| **NV NEVADA Total** | **843.72** | **714.09** | **846.44** | **460.26** | **2,864.51** |

**STATE: NY - NEW YORK**

| REGISTRANT ZIP CODE 3 | 1ST QUARTER | 2ND QUARTER | 3RD QUARTER | 4TH QUARTER | **TOTAL GRAMS** |
|---|---|---|---|---|---|
| 100 | 226.05 | 226.59 | 133.34 | 99.90 | **685.87** |

| | | | | | |
|---|---|---|---|---|---|
| 101 | 0 | 41.40 | 6.10 | 2.40 | **49.89** |
| 103 | 67.54 | 25.06 | 51.20 | 40.85 | **184.65** |
| 104 | 59.37 | 72.44 | 67.21 | 18.96 | **217.98** |
| 105 | 132.90 | 95.87 | 28.87 | 48.48 | **306.11** |
| 106 | 2.18 | 0 | 12.64 | 11.11 | **25.93** |
| 107 | 57.74 | 70.81 | 13.07 | 1.09 | **142.71** |
| 108 | 6.54 | 1.09 | 17.43 | 10.89 | **35.95** |
| 109 | 53.38 | 63.73 | 11.98 | 34.64 | **163.73** |
| 110 | 52.83 | 45.75 | 9.80 | 5.45 | **113.84** |
| 111 | 0 | 4.36 | 0 | 3.27 | **7.63** |
| 112 | 124.73 | 49.02 | 130.51 | 18.30 | **322.56** |
| 113 | 57.74 | 61.01 | 106.21 | 28.32 | **253.28** |
| 114 | 18.52 | 40.85 | 27.78 | 27.45 | **114.60** |
| 115 | 244.56 | 157.96 | 61.55 | 20.70 | **484.77** |
| 116 | 9.80 | 0 | 3.27 | 0 | **13.07** |
| 117 | 125.82 | 66.45 | 32.68 | 35.95 | **260.91** |
| 118 | 34.86 | 36.49 | 0 | 0 | **71.35** |
| 119 | 12.53 | 1.63 | 13.07 | 1.09 | **28.32** |
| 120 | 93.14 | 52.29 | 17.43 | 9.26 | **172.12** |
| 121 | 16.34 | 23.97 | 9.80 | 11.44 | **61.55** |
| 122 | 38.67 | 5.99 | 16.89 | 7.08 | **68.63** |
| 123 | 72.99 | 70.81 | 3.81 | 0.54 | **148.16** |
| 124 | 69.72 | 69.72 | 18.52 | 1.09 | **159.05** |
| 125 | 102.40 | 22.88 | 9.91 | 13.07 | **148.26** |
| 126 | 108.39 | 0 | 17.43 | 9.80 | **135.63** |
| 127 | 14.71 | 8.72 | 0 | 0 | **23.42** |
| 128 | 128.55 | 106.76 | 151.97 | 68.63 | **455.90** |
| 129 | 74.62 | 40.85 | 20.70 | 10.35 | **146.52** |
| 130 | 128.00 | 106.76 | 105.67 | 18.52 | **358.95** |
| 131 | 17.43 | 2.72 | 5.45 | 3.27 | **28.87** |
| 132 | 86.61 | 47.39 | 47.39 | 37.58 | **218.96** |
| 133 | 44.12 | 58.28 | 12.53 | 4.36 | **119.29** |
| 134 | 46.84 | 13.62 | 11.44 | 11.98 | **83.88** |
| 135 | 11.44 | 21.79 | 5.99 | 1.09 | **40.31** |
| 136 | 2.18 | 14.71 | 2.18 | 0.54 | **19.61** |
| 137 | 0 | 2.18 | 4.36 | 1.09 | **7.63** |
| 138 | 30.50 | 17.43 | 2.18 | 0 | **50.11** |
| 139 | 17.43 | 0.54 | 0 | 1.09 | **19.06** |
| 140 | 39.76 | 23.97 | 15.25 | 28.32 | **107.30** |
| 141 | 38.13 | 27.78 | 36.49 | 2.18 | **104.58** |

| | | | | | |
|---|---|---|---|---|---|
| 142 | 123.64 | 56.10 | 104.04 | 88.78 | **372.57** |
| 143 | 63.18 | 57.74 | 61.01 | 4.36 | **186.28** |
| 144 | 16.34 | 20.70 | 2.72 | 4.90 | **44.66** |
| 145 | 2.72 | 0 | 0.54 | 0.54 | **3.81** |
| 146 | 56.65 | 31.05 | 65.80 | 19.06 | **172.56** |
| 147 | 63.18 | 28.32 | 1.09 | 2.18 | **94.78** |
| 148 | 116.02 | 106.76 | 71.90 | 31.59 | **326.27** |
| 149 | 23.42 | 13.62 | 16.89 | 28.32 | **82.25** |
| **NY - NEW YORK - Total** | **2,934.23** | **2,113.93** | **1,566.09** | **829.89** | **7,444.14** |

**STATE: OH - OHIO**

| REGISTRANT ZIP CODE 3 | 1ST QUARTER | 2ND QUARTER | 3RD QUARTER | 4TH QUARTER | **TOTAL GRAMS** |
|---|---|---|---|---|---|
| 430 | 105.67 | 102.40 | 44.99 | 41.40 | **294.46** |
| 431 | 117.65 | 88.24 | 143.25 | 126.37 | **475.51** |
| 432 | 104.04 | 98.59 | 97.50 | 74.19 | **374.31** |
| 433 | 14.71 | 3.27 | 14.71 | 2.18 | **34.86** |
| 434 | 52.29 | 71.90 | 36.49 | 22.88 | **183.56** |
| 435 | 41.94 | 53.38 | 26.69 | 17.97 | **139.98** |
| 436 | 58.83 | 54.47 | 41.94 | 9.26 | **164.50** |
| 437 | 245.11 | 128.55 | 237.48 | 76.80 | **687.94** |
| 439 | 41.40 | 15.80 | 9.80 | 3.27 | **70.26** |
| 440 | 173.21 | 93.14 | 112.21 | 40.63 | **419.19** |
| 441 | 687.40 | 374.20 | 554.49 | 220.60 | **1,836.69** |
| 442 | 200.99 | 92.60 | 89.33 | 9.80 | **392.72** |
| 443 | 44.66 | 43.58 | 43.03 | 39.76 | **171.03** |
| 444 | 70.81 | 117.11 | 17.97 | 15.25 | **221.14** |
| 445 | 35.95 | 26.69 | 25.60 | 12.53 | **100.77** |
| 446 | 62.64 | 56.65 | 15.25 | 24.51 | **159.05** |
| 447 | 51.20 | 21.79 | 17.97 | 25.60 | **116.56** |
| 448 | 130.73 | 96.41 | 41.40 | 27.78 | **296.31** |
| 449 | 39.22 | 7.63 | 16.34 | 9.26 | **72.44** |
| 450 | 111.66 | 98.04 | 21.79 | 7.63 | **239.12** |
| 451 | 53.92 | 53.38 | 40.31 | 59.37 | **206.98** |
| 452 | 571.92 | 461.35 | 447.73 | 381.14 | **1,862.15** |
| 453 | 119.83 | 82.79 | 19.06 | 24.51 | **246.20** |
| 454 | 124.19 | 77.35 | 45.21 | 52.83 | **299.58** |
| 455 | 47.93 | 33.77 | 18.52 | 3.27 | **103.49** |
| 456 | 11.98 | 17.97 | 23.42 | 1.63 | **55.01** |
| 457 | 31.59 | 9.80 | 11.98 | 2.72 | **56.10** |
| 458 | 120.92 | 123.64 | 52.83 | 61.01 | **358.40** |
| **OH - OHIO Total** | **3,472.38** | **2,504.47** | **2,267.32** | **1,394.15** | **9,638.32** |

**STATE: OK - OKLAHOMA**

| REGISTRANT ZIP CODE 3 | 1ST QUARTER | 2ND QUARTER | 3RD QUARTER | 4TH QUARTER | TOTAL GRAMS |
|---|---|---|---|---|---|
| 730 | 696.11 | 495.12 | 423.77 | 94.23 | **1,709.23** |
| 731 | 993.51 | 890.56 | 612.23 | 212.97 | **2,709.28** |
| 734 | 32.68 | 66.45 | 21.79 | 4.36 | **125.28** |
| 735 | 463.53 | 211.34 | 168.85 | 238.57 | **1,082.29** |
| 736 | 24.51 | 23.42 | 27.23 | 19.06 | **94.23** |
| 737 | 115.47 | 83.34 | 74.62 | 20.15 | **293.59** |
| 738 | 16.34 | 7.63 | 3.81 | 0.54 | **28.32** |
| 739 | 38.13 | 5.99 | 5.99 | 14.16 | **64.27** |
| 740 | 486.95 | 332.80 | 208.07 | 117.11 | **1,144.93** |
| 741 | 334.98 | 280.51 | 212.43 | 114.38 | **942.31** |
| 743 | 101.86 | 54.47 | 23.42 | 17.43 | **197.18** |
| 744 | 175.93 | 66.45 | 58.28 | 43.03 | **343.70** |
| 745 | 21.24 | 17.43 | 17.43 | 3.27 | **59.37** |
| 746 | 46.84 | 5.45 | 5.99 | 3.27 | **61.55** |
| 747 | 104.58 | 80.61 | 92.60 | 8.72 | **286.51** |
| 748 | 145.43 | 65.36 | 143.25 | 61.55 | **415.60** |
| 749 | 32.68 | 27.23 | 27.78 | 21.24 | **108.94** |
| **OK - OKLAHOMA Total** | **3,830.79** | **2,714.18** | **2,127.55** | **994.05** | **9,666.57** |

**STATE: OR - OREGON**

| REGISTRANT ZIP CODE 3 | 1ST QUARTER | 2ND QUARTER | 3RD QUARTER | 4TH QUARTER | TOTAL GRAMS |
|---|---|---|---|---|---|
| 970 | 139.44 | 69.18 | 34.86 | 29.41 | **272.89** |
| 971 | 95.32 | 96.41 | 67.00 | 36.49 | **295.22** |
| 972 | 274.52 | 148.70 | 185.96 | 49.57 | **658.75** |
| 973 | 103.49 | 84.43 | 67.00 | 94.78 | **349.69** |
| 974 | 181.93 | 120.38 | 141.07 | 35.40 | **478.78** |
| 975 | 173.21 | 70.81 | 81.70 | 70.26 | **395.99** |
| 976 | 24.51 | 16.34 | 3.27 | 7.63 | **51.75** |
| 977 | 37.04 | 35.95 | 36.71 | 15.25 | **124.95** |
| 978 | 56.65 | 19.06 | 17.43 | 3.27 | **96.41** |
| 979 | 9.80 | 2.72 | 4.90 | 0 | **17.43** |
| **OR OREGON Total** | **1,095.91** | **663.97** | **639.90** | **342.06** | **2,741.85** |

**STATE: PA - PENNSYLVANIA**

| REGISTRANT ZIP CODE 3 | 1ST QUARTER | 2ND QUARTER | 3RD QUARTER | 4TH QUARTER | TOTAL GRAMS |
|---|---|---|---|---|---|
| 150 | 93.69 | 34.86 | 1.09 | 3.81 | **133.45** |
| 151 | 7.63 | 86.06 | 13.07 | 8.72 | **115.47** |
| 152 | 80.07 | 98.04 | 104.04 | 33.77 | **315.92** |
| 153 | 6.54 | 0 | 0 | 0.54 | **7.08** |
| 154 | 25.06 | 4.36 | 0 | 21.79 | **51.20** |

| | | | | | |
|---|---|---|---|---|---|
| 155 | 1.09 | 2.18 | 4.36 | 2.18 | **9.80** |
| 156 | 28.32 | 35.40 | 1.09 | 1.09 | **65.91** |
| 157 | 22.33 | 13.07 | 10.35 | 10.89 | **56.65** |
| 158 | 61.01 | 49.57 | 1.09 | 2.18 | **113.84** |
| 159 | 21.24 | 35.95 | 0 | 1.09 | **58.28** |
| 160 | 19.06 | 14.16 | 24.51 | 5.99 | **63.73** |
| 161 | 16.34 | 11.44 | 3.81 | 3.81 | **35.40** |
| 162 | 0 | 3.81 | 1.63 | 0 | **5.45** |
| 163 | 44.12 | 4.36 | 7.63 | 1.09 | **57.19** |
| 164 | 9.26 | 43.58 | 9.26 | 0.54 | **62.64** |
| 165 | 53.38 | 32.14 | 15.25 | 4.90 | **105.67** |
| 166 | 14.71 | 19.06 | 17.43 | 3.81 | **55.01** |
| 167 | 4.36 | 0 | 0 | 0 | **4.36** |
| 168 | 32.14 | 7.08 | 20.70 | 0 | **59.92** |
| 169 | 2.72 | 1.09 | 4.90 | 0 | **8.72** |
| 170 | 120.92 | 57.74 | 20.70 | 13.62 | **212.97** |
| 171 | 55.01 | 44.12 | 3.27 | 9.26 | **111.66** |
| 172 | 4.36 | 4.90 | 2.18 | 2.18 | **13.62** |
| 173 | 6.54 | 68.09 | 25.60 | 36.49 | **136.72** |
| 174 | 2.18 | 6.54 | 27.78 | 3.81 | **40.31** |
| 175 | 29.96 | 9.80 | 7.08 | 8.17 | **55.01** |
| 176 | 41.94 | 14.71 | 5.45 | 3.27 | **65.36** |
| 177 | 29.41 | 9.80 | 2.18 | 8.72 | **50.11** |
| 178 | 47.93 | 40.31 | 15.25 | 0 | **103.49** |
| 179 | 13.07 | 10.89 | 0 | 4.90 | **28.87** |
| 180 | 37.04 | 37.04 | 17.43 | 1.09 | **92.60** |
| 181 | 50.11 | 78.44 | 10.35 | 10.89 | **149.79** |
| 182 | 28.87 | 14.71 | 4.36 | 3.27 | **51.20** |
| 183 | 0.54 | 2.18 | 1.63 | 7.08 | **11.44** |
| 184 | 6.54 | 8.72 | 9.80 | 0 | **25.06** |
| 185 | 10.89 | 28.87 | 21.24 | 5.99 | **67.00** |
| 186 | 9.80 | 13.07 | 1.09 | 0.54 | **24.51** |
| 187 | 84.43 | 191.73 | 122.01 | 54.47 | **452.64** |
| 188 | 125.82 | 74.62 | 70.26 | 35.40 | **306.11** |
| 189 | 91.51 | 50.66 | 8.72 | 0 | **150.88** |
| 190 | 260.91 | 249.47 | 114.38 | 77.02 | **701.78** |
| 191 | 155.24 | 112.75 | 133.67 | 62.09 | **463.75** |
| 193 | 140.53 | 69.72 | 35.95 | 58.28 | **304.48** |
| 194 | 74.62 | 49.02 | 15.80 | 7.63 | **147.07** |
| 195 | 17.43 | 4.36 | 0 | 0 | **21.79** |

| | | | | | |
|---|---|---|---|---|---|
| 196 | 40.31 | 8.72 | 9.26 | 2.18 | **60.46** |
| **PA - PENNSYLVANIA - Total** | **2,028.96** | **1,757.16** | **925.64** | **522.57** | **5,234.34** |

**STATE: PR - PUERTO RICO**

| REGISTRANT ZIP CODE 3 | 1ST QUARTER | 2ND QUARTER | 3RD QUARTER | 4TH QUARTER | TOTAL GRAMS |
|---|---|---|---|---|---|
| 006 | 906.36 | 959.19 | 1,322.50 | 905.82 | **4,093.87** |
| 007 | 744.59 | 678.68 | 1,161.82 | 836.64 | **3,421.73** |
| 009 | 1,278.38 | 1,238.62 | 1,482.09 | 1,196.13 | **5,195.23** |
| **PR - PUERTO RICO - Total** | **2,929.33** | **2,876.49** | **3,966.41** | **2,938.59** | **12,710.83** |

**STATE: RI - RHODE ISLAND**

| REGISTRANT ZIP CODE 3 | 1ST QUARTER | 2ND QUARTER | 3RD QUARTER | 4TH QUARTER | TOTAL GRAMS |
|---|---|---|---|---|---|
| 028 | 23.42 | 51.75 | 14.16 | 8.72 | **98.04** |
| 029 | 19.06 | 24.51 | 26.15 | 100.22 | **169.94** |
| **RI RHODE ISLAND Total** | **42.49** | **76.26** | **40.31** | **108.94** | **267.99** |

**STATE: SC - SOUTH CAROLINA**

| REGISTRANT ZIP CODE 3 | 1ST QUARTER | 2ND QUARTER | 3RD QUARTER | 4TH QUARTER | TOTAL GRAMS |
|---|---|---|---|---|---|
| 290 | 404.70 | 230.40 | 74.08 | 17.43 | **726.61** |
| 291 | 297.40 | 257.64 | 103.49 | 42.49 | **701.01** |
| 292 | 352.96 | 269.62 | 135.08 | 89.33 | **846.99** |
| 293 | 209.16 | 122.01 | 70.26 | 9.80 | **411.24** |
| 294 | 523.34 | 260.91 | 240.10 | 244.41 | **1,268.75** |
| 295 | 355.68 | 242.39 | 153.06 | 56.65 | **807.77** |
| 296 | 457.54 | 252.74 | 281.60 | 132.90 | **1,124.78** |
| 297 | 67.54 | 50.66 | 9.26 | 10.35 | **137.81** |
| 298 | 142.71 | 126.37 | 30.50 | 21.79 | **321.37** |
| 299 | 80.07 | 17.97 | 22.33 | 14.16 | **134.54** |
| **SC - SOUTH CAROLINA Total** | **2,891.09** | **1,830.69** | **1,119.77** | **639.31** | **6,480.87** |

**STATE: SD - SOUTH DAKOTA**

| REGISTRANT ZIP CODE 3 | 1ST QUARTER | 2ND QUARTER | 3RD QUARTER | 4TH QUARTER | TOTAL GRAMS |
|---|---|---|---|---|---|
| 570 | 141.07 | 52.29 | 59.92 | 15.25 | **268.53** |
| 571 | 53.38 | 113.30 | 63.73 | 30.50 | **260.91** |
| 572 | 50.66 | 46.30 | 10.89 | 3.81 | **111.66** |
| 573 | 24.51 | 29.96 | 23.42 | 10.89 | **88.78** |
| 574 | 11.98 | 7.63 | 11.98 | 2.72 | **34.32** |
| 575 | 3.27 | 4.90 | 2.18 | 0 | **10.35** |
| 577 | 80.61 | 65.36 | 32.68 | 15.80 | **194.45** |
| **SD SOUTH DAKOTA - Total** | **365.49** | **319.73** | **204.80** | **78.98** | **969.00** |

**STATE: TN - TENNESSEE**

| REGISTRANT ZIP CODE 3 | 1ST QUARTER | 2ND QUARTER | 3RD QUARTER | 4TH QUARTER | TOTAL GRAMS |
|---|---|---|---|---|---|
| 370 | 535.43 | 311.02 | 178.77 | 92.71 | **1,117.92** |
| 371 | 484.77 | 326.27 | 202.08 | 199.90 | **1,213.02** |

| | | | | | |
|---|---|---|---|---|---|
| 372 | 380.74 | 187.37 | 141.62 | 123.10 | **832.83** |
| 373 | 370.39 | 253.82 | 85.52 | 101.86 | **811.58** |
| 374 | 156.33 | 135.08 | 90.42 | 63.18 | **445.01** |
| 376 | 129.64 | 118.20 | 75.17 | 24.51 | **347.51** |
| 377 | 118.74 | 106.76 | 23.42 | 21.24 | **270.17** |
| 378 | 138.90 | 88.24 | 104.04 | 51.75 | **382.92** |
| 379 | 93.14 | 138.90 | 76.80 | 63.73 | **372.57** |
| 380 | 172.12 | 95.32 | 33.77 | 54.47 | **355.68** |
| 381 | 380.74 | 281.06 | 198.27 | 185.74 | **1,045.80** |
| 382 | 63.18 | 19.61 | 17.97 | 10.89 | **111.66** |
| 383 | 199.90 | 131.81 | 124.73 | 67.54 | **523.99** |
| 384 | 192.82 | 84.97 | 58.83 | 49.02 | **385.64** |
| 385 | 98.04 | 46.30 | 50.11 | 27.23 | **221.69** |
| **TN TENNESSEE Total** | **3,514.87** | **2,324.73** | **1,461.51** | **1,136.87** | **8,437.97** |

**STATE: TX - TEXAS**

| REGISTRANT ZIP CODE 3 | 1ST QUARTER | 2ND QUARTER | 3RD QUARTER | 4TH QUARTER | **TOTAL GRAMS** |
|---|---|---|---|---|---|
| 750 | 408.52 | 395.99 | 395.44 | 330.08 | **1,530.03** |
| 751 | 160.68 | 92.05 | 78.98 | 26.15 | **357.86** |
| 752 | 265.81 | 149.79 | 284.33 | 157.96 | **857.88** |
| 753 | 5.45 | 6.54 | 6.54 | 4.36 | **22.88** |
| 754 | 103.49 | 65.91 | 173.76 | 31.05 | **374.20** |
| 755 | 56.65 | 38.13 | 40.85 | 30.50 | **166.13** |
| 756 | 60.94 | 103.19 | 190.16 | 58.83 | **413.11** |
| 757 | 142.71 | 49.02 | 158.50 | 70.81 | **421.04** |
| 758 | 13.62 | 6.54 | 12.53 | 8.72 | **41.40** |
| 759 | 104.04 | 60.46 | 59.81 | 34.32 | **258.62** |
| 760 | 232.58 | 187.37 | 233.67 | 145.43 | **799.06** |
| 761 | 125.82 | 151.97 | 111.66 | 94.78 | **484.23** |
| 762 | 109.48 | 121.47 | 80.07 | 46.84 | **357.86** |
| 763 | 65.36 | 38.67 | 72.99 | 17.43 | **194.45** |
| 764 | 53.38 | 20.15 | 40.31 | 21.79 | **135.63** |
| 765 | 50.66 | 87.15 | 56.10 | 15.80 | **209.70** |
| 766 | 15.80 | 8.72 | 13.62 | 0 | **38.13** |
| 767 | 41.40 | 47.93 | 52.29 | 86.06 | **227.68** |
| 768 | 33.77 | 19.61 | 42.49 | 13.62 | **109.48** |
| 769 | 67.00 | 67.54 | 52.29 | 28.32 | **215.15** |
| 770 | 1,516.41 | 963.55 | 1,313.79 | 1,234.81 | **5,028.56** |
| 773 | 303.94 | 228.77 | 175.27 | 196.63 | **904.61** |
| 774 | 177.02 | 198.27 | 123.10 | 167.22 | **665.61** |
| 775 | 267.44 | 195.54 | 136.72 | 111.66 | **711.36** |

| | | | | | |
|---|---|---|---|---|---|
| 776 | 44.66 | 40.31 | 15.25 | 16.89 | **117.11** |
| 777 | 72.44 | 92.60 | 71.90 | 59.92 | **296.85** |
| 778 | 142.16 | 117.65 | 59.37 | 33.23 | **352.41** |
| 779 | 28.87 | 35.95 | 33.77 | 29.41 | **128.00** |
| 780 | 133.45 | 144.34 | 138.90 | 96.41 | **513.10** |
| 781 | 70.26 | 52.83 | 58.28 | 47.39 | **228.77** |
| 782 | 597.52 | 429.21 | 525.62 | 417.78 | **1,970.13** |
| 783 | 15.25 | 3.27 | 16.89 | 16.34 | **51.75** |
| 784 | 115.47 | 65.36 | 104.04 | 92.60 | **377.47** |
| 785 | 661.80 | 553.95 | 607.33 | 434.12 | **2,257.19** |
| 786 | 142.16 | 155.24 | 131.16 | 95.32 | **523.88** |
| 787 | 190.10 | 149.79 | 187.92 | 106.21 | **634.02** |
| 788 | 64.27 | 47.93 | 74.08 | 48.48 | **234.76** |
| 789 | 25.60 | 15.25 | 16.89 | 4.36 | **62.09** |
| 790 | 34.32 | 11.98 | 13.07 | 8.17 | **67.54** |
| 791 | 61.55 | 51.75 | 63.73 | 23.97 | **200.99** |
| 792 | 2.72 | 0.54 | 8.72 | 8.72 | **20.70** |
| 793 | 78.44 | 40.31 | 33.77 | 48.48 | **200.99** |
| 794 | 83.34 | 81.70 | 106.21 | 94.23 | **365.49** |
| 795 | 36.49 | 22.88 | 17.43 | 27.23 | **104.04** |
| 796 | 58.83 | 52.83 | 79.52 | 51.75 | **242.93** |
| 797 | 243.48 | 229.31 | 213.52 | 210.25 | **896.56** |
| 798 | 2.18 | 1.09 | 0 | 0 | **3.27** |
| 799 | 130.73 | 98.04 | 101.86 | 107.30 | **437.93** |
| **TX - TEXAS Total** | **7,418.03** | **5,798.44** | **6,584.46** | **5,011.67** | **24,812.60** |

**STATE: UT - UTAH**

| REGISTRANT ZIP CODE 3 | 1ST QUARTER | 2ND QUARTER | 3RD QUARTER | 4TH QUARTER | **TOTAL GRAMS** |
|---|---|---|---|---|---|
| 840 | 440.11 | 332.80 | 280.73 | 251.21 | **1,304.85** |
| 841 | 269.08 | 214.61 | 196.63 | 226.59 | **906.90** |
| 843 | 33.23 | 27.23 | 6.54 | 9.80 | **76.80** |
| 844 | 50.66 | 34.32 | 50.11 | 22.33 | **157.41** |
| 845 | 71.90 | 39.22 | 15.80 | 26.15 | **153.06** |
| 846 | 166.67 | 93.14 | 101.64 | 76.80 | **438.26** |
| 847 | 193.91 | 142.71 | 62.09 | 94.99 | **493.70** |
| **UT - UTAH - Total** | **1,225.55** | **884.03** | **713.54** | **707.88** | **3,530.99** |

**STATE: VA - VIRGINIA**

| REGISTRANT ZIP CODE 3 | 1ST QUARTER | 2ND QUARTER | 3RD QUARTER | 4TH QUARTER | **TOTAL GRAMS** |
|---|---|---|---|---|---|
| 201 | 199.36 | 127.46 | 37.58 | 48.48 | **412.87** |
| 220 | 210.79 | 112.21 | 237.48 | 134.54 | **695.02** |
| 221 | 68.09 | 77.35 | 61.55 | 14.71 | **221.69** |

| | | | | | |
|---|---|---|---|---|---|
| 222 | 24.51 | 30.50 | 25.06 | 17.43 | **97.50** |
| 223 | 167.76 | 128.55 | 90.96 | 61.01 | **448.28** |
| 224 | 90.42 | 86.61 | 94.88 | 61.01 | **332.91** |
| 225 | 22.88 | 5.45 | 13.07 | 1.09 | **42.49** |
| 226 | 171.58 | 136.72 | 33.77 | 0 | **342.06** |
| 227 | 17.43 | 0 | 26.15 | 0 | **43.58** |
| 228 | 47.93 | 4.36 | 21.79 | 0 | **74.08** |
| 229 | 144.89 | 113.30 | 41.40 | 27.23 | **326.81** |
| 230 | 113.30 | 17.97 | 2.18 | 0 | **133.45** |
| 231 | 237.48 | 147.07 | 42.49 | 20.70 | **447.73** |
| 232 | 545.23 | 301.21 | 144.34 | 81.70 | **1,072.49** |
| 233 | 161.23 | 74.08 | 17.97 | 22.88 | **276.16** |
| 234 | 333.89 | 183.56 | 114.93 | 48.48 | **680.86** |
| 235 | 116.56 | 122.01 | 26.15 | 23.97 | **288.68** |
| 236 | 201.53 | 120.38 | 120.92 | 26.15 | **468.98** |
| 237 | 34.32 | 45.75 | 15.25 | 9.26 | **104.58** |
| 238 | 88.78 | 69.18 | 84.97 | 157.41 | **400.35** |
| 239 | 72.99 | 50.66 | 48.48 | 26.69 | **198.81** |
| 240 | 56.10 | 138.35 | 13.07 | 29.41 | **236.94** |
| 241 | 105.67 | 105.67 | 94.23 | 39.22 | **344.79** |
| 242 | 114.38 | 79.52 | 36.49 | 5.45 | **235.85** |
| 243 | 25.06 | 10.89 | 5.99 | 1.63 | **43.58** |
| 244 | 68.63 | 18.52 | 11.98 | 1.09 | **100.22** |
| 245 | 108.39 | 63.18 | 77.89 | 51.20 | **300.67** |
| **VA VIRGINIA Total** | **3,549.18** | **2,370.48** | **1,541.03** | **910.72** | **8,371.41** |

**STATE: VI - VIRGIN ISLANDS**

| REGISTRANT ZIP CODE 3 | 1ST QUARTER | 2ND QUARTER | 3RD QUARTER | 4TH QUARTER | **TOTAL GRAMS** |
|---|---|---|---|---|---|
| 008 | 61.55 | 72.44 | 24.51 | 2.18 | **160.68** |
| **VI - VIRGIN ISLANDS Total** | **61.55** | **72.44** | **24.51** | **2.18** | **160.68** |

**STATE: VT - VERMONT**

| REGISTRANT ZIP CODE 3 | 1ST QUARTER | 2ND QUARTER | 3RD QUARTER | 4TH QUARTER | **TOTAL GRAMS** |
|---|---|---|---|---|---|
| 050 | 0.54 | 1.09 | 0 | 2.18 | **3.81** |
| 051 | 0 | 0 | 1.09 | 0 | **1.09** |
| 052 | 0 | 0 | 0 | 1.09 | **1.09** |
| 054 | 137.26 | 177.02 | 105.67 | 133.45 | **553.40** |
| 056 | 4.36 | 0 | 0 | 0 | **4.36** |
| 057 | 24.51 | 77.35 | 31.05 | 17.43 | **150.33** |
| **VT VERMONT Total** | **166.67** | **255.46** | **137.81** | **154.15** | **714.09** |

**STATE: WA - WASHINGTON**

| REGISTRANT ZIP CODE 3 | 1ST QUARTER | 2ND QUARTER | 3RD QUARTER | 4TH QUARTER | **TOTAL GRAMS** |
|---|---|---|---|---|---|

| | | | | | |
|---|---|---|---|---|---|
| 980 | 354.59 | 208.62 | 136.39 | 138.35 | **837.95** |
| 981 | 195.54 | 88.24 | 40.85 | 34.86 | **359.49** |
| 982 | 376.38 | 342.06 | 184.10 | 135.08 | **1,037.63** |
| 983 | 318.10 | 139.44 | 14.71 | 33.99 | **506.23** |
| 984 | 60.46 | 93.69 | 21.79 | 11.98 | **187.92** |
| 985 | 167.76 | 173.21 | 80.07 | 63.73 | **484.77** |
| 986 | 93.69 | 89.33 | 92.60 | 19.61 | **295.22** |
| 988 | 46.84 | 49.57 | 31.59 | 28.32 | **156.33** |
| 989 | 61.55 | 41.94 | 20.70 | 12.53 | **136.72** |
| 990 | 15.25 | 4.36 | 0 | 2.18 | **21.79** |
| 991 | 68.09 | 77.35 | 27.78 | 11.44 | **184.65** |
| 992 | 123.64 | 52.83 | 37.58 | 70.81 | **284.87** |
| 993 | 73.53 | 74.08 | 87.15 | 46.30 | **281.06** |
| 994 | 26.15 | 10.89 | 17.43 | 24.51 | **78.98** |
| **WA WASHINGTON Total** | **1,981.57** | **1,445.60** | **792.74** | **633.69** | **4,853.60** |

**STATE: WI - WISCONSIN**

| REGISTRANT ZIP CODE 3 | 1ST QUARTER | 2ND QUARTER | 3RD QUARTER | 4TH QUARTER | TOTAL GRAMS |
|---|---|---|---|---|---|
| 530 | 87.15 | 69.18 | 80.61 | 53.92 | **290.86** |
| 531 | 121.47 | 37.58 | 50.11 | 26.15 | **235.31** |
| 532 | 84.43 | 31.05 | 42.49 | 40.85 | **198.81** |
| 534 | 2.18 | 0 | 0.54 | 2.18 | **4.90** |
| 535 | 83.88 | 49.57 | 59.37 | 23.42 | **216.24** |
| 537 | 55.01 | 62.09 | 69.18 | 36.49 | **222.78** |
| 538 | 4.36 | 0 | 5.45 | 0 | **9.80** |
| 539 | 2.72 | 2.18 | 5.99 | 1.63 | **12.53** |
| 540 | 2.18 | 0.54 | 1.09 | 0 | **3.81** |
| 541 | 7.08 | 14.16 | 42.49 | 26.15 | **89.87** |
| 542 | 39.76 | 49.57 | 45.75 | 29.96 | **165.04** |
| 543 | 96.41 | 110.03 | 99.68 | 115.47 | **421.59** |
| 544 | 86.06 | 62.09 | 93.69 | 44.12 | **285.96** |
| 545 | 1.63 | 0.54 | 0 | 0 | **2.18** |
| 546 | 17.43 | 13.62 | 23.97 | 2.18 | **57.19** |
| 547 | 24.51 | 10.89 | 4.90 | 5.99 | **46.30** |
| 548 | 10.35 | 6.54 | 6.54 | 7.08 | **30.50** |
| 549 | 29.96 | 39.76 | 13.62 | 11.98 | **95.32** |
| **WI - WISCONSIN Total** | **756.57** | **559.39** | **645.45** | **427.58** | **2,389.00** |

**STATE: WV - WEST VIRGINIA**

| REGISTRANT ZIP CODE 3 | 1ST QUARTER | 2ND QUARTER | 3RD QUARTER | 4TH QUARTER | TOTAL GRAMS |
|---|---|---|---|---|---|
| 247 | 15.80 | 6.54 | 5.45 | 8.17 | **35.95** |
| 248 | 14.16 | 17.43 | 9.80 | 21.79 | **63.18** |

| | | | | | |
|---|---|---|---|---|---|
| 249 | 10.89 | 8.72 | 13.62 | 6.54 | **39.76** |
| 251 | 10.89 | 9.80 | 2.18 | 0 | **22.88** |
| 252 | 0 | 0 | 4.36 | 0 | **4.36** |
| 253 | 28.87 | 25.60 | 21.24 | 16.89 | **92.60** |
| 254 | 13.07 | 7.08 | 0.54 | 1.63 | **22.33** |
| 255 | 8.72 | 6.54 | 8.72 | 4.36 | **28.32** |
| 256 | 9.26 | 0 | 0 | 0 | **9.26** |
| 257 | 101.31 | 32.68 | 64.82 | 26.15 | **224.96** |
| 258 | 11.98 | 7.63 | 7.63 | 8.72 | **35.95** |
| 259 | 1.09 | 1.63 | 2.18 | 1.09 | **5.99** |
| 260 | 38.13 | 16.89 | 1.09 | 7.30 | **63.40** |
| 261 | 19.06 | 17.43 | 21.79 | 10.35 | **68.63** |
| 262 | 5.99 | 0.54 | 3.27 | 2.18 | **11.98** |
| 263 | 13.07 | 4.36 | 16.34 | 14.16 | **47.93** |
| 264 | 1.09 | 6.54 | 2.72 | 0 | **10.35** |
| 265 | 16.89 | 17.97 | 17.43 | 7.63 | **59.92** |
| 266 | 0 | 5.99 | 0 | 21.79 | **27.78** |
| 267 | 4.36 | 4.36 | 0 | 0 | **8.72** |
| 268 | 0 | 2.72 | 0 | 1.09 | **3.81** |
| **WV WEST VIRGINIA Total** | **324.63** | **200.45** | **203.17** | **159.81** | **888.06** |

**STATE: WY - WYOMING**

| REGISTRANT ZIP CODE 3 | 1ST QUARTER | 2ND QUARTER | 3RD QUARTER | 4TH QUARTER | **TOTAL GRAMS** |
|---|---|---|---|---|---|
| 820 | 15.80 | 33.23 | 19.06 | 15.80 | **83.88** |
| 822 | 13.07 | 18.52 | 0 | 0 | **31.59** |
| 823 | 17.43 | 0 | 0 | 0 | **17.43** |
| 824 | 23.97 | 8.17 | 5.99 | 2.72 | **40.85** |
| 825 | 38.13 | 29.96 | 12.53 | 5.99 | **86.61** |
| 826 | 17.97 | 38.67 | 15.80 | 8.61 | **81.05** |
| 827 | 4.36 | 9.80 | 15.25 | 3.27 | **32.68** |
| 828 | 65.91 | 19.61 | 0.54 | 1.09 | **87.15** |
| 829 | 4.90 | 15.25 | 1.09 | 2.18 | **23.42** |
| 830 | 1.09 | 1.09 | 4.36 | 0 | **6.54** |
| 831 | 15.25 | 21.79 | 1.09 | 5.45 | **43.58** |
| **WY - WYOMING Total** | **217.88** | **196.09** | **75.71** | **45.10** | **534.77** |

**DRUG: 9250B - METHADONE**

**STATE: AK - ALASKA**

| REGISTRANT ZIP CODE 3 | 1ST QUARTER | 2ND QUARTER | 3RD QUARTER | 4TH QUARTER | **TOTAL GRAMS** |
|---|---|---|---|---|---|
| 995 | 5,199.86 | 5,864.85 | 5,656.63 | 5,387.15 | **22,108.49** |
| 996 | 2,874.60 | 2,938.82 | 3,102.41 | 3,006.79 | **11,922.62** |
| 997 | 971.05 | 976.24 | 994.30 | 550.06 | **3,491.65** |

| | | | | | |
|---|---|---|---|---|---|
| 998 | 96.15 | 101.51 | 94.81 | 65.29 | **357.76** |
| 999 | 38.91 | 40.25 | 41.14 | 41.77 | **162.07** |
| **AK ALASKA - Total** | **9,180.57** | **9,921.67** | **9,889.29** | **9,051.06** | **38,042.59** |

**STATE: AL - ALABAMA**

| REGISTRANT ZIP CODE 3 | 1ST QUARTER | 2ND QUARTER | 3RD QUARTER | 4TH QUARTER | **TOTAL GRAMS** |
|---|---|---|---|---|---|
| 350 | 12,501.12 | 9,960.04 | 12,969.34 | 13,231.66 | **48,662.16** |
| 351 | 855.49 | 832.24 | 835.37 | 822.40 | **3,345.50** |
| 352 | 10,964.99 | 17,044.22 | 10,769.11 | 12,594.58 | **51,372.90** |
| 354 | 4,601.06 | 3,434.14 | 3,889.84 | 3,526.26 | **15,451.30** |
| 355 | 4,918.31 | 5,832.83 | 5,829.70 | 4,598.11 | **21,178.94** |
| 356 | 3,017.71 | 3,011.98 | 2,087.89 | 2,576.95 | **10,694.52** |
| 357 | 868.91 | 832.69 | 511.60 | 851.02 | **3,064.21** |
| 358 | 13,980.10 | 8,322.03 | 13,978.40 | 11,811.54 | **48,092.07** |
| 359 | 3,485.12 | 2,321.24 | 3,306.78 | 2,650.55 | **11,763.69** |
| 360 | 364.92 | 355.08 | 329.59 | 328.24 | **1,377.82** |
| 361 | 2,351.82 | 3,471.61 | 1,277.65 | 2,381.79 | **9,482.88** |
| 362 | 4,281.94 | 4,117.28 | 3,224.76 | 3,630.82 | **15,254.80** |
| 363 | 2,329.73 | 2,329.46 | 2,313.81 | 2,267.30 | **9,240.31** |
| 364 | 235.50 | 227.62 | 234.33 | 224.94 | **922.39** |
| 365 | 2,794.55 | 2,807.97 | 2,403.70 | 2,805.29 | **10,811.51** |
| 366 | 12,152.30 | 13,626.45 | 14,287.59 | 10,864.46 | **50,930.80** |
| 367 | 185.14 | 176.64 | 177.36 | 180.67 | **719.81** |
| 368 | 514.73 | 538.16 | 514.46 | 497.55 | **2,064.90** |
| 369 | 3.58 | 8.50 | 7.16 | 8.94 | **28.17** |
| **AL ALABAMA - Total** | **80,407.01** | **79,250.19** | **78,948.42** | **75,853.08** | **314,458.70** |

**STATE: AR - ARKANSAS**

| REGISTRANT ZIP CODE 3 | 1ST QUARTER | 2ND QUARTER | 3RD QUARTER | 4TH QUARTER | **TOTAL GRAMS** |
|---|---|---|---|---|---|
| 716 | 111.44 | 105.54 | 105.54 | 114.48 | **437.00** |
| 717 | 212.06 | 208.22 | 190.95 | 190.24 | **801.47** |
| 718 | 1,908.65 | 1,532.11 | 2,246.73 | 1,887.63 | **7,575.12** |
| 719 | 201.24 | 235.23 | 232.81 | 211.53 | **880.81** |
| 720 | 1,312.53 | 1,262.62 | 1,281.05 | 1,208.33 | **5,064.54** |
| 721 | 1,602.05 | 1,577.27 | 1,544.72 | 2,019.11 | **6,743.15** |
| 722 | 277.00 | 674.11 | 1,122.11 | 264.47 | **2,337.69** |
| 723 | 147.13 | 148.02 | 196.77 | 256.69 | **748.61** |
| 724 | 2,258.81 | 2,226.16 | 2,239.13 | 2,210.51 | **8,934.61** |
| 725 | 194.53 | 191.40 | 182.01 | 172.17 | **740.12** |
| 726 | 216.44 | 200.35 | 220.92 | 216.00 | **853.70** |
| 727 | 3,183.17 | 4,124.79 | 3,128.07 | 3,000.26 | **13,436.30** |
| 728 | 118.51 | 115.38 | 104.20 | 86.31 | **424.39** |

| | | | | | |
|---|---|---|---|---|---|
| 729 | 555.42 | 452.12 | 464.19 | 407.85 | **1,879.58** |
| **AR ARKANSAS - Total** | **12,298.98** | **13,053.32** | **13,259.21** | **12,245.59** | **50,857.10** |

**STATE: AZ - ARIZONA**

| REGISTRANT ZIP CODE 3 | 1ST QUARTER | 2ND QUARTER | 3RD QUARTER | 4TH QUARTER | TOTAL GRAMS |
|---|---|---|---|---|---|
| 850 | 33,746.96 | 40,100.87 | 39,777.64 | 40,035.67 | **153,661.14** |
| 851 | 351.14 | 885.10 | 2,347.44 | 1,461.09 | **5,044.77** |
| 852 | 18,423.66 | 19,218.42 | 20,409.85 | 19,770.26 | **77,822.19** |
| 853 | 13,164.05 | 10,752.57 | 11,683.19 | 15,247.64 | **50,847.44** |
| 855 | 899.77 | 708.81 | 1,305.38 | 849.23 | **3,763.19** |
| 856 | 509.36 | 610.43 | 943.68 | 1,142.77 | **3,206.25** |
| 857 | 30,353.70 | 28,618.03 | 29,092.42 | 29,354.03 | **117,418.17** |
| 859 | 272.34 | 252.31 | 241.04 | 238.80 | **1,004.50** |
| 860 | 823.83 | 811.94 | 818.91 | 818.55 | **3,273.24** |
| 863 | 1,110.40 | 1,692.03 | 1,054.32 | 1,070.69 | **4,927.43** |
| 864 | 1,206.55 | 1,489.62 | 1,455.19 | 2,355.85 | **6,507.21** |
| 865 | 3.58 | 4.02 | 4.47 | 3.58 | **15.65** |
| **AZ ARIZONA - Total** | **100,865.33** | **105,144.14** | **109,133.53** | **112,348.18** | **427,491.18** |

**STATE: CA - CALIFORNIA**

| REGISTRANT ZIP CODE 3 | 1ST QUARTER | 2ND QUARTER | 3RD QUARTER | 4TH QUARTER | TOTAL GRAMS |
|---|---|---|---|---|---|
| 900 | 14,435.00 | 14,923.53 | 13,388.50 | 13,936.80 | **56,683.83** |
| 902 | 3,502.38 | 3,680.10 | 2,430.26 | 3,474.21 | **13,086.95** |
| 903 | 699.87 | 1,380.95 | 1,202.07 | 1,520.03 | **4,802.93** |
| 904 | 159.20 | 169.76 | 164.22 | 164.93 | **658.11** |
| 905 | 351.77 | 300.70 | 248.29 | 234.24 | **1,134.99** |
| 906 | 6,924.00 | 5,024.29 | 5,218.47 | 5,750.19 | **22,916.94** |
| 907 | 3,848.42 | 4,232.57 | 3,043.20 | 4,113.35 | **15,237.54** |
| 908 | 1,797.65 | 1,787.01 | 1,303.32 | 1,360.29 | **6,248.28** |
| 910 | 311.16 | 284.60 | 338.89 | 312.32 | **1,246.97** |
| 911 | 593.08 | 1,018.36 | 607.57 | 1,016.31 | **3,235.31** |
| 912 | 954.68 | 1,455.01 | 1,239.91 | 1,539.80 | **5,189.39** |
| 913 | 5,542.51 | 5,802.51 | 5,305.49 | 5,447.70 | **22,098.21** |
| 914 | 2,871.74 | 2,674.35 | 2,511.21 | 3,003.36 | **11,060.65** |
| 915 | 145.61 | 164.75 | 205.62 | 182.10 | **698.08** |
| 916 | 941.98 | 868.73 | 937.87 | 1,012.01 | **3,760.59** |
| 917 | 8,204.06 | 8,260.59 | 7,529.24 | 8,719.15 | **32,713.04** |
| 918 | 37.56 | 28.17 | 30.86 | 79.15 | **175.75** |
| 919 | 2,731.14 | 3,207.86 | 2,774.70 | 2,618.89 | **11,332.58** |
| 920 | 11,346.27 | 10,917.31 | 11,160.86 | 12,903.06 | **46,327.51** |
| 921 | 10,181.58 | 9,395.94 | 9,356.32 | 9,353.37 | **38,287.21** |
| 922 | 6,395.59 | 4,029.18 | 7,255.37 | 6,223.68 | **23,903.82** |

| | | | | | |
|---|---|---|---|---|---|
| 923 | 8,090.30 | 7,916.33 | 7,166.11 | 7,645.69 | **30,818.43** |
| 924 | 155.18 | 120.74 | 120.74 | 122.80 | **519.47** |
| 925 | 8,101.83 | 8,186.80 | 8,797.32 | 7,968.48 | **33,054.43** |
| 926 | 1,797.12 | 1,823.86 | 1,663.23 | 1,597.85 | **6,882.05** |
| 927 | 3,475.55 | 3,240.86 | 4,018.90 | 2,937.30 | **13,672.60** |
| 928 | 4,109.50 | 4,455.12 | 4,833.48 | 4,548.03 | **17,946.13** |
| 930 | 9,859.87 | 10,732.71 | 9,964.33 | 10,499.27 | **41,056.18** |
| 931 | 2,631.15 | 2,674.79 | 2,672.82 | 2,644.38 | **10,623.15** |
| 932 | 7,597.03 | 7,956.85 | 8,051.84 | 7,526.38 | **31,132.10** |
| 933 | 9,942.15 | 9,753.43 | 10,229.97 | 10,214.85 | **40,140.40** |
| 934 | 6,122.97 | 6,120.02 | 5,532.04 | 5,492.60 | **23,267.64** |
| 935 | 3,694.68 | 3,019.05 | 3,221.27 | 3,352.66 | **13,287.65** |
| 936 | 1,671.27 | 1,503.56 | 1,342.14 | 1,517.35 | **6,034.32** |
| 937 | 15,530.63 | 16,314.75 | 17,072.13 | 16,554.99 | **65,472.49** |
| 939 | 1,892.37 | 1,873.95 | 2,034.04 | 2,920.22 | **8,720.58** |
| 940 | 3,340.05 | 4,166.92 | 4,588.72 | 4,200.91 | **16,296.59** |
| 941 | 19,584.59 | 18,951.53 | 17,730.50 | 19,583.40 | **75,850.01** |
| 943 | 397.56 | 436.20 | 400.15 | 442.55 | **1,676.46** |
| 944 | 250.70 | 262.60 | 224.05 | 211.44 | **948.78** |
| 945 | 21,046.04 | 20,004.42 | 19,732.88 | 20,332.31 | **81,115.64** |
| 946 | 8,537.94 | 9,050.61 | 8,483.47 | 8,849.55 | **34,921.58** |
| 947 | 924.36 | 979.73 | 900.84 | 922.66 | **3,727.59** |
| 948 | 2,586.69 | 3,646.29 | 2,512.28 | 2,486.25 | **11,231.52** |
| 949 | 2,776.75 | 2,778.99 | 3,418.40 | 2,617.91 | **11,592.05** |
| 950 | 6,133.17 | 6,632.33 | 6,051.60 | 7,161.01 | **25,978.12** |
| 951 | 2,532.40 | 3,372.51 | 1,583.09 | 4,140.62 | **11,628.63** |
| 952 | 13,792.63 | 13,626.01 | 14,858.04 | 12,022.26 | **54,298.93** |
| 953 | 13,740.40 | 16,619.65 | 16,095.71 | 15,790.99 | **62,246.75** |
| 954 | 5,285.73 | 6,289.42 | 5,408.79 | 5,744.64 | **22,728.58** |
| 955 | 912.82 | 916.67 | 933.31 | 819.36 | **3,582.16** |
| 956 | 12,520.97 | 12,539.40 | 13,106.52 | 12,189.33 | **50,356.22** |
| 957 | 375.74 | 345.95 | 330.03 | 291.04 | **1,342.76** |
| 958 | 18,028.78 | 17,731.60 | 18,284.42 | 16,662.21 | **70,707.00** |
| 959 | 10,661.25 | 10,602.66 | 9,322.33 | 9,932.67 | **40,518.91** |
| 960 | 2,555.21 | 2,595.28 | 3,101.87 | 4,113.08 | **12,365.44** |
| 961 | 155.63 | 158.22 | 167.43 | 165.46 | **646.74** |
| **CA - CALIFORNIA - Total** | **312,786.25** | **317,006.09** | **310,207.00** | **317,187.44** | **1,257,186.79** |

**STATE: CO - COLORADO**

| REGISTRANT ZIP CODE 3 | 1ST QUARTER | 2ND QUARTER | 3RD QUARTER | 4TH QUARTER | **TOTAL GRAMS** |
|---|---|---|---|---|---|
| 800 | 8,616.38 | 9,933.83 | 9,307.47 | 8,377.39 | **36,235.07** |

| | | | | | |
|---|---|---|---|---|---|
| 801 | 3,970.06 | 3,220.47 | 3,407.31 | 4,271.21 | **14,869.04** |
| 802 | 18,719.79 | 19,129.34 | 19,643.62 | 19,579.13 | **77,071.88** |
| 803 | 785.28 | 917.21 | 1,055.03 | 1,039.83 | **3,797.35** |
| 804 | 145.52 | 171.46 | 171.72 | 162.24 | **650.94** |
| 805 | 2,709.85 | 3,362.41 | 2,713.97 | 3,019.14 | **11,805.36** |
| 806 | 1,165.76 | 1,567.97 | 1,523.52 | 1,676.82 | **5,934.08** |
| 807 | 38.91 | 41.86 | 44.27 | 52.32 | **177.36** |
| 808 | 164.39 | 137.11 | 138.01 | 163.94 | **603.45** |
| 809 | 4,385.33 | 5,071.70 | 5,069.46 | 5,729.53 | **20,256.01** |
| 810 | 4,467.89 | 4,733.52 | 4,940.22 | 5,317.66 | **19,459.28** |
| 811 | 579.57 | 579.21 | 572.86 | 593.52 | **2,325.17** |
| 812 | 128.88 | 133.27 | 126.92 | 135.59 | **524.66** |
| 813 | 282.63 | 485.66 | 476.27 | 457.93 | **1,702.49** |
| 814 | 49.19 | 41.14 | 48.30 | 41.14 | **179.77** |
| 815 | 1,910.44 | 2,759.22 | 1,938.25 | 1,896.04 | **8,503.96** |
| 816 | 115.74 | 113.14 | 106.34 | 106.25 | **441.48** |
| **CO - COLORADO - Total** | **48,235.62** | **52,398.51** | **51,283.54** | **52,619.69** | **204,537.35** |

**STATE: CT - CONNECTICUT**

| REGISTRANT ZIP CODE 3 | 1ST QUARTER | 2ND QUARTER | 3RD QUARTER | 4TH QUARTER | TOTAL GRAMS |
|---|---|---|---|---|---|
| 060 | 18,820.50 | 15,025.92 | 14,896.50 | 15,077.44 | **63,820.36** |
| 061 | 15,770.96 | 13,443.46 | 13,432.99 | 9,760.05 | **52,407.46** |
| 062 | 6,096.23 | 6,155.26 | 6,092.47 | 6,339.51 | **24,683.47** |
| 063 | 5,116.50 | 7,496.41 | 5,211.04 | 5,096.92 | **22,920.88** |
| 064 | 8,158.36 | 14,636.50 | 14,721.65 | 10,255.55 | **47,772.05** |
| 065 | 30,239.84 | 28,737.61 | 32,561.71 | 25,813.99 | **117,353.15** |
| 066 | 22,732.96 | 22,658.73 | 8,899.28 | 22,713.29 | **77,004.26** |
| 067 | 13,265.20 | 15,479.02 | 10,282.20 | 11,010.96 | **50,037.39** |
| 068 | 7,097.24 | 7,152.34 | 7,318.88 | 5,989.35 | **27,557.81** |
| 069 | 1,906.06 | 1,909.81 | 1,919.38 | 3,710.96 | **9,446.21** |
| **CT CONNECTICUT Total** | **129,203.86** | **132,695.06** | **115,336.10** | **115,768.01** | **493,003.03** |

**STATE: DC - DISTRICT OF COLUMBIA**

| REGISTRANT ZIP CODE 3 | 1ST QUARTER | 2ND QUARTER | 3RD QUARTER | 4TH QUARTER | TOTAL GRAMS |
|---|---|---|---|---|---|
| 200 | 9,930.13 | 8,553.66 | 9,259.26 | 8,872.85 | **36,615.91** |
| 204 | 864.88 | 396.22 | 855.05 | 808.54 | **2,924.69** |
| **DC - DISTRICT OF COLUMBIA** | **10,795.02** | **8,949.88** | **10,114.31** | **9,681.39** | **39,540.59** |

**STATE: DE - DELAWARE**

| REGISTRANT ZIP CODE 3 | 1ST QUARTER | 2ND QUARTER | 3RD QUARTER | 4TH QUARTER | TOTAL GRAMS |
|---|---|---|---|---|---|
| 197 | 13,143.66 | 12,515.52 | 12,215.09 | 14,211.03 | **52,085.29** |
| 198 | 7,280.33 | 5,812.17 | 6,861.21 | 7,657.85 | **27,611.56** |
| 199 | 15,828.64 | 15,346.29 | 14,143.06 | 17,099.05 | **62,417.05** |

| DE DELAWARE Total | 36,252.63 | 33,673.98 | 33,219.36 | 38,967.93 | 142,113.90 |
|---|---|---|---|---|---|

STATE: FL - FLORIDA

| REGISTRANT ZIP CODE 3 | 1ST QUARTER | 2ND QUARTER | 3RD QUARTER | 4TH QUARTER | TOTAL GRAMS |
|---|---|---|---|---|---|
| 320 | 6,992.33 | 8,192.88 | 7,840.40 | 7,178.99 | **30,204.60** |
| 321 | 10,397.04 | 13,013.97 | 7,832.17 | 10,858.46 | **42,101.64** |
| 322 | 26,204.92 | 20,979.69 | 24,617.23 | 20,689.47 | **92,491.31** |
| 323 | 5,105.86 | 4,647.39 | 4,100.11 | 4,034.46 | **17,887.82** |
| 324 | 4,320.94 | 4,288.20 | 4,266.29 | 6,113.67 | **18,989.10** |
| 325 | 13,020.76 | 11,264.97 | 12,489.49 | 12,219.65 | **48,994.87** |
| 326 | 3,601.48 | 5,078.40 | 4,094.74 | 3,537.80 | **16,312.42** |
| 327 | 1,681.47 | 1,505.81 | 1,633.62 | 1,621.64 | **6,442.54** |
| 328 | 15,151.37 | 18,634.57 | 10,729.41 | 17,786.00 | **62,301.35** |
| 329 | 5,352.09 | 6,754.06 | 5,204.07 | 7,424.68 | **24,734.90** |
| 330 | 6,645.12 | 7,406.26 | 5,933.27 | 5,170.08 | **25,154.73** |
| 331 | 2,119.10 | 2,459.15 | 2,346.06 | 2,624.06 | **9,548.37** |
| 333 | 3,109.65 | 4,734.33 | 3,047.13 | 2,628.82 | **13,519.93** |
| 334 | 9,029.42 | 10,993.96 | 8,015.43 | 9,565.26 | **37,604.08** |
| 335 | 1,557.51 | 1,432.38 | 1,464.76 | 1,513.50 | **5,968.15** |
| 336 | 12,727.70 | 12,407.12 | 10,644.07 | 12,506.20 | **48,285.09** |
| 337 | 21,212.93 | 25,207.50 | 21,566.94 | 18,687.15 | **86,674.52** |
| 338 | 2,857.88 | 2,830.06 | 1,560.73 | 3,267.87 | **10,516.54** |
| 339 | 13,740.04 | 13,391.58 | 16,593.45 | 14,351.99 | **58,077.06** |
| 341 | 3,589.76 | 2,820.49 | 4,694.35 | 3,188.18 | **14,292.78** |
| 342 | 13,043.30 | 13,168.97 | 14,532.75 | 13,833.24 | **54,578.26** |
| 344 | 9,595.48 | 11,632.75 | 9,214.47 | 6,802.54 | **37,245.23** |
| 346 | 15,636.80 | 16,919.28 | 18,322.23 | 16,554.00 | **67,432.30** |
| 347 | 3,836.35 | 6,002.94 | 4,495.16 | 4,547.76 | **18,882.22** |
| 349 | 2,621.40 | 3,121.01 | 2,618.80 | 2,734.90 | **11,096.11** |
| **FL FLORIDA - Total** | **213,150.70** | **228,887.74** | **207,857.13** | **209,440.36** | **859,335.92** |

STATE: GA - GEORGIA

| REGISTRANT ZIP CODE 3 | 1ST QUARTER | 2ND QUARTER | 3RD QUARTER | 4TH QUARTER | TOTAL GRAMS |
|---|---|---|---|---|---|
| 300 | 15,084.77 | 14,336.07 | 14,962.15 | 14,181.43 | **58,564.42** |
| 301 | 12,885.17 | 13,686.29 | 13,158.68 | 13,169.06 | **52,899.20** |
| 302 | 8,777.73 | 8,244.13 | 7,608.75 | 7,952.11 | **32,582.72** |
| 303 | 3,946.72 | 3,881.79 | 3,697.00 | 4,726.01 | **16,251.52** |
| 304 | 1,240.53 | 1,588.90 | 1,194.11 | 1,184.36 | **5,207.91** |
| 305 | 12,868.00 | 12,307.57 | 12,251.76 | 13,070.85 | **50,498.18** |
| 306 | 4,885.12 | 5,518.81 | 5,266.32 | 5,305.40 | **20,975.65** |
| 307 | 25,097.94 | 27,585.35 | 25,592.00 | 26,260.48 | **104,535.77** |
| 308 | 1,006.20 | 986.97 | 1,657.77 | 899.32 | **4,550.26** |

| | | | | | |
|---|---|---|---|---|---|
| 309 | 3,137.02 | 4,453.66 | 3,763.81 | 3,823.11 | **15,177.61** |
| 310 | 5,190.74 | 3,891.18 | 5,488.93 | 3,586.45 | **18,157.30** |
| 312 | 1,912.05 | 2,545.82 | 1,849.62 | 1,972.15 | **8,279.64** |
| 313 | 273.87 | 267.69 | 230.84 | 218.23 | **990.64** |
| 314 | 5,415.68 | 4,623.69 | 4,166.56 | 4,480.94 | **18,686.88** |
| 315 | 2,516.39 | 3,480.56 | 2,900.99 | 2,345.56 | **11,243.50** |
| 316 | 1,470.39 | 2,582.13 | 1,452.77 | 2,009.63 | **7,514.93** |
| 317 | 2,785.70 | 2,764.32 | 3,085.14 | 2,176.70 | **10,811.86** |
| 318 | 90.87 | 92.66 | 78.17 | 76.92 | **338.62** |
| 319 | 2,654.76 | 3,048.20 | 3,172.71 | 3,058.04 | **11,933.71** |
| 398 | 677.06 | 1,235.17 | 766.95 | 1,284.36 | **3,963.53** |
| **GA GEORGIA Total** | **111,916.72** | **117,120.96** | **112,345.05** | **111,781.13** | **453,163.86** |

**STATE: GU - GUAM**

| REGISTRANT ZIP CODE 3 | 1ST QUARTER | 2ND QUARTER | 3RD QUARTER | 4TH QUARTER | TOTAL GRAMS |
|---|---|---|---|---|---|
| 969 | 45.61 | 36.22 | 33.09 | 16.10 | **131.03** |
| **GU - GUAM - Total** | **45.61** | **36.22** | **33.09** | **16.10** | **131.03** |

**STATE: HI - HAWAII**

| REGISTRANT ZIP CODE 3 | 1ST QUARTER | 2ND QUARTER | 3RD QUARTER | 4TH QUARTER | TOTAL GRAMS |
|---|---|---|---|---|---|
| 967 | 4,070.86 | 3,147.75 | 3,747.98 | 2,985.60 | **13,952.19** |
| 968 | 3,575.63 | 4,145.01 | 4,798.46 | 3,612.39 | **16,131.49** |
| **HI - HAWAII - Total** | **7,646.49** | **7,292.76** | **8,546.44** | **6,597.99** | **30,083.68** |

**STATE: IA - IOWA**

| REGISTRANT ZIP CODE 3 | 1ST QUARTER | 2ND QUARTER | 3RD QUARTER | 4TH QUARTER | TOTAL GRAMS |
|---|---|---|---|---|---|
| 500 | 602.29 | 790.56 | 512.49 | 946.01 | **2,851.35** |
| 501 | 526.89 | 174.68 | 262.77 | 516.78 | **1,481.13** |
| 502 | 182.90 | 264.30 | 160.99 | 310.36 | **918.55** |
| 503 | 4,264.50 | 4,518.24 | 3,997.52 | 4,588.00 | **17,368.26** |
| 504 | 116.27 | 126.92 | 147.13 | 119.94 | **510.26** |
| 505 | 251.77 | 164.57 | 258.03 | 342.11 | **1,016.49** |
| 506 | 419.12 | 433.78 | 71.55 | 73.79 | **998.24** |
| 507 | 676.61 | 417.68 | 768.02 | 773.66 | **2,635.98** |
| 508 | 55.45 | 57.69 | 66.19 | 63.95 | **243.28** |
| 510 | 130.58 | 120.30 | 111.80 | 114.75 | **477.43** |
| 511 | 295.69 | 711.14 | 275.03 | 677.96 | **1,959.81** |
| 512 | 13.86 | 10.29 | 8.94 | 6.26 | **39.35** |
| 513 | 26.38 | 31.75 | 27.28 | 16.10 | **101.51** |
| 514 | 20.57 | 24.60 | 112.25 | 28.17 | **185.59** |
| 515 | 181.21 | 168.59 | 164.12 | 146.68 | **660.60** |
| 516 | 19.68 | 21.91 | 15.20 | 13.42 | **70.21** |
| 520 | 339.07 | 239.52 | 218.50 | 77.90 | **874.99** |

| | | | | | |
|---|---|---|---|---|---|
| 521 | 8.05 | 93.46 | 6.71 | 95.25 | **203.48** |
| 522 | 151.15 | 128.17 | 152.32 | 143.82 | **575.46** |
| 523 | 999.85 | 789.93 | 447.29 | 96.60 | **2,333.67** |
| 524 | 1,885.48 | 2,011.06 | 1,917.24 | 3,315.99 | **9,129.77** |
| 525 | 113.59 | 291.13 | 281.29 | 408.11 | **1,094.12** |
| 526 | 138.36 | 412.32 | 392.73 | 143.37 | **1,086.79** |
| 527 | 94.54 | 87.65 | 85.06 | 98.38 | **365.63** |
| 528 | 1,729.32 | 1,728.88 | 1,739.88 | 1,737.37 | **6,935.45** |
| **IA IOWA Total** | **13,243.20** | **13,819.11** | **12,200.33** | **14,854.73** | **54,117.37** |

**STATE: ID - IDAHO**

| REGISTRANT ZIP CODE 3 | 1ST QUARTER | 2ND QUARTER | 3RD QUARTER | 4TH QUARTER | TOTAL GRAMS |
|---|---|---|---|---|---|
| 832 | 662.75 | 661.95 | 627.51 | 603.27 | **2,555.48** |
| 833 | 218.23 | 224.05 | 252.04 | 213.31 | **907.64** |
| 834 | 296.58 | 306.69 | 306.51 | 290.05 | **1,199.84** |
| 835 | 279.50 | 285.31 | 280.84 | 276.82 | **1,122.47** |
| 836 | 3,896.81 | 3,571.07 | 3,423.41 | 3,392.64 | **14,283.93** |
| 837 | 4,681.29 | 5,235.37 | 4,533.53 | 4,704.45 | **19,154.65** |
| 838 | 902.90 | 821.60 | 829.91 | 797.00 | **3,351.41** |
| **ID IDAHO Total** | **10,938.06** | **11,106.03** | **10,253.76** | **10,277.55** | **42,575.41** |

**STATE: IL - ILLINOIS**

| REGISTRANT ZIP CODE 3 | 1ST QUARTER | 2ND QUARTER | 3RD QUARTER | 4TH QUARTER | TOTAL GRAMS |
|---|---|---|---|---|---|
| 600 | 9,161.07 | 9,369.02 | 9,084.96 | 9,387.98 | **37,003.03** |
| 601 | 6,327.08 | 8,375.97 | 6,818.73 | 7,363.68 | **28,885.45** |
| 602 | 4,408.86 | 4,035.44 | 3,984.82 | 3,871.86 | **16,300.98** |
| 603 | 40.25 | 54.11 | 47.40 | 54.11 | **195.87** |
| 604 | 7,818.84 | 8,456.55 | 6,930.88 | 8,418.63 | **31,624.91** |
| 605 | 6,153.47 | 6,063.85 | 6,829.91 | 5,981.30 | **25,028.53** |
| 606 | 55,528.73 | 56,189.70 | 57,370.13 | 58,056.76 | **227,145.31** |
| 607 | 53.57 | 399.71 | 62.07 | 53.57 | **568.93** |
| 608 | 3,824.01 | 3,644.68 | 3,774.82 | 4,080.70 | **15,324.20** |
| 609 | 1,380.42 | 740.56 | 1,377.11 | 1,438.37 | **4,936.46** |
| 610 | 732.07 | 1,059.69 | 724.46 | 1,139.47 | **3,655.68** |
| 611 | 2,817.63 | 2,847.59 | 2,847.59 | 2,723.72 | **11,236.53** |
| 612 | 766.95 | 867.57 | 896.19 | 936.44 | **3,467.14** |
| 613 | 922.57 | 623.84 | 741.28 | 1,135.44 | **3,423.14** |
| 614 | 111.35 | 116.27 | 113.14 | 123.43 | **464.19** |
| 615 | 119.85 | 111.35 | 126.92 | 120.30 | **478.41** |
| 616 | 8,527.70 | 7,663.47 | 7,137.94 | 7,450.81 | **30,779.92** |
| 617 | 1,436.32 | 1,834.06 | 1,593.91 | 1,283.73 | **6,148.02** |
| 618 | 1,332.66 | 3,129.86 | 2,457.72 | 1,644.89 | **8,565.13** |

| | | | | | |
|---|---|---|---|---|---|
| 619 | 36.22 | 31.75 | 43.83 | 34.26 | **146.06** |
| 620 | 3,866.67 | 4,513.68 | 3,527.51 | 4,649.72 | **16,557.58** |
| 622 | 783.14 | 1,335.79 | 775.09 | 1,002.62 | **3,896.63** |
| 623 | 481.90 | 294.08 | 295.42 | 331.02 | **1,402.42** |
| 624 | 137.74 | 128.35 | 114.93 | 105.99 | **487.00** |
| 625 | 1,889.87 | 2,104.97 | 2,297.62 | 2,308.45 | **8,600.91** |
| 626 | 118.96 | 110.46 | 105.54 | 118.51 | **453.46** |
| 627 | 3,036.94 | 3,793.78 | 3,272.16 | 4,234.09 | **14,336.96** |
| 628 | 185.14 | 168.15 | 190.95 | 172.62 | **716.86** |
| 629 | 402.75 | 415.27 | 923.11 | 958.62 | **2,699.75** |
| **IL ILLINOIS Total** | **122,402.70** | **128,479.56** | **124,466.14** | **129,181.06** | **504,529.46** |

**STATE: IN - INDIANA**

| REGISTRANT ZIP CODE 3 | 1ST QUARTER | 2ND QUARTER | 3RD QUARTER | 4TH QUARTER | TOTAL GRAMS |
|---|---|---|---|---|---|
| 460 | 441.21 | 406.50 | 417.24 | 335.49 | **1,600.44** |
| 461 | 2,807.07 | 2,783.02 | 3,563.65 | 3,311.52 | **12,465.25** |
| 462 | 17,932.88 | 17,148.60 | 13,495.11 | 18,274.38 | **66,850.96** |
| 463 | 3,535.47 | 2,800.90 | 3,802.54 | 2,829.34 | **12,968.26** |
| 464 | 6,261.25 | 6,727.23 | 6,954.23 | 7,592.11 | **27,534.82** |
| 465 | 1,170.95 | 1,166.21 | 1,082.76 | 1,107.45 | **4,527.36** |
| 466 | 4,069.07 | 3,566.42 | 4,465.74 | 4,465.29 | **16,566.52** |
| 467 | 757.83 | 754.61 | 702.46 | 658.28 | **2,873.17** |
| 468 | 6,192.56 | 7,521.46 | 6,981.42 | 7,244.01 | **27,939.45** |
| 469 | 7,854.08 | 7,560.36 | 8,174.37 | 8,978.43 | **32,567.25** |
| 470 | 4,708.12 | 5,378.47 | 5,436.16 | 4,694.26 | **20,217.02** |
| 471 | 12,620.07 | 10,444.18 | 10,851.40 | 10,810.70 | **44,726.35** |
| 472 | 308.93 | 301.41 | 278.34 | 228.97 | **1,117.64** |
| 473 | 6,799.77 | 5,734.27 | 5,688.56 | 6,164.12 | **24,386.71** |
| 474 | 900.21 | 1,397.95 | 1,386.77 | 1,801.77 | **5,486.70** |
| 475 | 221.01 | 213.76 | 204.28 | 210.99 | **850.04** |
| 476 | 194.98 | 199.90 | 193.10 | 195.87 | **783.85** |
| 477 | 4,065.41 | 3,735.64 | 3,034.52 | 3,862.47 | **14,698.03** |
| 478 | 1,668.95 | 887.24 | 1,808.48 | 992.78 | **5,357.46** |
| 479 | 1,191.88 | 1,904.80 | 1,921.35 | 1,976.27 | **6,994.30** |
| **IN INDIANA Total** | **83,701.69** | **80,632.93** | **80,442.47** | **85,734.50** | **330,511.59** |

**STATE: KS - KANSAS**

| REGISTRANT ZIP CODE 3 | 1ST QUARTER | 2ND QUARTER | 3RD QUARTER | 4TH QUARTER | TOTAL GRAMS |
|---|---|---|---|---|---|
| 660 | 1,247.42 | 1,580.58 | 1,197.06 | 1,449.46 | **5,474.53** |
| 661 | 3,518.48 | 3,497.10 | 3,215.99 | 3,679.56 | **13,911.14** |
| 662 | 1,748.73 | 1,794.08 | 1,499.01 | 1,707.05 | **6,748.87** |
| 664 | 80.05 | 88.10 | 63.06 | 63.06 | **294.26** |

| | | | | | |
|---|---|---|---|---|---|
| 665 | 120.74 | 110.46 | 119.67 | 112.25 | **463.12** |
| 666 | 1,724.40 | 1,837.99 | 1,902.66 | 1,604.82 | **7,069.87** |
| 667 | 298.73 | 281.29 | 291.13 | 276.37 | **1,147.52** |
| 668 | 90.78 | 69.32 | 69.32 | 71.10 | **300.52** |
| 669 | 80.50 | 73.79 | 67.08 | 52.77 | **274.13** |
| 670 | 450.78 | 467.77 | 454.80 | 408.56 | **1,781.91** |
| 671 | 203.92 | 183.80 | 163.32 | 151.15 | **702.19** |
| 672 | 11,821.28 | 10,323.35 | 9,260.80 | 9,641.51 | **41,046.94** |
| 673 | 113.59 | 99.28 | 109.12 | 93.46 | **415.45** |
| 674 | 180.67 | 176.20 | 169.04 | 166.81 | **692.71** |
| 675 | 160.99 | 170.83 | 170.38 | 170.83 | **673.04** |
| 676 | 31.75 | 67.97 | 59.92 | 40.70 | **200.35** |
| 677 | 29.96 | 34.88 | 22.36 | 26.38 | **113.59** |
| 678 | 129.69 | 119.40 | 123.87 | 127.54 | **500.51** |
| 679 | 26.38 | 33.54 | 37.56 | 36.22 | **133.71** |
| **KS - KANSAS - Total** | **22,058.86** | **21,009.73** | **18,996.16** | **19,879.62** | **81,944.36** |

**STATE: KY - KENTUCKY**

| REGISTRANT ZIP CODE 3 | 1ST QUARTER | 2ND QUARTER | 3RD QUARTER | 4TH QUARTER | TOTAL GRAMS |
|---|---|---|---|---|---|
| 400 | 487.18 | 645.76 | 563.02 | 645.31 | **2,341.27** |
| 401 | 160.99 | 155.18 | 160.54 | 165.46 | **642.18** |
| 402 | 12,860.85 | 9,251.94 | 12,474.02 | 10,748.54 | **45,335.35** |
| 403 | 1,584.25 | 1,563.41 | 1,780.75 | 2,010.16 | **6,938.58** |
| 404 | 1,330.24 | 1,868.40 | 1,793.54 | 1,204.31 | **6,196.49** |
| 405 | 3,975.65 | 4,266.40 | 4,441.83 | 4,792.20 | **17,476.07** |
| 406 | 1,618.42 | 1,584.52 | 1,622.89 | 1,598.65 | **6,424.48** |
| 407 | 1,719.48 | 1,511.54 | 1,716.80 | 1,683.26 | **6,631.08** |
| 408 | 57.69 | 48.74 | 55.01 | 45.17 | **206.61** |
| 409 | 120.74 | 124.77 | 120.74 | 100.17 | **466.43** |
| 410 | 7,688.17 | 10,048.85 | 10,022.29 | 10,890.66 | **38,649.98** |
| 411 | 3,146.50 | 4,925.91 | 5,454.05 | 5,175.45 | **18,701.90** |
| 412 | 855.05 | 1,205.65 | 1,226.22 | 857.73 | **4,144.65** |
| 413 | 164.12 | 139.97 | 142.21 | 130.14 | **576.44** |
| 414 | 72.89 | 71.55 | 84.52 | 74.68 | **303.65** |
| 415 | 1,168.53 | 1,173.90 | 1,165.40 | 1,149.30 | **4,657.14** |
| 416 | 127.45 | 135.05 | 126.11 | 128.79 | **517.41** |
| 417 | 2,528.47 | 2,136.72 | 2,315.60 | 1,970.36 | **8,951.16** |
| 418 | 24.60 | 31.30 | 28.62 | 33.09 | **117.61** |
| 420 | 1,476.48 | 1,890.22 | 1,363.78 | 1,861.96 | **6,592.44** |
| 421 | 2,502.98 | 1,696.05 | 2,217.66 | 1,677.80 | **8,094.50** |
| 422 | 1,694.44 | 1,151.09 | 1,686.84 | 1,700.70 | **6,233.07** |

| | | | | | |
|---|---|---|---|---|---|
| 423 | 706.58 | 1,233.38 | 698.53 | 689.58 | **3,328.06** |
| 424 | 272.34 | 245.96 | 257.59 | 268.77 | **1,044.66** |
| 425 | 1,130.52 | 1,273.63 | 1,533.00 | 2,352.27 | **6,289.42** |
| 426 | 60.82 | 63.95 | 60.82 | 52.77 | **238.36** |
| 427 | 1,635.41 | 1,613.94 | 2,339.75 | 877.85 | **6,466.96** |
| **KY - KENTUCKY - Total** | **49,170.84** | **50,057.80** | **55,452.14** | **52,885.16** | **207,565.94** |

**STATE: LA - LOUISIANA**

| REGISTRANT ZIP CODE 3 | 1ST QUARTER | 2ND QUARTER | 3RD QUARTER | 4TH QUARTER | TOTAL GRAMS |
|---|---|---|---|---|---|
| 700 | 7,754.27 | 6,233.43 | 7,211.28 | 7,070.14 | **28,269.12** |
| 701 | 4,533.53 | 5,904.83 | 4,423.70 | 5,824.60 | **20,686.67** |
| 703 | 131.03 | 158.76 | 119.85 | 130.58 | **540.22** |
| 704 | 3,845.65 | 3,821.50 | 3,799.59 | 2,746.70 | **14,213.45** |
| 705 | 3,135.41 | 3,916.67 | 3,073.70 | 3,970.96 | **14,096.73** |
| 706 | 1,898.36 | 1,853.64 | 1,649.72 | 1,972.15 | **7,373.88** |
| 707 | 339.42 | 330.48 | 381.91 | 313.93 | **1,365.75** |
| 708 | 4,173.00 | 3,893.23 | 3,790.47 | 3,782.95 | **15,639.66** |
| 710 | 301.23 | 354.18 | 294.08 | 276.37 | **1,225.86** |
| 711 | 3,598.53 | 4,170.14 | 3,600.93 | 3,659.65 | **15,029.25** |
| 712 | 883.13 | 886.35 | 912.20 | 886.53 | **3,568.21** |
| 713 | 3,675.54 | 3,652.73 | 4,650.43 | 2,487.33 | **14,466.03** |
| 714 | 313.49 | 362.23 | 342.56 | 305.88 | **1,324.16** |
| **LA - LOUISIANA - Total** | **34,582.60** | **35,538.18** | **34,250.40** | **33,427.79** | **137,798.97** |

**STATE: MA - MASSACHUSETTS**

| REGISTRANT ZIP CODE 3 | 1ST QUARTER | 2ND QUARTER | 3RD QUARTER | 4TH QUARTER | TOTAL GRAMS |
|---|---|---|---|---|---|
| 010 | 11,627.38 | 12,618.64 | 11,741.50 | 13,475.75 | **49,463.27** |
| 011 | 8,603.86 | 9,134.15 | 9,818.74 | 10,479.23 | **38,035.98** |
| 012 | 4,586.04 | 6,347.56 | 4,707.23 | 6,157.94 | **21,798.76** |
| 013 | 1,393.30 | 1,404.30 | 1,349.02 | 1,405.37 | **5,551.99** |
| 014 | 7,288.47 | 7,229.44 | 7,063.97 | 7,799.62 | **29,381.49** |
| 015 | 4,196.08 | 4,497.04 | 4,155.83 | 4,482.91 | **17,331.86** |
| 016 | 9,870.48 | 9,389.95 | 9,395.86 | 10,250.25 | **38,906.53** |
| 017 | 3,811.66 | 3,605.15 | 3,633.50 | 3,943.32 | **14,993.63** |
| 018 | 17,808.49 | 18,062.68 | 20,050.03 | 16,218.24 | **72,139.44** |
| 019 | 15,557.55 | 13,229.25 | 15,560.95 | 14,659.75 | **59,007.50** |
| 020 | 583.33 | 710.06 | 481.01 | 598.60 | **2,373.00** |
| 021 | 26,535.17 | 25,623.40 | 24,999.50 | 27,726.21 | **104,884.27** |
| 022 | 344.61 | 359.10 | 288.80 | 329.85 | **1,322.37** |
| 023 | 9,071.09 | 10,498.91 | 8,409.06 | 10,238.38 | **38,217.44** |
| 024 | 1,224.97 | 1,219.69 | 1,225.24 | 1,183.47 | **4,853.37** |
| 025 | 2,462.19 | 4,023.55 | 3,524.20 | 3,284.86 | **13,294.81** |

| | | | | | |
|---|---|---|---|---|---|
| 026 | 3,186.39 | 3,087.92 | 3,643.88 | 2,862.26 | **12,780.44** |
| 027 | 20,445.36 | 18,727.22 | 20,632.56 | 20,077.76 | **79,882.89** |
| **MA - MASSACHUSETTS - Total** | **148,596.41** | **149,768.00** | **150,680.87** | **155,173.78** | **604,219.06** |

**STATE: MD - MARYLAND**

| REGISTRANT ZIP CODE 3 | 1ST QUARTER | 2ND QUARTER | 3RD QUARTER | 4TH QUARTER | TOTAL GRAMS |
|---|---|---|---|---|---|
| 206 | 4,341.15 | 4,608.66 | 4,586.21 | 4,546.15 | **18,082.17** |
| 207 | 2,120.53 | 2,884.53 | 2,173.17 | 2,358.80 | **9,537.03** |
| 208 | 3,094.71 | 2,923.97 | 2,653.05 | 3,131.56 | **11,803.30** |
| 209 | 763.82 | 1,387.66 | 764.71 | 794.23 | **3,710.42** |
| 210 | 33,546.62 | 34,287.36 | 32,765.18 | 31,662.30 | **132,261.46** |
| 211 | 3,657.65 | 5,560.04 | 5,603.24 | 3,610.16 | **18,431.08** |
| 212 | 101,707.75 | 115,310.14 | 109,399.23 | 110,939.10 | **437,356.22** |
| 214 | 2,892.58 | 2,321.77 | 2,325.89 | 3,150.35 | **10,690.58** |
| 215 | 3,370.73 | 3,877.13 | 2,866.64 | 2,777.38 | **12,891.88** |
| 216 | 3,602.29 | 2,670.23 | 2,938.01 | 3,226.73 | **12,437.26** |
| 217 | 14,735.33 | 15,714.16 | 15,426.61 | 13,615.81 | **59,491.91** |
| 218 | 5,453.16 | 4,499.82 | 5,611.56 | 4,799.80 | **20,364.33** |
| 219 | 6,428.50 | 8,843.65 | 7,898.98 | 7,422.63 | **30,593.76** |
| **MD MARYLAND Total** | **185,714.81** | **204,889.13** | **195,012.49** | **192,034.97** | **777,651.39** |

**STATE: ME - MAINE**

| REGISTRANT ZIP CODE 3 | 1ST QUARTER | 2ND QUARTER | 3RD QUARTER | 4TH QUARTER | TOTAL GRAMS |
|---|---|---|---|---|---|
| 039 | 70.66 | 65.92 | 64.84 | 61.71 | **263.13** |
| 040 | 3,636.63 | 3,626.43 | 3,995.64 | 3,465.71 | **14,724.42** |
| 041 | 5,899.91 | 6,276.45 | 5,660.21 | 6,717.30 | **24,553.87** |
| 042 | 1,924.21 | 2,518.63 | 2,540.63 | 2,522.74 | **9,506.22** |
| 043 | 505.78 | 449.61 | 463.66 | 472.87 | **1,891.92** |
| 044 | 8,889.80 | 10,211.81 | 10,350.44 | 10,028.01 | **39,480.07** |
| 045 | 72.00 | 62.61 | 74.68 | 88.55 | **297.84** |
| 046 | 1,122.47 | 1,475.13 | 1,086.34 | 1,391.69 | **5,075.63** |
| 047 | 52.77 | 59.48 | 60.37 | 63.50 | **236.12** |
| 048 | 1,602.41 | 1,603.03 | 1,443.83 | 1,664.39 | **6,313.66** |
| 049 | 4,150.37 | 4,081.59 | 4,933.96 | 4,874.30 | **18,040.23** |
| **ME MAINE Total** | **27,927.01** | **30,430.71** | **30,674.61** | **31,350.78** | **120,383.11** |

**STATE: MI - MICHIGAN**

| REGISTRANT ZIP CODE 3 | 1ST QUARTER | 2ND QUARTER | 3RD QUARTER | 4TH QUARTER | TOTAL GRAMS |
|---|---|---|---|---|---|
| 480 | 16,215.01 | 15,975.67 | 17,303.58 | 15,533.17 | **65,027.44** |
| 481 | 10,984.04 | 9,822.93 | 11,180.36 | 9,669.63 | **41,656.95** |
| 482 | 15,494.12 | 16,108.79 | 15,461.21 | 18,761.51 | **65,825.62** |
| 483 | 12,654.24 | 12,430.19 | 12,120.19 | 9,510.69 | **46,715.32** |
| 484 | 858.62 | 874.72 | 876.06 | 797.36 | **3,406.77** |

| | | | | | |
|---|---|---|---|---|---|
| 485 | 9,769.08 | 8,480.25 | 9,684.12 | 8,795.08 | **36,728.54** |
| 486 | 5,248.43 | 5,201.56 | 4,572.17 | 5,889.71 | **20,911.88** |
| 487 | 388.26 | 372.07 | 347.47 | 340.77 | **1,448.57** |
| 488 | 4,502.95 | 5,821.47 | 5,439.03 | 5,323.65 | **21,087.09** |
| 489 | 6,703.39 | 8,111.22 | 7,941.03 | 8,505.03 | **31,260.68** |
| 490 | 10,844.87 | 9,649.86 | 8,918.33 | 11,864.48 | **41,277.54** |
| 491 | 334.95 | 305.71 | 300.07 | 281.83 | **1,222.56** |
| 492 | 4,208.60 | 4,006.46 | 4,014.51 | 3,889.30 | **16,118.88** |
| 493 | 726.25 | 626.97 | 669.55 | 560.43 | **2,583.21** |
| 494 | 9,640.65 | 9,358.29 | 8,860.73 | 9,271.08 | **37,130.75** |
| 495 | 8,784.35 | 10,050.64 | 7,854.26 | 8,403.51 | **35,092.77** |
| 496 | 1,902.48 | 1,826.63 | 1,960.52 | 1,735.67 | **7,425.31** |
| 497 | 4,339.18 | 3,830.00 | 4,697.48 | 4,005.48 | **16,872.14** |
| 498 | 475.37 | 457.93 | 456.59 | 397.29 | **1,787.19** |
| 499 | 68.42 | 64.84 | 58.14 | 73.79 | **265.19** |
| **MI - MICHIGAN Total** | **124,143.27** | **123,376.22** | **122,715.41** | **123,609.46** | **493,844.37** |

**STATE: MN - MINNESOTA**

| REGISTRANT ZIP CODE 3 | 1ST QUARTER | 2ND QUARTER | 3RD QUARTER | 4TH QUARTER | TOTAL GRAMS |
|---|---|---|---|---|---|
| 550 | 693.52 | 650.77 | 615.70 | 545.76 | **2,505.75** |
| 551 | 10,997.63 | 13,015.13 | 14,228.65 | 11,810.28 | **50,051.70** |
| 553 | 2,246.29 | 2,855.46 | 2,147.63 | 2,110.25 | **9,359.63** |
| 554 | 24,701.70 | 26,095.73 | 28,011.62 | 23,894.34 | **102,703.38** |
| 556 | 15.65 | 15.20 | 21.02 | 16.10 | **67.97** |
| 557 | 229.77 | 249.18 | 234.06 | 236.57 | **949.58** |
| 558 | 1,483.72 | 2,734.45 | 2,720.50 | 3,925.88 | **10,864.55** |
| 559 | 2,095.31 | 1,489.89 | 1,481.39 | 2,141.64 | **7,208.24** |
| 560 | 168.24 | 206.79 | 181.65 | 199.00 | **755.68** |
| 561 | 39.71 | 38.10 | 30.68 | 33.99 | **142.48** |
| 562 | 85.42 | 81.84 | 80.59 | 66.10 | **313.93** |
| 563 | 1,900.96 | 1,904.18 | 1,958.74 | 2,088.16 | **7,852.03** |
| 564 | 2,919.95 | 3,820.43 | 3,780.36 | 3,783.76 | **14,304.50** |
| 565 | 145.34 | 160.63 | 139.97 | 141.76 | **587.71** |
| 566 | 63.06 | 72.54 | 59.92 | 70.66 | **266.17** |
| 567 | 60.37 | 86.13 | 74.06 | 68.42 | **288.98** |
| **MN MINNESOTA Total** | **47,846.62** | **53,476.44** | **55,766.55** | **51,132.67** | **208,222.28** |

**STATE: MO - MISSOURI**

| REGISTRANT ZIP CODE 3 | 1ST QUARTER | 2ND QUARTER | 3RD QUARTER | 4TH QUARTER | TOTAL GRAMS |
|---|---|---|---|---|---|
| 630 | 4,391.06 | 4,435.42 | 4,992.45 | 5,084.04 | **18,902.97** |
| 631 | 9,630.18 | 10,337.68 | 11,428.98 | 8,884.43 | **40,281.27** |
| 633 | 2,073.67 | 1,629.60 | 2,509.60 | 1,757.85 | **7,970.71** |

| | | | | | |
|---|---|---|---|---|---|
| 634 | 189.61 | 175.75 | 204.82 | 209.29 | **779.47** |
| 635 | 27.28 | 23.25 | 25.49 | 13.95 | **89.98** |
| 636 | 192.74 | 175.30 | 170.83 | 136.84 | **675.72** |
| 637 | 1,514.31 | 1,516.54 | 2,013.92 | 550.23 | **5,595.01** |
| 638 | 150.08 | 113.59 | 109.12 | 103.75 | **476.54** |
| 639 | 902.72 | 802.72 | 1,028.56 | 608.01 | **3,342.02** |
| 640 | 761.58 | 752.19 | 763.01 | 730.01 | **3,006.79** |
| 641 | 4,188.02 | 4,753.19 | 4,699.36 | 4,444.62 | **18,085.18** |
| 644 | 59.48 | 45.17 | 53.75 | 37.12 | **195.52** |
| 645 | 2,125.09 | 1,418.61 | 1,915.80 | 1,573.70 | **7,033.20** |
| 646 | 61.71 | 54.11 | 49.64 | 51.88 | **217.34** |
| 647 | 113.59 | 119.85 | 119.40 | 120.74 | **473.58** |
| 648 | 1,355.46 | 1,155.56 | 2,036.55 | 1,131.42 | **5,678.99** |
| 650 | 105.54 | 92.57 | 84.97 | 93.91 | **376.99** |
| 651 | 88.10 | 77.81 | 74.68 | 75.58 | **316.17** |
| 652 | 1,428.63 | 1,226.85 | 1,107.18 | 1,353.85 | **5,116.50** |
| 653 | 80.50 | 75.58 | 79.15 | 65.29 | **300.52** |
| 654 | 80.94 | 66.63 | 84.52 | 78.71 | **310.80** |
| 655 | 143.55 | 160.54 | 148.47 | 125.66 | **578.23** |
| 656 | 317.15 | 249.18 | 284.87 | 244.26 | **1,095.46** |
| 657 | 1,142.15 | 1,348.76 | 1,270.76 | 1,224.25 | **4,985.92** |
| 658 | 3,763.01 | 2,989.62 | 3,639.76 | 3,406.32 | **13,798.71** |
| **MO MISSOURI - Total** | **34,886.15** | **33,796.08** | **38,895.65** | **32,105.72** | **139,683.60** |

**STATE: MS - MISSISSIPPI**

| REGISTRANT ZIP CODE 3 | 1ST QUARTER | 2ND QUARTER | 3RD QUARTER | 4TH QUARTER | TOTAL GRAMS |
|---|---|---|---|---|---|
| 386 | 1,753.47 | 1,599.63 | 2,023.04 | 2,024.47 | **7,400.62** |
| 387 | 177.99 | 144.45 | 160.54 | 132.82 | **615.79** |
| 388 | 91.94 | 86.31 | 76.83 | 61.18 | **316.26** |
| 389 | 66.63 | 47.40 | 53.22 | 55.90 | **223.15** |
| 390 | 174.86 | 154.73 | 159.65 | 149.81 | **639.05** |
| 391 | 134.43 | 118.06 | 126.11 | 131.48 | **510.08** |
| 392 | 1,624.59 | 1,485.60 | 1,771.18 | 1,606.16 | **6,487.53** |
| 393 | 923.92 | 920.34 | 931.52 | 915.87 | **3,691.64** |
| 394 | 200.35 | 198.56 | 183.35 | 165.46 | **747.72** |
| 395 | 2,594.48 | 4,339.45 | 2,532.31 | 2,662.18 | **12,128.42** |
| 396 | 130.58 | 110.01 | 121.64 | 104.64 | **466.88** |
| 397 | 68.87 | 88.72 | 84.88 | 75.85 | **318.32** |
| **MS MISSISSIPPI - Total** | **7,942.09** | **9,293.26** | **8,224.28** | **8,085.82** | **33,545.46** |

**STATE: MT - MONTANA**

| REGISTRANT ZIP CODE 3 | 1ST QUARTER | 2ND QUARTER | 3RD QUARTER | 4TH QUARTER | TOTAL GRAMS |
|---|---|---|---|---|---|

| | | | | | |
|---|---|---|---|---|---|
| 590 | 44.72 | 51.88 | 64.84 | 62.61 | **224.05** |
| 591 | 2,277.14 | 1,637.20 | 1,656.43 | 1,629.95 | **7,200.72** |
| 592 | 20.12 | 17.89 | 19.23 | 14.76 | **72.00** |
| 593 | 38.01 | 39.35 | 23.70 | 16.10 | **117.17** |
| 594 | 105.54 | 110.73 | 109.56 | 105.54 | **431.37** |
| 595 | 18.78 | 17.89 | 20.57 | 15.20 | **72.45** |
| 596 | 147.58 | 129.24 | 103.30 | 93.91 | **474.03** |
| 597 | 1,443.11 | 1,167.64 | 1,449.20 | 1,414.23 | **5,474.18** |
| 598 | 2,553.15 | 2,583.56 | 2,538.75 | 2,510.04 | **10,185.52** |
| 599 | 2,358.44 | 2,308.98 | 2,314.53 | 2,315.33 | **9,297.29** |
| **MT - MONTANA Total** | **9,006.61** | **8,064.36** | **8,300.12** | **8,177.68** | **33,548.77** |

**STATE: NC - NORTH CAROLINA**

| REGISTRANT ZIP CODE 3 | 1ST QUARTER | 2ND QUARTER | 3RD QUARTER | 4TH QUARTER | TOTAL GRAMS |
|---|---|---|---|---|---|
| 270 | 414.64 | 428.24 | 415.90 | 373.23 | **1,632.01** |
| 271 | 4,418.34 | 3,954.14 | 4,495.43 | 4,076.14 | **16,944.05** |
| 272 | 6,974.35 | 5,514.69 | 7,091.88 | 5,631.86 | **25,212.78** |
| 273 | 4,553.39 | 5,667.37 | 5,353.43 | 4,997.91 | **20,572.09** |
| 274 | 6,151.77 | 6,367.23 | 6,399.61 | 7,234.98 | **26,153.60** |
| 275 | 8,693.82 | 14,958.75 | 11,606.00 | 9,048.48 | **44,307.06** |
| 276 | 6,536.36 | 6,846.81 | 7,587.02 | 6,232.72 | **27,202.91** |
| 277 | 7,625.03 | 7,233.91 | 6,691.81 | 6,531.09 | **28,081.83** |
| 278 | 5,928.98 | 6,665.96 | 5,955.18 | 6,278.06 | **24,828.19** |
| 279 | 1,215.04 | 1,365.30 | 1,578.71 | 1,440.43 | **5,599.48** |
| 280 | 7,179.08 | 8,876.03 | 9,021.19 | 7,059.05 | **32,135.34** |
| 281 | 6,845.47 | 6,012.51 | 6,959.68 | 6,115.37 | **25,933.04** |
| 282 | 7,589.34 | 7,647.92 | 6,994.39 | 8,548.94 | **30,780.60** |
| 283 | 11,796.24 | 13,417.52 | 13,211.45 | 14,368.63 | **52,793.84** |
| 284 | 6,578.57 | 8,258.35 | 7,492.66 | 7,163.25 | **29,492.83** |
| 285 | 5,796.07 | 6,042.12 | 5,446.00 | 6,086.57 | **23,370.76** |
| 286 | 13,485.05 | 12,593.15 | 11,967.52 | 13,666.97 | **51,712.69** |
| 287 | 8,540.45 | 7,583.62 | 9,040.68 | 7,539.43 | **32,704.18** |
| 288 | 15,394.23 | 15,180.38 | 15,603.52 | 16,153.04 | **62,331.18** |
| 289 | 1,213.79 | 907.82 | 1,039.29 | 1,006.20 | **4,167.10** |
| **NC - NORTH CAROLINA Total** | **136,930.02** | **145,521.83** | **143,951.35** | **139,552.35** | **565,955.56** |

**STATE: ND - NORTH DAKOTA**

| REGISTRANT ZIP CODE 3 | 1ST QUARTER | 2ND QUARTER | 3RD QUARTER | 4TH QUARTER | TOTAL GRAMS |
|---|---|---|---|---|---|
| 580 | 68.42 | 55.90 | 47.05 | 42.04 | **213.40** |
| 581 | 2,213.91 | 2,205.05 | 2,508.79 | 2,823.26 | **9,751.02** |
| 582 | 175.57 | 177.09 | 145.34 | 152.50 | **650.50** |
| 583 | 19.95 | 20.57 | 17.89 | 18.78 | **77.19** |

| | | | | | |
|---|---|---|---|---|---|
| 584 | 20.12 | 36.67 | 29.96 | 25.04 | **111.80** |
| 585 | 1,054.41 | 1,438.28 | 1,431.67 | 1,433.54 | **5,357.90** |
| 586 | 44.27 | 44.27 | 38.01 | 25.94 | **152.50** |
| 587 | 1,117.46 | 1,103.24 | 1,110.84 | 1,454.29 | **4,785.84** |
| 588 | 44.27 | 46.51 | 59.92 | 54.20 | **204.91** |
| **ND NORTH DAKOTA Total** | **4,758.39** | **5,127.60** | **5,389.48** | **6,029.60** | **21,305.06** |

**STATE: NE - NEBRASKA**

| REGISTRANT ZIP CODE 3 | 1ST QUARTER | 2ND QUARTER | 3RD QUARTER | 4TH QUARTER | **TOTAL GRAMS** |
|---|---|---|---|---|---|
| 680 | 226.10 | 183.53 | 234.60 | 240.06 | **884.29** |
| 681 | 4,884.68 | 5,481.96 | 4,610.01 | 4,908.74 | **19,885.38** |
| 683 | 54.02 | 42.22 | 39.17 | 39.26 | **174.68** |
| 684 | 32.65 | 25.94 | 31.84 | 33.54 | **123.96** |
| 685 | 1,419.14 | 1,754.28 | 1,399.02 | 1,446.07 | **6,018.51** |
| 686 | 51.88 | 38.46 | 38.46 | 50.53 | **179.33** |
| 687 | 115.82 | 116.72 | 116.72 | 122.98 | **472.24** |
| 688 | 137.29 | 143.10 | 142.21 | 145.79 | **568.39** |
| 689 | 36.22 | 33.63 | 39.53 | 38.91 | **148.29** |
| 690 | 5.37 | 6.26 | 6.26 | 8.94 | **26.83** |
| 691 | 104.20 | 82.28 | 86.76 | 79.51 | **352.75** |
| 692 | 3.13 | 6.26 | 6.71 | 4.47 | **20.57** |
| 693 | 92.75 | 72.89 | 84.16 | 77.99 | **327.80** |
| **NE NEBRASKA Total** | **7,163.25** | **7,987.53** | **6,835.46** | **7,196.79** | **29,183.03** |

**STATE: NH - NEW HAMPSHIRE**

| REGISTRANT ZIP CODE 3 | 1ST QUARTER | 2ND QUARTER | 3RD QUARTER | 4TH QUARTER | **TOTAL GRAMS** |
|---|---|---|---|---|---|
| 030 | 3,149.99 | 3,166.27 | 3,674.91 | 3,159.74 | **13,150.90** |
| 031 | 10,349.37 | 14,455.02 | 10,338.28 | 11,923.78 | **47,066.46** |
| 032 | 2,331.70 | 1,563.68 | 1,635.86 | 1,814.74 | **7,345.98** |
| 033 | 3,962.64 | 5,761.19 | 3,992.33 | 3,963.53 | **17,679.69** |
| 034 | 3,260.54 | 1,663.14 | 2,741.34 | 2,374.18 | **10,039.19** |
| 035 | 166.81 | 153.84 | 187.82 | 144.89 | **653.36** |
| 036 | 33.09 | 30.41 | 33.54 | 29.52 | **126.56** |
| 037 | 2,956.17 | 2,742.18 | 2,736.11 | 3,469.39 | **11,903.85** |
| 038 | 6,129.50 | 6,159.64 | 6,074.14 | 5,575.33 | **23,938.62** |
| **NH - NEW HAMPSHIRE Total** | **32,339.80** | **35,695.37** | **31,414.33** | **32,455.10** | **131,904.60** |

**STATE: NJ - NEW JERSEY**

| REGISTRANT ZIP CODE 3 | 1ST QUARTER | 2ND QUARTER | 3RD QUARTER | 4TH QUARTER | **TOTAL GRAMS** |
|---|---|---|---|---|---|
| 070 | 8,453.69 | 7,571.81 | 6,274.75 | 8,424.80 | **30,725.05** |
| 071 | 11,047.27 | 10,962.12 | 13,533.79 | 10,956.76 | **46,499.95** |
| 072 | 1,870.19 | 944.49 | 966.85 | 1,859.01 | **5,640.53** |
| 073 | 5,485.09 | 6,415.17 | 6,498.71 | 7,342.49 | **25,741.46** |

| | | | | | |
|---|---|---|---|---|---|
| 074 | 490.58 | 509.63 | 501.22 | 493.98 | **1,995.41** |
| 075 | 3,619.64 | 4,322.19 | 3,541.82 | 3,774.28 | **15,257.93** |
| 076 | 2,508.08 | 2,481.16 | 2,438.04 | 2,319.98 | **9,747.26** |
| 077 | 11,421.22 | 11,947.84 | 17,567.45 | 17,359.41 | **58,295.92** |
| 078 | 3,811.58 | 2,872.28 | 2,927.73 | 3,849.41 | **13,460.99** |
| 079 | 228.88 | 216.71 | 295.87 | 214.21 | **955.67** |
| 080 | 4,096.89 | 3,227.80 | 4,404.65 | 5,176.16 | **16,905.50** |
| 081 | 25,173.42 | 26,698.47 | 27,769.42 | 26,710.27 | **106,351.58** |
| 082 | 9,440.75 | 10,772.60 | 9,786.35 | 11,074.19 | **41,073.89** |
| 083 | 7,947.19 | 7,870.27 | 9,520.89 | 8,559.86 | **33,898.21** |
| 084 | 135.05 | 134.16 | 139.53 | 141.32 | **550.06** |
| 085 | 385.04 | 339.78 | 401.59 | 361.87 | **1,488.28** |
| 086 | 6,516.24 | 6,250.25 | 6,486.37 | 6,748.52 | **26,001.37** |
| 087 | 12,861.02 | 11,640.17 | 11,535.08 | 14,307.00 | **50,343.27** |
| 088 | 10,204.93 | 9,392.63 | 5,534.64 | 4,197.60 | **29,329.79** |
| 089 | 3,306.06 | 3,995.20 | 3,703.80 | 3,660.24 | **14,665.30** |
| **NJ NEW JERSEY - Total** | **129,002.80** | **128,564.72** | **133,828.54** | **137,531.35** | **528,927.41** |

**STATE: NM - NEW MEXICO**

| REGISTRANT ZIP CODE 3 | 1ST QUARTER | 2ND QUARTER | 3RD QUARTER | 4TH QUARTER | **TOTAL GRAMS** |
|---|---|---|---|---|---|
| 870 | 1,948.00 | 1,367.09 | 1,513.77 | 2,087.98 | **6,916.84** |
| 871 | 34,533.59 | 37,614.44 | 36,093.06 | 38,912.39 | **147,153.49** |
| 873 | 24.60 | 40.70 | 25.94 | 27.28 | **118.51** |
| 874 | 1,687.73 | 806.75 | 1,515.20 | 1,688.18 | **5,697.86** |
| 875 | 6,323.23 | 7,732.09 | 7,538.99 | 6,482.43 | **28,076.74** |
| 877 | 21.47 | 23.25 | 33.09 | 28.17 | **105.99** |
| 878 | 16.10 | 14.31 | 15.20 | 16.99 | **62.61** |
| 879 | 11.18 | 5.37 | 2.68 | 1.79 | **21.02** |
| 880 | 1,852.66 | 2,309.16 | 2,357.91 | 1,930.74 | **8,450.47** |
| 881 | 28.17 | 23.25 | 26.38 | 29.07 | **106.88** |
| 882 | 1,369.33 | 1,350.10 | 1,355.64 | 2,005.69 | **6,080.76** |
| 883 | 131.92 | 102.86 | 107.78 | 123.87 | **466.43** |
| 884 | 4.02 | 3.13 | 1.79 | 2.68 | **11.63** |
| **NM NEW MEXICO Total** | **47,952.00** | **51,392.49** | **50,587.44** | **53,337.28** | **203,269.21** |

**STATE: NV - NEVADA**

| REGISTRANT ZIP CODE 3 | 1ST QUARTER | 2ND QUARTER | 3RD QUARTER | 4TH QUARTER | **TOTAL GRAMS** |
|---|---|---|---|---|---|
| 890 | 12,342.00 | 9,279.13 | 9,764.08 | 10,020.41 | **41,405.62** |
| 891 | 14,903.03 | 11,600.82 | 14,529.80 | 14,165.95 | **55,199.60** |
| 893 | 26.83 | 29.52 | 23.25 | 22.36 | **101.96** |
| 894 | 4,499.82 | 3,386.02 | 3,924.27 | 3,921.59 | **15,731.69** |
| 895 | 3,446.30 | 3,344.52 | 3,346.13 | 4,130.79 | **14,267.74** |

| | | | | | |
|---|---|---|---|---|---|
| 897 | 1,402.51 | 2,127.60 | 1,353.58 | 1,404.66 | **6,288.35** |
| 898 | 56.79 | 34.97 | 46.96 | 52.77 | **191.49** |
| **NV NEVADA Total** | **36,677.29** | **29,802.57** | **32,988.07** | **33,718.52** | **133,186.45** |

**STATE: NY - NEW YORK**

| REGISTRANT ZIP CODE 3 | 1ST QUARTER | 2ND QUARTER | 3RD QUARTER | 4TH QUARTER | TOTAL GRAMS |
|---|---|---|---|---|---|
| 100 | 83,839.62 | 74,705.92 | 80,042.27 | 83,063.46 | **321,651.28** |
| 101 | 1,347.86 | 1,429.52 | 1,204.58 | 1,351.08 | **5,333.04** |
| 102 | 21.82 | 20.12 | 11.63 | 6.26 | **59.84** |
| 103 | 11,252.63 | 12,534.12 | 12,516.59 | 11,083.40 | **47,386.74** |
| 104 | 66,133.55 | 59,203.91 | 65,977.47 | 61,854.74 | **253,169.67** |
| 105 | 6,031.65 | 5,991.94 | 6,561.23 | 6,557.11 | **25,141.94** |
| 106 | 115.74 | 127.72 | 142.93 | 117.61 | **503.99** |
| 107 | 6,600.49 | 4,779.32 | 3,924.27 | 4,782.98 | **20,087.06** |
| 108 | 1,832.18 | 1,202.97 | 1,292.86 | 1,933.25 | **6,261.25** |
| 109 | 3,408.29 | 3,252.04 | 2,927.28 | 3,287.28 | **12,874.89** |
| 110 | 2,978.93 | 3,134.80 | 3,250.96 | 3,256.03 | **12,620.72** |
| 111 | 123.78 | 110.46 | 132.37 | 110.91 | **477.52** |
| 112 | 37,164.02 | 37,409.44 | 37,790.98 | 37,559.43 | **149,923.87** |
| 113 | 7,519.40 | 7,415.65 | 7,621.00 | 7,730.48 | **30,286.53** |
| 114 | 6,491.91 | 6,232.36 | 6,447.01 | 7,280.42 | **26,451.70** |
| 115 | 4,387.21 | 6,362.14 | 2,566.48 | 4,432.29 | **17,748.12** |
| 116 | 101.07 | 72.89 | 98.92 | 80.94 | **353.82** |
| 117 | 10,947.72 | 7,983.95 | 10,770.01 | 9,628.04 | **39,329.72** |
| 118 | 118.96 | 98.74 | 110.46 | 109.83 | **437.99** |
| 119 | 3,215.37 | 2,472.75 | 2,801.89 | 3,155.89 | **11,645.89** |
| 120 | 1,117.11 | 1,686.84 | 1,736.03 | 1,724.05 | **6,264.02** |
| 121 | 1,229.08 | 1,169.43 | 1,487.12 | 1,602.76 | **5,488.40** |
| 122 | 7,147.96 | 7,237.48 | 6,014.93 | 8,073.48 | **28,473.85** |
| 123 | 921.68 | 873.38 | 929.28 | 1,068.36 | **3,792.70** |
| 124 | 2,629.18 | 2,757.70 | 3,099.10 | 2,788.92 | **11,274.90** |
| 125 | 6,148.11 | 4,743.72 | 6,183.43 | 5,286.08 | **22,361.34** |
| 126 | 3,462.13 | 3,983.48 | 3,572.32 | 4,557.86 | **15,575.80** |
| 127 | 296.94 | 223.60 | 239.25 | 182.82 | **942.61** |
| 128 | 615.17 | 589.95 | 608.55 | 572.06 | **2,385.72** |
| 129 | 1,118.45 | 1,108.88 | 1,077.30 | 1,012.91 | **4,317.54** |
| 130 | 1,928.59 | 1,725.57 | 1,785.40 | 2,151.84 | **7,591.40** |
| 131 | 999.49 | 1,340.71 | 1,608.13 | 1,258.87 | **5,207.20** |
| 132 | 8,697.95 | 10,414.66 | 8,969.94 | 10,533.89 | **38,616.44** |
| 133 | 188.72 | 180.22 | 189.34 | 159.47 | **717.76** |
| 134 | 2,008.38 | 2,007.93 | 2,279.11 | 2,279.38 | **8,574.79** |

| | | | | | |
|---|---|---|---|---|---|
| 135 | 723.84 | 733.14 | 962.82 | 959.24 | **3,379.04** |
| 136 | 1,145.73 | 995.02 | 1,085.44 | 1,283.46 | **4,509.65** |
| 137 | 142.21 | 166.36 | 188.27 | 169.04 | **665.88** |
| 138 | 371.62 | 419.92 | 335.40 | 330.93 | **1,457.87** |
| 139 | 1,254.13 | 2,499.40 | 2,549.22 | 2,444.48 | **8,747.23** |
| 140 | 1,688.27 | 1,943.78 | 2,029.39 | 2,316.23 | **7,977.67** |
| 141 | 531.27 | 669.01 | 1,113.17 | 580.91 | **2,894.37** |
| 142 | 18,278.77 | 19,352.66 | 17,852.84 | 17,743.08 | **73,227.36** |
| 143 | 1,464.13 | 1,432.56 | 1,423.80 | 1,427.55 | **5,748.04** |
| 144 | 593.52 | 562.04 | 610.25 | 570.09 | **2,335.90** |
| 145 | 335.76 | 296.14 | 296.94 | 263.40 | **1,192.24** |
| 146 | 6,489.47 | 6,421.11 | 6,443.71 | 7,138.77 | **26,493.06** |
| 147 | 310.63 | 294.17 | 291.84 | 296.23 | **1,192.86** |
| 148 | 510.61 | 480.92 | 481.28 | 469.29 | **1,942.10** |
| 149 | 118.96 | 121.64 | 131.03 | 125.22 | **496.84** |
| **NY - NEW YORK - Total** | **326,100.04** | **310,972.18** | **321,765.83** | **326,752.10** | **1,285,590.16** |

**STATE: OH - OHIO**

| REGISTRANT ZIP CODE 3 | 1ST QUARTER | 2ND QUARTER | 3RD QUARTER | 4TH QUARTER | TOTAL GRAMS |
|---|---|---|---|---|---|
| 430 | 1,101.10 | 1,021.58 | 1,073.10 | 1,002.09 | **4,197.87** |
| 431 | 624.38 | 652.64 | 600.77 | 617.23 | **2,495.02** |
| 432 | 12,123.76 | 13,297.60 | 14,663.87 | 15,666.02 | **55,751.26** |
| 433 | 487.45 | 492.46 | 470.45 | 450.87 | **1,901.23** |
| 434 | 235.05 | 251.42 | 206.96 | 125.93 | **819.36** |
| 435 | 193.91 | 194.26 | 207.41 | 199.63 | **795.21** |
| 436 | 6,291.03 | 6,421.08 | 6,752.36 | 7,938.96 | **27,403.43** |
| 437 | 931.43 | 915.15 | 1,075.34 | 1,076.23 | **3,998.15** |
| 438 | 62.88 | 62.16 | 57.24 | 58.14 | **240.41** |
| 439 | 331.46 | 290.59 | 294.70 | 226.73 | **1,143.49** |
| 440 | 1,074.62 | 1,069.52 | 1,053.96 | 1,193.13 | **4,391.24** |
| 441 | 8,040.92 | 7,681.29 | 8,518.62 | 7,848.36 | **32,089.19** |
| 442 | 2,291.01 | 1,984.05 | 2,669.69 | 2,441.71 | **9,386.46** |
| 443 | 3,693.60 | 4,100.47 | 3,772.40 | 3,997.97 | **15,564.44** |
| 444 | 1,277.38 | 1,185.44 | 1,214.60 | 1,215.67 | **4,893.08** |
| 445 | 5,102.55 | 5,982.91 | 6,398.18 | 6,668.38 | **24,152.02** |
| 446 | 477.52 | 480.29 | 514.73 | 569.29 | **2,041.83** |
| 447 | 1,544.09 | 1,666.18 | 1,565.92 | 1,601.24 | **6,377.43** |
| 448 | 481.90 | 464.73 | 392.55 | 308.93 | **1,648.11** |
| 449 | 107.33 | 139.53 | 90.69 | 195.61 | **533.15** |
| 450 | 3,145.96 | 2,499.49 | 2,418.64 | 1,973.30 | **10,037.39** |
| 451 | 577.78 | 742.35 | 1,367.09 | 1,986.73 | **4,673.96** |

| | | | | | |
|---|---|---|---|---|---|
| 452 | 13,795.37 | 13,783.69 | 14,347.73 | 14,259.89 | **56,186.67** |
| 453 | 712.84 | 680.46 | 687.26 | 650.77 | **2,731.32** |
| 454 | 5,600.91 | 6,035.32 | 5,194.41 | 6,197.66 | **23,028.30** |
| 455 | 133.27 | 165.11 | 155.18 | 142.57 | **596.12** |
| 456 | 2,038.52 | 3,424.30 | 3,406.86 | 3,313.30 | **12,182.98** |
| 457 | 282.18 | 346.13 | 251.15 | 294.97 | **1,174.44** |
| 458 | 699.96 | 819.99 | 488.34 | 1,391.87 | **3,400.15** |
| **OH - OHIO Total** | **73,460.16** | **76,850.17** | **79,910.20** | **83,613.15** | **313,833.69** |

**STATE: OK - OKLAHOMA**

| REGISTRANT ZIP CODE 3 | 1ST QUARTER | 2ND QUARTER | 3RD QUARTER | 4TH QUARTER | TOTAL GRAMS |
|---|---|---|---|---|---|
| 730 | 601.04 | 556.67 | 568.75 | 565.71 | **2,292.17** |
| 731 | 11,506.01 | 10,915.70 | 11,712.97 | 12,147.92 | **46,282.61** |
| 734 | 2,325.08 | 1,588.54 | 1,703.21 | 1,308.51 | **6,925.34** |
| 735 | 1,011.57 | 897.08 | 978.56 | 832.15 | **3,719.36** |
| 736 | 19.68 | 13.86 | 16.10 | 16.10 | **65.74** |
| 737 | 63.50 | 59.92 | 57.69 | 53.66 | **234.78** |
| 738 | 40.25 | 42.04 | 46.06 | 31.75 | **160.10** |
| 739 | 23.25 | 15.65 | 14.31 | 14.31 | **67.53** |
| 740 | 2,209.17 | 1,834.68 | 2,190.74 | 1,792.11 | **8,026.70** |
| 741 | 10,258.86 | 10,097.51 | 12,589.93 | 9,043.28 | **41,989.58** |
| 743 | 627.42 | 1,157.80 | 1,156.91 | 624.74 | **3,566.87** |
| 744 | 278.16 | 279.95 | 260.27 | 268.32 | **1,086.70** |
| 745 | 598.35 | 578.23 | 760.69 | 654.25 | **2,591.52** |
| 746 | 1,077.30 | 1,083.57 | 1,374.69 | 819.72 | **4,355.28** |
| 747 | 101.51 | 93.55 | 86.67 | 84.07 | **365.81** |
| 748 | 199.00 | 158.76 | 143.10 | 131.92 | **632.79** |
| 749 | 3,688.95 | 5,580.61 | 3,543.61 | 3,968.90 | **16,782.07** |
| **OK - OKLAHOMA Total** | **34,629.11** | **34,954.14** | **37,204.27** | **32,357.42** | **139,144.94** |

**STATE: OR - OREGON**

| REGISTRANT ZIP CODE 3 | 1ST QUARTER | 2ND QUARTER | 3RD QUARTER | 4TH QUARTER | TOTAL GRAMS |
|---|---|---|---|---|---|
| 970 | 1,682.19 | 1,611.53 | 1,596.06 | 1,536.76 | **6,426.53** |
| 971 | 1,011.48 | 988.76 | 1,024.98 | 954.32 | **3,979.54** |
| 972 | 33,477.43 | 35,237.14 | 35,054.70 | 32,493.17 | **136,262.44** |
| 973 | 9,562.57 | 9,424.29 | 9,510.60 | 9,770.43 | **38,267.89** |
| 974 | 7,788.35 | 7,343.65 | 8,288.49 | 8,353.70 | **31,774.19** |
| 975 | 6,677.32 | 5,839.36 | 6,376.09 | 7,373.43 | **26,266.20** |
| 976 | 223.15 | 184.07 | 175.12 | 183.35 | **765.70** |
| 977 | 1,173.27 | 1,451.61 | 1,094.30 | 1,436.59 | **5,155.77** |
| 978 | 285.31 | 631.89 | 1,008.79 | 969.44 | **2,895.44** |
| 979 | 98.83 | 76.74 | 63.95 | 55.01 | **294.53** |

| OR OREGON Total | 61,979.90 | 62,789.04 | 64,193.09 | 63,126.19 | 252,088.22 |
|---|---|---|---|---|---|

**STATE: PA - PENNSYLVANIA**

| REGISTRANT ZIP CODE 3 | 1ST QUARTER | 2ND QUARTER | 3RD QUARTER | 4TH QUARTER | TOTAL GRAMS |
|---|---|---|---|---|---|
| 150 | 4,672.97 | 4,213.88 | 4,309.49 | 4,791.12 | **17,987.46** |
| 151 | 376.54 | 370.37 | 392.91 | 407.76 | **1,547.58** |
| 152 | 22,927.05 | 23,248.85 | 21,661.47 | 21,491.98 | **89,329.36** |
| 153 | 4,844.07 | 4,260.74 | 4,413.95 | 3,488.88 | **17,007.64** |
| 154 | 3,083.89 | 3,057.06 | 3,065.73 | 3,844.13 | **13,050.82** |
| 155 | 64.84 | 66.19 | 58.40 | 77.81 | **267.25** |
| 156 | 7,612.24 | 7,177.11 | 7,183.55 | 7,402.50 | **29,375.41** |
| 157 | 895.12 | 864.88 | 841.09 | 806.93 | **3,408.02** |
| 158 | 122.26 | 94.36 | 120.92 | 78.71 | **416.25** |
| 159 | 3,040.16 | 3,011.44 | 3,734.66 | 3,016.54 | **12,802.80** |
| 160 | 4,723.06 | 4,531.84 | 3,839.21 | 4,613.40 | **17,707.51** |
| 161 | 7,362.97 | 4,876.81 | 7,402.59 | 6,435.03 | **26,077.39** |
| 162 | 206.16 | 151.60 | 113.95 | 81.30 | **553.01** |
| 163 | 1,522.00 | 1,505.63 | 1,525.49 | 1,536.40 | **6,089.52** |
| 164 | 79.15 | 72.89 | 87.20 | 92.57 | **331.82** |
| 165 | 4,193.30 | 4,097.34 | 3,685.73 | 3,881.70 | **15,858.07** |
| 166 | 5,881.40 | 5,734.45 | 5,173.57 | 4,701.50 | **21,490.91** |
| 167 | 132.82 | 128.79 | 137.83 | 112.25 | **511.69** |
| 168 | 3,002.05 | 3,081.66 | 2,468.99 | 2,570.95 | **11,123.65** |
| 169 | 73.52 | 57.69 | 67.17 | 57.60 | **255.98** |
| 170 | 6,156.69 | 7,019.07 | 6,205.88 | 6,520.62 | **25,902.27** |
| 171 | 4,641.85 | 4,747.03 | 4,647.30 | 5,522.03 | **19,558.20** |
| 172 | 378.87 | 346.76 | 312.32 | 293.54 | **1,331.49** |
| 173 | 1,741.84 | 2,305.32 | 1,789.52 | 2,281.61 | **8,118.29** |
| 174 | 3,929.55 | 3,919.71 | 3,917.83 | 4,802.03 | **16,569.12** |
| 175 | 309.10 | 281.29 | 236.12 | 312.32 | **1,138.84** |
| 176 | 5,596.17 | 5,597.07 | 4,701.86 | 5,596.62 | **21,491.72** |
| 177 | 3,858.08 | 4,496.15 | 3,822.58 | 4,456.53 | **16,633.34** |
| 178 | 920.87 | 933.31 | 927.31 | 875.26 | **3,656.75** |
| 179 | 135.95 | 111.62 | 104.38 | 135.05 | **487.00** |
| 180 | 6,146.59 | 6,514.27 | 5,202.19 | 6,541.10 | **24,404.15** |
| 181 | 411.87 | 401.41 | 323.68 | 370.64 | **1,507.60** |
| 182 | 134.16 | 138.63 | 118.51 | 125.22 | **516.52** |
| 183 | 3,691.37 | 3,645.13 | 2,504.32 | 2,495.38 | **12,336.19** |
| 184 | 213.58 | 198.47 | 193.91 | 193.19 | **799.15** |
| 185 | 2,788.65 | 2,757.88 | 3,605.77 | 2,801.98 | **11,954.28** |
| 186 | 364.47 | 315.10 | 322.61 | 290.68 | **1,292.86** |

| | | | | | |
|---|---|---|---|---|---|
| 187 | 5,788.91 | 5,392.61 | 5,215.07 | 5,864.67 | **22,261.26** |
| 188 | 126.56 | 123.87 | 85.86 | 88.55 | **424.84** |
| 189 | 7,372.18 | 5,519.79 | 6,677.68 | 6,431.54 | **26,001.19** |
| 190 | 15,061.79 | 15,702.16 | 13,858.37 | 16,458.57 | **61,080.88** |
| 191 | 41,645.56 | 46,643.30 | 39,458.26 | 42,251.39 | **169,998.52** |
| 193 | 3,804.51 | 4,695.78 | 3,859.69 | 4,794.16 | **17,154.14** |
| 194 | 6,666.50 | 6,007.68 | 5,603.95 | 5,423.82 | **23,701.96** |
| 195 | 155.18 | 133.00 | 136.22 | 139.35 | **563.74** |
| 196 | 2,419.21 | 4,576.16 | 4,546.95 | 4,866.70 | **16,409.02** |
| **PA - PENNSYLVANIA - Total** | **199,275.64** | **203,126.13** | **188,662.07** | **199,421.62** | **790,485.46** |

**STATE: PR - PUERTO RICO**

| REGISTRANT ZIP CODE 3 | 1ST QUARTER | 2ND QUARTER | 3RD QUARTER | 4TH QUARTER | TOTAL GRAMS |
|---|---|---|---|---|---|
| 006 | 1,086.25 | 1,547.31 | 1,613.05 | 1,079.99 | **5,326.60** |
| 007 | 4,472.00 | 4,483.18 | 5,368.19 | 4,472.00 | **18,795.37** |
| 009 | 15,096.13 | 15,520.52 | 17,210.76 | 13,346.68 | **61,174.10** |
| **PR - PUERTO RICO - Total** | **20,654.38** | **21,551.02** | **24,192.00** | **18,898.67** | **85,296.07** |

**STATE: RI - RHODE ISLAND**

| REGISTRANT ZIP CODE 3 | 1ST QUARTER | 2ND QUARTER | 3RD QUARTER | 4TH QUARTER | TOTAL GRAMS |
|---|---|---|---|---|---|
| 028 | 21,234.93 | 26,897.56 | 25,183.98 | 25,218.50 | **98,534.97** |
| 029 | 21,160.07 | 22,743.70 | 24,448.07 | 24,238.87 | **92,590.70** |
| **RI RHODE ISLAND Total** | **42,395.01** | **49,641.26** | **49,632.04** | **49,457.37** | **191,125.68** |

**STATE: SC - SOUTH CAROLINA**

| REGISTRANT ZIP CODE 3 | 1ST QUARTER | 2ND QUARTER | 3RD QUARTER | 4TH QUARTER | TOTAL GRAMS |
|---|---|---|---|---|---|
| 290 | 713.82 | 670.35 | 632.52 | 605.51 | **2,622.20** |
| 291 | 5,357.46 | 3,603.54 | 4,140.62 | 4,419.23 | **17,520.85** |
| 292 | 2,312.11 | 2,642.50 | 2,118.21 | 2,300.13 | **9,372.95** |
| 293 | 5,783.19 | 5,178.75 | 6,232.18 | 5,649.12 | **22,843.24** |
| 294 | 8,317.36 | 9,117.85 | 8,780.52 | 8,485.75 | **34,701.48** |
| 295 | 13,536.57 | 14,570.76 | 15,121.26 | 14,365.23 | **57,593.81** |
| 296 | 13,081.94 | 15,256.05 | 13,092.58 | 14,355.30 | **55,785.87** |
| 297 | 4,543.28 | 3,088.63 | 4,760.62 | 7,343.02 | **19,735.56** |
| 298 | 1,468.87 | 1,528.08 | 1,515.83 | 947.62 | **5,460.40** |
| 299 | 935.54 | 1,090.72 | 1,239.37 | 1,183.29 | **4,448.92** |
| **SC - SOUTH CAROLINA Total** | **56,050.15** | **56,747.24** | **57,633.72** | **59,654.20** | **230,085.30** |

**STATE: SD - SOUTH DAKOTA**

| REGISTRANT ZIP CODE 3 | 1ST QUARTER | 2ND QUARTER | 3RD QUARTER | 4TH QUARTER | TOTAL GRAMS |
|---|---|---|---|---|---|
| 570 | 265.64 | 253.12 | 233.71 | 219.49 | **971.94** |
| 571 | 1,302.69 | 1,176.05 | 772.05 | 1,308.51 | **4,559.29** |
| 572 | 32.65 | 28.62 | 24.77 | 25.49 | **111.53** |
| 573 | 71.55 | 71.55 | 75.13 | 72.89 | **291.13** |

| | | | | | |
|---|---|---|---|---|---|
| 574 | 16.55 | 10.73 | 7.16 | 8.50 | **42.93** |
| 575 | 20.57 | 17.44 | 24.77 | 17.08 | **79.87** |
| 576 | 5.37 | 5.81 | 0.89 | 0.45 | **12.52** |
| 577 | 261.16 | 565.26 | 413.57 | 446.48 | **1,686.48** |
| **SD SOUTH DAKOTA - Total** | **1,976.18** | **2,128.58** | **1,552.05** | **2,098.89** | **7,755.70** |

**STATE: TN - TENNESSEE**

| REGISTRANT ZIP CODE 3 | 1ST QUARTER | 2ND QUARTER | 3RD QUARTER | 4TH QUARTER | TOTAL GRAMS |
|---|---|---|---|---|---|
| 370 | 824.82 | 779.56 | 683.23 | 621.25 | **2,908.86** |
| 371 | 836.44 | 788.68 | 794.85 | 822.67 | **3,242.65** |
| 372 | 5,505.31 | 7,202.96 | 6,785.45 | 5,471.85 | **24,965.57** |
| 373 | 573.94 | 546.93 | 552.56 | 552.74 | **2,226.16** |
| 374 | 6,678.48 | 7,112.63 | 6,604.16 | 6,952.26 | **27,347.53** |
| 376 | 1,878.33 | 1,807.94 | 1,984.23 | 2,268.11 | **7,938.60** |
| 377 | 110.46 | 125.22 | 108.22 | 114.93 | **458.83** |
| 378 | 160.54 | 152.05 | 162.33 | 154.73 | **629.66** |
| 379 | 10,879.75 | 12,288.43 | 10,716.88 | 9,144.61 | **43,029.67** |
| 380 | 2,736.86 | 3,233.26 | 2,508.88 | 3,057.95 | **11,536.96** |
| 381 | 8,895.97 | 10,220.85 | 7,598.91 | 11,078.04 | **37,793.77** |
| 382 | 2,968.07 | 2,540.10 | 2,179.21 | 2,927.37 | **10,614.74** |
| 383 | 2,994.00 | 4,482.91 | 2,979.07 | 2,976.65 | **13,432.64** |
| 384 | 931.96 | 1,610.46 | 932.50 | 1,317.81 | **4,792.73** |
| 385 | 136.40 | 123.87 | 132.19 | 131.48 | **523.94** |
| **TN TENNESSEE Total** | **46,111.34** | **53,015.82** | **44,722.67** | **47,592.46** | **191,442.29** |

**STATE: TX - TEXAS**

| REGISTRANT ZIP CODE 3 | 1ST QUARTER | 2ND QUARTER | 3RD QUARTER | 4TH QUARTER | TOTAL GRAMS |
|---|---|---|---|---|---|
| 750 | 6,214.02 | 7,155.74 | 7,172.28 | 6,753.70 | **27,295.75** |
| 751 | 1,026.77 | 1,011.12 | 1,021.85 | 1,021.85 | **4,081.59** |
| 752 | 8,890.78 | 10,062.54 | 8,217.84 | 8,371.05 | **35,542.20** |
| 753 | 29.16 | 14.42 | 30.68 | 19.50 | **93.75** |
| 754 | 924.18 | 905.94 | 668.39 | 793.15 | **3,291.66** |
| 755 | 194.98 | 170.83 | 158.13 | 139.79 | **663.73** |
| 756 | 2,576.77 | 2,828.99 | 3,064.66 | 2,722.11 | **11,192.52** |
| 757 | 4,260.03 | 3,790.65 | 4,754.90 | 2,768.44 | **15,574.01** |
| 758 | 132.82 | 134.61 | 107.78 | 105.54 | **480.74** |
| 759 | 1,621.55 | 1,621.99 | 1,360.56 | 1,503.04 | **6,107.14** |
| 760 | 1,889.42 | 2,538.04 | 1,688.81 | 1,885.22 | **8,001.48** |
| 761 | 10,950.41 | 14,694.01 | 11,447.25 | 13,568.32 | **50,659.98** |
| 762 | 2,863.69 | 3,540.66 | 2,990.69 | 3,346.58 | **12,741.62** |
| 763 | 908.53 | 899.23 | 895.03 | 851.56 | **3,554.35** |
| 764 | 160.10 | 137.47 | 160.54 | 121.64 | **579.75** |

| | | | | | |
|---|---|---|---|---|---|
| 765 | 769.09 | 487.54 | 713.19 | 587.17 | **2,557.00** |
| 766 | 145.34 | 145.79 | 126.56 | 125.66 | **543.35** |
| 767 | 2,097.82 | 2,095.40 | 2,087.98 | 2,657.71 | **8,938.90** |
| 768 | 163.05 | 166.81 | 166.36 | 173.51 | **669.73** |
| 769 | 733.14 | 489.77 | 895.03 | 656.94 | **2,774.88** |
| 770 | 13,606.57 | 12,809.24 | 12,932.02 | 12,686.82 | **52,034.66** |
| 772 | 429.31 | 429.31 | 429.31 | 429.31 | **1,717.25** |
| 773 | 2,933.11 | 3,772.33 | 3,863.41 | 3,641.64 | **14,210.48** |
| 774 | 701.03 | 668.83 | 673.93 | 608.19 | **2,651.99** |
| 775 | 4,294.64 | 4,773.32 | 4,056.01 | 3,853.08 | **16,977.05** |
| 776 | 1,542.84 | 822.85 | 812.83 | 823.74 | **4,002.26** |
| 777 | 1,929.94 | 2,877.73 | 2,052.56 | 2,069.64 | **8,929.87** |
| 778 | 326.10 | 365.81 | 343.18 | 342.02 | **1,377.11** |
| 779 | 1,064.25 | 1,082.85 | 1,053.96 | 1,096.53 | **4,297.59** |
| 780 | 1,720.83 | 2,048.27 | 1,672.44 | 1,984.76 | **7,426.29** |
| 781 | 1,725.30 | 1,979.40 | 1,669.49 | 1,696.95 | **7,071.13** |
| 782 | 26,955.07 | 32,785.22 | 31,464.28 | 33,046.02 | **124,250.58** |
| 783 | 217.34 | 197.22 | 199.45 | 214.21 | **828.21** |
| 784 | 6,099.54 | 5,943.91 | 6,908.79 | 6,394.96 | **25,347.21** |
| 785 | 1,675.21 | 1,227.65 | 1,154.13 | 1,495.44 | **5,552.44** |
| 786 | 1,823.59 | 2,127.24 | 1,584.43 | 1,574.77 | **7,110.03** |
| 787 | 7,092.14 | 6,422.15 | 6,723.47 | 8,957.51 | **29,195.27** |
| 788 | 56.79 | 62.61 | 56.79 | 67.08 | **243.28** |
| 789 | 55.01 | 47.40 | 43.83 | 58.58 | **204.82** |
| 790 | 194.08 | 174.41 | 200.79 | 206.16 | **775.44** |
| 791 | 1,104.14 | 1,121.49 | 1,007.09 | 1,368.88 | **4,601.60** |
| 792 | 46.06 | 38.46 | 35.78 | 41.59 | **161.89** |
| 793 | 96.15 | 98.12 | 88.99 | 88.10 | **371.35** |
| 794 | 1,465.74 | 2,765.04 | 2,163.46 | 2,027.07 | **8,421.31** |
| 795 | 117.17 | 108.76 | 81.84 | 78.71 | **386.47** |
| 796 | 402.93 | 359.82 | 563.92 | 450.42 | **1,777.08** |
| 797 | 1,976.98 | 1,591.23 | 2,525.61 | 2,237.07 | **8,330.89** |
| 798 | 7.16 | 9.84 | 6.71 | 8.05 | **31.75** |
| 799 | 2,225.62 | 2,674.35 | 2,657.71 | 2,952.59 | **10,510.27** |
| **TX - TEXAS Total** | **128,436.28** | **142,276.36** | **134,754.71** | **138,672.36** | **544,139.71** |

**STATE: UT - UTAH**

| REGISTRANT ZIP CODE 3 | 1ST QUARTER | 2ND QUARTER | 3RD QUARTER | 4TH QUARTER | TOTAL GRAMS |
|---|---|---|---|---|---|
| 840 | 6,878.65 | 8,383.66 | 7,227.65 | 6,997.07 | **29,487.03** |
| 841 | 12,383.42 | 13,163.96 | 13,966.95 | 12,698.07 | **52,212.39** |
| 843 | 328.24 | 172.17 | 166.36 | 312.59 | **979.37** |

| | | | | | |
|---|---|---|---|---|---|
| 844 | 2,853.85 | 2,909.93 | 2,673.54 | 2,612.63 | **11,049.95** |
| 845 | 152.05 | 512.49 | 151.69 | 492.46 | **1,308.69** |
| 846 | 1,124.26 | 1,400.81 | 579.48 | 1,119.25 | **4,223.80** |
| 847 | 2,003.19 | 2,534.37 | 2,742.05 | 2,232.51 | **9,512.12** |
| **UT - UTAH - Total** | **25,723.66** | **29,077.39** | **27,507.72** | **26,464.58** | **108,773.35** |

**STATE: VA - VIRGINIA**

| REGISTRANT ZIP CODE 3 | 1ST QUARTER | 2ND QUARTER | 3RD QUARTER | 4TH QUARTER | **TOTAL GRAMS** |
|---|---|---|---|---|---|
| 201 | 2,646.62 | 2,803.94 | 3,077.99 | 2,564.07 | **11,092.62** |
| 220 | 1,344.82 | 1,703.83 | 2,111.59 | 1,343.12 | **6,503.36** |
| 221 | 325.38 | 357.13 | 372.25 | 290.77 | **1,345.54** |
| 222 | 83.63 | 79.06 | 74.24 | 84.97 | **321.89** |
| 223 | 446.04 | 734.30 | 433.78 | 467.23 | **2,081.36** |
| 224 | 3,966.84 | 4,658.75 | 3,922.93 | 4,732.54 | **17,281.06** |
| 225 | 120.39 | 105.63 | 116.72 | 96.24 | **438.97** |
| 226 | 6,635.55 | 6,438.88 | 5,752.24 | 7,109.32 | **25,935.99** |
| 227 | 149.81 | 131.92 | 130.58 | 159.20 | **571.52** |
| 228 | 3,878.66 | 3,881.78 | 3,873.54 | 3,881.94 | **15,515.91** |
| 229 | 5,037.44 | 4,019.88 | 5,172.40 | 4,496.60 | **18,726.32** |
| 230 | 4,018.99 | 2,613.44 | 3,028.08 | 3,740.83 | **13,401.33** |
| 231 | 966.58 | 1,423.17 | 1,226.85 | 1,095.55 | **4,712.15** |
| 232 | 16,979.56 | 22,103.93 | 18,148.36 | 19,000.45 | **76,232.31** |
| 233 | 4,279.61 | 3,779.47 | 4,128.73 | 3,760.06 | **15,947.87** |
| 234 | 8,197.53 | 7,806.14 | 7,947.28 | 9,568.01 | **33,518.97** |
| 235 | 1,301.17 | 790.52 | 1,291.52 | 1,291.31 | **4,674.51** |
| 236 | 4,271.83 | 5,185.28 | 4,970.63 | 5,312.91 | **19,740.66** |
| 237 | 1,269.24 | 723.57 | 1,292.23 | 1,294.73 | **4,579.78** |
| 238 | 1,582.01 | 3,294.52 | 2,045.31 | 3,776.78 | **10,698.63** |
| 239 | 111.80 | 117.88 | 101.96 | 78.89 | **410.53** |
| 240 | 4,874.03 | 3,940.99 | 4,288.83 | 5,159.61 | **18,263.46** |
| 241 | 820.16 | 798.97 | 787.34 | 684.31 | **3,090.78** |
| 242 | 561.24 | 563.02 | 571.97 | 464.64 | **2,160.87** |
| 243 | 6,520.80 | 7,067.91 | 6,433.78 | 7,450.80 | **27,473.28** |
| 244 | 822.40 | 1,078.20 | 873.47 | 1,203.33 | **3,977.40** |
| 245 | 4,285.25 | 4,846.40 | 5,955.45 | 4,895.86 | **19,982.95** |
| 246 | 3,791.36 | 3,719.81 | 3,846.81 | 3,743.96 | **15,101.94** |
| **VA VIRGINIA Total** | **89,288.76** | **94,768.34** | **91,976.86** | **97,748.01** | **373,781.97** |

**STATE: VI - VIRGIN ISLANDS**

| REGISTRANT ZIP CODE 3 | 1ST QUARTER | 2ND QUARTER | 3RD QUARTER | 4TH QUARTER | **TOTAL GRAMS** |
|---|---|---|---|---|---|
| 008 | 4.47 | 9.84 | 12.52 | 6.26 | **33.09** |
| **VI - VIRGIN ISLANDS Total** | **4.47** | **9.84** | **12.52** | **6.26** | **33.09** |

**STATE: VT - VERMONT**

| REGISTRANT ZIP CODE 3 | 1ST QUARTER | 2ND QUARTER | 3RD QUARTER | 4TH QUARTER | TOTAL GRAMS |
|---|---|---|---|---|---|
| 050 | 392.64 | 338.53 | 346.67 | 261.61 | **1,339.45** |
| 051 | 164.12 | 161.89 | 148.92 | 152.50 | **627.42** |
| 052 | 131.03 | 142.03 | 146.68 | 155.00 | **574.74** |
| 053 | 2,446.63 | 2,098.26 | 2,076.80 | 2,474.89 | **9,096.58** |
| 054 | 7,232.83 | 7,929.57 | 6,771.68 | 9,408.64 | **31,342.73** |
| 056 | 2,227.06 | 2,561.11 | 2,324.55 | 2,819.60 | **9,932.31** |
| 057 | 2,179.92 | 3,061.53 | 2,121.07 | 2,146.47 | **9,508.99** |
| 058 | 3,844.76 | 5,469.26 | 4,228.19 | 4,955.51 | **18,497.71** |
| **VT VERMONT Total** | **18,618.99** | **21,762.18** | **18,164.55** | **22,374.22** | **80,919.95** |

**STATE: WA - WASHINGTON**

| REGISTRANT ZIP CODE 3 | 1ST QUARTER | 2ND QUARTER | 3RD QUARTER | 4TH QUARTER | TOTAL GRAMS |
|---|---|---|---|---|---|
| 980 | 16,279.15 | 20,016.31 | 16,951.03 | 18,661.66 | **71,908.15** |
| 981 | 23,900.96 | 24,239.04 | 23,144.66 | 20,574.51 | **91,859.17** |
| 982 | 15,584.83 | 16,275.49 | 15,806.46 | 16,955.32 | **64,622.10** |
| 983 | 2,396.28 | 2,414.61 | 2,327.68 | 2,525.70 | **9,664.26** |
| 984 | 12,319.82 | 16,266.10 | 13,062.35 | 14,277.31 | **55,925.58** |
| 985 | 8,487.59 | 13,610.08 | 8,230.89 | 9,802.09 | **40,130.65** |
| 986 | 7,676.28 | 8,363.44 | 10,327.55 | 8,033.95 | **34,401.22** |
| 988 | 484.68 | 460.97 | 420.90 | 395.86 | **1,762.42** |
| 989 | 2,001.13 | 1,976.53 | 2,020.81 | 1,958.20 | **7,956.67** |
| 990 | 209.74 | 230.31 | 214.66 | 220.02 | **874.72** |
| 991 | 367.96 | 387.72 | 350.60 | 333.97 | **1,440.25** |
| 992 | 12,931.06 | 14,057.37 | 12,505.05 | 13,560.45 | **53,053.93** |
| 993 | 561.33 | 496.93 | 501.58 | 474.03 | **2,033.87** |
| 994 | 212.87 | 195.43 | 207.95 | 216.00 | **832.24** |
| **WA WASHINGTON Total** | **103,413.66** | **118,990.35** | **106,072.17** | **107,989.05** | **436,465.23** |

**STATE: WI - WISCONSIN**

| REGISTRANT ZIP CODE 3 | 1ST QUARTER | 2ND QUARTER | 3RD QUARTER | 4TH QUARTER | TOTAL GRAMS |
|---|---|---|---|---|---|
| 530 | 2,921.74 | 2,810.92 | 2,755.11 | 2,831.04 | **11,318.81** |
| 531 | 4,980.47 | 5,343.59 | 5,285.19 | 5,079.03 | **20,688.28** |
| 532 | 13,877.96 | 15,400.32 | 15,065.09 | 19,063.96 | **63,407.33** |
| 534 | 3,877.58 | 3,389.24 | 3,625.90 | 3,569.73 | **14,462.45** |
| 535 | 3,136.66 | 3,375.91 | 2,793.57 | 3,089.08 | **12,395.22** |
| 537 | 10,333.07 | 11,194.43 | 11,291.56 | 11,206.96 | **44,026.03** |
| 538 | 87.65 | 78.80 | 71.55 | 68.42 | **306.42** |
| 539 | 312.95 | 320.64 | 300.52 | 279.95 | **1,214.06** |
| 540 | 193.19 | 180.67 | 178.97 | 234.51 | **787.34** |
| 541 | 179.86 | 168.33 | 173.60 | 158.22 | **680.01** |

| | | | | | |
|---|---|---|---|---|---|
| 542 | 50.98 | 45.61 | 46.06 | 46.51 | **189.17** |
| 543 | 4,341.06 | 4,636.75 | 4,441.59 | 4,586.84 | **18,006.24** |
| 544 | 2,615.32 | 1,943.35 | 1,897.74 | 2,464.34 | **8,920.75** |
| 545 | 42.31 | 32.02 | 37.56 | 39.80 | **151.69** |
| 546 | 3,293.45 | 3,336.92 | 3,265.10 | 3,290.94 | **13,186.41** |
| 547 | 2,408.53 | 3,687.34 | 3,083.18 | 2,694.38 | **11,873.43** |
| 548 | 271.45 | 254.46 | 220.74 | 217.97 | **964.61** |
| 549 | 8,773.35 | 7,156.18 | 8,436.25 | 8,426.68 | **32,792.46** |
| **WI - WISCONSIN Total** | **61,697.57** | **63,355.48** | **62,969.27** | **67,348.36** | **255,370.69** |

**STATE: WV - WEST VIRGINIA**

| REGISTRANT ZIP CODE 3 | 1ST QUARTER | 2ND QUARTER | 3RD QUARTER | 4TH QUARTER | TOTAL GRAMS |
|---|---|---|---|---|---|
| 247 | 38.73 | 51.43 | 38.01 | 43.38 | **171.55** |
| 248 | 17.89 | 20.57 | 16.99 | 17.89 | **73.34** |
| 249 | 23.25 | 24.60 | 22.36 | 23.25 | **93.46** |
| 250 | 26.38 | 32.65 | 20.12 | 34.43 | **113.59** |
| 251 | 26.83 | 23.70 | 24.15 | 26.38 | **101.07** |
| 252 | 25.04 | 16.10 | 5.81 | 4.92 | **51.88** |
| 253 | 3,034.70 | 3,740.29 | 3,286.12 | 1,785.40 | **11,846.51** |
| 254 | 3,775.08 | 2,940.79 | 4,314.85 | 3,587.98 | **14,618.70** |
| 255 | 65.74 | 56.79 | 63.50 | 76.02 | **262.06** |
| 256 | 2,454.23 | 2,776.22 | 2,701.09 | 2,205.59 | **10,137.13** |
| 257 | 4,648.29 | 5,957.69 | 5,074.92 | 4,887.09 | **20,567.98** |
| 258 | 3,465.80 | 3,715.34 | 3,028.44 | 3,895.74 | **14,105.31** |
| 259 | 36.67 | 48.30 | 52.77 | 47.40 | **185.14** |
| 260 | 1,367.18 | 969.80 | 1,314.50 | 920.61 | **4,572.08** |
| 261 | 999.49 | 853.26 | 1,147.07 | 1,129.18 | **4,129.00** |
| 262 | 66.19 | 57.24 | 59.75 | 58.58 | **241.76** |
| 263 | 1,431.93 | 1,448.48 | 1,043.76 | 1,751.68 | **5,675.86** |
| 264 | 53.66 | 49.64 | 49.64 | 48.30 | **201.24** |
| 265 | 2,120.18 | 2,202.64 | 2,745.00 | 2,588.57 | **9,656.39** |
| 266 | 13.86 | 4.02 | 3.58 | 6.71 | **28.17** |
| 267 | 41.14 | 51.88 | 45.17 | 48.30 | **186.48** |
| 268 | 25.49 | 18.78 | 23.25 | 22.81 | **90.33** |
| **WV WEST VIRGINIA Total** | **23,757.77** | **25,060.19** | **25,080.85** | **23,210.22** | **97,109.03** |

**STATE: WY - WYOMING**

| REGISTRANT ZIP CODE 3 | 1ST QUARTER | 2ND QUARTER | 3RD QUARTER | 4TH QUARTER | TOTAL GRAMS |
|---|---|---|---|---|---|
| 820 | 116.72 | 137.74 | 117.17 | 103.30 | **474.93** |
| 822 | 35.78 | 26.83 | 28.17 | 24.15 | **114.93** |
| 823 | 9.39 | 9.39 | 11.63 | 9.84 | **40.25** |
| 824 | 42.48 | 45.61 | 52.32 | 29.07 | **169.49** |

| | | | | | |
|---|---|---|---|---|---|
| 825 | 82.28 | 95.70 | 77.01 | 80.76 | **335.76** |
| 826 | 189.61 | 183.35 | 160.99 | 154.82 | **688.78** |
| 827 | 25.49 | 21.47 | 26.83 | 28.62 | **102.41** |
| 828 | 67.97 | 46.06 | 52.32 | 51.88 | **218.23** |
| 829 | 201.33 | 212.87 | 199.00 | 202.13 | **815.34** |
| 830 | 23.70 | 22.81 | 37.12 | 18.78 | **102.41** |
| 831 | 34.88 | 32.20 | 31.30 | 25.04 | **123.43** |
| **WY - WYOMING Total** | **829.65** | **834.03** | **793.87** | **728.40** | **3,185.94** |

**DRUG: 9300 - MORPHINE**

**STATE: AK - ALASKA**

| REGISTRANT ZIP CODE 3 | 1ST QUARTER | 2ND QUARTER | 3RD QUARTER | 4TH QUARTER | TOTAL GRAMS |
|---|---|---|---|---|---|
| 995 | 3,637.91 | 3,466.35 | 3,350.98 | 3,054.81 | **13,510.05** |
| 996 | 3,014.00 | 2,866.68 | 2,919.59 | 2,653.83 | **11,454.10** |
| 997 | 917.75 | 782.65 | 770.47 | 669.05 | **3,139.92** |
| 998 | 383.51 | 336.42 | 392.90 | 297.65 | **1,410.48** |
| 999 | 119.30 | 152.60 | 134.13 | 123.18 | **529.22** |
| **AK ALASKA - Total** | **8,072.48** | **7,604.69** | **7,568.06** | **6,798.53** | **30,043.76** |

**STATE: AL - ALABAMA**

| REGISTRANT ZIP CODE 3 | 1ST QUARTER | 2ND QUARTER | 3RD QUARTER | 4TH QUARTER | TOTAL GRAMS |
|---|---|---|---|---|---|
| 350 | 6,304.90 | 5,976.95 | 5,961.10 | 5,705.68 | **23,948.64** |
| 351 | 3,420.31 | 3,101.57 | 2,998.45 | 2,927.10 | **12,447.43** |
| 352 | 5,235.38 | 5,001.49 | 4,747.26 | 4,311.21 | **19,295.34** |
| 354 | 3,262.49 | 3,182.10 | 3,392.99 | 3,105.46 | **12,943.05** |
| 355 | 1,713.72 | 1,807.25 | 1,856.46 | 1,833.45 | **7,210.88** |
| 356 | 3,666.72 | 3,700.75 | 3,455.79 | 3,552.19 | **14,375.44** |
| 357 | 2,092.66 | 1,856.54 | 1,964.62 | 1,815.37 | **7,729.19** |
| 358 | 3,298.21 | 2,897.45 | 2,903.54 | 2,698.33 | **11,797.53** |
| 359 | 4,841.08 | 4,668.99 | 4,532.05 | 4,464.99 | **18,507.11** |
| 360 | 2,855.81 | 3,113.34 | 2,888.57 | 2,775.34 | **11,633.06** |
| 361 | 2,486.91 | 2,328.94 | 2,215.95 | 2,154.29 | **9,186.08** |
| 362 | 1,922.80 | 1,919.51 | 1,914.73 | 1,949.52 | **7,706.56** |
| 363 | 1,809.13 | 1,705.37 | 1,766.79 | 1,683.70 | **6,964.99** |
| 364 | 882.24 | 782.67 | 847.48 | 887.65 | **3,400.04** |
| 365 | 3,714.94 | 3,821.23 | 3,717.91 | 3,435.51 | **14,689.59** |
| 366 | 3,605.50 | 3,229.94 | 3,264.64 | 3,280.52 | **13,380.60** |
| 367 | 524.92 | 505.74 | 529.84 | 491.58 | **2,052.09** |
| 368 | 3,266.98 | 3,093.32 | 2,954.01 | 2,744.81 | **12,059.12** |
| 369 | 114.86 | 130.13 | 86.55 | 117.67 | **449.21** |
| **AL ALABAMA - Total** | **55,019.57** | **52,823.27** | **51,998.73** | **49,934.38** | **209,775.95** |

**STATE: AR - ARKANSAS**

| REGISTRANT ZIP CODE 3 | 1ST QUARTER | 2ND QUARTER | 3RD QUARTER | 4TH QUARTER | TOTAL GRAMS |
|---|---|---|---|---|---|
| 716 | 963.64 | 925.21 | 871.61 | 806.25 | **3,566.72** |
| 717 | 964.55 | 843.22 | 881.72 | 792.80 | **3,482.29** |
| 718 | 731.94 | 643.75 | 652.86 | 632.31 | **2,660.85** |
| 719 | 2,141.53 | 2,127.01 | 1,945.45 | 1,932.95 | **8,146.94** |
| 720 | 3,440.83 | 3,284.22 | 3,204.24 | 2,931.64 | **12,860.94** |
| 721 | 3,597.05 | 3,394.91 | 3,403.98 | 3,131.37 | **13,527.31** |
| 722 | 2,052.86 | 1,883.57 | 2,072.31 | 1,790.33 | **7,799.06** |
| 723 | 1,022.90 | 900.58 | 991.59 | 994.31 | **3,909.38** |
| 724 | 2,207.22 | 2,128.80 | 2,194.24 | 2,024.56 | **8,554.81** |
| 725 | 1,078.88 | 1,158.33 | 1,079.71 | 1,299.80 | **4,616.72** |
| 726 | 1,896.59 | 1,754.30 | 1,851.86 | 1,696.01 | **7,198.76** |
| 727 | 5,615.49 | 4,901.67 | 4,783.76 | 4,550.19 | **19,851.10** |
| 728 | 1,084.14 | 1,074.18 | 930.07 | 952.38 | **4,040.78** |
| 729 | 2,798.20 | 2,518.11 | 2,434.85 | 2,373.10 | **10,124.26** |
| **AR ARKANSAS - Total** | **29,595.83** | **27,537.85** | **27,298.24** | **25,907.98** | **110,339.90** |

**STATE: AS - AMERICAN SAMOA**

| REGISTRANT ZIP CODE 3 | 1ST QUARTER | 2ND QUARTER | 3RD QUARTER | 4TH QUARTER | TOTAL GRAMS |
|---|---|---|---|---|---|
| 967 | 19.41 | 34.95 | 0.30 | 12.41 | **67.07** |
| **AS - AMERICAN SAMOA - Tota** | **19.41** | **34.95** | **0.30** | **12.41** | **67.07** |

**STATE: AZ - ARIZONA**

| REGISTRANT ZIP CODE 3 | 1ST QUARTER | 2ND QUARTER | 3RD QUARTER | 4TH QUARTER | TOTAL GRAMS |
|---|---|---|---|---|---|
| 850 | 16,691.40 | 15,269.31 | 14,916.32 | 14,905.91 | **61,782.93** |
| 851 | 4,184.23 | 3,723.30 | 3,491.52 | 3,584.31 | **14,983.36** |
| 852 | 23,036.57 | 21,229.95 | 19,323.37 | 20,885.57 | **84,475.47** |
| 853 | 14,955.55 | 13,942.98 | 13,678.35 | 13,197.01 | **55,773.89** |
| 855 | 2,394.86 | 2,262.54 | 2,269.77 | 2,071.63 | **8,998.80** |
| 856 | 4,243.66 | 3,837.49 | 3,819.00 | 3,500.31 | **15,400.46** |
| 857 | 23,930.27 | 22,339.21 | 20,916.83 | 20,114.76 | **87,301.06** |
| 859 | 2,275.74 | 2,305.83 | 1,967.01 | 2,004.97 | **8,553.54** |
| 860 | 1,888.80 | 1,446.49 | 1,476.21 | 1,425.65 | **6,237.16** |
| 863 | 5,668.48 | 5,238.40 | 5,025.91 | 4,580.20 | **20,512.98** |
| 864 | 4,826.17 | 4,971.87 | 5,073.75 | 4,866.13 | **19,737.93** |
| 865 | 163.10 | 114.00 | 163.65 | 105.83 | **546.59** |
| **AZ ARIZONA - Total** | **104,258.83** | **96,681.36** | **92,121.69** | **91,242.29** | **384,304.17** |

**STATE: CA - CALIFORNIA**

| REGISTRANT ZIP CODE 3 | 1ST QUARTER | 2ND QUARTER | 3RD QUARTER | 4TH QUARTER | TOTAL GRAMS |
|---|---|---|---|---|---|
| 900 | 5,032.27 | 5,130.13 | 5,093.20 | 5,017.89 | **20,273.49** |
| 902 | 3,584.59 | 3,215.52 | 3,226.09 | 3,313.06 | **13,339.26** |
| 903 | 190.03 | 172.45 | 202.65 | 181.80 | **746.92** |

| | | | | | |
|---|---|---|---|---|---|
| 904 | 797.62 | 760.68 | 782.63 | 792.38 | **3,133.31** |
| 905 | 1,909.14 | 1,605.40 | 1,688.88 | 1,799.67 | **7,003.10** |
| 906 | 5,537.89 | 4,637.65 | 5,478.27 | 5,275.65 | **20,929.47** |
| 907 | 3,580.37 | 3,274.63 | 3,218.46 | 3,173.32 | **13,246.79** |
| 908 | 3,395.37 | 3,300.98 | 3,162.53 | 3,200.03 | **13,058.91** |
| 910 | 1,730.44 | 1,727.81 | 1,778.92 | 1,491.67 | **6,728.83** |
| 911 | 1,036.46 | 1,048.60 | 974.48 | 1,068.83 | **4,128.38** |
| 912 | 2,543.02 | 1,856.19 | 2,160.30 | 2,165.74 | **8,725.25** |
| 913 | 9,009.73 | 8,444.00 | 8,274.35 | 8,231.46 | **33,959.54** |
| 914 | 3,272.42 | 2,421.05 | 2,565.07 | 2,093.43 | **10,351.97** |
| 915 | 902.93 | 1,047.99 | 1,406.82 | 1,165.92 | **4,523.66** |
| 916 | 383.40 | 317.88 | 378.79 | 386.61 | **1,466.67** |
| 917 | 11,519.23 | 11,514.19 | 11,639.47 | 10,987.94 | **45,660.82** |
| 918 | 185.93 | 154.63 | 186.07 | 169.70 | **696.32** |
| 919 | 3,195.36 | 2,944.03 | 2,928.77 | 2,958.67 | **12,026.82** |
| 920 | 14,941.43 | 14,915.29 | 14,225.27 | 13,260.52 | **57,342.51** |
| 921 | 9,816.26 | 9,080.43 | 9,787.01 | 8,311.29 | **36,994.99** |
| 922 | 7,136.21 | 7,006.89 | 6,879.39 | 6,291.84 | **27,314.33** |
| 923 | 13,497.94 | 13,147.52 | 12,692.91 | 12,291.56 | **51,629.93** |
| 924 | 1,467.55 | 1,296.75 | 1,227.53 | 1,134.41 | **5,126.24** |
| 925 | 11,520.28 | 11,044.41 | 10,375.86 | 10,628.81 | **43,569.36** |
| 926 | 9,368.57 | 9,368.17 | 9,043.06 | 8,676.69 | **36,456.50** |
| 927 | 2,402.87 | 2,240.94 | 2,023.65 | 2,089.93 | **8,757.39** |
| 928 | 8,798.05 | 8,543.94 | 8,911.52 | 8,310.03 | **34,563.55** |
| 930 | 5,109.22 | 4,719.96 | 4,491.79 | 4,173.05 | **18,494.01** |
| 931 | 752.64 | 764.02 | 714.03 | 738.11 | **2,968.79** |
| 932 | 5,320.94 | 5,134.86 | 5,392.83 | 4,764.58 | **20,613.21** |
| 933 | 6,632.78 | 6,446.39 | 6,430.44 | 6,215.35 | **25,724.96** |
| 934 | 6,197.44 | 6,099.78 | 5,923.47 | 6,299.25 | **24,519.95** |
| 935 | 7,212.16 | 6,841.84 | 6,462.39 | 6,338.11 | **26,854.50** |
| 936 | 4,623.91 | 4,345.49 | 4,380.54 | 4,077.50 | **17,427.44** |
| 937 | 6,852.10 | 7,239.41 | 6,913.27 | 6,650.48 | **27,655.25** |
| 939 | 2,440.81 | 2,300.43 | 2,280.09 | 2,159.66 | **9,180.99** |
| 940 | 6,170.53 | 6,099.57 | 5,613.81 | 5,936.94 | **23,820.85** |
| 941 | 10,259.64 | 9,044.88 | 9,051.87 | 8,496.85 | **36,853.25** |
| 943 | 1,342.47 | 1,190.54 | 1,190.35 | 1,315.84 | **5,039.20** |
| 944 | 926.85 | 917.16 | 831.62 | 778.18 | **3,453.80** |
| 945 | 25,692.45 | 24,792.03 | 22,529.74 | 22,105.60 | **95,119.82** |
| 946 | 2,513.80 | 2,455.05 | 2,461.86 | 2,353.95 | **9,784.65** |
| 947 | 2,227.78 | 1,447.60 | 1,557.04 | 1,908.42 | **7,140.84** |

| | | | | | |
|---|---|---|---|---|---|
| 948 | 866.84 | 970.05 | 875.92 | 779.35 | **3,492.16** |
| 949 | 4,207.02 | 3,945.33 | 3,732.83 | 3,783.47 | **15,668.65** |
| 950 | 4,337.59 | 4,130.03 | 4,134.54 | 3,590.58 | **16,192.74** |
| 951 | 2,902.38 | 2,443.57 | 2,310.32 | 2,097.90 | **9,754.17** |
| 952 | 5,392.99 | 5,356.17 | 4,731.34 | 4,956.75 | **20,437.25** |
| 953 | 16,078.97 | 15,128.87 | 15,276.56 | 14,417.76 | **60,902.16** |
| 954 | 8,140.60 | 7,535.48 | 7,557.62 | 7,339.85 | **30,573.56** |
| 955 | 3,473.57 | 3,311.18 | 3,340.22 | 2,971.57 | **13,096.54** |
| 956 | 18,863.38 | 18,134.76 | 17,874.50 | 16,858.08 | **71,730.73** |
| 957 | 2,364.12 | 2,442.67 | 2,439.12 | 2,261.35 | **9,507.25** |
| 958 | 10,962.87 | 11,176.38 | 10,177.51 | 9,399.25 | **41,716.01** |
| 959 | 7,720.62 | 7,182.51 | 6,837.33 | 6,010.04 | **27,750.50** |
| 960 | 5,100.66 | 5,315.83 | 5,353.37 | 5,110.27 | **20,880.14** |
| 961 | 1,269.39 | 1,104.72 | 1,037.31 | 1,084.41 | **4,495.83** |
| **CA - CALIFORNIA - Total** | **322,711.88** | **308,234.72** | **302,215.57** | **289,441.33** | **1,222,603.50** |

**STATE: CO - COLORADO**

| REGISTRANT ZIP CODE 3 | 1ST QUARTER | 2ND QUARTER | 3RD QUARTER | 4TH QUARTER | TOTAL GRAMS |
|---|---|---|---|---|---|
| 800 | 12,426.50 | 10,159.46 | 9,933.77 | 9,778.07 | **42,297.80** |
| 801 | 7,965.44 | 8,638.67 | 9,868.43 | 8,897.76 | **35,370.31** |
| 802 | 8,701.16 | 8,639.77 | 8,554.13 | 8,192.31 | **34,087.36** |
| 803 | 749.30 | 645.79 | 740.49 | 614.43 | **2,750.01** |
| 804 | 2,279.32 | 2,032.23 | 2,058.00 | 1,819.44 | **8,188.99** |
| 805 | 5,343.43 | 5,084.35 | 5,139.35 | 4,588.72 | **20,155.85** |
| 806 | 2,308.38 | 2,150.62 | 2,205.10 | 2,140.07 | **8,804.16** |
| 807 | 831.19 | 843.51 | 848.66 | 803.76 | **3,327.12** |
| 808 | 1,364.45 | 1,212.68 | 1,201.80 | 1,183.38 | **4,962.31** |
| 809 | 6,724.62 | 6,222.91 | 6,604.99 | 6,078.56 | **25,631.09** |
| 810 | 3,468.69 | 3,295.95 | 3,300.82 | 3,036.47 | **13,101.92** |
| 811 | 498.89 | 508.78 | 480.24 | 507.79 | **1,995.70** |
| 812 | 874.76 | 938.85 | 977.75 | 873.80 | **3,665.15** |
| 813 | 489.65 | 487.24 | 499.32 | 488.51 | **1,964.72** |
| 814 | 540.27 | 461.45 | 478.36 | 476.17 | **1,956.25** |
| 815 | 1,760.82 | 1,658.93 | 1,777.57 | 1,560.63 | **6,757.95** |
| 816 | 929.08 | 862.01 | 892.81 | 798.89 | **3,482.79** |
| **CO - COLORADO - Total** | **57,255.96** | **53,843.19** | **55,561.58** | **51,838.75** | **218,499.49** |

**STATE: CT - CONNECTICUT**

| REGISTRANT ZIP CODE 3 | 1ST QUARTER | 2ND QUARTER | 3RD QUARTER | 4TH QUARTER | TOTAL GRAMS |
|---|---|---|---|---|---|
| 060 | 5,009.34 | 4,902.18 | 4,660.83 | 4,550.22 | **19,122.55** |
| 061 | 1,382.44 | 1,574.24 | 1,253.30 | 1,371.57 | **5,581.55** |
| 062 | 1,269.84 | 1,181.80 | 1,259.26 | 1,155.55 | **4,866.45** |

| | | | | | |
|---|---|---|---|---|---|
| 063 | 1,646.02 | 1,661.49 | 1,721.66 | 1,564.12 | **6,593.28** |
| 064 | 8,626.17 | 8,724.72 | 8,523.35 | 8,418.71 | **34,292.95** |
| 065 | 2,851.96 | 2,814.60 | 3,176.94 | 2,930.79 | **11,774.29** |
| 066 | 1,132.96 | 948.35 | 1,167.99 | 1,139.22 | **4,388.52** |
| 067 | 2,401.89 | 2,325.87 | 2,479.22 | 2,329.29 | **9,536.28** |
| 068 | 2,845.37 | 2,644.26 | 2,725.02 | 2,485.79 | **10,700.43** |
| 069 | 233.94 | 203.00 | 216.15 | 202.14 | **855.23** |
| **CT CONNECTICUT Total** | **27,399.91** | **26,980.50** | **27,183.72** | **26,147.40** | **107,711.52** |

**STATE: DC - DISTRICT OF COLUMBIA**

| REGISTRANT ZIP CODE 3 | 1ST QUARTER | 2ND QUARTER | 3RD QUARTER | 4TH QUARTER | TOTAL GRAMS |
|---|---|---|---|---|---|
| 200 | 2,113.71 | 2,023.03 | 2,018.01 | 1,977.85 | **8,132.61** |
| 203 | 19.93 | 11.28 | 3.38 | 0 | **34.59** |
| 204 | 556.86 | 742.52 | 640.25 | 338.40 | **2,278.03** |
| 205 | 12.71 | 0 | 0 | 0 | **12.71** |
| **DC - DISTRICT OF COLUMBIA** | **2,703.21** | **2,776.84** | **2,661.65** | **2,316.25** | **10,457.95** |

**STATE: DE - DELAWARE**

| REGISTRANT ZIP CODE 3 | 1ST QUARTER | 2ND QUARTER | 3RD QUARTER | 4TH QUARTER | TOTAL GRAMS |
|---|---|---|---|---|---|
| 197 | 5,852.09 | 5,659.64 | 5,873.11 | 5,303.17 | **22,688.01** |
| 198 | 3,334.29 | 3,308.71 | 3,611.09 | 3,152.69 | **13,406.79** |
| 199 | 6,260.87 | 5,835.06 | 6,239.33 | 5,964.69 | **24,299.95** |
| **DE DELAWARE Total** | **15,447.25** | **14,803.41** | **15,723.54** | **14,420.55** | **60,394.75** |

**STATE: FL - FLORIDA**

| REGISTRANT ZIP CODE 3 | 1ST QUARTER | 2ND QUARTER | 3RD QUARTER | 4TH QUARTER | TOTAL GRAMS |
|---|---|---|---|---|---|
| 320 | 8,609.32 | 8,249.44 | 8,353.72 | 8,020.44 | **33,232.92** |
| 321 | 12,030.94 | 11,397.04 | 11,511.80 | 11,050.49 | **45,990.27** |
| 322 | 15,067.86 | 14,418.98 | 14,250.88 | 13,137.62 | **56,875.34** |
| 323 | 4,279.14 | 4,267.14 | 3,943.51 | 4,081.60 | **16,571.40** |
| 324 | 4,064.61 | 3,809.89 | 3,794.97 | 3,631.88 | **15,301.36** |
| 325 | 6,393.59 | 5,995.97 | 6,329.20 | 5,490.40 | **24,209.16** |
| 326 | 6,913.01 | 6,736.96 | 6,431.94 | 6,247.27 | **26,329.18** |
| 327 | 17,044.00 | 15,672.95 | 16,458.19 | 15,416.23 | **64,591.37** |
| 328 | 8,774.92 | 8,421.54 | 8,414.41 | 8,209.48 | **33,820.35** |
| 329 | 13,803.36 | 13,596.21 | 13,412.59 | 13,458.39 | **54,270.55** |
| 330 | 16,372.05 | 14,962.55 | 15,377.74 | 15,138.85 | **61,851.20** |
| 331 | 6,030.04 | 5,638.15 | 5,664.06 | 5,770.54 | **23,102.80** |
| 333 | 12,272.85 | 12,168.76 | 11,450.59 | 11,879.47 | **47,771.66** |
| 334 | 22,781.63 | 22,920.43 | 22,256.32 | 22,129.10 | **90,087.48** |
| 335 | 12,729.33 | 11,956.94 | 12,002.78 | 12,037.96 | **48,727.01** |
| 336 | 20,637.06 | 17,647.68 | 21,503.11 | 18,789.59 | **78,577.44** |
| 337 | 19,218.92 | 18,313.00 | 18,718.66 | 18,142.24 | **74,392.82** |

| | | | | | |
|---|---|---|---|---|---|
| 338 | 6,681.26 | 5,906.44 | 5,782.94 | 5,745.76 | **24,116.39** |
| 339 | 15,825.07 | 15,275.19 | 14,729.03 | 14,710.08 | **60,539.37** |
| 341 | 4,695.01 | 4,430.82 | 4,387.26 | 4,567.98 | **18,081.07** |
| 342 | 16,383.14 | 15,158.19 | 14,719.95 | 14,698.52 | **60,959.81** |
| 344 | 9,852.99 | 9,161.71 | 9,034.39 | 9,209.47 | **37,258.56** |
| 346 | 21,227.96 | 19,953.94 | 19,711.43 | 20,089.16 | **80,982.49** |
| 347 | 8,346.54 | 7,792.20 | 7,901.18 | 7,876.07 | **31,915.99** |
| 349 | 12,507.04 | 12,858.14 | 12,568.73 | 12,705.47 | **50,639.37** |
| **FL FLORIDA - Total** | **302,541.63** | **286,710.23** | **288,709.39** | **282,234.08** | **1,160,195.33** |

**STATE: GA - GEORGIA**

| REGISTRANT ZIP CODE 3 | 1ST QUARTER | 2ND QUARTER | 3RD QUARTER | 4TH QUARTER | TOTAL GRAMS |
|---|---|---|---|---|---|
| 300 | 18,898.58 | 17,800.85 | 17,972.22 | 23,184.23 | **77,855.89** |
| 301 | 12,530.93 | 12,783.59 | 13,058.00 | 12,555.17 | **50,927.68** |
| 302 | 8,627.79 | 8,744.64 | 8,374.34 | 7,607.80 | **33,354.56** |
| 303 | 5,010.08 | 4,526.47 | 4,612.29 | 4,435.87 | **18,584.71** |
| 304 | 1,817.68 | 1,828.21 | 1,770.57 | 1,754.60 | **7,171.06** |
| 305 | 7,171.22 | 7,127.48 | 7,207.48 | 6,978.34 | **28,484.52** |
| 306 | 3,469.38 | 3,106.07 | 3,289.00 | 3,252.73 | **13,117.18** |
| 307 | 2,698.10 | 2,353.15 | 2,363.97 | 2,147.04 | **9,562.26** |
| 308 | 1,646.21 | 1,704.13 | 1,709.73 | 1,707.80 | **6,767.87** |
| 309 | 3,390.95 | 3,136.43 | 3,613.47 | 2,948.61 | **13,089.46** |
| 310 | 4,964.15 | 5,023.49 | 4,920.86 | 4,738.44 | **19,646.95** |
| 312 | 2,392.86 | 2,042.49 | 1,861.49 | 1,700.86 | **7,997.70** |
| 313 | 1,813.58 | 1,723.54 | 1,723.80 | 1,624.87 | **6,885.78** |
| 314 | 2,149.30 | 2,121.12 | 2,149.67 | 1,906.45 | **8,326.54** |
| 315 | 3,076.41 | 3,176.15 | 3,329.86 | 2,998.01 | **12,580.43** |
| 316 | 2,800.27 | 2,888.91 | 2,752.47 | 2,734.53 | **11,176.19** |
| 317 | 2,854.45 | 2,934.66 | 2,765.57 | 2,650.88 | **11,205.56** |
| 318 | 447.74 | 328.25 | 361.22 | 328.26 | **1,465.48** |
| 319 | 2,854.77 | 2,757.25 | 2,918.14 | 2,199.63 | **10,729.79** |
| 398 | 754.56 | 681.68 | 787.67 | 739.49 | **2,963.40** |
| **GA GEORGIA Total** | **89,369.02** | **86,788.55** | **87,541.81** | **88,193.63** | **351,893.00** |

**STATE: GU - GUAM**

| REGISTRANT ZIP CODE 3 | 1ST QUARTER | 2ND QUARTER | 3RD QUARTER | 4TH QUARTER | TOTAL GRAMS |
|---|---|---|---|---|---|
| 969 | 740.46 | 874.34 | 726.91 | 589.27 | **2,930.98** |
| **GU - GUAM - Total** | **740.46** | **874.34** | **726.91** | **589.27** | **2,930.98** |

**STATE: HI - HAWAII**

| REGISTRANT ZIP CODE 3 | 1ST QUARTER | 2ND QUARTER | 3RD QUARTER | 4TH QUARTER | TOTAL GRAMS |
|---|---|---|---|---|---|
| 967 | 9,153.89 | 9,215.18 | 8,629.74 | 8,387.84 | **35,386.65** |
| 968 | 5,455.43 | 5,717.22 | 5,766.53 | 4,837.78 | **21,776.96** |

| HI - HAWAII - Total | 14,609.32 | 14,932.41 | 14,396.26 | 13,225.62 | 57,163.61 |
|---|---|---|---|---|---|

**STATE: IA - IOWA**

| REGISTRANT ZIP CODE 3 | 1ST QUARTER | 2ND QUARTER | 3RD QUARTER | 4TH QUARTER | TOTAL GRAMS |
|---|---|---|---|---|---|
| 500 | 1,734.82 | 1,794.77 | 1,854.86 | 1,677.82 | **7,062.26** |
| 501 | 1,945.62 | 1,724.42 | 1,637.00 | 1,629.60 | **6,936.64** |
| 502 | 1,575.71 | 1,575.58 | 1,471.39 | 1,409.26 | **6,031.94** |
| 503 | 4,110.39 | 3,925.39 | 4,332.84 | 3,310.84 | **15,679.46** |
| 504 | 856.76 | 879.63 | 851.46 | 798.85 | **3,386.70** |
| 505 | 1,162.74 | 1,127.39 | 1,096.15 | 1,177.72 | **4,564.00** |
| 506 | 906.01 | 802.92 | 773.94 | 766.30 | **3,249.17** |
| 507 | 600.15 | 709.72 | 764.40 | 726.61 | **2,800.88** |
| 508 | 351.78 | 383.67 | 384.35 | 534.25 | **1,654.04** |
| 510 | 474.59 | 476.79 | 439.60 | 372.84 | **1,763.81** |
| 511 | 865.55 | 848.79 | 1,114.22 | 889.81 | **3,718.36** |
| 512 | 328.91 | 371.49 | 255.95 | 293.19 | **1,249.53** |
| 513 | 306.33 | 291.06 | 338.72 | 300.19 | **1,236.29** |
| 514 | 416.07 | 413.73 | 505.21 | 495.98 | **1,830.98** |
| 515 | 985.18 | 871.91 | 916.79 | 825.25 | **3,599.13** |
| 516 | 152.92 | 151.85 | 154.21 | 152.24 | **611.22** |
| 520 | 1,077.36 | 1,129.99 | 1,089.47 | 943.80 | **4,240.62** |
| 521 | 252.39 | 273.83 | 366.92 | 299.29 | **1,192.43** |
| 522 | 1,517.58 | 1,543.57 | 1,443.22 | 1,298.77 | **5,803.14** |
| 523 | 740.03 | 748.92 | 658.08 | 613.98 | **2,761.02** |
| 524 | 1,252.44 | 1,153.61 | 1,093.39 | 1,018.24 | **4,517.68** |
| 525 | 1,200.36 | 1,154.77 | 1,242.74 | 1,100.40 | **4,698.26** |
| 526 | 1,003.24 | 907.61 | 947.66 | 998.67 | **3,857.18** |
| 527 | 902.07 | 905.79 | 1,002.42 | 908.74 | **3,719.02** |
| 528 | 1,067.32 | 1,053.74 | 1,050.33 | 1,117.86 | **4,289.25** |
| **IA IOWA Total** | **25,786.32** | **25,220.93** | **25,785.28** | **23,660.49** | **100,453.03** |

**STATE: ID - IDAHO**

| REGISTRANT ZIP CODE 3 | 1ST QUARTER | 2ND QUARTER | 3RD QUARTER | 4TH QUARTER | TOTAL GRAMS |
|---|---|---|---|---|---|
| 832 | 2,223.77 | 2,196.65 | 2,238.02 | 2,260.41 | **8,918.86** |
| 833 | 2,045.93 | 2,094.62 | 2,205.01 | 2,038.84 | **8,384.39** |
| 834 | 1,362.44 | 1,433.58 | 1,482.78 | 1,503.06 | **5,781.85** |
| 835 | 1,316.75 | 1,298.97 | 1,392.24 | 1,238.16 | **5,246.12** |
| 836 | 6,327.80 | 6,287.83 | 5,861.39 | 5,734.24 | **24,211.25** |
| 837 | 4,811.70 | 4,728.38 | 4,631.84 | 3,733.27 | **17,905.19** |
| 838 | 3,548.18 | 3,398.65 | 3,419.04 | 3,284.15 | **13,650.02** |
| **ID IDAHO Total** | **21,636.56** | **21,438.67** | **21,230.30** | **19,792.14** | **84,097.68** |

**STATE: IL - ILLINOIS**

| REGISTRANT ZIP CODE 3 | 1ST QUARTER | 2ND QUARTER | 3RD QUARTER | 4TH QUARTER | **TOTAL GRAMS** |
|---|---|---|---|---|---|
| 600 | 17,074.56 | 17,098.49 | 16,464.71 | 17,900.93 | **68,538.69** |
| 601 | 7,402.44 | 7,300.07 | 7,050.77 | 7,111.81 | **28,865.09** |
| 602 | 315.09 | 256.82 | 243.26 | 265.28 | **1,080.46** |
| 603 | 438.01 | 472.57 | 297.59 | 385.04 | **1,593.21** |
| 604 | 8,989.07 | 8,746.60 | 8,336.70 | 8,516.49 | **34,588.85** |
| 605 | 4,512.78 | 4,448.69 | 4,505.32 | 4,670.42 | **18,137.21** |
| 606 | 8,131.37 | 8,310.29 | 8,518.04 | 8,457.11 | **33,416.80** |
| 607 | 790.66 | 813.74 | 911.34 | 796.22 | **3,311.96** |
| 608 | 366.41 | 417.36 | 437.37 | 421.37 | **1,642.51** |
| 609 | 1,386.62 | 1,234.07 | 1,366.85 | 1,314.22 | **5,301.76** |
| 610 | 2,602.92 | 2,566.48 | 2,595.61 | 2,578.74 | **10,343.74** |
| 611 | 3,077.32 | 2,912.64 | 3,066.60 | 2,715.52 | **11,772.08** |
| 612 | 1,418.57 | 1,412.22 | 1,357.69 | 1,207.96 | **5,396.45** |
| 613 | 977.99 | 904.92 | 916.03 | 815.99 | **3,614.92** |
| 614 | 953.23 | 766.48 | 769.44 | 808.14 | **3,297.28** |
| 615 | 2,027.37 | 1,854.83 | 1,733.41 | 1,425.72 | **7,041.33** |
| 616 | 3,005.85 | 2,922.56 | 2,939.48 | 3,125.03 | **11,992.91** |
| 617 | 1,472.37 | 1,502.41 | 1,504.47 | 1,554.43 | **6,033.69** |
| 618 | 2,204.44 | 2,106.21 | 2,222.74 | 1,991.75 | **8,525.12** |
| 619 | 711.75 | 675.05 | 644.47 | 641.93 | **2,673.20** |
| 620 | 2,715.06 | 2,802.65 | 2,441.58 | 2,519.13 | **10,478.42** |
| 622 | 2,832.04 | 2,679.18 | 2,792.15 | 2,774.71 | **11,078.09** |
| 623 | 955.43 | 1,055.95 | 924.85 | 911.94 | **3,848.17** |
| 624 | 1,034.43 | 991.96 | 994.90 | 895.62 | **3,916.91** |
| 625 | 1,805.29 | 1,672.90 | 1,557.26 | 1,604.88 | **6,640.34** |
| 626 | 1,266.79 | 1,223.10 | 1,212.54 | 1,229.26 | **4,931.68** |
| 627 | 1,509.29 | 1,413.42 | 1,532.47 | 1,454.40 | **5,909.59** |
| 628 | 1,781.75 | 1,698.37 | 1,605.14 | 1,496.35 | **6,581.60** |
| 629 | 3,507.50 | 3,247.99 | 3,293.93 | 3,458.51 | **13,507.93** |
| **IL ILLINOIS Total** | **85,266.39** | **83,508.02** | **82,236.71** | **83,048.90** | **334,060.01** |

**STATE: IN - INDIANA**

| REGISTRANT ZIP CODE 3 | 1ST QUARTER | 2ND QUARTER | 3RD QUARTER | 4TH QUARTER | **TOTAL GRAMS** |
|---|---|---|---|---|---|
| 460 | 4,092.49 | 4,137.48 | 4,171.42 | 4,232.87 | **16,634.25** |
| 461 | 6,579.48 | 6,250.78 | 6,016.16 | 5,829.16 | **24,675.58** |
| 462 | 10,548.75 | 9,851.83 | 10,168.35 | 9,980.82 | **40,549.76** |
| 463 | 4,827.92 | 5,022.97 | 5,222.31 | 5,142.18 | **20,215.38** |
| 464 | 391.62 | 367.28 | 457.83 | 333.49 | **1,550.22** |
| 465 | 4,186.96 | 3,960.13 | 4,103.80 | 3,732.37 | **15,983.26** |
| 466 | 1,405.05 | 1,455.22 | 1,329.92 | 1,110.40 | **5,300.60** |

| | | | | | |
|---|---|---|---|---|---|
| 467 | 2,425.37 | 2,466.24 | 2,490.02 | 2,340.09 | **9,721.72** |
| 468 | 3,643.42 | 3,411.37 | 3,949.05 | 3,199.95 | **14,203.78** |
| 469 | 3,402.26 | 3,350.47 | 3,291.35 | 3,177.08 | **13,221.17** |
| 470 | 1,188.66 | 1,275.16 | 1,301.52 | 1,172.85 | **4,938.18** |
| 471 | 2,897.55 | 3,054.70 | 2,989.47 | 2,960.97 | **11,902.69** |
| 472 | 1,846.56 | 1,629.28 | 1,584.45 | 1,431.93 | **6,492.22** |
| 473 | 4,180.85 | 4,166.19 | 4,270.96 | 4,078.07 | **16,696.06** |
| 474 | 1,535.53 | 1,519.60 | 1,597.25 | 1,602.84 | **6,255.22** |
| 475 | 1,442.51 | 1,496.74 | 1,514.77 | 1,401.72 | **5,855.74** |
| 476 | 1,001.57 | 978.19 | 1,049.14 | 933.00 | **3,961.90** |
| 477 | 3,190.47 | 2,497.16 | 2,910.62 | 2,423.45 | **11,021.70** |
| 478 | 1,286.42 | 1,262.40 | 1,357.86 | 1,176.39 | **5,083.07** |
| 479 | 2,017.23 | 1,772.42 | 1,771.55 | 1,723.91 | **7,285.11** |
| **IN INDIANA Total** | **62,090.65** | **59,925.59** | **61,547.82** | **57,983.54** | **241,547.60** |

**STATE: KS - KANSAS**

| REGISTRANT ZIP CODE 3 | 1ST QUARTER | 2ND QUARTER | 3RD QUARTER | 4TH QUARTER | **TOTAL GRAMS** |
|---|---|---|---|---|---|
| 660 | 4,851.29 | 4,634.38 | 4,844.01 | 4,298.72 | **18,628.40** |
| 661 | 1,258.31 | 961.33 | 1,127.46 | 965.38 | **4,312.48** |
| 662 | 5,374.80 | 5,395.57 | 5,479.19 | 5,151.89 | **21,401.46** |
| 664 | 542.75 | 531.81 | 616.10 | 512.97 | **2,203.63** |
| 665 | 933.66 | 1,048.19 | 1,066.15 | 943.28 | **3,991.28** |
| 666 | 2,350.72 | 2,795.72 | 2,506.56 | 2,510.60 | **10,163.60** |
| 667 | 1,951.24 | 1,746.41 | 1,761.85 | 1,616.74 | **7,076.23** |
| 668 | 642.10 | 521.21 | 492.67 | 631.60 | **2,287.58** |
| 669 | 572.18 | 541.18 | 539.77 | 499.91 | **2,153.04** |
| 670 | 1,815.78 | 1,813.42 | 1,821.08 | 1,745.24 | **7,195.51** |
| 671 | 1,095.87 | 1,127.47 | 1,168.43 | 1,066.79 | **4,458.55** |
| 672 | 5,896.73 | 5,802.96 | 5,462.29 | 5,586.99 | **22,748.97** |
| 673 | 1,158.61 | 1,082.59 | 1,206.38 | 1,070.42 | **4,517.99** |
| 674 | 1,676.17 | 1,797.78 | 1,830.21 | 1,709.08 | **7,013.25** |
| 675 | 1,147.57 | 1,034.01 | 1,262.35 | 1,141.58 | **4,585.51** |
| 676 | 798.54 | 816.63 | 766.42 | 752.19 | **3,133.78** |
| 677 | 542.79 | 505.36 | 411.13 | 450.12 | **1,909.40** |
| 678 | 788.54 | 849.57 | 801.68 | 742.07 | **3,181.85** |
| 679 | 246.17 | 254.74 | 323.77 | 281.89 | **1,106.57** |
| **KS - KANSAS - Total** | **33,643.81** | **33,260.31** | **33,487.50** | **31,677.46** | **132,069.08** |

**STATE: KY - KENTUCKY**

| REGISTRANT ZIP CODE 3 | 1ST QUARTER | 2ND QUARTER | 3RD QUARTER | 4TH QUARTER | **TOTAL GRAMS** |
|---|---|---|---|---|---|
| 400 | 1,277.50 | 1,202.97 | 1,133.63 | 1,123.90 | **4,738.01** |
| 401 | 865.90 | 742.50 | 819.34 | 714.63 | **3,142.37** |

| | | | | | |
|---|---|---|---|---|---|
| 402 | 5,612.16 | 5,505.64 | 5,801.95 | 5,458.68 | **22,378.43** |
| 403 | 1,997.47 | 2,004.69 | 1,866.04 | 2,051.77 | **7,919.97** |
| 404 | 1,076.52 | 1,073.42 | 952.98 | 879.47 | **3,982.40** |
| 405 | 2,723.35 | 2,429.88 | 2,610.88 | 2,555.95 | **10,320.07** |
| 406 | 199.16 | 205.30 | 240.35 | 268.07 | **912.87** |
| 407 | 994.29 | 989.33 | 984.03 | 900.69 | **3,868.34** |
| 408 | 239.52 | 285.65 | 213.70 | 230.34 | **969.21** |
| 409 | 483.59 | 385.67 | 472.13 | 519.17 | **1,860.56** |
| 410 | 4,715.52 | 4,446.93 | 4,541.24 | 4,004.23 | **17,707.92** |
| 411 | 956.15 | 890.88 | 741.65 | 679.00 | **3,267.68** |
| 412 | 263.58 | 238.45 | 207.17 | 214.56 | **923.75** |
| 413 | 243.56 | 247.98 | 259.98 | 261.40 | **1,012.92** |
| 414 | 203.33 | 170.18 | 185.82 | 150.96 | **710.30** |
| 415 | 497.74 | 443.59 | 458.14 | 384.55 | **1,784.02** |
| 416 | 317.46 | 311.06 | 265.05 | 291.63 | **1,185.20** |
| 417 | 932.11 | 885.04 | 725.51 | 869.35 | **3,412.02** |
| 418 | 156.49 | 140.40 | 166.72 | 117.50 | **581.10** |
| 420 | 2,409.86 | 2,246.38 | 2,240.95 | 2,094.21 | **8,991.39** |
| 421 | 2,224.02 | 2,069.32 | 2,165.68 | 1,937.62 | **8,396.64** |
| 422 | 1,137.78 | 933.41 | 1,037.60 | 1,027.07 | **4,135.86** |
| 423 | 1,465.46 | 1,535.80 | 1,558.96 | 1,262.36 | **5,822.59** |
| 424 | 1,568.97 | 1,530.32 | 1,600.38 | 1,491.36 | **6,191.03** |
| 425 | 493.19 | 515.04 | 497.94 | 452.52 | **1,958.69** |
| 426 | 421.18 | 379.33 | 415.10 | 388.56 | **1,604.17** |
| 427 | 1,647.14 | 1,485.10 | 1,586.40 | 1,598.42 | **6,317.06** |
| **KY - KENTUCKY - Total** | **35,123.02** | **33,294.25** | **33,749.32** | **31,927.98** | **134,094.57** |

**STATE: LA - LOUISIANA**

| REGISTRANT ZIP CODE 3 | 1ST QUARTER | 2ND QUARTER | 3RD QUARTER | 4TH QUARTER | TOTAL GRAMS |
|---|---|---|---|---|---|
| 700 | 4,441.40 | 4,231.40 | 4,126.11 | 3,597.36 | **16,396.27** |
| 701 | 3,358.62 | 3,205.64 | 3,193.62 | 3,586.93 | **13,344.81** |
| 703 | 1,344.09 | 1,244.25 | 1,224.04 | 1,180.29 | **4,992.67** |
| 704 | 4,694.41 | 4,363.62 | 4,329.82 | 4,182.80 | **17,570.65** |
| 705 | 4,950.36 | 4,669.77 | 4,833.89 | 4,832.94 | **19,286.95** |
| 706 | 1,660.46 | 1,474.18 | 1,770.02 | 1,638.43 | **6,543.08** |
| 707 | 4,814.45 | 4,391.17 | 4,447.14 | 4,105.33 | **17,758.09** |
| 708 | 3,490.05 | 3,427.13 | 3,270.13 | 3,146.41 | **13,333.72** |
| 710 | 1,282.39 | 1,374.14 | 1,343.82 | 1,202.11 | **5,202.47** |
| 711 | 5,502.09 | 5,385.04 | 5,325.67 | 4,987.74 | **21,200.53** |
| 712 | 1,900.52 | 1,731.08 | 2,094.99 | 2,231.15 | **7,957.73** |
| 713 | 3,495.89 | 3,011.97 | 3,265.65 | 2,909.50 | **12,683.00** |

| | | | | | |
|---|---|---|---|---|---|
| 714 | 1,652.67 | 1,857.04 | 1,743.08 | 1,719.09 | **6,971.89** |
| **LA - LOUISIANA - Total** | **42,587.39** | **40,366.44** | **40,967.97** | **39,320.08** | **163,241.88** |

**STATE: MA - MASSACHUSETTS**

| REGISTRANT ZIP CODE 3 | 1ST QUARTER | 2ND QUARTER | 3RD QUARTER | 4TH QUARTER | TOTAL GRAMS |
|---|---|---|---|---|---|
| 010 | 6,043.45 | 6,009.43 | 5,880.63 | 5,708.80 | **23,642.31** |
| 011 | 2,607.21 | 2,382.90 | 2,628.87 | 2,443.45 | **10,062.43** |
| 012 | 976.73 | 946.82 | 934.34 | 988.90 | **3,846.80** |
| 013 | 1,094.61 | 1,122.08 | 1,106.87 | 1,103.94 | **4,427.49** |
| 014 | 1,866.59 | 1,817.72 | 1,925.39 | 1,851.92 | **7,461.63** |
| 015 | 2,486.42 | 2,449.94 | 2,442.81 | 2,281.35 | **9,660.51** |
| 016 | 1,597.75 | 1,759.68 | 1,528.61 | 1,509.94 | **6,395.98** |
| 017 | 2,958.00 | 2,973.89 | 2,930.11 | 3,019.33 | **11,881.32** |
| 018 | 6,895.53 | 7,121.36 | 7,108.62 | 6,506.36 | **27,631.87** |
| 019 | 2,967.91 | 2,898.43 | 2,861.16 | 2,581.95 | **11,309.45** |
| 020 | 1,765.09 | 1,647.22 | 1,725.77 | 1,616.15 | **6,754.23** |
| 021 | 5,896.55 | 5,696.53 | 5,895.63 | 5,731.80 | **23,220.50** |
| 022 | 1,001.39 | 917.79 | 771.43 | 785.36 | **3,475.97** |
| 023 | 3,810.32 | 3,800.83 | 3,820.70 | 3,204.31 | **14,636.15** |
| 024 | 1,052.72 | 1,168.79 | 885.42 | 923.95 | **4,030.87** |
| 025 | 931.48 | 892.46 | 935.44 | 1,051.03 | **3,810.40** |
| 026 | 1,200.68 | 1,308.08 | 1,459.78 | 1,494.59 | **5,463.13** |
| 027 | 7,027.14 | 7,090.60 | 6,972.72 | 7,437.09 | **28,527.55** |
| **MA - MASSACHUSETTS - Total** | **52,179.56** | **52,004.55** | **51,814.29** | **50,240.20** | **206,238.60** |

**STATE: MD - MARYLAND**

| REGISTRANT ZIP CODE 3 | 1ST QUARTER | 2ND QUARTER | 3RD QUARTER | 4TH QUARTER | TOTAL GRAMS |
|---|---|---|---|---|---|
| 206 | 3,676.14 | 3,694.47 | 3,250.45 | 3,331.89 | **13,952.96** |
| 207 | 4,348.94 | 3,915.55 | 3,881.05 | 3,562.49 | **15,708.02** |
| 208 | 4,390.34 | 4,387.82 | 4,451.28 | 4,127.84 | **17,357.29** |
| 209 | 787.79 | 612.28 | 594.87 | 579.71 | **2,574.65** |
| 210 | 8,687.63 | 8,618.50 | 8,839.24 | 8,567.81 | **34,713.18** |
| 211 | 4,029.17 | 3,494.59 | 3,630.17 | 3,521.54 | **14,675.47** |
| 212 | 12,448.11 | 12,368.96 | 12,135.79 | 11,814.54 | **48,767.40** |
| 214 | 662.52 | 682.06 | 738.56 | 693.38 | **2,776.52** |
| 215 | 1,271.68 | 1,235.93 | 1,372.15 | 1,159.51 | **5,039.27** |
| 216 | 2,096.91 | 1,924.74 | 1,856.48 | 1,869.62 | **7,747.76** |
| 217 | 6,044.87 | 5,803.54 | 5,795.11 | 5,518.31 | **23,161.84** |
| 218 | 1,683.31 | 1,513.84 | 1,866.83 | 1,609.41 | **6,673.39** |
| 219 | 2,392.39 | 2,222.09 | 2,113.07 | 2,129.10 | **8,856.65** |
| **MD MARYLAND Total** | **52,519.81** | **50,474.37** | **50,525.07** | **48,485.15** | **202,004.40** |

**STATE: ME - MAINE**

| REGISTRANT ZIP CODE 3 | 1ST QUARTER | 2ND QUARTER | 3RD QUARTER | 4TH QUARTER | TOTAL GRAMS |
|---|---|---|---|---|---|
| 039 | 254.15 | 254.10 | 228.73 | 197.25 | **934.22** |
| 040 | 3,400.30 | 3,369.77 | 3,391.33 | 2,974.88 | **13,136.27** |
| 041 | 806.95 | 799.32 | 987.20 | 974.63 | **3,568.10** |
| 042 | 1,511.23 | 1,523.30 | 1,383.35 | 1,245.68 | **5,663.56** |
| 043 | 1,949.76 | 2,091.72 | 2,245.81 | 2,173.80 | **8,461.08** |
| 044 | 947.19 | 1,030.30 | 1,111.28 | 1,079.99 | **4,168.77** |
| 045 | 311.08 | 311.49 | 408.02 | 377.85 | **1,408.44** |
| 046 | 438.22 | 515.13 | 514.32 | 451.15 | **1,918.83** |
| 047 | 476.52 | 480.02 | 616.22 | 440.84 | **2,013.61** |
| 048 | 412.27 | 385.72 | 368.01 | 379.99 | **1,546.00** |
| 049 | 2,337.67 | 2,369.02 | 2,399.00 | 2,110.58 | **9,216.27** |
| **ME MAINE Total** | **12,845.34** | **13,129.89** | **13,653.27** | **12,406.65** | **52,035.14** |

**STATE: MI - MICHIGAN**

| REGISTRANT ZIP CODE 3 | 1ST QUARTER | 2ND QUARTER | 3RD QUARTER | 4TH QUARTER | TOTAL GRAMS |
|---|---|---|---|---|---|
| 480 | 14,633.64 | 14,595.45 | 14,826.92 | 14,127.49 | **58,183.50** |
| 481 | 14,936.29 | 15,231.91 | 16,211.15 | 15,314.21 | **61,693.56** |
| 482 | 8,481.83 | 7,995.57 | 7,488.49 | 7,621.18 | **31,587.07** |
| 483 | 9,923.22 | 9,679.01 | 9,645.04 | 9,653.22 | **38,900.49** |
| 484 | 5,247.33 | 4,878.27 | 4,958.51 | 5,048.52 | **20,132.64** |
| 485 | 2,841.21 | 2,638.38 | 2,887.89 | 2,185.86 | **10,553.35** |
| 486 | 8,719.26 | 8,353.74 | 8,687.56 | 7,981.11 | **33,741.66** |
| 487 | 4,162.65 | 4,106.62 | 3,829.54 | 3,791.28 | **15,890.08** |
| 488 | 6,780.57 | 6,581.41 | 7,173.72 | 6,353.31 | **26,889.02** |
| 489 | 3,245.99 | 3,060.81 | 2,882.64 | 2,636.96 | **11,826.40** |
| 490 | 11,150.00 | 10,733.51 | 11,240.13 | 10,246.14 | **43,369.78** |
| 491 | 1,476.51 | 1,462.39 | 1,611.63 | 1,541.64 | **6,092.16** |
| 492 | 4,133.83 | 3,767.63 | 4,107.03 | 3,499.53 | **15,508.03** |
| 493 | 2,789.63 | 2,615.09 | 3,008.58 | 2,461.07 | **10,874.38** |
| 494 | 7,828.42 | 7,164.21 | 7,230.74 | 6,496.48 | **28,719.85** |
| 495 | 5,677.41 | 5,827.65 | 5,784.56 | 5,029.49 | **22,319.10** |
| 496 | 6,168.04 | 6,445.18 | 6,557.03 | 5,946.22 | **25,116.47** |
| 497 | 5,358.77 | 5,413.20 | 5,526.46 | 5,064.14 | **21,362.56** |
| 498 | 2,773.31 | 2,689.26 | 2,983.00 | 2,589.47 | **11,035.04** |
| 499 | 948.77 | 933.13 | 891.02 | 909.95 | **3,682.87** |
| **MI - MICHIGAN Total** | **127,276.68** | **124,172.43** | **127,531.64** | **118,497.27** | **497,478.02** |

**STATE: MN - MINNESOTA**

| REGISTRANT ZIP CODE 3 | 1ST QUARTER | 2ND QUARTER | 3RD QUARTER | 4TH QUARTER | TOTAL GRAMS |
|---|---|---|---|---|---|
| 550 | 3,940.38 | 3,685.18 | 3,680.61 | 3,540.86 | **14,847.03** |
| 551 | 5,214.37 | 4,656.63 | 4,568.59 | 4,506.29 | **18,945.89** |

| | | | | | |
|---|---|---|---|---|---|
| 553 | 4,456.33 | 4,025.61 | 4,157.90 | 3,869.70 | **16,509.54** |
| 554 | 8,923.02 | 8,405.40 | 8,117.87 | 8,113.36 | **33,559.66** |
| 556 | 137.73 | 141.79 | 152.24 | 135.00 | **566.76** |
| 557 | 1,304.49 | 1,321.76 | 1,236.28 | 1,164.78 | **5,027.31** |
| 558 | 1,023.45 | 1,054.65 | 954.40 | 981.25 | **4,013.75** |
| 559 | 1,353.09 | 1,509.67 | 1,316.09 | 1,356.23 | **5,535.08** |
| 560 | 1,211.74 | 1,285.03 | 1,307.36 | 1,236.15 | **5,040.28** |
| 561 | 731.75 | 635.45 | 617.96 | 632.00 | **2,617.17** |
| 562 | 749.25 | 726.21 | 742.58 | 753.44 | **2,971.47** |
| 563 | 2,735.97 | 2,624.08 | 3,016.81 | 2,796.12 | **11,172.97** |
| 564 | 1,192.11 | 1,181.67 | 1,152.82 | 1,052.54 | **4,579.14** |
| 565 | 984.39 | 1,062.49 | 1,139.63 | 975.08 | **4,161.60** |
| 566 | 606.82 | 508.92 | 655.78 | 562.54 | **2,334.06** |
| 567 | 441.65 | 455.55 | 429.52 | 414.66 | **1,741.39** |
| **MN MINNESOTA Total** | **35,006.53** | **33,280.09** | **33,246.45** | **32,090.00** | **133,623.08** |

**STATE: MO - MISSOURI**

| REGISTRANT ZIP CODE 3 | 1ST QUARTER | 2ND QUARTER | 3RD QUARTER | 4TH QUARTER | TOTAL GRAMS |
|---|---|---|---|---|---|
| 630 | 7,418.73 | 7,032.17 | 7,237.53 | 6,743.41 | **28,431.83** |
| 631 | 12,245.23 | 12,444.12 | 12,468.48 | 11,733.07 | **48,890.89** |
| 633 | 4,113.10 | 4,011.04 | 4,303.89 | 3,818.47 | **16,246.50** |
| 634 | 512.94 | 440.21 | 444.15 | 479.48 | **1,876.77** |
| 635 | 1,091.51 | 1,076.75 | 1,040.07 | 973.43 | **4,181.77** |
| 636 | 1,745.86 | 1,728.80 | 1,703.82 | 1,532.90 | **6,711.39** |
| 637 | 1,152.87 | 1,017.92 | 994.02 | 1,034.84 | **4,199.65** |
| 638 | 992.76 | 1,017.79 | 1,061.76 | 990.86 | **4,063.18** |
| 639 | 959.28 | 845.90 | 851.04 | 854.18 | **3,510.41** |
| 640 | 6,331.53 | 5,924.51 | 5,710.18 | 5,513.06 | **23,479.28** |
| 641 | 4,891.94 | 3,785.73 | 3,857.57 | 3,788.56 | **16,323.81** |
| 644 | 550.97 | 528.73 | 579.01 | 499.62 | **2,158.33** |
| 645 | 1,103.60 | 947.25 | 917.55 | 965.02 | **3,933.43** |
| 646 | 569.45 | 580.65 | 592.25 | 549.09 | **2,291.44** |
| 647 | 1,693.40 | 1,706.49 | 1,789.15 | 1,400.21 | **6,589.24** |
| 648 | 3,194.03 | 2,504.34 | 2,413.96 | 2,050.70 | **10,163.04** |
| 650 | 1,079.18 | 1,220.61 | 1,266.72 | 1,157.72 | **4,724.23** |
| 651 | 804.83 | 994.16 | 898.80 | 864.08 | **3,561.87** |
| 652 | 3,240.61 | 3,316.02 | 3,166.92 | 3,292.65 | **13,016.21** |
| 653 | 714.29 | 762.90 | 703.29 | 681.67 | **2,862.15** |
| 654 | 936.15 | 1,038.45 | 1,156.62 | 1,178.55 | **4,309.77** |
| 655 | 1,546.52 | 1,637.03 | 1,674.72 | 1,598.54 | **6,456.81** |
| 656 | 3,374.79 | 3,339.37 | 3,290.48 | 3,045.50 | **13,050.14** |

| | | | | | |
|---|---|---|---|---|---|
| 657 | 6,512.30 | 5,474.16 | 5,297.81 | 5,283.27 | **22,567.54** |
| 658 | 5,774.51 | 5,681.42 | 5,392.71 | 4,845.47 | **21,694.11** |
| **MO MISSOURI - Total** | **72,550.39** | **69,056.52** | **68,812.51** | **64,874.35** | **275,293.78** |

**STATE: MS - MISSISSIPPI**

| REGISTRANT ZIP CODE 3 | 1ST QUARTER | 2ND QUARTER | 3RD QUARTER | 4TH QUARTER | TOTAL GRAMS |
|---|---|---|---|---|---|
| 386 | 3,228.46 | 2,937.60 | 2,908.26 | 2,919.35 | **11,993.67** |
| 387 | 370.93 | 405.02 | 336.96 | 310.85 | **1,423.76** |
| 388 | 3,171.75 | 2,620.28 | 2,850.43 | 2,532.22 | **11,174.68** |
| 389 | 537.91 | 473.76 | 471.40 | 437.55 | **1,920.62** |
| 390 | 1,898.72 | 1,908.63 | 1,841.84 | 1,712.30 | **7,361.48** |
| 391 | 6,424.69 | 3,833.67 | 5,162.41 | 6,316.50 | **21,737.26** |
| 392 | 2,403.68 | 2,429.87 | 2,469.17 | 2,367.92 | **9,670.64** |
| 393 | 2,084.11 | 2,146.95 | 2,084.27 | 1,960.97 | **8,276.30** |
| 394 | 3,950.63 | 3,965.99 | 3,855.17 | 4,005.87 | **15,777.66** |
| 395 | 3,924.95 | 3,627.82 | 3,401.47 | 3,348.90 | **14,303.13** |
| 396 | 1,412.77 | 1,264.11 | 1,281.91 | 1,230.71 | **5,189.49** |
| 397 | 864.11 | 857.67 | 708.92 | 873.33 | **3,304.03** |
| **MS MISSISSIPPI - Total** | **30,272.71** | **26,471.37** | **27,372.20** | **28,016.47** | **112,132.74** |

**STATE: MT - MONTANA**

| REGISTRANT ZIP CODE 3 | 1ST QUARTER | 2ND QUARTER | 3RD QUARTER | 4TH QUARTER | TOTAL GRAMS |
|---|---|---|---|---|---|
| 590 | 831.55 | 771.13 | 763.61 | 787.74 | **3,154.03** |
| 591 | 2,532.74 | 2,450.70 | 2,489.47 | 2,261.02 | **9,733.93** |
| 592 | 488.28 | 468.55 | 428.17 | 399.19 | **1,784.19** |
| 593 | 483.30 | 451.75 | 413.52 | 365.95 | **1,714.53** |
| 594 | 1,491.69 | 1,409.66 | 1,502.41 | 1,298.05 | **5,701.81** |
| 595 | 256.42 | 248.63 | 234.53 | 256.32 | **995.90** |
| 596 | 1,243.11 | 1,262.29 | 1,256.76 | 1,051.37 | **4,813.52** |
| 597 | 1,591.79 | 1,722.81 | 1,716.02 | 1,736.73 | **6,767.35** |
| 598 | 2,161.72 | 2,169.72 | 2,195.13 | 2,072.49 | **8,599.05** |
| 599 | 2,073.77 | 2,185.88 | 2,148.88 | 2,209.48 | **8,618.01** |
| **MT - MONTANA Total** | **13,154.38** | **13,141.11** | **13,148.51** | **12,438.34** | **51,882.33** |

**STATE: NC - NORTH CAROLINA**

| REGISTRANT ZIP CODE 3 | 1ST QUARTER | 2ND QUARTER | 3RD QUARTER | 4TH QUARTER | TOTAL GRAMS |
|---|---|---|---|---|---|
| 270 | 3,167.12 | 2,936.97 | 2,913.30 | 2,720.09 | **11,737.49** |
| 271 | 2,434.71 | 2,579.63 | 2,775.14 | 2,437.61 | **10,227.09** |
| 272 | 6,465.55 | 6,295.91 | 6,085.17 | 5,431.79 | **24,278.43** |
| 273 | 4,741.66 | 4,320.82 | 4,028.73 | 3,490.16 | **16,581.38** |
| 274 | 3,027.13 | 2,541.10 | 2,513.88 | 2,254.14 | **10,336.24** |
| 275 | 10,993.26 | 15,225.66 | 14,892.15 | 9,253.77 | **50,364.84** |
| 276 | 3,681.72 | 3,378.40 | 3,120.09 | 3,054.91 | **13,235.13** |

| | | | | | |
|---|---|---|---|---|---|
| 277 | 4,013.75 | 3,595.37 | 3,754.21 | 4,027.85 | **15,391.18** |
| 278 | 6,666.56 | 6,242.67 | 6,280.93 | 5,740.62 | **24,930.78** |
| 279 | 2,487.84 | 2,720.12 | 2,573.28 | 1,947.64 | **9,728.88** |
| 280 | 7,759.80 | 7,552.91 | 7,414.00 | 7,197.47 | **29,924.17** |
| 281 | 6,491.03 | 6,225.77 | 6,485.09 | 5,924.25 | **25,126.15** |
| 282 | 4,181.79 | 4,068.27 | 4,669.41 | 4,859.95 | **17,779.42** |
| 283 | 8,128.36 | 7,809.41 | 7,624.70 | 6,364.37 | **29,926.84** |
| 284 | 5,756.14 | 5,869.31 | 5,853.67 | 5,079.23 | **22,558.35** |
| 285 | 5,907.15 | 6,195.57 | 6,926.74 | 6,149.83 | **25,179.31** |
| 286 | 10,302.11 | 9,713.99 | 9,908.33 | 8,664.35 | **38,588.78** |
| 287 | 6,140.22 | 5,824.11 | 6,113.00 | 5,551.57 | **23,628.90** |
| 288 | 2,446.54 | 2,647.59 | 2,481.35 | 2,423.63 | **9,999.11** |
| 289 | 816.34 | 731.43 | 745.95 | 719.33 | **3,013.04** |
| **NC - NORTH CAROLINA Total** | **105,608.79** | **106,475.01** | **107,159.13** | **93,292.58** | **412,535.51** |

**STATE: ND - NORTH DAKOTA**

| REGISTRANT ZIP CODE 3 | 1ST QUARTER | 2ND QUARTER | 3RD QUARTER | 4TH QUARTER | **TOTAL GRAMS** |
|---|---|---|---|---|---|
| 580 | 296.38 | 278.52 | 331.83 | 283.19 | **1,189.91** |
| 581 | 1,031.12 | 1,002.28 | 1,157.61 | 1,152.18 | **4,343.19** |
| 582 | 723.28 | 701.02 | 695.08 | 753.67 | **2,873.05** |
| 583 | 291.54 | 328.97 | 307.19 | 304.06 | **1,231.75** |
| 584 | 257.44 | 223.80 | 248.55 | 244.20 | **973.99** |
| 585 | 1,591.16 | 1,310.81 | 1,335.66 | 1,233.85 | **5,471.48** |
| 586 | 204.22 | 184.40 | 246.42 | 285.32 | **920.35** |
| 587 | 286.52 | 347.39 | 311.09 | 306.41 | **1,251.41** |
| 588 | 292.47 | 217.33 | 241.53 | 186.70 | **938.03** |
| **ND NORTH DAKOTA Total** | **4,974.12** | **4,594.51** | **4,874.95** | **4,749.57** | **19,193.15** |

**STATE: NE - NEBRASKA**

| REGISTRANT ZIP CODE 3 | 1ST QUARTER | 2ND QUARTER | 3RD QUARTER | 4TH QUARTER | **TOTAL GRAMS** |
|---|---|---|---|---|---|
| 680 | 1,959.66 | 1,865.12 | 1,927.90 | 1,849.52 | **7,602.20** |
| 681 | 4,967.98 | 5,093.97 | 4,986.29 | 4,486.21 | **19,534.45** |
| 683 | 474.92 | 569.00 | 453.56 | 483.49 | **1,980.98** |
| 684 | 484.68 | 496.82 | 505.31 | 408.93 | **1,895.73** |
| 685 | 2,046.82 | 2,269.23 | 1,926.01 | 2,084.66 | **8,326.72** |
| 686 | 363.08 | 343.07 | 369.55 | 380.76 | **1,456.46** |
| 687 | 840.01 | 805.24 | 839.54 | 848.92 | **3,333.71** |
| 688 | 1,455.84 | 1,403.32 | 1,626.96 | 1,519.37 | **6,005.48** |
| 689 | 529.50 | 502.34 | 595.37 | 554.29 | **2,181.49** |
| 690 | 289.44 | 349.92 | 305.88 | 279.14 | **1,224.38** |
| 691 | 652.06 | 600.61 | 593.53 | 424.01 | **2,270.21** |
| 692 | 22.26 | 21.20 | 29.25 | 46.17 | **118.88** |

| | | | | | |
|---|---|---|---|---|---|
| 693 | 875.65 | 777.09 | 904.60 | 779.98 | **3,337.32** |
| **NE NEBRASKA Total** | **14,961.89** | **15,096.92** | **15,063.74** | **14,145.46** | **59,268.01** |

**STATE: NH - NEW HAMPSHIRE**

| REGISTRANT ZIP CODE 3 | 1ST QUARTER | 2ND QUARTER | 3RD QUARTER | 4TH QUARTER | TOTAL GRAMS |
|---|---|---|---|---|---|
| 030 | 3,918.87 | 3,835.98 | 3,803.93 | 3,713.88 | **15,272.65** |
| 031 | 2,219.05 | 2,063.56 | 2,303.71 | 1,869.57 | **8,455.89** |
| 032 | 1,619.36 | 1,589.47 | 1,754.57 | 1,678.85 | **6,642.26** |
| 033 | 2,105.01 | 1,367.76 | 1,884.99 | 1,442.86 | **6,800.61** |
| 034 | 742.40 | 621.74 | 635.94 | 641.40 | **2,641.48** |
| 035 | 495.10 | 425.54 | 502.35 | 419.38 | **1,842.37** |
| 036 | 158.97 | 194.09 | 124.08 | 114.60 | **591.75** |
| 037 | 880.61 | 763.09 | 776.41 | 713.48 | **3,133.59** |
| 038 | 3,366.60 | 3,267.64 | 3,348.85 | 3,355.49 | **13,338.57** |
| **NH - NEW HAMPSHIRE Total** | **15,505.98** | **14,128.87** | **15,134.83** | **13,949.51** | **58,719.18** |

**STATE: NJ - NEW JERSEY**

| REGISTRANT ZIP CODE 3 | 1ST QUARTER | 2ND QUARTER | 3RD QUARTER | 4TH QUARTER | TOTAL GRAMS |
|---|---|---|---|---|---|
| 070 | 7,674.11 | 7,718.19 | 7,447.25 | 7,885.87 | **30,725.42** |
| 071 | 1,231.05 | 974.46 | 1,031.82 | 1,056.13 | **4,293.46** |
| 072 | 491.03 | 381.99 | 479.46 | 434.29 | **1,786.77** |
| 073 | 605.88 | 563.57 | 674.87 | 611.67 | **2,455.98** |
| 074 | 4,178.17 | 3,588.07 | 3,980.13 | 4,296.10 | **16,042.47** |
| 075 | 790.42 | 656.29 | 565.08 | 498.63 | **2,510.42** |
| 076 | 4,930.69 | 4,763.53 | 5,066.75 | 4,522.18 | **19,283.16** |
| 077 | 4,481.35 | 4,067.64 | 4,122.21 | 4,236.67 | **16,907.87** |
| 078 | 2,374.20 | 1,992.53 | 2,223.23 | 1,992.83 | **8,582.79** |
| 079 | 1,884.24 | 1,752.23 | 1,749.83 | 1,953.43 | **7,339.74** |
| 080 | 13,580.48 | 13,528.24 | 13,464.95 | 13,234.55 | **53,808.23** |
| 081 | 1,117.87 | 1,225.38 | 1,113.74 | 1,169.99 | **4,626.98** |
| 082 | 2,350.24 | 2,231.63 | 2,247.30 | 2,063.18 | **8,892.34** |
| 083 | 1,772.15 | 1,639.43 | 1,556.01 | 1,490.64 | **6,458.24** |
| 084 | 356.66 | 368.31 | 359.19 | 309.89 | **1,394.05** |
| 085 | 1,658.90 | 1,661.03 | 1,683.16 | 1,820.09 | **6,823.18** |
| 086 | 1,687.91 | 1,578.96 | 1,671.58 | 1,508.58 | **6,447.02** |
| 087 | 4,059.09 | 3,722.85 | 4,147.53 | 3,629.44 | **15,558.91** |
| 088 | 7,351.18 | 6,827.01 | 7,417.45 | 7,073.70 | **28,669.35** |
| 089 | 544.67 | 502.65 | 703.96 | 472.74 | **2,224.02** |
| **NJ NEW JERSEY - Total** | **63,120.29** | **59,743.99** | **61,705.49** | **60,260.62** | **244,830.39** |

**STATE: NM - NEW MEXICO**

| REGISTRANT ZIP CODE 3 | 1ST QUARTER | 2ND QUARTER | 3RD QUARTER | 4TH QUARTER | TOTAL GRAMS |
|---|---|---|---|---|---|
| 870 | 1,530.25 | 1,380.09 | 1,288.28 | 1,309.65 | **5,508.26** |

| | | | | | |
|---|---|---|---|---|---|
| 871 | 9,626.65 | 9,344.98 | 9,217.70 | 8,894.59 | **37,083.92** |
| 873 | 783.46 | 794.62 | 786.14 | 774.71 | **3,138.93** |
| 874 | 893.66 | 786.52 | 689.25 | 654.60 | **3,024.03** |
| 875 | 2,107.97 | 2,024.26 | 1,855.31 | 2,005.77 | **7,993.30** |
| 877 | 379.51 | 315.47 | 328.76 | 297.14 | **1,320.89** |
| 878 | 100.54 | 93.55 | 66.06 | 90.47 | **350.62** |
| 879 | 191.02 | 142.40 | 109.38 | 115.70 | **558.50** |
| 880 | 1,934.84 | 1,773.52 | 1,778.99 | 1,631.25 | **7,118.60** |
| 881 | 447.99 | 406.00 | 395.02 | 422.25 | **1,671.27** |
| 882 | 2,219.73 | 1,942.93 | 2,078.69 | 1,986.44 | **8,227.78** |
| 883 | 580.51 | 496.24 | 497.64 | 575.02 | **2,149.41** |
| 884 | 36.78 | 49.74 | 28.59 | 35.49 | **150.61** |
| **NM NEW MEXICO Total** | **20,832.92** | **19,550.31** | **19,119.81** | **18,793.07** | **78,296.12** |

**STATE: NV - NEVADA**

| REGISTRANT ZIP CODE 3 | 1ST QUARTER | 2ND QUARTER | 3RD QUARTER | 4TH QUARTER | **TOTAL GRAMS** |
|---|---|---|---|---|---|
| 890 | 10,127.49 | 10,119.07 | 9,544.71 | 9,919.98 | **39,711.25** |
| 891 | 22,511.08 | 20,873.13 | 20,360.32 | 19,721.98 | **83,466.50** |
| 893 | 56.32 | 61.13 | 66.51 | 40.80 | **224.77** |
| 894 | 3,269.72 | 3,077.15 | 2,660.98 | 2,595.58 | **11,603.43** |
| 895 | 3,797.89 | 3,485.47 | 3,676.86 | 3,418.87 | **14,379.10** |
| 897 | 1,505.42 | 987.94 | 1,013.48 | 948.17 | **4,455.02** |
| 898 | 333.99 | 242.16 | 270.58 | 239.06 | **1,085.80** |
| **NV NEVADA Total** | **41,601.91** | **38,846.06** | **37,593.44** | **36,884.44** | **154,925.85** |

**STATE: NY - NEW YORK**

| REGISTRANT ZIP CODE 3 | 1ST QUARTER | 2ND QUARTER | 3RD QUARTER | 4TH QUARTER | **TOTAL GRAMS** |
|---|---|---|---|---|---|
| 063 | 0 | 0 | 0.08 | 0 | **0.08** |
| 100 | 9,405.58 | 7,533.07 | 8,901.62 | 8,168.66 | **34,008.92** |
| 101 | 168.74 | 128.51 | 179.80 | 187.13 | **664.17** |
| 102 | 7.90 | 5.64 | 10.15 | 4.51 | **28.20** |
| 103 | 4,414.00 | 4,102.47 | 4,468.03 | 4,239.10 | **17,223.59** |
| 104 | 4,289.31 | 3,257.64 | 4,184.90 | 3,688.83 | **15,420.67** |
| 105 | 2,079.53 | 1,679.42 | 2,242.95 | 2,133.03 | **8,134.93** |
| 106 | 321.00 | 318.00 | 381.65 | 324.92 | **1,345.57** |
| 107 | 591.41 | 486.86 | 565.44 | 593.26 | **2,236.96** |
| 108 | 241.35 | 171.68 | 252.33 | 228.12 | **893.49** |
| 109 | 2,275.84 | 1,957.66 | 2,165.75 | 2,248.23 | **8,647.49** |
| 110 | 2,839.55 | 2,788.25 | 3,668.15 | 3,287.23 | **12,583.18** |
| 111 | 525.78 | 492.25 | 424.33 | 390.85 | **1,833.21** |
| 112 | 5,900.38 | 4,770.22 | 5,535.94 | 5,217.85 | **21,424.38** |
| 113 | 2,347.91 | 1,940.49 | 2,249.89 | 2,395.43 | **8,933.72** |

| | | | | | |
|---|---|---|---|---|---|
| 114 | 1,231.27 | 1,078.35 | 1,108.15 | 1,094.40 | **4,512.17** |
| 115 | 3,568.94 | 3,020.74 | 3,323.91 | 3,170.15 | **13,083.74** |
| 116 | 341.38 | 337.71 | 338.54 | 297.13 | **1,314.76** |
| 117 | 13,271.70 | 11,988.07 | 13,419.60 | 13,442.46 | **52,121.83** |
| 118 | 373.66 | 361.59 | 402.03 | 405.31 | **1,542.59** |
| 119 | 1,659.57 | 1,480.07 | 1,660.28 | 1,588.69 | **6,388.60** |
| 120 | 3,118.49 | 3,120.39 | 3,122.05 | 3,064.58 | **12,425.51** |
| 121 | 1,385.04 | 1,271.02 | 1,192.28 | 1,232.58 | **5,080.92** |
| 122 | 1,975.54 | 1,958.44 | 2,134.11 | 1,815.96 | **7,884.04** |
| 123 | 854.66 | 689.22 | 716.87 | 826.32 | **3,087.06** |
| 124 | 2,083.10 | 1,773.28 | 1,918.35 | 1,766.12 | **7,540.86** |
| 125 | 2,691.71 | 2,591.76 | 2,629.08 | 2,530.04 | **10,442.60** |
| 126 | 834.62 | 711.46 | 849.87 | 792.53 | **3,188.48** |
| 127 | 938.92 | 770.64 | 922.37 | 769.93 | **3,401.87** |
| 128 | 2,560.99 | 2,330.01 | 2,617.39 | 2,250.05 | **9,758.43** |
| 129 | 2,093.99 | 1,796.95 | 2,079.84 | 1,807.35 | **7,778.14** |
| 130 | 3,667.23 | 3,456.45 | 3,416.53 | 3,119.38 | **13,659.59** |
| 131 | 1,256.97 | 1,203.44 | 1,266.03 | 1,313.89 | **5,040.34** |
| 132 | 2,815.79 | 2,292.85 | 2,630.49 | 2,471.28 | **10,210.41** |
| 133 | 1,015.56 | 1,022.67 | 1,032.96 | 1,020.95 | **4,092.14** |
| 134 | 1,537.97 | 1,413.75 | 1,582.73 | 1,641.84 | **6,176.29** |
| 135 | 477.09 | 438.21 | 512.31 | 471.31 | **1,898.92** |
| 136 | 1,964.89 | 1,996.18 | 2,103.06 | 1,958.74 | **8,022.87** |
| 137 | 744.45 | 663.73 | 696.44 | 556.89 | **2,661.52** |
| 138 | 2,827.21 | 2,538.88 | 2,348.31 | 2,516.32 | **10,230.72** |
| 139 | 603.79 | 553.85 | 578.83 | 534.71 | **2,271.19** |
| 140 | 3,898.82 | 3,646.49 | 3,588.55 | 3,707.65 | **14,841.50** |
| 141 | 1,833.45 | 1,650.99 | 1,839.08 | 1,735.12 | **7,058.64** |
| 142 | 5,100.38 | 5,178.08 | 5,093.42 | 5,062.04 | **20,433.92** |
| 143 | 733.45 | 588.52 | 656.97 | 592.57 | **2,571.51** |
| 144 | 2,500.85 | 2,416.85 | 2,453.40 | 2,492.40 | **9,863.50** |
| 145 | 2,124.97 | 1,978.46 | 2,103.00 | 2,203.25 | **8,409.69** |
| 146 | 5,167.01 | 4,554.15 | 5,061.21 | 4,398.93 | **19,181.31** |
| 147 | 1,772.42 | 1,683.60 | 1,631.22 | 1,534.17 | **6,621.42** |
| 148 | 3,363.19 | 3,385.27 | 3,186.11 | 3,007.65 | **12,942.22** |
| 149 | 663.16 | 669.87 | 682.51 | 614.29 | **2,629.82** |
| **NY - NEW YORK - Total** | **118,460.51** | **106,244.15** | **116,128.85** | **110,914.13** | **451,747.65** |

**STATE: OH - OHIO**

| REGISTRANT ZIP CODE 3 | 1ST QUARTER | 2ND QUARTER | 3RD QUARTER | 4TH QUARTER | **TOTAL GRAMS** |
|---|---|---|---|---|---|
| 430 | 5,376.55 | 5,570.43 | 5,471.15 | 5,401.41 | **21,819.54** |

| | | | | | |
|---|---|---|---|---|---|
| 431 | 2,885.19 | 3,074.10 | 3,006.64 | 3,098.04 | **12,063.97** |
| 432 | 6,607.25 | 6,285.58 | 6,902.81 | 6,502.62 | **26,298.27** |
| 433 | 1,218.36 | 1,095.05 | 1,184.14 | 1,080.69 | **4,578.22** |
| 434 | 896.28 | 797.01 | 823.32 | 706.08 | **3,222.69** |
| 435 | 2,621.28 | 2,615.95 | 2,492.30 | 2,338.83 | **10,068.35** |
| 436 | 3,468.85 | 3,380.13 | 3,419.28 | 3,329.80 | **13,598.06** |
| 437 | 1,555.25 | 1,511.21 | 1,469.34 | 1,322.36 | **5,858.17** |
| 438 | 346.33 | 270.72 | 271.92 | 250.87 | **1,139.84** |
| 439 | 1,909.74 | 1,646.78 | 1,779.52 | 1,489.62 | **6,825.66** |
| 440 | 4,296.19 | 4,244.34 | 4,229.82 | 3,942.25 | **16,712.60** |
| 441 | 6,257.17 | 6,712.63 | 6,609.57 | 6,510.24 | **26,089.60** |
| 442 | 3,707.44 | 3,647.23 | 3,547.02 | 3,547.14 | **14,448.83** |
| 443 | 2,069.21 | 2,009.94 | 1,949.16 | 2,030.77 | **8,059.09** |
| 444 | 2,935.12 | 2,562.36 | 2,755.32 | 2,547.09 | **10,799.89** |
| 445 | 2,130.95 | 2,163.35 | 2,020.53 | 2,075.37 | **8,390.20** |
| 446 | 2,672.11 | 2,646.10 | 2,612.26 | 2,677.82 | **10,608.29** |
| 447 | 2,196.38 | 2,047.90 | 2,036.08 | 2,009.04 | **8,289.41** |
| 448 | 1,979.32 | 2,081.43 | 1,953.78 | 1,898.78 | **7,913.30** |
| 449 | 735.41 | 703.72 | 757.37 | 798.91 | **2,995.41** |
| 450 | 6,160.19 | 5,871.02 | 5,875.55 | 5,688.63 | **23,595.38** |
| 451 | 2,280.52 | 2,095.99 | 2,138.89 | 2,039.35 | **8,554.76** |
| 452 | 6,541.44 | 6,210.85 | 6,025.50 | 5,841.53 | **24,619.32** |
| 453 | 4,073.18 | 4,011.05 | 4,010.07 | 3,854.66 | **15,948.96** |
| 454 | 3,888.86 | 3,805.91 | 3,699.39 | 3,451.19 | **14,845.35** |
| 455 | 804.42 | 748.97 | 749.23 | 723.46 | **3,026.08** |
| 456 | 4,306.53 | 4,152.11 | 4,398.04 | 3,850.86 | **16,707.55** |
| 457 | 1,264.99 | 1,225.58 | 1,217.15 | 1,084.74 | **4,792.47** |
| 458 | 2,228.90 | 2,080.79 | 2,065.26 | 2,059.89 | **8,434.83** |
| **OH - OHIO Total** | **87,413.41** | **85,268.23** | **85,470.42** | **82,152.02** | **340,304.08** |

**STATE: OK - OKLAHOMA**

| REGISTRANT ZIP CODE 3 | 1ST QUARTER | 2ND QUARTER | 3RD QUARTER | 4TH QUARTER | **TOTAL GRAMS** |
|---|---|---|---|---|---|
| 730 | 9,133.53 | 8,615.78 | 8,493.13 | 8,092.25 | **34,334.69** |
| 731 | 13,354.94 | 13,050.75 | 12,769.43 | 12,707.98 | **51,883.11** |
| 734 | 1,414.51 | 1,345.14 | 1,342.52 | 1,268.27 | **5,370.44** |
| 735 | 3,392.38 | 3,179.79 | 3,077.91 | 3,001.57 | **12,651.65** |
| 736 | 663.67 | 548.01 | 627.64 | 582.69 | **2,422.02** |
| 737 | 906.86 | 854.89 | 935.01 | 861.37 | **3,558.13** |
| 738 | 180.90 | 170.28 | 198.84 | 201.72 | **751.73** |
| 739 | 30.00 | 36.99 | 48.65 | 39.74 | **155.39** |
| 740 | 9,070.26 | 9,130.18 | 8,132.86 | 8,320.63 | **34,653.93** |

| | | | | | |
|---|---|---|---|---|---|
| 741 | 5,561.62 | 5,527.24 | 4,991.21 | 5,178.56 | **21,258.63** |
| 743 | 1,253.52 | 1,122.71 | 1,081.49 | 1,074.87 | **4,532.59** |
| 744 | 3,350.55 | 3,163.25 | 2,778.82 | 2,595.79 | **11,888.42** |
| 745 | 1,179.20 | 1,070.95 | 1,061.38 | 923.51 | **4,235.04** |
| 746 | 647.06 | 619.63 | 663.60 | 561.47 | **2,491.76** |
| 747 | 801.21 | 824.98 | 688.71 | 730.28 | **3,045.18** |
| 748 | 1,954.50 | 1,873.74 | 1,885.82 | 1,811.12 | **7,525.18** |
| 749 | 831.85 | 826.48 | 759.89 | 700.30 | **3,118.51** |
| **OK - OKLAHOMA Total** | **53,726.55** | **51,960.79** | **49,536.92** | **48,652.13** | **203,876.39** |

**STATE: OR - OREGON**

| REGISTRANT ZIP CODE 3 | 1ST QUARTER | 2ND QUARTER | 3RD QUARTER | 4TH QUARTER | TOTAL GRAMS |
|---|---|---|---|---|---|
| 970 | 8,601.66 | 8,537.03 | 8,537.13 | 8,457.25 | **34,133.07** |
| 971 | 4,738.67 | 4,754.60 | 4,783.98 | 4,482.31 | **18,759.57** |
| 972 | 12,714.70 | 12,466.13 | 12,880.28 | 12,228.45 | **50,289.56** |
| 973 | 6,086.25 | 5,828.20 | 5,615.18 | 5,482.32 | **23,011.96** |
| 974 | 8,794.09 | 8,725.76 | 8,521.88 | 8,118.14 | **34,159.86** |
| 975 | 5,744.16 | 5,942.56 | 5,807.06 | 5,719.11 | **23,212.88** |
| 976 | 1,097.39 | 1,068.29 | 1,173.86 | 1,253.97 | **4,593.51** |
| 977 | 2,839.69 | 2,829.59 | 3,025.91 | 3,208.81 | **11,904.01** |
| 978 | 1,602.97 | 1,646.42 | 1,606.72 | 1,425.55 | **6,281.66** |
| 979 | 363.53 | 338.68 | 417.04 | 409.16 | **1,528.42** |
| **OR OREGON Total** | **52,583.12** | **52,137.26** | **52,369.04** | **50,785.07** | **207,874.49** |

**STATE: PA - PENNSYLVANIA**

| REGISTRANT ZIP CODE 3 | 1ST QUARTER | 2ND QUARTER | 3RD QUARTER | 4TH QUARTER | TOTAL GRAMS |
|---|---|---|---|---|---|
| 150 | 7,975.58 | 8,215.69 | 8,061.26 | 8,020.38 | **32,272.91** |
| 151 | 2,845.63 | 2,835.59 | 2,598.45 | 2,725.75 | **11,005.42** |
| 152 | 7,303.29 | 6,879.21 | 7,495.29 | 5,995.12 | **27,672.91** |
| 153 | 1,850.52 | 1,646.75 | 1,707.84 | 1,776.04 | **6,981.16** |
| 154 | 1,920.71 | 1,852.42 | 1,926.71 | 2,084.32 | **7,784.16** |
| 155 | 920.10 | 851.73 | 772.64 | 790.48 | **3,334.95** |
| 156 | 4,322.27 | 3,952.48 | 4,074.15 | 3,691.90 | **16,040.80** |
| 157 | 2,398.85 | 2,063.53 | 1,999.44 | 2,211.21 | **8,673.04** |
| 158 | 1,531.59 | 1,552.83 | 1,569.50 | 1,512.68 | **6,166.59** |
| 159 | 1,957.42 | 1,717.91 | 1,732.22 | 1,646.44 | **7,053.99** |
| 160 | 2,372.87 | 2,534.45 | 2,399.60 | 2,488.29 | **9,795.21** |
| 161 | 2,949.36 | 2,804.27 | 2,548.38 | 2,572.45 | **10,874.47** |
| 162 | 1,123.19 | 1,007.59 | 984.80 | 924.41 | **4,039.99** |
| 163 | 3,419.33 | 3,403.23 | 3,623.38 | 3,155.03 | **13,600.98** |
| 164 | 1,204.89 | 1,208.24 | 1,263.77 | 1,163.46 | **4,840.35** |
| 165 | 3,179.37 | 3,028.30 | 3,174.47 | 3,016.42 | **12,398.55** |

| | | | | | |
|---|---|---|---|---|---|
| 166 | 3,679.88 | 3,418.36 | 3,394.61 | 3,575.48 | **14,068.33** |
| 167 | 1,157.36 | 1,086.37 | 980.48 | 966.62 | **4,190.83** |
| 168 | 909.57 | 839.07 | 748.66 | 777.09 | **3,274.38** |
| 169 | 595.17 | 578.73 | 539.57 | 571.21 | **2,284.69** |
| 170 | 8,853.17 | 8,140.04 | 8,011.59 | 7,792.64 | **32,797.44** |
| 171 | 1,966.83 | 1,802.09 | 1,485.88 | 1,373.57 | **6,628.37** |
| 172 | 2,402.48 | 2,124.33 | 1,965.32 | 1,851.95 | **8,344.08** |
| 173 | 3,294.53 | 3,128.08 | 2,925.96 | 3,016.69 | **12,365.26** |
| 174 | 1,897.31 | 1,969.98 | 1,899.21 | 1,878.21 | **7,644.71** |
| 175 | 2,790.98 | 2,922.73 | 2,831.60 | 2,922.35 | **11,467.67** |
| 176 | 2,762.49 | 2,333.12 | 2,817.96 | 2,536.87 | **10,450.43** |
| 177 | 1,618.29 | 1,485.54 | 1,437.38 | 1,409.18 | **5,950.38** |
| 178 | 2,209.38 | 2,074.47 | 2,049.50 | 1,942.66 | **8,276.00** |
| 179 | 1,045.67 | 930.49 | 941.80 | 953.30 | **3,871.27** |
| 180 | 3,600.09 | 3,308.64 | 3,149.70 | 3,126.59 | **13,185.02** |
| 181 | 2,737.35 | 2,733.14 | 3,167.57 | 2,691.66 | **11,329.72** |
| 182 | 1,324.64 | 1,254.02 | 1,098.69 | 1,062.22 | **4,739.56** |
| 183 | 1,979.16 | 1,918.09 | 1,780.86 | 1,700.71 | **7,378.84** |
| 184 | 2,224.28 | 1,892.66 | 1,891.19 | 1,760.96 | **7,769.09** |
| 185 | 2,341.72 | 2,275.91 | 2,500.21 | 2,197.77 | **9,315.61** |
| 186 | 1,818.60 | 1,772.30 | 1,741.94 | 1,638.64 | **6,971.48** |
| 187 | 5,777.64 | 6,061.51 | 5,748.36 | 5,980.59 | **23,568.10** |
| 188 | 755.52 | 728.37 | 726.68 | 793.35 | **3,003.91** |
| 189 | 3,403.18 | 3,057.56 | 3,036.23 | 2,779.35 | **12,276.32** |
| 190 | 41,235.74 | 37,197.71 | 37,893.46 | 46,274.58 | **162,601.49** |
| 191 | 10,206.65 | 9,413.49 | 10,205.11 | 9,398.53 | **39,223.78** |
| 193 | 4,316.80 | 3,927.15 | 4,104.08 | 3,963.70 | **16,311.72** |
| 194 | 5,912.41 | 5,428.69 | 5,726.35 | 5,355.12 | **22,422.56** |
| 195 | 1,537.97 | 1,463.67 | 1,562.48 | 1,335.33 | **5,899.45** |
| 196 | 1,734.19 | 1,654.42 | 1,747.62 | 1,611.75 | **6,747.97** |
| **PA - PENNSYLVANIA - Total** | **173,364.04** | **162,474.94** | **164,041.94** | **167,013.02** | **666,893.94** |

**STATE: PR - PUERTO RICO**

| REGISTRANT ZIP CODE 3 | 1ST QUARTER | 2ND QUARTER | 3RD QUARTER | 4TH QUARTER | **TOTAL GRAMS** |
|---|---|---|---|---|---|
| 006 | 396.12 | 410.08 | 277.60 | 297.61 | **1,381.41** |
| 007 | 369.91 | 454.41 | 461.63 | 451.40 | **1,737.35** |
| 009 | 1,471.65 | 1,130.16 | 883.65 | 875.06 | **4,360.52** |
| **PR - PUERTO RICO - Total** | **2,237.68** | **1,994.64** | **1,622.88** | **1,624.07** | **7,479.28** |

**STATE: RI - RHODE ISLAND**

| REGISTRANT ZIP CODE 3 | 1ST QUARTER | 2ND QUARTER | 3RD QUARTER | 4TH QUARTER | **TOTAL GRAMS** |
|---|---|---|---|---|---|
| 028 | 5,588.94 | 5,459.29 | 5,382.26 | 5,424.62 | **21,855.11** |

| | | | | | |
|---|---|---|---|---|---|
| 029 | 3,241.12 | 3,087.34 | 2,936.76 | 3,228.52 | **12,493.73** |
| **RI RHODE ISLAND Total** | **8,830.06** | **8,546.62** | **8,319.02** | **8,653.14** | **34,348.84** |

**STATE: SC - SOUTH CAROLINA**

| REGISTRANT ZIP CODE 3 | 1ST QUARTER | 2ND QUARTER | 3RD QUARTER | 4TH QUARTER | TOTAL GRAMS |
|---|---|---|---|---|---|
| 290 | 4,921.46 | 4,860.36 | 4,571.66 | 4,755.98 | **19,109.47** |
| 291 | 4,085.13 | 4,063.59 | 4,133.61 | 3,884.66 | **16,166.99** |
| 292 | 4,187.82 | 4,127.90 | 4,026.71 | 4,165.53 | **16,507.97** |
| 293 | 4,575.73 | 4,775.25 | 4,553.51 | 4,307.24 | **18,211.73** |
| 294 | 10,910.20 | 10,244.75 | 10,284.77 | 9,822.89 | **41,262.62** |
| 295 | 8,204.52 | 8,190.38 | 8,240.86 | 7,812.06 | **32,447.82** |
| 296 | 10,500.49 | 9,913.41 | 9,757.65 | 9,510.92 | **39,682.46** |
| 297 | 2,239.00 | 2,251.74 | 2,322.75 | 2,417.27 | **9,230.76** |
| 298 | 2,236.49 | 2,159.34 | 2,212.05 | 1,852.31 | **8,460.20** |
| 299 | 2,254.76 | 2,272.45 | 2,303.53 | 2,157.78 | **8,988.51** |
| **SC - SOUTH CAROLINA Total** | **54,115.61** | **52,859.19** | **52,407.11** | **50,686.63** | **210,068.54** |

**STATE: SD - SOUTH DAKOTA**

| REGISTRANT ZIP CODE 3 | 1ST QUARTER | 2ND QUARTER | 3RD QUARTER | 4TH QUARTER | TOTAL GRAMS |
|---|---|---|---|---|---|
| 570 | 1,250.19 | 1,330.50 | 1,211.11 | 1,025.22 | **4,817.01** |
| 571 | 2,080.25 | 2,055.56 | 2,226.85 | 1,891.98 | **8,254.64** |
| 572 | 280.21 | 317.37 | 283.30 | 249.06 | **1,129.93** |
| 573 | 317.52 | 373.07 | 441.19 | 391.53 | **1,523.31** |
| 574 | 337.90 | 317.44 | 312.31 | 294.28 | **1,261.93** |
| 575 | 384.59 | 418.15 | 377.10 | 259.06 | **1,438.90** |
| 576 | 55.37 | 52.93 | 58.82 | 45.84 | **212.96** |
| 577 | 1,648.64 | 1,540.06 | 1,526.52 | 1,251.70 | **5,966.92** |
| **SD SOUTH DAKOTA - Total** | **6,354.66** | **6,405.08** | **6,437.19** | **5,408.67** | **24,605.60** |

**STATE: TN - TENNESSEE**

| REGISTRANT ZIP CODE 3 | 1ST QUARTER | 2ND QUARTER | 3RD QUARTER | 4TH QUARTER | TOTAL GRAMS |
|---|---|---|---|---|---|
| 370 | 10,576.08 | 10,233.08 | 9,978.69 | 9,438.34 | **40,226.19** |
| 371 | 10,803.66 | 10,701.68 | 10,663.37 | 10,112.36 | **42,281.07** |
| 372 | 6,620.49 | 6,128.44 | 6,823.19 | 6,100.71 | **25,672.83** |
| 373 | 8,024.89 | 7,903.85 | 7,497.30 | 7,170.84 | **30,596.89** |
| 374 | 2,479.64 | 2,286.01 | 2,249.77 | 2,107.60 | **9,123.02** |
| 376 | 7,023.40 | 7,470.37 | 7,286.52 | 6,977.86 | **28,758.15** |
| 377 | 5,853.64 | 5,916.80 | 5,676.88 | 5,805.82 | **23,253.13** |
| 378 | 9,035.69 | 9,160.07 | 8,692.77 | 8,437.07 | **35,325.60** |
| 379 | 6,393.44 | 6,115.25 | 6,290.65 | 5,825.68 | **24,625.02** |
| 380 | 3,231.26 | 3,151.29 | 3,113.89 | 3,009.41 | **12,505.85** |
| 381 | 35,649.04 | 30,130.38 | 33,652.86 | 45,286.12 | **144,718.41** |
| 382 | 1,720.68 | 1,443.61 | 1,418.56 | 1,379.28 | **5,962.12** |

| | | | | | |
|---|---|---|---|---|---|
| 383 | 4,716.33 | 4,370.40 | 4,080.14 | 3,962.80 | **17,129.67** |
| 384 | 3,379.93 | 3,064.77 | 2,775.55 | 2,607.58 | **11,827.83** |
| 385 | 5,515.77 | 5,347.37 | 4,993.85 | 4,708.81 | **20,565.80** |
| **TN TENNESSEE Total** | **121,023.94** | **113,423.36** | **115,194.01** | **122,930.27** | **472,571.57** |

**STATE: TX - TEXAS**

| REGISTRANT ZIP CODE 3 | 1ST QUARTER | 2ND QUARTER | 3RD QUARTER | 4TH QUARTER | TOTAL GRAMS |
|---|---|---|---|---|---|
| 750 | 12,950.46 | 12,489.34 | 12,083.23 | 12,157.78 | **49,680.81** |
| 751 | 4,644.57 | 4,293.18 | 4,119.27 | 4,022.62 | **17,079.64** |
| 752 | 6,288.71 | 6,247.29 | 6,860.35 | 5,821.45 | **25,217.80** |
| 753 | 173.16 | 151.46 | 130.08 | 213.37 | **668.07** |
| 754 | 3,317.47 | 3,116.59 | 3,362.72 | 2,679.63 | **12,476.41** |
| 755 | 1,078.34 | 907.92 | 994.47 | 983.30 | **3,964.02** |
| 756 | 2,740.37 | 2,959.14 | 2,704.47 | 2,504.84 | **10,908.82** |
| 757 | 3,535.61 | 3,191.71 | 3,106.23 | 2,975.30 | **12,808.85** |
| 758 | 632.83 | 573.54 | 644.16 | 621.86 | **2,472.38** |
| 759 | 1,869.45 | 1,788.51 | 1,794.06 | 1,612.04 | **7,064.06** |
| 760 | 8,853.41 | 8,148.12 | 7,966.87 | 7,820.38 | **32,788.79** |
| 761 | 10,651.93 | 9,628.49 | 9,537.60 | 9,179.17 | **38,997.19** |
| 762 | 3,132.02 | 3,397.86 | 3,028.24 | 3,137.96 | **12,696.09** |
| 763 | 1,924.70 | 1,808.95 | 1,789.45 | 1,721.66 | **7,244.75** |
| 764 | 837.34 | 819.82 | 764.53 | 712.74 | **3,134.43** |
| 765 | 3,329.97 | 2,958.32 | 2,762.36 | 2,713.75 | **11,764.41** |
| 766 | 961.14 | 847.14 | 934.50 | 827.34 | **3,570.12** |
| 767 | 2,169.14 | 2,136.69 | 2,065.16 | 1,913.87 | **8,284.86** |
| 768 | 775.67 | 781.94 | 894.94 | 929.49 | **3,382.03** |
| 769 | 947.58 | 835.64 | 856.93 | 870.68 | **3,510.84** |
| 770 | 10,905.13 | 11,184.87 | 11,176.60 | 11,670.78 | **44,937.38** |
| 773 | 8,831.79 | 7,795.02 | 7,664.78 | 7,064.35 | **31,355.93** |
| 774 | 4,917.25 | 4,828.34 | 4,643.01 | 4,436.20 | **18,824.79** |
| 775 | 9,092.10 | 8,862.21 | 8,089.82 | 8,021.25 | **34,065.38** |
| 776 | 2,113.03 | 1,817.02 | 1,828.66 | 1,859.35 | **7,618.06** |
| 777 | 1,634.53 | 1,870.07 | 2,183.79 | 1,826.02 | **7,514.41** |
| 778 | 1,585.75 | 1,560.00 | 1,723.83 | 1,677.66 | **6,547.24** |
| 779 | 1,804.88 | 1,831.65 | 1,835.11 | 1,712.08 | **7,183.72** |
| 780 | 2,262.85 | 2,307.81 | 2,396.47 | 2,146.91 | **9,114.04** |
| 781 | 2,389.33 | 2,096.85 | 2,179.98 | 2,203.81 | **8,869.97** |
| 782 | 11,410.53 | 10,603.01 | 10,716.70 | 10,352.09 | **43,082.34** |
| 783 | 1,312.10 | 1,162.27 | 1,208.95 | 1,303.91 | **4,987.24** |
| 784 | 2,400.56 | 2,266.50 | 2,264.87 | 2,135.38 | **9,067.31** |
| 785 | 2,384.79 | 2,115.31 | 2,135.54 | 1,935.81 | **8,571.44** |

| | | | | | |
|---|---|---|---|---|---|
| 786 | 6,444.92 | 5,743.83 | 5,938.39 | 5,786.70 | **23,913.84** |
| 787 | 5,254.24 | 5,037.38 | 4,929.19 | 4,475.27 | **19,696.07** |
| 788 | 367.76 | 318.74 | 345.45 | 307.01 | **1,338.95** |
| 789 | 386.11 | 360.35 | 372.09 | 342.25 | **1,460.79** |
| 790 | 1,165.90 | 1,165.42 | 1,293.16 | 1,306.49 | **4,930.98** |
| 791 | 3,145.85 | 3,543.94 | 3,086.30 | 2,608.87 | **12,384.95** |
| 792 | 243.04 | 264.67 | 273.73 | 290.01 | **1,071.44** |
| 793 | 577.49 | 610.31 | 635.64 | 564.04 | **2,387.47** |
| 794 | 3,198.00 | 2,988.10 | 2,951.49 | 2,799.29 | **11,936.88** |
| 795 | 419.05 | 467.19 | 454.98 | 356.70 | **1,697.92** |
| 796 | 1,264.94 | 1,192.66 | 1,359.42 | 1,121.76 | **4,938.78** |
| 797 | 2,156.79 | 2,002.88 | 1,707.91 | 1,863.26 | **7,730.84** |
| 798 | 73.45 | 61.69 | 78.85 | 72.39 | **286.38** |
| 799 | 2,919.18 | 2,453.06 | 2,684.78 | 2,754.02 | **10,811.04** |
| **TX - TEXAS Total** | **161,475.20** | **153,592.78** | **152,559.09** | **146,412.89** | **614,039.95** |

**STATE: UT - UTAH**

| REGISTRANT ZIP CODE 3 | 1ST QUARTER | 2ND QUARTER | 3RD QUARTER | 4TH QUARTER | **TOTAL GRAMS** |
|---|---|---|---|---|---|
| 840 | 14,997.04 | 14,681.11 | 14,455.06 | 14,421.58 | **58,554.79** |
| 841 | 10,123.10 | 9,841.11 | 10,051.36 | 9,557.57 | **39,573.14** |
| 843 | 1,463.66 | 1,544.76 | 1,615.35 | 1,473.00 | **6,096.77** |
| 844 | 3,354.66 | 3,488.19 | 3,654.90 | 3,419.15 | **13,916.90** |
| 845 | 1,156.06 | 1,068.86 | 994.67 | 966.56 | **4,186.15** |
| 846 | 2,702.08 | 2,514.78 | 2,552.35 | 2,707.60 | **10,476.82** |
| 847 | 4,759.40 | 4,439.19 | 4,481.78 | 4,367.97 | **18,048.34** |
| **UT - UTAH - Total** | **38,556.00** | **37,578.00** | **37,805.48** | **36,913.42** | **150,852.91** |

**STATE: VA - VIRGINIA**

| REGISTRANT ZIP CODE 3 | 1ST QUARTER | 2ND QUARTER | 3RD QUARTER | 4TH QUARTER | **TOTAL GRAMS** |
|---|---|---|---|---|---|
| 201 | 5,870.64 | 5,155.64 | 5,773.76 | 5,270.11 | **22,070.16** |
| 220 | 2,035.16 | 1,892.85 | 1,985.53 | 1,903.23 | **7,816.77** |
| 221 | 2,315.33 | 2,663.42 | 2,543.38 | 2,319.46 | **9,841.58** |
| 222 | 468.73 | 453.58 | 396.34 | 405.69 | **1,724.34** |
| 223 | 1,145.82 | 1,164.96 | 1,048.12 | 1,012.63 | **4,371.53** |
| 224 | 2,156.97 | 2,067.63 | 2,054.70 | 2,377.06 | **8,656.36** |
| 225 | 676.00 | 704.56 | 752.60 | 744.54 | **2,877.70** |
| 226 | 2,206.38 | 2,033.98 | 2,159.67 | 2,139.60 | **8,539.64** |
| 227 | 483.05 | 485.25 | 505.58 | 359.75 | **1,833.63** |
| 228 | 1,739.40 | 1,769.31 | 1,752.84 | 1,837.40 | **7,098.95** |
| 229 | 1,990.60 | 2,010.46 | 1,910.73 | 2,097.54 | **8,009.33** |
| 230 | 1,833.90 | 1,632.02 | 1,725.67 | 1,501.22 | **6,692.80** |
| 231 | 2,729.24 | 2,733.32 | 3,025.83 | 2,905.76 | **11,394.14** |

| | | | | | |
|---|---|---|---|---|---|
| 232 | 5,706.79 | 4,778.01 | 5,246.81 | 4,937.39 | **20,669.00** |
| 233 | 2,190.29 | 1,889.05 | 2,261.95 | 2,140.93 | **8,482.22** |
| 234 | 4,567.38 | 4,773.30 | 4,452.44 | 4,459.05 | **18,252.17** |
| 235 | 1,613.73 | 1,631.03 | 1,678.45 | 1,444.66 | **6,367.87** |
| 236 | 2,518.67 | 2,681.95 | 3,119.43 | 2,604.19 | **10,924.25** |
| 237 | 1,152.53 | 1,111.55 | 1,176.96 | 1,052.68 | **4,493.71** |
| 238 | 2,699.51 | 2,812.95 | 2,747.53 | 2,566.51 | **10,826.49** |
| 239 | 663.38 | 619.77 | 574.27 | 546.14 | **2,403.56** |
| 240 | 3,768.32 | 3,634.31 | 3,762.67 | 3,463.19 | **14,628.49** |
| 241 | 3,608.17 | 3,402.89 | 3,504.14 | 3,279.11 | **13,794.31** |
| 242 | 3,367.64 | 3,392.15 | 3,569.33 | 3,077.81 | **13,406.93** |
| 243 | 2,021.00 | 1,840.45 | 1,877.47 | 1,484.21 | **7,223.14** |
| 244 | 1,041.83 | 1,045.53 | 1,042.11 | 1,068.09 | **4,197.57** |
| 245 | 3,342.85 | 2,877.37 | 2,874.36 | 2,632.38 | **11,726.96** |
| 246 | 737.68 | 648.67 | 732.58 | 674.92 | **2,793.85** |
| **VA VIRGINIA Total** | **64,650.97** | **61,905.95** | **64,255.25** | **60,305.26** | **251,117.44** |

**STATE: VI - VIRGIN ISLANDS**

| REGISTRANT ZIP CODE 3 | 1ST QUARTER | 2ND QUARTER | 3RD QUARTER | 4TH QUARTER | TOTAL GRAMS |
|---|---|---|---|---|---|
| 008 | 105.70 | 80.54 | 73.02 | 66.97 | **326.22** |
| **VI - VIRGIN ISLANDS Total** | **105.70** | **80.54** | **73.02** | **66.97** | **326.22** |

**STATE: VT - VERMONT**

| REGISTRANT ZIP CODE 3 | 1ST QUARTER | 2ND QUARTER | 3RD QUARTER | 4TH QUARTER | TOTAL GRAMS |
|---|---|---|---|---|---|
| 050 | 864.17 | 1,038.45 | 718.33 | 685.67 | **3,306.62** |
| 051 | 475.81 | 391.74 | 439.58 | 385.31 | **1,692.43** |
| 052 | 379.60 | 335.68 | 345.55 | 397.73 | **1,458.57** |
| 053 | 555.28 | 463.03 | 500.29 | 480.60 | **1,999.20** |
| 054 | 3,007.40 | 3,161.78 | 3,093.50 | 3,241.11 | **12,503.78** |
| 056 | 546.49 | 589.80 | 684.07 | 588.05 | **2,408.41** |
| 057 | 1,296.42 | 1,249.14 | 1,402.24 | 1,196.81 | **5,144.61** |
| 058 | 685.05 | 676.92 | 781.23 | 674.74 | **2,817.94** |
| **VT VERMONT Total** | **7,810.23** | **7,906.55** | **7,964.77** | **7,650.01** | **31,331.55** |

**STATE: WA - WASHINGTON**

| REGISTRANT ZIP CODE 3 | 1ST QUARTER | 2ND QUARTER | 3RD QUARTER | 4TH QUARTER | TOTAL GRAMS |
|---|---|---|---|---|---|
| 980 | 11,267.42 | 11,323.57 | 11,122.54 | 11,046.83 | **44,760.36** |
| 981 | 7,780.72 | 7,754.76 | 7,759.92 | 7,393.79 | **30,689.19** |
| 982 | 9,542.93 | 9,366.16 | 9,222.54 | 9,349.15 | **37,480.78** |
| 983 | 9,925.09 | 9,680.87 | 9,818.83 | 9,218.68 | **38,643.47** |
| 984 | 5,089.67 | 4,759.26 | 4,943.32 | 4,858.04 | **19,650.28** |
| 985 | 5,433.26 | 5,241.07 | 5,249.10 | 5,319.92 | **21,243.36** |
| 986 | 11,058.56 | 10,848.08 | 10,248.05 | 10,246.53 | **42,401.21** |

| | | | | | |
|---|---|---|---|---|---|
| 988 | 2,495.14 | 2,392.42 | 2,498.44 | 2,350.26 | **9,736.26** |
| 989 | 2,557.05 | 2,533.03 | 2,533.89 | 2,468.11 | **10,092.07** |
| 990 | 1,443.20 | 1,334.07 | 1,368.07 | 1,303.46 | **5,448.80** |
| 991 | 1,879.05 | 1,605.35 | 1,713.23 | 1,649.28 | **6,846.90** |
| 992 | 7,761.64 | 7,649.65 | 8,169.45 | 7,479.43 | **31,060.17** |
| 993 | 4,018.13 | 3,968.05 | 3,711.06 | 3,642.32 | **15,339.57** |
| 994 | 985.98 | 1,013.51 | 948.06 | 1,120.43 | **4,067.99** |
| **WA WASHINGTON Total** | **81,237.84** | **79,469.85** | **79,306.50** | **77,446.24** | **317,460.42** |

**STATE: WI - WISCONSIN**

| REGISTRANT ZIP CODE 3 | 1ST QUARTER | 2ND QUARTER | 3RD QUARTER | 4TH QUARTER | TOTAL GRAMS |
|---|---|---|---|---|---|
| 530 | 6,571.84 | 6,407.81 | 6,279.03 | 5,922.98 | **25,181.66** |
| 531 | 6,045.80 | 5,835.52 | 5,898.70 | 5,719.60 | **23,499.62** |
| 532 | 8,751.38 | 7,924.71 | 7,884.52 | 7,778.36 | **32,338.97** |
| 534 | 1,276.45 | 1,188.80 | 1,181.73 | 1,253.06 | **4,900.04** |
| 535 | 5,998.55 | 5,789.36 | 5,695.98 | 5,610.10 | **23,093.99** |
| 537 | 4,782.86 | 4,462.84 | 4,343.84 | 4,498.72 | **18,088.26** |
| 538 | 562.26 | 630.83 | 546.44 | 570.08 | **2,309.61** |
| 539 | 2,875.03 | 2,826.92 | 2,808.72 | 2,725.82 | **11,236.49** |
| 540 | 716.23 | 732.96 | 839.28 | 748.26 | **3,036.73** |
| 541 | 1,189.01 | 1,224.18 | 1,131.29 | 1,068.17 | **4,612.66** |
| 542 | 896.56 | 1,062.39 | 1,041.40 | 861.76 | **3,862.11** |
| 543 | 1,581.17 | 1,661.87 | 1,817.10 | 1,570.32 | **6,630.46** |
| 544 | 2,926.63 | 2,792.46 | 2,927.17 | 2,740.88 | **11,387.14** |
| 545 | 832.88 | 832.88 | 923.28 | 792.02 | **3,381.06** |
| 546 | 2,662.20 | 2,710.87 | 2,642.59 | 2,542.75 | **10,558.41** |
| 547 | 2,200.30 | 2,177.67 | 2,313.77 | 2,100.73 | **8,792.47** |
| 548 | 1,778.57 | 1,798.66 | 1,768.85 | 1,743.01 | **7,089.09** |
| 549 | 4,577.93 | 4,714.55 | 4,959.16 | 4,364.88 | **18,616.51** |
| **WI - WISCONSIN Total** | **56,225.67** | **54,775.27** | **55,002.84** | **52,611.50** | **218,615.28** |

**STATE: WV - WEST VIRGINIA**

| REGISTRANT ZIP CODE 3 | 1ST QUARTER | 2ND QUARTER | 3RD QUARTER | 4TH QUARTER | TOTAL GRAMS |
|---|---|---|---|---|---|
| 247 | 592.01 | 607.84 | 580.20 | 577.02 | **2,357.08** |
| 248 | 278.30 | 262.54 | 257.78 | 267.60 | **1,066.22** |
| 249 | 282.66 | 295.42 | 254.61 | 261.57 | **1,094.26** |
| 250 | 435.56 | 473.38 | 477.07 | 524.90 | **1,910.91** |
| 251 | 236.31 | 213.04 | 209.05 | 223.39 | **881.79** |
| 252 | 353.31 | 346.07 | 311.63 | 271.11 | **1,282.11** |
| 253 | 1,015.84 | 854.93 | 974.94 | 880.06 | **3,725.77** |
| 254 | 2,683.71 | 2,353.53 | 2,606.48 | 2,305.42 | **9,949.14** |
| 255 | 795.05 | 798.58 | 958.97 | 844.76 | **3,397.35** |

| | | | | | |
|---|---|---|---|---|---|
| 256 | 347.23 | 250.58 | 326.76 | 224.31 | **1,148.88** |
| 257 | 958.26 | 742.48 | 858.14 | 747.04 | **3,305.92** |
| 258 | 858.77 | 703.00 | 983.37 | 666.07 | **3,211.22** |
| 259 | 210.11 | 262.82 | 221.39 | 183.11 | **877.43** |
| 260 | 1,550.29 | 1,475.55 | 1,643.91 | 1,679.13 | **6,348.88** |
| 261 | 861.34 | 775.31 | 779.70 | 665.64 | **3,081.99** |
| 262 | 578.51 | 555.32 | 597.17 | 535.32 | **2,266.32** |
| 263 | 1,387.17 | 1,302.56 | 1,169.23 | 1,291.37 | **5,150.33** |
| 264 | 314.84 | 377.05 | 340.34 | 365.58 | **1,397.81** |
| 265 | 1,365.63 | 1,132.43 | 1,322.91 | 1,285.45 | **5,106.42** |
| 266 | 300.92 | 278.87 | 289.07 | 281.13 | **1,149.99** |
| 267 | 439.65 | 535.27 | 505.34 | 452.09 | **1,932.36** |
| 268 | 317.85 | 305.89 | 357.37 | 277.89 | **1,259.00** |
| **WV WEST VIRGINIA Total** | **16,163.33** | **14,902.48** | **16,025.42** | **14,809.95** | **61,901.17** |

**STATE: WY - WYOMING**

| REGISTRANT ZIP CODE 3 | 1ST QUARTER | 2ND QUARTER | 3RD QUARTER | 4TH QUARTER | TOTAL GRAMS |
|---|---|---|---|---|---|
| 820 | 1,166.93 | 1,090.61 | 1,013.19 | 1,017.68 | **4,288.42** |
| 822 | 227.86 | 204.16 | 240.85 | 171.52 | **844.38** |
| 823 | 127.74 | 108.76 | 115.44 | 115.96 | **467.90** |
| 824 | 739.12 | 649.39 | 733.28 | 638.85 | **2,760.64** |
| 825 | 536.61 | 727.20 | 566.59 | 484.75 | **2,315.14** |
| 826 | 1,060.69 | 1,061.84 | 1,081.78 | 1,083.60 | **4,287.91** |
| 827 | 357.27 | 321.45 | 400.70 | 366.44 | **1,445.87** |
| 828 | 1,066.21 | 553.14 | 750.85 | 619.48 | **2,989.68** |
| 829 | 1,425.62 | 1,643.43 | 1,657.99 | 1,445.24 | **6,172.27** |
| 830 | 62.42 | 56.29 | 55.38 | 67.68 | **241.77** |
| 831 | 242.70 | 268.46 | 261.37 | 220.10 | **992.62** |
| **WY - WYOMING Total** | **7,013.17** | **6,684.72** | **6,877.41** | **6,231.29** | **26,806.60** |

**DRUG: 9652 - OXYMORPHONE**

**STATE: AK - ALASKA**

| REGISTRANT ZIP CODE 3 | 1ST QUARTER | 2ND QUARTER | 3RD QUARTER | 4TH QUARTER | TOTAL GRAMS |
|---|---|---|---|---|---|
| 995 | 27.74 | 19.36 | 12.85 | 12.58 | **72.53** |
| 996 | 25.87 | 24.98 | 34.79 | 44.61 | **130.25** |
| 997 | 8.39 | 7.14 | 4.10 | 2.14 | **21.77** |
| **AK ALASKA - Total** | **62.00** | **51.47** | **51.74** | **59.32** | **224.54** |

**STATE: AL - ALABAMA**

| REGISTRANT ZIP CODE 3 | 1ST QUARTER | 2ND QUARTER | 3RD QUARTER | 4TH QUARTER | TOTAL GRAMS |
|---|---|---|---|---|---|
| 350 | 93.67 | 108.35 | 81.81 | 84.70 | **368.53** |
| 351 | 106.03 | 107.50 | 52.59 | 44.07 | **310.18** |
| 352 | 66.82 | 39.25 | 45.99 | 45.45 | **197.51** |

| | | | | | |
|---|---|---|---|---|---|
| 354 | 57.59 | 61.06 | 59.28 | 55.98 | **233.91** |
| 355 | 61.42 | 60.22 | 58.48 | 49.02 | **229.14** |
| 356 | 112.49 | 90.95 | 87.02 | 87.92 | **378.38** |
| 357 | 76.45 | 70.57 | 46.75 | 52.81 | **246.58** |
| 358 | 113.03 | 102.86 | 85.55 | 83.86 | **385.30** |
| 359 | 314.64 | 261.47 | 234.22 | 205.58 | **1,015.92** |
| 360 | 22.75 | 22.93 | 23.82 | 23.82 | **93.31** |
| 361 | 20.25 | 20.70 | 15.88 | 13.56 | **70.39** |
| 362 | 73.87 | 68.33 | 61.55 | 52.41 | **256.17** |
| 363 | 58.88 | 57.09 | 59.15 | 51.56 | **226.68** |
| 364 | 16.77 | 20.70 | 14.10 | 9.81 | **61.38** |
| 365 | 104.42 | 98.04 | 96.93 | 84.48 | **383.87** |
| 366 | 111.16 | 109.10 | 121.50 | 95.72 | **437.49** |
| 367 | 4.15 | 7.49 | 3.12 | 4.10 | **18.87** |
| 368 | 143.58 | 143.09 | 135.33 | 135.60 | **557.61** |
| 369 | 9.10 | 11.24 | 3.75 | 0 | **24.09** |
| **AL ALABAMA - Total** | **1,567.06** | **1,460.95** | **1,286.81** | **1,180.47** | **5,495.29** |

**STATE: AR - ARKANSAS**

| REGISTRANT ZIP CODE 3 | 1ST QUARTER | 2ND QUARTER | 3RD QUARTER | 4TH QUARTER | **TOTAL GRAMS** |
|---|---|---|---|---|---|
| 716 | 5.53 | 5.89 | 3.57 | 4.46 | **19.45** |
| 717 | 2.68 | 8.03 | 4.01 | 8.74 | **23.46** |
| 719 | 12.76 | 8.65 | 4.10 | 7.40 | **32.92** |
| 720 | 55.44 | 45.99 | 35.55 | 40.59 | **177.57** |
| 721 | 21.05 | 20.47 | 8.43 | 10.93 | **60.89** |
| 722 | 25.25 | 23.95 | 26.90 | 20.96 | **97.06** |
| 723 | 10.08 | 4.28 | 5.35 | 0 | **19.72** |
| 724 | 4.28 | 6.42 | 4.82 | 2.14 | **17.66** |
| 725 | 8.65 | 3.75 | 2.41 | 5.98 | **20.79** |
| 726 | 1.16 | 1.34 | 0.45 | 2.14 | **5.08** |
| 727 | 105.27 | 66.91 | 43.09 | 35.42 | **250.68** |
| 729 | 38.54 | 29.17 | 20.07 | 24.62 | **112.40** |
| **AR ARKANSAS - Total** | **290.69** | **224.85** | **158.75** | **163.39** | **837.68** |

**STATE: AZ - ARIZONA**

| REGISTRANT ZIP CODE 3 | 1ST QUARTER | 2ND QUARTER | 3RD QUARTER | 4TH QUARTER | **TOTAL GRAMS** |
|---|---|---|---|---|---|
| 850 | 292.16 | 245.82 | 222.94 | 219.68 | **980.60** |
| 851 | 40.06 | 20.34 | 35.28 | 28.64 | **124.31** |
| 852 | 530.09 | 522.86 | 478.52 | 510.46 | **2,041.93** |
| 853 | 290.11 | 269.59 | 224.10 | 271.82 | **1,055.62** |
| 855 | 34.44 | 20.52 | 22.48 | 17.31 | **94.74** |
| 856 | 51.03 | 25.51 | 19.72 | 26.50 | **122.75** |

| | | | | | |
|---|---|---|---|---|---|
| 857 | 181.59 | 155.49 | 117.49 | 136.49 | **591.06** |
| 859 | 13.38 | 8.47 | 13.29 | 5.98 | **41.13** |
| 860 | 19.18 | 17.22 | 14.72 | 6.69 | **57.81** |
| 863 | 47.99 | 26.67 | 30.24 | 33.99 | **138.90** |
| 864 | 21.68 | 32.03 | 16.50 | 22.84 | **93.05** |
| **AZ ARIZONA - Total** | **1,521.70** | **1,344.53** | **1,195.28** | **1,280.39** | **5,341.89** |

**STATE: CA - CALIFORNIA**

| REGISTRANT ZIP CODE 3 | 1ST QUARTER | 2ND QUARTER | 3RD QUARTER | 4TH QUARTER | **TOTAL GRAMS** |
|---|---|---|---|---|---|
| 900 | 237.12 | 239.97 | 229.89 | 214.82 | **921.81** |
| 902 | 136.13 | 136.27 | 117.94 | 120.52 | **510.86** |
| 903 | 9.81 | 3.57 | 10.71 | 0 | **24.09** |
| 904 | 26.58 | 40.68 | 50.23 | 36.49 | **153.98** |
| 905 | 12.22 | 8.74 | 8.56 | 6.69 | **36.22** |
| 906 | 28.37 | 61.02 | 16.24 | 24.09 | **129.71** |
| 907 | 22.21 | 13.47 | 16.77 | 20.88 | **73.33** |
| 908 | 41.04 | 47.46 | 47.19 | 24.71 | **160.40** |
| 910 | 6.33 | 17.84 | 16.24 | 16.77 | **57.18** |
| 911 | 72.35 | 89.75 | 73.78 | 63.52 | **299.39** |
| 912 | 22.12 | 21.14 | 26.76 | 20.07 | **90.10** |
| 913 | 138.36 | 148.62 | 99.74 | 108.21 | **494.94** |
| 914 | 156.74 | 173.25 | 149.87 | 114.72 | **594.58** |
| 915 | 3.75 | 4.64 | 4.64 | 22.39 | **35.42** |
| 916 | 11.78 | 5.35 | 9.63 | 34.70 | **61.47** |
| 917 | 133.10 | 115.17 | 95.63 | 87.96 | **431.87** |
| 919 | 56.47 | 44.43 | 50.67 | 46.48 | **198.05** |
| 920 | 401.89 | 496.10 | 425.00 | 474.86 | **1,797.85** |
| 921 | 129.80 | 124.27 | 95.68 | 86.36 | **436.10** |
| 922 | 98.67 | 80.20 | 89.83 | 77.17 | **345.87** |
| 923 | 78.50 | 71.72 | 64.86 | 97.60 | **312.68** |
| 924 | 27.66 | 17.49 | 11.06 | 11.95 | **68.16** |
| 925 | 77.12 | 77.92 | 67.00 | 71.64 | **293.68** |
| 926 | 267.54 | 230.70 | 180.38 | 188.59 | **867.21** |
| 927 | 27.12 | 19.54 | 22.12 | 22.48 | **91.26** |
| 928 | 173.78 | 170.44 | 124.98 | 139.26 | **608.46** |
| 930 | 85.20 | 66.10 | 62.09 | 53.88 | **267.27** |
| 931 | 0.45 | 0 | 1.78 | 0.45 | **2.68** |
| 932 | 36.84 | 69.49 | 30.87 | 42.64 | **179.85** |
| 933 | 9.81 | 9.81 | 2.23 | 9.99 | **31.85** |
| 934 | 103.57 | 96.08 | 79.04 | 75.83 | **354.52** |
| 935 | 74.31 | 117.40 | 56.11 | 14.36 | **262.19** |

| | | | | | |
|---|---|---|---|---|---|
| 936 | 150.59 | 156.21 | 100.99 | 65.12 | **472.90** |
| 937 | 259.33 | 277.71 | 227.66 | 155.14 | **919.84** |
| 939 | 1.25 | 3.39 | 7.05 | 3.30 | **14.99** |
| 940 | 208.62 | 244.88 | 207.50 | 160.93 | **821.94** |
| 941 | 35.06 | 31.94 | 23.73 | 24.27 | **114.99** |
| 943 | 7.05 | 17.31 | 16.06 | 14.36 | **54.77** |
| 944 | 15.34 | 9.90 | 4.01 | 1.78 | **31.05** |
| 945 | 245.77 | 190.51 | 196.53 | 187.25 | **820.06** |
| 946 | 1.12 | 1.61 | 1.34 | 1.78 | **5.84** |
| 947 | 4.01 | 0.45 | 0 | 0 | **4.46** |
| 948 | 0 | 0 | 0 | 3.57 | **3.57** |
| 949 | 37.91 | 61.11 | 53.79 | 42.46 | **195.28** |
| 950 | 59.15 | 72.35 | 59.50 | 34.30 | **225.30** |
| 951 | 23.28 | 18.87 | 20.65 | 13.52 | **76.32** |
| 952 | 16.06 | 17.57 | 37.65 | 27.48 | **98.76** |
| 953 | 96.75 | 88.23 | 74.49 | 71.19 | **330.66** |
| 954 | 65.08 | 87.20 | 57.18 | 46.52 | **255.99** |
| 955 | 32.38 | 29.08 | 22.21 | 17.22 | **100.90** |
| 956 | 90.37 | 77.79 | 70.03 | 73.06 | **311.25** |
| 957 | 11.60 | 20.16 | 10.79 | 17.57 | **60.13** |
| 958 | 37.91 | 25.69 | 16.86 | 30.06 | **110.53** |
| 959 | 75.92 | 81.27 | 60.93 | 38.00 | **256.12** |
| 960 | 10.71 | 12.49 | 8.21 | 5.26 | **36.67** |
| 961 | 0.54 | 1.61 | 1.07 | 1.34 | **4.55** |
| **CA - CALIFORNIA - Total** | **4,192.56** | **4,345.95** | **3,615.77** | **3,365.58** | **15,519.86** |

**STATE: CO - COLORADO**

| REGISTRANT ZIP CODE 3 | 1ST QUARTER | 2ND QUARTER | 3RD QUARTER | 4TH QUARTER | TOTAL GRAMS |
|---|---|---|---|---|---|
| 800 | 158.79 | 175.70 | 164.19 | 182.43 | **681.12** |
| 801 | 193.32 | 208.62 | 213.12 | 163.39 | **778.45** |
| 802 | 203.76 | 213.88 | 143.27 | 172.76 | **733.66** |
| 803 | 25.34 | 15.43 | 18.82 | 15.79 | **75.38** |
| 804 | 15.08 | 10.48 | 19.09 | 8.74 | **53.39** |
| 805 | 96.53 | 109.55 | 91.80 | 95.68 | **393.55** |
| 806 | 65.03 | 65.75 | 59.68 | 68.20 | **258.66** |
| 807 | 14.81 | 10.44 | 7.72 | 8.30 | **41.26** |
| 808 | 20.88 | 17.31 | 17.84 | 14.63 | **70.65** |
| 809 | 125.56 | 121.95 | 110.44 | 94.74 | **452.70** |
| 810 | 38.54 | 48.44 | 57.45 | 56.83 | **201.26** |
| 811 | 12.85 | 4.55 | 5.35 | 7.14 | **29.89** |
| 812 | 9.99 | 12.13 | 13.92 | 8.30 | **44.34** |

| | | | | | |
|---|---|---|---|---|---|
| 813 | 2.23 | 0.89 | 2.68 | 1.78 | **7.58** |
| 814 | 1.34 | 1.34 | 0 | 0 | **2.68** |
| 815 | 6.07 | 1.61 | 0 | 2.50 | **10.17** |
| 816 | 4.01 | 3.75 | 3.12 | 0.45 | **11.33** |
| **CO - COLORADO - Total** | **994.11** | **1,021.81** | **928.50** | **901.65** | **3,846.07** |

**STATE: CT - CONNECTICUT**

| REGISTRANT ZIP CODE 3 | 1ST QUARTER | 2ND QUARTER | 3RD QUARTER | 4TH QUARTER | **TOTAL GRAMS** |
|---|---|---|---|---|---|
| 060 | 178.96 | 154.73 | 128.24 | 117.36 | **579.29** |
| 061 | 27.83 | 24.62 | 5.53 | 10.88 | **68.87** |
| 062 | 24.98 | 35.06 | 18.02 | 14.27 | **92.33** |
| 063 | 49.78 | 56.56 | 27.48 | 33.01 | **166.82** |
| 064 | 427.94 | 411.21 | 366.56 | 397.07 | **1,602.79** |
| 065 | 137.25 | 143.14 | 144.21 | 111.20 | **535.80** |
| 066 | 91.89 | 80.38 | 81.36 | 96.26 | **349.88** |
| 067 | 52.19 | 48.22 | 37.20 | 40.50 | **178.11** |
| 068 | 98.40 | 82.16 | 91.08 | 88.85 | **360.50** |
| 069 | 30.60 | 27.66 | 35.15 | 34.84 | **128.24** |
| **CT CONNECTICUT Total** | **1,119.81** | **1,063.74** | **934.83** | **944.24** | **4,062.62** |

**STATE: DC - DISTRICT OF COLUMBIA**

| REGISTRANT ZIP CODE 3 | 1ST QUARTER | 2ND QUARTER | 3RD QUARTER | 4TH QUARTER | **TOTAL GRAMS** |
|---|---|---|---|---|---|
| 200 | 56.78 | 62.27 | 42.82 | 31.89 | **193.76** |
| **DC - DISTRICT OF COLUMBIA** | **56.78** | **62.27** | **42.82** | **31.89** | **193.76** |

**STATE: DE - DELAWARE**

| REGISTRANT ZIP CODE 3 | 1ST QUARTER | 2ND QUARTER | 3RD QUARTER | 4TH QUARTER | **TOTAL GRAMS** |
|---|---|---|---|---|---|
| 197 | 436.55 | 480.49 | 427.85 | 364.38 | **1,709.26** |
| 198 | 278.07 | 332.04 | 183.24 | 197.60 | **990.94** |
| 199 | 475.44 | 461.88 | 393.86 | 381.42 | **1,712.61** |
| **DE DELAWARE Total** | **1,190.06** | **1,274.41** | **1,004.95** | **943.40** | **4,412.82** |

**STATE: FL - FLORIDA**

| REGISTRANT ZIP CODE 3 | 1ST QUARTER | 2ND QUARTER | 3RD QUARTER | 4TH QUARTER | **TOTAL GRAMS** |
|---|---|---|---|---|---|
| 320 | 300.19 | 319.28 | 288.82 | 183.64 | **1,091.93** |
| 321 | 130.02 | 121.59 | 105.71 | 85.33 | **442.66** |
| 322 | 535.71 | 571.79 | 513.49 | 436.91 | **2,057.90** |
| 323 | 246.98 | 250.23 | 251.08 | 208.75 | **957.04** |
| 324 | 207.37 | 193.76 | 186.98 | 133.19 | **721.31** |
| 325 | 65.03 | 62.80 | 65.39 | 53.79 | **247.02** |
| 326 | 199.47 | 193.27 | 199.38 | 172.53 | **764.66** |
| 327 | 404.83 | 389.40 | 318.35 | 282.84 | **1,395.42** |
| 328 | 220.21 | 225.79 | 189.35 | 192.60 | **827.96** |
| 329 | 549.85 | 551.09 | 507.43 | 395.24 | **2,003.61** |

| | | | | | |
|---|---|---|---|---|---|
| 330 | 441.01 | 520.50 | 382.31 | 311.34 | **1,655.16** |
| 331 | 129.27 | 103.75 | 85.20 | 79.58 | **397.79** |
| 333 | 248.67 | 292.30 | 272.63 | 228.11 | **1,041.71** |
| 334 | 778.98 | 807.71 | 690.49 | 540.84 | **2,818.01** |
| 335 | 314.69 | 295.29 | 262.81 | 244.79 | **1,117.58** |
| 336 | 345.69 | 358.13 | 320.49 | 303.00 | **1,327.31** |
| 337 | 383.60 | 390.20 | 417.32 | 388.87 | **1,580.00** |
| 338 | 164.59 | 158.57 | 154.33 | 85.64 | **563.14** |
| 339 | 217.00 | 190.42 | 178.60 | 123.87 | **709.89** |
| 341 | 89.03 | 79.22 | 77.26 | 88.32 | **333.82** |
| 342 | 598.55 | 617.51 | 506.80 | 427.67 | **2,150.54** |
| 344 | 133.46 | 103.39 | 94.03 | 68.16 | **399.04** |
| 346 | 741.51 | 905.57 | 834.56 | 831.44 | **3,313.08** |
| 347 | 185.51 | 190.91 | 174.99 | 149.03 | **700.43** |
| 349 | 537.00 | 602.61 | 489.14 | 445.25 | **2,074.00** |
| **FL FLORIDA - Total** | **8,168.25** | **8,495.11** | **7,566.93** | **6,460.72** | **30,691.01** |

**STATE: GA - GEORGIA**

| REGISTRANT ZIP CODE 3 | 1ST QUARTER | 2ND QUARTER | 3RD QUARTER | 4TH QUARTER | **TOTAL GRAMS** |
|---|---|---|---|---|---|
| 300 | 578.04 | 668.76 | 560.77 | 461.35 | **2,268.92** |
| 301 | 401.76 | 437.98 | 393.15 | 378.43 | **1,611.31** |
| 302 | 168.43 | 194.61 | 162.32 | 182.84 | **708.19** |
| 303 | 85.91 | 136.67 | 108.39 | 112.67 | **443.64** |
| 304 | 43.27 | 47.91 | 22.39 | 26.23 | **139.79** |
| 305 | 187.97 | 161.92 | 138.54 | 136.45 | **624.87** |
| 306 | 78.33 | 79.66 | 74.58 | 57.27 | **289.84** |
| 307 | 64.45 | 49.24 | 76.94 | 51.79 | **242.43** |
| 308 | 28.01 | 33.19 | 30.69 | 21.95 | **113.83** |
| 309 | 30.33 | 25.42 | 22.75 | 11.78 | **90.28** |
| 310 | 78.86 | 82.25 | 89.75 | 61.15 | **312.01** |
| 312 | 47.01 | 41.57 | 32.03 | 35.33 | **155.94** |
| 313 | 72.26 | 72.80 | 58.16 | 68.69 | **271.91** |
| 314 | 11.42 | 21.95 | 25.16 | 31.85 | **90.37** |
| 315 | 81.18 | 93.14 | 70.65 | 58.34 | **303.31** |
| 316 | 120.08 | 105.27 | 59.06 | 72.62 | **357.02** |
| 317 | 78.64 | 90.64 | 96.48 | 70.34 | **336.10** |
| 318 | 1.78 | 0 | 0 | 0 | **1.78** |
| 319 | 99.11 | 94.38 | 89.30 | 73.51 | **356.30** |
| 398 | 13.83 | 18.38 | 12.49 | 11.82 | **56.51** |
| **GA GEORGIA Total** | **2,270.66** | **2,455.73** | **2,123.60** | **1,924.39** | **8,774.38** |

**STATE: HI - HAWAII**

| REGISTRANT ZIP CODE 3 | 1ST QUARTER | 2ND QUARTER | 3RD QUARTER | 4TH QUARTER | TOTAL GRAMS |
|---|---|---|---|---|---|
| 967 | 121.41 | 123.91 | 99.11 | 93.22 | **437.66** |
| 968 | 81.89 | 86.09 | 65.12 | 55.31 | **288.42** |
| **HI - HAWAII - Total** | **203.31** | **210.00** | **164.24** | **148.53** | **726.08** |

**STATE: IA - IOWA**

| REGISTRANT ZIP CODE 3 | 1ST QUARTER | 2ND QUARTER | 3RD QUARTER | 4TH QUARTER | TOTAL GRAMS |
|---|---|---|---|---|---|
| 500 | 32.56 | 37.69 | 21.59 | 26.32 | **118.16** |
| 501 | 9.01 | 16.41 | 6.33 | 9.63 | **41.39** |
| 502 | 45.68 | 48.89 | 39.88 | 34.88 | **169.32** |
| 503 | 81.58 | 86.89 | 71.55 | 60.98 | **300.99** |
| 504 | 37.78 | 25.20 | 29.97 | 11.78 | **104.73** |
| 505 | 2.68 | 3.93 | 3.12 | 2.14 | **11.86** |
| 506 | 0 | 0 | 0 | 0.45 | **0.45** |
| 508 | 1.34 | 1.20 | 0.40 | 1.61 | **4.55** |
| 510 | 5.62 | 3.12 | 1.34 | 0 | **10.08** |
| 511 | 8.12 | 13.38 | 11.15 | 12.13 | **44.78** |
| 512 | 0.27 | 0 | 1.07 | 2.14 | **3.48** |
| 513 | 1.61 | 1.61 | 2.45 | 1.61 | **7.27** |
| 514 | 12.04 | 10.08 | 4.19 | 7.58 | **33.90** |
| 515 | 26.67 | 34.61 | 13.83 | 23.02 | **98.13** |
| 520 | 3.03 | 3.21 | 3.48 | 1.61 | **11.33** |
| 522 | 1.78 | 0 | 2.68 | 0 | **4.46** |
| 523 | 4.73 | 3.66 | 3.03 | 5.80 | **17.22** |
| 524 | 7.14 | 6.24 | 1.78 | 1.34 | **16.50** |
| 525 | 0 | 1.78 | 0 | 0 | **1.78** |
| 526 | 0 | 0.54 | 1.34 | 0.80 | **2.68** |
| 527 | 7.14 | 5.80 | 9.28 | 14.90 | **37.11** |
| 528 | 1.78 | 2.68 | 2.23 | 3.12 | **9.81** |
| **IA IOWA Total** | **290.56** | **306.93** | **230.70** | **221.82** | **1,050.00** |

**STATE: ID - IDAHO**

| REGISTRANT ZIP CODE 3 | 1ST QUARTER | 2ND QUARTER | 3RD QUARTER | 4TH QUARTER | TOTAL GRAMS |
|---|---|---|---|---|---|
| 832 | 44.43 | 49.15 | 53.62 | 43.71 | **190.91** |
| 833 | 27.74 | 23.95 | 34.08 | 28.99 | **114.77** |
| 834 | 34.66 | 22.84 | 27.30 | 25.34 | **110.13** |
| 835 | 18.02 | 19.27 | 19.67 | 19.80 | **76.77** |
| 836 | 235.16 | 241.36 | 208.84 | 166.47 | **851.82** |
| 837 | 173.25 | 170.84 | 153.17 | 156.39 | **653.64** |
| 838 | 40.41 | 36.75 | 37.91 | 39.07 | **154.15** |
| **ID IDAHO Total** | **573.66** | **564.16** | **534.59** | **479.77** | **2,152.19** |

**STATE: IL - ILLINOIS**

| REGISTRANT ZIP CODE 3 | 1ST QUARTER | 2ND QUARTER | 3RD QUARTER | 4TH QUARTER | TOTAL GRAMS |
|---|---|---|---|---|---|
| 600 | 467.28 | 540.26 | 441.23 | 467.10 | **1,915.87** |
| 601 | 238.64 | 182.43 | 181.45 | 167.09 | **769.61** |
| 602 | 33.77 | 25.96 | 32.52 | 23.28 | **115.53** |
| 603 | 21.50 | 22.39 | 15.88 | 15.25 | **75.03** |
| 604 | 280.57 | 268.39 | 223.74 | 224.36 | **997.06** |
| 605 | 187.34 | 186.40 | 162.36 | 146.57 | **682.68** |
| 606 | 119.10 | 131.23 | 103.22 | 106.52 | **460.06** |
| 607 | 5.71 | 6.60 | 6.16 | 3.93 | **22.39** |
| 608 | 5.98 | 1.07 | 0 | 0 | **7.05** |
| 609 | 7.85 | 6.24 | 6.07 | 10.71 | **30.87** |
| 610 | 19.85 | 28.50 | 16.73 | 15.57 | **80.65** |
| 611 | 11.78 | 7.67 | 9.10 | 2.77 | **31.31** |
| 612 | 2.01 | 4.24 | 2.68 | 5.58 | **14.50** |
| 613 | 39.07 | 23.37 | 31.85 | 22.84 | **117.13** |
| 614 | 3.57 | 5.35 | 5.35 | 4.46 | **18.73** |
| 615 | 12.76 | 6.96 | 8.21 | 8.92 | **36.84** |
| 616 | 5.35 | 7.49 | 5.08 | 12.49 | **30.42** |
| 617 | 14.45 | 9.28 | 12.49 | 11.24 | **47.46** |
| 618 | 26.94 | 21.23 | 20.43 | 4.37 | **72.97** |
| 619 | 2.77 | 2.59 | 2.59 | 1.69 | **9.63** |
| 620 | 20.61 | 14.90 | 15.43 | 10.79 | **61.73** |
| 622 | 30.82 | 12.58 | 11.64 | 11.11 | **66.15** |
| 623 | 14.27 | 9.01 | 24.84 | 17.93 | **66.06** |
| 624 | 10.88 | 18.73 | 7.76 | 5.71 | **43.09** |
| 625 | 8.03 | 6.96 | 19.27 | 15.88 | **50.14** |
| 626 | 17.13 | 11.60 | 0.45 | 3.75 | **32.92** |
| 627 | 30.60 | 28.64 | 18.64 | 18.20 | **96.08** |
| 628 | 0 | 0.89 | 0 | 0 | **0.89** |
| 629 | 28.99 | 23.82 | 20.43 | 26.05 | **99.29** |
| **IL ILLINOIS Total** | **1,667.60** | **1,614.79** | **1,405.59** | **1,364.15** | **6,052.14** |

**STATE: IN - INDIANA**

| REGISTRANT ZIP CODE 3 | 1ST QUARTER | 2ND QUARTER | 3RD QUARTER | 4TH QUARTER | TOTAL GRAMS |
|---|---|---|---|---|---|
| 460 | 123.82 | 110.80 | 94.74 | 86.71 | **416.08** |
| 461 | 287.26 | 316.78 | 201.30 | 128.95 | **934.30** |
| 462 | 253.62 | 269.37 | 218.88 | 271.64 | **1,013.51** |
| 463 | 169.77 | 135.06 | 122.04 | 107.36 | **534.23** |
| 464 | 1.07 | 4.28 | 3.48 | 1.34 | **10.17** |
| 465 | 145.77 | 122.66 | 147.37 | 107.41 | **523.22** |
| 466 | 17.66 | 4.46 | 13.02 | 8.30 | **43.45** |

| | | | | | |
|---|---|---|---|---|---|
| 467 | 29.35 | 24.98 | 26.32 | 21.41 | **102.06** |
| 468 | 49.51 | 45.94 | 50.14 | 36.40 | **181.99** |
| 469 | 32.74 | 39.43 | 36.89 | 22.21 | **131.27** |
| 470 | 43.36 | 38.90 | 30.33 | 25.34 | **137.92** |
| 471 | 64.59 | 56.83 | 53.70 | 51.83 | **226.95** |
| 472 | 30.15 | 39.52 | 49.24 | 21.32 | **140.24** |
| 473 | 64.28 | 78.19 | 62.94 | 37.65 | **243.05** |
| 474 | 69.90 | 80.38 | 87.52 | 85.51 | **323.30** |
| 475 | 45.41 | 37.74 | 52.37 | 47.64 | **183.15** |
| 476 | 57.45 | 31.49 | 12.67 | 16.15 | **117.76** |
| 477 | 115.79 | 116.51 | 113.56 | 90.82 | **436.68** |
| 478 | 93.76 | 92.24 | 66.82 | 58.34 | **311.16** |
| 479 | 33.72 | 40.86 | 41.57 | 25.78 | **141.93** |
| **IN INDIANA Total** | **1,728.98** | **1,686.43** | **1,484.90** | **1,252.11** | **6,152.41** |

**STATE: KS - KANSAS**

| REGISTRANT ZIP CODE 3 | 1ST QUARTER | 2ND QUARTER | 3RD QUARTER | 4TH QUARTER | **TOTAL GRAMS** |
|---|---|---|---|---|---|
| 660 | 245.95 | 231.50 | 217.85 | 187.56 | **882.87** |
| 661 | 61.38 | 37.11 | 28.10 | 44.61 | **171.19** |
| 662 | 96.84 | 92.51 | 91.53 | 79.26 | **360.14** |
| 664 | 18.29 | 15.52 | 10.97 | 12.40 | **57.18** |
| 665 | 1.87 | 0.54 | 0 | 0 | **2.41** |
| 666 | 17.80 | 19.94 | 36.22 | 16.68 | **90.64** |
| 667 | 34.26 | 22.79 | 15.83 | 12.00 | **84.88** |
| 668 | 7.67 | 5.35 | 5.00 | 5.89 | **23.91** |
| 669 | 4.46 | 5.35 | 7.58 | 1.78 | **19.18** |
| 670 | 27.03 | 33.54 | 27.57 | 33.99 | **122.13** |
| 671 | 10.71 | 15.17 | 12.49 | 14.10 | **52.46** |
| 672 | 54.51 | 74.31 | 57.36 | 55.85 | **242.03** |
| 673 | 0 | 0 | 1.61 | 0 | **1.61** |
| 674 | 23.28 | 21.14 | 23.73 | 18.73 | **86.89** |
| 675 | 22.12 | 22.75 | 30.33 | 25.07 | **100.27** |
| 676 | 15.34 | 21.05 | 19.80 | 19.45 | **75.65** |
| 678 | 11.69 | 19.36 | 8.30 | 17.22 | **56.56** |
| 679 | 3.75 | 0 | 0.89 | 0.45 | **5.08** |
| **KS - KANSAS - Total** | **656.94** | **637.94** | **595.16** | **545.03** | **2,435.08** |

**STATE: KY - KENTUCKY**

| REGISTRANT ZIP CODE 3 | 1ST QUARTER | 2ND QUARTER | 3RD QUARTER | 4TH QUARTER | **TOTAL GRAMS** |
|---|---|---|---|---|---|
| 400 | 18.38 | 9.81 | 14.81 | 6.60 | **49.60** |
| 401 | 6.42 | 9.63 | 7.76 | 4.46 | **28.28** |
| 402 | 137.92 | 166.56 | 134.44 | 144.65 | **583.57** |

| | | | | | |
|---|---|---|---|---|---|
| 403 | 128.73 | 157.68 | 127.17 | 121.68 | **535.26** |
| 404 | 58.21 | 61.24 | 58.92 | 68.74 | **247.11** |
| 405 | 108.26 | 108.39 | 95.28 | 153.04 | **464.96** |
| 406 | 7.67 | 8.39 | 3.61 | 8.47 | **28.15** |
| 407 | 201.26 | 241.45 | 259.73 | 245.82 | **948.26** |
| 408 | 2.32 | 1.96 | 2.23 | 8.47 | **14.99** |
| 409 | 124.72 | 140.51 | 163.12 | 182.52 | **610.87** |
| 410 | 82.83 | 73.02 | 77.57 | 93.14 | **326.55** |
| 411 | 53.57 | 48.17 | 33.63 | 55.85 | **191.22** |
| 412 | 8.30 | 9.19 | 6.16 | 7.76 | **31.40** |
| 413 | 4.01 | 2.41 | 2.41 | 2.77 | **11.60** |
| 414 | 6.42 | 3.48 | 1.34 | 2.27 | **13.52** |
| 415 | 73.06 | 61.06 | 56.02 | 56.20 | **246.35** |
| 416 | 95.10 | 86.36 | 112.18 | 116.51 | **410.14** |
| 417 | 9.32 | 13.56 | 10.04 | 13.74 | **46.66** |
| 420 | 28.81 | 26.94 | 31.40 | 24.53 | **111.69** |
| 421 | 76.41 | 68.02 | 56.78 | 51.30 | **252.51** |
| 422 | 57.05 | 68.25 | 38.40 | 39.88 | **203.58** |
| 423 | 12.31 | 7.58 | 12.13 | 9.19 | **41.22** |
| 424 | 22.48 | 19.18 | 22.66 | 22.93 | **87.25** |
| 425 | 69.94 | 75.61 | 54.86 | 45.59 | **246.00** |
| 426 | 9.05 | 7.72 | 13.11 | 11.24 | **41.13** |
| 427 | 81.67 | 86.09 | 46.48 | 51.30 | **265.53** |
| **KY - KENTUCKY - Total** | **1,484.23** | **1,562.25** | **1,442.26** | **1,548.64** | **6,037.38** |

**STATE: LA - LOUISIANA**

| REGISTRANT ZIP CODE 3 | 1ST QUARTER | 2ND QUARTER | 3RD QUARTER | 4TH QUARTER | **TOTAL GRAMS** |
|---|---|---|---|---|---|
| 700 | 400.64 | 384.81 | 304.12 | 289.44 | **1,379.01** |
| 701 | 189.84 | 179.13 | 139.97 | 111.33 | **620.28** |
| 703 | 123.73 | 115.62 | 82.07 | 65.57 | **386.99** |
| 704 | 449.71 | 489.72 | 413.49 | 431.06 | **1,783.98** |
| 705 | 193.23 | 214.59 | 150.85 | 173.25 | **731.92** |
| 706 | 35.68 | 26.50 | 24.27 | 19.45 | **105.89** |
| 707 | 871.49 | 958.87 | 933.05 | 836.34 | **3,599.76** |
| 708 | 466.43 | 494.76 | 359.52 | 381.64 | **1,702.35** |
| 710 | 20.25 | 36.40 | 14.54 | 20.52 | **91.71** |
| 711 | 201.12 | 194.03 | 181.10 | 142.20 | **718.45** |
| 712 | 16.41 | 14.81 | 10.71 | 9.37 | **51.30** |
| 713 | 28.55 | 17.40 | 19.98 | 15.43 | **81.36** |
| 714 | 4.46 | 2.32 | 4.73 | 4.28 | **15.79** |
| **LA - LOUISIANA - Total** | **3,001.56** | **3,128.95** | **2,638.39** | **2,499.89** | **11,268.78** |

**STATE: MA - MASSACHUSETTS**

| REGISTRANT ZIP CODE 3 | 1ST QUARTER | 2ND QUARTER | 3RD QUARTER | 4TH QUARTER | TOTAL GRAMS |
|---|---|---|---|---|---|
| 010 | 36.04 | 43.27 | 41.57 | 48.62 | **169.50** |
| 011 | 5.89 | 6.51 | 19.27 | 11.33 | **43.00** |
| 012 | 19.98 | 23.55 | 19.27 | 12.85 | **75.65** |
| 013 | 11.24 | 15.79 | 7.76 | 9.37 | **44.16** |
| 014 | 3.57 | 2.94 | 8.21 | 3.48 | **18.20** |
| 015 | 44.43 | 41.57 | 47.82 | 40.32 | **174.14** |
| 016 | 4.46 | 6.24 | 9.63 | 6.24 | **26.58** |
| 017 | 12.49 | 13.83 | 5.35 | 8.03 | **39.70** |
| 018 | 181.99 | 192.56 | 180.20 | 170.08 | **724.83** |
| 019 | 57.27 | 73.60 | 71.90 | 52.19 | **254.96** |
| 020 | 54.24 | 40.50 | 67.35 | 40.32 | **202.42** |
| 021 | 54.69 | 59.10 | 57.32 | 60.04 | **231.14** |
| 022 | 17.31 | 22.75 | 28.73 | 26.32 | **95.10** |
| 023 | 30.24 | 24.71 | 13.38 | 16.33 | **84.66** |
| 024 | 14.45 | 24.00 | 21.77 | 18.56 | **78.77** |
| 025 | 11.60 | 22.30 | 13.38 | 11.60 | **58.88** |
| 026 | 23.37 | 15.97 | 15.17 | 17.40 | **71.90** |
| 027 | 216.78 | 307.15 | 299.21 | 220.79 | **1,043.94** |
| **MA - MASSACHUSETTS - Total** | **800.04** | **936.35** | **927.29** | **773.85** | **3,437.53** |

**STATE: MD - MARYLAND**

| REGISTRANT ZIP CODE 3 | 1ST QUARTER | 2ND QUARTER | 3RD QUARTER | 4TH QUARTER | TOTAL GRAMS |
|---|---|---|---|---|---|
| 206 | 739.24 | 691.82 | 417.68 | 344.84 | **2,193.58** |
| 207 | 410.82 | 520.88 | 480.04 | 413.14 | **1,824.88** |
| 208 | 289.22 | 285.03 | 292.43 | 287.21 | **1,153.89** |
| 209 | 50.76 | 65.12 | 48.04 | 44.87 | **208.80** |
| 210 | 876.98 | 924.93 | 747.62 | 644.72 | **3,194.25** |
| 211 | 369.95 | 370.18 | 314.55 | 297.87 | **1,352.56** |
| 212 | 499.53 | 468.71 | 373.21 | 333.15 | **1,674.61** |
| 214 | 73.87 | 84.66 | 62.63 | 70.74 | **291.90** |
| 215 | 125.34 | 124.36 | 107.23 | 109.95 | **466.88** |
| 216 | 199.25 | 168.74 | 107.59 | 119.50 | **595.08** |
| 217 | 974.53 | 967.04 | 813.95 | 621.75 | **3,377.27** |
| 218 | 71.32 | 68.96 | 45.59 | 51.34 | **237.21** |
| 219 | 141.13 | 137.61 | 112.76 | 107.23 | **498.73** |
| **MD MARYLAND Total** | **4,821.94** | **4,878.03** | **3,923.32** | **3,446.33** | **17,069.62** |

**STATE: ME - MAINE**

| REGISTRANT ZIP CODE 3 | 1ST QUARTER | 2ND QUARTER | 3RD QUARTER | 4TH QUARTER | TOTAL GRAMS |
|---|---|---|---|---|---|
| 040 | 57.09 | 68.78 | 65.93 | 69.49 | **261.30** |

| | | | | | |
|---|---|---|---|---|---|
| 041 | 2.50 | 3.03 | 3.48 | 3.75 | **12.76** |
| 042 | 107.81 | 63.96 | 63.29 | 52.01 | **287.08** |
| 043 | 96.35 | 95.41 | 94.56 | 87.16 | **373.48** |
| 044 | 2.94 | 3.48 | 1.87 | 3.03 | **11.33** |
| 045 | 26.94 | 20.79 | 17.66 | 17.66 | **83.05** |
| 046 | 1.34 | 2.14 | 4.10 | 3.39 | **10.97** |
| 048 | 4.19 | 2.77 | 2.59 | 6.07 | **15.61** |
| 049 | 57.54 | 57.45 | 59.77 | 45.32 | **220.08** |
| **ME MAINE Total** | **356.71** | **317.81** | **313.26** | **287.88** | **1,275.66** |

**STATE: MI - MICHIGAN**

| REGISTRANT ZIP CODE 3 | 1ST QUARTER | 2ND QUARTER | 3RD QUARTER | 4TH QUARTER | TOTAL GRAMS |
|---|---|---|---|---|---|
| 480 | 1,529.42 | 1,620.01 | 1,265.27 | 1,131.50 | **5,546.19** |
| 481 | 1,702.48 | 1,822.96 | 1,586.15 | 1,454.03 | **6,565.63** |
| 482 | 2,557.47 | 2,940.45 | 2,523.22 | 2,138.01 | **10,159.15** |
| 483 | 665.24 | 771.13 | 721.62 | 663.28 | **2,821.27** |
| 484 | 69.58 | 81.18 | 83.41 | 59.77 | **293.95** |
| 485 | 54.33 | 69.23 | 64.77 | 47.99 | **236.32** |
| 486 | 79.00 | 77.92 | 55.62 | 54.06 | **266.60** |
| 487 | 32.29 | 27.39 | 26.58 | 27.48 | **113.74** |
| 488 | 103.08 | 115.08 | 95.59 | 111.65 | **425.40** |
| 489 | 42.82 | 36.40 | 28.37 | 35.68 | **143.27** |
| 490 | 189.88 | 186.36 | 189.57 | 168.29 | **734.11** |
| 491 | 34.79 | 31.58 | 17.13 | 27.12 | **110.62** |
| 492 | 61.55 | 65.30 | 49.07 | 29.89 | **205.81** |
| 493 | 36.40 | 30.96 | 28.90 | 21.32 | **117.58** |
| 494 | 48.62 | 46.66 | 24.71 | 24.09 | **144.07** |
| 495 | 46.12 | 38.27 | 39.88 | 42.55 | **166.82** |
| 496 | 90.64 | 81.45 | 89.83 | 95.99 | **357.91** |
| 497 | 62.80 | 68.07 | 52.72 | 40.14 | **223.74** |
| 498 | 1.96 | 1.61 | 10.08 | 0.89 | **14.54** |
| **MI - MICHIGAN Total** | **7,408.49** | **8,112.00** | **6,952.49** | **6,173.73** | **28,646.71** |

**STATE: MN - MINNESOTA**

| REGISTRANT ZIP CODE 3 | 1ST QUARTER | 2ND QUARTER | 3RD QUARTER | 4TH QUARTER | TOTAL GRAMS |
|---|---|---|---|---|---|
| 550 | 30.06 | 30.96 | 39.61 | 28.99 | **129.62** |
| 551 | 69.41 | 55.22 | 48.71 | 64.86 | **238.19** |
| 553 | 33.63 | 27.74 | 20.52 | 22.12 | **104.02** |
| 554 | 12.13 | 12.49 | 16.41 | 14.99 | **56.02** |
| 557 | 13.29 | 16.68 | 12.49 | 14.18 | **56.65** |
| 558 | 4.28 | 5.35 | 3.21 | 6.42 | **19.27** |
| 559 | 2.85 | 3.57 | 0.54 | 3.66 | **10.62** |

| | | | | | |
|---|---|---|---|---|---|
| 560 | 2.23 | 6.78 | 9.37 | 7.58 | **25.96** |
| 563 | 23.19 | 17.84 | 17.49 | 17.31 | **75.83** |
| 564 | 0 | 0.45 | 0.45 | 0 | **0.89** |
| 565 | 2.68 | 1.78 | 2.23 | 0 | **6.69** |
| 567 | 10.35 | 9.99 | 6.42 | 6.42 | **33.19** |
| **MN MINNESOTA Total** | **204.11** | **188.86** | **177.44** | **186.54** | **756.95** |

**STATE: MO - MISSOURI**

| REGISTRANT ZIP CODE 3 | 1ST QUARTER | 2ND QUARTER | 3RD QUARTER | 4TH QUARTER | TOTAL GRAMS |
|---|---|---|---|---|---|
| 630 | 88.27 | 99.29 | 84.84 | 67.44 | **339.85** |
| 631 | 487.58 | 667.74 | 511.71 | 571.39 | **2,238.41** |
| 633 | 92.69 | 125.43 | 95.99 | 78.68 | **392.79** |
| 634 | 26.76 | 26.41 | 23.82 | 20.79 | **97.77** |
| 635 | 2.68 | 3.21 | 19.54 | 7.85 | **33.28** |
| 636 | 30.11 | 25.60 | 27.74 | 18.64 | **102.10** |
| 637 | 8.12 | 7.81 | 7.23 | 5.89 | **29.04** |
| 638 | 5.80 | 5.35 | 2.68 | 6.69 | **20.52** |
| 639 | 1.61 | 3.75 | 1.07 | 0 | **6.42** |
| 640 | 100.76 | 89.70 | 71.99 | 84.79 | **347.25** |
| 641 | 115.71 | 74.18 | 76.77 | 44.69 | **311.34** |
| 644 | 12.49 | 4.01 | 8.47 | 11.60 | **36.58** |
| 645 | 20.70 | 14.45 | 23.19 | 12.22 | **70.57** |
| 646 | 13.56 | 12.89 | 12.85 | 11.78 | **51.07** |
| 647 | 40.06 | 26.09 | 24.62 | 24.49 | **115.26** |
| 648 | 13.92 | 9.28 | 0.45 | 3.57 | **27.21** |
| 650 | 40.41 | 38.45 | 42.20 | 43.67 | **164.73** |
| 651 | 20.52 | 27.03 | 13.20 | 24.35 | **85.11** |
| 652 | 23.64 | 35.64 | 43.98 | 34.26 | **137.52** |
| 653 | 1.78 | 0.89 | 10.71 | 9.01 | **22.39** |
| 654 | 8.83 | 9.14 | 11.06 | 13.25 | **42.29** |
| 655 | 7.23 | 5.71 | 3.84 | 5.17 | **21.95** |
| 656 | 29.97 | 27.83 | 11.86 | 10.44 | **80.11** |
| 657 | 105.62 | 75.20 | 58.34 | 49.78 | **288.95** |
| 658 | 73.51 | 49.15 | 23.60 | 15.88 | **162.14** |
| **MO MISSOURI - Total** | **1,372.32** | **1,464.25** | **1,211.74** | **1,176.32** | **5,224.63** |

**STATE: MS - MISSISSIPPI**

| REGISTRANT ZIP CODE 3 | 1ST QUARTER | 2ND QUARTER | 3RD QUARTER | 4TH QUARTER | TOTAL GRAMS |
|---|---|---|---|---|---|
| 386 | 38.81 | 36.13 | 43.62 | 36.93 | **155.49** |
| 387 | 0.80 | 3.75 | 2.27 | 1.20 | **8.03** |
| 388 | 134.57 | 155.14 | 135.29 | 126.10 | **551.09** |
| 389 | 10.44 | 9.63 | 5.53 | 8.03 | **33.63** |

| | | | | | |
|---|---|---|---|---|---|
| 390 | 33.36 | 27.03 | 28.90 | 10.53 | **99.83** |
| 391 | 24.80 | 32.20 | 31.94 | 32.20 | **121.15** |
| 392 | 40.59 | 50.14 | 45.94 | 50.05 | **186.72** |
| 393 | 28.06 | 20.52 | 19.45 | 28.90 | **96.93** |
| 394 | 66.91 | 46.66 | 35.24 | 35.33 | **184.13** |
| 395 | 659.35 | 570.14 | 432.71 | 438.73 | **2,100.94** |
| 396 | 27.12 | 17.57 | 10.62 | 18.47 | **73.78** |
| 397 | 17.75 | 10.62 | 10.97 | 10.35 | **49.69** |
| **MS MISSISSIPPI - Total** | **1,082.56** | **979.53** | **802.49** | **796.82** | **3,661.40** |

**STATE: MT - MONTANA**

| REGISTRANT ZIP CODE 3 | 1ST QUARTER | 2ND QUARTER | 3RD QUARTER | 4TH QUARTER | TOTAL GRAMS |
|---|---|---|---|---|---|
| 590 | 0.54 | 0 | 1.07 | 2.77 | **4.37** |
| 591 | 18.38 | 15.70 | 10.62 | 16.06 | **60.75** |
| 593 | 0 | 1.34 | 0 | 0 | **1.34** |
| 594 | 5.26 | 7.32 | 4.91 | 4.64 | **22.12** |
| 595 | 1.61 | 2.41 | 1.61 | 4.01 | **9.63** |
| 596 | 0 | 1.07 | 1.07 | 0.54 | **2.68** |
| 597 | 8.39 | 5.00 | 11.78 | 7.85 | **33.01** |
| 598 | 3.97 | 7.67 | 8.47 | 7.85 | **27.97** |
| 599 | 57.01 | 55.93 | 52.90 | 52.10 | **217.94** |
| **MT - MONTANA Total** | **95.14** | **96.44** | **92.42** | **95.81** | **379.81** |

**STATE: NC - NORTH CAROLINA**

| REGISTRANT ZIP CODE 3 | 1ST QUARTER | 2ND QUARTER | 3RD QUARTER | 4TH QUARTER | TOTAL GRAMS |
|---|---|---|---|---|---|
| 270 | 145.68 | 143.09 | 93.05 | 109.55 | **491.37** |
| 271 | 146.62 | 151.48 | 109.46 | 149.52 | **557.07** |
| 272 | 447.65 | 391.28 | 393.37 | 291.18 | **1,523.47** |
| 273 | 151.39 | 136.49 | 149.34 | 130.96 | **568.18** |
| 274 | 211.43 | 201.08 | 163.17 | 172.80 | **748.47** |
| 275 | 677.64 | 2,167.45 | 1,619.25 | 588.61 | **5,052.94** |
| 276 | 257.46 | 240.78 | 218.03 | 177.97 | **894.24** |
| 277 | 160.49 | 175.57 | 165.13 | 129.22 | **630.40** |
| 278 | 1,130.11 | 1,100.94 | 850.84 | 763.73 | **3,845.62** |
| 279 | 149.38 | 158.30 | 132.57 | 109.01 | **549.27** |
| 280 | 1,371.60 | 1,418.93 | 1,286.14 | 1,063.29 | **5,139.97** |
| 281 | 708.42 | 656.85 | 662.38 | 450.91 | **2,478.57** |
| 282 | 399.93 | 393.10 | 315.13 | 312.01 | **1,420.18** |
| 283 | 454.97 | 521.52 | 396.36 | 356.30 | **1,729.16** |
| 284 | 346.58 | 354.34 | 253.89 | 213.93 | **1,168.74** |
| 285 | 523.40 | 549.71 | 500.74 | 416.70 | **1,990.54** |
| 286 | 1,062.13 | 1,138.01 | 894.15 | 847.00 | **3,941.30** |

| | | | | | |
|---|---|---|---|---|---|
| 287 | 203.40 | 244.48 | 206.92 | 192.07 | **846.87** |
| 288 | 16.37 | 16.46 | 20.43 | 17.35 | **70.61** |
| 289 | 140.42 | 126.59 | 101.61 | 85.82 | **454.44** |
| **NC - NORTH CAROLINA Total** | **8,705.06** | **10,286.45** | **8,531.96** | **6,577.94** | **34,101.41** |

**STATE: ND - NORTH DAKOTA**

| REGISTRANT ZIP CODE 3 | 1ST QUARTER | 2ND QUARTER | 3RD QUARTER | 4TH QUARTER | TOTAL GRAMS |
|---|---|---|---|---|---|
| 580 | 4.28 | 0 | 0 | 0 | **4.28** |
| 581 | 2.14 | 3.84 | 3.12 | 3.57 | **12.67** |
| 582 | 0.89 | 0 | 0 | 0 | **0.89** |
| 585 | 28.81 | 38.09 | 32.65 | 33.01 | **132.57** |
| 586 | 0 | 4.46 | 0 | 4.46 | **8.92** |
| 587 | 19.63 | 14.99 | 5.17 | 8.03 | **47.82** |
| 588 | 3.21 | 0 | 0 | 0 | **3.21** |
| **ND NORTH DAKOTA Total** | **58.97** | **61.38** | **40.95** | **49.07** | **210.36** |

**STATE: NE - NEBRASKA**

| REGISTRANT ZIP CODE 3 | 1ST QUARTER | 2ND QUARTER | 3RD QUARTER | 4TH QUARTER | TOTAL GRAMS |
|---|---|---|---|---|---|
| 680 | 13.02 | 26.23 | 26.23 | 18.20 | **83.68** |
| 681 | 144.88 | 195.99 | 145.81 | 159.86 | **646.55** |
| 683 | 1.34 | 0 | 0 | 1.47 | **2.81** |
| 684 | 14.41 | 16.64 | 18.64 | 5.53 | **55.22** |
| 685 | 73.69 | 72.35 | 70.52 | 64.86 | **281.41** |
| 686 | 27.08 | 30.78 | 24.98 | 27.03 | **109.86** |
| 687 | 29.62 | 30.42 | 22.12 | 26.85 | **109.01** |
| 688 | 7.63 | 7.36 | 9.50 | 14.85 | **39.34** |
| 689 | 0.80 | 0 | 0 | 0.27 | **1.07** |
| 691 | 2.85 | 0 | 0 | 0 | **2.85** |
| 693 | 9.41 | 2.45 | 0.89 | 0 | **12.76** |
| **NE NEBRASKA Total** | **324.72** | **382.22** | **318.70** | **318.93** | **1,344.57** |

**STATE: NH - NEW HAMPSHIRE**

| REGISTRANT ZIP CODE 3 | 1ST QUARTER | 2ND QUARTER | 3RD QUARTER | 4TH QUARTER | TOTAL GRAMS |
|---|---|---|---|---|---|
| 030 | 94.29 | 76.90 | 64.23 | 76.99 | **312.41** |
| 031 | 30.02 | 31.67 | 21.86 | 16.15 | **99.69** |
| 032 | 31.22 | 18.02 | 15.61 | 26.67 | **91.53** |
| 033 | 49.47 | 47.73 | 39.79 | 32.03 | **169.01** |
| 034 | 4.46 | 4.46 | 1.78 | 4.91 | **15.61** |
| 035 | 9.10 | 4.82 | 0 | 0 | **13.92** |
| 038 | 87.87 | 79.49 | 63.34 | 42.91 | **273.61** |
| **NH - NEW HAMPSHIRE Total** | **306.44** | **263.08** | **206.61** | **199.65** | **975.78** |

**STATE: NJ - NEW JERSEY**

| REGISTRANT ZIP CODE 3 | 1ST QUARTER | 2ND QUARTER | 3RD QUARTER | 4TH QUARTER | TOTAL GRAMS |
|---|---|---|---|---|---|

| | | | | | |
|---|---|---|---|---|---|
| 070 | 442.26 | 395.82 | 383.29 | 391.59 | **1,612.96** |
| 071 | 53.88 | 31.22 | 46.03 | 21.19 | **152.33** |
| 072 | 13.92 | 9.37 | 17.84 | 16.06 | **57.18** |
| 073 | 7.14 | 9.46 | 21.77 | 10.88 | **49.24** |
| 074 | 157.10 | 149.69 | 160.22 | 174.94 | **641.96** |
| 075 | 56.38 | 40.14 | 40.06 | 63.43 | **200.01** |
| 076 | 201.70 | 192.34 | 213.66 | 193.14 | **800.84** |
| 077 | 252.15 | 283.87 | 224.54 | 265.76 | **1,026.32** |
| 078 | 77.08 | 78.15 | 57.81 | 60.57 | **273.61** |
| 079 | 88.85 | 57.63 | 66.10 | 37.83 | **250.41** |
| 080 | 576.21 | 570.81 | 478.88 | 491.41 | **2,117.31** |
| 081 | 32.65 | 32.47 | 25.25 | 18.29 | **108.66** |
| 082 | 50.67 | 45.32 | 50.05 | 45.14 | **191.18** |
| 083 | 32.29 | 37.11 | 30.51 | 34.08 | **133.99** |
| 084 | 2.41 | 2.14 | 5.08 | 1.61 | **11.24** |
| 085 | 95.28 | 103.66 | 103.31 | 106.16 | **408.40** |
| 086 | 27.39 | 26.94 | 40.64 | 30.51 | **125.47** |
| 087 | 181.10 | 168.07 | 182.08 | 165.04 | **696.28** |
| 088 | 274.10 | 285.03 | 339.40 | 302.91 | **1,201.44** |
| 089 | 30.87 | 14.63 | 24.98 | 22.12 | **92.60** |
| **NJ NEW JERSEY - Total** | **2,653.42** | **2,533.88** | **2,511.48** | **2,452.65** | **10,151.43** |

**STATE: NM - NEW MEXICO**

| REGISTRANT ZIP CODE 3 | 1ST QUARTER | 2ND QUARTER | 3RD QUARTER | 4TH QUARTER | TOTAL GRAMS |
|---|---|---|---|---|---|
| 870 | 50.40 | 38.18 | 38.18 | 36.31 | **163.08** |
| 871 | 175.97 | 183.33 | 167.89 | 116.87 | **644.05** |
| 873 | 13.38 | 8.30 | 12.31 | 1.07 | **35.06** |
| 874 | 7.18 | 4.68 | 4.19 | 0.94 | **16.99** |
| 875 | 43.45 | 40.32 | 27.61 | 33.36 | **144.74** |
| 877 | 4.82 | 4.91 | 6.78 | 8.30 | **24.80** |
| 879 | 0.89 | 3.12 | 2.68 | 1.78 | **8.47** |
| 880 | 13.65 | 15.43 | 13.11 | 12.31 | **54.51** |
| 881 | 0 | 0 | 3.57 | 0.45 | **4.01** |
| 882 | 16.59 | 8.03 | 6.69 | 11.91 | **43.22** |
| 883 | 17.31 | 19.89 | 13.20 | 16.68 | **67.09** |
| **NM NEW MEXICO Total** | **343.64** | **326.20** | **296.22** | **239.97** | **1,206.03** |

**STATE: NV - NEVADA**

| REGISTRANT ZIP CODE 3 | 1ST QUARTER | 2ND QUARTER | 3RD QUARTER | 4TH QUARTER | TOTAL GRAMS |
|---|---|---|---|---|---|
| 890 | 328.61 | 343.10 | 327.49 | 280.30 | **1,279.49** |
| 891 | 791.65 | 635.89 | 538.69 | 533.88 | **2,500.11** |
| 894 | 45.63 | 26.50 | 24.09 | 22.12 | **118.34** |

| | | | | | |
|---|---|---|---|---|---|
| 895 | 53.35 | 26.85 | 37.83 | 36.58 | **154.60** |
| 897 | 27.83 | 23.19 | 26.05 | 7.67 | **84.75** |
| 898 | 5.00 | 6.60 | 5.89 | 6.42 | **23.91** |
| **NV NEVADA Total** | **1,252.06** | **1,062.13** | **960.03** | **886.97** | **4,161.20** |

**STATE: NY - NEW YORK**

| REGISTRANT ZIP CODE 3 | 1ST QUARTER | 2ND QUARTER | 3RD QUARTER | 4TH QUARTER | TOTAL GRAMS |
|---|---|---|---|---|---|
| 100 | 653.11 | 585.57 | 500.65 | 479.86 | **2,219.19** |
| 101 | 20.34 | 17.31 | 8.74 | 17.57 | **63.96** |
| 103 | 484.10 | 511.22 | 485.66 | 545.70 | **2,026.67** |
| 104 | 356.93 | 236.05 | 248.27 | 221.87 | **1,063.12** |
| 105 | 208.93 | 172.49 | 169.32 | 149.38 | **700.12** |
| 106 | 17.31 | 9.46 | 5.71 | 3.75 | **36.22** |
| 107 | 39.70 | 46.39 | 41.22 | 51.92 | **179.22** |
| 108 | 41.04 | 26.41 | 36.58 | 24.53 | **128.55** |
| 109 | 405.10 | 331.28 | 265.58 | 256.92 | **1,258.89** |
| 110 | 96.44 | 82.25 | 65.93 | 100.45 | **345.06** |
| 111 | 80.47 | 85.64 | 28.90 | 76.81 | **271.82** |
| 112 | 950.09 | 864.13 | 755.03 | 650.70 | **3,219.95** |
| 113 | 215.22 | 208.66 | 227.89 | 237.03 | **888.80** |
| 114 | 248.99 | 112.40 | 131.76 | 81.45 | **574.60** |
| 115 | 264.60 | 265.53 | 189.35 | 189.21 | **908.69** |
| 116 | 42.46 | 44.61 | 57.27 | 47.28 | **191.62** |
| 117 | 745.84 | 705.65 | 625.54 | 627.01 | **2,704.04** |
| 118 | 9.01 | 15.97 | 7.67 | 11.51 | **44.16** |
| 119 | 75.38 | 62.13 | 64.19 | 50.98 | **252.69** |
| 120 | 86.00 | 87.69 | 68.69 | 87.43 | **329.81** |
| 121 | 31.76 | 22.30 | 24.71 | 15.34 | **94.12** |
| 122 | 20.79 | 20.96 | 16.50 | 19.27 | **77.52** |
| 123 | 9.81 | 18.47 | 18.02 | 22.84 | **69.14** |
| 124 | 182.26 | 179.36 | 149.11 | 184.49 | **695.21** |
| 125 | 297.60 | 287.70 | 291.72 | 266.83 | **1,143.85** |
| 126 | 129.35 | 89.39 | 130.16 | 140.86 | **489.76** |
| 127 | 88.67 | 85.73 | 40.14 | 42.11 | **256.66** |
| 128 | 161.34 | 174.54 | 155.14 | 142.16 | **633.17** |
| 129 | 207.41 | 205.81 | 183.82 | 145.28 | **742.32** |
| 130 | 279.99 | 264.91 | 262.99 | 211.83 | **1,019.71** |
| 131 | 83.50 | 66.77 | 55.40 | 60.22 | **265.89** |
| 132 | 63.07 | 36.93 | 63.16 | 60.04 | **223.20** |
| 133 | 52.90 | 54.95 | 46.03 | 28.99 | **182.88** |
| 134 | 33.05 | 50.63 | 39.03 | 38.85 | **161.56** |

| | | | | | |
|---|---|---|---|---|---|
| 135 | 8.92 | 16.06 | 10.26 | 8.30 | **43.53** |
| 136 | 120.75 | 100.54 | 98.80 | 94.38 | **414.47** |
| 137 | 58.16 | 50.76 | 53.08 | 42.95 | **204.96** |
| 138 | 132.39 | 119.54 | 97.24 | 137.74 | **486.91** |
| 139 | 133.37 | 124.80 | 118.74 | 66.28 | **443.20** |
| 140 | 429.99 | 389.09 | 308.85 | 381.73 | **1,509.66** |
| 141 | 173.02 | 172.44 | 150.85 | 181.81 | **678.13** |
| 142 | 454.44 | 505.82 | 455.19 | 483.21 | **1,898.66** |
| 143 | 193.76 | 205.14 | 159.64 | 205.23 | **763.77** |
| 144 | 89.12 | 78.42 | 56.29 | 48.35 | **272.18** |
| 145 | 45.41 | 48.44 | 23.28 | 46.48 | **163.61** |
| 146 | 55.31 | 51.21 | 41.66 | 53.04 | **201.21** |
| 147 | 61.69 | 67.93 | 52.81 | 62.80 | **245.24** |
| 148 | 147.60 | 167.76 | 120.79 | 123.78 | **559.93** |
| 149 | 14.81 | 14.45 | 10.48 | 17.04 | **56.78** |
| **NY - NEW YORK - Total** | **8,801.28** | **8,141.71** | **7,217.85** | **7,243.58** | **31,404.42** |

**STATE: OH - OHIO**

| REGISTRANT ZIP CODE 3 | 1ST QUARTER | 2ND QUARTER | 3RD QUARTER | 4TH QUARTER | TOTAL GRAMS |
|---|---|---|---|---|---|
| 430 | 145.68 | 134.66 | 121.50 | 117.04 | **518.89** |
| 431 | 70.07 | 85.37 | 70.21 | 76.54 | **302.20** |
| 432 | 65.79 | 80.82 | 85.69 | 90.33 | **322.62** |
| 433 | 28.41 | 29.80 | 40.32 | 34.61 | **133.15** |
| 434 | 55.76 | 71.90 | 75.25 | 41.66 | **244.57** |
| 435 | 57.54 | 49.51 | 40.14 | 62.80 | **210.00** |
| 436 | 120.57 | 111.78 | 120.57 | 155.00 | **507.92** |
| 437 | 13.92 | 8.47 | 7.67 | 9.90 | **39.97** |
| 438 | 16.06 | 17.84 | 12.85 | 15.17 | **61.91** |
| 439 | 129.76 | 142.25 | 128.73 | 46.12 | **446.85** |
| 440 | 112.09 | 102.19 | 79.58 | 110.13 | **403.99** |
| 441 | 61.02 | 80.60 | 37.20 | 43.53 | **222.36** |
| 442 | 106.52 | 100.36 | 84.75 | 84.66 | **376.29** |
| 443 | 43.89 | 36.93 | 32.12 | 32.65 | **145.59** |
| 444 | 99.34 | 78.91 | 56.11 | 58.08 | **292.43** |
| 445 | 84.97 | 68.91 | 74.18 | 40.64 | **268.70** |
| 446 | 134.39 | 129.67 | 105.13 | 114.46 | **483.65** |
| 447 | 108.75 | 83.59 | 89.83 | 67.62 | **349.79** |
| 448 | 137.38 | 105.18 | 69.05 | 44.92 | **356.53** |
| 449 | 22.30 | 9.81 | 10.26 | 9.81 | **52.19** |
| 450 | 289.40 | 381.91 | 278.34 | 309.92 | **1,259.56** |
| 451 | 128.77 | 136.67 | 136.49 | 109.73 | **511.66** |

| | | | | | |
|---|---|---|---|---|---|
| 452 | 142.20 | 131.54 | 107.68 | 109.15 | **490.57** |
| 453 | 102.73 | 104.24 | 80.51 | 103.04 | **390.52** |
| 454 | 142.74 | 127.30 | 104.82 | 116.91 | **491.77** |
| 455 | 28.86 | 30.87 | 18.07 | 19.85 | **97.64** |
| 456 | 671.39 | 548.02 | 448.77 | 413.76 | **2,081.94** |
| 457 | 29.17 | 25.51 | 28.10 | 41.93 | **124.72** |
| 458 | 15.43 | 21.68 | 9.01 | 11.15 | **57.27** |
| **OH - OHIO Total** | **3,164.90** | **3,036.31** | **2,552.92** | **2,491.10** | **11,245.23** |

**STATE: OK - OKLAHOMA**

| REGISTRANT ZIP CODE 3 | 1ST QUARTER | 2ND QUARTER | 3RD QUARTER | 4TH QUARTER | **TOTAL GRAMS** |
|---|---|---|---|---|---|
| 730 | 280.65 | 281.73 | 233.86 | 232.84 | **1,029.08** |
| 731 | 213.70 | 242.83 | 212.94 | 236.23 | **905.70** |
| 734 | 24.98 | 19.80 | 13.29 | 14.63 | **72.71** |
| 735 | 44.69 | 41.22 | 37.56 | 36.75 | **160.22** |
| 736 | 1.34 | 1.34 | 1.34 | 1.78 | **5.80** |
| 737 | 15.25 | 9.28 | 9.28 | 9.99 | **43.80** |
| 738 | 4.01 | 4.37 | 10.08 | 11.51 | **29.97** |
| 740 | 115.17 | 103.17 | 85.06 | 100.54 | **403.94** |
| 741 | 76.27 | 87.25 | 76.01 | 61.69 | **301.22** |
| 743 | 41.75 | 35.15 | 24.80 | 27.12 | **128.82** |
| 744 | 33.90 | 27.30 | 24.00 | 23.02 | **108.21** |
| 745 | 83.14 | 95.63 | 55.13 | 57.45 | **291.36** |
| 746 | 6.87 | 3.03 | 3.93 | 2.68 | **16.50** |
| 747 | 12.40 | 8.74 | 7.14 | 3.48 | **31.76** |
| 748 | 42.64 | 27.57 | 39.52 | 23.55 | **133.28** |
| 749 | 9.28 | 14.50 | 8.03 | 22.12 | **53.93** |
| **OK - OKLAHOMA Total** | **1,006.07** | **1,002.90** | **841.96** | **865.38** | **3,716.31** |

**STATE: OR - OREGON**

| REGISTRANT ZIP CODE 3 | 1ST QUARTER | 2ND QUARTER | 3RD QUARTER | 4TH QUARTER | **TOTAL GRAMS** |
|---|---|---|---|---|---|
| 970 | 135.06 | 119.68 | 117.58 | 124.89 | **497.21** |
| 971 | 14.90 | 14.45 | 20.61 | 19.18 | **69.14** |
| 972 | 121.95 | 122.22 | 99.47 | 102.86 | **446.50** |
| 973 | 76.59 | 77.48 | 60.35 | 70.25 | **284.67** |
| 974 | 209.47 | 199.79 | 185.11 | 132.21 | **726.57** |
| 975 | 41.57 | 46.66 | 35.42 | 32.29 | **155.94** |
| 976 | 2.41 | 1.07 | 1.61 | 3.21 | **8.30** |
| 977 | 61.47 | 45.85 | 36.49 | 54.15 | **197.96** |
| 978 | 36.04 | 33.63 | 18.02 | 17.22 | **104.91** |
| 979 | 8.74 | 8.92 | 5.53 | 9.63 | **32.83** |
| **OR OREGON Total** | **708.19** | **669.74** | **580.18** | **565.90** | **2,524.02** |

**STATE: PA - PENNSYLVANIA**

| REGISTRANT ZIP CODE 3 | 1ST QUARTER | 2ND QUARTER | 3RD QUARTER | 4TH QUARTER | TOTAL GRAMS |
|---|---|---|---|---|---|
| 150 | 615.01 | 583.88 | 443.73 | 415.58 | **2,058.21** |
| 151 | 403.50 | 360.90 | 312.10 | 351.93 | **1,428.43** |
| 152 | 960.79 | 1,085.91 | 879.83 | 849.64 | **3,776.17** |
| 153 | 245.73 | 231.86 | 188.14 | 139.84 | **805.57** |
| 154 | 733.13 | 657.21 | 569.78 | 455.24 | **2,415.36** |
| 155 | 89.43 | 71.28 | 62.85 | 65.79 | **289.35** |
| 156 | 722.02 | 765.29 | 609.57 | 538.92 | **2,635.80** |
| 157 | 159.24 | 198.54 | 188.55 | 166.78 | **713.10** |
| 158 | 164.86 | 185.29 | 147.24 | 157.59 | **654.98** |
| 159 | 380.39 | 319.06 | 253.09 | 242.29 | **1,194.83** |
| 160 | 249.97 | 311.92 | 214.42 | 202.19 | **978.50** |
| 161 | 583.30 | 520.18 | 436.91 | 331.86 | **1,872.25** |
| 162 | 139.97 | 120.03 | 99.65 | 105.09 | **464.74** |
| 163 | 329.63 | 312.55 | 283.33 | 287.66 | **1,213.17** |
| 164 | 63.83 | 77.17 | 63.52 | 64.54 | **269.06** |
| 165 | 98.62 | 98.40 | 104.96 | 88.72 | **390.70** |
| 166 | 215.22 | 242.87 | 194.08 | 165.62 | **817.79** |
| 167 | 68.69 | 49.07 | 62.71 | 53.62 | **234.09** |
| 168 | 29.84 | 31.31 | 21.05 | 28.10 | **110.31** |
| 169 | 1.78 | 1.43 | 0 | 0 | **3.21** |
| 170 | 307.51 | 252.73 | 218.30 | 194.66 | **973.19** |
| 171 | 65.21 | 65.61 | 30.42 | 50.58 | **211.83** |
| 172 | 185.20 | 194.79 | 102.95 | 90.77 | **573.71** |
| 173 | 189.21 | 178.15 | 174.58 | 137.38 | **679.33** |
| 174 | 109.01 | 106.20 | 99.11 | 82.39 | **396.72** |
| 175 | 120.43 | 107.85 | 121.01 | 83.95 | **433.25** |
| 176 | 87.78 | 67.49 | 101.70 | 110.84 | **367.81** |
| 177 | 29.13 | 22.44 | 18.78 | 25.87 | **96.21** |
| 178 | 13.74 | 11.69 | 13.43 | 5.49 | **44.34** |
| 179 | 10.97 | 6.07 | 10.44 | 10.08 | **37.56** |
| 180 | 51.43 | 40.77 | 44.16 | 35.77 | **172.13** |
| 181 | 29.53 | 33.54 | 35.06 | 21.72 | **119.85** |
| 182 | 40.86 | 45.05 | 10.71 | 39.97 | **136.58** |
| 183 | 10.35 | 5.53 | 6.87 | 11.86 | **34.61** |
| 184 | 55.04 | 57.27 | 79.58 | 68.69 | **260.58** |
| 185 | 19.45 | 13.02 | 11.60 | 18.11 | **62.18** |
| 186 | 41.22 | 38.36 | 32.74 | 37.11 | **149.43** |
| 187 | 370.49 | 498.59 | 439.85 | 493.11 | **1,802.04** |

| | | | | | |
|---|---|---|---|---|---|
| 188 | 15.61 | 19.18 | 5.17 | 14.99 | **54.95** |
| 189 | 93.31 | 84.08 | 93.94 | 74.40 | **345.73** |
| 190 | 559.35 | 583.75 | 533.92 | 500.96 | **2,177.97** |
| 191 | 165.75 | 160.22 | 152.37 | 110.44 | **588.79** |
| 193 | 168.92 | 155.54 | 150.85 | 125.34 | **600.65** |
| 194 | 110.04 | 112.63 | 77.92 | 107.68 | **408.27** |
| 195 | 34.79 | 31.58 | 21.32 | 29.08 | **116.78** |
| 196 | 46.12 | 50.05 | 29.44 | 42.11 | **167.71** |
| **PA - PENNSYLVANIA - Total** | **9,185.42** | **9,166.33** | **7,751.72** | **7,234.35** | **33,337.82** |

**STATE: RI - RHODE ISLAND**

| REGISTRANT ZIP CODE 3 | 1ST QUARTER | 2ND QUARTER | 3RD QUARTER | 4TH QUARTER | TOTAL GRAMS |
|---|---|---|---|---|---|
| 028 | 97.06 | 99.20 | 112.05 | 117.76 | **426.07** |
| 029 | 61.42 | 52.54 | 41.13 | 38.98 | **194.08** |
| **RI RHODE ISLAND Total** | **158.48** | **151.75** | **153.17** | **156.74** | **620.14** |

**STATE: SC - SOUTH CAROLINA**

| REGISTRANT ZIP CODE 3 | 1ST QUARTER | 2ND QUARTER | 3RD QUARTER | 4TH QUARTER | TOTAL GRAMS |
|---|---|---|---|---|---|
| 290 | 318.39 | 388.33 | 302.07 | 353.36 | **1,362.15** |
| 291 | 288.91 | 309.56 | 268.43 | 285.03 | **1,151.92** |
| 292 | 556.72 | 497.43 | 442.97 | 434.19 | **1,931.31** |
| 293 | 46.57 | 42.55 | 25.96 | 44.16 | **159.24** |
| 294 | 533.57 | 518.44 | 487.71 | 447.34 | **1,987.06** |
| 295 | 270.84 | 286.32 | 278.25 | 235.51 | **1,070.92** |
| 296 | 299.39 | 293.28 | 253.13 | 214.37 | **1,060.17** |
| 297 | 159.73 | 178.78 | 137.61 | 141.31 | **617.42** |
| 298 | 43.67 | 52.28 | 33.05 | 32.74 | **161.74** |
| 299 | 90.82 | 92.33 | 86.09 | 60.31 | **329.54** |
| **SC - SOUTH CAROLINA Total** | **2,608.59** | **2,659.31** | **2,315.27** | **2,248.32** | **9,831.48** |

**STATE: SD - SOUTH DAKOTA**

| REGISTRANT ZIP CODE 3 | 1ST QUARTER | 2ND QUARTER | 3RD QUARTER | 4TH QUARTER | TOTAL GRAMS |
|---|---|---|---|---|---|
| 570 | 32.47 | 26.67 | 27.92 | 22.21 | **109.28** |
| 571 | 65.03 | 69.58 | 51.38 | 48.08 | **234.09** |
| 573 | 0 | 0.89 | 0 | 0 | **0.89** |
| 574 | 4.46 | 7.14 | 7.14 | 7.14 | **25.87** |
| 575 | 17.57 | 8.70 | 13.34 | 8.12 | **47.73** |
| 576 | 4.28 | 1.07 | 0 | 4.46 | **9.81** |
| 577 | 104.46 | 118.11 | 100.81 | 96.75 | **420.13** |
| **SD SOUTH DAKOTA - Total** | **228.29** | **232.17** | **200.59** | **186.76** | **847.81** |

**STATE: TN - TENNESSEE**

| REGISTRANT ZIP CODE 3 | 1ST QUARTER | 2ND QUARTER | 3RD QUARTER | 4TH QUARTER | TOTAL GRAMS |
|---|---|---|---|---|---|
| 370 | 1,037.47 | 992.02 | 766.54 | 614.79 | **3,410.81** |

| | | | | | |
|---|---|---|---|---|---|
| 371 | 735.14 | 633.44 | 486.33 | 450.42 | **2,305.32** |
| 372 | 637.85 | 651.86 | 725.95 | 582.50 | **2,598.15** |
| 373 | 650.97 | 562.38 | 520.09 | 514.52 | **2,247.96** |
| 374 | 406.66 | 429.19 | 326.78 | 332.04 | **1,494.67** |
| 376 | 530.31 | 625.99 | 498.68 | 519.02 | **2,174.00** |
| 377 | 1,822.11 | 1,749.27 | 1,506.04 | 1,268.16 | **6,345.60** |
| 378 | 2,049.69 | 1,945.13 | 1,804.63 | 1,494.62 | **7,294.08** |
| 379 | 761.05 | 752.44 | 711.32 | 620.19 | **2,845.00** |
| 380 | 48.44 | 32.12 | 20.25 | 26.09 | **126.90** |
| 381 | 63.16 | 54.06 | 60.31 | 28.01 | **205.54** |
| 382 | 62.54 | 42.20 | 36.49 | 44.25 | **185.47** |
| 383 | 106.07 | 100.63 | 55.76 | 39.79 | **302.24** |
| 384 | 106.34 | 95.19 | 79.98 | 73.73 | **355.23** |
| 385 | 447.12 | 390.87 | 277.26 | 244.44 | **1,359.69** |
| **TN TENNESSEE Total** | **9,464.91** | **9,056.78** | **7,876.40** | **6,852.58** | **33,250.66** |

**STATE: TX - TEXAS**

| REGISTRANT ZIP CODE 3 | 1ST QUARTER | 2ND QUARTER | 3RD QUARTER | 4TH QUARTER | TOTAL GRAMS |
|---|---|---|---|---|---|
| 750 | 467.95 | 494.18 | 489.14 | 411.84 | **1,863.11** |
| 751 | 61.38 | 55.49 | 46.39 | 29.08 | **192.34** |
| 752 | 90.36 | 68.33 | 44.78 | 46.75 | **250.22** |
| 753 | 0 | 0 | 0.89 | 0 | **0.89** |
| 754 | 30.60 | 29.97 | 23.37 | 19.54 | **103.48** |
| 757 | 16.50 | 35.68 | 15.79 | 7.76 | **75.74** |
| 758 | 11.24 | 10.53 | 11.60 | 11.78 | **45.14** |
| 759 | 35.33 | 33.63 | 35.06 | 29.17 | **133.19** |
| 760 | 203.53 | 187.70 | 170.30 | 152.33 | **713.86** |
| 761 | 95.19 | 87.07 | 70.30 | 85.46 | **338.02** |
| 762 | 88.59 | 79.66 | 68.07 | 53.88 | **290.20** |
| 763 | 106.61 | 96.70 | 103.22 | 83.77 | **390.29** |
| 764 | 12.40 | 9.81 | 10.97 | 12.22 | **45.41** |
| 765 | 29.48 | 24.80 | 13.83 | 13.16 | **81.27** |
| 766 | 1.78 | 4.46 | 1.78 | 4.46 | **12.49** |
| 767 | 18.29 | 3.52 | 11.69 | 6.74 | **40.23** |
| 768 | 5.35 | 8.03 | 6.42 | 1.61 | **21.41** |
| 769 | 7.90 | 4.59 | 4.42 | 1.69 | **18.60** |
| 770 | 223.20 | 262.97 | 234.44 | 239.26 | **959.88** |
| 773 | 110.89 | 136.05 | 141.04 | 119.54 | **507.52** |
| 774 | 216.42 | 203.49 | 137.74 | 141.13 | **698.78** |
| 775 | 236.18 | 218.92 | 221.87 | 177.88 | **854.85** |
| 776 | 84.48 | 56.74 | 59.06 | 53.62 | **253.89** |

| | | | | | |
|---|---|---|---|---|---|
| 777 | 21.54 | 31.76 | 21.59 | 16.41 | **91.31** |
| 778 | 12.49 | 13.83 | 12.58 | 12.13 | **51.03** |
| 779 | 24.80 | 26.67 | 10.88 | 8.03 | **70.39** |
| 780 | 57.72 | 51.56 | 54.69 | 55.76 | **219.72** |
| 781 | 49.07 | 53.79 | 59.77 | 40.23 | **202.86** |
| 782 | 410.81 | 313.44 | 243.23 | 247.38 | **1,214.86** |
| 783 | 12.04 | 8.92 | 8.92 | 14.63 | **44.52** |
| 784 | 28.01 | 30.87 | 27.12 | 33.19 | **119.18** |
| 785 | 4.64 | 4.55 | 2.68 | 3.75 | **15.61** |
| 786 | 181.27 | 144.43 | 139.17 | 119.76 | **584.64** |
| 787 | 156.52 | 159.11 | 149.78 | 144.12 | **609.53** |
| 788 | 8.92 | 5.71 | 7.58 | 1.96 | **24.18** |
| 789 | 8.03 | 13.38 | 13.38 | 8.03 | **42.82** |
| 790 | 15.79 | 12.67 | 9.28 | 5.62 | **43.36** |
| 791 | 36.49 | 53.66 | 46.17 | 36.22 | **172.53** |
| 792 | 0 | 2.68 | 7.14 | 8.56 | **18.38** |
| 793 | 1.78 | 0 | 0 | 0 | **1.78** |
| 794 | 4.55 | 7.49 | 3.12 | 4.73 | **19.89** |
| 795 | 5.26 | 0.80 | 4.82 | 5.49 | **16.37** |
| 796 | 7.32 | 12.53 | 7.00 | 4.77 | **31.62** |
| 797 | 2.19 | 5.00 | 1.61 | 3.21 | **12.00** |
| 799 | 40.06 | 31.31 | 27.66 | 17.93 | **116.95** |
| **TX - TEXAS Total** | **3,242.95** | **3,096.50** | **2,780.32** | **2,494.58** | **11,614.35** |

**STATE: UT - UTAH**

| REGISTRANT ZIP CODE 3 | 1ST QUARTER | 2ND QUARTER | 3RD QUARTER | 4TH QUARTER | **TOTAL GRAMS** |
|---|---|---|---|---|---|
| 840 | 1,011.73 | 1,113.39 | 820.69 | 968.73 | **3,914.53** |
| 841 | 278.83 | 346.36 | 281.68 | 286.72 | **1,193.59** |
| 843 | 89.79 | 71.64 | 86.44 | 109.64 | **357.51** |
| 844 | 253.58 | 269.46 | 248.18 | 209.47 | **980.69** |
| 845 | 54.24 | 37.20 | 33.45 | 51.56 | **176.46** |
| 846 | 216.60 | 280.97 | 231.54 | 258.49 | **987.60** |
| 847 | 182.88 | 184.40 | 207.99 | 210.94 | **786.21** |
| **UT - UTAH - Total** | **2,087.65** | **2,303.40** | **1,909.99** | **2,095.54** | **8,396.58** |

**STATE: VA - VIRGINIA**

| REGISTRANT ZIP CODE 3 | 1ST QUARTER | 2ND QUARTER | 3RD QUARTER | 4TH QUARTER | **TOTAL GRAMS** |
|---|---|---|---|---|---|
| 201 | 608.41 | 628.26 | 576.70 | 555.69 | **2,369.06** |
| 220 | 299.03 | 322.76 | 388.78 | 409.43 | **1,420.00** |
| 221 | 326.64 | 446.94 | 341.67 | 154.51 | **1,269.77** |
| 222 | 81.27 | 136.98 | 122.04 | 176.28 | **516.57** |
| 223 | 104.46 | 97.15 | 82.25 | 70.48 | **354.34** |

| | | | | | |
|---|---|---|---|---|---|
| 224 | 568.09 | 488.74 | 300.10 | 249.07 | **1,606.00** |
| 225 | 59.06 | 108.57 | 86.09 | 72.62 | **326.33** |
| 226 | 186.58 | 167.76 | 141.31 | 144.56 | **640.22** |
| 227 | 27.03 | 36.58 | 23.28 | 18.47 | **105.36** |
| 228 | 18.11 | 13.02 | 9.28 | 13.92 | **54.33** |
| 229 | 75.47 | 65.93 | 57.36 | 48.53 | **247.29** |
| 230 | 76.32 | 76.50 | 25.65 | 35.28 | **213.75** |
| 231 | 115.48 | 130.11 | 111.74 | 137.12 | **494.45** |
| 232 | 122.93 | 145.68 | 126.14 | 109.55 | **504.30** |
| 233 | 193.41 | 196.89 | 194.48 | 226.59 | **811.36** |
| 234 | 242.74 | 286.23 | 276.24 | 223.92 | **1,029.13** |
| 235 | 89.21 | 118.07 | 118.11 | 111.20 | **436.59** |
| 236 | 81.31 | 76.90 | 88.01 | 93.31 | **339.53** |
| 237 | 74.45 | 84.57 | 66.19 | 57.72 | **282.93** |
| 238 | 135.33 | 155.58 | 96.30 | 74.22 | **461.44** |
| 239 | 25.87 | 14.63 | 8.47 | 4.01 | **52.99** |
| 240 | 131.41 | 129.71 | 104.55 | 100.50 | **466.17** |
| 241 | 74.00 | 68.87 | 76.81 | 67.62 | **287.30** |
| 242 | 269.15 | 315.09 | 293.95 | 231.41 | **1,109.59** |
| 243 | 95.01 | 50.14 | 36.58 | 41.22 | **222.94** |
| 244 | 17.75 | 22.66 | 20.52 | 22.93 | **83.86** |
| 245 | 126.32 | 120.52 | 85.91 | 83.77 | **416.52** |
| 246 | 53.70 | 52.37 | 16.86 | 38.00 | **160.93** |
| **VA VIRGINIA Total** | **4,278.56** | **4,557.20** | **3,875.37** | **3,571.92** | **16,283.06** |

**STATE: VT - VERMONT**

| REGISTRANT ZIP CODE 3 | 1ST QUARTER | 2ND QUARTER | 3RD QUARTER | 4TH QUARTER | **TOTAL GRAMS** |
|---|---|---|---|---|---|
| 050 | 3.57 | 3.57 | 1.78 | 0 | **8.92** |
| 051 | 0 | 0 | 4.46 | 5.35 | **9.81** |
| 052 | 18.73 | 15.17 | 7.14 | 8.92 | **49.96** |
| 054 | 67.89 | 78.33 | 61.64 | 57.72 | **265.58** |
| 056 | 4.28 | 12.85 | 7.49 | 7.49 | **32.12** |
| 057 | 5.35 | 8.03 | 9.63 | 11.24 | **34.26** |
| 058 | 2.85 | 3.12 | 3.48 | 4.46 | **13.92** |
| **VT VERMONT Total** | **102.68** | **121.06** | **95.63** | **95.19** | **414.56** |

**STATE: WA - WASHINGTON**

| REGISTRANT ZIP CODE 3 | 1ST QUARTER | 2ND QUARTER | 3RD QUARTER | 4TH QUARTER | **TOTAL GRAMS** |
|---|---|---|---|---|---|
| 980 | 269.15 | 276.42 | 226.68 | 234.27 | **1,006.51** |
| 981 | 79.40 | 59.59 | 47.28 | 50.05 | **236.32** |
| 982 | 195.95 | 189.17 | 147.37 | 163.43 | **695.93** |
| 983 | 87.60 | 88.59 | 78.59 | 59.28 | **314.06** |

| | | | | | |
|---|---|---|---|---|---|
| 984 | 40.72 | 62.40 | 61.87 | 59.10 | **224.10** |
| 985 | 106.70 | 103.53 | 92.47 | 124.09 | **426.78** |
| 986 | 171.91 | 156.21 | 174.90 | 164.50 | **667.51** |
| 988 | 17.40 | 15.25 | 19.54 | 6.87 | **59.06** |
| 989 | 35.33 | 51.38 | 38.09 | 30.51 | **155.31** |
| 990 | 4.91 | 5.31 | 10.35 | 10.21 | **30.78** |
| 991 | 2.59 | 0 | 0.40 | 0 | **2.99** |
| 992 | 105.80 | 94.92 | 79.93 | 90.28 | **370.94** |
| 993 | 63.16 | 74.80 | 50.45 | 59.32 | **247.74** |
| 994 | 7.14 | 9.81 | 2.68 | 8.03 | **27.66** |
| **WA WASHINGTON Total** | **1,187.74** | **1,187.39** | **1,030.60** | **1,059.95** | **4,465.67** |

**STATE: WI - WISCONSIN**

| REGISTRANT ZIP CODE 3 | 1ST QUARTER | 2ND QUARTER | 3RD QUARTER | 4TH QUARTER | TOTAL GRAMS |
|---|---|---|---|---|---|
| 530 | 91.17 | 80.29 | 106.78 | 69.41 | **347.65** |
| 531 | 121.82 | 97.55 | 88.14 | 96.17 | **403.68** |
| 532 | 57.45 | 79.71 | 63.25 | 48.35 | **248.76** |
| 534 | 48.17 | 42.20 | 48.17 | 34.61 | **173.16** |
| 535 | 16.55 | 18.64 | 17.66 | 10.79 | **63.65** |
| 537 | 2.41 | 2.94 | 4.01 | 3.57 | **12.94** |
| 539 | 22.84 | 24.71 | 21.05 | 9.37 | **77.97** |
| 540 | 8.83 | 8.65 | 12.40 | 6.16 | **36.04** |
| 541 | 10.53 | 10.97 | 5.35 | 4.46 | **31.31** |
| 542 | 14.99 | 16.86 | 10.08 | 9.01 | **50.94** |
| 543 | 9.05 | 9.32 | 10.17 | 6.24 | **34.79** |
| 544 | 61.69 | 62.80 | 51.83 | 75.52 | **251.84** |
| 545 | 7.14 | 4.82 | 2.94 | 4.82 | **19.72** |
| 547 | 0 | 0 | 0.54 | 0 | **0.54** |
| 548 | 18.64 | 22.21 | 19.54 | 20.34 | **80.74** |
| 549 | 104.82 | 100.27 | 105.80 | 77.88 | **388.78** |
| **WI - WISCONSIN Total** | **596.10** | **581.96** | **567.73** | **476.69** | **2,222.49** |

**STATE: WV - WEST VIRGINIA**

| REGISTRANT ZIP CODE 3 | 1ST QUARTER | 2ND QUARTER | 3RD QUARTER | 4TH QUARTER | TOTAL GRAMS |
|---|---|---|---|---|---|
| 247 | 28.50 | 33.77 | 57.27 | 29.13 | **148.67** |
| 248 | 85.11 | 51.38 | 17.13 | 14.81 | **168.43** |
| 249 | 27.03 | 15.70 | 12.67 | 19.63 | **75.03** |
| 250 | 15.97 | 13.65 | 16.41 | 22.30 | **68.33** |
| 251 | 6.87 | 9.63 | 7.14 | 3.21 | **26.85** |
| 252 | 4.46 | 3.57 | 1.78 | 1.78 | **11.60** |
| 253 | 72.35 | 87.60 | 86.09 | 77.79 | **323.83** |
| 254 | 417.32 | 355.77 | 241.31 | 202.06 | **1,216.47** |

| | | | | | |
|---|---|---|---|---|---|
| 255 | 105.13 | 99.60 | 66.95 | 63.03 | **334.72** |
| 256 | 15.97 | 12.40 | 16.50 | 24.27 | **69.14** |
| 257 | 14.99 | 28.41 | 6.69 | 27.21 | **77.30** |
| 258 | 8.12 | 7.76 | 1.96 | 0 | **17.84** |
| 259 | 11.60 | 10.17 | 14.10 | 2.94 | **38.81** |
| 260 | 115.93 | 121.33 | 85.24 | 65.84 | **388.33** |
| 261 | 18.11 | 16.95 | 16.86 | 7.76 | **59.68** |
| 263 | 6.07 | 7.49 | 5.71 | 6.78 | **26.05** |
| 264 | 4.82 | 3.39 | 4.64 | 3.39 | **16.24** |
| 265 | 28.28 | 24.18 | 24.18 | 17.31 | **93.94** |
| 267 | 44.87 | 38.98 | 35.86 | 21.68 | **141.40** |
| 268 | 0 | 0 | 0 | 0.45 | **0.45** |
| **WV WEST VIRGINIA Total** | **1,031.49** | **941.75** | **718.50** | **611.36** | **3,303.09** |

**STATE: WY - WYOMING**

| REGISTRANT ZIP CODE 3 | 1ST QUARTER | 2ND QUARTER | 3RD QUARTER | 4TH QUARTER | TOTAL GRAMS |
|---|---|---|---|---|---|
| 820 | 12.13 | 15.17 | 12.85 | 14.18 | **54.33** |
| 824 | 1.16 | 2.94 | 2.85 | 2.23 | **9.19** |
| 825 | 20.52 | 23.55 | 26.05 | 14.54 | **84.66** |
| 826 | 21.41 | 20.30 | 20.07 | 14.63 | **76.41** |
| 827 | 10.53 | 14.18 | 11.06 | 8.83 | **44.61** |
| 828 | 6.96 | 3.21 | 5.35 | 4.55 | **20.07** |
| 829 | 6.24 | 11.42 | 15.17 | 8.03 | **40.86** |
| 830 | 0 | 1.07 | 1.07 | 0 | **2.14** |
| 831 | 2.41 | 2.14 | 1.74 | 1.07 | **7.36** |
| **WY - WYOMING Total** | **81.36** | **93.98** | **96.21** | **68.07** | **339.62** |

**DRUG: 9780 - TAPENTADOL**

**STATE: AK - ALASKA**

| REGISTRANT ZIP CODE 3 | 1ST QUARTER | 2ND QUARTER | 3RD QUARTER | 4TH QUARTER | TOTAL GRAMS |
|---|---|---|---|---|---|
| 995 | 1,061.50 | 1,034.00 | 881.50 | 908.50 | **3,885.50** |
| 996 | 369.00 | 412.50 | 381.00 | 378.50 | **1,541.00** |
| 997 | 129.00 | 109.00 | 118.00 | 71.00 | **427.00** |
| **AK ALASKA - Total** | **1,559.50** | **1,555.50** | **1,380.50** | **1,358.00** | **5,853.50** |

**STATE: AL - ALABAMA**

| REGISTRANT ZIP CODE 3 | 1ST QUARTER | 2ND QUARTER | 3RD QUARTER | 4TH QUARTER | TOTAL GRAMS |
|---|---|---|---|---|---|
| 350 | 4,392.50 | 3,952.00 | 4,101.50 | 3,653.00 | **16,099.00** |
| 351 | 2,316.50 | 1,931.50 | 2,052.00 | 1,867.50 | **8,167.50** |
| 352 | 2,710.50 | 2,478.50 | 2,421.50 | 2,313.00 | **9,923.50** |
| 354 | 2,018.50 | 2,002.50 | 1,877.00 | 1,886.50 | **7,784.50** |
| 355 | 1,532.00 | 1,549.00 | 1,381.00 | 1,386.00 | **5,848.00** |
| 356 | 1,944.50 | 1,896.00 | 1,799.50 | 1,805.00 | **7,445.00** |

| | | | | | |
|---|---|---|---|---|---|
| 357 | 1,175.50 | 1,181.00 | 1,009.50 | 1,118.00 | **4,484.00** |
| 358 | 1,547.50 | 1,401.50 | 1,355.00 | 1,409.50 | **5,713.50** |
| 359 | 1,240.50 | 1,182.00 | 946.50 | 1,030.00 | **4,399.00** |
| 360 | 2,977.00 | 2,808.50 | 2,640.00 | 2,562.50 | **10,988.00** |
| 361 | 1,898.00 | 1,691.50 | 1,653.00 | 1,611.00 | **6,853.50** |
| 362 | 991.00 | 938.00 | 818.50 | 572.50 | **3,320.00** |
| 363 | 726.50 | 789.50 | 686.00 | 628.00 | **2,830.00** |
| 364 | 210.00 | 266.00 | 257.00 | 318.50 | **1,051.50** |
| 365 | 2,202.00 | 2,201.00 | 1,834.50 | 1,806.00 | **8,043.50** |
| 366 | 1,579.00 | 1,458.50 | 1,298.00 | 1,269.50 | **5,605.00** |
| 367 | 389.00 | 405.00 | 349.50 | 367.50 | **1,511.00** |
| 368 | 1,893.00 | 1,753.00 | 1,858.00 | 1,806.00 | **7,310.00** |
| 369 | 202.00 | 273.00 | 153.00 | 173.00 | **801.00** |
| **AL ALABAMA - Total** | **31,945.50** | **30,158.00** | **28,491.00** | **27,583.00** | **118,177.50** |

**STATE: AR - ARKANSAS**

| REGISTRANT ZIP CODE 3 | 1ST QUARTER | 2ND QUARTER | 3RD QUARTER | 4TH QUARTER | TOTAL GRAMS |
|---|---|---|---|---|---|
| 716 | 283.50 | 233.50 | 172.00 | 230.50 | **919.50** |
| 717 | 583.00 | 636.50 | 553.50 | 529.00 | **2,302.00** |
| 718 | 166.50 | 77.00 | 48.00 | 44.00 | **335.50** |
| 719 | 421.00 | 482.00 | 394.00 | 441.50 | **1,738.50** |
| 720 | 1,776.00 | 1,721.50 | 1,829.50 | 1,875.50 | **7,202.50** |
| 721 | 1,298.00 | 1,185.00 | 1,158.00 | 999.50 | **4,640.50** |
| 722 | 964.00 | 986.00 | 818.50 | 738.00 | **3,506.50** |
| 723 | 131.50 | 123.50 | 89.50 | 94.00 | **438.50** |
| 724 | 713.50 | 925.50 | 626.00 | 709.50 | **2,974.50** |
| 725 | 399.50 | 472.00 | 317.50 | 345.50 | **1,534.50** |
| 726 | 587.00 | 564.50 | 527.50 | 558.50 | **2,237.50** |
| 727 | 1,937.50 | 1,932.50 | 1,581.50 | 1,552.00 | **7,003.50** |
| 728 | 393.00 | 332.50 | 427.50 | 354.00 | **1,507.00** |
| 729 | 952.50 | 822.00 | 786.00 | 693.50 | **3,254.00** |
| **AR ARKANSAS - Total** | **10,606.50** | **10,494.00** | **9,329.00** | **9,165.00** | **39,594.50** |

**STATE: AZ - ARIZONA**

| REGISTRANT ZIP CODE 3 | 1ST QUARTER | 2ND QUARTER | 3RD QUARTER | 4TH QUARTER | TOTAL GRAMS |
|---|---|---|---|---|---|
| 850 | 3,694.00 | 3,907.00 | 3,443.00 | 3,443.50 | **14,487.50** |
| 851 | 677.00 | 557.50 | 510.00 | 650.00 | **2,394.50** |
| 852 | 9,302.00 | 9,694.50 | 8,528.00 | 9,121.50 | **36,646.00** |
| 853 | 4,290.00 | 4,328.00 | 3,870.00 | 4,032.50 | **16,520.50** |
| 855 | 238.00 | 187.50 | 153.50 | 184.00 | **763.00** |
| 856 | 1,005.00 | 907.50 | 845.50 | 814.50 | **3,572.50** |
| 857 | 3,896.00 | 3,554.50 | 3,307.50 | 3,043.00 | **13,801.00** |

| | | | | | |
|---|---|---|---|---|---|
| 859 | 85.50 | 70.00 | 74.00 | 74.50 | **304.00** |
| 860 | 151.50 | 208.00 | 116.00 | 192.50 | **668.00** |
| 863 | 347.50 | 410.50 | 423.50 | 396.00 | **1,577.50** |
| 864 | 121.00 | 95.00 | 140.50 | 106.00 | **462.50** |
| **AZ ARIZONA - Total** | **23,807.50** | **23,920.00** | **21,411.50** | **22,058.00** | **91,197.00** |

**STATE: CA - CALIFORNIA**

| REGISTRANT ZIP CODE 3 | 1ST QUARTER | 2ND QUARTER | 3RD QUARTER | 4TH QUARTER | **TOTAL GRAMS** |
|---|---|---|---|---|---|
| 900 | 1,522.50 | 1,480.50 | 1,348.50 | 1,262.50 | **5,614.00** |
| 902 | 1,556.50 | 1,457.00 | 1,316.50 | 1,318.50 | **5,648.50** |
| 903 | 78.50 | 40.00 | 62.50 | 77.50 | **258.50** |
| 904 | 990.50 | 1,111.00 | 856.00 | 823.00 | **3,780.50** |
| 905 | 372.00 | 475.50 | 530.00 | 350.50 | **1,728.00** |
| 906 | 675.50 | 614.00 | 575.50 | 456.00 | **2,321.00** |
| 907 | 957.00 | 737.50 | 675.50 | 667.00 | **3,037.00** |
| 908 | 476.00 | 456.50 | 419.50 | 335.00 | **1,687.00** |
| 910 | 313.00 | 317.00 | 208.00 | 185.50 | **1,023.50** |
| 911 | 461.00 | 450.00 | 584.00 | 488.50 | **1,983.50** |
| 912 | 163.00 | 168.50 | 137.50 | 150.00 | **619.00** |
| 913 | 3,627.50 | 3,474.50 | 3,056.00 | 3,023.50 | **13,181.50** |
| 914 | 981.00 | 773.50 | 812.50 | 627.00 | **3,194.00** |
| 915 | 121.00 | 152.00 | 70.00 | 110.50 | **453.50** |
| 916 | 112.50 | 80.00 | 57.50 | 82.50 | **332.50** |
| 917 | 3,641.00 | 4,134.50 | 3,730.50 | 3,724.50 | **15,230.50** |
| 918 | 0 | 5.00 | 22.50 | 10.00 | **37.50** |
| 919 | 1,198.50 | 1,367.00 | 1,163.00 | 965.00 | **4,693.50** |
| 920 | 6,214.50 | 6,234.00 | 6,418.50 | 5,811.50 | **24,678.50** |
| 921 | 2,510.00 | 2,440.00 | 2,331.50 | 2,362.00 | **9,643.50** |
| 922 | 1,609.00 | 1,692.50 | 1,679.50 | 1,636.00 | **6,617.00** |
| 923 | 2,356.00 | 2,308.00 | 1,937.00 | 2,269.50 | **8,870.50** |
| 924 | 251.50 | 245.00 | 233.00 | 142.00 | **871.50** |
| 925 | 4,667.50 | 4,693.50 | 4,746.00 | 4,711.00 | **18,818.00** |
| 926 | 4,734.50 | 4,579.00 | 4,167.00 | 4,501.00 | **17,981.50** |
| 927 | 817.50 | 815.50 | 675.00 | 786.50 | **3,094.50** |
| 928 | 2,821.00 | 2,556.50 | 2,624.50 | 2,504.00 | **10,506.00** |
| 930 | 2,268.00 | 2,039.50 | 1,950.00 | 1,845.50 | **8,103.00** |
| 931 | 296.50 | 202.00 | 243.00 | 284.50 | **1,026.00** |
| 932 | 2,313.00 | 2,271.00 | 2,632.50 | 2,394.50 | **9,611.00** |
| 933 | 1,017.00 | 991.00 | 1,016.50 | 889.50 | **3,914.00** |
| 934 | 1,337.00 | 1,168.00 | 1,249.00 | 1,288.00 | **5,042.00** |
| 935 | 1,059.50 | 976.50 | 1,114.50 | 933.00 | **4,083.50** |

| | | | | | |
|---|---|---|---|---|---|
| 936 | 1,709.50 | 1,657.50 | 1,804.00 | 1,644.50 | **6,815.50** |
| 937 | 2,902.00 | 2,725.00 | 2,597.00 | 2,650.50 | **10,874.50** |
| 939 | 616.50 | 700.00 | 529.00 | 675.50 | **2,521.00** |
| 940 | 997.50 | 1,007.50 | 1,207.00 | 1,152.00 | **4,364.00** |
| 941 | 489.50 | 493.50 | 541.50 | 587.00 | **2,111.50** |
| 943 | 5.00 | 0 | 5.00 | 0 | **10.00** |
| 944 | 151.00 | 236.50 | 207.00 | 262.00 | **856.50** |
| 945 | 2,836.00 | 2,584.50 | 2,173.50 | 2,261.00 | **9,855.00** |
| 946 | 246.50 | 239.50 | 200.00 | 210.50 | **896.50** |
| 947 | 64.00 | 32.00 | 10.00 | 37.50 | **143.50** |
| 948 | 5.00 | 3.00 | 5.00 | 6.00 | **19.00** |
| 949 | 268.00 | 253.00 | 274.00 | 274.50 | **1,069.50** |
| 950 | 464.00 | 461.00 | 400.00 | 509.00 | **1,834.00** |
| 951 | 354.00 | 281.00 | 369.00 | 220.00 | **1,224.00** |
| 952 | 273.50 | 317.00 | 301.00 | 237.00 | **1,128.50** |
| 953 | 1,358.00 | 1,319.00 | 1,157.00 | 1,124.50 | **4,958.50** |
| 954 | 403.50 | 413.50 | 466.00 | 366.00 | **1,649.00** |
| 955 | 9.00 | 9.00 | 18.00 | 3.00 | **39.00** |
| 956 | 1,793.00 | 1,972.00 | 1,956.00 | 2,047.50 | **7,768.50** |
| 957 | 476.00 | 570.00 | 510.50 | 443.00 | **1,999.50** |
| 958 | 1,433.00 | 1,272.00 | 1,060.50 | 1,100.00 | **4,865.50** |
| 959 | 1,010.50 | 1,095.00 | 936.00 | 973.00 | **4,014.50** |
| 960 | 466.00 | 460.00 | 517.50 | 422.50 | **1,866.00** |
| 961 | 6.00 | 21.00 | 0 | 13.00 | **40.00** |
| **CA - CALIFORNIA - Total** | **69,856.50** | **68,628.50** | **65,887.50** | **64,235.00** | **268,607.50** |

**STATE: CO - COLORADO**

| REGISTRANT ZIP CODE 3 | 1ST QUARTER | 2ND QUARTER | 3RD QUARTER | 4TH QUARTER | **TOTAL GRAMS** |
|---|---|---|---|---|---|
| 800 | 2,655.00 | 3,006.50 | 2,745.00 | 3,121.50 | **11,528.00** |
| 801 | 4,884.00 | 5,433.00 | 5,420.00 | 5,925.00 | **21,662.00** |
| 802 | 2,681.00 | 2,726.00 | 2,519.50 | 2,591.00 | **10,517.50** |
| 803 | 611.00 | 749.50 | 801.50 | 694.50 | **2,856.50** |
| 804 | 647.00 | 596.50 | 561.00 | 640.00 | **2,444.50** |
| 805 | 2,976.00 | 2,859.00 | 2,652.50 | 2,249.00 | **10,736.50** |
| 806 | 1,145.00 | 1,001.50 | 1,064.00 | 987.50 | **4,198.00** |
| 807 | 178.50 | 183.00 | 146.50 | 165.50 | **673.50** |
| 808 | 431.00 | 441.50 | 498.00 | 457.00 | **1,827.50** |
| 809 | 3,111.50 | 2,911.50 | 2,790.50 | 2,840.00 | **11,653.50** |
| 810 | 1,401.00 | 1,365.50 | 1,240.50 | 1,336.00 | **5,343.00** |
| 811 | 79.00 | 122.00 | 141.50 | 140.50 | **483.00** |
| 812 | 302.00 | 294.00 | 295.50 | 514.50 | **1,406.00** |

| | | | | | |
|---|---|---|---|---|---|
| 813 | 250.00 | 196.50 | 176.50 | 134.00 | **757.00** |
| 814 | 79.00 | 89.00 | 69.00 | 127.00 | **364.00** |
| 815 | 230.00 | 250.00 | 323.00 | 252.00 | **1,055.00** |
| 816 | 148.50 | 178.00 | 159.50 | 266.50 | **752.50** |
| **CO - COLORADO - Total** | **21,809.50** | **22,403.00** | **21,604.00** | **22,441.50** | **88,258.00** |

**STATE: CT - CONNECTICUT**

| REGISTRANT ZIP CODE 3 | 1ST QUARTER | 2ND QUARTER | 3RD QUARTER | 4TH QUARTER | TOTAL GRAMS |
|---|---|---|---|---|---|
| 060 | 1,099.50 | 1,167.50 | 1,164.00 | 924.50 | **4,355.50** |
| 061 | 394.00 | 285.00 | 309.50 | 232.50 | **1,221.00** |
| 062 | 451.00 | 433.50 | 419.00 | 331.50 | **1,635.00** |
| 063 | 928.00 | 882.00 | 651.00 | 616.00 | **3,077.00** |
| 064 | 2,026.00 | 1,893.50 | 1,879.50 | 1,838.50 | **7,637.50** |
| 065 | 679.50 | 596.00 | 677.50 | 515.00 | **2,468.00** |
| 066 | 286.00 | 265.50 | 245.50 | 265.50 | **1,062.50** |
| 067 | 616.00 | 623.50 | 557.50 | 648.00 | **2,445.00** |
| 068 | 1,102.50 | 951.50 | 1,015.50 | 930.00 | **3,999.50** |
| 069 | 130.00 | 116.00 | 124.00 | 107.00 | **477.00** |
| **CT CONNECTICUT Total** | **7,712.50** | **7,214.00** | **7,043.00** | **6,408.50** | **28,378.00** |

**STATE: DC - DISTRICT OF COLUMBIA**

| REGISTRANT ZIP CODE 3 | 1ST QUARTER | 2ND QUARTER | 3RD QUARTER | 4TH QUARTER | TOTAL GRAMS |
|---|---|---|---|---|---|
| 200 | 719.00 | 609.50 | 548.00 | 644.00 | **2,520.50** |
| 204 | 15.00 | 15.00 | 0 | 0 | **30.00** |
| **DC - DISTRICT OF COLUMBIA** | **734.00** | **624.50** | **548.00** | **644.00** | **2,550.50** |

**STATE: DE - DELAWARE**

| REGISTRANT ZIP CODE 3 | 1ST QUARTER | 2ND QUARTER | 3RD QUARTER | 4TH QUARTER | TOTAL GRAMS |
|---|---|---|---|---|---|
| 197 | 2,077.00 | 2,092.50 | 2,022.00 | 2,034.00 | **8,225.50** |
| 198 | 844.50 | 842.50 | 921.50 | 931.50 | **3,540.00** |
| 199 | 1,676.50 | 1,566.50 | 1,523.00 | 1,438.50 | **6,204.50** |
| **DE DELAWARE Total** | **4,598.00** | **4,501.50** | **4,466.50** | **4,404.00** | **17,970.00** |

**STATE: FL - FLORIDA**

| REGISTRANT ZIP CODE 3 | 1ST QUARTER | 2ND QUARTER | 3RD QUARTER | 4TH QUARTER | TOTAL GRAMS |
|---|---|---|---|---|---|
| 320 | 2,074.50 | 1,805.50 | 1,989.50 | 1,844.00 | **7,713.50** |
| 321 | 1,652.50 | 1,317.50 | 1,382.50 | 1,390.00 | **5,742.50** |
| 322 | 3,074.50 | 2,939.00 | 2,894.50 | 2,960.00 | **11,868.00** |
| 323 | 2,312.00 | 2,254.00 | 2,257.50 | 2,168.50 | **8,992.00** |
| 324 | 1,086.50 | 1,207.50 | 1,255.50 | 1,125.00 | **4,674.50** |
| 325 | 2,425.50 | 2,133.00 | 1,956.50 | 1,858.00 | **8,373.00** |
| 326 | 1,009.50 | 1,132.50 | 1,001.00 | 983.50 | **4,126.50** |
| 327 | 3,368.50 | 3,442.50 | 3,203.50 | 3,169.00 | **13,183.50** |
| 328 | 2,200.50 | 2,256.50 | 2,185.50 | 2,298.00 | **8,940.50** |

| | | | | | |
|---|---|---|---|---|---|
| 329 | 3,081.50 | 2,660.50 | 2,679.00 | 2,534.50 | **10,955.50** |
| 330 | 2,171.50 | 2,224.50 | 1,978.00 | 1,733.50 | **8,107.50** |
| 331 | 374.50 | 478.00 | 426.50 | 441.50 | **1,720.50** |
| 333 | 1,511.50 | 1,596.00 | 1,298.00 | 1,412.00 | **5,817.50** |
| 334 | 5,192.50 | 4,943.50 | 5,095.50 | 4,663.50 | **19,895.00** |
| 335 | 3,813.00 | 3,776.50 | 3,483.00 | 3,389.00 | **14,461.50** |
| 336 | 2,201.00 | 2,370.50 | 2,081.00 | 2,430.50 | **9,083.00** |
| 337 | 2,690.50 | 2,571.50 | 2,343.00 | 2,171.50 | **9,776.50** |
| 338 | 1,783.50 | 1,726.00 | 1,792.00 | 1,843.45 | **7,144.95** |
| 339 | 5,935.50 | 5,450.50 | 5,067.00 | 5,643.00 | **22,096.00** |
| 341 | 2,853.00 | 2,500.50 | 2,032.00 | 2,397.00 | **9,782.50** |
| 342 | 3,720.50 | 3,423.00 | 3,287.00 | 3,251.00 | **13,681.50** |
| 344 | 1,490.00 | 1,452.50 | 1,205.50 | 1,227.00 | **5,375.00** |
| 346 | 2,707.00 | 2,933.50 | 2,596.50 | 2,476.00 | **10,713.00** |
| 347 | 2,134.00 | 1,733.50 | 1,872.00 | 1,699.50 | **7,439.00** |
| 349 | 2,008.00 | 1,784.00 | 1,741.50 | 1,595.00 | **7,128.50** |
| **FL FLORIDA - Total** | **62,871.50** | **60,112.50** | **57,103.50** | **56,703.95** | **236,791.45** |

**STATE: GA - GEORGIA**

| REGISTRANT ZIP CODE 3 | 1ST QUARTER | 2ND QUARTER | 3RD QUARTER | 4TH QUARTER | **TOTAL GRAMS** |
|---|---|---|---|---|---|
| 300 | 7,730.50 | 7,492.00 | 7,760.50 | 7,249.50 | **30,232.50** |
| 301 | 6,062.00 | 5,644.00 | 5,469.00 | 5,404.50 | **22,579.50** |
| 302 | 3,720.50 | 3,737.50 | 3,407.50 | 3,469.00 | **14,334.50** |
| 303 | 2,290.00 | 2,126.50 | 2,078.50 | 2,114.00 | **8,609.00** |
| 304 | 2,073.00 | 2,176.00 | 2,166.00 | 2,153.50 | **8,568.50** |
| 305 | 2,706.00 | 2,910.00 | 3,115.00 | 2,663.50 | **11,394.50** |
| 306 | 1,403.50 | 1,334.50 | 1,395.00 | 1,425.50 | **5,558.50** |
| 307 | 838.50 | 718.00 | 782.00 | 735.50 | **3,074.00** |
| 308 | 445.50 | 452.50 | 584.00 | 466.00 | **1,948.00** |
| 309 | 740.50 | 643.50 | 615.00 | 648.50 | **2,647.50** |
| 310 | 2,699.00 | 2,452.50 | 2,418.00 | 2,378.00 | **9,947.50** |
| 312 | 608.00 | 515.00 | 619.50 | 581.50 | **2,324.00** |
| 313 | 3,242.50 | 3,088.00 | 2,853.00 | 2,962.50 | **12,146.00** |
| 314 | 1,576.00 | 1,575.00 | 1,418.00 | 1,519.00 | **6,088.00** |
| 315 | 5,140.00 | 4,540.50 | 4,931.50 | 5,108.50 | **19,720.50** |
| 316 | 986.50 | 1,059.00 | 854.50 | 879.00 | **3,779.00** |
| 317 | 1,166.50 | 1,079.50 | 1,101.00 | 1,113.50 | **4,460.50** |
| 318 | 192.00 | 145.00 | 146.00 | 86.00 | **569.00** |
| 319 | 620.00 | 508.00 | 450.50 | 466.00 | **2,044.50** |
| 398 | 226.00 | 185.00 | 175.50 | 210.00 | **796.50** |
| **GA GEORGIA Total** | **44,466.50** | **42,382.00** | **42,340.00** | **41,633.50** | **170,822.00** |

**STATE: HI - HAWAII**

| REGISTRANT ZIP CODE 3 | 1ST QUARTER | 2ND QUARTER | 3RD QUARTER | 4TH QUARTER | TOTAL GRAMS |
|---|---|---|---|---|---|
| 967 | 779.50 | 738.50 | 690.50 | 695.50 | **2,904.00** |
| 968 | 721.00 | 536.50 | 664.50 | 714.50 | **2,636.50** |
| **HI - HAWAII - Total** | **1,500.50** | **1,275.00** | **1,355.00** | **1,410.00** | **5,540.50** |

**STATE: IA - IOWA**

| REGISTRANT ZIP CODE 3 | 1ST QUARTER | 2ND QUARTER | 3RD QUARTER | 4TH QUARTER | TOTAL GRAMS |
|---|---|---|---|---|---|
| 500 | 1,129.00 | 867.00 | 817.00 | 775.50 | **3,588.50** |
| 501 | 588.50 | 574.50 | 568.00 | 559.50 | **2,290.50** |
| 502 | 1,039.50 | 1,090.00 | 1,059.50 | 1,126.50 | **4,315.50** |
| 503 | 1,577.00 | 1,659.50 | 1,381.00 | 1,441.50 | **6,059.00** |
| 504 | 34.00 | 24.00 | 20.00 | 55.00 | **133.00** |
| 505 | 187.00 | 154.50 | 134.50 | 176.50 | **652.50** |
| 506 | 379.00 | 326.00 | 307.00 | 281.50 | **1,293.50** |
| 507 | 278.00 | 176.50 | 219.50 | 277.00 | **951.00** |
| 508 | 88.50 | 128.50 | 136.50 | 190.00 | **543.50** |
| 510 | 203.50 | 281.00 | 262.00 | 310.00 | **1,056.50** |
| 511 | 446.00 | 248.50 | 307.50 | 380.50 | **1,382.50** |
| 512 | 122.50 | 72.50 | 105.50 | 83.00 | **383.50** |
| 513 | 182.50 | 194.50 | 234.00 | 207.00 | **818.00** |
| 514 | 57.00 | 35.00 | 40.00 | 40.00 | **172.00** |
| 515 | 477.50 | 447.50 | 366.00 | 397.00 | **1,688.00** |
| 516 | 165.50 | 144.00 | 126.50 | 188.00 | **624.00** |
| 520 | 348.00 | 354.00 | 365.00 | 313.00 | **1,380.00** |
| 521 | 35.00 | 20.00 | 50.00 | 20.00 | **125.00** |
| 522 | 78.00 | 56.00 | 12.00 | 47.00 | **193.00** |
| 523 | 145.00 | 192.00 | 157.00 | 107.50 | **601.50** |
| 524 | 156.50 | 108.00 | 141.00 | 173.00 | **578.50** |
| 525 | 228.50 | 207.00 | 194.00 | 190.00 | **819.50** |
| 526 | 0 | 5.00 | 7.50 | 15.00 | **27.50** |
| 527 | 85.50 | 108.50 | 113.50 | 110.00 | **417.50** |
| 528 | 129.50 | 84.00 | 44.00 | 108.00 | **365.50** |
| **IA IOWA Total** | **8,161.00** | **7,558.00** | **7,168.50** | **7,572.00** | **30,459.50** |

**STATE: ID - IDAHO**

| REGISTRANT ZIP CODE 3 | 1ST QUARTER | 2ND QUARTER | 3RD QUARTER | 4TH QUARTER | TOTAL GRAMS |
|---|---|---|---|---|---|
| 832 | 990.50 | 1,033.00 | 1,015.50 | 977.00 | **4,016.00** |
| 833 | 369.00 | 402.00 | 372.00 | 373.00 | **1,516.00** |
| 834 | 680.00 | 789.00 | 713.00 | 654.00 | **2,836.00** |
| 835 | 82.00 | 83.00 | 83.00 | 138.00 | **386.00** |
| 836 | 1,167.50 | 991.50 | 1,027.00 | 1,211.00 | **4,397.00** |

| | | | | | |
|---|---|---|---|---|---|
| 837 | 794.50 | 788.00 | 703.50 | 625.00 | **2,911.00** |
| 838 | 553.00 | 623.00 | 634.50 | 466.00 | **2,276.50** |
| **ID IDAHO Total** | **4,636.50** | **4,709.50** | **4,548.50** | **4,444.00** | **18,338.50** |

**STATE: IL - ILLINOIS**

| REGISTRANT ZIP CODE 3 | 1ST QUARTER | 2ND QUARTER | 3RD QUARTER | 4TH QUARTER | TOTAL GRAMS |
|---|---|---|---|---|---|
| 600 | 4,067.50 | 4,165.50 | 3,936.50 | 4,357.50 | **16,527.00** |
| 601 | 2,197.50 | 2,345.50 | 2,608.50 | 2,379.00 | **9,530.50** |
| 602 | 244.50 | 172.50 | 164.50 | 168.00 | **749.50** |
| 603 | 243.50 | 223.50 | 319.50 | 219.50 | **1,006.00** |
| 604 | 2,749.00 | 2,875.00 | 2,679.00 | 2,472.50 | **10,775.50** |
| 605 | 2,660.00 | 2,296.00 | 2,200.50 | 2,101.50 | **9,258.00** |
| 606 | 1,220.50 | 1,164.50 | 1,171.50 | 1,202.50 | **4,759.00** |
| 607 | 200.50 | 218.50 | 176.50 | 238.00 | **833.50** |
| 608 | 214.50 | 231.00 | 218.00 | 185.50 | **849.00** |
| 609 | 581.00 | 471.50 | 520.50 | 478.00 | **2,051.00** |
| 610 | 615.00 | 655.50 | 760.50 | 687.00 | **2,718.00** |
| 611 | 703.50 | 604.00 | 700.00 | 598.00 | **2,605.50** |
| 612 | 122.50 | 108.00 | 164.50 | 151.00 | **546.00** |
| 613 | 117.00 | 69.50 | 74.00 | 107.00 | **367.50** |
| 614 | 57.00 | 68.00 | 93.00 | 108.00 | **326.00** |
| 615 | 392.00 | 306.00 | 421.00 | 464.00 | **1,583.00** |
| 616 | 277.00 | 303.00 | 230.00 | 230.00 | **1,040.00** |
| 617 | 551.50 | 607.50 | 636.50 | 551.50 | **2,347.00** |
| 618 | 1,530.50 | 1,601.00 | 1,634.50 | 1,392.00 | **6,158.00** |
| 619 | 244.50 | 162.50 | 257.50 | 320.00 | **984.50** |
| 620 | 813.00 | 799.50 | 854.00 | 841.50 | **3,308.00** |
| 622 | 1,927.00 | 1,822.00 | 1,919.00 | 1,760.00 | **7,428.00** |
| 623 | 206.50 | 195.00 | 233.00 | 202.00 | **836.50** |
| 624 | 152.50 | 160.00 | 165.00 | 109.00 | **586.50** |
| 625 | 431.00 | 436.50 | 496.50 | 506.00 | **1,870.00** |
| 626 | 151.50 | 186.50 | 190.00 | 174.00 | **702.00** |
| 627 | 173.00 | 105.00 | 133.00 | 107.00 | **518.00** |
| 628 | 392.00 | 345.50 | 354.00 | 337.00 | **1,428.50** |
| 629 | 318.50 | 392.50 | 259.00 | 265.50 | **1,235.50** |
| **IL ILLINOIS Total** | **23,554.00** | **23,091.00** | **23,570.00** | **22,712.50** | **92,927.50** |

**STATE: IN - INDIANA**

| REGISTRANT ZIP CODE 3 | 1ST QUARTER | 2ND QUARTER | 3RD QUARTER | 4TH QUARTER | TOTAL GRAMS |
|---|---|---|---|---|---|
| 460 | 1,158.00 | 931.00 | 891.50 | 942.00 | **3,922.50** |
| 461 | 1,874.50 | 2,170.00 | 1,981.50 | 2,028.50 | **8,054.50** |
| 462 | 1,263.00 | 1,428.50 | 1,583.00 | 1,540.00 | **5,814.50** |

| | | | | | |
|---|---|---|---|---|---|
| 463 | 2,671.00 | 3,094.00 | 2,806.50 | 2,746.50 | **11,318.00** |
| 464 | 279.50 | 305.50 | 317.00 | 344.50 | **1,246.50** |
| 465 | 1,795.50 | 1,701.00 | 1,896.00 | 1,814.50 | **7,207.00** |
| 466 | 505.00 | 375.50 | 299.50 | 273.50 | **1,453.50** |
| 467 | 2,219.00 | 2,441.00 | 2,597.00 | 2,599.50 | **9,856.50** |
| 468 | 2,865.50 | 3,075.00 | 3,429.50 | 3,471.50 | **12,841.50** |
| 469 | 1,016.50 | 886.00 | 990.50 | 931.00 | **3,824.00** |
| 470 | 205.00 | 210.50 | 305.50 | 323.00 | **1,044.00** |
| 471 | 473.50 | 418.50 | 463.50 | 490.50 | **1,846.00** |
| 472 | 465.00 | 580.50 | 641.50 | 412.00 | **2,099.00** |
| 473 | 590.50 | 526.00 | 613.50 | 609.00 | **2,339.00** |
| 474 | 467.50 | 500.50 | 438.50 | 474.50 | **1,881.00** |
| 475 | 545.50 | 536.00 | 609.00 | 521.00 | **2,211.50** |
| 476 | 587.00 | 559.50 | 538.50 | 500.50 | **2,185.50** |
| 477 | 1,083.50 | 977.00 | 1,060.00 | 1,007.50 | **4,128.00** |
| 478 | 3,837.00 | 3,551.00 | 3,599.00 | 3,618.50 | **14,605.50** |
| 479 | 549.00 | 586.00 | 577.00 | 505.00 | **2,217.00** |
| **IN INDIANA Total** | **24,451.00** | **24,853.00** | **25,638.00** | **25,153.00** | **100,095.00** |

**STATE: KS - KANSAS**

| REGISTRANT ZIP CODE 3 | 1ST QUARTER | 2ND QUARTER | 3RD QUARTER | 4TH QUARTER | **TOTAL GRAMS** |
|---|---|---|---|---|---|
| 660 | 1,429.50 | 1,472.50 | 1,294.00 | 1,084.50 | **5,280.50** |
| 661 | 130.00 | 157.50 | 145.00 | 143.50 | **576.00** |
| 662 | 1,181.00 | 1,086.50 | 1,128.50 | 1,185.00 | **4,581.00** |
| 664 | 95.00 | 131.00 | 129.00 | 140.00 | **495.00** |
| 665 | 216.50 | 257.50 | 191.50 | 176.50 | **842.00** |
| 666 | 695.00 | 690.00 | 582.00 | 563.00 | **2,530.00** |
| 667 | 147.00 | 139.00 | 148.00 | 183.00 | **617.00** |
| 668 | 70.00 | 48.00 | 12.00 | 18.00 | **148.00** |
| 669 | 5.00 | 0 | 20.00 | 38.00 | **63.00** |
| 670 | 70.00 | 90.00 | 97.50 | 146.00 | **403.50** |
| 671 | 73.00 | 95.00 | 80.00 | 70.00 | **318.00** |
| 672 | 577.00 | 593.00 | 441.50 | 412.00 | **2,023.50** |
| 673 | 138.00 | 199.00 | 141.00 | 117.00 | **595.00** |
| 674 | 141.50 | 163.00 | 151.00 | 211.00 | **666.50** |
| 675 | 141.00 | 107.00 | 125.00 | 126.00 | **499.00** |
| 676 | 89.00 | 83.50 | 52.00 | 67.00 | **291.50** |
| 677 | 50.00 | 45.00 | 32.00 | 87.00 | **214.00** |
| 678 | 51.00 | 75.00 | 44.00 | 63.00 | **233.00** |
| 679 | 0 | 0 | 7.50 | 26.00 | **33.50** |
| **KS - KANSAS - Total** | **5,299.50** | **5,432.50** | **4,821.50** | **4,856.50** | **20,410.00** |

**STATE: KY - KENTUCKY**

| REGISTRANT ZIP CODE 3 | 1ST QUARTER | 2ND QUARTER | 3RD QUARTER | 4TH QUARTER | TOTAL GRAMS |
|---|---|---|---|---|---|
| 400 | 378.50 | 370.50 | 312.00 | 249.50 | **1,310.50** |
| 401 | 222.50 | 306.00 | 273.50 | 230.50 | **1,032.50** |
| 402 | 1,373.50 | 1,364.00 | 1,361.50 | 1,538.50 | **5,637.50** |
| 403 | 376.00 | 402.00 | 478.00 | 387.00 | **1,643.00** |
| 404 | 158.50 | 112.50 | 100.50 | 112.00 | **483.50** |
| 405 | 593.50 | 508.00 | 450.00 | 474.00 | **2,025.50** |
| 406 | 0 | 16.00 | 19.00 | 13.00 | **48.00** |
| 407 | 137.00 | 63.00 | 118.50 | 141.00 | **459.50** |
| 408 | 34.00 | 0 | 0 | 0 | **34.00** |
| 409 | 40.00 | 32.50 | 8.00 | 5.00 | **85.50** |
| 410 | 1,736.50 | 1,661.00 | 1,555.50 | 1,662.50 | **6,615.50** |
| 411 | 103.00 | 126.00 | 101.50 | 130.00 | **460.50** |
| 412 | 42.50 | 32.50 | 30.00 | 15.00 | **120.00** |
| 413 | 0 | 5.00 | 10.00 | 5.00 | **20.00** |
| 415 | 150.50 | 104.00 | 115.00 | 109.00 | **478.50** |
| 416 | 133.00 | 88.50 | 109.50 | 109.50 | **440.50** |
| 417 | 60.00 | 53.00 | 42.50 | 59.00 | **214.50** |
| 418 | 57.00 | 21.00 | 53.00 | 42.00 | **173.00** |
| 420 | 1,340.50 | 1,162.00 | 1,325.50 | 1,056.00 | **4,884.00** |
| 421 | 377.00 | 446.50 | 447.00 | 449.50 | **1,720.00** |
| 422 | 823.50 | 357.00 | 658.00 | 314.50 | **2,153.00** |
| 423 | 918.00 | 882.50 | 801.50 | 736.00 | **3,338.00** |
| 424 | 390.00 | 484.00 | 444.00 | 425.50 | **1,743.50** |
| 425 | 25.00 | 32.00 | 33.00 | 20.00 | **110.00** |
| 426 | 37.00 | 96.00 | 100.00 | 40.00 | **273.00** |
| 427 | 453.50 | 448.50 | 414.50 | 400.00 | **1,716.50** |
| **KY - KENTUCKY - Total** | **9,960.50** | **9,174.00** | **9,361.50** | **8,724.00** | **37,220.00** |

**STATE: LA - LOUISIANA**

| REGISTRANT ZIP CODE 3 | 1ST QUARTER | 2ND QUARTER | 3RD QUARTER | 4TH QUARTER | TOTAL GRAMS |
|---|---|---|---|---|---|
| 700 | 1,428.00 | 1,382.50 | 1,316.00 | 1,237.00 | **5,363.50** |
| 701 | 170.00 | 199.00 | 140.00 | 218.50 | **727.50** |
| 703 | 468.50 | 703.50 | 584.50 | 563.50 | **2,320.00** |
| 704 | 2,648.50 | 2,555.00 | 2,404.00 | 2,679.00 | **10,286.50** |
| 705 | 1,019.00 | 1,141.00 | 1,212.50 | 1,254.50 | **4,627.00** |
| 706 | 387.00 | 249.50 | 390.50 | 333.00 | **1,360.00** |
| 707 | 2,576.50 | 2,293.00 | 2,136.50 | 1,779.00 | **8,785.00** |
| 708 | 902.00 | 753.50 | 848.00 | 946.00 | **3,449.50** |
| 710 | 204.50 | 258.00 | 212.00 | 215.00 | **889.50** |

| | | | | | |
|---|---|---|---|---|---|
| 711 | 1,054.00 | 950.50 | 1,027.00 | 808.50 | **3,840.00** |
| 712 | 323.50 | 313.00 | 336.50 | 280.00 | **1,253.00** |
| 713 | 518.50 | 455.00 | 538.00 | 561.00 | **2,072.50** |
| 714 | 432.50 | 503.00 | 296.50 | 428.00 | **1,660.00** |
| **LA - LOUISIANA - Total** | **12,132.50** | **11,756.50** | **11,442.00** | **11,303.00** | **46,634.00** |

**STATE: MA - MASSACHUSETTS**

| REGISTRANT ZIP CODE 3 | 1ST QUARTER | 2ND QUARTER | 3RD QUARTER | 4TH QUARTER | **TOTAL GRAMS** |
|---|---|---|---|---|---|
| 010 | 954.50 | 888.50 | 974.50 | 953.50 | **3,771.00** |
| 011 | 191.50 | 202.50 | 144.00 | 149.00 | **687.00** |
| 012 | 60.00 | 60.00 | 105.00 | 60.00 | **285.00** |
| 013 | 128.00 | 48.00 | 30.00 | 25.00 | **231.00** |
| 014 | 257.00 | 190.00 | 261.00 | 236.00 | **944.00** |
| 015 | 467.00 | 456.50 | 441.00 | 537.50 | **1,902.00** |
| 016 | 136.00 | 179.50 | 250.00 | 209.00 | **774.50** |
| 017 | 555.50 | 598.00 | 574.50 | 536.00 | **2,264.00** |
| 018 | 7,703.00 | 7,650.50 | 8,014.50 | 7,565.50 | **30,933.50** |
| 019 | 1,178.50 | 1,383.50 | 1,326.00 | 1,091.00 | **4,979.00** |
| 020 | 320.50 | 244.50 | 322.00 | 238.00 | **1,125.00** |
| 021 | 639.50 | 737.50 | 761.50 | 642.50 | **2,781.00** |
| 022 | 96.50 | 107.50 | 96.00 | 110.00 | **410.00** |
| 023 | 450.00 | 567.00 | 623.00 | 644.50 | **2,284.50** |
| 024 | 303.00 | 350.00 | 387.00 | 279.00 | **1,319.00** |
| 025 | 75.00 | 44.00 | 53.00 | 111.50 | **283.50** |
| 026 | 214.50 | 342.50 | 304.50 | 251.50 | **1,113.00** |
| 027 | 1,774.50 | 1,554.00 | 1,609.00 | 1,686.50 | **6,624.00** |
| **MA - MASSACHUSETTS - Total** | **15,504.50** | **15,604.00** | **16,276.50** | **15,326.00** | **62,711.00** |

**STATE: MD - MARYLAND**

| REGISTRANT ZIP CODE 3 | 1ST QUARTER | 2ND QUARTER | 3RD QUARTER | 4TH QUARTER | **TOTAL GRAMS** |
|---|---|---|---|---|---|
| 206 | 1,649.50 | 1,712.00 | 1,644.00 | 1,577.50 | **6,583.00** |
| 207 | 1,597.00 | 1,562.50 | 1,766.00 | 1,683.50 | **6,609.00** |
| 208 | 2,554.00 | 3,060.50 | 2,634.00 | 2,576.00 | **10,824.50** |
| 209 | 352.00 | 244.00 | 266.00 | 255.50 | **1,117.50** |
| 210 | 4,607.00 | 4,636.00 | 4,573.50 | 5,033.00 | **18,849.50** |
| 211 | 2,261.50 | 2,201.00 | 2,272.00 | 2,030.50 | **8,765.00** |
| 212 | 2,957.50 | 2,747.50 | 2,709.50 | 2,838.50 | **11,253.00** |
| 214 | 1,068.50 | 949.00 | 914.50 | 706.50 | **3,638.50** |
| 215 | 326.00 | 534.50 | 573.00 | 688.50 | **2,122.00** |
| 216 | 1,259.50 | 1,214.50 | 1,239.50 | 1,044.00 | **4,757.50** |
| 217 | 3,566.50 | 3,663.00 | 3,542.00 | 3,499.50 | **14,271.00** |
| 218 | 872.50 | 790.50 | 746.50 | 669.50 | **3,079.00** |

| | | | | | |
|---|---|---|---|---|---|
| 219 | 285.00 | 266.00 | 270.50 | 217.50 | **1,039.00** |
| **MD MARYLAND Total** | **23,356.50** | **23,581.00** | **23,151.00** | **22,820.00** | **92,908.50** |

**STATE: ME - MAINE**

| REGISTRANT ZIP CODE 3 | 1ST QUARTER | 2ND QUARTER | 3RD QUARTER | 4TH QUARTER | TOTAL GRAMS |
|---|---|---|---|---|---|
| 039 | 90.00 | 92.00 | 133.00 | 129.00 | **444.00** |
| 040 | 232.00 | 309.50 | 308.00 | 277.00 | **1,126.50** |
| 041 | 21.00 | 34.00 | 22.00 | 28.00 | **105.00** |
| 042 | 215.00 | 280.00 | 216.00 | 181.00 | **892.00** |
| 043 | 59.00 | 63.50 | 95.50 | 74.00 | **292.00** |
| 044 | 70.00 | 67.00 | 67.00 | 92.50 | **296.50** |
| 045 | 83.50 | 62.00 | 74.00 | 79.00 | **298.50** |
| 046 | 30.00 | 32.50 | 68.50 | 17.50 | **148.50** |
| 047 | 170.00 | 129.00 | 150.00 | 151.00 | **600.00** |
| 048 | 27.00 | 34.50 | 45.00 | 27.00 | **133.50** |
| 049 | 186.00 | 145.00 | 187.00 | 142.00 | **660.00** |
| **ME MAINE Total** | **1,183.50** | **1,249.00** | **1,366.00** | **1,198.00** | **4,996.50** |

**STATE: MI - MICHIGAN**

| REGISTRANT ZIP CODE 3 | 1ST QUARTER | 2ND QUARTER | 3RD QUARTER | 4TH QUARTER | TOTAL GRAMS |
|---|---|---|---|---|---|
| 480 | 1,403.00 | 1,462.50 | 1,535.50 | 1,548.00 | **5,949.00** |
| 481 | 2,263.50 | 2,157.50 | 2,203.00 | 2,016.00 | **8,640.00** |
| 482 | 216.50 | 218.00 | 166.50 | 211.50 | **812.50** |
| 483 | 1,552.00 | 1,546.50 | 1,463.00 | 1,549.00 | **6,110.50** |
| 484 | 351.00 | 373.50 | 372.00 | 402.50 | **1,499.00** |
| 485 | 374.50 | 422.50 | 340.00 | 301.00 | **1,438.00** |
| 486 | 523.00 | 595.00 | 633.00 | 670.50 | **2,421.50** |
| 487 | 263.00 | 255.50 | 290.00 | 283.50 | **1,092.00** |
| 488 | 1,348.00 | 1,269.00 | 1,263.00 | 1,217.00 | **5,097.00** |
| 489 | 236.00 | 213.00 | 241.00 | 239.00 | **929.00** |
| 490 | 1,812.50 | 1,758.50 | 1,606.00 | 1,473.00 | **6,650.00** |
| 491 | 311.00 | 195.00 | 401.00 | 320.00 | **1,227.00** |
| 492 | 393.50 | 256.00 | 339.50 | 218.00 | **1,207.00** |
| 493 | 684.50 | 633.50 | 621.50 | 573.50 | **2,513.00** |
| 494 | 598.50 | 513.00 | 545.00 | 526.50 | **2,183.00** |
| 495 | 862.00 | 598.50 | 556.00 | 589.00 | **2,605.50** |
| 496 | 573.50 | 413.00 | 501.50 | 385.00 | **1,873.00** |
| 497 | 390.50 | 419.50 | 410.00 | 418.00 | **1,638.00** |
| 498 | 96.00 | 51.00 | 47.00 | 90.00 | **284.00** |
| 499 | 24.00 | 57.00 | 88.00 | 79.00 | **248.00** |
| **MI - MICHIGAN Total** | **14,276.50** | **13,408.00** | **13,622.50** | **13,110.00** | **54,417.00** |

**STATE: MN - MINNESOTA**

| REGISTRANT ZIP CODE 3 | 1ST QUARTER | 2ND QUARTER | 3RD QUARTER | 4TH QUARTER | TOTAL GRAMS |
|---|---|---|---|---|---|
| 550 | 356.00 | 387.50 | 507.00 | 492.50 | **1,743.00** |
| 551 | 658.50 | 746.00 | 590.00 | 615.50 | **2,610.00** |
| 553 | 404.00 | 435.00 | 317.00 | 258.50 | **1,414.50** |
| 554 | 621.50 | 633.50 | 602.50 | 486.50 | **2,344.00** |
| 557 | 0 | 0 | 15.00 | 8.00 | **23.00** |
| 558 | 96.00 | 130.00 | 96.00 | 103.00 | **425.00** |
| 559 | 145.00 | 110.00 | 95.50 | 62.50 | **413.00** |
| 560 | 132.00 | 140.50 | 99.00 | 123.50 | **495.00** |
| 561 | 235.00 | 210.00 | 210.00 | 215.00 | **870.00** |
| 562 | 63.00 | 62.50 | 37.50 | 60.00 | **223.00** |
| 563 | 267.50 | 199.00 | 161.50 | 172.00 | **800.00** |
| 564 | 48.00 | 51.00 | 129.00 | 74.00 | **302.00** |
| 565 | 126.50 | 150.50 | 138.00 | 145.00 | **560.00** |
| 566 | 86.50 | 75.00 | 33.00 | 48.00 | **242.50** |
| 567 | 202.00 | 226.50 | 155.00 | 152.00 | **735.50** |
| **MN MINNESOTA Total** | **3,441.50** | **3,557.00** | **3,186.00** | **3,016.00** | **13,200.50** |

**STATE: MO - MISSOURI**

| REGISTRANT ZIP CODE 3 | 1ST QUARTER | 2ND QUARTER | 3RD QUARTER | 4TH QUARTER | TOTAL GRAMS |
|---|---|---|---|---|---|
| 630 | 1,835.00 | 1,567.50 | 1,667.50 | 1,471.00 | **6,541.00** |
| 631 | 5,525.00 | 5,261.50 | 4,903.50 | 4,455.50 | **20,145.50** |
| 633 | 1,949.50 | 1,899.00 | 1,797.00 | 1,724.00 | **7,369.50** |
| 634 | 296.50 | 357.50 | 290.50 | 260.50 | **1,205.00** |
| 635 | 166.50 | 127.50 | 123.00 | 101.50 | **518.50** |
| 636 | 215.50 | 210.00 | 117.50 | 163.50 | **706.50** |
| 637 | 462.00 | 491.00 | 497.50 | 332.00 | **1,782.50** |
| 638 | 428.00 | 518.00 | 338.00 | 308.00 | **1,592.00** |
| 639 | 176.00 | 257.00 | 195.00 | 148.50 | **776.50** |
| 640 | 1,213.50 | 1,030.50 | 1,059.50 | 1,184.50 | **4,488.00** |
| 641 | 630.50 | 671.50 | 517.00 | 290.00 | **2,109.00** |
| 644 | 111.00 | 149.00 | 82.50 | 115.00 | **457.50** |
| 645 | 286.00 | 306.00 | 304.00 | 330.50 | **1,226.50** |
| 646 | 108.00 | 57.00 | 51.00 | 61.00 | **277.00** |
| 647 | 269.00 | 246.00 | 186.00 | 214.00 | **915.00** |
| 648 | 686.00 | 534.00 | 632.00 | 679.50 | **2,531.50** |
| 650 | 371.00 | 299.00 | 511.00 | 438.00 | **1,619.00** |
| 651 | 433.50 | 480.50 | 429.00 | 526.00 | **1,869.00** |
| 652 | 433.00 | 338.50 | 343.00 | 393.50 | **1,508.00** |
| 653 | 283.50 | 294.50 | 331.00 | 247.50 | **1,156.50** |
| 654 | 204.00 | 205.50 | 224.50 | 276.50 | **910.50** |

| | | | | | |
|---|---|---|---|---|---|
| 655 | 325.50 | 317.50 | 309.50 | 324.50 | **1,277.00** |
| 656 | 416.00 | 484.00 | 425.50 | 431.50 | **1,757.00** |
| 657 | 620.00 | 658.50 | 676.50 | 599.50 | **2,554.50** |
| 658 | 657.50 | 644.00 | 602.00 | 531.00 | **2,434.50** |
| **MO MISSOURI - Total** | **18,102.00** | **17,405.00** | **16,613.50** | **15,607.00** | **67,727.50** |

**STATE: MS - MISSISSIPPI**

| REGISTRANT ZIP CODE 3 | 1ST QUARTER | 2ND QUARTER | 3RD QUARTER | 4TH QUARTER | TOTAL GRAMS |
|---|---|---|---|---|---|
| 386 | 718.50 | 696.50 | 705.50 | 766.50 | **2,887.00** |
| 387 | 227.00 | 213.50 | 182.00 | 196.00 | **818.50** |
| 388 | 1,070.50 | 993.50 | 946.50 | 1,308.00 | **4,318.50** |
| 389 | 99.00 | 113.00 | 85.50 | 88.50 | **386.00** |
| 390 | 1,011.00 | 766.00 | 853.50 | 737.50 | **3,368.00** |
| 391 | 619.00 | 616.50 | 689.00 | 542.50 | **2,467.00** |
| 392 | 776.50 | 768.50 | 732.50 | 771.00 | **3,048.50** |
| 393 | 453.50 | 535.50 | 381.50 | 484.00 | **1,854.50** |
| 394 | 1,012.50 | 779.00 | 895.50 | 892.50 | **3,579.50** |
| 395 | 1,308.00 | 1,054.50 | 1,042.50 | 1,006.50 | **4,411.50** |
| 396 | 148.50 | 132.50 | 209.50 | 210.00 | **700.50** |
| 397 | 532.50 | 648.50 | 582.50 | 599.00 | **2,362.50** |
| **MS MISSISSIPPI - Total** | **7,976.50** | **7,317.50** | **7,306.00** | **7,602.00** | **30,202.00** |

**STATE: MT - MONTANA**

| REGISTRANT ZIP CODE 3 | 1ST QUARTER | 2ND QUARTER | 3RD QUARTER | 4TH QUARTER | TOTAL GRAMS |
|---|---|---|---|---|---|
| 590 | 116.00 | 114.00 | 120.00 | 134.00 | **484.00** |
| 591 | 359.50 | 352.00 | 296.00 | 275.00 | **1,282.50** |
| 593 | 64.00 | 61.00 | 46.00 | 45.00 | **216.00** |
| 594 | 157.00 | 121.00 | 104.00 | 132.00 | **514.00** |
| 595 | 73.00 | 73.50 | 73.00 | 36.00 | **255.50** |
| 596 | 96.00 | 97.00 | 133.00 | 106.00 | **432.00** |
| 597 | 293.50 | 190.00 | 316.50 | 242.00 | **1,042.00** |
| 598 | 245.00 | 223.50 | 223.00 | 228.50 | **920.00** |
| 599 | 80.00 | 80.50 | 116.00 | 94.50 | **371.00** |
| **MT - MONTANA Total** | **1,484.00** | **1,312.50** | **1,427.50** | **1,293.00** | **5,517.00** |

**STATE: NC - NORTH CAROLINA**

| REGISTRANT ZIP CODE 3 | 1ST QUARTER | 2ND QUARTER | 3RD QUARTER | 4TH QUARTER | TOTAL GRAMS |
|---|---|---|---|---|---|
| 270 | 3,686.50 | 3,733.50 | 3,626.00 | 3,758.00 | **14,804.00** |
| 271 | 4,425.50 | 3,915.50 | 3,832.50 | 3,625.50 | **15,799.00** |
| 272 | 6,031.50 | 6,035.50 | 5,561.50 | 4,724.00 | **22,352.50** |
| 273 | 3,954.00 | 3,894.00 | 3,433.50 | 3,265.00 | **14,546.50** |
| 274 | 3,056.50 | 2,493.50 | 2,821.00 | 2,576.50 | **10,947.50** |
| 275 | 10,551.00 | 14,176.50 | 9,776.00 | 9,399.90 | **43,903.40** |

| | | | | | |
|---|---|---|---|---|---|
| 276 | 3,579.00 | 3,435.50 | 3,320.50 | 3,546.50 | **13,881.50** |
| 277 | 2,200.00 | 2,027.50 | 2,351.50 | 2,014.50 | **8,593.50** |
| 278 | 3,899.00 | 3,621.00 | 3,768.50 | 3,541.50 | **14,830.00** |
| 279 | 1,730.00 | 1,456.00 | 1,396.50 | 1,253.00 | **5,835.50** |
| 280 | 4,562.00 | 4,690.50 | 4,103.50 | 4,087.00 | **17,443.00** |
| 281 | 4,238.50 | 4,036.50 | 4,271.00 | 4,069.50 | **16,615.50** |
| 282 | 2,800.00 | 2,493.00 | 2,671.50 | 2,405.00 | **10,369.50** |
| 283 | 5,511.50 | 5,712.50 | 5,751.00 | 5,265.00 | **22,240.00** |
| 284 | 4,534.00 | 4,537.50 | 4,291.00 | 3,687.50 | **17,050.00** |
| 285 | 6,068.50 | 6,420.50 | 7,432.50 | 5,698.00 | **25,619.50** |
| 286 | 6,383.00 | 5,551.00 | 5,737.00 | 5,090.00 | **22,761.00** |
| 287 | 3,143.00 | 3,182.00 | 2,906.50 | 3,056.00 | **12,287.50** |
| 288 | 881.50 | 983.50 | 922.00 | 1,104.00 | **3,891.00** |
| 289 | 99.00 | 107.00 | 128.00 | 90.00 | **424.00** |
| **NC - NORTH CAROLINA Total** | **81,334.00** | **82,502.50** | **78,101.50** | **72,256.40** | **314,194.40** |

**STATE: ND - NORTH DAKOTA**

| REGISTRANT ZIP CODE 3 | 1ST QUARTER | 2ND QUARTER | 3RD QUARTER | 4TH QUARTER | TOTAL GRAMS |
|---|---|---|---|---|---|
| 580 | 140.00 | 139.50 | 125.00 | 114.00 | **518.50** |
| 581 | 17.50 | 52.00 | 60.00 | 54.00 | **183.50** |
| 582 | 278.00 | 276.50 | 302.50 | 275.50 | **1,132.50** |
| 583 | 16.50 | 21.00 | 15.00 | 15.00 | **67.50** |
| 584 | 28.00 | 56.00 | 52.00 | 25.00 | **161.00** |
| 585 | 737.00 | 625.50 | 569.00 | 534.00 | **2,465.50** |
| 586 | 135.50 | 131.50 | 132.50 | 118.50 | **518.00** |
| 587 | 102.50 | 80.00 | 20.00 | 75.00 | **277.50** |
| 588 | 31.00 | 45.50 | 10.00 | 0 | **86.50** |
| **ND NORTH DAKOTA Total** | **1,486.00** | **1,427.50** | **1,286.00** | **1,211.00** | **5,410.50** |

**STATE: NE - NEBRASKA**

| REGISTRANT ZIP CODE 3 | 1ST QUARTER | 2ND QUARTER | 3RD QUARTER | 4TH QUARTER | TOTAL GRAMS |
|---|---|---|---|---|---|
| 680 | 956.50 | 995.00 | 1,058.00 | 1,099.50 | **4,109.00** |
| 681 | 4,142.50 | 3,876.50 | 3,741.00 | 3,960.00 | **15,720.00** |
| 683 | 161.50 | 121.00 | 164.00 | 222.00 | **668.50** |
| 684 | 182.00 | 157.00 | 125.00 | 170.00 | **634.00** |
| 685 | 1,166.50 | 1,145.00 | 1,162.50 | 1,229.00 | **4,703.00** |
| 686 | 434.00 | 387.00 | 514.00 | 403.50 | **1,738.50** |
| 687 | 289.50 | 215.50 | 271.50 | 243.50 | **1,020.00** |
| 688 | 642.00 | 656.00 | 631.50 | 629.50 | **2,559.00** |
| 689 | 276.50 | 166.00 | 117.00 | 154.00 | **713.50** |
| 690 | 54.00 | 40.00 | 46.00 | 43.50 | **183.50** |
| 691 | 220.00 | 280.50 | 276.50 | 240.50 | **1,017.50** |

| | | | | | |
|---|---|---|---|---|---|
| 692 | 0 | 7.50 | 22.50 | 7.50 | **37.50** |
| 693 | 12.00 | 14.00 | 29.00 | 52.00 | **107.00** |
| **NE NEBRASKA Total** | **8,537.00** | **8,061.00** | **8,158.50** | **8,454.50** | **33,211.00** |

**STATE: NH - NEW HAMPSHIRE**

| REGISTRANT ZIP CODE 3 | 1ST QUARTER | 2ND QUARTER | 3RD QUARTER | 4TH QUARTER | TOTAL GRAMS |
|---|---|---|---|---|---|
| 030 | 1,526.00 | 1,327.50 | 1,620.00 | 1,492.50 | **5,966.00** |
| 031 | 544.00 | 529.00 | 486.50 | 665.00 | **2,224.50** |
| 032 | 805.50 | 671.00 | 631.00 | 616.00 | **2,723.50** |
| 033 | 513.00 | 612.50 | 656.50 | 702.50 | **2,484.50** |
| 034 | 102.00 | 157.50 | 118.50 | 168.50 | **546.50** |
| 035 | 253.00 | 275.00 | 195.00 | 255.00 | **978.00** |
| 036 | 0 | 0 | 0 | 20.00 | **20.00** |
| 037 | 153.50 | 175.00 | 318.50 | 346.00 | **993.00** |
| 038 | 1,835.00 | 1,851.00 | 1,957.00 | 1,773.50 | **7,416.50** |
| **NH - NEW HAMPSHIRE Total** | **5,732.00** | **5,598.50** | **5,983.00** | **6,039.00** | **23,352.50** |

**STATE: NJ - NEW JERSEY**

| REGISTRANT ZIP CODE 3 | 1ST QUARTER | 2ND QUARTER | 3RD QUARTER | 4TH QUARTER | TOTAL GRAMS |
|---|---|---|---|---|---|
| 070 | 4,350.50 | 4,242.50 | 4,132.50 | 4,143.00 | **16,868.50** |
| 071 | 373.50 | 328.00 | 360.50 | 442.00 | **1,504.00** |
| 072 | 203.50 | 196.50 | 302.50 | 259.50 | **962.00** |
| 073 | 230.00 | 198.50 | 172.50 | 163.00 | **764.00** |
| 074 | 1,612.00 | 1,695.50 | 1,585.50 | 1,867.50 | **6,760.50** |
| 075 | 201.00 | 255.00 | 249.00 | 187.50 | **892.50** |
| 076 | 1,961.50 | 1,787.50 | 1,766.00 | 1,715.00 | **7,230.00** |
| 077 | 3,007.50 | 2,930.50 | 2,829.00 | 2,945.00 | **11,712.00** |
| 078 | 1,571.50 | 1,440.50 | 1,495.50 | 1,502.50 | **6,010.00** |
| 079 | 764.50 | 633.50 | 691.00 | 586.00 | **2,675.00** |
| 080 | 5,512.00 | 5,212.50 | 5,336.50 | 5,693.00 | **21,754.00** |
| 081 | 268.50 | 226.00 | 362.00 | 343.00 | **1,199.50** |
| 082 | 1,003.50 | 951.00 | 1,055.00 | 996.00 | **4,005.50** |
| 083 | 1,080.00 | 967.00 | 897.00 | 889.00 | **3,833.00** |
| 084 | 29.00 | 14.00 | 20.00 | 32.50 | **95.50** |
| 085 | 1,295.00 | 1,268.50 | 1,385.00 | 1,409.00 | **5,357.50** |
| 086 | 967.50 | 913.00 | 1,020.00 | 1,084.50 | **3,985.00** |
| 087 | 2,145.00 | 1,962.00 | 1,865.00 | 1,582.00 | **7,554.00** |
| 088 | 3,034.00 | 2,779.00 | 2,700.50 | 2,996.00 | **11,509.50** |
| 089 | 315.50 | 395.00 | 381.00 | 342.00 | **1,433.50** |
| **NJ NEW JERSEY - Total** | **29,925.50** | **28,396.00** | **28,606.00** | **29,178.00** | **116,105.50** |

**STATE: NM - NEW MEXICO**

| REGISTRANT ZIP CODE 3 | 1ST QUARTER | 2ND QUARTER | 3RD QUARTER | 4TH QUARTER | TOTAL GRAMS |
|---|---|---|---|---|---|

| | | | | | |
|---|---|---|---|---|---|
| 870 | 131.00 | 139.00 | 188.50 | 166.00 | **624.50** |
| 871 | 920.50 | 927.00 | 942.00 | 809.00 | **3,598.50** |
| 873 | 30.00 | 0 | 0 | 0 | **30.00** |
| 874 | 113.00 | 56.00 | 76.00 | 68.00 | **313.00** |
| 875 | 131.00 | 76.00 | 153.00 | 103.00 | **463.00** |
| 877 | 40.00 | 60.00 | 50.00 | 31.50 | **181.50** |
| 878 | 40.00 | 30.00 | 30.00 | 30.00 | **130.00** |
| 879 | 0 | 15.00 | 0 | 0 | **15.00** |
| 880 | 419.50 | 368.00 | 242.00 | 325.00 | **1,354.50** |
| 881 | 56.00 | 46.00 | 111.00 | 96.00 | **309.00** |
| 882 | 175.00 | 165.00 | 152.00 | 166.00 | **658.00** |
| 883 | 55.00 | 85.00 | 55.00 | 96.50 | **291.50** |
| 884 | 28.00 | 39.00 | 25.00 | 23.00 | **115.00** |
| **NM NEW MEXICO Total** | **2,139.00** | **2,006.00** | **2,024.50** | **1,914.00** | **8,083.50** |

**STATE: NV - NEVADA**

| REGISTRANT ZIP CODE 3 | 1ST QUARTER | 2ND QUARTER | 3RD QUARTER | 4TH QUARTER | TOTAL GRAMS |
|---|---|---|---|---|---|
| 890 | 2,176.00 | 2,001.00 | 1,825.00 | 2,023.00 | **8,025.00** |
| 891 | 3,848.50 | 3,675.50 | 3,159.50 | 3,189.50 | **13,873.00** |
| 893 | 0 | 5.00 | 0 | 0 | **5.00** |
| 894 | 488.00 | 444.00 | 363.50 | 386.00 | **1,681.50** |
| 895 | 429.00 | 517.00 | 387.50 | 396.50 | **1,730.00** |
| 897 | 86.00 | 118.00 | 149.50 | 72.00 | **425.50** |
| 898 | 109.00 | 87.00 | 125.00 | 63.50 | **384.50** |
| **NV NEVADA Total** | **7,136.50** | **6,847.50** | **6,010.00** | **6,130.50** | **26,124.50** |

**STATE: NY - NEW YORK**

| REGISTRANT ZIP CODE 3 | 1ST QUARTER | 2ND QUARTER | 3RD QUARTER | 4TH QUARTER | TOTAL GRAMS |
|---|---|---|---|---|---|
| 100 | 2,555.50 | 2,218.00 | 2,628.00 | 2,363.50 | **9,765.00** |
| 101 | 10.00 | 5.00 | 5.00 | 80.00 | **100.00** |
| 102 | 0 | 25.00 | 20.00 | 15.00 | **60.00** |
| 103 | 1,646.50 | 1,750.00 | 1,965.50 | 1,545.50 | **6,907.50** |
| 104 | 952.00 | 977.50 | 1,397.50 | 1,078.00 | **4,405.00** |
| 105 | 995.50 | 750.00 | 955.00 | 944.50 | **3,645.00** |
| 106 | 92.50 | 96.50 | 68.50 | 158.00 | **415.50** |
| 107 | 346.50 | 313.50 | 374.50 | 374.00 | **1,408.50** |
| 108 | 104.50 | 139.50 | 85.50 | 92.00 | **421.50** |
| 109 | 1,370.00 | 1,283.00 | 1,446.00 | 1,602.00 | **5,701.00** |
| 110 | 1,100.50 | 1,204.00 | 1,364.00 | 1,327.50 | **4,996.00** |
| 111 | 896.00 | 785.00 | 869.50 | 578.00 | **3,128.50** |
| 112 | 2,743.50 | 2,148.50 | 2,177.50 | 2,137.50 | **9,207.00** |
| 113 | 1,945.50 | 1,744.00 | 1,796.00 | 2,013.50 | **7,499.00** |

| | | | | | |
|---|---|---|---|---|---|
| 114 | 595.00 | 655.00 | 695.00 | 629.50 | **2,574.50** |
| 115 | 3,366.00 | 3,354.50 | 3,287.00 | 3,249.00 | **13,256.50** |
| 116 | 95.50 | 44.00 | 84.00 | 43.00 | **266.50** |
| 117 | 10,861.00 | 10,688.50 | 11,392.50 | 10,869.00 | **43,811.00** |
| 118 | 714.50 | 671.00 | 591.50 | 572.00 | **2,549.00** |
| 119 | 1,186.50 | 1,153.00 | 1,116.00 | 1,003.00 | **4,458.50** |
| 120 | 498.00 | 564.50 | 489.50 | 438.00 | **1,990.00** |
| 121 | 380.00 | 297.00 | 272.00 | 417.00 | **1,366.00** |
| 122 | 864.00 | 855.50 | 739.00 | 662.00 | **3,120.50** |
| 123 | 205.00 | 257.50 | 121.50 | 203.00 | **787.00** |
| 124 | 412.50 | 383.00 | 432.00 | 456.00 | **1,683.50** |
| 125 | 1,423.50 | 1,225.50 | 1,492.00 | 1,453.00 | **5,594.00** |
| 126 | 202.00 | 158.00 | 200.00 | 188.00 | **748.00** |
| 127 | 280.00 | 386.50 | 432.50 | 325.00 | **1,424.00** |
| 128 | 984.50 | 946.50 | 932.50 | 909.50 | **3,773.00** |
| 129 | 596.00 | 508.00 | 544.00 | 411.50 | **2,059.50** |
| 130 | 3,779.50 | 3,633.50 | 3,475.00 | 3,330.00 | **14,218.00** |
| 131 | 744.50 | 672.50 | 635.00 | 623.00 | **2,675.00** |
| 132 | 1,528.00 | 1,151.50 | 1,124.00 | 1,148.00 | **4,951.50** |
| 133 | 1,106.00 | 1,086.50 | 992.50 | 971.50 | **4,156.50** |
| 134 | 1,465.50 | 1,315.00 | 1,235.50 | 1,275.00 | **5,291.00** |
| 135 | 462.00 | 455.50 | 467.50 | 459.00 | **1,844.00** |
| 136 | 2,421.50 | 2,352.50 | 2,095.50 | 2,119.50 | **8,989.00** |
| 137 | 743.00 | 703.50 | 658.00 | 715.00 | **2,819.50** |
| 138 | 1,189.50 | 1,281.50 | 1,106.00 | 1,172.50 | **4,749.50** |
| 139 | 947.00 | 967.50 | 826.50 | 693.50 | **3,434.50** |
| 140 | 3,641.50 | 3,547.00 | 3,198.50 | 3,447.00 | **13,834.00** |
| 141 | 1,495.50 | 1,546.50 | 1,383.50 | 1,349.00 | **5,774.50** |
| 142 | 4,523.50 | 3,944.50 | 3,732.00 | 3,760.50 | **15,960.50** |
| 143 | 455.00 | 498.50 | 318.00 | 252.00 | **1,523.50** |
| 144 | 1,306.00 | 1,427.00 | 1,481.50 | 1,506.00 | **5,720.50** |
| 145 | 1,336.00 | 1,107.50 | 1,153.50 | 1,111.00 | **4,708.00** |
| 146 | 1,773.00 | 1,611.00 | 1,868.50 | 1,513.00 | **6,765.50** |
| 147 | 579.00 | 539.00 | 505.50 | 406.00 | **2,029.50** |
| 148 | 1,459.00 | 1,403.00 | 1,454.50 | 1,250.00 | **5,566.50** |
| 149 | 144.50 | 141.50 | 136.50 | 160.00 | **582.50** |
| **NY - NEW YORK - Total** | **68,522.00** | **64,972.50** | **65,821.00** | **63,399.00** | **262,714.50** |

**STATE: OH - OHIO**

| REGISTRANT ZIP CODE 3 | 1ST QUARTER | 2ND QUARTER | 3RD QUARTER | 4TH QUARTER | **TOTAL GRAMS** |
|---|---|---|---|---|---|
| 430 | 2,331.50 | 2,277.00 | 2,151.50 | 2,314.50 | **9,074.50** |

| | | | | | |
|---|---|---|---|---|---|
| 431 | 1,785.00 | 1,627.50 | 1,506.50 | 1,605.50 | **6,524.50** |
| 432 | 2,508.50 | 2,227.00 | 2,126.50 | 1,912.50 | **8,774.50** |
| 433 | 425.00 | 443.50 | 438.00 | 486.00 | **1,792.50** |
| 434 | 600.50 | 614.00 | 648.00 | 477.50 | **2,340.00** |
| 435 | 2,044.50 | 2,094.00 | 1,967.50 | 1,904.50 | **8,010.50** |
| 436 | 3,025.00 | 2,749.00 | 2,495.00 | 2,373.00 | **10,642.00** |
| 437 | 236.00 | 262.00 | 178.50 | 166.50 | **843.00** |
| 438 | 174.00 | 206.00 | 141.00 | 162.00 | **683.00** |
| 439 | 630.00 | 673.50 | 647.50 | 483.00 | **2,434.00** |
| 440 | 3,575.50 | 3,475.00 | 3,204.50 | 3,171.00 | **13,426.00** |
| 441 | 2,174.50 | 2,259.00 | 2,161.50 | 2,265.50 | **8,860.50** |
| 442 | 2,100.50 | 2,119.00 | 2,080.00 | 2,246.50 | **8,546.00** |
| 443 | 706.50 | 614.00 | 594.50 | 577.50 | **2,492.50** |
| 444 | 958.50 | 939.50 | 779.00 | 831.00 | **3,508.00** |
| 445 | 539.50 | 540.00 | 399.50 | 472.50 | **1,951.50** |
| 446 | 2,374.50 | 2,364.00 | 2,324.00 | 2,679.50 | **9,742.00** |
| 447 | 1,199.00 | 1,167.00 | 982.50 | 1,077.50 | **4,426.00** |
| 448 | 447.00 | 377.50 | 315.50 | 264.00 | **1,404.00** |
| 449 | 194.00 | 182.00 | 203.00 | 215.00 | **794.00** |
| 450 | 2,592.00 | 2,624.50 | 2,442.50 | 2,253.50 | **9,912.50** |
| 451 | 869.00 | 848.00 | 747.50 | 698.00 | **3,162.50** |
| 452 | 2,185.00 | 2,398.50 | 1,938.50 | 2,058.50 | **8,580.50** |
| 453 | 1,235.50 | 1,234.00 | 1,150.50 | 1,263.00 | **4,883.00** |
| 454 | 1,386.50 | 1,261.50 | 1,048.00 | 1,097.50 | **4,793.50** |
| 455 | 483.00 | 579.50 | 530.50 | 676.50 | **2,269.50** |
| 456 | 859.50 | 823.00 | 808.00 | 908.50 | **3,399.00** |
| 457 | 145.00 | 131.00 | 211.00 | 116.00 | **603.00** |
| 458 | 647.50 | 624.50 | 510.00 | 719.00 | **2,501.00** |
| **OH - OHIO Total** | **38,432.50** | **37,735.00** | **34,730.50** | **35,475.50** | **146,373.50** |

**STATE: OK - OKLAHOMA**

| REGISTRANT ZIP CODE 3 | 1ST QUARTER | 2ND QUARTER | 3RD QUARTER | 4TH QUARTER | TOTAL GRAMS |
|---|---|---|---|---|---|
| 730 | 2,593.50 | 2,389.00 | 2,386.00 | 2,191.00 | **9,559.50** |
| 731 | 1,544.50 | 1,363.00 | 1,302.50 | 1,237.50 | **5,447.50** |
| 734 | 426.00 | 314.50 | 402.50 | 300.00 | **1,443.00** |
| 735 | 494.00 | 517.50 | 481.50 | 403.00 | **1,896.00** |
| 736 | 103.00 | 99.00 | 99.00 | 93.00 | **394.00** |
| 737 | 87.00 | 75.00 | 68.00 | 62.00 | **292.00** |
| 738 | 21.00 | 21.00 | 26.00 | 18.00 | **86.00** |
| 739 | 15.00 | 0 | 10.00 | 0 | **25.00** |
| 740 | 1,818.50 | 1,867.00 | 1,908.00 | 1,947.50 | **7,541.00** |

| | | | | | |
|---|---|---|---|---|---|
| 741 | 740.00 | 742.00 | 692.50 | 640.00 | **2,814.50** |
| 743 | 68.00 | 73.00 | 162.50 | 109.00 | **412.50** |
| 744 | 478.50 | 539.50 | 632.50 | 515.00 | **2,165.50** |
| 745 | 150.50 | 98.50 | 129.00 | 51.50 | **429.50** |
| 746 | 42.00 | 45.00 | 36.00 | 30.00 | **153.00** |
| 747 | 20.00 | 64.50 | 40.00 | 49.00 | **173.50** |
| 748 | 362.00 | 402.00 | 237.00 | 171.00 | **1,172.00** |
| 749 | 47.00 | 57.00 | 54.00 | 34.00 | **192.00** |
| **OK - OKLAHOMA Total** | **9,010.50** | **8,667.50** | **8,667.00** | **7,851.50** | **34,196.50** |

**STATE: OR - OREGON**

| REGISTRANT ZIP CODE 3 | 1ST QUARTER | 2ND QUARTER | 3RD QUARTER | 4TH QUARTER | TOTAL GRAMS |
|---|---|---|---|---|---|
| 970 | 1,163.00 | 1,134.50 | 1,122.50 | 1,055.00 | **4,475.00** |
| 971 | 154.00 | 154.00 | 130.00 | 112.00 | **550.00** |
| 972 | 543.00 | 650.00 | 663.00 | 572.00 | **2,428.00** |
| 973 | 799.50 | 820.00 | 855.50 | 834.50 | **3,309.50** |
| 974 | 789.00 | 722.50 | 697.50 | 766.50 | **2,975.50** |
| 975 | 847.00 | 1,146.50 | 1,010.00 | 745.50 | **3,749.00** |
| 976 | 83.00 | 136.00 | 80.00 | 103.00 | **402.00** |
| 977 | 283.00 | 311.00 | 260.00 | 208.50 | **1,062.50** |
| 978 | 93.50 | 92.00 | 33.00 | 65.50 | **284.00** |
| 979 | 12.00 | 15.00 | 6.00 | 11.00 | **44.00** |
| **OR OREGON Total** | **4,767.00** | **5,181.50** | **4,857.50** | **4,473.50** | **19,279.50** |

**STATE: PA - PENNSYLVANIA**

| REGISTRANT ZIP CODE 3 | 1ST QUARTER | 2ND QUARTER | 3RD QUARTER | 4TH QUARTER | TOTAL GRAMS |
|---|---|---|---|---|---|
| 150 | 1,098.00 | 1,083.00 | 963.50 | 878.00 | **4,022.50** |
| 151 | 518.00 | 624.00 | 713.00 | 694.00 | **2,549.00** |
| 152 | 454.00 | 505.00 | 546.50 | 468.50 | **1,974.00** |
| 153 | 454.50 | 407.50 | 374.50 | 531.00 | **1,767.50** |
| 154 | 116.00 | 123.00 | 139.50 | 185.00 | **563.50** |
| 155 | 223.50 | 270.00 | 211.50 | 273.00 | **978.00** |
| 156 | 319.00 | 294.00 | 311.50 | 290.00 | **1,214.50** |
| 157 | 99.50 | 94.00 | 119.50 | 201.00 | **514.00** |
| 158 | 107.00 | 134.00 | 157.00 | 159.00 | **557.00** |
| 159 | 316.50 | 254.00 | 212.00 | 194.00 | **976.50** |
| 160 | 240.50 | 244.00 | 231.50 | 227.50 | **943.50** |
| 161 | 426.00 | 512.50 | 490.00 | 383.50 | **1,812.00** |
| 162 | 229.50 | 267.00 | 240.50 | 255.50 | **992.50** |
| 163 | 450.50 | 413.50 | 395.00 | 446.50 | **1,705.50** |
| 164 | 237.50 | 241.00 | 211.00 | 293.00 | **982.50** |
| 165 | 650.50 | 740.00 | 632.00 | 639.00 | **2,661.50** |

| | | | | | |
|---|---|---|---|---|---|
| 166 | 606.00 | 551.50 | 454.50 | 566.50 | **2,178.50** |
| 167 | 45.00 | 40.00 | 60.00 | 33.00 | **178.00** |
| 168 | 542.00 | 587.00 | 518.50 | 518.50 | **2,166.00** |
| 169 | 75.00 | 50.00 | 50.00 | 83.00 | **258.00** |
| 170 | 1,819.00 | 1,507.50 | 1,649.50 | 1,591.00 | **6,567.00** |
| 171 | 509.50 | 544.00 | 476.50 | 594.50 | **2,124.50** |
| 172 | 948.50 | 741.00 | 767.00 | 736.50 | **3,193.00** |
| 173 | 1,669.50 | 1,827.50 | 1,859.50 | 1,868.00 | **7,224.50** |
| 174 | 551.50 | 578.00 | 417.00 | 422.00 | **1,968.50** |
| 175 | 328.50 | 415.00 | 369.50 | 331.00 | **1,444.00** |
| 176 | 386.50 | 388.00 | 402.00 | 401.50 | **1,578.00** |
| 177 | 147.00 | 102.00 | 102.00 | 116.00 | **467.00** |
| 178 | 230.50 | 162.50 | 112.50 | 107.00 | **612.50** |
| 179 | 248.00 | 211.00 | 183.00 | 211.50 | **853.50** |
| 180 | 3,922.50 | 3,632.50 | 3,488.00 | 3,171.50 | **14,214.50** |
| 181 | 1,574.00 | 1,810.00 | 1,831.50 | 2,297.50 | **7,513.00** |
| 182 | 390.00 | 333.00 | 247.00 | 397.00 | **1,367.00** |
| 183 | 1,854.00 | 2,082.00 | 1,884.00 | 1,663.50 | **7,483.50** |
| 184 | 830.50 | 848.00 | 940.50 | 816.50 | **3,435.50** |
| 185 | 569.50 | 443.00 | 556.50 | 499.50 | **2,068.50** |
| 186 | 300.50 | 272.00 | 228.50 | 307.00 | **1,108.00** |
| 187 | 5,598.00 | 5,678.50 | 5,486.00 | 5,285.50 | **22,048.00** |
| 188 | 91.00 | 86.00 | 67.00 | 98.00 | **342.00** |
| 189 | 1,482.00 | 1,496.00 | 1,372.00 | 1,393.00 | **5,743.00** |
| 190 | 6,124.00 | 5,998.50 | 5,640.50 | 5,906.50 | **23,669.50** |
| 191 | 1,516.00 | 1,636.00 | 1,462.50 | 1,538.00 | **6,152.50** |
| 193 | 1,930.00 | 1,710.00 | 1,861.50 | 1,713.50 | **7,215.00** |
| 194 | 1,799.00 | 1,711.50 | 1,778.50 | 1,697.50 | **6,986.50** |
| 195 | 860.50 | 945.50 | 935.50 | 865.00 | **3,606.50** |
| 196 | 857.00 | 850.50 | 868.00 | 780.50 | **3,356.00** |
| **PA - PENNSYLVANIA - Total** | **43,745.50** | **43,444.50** | **42,017.50** | **42,128.50** | **171,336.00** |

**STATE: PR - PUERTO RICO**

| REGISTRANT ZIP CODE 3 | 1ST QUARTER | 2ND QUARTER | 3RD QUARTER | 4TH QUARTER | **TOTAL GRAMS** |
|---|---|---|---|---|---|
| 006 | 45.00 | 55.00 | 102.50 | 25.00 | **227.50** |
| 007 | 20.00 | 50.00 | 22.50 | 6.00 | **98.50** |
| 009 | 344.00 | 392.00 | 302.50 | 229.00 | **1,267.50** |
| **PR - PUERTO RICO - Total** | **409.00** | **497.00** | **427.50** | **260.00** | **1,593.50** |

**STATE: RI - RHODE ISLAND**

| REGISTRANT ZIP CODE 3 | 1ST QUARTER | 2ND QUARTER | 3RD QUARTER | 4TH QUARTER | **TOTAL GRAMS** |
|---|---|---|---|---|---|
| 028 | 1,432.00 | 1,373.50 | 1,416.00 | 1,189.00 | **5,410.50** |

| | | | | | |
|---|---|---|---|---|---|
| 029 | 429.00 | 384.00 | 476.50 | 427.00 | **1,716.50** |
| **RI RHODE ISLAND Total** | **1,861.00** | **1,757.50** | **1,892.50** | **1,616.00** | **7,127.00** |

**STATE: SC - SOUTH CAROLINA**

| REGISTRANT ZIP CODE 3 | 1ST QUARTER | 2ND QUARTER | 3RD QUARTER | 4TH QUARTER | TOTAL GRAMS |
|---|---|---|---|---|---|
| 290 | 2,728.50 | 2,503.50 | 2,399.50 | 2,315.00 | **9,946.50** |
| 291 | 2,497.50 | 2,375.00 | 2,362.00 | 2,448.00 | **9,682.50** |
| 292 | 2,915.50 | 2,848.00 | 2,599.50 | 2,632.00 | **10,995.00** |
| 293 | 2,267.00 | 2,096.00 | 2,183.50 | 2,122.00 | **8,668.50** |
| 294 | 3,195.50 | 2,941.00 | 2,658.00 | 2,956.00 | **11,750.50** |
| 295 | 3,779.00 | 3,736.50 | 3,677.50 | 3,228.50 | **14,421.50** |
| 296 | 10,039.00 | 9,391.50 | 9,613.50 | 9,297.00 | **38,341.00** |
| 297 | 1,490.00 | 1,347.50 | 1,282.00 | 1,112.50 | **5,232.00** |
| 298 | 1,392.50 | 1,577.50 | 1,712.00 | 1,406.50 | **6,088.50** |
| 299 | 1,506.00 | 1,446.00 | 1,408.50 | 1,516.50 | **5,877.00** |
| **SC - SOUTH CAROLINA Total** | **31,810.50** | **30,262.50** | **29,896.00** | **29,034.00** | **121,003.00** |

**STATE: SD - SOUTH DAKOTA**

| REGISTRANT ZIP CODE 3 | 1ST QUARTER | 2ND QUARTER | 3RD QUARTER | 4TH QUARTER | TOTAL GRAMS |
|---|---|---|---|---|---|
| 570 | 747.00 | 667.50 | 535.50 | 557.50 | **2,507.50** |
| 571 | 819.00 | 855.50 | 976.00 | 897.00 | **3,547.50** |
| 572 | 149.50 | 113.00 | 88.00 | 99.50 | **450.00** |
| 573 | 239.00 | 231.00 | 224.50 | 215.00 | **909.50** |
| 574 | 458.50 | 534.00 | 421.00 | 420.00 | **1,833.50** |
| 575 | 70.00 | 85.00 | 113.00 | 103.00 | **371.00** |
| 576 | 85.00 | 70.00 | 68.00 | 85.00 | **308.00** |
| 577 | 531.00 | 494.50 | 488.50 | 449.50 | **1,963.50** |
| **SD SOUTH DAKOTA - Total** | **3,099.00** | **3,050.50** | **2,914.50** | **2,826.50** | **11,890.50** |

**STATE: TN - TENNESSEE**

| REGISTRANT ZIP CODE 3 | 1ST QUARTER | 2ND QUARTER | 3RD QUARTER | 4TH QUARTER | TOTAL GRAMS |
|---|---|---|---|---|---|
| 370 | 4,173.50 | 4,057.00 | 3,913.00 | 4,184.50 | **16,328.00** |
| 371 | 4,189.50 | 3,883.50 | 4,033.00 | 4,250.00 | **16,356.00** |
| 372 | 1,615.00 | 1,591.00 | 1,445.00 | 1,394.50 | **6,045.50** |
| 373 | 1,977.00 | 2,166.50 | 1,751.50 | 1,922.00 | **7,817.00** |
| 374 | 1,248.50 | 1,203.50 | 1,077.00 | 1,259.00 | **4,788.00** |
| 376 | 2,053.00 | 2,451.50 | 2,656.50 | 2,557.00 | **9,718.00** |
| 377 | 736.50 | 770.50 | 678.00 | 636.50 | **2,821.50** |
| 378 | 1,086.00 | 1,252.00 | 1,210.50 | 1,264.50 | **4,813.00** |
| 379 | 672.50 | 614.50 | 782.00 | 814.50 | **2,883.50** |
| 380 | 601.00 | 670.00 | 782.00 | 882.00 | **2,935.00** |
| 381 | 944.00 | 1,082.00 | 1,169.50 | 1,105.50 | **4,301.00** |
| 382 | 519.00 | 448.50 | 411.50 | 317.50 | **1,696.50** |

| | | | | | |
|---|---|---|---|---|---|
| 383 | 1,071.50 | 883.00 | 757.00 | 778.50 | **3,490.00** |
| 384 | 528.50 | 497.00 | 584.00 | 609.00 | **2,218.50** |
| 385 | 538.50 | 536.50 | 673.00 | 561.50 | **2,309.50** |
| **TN TENNESSEE Total** | **21,954.00** | **22,107.00** | **21,923.50** | **22,536.50** | **88,521.00** |

**STATE: TX - TEXAS**

| REGISTRANT ZIP CODE 3 | 1ST QUARTER | 2ND QUARTER | 3RD QUARTER | 4TH QUARTER | TOTAL GRAMS |
|---|---|---|---|---|---|
| 750 | 9,469.00 | 9,353.50 | 9,151.00 | 8,116.00 | **36,089.50** |
| 751 | 2,498.00 | 2,877.00 | 2,731.50 | 2,445.00 | **10,551.50** |
| 752 | 2,276.00 | 2,142.50 | 1,962.50 | 2,009.50 | **8,390.50** |
| 753 | 10.00 | 0 | 0 | 0 | **10.00** |
| 754 | 862.50 | 715.00 | 787.50 | 704.00 | **3,069.00** |
| 755 | 22.00 | 17.00 | 90.00 | 30.00 | **159.00** |
| 756 | 339.00 | 335.50 | 332.50 | 234.00 | **1,241.00** |
| 757 | 703.50 | 645.00 | 600.50 | 614.50 | **2,563.50** |
| 758 | 204.00 | 206.00 | 219.50 | 232.50 | **862.00** |
| 759 | 246.00 | 187.00 | 199.50 | 178.00 | **810.50** |
| 760 | 4,349.00 | 3,700.50 | 3,764.50 | 3,362.00 | **15,176.00** |
| 761 | 2,760.50 | 2,649.50 | 2,479.50 | 2,569.00 | **10,458.50** |
| 762 | 1,562.00 | 1,704.50 | 1,908.00 | 1,692.00 | **6,866.50** |
| 763 | 781.50 | 746.50 | 756.50 | 668.00 | **2,952.50** |
| 764 | 167.00 | 116.50 | 44.50 | 118.50 | **446.50** |
| 765 | 1,174.50 | 1,215.00 | 1,216.00 | 1,095.00 | **4,700.50** |
| 766 | 163.00 | 140.50 | 112.50 | 127.00 | **543.00** |
| 767 | 292.00 | 257.00 | 244.00 | 212.00 | **1,005.00** |
| 768 | 24.00 | 18.00 | 33.00 | 32.00 | **107.00** |
| 769 | 313.00 | 236.00 | 166.00 | 170.00 | **885.00** |
| 770 | 3,206.00 | 3,142.00 | 3,196.50 | 2,970.50 | **12,515.00** |
| 773 | 3,367.00 | 3,168.50 | 3,240.50 | 2,856.50 | **12,632.50** |
| 774 | 3,010.50 | 3,352.00 | 3,684.50 | 3,848.00 | **13,895.00** |
| 775 | 3,023.50 | 3,113.50 | 3,058.50 | 2,921.50 | **12,117.00** |
| 776 | 533.00 | 435.50 | 468.00 | 476.50 | **1,913.00** |
| 777 | 209.50 | 229.50 | 199.00 | 207.00 | **845.00** |
| 778 | 300.00 | 238.00 | 284.50 | 287.00 | **1,109.50** |
| 779 | 233.50 | 234.00 | 226.00 | 413.00 | **1,106.50** |
| 780 | 868.00 | 767.00 | 809.50 | 779.00 | **3,223.50** |
| 781 | 1,317.50 | 1,518.00 | 1,078.00 | 1,169.50 | **5,083.00** |
| 782 | 5,948.50 | 5,975.00 | 5,404.50 | 4,935.50 | **22,263.50** |
| 783 | 293.00 | 292.50 | 435.50 | 409.00 | **1,430.00** |
| 784 | 829.00 | 838.50 | 872.50 | 938.00 | **3,478.00** |
| 785 | 327.50 | 259.50 | 226.00 | 328.50 | **1,141.50** |

| | | | | | |
|---|---|---|---|---|---|
| 786 | 2,698.00 | 2,926.00 | 2,799.00 | 2,658.50 | **11,081.50** |
| 787 | 2,979.50 | 3,152.50 | 2,935.00 | 3,012.50 | **12,079.50** |
| 788 | 65.00 | 76.00 | 61.00 | 114.50 | **316.50** |
| 789 | 143.00 | 185.50 | 199.00 | 153.00 | **680.50** |
| 790 | 223.00 | 207.00 | 214.00 | 300.00 | **944.00** |
| 791 | 638.00 | 672.00 | 739.50 | 712.50 | **2,762.00** |
| 792 | 0 | 3.00 | 0 | 0 | **3.00** |
| 793 | 73.00 | 45.00 | 30.00 | 96.00 | **244.00** |
| 794 | 732.50 | 771.00 | 867.50 | 717.50 | **3,088.50** |
| 795 | 0 | 0 | 0 | 10.00 | **10.00** |
| 796 | 39.00 | 96.50 | 75.50 | 60.00 | **271.00** |
| 797 | 133.00 | 141.50 | 102.00 | 203.50 | **580.00** |
| 798 | 0 | 9.00 | 5.00 | 40.00 | **54.00** |
| 799 | 2,014.00 | 1,616.00 | 1,687.00 | 1,540.50 | **6,857.50** |
| **TX - TEXAS Total** | **61,421.00** | **60,727.00** | **59,697.00** | **56,767.00** | **238,612.00** |

**STATE: UT - UTAH**

| REGISTRANT ZIP CODE 3 | 1ST QUARTER | 2ND QUARTER | 3RD QUARTER | 4TH QUARTER | TOTAL GRAMS |
|---|---|---|---|---|---|
| 840 | 3,861.50 | 4,204.00 | 4,050.00 | 3,659.50 | **15,775.00** |
| 841 | 1,138.50 | 1,121.50 | 984.00 | 1,138.50 | **4,382.50** |
| 843 | 285.00 | 354.50 | 236.00 | 275.00 | **1,150.50** |
| 844 | 554.50 | 662.00 | 747.00 | 785.00 | **2,748.50** |
| 845 | 599.50 | 596.00 | 665.50 | 585.50 | **2,446.50** |
| 846 | 712.00 | 772.00 | 781.00 | 814.00 | **3,079.00** |
| 847 | 1,337.00 | 1,352.00 | 1,283.00 | 1,350.00 | **5,322.00** |
| **UT - UTAH - Total** | **8,488.00** | **9,062.00** | **8,746.50** | **8,607.50** | **34,904.00** |

**STATE: VA - VIRGINIA**

| REGISTRANT ZIP CODE 3 | 1ST QUARTER | 2ND QUARTER | 3RD QUARTER | 4TH QUARTER | TOTAL GRAMS |
|---|---|---|---|---|---|
| 201 | 3,193.00 | 3,070.50 | 3,007.50 | 2,583.50 | **11,854.50** |
| 220 | 1,286.00 | 1,514.50 | 1,315.50 | 1,312.50 | **5,428.50** |
| 221 | 1,128.50 | 1,354.50 | 1,325.00 | 1,261.50 | **5,069.50** |
| 222 | 884.50 | 717.00 | 867.50 | 941.00 | **3,410.00** |
| 223 | 928.50 | 834.50 | 946.50 | 850.00 | **3,559.50** |
| 224 | 1,022.50 | 839.00 | 995.50 | 981.50 | **3,838.50** |
| 225 | 405.50 | 527.50 | 457.50 | 547.00 | **1,937.50** |
| 226 | 969.50 | 1,025.00 | 1,020.00 | 975.50 | **3,990.00** |
| 227 | 214.50 | 186.50 | 173.50 | 154.00 | **728.50** |
| 228 | 659.00 | 812.00 | 693.50 | 573.00 | **2,737.50** |
| 229 | 258.00 | 362.50 | 284.50 | 292.50 | **1,197.50** |
| 230 | 252.50 | 298.50 | 227.50 | 142.50 | **921.00** |
| 231 | 1,124.00 | 819.50 | 778.00 | 792.00 | **3,513.50** |

| | | | | | |
|---|---|---|---|---|---|
| 232 | 1,254.00 | 1,022.50 | 977.50 | 884.50 | **4,138.50** |
| 233 | 1,342.50 | 1,361.00 | 1,352.00 | 1,524.50 | **5,580.00** |
| 234 | 3,528.50 | 3,855.50 | 3,812.00 | 3,471.50 | **14,667.50** |
| 235 | 829.00 | 766.50 | 756.50 | 839.50 | **3,191.50** |
| 236 | 1,048.50 | 1,008.00 | 1,041.50 | 1,151.50 | **4,249.50** |
| 237 | 496.00 | 355.50 | 603.00 | 492.50 | **1,947.00** |
| 238 | 750.50 | 845.00 | 765.50 | 836.00 | **3,197.00** |
| 239 | 224.00 | 264.50 | 195.50 | 157.00 | **841.00** |
| 240 | 1,074.00 | 1,084.00 | 1,151.00 | 1,292.50 | **4,601.50** |
| 241 | 1,016.50 | 1,078.00 | 962.00 | 972.50 | **4,029.00** |
| 242 | 821.50 | 938.00 | 993.50 | 765.50 | **3,518.50** |
| 243 | 855.00 | 825.00 | 712.50 | 803.00 | **3,195.50** |
| 244 | 375.00 | 434.50 | 357.00 | 345.50 | **1,512.00** |
| 245 | 1,200.00 | 1,153.50 | 1,125.00 | 1,048.50 | **4,527.00** |
| 246 | 167.50 | 229.00 | 312.50 | 173.50 | **882.50** |
| **VA VIRGINIA Total** | **27,308.50** | **27,582.00** | **27,209.00** | **26,164.50** | **108,264.00** |

**STATE: VI - VIRGIN ISLANDS**

| REGISTRANT ZIP CODE 3 | 1ST QUARTER | 2ND QUARTER | 3RD QUARTER | 4TH QUARTER | **TOTAL GRAMS** |
|---|---|---|---|---|---|
| 008 | 0 | 5.00 | 0 | 0 | **5.00** |
| **VI - VIRGIN ISLANDS Total** | **0.00** | **5.00** | **0.00** | **0.00** | **5.00** |

**STATE: VT - VERMONT**

| REGISTRANT ZIP CODE 3 | 1ST QUARTER | 2ND QUARTER | 3RD QUARTER | 4TH QUARTER | **TOTAL GRAMS** |
|---|---|---|---|---|---|
| 050 | 0 | 0 | 0 | 26.00 | **26.00** |
| 051 | 24.00 | 19.00 | 32.00 | 54.00 | **129.00** |
| 052 | 160.00 | 128.00 | 163.00 | 167.00 | **618.00** |
| 054 | 207.50 | 209.00 | 223.50 | 248.00 | **888.00** |
| 056 | 91.00 | 96.00 | 66.00 | 17.00 | **270.00** |
| 057 | 298.00 | 198.00 | 199.00 | 226.00 | **921.00** |
| 058 | 30.00 | 13.00 | 60.00 | 42.00 | **145.00** |
| **VT VERMONT Total** | **810.50** | **663.00** | **743.50** | **780.00** | **2,997.00** |

**STATE: WA - WASHINGTON**

| REGISTRANT ZIP CODE 3 | 1ST QUARTER | 2ND QUARTER | 3RD QUARTER | 4TH QUARTER | **TOTAL GRAMS** |
|---|---|---|---|---|---|
| 980 | 1,670.00 | 1,546.50 | 1,494.00 | 1,487.50 | **6,198.00** |
| 981 | 905.00 | 923.00 | 993.00 | 1,014.50 | **3,835.50** |
| 982 | 735.50 | 613.50 | 589.50 | 751.50 | **2,690.00** |
| 983 | 1,519.50 | 1,497.50 | 1,764.00 | 1,717.00 | **6,498.00** |
| 984 | 510.50 | 568.00 | 464.00 | 474.50 | **2,017.00** |
| 985 | 584.00 | 554.00 | 568.00 | 588.50 | **2,294.50** |
| 986 | 1,240.00 | 1,140.00 | 1,030.00 | 1,119.00 | **4,529.00** |
| 988 | 399.00 | 508.00 | 424.00 | 319.00 | **1,650.00** |

| | | | | | |
|---|---|---|---|---|---|
| 989 | 290.50 | 280.50 | 352.00 | 422.50 | **1,345.50** |
| 990 | 591.50 | 473.00 | 449.50 | 491.50 | **2,005.50** |
| 991 | 94.00 | 74.00 | 78.00 | 73.00 | **319.00** |
| 992 | 3,127.00 | 2,917.00 | 3,051.50 | 3,004.00 | **12,099.50** |
| 993 | 827.00 | 919.00 | 800.50 | 844.50 | **3,391.00** |
| 994 | 59.00 | 47.00 | 60.50 | 55.00 | **221.50** |
| **WA WASHINGTON Total** | **12,552.50** | **12,061.00** | **12,118.50** | **12,362.00** | **49,094.00** |

**STATE: WI - WISCONSIN**

| REGISTRANT ZIP CODE 3 | 1ST QUARTER | 2ND QUARTER | 3RD QUARTER | 4TH QUARTER | TOTAL GRAMS |
|---|---|---|---|---|---|
| 530 | 1,736.00 | 1,552.00 | 1,146.50 | 1,154.50 | **5,589.00** |
| 531 | 1,408.00 | 1,208.50 | 1,046.50 | 1,192.00 | **4,855.00** |
| 532 | 1,435.00 | 1,258.50 | 1,310.00 | 1,099.00 | **5,102.50** |
| 534 | 461.00 | 507.50 | 484.00 | 530.00 | **1,982.50** |
| 535 | 598.00 | 543.00 | 558.00 | 443.50 | **2,142.50** |
| 537 | 173.50 | 159.50 | 140.50 | 117.50 | **591.00** |
| 538 | 0 | 20.00 | 0 | 0 | **20.00** |
| 539 | 216.00 | 125.50 | 147.50 | 140.00 | **629.00** |
| 540 | 193.00 | 144.50 | 155.50 | 168.00 | **661.00** |
| 541 | 474.50 | 405.50 | 348.00 | 348.00 | **1,576.00** |
| 542 | 209.00 | 198.00 | 162.00 | 111.00 | **680.00** |
| 543 | 427.50 | 515.50 | 349.50 | 441.50 | **1,734.00** |
| 544 | 570.50 | 520.00 | 631.00 | 603.00 | **2,324.50** |
| 545 | 163.00 | 161.00 | 121.50 | 141.50 | **587.00** |
| 546 | 9.00 | 38.00 | 51.00 | 23.00 | **121.00** |
| 547 | 107.50 | 142.50 | 108.00 | 118.00 | **476.00** |
| 548 | 65.00 | 94.00 | 138.50 | 91.00 | **388.50** |
| 549 | 1,103.00 | 1,098.50 | 984.50 | 889.00 | **4,075.00** |
| **WI - WISCONSIN Total** | **9,349.50** | **8,692.00** | **7,882.50** | **7,610.50** | **33,534.50** |

**STATE: WV - WEST VIRGINIA**

| REGISTRANT ZIP CODE 3 | 1ST QUARTER | 2ND QUARTER | 3RD QUARTER | 4TH QUARTER | TOTAL GRAMS |
|---|---|---|---|---|---|
| 247 | 120.00 | 138.00 | 167.00 | 155.00 | **580.00** |
| 248 | 0 | 22.50 | 12.00 | 6.00 | **40.50** |
| 249 | 63.00 | 45.00 | 41.00 | 36.00 | **185.00** |
| 250 | 71.00 | 85.00 | 120.00 | 107.50 | **383.50** |
| 251 | 80.00 | 105.00 | 85.00 | 110.00 | **380.00** |
| 252 | 45.00 | 37.50 | 30.00 | 29.00 | **141.50** |
| 253 | 75.50 | 101.50 | 85.00 | 85.00 | **347.00** |
| 254 | 856.00 | 923.50 | 1,014.00 | 948.00 | **3,741.50** |
| 255 | 374.50 | 370.50 | 310.00 | 349.50 | **1,404.50** |
| 256 | 22.00 | 40.00 | 34.00 | 22.00 | **118.00** |

| | | | | | |
|---|---|---|---|---|---|
| 257 | 307.00 | 324.00 | 260.50 | 264.50 | **1,156.00** |
| 258 | 18.00 | 24.00 | 30.00 | 40.00 | **112.00** |
| 259 | 10.00 | 22.50 | 5.00 | 10.00 | **47.50** |
| 260 | 843.50 | 873.50 | 948.00 | 979.50 | **3,644.50** |
| 261 | 233.00 | 199.50 | 171.00 | 215.00 | **818.50** |
| 262 | 0 | 54.00 | 18.00 | 0 | **72.00** |
| 263 | 0 | 0 | 6.00 | 12.00 | **18.00** |
| 265 | 155.00 | 195.50 | 93.00 | 130.00 | **573.50** |
| 266 | 42.00 | 57.00 | 59.50 | 20.00 | **178.50** |
| 267 | 37.50 | 104.00 | 129.00 | 154.50 | **425.00** |
| 268 | 51.00 | 37.00 | 5.00 | 3.00 | **96.00** |
| **WV WEST VIRGINIA Total** | **3,404.00** | **3,759.50** | **3,623.00** | **3,676.50** | **14,463.00** |

**STATE: WY - WYOMING**

| REGISTRANT ZIP CODE 3 | 1ST QUARTER | 2ND QUARTER | 3RD QUARTER | 4TH QUARTER | TOTAL GRAMS |
|---|---|---|---|---|---|
| 820 | 773.50 | 697.00 | 557.00 | 543.00 | **2,570.50** |
| 822 | 36.00 | 65.00 | 39.00 | 54.00 | **194.00** |
| 823 | 20.00 | 6.00 | 0 | 0 | **26.00** |
| 824 | 98.00 | 32.00 | 30.00 | 41.00 | **201.00** |
| 826 | 128.50 | 209.50 | 156.00 | 140.00 | **634.00** |
| 827 | 108.50 | 176.50 | 151.00 | 175.50 | **611.50** |
| 828 | 55.00 | 78.00 | 64.00 | 88.00 | **285.00** |
| 829 | 60.00 | 15.00 | 135.00 | 18.00 | **228.00** |
| 830 | 0 | 0 | 5.00 | 0 | **5.00** |
| 831 | 5.00 | 10.00 | 21.00 | 0 | **36.00** |
| **WY - WYOMING Total** | **1,284.50** | **1,289.00** | **1,158.00** | **1,059.50** | **4,791.00** |

**DRUG: 9801 - FENTANYL BASE**

**STATE: AK - ALASKA**

| REGISTRANT ZIP CODE 3 | 1ST QUARTER | 2ND QUARTER | 3RD QUARTER | 4TH QUARTER | TOTAL GRAMS |
|---|---|---|---|---|---|
| 995 | 55.83 | 49.52 | 44.65 | 38.01 | **188.01** |
| 996 | 45.41 | 42.99 | 39.91 | 37.32 | **165.63** |
| 997 | 14.56 | 11.20 | 14.06 | 12.22 | **52.05** |
| 998 | 8.48 | 7.84 | 8.98 | 8.43 | **33.75** |
| 999 | 3.98 | 2.73 | 3.84 | 3.76 | **14.32** |
| **AK ALASKA - Total** | **128.28** | **114.28** | **111.45** | **99.74** | **453.75** |

**STATE: AL - ALABAMA**

| REGISTRANT ZIP CODE 3 | 1ST QUARTER | 2ND QUARTER | 3RD QUARTER | 4TH QUARTER | TOTAL GRAMS |
|---|---|---|---|---|---|
| 350 | 90.55 | 93.85 | 87.47 | 82.81 | **354.68** |
| 351 | 56.08 | 52.69 | 49.27 | 48.65 | **206.69** |
| 352 | 99.34 | 87.53 | 84.82 | 87.99 | **359.68** |
| 354 | 54.73 | 63.39 | 59.35 | 57.46 | **234.93** |

| | | | | | |
|---|---|---|---|---|---|
| 355 | 34.83 | 31.37 | 34.23 | 27.61 | **128.04** |
| 356 | 55.67 | 54.09 | 53.17 | 49.53 | **212.47** |
| 357 | 17.92 | 15.39 | 15.45 | 15.74 | **64.50** |
| 358 | 46.03 | 39.62 | 39.39 | 40.12 | **165.17** |
| 359 | 68.65 | 66.69 | 63.51 | 58.33 | **257.17** |
| 360 | 44.69 | 40.58 | 39.46 | 37.22 | **161.95** |
| 361 | 59.81 | 62.27 | 58.81 | 65.88 | **246.77** |
| 362 | 29.02 | 25.53 | 25.15 | 24.22 | **103.93** |
| 363 | 61.26 | 58.03 | 60.32 | 50.72 | **230.33** |
| 364 | 23.07 | 20.52 | 18.70 | 18.19 | **80.47** |
| 365 | 60.83 | 57.32 | 55.36 | 54.09 | **227.60** |
| 366 | 43.80 | 40.60 | 38.91 | 38.82 | **162.13** |
| 367 | 20.14 | 17.52 | 16.96 | 14.17 | **68.80** |
| 368 | 43.60 | 41.88 | 39.01 | 39.41 | **163.90** |
| 369 | 3.49 | 3.71 | 3.43 | 2.66 | **13.29** |
| **AL ALABAMA - Total** | **913.51** | **872.60** | **842.78** | **813.62** | **3,442.51** |

**STATE: AR - ARKANSAS**

| REGISTRANT ZIP CODE 3 | 1ST QUARTER | 2ND QUARTER | 3RD QUARTER | 4TH QUARTER | **TOTAL GRAMS** |
|---|---|---|---|---|---|
| 716 | 19.88 | 18.49 | 18.66 | 18.90 | **75.93** |
| 717 | 20.97 | 20.21 | 17.95 | 17.99 | **77.13** |
| 718 | 14.77 | 14.90 | 13.65 | 12.54 | **55.86** |
| 719 | 43.99 | 45.16 | 38.53 | 40.87 | **168.55** |
| 720 | 50.89 | 46.21 | 41.55 | 38.90 | **177.55** |
| 721 | 70.96 | 69.62 | 67.25 | 65.77 | **273.60** |
| 722 | 46.29 | 52.15 | 43.54 | 47.06 | **189.03** |
| 723 | 12.82 | 12.43 | 14.92 | 15.69 | **55.87** |
| 724 | 37.81 | 34.68 | 38.46 | 35.21 | **146.16** |
| 725 | 31.17 | 25.16 | 25.76 | 21.40 | **103.49** |
| 726 | 25.73 | 24.99 | 26.12 | 21.89 | **98.73** |
| 727 | 78.55 | 72.92 | 69.15 | 64.15 | **284.78** |
| 728 | 16.07 | 15.45 | 15.95 | 15.87 | **63.33** |
| 729 | 37.56 | 31.18 | 34.56 | 33.32 | **136.63** |
| **AR ARKANSAS - Total** | **507.47** | **483.56** | **466.04** | **449.57** | **1,906.65** |

**STATE: AS - AMERICAN SAMOA**

| REGISTRANT ZIP CODE 3 | 1ST QUARTER | 2ND QUARTER | 3RD QUARTER | 4TH QUARTER | **TOTAL GRAMS** |
|---|---|---|---|---|---|
| 967 | 0.48 | 0.13 | 0 | 0.04 | **0.65** |
| **AS - AMERICAN SAMOA - Tota** | **0.48** | **0.13** | **0.00** | **0.04** | **0.65** |

**STATE: AZ - ARIZONA**

| REGISTRANT ZIP CODE 3 | 1ST QUARTER | 2ND QUARTER | 3RD QUARTER | 4TH QUARTER | **TOTAL GRAMS** |
|---|---|---|---|---|---|
| 850 | 157.36 | 153.75 | 150.75 | 133.72 | **595.57** |

| | | | | | |
|---|---|---|---|---|---|
| 851 | 33.04 | 25.09 | 24.02 | 24.99 | **107.14** |
| 852 | 287.25 | 273.56 | 257.17 | 244.43 | **1,062.41** |
| 853 | 161.66 | 135.42 | 126.64 | 129.70 | **553.43** |
| 855 | 10.74 | 10.95 | 10.37 | 11.42 | **43.48** |
| 856 | 38.06 | 33.78 | 31.08 | 30.52 | **133.44** |
| 857 | 169.60 | 157.12 | 142.39 | 140.79 | **609.90** |
| 859 | 13.14 | 11.65 | 11.29 | 8.63 | **44.71** |
| 860 | 14.52 | 12.04 | 12.78 | 12.06 | **51.40** |
| 863 | 35.21 | 34.39 | 31.60 | 31.23 | **132.43** |
| 864 | 32.43 | 28.80 | 27.44 | 32.05 | **120.71** |
| 865 | 0.74 | 0.30 | 0.87 | 0.53 | **2.44** |
| **AZ ARIZONA - Total** | **953.74** | **876.85** | **826.41** | **800.08** | **3,457.08** |

**STATE: CA - CALIFORNIA**

| REGISTRANT ZIP CODE 3 | 1ST QUARTER | 2ND QUARTER | 3RD QUARTER | 4TH QUARTER | TOTAL GRAMS |
|---|---|---|---|---|---|
| 900 | 117.82 | 112.67 | 108.85 | 107.22 | **446.56** |
| 902 | 62.92 | 60.43 | 59.08 | 58.37 | **240.81** |
| 903 | 3.68 | 3.99 | 3.57 | 3.66 | **14.89** |
| 904 | 69.42 | 52.07 | 52.21 | 51.17 | **224.87** |
| 905 | 29.82 | 33.14 | 35.89 | 31.74 | **130.59** |
| 906 | 55.85 | 54.13 | 54.40 | 57.99 | **222.37** |
| 907 | 50.26 | 46.90 | 39.72 | 41.29 | **178.17** |
| 908 | 112.67 | 111.22 | 103.82 | 103.85 | **431.55** |
| 910 | 33.97 | 33.89 | 36.08 | 32.37 | **136.32** |
| 911 | 16.80 | 16.54 | 14.63 | 13.72 | **61.70** |
| 912 | 27.41 | 24.07 | 25.99 | 26.78 | **104.24** |
| 913 | 125.47 | 126.78 | 113.26 | 110.78 | **476.29** |
| 914 | 58.85 | 47.44 | 43.85 | 39.67 | **189.82** |
| 915 | 8.88 | 11.80 | 9.97 | 12.27 | **42.92** |
| 916 | 2.99 | 4.29 | 5.06 | 4.57 | **16.91** |
| 917 | 230.52 | 192.46 | 212.96 | 171.64 | **807.58** |
| 918 | 4.12 | 2.66 | 3.22 | 3.67 | **13.66** |
| 919 | 27.66 | 23.81 | 23.78 | 19.63 | **94.88** |
| 920 | 195.41 | 186.81 | 178.62 | 166.98 | **727.82** |
| 921 | 226.83 | 210.62 | 178.93 | 193.17 | **809.55** |
| 922 | 128.33 | 134.97 | 113.22 | 106.86 | **483.38** |
| 923 | 150.41 | 141.05 | 140.49 | 136.43 | **568.38** |
| 924 | 21.35 | 18.01 | 17.35 | 14.40 | **71.10** |
| 925 | 188.69 | 217.76 | 156.86 | 177.30 | **740.60** |
| 926 | 181.34 | 177.70 | 169.60 | 173.05 | **701.70** |
| 927 | 33.07 | 29.00 | 27.35 | 25.04 | **114.46** |

| | | | | | |
|---|---|---|---|---|---|
| 928 | 149.67 | 136.33 | 140.25 | 134.70 | **560.96** |
| 930 | 69.79 | 66.84 | 63.22 | 61.87 | **261.73** |
| 931 | 13.21 | 17.24 | 12.41 | 16.57 | **59.43** |
| 932 | 68.52 | 62.61 | 71.57 | 63.57 | **266.27** |
| 933 | 78.37 | 75.26 | 73.46 | 72.02 | **299.12** |
| 934 | 78.78 | 69.67 | 61.65 | 63.23 | **273.34** |
| 935 | 56.89 | 53.57 | 49.88 | 47.78 | **208.13** |
| 936 | 41.74 | 37.84 | 35.52 | 33.98 | **149.08** |
| 937 | 76.89 | 65.45 | 67.54 | 63.55 | **273.44** |
| 939 | 19.03 | 19.73 | 15.96 | 18.68 | **73.40** |
| 940 | 78.25 | 70.66 | 75.12 | 70.40 | **294.43** |
| 941 | 88.53 | 94.69 | 86.89 | 71.11 | **341.22** |
| 943 | 24.26 | 25.88 | 21.88 | 28.76 | **100.79** |
| 944 | 11.04 | 12.39 | 9.89 | 8.69 | **42.01** |
| 945 | 272.26 | 242.11 | 244.83 | 230.64 | **989.84** |
| 946 | 32.26 | 34.76 | 30.26 | 26.17 | **123.44** |
| 947 | 11.18 | 10.48 | 10.84 | 10.81 | **43.30** |
| 948 | 8.53 | 7.36 | 5.21 | 4.76 | **25.86** |
| 949 | 37.40 | 32.18 | 33.63 | 31.03 | **134.23** |
| 950 | 70.15 | 61.53 | 62.30 | 55.69 | **249.67** |
| 951 | 43.72 | 42.16 | 42.56 | 39.60 | **168.05** |
| 952 | 67.50 | 63.14 | 60.57 | 62.91 | **254.12** |
| 953 | 159.76 | 149.29 | 137.50 | 129.47 | **576.03** |
| 954 | 73.71 | 68.18 | 63.01 | 58.65 | **263.55** |
| 955 | 31.03 | 28.27 | 25.34 | 24.67 | **109.31** |
| 956 | 200.99 | 180.98 | 173.89 | 169.33 | **725.19** |
| 957 | 26.79 | 25.89 | 24.27 | 19.65 | **96.61** |
| 958 | 109.25 | 103.76 | 101.65 | 90.64 | **405.30** |
| 959 | 95.33 | 86.95 | 75.74 | 64.18 | **322.20** |
| 960 | 52.56 | 47.65 | 44.96 | 41.54 | **186.70** |
| 961 | 9.54 | 9.26 | 8.44 | 7.97 | **35.21** |
| **CA - CALIFORNIA - Total** | **4,321.45** | **4,076.32** | **3,859.00** | **3,706.27** | **15,963.04** |

**STATE: CO - COLORADO**

| REGISTRANT ZIP CODE 3 | 1ST QUARTER | 2ND QUARTER | 3RD QUARTER | 4TH QUARTER | **TOTAL GRAMS** |
|---|---|---|---|---|---|
| 800 | 149.14 | 129.38 | 131.59 | 126.91 | **537.02** |
| 801 | 116.76 | 114.02 | 125.36 | 104.18 | **460.32** |
| 802 | 109.39 | 109.57 | 99.97 | 99.89 | **418.82** |
| 803 | 19.20 | 16.08 | 17.77 | 16.60 | **69.65** |
| 804 | 30.17 | 27.14 | 25.31 | 28.94 | **111.56** |
| 805 | 83.90 | 85.44 | 82.23 | 75.92 | **327.49** |

| | | | | | |
|---|---|---|---|---|---|
| 806 | 27.90 | 24.80 | 22.62 | 23.02 | **98.34** |
| 807 | 14.60 | 14.29 | 13.61 | 12.15 | **54.66** |
| 808 | 15.87 | 16.60 | 15.71 | 16.35 | **64.53** |
| 809 | 94.57 | 86.65 | 86.61 | 75.78 | **343.61** |
| 810 | 41.65 | 40.88 | 43.55 | 39.07 | **165.14** |
| 811 | 8.90 | 9.60 | 11.67 | 11.17 | **41.33** |
| 812 | 14.79 | 13.78 | 13.15 | 15.53 | **57.24** |
| 813 | 8.01 | 9.59 | 9.87 | 8.03 | **35.50** |
| 814 | 8.00 | 7.18 | 7.61 | 7.14 | **29.93** |
| 815 | 27.70 | 25.55 | 26.04 | 24.86 | **104.15** |
| 816 | 12.80 | 13.03 | 11.99 | 12.76 | **50.58** |
| **CO - COLORADO - Total** | **783.34** | **743.57** | **744.66** | **698.29** | **2,969.87** |

**STATE: CT - CONNECTICUT**

| REGISTRANT ZIP CODE 3 | 1ST QUARTER | 2ND QUARTER | 3RD QUARTER | 4TH QUARTER | TOTAL GRAMS |
|---|---|---|---|---|---|
| 060 | 106.35 | 98.25 | 96.37 | 92.06 | **393.04** |
| 061 | 32.17 | 28.60 | 26.03 | 26.54 | **113.34** |
| 062 | 28.12 | 24.73 | 26.21 | 25.21 | **104.27** |
| 063 | 49.22 | 43.52 | 41.46 | 42.11 | **176.31** |
| 064 | 159.50 | 154.13 | 167.66 | 138.24 | **619.53** |
| 065 | 61.57 | 62.43 | 56.96 | 59.91 | **240.87** |
| 066 | 23.08 | 21.36 | 23.15 | 24.65 | **92.25** |
| 067 | 61.92 | 59.88 | 56.12 | 56.42 | **234.34** |
| 068 | 57.21 | 56.91 | 55.65 | 50.45 | **220.22** |
| 069 | 8.36 | 8.86 | 7.68 | 7.40 | **32.30** |
| **CT CONNECTICUT Total** | **587.49** | **558.68** | **557.30** | **522.99** | **2,226.46** |

**STATE: DC - DISTRICT OF COLUMBIA**

| REGISTRANT ZIP CODE 3 | 1ST QUARTER | 2ND QUARTER | 3RD QUARTER | 4TH QUARTER | TOTAL GRAMS |
|---|---|---|---|---|---|
| 200 | 57.25 | 50.97 | 46.30 | 47.09 | **201.61** |
| 203 | 0.03 | 0 | 0 | 0 | **0.03** |
| 204 | 3.49 | 2.00 | 5.92 | 1.68 | **13.09** |
| 205 | 0.02 | 0 | 0 | 0 | **0.02** |
| **DC - DISTRICT OF COLUMBIA** | **60.79** | **52.98** | **52.22** | **48.76** | **214.74** |

**STATE: DE - DELAWARE**

| REGISTRANT ZIP CODE 3 | 1ST QUARTER | 2ND QUARTER | 3RD QUARTER | 4TH QUARTER | TOTAL GRAMS |
|---|---|---|---|---|---|
| 197 | 69.91 | 70.12 | 68.65 | 67.47 | **276.16** |
| 198 | 36.61 | 35.31 | 34.03 | 27.00 | **132.95** |
| 199 | 67.52 | 62.64 | 58.01 | 54.16 | **242.32** |
| **DE DELAWARE Total** | **174.04** | **168.08** | **160.69** | **148.63** | **651.43** |

**STATE: FL - FLORIDA**

| REGISTRANT ZIP CODE 3 | 1ST QUARTER | 2ND QUARTER | 3RD QUARTER | 4TH QUARTER | TOTAL GRAMS |
|---|---|---|---|---|---|

| | | | | | |
|---|---|---|---|---|---|
| 320 | 116.97 | 108.57 | 105.01 | 106.60 | **437.14** |
| 321 | 155.07 | 137.49 | 123.73 | 119.77 | **536.06** |
| 322 | 211.64 | 202.72 | 192.58 | 179.97 | **786.91** |
| 323 | 65.42 | 65.39 | 62.82 | 62.25 | **255.88** |
| 324 | 84.08 | 81.09 | 76.13 | 72.44 | **313.75** |
| 325 | 90.56 | 81.30 | 85.24 | 80.09 | **337.19** |
| 326 | 82.39 | 86.74 | 75.07 | 72.68 | **316.89** |
| 327 | 209.37 | 184.11 | 175.19 | 167.60 | **736.27** |
| 328 | 115.92 | 110.29 | 106.57 | 112.26 | **445.03** |
| 329 | 211.63 | 192.36 | 185.59 | 182.58 | **772.16** |
| 330 | 167.66 | 164.46 | 158.78 | 144.85 | **635.75** |
| 331 | 142.82 | 127.34 | 126.03 | 117.05 | **513.24** |
| 333 | 167.68 | 153.44 | 147.62 | 140.38 | **609.12** |
| 334 | 323.89 | 318.13 | 296.64 | 296.20 | **1,234.85** |
| 335 | 120.05 | 112.97 | 109.76 | 109.14 | **451.93** |
| 336 | 439.52 | 268.98 | 276.20 | 236.30 | **1,221.01** |
| 337 | 173.09 | 156.37 | 153.56 | 143.71 | **626.74** |
| 338 | 101.32 | 91.14 | 85.80 | 82.14 | **360.40** |
| 339 | 175.79 | 164.63 | 157.17 | 165.36 | **662.95** |
| 341 | 69.94 | 61.43 | 54.28 | 61.38 | **247.03** |
| 342 | 184.08 | 167.72 | 152.11 | 151.53 | **655.44** |
| 344 | 80.01 | 70.32 | 63.41 | 69.54 | **283.28** |
| 346 | 178.11 | 162.84 | 156.21 | 162.64 | **659.80** |
| 347 | 82.08 | 75.75 | 74.07 | 75.64 | **307.54** |
| 349 | 129.74 | 128.05 | 113.75 | 115.01 | **486.54** |
| **FL FLORIDA - Total** | **3,878.80** | **3,473.63** | **3,313.34** | **3,227.11** | **13,892.88** |

**STATE: GA - GEORGIA**

| REGISTRANT ZIP CODE 3 | 1ST QUARTER | 2ND QUARTER | 3RD QUARTER | 4TH QUARTER | **TOTAL GRAMS** |
|---|---|---|---|---|---|
| 300 | 372.02 | 347.44 | 330.09 | 320.26 | **1,369.81** |
| 301 | 230.20 | 212.54 | 201.89 | 200.04 | **844.67** |
| 302 | 166.30 | 159.01 | 150.31 | 140.84 | **616.46** |
| 303 | 147.94 | 132.95 | 139.87 | 134.54 | **555.31** |
| 304 | 38.40 | 37.06 | 39.36 | 36.86 | **151.68** |
| 305 | 155.48 | 142.67 | 139.86 | 134.35 | **572.36** |
| 306 | 70.30 | 68.83 | 67.90 | 69.22 | **276.24** |
| 307 | 40.54 | 35.92 | 34.52 | 32.77 | **143.74** |
| 308 | 37.84 | 35.83 | 35.14 | 33.78 | **142.60** |
| 309 | 66.38 | 71.52 | 68.74 | 59.34 | **265.98** |
| 310 | 118.06 | 115.58 | 114.12 | 105.55 | **453.31** |
| 312 | 43.12 | 41.07 | 40.26 | 36.33 | **160.78** |

| | | | | | |
|---|---|---|---|---|---|
| 313 | 26.10 | 25.97 | 26.71 | 24.35 | **103.13** |
| 314 | 40.55 | 42.87 | 45.56 | 46.12 | **175.10** |
| 315 | 75.81 | 69.84 | 74.56 | 68.53 | **288.74** |
| 316 | 54.50 | 49.94 | 49.18 | 46.60 | **200.22** |
| 317 | 92.93 | 86.10 | 86.67 | 83.78 | **349.48** |
| 318 | 4.83 | 4.31 | 4.34 | 3.73 | **17.20** |
| 319 | 34.96 | 27.23 | 27.57 | 29.62 | **119.38** |
| 398 | 23.86 | 23.76 | 25.02 | 23.00 | **95.64** |
| **GA GEORGIA Total** | **1,840.13** | **1,730.42** | **1,701.67** | **1,629.59** | **6,901.81** |

**STATE: GU - GUAM**

| REGISTRANT ZIP CODE 3 | 1ST QUARTER | 2ND QUARTER | 3RD QUARTER | 4TH QUARTER | TOTAL GRAMS |
|---|---|---|---|---|---|
| 969 | 7.39 | 6.87 | 6.02 | 7.28 | **27.56** |
| **GU - GUAM - Total** | **7.39** | **6.87** | **6.02** | **7.28** | **27.56** |

**STATE: HI - HAWAII**

| REGISTRANT ZIP CODE 3 | 1ST QUARTER | 2ND QUARTER | 3RD QUARTER | 4TH QUARTER | TOTAL GRAMS |
|---|---|---|---|---|---|
| 967 | 77.15 | 74.85 | 69.45 | 66.86 | **288.30** |
| 968 | 55.61 | 49.58 | 51.46 | 45.11 | **201.76** |
| **HI - HAWAII - Total** | **132.76** | **124.43** | **120.91** | **111.96** | **490.07** |

**STATE: IA - IOWA**

| REGISTRANT ZIP CODE 3 | 1ST QUARTER | 2ND QUARTER | 3RD QUARTER | 4TH QUARTER | TOTAL GRAMS |
|---|---|---|---|---|---|
| 500 | 51.66 | 51.29 | 51.16 | 48.90 | **203.00** |
| 501 | 50.70 | 50.92 | 47.99 | 44.58 | **194.19** |
| 502 | 49.21 | 48.31 | 48.33 | 48.83 | **194.68** |
| 503 | 66.02 | 64.90 | 61.12 | 60.59 | **252.62** |
| 504 | 25.25 | 26.36 | 27.29 | 25.56 | **104.46** |
| 505 | 32.07 | 35.66 | 32.09 | 32.39 | **132.21** |
| 506 | 34.22 | 35.09 | 33.10 | 33.90 | **136.32** |
| 507 | 12.93 | 14.37 | 11.68 | 14.85 | **53.83** |
| 508 | 7.50 | 6.53 | 5.40 | 5.54 | **24.97** |
| 510 | 6.90 | 7.70 | 7.78 | 7.15 | **29.54** |
| 511 | 16.76 | 15.19 | 15.50 | 18.09 | **65.54** |
| 512 | 7.90 | 7.77 | 6.09 | 5.69 | **27.45** |
| 513 | 9.40 | 7.95 | 8.13 | 7.24 | **32.72** |
| 514 | 17.13 | 16.65 | 16.55 | 19.28 | **69.61** |
| 515 | 21.69 | 20.45 | 17.53 | 16.54 | **76.21** |
| 516 | 5.65 | 5.28 | 5.47 | 4.86 | **21.27** |
| 520 | 30.44 | 25.33 | 26.81 | 25.05 | **107.64** |
| 521 | 11.34 | 10.91 | 10.70 | 11.46 | **44.42** |
| 522 | 29.53 | 28.14 | 28.06 | 25.97 | **111.70** |
| 523 | 18.97 | 15.40 | 13.09 | 11.90 | **59.36** |

| | | | | | |
|---|---|---|---|---|---|
| 524 | 22.93 | 21.20 | 20.81 | 19.10 | **84.04** |
| 525 | 25.72 | 25.87 | 28.31 | 25.16 | **105.06** |
| 526 | 20.97 | 21.28 | 19.48 | 19.56 | **81.29** |
| 527 | 15.69 | 13.61 | 13.21 | 12.25 | **54.76** |
| 528 | 24.40 | 22.74 | 21.71 | 21.42 | **90.27** |
| **IA IOWA Total** | **615.00** | **598.90** | **577.40** | **565.87** | **2,357.16** |

**STATE: ID - IDAHO**

| REGISTRANT ZIP CODE 3 | 1ST QUARTER | 2ND QUARTER | 3RD QUARTER | 4TH QUARTER | **TOTAL GRAMS** |
|---|---|---|---|---|---|
| 832 | 45.50 | 40.85 | 43.08 | 43.45 | **172.88** |
| 833 | 37.90 | 34.59 | 36.43 | 32.28 | **141.20** |
| 834 | 46.76 | 46.40 | 45.75 | 46.14 | **185.05** |
| 835 | 28.63 | 29.07 | 28.25 | 26.04 | **112.00** |
| 836 | 108.03 | 102.21 | 94.70 | 90.30 | **395.24** |
| 837 | 82.76 | 77.12 | 71.52 | 72.41 | **303.81** |
| 838 | 57.19 | 53.36 | 59.57 | 50.17 | **220.29** |
| **ID IDAHO Total** | **406.77** | **383.61** | **379.31** | **360.79** | **1,530.47** |

**STATE: IL - ILLINOIS**

| REGISTRANT ZIP CODE 3 | 1ST QUARTER | 2ND QUARTER | 3RD QUARTER | 4TH QUARTER | **TOTAL GRAMS** |
|---|---|---|---|---|---|
| 600 | 301.12 | 304.35 | 293.13 | 291.86 | **1,190.45** |
| 601 | 183.54 | 180.34 | 175.34 | 160.62 | **699.84** |
| 602 | 9.39 | 8.44 | 7.27 | 7.21 | **32.31** |
| 603 | 12.93 | 8.67 | 7.05 | 8.38 | **37.03** |
| 604 | 216.79 | 201.26 | 196.16 | 191.34 | **805.54** |
| 605 | 126.71 | 121.27 | 124.20 | 118.66 | **490.84** |
| 606 | 186.75 | 184.82 | 177.31 | 171.92 | **720.80** |
| 607 | 18.74 | 20.17 | 23.20 | 20.14 | **82.25** |
| 608 | 9.88 | 10.60 | 7.01 | 9.67 | **37.15** |
| 609 | 31.74 | 25.47 | 25.49 | 25.31 | **108.01** |
| 610 | 74.32 | 74.43 | 71.97 | 70.45 | **291.17** |
| 611 | 69.79 | 71.45 | 68.48 | 67.96 | **277.69** |
| 612 | 22.93 | 21.16 | 18.38 | 15.62 | **78.09** |
| 613 | 21.06 | 21.68 | 19.94 | 15.98 | **78.66** |
| 614 | 27.88 | 25.27 | 24.82 | 22.56 | **100.53** |
| 615 | 40.96 | 38.23 | 34.66 | 31.08 | **144.94** |
| 616 | 83.62 | 82.46 | 86.09 | 87.09 | **339.26** |
| 617 | 56.95 | 54.94 | 53.53 | 51.27 | **216.69** |
| 618 | 45.36 | 45.84 | 49.02 | 44.04 | **184.25** |
| 619 | 19.04 | 18.37 | 17.34 | 16.87 | **71.61** |
| 620 | 66.00 | 62.02 | 58.93 | 54.29 | **241.24** |
| 622 | 56.51 | 56.45 | 53.06 | 54.79 | **220.81** |

| | | | | | |
|---|---|---|---|---|---|
| 623 | 31.89 | 33.09 | 34.41 | 30.70 | **130.08** |
| 624 | 26.84 | 24.31 | 24.56 | 25.47 | **101.19** |
| 625 | 49.39 | 45.32 | 40.67 | 40.76 | **176.15** |
| 626 | 33.61 | 30.53 | 32.48 | 27.82 | **124.44** |
| 627 | 48.46 | 45.67 | 45.50 | 43.24 | **182.86** |
| 628 | 33.99 | 31.20 | 32.42 | 29.19 | **126.81** |
| 629 | 51.33 | 52.45 | 49.20 | 52.05 | **205.03** |
| **IL ILLINOIS Total** | **1,957.48** | **1,900.26** | **1,851.64** | **1,786.33** | **7,495.72** |

**STATE: IN - INDIANA**

| REGISTRANT ZIP CODE 3 | 1ST QUARTER | 2ND QUARTER | 3RD QUARTER | 4TH QUARTER | TOTAL GRAMS |
|---|---|---|---|---|---|
| 460 | 82.93 | 84.50 | 84.37 | 82.67 | **334.48** |
| 461 | 103.37 | 105.77 | 92.61 | 91.24 | **392.99** |
| 462 | 222.31 | 194.24 | 193.56 | 188.61 | **798.72** |
| 463 | 121.71 | 123.18 | 113.65 | 116.70 | **475.24** |
| 464 | 9.06 | 6.86 | 6.87 | 7.81 | **30.61** |
| 465 | 82.31 | 82.68 | 77.75 | 76.40 | **319.13** |
| 466 | 24.61 | 23.27 | 19.99 | 19.53 | **87.41** |
| 467 | 42.98 | 42.52 | 47.03 | 44.98 | **177.51** |
| 468 | 71.66 | 65.46 | 67.86 | 61.03 | **266.00** |
| 469 | 54.81 | 50.91 | 52.37 | 48.58 | **206.68** |
| 470 | 32.53 | 33.20 | 32.85 | 25.04 | **123.61** |
| 471 | 58.54 | 52.32 | 53.86 | 50.46 | **215.17** |
| 472 | 30.23 | 27.71 | 27.86 | 24.22 | **110.01** |
| 473 | 68.60 | 69.32 | 63.85 | 57.45 | **259.23** |
| 474 | 44.47 | 44.25 | 41.98 | 38.86 | **169.55** |
| 475 | 33.02 | 38.88 | 32.82 | 29.41 | **134.13** |
| 476 | 11.47 | 13.18 | 12.10 | 12.12 | **48.88** |
| 477 | 47.46 | 36.01 | 48.92 | 49.40 | **181.80** |
| 478 | 22.17 | 21.51 | 21.78 | 21.47 | **86.94** |
| 479 | 30.14 | 26.96 | 26.36 | 29.19 | **112.65** |
| **IN INDIANA Total** | **1,194.38** | **1,142.72** | **1,118.46** | **1,075.17** | **4,530.74** |

**STATE: KS - KANSAS**

| REGISTRANT ZIP CODE 3 | 1ST QUARTER | 2ND QUARTER | 3RD QUARTER | 4TH QUARTER | TOTAL GRAMS |
|---|---|---|---|---|---|
| 660 | 102.64 | 98.33 | 96.84 | 86.31 | **384.12** |
| 661 | 27.60 | 28.59 | 25.80 | 24.66 | **106.65** |
| 662 | 113.78 | 107.23 | 94.44 | 92.08 | **407.53** |
| 664 | 17.48 | 15.84 | 17.52 | 14.12 | **64.96** |
| 665 | 32.93 | 29.24 | 28.55 | 28.01 | **118.72** |
| 666 | 43.75 | 44.99 | 40.54 | 42.40 | **171.68** |
| 667 | 41.68 | 38.43 | 40.71 | 38.05 | **158.87** |

| | | | | | |
|---|---|---|---|---|---|
| 668 | 5.43 | 5.70 | 4.89 | 4.56 | **20.58** |
| 669 | 12.46 | 12.62 | 12.66 | 11.99 | **49.74** |
| 670 | 41.83 | 37.41 | 36.50 | 35.71 | **151.46** |
| 671 | 18.39 | 17.84 | 18.73 | 17.45 | **72.41** |
| 672 | 78.41 | 79.21 | 74.22 | 69.38 | **301.22** |
| 673 | 14.84 | 12.76 | 13.03 | 11.70 | **52.32** |
| 674 | 53.69 | 54.04 | 50.52 | 49.05 | **207.30** |
| 675 | 26.81 | 27.34 | 25.23 | 24.53 | **103.91** |
| 676 | 23.37 | 25.86 | 23.54 | 22.90 | **95.67** |
| 677 | 13.92 | 14.23 | 11.41 | 12.06 | **51.62** |
| 678 | 38.34 | 38.36 | 34.85 | 32.29 | **143.84** |
| 679 | 8.50 | 8.87 | 11.02 | 10.90 | **39.30** |
| **KS - KANSAS - Total** | **715.84** | **696.92** | **661.01** | **628.16** | **2,701.93** |

**STATE: KY - KENTUCKY**

| REGISTRANT ZIP CODE 3 | 1ST QUARTER | 2ND QUARTER | 3RD QUARTER | 4TH QUARTER | TOTAL GRAMS |
|---|---|---|---|---|---|
| 400 | 31.21 | 30.53 | 29.42 | 29.74 | **120.89** |
| 401 | 15.87 | 15.34 | 16.46 | 14.50 | **62.17** |
| 402 | 134.59 | 123.34 | 120.08 | 110.88 | **488.89** |
| 403 | 45.35 | 43.09 | 41.10 | 42.78 | **172.32** |
| 404 | 26.33 | 23.08 | 22.06 | 21.23 | **92.71** |
| 405 | 53.29 | 53.46 | 46.24 | 49.79 | **202.79** |
| 406 | 5.45 | 5.44 | 5.59 | 5.09 | **21.57** |
| 407 | 24.63 | 24.15 | 23.16 | 23.29 | **95.23** |
| 408 | 4.62 | 3.82 | 3.66 | 3.24 | **15.35** |
| 409 | 8.22 | 8.44 | 7.51 | 7.90 | **32.07** |
| 410 | 86.46 | 86.25 | 71.60 | 73.33 | **317.64** |
| 411 | 20.24 | 18.08 | 18.03 | 16.38 | **72.73** |
| 412 | 8.06 | 8.64 | 7.59 | 7.32 | **31.61** |
| 413 | 6.42 | 5.71 | 7.38 | 6.85 | **26.36** |
| 414 | 1.72 | 1.94 | 2.06 | 1.71 | **7.43** |
| 415 | 10.87 | 11.76 | 11.05 | 9.96 | **43.64** |
| 416 | 8.78 | 8.92 | 9.55 | 8.02 | **35.28** |
| 417 | 28.53 | 28.19 | 25.67 | 25.59 | **107.98** |
| 418 | 2.60 | 1.86 | 2.05 | 1.61 | **8.12** |
| 420 | 38.06 | 36.75 | 35.71 | 30.68 | **141.20** |
| 421 | 41.13 | 40.65 | 41.60 | 37.56 | **160.94** |
| 422 | 14.83 | 13.71 | 14.15 | 14.62 | **57.30** |
| 423 | 21.08 | 18.97 | 20.15 | 17.58 | **77.78** |
| 424 | 22.92 | 20.84 | 19.06 | 20.54 | **83.36** |
| 425 | 13.92 | 14.14 | 13.66 | 12.37 | **54.09** |

| | | | | | |
|---|---|---|---|---|---|
| 426 | 10.97 | 11.22 | 9.58 | 7.82 | **39.58** |
| 427 | 39.88 | 32.95 | 31.78 | 31.30 | **135.92** |
| **KY - KENTUCKY - Total** | **726.03** | **691.27** | **655.97** | **631.69** | **2,704.96** |

**STATE: LA - LOUISIANA**

| REGISTRANT ZIP CODE 3 | 1ST QUARTER | 2ND QUARTER | 3RD QUARTER | 4TH QUARTER | TOTAL GRAMS |
|---|---|---|---|---|---|
| 700 | 61.79 | 58.29 | 61.07 | 59.25 | **240.39** |
| 701 | 43.82 | 38.21 | 42.19 | 42.63 | **166.85** |
| 703 | 43.88 | 41.78 | 36.47 | 35.64 | **157.77** |
| 704 | 77.29 | 74.14 | 68.97 | 70.09 | **290.48** |
| 705 | 133.60 | 125.53 | 124.17 | 111.64 | **494.94** |
| 706 | 28.26 | 28.61 | 28.41 | 27.44 | **112.72** |
| 707 | 77.97 | 65.55 | 73.45 | 60.53 | **277.50** |
| 708 | 63.63 | 58.92 | 61.22 | 55.70 | **239.48** |
| 710 | 21.77 | 21.78 | 19.90 | 19.10 | **82.55** |
| 711 | 61.34 | 55.45 | 52.83 | 50.99 | **220.61** |
| 712 | 77.49 | 77.73 | 77.29 | 72.25 | **304.75** |
| 713 | 50.98 | 45.03 | 41.32 | 44.00 | **181.34** |
| 714 | 24.73 | 25.27 | 20.63 | 22.65 | **93.27** |
| **LA - LOUISIANA - Total** | **766.54** | **716.30** | **707.91** | **671.91** | **2,862.66** |

**STATE: MA - MASSACHUSETTS**

| REGISTRANT ZIP CODE 3 | 1ST QUARTER | 2ND QUARTER | 3RD QUARTER | 4TH QUARTER | TOTAL GRAMS |
|---|---|---|---|---|---|
| 010 | 98.44 | 95.56 | 88.02 | 85.55 | **367.56** |
| 011 | 26.05 | 26.72 | 27.12 | 27.59 | **107.47** |
| 012 | 20.33 | 20.66 | 21.48 | 21.22 | **83.69** |
| 013 | 11.28 | 10.32 | 11.63 | 11.57 | **44.81** |
| 014 | 25.95 | 24.25 | 23.63 | 22.28 | **96.12** |
| 015 | 39.80 | 42.02 | 40.08 | 37.19 | **159.09** |
| 016 | 30.23 | 30.48 | 27.62 | 28.78 | **117.11** |
| 017 | 52.63 | 52.17 | 47.89 | 52.45 | **205.15** |
| 018 | 123.11 | 121.49 | 109.61 | 109.88 | **464.09** |
| 019 | 56.80 | 53.48 | 50.77 | 47.75 | **208.80** |
| 020 | 38.80 | 35.56 | 38.54 | 34.88 | **147.78** |
| 021 | 125.75 | 117.75 | 114.27 | 117.69 | **475.46** |
| 022 | 20.62 | 15.66 | 17.51 | 17.96 | **71.76** |
| 023 | 64.06 | 61.48 | 57.00 | 51.84 | **234.39** |
| 024 | 29.96 | 29.95 | 27.33 | 22.43 | **109.66** |
| 025 | 18.22 | 15.55 | 14.89 | 14.68 | **63.35** |
| 026 | 17.76 | 16.65 | 18.54 | 18.11 | **71.07** |
| 027 | 119.82 | 115.20 | 112.83 | 124.51 | **472.36** |
| **MA - MASSACHUSETTS - Total** | **919.63** | **884.96** | **848.75** | **846.38** | **3,499.72** |

STATE: MD - MARYLAND

| REGISTRANT ZIP CODE 3 | 1ST QUARTER | 2ND QUARTER | 3RD QUARTER | 4TH QUARTER | TOTAL GRAMS |
|---|---|---|---|---|---|
| 206 | 53.49 | 49.58 | 45.11 | 39.15 | **187.34** |
| 207 | 72.07 | 61.60 | 59.99 | 52.38 | **246.03** |
| 208 | 96.00 | 82.24 | 84.27 | 78.33 | **340.83** |
| 209 | 21.73 | 21.78 | 24.71 | 21.35 | **89.57** |
| 210 | 120.33 | 113.69 | 112.12 | 109.70 | **455.85** |
| 211 | 60.03 | 60.99 | 52.62 | 46.00 | **219.63** |
| 212 | 178.92 | 167.68 | 164.91 | 155.44 | **666.95** |
| 214 | 14.67 | 14.06 | 12.95 | 14.17 | **55.84** |
| 215 | 18.66 | 20.76 | 19.71 | 16.69 | **75.82** |
| 216 | 24.22 | 24.30 | 23.99 | 21.98 | **94.49** |
| 217 | 114.00 | 101.98 | 101.38 | 97.19 | **414.54** |
| 218 | 28.72 | 27.95 | 27.12 | 24.58 | **108.37** |
| 219 | 14.01 | 12.58 | 12.41 | 11.79 | **50.79** |
| **MD MARYLAND Total** | **816.84** | **759.18** | **741.28** | **688.74** | **3,006.05** |

STATE: ME - MAINE

| REGISTRANT ZIP CODE 3 | 1ST QUARTER | 2ND QUARTER | 3RD QUARTER | 4TH QUARTER | TOTAL GRAMS |
|---|---|---|---|---|---|
| 039 | 2.42 | 3.64 | 3.93 | 2.94 | **12.93** |
| 040 | 65.20 | 69.86 | 67.71 | 60.96 | **263.73** |
| 041 | 29.33 | 27.21 | 28.61 | 26.63 | **111.77** |
| 042 | 27.98 | 28.29 | 29.62 | 27.59 | **113.48** |
| 043 | 24.03 | 25.40 | 29.19 | 26.85 | **105.47** |
| 044 | 26.62 | 26.20 | 29.04 | 28.73 | **110.59** |
| 045 | 4.56 | 5.18 | 5.10 | 6.51 | **21.36** |
| 046 | 11.24 | 9.34 | 8.73 | 9.35 | **38.66** |
| 047 | 12.65 | 11.86 | 13.85 | 12.07 | **50.43** |
| 048 | 3.89 | 3.15 | 3.74 | 4.54 | **15.31** |
| 049 | 27.22 | 27.60 | 27.17 | 24.12 | **106.12** |
| **ME MAINE Total** | **235.14** | **237.74** | **246.68** | **230.30** | **949.85** |

STATE: MI - MICHIGAN

| REGISTRANT ZIP CODE 3 | 1ST QUARTER | 2ND QUARTER | 3RD QUARTER | 4TH QUARTER | TOTAL GRAMS |
|---|---|---|---|---|---|
| 480 | 206.69 | 199.18 | 194.18 | 186.04 | **786.09** |
| 481 | 250.28 | 242.38 | 232.35 | 220.38 | **945.40** |
| 482 | 63.44 | 56.64 | 54.29 | 55.78 | **230.15** |
| 483 | 169.66 | 149.30 | 161.02 | 160.01 | **639.99** |
| 484 | 73.43 | 69.20 | 66.22 | 66.71 | **275.56** |
| 485 | 38.86 | 40.72 | 32.98 | 30.97 | **143.54** |
| 486 | 133.11 | 118.01 | 120.85 | 108.34 | **480.30** |
| 487 | 56.46 | 56.79 | 53.91 | 49.32 | **216.48** |

| | | | | | |
|---|---|---|---|---|---|
| 488 | 86.95 | 87.30 | 80.02 | 73.38 | **327.65** |
| 489 | 38.88 | 36.45 | 35.71 | 34.93 | **145.96** |
| 490 | 204.59 | 199.45 | 200.16 | 191.42 | **795.61** |
| 491 | 21.17 | 20.92 | 23.29 | 20.15 | **85.54** |
| 492 | 59.35 | 52.34 | 50.98 | 45.40 | **208.08** |
| 493 | 31.73 | 29.14 | 29.23 | 25.02 | **115.11** |
| 494 | 107.54 | 105.10 | 101.42 | 92.69 | **406.75** |
| 495 | 108.66 | 105.98 | 106.43 | 97.53 | **418.60** |
| 496 | 87.88 | 84.11 | 82.05 | 75.07 | **329.11** |
| 497 | 58.01 | 58.24 | 57.20 | 52.52 | **225.98** |
| 498 | 50.18 | 48.77 | 48.84 | 41.50 | **189.29** |
| 499 | 17.23 | 15.81 | 16.03 | 15.72 | **64.79** |
| **MI - MICHIGAN Total** | **1,864.11** | **1,775.82** | **1,747.16** | **1,642.86** | **7,029.96** |

**STATE: MN - MINNESOTA**

| REGISTRANT ZIP CODE 3 | 1ST QUARTER | 2ND QUARTER | 3RD QUARTER | 4TH QUARTER | **TOTAL GRAMS** |
|---|---|---|---|---|---|
| 550 | 66.73 | 67.89 | 62.83 | 59.94 | **257.39** |
| 551 | 99.04 | 91.04 | 84.75 | 85.31 | **360.14** |
| 553 | 83.05 | 79.38 | 70.91 | 67.60 | **300.94** |
| 554 | 189.67 | 139.28 | 166.83 | 151.86 | **647.64** |
| 556 | 1.33 | 2.01 | 1.69 | 1.39 | **6.42** |
| 557 | 24.32 | 24.19 | 26.57 | 25.03 | **100.11** |
| 558 | 18.39 | 18.97 | 17.64 | 16.32 | **71.32** |
| 559 | 37.85 | 43.06 | 37.11 | 42.33 | **160.35** |
| 560 | 28.50 | 28.69 | 25.77 | 25.98 | **108.94** |
| 561 | 11.59 | 11.63 | 10.46 | 10.40 | **44.08** |
| 562 | 23.15 | 20.14 | 22.83 | 22.45 | **88.57** |
| 563 | 46.51 | 42.85 | 43.62 | 43.01 | **175.98** |
| 564 | 20.11 | 20.99 | 22.27 | 17.58 | **80.95** |
| 565 | 22.43 | 20.19 | 16.57 | 14.09 | **73.28** |
| 566 | 8.82 | 9.49 | 9.22 | 10.06 | **37.60** |
| 567 | 19.73 | 19.27 | 20.39 | 20.81 | **80.19** |
| **MN MINNESOTA Total** | **701.21** | **639.08** | **639.46** | **614.14** | **2,593.89** |

**STATE: MO - MISSOURI**

| REGISTRANT ZIP CODE 3 | 1ST QUARTER | 2ND QUARTER | 3RD QUARTER | 4TH QUARTER | **TOTAL GRAMS** |
|---|---|---|---|---|---|
| 630 | 114.93 | 112.61 | 105.54 | 103.39 | **436.47** |
| 631 | 243.10 | 250.56 | 228.74 | 246.51 | **968.92** |
| 633 | 59.97 | 60.02 | 61.93 | 55.11 | **237.03** |
| 634 | 22.88 | 20.81 | 19.66 | 19.63 | **82.98** |
| 635 | 20.67 | 19.68 | 17.99 | 20.18 | **78.51** |
| 636 | 30.00 | 30.47 | 27.47 | 23.92 | **111.86** |

| | | | | | |
|---|---|---|---|---|---|
| 637 | 20.80 | 18.62 | 15.49 | 16.77 | **71.68** |
| 638 | 27.19 | 23.21 | 23.47 | 20.85 | **94.72** |
| 639 | 10.72 | 8.93 | 7.75 | 8.24 | **35.64** |
| 640 | 144.40 | 139.44 | 133.98 | 124.21 | **542.03** |
| 641 | 107.58 | 91.45 | 85.81 | 86.06 | **370.90** |
| 644 | 18.14 | 17.99 | 18.23 | 16.09 | **70.46** |
| 645 | 23.13 | 23.08 | 23.98 | 23.51 | **93.71** |
| 646 | 14.68 | 12.33 | 14.10 | 11.28 | **52.39** |
| 647 | 31.84 | 33.05 | 30.61 | 28.23 | **123.73** |
| 648 | 56.90 | 47.30 | 45.76 | 44.24 | **194.21** |
| 650 | 30.08 | 33.62 | 30.28 | 28.59 | **122.57** |
| 651 | 19.58 | 20.34 | 19.12 | 19.98 | **79.03** |
| 652 | 60.98 | 59.31 | 59.01 | 56.43 | **235.73** |
| 653 | 20.94 | 21.81 | 21.78 | 20.29 | **84.82** |
| 654 | 23.78 | 25.41 | 24.78 | 22.25 | **96.22** |
| 655 | 25.73 | 24.00 | 24.27 | 21.64 | **95.64** |
| 656 | 61.57 | 57.90 | 58.93 | 55.79 | **234.18** |
| 657 | 89.13 | 87.95 | 80.65 | 78.73 | **336.46** |
| 658 | 90.21 | 89.55 | 80.19 | 71.19 | **331.13** |
| **MO MISSOURI - Total** | **1,368.95** | **1,329.44** | **1,259.52** | **1,223.10** | **5,181.01** |

**STATE: MS - MISSISSIPPI**

| REGISTRANT ZIP CODE 3 | 1ST QUARTER | 2ND QUARTER | 3RD QUARTER | 4TH QUARTER | TOTAL GRAMS |
|---|---|---|---|---|---|
| 386 | 71.67 | 69.44 | 63.44 | 61.12 | **265.67** |
| 387 | 14.40 | 14.57 | 15.48 | 14.02 | **58.47** |
| 388 | 67.12 | 62.83 | 62.31 | 58.32 | **250.58** |
| 389 | 16.41 | 15.87 | 14.96 | 15.30 | **62.54** |
| 390 | 42.37 | 40.11 | 39.15 | 37.05 | **158.69** |
| 391 | 311.81 | 295.89 | 365.25 | 349.95 | **1,322.90** |
| 392 | 69.58 | 66.17 | 62.39 | 58.74 | **256.87** |
| 393 | 47.19 | 46.49 | 39.94 | 42.28 | **175.91** |
| 394 | 89.70 | 81.24 | 80.35 | 81.66 | **332.94** |
| 395 | 60.85 | 58.52 | 57.30 | 54.20 | **230.87** |
| 396 | 29.80 | 27.45 | 30.22 | 26.83 | **114.30** |
| 397 | 26.45 | 24.13 | 26.19 | 26.01 | **102.78** |
| **MS MISSISSIPPI - Total** | **847.35** | **802.70** | **856.99** | **825.48** | **3,332.52** |

**STATE: MT - MONTANA**

| REGISTRANT ZIP CODE 3 | 1ST QUARTER | 2ND QUARTER | 3RD QUARTER | 4TH QUARTER | TOTAL GRAMS |
|---|---|---|---|---|---|
| 590 | 10.80 | 11.03 | 9.93 | 9.91 | **41.68** |
| 591 | 44.09 | 41.25 | 41.57 | 38.43 | **165.34** |
| 592 | 9.17 | 7.97 | 9.37 | 9.49 | **36.00** |

| | | | | | |
|---|---|---|---|---|---|
| 593 | 9.30 | 10.06 | 10.08 | 9.71 | **39.15** |
| 594 | 33.73 | 31.56 | 34.16 | 30.87 | **130.32** |
| 595 | 7.85 | 8.09 | 8.34 | 8.98 | **33.26** |
| 596 | 19.42 | 20.71 | 18.87 | 18.30 | **77.31** |
| 597 | 24.98 | 22.55 | 26.93 | 22.28 | **96.74** |
| 598 | 39.06 | 36.58 | 35.16 | 35.02 | **145.82** |
| 599 | 26.35 | 23.78 | 21.94 | 23.64 | **95.71** |
| **MT - MONTANA Total** | **224.75** | **213.58** | **216.36** | **206.62** | **861.32** |

**STATE: NC - NORTH CAROLINA**

| REGISTRANT ZIP CODE 3 | 1ST QUARTER | 2ND QUARTER | 3RD QUARTER | 4TH QUARTER | TOTAL GRAMS |
|---|---|---|---|---|---|
| 270 | 40.49 | 40.66 | 40.01 | 36.93 | **158.09** |
| 271 | 59.01 | 59.62 | 56.65 | 61.43 | **236.71** |
| 272 | 115.34 | 100.12 | 104.00 | 92.18 | **411.64** |
| 273 | 63.28 | 64.12 | 60.84 | 51.77 | **240.01** |
| 274 | 46.35 | 44.23 | 41.76 | 39.59 | **171.93** |
| 275 | 194.66 | 183.26 | 189.52 | 162.38 | **729.82** |
| 276 | 92.02 | 80.44 | 81.08 | 80.83 | **334.36** |
| 277 | 64.48 | 60.95 | 52.16 | 49.47 | **227.05** |
| 278 | 142.49 | 130.01 | 122.98 | 112.41 | **507.89** |
| 279 | 54.12 | 51.40 | 47.50 | 34.30 | **187.32** |
| 280 | 157.00 | 141.69 | 137.70 | 129.34 | **565.73** |
| 281 | 111.03 | 106.08 | 102.94 | 99.13 | **419.17** |
| 282 | 86.54 | 90.38 | 94.09 | 94.62 | **365.63** |
| 283 | 150.74 | 133.65 | 124.81 | 106.78 | **515.98** |
| 284 | 119.19 | 111.35 | 109.70 | 94.49 | **434.73** |
| 285 | 127.47 | 123.63 | 121.99 | 110.16 | **483.24** |
| 286 | 191.52 | 172.48 | 157.64 | 151.86 | **673.51** |
| 287 | 87.92 | 85.66 | 86.97 | 86.29 | **346.84** |
| 288 | 36.40 | 40.50 | 37.53 | 34.98 | **149.40** |
| 289 | 11.33 | 10.67 | 10.27 | 10.78 | **43.05** |
| **NC - NORTH CAROLINA Total** | **1,951.39** | **1,830.89** | **1,780.13** | **1,639.72** | **7,202.12** |

**STATE: ND - NORTH DAKOTA**

| REGISTRANT ZIP CODE 3 | 1ST QUARTER | 2ND QUARTER | 3RD QUARTER | 4TH QUARTER | TOTAL GRAMS |
|---|---|---|---|---|---|
| 580 | 5.71 | 5.80 | 5.20 | 5.06 | **21.78** |
| 581 | 32.26 | 27.43 | 25.79 | 27.57 | **113.05** |
| 582 | 30.34 | 28.10 | 25.96 | 25.56 | **109.96** |
| 583 | 12.58 | 13.13 | 11.33 | 11.73 | **48.76** |
| 584 | 8.83 | 8.61 | 7.93 | 7.75 | **33.11** |
| 585 | 42.30 | 35.57 | 35.84 | 37.00 | **150.71** |
| 586 | 11.12 | 9.70 | 11.03 | 9.90 | **41.75** |

| | | | | | |
|---|---|---|---|---|---|
| 587 | 12.14 | 11.68 | 13.67 | 13.15 | **50.63** |
| 588 | 9.41 | 10.09 | 9.35 | 8.97 | **37.83** |
| **ND NORTH DAKOTA Total** | **164.67** | **150.10** | **146.10** | **146.69** | **607.57** |

**STATE: NE - NEBRASKA**

| REGISTRANT ZIP CODE 3 | 1ST QUARTER | 2ND QUARTER | 3RD QUARTER | 4TH QUARTER | TOTAL GRAMS |
|---|---|---|---|---|---|
| 680 | 48.18 | 43.39 | 41.51 | 43.06 | **176.14** |
| 681 | 86.71 | 90.46 | 78.90 | 74.70 | **330.77** |
| 683 | 17.34 | 16.27 | 16.31 | 17.52 | **67.44** |
| 684 | 12.55 | 12.24 | 10.91 | 9.77 | **45.46** |
| 685 | 41.88 | 40.35 | 38.42 | 35.30 | **155.96** |
| 686 | 12.43 | 12.20 | 9.38 | 10.38 | **44.40** |
| 687 | 26.26 | 24.73 | 24.75 | 24.05 | **99.80** |
| 688 | 42.88 | 45.77 | 45.20 | 44.99 | **178.84** |
| 689 | 21.13 | 19.99 | 21.50 | 22.96 | **85.57** |
| 690 | 14.55 | 14.63 | 14.22 | 13.36 | **56.75** |
| 691 | 21.38 | 22.12 | 22.00 | 20.41 | **85.91** |
| 692 | 0.62 | 0.80 | 0.65 | 0.98 | **3.05** |
| 693 | 23.16 | 23.71 | 24.30 | 23.54 | **94.71** |
| **NE NEBRASKA Total** | **369.05** | **366.68** | **348.05** | **341.00** | **1,424.79** |

**STATE: NH - NEW HAMPSHIRE**

| REGISTRANT ZIP CODE 3 | 1ST QUARTER | 2ND QUARTER | 3RD QUARTER | 4TH QUARTER | TOTAL GRAMS |
|---|---|---|---|---|---|
| 030 | 65.71 | 65.08 | 62.62 | 62.66 | **256.08** |
| 031 | 30.56 | 27.86 | 29.78 | 29.29 | **117.49** |
| 032 | 15.78 | 18.19 | 19.05 | 18.73 | **71.75** |
| 033 | 35.45 | 28.99 | 31.32 | 30.18 | **125.94** |
| 034 | 7.25 | 7.09 | 8.13 | 5.93 | **28.40** |
| 035 | 8.91 | 7.85 | 7.81 | 7.18 | **31.74** |
| 036 | 0.57 | 0.42 | 0.95 | 0.86 | **2.80** |
| 037 | 16.11 | 13.65 | 15.23 | 13.94 | **58.93** |
| 038 | 49.69 | 45.60 | 46.40 | 44.91 | **186.60** |
| **NH - NEW HAMPSHIRE Total** | **230.03** | **214.73** | **221.28** | **213.68** | **879.73** |

**STATE: NJ - NEW JERSEY**

| REGISTRANT ZIP CODE 3 | 1ST QUARTER | 2ND QUARTER | 3RD QUARTER | 4TH QUARTER | TOTAL GRAMS |
|---|---|---|---|---|---|
| 070 | 238.74 | 213.98 | 202.95 | 199.11 | **854.78** |
| 071 | 26.45 | 26.47 | 25.21 | 26.19 | **104.31** |
| 072 | 10.64 | 10.19 | 11.91 | 9.19 | **41.93** |
| 073 | 9.70 | 6.64 | 8.98 | 8.44 | **33.76** |
| 074 | 109.81 | 101.87 | 120.28 | 86.85 | **418.82** |
| 075 | 16.92 | 15.32 | 12.88 | 12.49 | **57.61** |
| 076 | 163.34 | 162.38 | 155.84 | 148.90 | **630.45** |

| | | | | | |
|---|---|---|---|---|---|
| 077 | 112.54 | 99.64 | 106.93 | 95.98 | **415.09** |
| 078 | 50.78 | 49.15 | 46.76 | 48.67 | **195.36** |
| 079 | 38.53 | 38.70 | 47.70 | 46.90 | **171.83** |
| 080 | 216.75 | 204.55 | 196.33 | 194.03 | **811.65** |
| 081 | 18.11 | 16.68 | 18.79 | 16.63 | **70.21** |
| 082 | 35.70 | 39.18 | 36.95 | 34.10 | **145.94** |
| 083 | 18.92 | 17.27 | 15.84 | 15.94 | **67.97** |
| 084 | 8.54 | 8.19 | 6.95 | 7.40 | **31.08** |
| 085 | 42.42 | 36.60 | 34.94 | 37.84 | **151.80** |
| 086 | 41.87 | 35.52 | 34.96 | 35.72 | **148.06** |
| 087 | 85.37 | 82.38 | 75.88 | 76.18 | **319.81** |
| 088 | 194.11 | 173.36 | 163.01 | 155.17 | **685.64** |
| 089 | 19.93 | 21.63 | 22.35 | 23.79 | **87.70** |
| **NJ NEW JERSEY - Total** | **1,459.17** | **1,359.68** | **1,345.44** | **1,279.52** | **5,443.80** |

**STATE: NM - NEW MEXICO**

| REGISTRANT ZIP CODE 3 | 1ST QUARTER | 2ND QUARTER | 3RD QUARTER | 4TH QUARTER | **TOTAL GRAMS** |
|---|---|---|---|---|---|
| 870 | 14.96 | 13.38 | 15.57 | 11.71 | **55.62** |
| 871 | 128.66 | 117.76 | 110.99 | 100.74 | **458.15** |
| 873 | 6.78 | 5.24 | 6.32 | 6.36 | **24.70** |
| 874 | 20.17 | 21.27 | 19.69 | 18.66 | **79.78** |
| 875 | 37.09 | 33.17 | 31.80 | 36.27 | **138.33** |
| 877 | 2.72 | 3.87 | 4.77 | 3.98 | **15.34** |
| 878 | 0.80 | 0.73 | 0.64 | 0.47 | **2.64** |
| 879 | 1.24 | 1.05 | 1.24 | 1.13 | **4.67** |
| 880 | 18.03 | 19.90 | 18.94 | 18.78 | **75.65** |
| 881 | 7.63 | 6.80 | 6.69 | 6.05 | **27.16** |
| 882 | 32.04 | 31.71 | 29.15 | 30.96 | **123.85** |
| 883 | 9.21 | 9.35 | 11.38 | 9.10 | **39.05** |
| 884 | 2.72 | 2.90 | 3.84 | 2.86 | **12.32** |
| **NM NEW MEXICO Total** | **282.04** | **267.12** | **261.01** | **247.09** | **1,057.26** |

**STATE: NV - NEVADA**

| REGISTRANT ZIP CODE 3 | 1ST QUARTER | 2ND QUARTER | 3RD QUARTER | 4TH QUARTER | **TOTAL GRAMS** |
|---|---|---|---|---|---|
| 890 | 91.88 | 76.08 | 72.49 | 72.29 | **312.73** |
| 891 | 177.34 | 167.61 | 159.80 | 153.02 | **657.78** |
| 893 | 1.17 | 0.92 | 1.17 | 0.91 | **4.16** |
| 894 | 22.68 | 23.85 | 21.34 | 20.29 | **88.16** |
| 895 | 43.50 | 40.70 | 36.80 | 30.13 | **151.13** |
| 897 | 9.52 | 10.97 | 7.14 | 7.90 | **35.53** |
| 898 | 8.50 | 6.96 | 8.90 | 7.31 | **31.68** |
| **NV NEVADA Total** | **354.61** | **327.08** | **307.64** | **291.84** | **1,281.17** |

**STATE: NY - NEW YORK**

| REGISTRANT ZIP CODE 3 | 1ST QUARTER | 2ND QUARTER | 3RD QUARTER | 4TH QUARTER | **TOTAL GRAMS** |
|---|---|---|---|---|---|
| 100 | 262.05 | 232.08 | 251.76 | 254.28 | **1,000.16** |
| 101 | 2.88 | 3.53 | 4.59 | 3.28 | **14.28** |
| 102 | 0.55 | 0.09 | 0.06 | 0.12 | **0.82** |
| 103 | 98.80 | 90.37 | 87.60 | 79.40 | **356.16** |
| 104 | 100.71 | 88.33 | 93.43 | 86.47 | **368.95** |
| 105 | 81.59 | 71.42 | 66.63 | 68.03 | **287.68** |
| 106 | 12.31 | 11.25 | 10.47 | 12.91 | **46.93** |
| 107 | 22.87 | 19.20 | 21.61 | 23.49 | **87.18** |
| 108 | 5.93 | 4.20 | 4.46 | 4.37 | **18.97** |
| 109 | 81.07 | 68.18 | 73.07 | 70.88 | **293.21** |
| 110 | 67.11 | 57.36 | 62.75 | 58.40 | **245.62** |
| 111 | 13.39 | 10.88 | 10.49 | 10.41 | **45.16** |
| 112 | 146.07 | 125.23 | 131.77 | 126.12 | **529.19** |
| 113 | 65.93 | 60.38 | 64.12 | 60.58 | **251.01** |
| 114 | 35.17 | 31.69 | 32.30 | 32.67 | **131.82** |
| 115 | 103.54 | 87.65 | 98.65 | 91.00 | **380.84** |
| 116 | 6.44 | 6.18 | 5.86 | 6.69 | **25.17** |
| 117 | 315.32 | 293.70 | 292.51 | 287.48 | **1,189.01** |
| 118 | 10.25 | 11.65 | 11.44 | 10.82 | **44.15** |
| 119 | 38.56 | 34.20 | 35.61 | 33.58 | **141.96** |
| 120 | 72.61 | 63.69 | 63.46 | 56.90 | **256.66** |
| 121 | 34.15 | 31.82 | 32.69 | 29.70 | **128.35** |
| 122 | 50.02 | 53.02 | 47.63 | 44.48 | **195.16** |
| 123 | 24.54 | 23.35 | 22.37 | 24.69 | **94.95** |
| 124 | 36.62 | 35.36 | 36.33 | 33.25 | **141.57** |
| 125 | 65.98 | 55.27 | 59.95 | 53.79 | **234.99** |
| 126 | 24.96 | 23.15 | 24.81 | 20.55 | **93.48** |
| 127 | 14.85 | 12.88 | 14.85 | 10.97 | **53.55** |
| 128 | 70.95 | 62.05 | 58.74 | 51.96 | **243.70** |
| 129 | 33.22 | 27.68 | 28.54 | 25.93 | **115.38** |
| 130 | 68.70 | 65.52 | 64.76 | 59.91 | **258.90** |
| 131 | 18.43 | 18.39 | 17.05 | 16.45 | **70.31** |
| 132 | 64.57 | 61.59 | 57.42 | 56.52 | **240.10** |
| 133 | 25.24 | 24.93 | 25.83 | 20.03 | **96.03** |
| 134 | 41.22 | 37.63 | 39.11 | 34.42 | **152.38** |
| 135 | 11.11 | 10.58 | 12.08 | 10.52 | **44.28** |
| 136 | 45.31 | 38.22 | 39.49 | 32.42 | **155.44** |
| 137 | 20.57 | 20.17 | 19.68 | 17.13 | **77.55** |

| | | | | | |
|---|---|---|---|---|---|
| 138 | 55.58 | 54.18 | 46.41 | 44.85 | **201.03** |
| 139 | 20.87 | 19.13 | 16.77 | 15.09 | **71.86** |
| 140 | 67.35 | 66.16 | 58.48 | 55.80 | **247.79** |
| 141 | 33.36 | 32.94 | 31.70 | 27.12 | **125.12** |
| 142 | 99.51 | 93.00 | 90.62 | 86.77 | **369.90** |
| 143 | 11.16 | 7.73 | 8.50 | 8.20 | **35.60** |
| 144 | 53.96 | 53.41 | 52.74 | 52.28 | **212.38** |
| 145 | 47.30 | 42.20 | 39.74 | 37.10 | **166.34** |
| 146 | 103.15 | 99.67 | 95.83 | 95.18 | **393.82** |
| 147 | 35.11 | 32.12 | 28.85 | 25.32 | **121.40** |
| 148 | 59.04 | 58.49 | 53.91 | 48.65 | **220.10** |
| 149 | 17.40 | 14.30 | 13.64 | 13.73 | **59.07** |
| **NY - NEW YORK - Total** | **2,797.38** | **2,546.22** | **2,561.17** | **2,430.71** | **10,335.48** |

**STATE: OH - OHIO**

| REGISTRANT ZIP CODE 3 | 1ST QUARTER | 2ND QUARTER | 3RD QUARTER | 4TH QUARTER | **TOTAL GRAMS** |
|---|---|---|---|---|---|
| 430 | 88.10 | 86.03 | 79.03 | 77.51 | **330.68** |
| 431 | 50.12 | 48.42 | 47.24 | 43.53 | **189.31** |
| 432 | 110.43 | 108.08 | 101.31 | 100.94 | **420.77** |
| 433 | 21.76 | 20.57 | 19.54 | 20.50 | **82.37** |
| 434 | 14.01 | 13.76 | 15.21 | 13.21 | **56.19** |
| 435 | 51.25 | 47.92 | 43.43 | 42.56 | **185.16** |
| 436 | 79.84 | 84.96 | 78.24 | 72.07 | **315.12** |
| 437 | 22.40 | 21.98 | 22.29 | 20.23 | **86.91** |
| 438 | 2.42 | 2.70 | 2.32 | 2.05 | **9.50** |
| 439 | 24.81 | 21.49 | 19.93 | 16.69 | **82.91** |
| 440 | 71.74 | 70.99 | 65.81 | 57.92 | **266.47** |
| 441 | 135.03 | 131.77 | 126.36 | 117.97 | **511.13** |
| 442 | 45.34 | 44.92 | 38.99 | 42.16 | **171.41** |
| 443 | 23.43 | 21.58 | 21.77 | 22.10 | **88.89** |
| 444 | 29.26 | 29.65 | 27.56 | 28.46 | **114.93** |
| 445 | 35.24 | 35.52 | 34.45 | 36.94 | **142.15** |
| 446 | 42.35 | 40.10 | 41.63 | 38.04 | **162.13** |
| 447 | 35.63 | 33.62 | 34.06 | 30.63 | **133.95** |
| 448 | 30.89 | 30.17 | 30.10 | 28.92 | **120.08** |
| 449 | 13.23 | 13.36 | 11.69 | 11.16 | **49.44** |
| 450 | 93.66 | 94.19 | 88.52 | 86.71 | **363.09** |
| 451 | 26.68 | 24.77 | 23.69 | 21.35 | **96.49** |
| 452 | 110.06 | 107.93 | 103.78 | 89.85 | **411.62** |
| 453 | 79.08 | 77.22 | 72.05 | 72.18 | **300.54** |
| 454 | 66.14 | 63.69 | 57.81 | 57.58 | **245.23** |

| | | | | | |
|---|---|---|---|---|---|
| 455 | 13.16 | 12.19 | 11.52 | 11.95 | **48.83** |
| 456 | 55.37 | 53.43 | 54.80 | 57.42 | **221.02** |
| 457 | 16.95 | 16.17 | 16.05 | 14.69 | **63.87** |
| 458 | 34.74 | 31.77 | 33.28 | 32.04 | **131.84** |
| **OH - OHIO Total** | **1,423.15** | **1,388.98** | **1,322.48** | **1,267.39** | **5,402.00** |

**STATE: OK - OKLAHOMA**

| REGISTRANT ZIP CODE 3 | 1ST QUARTER | 2ND QUARTER | 3RD QUARTER | 4TH QUARTER | TOTAL GRAMS |
|---|---|---|---|---|---|
| 730 | 118.68 | 113.95 | 114.25 | 108.38 | **455.26** |
| 731 | 152.09 | 143.67 | 134.94 | 135.44 | **566.13** |
| 734 | 21.53 | 20.85 | 20.33 | 18.05 | **80.76** |
| 735 | 62.10 | 60.85 | 62.60 | 55.12 | **240.66** |
| 736 | 19.60 | 18.06 | 19.66 | 17.79 | **75.11** |
| 737 | 22.00 | 22.97 | 21.38 | 18.82 | **85.17** |
| 738 | 6.27 | 5.86 | 5.17 | 4.94 | **22.24** |
| 739 | 3.91 | 2.69 | 3.28 | 3.06 | **12.94** |
| 740 | 101.23 | 93.40 | 88.73 | 85.60 | **368.97** |
| 741 | 89.38 | 88.17 | 81.57 | 79.11 | **338.23** |
| 743 | 21.94 | 18.96 | 17.77 | 15.62 | **74.29** |
| 744 | 40.20 | 41.61 | 35.70 | 32.94 | **150.45** |
| 745 | 22.73 | 21.51 | 20.16 | 19.01 | **83.41** |
| 746 | 13.78 | 15.69 | 12.62 | 12.61 | **54.70** |
| 747 | 19.25 | 14.45 | 14.12 | 12.87 | **60.69** |
| 748 | 32.89 | 31.76 | 31.58 | 30.70 | **126.93** |
| 749 | 10.26 | 9.61 | 9.55 | 7.77 | **37.19** |
| **OK - OKLAHOMA Total** | **757.84** | **724.05** | **693.42** | **657.84** | **2,833.15** |

**STATE: OR - OREGON**

| REGISTRANT ZIP CODE 3 | 1ST QUARTER | 2ND QUARTER | 3RD QUARTER | 4TH QUARTER | TOTAL GRAMS |
|---|---|---|---|---|---|
| 970 | 91.87 | 82.19 | 81.94 | 81.84 | **337.84** |
| 971 | 46.97 | 46.90 | 43.60 | 43.17 | **180.64** |
| 972 | 134.36 | 123.46 | 116.73 | 118.85 | **493.40** |
| 973 | 77.76 | 76.10 | 71.26 | 69.86 | **294.98** |
| 974 | 93.17 | 86.27 | 83.26 | 82.09 | **344.79** |
| 975 | 53.65 | 55.78 | 53.11 | 52.84 | **215.38** |
| 976 | 8.30 | 7.77 | 9.67 | 9.12 | **34.86** |
| 977 | 37.44 | 36.97 | 37.95 | 33.51 | **145.88** |
| 978 | 30.45 | 28.40 | 26.85 | 24.86 | **110.56** |
| 979 | 9.26 | 9.01 | 8.04 | 6.99 | **33.29** |
| **OR OREGON Total** | **583.22** | **552.85** | **532.42** | **523.14** | **2,191.63** |

**STATE: PA - PENNSYLVANIA**

| REGISTRANT ZIP CODE 3 | 1ST QUARTER | 2ND QUARTER | 3RD QUARTER | 4TH QUARTER | TOTAL GRAMS |
|---|---|---|---|---|---|

| | | | | | |
|---|---|---|---|---|---|
| 150 | 122.98 | 121.38 | 113.08 | 110.79 | **468.22** |
| 151 | 74.06 | 71.17 | 64.96 | 59.86 | **270.05** |
| 152 | 153.56 | 158.82 | 154.68 | 150.88 | **617.94** |
| 153 | 30.46 | 29.08 | 26.37 | 28.66 | **114.57** |
| 154 | 28.93 | 27.26 | 24.64 | 22.80 | **103.64** |
| 155 | 21.12 | 19.87 | 17.38 | 16.24 | **74.61** |
| 156 | 70.53 | 70.99 | 63.49 | 64.63 | **269.65** |
| 157 | 45.97 | 41.79 | 40.47 | 42.32 | **170.55** |
| 158 | 40.79 | 38.37 | 39.77 | 38.52 | **157.44** |
| 159 | 43.90 | 37.38 | 37.05 | 38.27 | **156.60** |
| 160 | 60.27 | 67.62 | 62.00 | 60.59 | **250.49** |
| 161 | 46.54 | 44.90 | 44.28 | 40.45 | **176.17** |
| 162 | 23.48 | 22.37 | 20.96 | 17.37 | **84.18** |
| 163 | 57.47 | 52.84 | 52.72 | 47.42 | **210.46** |
| 164 | 18.77 | 16.31 | 19.02 | 17.81 | **71.92** |
| 165 | 47.43 | 46.30 | 45.62 | 40.85 | **180.20** |
| 166 | 66.63 | 62.72 | 58.76 | 57.00 | **245.10** |
| 167 | 10.91 | 8.66 | 7.98 | 7.42 | **34.97** |
| 168 | 22.53 | 21.06 | 22.18 | 20.49 | **86.25** |
| 169 | 12.69 | 11.20 | 11.09 | 9.38 | **44.36** |
| 170 | 141.37 | 123.12 | 111.29 | 116.87 | **492.65** |
| 171 | 25.59 | 19.79 | 18.49 | 20.71 | **84.59** |
| 172 | 39.83 | 34.18 | 31.88 | 30.88 | **136.76** |
| 173 | 65.22 | 60.70 | 58.21 | 56.34 | **240.48** |
| 174 | 36.47 | 36.40 | 34.91 | 35.26 | **143.04** |
| 175 | 42.10 | 43.37 | 41.34 | 38.43 | **165.24** |
| 176 | 42.37 | 35.56 | 34.28 | 34.19 | **146.40** |
| 177 | 52.22 | 44.50 | 44.65 | 47.57 | **188.94** |
| 178 | 70.69 | 63.99 | 60.30 | 56.59 | **251.57** |
| 179 | 24.42 | 24.02 | 22.29 | 22.50 | **93.23** |
| 180 | 91.05 | 89.27 | 79.65 | 83.82 | **343.79** |
| 181 | 59.66 | 64.58 | 53.64 | 54.66 | **232.55** |
| 182 | 32.14 | 32.38 | 26.41 | 23.01 | **113.93** |
| 183 | 35.52 | 32.16 | 26.57 | 27.96 | **122.21** |
| 184 | 40.47 | 39.70 | 41.08 | 40.90 | **162.16** |
| 185 | 55.67 | 43.89 | 44.61 | 37.32 | **181.49** |
| 186 | 46.38 | 41.70 | 41.11 | 39.62 | **168.81** |
| 187 | 174.44 | 194.06 | 169.28 | 170.50 | **708.28** |
| 188 | 20.00 | 17.37 | 17.54 | 16.08 | **70.99** |
| 189 | 78.44 | 70.66 | 65.64 | 64.89 | **279.63** |

| | | | | | |
|---|---|---|---|---|---|
| 190 | 533.73 | 461.69 | 434.17 | 722.41 | **2,152.01** |
| 191 | 212.19 | 190.94 | 184.06 | 183.95 | **771.15** |
| 193 | 66.19 | 60.47 | 56.57 | 53.06 | **236.30** |
| 194 | 104.85 | 106.16 | 103.00 | 97.92 | **411.93** |
| 195 | 30.92 | 31.28 | 31.07 | 25.05 | **118.32** |
| 196 | 34.21 | 30.45 | 27.56 | 26.24 | **118.46** |
| **PA - PENNSYLVANIA - Total** | **3,155.19** | **2,962.49** | **2,786.12** | **3,018.48** | **11,922.28** |

**STATE: PR - PUERTO RICO**

| REGISTRANT ZIP CODE 3 | 1ST QUARTER | 2ND QUARTER | 3RD QUARTER | 4TH QUARTER | TOTAL GRAMS |
|---|---|---|---|---|---|
| 006 | 40.42 | 44.08 | 39.50 | 47.40 | **171.40** |
| 007 | 60.80 | 61.42 | 45.51 | 57.98 | **225.71** |
| 009 | 63.50 | 69.15 | 54.62 | 68.09 | **255.36** |
| **PR - PUERTO RICO - Total** | **164.72** | **174.65** | **139.62** | **173.47** | **652.46** |

**STATE: RI - RHODE ISLAND**

| REGISTRANT ZIP CODE 3 | 1ST QUARTER | 2ND QUARTER | 3RD QUARTER | 4TH QUARTER | TOTAL GRAMS |
|---|---|---|---|---|---|
| 028 | 75.46 | 70.50 | 72.90 | 67.60 | **286.47** |
| 029 | 42.43 | 40.87 | 37.47 | 42.02 | **162.78** |
| **RI RHODE ISLAND Total** | **117.89** | **111.37** | **110.37** | **109.62** | **449.24** |

**STATE: SC - SOUTH CAROLINA**

| REGISTRANT ZIP CODE 3 | 1ST QUARTER | 2ND QUARTER | 3RD QUARTER | 4TH QUARTER | TOTAL GRAMS |
|---|---|---|---|---|---|
| 290 | 55.66 | 55.64 | 55.03 | 49.97 | **216.31** |
| 291 | 71.58 | 66.93 | 70.78 | 60.40 | **269.69** |
| 292 | 69.98 | 63.31 | 60.36 | 54.83 | **248.46** |
| 293 | 61.87 | 56.85 | 54.39 | 53.20 | **226.32** |
| 294 | 141.92 | 128.25 | 119.33 | 121.52 | **511.02** |
| 295 | 156.52 | 145.81 | 139.36 | 131.13 | **572.82** |
| 296 | 167.08 | 151.78 | 145.81 | 140.93 | **605.59** |
| 297 | 38.66 | 32.28 | 32.21 | 32.06 | **135.22** |
| 298 | 56.60 | 52.40 | 52.94 | 51.08 | **213.02** |
| 299 | 50.27 | 46.45 | 45.47 | 39.48 | **181.67** |
| **SC - SOUTH CAROLINA Total** | **870.13** | **799.71** | **775.69** | **734.59** | **3,180.12** |

**STATE: SD - SOUTH DAKOTA**

| REGISTRANT ZIP CODE 3 | 1ST QUARTER | 2ND QUARTER | 3RD QUARTER | 4TH QUARTER | TOTAL GRAMS |
|---|---|---|---|---|---|
| 570 | 39.62 | 34.05 | 35.58 | 32.57 | **141.83** |
| 571 | 76.81 | 68.24 | 66.26 | 67.35 | **278.66** |
| 572 | 12.38 | 10.58 | 10.00 | 9.76 | **42.73** |
| 573 | 16.64 | 16.42 | 16.59 | 14.96 | **64.61** |
| 574 | 10.58 | 10.61 | 9.81 | 8.44 | **39.45** |
| 575 | 14.24 | 14.26 | 14.46 | 14.08 | **57.04** |
| 576 | 1.98 | 2.16 | 1.89 | 1.98 | **8.00** |

| | | | | | |
|---|---|---|---|---|---|
| 577 | 19.86 | 18.53 | 20.82 | 15.84 | **75.04** |
| **SD SOUTH DAKOTA - Total** | **192.11** | **174.83** | **175.41** | **164.99** | **707.34** |

**STATE: TN - TENNESSEE**

| REGISTRANT ZIP CODE 3 | 1ST QUARTER | 2ND QUARTER | 3RD QUARTER | 4TH QUARTER | TOTAL GRAMS |
|---|---|---|---|---|---|
| 370 | 111.08 | 112.63 | 94.69 | 89.84 | **408.24** |
| 371 | 101.12 | 94.79 | 90.30 | 83.25 | **369.46** |
| 372 | 93.98 | 88.70 | 96.31 | 95.52 | **374.50** |
| 373 | 77.39 | 77.45 | 78.12 | 70.93 | **303.89** |
| 374 | 35.39 | 36.11 | 30.93 | 33.53 | **135.95** |
| 376 | 83.68 | 80.70 | 74.42 | 74.30 | **313.10** |
| 377 | 43.05 | 40.92 | 37.47 | 39.43 | **160.87** |
| 378 | 84.76 | 80.75 | 81.21 | 77.75 | **324.48** |
| 379 | 65.85 | 69.48 | 63.63 | 68.45 | **267.41** |
| 380 | 59.14 | 54.30 | 52.52 | 50.47 | **216.43** |
| 381 | 398.05 | 367.06 | 385.22 | 753.70 | **1,904.03** |
| 382 | 17.01 | 15.89 | 14.96 | 14.86 | **62.72** |
| 383 | 49.41 | 49.75 | 45.35 | 46.35 | **190.86** |
| 384 | 26.50 | 25.11 | 21.72 | 19.67 | **93.00** |
| 385 | 51.24 | 46.92 | 42.39 | 44.06 | **184.61** |
| **TN TENNESSEE Total** | **1,297.64** | **1,240.55** | **1,209.23** | **1,562.13** | **5,309.56** |

**STATE: TX - TEXAS**

| REGISTRANT ZIP CODE 3 | 1ST QUARTER | 2ND QUARTER | 3RD QUARTER | 4TH QUARTER | TOTAL GRAMS |
|---|---|---|---|---|---|
| 750 | 331.94 | 315.28 | 296.44 | 293.06 | **1,236.72** |
| 751 | 92.64 | 94.75 | 86.96 | 83.71 | **358.07** |
| 752 | 128.98 | 141.07 | 141.91 | 153.10 | **565.06** |
| 753 | 10.13 | 11.69 | 10.10 | 9.36 | **41.28** |
| 754 | 85.33 | 82.23 | 74.04 | 73.92 | **315.53** |
| 755 | 20.95 | 19.56 | 20.10 | 21.67 | **82.28** |
| 756 | 106.23 | 103.00 | 53.23 | 69.55 | **332.01** |
| 757 | 90.75 | 95.97 | 87.90 | 84.70 | **359.32** |
| 758 | 12.45 | 11.16 | 12.53 | 10.48 | **46.61** |
| 759 | 36.33 | 33.21 | 32.39 | 30.78 | **132.71** |
| 760 | 191.24 | 185.41 | 174.81 | 168.20 | **719.65** |
| 761 | 180.73 | 181.02 | 168.46 | 162.22 | **692.43** |
| 762 | 90.54 | 84.08 | 78.64 | 73.73 | **326.99** |
| 763 | 51.87 | 48.44 | 40.36 | 40.00 | **180.67** |
| 764 | 30.09 | 28.21 | 29.06 | 27.97 | **115.32** |
| 765 | 52.48 | 47.34 | 47.49 | 41.09 | **188.40** |
| 766 | 15.75 | 15.95 | 13.82 | 10.59 | **56.12** |
| 767 | 26.91 | 25.93 | 22.72 | 22.72 | **98.28** |

| | | | | | |
|---|---|---|---|---|---|
| 768 | 25.39 | 28.33 | 27.56 | 24.56 | **105.84** |
| 769 | 15.68 | 15.38 | 15.13 | 14.94 | **61.13** |
| 770 | 242.14 | 232.58 | 224.76 | 232.87 | **932.35** |
| 772 | 0 | 0 | 0.02 | 0 | **0.02** |
| 773 | 157.76 | 150.12 | 146.92 | 140.57 | **595.37** |
| 774 | 109.89 | 110.17 | 101.01 | 99.48 | **420.55** |
| 775 | 149.54 | 144.88 | 144.30 | 137.76 | **576.49** |
| 776 | 38.21 | 36.01 | 37.64 | 35.35 | **147.21** |
| 777 | 21.93 | 22.27 | 23.02 | 21.82 | **89.03** |
| 778 | 41.94 | 39.53 | 41.08 | 41.67 | **164.21** |
| 779 | 35.54 | 33.09 | 33.04 | 33.92 | **135.60** |
| 780 | 49.98 | 56.67 | 51.62 | 49.91 | **208.18** |
| 781 | 47.45 | 43.62 | 43.54 | 43.57 | **178.18** |
| 782 | 254.27 | 236.31 | 236.47 | 227.27 | **954.32** |
| 783 | 26.71 | 25.93 | 22.74 | 25.80 | **101.19** |
| 784 | 55.17 | 56.28 | 52.58 | 51.01 | **215.05** |
| 785 | 92.67 | 86.89 | 88.54 | 85.50 | **353.59** |
| 786 | 116.88 | 106.96 | 101.42 | 98.54 | **423.79** |
| 787 | 110.84 | 118.04 | 102.26 | 97.56 | **428.70** |
| 788 | 9.24 | 6.71 | 7.14 | 6.52 | **29.61** |
| 789 | 5.79 | 5.44 | 5.73 | 5.55 | **22.51** |
| 790 | 36.76 | 34.97 | 32.69 | 32.40 | **136.83** |
| 791 | 63.80 | 61.67 | 60.33 | 57.87 | **243.67** |
| 792 | 6.98 | 6.16 | 5.96 | 6.34 | **25.43** |
| 793 | 21.98 | 20.99 | 16.08 | 15.19 | **74.24** |
| 794 | 74.09 | 70.73 | 67.56 | 69.15 | **281.53** |
| 795 | 21.23 | 20.99 | 21.43 | 18.51 | **82.16** |
| 796 | 45.46 | 43.64 | 43.74 | 41.89 | **174.73** |
| 797 | 27.57 | 24.04 | 25.08 | 23.84 | **100.52** |
| 798 | 4.11 | 2.34 | 2.44 | 2.30 | **11.19** |
| 799 | 80.10 | 73.11 | 77.81 | 68.86 | **299.87** |
| **TX - TEXAS Total** | **3,544.43** | **3,438.13** | **3,250.61** | **3,187.39** | **13,420.57** |

**STATE: UT - UTAH**

| REGISTRANT ZIP CODE 3 | 1ST QUARTER | 2ND QUARTER | 3RD QUARTER | 4TH QUARTER | **TOTAL GRAMS** |
|---|---|---|---|---|---|
| 840 | 220.65 | 196.59 | 206.45 | 192.42 | **816.11** |
| 841 | 134.89 | 127.15 | 124.33 | 124.36 | **510.73** |
| 843 | 31.95 | 32.60 | 37.34 | 32.75 | **134.63** |
| 844 | 42.85 | 43.93 | 41.58 | 40.09 | **168.45** |
| 845 | 19.44 | 18.65 | 19.48 | 18.30 | **75.87** |
| 846 | 56.84 | 55.05 | 57.60 | 52.86 | **222.35** |

| | | | | | |
|---|---|---|---|---|---|
| 847 | 84.50 | 87.68 | 81.87 | 82.83 | **336.88** |
| **UT - UTAH - Total** | **591.12** | **561.64** | **568.64** | **543.61** | **2,265.01** |

**STATE: VA - VIRGINIA**

| REGISTRANT ZIP CODE 3 | 1ST QUARTER | 2ND QUARTER | 3RD QUARTER | 4TH QUARTER | TOTAL GRAMS |
|---|---|---|---|---|---|
| 201 | 77.13 | 72.37 | 74.01 | 68.96 | **292.46** |
| 220 | 38.73 | 38.85 | 34.96 | 38.03 | **150.57** |
| 221 | 26.83 | 24.59 | 26.12 | 25.76 | **103.30** |
| 222 | 9.56 | 9.57 | 8.17 | 7.36 | **34.66** |
| 223 | 16.31 | 19.02 | 15.37 | 16.19 | **66.90** |
| 224 | 29.11 | 27.59 | 25.43 | 25.58 | **107.71** |
| 225 | 11.47 | 11.04 | 10.34 | 9.69 | **42.54** |
| 226 | 30.82 | 33.08 | 32.50 | 29.48 | **125.89** |
| 227 | 7.72 | 8.96 | 8.36 | 8.15 | **33.19** |
| 228 | 34.83 | 33.31 | 31.20 | 30.84 | **130.18** |
| 229 | 49.04 | 48.65 | 47.78 | 45.38 | **190.85** |
| 230 | 38.34 | 38.75 | 40.16 | 33.44 | **150.69** |
| 231 | 52.15 | 54.03 | 47.46 | 51.17 | **204.82** |
| 232 | 109.32 | 92.55 | 103.36 | 88.42 | **393.65** |
| 233 | 47.70 | 44.72 | 45.23 | 43.58 | **181.23** |
| 234 | 93.51 | 87.13 | 89.43 | 84.37 | **354.43** |
| 235 | 35.63 | 31.53 | 30.96 | 29.85 | **127.96** |
| 236 | 47.43 | 50.78 | 46.46 | 45.91 | **190.58** |
| 237 | 22.94 | 25.33 | 24.34 | 16.75 | **89.35** |
| 238 | 35.52 | 30.51 | 31.14 | 32.32 | **129.48** |
| 239 | 12.89 | 9.66 | 8.91 | 8.54 | **40.00** |
| 240 | 71.04 | 67.90 | 68.33 | 68.31 | **275.57** |
| 241 | 48.83 | 46.78 | 45.36 | 46.35 | **187.33** |
| 242 | 54.86 | 53.89 | 48.19 | 45.97 | **202.91** |
| 243 | 27.00 | 25.76 | 24.88 | 25.47 | **103.10** |
| 244 | 31.21 | 32.16 | 28.72 | 28.60 | **120.70** |
| 245 | 62.26 | 58.67 | 56.69 | 49.26 | **226.88** |
| 246 | 15.99 | 17.47 | 14.03 | 13.36 | **60.85** |
| **VA VIRGINIA Total** | **1,138.16** | **1,094.65** | **1,067.88** | **1,017.08** | **4,317.77** |

**STATE: VI - VIRGIN ISLANDS**

| REGISTRANT ZIP CODE 3 | 1ST QUARTER | 2ND QUARTER | 3RD QUARTER | 4TH QUARTER | TOTAL GRAMS |
|---|---|---|---|---|---|
| 008 | 3.11 | 4.27 | 2.46 | 2.59 | **12.43** |
| **VI - VIRGIN ISLANDS Total** | **3.11** | **4.27** | **2.46** | **2.59** | **12.43** |

**STATE: VT - VERMONT**

| REGISTRANT ZIP CODE 3 | 1ST QUARTER | 2ND QUARTER | 3RD QUARTER | 4TH QUARTER | TOTAL GRAMS |
|---|---|---|---|---|---|
| 050 | 6.50 | 5.83 | 5.70 | 6.39 | **24.42** |

| | | | | | |
|---|---|---|---|---|---|
| 051 | 4.37 | 4.26 | 4.13 | 4.46 | **17.21** |
| 052 | 7.90 | 7.25 | 9.00 | 8.20 | **32.35** |
| 053 | 4.26 | 3.40 | 2.70 | 2.71 | **13.07** |
| 054 | 47.79 | 42.92 | 43.85 | 45.82 | **180.38** |
| 056 | 16.88 | 16.71 | 16.81 | 14.97 | **65.37** |
| 057 | 19.58 | 17.47 | 21.87 | 16.33 | **75.25** |
| 058 | 11.93 | 11.07 | 11.94 | 11.79 | **46.73** |
| **VT VERMONT Total** | **119.22** | **108.92** | **115.99** | **110.66** | **454.79** |

**STATE: WA - WASHINGTON**

| REGISTRANT ZIP CODE 3 | 1ST QUARTER | 2ND QUARTER | 3RD QUARTER | 4TH QUARTER | TOTAL GRAMS |
|---|---|---|---|---|---|
| 980 | 129.37 | 128.01 | 119.45 | 117.85 | **494.67** |
| 981 | 131.56 | 120.28 | 116.41 | 108.59 | **476.84** |
| 982 | 110.78 | 100.77 | 97.22 | 88.57 | **397.34** |
| 983 | 98.14 | 98.93 | 95.39 | 95.07 | **387.53** |
| 984 | 52.93 | 54.56 | 52.49 | 48.17 | **208.15** |
| 985 | 57.81 | 49.92 | 51.30 | 51.56 | **210.59** |
| 986 | 124.34 | 122.67 | 112.14 | 108.81 | **467.96** |
| 988 | 27.58 | 24.60 | 23.27 | 22.60 | **98.06** |
| 989 | 37.50 | 43.02 | 39.21 | 35.66 | **155.38** |
| 990 | 13.04 | 14.15 | 13.43 | 13.28 | **53.91** |
| 991 | 22.02 | 20.81 | 20.44 | 20.94 | **84.21** |
| 992 | 114.88 | 109.40 | 103.33 | 93.90 | **421.50** |
| 993 | 77.85 | 72.31 | 70.35 | 67.88 | **288.40** |
| 994 | 18.65 | 18.55 | 16.11 | 16.98 | **70.29** |
| **WA WASHINGTON Total** | **1,016.45** | **977.97** | **930.54** | **889.86** | **3,814.82** |

**STATE: WI - WISCONSIN**

| REGISTRANT ZIP CODE 3 | 1ST QUARTER | 2ND QUARTER | 3RD QUARTER | 4TH QUARTER | TOTAL GRAMS |
|---|---|---|---|---|---|
| 530 | 85.54 | 79.88 | 80.65 | 73.43 | **319.49** |
| 531 | 89.24 | 84.25 | 79.33 | 82.65 | **335.47** |
| 532 | 129.51 | 134.96 | 125.28 | 123.42 | **513.18** |
| 534 | 14.51 | 12.76 | 11.97 | 12.10 | **51.34** |
| 535 | 86.09 | 78.73 | 69.47 | 68.55 | **302.85** |
| 537 | 57.25 | 55.75 | 55.08 | 52.25 | **220.33** |
| 538 | 14.21 | 13.59 | 13.34 | 13.02 | **54.17** |
| 539 | 41.79 | 40.39 | 40.18 | 35.06 | **157.43** |
| 540 | 12.50 | 12.97 | 11.36 | 11.14 | **47.97** |
| 541 | 24.19 | 24.14 | 23.99 | 24.49 | **96.81** |
| 542 | 17.21 | 16.88 | 15.92 | 14.23 | **64.24** |
| 543 | 33.23 | 37.46 | 32.09 | 32.43 | **135.21** |
| 544 | 47.37 | 43.65 | 36.97 | 37.39 | **165.38** |

| | | | | | |
|---|---|---|---|---|---|
| 545 | 10.72 | 10.07 | 8.50 | 6.76 | **36.05** |
| 546 | 49.34 | 50.14 | 47.22 | 46.95 | **193.66** |
| 547 | 47.61 | 48.29 | 51.08 | 44.12 | **191.10** |
| 548 | 26.56 | 27.49 | 27.55 | 24.88 | **106.48** |
| 549 | 115.12 | 122.75 | 112.52 | 103.00 | **453.38** |
| **WI - WISCONSIN Total** | **901.99** | **894.16** | **842.53** | **805.86** | **3,444.54** |

**STATE: WV - WEST VIRGINIA**

| REGISTRANT ZIP CODE 3 | 1ST QUARTER | 2ND QUARTER | 3RD QUARTER | 4TH QUARTER | TOTAL GRAMS |
|---|---|---|---|---|---|
| 247 | 11.19 | 10.52 | 10.58 | 9.16 | **41.44** |
| 248 | 4.45 | 4.77 | 2.90 | 3.24 | **15.36** |
| 249 | 4.56 | 4.25 | 2.91 | 3.01 | **14.73** |
| 250 | 12.46 | 11.17 | 9.61 | 11.24 | **44.48** |
| 251 | 8.55 | 9.07 | 7.57 | 6.77 | **31.96** |
| 252 | 7.95 | 5.84 | 4.80 | 4.78 | **23.37** |
| 253 | 37.90 | 36.87 | 36.30 | 39.50 | **150.57** |
| 254 | 45.67 | 42.45 | 41.77 | 38.80 | **168.70** |
| 255 | 19.27 | 18.49 | 17.87 | 18.67 | **74.30** |
| 256 | 8.01 | 7.69 | 7.12 | 7.20 | **30.01** |
| 257 | 30.73 | 29.07 | 36.62 | 28.02 | **124.43** |
| 258 | 12.81 | 13.13 | 13.21 | 14.03 | **53.19** |
| 259 | 6.18 | 4.66 | 4.59 | 3.92 | **19.35** |
| 260 | 26.20 | 23.61 | 22.12 | 19.25 | **91.17** |
| 261 | 15.04 | 12.87 | 13.24 | 10.97 | **52.12** |
| 262 | 13.90 | 12.10 | 9.17 | 10.87 | **46.04** |
| 263 | 20.15 | 20.03 | 19.23 | 18.50 | **77.92** |
| 264 | 7.66 | 7.45 | 6.60 | 6.74 | **28.45** |
| 265 | 40.92 | 30.42 | 36.07 | 32.02 | **139.44** |
| 266 | 4.90 | 4.46 | 4.91 | 4.32 | **18.58** |
| 267 | 12.99 | 12.01 | 12.74 | 12.92 | **50.66** |
| 268 | 3.70 | 2.54 | 3.26 | 3.00 | **12.50** |
| **WV WEST VIRGINIA Total** | **355.17** | **323.47** | **323.19** | **306.92** | **1,308.75** |

**STATE: WY - WYOMING**

| REGISTRANT ZIP CODE 3 | 1ST QUARTER | 2ND QUARTER | 3RD QUARTER | 4TH QUARTER | TOTAL GRAMS |
|---|---|---|---|---|---|
| 820 | 17.00 | 16.12 | 16.19 | 15.18 | **64.50** |
| 822 | 5.69 | 4.16 | 3.39 | 3.17 | **16.42** |
| 823 | 0.49 | 0.79 | 1.56 | 0.97 | **3.81** |
| 824 | 14.87 | 12.79 | 14.88 | 14.07 | **56.61** |
| 825 | 4.43 | 4.95 | 4.47 | 4.96 | **18.81** |
| 826 | 17.79 | 17.49 | 17.51 | 14.52 | **67.31** |
| 827 | 4.50 | 3.64 | 2.93 | 3.04 | **14.11** |

| | | | | | |
|---|---|---|---|---|---|
| 828 | 5.18 | 5.72 | 5.12 | 4.93 | **20.95** |
| 829 | 19.96 | 19.38 | 19.86 | 21.85 | **81.05** |
| 830 | 4.28 | 3.40 | 3.93 | 3.32 | **14.93** |
| 831 | 3.97 | 4.77 | 4.35 | 4.53 | **17.62** |
| **WY - WYOMING Total** | **98.18** | **93.21** | **94.20** | **90.56** | **376.15** |

**DRUG: 1100 - AMPHETAMINE**

| STATE | 1ST QUARTER | 2ND QUARTER | 3RD QUARTER | 4TH QUARTER | TOTAL GRAMS |
|---|---|---|---|---|---|
| ALABAMA | 135,557.55 | 138,115.57 | 137,639.22 | 137,750.28 | 549,062.62 |
| ALASKA | 8,408.96 | 8,675.49 | 8,755.54 | 8,750.60 | 34,590.59 |
| AMERICAN SAMOA | 3.75 | 0 | 0 | 0 | 3.75 |
| ARIZONA | 92,745.26 | 95,104.00 | 95,879.30 | 101,190.12 | 384,918.68 |
| ARKANSAS | 54,804.47 | 54,426.63 | 55,052.16 | 56,544.45 | 220,827.70 |
| CALIFORNIA | 308,659.71 | 315,354.24 | 319,527.54 | 322,716.52 | 1,266,258.02 |
| COLORADO | 89,568.67 | 93,525.58 | 95,621.16 | 99,986.74 | 378,702.16 |
| CONNECTICUT | 56,694.65 | 60,157.98 | 59,588.28 | 63,488.75 | 239,929.66 |
| DELAWARE | 21,191.28 | 21,702.52 | 21,954.03 | 22,230.26 | 87,078.09 |
| DISTRICT OF COLUMBIA | 12,465.18 | 13,052.50 | 13,214.31 | 12,972.12 | 51,704.11 |
| FLORIDA | 332,225.20 | 343,548.71 | 327,059.59 | 360,147.70 | 1,362,981.20 |
| GEORGIA | 200,316.71 | 205,396.82 | 207,456.66 | 211,504.15 | 824,674.34 |
| GUAM | 422.85 | 432.20 | 452.03 | 408.34 | 1,715.41 |
| HAWAII | 10,566.38 | 10,703.49 | 10,879.15 | 10,914.45 | 43,063.47 |
| IDAHO | 29,552.81 | 30,144.15 | 30,958.45 | 31,282.85 | 121,938.25 |
| ILLINOIS | 201,842.70 | 209,900.70 | 215,539.49 | 217,804.48 | 845,087.37 |
| INDIANA | 137,705.23 | 141,812.97 | 150,867.28 | 144,113.02 | 574,498.49 |
| IOWA | 53,903.62 | 54,226.56 | 54,893.88 | 56,174.78 | 219,198.84 |
| KANSAS | 64,919.88 | 66,345.39 | 67,552.57 | 69,247.62 | 268,065.45 |
| KENTUCKY | 73,890.93 | 72,951.47 | 76,156.50 | 77,022.51 | 300,021.42 |
| LOUISIANA | 173,158.96 | 172,307.65 | 172,140.33 | 179,444.24 | 697,051.17 |
| MAINE | 24,795.54 | 26,360.02 | 27,057.58 | 26,795.10 | 105,008.24 |
| MARYLAND | 98,522.40 | 101,683.07 | 101,932.68 | 105,036.34 | 407,174.51 |
| MASSACHUSETTS | 180,345.49 | 193,594.89 | 188,892.05 | 196,777.68 | 759,610.12 |
| MICHIGAN | 246,308.62 | 248,975.11 | 268,475.31 | 247,333.12 | 1,011,092.15 |
| MINNESOTA | 112,676.93 | 115,733.08 | 116,320.27 | 120,667.86 | 465,398.14 |
| MISSISSIPPI | 63,631.03 | 61,764.81 | 63,080.31 | 64,777.45 | 253,253.60 |
| MISSOURI | 152,401.88 | 152,864.74 | 150,240.80 | 153,406.12 | 608,913.54 |
| MONTANA | 15,607.82 | 15,775.09 | 16,427.60 | 16,409.41 | 64,219.93 |
| NEBRASKA | 28,548.26 | 29,131.35 | 29,935.52 | 30,637.56 | 118,252.69 |
| NEVADA | 27,965.72 | 27,921.14 | 28,386.39 | 30,092.64 | 114,365.89 |
| NEW HAMPSHIRE | 41,145.99 | 43,248.82 | 43,741.04 | 44,492.27 | 172,628.12 |
| NEW JERSEY | 113,455.51 | 114,175.15 | 117,889.28 | 122,312.45 | 467,832.40 |
| NEW MEXICO | 15,978.33 | 15,913.78 | 15,937.83 | 16,548.06 | 64,378.00 |
| NEW YORK | 258,552.62 | 260,132.23 | 265,551.13 | 277,775.49 | 1,062,011.46 |
| NORTH CAROLINA | 228,398.99 | 235,252.31 | 236,905.59 | 232,885.37 | 933,442.27 |
| NORTH DAKOTA | 9,886.22 | 9,602.26 | 10,043.39 | 10,218.65 | 39,750.51 |
| OHIO | 207,450.24 | 209,989.32 | 219,576.63 | 221,604.47 | 858,620.65 |
| OKLAHOMA | 55,428.77 | 55,951.98 | 57,125.39 | 57,711.93 | 226,218.08 |

| | | | | | |
|---|---|---|---|---|---|
| OREGON | 64,547.54 | 67,166.78 | 67,787.61 | 71,150.45 | 270,652.38 |
| PENNSYLVANIA | 218,795.65 | 219,534.66 | 224,398.25 | 232,492.82 | 895,221.38 |
| PUERTO RICO | 9,381.13 | 9,405.29 | 7,192.96 | 9,928.22 | 35,907.60 |
| RHODE ISLAND | 37,201.65 | 40,273.42 | 39,544.00 | 40,966.39 | 157,985.45 |
| SOUTH CAROLINA | 141,288.30 | 148,015.69 | 146,656.89 | 148,996.55 | 584,957.42 |
| SOUTH DAKOTA | 14,516.62 | 15,133.45 | 14,741.20 | 15,335.62 | 59,726.90 |
| TENNESSEE | 136,353.89 | 140,168.83 | 141,245.65 | 143,514.98 | 561,283.36 |
| TEXAS | 464,484.94 | 472,807.55 | 479,642.83 | 501,915.77 | 1,918,851.09 |
| UTAH | 91,895.15 | 94,507.39 | 97,176.75 | 101,148.36 | 384,727.64 |
| VERMONT | 10,094.12 | 10,646.01 | 10,770.11 | 10,768.26 | 42,278.50 |
| VIRGIN ISLANDS | 335.93 | 426.39 | 317.39 | 380.70 | 1,460.40 |
| VIRGINIA | 182,704.81 | 192,732.07 | 192,152.37 | 193,463.51 | 761,052.77 |
| WASHINGTON | 117,614.32 | 123,148.03 | 122,438.52 | 127,239.79 | 490,440.66 |
| WEST VIRGINIA | 24,232.75 | 23,578.42 | 24,089.83 | 24,343.76 | 96,244.76 |
| WISCONSIN | 125,044.43 | 127,323.46 | 128,286.26 | 128,124.25 | 508,778.40 |
| WYOMING | 6,986.48 | 7,170.96 | 7,449.29 | 7,434.14 | 29,040.88 |
| **1100 - AMPHETAMINE Total** | **5,585,182.82** | **5,717,992.18** | **5,782,596.17** | **5,916,379.52** | **23,002,150.68** |

**DRUG: 1105 - METHAMPHETAMINE**

| STATE | 1ST QUARTER | 2ND QUARTER | 3RD QUARTER | 4TH QUARTER | TOTAL GRAMS |
|---|---|---|---|---|---|
| ALABAMA | 28.53 | 32.54 | 30.93 | 30.53 | 122.54 |
| ALASKA | 0.40 | 0 | 2.01 | 3.21 | 5.62 |
| ARIZONA | 26.12 | 16.07 | 21.70 | 18.48 | 82.37 |
| ARKANSAS | 3.62 | 2.81 | 1.61 | 0.80 | 8.84 |
| CALIFORNIA | 240.65 | 237.04 | 226.58 | 252.29 | 956.56 |
| COLORADO | 49.82 | 46.61 | 51.82 | 43.79 | 192.04 |
| CONNECTICUT | 24.91 | 22.90 | 20.09 | 20.89 | 88.79 |
| DELAWARE | 4.42 | 2.81 | 5.22 | 4.02 | 16.47 |
| DISTRICT OF COLUMBIA | 4.82 | 4.42 | 4.82 | 6.03 | 20.09 |
| FLORIDA | 56.25 | 46.61 | 54.24 | 54.64 | 211.74 |
| GEORGIA | 24.51 | 24.51 | 19.69 | 27.72 | 96.42 |
| HAWAII | 5.63 | 4.42 | 3.62 | 4.82 | 18.48 |
| IDAHO | 11.25 | 11.65 | 13.66 | 16.07 | 52.62 |
| ILLINOIS | 62.67 | 56.65 | 53.83 | 55.44 | 228.59 |
| INDIANA | 10.05 | 18.88 | 11.65 | 30.13 | 70.71 |
| IOWA | 13.66 | 13.26 | 12.45 | 16.87 | 56.25 |
| KANSAS | 13.26 | 13.26 | 12.86 | 13.66 | 53.03 |
| KENTUCKY | 12.46 | 10.85 | 11.65 | 4.82 | 39.77 |
| LOUISIANA | 20.49 | 12.86 | 9.24 | 10.85 | 53.44 |
| MAINE | 13.26 | 13.66 | 8.84 | 10.04 | 45.80 |
| MARYLAND | 32.14 | 28.12 | 41.38 | 30.93 | 132.58 |

| | | | | | |
|---|---|---|---|---|---|
| MASSACHUSETTS | 24.91 | 24.11 | 22.50 | 26.52 | 98.04 |
| MICHIGAN | 22.10 | 23.30 | 23.30 | 16.87 | 85.58 |
| MINNESOTA | 46.21 | 52.23 | 55.04 | 61.07 | 214.55 |
| MISSISSIPPI | 8.03 | 7.23 | 7.63 | 6.43 | 29.33 |
| MISSOURI | 61.87 | 47.00 | 52.23 | 42.99 | 204.08 |
| NEBRASKA | 5.22 | 6.03 | 6.03 | 5.62 | 22.90 |
| NEVADA | 11.25 | 7.63 | 10.85 | 10.04 | 39.78 |
| NEW HAMPSHIRE | 2.81 | 8.84 | 6.43 | 6.03 | 24.10 |
| NEW JERSEY | 33.75 | 30.54 | 32.14 | 44.19 | 140.63 |
| NEW MEXICO | 5.63 | 4.82 | 7.63 | 10.45 | 28.52 |
| NEW YORK | 82.37 | 77.54 | 93.61 | 80.35 | 333.87 |
| NORTH CAROLINA | 45.41 | 47.01 | 32.54 | 50.22 | 175.18 |
| NORTH DAKOTA | 7.23 | 6.83 | 13.26 | 5.22 | 32.54 |
| OHIO | 45.00 | 31.74 | 33.75 | 30.94 | 141.43 |
| OKLAHOMA | 8.44 | 10.85 | 8.44 | 8.84 | 36.56 |
| OREGON | 38.57 | 34.95 | 36.96 | 51.02 | 161.50 |
| PENNSYLVANIA | 85.18 | 104.47 | 94.82 | 109.69 | 394.15 |
| RHODE ISLAND | 0.80 | 1.21 | 3.21 | 2.81 | 8.04 |
| SOUTH CAROLINA | 12.46 | 12.05 | 11.65 | 13.66 | 49.82 |
| SOUTH DAKOTA | 5.22 | 4.82 | 6.83 | 6.43 | 23.30 |
| TENNESSEE | 10.05 | 14.46 | 16.87 | 14.06 | 55.45 |
| TEXAS | 83.97 | 87.19 | 82.76 | 95.21 | 349.13 |
| UTAH | 26.11 | 21.70 | 30.93 | 25.31 | 104.05 |
| VERMONT | 1.61 | 0.80 | 2.41 | 2.41 | 7.23 |
| VIRGINIA | 32.95 | 32.95 | 30.54 | 29.73 | 126.16 |
| WASHINGTON | 83.96 | 68.30 | 89.98 | 96.41 | 338.65 |
| WEST VIRGINIA | 4.82 | 3.21 | 3.21 | 2.81 | 14.06 |
| WISCONSIN | 26.92 | 25.71 | 26.11 | 26.92 | 105.66 |
| WYOMING | 4.42 | 4.82 | 4.42 | 6.03 | 19.68 |
| **1105 METHAMPHETAMINE** | **1,486.16** | **1,422.29** | **1,463.97** | **1,544.31** | **5,916.73** |

**DRUG: 1205 - LISDEXAMFETAMINE**

| STATE | 1ST QUARTER | 2ND QUARTER | 3RD QUARTER | 4TH QUARTER | TOTAL GRAMS |
|---|---|---|---|---|---|
| ALABAMA | 72,684.51 | 69,709.03 | 70,744.99 | 73,423.90 | 286,562.44 |
| ALASKA | 2,622.26 | 2,924.61 | 2,926.34 | 3,218.28 | 11,691.49 |
| ARIZONA | 38,891.68 | 39,287.10 | 39,838.61 | 42,403.06 | 160,420.44 |
| ARKANSAS | 29,728.85 | 28,297.42 | 28,389.91 | 30,014.95 | 116,431.13 |
| CALIFORNIA | 113,460.22 | 115,698.62 | 114,465.53 | 117,817.94 | 461,442.31 |
| COLORADO | 29,099.82 | 29,420.09 | 30,228.85 | 31,315.10 | 120,063.85 |
| CONNECTICUT | 35,787.28 | 36,530.14 | 35,242.13 | 37,703.10 | 145,262.66 |
| DELAWARE | 13,714.27 | 13,388.22 | 13,373.77 | 13,981.93 | 54,458.18 |

| | | | | | |
|---|---|---|---|---|---|
| DISTRICT OF COLUMBIA | 5,876.39 | 6,140.00 | 6,428.47 | 6,477.03 | 24,921.89 |
| FLORIDA | 138,109.16 | 134,669.84 | 130,337.96 | 140,054.55 | 543,171.52 |
| GEORGIA | 97,874.64 | 93,923.91 | 96,653.12 | 100,440.25 | 388,891.92 |
| GUAM | 11.56 | 25.44 | 0.58 | 19.08 | 56.65 |
| HAWAII | 1,479.36 | 1,633.13 | 1,718.69 | 1,922.76 | 6,753.94 |
| IDAHO | 9,716.70 | 10,184.97 | 9,972.80 | 10,588.48 | 40,462.95 |
| ILLINOIS | 71,653.18 | 72,790.88 | 73,851.70 | 76,946.27 | 295,242.03 |
| INDIANA | 81,266.41 | 79,897.47 | 81,195.30 | 83,712.93 | 326,072.10 |
| IOWA | 33,337.29 | 33,284.11 | 33,194.50 | 34,899.90 | 134,715.80 |
| KANSAS | 31,302.38 | 32,704.85 | 32,827.41 | 34,045.47 | 130,880.11 |
| KENTUCKY | 40,799.41 | 39,342.60 | 38,911.91 | 40,029.96 | 159,083.87 |
| LOUISIANA | 90,049.48 | 83,730.27 | 86,894.21 | 91,658.33 | 352,332.29 |
| MAINE | 16,483.94 | 17,021.58 | 16,996.14 | 17,354.56 | 67,856.22 |
| MARYLAND | 56,908.22 | 57,783.41 | 56,306.36 | 59,948.39 | 230,946.38 |
| MASSACHUSETTS | 38,428.62 | 39,759.98 | 39,760.56 | 42,471.27 | 160,420.43 |
| MICHIGAN | 61,179.17 | 62,786.86 | 61,346.24 | 65,791.83 | 251,104.09 |
| MINNESOTA | 31,096.00 | 31,757.35 | 32,086.86 | 35,199.35 | 130,139.56 |
| MISSISSIPPI | 37,425.62 | 34,825.32 | 35,721.38 | 37,859.19 | 145,831.51 |
| MISSOURI | 73,131.96 | 69,654.11 | 67,176.95 | 69,524.62 | 279,487.65 |
| MONTANA | 6,535.42 | 6,552.19 | 6,547.56 | 6,823.89 | 26,459.06 |
| NEBRASKA | 17,133.19 | 17,519.74 | 17,420.83 | 18,935.75 | 71,009.51 |
| NEVADA | 8,184.74 | 7,959.28 | 7,621.09 | 8,333.89 | 32,099.00 |
| NEW HAMPSHIRE | 14,355.38 | 14,406.25 | 14,386.60 | 15,043.32 | 58,191.55 |
| NEW JERSEY | 47,861.56 | 47,807.14 | 47,111.10 | 49,452.99 | 192,232.79 |
| NEW MEXICO | 3,850.15 | 3,801.01 | 3,665.15 | 3,876.16 | 15,192.47 |
| NEW YORK | 87,447.45 | 87,654.99 | 88,900.22 | 94,126.82 | 358,129.48 |
| NORTH CAROLINA | 114,135.44 | 111,261.13 | 111,682.56 | 113,616.31 | 450,695.43 |
| NORTH DAKOTA | 6,149.83 | 5,969.46 | 6,063.69 | 6,451.60 | 24,634.58 |
| OHIO | 94,558.20 | 93,159.66 | 93,034.79 | 97,678.67 | 378,431.31 |
| OKLAHOMA | 37,576.73 | 36,697.79 | 37,186.28 | 39,447.23 | 150,908.03 |
| OREGON | 15,220.79 | 15,938.22 | 15,794.85 | 16,934.28 | 63,888.14 |
| PENNSYLVANIA | 90,622.38 | 92,042.19 | 92,470.56 | 98,639.47 | 373,774.60 |
| PUERTO RICO | 2,332.63 | 2,055.15 | 1,990.98 | 2,336.10 | 8,714.86 |
| RHODE ISLAND | 7,494.49 | 7,475.99 | 7,502.58 | 8,224.63 | 30,697.69 |
| SOUTH CAROLINA | 62,989.23 | 61,473.16 | 59,981.69 | 64,524.76 | 248,968.84 |
| SOUTH DAKOTA | 9,635.77 | 9,488.36 | 9,736.36 | 10,025.41 | 38,885.90 |
| TENNESSEE | 67,045.62 | 64,708.31 | 66,422.87 | 68,907.29 | 267,084.09 |
| TEXAS | 264,408.18 | 257,403.65 | 253,117.37 | 269,220.59 | 1,044,149.80 |
| UTAH | 22,315.83 | 22,767.67 | 23,122.08 | 24,415.50 | 92,621.09 |
| VERMONT | 7,209.49 | 7,203.70 | 7,534.96 | 7,440.15 | 29,388.29 |

| | | | | | |
|---|---|---|---|---|---|
| VIRGIN ISLANDS | 83.25 | 66.48 | 67.64 | 106.95 | 324.31 |
| VIRGINIA | 68,507.16 | 69,717.70 | 68,447.62 | 72,278.11 | 278,950.59 |
| WASHINGTON | 35,562.52 | 37,563.20 | 37,517.53 | 39,642.05 | 150,285.30 |
| WEST VIRGINIA | 18,935.67 | 18,526.95 | 18,026.89 | 19,354.79 | 74,844.29 |
| WISCONSIN | 74,019.92 | 76,135.19 | 75,204.45 | 79,086.97 | 304,446.54 |
| WYOMING | 2,639.03 | 2,629.20 | 2,808.41 | 2,773.15 | 10,849.78 |
| **1205 - LISDEXAMFETAMINE T** | **2,438,928.43** | **2,415,155.03** | **2,409,957.98** | **2,536,519.30** | **9,800,560.75** |

**DRUG: 1724 - METHYLPHENIDATE (DL;D;L;ISOMERS)**

| STATE | 1ST QUARTER | 2ND QUARTER | 3RD QUARTER | 4TH QUARTER | TOTAL GRAMS |
|---|---|---|---|---|---|
| ALABAMA | 74,208.96 | 68,629.22 | 71,699.82 | 76,268.23 | 290,806.23 |
| ALASKA | 7,834.35 | 7,957.58 | 8,027.40 | 8,627.62 | 32,446.95 |
| AMERICAN SAMOA | 8.65 | 14.70 | 2.59 | 9.08 | 35.02 |
| ARIZONA | 75,978.71 | 77,692.53 | 73,482.60 | 79,202.24 | 306,356.07 |
| ARKANSAS | 40,141.16 | 36,699.49 | 37,562.60 | 39,549.34 | 153,952.59 |
| CALIFORNIA | 261,691.50 | 262,117.30 | 251,569.47 | 272,242.86 | 1,047,621.13 |
| COLORADO | 62,218.53 | 61,781.15 | 61,665.86 | 67,819.66 | 253,485.20 |
| CONNECTICUT | 54,834.76 | 54,580.53 | 50,871.75 | 56,402.44 | 216,689.47 |
| DELAWARE | 18,055.71 | 17,333.55 | 16,502.44 | 18,279.70 | 70,171.41 |
| DISTRICT OF COLUMBIA | 8,338.02 | 8,275.83 | 8,253.54 | 8,451.18 | 33,318.56 |
| FLORIDA | 197,331.46 | 192,264.30 | 182,441.17 | 205,502.62 | 777,539.54 |
| GEORGIA | 167,013.09 | 149,196.94 | 155,093.66 | 167,254.80 | 638,558.48 |
| GUAM | 343.40 | 453.68 | 405.68 | 319.84 | 1,522.60 |
| HAWAII | 10,536.94 | 10,767.34 | 10,532.17 | 10,927.84 | 42,764.29 |
| IDAHO | 28,402.19 | 28,027.75 | 26,769.29 | 29,774.01 | 112,973.23 |
| ILLINOIS | 170,894.16 | 164,021.96 | 161,253.04 | 172,582.29 | 668,751.45 |
| INDIANA | 116,194.08 | 110,181.12 | 113,369.27 | 118,968.14 | 458,712.61 |
| IOWA | 78,862.48 | 77,803.95 | 75,910.86 | 83,152.00 | 315,729.29 |
| KANSAS | 49,049.62 | 47,884.49 | 47,608.23 | 54,482.90 | 199,025.25 |
| KENTUCKY | 75,259.04 | 69,633.34 | 71,739.04 | 77,172.91 | 293,804.33 |
| LOUISIANA | 85,808.99 | 74,389.33 | 77,222.99 | 85,274.21 | 322,695.52 |
| MAINE | 34,787.81 | 34,795.32 | 35,411.59 | 35,487.40 | 140,482.12 |
| MARYLAND | 100,701.31 | 100,058.63 | 93,025.07 | 103,751.13 | 397,536.14 |
| MASSACHUSETTS | 143,883.34 | 144,626.21 | 132,815.51 | 148,819.79 | 570,144.86 |
| MICHIGAN | 175,177.52 | 171,263.53 | 173,286.83 | 171,856.27 | 691,584.16 |
| MINNESOTA | 101,823.36 | 102,402.43 | 95,584.23 | 105,601.12 | 405,411.14 |
| MISSISSIPPI | 44,107.88 | 39,040.08 | 40,925.18 | 46,561.13 | 170,634.28 |
| MISSOURI | 142,286.74 | 138,281.95 | 132,480.98 | 146,659.18 | 559,708.85 |
| MONTANA | 16,978.65 | 17,142.74 | 16,376.55 | 17,601.06 | 68,098.99 |
| NEBRASKA | 31,299.78 | 30,116.62 | 30,183.50 | 32,594.14 | 124,194.04 |
| NEVADA | 14,904.21 | 14,868.03 | 14,260.64 | 15,604.73 | 59,637.61 |

| | | | | | |
|---|---|---|---|---|---|
| NEW HAMPSHIRE | 34,101.02 | 34,236.64 | 33,130.18 | 35,214.16 | 136,682.00 |
| NEW JERSEY | 92,757.36 | 89,496.56 | 83,250.81 | 94,696.34 | 360,201.08 |
| NEW MEXICO | 18,155.83 | 17,983.96 | 16,990.89 | 18,571.10 | 71,701.78 |
| NEW YORK | 228,576.00 | 224,969.15 | 211,201.46 | 234,878.10 | 899,624.71 |
| NORTH CAROLINA | 197,573.85 | 187,942.92 | 179,321.06 | 194,028.48 | 758,866.32 |
| NORTH DAKOTA | 15,323.39 | 14,838.10 | 14,545.69 | 15,733.46 | 60,440.63 |
| OHIO | 200,848.71 | 188,998.68 | 188,875.03 | 206,729.24 | 785,451.66 |
| OKLAHOMA | 47,468.47 | 44,301.03 | 44,900.09 | 48,747.77 | 185,417.36 |
| OREGON | 63,305.28 | 63,665.64 | 60,005.98 | 67,126.13 | 254,103.03 |
| PENNSYLVANIA | 220,220.47 | 217,694.25 | 210,833.55 | 229,783.27 | 878,531.54 |
| PUERTO RICO | 15,514.70 | 13,233.10 | 9,923.32 | 14,715.52 | 53,386.65 |
| RHODE ISLAND | 19,001.03 | 18,563.64 | 16,917.74 | 19,427.80 | 73,910.21 |
| SOUTH CAROLINA | 91,374.42 | 85,557.76 | 81,605.14 | 91,849.67 | 350,387.00 |
| SOUTH DAKOTA | 19,802.18 | 19,176.45 | 19,112.72 | 20,697.54 | 78,788.90 |
| TENNESSEE | 83,715.64 | 78,424.14 | 80,371.73 | 86,496.07 | 329,007.58 |
| TEXAS | 337,184.79 | 312,167.22 | 305,926.76 | 348,451.01 | 1,303,729.79 |
| UTAH | 47,063.52 | 46,964.69 | 46,352.63 | 52,378.27 | 192,759.11 |
| VERMONT | 18,409.16 | 18,788.31 | 18,576.26 | 18,672.00 | 74,445.73 |
| VIRGIN ISLANDS | 95.20 | 155.20 | 84.26 | 135.00 | 469.65 |
| VIRGINIA | 135,278.76 | 131,574.81 | 126,676.22 | 137,371.32 | 530,901.12 |
| WASHINGTON | 112,592.41 | 117,343.15 | 108,824.74 | 120,376.59 | 459,136.89 |
| WEST VIRGINIA | 30,113.03 | 28,193.39 | 27,842.85 | 30,456.46 | 116,605.74 |
| WISCONSIN | 103,286.79 | 101,930.14 | 98,829.22 | 107,126.30 | 411,172.45 |
| WYOMING | 7,790.88 | 7,709.43 | 7,298.82 | 7,828.86 | 30,627.99 |
| **1724 - METHYLPHENIDATE (DI** | **4,528,509.32** | **4,376,209.98** | **4,257,730.70** | **4,658,290.32** | **17,820,740.31** |

**DRUG: 9050 - CODEINE**

| STATE | 1ST QUARTER | 2ND QUARTER | 3RD QUARTER | 4TH QUARTER | TOTAL GRAMS |
|---|---|---|---|---|---|
| ALABAMA | 45,544.99 | 45,702.76 | 42,737.56 | 43,516.47 | 177,501.78 |
| ALASKA | 5,081.99 | 4,825.72 | 5,295.10 | 4,819.05 | 20,021.86 |
| AMERICAN SAMOA | 274.05 | 293.94 | 243.11 | 388.98 | 1,200.08 |
| ARIZONA | 50,760.76 | 49,370.61 | 44,786.56 | 45,663.09 | 190,581.01 |
| ARKANSAS | 53,128.12 | 50,059.78 | 50,168.75 | 48,345.43 | 201,702.07 |
| CALIFORNIA | 347,633.47 | 328,684.36 | 325,344.88 | 322,475.55 | 1,324,138.27 |
| COLORADO | 16,900.66 | 16,906.46 | 16,549.95 | 16,913.43 | 67,270.50 |
| CONNECTICUT | 16,008.51 | 18,575.55 | 16,263.14 | 16,334.85 | 67,182.04 |
| DELAWARE | 6,882.16 | 6,966.43 | 6,868.92 | 6,470.13 | 27,187.64 |
| DISTRICT OF COLUMBIA | 8,331.51 | 8,824.41 | 7,923.30 | 8,012.24 | 33,091.45 |
| FLORIDA | 130,853.28 | 127,977.69 | 125,752.44 | 127,042.54 | 511,625.95 |
| GEORGIA | 78,186.36 | 81,419.32 | 78,732.14 | 77,469.58 | 315,807.40 |
| GUAM | 616.62 | 799.46 | 846.55 | 596.73 | 2,859.35 |

| | | | | | |
|---|---|---|---|---|---|
| HAWAII | 10,649.05 | 9,722.89 | 9,723.51 | 10,010.72 | 40,106.16 |
| IDAHO | 10,844.89 | 11,229.46 | 11,218.81 | 11,609.51 | 44,902.67 |
| ILLINOIS | 125,405.08 | 125,834.10 | 125,313.89 | 123,345.23 | 499,898.30 |
| INDIANA | 38,977.83 | 38,201.85 | 36,552.92 | 34,163.06 | 147,895.67 |
| IOWA | 14,177.70 | 14,301.18 | 14,045.63 | 13,401.78 | 55,926.30 |
| KANSAS | 18,992.79 | 19,153.00 | 20,524.53 | 20,443.61 | 79,113.93 |
| KENTUCKY | 37,144.49 | 36,462.58 | 34,817.59 | 35,296.99 | 143,721.65 |
| LOUISIANA | 34,558.38 | 34,328.82 | 34,239.63 | 32,623.23 | 135,750.06 |
| MAINE | 7,414.71 | 7,652.23 | 7,877.92 | 7,564.99 | 30,509.85 |
| MARYLAND | 42,941.25 | 44,491.61 | 44,412.52 | 43,487.14 | 175,332.52 |
| MASSACHUSETTS | 33,244.30 | 39,146.96 | 34,102.87 | 35,214.96 | 141,709.08 |
| MICHIGAN | 168,290.24 | 169,054.48 | 175,662.59 | 162,233.42 | 675,240.73 |
| MINNESOTA | 26,444.88 | 26,128.86 | 25,742.67 | 25,485.33 | 103,801.74 |
| MISSISSIPPI | 20,368.82 | 19,509.49 | 18,246.43 | 17,419.68 | 75,544.42 |
| MISSOURI | 55,502.10 | 56,546.13 | 56,785.45 | 57,392.58 | 226,226.25 |
| MONTANA | 7,629.17 | 7,519.49 | 7,983.60 | 7,141.01 | 30,273.27 |
| NEBRASKA | 8,518.47 | 8,623.59 | 8,712.76 | 7,906.48 | 33,761.30 |
| NEVADA | 16,427.91 | 16,530.27 | 16,490.78 | 16,741.99 | 66,190.95 |
| NEW HAMPSHIRE | 6,119.31 | 6,346.95 | 5,450.60 | 5,959.51 | 23,876.38 |
| NEW JERSEY | 59,848.83 | 59,236.44 | 59,188.83 | 58,850.32 | 237,124.43 |
| NEW MEXICO | 14,953.48 | 14,617.48 | 13,794.09 | 14,386.26 | 57,751.31 |
| NEW YORK | 106,998.32 | 103,273.09 | 104,859.32 | 105,387.82 | 420,518.55 |
| NORTH CAROLINA | 36,757.08 | 38,943.79 | 37,595.58 | 35,359.60 | 148,656.06 |
| NORTH DAKOTA | 4,048.00 | 4,206.17 | 3,891.61 | 3,737.96 | 15,883.73 |
| OHIO | 61,303.89 | 60,334.80 | 60,340.45 | 60,472.55 | 242,451.69 |
| OKLAHOMA | 71,757.05 | 73,702.57 | 71,506.08 | 71,129.59 | 288,095.29 |
| OREGON | 26,199.06 | 26,665.08 | 26,103.10 | 24,565.28 | 103,532.51 |
| PENNSYLVANIA | 94,013.24 | 91,044.89 | 90,173.31 | 91,456.44 | 366,687.88 |
| PUERTO RICO | 9,967.55 | 9,569.73 | 8,232.87 | 9,455.91 | 37,226.06 |
| RHODE ISLAND | 4,477.25 | 4,802.65 | 4,450.66 | 4,601.16 | 18,331.72 |
| SOUTH CAROLINA | 58,678.76 | 60,022.03 | 58,849.03 | 55,283.77 | 232,833.58 |
| SOUTH DAKOTA | 4,787.72 | 4,692.60 | 5,010.99 | 4,847.90 | 19,339.21 |
| TENNESSEE | 57,868.48 | 52,087.86 | 56,738.81 | 51,854.44 | 218,549.59 |
| TEXAS | 823,197.09 | 849,402.09 | 839,948.89 | 810,837.80 | 3,323,385.87 |
| UTAH | 12,616.96 | 13,172.67 | 12,506.58 | 12,425.58 | 50,721.79 |
| VERMONT | 2,506.12 | 2,351.66 | 2,258.02 | 2,281.78 | 9,397.58 |
| VIRGIN ISLANDS | 580.22 | 563.65 | 707.38 | 532.63 | 2,383.89 |
| VIRGINIA | 39,397.47 | 42,307.87 | 39,888.97 | 39,900.74 | 161,495.05 |
| WASHINGTON | 45,286.41 | 45,742.94 | 44,598.58 | 43,452.50 | 179,080.43 |
| WEST VIRGINIA | 35,997.92 | 36,377.77 | 37,500.65 | 34,422.85 | 144,299.19 |

| | | | | | |
|---|---|---|---|---|---|
| WISCONSIN | 36,752.57 | 36,217.11 | 37,027.55 | 35,469.33 | 145,466.57 |
| WYOMING | 3,068.62 | 3,377.20 | 3,333.91 | 3,042.42 | 12,822.15 |
| **9050 - CODEINE Total** | **3,054,915.94** | **3,063,902.59** | **3,027,922.34** | **2,959,243.91** | **12,105,984.78** |

**DRUG: 9064 - BUPRENORPHINE**

| STATE | 1ST QUARTER | 2ND QUARTER | 3RD QUARTER | 4TH QUARTER | TOTAL GRAMS |
|---|---|---|---|---|---|
| ALABAMA | 31,241.60 | 32,965.99 | 33,438.34 | 33,794.65 | 131,440.58 |
| ALASKA | 2,656.71 | 2,593.91 | 2,616.65 | 2,620.06 | 10,487.33 |
| ARIZONA | 13,164.25 | 13,681.46 | 14,475.12 | 15,061.49 | 56,382.32 |
| ARKANSAS | 8,143.68 | 8,688.79 | 9,046.96 | 9,409.73 | 35,289.16 |
| CALIFORNIA | 47,782.46 | 49,845.50 | 53,089.05 | 55,354.57 | 206,071.57 |
| COLORADO | 9,953.90 | 10,460.96 | 10,687.08 | 11,557.72 | 42,659.66 |
| CONNECTICUT | 17,914.95 | 17,547.79 | 18,797.14 | 18,856.96 | 73,116.83 |
| DELAWARE | 4,324.36 | 4,166.79 | 4,387.13 | 4,498.12 | 17,376.39 |
| DISTRICT OF COLUMBIA | 3,375.68 | 3,583.36 | 3,607.90 | 3,373.91 | 13,940.85 |
| FLORIDA | 55,849.42 | 57,961.54 | 60,680.34 | 60,429.93 | 234,921.23 |
| GEORGIA | 19,142.31 | 19,491.18 | 20,176.14 | 20,330.75 | 79,140.38 |
| GUAM | 2.66 | 4.34 | 12.48 | 0.83 | 20.31 |
| HAWAII | 2,177.32 | 2,219.76 | 2,264.99 | 2,217.78 | 8,879.84 |
| IDAHO | 4,619.70 | 5,040.41 | 5,021.35 | 5,240.96 | 19,922.41 |
| ILLINOIS | 18,517.89 | 19,586.80 | 19,976.88 | 20,339.60 | 78,421.17 |
| INDIANA | 27,911.85 | 29,135.06 | 31,367.65 | 32,645.30 | 121,059.87 |
| IOWA | 2,367.86 | 2,515.46 | 2,718.40 | 2,578.79 | 10,180.51 |
| KANSAS | 3,025.98 | 3,136.68 | 3,242.35 | 3,318.77 | 12,723.79 |
| KENTUCKY | 42,326.54 | 44,182.94 | 45,809.89 | 47,330.66 | 179,650.03 |
| LOUISIANA | 22,486.90 | 23,727.54 | 24,111.89 | 24,468.96 | 94,795.28 |
| MAINE | 12,538.63 | 13,365.76 | 14,059.11 | 14,399.38 | 54,362.88 |
| MARYLAND | 32,061.26 | 32,450.53 | 34,275.99 | 35,149.01 | 133,936.79 |
| MASSACHUSETTS | 47,002.35 | 46,390.84 | 48,569.65 | 50,227.99 | 192,190.83 |
| MICHIGAN | 32,853.28 | 32,754.31 | 33,934.97 | 35,152.61 | 134,695.17 |
| MINNESOTA | 10,254.75 | 10,770.22 | 11,024.75 | 11,601.55 | 43,651.27 |
| MISSISSIPPI | 11,247.01 | 11,610.13 | 11,933.86 | 12,162.88 | 46,953.89 |
| MISSOURI | 16,050.86 | 17,473.83 | 18,648.66 | 18,561.92 | 70,735.27 |
| MONTANA | 3,434.05 | 3,767.57 | 3,987.40 | 4,122.69 | 15,311.71 |
| NEBRASKA | 1,706.50 | 1,690.42 | 1,954.24 | 2,038.30 | 7,389.45 |
| NEVADA | 4,329.44 | 4,347.38 | 4,504.95 | 4,675.48 | 17,857.25 |
| NEW HAMPSHIRE | 9,573.43 | 9,914.33 | 10,364.65 | 10,594.53 | 40,446.94 |
| NEW JERSEY | 29,225.78 | 29,542.76 | 31,614.77 | 32,521.75 | 122,905.06 |
| NEW MEXICO | 9,151.52 | 9,390.80 | 9,296.73 | 9,621.18 | 37,460.23 |
| NEW YORK | 68,607.00 | 67,157.11 | 71,887.74 | 75,627.52 | 283,279.37 |
| NORTH CAROLINA | 51,440.53 | 52,772.36 | 56,553.60 | 53,559.03 | 214,325.52 |

| | | | | | |
|---|---|---|---|---|---|
| NORTH DAKOTA | 1,311.83 | 1,453.93 | 1,597.52 | 1,636.04 | 5,999.33 |
| OHIO | 59,742.45 | 60,549.65 | 61,323.32 | 62,926.34 | 244,541.76 |
| OKLAHOMA | 16,205.87 | 16,768.47 | 17,901.95 | 18,567.63 | 69,443.93 |
| OREGON | 13,827.45 | 15,087.87 | 15,393.72 | 16,339.10 | 60,648.14 |
| PENNSYLVANIA | 79,677.19 | 80,879.27 | 83,801.84 | 87,174.14 | 331,532.43 |
| PUERTO RICO | 2,149.24 | 2,240.60 | 2,084.30 | 2,286.09 | 8,760.22 |
| RHODE ISLAND | 8,707.91 | 8,442.62 | 9,075.37 | 9,119.39 | 35,345.29 |
| SOUTH CAROLINA | 15,021.60 | 15,989.45 | 16,820.07 | 16,886.39 | 64,717.50 |
| SOUTH DAKOTA | 780.73 | 871.68 | 1,050.30 | 1,125.97 | 3,828.67 |
| TENNESSEE | 39,141.41 | 43,115.01 | 43,684.62 | 44,409.10 | 170,350.15 |
| TEXAS | 30,619.84 | 31,251.33 | 32,820.21 | 32,252.14 | 126,943.52 |
| UTAH | 11,093.22 | 11,022.38 | 11,281.12 | 11,947.19 | 45,343.90 |
| VERMONT | 7,478.16 | 7,492.76 | 7,634.51 | 7,976.57 | 30,581.99 |
| VIRGIN ISLANDS | 17.74 | 19.02 | 23.63 | 23.79 | 84.19 |
| VIRGINIA | 24,281.03 | 25,656.95 | 27,200.65 | 26,983.25 | 104,121.87 |
| WASHINGTON | 27,921.27 | 29,324.00 | 31,006.99 | 33,369.27 | 121,621.53 |
| WEST VIRGINIA | 18,084.61 | 19,110.25 | 19,779.77 | 20,117.97 | 77,092.60 |
| WISCONSIN | 13,433.94 | 13,878.31 | 14,583.69 | 14,968.85 | 56,864.79 |
| WYOMING | 1,429.05 | 1,479.57 | 1,487.94 | 1,571.94 | 5,968.49 |
| **9064 - BUPRENORPHINE Tota** | **1,047,386.94** | **1,078,567.72** | **1,126,690.29** | **1,153,126.51** | **4,405,771.45** |

**DRUG: 9143 - OXYCODONE**

| STATE | 1ST QUARTER | 2ND QUARTER | 3RD QUARTER | 4TH QUARTER | TOTAL GRAMS |
|---|---|---|---|---|---|
| ALABAMA | 175,022.03 | 172,748.95 | 170,532.67 | 170,696.51 | 689,000.17 |
| ALASKA | 23,379.75 | 22,660.91 | 22,660.05 | 20,805.64 | 89,506.34 |
| AMERICAN SAMOA | 10.76 | 15.69 | 6.72 | 21.96 | 55.13 |
| ARIZONA | 300,846.19 | 291,346.21 | 286,808.81 | 285,485.97 | 1,164,487.18 |
| ARKANSAS | 94,302.17 | 89,526.78 | 87,045.18 | 83,685.79 | 354,559.92 |
| CALIFORNIA | 614,695.39 | 608,555.76 | 587,643.27 | 576,448.13 | 2,387,342.55 |
| COLORADO | 167,623.81 | 166,628.71 | 165,933.81 | 163,506.36 | 663,692.69 |
| CONNECTICUT | 128,530.58 | 125,908.35 | 125,426.03 | 124,395.80 | 504,260.77 |
| DELAWARE | 53,528.55 | 52,330.07 | 51,549.11 | 48,239.45 | 205,647.19 |
| DISTRICT OF COLUMBIA | 11,681.27 | 11,037.41 | 11,119.13 | 10,366.42 | 44,204.22 |
| FLORIDA | 843,974.72 | 837,906.70 | 842,693.23 | 855,486.60 | 3,380,061.24 |
| GEORGIA | 363,505.56 | 360,724.91 | 360,539.31 | 369,182.95 | 1,453,952.73 |
| GUAM | 732.22 | 762.11 | 724.64 | 537.54 | 2,756.52 |
| HAWAII | 31,225.04 | 32,086.00 | 31,206.51 | 29,133.83 | 123,651.39 |
| IDAHO | 47,110.32 | 48,187.10 | 47,426.83 | 46,506.74 | 189,231.00 |
| ILLINOIS | 116,449.46 | 114,456.05 | 114,361.40 | 111,833.05 | 457,099.97 |
| INDIANA | 177,046.26 | 175,204.12 | 176,454.16 | 172,030.16 | 700,734.70 |
| IOWA | 38,867.65 | 38,839.37 | 37,871.48 | 37,256.51 | 152,835.01 |

| | | | | | |
|---|---|---|---|---|---|
| KANSAS | 92,936.22 | 91,991.63 | 90,336.94 | 88,960.78 | 364,225.57 |
| KENTUCKY | 168,229.50 | 164,883.93 | 165,536.85 | 163,699.89 | 662,350.17 |
| LOUISIANA | 148,661.39 | 146,123.87 | 145,896.69 | 142,526.39 | 583,208.34 |
| MAINE | 40,797.41 | 40,767.34 | 41,511.33 | 38,729.16 | 161,805.24 |
| MARYLAND | 243,673.71 | 242,105.60 | 239,600.87 | 239,593.58 | 964,973.75 |
| MASSACHUSETTS | 187,297.80 | 183,742.77 | 182,258.72 | 179,521.34 | 732,820.63 |
| MICHIGAN | 198,515.69 | 197,594.33 | 200,135.47 | 191,767.23 | 788,012.72 |
| MINNESOTA | 102,521.74 | 102,847.14 | 98,366.26 | 97,229.24 | 400,964.38 |
| MISSISSIPPI | 80,350.69 | 77,636.97 | 79,207.80 | 78,702.24 | 315,897.71 |
| MISSOURI | 201,805.63 | 195,316.33 | 190,698.98 | 186,448.70 | 774,269.64 |
| MONTANA | 27,919.67 | 28,055.69 | 27,645.84 | 26,600.23 | 110,221.43 |
| NEBRASKA | 31,251.94 | 30,250.71 | 29,891.34 | 29,349.56 | 120,743.56 |
| NEVADA | 135,635.35 | 135,627.73 | 132,387.63 | 134,429.04 | 538,079.75 |
| NEW HAMPSHIRE | 45,643.45 | 44,674.12 | 44,999.46 | 43,444.69 | 178,761.72 |
| NEW JERSEY | 361,611.68 | 348,469.38 | 357,651.24 | 354,560.93 | 1,422,293.24 |
| NEW MEXICO | 70,771.16 | 68,452.51 | 66,088.46 | 65,423.12 | 270,735.26 |
| NEW YORK | 552,548.94 | 502,178.55 | 517,455.44 | 509,243.36 | 2,081,426.29 |
| NORTH CAROLINA | 490,528.39 | 482,506.89 | 484,374.30 | 432,490.46 | 1,889,900.04 |
| NORTH DAKOTA | 11,573.09 | 11,115.35 | 11,045.22 | 10,885.71 | 44,619.37 |
| OHIO | 343,416.78 | 337,971.12 | 334,361.12 | 322,699.86 | 1,338,448.88 |
| OKLAHOMA | 143,372.51 | 140,933.98 | 141,637.92 | 138,811.54 | 564,755.96 |
| OREGON | 127,621.47 | 127,966.22 | 125,690.66 | 124,141.38 | 505,419.73 |
| PENNSYLVANIA | 528,758.98 | 513,827.35 | 504,837.17 | 498,701.92 | 2,046,125.42 |
| PUERTO RICO | 12,721.42 | 11,584.08 | 10,815.56 | 11,903.32 | 47,024.38 |
| RHODE ISLAND | 29,761.19 | 29,266.63 | 29,504.01 | 29,742.33 | 118,274.17 |
| SOUTH CAROLINA | 176,489.52 | 171,678.21 | 167,890.84 | 163,526.79 | 679,585.37 |
| SOUTH DAKOTA | 15,193.18 | 12,259.99 | 13,136.02 | 11,099.21 | 51,688.40 |
| TENNESSEE | 353,235.60 | 347,402.51 | 339,710.06 | 343,453.96 | 1,383,802.14 |
| TEXAS | 262,056.40 | 256,724.63 | 261,959.88 | 237,872.95 | 1,018,613.87 |
| UTAH | 132,807.19 | 132,809.31 | 133,035.40 | 133,645.24 | 532,297.13 |
| VERMONT | 16,590.29 | 16,923.99 | 16,798.31 | 16,170.95 | 66,483.54 |
| VIRGIN ISLANDS | 440.18 | 457.44 | 420.68 | 381.01 | 1,699.32 |
| VIRGINIA | 233,617.07 | 233,738.89 | 231,651.51 | 224,094.34 | 923,101.80 |
| WASHINGTON | 218,406.95 | 218,136.83 | 216,311.43 | 214,444.80 | 867,300.00 |
| WEST VIRGINIA | 57,254.62 | 54,597.23 | 53,856.88 | 51,427.06 | 217,135.79 |
| WISCONSIN | 135,556.37 | 132,151.39 | 126,740.56 | 124,370.98 | 518,819.30 |
| WYOMING | 20,484.18 | 20,197.49 | 20,251.33 | 19,362.65 | 80,295.64 |
| **9143 - OXYCODONE Total** | **9,188,589.09** | **9,001,893.35** | **8,953,700.53** | **8,785,075.39** | **35,929,258.36** |

**DRUG: 9150 - HYDROMORPHONE**

| STATE | 1ST QUARTER | 2ND QUARTER | 3RD QUARTER | 4TH QUARTER | TOTAL GRAMS |
|---|---|---|---|---|---|

| | | | | | |
|---|---|---|---|---|---|
| ALABAMA | 2,262.18 | 2,304.41 | 2,119.89 | 1,935.05 | 8,621.53 |
| ALASKA | 566.64 | 496.58 | 487.00 | 425.49 | 1,975.71 |
| AMERICAN SAMOA | 2.84 | 2.13 | 1.46 | 1.33 | 7.76 |
| ARIZONA | 4,881.95 | 4,405.59 | 4,486.70 | 4,643.83 | 18,418.08 |
| ARKANSAS | 2,097.34 | 2,021.15 | 2,085.85 | 1,797.70 | 8,002.04 |
| CALIFORNIA | 25,774.72 | 25,502.79 | 25,024.81 | 24,512.55 | 100,814.87 |
| COLORADO | 5,010.30 | 4,976.28 | 5,051.22 | 5,070.39 | 20,108.19 |
| CONNECTICUT | 4,448.49 | 4,347.28 | 4,426.13 | 4,758.33 | 17,980.22 |
| DELAWARE | 1,255.46 | 1,202.64 | 1,170.35 | 1,159.06 | 4,787.51 |
| DISTRICT OF COLUMBIA | 401.31 | 383.28 | 375.29 | 344.43 | 1,504.31 |
| FLORIDA | 34,428.22 | 32,328.92 | 33,411.95 | 31,753.69 | 131,922.78 |
| GEORGIA | 6,028.73 | 5,822.72 | 5,648.38 | 5,774.32 | 23,274.15 |
| GUAM | 26.17 | 21.07 | 12.50 | 12.86 | 72.61 |
| HAWAII | 481.52 | 505.51 | 503.65 | 515.39 | 2,006.06 |
| IDAHO | 1,340.15 | 1,350.69 | 1,344.70 | 1,254.71 | 5,290.25 |
| ILLINOIS | 6,788.14 | 6,958.96 | 7,056.39 | 6,699.29 | 27,502.77 |
| INDIANA | 4,260.94 | 4,462.79 | 4,117.66 | 3,989.31 | 16,830.70 |
| IOWA | 2,057.58 | 1,965.70 | 1,892.79 | 1,897.33 | 7,813.40 |
| KANSAS | 2,474.96 | 2,445.73 | 2,441.01 | 2,441.07 | 9,802.77 |
| KENTUCKY | 2,214.27 | 2,136.96 | 2,354.21 | 2,036.10 | 8,741.55 |
| LOUISIANA | 2,444.77 | 2,404.35 | 2,448.31 | 1,967.88 | 9,265.32 |
| MAINE | 1,296.89 | 1,223.50 | 1,445.67 | 1,334.98 | 5,301.05 |
| MARYLAND | 5,422.12 | 5,520.79 | 5,477.11 | 5,437.84 | 21,857.86 |
| MASSACHUSETTS | 5,650.22 | 5,725.52 | 5,536.52 | 6,286.38 | 23,198.64 |
| MICHIGAN | 5,721.03 | 5,600.44 | 5,700.56 | 5,894.62 | 22,916.66 |
| MINNESOTA | 4,314.55 | 4,455.81 | 4,545.47 | 4,406.20 | 17,722.03 |
| MISSISSIPPI | 5,104.46 | 5,032.64 | 5,077.58 | 6,591.99 | 21,806.67 |
| MISSOURI | 4,031.12 | 3,815.62 | 3,888.47 | 3,735.92 | 15,471.13 |
| MONTANA | 1,322.67 | 1,374.96 | 1,237.59 | 1,309.68 | 5,244.91 |
| NEBRASKA | 888.34 | 852.69 | 817.15 | 844.17 | 3,402.35 |
| NEVADA | 2,169.78 | 2,103.50 | 2,079.67 | 1,830.20 | 8,183.15 |
| NEW HAMPSHIRE | 1,512.86 | 1,492.60 | 1,567.57 | 1,486.64 | 6,059.66 |
| NEW JERSEY | 6,706.35 | 6,523.72 | 6,853.15 | 6,457.52 | 26,540.73 |
| NEW MEXICO | 1,028.30 | 902.27 | 933.04 | 966.50 | 3,830.12 |
| NEW YORK | 14,393.39 | 13,529.97 | 14,167.94 | 14,033.12 | 56,124.42 |
| NORTH CAROLINA | 8,392.23 | 8,138.22 | 8,364.25 | 8,038.01 | 32,932.71 |
| NORTH DAKOTA | 575.55 | 476.22 | 490.15 | 471.55 | 2,013.46 |
| OHIO | 5,943.59 | 6,069.12 | 6,434.05 | 5,996.69 | 24,443.46 |
| OKLAHOMA | 2,512.41 | 2,536.40 | 2,427.80 | 2,602.31 | 10,078.91 |
| OREGON | 3,467.73 | 3,462.56 | 3,605.16 | 3,525.82 | 14,061.27 |

| | | | | | |
|---|---|---|---|---|---|
| PENNSYLVANIA | 10,635.89 | 10,145.37 | 9,845.17 | 10,960.10 | 41,586.54 |
| PUERTO RICO | 54.54 | 53.91 | 50.34 | 48.21 | 207.00 |
| RHODE ISLAND | 606.21 | 612.59 | 595.02 | 942.16 | 2,755.98 |
| SOUTH CAROLINA | 4,198.02 | 4,309.00 | 4,082.37 | 3,929.24 | 16,518.63 |
| SOUTH DAKOTA | 372.11 | 405.41 | 424.32 | 421.05 | 1,622.88 |
| TENNESSEE | 5,160.77 | 4,800.19 | 4,669.36 | 5,977.27 | 20,607.60 |
| TEXAS | 16,038.37 | 17,803.44 | 16,722.72 | 16,832.13 | 67,396.65 |
| UTAH | 2,247.51 | 2,250.85 | 2,436.85 | 2,374.80 | 9,310.02 |
| VERMONT | 864.97 | 952.96 | 996.56 | 865.62 | 3,680.11 |
| VIRGIN ISLANDS | 12.57 | 20.44 | 7.91 | 19.95 | 60.87 |
| VIRGINIA | 7,917.08 | 7,702.34 | 7,749.94 | 7,539.10 | 30,908.45 |
| WASHINGTON | 7,358.68 | 7,777.55 | 7,465.50 | 7,697.44 | 30,299.17 |
| WEST VIRGINIA | 967.65 | 997.47 | 961.53 | 929.70 | 3,856.36 |
| WISCONSIN | 2,735.25 | 2,644.19 | 2,611.82 | 2,391.55 | 10,382.81 |
| WYOMING | 459.72 | 454.05 | 573.50 | 609.31 | 2,096.58 |
| **9150 - HYDROMORPHONE To** | **249,329.64** | **245,811.82** | **246,302.04** | **245,777.89** | **987,221.39** |

**DRUG: 9193 - HYDROCODONE**

| STATE | 1ST QUARTER | 2ND QUARTER | 3RD QUARTER | 4TH QUARTER | TOTAL GRAMS |
|---|---|---|---|---|---|
| ALABAMA | 194,106.07 | 190,545.56 | 188,902.31 | 185,746.59 | 759,300.53 |
| ALASKA | 8,281.17 | 8,237.70 | 8,196.46 | 7,936.52 | 32,651.84 |
| AMERICAN SAMOA | 0 | 0.61 | 0 | 3.03 | 3.63 |
| ARIZONA | 77,974.44 | 76,192.40 | 74,290.92 | 75,882.68 | 304,340.44 |
| ARKANSAS | 103,763.17 | 102,308.99 | 101,747.15 | 100,169.23 | 407,988.54 |
| CALIFORNIA | 685,151.80 | 676,648.13 | 660,469.26 | 649,564.43 | 2,671,833.62 |
| COLORADO | 48,497.32 | 47,501.70 | 47,288.42 | 46,260.08 | 189,547.52 |
| CONNECTICUT | 15,243.64 | 14,682.74 | 14,380.04 | 14,529.80 | 58,836.21 |
| DELAWARE | 4,457.11 | 4,711.74 | 4,430.76 | 4,179.32 | 17,778.93 |
| DISTRICT OF COLUMBIA | 1,058.09 | 969.80 | 952.07 | 905.09 | 3,885.04 |
| FLORIDA | 272,128.93 | 261,936.03 | 258,746.21 | 260,571.08 | 1,053,382.25 |
| GEORGIA | 194,375.53 | 190,516.79 | 188,817.10 | 187,944.01 | 761,653.42 |
| GUAM | 126.82 | 100.90 | 120.90 | 98.30 | 446.92 |
| HAWAII | 13,283.15 | 12,928.29 | 13,111.38 | 12,806.35 | 52,129.17 |
| IDAHO | 45,282.27 | 45,411.26 | 45,485.77 | 44,355.48 | 180,534.78 |
| ILLINOIS | 229,781.79 | 227,885.18 | 226,291.32 | 221,173.29 | 905,131.57 |
| INDIANA | 177,190.90 | 175,696.23 | 175,207.32 | 168,705.29 | 696,799.74 |
| IOWA | 41,749.61 | 41,159.53 | 40,514.33 | 39,793.18 | 163,216.64 |
| KANSAS | 62,624.21 | 62,005.83 | 60,917.79 | 60,725.79 | 246,273.62 |
| KENTUCKY | 169,500.70 | 165,654.30 | 165,755.86 | 162,808.39 | 663,719.26 |
| LOUISIANA | 126,704.27 | 124,180.25 | 121,324.53 | 117,536.67 | 489,745.71 |
| MAINE | 13,701.99 | 13,535.58 | 13,756.03 | 12,813.19 | 53,806.80 |

| | | | | | |
|---|---|---|---|---|---|
| MARYLAND | 25,776.10 | 25,202.59 | 24,998.90 | 24,552.96 | 100,530.54 |
| MASSACHUSETTS | 22,840.66 | 22,362.53 | 22,065.87 | 21,898.59 | 89,167.65 |
| MICHIGAN | 260,080.32 | 257,619.24 | 261,351.26 | 240,021.77 | 1,019,072.59 |
| MINNESOTA | 32,501.64 | 32,384.94 | 31,331.28 | 30,766.76 | 126,984.62 |
| MISSISSIPPI | 79,227.87 | 77,074.67 | 75,799.54 | 75,352.30 | 307,454.38 |
| MISSOURI | 131,657.60 | 129,704.10 | 127,779.91 | 126,760.46 | 515,902.06 |
| MONTANA | 20,423.48 | 20,380.14 | 20,089.33 | 19,477.54 | 80,370.49 |
| NEBRASKA | 23,970.08 | 23,601.67 | 22,962.70 | 22,865.16 | 93,399.61 |
| NEVADA | 68,989.24 | 67,908.27 | 66,175.94 | 67,077.48 | 270,150.93 |
| NEW HAMPSHIRE | 7,385.84 | 7,040.25 | 7,020.17 | 6,942.55 | 28,388.81 |
| NEW JERSEY | 26,213.06 | 24,889.34 | 25,114.73 | 25,175.95 | 101,393.08 |
| NEW MEXICO | 27,759.73 | 27,282.16 | 26,176.86 | 26,194.58 | 107,413.33 |
| NEW YORK | 131,085.68 | 125,678.80 | 125,256.94 | 123,726.07 | 505,747.50 |
| NORTH CAROLINA | 158,256.39 | 149,012.63 | 147,565.69 | 140,857.33 | 595,692.04 |
| NORTH DAKOTA | 8,050.44 | 7,554.84 | 7,579.90 | 7,390.71 | 30,575.89 |
| OHIO | 131,646.17 | 130,711.60 | 129,633.27 | 126,375.80 | 518,366.85 |
| OKLAHOMA | 132,525.66 | 131,310.16 | 131,300.83 | 129,061.11 | 524,197.75 |
| OREGON | 73,346.36 | 72,690.66 | 71,818.44 | 71,077.51 | 288,932.98 |
| PENNSYLVANIA | 116,461.12 | 114,895.96 | 110,216.30 | 113,416.28 | 454,989.65 |
| PUERTO RICO | 567.72 | 406.49 | 342.72 | 405.56 | 1,722.49 |
| RHODE ISLAND | 7,681.15 | 7,595.34 | 7,559.00 | 7,545.55 | 30,381.05 |
| SOUTH CAROLINA | 104,035.86 | 100,915.12 | 97,637.89 | 95,491.00 | 398,079.87 |
| SOUTH DAKOTA | 10,375.49 | 10,114.80 | 10,304.10 | 9,918.48 | 40,712.87 |
| TENNESSEE | 185,650.62 | 183,346.41 | 179,318.75 | 186,724.50 | 735,040.29 |
| TEXAS | 525,382.04 | 511,151.94 | 504,238.60 | 493,467.49 | 2,034,240.07 |
| UTAH | 45,932.81 | 46,109.33 | 45,311.30 | 45,939.13 | 183,292.58 |
| VERMONT | 3,660.15 | 3,686.74 | 3,683.83 | 3,559.26 | 14,589.98 |
| VIRGIN ISLANDS | 256.51 | 186.01 | 181.09 | 200.01 | 823.62 |
| VIRGINIA | 80,239.56 | 78,298.17 | 76,038.87 | 73,872.10 | 308,448.70 |
| WASHINGTON | 100,680.42 | 99,229.81 | 97,950.74 | 96,731.38 | 394,592.36 |
| WEST VIRGINIA | 40,550.69 | 39,007.70 | 38,559.69 | 36,519.54 | 154,637.62 |
| WISCONSIN | 60,237.95 | 60,523.54 | 59,485.20 | 58,072.15 | 238,318.84 |
| WYOMING | 8,595.68 | 8,599.90 | 8,811.08 | 8,337.73 | 34,344.39 |
| **9193 - HYDROCODONE Total** | **5,137,027.07** | **5,039,285.38** | **4,974,366.65** | **4,890,282.54** | **20,040,961.64** |

**DRUG: 9230 - MEPERIDINE (PETHIDINE)**

| STATE | 1ST QUARTER | 2ND QUARTER | 3RD QUARTER | 4TH QUARTER | TOTAL GRAMS |
|---|---|---|---|---|---|
| ALABAMA | 4,597.71 | 3,458.22 | 2,543.47 | 1,813.92 | 12,413.32 |
| ALASKA | 356.77 | 237.48 | 74.08 | 93.69 | 762.02 |
| ARIZONA | 3,253.35 | 2,749.95 | 2,565.74 | 1,896.25 | 10,465.30 |
| ARKANSAS | 4,559.03 | 2,939.13 | 2,173.09 | 1,440.15 | 11,111.41 |

| | | | | | |
|---|---|---|---|---|---|
| CALIFORNIA | 10,796.25 | 8,817.40 | 10,360.50 | 7,031.92 | 37,006.07 |
| COLORADO | 1,061.60 | 805.05 | 366.79 | 266.35 | 2,499.79 |
| CONNECTICUT | 644.37 | 454.81 | 76.47 | 58.28 | 1,233.94 |
| DELAWARE | 92.05 | 133.45 | 28.54 | 31.05 | 285.09 |
| DISTRICT OF COLUMBIA | 121.47 | 95.87 | 154.15 | 69.72 | 441.20 |
| FLORIDA | 4,885.85 | 3,935.26 | 2,949.16 | 2,157.29 | 13,927.55 |
| GEORGIA | 6,059.10 | 4,044.85 | 2,933.80 | 1,731.89 | 14,769.64 |
| GUAM | 14.16 | 16.34 | 34.86 | 19.61 | 84.97 |
| HAWAII | 230.40 | 166.13 | 183.56 | 72.44 | 652.54 |
| IDAHO | 599.16 | 418.86 | 255.68 | 155.45 | 1,429.15 |
| ILLINOIS | 1,624.80 | 1,414.55 | 1,162.36 | 640.23 | 4,841.95 |
| INDIANA | 1,973.40 | 1,387.86 | 1,410.74 | 1,138.07 | 5,910.08 |
| IOWA | 1,502.79 | 1,097.55 | 1,154.74 | 965.95 | 4,721.02 |
| KANSAS | 1,128.59 | 733.69 | 552.86 | 381.28 | 2,796.43 |
| KENTUCKY | 1,217.92 | 931.96 | 947.65 | 597.20 | 3,694.72 |
| LOUISIANA | 3,743.09 | 2,706.01 | 2,035.93 | 1,503.66 | 9,988.70 |
| MAINE | 184.10 | 88.24 | 93.36 | 63.73 | 429.43 |
| MARYLAND | 1,374.25 | 1,253.33 | 937.95 | 458.08 | 4,023.61 |
| MASSACHUSETTS | 1,287.10 | 878.04 | 455.47 | 349.25 | 2,969.85 |
| MICHIGAN | 1,734.83 | 1,385.14 | 958.65 | 556.67 | 4,635.29 |
| MINNESOTA | 390.54 | 340.43 | 264.17 | 164.50 | 1,159.64 |
| MISSISSIPPI | 2,531.16 | 1,768.82 | 1,572.40 | 1,275.44 | 7,147.83 |
| MISSOURI | 2,013.17 | 1,519.68 | 1,312.15 | 566.48 | 5,411.47 |
| MONTANA | 294.13 | 212.97 | 172.67 | 111.66 | 791.43 |
| NEBRASKA | 439.02 | 206.98 | 263.08 | 148.16 | 1,057.24 |
| NEVADA | 843.72 | 714.09 | 846.44 | 460.26 | 2,864.51 |
| NEW HAMPSHIRE | 416.14 | 243.91 | 131.95 | 54.01 | 846.01 |
| NEW JERSEY | 1,241.89 | 860.61 | 555.47 | 416.58 | 3,074.54 |
| NEW MEXICO | 590.44 | 477.15 | 357.32 | 244.02 | 1,668.92 |
| NEW YORK | 2,934.23 | 2,113.93 | 1,566.09 | 829.89 | 7,444.14 |
| NORTH CAROLINA | 3,378.15 | 2,074.17 | 1,053.43 | 814.20 | 7,319.95 |
| NORTH DAKOTA | 154.15 | 136.17 | 79.74 | 42.49 | 412.55 |
| OHIO | 3,472.38 | 2,504.47 | 2,267.32 | 1,394.15 | 9,638.32 |
| OKLAHOMA | 3,830.79 | 2,714.18 | 2,127.55 | 994.05 | 9,666.57 |
| OREGON | 1,095.91 | 663.97 | 639.90 | 342.06 | 2,741.85 |
| PENNSYLVANIA | 2,028.96 | 1,757.16 | 925.64 | 522.57 | 5,234.34 |
| PUERTO RICO | 2,929.33 | 2,876.49 | 3,966.41 | 2,938.59 | 12,710.83 |
| RHODE ISLAND | 42.49 | 76.26 | 40.31 | 108.94 | 267.99 |
| SOUTH CAROLINA | 2,891.09 | 1,830.69 | 1,119.77 | 639.31 | 6,480.87 |
| SOUTH DAKOTA | 365.49 | 319.73 | 204.80 | 78.98 | 969.00 |

| | | | | | |
|---|---|---|---|---|---|
| TENNESSEE | 3,514.87 | 2,324.73 | 1,461.51 | 1,136.87 | 8,437.97 |
| TEXAS | 7,418.03 | 5,798.44 | 6,584.46 | 5,011.67 | 24,812.60 |
| UTAH | 1,225.55 | 884.03 | 713.54 | 707.88 | 3,530.99 |
| VERMONT | 166.67 | 255.46 | 137.81 | 154.15 | 714.09 |
| VIRGIN ISLANDS | 61.55 | 72.44 | 24.51 | 2.18 | 160.68 |
| VIRGINIA | 3,549.18 | 2,370.48 | 1,541.03 | 910.72 | 8,371.41 |
| WASHINGTON | 1,981.57 | 1,445.60 | 792.74 | 633.69 | 4,853.60 |
| WEST VIRGINIA | 324.63 | 200.45 | 203.17 | 159.81 | 888.06 |
| WISCONSIN | 756.57 | 559.39 | 645.45 | 427.58 | 2,389.00 |
| WYOMING | 217.88 | 196.09 | 75.71 | 45.10 | 534.77 |
| **9230 - MEPERIDINE (PETHIDIN** | **104,141.83** | **77,668.15** | **66,056.12** | **44,828.10** | **292,694.19** |

**DRUG: 9250B - METHADONE**

| STATE | 1ST QUARTER | 2ND QUARTER | 3RD QUARTER | 4TH QUARTER | TOTAL GRAMS |
|---|---|---|---|---|---|
| ALABAMA | 80,407.01 | 79,250.19 | 78,948.42 | 75,853.08 | 314,458.70 |
| ALASKA | 9,180.57 | 9,921.67 | 9,889.29 | 9,051.06 | 38,042.59 |
| ARIZONA | 100,865.33 | 105,144.14 | 109,133.53 | 112,348.18 | 427,491.18 |
| ARKANSAS | 12,298.98 | 13,053.32 | 13,259.21 | 12,245.59 | 50,857.10 |
| CALIFORNIA | 312,786.25 | 317,006.09 | 310,207.00 | 317,187.44 | 1,257,186.79 |
| COLORADO | 48,235.62 | 52,398.51 | 51,283.54 | 52,619.69 | 204,537.35 |
| CONNECTICUT | 129,203.86 | 132,695.06 | 115,336.10 | 115,768.01 | 493,003.03 |
| DELAWARE | 36,252.63 | 33,673.98 | 33,219.36 | 38,967.93 | 142,113.90 |
| DISTRICT OF COLUMBIA | 10,795.02 | 8,949.88 | 10,114.31 | 9,681.39 | 39,540.59 |
| FLORIDA | 213,150.70 | 228,887.74 | 207,857.13 | 209,440.36 | 859,335.92 |
| GEORGIA | 111,916.72 | 117,120.96 | 112,345.05 | 111,781.13 | 453,163.86 |
| GUAM | 45.61 | 36.22 | 33.09 | 16.10 | 131.03 |
| HAWAII | 7,646.49 | 7,292.76 | 8,546.44 | 6,597.99 | 30,083.68 |
| IDAHO | 10,938.06 | 11,106.03 | 10,253.76 | 10,277.55 | 42,575.41 |
| ILLINOIS | 122,402.70 | 128,479.56 | 124,466.14 | 129,181.06 | 504,529.46 |
| INDIANA | 83,701.69 | 80,632.93 | 80,442.47 | 85,734.50 | 330,511.59 |
| IOWA | 13,243.20 | 13,819.11 | 12,200.33 | 14,854.73 | 54,117.37 |
| KANSAS | 22,058.86 | 21,009.73 | 18,996.16 | 19,879.62 | 81,944.36 |
| KENTUCKY | 49,170.84 | 50,057.80 | 55,452.14 | 52,885.16 | 207,565.94 |
| LOUISIANA | 34,582.60 | 35,538.18 | 34,250.40 | 33,427.79 | 137,798.97 |
| MAINE | 27,927.01 | 30,430.71 | 30,674.61 | 31,350.78 | 120,383.11 |
| MARYLAND | 185,714.81 | 204,889.13 | 195,012.49 | 192,034.97 | 777,651.39 |
| MASSACHUSETTS | 148,596.41 | 149,768.00 | 150,680.87 | 155,173.78 | 604,219.06 |
| MICHIGAN | 124,143.27 | 123,376.22 | 122,715.41 | 123,609.46 | 493,844.37 |
| MINNESOTA | 47,846.62 | 53,476.44 | 55,766.55 | 51,132.67 | 208,222.28 |
| MISSISSIPPI | 7,942.09 | 9,293.26 | 8,224.28 | 8,085.82 | 33,545.46 |
| MISSOURI | 34,886.15 | 33,796.08 | 38,895.65 | 32,105.72 | 139,683.60 |

| | | | | | |
|---|---|---|---|---|---|
| MONTANA | 9,006.61 | 8,064.36 | 8,300.12 | 8,177.68 | 33,548.77 |
| NEBRASKA | 7,163.25 | 7,987.53 | 6,835.46 | 7,196.79 | 29,183.03 |
| NEVADA | 36,677.29 | 29,802.57 | 32,988.07 | 33,718.52 | 133,186.45 |
| NEW HAMPSHIRE | 32,339.80 | 35,695.37 | 31,414.33 | 32,455.10 | 131,904.60 |
| NEW JERSEY | 129,002.80 | 128,564.72 | 133,828.54 | 137,531.35 | 528,927.41 |
| NEW MEXICO | 47,952.00 | 51,392.49 | 50,587.44 | 53,337.28 | 203,269.21 |
| NEW YORK | 326,100.04 | 310,972.18 | 321,765.83 | 326,752.10 | 1,285,590.16 |
| NORTH CAROLINA | 136,930.02 | 145,521.83 | 143,951.35 | 139,552.35 | 565,955.56 |
| NORTH DAKOTA | 4,758.39 | 5,127.60 | 5,389.48 | 6,029.60 | 21,305.06 |
| OHIO | 73,460.16 | 76,850.17 | 79,910.20 | 83,613.15 | 313,833.69 |
| OKLAHOMA | 34,629.11 | 34,954.14 | 37,204.27 | 32,357.42 | 139,144.94 |
| OREGON | 61,979.90 | 62,789.04 | 64,193.09 | 63,126.19 | 252,088.22 |
| PENNSYLVANIA | 199,275.64 | 203,126.13 | 188,662.07 | 199,421.62 | 790,485.46 |
| PUERTO RICO | 20,654.38 | 21,551.02 | 24,192.00 | 18,898.67 | 85,296.07 |
| RHODE ISLAND | 42,395.01 | 49,641.26 | 49,632.04 | 49,457.37 | 191,125.68 |
| SOUTH CAROLINA | 56,050.15 | 56,747.24 | 57,633.72 | 59,654.20 | 230,085.30 |
| SOUTH DAKOTA | 1,976.18 | 2,128.58 | 1,552.05 | 2,098.89 | 7,755.70 |
| TENNESSEE | 46,111.34 | 53,015.82 | 44,722.67 | 47,592.46 | 191,442.29 |
| TEXAS | 128,436.28 | 142,276.36 | 134,754.71 | 138,672.36 | 544,139.71 |
| UTAH | 25,723.66 | 29,077.39 | 27,507.72 | 26,464.58 | 108,773.35 |
| VERMONT | 18,618.99 | 21,762.18 | 18,164.55 | 22,374.22 | 80,919.95 |
| VIRGIN ISLANDS | 4.47 | 9.84 | 12.52 | 6.26 | 33.09 |
| VIRGINIA | 89,288.76 | 94,768.34 | 91,976.86 | 97,748.01 | 373,781.97 |
| WASHINGTON | 103,413.66 | 118,990.35 | 106,072.17 | 107,989.05 | 436,465.23 |
| WEST VIRGINIA | 23,757.77 | 25,060.19 | 25,080.85 | 23,210.22 | 97,109.03 |
| WISCONSIN | 61,697.57 | 63,355.48 | 62,969.27 | 67,348.36 | 255,370.69 |
| WYOMING | 829.65 | 834.03 | 793.87 | 728.40 | 3,185.94 |
| **9250B METHADONE Total** | **3,704,171.99** | **3,831,169.93** | **3,748,298.99** | **3,796,803.76** | **15,080,444.66** |

**DRUG: 9300 - MORPHINE**

| STATE | 1ST QUARTER | 2ND QUARTER | 3RD QUARTER | 4TH QUARTER | TOTAL GRAMS |
|---|---|---|---|---|---|
| ALABAMA | 55,019.57 | 52,823.27 | 51,998.73 | 49,934.38 | 209,775.95 |
| ALASKA | 8,072.48 | 7,604.69 | 7,568.06 | 6,798.53 | 30,043.76 |
| AMERICAN SAMOA | 19.41 | 34.95 | 0.30 | 12.41 | 67.07 |
| ARIZONA | 104,258.83 | 96,681.36 | 92,121.69 | 91,242.29 | 384,304.17 |
| ARKANSAS | 29,595.83 | 27,537.85 | 27,298.24 | 25,907.98 | 110,339.90 |
| CALIFORNIA | 322,711.88 | 308,234.72 | 302,215.57 | 289,441.33 | 1,222,603.50 |
| COLORADO | 57,255.96 | 53,843.19 | 55,561.58 | 51,838.75 | 218,499.49 |
| CONNECTICUT | 27,399.91 | 26,980.50 | 27,183.72 | 26,147.40 | 107,711.52 |
| DELAWARE | 15,447.25 | 14,803.41 | 15,723.54 | 14,420.55 | 60,394.75 |
| DISTRICT OF COLUMBIA | 2,703.21 | 2,776.84 | 2,661.65 | 2,316.25 | 10,457.95 |

| | | | | | |
|---|---|---|---|---|---|
| FLORIDA | 302,541.63 | 286,710.23 | 288,709.39 | 282,234.08 | 1,160,195.33 |
| GEORGIA | 89,369.02 | 86,788.55 | 87,541.81 | 88,193.63 | 351,893.00 |
| GUAM | 740.46 | 874.34 | 726.91 | 589.27 | 2,930.98 |
| HAWAII | 14,609.32 | 14,932.41 | 14,396.26 | 13,225.62 | 57,163.61 |
| IDAHO | 21,636.56 | 21,438.67 | 21,230.30 | 19,792.14 | 84,097.68 |
| ILLINOIS | 85,266.39 | 83,508.02 | 82,236.71 | 83,048.90 | 334,060.01 |
| INDIANA | 62,090.65 | 59,925.59 | 61,547.82 | 57,983.54 | 241,547.60 |
| IOWA | 25,786.32 | 25,220.93 | 25,785.28 | 23,660.49 | 100,453.03 |
| KANSAS | 33,643.81 | 33,260.31 | 33,487.50 | 31,677.46 | 132,069.08 |
| KENTUCKY | 35,123.02 | 33,294.25 | 33,749.32 | 31,927.98 | 134,094.57 |
| LOUISIANA | 42,587.39 | 40,366.44 | 40,967.97 | 39,320.08 | 163,241.88 |
| MAINE | 12,845.34 | 13,129.89 | 13,653.27 | 12,406.65 | 52,035.14 |
| MARYLAND | 52,519.81 | 50,474.37 | 50,525.07 | 48,485.15 | 202,004.40 |
| MASSACHUSETTS | 52,179.56 | 52,004.55 | 51,814.29 | 50,240.20 | 206,238.60 |
| MICHIGAN | 127,276.68 | 124,172.43 | 127,531.64 | 118,497.27 | 497,478.02 |
| MINNESOTA | 35,006.53 | 33,280.09 | 33,246.45 | 32,090.00 | 133,623.08 |
| MISSISSIPPI | 30,272.71 | 26,471.37 | 27,372.20 | 28,016.47 | 112,132.74 |
| MISSOURI | 72,550.39 | 69,056.52 | 68,812.51 | 64,874.35 | 275,293.78 |
| MONTANA | 13,154.38 | 13,141.11 | 13,148.51 | 12,438.34 | 51,882.33 |
| NEBRASKA | 14,961.89 | 15,096.92 | 15,063.74 | 14,145.46 | 59,268.01 |
| NEVADA | 41,601.91 | 38,846.06 | 37,593.44 | 36,884.44 | 154,925.85 |
| NEW HAMPSHIRE | 15,505.98 | 14,128.87 | 15,134.83 | 13,949.51 | 58,719.18 |
| NEW JERSEY | 63,120.29 | 59,743.99 | 61,705.49 | 60,260.62 | 244,830.39 |
| NEW MEXICO | 20,832.92 | 19,550.31 | 19,119.81 | 18,793.07 | 78,296.12 |
| NEW YORK | 118,460.51 | 106,244.15 | 116,128.85 | 110,914.13 | 451,747.65 |
| NORTH CAROLINA | 105,608.79 | 106,475.01 | 107,159.13 | 93,292.58 | 412,535.51 |
| NORTH DAKOTA | 4,974.12 | 4,594.51 | 4,874.95 | 4,749.57 | 19,193.15 |
| OHIO | 87,413.41 | 85,268.23 | 85,470.42 | 82,152.02 | 340,304.08 |
| OKLAHOMA | 53,726.55 | 51,960.79 | 49,536.92 | 48,652.13 | 203,876.39 |
| OREGON | 52,583.12 | 52,137.26 | 52,369.04 | 50,785.07 | 207,874.49 |
| PENNSYLVANIA | 173,364.04 | 162,474.94 | 164,041.94 | 167,013.02 | 666,893.94 |
| PUERTO RICO | 2,237.68 | 1,994.64 | 1,622.88 | 1,624.07 | 7,479.28 |
| RHODE ISLAND | 8,830.06 | 8,546.62 | 8,319.02 | 8,653.14 | 34,348.84 |
| SOUTH CAROLINA | 54,115.61 | 52,859.19 | 52,407.11 | 50,686.63 | 210,068.54 |
| SOUTH DAKOTA | 6,354.66 | 6,405.08 | 6,437.19 | 5,408.67 | 24,605.60 |
| TENNESSEE | 121,023.94 | 113,423.36 | 115,194.01 | 122,930.27 | 472,571.57 |
| TEXAS | 161,475.20 | 153,592.78 | 152,559.09 | 146,412.89 | 614,039.95 |
| UTAH | 38,556.00 | 37,578.00 | 37,805.48 | 36,913.42 | 150,852.91 |
| VERMONT | 7,810.23 | 7,906.55 | 7,964.77 | 7,650.01 | 31,331.55 |
| VIRGIN ISLANDS | 105.70 | 80.54 | 73.02 | 66.97 | 326.22 |

| | | | | | |
|---|---|---|---|---|---|
| VIRGINIA | 64,650.97 | 61,905.95 | 64,255.25 | 60,305.26 | 251,117.44 |
| WASHINGTON | 81,237.84 | 79,469.85 | 79,306.50 | 77,446.24 | 317,460.42 |
| WEST VIRGINIA | 16,163.33 | 14,902.48 | 16,025.42 | 14,809.95 | 61,901.17 |
| WISCONSIN | 56,225.67 | 54,775.27 | 55,002.84 | 52,611.50 | 218,615.28 |
| WYOMING | 7,013.17 | 6,684.72 | 6,877.41 | 6,231.29 | 26,806.60 |
| **9300 - MORPHINE Total** | **3,109,637.88** | **2,976,016.91** | **2,990,864.84** | **2,890,103.41** | **11,966,623.04** |

**DRUG: 9652 - OXYMORPHONE**

| STATE | 1ST QUARTER | 2ND QUARTER | 3RD QUARTER | 4TH QUARTER | TOTAL GRAMS |
|---|---|---|---|---|---|
| ALABAMA | 1,567.06 | 1,460.95 | 1,286.81 | 1,180.47 | 5,495.29 |
| ALASKA | 62.00 | 51.47 | 51.74 | 59.32 | 224.54 |
| ARIZONA | 1,521.70 | 1,344.53 | 1,195.28 | 1,280.39 | 5,341.89 |
| ARKANSAS | 290.69 | 224.85 | 158.75 | 163.39 | 837.68 |
| CALIFORNIA | 4,192.56 | 4,345.95 | 3,615.77 | 3,365.58 | 15,519.86 |
| COLORADO | 994.11 | 1,021.81 | 928.50 | 901.65 | 3,846.07 |
| CONNECTICUT | 1,119.81 | 1,063.74 | 934.83 | 944.24 | 4,062.62 |
| DELAWARE | 1,190.06 | 1,274.41 | 1,004.95 | 943.40 | 4,412.82 |
| DISTRICT OF COLUMBIA | 56.78 | 62.27 | 42.82 | 31.89 | 193.76 |
| FLORIDA | 8,168.25 | 8,495.11 | 7,566.93 | 6,460.72 | 30,691.01 |
| GEORGIA | 2,270.66 | 2,455.73 | 2,123.60 | 1,924.39 | 8,774.38 |
| HAWAII | 203.31 | 210.00 | 164.24 | 148.53 | 726.08 |
| IDAHO | 573.66 | 564.16 | 534.59 | 479.77 | 2,152.19 |
| ILLINOIS | 1,667.60 | 1,614.79 | 1,405.59 | 1,364.15 | 6,052.14 |
| INDIANA | 1,728.98 | 1,686.43 | 1,484.90 | 1,252.11 | 6,152.41 |
| IOWA | 290.56 | 306.93 | 230.70 | 221.82 | 1,050.00 |
| KANSAS | 656.94 | 637.94 | 595.16 | 545.03 | 2,435.08 |
| KENTUCKY | 1,484.23 | 1,562.25 | 1,442.26 | 1,548.64 | 6,037.38 |
| LOUISIANA | 3,001.56 | 3,128.95 | 2,638.39 | 2,499.89 | 11,268.78 |
| MAINE | 356.71 | 317.81 | 313.26 | 287.88 | 1,275.66 |
| MARYLAND | 4,821.94 | 4,878.03 | 3,923.32 | 3,446.33 | 17,069.62 |
| MASSACHUSETTS | 800.04 | 936.35 | 927.29 | 773.85 | 3,437.53 |
| MICHIGAN | 7,408.49 | 8,112.00 | 6,952.49 | 6,173.73 | 28,646.71 |
| MINNESOTA | 204.11 | 188.86 | 177.44 | 186.54 | 756.95 |
| MISSISSIPPI | 1,082.56 | 979.53 | 802.49 | 796.82 | 3,661.40 |
| MISSOURI | 1,372.32 | 1,464.25 | 1,211.74 | 1,176.32 | 5,224.63 |
| MONTANA | 95.14 | 96.44 | 92.42 | 95.81 | 379.81 |
| NEBRASKA | 324.72 | 382.22 | 318.70 | 318.93 | 1,344.57 |
| NEVADA | 1,252.06 | 1,062.13 | 960.03 | 886.97 | 4,161.20 |
| NEW HAMPSHIRE | 306.44 | 263.08 | 206.61 | 199.65 | 975.78 |
| NEW JERSEY | 2,653.42 | 2,533.88 | 2,511.48 | 2,452.65 | 10,151.43 |
| NEW MEXICO | 343.64 | 326.20 | 296.22 | 239.97 | 1,206.03 |

| | | | | | |
|---|---|---|---|---|---|
| NEW YORK | 8,801.28 | 8,141.71 | 7,217.85 | 7,243.58 | 31,404.42 |
| NORTH CAROLINA | 8,705.06 | 10,286.45 | 8,531.96 | 6,577.94 | 34,101.41 |
| NORTH DAKOTA | 58.97 | 61.38 | 40.95 | 49.07 | 210.36 |
| OHIO | 3,164.90 | 3,036.31 | 2,552.92 | 2,491.10 | 11,245.23 |
| OKLAHOMA | 1,006.07 | 1,002.90 | 841.96 | 865.38 | 3,716.31 |
| OREGON | 708.19 | 669.74 | 580.18 | 565.90 | 2,524.02 |
| PENNSYLVANIA | 9,185.42 | 9,166.33 | 7,751.72 | 7,234.35 | 33,337.82 |
| RHODE ISLAND | 158.48 | 151.75 | 153.17 | 156.74 | 620.14 |
| SOUTH CAROLINA | 2,608.59 | 2,659.31 | 2,315.27 | 2,248.32 | 9,831.48 |
| SOUTH DAKOTA | 228.29 | 232.17 | 200.59 | 186.76 | 847.81 |
| TENNESSEE | 9,464.91 | 9,056.78 | 7,876.40 | 6,852.58 | 33,250.66 |
| TEXAS | 3,242.95 | 3,096.50 | 2,780.32 | 2,494.58 | 11,614.35 |
| UTAH | 2,087.65 | 2,303.40 | 1,909.99 | 2,095.54 | 8,396.58 |
| VERMONT | 102.68 | 121.06 | 95.63 | 95.19 | 414.56 |
| VIRGINIA | 4,278.56 | 4,557.20 | 3,875.37 | 3,571.92 | 16,283.06 |
| WASHINGTON | 1,187.74 | 1,187.39 | 1,030.60 | 1,059.95 | 4,465.67 |
| WEST VIRGINIA | 1,031.49 | 941.75 | 718.50 | 611.36 | 3,303.09 |
| WISCONSIN | 596.10 | 581.96 | 567.73 | 476.69 | 2,222.49 |
| WYOMING | 81.36 | 93.98 | 96.21 | 68.07 | 339.62 |
| **9652 - OXYMORPHONE Total** | **108,760.80** | **110,401.08** | **95,236.63** | **87,295.87** | **401,694.39** |

**DRUG: 9780 - TAPENTADOL**

| STATE | 1ST QUARTER | 2ND QUARTER | 3RD QUARTER | 4TH QUARTER | TOTAL GRAMS |
|---|---|---|---|---|---|
| ALABAMA | 31,945.50 | 30,158.00 | 28,491.00 | 27,583.00 | 118,177.50 |
| ALASKA | 1,559.50 | 1,555.50 | 1,380.50 | 1,358.00 | 5,853.50 |
| ARIZONA | 23,807.50 | 23,920.00 | 21,411.50 | 22,058.00 | 91,197.00 |
| ARKANSAS | 10,606.50 | 10,494.00 | 9,329.00 | 9,165.00 | 39,594.50 |
| CALIFORNIA | 69,856.50 | 68,628.50 | 65,887.50 | 64,235.00 | 268,607.50 |
| COLORADO | 21,809.50 | 22,403.00 | 21,604.00 | 22,441.50 | 88,258.00 |
| CONNECTICUT | 7,712.50 | 7,214.00 | 7,043.00 | 6,408.50 | 28,378.00 |
| DELAWARE | 4,598.00 | 4,501.50 | 4,466.50 | 4,404.00 | 17,970.00 |
| DISTRICT OF COLUMBIA | 734.00 | 624.50 | 548.00 | 644.00 | 2,550.50 |
| FLORIDA | 62,871.50 | 60,112.50 | 57,103.50 | 56,703.95 | 236,791.45 |
| GEORGIA | 44,466.50 | 42,382.00 | 42,340.00 | 41,633.50 | 170,822.00 |
| HAWAII | 1,500.50 | 1,275.00 | 1,355.00 | 1,410.00 | 5,540.50 |
| IDAHO | 4,636.50 | 4,709.50 | 4,548.50 | 4,444.00 | 18,338.50 |
| ILLINOIS | 23,554.00 | 23,091.00 | 23,570.00 | 22,712.50 | 92,927.50 |
| INDIANA | 24,451.00 | 24,853.00 | 25,638.00 | 25,153.00 | 100,095.00 |
| IOWA | 8,161.00 | 7,558.00 | 7,168.50 | 7,572.00 | 30,459.50 |
| KANSAS | 5,299.50 | 5,432.50 | 4,821.50 | 4,856.50 | 20,410.00 |
| KENTUCKY | 9,960.50 | 9,174.00 | 9,361.50 | 8,724.00 | 37,220.00 |

| | | | | | |
|---|---|---|---|---|---|
| LOUISIANA | 12,132.50 | 11,756.50 | 11,442.00 | 11,303.00 | 46,634.00 |
| MAINE | 1,183.50 | 1,249.00 | 1,366.00 | 1,198.00 | 4,996.50 |
| MARYLAND | 23,356.50 | 23,581.00 | 23,151.00 | 22,820.00 | 92,908.50 |
| MASSACHUSETTS | 15,504.50 | 15,604.00 | 16,276.50 | 15,326.00 | 62,711.00 |
| MICHIGAN | 14,276.50 | 13,408.00 | 13,622.50 | 13,110.00 | 54,417.00 |
| MINNESOTA | 3,441.50 | 3,557.00 | 3,186.00 | 3,016.00 | 13,200.50 |
| MISSISSIPPI | 7,976.50 | 7,317.50 | 7,306.00 | 7,602.00 | 30,202.00 |
| MISSOURI | 18,102.00 | 17,405.00 | 16,613.50 | 15,607.00 | 67,727.50 |
| MONTANA | 1,484.00 | 1,312.50 | 1,427.50 | 1,293.00 | 5,517.00 |
| NEBRASKA | 8,537.00 | 8,061.00 | 8,158.50 | 8,454.50 | 33,211.00 |
| NEVADA | 7,136.50 | 6,847.50 | 6,010.00 | 6,130.50 | 26,124.50 |
| NEW HAMPSHIRE | 5,732.00 | 5,598.50 | 5,983.00 | 6,039.00 | 23,352.50 |
| NEW JERSEY | 29,925.50 | 28,396.00 | 28,606.00 | 29,178.00 | 116,105.50 |
| NEW MEXICO | 2,139.00 | 2,006.00 | 2,024.50 | 1,914.00 | 8,083.50 |
| NEW YORK | 68,522.00 | 64,972.50 | 65,821.00 | 63,399.00 | 262,714.50 |
| NORTH CAROLINA | 81,334.00 | 82,502.50 | 78,101.50 | 72,256.40 | 314,194.40 |
| NORTH DAKOTA | 1,486.00 | 1,427.50 | 1,286.00 | 1,211.00 | 5,410.50 |
| OHIO | 38,432.50 | 37,735.00 | 34,730.50 | 35,475.50 | 146,373.50 |
| OKLAHOMA | 9,010.50 | 8,667.50 | 8,667.00 | 7,851.50 | 34,196.50 |
| OREGON | 4,767.00 | 5,181.50 | 4,857.50 | 4,473.50 | 19,279.50 |
| PENNSYLVANIA | 43,745.50 | 43,444.50 | 42,017.50 | 42,128.50 | 171,336.00 |
| PUERTO RICO | 409.00 | 497.00 | 427.50 | 260.00 | 1,593.50 |
| RHODE ISLAND | 1,861.00 | 1,757.50 | 1,892.50 | 1,616.00 | 7,127.00 |
| SOUTH CAROLINA | 31,810.50 | 30,262.50 | 29,896.00 | 29,034.00 | 121,003.00 |
| SOUTH DAKOTA | 3,099.00 | 3,050.50 | 2,914.50 | 2,826.50 | 11,890.50 |
| TENNESSEE | 21,954.00 | 22,107.00 | 21,923.50 | 22,536.50 | 88,521.00 |
| TEXAS | 61,421.00 | 60,727.00 | 59,697.00 | 56,767.00 | 238,612.00 |
| UTAH | 8,488.00 | 9,062.00 | 8,746.50 | 8,607.50 | 34,904.00 |
| VERMONT | 810.50 | 663.00 | 743.50 | 780.00 | 2,997.00 |
| VIRGIN ISLANDS | 0 | 5.00 | 0 | 0 | 5.00 |
| VIRGINIA | 27,308.50 | 27,582.00 | 27,209.00 | 26,164.50 | 108,264.00 |
| WASHINGTON | 12,552.50 | 12,061.00 | 12,118.50 | 12,362.00 | 49,094.00 |
| WEST VIRGINIA | 3,404.00 | 3,759.50 | 3,623.00 | 3,676.50 | 14,463.00 |
| WISCONSIN | 9,349.50 | 8,692.00 | 7,882.50 | 7,610.50 | 33,534.50 |
| WYOMING | 1,284.50 | 1,289.00 | 1,158.00 | 1,059.50 | 4,791.00 |
| **9780 - TAPENTADOL Total** | **939,507.50** | **919,632.00** | **894,953.50** | **874,593.35** | **3,628,686.35** |

**DRUG: 9801 - FENTANYL BASE**

| STATE | 1ST QUARTER | 2ND QUARTER | 3RD QUARTER | 4TH QUARTER | TOTAL GRAMS |
|---|---|---|---|---|---|
| ALABAMA | 913.51 | 872.60 | 842.78 | 813.62 | 3,442.51 |
| ALASKA | 128.28 | 114.28 | 111.45 | 99.74 | 453.75 |

| | | | | | |
|---|---|---|---|---|---|
| AMERICAN SAMOA | 0.48 | 0.13 | 0 | 0.04 | 0.65 |
| ARIZONA | 953.74 | 876.85 | 826.41 | 800.08 | 3,457.08 |
| ARKANSAS | 507.47 | 483.56 | 466.04 | 449.57 | 1,906.65 |
| CALIFORNIA | 4,321.45 | 4,076.32 | 3,859.00 | 3,706.27 | 15,963.04 |
| COLORADO | 783.34 | 743.57 | 744.66 | 698.29 | 2,969.87 |
| CONNECTICUT | 587.49 | 558.68 | 557.30 | 522.99 | 2,226.46 |
| DELAWARE | 174.04 | 168.08 | 160.69 | 148.63 | 651.43 |
| DISTRICT OF COLUMBIA | 60.79 | 52.98 | 52.22 | 48.76 | 214.74 |
| FLORIDA | 3,878.80 | 3,473.63 | 3,313.34 | 3,227.11 | 13,892.88 |
| GEORGIA | 1,840.13 | 1,730.42 | 1,701.67 | 1,629.59 | 6,901.81 |
| GUAM | 7.39 | 6.87 | 6.02 | 7.28 | 27.56 |
| HAWAII | 132.76 | 124.43 | 120.91 | 111.96 | 490.07 |
| IDAHO | 406.77 | 383.61 | 379.31 | 360.79 | 1,530.47 |
| ILLINOIS | 1,957.48 | 1,900.26 | 1,851.64 | 1,786.33 | 7,495.72 |
| INDIANA | 1,194.38 | 1,142.72 | 1,118.46 | 1,075.17 | 4,530.74 |
| IOWA | 615.00 | 598.90 | 577.40 | 565.87 | 2,357.16 |
| KANSAS | 715.84 | 696.92 | 661.01 | 628.16 | 2,701.93 |
| KENTUCKY | 726.03 | 691.27 | 655.97 | 631.69 | 2,704.96 |
| LOUISIANA | 766.54 | 716.30 | 707.91 | 671.91 | 2,862.66 |
| MAINE | 235.14 | 237.74 | 246.68 | 230.30 | 949.85 |
| MARYLAND | 816.84 | 759.18 | 741.28 | 688.74 | 3,006.05 |
| MASSACHUSETTS | 919.63 | 884.96 | 848.75 | 846.38 | 3,499.72 |
| MICHIGAN | 1,864.11 | 1,775.82 | 1,747.16 | 1,642.86 | 7,029.96 |
| MINNESOTA | 701.21 | 639.08 | 639.46 | 614.14 | 2,593.89 |
| MISSISSIPPI | 847.35 | 802.70 | 856.99 | 825.48 | 3,332.52 |
| MISSOURI | 1,368.95 | 1,329.44 | 1,259.52 | 1,223.10 | 5,181.01 |
| MONTANA | 224.75 | 213.58 | 216.36 | 206.62 | 861.32 |
| NEBRASKA | 369.05 | 366.68 | 348.05 | 341.00 | 1,424.79 |
| NEVADA | 354.61 | 327.08 | 307.64 | 291.84 | 1,281.17 |
| NEW HAMPSHIRE | 230.03 | 214.73 | 221.28 | 213.68 | 879.73 |
| NEW JERSEY | 1,459.17 | 1,359.68 | 1,345.44 | 1,279.52 | 5,443.80 |
| NEW MEXICO | 282.04 | 267.12 | 261.01 | 247.09 | 1,057.26 |
| NEW YORK | 2,797.38 | 2,546.22 | 2,561.17 | 2,430.71 | 10,335.48 |
| NORTH CAROLINA | 1,951.39 | 1,830.89 | 1,780.13 | 1,639.72 | 7,202.12 |
| NORTH DAKOTA | 164.67 | 150.10 | 146.10 | 146.69 | 607.57 |
| OHIO | 1,423.15 | 1,388.98 | 1,322.48 | 1,267.39 | 5,402.00 |
| OKLAHOMA | 757.84 | 724.05 | 693.42 | 657.84 | 2,833.15 |
| OREGON | 583.22 | 552.85 | 532.42 | 523.14 | 2,191.63 |
| PENNSYLVANIA | 3,155.19 | 2,962.49 | 2,786.12 | 3,018.48 | 11,922.28 |
| PUERTO RICO | 164.72 | 174.65 | 139.62 | 173.47 | 652.46 |

| | | | | | |
|---|---|---|---|---|---|
| RHODE ISLAND | 117.89 | 111.37 | 110.37 | 109.62 | 449.24 |
| SOUTH CAROLINA | 870.13 | 799.71 | 775.69 | 734.59 | 3,180.12 |
| SOUTH DAKOTA | 192.11 | 174.83 | 175.41 | 164.99 | 707.34 |
| TENNESSEE | 1,297.64 | 1,240.55 | 1,209.23 | 1,562.13 | 5,309.56 |
| TEXAS | 3,544.43 | 3,438.13 | 3,250.61 | 3,187.39 | 13,420.57 |
| UTAH | 591.12 | 561.64 | 568.64 | 543.61 | 2,265.01 |
| VERMONT | 119.22 | 108.92 | 115.99 | 110.66 | 454.79 |
| VIRGIN ISLANDS | 3.11 | 4.27 | 2.46 | 2.59 | 12.43 |
| VIRGINIA | 1,138.16 | 1,094.65 | 1,067.88 | 1,017.08 | 4,317.77 |
| WASHINGTON | 1,016.45 | 977.97 | 930.54 | 889.86 | 3,814.82 |
| WEST VIRGINIA | 355.17 | 323.47 | 323.19 | 306.92 | 1,308.75 |
| WISCONSIN | 901.99 | 894.16 | 842.53 | 805.86 | 3,444.54 |
| WYOMING | 98.18 | 93.21 | 94.20 | 90.56 | 376.15 |
| **9801 - FENTANYL BASE Total** | **51,587.79** | **48,743.30** | **47,182.03** | **46,017.90** | **193,531.02** |

**DRUG: 1100 - AMPHETAMINE**

| STATE NAME | 1ST QUARTER | 2ND QUARTER | 3RD QUARTER | 4TH QUARTER | TOTAL GRAMS |
|---|---|---|---|---|---|
| ALABAMA | 2,836.09 | 2,889.61 | 2,879.64 | 2,881.96 | 11,487.30 |
| ALASKA | 1,183.98 | 1,221.50 | 1,232.77 | 1,232.08 | 4,870.33 |
| AMERICAN SAMOA | 5.72 | 0.00 | 0.00 | 0.00 | 5.72 |
| ARIZONA | 1,450.95 | 1,487.86 | 1,499.99 | 1,583.07 | 6,021.87 |
| ARKANSAS | 1,879.49 | 1,866.53 | 1,887.99 | 1,939.16 | 7,573.18 |
| CALIFORNIA | 828.53 | 846.50 | 857.70 | 866.26 | 3,398.99 |
| COLORADO | 1,780.97 | 1,859.65 | 1,901.32 | 1,988.13 | 7,530.07 |
| CONNECTICUT | 1,586.26 | 1,683.17 | 1,667.23 | 1,776.36 | 6,713.01 |
| DELAWARE | 2,360.00 | 2,416.94 | 2,444.95 | 2,475.71 | 9,697.60 |
| DISTRICT OF COLUMBIA | 2,071.58 | 2,169.19 | 2,196.08 | 2,155.83 | 8,592.68 |
| FLORIDA | 1,767.03 | 1,827.26 | 1,739.56 | 1,915.55 | 7,249.39 |
| GEORGIA | 2,067.75 | 2,120.19 | 2,141.45 | 2,183.23 | 8,512.63 |
| GUAM | 236.98 | 242.22 | 253.34 | 228.85 | 961.39 |
| HAWAII | 776.77 | 786.85 | 799.76 | 802.36 | 3,165.73 |
| IDAHO | 1,885.25 | 1,922.97 | 1,974.92 | 1,995.61 | 7,778.75 |
| ILLINOIS | 1,573.13 | 1,635.93 | 1,679.88 | 1,697.54 | 6,586.48 |
| INDIANA | 2,123.83 | 2,187.19 | 2,326.83 | 2,222.66 | 8,860.52 |
| IOWA | 1,769.45 | 1,780.05 | 1,801.95 | 1,844.00 | 7,195.45 |
| KANSAS | 2,275.40 | 2,325.36 | 2,367.68 | 2,427.09 | 9,395.53 |
| KENTUCKY | 1,702.80 | 1,681.15 | 1,755.01 | 1,774.97 | 6,913.94 |
| LOUISIANA | 3,819.65 | 3,800.87 | 3,797.18 | 3,958.29 | 15,376.00 |
| MAINE | 1,866.63 | 1,984.40 | 2,036.91 | 2,017.15 | 7,905.10 |
| MARYLAND | 1,706.44 | 1,761.19 | 1,765.51 | 1,819.27 | 7,052.41 |
| MASSACHUSETTS | 2,754.36 | 2,956.72 | 2,884.89 | 3,005.33 | 11,601.30 |
| MICHIGAN | 2,492.08 | 2,519.06 | 2,716.36 | 2,502.45 | 10,229.96 |
| MINNESOTA | 2,124.41 | 2,182.03 | 2,193.10 | 2,275.07 | 8,774.60 |
| MISSISSIPPI | 2,144.41 | 2,081.52 | 2,125.85 | 2,183.05 | 8,534.82 |
| MISSOURI | 2,544.73 | 2,552.46 | 2,508.64 | 2,561.50 | 10,167.32 |
| MONTANA | 1,577.48 | 1,594.39 | 1,660.34 | 1,658.50 | 6,490.70 |
| NEBRASKA | 1,563.14 | 1,595.07 | 1,639.10 | 1,677.54 | 6,474.84 |
| NEVADA | 1,035.56 | 1,033.91 | 1,051.13 | 1,114.31 | 4,234.91 |
| NEW HAMPSHIRE | 3,125.48 | 3,285.21 | 3,322.60 | 3,379.66 | 13,112.95 |
| NEW JERSEY | 1,290.46 | 1,298.64 | 1,340.89 | 1,391.20 | 5,321.18 |
| NEW MEXICO | 775.96 | 772.82 | 773.99 | 803.62 | 3,126.39 |
| NEW YORK | 1,334.25 | 1,342.40 | 1,370.37 | 1,433.45 | 5,480.47 |
| NORTH CAROLINA | 2,395.25 | 2,467.13 | 2,484.46 | 2,442.30 | 9,789.15 |
| NORTH DAKOTA | 1,469.87 | 1,427.65 | 1,493.24 | 1,519.30 | 5,910.06 |
| OHIO | 1,798.21 | 1,820.22 | 1,903.32 | 1,920.90 | 7,442.64 |

| | | | | | |
|---|---|---|---|---|---|
| OKLAHOMA | 1,477.57 | 1,491.52 | 1,522.80 | 1,538.43 | 6,030.31 |
| OREGON | 1,684.84 | 1,753.21 | 1,769.42 | 1,857.19 | 7,064.66 |
| PENNSYLVANIA | 1,722.48 | 1,728.30 | 1,766.58 | 1,830.31 | 7,047.67 |
| PUERTO RICO | 251.79 | 252.44 | 193.06 | 266.47 | 963.76 |
| RHODE ISLAND | 3,534.37 | 3,826.21 | 3,756.91 | 3,892.05 | 15,009.54 |
| SOUTH CAROLINA | 3,054.64 | 3,200.09 | 3,170.71 | 3,221.29 | 12,646.73 |
| SOUTH DAKOTA | 1,782.97 | 1,858.74 | 1,810.56 | 1,883.57 | 7,335.83 |
| TENNESSEE | 2,148.62 | 2,208.74 | 2,225.71 | 2,261.47 | 8,844.53 |
| TEXAS | 1,847.18 | 1,880.28 | 1,907.47 | 1,996.04 | 7,630.97 |
| UTAH | 3,324.85 | 3,419.37 | 3,515.95 | 3,659.64 | 13,919.81 |
| VERMONT | 1,613.15 | 1,701.35 | 1,721.18 | 1,720.88 | 6,756.55 |
| VIRGIN ISLANDS | 305.88 | 388.24 | 288.99 | 346.64 | 1,329.75 |
| VIRGINIA | 2,283.52 | 2,408.84 | 2,401.60 | 2,417.98 | 9,511.94 |
| WASHINGTON | 1,749.03 | 1,831.32 | 1,820.77 | 1,892.17 | 7,293.30 |
| WEST VIRGINIA | 1,307.76 | 1,272.45 | 1,300.05 | 1,313.75 | 5,194.01 |
| WISCONSIN | 2,198.78 | 2,238.86 | 2,255.79 | 2,252.94 | 8,946.36 |
| WYOMING | 1,239.56 | 1,272.29 | 1,321.67 | 1,318.99 | 5,152.51 |
| **1100 - AMPHETAMINE Total** | **1,785.40** | **1,827.86** | **1,848.51** | **1,891.27** | **7,353.04** |

**DRUG: 1105 - METHAMPHETAMINE**

| STATE NAME | 1ST QUARTER | 2ND QUARTER | 3RD QUARTER | 4TH QUARTER | TOTAL GRAMS |
|---|---|---|---|---|---|
| ALABAMA | 0.60 | 0.68 | 0.65 | 0.64 | 2.56 |
| ALASKA | 0.06 | 0.00 | 0.28 | 0.45 | 0.79 |
| ARIZONA | 0.41 | 0.25 | 0.34 | 0.29 | 1.29 |
| ARKANSAS | 0.12 | 0.10 | 0.06 | 0.03 | 0.30 |
| CALIFORNIA | 0.65 | 0.64 | 0.61 | 0.68 | 2.57 |
| COLORADO | 0.99 | 0.93 | 1.03 | 0.87 | 3.82 |
| CONNECTICUT | 0.70 | 0.64 | 0.56 | 0.58 | 2.48 |
| DELAWARE | 0.49 | 0.31 | 0.58 | 0.45 | 1.83 |
| DISTRICT OF COLUMBIA | 0.80 | 0.73 | 0.80 | 1.00 | 3.34 |
| FLORIDA | 0.30 | 0.25 | 0.29 | 0.29 | 1.13 |
| GEORGIA | 0.25 | 0.25 | 0.20 | 0.29 | 1.00 |
| HAWAII | 0.41 | 0.32 | 0.27 | 0.35 | 1.36 |
| IDAHO | 0.72 | 0.74 | 0.87 | 1.03 | 3.36 |
| ILLINOIS | 0.49 | 0.44 | 0.42 | 0.43 | 1.78 |
| INDIANA | 0.15 | 0.29 | 0.18 | 0.46 | 1.09 |
| IOWA | 0.45 | 0.44 | 0.41 | 0.55 | 1.85 |
| KANSAS | 0.46 | 0.46 | 0.45 | 0.48 | 1.86 |
| KENTUCKY | 0.29 | 0.25 | 0.27 | 0.11 | 0.92 |
| LOUISIANA | 0.45 | 0.28 | 0.20 | 0.24 | 1.18 |
| MAINE | 1.00 | 1.03 | 0.67 | 0.76 | 3.45 |
| MARYLAND | 0.56 | 0.49 | 0.72 | 0.54 | 2.30 |

| | | | | | |
|---|---|---|---|---|---|
| MASSACHUSETTS | 0.38 | 0.37 | 0.34 | 0.41 | 1.50 |
| MICHIGAN | 0.22 | 0.24 | 0.24 | 0.17 | 0.87 |
| MINNESOTA | 0.87 | 0.98 | 1.04 | 1.15 | 4.05 |
| MISSISSIPPI | 0.27 | 0.24 | 0.26 | 0.22 | 0.99 |
| MISSOURI | 1.03 | 0.78 | 0.87 | 0.72 | 3.41 |
| NEBRASKA | 0.29 | 0.33 | 0.33 | 0.31 | 1.25 |
| NEVADA | 0.42 | 0.28 | 0.40 | 0.37 | 1.47 |
| NEW HAMPSHIRE | 0.21 | 0.67 | 0.49 | 0.46 | 1.83 |
| NEW JERSEY | 0.38 | 0.35 | 0.37 | 0.50 | 1.60 |
| NEW MEXICO | 0.27 | 0.23 | 0.37 | 0.51 | 1.39 |
| NEW YORK | 0.43 | 0.40 | 0.48 | 0.41 | 1.72 |
| NORTH CAROLINA | 0.48 | 0.49 | 0.34 | 0.53 | 1.84 |
| NORTH DAKOTA | 1.08 | 1.02 | 1.97 | 0.78 | 4.84 |
| OHIO | 0.39 | 0.28 | 0.29 | 0.27 | 1.23 |
| OKLAHOMA | 0.22 | 0.29 | 0.22 | 0.24 | 0.97 |
| OREGON | 1.01 | 0.91 | 0.96 | 1.33 | 4.22 |
| PENNSYLVANIA | 0.67 | 0.82 | 0.75 | 0.86 | 3.10 |
| RHODE ISLAND | 0.08 | 0.11 | 0.31 | 0.27 | 0.76 |
| SOUTH CAROLINA | 0.27 | 0.26 | 0.25 | 0.30 | 1.08 |
| SOUTH DAKOTA | 0.64 | 0.59 | 0.84 | 0.79 | 2.86 |
| TENNESSEE | 0.16 | 0.23 | 0.27 | 0.22 | 0.87 |
| TEXAS | 0.33 | 0.35 | 0.33 | 0.38 | 1.39 |
| UTAH | 0.94 | 0.78 | 1.12 | 0.92 | 3.76 |
| VERMONT | 0.26 | 0.13 | 0.39 | 0.39 | 1.16 |
| VIRGINIA | 0.41 | 0.41 | 0.38 | 0.37 | 1.58 |
| WASHINGTON | 1.25 | 1.02 | 1.34 | 1.43 | 5.04 |
| WEST VIRGINIA | 0.26 | 0.17 | 0.17 | 0.15 | 0.76 |
| WISCONSIN | 0.47 | 0.45 | 0.46 | 0.47 | 1.86 |
| WYOMING | 0.78 | 0.86 | 0.78 | 1.07 | 3.49 |
| **1105 - METHAMPHETAMINE Total** | **0.48** | **0.45** | **0.47** | **0.49** | **1.89** |

**DRUG: 1205 - LISDEXAMFETAMINE**

| STATE NAME | 1ST QUARTER | 2ND QUARTER | 3RD QUARTER | 4TH QUARTER | TOTAL GRAMS |
|---|---|---|---|---|---|
| ALABAMA | 1,520.68 | 1,458.43 | 1,480.10 | 1,536.15 | 5,995.36 |
| ALASKA | 369.21 | 411.78 | 412.03 | 453.13 | 1,646.15 |
| ARIZONA | 608.44 | 614.63 | 623.26 | 663.38 | 2,509.70 |
| ARKANSAS | 1,019.54 | 970.45 | 973.62 | 1,029.35 | 3,992.95 |
| CALIFORNIA | 304.56 | 310.57 | 307.26 | 316.26 | 1,238.64 |
| COLORADO | 578.62 | 584.99 | 601.07 | 622.67 | 2,387.34 |
| CONNECTICUT | 1,001.30 | 1,022.08 | 986.04 | 1,054.90 | 4,064.32 |
| DELAWARE | 1,527.31 | 1,491.00 | 1,489.39 | 1,557.12 | 6,064.83 |
| DISTRICT OF COLUMBIA | 976.59 | 1,020.40 | 1,068.34 | 1,076.41 | 4,141.75 |

| | | | | | |
|---|---|---|---|---|---|
| FLORIDA | 734.57 | 716.28 | 693.24 | 744.92 | 2,889.01 |
| GEORGIA | 1,010.30 | 969.52 | 997.69 | 1,036.79 | 4,014.30 |
| GUAM | 6.48 | 14.26 | 0.32 | 10.69 | 31.75 |
| HAWAII | 108.75 | 120.06 | 126.35 | 141.35 | 496.50 |
| IDAHO | 619.85 | 649.72 | 636.19 | 675.47 | 2,581.23 |
| ILLINOIS | 558.45 | 567.32 | 575.59 | 599.71 | 2,301.07 |
| INDIANA | 1,253.38 | 1,232.26 | 1,252.28 | 1,291.11 | 5,029.03 |
| IOWA | 1,094.33 | 1,092.59 | 1,089.65 | 1,145.63 | 4,422.20 |
| KANSAS | 1,097.13 | 1,146.28 | 1,150.58 | 1,193.27 | 4,587.27 |
| KENTUCKY | 940.22 | 906.64 | 896.72 | 922.48 | 3,666.06 |
| LOUISIANA | 1,986.37 | 1,846.98 | 1,916.77 | 2,021.86 | 7,771.97 |
| MAINE | 1,240.92 | 1,281.40 | 1,279.48 | 1,306.46 | 5,108.27 |
| MARYLAND | 985.67 | 1,000.83 | 975.25 | 1,038.33 | 4,000.07 |
| MASSACHUSETTS | 586.91 | 607.24 | 607.25 | 648.65 | 2,450.05 |
| MICHIGAN | 618.99 | 635.26 | 620.68 | 665.66 | 2,540.60 |
| MINNESOTA | 586.28 | 598.75 | 604.96 | 663.65 | 2,453.65 |
| MISSISSIPPI | 1,261.27 | 1,173.64 | 1,203.84 | 1,275.88 | 4,914.62 |
| MISSOURI | 1,221.12 | 1,163.05 | 1,121.69 | 1,160.89 | 4,666.74 |
| MONTANA | 660.53 | 662.23 | 661.76 | 689.69 | 2,674.21 |
| NEBRASKA | 938.12 | 959.28 | 953.86 | 1,036.81 | 3,888.08 |
| NEVADA | 303.08 | 294.73 | 282.21 | 308.60 | 1,188.61 |
| NEW HAMPSHIRE | 1,090.44 | 1,094.31 | 1,092.82 | 1,142.70 | 4,420.27 |
| NEW JERSEY | 544.38 | 543.76 | 535.85 | 562.48 | 2,186.48 |
| NEW MEXICO | 186.97 | 184.59 | 177.99 | 188.24 | 737.79 |
| NEW YORK | 451.27 | 452.34 | 458.77 | 485.74 | 1,848.11 |
| NORTH CAROLINA | 1,196.95 | 1,166.81 | 1,171.23 | 1,191.51 | 4,726.51 |
| NORTH DAKOTA | 914.35 | 887.53 | 901.54 | 959.22 | 3,662.64 |
| OHIO | 819.64 | 807.52 | 806.44 | 846.69 | 3,280.29 |
| OKLAHOMA | 1,001.69 | 978.26 | 991.28 | 1,051.55 | 4,022.76 |
| OREGON | 397.30 | 416.02 | 412.28 | 442.02 | 1,667.63 |
| PENNSYLVANIA | 713.43 | 724.61 | 727.98 | 776.54 | 2,942.56 |
| PUERTO RICO | 62.61 | 55.16 | 53.44 | 62.70 | 233.91 |
| RHODE ISLAND | 712.02 | 710.26 | 712.79 | 781.39 | 2,916.46 |
| SOUTH CAROLINA | 1,361.82 | 1,329.04 | 1,296.80 | 1,395.02 | 5,382.69 |
| SOUTH DAKOTA | 1,183.49 | 1,165.39 | 1,195.85 | 1,231.35 | 4,776.08 |
| TENNESSEE | 1,056.48 | 1,019.65 | 1,046.67 | 1,085.82 | 4,208.63 |
| TEXAS | 1,051.51 | 1,023.65 | 1,006.61 | 1,070.65 | 4,152.42 |
| UTAH | 807.41 | 823.76 | 836.58 | 883.38 | 3,351.12 |
| VERMONT | 1,152.15 | 1,151.23 | 1,204.17 | 1,189.01 | 4,696.56 |
| VIRGIN ISLANDS | 75.80 | 60.53 | 61.59 | 97.38 | 295.30 |
| VIRGINIA | 856.23 | 871.36 | 855.49 | 903.36 | 3,486.44 |

| | | | | | |
|---|---|---|---|---|---|
| WASHINGTON | 528.85 | 558.60 | 557.92 | 589.51 | 2,234.88 |
| WEST VIRGINIA | 1,021.90 | 999.84 | 972.85 | 1,044.51 | 4,039.10 |
| WISCONSIN | 1,301.57 | 1,338.76 | 1,322.40 | 1,390.67 | 5,353.39 |
| WYOMING | 468.22 | 466.48 | 498.28 | 492.02 | 1,925.00 |
| **1205 LISDEXAMFETAMINE Total** | **779.65** | **772.05** | **770.38** | **810.84** | **3,132.92** |

**DRUG: 1724 - METHYLPHENIDATE (DL;D;L;ISOMERS)**

| STATE NAME | 1ST QUARTER | 2ND QUARTER | 3RD QUARTER | 4TH QUARTER | TOTAL GRAMS |
|---|---|---|---|---|---|
| ALABAMA | 1,552.57 | 1,435.84 | 1,500.08 | 1,595.66 | 6,084.15 |
| ALASKA | 1,103.07 | 1,120.42 | 1,130.25 | 1,214.76 | 4,568.51 |
| AMERICAN SAMOA | 13.18 | 22.40 | 3.95 | 13.83 | 53.36 |
| ARIZONA | 1,188.65 | 1,215.46 | 1,149.60 | 1,239.08 | 4,792.79 |
| ARKANSAS | 1,376.62 | 1,258.59 | 1,288.19 | 1,356.33 | 5,279.73 |
| CALIFORNIA | 702.45 | 703.60 | 675.28 | 730.78 | 2,812.11 |
| COLORADO | 1,237.15 | 1,228.45 | 1,226.16 | 1,348.52 | 5,040.27 |
| CONNECTICUT | 1,534.23 | 1,527.11 | 1,423.35 | 1,578.09 | 6,062.78 |
| DELAWARE | 2,010.81 | 1,930.38 | 1,837.82 | 2,035.75 | 7,814.76 |
| DISTRICT OF COLUMBIA | 1,385.69 | 1,375.36 | 1,371.65 | 1,404.50 | 5,537.19 |
| FLORIDA | 1,049.56 | 1,022.61 | 970.36 | 1,093.02 | 4,135.56 |
| GEORGIA | 1,723.98 | 1,540.07 | 1,600.94 | 1,726.47 | 6,591.47 |
| GUAM | 192.45 | 254.26 | 227.36 | 179.25 | 853.33 |
| HAWAII | 774.60 | 791.54 | 774.25 | 803.34 | 3,143.74 |
| IDAHO | 1,811.85 | 1,787.96 | 1,707.68 | 1,899.36 | 7,206.85 |
| ILLINOIS | 1,331.92 | 1,278.36 | 1,256.78 | 1,345.08 | 5,212.15 |
| INDIANA | 1,792.07 | 1,699.33 | 1,748.50 | 1,834.85 | 7,074.75 |
| IOWA | 2,588.75 | 2,554.00 | 2,491.86 | 2,729.56 | 10,364.17 |
| KANSAS | 1,719.16 | 1,678.32 | 1,668.64 | 1,909.59 | 6,975.71 |
| KENTUCKY | 1,734.33 | 1,604.69 | 1,653.21 | 1,778.44 | 6,770.67 |
| LOUISIANA | 1,892.83 | 1,640.93 | 1,703.43 | 1,881.03 | 7,118.22 |
| MAINE | 2,618.85 | 2,619.42 | 2,665.81 | 2,671.52 | 10,575.60 |
| MARYLAND | 1,744.18 | 1,733.05 | 1,611.23 | 1,797.01 | 6,885.47 |
| MASSACHUSETTS | 2,197.49 | 2,208.83 | 2,028.45 | 2,272.88 | 8,707.65 |
| MICHIGAN | 1,772.40 | 1,732.80 | 1,753.27 | 1,738.80 | 6,997.26 |
| MINNESOTA | 1,919.77 | 1,930.69 | 1,802.14 | 1,991.00 | 7,643.61 |
| MISSISSIPPI | 1,486.47 | 1,315.68 | 1,379.21 | 1,569.14 | 5,750.50 |
| MISSOURI | 2,375.83 | 2,308.96 | 2,212.10 | 2,448.84 | 9,345.73 |
| MONTANA | 1,716.03 | 1,732.61 | 1,655.18 | 1,778.94 | 6,882.75 |
| NEBRASKA | 1,713.80 | 1,649.01 | 1,652.68 | 1,784.67 | 6,800.16 |
| NEVADA | 551.90 | 550.56 | 528.06 | 577.84 | 2,208.35 |
| NEW HAMPSHIRE | 2,590.34 | 2,600.64 | 2,516.59 | 2,674.89 | 10,382.46 |
| NEW JERSEY | 1,055.03 | 1,017.94 | 946.90 | 1,077.09 | 4,096.97 |
| NEW MEXICO | 881.70 | 873.36 | 825.13 | 901.87 | 3,482.06 |

| | | | | | |
|---|---|---|---|---|---|
| NEW YORK | 1,179.56 | 1,160.95 | 1,089.90 | 1,212.08 | 4,642.48 |
| NORTH CAROLINA | 2,071.99 | 1,970.98 | 1,880.57 | 2,034.80 | 7,958.34 |
| NORTH DAKOTA | 2,278.26 | 2,206.11 | 2,162.63 | 2,339.23 | 8,986.24 |
| OHIO | 1,740.98 | 1,638.27 | 1,637.19 | 1,791.96 | 6,808.40 |
| OKLAHOMA | 1,265.37 | 1,180.94 | 1,196.90 | 1,299.47 | 4,942.68 |
| OREGON | 1,652.42 | 1,661.82 | 1,566.30 | 1,752.15 | 6,632.68 |
| PENNSYLVANIA | 1,733.69 | 1,713.81 | 1,659.80 | 1,808.98 | 6,916.28 |
| PUERTO RICO | 416.41 | 355.18 | 266.34 | 394.96 | 1,432.90 |
| RHODE ISLAND | 1,805.21 | 1,763.65 | 1,607.28 | 1,845.75 | 7,021.90 |
| SOUTH CAROLINA | 1,975.51 | 1,849.75 | 1,764.30 | 1,985.78 | 7,575.34 |
| SOUTH DAKOTA | 2,432.16 | 2,355.31 | 2,347.48 | 2,542.13 | 9,677.09 |
| TENNESSEE | 1,319.17 | 1,235.78 | 1,266.47 | 1,362.98 | 5,184.40 |
| TEXAS | 1,340.93 | 1,241.44 | 1,216.62 | 1,385.74 | 5,184.73 |
| UTAH | 1,702.80 | 1,699.23 | 1,677.08 | 1,895.10 | 6,974.21 |
| VERMONT | 2,941.98 | 3,002.57 | 2,968.68 | 2,983.98 | 11,897.21 |
| VIRGIN ISLANDS | 86.68 | 141.32 | 76.72 | 122.92 | 427.64 |
| VIRGINIA | 1,690.77 | 1,644.47 | 1,583.25 | 1,716.92 | 6,635.41 |
| WASHINGTON | 1,674.35 | 1,745.00 | 1,618.32 | 1,790.11 | 6,827.78 |
| WEST VIRGINIA | 1,625.10 | 1,521.50 | 1,502.59 | 1,643.64 | 6,292.83 |
| WISCONSIN | 1,816.20 | 1,792.34 | 1,737.81 | 1,883.71 | 7,230.06 |
| WYOMING | 1,382.28 | 1,367.83 | 1,294.98 | 1,389.02 | 5,434.10 |
| **1724 - METHYLPHENIDATE (DL;D;L;ISOMERS) Total** | **1,447.62** | **1,398.93** | **1,361.06** | **1,489.10** | **5,696.71** |

**DRUG: 9050 - CODEINE**

| STATE NAME | 1ST QUARTER | 2ND QUARTER | 3RD QUARTER | 4TH QUARTER | TOTAL GRAMS |
|---|---|---|---|---|---|
| ALABAMA | 952.88 | 956.18 | 894.14 | 910.44 | 3,713.63 |
| ALASKA | 715.54 | 679.46 | 745.55 | 678.52 | 2,819.06 |
| AMERICAN SAMOA | 417.58 | 447.89 | 370.44 | 592.70 | 1,828.62 |
| ARIZONA | 794.13 | 772.38 | 700.66 | 714.38 | 2,981.55 |
| ARKANSAS | 1,822.00 | 1,716.78 | 1,720.51 | 1,657.98 | 6,917.28 |
| CALIFORNIA | 933.15 | 882.28 | 873.32 | 865.61 | 3,554.36 |
| COLORADO | 336.05 | 336.17 | 329.08 | 336.30 | 1,337.60 |
| CONNECTICUT | 447.90 | 519.73 | 455.03 | 457.03 | 1,879.69 |
| DELAWARE | 766.44 | 775.83 | 764.97 | 720.56 | 3,027.80 |
| DISTRICT OF COLUMBIA | 1,384.61 | 1,466.52 | 1,316.77 | 1,331.55 | 5,499.45 |
| FLORIDA | 695.98 | 680.68 | 668.85 | 675.71 | 2,721.22 |
| GEORGIA | 807.07 | 840.44 | 812.71 | 799.67 | 3,259.90 |
| GUAM | 345.58 | 448.05 | 474.44 | 334.43 | 1,602.51 |
| HAWAII | 782.84 | 714.76 | 714.81 | 735.92 | 2,948.33 |
| IDAHO | 691.82 | 716.36 | 715.68 | 740.60 | 2,864.45 |
| ILLINOIS | 977.39 | 980.73 | 976.68 | 961.33 | 3,896.13 |
| INDIANA | 601.16 | 589.19 | 563.76 | 526.90 | 2,281.00 |

| | | | | | |
|---|---|---|---|---|---|
| IOWA | 465.40 | 469.45 | 461.06 | 439.93 | 1,835.84 |
| KANSAS | 665.69 | 671.30 | 719.37 | 716.54 | 2,772.89 |
| KENTUCKY | 855.99 | 840.27 | 802.37 | 813.41 | 3,312.04 |
| LOUISIANA | 762.31 | 757.25 | 755.28 | 719.62 | 2,994.46 |
| MAINE | 558.19 | 576.07 | 593.06 | 569.50 | 2,296.80 |
| MARYLAND | 743.76 | 770.61 | 769.24 | 753.21 | 3,036.82 |
| MASSACHUSETTS | 507.73 | 597.88 | 520.84 | 537.83 | 2,164.28 |
| MICHIGAN | 1,702.72 | 1,710.45 | 1,777.31 | 1,641.43 | 6,831.90 |
| MINNESOTA | 498.59 | 492.63 | 485.35 | 480.50 | 1,957.07 |
| MISSISSIPPI | 686.44 | 657.48 | 614.92 | 587.06 | 2,545.90 |
| MISSOURI | 926.75 | 944.18 | 948.17 | 958.31 | 3,777.41 |
| MONTANA | 771.08 | 759.99 | 806.90 | 721.74 | 3,059.71 |
| NEBRASKA | 466.42 | 472.18 | 477.06 | 432.91 | 1,848.58 |
| NEVADA | 608.32 | 612.11 | 610.65 | 619.95 | 2,451.02 |
| NEW HAMPSHIRE | 464.83 | 482.12 | 414.03 | 452.69 | 1,813.67 |
| NEW JERSEY | 680.73 | 673.76 | 673.22 | 669.37 | 2,697.08 |
| NEW MEXICO | 726.19 | 709.87 | 669.88 | 698.64 | 2,804.58 |
| NEW YORK | 552.16 | 532.94 | 541.12 | 543.85 | 2,170.07 |
| NORTH CAROLINA | 385.48 | 408.41 | 394.27 | 370.82 | 1,558.98 |
| NORTH DAKOTA | 601.85 | 625.37 | 578.60 | 555.76 | 2,361.57 |
| OHIO | 531.39 | 522.99 | 523.04 | 524.18 | 2,101.60 |
| OKLAHOMA | 1,912.83 | 1,964.69 | 1,906.14 | 1,896.11 | 7,679.77 |
| OREGON | 683.86 | 696.02 | 681.35 | 641.21 | 2,702.44 |
| PENNSYLVANIA | 740.12 | 716.75 | 709.89 | 719.99 | 2,886.77 |
| PUERTO RICO | 267.53 | 256.85 | 220.97 | 253.80 | 999.15 |
| RHODE ISLAND | 425.37 | 456.28 | 422.84 | 437.14 | 1,741.62 |
| SOUTH CAROLINA | 1,268.63 | 1,297.67 | 1,272.31 | 1,195.23 | 5,033.84 |
| SOUTH DAKOTA | 588.04 | 576.36 | 615.47 | 595.43 | 2,375.30 |
| TENNESSEE | 911.87 | 820.78 | 894.07 | 817.11 | 3,443.84 |
| TEXAS | 3,273.73 | 3,377.94 | 3,340.35 | 3,224.58 | 13,216.59 |
| UTAH | 456.49 | 476.60 | 452.50 | 449.57 | 1,835.16 |
| VERMONT | 400.50 | 375.82 | 360.86 | 364.65 | 1,501.83 |
| VIRGIN ISLANDS | 528.32 | 513.22 | 644.10 | 484.98 | 2,170.62 |
| VIRGINIA | 492.41 | 528.78 | 498.55 | 498.70 | 2,018.43 |
| WASHINGTON | 673.45 | 680.24 | 663.22 | 646.18 | 2,663.09 |
| WEST VIRGINIA | 1,942.69 | 1,963.19 | 2,023.79 | 1,857.69 | 7,787.35 |
| WISCONSIN | 646.26 | 636.84 | 651.09 | 623.69 | 2,557.89 |
| WYOMING | 544.44 | 599.19 | 591.51 | 539.79 | 2,274.94 |
| **9050 - CODEINE Total** | **976.56** | **979.43** | **967.93** | **945.97** | **3,869.89** |

**DRUG: 9064 - BUPRENORPHINE**

| STATE NAME | 1ST QUARTER | 2ND QUARTER | 3RD QUARTER | 4TH QUARTER | TOTAL GRAMS |
|---|---|---|---|---|---|

| | | | | | |
|---|---|---|---|---|---|
| ALABAMA | 653.63 | 689.70 | 699.59 | 707.04 | 2,749.95 |
| ALASKA | 374.06 | 365.22 | 368.42 | 368.90 | 1,476.61 |
| ARIZONA | 205.95 | 214.04 | 226.46 | 235.63 | 882.07 |
| ARKANSAS | 279.28 | 297.98 | 310.26 | 322.70 | 1,210.22 |
| CALIFORNIA | 128.26 | 133.80 | 142.51 | 148.59 | 553.15 |
| COLORADO | 197.92 | 208.00 | 212.50 | 229.81 | 848.24 |
| CONNECTICUT | 501.24 | 490.97 | 525.93 | 527.60 | 2,045.74 |
| DELAWARE | 481.59 | 464.04 | 488.58 | 500.94 | 1,935.15 |
| DISTRICT OF COLUMBIA | 561.00 | 595.52 | 599.59 | 560.71 | 2,316.82 |
| FLORIDA | 297.05 | 308.28 | 322.75 | 321.41 | 1,249.49 |
| GEORGIA | 197.59 | 201.20 | 208.27 | 209.86 | 816.92 |
| GUAM | 1.49 | 2.43 | 7.00 | 0.47 | 11.38 |
| HAWAII | 160.06 | 163.18 | 166.51 | 163.04 | 652.79 |
| IDAHO | 294.70 | 321.54 | 320.32 | 334.33 | 1,270.90 |
| ILLINOIS | 144.33 | 152.66 | 155.70 | 158.52 | 611.20 |
| INDIANA | 430.49 | 449.35 | 483.78 | 503.49 | 1,867.11 |
| IOWA | 77.73 | 82.57 | 89.23 | 84.65 | 334.19 |
| KANSAS | 106.06 | 109.94 | 113.64 | 116.32 | 445.96 |
| KENTUCKY | 975.41 | 1,018.19 | 1,055.68 | 1,090.73 | 4,140.01 |
| LOUISIANA | 496.03 | 523.40 | 531.88 | 539.75 | 2,091.05 |
| MAINE | 943.92 | 1,006.18 | 1,058.38 | 1,084.00 | 4,092.48 |
| MARYLAND | 555.31 | 562.05 | 593.67 | 608.79 | 2,319.83 |
| MASSACHUSETTS | 717.85 | 708.51 | 741.79 | 767.12 | 2,935.27 |
| MICHIGAN | 332.40 | 331.40 | 343.34 | 355.66 | 1,362.81 |
| MINNESOTA | 193.34 | 203.06 | 207.86 | 218.74 | 823.00 |
| MISSISSIPPI | 379.03 | 391.27 | 402.18 | 409.90 | 1,582.38 |
| MISSOURI | 268.01 | 291.77 | 311.39 | 309.94 | 1,181.10 |
| MONTANA | 347.08 | 380.79 | 403.01 | 416.68 | 1,547.55 |
| NEBRASKA | 93.44 | 92.56 | 107.00 | 111.61 | 404.60 |
| NEVADA | 160.32 | 160.98 | 166.82 | 173.13 | 661.24 |
| NEW HAMPSHIRE | 727.20 | 753.10 | 787.31 | 804.77 | 3,072.38 |
| NEW JERSEY | 332.42 | 336.02 | 359.59 | 369.91 | 1,397.94 |
| NEW MEXICO | 444.43 | 456.05 | 451.48 | 467.23 | 1,819.18 |
| NEW YORK | 354.04 | 346.56 | 370.97 | 390.27 | 1,461.85 |
| NORTH CAROLINA | 539.46 | 553.43 | 593.09 | 561.68 | 2,247.66 |
| NORTH DAKOTA | 195.04 | 216.17 | 237.52 | 243.24 | 891.97 |
| OHIO | 517.86 | 524.85 | 531.56 | 545.45 | 2,119.72 |
| OKLAHOMA | 432.00 | 447.00 | 477.21 | 494.96 | 1,851.17 |
| OREGON | 360.93 | 393.83 | 401.81 | 426.49 | 1,583.06 |
| PENNSYLVANIA | 627.26 | 636.73 | 659.73 | 686.28 | 2,610.00 |
| PUERTO RICO | 57.69 | 60.14 | 55.94 | 61.36 | 235.12 |

| | | | | | |
|---|---|---|---|---|---|
| RHODE ISLAND | 827.30 | 802.10 | 862.21 | 866.39 | 3,358.01 |
| SOUTH CAROLINA | 324.77 | 345.69 | 363.65 | 365.08 | 1,399.19 |
| SOUTH DAKOTA | 95.89 | 107.06 | 129.00 | 138.29 | 470.25 |
| TENNESSEE | 616.78 | 679.39 | 688.37 | 699.79 | 2,684.33 |
| TEXAS | 121.77 | 124.28 | 130.52 | 128.26 | 504.83 |
| UTAH | 401.36 | 398.80 | 408.16 | 432.26 | 1,640.59 |
| VERMONT | 1,195.09 | 1,197.42 | 1,220.07 | 1,274.74 | 4,887.32 |
| VIRGIN ISLANDS | 16.16 | 17.32 | 21.52 | 21.66 | 76.66 |
| VIRGINIA | 303.47 | 320.67 | 339.96 | 337.25 | 1,301.36 |
| WASHINGTON | 415.21 | 436.07 | 461.10 | 496.23 | 1,808.62 |
| WEST VIRGINIA | 975.97 | 1,031.32 | 1,067.45 | 1,085.70 | 4,160.43 |
| WISCONSIN | 236.22 | 244.04 | 256.44 | 263.21 | 999.91 |
| WYOMING | 253.55 | 262.51 | 263.99 | 278.90 | 1,058.95 |
| **9064 BUPRENORPHINE Total** | **334.82** | **344.78** | **360.17** | **368.62** | **1,408.38** |

**DRUG: 9143 - OXYCODONE**

| STATE NAME | 1ST QUARTER | 2ND QUARTER | 3RD QUARTER | 4TH QUARTER | TOTAL GRAMS |
|---|---|---|---|---|---|
| ALABAMA | 3,661.75 | 3,614.19 | 3,567.83 | 3,571.25 | 14,415.03 |
| ALASKA | 3,291.85 | 3,190.64 | 3,190.52 | 2,929.42 | 12,602.43 |
| AMERICAN SAMOA | 16.39 | 23.91 | 10.25 | 33.47 | 84.01 |
| ARIZONA | 4,706.59 | 4,557.97 | 4,486.98 | 4,466.29 | 18,217.84 |
| ARKANSAS | 3,234.05 | 3,070.28 | 2,985.17 | 2,869.96 | 12,159.46 |
| CALIFORNIA | 1,650.01 | 1,633.53 | 1,577.40 | 1,547.35 | 6,408.29 |
| COLORADO | 3,333.01 | 3,313.23 | 3,299.41 | 3,251.14 | 13,196.80 |
| CONNECTICUT | 3,596.17 | 3,522.80 | 3,509.31 | 3,480.48 | 14,108.76 |
| DELAWARE | 5,961.30 | 5,827.83 | 5,740.86 | 5,372.27 | 22,902.26 |
| DISTRICT OF COLUMBIA | 1,941.30 | 1,834.30 | 1,847.88 | 1,722.79 | 7,346.27 |
| FLORIDA | 4,488.91 | 4,456.64 | 4,482.10 | 4,550.14 | 17,977.80 |
| GEORGIA | 3,752.26 | 3,723.55 | 3,721.64 | 3,810.86 | 15,008.31 |
| GUAM | 410.37 | 427.12 | 406.12 | 301.26 | 1,544.87 |
| HAWAII | 2,295.45 | 2,358.74 | 2,294.09 | 2,141.72 | 9,090.00 |
| IDAHO | 3,005.29 | 3,073.98 | 3,025.48 | 2,966.78 | 12,071.52 |
| ILLINOIS | 907.59 | 892.05 | 891.32 | 871.61 | 3,562.57 |
| INDIANA | 2,730.59 | 2,702.18 | 2,721.46 | 2,653.23 | 10,807.47 |
| IOWA | 1,275.87 | 1,274.95 | 1,243.17 | 1,222.99 | 5,016.98 |
| KANSAS | 3,257.36 | 3,224.25 | 3,166.25 | 3,118.02 | 12,765.88 |
| KENTUCKY | 3,876.82 | 3,799.72 | 3,814.77 | 3,772.44 | 15,263.75 |
| LOUISIANA | 3,279.27 | 3,223.29 | 3,218.28 | 3,143.94 | 12,864.78 |
| MAINE | 3,071.26 | 3,069.00 | 3,125.00 | 2,915.56 | 12,180.82 |
| MARYLAND | 4,220.52 | 4,193.36 | 4,149.97 | 4,149.85 | 16,713.69 |
| MASSACHUSETTS | 2,860.54 | 2,806.25 | 2,783.58 | 2,741.78 | 11,192.15 |
| MICHIGAN | 2,008.53 | 1,999.21 | 2,024.92 | 1,940.25 | 7,972.90 |

| | | | | | |
|---|---|---|---|---|---|
| MINNESOTA | 1,932.94 | 1,939.08 | 1,854.59 | 1,833.16 | 7,559.77 |
| MISSISSIPPI | 2,707.87 | 2,616.42 | 2,669.36 | 2,652.32 | 10,645.98 |
| MISSOURI | 3,369.65 | 3,261.29 | 3,184.19 | 3,113.22 | 12,928.35 |
| MONTANA | 2,821.84 | 2,835.58 | 2,794.16 | 2,688.48 | 11,140.06 |
| NEBRASKA | 1,711.18 | 1,656.36 | 1,636.68 | 1,607.01 | 6,611.23 |
| NEVADA | 5,022.51 | 5,022.22 | 4,902.25 | 4,977.84 | 19,924.81 |
| NEW HAMPSHIRE | 3,467.11 | 3,393.48 | 3,418.19 | 3,300.09 | 13,578.87 |
| NEW JERSEY | 4,113.01 | 3,963.53 | 4,067.97 | 4,032.82 | 16,177.32 |
| NEW MEXICO | 3,436.86 | 3,324.26 | 3,209.46 | 3,177.15 | 13,147.73 |
| NEW YORK | 2,851.41 | 2,591.47 | 2,670.31 | 2,627.93 | 10,741.13 |
| NORTH CAROLINA | 5,144.24 | 5,060.12 | 5,079.70 | 4,535.59 | 19,819.66 |
| NORTH DAKOTA | 1,720.67 | 1,652.62 | 1,642.19 | 1,618.47 | 6,633.95 |
| OHIO | 2,976.78 | 2,929.58 | 2,898.29 | 2,797.21 | 11,601.86 |
| OKLAHOMA | 3,821.89 | 3,756.89 | 3,775.65 | 3,700.31 | 15,054.74 |
| OREGON | 3,331.22 | 3,340.22 | 3,280.82 | 3,240.38 | 13,192.64 |
| PENNSYLVANIA | 4,162.68 | 4,045.13 | 3,974.35 | 3,926.05 | 16,108.21 |
| PUERTO RICO | 341.44 | 310.92 | 290.29 | 319.48 | 1,262.13 |
| RHODE ISLAND | 2,827.49 | 2,780.50 | 2,803.05 | 2,825.69 | 11,236.74 |
| SOUTH CAROLINA | 3,815.69 | 3,711.67 | 3,629.79 | 3,535.44 | 14,692.58 |
| SOUTH DAKOTA | 1,866.07 | 1,505.81 | 1,613.40 | 1,363.24 | 6,348.52 |
| TENNESSEE | 5,566.18 | 5,474.26 | 5,353.05 | 5,412.04 | 21,805.53 |
| TEXAS | 1,042.16 | 1,020.95 | 1,041.77 | 945.98 | 4,050.87 |
| UTAH | 4,805.09 | 4,805.17 | 4,813.35 | 4,835.41 | 19,259.02 |
| VERMONT | 2,651.30 | 2,704.63 | 2,684.55 | 2,584.29 | 10,624.77 |
| VIRGIN ISLANDS | 400.80 | 416.52 | 383.05 | 346.93 | 1,547.29 |
| VIRGINIA | 2,919.84 | 2,921.36 | 2,895.27 | 2,800.82 | 11,537.30 |
| WASHINGTON | 3,247.91 | 3,243.89 | 3,216.75 | 3,188.99 | 12,897.54 |
| WEST VIRGINIA | 3,089.84 | 2,946.43 | 2,906.48 | 2,775.35 | 11,718.11 |
| WISCONSIN | 2,383.62 | 2,323.75 | 2,228.61 | 2,186.94 | 9,122.92 |
| WYOMING | 3,634.36 | 3,583.49 | 3,593.04 | 3,435.37 | 14,246.26 |
| **9143 - OXYCODONE Total** | **2,937.29** | **2,877.61** | **2,862.21** | **2,808.30** | **11,485.41** |

**DRUG: 9150 - HYDROMORPHONE**

| STATE NAME | 1ST QUARTER | 2ND QUARTER | 3RD QUARTER | 4TH QUARTER | TOTAL GRAMS |
|---|---|---|---|---|---|
| ALABAMA | 47.33 | 48.21 | 44.35 | 40.48 | 180.38 |
| ALASKA | 79.78 | 69.92 | 68.57 | 59.91 | 278.18 |
| AMERICAN SAMOA | 4.32 | 3.24 | 2.23 | 2.03 | 11.82 |
| ARIZONA | 76.38 | 68.92 | 70.19 | 72.65 | 288.14 |
| ARKANSAS | 71.93 | 69.31 | 71.53 | 61.65 | 274.43 |
| CALIFORNIA | 69.19 | 68.46 | 67.17 | 65.80 | 270.62 |
| COLORADO | 99.62 | 98.95 | 100.44 | 100.82 | 399.83 |
| CONNECTICUT | 124.46 | 121.63 | 123.84 | 133.13 | 503.07 |

| | | | | | |
|---|---|---|---|---|---|
| DELAWARE | 139.82 | 133.93 | 130.34 | 129.08 | 533.17 |
| DISTRICT OF COLUMBIA | 66.69 | 63.70 | 62.37 | 57.24 | 250.00 |
| FLORIDA | 183.12 | 171.95 | 177.71 | 168.89 | 701.67 |
| GEORGIA | 62.23 | 60.10 | 58.30 | 59.60 | 240.25 |
| GUAM | 14.67 | 11.81 | 7.00 | 7.21 | 40.69 |
| HAWAII | 35.40 | 37.16 | 37.02 | 37.89 | 147.47 |
| IDAHO | 85.49 | 86.16 | 85.78 | 80.04 | 337.48 |
| ILLINOIS | 52.91 | 54.24 | 55.00 | 52.21 | 214.35 |
| INDIANA | 65.72 | 68.83 | 63.51 | 61.53 | 259.58 |
| IOWA | 67.54 | 64.53 | 62.13 | 62.28 | 256.48 |
| KANSAS | 86.75 | 85.72 | 85.56 | 85.56 | 343.58 |
| KENTUCKY | 51.03 | 49.25 | 54.25 | 46.92 | 201.45 |
| LOUISIANA | 53.93 | 53.04 | 54.01 | 43.41 | 204.38 |
| MAINE | 97.63 | 92.11 | 108.83 | 100.50 | 399.07 |
| MARYLAND | 93.91 | 95.62 | 94.87 | 94.19 | 378.59 |
| MASSACHUSETTS | 86.29 | 87.44 | 84.56 | 96.01 | 354.31 |
| MICHIGAN | 57.88 | 56.66 | 57.68 | 59.64 | 231.86 |
| MINNESOTA | 81.35 | 84.01 | 85.70 | 83.07 | 334.13 |
| MISSISSIPPI | 172.02 | 169.60 | 171.12 | 222.15 | 734.90 |
| MISSOURI | 67.31 | 63.71 | 64.93 | 62.38 | 258.33 |
| MONTANA | 133.68 | 138.97 | 125.08 | 132.37 | 530.10 |
| NEBRASKA | 48.64 | 46.69 | 44.74 | 46.22 | 186.29 |
| NEVADA | 80.35 | 77.89 | 77.01 | 67.77 | 303.02 |
| NEW HAMPSHIRE | 114.92 | 113.38 | 119.07 | 112.93 | 460.30 |
| NEW JERSEY | 76.28 | 74.20 | 77.95 | 73.45 | 301.88 |
| NEW MEXICO | 49.94 | 43.82 | 45.31 | 46.94 | 186.00 |
| NEW YORK | 74.28 | 69.82 | 73.11 | 72.42 | 289.63 |
| NORTH CAROLINA | 88.01 | 85.35 | 87.72 | 84.30 | 345.37 |
| NORTH DAKOTA | 85.57 | 70.80 | 72.87 | 70.11 | 299.36 |
| OHIO | 51.52 | 52.61 | 55.77 | 51.98 | 211.88 |
| OKLAHOMA | 66.97 | 67.61 | 64.72 | 69.37 | 268.67 |
| OREGON | 90.52 | 90.38 | 94.10 | 92.03 | 367.03 |
| PENNSYLVANIA | 83.73 | 79.87 | 77.51 | 86.28 | 327.39 |
| PUERTO RICO | 1.46 | 1.45 | 1.35 | 1.29 | 5.56 |
| RHODE ISLAND | 57.59 | 58.20 | 56.53 | 89.51 | 261.83 |
| SOUTH CAROLINA | 90.76 | 93.16 | 88.26 | 84.95 | 357.13 |
| SOUTH DAKOTA | 45.70 | 49.79 | 52.12 | 51.71 | 199.33 |
| TENNESSEE | 81.32 | 75.64 | 73.58 | 94.19 | 324.73 |
| TEXAS | 63.78 | 70.80 | 66.50 | 66.94 | 268.03 |
| UTAH | 81.32 | 81.44 | 88.17 | 85.92 | 336.85 |
| VERMONT | 138.23 | 152.29 | 159.26 | 138.34 | 588.12 |

| | | | | | |
|---|---|---|---|---|---|
| VIRGIN ISLANDS | 11.45 | 18.61 | 7.20 | 18.16 | 55.42 |
| VIRGINIA | 98.95 | 96.27 | 96.86 | 94.23 | 386.31 |
| WASHINGTON | 109.43 | 115.66 | 111.02 | 114.47 | 450.58 |
| WEST VIRGINIA | 52.22 | 53.83 | 51.89 | 50.17 | 208.12 |
| WISCONSIN | 48.10 | 46.50 | 45.93 | 42.05 | 182.57 |
| WYOMING | 81.57 | 80.56 | 101.75 | 108.11 | 371.98 |
| **9150 HYDROMORPHONE Total** | **79.70** | **78.58** | **78.73** | **78.57** | **315.58** |

**DRUG: 9193 - HYDROCODONE**

| STATE NAME | 1ST QUARTER | 2ND QUARTER | 3RD QUARTER | 4TH QUARTER | TOTAL GRAMS |
|---|---|---|---|---|---|
| ALABAMA | 4,061.02 | 3,986.53 | 3,952.15 | 3,886.13 | 15,885.83 |
| ALASKA | 1,165.98 | 1,159.86 | 1,154.06 | 1,117.46 | 4,597.35 |
| AMERICAN SAMOA | 0.00 | 0.92 | 0.00 | 4.61 | 5.53 |
| ARIZONA | 1,219.87 | 1,191.99 | 1,162.25 | 1,187.15 | 4,761.26 |
| ARKANSAS | 3,558.51 | 3,508.64 | 3,489.37 | 3,435.26 | 13,991.77 |
| CALIFORNIA | 1,839.14 | 1,816.31 | 1,772.88 | 1,743.61 | 7,171.95 |
| COLORADO | 964.32 | 944.52 | 940.28 | 919.83 | 3,768.94 |
| CONNECTICUT | 426.50 | 410.81 | 402.34 | 406.53 | 1,646.18 |
| DELAWARE | 496.37 | 524.73 | 493.44 | 465.44 | 1,979.98 |
| DISTRICT OF COLUMBIA | 175.84 | 161.17 | 158.22 | 150.42 | 645.65 |
| FLORIDA | 1,447.39 | 1,393.18 | 1,376.21 | 1,385.92 | 5,602.71 |
| GEORGIA | 2,006.43 | 1,966.59 | 1,949.05 | 1,940.04 | 7,862.10 |
| GUAM | 71.07 | 56.55 | 67.76 | 55.09 | 250.47 |
| HAWAII | 976.49 | 950.40 | 963.86 | 941.43 | 3,832.18 |
| IDAHO | 2,888.67 | 2,896.90 | 2,901.65 | 2,829.55 | 11,516.77 |
| ILLINOIS | 1,790.88 | 1,776.10 | 1,763.68 | 1,723.79 | 7,054.46 |
| INDIANA | 2,732.82 | 2,709.77 | 2,702.23 | 2,601.95 | 10,746.78 |
| IOWA | 1,370.48 | 1,351.11 | 1,329.93 | 1,306.26 | 5,357.77 |
| KANSAS | 2,194.94 | 2,173.27 | 2,135.13 | 2,128.40 | 8,631.74 |
| KENTUCKY | 3,906.12 | 3,817.48 | 3,819.82 | 3,751.89 | 15,295.30 |
| LOUISIANA | 2,794.92 | 2,739.25 | 2,676.25 | 2,592.70 | 10,803.12 |
| MAINE | 1,031.50 | 1,018.97 | 1,035.56 | 964.59 | 4,050.62 |
| MARYLAND | 446.45 | 436.52 | 432.99 | 425.27 | 1,741.23 |
| MASSACHUSETTS | 348.84 | 341.54 | 337.01 | 334.45 | 1,361.83 |
| MICHIGAN | 2,631.42 | 2,606.52 | 2,644.28 | 2,428.48 | 10,310.70 |
| MINNESOTA | 612.78 | 610.58 | 590.72 | 580.08 | 2,394.16 |
| MISSISSIPPI | 2,670.04 | 2,597.47 | 2,554.50 | 2,539.43 | 10,361.43 |
| MISSOURI | 2,198.35 | 2,165.73 | 2,133.60 | 2,116.58 | 8,614.27 |
| MONTANA | 2,064.20 | 2,059.82 | 2,030.43 | 1,968.59 | 8,123.03 |
| NEBRASKA | 1,312.46 | 1,292.29 | 1,257.31 | 1,251.97 | 5,114.03 |
| NEVADA | 2,554.64 | 2,514.61 | 2,450.46 | 2,483.84 | 10,003.55 |
| NEW HAMPSHIRE | 561.03 | 534.78 | 533.26 | 527.36 | 2,156.43 |

| | | | | | |
|---|---|---|---|---|---|
| NEW JERSEY | 298.15 | 283.09 | 285.66 | 286.35 | 1,153.26 |
| NEW MEXICO | 1,348.10 | 1,324.90 | 1,271.23 | 1,272.09 | 5,216.32 |
| NEW YORK | 676.46 | 648.56 | 646.38 | 638.48 | 2,609.89 |
| NORTH CAROLINA | 1,659.66 | 1,562.72 | 1,547.54 | 1,477.19 | 6,247.11 |
| NORTH DAKOTA | 1,196.93 | 1,123.24 | 1,126.97 | 1,098.84 | 4,545.99 |
| OHIO | 1,141.13 | 1,133.03 | 1,123.68 | 1,095.44 | 4,493.27 |
| OKLAHOMA | 3,532.74 | 3,500.34 | 3,500.09 | 3,440.39 | 13,973.57 |
| OREGON | 1,914.51 | 1,897.40 | 1,874.63 | 1,855.29 | 7,541.83 |
| PENNSYLVANIA | 916.84 | 904.52 | 867.68 | 892.87 | 3,581.92 |
| PUERTO RICO | 15.24 | 10.91 | 9.20 | 10.89 | 46.23 |
| RHODE ISLAND | 729.75 | 721.60 | 718.15 | 716.87 | 2,886.38 |
| SOUTH CAROLINA | 2,249.25 | 2,181.78 | 2,110.92 | 2,064.51 | 8,606.45 |
| SOUTH DAKOTA | 1,274.35 | 1,242.33 | 1,265.58 | 1,218.22 | 5,000.47 |
| TENNESSEE | 2,925.43 | 2,889.12 | 2,825.65 | 2,942.35 | 11,582.54 |
| TEXAS | 2,089.36 | 2,032.77 | 2,005.28 | 1,962.44 | 8,089.86 |
| UTAH | 1,661.89 | 1,668.28 | 1,639.41 | 1,662.12 | 6,631.70 |
| VERMONT | 584.93 | 589.18 | 588.72 | 568.81 | 2,331.63 |
| VIRGIN ISLANDS | 233.56 | 169.37 | 164.89 | 182.12 | 749.94 |
| VIRGINIA | 1,002.87 | 978.60 | 950.36 | 923.28 | 3,855.12 |
| WASHINGTON | 1,497.21 | 1,475.64 | 1,456.62 | 1,438.48 | 5,867.95 |
| WEST VIRGINIA | 2,188.39 | 2,105.12 | 2,080.94 | 1,970.84 | 8,345.28 |
| WISCONSIN | 1,059.22 | 1,064.25 | 1,045.99 | 1,021.14 | 4,190.60 |
| WYOMING | 1,525.07 | 1,525.82 | 1,563.28 | 1,479.30 | 6,093.47 |
| **9193 HYDROCODONE Total** | **1,642.14** | **1,610.89** | **1,590.14** | **1,563.26** | **6,406.44** |

**DRUG: 9230 - MEPERIDINE (PETHIDINE)**

| STATE NAME | 1ST QUARTER | 2ND QUARTER | 3RD QUARTER | 4TH QUARTER | TOTAL GRAMS |
|---|---|---|---|---|---|
| ALABAMA | 96.19 | 72.35 | 53.21 | 37.95 | 259.71 |
| ALASKA | 50.23 | 33.44 | 10.43 | 13.19 | 107.29 |
| ARIZONA | 50.90 | 43.02 | 40.14 | 29.67 | 163.72 |
| ARKANSAS | 156.35 | 100.80 | 74.52 | 49.39 | 381.06 |
| CALIFORNIA | 28.98 | 23.67 | 27.81 | 18.88 | 99.33 |
| COLORADO | 21.11 | 16.01 | 7.29 | 5.30 | 49.71 |
| CONNECTICUT | 18.03 | 12.73 | 2.14 | 1.63 | 34.52 |
| DELAWARE | 10.25 | 14.86 | 3.18 | 3.46 | 31.75 |
| DISTRICT OF COLUMBIA | 20.19 | 15.93 | 25.62 | 11.59 | 73.32 |
| FLORIDA | 25.99 | 20.93 | 15.69 | 11.47 | 74.08 |
| GEORGIA | 62.54 | 41.75 | 30.28 | 17.88 | 152.46 |
| GUAM | 7.94 | 9.16 | 19.54 | 10.99 | 47.62 |
| HAWAII | 16.94 | 12.21 | 13.49 | 5.33 | 47.97 |
| IDAHO | 38.22 | 26.72 | 16.31 | 9.92 | 91.17 |
| ILLINOIS | 12.66 | 11.02 | 9.06 | 4.99 | 37.74 |

| | | | | | |
|---|---|---|---|---|---|
| INDIANA | 30.44 | 21.41 | 21.76 | 17.55 | 91.15 |
| IOWA | 49.33 | 36.03 | 37.91 | 31.71 | 154.97 |
| KANSAS | 39.56 | 25.72 | 19.38 | 13.36 | 98.01 |
| KENTUCKY | 28.07 | 21.48 | 21.84 | 13.76 | 85.14 |
| LOUISIANA | 82.57 | 59.69 | 44.91 | 33.17 | 220.34 |
| MAINE | 13.86 | 6.64 | 7.03 | 4.80 | 32.33 |
| MARYLAND | 23.80 | 21.71 | 16.25 | 7.93 | 69.69 |
| MASSACHUSETTS | 19.66 | 13.41 | 6.96 | 5.33 | 45.36 |
| MICHIGAN | 17.55 | 14.01 | 9.70 | 5.63 | 46.90 |
| MINNESOTA | 7.36 | 6.42 | 4.98 | 3.10 | 21.86 |
| MISSISSIPPI | 85.30 | 59.61 | 52.99 | 42.98 | 240.89 |
| MISSOURI | 33.61 | 25.37 | 21.91 | 9.46 | 90.36 |
| MONTANA | 29.73 | 21.53 | 17.45 | 11.29 | 79.99 |
| NEBRASKA | 24.04 | 11.33 | 14.40 | 8.11 | 57.89 |
| NEVADA | 31.24 | 26.44 | 31.34 | 17.04 | 106.07 |
| NEW HAMPSHIRE | 31.61 | 18.53 | 10.02 | 4.10 | 64.26 |
| NEW JERSEY | 14.13 | 9.79 | 6.32 | 4.74 | 34.97 |
| NEW MEXICO | 28.67 | 23.17 | 17.35 | 11.85 | 81.05 |
| NEW YORK | 15.14 | 10.91 | 8.08 | 4.28 | 38.42 |
| NORTH CAROLINA | 35.43 | 21.75 | 11.05 | 8.54 | 76.77 |
| NORTH DAKOTA | 22.92 | 20.25 | 11.86 | 6.32 | 61.34 |
| OHIO | 30.10 | 21.71 | 19.65 | 12.08 | 83.55 |
| OKLAHOMA | 102.12 | 72.35 | 56.71 | 26.50 | 257.68 |
| OREGON | 28.61 | 17.33 | 16.70 | 8.93 | 71.57 |
| PENNSYLVANIA | 15.97 | 13.83 | 7.29 | 4.11 | 41.21 |
| PUERTO RICO | 78.62 | 77.20 | 106.46 | 78.87 | 341.16 |
| RHODE ISLAND | 4.04 | 7.24 | 3.83 | 10.35 | 25.46 |
| SOUTH CAROLINA | 62.51 | 39.58 | 24.21 | 13.82 | 140.12 |
| SOUTH DAKOTA | 44.89 | 39.27 | 25.15 | 9.70 | 119.02 |
| TENNESSEE | 55.39 | 36.63 | 23.03 | 17.91 | 132.96 |
| TEXAS | 29.50 | 23.06 | 26.19 | 19.93 | 98.68 |
| UTAH | 44.34 | 31.98 | 25.82 | 25.61 | 127.75 |
| VERMONT | 26.64 | 40.82 | 22.02 | 24.63 | 114.12 |
| VIRGIN ISLANDS | 56.04 | 65.96 | 22.32 | 1.98 | 146.31 |
| VIRGINIA | 44.36 | 29.63 | 19.26 | 11.38 | 104.63 |
| WASHINGTON | 29.47 | 21.50 | 11.79 | 9.42 | 72.18 |
| WEST VIRGINIA | 17.52 | 10.82 | 10.96 | 8.62 | 47.93 |
| WISCONSIN | 13.30 | 9.84 | 11.35 | 7.52 | 42.01 |
| WYOMING | 38.66 | 34.79 | 13.43 | 8.00 | 94.88 |
| **9230 MEPERIDINE (PETHIDINE) Total** | **33.29** | **24.83** | **21.12** | **14.33** | **93.56** |

**DRUG: 9250B - METHADONE**

DATE RANGE: 01/01/2019 TO 12/31/2019

ARCOS 3 - REPORT 03 by GRAMS
QUARTERLY DRUG DISTRIBUTION BY STATE PER 100K POPULATION

| STATE NAME | 1ST QUARTER | 2ND QUARTER | 3RD QUARTER | 4TH QUARTER | TOTAL GRAMS |
|---|---|---|---|---|---|
| ALABAMA | 1,682.25 | 1,658.05 | 1,651.73 | 1,586.97 | 6,579.00 |
| ALASKA | 1,292.62 | 1,396.96 | 1,392.40 | 1,274.38 | 5,356.37 |
| ARIZONA | 1,577.99 | 1,644.93 | 1,707.34 | 1,757.63 | 6,687.89 |
| ARKANSAS | 421.79 | 447.66 | 454.72 | 419.96 | 1,744.12 |
| CALIFORNIA | 839.61 | 850.93 | 832.68 | 851.42 | 3,374.64 |
| COLORADO | 959.11 | 1,041.89 | 1,019.72 | 1,046.28 | 4,067.00 |
| CONNECTICUT | 3,615.01 | 3,712.69 | 3,227.00 | 3,239.08 | 13,793.78 |
| DELAWARE | 4,037.34 | 3,750.16 | 3,699.53 | 4,339.73 | 15,826.76 |
| DISTRICT OF COLUMBIA | 1,794.02 | 1,487.38 | 1,680.89 | 1,608.94 | 6,571.23 |
| FLORIDA | 1,133.70 | 1,217.40 | 1,105.55 | 1,113.97 | 4,570.62 |
| GEORGIA | 1,155.25 | 1,208.97 | 1,159.67 | 1,153.85 | 4,677.75 |
| GUAM | 25.56 | 20.30 | 18.55 | 9.02 | 73.43 |
| HAWAII | 562.12 | 536.11 | 628.28 | 485.04 | 2,211.55 |
| IDAHO | 697.77 | 708.48 | 654.11 | 655.63 | 2,715.99 |
| ILLINOIS | 953.99 | 1,001.35 | 970.07 | 1,006.82 | 3,932.23 |
| INDIANA | 1,290.94 | 1,243.61 | 1,240.67 | 1,322.29 | 5,097.50 |
| IOWA | 434.72 | 453.63 | 400.49 | 487.62 | 1,776.46 |
| KANSAS | 773.15 | 736.38 | 665.80 | 696.77 | 2,872.10 |
| KENTUCKY | 1,133.13 | 1,153.57 | 1,277.89 | 1,218.73 | 4,783.32 |
| LOUISIANA | 762.85 | 783.92 | 755.52 | 737.37 | 3,039.66 |
| MAINE | 2,102.37 | 2,290.85 | 2,309.21 | 2,360.11 | 9,062.53 |
| MARYLAND | 3,216.65 | 3,548.75 | 3,377.69 | 3,326.11 | 13,469.20 |
| MASSACHUSETTS | 2,269.47 | 2,287.36 | 2,301.30 | 2,369.92 | 9,228.06 |
| MICHIGAN | 1,256.05 | 1,248.29 | 1,241.60 | 1,250.65 | 4,996.58 |
| MINNESOTA | 902.10 | 1,008.24 | 1,051.42 | 964.05 | 3,925.81 |
| MISSISSIPPI | 267.65 | 313.19 | 277.16 | 272.50 | 1,130.51 |
| MISSOURI | 582.51 | 564.31 | 649.46 | 536.08 | 2,332.36 |
| MONTANA | 910.30 | 815.06 | 838.89 | 826.52 | 3,390.77 |
| NEBRASKA | 392.22 | 437.35 | 374.27 | 394.06 | 1,597.90 |
| NEVADA | 1,358.14 | 1,103.57 | 1,221.53 | 1,248.58 | 4,931.82 |
| NEW HAMPSHIRE | 2,456.55 | 2,711.45 | 2,386.26 | 2,465.31 | 10,019.57 |
| NEW JERSEY | 1,467.29 | 1,462.31 | 1,522.18 | 1,564.30 | 6,016.08 |
| NEW MEXICO | 2,328.70 | 2,495.78 | 2,456.68 | 2,590.22 | 9,871.37 |
| NEW YORK | 1,682.83 | 1,604.76 | 1,660.46 | 1,686.19 | 6,634.24 |
| NORTH CAROLINA | 1,436.01 | 1,526.11 | 1,509.64 | 1,463.51 | 5,935.26 |
| NORTH DAKOTA | 707.47 | 762.36 | 801.30 | 896.47 | 3,167.61 |
| OHIO | 636.76 | 666.15 | 692.67 | 724.77 | 2,720.35 |
| OKLAHOMA | 923.11 | 931.77 | 991.76 | 862.55 | 3,709.20 |
| OREGON | 1,617.82 | 1,638.94 | 1,675.59 | 1,647.74 | 6,580.09 |
| PENNSYLVANIA | 1,568.81 | 1,599.12 | 1,485.25 | 1,569.95 | 6,223.13 |

| | | | | | |
|---|---|---|---|---|---|
| PUERTO RICO | 554.36 | 578.43 | 649.31 | 507.24 | 2,289.34 |
| RHODE ISLAND | 4,027.77 | 4,716.21 | 4,715.33 | 4,698.74 | 18,158.05 |
| SOUTH CAROLINA | 1,211.80 | 1,226.87 | 1,246.04 | 1,289.72 | 4,974.43 |
| SOUTH DAKOTA | 242.72 | 261.44 | 190.63 | 257.79 | 952.58 |
| TENNESSEE | 726.61 | 835.41 | 704.73 | 749.95 | 3,016.69 |
| TEXAS | 510.77 | 565.81 | 535.90 | 551.48 | 2,163.96 |
| UTAH | 930.71 | 1,052.05 | 995.26 | 957.51 | 3,935.52 |
| VERMONT | 2,975.51 | 3,477.83 | 2,902.89 | 3,575.64 | 12,931.86 |
| VIRGIN ISLANDS | 4.07 | 8.96 | 11.40 | 5.70 | 30.13 |
| VIRGINIA | 1,115.97 | 1,184.45 | 1,149.56 | 1,221.69 | 4,671.68 |
| WASHINGTON | 1,537.85 | 1,769.49 | 1,577.39 | 1,605.89 | 6,490.63 |
| WEST VIRGINIA | 1,282.13 | 1,352.42 | 1,353.53 | 1,252.58 | 5,240.66 |
| WISCONSIN | 1,084.89 | 1,114.04 | 1,107.25 | 1,184.25 | 4,490.44 |
| WYOMING | 147.20 | 147.98 | 140.85 | 129.23 | 565.26 |
| **9250B METHADONE Total** | **1,184.10** | **1,224.70** | **1,198.21** | **1,213.71** | **4,820.73** |

**DRUG: 9300 - MORPHINE**

| STATE NAME | 1ST QUARTER | 2ND QUARTER | 3RD QUARTER | 4TH QUARTER | TOTAL GRAMS |
|---|---|---|---|---|---|
| ALABAMA | 1,151.10 | 1,105.15 | 1,087.90 | 1,044.71 | 4,388.86 |
| ALASKA | 1,136.60 | 1,070.73 | 1,065.58 | 957.23 | 4,230.14 |
| AMERICAN SAMOA | 29.58 | 53.26 | 0.46 | 18.91 | 102.20 |
| ARIZONA | 1,631.08 | 1,512.53 | 1,441.20 | 1,427.44 | 6,012.25 |
| ARKANSAS | 1,014.97 | 944.40 | 936.18 | 888.50 | 3,784.05 |
| CALIFORNIA | 866.25 | 827.39 | 811.23 | 776.94 | 3,281.81 |
| COLORADO | 1,138.47 | 1,070.61 | 1,104.78 | 1,030.76 | 4,344.62 |
| CONNECTICUT | 766.62 | 754.89 | 760.58 | 731.58 | 3,013.67 |
| DELAWARE | 1,720.31 | 1,648.61 | 1,751.08 | 1,605.97 | 6,725.97 |
| DISTRICT OF COLUMBIA | 449.24 | 461.48 | 442.34 | 384.94 | 1,738.00 |
| FLORIDA | 1,609.15 | 1,524.95 | 1,535.58 | 1,501.14 | 6,170.82 |
| GEORGIA | 922.50 | 895.87 | 903.64 | 910.37 | 3,632.39 |
| GUAM | 414.99 | 490.02 | 407.39 | 330.25 | 1,642.65 |
| HAWAII | 1,073.98 | 1,097.73 | 1,058.31 | 972.26 | 4,202.28 |
| IDAHO | 1,380.25 | 1,367.63 | 1,354.33 | 1,262.59 | 5,364.80 |
| ILLINOIS | 664.55 | 650.85 | 640.94 | 647.27 | 2,603.61 |
| INDIANA | 957.63 | 924.24 | 949.26 | 894.28 | 3,725.40 |
| IOWA | 846.46 | 827.91 | 846.43 | 776.68 | 3,297.48 |
| KANSAS | 1,179.19 | 1,165.75 | 1,173.72 | 1,110.28 | 4,628.94 |
| KENTUCKY | 809.40 | 767.26 | 777.75 | 735.77 | 3,090.19 |
| LOUISIANA | 939.42 | 890.43 | 903.70 | 867.35 | 3,600.89 |
| MAINE | 967.01 | 988.43 | 1,027.83 | 933.98 | 3,917.24 |
| MARYLAND | 909.66 | 874.23 | 875.11 | 839.78 | 3,498.79 |
| MASSACHUSETTS | 796.92 | 794.25 | 791.34 | 767.30 | 3,149.82 |

| | | | | | |
|---|---|---|---|---|---|
| MICHIGAN | 1,287.75 | 1,256.34 | 1,290.33 | 1,198.92 | 5,033.35 |
| MINNESOTA | 660.01 | 627.46 | 626.83 | 605.02 | 2,519.32 |
| MISSISSIPPI | 1,020.21 | 892.10 | 922.46 | 944.17 | 3,778.95 |
| MISSOURI | 1,211.41 | 1,153.07 | 1,149.00 | 1,083.24 | 4,596.71 |
| MONTANA | 1,329.51 | 1,328.17 | 1,328.92 | 1,257.14 | 5,243.74 |
| NEBRASKA | 819.23 | 826.62 | 824.80 | 774.52 | 3,245.18 |
| NEVADA | 1,540.50 | 1,438.45 | 1,392.07 | 1,365.81 | 5,736.82 |
| NEW HAMPSHIRE | 1,177.84 | 1,073.24 | 1,149.65 | 1,059.61 | 4,460.35 |
| NEW JERSEY | 717.94 | 679.53 | 701.85 | 685.41 | 2,784.73 |
| NEW MEXICO | 1,011.71 | 949.42 | 928.52 | 912.65 | 3,802.30 |
| NEW YORK | 611.31 | 548.27 | 599.28 | 572.37 | 2,331.23 |
| NORTH CAROLINA | 1,107.53 | 1,116.62 | 1,123.79 | 978.37 | 4,326.32 |
| NORTH DAKOTA | 739.55 | 683.11 | 724.80 | 706.16 | 2,853.61 |
| OHIO | 757.71 | 739.12 | 740.87 | 712.10 | 2,949.80 |
| OKLAHOMA | 1,432.19 | 1,385.12 | 1,320.51 | 1,296.92 | 5,434.75 |
| OREGON | 1,372.54 | 1,360.90 | 1,366.95 | 1,325.61 | 5,426.01 |
| PENNSYLVANIA | 1,364.82 | 1,279.09 | 1,291.43 | 1,314.82 | 5,250.15 |
| PUERTO RICO | 60.06 | 53.54 | 43.56 | 43.59 | 200.74 |
| RHODE ISLAND | 838.91 | 811.98 | 790.36 | 822.10 | 3,263.34 |
| SOUTH CAROLINA | 1,169.98 | 1,142.81 | 1,133.04 | 1,095.84 | 4,541.66 |
| SOUTH DAKOTA | 780.50 | 786.69 | 790.64 | 664.31 | 3,022.13 |
| TENNESSEE | 1,907.06 | 1,787.29 | 1,815.19 | 1,937.10 | 7,446.64 |
| TEXAS | 642.16 | 610.81 | 606.70 | 582.26 | 2,441.94 |
| UTAH | 1,394.99 | 1,359.61 | 1,367.84 | 1,335.56 | 5,458.00 |
| VERMONT | 1,248.16 | 1,263.55 | 1,272.85 | 1,222.55 | 5,007.11 |
| VIRGIN ISLANDS | 96.24 | 73.33 | 66.49 | 60.97 | 297.04 |
| VIRGINIA | 808.03 | 773.73 | 803.09 | 753.72 | 3,138.57 |
| WASHINGTON | 1,208.08 | 1,181.79 | 1,179.36 | 1,151.70 | 4,720.92 |
| WEST VIRGINIA | 872.28 | 804.24 | 864.84 | 799.24 | 3,340.60 |
| WISCONSIN | 988.67 | 963.17 | 967.17 | 925.12 | 3,844.13 |
| WYOMING | 1,244.30 | 1,186.02 | 1,220.21 | 1,105.57 | 4,756.10 |
| **9300 - MORPHINE Total** | **994.05** | **951.34** | **956.08** | **923.87** | **3,825.34** |

**DRUG: 9652 - OXYMORPHONE**

| STATE NAME | 1ST QUARTER | 2ND QUARTER | 3RD QUARTER | 4TH QUARTER | TOTAL GRAMS |
|---|---|---|---|---|---|
| ALABAMA | 32.79 | 30.57 | 26.92 | 24.70 | 114.97 |
| ALASKA | 8.73 | 7.25 | 7.29 | 8.35 | 31.62 |
| ARIZONA | 23.81 | 21.03 | 18.70 | 20.03 | 83.57 |
| ARKANSAS | 9.97 | 7.71 | 5.44 | 5.60 | 28.73 |
| CALIFORNIA | 11.25 | 11.67 | 9.71 | 9.03 | 41.66 |
| COLORADO | 19.77 | 20.32 | 18.46 | 17.93 | 76.47 |
| CONNECTICUT | 31.33 | 29.76 | 26.16 | 26.42 | 113.67 |

| | | | | | |
|---|---|---|---|---|---|
| DELAWARE | 132.53 | 141.93 | 111.92 | 105.06 | 491.44 |
| DISTRICT OF COLUMBIA | 9.44 | 10.35 | 7.12 | 5.30 | 32.20 |
| FLORIDA | 43.45 | 45.18 | 40.25 | 34.36 | 163.24 |
| GEORGIA | 23.44 | 25.35 | 21.92 | 19.86 | 90.57 |
| HAWAII | 14.95 | 15.44 | 12.07 | 10.92 | 53.38 |
| IDAHO | 36.60 | 35.99 | 34.10 | 30.61 | 137.29 |
| ILLINOIS | 13.00 | 12.59 | 10.95 | 10.63 | 47.17 |
| INDIANA | 26.67 | 26.01 | 22.90 | 19.31 | 94.89 |
| IOWA | 9.54 | 10.08 | 7.57 | 7.28 | 34.47 |
| KANSAS | 23.03 | 22.36 | 20.86 | 19.10 | 85.35 |
| KENTUCKY | 34.20 | 36.00 | 33.24 | 35.69 | 139.13 |
| LOUISIANA | 66.21 | 69.02 | 58.20 | 55.14 | 248.57 |
| MAINE | 26.85 | 23.93 | 23.58 | 21.67 | 96.03 |
| MARYLAND | 83.52 | 84.49 | 67.95 | 59.69 | 295.65 |
| MASSACHUSETTS | 12.22 | 14.30 | 14.16 | 11.82 | 52.50 |
| MICHIGAN | 74.96 | 82.08 | 70.34 | 62.46 | 289.84 |
| MINNESOTA | 3.85 | 3.56 | 3.35 | 3.52 | 14.27 |
| MISSISSIPPI | 36.48 | 33.01 | 27.04 | 26.85 | 123.39 |
| MISSOURI | 22.91 | 24.45 | 20.23 | 19.64 | 87.24 |
| MONTANA | 9.62 | 9.75 | 9.34 | 9.68 | 38.39 |
| NEBRASKA | 17.78 | 20.93 | 17.45 | 17.46 | 73.62 |
| NEVADA | 46.36 | 39.33 | 35.55 | 32.84 | 154.09 |
| NEW HAMPSHIRE | 23.28 | 19.98 | 15.69 | 15.17 | 74.12 |
| NEW JERSEY | 30.18 | 28.82 | 28.57 | 27.90 | 115.46 |
| NEW MEXICO | 16.69 | 15.84 | 14.39 | 11.65 | 58.57 |
| NEW YORK | 45.42 | 42.01 | 37.25 | 37.38 | 162.06 |
| NORTH CAROLINA | 91.29 | 107.88 | 89.48 | 68.98 | 357.63 |
| NORTH DAKOTA | 8.77 | 9.13 | 6.09 | 7.29 | 31.28 |
| OHIO | 27.43 | 26.32 | 22.13 | 21.59 | 97.48 |
| OKLAHOMA | 26.82 | 26.73 | 22.44 | 23.07 | 99.07 |
| OREGON | 18.49 | 17.48 | 15.14 | 14.77 | 65.88 |
| PENNSYLVANIA | 72.31 | 72.16 | 61.03 | 56.95 | 262.45 |
| RHODE ISLAND | 15.06 | 14.42 | 14.55 | 14.89 | 58.92 |
| SOUTH CAROLINA | 56.40 | 57.49 | 50.06 | 48.61 | 212.56 |
| SOUTH DAKOTA | 28.04 | 28.52 | 24.64 | 22.94 | 104.13 |
| TENNESSEE | 149.15 | 142.71 | 124.11 | 107.98 | 523.95 |
| TEXAS | 12.90 | 12.31 | 11.06 | 9.92 | 46.19 |
| UTAH | 75.53 | 83.34 | 69.11 | 75.82 | 303.80 |
| VERMONT | 16.41 | 19.35 | 15.28 | 15.21 | 66.25 |
| VIRGINIA | 53.48 | 56.96 | 48.44 | 44.64 | 203.51 |
| WASHINGTON | 17.66 | 17.66 | 15.33 | 15.76 | 66.41 |

| | | | | | |
|---|---|---|---|---|---|
| WEST VIRGINIA | 55.67 | 50.82 | 38.77 | 32.99 | 178.26 |
| WISCONSIN | 10.48 | 10.23 | 9.98 | 8.38 | 39.08 |
| WYOMING | 14.44 | 16.67 | 17.07 | 12.08 | 60.26 |
| **9652 - OXYMORPHONE Total** | **34.77** | **35.29** | **30.44** | **27.91** | **128.41** |

**DRUG: 9780 - TAPENTADOL**

| STATE NAME | 1ST QUARTER | 2ND QUARTER | 3RD QUARTER | 4TH QUARTER | TOTAL GRAMS |
|---|---|---|---|---|---|
| ALABAMA | 668.35 | 630.96 | 596.08 | 577.08 | 2,472.47 |
| ALASKA | 219.58 | 219.01 | 194.37 | 191.21 | 824.17 |
| ARIZONA | 372.46 | 374.22 | 334.97 | 345.09 | 1,426.73 |
| ARKANSAS | 363.74 | 359.89 | 319.93 | 314.31 | 1,357.87 |
| CALIFORNIA | 187.51 | 184.22 | 176.86 | 172.42 | 721.02 |
| COLORADO | 433.66 | 445.46 | 429.57 | 446.22 | 1,754.91 |
| CONNECTICUT | 215.79 | 201.84 | 197.06 | 179.30 | 793.99 |
| DELAWARE | 512.06 | 501.32 | 497.42 | 490.46 | 2,001.26 |
| DISTRICT OF COLUMBIA | 121.98 | 103.79 | 91.07 | 107.03 | 423.87 |
| FLORIDA | 334.40 | 319.73 | 303.72 | 301.60 | 1,259.44 |
| GEORGIA | 459.00 | 437.48 | 437.05 | 429.76 | 1,763.30 |
| HAWAII | 110.31 | 93.73 | 99.61 | 103.65 | 407.30 |
| IDAHO | 295.77 | 300.43 | 290.16 | 283.49 | 1,169.86 |
| ILLINOIS | 183.58 | 179.97 | 183.70 | 177.02 | 724.26 |
| INDIANA | 377.11 | 383.31 | 395.42 | 387.94 | 1,543.77 |
| IOWA | 267.89 | 248.10 | 235.31 | 248.56 | 999.87 |
| KANSAS | 185.74 | 190.41 | 168.99 | 170.22 | 715.36 |
| KENTUCKY | 229.54 | 211.41 | 215.73 | 201.04 | 857.73 |
| LOUISIANA | 267.63 | 259.33 | 252.39 | 249.33 | 1,028.68 |
| MAINE | 89.09 | 94.03 | 102.83 | 90.19 | 376.14 |
| MARYLAND | 404.54 | 408.43 | 400.98 | 395.25 | 1,609.21 |
| MASSACHUSETTS | 236.80 | 238.32 | 248.59 | 234.07 | 957.77 |
| MICHIGAN | 144.45 | 135.66 | 137.83 | 132.64 | 550.58 |
| MINNESOTA | 64.89 | 67.06 | 60.07 | 56.86 | 248.88 |
| MISSISSIPPI | 268.81 | 246.60 | 246.22 | 256.19 | 1,017.83 |
| MISSOURI | 302.26 | 290.62 | 277.40 | 260.60 | 1,130.88 |
| MONTANA | 149.99 | 132.65 | 144.28 | 130.68 | 557.60 |
| NEBRASKA | 467.44 | 441.37 | 446.71 | 462.92 | 1,818.44 |
| NEVADA | 264.26 | 253.56 | 222.55 | 227.01 | 967.38 |
| NEW HAMPSHIRE | 435.41 | 425.27 | 454.47 | 458.73 | 1,773.87 |
| NEW JERSEY | 340.38 | 322.98 | 325.37 | 331.87 | 1,320.60 |
| NEW MEXICO | 103.88 | 97.42 | 98.32 | 92.95 | 392.56 |
| NEW YORK | 353.61 | 335.29 | 339.67 | 327.17 | 1,355.73 |
| NORTH CAROLINA | 852.96 | 865.22 | 819.06 | 757.76 | 3,295.00 |
| NORTH DAKOTA | 220.94 | 212.24 | 191.20 | 180.05 | 804.43 |

| | | | | | |
|---|---|---|---|---|---|
| OHIO | 333.14 | 327.09 | 301.05 | 307.51 | 1,268.79 |
| OKLAHOMA | 240.19 | 231.05 | 231.04 | 209.30 | 911.58 |
| OREGON | 124.43 | 135.25 | 126.79 | 116.77 | 503.24 |
| PENNSYLVANIA | 344.39 | 342.02 | 330.78 | 331.66 | 1,348.85 |
| PUERTO RICO | 10.98 | 13.34 | 11.47 | 6.98 | 42.77 |
| RHODE ISLAND | 176.81 | 166.97 | 179.80 | 153.53 | 677.11 |
| SOUTH CAROLINA | 687.74 | 654.27 | 646.35 | 627.71 | 2,616.08 |
| SOUTH DAKOTA | 380.63 | 374.67 | 357.97 | 347.16 | 1,460.43 |
| TENNESSEE | 345.94 | 348.36 | 345.46 | 355.12 | 1,394.89 |
| TEXAS | 244.26 | 241.50 | 237.41 | 225.75 | 948.92 |
| UTAH | 307.10 | 327.87 | 316.46 | 311.43 | 1,262.86 |
| VERMONT | 129.53 | 105.95 | 118.82 | 124.65 | 478.95 |
| VIRGIN ISLANDS | 0.00 | 4.55 | 0.00 | 0.00 | 4.55 |
| VIRGINIA | 341.31 | 344.73 | 340.07 | 327.01 | 1,353.13 |
| WASHINGTON | 186.67 | 179.36 | 180.21 | 183.83 | 730.07 |
| WEST VIRGINIA | 183.70 | 202.89 | 195.52 | 198.41 | 780.52 |
| WISCONSIN | 164.40 | 152.84 | 138.61 | 133.82 | 589.67 |
| WYOMING | 227.90 | 228.70 | 205.46 | 187.98 | 850.03 |
| **9780 - TAPENTADOL Total** | **300.33** | **293.98** | **286.09** | **279.58** | **1,159.97** |

**DRUG: 9801 - FENTANYL BASE**

| STATE NAME | 1ST QUARTER | 2ND QUARTER | 3RD QUARTER | 4TH QUARTER | TOTAL GRAMS |
|---|---|---|---|---|---|
| ALABAMA | 19.11 | 18.26 | 17.63 | 17.02 | 72.02 |
| ALASKA | 18.06 | 16.09 | 15.69 | 14.04 | 63.89 |
| AMERICAN SAMOA | 0.73 | 0.20 | 0.00 | 0.06 | 0.99 |
| ARIZONA | 14.92 | 13.72 | 12.93 | 12.52 | 54.08 |
| ARKANSAS | 17.40 | 16.58 | 15.98 | 15.42 | 65.39 |
| CALIFORNIA | 11.60 | 10.94 | 10.36 | 9.95 | 42.85 |
| COLORADO | 15.58 | 14.79 | 14.81 | 13.88 | 59.05 |
| CONNECTICUT | 16.44 | 15.63 | 15.59 | 14.63 | 62.29 |
| DELAWARE | 19.38 | 18.72 | 17.90 | 16.55 | 72.55 |
| DISTRICT OF COLUMBIA | 10.10 | 8.80 | 8.68 | 8.10 | 35.69 |
| FLORIDA | 20.63 | 18.48 | 17.62 | 17.16 | 73.89 |
| GEORGIA | 18.99 | 17.86 | 17.57 | 16.82 | 71.24 |
| GUAM | 4.14 | 3.85 | 3.38 | 4.08 | 15.45 |
| HAWAII | 9.76 | 9.15 | 8.89 | 8.23 | 36.03 |
| IDAHO | 25.95 | 24.47 | 24.20 | 23.02 | 97.63 |
| ILLINOIS | 15.26 | 14.81 | 14.43 | 13.92 | 58.42 |
| INDIANA | 18.42 | 17.62 | 17.25 | 16.58 | 69.88 |
| IOWA | 20.19 | 19.66 | 18.95 | 18.58 | 77.38 |
| KANSAS | 25.09 | 24.43 | 23.17 | 22.02 | 94.70 |
| KENTUCKY | 16.73 | 15.93 | 15.12 | 14.56 | 62.34 |

| | | | | | |
|---|---|---|---|---|---|
| LOUISIANA | 16.91 | 15.80 | 15.62 | 14.82 | 63.15 |
| MAINE | 17.70 | 17.90 | 18.57 | 17.34 | 71.51 |
| MARYLAND | 14.15 | 13.15 | 12.84 | 11.93 | 52.07 |
| MASSACHUSETTS | 14.05 | 13.52 | 12.96 | 12.93 | 53.45 |
| MICHIGAN | 18.86 | 17.97 | 17.68 | 16.62 | 71.13 |
| MINNESOTA | 13.22 | 12.05 | 12.06 | 11.58 | 48.91 |
| MISSISSIPPI | 28.56 | 27.05 | 28.88 | 27.82 | 112.31 |
| MISSOURI | 22.86 | 22.20 | 21.03 | 20.42 | 86.51 |
| MONTANA | 22.72 | 21.59 | 21.87 | 20.88 | 87.05 |
| NEBRASKA | 20.21 | 20.08 | 19.06 | 18.67 | 78.01 |
| NEVADA | 13.13 | 12.11 | 11.39 | 10.81 | 47.44 |
| NEW HAMPSHIRE | 17.47 | 16.31 | 16.81 | 16.23 | 66.82 |
| NEW JERSEY | 16.60 | 15.47 | 15.30 | 14.55 | 61.92 |
| NEW MEXICO | 13.70 | 12.97 | 12.68 | 12.00 | 51.34 |
| NEW YORK | 14.44 | 13.14 | 13.22 | 12.54 | 53.34 |
| NORTH CAROLINA | 20.46 | 19.20 | 18.67 | 17.20 | 75.53 |
| NORTH DAKOTA | 24.48 | 22.32 | 21.72 | 21.81 | 90.33 |
| OHIO | 12.34 | 12.04 | 11.46 | 10.99 | 46.83 |
| OKLAHOMA | 20.20 | 19.30 | 18.48 | 17.54 | 75.52 |
| OREGON | 15.22 | 14.43 | 13.90 | 13.66 | 57.21 |
| PENNSYLVANIA | 24.84 | 23.32 | 21.93 | 23.76 | 93.86 |
| PUERTO RICO | 4.42 | 4.69 | 3.75 | 4.66 | 17.51 |
| RHODE ISLAND | 11.20 | 10.58 | 10.49 | 10.41 | 42.68 |
| SOUTH CAROLINA | 18.81 | 17.29 | 16.77 | 15.88 | 68.75 |
| SOUTH DAKOTA | 23.60 | 21.47 | 21.54 | 20.26 | 86.88 |
| TENNESSEE | 20.45 | 19.55 | 19.05 | 24.62 | 83.67 |
| TEXAS | 14.10 | 13.67 | 12.93 | 12.68 | 53.37 |
| UTAH | 21.39 | 20.32 | 20.57 | 19.67 | 81.95 |
| VERMONT | 19.05 | 17.41 | 18.54 | 17.68 | 72.68 |
| VIRGIN ISLANDS | 2.83 | 3.89 | 2.24 | 2.36 | 11.32 |
| VIRGINIA | 14.23 | 13.68 | 13.35 | 12.71 | 53.97 |
| WASHINGTON | 15.12 | 14.54 | 13.84 | 13.23 | 56.73 |
| WEST VIRGINIA | 19.17 | 17.46 | 17.44 | 16.56 | 70.63 |
| WISCONSIN | 15.86 | 15.72 | 14.82 | 14.17 | 60.57 |
| WYOMING | 17.42 | 16.54 | 16.71 | 16.07 | 66.74 |
| **9801 - FENTANYL BASE Total** | **16.49** | **15.58** | **15.08** | **14.71** | **61.87** |

**DRUG: 1100 - AMPHETAMINE**

| RANK | STATE NAME | STATE POPULATION | TOTAL GRAMS | TOTAL GRAMS/100K POPULATION |
|---|---|---|---|---|
| 1 | LOUISIANA | 4,533,372 | 697,051.17 | 15,376.00 |
| 2 | RHODE ISLAND | 1,052,567 | 157,985.45 | 15,009.54 |
| 3 | UTAH | 2,763,885 | 384,727.64 | 13,919.81 |
| 4 | NEW HAMPSHIRE | 1,316,470 | 172,628.12 | 13,112.95 |
| 5 | SOUTH CAROLINA | 4,625,364 | 584,957.42 | 12,646.73 |
| 6 | MASSACHUSETTS | 6,547,629 | 759,610.12 | 11,601.30 |
| 7 | ALABAMA | 4,779,736 | 549,062.62 | 11,487.30 |
| 8 | MICHIGAN | 9,883,640 | 1,011,092.15 | 10,229.96 |
| 9 | MISSOURI | 5,988,927 | 608,913.54 | 10,167.32 |
| 10 | NORTH CAROLINA | 9,535,483 | 933,442.27 | 9,789.15 |
| 11 | DELAWARE | 897,934 | 87,078.09 | 9,697.60 |
| 12 | VIRGINIA | 8,001,024 | 761,052.77 | 9,511.94 |
| 13 | KANSAS | 2,853,118 | 268,065.45 | 9,395.53 |
| 14 | WISCONSIN | 5,686,986 | 508,778.40 | 8,946.36 |
| 15 | INDIANA | 6,483,802 | 574,498.49 | 8,860.52 |
| 16 | TENNESSEE | 6,346,105 | 561,283.36 | 8,844.53 |
| 17 | MINNESOTA | 5,303,925 | 465,398.14 | 8,774.60 |
| 18 | DISTRICT OF COLUMBIA | 601,723 | 51,704.11 | 8,592.68 |
| 19 | MISSISSIPPI | 2,967,297 | 253,253.60 | 8,534.82 |
| 20 | GEORGIA | 9,687,653 | 824,674.34 | 8,512.63 |
| 21 | MAINE | 1,328,361 | 105,008.24 | 7,905.10 |
| 22 | IDAHO | 1,567,582 | 121,938.25 | 7,778.75 |
| 23 | TEXAS | 25,145,561 | 1,918,851.09 | 7,630.97 |
| 24 | ARKANSAS | 2,915,918 | 220,827.70 | 7,573.18 |
| 25 | COLORADO | 5,029,196 | 378,702.16 | 7,530.07 |
| 26 | OHIO | 11,536,504 | 858,620.65 | 7,442.64 |
| 27 | SOUTH DAKOTA | 814,180 | 59,726.90 | 7,335.83 |
| 28 | WASHINGTON | 6,724,540 | 490,440.66 | 7,293.30 |
| 29 | FLORIDA | 18,801,310 | 1,362,981.20 | 7,249.39 |
| 30 | IOWA | 3,046,355 | 219,198.84 | 7,195.45 |
| 31 | OREGON | 3,831,074 | 270,652.38 | 7,064.66 |
| 32 | MARYLAND | 5,773,552 | 407,174.51 | 7,052.41 |
| 33 | PENNSYLVANIA | 12,702,379 | 895,221.38 | 7,047.67 |
| 34 | KENTUCKY | 4,339,367 | 300,021.42 | 6,913.94 |
| 35 | VERMONT | 625,741 | 42,278.50 | 6,756.55 |
| 36 | CONNECTICUT | 3,574,097 | 239,929.66 | 6,713.01 |
| 37 | ILLINOIS | 12,830,632 | 845,087.37 | 6,586.48 |
| 38 | MONTANA | 989,415 | 64,219.93 | 6,490.70 |
| 39 | NEBRASKA | 1,826,341 | 118,252.69 | 6,474.84 |
| 40 | OKLAHOMA | 3,751,351 | 226,218.08 | 6,030.31 |
| 41 | ARIZONA | 6,392,017 | 384,918.68 | 6,021.87 |
| 42 | NORTH DAKOTA | 672,591 | 39,750.51 | 5,910.06 |
| 43 | NEW YORK | 19,378,102 | 1,062,011.46 | 5,480.47 |

| | | | | |
|---|---|---|---|---|
| 44 | NEW JERSEY | 8,791,894 | 467,832.40 | 5,321.18 |
| 45 | WEST VIRGINIA | 1,852,994 | 96,244.76 | 5,194.01 |
| 46 | WYOMING | 563,626 | 29,040.88 | 5,152.51 |
| 47 | ALASKA | 710,231 | 34,590.59 | 4,870.33 |
| 48 | NEVADA | 2,700,551 | 114,365.89 | 4,234.91 |
| 49 | CALIFORNIA | 37,253,956 | 1,266,258.02 | 3,398.99 |
| 50 | HAWAII | 1,360,301 | 43,063.47 | 3,165.73 |
| 51 | NEW MEXICO | 2,059,179 | 64,378.00 | 3,126.39 |
| 52 | VIRGIN ISLANDS | 109,825 | 1,460.40 | 1,329.75 |
| 53 | PUERTO RICO | 3,725,789 | 35,907.60 | 963.76 |
| 54 | GUAM | 178,430 | 1,715.41 | 961.39 |
| 55 | AMERICAN SAMOA | 65,628 | 3.75 | 5.72 |
| **1100 - AMPHETAMINE Total** | | **312,825,210** | 23,002,150.68 | **7,353.04** |

**DRUG: 1105 - METHAMPHETAMINE**

| RANK | STATE NAME | STATE POPULATION | TOTAL GRAMS | TOTAL GRAMS/100K POPULATION |
|---|---|---|---|---|
| 1 | WASHINGTON | 6,724,540 | 338.65 | 5.04 |
| 2 | NORTH DAKOTA | 672,591 | 32.54 | 4.84 |
| 3 | OREGON | 3,831,074 | 161.50 | 4.22 |
| 4 | MINNESOTA | 5,303,925 | 214.55 | 4.05 |
| 5 | COLORADO | 5,029,196 | 192.04 | 3.82 |
| 6 | UTAH | 2,763,885 | 104.05 | 3.76 |
| 7 | WYOMING | 563,626 | 19.68 | 3.49 |
| 8 | MAINE | 1,328,361 | 45.80 | 3.45 |
| 9 | MISSOURI | 5,988,927 | 204.08 | 3.41 |
| 10 | IDAHO | 1,567,582 | 52.62 | 3.36 |
| 11 | DISTRICT OF COLUMBIA | 601,723 | 20.09 | 3.34 |
| 12 | PENNSYLVANIA | 12,702,379 | 394.15 | 3.10 |
| 13 | SOUTH DAKOTA | 814,180 | 23.30 | 2.86 |
| 14 | CALIFORNIA | 37,253,956 | 956.56 | 2.57 |
| 15 | ALABAMA | 4,779,736 | 122.54 | 2.56 |
| 16 | CONNECTICUT | 3,574,097 | 88.79 | 2.48 |
| 17 | MARYLAND | 5,773,552 | 132.58 | 2.30 |
| 18 | KANSAS | 2,853,118 | 53.03 | 1.86 |
| 19 | WISCONSIN | 5,686,986 | 105.66 | 1.86 |
| 20 | IOWA | 3,046,355 | 56.25 | 1.85 |
| 21 | NORTH CAROLINA | 9,535,483 | 175.18 | 1.84 |
| 22 | DELAWARE | 897,934 | 16.47 | 1.83 |
| 23 | NEW HAMPSHIRE | 1,316,470 | 24.10 | 1.83 |
| 24 | ILLINOIS | 12,830,632 | 228.59 | 1.78 |

| | | | | |
|---|---|---|---|---|
| 25 | NEW YORK | 19,378,102 | 333.87 | 1.72 |
| 26 | NEW JERSEY | 8,791,894 | 140.63 | 1.60 |
| 27 | VIRGINIA | 8,001,024 | 126.16 | 1.58 |
| 28 | MASSACHUSETTS | 6,547,629 | 98.04 | 1.50 |
| 29 | NEVADA | 2,700,551 | 39.78 | 1.47 |
| 30 | TEXAS | 25,145,561 | 349.13 | 1.39 |
| 31 | NEW MEXICO | 2,059,179 | 28.52 | 1.39 |
| 32 | HAWAII | 1,360,301 | 18.48 | 1.36 |
| 33 | ARIZONA | 6,392,017 | 82.37 | 1.29 |
| 34 | NEBRASKA | 1,826,341 | 22.90 | 1.25 |
| 35 | OHIO | 11,536,504 | 141.43 | 1.23 |
| 36 | LOUISIANA | 4,533,372 | 53.44 | 1.18 |
| 37 | VERMONT | 625,741 | 7.23 | 1.16 |
| 38 | FLORIDA | 18,801,310 | 211.74 | 1.13 |
| 39 | INDIANA | 6,483,802 | 70.71 | 1.09 |
| 40 | SOUTH CAROLINA | 4,625,364 | 49.82 | 1.08 |
| 41 | GEORGIA | 9,687,653 | 96.42 | 1.00 |
| 42 | MISSISSIPPI | 2,967,297 | 29.33 | 0.99 |
| 43 | OKLAHOMA | 3,751,351 | 36.56 | 0.97 |
| 44 | KENTUCKY | 4,339,367 | 39.77 | 0.92 |
| 45 | TENNESSEE | 6,346,105 | 55.45 | 0.87 |
| 46 | MICHIGAN | 9,883,640 | 85.58 | 0.87 |
| 47 | ALASKA | 710,231 | 5.62 | 0.79 |
| 48 | RHODE ISLAND | 1,052,567 | 8.04 | 0.76 |
| 49 | WEST VIRGINIA | 1,852,994 | 14.06 | 0.76 |
| 50 | ARKANSAS | 2,915,918 | 8.84 | 0.30 |
| **1105 - METHAMPHETAMINE Total** | | **312,825,210** | 5,916.73 | **1.89** |

**DRUG: 1205 - LISDEXAMFETAMINE**

| RANK | STATE NAME | STATE POPULATION | TOTAL GRAMS | TOTAL GRAMS/100K POPULATION |
|---|---|---|---|---|
| 1 | LOUISIANA | 4,533,372 | 352,332.29 | 7,771.97 |
| 2 | DELAWARE | 897,934 | 54,458.18 | 6,064.83 |
| 3 | ALABAMA | 4,779,736 | 286,562.44 | 5,995.36 |
| 4 | SOUTH CAROLINA | 4,625,364 | 248,968.84 | 5,382.69 |
| 5 | WISCONSIN | 5,686,986 | 304,446.54 | 5,353.39 |
| 6 | MAINE | 1,328,361 | 67,856.22 | 5,108.27 |
| 7 | INDIANA | 6,483,802 | 326,072.10 | 5,029.03 |
| 8 | MISSISSIPPI | 2,967,297 | 145,831.51 | 4,914.62 |
| 9 | SOUTH DAKOTA | 814,180 | 38,885.90 | 4,776.08 |
| 10 | NORTH CAROLINA | 9,535,483 | 450,695.43 | 4,726.51 |

| | | | | | |
|---|---|---|---|---|---|
| 11 | VERMONT | | 625,741 | 29,388.29 | 4,696.56 |
| 12 | MISSOURI | | 5,988,927 | 279,487.65 | 4,666.74 |
| 13 | KANSAS | | 2,853,118 | 130,880.11 | 4,587.27 |
| 14 | IOWA | | 3,046,355 | 134,715.80 | 4,422.20 |
| 15 | NEW HAMPSHIRE | | 1,316,470 | 58,191.55 | 4,420.27 |
| 16 | TENNESSEE | | 6,346,105 | 267,084.09 | 4,208.63 |
| 17 | TEXAS | | 25,145,561 | 1,044,149.80 | 4,152.42 |
| 18 | DISTRICT OF COLUMBIA | | 601,723 | 24,921.89 | 4,141.75 |
| 19 | CONNECTICUT | | 3,574,097 | 145,262.66 | 4,064.32 |
| 20 | WEST VIRGINIA | | 1,852,994 | 74,844.29 | 4,039.10 |
| 21 | OKLAHOMA | | 3,751,351 | 150,908.03 | 4,022.76 |
| 22 | GEORGIA | | 9,687,653 | 388,891.92 | 4,014.30 |
| 23 | MARYLAND | | 5,773,552 | 230,946.38 | 4,000.07 |
| 24 | ARKANSAS | | 2,915,918 | 116,431.13 | 3,992.95 |
| 25 | NEBRASKA | | 1,826,341 | 71,009.51 | 3,888.08 |
| 26 | KENTUCKY | | 4,339,367 | 159,083.87 | 3,666.06 |
| 27 | NORTH DAKOTA | | 672,591 | 24,634.58 | 3,662.64 |
| 28 | VIRGINIA | | 8,001,024 | 278,950.59 | 3,486.44 |
| 29 | UTAH | | 2,763,885 | 92,621.09 | 3,351.12 |
| 30 | OHIO | | 11,536,504 | 378,431.31 | 3,280.29 |
| 31 | PENNSYLVANIA | | 12,702,379 | 373,774.60 | 2,942.56 |
| 32 | RHODE ISLAND | | 1,052,567 | 30,697.69 | 2,916.46 |
| 33 | FLORIDA | | 18,801,310 | 543,171.52 | 2,889.01 |
| 34 | MONTANA | | 989,415 | 26,459.06 | 2,674.21 |
| 35 | IDAHO | | 1,567,582 | 40,462.95 | 2,581.23 |
| 36 | MICHIGAN | | 9,883,640 | 251,104.09 | 2,540.60 |
| 37 | ARIZONA | | 6,392,017 | 160,420.44 | 2,509.70 |
| 38 | MINNESOTA | | 5,303,925 | 130,139.56 | 2,453.65 |
| 39 | MASSACHUSETTS | | 6,547,629 | 160,420.43 | 2,450.05 |
| 40 | COLORADO | | 5,029,196 | 120,063.85 | 2,387.34 |
| 41 | ILLINOIS | | 12,830,632 | 295,242.03 | 2,301.07 |
| 42 | WASHINGTON | | 6,724,540 | 150,285.30 | 2,234.88 |
| 43 | NEW JERSEY | | 8,791,894 | 192,232.79 | 2,186.48 |
| 44 | WYOMING | | 563,626 | 10,849.78 | 1,925.00 |
| 45 | NEW YORK | | 19,378,102 | 358,129.48 | 1,848.11 |
| 46 | OREGON | | 3,831,074 | 63,888.14 | 1,667.63 |
| 47 | ALASKA | | 710,231 | 11,691.49 | 1,646.15 |
| 48 | CALIFORNIA | | 37,253,956 | 461,442.31 | 1,238.64 |
| 49 | NEVADA | | 2,700,551 | 32,099.00 | 1,188.61 |

| | | | | |
|---|---|---|---|---|
| 50 | NEW MEXICO | 2,059,179 | 15,192.47 | 737.79 |
| 51 | HAWAII | 1,360,301 | 6,753.94 | 496.50 |
| 52 | VIRGIN ISLANDS | 109,825 | 324.31 | 295.30 |
| 53 | PUERTO RICO | 3,725,789 | 8,714.86 | 233.91 |
| 54 | GUAM | 178,430 | 56.65 | 31.75 |
| **1205 - LISDEXAMFETAMINE Total** | | **312,825,210** | 9,800,560.75 | **3,132.92** |

**DRUG: 1724 - METHYLPHENIDATE (DL;D;L;ISOMERS)**

| RANK | STATE NAME | STATE POPULATION | TOTAL GRAMS | TOTAL GRAMS/100K POPULATION |
|---|---|---|---|---|
| 1 | VERMONT | 625,741 | 74,445.73 | 11,897.21 |
| 2 | MAINE | 1,328,361 | 140,482.12 | 10,575.60 |
| 3 | NEW HAMPSHIRE | 1,316,470 | 136,682.00 | 10,382.46 |
| 4 | IOWA | 3,046,355 | 315,729.29 | 10,364.17 |
| 5 | SOUTH DAKOTA | 814,180 | 78,788.90 | 9,677.09 |
| 6 | MISSOURI | 5,988,927 | 559,708.85 | 9,345.73 |
| 7 | NORTH DAKOTA | 672,591 | 60,440.63 | 8,986.24 |
| 8 | MASSACHUSETTS | 6,547,629 | 570,144.86 | 8,707.65 |
| 9 | NORTH CAROLINA | 9,535,483 | 758,866.32 | 7,958.34 |
| 10 | DELAWARE | 897,934 | 70,171.41 | 7,814.76 |
| 11 | MINNESOTA | 5,303,925 | 405,411.14 | 7,643.61 |
| 12 | SOUTH CAROLINA | 4,625,364 | 350,387.00 | 7,575.34 |
| 13 | WISCONSIN | 5,686,986 | 411,172.45 | 7,230.06 |
| 14 | IDAHO | 1,567,582 | 112,973.23 | 7,206.85 |
| 15 | LOUISIANA | 4,533,372 | 322,695.52 | 7,118.22 |
| 16 | INDIANA | 6,483,802 | 458,712.61 | 7,074.75 |
| 17 | RHODE ISLAND | 1,052,567 | 73,910.21 | 7,021.90 |
| 18 | MICHIGAN | 9,883,640 | 691,584.16 | 6,997.26 |
| 19 | KANSAS | 2,853,118 | 199,025.25 | 6,975.71 |
| 20 | UTAH | 2,763,885 | 192,759.11 | 6,974.21 |
| 21 | PENNSYLVANIA | 12,702,379 | 878,531.54 | 6,916.28 |
| 22 | MARYLAND | 5,773,552 | 397,536.14 | 6,885.47 |
| 23 | MONTANA | 989,415 | 68,098.99 | 6,882.75 |
| 24 | WASHINGTON | 6,724,540 | 459,136.89 | 6,827.78 |
| 25 | OHIO | 11,536,504 | 785,451.66 | 6,808.40 |
| 26 | NEBRASKA | 1,826,341 | 124,194.04 | 6,800.16 |
| 27 | KENTUCKY | 4,339,367 | 293,804.33 | 6,770.67 |
| 28 | VIRGINIA | 8,001,024 | 530,901.12 | 6,635.41 |
| 29 | OREGON | 3,831,074 | 254,103.03 | 6,632.68 |
| 30 | GEORGIA | 9,687,653 | 638,558.48 | 6,591.47 |
| 31 | WEST VIRGINIA | 1,852,994 | 116,605.74 | 6,292.83 |

| | | | | |
|---|---|---|---|---|
| 32 | ALABAMA | 4,779,736 | 290,806.23 | 6,084.15 |
| 33 | CONNECTICUT | 3,574,097 | 216,689.47 | 6,062.78 |
| 34 | MISSISSIPPI | 2,967,297 | 170,634.28 | 5,750.50 |
| 35 | DISTRICT OF COLUMBIA | 601,723 | 33,318.56 | 5,537.19 |
| 36 | WYOMING | 563,626 | 30,627.99 | 5,434.10 |
| 37 | ARKANSAS | 2,915,918 | 153,952.59 | 5,279.73 |
| 38 | ILLINOIS | 12,830,632 | 668,751.45 | 5,212.15 |
| 39 | TEXAS | 25,145,561 | 1,303,729.79 | 5,184.73 |
| 40 | TENNESSEE | 6,346,105 | 329,007.58 | 5,184.40 |
| 41 | COLORADO | 5,029,196 | 253,485.20 | 5,040.27 |
| 42 | OKLAHOMA | 3,751,351 | 185,417.36 | 4,942.68 |
| 43 | ARIZONA | 6,392,017 | 306,356.07 | 4,792.79 |
| 44 | NEW YORK | 19,378,102 | 899,624.71 | 4,642.48 |
| 45 | ALASKA | 710,231 | 32,446.95 | 4,568.51 |
| 46 | FLORIDA | 18,801,310 | 777,539.54 | 4,135.56 |
| 47 | NEW JERSEY | 8,791,894 | 360,201.08 | 4,096.97 |
| 48 | NEW MEXICO | 2,059,179 | 71,701.78 | 3,482.06 |
| 49 | HAWAII | 1,360,301 | 42,764.29 | 3,143.74 |
| 50 | CALIFORNIA | 37,253,956 | 1,047,621.13 | 2,812.11 |
| 51 | NEVADA | 2,700,551 | 59,637.61 | 2,208.35 |
| 52 | PUERTO RICO | 3,725,789 | 53,386.65 | 1,432.90 |
| 53 | GUAM | 178,430 | 1,522.60 | 853.33 |
| 54 | VIRGIN ISLANDS | 109,825 | 469.65 | 427.64 |
| 55 | AMERICAN SAMOA | 65,628 | 35.02 | 53.36 |
| **1724 - METHYLPHENIDATE (DL;D;L;ISOMERS) Total** | | **312,825,210** | 17,820,740.31 | **5,696.71** |

**DRUG: 9050 - CODEINE**

| RANK | STATE NAME | STATE POPULATION | TOTAL GRAMS | TOTAL GRAMS/100K POPULATION |
|---|---|---|---|---|
| 1 | TEXAS | 25,145,561 | 3,323,385.87 | 13,216.59 |
| 2 | WEST VIRGINIA | 1,852,994 | 144,299.19 | 7,787.35 |
| 3 | OKLAHOMA | 3,751,351 | 288,095.29 | 7,679.77 |
| 4 | ARKANSAS | 2,915,918 | 201,702.07 | 6,917.28 |
| 5 | MICHIGAN | 9,883,640 | 675,240.73 | 6,831.90 |
| 6 | DISTRICT OF COLUMBIA | 601,723 | 33,091.45 | 5,499.45 |
| 7 | SOUTH CAROLINA | 4,625,364 | 232,833.58 | 5,033.84 |
| 8 | ILLINOIS | 12,830,632 | 499,898.30 | 3,896.13 |
| 9 | MISSOURI | 5,988,927 | 226,226.25 | 3,777.41 |
| 10 | ALABAMA | 4,779,736 | 177,501.78 | 3,713.63 |
| 11 | CALIFORNIA | 37,253,956 | 1,324,138.27 | 3,554.36 |
| 12 | TENNESSEE | 6,346,105 | 218,549.59 | 3,443.84 |

| | | | | | |
|---|---|---|---|---|---|
| 13 | KENTUCKY | | 4,339,367 | 143,721.65 | 3,312.04 |
| 14 | GEORGIA | | 9,687,653 | 315,807.40 | 3,259.90 |
| 15 | MONTANA | | 989,415 | 30,273.27 | 3,059.71 |
| 16 | MARYLAND | | 5,773,552 | 175,332.52 | 3,036.82 |
| 17 | DELAWARE | | 897,934 | 27,187.64 | 3,027.80 |
| 18 | LOUISIANA | | 4,533,372 | 135,750.06 | 2,994.46 |
| 19 | ARIZONA | | 6,392,017 | 190,581.01 | 2,981.55 |
| 20 | HAWAII | | 1,360,301 | 40,106.16 | 2,948.33 |
| 21 | PENNSYLVANIA | | 12,702,379 | 366,687.88 | 2,886.77 |
| 22 | IDAHO | | 1,567,582 | 44,902.67 | 2,864.45 |
| 23 | ALASKA | | 710,231 | 20,021.86 | 2,819.06 |
| 24 | NEW MEXICO | | 2,059,179 | 57,751.31 | 2,804.58 |
| 25 | KANSAS | | 2,853,118 | 79,113.93 | 2,772.89 |
| 26 | FLORIDA | | 18,801,310 | 511,625.95 | 2,721.22 |
| 27 | OREGON | | 3,831,074 | 103,532.51 | 2,702.44 |
| 28 | NEW JERSEY | | 8,791,894 | 237,124.43 | 2,697.08 |
| 29 | WASHINGTON | | 6,724,540 | 179,080.43 | 2,663.09 |
| 30 | WISCONSIN | | 5,686,986 | 145,466.57 | 2,557.89 |
| 31 | MISSISSIPPI | | 2,967,297 | 75,544.42 | 2,545.90 |
| 32 | NEVADA | | 2,700,551 | 66,190.95 | 2,451.02 |
| 33 | SOUTH DAKOTA | | 814,180 | 19,339.21 | 2,375.30 |
| 34 | NORTH DAKOTA | | 672,591 | 15,883.73 | 2,361.57 |
| 35 | MAINE | | 1,328,361 | 30,509.85 | 2,296.80 |
| 36 | INDIANA | | 6,483,802 | 147,895.67 | 2,281.00 |
| 37 | WYOMING | | 563,626 | 12,822.15 | 2,274.94 |
| 38 | VIRGIN ISLANDS | | 109,825 | 2,383.89 | 2,170.62 |
| 39 | NEW YORK | | 19,378,102 | 420,518.55 | 2,170.07 |
| 40 | MASSACHUSETTS | | 6,547,629 | 141,709.08 | 2,164.28 |
| 41 | OHIO | | 11,536,504 | 242,451.69 | 2,101.60 |
| 42 | VIRGINIA | | 8,001,024 | 161,495.05 | 2,018.43 |
| 43 | MINNESOTA | | 5,303,925 | 103,801.74 | 1,957.07 |
| 44 | CONNECTICUT | | 3,574,097 | 67,182.04 | 1,879.69 |
| 45 | NEBRASKA | | 1,826,341 | 33,761.30 | 1,848.58 |
| 46 | IOWA | | 3,046,355 | 55,926.30 | 1,835.84 |
| 47 | UTAH | | 2,763,885 | 50,721.79 | 1,835.16 |
| 48 | AMERICAN SAMOA | | 65,628 | 1,200.08 | 1,828.62 |
| 49 | NEW HAMPSHIRE | | 1,316,470 | 23,876.38 | 1,813.67 |
| 50 | RHODE ISLAND | | 1,052,567 | 18,331.72 | 1,741.62 |
| 51 | GUAM | | 178,430 | 2,859.35 | 1,602.51 |

| | | | | |
|---|---|---|---|---|
| 52 | NORTH CAROLINA | 9,535,483 | 148,656.06 | 1,558.98 |
| 53 | VERMONT | 625,741 | 9,397.58 | 1,501.83 |
| 54 | COLORADO | 5,029,196 | 67,270.50 | 1,337.60 |
| 55 | PUERTO RICO | 3,725,789 | 37,226.06 | 999.15 |
| **9050 - CODEINE Total** | | **312,825,210** | 12,105,984.78 | **3,869.89** |

**DRUG: 9064 - BUPRENORPHINE**

| RANK | STATE NAME | STATE POPULATION | TOTAL GRAMS | TOTAL GRAMS/100K POPULATION |
|---|---|---|---|---|
| 1 | VERMONT | 625,741 | 30,581.99 | 4,887.32 |
| 2 | WEST VIRGINIA | 1,852,994 | 77,092.60 | 4,160.43 |
| 3 | KENTUCKY | 4,339,367 | 179,650.03 | 4,140.01 |
| 4 | MAINE | 1,328,361 | 54,362.88 | 4,092.48 |
| 5 | RHODE ISLAND | 1,052,567 | 35,345.29 | 3,358.01 |
| 6 | NEW HAMPSHIRE | 1,316,470 | 40,446.94 | 3,072.38 |
| 7 | MASSACHUSETTS | 6,547,629 | 192,190.83 | 2,935.27 |
| 8 | ALABAMA | 4,779,736 | 131,440.58 | 2,749.95 |
| 9 | TENNESSEE | 6,346,105 | 170,350.15 | 2,684.33 |
| 10 | PENNSYLVANIA | 12,702,379 | 331,532.43 | 2,610.00 |
| 11 | MARYLAND | 5,773,552 | 133,936.79 | 2,319.83 |
| 12 | DISTRICT OF COLUMBIA | 601,723 | 13,940.85 | 2,316.82 |
| 13 | NORTH CAROLINA | 9,535,483 | 214,325.52 | 2,247.66 |
| 14 | OHIO | 11,536,504 | 244,541.76 | 2,119.72 |
| 15 | LOUISIANA | 4,533,372 | 94,795.28 | 2,091.05 |
| 16 | CONNECTICUT | 3,574,097 | 73,116.83 | 2,045.74 |
| 17 | DELAWARE | 897,934 | 17,376.39 | 1,935.15 |
| 18 | INDIANA | 6,483,802 | 121,059.87 | 1,867.11 |
| 19 | OKLAHOMA | 3,751,351 | 69,443.93 | 1,851.17 |
| 20 | NEW MEXICO | 2,059,179 | 37,460.23 | 1,819.18 |
| 21 | WASHINGTON | 6,724,540 | 121,621.53 | 1,808.62 |
| 22 | UTAH | 2,763,885 | 45,343.90 | 1,640.59 |
| 23 | OREGON | 3,831,074 | 60,648.14 | 1,583.06 |
| 24 | MISSISSIPPI | 2,967,297 | 46,953.89 | 1,582.38 |
| 25 | MONTANA | 989,415 | 15,311.71 | 1,547.55 |
| 26 | ALASKA | 710,231 | 10,487.33 | 1,476.61 |
| 27 | NEW YORK | 19,378,102 | 283,279.37 | 1,461.85 |
| 28 | SOUTH CAROLINA | 4,625,364 | 64,717.50 | 1,399.19 |
| 29 | NEW JERSEY | 8,791,894 | 122,905.06 | 1,397.94 |
| 30 | MICHIGAN | 9,883,640 | 134,695.17 | 1,362.81 |
| 31 | VIRGINIA | 8,001,024 | 104,121.87 | 1,301.36 |
| 32 | IDAHO | 1,567,582 | 19,922.41 | 1,270.90 |

| | | | | |
|---|---|---|---|---|
| 33 | FLORIDA | 18,801,310 | 234,921.23 | 1,249.49 |
| 34 | ARKANSAS | 2,915,918 | 35,289.16 | 1,210.22 |
| 35 | MISSOURI | 5,988,927 | 70,735.27 | 1,181.10 |
| 36 | WYOMING | 563,626 | 5,968.49 | 1,058.95 |
| 37 | WISCONSIN | 5,686,986 | 56,864.79 | 999.91 |
| 38 | NORTH DAKOTA | 672,591 | 5,999.33 | 891.97 |
| 39 | ARIZONA | 6,392,017 | 56,382.32 | 882.07 |
| 40 | COLORADO | 5,029,196 | 42,659.66 | 848.24 |
| 41 | MINNESOTA | 5,303,925 | 43,651.27 | 823.00 |
| 42 | GEORGIA | 9,687,653 | 79,140.38 | 816.92 |
| 43 | NEVADA | 2,700,551 | 17,857.25 | 661.24 |
| 44 | HAWAII | 1,360,301 | 8,879.84 | 652.79 |
| 45 | ILLINOIS | 12,830,632 | 78,421.17 | 611.20 |
| 46 | CALIFORNIA | 37,253,956 | 206,071.57 | 553.15 |
| 47 | TEXAS | 25,145,561 | 126,943.52 | 504.83 |
| 48 | SOUTH DAKOTA | 814,180 | 3,828.67 | 470.25 |
| 49 | KANSAS | 2,853,118 | 12,723.79 | 445.96 |
| 50 | NEBRASKA | 1,826,341 | 7,389.45 | 404.60 |
| 51 | IOWA | 3,046,355 | 10,180.51 | 334.19 |
| 52 | PUERTO RICO | 3,725,789 | 8,760.22 | 235.12 |
| 53 | VIRGIN ISLANDS | 109,825 | 84.19 | 76.66 |
| 54 | GUAM | 178,430 | 20.31 | 11.38 |
| **9064 BUPRENORPHINE Total** | | **312,825,210** | 4,405,771.45 | **1,408.38** |

**DRUG: 9143 - OXYCODONE**

| RANK | STATE NAME | STATE POPULATION | TOTAL GRAMS | TOTAL GRAMS/100K POPULATION |
|---|---|---|---|---|
| 1 | DELAWARE | 897,934 | 205,647.19 | 22,902.26 |
| 2 | TENNESSEE | 6,346,105 | 1,383,802.14 | 21,805.53 |
| 3 | NEVADA | 2,700,551 | 538,079.75 | 19,924.81 |
| 4 | NORTH CAROLINA | 9,535,483 | 1,889,900.04 | 19,819.66 |
| 5 | UTAH | 2,763,885 | 532,297.13 | 19,259.02 |
| 6 | ARIZONA | 6,392,017 | 1,164,487.18 | 18,217.84 |
| 7 | FLORIDA | 18,801,310 | 3,380,061.24 | 17,977.80 |
| 8 | MARYLAND | 5,773,552 | 964,973.75 | 16,713.69 |
| 9 | NEW JERSEY | 8,791,894 | 1,422,293.24 | 16,177.32 |
| 10 | PENNSYLVANIA | 12,702,379 | 2,046,125.42 | 16,108.21 |
| 11 | KENTUCKY | 4,339,367 | 662,350.17 | 15,263.75 |
| 12 | OKLAHOMA | 3,751,351 | 564,755.96 | 15,054.74 |
| 13 | GEORGIA | 9,687,653 | 1,453,952.73 | 15,008.31 |
| 14 | SOUTH CAROLINA | 4,625,364 | 679,585.37 | 14,692.58 |

| | | | | | |
|---|---|---|---|---|---|
| 15 | ALABAMA | | 4,779,736 | 689,000.17 | 14,415.03 |
| 16 | WYOMING | | 563,626 | 80,295.64 | 14,246.26 |
| 17 | CONNECTICUT | | 3,574,097 | 504,260.77 | 14,108.76 |
| 18 | NEW HAMPSHIRE | | 1,316,470 | 178,761.72 | 13,578.87 |
| 19 | COLORADO | | 5,029,196 | 663,692.69 | 13,196.80 |
| 20 | OREGON | | 3,831,074 | 505,419.73 | 13,192.64 |
| 21 | NEW MEXICO | | 2,059,179 | 270,735.26 | 13,147.73 |
| 22 | MISSOURI | | 5,988,927 | 774,269.64 | 12,928.35 |
| 23 | WASHINGTON | | 6,724,540 | 867,300.00 | 12,897.54 |
| 24 | LOUISIANA | | 4,533,372 | 583,208.34 | 12,864.78 |
| 25 | KANSAS | | 2,853,118 | 364,225.57 | 12,765.88 |
| 26 | ALASKA | | 710,231 | 89,506.34 | 12,602.43 |
| 27 | MAINE | | 1,328,361 | 161,805.24 | 12,180.82 |
| 28 | ARKANSAS | | 2,915,918 | 354,559.92 | 12,159.46 |
| 29 | IDAHO | | 1,567,582 | 189,231.00 | 12,071.52 |
| 30 | WEST VIRGINIA | | 1,852,994 | 217,135.79 | 11,718.11 |
| 31 | OHIO | | 11,536,504 | 1,338,448.88 | 11,601.86 |
| 32 | VIRGINIA | | 8,001,024 | 923,101.80 | 11,537.30 |
| 33 | RHODE ISLAND | | 1,052,567 | 118,274.17 | 11,236.74 |
| 34 | MASSACHUSETTS | | 6,547,629 | 732,820.63 | 11,192.15 |
| 35 | MONTANA | | 989,415 | 110,221.43 | 11,140.06 |
| 36 | INDIANA | | 6,483,802 | 700,734.70 | 10,807.47 |
| 37 | NEW YORK | | 19,378,102 | 2,081,426.29 | 10,741.13 |
| 38 | MISSISSIPPI | | 2,967,297 | 315,897.71 | 10,645.98 |
| 39 | VERMONT | | 625,741 | 66,483.54 | 10,624.77 |
| 40 | WISCONSIN | | 5,686,986 | 518,819.30 | 9,122.92 |
| 41 | HAWAII | | 1,360,301 | 123,651.39 | 9,090.00 |
| 42 | MICHIGAN | | 9,883,640 | 788,012.72 | 7,972.90 |
| 43 | MINNESOTA | | 5,303,925 | 400,964.38 | 7,559.77 |
| 44 | DISTRICT OF COLUMBIA | | 601,723 | 44,204.22 | 7,346.27 |
| 45 | NORTH DAKOTA | | 672,591 | 44,619.37 | 6,633.95 |
| 46 | NEBRASKA | | 1,826,341 | 120,743.56 | 6,611.23 |
| 47 | CALIFORNIA | | 37,253,956 | 2,387,342.55 | 6,408.29 |
| 48 | SOUTH DAKOTA | | 814,180 | 51,688.40 | 6,348.52 |
| 49 | IOWA | | 3,046,355 | 152,835.01 | 5,016.98 |
| 50 | TEXAS | | 25,145,561 | 1,018,613.87 | 4,050.87 |
| 51 | ILLINOIS | | 12,830,632 | 457,099.97 | 3,562.57 |
| 52 | VIRGIN ISLANDS | | 109,825 | 1,699.32 | 1,547.29 |
| 53 | GUAM | | 178,430 | 2,756.52 | 1,544.87 |

| | | | | |
|---|---|---|---|---|
| 54 | PUERTO RICO | 3,725,789 | 47,024.38 | 1,262.13 |
| 55 | AMERICAN SAMOA | 65,628 | 55.13 | 84.01 |
| **9143 - OXYCODONE Total** | | **312,825,210** | 35,929,258.36 | **11,485.41** |

**DRUG: 9150 - HYDROMORPHONE**

| RANK | STATE NAME | STATE POPULATION | TOTAL GRAMS | TOTAL GRAMS/100K POPULATION |
|---|---|---|---|---|
| 1 | MISSISSIPPI | 2,967,297 | 21,806.67 | 734.90 |
| 2 | FLORIDA | 18,801,310 | 131,922.78 | 701.67 |
| 3 | VERMONT | 625,741 | 3,680.11 | 588.12 |
| 4 | DELAWARE | 897,934 | 4,787.51 | 533.17 |
| 5 | MONTANA | 989,415 | 5,244.91 | 530.10 |
| 6 | CONNECTICUT | 3,574,097 | 17,980.22 | 503.07 |
| 7 | NEW HAMPSHIRE | 1,316,470 | 6,059.66 | 460.30 |
| 8 | WASHINGTON | 6,724,540 | 30,299.17 | 450.58 |
| 9 | COLORADO | 5,029,196 | 20,108.19 | 399.83 |
| 10 | MAINE | 1,328,361 | 5,301.05 | 399.07 |
| 11 | VIRGINIA | 8,001,024 | 30,908.45 | 386.31 |
| 12 | MARYLAND | 5,773,552 | 21,857.86 | 378.59 |
| 13 | WYOMING | 563,626 | 2,096.58 | 371.98 |
| 14 | OREGON | 3,831,074 | 14,061.27 | 367.03 |
| 15 | SOUTH CAROLINA | 4,625,364 | 16,518.63 | 357.13 |
| 16 | MASSACHUSETTS | 6,547,629 | 23,198.64 | 354.31 |
| 17 | NORTH CAROLINA | 9,535,483 | 32,932.71 | 345.37 |
| 18 | KANSAS | 2,853,118 | 9,802.77 | 343.58 |
| 19 | IDAHO | 1,567,582 | 5,290.25 | 337.48 |
| 20 | UTAH | 2,763,885 | 9,310.02 | 336.85 |
| 21 | MINNESOTA | 5,303,925 | 17,722.03 | 334.13 |
| 22 | PENNSYLVANIA | 12,702,379 | 41,586.54 | 327.39 |
| 23 | TENNESSEE | 6,346,105 | 20,607.60 | 324.73 |
| 24 | NEVADA | 2,700,551 | 8,183.15 | 303.02 |
| 25 | NEW JERSEY | 8,791,894 | 26,540.73 | 301.88 |
| 26 | NORTH DAKOTA | 672,591 | 2,013.46 | 299.36 |
| 27 | NEW YORK | 19,378,102 | 56,124.42 | 289.63 |
| 28 | ARIZONA | 6,392,017 | 18,418.08 | 288.14 |
| 29 | ALASKA | 710,231 | 1,975.71 | 278.18 |
| 30 | ARKANSAS | 2,915,918 | 8,002.04 | 274.43 |
| 31 | CALIFORNIA | 37,253,956 | 100,814.87 | 270.62 |
| 32 | OKLAHOMA | 3,751,351 | 10,078.91 | 268.67 |
| 33 | TEXAS | 25,145,561 | 67,396.65 | 268.03 |
| 34 | RHODE ISLAND | 1,052,567 | 2,755.98 | 261.83 |

| | | | | |
|---|---|---|---|---|
| 35 | INDIANA | 6,483,802 | 16,830.70 | 259.58 |
| 36 | MISSOURI | 5,988,927 | 15,471.13 | 258.33 |
| 37 | IOWA | 3,046,355 | 7,813.40 | 256.48 |
| 38 | DISTRICT OF COLUMBIA | 601,723 | 1,504.31 | 250.00 |
| 39 | GEORGIA | 9,687,653 | 23,274.15 | 240.25 |
| 40 | MICHIGAN | 9,883,640 | 22,916.66 | 231.86 |
| 41 | ILLINOIS | 12,830,632 | 27,502.77 | 214.35 |
| 42 | OHIO | 11,536,504 | 24,443.46 | 211.88 |
| 43 | WEST VIRGINIA | 1,852,994 | 3,856.36 | 208.12 |
| 44 | LOUISIANA | 4,533,372 | 9,265.32 | 204.38 |
| 45 | KENTUCKY | 4,339,367 | 8,741.55 | 201.45 |
| 46 | SOUTH DAKOTA | 814,180 | 1,622.88 | 199.33 |
| 47 | NEBRASKA | 1,826,341 | 3,402.35 | 186.29 |
| 48 | NEW MEXICO | 2,059,179 | 3,830.12 | 186.00 |
| 49 | WISCONSIN | 5,686,986 | 10,382.81 | 182.57 |
| 50 | ALABAMA | 4,779,736 | 8,621.53 | 180.38 |
| 51 | HAWAII | 1,360,301 | 2,006.06 | 147.47 |
| 52 | VIRGIN ISLANDS | 109,825 | 60.87 | 55.42 |
| 53 | GUAM | 178,430 | 72.61 | 40.69 |
| 54 | AMERICAN SAMOA | 65,628 | 7.76 | 11.82 |
| 55 | PUERTO RICO | 3,725,789 | 207.00 | 5.56 |
| **9150 - HYDROMORPHONE Total** | | **312,825,210** | 987,221.39 | **315.58** |

**DRUG: 9193 - HYDROCODONE**

| RANK | STATE NAME | STATE POPULATION | TOTAL GRAMS | TOTAL GRAMS/100K POPULATION |
|---|---|---|---|---|
| 1 | ALABAMA | 4,779,736 | 759,300.53 | 15,885.83 |
| 2 | KENTUCKY | 4,339,367 | 663,719.26 | 15,295.30 |
| 3 | ARKANSAS | 2,915,918 | 407,988.54 | 13,991.77 |
| 4 | OKLAHOMA | 3,751,351 | 524,197.75 | 13,973.57 |
| 5 | TENNESSEE | 6,346,105 | 735,040.29 | 11,582.54 |
| 6 | IDAHO | 1,567,582 | 180,534.78 | 11,516.77 |
| 7 | LOUISIANA | 4,533,372 | 489,745.71 | 10,803.12 |
| 8 | INDIANA | 6,483,802 | 696,799.74 | 10,746.78 |
| 9 | MISSISSIPPI | 2,967,297 | 307,454.38 | 10,361.43 |
| 10 | MICHIGAN | 9,883,640 | 1,019,072.59 | 10,310.70 |
| 11 | NEVADA | 2,700,551 | 270,150.93 | 10,003.55 |
| 12 | KANSAS | 2,853,118 | 246,273.62 | 8,631.74 |
| 13 | MISSOURI | 5,988,927 | 515,902.06 | 8,614.27 |
| 14 | SOUTH CAROLINA | 4,625,364 | 398,079.87 | 8,606.45 |
| 15 | WEST VIRGINIA | 1,852,994 | 154,637.62 | 8,345.28 |

| | | | | | |
|---|---|---|---|---|---|
| 16 | MONTANA | | 989,415 | 80,370.49 | 8,123.03 |
| 17 | TEXAS | | 25,145,561 | 2,034,240.07 | 8,089.86 |
| 18 | GEORGIA | | 9,687,653 | 761,653.42 | 7,862.10 |
| 19 | OREGON | | 3,831,074 | 288,932.98 | 7,541.83 |
| 20 | CALIFORNIA | | 37,253,956 | 2,671,833.62 | 7,171.95 |
| 21 | ILLINOIS | | 12,830,632 | 905,131.57 | 7,054.46 |
| 22 | UTAH | | 2,763,885 | 183,292.58 | 6,631.70 |
| 23 | NORTH CAROLINA | | 9,535,483 | 595,692.04 | 6,247.11 |
| 24 | WYOMING | | 563,626 | 34,344.39 | 6,093.47 |
| 25 | WASHINGTON | | 6,724,540 | 394,592.36 | 5,867.95 |
| 26 | FLORIDA | | 18,801,310 | 1,053,382.25 | 5,602.71 |
| 27 | IOWA | | 3,046,355 | 163,216.64 | 5,357.77 |
| 28 | NEW MEXICO | | 2,059,179 | 107,413.33 | 5,216.32 |
| 29 | NEBRASKA | | 1,826,341 | 93,399.61 | 5,114.03 |
| 30 | SOUTH DAKOTA | | 814,180 | 40,712.87 | 5,000.47 |
| 31 | ARIZONA | | 6,392,017 | 304,340.44 | 4,761.26 |
| 32 | ALASKA | | 710,231 | 32,651.84 | 4,597.35 |
| 33 | NORTH DAKOTA | | 672,591 | 30,575.89 | 4,545.99 |
| 34 | OHIO | | 11,536,504 | 518,366.85 | 4,493.27 |
| 35 | WISCONSIN | | 5,686,986 | 238,318.84 | 4,190.60 |
| 36 | MAINE | | 1,328,361 | 53,806.80 | 4,050.62 |
| 37 | VIRGINIA | | 8,001,024 | 308,448.70 | 3,855.12 |
| 38 | HAWAII | | 1,360,301 | 52,129.17 | 3,832.18 |
| 39 | COLORADO | | 5,029,196 | 189,547.52 | 3,768.94 |
| 40 | PENNSYLVANIA | | 12,702,379 | 454,989.65 | 3,581.92 |
| 41 | RHODE ISLAND | | 1,052,567 | 30,381.05 | 2,886.38 |
| 42 | NEW YORK | | 19,378,102 | 505,747.50 | 2,609.89 |
| 43 | MINNESOTA | | 5,303,925 | 126,984.62 | 2,394.16 |
| 44 | VERMONT | | 625,741 | 14,589.98 | 2,331.63 |
| 45 | NEW HAMPSHIRE | | 1,316,470 | 28,388.81 | 2,156.43 |
| 46 | DELAWARE | | 897,934 | 17,778.93 | 1,979.98 |
| 47 | MARYLAND | | 5,773,552 | 100,530.54 | 1,741.23 |
| 48 | CONNECTICUT | | 3,574,097 | 58,836.21 | 1,646.18 |
| 49 | MASSACHUSETTS | | 6,547,629 | 89,167.65 | 1,361.83 |
| 50 | NEW JERSEY | | 8,791,894 | 101,393.08 | 1,153.26 |
| 51 | VIRGIN ISLANDS | | 109,825 | 823.62 | 749.94 |
| 52 | DISTRICT OF COLUMBIA | | 601,723 | 3,885.04 | 645.65 |
| 53 | GUAM | | 178,430 | 446.92 | 250.47 |
| 54 | PUERTO RICO | | 3,725,789 | 1,722.49 | 46.23 |

| | | | | |
|---|---|---|---|---|
| 55 | AMERICAN SAMOA | 65,628 | 3.63 | 5.53 |
| **9193 - HYDROCODONE Total** | | **312,825,210** | 20,040,961.64 | **6,406.44** |

**DRUG: 9230 - MEPERIDINE (PETHIDINE)**

| RANK | STATE NAME | STATE POPULATION | TOTAL GRAMS | TOTAL GRAMS/100K POPULATION |
|---|---|---|---|---|
| 1 | ARKANSAS | 2,915,918 | 11,111.41 | 381.06 |
| 2 | PUERTO RICO | 3,725,789 | 12,710.83 | 341.16 |
| 3 | ALABAMA | 4,779,736 | 12,413.32 | 259.71 |
| 4 | OKLAHOMA | 3,751,351 | 9,666.57 | 257.68 |
| 5 | MISSISSIPPI | 2,967,297 | 7,147.83 | 240.89 |
| 6 | LOUISIANA | 4,533,372 | 9,988.70 | 220.34 |
| 7 | ARIZONA | 6,392,017 | 10,465.30 | 163.72 |
| 8 | IOWA | 3,046,355 | 4,721.02 | 154.97 |
| 9 | GEORGIA | 9,687,653 | 14,769.64 | 152.46 |
| 10 | VIRGIN ISLANDS | 109,825 | 160.68 | 146.31 |
| 11 | SOUTH CAROLINA | 4,625,364 | 6,480.87 | 140.12 |
| 12 | TENNESSEE | 6,346,105 | 8,437.97 | 132.96 |
| 13 | UTAH | 2,763,885 | 3,530.99 | 127.75 |
| 14 | SOUTH DAKOTA | 814,180 | 969.00 | 119.02 |
| 15 | VERMONT | 625,741 | 714.09 | 114.12 |
| 16 | ALASKA | 710,231 | 762.02 | 107.29 |
| 17 | NEVADA | 2,700,551 | 2,864.51 | 106.07 |
| 18 | VIRGINIA | 8,001,024 | 8,371.41 | 104.63 |
| 19 | CALIFORNIA | 37,253,956 | 37,006.07 | 99.33 |
| 20 | TEXAS | 25,145,561 | 24,812.60 | 98.68 |
| 21 | KANSAS | 2,853,118 | 2,796.43 | 98.01 |
| 22 | WYOMING | 563,626 | 534.77 | 94.88 |
| 23 | IDAHO | 1,567,582 | 1,429.15 | 91.17 |
| 24 | INDIANA | 6,483,802 | 5,910.08 | 91.15 |
| 25 | MISSOURI | 5,988,927 | 5,411.47 | 90.36 |
| 26 | KENTUCKY | 4,339,367 | 3,694.72 | 85.14 |
| 27 | OHIO | 11,536,504 | 9,638.32 | 83.55 |
| 28 | NEW MEXICO | 2,059,179 | 1,668.92 | 81.05 |
| 29 | MONTANA | 989,415 | 791.43 | 79.99 |
| 30 | NORTH CAROLINA | 9,535,483 | 7,319.95 | 76.77 |
| 31 | FLORIDA | 18,801,310 | 13,927.55 | 74.08 |
| 32 | DISTRICT OF COLUMBIA | 601,723 | 441.20 | 73.32 |
| 33 | WASHINGTON | 6,724,540 | 4,853.60 | 72.18 |
| 34 | OREGON | 3,831,074 | 2,741.85 | 71.57 |
| 35 | MARYLAND | 5,773,552 | 4,023.61 | 69.69 |

| | | | | |
|---|---|---|---|---|
| 36 | NEW HAMPSHIRE | 1,316,470 | 846.01 | 64.26 |
| 37 | NORTH DAKOTA | 672,591 | 412.55 | 61.34 |
| 38 | NEBRASKA | 1,826,341 | 1,057.24 | 57.89 |
| 39 | COLORADO | 5,029,196 | 2,499.79 | 49.71 |
| 40 | HAWAII | 1,360,301 | 652.54 | 47.97 |
| 41 | WEST VIRGINIA | 1,852,994 | 888.06 | 47.93 |
| 42 | GUAM | 178,430 | 84.97 | 47.62 |
| 43 | MICHIGAN | 9,883,640 | 4,635.29 | 46.90 |
| 44 | MASSACHUSETTS | 6,547,629 | 2,969.85 | 45.36 |
| 45 | WISCONSIN | 5,686,986 | 2,389.00 | 42.01 |
| 46 | PENNSYLVANIA | 12,702,379 | 5,234.34 | 41.21 |
| 47 | NEW YORK | 19,378,102 | 7,444.14 | 38.42 |
| 48 | ILLINOIS | 12,830,632 | 4,841.95 | 37.74 |
| 49 | NEW JERSEY | 8,791,894 | 3,074.54 | 34.97 |
| 50 | CONNECTICUT | 3,574,097 | 1,233.94 | 34.52 |
| 51 | MAINE | 1,328,361 | 429.43 | 32.33 |
| 52 | DELAWARE | 897,934 | 285.09 | 31.75 |
| 53 | RHODE ISLAND | 1,052,567 | 267.99 | 25.46 |
| 54 | MINNESOTA | 5,303,925 | 1,159.64 | 21.86 |
| **9230 - MEPERIDINE (PETHIDINE) Total** | | **312,825,210** | 292,694.19 | **93.56** |

**DRUG: 9250B - METHADONE**

| RANK | STATE NAME | STATE POPULATION | TOTAL GRAMS | TOTAL GRAMS/100K POPULATION |
|---|---|---|---|---|
| 1 | RHODE ISLAND | 1,052,567 | 191,125.68 | 18,158.05 |
| 2 | DELAWARE | 897,934 | 142,113.90 | 15,826.76 |
| 3 | CONNECTICUT | 3,574,097 | 493,003.03 | 13,793.78 |
| 4 | MARYLAND | 5,773,552 | 777,651.39 | 13,469.20 |
| 5 | VERMONT | 625,741 | 80,919.95 | 12,931.86 |
| 6 | NEW HAMPSHIRE | 1,316,470 | 131,904.60 | 10,019.57 |
| 7 | NEW MEXICO | 2,059,179 | 203,269.21 | 9,871.37 |
| 8 | MASSACHUSETTS | 6,547,629 | 604,219.06 | 9,228.06 |
| 9 | MAINE | 1,328,361 | 120,383.11 | 9,062.53 |
| 10 | ARIZONA | 6,392,017 | 427,491.18 | 6,687.89 |
| 11 | NEW YORK | 19,378,102 | 1,285,590.16 | 6,634.24 |
| 12 | OREGON | 3,831,074 | 252,088.22 | 6,580.09 |
| 13 | ALABAMA | 4,779,736 | 314,458.70 | 6,579.00 |
| 14 | DISTRICT OF COLUMBIA | 601,723 | 39,540.59 | 6,571.23 |
| 15 | WASHINGTON | 6,724,540 | 436,465.23 | 6,490.63 |
| 16 | PENNSYLVANIA | 12,702,379 | 790,485.46 | 6,223.13 |
| 17 | NEW JERSEY | 8,791,894 | 528,927.41 | 6,016.08 |

| | | | | |
|---|---|---|---|---|
| 18 | NORTH CAROLINA | 9,535,483 | 565,955.56 | 5,935.26 |
| 19 | ALASKA | 710,231 | 38,042.59 | 5,356.37 |
| 20 | WEST VIRGINIA | 1,852,994 | 97,109.03 | 5,240.66 |
| 21 | INDIANA | 6,483,802 | 330,511.59 | 5,097.50 |
| 22 | MICHIGAN | 9,883,640 | 493,844.37 | 4,996.58 |
| 23 | SOUTH CAROLINA | 4,625,364 | 230,085.30 | 4,974.43 |
| 24 | NEVADA | 2,700,551 | 133,186.45 | 4,931.82 |
| 25 | KENTUCKY | 4,339,367 | 207,565.94 | 4,783.32 |
| 26 | GEORGIA | 9,687,653 | 453,163.86 | 4,677.75 |
| 27 | VIRGINIA | 8,001,024 | 373,781.97 | 4,671.68 |
| 28 | FLORIDA | 18,801,310 | 859,335.92 | 4,570.62 |
| 29 | WISCONSIN | 5,686,986 | 255,370.69 | 4,490.44 |
| 30 | COLORADO | 5,029,196 | 204,537.35 | 4,067.00 |
| 31 | UTAH | 2,763,885 | 108,773.35 | 3,935.52 |
| 32 | ILLINOIS | 12,830,632 | 504,529.46 | 3,932.23 |
| 33 | MINNESOTA | 5,303,925 | 208,222.28 | 3,925.81 |
| 34 | OKLAHOMA | 3,751,351 | 139,144.94 | 3,709.20 |
| 35 | MONTANA | 989,415 | 33,548.77 | 3,390.77 |
| 36 | CALIFORNIA | 37,253,956 | 1,257,186.79 | 3,374.64 |
| 37 | NORTH DAKOTA | 672,591 | 21,305.06 | 3,167.61 |
| 38 | LOUISIANA | 4,533,372 | 137,798.97 | 3,039.66 |
| 39 | TENNESSEE | 6,346,105 | 191,442.29 | 3,016.69 |
| 40 | KANSAS | 2,853,118 | 81,944.36 | 2,872.10 |
| 41 | OHIO | 11,536,504 | 313,833.69 | 2,720.35 |
| 42 | IDAHO | 1,567,582 | 42,575.41 | 2,715.99 |
| 43 | MISSOURI | 5,988,927 | 139,683.60 | 2,332.36 |
| 44 | PUERTO RICO | 3,725,789 | 85,296.07 | 2,289.34 |
| 45 | HAWAII | 1,360,301 | 30,083.68 | 2,211.55 |
| 46 | TEXAS | 25,145,561 | 544,139.71 | 2,163.96 |
| 47 | IOWA | 3,046,355 | 54,117.37 | 1,776.46 |
| 48 | ARKANSAS | 2,915,918 | 50,857.10 | 1,744.12 |
| 49 | NEBRASKA | 1,826,341 | 29,183.03 | 1,597.90 |
| 50 | MISSISSIPPI | 2,967,297 | 33,545.46 | 1,130.51 |
| 51 | SOUTH DAKOTA | 814,180 | 7,755.70 | 952.58 |
| 52 | WYOMING | 563,626 | 3,185.94 | 565.26 |
| 53 | GUAM | 178,430 | 131.03 | 73.43 |
| 54 | VIRGIN ISLANDS | 109,825 | 33.09 | 30.13 |
| **9250B METHADONE Total** | | **312,825,210** | 15,080,444.66 | **4,820.73** |

**DRUG: 9300 - MORPHINE**

| RANK | STATE NAME | STATE POPULATION | TOTAL GRAMS | TOTAL GRAMS/100K POPULATION |
|---|---|---|---|---|
| 1 | TENNESSEE | 6,346,105 | 472,571.57 | 7,446.64 |
| 2 | DELAWARE | 897,934 | 60,394.75 | 6,725.97 |
| 3 | FLORIDA | 18,801,310 | 1,160,195.33 | 6,170.82 |
| 4 | ARIZONA | 6,392,017 | 384,304.17 | 6,012.25 |
| 5 | NEVADA | 2,700,551 | 154,925.85 | 5,736.82 |
| 6 | UTAH | 2,763,885 | 150,852.91 | 5,458.00 |
| 7 | OKLAHOMA | 3,751,351 | 203,876.39 | 5,434.75 |
| 8 | OREGON | 3,831,074 | 207,874.49 | 5,426.01 |
| 9 | IDAHO | 1,567,582 | 84,097.68 | 5,364.80 |
| 10 | PENNSYLVANIA | 12,702,379 | 666,893.94 | 5,250.15 |
| 11 | MONTANA | 989,415 | 51,882.33 | 5,243.74 |
| 12 | MICHIGAN | 9,883,640 | 497,478.02 | 5,033.35 |
| 13 | VERMONT | 625,741 | 31,331.55 | 5,007.11 |
| 14 | WYOMING | 563,626 | 26,806.60 | 4,756.10 |
| 15 | WASHINGTON | 6,724,540 | 317,460.42 | 4,720.92 |
| 16 | KANSAS | 2,853,118 | 132,069.08 | 4,628.94 |
| 17 | MISSOURI | 5,988,927 | 275,293.78 | 4,596.71 |
| 18 | SOUTH CAROLINA | 4,625,364 | 210,068.54 | 4,541.66 |
| 19 | NEW HAMPSHIRE | 1,316,470 | 58,719.18 | 4,460.35 |
| 20 | ALABAMA | 4,779,736 | 209,775.95 | 4,388.86 |
| 21 | COLORADO | 5,029,196 | 218,499.49 | 4,344.62 |
| 22 | NORTH CAROLINA | 9,535,483 | 412,535.51 | 4,326.32 |
| 23 | ALASKA | 710,231 | 30,043.76 | 4,230.14 |
| 24 | HAWAII | 1,360,301 | 57,163.61 | 4,202.28 |
| 25 | MAINE | 1,328,361 | 52,035.14 | 3,917.24 |
| 26 | WISCONSIN | 5,686,986 | 218,615.28 | 3,844.13 |
| 27 | NEW MEXICO | 2,059,179 | 78,296.12 | 3,802.30 |
| 28 | ARKANSAS | 2,915,918 | 110,339.90 | 3,784.05 |
| 29 | MISSISSIPPI | 2,967,297 | 112,132.74 | 3,778.95 |
| 30 | INDIANA | 6,483,802 | 241,547.60 | 3,725.40 |
| 31 | GEORGIA | 9,687,653 | 351,893.00 | 3,632.39 |
| 32 | LOUISIANA | 4,533,372 | 163,241.88 | 3,600.89 |
| 33 | MARYLAND | 5,773,552 | 202,004.40 | 3,498.79 |
| 34 | WEST VIRGINIA | 1,852,994 | 61,901.17 | 3,340.60 |
| 35 | IOWA | 3,046,355 | 100,453.03 | 3,297.48 |
| 36 | CALIFORNIA | 37,253,956 | 1,222,603.50 | 3,281.81 |
| 37 | RHODE ISLAND | 1,052,567 | 34,348.84 | 3,263.34 |
| 38 | NEBRASKA | 1,826,341 | 59,268.01 | 3,245.18 |

| | | | | |
|---|---|---|---|---|
| 39 | MASSACHUSETTS | 6,547,629 | 206,238.60 | 3,149.82 |
| 40 | VIRGINIA | 8,001,024 | 251,117.44 | 3,138.57 |
| 41 | KENTUCKY | 4,339,367 | 134,094.57 | 3,090.19 |
| 42 | SOUTH DAKOTA | 814,180 | 24,605.60 | 3,022.13 |
| 43 | CONNECTICUT | 3,574,097 | 107,711.52 | 3,013.67 |
| 44 | OHIO | 11,536,504 | 340,304.08 | 2,949.80 |
| 45 | NORTH DAKOTA | 672,591 | 19,193.15 | 2,853.61 |
| 46 | NEW JERSEY | 8,791,894 | 244,830.39 | 2,784.73 |
| 47 | ILLINOIS | 12,830,632 | 334,060.01 | 2,603.61 |
| 48 | MINNESOTA | 5,303,925 | 133,623.08 | 2,519.32 |
| 49 | TEXAS | 25,145,561 | 614,039.95 | 2,441.94 |
| 50 | NEW YORK | 19,378,102 | 451,747.65 | 2,331.23 |
| 51 | DISTRICT OF COLUMBIA | 601,723 | 10,457.95 | 1,738.00 |
| 52 | GUAM | 178,430 | 2,930.98 | 1,642.65 |
| 53 | VIRGIN ISLANDS | 109,825 | 326.22 | 297.04 |
| 54 | PUERTO RICO | 3,725,789 | 7,479.28 | 200.74 |
| 55 | AMERICAN SAMOA | 65,628 | 67.07 | 102.20 |
| **9300 - MORPHINE Total** | | **312,825,210** | 11,966,623.04 | **3,825.34** |

**DRUG: 9652 - OXYMORPHONE**

| RANK | STATE NAME | STATE POPULATION | TOTAL GRAMS | TOTAL GRAMS/100K POPULATION |
|---|---|---|---|---|
| 1 | TENNESSEE | 6,346,105 | 33,250.66 | 523.95 |
| 2 | DELAWARE | 897,934 | 4,412.82 | 491.44 |
| 3 | NORTH CAROLINA | 9,535,483 | 34,101.41 | 357.63 |
| 4 | UTAH | 2,763,885 | 8,396.58 | 303.80 |
| 5 | MARYLAND | 5,773,552 | 17,069.62 | 295.65 |
| 6 | MICHIGAN | 9,883,640 | 28,646.71 | 289.84 |
| 7 | PENNSYLVANIA | 12,702,379 | 33,337.82 | 262.45 |
| 8 | LOUISIANA | 4,533,372 | 11,268.78 | 248.57 |
| 9 | SOUTH CAROLINA | 4,625,364 | 9,831.48 | 212.56 |
| 10 | VIRGINIA | 8,001,024 | 16,283.06 | 203.51 |
| 11 | WEST VIRGINIA | 1,852,994 | 3,303.09 | 178.26 |
| 12 | FLORIDA | 18,801,310 | 30,691.01 | 163.24 |
| 13 | NEW YORK | 19,378,102 | 31,404.42 | 162.06 |
| 14 | NEVADA | 2,700,551 | 4,161.20 | 154.09 |
| 15 | KENTUCKY | 4,339,367 | 6,037.38 | 139.13 |
| 16 | IDAHO | 1,567,582 | 2,152.19 | 137.29 |
| 17 | MISSISSIPPI | 2,967,297 | 3,661.40 | 123.39 |
| 18 | NEW JERSEY | 8,791,894 | 10,151.43 | 115.46 |
| 19 | ALABAMA | 4,779,736 | 5,495.29 | 114.97 |

| | | | | |
|---|---|---|---|---|
| 20 | CONNECTICUT | 3,574,097 | 4,062.62 | 113.67 |
| 21 | SOUTH DAKOTA | 814,180 | 847.81 | 104.13 |
| 22 | OKLAHOMA | 3,751,351 | 3,716.31 | 99.07 |
| 23 | OHIO | 11,536,504 | 11,245.23 | 97.48 |
| 24 | MAINE | 1,328,361 | 1,275.66 | 96.03 |
| 25 | INDIANA | 6,483,802 | 6,152.41 | 94.89 |
| 26 | GEORGIA | 9,687,653 | 8,774.38 | 90.57 |
| 27 | MISSOURI | 5,988,927 | 5,224.63 | 87.24 |
| 28 | KANSAS | 2,853,118 | 2,435.08 | 85.35 |
| 29 | ARIZONA | 6,392,017 | 5,341.89 | 83.57 |
| 30 | COLORADO | 5,029,196 | 3,846.07 | 76.47 |
| 31 | NEW HAMPSHIRE | 1,316,470 | 975.78 | 74.12 |
| 32 | NEBRASKA | 1,826,341 | 1,344.57 | 73.62 |
| 33 | WASHINGTON | 6,724,540 | 4,465.67 | 66.41 |
| 34 | VERMONT | 625,741 | 414.56 | 66.25 |
| 35 | OREGON | 3,831,074 | 2,524.02 | 65.88 |
| 36 | WYOMING | 563,626 | 339.62 | 60.26 |
| 37 | RHODE ISLAND | 1,052,567 | 620.14 | 58.92 |
| 38 | NEW MEXICO | 2,059,179 | 1,206.03 | 58.57 |
| 39 | HAWAII | 1,360,301 | 726.08 | 53.38 |
| 40 | MASSACHUSETTS | 6,547,629 | 3,437.53 | 52.50 |
| 41 | ILLINOIS | 12,830,632 | 6,052.14 | 47.17 |
| 42 | TEXAS | 25,145,561 | 11,614.35 | 46.19 |
| 43 | CALIFORNIA | 37,253,956 | 15,519.86 | 41.66 |
| 44 | WISCONSIN | 5,686,986 | 2,222.49 | 39.08 |
| 45 | MONTANA | 989,415 | 379.81 | 38.39 |
| 46 | IOWA | 3,046,355 | 1,050.00 | 34.47 |
| 47 | DISTRICT OF COLUMBIA | 601,723 | 193.76 | 32.20 |
| 48 | ALASKA | 710,231 | 224.54 | 31.62 |
| 49 | NORTH DAKOTA | 672,591 | 210.36 | 31.28 |
| 50 | ARKANSAS | 2,915,918 | 837.68 | 28.73 |
| 51 | MINNESOTA | 5,303,925 | 756.95 | 14.27 |
| **9652 - OXYMORPHONE - Total** | | **312,825,210** | 401,694.39 | **128.41** |

**DRUG: 9780 - TAPENTADOL**

| RANK | STATE NAME | STATE POPULATION | TOTAL GRAMS | TOTAL GRAMS/100K POPULATION |
|---|---|---|---|---|
| 1 | NORTH CAROLINA | 9,535,483 | 314,194.40 | 3,295.00 |
| 2 | SOUTH CAROLINA | 4,625,364 | 121,003.00 | 2,616.08 |
| 3 | ALABAMA | 4,779,736 | 118,177.50 | 2,472.47 |
| 4 | DELAWARE | 897,934 | 17,970.00 | 2,001.26 |

| | | | | | |
|---|---|---|---|---|---|
| 5 | NEBRASKA | | 1,826,341 | 33,211.00 | 1,818.44 |
| 6 | NEW HAMPSHIRE | | 1,316,470 | 23,352.50 | 1,773.87 |
| 7 | GEORGIA | | 9,687,653 | 170,822.00 | 1,763.30 |
| 8 | COLORADO | | 5,029,196 | 88,258.00 | 1,754.91 |
| 9 | MARYLAND | | 5,773,552 | 92,908.50 | 1,609.21 |
| 10 | INDIANA | | 6,483,802 | 100,095.00 | 1,543.77 |
| 11 | SOUTH DAKOTA | | 814,180 | 11,890.50 | 1,460.43 |
| 12 | ARIZONA | | 6,392,017 | 91,197.00 | 1,426.73 |
| 13 | TENNESSEE | | 6,346,105 | 88,521.00 | 1,394.89 |
| 14 | ARKANSAS | | 2,915,918 | 39,594.50 | 1,357.87 |
| 15 | NEW YORK | | 19,378,102 | 262,714.50 | 1,355.73 |
| 16 | VIRGINIA | | 8,001,024 | 108,264.00 | 1,353.13 |
| 17 | PENNSYLVANIA | | 12,702,379 | 171,336.00 | 1,348.85 |
| 18 | NEW JERSEY | | 8,791,894 | 116,105.50 | 1,320.60 |
| 19 | OHIO | | 11,536,504 | 146,373.50 | 1,268.79 |
| 20 | UTAH | | 2,763,885 | 34,904.00 | 1,262.86 |
| 21 | FLORIDA | | 18,801,310 | 236,791.45 | 1,259.44 |
| 22 | IDAHO | | 1,567,582 | 18,338.50 | 1,169.86 |
| 23 | MISSOURI | | 5,988,927 | 67,727.50 | 1,130.88 |
| 24 | LOUISIANA | | 4,533,372 | 46,634.00 | 1,028.68 |
| 25 | MISSISSIPPI | | 2,967,297 | 30,202.00 | 1,017.83 |
| 26 | IOWA | | 3,046,355 | 30,459.50 | 999.87 |
| 27 | NEVADA | | 2,700,551 | 26,124.50 | 967.38 |
| 28 | MASSACHUSETTS | | 6,547,629 | 62,711.00 | 957.77 |
| 29 | TEXAS | | 25,145,561 | 238,612.00 | 948.92 |
| 30 | OKLAHOMA | | 3,751,351 | 34,196.50 | 911.58 |
| 31 | KENTUCKY | | 4,339,367 | 37,220.00 | 857.73 |
| 32 | WYOMING | | 563,626 | 4,791.00 | 850.03 |
| 33 | ALASKA | | 710,231 | 5,853.50 | 824.17 |
| 34 | NORTH DAKOTA | | 672,591 | 5,410.50 | 804.43 |
| 35 | CONNECTICUT | | 3,574,097 | 28,378.00 | 793.99 |
| 36 | WEST VIRGINIA | | 1,852,994 | 14,463.00 | 780.52 |
| 37 | WASHINGTON | | 6,724,540 | 49,094.00 | 730.07 |
| 38 | ILLINOIS | | 12,830,632 | 92,927.50 | 724.26 |
| 39 | CALIFORNIA | | 37,253,956 | 268,607.50 | 721.02 |
| 40 | KANSAS | | 2,853,118 | 20,410.00 | 715.36 |
| 41 | RHODE ISLAND | | 1,052,567 | 7,127.00 | 677.11 |
| 42 | WISCONSIN | | 5,686,986 | 33,534.50 | 589.67 |
| 43 | MONTANA | | 989,415 | 5,517.00 | 557.60 |

| | | | | |
|---|---|---|---|---|
| 44 | MICHIGAN | 9,883,640 | 54,417.00 | 550.58 |
| 45 | OREGON | 3,831,074 | 19,279.50 | 503.24 |
| 46 | VERMONT | 625,741 | 2,997.00 | 478.95 |
| 47 | DISTRICT OF COLUMBIA | 601,723 | 2,550.50 | 423.87 |
| 48 | HAWAII | 1,360,301 | 5,540.50 | 407.30 |
| 49 | NEW MEXICO | 2,059,179 | 8,083.50 | 392.56 |
| 50 | MAINE | 1,328,361 | 4,996.50 | 376.14 |
| 51 | MINNESOTA | 5,303,925 | 13,200.50 | 248.88 |
| 52 | PUERTO RICO | 3,725,789 | 1,593.50 | 42.77 |
| 53 | VIRGIN ISLANDS | 109,825 | 5.00 | 4.55 |
| **9780 - TAPENTADOL Total** | | **312,825,210** | 3,628,686.35 | **1,159.97** |

**DRUG: 9801 - FENTANYL BASE**

| RANK | STATE NAME | STATE POPULATION | TOTAL GRAMS | TOTAL GRAMS/100K POPULATION |
|---|---|---|---|---|
| 1 | MISSISSIPPI | 2,967,297 | 3,332.52 | 112.31 |
| 2 | IDAHO | 1,567,582 | 1,530.47 | 97.63 |
| 3 | KANSAS | 2,853,118 | 2,701.93 | 94.70 |
| 4 | PENNSYLVANIA | 12,702,379 | 11,922.28 | 93.86 |
| 5 | NORTH DAKOTA | 672,591 | 607.57 | 90.33 |
| 6 | MONTANA | 989,415 | 861.32 | 87.05 |
| 7 | SOUTH DAKOTA | 814,180 | 707.34 | 86.88 |
| 8 | MISSOURI | 5,988,927 | 5,181.01 | 86.51 |
| 9 | TENNESSEE | 6,346,105 | 5,309.56 | 83.67 |
| 10 | UTAH | 2,763,885 | 2,265.01 | 81.95 |
| 11 | NEBRASKA | 1,826,341 | 1,424.79 | 78.01 |
| 12 | IOWA | 3,046,355 | 2,357.16 | 77.38 |
| 13 | NORTH CAROLINA | 9,535,483 | 7,202.12 | 75.53 |
| 14 | OKLAHOMA | 3,751,351 | 2,833.15 | 75.52 |
| 15 | FLORIDA | 18,801,310 | 13,892.88 | 73.89 |
| 16 | VERMONT | 625,741 | 454.79 | 72.68 |
| 17 | DELAWARE | 897,934 | 651.43 | 72.55 |
| 18 | ALABAMA | 4,779,736 | 3,442.51 | 72.02 |
| 19 | MAINE | 1,328,361 | 949.85 | 71.51 |
| 20 | GEORGIA | 9,687,653 | 6,901.81 | 71.24 |
| 21 | MICHIGAN | 9,883,640 | 7,029.96 | 71.13 |
| 22 | WEST VIRGINIA | 1,852,994 | 1,308.75 | 70.63 |
| 23 | INDIANA | 6,483,802 | 4,530.74 | 69.88 |
| 24 | SOUTH CAROLINA | 4,625,364 | 3,180.12 | 68.75 |
| 25 | NEW HAMPSHIRE | 1,316,470 | 879.73 | 66.82 |
| 26 | WYOMING | 563,626 | 376.15 | 66.74 |

| | | | | | |
|---|---|---|---|---|---|
| 27 | ARKANSAS | | 2,915,918 | 1,906.65 | 65.39 |
| 28 | ALASKA | | 710,231 | 453.75 | 63.89 |
| 29 | LOUISIANA | | 4,533,372 | 2,862.66 | 63.15 |
| 30 | KENTUCKY | | 4,339,367 | 2,704.96 | 62.34 |
| 31 | CONNECTICUT | | 3,574,097 | 2,226.46 | 62.29 |
| 32 | NEW JERSEY | | 8,791,894 | 5,443.80 | 61.92 |
| 33 | WISCONSIN | | 5,686,986 | 3,444.54 | 60.57 |
| 34 | COLORADO | | 5,029,196 | 2,969.87 | 59.05 |
| 35 | ILLINOIS | | 12,830,632 | 7,495.72 | 58.42 |
| 36 | OREGON | | 3,831,074 | 2,191.63 | 57.21 |
| 37 | WASHINGTON | | 6,724,540 | 3,814.82 | 56.73 |
| 38 | ARIZONA | | 6,392,017 | 3,457.08 | 54.08 |
| 39 | VIRGINIA | | 8,001,024 | 4,317.77 | 53.97 |
| 40 | MASSACHUSETTS | | 6,547,629 | 3,499.72 | 53.45 |
| 41 | TEXAS | | 25,145,561 | 13,420.57 | 53.37 |
| 42 | NEW YORK | | 19,378,102 | 10,335.48 | 53.34 |
| 43 | MARYLAND | | 5,773,552 | 3,006.05 | 52.07 |
| 44 | NEW MEXICO | | 2,059,179 | 1,057.26 | 51.34 |
| 45 | MINNESOTA | | 5,303,925 | 2,593.89 | 48.91 |
| 46 | NEVADA | | 2,700,551 | 1,281.17 | 47.44 |
| 47 | OHIO | | 11,536,504 | 5,402.00 | 46.83 |
| 48 | CALIFORNIA | | 37,253,956 | 15,963.04 | 42.85 |
| 49 | RHODE ISLAND | | 1,052,567 | 449.24 | 42.68 |
| 50 | HAWAII | | 1,360,301 | 490.07 | 36.03 |
| 51 | DISTRICT OF COLUMBIA | | 601,723 | 214.74 | 35.69 |
| 52 | PUERTO RICO | | 3,725,789 | 652.46 | 17.51 |
| 53 | GUAM | | 178,430 | 27.56 | 15.45 |
| 54 | VIRGIN ISLANDS | | 109,825 | 12.43 | 11.32 |
| 55 | AMERICAN SAMOA | | 65,628 | 0.65 | 0.99 |
| **9801 - FENTANYL BASE Total** | | | **312,825,210** | 193,531.02 | **61.87** |

**STATE: AK - ALASKA**

**BUSINESS ACTIVITY: A - PHARMACIES**

| DRUG NAME | DRUG CODE | NUMBER OF REGISTRANT SOLD TO | TOTAL GRAMS | AVG GRAMS |
|---|---|---|---|---|
| AMPHETAMINE | 1100 | 105 | **28,717.50** | 273.50 |
| METHAMPHETAMINE | 1105 | 3 | **5.62** | 1.87 |
| LISDEXAMFETAMINE | 1205 | 100 | **10,478.64** | 104.79 |
| METHYLPHENIDATE (DL;D;L;ISOMERS) | 1724 | 107 | **25,352.45** | 236.94 |
| CODEINE | 9050 | 104 | **14,493.69** | 139.36 |
| BUPRENORPHINE | 9064 | 103 | **9,169.48** | 89.02 |
| OXYCODONE | 9143 | 108 | **78,618.17** | 727.95 |
| HYDROMORPHONE | 9150 | 98 | **1,589.50** | 16.22 |
| HYDROCODONE | 9193 | 110 | **28,945.63** | 263.14 |
| MEPERIDINE (PETHIDINE) | 9230 | 24 | **601.34** | 25.06 |
| METHADONE | 9250B | 87 | **7,801.67** | 89.67 |
| MORPHINE | 9300 | 108 | **24,881.68** | 230.39 |
| OXYMORPHONE | 9652 | 15 | **203.67** | 13.58 |
| TAPENTADOL | 9780 | 46 | **5,492.50** | 119.40 |
| FENTANYL BASE | 9801 | 97 | **347.59** | 3.58 |

**BUSINESS ACTIVITY: B - HOSPITALS**

| DRUG NAME | DRUG CODE | NUMBER OF REGISTRANT SOLD TO | TOTAL GRAMS | AVG GRAMS |
|---|---|---|---|---|
| AMPHETAMINE | 1100 | 30 | **5,873.09** | 195.77 |
| LISDEXAMFETAMINE | 1205 | 19 | **1,212.85** | 63.83 |
| METHYLPHENIDATE (DL;D;L;ISOMERS) | 1724 | 32 | **7,094.50** | 221.70 |
| CODEINE | 9050 | 36 | **5,308.66** | 147.46 |
| BUPRENORPHINE | 9064 | 35 | **1,249.09** | 35.69 |
| OXYCODONE | 9143 | 50 | **10,859.08** | 217.18 |
| HYDROMORPHONE | 9150 | 44 | **361.23** | 8.21 |
| HYDROCODONE | 9193 | 53 | **3,617.56** | 68.26 |
| MEPERIDINE (PETHIDINE) | 9230 | 28 | **131.81** | 4.71 |
| METHADONE | 9250B | 25 | **995.91** | 39.84 |
| MORPHINE | 9300 | 48 | **5,130.69** | 106.89 |
| OXYMORPHONE | 9652 | 1 | **20.88** | 20.88 |
| TAPENTADOL | 9780 | 4 | **361.00** | 90.25 |
| FENTANYL BASE | 9801 | 53 | **100.32** | 1.89 |

**BUSINESS ACTIVITY: C - PRACTITIONERS**

| DRUG NAME | DRUG CODE | NUMBER OF REGISTRANT SOLD TO | TOTAL GRAMS | AVG GRAMS |
|---|---|---|---|---|
| CODEINE | 9050 | 12 | **191.89** | 15.99 |
| BUPRENORPHINE | 9064 | 59 | **4.52** | 0.08 |
| OXYCODONE | 9143 | 5 | **28.19** | 5.64 |
| HYDROMORPHONE | 9150 | 48 | **24.98** | 0.52 |
| HYDROCODONE | 9193 | 37 | **79.87** | 2.16 |
| MEPERIDINE (PETHIDINE) | 9230 | 11 | **28.87** | 2.62 |
| METHADONE | 9250B | 7 | **51.79** | 7.40 |
| MORPHINE | 9300 | 25 | **29.89** | 1.20 |
| FENTANYL BASE | 9801 | 83 | **5.83** | 0.07 |

**BUSINESS ACTIVITY: M - MID-LEVEL PRACTITIONERS**

| DRUG NAME | DRUG CODE | NUMBER OF REGISTRANT SOLD TO | TOTAL GRAMS | AVG GRAMS |
|---|---|---|---|---|
| CODEINE | 9050 | 5 | **27.63** | 5.53 |
| BUPRENORPHINE | 9064 | 1 | **3.93** | 3.93 |
| OXYCODONE | 9143 | 2 | **0.90** | 0.45 |
| HYDROCODONE | 9193 | 4 | **8.78** | 2.19 |
| MORPHINE | 9300 | 5 | **1.50** | 0.30 |
| FENTANYL BASE | 9801 | 5 | **0.02** | 0.00 |

**BUSINESS ACTIVITY: NTP - NARCOTIC TREATMENT PROGRAMS**

| DRUG NAME | DRUG CODE | NUMBER OF REGISTRANT SOLD TO | TOTAL GRAMS | AVG GRAMS |
|---|---|---|---|---|
| BUPRENORPHINE | 9064 | 2 | **60.30** | 30.15 |
| METHADONE | 9250B | 4 | **29,193.22** | 7,298.30 |

**STATE: AL - ALABAMA**

**BUSINESS ACTIVITY: A - PHARMACIES**

| DRUG NAME | DRUG CODE | NUMBER OF REGISTRANT SOLD TO | TOTAL GRAMS | AVG GRAMS |
|---|---|---|---|---|
| AMPHETAMINE | 1100 | 1,291 | **540,607.57** | 418.75 |
| METHAMPHETAMINE | 1105 | 37 | **122.54** | 3.31 |
| LISDEXAMFETAMINE | 1205 | 1,260 | **283,478.85** | 224.98 |
| METHYLPHENIDATE (DL;D;L;ISOMERS) | 1724 | 1,276 | **283,883.93** | 222.48 |
| CODEINE | 9050 | 1,258 | **172,641.60** | 137.23 |
| BUPRENORPHINE | 9064 | 1,231 | **129,123.23** | 104.89 |
| OXYCODONE | 9143 | 1,296 | **659,430.54** | 508.82 |
| HYDROMORPHONE | 9150 | 979 | **6,643.49** | 6.79 |
| HYDROCODONE | 9193 | 1,313 | **728,292.67** | 554.68 |
| MEPERIDINE (PETHIDINE) | 9230 | 342 | **4,817.43** | 14.09 |
| METHADONE | 9250B | 1,023 | **44,923.57** | 43.91 |
| MORPHINE | 9300 | 1,256 | **193,196.85** | 153.82 |
| OXYMORPHONE | 9652 | 416 | **5,461.84** | 13.13 |
| TAPENTADOL | 9780 | 890 | **117,302.00** | 131.80 |
| FENTANYL BASE | 9801 | 1,190 | **3,024.88** | 2.54 |

**BUSINESS ACTIVITY: B - HOSPITALS**

| DRUG NAME | DRUG CODE | NUMBER OF REGISTRANT SOLD TO | TOTAL GRAMS | AVG GRAMS |
|---|---|---|---|---|
| AMPHETAMINE | 1100 | 62 | **8,455.06** | 136.37 |
| LISDEXAMFETAMINE | 1205 | 26 | **3,083.59** | 118.60 |
| METHYLPHENIDATE (DL;D;L;ISOMERS) | 1724 | 82 | **6,922.30** | 84.42 |
| CODEINE | 9050 | 114 | **3,182.78** | 27.92 |
| BUPRENORPHINE | 9064 | 103 | **1,529.51** | 14.85 |
| OXYCODONE | 9143 | 138 | **29,192.43** | 211.54 |
| HYDROMORPHONE | 9150 | 123 | **1,928.49** | 15.68 |
| HYDROCODONE | 9193 | 147 | **30,375.16** | 206.63 |

| | | | | |
|---|---|---|---|---|
| MEPERIDINE (PETHIDINE) | 9230 | 109 | **6,530.80** | 59.92 |
| METHADONE | 9250B | 103 | **2,196.83** | 21.33 |
| MORPHINE | 9300 | 144 | **16,275.39** | 113.02 |
| OXYMORPHONE | 9652 | 6 | **33.45** | 5.58 |
| TAPENTADOL | 9780 | 25 | **875.50** | 35.02 |
| FENTANYL BASE | 9801 | 128 | **379.64** | 2.97 |

**BUSINESS ACTIVITY: C - PRACTITIONERS**

| DRUG NAME | DRUG CODE | NUMBER OF REGISTRANT SOLD TO | TOTAL GRAMS | AVG GRAMS |
|---|---|---|---|---|
| CODEINE | 9050 | 88 | **1,669.10** | 18.97 |
| BUPRENORPHINE | 9064 | 321 | **34.67** | 0.11 |
| OXYCODONE | 9143 | 30 | **377.20** | 12.57 |
| HYDROMORPHONE | 9150 | 99 | **48.84** | 0.49 |
| HYDROCODONE | 9193 | 118 | **632.70** | 5.36 |
| MEPERIDINE (PETHIDINE) | 9230 | 84 | **1,065.08** | 12.68 |
| METHADONE | 9250B | 4 | **3.04** | 0.76 |
| MORPHINE | 9300 | 105 | **303.34** | 2.89 |
| FENTANYL BASE | 9801 | 239 | **37.82** | 0.16 |

**BUSINESS ACTIVITY: D - TEACHING INSTITUTIONS**

| DRUG NAME | DRUG CODE | NUMBER OF REGISTRANT SOLD TO | TOTAL GRAMS | AVG GRAMS |
|---|---|---|---|---|
| BUPRENORPHINE | 9064 | 1 | **0.02** | 0.02 |
| HYDROMORPHONE | 9150 | 1 | **0.71** | 0.71 |
| MORPHINE | 9300 | 1 | **0.38** | 0.38 |
| FENTANYL BASE | 9801 | 1 | **0.16** | 0.16 |

**BUSINESS ACTIVITY: M - MID-LEVEL PRACTITIONERS**

| DRUG NAME | DRUG CODE | NUMBER OF REGISTRANT SOLD TO | TOTAL GRAMS | AVG GRAMS |
|---|---|---|---|---|
| CODEINE | 9050 | 13 | **8.31** | 0.64 |
| BUPRENORPHINE | 9064 | 2 | **12.92** | 6.46 |

**BUSINESS ACTIVITY: NTP - NARCOTIC TREATMENT PROGRAMS**

| DRUG NAME | DRUG CODE | NUMBER OF REGISTRANT SOLD TO | TOTAL GRAMS | AVG GRAMS |
|---|---|---|---|---|
| BUPRENORPHINE | 9064 | 11 | **740.24** | 67.29 |
| METHADONE | 9250B | 22 | **267,335.27** | 12,151.60 |

**STATE: AR - ARKANSAS**

**BUSINESS ACTIVITY: A - PHARMACIES**

| DRUG NAME | DRUG CODE | NUMBER OF REGISTRANT SOLD TO | TOTAL GRAMS | AVG GRAMS |
|---|---|---|---|---|
| AMPHETAMINE | 1100 | 744 | **216,186.85** | 290.57 |
| METHAMPHETAMINE | 1105 | 6 | **8.84** | 1.47 |
| LISDEXAMFETAMINE | 1205 | 736 | **115,157.52** | 156.46 |
| METHYLPHENIDATE (DL;D;L;ISOMERS) | 1724 | 742 | **150,012.53** | 202.17 |
| CODEINE | 9050 | 751 | **197,829.48** | 263.42 |
| BUPRENORPHINE | 9064 | 711 | **34,115.47** | 47.98 |

| | | | | |
|---|---|---|---|---|
| OXYCODONE | 9143 | 750 | **340,002.58** | 453.34 |
| HYDROMORPHONE | 9150 | 672 | **6,583.36** | 9.80 |
| HYDROCODONE | 9193 | 756 | **394,305.24** | 521.57 |
| MEPERIDINE (PETHIDINE) | 9230 | 428 | **7,260.47** | 16.96 |
| METHADONE | 9250B | 619 | **16,018.61** | 25.88 |
| MORPHINE | 9300 | 740 | **101,752.88** | 137.50 |
| OXYMORPHONE | 9652 | 124 | **833.76** | 6.72 |
| TAPENTADOL | 9780 | 462 | **38,762.50** | 83.90 |
| FENTANYL BASE | 9801 | 699 | **1,640.21** | 2.35 |

**BUSINESS ACTIVITY: B - HOSPITALS**

| DRUG NAME | DRUG CODE | NUMBER OF REGISTRANT SOLD TO | TOTAL GRAMS | AVG GRAMS |
|---|---|---|---|---|
| AMPHETAMINE | 1100 | 48 | **4,640.85** | 96.68 |
| LISDEXAMFETAMINE | 1205 | 26 | **1,272.40** | 48.94 |
| METHYLPHENIDATE (DL;D;L;ISOMERS) | 1724 | 71 | **3,940.06** | 55.49 |
| CODEINE | 9050 | 90 | **3,465.27** | 38.50 |
| BUPRENORPHINE | 9064 | 55 | **1,147.77** | 20.87 |
| OXYCODONE | 9143 | 124 | **14,527.30** | 117.16 |
| HYDROMORPHONE | 9150 | 108 | **1,391.58** | 12.89 |
| HYDROCODONE | 9193 | 122 | **13,487.57** | 110.55 |
| MEPERIDINE (PETHIDINE) | 9230 | 85 | **3,426.08** | 40.31 |
| METHADONE | 9250B | 85 | **1,490.79** | 17.54 |
| MORPHINE | 9300 | 121 | **8,541.79** | 70.59 |
| OXYMORPHONE | 9652 | 3 | **3.93** | 1.31 |
| TAPENTADOL | 9780 | 19 | **832.00** | 43.79 |
| FENTANYL BASE | 9801 | 121 | **255.68** | 2.11 |

**BUSINESS ACTIVITY: C - PRACTITIONERS**

| DRUG NAME | DRUG CODE | NUMBER OF REGISTRANT SOLD TO | TOTAL GRAMS | AVG GRAMS |
|---|---|---|---|---|
| LISDEXAMFETAMINE | 1205 | 1 | **1.21** | 1.21 |
| CODEINE | 9050 | 39 | **390.57** | 10.01 |
| BUPRENORPHINE | 9064 | 166 | **16.05** | 0.10 |
| OXYCODONE | 9143 | 6 | **30.03** | 5.01 |
| HYDROMORPHONE | 9150 | 66 | **27.09** | 0.41 |
| HYDROCODONE | 9193 | 72 | **195.73** | 2.72 |
| MEPERIDINE (PETHIDINE) | 9230 | 36 | **424.86** | 11.80 |
| METHADONE | 9250B | 1 | **0.89** | 0.89 |
| MORPHINE | 9300 | 56 | **45.24** | 0.81 |
| FENTANYL BASE | 9801 | 148 | **10.76** | 0.07 |

**BUSINESS ACTIVITY: M - MID-LEVEL PRACTITIONERS**

| DRUG NAME | DRUG CODE | NUMBER OF REGISTRANT SOLD TO | TOTAL GRAMS | AVG GRAMS |
|---|---|---|---|---|
| CODEINE | 9050 | 20 | **16.76** | 0.84 |

| | | | | |
|---|---|---|---|---|
| BUPRENORPHINE | 9064 | 1 | **0.75** | 0.75 |

**BUSINESS ACTIVITY: NTP - NARCOTIC TREATMENT PROGRAMS**

| DRUG NAME | DRUG CODE | NUMBER OF REGISTRANT SOLD TO | TOTAL GRAMS | AVG GRAMS |
|---|---|---|---|---|
| BUPRENORPHINE | 9064 | 2 | **9.12** | 4.56 |
| METHADONE | 9250B | 7 | **33,346.81** | 4,763.83 |

**STATE: AS - AMERICAN SAMOA**

**BUSINESS ACTIVITY: B - HOSPITALS**

| DRUG NAME | DRUG CODE | NUMBER OF REGISTRANT SOLD TO | TOTAL GRAMS | AVG GRAMS |
|---|---|---|---|---|
| AMPHETAMINE | 1100 | 1 | **3.75** | 3.75 |
| METHYLPHENIDATE (DL;D;L;ISOMERS) | 1724 | 1 | **35.02** | 35.02 |
| CODEINE | 9050 | 1 | **1,200.08** | 1,200.08 |
| OXYCODONE | 9143 | 1 | **55.13** | 55.13 |
| HYDROMORPHONE | 9150 | 1 | **7.76** | 7.76 |
| HYDROCODONE | 9193 | 1 | **3.63** | 3.63 |
| MORPHINE | 9300 | 1 | **67.07** | 67.07 |
| FENTANYL BASE | 9801 | 1 | **0.65** | 0.65 |

**STATE: AZ - ARIZONA**

**BUSINESS ACTIVITY: A - PHARMACIES**

| DRUG NAME | DRUG CODE | NUMBER OF REGISTRANT SOLD TO | TOTAL GRAMS | AVG GRAMS |
|---|---|---|---|---|
| AMPHETAMINE | 1100 | 1,105 | **375,929.86** | 340.21 |
| METHAMPHETAMINE | 1105 | 32 | **82.37** | 2.57 |
| LISDEXAMFETAMINE | 1205 | 1,056 | **158,732.96** | 150.32 |
| METHYLPHENIDATE (DL;D;L;ISOMERS) | 1724 | 1,098 | **292,361.96** | 266.27 |
| CODEINE | 9050 | 1,080 | **176,671.52** | 163.58 |
| BUPRENORPHINE | 9064 | 1,090 | **51,009.75** | 46.80 |
| OXYCODONE | 9143 | 1,120 | **1,116,207.56** | 996.61 |
| HYDROMORPHONE | 9150 | 988 | **16,164.06** | 16.36 |
| HYDROCODONE | 9193 | 1,124 | **290,208.65** | 258.19 |
| MEPERIDINE (PETHIDINE) | 9230 | 191 | **4,294.32** | 22.48 |
| METHADONE | 9250B | 841 | **26,500.00** | 31.51 |
| MORPHINE | 9300 | 1,106 | **352,959.41** | 319.13 |
| OXYMORPHONE | 9652 | 412 | **5,336.01** | 12.95 |
| TAPENTADOL | 9780 | 730 | **90,471.50** | 123.93 |
| FENTANYL BASE | 9801 | 986 | **2,951.31** | 2.99 |

**BUSINESS ACTIVITY: B - HOSPITALS**

| DRUG NAME | DRUG CODE | NUMBER OF REGISTRANT SOLD TO | TOTAL GRAMS | AVG GRAMS |
|---|---|---|---|---|
| AMPHETAMINE | 1100 | 96 | **8,988.82** | 93.63 |
| LISDEXAMFETAMINE | 1205 | 38 | **1,687.47** | 44.41 |
| METHYLPHENIDATE (DL;D;L;ISOMERS) | 1724 | 112 | **13,994.11** | 124.95 |
| CODEINE | 9050 | 130 | **10,950.28** | 84.23 |

| | | | | |
|---|---|---|---|---|
| BUPRENORPHINE | 9064 | 88 | **3,630.72** | 41.26 |
| OXYCODONE | 9143 | 162 | **48,050.30** | 296.61 |
| HYDROMORPHONE | 9150 | 123 | **2,069.19** | 16.82 |
| HYDROCODONE | 9193 | 154 | **13,332.38** | 86.57 |
| MEPERIDINE (PETHIDINE) | 9230 | 86 | **3,305.16** | 38.43 |
| METHADONE | 9250B | 112 | **3,828.30** | 34.18 |
| MORPHINE | 9300 | 157 | **31,144.11** | 198.37 |
| OXYMORPHONE | 9652 | 6 | **5.89** | 0.98 |
| TAPENTADOL | 9780 | 11 | **725.50** | 65.95 |
| FENTANYL BASE | 9801 | 124 | **452.95** | 3.65 |

**BUSINESS ACTIVITY: C - PRACTITIONERS**

| DRUG NAME | DRUG CODE | NUMBER OF REGISTRANT SOLD TO | TOTAL GRAMS | AVG GRAMS |
|---|---|---|---|---|
| CODEINE | 9050 | 84 | **2,723.62** | 32.42 |
| BUPRENORPHINE | 9064 | 678 | **102.47** | 0.15 |
| OXYCODONE | 9143 | 101 | **228.88** | 2.27 |
| HYDROMORPHONE | 9150 | 408 | **184.83** | 0.45 |
| HYDROCODONE | 9193 | 286 | **799.40** | 2.80 |
| MEPERIDINE (PETHIDINE) | 9230 | 297 | **2,865.82** | 9.65 |
| METHADONE | 9250B | 16 | **102.50** | 6.41 |
| MORPHINE | 9300 | 234 | **200.27** | 0.86 |
| FENTANYL BASE | 9801 | 750 | **52.71** | 0.07 |

**BUSINESS ACTIVITY: M - MID-LEVEL PRACTITIONERS**

| DRUG NAME | DRUG CODE | NUMBER OF REGISTRANT SOLD TO | TOTAL GRAMS | AVG GRAMS |
|---|---|---|---|---|
| CODEINE | 9050 | 12 | **235.59** | 19.63 |
| BUPRENORPHINE | 9064 | 3 | **4.62** | 1.54 |
| OXYCODONE | 9143 | 1 | **0.45** | 0.45 |
| MORPHINE | 9300 | 1 | **0.38** | 0.38 |
| FENTANYL BASE | 9801 | 3 | **0.12** | 0.04 |

**BUSINESS ACTIVITY: NTP - NARCOTIC TREATMENT PROGRAMS**

| DRUG NAME | DRUG CODE | NUMBER OF REGISTRANT SOLD TO | TOTAL GRAMS | AVG GRAMS |
|---|---|---|---|---|
| BUPRENORPHINE | 9064 | 28 | **1,634.77** | 58.38 |
| METHADONE | 9250B | 61 | **397,060.38** | 6,509.19 |

**STATE: CA - CALIFORNIA**

**BUSINESS ACTIVITY: A - PHARMACIES**

| DRUG NAME | DRUG CODE | NUMBER OF REGISTRANT SOLD TO | TOTAL GRAMS | AVG GRAMS |
|---|---|---|---|---|
| AMPHETAMINE | 1100 | 5,438 | **1,232,609.22** | 226.67 |
| METHAMPHETAMINE | 1105 | 254 | **946.52** | 3.73 |
| LISDEXAMFETAMINE | 1205 | 4,717 | **456,272.94** | 96.73 |
| METHYLPHENIDATE (DL;D;L;ISOMERS) | 1724 | 5,293 | **1,014,718.85** | 191.71 |
| CODEINE | 9050 | 6,003 | **1,265,091.37** | 210.74 |

| | | | | |
|---|---|---|---|---|
| BUPRENORPHINE | 9064 | 5,228 | **193,350.76** | 36.98 |
| OXYCODONE | 9143 | 5,709 | **2,270,187.83** | 397.65 |
| HYDROMORPHONE | 9150 | 4,781 | **83,882.02** | 17.54 |
| HYDROCODONE | 9193 | 6,010 | **2,553,110.62** | 424.81 |
| MEPERIDINE (PETHIDINE) | 9230 | 457 | **11,478.42** | 25.12 |
| METHADONE | 9250B | 4,346 | **209,440.93** | 48.19 |
| MORPHINE | 9300 | 5,326 | **1,122,423.11** | 210.74 |
| OXYMORPHONE | 9652 | 942 | **15,443.41** | 16.39 |
| TAPENTADOL | 9780 | 2,557 | **265,990.00** | 104.02 |
| FENTANYL BASE | 9801 | 4,504 | **13,365.01** | 2.97 |

**BUSINESS ACTIVITY: B - HOSPITALS**

| DRUG NAME | DRUG CODE | NUMBER OF REGISTRANT SOLD TO | TOTAL GRAMS | AVG GRAMS |
|---|---|---|---|---|
| AMPHETAMINE | 1100 | 263 | **32,872.29** | 124.99 |
| METHAMPHETAMINE | 1105 | 3 | **10.04** | 3.35 |
| LISDEXAMFETAMINE | 1205 | 100 | **5,169.37** | 51.69 |
| METHYLPHENIDATE (DL;D;L;ISOMERS) | 1724 | 404 | **32,724.58** | 81.00 |
| CODEINE | 9050 | 572 | **42,913.09** | 75.02 |
| BUPRENORPHINE | 9064 | 456 | **7,537.75** | 16.53 |
| OXYCODONE | 9143 | 679 | **116,613.94** | 171.74 |
| HYDROMORPHONE | 9150 | 705 | **15,924.10** | 22.59 |
| HYDROCODONE | 9193 | 754 | **109,774.70** | 145.59 |
| MEPERIDINE (PETHIDINE) | 9230 | 577 | **18,042.77** | 31.27 |
| METHADONE | 9250B | 550 | **24,439.02** | 44.43 |
| MORPHINE | 9300 | 747 | **99,617.13** | 133.36 |
| OXYMORPHONE | 9652 | 20 | **76.45** | 3.82 |
| TAPENTADOL | 9780 | 63 | **2,617.50** | 41.55 |
| FENTANYL BASE | 9801 | 850 | **2,089.42** | 2.46 |

**BUSINESS ACTIVITY: C - PRACTITIONERS**

| DRUG NAME | DRUG CODE | NUMBER OF REGISTRANT SOLD TO | TOTAL GRAMS | AVG GRAMS |
|---|---|---|---|---|
| AMPHETAMINE | 1100 | 14 | **776.51** | 55.46 |
| METHYLPHENIDATE (DL;D;L;ISOMERS) | 1724 | 4 | **175.11** | 43.78 |
| CODEINE | 9050 | 499 | **15,829.29** | 31.72 |
| BUPRENORPHINE | 9064 | 3,017 | **1,028.16** | 0.34 |
| OXYCODONE | 9143 | 197 | **540.32** | 2.74 |
| HYDROMORPHONE | 9150 | 1,597 | **1,008.18** | 0.63 |
| HYDROCODONE | 9193 | 1,932 | **8,948.30** | 4.63 |
| MEPERIDINE (PETHIDINE) | 9230 | 637 | **7,484.88** | 11.75 |
| METHADONE | 9250B | 150 | **398.54** | 2.66 |
| MORPHINE | 9300 | 725 | **561.95** | 0.78 |
| FENTANYL BASE | 9801 | 2,437 | **508.60** | 0.21 |

**BUSINESS ACTIVITY: D - TEACHING INSTITUTIONS**

| DRUG NAME | DRUG CODE | NUMBER OF REGISTRANT SOLD TO | TOTAL GRAMS | AVG GRAMS |
|---|---|---|---|---|
| METHYLPHENIDATE (DL;D;L;ISOMERS) | 1724 | 1 | **2.59** | 2.59 |
| BUPRENORPHINE | 9064 | 7 | **0.28** | 0.04 |
| OXYCODONE | 9143 | 1 | **0.45** | 0.45 |
| HYDROMORPHONE | 9150 | 2 | **0.57** | 0.28 |
| MORPHINE | 9300 | 2 | **1.32** | 0.66 |

**BUSINESS ACTIVITY: M - MID-LEVEL PRACTITIONERS**

| DRUG NAME | DRUG CODE | NUMBER OF REGISTRANT SOLD TO | TOTAL GRAMS | AVG GRAMS |
|---|---|---|---|---|
| CODEINE | 9050 | 25 | **304.51** | 12.18 |
| BUPRENORPHINE | 9064 | 5 | **9.29** | 1.86 |

**BUSINESS ACTIVITY: NTP - NARCOTIC TREATMENT PROGRAMS**

| DRUG NAME | DRUG CODE | NUMBER OF REGISTRANT SOLD TO | TOTAL GRAMS | AVG GRAMS |
|---|---|---|---|---|
| BUPRENORPHINE | 9064 | 119 | **4,145.33** | 34.83 |
| METHADONE | 9250B | 158 | **1,022,908.29** | 6,474.10 |

**STATE: CO - COLORADO**

**BUSINESS ACTIVITY: A - PHARMACIES**

| DRUG NAME | DRUG CODE | NUMBER OF REGISTRANT SOLD TO | TOTAL GRAMS | AVG GRAMS |
|---|---|---|---|---|
| AMPHETAMINE | 1100 | 795 | **329,733.86** | 414.76 |
| METHAMPHETAMINE | 1105 | 71 | **171.55** | 2.42 |
| LISDEXAMFETAMINE | 1205 | 758 | **113,061.33** | 149.16 |
| METHYLPHENIDATE (DL;D;L;ISOMERS) | 1724 | 794 | **210,856.06** | 265.56 |
| CODEINE | 9050 | 768 | **55,890.70** | 72.77 |
| BUPRENORPHINE | 9064 | 779 | **36,158.62** | 46.42 |
| OXYCODONE | 9143 | 802 | **567,366.79** | 707.44 |
| HYDROMORPHONE | 9150 | 773 | **15,885.22** | 20.55 |
| HYDROCODONE | 9193 | 807 | **162,150.78** | 200.93 |
| MEPERIDINE (PETHIDINE) | 9230 | 101 | **1,595.39** | 15.80 |
| METHADONE | 9250B | 628 | **20,357.80** | 32.42 |
| MORPHINE | 9300 | 787 | **178,146.33** | 226.36 |
| OXYMORPHONE | 9652 | 310 | **3,662.83** | 11.82 |
| TAPENTADOL | 9780 | 605 | **84,185.50** | 139.15 |
| FENTANYL BASE | 9801 | 739 | **2,299.29** | 3.11 |

**BUSINESS ACTIVITY: B - HOSPITALS**

| DRUG NAME | DRUG CODE | NUMBER OF REGISTRANT SOLD TO | TOTAL GRAMS | AVG GRAMS |
|---|---|---|---|---|
| AMPHETAMINE | 1100 | 121 | **48,968.30** | 404.70 |
| METHAMPHETAMINE | 1105 | 5 | **20.49** | 4.10 |
| LISDEXAMFETAMINE | 1205 | 75 | **7,002.53** | 93.37 |
| METHYLPHENIDATE (DL;D;L;ISOMERS) | 1724 | 136 | **42,617.89** | 313.37 |
| CODEINE | 9050 | 145 | **10,217.55** | 70.47 |

| | | | | |
|---|---|---|---|---|
| BUPRENORPHINE | 9064 | 137 | **4,972.13** | 36.29 |
| OXYCODONE | 9143 | 209 | **95,960.56** | 459.14 |
| HYDROMORPHONE | 9150 | 194 | **3,943.44** | 20.33 |
| HYDROCODONE | 9193 | 218 | **26,623.90** | 122.13 |
| MEPERIDINE (PETHIDINE) | 9230 | 90 | **733.15** | 8.15 |
| METHADONE | 9250B | 135 | **5,376.66** | 39.83 |
| MORPHINE | 9300 | 204 | **40,105.43** | 196.60 |
| OXYMORPHONE | 9652 | 35 | **183.24** | 5.24 |
| TAPENTADOL | 9780 | 56 | **4,072.50** | 72.72 |
| FENTANYL BASE | 9801 | 208 | **612.06** | 2.94 |

**BUSINESS ACTIVITY: C - PRACTITIONERS**

| DRUG NAME | DRUG CODE | NUMBER OF REGISTRANT SOLD TO | TOTAL GRAMS | AVG GRAMS |
|---|---|---|---|---|
| METHYLPHENIDATE (DL;D;L;ISOMERS) | 1724 | 2 | **11.24** | 5.62 |
| CODEINE | 9050 | 58 | **1,152.06** | 19.86 |
| BUPRENORPHINE | 9064 | 808 | **571.43** | 0.71 |
| OXYCODONE | 9143 | 99 | **365.34** | 3.69 |
| HYDROMORPHONE | 9150 | 520 | **279.07** | 0.54 |
| HYDROCODONE | 9193 | 311 | **772.85** | 2.49 |
| MEPERIDINE (PETHIDINE) | 9230 | 67 | **171.25** | 2.56 |
| METHADONE | 9250B | 31 | **48.12** | 1.55 |
| MORPHINE | 9300 | 217 | **239.24** | 1.10 |
| FENTANYL BASE | 9801 | 528 | **56.68** | 0.11 |

**BUSINESS ACTIVITY: M - MID-LEVEL PRACTITIONERS**

| DRUG NAME | DRUG CODE | NUMBER OF REGISTRANT SOLD TO | TOTAL GRAMS | AVG GRAMS |
|---|---|---|---|---|
| CODEINE | 9050 | 12 | **10.19** | 0.85 |
| BUPRENORPHINE | 9064 | 4 | **7.55** | 1.89 |
| HYDROMORPHONE | 9150 | 7 | **0.45** | 0.06 |
| MORPHINE | 9300 | 21 | **8.48** | 0.40 |
| FENTANYL BASE | 9801 | 53 | **1.84** | 0.03 |

**BUSINESS ACTIVITY: NTP - NARCOTIC TREATMENT PROGRAMS**

| DRUG NAME | DRUG CODE | NUMBER OF REGISTRANT SOLD TO | TOTAL GRAMS | AVG GRAMS |
|---|---|---|---|---|
| BUPRENORPHINE | 9064 | 15 | **949.92** | 63.33 |
| METHADONE | 9250B | 25 | **178,754.78** | 7,150.19 |

**STATE: CT - CONNECTICUT**

**BUSINESS ACTIVITY: A - PHARMACIES**

| DRUG NAME | DRUG CODE | NUMBER OF REGISTRANT SOLD TO | TOTAL GRAMS | AVG GRAMS |
|---|---|---|---|---|
| AMPHETAMINE | 1100 | 686 | **234,963.82** | 342.51 |
| METHAMPHETAMINE | 1105 | 23 | **77.14** | 3.35 |
| LISDEXAMFETAMINE | 1205 | 673 | **143,372.85** | 213.04 |
| METHYLPHENIDATE (DL;D;L;ISOMERS) | 1724 | 689 | **211,889.77** | 307.53 |

| | | | | |
|---|---|---|---|---|
| CODEINE | 9050 | 677 | **65,149.31** | 96.23 |
| BUPRENORPHINE | 9064 | 679 | **70,037.36** | 103.15 |
| OXYCODONE | 9143 | 695 | **487,524.08** | 701.47 |
| HYDROMORPHONE | 9150 | 670 | **15,151.91** | 22.61 |
| HYDROCODONE | 9193 | 688 | **57,055.92** | 82.93 |
| MEPERIDINE (PETHIDINE) | 9230 | 51 | **943.94** | 18.51 |
| METHADONE | 9250B | 494 | **16,271.91** | 32.94 |
| MORPHINE | 9300 | 662 | **98,732.51** | 149.14 |
| OXYMORPHONE | 9652 | 240 | **4,055.67** | 16.90 |
| TAPENTADOL | 9780 | 352 | **28,089.00** | 79.80 |
| FENTANYL BASE | 9801 | 614 | **1,954.98** | 3.18 |

**BUSINESS ACTIVITY: B - HOSPITALS**

| DRUG NAME | DRUG CODE | NUMBER OF REGISTRANT SOLD TO | TOTAL GRAMS | AVG GRAMS |
|---|---|---|---|---|
| AMPHETAMINE | 1100 | 43 | **4,964.51** | 115.45 |
| METHAMPHETAMINE | 1105 | 1 | **11.65** | 11.65 |
| LISDEXAMFETAMINE | 1205 | 24 | **1,889.81** | 78.74 |
| METHYLPHENIDATE (DL;D;L;ISOMERS) | 1724 | 49 | **4,799.70** | 97.95 |
| CODEINE | 9050 | 63 | **1,899.06** | 30.14 |
| BUPRENORPHINE | 9064 | 65 | **2,917.43** | 44.88 |
| OXYCODONE | 9143 | 76 | **16,353.19** | 215.17 |
| HYDROMORPHONE | 9150 | 66 | **2,749.15** | 41.65 |
| HYDROCODONE | 9193 | 58 | **1,518.14** | 26.17 |
| MEPERIDINE (PETHIDINE) | 9230 | 42 | **231.49** | 5.51 |
| METHADONE | 9250B | 51 | **3,750.13** | 73.53 |
| MORPHINE | 9300 | 63 | **8,860.58** | 140.64 |
| OXYMORPHONE | 9652 | 4 | **6.96** | 1.74 |
| TAPENTADOL | 9780 | 7 | **289.00** | 41.29 |
| FENTANYL BASE | 9801 | 70 | **243.71** | 3.48 |

**BUSINESS ACTIVITY: C - PRACTITIONERS**

| DRUG NAME | DRUG CODE | NUMBER OF REGISTRANT SOLD TO | TOTAL GRAMS | AVG GRAMS |
|---|---|---|---|---|
| AMPHETAMINE | 1100 | 1 | **1.32** | 1.32 |
| CODEINE | 9050 | 28 | **132.85** | 4.74 |
| BUPRENORPHINE | 9064 | 325 | **32.35** | 0.10 |
| OXYCODONE | 9143 | 39 | **382.60** | 9.81 |
| HYDROMORPHONE | 9150 | 155 | **79.16** | 0.51 |
| HYDROCODONE | 9193 | 86 | **262.15** | 3.05 |
| MEPERIDINE (PETHIDINE) | 9230 | 18 | **58.50** | 3.25 |
| METHADONE | 9250B | 17 | **84.79** | 4.99 |
| MORPHINE | 9300 | 105 | **117.12** | 1.12 |
| FENTANYL BASE | 9801 | 221 | **27.78** | 0.13 |

**BUSINESS ACTIVITY: M - MID-LEVEL PRACTITIONERS**

| DRUG NAME | DRUG CODE | NUMBER OF REGISTRANT SOLD TO | TOTAL GRAMS | AVG GRAMS |
|---|---|---|---|---|
| CODEINE | 9050 | 2 | **0.82** | 0.41 |
| BUPRENORPHINE | 9064 | 1 | **0.94** | 0.94 |
| OXYCODONE | 9143 | 1 | **0.90** | 0.90 |
| MORPHINE | 9300 | 1 | **1.32** | 1.32 |

**BUSINESS ACTIVITY: NTP - NARCOTIC TREATMENT PROGRAMS**

| DRUG NAME | DRUG CODE | NUMBER OF REGISTRANT SOLD TO | TOTAL GRAMS | AVG GRAMS |
|---|---|---|---|---|
| BUPRENORPHINE | 9064 | 6 | **128.76** | 21.46 |
| METHADONE | 9250B | 43 | **472,896.20** | 10,997.59 |

**STATE: DC - DISTRICT OF COLUMBIA**

**BUSINESS ACTIVITY: A - PHARMACIES**

| DRUG NAME | DRUG CODE | NUMBER OF REGISTRANT SOLD TO | TOTAL GRAMS | AVG GRAMS |
|---|---|---|---|---|
| AMPHETAMINE | 1100 | 145 | **48,812.89** | 336.64 |
| METHAMPHETAMINE | 1105 | 6 | **20.09** | 3.35 |
| LISDEXAMFETAMINE | 1205 | 133 | **24,506.82** | 184.26 |
| METHYLPHENIDATE (DL;D;L;ISOMERS) | 1724 | 140 | **30,168.63** | 215.49 |
| CODEINE | 9050 | 153 | **31,997.79** | 209.14 |
| BUPRENORPHINE | 9064 | 141 | **13,498.10** | 95.73 |
| OXYCODONE | 9143 | 147 | **37,130.91** | 252.59 |
| HYDROMORPHONE | 9150 | 103 | **753.83** | 7.32 |
| HYDROCODONE | 9193 | 142 | **3,503.45** | 24.67 |
| MEPERIDINE (PETHIDINE) | 9230 | 6 | **77.89** | 12.98 |
| METHADONE | 9250B | 44 | **886.26** | 20.14 |
| MORPHINE | 9300 | 102 | **6,952.45** | 68.16 |
| OXYMORPHONE | 9652 | 18 | **190.20** | 10.57 |
| TAPENTADOL | 9780 | 43 | **2,342.00** | 54.47 |
| FENTANYL BASE | 9801 | 77 | **116.86** | 1.52 |

**BUSINESS ACTIVITY: B - HOSPITALS**

| DRUG NAME | DRUG CODE | NUMBER OF REGISTRANT SOLD TO | TOTAL GRAMS | AVG GRAMS |
|---|---|---|---|---|
| AMPHETAMINE | 1100 | 10 | **2,888.73** | 288.87 |
| LISDEXAMFETAMINE | 1205 | 6 | **415.08** | 69.18 |
| METHYLPHENIDATE (DL;D;L;ISOMERS) | 1724 | 14 | **3,142.67** | 224.48 |
| CODEINE | 9050 | 17 | **1,088.95** | 64.06 |
| BUPRENORPHINE | 9064 | 23 | **437.22** | 19.01 |
| OXYCODONE | 9143 | 21 | **7,070.18** | 336.68 |
| HYDROMORPHONE | 9150 | 26 | **749.74** | 28.84 |
| HYDROCODONE | 9193 | 23 | **379.09** | 16.48 |
| MEPERIDINE (PETHIDINE) | 9230 | 7 | **356.77** | 50.97 |
| METHADONE | 9250B | 14 | **794.38** | 56.74 |

| | | | | |
|---|---|---|---|---|
| MORPHINE | 9300 | 21 | **3,492.71** | 166.32 |
| OXYMORPHONE | 9652 | 2 | **3.57** | 1.78 |
| TAPENTADOL | 9780 | 3 | **208.50** | 69.50 |
| FENTANYL BASE | 9801 | 22 | **97.40** | 4.43 |

**BUSINESS ACTIVITY: C - PRACTITIONERS**

| DRUG NAME | DRUG CODE | NUMBER OF REGISTRANT SOLD TO | TOTAL GRAMS | AVG GRAMS |
|---|---|---|---|---|
| AMPHETAMINE | 1100 | 2 | **2.50** | 1.25 |
| METHYLPHENIDATE (DL;D;L;ISOMERS) | 1724 | 3 | **6.83** | 2.28 |
| CODEINE | 9050 | 3 | **4.71** | 1.57 |
| BUPRENORPHINE | 9064 | 19 | **1.92** | 0.10 |
| OXYCODONE | 9143 | 4 | **3.14** | 0.78 |
| HYDROMORPHONE | 9150 | 4 | **0.74** | 0.18 |
| HYDROCODONE | 9193 | 6 | **2.51** | 0.42 |
| MEPERIDINE (PETHIDINE) | 9230 | 3 | **6.54** | 2.18 |
| MORPHINE | 9300 | 3 | **12.78** | 4.26 |
| FENTANYL BASE | 9801 | 17 | **0.41** | 0.02 |

**BUSINESS ACTIVITY: D - TEACHING INSTITUTIONS**

| DRUG NAME | DRUG CODE | NUMBER OF REGISTRANT SOLD TO | TOTAL GRAMS | AVG GRAMS |
|---|---|---|---|---|
| BUPRENORPHINE | 9064 | 1 | **0.01** | 0.01 |

**BUSINESS ACTIVITY: M - MID-LEVEL PRACTITIONERS**

| DRUG NAME | DRUG CODE | NUMBER OF REGISTRANT SOLD TO | TOTAL GRAMS | AVG GRAMS |
|---|---|---|---|---|
| METHYLPHENIDATE (DL;D;L;ISOMERS) | 1724 | 1 | **0.43** | 0.43 |
| FENTANYL BASE | 9801 | 1 | **0.08** | 0.08 |

**BUSINESS ACTIVITY: NTP - NARCOTIC TREATMENT PROGRAMS**

| DRUG NAME | DRUG CODE | NUMBER OF REGISTRANT SOLD TO | TOTAL GRAMS | AVG GRAMS |
|---|---|---|---|---|
| BUPRENORPHINE | 9064 | 1 | **3.60** | 3.60 |
| METHADONE | 9250B | 6 | **37,859.95** | 6,309.99 |

**STATE: DE - DELAWARE**

**BUSINESS ACTIVITY: A - PHARMACIES**

| DRUG NAME | DRUG CODE | NUMBER OF REGISTRANT SOLD TO | TOTAL GRAMS | AVG GRAMS |
|---|---|---|---|---|
| AMPHETAMINE | 1100 | 203 | **84,680.27** | 417.14 |
| METHAMPHETAMINE | 1105 | 3 | **16.47** | 5.49 |
| LISDEXAMFETAMINE | 1205 | 198 | **53,861.58** | 272.03 |
| METHYLPHENIDATE (DL;D;L;ISOMERS) | 1724 | 201 | **68,373.15** | 340.16 |
| CODEINE | 9050 | 195 | **25,737.74** | 131.99 |
| BUPRENORPHINE | 9064 | 198 | **14,161.07** | 71.52 |
| OXYCODONE | 9143 | 203 | **197,417.16** | 972.50 |
| HYDROMORPHONE | 9150 | 197 | **4,371.45** | 22.19 |
| HYDROCODONE | 9193 | 197 | **16,740.62** | 84.98 |
| MEPERIDINE (PETHIDINE) | 9230 | 7 | **75.17** | 10.74 |

| | | | | |
|---|---|---|---|---|
| METHADONE | 9250B | 150 | **6,678.13** | 44.52 |
| MORPHINE | 9300 | 198 | **56,281.71** | 284.25 |
| OXYMORPHONE | 9652 | 116 | **4,399.88** | 37.93 |
| TAPENTADOL | 9780 | 143 | **17,651.00** | 123.43 |
| FENTANYL BASE | 9801 | 184 | **571.99** | 3.11 |

**BUSINESS ACTIVITY: B - HOSPITALS**

| DRUG NAME | DRUG CODE | NUMBER OF REGISTRANT SOLD TO | TOTAL GRAMS | AVG GRAMS |
|---|---|---|---|---|
| AMPHETAMINE | 1100 | 15 | **2,397.82** | 159.85 |
| LISDEXAMFETAMINE | 1205 | 9 | **596.60** | 66.29 |
| METHYLPHENIDATE (DL;D;L;ISOMERS) | 1724 | 18 | **1,798.26** | 99.90 |
| CODEINE | 9050 | 18 | **915.72** | 50.87 |
| BUPRENORPHINE | 9064 | 20 | **966.07** | 48.30 |
| OXYCODONE | 9143 | 29 | **8,228.24** | 283.73 |
| HYDROMORPHONE | 9150 | 27 | **402.84** | 14.92 |
| HYDROCODONE | 9193 | 27 | **964.82** | 35.73 |
| MEPERIDINE (PETHIDINE) | 9230 | 11 | **206.98** | 18.82 |
| METHADONE | 9250B | 18 | **1,111.38** | 61.74 |
| MORPHINE | 9300 | 28 | **4,107.94** | 146.71 |
| OXYMORPHONE | 9652 | 3 | **12.94** | 4.31 |
| TAPENTADOL | 9780 | 4 | **319.00** | 79.75 |
| FENTANYL BASE | 9801 | 34 | **76.33** | 2.25 |

**BUSINESS ACTIVITY: C - PRACTITIONERS**

| DRUG NAME | DRUG CODE | NUMBER OF REGISTRANT SOLD TO | TOTAL GRAMS | AVG GRAMS |
|---|---|---|---|---|
| CODEINE | 9050 | 14 | **531.96** | 38.00 |
| BUPRENORPHINE | 9064 | 73 | **6.39** | 0.09 |
| OXYCODONE | 9143 | 3 | **1.79** | 0.60 |
| HYDROMORPHONE | 9150 | 40 | **13.22** | 0.33 |
| HYDROCODONE | 9193 | 29 | **73.48** | 2.53 |
| MEPERIDINE (PETHIDINE) | 9230 | 2 | **2.94** | 1.47 |
| METHADONE | 9250B | 4 | **7.87** | 1.97 |
| MORPHINE | 9300 | 10 | **5.10** | 0.51 |
| FENTANYL BASE | 9801 | 52 | **3.11** | 0.06 |

**BUSINESS ACTIVITY: M - MID-LEVEL PRACTITIONERS**

| DRUG NAME | DRUG CODE | NUMBER OF REGISTRANT SOLD TO | TOTAL GRAMS | AVG GRAMS |
|---|---|---|---|---|
| CODEINE | 9050 | 1 | **2.21** | 2.21 |

**BUSINESS ACTIVITY: NTP - NARCOTIC TREATMENT PROGRAMS**

| DRUG NAME | DRUG CODE | NUMBER OF REGISTRANT SOLD TO | TOTAL GRAMS | AVG GRAMS |
|---|---|---|---|---|
| BUPRENORPHINE | 9064 | 16 | **2,242.86** | 140.18 |
| METHADONE | 9250B | 15 | **134,316.52** | 8,954.43 |

**STATE: FL - FLORIDA**

**BUSINESS ACTIVITY: A - PHARMACIES**

| DRUG NAME | DRUG CODE | NUMBER OF REGISTRANT SOLD TO | TOTAL GRAMS | AVG GRAMS |
|---|---|---|---|---|
| AMPHETAMINE | 1100 | 4,340 | **1,331,091.16** | 306.70 |
| METHAMPHETAMINE | 1105 | 88 | **206.91** | 2.35 |
| LISDEXAMFETAMINE | 1205 | 3,969 | **535,528.31** | 134.93 |
| METHYLPHENIDATE (DL;D;L;ISOMERS) | 1724 | 4,171 | **750,185.21** | 179.86 |
| CODEINE | 9050 | 4,287 | **496,690.57** | 115.86 |
| BUPRENORPHINE | 9064 | 4,054 | **223,336.58** | 55.09 |
| OXYCODONE | 9143 | 4,485 | **3,270,159.11** | 729.13 |
| HYDROMORPHONE | 9150 | 3,832 | **123,364.20** | 32.19 |
| HYDROCODONE | 9193 | 4,413 | **1,009,290.04** | 228.71 |
| MEPERIDINE (PETHIDINE) | 9230 | 387 | **6,321.53** | 16.33 |
| METHADONE | 9250B | 3,175 | **144,987.07** | 45.67 |
| MORPHINE | 9300 | 4,126 | **1,095,511.63** | 265.51 |
| OXYMORPHONE | 9652 | 1,429 | **30,665.67** | 21.46 |
| TAPENTADOL | 9780 | 2,375 | **233,293.50** | 98.23 |
| FENTANYL BASE | 9801 | 3,771 | **12,240.64** | 3.25 |

**BUSINESS ACTIVITY: B - HOSPITALS**

| DRUG NAME | DRUG CODE | NUMBER OF REGISTRANT SOLD TO | TOTAL GRAMS | AVG GRAMS |
|---|---|---|---|---|
| AMPHETAMINE | 1100 | 194 | **31,771.72** | 163.77 |
| METHAMPHETAMINE | 1105 | 1 | **4.82** | 4.82 |
| LISDEXAMFETAMINE | 1205 | 89 | **7,628.03** | 85.71 |
| METHYLPHENIDATE (DL;D;L;ISOMERS) | 1724 | 274 | **27,135.56** | 99.03 |
| CODEINE | 9050 | 382 | **12,977.76** | 33.97 |
| BUPRENORPHINE | 9064 | 307 | **8,293.80** | 27.02 |
| OXYCODONE | 9143 | 595 | **109,767.49** | 184.48 |
| HYDROMORPHONE | 9150 | 595 | **8,236.64** | 13.84 |
| HYDROCODONE | 9193 | 575 | **40,844.54** | 71.03 |
| MEPERIDINE (PETHIDINE) | 9230 | 422 | **4,864.82** | 11.53 |
| METHADONE | 9250B | 321 | **7,771.33** | 24.21 |
| MORPHINE | 9300 | 614 | **63,902.91** | 104.08 |
| OXYMORPHONE | 9652 | 13 | **25.34** | 1.95 |
| TAPENTADOL | 9780 | 70 | **3,497.95** | 49.97 |
| FENTANYL BASE | 9801 | 816 | **1,575.54** | 1.93 |

**BUSINESS ACTIVITY: C - PRACTITIONERS**

| DRUG NAME | DRUG CODE | NUMBER OF REGISTRANT SOLD TO | TOTAL GRAMS | AVG GRAMS |
|---|---|---|---|---|
| AMPHETAMINE | 1100 | 9 | **118.32** | 13.15 |
| LISDEXAMFETAMINE | 1205 | 1 | **15.18** | 15.18 |
| METHYLPHENIDATE (DL;D;L;ISOMERS) | 1724 | 3 | **218.77** | 72.92 |
| CODEINE | 9050 | 147 | **1,951.73** | 13.28 |

| | | | | |
|---|---|---|---|---|
| BUPRENORPHINE | 9064 | 2,168 | **1,068.73** | 0.49 |
| OXYCODONE | 9143 | 73 | **134.64** | 1.84 |
| HYDROMORPHONE | 9150 | 709 | **321.94** | 0.45 |
| HYDROCODONE | 9193 | 837 | **3,247.66** | 3.88 |
| MEPERIDINE (PETHIDINE) | 9230 | 203 | **2,741.19** | 13.50 |
| METHADONE | 9250B | 38 | **95.88** | 2.52 |
| MORPHINE | 9300 | 339 | **780.79** | 2.30 |
| FENTANYL BASE | 9801 | 1,203 | **76.69** | 0.06 |

**BUSINESS ACTIVITY: M - MID-LEVEL PRACTITIONERS**

| DRUG NAME | DRUG CODE | NUMBER OF REGISTRANT SOLD TO | TOTAL GRAMS | AVG GRAMS |
|---|---|---|---|---|
| CODEINE | 9050 | 6 | **5.88** | 0.98 |
| BUPRENORPHINE | 9064 | 3 | **1.79** | 0.60 |

**BUSINESS ACTIVITY: NTP - NARCOTIC TREATMENT PROGRAMS**

| DRUG NAME | DRUG CODE | NUMBER OF REGISTRANT SOLD TO | TOTAL GRAMS | AVG GRAMS |
|---|---|---|---|---|
| BUPRENORPHINE | 9064 | 57 | **2,220.33** | 38.95 |
| METHADONE | 9250B | 61 | **706,481.64** | 11,581.67 |

**STATE: GA - GEORGIA**

**BUSINESS ACTIVITY: A - PHARMACIES**

| DRUG NAME | DRUG CODE | NUMBER OF REGISTRANT SOLD TO | TOTAL GRAMS | AVG GRAMS |
|---|---|---|---|---|
| AMPHETAMINE | 1100 | 2,140 | **807,712.51** | 377.44 |
| METHAMPHETAMINE | 1105 | 37 | **87.99** | 2.38 |
| LISDEXAMFETAMINE | 1205 | 2,038 | **382,652.48** | 187.76 |
| METHYLPHENIDATE (DL;D;L;ISOMERS) | 1724 | 2,125 | **618,936.05** | 291.26 |
| CODEINE | 9050 | 2,130 | **304,361.50** | 142.89 |
| BUPRENORPHINE | 9064 | 2,044 | **74,469.44** | 36.43 |
| OXYCODONE | 9143 | 2,152 | **1,398,862.63** | 650.03 |
| HYDROMORPHONE | 9150 | 1,841 | **19,351.96** | 10.51 |
| HYDROCODONE | 9193 | 2,168 | **728,470.02** | 336.01 |
| MEPERIDINE (PETHIDINE) | 9230 | 572 | **8,512.92** | 14.88 |
| METHADONE | 9250B | 1,671 | **64,604.57** | 38.66 |
| MORPHINE | 9300 | 2,065 | **324,209.85** | 157.00 |
| OXYMORPHONE | 9652 | 588 | **8,767.96** | 14.91 |
| TAPENTADOL | 9780 | 1,397 | **167,480.00** | 119.89 |
| FENTANYL BASE | 9801 | 1,962 | **5,840.83** | 2.98 |

**BUSINESS ACTIVITY: B - HOSPITALS**

| DRUG NAME | DRUG CODE | NUMBER OF REGISTRANT SOLD TO | TOTAL GRAMS | AVG GRAMS |
|---|---|---|---|---|
| AMPHETAMINE | 1100 | 104 | **16,939.11** | 162.88 |
| METHAMPHETAMINE | 1105 | 2 | **8.44** | 4.22 |
| LISDEXAMFETAMINE | 1205 | 57 | **6,239.43** | 109.46 |
| METHYLPHENIDATE (DL;D;L;ISOMERS) | 1724 | 135 | **19,622.00** | 145.35 |

| | | | | |
|---|---|---|---|---|
| CODEINE | 9050 | 180 | **9,674.14** | 53.75 |
| BUPRENORPHINE | 9064 | 116 | **2,047.50** | 17.65 |
| OXYCODONE | 9143 | 223 | **53,955.62** | 241.95 |
| HYDROMORPHONE | 9150 | 195 | **3,728.51** | 19.12 |
| HYDROCODONE | 9193 | 220 | **31,196.87** | 141.80 |
| MEPERIDINE (PETHIDINE) | 9230 | 157 | **4,598.25** | 29.29 |
| METHADONE | 9250B | 157 | **3,080.40** | 19.62 |
| MORPHINE | 9300 | 211 | **27,250.41** | 129.15 |
| OXYMORPHONE | 9652 | 7 | **6.42** | 0.92 |
| TAPENTADOL | 9780 | 50 | **3,342.00** | 66.84 |
| FENTANYL BASE | 9801 | 201 | **1,005.05** | 5.00 |

**BUSINESS ACTIVITY: C - PRACTITIONERS**

| DRUG NAME | DRUG CODE | NUMBER OF REGISTRANT SOLD TO | TOTAL GRAMS | AVG GRAMS |
|---|---|---|---|---|
| AMPHETAMINE | 1100 | 4 | **22.72** | 5.68 |
| METHYLPHENIDATE (DL;D;L;ISOMERS) | 1724 | 1 | **0.43** | 0.43 |
| CODEINE | 9050 | 119 | **1,762.94** | 14.81 |
| BUPRENORPHINE | 9064 | 831 | **846.20** | 1.02 |
| OXYCODONE | 9143 | 110 | **1,134.48** | 10.31 |
| HYDROMORPHONE | 9150 | 409 | **193.67** | 0.47 |
| HYDROCODONE | 9193 | 419 | **1,986.53** | 4.74 |
| MEPERIDINE (PETHIDINE) | 9230 | 178 | **1,658.46** | 9.32 |
| METHADONE | 9250B | 8 | **55.10** | 6.89 |
| MORPHINE | 9300 | 246 | **432.75** | 1.76 |
| FENTANYL BASE | 9801 | 827 | **55.92** | 0.07 |

**BUSINESS ACTIVITY: M - MID-LEVEL PRACTITIONERS**

| DRUG NAME | DRUG CODE | NUMBER OF REGISTRANT SOLD TO | TOTAL GRAMS | AVG GRAMS |
|---|---|---|---|---|
| CODEINE | 9050 | 14 | **8.82** | 0.63 |
| BUPRENORPHINE | 9064 | 2 | **30.60** | 15.30 |

**BUSINESS ACTIVITY: NTP - NARCOTIC TREATMENT PROGRAMS**

| DRUG NAME | DRUG CODE | NUMBER OF REGISTRANT SOLD TO | TOTAL GRAMS | AVG GRAMS |
|---|---|---|---|---|
| BUPRENORPHINE | 9064 | 37 | **1,746.65** | 47.21 |
| METHADONE | 9250B | 72 | **385,423.79** | 5,353.11 |

**STATE: GU - GUAM**

**BUSINESS ACTIVITY: A - PHARMACIES**

| DRUG NAME | DRUG CODE | NUMBER OF REGISTRANT SOLD TO | TOTAL GRAMS | AVG GRAMS |
|---|---|---|---|---|
| AMPHETAMINE | 1100 | 14 | **1,012.46** | 72.32 |
| LISDEXAMFETAMINE | 1205 | 4 | **27.17** | 6.79 |
| METHYLPHENIDATE (DL;D;L;ISOMERS) | 1724 | 13 | **654.80** | 50.37 |
| CODEINE | 9050 | 20 | **2,572.56** | 128.63 |
| BUPRENORPHINE | 9064 | 4 | **20.16** | 5.04 |

| | | | | |
|---|---|---|---|---|
| OXYCODONE | 9143 | 15 | **2,098.26** | 139.88 |
| HYDROMORPHONE | 9150 | 6 | **47.17** | 7.86 |
| HYDROCODONE | 9193 | 14 | **299.36** | 21.38 |
| MEPERIDINE (PETHIDINE) | 9230 | 1 | **2.18** | 2.18 |
| METHADONE | 9250B | 3 | **101.96** | 33.99 |
| MORPHINE | 9300 | 10 | **2,464.23** | 246.42 |
| FENTANYL BASE | 9801 | 7 | **12.74** | 1.82 |

**BUSINESS ACTIVITY: B - HOSPITALS**

| DRUG NAME | DRUG CODE | NUMBER OF REGISTRANT SOLD TO | TOTAL GRAMS | AVG GRAMS |
|---|---|---|---|---|
| AMPHETAMINE | 1100 | 4 | **702.95** | 175.74 |
| LISDEXAMFETAMINE | 1205 | 1 | **29.48** | 29.48 |
| METHYLPHENIDATE (DL;D;L;ISOMERS) | 1724 | 4 | **867.79** | 216.95 |
| CODEINE | 9050 | 6 | **284.59** | 47.43 |
| BUPRENORPHINE | 9064 | 2 | **0.01** | 0.01 |
| OXYCODONE | 9143 | 7 | **658.26** | 94.04 |
| HYDROMORPHONE | 9150 | 6 | **23.13** | 3.86 |
| HYDROCODONE | 9193 | 7 | **136.68** | 19.53 |
| MEPERIDINE (PETHIDINE) | 9230 | 3 | **13.07** | 4.36 |
| METHADONE | 9250B | 3 | **29.07** | 9.69 |
| MORPHINE | 9300 | 7 | **464.61** | 66.37 |
| FENTANYL BASE | 9801 | 7 | **14.52** | 2.07 |

**BUSINESS ACTIVITY: C - PRACTITIONERS**

| DRUG NAME | DRUG CODE | NUMBER OF REGISTRANT SOLD TO | TOTAL GRAMS | AVG GRAMS |
|---|---|---|---|---|
| CODEINE | 9050 | 1 | **2.21** | 2.21 |
| BUPRENORPHINE | 9064 | 3 | **0.14** | 0.05 |
| HYDROMORPHONE | 9150 | 2 | **2.31** | 1.15 |
| HYDROCODONE | 9193 | 4 | **10.88** | 2.72 |
| MEPERIDINE (PETHIDINE) | 9230 | 3 | **69.72** | 23.24 |
| MORPHINE | 9300 | 3 | **2.14** | 0.71 |
| FENTANYL BASE | 9801 | 5 | **0.31** | 0.06 |

**STATE: HI - HAWAII**

**BUSINESS ACTIVITY: A - PHARMACIES**

| DRUG NAME | DRUG CODE | NUMBER OF REGISTRANT SOLD TO | TOTAL GRAMS | AVG GRAMS |
|---|---|---|---|---|
| AMPHETAMINE | 1100 | 213 | **34,485.41** | 161.90 |
| METHAMPHETAMINE | 1105 | 6 | **18.48** | 3.08 |
| LISDEXAMFETAMINE | 1205 | 145 | **5,370.55** | 37.04 |
| METHYLPHENIDATE (DL;D;L;ISOMERS) | 1724 | 213 | **32,327.65** | 151.77 |
| CODEINE | 9050 | 228 | **35,582.78** | 156.06 |
| BUPRENORPHINE | 9064 | 202 | **8,550.95** | 42.33 |
| OXYCODONE | 9143 | 228 | **109,004.53** | 478.09 |

| | | | | |
|---|---|---|---|---|
| HYDROMORPHONE | 9150 | 134 | **1,572.17** | 11.73 |
| HYDROCODONE | 9193 | 229 | **46,895.14** | 204.78 |
| MEPERIDINE (PETHIDINE) | 9230 | 21 | **313.74** | 14.94 |
| METHADONE | 9250B | 143 | **8,510.84** | 59.52 |
| MORPHINE | 9300 | 197 | **51,330.77** | 260.56 |
| OXYMORPHONE | 9652 | 33 | **491.99** | 14.91 |
| TAPENTADOL | 9780 | 54 | **5,004.50** | 92.68 |
| FENTANYL BASE | 9801 | 164 | **377.62** | 2.30 |

**BUSINESS ACTIVITY: B - HOSPITALS**

| DRUG NAME | DRUG CODE | NUMBER OF REGISTRANT SOLD TO | TOTAL GRAMS | AVG GRAMS |
|---|---|---|---|---|
| AMPHETAMINE | 1100 | 16 | **8,555.54** | 534.72 |
| LISDEXAMFETAMINE | 1205 | 11 | **1,383.39** | 125.76 |
| METHYLPHENIDATE (DL;D;L;ISOMERS) | 1724 | 22 | **10,436.65** | 474.39 |
| CODEINE | 9050 | 33 | **4,010.34** | 121.53 |
| BUPRENORPHINE | 9064 | 23 | **323.35** | 14.06 |
| OXYCODONE | 9143 | 39 | **13,154.73** | 337.30 |
| HYDROMORPHONE | 9150 | 38 | **402.21** | 10.58 |
| HYDROCODONE | 9193 | 36 | **3,807.01** | 105.75 |
| MEPERIDINE (PETHIDINE) | 9230 | 21 | **219.51** | 10.45 |
| METHADONE | 9250B | 26 | **808.81** | 31.11 |
| MORPHINE | 9300 | 38 | **5,597.60** | 147.31 |
| OXYMORPHONE | 9652 | 3 | **233.28** | 77.76 |
| TAPENTADOL | 9780 | 8 | **536.00** | 67.00 |
| FENTANYL BASE | 9801 | 43 | **108.29** | 2.52 |

**BUSINESS ACTIVITY: C - PRACTITIONERS**

| DRUG NAME | DRUG CODE | NUMBER OF REGISTRANT SOLD TO | TOTAL GRAMS | AVG GRAMS |
|---|---|---|---|---|
| AMPHETAMINE | 1100 | 1 | **22.52** | 22.52 |
| CODEINE | 9050 | 16 | **513.04** | 32.07 |
| BUPRENORPHINE | 9064 | 93 | **5.55** | 0.06 |
| OXYCODONE | 9143 | 16 | **1,492.13** | 93.26 |
| HYDROMORPHONE | 9150 | 50 | **31.68** | 0.63 |
| HYDROCODONE | 9193 | 50 | **1,427.02** | 28.54 |
| MEPERIDINE (PETHIDINE) | 9230 | 16 | **119.29** | 7.46 |
| METHADONE | 9250B | 3 | **6.80** | 2.27 |
| MORPHINE | 9300 | 21 | **234.50** | 11.17 |
| OXYMORPHONE | 9652 | 1 | **0.80** | 0.80 |
| FENTANYL BASE | 9801 | 53 | **4.13** | 0.08 |

**BUSINESS ACTIVITY: M - MID-LEVEL PRACTITIONERS**

| DRUG NAME | DRUG CODE | NUMBER OF REGISTRANT SOLD TO | TOTAL GRAMS | AVG GRAMS |
|---|---|---|---|---|
| MORPHINE | 9300 | 1 | **0.75** | 0.75 |

| | | | | |
|---|---|---|---|---|
| FENTANYL BASE | 9801 | 1 | **0.02** | 0.02 |

**BUSINESS ACTIVITY: NTP - NARCOTIC TREATMENT PROGRAMS**

| DRUG NAME | DRUG CODE | NUMBER OF REGISTRANT SOLD TO | TOTAL GRAMS | AVG GRAMS |
|---|---|---|---|---|
| METHADONE | 9250B | 4 | **20,757.24** | 5,189.31 |

**STATE: IA - IOWA**

**BUSINESS ACTIVITY: A - PHARMACIES**

| DRUG NAME | DRUG CODE | NUMBER OF REGISTRANT SOLD TO | TOTAL GRAMS | AVG GRAMS |
|---|---|---|---|---|
| AMPHETAMINE | 1100 | 671 | **213,536.73** | 318.24 |
| METHAMPHETAMINE | 1105 | 14 | **56.25** | 4.02 |
| LISDEXAMFETAMINE | 1205 | 658 | **132,795.93** | 201.82 |
| METHYLPHENIDATE (DL;D;L;ISOMERS) | 1724 | 677 | **309,062.00** | 456.52 |
| CODEINE | 9050 | 659 | **53,768.92** | 81.59 |
| BUPRENORPHINE | 9064 | 574 | **8,572.71** | 14.94 |
| OXYCODONE | 9143 | 676 | **140,959.77** | 208.52 |
| HYDROMORPHONE | 9150 | 611 | **6,293.54** | 10.30 |
| HYDROCODONE | 9193 | 687 | **152,723.50** | 222.30 |
| MEPERIDINE (PETHIDINE) | 9230 | 66 | **1,074.67** | 16.28 |
| METHADONE | 9250B | 478 | **11,022.59** | 23.06 |
| MORPHINE | 9300 | 669 | **89,819.38** | 134.26 |
| OXYMORPHONE | 9652 | 104 | **1,033.05** | 9.93 |
| TAPENTADOL | 9780 | 359 | **29,887.00** | 83.25 |
| FENTANYL BASE | 9801 | 650 | **2,094.77** | 3.22 |

**BUSINESS ACTIVITY: B - HOSPITALS**

| DRUG NAME | DRUG CODE | NUMBER OF REGISTRANT SOLD TO | TOTAL GRAMS | AVG GRAMS |
|---|---|---|---|---|
| AMPHETAMINE | 1100 | 53 | **5,662.10** | 106.83 |
| LISDEXAMFETAMINE | 1205 | 38 | **1,919.87** | 50.52 |
| METHYLPHENIDATE (DL;D;L;ISOMERS) | 1724 | 79 | **6,667.29** | 84.40 |
| CODEINE | 9050 | 102 | **1,879.75** | 18.43 |
| BUPRENORPHINE | 9064 | 62 | **675.91** | 10.90 |
| OXYCODONE | 9143 | 145 | **11,756.05** | 81.08 |
| HYDROMORPHONE | 9150 | 137 | **1,489.22** | 10.87 |
| HYDROCODONE | 9193 | 145 | **10,155.45** | 70.04 |
| MEPERIDINE (PETHIDINE) | 9230 | 94 | **2,598.16** | 27.64 |
| METHADONE | 9250B | 84 | **859.43** | 10.23 |
| MORPHINE | 9300 | 144 | **10,585.93** | 73.51 |
| OXYMORPHONE | 9652 | 6 | **16.95** | 2.82 |
| TAPENTADOL | 9780 | 27 | **572.50** | 21.20 |
| FENTANYL BASE | 9801 | 149 | **239.08** | 1.60 |

**BUSINESS ACTIVITY: C - PRACTITIONERS**

| DRUG NAME | DRUG CODE | NUMBER OF REGISTRANT SOLD TO | TOTAL GRAMS | AVG GRAMS |
|---|---|---|---|---|

| | | | | |
|---|---|---|---|---|
| CODEINE | 9050 | 13 | **277.63** | 21.36 |
| BUPRENORPHINE | 9064 | 287 | **28.08** | 0.10 |
| OXYCODONE | 9143 | 6 | **119.19** | 19.86 |
| HYDROMORPHONE | 9150 | 86 | **30.46** | 0.35 |
| HYDROCODONE | 9193 | 114 | **337.70** | 2.96 |
| MEPERIDINE (PETHIDINE) | 9230 | 23 | **1,048.20** | 45.57 |
| MORPHINE | 9300 | 61 | **47.71** | 0.78 |
| FENTANYL BASE | 9801 | 106 | **23.31** | 0.22 |

**BUSINESS ACTIVITY: D - TEACHING INSTITUTIONS**

| DRUG NAME | DRUG CODE | NUMBER OF REGISTRANT SOLD TO | TOTAL GRAMS | AVG GRAMS |
|---|---|---|---|---|
| BUPRENORPHINE | 9064 | 1 | **0.00** | 0.00 |
| HYDROMORPHONE | 9150 | 2 | **0.18** | 0.09 |

**BUSINESS ACTIVITY: M - MID-LEVEL PRACTITIONERS**

| DRUG NAME | DRUG CODE | NUMBER OF REGISTRANT SOLD TO | TOTAL GRAMS | AVG GRAMS |
|---|---|---|---|---|
| BUPRENORPHINE | 9064 | 1 | **0.01** | 0.01 |

**BUSINESS ACTIVITY: NTP - NARCOTIC TREATMENT PROGRAMS**

| DRUG NAME | DRUG CODE | NUMBER OF REGISTRANT SOLD TO | TOTAL GRAMS | AVG GRAMS |
|---|---|---|---|---|
| BUPRENORPHINE | 9064 | 16 | **903.79** | 56.49 |
| METHADONE | 9250B | 20 | **42,235.36** | 2,111.77 |

**STATE: ID - IDAHO**

**BUSINESS ACTIVITY: A - PHARMACIES**

| DRUG NAME | DRUG CODE | NUMBER OF REGISTRANT SOLD TO | TOTAL GRAMS | AVG GRAMS |
|---|---|---|---|---|
| AMPHETAMINE | 1100 | 317 | **120,467.33** | 380.02 |
| METHAMPHETAMINE | 1105 | 13 | **52.62** | 4.05 |
| LISDEXAMFETAMINE | 1205 | 304 | **40,226.51** | 132.32 |
| METHYLPHENIDATE (DL;D;L;ISOMERS) | 1724 | 313 | **111,066.33** | 354.84 |
| CODEINE | 9050 | 308 | **39,206.02** | 127.29 |
| BUPRENORPHINE | 9064 | 314 | **19,116.34** | 60.88 |
| OXYCODONE | 9143 | 318 | **179,725.05** | 565.17 |
| HYDROMORPHONE | 9150 | 299 | **4,738.47** | 15.85 |
| HYDROCODONE | 9193 | 323 | **172,387.76** | 533.71 |
| MEPERIDINE (PETHIDINE) | 9230 | 67 | **877.71** | 13.10 |
| METHADONE | 9250B | 294 | **16,072.73** | 54.67 |
| MORPHINE | 9300 | 315 | **77,293.16** | 245.38 |
| OXYMORPHONE | 9652 | 118 | **2,121.86** | 17.98 |
| TAPENTADOL | 9780 | 189 | **18,160.50** | 96.09 |
| FENTANYL BASE | 9801 | 305 | **1,371.65** | 4.50 |

**BUSINESS ACTIVITY: B - HOSPITALS**

| DRUG NAME | DRUG CODE | NUMBER OF REGISTRANT SOLD TO | TOTAL GRAMS | AVG GRAMS |
|---|---|---|---|---|
| AMPHETAMINE | 1100 | 22 | **1,468.47** | 66.75 |

| | | | | |
|---|---|---|---|---|
| LISDEXAMFETAMINE | 1205 | 13 | **236.44** | 18.19 |
| METHYLPHENIDATE (DL;D;L;ISOMERS) | 1724 | 34 | **1,906.90** | 56.09 |
| CODEINE | 9050 | 37 | **1,268.75** | 34.29 |
| BUPRENORPHINE | 9064 | 38 | **596.08** | 15.69 |
| OXYCODONE | 9143 | 59 | **9,450.63** | 160.18 |
| HYDROMORPHONE | 9150 | 54 | **501.64** | 9.29 |
| HYDROCODONE | 9193 | 62 | **7,890.65** | 127.27 |
| MEPERIDINE (PETHIDINE) | 9230 | 38 | **448.06** | 11.79 |
| METHADONE | 9250B | 41 | **908.17** | 22.15 |
| MORPHINE | 9300 | 58 | **6,666.13** | 114.93 |
| OXYMORPHONE | 9652 | 3 | **30.33** | 10.11 |
| TAPENTADOL | 9780 | 9 | **178.00** | 19.78 |
| FENTANYL BASE | 9801 | 58 | **139.20** | 2.40 |

**BUSINESS ACTIVITY: C - PRACTITIONERS**

| DRUG NAME | DRUG CODE | NUMBER OF REGISTRANT SOLD TO | TOTAL GRAMS | AVG GRAMS |
|---|---|---|---|---|
| AMPHETAMINE | 1100 | 1 | **2.44** | 2.44 |
| CODEINE | 9050 | 18 | **4,380.16** | 243.34 |
| BUPRENORPHINE | 9064 | 143 | **14.19** | 0.10 |
| OXYCODONE | 9143 | 15 | **55.31** | 3.69 |
| HYDROMORPHONE | 9150 | 79 | **50.09** | 0.63 |
| HYDROCODONE | 9193 | 87 | **255.77** | 2.94 |
| MEPERIDINE (PETHIDINE) | 9230 | 32 | **101.75** | 3.18 |
| METHADONE | 9250B | 5 | **14.67** | 2.93 |
| MORPHINE | 9300 | 65 | **138.35** | 2.13 |
| FENTANYL BASE | 9801 | 139 | **19.35** | 0.14 |

**BUSINESS ACTIVITY: D - TEACHING INSTITUTIONS**

| DRUG NAME | DRUG CODE | NUMBER OF REGISTRANT SOLD TO | TOTAL GRAMS | AVG GRAMS |
|---|---|---|---|---|
| BUPRENORPHINE | 9064 | 1 | **0.00** | 0.00 |

**BUSINESS ACTIVITY: M - MID-LEVEL PRACTITIONERS**

| DRUG NAME | DRUG CODE | NUMBER OF REGISTRANT SOLD TO | TOTAL GRAMS | AVG GRAMS |
|---|---|---|---|---|
| CODEINE | 9050 | 2 | **47.74** | 23.87 |
| BUPRENORPHINE | 9064 | 1 | **0.01** | 0.01 |
| HYDROMORPHONE | 9150 | 1 | **0.04** | 0.04 |
| HYDROCODONE | 9193 | 1 | **0.61** | 0.61 |
| MEPERIDINE (PETHIDINE) | 9230 | 1 | **1.63** | 1.63 |
| MORPHINE | 9300 | 1 | **0.04** | 0.04 |
| FENTANYL BASE | 9801 | 3 | **0.27** | 0.09 |

**BUSINESS ACTIVITY: NTP - NARCOTIC TREATMENT PROGRAMS**

| DRUG NAME | DRUG CODE | NUMBER OF REGISTRANT SOLD TO | TOTAL GRAMS | AVG GRAMS |
|---|---|---|---|---|
| BUPRENORPHINE | 9064 | 4 | **195.79** | 48.95 |

| | | | | |
|---|---|---|---|---|
| METHADONE | 9250B | 4 | **25,579.84** | 6,394.96 |

**STATE: IL - ILLINOIS**

**BUSINESS ACTIVITY: A - PHARMACIES**

| DRUG NAME | DRUG CODE | NUMBER OF REGISTRANT SOLD TO | TOTAL GRAMS | AVG GRAMS |
|---|---|---|---|---|
| AMPHETAMINE | 1100 | 2,058 | **834,180.59** | 405.34 |
| METHAMPHETAMINE | 1105 | 68 | **228.59** | 3.36 |
| LISDEXAMFETAMINE | 1205 | 1,887 | **292,796.09** | 155.16 |
| METHYLPHENIDATE (DL;D;L;ISOMERS) | 1724 | 2,042 | **657,692.68** | 322.08 |
| CODEINE | 9050 | 2,126 | **487,482.23** | 229.30 |
| BUPRENORPHINE | 9064 | 1,927 | **73,355.43** | 38.07 |
| OXYCODONE | 9143 | 2,028 | **433,387.37** | 213.70 |
| HYDROMORPHONE | 9150 | 1,697 | **22,916.38** | 13.50 |
| HYDROCODONE | 9193 | 2,128 | **856,883.24** | 402.67 |
| MEPERIDINE (PETHIDINE) | 9230 | 110 | **1,839.95** | 16.73 |
| METHADONE | 9250B | 1,307 | **33,658.51** | 25.75 |
| MORPHINE | 9300 | 1,912 | **305,606.45** | 159.84 |
| OXYMORPHONE | 9652 | 517 | **5,897.54** | 11.41 |
| TAPENTADOL | 9780 | 996 | **91,339.00** | 91.71 |
| FENTANYL BASE | 9801 | 1,855 | **6,557.04** | 3.53 |

**BUSINESS ACTIVITY: B - HOSPITALS**

| DRUG NAME | DRUG CODE | NUMBER OF REGISTRANT SOLD TO | TOTAL GRAMS | AVG GRAMS |
|---|---|---|---|---|
| AMPHETAMINE | 1100 | 143 | **10,905.53** | 76.26 |
| LISDEXAMFETAMINE | 1205 | 57 | **2,445.94** | 42.91 |
| METHYLPHENIDATE (DL;D;L;ISOMERS) | 1724 | 193 | **11,046.32** | 57.23 |
| CODEINE | 9050 | 223 | **9,283.37** | 41.63 |
| BUPRENORPHINE | 9064 | 163 | **2,954.90** | 18.13 |
| OXYCODONE | 9143 | 228 | **23,566.91** | 103.36 |
| HYDROMORPHONE | 9150 | 233 | **4,310.42** | 18.50 |
| HYDROCODONE | 9193 | 252 | **45,833.54** | 181.88 |
| MEPERIDINE (PETHIDINE) | 9230 | 154 | **2,396.08** | 15.56 |
| METHADONE | 9250B | 191 | **5,270.55** | 27.59 |
| MORPHINE | 9300 | 238 | **28,229.05** | 118.61 |
| OXYMORPHONE | 9652 | 16 | **154.60** | 9.66 |
| TAPENTADOL | 9780 | 44 | **1,588.50** | 36.10 |
| FENTANYL BASE | 9801 | 233 | **884.07** | 3.79 |

**BUSINESS ACTIVITY: C - PRACTITIONERS**

| DRUG NAME | DRUG CODE | NUMBER OF REGISTRANT SOLD TO | TOTAL GRAMS | AVG GRAMS |
|---|---|---|---|---|
| AMPHETAMINE | 1100 | 1 | **1.25** | 1.25 |
| METHYLPHENIDATE (DL;D;L;ISOMERS) | 1724 | 1 | **12.45** | 12.45 |
| CODEINE | 9050 | 187 | **3,079.88** | 16.47 |

| | | | | |
|---|---|---|---|---|
| BUPRENORPHINE | 9064 | 912 | **81.21** | 0.09 |
| OXYCODONE | 9143 | 41 | **145.68** | 3.55 |
| HYDROMORPHONE | 9150 | 441 | **264.53** | 0.60 |
| HYDROCODONE | 9193 | 493 | **2,396.85** | 4.86 |
| MEPERIDINE (PETHIDINE) | 9230 | 102 | **603.73** | 5.92 |
| METHADONE | 9250B | 28 | **53.22** | 1.90 |
| MORPHINE | 9300 | 183 | **217.59** | 1.19 |
| FENTANYL BASE | 9801 | 622 | **52.23** | 0.08 |

**BUSINESS ACTIVITY: D - TEACHING INSTITUTIONS**

| DRUG NAME | DRUG CODE | NUMBER OF REGISTRANT SOLD TO | TOTAL GRAMS | AVG GRAMS |
|---|---|---|---|---|
| CODEINE | 9050 | 1 | **45.45** | 45.45 |
| BUPRENORPHINE | 9064 | 2 | **0.77** | 0.38 |
| HYDROMORPHONE | 9150 | 2 | **11.44** | 5.72 |
| HYDROCODONE | 9193 | 1 | **17.94** | 17.94 |
| MEPERIDINE (PETHIDINE) | 9230 | 1 | **2.18** | 2.18 |
| METHADONE | 9250B | 1 | **12.88** | 12.88 |
| MORPHINE | 9300 | 1 | **6.92** | 6.92 |
| FENTANYL BASE | 9801 | 2 | **2.39** | 1.19 |

**BUSINESS ACTIVITY: M - MID-LEVEL PRACTITIONERS**

| DRUG NAME | DRUG CODE | NUMBER OF REGISTRANT SOLD TO | TOTAL GRAMS | AVG GRAMS |
|---|---|---|---|---|
| CODEINE | 9050 | 4 | **7.36** | 1.84 |

**BUSINESS ACTIVITY: NTP - NARCOTIC TREATMENT PROGRAMS**

| DRUG NAME | DRUG CODE | NUMBER OF REGISTRANT SOLD TO | TOTAL GRAMS | AVG GRAMS |
|---|---|---|---|---|
| BUPRENORPHINE | 9064 | 14 | **2,028.86** | 144.92 |
| METHADONE | 9250B | 91 | **465,534.31** | 5,115.76 |

**STATE: IN - INDIANA**

**BUSINESS ACTIVITY: A - PHARMACIES**

| DRUG NAME | DRUG CODE | NUMBER OF REGISTRANT SOLD TO | TOTAL GRAMS | AVG GRAMS |
|---|---|---|---|---|
| AMPHETAMINE | 1100 | 1,133 | **565,018.15** | 498.69 |
| METHAMPHETAMINE | 1105 | 27 | **70.71** | 2.62 |
| LISDEXAMFETAMINE | 1205 | 1,120 | **320,936.26** | 286.55 |
| METHYLPHENIDATE (DL;D;L;ISOMERS) | 1724 | 1,138 | **450,068.68** | 395.49 |
| CODEINE | 9050 | 1,109 | **143,964.50** | 129.81 |
| BUPRENORPHINE | 9064 | 1,105 | **117,226.33** | 106.09 |
| OXYCODONE | 9143 | 1,140 | **670,197.56** | 587.89 |
| HYDROMORPHONE | 9150 | 982 | **13,996.37** | 14.25 |
| HYDROCODONE | 9193 | 1,156 | **663,368.90** | 573.85 |
| MEPERIDINE (PETHIDINE) | 9230 | 106 | **1,283.83** | 12.11 |
| METHADONE | 9250B | 943 | **39,178.66** | 41.55 |
| MORPHINE | 9300 | 1,111 | **223,105.13** | 200.81 |

| | | | | |
|---|---|---|---|---|
| OXYMORPHONE | 9652 | 410 | **6,107.72** | 14.90 |
| TAPENTADOL | 9780 | 706 | **99,218.50** | 140.54 |
| FENTANYL BASE | 9801 | 1,080 | **3,970.15** | 3.68 |

**BUSINESS ACTIVITY: B - HOSPITALS**

| DRUG NAME | DRUG CODE | NUMBER OF REGISTRANT SOLD TO | TOTAL GRAMS | AVG GRAMS |
|---|---|---|---|---|
| AMPHETAMINE | 1100 | 125 | **9,480.34** | 75.84 |
| LISDEXAMFETAMINE | 1205 | 77 | **5,135.84** | 66.70 |
| METHYLPHENIDATE (DL;D;L;ISOMERS) | 1724 | 144 | **8,643.93** | 60.03 |
| CODEINE | 9050 | 158 | **3,628.28** | 22.96 |
| BUPRENORPHINE | 9064 | 141 | **2,851.07** | 20.22 |
| OXYCODONE | 9143 | 247 | **30,513.46** | 123.54 |
| HYDROMORPHONE | 9150 | 245 | **2,705.71** | 11.04 |
| HYDROCODONE | 9193 | 275 | **32,739.65** | 119.05 |
| MEPERIDINE (PETHIDINE) | 9230 | 196 | **4,514.91** | 23.04 |
| METHADONE | 9250B | 152 | **4,499.21** | 29.60 |
| MORPHINE | 9300 | 259 | **18,340.39** | 70.81 |
| OXYMORPHONE | 9652 | 12 | **44.69** | 3.72 |
| TAPENTADOL | 9780 | 31 | **876.50** | 28.27 |
| FENTANYL BASE | 9801 | 299 | **544.40** | 1.82 |

**BUSINESS ACTIVITY: C - PRACTITIONERS**

| DRUG NAME | DRUG CODE | NUMBER OF REGISTRANT SOLD TO | TOTAL GRAMS | AVG GRAMS |
|---|---|---|---|---|
| CODEINE | 9050 | 31 | **301.42** | 9.72 |
| BUPRENORPHINE | 9064 | 497 | **121.00** | 0.24 |
| OXYCODONE | 9143 | 14 | **23.67** | 1.69 |
| HYDROMORPHONE | 9150 | 197 | **128.62** | 0.65 |
| HYDROCODONE | 9193 | 240 | **690.89** | 2.88 |
| MEPERIDINE (PETHIDINE) | 9230 | 40 | **111.33** | 2.78 |
| METHADONE | 9250B | 3 | **3.22** | 1.07 |
| MORPHINE | 9300 | 101 | **102.08** | 1.01 |
| FENTANYL BASE | 9801 | 224 | **16.19** | 0.07 |

**BUSINESS ACTIVITY: M - MID-LEVEL PRACTITIONERS**

| DRUG NAME | DRUG CODE | NUMBER OF REGISTRANT SOLD TO | TOTAL GRAMS | AVG GRAMS |
|---|---|---|---|---|
| CODEINE | 9050 | 2 | **1.47** | 0.74 |
| HYDROCODONE | 9193 | 1 | **0.30** | 0.30 |

**BUSINESS ACTIVITY: NTP - NARCOTIC TREATMENT PROGRAMS**

| DRUG NAME | DRUG CODE | NUMBER OF REGISTRANT SOLD TO | TOTAL GRAMS | AVG GRAMS |
|---|---|---|---|---|
| BUPRENORPHINE | 9064 | 10 | **861.46** | 86.15 |
| METHADONE | 9250B | 20 | **286,830.50** | 14,341.53 |

**STATE: KS - KANSAS**

**BUSINESS ACTIVITY: A - PHARMACIES**

| DRUG NAME | DRUG CODE | NUMBER OF REGISTRANT SOLD TO | TOTAL GRAMS | AVG GRAMS |
|---|---|---|---|---|
| AMPHETAMINE | 1100 | 612 | **258,982.85** | 423.17 |
| METHAMPHETAMINE | 1105 | 18 | **51.42** | 2.86 |
| LISDEXAMFETAMINE | 1205 | 605 | **128,854.44** | 212.98 |
| METHYLPHENIDATE (DL;D;L;ISOMERS) | 1724 | 611 | **188,645.58** | 308.75 |
| CODEINE | 9050 | 606 | **74,995.22** | 123.75 |
| BUPRENORPHINE | 9064 | 533 | **11,609.00** | 21.78 |
| OXYCODONE | 9143 | 616 | **339,544.00** | 551.21 |
| HYDROMORPHONE | 9150 | 557 | **8,470.15** | 15.21 |
| HYDROCODONE | 9193 | 619 | **232,926.12** | 376.29 |
| MEPERIDINE (PETHIDINE) | 9230 | 101 | **1,535.47** | 15.20 |
| METHADONE | 9250B | 483 | **18,469.81** | 38.24 |
| MORPHINE | 9300 | 605 | **121,233.24** | 200.39 |
| OXYMORPHONE | 9652 | 159 | **2,271.78** | 14.29 |
| TAPENTADOL | 9780 | 259 | **19,594.50** | 75.65 |
| FENTANYL BASE | 9801 | 585 | **2,381.36** | 4.07 |

**BUSINESS ACTIVITY: B - HOSPITALS**

| DRUG NAME | DRUG CODE | NUMBER OF REGISTRANT SOLD TO | TOTAL GRAMS | AVG GRAMS |
|---|---|---|---|---|
| AMPHETAMINE | 1100 | 52 | **9,082.60** | 174.67 |
| METHAMPHETAMINE | 1105 | 1 | **1.61** | 1.61 |
| LISDEXAMFETAMINE | 1205 | 29 | **2,025.66** | 69.85 |
| METHYLPHENIDATE (DL;D;L;ISOMERS) | 1724 | 84 | **8,418.31** | 100.22 |
| CODEINE | 9050 | 138 | **3,479.84** | 25.22 |
| BUPRENORPHINE | 9064 | 43 | **769.07** | 17.89 |
| OXYCODONE | 9143 | 199 | **24,669.91** | 123.97 |
| HYDROMORPHONE | 9150 | 171 | **1,271.42** | 7.44 |
| HYDROCODONE | 9193 | 211 | **13,164.81** | 62.39 |
| MEPERIDINE (PETHIDINE) | 9230 | 127 | **1,209.75** | 9.53 |
| METHADONE | 9250B | 89 | **1,303.11** | 14.64 |
| MORPHINE | 9300 | 202 | **10,793.74** | 53.43 |
| OXYMORPHONE | 9652 | 10 | **163.30** | 16.33 |
| TAPENTADOL | 9780 | 19 | **815.50** | 42.92 |
| FENTANYL BASE | 9801 | 208 | **310.70** | 1.49 |

**BUSINESS ACTIVITY: C - PRACTITIONERS**

| DRUG NAME | DRUG CODE | NUMBER OF REGISTRANT SOLD TO | TOTAL GRAMS | AVG GRAMS |
|---|---|---|---|---|
| METHYLPHENIDATE (DL;D;L;ISOMERS) | 1724 | 1 | **1,961.36** | 1,961.36 |
| CODEINE | 9050 | 27 | **638.87** | 23.66 |
| BUPRENORPHINE | 9064 | 284 | **24.89** | 0.09 |
| OXYCODONE | 9143 | 4 | **11.65** | 2.91 |
| HYDROMORPHONE | 9150 | 117 | **60.48** | 0.52 |

| | | | | |
|---|---|---|---|---|
| HYDROCODONE | 9193 | 96 | **182.69** | 1.90 |
| MEPERIDINE (PETHIDINE) | 9230 | 10 | **51.20** | 5.12 |
| METHADONE | 9250B | 2 | **7.51** | 3.76 |
| MORPHINE | 9300 | 33 | **35.45** | 1.07 |
| FENTANYL BASE | 9801 | 103 | **9.31** | 0.09 |

**BUSINESS ACTIVITY: D - TEACHING INSTITUTIONS**

| DRUG NAME | DRUG CODE | NUMBER OF REGISTRANT SOLD TO | TOTAL GRAMS | AVG GRAMS |
|---|---|---|---|---|
| BUPRENORPHINE | 9064 | 1 | **0.00** | 0.00 |

**BUSINESS ACTIVITY: M - MID-LEVEL PRACTITIONERS**

| DRUG NAME | DRUG CODE | NUMBER OF REGISTRANT SOLD TO | TOTAL GRAMS | AVG GRAMS |
|---|---|---|---|---|
| HYDROMORPHONE | 9150 | 14 | **0.73** | 0.05 |
| MORPHINE | 9300 | 21 | **6.66** | 0.32 |
| FENTANYL BASE | 9801 | 37 | **0.54** | 0.01 |

**BUSINESS ACTIVITY: NTP - NARCOTIC TREATMENT PROGRAMS**

| DRUG NAME | DRUG CODE | NUMBER OF REGISTRANT SOLD TO | TOTAL GRAMS | AVG GRAMS |
|---|---|---|---|---|
| BUPRENORPHINE | 9064 | 6 | **320.82** | 53.47 |
| METHADONE | 9250B | 9 | **62,163.93** | 6,907.10 |

**STATE: KY - KENTUCKY**

**BUSINESS ACTIVITY: A - PHARMACIES**

| DRUG NAME | DRUG CODE | NUMBER OF REGISTRANT SOLD TO | TOTAL GRAMS | AVG GRAMS |
|---|---|---|---|---|
| AMPHETAMINE | 1100 | 1,092 | **292,529.32** | 267.88 |
| METHAMPHETAMINE | 1105 | 18 | **39.77** | 2.21 |
| LISDEXAMFETAMINE | 1205 | 1,040 | **157,531.09** | 151.47 |
| METHYLPHENIDATE (DL;D;L;ISOMERS) | 1724 | 1,094 | **286,618.91** | 261.99 |
| CODEINE | 9050 | 1,086 | **140,240.00** | 129.13 |
| BUPRENORPHINE | 9064 | 1,017 | **176,940.40** | 173.98 |
| OXYCODONE | 9143 | 1,105 | **634,779.80** | 574.46 |
| HYDROMORPHONE | 9150 | 757 | **6,819.66** | 9.01 |
| HYDROCODONE | 9193 | 1,112 | **640,816.17** | 576.27 |
| MEPERIDINE (PETHIDINE) | 9230 | 109 | **1,312.37** | 12.04 |
| METHADONE | 9250B | 841 | **30,966.90** | 36.82 |
| MORPHINE | 9300 | 1,057 | **120,018.29** | 113.55 |
| OXYMORPHONE | 9652 | 365 | **6,019.44** | 16.49 |
| TAPENTADOL | 9780 | 430 | **35,980.00** | 83.67 |
| FENTANYL BASE | 9801 | 993 | **2,298.81** | 2.32 |

**BUSINESS ACTIVITY: B - HOSPITALS**

| DRUG NAME | DRUG CODE | NUMBER OF REGISTRANT SOLD TO | TOTAL GRAMS | AVG GRAMS |
|---|---|---|---|---|
| AMPHETAMINE | 1100 | 56 | **7,492.09** | 133.79 |
| LISDEXAMFETAMINE | 1205 | 30 | **1,552.78** | 51.76 |
| METHYLPHENIDATE (DL;D;L;ISOMERS) | 1724 | 86 | **7,185.42** | 83.55 |

| | | | | |
|---|---|---|---|---|
| CODEINE | 9050 | 133 | **3,103.36** | 23.33 |
| BUPRENORPHINE | 9064 | 102 | **2,169.09** | 21.27 |
| OXYCODONE | 9143 | 152 | **27,553.92** | 181.28 |
| HYDROMORPHONE | 9150 | 138 | **1,777.28** | 12.88 |
| HYDROCODONE | 9193 | 153 | **22,300.19** | 145.75 |
| MEPERIDINE (PETHIDINE) | 9230 | 108 | **2,153.15** | 19.94 |
| METHADONE | 9250B | 108 | **2,469.65** | 22.87 |
| MORPHINE | 9300 | 147 | **13,619.33** | 92.65 |
| OXYMORPHONE | 9652 | 13 | **17.93** | 1.38 |
| TAPENTADOL | 9780 | 19 | **1,240.00** | 65.26 |
| FENTANYL BASE | 9801 | 147 | **384.55** | 2.62 |

**BUSINESS ACTIVITY: C - PRACTITIONERS**

| DRUG NAME | DRUG CODE | NUMBER OF REGISTRANT SOLD TO | TOTAL GRAMS | AVG GRAMS |
|---|---|---|---|---|
| CODEINE | 9050 | 11 | **378.29** | 34.39 |
| BUPRENORPHINE | 9064 | 290 | **141.51** | 0.49 |
| OXYCODONE | 9143 | 13 | **16.45** | 1.27 |
| HYDROMORPHONE | 9150 | 101 | **144.54** | 1.43 |
| HYDROCODONE | 9193 | 131 | **602.90** | 4.60 |
| MEPERIDINE (PETHIDINE) | 9230 | 40 | **229.20** | 5.73 |
| MORPHINE | 9300 | 97 | **456.94** | 4.71 |
| FENTANYL BASE | 9801 | 302 | **21.61** | 0.07 |

**BUSINESS ACTIVITY: D - TEACHING INSTITUTIONS**

| DRUG NAME | DRUG CODE | NUMBER OF REGISTRANT SOLD TO | TOTAL GRAMS | AVG GRAMS |
|---|---|---|---|---|
| HYDROMORPHONE | 9150 | 1 | **0.07** | 0.07 |

**BUSINESS ACTIVITY: NTP - NARCOTIC TREATMENT PROGRAMS**

| DRUG NAME | DRUG CODE | NUMBER OF REGISTRANT SOLD TO | TOTAL GRAMS | AVG GRAMS |
|---|---|---|---|---|
| BUPRENORPHINE | 9064 | 18 | **399.04** | 22.17 |
| METHADONE | 9250B | 28 | **174,129.39** | 6,218.91 |

**STATE: LA - LOUISIANA**

**BUSINESS ACTIVITY: A - PHARMACIES**

| DRUG NAME | DRUG CODE | NUMBER OF REGISTRANT SOLD TO | TOTAL GRAMS | AVG GRAMS |
|---|---|---|---|---|
| AMPHETAMINE | 1100 | 1,132 | **688,818.31** | 608.50 |
| METHAMPHETAMINE | 1105 | 15 | **49.42** | 3.29 |
| LISDEXAMFETAMINE | 1205 | 1,101 | **350,105.45** | 317.99 |
| METHYLPHENIDATE (DL;D;L;ISOMERS) | 1724 | 1,121 | **318,229.56** | 283.88 |
| CODEINE | 9050 | 1,109 | **132,162.20** | 119.17 |
| BUPRENORPHINE | 9064 | 989 | **93,180.06** | 94.22 |
| OXYCODONE | 9143 | 1,129 | **559,603.42** | 495.66 |
| HYDROMORPHONE | 9150 | 836 | **7,457.04** | 8.92 |
| HYDROCODONE | 9193 | 1,141 | **462,435.94** | 405.29 |

| | | | | |
|---|---|---|---|---|
| MEPERIDINE (PETHIDINE) | 9230 | 409 | **5,517.14** | 13.49 |
| METHADONE | 9250B | 642 | **22,275.75** | 34.70 |
| MORPHINE | 9300 | 1,025 | **150,526.45** | 146.86 |
| OXYMORPHONE | 9652 | 351 | **11,249.16** | 32.05 |
| TAPENTADOL | 9780 | 523 | **46,225.00** | 88.38 |
| FENTANYL BASE | 9801 | 950 | **2,535.93** | 2.67 |

**BUSINESS ACTIVITY: B - HOSPITALS**

| DRUG NAME | DRUG CODE | NUMBER OF REGISTRANT SOLD TO | TOTAL GRAMS | AVG GRAMS |
|---|---|---|---|---|
| AMPHETAMINE | 1100 | 57 | **8,232.86** | 144.44 |
| LISDEXAMFETAMINE | 1205 | 28 | **2,226.84** | 79.53 |
| METHYLPHENIDATE (DL;D;L;ISOMERS) | 1724 | 105 | **4,465.96** | 42.53 |
| CODEINE | 9050 | 129 | **2,528.73** | 19.60 |
| BUPRENORPHINE | 9064 | 95 | **1,152.63** | 12.13 |
| OXYCODONE | 9143 | 196 | **23,580.98** | 120.31 |
| HYDROMORPHONE | 9150 | 181 | **1,766.43** | 9.76 |
| HYDROCODONE | 9193 | 203 | **26,801.46** | 132.03 |
| MEPERIDINE (PETHIDINE) | 9230 | 140 | **3,769.24** | 26.92 |
| METHADONE | 9250B | 125 | **1,765.29** | 14.12 |
| MORPHINE | 9300 | 193 | **12,663.02** | 65.61 |
| OXYMORPHONE | 9652 | 9 | **19.63** | 2.18 |
| TAPENTADOL | 9780 | 12 | **409.00** | 34.08 |
| FENTANYL BASE | 9801 | 191 | **306.32** | 1.60 |

**BUSINESS ACTIVITY: C - PRACTITIONERS**

| DRUG NAME | DRUG CODE | NUMBER OF REGISTRANT SOLD TO | TOTAL GRAMS | AVG GRAMS |
|---|---|---|---|---|
| METHAMPHETAMINE | 1105 | 1 | **4.02** | 4.02 |
| CODEINE | 9050 | 57 | **1,054.43** | 18.50 |
| BUPRENORPHINE | 9064 | 358 | **18.14** | 0.05 |
| OXYCODONE | 9143 | 18 | **23.94** | 1.33 |
| HYDROMORPHONE | 9150 | 110 | **41.85** | 0.38 |
| HYDROCODONE | 9193 | 202 | **508.31** | 2.52 |
| MEPERIDINE (PETHIDINE) | 9230 | 90 | **702.32** | 7.80 |
| METHADONE | 9250B | 4 | **9.48** | 2.37 |
| MORPHINE | 9300 | 105 | **52.41** | 0.50 |
| FENTANYL BASE | 9801 | 257 | **20.36** | 0.08 |

**BUSINESS ACTIVITY: D - TEACHING INSTITUTIONS**

| DRUG NAME | DRUG CODE | NUMBER OF REGISTRANT SOLD TO | TOTAL GRAMS | AVG GRAMS |
|---|---|---|---|---|
| FENTANYL BASE | 9801 | 1 | **0.05** | 0.05 |

**BUSINESS ACTIVITY: M - MID-LEVEL PRACTITIONERS**

| DRUG NAME | DRUG CODE | NUMBER OF REGISTRANT SOLD TO | TOTAL GRAMS | AVG GRAMS |
|---|---|---|---|---|
| CODEINE | 9050 | 6 | **4.70** | 0.78 |

**BUSINESS ACTIVITY: NTP - NARCOTIC TREATMENT PROGRAMS**

| DRUG NAME | DRUG CODE | NUMBER OF REGISTRANT SOLD TO | TOTAL GRAMS | AVG GRAMS |
|---|---|---|---|---|
| BUPRENORPHINE | 9064 | 9 | **444.45** | 49.38 |
| METHADONE | 9250B | 10 | **113,748.45** | 11,374.85 |

**STATE: MA - MASSACHUSETTS**

**BUSINESS ACTIVITY: A - PHARMACIES**

| DRUG NAME | DRUG CODE | NUMBER OF REGISTRANT SOLD TO | TOTAL GRAMS | AVG GRAMS |
|---|---|---|---|---|
| AMPHETAMINE | 1100 | 1,110 | **731,990.68** | 659.45 |
| METHAMPHETAMINE | 1105 | 31 | **98.04** | 3.16 |
| LISDEXAMFETAMINE | 1205 | 1,067 | **156,360.44** | 146.54 |
| METHYLPHENIDATE (DL;D;L;ISOMERS) | 1724 | 1,101 | **545,999.47** | 495.91 |
| CODEINE | 9050 | 1,082 | **135,009.91** | 124.78 |
| BUPRENORPHINE | 9064 | 1,104 | **179,921.07** | 162.97 |
| OXYCODONE | 9143 | 1,112 | **678,375.15** | 610.05 |
| HYDROMORPHONE | 9150 | 1,048 | **16,515.18** | 15.76 |
| HYDROCODONE | 9193 | 1,102 | **85,923.62** | 77.97 |
| MEPERIDINE (PETHIDINE) | 9230 | 88 | **1,934.19** | 21.98 |
| METHADONE | 9250B | 895 | **50,399.17** | 56.31 |
| MORPHINE | 9300 | 1,042 | **184,269.04** | 176.84 |
| OXYMORPHONE | 9652 | 142 | **3,068.20** | 21.61 |
| TAPENTADOL | 9780 | 427 | **60,964.50** | 142.77 |
| FENTANYL BASE | 9801 | 940 | **2,925.40** | 3.11 |

**BUSINESS ACTIVITY: B - HOSPITALS**

| DRUG NAME | DRUG CODE | NUMBER OF REGISTRANT SOLD TO | TOTAL GRAMS | AVG GRAMS |
|---|---|---|---|---|
| AMPHETAMINE | 1100 | 151 | **27,618.64** | 182.90 |
| LISDEXAMFETAMINE | 1205 | 80 | **4,059.99** | 50.75 |
| METHYLPHENIDATE (DL;D;L;ISOMERS) | 1724 | 163 | **24,145.39** | 148.13 |
| CODEINE | 9050 | 150 | **6,318.29** | 42.12 |
| BUPRENORPHINE | 9064 | 168 | **11,294.15** | 67.23 |
| OXYCODONE | 9143 | 200 | **54,413.29** | 272.07 |
| HYDROMORPHONE | 9150 | 169 | **6,583.53** | 38.96 |
| HYDROCODONE | 9193 | 162 | **2,844.20** | 17.56 |
| MEPERIDINE (PETHIDINE) | 9230 | 88 | **954.84** | 10.85 |
| METHADONE | 9250B | 162 | **12,400.48** | 76.55 |
| MORPHINE | 9300 | 192 | **21,927.49** | 114.21 |
| OXYMORPHONE | 9652 | 17 | **369.33** | 21.73 |
| TAPENTADOL | 9780 | 18 | **1,746.50** | 97.03 |
| FENTANYL BASE | 9801 | 182 | **550.74** | 3.03 |

**BUSINESS ACTIVITY: C - PRACTITIONERS**

| DRUG NAME | DRUG CODE | NUMBER OF REGISTRANT SOLD TO | TOTAL GRAMS | AVG GRAMS |
|---|---|---|---|---|

| | | | | |
|---|---|---|---|---|
| AMPHETAMINE | 1100 | 1 | **0.80** | 0.80 |
| CODEINE | 9050 | 21 | **380.88** | 18.14 |
| BUPRENORPHINE | 9064 | 595 | **101.56** | 0.17 |
| OXYCODONE | 9143 | 22 | **29.49** | 1.34 |
| HYDROMORPHONE | 9150 | 245 | **99.93** | 0.41 |
| HYDROCODONE | 9193 | 132 | **398.93** | 3.02 |
| MEPERIDINE (PETHIDINE) | 9230 | 19 | **80.83** | 4.25 |
| METHADONE | 9250B | 36 | **114.66** | 3.19 |
| MORPHINE | 9300 | 82 | **42.07** | 0.51 |
| FENTANYL BASE | 9801 | 265 | **23.59** | 0.09 |

**BUSINESS ACTIVITY: M - MID-LEVEL PRACTITIONERS**

| DRUG NAME | DRUG CODE | NUMBER OF REGISTRANT SOLD TO | TOTAL GRAMS | AVG GRAMS |
|---|---|---|---|---|
| OXYCODONE | 9143 | 1 | **2.69** | 2.69 |
| HYDROCODONE | 9193 | 1 | **0.91** | 0.91 |

**BUSINESS ACTIVITY: NTP - NARCOTIC TREATMENT PROGRAMS**

| DRUG NAME | DRUG CODE | NUMBER OF REGISTRANT SOLD TO | TOTAL GRAMS | AVG GRAMS |
|---|---|---|---|---|
| BUPRENORPHINE | 9064 | 37 | **874.04** | 23.62 |
| METHADONE | 9250B | 77 | **541,304.74** | 7,029.93 |

**STATE: MD - MARYLAND**

**BUSINESS ACTIVITY: A - PHARMACIES**

| DRUG NAME | DRUG CODE | NUMBER OF REGISTRANT SOLD TO | TOTAL GRAMS | AVG GRAMS |
|---|---|---|---|---|
| AMPHETAMINE | 1100 | 1,218 | **397,011.97** | 325.95 |
| METHAMPHETAMINE | 1105 | 29 | **132.58** | 4.57 |
| LISDEXAMFETAMINE | 1205 | 1,172 | **228,903.90** | 195.31 |
| METHYLPHENIDATE (DL;D;L;ISOMERS) | 1724 | 1,206 | **381,956.31** | 316.71 |
| CODEINE | 9050 | 1,209 | **165,147.76** | 136.60 |
| BUPRENORPHINE | 9064 | 1,191 | **123,138.49** | 103.39 |
| OXYCODONE | 9143 | 1,236 | **919,403.43** | 743.85 |
| HYDROMORPHONE | 9150 | 1,107 | **17,906.50** | 16.18 |
| HYDROCODONE | 9193 | 1,207 | **96,726.65** | 80.14 |
| MEPERIDINE (PETHIDINE) | 9230 | 129 | **2,695.66** | 20.90 |
| METHADONE | 9250B | 881 | **31,296.40** | 35.52 |
| MORPHINE | 9300 | 1,153 | **187,374.83** | 162.51 |
| OXYMORPHONE | 9652 | 680 | **16,833.93** | 24.76 |
| TAPENTADOL | 9780 | 733 | **89,659.50** | 122.32 |
| FENTANYL BASE | 9801 | 1,021 | **2,587.14** | 2.53 |

**BUSINESS ACTIVITY: B - HOSPITALS**

| DRUG NAME | DRUG CODE | NUMBER OF REGISTRANT SOLD TO | TOTAL GRAMS | AVG GRAMS |
|---|---|---|---|---|
| AMPHETAMINE | 1100 | 55 | **10,161.43** | 184.75 |
| LISDEXAMFETAMINE | 1205 | 27 | **2,042.49** | 75.65 |

| | | | | |
|---|---|---|---|---|
| METHYLPHENIDATE (DL;D;L;ISOMERS) | 1724 | 65 | **15,500.18** | 238.46 |
| CODEINE | 9050 | 82 | **3,725.39** | 45.43 |
| BUPRENORPHINE | 9064 | 84 | **2,272.26** | 27.05 |
| OXYCODONE | 9143 | 138 | **43,048.13** | 311.94 |
| HYDROMORPHONE | 9150 | 130 | **3,822.34** | 29.40 |
| HYDROCODONE | 9193 | 119 | **2,537.46** | 21.32 |
| MEPERIDINE (PETHIDINE) | 9230 | 74 | **900.37** | 12.17 |
| METHADONE | 9250B | 68 | **7,554.83** | 111.10 |
| MORPHINE | 9300 | 115 | **13,961.62** | 121.41 |
| OXYMORPHONE | 9652 | 5 | **144.30** | 28.86 |
| TAPENTADOL | 9780 | 17 | **2,782.50** | 163.68 |
| FENTANYL BASE | 9801 | 212 | **377.45** | 1.78 |

**BUSINESS ACTIVITY: C - PRACTITIONERS**

| DRUG NAME | DRUG CODE | NUMBER OF REGISTRANT SOLD TO | TOTAL GRAMS | AVG GRAMS |
|---|---|---|---|---|
| AMPHETAMINE | 1100 | 1 | **1.10** | 1.10 |
| METHYLPHENIDATE (DL;D;L;ISOMERS) | 1724 | 4 | **79.65** | 19.91 |
| CODEINE | 9050 | 101 | **6,456.15** | 63.92 |
| BUPRENORPHINE | 9064 | 565 | **144.64** | 0.26 |
| OXYCODONE | 9143 | 59 | **2,522.19** | 42.75 |
| HYDROMORPHONE | 9150 | 257 | **129.02** | 0.50 |
| HYDROCODONE | 9193 | 229 | **1,266.44** | 5.53 |
| MEPERIDINE (PETHIDINE) | 9230 | 45 | **427.58** | 9.50 |
| METHADONE | 9250B | 21 | **92.84** | 4.42 |
| MORPHINE | 9300 | 110 | **667.95** | 6.07 |
| OXYMORPHONE | 9652 | 6 | **91.40** | 15.23 |
| TAPENTADOL | 9780 | 8 | **466.50** | 58.31 |
| FENTANYL BASE | 9801 | 351 | **41.44** | 0.12 |

**BUSINESS ACTIVITY: D - TEACHING INSTITUTIONS**

| DRUG NAME | DRUG CODE | NUMBER OF REGISTRANT SOLD TO | TOTAL GRAMS | AVG GRAMS |
|---|---|---|---|---|
| FENTANYL BASE | 9801 | 1 | **0.02** | 0.02 |

**BUSINESS ACTIVITY: M - MID-LEVEL PRACTITIONERS**

| DRUG NAME | DRUG CODE | NUMBER OF REGISTRANT SOLD TO | TOTAL GRAMS | AVG GRAMS |
|---|---|---|---|---|
| CODEINE | 9050 | 5 | **3.23** | 0.65 |
| BUPRENORPHINE | 9064 | 4 | **4.74** | 1.19 |

**BUSINESS ACTIVITY: NTP - NARCOTIC TREATMENT PROGRAMS**

| DRUG NAME | DRUG CODE | NUMBER OF REGISTRANT SOLD TO | TOTAL GRAMS | AVG GRAMS |
|---|---|---|---|---|
| BUPRENORPHINE | 9064 | 61 | **8,376.66** | 137.32 |
| METHADONE | 9250B | 86 | **738,707.32** | 8,589.62 |

**STATE: ME - MAINE**

**BUSINESS ACTIVITY: A - PHARMACIES**

| DRUG NAME | DRUG CODE | NUMBER OF REGISTRANT SOLD TO | TOTAL GRAMS | AVG GRAMS |
|---|---|---|---|---|
| AMPHETAMINE | 1100 | 297 | **103,327.78** | 347.90 |
| METHAMPHETAMINE | 1105 | 5 | **45.80** | 9.16 |
| LISDEXAMFETAMINE | 1205 | 294 | **67,447.51** | 229.41 |
| METHYLPHENIDATE (DL;D;L;ISOMERS) | 1724 | 297 | **138,197.70** | 465.31 |
| CODEINE | 9050 | 293 | **29,539.29** | 100.82 |
| BUPRENORPHINE | 9064 | 302 | **52,430.17** | 173.61 |
| OXYCODONE | 9143 | 298 | **153,877.00** | 516.37 |
| HYDROMORPHONE | 9150 | 283 | **4,450.23** | 15.73 |
| HYDROCODONE | 9193 | 300 | **50,897.74** | 169.66 |
| MEPERIDINE (PETHIDINE) | 9230 | 27 | **298.49** | 11.06 |
| METHADONE | 9250B | 277 | **12,775.43** | 46.12 |
| MORPHINE | 9300 | 298 | **48,086.21** | 161.36 |
| OXYMORPHONE | 9652 | 90 | **1,275.12** | 14.17 |
| TAPENTADOL | 9780 | 93 | **4,969.50** | 53.44 |
| FENTANYL BASE | 9801 | 289 | **838.93** | 2.90 |

**BUSINESS ACTIVITY: B - HOSPITALS**

| DRUG NAME | DRUG CODE | NUMBER OF REGISTRANT SOLD TO | TOTAL GRAMS | AVG GRAMS |
|---|---|---|---|---|
| AMPHETAMINE | 1100 | 24 | **1,676.29** | 69.85 |
| LISDEXAMFETAMINE | 1205 | 14 | **408.72** | 29.19 |
| METHYLPHENIDATE (DL;D;L;ISOMERS) | 1724 | 38 | **2,284.42** | 60.12 |
| CODEINE | 9050 | 36 | **675.60** | 18.77 |
| BUPRENORPHINE | 9064 | 48 | **1,517.78** | 31.62 |
| OXYCODONE | 9143 | 49 | **7,907.17** | 161.37 |
| HYDROMORPHONE | 9150 | 43 | **813.12** | 18.91 |
| HYDROCODONE | 9193 | 46 | **2,805.09** | 60.98 |
| MEPERIDINE (PETHIDINE) | 9230 | 30 | **113.84** | 3.79 |
| METHADONE | 9250B | 39 | **1,074.89** | 27.56 |
| MORPHINE | 9300 | 48 | **3,933.39** | 81.95 |
| OXYMORPHONE | 9652 | 1 | **0.54** | 0.54 |
| TAPENTADOL | 9780 | 1 | **27.00** | 27.00 |
| FENTANYL BASE | 9801 | 50 | **106.31** | 2.13 |

**BUSINESS ACTIVITY: C - PRACTITIONERS**

| DRUG NAME | DRUG CODE | NUMBER OF REGISTRANT SOLD TO | TOTAL GRAMS | AVG GRAMS |
|---|---|---|---|---|
| AMPHETAMINE | 1100 | 1 | **4.17** | 4.17 |
| CODEINE | 9050 | 7 | **231.75** | 33.11 |
| BUPRENORPHINE | 9064 | 171 | **32.18** | 0.19 |
| OXYCODONE | 9143 | 5 | **21.07** | 4.21 |
| HYDROMORPHONE | 9150 | 81 | **37.70** | 0.47 |
| HYDROCODONE | 9193 | 55 | **103.97** | 1.89 |

| | | | | |
|---|---|---|---|---|
| MEPERIDINE (PETHIDINE) | 9230 | 4 | **17.10** | 4.28 |
| METHADONE | 9250B | 3 | **33.90** | 11.30 |
| MORPHINE | 9300 | 27 | **15.54** | 0.58 |
| FENTANYL BASE | 9801 | 48 | **4.61** | 0.10 |

**BUSINESS ACTIVITY: D - TEACHING INSTITUTIONS**

| DRUG NAME | DRUG CODE | NUMBER OF REGISTRANT SOLD TO | TOTAL GRAMS | AVG GRAMS |
|---|---|---|---|---|
| BUPRENORPHINE | 9064 | 1 | **0.01** | 0.01 |

**BUSINESS ACTIVITY: M - MID-LEVEL PRACTITIONERS**

| DRUG NAME | DRUG CODE | NUMBER OF REGISTRANT SOLD TO | TOTAL GRAMS | AVG GRAMS |
|---|---|---|---|---|
| CODEINE | 9050 | 4 | **63.21** | 15.80 |
| BUPRENORPHINE | 9064 | 5 | **40.44** | 8.09 |
| FENTANYL BASE | 9801 | 1 | **0.00** | 0.00 |

**BUSINESS ACTIVITY: NTP - NARCOTIC TREATMENT PROGRAMS**

| DRUG NAME | DRUG CODE | NUMBER OF REGISTRANT SOLD TO | TOTAL GRAMS | AVG GRAMS |
|---|---|---|---|---|
| BUPRENORPHINE | 9064 | 5 | **342.30** | 68.46 |
| METHADONE | 9250B | 10 | **106,498.89** | 10,649.89 |

**STATE: MI - MICHIGAN**

**BUSINESS ACTIVITY: A - PHARMACIES**

| DRUG NAME | DRUG CODE | NUMBER OF REGISTRANT SOLD TO | TOTAL GRAMS | AVG GRAMS |
|---|---|---|---|---|
| AMPHETAMINE | 1100 | 2,326 | **991,450.26** | 426.25 |
| METHAMPHETAMINE | 1105 | 31 | **85.58** | 2.76 |
| LISDEXAMFETAMINE | 1205 | 2,088 | **247,021.55** | 118.31 |
| METHYLPHENIDATE (DL;D;L;ISOMERS) | 1724 | 2,236 | **675,756.21** | 302.22 |
| CODEINE | 9050 | 2,350 | **658,710.95** | 280.30 |
| BUPRENORPHINE | 9064 | 2,185 | **130,663.29** | 59.80 |
| OXYCODONE | 9143 | 2,353 | **741,723.00** | 315.22 |
| HYDROMORPHONE | 9150 | 1,715 | **18,917.58** | 11.03 |
| HYDROCODONE | 9193 | 2,393 | **973,247.33** | 406.71 |
| MEPERIDINE (PETHIDINE) | 9230 | 121 | **2,611.78** | 21.58 |
| METHADONE | 9250B | 1,722 | **81,027.01** | 47.05 |
| MORPHINE | 9300 | 2,199 | **461,831.18** | 210.02 |
| OXYMORPHONE | 9652 | 683 | **28,394.38** | 41.57 |
| TAPENTADOL | 9780 | 711 | **53,314.00** | 74.98 |
| FENTANYL BASE | 9801 | 1,969 | **6,137.22** | 3.12 |

**BUSINESS ACTIVITY: B - HOSPITALS**

| DRUG NAME | DRUG CODE | NUMBER OF REGISTRANT SOLD TO | TOTAL GRAMS | AVG GRAMS |
|---|---|---|---|---|
| AMPHETAMINE | 1100 | 112 | **19,635.25** | 175.31 |
| LISDEXAMFETAMINE | 1205 | 62 | **4,082.54** | 65.85 |
| METHYLPHENIDATE (DL;D;L;ISOMERS) | 1724 | 156 | **15,827.95** | 101.46 |
| CODEINE | 9050 | 182 | **14,036.23** | 77.12 |

| | | | | |
|---|---|---|---|---|
| BUPRENORPHINE | 9064 | 132 | **3,300.24** | 25.00 |
| OXYCODONE | 9143 | 185 | **45,792.70** | 247.53 |
| HYDROMORPHONE | 9150 | 182 | **3,792.46** | 20.84 |
| HYDROCODONE | 9193 | 197 | **44,728.12** | 227.05 |
| MEPERIDINE (PETHIDINE) | 9230 | 107 | **1,550.18** | 14.49 |
| METHADONE | 9250B | 156 | **6,201.84** | 39.76 |
| MORPHINE | 9300 | 190 | **35,322.62** | 185.91 |
| OXYMORPHONE | 9652 | 13 | **252.33** | 19.41 |
| TAPENTADOL | 9780 | 20 | **1,103.00** | 55.15 |
| FENTANYL BASE | 9801 | 187 | **826.95** | 4.42 |

**BUSINESS ACTIVITY: C - PRACTITIONERS**

| DRUG NAME | DRUG CODE | NUMBER OF REGISTRANT SOLD TO | TOTAL GRAMS | AVG GRAMS |
|---|---|---|---|---|
| AMPHETAMINE | 1100 | 3 | **6.64** | 2.21 |
| CODEINE | 9050 | 102 | **1,984.25** | 19.45 |
| BUPRENORPHINE | 9064 | 784 | **479.56** | 0.61 |
| OXYCODONE | 9143 | 46 | **497.02** | 10.80 |
| HYDROMORPHONE | 9150 | 408 | **206.62** | 0.51 |
| HYDROCODONE | 9193 | 249 | **1,078.19** | 4.33 |
| MEPERIDINE (PETHIDINE) | 9230 | 80 | **473.33** | 5.92 |
| METHADONE | 9250B | 9 | **70.39** | 7.82 |
| MORPHINE | 9300 | 200 | **321.48** | 1.61 |
| FENTANYL BASE | 9801 | 517 | **63.86** | 0.12 |

**BUSINESS ACTIVITY: D - TEACHING INSTITUTIONS**

| DRUG NAME | DRUG CODE | NUMBER OF REGISTRANT SOLD TO | TOTAL GRAMS | AVG GRAMS |
|---|---|---|---|---|
| CODEINE | 9050 | 1 | **229.65** | 229.65 |
| BUPRENORPHINE | 9064 | 1 | **1.35** | 1.35 |
| HYDROCODONE | 9193 | 1 | **18.94** | 18.94 |
| METHADONE | 9250B | 1 | **44.36** | 44.36 |
| MORPHINE | 9300 | 1 | **2.74** | 2.74 |
| FENTANYL BASE | 9801 | 1 | **1.94** | 1.94 |

**BUSINESS ACTIVITY: M - MID-LEVEL PRACTITIONERS**

| DRUG NAME | DRUG CODE | NUMBER OF REGISTRANT SOLD TO | TOTAL GRAMS | AVG GRAMS |
|---|---|---|---|---|
| CODEINE | 9050 | 3 | **279.65** | 93.22 |
| BUPRENORPHINE | 9064 | 4 | **4.93** | 1.23 |

**BUSINESS ACTIVITY: NTP - NARCOTIC TREATMENT PROGRAMS**

| DRUG NAME | DRUG CODE | NUMBER OF REGISTRANT SOLD TO | TOTAL GRAMS | AVG GRAMS |
|---|---|---|---|---|
| BUPRENORPHINE | 9064 | 13 | **245.80** | 18.91 |
| METHADONE | 9250B | 47 | **406,500.78** | 8,648.95 |

**STATE: MN - MINNESOTA**

**BUSINESS ACTIVITY: A - PHARMACIES**

| DRUG NAME | DRUG CODE | NUMBER OF REGISTRANT SOLD TO | TOTAL GRAMS | AVG GRAMS |
|---|---|---|---|---|
| AMPHETAMINE | 1100 | 990 | **457,844.30** | 462.47 |
| METHAMPHETAMINE | 1105 | 34 | **171.15** | 5.03 |
| LISDEXAMFETAMINE | 1205 | 973 | **128,499.49** | 132.07 |
| METHYLPHENIDATE (DL;D;L;ISOMERS) | 1724 | 992 | **397,664.57** | 400.87 |
| CODEINE | 9050 | 980 | **98,890.92** | 100.91 |
| BUPRENORPHINE | 9064 | 938 | **39,528.54** | 42.14 |
| OXYCODONE | 9143 | 1,002 | **371,441.46** | 370.70 |
| HYDROMORPHONE | 9150 | 930 | **13,887.59** | 14.93 |
| HYDROCODONE | 9193 | 1,005 | **117,634.25** | 117.05 |
| MEPERIDINE (PETHIDINE) | 9230 | 51 | **444.47** | 8.72 |
| METHADONE | 9250B | 799 | **20,522.19** | 25.68 |
| MORPHINE | 9300 | 972 | **119,080.08** | 122.51 |
| OXYMORPHONE | 9652 | 88 | **730.27** | 8.30 |
| TAPENTADOL | 9780 | 247 | **12,994.00** | 52.61 |
| FENTANYL BASE | 9801 | 894 | **2,170.13** | 2.43 |

**BUSINESS ACTIVITY: B - HOSPITALS**

| DRUG NAME | DRUG CODE | NUMBER OF REGISTRANT SOLD TO | TOTAL GRAMS | AVG GRAMS |
|---|---|---|---|---|
| AMPHETAMINE | 1100 | 82 | **7,550.04** | 92.07 |
| METHAMPHETAMINE | 1105 | 3 | **43.40** | 14.47 |
| LISDEXAMFETAMINE | 1205 | 47 | **1,640.07** | 34.90 |
| METHYLPHENIDATE (DL;D;L;ISOMERS) | 1724 | 96 | **7,731.44** | 80.54 |
| CODEINE | 9050 | 124 | **3,851.09** | 31.06 |
| BUPRENORPHINE | 9064 | 86 | **2,131.68** | 24.79 |
| OXYCODONE | 9143 | 147 | **29,304.49** | 199.35 |
| HYDROMORPHONE | 9150 | 147 | **3,700.27** | 25.17 |
| HYDROCODONE | 9193 | 146 | **8,759.54** | 60.00 |
| MEPERIDINE (PETHIDINE) | 9230 | 88 | **516.36** | 5.87 |
| METHADONE | 9250B | 103 | **2,906.23** | 28.22 |
| MORPHINE | 9300 | 151 | **14,437.83** | 95.61 |
| OXYMORPHONE | 9652 | 3 | **26.67** | 8.89 |
| TAPENTADOL | 9780 | 11 | **206.50** | 18.77 |
| FENTANYL BASE | 9801 | 144 | **383.34** | 2.66 |

**BUSINESS ACTIVITY: C - PRACTITIONERS**

| DRUG NAME | DRUG CODE | NUMBER OF REGISTRANT SOLD TO | TOTAL GRAMS | AVG GRAMS |
|---|---|---|---|---|
| AMPHETAMINE | 1100 | 1 | **3.80** | 3.80 |
| METHYLPHENIDATE (DL;D;L;ISOMERS) | 1724 | 1 | **15.13** | 15.13 |
| CODEINE | 9050 | 41 | **1,059.73** | 25.85 |
| BUPRENORPHINE | 9064 | 578 | **94.08** | 0.16 |
| OXYCODONE | 9143 | 61 | **218.43** | 3.58 |

| | | | | |
|---|---|---|---|---|
| HYDROMORPHONE | 9150 | 356 | **134.16** | 0.38 |
| HYDROCODONE | 9193 | 249 | **590.83** | 2.37 |
| MEPERIDINE (PETHIDINE) | 9230 | 42 | **198.81** | 4.73 |
| METHADONE | 9250B | 2 | **5.90** | 2.95 |
| MORPHINE | 9300 | 134 | **105.17** | 0.78 |
| FENTANYL BASE | 9801 | 330 | **40.42** | 0.12 |

**BUSINESS ACTIVITY: M - MID-LEVEL PRACTITIONERS**

| DRUG NAME | DRUG CODE | NUMBER OF REGISTRANT SOLD TO | TOTAL GRAMS | AVG GRAMS |
|---|---|---|---|---|
| BUPRENORPHINE | 9064 | 3 | **4.52** | 1.51 |
| FENTANYL BASE | 9801 | 1 | **0.01** | 0.01 |

**BUSINESS ACTIVITY: NTP - NARCOTIC TREATMENT PROGRAMS**

| DRUG NAME | DRUG CODE | NUMBER OF REGISTRANT SOLD TO | TOTAL GRAMS | AVG GRAMS |
|---|---|---|---|---|
| BUPRENORPHINE | 9064 | 15 | **1,892.45** | 126.16 |
| METHADONE | 9250B | 16 | **184,787.96** | 11,549.25 |

**STATE: MO - MISSOURI**

**BUSINESS ACTIVITY: A - PHARMACIES**

| DRUG NAME | DRUG CODE | NUMBER OF REGISTRANT SOLD TO | TOTAL GRAMS | AVG GRAMS |
|---|---|---|---|---|
| AMPHETAMINE | 1100 | 1,253 | **599,592.08** | 478.53 |
| METHAMPHETAMINE | 1105 | 24 | **204.08** | 8.50 |
| LISDEXAMFETAMINE | 1205 | 1,220 | **276,779.25** | 226.87 |
| METHYLPHENIDATE (DL;D;L;ISOMERS) | 1724 | 1,253 | **551,396.18** | 440.06 |
| CODEINE | 9050 | 1,244 | **217,933.63** | 175.19 |
| BUPRENORPHINE | 9064 | 1,200 | **67,975.16** | 56.65 |
| OXYCODONE | 9143 | 1,262 | **734,738.90** | 582.20 |
| HYDROMORPHONE | 9150 | 1,064 | **12,921.95** | 12.14 |
| HYDROCODONE | 9193 | 1,264 | **495,045.28** | 391.65 |
| MEPERIDINE (PETHIDINE) | 9230 | 152 | **3,292.09** | 21.66 |
| METHADONE | 9250B | 1,012 | **30,670.23** | 30.31 |
| MORPHINE | 9300 | 1,229 | **253,508.42** | 206.27 |
| OXYMORPHONE | 9652 | 355 | **5,208.39** | 14.67 |
| TAPENTADOL | 9780 | 675 | **66,677.50** | 98.78 |
| FENTANYL BASE | 9801 | 1,173 | **4,603.88** | 3.92 |

**BUSINESS ACTIVITY: B - HOSPITALS**

| DRUG NAME | DRUG CODE | NUMBER OF REGISTRANT SOLD TO | TOTAL GRAMS | AVG GRAMS |
|---|---|---|---|---|
| AMPHETAMINE | 1100 | 94 | **9,321.46** | 99.16 |
| LISDEXAMFETAMINE | 1205 | 48 | **2,708.40** | 56.42 |
| METHYLPHENIDATE (DL;D;L;ISOMERS) | 1724 | 124 | **8,312.67** | 67.04 |
| CODEINE | 9050 | 167 | **4,737.23** | 28.37 |
| BUPRENORPHINE | 9064 | 104 | **1,670.46** | 16.06 |
| OXYCODONE | 9143 | 223 | **39,412.53** | 176.74 |

| | | | | |
|---|---|---|---|---|
| HYDROMORPHONE | 9150 | 180 | **2,438.36** | 13.55 |
| HYDROCODONE | 9193 | 230 | **19,638.15** | 85.38 |
| MEPERIDINE (PETHIDINE) | 9230 | 148 | **1,988.11** | 13.43 |
| METHADONE | 9250B | 120 | **2,763.13** | 23.03 |
| MORPHINE | 9300 | 218 | **21,699.42** | 99.54 |
| OXYMORPHONE | 9652 | 14 | **16.24** | 1.16 |
| TAPENTADOL | 9780 | 30 | **1,050.00** | 35.00 |
| FENTANYL BASE | 9801 | 251 | **563.24** | 2.24 |

**BUSINESS ACTIVITY: C - PRACTITIONERS**

| DRUG NAME | DRUG CODE | NUMBER OF REGISTRANT SOLD TO | TOTAL GRAMS | AVG GRAMS |
|---|---|---|---|---|
| CODEINE | 9050 | 63 | **3,376.37** | 53.59 |
| BUPRENORPHINE | 9064 | 503 | **102.14** | 0.20 |
| OXYCODONE | 9143 | 11 | **118.21** | 10.75 |
| HYDROMORPHONE | 9150 | 163 | **101.40** | 0.62 |
| HYDROCODONE | 9193 | 216 | **1,202.20** | 5.57 |
| MEPERIDINE (PETHIDINE) | 9230 | 15 | **130.18** | 8.68 |
| METHADONE | 9250B | 7 | **25.04** | 3.58 |
| MORPHINE | 9300 | 110 | **65.13** | 0.59 |
| FENTANYL BASE | 9801 | 161 | **11.56** | 0.07 |

**BUSINESS ACTIVITY: D - TEACHING INSTITUTIONS**

| DRUG NAME | DRUG CODE | NUMBER OF REGISTRANT SOLD TO | TOTAL GRAMS | AVG GRAMS |
|---|---|---|---|---|
| CODEINE | 9050 | 1 | **179.02** | 179.02 |
| BUPRENORPHINE | 9064 | 1 | **1.24** | 1.24 |
| HYDROMORPHONE | 9150 | 2 | **9.25** | 4.62 |
| HYDROCODONE | 9193 | 1 | **16.43** | 16.43 |
| MORPHINE | 9300 | 2 | **5.01** | 2.51 |
| FENTANYL BASE | 9801 | 3 | **1.73** | 0.58 |

**BUSINESS ACTIVITY: M - MID-LEVEL PRACTITIONERS**

| DRUG NAME | DRUG CODE | NUMBER OF REGISTRANT SOLD TO | TOTAL GRAMS | AVG GRAMS |
|---|---|---|---|---|
| HYDROMORPHONE | 9150 | 3 | **0.18** | 0.06 |
| MEPERIDINE (PETHIDINE) | 9230 | 1 | **1.09** | 1.09 |
| MORPHINE | 9300 | 29 | **15.79** | 0.54 |
| FENTANYL BASE | 9801 | 51 | **0.62** | 0.01 |

**BUSINESS ACTIVITY: NTP - NARCOTIC TREATMENT PROGRAMS**

| DRUG NAME | DRUG CODE | NUMBER OF REGISTRANT SOLD TO | TOTAL GRAMS | AVG GRAMS |
|---|---|---|---|---|
| BUPRENORPHINE | 9064 | 16 | **986.27** | 61.64 |
| METHADONE | 9250B | 16 | **106,225.20** | 6,639.08 |

**STATE: MS - MISSISSIPPI**

**BUSINESS ACTIVITY: A - PHARMACIES**

| DRUG NAME | DRUG CODE | NUMBER OF REGISTRANT SOLD TO | TOTAL GRAMS | AVG GRAMS |
|---|---|---|---|---|

| | | | | |
|---|---|---|---|---|
| AMPHETAMINE | 1100 | 705 | **245,599.41** | 348.37 |
| METHAMPHETAMINE | 1105 | 5 | **29.33** | 5.87 |
| LISDEXAMFETAMINE | 1205 | 680 | **141,669.76** | 208.34 |
| METHYLPHENIDATE (DL;D;L;ISOMERS) | 1724 | 694 | **165,078.71** | 237.87 |
| CODEINE | 9050 | 699 | **71,263.50** | 101.95 |
| BUPRENORPHINE | 9064 | 638 | **45,388.46** | 71.14 |
| OXYCODONE | 9143 | 706 | **303,009.80** | 429.19 |
| HYDROMORPHONE | 9150 | 512 | **20,550.75** | 40.14 |
| HYDROCODONE | 9193 | 722 | **293,029.23** | 405.86 |
| MEPERIDINE (PETHIDINE) | 9230 | 208 | **3,299.06** | 15.86 |
| METHADONE | 9250B | 352 | **6,761.75** | 19.21 |
| MORPHINE | 9300 | 670 | **104,590.61** | 156.11 |
| OXYMORPHONE | 9652 | 152 | **3,640.35** | 23.95 |
| TAPENTADOL | 9780 | 381 | **28,932.00** | 75.94 |
| FENTANYL BASE | 9801 | 658 | **3,051.45** | 4.64 |

**BUSINESS ACTIVITY: B - HOSPITALS**

| DRUG NAME | DRUG CODE | NUMBER OF REGISTRANT SOLD TO | TOTAL GRAMS | AVG GRAMS |
|---|---|---|---|---|
| AMPHETAMINE | 1100 | 51 | **7,654.19** | 150.08 |
| LISDEXAMFETAMINE | 1205 | 35 | **4,161.74** | 118.91 |
| METHYLPHENIDATE (DL;D;L;ISOMERS) | 1724 | 71 | **5,555.56** | 78.25 |
| CODEINE | 9050 | 93 | **3,506.06** | 37.70 |
| BUPRENORPHINE | 9064 | 62 | **1,360.33** | 21.94 |
| OXYCODONE | 9143 | 133 | **12,863.04** | 96.71 |
| HYDROMORPHONE | 9150 | 123 | **1,237.91** | 10.06 |
| HYDROCODONE | 9193 | 147 | **14,338.51** | 97.54 |
| MEPERIDINE (PETHIDINE) | 9230 | 95 | **3,525.76** | 37.11 |
| METHADONE | 9250B | 56 | **309.28** | 5.52 |
| MORPHINE | 9300 | 133 | **7,495.58** | 56.36 |
| OXYMORPHONE | 9652 | 5 | **21.05** | 4.21 |
| TAPENTADOL | 9780 | 14 | **1,270.00** | 90.71 |
| FENTANYL BASE | 9801 | 135 | **274.85** | 2.04 |

**BUSINESS ACTIVITY: C - PRACTITIONERS**

| DRUG NAME | DRUG CODE | NUMBER OF REGISTRANT SOLD TO | TOTAL GRAMS | AVG GRAMS |
|---|---|---|---|---|
| CODEINE | 9050 | 11 | **771.91** | 70.17 |
| BUPRENORPHINE | 9064 | 112 | **4.81** | 0.04 |
| OXYCODONE | 9143 | 14 | **24.88** | 1.78 |
| HYDROMORPHONE | 9150 | 38 | **17.94** | 0.47 |
| HYDROCODONE | 9193 | 43 | **86.65** | 2.02 |
| MEPERIDINE (PETHIDINE) | 9230 | 37 | **323.00** | 8.73 |
| METHADONE | 9250B | 1 | **0.18** | 0.18 |

| | | | | |
|---|---|---|---|---|
| MORPHINE | 9300 | 45 | **46.55** | 1.03 |
| FENTANYL BASE | 9801 | 104 | **6.22** | 0.06 |

**BUSINESS ACTIVITY: D - TEACHING INSTITUTIONS**

| DRUG NAME | DRUG CODE | NUMBER OF REGISTRANT SOLD TO | TOTAL GRAMS | AVG GRAMS |
|---|---|---|---|---|
| BUPRENORPHINE | 9064 | 1 | **0.00** | 0.00 |
| HYDROMORPHONE | 9150 | 1 | **0.07** | 0.07 |

**BUSINESS ACTIVITY: M - MID-LEVEL PRACTITIONERS**

| DRUG NAME | DRUG CODE | NUMBER OF REGISTRANT SOLD TO | TOTAL GRAMS | AVG GRAMS |
|---|---|---|---|---|
| CODEINE | 9050 | 2 | **2.94** | 1.47 |

**BUSINESS ACTIVITY: NTP - NARCOTIC TREATMENT PROGRAMS**

| DRUG NAME | DRUG CODE | NUMBER OF REGISTRANT SOLD TO | TOTAL GRAMS | AVG GRAMS |
|---|---|---|---|---|
| BUPRENORPHINE | 9064 | 4 | **200.29** | 50.07 |
| METHADONE | 9250B | 5 | **26,474.24** | 5,294.85 |

**STATE: MT - MONTANA**

**BUSINESS ACTIVITY: A - PHARMACIES**

| DRUG NAME | DRUG CODE | NUMBER OF REGISTRANT SOLD TO | TOTAL GRAMS | AVG GRAMS |
|---|---|---|---|---|
| AMPHETAMINE | 1100 | 228 | **60,231.35** | 264.17 |
| LISDEXAMFETAMINE | 1205 | 223 | **24,814.94** | 111.28 |
| METHYLPHENIDATE (DL;D;L;ISOMERS) | 1724 | 230 | **62,659.77** | 272.43 |
| CODEINE | 9050 | 229 | **26,702.50** | 116.60 |
| BUPRENORPHINE | 9064 | 218 | **13,054.81** | 59.88 |
| OXYCODONE | 9143 | 230 | **99,092.54** | 430.84 |
| HYDROMORPHONE | 9150 | 218 | **4,463.88** | 20.48 |
| HYDROCODONE | 9193 | 231 | **69,975.18** | 302.92 |
| MEPERIDINE (PETHIDINE) | 9230 | 39 | **305.57** | 7.84 |
| METHADONE | 9250B | 190 | **4,928.14** | 25.94 |
| MORPHINE | 9300 | 227 | **44,399.61** | 195.59 |
| OXYMORPHONE | 9652 | 43 | **379.81** | 8.83 |
| TAPENTADOL | 9780 | 92 | **5,382.00** | 58.50 |
| FENTANYL BASE | 9801 | 217 | **681.56** | 3.14 |

**BUSINESS ACTIVITY: B - HOSPITALS**

| DRUG NAME | DRUG CODE | NUMBER OF REGISTRANT SOLD TO | TOTAL GRAMS | AVG GRAMS |
|---|---|---|---|---|
| AMPHETAMINE | 1100 | 38 | **3,988.58** | 104.96 |
| LISDEXAMFETAMINE | 1205 | 21 | **1,644.12** | 78.29 |
| METHYLPHENIDATE (DL;D;L;ISOMERS) | 1724 | 46 | **5,439.23** | 118.24 |
| CODEINE | 9050 | 53 | **3,188.50** | 60.16 |
| BUPRENORPHINE | 9064 | 31 | **1,208.01** | 38.97 |
| OXYCODONE | 9143 | 87 | **11,123.96** | 127.86 |
| HYDROMORPHONE | 9150 | 72 | **750.88** | 10.43 |
| HYDROCODONE | 9193 | 89 | **10,252.10** | 115.19 |

| | | | | |
|---|---|---|---|---|
| MEPERIDINE (PETHIDINE) | 9230 | 42 | **471.70** | 11.23 |
| METHADONE | 9250B | 34 | **523.05** | 15.38 |
| MORPHINE | 9300 | 85 | **7,440.09** | 87.53 |
| TAPENTADOL | 9780 | 7 | **135.00** | 19.29 |
| FENTANYL BASE | 9801 | 86 | **173.51** | 2.02 |

**BUSINESS ACTIVITY: C - PRACTITIONERS**

| DRUG NAME | DRUG CODE | NUMBER OF REGISTRANT SOLD TO | TOTAL GRAMS | AVG GRAMS |
|---|---|---|---|---|
| CODEINE | 9050 | 16 | **382.28** | 23.89 |
| BUPRENORPHINE | 9064 | 135 | **6.72** | 0.05 |
| OXYCODONE | 9143 | 4 | **4.93** | 1.23 |
| HYDROMORPHONE | 9150 | 71 | **30.14** | 0.42 |
| HYDROCODONE | 9193 | 68 | **143.21** | 2.11 |
| MEPERIDINE (PETHIDINE) | 9230 | 7 | **14.16** | 2.02 |
| METHADONE | 9250B | 3 | **13.42** | 4.47 |
| MORPHINE | 9300 | 52 | **42.19** | 0.81 |
| FENTANYL BASE | 9801 | 78 | **6.23** | 0.08 |

**BUSINESS ACTIVITY: M - MID-LEVEL PRACTITIONERS**

| DRUG NAME | DRUG CODE | NUMBER OF REGISTRANT SOLD TO | TOTAL GRAMS | AVG GRAMS |
|---|---|---|---|---|
| BUPRENORPHINE | 9064 | 2 | **1.38** | 0.69 |
| MORPHINE | 9300 | 3 | **0.45** | 0.15 |
| FENTANYL BASE | 9801 | 4 | **0.01** | 0.00 |

**BUSINESS ACTIVITY: NTP - NARCOTIC TREATMENT PROGRAMS**

| DRUG NAME | DRUG CODE | NUMBER OF REGISTRANT SOLD TO | TOTAL GRAMS | AVG GRAMS |
|---|---|---|---|---|
| BUPRENORPHINE | 9064 | 4 | **1,040.78** | 260.20 |
| METHADONE | 9250B | 4 | **28,084.16** | 7,021.04 |

**STATE: NC - NORTH CAROLINA**

**BUSINESS ACTIVITY: A - PHARMACIES**

| DRUG NAME | DRUG CODE | NUMBER OF REGISTRANT SOLD TO | TOTAL GRAMS | AVG GRAMS |
|---|---|---|---|---|
| AMPHETAMINE | 1100 | 2,035 | **901,955.12** | 443.22 |
| METHAMPHETAMINE | 1105 | 53 | **175.18** | 3.31 |
| LISDEXAMFETAMINE | 1205 | 1,992 | **440,554.98** | 221.16 |
| METHYLPHENIDATE (DL;D;L;ISOMERS) | 1724 | 2,021 | **726,880.58** | 359.66 |
| CODEINE | 9050 | 1,939 | **142,564.77** | 73.52 |
| BUPRENORPHINE | 9064 | 2,028 | **195,227.83** | 96.27 |
| OXYCODONE | 9143 | 2,043 | **1,799,379.53** | 880.75 |
| HYDROMORPHONE | 9150 | 1,875 | **27,255.20** | 14.54 |
| HYDROCODONE | 9193 | 2,054 | **563,031.67** | 274.11 |
| MEPERIDINE (PETHIDINE) | 9230 | 346 | **5,757.89** | 16.64 |
| METHADONE | 9250B | 1,747 | **73,675.57** | 42.17 |
| MORPHINE | 9300 | 1,980 | **367,692.30** | 185.70 |

| | | | | |
|---|---|---|---|---|
| OXYMORPHONE | 9652 | 1,045 | **31,117.21** | 29.78 |
| TAPENTADOL | 9780 | 1,663 | **299,665.50** | 180.20 |
| FENTANYL BASE | 9801 | 1,880 | **6,279.17** | 3.34 |

**BUSINESS ACTIVITY: B - HOSPITALS**

| DRUG NAME | DRUG CODE | NUMBER OF REGISTRANT SOLD TO | TOTAL GRAMS | AVG GRAMS |
|---|---|---|---|---|
| AMPHETAMINE | 1100 | 152 | **31,487.15** | 207.15 |
| LISDEXAMFETAMINE | 1205 | 81 | **10,140.45** | 125.19 |
| METHYLPHENIDATE (DL;D;L;ISOMERS) | 1724 | 176 | **31,866.19** | 181.06 |
| CODEINE | 9050 | 163 | **5,643.68** | 34.62 |
| BUPRENORPHINE | 9064 | 314 | **5,324.87** | 16.96 |
| OXYCODONE | 9143 | 268 | **90,499.67** | 337.69 |
| HYDROMORPHONE | 9150 | 333 | **5,530.52** | 16.61 |
| HYDROCODONE | 9193 | 317 | **32,184.31** | 101.53 |
| MEPERIDINE (PETHIDINE) | 9230 | 144 | **1,266.94** | 8.80 |
| METHADONE | 9250B | 189 | **17,804.64** | 94.20 |
| MORPHINE | 9300 | 310 | **44,647.40** | 144.02 |
| OXYMORPHONE | 9652 | 53 | **2,984.20** | 56.31 |
| TAPENTADOL | 9780 | 90 | **14,528.90** | 161.43 |
| FENTANYL BASE | 9801 | 342 | **891.38** | 2.61 |

**BUSINESS ACTIVITY: C - PRACTITIONERS**

| DRUG NAME | DRUG CODE | NUMBER OF REGISTRANT SOLD TO | TOTAL GRAMS | AVG GRAMS |
|---|---|---|---|---|
| METHYLPHENIDATE (DL;D;L;ISOMERS) | 1724 | 1 | **119.54** | 119.54 |
| CODEINE | 9050 | 50 | **436.72** | 8.73 |
| BUPRENORPHINE | 9064 | 996 | **300.57** | 0.30 |
| OXYCODONE | 9143 | 14 | **19.95** | 1.42 |
| HYDROMORPHONE | 9150 | 339 | **145.17** | 0.43 |
| HYDROCODONE | 9193 | 227 | **476.06** | 2.10 |
| MEPERIDINE (PETHIDINE) | 9230 | 59 | **295.11** | 5.00 |
| METHADONE | 9250B | 5 | **32.91** | 6.58 |
| MORPHINE | 9300 | 136 | **152.21** | 1.12 |
| FENTANYL BASE | 9801 | 424 | **29.40** | 0.07 |

**BUSINESS ACTIVITY: D - TEACHING INSTITUTIONS**

| DRUG NAME | DRUG CODE | NUMBER OF REGISTRANT SOLD TO | TOTAL GRAMS | AVG GRAMS |
|---|---|---|---|---|
| BUPRENORPHINE | 9064 | 5 | **0.05** | 0.01 |
| HYDROMORPHONE | 9150 | 2 | **0.51** | 0.26 |
| MORPHINE | 9300 | 1 | **0.19** | 0.19 |
| FENTANYL BASE | 9801 | 1 | **0.06** | 0.06 |

**BUSINESS ACTIVITY: M - MID-LEVEL PRACTITIONERS**

| DRUG NAME | DRUG CODE | NUMBER OF REGISTRANT SOLD TO | TOTAL GRAMS | AVG GRAMS |
|---|---|---|---|---|
| CODEINE | 9050 | 16 | **10.88** | 0.68 |

| | | | | |
|---|---|---|---|---|
| BUPRENORPHINE | 9064 | 4 | **25.10** | 6.28 |
| OXYCODONE | 9143 | 1 | **0.90** | 0.90 |
| HYDROMORPHONE | 9150 | 8 | **1.30** | 0.16 |
| MORPHINE | 9300 | 61 | **43.41** | 0.71 |
| FENTANYL BASE | 9801 | 83 | **2.11** | 0.03 |

**BUSINESS ACTIVITY: NTP - NARCOTIC TREATMENT PROGRAMS**

| DRUG NAME | DRUG CODE | NUMBER OF REGISTRANT SOLD TO | TOTAL GRAMS | AVG GRAMS |
|---|---|---|---|---|
| BUPRENORPHINE | 9064 | 67 | **13,447.09** | 200.70 |
| METHADONE | 9250B | 77 | **474,442.44** | 6,161.59 |

**STATE: ND - NORTH DAKOTA**

**BUSINESS ACTIVITY: A - PHARMACIES**

| DRUG NAME | DRUG CODE | NUMBER OF REGISTRANT SOLD TO | TOTAL GRAMS | AVG GRAMS |
|---|---|---|---|---|
| AMPHETAMINE | 1100 | 174 | **36,895.86** | 212.05 |
| METHAMPHETAMINE | 1105 | 4 | **19.68** | 4.92 |
| LISDEXAMFETAMINE | 1205 | 171 | **23,232.68** | 135.86 |
| METHYLPHENIDATE (DL;D;L;ISOMERS) | 1724 | 174 | **55,221.08** | 317.36 |
| CODEINE | 9050 | 169 | **13,352.69** | 79.01 |
| BUPRENORPHINE | 9064 | 142 | **5,281.10** | 37.19 |
| OXYCODONE | 9143 | 173 | **37,633.08** | 217.53 |
| HYDROMORPHONE | 9150 | 153 | **1,472.34** | 9.62 |
| HYDROCODONE | 9193 | 176 | **24,918.87** | 141.58 |
| MEPERIDINE (PETHIDINE) | 9230 | 23 | **240.21** | 10.44 |
| METHADONE | 9250B | 121 | **2,482.76** | 20.52 |
| MORPHINE | 9300 | 165 | **14,925.67** | 90.46 |
| OXYMORPHONE | 9652 | 15 | **204.83** | 13.66 |
| TAPENTADOL | 9780 | 64 | **4,710.50** | 73.60 |
| FENTANYL BASE | 9801 | 165 | **468.43** | 2.84 |

**BUSINESS ACTIVITY: B - HOSPITALS**

| DRUG NAME | DRUG CODE | NUMBER OF REGISTRANT SOLD TO | TOTAL GRAMS | AVG GRAMS |
|---|---|---|---|---|
| AMPHETAMINE | 1100 | 22 | **2,854.65** | 129.76 |
| METHAMPHETAMINE | 1105 | 1 | **12.86** | 12.86 |
| LISDEXAMFETAMINE | 1205 | 18 | **1,400.16** | 77.79 |
| METHYLPHENIDATE (DL;D;L;ISOMERS) | 1724 | 28 | **5,214.36** | 186.23 |
| CODEINE | 9050 | 46 | **2,434.27** | 52.92 |
| BUPRENORPHINE | 9064 | 20 | **548.97** | 27.45 |
| OXYCODONE | 9143 | 51 | **6,976.61** | 136.80 |
| HYDROMORPHONE | 9150 | 52 | **529.56** | 10.18 |
| HYDROCODONE | 9193 | 55 | **5,575.13** | 101.37 |
| MEPERIDINE (PETHIDINE) | 9230 | 21 | **125.28** | 5.97 |
| METHADONE | 9250B | 22 | **405.97** | 18.45 |

| | | | | |
|---|---|---|---|---|
| MORPHINE | 9300 | 56 | **4,212.36** | 75.22 |
| OXYMORPHONE | 9652 | 1 | **5.53** | 5.53 |
| TAPENTADOL | 9780 | 6 | **700.00** | 116.67 |
| FENTANYL BASE | 9801 | 54 | **134.48** | 2.49 |

**BUSINESS ACTIVITY: C - PRACTITIONERS**

| DRUG NAME | DRUG CODE | NUMBER OF REGISTRANT SOLD TO | TOTAL GRAMS | AVG GRAMS |
|---|---|---|---|---|
| CODEINE | 9050 | 6 | **96.77** | 16.13 |
| BUPRENORPHINE | 9064 | 66 | **30.42** | 0.46 |
| OXYCODONE | 9143 | 5 | **9.68** | 1.94 |
| HYDROMORPHONE | 9150 | 30 | **11.57** | 0.39 |
| HYDROCODONE | 9193 | 24 | **81.88** | 3.41 |
| MEPERIDINE (PETHIDINE) | 9230 | 14 | **47.06** | 3.36 |
| METHADONE | 9250B | 2 | **26.12** | 13.06 |
| MORPHINE | 9300 | 22 | **55.12** | 2.51 |
| FENTANYL BASE | 9801 | 45 | **4.65** | 0.10 |

**BUSINESS ACTIVITY: M - MID-LEVEL PRACTITIONERS**

| DRUG NAME | DRUG CODE | NUMBER OF REGISTRANT SOLD TO | TOTAL GRAMS | AVG GRAMS |
|---|---|---|---|---|
| LISDEXAMFETAMINE | 1205 | 1 | **1.73** | 1.73 |
| METHYLPHENIDATE (DL;D;L;ISOMERS) | 1724 | 1 | **5.19** | 5.19 |
| BUPRENORPHINE | 9064 | 2 | **138.84** | 69.42 |
| METHADONE | 9250B | 1 | **1.34** | 1.34 |

**BUSINESS ACTIVITY: NTP - NARCOTIC TREATMENT PROGRAMS**

| DRUG NAME | DRUG CODE | NUMBER OF REGISTRANT SOLD TO | TOTAL GRAMS | AVG GRAMS |
|---|---|---|---|---|
| METHADONE | 9250B | 3 | **18,388.86** | 6,129.62 |

**STATE: NE - NEBRASKA**

**BUSINESS ACTIVITY: A - PHARMACIES**

| DRUG NAME | DRUG CODE | NUMBER OF REGISTRANT SOLD TO | TOTAL GRAMS | AVG GRAMS |
|---|---|---|---|---|
| AMPHETAMINE | 1100 | 417 | **114,325.32** | 274.16 |
| METHAMPHETAMINE | 1105 | 12 | **22.90** | 1.91 |
| LISDEXAMFETAMINE | 1205 | 411 | **69,961.08** | 170.22 |
| METHYLPHENIDATE (DL;D;L;ISOMERS) | 1724 | 422 | **119,605.39** | 283.43 |
| CODEINE | 9050 | 403 | **31,078.98** | 77.12 |
| BUPRENORPHINE | 9064 | 382 | **7,014.00** | 18.36 |
| OXYCODONE | 9143 | 429 | **112,545.72** | 262.34 |
| HYDROMORPHONE | 9150 | 361 | **2,812.91** | 7.79 |
| HYDROCODONE | 9193 | 431 | **88,134.61** | 204.49 |
| MEPERIDINE (PETHIDINE) | 9230 | 42 | **441.20** | 10.50 |
| METHADONE | 9250B | 307 | **7,133.20** | 23.24 |
| MORPHINE | 9300 | 421 | **52,495.78** | 124.69 |
| OXYMORPHONE | 9652 | 115 | **1,308.35** | 11.38 |

| | | | | |
|---|---|---|---|---|
| TAPENTADOL | 9780 | 289 | **32,117.00** | 111.13 |
| FENTANYL BASE | 9801 | 389 | **1,235.25** | 3.18 |

**BUSINESS ACTIVITY: B - HOSPITALS**

| DRUG NAME | DRUG CODE | NUMBER OF REGISTRANT SOLD TO | TOTAL GRAMS | AVG GRAMS |
|---|---|---|---|---|
| AMPHETAMINE | 1100 | 39 | **3,926.21** | 100.67 |
| LISDEXAMFETAMINE | 1205 | 28 | **1,035.38** | 36.98 |
| METHYLPHENIDATE (DL;D;L;ISOMERS) | 1724 | 56 | **4,588.65** | 81.94 |
| CODEINE | 9050 | 79 | **1,615.00** | 20.44 |
| BUPRENORPHINE | 9064 | 43 | **360.49** | 8.38 |
| OXYCODONE | 9143 | 121 | **8,111.82** | 67.04 |
| HYDROMORPHONE | 9150 | 109 | **546.85** | 5.02 |
| HYDROCODONE | 9193 | 127 | **5,161.41** | 40.64 |
| MEPERIDINE (PETHIDINE) | 9230 | 92 | **556.13** | 6.04 |
| METHADONE | 9250B | 53 | **781.36** | 14.74 |
| MORPHINE | 9300 | 124 | **6,736.22** | 54.32 |
| OXYMORPHONE | 9652 | 5 | **36.22** | 7.24 |
| TAPENTADOL | 9780 | 40 | **1,019.00** | 25.48 |
| FENTANYL BASE | 9801 | 132 | **179.46** | 1.36 |

**BUSINESS ACTIVITY: C - PRACTITIONERS**

| DRUG NAME | DRUG CODE | NUMBER OF REGISTRANT SOLD TO | TOTAL GRAMS | AVG GRAMS |
|---|---|---|---|---|
| AMPHETAMINE | 1100 | 1 | **1.16** | 1.16 |
| LISDEXAMFETAMINE | 1205 | 1 | **13.05** | 13.05 |
| CODEINE | 9050 | 24 | **1,066.44** | 44.44 |
| BUPRENORPHINE | 9064 | 173 | **14.67** | 0.08 |
| OXYCODONE | 9143 | 7 | **86.02** | 12.29 |
| HYDROMORPHONE | 9150 | 83 | **39.65** | 0.48 |
| HYDROCODONE | 9193 | 51 | **103.59** | 2.03 |
| MEPERIDINE (PETHIDINE) | 9230 | 17 | **59.92** | 3.52 |
| METHADONE | 9250B | 2 | **3.22** | 1.61 |
| MORPHINE | 9300 | 37 | **30.02** | 0.81 |
| TAPENTADOL | 9780 | 1 | **75.00** | 75.00 |
| FENTANYL BASE | 9801 | 94 | **9.47** | 0.10 |

**BUSINESS ACTIVITY: D - TEACHING INSTITUTIONS**

| DRUG NAME | DRUG CODE | NUMBER OF REGISTRANT SOLD TO | TOTAL GRAMS | AVG GRAMS |
|---|---|---|---|---|
| HYDROMORPHONE | 9150 | 1 | **2.66** | 2.66 |

**BUSINESS ACTIVITY: M - MID-LEVEL PRACTITIONERS**

| DRUG NAME | DRUG CODE | NUMBER OF REGISTRANT SOLD TO | TOTAL GRAMS | AVG GRAMS |
|---|---|---|---|---|
| CODEINE | 9050 | 2 | **0.88** | 0.44 |
| HYDROMORPHONE | 9150 | 3 | **0.28** | 0.09 |
| MORPHINE | 9300 | 17 | **5.99** | 0.35 |

| | | | | |
|---|---|---|---|---|
| FENTANYL BASE | 9801 | 28 | **0.62** | 0.02 |

**BUSINESS ACTIVITY: NTP - NARCOTIC TREATMENT PROGRAMS**

| DRUG NAME | DRUG CODE | NUMBER OF REGISTRANT SOLD TO | TOTAL GRAMS | AVG GRAMS |
|---|---|---|---|---|
| BUPRENORPHINE | 9064 | 1 | **0.30** | 0.30 |
| METHADONE | 9250B | 3 | **21,265.25** | 7,088.42 |

**STATE: NH - NEW HAMPSHIRE**

**BUSINESS ACTIVITY: A - PHARMACIES**

| DRUG NAME | DRUG CODE | NUMBER OF REGISTRANT SOLD TO | TOTAL GRAMS | AVG GRAMS |
|---|---|---|---|---|
| AMPHETAMINE | 1100 | 265 | **169,639.13** | 640.15 |
| METHAMPHETAMINE | 1105 | 7 | **24.10** | 3.44 |
| LISDEXAMFETAMINE | 1205 | 265 | **57,767.22** | 217.99 |
| METHYLPHENIDATE (DL;D;L;ISOMERS) | 1724 | 265 | **134,584.44** | 507.87 |
| CODEINE | 9050 | 257 | **23,399.05** | 91.05 |
| BUPRENORPHINE | 9064 | 266 | **38,997.64** | 146.61 |
| OXYCODONE | 9143 | 264 | **170,486.96** | 645.78 |
| HYDROMORPHONE | 9150 | 258 | **5,298.12** | 20.54 |
| HYDROCODONE | 9193 | 265 | **27,442.37** | 103.56 |
| MEPERIDINE (PETHIDINE) | 9230 | 22 | **616.59** | 28.03 |
| METHADONE | 9250B | 241 | **14,280.26** | 59.25 |
| MORPHINE | 9300 | 260 | **54,811.65** | 210.81 |
| OXYMORPHONE | 9652 | 80 | **975.78** | 12.20 |
| TAPENTADOL | 9780 | 179 | **23,229.50** | 129.77 |
| FENTANYL BASE | 9801 | 251 | **767.43** | 3.06 |

**BUSINESS ACTIVITY: B - HOSPITALS**

| DRUG NAME | DRUG CODE | NUMBER OF REGISTRANT SOLD TO | TOTAL GRAMS | AVG GRAMS |
|---|---|---|---|---|
| AMPHETAMINE | 1100 | 23 | **2,988.98** | 129.96 |
| LISDEXAMFETAMINE | 1205 | 13 | **424.33** | 32.64 |
| METHYLPHENIDATE (DL;D;L;ISOMERS) | 1724 | 35 | **2,097.56** | 59.93 |
| CODEINE | 9050 | 29 | **464.53** | 16.02 |
| BUPRENORPHINE | 9064 | 32 | **882.85** | 27.59 |
| OXYCODONE | 9143 | 44 | **8,267.45** | 187.90 |
| HYDROMORPHONE | 9150 | 40 | **731.72** | 18.29 |
| HYDROCODONE | 9193 | 44 | **937.77** | 21.31 |
| MEPERIDINE (PETHIDINE) | 9230 | 27 | **204.80** | 7.59 |
| METHADONE | 9250B | 33 | **1,184.38** | 35.89 |
| MORPHINE | 9300 | 41 | **3,886.92** | 94.80 |
| TAPENTADOL | 9780 | 1 | **123.00** | 123.00 |
| FENTANYL BASE | 9801 | 50 | **103.07** | 2.06 |

**BUSINESS ACTIVITY: C - PRACTITIONERS**

| DRUG NAME | DRUG CODE | NUMBER OF REGISTRANT SOLD TO | TOTAL GRAMS | AVG GRAMS |
|---|---|---|---|---|

| | | | | |
|---|---|---|---|---|
| CODEINE | 9050 | 1 | **8.37** | 8.37 |
| BUPRENORPHINE | 9064 | 193 | **12.45** | 0.06 |
| OXYCODONE | 9143 | 6 | **7.31** | 1.22 |
| HYDROMORPHONE | 9150 | 87 | **29.83** | 0.34 |
| HYDROCODONE | 9193 | 15 | **8.66** | 0.58 |
| MEPERIDINE (PETHIDINE) | 9230 | 6 | **24.62** | 4.10 |
| METHADONE | 9250B | 5 | **8.77** | 1.75 |
| MORPHINE | 9300 | 34 | **20.60** | 0.61 |
| FENTANYL BASE | 9801 | 91 | **9.23** | 0.10 |

**BUSINESS ACTIVITY: M - MID-LEVEL PRACTITIONERS**

| DRUG NAME | DRUG CODE | NUMBER OF REGISTRANT SOLD TO | TOTAL GRAMS | AVG GRAMS |
|---|---|---|---|---|
| CODEINE | 9050 | 1 | **4.42** | 4.42 |
| BUPRENORPHINE | 9064 | 2 | **2.29** | 1.15 |

**BUSINESS ACTIVITY: NTP - NARCOTIC TREATMENT PROGRAMS**

| DRUG NAME | DRUG CODE | NUMBER OF REGISTRANT SOLD TO | TOTAL GRAMS | AVG GRAMS |
|---|---|---|---|---|
| BUPRENORPHINE | 9064 | 5 | **551.70** | 110.34 |
| METHADONE | 9250B | 9 | **116,431.20** | 12,936.80 |

**STATE: NJ - NEW JERSEY**

**BUSINESS ACTIVITY: A - PHARMACIES**

| DRUG NAME | DRUG CODE | NUMBER OF REGISTRANT SOLD TO | TOTAL GRAMS | AVG GRAMS |
|---|---|---|---|---|
| AMPHETAMINE | 1100 | 1,974 | **463,948.34** | 235.03 |
| METHAMPHETAMINE | 1105 | 58 | **140.63** | 2.42 |
| LISDEXAMFETAMINE | 1205 | 1,758 | **190,771.84** | 108.52 |
| METHYLPHENIDATE (DL;D;L;ISOMERS) | 1724 | 1,944 | **356,315.38** | 183.29 |
| CODEINE | 9050 | 1,989 | **227,810.42** | 114.54 |
| BUPRENORPHINE | 9064 | 1,856 | **117,671.02** | 63.40 |
| OXYCODONE | 9143 | 2,038 | **1,390,724.47** | 682.40 |
| HYDROMORPHONE | 9150 | 1,666 | **22,643.23** | 13.59 |
| HYDROCODONE | 9193 | 1,949 | **99,393.07** | 51.00 |
| MEPERIDINE (PETHIDINE) | 9230 | 84 | **2,366.89** | 28.18 |
| METHADONE | 9250B | 1,077 | **30,792.58** | 28.59 |
| MORPHINE | 9300 | 1,760 | **232,635.48** | 132.18 |
| OXYMORPHONE | 9652 | 542 | **10,135.64** | 18.70 |
| TAPENTADOL | 9780 | 1,074 | **114,831.50** | 106.92 |
| FENTANYL BASE | 9801 | 1,635 | **4,821.72** | 2.95 |

**BUSINESS ACTIVITY: B - HOSPITALS**

| DRUG NAME | DRUG CODE | NUMBER OF REGISTRANT SOLD TO | TOTAL GRAMS | AVG GRAMS |
|---|---|---|---|---|
| AMPHETAMINE | 1100 | 58 | **3,874.08** | 66.79 |
| LISDEXAMFETAMINE | 1205 | 26 | **1,460.95** | 56.19 |
| METHYLPHENIDATE (DL;D;L;ISOMERS) | 1724 | 104 | **3,876.62** | 37.28 |

| | | | | |
|---|---|---|---|---|
| CODEINE | 9050 | 103 | **3,629.53** | 35.24 |
| BUPRENORPHINE | 9064 | 97 | **1,348.99** | 13.91 |
| OXYCODONE | 9143 | 129 | **31,415.56** | 243.53 |
| HYDROMORPHONE | 9150 | 114 | **3,772.39** | 33.09 |
| HYDROCODONE | 9193 | 91 | **1,118.37** | 12.29 |
| MEPERIDINE (PETHIDINE) | 9230 | 63 | **470.61** | 7.47 |
| METHADONE | 9250B | 105 | **4,503.66** | 42.89 |
| MORPHINE | 9300 | 120 | **11,877.17** | 98.98 |
| OXYMORPHONE | 9652 | 2 | **15.79** | 7.90 |
| TAPENTADOL | 9780 | 32 | **1,274.00** | 39.81 |
| FENTANYL BASE | 9801 | 111 | **569.09** | 5.13 |

**BUSINESS ACTIVITY: C - PRACTITIONERS**

| DRUG NAME | DRUG CODE | NUMBER OF REGISTRANT SOLD TO | TOTAL GRAMS | AVG GRAMS |
|---|---|---|---|---|
| AMPHETAMINE | 1100 | 4 | **9.98** | 2.49 |
| METHYLPHENIDATE (DL;D;L;ISOMERS) | 1724 | 1 | **9.08** | 9.08 |
| CODEINE | 9050 | 197 | **5,677.12** | 28.82 |
| BUPRENORPHINE | 9064 | 629 | **1,684.99** | 2.68 |
| OXYCODONE | 9143 | 107 | **153.21** | 1.43 |
| HYDROMORPHONE | 9150 | 297 | **125.11** | 0.42 |
| HYDROCODONE | 9193 | 277 | **881.65** | 3.18 |
| MEPERIDINE (PETHIDINE) | 9230 | 85 | **237.05** | 2.79 |
| METHADONE | 9250B | 34 | **454.53** | 13.37 |
| MORPHINE | 9300 | 144 | **317.74** | 2.21 |
| FENTANYL BASE | 9801 | 479 | **53.00** | 0.11 |

**BUSINESS ACTIVITY: D - TEACHING INSTITUTIONS**

| DRUG NAME | DRUG CODE | NUMBER OF REGISTRANT SOLD TO | TOTAL GRAMS | AVG GRAMS |
|---|---|---|---|---|
| BUPRENORPHINE | 9064 | 2 | **0.02** | 0.01 |

**BUSINESS ACTIVITY: M - MID-LEVEL PRACTITIONERS**

| DRUG NAME | DRUG CODE | NUMBER OF REGISTRANT SOLD TO | TOTAL GRAMS | AVG GRAMS |
|---|---|---|---|---|
| CODEINE | 9050 | 7 | **7.36** | 1.05 |
| BUPRENORPHINE | 9064 | 2 | **11.54** | 5.77 |

**BUSINESS ACTIVITY: NTP - NARCOTIC TREATMENT PROGRAMS**

| DRUG NAME | DRUG CODE | NUMBER OF REGISTRANT SOLD TO | TOTAL GRAMS | AVG GRAMS |
|---|---|---|---|---|
| BUPRENORPHINE | 9064 | 28 | **2,188.51** | 78.16 |
| METHADONE | 9250B | 37 | **493,176.63** | 13,329.10 |

**STATE: NM - NEW MEXICO**

**BUSINESS ACTIVITY: A - PHARMACIES**

| DRUG NAME | DRUG CODE | NUMBER OF REGISTRANT SOLD TO | TOTAL GRAMS | AVG GRAMS |
|---|---|---|---|---|
| AMPHETAMINE | 1100 | 300 | **61,789.17** | 205.96 |
| METHAMPHETAMINE | 1105 | 16 | **28.52** | 1.78 |

| | | | | |
|---|---|---|---|---|
| LISDEXAMFETAMINE | 1205 | 267 | **14,860.64** | 55.66 |
| METHYLPHENIDATE (DL;D;L;ISOMERS) | 1724 | 308 | **65,797.55** | 213.63 |
| CODEINE | 9050 | 307 | **50,445.45** | 164.32 |
| BUPRENORPHINE | 9064 | 299 | **35,100.09** | 117.39 |
| OXYCODONE | 9143 | 310 | **250,414.69** | 807.79 |
| HYDROMORPHONE | 9150 | 269 | **3,221.74** | 11.98 |
| HYDROCODONE | 9193 | 310 | **98,584.98** | 318.02 |
| MEPERIDINE (PETHIDINE) | 9230 | 67 | **1,228.82** | 18.34 |
| METHADONE | 9250B | 244 | **6,009.21** | 24.63 |
| MORPHINE | 9300 | 305 | **67,277.09** | 220.58 |
| OXYMORPHONE | 9652 | 117 | **1,197.11** | 10.23 |
| TAPENTADOL | 9780 | 126 | **8,006.50** | 63.54 |
| FENTANYL BASE | 9801 | 284 | **840.09** | 2.96 |

**BUSINESS ACTIVITY: B - HOSPITALS**

| DRUG NAME | DRUG CODE | NUMBER OF REGISTRANT SOLD TO | TOTAL GRAMS | AVG GRAMS |
|---|---|---|---|---|
| AMPHETAMINE | 1100 | 43 | **2,584.32** | 60.10 |
| LISDEXAMFETAMINE | 1205 | 12 | **331.83** | 27.65 |
| METHYLPHENIDATE (DL;D;L;ISOMERS) | 1724 | 56 | **5,904.23** | 105.43 |
| CODEINE | 9050 | 59 | **7,036.44** | 119.26 |
| BUPRENORPHINE | 9064 | 62 | **1,902.16** | 30.68 |
| OXYCODONE | 9143 | 90 | **20,308.46** | 225.65 |
| HYDROMORPHONE | 9150 | 61 | **564.08** | 9.25 |
| HYDROCODONE | 9193 | 93 | **8,745.71** | 94.04 |
| MEPERIDINE (PETHIDINE) | 9230 | 46 | **426.49** | 9.27 |
| METHADONE | 9250B | 49 | **1,904.45** | 38.87 |
| MORPHINE | 9300 | 91 | **10,991.46** | 120.79 |
| OXYMORPHONE | 9652 | 4 | **8.92** | 2.23 |
| TAPENTADOL | 9780 | 4 | **77.00** | 19.25 |
| FENTANYL BASE | 9801 | 100 | **204.76** | 2.05 |

**BUSINESS ACTIVITY: C - PRACTITIONERS**

| DRUG NAME | DRUG CODE | NUMBER OF REGISTRANT SOLD TO | TOTAL GRAMS | AVG GRAMS |
|---|---|---|---|---|
| CODEINE | 9050 | 8 | **103.82** | 12.98 |
| BUPRENORPHINE | 9064 | 143 | **11.41** | 0.08 |
| OXYCODONE | 9143 | 3 | **12.10** | 4.03 |
| HYDROMORPHONE | 9150 | 69 | **42.10** | 0.61 |
| HYDROCODONE | 9193 | 40 | **80.40** | 2.01 |
| MEPERIDINE (PETHIDINE) | 9230 | 4 | **13.62** | 3.40 |
| METHADONE | 9250B | 2 | **0.72** | 0.36 |
| MORPHINE | 9300 | 26 | **19.71** | 0.76 |
| FENTANYL BASE | 9801 | 92 | **11.28** | 0.12 |

**BUSINESS ACTIVITY: D - TEACHING INSTITUTIONS**

| DRUG NAME | DRUG CODE | NUMBER OF REGISTRANT SOLD TO | TOTAL GRAMS | AVG GRAMS |
|---|---|---|---|---|
| BUPRENORPHINE | 9064 | 1 | **0.01** | 0.01 |
| FENTANYL BASE | 9801 | 1 | **0.02** | 0.02 |

**BUSINESS ACTIVITY: M - MID-LEVEL PRACTITIONERS**

| DRUG NAME | DRUG CODE | NUMBER OF REGISTRANT SOLD TO | TOTAL GRAMS | AVG GRAMS |
|---|---|---|---|---|
| AMPHETAMINE | 1100 | 1 | **4.50** | 4.50 |
| CODEINE | 9050 | 3 | **165.61** | 55.20 |
| BUPRENORPHINE | 9064 | 6 | **1.10** | 0.18 |
| HYDROMORPHONE | 9150 | 3 | **2.21** | 0.74 |
| HYDROCODONE | 9193 | 3 | **2.24** | 0.75 |
| MORPHINE | 9300 | 19 | **7.86** | 0.41 |
| FENTANYL BASE | 9801 | 45 | **1.12** | 0.02 |

**BUSINESS ACTIVITY: NTP - NARCOTIC TREATMENT PROGRAMS**

| DRUG NAME | DRUG CODE | NUMBER OF REGISTRANT SOLD TO | TOTAL GRAMS | AVG GRAMS |
|---|---|---|---|---|
| BUPRENORPHINE | 9064 | 4 | **445.47** | 111.37 |
| METHADONE | 9250B | 17 | **195,354.85** | 11,491.46 |

**STATE: NV - NEVADA**

**BUSINESS ACTIVITY: A - PHARMACIES**

| DRUG NAME | DRUG CODE | NUMBER OF REGISTRANT SOLD TO | TOTAL GRAMS | AVG GRAMS |
|---|---|---|---|---|
| AMPHETAMINE | 1100 | 453 | **110,813.01** | 244.62 |
| METHAMPHETAMINE | 1105 | 12 | **39.78** | 3.31 |
| LISDEXAMFETAMINE | 1205 | 414 | **31,582.76** | 76.29 |
| METHYLPHENIDATE (DL;D;L;ISOMERS) | 1724 | 428 | **55,529.40** | 129.74 |
| CODEINE | 9050 | 445 | **63,685.83** | 143.11 |
| BUPRENORPHINE | 9064 | 437 | **16,879.31** | 38.63 |
| OXYCODONE | 9143 | 462 | **509,332.05** | 1,102.45 |
| HYDROMORPHONE | 9150 | 420 | **6,515.94** | 15.51 |
| HYDROCODONE | 9193 | 460 | **251,326.01** | 546.36 |
| MEPERIDINE (PETHIDINE) | 9230 | 49 | **815.40** | 16.64 |
| METHADONE | 9250B | 415 | **26,279.44** | 63.32 |
| MORPHINE | 9300 | 450 | **139,435.53** | 309.86 |
| OXYMORPHONE | 9652 | 222 | **4,147.73** | 18.68 |
| TAPENTADOL | 9780 | 262 | **25,708.00** | 98.12 |
| FENTANYL BASE | 9801 | 393 | **987.18** | 2.51 |

**BUSINESS ACTIVITY: B - HOSPITALS**

| DRUG NAME | DRUG CODE | NUMBER OF REGISTRANT SOLD TO | TOTAL GRAMS | AVG GRAMS |
|---|---|---|---|---|
| AMPHETAMINE | 1100 | 38 | **3,535.38** | 93.04 |
| LISDEXAMFETAMINE | 1205 | 13 | **516.24** | 39.71 |
| METHYLPHENIDATE (DL;D;L;ISOMERS) | 1724 | 48 | **4,090.61** | 85.22 |

| | | | | |
|---|---|---|---|---|
| CODEINE | 9050 | 45 | **2,326.01** | 51.69 |
| BUPRENORPHINE | 9064 | 41 | **772.00** | 18.83 |
| OXYCODONE | 9143 | 94 | **28,667.06** | 304.97 |
| HYDROMORPHONE | 9150 | 81 | **1,508.12** | 18.62 |
| HYDROCODONE | 9193 | 99 | **18,455.69** | 186.42 |
| MEPERIDINE (PETHIDINE) | 9230 | 73 | **1,342.98** | 18.40 |
| METHADONE | 9250B | 49 | **1,528.44** | 31.19 |
| MORPHINE | 9300 | 92 | **15,398.24** | 167.37 |
| OXYMORPHONE | 9652 | 6 | **13.47** | 2.25 |
| TAPENTADOL | 9780 | 10 | **416.50** | 41.65 |
| FENTANYL BASE | 9801 | 122 | **264.88** | 2.17 |

**BUSINESS ACTIVITY: C - PRACTITIONERS**

| DRUG NAME | DRUG CODE | NUMBER OF REGISTRANT SOLD TO | TOTAL GRAMS | AVG GRAMS |
|---|---|---|---|---|
| AMPHETAMINE | 1100 | 1 | **17.50** | 17.50 |
| METHYLPHENIDATE (DL;D;L;ISOMERS) | 1724 | 1 | **17.60** | 17.60 |
| CODEINE | 9050 | 17 | **143.97** | 8.47 |
| BUPRENORPHINE | 9064 | 244 | **65.93** | 0.27 |
| OXYCODONE | 9143 | 12 | **80.64** | 6.72 |
| HYDROMORPHONE | 9150 | 131 | **159.08** | 1.21 |
| HYDROCODONE | 9193 | 109 | **369.23** | 3.39 |
| MEPERIDINE (PETHIDINE) | 9230 | 55 | **703.95** | 12.80 |
| METHADONE | 9250B | 10 | **24.51** | 2.45 |
| MORPHINE | 9300 | 69 | **91.71** | 1.33 |
| FENTANYL BASE | 9801 | 179 | **29.10** | 0.16 |

**BUSINESS ACTIVITY: D - TEACHING INSTITUTIONS**

| DRUG NAME | DRUG CODE | NUMBER OF REGISTRANT SOLD TO | TOTAL GRAMS | AVG GRAMS |
|---|---|---|---|---|
| MEPERIDINE (PETHIDINE) | 9230 | 1 | **2.18** | 2.18 |
| MORPHINE | 9300 | 1 | **0.19** | 0.19 |
| FENTANYL BASE | 9801 | 1 | **0.00** | 0.00 |

**BUSINESS ACTIVITY: M - MID-LEVEL PRACTITIONERS**

| DRUG NAME | DRUG CODE | NUMBER OF REGISTRANT SOLD TO | TOTAL GRAMS | AVG GRAMS |
|---|---|---|---|---|
| CODEINE | 9050 | 2 | **35.14** | 17.57 |
| BUPRENORPHINE | 9064 | 2 | **34.20** | 17.10 |
| MORPHINE | 9300 | 1 | **0.19** | 0.19 |

**BUSINESS ACTIVITY: NTP - NARCOTIC TREATMENT PROGRAMS**

| DRUG NAME | DRUG CODE | NUMBER OF REGISTRANT SOLD TO | TOTAL GRAMS | AVG GRAMS |
|---|---|---|---|---|
| BUPRENORPHINE | 9064 | 8 | **105.81** | 13.23 |
| METHADONE | 9250B | 14 | **105,354.06** | 7,525.29 |

**STATE: NY - NEW YORK**

**BUSINESS ACTIVITY: A - PHARMACIES**

| DRUG NAME | DRUG CODE | NUMBER OF REGISTRANT SOLD TO | TOTAL GRAMS | AVG GRAMS |
|---|---|---|---|---|
| AMPHETAMINE | 1100 | 4,490 | **1,052,346.50** | 234.38 |
| METHAMPHETAMINE | 1105 | 95 | **329.85** | 3.47 |
| LISDEXAMFETAMINE | 1205 | 3,593 | **356,543.18** | 99.23 |
| METHYLPHENIDATE (DL;D;L;ISOMERS) | 1724 | 4,343 | **889,544.67** | 204.82 |
| CODEINE | 9050 | 4,657 | **407,357.37** | 87.47 |
| BUPRENORPHINE | 9064 | 4,015 | **273,887.77** | 68.22 |
| OXYCODONE | 9143 | 4,869 | **2,013,761.79** | 413.59 |
| HYDROMORPHONE | 9150 | 3,642 | **44,851.26** | 12.32 |
| HYDROCODONE | 9193 | 4,445 | **485,905.65** | 109.32 |
| MEPERIDINE (PETHIDINE) | 9230 | 179 | **4,147.90** | 23.17 |
| METHADONE | 9250B | 2,894 | **99,718.25** | 34.46 |
| MORPHINE | 9300 | 3,845 | **409,831.37** | 106.59 |
| OXYMORPHONE | 9652 | 1,369 | **31,246.87** | 22.82 |
| TAPENTADOL | 9780 | 2,162 | **257,892.50** | 119.28 |
| FENTANYL BASE | 9801 | 3,562 | **8,567.34** | 2.41 |

**BUSINESS ACTIVITY: B - HOSPITALS**

| DRUG NAME | DRUG CODE | NUMBER OF REGISTRANT SOLD TO | TOTAL GRAMS | AVG GRAMS |
|---|---|---|---|---|
| AMPHETAMINE | 1100 | 186 | **9,661.26** | 51.94 |
| LISDEXAMFETAMINE | 1205 | 87 | **1,586.31** | 18.23 |
| METHYLPHENIDATE (DL;D;L;ISOMERS) | 1724 | 245 | **10,062.13** | 41.07 |
| CODEINE | 9050 | 261 | **9,425.71** | 36.11 |
| BUPRENORPHINE | 9064 | 241 | **5,820.34** | 24.15 |
| OXYCODONE | 9143 | 324 | **67,139.88** | 207.22 |
| HYDROMORPHONE | 9150 | 273 | **10,991.57** | 40.26 |
| HYDROCODONE | 9193 | 240 | **18,386.84** | 76.61 |
| MEPERIDINE (PETHIDINE) | 9230 | 173 | **2,261.54** | 13.07 |
| METHADONE | 9250B | 253 | **24,841.16** | 98.19 |
| MORPHINE | 9300 | 299 | **41,368.17** | 138.36 |
| OXYMORPHONE | 9652 | 24 | **157.54** | 6.56 |
| TAPENTADOL | 9780 | 78 | **4,822.00** | 61.82 |
| FENTANYL BASE | 9801 | 279 | **1,639.05** | 5.87 |

**BUSINESS ACTIVITY: C - PRACTITIONERS**

| DRUG NAME | DRUG CODE | NUMBER OF REGISTRANT SOLD TO | TOTAL GRAMS | AVG GRAMS |
|---|---|---|---|---|
| AMPHETAMINE | 1100 | 1 | **3.70** | 3.70 |
| METHAMPHETAMINE | 1105 | 1 | **4.02** | 4.02 |
| METHYLPHENIDATE (DL;D;L;ISOMERS) | 1724 | 1 | **17.90** | 17.90 |
| CODEINE | 9050 | 113 | **3,725.52** | 32.97 |
| BUPRENORPHINE | 9064 | 1,164 | **269.09** | 0.23 |
| OXYCODONE | 9143 | 144 | **524.63** | 3.64 |

| | | | | |
|---|---|---|---|---|
| HYDROMORPHONE | 9150 | 621 | **281.23** | 0.45 |
| HYDROCODONE | 9193 | 401 | **1,453.80** | 3.63 |
| MEPERIDINE (PETHIDINE) | 9230 | 148 | **1,034.69** | 6.99 |
| METHADONE | 9250B | 52 | **109.30** | 2.10 |
| MORPHINE | 9300 | 327 | **547.36** | 1.67 |
| FENTANYL BASE | 9801 | 1,059 | **128.64** | 0.12 |

**BUSINESS ACTIVITY: D - TEACHING INSTITUTIONS**

| DRUG NAME | DRUG CODE | NUMBER OF REGISTRANT SOLD TO | TOTAL GRAMS | AVG GRAMS |
|---|---|---|---|---|
| BUPRENORPHINE | 9064 | 8 | **0.10** | 0.01 |
| HYDROMORPHONE | 9150 | 2 | **0.05** | 0.03 |
| MORPHINE | 9300 | 1 | **0.75** | 0.75 |
| FENTANYL BASE | 9801 | 1 | **0.00** | 0.00 |

**BUSINESS ACTIVITY: M - MID-LEVEL PRACTITIONERS**

| DRUG NAME | DRUG CODE | NUMBER OF REGISTRANT SOLD TO | TOTAL GRAMS | AVG GRAMS |
|---|---|---|---|---|
| CODEINE | 9050 | 2 | **9.95** | 4.97 |
| BUPRENORPHINE | 9064 | 8 | **180.06** | 22.51 |
| HYDROMORPHONE | 9150 | 3 | **0.31** | 0.10 |
| HYDROCODONE | 9193 | 1 | **1.21** | 1.21 |
| METHADONE | 9250B | 1 | **51.43** | 51.43 |
| FENTANYL BASE | 9801 | 2 | **0.44** | 0.22 |

**BUSINESS ACTIVITY: NTP - NARCOTIC TREATMENT PROGRAMS**

| DRUG NAME | DRUG CODE | NUMBER OF REGISTRANT SOLD TO | TOTAL GRAMS | AVG GRAMS |
|---|---|---|---|---|
| BUPRENORPHINE | 9064 | 54 | **3,122.01** | 57.81 |
| METHADONE | 9250B | 123 | **1,160,870.02** | 9,437.97 |

**STATE: OH - OHIO**

**BUSINESS ACTIVITY: A - PHARMACIES**

| DRUG NAME | DRUG CODE | NUMBER OF REGISTRANT SOLD TO | TOTAL GRAMS | AVG GRAMS |
|---|---|---|---|---|
| AMPHETAMINE | 1100 | 2,219 | **842,065.86** | 379.48 |
| METHAMPHETAMINE | 1105 | 46 | **139.02** | 3.02 |
| LISDEXAMFETAMINE | 1205 | 2,155 | **374,224.36** | 173.65 |
| METHYLPHENIDATE (DL;D;L;ISOMERS) | 1724 | 2,208 | **770,228.36** | 348.84 |
| CODEINE | 9050 | 2,152 | **235,974.32** | 109.65 |
| BUPRENORPHINE | 9064 | 2,131 | **216,460.27** | 101.58 |
| OXYCODONE | 9143 | 2,218 | **1,263,873.24** | 569.83 |
| HYDROMORPHONE | 9150 | 1,805 | **18,943.63** | 10.50 |
| HYDROCODONE | 9193 | 2,223 | **498,658.81** | 224.32 |
| MEPERIDINE (PETHIDINE) | 9230 | 172 | **3,037.18** | 17.66 |
| METHADONE | 9250B | 1,825 | **55,969.58** | 30.67 |
| MORPHINE | 9300 | 2,111 | **309,828.50** | 146.77 |
| OXYMORPHONE | 9652 | 771 | **11,092.64** | 14.39 |

| | | | | |
|---|---|---|---|---|
| TAPENTADOL | 9780 | 1,385 | **143,484.50** | 103.60 |
| FENTANYL BASE | 9801 | 1,939 | **4,465.11** | 2.30 |

**BUSINESS ACTIVITY: B - HOSPITALS**

| DRUG NAME | DRUG CODE | NUMBER OF REGISTRANT SOLD TO | TOTAL GRAMS | AVG GRAMS |
|---|---|---|---|---|
| AMPHETAMINE | 1100 | 168 | **16,548.45** | 98.50 |
| METHAMPHETAMINE | 1105 | 1 | **2.41** | 2.41 |
| LISDEXAMFETAMINE | 1205 | 71 | **4,204.52** | 59.22 |
| METHYLPHENIDATE (DL;D;L;ISOMERS) | 1724 | 215 | **15,214.99** | 70.77 |
| CODEINE | 9050 | 250 | **5,674.65** | 22.70 |
| BUPRENORPHINE | 9064 | 259 | **9,323.11** | 36.00 |
| OXYCODONE | 9143 | 388 | **74,512.44** | 192.04 |
| HYDROMORPHONE | 9150 | 376 | **5,339.30** | 14.20 |
| HYDROCODONE | 9193 | 383 | **18,978.00** | 49.55 |
| MEPERIDINE (PETHIDINE) | 9230 | 263 | **6,193.50** | 23.55 |
| METHADONE | 9250B | 253 | **9,083.25** | 35.90 |
| MORPHINE | 9300 | 390 | **30,280.23** | 77.64 |
| OXYMORPHONE | 9652 | 34 | **152.59** | 4.49 |
| TAPENTADOL | 9780 | 62 | **2,889.00** | 46.60 |
| FENTANYL BASE | 9801 | 456 | **909.82** | 2.00 |

**BUSINESS ACTIVITY: C - PRACTITIONERS**

| DRUG NAME | DRUG CODE | NUMBER OF REGISTRANT SOLD TO | TOTAL GRAMS | AVG GRAMS |
|---|---|---|---|---|
| AMPHETAMINE | 1100 | 1 | **6.34** | 6.34 |
| LISDEXAMFETAMINE | 1205 | 1 | **2.43** | 2.43 |
| METHYLPHENIDATE (DL;D;L;ISOMERS) | 1724 | 1 | **8.30** | 8.30 |
| CODEINE | 9050 | 20 | **802.73** | 40.14 |
| BUPRENORPHINE | 9064 | 940 | **210.34** | 0.22 |
| OXYCODONE | 9143 | 19 | **63.20** | 3.33 |
| HYDROMORPHONE | 9150 | 347 | **160.12** | 0.46 |
| HYDROCODONE | 9193 | 259 | **730.04** | 2.82 |
| MEPERIDINE (PETHIDINE) | 9230 | 43 | **407.64** | 9.48 |
| METHADONE | 9250B | 10 | **23.79** | 2.38 |
| MORPHINE | 9300 | 131 | **181.82** | 1.39 |
| FENTANYL BASE | 9801 | 349 | **25.89** | 0.07 |

**BUSINESS ACTIVITY: D - TEACHING INSTITUTIONS**

| DRUG NAME | DRUG CODE | NUMBER OF REGISTRANT SOLD TO | TOTAL GRAMS | AVG GRAMS |
|---|---|---|---|---|
| BUPRENORPHINE | 9064 | 5 | **0.09** | 0.02 |
| HYDROMORPHONE | 9150 | 2 | **0.11** | 0.05 |
| MORPHINE | 9300 | 1 | **0.08** | 0.08 |
| FENTANYL BASE | 9801 | 2 | **0.26** | 0.13 |

**BUSINESS ACTIVITY: M - MID-LEVEL PRACTITIONERS**

| DRUG NAME | DRUG CODE | NUMBER OF REGISTRANT SOLD TO | TOTAL GRAMS | AVG GRAMS |
|---|---|---|---|---|
| BUPRENORPHINE | 9064 | 7 | **22.01** | 3.14 |
| HYDROMORPHONE | 9150 | 4 | **0.30** | 0.08 |
| MORPHINE | 9300 | 39 | **13.46** | 0.35 |
| FENTANYL BASE | 9801 | 102 | **0.91** | 0.01 |

**BUSINESS ACTIVITY: NTP - NARCOTIC TREATMENT PROGRAMS**

| DRUG NAME | DRUG CODE | NUMBER OF REGISTRANT SOLD TO | TOTAL GRAMS | AVG GRAMS |
|---|---|---|---|---|
| BUPRENORPHINE | 9064 | 48 | **18,525.94** | 385.96 |
| METHADONE | 9250B | 45 | **248,757.06** | 5,527.93 |

**STATE: OK - OKLAHOMA**

**BUSINESS ACTIVITY: A - PHARMACIES**

| DRUG NAME | DRUG CODE | NUMBER OF REGISTRANT SOLD TO | TOTAL GRAMS | AVG GRAMS |
|---|---|---|---|---|
| AMPHETAMINE | 1100 | 855 | **218,504.47** | 255.56 |
| METHAMPHETAMINE | 1105 | 10 | **36.16** | 3.62 |
| LISDEXAMFETAMINE | 1205 | 834 | **145,927.47** | 174.97 |
| METHYLPHENIDATE (DL;D;L;ISOMERS) | 1724 | 853 | **176,873.27** | 207.35 |
| CODEINE | 9050 | 879 | **268,481.29** | 305.44 |
| BUPRENORPHINE | 9064 | 859 | **65,533.34** | 76.29 |
| OXYCODONE | 9143 | 865 | **539,966.89** | 624.24 |
| HYDROMORPHONE | 9150 | 725 | **8,601.57** | 11.86 |
| HYDROCODONE | 9193 | 883 | **493,497.79** | 558.89 |
| MEPERIDINE (PETHIDINE) | 9230 | 325 | **6,629.94** | 20.40 |
| METHADONE | 9250B | 621 | **14,782.55** | 23.80 |
| MORPHINE | 9300 | 849 | **185,729.67** | 218.76 |
| OXYMORPHONE | 9652 | 283 | **3,679.47** | 13.00 |
| TAPENTADOL | 9780 | 440 | **33,885.50** | 77.01 |
| FENTANYL BASE | 9801 | 818 | **2,461.93** | 3.01 |

**BUSINESS ACTIVITY: B - HOSPITALS**

| DRUG NAME | DRUG CODE | NUMBER OF REGISTRANT SOLD TO | TOTAL GRAMS | AVG GRAMS |
|---|---|---|---|---|
| AMPHETAMINE | 1100 | 73 | **7,713.61** | 105.67 |
| METHAMPHETAMINE | 1105 | 1 | **0.40** | 0.40 |
| LISDEXAMFETAMINE | 1205 | 45 | **4,975.71** | 110.57 |
| METHYLPHENIDATE (DL;D;L;ISOMERS) | 1724 | 98 | **8,544.08** | 87.18 |
| CODEINE | 9050 | 148 | **18,818.06** | 127.15 |
| BUPRENORPHINE | 9064 | 89 | **2,030.96** | 22.82 |
| OXYCODONE | 9143 | 194 | **24,581.82** | 126.71 |
| HYDROMORPHONE | 9150 | 163 | **1,434.89** | 8.80 |
| HYDROCODONE | 9193 | 201 | **30,163.18** | 150.07 |
| MEPERIDINE (PETHIDINE) | 9230 | 132 | **2,860.70** | 21.67 |
| METHADONE | 9250B | 87 | **1,230.78** | 14.15 |

| | | | | |
|---|---|---|---|---|
| MORPHINE | 9300 | 197 | **17,566.60** | 89.17 |
| OXYMORPHONE | 9652 | 9 | **36.84** | 4.09 |
| TAPENTADOL | 9780 | 16 | **311.00** | 19.44 |
| FENTANYL BASE | 9801 | 199 | **350.00** | 1.76 |

**BUSINESS ACTIVITY: C - PRACTITIONERS**

| DRUG NAME | DRUG CODE | NUMBER OF REGISTRANT SOLD TO | TOTAL GRAMS | AVG GRAMS |
|---|---|---|---|---|
| LISDEXAMFETAMINE | 1205 | 2 | **4.86** | 2.43 |
| CODEINE | 9050 | 44 | **783.59** | 17.81 |
| BUPRENORPHINE | 9064 | 284 | **696.54** | 2.45 |
| OXYCODONE | 9143 | 11 | **207.25** | 18.84 |
| HYDROMORPHONE | 9150 | 81 | **37.57** | 0.46 |
| HYDROCODONE | 9193 | 59 | **526.99** | 8.93 |
| MEPERIDINE (PETHIDINE) | 9230 | 28 | **175.93** | 6.28 |
| METHADONE | 9250B | 6 | **38.46** | 6.41 |
| MORPHINE | 9300 | 54 | **514.86** | 9.53 |
| FENTANYL BASE | 9801 | 161 | **18.43** | 0.11 |

**BUSINESS ACTIVITY: D - TEACHING INSTITUTIONS**

| DRUG NAME | DRUG CODE | NUMBER OF REGISTRANT SOLD TO | TOTAL GRAMS | AVG GRAMS |
|---|---|---|---|---|
| BUPRENORPHINE | 9064 | 1 | **0.26** | 0.26 |
| HYDROMORPHONE | 9150 | 1 | **4.22** | 4.22 |
| HYDROCODONE | 9193 | 1 | **9.79** | 9.79 |
| METHADONE | 9250B | 1 | **39.80** | 39.80 |
| MORPHINE | 9300 | 1 | **53.96** | 53.96 |
| FENTANYL BASE | 9801 | 1 | **1.24** | 1.24 |

**BUSINESS ACTIVITY: M - MID-LEVEL PRACTITIONERS**

| DRUG NAME | DRUG CODE | NUMBER OF REGISTRANT SOLD TO | TOTAL GRAMS | AVG GRAMS |
|---|---|---|---|---|
| CODEINE | 9050 | 12 | **12.35** | 1.03 |
| HYDROMORPHONE | 9150 | 9 | **0.66** | 0.07 |
| MORPHINE | 9300 | 27 | **11.30** | 0.42 |
| FENTANYL BASE | 9801 | 51 | **1.56** | 0.03 |

**BUSINESS ACTIVITY: NTP - NARCOTIC TREATMENT PROGRAMS**

| DRUG NAME | DRUG CODE | NUMBER OF REGISTRANT SOLD TO | TOTAL GRAMS | AVG GRAMS |
|---|---|---|---|---|
| BUPRENORPHINE | 9064 | 12 | **1,182.83** | 98.57 |
| METHADONE | 9250B | 19 | **123,053.34** | 6,476.49 |

**STATE: OR - OREGON**

**BUSINESS ACTIVITY: A - PHARMACIES**

| DRUG NAME | DRUG CODE | NUMBER OF REGISTRANT SOLD TO | TOTAL GRAMS | AVG GRAMS |
|---|---|---|---|---|
| AMPHETAMINE | 1100 | 663 | **266,981.15** | 402.69 |
| METHAMPHETAMINE | 1105 | 37 | **160.69** | 4.34 |
| LISDEXAMFETAMINE | 1205 | 624 | **63,425.09** | 101.64 |

| | | | | |
|---|---|---|---|---|
| METHYLPHENIDATE (DL;D;L;ISOMERS) | 1724 | 662 | **249,524.83** | 376.93 |
| CODEINE | 9050 | 653 | **95,828.98** | 146.75 |
| BUPRENORPHINE | 9064 | 666 | **53,487.84** | 80.31 |
| OXYCODONE | 9143 | 679 | **477,685.36** | 703.51 |
| HYDROMORPHONE | 9150 | 646 | **11,905.70** | 18.43 |
| HYDROCODONE | 9193 | 674 | **272,792.11** | 404.74 |
| MEPERIDINE (PETHIDINE) | 9230 | 86 | **1,821.44** | 21.18 |
| METHADONE | 9250B | 605 | **37,013.49** | 61.18 |
| MORPHINE | 9300 | 668 | **192,768.83** | 288.58 |
| OXYMORPHONE | 9652 | 212 | **2,520.00** | 11.89 |
| TAPENTADOL | 9780 | 244 | **19,102.00** | 78.29 |
| FENTANYL BASE | 9801 | 614 | **1,917.64** | 3.12 |

**BUSINESS ACTIVITY: B - HOSPITALS**

| DRUG NAME | DRUG CODE | NUMBER OF REGISTRANT SOLD TO | TOTAL GRAMS | AVG GRAMS |
|---|---|---|---|---|
| AMPHETAMINE | 1100 | 50 | **3,664.62** | 73.29 |
| METHAMPHETAMINE | 1105 | 1 | **0.80** | 0.80 |
| LISDEXAMFETAMINE | 1205 | 20 | **463.06** | 23.15 |
| METHYLPHENIDATE (DL;D;L;ISOMERS) | 1724 | 56 | **4,575.09** | 81.70 |
| CODEINE | 9050 | 79 | **2,917.39** | 36.93 |
| BUPRENORPHINE | 9064 | 73 | **1,538.50** | 21.08 |
| OXYCODONE | 9143 | 93 | **27,593.25** | 296.70 |
| HYDROMORPHONE | 9150 | 77 | **2,036.77** | 26.45 |
| HYDROCODONE | 9193 | 97 | **15,461.98** | 159.40 |
| MEPERIDINE (PETHIDINE) | 9230 | 54 | **541.42** | 10.03 |
| METHADONE | 9250B | 69 | **2,808.84** | 40.71 |
| MORPHINE | 9300 | 93 | **14,949.60** | 160.75 |
| OXYMORPHONE | 9652 | 2 | **4.01** | 2.01 |
| TAPENTADOL | 9780 | 5 | **177.50** | 35.50 |
| FENTANYL BASE | 9801 | 81 | **241.35** | 2.98 |

**BUSINESS ACTIVITY: C - PRACTITIONERS**

| DRUG NAME | DRUG CODE | NUMBER OF REGISTRANT SOLD TO | TOTAL GRAMS | AVG GRAMS |
|---|---|---|---|---|
| AMPHETAMINE | 1100 | 1 | **6.60** | 6.60 |
| METHYLPHENIDATE (DL;D;L;ISOMERS) | 1724 | 1 | **3.11** | 3.11 |
| CODEINE | 9050 | 71 | **4,749.01** | 66.89 |
| BUPRENORPHINE | 9064 | 604 | **212.14** | 0.35 |
| OXYCODONE | 9143 | 61 | **141.12** | 2.31 |
| HYDROMORPHONE | 9150 | 348 | **118.46** | 0.34 |
| HYDROCODONE | 9193 | 306 | **670.41** | 2.19 |
| MEPERIDINE (PETHIDINE) | 9230 | 77 | **374.64** | 4.87 |
| METHADONE | 9250B | 21 | **47.58** | 2.27 |

| | | | | |
|---|---|---|---|---|
| MORPHINE | 9300 | 141 | **156.06** | 1.11 |
| FENTANYL BASE | 9801 | 374 | **32.62** | 0.09 |

**BUSINESS ACTIVITY: D - TEACHING INSTITUTIONS**

| DRUG NAME | DRUG CODE | NUMBER OF REGISTRANT SOLD TO | TOTAL GRAMS | AVG GRAMS |
|---|---|---|---|---|
| BUPRENORPHINE | 9064 | 1 | **0.02** | 0.02 |
| HYDROMORPHONE | 9150 | 1 | **0.07** | 0.07 |
| FENTANYL BASE | 9801 | 1 | **0.01** | 0.01 |

**BUSINESS ACTIVITY: M - MID-LEVEL PRACTITIONERS**

| DRUG NAME | DRUG CODE | NUMBER OF REGISTRANT SOLD TO | TOTAL GRAMS | AVG GRAMS |
|---|---|---|---|---|
| CODEINE | 9050 | 2 | **37.13** | 18.56 |
| BUPRENORPHINE | 9064 | 9 | **210.21** | 23.36 |
| HYDROMORPHONE | 9150 | 3 | **0.27** | 0.09 |
| HYDROCODONE | 9193 | 6 | **8.48** | 1.41 |
| MEPERIDINE (PETHIDINE) | 9230 | 1 | **4.36** | 4.36 |

**BUSINESS ACTIVITY: NTP - NARCOTIC TREATMENT PROGRAMS**

| DRUG NAME | DRUG CODE | NUMBER OF REGISTRANT SOLD TO | TOTAL GRAMS | AVG GRAMS |
|---|---|---|---|---|
| BUPRENORPHINE | 9064 | 18 | **5,199.43** | 288.86 |
| METHADONE | 9250B | 20 | **212,218.31** | 10,610.92 |

**STATE: PA - PENNSYLVANIA**

**BUSINESS ACTIVITY: A - PHARMACIES**

| DRUG NAME | DRUG CODE | NUMBER OF REGISTRANT SOLD TO | TOTAL GRAMS | AVG GRAMS |
|---|---|---|---|---|
| AMPHETAMINE | 1100 | 2,933 | **881,761.23** | 300.63 |
| METHAMPHETAMINE | 1105 | 61 | **394.15** | 6.46 |
| LISDEXAMFETAMINE | 1205 | 2,685 | **369,741.78** | 137.71 |
| METHYLPHENIDATE (DL;D;L;ISOMERS) | 1724 | 2,887 | **865,385.53** | 299.75 |
| CODEINE | 9050 | 2,913 | **346,534.19** | 118.96 |
| BUPRENORPHINE | 9064 | 2,839 | **319,148.65** | 112.42 |
| OXYCODONE | 9143 | 3,001 | **1,949,879.93** | 649.74 |
| HYDROMORPHONE | 9150 | 2,435 | **34,379.58** | 14.12 |
| HYDROCODONE | 9193 | 2,958 | **433,756.31** | 146.64 |
| MEPERIDINE (PETHIDINE) | 9230 | 189 | **3,575.76** | 18.92 |
| METHADONE | 9250B | 2,104 | **77,799.56** | 36.98 |
| MORPHINE | 9300 | 2,849 | **619,003.08** | 217.27 |
| OXYMORPHONE | 9652 | 1,216 | **32,943.78** | 27.09 |
| TAPENTADOL | 9780 | 1,374 | **170,169.00** | 123.85 |
| FENTANYL BASE | 9801 | 2,690 | **10,676.17** | 3.97 |

**BUSINESS ACTIVITY: B - HOSPITALS**

| DRUG NAME | DRUG CODE | NUMBER OF REGISTRANT SOLD TO | TOTAL GRAMS | AVG GRAMS |
|---|---|---|---|---|
| AMPHETAMINE | 1100 | 170 | **13,457.74** | 79.16 |
| LISDEXAMFETAMINE | 1205 | 63 | **4,032.83** | 64.01 |

| | | | | |
|---|---|---|---|---|
| METHYLPHENIDATE (DL;D;L;ISOMERS) | 1724 | 227 | **13,140.31** | 57.89 |
| CODEINE | 9050 | 242 | **10,243.97** | 42.33 |
| BUPRENORPHINE | 9064 | 233 | **10,179.69** | 43.69 |
| OXYCODONE | 9143 | 319 | **96,024.98** | 301.02 |
| HYDROMORPHONE | 9150 | 285 | **6,927.70** | 24.31 |
| HYDROCODONE | 9193 | 271 | **20,137.49** | 74.31 |
| MEPERIDINE (PETHIDINE) | 9230 | 184 | **1,334.48** | 7.25 |
| METHADONE | 9250B | 227 | **13,159.11** | 57.97 |
| MORPHINE | 9300 | 312 | **47,757.33** | 153.07 |
| OXYMORPHONE | 9652 | 49 | **394.04** | 8.04 |
| TAPENTADOL | 9780 | 47 | **1,167.00** | 24.83 |
| FENTANYL BASE | 9801 | 343 | **1,192.31** | 3.48 |

**BUSINESS ACTIVITY: C - PRACTITIONERS**

| DRUG NAME | DRUG CODE | NUMBER OF REGISTRANT SOLD TO | TOTAL GRAMS | AVG GRAMS |
|---|---|---|---|---|
| AMPHETAMINE | 1100 | 1 | **1.16** | 1.16 |
| METHYLPHENIDATE (DL;D;L;ISOMERS) | 1724 | 2 | **5.62** | 2.81 |
| CODEINE | 9050 | 153 | **9,466.88** | 61.88 |
| BUPRENORPHINE | 9064 | 1,111 | **931.95** | 0.84 |
| OXYCODONE | 9143 | 56 | **220.51** | 3.94 |
| HYDROMORPHONE | 9150 | 436 | **278.91** | 0.64 |
| HYDROCODONE | 9193 | 383 | **1,043.78** | 2.73 |
| MEPERIDINE (PETHIDINE) | 9230 | 77 | **324.09** | 4.21 |
| METHADONE | 9250B | 50 | **599.43** | 11.99 |
| MORPHINE | 9300 | 175 | **91.52** | 0.52 |
| FENTANYL BASE | 9801 | 573 | **52.39** | 0.09 |

**BUSINESS ACTIVITY: D - TEACHING INSTITUTIONS**

| DRUG NAME | DRUG CODE | NUMBER OF REGISTRANT SOLD TO | TOTAL GRAMS | AVG GRAMS |
|---|---|---|---|---|
| AMPHETAMINE | 1100 | 1 | **1.25** | 1.25 |
| METHYLPHENIDATE (DL;D;L;ISOMERS) | 1724 | 1 | **0.09** | 0.09 |
| CODEINE | 9050 | 1 | **432.99** | 432.99 |
| BUPRENORPHINE | 9064 | 2 | **1.20** | 0.60 |
| HYDROMORPHONE | 9150 | 1 | **0.34** | 0.34 |
| HYDROCODONE | 9193 | 2 | **52.07** | 26.03 |
| METHADONE | 9250B | 1 | **32.38** | 32.38 |
| MORPHINE | 9300 | 2 | **42.00** | 21.00 |
| FENTANYL BASE | 9801 | 3 | **1.41** | 0.47 |

**BUSINESS ACTIVITY: M - MID-LEVEL PRACTITIONERS**

| DRUG NAME | DRUG CODE | NUMBER OF REGISTRANT SOLD TO | TOTAL GRAMS | AVG GRAMS |
|---|---|---|---|---|
| CODEINE | 9050 | 10 | **9.85** | 0.99 |
| BUPRENORPHINE | 9064 | 1 | **7.68** | 7.68 |

**BUSINESS ACTIVITY: NTP - NARCOTIC TREATMENT PROGRAMS**

| DRUG NAME | DRUG CODE | NUMBER OF REGISTRANT SOLD TO | TOTAL GRAMS | AVG GRAMS |
|---|---|---|---|---|
| BUPRENORPHINE | 9064 | 22 | **1,263.27** | 57.42 |
| METHADONE | 9250B | 89 | **698,894.98** | 7,852.75 |

**STATE: PR - PUERTO RICO**

**BUSINESS ACTIVITY: A - PHARMACIES**

| DRUG NAME | DRUG CODE | NUMBER OF REGISTRANT SOLD TO | TOTAL GRAMS | AVG GRAMS |
|---|---|---|---|---|
| AMPHETAMINE | 1100 | 661 | **35,776.66** | 54.13 |
| LISDEXAMFETAMINE | 1205 | 282 | **8,711.97** | 30.89 |
| METHYLPHENIDATE (DL;D;L;ISOMERS) | 1724 | 695 | **53,186.00** | 76.53 |
| CODEINE | 9050 | 993 | **34,461.23** | 34.70 |
| BUPRENORPHINE | 9064 | 263 | **8,032.02** | 30.54 |
| OXYCODONE | 9143 | 798 | **42,446.32** | 53.19 |
| HYDROMORPHONE | 9150 | 21 | **134.05** | 6.38 |
| HYDROCODONE | 9193 | 443 | **1,572.19** | 3.55 |
| MEPERIDINE (PETHIDINE) | 9230 | 626 | **4,324.82** | 6.91 |
| METHADONE | 9250B | 12 | **155.45** | 12.95 |
| MORPHINE | 9300 | 137 | **4,237.86** | 30.93 |
| TAPENTADOL | 9780 | 25 | **1,022.00** | 40.88 |
| FENTANYL BASE | 9801 | 740 | **525.37** | 0.71 |

**BUSINESS ACTIVITY: B - HOSPITALS**

| DRUG NAME | DRUG CODE | NUMBER OF REGISTRANT SOLD TO | TOTAL GRAMS | AVG GRAMS |
|---|---|---|---|---|
| AMPHETAMINE | 1100 | 8 | **130.95** | 16.37 |
| LISDEXAMFETAMINE | 1205 | 1 | **2.89** | 2.89 |
| METHYLPHENIDATE (DL;D;L;ISOMERS) | 1724 | 14 | **200.65** | 14.33 |
| CODEINE | 9050 | 74 | **2,764.84** | 37.36 |
| BUPRENORPHINE | 9064 | 21 | **721.51** | 34.36 |
| OXYCODONE | 9143 | 68 | **4,578.07** | 67.32 |
| HYDROMORPHONE | 9150 | 4 | **72.95** | 18.24 |
| HYDROCODONE | 9193 | 23 | **150.31** | 6.54 |
| MEPERIDINE (PETHIDINE) | 9230 | 93 | **8,347.34** | 89.76 |
| METHADONE | 9250B | 2 | **101.07** | 50.53 |
| MORPHINE | 9300 | 84 | **3,239.09** | 38.56 |
| TAPENTADOL | 9780 | 4 | **571.50** | 142.88 |
| FENTANYL BASE | 9801 | 83 | **125.34** | 1.51 |

**BUSINESS ACTIVITY: C - PRACTITIONERS**

| DRUG NAME | DRUG CODE | NUMBER OF REGISTRANT SOLD TO | TOTAL GRAMS | AVG GRAMS |
|---|---|---|---|---|
| BUPRENORPHINE | 9064 | 15 | **6.68** | 0.45 |
| MEPERIDINE (PETHIDINE) | 9230 | 3 | **38.67** | 12.89 |
| MORPHINE | 9300 | 3 | **2.33** | 0.78 |

| | | | | |
|---|---|---|---|---|
| FENTANYL BASE | 9801 | 9 | **1.76** | 0.20 |

**BUSINESS ACTIVITY: NTP - NARCOTIC TREATMENT PROGRAMS**

| DRUG NAME | DRUG CODE | NUMBER OF REGISTRANT SOLD TO | TOTAL GRAMS | AVG GRAMS |
|---|---|---|---|---|
| METHADONE | 9250B | 5 | **85,039.55** | 17,007.91 |

**STATE: RI - RHODE ISLAND**

**BUSINESS ACTIVITY: A - PHARMACIES**

| DRUG NAME | DRUG CODE | NUMBER OF REGISTRANT SOLD TO | TOTAL GRAMS | AVG GRAMS |
|---|---|---|---|---|
| AMPHETAMINE | 1100 | 189 | **155,140.27** | 820.85 |
| METHAMPHETAMINE | 1105 | 3 | **8.04** | 2.68 |
| LISDEXAMFETAMINE | 1205 | 176 | **30,273.94** | 172.01 |
| METHYLPHENIDATE (DL;D;L;ISOMERS) | 1724 | 185 | **71,366.09** | 385.76 |
| CODEINE | 9050 | 178 | **17,887.56** | 100.49 |
| BUPRENORPHINE | 9064 | 190 | **33,796.61** | 177.88 |
| OXYCODONE | 9143 | 191 | **110,670.16** | 579.42 |
| HYDROMORPHONE | 9150 | 149 | **2,285.40** | 15.34 |
| HYDROCODONE | 9193 | 189 | **28,739.73** | 152.06 |
| MEPERIDINE (PETHIDINE) | 9230 | 8 | **97.50** | 12.19 |
| METHADONE | 9250B | 149 | **5,000.77** | 33.56 |
| MORPHINE | 9300 | 183 | **30,106.02** | 164.51 |
| OXYMORPHONE | 9652 | 48 | **619.25** | 12.90 |
| TAPENTADOL | 9780 | 85 | **7,122.00** | 83.79 |
| FENTANYL BASE | 9801 | 161 | **377.80** | 2.35 |

**BUSINESS ACTIVITY: B - HOSPITALS**

| DRUG NAME | DRUG CODE | NUMBER OF REGISTRANT SOLD TO | TOTAL GRAMS | AVG GRAMS |
|---|---|---|---|---|
| AMPHETAMINE | 1100 | 16 | **2,845.18** | 177.82 |
| LISDEXAMFETAMINE | 1205 | 9 | **423.75** | 47.08 |
| METHYLPHENIDATE (DL;D;L;ISOMERS) | 1724 | 19 | **2,544.11** | 133.90 |
| CODEINE | 9050 | 17 | **439.37** | 25.85 |
| BUPRENORPHINE | 9064 | 15 | **827.41** | 55.16 |
| OXYCODONE | 9143 | 20 | **7,595.04** | 379.75 |
| HYDROMORPHONE | 9150 | 16 | **453.69** | 28.36 |
| HYDROCODONE | 9193 | 20 | **1,563.35** | 78.17 |
| MEPERIDINE (PETHIDINE) | 9230 | 12 | **154.69** | 12.89 |
| METHADONE | 9250B | 14 | **1,515.47** | 108.25 |
| MORPHINE | 9300 | 19 | **4,236.85** | 222.99 |
| OXYMORPHONE | 9652 | 1 | **0.89** | 0.89 |
| TAPENTADOL | 9780 | 1 | **5.00** | 5.00 |
| FENTANYL BASE | 9801 | 15 | **64.15** | 4.28 |

**BUSINESS ACTIVITY: C - PRACTITIONERS**

| DRUG NAME | DRUG CODE | NUMBER OF REGISTRANT SOLD TO | TOTAL GRAMS | AVG GRAMS |
|---|---|---|---|---|

| | | | | |
|---|---|---|---|---|
| CODEINE | 9050 | 1 | **4.78** | 4.78 |
| BUPRENORPHINE | 9064 | 86 | **12.88** | 0.15 |
| OXYCODONE | 9143 | 3 | **8.97** | 2.99 |
| HYDROMORPHONE | 9150 | 38 | **16.89** | 0.44 |
| HYDROCODONE | 9193 | 22 | **77.96** | 3.54 |
| MEPERIDINE (PETHIDINE) | 9230 | 7 | **15.80** | 2.26 |
| METHADONE | 9250B | 3 | **28.62** | 9.54 |
| MORPHINE | 9300 | 9 | **5.98** | 0.66 |
| FENTANYL BASE | 9801 | 39 | **7.30** | 0.19 |

**BUSINESS ACTIVITY: M - MID-LEVEL PRACTITIONERS**

| DRUG NAME | DRUG CODE | NUMBER OF REGISTRANT SOLD TO | TOTAL GRAMS | AVG GRAMS |
|---|---|---|---|---|
| BUPRENORPHINE | 9064 | 1 | **1.26** | 1.26 |

**BUSINESS ACTIVITY: NTP - NARCOTIC TREATMENT PROGRAMS**

| DRUG NAME | DRUG CODE | NUMBER OF REGISTRANT SOLD TO | TOTAL GRAMS | AVG GRAMS |
|---|---|---|---|---|
| BUPRENORPHINE | 9064 | 13 | **707.13** | 54.39 |
| METHADONE | 9250B | 22 | **184,580.82** | 8,390.04 |

**STATE: SC - SOUTH CAROLINA**

**BUSINESS ACTIVITY: A - PHARMACIES**

| DRUG NAME | DRUG CODE | NUMBER OF REGISTRANT SOLD TO | TOTAL GRAMS | AVG GRAMS |
|---|---|---|---|---|
| AMPHETAMINE | 1100 | 1,061 | **575,849.77** | 542.74 |
| METHAMPHETAMINE | 1105 | 17 | **32.95** | 1.94 |
| LISDEXAMFETAMINE | 1205 | 1,044 | **247,417.55** | 236.99 |
| METHYLPHENIDATE (DL;D;L;ISOMERS) | 1724 | 1,060 | **342,149.84** | 322.78 |
| CODEINE | 9050 | 1,048 | **229,053.31** | 218.56 |
| BUPRENORPHINE | 9064 | 1,024 | **61,717.42** | 60.27 |
| OXYCODONE | 9143 | 1,065 | **655,799.77** | 615.77 |
| HYDROMORPHONE | 9150 | 972 | **14,003.19** | 14.41 |
| HYDROCODONE | 9193 | 1,080 | **385,822.56** | 357.24 |
| MEPERIDINE (PETHIDINE) | 9230 | 304 | **4,617.32** | 15.19 |
| METHADONE | 9250B | 816 | **26,184.99** | 32.09 |
| MORPHINE | 9300 | 1,040 | **194,525.73** | 187.04 |
| OXYMORPHONE | 9652 | 466 | **9,826.48** | 21.09 |
| TAPENTADOL | 9780 | 797 | **120,110.00** | 150.70 |
| FENTANYL BASE | 9801 | 980 | **2,818.59** | 2.88 |

**BUSINESS ACTIVITY: B - HOSPITALS**

| DRUG NAME | DRUG CODE | NUMBER OF REGISTRANT SOLD TO | TOTAL GRAMS | AVG GRAMS |
|---|---|---|---|---|
| AMPHETAMINE | 1100 | 61 | **9,101.12** | 149.20 |
| METHAMPHETAMINE | 1105 | 1 | **16.88** | 16.88 |
| LISDEXAMFETAMINE | 1205 | 25 | **1,544.61** | 61.78 |
| METHYLPHENIDATE (DL;D;L;ISOMERS) | 1724 | 77 | **8,237.16** | 106.98 |

| | | | | |
|---|---|---|---|---|
| CODEINE | 9050 | 92 | **3,360.44** | 36.53 |
| BUPRENORPHINE | 9064 | 77 | **1,304.20** | 16.94 |
| OXYCODONE | 9143 | 148 | **23,767.22** | 160.59 |
| HYDROMORPHONE | 9150 | 148 | **2,422.42** | 16.37 |
| HYDROCODONE | 9193 | 152 | **12,105.48** | 79.64 |
| MEPERIDINE (PETHIDINE) | 9230 | 99 | **1,571.60** | 15.87 |
| METHADONE | 9250B | 91 | **1,772.00** | 19.47 |
| MORPHINE | 9300 | 154 | **15,500.38** | 100.65 |
| OXYMORPHONE | 9652 | 2 | **5.00** | 2.50 |
| TAPENTADOL | 9780 | 25 | **893.00** | 35.72 |
| FENTANYL BASE | 9801 | 178 | **351.22** | 1.97 |

**BUSINESS ACTIVITY: C - PRACTITIONERS**

| DRUG NAME | DRUG CODE | NUMBER OF REGISTRANT SOLD TO | TOTAL GRAMS | AVG GRAMS |
|---|---|---|---|---|
| AMPHETAMINE | 1100 | 2 | **6.54** | 3.27 |
| LISDEXAMFETAMINE | 1205 | 1 | **6.68** | 6.68 |
| CODEINE | 9050 | 17 | **419.84** | 24.70 |
| BUPRENORPHINE | 9064 | 417 | **71.55** | 0.17 |
| OXYCODONE | 9143 | 3 | **18.38** | 6.13 |
| HYDROMORPHONE | 9150 | 149 | **93.02** | 0.62 |
| HYDROCODONE | 9193 | 69 | **151.83** | 2.20 |
| MEPERIDINE (PETHIDINE) | 9230 | 40 | **291.95** | 7.30 |
| METHADONE | 9250B | 1 | **1.07** | 1.07 |
| MORPHINE | 9300 | 56 | **30.96** | 0.55 |
| FENTANYL BASE | 9801 | 230 | **9.46** | 0.04 |

**BUSINESS ACTIVITY: D - TEACHING INSTITUTIONS**

| DRUG NAME | DRUG CODE | NUMBER OF REGISTRANT SOLD TO | TOTAL GRAMS | AVG GRAMS |
|---|---|---|---|---|
| BUPRENORPHINE | 9064 | 1 | **0.01** | 0.01 |

**BUSINESS ACTIVITY: M - MID-LEVEL PRACTITIONERS**

| DRUG NAME | DRUG CODE | NUMBER OF REGISTRANT SOLD TO | TOTAL GRAMS | AVG GRAMS |
|---|---|---|---|---|
| MORPHINE | 9300 | 30 | **11.47** | 0.38 |
| FENTANYL BASE | 9801 | 65 | **0.85** | 0.01 |

**BUSINESS ACTIVITY: NTP - NARCOTIC TREATMENT PROGRAMS**

| DRUG NAME | DRUG CODE | NUMBER OF REGISTRANT SOLD TO | TOTAL GRAMS | AVG GRAMS |
|---|---|---|---|---|
| BUPRENORPHINE | 9064 | 16 | **1,624.32** | 101.52 |
| METHADONE | 9250B | 22 | **202,127.24** | 9,187.60 |

**STATE: SD - SOUTH DAKOTA**

**BUSINESS ACTIVITY: A - PHARMACIES**

| DRUG NAME | DRUG CODE | NUMBER OF REGISTRANT SOLD TO | TOTAL GRAMS | AVG GRAMS |
|---|---|---|---|---|
| AMPHETAMINE | 1100 | 183 | **56,978.67** | 311.36 |
| METHAMPHETAMINE | 1105 | 3 | **23.30** | 7.77 |

| | | | | |
|---|---|---|---|---|
| LISDEXAMFETAMINE | 1205 | 177 | **38,097.37** | 215.24 |
| METHYLPHENIDATE (DL;D;L;ISOMERS) | 1724 | 181 | **75,224.52** | 415.61 |
| CODEINE | 9050 | 182 | **17,618.85** | 96.81 |
| BUPRENORPHINE | 9064 | 150 | **3,414.03** | 22.76 |
| OXYCODONE | 9143 | 183 | **46,806.13** | 255.77 |
| HYDROMORPHONE | 9150 | 159 | **1,221.92** | 7.69 |
| HYDROCODONE | 9193 | 187 | **33,875.39** | 181.15 |
| MEPERIDINE (PETHIDINE) | 9230 | 23 | **507.10** | 22.05 |
| METHADONE | 9250B | 132 | **3,252.75** | 24.64 |
| MORPHINE | 9300 | 179 | **19,741.08** | 110.29 |
| OXYMORPHONE | 9652 | 47 | **836.92** | 17.81 |
| TAPENTADOL | 9780 | 120 | **11,344.00** | 94.53 |
| FENTANYL BASE | 9801 | 179 | **592.54** | 3.31 |

**BUSINESS ACTIVITY: B - HOSPITALS**

| DRUG NAME | DRUG CODE | NUMBER OF REGISTRANT SOLD TO | TOTAL GRAMS | AVG GRAMS |
|---|---|---|---|---|
| AMPHETAMINE | 1100 | 28 | **2,748.23** | 98.15 |
| LISDEXAMFETAMINE | 1205 | 16 | **788.53** | 49.28 |
| METHYLPHENIDATE (DL;D;L;ISOMERS) | 1724 | 35 | **3,563.51** | 101.81 |
| CODEINE | 9050 | 53 | **1,654.06** | 31.21 |
| BUPRENORPHINE | 9064 | 20 | **412.43** | 20.62 |
| OXYCODONE | 9143 | 68 | **4,875.99** | 71.71 |
| HYDROMORPHONE | 9150 | 65 | **348.87** | 5.37 |
| HYDROCODONE | 9193 | 71 | **6,777.00** | 95.45 |
| MEPERIDINE (PETHIDINE) | 9230 | 30 | **349.14** | 11.64 |
| METHADONE | 9250B | 27 | **1,025.34** | 37.98 |
| MORPHINE | 9300 | 75 | **4,810.27** | 64.14 |
| OXYMORPHONE | 9652 | 4 | **10.88** | 2.72 |
| TAPENTADOL | 9780 | 13 | **546.50** | 42.04 |
| FENTANYL BASE | 9801 | 73 | **109.48** | 1.50 |

**BUSINESS ACTIVITY: C - PRACTITIONERS**

| DRUG NAME | DRUG CODE | NUMBER OF REGISTRANT SOLD TO | TOTAL GRAMS | AVG GRAMS |
|---|---|---|---|---|
| METHYLPHENIDATE (DL;D;L;ISOMERS) | 1724 | 1 | **0.86** | 0.86 |
| CODEINE | 9050 | 3 | **66.30** | 22.10 |
| BUPRENORPHINE | 9064 | 64 | **2.21** | 0.03 |
| OXYCODONE | 9143 | 4 | **6.28** | 1.57 |
| HYDROMORPHONE | 9150 | 33 | **52.09** | 1.58 |
| HYDROCODONE | 9193 | 23 | **60.47** | 2.63 |
| MEPERIDINE (PETHIDINE) | 9230 | 8 | **112.75** | 14.09 |
| METHADONE | 9250B | 1 | **0.18** | 0.18 |
| MORPHINE | 9300 | 17 | **53.87** | 3.17 |

| | | | | |
|---|---|---|---|---|
| FENTANYL BASE | 9801 | 52 | **5.24** | 0.10 |

**BUSINESS ACTIVITY: M - MID-LEVEL PRACTITIONERS**

| DRUG NAME | DRUG CODE | NUMBER OF REGISTRANT SOLD TO | TOTAL GRAMS | AVG GRAMS |
|---|---|---|---|---|
| MORPHINE | 9300 | 2 | **0.38** | 0.19 |
| FENTANYL BASE | 9801 | 7 | **0.08** | 0.01 |

**BUSINESS ACTIVITY: NTP - NARCOTIC TREATMENT PROGRAMS**

| DRUG NAME | DRUG CODE | NUMBER OF REGISTRANT SOLD TO | TOTAL GRAMS | AVG GRAMS |
|---|---|---|---|---|
| METHADONE | 9250B | 1 | **3,477.43** | 3,477.43 |

**STATE: TN - TENNESSEE**

**BUSINESS ACTIVITY: A - PHARMACIES**

| DRUG NAME | DRUG CODE | NUMBER OF REGISTRANT SOLD TO | TOTAL GRAMS | AVG GRAMS |
|---|---|---|---|---|
| AMPHETAMINE | 1100 | 1,458 | **553,941.31** | 379.93 |
| METHAMPHETAMINE | 1105 | 19 | **54.24** | 2.85 |
| LISDEXAMFETAMINE | 1205 | 1,401 | **263,511.85** | 188.09 |
| METHYLPHENIDATE (DL;D;L;ISOMERS) | 1724 | 1,446 | **323,631.77** | 223.81 |
| CODEINE | 9050 | 1,388 | **213,161.29** | 153.57 |
| BUPRENORPHINE | 9064 | 1,376 | **165,619.66** | 120.36 |
| OXYCODONE | 9143 | 1,474 | **1,324,350.70** | 898.47 |
| HYDROMORPHONE | 9150 | 1,179 | **17,492.81** | 14.84 |
| HYDROCODONE | 9193 | 1,480 | **699,977.77** | 472.96 |
| MEPERIDINE (PETHIDINE) | 9230 | 426 | **6,011.72** | 14.11 |
| METHADONE | 9250B | 873 | **29,810.71** | 34.15 |
| MORPHINE | 9300 | 1,445 | **448,625.13** | 310.47 |
| OXYMORPHONE | 9652 | 766 | **33,180.50** | 43.32 |
| TAPENTADOL | 9780 | 1,011 | **86,444.00** | 85.50 |
| FENTANYL BASE | 9801 | 1,340 | **4,734.62** | 3.53 |

**BUSINESS ACTIVITY: B - HOSPITALS**

| DRUG NAME | DRUG CODE | NUMBER OF REGISTRANT SOLD TO | TOTAL GRAMS | AVG GRAMS |
|---|---|---|---|---|
| AMPHETAMINE | 1100 | 82 | **6,244.44** | 76.15 |
| METHAMPHETAMINE | 1105 | 1 | **1.21** | 1.21 |
| LISDEXAMFETAMINE | 1205 | 40 | **2,818.82** | 70.47 |
| METHYLPHENIDATE (DL;D;L;ISOMERS) | 1724 | 103 | **4,881.26** | 47.39 |
| CODEINE | 9050 | 130 | **2,712.98** | 20.87 |
| BUPRENORPHINE | 9064 | 135 | **3,655.55** | 27.08 |
| OXYCODONE | 9143 | 184 | **58,953.49** | 320.40 |
| HYDROMORPHONE | 9150 | 172 | **2,956.64** | 17.19 |
| HYDROCODONE | 9193 | 193 | **34,092.59** | 176.65 |
| MEPERIDINE (PETHIDINE) | 9230 | 118 | **1,785.49** | 15.13 |
| METHADONE | 9250B | 123 | **1,771.81** | 14.40 |
| MORPHINE | 9300 | 185 | **23,797.52** | 128.64 |

| | | | | |
|---|---|---|---|---|
| OXYMORPHONE | 9652 | 14 | **70.16** | 5.01 |
| TAPENTADOL | 9780 | 33 | **2,072.00** | 62.79 |
| FENTANYL BASE | 9801 | 173 | **536.17** | 3.10 |

**BUSINESS ACTIVITY: C - PRACTITIONERS**

| DRUG NAME | DRUG CODE | NUMBER OF REGISTRANT SOLD TO | TOTAL GRAMS | AVG GRAMS |
|---|---|---|---|---|
| AMPHETAMINE | 1100 | 9 | **1,096.35** | 121.82 |
| LISDEXAMFETAMINE | 1205 | 6 | **753.42** | 125.57 |
| METHYLPHENIDATE (DL;D;L;ISOMERS) | 1724 | 2 | **494.56** | 247.28 |
| CODEINE | 9050 | 71 | **2,617.89** | 36.87 |
| BUPRENORPHINE | 9064 | 580 | **107.81** | 0.19 |
| OXYCODONE | 9143 | 71 | **497.95** | 7.01 |
| HYDROMORPHONE | 9150 | 260 | **157.82** | 0.61 |
| HYDROCODONE | 9193 | 262 | **968.72** | 3.70 |
| MEPERIDINE (PETHIDINE) | 9230 | 121 | **629.33** | 5.20 |
| METHADONE | 9250B | 9 | **16.19** | 1.80 |
| MORPHINE | 9300 | 139 | **127.42** | 0.92 |
| TAPENTADOL | 9780 | 1 | **5.00** | 5.00 |
| FENTANYL BASE | 9801 | 418 | **37.34** | 0.09 |

**BUSINESS ACTIVITY: D - TEACHING INSTITUTIONS**

| DRUG NAME | DRUG CODE | NUMBER OF REGISTRANT SOLD TO | TOTAL GRAMS | AVG GRAMS |
|---|---|---|---|---|
| BUPRENORPHINE | 9064 | 2 | **0.13** | 0.06 |
| FENTANYL BASE | 9801 | 1 | **0.01** | 0.01 |

**BUSINESS ACTIVITY: M - MID-LEVEL PRACTITIONERS**

| DRUG NAME | DRUG CODE | NUMBER OF REGISTRANT SOLD TO | TOTAL GRAMS | AVG GRAMS |
|---|---|---|---|---|
| AMPHETAMINE | 1100 | 1 | **1.26** | 1.26 |
| CODEINE | 9050 | 27 | **57.42** | 2.13 |
| HYDROMORPHONE | 9150 | 2 | **0.34** | 0.17 |
| HYDROCODONE | 9193 | 1 | **1.21** | 1.21 |
| MEPERIDINE (PETHIDINE) | 9230 | 4 | **11.44** | 2.86 |
| MORPHINE | 9300 | 58 | **21.51** | 0.37 |
| FENTANYL BASE | 9801 | 86 | **1.43** | 0.02 |

**BUSINESS ACTIVITY: NTP - NARCOTIC TREATMENT PROGRAMS**

| DRUG NAME | DRUG CODE | NUMBER OF REGISTRANT SOLD TO | TOTAL GRAMS | AVG GRAMS |
|---|---|---|---|---|
| BUPRENORPHINE | 9064 | 13 | **967.00** | 74.38 |
| METHADONE | 9250B | 13 | **159,843.59** | 12,295.66 |

**STATE: TX - TEXAS**

**BUSINESS ACTIVITY: A - PHARMACIES**

| DRUG NAME | DRUG CODE | NUMBER OF REGISTRANT SOLD TO | TOTAL GRAMS | AVG GRAMS |
|---|---|---|---|---|
| AMPHETAMINE | 1100 | 4,605 | **1,881,404.93** | 408.56 |
| METHAMPHETAMINE | 1105 | 133 | **349.13** | 2.63 |

| | | | | |
|---|---|---|---|---|
| LISDEXAMFETAMINE | 1205 | 4,386 | **1,034,493.76** | 235.86 |
| METHYLPHENIDATE (DL;D;L;ISOMERS) | 1724 | 4,551 | **1,258,688.47** | 276.57 |
| CODEINE | 9050 | 4,963 | **3,221,756.00** | 649.15 |
| BUPRENORPHINE | 9064 | 4,238 | **119,696.74** | 28.24 |
| OXYCODONE | 9143 | 4,535 | **970,662.35** | 214.04 |
| HYDROMORPHONE | 9150 | 3,834 | **56,115.11** | 14.64 |
| HYDROCODONE | 9193 | 4,949 | **1,925,741.20** | 389.12 |
| MEPERIDINE (PETHIDINE) | 9230 | 500 | **7,961.87** | 15.92 |
| METHADONE | 9250B | 3,239 | **85,021.93** | 26.25 |
| MORPHINE | 9300 | 4,307 | **543,446.43** | 126.18 |
| OXYMORPHONE | 9652 | 920 | **11,263.07** | 12.24 |
| TAPENTADOL | 9780 | 2,380 | **235,286.00** | 98.86 |
| FENTANYL BASE | 9801 | 4,091 | **11,233.64** | 2.75 |

**BUSINESS ACTIVITY: B - HOSPITALS**

| DRUG NAME | DRUG CODE | NUMBER OF REGISTRANT SOLD TO | TOTAL GRAMS | AVG GRAMS |
|---|---|---|---|---|
| AMPHETAMINE | 1100 | 269 | **37,425.62** | 139.13 |
| LISDEXAMFETAMINE | 1205 | 122 | **9,647.33** | 79.08 |
| METHYLPHENIDATE (DL;D;L;ISOMERS) | 1724 | 412 | **44,999.03** | 109.22 |
| CODEINE | 9050 | 919 | **98,818.70** | 107.53 |
| BUPRENORPHINE | 9064 | 338 | **4,877.22** | 14.43 |
| OXYCODONE | 9143 | 746 | **47,908.72** | 64.22 |
| HYDROMORPHONE | 9150 | 1,088 | **10,971.80** | 10.08 |
| HYDROCODONE | 9193 | 1,211 | **106,574.37** | 88.01 |
| MEPERIDINE (PETHIDINE) | 9230 | 697 | **14,201.09** | 20.37 |
| METHADONE | 9250B | 522 | **8,725.04** | 16.71 |
| MORPHINE | 9300 | 1,241 | **70,256.63** | 56.61 |
| OXYMORPHONE | 9652 | 28 | **346.82** | 12.39 |
| TAPENTADOL | 9780 | 90 | **3,316.00** | 36.84 |
| FENTANYL BASE | 9801 | 1,303 | **2,090.70** | 1.60 |

**BUSINESS ACTIVITY: C - PRACTITIONERS**

| DRUG NAME | DRUG CODE | NUMBER OF REGISTRANT SOLD TO | TOTAL GRAMS | AVG GRAMS |
|---|---|---|---|---|
| AMPHETAMINE | 1100 | 6 | **20.54** | 3.42 |
| LISDEXAMFETAMINE | 1205 | 2 | **8.70** | 4.35 |
| METHYLPHENIDATE (DL;D;L;ISOMERS) | 1724 | 1 | **42.28** | 42.28 |
| CODEINE | 9050 | 302 | **2,724.87** | 9.02 |
| BUPRENORPHINE | 9064 | 2,118 | **564.33** | 0.27 |
| OXYCODONE | 9143 | 26 | **42.81** | 1.65 |
| HYDROMORPHONE | 9150 | 736 | **302.73** | 0.41 |
| HYDROCODONE | 9193 | 839 | **1,920.92** | 2.29 |
| MEPERIDINE (PETHIDINE) | 9230 | 364 | **2,643.10** | 7.26 |

| | | | | |
|---|---|---|---|---|
| METHADONE | 9250B | 38 | **93.73** | 2.47 |
| MORPHINE | 9300 | 404 | **245.99** | 0.61 |
| OXYMORPHONE | 9652 | 1 | **4.46** | 4.46 |
| TAPENTADOL | 9780 | 1 | **10.00** | 10.00 |
| FENTANYL BASE | 9801 | 1,366 | **71.82** | 0.05 |

**BUSINESS ACTIVITY: D - TEACHING INSTITUTIONS**

| DRUG NAME | DRUG CODE | NUMBER OF REGISTRANT SOLD TO | TOTAL GRAMS | AVG GRAMS |
|---|---|---|---|---|
| BUPRENORPHINE | 9064 | 3 | **0.81** | 0.27 |
| HYDROMORPHONE | 9150 | 1 | **5.71** | 5.71 |
| HYDROCODONE | 9193 | 1 | **2.37** | 2.37 |
| MEPERIDINE (PETHIDINE) | 9230 | 1 | **6.54** | 6.54 |
| METHADONE | 9250B | 1 | **22.72** | 22.72 |
| MORPHINE | 9300 | 2 | **7.52** | 3.76 |
| FENTANYL BASE | 9801 | 3 | **3.62** | 1.21 |

**BUSINESS ACTIVITY: M - MID-LEVEL PRACTITIONERS**

| DRUG NAME | DRUG CODE | NUMBER OF REGISTRANT SOLD TO | TOTAL GRAMS | AVG GRAMS |
|---|---|---|---|---|
| CODEINE | 9050 | 91 | **86.30** | 0.95 |
| HYDROMORPHONE | 9150 | 17 | **1.29** | 0.08 |
| HYDROCODONE | 9193 | 1 | **1.21** | 1.21 |
| MORPHINE | 9300 | 209 | **83.39** | 0.40 |
| FENTANYL BASE | 9801 | 455 | **20.79** | 0.05 |

**BUSINESS ACTIVITY: NTP - NARCOTIC TREATMENT PROGRAMS**

| DRUG NAME | DRUG CODE | NUMBER OF REGISTRANT SOLD TO | TOTAL GRAMS | AVG GRAMS |
|---|---|---|---|---|
| BUPRENORPHINE | 9064 | 45 | **1,804.43** | 40.10 |
| METHADONE | 9250B | 97 | **450,276.29** | 4,642.02 |

**STATE: UT - UTAH**

**BUSINESS ACTIVITY: A - PHARMACIES**

| DRUG NAME | DRUG CODE | NUMBER OF REGISTRANT SOLD TO | TOTAL GRAMS | AVG GRAMS |
|---|---|---|---|---|
| AMPHETAMINE | 1100 | 527 | **380,730.92** | 722.45 |
| METHAMPHETAMINE | 1105 | 25 | **104.05** | 4.16 |
| LISDEXAMFETAMINE | 1205 | 513 | **91,795.92** | 178.94 |
| METHYLPHENIDATE (DL;D;L;ISOMERS) | 1724 | 522 | **188,899.72** | 361.88 |
| CODEINE | 9050 | 508 | **49,074.19** | 96.60 |
| BUPRENORPHINE | 9064 | 521 | **43,762.74** | 84.00 |
| OXYCODONE | 9143 | 533 | **511,155.79** | 959.02 |
| HYDROMORPHONE | 9150 | 495 | **8,029.05** | 16.22 |
| HYDROCODONE | 9193 | 532 | **174,278.65** | 327.59 |
| MEPERIDINE (PETHIDINE) | 9230 | 129 | **2,370.26** | 18.37 |
| METHADONE | 9250B | 476 | **25,598.62** | 53.78 |
| MORPHINE | 9300 | 519 | **140,411.16** | 270.54 |

| | | | | |
|---|---|---|---|---|
| OXYMORPHONE | 9652 | 330 | **8,347.34** | 25.29 |
| TAPENTADOL | 9780 | 325 | **32,726.50** | 100.70 |
| FENTANYL BASE | 9801 | 498 | **2,071.50** | 4.16 |

**BUSINESS ACTIVITY: B - HOSPITALS**

| DRUG NAME | DRUG CODE | NUMBER OF REGISTRANT SOLD TO | TOTAL GRAMS | AVG GRAMS |
|---|---|---|---|---|
| AMPHETAMINE | 1100 | 32 | **3,994.82** | 124.84 |
| LISDEXAMFETAMINE | 1205 | 16 | **818.01** | 51.13 |
| METHYLPHENIDATE (DL;D;L;ISOMERS) | 1724 | 45 | **3,859.38** | 85.76 |
| CODEINE | 9050 | 35 | **1,231.64** | 35.19 |
| BUPRENORPHINE | 9064 | 48 | **1,226.60** | 25.55 |
| OXYCODONE | 9143 | 91 | **21,127.22** | 232.17 |
| HYDROMORPHONE | 9150 | 85 | **1,232.27** | 14.50 |
| HYDROCODONE | 9193 | 115 | **8,721.55** | 75.84 |
| MEPERIDINE (PETHIDINE) | 9230 | 70 | **990.79** | 14.15 |
| METHADONE | 9250B | 58 | **1,113.89** | 19.20 |
| MORPHINE | 9300 | 105 | **10,396.74** | 99.02 |
| OXYMORPHONE | 9652 | 11 | **49.24** | 4.48 |
| TAPENTADOL | 9780 | 16 | **2,177.50** | 136.09 |
| FENTANYL BASE | 9801 | 110 | **179.48** | 1.63 |

**BUSINESS ACTIVITY: C - PRACTITIONERS**

| DRUG NAME | DRUG CODE | NUMBER OF REGISTRANT SOLD TO | TOTAL GRAMS | AVG GRAMS |
|---|---|---|---|---|
| AMPHETAMINE | 1100 | 1 | **1.90** | 1.90 |
| LISDEXAMFETAMINE | 1205 | 2 | **7.15** | 3.58 |
| CODEINE | 9050 | 10 | **415.95** | 41.60 |
| BUPRENORPHINE | 9064 | 200 | **38.66** | 0.19 |
| OXYCODONE | 9143 | 12 | **14.12** | 1.18 |
| HYDROMORPHONE | 9150 | 96 | **48.66** | 0.51 |
| HYDROCODONE | 9193 | 78 | **292.38** | 3.75 |
| MEPERIDINE (PETHIDINE) | 9230 | 19 | **113.30** | 5.96 |
| METHADONE | 9250B | 5 | **46.15** | 9.23 |
| MORPHINE | 9300 | 76 | **45.01** | 0.59 |
| FENTANYL BASE | 9801 | 183 | **13.52** | 0.07 |

**BUSINESS ACTIVITY: M - MID-LEVEL PRACTITIONERS**

| DRUG NAME | DRUG CODE | NUMBER OF REGISTRANT SOLD TO | TOTAL GRAMS | AVG GRAMS |
|---|---|---|---|---|
| BUPRENORPHINE | 9064 | 1 | **1.26** | 1.26 |
| HYDROMORPHONE | 9150 | 1 | **0.04** | 0.04 |
| MEPERIDINE (PETHIDINE) | 9230 | 2 | **56.65** | 28.32 |
| FENTANYL BASE | 9801 | 12 | **0.52** | 0.04 |

**BUSINESS ACTIVITY: NTP - NARCOTIC TREATMENT PROGRAMS**

| DRUG NAME | DRUG CODE | NUMBER OF REGISTRANT SOLD TO | TOTAL GRAMS | AVG GRAMS |
|---|---|---|---|---|

| | | | | |
|---|---|---|---|---|
| BUPRENORPHINE | 9064 | 11 | **314.66** | 28.61 |
| METHADONE | 9250B | 16 | **82,014.69** | 5,125.92 |

**STATE: VA - VIRGINIA**

**BUSINESS ACTIVITY: A - PHARMACIES**

| DRUG NAME | DRUG CODE | NUMBER OF REGISTRANT SOLD TO | TOTAL GRAMS | AVG GRAMS |
|---|---|---|---|---|
| AMPHETAMINE | 1100 | 1,534 | **730,667.71** | 476.32 |
| METHAMPHETAMINE | 1105 | 45 | **126.16** | 2.80 |
| LISDEXAMFETAMINE | 1205 | 1,476 | **271,628.38** | 184.03 |
| METHYLPHENIDATE (DL;D;L;ISOMERS) | 1724 | 1,524 | **498,447.72** | 327.07 |
| CODEINE | 9050 | 1,484 | **152,634.25** | 102.85 |
| BUPRENORPHINE | 9064 | 1,464 | **97,486.76** | 66.59 |
| OXYCODONE | 9143 | 1,535 | **870,607.00** | 567.17 |
| HYDROMORPHONE | 9150 | 1,432 | **26,395.94** | 18.43 |
| HYDROCODONE | 9193 | 1,543 | **294,155.57** | 190.64 |
| MEPERIDINE (PETHIDINE) | 9230 | 295 | **5,278.57** | 17.89 |
| METHADONE | 9250B | 1,161 | **43,209.72** | 37.22 |
| MORPHINE | 9300 | 1,466 | **229,762.71** | 156.73 |
| OXYMORPHONE | 9652 | 708 | **16,217.75** | 22.91 |
| TAPENTADOL | 9780 | 938 | **105,873.00** | 112.87 |
| FENTANYL BASE | 9801 | 1,366 | **3,697.49** | 2.71 |

**BUSINESS ACTIVITY: B - HOSPITALS**

| DRUG NAME | DRUG CODE | NUMBER OF REGISTRANT SOLD TO | TOTAL GRAMS | AVG GRAMS |
|---|---|---|---|---|
| AMPHETAMINE | 1100 | 97 | **30,383.89** | 313.24 |
| LISDEXAMFETAMINE | 1205 | 48 | **7,322.21** | 152.55 |
| METHYLPHENIDATE (DL;D;L;ISOMERS) | 1724 | 121 | **32,453.40** | 268.21 |
| CODEINE | 9050 | 125 | **5,005.22** | 40.04 |
| BUPRENORPHINE | 9064 | 121 | **2,593.17** | 21.43 |
| OXYCODONE | 9143 | 180 | **52,307.44** | 290.60 |
| HYDROMORPHONE | 9150 | 175 | **4,365.32** | 24.94 |
| HYDROCODONE | 9193 | 178 | **13,624.92** | 76.54 |
| MEPERIDINE (PETHIDINE) | 9230 | 104 | **2,301.85** | 22.13 |
| METHADONE | 9250B | 117 | **3,380.88** | 28.90 |
| MORPHINE | 9300 | 177 | **21,206.16** | 119.81 |
| OXYMORPHONE | 9652 | 11 | **52.19** | 4.74 |
| TAPENTADOL | 9780 | 39 | **2,289.00** | 58.69 |
| FENTANYL BASE | 9801 | 192 | **589.45** | 3.07 |

**BUSINESS ACTIVITY: C - PRACTITIONERS**

| DRUG NAME | DRUG CODE | NUMBER OF REGISTRANT SOLD TO | TOTAL GRAMS | AVG GRAMS |
|---|---|---|---|---|
| AMPHETAMINE | 1100 | 1 | **1.16** | 1.16 |
| CODEINE | 9050 | 112 | **3,459.28** | 30.89 |

| | | | | |
|---|---|---|---|---|
| BUPRENORPHINE | 9064 | 858 | **89.55** | 0.10 |
| OXYCODONE | 9143 | 27 | **187.37** | 6.94 |
| HYDROMORPHONE | 9150 | 356 | **146.19** | 0.41 |
| HYDROCODONE | 9193 | 182 | **667.03** | 3.66 |
| MEPERIDINE (PETHIDINE) | 9230 | 85 | **791.00** | 9.31 |
| METHADONE | 9250B | 25 | **70.30** | 2.81 |
| MORPHINE | 9300 | 140 | **132.82** | 0.95 |
| OXYMORPHONE | 9652 | 1 | **13.11** | 13.11 |
| TAPENTADOL | 9780 | 1 | **102.00** | 102.00 |
| FENTANYL BASE | 9801 | 433 | **29.52** | 0.07 |

**BUSINESS ACTIVITY: D - TEACHING INSTITUTIONS**

| DRUG NAME | DRUG CODE | NUMBER OF REGISTRANT SOLD TO | TOTAL GRAMS | AVG GRAMS |
|---|---|---|---|---|
| CODEINE | 9050 | 1 | **376.15** | 376.15 |
| BUPRENORPHINE | 9064 | 2 | **0.72** | 0.36 |
| HYDROMORPHONE | 9150 | 1 | **1.00** | 1.00 |
| HYDROCODONE | 9193 | 1 | **1.18** | 1.18 |
| METHADONE | 9250B | 1 | **5.01** | 5.01 |
| MORPHINE | 9300 | 1 | **11.77** | 11.77 |
| FENTANYL BASE | 9801 | 1 | **1.10** | 1.10 |

**BUSINESS ACTIVITY: M - MID-LEVEL PRACTITIONERS**

| DRUG NAME | DRUG CODE | NUMBER OF REGISTRANT SOLD TO | TOTAL GRAMS | AVG GRAMS |
|---|---|---|---|---|
| CODEINE | 9050 | 17 | **20.16** | 1.19 |
| BUPRENORPHINE | 9064 | 1 | **0.01** | 0.01 |
| MORPHINE | 9300 | 11 | **3.99** | 0.36 |
| FENTANYL BASE | 9801 | 13 | **0.23** | 0.02 |

**BUSINESS ACTIVITY: NTP - NARCOTIC TREATMENT PROGRAMS**

| DRUG NAME | DRUG CODE | NUMBER OF REGISTRANT SOLD TO | TOTAL GRAMS | AVG GRAMS |
|---|---|---|---|---|
| BUPRENORPHINE | 9064 | 32 | **3,951.66** | 123.49 |
| METHADONE | 9250B | 42 | **327,116.07** | 7,788.48 |

**STATE: VI - VIRGIN ISLANDS**

**BUSINESS ACTIVITY: A - PHARMACIES**

| DRUG NAME | DRUG CODE | NUMBER OF REGISTRANT SOLD TO | TOTAL GRAMS | AVG GRAMS |
|---|---|---|---|---|
| AMPHETAMINE | 1100 | 20 | **1,460.40** | 73.02 |
| LISDEXAMFETAMINE | 1205 | 13 | **324.31** | 24.95 |
| METHYLPHENIDATE (DL;D;L;ISOMERS) | 1724 | 17 | **469.22** | 27.60 |
| CODEINE | 9050 | 22 | **2,324.21** | 105.65 |
| BUPRENORPHINE | 9064 | 14 | **83.98** | 6.00 |
| OXYCODONE | 9143 | 22 | **1,681.39** | 76.43 |
| HYDROMORPHONE | 9150 | 12 | **45.22** | 3.77 |
| HYDROCODONE | 9193 | 22 | **794.06** | 36.09 |

| | | | | |
|---|---|---|---|---|
| MEPERIDINE (PETHIDINE) | 9230 | 6 | **104.58** | 17.43 |
| METHADONE | 9250B | 4 | **33.09** | 8.27 |
| MORPHINE | 9300 | 12 | **295.80** | 24.65 |
| TAPENTADOL | 9780 | 1 | **5.00** | 5.00 |
| FENTANYL BASE | 9801 | 11 | **10.12** | 0.92 |

**BUSINESS ACTIVITY: B - HOSPITALS**

| DRUG NAME | DRUG CODE | NUMBER OF REGISTRANT SOLD TO | TOTAL GRAMS | AVG GRAMS |
|---|---|---|---|---|
| METHYLPHENIDATE (DL;D;L;ISOMERS) | 1724 | 1 | **0.43** | 0.43 |
| CODEINE | 9050 | 2 | **59.67** | 29.84 |
| OXYCODONE | 9143 | 2 | **17.93** | 8.97 |
| HYDROMORPHONE | 9150 | 2 | **15.42** | 7.71 |
| HYDROCODONE | 9193 | 2 | **29.57** | 14.78 |
| MEPERIDINE (PETHIDINE) | 9230 | 2 | **56.10** | 28.05 |
| MORPHINE | 9300 | 3 | **29.87** | 9.96 |
| FENTANYL BASE | 9801 | 3 | **2.25** | 0.75 |

**BUSINESS ACTIVITY: C - PRACTITIONERS**

| DRUG NAME | DRUG CODE | NUMBER OF REGISTRANT SOLD TO | TOTAL GRAMS | AVG GRAMS |
|---|---|---|---|---|
| BUPRENORPHINE | 9064 | 5 | **0.21** | 0.04 |
| HYDROMORPHONE | 9150 | 2 | **0.23** | 0.12 |
| MORPHINE | 9300 | 1 | **0.56** | 0.56 |
| FENTANYL BASE | 9801 | 2 | **0.07** | 0.03 |

**STATE: VT - VERMONT**

**BUSINESS ACTIVITY: A - PHARMACIES**

| DRUG NAME | DRUG CODE | NUMBER OF REGISTRANT SOLD TO | TOTAL GRAMS | AVG GRAMS |
|---|---|---|---|---|
| AMPHETAMINE | 1100 | 141 | **41,144.36** | 291.80 |
| METHAMPHETAMINE | 1105 | 5 | **7.23** | 1.45 |
| LISDEXAMFETAMINE | 1205 | 141 | **28,977.26** | 205.51 |
| METHYLPHENIDATE (DL;D;L;ISOMERS) | 1724 | 141 | **72,085.59** | 511.25 |
| CODEINE | 9050 | 136 | **9,082.14** | 66.78 |
| BUPRENORPHINE | 9064 | 138 | **25,748.91** | 186.59 |
| OXYCODONE | 9143 | 140 | **61,805.83** | 441.47 |
| HYDROMORPHONE | 9150 | 139 | **3,094.45** | 22.26 |
| HYDROCODONE | 9193 | 141 | **13,781.22** | 97.74 |
| MEPERIDINE (PETHIDINE) | 9230 | 14 | **192.27** | 13.73 |
| METHADONE | 9250B | 134 | **9,807.19** | 73.19 |
| MORPHINE | 9300 | 141 | **28,074.94** | 199.11 |
| OXYMORPHONE | 9652 | 19 | **411.88** | 21.68 |
| TAPENTADOL | 9780 | 39 | **2,994.00** | 76.77 |
| FENTANYL BASE | 9801 | 135 | **417.14** | 3.09 |

**BUSINESS ACTIVITY: B - HOSPITALS**

| DRUG NAME | DRUG CODE | NUMBER OF REGISTRANT SOLD TO | TOTAL GRAMS | AVG GRAMS |
|---|---|---|---|---|
| AMPHETAMINE | 1100 | 12 | **1,134.15** | 94.51 |
| LISDEXAMFETAMINE | 1205 | 6 | **411.03** | 68.50 |
| METHYLPHENIDATE (DL;D;L;ISOMERS) | 1724 | 15 | **2,360.14** | 157.34 |
| CODEINE | 9050 | 13 | **269.69** | 20.75 |
| BUPRENORPHINE | 9064 | 17 | **1,195.09** | 70.30 |
| OXYCODONE | 9143 | 17 | **4,675.29** | 275.02 |
| HYDROMORPHONE | 9150 | 15 | **566.21** | 37.75 |
| HYDROCODONE | 9193 | 17 | **779.99** | 45.88 |
| MEPERIDINE (PETHIDINE) | 9230 | 8 | **518.54** | 64.82 |
| METHADONE | 9250B | 16 | **600.77** | 37.55 |
| MORPHINE | 9300 | 17 | **3,243.17** | 190.77 |
| OXYMORPHONE | 9652 | 1 | **2.68** | 2.68 |
| TAPENTADOL | 9780 | 1 | **3.00** | 3.00 |
| FENTANYL BASE | 9801 | 17 | **36.67** | 2.16 |

**BUSINESS ACTIVITY: C - PRACTITIONERS**

| DRUG NAME | DRUG CODE | NUMBER OF REGISTRANT SOLD TO | TOTAL GRAMS | AVG GRAMS |
|---|---|---|---|---|
| CODEINE | 9050 | 5 | **39.12** | 7.82 |
| BUPRENORPHINE | 9064 | 102 | **8.83** | 0.09 |
| OXYCODONE | 9143 | 2 | **2.42** | 1.21 |
| HYDROMORPHONE | 9150 | 48 | **19.44** | 0.41 |
| HYDROCODONE | 9193 | 22 | **28.77** | 1.31 |
| MEPERIDINE (PETHIDINE) | 9230 | 2 | **3.27** | 1.63 |
| METHADONE | 9250B | 1 | **2.86** | 2.86 |
| MORPHINE | 9300 | 19 | **13.44** | 0.71 |
| FENTANYL BASE | 9801 | 26 | **0.99** | 0.04 |

**BUSINESS ACTIVITY: D - TEACHING INSTITUTIONS**

| DRUG NAME | DRUG CODE | NUMBER OF REGISTRANT SOLD TO | TOTAL GRAMS | AVG GRAMS |
|---|---|---|---|---|
| BUPRENORPHINE | 9064 | 1 | **0.01** | 0.01 |

**BUSINESS ACTIVITY: M - MID-LEVEL PRACTITIONERS**

| DRUG NAME | DRUG CODE | NUMBER OF REGISTRANT SOLD TO | TOTAL GRAMS | AVG GRAMS |
|---|---|---|---|---|
| CODEINE | 9050 | 2 | **6.63** | 3.32 |

**BUSINESS ACTIVITY: NTP - NARCOTIC TREATMENT PROGRAMS**

| DRUG NAME | DRUG CODE | NUMBER OF REGISTRANT SOLD TO | TOTAL GRAMS | AVG GRAMS |
|---|---|---|---|---|
| BUPRENORPHINE | 9064 | 9 | **3,629.16** | 403.24 |
| METHADONE | 9250B | 9 | **70,509.13** | 7,834.35 |

**STATE: WA - WASHINGTON**

**BUSINESS ACTIVITY: A - PHARMACIES**

| DRUG NAME | DRUG CODE | NUMBER OF REGISTRANT SOLD TO | TOTAL GRAMS | AVG GRAMS |
|---|---|---|---|---|
| AMPHETAMINE | 1100 | 1,189 | **475,361.73** | 399.80 |

| | | | | |
|---|---|---|---|---|
| METHAMPHETAMINE | 1105 | 86 | **338.65** | 3.94 |
| LISDEXAMFETAMINE | 1205 | 1,125 | **146,948.40** | 130.62 |
| METHYLPHENIDATE (DL;D;L;ISOMERS) | 1724 | 1,190 | **436,996.06** | 367.22 |
| CODEINE | 9050 | 1,189 | **167,206.16** | 140.63 |
| BUPRENORPHINE | 9064 | 1,201 | **114,080.47** | 94.99 |
| OXYCODONE | 9143 | 1,213 | **809,555.24** | 667.40 |
| HYDROMORPHONE | 9150 | 1,140 | **25,307.09** | 22.20 |
| HYDROCODONE | 9193 | 1,223 | **370,706.40** | 303.11 |
| MEPERIDINE (PETHIDINE) | 9230 | 187 | **3,235.99** | 17.30 |
| METHADONE | 9250B | 1,106 | **65,832.85** | 59.52 |
| MORPHINE | 9300 | 1,182 | **291,429.31** | 246.56 |
| OXYMORPHONE | 9652 | 411 | **4,437.39** | 10.80 |
| TAPENTADOL | 9780 | 563 | **46,903.50** | 83.31 |
| FENTANYL BASE | 9801 | 1,072 | **3,266.28** | 3.05 |

**BUSINESS ACTIVITY: B - HOSPITALS**

| DRUG NAME | DRUG CODE | NUMBER OF REGISTRANT SOLD TO | TOTAL GRAMS | AVG GRAMS |
|---|---|---|---|---|
| AMPHETAMINE | 1100 | 95 | **15,078.93** | 158.73 |
| LISDEXAMFETAMINE | 1205 | 41 | **3,334.48** | 81.33 |
| METHYLPHENIDATE (DL;D;L;ISOMERS) | 1724 | 111 | **22,122.02** | 199.30 |
| CODEINE | 9050 | 153 | **6,043.89** | 39.50 |
| BUPRENORPHINE | 9064 | 154 | **4,648.72** | 30.19 |
| OXYCODONE | 9143 | 223 | **57,686.61** | 258.68 |
| HYDROMORPHONE | 9150 | 199 | **4,687.58** | 23.56 |
| HYDROCODONE | 9193 | 209 | **22,698.97** | 108.61 |
| MEPERIDINE (PETHIDINE) | 9230 | 140 | **1,120.42** | 8.00 |
| METHADONE | 9250B | 126 | **7,270.76** | 57.70 |
| MORPHINE | 9300 | 221 | **25,715.34** | 116.36 |
| OXYMORPHONE | 9652 | 9 | **28.28** | 3.14 |
| TAPENTADOL | 9780 | 15 | **2,190.50** | 146.03 |
| FENTANYL BASE | 9801 | 220 | **513.49** | 2.33 |

**BUSINESS ACTIVITY: C - PRACTITIONERS**

| DRUG NAME | DRUG CODE | NUMBER OF REGISTRANT SOLD TO | TOTAL GRAMS | AVG GRAMS |
|---|---|---|---|---|
| LISDEXAMFETAMINE | 1205 | 1 | **2.43** | 2.43 |
| METHYLPHENIDATE (DL;D;L;ISOMERS) | 1724 | 2 | **18.81** | 9.40 |
| CODEINE | 9050 | 113 | **5,822.42** | 51.53 |
| BUPRENORPHINE | 9064 | 881 | **526.21** | 0.60 |
| OXYCODONE | 9143 | 31 | **55.01** | 1.77 |
| HYDROMORPHONE | 9150 | 482 | **304.04** | 0.63 |
| HYDROCODONE | 9193 | 365 | **1,185.23** | 3.25 |
| MEPERIDINE (PETHIDINE) | 9230 | 95 | **484.12** | 5.10 |

| | | | | |
|---|---|---|---|---|
| METHADONE | 9250B | 44 | **111.80** | 2.54 |
| MORPHINE | 9300 | 240 | **314.63** | 1.31 |
| FENTANYL BASE | 9801 | 630 | **34.70** | 0.06 |

**BUSINESS ACTIVITY: D - TEACHING INSTITUTIONS**

| DRUG NAME | DRUG CODE | NUMBER OF REGISTRANT SOLD TO | TOTAL GRAMS | AVG GRAMS |
|---|---|---|---|---|
| HYDROMORPHONE | 9150 | 1 | **0.04** | 0.04 |

**BUSINESS ACTIVITY: M - MID-LEVEL PRACTITIONERS**

| DRUG NAME | DRUG CODE | NUMBER OF REGISTRANT SOLD TO | TOTAL GRAMS | AVG GRAMS |
|---|---|---|---|---|
| CODEINE | 9050 | 4 | **7.96** | 1.99 |
| BUPRENORPHINE | 9064 | 11 | **329.10** | 29.92 |
| OXYCODONE | 9143 | 1 | **3.14** | 3.14 |
| HYDROMORPHONE | 9150 | 4 | **0.43** | 0.11 |
| HYDROCODONE | 9193 | 1 | **1.76** | 1.76 |
| MEPERIDINE (PETHIDINE) | 9230 | 1 | **13.07** | 13.07 |
| MORPHINE | 9300 | 1 | **1.13** | 1.13 |
| FENTANYL BASE | 9801 | 16 | **0.34** | 0.02 |

**BUSINESS ACTIVITY: NTP - NARCOTIC TREATMENT PROGRAMS**

| DRUG NAME | DRUG CODE | NUMBER OF REGISTRANT SOLD TO | TOTAL GRAMS | AVG GRAMS |
|---|---|---|---|---|
| BUPRENORPHINE | 9064 | 20 | **2,037.03** | 101.85 |
| METHADONE | 9250B | 30 | **363,249.83** | 12,108.33 |

**STATE: WI - WISCONSIN**

**BUSINESS ACTIVITY: A - PHARMACIES**

| DRUG NAME | DRUG CODE | NUMBER OF REGISTRANT SOLD TO | TOTAL GRAMS | AVG GRAMS |
|---|---|---|---|---|
| AMPHETAMINE | 1100 | 1,045 | **497,482.78** | 476.06 |
| METHAMPHETAMINE | 1105 | 24 | **104.06** | 4.34 |
| LISDEXAMFETAMINE | 1205 | 1,031 | **301,369.31** | 292.31 |
| METHYLPHENIDATE (DL;D;L;ISOMERS) | 1724 | 1,045 | **402,969.83** | 385.62 |
| CODEINE | 9050 | 1,047 | **140,904.32** | 134.58 |
| BUPRENORPHINE | 9064 | 1,017 | **51,090.67** | 50.24 |
| OXYCODONE | 9143 | 1,056 | **484,643.96** | 458.94 |
| HYDROMORPHONE | 9150 | 859 | **7,731.29** | 9.00 |
| HYDROCODONE | 9193 | 1,058 | **226,289.08** | 213.88 |
| MEPERIDINE (PETHIDINE) | 9230 | 49 | **558.30** | 11.39 |
| METHADONE | 9250B | 764 | **19,041.24** | 24.92 |
| MORPHINE | 9300 | 1,039 | **198,381.09** | 190.93 |
| OXYMORPHONE | 9652 | 239 | **2,220.53** | 9.29 |
| TAPENTADOL | 9780 | 506 | **33,216.50** | 65.65 |
| FENTANYL BASE | 9801 | 966 | **2,945.93** | 3.05 |

**BUSINESS ACTIVITY: B - HOSPITALS**

| DRUG NAME | DRUG CODE | NUMBER OF REGISTRANT SOLD TO | TOTAL GRAMS | AVG GRAMS |
|---|---|---|---|---|

| | | | | |
|---|---|---|---|---|
| AMPHETAMINE | 1100 | 103 | **11,288.66** | 109.60 |
| METHAMPHETAMINE | 1105 | 1 | **1.61** | 1.61 |
| LISDEXAMFETAMINE | 1205 | 64 | **3,077.23** | 48.08 |
| METHYLPHENIDATE (DL;D;L;ISOMERS) | 1724 | 127 | **8,202.63** | 64.59 |
| CODEINE | 9050 | 172 | **4,080.44** | 23.72 |
| BUPRENORPHINE | 9064 | 115 | **2,959.90** | 25.74 |
| OXYCODONE | 9143 | 172 | **33,615.03** | 195.44 |
| HYDROMORPHONE | 9150 | 160 | **2,423.66** | 15.15 |
| HYDROCODONE | 9193 | 170 | **11,425.49** | 67.21 |
| MEPERIDINE (PETHIDINE) | 9230 | 98 | **1,559.99** | 15.92 |
| METHADONE | 9250B | 131 | **2,228.12** | 17.01 |
| MORPHINE | 9300 | 165 | **20,060.98** | 121.58 |
| OXYMORPHONE | 9652 | 3 | **1.96** | 0.65 |
| TAPENTADOL | 9780 | 15 | **318.00** | 21.20 |
| FENTANYL BASE | 9801 | 160 | **469.06** | 2.93 |

**BUSINESS ACTIVITY: C - PRACTITIONERS**

| DRUG NAME | DRUG CODE | NUMBER OF REGISTRANT SOLD TO | TOTAL GRAMS | AVG GRAMS |
|---|---|---|---|---|
| AMPHETAMINE | 1100 | 1 | **6.96** | 6.96 |
| CODEINE | 9050 | 44 | **465.90** | 10.59 |
| BUPRENORPHINE | 9064 | 578 | **259.13** | 0.45 |
| OXYCODONE | 9143 | 53 | **560.31** | 10.57 |
| HYDROMORPHONE | 9150 | 388 | **227.87** | 0.59 |
| HYDROCODONE | 9193 | 238 | **604.27** | 2.54 |
| MEPERIDINE (PETHIDINE) | 9230 | 47 | **270.71** | 5.76 |
| METHADONE | 9250B | 20 | **36.85** | 1.84 |
| MORPHINE | 9300 | 143 | **173.21** | 1.21 |
| FENTANYL BASE | 9801 | 320 | **29.54** | 0.09 |

**BUSINESS ACTIVITY: M - MID-LEVEL PRACTITIONERS**

| DRUG NAME | DRUG CODE | NUMBER OF REGISTRANT SOLD TO | TOTAL GRAMS | AVG GRAMS |
|---|---|---|---|---|
| CODEINE | 9050 | 3 | **15.91** | 5.30 |
| FENTANYL BASE | 9801 | 2 | **0.01** | 0.00 |

**BUSINESS ACTIVITY: NTP - NARCOTIC TREATMENT PROGRAMS**

| DRUG NAME | DRUG CODE | NUMBER OF REGISTRANT SOLD TO | TOTAL GRAMS | AVG GRAMS |
|---|---|---|---|---|
| BUPRENORPHINE | 9064 | 17 | **2,555.10** | 150.30 |
| METHADONE | 9250B | 21 | **234,064.48** | 11,145.93 |

**STATE: WV - WEST VIRGINIA**

**BUSINESS ACTIVITY: A - PHARMACIES**

| DRUG NAME | DRUG CODE | NUMBER OF REGISTRANT SOLD TO | TOTAL GRAMS | AVG GRAMS |
|---|---|---|---|---|
| AMPHETAMINE | 1100 | 545 | **94,054.25** | 172.58 |
| METHAMPHETAMINE | 1105 | 3 | **14.06** | 4.69 |

| | | | | |
|---|---|---|---|---|
| LISDEXAMFETAMINE | 1205 | 531 | **74,071.37** | 139.49 |
| METHYLPHENIDATE (DL;D;L;ISOMERS) | 1724 | 545 | **114,408.55** | 209.92 |
| CODEINE | 9050 | 549 | **142,415.86** | 259.41 |
| BUPRENORPHINE | 9064 | 498 | **73,430.28** | 147.45 |
| OXYCODONE | 9143 | 551 | **203,969.20** | 370.18 |
| HYDROMORPHONE | 9150 | 352 | **3,097.96** | 8.80 |
| HYDROCODONE | 9193 | 550 | **141,944.65** | 258.08 |
| MEPERIDINE (PETHIDINE) | 9230 | 23 | **161.77** | 7.03 |
| METHADONE | 9250B | 301 | **7,790.85** | 25.88 |
| MORPHINE | 9300 | 525 | **54,820.15** | 104.42 |
| OXYMORPHONE | 9652 | 143 | **3,282.97** | 22.96 |
| TAPENTADOL | 9780 | 169 | **14,277.00** | 84.48 |
| FENTANYL BASE | 9801 | 488 | **1,131.74** | 2.32 |

**BUSINESS ACTIVITY: B - HOSPITALS**

| DRUG NAME | DRUG CODE | NUMBER OF REGISTRANT SOLD TO | TOTAL GRAMS | AVG GRAMS |
|---|---|---|---|---|
| AMPHETAMINE | 1100 | 33 | **2,190.52** | 66.38 |
| LISDEXAMFETAMINE | 1205 | 19 | **772.92** | 40.68 |
| METHYLPHENIDATE (DL;D;L;ISOMERS) | 1724 | 48 | **2,197.19** | 45.77 |
| CODEINE | 9050 | 70 | **1,830.03** | 26.14 |
| BUPRENORPHINE | 9064 | 61 | **3,048.71** | 49.98 |
| OXYCODONE | 9143 | 80 | **13,166.14** | 164.58 |
| HYDROMORPHONE | 9150 | 67 | **733.72** | 10.95 |
| HYDROCODONE | 9193 | 78 | **12,569.17** | 161.14 |
| MEPERIDINE (PETHIDINE) | 9230 | 50 | **507.32** | 10.15 |
| METHADONE | 9250B | 46 | **701.03** | 15.24 |
| MORPHINE | 9300 | 76 | **7,049.14** | 92.75 |
| OXYMORPHONE | 9652 | 6 | **20.12** | 3.35 |
| TAPENTADOL | 9780 | 9 | **186.00** | 20.67 |
| FENTANYL BASE | 9801 | 76 | **172.66** | 2.27 |

**BUSINESS ACTIVITY: C - PRACTITIONERS**

| DRUG NAME | DRUG CODE | NUMBER OF REGISTRANT SOLD TO | TOTAL GRAMS | AVG GRAMS |
|---|---|---|---|---|
| CODEINE | 9050 | 19 | **49.77** | 2.62 |
| BUPRENORPHINE | 9064 | 112 | **11.15** | 0.10 |
| OXYCODONE | 9143 | 1 | **0.45** | 0.45 |
| HYDROMORPHONE | 9150 | 39 | **24.64** | 0.63 |
| HYDROCODONE | 9193 | 42 | **123.79** | 2.95 |
| MEPERIDINE (PETHIDINE) | 9230 | 5 | **218.96** | 43.79 |
| MORPHINE | 9300 | 29 | **17.69** | 0.61 |
| FENTANYL BASE | 9801 | 48 | **3.85** | 0.08 |

**BUSINESS ACTIVITY: D - TEACHING INSTITUTIONS**

| DRUG NAME | DRUG CODE | NUMBER OF REGISTRANT SOLD TO | TOTAL GRAMS | AVG GRAMS |
|---|---|---|---|---|
| HYDROMORPHONE | 9150 | 1 | **0.04** | 0.04 |
| MORPHINE | 9300 | 1 | **0.19** | 0.19 |

**BUSINESS ACTIVITY: M - MID-LEVEL PRACTITIONERS**

| DRUG NAME | DRUG CODE | NUMBER OF REGISTRANT SOLD TO | TOTAL GRAMS | AVG GRAMS |
|---|---|---|---|---|
| CODEINE | 9050 | 6 | **3.53** | 0.59 |
| MORPHINE | 9300 | 35 | **14.00** | 0.40 |
| FENTANYL BASE | 9801 | 47 | **0.50** | 0.01 |

**BUSINESS ACTIVITY: NTP - NARCOTIC TREATMENT PROGRAMS**

| DRUG NAME | DRUG CODE | NUMBER OF REGISTRANT SOLD TO | TOTAL GRAMS | AVG GRAMS |
|---|---|---|---|---|
| BUPRENORPHINE | 9064 | 7 | **602.46** | 86.07 |
| METHADONE | 9250B | 9 | **88,617.15** | 9,846.35 |

**STATE: WY - WYOMING**

**BUSINESS ACTIVITY: A - PHARMACIES**

| DRUG NAME | DRUG CODE | NUMBER OF REGISTRANT SOLD TO | TOTAL GRAMS | AVG GRAMS |
|---|---|---|---|---|
| AMPHETAMINE | 1100 | 109 | **27,362.38** | 251.03 |
| METHAMPHETAMINE | 1105 | 5 | **19.68** | 3.94 |
| LISDEXAMFETAMINE | 1205 | 106 | **10,310.99** | 97.27 |
| METHYLPHENIDATE (DL;D;L;ISOMERS) | 1724 | 107 | **28,701.10** | 268.23 |
| CODEINE | 9050 | 111 | **11,465.95** | 103.30 |
| BUPRENORPHINE | 9064 | 109 | **5,531.93** | 50.75 |
| OXYCODONE | 9143 | 114 | **74,732.28** | 655.55 |
| HYDROMORPHONE | 9150 | 108 | **1,830.91** | 16.95 |
| HYDROCODONE | 9193 | 114 | **30,254.46** | 265.39 |
| MEPERIDINE (PETHIDINE) | 9230 | 23 | **370.39** | 16.10 |
| METHADONE | 9250B | 97 | **2,898.84** | 29.88 |
| MORPHINE | 9300 | 110 | **21,971.69** | 199.74 |
| OXYMORPHONE | 9652 | 29 | **337.84** | 11.65 |
| TAPENTADOL | 9780 | 52 | **4,766.00** | 91.65 |
| FENTANYL BASE | 9801 | 106 | **328.60** | 3.10 |

**BUSINESS ACTIVITY: B - HOSPITALS**

| DRUG NAME | DRUG CODE | NUMBER OF REGISTRANT SOLD TO | TOTAL GRAMS | AVG GRAMS |
|---|---|---|---|---|
| AMPHETAMINE | 1100 | 16 | **1,678.49** | 104.91 |
| LISDEXAMFETAMINE | 1205 | 10 | **538.79** | 53.88 |
| METHYLPHENIDATE (DL;D;L;ISOMERS) | 1724 | 22 | **1,926.89** | 87.59 |
| CODEINE | 9050 | 32 | **1,228.01** | 38.38 |
| BUPRENORPHINE | 9064 | 16 | **431.36** | 26.96 |
| OXYCODONE | 9143 | 46 | **5,550.26** | 120.66 |
| HYDROMORPHONE | 9150 | 40 | **243.13** | 6.08 |
| HYDROCODONE | 9193 | 46 | **4,003.24** | 87.03 |

| | | | | |
|---|---|---|---|---|
| MEPERIDINE (PETHIDINE) | 9230 | 28 | **157.41** | 5.62 |
| METHADONE | 9250B | 16 | **287.10** | 17.94 |
| MORPHINE | 9300 | 44 | **4,797.79** | 109.04 |
| OXYMORPHONE | 9652 | 1 | **1.78** | 1.78 |
| TAPENTADOL | 9780 | 3 | **25.00** | 8.33 |
| FENTANYL BASE | 9801 | 47 | **42.17** | 0.90 |

**BUSINESS ACTIVITY: C - PRACTITIONERS**

| DRUG NAME | DRUG CODE | NUMBER OF REGISTRANT SOLD TO | TOTAL GRAMS | AVG GRAMS |
|---|---|---|---|---|
| CODEINE | 9050 | 7 | **125.53** | 17.93 |
| BUPRENORPHINE | 9064 | 64 | **5.09** | 0.08 |
| OXYCODONE | 9143 | 1 | **13.11** | 13.11 |
| HYDROMORPHONE | 9150 | 32 | **22.54** | 0.70 |
| HYDROCODONE | 9193 | 21 | **86.68** | 4.13 |
| MEPERIDINE (PETHIDINE) | 9230 | 4 | **6.97** | 1.74 |
| MORPHINE | 9300 | 24 | **37.13** | 1.55 |
| FENTANYL BASE | 9801 | 38 | **5.38** | 0.14 |

**BUSINESS ACTIVITY: D - TEACHING INSTITUTIONS**

| DRUG NAME | DRUG CODE | NUMBER OF REGISTRANT SOLD TO | TOTAL GRAMS | AVG GRAMS |
|---|---|---|---|---|
| BUPRENORPHINE | 9064 | 1 | **0.00** | 0.00 |

**BUSINESS ACTIVITY: M - MID-LEVEL PRACTITIONERS**

| DRUG NAME | DRUG CODE | NUMBER OF REGISTRANT SOLD TO | TOTAL GRAMS | AVG GRAMS |
|---|---|---|---|---|
| CODEINE | 9050 | 2 | **2.65** | 1.33 |
| BUPRENORPHINE | 9064 | 1 | **0.12** | 0.12 |

**ENTIRE UNITED STATES**

**BUSINESS ACTIVITY: A - PHARMACIES**

| DRUG NAME | DRUG CODE | NUMBER OF REGISTRANT SOLD TO | **TOTAL GRAMS** | AVG GRAMS |
|---|---|---|---|---|
| AMPHETAMINE | 1100 | 62,406 | **22,455,505.83** | 359.83 |
| METHAMPHETAMINE | 1105 | 1,677 | **5,772.08** | 3.44 |
| LISDEXAMFETAMINE | 1205 | 57,790 | **9,663,770.09** | 167.22 |
| METHYLPHENIDATE (DL;D;L;ISOMERS) | 1724 | 61,591 | **17,262,528.67** | 280.28 |
| CODEINE | 9050 | 63,304 | **11,635,286.84** | 183.80 |
| BUPRENORPHINE | 9064 | 58,822 | **4,157,282.31** | 70.68 |
| OXYCODONE | 9143 | 63,682 | **34,178,438.20** | 536.70 |
| HYDROMORPHONE | 9150 | 52,796 | **830,351.22** | 15.73 |
| HYDROCODONE | 9193 | 63,700 | **19,048,664.24** | 299.04 |
| MEPERIDINE (PETHIDINE) | 9230 | 8,568 | **141,088.83** | 16.47 |
| METHADONE | 9250B | 45,805 | **1,746,684.03** | 38.13 |
| MORPHINE | 9300 | 59,115 | **10,951,849.52** | 185.26 |
| OXYMORPHONE | 9652 | 19,013 | **395,295.22** | 20.79 |
| TAPENTADOL | 9780 | 32,748 | **3,554,253.00** | 108.53 |
| FENTANYL BASE | 9801 | 55,326 | **166,588.18** | 3.01 |

**BUSINESS ACTIVITY: B - HOSPITALS**

| DRUG NAME | DRUG CODE | NUMBER OF REGISTRANT SOLD TO | **TOTAL GRAMS** | AVG GRAMS |
|---|---|---|---|---|
| AMPHETAMINE | 1100 | 3,996 | **544,493.83** | 136.26 |
| METHAMPHETAMINE | 1105 | 23 | **136.62** | 5.94 |
| LISDEXAMFETAMINE | 1205 | 1,981 | **135,973.81** | 68.64 |
| METHYLPHENIDATE (DL;D;L;ISOMERS) | 1724 | 5,332 | **554,984.68** | 104.09 |
| CODEINE | 9050 | 6,985 | **373,026.97** | 53.40 |
| BUPRENORPHINE | 9064 | 5,431 | **138,910.80** | 25.58 |
| OXYCODONE | 9143 | 8,901 | **1,739,327.08** | 195.41 |
| HYDROMORPHONE | 9150 | 8,656 | **150,286.17** | 17.36 |
| HYDROCODONE | 9193 | 9,335 | **951,272.83** | 101.90 |
| MEPERIDINE (PETHIDINE) | 9230 | 5,988 | **120,977.85** | 20.20 |
| METHADONE | 9250B | 5,806 | **216,211.45** | 37.24 |
| MORPHINE | 9300 | 9,468 | **1,005,685.62** | 106.22 |
| OXYMORPHONE | 9652 | 510 | **6,289.40** | 12.33 |
| TAPENTADOL | 9780 | 1,254 | **73,774.85** | 58.83 |
| FENTANYL BASE | 9801 | 10,128 | **25,038.18** | 2.47 |

**BUSINESS ACTIVITY: C - PRACTITIONERS**

| DRUG NAME | DRUG CODE | NUMBER OF REGISTRANT SOLD TO | **TOTAL GRAMS** | AVG GRAMS |
|---|---|---|---|---|
| AMPHETAMINE | 1100 | 70 | **2,144.00** | 30.63 |
| METHAMPHETAMINE | 1105 | 2 | **8.03** | 4.02 |
| LISDEXAMFETAMINE | 1205 | 18 | **815.11** | 45.28 |
| METHYLPHENIDATE (DL;D;L;ISOMERS) | 1724 | 34 | **3,218.66** | 94.67 |
| CODEINE | 9050 | 3,218 | **94,882.57** | 29.48 |
| BUPRENORPHINE | 9064 | 27,308 | **11,258.05** | 0.41 |

| | | | | |
|---|---|---|---|---|
| OXYCODONE | 9143 | 1,695 | **11,483.66** | 6.78 |
| HYDROMORPHONE | 9150 | 12,373 | **6,538.15** | 0.53 |
| HYDROCODONE | 9193 | 11,094 | **40,879.15** | 3.68 |
| MEPERIDINE (PETHIDINE) | 9230 | 3,553 | **30,528.37** | 8.59 |
| METHADONE | 9250B | 745 | **3,076.83** | 4.13 |
| MORPHINE | 9300 | 6,088 | **8,701.46** | 1.43 |
| OXYMORPHONE | 9652 | 9 | **109.77** | 12.20 |
| TAPENTADOL | 9780 | 12 | **658.50** | 54.88 |
| FENTANYL BASE | 9801 | 17,873 | **1,855.62** | 0.10 |

**BUSINESS ACTIVITY: D - TEACHING INSTITUTIONS**

| DRUG NAME | DRUG CODE | NUMBER OF REGISTRANT SOLD TO | **TOTAL GRAMS** | AVG GRAMS |
|---|---|---|---|---|
| AMPHETAMINE | 1100 | 1 | **1.25** | 1.25 |
| METHYLPHENIDATE (DL;D;L;ISOMERS) | 1724 | 2 | **2.68** | 1.34 |
| CODEINE | 9050 | 5 | **1,263.26** | 252.65 |
| BUPRENORPHINE | 9064 | 53 | **7.11** | 0.13 |
| OXYCODONE | 9143 | 1 | **0.45** | 0.45 |
| HYDROMORPHONE | 9150 | 25 | **37.02** | 1.48 |
| HYDROCODONE | 9193 | 8 | **118.73** | 14.84 |
| MEPERIDINE (PETHIDINE) | 9230 | 3 | **10.89** | 3.63 |
| METHADONE | 9250B | 6 | **157.15** | 26.19 |
| MORPHINE | 9300 | 18 | **133.00** | 7.39 |
| FENTANYL BASE | 9801 | 25 | **14.01** | 0.56 |

**BUSINESS ACTIVITY: M - MID-LEVEL PRACTITIONERS**

| DRUG NAME | DRUG CODE | NUMBER OF REGISTRANT SOLD TO | **TOTAL GRAMS** | AVG GRAMS |
|---|---|---|---|---|
| AMPHETAMINE | 1100 | 2 | **5.77** | 2.88 |
| LISDEXAMFETAMINE | 1205 | 1 | **1.73** | 1.73 |
| METHYLPHENIDATE (DL;D;L;ISOMERS) | 1724 | 2 | **5.62** | 2.81 |
| CODEINE | 9050 | 347 | **1,525.15** | 4.40 |
| BUPRENORPHINE | 9064 | 100 | **1,093.19** | 10.93 |
| OXYCODONE | 9143 | 7 | **8.97** | 1.28 |
| HYDROMORPHONE | 9150 | 82 | **8.83** | 0.11 |
| HYDROCODONE | 9193 | 20 | **26.69** | 1.33 |
| MEPERIDINE (PETHIDINE) | 9230 | 10 | **88.24** | 8.82 |
| METHADONE | 9250B | 2 | **52.77** | 26.38 |
| MORPHINE | 9300 | 593 | **253.43** | 0.43 |
| FENTANYL BASE | 9801 | 1,174 | **35.02** | 0.03 |

**BUSINESS ACTIVITY: NTP - NARCOTIC TREATMENT PROGRAMS**

| DRUG NAME | DRUG CODE | NUMBER OF REGISTRANT SOLD TO | **TOTAL GRAMS** | AVG GRAMS |
|---|---|---|---|---|

| | | | | |
|---|---|---|---|---|
| BUPRENORPHINE | 9064 | 976 | **97,220.00** | 99.61 |
| METHADONE | 9250B | 1,664 | **13,114,262.44** | 7,881.17 |

Apr 8, 2020 11:12:25 AM
